# Early Epigenetic Biomarkers for Perinatal Suicidal Ideation: DNA Methylation Signatures Across the Peripartum Period

**DOI:** 10.64898/2026.03.30.26349727

**Authors:** Emma Simpson-Wade, Julien Dubreucq, Joëlle Rüegg, Alkistis Skalkidou, Marie E. Gaine

**Affiliations:** Molecular Medicine Program, University of Iowa Carver College of Medicine, Iowa City, IA; Psychiatry B and Perinatal Psychiatry, Clermont Ferrand University Hospital; Clermont Auvergne University, Inserm Inserm U-1107, Neuro-Dol; Department of Organismal Biology, Uppsala University, Uppsala, Sweden; Department of Women’s and Children’s Health, Uppsala University, Uppsala, Sweden; Pharmaceutical Sciences and Experimental Therapeutics, University of Iowa, Iowa City, IA; Iowa Neuroscience Institute, University of Iowa, Iowa City, IA

**Author notes:** Correspondence: Marie E. Gaine.

## Abstract

Mental health conditions, including perinatal suicidality, remains a significant health burden representing a leading cause of maternal mortality in the United States. Although the etiology of perinatal suicidal ideation (SI) is not well understood, DNA methylation may provide meaningful mechanistic insights and/or serve as clinical biomarkers during the peripartum period. Using data provided by the Swedish BASIC cohort, we performed a retrospective analysis of DNA methylation changes associated with perinatal SI at three perinatal timepoints (17- and 38-weeks gestation and 8 weeks post-partum) through a targeted and genome-wide approach. Targeted analysis of *a priori* genes revealed 1, 10, and 4 significantly differentially methylated probes at each timepoint and implicated genes associated with the hypothalamic-pituitary-adrenal axis. Genome-wide results identified 465, 2,880, and 510 differentially methylated probes and 7, 25, and 12 differentially methylated regions at each timepoint. Pathway analysis at 38-weeks gestation identified vitamin digestion and absorption as the top term differentially methylated in perinatal SI. Additionally, genes implicated in estrogen and oxytocin signaling were also significantly differentially methylated. Post-partum ideation-risk was successfully predicted using the top ten genome-wide differentially methylated probes at 17 weeks (AUC=66.9%), with prediction accuracy highest when DNA methylation and depression severity were combined (AUC=93.2%). Furthermore, the prediction accuracy for identifying novel SI in the post-partum period increased to 86.2% with 17-week biomarkers. Our results deliver novel insights regarding the role of DNA methylation and perinatal SI, with biomarkers providing both mechanistic insights and clinical usefulness, contributing to the field of perinatal psychiatry and epigenetics.

## 1. Introduction

Mental health conditions remain a leading cause of maternal mortality in the United States (1, 2), with approximately 6.5% of maternal deaths resulting from suicide, while other studies indicate significantly higher incidence (3–5). Notably, suicide attempts, non-suicidal self-injury (NSSI), and suicidal ideation (SI) remain excluded from standard measures of maternal morbidity (6) and underrepresented in most maternal mortality studies.

At present, the etiology of perinatal SI is not well understood, although it is likely attributed to the complex interplay between psychosocial, clinical, and biological factors (7, 8). Diagnosis of major depressive disorder (MDD), discontinuation/lack of psychopharmacological medication, young age, and marital status are the most well-documented psychosocial factors identified (4, 7, 9–12). The biological risk factors are less understood due to the limited data regarding perinatal-related SI, but may involve factors such as hormone dysregulation, inflammation, and/or alterations to neurotransmission (7, 13–15).

Currently, various psychiatric screening surveys such as the Edinburgh Postnatal Depression Scale (EPDS), are implemented in an effort to identify at risk individuals for depressive symptoms including passive SI and NSSI; however, screening can be difficult due to limited mental health training among obstetric clinicians and reliance of patient self-disclosure (6), which is often reduced due to the continued stigma surrounding mental health (4). Therefore, establishing biological markers associated with SI is critical to enhance screening for mental health changes during the perinatal period; ultimately reducing maternal mortality rates through precision prevention approaches and improved perinatal outcomes.

Over the past decade, advances to our understanding of epigenetic processes have unlocked novel mechanisms relating to mental health disorders, including suicidality (16–21). Studies implementing candidate gene approaches, identified DNA methylation (DNAm) changes in hypothalamic-pituitary-adrenal (HPA) -axis genes, such as *NR3C1, FKBP5,* and *CRH* (21), and *BDNF* (21, 22) to be significantly correlated with SI severity in individuals with MDD. Notably, de novo DNAm has been previously linked to changes in brain-related genes, with one study showing a positive correlation between *DNMT3B* expression and *GABRA1* promoter methylation in brains of individuals who died by suicide (23), suggesting psychiatric phenotypes may be a consequence of aberrant DNAm patterns. Recent studies have also identified dynamic alterations to the epigenome associated with pregnancy alone, with longitudinal studies exhibiting DNAm changes highly relevant to the physiological demands of pregnancy, such as immune regulation (24, 25). It is reasonable, therefore, to hypothesize that DNAm would dynamically regulate genes associated with maternal behavior, and aberrant DNAm patterns of these genes may lead to increased psychiatric risk during the peripartum period (18, 26–28); however, the specific targets of this epigenetic regulation have not been elucidated.

In this study, we leveraged DNAm and psychiatric data from a large perinatal cohort to understand the inherent DNAm patterns associated with perinatal SI throughout the peripartum period. We hypothesized that DNAm changes would correlate with SI across multiple timepoints throughout pregnancy and the post-partum period. We implemented both candidate gene and genome-wide approaches, to ascertain brain-relevant mechanisms as well as identify peripheral biomarkers independent of bias. Genome-wide DNAm results were used to determine differentially methylated regions (DMRs) associated with perinatal SI and assess changes to biological pathways. Finally, to demonstrate the clinical applicability of our epigenetic biomarkers, we performed risk-prediction modeling of perinatal SI based on DNAm and depression severity through independent and combined paradigms across multiple timepoints.

## 2. Materials and Methods

### 2.1 Data source

Data for this study were provided by the “Biology, Affect, Stress, Imaging, and Cognition during Pregnancy and the Puerperium” (BASIC) cohort. Sample recruitment and detailed information regarding informed consent and protection of human rights have been previously described (29). Briefly, the BASIC cohort was a population-based, prospective cohort study developed by the Department of Obstetrics and Gynecology at Uppsala University Hospital to better understand the biological processes associated with post-partum depression (PPD). Between September 2009 to November 2018, pregnant people were recruited for participation from baseline (gestational weeks 16-18) through one-year post-partum. Exclusion criteria included adolescence (<18 years of age), language insufficiency, protected identity, non-viable pregnancy, and known blood-borne pathogens. Psychological data were acquired primarily through self-reported surveys and questionnaires taken at multiple timepoints throughout the perinatal period. Surveys included sociodemographic variables, health, and lifestyle information. Maternal blood was collected at three timepoints (17- and 38-weeks gestation (17w; 38w) and 8-weeks post-partum (8pp)). Participation in each modality of the study was optional and written consent was previously provided for each item.

### 2.2 Current study cohort & design

For this sub-study, we used the original BASIC dataset to analyze perinatal SI in a case-control design using the EPDS at three timepoints (17w, 38w, and 8pp). The EPDS is a 10-item self-screening tool used to identify individuals at risk of perinatal depression. Each item assesses behavior over the previous 7 days using a sliding-scale (0–4) based on severity, with a total score ranging from 0-30. Item 10 of the EPDS has been previously established as an informal screen for passive SI (30, 31). Using the calculated total score (continuous variable), we classified pregnant individuals into their respective categories: 1) non-SI controls (NC): total EPDS score 0-4 and a negative response to question 10 (0) or 2) perinatal SI: a positive response to question 10 (1–3). Individuals without DNAm data or without corresponding EPDS responses/covariate data were excluded from analyses resulting in groups of NC (*n* = 184, 103, 127) and SI (*n* = 40, 24, 31) at each timepoint respectively. At all timepoints, demographic variables were separately examined for normality using independent samples *t*-test for continuous variables or Fisher’s Exact Test for categorical variables **(Table 1).**

**Table 1.**
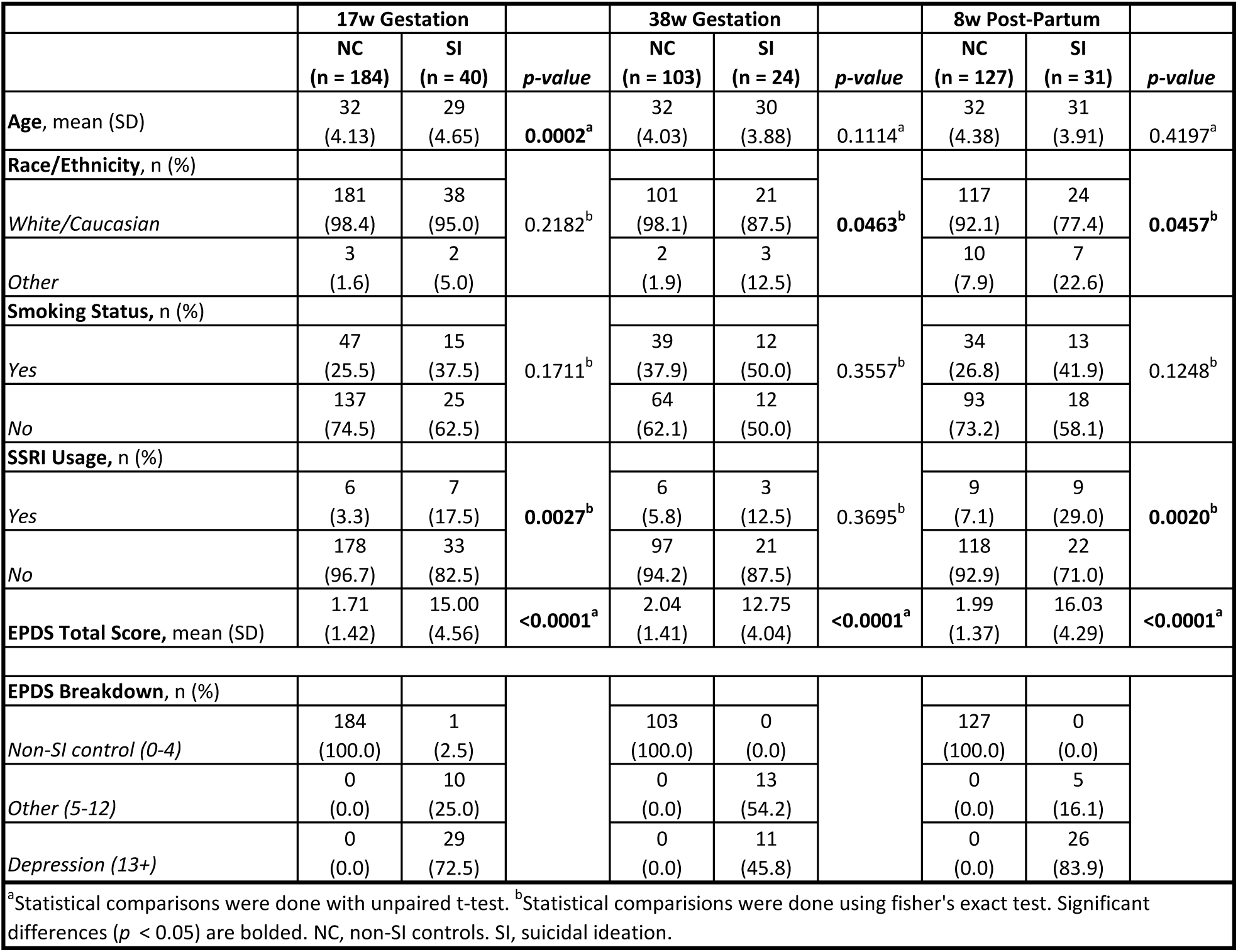
Demographic Information for Primary Cohorts (SI v NC). ^a^Statistical comparisons were done with unpaired t-test. ^b^Statistical comparisions were done using fisher’s exact test. Significant differences (*p* < 0.05) are bolded. *NC*, non-SI controls. *SI*, suicidal ideation.

### 2.3. DNAm array

Peripheral blood samples were previously collected in heparin tubes via venipuncture. Following centrifugation, buffy coats were aliquoted and stored in −80°C. DNA was extracted from buffy coats with Chemagen’s magnetic bead technology using Hamilton ChemagicSTAR®. Genomic DNA (250 ng) was bisulfite-converted using the EZ DNA Methylation Kit (Zymo Research, CA, USA) and DNAm patterns identified using the Infinium Methylation EPIC v2.0 BeadChip (Illumina, CA, USA) per manufacturer’s protocol.

### 2.4 Data pre-processing, quality control, and normalization

Data pre-processing and normalization of Illumina EPIC v2.0 data was performed in R version 4.3.1 following the *ChAMP* pipeline (ChAMP v2.29 and ChAMPdata v2.31) (32). Poor quality probes (detection *p-value* < 0.1), probes with less than 3 beads in >1% of samples, polymorphic probes (33), and CpG probes located on sex chromosomes were removed prior to analysis. Correction for technical variation and batch effects was achieved using *FunctionalNormalization* and *Combat* methods (34, 35). To account for cell-type heterogeneity, white blood cell count proportions (CD8+ T cells, CD4+ T cells, natural killer cells, B cells, monocytes, and granulocytes) were estimated using *RefbaseEWAS* package (36). Quality controls were conducted separately for each DNAm analysis.

## 3. Modeling

### 3.1 Targeted analysis of behavior-related genes

Initially, an *a priori* analysis was performed based on a thorough literature search of genes relevant to maternal behavior **(Supplementary Table 1)**. This was to test the hypothesis that behavior-related genes will be differentially methylated in pregnant people with SI. At each timepoint, 15 genes (∼360 CpG sites) were investigated for differential DNAm between SI and NC. Following pre-processing, individual beta values for each targeted CpG site were extracted and combined into a data matrix and log-transformed to M-values. Targeted CpGs were investigated with linear models adjusted for age (at partus), ethnicity, smoking status prior to pregnancy, and usage of serotonin selective reuptake inhibitors (SSRIs) using *limma* (37). SSRI usage was acquired through self-report surveys taken at 17w- and 32-weeks gestation and 6-weeks post-partum. Targeted results for each timepoint were independently adjusted for multiple comparisons using a Bonferroni-corrected significance threshold of *p* = 1.39 ×10^-4^.

### 3.2 Genome-wide analysis

We also performed a genome-wide analysis to identify novel differentially methylated probes (DMPs). Following pre-processing, all available probes (∼870,000 CpGs) were log-transformed from beta-values to M-values and similarly interrogated for differential DNAm with linear models adjusted for age (at partus), ethnicity, smoking status prior to pregnancy, and usage of SSRIs using *limma* (37). Results for each timepoint were annotated to UCSC Genome Browser hg38 reference genome (38) and independently adjusted for multiple comparisons using Bonferroni correction. This resulted in a Bonferroni-corrected significance threshold of *p* = 5.75×10^-8^, 5.74×10^-8^, and 5.75×10^-8^ for each timepoint respectively.

### 3.3 Identification of differentially methylated regions

At all timepoints using genome-wide analyses, differentially methylated regions (DMRs) were independently identified using *DMRcate* packages in R (39). Briefly, DMPs for each timepoint were identified using the cpg.annotate function with arguments epicv2Filter and epicv2Remap set to “mean” and “TRUE” respectively. These DMPs were subsequently used for DMR interrogation using the following parameters: FDR corrected *p-*value ≤ 0.05 and sliding window of 1000 base pairs. DMRs with a minimum of three probes, a Fisher’s combined probability ≤ 0.01, and a mean differential of 5% were considered significant.

### 3.4 Pathway enrichment analyses

The missMethyl package (40) was used to test gene ontology (GO) and Kyoto Encyclopedia of Genes and Genomes (KEGG) pathway enrichment from the unfiltered DMR results (FDR < 0.05). For both tests, prior probabilities argument was used to take into account potential bias of significant differences due to the number of probes per gene. Pathways with an FDR corrected *p-*value ≤ 0.05 were considered significant.

### 3.5 Clinical-predictive modeling of perinatal SI

Genome-wide DNAm beta-values and EPDS data (total scores and endorsement of SI) were tested for perinatal SI-risk prediction across timepoints (1) 17w – 38w gestation, 2) 38w gestation – 8w pp, and 3) 17w gestation – 8w pp) in an independent manner. Importantly, all individuals with DNAm data at timepoint “1” and EPDS data from timepoints “1” and “2” were used in our predictive model, regardless of psychiatric groupings or overlap with the primary DNAm analyses. This allowed us to determine prediction accuracy of our DNAm biomarkers for dynamic depression-like symptoms as well as both transient and enduring SI. This resulted in *n* = 102, 55, and 107 individuals for each respective model. Generalized linear models were fitted to identify risk-prediction value of perinatal SI based on (i) DNAm (beta-values) from the top 10 genome-wide Bonferroni-significant CpGs at timepoint “1”, (ii) depression severity (EPDS total score) alone, and (iii) DNAm and depression severity combined. Logistic form was used in our generalized linear models where SI endorsement at timepoint “2” was the outcome and individual DNAm and EPDS data were included as independent, additive predictors. For example, to test SI-risk at 8w pp from 17w gestation using model iii, we leveraged the top 10 CpGs from the 17w SI v NC genome-wide analysis and EPDS responses at 17w gestation and 8w pp in the form below:

8w SI_Outcome ∼ EPDS_17w + cg11693709_TC21 + cg21814633_TC21 + cg07466643_BC21 + cg20820280_TC21 + cg01428240_TC21 + cg11093223_BC21 + cg01376622_TC21 + cg11219690_TC21 + cg17283823_BC21 + cg10629173_BC21.

Area under the curve (AUC) of the receiver operating characteristic (ROC) was used to evaluate the predictive performance of each model using *pROC* package in R (41).

## 4. Results

### 4.1 Demographics

For our primary analyses we included *n* = 184, 103, and 127 NC and *n* = 40, 24, 31 SI individuals at each timepoint **(Table 1).** Participants primarily self-identified as White/Caucasian (77.4-98.4%) with average ages between 29-32 years old. Smoking status prior to pregnancy was moderate (25.5-50%) but did not differ between SI and NC individuals at any timepoint. SSRI usage was self-disclosed in pregnancy between 3.3-17.5% whereas 7.1% of NC and 29.0% of SI individuals endorsed medication use at 8w pp. Average EPDS scores ranged from 1.71-2.04 for NC and 12.75-16.03 for individuals with SI. Examination of total EPDS scores revealed clinically significant depression criteria (EPDS ≥ 13) was met in 72.5%, 45.8%, and 83.9% of participants who endorsed SI at each timepoint, respectively.

The test cohorts for our predictive modeling analyses are outlined in **Supplementary Table 2.** For each model, *n* = 102, 55, and 107 individuals were independently tested for biomarker prediction. Model cohorts overlapped with primary cohorts 47.1%, 47.3%, and 52.3% respectively. For example, *n* = 48 individuals in 17w-38w gestation model were also used in 17w SI v NC primary analysis. Thus, participant demographics for modeling were similar to primary cohorts with average ages between 29-34 years and >95% Caucasian. Average EPDS scores at model baseline ranged from 5.31-6.26 for individuals without SI (SI Null) and 12.50-13.71 for individuals with SI.

### 4.2 Targeted analysis DMPs associated with perinatal SI

At each timepoint, 15 genes (∼360 CpGs, **Supplementary Table 1**) were investigated for differential DNAm between SI and NC based on previous associations with non-pregnancy related suicidality (ideation, attempt, or death) or maternal behavior (21, 23, 24, 26, 28, 42–46). Following Bonferroni correction (*p*-value = 1.39×10^-4^), one, ten, and four CpGs remained significantly differentially methylated between SI and NC at each respective timepoint **(Figure 1)**; with no overlap between timepoints. The top CpGs for each timepoint were cg27399558_TC21, cg20801491_TC21, and cg12067298_BC21, annotated to *COMT, CRHBP, and BDNF* respectively. Although 38w gestation resulted in four CpGs significantly hypomethylated in perinatal SI, the majority of significant CpG sites at all timepoints reflected a hypermethylated status in individuals with perinatal SI (73.3%). For all significant targeted CpG site results, see **Supplementary Table 3.**

**Figure 1.**
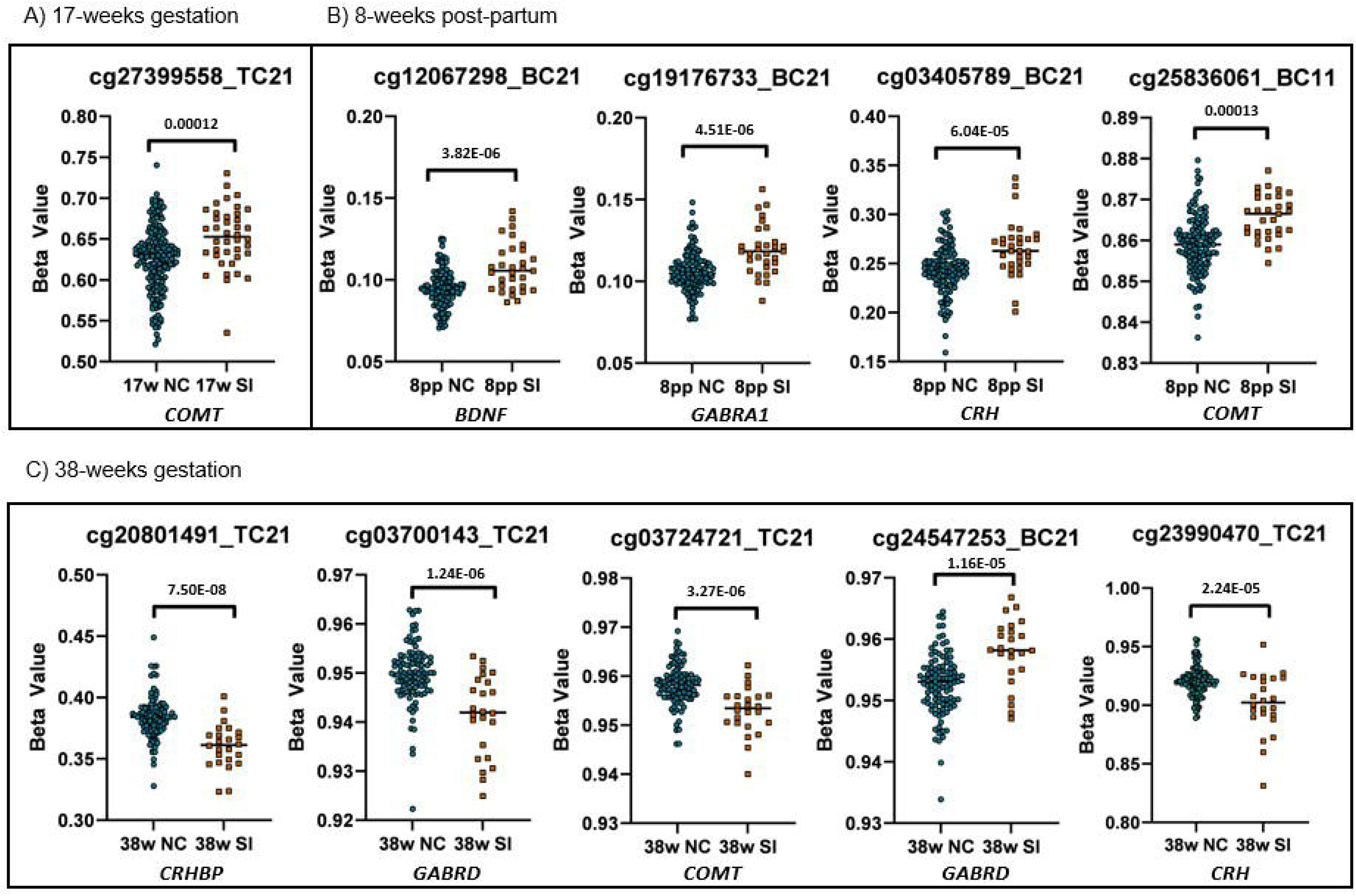
Targeted Significantly Differentially Methylated Probe Results. Individual CpG plots with significant DNA methylation (β-value) changes between perinatal SI and NC individuals at **A)** 17-weeks gestation (*n* = 224), **B)** 8-weeks pp (*n* = 127), and **C)** 38-weeks gestation (*n* = 158) (Bonferroni ≤ 0.000139). At 38w gestation, 10 CpGs remained significant following Bonferroni correction, with the top five significant CpGs displayed here. Unadjusted *p*-values displayed. *SI*, suicidal ideation. *NC*, non-SI controls.

### 4.3 Genome-wide DMPs associated with perinatal SI

Adjusted linear models using Bonferroni correction identified 465, 2,880, and 510 DMPs between SI and NC at each timepoint with the top five CpGs shown in **Table 2**. The top CpGs for our genome-wide analysis were cg11693709_TC21, cg11712199_TC21, and cg16386488_TC21, associated with *BUB1-PAK6*, *CSRNP3*, and *ENSG00000310033* respectively. Similar to our targeted analysis, 17w gestation and 8w post-partum showed an overall hypermethylated status in perinatal SI with only one and two CpGs hypomethylated in the top 50 hits respectively. At 38w gestation, perinatal SI showed an opposite trend where 78% of the top 50 CpGs were hypomethylated in perinatal SI vs NC. The complete list of significant genome-wide DMP results can be found in **Supplementary Table 4.**

**Table 2.**
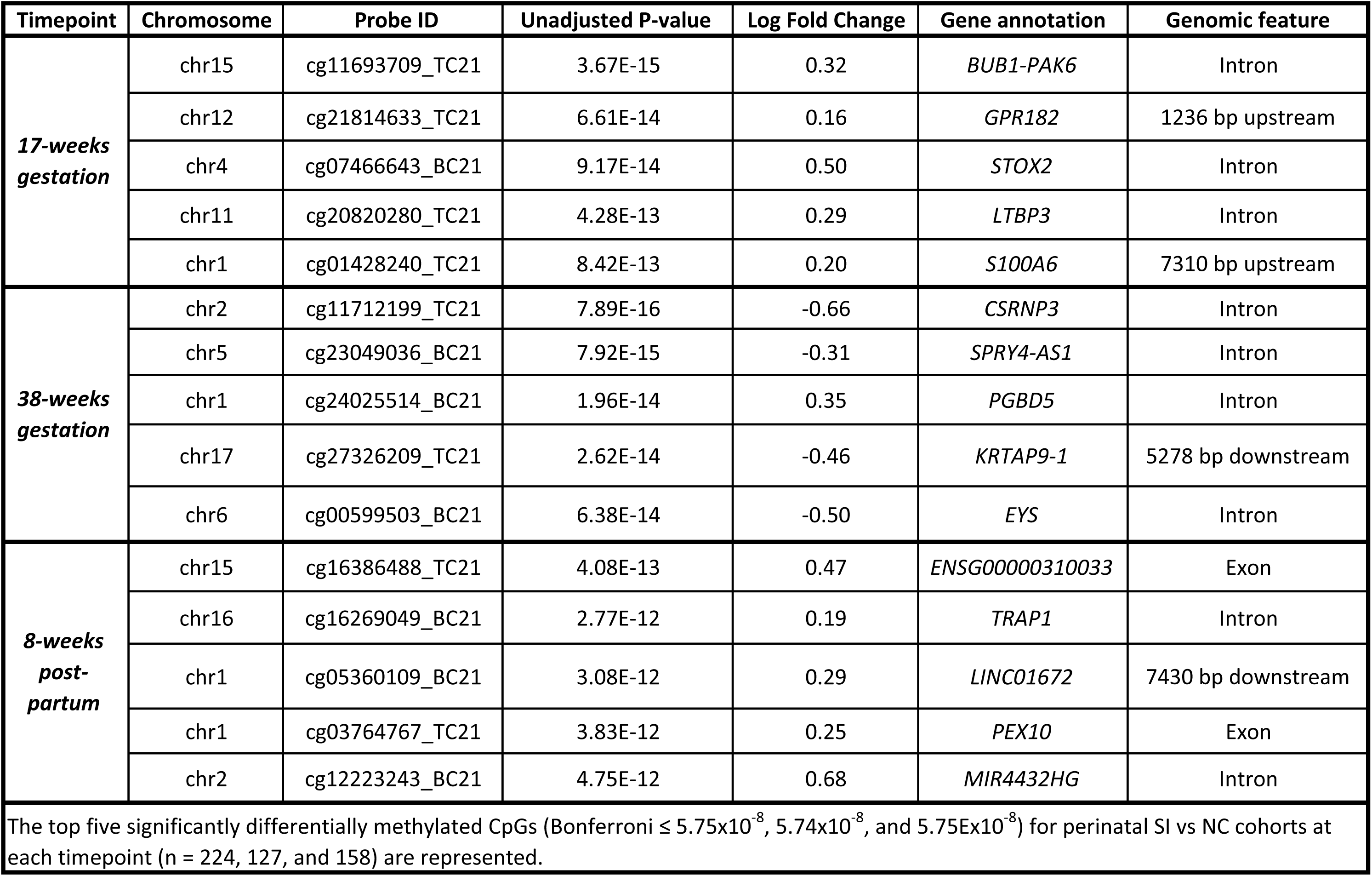
Genome-wide Significantly Differentially Methylated Probes. The top five significantly differentially methylated CpGs (Bonferroni ≤ 5.75×10^-8^, 5.74×10^-8^, and 5.75E×10^-8^) between perinatal SI and NC cohorts at each timepoint (n = 224, 127, and 158) are represented.

| Timepoint | Chromosome | Probe ID | Unadjusted P-value | Log Fold Change | Gene annotation | Genomic feature |
| --- | --- | --- | --- | --- | --- | --- |
| **17-weeks**<br>**gestation** | chr15 | cg11693709_TC21 | 3.67E-15 | 0.32 | *BUB1-PAK6* | Intron |
|  | chr12 | cg21814633_TC21 | 6.61E-14 | 0.16 | *GPR182* | 1236 bp upstream |
|  | chr4 | cg07466643_BC21 | 9.17E-14 | 0.50 | *STOX2* | Intron |
|  | chr11 | cg20820280_TC21 | 4.28E-13 | 0.29 | *LTBP3* | Intron |
|  | chr1 | cg01428240_TC21 | 8.42E-13 | 0.20 | *S100A6* | 7310 bp upstream |
| **38-weeks**<br>**gestation** | chr2 | cg11712199_TC21 | 7.89E-16 | -0.66 | *CSRNP3* | Intron |
|  | chr5 | cg23049036_BC21 | 7.92E-15 | -0.31 | *SPRY4-AS1* | Intron |
|  | chr1 | cg24025514_BC21 | 1.96E-14 | 0.35 | *PGBD5* | Intron |
|  | chr17 | cg27326209_TC21 | 2.62E-14 | -0.46 | *KRTAP9-1* | 5278 bp downstream |
|  | chr6 | cg00599503_BC21 | 6.38E-14 | -0.50 | *EYS* | Intron |
| **8-weeks**<br>**post-partum** | chr15 | cg16386488_TC21 | 4.08E-13 | 0.47 | *ENSG00000310033* | Exon |
|  | chr16 | cg16269049_BC21 | 2.77E-12 | 0.19 | *TRAP1* | Intron |
|  | chr1 | cg05360109_BC21 | 3.08E-12 | 0.29 | *LINC01672* | 7430 bp downstream |
|  | chr1 | cg03764767_TC21 | 3.83E-12 | 0.25 | *PEX10* | Exon |
|  | chr2 | cg12223243_BC21 | 4.75E-12 | 0.68 | *MIR4432HG* | Intron |
| The top five significantly differentially methylated CpGs (Bonferroni $\leq 5.75 \times 10^{-8}$, $5.74 \times 10^{-8}$, and $5.75 \times 10^{-8}$) for perinatal SI vs NC cohorts at each timepoint (n = 224, 127, and 158) are represented. | | | | | | |

### 4.4. DMRs associated with perinatal SI

Our DMR analysis resulted in 7, 25, and 12 DMRs that remained significant following our pre-determined criteria (minimum of 3 probes, a Fisher’s probability ≤ 0.01, and ±5% mean methylation difference). The top DMRs by timepoint were annotated to *DUSP22, MPL*, and *MDGA1* respectively **(Table 3).** Only one DMR (chr2: 238644242-238644765; hg38) was shared across all timepoints. This DMR included 3 CpGs annotated to *LINC01937.* The significant results from the DMR analysis for each timepoint are in **Supplementary Table 5.**

**Table 3.** Genome-wide Differentially Methylated Regions. The top five differentially methylated regions between SI and NC and from each timepoint (*n* = 224, 127, and 158) analysis are represented. *NC*, non-SI controls. *SI*, suicidal ideation. *Minimum smoothed FDR*, minimum false discovery rate of the smoothed estimate. *Stouffer*, *p*-value from Stouffer’s test. *HMFDR*, *p*-value from harmonic mean test. *Fisher, p*-value of Fisher’s combined probability test. *Mean differential*, mean differential across the DMR (logit2-space).

| Timepoint | Chromosome | Start | End | Width | *n* CpGs | Minimum smoothed FDR | Stouffer | HMFDR | Fisher | Mean differential | Overlapping genes |
| --- | --- | --- | --- | --- | --- | --- | --- | --- | --- | --- | --- |
| **17-weeks gestation** | chr6 | 291124 | 293285 | 2162 | 11 | 1.18E-09 | 1.47E-13 | 0.003076 | 8.15E-12 | -0.075359296 | DUSP22, AL365272.1 |
|  | chr7 | 39130897 | 39131513 | 617 | 6 | 2.71E-10 | 1.05E-12 | 0.000532 | 1.97E-11 | -0.065721821 | POU6F2 |
|  | chr1 | 18784240 | 18784595 | 356 | 4 | 4.50E-09 | 1.35E-09 | 0.000111 | 1.86E-09 | -0.08903495 | Intergenic |
|  | chr4 | 48944587 | 48944782 | 196 | 3 | 3.65E-08 | 2.68E-09 | 8.45E-05 | 7.49E-09 | -0.051990476 | Intergenic |
|  | chr2 | 238644242 | 238644765 | 524 | 3 | 7.94E-08 | 7.32E-08 | 6.84E-05 | 5.22E-08 | -0.084723121 | LINC01937 |
| **38-weeks gestation** | chr1 | 43348221 | 43349364 | 1144 | 10 | 9.86E-73 | 3.17E-46 | 9.87E-09 | 1.14E-56 | -0.063786843 | MPL, AL139289.2 |
|  | chr15 | 67063159 | 67065023 | 1865 | 7 | 1.06E-17 | 9.10E-15 | 4.89E-05 | 4.83E-18 | -0.075684347 | SMAD3, AC087482.1 |
|  | chr2 | 239318494 | 239319523 | 1030 | 4 | 3.28E-22 | 1.25E-15 | 9.86E-07 | 6.08E-17 | -0.098462884 | HDAC4 |
|  | chr2 | 238644242 | 238644765 | 524 | 3 | 4.03E-20 | 1.19E-15 | 6.25E-08 | 1.74E-16 | -0.126609721 | LINC01937 |
|  | chr1 | 100538078 | 100539565 | 1488 | 7 | 2.38E-17 | 1.14E-12 | 2.51E-05 | 4.85E-16 | -0.065265706 | GPR88 |
| **8-weeks post-partum** | chr6 | 37648634 | 37650738 | 2105 | 14 | 7.69E-23 | 5.21E-30 | 8.61E-05 | 6.52E-28 | 0.055321135 | MDGA1 |
|  | chr10 | 70494558 | 70495313 | 756 | 4 | 3.00E-15 | 8.16E-18 | 2.05E-05 | 1.19E-16 | 0.060656724 | PALD1 |
|  | chr18 | 49723780 | 49724526 | 747 | 3 | 5.01E-11 | 1.36E-10 | 1.45E-05 | 1.45E-10 | 0.052991912 | Intergenic |
|  | chr21 | 46184138 | 46185260 | 1123 | 7 | 8.26E-08 | 9.73E-12 | 0.001934 | 2.73E-10 | -0.053515836 | AP001468.1, SPATC1L |
|  | chr6 | 2952793 | 2953208 | 416 | 3 | 1.07E-07 | 2.69E-08 | 0.000136 | 7.08E-08 | 0.058247703 | SERPINB6 |

### 4.5 Pathway enrichment analyses at each timepoint

GO and KEGG pathway enrichment analyses were conducted using all DMRs that met FDR significance (≤ 0.05), regardless of our applied criteria. At 38w gestation and 8w post-partum, 427 and 14 GO processes were found to be significantly enriched (FDR ≤ 0.05) between perinatal SI and NC **(Supplementary Table 6).** Upon examination of the top 20 GO terms, regardless of significance, we noted 38w gestation and 8w post-partum shared the most similarities, with eight processes similarly enriched at each timepoint between SI vs NC. In addition, both timepoints implicated “anatomical structure development” as the top GO enriched trait and included dysregulation of pathways associated with developmental process. In contrast, GO term “mRNA base-pairing translational repressor” was the only nominally dysregulated pathway identified at 17w gestation (FDR = 0.05). KEGG pathway enrichment was only statistically significant at 38w gestation and identified 10 KEGG traits (FDR ≤ 0.05) differentiating perinatal SI and NC **(Figure 2; Supplementary Table 7).** Signaling cascades such as MAPK, PI3K-AKT, and mTOR pathways were all implicated in KEGG analysis, as well hormone-specific processes such as Oxytocin and Estrogen signaling. “Vitamin Digestion and Absorption” was the most significantly dysregulated pathway and hypermethylated in individuals with perinatal SI. Of note, GO and KEGG pathway analyses using DMPs failed to meet significance at all timepoints.

**Figure 2.**
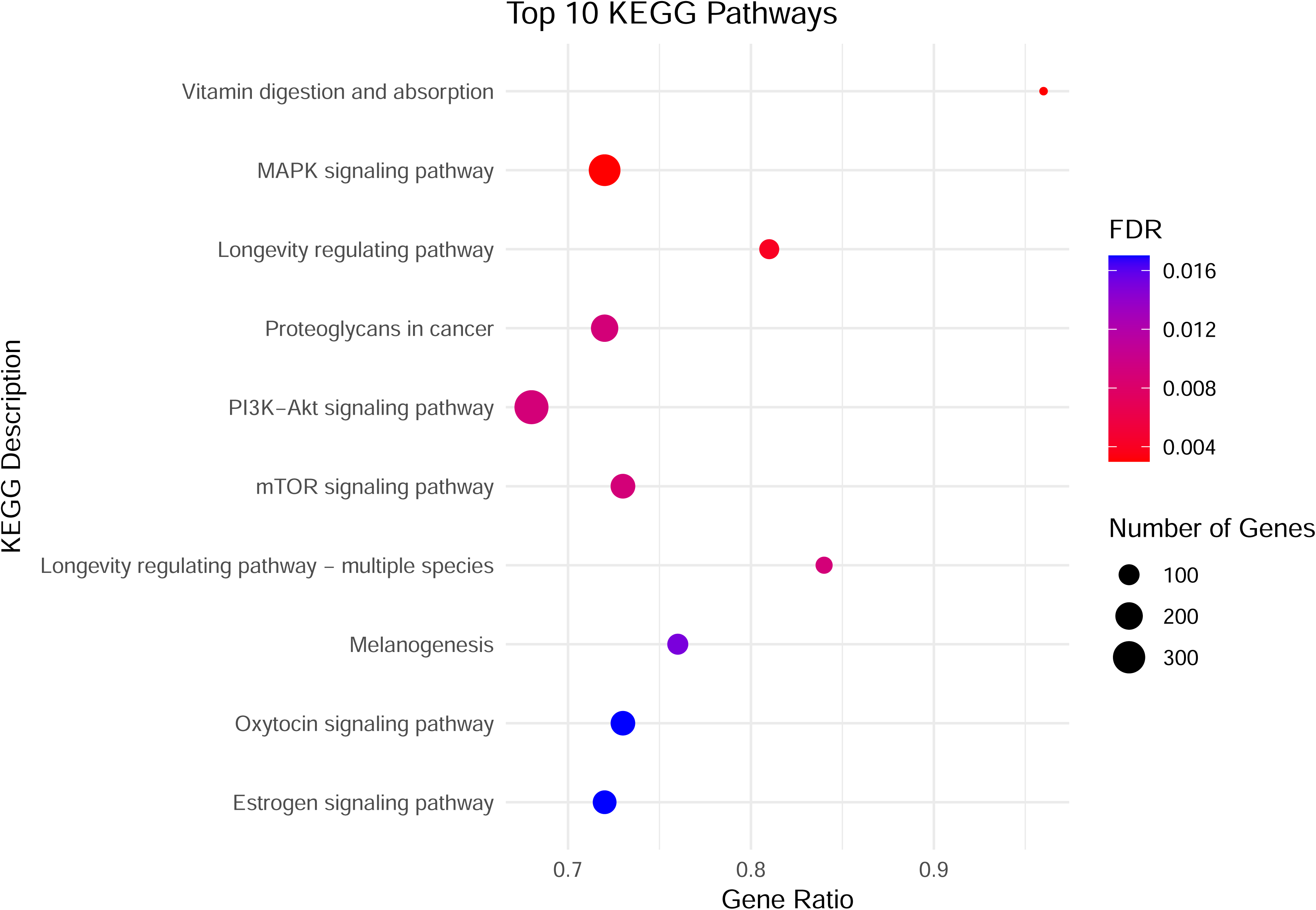
KEGG Pathway Results at 38-weeks Gestation. All significant KEGG pathways (enriched) ordered by -log of the adjusted *p*-value (FDR). Gene ratio is the proportion of differentially methylated genes in 38-weeks SI vs NC compared to the total number of genes (size of point) assigned to that pathway.

### 4.6 Perinatal DNAm biomarkers are predictive of SI-risk

Performance evaluation of DNAm changes as biomarkers for perinatal SI-risk prediction was conducted to verify clinical applicability. When predicting SI at 38w gestation using 17w gestation classifiers, AUC was 100% based on 17w DNAm data alone, 53% for 17w depression severity alone, and 100% for combined classification. We conducted two models to predict SI risk at 8w pp with classifiers from both 17w and 38w gestation. Using 38w gestation classifiers, AUC was 92.6% for DNAm data alone, 89.2% for depression severity alone, and 100% for combined classification. 17w gestation classifiers resulted in slightly reduced but effective prediction capability for 8w pp SI-prediction, with AUC of 66.9% for DNAm data alone, 88% for depression severity alone, and 93.2% for combined classifiers. To determine if DNAm biomarkers withstood identification of novel SI, we repeated our 17w gestation to 8w pp prediction analysis only with SI-Null individuals at 17w gestation (*n* = 100). This yielded improved prediction accuracy for our DNAm biomarkers with AUC showing 86.2% for DNAm data alone, 82.6% for depression severity alone, and 94.9% for combined classification **(Figure 3A-D).**

**Figure 3.**
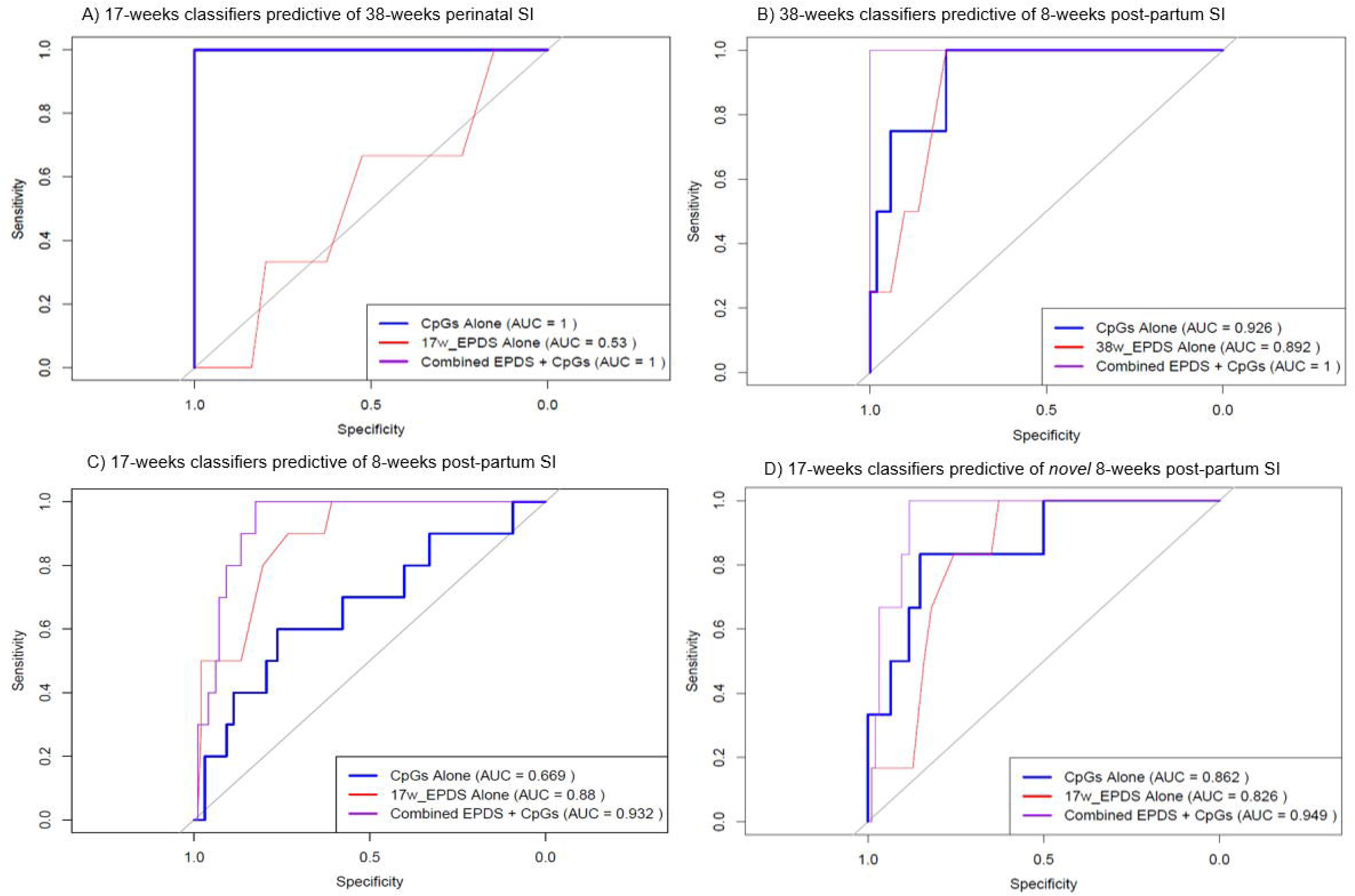
Clinical-Epigenetic Models Predictive of Perinatal SI Risk. AUC of the ROC curves presented for classification of SI risk at **A)** 38-weeks gestation based on 17-weeks classifiers (*n* = 102), **B)** 8-weeks pp based on 17-weeks classifiers (*n* = 107), **C)** 8-weeks pp based on 38-weeks classifiers (*n* = 55) and **D)** *novel* SI at 8-weeks pp based on 17-weeks gestation (n = 100). Classifiers for each prediction model included the beta-values of the top 10 significantly differentially methylated CpG sites from the SI v NC whole-genome analysis (CpGs Alone), depression severity based on EPDS total score (EPDS Alone), and the beta-values and depression severity combined (Combined EPDS + CpGs) from each classification timepoint. *NC*, non-SI controls. *SI,* suicidal ideation. *AUC,* area under the curve. *ROC,* receiver operating characteristic. *EPDS,* Edinburgh Postnatal Depression Scale.

## 5. Discussion

This study provides evidence of distinct DNAm biomarkers associated with perinatal SI, producing meaningful biological targets for therapeutic application and resulting in clinically relevant methods for risk prediction. Our findings show significant DNAm changes across numerous CpG loci in pregnant individuals who endorsed SI on EPDS questionnaires, and although no common loci across time points were identified, we noted more hypermethylated CpG sites associated with perinatal SI at 17w gestation and 8w pp. Our initial targeted analyses identified several brain-relevant genes (*COMT, CRHBP, GABRD, CRH, NR3C1, BDNF,* and *GABRA1)* that were altered in perinatal SI, which suggests a role for stress response as a driver of this phenotype (48, 49). HPA axis dysregulation has been increasingly associated with psychiatric disorders (50, 51) and is one of the most well-documented biological risk factors for PPD (47, 49). When experiencing stress, secretion of corticotropin-releasing hormone (*CRH)* from the hypothalamus will activate the pituitary gland to produce adrenocorticotropic hormone (*ACTH),* which in turn promotes glucocorticoid release, such as cortisol, that functions to regulate immune, metabolic, and endocrine systems (52). Cortisol initiates stress-response signaling by binding *NR3C1*, or glucocorticoid receptor (GR), resulting in GR nuclear translocation where it mediates transcription of stress-response genes while also providing negative feedback through inhibition of further hormone secretion. Similarly, corticotropin-releasing hormone binding protein *(CRHBP)*, works as a negative regulator by binding to and inactivating *CRH*, reducing stress-response signaling. Notably, the association between stress and epigenetic modifications are well-documented with DNAm providing a potential mechanism for impaired stress response (52–55). For example, maternal stress, depression, and anxiety were all significantly associated with hypermethylation of *NR3C1* (56–58). Our targeted analysis highlights a role for impaired stress response as a potential driver of perinatal SI. From our *a priori* candidate genes, we noted several related to the HPA axis, or its regulation, were differentially methylated across all timepoints. Epigenetic alterations to HPA-axis genes have also been identified in cases of non-pregnancy related suicidality (21, 43, 59) as well, with similar promoter-related methylation patterns of *NR3C1* (cg03906910_TC21; cg15645634_TC11) and *CRH* (cg23990470_TC21) found in our study. Interestingly, our study revealed hypomethylation of *CRHBP* (cg20801491_TC21) to be associated with perinatal SI, a deviation from Roy et al. 2017 where hypermethylation of CpG islands near the *CRHBP* transcription start site (TSS) was identified in MDD individuals with SI (21). This apparent departure of *CRHBP* methylation could be explained by our cohort background and the specific requirements surrounding fetal development. Specifically, *CRHBP* is imperative for regulating the increased *CRH* levels from placental secretion and ensuring appropriate stress response (60). Therefore, the hypomethylation of *CRHBP* found in our study may instead be an attempt to mitigate the primary stress-response dysfunction seen in perinatal SI, rather than a significant driving factor of this phenotype. However, it is important to note that while both results are reflective of TSS regions, direct comparison of CpG probes are not available due to methodological differences between the current study and Roy et al. 2017. Similarly, other HPA-axis related genes showing strong associations with non-pregnancy related suicidality, such as *FKBP5 and SKA2* (21, 59), were not significantly differentially methylated in perinatal SI. This suggests that the epigenetic differences present in the current study, along with any proposed HPA-axis dysregulation, may be female and/or perinatal-specific. Roy et al. 2017, among others, also identified hypermethylation of brain-derived neurotrophic factor (*BDNF),* a critical growth factor for neuronal plasticity and a known regulator of the HPA-axis, as a significant biomarker of suicide risk (22, 61). Our data supports previous correlations, with significant hypermethylation of *BDNF* loci (cg12067298_BC21; cg11718030_TC21) identified in women with perinatal SI compared to NC. Furthermore, *BDNF* dysfunction may play a more significant role in the onset of post-partum mood disorders, with previous studies reporting lower levels of BDNF in individuals with PPD compared to controls (62), a finding that is supported in our 8w pp results with *BDNF* TSS hypermethylation as the most significantly differentially methylated CpG.

In addition to HPA-axis dysregulation, our study supports a role for increased neuronal excitation associated with perinatal SI as evidenced by hypermethylation of CpGs associated with *COMT* (cg27399558_TC21; cg25836061_BC11) and *GABRA1* (cg02065387_BC21; cg24523000_TC21; cg19176733_BC21) TSS. *COMT* encodes an enzyme vital to degradation of catecholamines, such as dopamine, and is crucial for proper brain functioning (63, 64). When considering evolutionary advantage, promoter hypermethylation of *COMT* may be beneficial during the peripartum, as increased dopamine availability would reinforce rewarding affects associated with parental care (65). However, *COMT* promoter hypermethylation, and subsequent reduction in activity, may also heighten epinephrine and norepinephrine, neurotransmitters highly involved in acute stress response, and thus may consequently contribute to heightened risk of SI in the peripartum. *COMT* DNAm has been shown to correlate with psychiatric disorders such as MDD, schizophrenia, bipolar disorder, and suicide completion; however, hypomethylation of *COMT* promoter was primarily identified in these individuals (66, 67). This deviation may again highlight the unique nuances associated with perinatal-related SI, which is further supported by more recent studies examining sex-specific differences associated with *COMT* polymorphisms and psychiatric risk (68). Moreover, GABAergic dysfunction has been increasingly associated with neuropsychiatric phenotypes, including PPD (69, 70). Recently, a study evaluating the pharmacological effects of an α1 receptor agonist in PPD-like mice identified reduced expression of the α1 subunit of the gamma-aminobutyric acid receptor (gabra1) in these mice, and following drug treatment, gabra1 expression and behavioral phenotypes were attenuated (70). In addition, *gabra1* loss of function mutations have previously been shown to reduce and/or abolish GABAergic signaling either through lowered gating channel efficacy or receptor surface expression (71, 72). Consequently, hypermethylation of *GABRA1* promoter CpGs in our study may result in heightened perinatal SI due to reduced GABAergic neurotransmission and increased neuronal excitation.

Examination of our genome-wide results show notable temporal patterns associated with perinatal SI, with predicted RNA genes becoming increasingly relevant especially at 8w pp. *BUB1-PAK6*, the top 17w gestation DMP, is a read-through transcript that produces a serine/threonine protein kinase which notably interacts with androgen receptor (AR) to regulate gene transcription (73). Among altered gene regulation, reduced AR signaling due to increased PAK6 activity has also been thought to impact dopaminergic transmission (74, 75), potentially generating additive behavioral dysfunction. At 38w, perinatal SI was most associated with DNAm changes in *CSRNP3,* a gene predicted to exhibit DNA-binding transcription factor activity and thereby positive regulation of genes transcribed by RNA polymerase II. Previous studies reflecting on stress show slightly confounding results with one showing *CSRNP3* promoter hypermethylation associated with early life stress (76); while another found increased csrnp3 levels in the rostral posterior hypothalamic (rPH) region of rats undergoing homotypic stress (77). As our results showed altered DNAm of CpGs located within intronic regions of their respective gene, our ability to draw inferences on transcriptional consequences is limited due to intronic DNAm capable of enhancing (78) or decreasing gene expression (79), as well as resulting in alternative splicing (80). Nevertheless, our results suggest a link between gene regulation and perinatal SI. Notably, three out of the top five significant DMPs were annotated to lncRNAs at 8w pp. As lncRNAs have been established as key regulators of gene expression through alterations to RNA processing and interactions with DNA and proteins (81), this finding could suggest additional molecular remodeling may be influencing SI following parturition.

The timepoint with the most DNAm changes associated with SI was at 38w gestation, suggesting increased epigenetic modifications, and potential for signaling dysregulation, as pregnancy progresses. Indeed, a recent report by Fradin et al. 2023 showed significant DNAm changes in maternal blood from early to late pregnancy in genes thought to be important for maternal adaptation to embryonic development and labor (82). Interestingly, this study also revealed significant hypomethylation of arginine vasopressin (*AVP)* a crucial neuropeptide for socio-emotional regulation, during late-stage pregnancy, which may implicate a role for DNAm in maternal adaptation to care (82). Similarly, the increased volume of DNAm changes associated with SI at 38w in the current study could possibly be associated with dysregulation of an evolutionary response to “prime” maternal behavior in such a way that maximizes offspring survivability (83–87). When properly maintained, this molecular reprogramming, influenced by hormonal fluctuations (83), is thought to improve environmental responsiveness (85, 88) while also providing rewarding effects associated with caregiving (65). Although evolutionarily advantageous, hormonal imbalances may result in improper reprogramming and consequently, lead to maladaptive phenotypes (83) such as SI. The significant dysregulation of Oxytocin and Estrogen signaling found in our 38w KEGG Pathway results provides additional evidence supporting a role for hormone dysregulation in the onset of perinatal SI (89, 90). However, future efforts to identify specific correlations between perinatal hormone levels and DNAm will be required to establish this.

One surprising finding was the lack of commonly shared CpGs associated with SI across each perinatal timepoint in both the targeted and genome-wide analyses. This distinction in DNAm differences may be reflective of intrapersonal DNAm trends throughout pregnancy due to the primarily independent sample populations making up each timepoint. Therefore, future studies surveilling individual DNAm trajectories throughout the peripartum period and its association with behavioral phenotypes would be required to parse out this discrepancy. However, in an alternative effort to gauge the clinical efficacy of our genome-wide DNAm biomarkers, we inspected the AUC of the ROC classification curves for “future” perinatal SI prediction-risk in individuals with overlapping timepoints. All three risk-prediction models identified significant prediction accuracy for the top 10 genome-wide DMPs alone, with two out of three models showing better performance of DNAm alone at predicting individuals with perinatal SI than depression severity based on total EPDS scores. This suggests that DNAm patterns early in pregnancy informs SI risk in later trimesters. Furthermore, when examining prediction accuracy of novel SI at 8w pp (*i.e.* individuals who were SI null at 17w and later transitioned), our 17w gestation to 8w pp model showed similar albeit improved predictive capacity for our DNAm biomarkers. The finding that epigenetic biomarkers did not overlap across timepoints, although explanatory power was still achieved when leveraging DNAm in our ROC approach, might also suggest that these DNAm modifications are occurring *prior* to SI. Importantly, this method has been similarly established in cases of non-pregnancy related suicidality (91) and PPD (92), further highlighting the usefulness of DNAm in risk-prediction.

There are several key limitations to consider in the interpretation of our findings. While the EPDS is a highly robust screening tool with question 10 used as a standard clinical measure for perinatal SI (30, 31) we cannot guarantee the distinction between SI and thoughts of NSSI. Furthermore, while our cohorts were exempt from NSSI and suicide attempt in the year prior to conception, we did not include historical psychiatric behavior and diagnoses from individuals as covariates in our analyses. In addition, we acknowledge several of the DMP findings in this study resulted in minimal variance in DNA methylation between groups; however, we cannot definitively negate their biological implications in the current study. Access to individuals with both DNAm data and clinical data at all timepoints also hindered our ability to track longitudinal changes across the peripartum period and likely contributed to the independent biomarker results at each timepoint. Our use of AUC analyses as prediction risk-assessment was an attempt to reduce this limitation and provide clarity of the usefulness of these epigenetic biomarkers. While these results indicate significant prediction accuracy of our DNAm biomarkers, the strikingly high AUC seen in our 17w-38w gestation model may throw doubt on our model, reflecting small sample sizes with few representative individuals with SI. It is of note, however, that our model withstood identification of individuals who “switched” between psychiatric phenotypes across timepoints and remained effective at predicting novel post-partum SI, reflecting its usefulness in clinical application. Interestingly, when compared to similar risk-prediction models, smaller sample populations instead hindered model prediction accuracy (91), suggesting our model’s AUC scores may also be reflective of the relatively short time periods we are predicting for. However, repetition in larger cohorts will be necessary to ensure prediction accuracy remains effective.

In summary, we provide evidence of biomarkers associated with SI at multiple perinatal timepoints, suggesting altered stress response, hormone signaling, and developmental processes. DNAm changes found in this study provide both mechanistic insights, through identification of brain-relevant genes, and clinical usefulness, as evidenced by risk-prediction modeling, contributing to the field of perinatal psychiatry and epigenetics. Although future longitudinal work is required, we propose that DNAm modifications may occur prior to the appearance of perinatal SI, thus reflecting an important role in risk-prediction, and when implemented with readily available screening tools has the potential to reduce suicide-related maternal mortality.

## Acknowledgements

This work was supported by Roy J. Carver Charitable Trust Early Stage Investigator Award (MEG); Environmental Health Sciences Center (EHSRC) awards (NIH P30 ES005605; MEG); the University of Iowa Hawkeye Intellectual and Developmental Disabilities Research Center (HAWK-IDDRC, P50 HD103556); Pharmacological Sciences Training Program Fellowship (NIH T32 GM144636; ESW); the European Union’s Horizon Europe Research and Innovation Program (101057604; AS & JR); Swedish Research Council VR (2024-03199; AS); Swedish Research Council for Health, Working Life and Welfare Grants (2025-02047, 2025-02053; AS), and the Swedish Brain Foundation (2024-0138, AS).

## Author Contributions

**ESW:** Conceptualization, Investigation, Methodology, Formal Analysis, Software, Writing-original draft, Writing-review and editing, Visualization. **JD:** Writing-review and editing. **JR:** Writing-review and editing. **AS:** Conceptualization, Data curation, Writing-review and editing. **MEG:** Conceptualization, Supervision, Funding Acquisition, Writing-review and editing, original draft. All authors edited the manuscript.

## Data Availability Statement

Data available on request from the authors.

