## Supplementary Table S3 for "Early Epigenetic Biomarkers for Perinatal Suicidal Ideation: DNA Methylation Signatures Across the Peripartum Period"

**Supplementary Table S3.1. Significant Target CpGs in Perinatal SI v NC cohort at 17w gestation.**

| Probe ID | Chr | Position | Gene Feature | Gene Annotation | Log FC | *p* -value | FDR |
|---|---|---|---|---|---|---|---|
| cg27399558_TC21 | chr22 | 19941093 | TSS1500;TSS150( | COMT;COMT | 0.178628 | 0.000118 | 0.030693 |
| cg26840770_BC21 | chr11 | 27701743 | TSS1500;TSS150( | BDNF;BDNF;BDNF;B | -0.20348 | 0.000249 | 0.030693 |
| cg09926649_BC21 | chr22 | 19950573 | TSS1500;TSS150( | COMT;COMT | 0.185996 | 0.000256 | 0.030693 |
| cg11514288_BC21 | chr13 | 46897062 | TSS200;TSS200 | HTR2A;HTR2A | -0.17655 | 0.00069 | 0.03976 |
| cg15825127_BC11 | chr1 | 2025634 | exon_4 | GABRD | 0.11549 | 0.000713 | 0.03976 |
| cg02613510_TC21 | chr11 | 27702242 | TSS1500;TSS150( | BDNF;BDNF;BDNF;B | -0.22564 | 0.000781 | 0.03976 |
| cg17344495_BC21 | chr13 | 46831761 | 3UTR;exon_4;3U | HTR2A;HTR2A;HTR2 | -0.17418 | 0.000826 | 0.03976 |
| cg03710862_TC21 | chr3 | 8770042 | TSS1500;TSS150( | OXTR;OXTR;OXTR;O | -0.23592 | 0.000884 | 0.03976 |

**Supplementary Table S3.2. Significant Target CpGs in Perinatal SI v NC cohort at 38w gestation.**

| Probe ID | Chr | Position | Gene Feature | Gene Annotation | Log FC | *p* -value | FDR |
|---|---|---|---|---|---|---|---|
| cg20801491_TC21 | chr5 | 76952695 | TSS1500 | CRHBP | -0.14983 | 7.50E-08 | 2.71E-05 |
| cg03700143_TC21 | chr1 | 2030323 | exon_9;3UTR | GABRD;GABRD | -0.22385 | 1.24E-06 | 0.000225 |
| cg03724721_TC21 | chr22 | 19951538 | 5UTR;exon_1 | COMT;COMT | -0.17498 | 3.27E-06 | 0.000393 |
| cg24547253_BC21 | chr1 | 2030643 | exon_9;3UTR | GABRD;GABRD | 0.179303 | 1.16E-05 | 0.00105 |
| cg23990470_TC21 | chr8 | 66179847 | TSS1500 | CRH | -0.32186 | 2.24E-05 | 0.001566 |
| cg03906910_TC21 | chr5 | 143434823 | 5UTR;exon_1;TS! | NR3C1;NR3C1;NR3C | 0.173252 | 2.60E-05 | 0.001566 |
| cg11718030_TC21 | chr11 | 27722816 | TSS1500 | BDNF | 0.121174 | 3.32E-05 | 0.001711 |
| cg15645634_TC11 | chr5 | 143404074 | 5UTR;exon_1;TS! | NR3C1;NR3C1;NR3C | 0.236069 | 3.87E-05 | 0.001747 |
| cg02065387_BC21 | chr5 | 161847301 | 5UTR;exon_1;TS! | GABRA1;GABRA1;G/ | 0.212263 | 4.64E-05 | 0.001861 |
| cg24523000_TC21 | chr5 | 161846833 | TSS1500;TSS150( | GABRA1;GABRA1;G/ | 0.191682 | 6.19E-05 | 0.002236 |
| cg02527472_TC21 | chr11 | 27721801 | exon_1;5UTR;TS! | BDNF;BDNF;BDNF;B | 0.205729 | 0.000316 | 0.010387 |
| cg15264052_BC21 | chr11 | 27506464 | TSS1500;TSS150( | BDNF-AS;BDNF-AS;B | 0.116153 | 0.000374 | 0.010684 |
| cg01086446_BC21 | chr5 | 161846847 | TSS1500;TSS150( | GABRA1;GABRA1;G/ | 0.164456 | 0.000385 | 0.010684 |
| cg25085537_BC21 | chr3 | 8770053 | TSS1500;TSS150( | OXTR;OXTR;OXTR;O | 0.201525 | 0.000457 | 0.011371 |
| cg00889627_TC11 | chr1 | 2028191 | exon_6 | GABRD | 0.153731 | 0.000472 | 0.011371 |
| cg24924243_TC11 | chr11 | 27506883 | exon_1;exon_1;e | BDNF-AS;BDNF-AS;B | -0.20284 | 0.000568 | 0.012673 |
| cg03747251_BC21 | chr11 | 27701175 | TSS1500;TSS150( | BDNF;BDNF;BDNF;B | 0.264409 | 0.000601 | 0.012673 |
| cg23448729_TC11 | chr5 | 63960458 | exon_1 | HTR1A | 0.191968 | 0.000632 | 0.012673 |
| cg19035496_BC21 | chr8 | 66178557 | TSS200 | CRH | 0.155211 | 0.000689 | 0.013097 |
| cg25412831_BC11 | chr11 | 27720591 | 5UTR;exon_1;5U | BDNF;BDNF;BDNF;B | -0.13841 | 0.000758 | 0.013679 |
| cg18117895_TC11 | chr11 | 27700519 | TSS1500;TSS200; | BDNF;BDNF;BDNF;B | -0.15591 | 0.000806 | 0.013864 |
| cg27068143_BC21 | chr13 | 46897129 | TSS200;TSS200 | HTR2A;HTR2A | -0.14306 | 0.001017 | 0.016695 |
| cg03257388_BC21 | chr3 | 8767527 | exon_3;exon_4;e | OXTR;OXTR;OXTR;O | -0.21525 | 0.001183 | 0.018513 |
| cg13648501_TC21 | chr5 | 143405693 | TSS1500;TSS150( | NR3C1;NR3C1;NR3C | 0.184164 | 0.001273 | 0.018513 |
| cg21919834_TC21 | chr22 | 19961310 | TSS1500 | COMT | -0.14822 | 0.001327 | 0.018513 |
| cg25913647_TC21 | chr5 | 76952306 | TSS1500 | CRHBP | 0.135307 | 0.001333 | 0.018513 |
| cg01515809_BC11 | chr17 | 59155035 | exon_1;TSS1500 | SKA2;PRR11 | 0.145087 | 0.001559 | 0.020844 |
| cg14483142_TC21 | chr3 | 8770072 | TSS1500;TSS150( | OXTR;OXTR;OXTR;O | 0.23113 | 0.002273 | 0.029306 |
| cg01294526_TC21 | chr5 | 143436852 | TSS1500;TSS150( | NR3C1;NR3C1;NR3C | 0.142667 | 0.002461 | 0.030632 |
| cg26609236_TC21 | chr1 | 2017915 | TSS1500 | GABRD | 0.123069 | 0.002821 | 0.033668 |
| cg16327961_BC21 | chr12 | 72031260 | exon_10 | TPH2 | -0.13991 | 0.002891 | 0.033668 |
| cg27569822_BC11 | chr17 | 30236101 | TSS1500 | SLC6A4 | 0.115833 | 0.003117 | 0.035159 |
| cg00850109_BC21 | chr5 | 161847838 | TSS1500;TSS150( | GABRA1;GABRA1 | -0.21353 | 0.003475 | 0.037874 |
| cg17036624_BC21 | chr3 | 8769915 | TSS1500;TSS150( | OXTR;OXTR;OXTR;O | 0.113902 | 0.003649 | 0.037874 |
| cg26720913_BC21 | chr5 | 143435369 | 5UTR;exon_1;5U | NR3C1;NR3C1;NR3C | 0.249752 | 0.003694 | 0.037874 |
| cg25457956_BC21 | chr11 | 27722117 | TSS200;TSS1500; | BDNF;BDNF;BDNF;B | 0.203759 | 0.003777 | 0.037874 |
| cg18640030_BC21 | chr8 | 66178517 | TSS200 | CRH | -0.21638 | 0.004123 | 0.039426 |
| cg12296752_BC11 | chr11 | 27659664 | TSS200;TSS200 | BDNF;BDNF | -0.12714 | 0.004255 | 0.039426 |
| cg11171527_BC21 | chr3 | 8768520 | 5UTR;exon_2;5U | OXTR;OXTR;OXTR;O | -0.24066 | 0.004259 | 0.039426 |
| cg24650785_TC21 | chr11 | 27720369 | 5UTR;exon_1;TS! | BDNF;BDNF;BDNF | 0.169467 | 0.005139 | 0.046376 |
| cg03167496_TC21 | chr11 | 27722072 | TSS200;TSS1500; | BDNF;BDNF;BDNF;B | 0.091764 | 0.005448 | 0.047418 |
| cg00629244_TC11 | chr5 | 143403814 | TSS200;TSS1500; | NR3C1;NR3C1;NR3C | 0.15978 | 0.005522 | 0.047418 |
| cg01294490_BC21 | chr6 | 35689129 | TSS1500;TSS150( | FKBP5;FKBP5;FKBP5 | -0.15372 | 0.005731 | 0.047418 |
| cg00247334_BC21 | chr3 | 8769857 | TSS1500;TSS150( | OXTR;OXTR;OXTR;O | 0.15176 | 0.00578 | 0.047418 |
| cg02834846_TC11 | chr8 | 66179232 | TSS1500 | CRH | -0.09366 | 0.006002 | 0.048145 |
| cg06841846_BC11 | chr17 | 30237076 | TSS1500 | SLC6A4 | 0.122378 | 0.006408 | 0.048412 |
| cg09353063_BC21 | chr3 | 8769406 | 5UTR;exon_1;5U | OXTR;OXTR;OXTR;O | 0.106859 | 0.006493 | 0.048412 |
| cg11808581_BC11 | chr1 | 2019286 | TSS200 | GABRD | -0.12017 | 0.006499 | 0.048412 |
| cg27193031_TC12 | chr11 | 27699541 | exon_1;5UTR | BDNF;BDNF | 0.05611 | 0.006585 | 0.048412 |

| | | | | | | | |
|---|---|---|---|---|---|---|---|
| cg08652028_BC21 | chr5 | 143434671 | 5UTR;exon_1;5U | NR3C1;NR3C1;NR3C | 0.090595 | 0.006705 | 0.048412 |
| cg23619332_TC21 | chr11 | 27700513 | TSS1500;TSS200; | BDNF;BDNF;BDNF;B | 0.110048 | 0.00694 | 0.049123 |

**Supplementary Table S3.3. Significant Target CpGs in Perinatal SI v NC cohort at 8w post-partum.**

| Probe ID | Chr | Position | Gene Feature | Gene Annotation | Log FC | *p*-value | FDR |
|---|---|---|---|---|---|---|---|
| cg12067298_BC21 | chr11 | 27702111 | TSS1500;TSS1500;TS | BDNF;BDNF;BDNF;B | 0.206631 | 3.82E-06 | 0.000815 |
| cg19176733_BC21 | chr5 | 161846971 | TSS1500;TSS1500;TS | GABRA1;GABRA1;G | 0.195603 | 4.51E-06 | 0.000815 |
| cg03405789_BC21 | chr8 | 66176660 | 3UTR;exon_2 | CRH;CRH | 0.179194 | 6.04E-05 | 0.007264 |
| cg25836061_BC11 | chr22 | 19951505 | TSS200 | COMT | 0.076063 | 0.000125 | 0.011324 |
