## Supplementary Table S4 for "Early Epigenetic Biomarkers for Perinatal Suicidal Ideation: DNA Methylation Signatures Across the Peripartum Period"

**Supplementary Table S4.1. Significant Genome-wide CpGs in Perinatal SI v NC cohort at 17w gestation.**

| Probe ID | Chr | Position | Gene Fea | Gene Ann | Log FC | *p* -value | FDR |
|---|---|---|---|---|---|---|---|
| cg11693709_TC21 | chr15 | 40249818 | | | 0.322964851 | 3.67E-15 | 3.19E-09 |
| cg21814633_TC21 | chr12 | 56993255 | TSS1500 | GPR182 | 0.161280115 | 6.61E-14 | 2.66E-08 |
| cg07466643_BC21 | chr4 | 183876448 | | | 0.495868898 | 9.17E-14 | 2.66E-08 |
| cg20820280_TC21 | chr11 | 65542153 | | | 0.289688003 | 4.28E-13 | 9.31E-08 |
| cg01428240_TC21 | chr1 | 153527289 | | | 0.204247178 | 8.42E-13 | 1.46E-07 |
| cg11093223_BC21 | chr9 | 34363684 | | | 0.159469971 | 1.27E-12 | 1.85E-07 |
| cg01376622_TC21 | chr12 | 7480045 | | | 0.253599407 | 1.60E-12 | 1.99E-07 |
| cg11219690_TC21 | chr22 | 23306067 | | | 0.307955211 | 2.71E-12 | 2.91E-07 |
| cg17283823_BC21 | chr12 | 11018653 | | | 0.28300112 | 3.01E-12 | 2.91E-07 |
| cg10629173_BC21 | chr10 | 5534471 | | | 0.197418183 | 1.09E-11 | 7.98E-07 |
| cg17288956_TC11 | chr12 | 11707195 | | | 0.16197169 | 1.10E-11 | 7.98E-07 |
| cg09743439_TC21 | chr6 | 79422014 | | | 0.326430594 | 1.10E-11 | 7.98E-07 |
| cg19721554_BC21 | chr5 | 72443538 | 3UTR;exo | ZNF366;ZN | 0.305475206 | 1.34E-11 | 8.94E-07 |
| cg03510569_TC21 | chr2 | 130988231 | | | 0.30214536 | 1.57E-11 | 9.74E-07 |
| cg02030542_BC21 | chr16 | 70688551 | | | 0.176721983 | 2.03E-11 | 1.08E-06 |
| cg23695907_BC21 | chr2 | 10101443 | | | 0.22702226 | 2.14E-11 | 1.08E-06 |
| cg24526299_BC21 | chr19 | 40241976 | exon_6;e | AKT2;AKT2 | 0.211909402 | 2.28E-11 | 1.08E-06 |
| cg17255391_BC21 | chr12 | 8038773 | | | 0.244331132 | 2.45E-11 | 1.08E-06 |
| cg10819493_BC21 | chr11 | 70281474 | | | 0.208896854 | 2.48E-11 | 1.08E-06 |
| cg16397176_BC21 | chr5 | 111563616 | | | 0.248234225 | 2.57E-11 | 1.08E-06 |
| cg06127719_BC21 | chr18 | 11150785 | TSS1500;T | PIEZO2;PIE | 0.280549872 | 2.62E-11 | 1.08E-06 |
| cg00654016_BC21 | chr1 | 41504163 | | | 0.177567057 | 3.71E-11 | 1.47E-06 |
| cg13721556_BC21 | chr9 | 68714833 | | | 0.174717515 | 3.95E-11 | 1.50E-06 |
| cg23924005_BC21 | chr19 | 2728759 | | | 0.211232991 | 4.32E-11 | 1.56E-06 |
| cg12585732_TC21 | chr10 | 79160820 | | | 0.187449135 | 5.30E-11 | 1.84E-06 |
| cg25370833_TC21 | chr6 | 149630240 | | | 0.427120898 | 6.20E-11 | 2.08E-06 |
| cg04839975_TC21 | chr3 | 193938829 | | | 0.308706183 | 6.63E-11 | 2.14E-06 |
| cg22737154_BC21 | chr2 | 64404480 | | | 0.219562491 | 8.14E-11 | 2.40E-06 |
| cg15062874_BC21 | chr14 | 23147422 | | | 0.270102158 | 8.31E-11 | 2.40E-06 |
| cg22755115_TC21 | chr17 | 59381937 | | | 0.136703961 | 8.52E-11 | 2.40E-06 |
| cg26156715_BC21 | chr5 | 65588685 | | | 0.386478389 | 8.55E-11 | 2.40E-06 |
| cg00190178_BC21 | chr1 | 9360959 | | | 0.230675454 | 9.43E-11 | 2.56E-06 |
| cg21811262_BC21 | chr16 | 81501128 | | | 0.258967948 | 9.97E-11 | 2.63E-06 |
| cg14394389_TC21 | chr9 | 133419355 | TSS1500;T | REXO4;REX | 0.200470174 | 1.07E-10 | 2.74E-06 |
| cg25093475_TC21 | chr20 | 25039789 | | | 0.153151184 | 1.12E-10 | 2.77E-06 |
| cg00080260_BC21 | chr1 | 3198445 | | | 0.264012657 | 1.34E-10 | 3.15E-06 |
| cg01048287_TC21 | chr8 | 29905341 | | | 0.265780185 | 1.37E-10 | 3.15E-06 |
| cg13762474_BC11 | chr15 | 42079610 | exon_13 | PLA2G4D | -0.31657135 | 1.38E-10 | 3.15E-06 |
| cg19270739_BC21 | chr1 | 1433466 | | | 0.205073142 | 1.55E-10 | 3.45E-06 |
| cg15993296_BC21 | chr19 | 7848988 | exon_4;e | EVI5L;EVI5 | 0.233675197 | 1.72E-10 | 3.74E-06 |
| cg18402315_TC21 | chr4 | 6834969 | | | 0.295117742 | 2.04E-10 | 4.33E-06 |
| cg12833422_TC21 | chr11 | 118295334 | | | 0.295188981 | 2.23E-10 | 4.62E-06 |
| cg10585061_TC21 | chr8 | 121784653 | | | 0.22064448 | 2.41E-10 | 4.83E-06 |

| | | | | | | | |
|---|---|---|---|---|---|---|---|
| cg17594864_TC21 | chr12 | 50241149 | | | 0.259152182 | 2.49E-10 | 4.83E-06 |
| cg21161367_TC21 | chr1 | 107932105 | | | 0.402777682 | 2.50E-10 | 4.83E-06 |
| cg15404463_BC21 | chr9 | 87509819 | | | 0.240927052 | 2.57E-10 | 4.85E-06 |
| cg22654923_BC21 | chr6 | 27162243 | | | 0.276541661 | 2.63E-10 | 4.86E-06 |
| cg23007535_TC21 | chr3 | 47108394 | | | 0.49755678 | 2.68E-10 | 4.86E-06 |
| cg12262372_TC21 | chr1 | 61076539 | TSS1500 | NFIA | 0.220879311 | 2.78E-10 | 4.93E-06 |
| cg13886001_TC21 | chr15 | 63912121 | exon_10; | DAPK2;DA | 0.174965924 | 2.96E-10 | 5.11E-06 |
| cg24855446_TC21 | chr3 | 105465262 | | | 0.26231525 | 3.04E-10 | 5.11E-06 |
| cg08172999_TC11 | chr2 | 199910999 | TSS1500 | C2orf69 | -0.536798112 | 3.09E-10 | 5.11E-06 |
| cg19872661_BC21 | chr5 | 144536383 | | | 0.300790977 | 3.12E-10 | 5.11E-06 |
| cg06822989_BC21 | chr5 | 24832176 | | | 0.216057879 | 3.20E-10 | 5.16E-06 |
| cg26660461_BC21 | chr1 | 94690001 | | | 0.249200826 | 3.44E-10 | 5.44E-06 |
| cg22521947_BC21 | chr16 | 56404419 | | | 0.400392287 | 3.62E-10 | 5.62E-06 |
| cg06665786_BC21 | chr1 | 8198053 | | | 0.188801418 | 3.75E-10 | 5.72E-06 |
| cg19483242_BC21 | chr16 | 57633138 | | | 0.123360882 | 3.92E-10 | 5.85E-06 |
| cg02915831_TC21 | chr2 | 64679842 | | | 0.351737953 | 4.04E-10 | 5.85E-06 |
| cg02676101_BC21 | chr2 | 36655099 | | | 0.267065876 | 4.04E-10 | 5.85E-06 |
| cg02415808_BC21 | chr3 | 46207017 | | | 0.293975918 | 4.24E-10 | 6.03E-06 |
| cg03275872_BC21 | chr2 | 79280142 | | | 0.535891254 | 4.32E-10 | 6.03E-06 |
| cg22649617_BC21 | chr22 | 39636189 | | | 0.30770216 | 4.37E-10 | 6.03E-06 |
| cg14285115_TC21 | chr9 | 127966215 | | | 0.122314349 | 4.56E-10 | 6.10E-06 |
| cg09050062_TC11 | chr8 | 143952167 | TSS1500 | PLEC | -0.308502299 | 4.56E-10 | 6.10E-06 |
| cg13066220_BC21 | chr8 | 123022506 | | | 0.169232899 | 4.90E-10 | 6.30E-06 |
| cg14209730_BC21 | chr2 | 64405502 | | | 0.163947071 | 4.92E-10 | 6.30E-06 |
| cg04730839_TC21 | chr6 | 43018146 | exon_5;e | KLHDC3;KL | 0.349691942 | 4.94E-10 | 6.30E-06 |
| cg09977284_BC21 | chr6 | 113904007 | TSS1500 | LINC0288C | 0.247155314 | 5.04E-10 | 6.30E-06 |
| cg12421814_TC11 | chr4 | 3447643 | | | 0.102600126 | 5.07E-10 | 6.30E-06 |
| cg20294280_BC21 | chr6 | 30190606 | | | 0.391673796 | 5.20E-10 | 6.31E-06 |
| cg19872136_BC21 | chr1 | 8699861 | | | 0.350489895 | 5.28E-10 | 6.31E-06 |
| cg18338984_TC21 | chr7 | 2607321 | exon_21; | IQCE;IQCE_ | 0.269265097 | 5.31E-10 | 6.31E-06 |
| cg26346574_TC21 | chr5 | 120615832 | | | 0.320022153 | 5.37E-10 | 6.31E-06 |
| cg12904050_BC21 | chr6 | 161379953 | | | 0.161474257 | 5.48E-10 | 6.35E-06 |
| cg22982489_BC21 | chr1 | 8198286 | | | 0.22457422 | 5.55E-10 | 6.35E-06 |
| cg13491481_BC11 | chr3 | 62875467 | TSS200;T: | CADPS;CAI | 0.378486824 | 5.87E-10 | 6.63E-06 |
| cg24562233_BC21 | chr19 | 42278181 | | | 0.107801342 | 6.23E-10 | 6.94E-06 |
| cg21777154_BC21 | chr1 | 184427170 | | | 0.163175205 | 6.30E-10 | 6.94E-06 |
| cg20576068_BC21 | chr8 | 24039468 | | | 0.199877037 | 6.50E-10 | 7.07E-06 |
| cg08018499_BC21 | chr17 | 50115427 | | | 0.167289431 | 6.93E-10 | 7.44E-06 |
| cg07848273_BC21 | chr9 | 134441790 | | | 0.207601726 | 7.23E-10 | 7.58E-06 |
| cg09181319_BC21 | chr6 | 13558903 | | | 0.277414867 | 7.28E-10 | 7.58E-06 |
| cg00247963_BC21 | chr16 | 20851829 | | | 0.146064667 | 7.32E-10 | 7.58E-06 |
| cg13794588_BC11 | chr9 | 134880698 | TSS200;T: | FCN2;FCN2 | 0.160685677 | 7.53E-10 | 7.71E-06 |
| cg14587890_BC21 | chr10 | 4280152 | | | -0.324202777 | 7.68E-10 | 7.77E-06 |
| cg17559990_TC21 | chr12 | 47753617 | | | 0.195253269 | 7.83E-10 | 7.83E-06 |
| cg18203859_TC21 | chr1 | 202966193 | | | 0.112246551 | 9.08E-10 | 8.97E-06 |
| cg11718989_BC21 | chr5 | 157563915 | | | 0.271229628 | 9.32E-10 | 9.11E-06 |
| cg23167654_BC21 | chr18 | 3062989 | | | 0.318539295 | 9.52E-10 | 9.20E-06 |

| | | | | | | | |
|---|---|---|---|---|---|---|---|
| cg21920275_TC21 | chr16 | 87849930 | | | 0.167129674 | 1.00E-09 | 9.59E-06 |
| cg16761702_TC21 | chr20 | 44435796 | | | -0.443945479 | 1.03E-09 | 9.75E-06 |
| cg05912614_BC21 | chr3 | 184144711 | | | 0.1319561 | 1.05E-09 | 9.81E-06 |
| cg25787078_BC21 | chr21 | 32485061 | | | 0.204092498 | 1.07E-09 | 9.87E-06 |
| cg25927699_BC21 | chr21 | 43763031 | | | 0.228204283 | 1.08E-09 | 9.87E-06 |
| cg19048275_TC21 | chr13 | 98454564 | exon_28; | FARP1;FAF | 0.220914588 | 1.09E-09 | 9.87E-06 |
| cg16147729_TC21 | chr10 | 119424261 | | | 0.19528949 | 1.15E-09 | 1.03E-05 |
| cg21325822_BC21 | chr3 | 154779802 | | | 0.167010589 | 1.16E-09 | 1.03E-05 |
| cg10454368_BC21 | chr6 | 30343849 | TSS1500; | RPP21;RPF | 0.372935392 | 1.22E-09 | 1.06E-05 |
| cg22687206_TC21 | chr16 | 53504580 | TSS1500; | AKTIP;AKT | 0.307262149 | 1.22E-09 | 1.06E-05 |
| cg02432487_TC21 | chr2 | 241901202 | TSS1500 | FAM240C | 0.263149785 | 1.28E-09 | 1.10E-05 |
| cg09476232_BC21 | chr22 | 19664037 | | | 0.199953389 | 1.33E-09 | 1.13E-05 |
| cg01885534_TC21 | chr1 | 207812168 | | | 0.234946029 | 1.36E-09 | 1.14E-05 |
| cg16181099_BC21 | chr10 | 91602895 | | | 0.3518764 | 1.36E-09 | 1.14E-05 |
| cg00654018_BC21 | chr1 | 41504306 | | | 0.245012487 | 1.39E-09 | 1.14E-05 |
| cg02587451_BC21 | chr2 | 27129344 | exon_4 | ABHD1 | 0.143396782 | 1.40E-09 | 1.14E-05 |
| cg26467181_BC21 | chr9 | 32738541 | | | 0.397437677 | 1.41E-09 | 1.14E-05 |
| cg26500080_TC11 | chr17 | 6435234 | TSS200;T | AIPL1;AIPL | 0.25125039 | 1.42E-09 | 1.14E-05 |
| cg27562582_TC21 | chr3 | 10028611 | | | 0.255472793 | 1.45E-09 | 1.16E-05 |
| cg12207127_BC21 | chr10 | 79119487 | | | 0.155864253 | 1.47E-09 | 1.16E-05 |
| cg06085518_TC11 | chr6 | 113904015 | TSS1500 | LINC0288C | 0.284264341 | 1.47E-09 | 1.16E-05 |
| cg06379883_BC21 | chr2 | 222557086 | | | 0.182672008 | 1.50E-09 | 1.16E-05 |
| cg09985873_TC21 | chr6 | 113903970 | TSS1500 | LINC0288C | 0.30877012 | 1.55E-09 | 1.18E-05 |
| cg19015511_BC21 | chr17 | 3794885 | | | 0.220202459 | 1.55E-09 | 1.18E-05 |
| cg10326339_BC21 | chr1 | 21254877 | | | 0.297795198 | 1.56E-09 | 1.18E-05 |
| cg22359668_BC21 | chr1 | 246900735 | | | 0.29059604 | 1.58E-09 | 1.18E-05 |
| cg23244540_BC21 | chr2 | 126774812 | | | 0.290651671 | 1.63E-09 | 1.21E-05 |
| cg03931540_BC21 | chr2 | 190601382 | | | 0.263076496 | 1.65E-09 | 1.22E-05 |
| cg14798125_TC21 | chr1 | 174449077 | exon_1 | GPR52 | 0.381313582 | 1.70E-09 | 1.24E-05 |
| cg21041051_BC21 | chr15 | 41892733 | TSS200 | LOC10537 | 0.178832823 | 1.73E-09 | 1.25E-05 |
| cg03282889_TC21 | chr3 | 125988076 | | | 0.496091194 | 1.84E-09 | 1.32E-05 |
| cg16537044_BC11 | chr7 | 151187761 | exon_1;5 | ASB10;ASE | 0.176840619 | 1.87E-09 | 1.32E-05 |
| cg10456327_BC21 | chr10 | 99733710 | | | 0.264037066 | 1.87E-09 | 1.32E-05 |
| cg01635063_BC11 | chr19 | 728385 | | | 0.287762536 | 1.88E-09 | 1.32E-05 |
| cg25136988_TC11 | chr6 | 31681846 | TSS1500 | LY6G5C | 0.177942082 | 1.89E-09 | 1.32E-05 |
| cg13072943_BC21 | chr6 | 166597823 | | | 0.193551279 | 1.93E-09 | 1.33E-05 |
| cg05768419_TC21 | chr1 | 174448829 | exon_1 | GPR52 | 0.415789793 | 2.03E-09 | 1.39E-05 |
| cg18052824_TC21 | chr12 | 108577365 | | | 0.177902323 | 2.06E-09 | 1.40E-05 |
| cg13835272_TC21 | chr7 | 40447553 | | | 0.18854621 | 2.13E-09 | 1.44E-05 |
| cg26954905_TC21 | chr5 | 169945278 | | | 0.289217087 | 2.16E-09 | 1.45E-05 |
| cg08294136_BC11 | chr5 | 39760698 | | | 0.260814631 | 2.29E-09 | 1.52E-05 |
| cg24820828_BC21 | chr3 | 133587370 | exon_5;3 | CDV3;CDV | 0.321647684 | 2.40E-09 | 1.57E-05 |
| cg24526986_TC21 | chr19 | 40281005 | | | 0.102744685 | 2.41E-09 | 1.57E-05 |
| cg13085630_TC21 | chr5 | 172904794 | | | 0.188188366 | 2.45E-09 | 1.59E-05 |
| cg08650597_BC21 | chr15 | 100902501 | | | 0.169148418 | 2.49E-09 | 1.61E-05 |
| cg24363719_TC21 | chr21 | 31701856 | exon_5;e | SCAF4;SCA | 0.246680139 | 2.54E-09 | 1.62E-05 |
| cg03240624_TC21 | chr7 | 106355050 | | | 0.276741288 | 2.55E-09 | 1.62E-05 |

| | | | | | | | |
|---|---|---|---|---|---|---|---|
| cg06311141_TC21 | chr4 | 17097640 | | | 0.270643886 | 2.81E-09 | 1.77E-05 |
| cg10659811_BC21 | chr12 | 54439618 | | | 0.11667996 | 2.89E-09 | 1.81E-05 |
| cg08432509_BC21 | chr20 | 63686415 | | | 0.234317466 | 2.94E-09 | 1.82E-05 |
| cg09349790_BC21 | chr13 | 99655420 | | | 0.113650944 | 2.96E-09 | 1.82E-05 |
| cg06725204_BC21 | chr7 | 129388057 | TSS200 | AHCYL2 | 0.316229246 | 2.99E-09 | 1.82E-05 |
| cg01279538_TC21 | chr11 | 1025794 | | | 0.150363749 | 3.00E-09 | 1.82E-05 |
| cg07328962_BC21 | chr1 | 236516769 | TSS1500 | LGALS8 | 0.197496877 | 3.07E-09 | 1.85E-05 |
| cg18368368_TC21 | chr1 | 234691978 | | | 0.132332876 | 3.12E-09 | 1.86E-05 |
| cg20039929_TC21 | chr14 | 104682057 | | | 0.327190132 | 3.12E-09 | 1.86E-05 |
| cg27052234_BC11 | chr1 | 153533474 | | | 0.161791216 | 3.17E-09 | 1.88E-05 |
| cg27090964_TC21 | chr18 | 25042949 | | | 0.309938224 | 3.28E-09 | 1.93E-05 |
| cg22693806_TC21 | chr1 | 170662706 | TSS1500;T | PRRX1;PRF | -0.263260899 | 3.38E-09 | 1.97E-05 |
| cg12158309_BC21 | chr7 | 116433161 | | | 0.271192731 | 3.39E-09 | 1.97E-05 |
| cg15682223_BC21 | chr8 | 20274641 | TSS1500;T | LZTS1-AS1 | 0.421804951 | 3.44E-09 | 1.97E-05 |
| cg25440894_BC21 | chr22 | 28986614 | | | 0.160379279 | 3.45E-09 | 1.97E-05 |
| cg11287725_BC21 | chr9 | 134323889 | | | 0.212920713 | 3.47E-09 | 1.97E-05 |
| cg09784847_TC11 | chr2 | 74465423 | TSS200;TS | MOGS;MC | 0.323614231 | 3.49E-09 | 1.97E-05 |
| cg04796653_BC21 | chr8 | 134754607 | | | 0.195210194 | 3.71E-09 | 2.07E-05 |
| cg11865226_TC21 | chr4 | 183912666 | | | 0.233250784 | 3.75E-09 | 2.07E-05 |
| cg15208139_BC21 | chr11 | 36037175 | | | 0.170002446 | 3.76E-09 | 2.07E-05 |
| cg09427812_BC21 | chr6 | 134540103 | TSS200;TS | LOC10192 | 0.123682018 | 3.77E-09 | 2.07E-05 |
| cg00032661_BC21 | chr3 | 46088029 | | | 0.296106737 | 3.82E-09 | 2.09E-05 |
| cg12841013_BC21 | chr11 | 13884309 | | | 0.324868114 | 3.86E-09 | 2.09E-05 |
| cg24065136_TC21 | chr7 | 152139237 | exon_57 | KMT2C | 0.363852724 | 3.86E-09 | 2.09E-05 |
| cg14690916_BC21 | chr10 | 14182878 | | | 0.128597875 | 4.01E-09 | 2.15E-05 |
| cg11314927_TC21 | chr7 | 81367587 | | | 0.353437276 | 4.12E-09 | 2.20E-05 |
| cg04375036_TC21 | chr12 | 110744014 | TSS1500;T | PPP1CC;PF | 0.23814807 | 4.18E-09 | 2.21E-05 |
| cg26717280_TC21 | chr5 | 176852816 | | | 0.114346452 | 4.20E-09 | 2.22E-05 |
| cg23135828_TC21 | chr1 | 202296022 | | | 0.160676564 | 4.35E-09 | 2.28E-05 |
| cg22204103_TC21 | chr4 | 41538098 | TSS200;TS | LIMCH1;LI | 0.173634915 | 4.42E-09 | 2.30E-05 |
| cg25387658_BC21 | chr20 | 50917194 | | | 0.125722419 | 4.47E-09 | 2.32E-05 |
| cg14425470_BC21 | chr9 | 134652940 | TSS200 | COL5A1-AS | 0.170679509 | 4.55E-09 | 2.34E-05 |
| cg07544511_BC21 | chr15 | 75455742 | 5UTR;exo | SIN3A;SIN3 | -0.345914513 | 4.61E-09 | 2.36E-05 |
| cg26451735_TC11 | chr15 | 74202559 | exon_1;5 | STRA6;STR | 0.370377216 | 4.68E-09 | 2.37E-05 |
| cg00533891_BC21 | chr10 | 79159485 | | | 0.357422755 | 4.69E-09 | 2.37E-05 |
| cg25170409_BC21 | chr1 | 152248588 | | | 0.474922978 | 4.74E-09 | 2.38E-05 |
| cg13278478_BC21 | chr7 | 56093218 | TSS1500;T | PHKG1;PH | 0.359799166 | 4.83E-09 | 2.41E-05 |
| cg25214048_BC11 | chr20 | 36538170 | | | 0.151501485 | 4.85E-09 | 2.41E-05 |
| cg01616145_BC21 | chr2 | 88057421 | | | 0.267333795 | 4.98E-09 | 2.46E-05 |
| cg15251319_BC11 | chr3 | 195115573 | | | 0.149507029 | 5.14E-09 | 2.52E-05 |
| cg22413085_BC21 | chr17 | 31565048 | | | 0.22710577 | 5.16E-09 | 2.52E-05 |
| cg08188318_BC21 | chr6 | 35017176 | | | 0.215028489 | 5.34E-09 | 2.58E-05 |
| cg13789629_BC21 | chr5 | 16438154 | | | 0.247963857 | 5.37E-09 | 2.58E-05 |
| cg13777501_BC21 | chr9 | 75962336 | | | 0.142571921 | 5.38E-09 | 2.58E-05 |
| cg26531901_BC11 | chr17 | 35752930 | exon_1;5 | GAS2L2;GA | 0.209771779 | 5.41E-09 | 2.59E-05 |
| cg16506513_TC21 | chr8 | 90642357 | | | 0.256851179 | 5.46E-09 | 2.60E-05 |
| cg13519106_TC21 | chr9 | 6654943 | | | 0.284175838 | 5.81E-09 | 2.74E-05 |

| | | | | | | | |
|---|---|---|---|---|---|---|---|
| cg17478888_BC21 | chr16 | 84091596 | | | 0.149676236 | 5.89E-09 | 2.77E-05 |
| cg12939515_BC11 | chr1 | 89820994 | TSS1500; | LRRC8D;LF | -0.286370368 | 5.92E-09 | 2.77E-05 |
| cg00854187_TC21 | chr13 | 27466502 | | | 0.219997858 | 6.00E-09 | 2.79E-05 |
| cg22515654_BC21 | chr1 | 10530615 | | | 0.221880362 | 6.04E-09 | 2.80E-05 |
| cg26732544_BC21 | chr10 | 31019163 | | | 0.265944089 | 6.09E-09 | 2.80E-05 |
| cg01092932_BC21 | chr1 | 198858931 | exon_1 | MIR181B1 | 0.294670257 | 6.22E-09 | 2.85E-05 |
| cg25802611_BC21 | chr12 | 119835221 | | | 0.304024519 | 6.39E-09 | 2.91E-05 |
| cg26715240_BC21 | chr12 | 677037 | | | 0.420082673 | 6.50E-09 | 2.93E-05 |
| cg06264922_TC11 | chr11 | 1298418 | | | 0.130679658 | 6.53E-09 | 2.93E-05 |
| cg07279963_TC11 | chr3 | 61560852 | TSS1500; | PTPRG;PTF | -0.240190591 | 6.54E-09 | 2.93E-05 |
| cg03225003_BC21 | chr1 | 5882945 | | | 0.27112822 | 6.77E-09 | 3.02E-05 |
| cg03537163_BC11 | chr19 | 814520 | exon_7;e | PLPPR3;PL | 0.231081858 | 6.92E-09 | 3.07E-05 |
| cg04905779_TC21 | chr20 | 63733971 | | | 0.217177497 | 7.02E-09 | 3.10E-05 |
| cg09575472_BC21 | chr5 | 174618106 | | | 0.132248121 | 7.24E-09 | 3.17E-05 |
| cg20681068_TC21 | chr20 | 3408888 | TSS1500 | DNAAF9 | -0.378341705 | 7.28E-09 | 3.17E-05 |
| cg10079354_BC21 | chr6 | 128640752 | | | 0.253209934 | 7.30E-09 | 3.17E-05 |
| cg00263902_BC21 | chr20 | 48932525 | | | 0.192608303 | 7.52E-09 | 3.25E-05 |
| cg13864940_BC21 | chr12 | 3220238 | | | 0.272991563 | 7.58E-09 | 3.26E-05 |
| cg22290393_BC21 | chr17 | 19364953 | | | -0.269861267 | 7.63E-09 | 3.27E-05 |
| cg22176903_TC21 | chr16 | 57810067 | TSS1500 | LOC38828 | 0.188225817 | 7.67E-09 | 3.27E-05 |
| cg11886561_BC21 | chr7 | 149777179 | | | 0.154057009 | 8.02E-09 | 3.40E-05 |
| cg22510036_BC21 | chr17 | 39649336 | | | 0.223858371 | 8.14E-09 | 3.44E-05 |
| cg17510321_TC21 | chr15 | 90855382 | | | 0.182480738 | 8.19E-09 | 3.44E-05 |
| cg05118252_BC21 | chr3 | 63863373 | TSS1500; | ATXN7;AT | -0.345369652 | 8.22E-09 | 3.44E-05 |
| cg07546579_BC21 | chr16 | 87173723 | | | 0.187907859 | 8.49E-09 | 3.53E-05 |
| cg13897833_BC21 | chr13 | 73988607 | | | 0.196993691 | 8.58E-09 | 3.54E-05 |
| cg06447341_TC21 | chr1 | 31987329 | | | 0.103951077 | 8.59E-09 | 3.54E-05 |
| cg06046647_TC21 | chr4 | 158208862 | | | 0.26145561 | 8.78E-09 | 3.60E-05 |
| cg23851780_BC21 | chr19 | 801569 | | | 0.156454821 | 9.03E-09 | 3.67E-05 |
| cg10553506_BC21 | chr7 | 2007249 | | | 0.219692605 | 9.07E-09 | 3.67E-05 |
| cg18603250_TC21 | chr11 | 94767886 | TSS1500; | AMOTL1;A | 0.171833227 | 9.08E-09 | 3.67E-05 |
| cg14137606_BC21 | chr1 | 46569869 | exon_8 | MKNK1-AS | 0.302183651 | 9.18E-09 | 3.69E-05 |
| cg02425407_BC21 | chr1 | 9842929 | | | 0.111146146 | 9.24E-09 | 3.70E-05 |
| cg06264518_TC21 | chr1 | 112360607 | | | 0.177521171 | 9.26E-09 | 3.70E-05 |
| cg21741807_BC21 | chr10 | 133278067 | TSS1500; | ADAM8;AI | 0.145173743 | 9.58E-09 | 3.80E-05 |
| cg08889109_BC21 | chr7 | 141665004 | exon_7 | DENND11 | 0.163192537 | 9.80E-09 | 3.87E-05 |
| cg06636342_BC21 | chr19 | 13995301 | | | 0.192917234 | 9.91E-09 | 3.89E-05 |
| cg14706381_TC21 | chr10 | 14478742 | | | 0.197483028 | 1.00E-08 | 3.89E-05 |
| cg09694675_TC21 | chr12 | 51440285 | | | 0.247898758 | 1.01E-08 | 3.89E-05 |
| cg17330838_TC11 | chr11 | 57499628 | | | 0.162051378 | 1.01E-08 | 3.89E-05 |
| cg25540925_BC21 | chr12 | 132071090 | | | 0.345775118 | 1.01E-08 | 3.89E-05 |
| cg03459735_BC21 | chr2 | 126983556 | | | 0.241050652 | 1.01E-08 | 3.89E-05 |
| cg23234602_TC11 | chr5 | 181001536 | | | 0.376421141 | 1.02E-08 | 3.90E-05 |
| cg19628958_TC21 | chr1 | 112360574 | | | 0.143516759 | 1.04E-08 | 3.98E-05 |
| cg00653897_BC21 | chr1 | 41497015 | | | 0.190066811 | 1.05E-08 | 3.99E-05 |
| cg21367940_BC21 | chr5 | 16758113 | | | 0.199344172 | 1.06E-08 | 4.01E-05 |
| cg10583769_TC21 | chr15 | 71189122 | TSS200;T | THSD4-AS | 0.179995979 | 1.07E-08 | 4.03E-05 |

| | | | | | | | |
|---|---|---|---|---|---|---|---|
| cg18208380_BC21 | chr20 | 62667599 | TSS1500 | SLCO4A1-/ | 0.261790279 | 1.08E-08 | 4.04E-05 |
| cg26899988_TC21 | chr11 | 128451213 | | | 0.276577461 | 1.09E-08 | 4.08E-05 |
| cg13807341_BC21 | chr11 | 118575269 | | | -0.240691976 | 1.11E-08 | 4.12E-05 |
| cg06601581_TC11 | chr16 | 85371048 | | | 0.255592864 | 1.15E-08 | 4.24E-05 |
| cg12702344_TC21 | chr20 | 57974559 | | | 0.188273827 | 1.16E-08 | 4.27E-05 |
| cg17830061_BC11 | chr5 | 128377836 | exon_13 | FBN2 | 0.249753819 | 1.18E-08 | 4.32E-05 |
| cg07187971_TC21 | chr18 | 62813785 | | | 0.362394277 | 1.18E-08 | 4.33E-05 |
| cg08142776_BC21 | chr5 | 173824379 | | | 0.152256218 | 1.23E-08 | 4.46E-05 |
| cg00206266_TC21 | chr20 | 44930205 | | | 0.267696557 | 1.26E-08 | 4.54E-05 |
| cg26197687_TC21 | chr22 | 29343418 | | | 0.166909781 | 1.26E-08 | 4.54E-05 |
| cg11041314_BC21 | chr5 | 139642065 | | | 0.179689076 | 1.26E-08 | 4.54E-05 |
| cg10304765_BC21 | chr6 | 155945621 | TSS1500 | MIR1202 | 0.126855986 | 1.27E-08 | 4.54E-05 |
| cg11538410_TC21 | chr8 | 141131356 | | | 0.176049469 | 1.29E-08 | 4.57E-05 |
| cg24158174_TC21 | chr19 | 13128821 | | | 0.092823537 | 1.29E-08 | 4.57E-05 |
| cg20066640_TC21 | chr5 | 80952984 | | | 0.234558772 | 1.30E-08 | 4.58E-05 |
| cg19487251_TC21 | chr14 | 51019321 | | | -0.413218136 | 1.30E-08 | 4.58E-05 |
| cg08564636_TC21 | chr6 | 165910904 | | | 0.309559672 | 1.31E-08 | 4.59E-05 |
| cg01705036_TC21 | chr7 | 995782 | | | 0.196194325 | 1.32E-08 | 4.59E-05 |
| cg26150489_BC21 | chr11 | 62508434 | | | 0.176089355 | 1.32E-08 | 4.59E-05 |
| cg13484663_BC21 | chr9 | 20356634 | | | 0.172927097 | 1.33E-08 | 4.60E-05 |
| cg17277833_TC21 | chr2 | 43331405 | | | 0.204483999 | 1.33E-08 | 4.60E-05 |
| cg14357604_BC21 | chr9 | 131421313 | | | 0.180841897 | 1.34E-08 | 4.62E-05 |
| cg21292007_TC21 | chr17 | 44905664 | 3UTR;exo | GFAP;GFAI | 0.134564862 | 1.35E-08 | 4.63E-05 |
| cg04158698_BC21 | chr1 | 201553913 | | | 0.146947346 | 1.36E-08 | 4.64E-05 |
| cg10193870_BC11 | chr14 | 72671298 | exon_10; | DPF3;DPF3 | 0.340940993 | 1.37E-08 | 4.64E-05 |
| cg08986751_BC21 | chr14 | 23041684 | TSS1500 | PSMB11 | 0.171636531 | 1.38E-08 | 4.66E-05 |
| cg07207982_TC21 | chr6 | 35017153 | | | 0.271234485 | 1.38E-08 | 4.66E-05 |
| cg07390242_TC21 | chr13 | 51644865 | | | 0.217341573 | 1.39E-08 | 4.66E-05 |
| cg21025449_TC21 | chr3 | 132654150 | TSS1500 | UBA5 | 0.231833279 | 1.40E-08 | 4.66E-05 |
| cg10524033_TC21 | chr4 | 133149278 | TSS200;T! | PCDH10;P( | -0.255366265 | 1.40E-08 | 4.66E-05 |
| cg07140337_BC21 | chr1 | 222544201 | | | 0.344014302 | 1.41E-08 | 4.68E-05 |
| cg01432214_BC21 | chr5 | 115297735 | TSS1500;1 | CCDC112;( | 0.215709601 | 1.49E-08 | 4.94E-05 |
| cg16445774_BC21 | chr11 | 64138447 | | | 0.115141476 | 1.51E-08 | 4.97E-05 |
| cg06283304_BC21 | chr12 | 117189051 | | | 0.236219365 | 1.51E-08 | 4.97E-05 |
| cg08796342_TC21 | chr14 | 91867685 | TSS200 | TC2N | 0.109322053 | 1.53E-08 | 5.01E-05 |
| cg17559991_BC21 | chr12 | 47753692 | | | 0.151164553 | 1.54E-08 | 5.01E-05 |
| cg03216879_BC21 | chr10 | 79160914 | | | 0.23834822 | 1.54E-08 | 5.01E-05 |
| cg07238669_BC21 | chr7 | 7420479 | | | -0.408761908 | 1.60E-08 | 5.16E-05 |
| cg09844227_BC21 | chr13 | 51895806 | | | 0.179539484 | 1.61E-08 | 5.18E-05 |
| cg21610839_BC22 | chr3 | 23819821 | | | 0.18914184 | 1.64E-08 | 5.25E-05 |
| cg24156299_TC21 | chr19 | 13054149 | | | 0.17765297 | 1.64E-08 | 5.25E-05 |
| cg10335988_TC21 | chr17 | 49266498 | | | 0.181792145 | 1.68E-08 | 5.35E-05 |
| cg10721256_BC21 | chr16 | 67526653 | | | 0.230313299 | 1.69E-08 | 5.35E-05 |
| cg07876162_TC11 | chr19 | 728176 | | | 0.341441207 | 1.69E-08 | 5.35E-05 |
| cg01819046_BC21 | chr2 | 588708 | | | 0.179645579 | 1.70E-08 | 5.37E-05 |
| cg12473666_BC21 | chr11 | 57600619 | | | 0.251094152 | 1.73E-08 | 5.40E-05 |
| cg06174795_BC21 | chr3 | 188037725 | | | 0.328838496 | 1.73E-08 | 5.40E-05 |

| | | | | | | | |
|---|---|---|---|---|---|---|---|
| cg11266339_TC21 | chr12 | 131947482 | | | 0.103391739 | 1.73E-08 | 5.40E-05 |
| cg16850067_TC21 | chr9 | 114348724 | | | 0.215768942 | 1.74E-08 | 5.41E-05 |
| cg09070662_BC21 | chr15 | 65368384 | | | 0.218154731 | 1.77E-08 | 5.47E-05 |
| cg25240047_BC11 | chr5 | 33892138 | TSS200;T! | ADAMTS1. | 0.281811825 | 1.80E-08 | 5.54E-05 |
| cg05500898_TC21 | chr11 | 93775492 | | | 0.175931246 | 1.80E-08 | 5.54E-05 |
| cg15158608_TC21 | chr17 | 40015364 | TSS200;T! | CSF3;CSF3 | 0.167602612 | 1.81E-08 | 5.54E-05 |
| cg04842115_TC21 | chr3 | 46923796 | | | 0.189194499 | 1.84E-08 | 5.61E-05 |
| cg13095372_BC21 | chr2 | 46697990 | TSS1500; | SOCS5;SO( | 0.203121496 | 1.86E-08 | 5.65E-05 |
| cg05673892_TC21 | chr3 | 37992443 | | | 0.209278889 | 1.87E-08 | 5.68E-05 |
| cg05130679_TC11 | chr11 | 94769658 | | | 0.166929013 | 1.88E-08 | 5.69E-05 |
| cg13917812_TC21 | chr22 | 27757753 | | | 0.225332682 | 1.91E-08 | 5.72E-05 |
| cg22619628_BC21 | chr3 | 53883414 | TSS1500 | ACTR8 | 0.190617199 | 1.91E-08 | 5.72E-05 |
| cg22918476_TC21 | chr11 | 1830429 | | | 0.307531731 | 1.92E-08 | 5.72E-05 |
| cg14674306_TC21 | chr17 | 82603585 | exon_9;3 | FOXK2;FO) | 0.181503658 | 1.95E-08 | 5.80E-05 |
| cg01287505_BC21 | chr17 | 58529829 | TSS1500; | SEPTIN4;SI | 0.277512875 | 1.96E-08 | 5.82E-05 |
| cg18099734_TC21 | chr13 | 111371353 | | | 0.252776592 | 1.97E-08 | 5.82E-05 |
| cg26302238_BC21 | chr9 | 123221656 | | | 0.473782901 | 1.98E-08 | 5.84E-05 |
| cg14297340_TC21 | chr9 | 70256427 | | | 0.292245391 | 1.99E-08 | 5.84E-05 |
| cg01682917_TC11 | chr13 | 114100469 | | | 0.152586771 | 1.99E-08 | 5.84E-05 |
| cg09731420_BC21 | chr5 | 138283579 | | | 0.279725293 | 2.04E-08 | 5.96E-05 |
| cg10285815_BC21 | chr18 | 48552723 | | | 0.242784666 | 2.06E-08 | 5.99E-05 |
| cg25790453_BC21 | chr13 | 112979276 | TSS200 | MCF2L | 0.290173461 | 2.07E-08 | 6.00E-05 |
| cg18146221_TC21 | chr2 | 181497186 | | | 0.326743453 | 2.09E-08 | 6.04E-05 |
| cg00875413_BC21 | chr1 | 64343852 | | | 0.311005046 | 2.10E-08 | 6.04E-05 |
| cg03557803_BC21 | chr13 | 40987702 | | | 0.214195998 | 2.11E-08 | 6.04E-05 |
| cg24603297_TC21 | chr5 | 172901218 | | | 0.173401207 | 2.12E-08 | 6.04E-05 |
| cg14042143_BC22 | chr7 | 2607148 | exon_21; | IQCE;IQCE | 0.406759184 | 2.12E-08 | 6.04E-05 |
| cg08304863_BC21 | chr20 | 62692919 | | | 0.293795731 | 2.14E-08 | 6.06E-05 |
| cg07756014_TC21 | chr9 | 123170132 | | | 0.252804625 | 2.14E-08 | 6.06E-05 |
| cg06038342_BC21 | chr13 | 114099333 | | | 0.866566357 | 2.15E-08 | 6.06E-05 |
| cg01890741_BC21 | chr1 | 37948118 | TSS1500; | INPP5B;IN | 0.163601911 | 2.17E-08 | 6.10E-05 |
| cg06306796_BC21 | chr4 | 16586293 | | | 0.160762211 | 2.18E-08 | 6.12E-05 |
| cg18276409_TC21 | chr12 | 124393051 | | | 0.159399144 | 2.20E-08 | 6.12E-05 |
| cg17007339_TC21 | chr11 | 85719586 | TSS1500; | SYTL2;SYT | 0.260876503 | 2.20E-08 | 6.12E-05 |
| cg04657831_TC11 | chr21 | 45288036 | TSS200;T! | LINC0020! | -0.263690625 | 2.20E-08 | 6.12E-05 |
| cg17741068_BC21 | chr1 | 41509330 | 3UTR;exo | HIVEP3;HI' | 0.212970943 | 2.21E-08 | 6.12E-05 |
| cg01166384_BC21 | chr9 | 136078756 | | | 0.120906258 | 2.22E-08 | 6.14E-05 |
| cg25397498_BC21 | chr20 | 51619007 | exon_20 | ATP9A | 0.267576836 | 2.24E-08 | 6.15E-05 |
| cg08811097_BC21 | chr16 | 47004908 | | | 0.25706017 | 2.27E-08 | 6.23E-05 |
| cg04679323_TC21 | chr2 | 45639574 | | | 0.124687185 | 2.34E-08 | 6.39E-05 |
| cg24611115_BC21 | chr3 | 33110953 | | | -0.353983463 | 2.38E-08 | 6.45E-05 |
| cg14730984_TC21 | chr15 | 70421253 | | | 0.114074616 | 2.38E-08 | 6.45E-05 |
| cg14270760_BC21 | chr2 | 120820590 | | | 0.401902148 | 2.38E-08 | 6.45E-05 |
| cg01088387_BC21 | chr11 | 78504399 | | | 0.293291647 | 2.39E-08 | 6.45E-05 |
| cg12200866_TC21 | chr3 | 121431868 | 3UTR;exo | POLQ;POL | 0.234995791 | 2.41E-08 | 6.48E-05 |
| cg05003599_TC11 | chr7 | 100589776 | exon_1;e | FBXO24;P( | 0.225852911 | 2.41E-08 | 6.48E-05 |
| cg22728019_TC21 | chr6 | 106078221 | | | 0.262484481 | 2.43E-08 | 6.50E-05 |

| | | | | | | | |
|---|---|---|---|---|---|---|---|
| cg26995224_TC21 | chr12 | 121536241 | | | 0.348916651 | 2.49E-08 | 6.62E-05 |
| cg18554180_BC21 | chr13 | 28483689 | | | 0.310956103 | 2.49E-08 | 6.62E-05 |
| cg00873965_TC21 | chr1 | 64172090 | | | -0.661502214 | 2.50E-08 | 6.62E-05 |
| cg16561057_TC21 | chr7 | 157851863 | | | -0.479213277 | 2.54E-08 | 6.71E-05 |
| cg01450021_TC21 | chr1 | 154972650 | | | 0.116120975 | 2.55E-08 | 6.71E-05 |
| cg11299350_TC21 | chr6 | 68126581 | | | 0.173135098 | 2.56E-08 | 6.72E-05 |
| cg03580279_TC21 | chr1 | 2084535 | | | 0.175834683 | 2.56E-08 | 6.72E-05 |
| cg26785800_TC21 | chr1 | 27528534 | | | 0.196494368 | 2.62E-08 | 6.83E-05 |
| cg15005441_TC21 | chr2 | 20353326 | TSS1500;` | PUM2;PUI | 0.245029185 | 2.62E-08 | 6.83E-05 |
| cg00617135_TC21 | chr13 | 98301017 | | | -0.236881295 | 2.64E-08 | 6.85E-05 |
| cg15233131_TC21 | chr19 | 817832 | | | 0.15578554 | 2.65E-08 | 6.86E-05 |
| cg11135409_TC21 | chr1 | 184545623 | | | 0.306351121 | 2.67E-08 | 6.87E-05 |
| cg12584528_TC21 | chr17 | 81962796 | | | 0.170998525 | 2.67E-08 | 6.87E-05 |
| cg15970074_TC21 | chr8 | 143826052 | | | 0.094690369 | 2.70E-08 | 6.92E-05 |
| cg02674129_BC21 | chr2 | 36439460 | | | 0.271555006 | 2.71E-08 | 6.94E-05 |
| cg26440820_BC21 | chr7 | 148976649 | | | 0.124037616 | 2.73E-08 | 6.95E-05 |
| cg12648691_TC21 | chr13 | 113365369 | TSS1500;` | GRTP1;GR` | -0.223804468 | 2.81E-08 | 7.14E-05 |
| cg04771543_BC21 | chr3 | 155731428 | | | 0.305400153 | 2.82E-08 | 7.16E-05 |
| cg13882251_TC21 | chr19 | 40266359 | TSS1500 | AKT2 | 0.227800669 | 2.84E-08 | 7.17E-05 |
| cg02671171_TC21 | chr17 | 353296 | TSS1500;` | RPH3AL;RI | 0.328449618 | 2.84E-08 | 7.17E-05 |
| cg03252549_BC21 | chr2 | 98705213 | | | -0.200822313 | 2.87E-08 | 7.21E-05 |
| cg17192147_BC21 | chr11 | 102607284 | | | 0.252037086 | 2.90E-08 | 7.26E-05 |
| cg08423424_TC21 | chr2 | 234989006 | | | 0.175643036 | 2.91E-08 | 7.26E-05 |
| cg23630380_BC21 | chr17 | 38573161 | | | 0.158342476 | 2.93E-08 | 7.26E-05 |
| cg19121470_BC21 | chr5 | 40784881 | | | 0.227727315 | 2.93E-08 | 7.26E-05 |
| cg12173308_BC21 | chr7 | 47351226 | | | 0.258501721 | 2.93E-08 | 7.26E-05 |
| cg12235672_BC21 | chr16 | 8956453 | TSS200 | USP7 | 0.124515919 | 2.94E-08 | 7.26E-05 |
| cg21151899_BC21 | chr22 | 41941653 | TSS1500 | CENPM | 0.202656348 | 2.95E-08 | 7.26E-05 |
| cg20784813_TC21 | chr1 | 21672613 | | | 0.138334299 | 2.96E-08 | 7.26E-05 |
| cg13110988_TC21 | chr10 | 85635937 | | | 0.328036242 | 2.99E-08 | 7.33E-05 |
| cg10837110_BC11 | chr1 | 2026272 | | | 0.282457619 | 3.06E-08 | 7.47E-05 |
| cg03633040_BC21 | chr21 | 46137385 | | | 0.274472873 | 3.07E-08 | 7.48E-05 |
| cg10728716_BC21 | chr1 | 19078550 | | | 0.281483768 | 3.13E-08 | 7.61E-05 |
| cg26466027_BC21 | chr3 | 45948252 | exon_2;3 | CXCR6;CX( | 0.551393526 | 3.17E-08 | 7.67E-05 |
| cg15061576_BC21 | chr10 | 53030160 | | | 0.216425402 | 3.17E-08 | 7.67E-05 |
| cg03478916_BC21 | chr8 | 22563516 | | | 0.184747027 | 3.22E-08 | 7.77E-05 |
| cg20544389_BC21 | chr15 | 67244256 | | | 0.335022047 | 3.23E-08 | 7.77E-05 |
| cg15616183_BC21 | chr14 | 55587662 | | | 0.17408153 | 3.27E-08 | 7.81E-05 |
| cg22936466_BC21 | chr17 | 74768385 | | | 0.166558256 | 3.28E-08 | 7.81E-05 |
| cg14731091_TC21 | chr4 | 38114516 | | | 0.122338214 | 3.29E-08 | 7.81E-05 |
| cg15350671_TC21 | chr14 | 103801726 | | | 0.12166415 | 3.29E-08 | 7.81E-05 |
| cg16613029_BC21 | chr16 | 8958905 | | | 0.116948911 | 3.33E-08 | 7.90E-05 |
| cg26089783_BC21 | chr22 | 21018029 | exon_3;5 | P2RX6;P2F | 0.134210298 | 3.35E-08 | 7.91E-05 |
| cg01208787_BC21 | chr7 | 29486693 | | | 0.278476766 | 3.36E-08 | 7.91E-05 |
| cg13229078_TC11 | chr1 | 234863022 | | | 0.149803299 | 3.37E-08 | 7.92E-05 |
| cg05441738_BC21 | chr3 | 124352074 | | | -0.534948217 | 3.40E-08 | 7.96E-05 |
| cg06127256_TC11 | chr16 | 73172689 | | | 0.200641101 | 3.40E-08 | 7.96E-05 |

| | | | | | | | |
|---|---|---|---|---|---|---|---|
| cg01241697_TC21 | chr19 | 8259867 | | | 0.225643343 | 3.43E-08 | 8.00E-05 |
| cg06252795_BC21 | chr2 | 16854881 | | | 0.218023159 | 3.45E-08 | 8.01E-05 |
| cg22670441_BC21 | chr17 | 50266718 | | | 0.106402959 | 3.52E-08 | 8.13E-05 |
| cg24446898_TC21 | chr19 | 35605081 | | | 0.175409982 | 3.52E-08 | 8.13E-05 |
| cg01317045_BC21 | chr16 | 57815751 | | | 0.195715703 | 3.53E-08 | 8.13E-05 |
| cg02592957_BC21 | chr3 | 190281676 | | | 0.186546569 | 3.57E-08 | 8.20E-05 |
| cg05085844_TC21 | chr7 | 911907 | | | 0.427457251 | 3.57E-08 | 8.20E-05 |
| cg01025296_BC21 | chr2 | 199954053 | TSS1500;T | MAIP1;MA | 0.166841043 | 3.59E-08 | 8.21E-05 |
| cg04640684_TC21 | chr8 | 106751306 | exon_16; | OXR1;OXR | 0.297892029 | 3.62E-08 | 8.22E-05 |
| cg09353140_BC21 | chr10 | 35398447 | | | 0.202090128 | 3.64E-08 | 8.22E-05 |
| cg04647790_TC21 | chr13 | 51845129 | 5UTR;exo | TMEM272 | 0.168980006 | 3.64E-08 | 8.22E-05 |
| cg23796525_BC21 | chr17 | 41822601 | exon_10; | FKBP10;FK | 0.123810554 | 3.64E-08 | 8.22E-05 |
| cg06597454_BC21 | chr2 | 168796874 | | | 0.171424226 | 3.64E-08 | 8.22E-05 |
| cg09797698_BC21 | chr2 | 174761219 | | | 0.169620427 | 3.65E-08 | 8.23E-05 |
| cg05418947_TC21 | chr13 | 106151304 | | | 0.233301021 | 3.67E-08 | 8.25E-05 |
| cg11623476_BC11 | chr7 | 2526393 | exon_6;e | LFNG;LFNG | 0.19771248 | 3.68E-08 | 8.26E-05 |
| cg19758448_BC21 | chr17 | 39672043 | 3UTR;exo | PGAP3;PG. | 0.25222636 | 3.71E-08 | 8.29E-05 |
| cg15596811_BC21 | chr10 | 113666121 | | | 0.205420199 | 3.73E-08 | 8.31E-05 |
| cg06851855_BC21 | chr4 | 94955618 | | | -0.205717583 | 3.81E-08 | 8.47E-05 |
| cg14766884_BC21 | chr19 | 2784735 | TSS1500;T | SGTA;THO | 0.27861872 | 3.85E-08 | 8.51E-05 |
| cg00418068_BC21 | chr1 | 33429904 | | | 0.324400909 | 3.85E-08 | 8.51E-05 |
| cg09615693_TC21 | chr12 | 94177809 | | | 0.322345813 | 3.87E-08 | 8.51E-05 |
| cg09868274_TC21 | chr12 | 109096302 | TSS1500 | UNG | 0.221840738 | 3.88E-08 | 8.51E-05 |
| cg23737927_TC12 | chr12 | 5039987 | | | 0.213740884 | 3.89E-08 | 8.51E-05 |
| cg20796101_BC21 | chr15 | 90645809 | | | 0.228686483 | 3.89E-08 | 8.51E-05 |
| cg08567670_BC21 | chr12 | 113243861 | TSS1500;T | TPCN1;TPC | 0.13865965 | 3.90E-08 | 8.51E-05 |
| cg15823423_TC21 | chr11 | 64117052 | exon_3;e | FLRT1;FLRT | 0.336838032 | 3.90E-08 | 8.51E-05 |
| cg15457922_BC21 | chr10 | 99331592 | | | 0.177779578 | 3.92E-08 | 8.53E-05 |
| cg22796704_BC21 | chr10 | 48465491 | | | 0.189269362 | 3.95E-08 | 8.56E-05 |
| cg13937852_BC21 | chr22 | 45477095 | | | 0.226759064 | 3.96E-08 | 8.56E-05 |
| cg15383740_BC21 | chr8 | 129726695 | | | 0.174326949 | 3.99E-08 | 8.61E-05 |
| cg27042520_TC21 | chr19 | 33499555 | | | 0.20423636 | 4.00E-08 | 8.61E-05 |
| cg10603235_TC21 | chr22 | 42940043 | | | 0.209736032 | 4.04E-08 | 8.69E-05 |
| cg21336878_BC21 | chr4 | 4284953 | | | 0.156723119 | 4.09E-08 | 8.74E-05 |
| cg01312837_BC21 | chr16 | 3873771 | | | 0.124794699 | 4.09E-08 | 8.74E-05 |
| cg25283465_TC11 | chr1 | 110780719 | | | 0.262601219 | 4.10E-08 | 8.74E-05 |
| cg00761968_BC21 | chr13 | 52740007 | TSS200;TS | CNMD;CNI | 0.168812272 | 4.14E-08 | 8.80E-05 |
| cg15603964_TC21 | chr1 | 19084244 | | | -0.326793026 | 4.16E-08 | 8.82E-05 |
| cg07738859_BC21 | chr7 | 1982740 | | | 0.291859819 | 4.17E-08 | 8.82E-05 |
| cg26466202_BC21 | chr22 | 46043963 | | | 0.151290799 | 4.26E-08 | 9.00E-05 |
| cg09319617_TC11 | chr1 | 175596235 | | | 0.190580502 | 4.29E-08 | 9.00E-05 |
| cg06440430_BC21 | chr16 | 27521357 | | | 0.183241127 | 4.29E-08 | 9.00E-05 |
| cg02540858_BC21 | chr16 | 84240956 | TSS1500 | KCNG4 | 0.2704737 | 4.29E-08 | 9.00E-05 |
| cg12371954_BC21 | chr5 | 154466504 | | | 0.281623436 | 4.32E-08 | 9.03E-05 |
| cg05897803_BC21 | chr6 | 39322702 | exon_1;5 | KCNK16;K( | 0.19689029 | 4.33E-08 | 9.03E-05 |
| cg11626857_TC21 | chr8 | 80703027 | | | 0.223887951 | 4.34E-08 | 9.04E-05 |
| cg14188840_BC11 | chr7 | 27180779 | TSS1500;T | HOXA10;H | 0.182028933 | 4.37E-08 | 9.06E-05 |

| | | | | | | | |
|---|---|---|---|---|---|---|---|
| cg03714754_BC21 | chr15 | 68700734 | | | 0.186001608 | 4.38E-08 | 9.06E-05 |
| cg02999604_BC21 | chr5 | 134568542 | | | 0.120051431 | 4.38E-08 | 9.06E-05 |
| cg02184744_BC21 | chr16 | 89414366 | | | 0.371282584 | 4.40E-08 | 9.06E-05 |
| cg16899859_TC21 | chr4 | 34668533 | | | 0.285647783 | 4.47E-08 | 9.18E-05 |
| cg15108134_TC21 | chr2 | 73270522 | | | 0.28720797 | 4.47E-08 | 9.18E-05 |
| cg22641406_BC21 | chr22 | 45299139 | | | 0.10423075 | 4.50E-08 | 9.21E-05 |
| cg18099923_TC21 | chr12 | 111773256 | | | 0.177313668 | 4.56E-08 | 9.27E-05 |
| cg10992481_BC21 | chr6 | 3427999 | | | 0.184621094 | 4.56E-08 | 9.27E-05 |
| cg23823134_TC21 | chr17 | 39200301 | 5UTR;exo | RPL19;RPL | -0.408244069 | 4.57E-08 | 9.27E-05 |
| cg05407171_TC21 | chr2 | 137963863 | TSS1500;' | HNMT;HNI | 0.188737415 | 4.57E-08 | 9.27E-05 |
| cg21868776_BC21 | chr16 | 85377155 | | | 0.140545058 | 4.61E-08 | 9.33E-05 |
| cg22422251_TC21 | chr5 | 113034494 | | | 0.348385109 | 4.63E-08 | 9.34E-05 |
| cg17398495_BC21 | chr12 | 25976732 | | | 0.242458771 | 4.68E-08 | 9.39E-05 |
| cg15712177_BC11 | chr1 | 24909805 | | | 0.230434685 | 4.68E-08 | 9.39E-05 |
| cg21044433_TC21 | chr2 | 101255701 | | | 0.104850063 | 4.69E-08 | 9.39E-05 |
| cg00471198_TC21 | chr1 | 27529083 | | | 0.22062266 | 4.70E-08 | 9.39E-05 |
| cg02301040_TC21 | chr16 | 70401308 | | | 0.153415288 | 4.71E-08 | 9.39E-05 |
| cg16745596_TC21 | chr19 | 39205161 | TSS1500 | SYCN | 0.140263631 | 4.74E-08 | 9.44E-05 |
| cg03630479_BC21 | chr16 | 2233363 | | | 0.30190575 | 4.78E-08 | 9.48E-05 |
| cg25284762_BC21 | chr11 | 57500206 | | | 0.179189866 | 4.79E-08 | 9.48E-05 |
| cg25493687_BC21 | chr8 | 27430697 | | | 0.222425661 | 4.87E-08 | 9.63E-05 |
| cg24270783_BC21 | chr19 | 18645259 | | | 0.203557651 | 4.99E-08 | 9.85E-05 |
| cg02843201_BC21 | chr16 | 22092275 | TSS1500 | VWA3A | 0.135302436 | 5.07E-08 | 9.97E-05 |
| cg17167911_TC21 | chr12 | 1631172 | | | 0.415491238 | 5.09E-08 | 9.99E-05 |
| cg12929160_TC21 | chr13 | 76456509 | | | 0.149144972 | 5.15E-08 | 0.00010057 |
| cg01113530_BC21 | chr10 | 44956016 | | | 0.271301047 | 5.15E-08 | 0.00010057 |
| cg00087983_BC21 | chr17 | 82275127 | TSS1500;' | CSNK1D;C! | 0.203436394 | 5.16E-08 | 0.00010057 |
| cg16343710_TC21 | chr1 | 24573202 | | | 0.152687316 | 5.18E-08 | 0.00010086 |
| cg25592854_BC11 | chr20 | 64057272 | 5UTR;exo | TCEA2;TCE | 0.161256244 | 5.23E-08 | 0.00010117 |
| cg16163558_TC21 | chr5 | 148637235 | TSS200;T! | HTR4;HTR | 0.378679964 | 5.23E-08 | 0.00010117 |
| cg26060605_BC21 | chr22 | 19664190 | | | 0.24545815 | 5.24E-08 | 0.00010117 |
| cg05994140_BC21 | chr2 | 213239167 | | | 0.146080568 | 5.25E-08 | 0.00010117 |
| cg17082959_BC21 | chr1 | 209208432 | | | 0.163439235 | 5.35E-08 | 0.00010287 |
| cg19066120_BC21 | chr19 | 6376457 | TSS1500 | PSPN | 0.145210963 | 5.45E-08 | 0.00010463 |
| cg09615411_TC21 | chr8 | 124752195 | | | -0.62627634 | 5.48E-08 | 0.00010484 |
| cg10641688_TC21 | chr12 | 57091995 | exon_2;e | NAB2;NAB | 0.144128003 | 5.49E-08 | 0.00010484 |
| cg07559029_BC21 | chr11 | 118180682 | | | 0.243568425 | 5.50E-08 | 0.00010484 |
| cg15962969_TC21 | chr6 | 138545750 | | | 0.233611351 | 5.57E-08 | 0.0001059 |
| cg24190664_TC21 | chr17 | 73262485 | TSS1500 | CPSF4L | 0.15644065 | 5.60E-08 | 0.0001059 |
| cg05272917_BC21 | chr3 | 47917751 | | | 0.310065666 | 5.60E-08 | 0.0001059 |
| cg19999362_BC21 | chr22 | 23250663 | | | 0.143780439 | 5.61E-08 | 0.0001059 |
| cg12993543_TC21 | chr12 | 108391128 | | | 0.261845337 | 5.61E-08 | 0.0001059 |
| cg12798017_BC21 | chr4 | 107871532 | | | 0.33001047 | 5.62E-08 | 0.0001059 |
| cg01642987_TC21 | chr1 | 7616794 | | | 0.185723428 | 5.64E-08 | 0.00010594 |
| cg19042940_BC21 | chr13 | 98106241 | | | 0.442679923 | 5.74E-08 | 0.00010743 |
| cg11883790_TC21 | chr3 | 195156377 | | | 0.150380557 | 5.74E-08 | 0.00010743 |

**Supplementary Table S4.2. Significant Genome-wide CpGs in Perinatal SI v NC cohort at 38w gestation.**

| Probe ID | Chr | Position | Gene Featu | Gene Anno | Log FC | *p* -value | FDR |
|---|---|---|---|---|---|---|---|
| cg11712199_TC21 | chr2 | 165572734 | | | -0.662843 | 7.89E-16 | 6.88E-10 |
| cg23049036_BC21 | chr5 | 142587196 | | | -0.307858 | 7.92E-15 | 3.45E-09 |
| cg24025514_BC21 | chr1 | 230377633 | | | 0.351018 | 1.96E-14 | 5.69E-09 |
| cg27326209_TC21 | chr17 | 41195917 | | | -0.463637 | 2.62E-14 | 5.71E-09 |
| cg00599503_BC21 | chr6 | 64734757 | | | -0.499967 | 6.38E-14 | 8.20E-09 |
| cg13611870_BC21 | chr6 | 44290050 | | | -0.184315 | 7.29E-14 | 8.20E-09 |
| cg10553141_BC21 | chr7 | 1994971 | | | 0.326814 | 7.41E-14 | 8.20E-09 |
| cg13480504_BC21 | chr8 | 10525446 | TSS200;TSS | PRSS55;PRS | -0.214155 | 7.59E-14 | 8.20E-09 |
| cg15029283_TC21 | chr10 | 48708686 | | | -0.499907 | 8.47E-14 | 8.20E-09 |
| cg26363309_BC21 | chr13 | 29023910 | | | -0.350271 | 1.04E-13 | 9.05E-09 |
| cg11083325_BC21 | chr19 | 48741411 | exon_8;exo | IZUMO1;IZ | -0.280384 | 1.30E-13 | 1.03E-08 |
| cg19264824_TC21 | chr1 | 165367778 | | | 0.36648 | 1.65E-13 | 1.12E-08 |
| cg21300659_BC21 | chr15 | 59701916 | | | 0.434027 | 1.78E-13 | 1.12E-08 |
| cg24435439_BC21 | chr22 | 38844654 | TSS1500 | NPTXR | -0.240689 | 1.84E-13 | 1.12E-08 |
| cg08957596_TC21 | chr5 | 177400739 | TSS200 | PFN3 | -0.492566 | 2.01E-13 | 1.12E-08 |
| cg26981929_BC21 | chr10 | 23886569 | | | -0.461284 | 2.06E-13 | 1.12E-08 |
| cg24158959_TC21 | chr19 | 38613936 | exon_8;exo | MAP4K1;M | 0.279043 | 2.26E-13 | 1.16E-08 |
| cg04233789_BC21 | chr11 | 70688356 | | | -0.235746 | 2.73E-13 | 1.32E-08 |
| cg11522683_BC21 | chr6 | 37533652 | | | -0.324151 | 3.18E-13 | 1.46E-08 |
| cg08725491_TC21 | chr1 | 199226958 | | | -0.626834 | 4.07E-13 | 1.75E-08 |
| cg21169049_BC21 | chr16 | 13928943 | | | -0.463387 | 4.27E-13 | 1.75E-08 |
| cg04011062_TC21 | chr7 | 143027287 | TSS200 | OR9A2 | 0.361826 | 4.42E-13 | 1.75E-08 |
| cg16810626_BC21 | chr8 | 11736494 | | | -0.46152 | 4.84E-13 | 1.83E-08 |
| cg16113168_TC21 | chr14 | 54490683 | | | 0.357061 | 5.08E-13 | 1.84E-08 |
| cg01875206_BC21 | chr1 | 206603719 | | | -0.408416 | 6.21E-13 | 2.13E-08 |
| cg06126386_BC21 | chr4 | 2281656 | | | -0.156806 | 6.37E-13 | 2.13E-08 |
| cg10782081_TC21 | chr1 | 51088772 | | | -0.381448 | 6.67E-13 | 2.15E-08 |
| cg03770410_BC21 | chr20 | 59191277 | exon_4;exo | ZNF831;ZN | -0.222337 | 7.10E-13 | 2.21E-08 |
| cg03089778_BC21 | chr14 | 91277759 | | | -0.319077 | 7.52E-13 | 2.26E-08 |
| cg22852824_TC21 | chr12 | 19212865 | | | -0.394997 | 8.18E-13 | 2.32E-08 |
| cg10191248_TC21 | chr10 | 132627581 | | | -0.330318 | 8.26E-13 | 2.32E-08 |
| cg05413199_BC21 | chr16 | 67396551 | | | -0.397637 | 9.71E-13 | 2.64E-08 |
| cg27281030_BC11 | chr19 | 53809627 | exon_3;exo | NLRP12;NL | -0.361709 | 1.01E-12 | 2.66E-08 |
| cg12706801_BC21 | chr11 | 66688769 | exon_31 | SPTBN2 | 0.249264 | 1.08E-12 | 2.68E-08 |
| cg23208557_BC21 | chr4 | 123462216 | | | 0.22396 | 1.08E-12 | 2.68E-08 |
| cg24076840_BC21 | chr14 | 61768487 | | | -0.194782 | 1.11E-12 | 2.69E-08 |
| cg26773506_TC21 | chr11 | 118359296 | TSS1500;TS | UBE4A;UBE | -0.202913 | 1.29E-12 | 3.03E-08 |
| cg19861048_TC21 | chr1 | 1280967 | | | -0.231365 | 1.42E-12 | 3.21E-08 |
| cg20751715_BC21 | chr6 | 149406553 | | | -0.179621 | 1.44E-12 | 3.21E-08 |
| cg19949417_BC21 | chr14 | 99828898 | | | -0.293197 | 1.53E-12 | 3.33E-08 |
| cg23859430_TC21 | chr14 | 101632642 | | | -0.229492 | 1.63E-12 | 3.47E-08 |
| cg06847624_TC11 | chr5 | 177400670 | TSS200 | PFN3 | -0.494435 | 1.68E-12 | 3.48E-08 |
| cg14838738_TC21 | chr3 | 101646449 | | | 0.449342 | 1.91E-12 | 3.87E-08 |
| cg27426468_TC21 | chr13 | 95894604 | exon_24 | UGGT2 | -0.387913 | 2.02E-12 | 3.95E-08 |

| | | | | | | | |
|---|---|---|---|---|---|---|---|
| cg05334993_BC21 | chr15 | 60596022 | | | -0.38719 | 2.08E-12 | 3.95E-08 |
| cg02740307_BC21 | chr2 | 43461743 | | | -0.300082 | 2.09E-12 | 3.95E-08 |
| cg13924954_BC11 | chr19 | 53724022 | TSS1500 | MIR516B2 | -0.3096 | 2.25E-12 | 4.17E-08 |
| cg03411046_BC21 | chr2 | 120847986 | | | 0.289245 | 2.37E-12 | 4.22E-08 |
| cg09422187_TC21 | chr2 | 66461724 | | | -0.282986 | 2.37E-12 | 4.22E-08 |
| cg20482594_BC21 | chr15 | 62254421 | | | -0.240933 | 2.75E-12 | 4.79E-08 |
| cg09922473_BC21 | chr6 | 106910661 | | | 0.37433 | 2.80E-12 | 4.79E-08 |
| cg15123573_TC21 | chr5 | 177400081 | | | -0.282248 | 3.10E-12 | 4.97E-08 |
| cg23870409_BC21 | chr19 | 1239803 | | | 0.149181 | 3.12E-12 | 4.97E-08 |
| cg05394840_TC21 | chr6 | 117910558 | | | -0.311214 | 3.14E-12 | 4.97E-08 |
| cg05735373_BC21 | chr9 | 132481417 | | | -0.204158 | 3.21E-12 | 4.97E-08 |
| cg02510185_TC11 | chr16 | 85088763 | exon_13;3U | KIAA0513;K | -0.242071 | 3.22E-12 | 4.97E-08 |
| cg26624673_TC21 | chr8 | 66436857 | | | -0.107748 | 3.25E-12 | 4.97E-08 |
| cg17840363_BC21 | chr4 | 70914630 | | | 0.461636 | 3.49E-12 | 5.24E-08 |
| cg16316624_TC11 | chr11 | 61392177 | TSS1500;TS | TMEM216; | -0.345649 | 3.56E-12 | 5.25E-08 |
| cg09034922_TC21 | chr11 | 44576811 | | | -0.285561 | 3.84E-12 | 5.58E-08 |
| cg09303171_BC21 | chr10 | 15672771 | | | -0.25904 | 3.98E-12 | 5.68E-08 |
| cg16288834_BC21 | chr17 | 35073258 | | | -0.350388 | 4.20E-12 | 5.81E-08 |
| cg06998181_BC21 | chr11 | 57600570 | | | 0.265136 | 4.26E-12 | 5.81E-08 |
| cg26576712_TC21 | chr5 | 77640157 | TSS1500 | OTP | -0.202569 | 4.26E-12 | 5.81E-08 |
| cg08706567_TC21 | chr1 | 43349312 | exon_10 | MPL | -0.603588 | 4.49E-12 | 5.98E-08 |
| cg03296810_TC21 | chr15 | 88283223 | | | 0.268678 | 4.54E-12 | 5.98E-08 |
| cg10713002_TC21 | chr8 | 143822877 | | | 0.237087 | 4.64E-12 | 5.98E-08 |
| cg14660128_BC21 | chr10 | 11270726 | exon_7;exo | CELF2;CELF | -0.243802 | 4.67E-12 | 5.98E-08 |
| cg08716773_TC21 | chr3 | 137971466 | | | -0.258093 | 4.78E-12 | 6.03E-08 |
| cg21386414_TC21 | chr1 | 26094858 | | | -0.294275 | 5.16E-12 | 6.37E-08 |
| cg22130008_TC21 | chr4 | 154627380 | | | -0.412511 | 5.19E-12 | 6.37E-08 |
| cg10033694_BC21 | chr2 | 138515008 | | | 0.323309 | 5.61E-12 | 6.71E-08 |
| cg13393612_TC11 | chr7 | 101917011 | | | -0.229397 | 5.63E-12 | 6.71E-08 |
| cg00885506_BC21 | chr9 | 113340512 | TSS1500;TS | WDR31;WI | -0.436179 | 6.01E-12 | 6.98E-08 |
| cg00837209_BC21 | chr11 | 132099835 | | | -0.27961 | 6.03E-12 | 6.98E-08 |
| cg13027778_BC21 | chr8 | 117863813 | | | -0.588916 | 6.09E-12 | 6.98E-08 |
| cg01060989_TC21 | chr1 | 221772472 | | | -0.273852 | 6.76E-12 | 7.64E-08 |
| cg11848350_TC21 | chr9 | 84075652 | exon_3 | LOC10192 | -0.34492 | 7.01E-12 | 7.64E-08 |
| cg20166027_TC21 | chr10 | 46019369 | 5UTR;exon | NCOA4;NC | -0.187637 | 7.03E-12 | 7.64E-08 |
| cg20744302_TC21 | chr16 | 58678958 | | | 0.200515 | 7.08E-12 | 7.64E-08 |
| cg08594916_BC21 | chr13 | 98278575 | | | -0.423437 | 7.11E-12 | 7.64E-08 |
| cg18187629_BC21 | chr18 | 57750595 | | | -0.230138 | 7.72E-12 | 8.21E-08 |
| cg02441711_TC21 | chr17 | 82333229 | | | -0.402601 | 8.01E-12 | 8.22E-08 |
| cg00673776_BC21 | chr1 | 43338500 | | | 0.346767 | 8.01E-12 | 8.22E-08 |
| cg05220114_BC21 | chr19 | 57712325 | | | -0.245789 | 8.02E-12 | 8.22E-08 |
| cg07460605_TC21 | chr4 | 55271733 | | | -0.481771 | 8.24E-12 | 8.35E-08 |
| cg01128522_BC21 | chr1 | 101081420 | | | 0.353884 | 8.83E-12 | 8.76E-08 |
| cg13702833_BC21 | chr4 | 24321621 | | | 0.385125 | 8.85E-12 | 8.76E-08 |
| cg16175661_BC21 | chr11 | 32713490 | | | -0.228204 | 9.00E-12 | 8.81E-08 |
| cg22572859_TC21 | chr16 | 48099618 | | | -0.306835 | 9.25E-12 | 8.88E-08 |
| cg26492771_BC21 | chr1 | 1899127 | | | 0.224179 | 9.27E-12 | 8.88E-08 |

| | | | | | | | |
|---|---|---|---|---|---|---|---|
| cg14387705_BC21 | chr5 | 174329769 | | | -0.186311 | 9.60E-12 | 9.00E-08 |
| cg11032640_TC21 | chr2 | 150256144 | | | -0.322347 | 9.61E-12 | 9.00E-08 |
| cg21967741_BC21 | chr16 | 89467676 | | | -0.289385 | 9.81E-12 | 9.00E-08 |
| cg04187545_TC21 | chr8 | 56074491 | exon_1;5U | RPS20;RPS: | -0.42461 | 9.82E-12 | 9.00E-08 |
| cg01402552_BC21 | chr13 | 86945788 | | | -0.316259 | 9.93E-12 | 9.01E-08 |
| cg12861636_BC21 | chr12 | 92224946 | | | -0.272692 | 1.01E-11 | 9.05E-08 |
| cg00883565_BC21 | chr10 | 132037912 | | | -0.238303 | 1.02E-11 | 9.05E-08 |
| cg11675750_BC21 | chr7 | 128298705 | 3UTR;exon | RBM28;RBI | 0.433641 | 1.04E-11 | 9.05E-08 |
| cg00757033_TC21 | chr12 | 89526873 | TSS1500;TS | POC1B-GAI | -0.359979 | 1.04E-11 | 9.05E-08 |
| cg21247398_BC21 | chr7 | 151878192 | TSS1500 | PRKAG2 | -0.314194 | 1.05E-11 | 9.09E-08 |
| cg26065145_BC21 | chr22 | 19823860 | | | 0.354531 | 1.09E-11 | 9.34E-08 |
| cg03634007_TC21 | chr12 | 124722990 | | | 0.321977 | 1.11E-11 | 9.37E-08 |
| cg17728281_TC21 | chr12 | 186221 | | | 0.306872 | 1.13E-11 | 9.48E-08 |
| cg13546858_BC21 | chr1 | 933966 | | | -0.608187 | 1.15E-11 | 9.57E-08 |
| cg06265999_TC21 | chr4 | 39546805 | 3UTR;exon | SMIM14;SI | 0.349223 | 1.18E-11 | 9.68E-08 |
| cg19309499_BC11 | chr8 | 1200488 | | | -0.199052 | 1.19E-11 | 9.68E-08 |
| cg08845921_TC21 | chr4 | 99090059 | TSS1500 | ADH5 | 0.214861 | 1.20E-11 | 9.71E-08 |
| cg24785605_BC11 | chr19 | 54431839 | | | -0.294025 | 1.23E-11 | 9.83E-08 |
| cg16909583_BC21 | chr21 | 43675275 | | | 0.360816 | 1.24E-11 | 9.83E-08 |
| cg16178299_TC21 | chr19 | 36246667 | TSS1500 | ZNF565 | 0.507395 | 1.25E-11 | 9.85E-08 |
| cg13931250_TC21 | chr1 | 179808237 | | | 0.331114 | 1.31E-11 | 1.02E-07 |
| cg07079735_BC21 | chr5 | 66054012 | exon_21;e | ERBIN;ERBI | -0.421152 | 1.34E-11 | 1.03E-07 |
| cg22402261_BC21 | chr1 | 35561010 | exon_4;exc | NCDN;NCD | -0.23683 | 1.36E-11 | 1.04E-07 |
| cg15756996_BC21 | chr6 | 107690240 | | | -0.212739 | 1.43E-11 | 1.08E-07 |
| cg13069677_TC21 | chr8 | 123276251 | TSS1500 | ZHX1 | 0.347772 | 1.46E-11 | 1.10E-07 |
| cg08717401_BC21 | chr10 | 24098215 | | | -0.317625 | 1.55E-11 | 1.15E-07 |
| cg17727502_BC21 | chr1 | 3728557 | | | -0.220046 | 1.59E-11 | 1.15E-07 |
| cg12603131_BC21 | chr22 | 38828496 | exon_2 | NPTXR | -0.297545 | 1.59E-11 | 1.15E-07 |
| cg00673940_BC21 | chr1 | 43349309 | exon_10 | MPL | -0.555713 | 1.59E-11 | 1.15E-07 |
| cg01430218_TC11 | chr7 | 1229545 | | | -0.221898 | 1.59E-11 | 1.15E-07 |
| cg04766005_TC21 | chr10 | 132066259 | | | 0.203797 | 1.61E-11 | 1.15E-07 |
| cg05130679_TC11 | chr11 | 94769658 | | | -0.213271 | 1.63E-11 | 1.15E-07 |
| cg20020481_BC21 | chr19 | 9945765 | | | -0.283142 | 1.63E-11 | 1.15E-07 |
| cg13830226_BC21 | chr9 | 82475760 | | | -0.473255 | 1.69E-11 | 1.18E-07 |
| cg23714263_BC21 | chr2 | 218245650 | | | 0.248324 | 1.77E-11 | 1.22E-07 |
| cg11687966_BC21 | chr1 | 204153193 | TSS1500;TS | ETNK2;ETN | -0.362435 | 1.78E-11 | 1.22E-07 |
| cg23854008_BC21 | chr11 | 3381544 | | | 0.262568 | 1.93E-11 | 1.30E-07 |
| cg13617603_TC21 | chr6 | 32875278 | | | -0.29637 | 1.93E-11 | 1.30E-07 |
| cg26229043_BC21 | chr10 | 24118548 | | | -0.380996 | 1.95E-11 | 1.30E-07 |
| cg02828261_BC21 | chr12 | 52137988 | | | -0.261347 | 1.97E-11 | 1.31E-07 |
| cg05819073_BC21 | chr4 | 109068049 | | | -0.362388 | 2.01E-11 | 1.33E-07 |
| cg00526445_BC21 | chr1 | 155200526 | exon_13;e | THBS3;THB | -0.220051 | 2.05E-11 | 1.34E-07 |
| cg07865091_BC21 | chr1 | 43348635 | | | -0.484755 | 2.08E-11 | 1.35E-07 |
| cg22598810_BC21 | chr1 | 6115807 | | | -0.35525 | 2.19E-11 | 1.41E-07 |
| cg22949004_TC21 | chr19 | 49909739 | exon_2;exc | NUP62;NU | -0.260079 | 2.20E-11 | 1.41E-07 |
| cg24121733_TC11 | chr19 | 7889099 | | | -0.170649 | 2.28E-11 | 1.44E-07 |
| cg25937550_BC21 | chr4 | 76581559 | | | -0.290948 | 2.30E-11 | 1.44E-07 |

| | | | | | | | |
|---|---|---|---|---|---|---|---|
| cg23512079_BC21 | chr2 | 31126891 | | | -0.2092 | 2.33E-11 | 1.44E-07 |
| cg21545859_BC21 | chr3 | 5026352 | | | -0.231385 | 2.33E-11 | 1.44E-07 |
| cg05334611_BC11 | chr19 | 40279431 | | | 0.188875 | 2.35E-11 | 1.44E-07 |
| cg26474010_BC21 | chr16 | 87436550 | | | -0.214088 | 2.37E-11 | 1.44E-07 |
| cg06120122_TC21 | chr11 | 22180375 | | | -0.279433 | 2.37E-11 | 1.44E-07 |
| cg13540060_TC21 | chr6 | 141471908 | | | -0.196211 | 2.43E-11 | 1.47E-07 |
| cg00863890_TC21 | chr16 | 80277521 | | | -0.196725 | 2.46E-11 | 1.48E-07 |
| cg16658020_TC21 | chr17 | 41047389 | TSS200 | KRTAP2-1 | -0.346429 | 2.48E-11 | 1.48E-07 |
| cg03127182_BC21 | chr1 | 26094617 | | | -0.256943 | 2.52E-11 | 1.49E-07 |
| cg12053563_BC21 | chr14 | 29168374 | | | -0.265075 | 2.53E-11 | 1.49E-07 |
| cg17228637_BC21 | chr12 | 6279846 | | | 0.257585 | 2.55E-11 | 1.49E-07 |
| cg27602795_TC21 | chr6 | 34146157 | TSS200;TSS | GRM4;GRM | -0.262516 | 2.56E-11 | 1.49E-07 |
| cg15793395_BC21 | chr3 | 197782356 | exon_10;3U | FYTTD1;FYT | -0.301743 | 2.63E-11 | 1.51E-07 |
| cg08526880_TC21 | chr3 | 128568517 | | | -0.166 | 2.67E-11 | 1.53E-07 |
| cg05069228_BC21 | chr3 | 61807949 | | | -0.351594 | 2.71E-11 | 1.54E-07 |
| cg07556625_BC21 | chr5 | 567723 | | | -0.217184 | 2.71E-11 | 1.54E-07 |
| cg13393721_TC21 | chr1 | 43349364 | | | -0.764162 | 2.78E-11 | 1.56E-07 |
| cg11201398_TC21 | chr15 | 47599307 | | | -0.270755 | 2.80E-11 | 1.57E-07 |
| cg09943102_TC21 | chr7 | 73743458 | | | -0.347172 | 2.82E-11 | 1.57E-07 |
| cg15976283_BC21 | chr2 | 237133708 | | | -0.364661 | 2.85E-11 | 1.57E-07 |
| cg02958004_BC11 | chr18 | 22418006 | TSS200 | CTAGE1 | -0.263035 | 2.87E-11 | 1.57E-07 |
| cg25476599_TC21 | chr2 | 219042380 | TSS1500;TS | CFAP65;CF | 0.325093 | 2.90E-11 | 1.58E-07 |
| cg01287088_TC11 | chr5 | 177400391 | exon_1 | PFN3 | -0.353438 | 2.92E-11 | 1.58E-07 |
| cg26697065_TC21 | chr16 | 30445058 | exon_1 | SEPHS2 | 0.371835 | 2.96E-11 | 1.59E-07 |
| cg17087405_BC21 | chr11 | 130102771 | | | 0.257501 | 3.01E-11 | 1.60E-07 |
| cg26486411_BC21 | chr22 | 46857220 | | | 0.364059 | 3.01E-11 | 1.60E-07 |
| cg16087263_TC21 | chr1 | 20138725 | TSS1500;TS | PLA2G2F;P | -0.245397 | 3.03E-11 | 1.60E-07 |
| cg20731581_TC21 | chr13 | 98753015 | TSS1500 | SLC15A1 | 0.327264 | 3.10E-11 | 1.63E-07 |
| cg24603113_TC11 | chr7 | 1065685 | | | 0.321704 | 3.16E-11 | 1.65E-07 |
| cg10417662_BC21 | chr6 | 166098399 | exon_2 | LOC729681 | -0.4346 | 3.31E-11 | 1.72E-07 |
| cg17566938_TC21 | chr12 | 48336849 | 3UTR;exon | ZNF641;ZN | -0.233533 | 3.40E-11 | 1.75E-07 |
| cg10824065_BC21 | chr7 | 29240041 | | | 0.249792 | 3.42E-11 | 1.75E-07 |
| cg19077019_BC21 | chr20 | 1994315 | TSS1500;TS | PDYN;PDYN | -0.186386 | 3.50E-11 | 1.78E-07 |
| cg16502517_TC21 | chr3 | 46677237 | | | -0.16005 | 3.51E-11 | 1.78E-07 |
| cg26271763_BC21 | chr22 | 35299019 | TSS1500;TS | TOM1;TOM | -0.209544 | 3.53E-11 | 1.78E-07 |
| cg26376030_BC11 | chr11 | 646373 | | | 0.30786 | 3.58E-11 | 1.79E-07 |
| cg00752478_TC21 | chr16 | 67396456 | | | -0.198564 | 3.62E-11 | 1.80E-07 |
| cg15156944_BC11 | chr10 | 67852851 | | | 0.751517 | 3.69E-11 | 1.83E-07 |
| cg08532042_BC21 | chr10 | 71074228 | | | -0.376184 | 3.87E-11 | 1.90E-07 |
| cg16348233_BC21 | chr10 | 62305026 | | | -0.311674 | 3.97E-11 | 1.94E-07 |
| cg04302775_BC21 | chr22 | 48644786 | | | -0.328483 | 4.03E-11 | 1.95E-07 |
| cg09134668_TC21 | chr5 | 178564669 | | | -0.370576 | 4.04E-11 | 1.95E-07 |
| cg21877680_BC11 | chr8 | 43277364 | | | -0.420967 | 4.06E-11 | 1.96E-07 |
| cg05739937_BC21 | chr3 | 160320708 | | | -0.377164 | 4.10E-11 | 1.96E-07 |
| cg18682454_BC21 | chr13 | 44160636 | exon_2 | SMIM2 | -0.223512 | 4.12E-11 | 1.96E-07 |
| cg13235717_TC21 | chr1 | 26862014 | TSS1500 | SFN | -0.15108 | 4.23E-11 | 1.98E-07 |
| cg05250138_TC21 | chr11 | 134562929 | | | -0.210599 | 4.24E-11 | 1.98E-07 |

| | | | | | | | |
|---|---|---|---|---|---|---|---|
| cg05901920_BC21 | chr1 | 217078137 | 5UTR;exon_ | ESRRG;ESR | 0.30065 | 4.26E-11 | 1.98E-07 |
| cg26074430_BC21 | chr6 | 33196680 | | | 0.308998 | 4.27E-11 | 1.98E-07 |
| cg08524618_TC21 | chr4 | 148814208 | | | -0.412558 | 4.27E-11 | 1.98E-07 |
| cg17705814_BC21 | chr20 | 37521733 | TSS1500;TS | BLCAP;BLC | -0.222999 | 4.31E-11 | 1.99E-07 |
| cg02143055_BC21 | chr1 | 234863640 | | | -0.418135 | 4.34E-11 | 1.99E-07 |
| cg05722870_TC21 | chr10 | 118764053 | | | -0.242436 | 4.38E-11 | 1.99E-07 |
| cg07241573_BC21 | chr18 | 77248743 | TSS1500 | GALR1 | 0.38155 | 4.40E-11 | 1.99E-07 |
| cg07823663_TC21 | chr10 | 17006762 | | | 0.361938 | 4.43E-11 | 1.99E-07 |
| cg23964342_BC21 | chr1 | 93039027 | | | 0.272131 | 4.43E-11 | 1.99E-07 |
| cg13419814_BC21 | chr20 | 2664408 | TSS200;TSS | IDH3B;IDH3 | -0.384702 | 4.46E-11 | 1.99E-07 |
| cg10582687_BC11 | chr8 | 47179801 | | | -0.226509 | 4.51E-11 | 2.00E-07 |
| cg20888995_BC21 | chr3 | 56788031 | | | 0.2482 | 4.59E-11 | 2.03E-07 |
| cg27208169_TC21 | chr17 | 17700270 | | | -0.190562 | 4.67E-11 | 2.05E-07 |
| cg21766218_TC21 | chr11 | 36295172 | TSS1500 | PRR5L | 0.189792 | 4.68E-11 | 2.05E-07 |
| cg08805287_BC21 | chr2 | 3754460 | | | -0.250382 | 4.74E-11 | 2.06E-07 |
| cg16313807_TC11 | chr18 | 79963226 | TSS1500 | HSBP1L1 | -0.281899 | 4.76E-11 | 2.06E-07 |
| cg21007939_TC21 | chr6 | 167666985 | exon_3 | LINC02538 | -0.31037 | 4.80E-11 | 2.07E-07 |
| cg23901655_BC21 | chr19 | 2045013 | | | 0.357386 | 4.96E-11 | 2.13E-07 |
| cg10484096_BC21 | chr6 | 170162660 | exon_2 | LOC10272 | 0.411919 | 5.06E-11 | 2.15E-07 |
| cg10328844_TC21 | chr15 | 97874501 | TSS200;TSS | LINC00923 | 0.263635 | 5.07E-11 | 2.15E-07 |
| cg27438218_TC21 | chr10 | 117555422 | | | 0.462912 | 5.12E-11 | 2.16E-07 |
| cg12881469_TC21 | chr7 | 101598738 | | | -0.281755 | 5.17E-11 | 2.18E-07 |
| cg00531789_BC21 | chr6 | 36959472 | | | -0.485983 | 5.34E-11 | 2.24E-07 |
| cg06195227_BC21 | chr4 | 144580747 | | | -0.227829 | 5.44E-11 | 2.26E-07 |
| cg22561623_BC21 | chr17 | 43077600 | | | -0.158345 | 5.46E-11 | 2.26E-07 |
| cg12159502_BC21 | chr10 | 67883789 | TSS1500;TS | SIRT1;SIRT | -0.516615 | 5.60E-11 | 2.31E-07 |
| cg01150955_BC21 | chr4 | 83686184 | | | -0.368579 | 5.62E-11 | 2.31E-07 |
| cg01534262_BC21 | chr4 | 184932406 | | | 0.24648 | 5.74E-11 | 2.33E-07 |
| cg09247657_BC21 | chr14 | 99795812 | | | 0.457288 | 5.80E-11 | 2.33E-07 |
| cg12845952_BC21 | chr3 | 167380331 | 5UTR;exon_ | ZBBX;ZBBX | -0.370417 | 5.82E-11 | 2.33E-07 |
| cg09115440_BC21 | chr1 | 233326829 | TSS1500 | MAP3K21 | -0.237515 | 5.84E-11 | 2.33E-07 |
| cg18645316_TC21 | chr10 | 71622238 | | | -0.324506 | 5.89E-11 | 2.33E-07 |
| cg14008004_BC21 | chr9 | 98551677 | | | 0.26374 | 5.90E-11 | 2.33E-07 |
| cg04711862_BC21 | chr3 | 32019059 | | | -0.1803 | 5.93E-11 | 2.33E-07 |
| cg11449021_BC21 | chr17 | 42768102 | | | -0.19825 | 5.94E-11 | 2.33E-07 |
| cg05628927_BC21 | chr16 | 15592594 | | | 0.20978 | 5.98E-11 | 2.33E-07 |
| cg24247537_BC11 | chr11 | 457278 | | | 0.262368 | 5.99E-11 | 2.33E-07 |
| cg16116203_BC21 | chr12 | 6767756 | | | -0.343334 | 6.03E-11 | 2.33E-07 |
| cg14370713_TC21 | chr9 | 109255705 | | | -0.177353 | 6.04E-11 | 2.33E-07 |
| cg13025126_BC21 | chr8 | 117460792 | | | -0.343237 | 6.04E-11 | 2.33E-07 |
| cg00516250_BC21 | chr1 | 30817370 | | | -0.266217 | 6.04E-11 | 2.33E-07 |
| cg26286393_TC21 | chr7 | 92756076 | | | -0.312096 | 6.09E-11 | 2.34E-07 |
| cg04745574_BC21 | chr15 | 70024706 | | | -0.383259 | 6.21E-11 | 2.35E-07 |
| cg09137243_BC21 | chr15 | 69288498 | | | -0.238584 | 6.21E-11 | 2.35E-07 |
| cg09865698_BC21 | chr11 | 119726761 | | | -0.140733 | 6.24E-11 | 2.35E-07 |
| cg20508648_BC21 | chr15 | 64567036 | | | -0.338698 | 6.25E-11 | 2.35E-07 |
| cg11550599_TC21 | chr13 | 113321653 | exon_8 | LAMP1 | 0.307381 | 6.25E-11 | 2.35E-07 |

| | | | | | | | |
|---|---|---|---|---|---|---|---|
| cg12975201_BC21 | chr3 | 26646637 | | | -0.666388 | 6.30E-11 | 2.36E-07 |
| cg25488751_TC21 | chr20 | 59159747 | 5UTR;exon_ | ZNF831;ZN | 0.238755 | 6.41E-11 | 2.39E-07 |
| cg04112961_BC11 | chr1 | 7954513 | | | -0.448197 | 6.52E-11 | 2.42E-07 |
| cg25008182_TC21 | chr3 | 182405915 | | | -0.531847 | 6.59E-11 | 2.42E-07 |
| cg15012214_TC21 | chr3 | 56776292 | TSS1500;TS | ARHGEF3;A | 0.231911 | 6.59E-11 | 2.42E-07 |
| cg00178749_BC21 | chr2 | 11541733 | TSS1500;TS | MIR4429;G | -0.178425 | 6.77E-11 | 2.48E-07 |
| cg07599644_BC21 | chr11 | 33672320 | exon_21;3U | KIAA1549L | -0.272265 | 6.94E-11 | 2.53E-07 |
| cg00527391_BC21 | chr18 | 42192464 | | | -0.330892 | 7.00E-11 | 2.54E-07 |
| cg10584271_BC21 | chr3 | 52780627 | | | -0.281033 | 7.05E-11 | 2.55E-07 |
| cg05989861_TC21 | chr2 | 95274283 | TSS200;TSS | PROM2;PR | 0.25329 | 7.16E-11 | 2.58E-07 |
| cg27390775_BC21 | chr20 | 45827845 | TSS1500 | TNNC2 | -0.296483 | 7.52E-11 | 2.69E-07 |
| cg24200022_BC21 | chr5 | 1850761 | | | 0.267014 | 7.58E-11 | 2.69E-07 |
| cg09451572_TC11 | chr22 | 46364223 | exon_34;ex | CELSR1;CEL | -0.126504 | 7.62E-11 | 2.69E-07 |
| cg04704382_BC21 | chr3 | 129892911 | TSS1500 | TMCC1-DT | 0.288242 | 7.63E-11 | 2.69E-07 |
| cg03887755_BC21 | chr10 | 131998083 | | | -0.212747 | 7.63E-11 | 2.69E-07 |
| cg22491440_TC21 | chr1 | 17984862 | | | 0.402243 | 7.85E-11 | 2.76E-07 |
| cg06570734_TC21 | chr15 | 88580727 | | | -0.168408 | 7.92E-11 | 2.76E-07 |
| cg27255235_TC21 | chr12 | 31729997 | TSS1500;TS | AMN1;AMI | 0.180161 | 7.93E-11 | 2.76E-07 |
| cg13554229_BC21 | chr11 | 7655127 | | | -0.473692 | 8.08E-11 | 2.80E-07 |
| cg23603376_TC21 | chr7 | 19107759 | | | -0.29532 | 8.15E-11 | 2.82E-07 |
| cg16929850_BC21 | chr10 | 18700168 | | | -0.25801 | 8.21E-11 | 2.82E-07 |
| cg08474468_TC21 | chr5 | 125317295 | | | -0.287792 | 8.22E-11 | 2.82E-07 |
| cg20515751_TC21 | chr11 | 94543345 | TSS1500 | FUT4 | -0.16617 | 8.29E-11 | 2.83E-07 |
| cg13912631_BC21 | chr9 | 133199892 | | | -0.301836 | 8.34E-11 | 2.83E-07 |
| cg11984586_BC21 | chr3 | 57666199 | exon_4 | DENND6A | -0.342201 | 8.36E-11 | 2.83E-07 |
| cg07129714_TC21 | chr3 | 154326844 | | | -0.307298 | 8.41E-11 | 2.84E-07 |
| cg04729852_BC21 | chr4 | 56320570 | | | 0.22044 | 8.45E-11 | 2.84E-07 |
| cg13831216_TC21 | chr7 | 70313653 | | | -0.257062 | 8.61E-11 | 2.89E-07 |
| cg16862127_TC21 | chr18 | 6219349 | | | 0.230723 | 8.76E-11 | 2.92E-07 |
| cg09119349_BC21 | chr19 | 45591611 | exon_2 | GPR4 | -0.286385 | 8.78E-11 | 2.92E-07 |
| cg18813194_TC21 | chr12 | 50105525 | | | -0.274097 | 9.05E-11 | 2.99E-07 |
| cg25806249_TC21 | chr13 | 23723615 | | | -0.205655 | 9.05E-11 | 2.99E-07 |
| cg09208020_TC21 | chr14 | 22470082 | | | 0.202424 | 9.26E-11 | 3.04E-07 |
| cg01297323_BC21 | chr17 | 74437939 | | | -0.291815 | 9.34E-11 | 3.06E-07 |
| cg09922480_TC21 | chr6 | 106911426 | | | 0.238651 | 9.42E-11 | 3.07E-07 |
| cg19656689_TC21 | chr12 | 70719749 | | | -0.38548 | 9.48E-11 | 3.08E-07 |
| cg08174558_TC21 | chr5 | 79235265 | TSS1500 | JMY | 0.490083 | 1.01E-10 | 3.27E-07 |
| cg05970234_BC21 | chr18 | 63126588 | 3UTR;exon_ | BCL2;BCL2 | 0.215495 | 1.03E-10 | 3.31E-07 |
| cg05804525_TC21 | chr9 | 96501839 | 3UTR;exon_ | CDC14B;CD | 0.323606 | 1.05E-10 | 3.36E-07 |
| cg08153693_TC21 | chr8 | 63768078 | TSS1500 | LINC01289 | -0.259479 | 1.06E-10 | 3.40E-07 |
| cg04999902_BC21 | chr17 | 49500492 | | | -0.134508 | 1.08E-10 | 3.44E-07 |
| cg11786848_BC21 | chr13 | 70961041 | | | 0.39177 | 1.08E-10 | 3.44E-07 |
| cg20061862_TC21 | chr15 | 55196768 | | | -0.307259 | 1.10E-10 | 3.47E-07 |
| cg14965300_BC21 | chr10 | 14909153 | exon_14;ex | DCLRE1C;D | -0.299694 | 1.11E-10 | 3.50E-07 |
| cg08911237_TC21 | chr12 | 104356640 | | | 0.251727 | 1.11E-10 | 3.50E-07 |
| cg16082094_TC21 | chr11 | 18651249 | | | -0.30091 | 1.13E-10 | 3.55E-07 |
| cg20953257_TC21 | chr19 | 11178673 | exon_5;exo | KANK2;KAN | -0.189322 | 1.15E-10 | 3.60E-07 |

| | | | | | | | |
|---|---|---|---|---|---|---|---|
| cg05981178_TC21 | chr3 | 190941839 | | | -0.273684 | 1.16E-10 | 3.62E-07 |
| cg07917031_BC21 | chr1 | 232435589 | | | 0.23334 | 1.18E-10 | 3.66E-07 |
| cg12031808_BC21 | chr5 | 133114132 | | | -0.280221 | 1.18E-10 | 3.66E-07 |
| cg04073614_TC21 | chr13 | 97980018 | | | 0.266206 | 1.19E-10 | 3.67E-07 |
| cg15911859_TC11 | chr22 | 45414162 | TSS1500;TS | SMC1B;SM | -0.624533 | 1.20E-10 | 3.67E-07 |
| cg15847600_BC21 | chr8 | 142913584 | | | -0.265126 | 1.20E-10 | 3.67E-07 |
| cg24260907_TC21 | chr4 | 119519687 | | | 0.407574 | 1.21E-10 | 3.67E-07 |
| cg09368485_BC21 | chr6 | 30006245 | | | -0.32173 | 1.21E-10 | 3.68E-07 |
| cg09124636_TC21 | chr14 | 23305262 | | | 0.444086 | 1.22E-10 | 3.68E-07 |
| cg19514045_TC21 | chr6 | 149865144 | TSS1500 | LRP11 | -0.418846 | 1.22E-10 | 3.68E-07 |
| cg01991597_BC21 | chr1 | 220777163 | | | 0.279864 | 1.23E-10 | 3.68E-07 |
| cg08203845_BC21 | chr1 | 184408212 | | | 0.208678 | 1.25E-10 | 3.74E-07 |
| cg18457394_BC21 | chr2 | 234462482 | | | -0.14753 | 1.26E-10 | 3.77E-07 |
| cg13843727_BC11 | chr22 | 41205348 | 5UTR;exon_ | L3MBTL2;L | 0.224711 | 1.30E-10 | 3.85E-07 |
| cg13364311_TC21 | chr2 | 221468569 | | | -0.205604 | 1.31E-10 | 3.87E-07 |
| cg08528513_BC21 | chr5 | 132745651 | | | 0.26162 | 1.38E-10 | 4.07E-07 |
| cg16563487_TC21 | chr11 | 69979173 | | | -0.377618 | 1.38E-10 | 4.07E-07 |
| cg23826829_TC21 | chr2 | 72182783 | | | -0.305098 | 1.40E-10 | 4.11E-07 |
| cg20603480_BC21 | chr9 | 95426500 | | | 0.315094 | 1.41E-10 | 4.11E-07 |
| cg02084211_BC21 | chr1 | 180187017 | | | -0.475029 | 1.42E-10 | 4.13E-07 |
| cg25488108_TC21 | chr20 | 59105950 | | | -0.229869 | 1.42E-10 | 4.13E-07 |
| cg06181703_BC11 | chr1 | 1093498 | | | -0.234712 | 1.43E-10 | 4.15E-07 |
| cg10354474_BC21 | chr3 | 50313329 | TSS1500 | HYAL1 | -0.214451 | 1.44E-10 | 4.15E-07 |
| cg10416206_BC11 | chr2 | 222298078 | TSS200 | CCDC140 | -0.300331 | 1.45E-10 | 4.18E-07 |
| cg26775604_BC21 | chr6 | 26235443 | TSS1500 | H1-3 | -0.343005 | 1.46E-10 | 4.18E-07 |
| cg09918587_BC21 | chr6 | 106509559 | | | -0.282733 | 1.46E-10 | 4.18E-07 |
| cg19311244_TC21 | chr4 | 76420759 | | | -0.315799 | 1.47E-10 | 4.18E-07 |
| cg27543493_BC11 | chr17 | 39980844 | 5UTR;exon_ | PSMD3;PSI | -0.291787 | 1.50E-10 | 4.26E-07 |
| cg01902489_BC21 | chr8 | 86125139 | | | -0.221276 | 1.51E-10 | 4.27E-07 |
| cg06715007_TC21 | chr13 | 44650578 | | | -0.246899 | 1.52E-10 | 4.27E-07 |
| cg06664486_TC21 | chr6 | 170064153 | | | -0.142576 | 1.53E-10 | 4.27E-07 |
| cg11886879_TC21 | chr18 | 58765259 | | | 0.41894 | 1.53E-10 | 4.27E-07 |
| cg15346317_TC11 | chr8 | 119856105 | TSS1500 | DSCC1 | -0.376256 | 1.53E-10 | 4.27E-07 |
| cg19254026_BC21 | chr2 | 189738502 | | | 0.367902 | 1.54E-10 | 4.28E-07 |
| cg11638553_BC21 | chr7 | 137920259 | | | 0.275722 | 1.54E-10 | 4.28E-07 |
| cg10262997_TC21 | chr11 | 18633339 | | | -0.261442 | 1.58E-10 | 4.35E-07 |
| cg10300540_TC21 | chr15 | 85627824 | | | -0.291758 | 1.58E-10 | 4.35E-07 |
| cg04104144_TC21 | chr2 | 212956901 | | | 0.182071 | 1.58E-10 | 4.35E-07 |
| cg05274464_TC11 | chr16 | 71895555 | 5UTR;exon_ | IST1;IST1;IS | 0.255252 | 1.62E-10 | 4.43E-07 |
| cg05433098_TC22 | chr3 | 123365823 | | | -0.174237 | 1.62E-10 | 4.43E-07 |
| cg00252418_BC21 | chr7 | 154748615 | | | -0.232777 | 1.65E-10 | 4.49E-07 |
| cg05729088_BC21 | chr20 | 13107477 | | | -0.231425 | 1.67E-10 | 4.54E-07 |
| cg26840437_TC21 | chr5 | 1850957 | | | 0.462119 | 1.70E-10 | 4.56E-07 |
| cg27426044_TC21 | chr1 | 204870736 | TSS200;TSS | NFASC;NFA | -0.217541 | 1.70E-10 | 4.56E-07 |
| cg10054971_TC21 | chr17 | 8332793 | | | -0.192815 | 1.70E-10 | 4.56E-07 |
| cg17343184_BC21 | chr2 | 188969543 | | | 0.347386 | 1.72E-10 | 4.59E-07 |
| cg21789280_TC21 | chr1 | 22024932 | exon_2;TSS | LINC01635_ | -0.277694 | 1.72E-10 | 4.59E-07 |

| | | | | | | | |
|---|---|---|---|---|---|---|---|
| cg20483325_TC21 | chr12 | 29844873 | | | -0.24269 | 1.73E-10 | 4.60E-07 |
| cg09795209_BC21 | chr6 | 87628706 | | | 0.591743 | 1.73E-10 | 4.60E-07 |
| cg08889243_TC21 | chr1 | 57427158 | | | 0.372208 | 1.75E-10 | 4.63E-07 |
| cg25490949_TC21 | chr9 | 16058432 | | | -0.154487 | 1.78E-10 | 4.69E-07 |
| cg01108633_TC21 | chr7 | 97270527 | | | -0.273918 | 1.81E-10 | 4.77E-07 |
| cg19051042_TC22 | chr17 | 41524067 | | | -0.217831 | 1.85E-10 | 4.81E-07 |
| cg17063565_TC21 | chr18 | 27298453 | | | -0.427753 | 1.85E-10 | 4.81E-07 |
| cg10439075_BC21 | chr15 | 34977820 | | | -0.282186 | 1.85E-10 | 4.81E-07 |
| cg21530280_BC21 | chr4 | 173530243 | TSS1500;TS | HAND2-AS: | -0.337352 | 1.85E-10 | 4.81E-07 |
| cg19090574_BC21 | chr1 | 205271782 | | | -0.219082 | 1.87E-10 | 4.84E-07 |
| cg09644892_TC21 | chr6 | 63574436 | | | -0.215122 | 1.87E-10 | 4.84E-07 |
| cg26145670_TC21 | chr6 | 28586720 | exon_1;5U | ZBED9;ZBE | -0.171779 | 1.88E-10 | 4.86E-07 |
| cg24834300_BC21 | chr18 | 43396235 | | | -0.349986 | 1.89E-10 | 4.86E-07 |
| cg23646888_BC21 | chr3 | 113210825 | TSS1500;TS | BOC;BOC;B | -0.162501 | 1.90E-10 | 4.86E-07 |
| cg12372706_TC21 | chr2 | 106975914 | | | 0.345372 | 1.92E-10 | 4.91E-07 |
| cg00282582_TC21 | chr1 | 15407276 | | | -0.25169 | 1.96E-10 | 4.98E-07 |
| cg18929251_BC21 | chr7 | 108104122 | | | -0.247246 | 1.97E-10 | 4.98E-07 |
| cg20054983_TC21 | chr11 | 124626465 | | | -0.243979 | 1.98E-10 | 4.98E-07 |
| cg08917665_TC21 | chr12 | 9708440 | | | -0.221858 | 1.98E-10 | 4.98E-07 |
| cg07860918_TC21 | chr16 | 90026347 | | | -0.344673 | 1.98E-10 | 4.98E-07 |
| cg18125814_TC21 | chr11 | 49433662 | | | -0.302728 | 1.99E-10 | 4.98E-07 |
| cg25712987_BC21 | chr3 | 148741450 | exon_3;exc | AGTR1;AGT | -0.343154 | 2.00E-10 | 4.98E-07 |
| cg10084554_TC21 | chr10 | 126302590 | | | -0.197626 | 2.00E-10 | 4.98E-07 |
| cg09332650_TC21 | chr19 | 49616743 | | | -0.273692 | 2.01E-10 | 4.98E-07 |
| cg16975250_TC21 | chr11 | 119417701 | 3UTR;exon | THY1;THY1 | -0.143108 | 2.02E-10 | 4.98E-07 |
| cg17056731_BC21 | chr6 | 166440395 | | | 0.632907 | 2.02E-10 | 4.98E-07 |
| cg06595320_BC21 | chr3 | 27712474 | | | -0.279789 | 2.02E-10 | 4.98E-07 |
| cg08802535_TC21 | chr11 | 26231764 | | | -0.266933 | 2.03E-10 | 4.98E-07 |
| cg17277615_TC21 | chr10 | 22554918 | | | -0.271755 | 2.03E-10 | 4.98E-07 |
| cg11550865_BC21 | chr15 | 59209209 | | | 0.269648 | 2.05E-10 | 5.01E-07 |
| cg13081156_TC21 | chr5 | 134945306 | | | -0.305764 | 2.05E-10 | 5.01E-07 |
| cg06240320_TC21 | chr10 | 48944654 | | | -0.300254 | 2.06E-10 | 5.01E-07 |
| cg09618773_TC21 | chr8 | 1099167 | | | -0.280046 | 2.08E-10 | 5.05E-07 |
| cg09636214_TC21 | chr14 | 73245783 | exon_1 | PAPLN-AS1 | -0.579198 | 2.09E-10 | 5.07E-07 |
| cg00470471_BC21 | chr1 | 27492888 | | | 0.243214 | 2.15E-10 | 5.18E-07 |
| cg23049123_TC11 | chr6 | 128027739 | TSS200 | PTPRK-AS1 | -0.23291 | 2.15E-10 | 5.18E-07 |
| cg00123478_BC21 | chr4 | 10456541 | | | -0.227568 | 2.17E-10 | 5.20E-07 |
| cg23255231_TC21 | chr3 | 111609744 | | | -0.302654 | 2.20E-10 | 5.26E-07 |
| cg00606578_TC21 | chr6 | 30219747 | | | -0.402973 | 2.22E-10 | 5.29E-07 |
| cg11260439_TC21 | chr17 | 41540336 | | | -0.154538 | 2.25E-10 | 5.36E-07 |
| cg15028458_TC11 | chr3 | 58874564 | | | 0.215883 | 2.27E-10 | 5.38E-07 |
| cg20748675_TC21 | chr5 | 50853569 | | | 0.263269 | 2.28E-10 | 5.38E-07 |
| cg13082558_BC21 | chr8 | 124596661 | | | 0.217369 | 2.28E-10 | 5.38E-07 |
| cg15229836_BC21 | chr2 | 238644739 | | | -1.387998 | 2.29E-10 | 5.39E-07 |
| cg21814602_TC21 | chr7 | 67820218 | | | -0.332014 | 2.30E-10 | 5.39E-07 |
| cg13588196_BC21 | chr9 | 34620938 | TSS1500;TS | DCTN3;DCT | -0.295299 | 2.31E-10 | 5.39E-07 |
| cg25806481_BC21 | chr21 | 33971798 | | | -0.291043 | 2.31E-10 | 5.39E-07 |

| | | | | | | | |
|---|---|---|---|---|---|---|---|
| cg21251385_TC12 | chr20 | 53660511 | | | 0.330869 | 2.32E-10 | 5.39E-07 |
| cg11839415_BC11 | chr1 | 43349093 | | | -0.554083 | 2.32E-10 | 5.39E-07 |
| cg04071866_BC21 | chr16 | 4650736 | | | -0.124125 | 2.34E-10 | 5.42E-07 |
| cg01008680_TC21 | chr22 | 39291339 | | | -0.263942 | 2.35E-10 | 5.42E-07 |
| cg26040401_BC21 | chr18 | 31576700 | | | 0.338438 | 2.38E-10 | 5.48E-07 |
| cg12434134_BC21 | chr7 | 4727775 | | | -0.187488 | 2.40E-10 | 5.51E-07 |
| cg01921037_TC21 | chr17 | 39029732 | TSS200 | LRRC37A11 | 0.198654 | 2.40E-10 | 5.51E-07 |
| cg07650831_BC21 | chr7 | 158879672 | | | 0.361 | 2.45E-10 | 5.59E-07 |
| cg01856541_BC21 | chr1 | 204972091 | | | 0.312362 | 2.47E-10 | 5.60E-07 |
| cg18637510_TC21 | chr5 | 99771767 | | | -0.298289 | 2.47E-10 | 5.60E-07 |
| cg24208317_BC21 | chr1 | 152510133 | TSS1500 | LCE5A | -0.186625 | 2.47E-10 | 5.60E-07 |
| cg04467832_TC21 | chr6 | 168625812 | | | -0.256204 | 2.48E-10 | 5.60E-07 |
| cg04342062_BC21 | chr2 | 236567555 | | | -0.158828 | 2.48E-10 | 5.60E-07 |
| cg09939441_TC21 | chr10 | 11603009 | | | -0.416767 | 2.49E-10 | 5.60E-07 |
| cg03414830_BC21 | chr19 | 18588593 | TSS1500;TS | REX1BD;RE | 0.242381 | 2.49E-10 | 5.60E-07 |
| cg26792783_TC21 | chr13 | 43876018 | | | -0.302021 | 2.51E-10 | 5.63E-07 |
| cg00080816_TC21 | chr6 | 159990048 | | | -0.194222 | 2.54E-10 | 5.67E-07 |
| cg19656064_BC21 | chr16 | 77432668 | | | 0.363864 | 2.56E-10 | 5.71E-07 |
| cg11794015_TC21 | chr5 | 149296574 | | | 0.264907 | 2.58E-10 | 5.74E-07 |
| cg04854098_TC21 | chr4 | 98566346 | | | -0.246804 | 2.60E-10 | 5.75E-07 |
| cg00788235_BC21 | chr11 | 118400176 | TSS1500;TS | ATP5MG;A | 0.30119 | 2.60E-10 | 5.75E-07 |
| cg14275216_BC21 | chr9 | 127494560 | | | -0.200208 | 2.62E-10 | 5.77E-07 |
| cg08559141_BC21 | chr20 | 36831553 | | | -0.225652 | 2.65E-10 | 5.84E-07 |
| cg04177244_BC21 | chr10 | 71214137 | | | -0.177647 | 2.67E-10 | 5.85E-07 |
| cg00642460_BC11 | chr5 | 177400696 | TSS200 | PFN3 | -0.44745 | 2.70E-10 | 5.92E-07 |
| cg27440866_BC21 | chr2 | 177333034 | | | -0.170128 | 2.76E-10 | 6.02E-07 |
| cg07595691_BC21 | chr11 | 15615896 | | | -0.120603 | 2.77E-10 | 6.04E-07 |
| cg15362075_TC21 | chr5 | 66595234 | TSS1500;TS | MAST4;MA | -0.412233 | 2.78E-10 | 6.04E-07 |
| cg25027454_BC21 | chr20 | 18225153 | | | -0.122313 | 2.83E-10 | 6.13E-07 |
| cg22743002_TC21 | chr6 | 34155438 | exon_1;5U | GRM4;GRN | -0.182819 | 2.85E-10 | 6.16E-07 |
| cg11085304_TC21 | chr10 | 11005623 | TSS200;TSS | CELF2;CELF | -0.27283 | 2.87E-10 | 6.16E-07 |
| cg18744117_TC21 | chr16 | 66832195 | TSS1500;TS | NAE1;NAE1 | -0.144298 | 2.88E-10 | 6.16E-07 |
| cg18548246_BC21 | chr21 | 32728817 | exon_1;exo | PAXBP1-AS | -0.237218 | 2.88E-10 | 6.16E-07 |
| cg16143893_TC21 | chr16 | 55566146 | TSS1500 | CAPNS2 | -0.457437 | 2.88E-10 | 6.16E-07 |
| cg25034558_TC21 | chr1 | 225740875 | | | 0.194378 | 2.88E-10 | 6.16E-07 |
| cg08952060_BC21 | chr5 | 177097385 | exon_17;ex | FGFR4;FGF | -0.162701 | 2.90E-10 | 6.17E-07 |
| cg12220237_BC21 | chr5 | 41009382 | exon_32 | MROH2B | -0.168549 | 2.91E-10 | 6.17E-07 |
| cg27480427_BC11 | chr7 | 35538900 | | | -0.164008 | 2.96E-10 | 6.24E-07 |
| cg21941957_TC21 | chr15 | 69282712 | | | 0.242747 | 2.96E-10 | 6.24E-07 |
| cg03366574_TC11 | chr7 | 2724965 | exon_6;3U | AMZ1;AMZ | 0.320534 | 2.97E-10 | 6.24E-07 |
| cg04373118_BC21 | chr2 | 238686842 | | | -0.490729 | 3.00E-10 | 6.24E-07 |
| cg19337279_BC21 | chr1 | 43338646 | exon_3 | MPL | 0.415639 | 3.00E-10 | 6.24E-07 |
| cg08542879_BC21 | chr1 | 203493078 | TSS1500 | OPTC | -0.141951 | 3.00E-10 | 6.24E-07 |
| cg26811832_TC21 | chr13 | 67228523 | exon_2;5U | PCDH9;PCD | 0.287619 | 3.01E-10 | 6.24E-07 |
| cg09371284_TC21 | chr1 | 210366879 | | | -0.306937 | 3.01E-10 | 6.24E-07 |
| cg12287196_BC21 | chr1 | 178894534 | | | -0.518063 | 3.01E-10 | 6.24E-07 |
| cg05980569_BC21 | chr11 | 6357147 | | | -0.159508 | 3.02E-10 | 6.24E-07 |

| | | | | | | | |
|---|---|---|---|---|---|---|---|
| cg17379860_TC21 | chr11 | 61392130 | TSS1500;TS | TMEM216; | -0.202437 | 3.02E-10 | 6.24E-07 |
| cg04851106_BC21 | chr19 | 35665773 | TSS1500;TS | UPK1A;UPK | -0.14785 | 3.03E-10 | 6.24E-07 |
| cg10582401_BC21 | chr7 | 3246821 | | | -0.302863 | 3.04E-10 | 6.24E-07 |
| cg01622304_TC21 | chr13 | 27923264 | | | -0.322618 | 3.04E-10 | 6.24E-07 |
| cg26710839_TC21 | chr11 | 107771383 | | | 0.336392 | 3.09E-10 | 6.34E-07 |
| cg17164077_BC21 | chr5 | 34950079 | | | -0.22024 | 3.11E-10 | 6.36E-07 |
| cg27634164_BC21 | chr5 | 117415439 | TSS200 | LINC00992 | 0.349563 | 3.12E-10 | 6.36E-07 |
| cg11926656_BC21 | chr16 | 52709576 | | | -0.157146 | 3.16E-10 | 6.42E-07 |
| cg16844420_TC21 | chr2 | 241153351 | | | -0.347026 | 3.18E-10 | 6.44E-07 |
| cg06077968_BC21 | chr4 | 473183 | | | -0.226001 | 3.18E-10 | 6.44E-07 |
| cg02801685_BC21 | chr5 | 132767909 | | | -0.231511 | 3.19E-10 | 6.44E-07 |
| cg15576669_TC21 | chr11 | 44764218 | | | -0.204598 | 3.21E-10 | 6.48E-07 |
| cg17282018_BC21 | chr12 | 10719725 | | | -0.249399 | 3.25E-10 | 6.54E-07 |
| cg24972504_BC21 | chr20 | 11219763 | | | -0.516329 | 3.26E-10 | 6.54E-07 |
| cg03028229_TC21 | chr17 | 55459022 | | | -0.190964 | 3.27E-10 | 6.55E-07 |
| cg20552263_BC11 | chr7 | 156127761 | | | -0.215133 | 3.29E-10 | 6.58E-07 |
| cg02394812_TC21 | chr15 | 43800290 | TSS1500;TS | HYPK;HYPK | -0.224297 | 3.31E-10 | 6.59E-07 |
| cg25383605_TC21 | chr6 | 155216913 | TSS200 | TIAM2 | -0.177157 | 3.32E-10 | 6.59E-07 |
| cg02848279_TC21 | chr2 | 56132404 | | | 0.347922 | 3.34E-10 | 6.61E-07 |
| cg17709512_BC21 | chr19 | 36800766 | | | 0.295111 | 3.34E-10 | 6.61E-07 |
| cg17602636_TC21 | chr12 | 50882863 | | | -0.347352 | 3.38E-10 | 6.68E-07 |
| cg24890423_BC21 | chr20 | 2497495 | | | -0.18169 | 3.39E-10 | 6.68E-07 |
| cg13560436_BC21 | chr5 | 56952760 | TSS1500;TS | MIER3;MIE | -0.198343 | 3.40E-10 | 6.69E-07 |
| cg22784595_BC21 | chr10 | 7413752 | | | -0.278955 | 3.41E-10 | 6.70E-07 |
| cg19059495_BC21 | chr6 | 30127718 | | | -0.349227 | 3.43E-10 | 6.71E-07 |
| cg05882344_BC21 | chr8 | 41307285 | | | -0.416565 | 3.44E-10 | 6.72E-07 |
| cg00108098_TC21 | chr3 | 122918817 | | | -0.178868 | 3.50E-10 | 6.82E-07 |
| cg20995327_BC21 | chr14 | 99218507 | | | -0.16641 | 3.54E-10 | 6.87E-07 |
| cg08956810_BC21 | chr5 | 177366317 | exon_3;exo | RGS14;RGS | -0.172327 | 3.55E-10 | 6.87E-07 |
| cg07833462_BC21 | chr12 | 11563910 | exon_8 | LINC01252 | -0.319799 | 3.55E-10 | 6.87E-07 |
| cg11696305_BC21 | chr2 | 85611384 | TSS1500;TS | USP39;USP | -0.15045 | 3.61E-10 | 6.98E-07 |
| cg10540467_BC21 | chr2 | 32246932 | | | -0.175429 | 3.63E-10 | 7.00E-07 |
| cg21489303_BC21 | chr19 | 57477814 | TSS1500;TS | ZNF772;ZN | -0.375997 | 3.64E-10 | 7.00E-07 |
| cg25330143_BC21 | chr2 | 219434412 | TSS1500 | SPEG | -0.212268 | 3.69E-10 | 7.06E-07 |
| cg20948699_BC21 | chr8 | 101393288 | | | 0.265507 | 3.69E-10 | 7.06E-07 |
| cg04263284_BC21 | chr2 | 231004463 | | | -0.33073 | 3.72E-10 | 7.10E-07 |
| cg26572392_TC21 | chr10 | 24208014 | TSS1500;TS | KIAA1217;I | 0.179256 | 3.73E-10 | 7.11E-07 |
| cg09843855_TC21 | chr10 | 132626156 | | | -0.254416 | 3.76E-10 | 7.16E-07 |
| cg25227449_TC21 | chr6 | 136901696 | | | 0.267265 | 3.78E-10 | 7.17E-07 |
| cg01528504_BC21 | chr22 | 49674788 | | | 0.238039 | 3.80E-10 | 7.19E-07 |
| cg11383115_TC21 | chr7 | 92608996 | 3UTR;exon_ | CDK6;CDK6 | 0.310478 | 3.80E-10 | 7.19E-07 |
| cg13852833_BC21 | chr12 | 62650715 | | | 0.336431 | 3.90E-10 | 7.35E-07 |
| cg07237196_TC21 | chr2 | 159027896 | | | -0.20169 | 3.93E-10 | 7.37E-07 |
| cg16408528_TC21 | chr14 | 39174952 | TSS1500 | PNN | -0.335466 | 3.94E-10 | 7.37E-07 |
| cg04890478_TC21 | chr15 | 47961587 | | | 0.352017 | 3.95E-10 | 7.37E-07 |
| cg02072717_TC21 | chr1 | 228375704 | exon_98;ex | OBSCN;OBS | -0.347719 | 3.95E-10 | 7.37E-07 |
| cg25313558_BC21 | chr14 | 58019619 | | | -0.228678 | 3.95E-10 | 7.37E-07 |

| | | | | | | | |
|---|---|---|---|---|---|---|---|
| cg23845936_BC21 | chr19 | 7140619 | | | -0.415222 | 3.96E-10 | 7.37E-07 |
| cg20531262_TC21 | chr8 | 9963432 | | | 0.493927 | 3.97E-10 | 7.37E-07 |
| cg19107504_TC21 | chr10 | 86707761 | | | -0.159841 | 4.03E-10 | 7.46E-07 |
| cg13857881_BC21 | chr3 | 25472383 | | | 0.288386 | 4.04E-10 | 7.47E-07 |
| cg12204155_TC21 | chr10 | 113819684 | | | -0.516169 | 4.07E-10 | 7.50E-07 |
| cg04712102_BC21 | chr10 | 267044 | | | 0.399621 | 4.07E-10 | 7.50E-07 |
| cg10189885_BC21 | chr2 | 197311709 | TSS1500;TS | ANKRD44;A | 0.168303 | 4.08E-10 | 7.50E-07 |
| cg17970299_TC11 | chr12 | 54379020 | | | -0.438661 | 4.16E-10 | 7.63E-07 |
| cg02417862_TC21 | chr6 | 160991220 | TSS1500;TS | MAP3K4;M | -0.558495 | 4.17E-10 | 7.63E-07 |
| cg13247077_TC21 | chr8 | 125039498 | TSS1500 | WASHC5-A | -0.465943 | 4.20E-10 | 7.67E-07 |
| cg20260841_TC21 | chr4 | 55553943 | | | -0.1825 | 4.26E-10 | 7.76E-07 |
| cg15238183_BC21 | chr7 | 139648090 | | | -0.204774 | 4.27E-10 | 7.76E-07 |
| cg03923353_BC21 | chr1 | 3982431 | | | 0.318479 | 4.28E-10 | 7.77E-07 |
| cg19120015_BC21 | chr13 | 106374866 | | | -0.239277 | 4.30E-10 | 7.79E-07 |
| cg00721062_BC21 | chr3 | 46756438 | exon_2 | PRSS43P | -0.425045 | 4.32E-10 | 7.80E-07 |
| cg13832201_TC21 | chr17 | 19379208 | TSS1500 | B9D1 | -0.207201 | 4.33E-10 | 7.80E-07 |
| cg05964971_BC21 | chr16 | 16195208 | | | -0.240568 | 4.34E-10 | 7.80E-07 |
| cg27305266_BC21 | chr11 | 70656026 | | | -0.22234 | 4.34E-10 | 7.80E-07 |
| cg21674021_TC21 | chr1 | 87212645 | TSS200 | LINC02801 | -0.277102 | 4.40E-10 | 7.85E-07 |
| cg06961812_BC21 | chr19 | 35812583 | | | -0.473771 | 4.40E-10 | 7.85E-07 |
| cg27241520_BC21 | chr17 | 81702126 | TSS1500 | MRPL12 | -0.30818 | 4.40E-10 | 7.85E-07 |
| cg04235562_TC21 | chr20 | 59472228 | | | -0.220389 | 4.42E-10 | 7.88E-07 |
| cg13859739_TC21 | chr1 | 5877318 | | | -0.312975 | 4.44E-10 | 7.90E-07 |
| cg18667130_BC21 | chr10 | 71822213 | | | 0.285123 | 4.46E-10 | 7.91E-07 |
| cg13534221_BC21 | chr3 | 19349464 | | | -0.27555 | 4.47E-10 | 7.91E-07 |
| cg15264020_TC11 | chr3 | 61688536 | | | -0.325892 | 4.50E-10 | 7.95E-07 |
| cg09104006_BC21 | chr12 | 29380524 | | | -0.426353 | 4.52E-10 | 7.97E-07 |
| cg09775695_TC11 | chr2 | 235384691 | | | -0.261711 | 4.55E-10 | 8.00E-07 |
| cg20435387_TC21 | chr6 | 143570378 | TSS1500 | PHACTR2-A | 0.474361 | 4.55E-10 | 8.00E-07 |
| cg01451195_BC21 | chr1 | 155026516 | | | -0.212522 | 4.59E-10 | 8.04E-07 |
| cg04039144_BC21 | chr16 | 56532919 | | | -0.155471 | 4.61E-10 | 8.06E-07 |
| cg07513607_TC21 | chr8 | 138831016 | | | 0.295252 | 4.62E-10 | 8.07E-07 |
| cg06621861_TC21 | chr2 | 25664820 | | | -0.227635 | 4.64E-10 | 8.08E-07 |
| cg08977842_TC21 | chr17 | 83050621 | | | -0.210784 | 4.73E-10 | 8.22E-07 |
| cg03796654_BC21 | chr2 | 171574162 | | | -0.177123 | 4.76E-10 | 8.24E-07 |
| cg04352391_BC21 | chr9 | 4836601 | | | -0.330403 | 4.76E-10 | 8.24E-07 |
| cg19651341_BC21 | chr4 | 8010611 | | | -0.260966 | 4.77E-10 | 8.24E-07 |
| cg05290450_BC21 | chr2 | 3269144 | | | -0.2405 | 4.78E-10 | 8.24E-07 |
| cg08140682_TC21 | chr9 | 15446301 | | | -0.133577 | 4.81E-10 | 8.28E-07 |
| cg06584287_TC21 | chr4 | 55101969 | exon_15 | KDR | -0.245567 | 4.83E-10 | 8.29E-07 |
| cg24352731_BC21 | chr19 | 28710521 | | | 0.411593 | 4.84E-10 | 8.29E-07 |
| cg12085226_BC21 | chr10 | 93397985 | | | -0.30138 | 4.90E-10 | 8.38E-07 |
| cg17882580_BC21 | chr7 | 47275465 | 3UTR;exon | TNS3;TNS3 | -0.366059 | 4.94E-10 | 8.42E-07 |
| cg03871754_BC21 | chr17 | 81346852 | | | -0.146902 | 4.95E-10 | 8.42E-07 |
| cg26890284_TC21 | chr6 | 116876382 | TSS1500 | RFX6 | -0.284358 | 4.95E-10 | 8.42E-07 |
| cg16592371_BC21 | chr15 | 66348206 | TSS1500 | SCARNA14 | -0.318863 | 4.96E-10 | 8.42E-07 |
| cg27483694_TC11 | chr16 | 88870900 | | | 0.423638 | 4.97E-10 | 8.43E-07 |

| | | | | | | | |
|---|---|---|---|---|---|---|---|
| cg22811734_TC21 | chr17 | 64087603 | | | 0.183757 | 5.00E-10 | 8.44E-07 |
| cg23506143_BC11 | chr5 | 56465447 | | | 0.178681 | 5.00E-10 | 8.44E-07 |
| cg02253587_TC21 | chr9 | 109816344 | | | -0.140085 | 5.14E-10 | 8.66E-07 |
| cg08749132_BC21 | chr9 | 3331995 | | | 0.228987 | 5.16E-10 | 8.67E-07 |
| cg05561235_BC21 | chr2 | 40453471 | TSS1500;TS | SLC8A1;SL( | -0.179165 | 5.20E-10 | 8.73E-07 |
| cg09385972_TC21 | chr16 | 722568 | exon_4;3U | ANTKMT;A | -0.167153 | 5.21E-10 | 8.73E-07 |
| cg05659486_BC21 | chr6 | 53927891 | | | -0.201142 | 5.23E-10 | 8.74E-07 |
| cg10708271_TC21 | chr3 | 112974050 | | | -0.259118 | 5.28E-10 | 8.81E-07 |
| cg07315493_TC11 | chr16 | 1558793 | | | -0.199571 | 5.37E-10 | 8.95E-07 |
| cg14792709_BC21 | chr10 | 23280382 | | | -0.245187 | 5.41E-10 | 8.99E-07 |
| cg08353847_BC21 | chr10 | 6926205 | | | 0.484214 | 5.43E-10 | 9.00E-07 |
| cg14438391_TC11 | chr9 | 135253165 | | | -0.251353 | 5.44E-10 | 9.00E-07 |
| cg02604754_BC21 | chr1 | 34870633 | | | -0.176208 | 5.45E-10 | 9.00E-07 |
| cg19853112_TC21 | chr11 | 112482141 | TSS200 | LINC02763 | -0.238728 | 5.45E-10 | 9.00E-07 |
| cg17895630_BC21 | chr12 | 89324531 | | | -0.184137 | 5.47E-10 | 9.00E-07 |
| cg03177862_BC21 | chr2 | 96550915 | | | -0.205032 | 5.48E-10 | 9.01E-07 |
| cg25194512_BC21 | chr10 | 120826075 | | | -0.338552 | 5.51E-10 | 9.03E-07 |
| cg01379297_BC21 | chr3 | 10767196 | | | -0.205714 | 5.52E-10 | 9.04E-07 |
| cg09495769_TC21 | chr7 | 27185723 | exon_1;TSS | HOXA11-AS | -0.297063 | 5.58E-10 | 9.13E-07 |
| cg12860635_BC21 | chr6 | 30127363 | | | -0.376405 | 5.61E-10 | 9.16E-07 |
| cg19549245_BC21 | chr14 | 58295807 | | | -0.171513 | 5.63E-10 | 9.16E-07 |
| cg00512998_BC21 | chr1 | 222011831 | | | -0.21238 | 5.67E-10 | 9.22E-07 |
| cg06674311_BC21 | chr8 | 57828390 | | | -0.266614 | 5.68E-10 | 9.22E-07 |
| cg07153053_BC21 | chr10 | 89224915 | | | -0.280425 | 5.72E-10 | 9.26E-07 |
| cg15687567_BC21 | chr12 | 43798022 | | | 0.239719 | 5.81E-10 | 9.39E-07 |
| cg05910682_BC21 | chr1 | 182863910 | | | -0.180393 | 5.82E-10 | 9.39E-07 |
| cg21871952_BC11 | chr1 | 111271524 | | | -0.234967 | 5.83E-10 | 9.39E-07 |
| cg15691344_BC11 | chr10 | 122496168 | | | -0.454036 | 5.85E-10 | 9.40E-07 |
| cg24927974_BC21 | chr7 | 35038657 | TSS1500;TS | DPY19L1;D | -0.232344 | 5.88E-10 | 9.41E-07 |
| cg05386208_TC21 | chr6 | 89887015 | | | -0.26762 | 5.88E-10 | 9.41E-07 |
| cg08650733_BC21 | chr10 | 73466600 | | | -0.260986 | 5.91E-10 | 9.45E-07 |
| cg06080616_TC21 | chr6 | 10116196 | | | -0.432201 | 6.08E-10 | 9.71E-07 |
| cg24729635_BC21 | chr16 | 8905485 | | | -0.212431 | 6.12E-10 | 9.72E-07 |
| cg08419479_TC11 | chr1 | 181191346 | | | -0.224788 | 6.13E-10 | 9.72E-07 |
| cg19395212_BC21 | chr2 | 41871192 | | | -0.183237 | 6.14E-10 | 9.72E-07 |
| cg08335107_TC21 | chr2 | 236732807 | | | -0.327297 | 6.14E-10 | 9.72E-07 |
| cg06660935_BC21 | chr9 | 84781865 | | | 0.284334 | 6.15E-10 | 9.73E-07 |
| cg20879166_TC21 | chr15 | 98916868 | exon_10;e) | IGF1R;IGF1 | 0.224403 | 6.21E-10 | 9.80E-07 |
| cg06534724_TC21 | chr12 | 12465657 | exon_4;exo | BORCS5;BC | 0.261684 | 6.23E-10 | 9.81E-07 |
| cg20572472_BC21 | chr1 | 44290626 | | | -0.237524 | 6.24E-10 | 9.81E-07 |
| cg15607522_BC21 | chr10 | 114648808 | | | 0.39245 | 6.25E-10 | 9.81E-07 |
| cg16717793_TC21 | chr12 | 87337144 | | | -0.394205 | 6.27E-10 | 9.82E-07 |
| cg15613761_BC21 | chr19 | 45901452 | exon_2 | MYPOP | -0.187292 | 6.28E-10 | 9.82E-07 |
| cg16327529_BC21 | chr12 | 111761040 | | | 0.311434 | 6.29E-10 | 9.83E-07 |
| cg01738638_TC21 | chr11 | 130068706 | TSS1500;TS | APLP2;APLI | -0.166195 | 6.32E-10 | 9.83E-07 |
| cg14603375_BC21 | chr19 | 57320181 | TSS1500 | ZNF543 | -0.303074 | 6.32E-10 | 9.83E-07 |
| cg15529239_BC21 | chr6 | 4649796 | | | 0.297326 | 6.35E-10 | 9.86E-07 |

| | | | | | | | |
|---|---|---|---|---|---|---|---|
| cg23525203_BC21 | chr18 | 48345687 | | | -0.1293 | 6.36E-10 | 9.86E-07 |
| cg17738121_BC21 | chr18 | 59502103 | | | -0.17435 | 6.46E-10 | 9.99E-07 |
| cg24185852_BC21 | chr5 | 169263814 | TSS200 | MIR585 | -0.115538 | 6.49E-10 | 1.00E-06 |
| cg16102939_BC11 | chr14 | 103942507 | 5UTR;exon | RD3L;RD3L | -0.194019 | 6.55E-10 | 1.01E-06 |
| cg13569079_TC21 | chr1 | 11701640 | | | 0.310569 | 6.56E-10 | 1.01E-06 |
| cg24436598_BC21 | chr11 | 62238612 | | | -0.314714 | 6.57E-10 | 1.01E-06 |
| cg26697125_TC21 | chr2 | 127811939 | TSS1500;TS | WDR33;WI | -0.292154 | 6.58E-10 | 1.01E-06 |
| cg07681487_TC21 | chr18 | 13482421 | | | 0.242357 | 6.60E-10 | 1.01E-06 |
| cg03060745_TC21 | chr13 | 30956910 | 5UTR;exon | TEX26;TEX2 | 0.335573 | 6.60E-10 | 1.01E-06 |
| cg09487449_TC11 | chr19 | 47480406 | exon_5;exo | KPTN;KPTN | -0.167655 | 6.62E-10 | 1.01E-06 |
| cg17810966_TC21 | chr4 | 94539620 | | | -0.294193 | 6.68E-10 | 1.02E-06 |
| cg00366392_TC21 | chr1 | 20788411 | TSS1500;TS | HP1BP3;HF | -0.195251 | 6.71E-10 | 1.02E-06 |
| cg17034390_BC11 | chr6 | 30127549 | | | -0.317377 | 6.75E-10 | 1.02E-06 |
| cg16567330_TC21 | chr6 | 106129683 | | | 0.204528 | 6.78E-10 | 1.03E-06 |
| cg09549067_TC21 | chr3 | 97252434 | | | -0.456436 | 6.78E-10 | 1.03E-06 |
| cg17186087_TC21 | chr2 | 231900706 | | | -0.177021 | 6.80E-10 | 1.03E-06 |
| cg17753090_BC21 | chr19 | 32599911 | | | -0.204878 | 6.88E-10 | 1.04E-06 |
| cg16938497_TC21 | chr11 | 116832634 | | | -0.165158 | 6.88E-10 | 1.04E-06 |
| cg27135218_TC21 | chr10 | 71240748 | | | 0.258159 | 6.91E-10 | 1.04E-06 |
| cg13266286_BC21 | chr5 | 104179239 | | | -0.353321 | 6.92E-10 | 1.04E-06 |
| cg14426343_BC21 | chr9 | 134698163 | | | -0.237322 | 6.95E-10 | 1.04E-06 |
| cg17906339_TC21 | chr1 | 204214401 | TSS1500 | GOLT1A | -0.134691 | 6.97E-10 | 1.04E-06 |
| cg11462194_TC21 | chr7 | 100560720 | | | 0.339568 | 6.99E-10 | 1.04E-06 |
| cg11032439_BC21 | chr12 | 106170430 | | | -0.333651 | 7.04E-10 | 1.04E-06 |
| cg01359962_BC21 | chr3 | 43106510 | TSS1500 | POMGNT2 | -0.228032 | 7.04E-10 | 1.04E-06 |
| cg14620697_BC21 | chr10 | 7132476 | | | -0.208476 | 7.05E-10 | 1.04E-06 |
| cg08645488_BC21 | chr14 | 64796842 | | | -0.204024 | 7.05E-10 | 1.04E-06 |
| cg05379059_TC21 | chr7 | 46786274 | | | -0.284619 | 7.09E-10 | 1.05E-06 |
| cg19928597_BC11 | chr7 | 73743053 | | | -0.354595 | 7.11E-10 | 1.05E-06 |
| cg02923490_TC21 | chr6 | 37904243 | | | 0.384549 | 7.14E-10 | 1.05E-06 |
| cg17971587_TC11 | chr11 | 120137615 | exon_1 | TRIM29 | 0.195162 | 7.17E-10 | 1.05E-06 |
| cg04880091_BC11 | chr8 | 43277308 | | | -0.49106 | 7.20E-10 | 1.06E-06 |
| cg19701538_BC21 | chr16 | 17381095 | | | 0.17798 | 7.21E-10 | 1.06E-06 |
| cg04178523_BC21 | chr19 | 12056480 | | | -0.243483 | 7.22E-10 | 1.06E-06 |
| cg21020487_BC21 | chr1 | 8701684 | | | 0.497322 | 7.25E-10 | 1.06E-06 |
| cg00673906_BC21 | chr1 | 43348932 | exon_9 | MPL | -0.367473 | 7.25E-10 | 1.06E-06 |
| cg01577678_TC21 | chr16 | 1050847 | | | -0.387088 | 7.31E-10 | 1.06E-06 |
| cg23093590_TC21 | chr15 | 42491174 | TSS200;TSS | ZNF106;ZN | -0.455625 | 7.32E-10 | 1.06E-06 |
| cg14722701_BC21 | chr12 | 28127289 | | | -0.319163 | 7.32E-10 | 1.06E-06 |
| cg26194641_TC21 | chr10 | 97611584 | exon_3;exo | HOGA1;HO | -0.313888 | 7.32E-10 | 1.06E-06 |
| cg12513061_TC21 | chr3 | 184360928 | TSS1500;TS | POLR2H;PC | -0.188561 | 7.35E-10 | 1.06E-06 |
| cg25290970_TC21 | chr21 | 35482447 | | | -0.340662 | 7.35E-10 | 1.06E-06 |
| cg16571889_BC21 | chr11 | 70424829 | | | -0.339981 | 7.38E-10 | 1.06E-06 |
| cg02827340_BC21 | chr2 | 221156394 | | | -0.30402 | 7.38E-10 | 1.06E-06 |
| cg05994850_BC21 | chr6 | 30127564 | | | -0.307097 | 7.43E-10 | 1.07E-06 |
| cg17241935_TC21 | chr19 | 45601310 | | | -0.137214 | 7.44E-10 | 1.07E-06 |
| cg04359635_TC21 | chr5 | 139390286 | TSS1500 | MZB1 | 0.15224 | 7.54E-10 | 1.08E-06 |

| | | | | | | | |
|---|---|---|---|---|---|---|---|
| cg21204639_BC21 | chr14 | 68723967 | | | -0.246939 | 7.59E-10 | 1.08E-06 |
| cg16191982_TC21 | chr19 | 16495667 | TSS1500;TS | C19orf44;C | -0.332778 | 7.59E-10 | 1.08E-06 |
| cg07719343_BC21 | chr3 | 196537816 | | | -0.354446 | 7.60E-10 | 1.08E-06 |
| cg15201733_TC21 | chr10 | 71303371 | | | -0.177186 | 7.60E-10 | 1.08E-06 |
| cg10086720_BC21 | chr6 | 33201482 | exon_2;exo | SLC39A7;SI | -0.294346 | 7.62E-10 | 1.08E-06 |
| cg21317867_TC21 | chr19 | 47992558 | TSS1500 | BSPH1 | -0.244213 | 7.65E-10 | 1.09E-06 |
| cg12302527_TC11 | chr1 | 3544817 | | | 0.175317 | 7.67E-10 | 1.09E-06 |
| cg00673938_BC11 | chr1 | 43349236 | | | -0.544343 | 7.67E-10 | 1.09E-06 |
| cg25940239_BC21 | chr8 | 132127953 | 3UTR;exon_ | KCNQ3;KCI | -0.386118 | 7.69E-10 | 1.09E-06 |
| cg19853845_TC21 | chr10 | 93417368 | | | -0.203916 | 7.74E-10 | 1.09E-06 |
| cg08999896_TC21 | chr5 | 179258786 | | | -0.208878 | 7.76E-10 | 1.09E-06 |
| cg05223897_TC21 | chr7 | 66377450 | exon_5 | LINC00174 | -0.185587 | 7.81E-10 | 1.10E-06 |
| cg02621993_BC11 | chr7 | 4834654 | exon_4 | RADIL | -0.20725 | 7.84E-10 | 1.10E-06 |
| cg10640362_BC21 | chr5 | 168600045 | | | 0.196936 | 7.88E-10 | 1.10E-06 |
| cg15362317_TC21 | chr8 | 143550305 | | | -0.167158 | 7.91E-10 | 1.11E-06 |
| cg17916960_TC21 | chr15 | 79154958 | | | -0.64358 | 7.95E-10 | 1.11E-06 |
| cg06382922_BC21 | chr11 | 11502722 | | | 0.22118 | 7.96E-10 | 1.11E-06 |
| cg12047780_BC21 | chr8 | 718961 | | | -0.320155 | 8.02E-10 | 1.11E-06 |
| cg08663909_TC21 | chr5 | 1850541 | | | 0.337673 | 8.02E-10 | 1.11E-06 |
| cg19130630_TC21 | chr7 | 47666876 | | | -0.217525 | 8.04E-10 | 1.12E-06 |
| cg25644759_TC21 | chr11 | 2997130 | | | 0.196752 | 8.16E-10 | 1.13E-06 |
| cg18037457_TC21 | chr3 | 113890421 | | | -0.171547 | 8.19E-10 | 1.13E-06 |
| cg22657492_TC11 | chr5 | 138257799 | exon_4 | GFRA3 | -0.242091 | 8.24E-10 | 1.14E-06 |
| cg06520831_TC21 | chr2 | 210324545 | TSS200;TSS | LANCL1-AS | 0.437906 | 8.26E-10 | 1.14E-06 |
| cg12196901_BC21 | chr17 | 28303055 | | | -0.309848 | 8.27E-10 | 1.14E-06 |
| cg10433547_TC21 | chr16 | 1436535 | | | 0.221648 | 8.29E-10 | 1.14E-06 |
| cg02356917_TC21 | chr20 | 62831485 | | | -0.212018 | 8.32E-10 | 1.14E-06 |
| cg02976459_BC21 | chr11 | 61391998 | TSS1500;TS | TMEM216; | -0.34623 | 8.35E-10 | 1.14E-06 |
| cg19818911_TC21 | chr4 | 76402001 | | | -0.190124 | 8.37E-10 | 1.14E-06 |
| cg17534540_TC11 | chr3 | 129305869 | exon_7;3U | HMCES;HM | -0.139411 | 8.39E-10 | 1.15E-06 |
| cg18603580_BC21 | chr8 | 104665673 | | | 0.393816 | 8.51E-10 | 1.16E-06 |
| cg01384173_TC21 | chr19 | 17453539 | | | -0.314729 | 8.52E-10 | 1.16E-06 |
| cg08030248_TC21 | chr2 | 38438614 | | | 0.280176 | 8.56E-10 | 1.16E-06 |
| cg02330587_TC21 | chr12 | 11162254 | | | -0.467573 | 8.67E-10 | 1.18E-06 |
| cg20185615_BC21 | chr7 | 2277271 | | | -0.510265 | 8.76E-10 | 1.19E-06 |
| cg17825846_BC21 | chr16 | 23803925 | | | -0.19951 | 8.76E-10 | 1.19E-06 |
| cg00399673_BC21 | chr10 | 122089399 | | | -0.379706 | 8.78E-10 | 1.19E-06 |
| cg17515932_BC21 | chr7 | 156397380 | | | -0.316289 | 8.79E-10 | 1.19E-06 |
| cg25149423_BC21 | chr16 | 11169054 | | | 0.319696 | 8.84E-10 | 1.19E-06 |
| cg13818205_TC21 | chr2 | 70705149 | | | -0.231993 | 8.87E-10 | 1.19E-06 |
| cg05745631_BC11 | chr6 | 30127401 | | | -0.271522 | 8.88E-10 | 1.19E-06 |
| cg27405177_TC21 | chr10 | 133113306 | | | -0.542244 | 8.88E-10 | 1.19E-06 |
| cg23518655_BC21 | chr20 | 63042935 | | | -0.338028 | 8.94E-10 | 1.19E-06 |
| cg15113457_TC21 | chr4 | 127163059 | | | -0.237137 | 8.94E-10 | 1.19E-06 |
| cg01219345_TC21 | chr19 | 56956005 | | | -0.254874 | 8.96E-10 | 1.19E-06 |
| cg18990588_BC21 | chr7 | 175713 | exon_2 | LOC10537 | -0.208914 | 8.96E-10 | 1.19E-06 |
| cg02178151_TC21 | chr14 | 24953880 | | | -0.230009 | 9.01E-10 | 1.19E-06 |

| | | | | | | | |
|---|---|---|---|---|---|---|---|
| cg25875824_TC11 | chr1 | 28736919 | TSS200;TSS | YTHDF2;YT | -0.268845 | 9.02E-10 | 1.19E-06 |
| cg10605590_BC21 | chr20 | 38634074 | | | -0.312799 | 9.02E-10 | 1.19E-06 |
| cg08819022_BC11 | chr22 | 18048681 | | | 0.347665 | 9.02E-10 | 1.19E-06 |
| cg07961559_BC21 | chr5 | 50751498 | | | 0.249357 | 9.06E-10 | 1.20E-06 |
| cg26684225_BC21 | chr10 | 131997955 | | | -0.193625 | 9.14E-10 | 1.21E-06 |
| cg07146298_TC21 | chr8 | 79621011 | | | -0.278575 | 9.17E-10 | 1.21E-06 |
| cg06618957_TC21 | chr4 | 52017743 | | | -0.421392 | 9.18E-10 | 1.21E-06 |
| cg13327513_TC21 | chr3 | 128462965 | exon_3;TSS | DNAJB8;DN | 0.181651 | 9.24E-10 | 1.21E-06 |
| cg05554722_BC21 | chr14 | 91884835 | | | -0.233334 | 9.25E-10 | 1.21E-06 |
| cg13358134_BC21 | chr9 | 93263131 | | | 0.242176 | 9.27E-10 | 1.21E-06 |
| cg27509052_TC11 | chr8 | 1062324 | | | -0.26361 | 9.29E-10 | 1.21E-06 |
| cg24103098_TC21 | chr7 | 45460517 | | | -0.153778 | 9.31E-10 | 1.22E-06 |
| cg16276982_TC11 | chr15 | 29675828 | TSS1500;TS | FAM189A1 | -0.392635 | 9.38E-10 | 1.22E-06 |
| cg00127991_TC21 | chr10 | 83724688 | | | 0.206104 | 9.47E-10 | 1.23E-06 |
| cg05757376_BC21 | chr13 | 35941932 | | | -0.443056 | 9.48E-10 | 1.23E-06 |
| cg16642517_BC21 | chr4 | 40438128 | exon_4;exo | RBM47;RBI | -0.451285 | 9.54E-10 | 1.24E-06 |
| cg23471798_BC21 | chr3 | 110392033 | | | -0.384999 | 9.57E-10 | 1.24E-06 |
| cg05603938_TC11 | chr12 | 6851257 | 5UTR;exon | CDCA3;CD( | 0.195823 | 9.61E-10 | 1.24E-06 |
| cg26271879_TC21 | chr22 | 35301648 | | | -0.321222 | 9.61E-10 | 1.24E-06 |
| cg14621323_BC11 | chr5 | 177400160 | 3UTR;exon | PFN3;PFN3 | -0.370363 | 9.62E-10 | 1.24E-06 |
| cg03257557_TC21 | chr12 | 121085974 | | | -0.184069 | 9.68E-10 | 1.25E-06 |
| cg22389370_BC11 | chr11 | 78574851 | 5UTR;exon | NARS2;NAF | 0.386177 | 9.69E-10 | 1.25E-06 |
| cg18094830_BC21 | chr19 | 11074203 | | | -0.258261 | 9.71E-10 | 1.25E-06 |
| cg11143475_TC21 | chr2 | 65027785 | | | 0.20348 | 9.71E-10 | 1.25E-06 |
| cg17801352_TC21 | chr2 | 1745304 | TSS1500 | PXDN | -0.255904 | 9.76E-10 | 1.25E-06 |
| cg12924849_BC21 | chr8 | 101764104 | | | -0.259908 | 9.77E-10 | 1.25E-06 |
| cg19905126_BC21 | chr10 | 21765045 | | | -0.227165 | 9.81E-10 | 1.25E-06 |
| cg06954658_TC21 | chr18 | 78980093 | 5UTR;exon | SALL3;SALL | -0.376765 | 9.83E-10 | 1.25E-06 |
| cg00493490_TC21 | chr2 | 17912082 | | | 0.221905 | 9.93E-10 | 1.26E-06 |
| cg16690253_BC21 | chr3 | 127414320 | | | 0.153899 | 9.94E-10 | 1.26E-06 |
| cg26414822_BC21 | chr10 | 100866419 | | | -0.137271 | 9.94E-10 | 1.26E-06 |
| cg02075828_BC21 | chr20 | 59351306 | | | -0.231461 | 1.00E-09 | 1.27E-06 |
| cg08958140_TC21 | chr14 | 57058293 | | | 0.282266 | 1.00E-09 | 1.27E-06 |
| cg10251070_BC21 | chr21 | 43362078 | exon_1 | LINC01679 | 0.186047 | 1.00E-09 | 1.27E-06 |
| cg19908990_TC21 | chr14 | 96270044 | | | -0.179657 | 1.01E-09 | 1.28E-06 |
| cg10649356_TC21 | chr22 | 37210737 | TSS200 | SSTR3 | 0.358247 | 1.02E-09 | 1.28E-06 |
| cg14772412_TC21 | chr4 | 177431024 | 3UTR;exon | AGA;AGA;A | 0.27945 | 1.02E-09 | 1.28E-06 |
| cg14580085_BC21 | chr2 | 238644765 | | | -1.03479 | 1.02E-09 | 1.28E-06 |
| cg13348338_BC21 | chr19 | 10640583 | | | 0.35557 | 1.02E-09 | 1.28E-06 |
| cg21457573_BC21 | chr1 | 6762335 | | | 0.255806 | 1.02E-09 | 1.28E-06 |
| cg04108406_BC21 | chr4 | 24965787 | | | 0.236656 | 1.03E-09 | 1.29E-06 |
| cg15653143_TC21 | chr11 | 61055218 | | | -0.219172 | 1.03E-09 | 1.29E-06 |
| cg04585717_TC21 | chr15 | 79058417 | exon_3;exo | RASGRF1;R | -0.245131 | 1.03E-09 | 1.29E-06 |
| cg14091763_BC21 | chr9 | 109329964 | | | -0.250941 | 1.03E-09 | 1.29E-06 |
| cg01857595_BC21 | chr14 | 37546983 | exon_13;3U | MIPOL1;MI | -0.23107 | 1.05E-09 | 1.31E-06 |
| cg12095600_TC21 | chr10 | 122162995 | TSS1500;TS | TACC2;TAC | -0.23153 | 1.05E-09 | 1.31E-06 |
| cg12133554_BC21 | chr4 | 305520 | TSS200 | ZNF732 | -0.195099 | 1.06E-09 | 1.31E-06 |

| | | | | | | | |
|---|---|---|---|---|---|---|---|
| cg06052716_BC21 | chr7 | 1240971 | | | -0.165715 | 1.07E-09 | 1.32E-06 |
| cg00968865_TC21 | chr7 | 13906545 | exon_10;e | ETV1;ETV1 | -0.199515 | 1.07E-09 | 1.32E-06 |
| cg03037450_TC21 | chr2 | 78087302 | TSS1500 | LOC10192 | -0.344855 | 1.07E-09 | 1.32E-06 |
| cg00554858_BC21 | chr5 | 96656187 | | | 0.286188 | 1.08E-09 | 1.34E-06 |
| cg11679196_TC21 | chr5 | 150374575 | | | -0.175611 | 1.09E-09 | 1.34E-06 |
| cg00810956_BC11 | chr3 | 27730275 | | | 0.204131 | 1.09E-09 | 1.34E-06 |
| cg01227835_TC21 | chr7 | 126377778 | | | -0.259508 | 1.09E-09 | 1.34E-06 |
| cg05915202_TC21 | chr7 | 126377789 | | | -0.28568 | 1.09E-09 | 1.34E-06 |
| cg09000015_TC21 | chr18 | 79000253 | | | -0.251506 | 1.09E-09 | 1.34E-06 |
| cg00403251_BC21 | chr9 | 127223918 | | | -0.220082 | 1.10E-09 | 1.35E-06 |
| cg15999067_BC21 | chr7 | 78040874 | | | 0.248248 | 1.11E-09 | 1.35E-06 |
| cg17942763_TC21 | chr1 | 2232033 | | | -0.158632 | 1.11E-09 | 1.35E-06 |
| cg04226884_BC21 | chr19 | 8424219 | | | -0.290924 | 1.12E-09 | 1.36E-06 |
| cg24686131_BC21 | chr19 | 49058742 | TSS1500;TS | CGB7;CGB7 | -0.222611 | 1.12E-09 | 1.36E-06 |
| cg01315092_BC11 | chr2 | 120222799 | exon_1 | TMEM185 | -0.272691 | 1.12E-09 | 1.36E-06 |
| cg26320010_BC21 | chr2 | 178259185 | | | -0.318031 | 1.12E-09 | 1.36E-06 |
| cg21026868_TC21 | chr1 | 219568810 | | | -0.209357 | 1.13E-09 | 1.37E-06 |
| cg09213428_BC21 | chr6 | 16332296 | | | 0.448839 | 1.13E-09 | 1.37E-06 |
| cg16664667_BC11 | chr10 | 7408851 | TSS200 | SFMBT2 | 0.399245 | 1.14E-09 | 1.37E-06 |
| cg21641924_BC21 | chr16 | 67506949 | | | 0.283135 | 1.14E-09 | 1.37E-06 |
| cg27269899_BC21 | chr11 | 72041184 | | | 0.268815 | 1.14E-09 | 1.37E-06 |
| cg05158074_TC21 | chr12 | 110846781 | TSS1500;TS | CCDC63;CC | -0.372254 | 1.14E-09 | 1.38E-06 |
| cg24350090_BC21 | chr11 | 115054782 | | | 0.347421 | 1.15E-09 | 1.38E-06 |
| cg04325291_BC21 | chr1 | 203554515 | | | -0.145462 | 1.15E-09 | 1.38E-06 |
| cg10657259_TC21 | chr6 | 168067718 | TSS200 | FRMD1 | 0.197548 | 1.16E-09 | 1.39E-06 |
| cg05300996_TC21 | chr3 | 120095846 | TSS1500;TS | GSK3B;GSK | 0.312955 | 1.16E-09 | 1.39E-06 |
| cg16306925_TC21 | chr19 | 53865704 | 5UTR;exon | MYADM;M | 0.181014 | 1.16E-09 | 1.39E-06 |
| cg19597318_BC11 | chr1 | 204870478 | TSS1500;TS | NFASC;NFA | -0.333579 | 1.16E-09 | 1.39E-06 |
| cg22533535_TC21 | chr17 | 41523608 | | | -0.118277 | 1.17E-09 | 1.39E-06 |
| cg06202984_BC21 | chr9 | 95317621 | 5UTR;exon | FANCC;FAN | 0.36682 | 1.17E-09 | 1.39E-06 |
| cg24986879_TC21 | chr20 | 13344782 | | | -0.231686 | 1.17E-09 | 1.39E-06 |
| cg16313267_BC21 | chr11 | 48101364 | | | -0.188557 | 1.17E-09 | 1.39E-06 |
| cg00362214_BC21 | chr3 | 193688978 | | | 0.268315 | 1.18E-09 | 1.39E-06 |
| cg12237293_BC21 | chr22 | 41261332 | | | 0.2775 | 1.18E-09 | 1.39E-06 |
| cg23696712_BC21 | chr12 | 52834357 | TSS200 | KRT79 | -0.191344 | 1.18E-09 | 1.40E-06 |
| cg12687449_BC21 | chr14 | 57060826 | | | -0.20046 | 1.19E-09 | 1.40E-06 |
| cg05484149_TC21 | chr2 | 31919983 | | | -0.387569 | 1.19E-09 | 1.40E-06 |
| cg13971025_TC12 | chr9 | 135104424 | | | 0.297813 | 1.19E-09 | 1.40E-06 |
| cg26481465_TC21 | chr12 | 105775473 | | | -0.239813 | 1.19E-09 | 1.40E-06 |
| cg04464316_BC21 | chr13 | 34748316 | | | -0.26425 | 1.20E-09 | 1.40E-06 |
| cg19717060_BC21 | chr17 | 72556939 | | | -0.300747 | 1.20E-09 | 1.40E-06 |
| cg06491878_BC21 | chr2 | 27059711 | | | -0.35245 | 1.20E-09 | 1.41E-06 |
| cg26890646_TC21 | chr12 | 108668249 | | | 0.225882 | 1.21E-09 | 1.41E-06 |
| cg14640509_BC21 | chr2 | 43596569 | TSS1500;TS | THADA;TH | 0.4786 | 1.21E-09 | 1.42E-06 |
| cg18139462_BC21 | chr19 | 35758311 | TSS1500 | HSPB6 | 0.280248 | 1.22E-09 | 1.43E-06 |
| cg03678547_BC21 | chr3 | 50540003 | | | -0.150911 | 1.23E-09 | 1.43E-06 |
| cg18048309_BC22 | chr8 | 144562303 | | | -0.316359 | 1.23E-09 | 1.44E-06 |

| | | | | | | | |
|---|---|---|---|---|---|---|---|
| cg05742863_BC11 | chr22 | 45413438 | TSS1500 | RIBC2 | -0.276812 | 1.24E-09 | 1.44E-06 |
| cg11236430_BC21 | chr6 | 36962067 | | | -0.353017 | 1.24E-09 | 1.44E-06 |
| cg10176692_TC21 | chr11 | 48216744 | TSS200 | OR4B1 | -0.607749 | 1.24E-09 | 1.44E-06 |
| cg10455757_TC21 | chr9 | 35111427 | 5UTR;exon | FAM214B;I | 0.172543 | 1.24E-09 | 1.44E-06 |
| cg18931815_TC21 | chr1 | 36100171 | exon_3 | COL8A2 | -0.26331 | 1.25E-09 | 1.44E-06 |
| cg12176605_BC21 | chr3 | 40476647 | TSS1500;TS | ZNF619;ZN | -0.260412 | 1.25E-09 | 1.44E-06 |
| cg02269978_BC21 | chr6 | 32879808 | | | -0.243625 | 1.25E-09 | 1.44E-06 |
| cg20942702_TC21 | chr16 | 713037 | | | 0.312942 | 1.25E-09 | 1.44E-06 |
| cg21175069_TC21 | chr2 | 64622921 | | | 0.243682 | 1.26E-09 | 1.45E-06 |
| cg08363783_TC21 | chr22 | 22884315 | | | -0.202294 | 1.26E-09 | 1.45E-06 |
| cg11646018_BC21 | chr8 | 84900414 | | | -0.27715 | 1.27E-09 | 1.45E-06 |
| cg27568504_BC21 | chr10 | 131260638 | | | 0.304851 | 1.27E-09 | 1.45E-06 |
| cg05696436_BC21 | chr16 | 1323785 | | | 0.238835 | 1.27E-09 | 1.45E-06 |
| cg04304036_BC21 | chr16 | 68300716 | exon_12;3I | SLC7A6;SLC | -0.205754 | 1.27E-09 | 1.45E-06 |
| cg18530790_TC21 | chr17 | 48705241 | | | -0.124626 | 1.27E-09 | 1.45E-06 |
| cg15301881_BC11 | chr10 | 132136296 | | | -0.154793 | 1.28E-09 | 1.46E-06 |
| cg13910191_TC21 | chr1 | 19890334 | | | 0.302604 | 1.29E-09 | 1.47E-06 |
| cg12137686_BC21 | chr10 | 52301356 | exon_4 | PRKG1-AS1 | 0.308407 | 1.29E-09 | 1.47E-06 |
| cg14080227_BC21 | chr9 | 38363160 | | | -0.233747 | 1.30E-09 | 1.47E-06 |
| cg00684861_BC21 | chr9 | 123425517 | | | 0.21616 | 1.30E-09 | 1.47E-06 |
| cg24217567_BC21 | chr2 | 207771504 | | | 0.274542 | 1.30E-09 | 1.47E-06 |
| cg15434747_BC21 | chr3 | 46607966 | | | -0.142456 | 1.30E-09 | 1.47E-06 |
| cg00698575_BC21 | chr9 | 137494152 | | | 0.173885 | 1.31E-09 | 1.48E-06 |
| cg00471159_TC21 | chr7 | 108001351 | | | -0.179276 | 1.32E-09 | 1.48E-06 |
| cg12008002_BC21 | chr2 | 96290504 | exon_20 | SNRNP200 | 0.205913 | 1.32E-09 | 1.49E-06 |
| cg08189615_BC11 | chr2 | 218292396 | | | -0.435361 | 1.33E-09 | 1.49E-06 |
| cg26786174_BC21 | chr7 | 55592934 | | | -0.268069 | 1.33E-09 | 1.49E-06 |
| cg17530503_TC21 | chr2 | 109204602 | | | -0.189495 | 1.33E-09 | 1.49E-06 |
| cg14210736_BC21 | chr13 | 39857496 | | | -0.23091 | 1.33E-09 | 1.49E-06 |
| cg27634071_TC21 | chr22 | 45413859 | 5UTR;exon | RIBC2;RIBC | -0.483132 | 1.34E-09 | 1.50E-06 |
| cg20213769_TC21 | chr16 | 2236286 | TSS200;TSS | DNASE1L2; | -0.131667 | 1.34E-09 | 1.50E-06 |
| cg12261506_TC21 | chr11 | 88576810 | | | -0.415083 | 1.35E-09 | 1.50E-06 |
| cg10577719_BC21 | chr7 | 2934545 | exon_11;e | CARD11;CA | 0.203619 | 1.36E-09 | 1.51E-06 |
| cg10070756_BC21 | chr9 | 126480364 | | | -0.300791 | 1.36E-09 | 1.51E-06 |
| cg25124064_TC21 | chr12 | 110613291 | TSS1500;TS | TCTN1;TCT | -0.297496 | 1.36E-09 | 1.51E-06 |
| cg08529194_BC21 | chr5 | 132787437 | | | -0.225185 | 1.37E-09 | 1.52E-06 |
| cg18728631_BC21 | chr14 | 20332593 | | | -0.16792 | 1.39E-09 | 1.54E-06 |
| cg04773730_BC21 | chr6 | 97669665 | | | 0.428209 | 1.39E-09 | 1.54E-06 |
| cg26644853_BC21 | chr16 | 57372345 | TSS200;TSS | CX3CL1;CX | -0.231673 | 1.40E-09 | 1.55E-06 |
| cg04373548_TC21 | chr6 | 28673952 | | | -0.369659 | 1.42E-09 | 1.57E-06 |
| cg10154780_TC11 | chr5 | 166962849 | | | -0.255026 | 1.42E-09 | 1.57E-06 |
| cg02645710_TC11 | chr12 | 85036307 | TSS200;TSS | LRRIQ1;TSF | 0.422198 | 1.42E-09 | 1.57E-06 |
| cg01349368_BC21 | chr12 | 2891737 | | | -0.253696 | 1.43E-09 | 1.57E-06 |
| cg04055053_TC21 | chr10 | 509193 | | | -0.307652 | 1.44E-09 | 1.58E-06 |
| cg26177192_TC21 | chr6 | 35464658 | | | -0.2592 | 1.44E-09 | 1.58E-06 |
| cg21685099_TC21 | chr16 | 70267905 | | | -0.140064 | 1.44E-09 | 1.58E-06 |
| cg10892799_TC21 | chr3 | 66407343 | TSS1500 | LRIG1 | -0.245772 | 1.44E-09 | 1.58E-06 |

| | | | | | | | |
|---|---|---|---|---|---|---|---|
| cg25589139_BC21 | chr18 | 46462076 | exon_8;3U | RNF165;RN | -0.178509 | 1.45E-09 | 1.59E-06 |
| cg09702434_BC21 | chr21 | 31249762 | | | -0.17556 | 1.46E-09 | 1.59E-06 |
| cg19012054_BC11 | chr9 | 124862084 | TSS200;TSS | ARPC5L;RP | -0.286014 | 1.46E-09 | 1.59E-06 |
| cg08816573_BC21 | chr1 | 16156760 | TSS1500;TS | EPHA2;EPH | -0.113346 | 1.47E-09 | 1.60E-06 |
| cg06236450_TC21 | chr13 | 26956526 | | | -0.304711 | 1.47E-09 | 1.60E-06 |
| cg26719062_TC21 | chr6 | 30127572 | | | -0.258825 | 1.47E-09 | 1.60E-06 |
| cg08943293_TC21 | chr1 | 181480563 | | | -0.157009 | 1.47E-09 | 1.60E-06 |
| cg25936482_TC21 | chr5 | 15928577 | | | -0.21337 | 1.48E-09 | 1.60E-06 |
| cg01145131_BC21 | chr12 | 15222219 | TSS1500;TS | RERG;RERG | -0.192764 | 1.48E-09 | 1.60E-06 |
| cg26643325_TC21 | chr10 | 43203819 | | | -0.342139 | 1.50E-09 | 1.62E-06 |
| cg00153057_TC21 | chr18 | 78448271 | | | 0.556006 | 1.50E-09 | 1.62E-06 |
| cg00356912_BC21 | chr1 | 20184228 | | | -0.343616 | 1.51E-09 | 1.62E-06 |
| cg19145215_TC21 | chr13 | 109297557 | exon_5;exo | LINC00370 | -0.300374 | 1.51E-09 | 1.63E-06 |
| cg01614041_BC11 | chr6 | 34155637 | TSS200 | GRM4 | -0.22968 | 1.51E-09 | 1.63E-06 |
| cg10208942_TC21 | chr17 | 82234878 | TSS1500;TS | SLC16A3;SL | -0.108121 | 1.52E-09 | 1.63E-06 |
| cg13007105_BC21 | chr2 | 11568190 | | | 0.335551 | 1.52E-09 | 1.63E-06 |
| cg08745810_TC21 | chr17 | 6304250 | | | -0.278604 | 1.52E-09 | 1.63E-06 |
| cg25322198_BC21 | chr20 | 46342463 | | | -0.133284 | 1.53E-09 | 1.64E-06 |
| cg13305245_BC21 | chr8 | 32514377 | | | -0.410822 | 1.53E-09 | 1.64E-06 |
| cg05185749_TC21 | chr9 | 114930479 | exon_1;5U | TNFSF8;TN | 0.53588 | 1.54E-09 | 1.64E-06 |
| cg11489779_TC21 | chr12 | 103587161 | TSS200 | STAB2 | -0.184153 | 1.54E-09 | 1.64E-06 |
| cg02929044_BC21 | chr5 | 61265824 | TSS1500 | LINC02057 | -0.43663 | 1.60E-09 | 1.70E-06 |
| cg09229850_BC21 | chr17 | 57762040 | | | -0.203228 | 1.60E-09 | 1.71E-06 |
| cg05272288_BC21 | chr3 | 49896092 | exon_9;exo | MST1R;MS | -0.244417 | 1.61E-09 | 1.71E-06 |
| cg20569034_BC21 | chr12 | 57727767 | exon_16;e | AGAP2;AG | 0.214402 | 1.62E-09 | 1.72E-06 |
| cg06205661_TC21 | chr3 | 115246132 | | | -0.363925 | 1.62E-09 | 1.72E-06 |
| cg26689578_TC21 | chr7 | 74571243 | | | -0.146538 | 1.63E-09 | 1.72E-06 |
| cg24878483_BC21 | chr2 | 141037751 | | | -0.298771 | 1.65E-09 | 1.74E-06 |
| cg26233832_TC21 | chr7 | 142963886 | | | -0.207315 | 1.65E-09 | 1.75E-06 |
| cg04979798_BC21 | chr8 | 92100202 | TSS1500 | RUNX1T1 | -0.282098 | 1.66E-09 | 1.75E-06 |
| cg07962822_BC21 | chr5 | 50927755 | | | -0.142245 | 1.67E-09 | 1.76E-06 |
| cg22744810_BC21 | chr17 | 58577905 | | | -0.137333 | 1.67E-09 | 1.76E-06 |
| cg26991336_TC21 | chr1 | 224069314 | | | -0.332839 | 1.68E-09 | 1.76E-06 |
| cg03704673_BC11 | chr17 | 4788992 | 5UTR;exon | GLTPD2;GL | -0.306146 | 1.68E-09 | 1.76E-06 |
| cg24656924_BC21 | chr20 | 37294890 | | | 0.319647 | 1.69E-09 | 1.77E-06 |
| cg06210630_BC21 | chr1 | 160115643 | TSS200 | ATP1A2 | -0.219564 | 1.69E-09 | 1.77E-06 |
| cg16517419_TC21 | chr5 | 3980333 | | | -0.200495 | 1.70E-09 | 1.77E-06 |
| cg23435684_TC21 | chr18 | 35939501 | | | -0.164182 | 1.71E-09 | 1.78E-06 |
| cg00949446_BC21 | chr3 | 194351337 | TSS200;TSS | CPN2;CPN2 | -0.173241 | 1.72E-09 | 1.79E-06 |
| cg26118553_TC21 | chr2 | 21022920 | exon_18 | APOB | -0.286897 | 1.72E-09 | 1.79E-06 |
| cg15415893_TC21 | chr19 | 56824725 | exon_3;5U | PEG3;PEG3 | -0.246479 | 1.73E-09 | 1.80E-06 |
| cg03185964_TC21 | chr12 | 124294592 | TSS1500 | RFLNA | -0.244414 | 1.76E-09 | 1.82E-06 |
| cg00743158_BC21 | chr14 | 100827298 | | | -0.138749 | 1.76E-09 | 1.83E-06 |
| cg00105116_BC21 | chr20 | 50320663 | | | -0.235002 | 1.76E-09 | 1.83E-06 |
| cg25277981_TC21 | chr20 | 42771518 | exon_5;exo | PTPRT;PTP | -0.251671 | 1.76E-09 | 1.83E-06 |
| cg21883463_BC21 | chr16 | 85987577 | | | -0.207917 | 1.77E-09 | 1.83E-06 |
| cg02853553_BC21 | chr7 | 44218455 | 3UTR;exon | CAMK2B;C | -0.289588 | 1.77E-09 | 1.83E-06 |

| | | | | | | | |
|---|---|---|---|---|---|---|---|
| cg13061702_TC21 | chr3 | 127028761 | | | -0.234516 | 1.78E-09 | 1.84E-06 |
| cg00079375_TC21 | chr15 | 78833493 | | | 0.258381 | 1.81E-09 | 1.86E-06 |
| cg03786743_TC21 | chr4 | 52861614 | TSS1500 | RASL11B | -0.277658 | 1.81E-09 | 1.86E-06 |
| cg08354106_BC21 | chr4 | 7960255 | | | -0.300903 | 1.81E-09 | 1.87E-06 |
| cg00088126_TC21 | chr1 | 3473745 | | | -0.137475 | 1.82E-09 | 1.87E-06 |
| cg10484450_TC11 | chr14 | 67515435 | TSS1500;TS | TMEM229I | -0.326228 | 1.82E-09 | 1.87E-06 |
| cg07959593_BC21 | chr2 | 240235111 | | | -0.243143 | 1.83E-09 | 1.87E-06 |
| cg03972135_TC21 | chr6 | 170248888 | | | 0.894522 | 1.83E-09 | 1.87E-06 |
| cg01607530_BC21 | chr11 | 1639762 | | | -0.228365 | 1.84E-09 | 1.88E-06 |
| cg23847472_TC21 | chr11 | 133913400 | | | -0.178283 | 1.85E-09 | 1.88E-06 |
| cg10834305_BC21 | chr7 | 30278360 | | | -0.18451 | 1.85E-09 | 1.88E-06 |
| cg07643840_BC21 | chr5 | 6453042 | | | -0.251944 | 1.85E-09 | 1.88E-06 |
| cg15281736_BC21 | chr9 | 17018196 | | | -0.217581 | 1.85E-09 | 1.88E-06 |
| cg13295459_TC21 | chr20 | 45236291 | | | -0.199191 | 1.85E-09 | 1.88E-06 |
| cg18182685_TC21 | chr16 | 88730987 | TSS200 | LOC10028 | -0.237107 | 1.87E-09 | 1.90E-06 |
| cg09308008_BC21 | chr6 | 143747626 | | | 0.295207 | 1.88E-09 | 1.90E-06 |
| cg06310633_TC21 | chr8 | 33812839 | | | -0.687886 | 1.88E-09 | 1.90E-06 |
| cg26645494_BC11 | chr7 | 44113901 | exon_21 | AEBP1 | -0.210391 | 1.88E-09 | 1.90E-06 |
| cg05479250_TC21 | chr1 | 227219445 | | | 0.399872 | 1.88E-09 | 1.90E-06 |
| cg21230309_BC21 | chr3 | 52827558 | | | -0.210667 | 1.88E-09 | 1.90E-06 |
| cg06112967_TC21 | chr2 | 230221010 | TSS1500;TS | SP110;SP1 | 0.220133 | 1.89E-09 | 1.91E-06 |
| cg17465304_TC21 | chr9 | 114099162 | exon_2;TSS | KIF12;KIF1 | -0.397859 | 1.90E-09 | 1.91E-06 |
| cg04868382_BC21 | chr3 | 48660903 | exon_1 | CELSR3 | -0.520686 | 1.91E-09 | 1.92E-06 |
| cg12714054_TC21 | chr8 | 71854435 | | | 0.228274 | 1.91E-09 | 1.92E-06 |
| cg20436376_BC21 | chr1 | 54479763 | | | -0.333481 | 1.91E-09 | 1.92E-06 |
| cg22579828_BC21 | chr20 | 43727016 | TSS200;TSS | GTSF1L;GT | -0.206151 | 1.93E-09 | 1.93E-06 |
| cg23357588_TC21 | chr1 | 168295863 | | | -0.272057 | 1.93E-09 | 1.94E-06 |
| cg16730459_BC21 | chr8 | 126472805 | | | -0.224364 | 1.94E-09 | 1.94E-06 |
| cg19476241_BC21 | chr3 | 12885908 | TSS1500 | LINC02022 | 0.217091 | 1.94E-09 | 1.94E-06 |
| cg10328573_BC21 | chr17 | 41571969 | exon_1 | KRT9 | -0.117705 | 1.94E-09 | 1.94E-06 |
| cg25224813_BC21 | chr4 | 156792451 | | | -0.302793 | 1.95E-09 | 1.94E-06 |
| cg14178428_TC21 | chr1 | 68384108 | | | -0.325683 | 1.95E-09 | 1.94E-06 |
| cg04743916_TC21 | chr12 | 122897443 | TSS1500 | VPS37B | -0.159911 | 1.96E-09 | 1.95E-06 |
| cg10557282_TC21 | chr3 | 181056668 | TSS200;TSS | SOX2-OT;S | -0.434341 | 1.97E-09 | 1.95E-06 |
| cg10437565_TC21 | chr20 | 40650708 | | | -0.18822 | 1.97E-09 | 1.95E-06 |
| cg02619699_TC21 | chr2 | 70686040 | | | -0.183874 | 1.97E-09 | 1.95E-06 |
| cg16038095_TC21 | chr11 | 13313312 | | | 0.133018 | 1.97E-09 | 1.95E-06 |
| cg02722672_BC11 | chr4 | 2965379 | | | -0.148484 | 1.97E-09 | 1.95E-06 |
| cg19557440_BC21 | chr14 | 59263587 | exon_2;exo | DAAM1;DA | -0.272756 | 1.98E-09 | 1.95E-06 |
| cg17205357_BC21 | chr2 | 20677662 | exon_2;3U | GDF7;GDF | -0.192762 | 1.98E-09 | 1.96E-06 |
| cg07020846_TC21 | chr15 | 70098024 | TSS1500;TS | TLE3;TLE3; | 0.375358 | 1.99E-09 | 1.96E-06 |
| cg17308545_BC21 | chr8 | 92138603 | | | -0.198062 | 1.99E-09 | 1.96E-06 |
| cg14533953_TC21 | chr4 | 37873404 | | | 0.405602 | 2.00E-09 | 1.96E-06 |
| cg00468175_BC21 | chr1 | 27357092 | exon_23;e | MAP3K6;M | 0.368176 | 2.00E-09 | 1.96E-06 |
| cg21650557_BC21 | chr3 | 157865165 | | | -0.219346 | 2.00E-09 | 1.96E-06 |
| cg16615829_TC11 | chr19 | 35755494 | 3UTR;exon | HSPB6;HSP | -0.234243 | 2.00E-09 | 1.96E-06 |
| cg24878478_BC21 | chr19 | 11208700 | exon_40;e | DOCK6;DO | -0.246272 | 2.01E-09 | 1.96E-06 |

| | | | | | | | |
|---|---|---|---|---|---|---|---|
| cg14058955_BC21 | chr20 | 24815553 | | | 0.22955 | 2.01E-09 | 1.96E-06 |
| cg05896465_BC21 | chr22 | 45164811 | TSS1500 | NUP50-DT | 0.209239 | 2.01E-09 | 1.97E-06 |
| cg13959721_TC21 | chr1 | 97632989 | | | 0.309925 | 2.03E-09 | 1.98E-06 |
| cg23973304_BC21 | chr19 | 4390307 | | | -0.270664 | 2.04E-09 | 1.99E-06 |
| cg09580974_BC22 | chr1 | 219833616 | | | 0.253083 | 2.04E-09 | 1.99E-06 |
| cg06354558_TC21 | chr8 | 144286315 | | | -0.193088 | 2.05E-09 | 1.99E-06 |
| cg13391389_TC21 | chr11 | 8767790 | TSS1500 | LOC10272 | 0.284291 | 2.06E-09 | 2.00E-06 |
| cg25395069_BC21 | chr20 | 51470300 | | | -0.177616 | 2.06E-09 | 2.00E-06 |
| cg16952407_BC21 | chr1 | 945889 | | | 0.169947 | 2.07E-09 | 2.01E-06 |
| cg07414162_BC21 | chr3 | 124427372 | | | 0.240014 | 2.07E-09 | 2.01E-06 |
| cg17206001_BC21 | chr11 | 118348886 | | | -0.268061 | 2.08E-09 | 2.01E-06 |
| cg24203562_TC21 | chr2 | 238333228 | | | 0.323282 | 2.10E-09 | 2.03E-06 |
| cg09606807_BC21 | chr15 | 96036679 | | | 0.262799 | 2.13E-09 | 2.06E-06 |
| cg02582780_BC21 | chr11 | 109122264 | | | 0.286072 | 2.14E-09 | 2.06E-06 |
| cg02344701_TC21 | chr22 | 39133118 | 3UTR;exon | CBX7;CBX7 | -0.149659 | 2.15E-09 | 2.07E-06 |
| cg12556057_BC21 | chr13 | 23268638 | | | -0.219886 | 2.16E-09 | 2.08E-06 |
| cg16620515_TC21 | chr11 | 73978397 | exon_4;5U | UCP2;UCP2 | -0.102921 | 2.16E-09 | 2.08E-06 |
| cg25279995_BC21 | chr20 | 43014068 | | | 0.2918 | 2.17E-09 | 2.08E-06 |
| cg01710886_TC21 | chr4 | 2817980 | TSS1500;TS | SH3BP2;SH | -0.471669 | 2.18E-09 | 2.09E-06 |
| cg03041617_TC21 | chr7 | 2655776 | | | -0.295195 | 2.18E-09 | 2.09E-06 |
| cg19088141_BC21 | chr1 | 112513529 | | | -0.290683 | 2.19E-09 | 2.09E-06 |
| cg27605438_BC21 | chr6 | 104537416 | | | -0.301492 | 2.19E-09 | 2.09E-06 |
| cg21497549_BC21 | chr9 | 133115910 | | | -0.15345 | 2.20E-09 | 2.10E-06 |
| cg13090456_TC21 | chr18 | 79719525 | | | -0.14753 | 2.20E-09 | 2.10E-06 |
| cg09800781_BC21 | chr10 | 102493258 | | | -0.159976 | 2.20E-09 | 2.10E-06 |
| cg19435731_TC21 | chr11 | 110311832 | | | 0.319348 | 2.24E-09 | 2.13E-06 |
| cg27574066_TC21 | chr17 | 28360359 | TSS1500 | MIR4723 | -0.218363 | 2.24E-09 | 2.13E-06 |
| cg16537371_BC21 | chr11 | 131944541 | | | -0.175052 | 2.26E-09 | 2.14E-06 |
| cg13022326_BC21 | chr19 | 29617061 | exon_7;3U | POP4;POP4 | -0.332107 | 2.26E-09 | 2.14E-06 |
| cg26069973_BC21 | chr7 | 80450897 | | | 0.422 | 2.28E-09 | 2.16E-06 |
| cg23792813_BC21 | chr2 | 28990876 | | | -0.205228 | 2.29E-09 | 2.17E-06 |
| cg21656937_TC21 | chr12 | 53692878 | | | -0.235465 | 2.29E-09 | 2.17E-06 |
| cg03415883_BC21 | chr10 | 100520151 | TSS1500 | SEC31B | -0.200824 | 2.30E-09 | 2.17E-06 |
| cg05014691_BC21 | chr17 | 19716939 | TSS1500;TS | SLC47A2;SL | -0.29978 | 2.30E-09 | 2.17E-06 |
| cg14683191_BC21 | chr3 | 54654546 | | | 0.228226 | 2.31E-09 | 2.17E-06 |
| cg21568131_BC21 | chr17 | 19555232 | exon_7 | SLC47A1 | 0.202037 | 2.31E-09 | 2.18E-06 |
| cg22827729_TC21 | chr6 | 4427270 | | | 0.258364 | 2.33E-09 | 2.19E-06 |
| cg17213087_TC21 | chr15 | 40944997 | | | -0.275978 | 2.34E-09 | 2.19E-06 |
| cg14165142_TC21 | chr20 | 3798008 | | | -0.130166 | 2.34E-09 | 2.19E-06 |
| cg20332679_BC21 | chr8 | 10825678 | TSS1500 | MIR1322 | -0.236258 | 2.34E-09 | 2.19E-06 |
| cg14531576_BC21 | chr1 | 186126625 | | | -0.315876 | 2.34E-09 | 2.19E-06 |
| cg01130153_BC21 | chr18 | 74408450 | TSS1500 | LINC01922 | -0.424157 | 2.34E-09 | 2.19E-06 |
| cg26591221_TC11 | chr16 | 88875927 | 3UTR;exon | CBFA2T3;C | -0.219565 | 2.35E-09 | 2.19E-06 |
| cg11577339_BC21 | chr1 | 7561678 | | | -0.126249 | 2.35E-09 | 2.19E-06 |
| cg00871124_BC21 | chr7 | 6524221 | | | -0.234264 | 2.36E-09 | 2.20E-06 |
| cg13834746_TC21 | chr13 | 86563051 | | | -0.373781 | 2.39E-09 | 2.22E-06 |
| cg05381232_BC21 | chr12 | 11664751 | | | 0.130341 | 2.40E-09 | 2.23E-06 |

| | | | | | | | |
|---|---|---|---|---|---|---|---|
| cg10765346_BC21 | chr7 | 23205676 | | | -0.370151 | 2.40E-09 | 2.23E-06 |
| cg11994854_BC21 | chr2 | 217142845 | | | -0.278357 | 2.40E-09 | 2.23E-06 |
| cg05706231_BC21 | chr2 | 96632631 | | | -0.229644 | 2.42E-09 | 2.24E-06 |
| cg22329579_BC21 | chr17 | 36464506 | | | -0.345872 | 2.43E-09 | 2.25E-06 |
| cg13246103_TC21 | chr2 | 231388908 | | | 0.191822 | 2.43E-09 | 2.25E-06 |
| cg24500049_TC21 | chr19 | 38711204 | | | -0.200888 | 2.45E-09 | 2.27E-06 |
| cg15059239_TC11 | chr5 | 121851876 | TSS200 | FTMT | -0.292328 | 2.46E-09 | 2.27E-06 |
| cg21354353_TC21 | chr6 | 12332026 | | | -0.186149 | 2.46E-09 | 2.27E-06 |
| cg01529671_TC21 | chr9 | 30996789 | | | 0.24393 | 2.46E-09 | 2.27E-06 |
| cg05944800_TC21 | chr2 | 118087970 | TSS1500;TS | INSIG2;INSI | -0.396886 | 2.47E-09 | 2.27E-06 |
| cg17266282_BC11 | chr6 | 111973329 | | | 0.260142 | 2.48E-09 | 2.27E-06 |
| cg13478953_BC21 | chr1 | 156596639 | | | -0.120191 | 2.48E-09 | 2.27E-06 |
| cg26157451_TC21 | chr15 | 89003871 | | | -0.261101 | 2.49E-09 | 2.28E-06 |
| cg22963915_BC11 | chr19 | 3785857 | | | -0.37396 | 2.49E-09 | 2.28E-06 |
| cg19254434_TC21 | chr1 | 60733025 | | | -0.248468 | 2.50E-09 | 2.29E-06 |
| cg14626898_TC21 | chr8 | 24932727 | | | -0.190213 | 2.50E-09 | 2.29E-06 |
| cg19207803_BC21 | chr4 | 1806092 | exon_14;e | FGFR3;FGF | -0.188796 | 2.51E-09 | 2.29E-06 |
| cg17677381_TC21 | chr7 | 85123069 | | | -0.347868 | 2.52E-09 | 2.29E-06 |
| cg03680932_BC11 | chr6 | 16306452 | exon_9;exo | ATXN1;ATX | -0.281562 | 2.52E-09 | 2.30E-06 |
| cg02224142_BC21 | chr8 | 141338762 | | | 0.283035 | 2.54E-09 | 2.31E-06 |
| cg21786034_TC21 | chr22 | 21983981 | TSS1500;TS | TOP3B;TOP | -0.161714 | 2.54E-09 | 2.31E-06 |
| cg17129435_BC21 | chr1 | 155169888 | | | -0.201125 | 2.54E-09 | 2.31E-06 |
| cg23199006_BC21 | chr12 | 3989261 | | | -0.141762 | 2.54E-09 | 2.31E-06 |
| cg22518656_BC21 | chr17 | 40172249 | | | 0.21415 | 2.54E-09 | 2.31E-06 |
| cg21884335_TC21 | chr16 | 86044305 | | | -0.224529 | 2.55E-09 | 2.31E-06 |
| cg12251898_BC21 | chr8 | 18804875 | exon_4;exo | PSD3;PSD3 | -0.24543 | 2.56E-09 | 2.32E-06 |
| cg16824480_TC21 | chr17 | 76193850 | | | -0.148759 | 2.57E-09 | 2.32E-06 |
| cg12227105_TC21 | chr1 | 150895490 | | | 0.352295 | 2.59E-09 | 2.33E-06 |
| cg03958112_TC21 | chr10 | 43376142 | | | -0.209959 | 2.60E-09 | 2.35E-06 |
| cg24624629_TC11 | chr17 | 41047369 | TSS200 | KRTAP2-1 | -0.272159 | 2.61E-09 | 2.35E-06 |
| cg01929087_BC21 | chr20 | 35604579 | exon_5 | FER1L4 | -0.149535 | 2.61E-09 | 2.35E-06 |
| cg18859219_BC21 | chr20 | 56616710 | | | -0.254243 | 2.62E-09 | 2.36E-06 |
| cg19722516_BC21 | chr14 | 76264802 | | | -0.218596 | 2.64E-09 | 2.37E-06 |
| cg22893593_BC21 | chr12 | 122197545 | | | 0.151367 | 2.64E-09 | 2.37E-06 |
| cg14947466_BC21 | chr11 | 2562475 | | | -0.220133 | 2.65E-09 | 2.37E-06 |
| cg02830023_BC21 | chr6 | 15575542 | | | -0.183271 | 2.65E-09 | 2.37E-06 |
| cg00645217_TC21 | chr1 | 40784966 | | | -0.127938 | 2.66E-09 | 2.37E-06 |
| cg26882454_BC21 | chr16 | 3364715 | | | -0.372987 | 2.66E-09 | 2.37E-06 |
| cg15901029_TC21 | chr2 | 224242664 | | | -0.29982 | 2.67E-09 | 2.38E-06 |
| cg20285761_TC21 | chr11 | 60783964 | TSS1500 | MS4A10 | -0.30882 | 2.67E-09 | 2.38E-06 |
| cg00938819_TC21 | chr9 | 124045232 | | | -0.143092 | 2.67E-09 | 2.38E-06 |
| cg11333117_BC21 | chr1 | 212284398 | TSS1500 | PPP2R5A | -0.226522 | 2.68E-09 | 2.38E-06 |
| cg21558323_BC11 | chr11 | 66267958 | TSS1500 | RAB1B | 0.328696 | 2.68E-09 | 2.38E-06 |
| cg12601856_TC21 | chr6 | 158313053 | 5UTR;exon | TULP4;TUL | -0.252074 | 2.68E-09 | 2.38E-06 |
| cg24279623_TC21 | chr8 | 6407469 | TSS1500 | MCPH1-DT | -0.20498 | 2.68E-09 | 2.38E-06 |
| cg26750002_TC21 | chr19 | 48741417 | exon_8;exo | IZUMO1;IZ | -0.273341 | 2.71E-09 | 2.40E-06 |
| cg15752247_BC21 | chr1 | 85307876 | | | 0.214243 | 2.72E-09 | 2.41E-06 |

| | | | | | | | |
|---|---|---|---|---|---|---|---|
| cg15627159_BC21 | chr11 | 117262705 | | | -0.360367 | 2.72E-09 | 2.41E-06 |
| cg14798397_TC21 | chr10 | 23968615 | | | 0.493503 | 2.72E-09 | 2.41E-06 |
| cg03032512_TC21 | chr17 | 79034336 | 5UTR;exon | C1QTNF1;C | -0.147332 | 2.73E-09 | 2.41E-06 |
| cg11000227_BC21 | chr7 | 48839298 | | | 0.18895 | 2.73E-09 | 2.41E-06 |
| cg13534497_TC21 | chr1 | 109904630 | | | -0.188052 | 2.74E-09 | 2.41E-06 |
| cg26228696_BC11 | chr1 | 150876713 | TSS200;TSS | ARNT;ARNT | 0.23842 | 2.75E-09 | 2.42E-06 |
| cg16726195_BC21 | chr6 | 37537248 | | | -0.27662 | 2.76E-09 | 2.43E-06 |
| cg10312841_TC21 | chr16 | 85179221 | | | 0.203679 | 2.77E-09 | 2.43E-06 |
| cg11697861_BC22 | chr19 | 15422798 | 3UTR;exon | WIZ;WIZ;W | -0.164441 | 2.78E-09 | 2.44E-06 |
| cg17469604_BC21 | chr2 | 128426813 | | | 0.414212 | 2.78E-09 | 2.44E-06 |
| cg02628202_TC21 | chr14 | 92748862 | TSS1500;TS | LGMN;LGM | -0.28606 | 2.79E-09 | 2.44E-06 |
| cg24610231_TC21 | chr15 | 29675416 | exon_1;TSS | LOC10013( | -0.35022 | 2.79E-09 | 2.44E-06 |
| cg23647640_TC21 | chr8 | 141433918 | exon_28;e› | MROH5;MI | -0.21786 | 2.80E-09 | 2.44E-06 |
| cg25875110_BC21 | chr10 | 59719478 | | | -0.247089 | 2.81E-09 | 2.45E-06 |
| cg07454951_TC21 | chr2 | 147323402 | | | -0.32683 | 2.81E-09 | 2.45E-06 |
| cg05490540_BC21 | chr3 | 128434166 | | | 0.191045 | 2.82E-09 | 2.46E-06 |
| cg08719267_BC21 | chr4 | 127641828 | | | -0.280155 | 2.83E-09 | 2.46E-06 |
| cg02800817_TC21 | chr6 | 30185557 | 3UTR;exon | TRIM26;TR | 0.364403 | 2.83E-09 | 2.46E-06 |
| cg14313414_TC21 | chr10 | 76977169 | | | -0.201037 | 2.84E-09 | 2.46E-06 |
| cg16156543_TC21 | chr6 | 169174019 | | | -0.221676 | 2.84E-09 | 2.46E-06 |
| cg05557384_TC21 | chr3 | 135514303 | | | -0.450763 | 2.85E-09 | 2.47E-06 |
| cg16316382_TC21 | chr10 | 93612312 | TSS1500 | PDE6C | -0.324237 | 2.85E-09 | 2.47E-06 |
| cg25363306_TC21 | chr20 | 49385774 | | | -0.456264 | 2.86E-09 | 2.48E-06 |
| cg17211793_BC21 | chr16 | 19065915 | | | 0.349835 | 2.86E-09 | 2.48E-06 |
| cg00667680_BC21 | chr8 | 1965103 | | | 0.126904 | 2.87E-09 | 2.48E-06 |
| cg24178000_BC21 | chr14 | 104723486 | TSS1500 | ADSS1 | -0.13396 | 2.88E-09 | 2.48E-06 |
| cg19313574_BC21 | chr1 | 165540012 | | | -0.24197 | 2.88E-09 | 2.48E-06 |
| cg03104501_TC11 | chr2 | 219002099 | TSS200;TSS | LOC10012S | 0.327455 | 2.88E-09 | 2.48E-06 |
| cg14983109_TC21 | chr10 | 43586023 | | | 0.448024 | 2.92E-09 | 2.50E-06 |
| cg07834691_TC21 | chr3 | 156865776 | | | -0.391192 | 2.92E-09 | 2.50E-06 |
| cg22934764_TC21 | chr17 | 74669443 | | | -0.18249 | 2.94E-09 | 2.52E-06 |
| cg24857016_BC21 | chr19 | 58397083 | | | 0.326729 | 2.94E-09 | 2.52E-06 |
| cg04213847_BC21 | chr6 | 136307813 | | | -0.311792 | 2.94E-09 | 2.52E-06 |
| cg09475796_TC21 | chr6 | 57206326 | | | 0.266677 | 2.94E-09 | 2.52E-06 |
| cg15680910_TC21 | chr10 | 121734810 | | | -0.156867 | 2.95E-09 | 2.52E-06 |
| cg04322572_TC11 | chr10 | 42952170 | | | -0.269478 | 2.99E-09 | 2.55E-06 |
| cg02725313_TC21 | chr20 | 31484110 | | | -0.189156 | 3.00E-09 | 2.56E-06 |
| cg17460204_BC21 | chr17 | 83087987 | | | -0.199608 | 3.02E-09 | 2.57E-06 |
| cg18892020_TC21 | chr19 | 10361775 | exon_13;e› | TYK2;TYK2; | -0.252045 | 3.02E-09 | 2.57E-06 |
| cg10336039_BC11 | chr17 | 78838907 | | | 0.222137 | 3.03E-09 | 2.58E-06 |
| cg00404788_TC21 | chr9 | 69517257 | exon_2;5U | APBA1;APE | 0.315166 | 3.04E-09 | 2.59E-06 |
| cg16329304_BC21 | chr17 | 4600326 | | | -0.155103 | 3.07E-09 | 2.61E-06 |
| cg06163186_BC21 | chr19 | 3610338 | | | -0.19062 | 3.07E-09 | 2.61E-06 |
| cg24071177_TC22 | chr20 | 36366947 | | | -0.184934 | 3.08E-09 | 2.61E-06 |
| cg12218126_TC11 | chr19 | 42408334 | exon_2 | LOC10193( | 0.245844 | 3.09E-09 | 2.62E-06 |
| cg19353410_BC21 | chr12 | 119365691 | | | 0.232913 | 3.10E-09 | 2.62E-06 |
| cg10403145_TC21 | chr13 | 99200596 | exon_1;TSS | UBAC2-AS1 | 0.357121 | 3.10E-09 | 2.62E-06 |

| | | | | | | | |
|---|---|---|---|---|---|---|---|
| cg05809029_BC21 | chr5 | 506505 | | | -0.410368 | 3.10E-09 | 2.62E-06 |
| cg19051042_TC21 | chr17 | 41524067 | | | -0.19625 | 3.11E-09 | 2.62E-06 |
| cg02663382_BC21 | chr19 | 37817818 | | | -0.410473 | 3.15E-09 | 2.65E-06 |
| cg16900185_BC21 | chr6 | 118810725 | | | 0.479856 | 3.15E-09 | 2.65E-06 |
| cg13722294_TC21 | chr9 | 68810341 | | | 0.314964 | 3.15E-09 | 2.65E-06 |
| cg00694777_BC21 | chr7 | 967204 | 3UTR;exon_ | COX19;CO) | -0.260418 | 3.16E-09 | 2.66E-06 |
| cg08403496_TC21 | chr5 | 114669544 | | | -0.37851 | 3.18E-09 | 2.66E-06 |
| cg15145672_BC21 | chr3 | 15333027 | TSS1500 | SH3BP5 | -0.20555 | 3.18E-09 | 2.66E-06 |
| cg00531320_BC21 | chr1 | 31881027 | | | 0.231322 | 3.19E-09 | 2.67E-06 |
| cg00114452_TC21 | chr15 | 31102780 | TSS1500;TS | TRPM1;TRI | -0.203917 | 3.19E-09 | 2.67E-06 |
| cg14657377_BC21 | chr10 | 11036729 | | | -0.466273 | 3.21E-09 | 2.68E-06 |
| cg16034426_BC21 | chr2 | 44291726 | | | -0.148763 | 3.22E-09 | 2.69E-06 |
| cg03655692_BC21 | chr8 | 117624257 | | | -0.159305 | 3.23E-09 | 2.70E-06 |
| cg09573585_BC21 | chr10 | 86968371 | TSS200 | ADIRF | -0.277995 | 3.24E-09 | 2.70E-06 |
| cg22124648_TC21 | chr1 | 27615355 | | | -0.201741 | 3.26E-09 | 2.72E-06 |
| cg05099035_TC21 | chr17 | 62737672 | exon_3 | LOC105371 | -0.126787 | 3.26E-09 | 2.72E-06 |
| cg01317870_TC21 | chr5 | 69518631 | | | -0.305326 | 3.27E-09 | 2.72E-06 |
| cg03662398_BC21 | chr17 | 55335467 | | | 0.175959 | 3.28E-09 | 2.72E-06 |
| cg01687663_BC21 | chr2 | 218858338 | TSS1500 | WNT6 | 0.327876 | 3.29E-09 | 2.73E-06 |
| cg12824973_BC21 | chr13 | 60148950 | | | -0.608774 | 3.30E-09 | 2.73E-06 |
| cg27141509_TC21 | chr10 | 5844148 | | | -0.185355 | 3.30E-09 | 2.73E-06 |
| cg18232089_BC21 | chr5 | 131271021 | | | 0.234771 | 3.32E-09 | 2.74E-06 |
| cg01840020_BC21 | chr17 | 4745654 | | | -0.19873 | 3.32E-09 | 2.75E-06 |
| cg14392588_TC21 | chr9 | 133346858 | TSS1500 | RPL7A | -0.1433 | 3.34E-09 | 2.75E-06 |
| cg00088620_BC21 | chr16 | 73191099 | | | -0.198111 | 3.34E-09 | 2.76E-06 |
| cg08851848_TC21 | chr3 | 66336529 | | | -0.161019 | 3.36E-09 | 2.77E-06 |
| cg01665908_BC21 | chr1 | 180187002 | | | -0.501355 | 3.38E-09 | 2.79E-06 |
| cg14227785_BC21 | chr8 | 141771502 | | | 0.431525 | 3.39E-09 | 2.79E-06 |
| cg23902177_BC21 | chr7 | 87416194 | | | 0.296859 | 3.40E-09 | 2.80E-06 |
| cg11794617_BC21 | chr7 | 140026284 | exon_11;e) | PARP12;PA | -0.172543 | 3.41E-09 | 2.80E-06 |
| cg26917745_BC21 | chr7 | 30900108 | | | -0.161066 | 3.41E-09 | 2.80E-06 |
| cg15196611_TC21 | chr4 | 40613138 | | | 0.364286 | 3.42E-09 | 2.80E-06 |
| cg00111006_BC21 | chr1 | 232092260 | | | -0.268676 | 3.43E-09 | 2.81E-06 |
| cg03564270_BC21 | chr6 | 81737749 | | | -0.330225 | 3.43E-09 | 2.81E-06 |
| cg13353015_TC11 | chr1 | 34081653 | | | -0.123015 | 3.43E-09 | 2.81E-06 |
| cg05359525_TC21 | chr1 | 75357257 | | | -0.250859 | 3.45E-09 | 2.81E-06 |
| cg09544728_TC11 | chr7 | 561866 | TSS200 | PRKAR1B-A | -0.226482 | 3.45E-09 | 2.81E-06 |
| cg21664764_TC21 | chr8 | 59307658 | | | 0.206895 | 3.48E-09 | 2.84E-06 |
| cg21129604_BC21 | chr14 | 95212345 | | | -0.153939 | 3.49E-09 | 2.84E-06 |
| cg10512089_BC21 | chr11 | 57641278 | exon_1 | MIR130A | 0.150522 | 3.49E-09 | 2.84E-06 |
| cg21950166_BC21 | chr1 | 26862858 | TSS1500 | SFN | -0.141808 | 3.50E-09 | 2.85E-06 |
| cg02019549_BC21 | chr6 | 119280960 | | | 0.216782 | 3.53E-09 | 2.86E-06 |
| cg04940310_BC21 | chr2 | 47748023 | | | 0.271655 | 3.53E-09 | 2.86E-06 |
| cg05956871_TC21 | chr19 | 35084558 | | | -0.216007 | 3.54E-09 | 2.87E-06 |
| cg19154320_BC21 | chr1 | 180244134 | | | -0.212031 | 3.54E-09 | 2.87E-06 |
| cg26395239_BC21 | chr22 | 42293165 | | | 0.288085 | 3.55E-09 | 2.87E-06 |
| cg18718397_TC21 | chr21 | 37455560 | | | -0.243794 | 3.55E-09 | 2.87E-06 |

| | | | | | | | |
|---|---|---|---|---|---|---|---|
| cg12163365_BC21 | chr8 | 139453339 | | | 0.266953 | 3.57E-09 | 2.88E-06 |
| cg08406615_BC21 | chr14 | 23452977 | | | -0.440927 | 3.59E-09 | 2.90E-06 |
| cg04912778_BC21 | chr10 | 77197190 | | | -0.261094 | 3.59E-09 | 2.90E-06 |
| cg13571802_TC21 | chr3 | 44874981 | | | -0.313116 | 3.60E-09 | 2.90E-06 |
| cg26257427_TC21 | chr22 | 33726490 | | | -0.195591 | 3.63E-09 | 2.92E-06 |
| cg20022216_BC11 | chr4 | 39698689 | | | 0.288814 | 3.65E-09 | 2.93E-06 |
| cg03154914_BC21 | chr9 | 131490178 | TSS1500 | SNORD62B | -0.191373 | 3.67E-09 | 2.94E-06 |
| cg01768056_BC21 | chr19 | 19366861 | | | -0.190863 | 3.67E-09 | 2.94E-06 |
| cg06394961_BC21 | chr10 | 23918579 | | | -0.381954 | 3.67E-09 | 2.94E-06 |
| cg20969377_BC21 | chr16 | 1431602 | | | 0.211956 | 3.69E-09 | 2.95E-06 |
| cg08138586_TC21 | chr2 | 204547063 | | | -0.341535 | 3.69E-09 | 2.96E-06 |
| cg03558688_TC21 | chr10 | 104300167 | exon_6;3U | GSTO2;GST | -0.234494 | 3.70E-09 | 2.96E-06 |
| cg18699466_BC21 | chr6 | 45447903 | | | 0.209138 | 3.71E-09 | 2.96E-06 |
| cg15724252_TC21 | chr8 | 96261108 | TSS1500;TS | PTDSS1;PTI | -0.341881 | 3.71E-09 | 2.96E-06 |
| cg14884081_BC21 | chr10 | 74224235 | | | -0.291992 | 3.72E-09 | 2.96E-06 |
| cg11415496_BC21 | chr4 | 76434901 | TSS1500 | SHROOM3 | -0.342531 | 3.72E-09 | 2.96E-06 |
| cg15921713_TC21 | chr7 | 16563348 | | | 0.542118 | 3.72E-09 | 2.96E-06 |
| cg08639799_BC21 | chr1 | 1939723 | exon_24 | CFAP74 | -0.200002 | 3.74E-09 | 2.97E-06 |
| cg03654618_BC21 | chr4 | 3206627 | exon_43;e | HTT;HTT | 0.387579 | 3.76E-09 | 2.98E-06 |
| cg10411143_TC21 | chr21 | 25177145 | | | -0.292755 | 3.77E-09 | 2.99E-06 |
| cg24853741_BC21 | chr20 | 17620198 | | | -0.29749 | 3.77E-09 | 2.99E-06 |
| cg17495466_TC21 | chr1 | 165543743 | TSS1500 | LRRC52 | -0.198556 | 3.78E-09 | 2.99E-06 |
| cg26568378_BC21 | chr14 | 103685378 | | | -0.333191 | 3.78E-09 | 2.99E-06 |
| cg07758667_BC21 | chr8 | 47439462 | | | 0.308247 | 3.81E-09 | 3.01E-06 |
| cg07287131_TC21 | chr12 | 90408039 | | | -0.280687 | 3.82E-09 | 3.01E-06 |
| cg20678800_BC21 | chr1 | 115314810 | | | -0.156237 | 3.82E-09 | 3.01E-06 |
| cg18088111_TC21 | chr14 | 24843507 | | | 0.250662 | 3.83E-09 | 3.02E-06 |
| cg16860070_TC21 | chr8 | 88542409 | | | -0.165177 | 3.83E-09 | 3.02E-06 |
| cg18824556_TC21 | chr12 | 21606155 | TSS1500 | GYS2 | -0.418961 | 3.84E-09 | 3.02E-06 |
| cg20201475_TC11 | chr3 | 169149519 | | | 0.338379 | 3.85E-09 | 3.03E-06 |
| cg15156391_BC21 | chr1 | 9959054 | | | 0.18683 | 3.85E-09 | 3.03E-06 |
| cg09604586_BC11 | chr19 | 3006683 | | | -0.364248 | 3.87E-09 | 3.04E-06 |
| cg25589092_BC21 | chr1 | 94324861 | | | -0.266826 | 3.87E-09 | 3.04E-06 |
| cg21460729_BC21 | chr7 | 149734176 | 3UTR;3UTF | KRBA1;KRB | -0.13178 | 3.88E-09 | 3.04E-06 |
| cg15690598_TC21 | chr8 | 133794372 | | | -0.169812 | 3.89E-09 | 3.05E-06 |
| cg08053057_BC21 | chr2 | 65231995 | | | 0.256328 | 3.91E-09 | 3.06E-06 |
| cg11085097_BC21 | chr19 | 9139970 | TSS1500;TS | ZNF317;ZN | -0.415617 | 3.93E-09 | 3.07E-06 |
| cg08822227_TC21 | chr4 | 2818741 | TSS200 | SH3BP2 | -0.65606 | 3.95E-09 | 3.09E-06 |
| cg16339560_TC21 | chr7 | 30994399 | | | -0.249814 | 3.97E-09 | 3.09E-06 |
| cg17996892_TC21 | chr17 | 28505566 | TSS1500 | FOXN1 | -0.152942 | 3.98E-09 | 3.10E-06 |
| cg11383231_BC21 | chr7 | 92631207 | | | -0.152459 | 3.98E-09 | 3.10E-06 |
| cg21523688_BC21 | chr15 | 45026839 | | | -0.389688 | 3.99E-09 | 3.10E-06 |
| cg23850272_TC21 | chr19 | 44357089 | TSS1500;TS | ZNF112;ZN | -0.441475 | 3.99E-09 | 3.10E-06 |
| cg25107893_TC11 | chr1 | 976221 | exon_4;exo | PERM1;PEF | 0.316831 | 4.00E-09 | 3.10E-06 |
| cg12729800_TC21 | chr17 | 27879818 | | | -0.19518 | 4.00E-09 | 3.10E-06 |
| cg04716048_TC21 | chr6 | 3732563 | | | -0.274846 | 4.01E-09 | 3.11E-06 |
| cg24691324_BC21 | chr4 | 113781941 | | | -0.383891 | 4.01E-09 | 3.11E-06 |

| | | | | | | | |
|---|---|---|---|---|---|---|---|
| cg01661846_TC21 | chr1 | 179806781 | | | 0.263084 | 4.03E-09 | 3.12E-06 |
| cg07636688_BC11 | chr22 | 45413521 | exon_1;exo | SMC1B;SM | -0.388931 | 4.09E-09 | 3.16E-06 |
| cg20704654_TC11 | chr20 | 31484315 | exon_5;TSS | REM1;LINC | -0.325543 | 4.09E-09 | 3.16E-06 |
| cg02340541_TC21 | chr11 | 68684231 | TSS1500 | GAL | -0.29344 | 4.09E-09 | 3.16E-06 |
| cg08070975_BC21 | chr5 | 66239170 | | | -0.147813 | 4.11E-09 | 3.16E-06 |
| cg10127249_TC21 | chr19 | 43607975 | exon_3 | ZNF428 | 0.206497 | 4.11E-09 | 3.16E-06 |
| cg04451579_BC21 | chr7 | 32020094 | | | -0.31456 | 4.11E-09 | 3.16E-06 |
| cg06779393_BC21 | chr1 | 159714547 | exon_1;5U | CRP;CRP;CI | -0.222474 | 4.13E-09 | 3.17E-06 |
| cg02302816_BC21 | chr2 | 31894119 | | | 0.316966 | 4.13E-09 | 3.17E-06 |
| cg21874525_BC21 | chr16 | 85594811 | | | -0.248016 | 4.15E-09 | 3.18E-06 |
| cg23647325_TC21 | chr4 | 8014342 | | | 0.304665 | 4.16E-09 | 3.19E-06 |
| cg12670990_BC21 | chr17 | 44905899 | 3UTR;exon | GFAP;GFAF | -0.195628 | 4.18E-09 | 3.20E-06 |
| cg23047434_TC21 | chr12 | 54016707 | TSS200;TSS | HOXC6;HO | -0.299755 | 4.18E-09 | 3.20E-06 |
| cg11265216_TC21 | chr1 | 221458614 | | | 0.265172 | 4.19E-09 | 3.20E-06 |
| cg01586506_BC11 | chr22 | 37983499 | exon_2 | SOX10 | -0.296013 | 4.19E-09 | 3.20E-06 |
| cg01523712_TC21 | chr20 | 44809217 | | | 0.286438 | 4.21E-09 | 3.21E-06 |
| cg24661236_TC11 | chr6 | 36962286 | | | -0.380067 | 4.22E-09 | 3.22E-06 |
| cg06360940_BC21 | chr3 | 171692394 | exon_13;ex | PLD1;PLD1 | -0.387244 | 4.23E-09 | 3.22E-06 |
| cg13352599_BC21 | chr9 | 3019704 | | | -0.366394 | 4.24E-09 | 3.23E-06 |
| cg19499720_BC21 | chr12 | 112524940 | | | -0.1604 | 4.25E-09 | 3.24E-06 |
| cg23450578_TC21 | chr4 | 1949151 | | | 0.211477 | 4.31E-09 | 3.27E-06 |
| cg02524833_BC21 | chr20 | 62379431 | | | 0.322038 | 4.31E-09 | 3.27E-06 |
| cg08871137_BC21 | chr5 | 50970560 | TSS1500 | LINC02106 | -0.337421 | 4.31E-09 | 3.27E-06 |
| cg13153721_BC21 | chr10 | 122560511 | TSS1500;TS | DMBT1;DM | -0.250534 | 4.31E-09 | 3.27E-06 |
| cg24591413_BC21 | chr19 | 44746906 | | | -0.121038 | 4.33E-09 | 3.28E-06 |
| cg23045001_TC21 | chr17 | 79841649 | | | -0.174704 | 4.34E-09 | 3.29E-06 |
| cg05457684_TC21 | chr11 | 2898578 | TSS1500 | SLC22A18 | 0.180184 | 4.35E-09 | 3.29E-06 |
| cg02843293_BC21 | chr11 | 61273941 | | | -0.143488 | 4.38E-09 | 3.30E-06 |
| cg17622922_TC11 | chr11 | 119317139 | TSS200 | MCAM | 0.101976 | 4.38E-09 | 3.30E-06 |
| cg14041778_TC21 | chr1 | 33472609 | TSS200;TSS | ZSCAN20;Z | -0.352856 | 4.39E-09 | 3.30E-06 |
| cg08736982_BC21 | chr7 | 138917279 | exon_2;exo | KIAA1549;I | -0.219204 | 4.39E-09 | 3.30E-06 |
| cg20446641_TC21 | chr11 | 66475606 | exon_7;exo | PELI3;PELI3 | -0.168562 | 4.39E-09 | 3.30E-06 |
| cg02177087_BC21 | chr16 | 19698667 | | | -0.210116 | 4.39E-09 | 3.30E-06 |
| cg18473733_BC21 | chr19 | 16326551 | | | -0.126401 | 4.39E-09 | 3.30E-06 |
| cg15433443_BC21 | chr8 | 80197655 | | | -0.245005 | 4.40E-09 | 3.31E-06 |
| cg27487996_BC21 | chr11 | 80502826 | | | -0.253611 | 4.41E-09 | 3.31E-06 |
| cg14465410_BC21 | chr3 | 196315984 | | | 0.273471 | 4.41E-09 | 3.31E-06 |
| cg04770591_TC21 | chr3 | 39106427 | TSS1500;TS | TTC21A;TT | -0.113527 | 4.42E-09 | 3.31E-06 |
| cg17009717_TC21 | chr2 | 112542131 | 5UTR;exon | POLR1B;PC | -0.280059 | 4.43E-09 | 3.31E-06 |
| cg26474827_BC21 | chr5 | 14945169 | | | -0.181608 | 4.43E-09 | 3.31E-06 |
| cg11648289_BC21 | chr7 | 155456979 | TSS1500 | EN2 | -0.232185 | 4.44E-09 | 3.31E-06 |
| cg26675664_TC11 | chr4 | 1415148 | | | 0.328655 | 4.46E-09 | 3.32E-06 |
| cg00157043_BC21 | chr11 | 35943357 | TSS1500;TS | LDLRAD3;L | 0.243882 | 4.46E-09 | 3.32E-06 |
| cg19719703_TC21 | chr13 | 113220844 | | | 0.247008 | 4.46E-09 | 3.32E-06 |
| cg15196156_TC21 | chr1 | 39922723 | | | -0.305185 | 4.46E-09 | 3.32E-06 |
| cg19849738_BC21 | chr14 | 91365080 | | | -0.22776 | 4.48E-09 | 3.34E-06 |
| cg22187733_BC21 | chr19 | 49961524 | TSS1500;TS | SIGLEC11;S | -0.269906 | 4.51E-09 | 3.35E-06 |

| | | | | | | | |
|---|---|---|---|---|---|---|---|
| cg19936757_TC21 | chr11 | 2430988 | | | -0.113898 | 4.51E-09 | 3.35E-06 |
| cg05437857_TC21 | chr6 | 148935799 | | | -0.227616 | 4.52E-09 | 3.35E-06 |
| cg13327949_BC21 | chr4 | 179465983 | | | -0.26329 | 4.52E-09 | 3.35E-06 |
| cg10623221_TC21 | chr3 | 173401422 | | | -0.152732 | 4.52E-09 | 3.35E-06 |
| cg16405983_TC21 | chr17 | 8673829 | | | -0.246113 | 4.52E-09 | 3.35E-06 |
| cg09355086_TC21 | chr6 | 30703531 | exon_11 | MDC1 | -0.141171 | 4.53E-09 | 3.35E-06 |
| cg24551923_TC21 | chr19 | 41753185 | | | -0.156524 | 4.54E-09 | 3.35E-06 |
| cg04363755_BC21 | chr1 | 20107530 | | | -0.201345 | 4.54E-09 | 3.35E-06 |
| cg12522173_BC21 | chr22 | 42379293 | | | 0.315889 | 4.57E-09 | 3.37E-06 |
| cg25362014_TC21 | chr20 | 49290455 | | | -0.200766 | 4.59E-09 | 3.38E-06 |
| cg13150989_TC21 | chr8 | 133236407 | | | 0.321314 | 4.61E-09 | 3.39E-06 |
| cg26624067_TC21 | chr5 | 71719386 | exon_1 | CARTPT | 0.356789 | 4.63E-09 | 3.41E-06 |
| cg12428229_BC21 | chr10 | 131734238 | | | -0.258775 | 4.63E-09 | 3.41E-06 |
| cg26391564_TC21 | chr7 | 122260854 | | | 0.289602 | 4.67E-09 | 3.43E-06 |
| cg12578844_TC11 | chr1 | 23019719 | exon_1;exc | KDM1A;KD | -0.26381 | 4.70E-09 | 3.44E-06 |
| cg22019970_TC21 | chr1 | 25998707 | TSS1500 | PAFAH2 | -0.21582 | 4.71E-09 | 3.44E-06 |
| cg05506600_TC21 | chr15 | 72155626 | | | -0.244227 | 4.71E-09 | 3.44E-06 |
| cg08950930_BC21 | chr16 | 1148408 | | | -0.258144 | 4.71E-09 | 3.44E-06 |
| cg13838832_TC21 | chr9 | 132758240 | | | -0.386779 | 4.71E-09 | 3.44E-06 |
| cg13609889_BC21 | chr3 | 178337838 | | | -0.435579 | 4.71E-09 | 3.44E-06 |
| cg20878902_TC21 | chr15 | 98899545 | exon_5;exc | IGF1R;IGF1 | -0.135769 | 4.74E-09 | 3.46E-06 |
| cg21915639_BC11 | chr1 | 46303636 | TSS200;TSS | UQCRH;UQ | -0.441864 | 4.74E-09 | 3.46E-06 |
| cg15476229_BC21 | chr15 | 38695109 | TSS1500 | LINC02694 | -0.339038 | 4.75E-09 | 3.46E-06 |
| cg01702225_TC21 | chr7 | 55016731 | | | -0.225555 | 4.75E-09 | 3.46E-06 |
| cg06321056_BC21 | chr2 | 68837340 | | | -0.591459 | 4.76E-09 | 3.46E-06 |
| cg02878043_BC21 | chr14 | 34035844 | | | -0.203455 | 4.77E-09 | 3.47E-06 |
| cg06581698_BC21 | chr4 | 54705653 | | | -0.135051 | 4.78E-09 | 3.47E-06 |
| cg02167021_TC21 | chr3 | 121660944 | TSS200;TSS | HCLS1;HCL | -0.467314 | 4.79E-09 | 3.48E-06 |
| cg12457188_BC21 | chr8 | 40054934 | | | -0.36395 | 4.80E-09 | 3.48E-06 |
| cg24570094_TC21 | chr16 | 30030122 | TSS200 | TLCD3B | 0.267517 | 4.80E-09 | 3.48E-06 |
| cg19720714_BC21 | chr17 | 3871000 | | | -0.206106 | 4.84E-09 | 3.50E-06 |
| cg20089491_TC21 | chr12 | 91863701 | | | 0.496198 | 4.85E-09 | 3.51E-06 |
| cg11524692_BC21 | chr3 | 193970069 | | | -0.238828 | 4.86E-09 | 3.51E-06 |
| cg02775263_BC21 | chr18 | 36644101 | | | -0.250945 | 4.87E-09 | 3.52E-06 |
| cg25727569_BC21 | chr3 | 53811260 | exon_49;e› | CACNA1D;( | -0.178368 | 4.87E-09 | 3.52E-06 |
| cg15700636_BC21 | chr11 | 57388577 | exon_4;exc | PRG2;PRG2 | -0.148717 | 4.88E-09 | 3.52E-06 |
| cg09572697_BC21 | chr6 | 168049404 | | | -0.149016 | 4.89E-09 | 3.52E-06 |
| cg04090021_TC21 | chr1 | 11398572 | | | 0.196506 | 4.90E-09 | 3.53E-06 |
| cg02700954_TC21 | chr2 | 39095075 | | | 0.475634 | 4.91E-09 | 3.53E-06 |
| cg21194066_TC21 | chr11 | 67285038 | | | -0.13548 | 4.92E-09 | 3.54E-06 |
| cg13971182_TC21 | chr9 | 95510311 | TSS1500;TS | PTCH1;PTC | -0.339294 | 4.93E-09 | 3.54E-06 |
| cg09082664_BC11 | chr4 | 23889078 | | | -0.363635 | 4.93E-09 | 3.54E-06 |
| cg14889801_BC21 | chr21 | 46653827 | exon_7;3U | PRMT2;PRI | -0.35272 | 4.96E-09 | 3.55E-06 |
| cg21412374_BC21 | chr8 | 143669268 | | | -0.157687 | 4.96E-09 | 3.55E-06 |
| cg13790051_TC21 | chr4 | 119683822 | | | -0.191788 | 4.98E-09 | 3.56E-06 |
| cg04936620_BC21 | chr19 | 18017957 | | | 0.264347 | 4.98E-09 | 3.56E-06 |
| cg17566926_BC21 | chr12 | 48335028 | 3UTR;exon | ZNF641;ZN | -0.270368 | 4.99E-09 | 3.56E-06 |

| | | | | | | | |
|---|---|---|---|---|---|---|---|
| cg13037895_TC21 | chr7 | 2222657 | exon_5;exc | MAD1L1;M | -0.151535 | 5.00E-09 | 3.57E-06 |
| cg05112200_BC21 | chr13 | 75374175 | | | -0.331426 | 5.04E-09 | 3.59E-06 |
| cg04309287_BC21 | chr2 | 234290764 | | | 0.269055 | 5.05E-09 | 3.60E-06 |
| cg19870319_BC21 | chr14 | 93009726 | | | -0.460287 | 5.05E-09 | 3.60E-06 |
| cg13448406_TC21 | chr9 | 33000170 | | | -0.372513 | 5.10E-09 | 3.62E-06 |
| cg12540553_TC21 | chr7 | 55475506 | | | -0.136662 | 5.10E-09 | 3.62E-06 |
| cg26648306_TC21 | chr15 | 40242237 | | | -0.173872 | 5.10E-09 | 3.62E-06 |
| cg09825411_BC21 | chr19 | 50769849 | TSS1500 | GPR32 | 0.369033 | 5.12E-09 | 3.63E-06 |
| cg21806649_BC21 | chr1 | 111629122 | | | -0.326228 | 5.14E-09 | 3.65E-06 |
| cg13784474_BC21 | chr4 | 3249378 | exon_1;exc | MSANTD1; | -0.22939 | 5.16E-09 | 3.65E-06 |
| cg23547340_BC21 | chr3 | 120595072 | TSS1500;TS | NDUFB4;NI | -0.369147 | 5.16E-09 | 3.66E-06 |
| cg19776080_BC21 | chr21 | 40847295 | TSS200;TSS | DSCAM;DS | -0.189452 | 5.19E-09 | 3.67E-06 |
| cg03463735_TC21 | chr2 | 66704050 | TSS1500;TS | LINC01797 | -0.358832 | 5.20E-09 | 3.68E-06 |
| cg23669584_TC21 | chr10 | 35174312 | TSS1500;TS | CREM;CREI | -0.339346 | 5.22E-09 | 3.69E-06 |
| cg16603004_BC21 | chr11 | 72573740 | | | -0.122024 | 5.22E-09 | 3.69E-06 |
| cg03403240_BC21 | chr3 | 194842387 | | | -0.269189 | 5.25E-09 | 3.70E-06 |
| cg09832244_BC11 | chr12 | 6620434 | exon_2;exc | LPAR5;LPAI | -0.138267 | 5.25E-09 | 3.70E-06 |
| cg19219106_BC21 | chr13 | 46600344 | | | 0.303906 | 5.26E-09 | 3.71E-06 |
| cg22971501_BC21 | chr19 | 11088800 | TSS1500;TS | LDLR;LDLR; | -0.335927 | 5.27E-09 | 3.71E-06 |
| cg08836623_BC21 | chr3 | 194537855 | | | 0.226509 | 5.27E-09 | 3.71E-06 |
| cg12640305_TC21 | chr17 | 82320403 | | | -0.202956 | 5.30E-09 | 3.72E-06 |
| cg20614337_BC21 | chr15 | 74051689 | | | -0.132441 | 5.31E-09 | 3.73E-06 |
| cg13687885_TC21 | chr6 | 75403002 | exon_3 | LOC10192ε | -0.193083 | 5.31E-09 | 3.73E-06 |
| cg14243899_TC21 | chr17 | 76584315 | | | 0.336603 | 5.35E-09 | 3.75E-06 |
| cg10114631_BC21 | chr8 | 95305240 | | | -0.169442 | 5.35E-09 | 3.75E-06 |
| cg21190363_TC21 | chr14 | 104737069 | | | 0.319494 | 5.36E-09 | 3.75E-06 |
| cg02003175_BC21 | chr7 | 2857763 | | | -0.217614 | 5.36E-09 | 3.75E-06 |
| cg03905795_TC21 | chr11 | 70824775 | | | 0.40078 | 5.37E-09 | 3.75E-06 |
| cg04770218_TC21 | chr6 | 32169113 | 3UTR;exon | AGPAT1;AC | -0.285367 | 5.37E-09 | 3.75E-06 |
| cg06308522_BC21 | chr1 | 225581082 | | | -0.235753 | 5.37E-09 | 3.75E-06 |
| cg12993255_BC21 | chr2 | 70949157 | TSS1500;TS | ATP6V1B1- | -0.389512 | 5.38E-09 | 3.75E-06 |
| cg11804021_TC21 | chr19 | 39858850 | | | 0.64561 | 5.39E-09 | 3.75E-06 |
| cg01329532_TC21 | chr2 | 23002356 | | | 0.243861 | 5.41E-09 | 3.76E-06 |
| cg25212335_BC21 | chr20 | 36435793 | | | 0.187136 | 5.41E-09 | 3.76E-06 |
| cg10422983_TC21 | chr6 | 166440323 | | | 0.359491 | 5.43E-09 | 3.77E-06 |
| cg14569784_BC21 | chr10 | 2979499 | | | 0.140033 | 5.43E-09 | 3.77E-06 |
| cg02617070_BC21 | chr2 | 29749093 | | | -0.222733 | 5.44E-09 | 3.77E-06 |
| cg23376746_BC21 | chr12 | 1027449 | TSS1500 | ERC1 | -0.096743 | 5.46E-09 | 3.78E-06 |
| cg07563385_BC21 | chr17 | 80068780 | exon_11;3l | CCDC40;CC | 0.191712 | 5.47E-09 | 3.79E-06 |
| cg19635650_BC21 | chr14 | 68401554 | | | 0.645361 | 5.48E-09 | 3.79E-06 |
| cg25657433_BC21 | chr10 | 123377004 | | | -0.318107 | 5.48E-09 | 3.79E-06 |
| cg02630477_BC11 | chr1 | 39922551 | | | -0.227245 | 5.48E-09 | 3.79E-06 |
| cg09661228_BC21 | chr1 | 161962422 | exon_16;3l | ATF6;ATF6 | 0.546318 | 5.49E-09 | 3.79E-06 |
| cg21538445_BC21 | chr16 | 56370989 | | | -0.252479 | 5.49E-09 | 3.79E-06 |
| cg26992245_BC21 | chr8 | 29991063 | | | 0.268624 | 5.52E-09 | 3.80E-06 |
| cg15328351_TC21 | chr10 | 84566922 | | | -0.19724 | 5.52E-09 | 3.80E-06 |
| cg15915766_BC21 | chr7 | 128789638 | | | -0.236892 | 5.52E-09 | 3.80E-06 |

| | | | | | | | |
|---|---|---|---|---|---|---|---|
| cg05803473_TC21 | chr3 | 170469121 | | | -0.234905 | 5.53E-09 | 3.80E-06 |
| cg12175311_BC21 | chr15 | 99997887 | | | 0.33905 | 5.56E-09 | 3.81E-06 |
| cg14068702_TC21 | chr5 | 107610945 | | | -0.373102 | 5.56E-09 | 3.81E-06 |
| cg12225812_BC21 | chr8 | 48217699 | | | -0.263947 | 5.56E-09 | 3.82E-06 |
| cg24260207_BC21 | chr1 | 59045930 | | | -0.261671 | 5.57E-09 | 3.82E-06 |
| cg08497322_BC21 | chr9 | 113742916 | | | 0.2913 | 5.59E-09 | 3.82E-06 |
| cg13051969_TC21 | chr2 | 170927108 | TSS1500 | GORASP2 | 0.219976 | 5.59E-09 | 3.82E-06 |
| cg09556447_BC11 | chr6 | 36959324 | exon_3;exc | PI16;PI16 | -0.293614 | 5.59E-09 | 3.82E-06 |
| cg11922102_BC21 | chr7 | 151992521 | | | -0.257638 | 5.59E-09 | 3.82E-06 |
| cg24810239_TC21 | chr19 | 55564361 | | | -0.152762 | 5.60E-09 | 3.82E-06 |
| cg10606176_BC21 | chr4 | 128560942 | | | 0.331807 | 5.61E-09 | 3.83E-06 |
| cg11740770_BC21 | chr9 | 111287326 | | | 0.308175 | 5.62E-09 | 3.83E-06 |
| cg16268546_TC11 | chr8 | 1703031 | exon_15;3l | DLGAP2;DL | -0.180885 | 5.62E-09 | 3.83E-06 |
| cg13492553_BC21 | chr16 | 78592110 | | | -0.414444 | 5.64E-09 | 3.84E-06 |
| cg13838664_BC21 | chr5 | 149394982 | | | -0.232726 | 5.66E-09 | 3.85E-06 |
| cg11009880_TC21 | chr8 | 67207872 | | | 0.261344 | 5.66E-09 | 3.85E-06 |
| cg08364956_BC21 | chr5 | 178980121 | 3UTR;exon | GRM6;GRN | -0.351166 | 5.68E-09 | 3.85E-06 |
| cg08838695_BC21 | chr5 | 167599551 | | | 0.302322 | 5.68E-09 | 3.86E-06 |
| cg16723994_TC21 | chr17 | 58324262 | TSS1500 | TSPOAP1-A | -0.135525 | 5.70E-09 | 3.87E-06 |
| cg10919470_BC21 | chr8 | 54393124 | | | 0.19016 | 5.73E-09 | 3.88E-06 |
| cg18241130_TC11 | chr17 | 13024657 | | | -0.299663 | 5.74E-09 | 3.89E-06 |
| cg17759235_TC21 | chr7 | 100367422 | exon_5;3U | PILRB;PILRI | 0.233276 | 5.76E-09 | 3.90E-06 |
| cg12162189_BC21 | chr10 | 122021697 | | | -0.240861 | 5.80E-09 | 3.92E-06 |
| cg20751313_BC21 | chr19 | 37412781 | exon_5;exc | ZNF569;ZN | -0.305783 | 5.80E-09 | 3.92E-06 |
| cg13030329_TC21 | chr8 | 118120939 | | | -0.162284 | 5.82E-09 | 3.92E-06 |
| cg07188233_TC11 | chr14 | 92106803 | TSS1500;TS | ATXN3;ATX | 0.199824 | 5.82E-09 | 3.93E-06 |
| cg03841251_TC21 | chr6 | 165853094 | | | -0.30323 | 5.84E-09 | 3.94E-06 |
| cg20142209_BC21 | chr15 | 26096415 | | | -0.19298 | 5.85E-09 | 3.94E-06 |
| cg07179033_TC21 | chr6 | 30071771 | | | -0.432771 | 5.86E-09 | 3.94E-06 |
| cg04133413_TC21 | chr2 | 216766217 | | | -1.076783 | 5.86E-09 | 3.94E-06 |
| cg14591622_BC21 | chr16 | 67209576 | | | -0.334018 | 5.87E-09 | 3.94E-06 |
| cg10676762_BC21 | chr11 | 103433671 | | | -0.26914 | 5.87E-09 | 3.94E-06 |
| cg03698489_BC21 | chr16 | 81960730 | exon_33;3l | PLCG2;PLC | -0.545916 | 5.88E-09 | 3.94E-06 |
| cg15570211_TC21 | chr5 | 138559813 | TSS1500 | SNORD63B | 0.400825 | 5.89E-09 | 3.94E-06 |
| cg14684743_BC21 | chr10 | 12832207 | exon_11;3l | CAMK1D;C | 0.234427 | 5.93E-09 | 3.97E-06 |
| cg12823233_BC21 | chr7 | 2277241 | | | -0.522377 | 5.94E-09 | 3.98E-06 |
| cg06856216_BC21 | chr18 | 2907324 | | | -0.239877 | 5.95E-09 | 3.98E-06 |
| cg18271682_TC21 | chr19 | 29617198 | exon_7;3U | POP4;POP4 | -0.267505 | 5.99E-09 | 4.00E-06 |
| cg25111284_BC21 | chr3 | 66453946 | | | -0.186351 | 6.02E-09 | 4.02E-06 |
| cg17291585_TC21 | chr12 | 11947726 | | | 0.12377 | 6.04E-09 | 4.03E-06 |
| cg01312388_BC21 | chr17 | 76656166 | | | 0.3715 | 6.04E-09 | 4.03E-06 |
| cg02201239_BC21 | chr11 | 134416581 | | | -0.165577 | 6.06E-09 | 4.04E-06 |
| cg02827075_TC11 | chr19 | 45591425 | exon_2 | GPR4 | -0.261081 | 6.08E-09 | 4.05E-06 |
| cg16238279_BC21 | chr6 | 12064204 | | | -0.313283 | 6.09E-09 | 4.05E-06 |
| cg04319959_TC21 | chr2 | 235142775 | | | -0.281072 | 6.10E-09 | 4.05E-06 |
| cg05752133_TC21 | chr22 | 36017815 | | | -0.191608 | 6.11E-09 | 4.05E-06 |
| cg03157531_BC11 | chr10 | 131981502 | | | 0.307699 | 6.11E-09 | 4.05E-06 |

| | | | | | | | |
|---|---|---|---|---|---|---|---|
| cg11543011_TC21 | chr15 | 52349527 | | | -0.357762 | 6.12E-09 | 4.05E-06 |
| cg08018809_BC21 | chr2 | 58038538 | | | 0.305292 | 6.12E-09 | 4.05E-06 |
| cg03001566_BC21 | chr8 | 128819034 | | | -0.192359 | 6.12E-09 | 4.05E-06 |
| cg17794358_BC21 | chr6 | 32129480 | exon_2;TSS | FKBPL;ATF6 | -0.301778 | 6.15E-09 | 4.07E-06 |
| cg10125068_BC21 | chr6 | 134814569 | | | -0.281547 | 6.16E-09 | 4.07E-06 |
| cg08629469_BC21 | chr5 | 141442308 | | | -0.245096 | 6.17E-09 | 4.08E-06 |
| cg24035776_BC21 | chr21 | 37044198 | | | 0.110738 | 6.19E-09 | 4.08E-06 |
| cg11810204_TC21 | chr5 | 76088497 | | | 0.318687 | 6.19E-09 | 4.08E-06 |
| cg20707970_TC11 | chr3 | 126395221 | TSS1500;TS | CFAP100-D | -0.149099 | 6.19E-09 | 4.08E-06 |
| cg00215887_BC21 | chr20 | 16729402 | TSS1500;TS | SNRPB2;SN | -0.202334 | 6.21E-09 | 4.08E-06 |
| cg17585880_BC21 | chr17 | 18146436 | | | -0.231585 | 6.21E-09 | 4.08E-06 |
| cg14856577_TC21 | chr5 | 135953479 | | | 0.1958 | 6.22E-09 | 4.09E-06 |
| cg15998962_TC21 | chr2 | 25342590 | 5UTR;exon | DNMT3A;D | 0.428643 | 6.23E-09 | 4.09E-06 |
| cg03454705_TC11 | chr17 | 82577552 | | | -0.174432 | 6.24E-09 | 4.10E-06 |
| cg18574298_TC21 | chr21 | 45595484 | | | -0.294841 | 6.25E-09 | 4.10E-06 |
| cg26376621_TC21 | chr3 | 4812852 | | | 0.229717 | 6.25E-09 | 4.10E-06 |
| cg16449328_BC21 | chr10 | 112989561 | | | -0.209928 | 6.25E-09 | 4.10E-06 |
| cg07347147_TC21 | chr2 | 68837228 | | | -0.190775 | 6.26E-09 | 4.10E-06 |
| cg01752072_TC21 | chr1 | 37890624 | 5UTR;exon | INPP5B;INF | 0.248522 | 6.27E-09 | 4.10E-06 |
| cg07893663_BC21 | chr5 | 38551243 | | | -0.265123 | 6.30E-09 | 4.11E-06 |
| cg07467482_TC21 | chr7 | 154308981 | | | -0.363123 | 6.31E-09 | 4.12E-06 |
| cg19751824_BC21 | chr11 | 125465039 | | | -0.232213 | 6.32E-09 | 4.12E-06 |
| cg22425466_BC21 | chr9 | 137221535 | exon_2;5U | RNF208;RN | -0.162088 | 6.35E-09 | 4.14E-06 |
| cg01440556_TC21 | chr2 | 241225894 | exon_25;3I | ANO7;ANO | -0.37945 | 6.36E-09 | 4.15E-06 |
| cg16554145_BC21 | chr1 | 241627648 | | | 0.270469 | 6.38E-09 | 4.15E-06 |
| cg04875514_TC21 | chr12 | 53103363 | TSS200 | SOAT2 | 0.113459 | 6.39E-09 | 4.15E-06 |
| cg17264915_BC21 | chr12 | 8823769 | exon_3 | A2ML1 | -0.139196 | 6.39E-09 | 4.15E-06 |
| cg04558587_TC21 | chr9 | 33287323 | | | -0.216038 | 6.39E-09 | 4.15E-06 |
| cg00003937_TC21 | chr1 | 19305249 | | | 0.35493 | 6.41E-09 | 4.16E-06 |
| cg22776856_TC21 | chr16 | 3022571 | TSS1500;TS | THOC6;THO | -0.209941 | 6.42E-09 | 4.16E-06 |
| cg18433015_TC21 | chr15 | 28930785 | | | -0.138643 | 6.42E-09 | 4.16E-06 |
| cg27391396_TC11 | chr19 | 13865926 | | | -0.293904 | 6.42E-09 | 4.16E-06 |
| cg07912161_TC21 | chr6 | 36962062 | | | -0.42237 | 6.43E-09 | 4.16E-06 |
| cg07042095_TC21 | chr2 | 73823843 | | | 0.275112 | 6.43E-09 | 4.16E-06 |
| cg17303874_BC21 | chr12 | 12921326 | | | -0.139931 | 6.46E-09 | 4.17E-06 |
| cg16459199_TC21 | chr2 | 180695805 | | | -0.374148 | 6.47E-09 | 4.18E-06 |
| cg27154217_BC21 | chr5 | 50727293 | | | 0.577656 | 6.49E-09 | 4.19E-06 |
| cg10541053_TC21 | chr1 | 52351607 | 3UTR;exon | CC2D1B;CC | 0.34249 | 6.50E-09 | 4.19E-06 |
| cg13658458_TC21 | chr14 | 91254474 | TSS1500;TS | GPR68;GPF | -0.1529 | 6.50E-09 | 4.19E-06 |
| cg26378982_TC11 | chr2 | 10913811 | exon_1 | KCNF1 | -0.233874 | 6.52E-09 | 4.19E-06 |
| cg09786420_BC21 | chr1 | 154610913 | | | -0.182421 | 6.52E-09 | 4.19E-06 |
| cg17357334_BC21 | chr13 | 78994542 | | | 0.362721 | 6.57E-09 | 4.22E-06 |
| cg06489436_TC21 | chr5 | 154630598 | | | -0.144849 | 6.59E-09 | 4.23E-06 |
| cg16747321_BC21 | chr17 | 43766780 | 3UTR;exon | DUSP3;DUS | -0.14585 | 6.60E-09 | 4.23E-06 |
| cg18392348_BC21 | chr21 | 42315650 | TSS1500 | TFF3 | -0.197797 | 6.60E-09 | 4.23E-06 |
| cg25292163_TC21 | chr18 | 70555090 | | | 0.21966 | 6.65E-09 | 4.26E-06 |
| cg04692435_BC21 | chr1 | 115655119 | | | -0.185883 | 6.68E-09 | 4.28E-06 |

| | | | | | | | |
|---|---|---|---|---|---|---|---|
| cg26875073_BC11 | chr15 | 24955343 | TSS1500 | SNRPN | -0.148701 | 6.69E-09 | 4.28E-06 |
| cg02998056_BC11 | chr2 | 73269559 | exon_6;exc | FBXO41;FB | -0.363751 | 6.71E-09 | 4.29E-06 |
| cg06225927_BC21 | chr4 | 8030109 | | | -0.245593 | 6.71E-09 | 4.29E-06 |
| cg00413961_BC21 | chr1 | 23905913 | | | -0.279859 | 6.72E-09 | 4.29E-06 |
| cg25673717_BC21 | chr4 | 83524264 | | | -0.355063 | 6.74E-09 | 4.30E-06 |
| cg08981212_TC21 | chr13 | 35696714 | TSS1500;TS | LINC00445 | -0.208639 | 6.75E-09 | 4.30E-06 |
| cg13147462_BC21 | chr8 | 96233765 | | | -0.452664 | 6.76E-09 | 4.31E-06 |
| cg20199629_TC21 | chr17 | 50130144 | 5UTR;exon | SAMD14;SA | 0.29471 | 6.77E-09 | 4.31E-06 |
| cg03647734_BC21 | chr2 | 150470886 | | | 0.270469 | 6.85E-09 | 4.36E-06 |
| cg23628300_TC21 | chr17 | 60092800 | | | -0.281064 | 6.86E-09 | 4.36E-06 |
| cg15997518_BC21 | chr1 | 81349189 | | | -0.197002 | 6.87E-09 | 4.36E-06 |
| cg12441067_BC21 | chr3 | 42853010 | | | -0.122987 | 6.87E-09 | 4.36E-06 |
| cg08215187_BC21 | chr6 | 18308071 | | | -0.292897 | 6.89E-09 | 4.37E-06 |
| cg00122555_BC21 | chr2 | 128406005 | | | -0.100658 | 6.94E-09 | 4.40E-06 |
| cg01872024_BC21 | chr11 | 94769638 | | | -0.156275 | 6.94E-09 | 4.40E-06 |
| cg11081272_TC21 | chr11 | 78419190 | TSS1500 | GAB2 | -0.475137 | 6.96E-09 | 4.41E-06 |
| cg05092932_TC12 | chr10 | 71377594 | | | -0.595725 | 6.97E-09 | 4.41E-06 |
| cg18030306_BC21 | chr15 | 85297158 | | | -0.526432 | 6.97E-09 | 4.41E-06 |
| cg07965640_BC21 | chr2 | 213285011 | TSS1500;TS | SPAG16-DT | -0.211381 | 6.98E-09 | 4.41E-06 |
| cg17336354_TC21 | chr3 | 34257413 | | | -0.151709 | 6.99E-09 | 4.41E-06 |
| cg21789877_TC21 | chr1 | 61973195 | | | -0.237796 | 6.99E-09 | 4.41E-06 |
| cg12626882_TC21 | chr17 | 11883095 | 5UTR;exon | DNAH9;DN | -0.217074 | 7.00E-09 | 4.41E-06 |
| cg08337541_BC21 | chr5 | 104190058 | | | -0.263641 | 7.03E-09 | 4.43E-06 |
| cg20797409_BC21 | chr1 | 42315592 | | | 0.233411 | 7.04E-09 | 4.43E-06 |
| cg11180296_TC21 | chr7 | 69374344 | | | 0.246224 | 7.05E-09 | 4.43E-06 |
| cg08170023_TC21 | chr10 | 43203894 | | | -0.324613 | 7.06E-09 | 4.44E-06 |
| cg21306849_TC21 | chr15 | 89479518 | | | -0.147257 | 7.08E-09 | 4.44E-06 |
| cg10951933_BC21 | chr12 | 1861004 | | | 0.228871 | 7.10E-09 | 4.46E-06 |
| cg13698860_TC21 | chr22 | 43129673 | exon_5;3U | BIK;BIK | -0.128727 | 7.11E-09 | 4.46E-06 |
| cg14800608_TC21 | chr10 | 24242677 | | | -0.167857 | 7.11E-09 | 4.46E-06 |
| cg23737190_TC11 | chr6 | 36962245 | | | -0.383048 | 7.15E-09 | 4.47E-06 |
| cg16579752_TC21 | chr11 | 70827107 | TSS1500 | SHANK2 | 0.449417 | 7.15E-09 | 4.47E-06 |
| cg16604729_BC21 | chr11 | 72690751 | | | 0.222142 | 7.16E-09 | 4.48E-06 |
| cg09228402_BC21 | chr1 | 3907755 | | | -0.283928 | 7.16E-09 | 4.48E-06 |
| cg00230393_TC21 | chr10 | 47310182 | exon_2 | GDF10 | 0.281086 | 7.18E-09 | 4.48E-06 |
| cg05493580_BC21 | chr10 | 23280735 | | | -0.276544 | 7.19E-09 | 4.48E-06 |
| cg13654525_TC21 | chr11 | 65593038 | 3UTR;exon | KCNK7;KCN | -0.234795 | 7.20E-09 | 4.48E-06 |
| cg04648412_BC21 | chr5 | 174336783 | | | -0.239939 | 7.20E-09 | 4.48E-06 |
| cg20942867_TC11 | chr1 | 27099653 | 3UTR;exon | SLC9A1;SLC | 0.164715 | 7.20E-09 | 4.48E-06 |
| cg14875529_BC21 | chr7 | 778044 | | | 0.256127 | 7.21E-09 | 4.48E-06 |
| cg08519191_TC21 | chr5 | 138458018 | | | -0.194153 | 7.22E-09 | 4.49E-06 |
| cg15144237_BC21 | chr2 | 16400125 | | | -0.184239 | 7.26E-09 | 4.51E-06 |
| cg16923709_TC21 | chr10 | 128991117 | | | -0.1982 | 7.27E-09 | 4.51E-06 |
| cg20012915_BC21 | chr11 | 7673951 | 5UTR;exon | CYB5R2;CY | 0.237094 | 7.28E-09 | 4.52E-06 |
| cg15057798_TC21 | chr14 | 96638693 | | | -0.171793 | 7.29E-09 | 4.52E-06 |
| cg04057432_BC21 | chr2 | 206423497 | | | 0.222206 | 7.29E-09 | 4.52E-06 |
| cg05062944_BC21 | chr2 | 226956710 | | | -0.172525 | 7.30E-09 | 4.52E-06 |

| | | | | | | | |
|---|---|---|---|---|---|---|---|
| cg20270653_TC21 | chr7 | 71497735 | | | -0.258329 | 7.30E-09 | 4.52E-06 |
| cg09684021_BC21 | chr19 | 7215307 | | | -0.119658 | 7.32E-09 | 4.53E-06 |
| cg04554196_TC21 | chr2 | 9007863 | | | 0.254169 | 7.35E-09 | 4.54E-06 |
| cg16751781_BC21 | chr15 | 78566247 | | | 0.323147 | 7.35E-09 | 4.54E-06 |
| cg10784917_TC21 | chr7 | 25297105 | | | 0.170905 | 7.38E-09 | 4.55E-06 |
| cg23271660_BC21 | chr10 | 69121261 | | | -0.347382 | 7.38E-09 | 4.55E-06 |
| cg06922943_TC21 | chr19 | 54163660 | | | -0.358241 | 7.38E-09 | 4.55E-06 |
| cg24370079_TC11 | chr5 | 180815568 | 5UTR;exon | MGAT1;MG | 0.124918 | 7.39E-09 | 4.55E-06 |
| cg25802503_TC21 | chr21 | 33644210 | | | -0.212069 | 7.39E-09 | 4.55E-06 |
| cg18842260_BC21 | chr13 | 67117537 | | | -0.25457 | 7.40E-09 | 4.55E-06 |
| cg05425310_BC21 | chr6 | 56808050 | | | 0.226952 | 7.40E-09 | 4.55E-06 |
| cg26383056_TC21 | chr22 | 41597353 | | | -0.153501 | 7.41E-09 | 4.55E-06 |
| cg01748263_TC21 | chr3 | 193379572 | TSS1500 | ATP13A5 | -0.332015 | 7.42E-09 | 4.55E-06 |
| cg18801197_TC11 | chr3 | 122516271 | TSS1500;TS | KPNA1;KPN | -0.195866 | 7.45E-09 | 4.57E-06 |
| cg01238590_BC21 | chr4 | 154785723 | | | -0.275317 | 7.49E-09 | 4.59E-06 |
| cg05517791_TC21 | chr7 | 73743021 | | | -0.264181 | 7.49E-09 | 4.59E-06 |
| cg13844930_TC21 | chr12 | 1279342 | | | 0.225455 | 7.50E-09 | 4.59E-06 |
| cg19671311_BC21 | chr12 | 122112216 | | | 0.105026 | 7.52E-09 | 4.60E-06 |
| cg25595166_BC21 | chr20 | 64147372 | | | -0.128995 | 7.53E-09 | 4.60E-06 |
| cg19367330_TC21 | chr14 | 34332171 | | | -0.223474 | 7.57E-09 | 4.62E-06 |
| cg16071782_BC21 | chr14 | 96565623 | exon_22;3U | PAPOLA;PA | 0.211566 | 7.60E-09 | 4.63E-06 |
| cg21608694_TC21 | chr16 | 64320723 | | | -0.308175 | 7.60E-09 | 4.63E-06 |
| cg16385933_TC11 | chr10 | 110872034 | TSS1500 | PDCD4-AS1 | 0.296782 | 7.63E-09 | 4.65E-06 |
| cg25700862_TC21 | chr13 | 30934234 | | | -0.235264 | 7.66E-09 | 4.67E-06 |
| cg06332752_TC21 | chr10 | 96257408 | | | -0.170877 | 7.68E-09 | 4.67E-06 |
| cg01179542_TC21 | chr1 | 27103798 | | | -0.205803 | 7.68E-09 | 4.67E-06 |
| cg03460315_BC21 | chr4 | 110089235 | | | -0.325748 | 7.69E-09 | 4.67E-06 |
| cg13049556_TC21 | chr17 | 73370062 | | | 0.351763 | 7.70E-09 | 4.67E-06 |
| cg06342931_BC21 | chr13 | 75815529 | | | -0.182094 | 7.70E-09 | 4.67E-06 |
| cg26411015_TC11 | chr15 | 83208357 | TSS1500 | HDGFL3 | 0.189279 | 7.72E-09 | 4.67E-06 |
| cg22449114_TC21 | chr20 | 609599 | | | -0.197316 | 7.72E-09 | 4.67E-06 |
| cg25549238_TC21 | chr6 | 157940936 | exon_17 | SNX9 | -0.302167 | 7.72E-09 | 4.67E-06 |
| cg21995171_TC21 | chr2 | 3105607 | | | -0.265966 | 7.73E-09 | 4.67E-06 |
| cg03104792_BC21 | chr2 | 86809959 | | | -0.253592 | 7.74E-09 | 4.68E-06 |
| cg00260259_BC21 | chr17 | 52057498 | | | -0.231014 | 7.74E-09 | 4.68E-06 |
| cg03277377_BC21 | chr13 | 29580370 | | | 0.178024 | 7.76E-09 | 4.68E-06 |
| cg01018437_TC21 | chr12 | 49951982 | exon_3;exo | AQP5-AS1; | -0.243869 | 7.76E-09 | 4.68E-06 |
| cg11989689_BC21 | chr1 | 31651559 | | | -0.265323 | 7.76E-09 | 4.68E-06 |
| cg09502182_BC21 | chr20 | 16215310 | | | -0.207537 | 7.76E-09 | 4.68E-06 |
| cg25513668_BC21 | chr16 | 78098933 | TSS1500;TS | WWOX;WV | -0.244288 | 7.78E-09 | 4.68E-06 |
| cg19942385_BC21 | chr1 | 39168624 | | | -0.347596 | 7.79E-09 | 4.69E-06 |
| cg19774236_TC21 | chr17 | 8015704 | | | -0.192732 | 7.80E-09 | 4.69E-06 |
| cg09318162_TC21 | chr6 | 32880065 | | | -0.247169 | 7.80E-09 | 4.69E-06 |
| cg16616587_BC21 | chr12 | 77894979 | | | -0.21562 | 7.81E-09 | 4.69E-06 |
| cg27502952_BC21 | chr2 | 241411986 | | | -0.344592 | 7.81E-09 | 4.69E-06 |
| cg27554147_BC21 | chr11 | 66021391 | | | -0.113981 | 7.83E-09 | 4.69E-06 |
| cg05301609_TC21 | chr17 | 19674404 | | | -0.34175 | 7.83E-09 | 4.69E-06 |

| | | | | | | | |
|---|---|---|---|---|---|---|---|
| cg23129055_BC21 | chr1 | 54413721 | TSS1500;TS | SSBP3;SSBI | -0.210328 | 7.85E-09 | 4.70E-06 |
| cg25515317_TC21 | chr7 | 116524054 | TSS1500;TS | CAV1;CAV1 | 0.3427 | 7.86E-09 | 4.70E-06 |
| cg11389793_BC21 | chr6 | 74932608 | | | -0.598538 | 7.86E-09 | 4.70E-06 |
| cg21250061_TC21 | chr9 | 37463347 | | | -0.43783 | 7.88E-09 | 4.71E-06 |
| cg14301508_BC21 | chr9 | 128733450 | exon_15;e | ZER1;ZER1; | -0.107213 | 7.90E-09 | 4.72E-06 |
| cg14086033_TC21 | chr11 | 61792349 | TSS1500 | FEN1 | -0.320926 | 7.91E-09 | 4.72E-06 |
| cg06322979_BC21 | chr16 | 81717786 | | | -0.505928 | 7.91E-09 | 4.72E-06 |
| cg13324082_BC21 | chr7 | 158843737 | | | -0.113199 | 7.92E-09 | 4.72E-06 |
| cg19871173_BC21 | chr1 | 229862095 | | | -0.254538 | 7.95E-09 | 4.73E-06 |
| cg06465731_TC21 | chr10 | 88064934 | | | -0.244009 | 7.95E-09 | 4.73E-06 |
| cg14318583_BC21 | chr4 | 7611767 | | | -0.263848 | 7.96E-09 | 4.73E-06 |
| cg07271319_BC21 | chr1 | 78648310 | TSS1500;TS | IFI44;IFI44 | -0.185997 | 7.98E-09 | 4.74E-06 |
| cg19909613_TC21 | chr8 | 108474924 | | | 0.279179 | 7.99E-09 | 4.75E-06 |
| cg23701962_BC21 | chr1 | 245715480 | | | -0.176358 | 8.04E-09 | 4.77E-06 |
| cg19189201_TC21 | chr15 | 98421554 | TSS1500 | LINC02351 | -0.229818 | 8.04E-09 | 4.77E-06 |
| cg00825491_BC21 | chr18 | 48922279 | | | -0.127374 | 8.07E-09 | 4.78E-06 |
| cg22110918_BC11 | chr8 | 90985429 | TSS200;TSS | C8orf88;C8 | -0.394264 | 8.07E-09 | 4.78E-06 |
| cg05259872_BC21 | chr4 | 168631159 | TSS1500 | PALLD | -0.180824 | 8.08E-09 | 4.78E-06 |
| cg09355028_BC21 | chr14 | 103069775 | | | -0.222191 | 8.08E-09 | 4.78E-06 |
| cg24637639_BC21 | chr10 | 15316755 | | | 0.297457 | 8.09E-09 | 4.78E-06 |
| cg18062379_TC21 | chr12 | 109278127 | 3UTR;exon | FOXN4;FO) | 0.173652 | 8.09E-09 | 4.78E-06 |
| cg21114878_TC21 | chr1 | 239399120 | | | -0.133315 | 8.10E-09 | 4.78E-06 |
| cg18254784_BC21 | chr20 | 34495336 | | | -0.503726 | 8.11E-09 | 4.79E-06 |
| cg21655740_TC21 | chr8 | 52033308 | | | 0.256535 | 8.13E-09 | 4.79E-06 |
| cg09160811_BC21 | chr4 | 151674407 | | | -0.271422 | 8.14E-09 | 4.80E-06 |
| cg11697038_TC21 | chr1 | 27604773 | TSS1500;TS | AHDC1;AHI | -0.201922 | 8.17E-09 | 4.81E-06 |
| cg02315376_TC21 | chr13 | 27403091 | | | -0.389552 | 8.17E-09 | 4.81E-06 |
| cg21021448_BC21 | chr8 | 93706148 | exon_6;exc | CIBAR1;CIB | 0.199052 | 8.20E-09 | 4.82E-06 |
| cg24640561_BC21 | chr13 | 108326790 | | | 0.153851 | 8.26E-09 | 4.85E-06 |
| cg16070857_TC21 | chr11 | 17565031 | | | -0.328288 | 8.26E-09 | 4.85E-06 |
| cg08533268_BC21 | chr3 | 48327687 | | | 0.205128 | 8.26E-09 | 4.85E-06 |
| cg25900312_TC21 | chr12 | 109766430 | | | -0.195891 | 8.27E-09 | 4.85E-06 |
| cg06991890_BC11 | chr16 | 30394970 | TSS1500;TS | ZNF48;ZNF | 0.224223 | 8.27E-09 | 4.85E-06 |
| cg02198653_TC21 | chr19 | 35714016 | | | -0.234209 | 8.30E-09 | 4.85E-06 |
| cg26992204_TC21 | chr5 | 1147343 | | | -0.348362 | 8.30E-09 | 4.85E-06 |
| cg21738177_TC21 | chr2 | 170052142 | | | -0.366502 | 8.33E-09 | 4.87E-06 |
| cg00574649_TC21 | chr19 | 45250411 | TSS1500;TS | MARK4;MA | -0.193176 | 8.33E-09 | 4.87E-06 |
| cg23853915_BC21 | chr19 | 850485 | | | -0.117162 | 8.35E-09 | 4.87E-06 |
| cg08761926_TC21 | chr10 | 131831765 | | | -0.218695 | 8.35E-09 | 4.87E-06 |
| cg05415135_TC21 | chr14 | 22691590 | | | -0.179767 | 8.37E-09 | 4.88E-06 |
| cg18982477_TC21 | chr4 | 37642476 | | | 0.290771 | 8.38E-09 | 4.89E-06 |
| cg05626927_BC21 | chr4 | 14360615 | | | -0.158588 | 8.39E-09 | 4.89E-06 |
| cg02303651_BC21 | chr7 | 158956953 | | | -0.282702 | 8.41E-09 | 4.89E-06 |
| cg17737681_BC11 | chr2 | 172087408 | | | 0.204742 | 8.41E-09 | 4.89E-06 |
| cg04114405_BC21 | chr15 | 49046959 | TSS1500;TS | SECISBP2L; | 0.343768 | 8.43E-09 | 4.90E-06 |
| cg25936595_TC21 | chr19 | 3772271 | TSS200;TSS | RAX2;RAX2 | -0.178965 | 8.44E-09 | 4.90E-06 |
| cg06399596_TC21 | chr6 | 1595441 | | | 0.239583 | 8.47E-09 | 4.92E-06 |

| | | | | | | | |
|---|---|---|---|---|---|---|---|
| cg04315843_BC21 | chr2 | 234881810 | | | -0.347582 | 8.50E-09 | 4.93E-06 |
| cg16594066_TC11 | chr16 | 30558405 | TSS200;TSS | ZNF764;ZN | -0.324927 | 8.51E-09 | 4.93E-06 |
| cg16284279_BC21 | chr6 | 77034235 | | | 0.4876 | 8.52E-09 | 4.93E-06 |
| cg23176632_BC21 | chr12 | 262973 | TSS200;TSS | SLC6A13;SI | -0.172447 | 8.52E-09 | 4.93E-06 |
| cg07642551_BC21 | chr3 | 45927233 | 3UTR;exon | FYCO1;FYC | -0.147663 | 8.53E-09 | 4.94E-06 |
| cg03604731_BC21 | chr2 | 96159061 | | | -0.182767 | 8.55E-09 | 4.94E-06 |
| cg03867877_BC21 | chr8 | 29270786 | | | -0.111881 | 8.58E-09 | 4.96E-06 |
| cg18088032_TC21 | chr14 | 21090497 | 3UTR;exon | ZNF219;ZN | -0.247664 | 8.62E-09 | 4.97E-06 |
| cg14184866_TC21 | chr9 | 79570222 | | | -0.227477 | 8.65E-09 | 4.99E-06 |
| cg24922733_TC21 | chr20 | 4845146 | | | 0.297103 | 8.67E-09 | 5.00E-06 |
| cg03031182_BC21 | chr21 | 45528788 | | | 0.243116 | 8.68E-09 | 5.00E-06 |
| cg12652115_BC21 | chr1 | 180248298 | exon_2 | LHX4 | -0.258219 | 8.69E-09 | 5.01E-06 |
| cg13570060_BC21 | chr9 | 33240073 | TSS200 | SPINK4 | -0.191268 | 8.72E-09 | 5.02E-06 |
| cg11682661_BC21 | chr20 | 44700558 | | | 0.238565 | 8.73E-09 | 5.02E-06 |
| cg02804707_TC21 | chr11 | 14904293 | | | -0.239526 | 8.73E-09 | 5.02E-06 |
| cg15677375_TC11 | chr20 | 5121877 | | | 0.197211 | 8.80E-09 | 5.05E-06 |
| cg16135310_TC21 | chr1 | 84001722 | | | -0.221618 | 8.82E-09 | 5.06E-06 |
| cg07701678_BC21 | chr1 | 229221431 | | | 0.197169 | 8.82E-09 | 5.06E-06 |
| cg21442603_BC21 | chr2 | 120918887 | | | 0.190774 | 8.82E-09 | 5.06E-06 |
| cg04723239_BC21 | chr11 | 67048799 | exon_8;3U | SYT12;SYT1 | -0.208566 | 8.84E-09 | 5.06E-06 |
| cg21837850_TC21 | chr16 | 83903990 | | | 0.266204 | 8.85E-09 | 5.07E-06 |
| cg17199683_BC21 | chr2 | 112520405 | exon_6;exo | TTL;TTL | 0.326173 | 8.86E-09 | 5.07E-06 |
| cg27131881_TC21 | chr22 | 45424469 | | | -0.17157 | 8.89E-09 | 5.08E-06 |
| cg01669540_BC21 | chr16 | 67850570 | | | -0.325086 | 8.91E-09 | 5.09E-06 |
| cg23286346_BC21 | chr12 | 76927372 | | | -0.180998 | 8.92E-09 | 5.09E-06 |
| cg07869795_BC21 | chr22 | 45413494 | exon_1;exo | SMC1B;SM | -0.242463 | 8.96E-09 | 5.11E-06 |
| cg20705772_BC21 | chr4 | 57165032 | | | 0.281368 | 8.97E-09 | 5.11E-06 |
| cg24603102_BC21 | chr8 | 117998934 | | | -0.356407 | 8.99E-09 | 5.12E-06 |
| cg00627223_BC21 | chr1 | 39454172 | | | -0.391353 | 8.99E-09 | 5.12E-06 |
| cg24172324_BC21 | chr2 | 231393652 | | | 0.193647 | 9.01E-09 | 5.12E-06 |
| cg14012310_TC21 | chr13 | 78607812 | | | -0.133267 | 9.01E-09 | 5.12E-06 |
| cg00005077_BC11 | chr10 | 45672788 | 5UTR;exon | ZFAND4;ZF | -0.255091 | 9.02E-09 | 5.13E-06 |
| cg03760670_BC21 | chr3 | 172078756 | | | -0.318742 | 9.05E-09 | 5.14E-06 |
| cg08884591_TC21 | chr11 | 62269719 | TSS1500 | SCGB2A2 | -0.339746 | 9.08E-09 | 5.15E-06 |
| cg11126410_BC11 | chr3 | 129122348 | TSS1500;TS | RAB43;RAE | -0.238445 | 9.10E-09 | 5.16E-06 |
| cg06333196_TC21 | chr4 | 20305956 | | | -0.396547 | 9.14E-09 | 5.18E-06 |
| cg24795721_TC21 | chr9 | 112037326 | | | -0.3048 | 9.20E-09 | 5.21E-06 |
| cg13411813_TC21 | chr13 | 96997727 | | | -0.22994 | 9.22E-09 | 5.22E-06 |
| cg05842470_BC21 | chr11 | 60346254 | | | -0.161532 | 9.22E-09 | 5.22E-06 |
| cg19695822_BC21 | chr14 | 51243562 | | | 0.386714 | 9.24E-09 | 5.22E-06 |
| cg04395776_BC21 | chr2 | 43371997 | | | -0.390963 | 9.24E-09 | 5.22E-06 |
| cg04559409_BC21 | chr6 | 132598672 | | | -0.344414 | 9.25E-09 | 5.22E-06 |
| cg19472302_BC21 | chr3 | 195367049 | | | 0.379426 | 9.26E-09 | 5.23E-06 |
| cg19700955_TC21 | chr14 | 101374410 | | | 0.238253 | 9.29E-09 | 5.23E-06 |
| cg18042004_TC11 | chr22 | 41544653 | TSS200;TSS | POLR3H;PC | 0.319972 | 9.29E-09 | 5.23E-06 |
| cg18017265_TC21 | chr2 | 23384383 | TSS1500 | KLHL29 | 0.283776 | 9.33E-09 | 5.25E-06 |
| cg15853550_TC21 | chr10 | 133162297 | | | -0.308124 | 9.36E-09 | 5.27E-06 |

| | | | | | | | |
|---|---|---|---|---|---|---|---|
| cg19919383_BC21 | chr3 | 17005342 | | | 0.265741 | 9.37E-09 | 5.27E-06 |
| cg23153655_TC21 | chr2 | 10080689 | TSS1500 | CYS1 | 0.278075 | 9.42E-09 | 5.30E-06 |
| cg06566158_TC21 | chr17 | 3571656 | | | -0.229075 | 9.43E-09 | 5.30E-06 |
| cg09440866_BC21 | chr12 | 113576705 | | | 0.266677 | 9.46E-09 | 5.31E-06 |
| cg04560534_TC21 | chr1 | 40092169 | exon_3;exo | PPT1;PPT1 | 0.2955 | 9.48E-09 | 5.31E-06 |
| cg06705597_TC21 | chr4 | 74375748 | | | 0.382589 | 9.48E-09 | 5.31E-06 |
| cg06842332_BC21 | chr4 | 93666395 | | | -0.304713 | 9.48E-09 | 5.31E-06 |
| cg21096444_BC21 | chr1 | 215189316 | | | -0.349445 | 9.49E-09 | 5.31E-06 |
| cg18241780_BC21 | chr12 | 118182411 | | | -0.264041 | 9.49E-09 | 5.31E-06 |
| cg03179401_TC21 | chr6 | 108549970 | | | -0.27104 | 9.53E-09 | 5.33E-06 |
| cg07719203_BC21 | chr5 | 14901858 | | | -0.289981 | 9.55E-09 | 5.34E-06 |
| cg13050716_BC21 | chr3 | 127697737 | | | -0.16995 | 9.56E-09 | 5.34E-06 |
| cg26463516_TC21 | chr19 | 35510240 | exon_5;exo | DMKN;DM | 0.237131 | 9.59E-09 | 5.35E-06 |
| cg04589975_TC11 | chr2 | 70191054 | TSS200;TSS | C2orf42;C2 | -0.48393 | 9.60E-09 | 5.35E-06 |
| cg18559785_TC21 | chr19 | 42411220 | | | -0.188479 | 9.60E-09 | 5.35E-06 |
| cg02388568_TC21 | chr6 | 73462016 | exon_1;exo | MTO1;MTO | -0.259505 | 9.60E-09 | 5.35E-06 |
| cg25141441_BC21 | chr15 | 60538677 | | | -0.266884 | 9.62E-09 | 5.35E-06 |
| cg13384396_TC21 | chr3 | 123448830 | exon_1;5U | ADCY5;ADC | -0.213332 | 9.63E-09 | 5.36E-06 |
| cg25066868_TC21 | chr11 | 1942753 | | | 0.183501 | 9.67E-09 | 5.37E-06 |
| cg10623785_TC21 | chr1 | 183804741 | TSS1500;TS | RGL1;RGL1 | -0.244844 | 9.68E-09 | 5.38E-06 |
| cg18692382_TC11 | chr19 | 39834258 | TSS200;TSS | DYRK1B;DY | 0.493627 | 9.74E-09 | 5.40E-06 |
| cg22025064_BC21 | chr1 | 21814907 | | | -0.180522 | 9.74E-09 | 5.40E-06 |
| cg20441066_BC21 | chr6 | 146316441 | | | -0.286597 | 9.74E-09 | 5.40E-06 |
| cg09726316_TC21 | chr14 | 105588547 | | | -0.280524 | 9.79E-09 | 5.42E-06 |
| cg16162350_BC21 | chr3 | 169149624 | | | -0.319706 | 9.79E-09 | 5.42E-06 |
| cg02725398_TC21 | chr3 | 9946460 | exon_4;exo | PRRT3;PRR | -0.484475 | 9.79E-09 | 5.42E-06 |
| cg25658377_TC21 | chr21 | 15246579 | | | -0.248737 | 9.80E-09 | 5.42E-06 |
| cg01514490_TC21 | chr10 | 7413376 | | | -0.228808 | 9.81E-09 | 5.42E-06 |
| cg09055594_TC21 | chr6 | 1995916 | | | 0.487958 | 9.84E-09 | 5.43E-06 |
| cg10620929_BC21 | chr7 | 5435799 | | | -0.228388 | 9.85E-09 | 5.44E-06 |
| cg27392956_TC21 | chr20 | 64256109 | | | -0.328763 | 9.87E-09 | 5.45E-06 |
| cg05726450_BC21 | chr17 | 8868630 | TSS1500;TS | PIK3R6;PIK | 0.14994 | 9.88E-09 | 5.45E-06 |
| cg17791346_TC21 | chr3 | 78046957 | | | -0.190545 | 9.90E-09 | 5.46E-06 |
| cg13778339_TC21 | chr9 | 87702510 | | | -0.33778 | 9.93E-09 | 5.47E-06 |
| cg07943959_BC21 | chr1 | 145885902 | TSS200;TSS | ANKRD35;A | -0.265167 | 9.97E-09 | 5.48E-06 |
| cg05856099_BC21 | chr9 | 90961268 | | | 0.145976 | 9.97E-09 | 5.48E-06 |
| cg14465144_BC21 | chr9 | 136268684 | | | 0.23418 | 9.98E-09 | 5.48E-06 |
| cg06715167_BC21 | chr2 | 235535377 | | | -0.37346 | 9.98E-09 | 5.48E-06 |
| cg15470519_BC21 | chr13 | 101700726 | | | -0.322714 | 9.99E-09 | 5.48E-06 |
| cg18785679_TC21 | chr10 | 133219000 | | | -0.204989 | 9.99E-09 | 5.48E-06 |
| cg14620572_BC11 | chr9 | 124453390 | exon_6 | ADGRD2 | 0.193831 | 1.01E-08 | 5.52E-06 |
| cg23269864_TC21 | chr1 | 8926011 | | | -0.223811 | 1.01E-08 | 5.52E-06 |
| cg03070533_TC11 | chr22 | 35257401 | TSS200;TSS | HMGXB4;H | 0.289389 | 1.01E-08 | 5.55E-06 |
| cg16920832_BC21 | chr5 | 150054682 | | | 0.261573 | 1.02E-08 | 5.56E-06 |
| cg07503367_TC21 | chr7 | 11833119 | TSS1500 | THSD7A | -0.450843 | 1.02E-08 | 5.56E-06 |
| cg05074859_BC21 | chr15 | 31317527 | | | 0.112654 | 1.02E-08 | 5.57E-06 |
| cg03173723_TC21 | chr12 | 52651029 | | | -0.213883 | 1.02E-08 | 5.58E-06 |

| | | | | | | | |
|---|---|---|---|---|---|---|---|
| cg21007215_BC21 | chr12 | 127596837 | | | 0.237133 | 1.02E-08 | 5.59E-06 |
| cg11049752_BC21 | chr7 | 55158096 | | | -0.144861 | 1.02E-08 | 5.59E-06 |
| cg20241156_BC21 | chr9 | 69723582 | exon_6;exo | PTAR1;PTA | 0.156905 | 1.02E-08 | 5.59E-06 |
| cg14915777_BC21 | chr10 | 35822550 | | | -0.194981 | 1.03E-08 | 5.60E-06 |
| cg16310668_BC21 | chr5 | 77118238 | | | -0.209487 | 1.03E-08 | 5.61E-06 |
| cg14208151_TC21 | chr3 | 123758042 | | | 0.195772 | 1.03E-08 | 5.61E-06 |
| cg10751207_BC21 | chr18 | 79356201 | | | -0.291064 | 1.04E-08 | 5.67E-06 |
| cg12155257_BC21 | chr17 | 1896248 | | | 0.296704 | 1.04E-08 | 5.67E-06 |
| cg02302721_TC21 | chr3 | 125399144 | | | -0.145449 | 1.04E-08 | 5.67E-06 |
| cg17253057_TC21 | chr19 | 49750731 | | | 0.217121 | 1.05E-08 | 5.68E-06 |
| cg27087228_BC21 | chr6 | 166908180 | | | -0.186394 | 1.05E-08 | 5.69E-06 |
| cg21047793_TC21 | chr5 | 69531426 | | | -0.231003 | 1.05E-08 | 5.70E-06 |
| cg12365876_TC21 | chr8 | 29287121 | | | 0.191245 | 1.05E-08 | 5.71E-06 |
| cg12697306_BC21 | chr2 | 241652989 | | | -0.168182 | 1.06E-08 | 5.71E-06 |
| cg12404940_TC21 | chr8 | 33806108 | | | -0.247512 | 1.06E-08 | 5.71E-06 |
| cg09085614_TC21 | chr5 | 96584046 | | | 0.224843 | 1.06E-08 | 5.71E-06 |
| cg13561163_BC21 | chr14 | 102242094 | TSS1500;TS | MOK;MOK; | -0.235005 | 1.06E-08 | 5.71E-06 |
| cg23507713_BC21 | chr16 | 49452882 | | | -0.215177 | 1.06E-08 | 5.71E-06 |
| cg13971025_TC13 | chr9 | 135104424 | | | 0.278744 | 1.06E-08 | 5.71E-06 |
| cg24276239_TC11 | chr15 | 74906985 | TSS200;TSS | FAM219B;F | 0.263703 | 1.06E-08 | 5.74E-06 |
| cg13865934_TC21 | chr1 | 2331035 | | | -0.207786 | 1.06E-08 | 5.74E-06 |
| cg20750472_TC21 | chr19 | 52496267 | | | -0.211058 | 1.07E-08 | 5.74E-06 |
| cg04587581_TC21 | chr16 | 88866774 | TSS200;TSS | PABPN1L;P | -0.190235 | 1.07E-08 | 5.74E-06 |
| cg17520314_BC11 | chr17 | 81637181 | TSS200;TSS | TSPAN10;T | 0.340828 | 1.07E-08 | 5.74E-06 |
| cg07648498_TC11 | chr16 | 89816777 | TSS200;TSS | FANCA;FAN | -0.27298 | 1.07E-08 | 5.74E-06 |
| cg10557174_BC21 | chr13 | 28592330 | | | 0.699931 | 1.07E-08 | 5.74E-06 |
| cg09783609_BC21 | chr11 | 36300817 | | | -0.140462 | 1.07E-08 | 5.74E-06 |
| cg16287656_BC21 | chr11 | 46382671 | exon_4;exo | MDK;MDK; | -0.173961 | 1.07E-08 | 5.76E-06 |
| cg15474999_BC21 | chr19 | 17558399 | | | 0.313021 | 1.07E-08 | 5.76E-06 |
| cg17332603_BC11 | chr4 | 7324849 | | | 0.14792 | 1.07E-08 | 5.76E-06 |
| cg08483594_BC21 | chr5 | 126679983 | | | -0.164972 | 1.08E-08 | 5.78E-06 |
| cg15262006_BC21 | chr14 | 70783798 | TSS1500;TS | MAP3K9;M | -0.257283 | 1.08E-08 | 5.80E-06 |
| cg04273745_TC21 | chr15 | 24675193 | TSS1500 | NPAP1 | -0.340889 | 1.08E-08 | 5.80E-06 |
| cg15343302_BC21 | chr1 | 75135651 | | | -0.237928 | 1.09E-08 | 5.82E-06 |
| cg12467559_BC21 | chr9 | 114421419 | | | -0.191839 | 1.10E-08 | 5.86E-06 |
| cg03954442_BC11 | chr15 | 31391940 | | | 0.256699 | 1.10E-08 | 5.89E-06 |
| cg05394663_BC11 | chr1 | 228061084 | exon_4;3U | WNT3A;WI | -0.266547 | 1.11E-08 | 5.94E-06 |
| cg12898290_BC21 | chr2 | 21270765 | | | 0.176503 | 1.11E-08 | 5.94E-06 |
| cg02836135_BC21 | chr6 | 107730889 | | | -0.135257 | 1.11E-08 | 5.94E-06 |
| cg23794157_BC21 | chr18 | 78683654 | | | -0.308325 | 1.12E-08 | 5.94E-06 |
| cg27509510_TC21 | chr2 | 25977132 | | | 0.222219 | 1.12E-08 | 5.97E-06 |
| cg11933951_TC21 | chr11 | 83286753 | TSS1500;TS | CCDC90B;C | -0.28929 | 1.12E-08 | 5.97E-06 |
| cg14653151_BC21 | chr8 | 79351279 | | | -0.230583 | 1.12E-08 | 5.97E-06 |
| cg13806603_BC21 | chr8 | 54213892 | | | -0.404198 | 1.13E-08 | 6.00E-06 |
| cg09527343_TC21 | chr8 | 57740655 | | | -0.307565 | 1.13E-08 | 6.00E-06 |
| cg25214146_BC21 | chr2 | 176575800 | | | -0.214005 | 1.13E-08 | 6.02E-06 |
| cg21486532_BC21 | chr2 | 241795720 | | | -0.336219 | 1.14E-08 | 6.03E-06 |

| | | | | | | | |
|---|---|---|---|---|---|---|---|
| cg23807113_TC21 | chr3 | 182371136 | | | -0.377275 | 1.14E-08 | 6.03E-06 |
| cg05285223_BC21 | chr3 | 69916336 | | | -0.425301 | 1.14E-08 | 6.03E-06 |
| cg09333298_BC21 | chr10 | 132113912 | | | -0.349462 | 1.14E-08 | 6.06E-06 |
| cg11620770_BC21 | chr17 | 57542109 | | | 0.265495 | 1.15E-08 | 6.08E-06 |
| cg01282402_BC21 | chr4 | 74503630 | | | -0.277837 | 1.15E-08 | 6.09E-06 |
| cg04110104_BC21 | chr11 | 39862300 | | | -0.257328 | 1.15E-08 | 6.09E-06 |
| cg04775302_BC21 | chr1 | 90840120 | | | 0.344292 | 1.15E-08 | 6.09E-06 |
| cg22581200_BC21 | chr15 | 60374376 | | | -0.189815 | 1.15E-08 | 6.09E-06 |
| cg15388804_TC21 | chr14 | 36523874 | | | 0.251467 | 1.15E-08 | 6.09E-06 |
| cg12672779_BC21 | chr5 | 143810819 | TSS1500 | HMHB1 | 0.243687 | 1.15E-08 | 6.09E-06 |
| cg22123285_TC21 | chr1 | 109269551 | exon_21 | CELSR2 | -0.311356 | 1.16E-08 | 6.09E-06 |
| cg05708566_BC21 | chr9 | 137187697 | exon_2;TSS | ANAPC2;SS | 0.185962 | 1.16E-08 | 6.10E-06 |
| cg02486750_TC21 | chr8 | 104618668 | | | -0.167438 | 1.16E-08 | 6.10E-06 |
| cg12906748_BC11 | chr11 | 8933029 | TSS200 | C11orf16 | -0.163105 | 1.16E-08 | 6.10E-06 |
| cg02927327_BC21 | chr6 | 35297982 | | | 0.263297 | 1.16E-08 | 6.10E-06 |
| cg12877860_BC21 | chr9 | 114504300 | exon_1;exo | WHRN;WH | -0.144263 | 1.16E-08 | 6.12E-06 |
| cg16932880_BC21 | chr2 | 64769007 | | | 0.152443 | 1.17E-08 | 6.13E-06 |
| cg05612205_TC21 | chr4 | 158201863 | exon_6;exo | GASK1B-AS | 0.166876 | 1.17E-08 | 6.13E-06 |
| cg13389669_BC21 | chr11 | 73403970 | 3UTR;exon | FAM168A;I | -0.183636 | 1.17E-08 | 6.13E-06 |
| cg23545793_BC21 | chr7 | 98329234 | | | -0.258447 | 1.17E-08 | 6.13E-06 |
| cg22536963_TC21 | chr1 | 40221564 | | | 0.28886 | 1.17E-08 | 6.13E-06 |
| cg02118194_TC21 | chr19 | 45901280 | exon_2 | MYPOP | -0.116798 | 1.17E-08 | 6.13E-06 |
| cg23787453_BC21 | chr8 | 126090581 | | | -0.296443 | 1.17E-08 | 6.14E-06 |
| cg17772680_BC21 | chr12 | 133076830 | | | 0.51428 | 1.18E-08 | 6.14E-06 |
| cg17628730_BC21 | chr8 | 37781178 | | | 0.151691 | 1.18E-08 | 6.16E-06 |
| cg25146415_BC21 | chr20 | 31833806 | exon_13;3U | MYLK2;MY | -0.155481 | 1.18E-08 | 6.16E-06 |
| cg13617225_TC21 | chr9 | 129494283 | | | -0.182259 | 1.18E-08 | 6.18E-06 |
| cg11110934_TC21 | chr6 | 161435345 | | | -0.373681 | 1.19E-08 | 6.19E-06 |
| cg10865262_TC21 | chr11 | 10398492 | | | -0.101813 | 1.19E-08 | 6.19E-06 |
| cg14406501_BC21 | chr17 | 80544380 | TSS1500;TS | RPTOR;RPT | -0.360499 | 1.19E-08 | 6.19E-06 |
| cg03515060_TC21 | chr21 | 45285912 | exon_2;exo | POFUT2;PO | -0.208161 | 1.19E-08 | 6.19E-06 |
| cg25839195_TC21 | chr5 | 4946770 | | | -0.230306 | 1.19E-08 | 6.20E-06 |
| cg03770575_BC21 | chr12 | 24261405 | | | -0.332931 | 1.19E-08 | 6.20E-06 |
| cg23351190_TC21 | chr4 | 7797974 | | | -0.329157 | 1.19E-08 | 6.21E-06 |
| cg04448477_TC21 | chr4 | 54228981 | TSS1500;TS | PDGFRA;PD | -0.203722 | 1.20E-08 | 6.22E-06 |
| cg02978894_TC21 | chr2 | 161623337 | TSS1500;TS | SLC4A10;SL | -0.406572 | 1.20E-08 | 6.22E-06 |
| cg14250330_BC21 | chr9 | 38734800 | | | -0.279465 | 1.20E-08 | 6.22E-06 |
| cg26003388_TC11 | chr17 | 78133452 | exon_6;TSS | TMC8;TMC | -0.154571 | 1.20E-08 | 6.22E-06 |
| cg08875198_BC11 | chr21 | 32277335 | | | -0.187717 | 1.20E-08 | 6.22E-06 |
| cg05508360_BC21 | chr19 | 34964202 | exon_1;5U | ZNF792;ZN | 0.287025 | 1.20E-08 | 6.23E-06 |
| cg10813469_BC21 | chr7 | 28062229 | | | -0.253559 | 1.21E-08 | 6.24E-06 |
| cg02694225_TC21 | chr2 | 21636471 | | | -0.276314 | 1.21E-08 | 6.25E-06 |
| cg21800290_BC21 | chr1 | 155200186 | | | -0.156107 | 1.21E-08 | 6.25E-06 |
| cg09958401_TC21 | chr5 | 58411703 | | | -0.240777 | 1.21E-08 | 6.25E-06 |
| cg02122105_BC21 | chr1 | 232743138 | TSS1500 | LINC01744 | -0.312165 | 1.22E-08 | 6.31E-06 |
| cg11714602_BC21 | chr5 | 132623571 | | | -0.289101 | 1.22E-08 | 6.31E-06 |
| cg02147681_BC11 | chr17 | 17700523 | | | -0.224257 | 1.23E-08 | 6.35E-06 |

| | | | | | | | |
|---|---|---|---|---|---|---|---|
| cg17844831_TC21 | chr17 | 17700832 | | | -0.235366 | 1.23E-08 | 6.35E-06 |
| cg11479256_TC21 | chr20 | 46244276 | | | -0.175465 | 1.23E-08 | 6.35E-06 |
| cg26281051_TC21 | chr20 | 227162 | TSS200 | DEFB129 | -0.306492 | 1.23E-08 | 6.36E-06 |
| cg17614731_BC21 | chr22 | 25068869 | TSS1500 | KIAA1671 | -0.138302 | 1.24E-08 | 6.36E-06 |
| cg17272224_BC21 | chr2 | 45010746 | TSS1500 | SIX2 | -0.148794 | 1.24E-08 | 6.36E-06 |
| cg20587213_BC21 | chr17 | 41522748 | | | -0.207595 | 1.24E-08 | 6.38E-06 |
| cg11664516_TC21 | chr15 | 62739361 | exon_30;e› | TLN2;TLN2 | -0.250705 | 1.24E-08 | 6.39E-06 |
| cg18158855_BC21 | chr20 | 46574466 | | | 0.662792 | 1.25E-08 | 6.39E-06 |
| cg05946089_TC21 | chr3 | 186722480 | exon_3;exo | KNG1;KNG | -0.178801 | 1.25E-08 | 6.40E-06 |
| cg18924706_TC21 | chr2 | 11671587 | TSS1500 | NTSR2 | -0.129193 | 1.25E-08 | 6.40E-06 |
| cg18188129_BC21 | chr12 | 119365540 | | | 0.208934 | 1.25E-08 | 6.40E-06 |
| cg16570210_TC21 | chr13 | 79389672 | | | 0.324012 | 1.25E-08 | 6.40E-06 |
| cg19781199_TC21 | chr18 | 58998848 | | | -0.297411 | 1.25E-08 | 6.40E-06 |
| cg04466022_TC11 | chr6 | 37650347 | exon_8 | MDGA1 | -0.283616 | 1.25E-08 | 6.40E-06 |
| cg13001732_TC21 | chr2 | 237257025 | | | 0.247255 | 1.25E-08 | 6.40E-06 |
| cg04770593_BC21 | chr2 | 55689658 | | | -0.275614 | 1.26E-08 | 6.42E-06 |
| cg07172248_TC21 | chr18 | 28189573 | | | 0.36537 | 1.26E-08 | 6.42E-06 |
| cg04314597_BC21 | chr14 | 92093471 | | | -0.244771 | 1.26E-08 | 6.42E-06 |
| cg18872311_TC21 | chr3 | 177531224 | | | -0.122033 | 1.26E-08 | 6.42E-06 |
| cg02347312_BC21 | chr13 | 113044292 | TSS1500;TS | MCF2L;MC | -0.236649 | 1.26E-08 | 6.42E-06 |
| cg10168286_TC21 | chr14 | 102738449 | | | -0.154495 | 1.26E-08 | 6.42E-06 |
| cg02049405_TC11 | chr6 | 30127488 | | | -0.246443 | 1.26E-08 | 6.42E-06 |
| cg18022921_TC21 | chr6 | 43769236 | TSS1500;TS | VEGFA;VEG | -0.130291 | 1.26E-08 | 6.42E-06 |
| cg09362240_TC21 | chr6 | 151147242 | | | 0.217495 | 1.26E-08 | 6.42E-06 |
| cg02400967_BC21 | chr2 | 71691124 | | | -0.250095 | 1.26E-08 | 6.42E-06 |
| cg12197690_BC21 | chr8 | 29671297 | | | 0.359291 | 1.26E-08 | 6.42E-06 |
| cg15344028_TC21 | chr2 | 203936787 | 5UTR;exon | ICOS;ICOS | -0.23826 | 1.27E-08 | 6.42E-06 |
| cg10875261_BC12 | chr2 | 95525606 | | | -0.213418 | 1.27E-08 | 6.42E-06 |
| cg04361498_BC21 | chr1 | 16430995 | | | -0.193346 | 1.27E-08 | 6.43E-06 |
| cg13869600_TC21 | chr10 | 43079694 | | | 0.176623 | 1.27E-08 | 6.43E-06 |
| cg16543461_BC21 | chr11 | 68912679 | | | -0.412368 | 1.28E-08 | 6.46E-06 |
| cg08057475_TC21 | chr15 | 45511143 | exon_1;exo | HMGN2P4( | -0.248359 | 1.28E-08 | 6.50E-06 |
| cg14717666_BC11 | chr4 | 118941837 | | | -0.211258 | 1.29E-08 | 6.51E-06 |
| cg13087191_TC21 | chr4 | 168183606 | | | -0.342928 | 1.29E-08 | 6.51E-06 |
| cg18432414_BC11 | chr17 | 8799168 | 5UTR;exon | MFSD6L;M | -0.281885 | 1.29E-08 | 6.52E-06 |
| cg01080888_TC21 | chr14 | 35480198 | | | -0.320265 | 1.29E-08 | 6.52E-06 |
| cg20791674_BC21 | chr8 | 141176394 | | | -0.302125 | 1.29E-08 | 6.52E-06 |
| cg25110341_TC21 | chr5 | 122500631 | | | -0.254569 | 1.29E-08 | 6.52E-06 |
| cg06824199_BC21 | chr1 | 46692137 | | | -0.2547 | 1.29E-08 | 6.52E-06 |
| cg01813978_TC21 | chr10 | 62236822 | | | -0.188098 | 1.30E-08 | 6.53E-06 |
| cg16233311_TC21 | chr2 | 174334531 | TSS1500 | SP9 | -0.171951 | 1.30E-08 | 6.53E-06 |
| cg19607123_BC21 | chr14 | 64946825 | 3UTR;exon | RAB15;RAE | 0.135255 | 1.30E-08 | 6.53E-06 |
| cg06754923_TC21 | chr5 | 157144034 | TSS1500;TS | MED7;MEI | -0.167968 | 1.30E-08 | 6.53E-06 |
| cg09400472_BC21 | chr1 | 36954373 | | | -0.180608 | 1.31E-08 | 6.55E-06 |
| cg07461772_TC11 | chr16 | 68448906 | TSS1500 | SMPD3 | -0.259366 | 1.31E-08 | 6.55E-06 |
| cg21217064_BC21 | chr11 | 44923758 | | | -0.110201 | 1.31E-08 | 6.55E-06 |
| cg10417176_BC21 | chr6 | 75752772 | | | -0.302949 | 1.31E-08 | 6.55E-06 |

| | | | | | | | |
|---|---|---|---|---|---|---|---|
| cg06642509_TC21 | chr5 | 1850829 | | | 0.33098 | 1.31E-08 | 6.55E-06 |
| cg04338863_BC21 | chr8 | 33813101 | | | -0.308406 | 1.31E-08 | 6.55E-06 |
| cg24382884_BC21 | chr9 | 133657079 | exon_11 | DBH | -0.142145 | 1.31E-08 | 6.55E-06 |
| cg13901857_TC21 | chr1 | 3485133 | | | -0.210349 | 1.31E-08 | 6.55E-06 |
| cg08940009_BC21 | chr21 | 46313055 | | | -0.317693 | 1.31E-08 | 6.55E-06 |
| cg13179592_TC21 | chr14 | 73287178 | exon_9;exo | NUMB;NUI | 0.316933 | 1.31E-08 | 6.55E-06 |
| cg15606779_BC21 | chr3 | 196198933 | | | -0.205414 | 1.31E-08 | 6.55E-06 |
| cg12682432_BC21 | chr5 | 2587802 | | | -0.311126 | 1.31E-08 | 6.55E-06 |
| cg26752179_BC21 | chr12 | 118649763 | | | -0.479502 | 1.31E-08 | 6.55E-06 |
| cg18902565_BC21 | chr22 | 49460989 | | | 0.261851 | 1.31E-08 | 6.55E-06 |
| cg06420033_BC21 | chr8 | 38106402 | 5UTR;exon | ASH2L;ASH | -0.337547 | 1.31E-08 | 6.55E-06 |
| cg22659094_TC21 | chr12 | 119444691 | | | -0.225543 | 1.32E-08 | 6.56E-06 |
| cg06864796_BC21 | chr17 | 49995469 | TSS1500 | DLX3 | -0.335305 | 1.32E-08 | 6.56E-06 |
| cg00936699_TC21 | chr22 | 20114307 | | | -0.167671 | 1.32E-08 | 6.56E-06 |
| cg24816286_BC21 | chr8 | 53841375 | | | -0.121576 | 1.32E-08 | 6.57E-06 |
| cg02156526_BC21 | chr6 | 24934041 | | | -0.136307 | 1.32E-08 | 6.57E-06 |
| cg11619277_TC21 | chr16 | 55752558 | | | -0.241321 | 1.32E-08 | 6.57E-06 |
| cg03008655_BC21 | chr7 | 158109513 | | | -0.1395 | 1.32E-08 | 6.57E-06 |
| cg04140971_TC11 | chr11 | 64149219 | | | -0.188219 | 1.32E-08 | 6.57E-06 |
| cg18809853_BC21 | chr13 | 61319408 | | | 0.2882 | 1.33E-08 | 6.58E-06 |
| cg22088096_TC21 | chr1 | 31577325 | exon_2;exo | TINAGL1;TI | -0.283649 | 1.33E-08 | 6.59E-06 |
| cg01052614_TC11 | chr8 | 142538222 | | | -0.100187 | 1.34E-08 | 6.62E-06 |
| cg01162866_BC21 | chr22 | 41845478 | | | -0.231164 | 1.34E-08 | 6.62E-06 |
| cg19747271_BC21 | chr6 | 28585613 | | | -0.195903 | 1.34E-08 | 6.63E-06 |
| cg02293358_TC21 | chr14 | 91888798 | | | 0.307659 | 1.34E-08 | 6.63E-06 |
| cg27185298_BC21 | chr6 | 30917428 | | | -0.18298 | 1.34E-08 | 6.63E-06 |
| cg24282430_BC21 | chr19 | 1049134 | | | 0.250016 | 1.35E-08 | 6.65E-06 |
| cg00766670_TC21 | chr14 | 93160749 | | | -0.147996 | 1.35E-08 | 6.67E-06 |
| cg24219095_BC21 | chr19 | 16414347 | | | -0.356564 | 1.36E-08 | 6.69E-06 |
| cg05204167_TC21 | chr4 | 3369341 | TSS1500;TS | RGS12;RGS | 0.247617 | 1.36E-08 | 6.69E-06 |
| cg04236827_BC21 | chr16 | 50740748 | TSS1500;TS | CYLD;CYLD | -0.146545 | 1.36E-08 | 6.71E-06 |
| cg03759077_BC21 | chr2 | 152719085 | 5UTR;exon | ARL6IP6;AI | 0.504655 | 1.36E-08 | 6.71E-06 |
| cg11502736_TC21 | chr1 | 179954710 | TSS200 | CEP350 | 0.235693 | 1.37E-08 | 6.73E-06 |
| cg23348497_TC21 | chr18 | 23885075 | | | 0.282763 | 1.37E-08 | 6.73E-06 |
| cg17047253_BC21 | chr10 | 98463209 | | | 0.2326 | 1.37E-08 | 6.76E-06 |
| cg04444703_BC21 | chr14 | 73276645 | exon_13;e> | NUMB;NUI | -0.24751 | 1.38E-08 | 6.76E-06 |
| cg26313301_TC21 | chr19 | 11108939 | | | 0.232423 | 1.38E-08 | 6.76E-06 |
| cg00521620_BC21 | chr6 | 29466639 | | | -0.421861 | 1.38E-08 | 6.77E-06 |
| cg18013874_BC21 | chr12 | 104358441 | | | -0.209211 | 1.38E-08 | 6.78E-06 |
| cg24047538_TC21 | chr7 | 29955300 | exon_3;exo | SCRN1;SCR | -0.34147 | 1.39E-08 | 6.79E-06 |
| cg19041421_TC21 | chr4 | 13657181 | | | -0.44116 | 1.39E-08 | 6.79E-06 |
| cg26485445_TC21 | chr22 | 46798869 | | | -0.203497 | 1.39E-08 | 6.79E-06 |
| cg18839354_BC21 | chr4 | 39457749 | TSS1500;TS | LIAS;LIAS;L | 0.182504 | 1.40E-08 | 6.84E-06 |
| cg15492379_BC21 | chr2 | 158735108 | TSS200 | PKP4-AS1 | -0.226677 | 1.40E-08 | 6.84E-06 |
| cg14649479_TC21 | chr8 | 24905803 | | | -0.244878 | 1.40E-08 | 6.84E-06 |
| cg13680579_TC21 | chr10 | 69833446 | | | -0.146815 | 1.40E-08 | 6.84E-06 |
| cg13851625_BC21 | chr7 | 26636881 | TSS1500;TS | LINC02860 | -0.358776 | 1.40E-08 | 6.84E-06 |

| | | | | | | | |
|---|---|---|---|---|---|---|---|
| cg14737671_BC21 | chr8 | 37775787 | | | -0.313784 | 1.40E-08 | 6.86E-06 |
| cg06398439_TC21 | chr4 | 28686632 | | | -0.171846 | 1.40E-08 | 6.86E-06 |
| cg03416228_TC21 | chr11 | 65592850 | 3UTR;exon | KCNK7;KCN | -0.270349 | 1.41E-08 | 6.86E-06 |
| cg02240722_BC21 | chr12 | 25885991 | | | -0.212465 | 1.41E-08 | 6.86E-06 |
| cg00579960_BC21 | chr21 | 31730280 | | | -0.204648 | 1.41E-08 | 6.86E-06 |
| cg09473396_BC21 | chr3 | 65146304 | | | -0.211573 | 1.41E-08 | 6.87E-06 |
| cg24262996_TC21 | chr5 | 171365551 | TSS1500 | SNORA70J | -0.151805 | 1.41E-08 | 6.88E-06 |
| cg26730543_TC21 | chr6 | 30071823 | | | -0.265678 | 1.42E-08 | 6.90E-06 |
| cg01440638_BC21 | chr22 | 20113416 | | | 0.24945 | 1.42E-08 | 6.92E-06 |
| cg10653520_BC21 | chr7 | 7113818 | | | -0.150943 | 1.43E-08 | 6.96E-06 |
| cg10507773_BC21 | chr7 | 534458 | | | 0.155946 | 1.43E-08 | 6.96E-06 |
| cg25475366_BC21 | chr8 | 143929702 | exon_24;e | PLEC;PLEC; | -0.184713 | 1.44E-08 | 6.96E-06 |
| cg26401906_TC11 | chr22 | 42616489 | | | -0.22173 | 1.44E-08 | 6.96E-06 |
| cg22027433_BC21 | chr17 | 64130689 | TSS1500 | ERN1 | -0.286392 | 1.44E-08 | 6.97E-06 |
| cg27477582_BC21 | chr10 | 10931542 | | | -0.426038 | 1.44E-08 | 6.98E-06 |
| cg01351177_BC11 | chr2 | 88691631 | TSS200 | RPIA | 0.205313 | 1.44E-08 | 6.99E-06 |
| cg18348862_BC21 | chr12 | 130212945 | | | -0.510636 | 1.44E-08 | 6.99E-06 |
| cg08865560_TC21 | chr7 | 142308489 | | | 0.20459 | 1.45E-08 | 6.99E-06 |
| cg21351992_TC21 | chr3 | 25577775 | | | -0.179612 | 1.45E-08 | 6.99E-06 |
| cg27332136_TC21 | chr20 | 57421473 | | | -0.250326 | 1.45E-08 | 6.99E-06 |
| cg24032246_BC21 | chr19 | 6829021 | | | -0.189494 | 1.45E-08 | 6.99E-06 |
| cg12693152_BC21 | chr18 | 36519069 | | | -0.194368 | 1.45E-08 | 6.99E-06 |
| cg23906001_BC21 | chr7 | 122319760 | 3UTR;exon | CADPS2;CA | -0.226551 | 1.45E-08 | 6.99E-06 |
| cg22200062_BC21 | chr14 | 74898725 | | | -0.420443 | 1.45E-08 | 6.99E-06 |
| cg04659190_BC21 | chr6 | 7446734 | | | -0.103722 | 1.46E-08 | 7.01E-06 |
| cg17520932_BC21 | chr7 | 120858576 | TSS1500 | TSPAN12 | -0.202948 | 1.46E-08 | 7.01E-06 |
| cg12291059_BC11 | chr18 | 22417961 | TSS200 | CTAGE1 | -0.263698 | 1.46E-08 | 7.01E-06 |
| cg02633838_BC21 | chr2 | 95121289 | | | -0.302129 | 1.46E-08 | 7.02E-06 |
| cg16796782_BC21 | chr10 | 126070762 | | | -0.225556 | 1.46E-08 | 7.02E-06 |
| cg22727668_BC21 | chr17 | 57243666 | | | -0.228787 | 1.46E-08 | 7.03E-06 |
| cg19294507_TC21 | chr11 | 68226184 | | | -0.141088 | 1.47E-08 | 7.06E-06 |
| cg09457671_BC21 | chr6 | 38614998 | | | -0.242208 | 1.47E-08 | 7.06E-06 |
| cg03623648_BC21 | chr2 | 12705530 | | | -0.297834 | 1.47E-08 | 7.07E-06 |
| cg05726002_BC21 | chr12 | 114903373 | | | 0.284417 | 1.47E-08 | 7.07E-06 |
| cg08028091_TC11 | chr10 | 43648751 | 5UTR;exon | ZNF32;ZNF | 0.419722 | 1.48E-08 | 7.09E-06 |
| cg07472806_BC21 | chr14 | 100966550 | TSS1500 | SNORD114 | -0.319437 | 1.48E-08 | 7.09E-06 |
| cg27310748_BC11 | chr19 | 3870880 | | | 0.294186 | 1.48E-08 | 7.10E-06 |
| cg14003265_BC21 | chr9 | 136902047 | | | -0.169023 | 1.49E-08 | 7.11E-06 |
| cg22976366_BC21 | chr17 | 32281348 | | | -0.190991 | 1.49E-08 | 7.11E-06 |
| cg06705595_TC21 | chr4 | 74375636 | | | -0.296308 | 1.49E-08 | 7.14E-06 |
| cg13173118_BC21 | chr11 | 115523458 | | | 0.21162 | 1.50E-08 | 7.15E-06 |
| cg13656701_BC21 | chr9 | 130915105 | | | -0.196562 | 1.50E-08 | 7.15E-06 |
| cg25429204_BC21 | chr19 | 42327756 | | | -0.300388 | 1.50E-08 | 7.16E-06 |
| cg19655486_BC21 | chr3 | 50268609 | TSS1500;TS | SEMA3B;SE | -0.262677 | 1.50E-08 | 7.17E-06 |
| cg00693946_BC21 | chr1 | 197360270 | | | -0.227855 | 1.51E-08 | 7.18E-06 |
| cg19354180_TC11 | chr14 | 91258204 | | | -0.173387 | 1.51E-08 | 7.18E-06 |
| cg15135704_BC11 | chr1 | 15876356 | exon_3 | SPEN | -0.214117 | 1.51E-08 | 7.19E-06 |

| | | | | | | | |
|---|---|---|---|---|---|---|---|
| cg09071040_TC21 | chr12 | 48964907 | | | 0.26926 | 1.51E-08 | 7.19E-06 |
| cg06545942_BC21 | chr2 | 127017267 | | | -0.211145 | 1.51E-08 | 7.19E-06 |
| cg22670630_TC21 | chr3 | 50313333 | TSS1500 | HYAL1 | -0.217199 | 1.51E-08 | 7.19E-06 |
| cg23470017_TC21 | chr11 | 128828452 | | | -0.434978 | 1.52E-08 | 7.19E-06 |
| cg20285483_TO11 | chr15 | 40872026 | | | 0.182499 | 1.52E-08 | 7.19E-06 |
| cg25674613_BC11 | chr8 | 1325642 | | | -0.21335 | 1.53E-08 | 7.24E-06 |
| cg09413498_TC21 | chr8 | 57261042 | TSS200 | LOC28617 | -0.226274 | 1.53E-08 | 7.25E-06 |
| cg14262766_BC21 | chr9 | 126616766 | | | 0.347138 | 1.53E-08 | 7.25E-06 |
| cg20718736_TC21 | chr11 | 8250714 | | | -0.225665 | 1.53E-08 | 7.25E-06 |
| cg16212902_BC21 | chr11 | 36745457 | | | 0.321011 | 1.53E-08 | 7.26E-06 |
| cg22369918_TC21 | chr17 | 28427350 | | | -0.122361 | 1.54E-08 | 7.28E-06 |
| cg16562689_BC21 | chr11 | 69936966 | | | 0.202747 | 1.54E-08 | 7.28E-06 |
| cg03954295_TC11 | chr19 | 34172265 | TSS1500;TS | LSM14A;LS | -0.45272 | 1.55E-08 | 7.31E-06 |
| cg03245326_BC21 | chr7 | 142835229 | | | -0.803432 | 1.55E-08 | 7.31E-06 |
| cg23965429_BC11 | chr14 | 99274661 | | | 0.116664 | 1.55E-08 | 7.33E-06 |
| cg26898121_BC21 | chr6 | 123741865 | | | -0.289594 | 1.55E-08 | 7.33E-06 |
| cg04933503_TC21 | chr3 | 53108958 | | | -0.447913 | 1.55E-08 | 7.33E-06 |
| cg11869406_BC21 | chr7 | 148717561 | | | -0.283567 | 1.56E-08 | 7.33E-06 |
| cg00193695_BC21 | chr18 | 78426879 | | | -0.145375 | 1.56E-08 | 7.33E-06 |
| cg05754819_TC21 | chr10 | 77638186 | TSS1500;TS | KCNMA1;K | -0.27665 | 1.56E-08 | 7.33E-06 |
| cg10011655_BC21 | chr17 | 76354885 | TSS1500;TS | PRPSAP1;P | -0.199559 | 1.56E-08 | 7.34E-06 |
| cg04229851_TC21 | chr4 | 7433027 | 3UTR;exon | PSAPL1;PS | -0.27486 | 1.56E-08 | 7.34E-06 |
| cg17012076_BC21 | chr11 | 122851760 | exon_3 | CRTAM | -0.140905 | 1.56E-08 | 7.34E-06 |
| cg17726655_BC21 | chr19 | 42005708 | | | -0.245888 | 1.57E-08 | 7.35E-06 |
| cg02611502_TC21 | chr2 | 29106895 | | | -0.359935 | 1.57E-08 | 7.35E-06 |
| cg16223109_BC11 | chr11 | 38648623 | TSS1500 | LINC01493 | -0.209998 | 1.57E-08 | 7.35E-06 |
| cg19111712_TC11 | chr11 | 1279835 | | | -0.151408 | 1.57E-08 | 7.36E-06 |
| cg13396410_BC21 | chr9 | 8376274 | | | -0.283586 | 1.57E-08 | 7.36E-06 |
| cg20910408_BC11 | chr7 | 44761990 | | | -0.200762 | 1.57E-08 | 7.37E-06 |
| cg13544811_BC21 | chr3 | 141387587 | 5UTR;exon | ZBTB38;ZB | -0.369878 | 1.58E-08 | 7.39E-06 |
| cg15377933_TC21 | chr17 | 79986208 | | | -0.335236 | 1.58E-08 | 7.39E-06 |
| cg04094885_BC21 | chr17 | 39867577 | TSS1500;TS | ZPBP2;ZPB | -0.271368 | 1.58E-08 | 7.39E-06 |
| cg01337514_TC21 | chr1 | 6450966 | | | -0.142316 | 1.59E-08 | 7.42E-06 |
| cg04863568_TC11 | chr16 | 28925181 | exon_1;5U | RABEP2;RA | 0.328641 | 1.59E-08 | 7.43E-06 |
| cg08058681_TC21 | chr5 | 64632542 | | | -0.11238 | 1.59E-08 | 7.45E-06 |
| cg18839315_TC21 | chr12 | 15176053 | | | -0.135209 | 1.60E-08 | 7.45E-06 |
| cg07573461_BC11 | chr1 | 16725160 | | | -0.18984 | 1.60E-08 | 7.47E-06 |
| cg24728778_BC21 | chr19 | 50938204 | | | -0.390986 | 1.61E-08 | 7.50E-06 |
| cg02534744_TC21 | chr19 | 863226 | exon_5;exc | CFD;CFD | -0.359914 | 1.61E-08 | 7.51E-06 |
| cg10267181_BC21 | chr11 | 8312275 | | | 0.205121 | 1.61E-08 | 7.52E-06 |
| cg17779400_TC21 | chr19 | 34484476 | | | -0.194372 | 1.61E-08 | 7.52E-06 |
| cg16389209_BC21 | chr10 | 62049362 | 5UTR;exon | ARID5B;AR | -0.186593 | 1.62E-08 | 7.52E-06 |
| cg14622830_BC21 | chr10 | 7301330 | | | -0.232354 | 1.62E-08 | 7.52E-06 |
| cg08097657_TC21 | chr3 | 50276565 | exon_18;e | SEMA3B;SE | -0.175976 | 1.62E-08 | 7.53E-06 |
| cg05558062_BC21 | chr3 | 135627426 | | | 0.265254 | 1.62E-08 | 7.53E-06 |
| cg23919660_BC21 | chr6 | 164108219 | | | -0.295358 | 1.63E-08 | 7.57E-06 |
| cg23506322_TC21 | chr2 | 185738590 | exon_1;exc | FSIP2-AS2; | -0.227858 | 1.63E-08 | 7.59E-06 |

| | | | | | | | |
|---|---|---|---|---|---|---|---|
| cg06529080_TC21 | chr7 | 101943481 | | | -0.580564 | 1.64E-08 | 7.60E-06 |
| cg23999833_BC21 | chr15 | 100772699 | | | -0.20906 | 1.65E-08 | 7.64E-06 |
| cg24827234_TC21 | chr11 | 115661842 | | | -0.174714 | 1.65E-08 | 7.64E-06 |
| cg02366262_BC21 | chr11 | 121427007 | | | -0.134479 | 1.65E-08 | 7.64E-06 |
| cg00561562_BC11 | chr4 | 143514585 | exon_1 | SMARCA5-/ | 0.13829 | 1.65E-08 | 7.64E-06 |
| cg24275466_TC21 | chr14 | 90812409 | | | -0.185176 | 1.65E-08 | 7.64E-06 |
| cg22006449_TC21 | chr11 | 111748188 | | | 0.307779 | 1.65E-08 | 7.64E-06 |
| cg13258599_BC21 | chr7 | 151392675 | | | -0.185035 | 1.66E-08 | 7.65E-06 |
| cg08173263_TC21 | chr19 | 14166099 | | | 0.242493 | 1.66E-08 | 7.66E-06 |
| cg20434511_BC21 | chr2 | 239319280 | | | -0.569577 | 1.66E-08 | 7.67E-06 |
| cg27150596_BC21 | chr22 | 32853020 | | | 0.284302 | 1.67E-08 | 7.69E-06 |
| cg21623186_BC21 | chr14 | 104517038 | | | -0.451476 | 1.67E-08 | 7.69E-06 |
| cg03340119_BC11 | chr1 | 246789206 | TSS1500 | LINC01341 | 0.332437 | 1.67E-08 | 7.69E-06 |
| cg05056583_BC21 | chr3 | 124318078 | | | 0.184466 | 1.67E-08 | 7.69E-06 |
| cg09684845_BC21 | chr13 | 51922665 | | | -0.126922 | 1.67E-08 | 7.70E-06 |
| cg22027689_BC21 | chr3 | 42653651 | TSS200 | ZBTB47 | -0.135601 | 1.68E-08 | 7.71E-06 |
| cg10741568_BC21 | chr13 | 48972001 | | | -0.226603 | 1.68E-08 | 7.72E-06 |
| cg02917889_TC21 | chr2 | 131034164 | TSS1500;TS | SMIM39;AI | -0.127063 | 1.68E-08 | 7.72E-06 |
| cg23995249_BC21 | chr3 | 132721556 | TSS200;TSS | NPHP3-AS1 | -0.156077 | 1.68E-08 | 7.72E-06 |
| cg19999342_BC21 | chr4 | 8066491 | | | 0.311107 | 1.68E-08 | 7.72E-06 |
| cg05728902_TC21 | chr12 | 57455246 | TSS200 | INHBE | -0.116639 | 1.69E-08 | 7.74E-06 |
| cg01327552_TC11 | chr6 | 166862321 | exon_1;5U | RPS6KA2;R | -0.30871 | 1.69E-08 | 7.74E-06 |
| cg02440468_BC21 | chr2 | 11544767 | | | -0.158894 | 1.69E-08 | 7.75E-06 |
| cg18210562_TC21 | chr8 | 22063334 | | | -0.136865 | 1.69E-08 | 7.75E-06 |
| cg05871806_TC21 | chr14 | 75699197 | exon_7 | TTLL5 | 0.396004 | 1.70E-08 | 7.77E-06 |
| cg22979448_BC21 | chr17 | 76715650 | 3UTR;exon | JMJD6;JMJ | 0.156887 | 1.70E-08 | 7.77E-06 |
| cg23907108_BC21 | chr11 | 64638521 | | | -0.219661 | 1.70E-08 | 7.77E-06 |
| cg27615154_TC21 | chr7 | 30770017 | TSS1500 | MINDY4 | -0.217738 | 1.70E-08 | 7.77E-06 |
| cg12513231_BC21 | chr17 | 28222321 | | | -0.194433 | 1.70E-08 | 7.77E-06 |
| cg23516370_TC21 | chr7 | 52120136 | | | -0.360114 | 1.70E-08 | 7.77E-06 |
| cg05636964_TC21 | chr11 | 59648800 | | | -0.308232 | 1.70E-08 | 7.78E-06 |
| cg23733975_BC21 | chr18 | 74248934 | | | 0.293503 | 1.71E-08 | 7.79E-06 |
| cg17287129_TC21 | chr12 | 11543960 | | | 0.185388 | 1.71E-08 | 7.79E-06 |
| cg03565777_TC11 | chr12 | 124543793 | | | 0.366458 | 1.71E-08 | 7.79E-06 |
| cg23580000_BC21 | chr16 | 50288245 | exon_2;exo | ADCY7;AD( | 0.15132 | 1.71E-08 | 7.79E-06 |
| cg07097098_BC21 | chr19 | 55210270 | TSS1500;TS | PTPRH;PTP | -0.200063 | 1.72E-08 | 7.83E-06 |
| cg14629755_TC21 | chr17 | 4786598 | TSS1500;TS | VMO1;VM( | -0.349402 | 1.72E-08 | 7.85E-06 |
| cg14045264_TC21 | chr2 | 207238672 | TSS1500 | MYOSLID | 0.189116 | 1.73E-08 | 7.88E-06 |
| cg00672093_BC21 | chr2 | 138897274 | TSS200 | YY1P2 | -0.210881 | 1.74E-08 | 7.89E-06 |
| cg27317004_BC21 | chr1 | 156928003 | exon_9 | LRRC71 | 0.208883 | 1.74E-08 | 7.89E-06 |
| cg23011561_TC21 | chr2 | 205273790 | | | 0.383472 | 1.74E-08 | 7.89E-06 |
| cg11788260_BC21 | chr1 | 223756884 | | | -0.211467 | 1.74E-08 | 7.90E-06 |
| cg18452347_TC21 | chr8 | 226533 | | | -0.349252 | 1.74E-08 | 7.91E-06 |
| cg12688068_BC21 | chr4 | 58565219 | | | -0.136857 | 1.74E-08 | 7.91E-06 |
| cg14405013_TC21 | chr13 | 32525633 | | | -0.136018 | 1.74E-08 | 7.91E-06 |
| cg09810593_BC21 | chr19 | 48740869 | 3UTR;exon | IZUMO1;IZ | -0.228999 | 1.75E-08 | 7.91E-06 |
| cg06200996_BC21 | chr12 | 129772361 | | | -0.464329 | 1.75E-08 | 7.93E-06 |

| | | | | | | | |
|---|---|---|---|---|---|---|---|
| cg07742814_BC21 | chr11 | 487269 | | | -0.27733 | 1.75E-08 | 7.93E-06 |
| cg24276305_BC21 | chr19 | 18856134 | | | -0.166038 | 1.76E-08 | 7.94E-06 |
| cg26718385_TC21 | chr1 | 163470030 | | | -0.351446 | 1.76E-08 | 7.94E-06 |
| cg04972856_TC21 | chr6 | 87322333 | TSS1500;TS | SMIM8;SM | -0.264762 | 1.76E-08 | 7.94E-06 |
| cg14517589_BC21 | chr7 | 1438090 | | | -0.193867 | 1.76E-08 | 7.96E-06 |
| cg02798703_TC21 | chr9 | 108855192 | exon_1 | ACTL7B | -0.215225 | 1.76E-08 | 7.97E-06 |
| cg16595288_TC21 | chr10 | 43203483 | | | -0.224695 | 1.77E-08 | 7.99E-06 |
| cg04590610_BC21 | chr19 | 43799706 | exon_2;exo | LYPD5;LYPI | -0.287325 | 1.77E-08 | 7.99E-06 |
| cg01979157_BC11 | chr1 | 2229574 | exon_1 | SKI | -0.20438 | 1.77E-08 | 7.99E-06 |
| cg10221624_TC21 | chr2 | 159922704 | | | -0.144512 | 1.77E-08 | 7.99E-06 |
| cg12669659_BC21 | chr11 | 59706233 | | | -0.182702 | 1.77E-08 | 7.99E-06 |
| cg15008743_BC21 | chr19 | 52638853 | TSS1500;TS | ZNF83;ZNF | -0.707123 | 1.78E-08 | 8.01E-06 |
| cg10106694_BC21 | chr6 | 132503497 | exon_2;exo | STX7;STX7; | 0.293405 | 1.78E-08 | 8.01E-06 |
| cg00710738_BC21 | chr11 | 90488600 | | | 0.180803 | 1.78E-08 | 8.02E-06 |
| cg24273709_TC21 | chr12 | 55914706 | | | 0.237734 | 1.78E-08 | 8.02E-06 |
| cg18403454_TC21 | chr6 | 162177865 | | | -0.207724 | 1.79E-08 | 8.02E-06 |
| cg02931873_TC21 | chr9 | 110427704 | exon_36 | SVEP1 | -0.296178 | 1.79E-08 | 8.02E-06 |
| cg05092777_TC21 | chr7 | 131683889 | | | -0.139673 | 1.79E-08 | 8.03E-06 |
| cg19549295_BC21 | chr7 | 4611443 | | | 0.266703 | 1.80E-08 | 8.09E-06 |
| cg06629999_BC21 | chr22 | 49324267 | | | -0.214268 | 1.81E-08 | 8.10E-06 |
| cg22326588_BC11 | chr12 | 111766800 | TSS200;TSS | ALDH2;ALD | 0.327194 | 1.81E-08 | 8.10E-06 |
| cg08941317_TC21 | chr11 | 83192826 | TSS1500;TS | ANKRD42;A | 0.206178 | 1.81E-08 | 8.10E-06 |
| cg05297931_BC21 | chr6 | 13429609 | | | -0.239973 | 1.82E-08 | 8.15E-06 |
| cg22970086_TC21 | chr17 | 76277269 | exon_16;ex | QRICH2;QR | -0.15217 | 1.83E-08 | 8.19E-06 |
| cg06458732_TC11 | chr2 | 26345596 | TSS1500;TS | SELENOI;SE | -0.396264 | 1.84E-08 | 8.23E-06 |
| cg03684245_BC21 | chr6 | 1228715 | | | -0.211621 | 1.84E-08 | 8.23E-06 |
| cg16323491_BC11 | chr10 | 72272972 | TSS1500 | DDIT4 | 0.212304 | 1.84E-08 | 8.23E-06 |
| cg08999334_TC21 | chr5 | 100698523 | | | -0.343139 | 1.84E-08 | 8.23E-06 |
| cg26122076_BC21 | chr1 | 19762415 | | | -0.172091 | 1.85E-08 | 8.24E-06 |
| cg04802442_TC21 | chr14 | 89367232 | | | -0.124643 | 1.85E-08 | 8.24E-06 |
| cg16388639_TC21 | chr9 | 96668199 | | | -0.26179 | 1.85E-08 | 8.24E-06 |
| cg21764456_BC21 | chr16 | 10683220 | | | -0.606148 | 1.85E-08 | 8.26E-06 |
| cg13539115_TC11 | chr3 | 52278844 | TSS200 | WDR82 | 0.216196 | 1.86E-08 | 8.28E-06 |
| cg12924799_TC21 | chr8 | 101757129 | | | -0.226334 | 1.86E-08 | 8.28E-06 |
| cg07481277_BC21 | chr4 | 184984186 | | | -0.273836 | 1.86E-08 | 8.28E-06 |
| cg03174507_TC21 | chr10 | 21500653 | | | -0.153006 | 1.86E-08 | 8.28E-06 |
| cg23613051_TC21 | chr4 | 2818701 | TSS200 | SH3BP2 | -0.661037 | 1.86E-08 | 8.28E-06 |
| cg16688330_BC21 | chr9 | 82882491 | | | -0.228635 | 1.87E-08 | 8.28E-06 |
| cg26824721_TC21 | chr8 | 8665215 | | | -0.311748 | 1.87E-08 | 8.29E-06 |
| cg27000690_BC21 | chr6 | 34469450 | | | -0.158679 | 1.87E-08 | 8.29E-06 |
| cg03173936_TC21 | chr2 | 106994766 | | | -0.225371 | 1.87E-08 | 8.30E-06 |
| cg05152382_TC21 | chr3 | 79247333 | | | 0.213386 | 1.88E-08 | 8.32E-06 |
| cg07517370_BC21 | chr10 | 122162991 | TSS1500;TS | TACC2;TAC | -0.213603 | 1.88E-08 | 8.32E-06 |
| cg10790672_BC11 | chr5 | 79088642 | exon_7;3U | BHMT2;BH | 0.137265 | 1.88E-08 | 8.34E-06 |
| cg12598007_BC21 | chr11 | 68839601 | | | 0.231653 | 1.89E-08 | 8.35E-06 |
| cg04030848_TC21 | chr22 | 30564894 | 5UTR;exon | GAL3ST1;G | -0.167616 | 1.89E-08 | 8.35E-06 |
| cg18274065_BC21 | chr18 | 74292598 | TSS1500;TS | CYB5A;CYB | -0.212411 | 1.89E-08 | 8.35E-06 |

| | | | | | | | |
|---|---|---|---|---|---|---|---|
| cg06600697_TC21 | chr9 | 32782403 | TSS1500 | TMEM215 | -0.298289 | 1.89E-08 | 8.35E-06 |
| cg25130977_TC21 | chr9 | 2858119 | | | -0.381944 | 1.89E-08 | 8.35E-06 |
| cg15819551_BC21 | chr14 | 75935360 | | | 0.182305 | 1.89E-08 | 8.35E-06 |
| cg17747948_TC21 | chr11 | 73665312 | | | -0.117671 | 1.90E-08 | 8.37E-06 |
| cg09152354_BC21 | chr6 | 10675294 | | | -0.305383 | 1.90E-08 | 8.37E-06 |
| cg08830588_BC21 | chr12 | 49545757 | | | -0.12413 | 1.90E-08 | 8.37E-06 |
| cg26558849_BC21 | chr22 | 50361338 | | | 0.205 | 1.90E-08 | 8.37E-06 |
| cg20911446_TC21 | chr22 | 38290143 | | | -0.137104 | 1.90E-08 | 8.37E-06 |
| cg06470438_BC21 | chr2 | 238922671 | | | -0.166743 | 1.90E-08 | 8.38E-06 |
| cg07291065_BC21 | chr1 | 235842609 | | | 0.547682 | 1.91E-08 | 8.38E-06 |
| cg26712080_TC11 | chr3 | 55485560 | TSS1500;TS | WNT5A;WI | -0.278857 | 1.91E-08 | 8.38E-06 |
| cg06221509_TC21 | chr20 | 47983341 | exon_1 | LINC01523 | -0.249978 | 1.91E-08 | 8.39E-06 |
| cg20709417_TC21 | chr13 | 52616506 | TSS1500;TS | HNRNPA1L | 0.200241 | 1.91E-08 | 8.39E-06 |
| cg22202837_BC21 | chr12 | 110500464 | | | -0.187799 | 1.91E-08 | 8.39E-06 |
| cg08273518_BC21 | chr5 | 82310071 | | | 0.201241 | 1.91E-08 | 8.39E-06 |
| cg17874490_BC21 | chr17 | 41500882 | | | -0.207923 | 1.91E-08 | 8.39E-06 |
| cg03383322_BC21 | chr3 | 123375803 | | | -0.129869 | 1.92E-08 | 8.39E-06 |
| cg06427395_TC21 | chr12 | 44476658 | | | -0.169583 | 1.92E-08 | 8.39E-06 |
| cg08663213_TC21 | chr17 | 29084579 | | | 0.391336 | 1.92E-08 | 8.42E-06 |
| cg19689427_TC21 | chr5 | 141339441 | exon_1;exo | PCDHGA2;I | -0.335256 | 1.93E-08 | 8.43E-06 |
| cg04566608_BC21 | chr19 | 15194627 | | | -0.179644 | 1.93E-08 | 8.44E-06 |
| cg26702212_BC21 | chr1 | 1069790 | TSS1500;TS | LOC10537 | -0.151821 | 1.93E-08 | 8.44E-06 |
| cg24420089_BC11 | chr11 | 457304 | | | 0.104129 | 1.93E-08 | 8.44E-06 |
| cg24229553_TC21 | chr8 | 140295146 | | | 0.211641 | 1.93E-08 | 8.44E-06 |
| cg21116900_BC21 | chr12 | 100356982 | TSS200;TSS | SLC17A8;SI | -0.204292 | 1.94E-08 | 8.47E-06 |
| cg23005321_TC21 | chr12 | 96401778 | TSS1500;TS | CDK17;CDK | -0.17224 | 1.94E-08 | 8.48E-06 |
| cg20907483_BC21 | chr15 | 101105759 | | | 0.247532 | 1.95E-08 | 8.48E-06 |
| cg02849754_TC21 | chr20 | 41999337 | | | 0.569191 | 1.95E-08 | 8.48E-06 |
| cg00478710_BC21 | chr3 | 133674121 | | | -0.238457 | 1.95E-08 | 8.48E-06 |
| cg24010825_BC21 | chr3 | 89257335 | | | -0.201358 | 1.96E-08 | 8.52E-06 |
| cg25981934_BC21 | chr17 | 4396393 | | | -0.147258 | 1.96E-08 | 8.52E-06 |
| cg13215559_BC21 | chr8 | 140361445 | | | 0.359447 | 1.96E-08 | 8.53E-06 |
| cg19944546_TC21 | chr14 | 99575442 | | | -0.205151 | 1.96E-08 | 8.53E-06 |
| cg26590607_BC21 | chr15 | 72155656 | | | -0.261598 | 1.98E-08 | 8.58E-06 |
| cg03654668_TC21 | chr10 | 14509050 | | | -0.261093 | 1.98E-08 | 8.59E-06 |
| cg16251237_BC21 | chr3 | 13260248 | | | 0.175156 | 1.98E-08 | 8.59E-06 |
| cg02135112_TC21 | chr2 | 170409028 | | | -0.332686 | 1.98E-08 | 8.59E-06 |
| cg22002205_TC21 | chr10 | 126216622 | | | -0.195905 | 1.98E-08 | 8.59E-06 |
| cg05163268_TC21 | chr5 | 180689385 | | | -0.124253 | 1.99E-08 | 8.64E-06 |
| cg11579758_TC11 | chr4 | 1092849 | | | -0.142612 | 2.00E-08 | 8.65E-06 |
| cg01416521_TC11 | chr19 | 55536265 | exon_2 | SBK2 | -0.146247 | 2.00E-08 | 8.65E-06 |
| cg14798919_TC11 | chr19 | 38849991 | TSS200;TSS | HNRNPL;HI | -0.385401 | 2.00E-08 | 8.65E-06 |
| cg00711383_TC21 | chr11 | 65626753 | | | -0.192007 | 2.00E-08 | 8.66E-06 |
| cg26089653_BC21 | chr22 | 21012110 | | | -0.172914 | 2.00E-08 | 8.66E-06 |
| cg22512861_TC21 | chr5 | 133998603 | | | -0.274667 | 2.01E-08 | 8.67E-06 |
| cg26507629_BC21 | chr16 | 49598992 | | | -0.251848 | 2.01E-08 | 8.67E-06 |
| cg02310457_BC21 | chr9 | 22682694 | exon_3 | LINC01239 | -0.318673 | 2.01E-08 | 8.69E-06 |

| | | | | | | | |
|---|---|---|---|---|---|---|---|
| cg07597598_BC21 | chr2 | 190699441 | | | -0.267694 | 2.02E-08 | 8.70E-06 |
| cg13135455_BC21 | chr2 | 240920901 | | | -0.173478 | 2.02E-08 | 8.71E-06 |
| cg06243914_TC21 | chr2 | 120476597 | | | -0.268214 | 2.02E-08 | 8.71E-06 |
| cg12841600_BC21 | chr6 | 30552502 | exon_8 | GNL1 | 0.153136 | 2.03E-08 | 8.74E-06 |
| cg15035716_TC21 | chr1 | 234742786 | | | 0.27165 | 2.03E-08 | 8.74E-06 |
| cg01905690_TC21 | chr14 | 56057670 | | | -0.353962 | 2.03E-08 | 8.74E-06 |
| cg25266899_BC21 | chr2 | 218431027 | | | -0.242582 | 2.03E-08 | 8.75E-06 |
| cg14836839_BC21 | chr17 | 44628716 | | | -0.42003 | 2.04E-08 | 8.77E-06 |
| cg24198381_TC11 | chr10 | 2266757 | | | -0.223608 | 2.04E-08 | 8.77E-06 |
| cg02102141_BC22 | chr1 | 230849455 | | | -0.188819 | 2.04E-08 | 8.78E-06 |
| cg01391052_BC21 | chr1 | 150233695 | | | 0.249614 | 2.05E-08 | 8.79E-06 |
| cg03050188_BC21 | chr2 | 11391480 | | | -0.16045 | 2.05E-08 | 8.79E-06 |
| cg02643214_BC21 | chr16 | 86275902 | | | -0.408214 | 2.05E-08 | 8.80E-06 |
| cg20936443_TC21 | chr3 | 158799673 | | | -0.324335 | 2.05E-08 | 8.80E-06 |
| cg25988219_TC21 | chr20 | 3044000 | | | -0.157629 | 2.05E-08 | 8.80E-06 |
| cg18242400_BC21 | chr4 | 61518465 | | | -0.297888 | 2.07E-08 | 8.85E-06 |
| cg12125104_BC21 | chr2 | 98878631 | | | -0.334752 | 2.07E-08 | 8.85E-06 |
| cg18157298_TC21 | chr9 | 137455253 | exon_6;exo | NSMF;NSM | -0.135794 | 2.07E-08 | 8.86E-06 |
| cg17948453_BC21 | chr20 | 36378136 | | | -0.221766 | 2.07E-08 | 8.86E-06 |
| cg24985874_BC21 | chr21 | 23386037 | TSS1500 | D21S2088E | -0.1862 | 2.07E-08 | 8.86E-06 |
| cg19154669_BC21 | chr13 | 110139881 | | | -0.438582 | 2.08E-08 | 8.90E-06 |
| cg03610970_TC21 | chr12 | 6619852 | 3UTR;exon_ | LPAR5;LPA | -0.351438 | 2.08E-08 | 8.90E-06 |
| cg00499277_BC21 | chr14 | 23408307 | TSS200 | MYH6 | -0.148525 | 2.09E-08 | 8.90E-06 |
| cg14014506_BC21 | chr9 | 114099124 | exon_2;TSS | KIF12;KIF1 | -0.232634 | 2.09E-08 | 8.91E-06 |
| cg03422137_BC21 | chr15 | 93982452 | | | -0.279225 | 2.09E-08 | 8.91E-06 |
| cg08644421_BC21 | chr5 | 142587485 | | | -0.27126 | 2.10E-08 | 8.94E-06 |
| cg23202887_TC21 | chr14 | 99225035 | | | -0.11749 | 2.10E-08 | 8.94E-06 |
| cg15960579_BC21 | chr16 | 77971049 | | | 0.25497 | 2.10E-08 | 8.96E-06 |
| cg08744877_BC21 | chr17 | 59848439 | | | -0.351306 | 2.11E-08 | 8.96E-06 |
| cg01147550_TC11 | chr17 | 49132665 | TSS200;TSS | B4GALNT2; | -0.316989 | 2.11E-08 | 8.98E-06 |
| cg03753813_BC21 | chr1 | 115545880 | | | -0.213906 | 2.11E-08 | 8.98E-06 |
| cg15826485_TC21 | chr14 | 21384296 | TSS1500 | SUPT16H | -0.275069 | 2.11E-08 | 8.98E-06 |
| cg25066165_TC21 | chr10 | 80529071 | | | 0.165912 | 2.11E-08 | 8.98E-06 |
| cg25097225_BC21 | chr20 | 25282552 | | | -0.619163 | 2.12E-08 | 8.98E-06 |
| cg03774025_TC11 | chr9 | 96383802 | TSS200;TSS | SLC35D2;SI | 0.20834 | 2.12E-08 | 8.99E-06 |
| cg16080512_TC21 | chr19 | 42076389 | | | -0.336595 | 2.13E-08 | 9.02E-06 |
| cg03783039_BC21 | chr4 | 183480567 | | | 0.226089 | 2.13E-08 | 9.02E-06 |
| cg14304756_BC11 | chr9 | 128883700 | TSS1500;TS | KYAT1;KYA | -0.174214 | 2.13E-08 | 9.03E-06 |
| cg11353357_TC21 | chr21 | 36060618 | TSS200;TSS | SETD4;SETI | -0.441815 | 2.14E-08 | 9.05E-06 |
| cg15380263_BC21 | chr5 | 133612824 | TSS1500 | FSTL4 | 0.301514 | 2.14E-08 | 9.05E-06 |
| cg18381315_BC21 | chr4 | 139656887 | | | -0.311675 | 2.14E-08 | 9.06E-06 |
| cg20712742_BC21 | chr11 | 57667733 | TSS1500 | ZDHHC5 | 0.197945 | 2.14E-08 | 9.06E-06 |
| cg10959672_BC21 | chr1 | 35506526 | | | -0.137501 | 2.14E-08 | 9.06E-06 |
| cg25642126_BC21 | chr3 | 36856914 | exon_13;ex | TRANK1;TR | -0.323797 | 2.15E-08 | 9.07E-06 |
| cg20451043_BC21 | chr15 | 58979728 | | | -0.129261 | 2.15E-08 | 9.08E-06 |
| cg19523145_BC21 | chr2 | 210955886 | | | -0.376444 | 2.16E-08 | 9.10E-06 |
| cg16166442_BC21 | chr20 | 2868226 | | | 0.322822 | 2.16E-08 | 9.10E-06 |

| | | | | | | | |
|---|---|---|---|---|---|---|---|
| cg19971915_TC21 | chr14 | 101183596 | | | -0.213891 | 2.16E-08 | 9.11E-06 |
| cg12233314_BC21 | chr8 | 16914647 | | | -0.27424 | 2.16E-08 | 9.11E-06 |
| cg06837552_TC22 | chr11 | 68859277 | | | 0.336825 | 2.16E-08 | 9.11E-06 |
| cg02749623_TC21 | chr21 | 44305236 | | | -0.15719 | 2.16E-08 | 9.11E-06 |
| cg09538263_BC21 | chr12 | 7618494 | | | 0.242841 | 2.16E-08 | 9.11E-06 |
| cg13273372_BC21 | chr8 | 143269437 | exon_2 | GLI4 | -0.174343 | 2.17E-08 | 9.11E-06 |
| cg07299810_TC21 | chr7 | 30465653 | | | 0.265259 | 2.18E-08 | 9.15E-06 |
| cg26065792_TC11 | chr5 | 4023638 | | | -0.219844 | 2.18E-08 | 9.17E-06 |
| cg17768691_BC21 | chr12 | 14568014 | TSS200 | PLBD1 | -0.141933 | 2.18E-08 | 9.18E-06 |
| cg15614897_BC21 | chr10 | 115657254 | | | -0.30224 | 2.19E-08 | 9.19E-06 |
| cg27041868_BC21 | chr7 | 5501935 | exon_3;exo | FBXL18;FB) | -0.243352 | 2.19E-08 | 9.19E-06 |
| cg08889585_TC11 | chr1 | 3857258 | TSS1500;TS | DFFB;DFFB | -0.354636 | 2.19E-08 | 9.19E-06 |
| cg22230482_BC21 | chr17 | 15399392 | | | -0.199371 | 2.19E-08 | 9.19E-06 |
| cg26988432_TC21 | chr17 | 48100634 | | | -0.097712 | 2.20E-08 | 9.22E-06 |
| cg06891424_BC21 | chr19 | 38492469 | | | -0.249899 | 2.21E-08 | 9.25E-06 |
| cg03807469_BC11 | chr9 | 132670201 | exon_1;TSS | GTF3C4;GT | 0.230918 | 2.21E-08 | 9.25E-06 |
| cg17748182_BC21 | chr1 | 1342061 | exon_4;exo | DVL1;DVL1 | -0.154476 | 2.21E-08 | 9.26E-06 |
| cg18047214_BC21 | chr17 | 75952021 | | | 0.28026 | 2.21E-08 | 9.26E-06 |
| cg12915752_TC21 | chr8 | 100871952 | | | -0.252529 | 2.22E-08 | 9.27E-06 |
| cg10187773_TC21 | chr5 | 100424949 | | | -0.222507 | 2.22E-08 | 9.27E-06 |
| cg18359780_BC21 | chr3 | 24963272 | | | -0.240728 | 2.22E-08 | 9.27E-06 |
| cg22704351_TC21 | chr2 | 240986131 | | | 0.152276 | 2.24E-08 | 9.34E-06 |
| cg22046535_BC21 | chr2 | 1621553 | | | -0.224012 | 2.24E-08 | 9.34E-06 |
| cg14669616_BC21 | chr6 | 151765412 | | | -0.391542 | 2.24E-08 | 9.35E-06 |
| cg25002149_TC21 | chr20 | 15497005 | | | -0.123401 | 2.24E-08 | 9.35E-06 |
| cg13971484_BC21 | chr9 | 131478575 | exon_18;e) | PRRC2B;PR | -0.144503 | 2.24E-08 | 9.35E-06 |
| cg21331845_TC21 | chr3 | 150775283 | | | -0.282347 | 2.25E-08 | 9.38E-06 |
| cg09597496_TC21 | chr3 | 52455402 | TSS1500;TS | NISCH;NISC | -0.221935 | 2.25E-08 | 9.38E-06 |
| cg02476671_TC21 | chr4 | 2429129 | | | -0.30891 | 2.26E-08 | 9.41E-06 |
| cg23912737_TC21 | chr19 | 2342175 | | | 0.444288 | 2.26E-08 | 9.42E-06 |
| cg24398176_BC21 | chr1 | 33082214 | 5UTR;exon_ | AZIN2;AZIN | -0.15735 | 2.27E-08 | 9.42E-06 |
| cg26211101_TC21 | chr22 | 30308487 | | | 0.121424 | 2.27E-08 | 9.43E-06 |
| cg08123444_BC11 | chr2 | 9692972 | | | 0.253232 | 2.27E-08 | 9.43E-06 |
| cg02133748_TC21 | chr8 | 66369657 | | | -0.272225 | 2.27E-08 | 9.43E-06 |
| cg25335944_BC21 | chr4 | 2817989 | TSS1500;TS | SH3BP2;SH | -0.401492 | 2.28E-08 | 9.44E-06 |
| cg02445907_BC21 | chr6 | 36326627 | exon_5 | BNIP5 | -0.11017 | 2.28E-08 | 9.44E-06 |
| cg15256315_BC21 | chr8 | 134779489 | | | 0.417217 | 2.28E-08 | 9.45E-06 |
| cg25849994_TC21 | chr22 | 38318130 | TSS1500;TS | CSNK1E;CS | -0.197991 | 2.29E-08 | 9.48E-06 |
| cg18887918_BC21 | chr6 | 42567961 | | | -0.095083 | 2.29E-08 | 9.49E-06 |
| cg27111836_TC21 | chr22 | 33657871 | | | -0.257056 | 2.29E-08 | 9.49E-06 |
| cg26129675_TC21 | chr3 | 63545055 | | | -0.279229 | 2.29E-08 | 9.49E-06 |
| cg18823249_TC21 | chr12 | 62145897 | | | -0.295788 | 2.31E-08 | 9.55E-06 |
| cg07068045_BC12 | chr11 | 69497434 | | | -0.333371 | 2.31E-08 | 9.55E-06 |
| cg16210378_BC21 | chr5 | 14901687 | | | -0.209083 | 2.32E-08 | 9.58E-06 |
| cg23685577_BC21 | chr21 | 30562159 | TSS1500 | KRTAP19-7 | -0.320509 | 2.33E-08 | 9.62E-06 |
| cg01603919_BC21 | chr19 | 17476624 | | | -0.162881 | 2.33E-08 | 9.64E-06 |
| cg24614955_BC21 | chr2 | 96641842 | TSS1500 | FER1L5 | -0.326103 | 2.34E-08 | 9.64E-06 |

| | | | | | | | |
|---|---|---|---|---|---|---|---|
| cg09439817_BC21 | chr17 | 47855700 | TSS200 | SP6 | -0.220331 | 2.34E-08 | 9.64E-06 |
| cg24458478_TC11 | chr2 | 112645674 | TSS1500;TS | SLC20A1;SI | -0.254339 | 2.34E-08 | 9.65E-06 |
| cg14520214_TC11 | chr7 | 1452195 | | | -0.144438 | 2.34E-08 | 9.65E-06 |
| cg13676800_BC21 | chr9 | 87107517 | | | -0.171596 | 2.35E-08 | 9.66E-06 |
| cg11072498_TC21 | chr2 | 86808486 | TSS1500;TS | CD8A;CD8/ | -0.315372 | 2.35E-08 | 9.66E-06 |
| cg13916402_TC21 | chr6 | 54515289 | | | -0.171395 | 2.35E-08 | 9.67E-06 |
| cg20635265_BC21 | chr6 | 11225420 | | | -0.296533 | 2.35E-08 | 9.67E-06 |
| cg22872992_BC21 | chr20 | 52863890 | | | -0.314567 | 2.35E-08 | 9.67E-06 |
| cg18626176_BC11 | chr13 | 36870973 | | | -0.182924 | 2.36E-08 | 9.67E-06 |
| cg03334077_TC21 | chr15 | 99056980 | | | -0.207952 | 2.36E-08 | 9.67E-06 |
| cg00533483_TC21 | chr14 | 75719741 | exon_11 | TTLL5 | -0.204121 | 2.36E-08 | 9.68E-06 |
| cg24709209_TC21 | chr19 | 49997459 | | | -0.276949 | 2.37E-08 | 9.72E-06 |
| cg14075393_TC21 | chr13 | 25911432 | | | -0.345112 | 2.37E-08 | 9.72E-06 |
| cg20167003_BC21 | chr15 | 91069191 | | | -0.273848 | 2.38E-08 | 9.73E-06 |
| cg13522462_BC21 | chr19 | 44072721 | | | -0.307425 | 2.38E-08 | 9.74E-06 |
| cg17401067_TC21 | chr22 | 41444288 | | | -0.347645 | 2.38E-08 | 9.74E-06 |
| cg01955962_TC21 | chr15 | 72797195 | | | 0.234328 | 2.38E-08 | 9.76E-06 |
| cg17339145_BC21 | chr3 | 46750533 | | | -0.264695 | 2.40E-08 | 9.80E-06 |
| cg22730006_BC21 | chr17 | 57421279 | | | 0.174849 | 2.40E-08 | 9.80E-06 |
| cg17885794_BC21 | chr1 | 176552057 | | | 0.233567 | 2.40E-08 | 9.81E-06 |
| cg00549574_TC11 | chr17 | 76354861 | TSS1500;TS | PRPSAP1;P | -0.205528 | 2.41E-08 | 9.83E-06 |
| cg11557071_BC21 | chr3 | 44624490 | TSS1500;TS | ZNF197;ZN | -0.41756 | 2.42E-08 | 9.88E-06 |
| cg00617691_TC21 | chr10 | 697991 | | | -0.154796 | 2.42E-08 | 9.88E-06 |
| cg24405068_TC21 | chr6 | 137045199 | TSS1500;TS | IL20RA;IL2( | 0.369277 | 2.43E-08 | 9.90E-06 |
| cg20513745_BC21 | chr15 | 64893471 | | | 0.146537 | 2.43E-08 | 9.90E-06 |
| cg07871104_TC21 | chr15 | 45921744 | | | -0.172159 | 2.44E-08 | 9.92E-06 |
| cg15294119_BC21 | chr18 | 58636208 | | | -0.268171 | 2.44E-08 | 9.94E-06 |
| cg24943150_BC21 | chr18 | 3924605 | | | -0.258049 | 2.44E-08 | 9.95E-06 |
| cg14666720_BC21 | chr2 | 99790091 | | | -0.18768 | 2.45E-08 | 9.98E-06 |
| cg19095448_TC21 | chr13 | 102999889 | | | 0.170599 | 2.45E-08 | 9.98E-06 |
| cg01910094_TC21 | chr1 | 210752046 | | | 0.206797 | 2.46E-08 | 9.99E-06 |
| cg22745781_TC11 | chr7 | 16421619 | TSS200;TSS | CRPPA;CRP | 0.167352 | 2.46E-08 | 9.99E-06 |
| cg06215859_BC11 | chr16 | 88919301 | | | -0.164121 | 2.46E-08 | 1.00E-05 |
| cg00878641_BC21 | chr5 | 177308686 | | | 0.194666 | 2.46E-08 | 1.00E-05 |
| cg17615919_BC21 | chr9 | 133602925 | | | 0.326564 | 2.47E-08 | 1.00E-05 |
| cg14438453_BC21 | chr6 | 39818790 | | | 0.117877 | 2.47E-08 | 1.00E-05 |
| cg14015192_BC21 | chr9 | 99183343 | | | 0.313693 | 2.48E-08 | 1.00E-05 |
| cg25543768_TC21 | chr1 | 210025218 | | | -0.195126 | 2.48E-08 | 1.00E-05 |
| cg01891940_BC11 | chr14 | 39347298 | | | 0.112397 | 2.48E-08 | 1.01E-05 |
| cg22327338_TC21 | chr7 | 2166056 | | | -0.144484 | 2.48E-08 | 1.01E-05 |
| cg19892584_BC21 | chr1 | 63363148 | | | -0.283098 | 2.48E-08 | 1.01E-05 |
| cg20738928_BC21 | chr2 | 99255525 | | | -0.352292 | 2.49E-08 | 1.01E-05 |
| cg17617491_BC21 | chr19 | 2607728 | | | -0.10785 | 2.49E-08 | 1.01E-05 |
| cg15520012_BC21 | chr10 | 103811317 | | | -0.413692 | 2.49E-08 | 1.01E-05 |
| cg10721333_TC21 | chr7 | 17402416 | | | 0.181223 | 2.49E-08 | 1.01E-05 |
| cg01434144_BC21 | chr6 | 147142843 | | | 0.190453 | 2.49E-08 | 1.01E-05 |
| cg03414367_BC21 | chr2 | 121055682 | | | -0.139953 | 2.50E-08 | 1.01E-05 |

| | | | | | | | |
|---|---|---|---|---|---|---|---|
| cg18399898_BC21 | chr10 | 89331924 | TSS1500;TS | IFIT3;IFIT3 | 0.274157 | 2.50E-08 | 1.01E-05 |
| cg24808897_BC21 | chr7 | 30854145 | | | -0.117131 | 2.50E-08 | 1.01E-05 |
| cg00007076_TC21 | chr8 | 66430365 | exon_1;3U | RRS1;RRS1 | -0.177003 | 2.50E-08 | 1.01E-05 |
| cg15145715_BC21 | chr1 | 246689888 | | | -0.207491 | 2.50E-08 | 1.01E-05 |
| cg01413404_BC21 | chr4 | 2817970 | TSS1500;TS | SH3BP2;SH | -0.21175 | 2.51E-08 | 1.01E-05 |
| cg15117913_BC21 | chr10 | 62091295 | exon_7;exc | ARID5B;AR | 0.257658 | 2.51E-08 | 1.01E-05 |
| cg07762474_BC21 | chr16 | 30821064 | | | -0.431874 | 2.51E-08 | 1.01E-05 |
| cg07760720_BC21 | chr5 | 140700427 | TSS200 | ZMAT2 | -0.312537 | 2.51E-08 | 1.01E-05 |
| cg10815770_BC21 | chr10 | 35306999 | | | 0.148218 | 2.51E-08 | 1.01E-05 |
| cg07119672_TC21 | chr5 | 176662802 | | | -0.128968 | 2.51E-08 | 1.01E-05 |
| cg11889371_BC21 | chr2 | 6826267 | | | -0.156403 | 2.52E-08 | 1.01E-05 |
| cg15345674_BC21 | chr10 | 86684644 | | | -0.176988 | 2.52E-08 | 1.01E-05 |
| cg10032604_BC21 | chr9 | 134594976 | | | -0.184149 | 2.52E-08 | 1.01E-05 |
| cg10667201_TC11 | chr13 | 39655885 | | | -0.227683 | 2.52E-08 | 1.01E-05 |
| cg09576488_BC21 | chr15 | 37984450 | | | 0.277067 | 2.52E-08 | 1.01E-05 |
| cg03293507_BC11 | chr6 | 30071198 | exon_4;exc | RNF39;RNF | -0.309087 | 2.52E-08 | 1.01E-05 |
| cg16063999_TC21 | chr11 | 17000208 | | | -0.183197 | 2.53E-08 | 1.01E-05 |
| cg26226576_TC21 | chr1 | 234585831 | | | -0.281549 | 2.53E-08 | 1.01E-05 |
| cg20983112_BC21 | chr11 | 75769201 | TSS1500 | DGAT2-DT | -0.265619 | 2.54E-08 | 1.02E-05 |
| cg18375707_TC21 | chr11 | 64267487 | exon_31;ex | PLCB3;PLCI | -0.317922 | 2.54E-08 | 1.02E-05 |
| cg24875899_BC21 | chr7 | 29609872 | | | -0.418323 | 2.54E-08 | 1.02E-05 |
| cg05210612_TC21 | chr2 | 41854516 | | | -0.163023 | 2.54E-08 | 1.02E-05 |
| cg01292722_BC21 | chr3 | 150972642 | exon_1;exc | CLRN1;CLR | -0.225575 | 2.55E-08 | 1.02E-05 |
| cg00608183_BC21 | chr2 | 9144763 | | | -0.19566 | 2.55E-08 | 1.02E-05 |
| cg13194590_TC21 | chr8 | 138700641 | | | -0.158135 | 2.56E-08 | 1.02E-05 |
| cg17800666_BC21 | chr7 | 74042556 | | | 0.309 | 2.56E-08 | 1.02E-05 |
| cg15118627_TC21 | chr4 | 87346219 | | | -0.272044 | 2.57E-08 | 1.02E-05 |
| cg18575811_BC11 | chr6 | 39229648 | TSS200 | KCNK5 | 0.332975 | 2.57E-08 | 1.02E-05 |
| cg09666988_BC21 | chr4 | 82515846 | | | 0.212608 | 2.57E-08 | 1.02E-05 |
| cg22399868_BC21 | chr11 | 19447694 | | | -0.273776 | 2.57E-08 | 1.02E-05 |
| cg03506316_TC21 | chr4 | 7923812 | | | -0.211647 | 2.57E-08 | 1.02E-05 |
| cg24836097_TC21 | chr4 | 4290840 | TSS1500;TS | LYAR;LYAR | -0.253816 | 2.57E-08 | 1.02E-05 |
| cg04958522_TC21 | chr10 | 33876435 | | | -0.246907 | 2.58E-08 | 1.02E-05 |
| cg09880955_BC21 | chr14 | 28771698 | TSS1500;TS | LINC01551 | -0.3138 | 2.58E-08 | 1.03E-05 |
| cg03778035_TC11 | chr6 | 32854215 | 5UTR;exon | PSMB9;PSN | 0.224064 | 2.60E-08 | 1.03E-05 |
| cg04846648_TC11 | chr14 | 104724253 | 5UTR;exon | ADSS1;ADS | 0.275993 | 2.60E-08 | 1.03E-05 |
| cg13237711_BC21 | chr8 | 141560497 | | | 0.276318 | 2.60E-08 | 1.03E-05 |
| cg16009311_TC11 | chr8 | 118074523 | | | -0.221492 | 2.60E-08 | 1.03E-05 |
| cg09600520_TC21 | chr6 | 4706010 | TSS200 | CDYL | -0.204368 | 2.60E-08 | 1.03E-05 |
| cg19438683_BC21 | chr1 | 47423701 | | | 0.268497 | 2.61E-08 | 1.03E-05 |
| cg10521706_BC21 | chr2 | 19356825 | | | -0.16036 | 2.61E-08 | 1.03E-05 |
| cg17600662_TC11 | chr17 | 3049473 | | | 0.250427 | 2.61E-08 | 1.03E-05 |
| cg17461670_BC21 | chr17 | 68930372 | | | 0.391502 | 2.61E-08 | 1.03E-05 |
| cg24664957_BC21 | chr12 | 122896456 | TSS1500 | VPS37B | -0.19195 | 2.61E-08 | 1.03E-05 |
| cg06622582_BC21 | chr8 | 17313156 | | | 0.327971 | 2.62E-08 | 1.04E-05 |
| cg25374696_TC21 | chr20 | 50138784 | | | -0.219332 | 2.63E-08 | 1.04E-05 |
| cg10836508_TC21 | chr16 | 68222569 | | | 0.4023 | 2.63E-08 | 1.04E-05 |

| | | | | | | | |
|---|---|---|---|---|---|---|---|
| cg22935728_BC21 | chr17 | 74736739 | TSS1500;TS | RAB37;RAB | 0.231136 | 2.63E-08 | 1.04E-05 |
| cg16621078_TC21 | chr11 | 74019924 | | | -0.123828 | 2.63E-08 | 1.04E-05 |
| cg00576433_TC21 | chr2 | 231393203 | | | 0.266661 | 2.64E-08 | 1.04E-05 |
| cg01472046_TC21 | chr1 | 156251577 | | | 0.112225 | 2.64E-08 | 1.04E-05 |
| cg14922116_BC21 | chr8 | 135578221 | | | -0.374279 | 2.64E-08 | 1.04E-05 |
| cg02922289_TC21 | chr7 | 44210888 | | | 0.317938 | 2.64E-08 | 1.04E-05 |
| cg23849483_TC11 | chr19 | 15113733 | exon_8;exo | SYDE1;SYD | 0.144786 | 2.64E-08 | 1.04E-05 |
| cg22626548_TC11 | chr5 | 2253015 | | | -0.188159 | 2.65E-08 | 1.04E-05 |
| cg26973137_TC21 | chr6 | 39698994 | | | 0.292489 | 2.65E-08 | 1.04E-05 |
| cg15276037_BC21 | chr5 | 80711340 | | | 0.242264 | 2.65E-08 | 1.04E-05 |
| cg21933812_TC21 | chr1 | 203272894 | | | -0.142503 | 2.65E-08 | 1.04E-05 |
| cg22663985_BC21 | chr17 | 49901574 | | | -0.201992 | 2.66E-08 | 1.05E-05 |
| cg27046460_BC21 | chr1 | 230701634 | | | -0.182288 | 2.67E-08 | 1.05E-05 |
| cg06468920_BC21 | chr17 | 63535906 | exon_6;exo | KCNH6;KCN | -0.246296 | 2.67E-08 | 1.05E-05 |
| cg16207671_BC21 | chr18 | 31105836 | | | 0.25494 | 2.67E-08 | 1.05E-05 |
| cg05913921_BC21 | chr1 | 245990816 | | | 0.470178 | 2.67E-08 | 1.05E-05 |
| cg17541584_BC21 | chr12 | 45218040 | | | -0.372149 | 2.68E-08 | 1.05E-05 |
| cg03625007_TC21 | chr2 | 196613343 | | | -0.183197 | 2.68E-08 | 1.05E-05 |
| cg17629593_BC21 | chr12 | 52977611 | | | -0.351471 | 2.68E-08 | 1.05E-05 |
| cg17507887_TC11 | chr6 | 50818798 | TSS200 | TFAP2B | -0.183595 | 2.68E-08 | 1.05E-05 |
| cg01308419_TC11 | chr7 | 6526260 | exon_4;exo | GRID2IP;GR | -0.182288 | 2.68E-08 | 1.05E-05 |
| cg22846767_TC21 | chr5 | 157116642 | | | -0.214062 | 2.68E-08 | 1.05E-05 |
| cg09079970_BC21 | chr6 | 3787616 | | | -0.325929 | 2.69E-08 | 1.05E-05 |
| cg04372929_TC21 | chr20 | 44717550 | | | -0.165668 | 2.69E-08 | 1.05E-05 |
| cg02287146_BC21 | chr5 | 114331143 | | | -0.255507 | 2.69E-08 | 1.05E-05 |
| cg03644585_TC21 | chr7 | 845188 | | | -0.215561 | 2.70E-08 | 1.05E-05 |
| cg03564201_TC21 | chr3 | 42906677 | | | -0.176278 | 2.70E-08 | 1.05E-05 |
| cg05947727_TC21 | chr15 | 69461957 | | | -0.271067 | 2.71E-08 | 1.06E-05 |
| cg21185075_BC21 | chr16 | 46704811 | 3UTR;exon | MYLK3;MY | -0.343128 | 2.71E-08 | 1.06E-05 |
| cg05058657_TC21 | chr9 | 126508817 | | | -0.18891 | 2.71E-08 | 1.06E-05 |
| cg24882332_TC21 | chr17 | 38907396 | | | -0.117796 | 2.72E-08 | 1.06E-05 |
| cg05053440_BC11 | chr1 | 227560741 | | | -0.19832 | 2.72E-08 | 1.06E-05 |
| cg08692187_TC21 | chr20 | 33133156 | | | -0.199696 | 2.72E-08 | 1.06E-05 |
| cg09147213_TC21 | chr5 | 79514779 | TSS1500;TS | HOMER1;H | -0.269614 | 2.73E-08 | 1.06E-05 |
| cg14996143_TC21 | chr2 | 172080600 | exon_10;3U | METAP1D;I | -0.204469 | 2.74E-08 | 1.06E-05 |
| cg00140057_TC21 | chr11 | 134401896 | | | -0.17164 | 2.74E-08 | 1.06E-05 |
| cg14350294_BC21 | chr18 | 60663420 | | | -0.234117 | 2.74E-08 | 1.06E-05 |
| cg14358282_TC11 | chr2 | 219001906 | TSS1500;TS | LOC10012S | 0.19231 | 2.74E-08 | 1.06E-05 |
| cg26113185_TC21 | chr22 | 23012117 | | | -0.159778 | 2.76E-08 | 1.07E-05 |
| cg06831584_BC21 | chr2 | 241310002 | | | 0.347186 | 2.76E-08 | 1.07E-05 |
| cg02659402_TC21 | chr1 | 236762577 | exon_21;ex | ACTN2;ACT | -0.385582 | 2.76E-08 | 1.07E-05 |
| cg13901892_BC21 | chr2 | 70780457 | | | -0.239051 | 2.76E-08 | 1.07E-05 |
| cg08929188_TC21 | chr11 | 14973776 | TSS1500;TS | CALCA;CAL | -0.260728 | 2.77E-08 | 1.07E-05 |
| cg15954792_TC21 | chr2 | 98370508 | | | -0.178416 | 2.77E-08 | 1.07E-05 |
| cg01328190_BC21 | chr16 | 14435857 | 3UTR;exon | PARN;PARN | 0.347139 | 2.77E-08 | 1.07E-05 |
| cg07742235_TC11 | chr11 | 69493984 | | | -0.226664 | 2.78E-08 | 1.07E-05 |
| cg21126847_BC21 | chr15 | 55258548 | | | 0.163427 | 2.78E-08 | 1.07E-05 |

| | | | | | | | |
|---|---|---|---|---|---|---|---|
| cg11995228_BC21 | chr19 | 50288592 | | | -0.173527 | 2.78E-08 | 1.07E-05 |
| cg23412875_BC11 | chr16 | 3283539 | 5UTR;exon_ | ZNF263;ZN | 0.311505 | 2.78E-08 | 1.07E-05 |
| cg22401907_BC11 | chr6 | 16353011 | | | -0.191026 | 2.78E-08 | 1.07E-05 |
| cg17121140_TC11 | chr13 | 111066656 | | | -0.26263 | 2.78E-08 | 1.07E-05 |
| cg27158519_BC21 | chr6 | 113259001 | | | -0.202601 | 2.79E-08 | 1.08E-05 |
| cg27188696_TC21 | chr2 | 173003071 | | | 0.423379 | 2.79E-08 | 1.08E-05 |
| cg05513593_TC21 | chr3 | 72397861 | | | 0.171213 | 2.79E-08 | 1.08E-05 |
| cg19223274_TC11 | chr6 | 143060861 | TSS200;TSS | AIG1;AIG1; | 0.364012 | 2.79E-08 | 1.08E-05 |
| cg03284768_BC21 | chr11 | 9276772 | | | 0.303657 | 2.80E-08 | 1.08E-05 |
| cg20373012_TC21 | chr11 | 122653235 | | | -0.281067 | 2.80E-08 | 1.08E-05 |
| cg11135756_TC21 | chr7 | 65731795 | | | -0.380857 | 2.80E-08 | 1.08E-05 |
| cg02400773_TC21 | chr17 | 13598987 | | | -0.330716 | 2.81E-08 | 1.08E-05 |
| cg22723129_BC21 | chr17 | 56885167 | TSS1500 | MTVR2 | -0.387153 | 2.81E-08 | 1.08E-05 |
| cg18971251_TC21 | chr12 | 120925937 | | | -0.263924 | 2.81E-08 | 1.08E-05 |
| cg08333515_TC22 | chr5 | 103582586 | | | -0.3012 | 2.82E-08 | 1.08E-05 |
| cg11978330_BC21 | chr7 | 156112957 | | | 0.153393 | 2.82E-08 | 1.08E-05 |
| cg02052527_BC21 | chr18 | 63995121 | | | -0.357115 | 2.83E-08 | 1.09E-05 |
| cg00735141_BC21 | chr1 | 48498510 | | | -0.277751 | 2.83E-08 | 1.09E-05 |
| cg08490624_BC21 | chr8 | 66436653 | | | -0.225668 | 2.83E-08 | 1.09E-05 |
| cg24341612_BC21 | chr20 | 18463244 | | | -0.531934 | 2.84E-08 | 1.09E-05 |
| cg15260400_BC21 | chr4 | 83116109 | TSS1500;TS | PLAC8;PLA | -0.124814 | 2.84E-08 | 1.09E-05 |
| cg14535045_BC21 | chr14 | 89626965 | | | 0.324234 | 2.84E-08 | 1.09E-05 |
| cg01450481_TC21 | chr1 | 154995861 | | | -0.215512 | 2.86E-08 | 1.09E-05 |
| cg15123289_BC11 | chr10 | 62815470 | | | 0.281681 | 2.86E-08 | 1.09E-05 |
| cg06665333_TC11 | chr16 | 87840231 | | | -0.170447 | 2.87E-08 | 1.10E-05 |
| cg09503767_BC21 | chr9 | 130185398 | | | -0.204459 | 2.88E-08 | 1.10E-05 |
| cg15337497_BC21 | chr5 | 58422471 | | | 0.214064 | 2.88E-08 | 1.10E-05 |
| cg12804006_BC21 | chr13 | 24688522 | exon_4;exc | ATP12A;AT | -0.208508 | 2.88E-08 | 1.10E-05 |
| cg19547516_TC21 | chr3 | 70748995 | | | -0.34021 | 2.90E-08 | 1.11E-05 |
| cg18704869_BC21 | chr14 | 104095053 | | | -0.198529 | 2.91E-08 | 1.11E-05 |
| cg06948784_TC11 | chr1 | 16535649 | exon_1 | LINC01783 | -0.312999 | 2.91E-08 | 1.11E-05 |
| cg10787865_TC21 | chr1 | 226444900 | | | -0.12186 | 2.91E-08 | 1.11E-05 |
| cg05555283_BC21 | chr14 | 104643630 | | | -0.247425 | 2.92E-08 | 1.11E-05 |
| cg19309844_TC21 | chr4 | 128683346 | | | -0.249366 | 2.92E-08 | 1.11E-05 |
| cg03182007_TC21 | chr18 | 26290486 | | | -0.134071 | 2.92E-08 | 1.11E-05 |
| cg17644569_BC11 | chr17 | 47229977 | | | 0.310377 | 2.92E-08 | 1.11E-05 |
| cg21527370_BC21 | chr6 | 109429387 | | | -0.130462 | 2.93E-08 | 1.11E-05 |
| cg23198793_BC11 | chr15 | 42402208 | TSS200 | CAPN3 | 0.129098 | 2.93E-08 | 1.11E-05 |
| cg06284586_BC11 | chr2 | 237387870 | exon_4;exc | COL6A3;CC | -0.169078 | 2.93E-08 | 1.11E-05 |
| cg06028875_TC21 | chr6 | 30074518 | | | -0.133664 | 2.93E-08 | 1.11E-05 |
| cg09579623_TC21 | chr6 | 150139983 | | | 0.261894 | 2.93E-08 | 1.11E-05 |
| cg03573068_BC21 | chr11 | 20598741 | TSS1500;TS | SLC6A5;SL | 0.377709 | 2.94E-08 | 1.12E-05 |
| cg03751759_BC21 | chr1 | 107153180 | | | -0.241698 | 2.94E-08 | 1.12E-05 |
| cg14639336_BC21 | chr10 | 8642894 | | | -0.151276 | 2.95E-08 | 1.12E-05 |
| cg14386401_TC21 | chr9 | 133055249 | | | 0.194487 | 2.95E-08 | 1.12E-05 |
| cg02067882_TC21 | chr2 | 220619784 | | | -0.175081 | 2.96E-08 | 1.12E-05 |
| cg19611374_BC21 | chr8 | 33538959 | | | -0.203725 | 2.96E-08 | 1.12E-05 |

| | | | | | | | |
|---|---|---|---|---|---|---|---|
| cg22369019_TC21 | chr17 | 28372166 | exon_1 | SARM1 | -0.249317 | 2.97E-08 | 1.12E-05 |
| cg20285263_TC21 | chr6 | 41424665 | | | 0.260277 | 2.97E-08 | 1.12E-05 |
| cg04941897_TC21 | chr1 | 43492659 | | | -0.192206 | 2.97E-08 | 1.12E-05 |
| cg05810879_TC21 | chr4 | 6965488 | | | 0.277934 | 2.97E-08 | 1.12E-05 |
| cg16922763_BC11 | chr13 | 111066825 | | | -0.261512 | 2.99E-08 | 1.13E-05 |
| cg20283224_TC21 | chr2 | 54866904 | | | -0.172584 | 3.00E-08 | 1.14E-05 |
| cg10309957_BC21 | chr1 | 161765444 | TSS1500 | ATF6 | -0.410333 | 3.02E-08 | 1.14E-05 |
| cg10211698_BC21 | chr15 | 55907889 | | | 0.276616 | 3.02E-08 | 1.14E-05 |
| cg02624558_BC11 | chr1 | 202808483 | exon_1;5U | KDM5B;KD | -0.28089 | 3.02E-08 | 1.14E-05 |
| cg14893069_TC21 | chr10 | 33684263 | | | -0.206431 | 3.03E-08 | 1.14E-05 |
| cg06851151_BC21 | chr5 | 134100677 | | | -0.082366 | 3.03E-08 | 1.14E-05 |
| cg25838091_TC21 | chr10 | 49679888 | | | -0.145009 | 3.03E-08 | 1.14E-05 |
| cg09317199_TC21 | chr6 | 27295365 | | | -0.260936 | 3.04E-08 | 1.15E-05 |
| cg17401974_TC21 | chr8 | 76810949 | | | -0.392477 | 3.05E-08 | 1.15E-05 |
| cg25096066_BC21 | chr20 | 25227411 | exon_14;3 | ENTPD6;EN | -0.383352 | 3.05E-08 | 1.15E-05 |
| cg00638021_BC21 | chr17 | 50189701 | exon_38 | COL1A1 | -0.191411 | 3.05E-08 | 1.15E-05 |
| cg14301513_TC21 | chr9 | 128733581 | | | -0.181747 | 3.06E-08 | 1.15E-05 |
| cg02498063_TC21 | chr12 | 119334772 | 5UTR;exon | CCDC60;CC | -0.227245 | 3.06E-08 | 1.15E-05 |
| cg16916261_BC21 | chr11 | 114285541 | | | -0.281759 | 3.06E-08 | 1.15E-05 |
| cg08352530_BC21 | chr7 | 6410350 | exon_12;e | DAGLB;DA | 0.151697 | 3.07E-08 | 1.15E-05 |
| cg14200859_BC21 | chr9 | 126932917 | | | -0.350965 | 3.07E-08 | 1.15E-05 |
| cg09984392_TC11 | chr8 | 124999542 | exon_1;TS | SQLE;SQLE | -0.285325 | 3.07E-08 | 1.15E-05 |
| cg14698932_TC21 | chr13 | 33514501 | | | -0.099006 | 3.07E-08 | 1.15E-05 |
| cg00135728_BC21 | chr9 | 98174196 | TSS1500 | CORO2A | 0.284493 | 3.08E-08 | 1.15E-05 |
| cg01361849_BC11 | chr11 | 65181005 | exon_2;TS | LOC72897 | -0.245581 | 3.08E-08 | 1.15E-05 |
| cg15391057_BC21 | chr3 | 37975697 | | | -0.265507 | 3.09E-08 | 1.16E-05 |
| cg09450907_BC21 | chr2 | 19361326 | | | -0.244993 | 3.10E-08 | 1.16E-05 |
| cg01854277_BC21 | chr11 | 45599628 | | | -0.410532 | 3.11E-08 | 1.16E-05 |
| cg26816919_TC21 | chr12 | 121040012 | TSS1500;T | OASL;OASL | -0.250188 | 3.11E-08 | 1.16E-05 |
| cg07796220_TC21 | chr17 | 8799051 | exon_1 | MFSD6L | -0.256024 | 3.11E-08 | 1.16E-05 |
| cg04958800_BC21 | chr7 | 127244461 | TSS1500 | GRM8 | -0.245917 | 3.12E-08 | 1.17E-05 |
| cg16846489_TC11 | chr9 | 19049462 | 5UTR;exon | RRAGA;RR | -0.367217 | 3.12E-08 | 1.17E-05 |
| cg12954779_BC21 | chr11 | 24687550 | | | -0.264074 | 3.12E-08 | 1.17E-05 |
| cg04801236_TC11 | chr12 | 107093231 | exon_1;5U | CRY1;CRY1 | -0.308362 | 3.12E-08 | 1.17E-05 |
| cg00659250_BC21 | chr7 | 73767906 | | | -0.208753 | 3.13E-08 | 1.17E-05 |
| cg10067427_BC21 | chr15 | 81324203 | TSS200;TS | TMC3-AS1; | 0.290213 | 3.13E-08 | 1.17E-05 |
| cg20084692_TC21 | chr7 | 139636218 | | | 0.24852 | 3.13E-08 | 1.17E-05 |
| cg08127916_TC21 | chr11 | 122773281 | | | 0.175717 | 3.13E-08 | 1.17E-05 |
| cg23345084_BC21 | chr2 | 39664201 | TSS1500 | TMEM178 | -0.331208 | 3.13E-08 | 1.17E-05 |
| cg26976693_BC21 | chr17 | 8899529 | | | -0.184315 | 3.14E-08 | 1.17E-05 |
| cg07686441_TC21 | chr15 | 52278320 | TSS1500 | MIR1266 | 0.232171 | 3.14E-08 | 1.17E-05 |
| cg11025743_TC21 | chr2 | 219439716 | | | -0.182772 | 3.15E-08 | 1.17E-05 |
| cg14987745_TC21 | chr12 | 103966244 | | | 0.656468 | 3.15E-08 | 1.17E-05 |
| cg15475055_TC11 | chr10 | 35090417 | TSS200;TS | CUL2;CUL2 | 0.311895 | 3.15E-08 | 1.17E-05 |
| cg11180966_TC21 | chr11 | 755608 | | | 0.226405 | 3.15E-08 | 1.17E-05 |
| cg14299508_BC21 | chr8 | 104895462 | | | -0.299227 | 3.16E-08 | 1.17E-05 |
| cg21534623_TC11 | chr1 | 64966672 | TSS200;TS | JAK1;JAK1; | 0.269195 | 3.17E-08 | 1.18E-05 |

| | | | | | | | |
|---|---|---|---|---|---|---|---|
| cg03770138_BC21 | chr9 | 133134264 | | | -0.119663 | 3.17E-08 | 1.18E-05 |
| cg22507734_TC21 | chr17 | 39511596 | exon_8;exo | CDK12;CDK | 0.270281 | 3.17E-08 | 1.18E-05 |
| cg15434749_TC21 | chr8 | 6391844 | | | -0.299125 | 3.17E-08 | 1.18E-05 |
| cg09992776_BC21 | chr15 | 39472357 | | | -0.225779 | 3.18E-08 | 1.18E-05 |
| cg00857137_TC21 | chr19 | 3006722 | | | -0.379864 | 3.18E-08 | 1.18E-05 |
| cg07813377_TC21 | chr12 | 1083259 | exon_2;exo | ERC1;ERC1 | -0.22609 | 3.19E-08 | 1.18E-05 |
| cg00259249_TC21 | chr1 | 34781125 | TSS1500;TS | GJB3;GJB3 | -0.223284 | 3.19E-08 | 1.18E-05 |
| cg10695105_BC11 | chr12 | 49549383 | | | -0.189828 | 3.19E-08 | 1.18E-05 |
| cg16246573_BC11 | chr2 | 240591974 | exon_4;exo | CAPN10;CA | -0.177966 | 3.21E-08 | 1.19E-05 |
| cg04775627_BC21 | chr2 | 42031184 | | | -0.221032 | 3.21E-08 | 1.19E-05 |
| cg01028287_BC21 | chr7 | 44235957 | | | -0.182401 | 3.22E-08 | 1.19E-05 |
| cg11631265_BC11 | chr8 | 143512793 | | | 0.248337 | 3.22E-08 | 1.19E-05 |
| cg00454305_BC21 | chr16 | 1379904 | TSS1500;TS | UNKL;UNKI | -0.217738 | 3.22E-08 | 1.19E-05 |
| cg17566923_BC11 | chr12 | 48334835 | 3UTR;exon_ | ZNF641;ZN | -0.168429 | 3.22E-08 | 1.19E-05 |
| cg11390645_BC11 | chr19 | 8308398 | TSS200;TSS | CD320;CD3 | -0.519819 | 3.23E-08 | 1.19E-05 |
| cg03405128_TC21 | chr4 | 76420688 | | | -0.189102 | 3.23E-08 | 1.19E-05 |
| cg07570439_BC21 | chr13 | 36207705 | | | -0.407673 | 3.23E-08 | 1.19E-05 |
| cg10623227_BC21 | chr2 | 172428697 | | | -0.187878 | 3.23E-08 | 1.19E-05 |
| cg08594102_BC21 | chr9 | 117247962 | | | 0.197685 | 3.23E-08 | 1.19E-05 |
| cg23815246_TC11 | chr6 | 81753716 | TSS1500 | TENT5A | 0.160363 | 3.23E-08 | 1.19E-05 |
| cg06361845_BC21 | chr9 | 133199499 | | | -0.389171 | 3.23E-08 | 1.19E-05 |
| cg03528118_TC21 | chr3 | 133533169 | | | -0.255739 | 3.24E-08 | 1.19E-05 |
| cg25191305_BC21 | chr7 | 48537670 | | | 0.19765 | 3.24E-08 | 1.19E-05 |
| cg02207638_TC11 | chr20 | 35292246 | exon_1 | FAM83C | -0.195234 | 3.25E-08 | 1.19E-05 |
| cg24505073_BC21 | chr18 | 31103831 | TSS1500;TS | DSC2;DSC2 | -0.238857 | 3.25E-08 | 1.19E-05 |
| cg23411206_TC21 | chr8 | 94668174 | exon_10;ex | ESRP1;ESRI | -0.192551 | 3.26E-08 | 1.20E-05 |
| cg19773376_TC11 | chr6 | 34359101 | | | 0.257717 | 3.26E-08 | 1.20E-05 |
| cg21624833_TC21 | chr8 | 48199151 | | | -0.291496 | 3.27E-08 | 1.20E-05 |
| cg16290299_TC21 | chr2 | 85669139 | TSS1500;TS | SFTPB;SFTF | 0.202097 | 3.28E-08 | 1.20E-05 |
| cg12847548_BC21 | chr11 | 74468163 | TSS1500 | KCNE3 | -0.111117 | 3.29E-08 | 1.21E-05 |
| cg15312323_BC21 | chr17 | 41161442 | TSS1500 | KRTAP4-4 | -0.357113 | 3.30E-08 | 1.21E-05 |
| cg03862319_BC21 | chr12 | 39323140 | | | 0.247953 | 3.31E-08 | 1.21E-05 |
| cg16619764_TC21 | chr1 | 115981769 | | | 0.18492 | 3.32E-08 | 1.21E-05 |
| cg16718760_TC21 | chr3 | 55687039 | | | -0.141908 | 3.32E-08 | 1.22E-05 |
| cg16533864_BC21 | chr9 | 93504738 | | | -0.118326 | 3.33E-08 | 1.22E-05 |
| cg09170311_BC21 | chr11 | 111515053 | TSS1500 | BTG4 | -0.142312 | 3.33E-08 | 1.22E-05 |
| cg23522493_TC21 | chr18 | 48085176 | | | -0.154994 | 3.33E-08 | 1.22E-05 |
| cg10753073_BC21 | chr1 | 64203916 | 5UTR;exon_ | UBE2U;UBI | -0.619512 | 3.33E-08 | 1.22E-05 |
| cg00265412_BC21 | chr5 | 16740948 | | | -0.203294 | 3.33E-08 | 1.22E-05 |
| cg15838493_BC21 | chr17 | 42098985 | TSS1500;TS | ZNF385C;ZI | 0.240557 | 3.33E-08 | 1.22E-05 |
| cg21731088_BC21 | chr18 | 52499403 | | | -0.201553 | 3.34E-08 | 1.22E-05 |
| cg26353448_BC21 | chr1 | 248360934 | TSS1500 | OR2T4 | -0.340152 | 3.35E-08 | 1.22E-05 |
| cg21776667_BC21 | chr6 | 143960315 | | | -0.352325 | 3.36E-08 | 1.22E-05 |
| cg20011248_TC11 | chr1 | 227559567 | | | -0.215153 | 3.36E-08 | 1.22E-05 |
| cg15128785_TC21 | chr22 | 41834875 | TSS1500 | SREBF2-AS: | -0.230111 | 3.36E-08 | 1.23E-05 |
| cg06334805_TC21 | chr4 | 20542526 | exon_21;ex | SLIT2;SLIT2 | -0.235748 | 3.37E-08 | 1.23E-05 |
| cg07991621_TC21 | chr4 | 2818752 | TSS200 | SH3BP2 | -0.631885 | 3.37E-08 | 1.23E-05 |

| | | | | | | | |
|---|---|---|---|---|---|---|---|
| cg08949533_TC21 | chr22 | 47614288 | | | 0.314266 | 3.37E-08 | 1.23E-05 |
| cg04471434_BC21 | chr4 | 159002795 | | | -0.364126 | 3.37E-08 | 1.23E-05 |
| cg18180155_BC21 | chr22 | 45413573 | exon_1;5U | SMC1B;SM | -0.410473 | 3.38E-08 | 1.23E-05 |
| cg04138155_BC11 | chr1 | 243255948 | TSS200;TSS | SDCCAG8;S | 0.21076 | 3.39E-08 | 1.23E-05 |
| cg06811801_BC21 | chr6 | 87318980 | | | -0.237986 | 3.39E-08 | 1.23E-05 |
| cg23439213_BC21 | chr18 | 36296164 | | | -0.267339 | 3.39E-08 | 1.23E-05 |
| cg16493726_TC21 | chr6 | 3963139 | | | -0.207163 | 3.39E-08 | 1.23E-05 |
| cg10441983_TC11 | chr4 | 1368605 | | | -0.138068 | 3.40E-08 | 1.23E-05 |
| cg15921461_BC21 | chr7 | 158581100 | | | -0.296239 | 3.41E-08 | 1.24E-05 |
| cg13656835_TC11 | chr11 | 65888998 | TSS1500;TS | FIBP;FIBP | -0.319157 | 3.42E-08 | 1.24E-05 |
| cg09247684_TC21 | chr14 | 58350964 | | | 0.311228 | 3.42E-08 | 1.24E-05 |
| cg13363748_BC21 | chr6 | 143540046 | | | 0.296545 | 3.42E-08 | 1.24E-05 |
| cg11119681_BC21 | chr20 | 38218239 | | | -0.108491 | 3.42E-08 | 1.24E-05 |
| cg00367546_BC21 | chr15 | 88621815 | | | -0.376054 | 3.42E-08 | 1.24E-05 |
| cg02130178_TC21 | chr12 | 14245351 | | | -0.253116 | 3.43E-08 | 1.24E-05 |
| cg00760646_TC21 | chr17 | 4740570 | exon_2;exo | ZMYND15;: | -0.203989 | 3.43E-08 | 1.24E-05 |
| cg26599122_BC21 | chr1 | 20187241 | TSS1500;TS | PLA2G2C;P | -0.141674 | 3.44E-08 | 1.24E-05 |
| cg11826104_TC21 | chr15 | 25405464 | exon_5;exo | UBE3A;UBE | -0.13044 | 3.44E-08 | 1.24E-05 |
| cg03844115_BC21 | chr10 | 1413132 | | | -0.231348 | 3.44E-08 | 1.24E-05 |
| cg03098814_BC11 | chr6 | 24911080 | TSS1500;TS | RIPOR2;RIF | 0.295503 | 3.45E-08 | 1.25E-05 |
| cg10112249_TC21 | chr1 | 30058198 | | | -0.196039 | 3.45E-08 | 1.25E-05 |
| cg24839322_BC21 | chr2 | 222714875 | | | -0.293638 | 3.46E-08 | 1.25E-05 |
| cg24036396_TC21 | chr19 | 7123103 | | | -0.18134 | 3.46E-08 | 1.25E-05 |
| cg04917275_BC11 | chr2 | 593391 | | | -0.270112 | 3.46E-08 | 1.25E-05 |
| cg02231734_TC21 | chr20 | 34714827 | 3UTR;exon | NCOA6;NC | -0.369592 | 3.46E-08 | 1.25E-05 |
| cg18468604_TC11 | chr17 | 18183934 | 5UTR;exon | ALKBH5;AL | 0.206847 | 3.46E-08 | 1.25E-05 |
| cg18909711_TC21 | chr5 | 64689455 | TSS1500 | SHISAL2B | -0.276148 | 3.48E-08 | 1.25E-05 |
| cg17059228_TC21 | chr10 | 32395672 | | | -0.336599 | 3.48E-08 | 1.25E-05 |
| cg14417498_TC11 | chr1 | 165352962 | | | 0.215633 | 3.48E-08 | 1.25E-05 |
| cg18158670_BC21 | chr15 | 98889996 | | | -0.263975 | 3.48E-08 | 1.25E-05 |
| cg04360009_BC21 | chr10 | 100655148 | | | 0.1401 | 3.49E-08 | 1.25E-05 |
| cg11231143_TC21 | chr17 | 28227219 | | | -0.216603 | 3.49E-08 | 1.25E-05 |
| cg06155794_BC21 | chr4 | 3706093 | | | -0.129521 | 3.49E-08 | 1.25E-05 |
| cg17788761_TC21 | chr1 | 77874894 | exon_16;e | MIGA1;MI | -0.297933 | 3.50E-08 | 1.26E-05 |
| cg19081437_BC21 | chr17 | 48578434 | TSS200 | HOXB4 | 0.277098 | 3.51E-08 | 1.26E-05 |
| cg18064706_BC11 | chr7 | 157853964 | TSS1500 | PTPRN2-AS | -0.197481 | 3.52E-08 | 1.26E-05 |
| cg10440011_TC11 | chr17 | 2029889 | TSS1500;TS | DPH1;DPH: | -0.169286 | 3.53E-08 | 1.27E-05 |
| cg00298027_TC21 | chr1 | 207096407 | | | 0.198205 | 3.53E-08 | 1.27E-05 |
| cg02384525_BC21 | chr2 | 71668972 | | | -0.293293 | 3.53E-08 | 1.27E-05 |
| cg11258616_TC21 | chr7 | 75106140 | | | -0.285717 | 3.54E-08 | 1.27E-05 |
| cg03369790_TC21 | chr6 | 170207423 | | | -0.115847 | 3.54E-08 | 1.27E-05 |
| cg17588475_BC21 | chr12 | 49821802 | | | -0.261034 | 3.54E-08 | 1.27E-05 |
| cg25148341_TC21 | chr5 | 14849826 | | | 0.181375 | 3.55E-08 | 1.27E-05 |
| cg17609498_BC21 | chr12 | 97557552 | | | -0.284513 | 3.55E-08 | 1.27E-05 |
| cg21279804_TC21 | chr2 | 41695436 | | | -0.205133 | 3.55E-08 | 1.27E-05 |
| cg15878199_BC21 | chr5 | 175468513 | | | -0.309104 | 3.56E-08 | 1.27E-05 |
| cg13804685_BC21 | chr14 | 23876751 | | | 0.233957 | 3.57E-08 | 1.27E-05 |

| | | | | | | | |
|---|---|---|---|---|---|---|---|
| cg00210007_BC21 | chr1 | 217078224 | 5UTR;exon | ESRRG;ESR | 0.217369 | 3.57E-08 | 1.27E-05 |
| cg07759647_BC21 | chr3 | 71200527 | | | 0.409311 | 3.57E-08 | 1.27E-05 |
| cg14599019_BC21 | chr19 | 39413184 | 5UTR;exon | PLEKHG2;P | -0.306536 | 3.58E-08 | 1.27E-05 |
| cg17866225_TC21 | chr18 | 58454930 | | | -0.229491 | 3.59E-08 | 1.28E-05 |
| cg26585864_TC21 | chr5 | 149406955 | exon_1;exc | CARMN;CA | -0.174475 | 3.60E-08 | 1.28E-05 |
| cg10656587_BC21 | chr7 | 7554758 | | | -0.230888 | 3.62E-08 | 1.29E-05 |
| cg07932504_BC21 | chr11 | 126102048 | | | -0.256613 | 3.62E-08 | 1.29E-05 |
| cg19503629_BC21 | chr14 | 52918000 | | | 0.154285 | 3.62E-08 | 1.29E-05 |
| cg04315226_BC21 | chr14 | 75077592 | exon_3;3U | ZC2HC1C;Z | 0.184535 | 3.64E-08 | 1.29E-05 |
| cg22895857_TC21 | chr16 | 62225931 | | | -0.185453 | 3.65E-08 | 1.30E-05 |
| cg13520744_BC21 | chr22 | 48642186 | | | -0.200434 | 3.66E-08 | 1.30E-05 |
| cg04011911_BC21 | chr1 | 226363344 | | | -0.188314 | 3.66E-08 | 1.30E-05 |
| cg25035939_TC21 | chr1 | 245510985 | | | 0.199922 | 3.67E-08 | 1.30E-05 |
| cg11414782_TC21 | chr7 | 97398635 | | | -0.310775 | 3.67E-08 | 1.30E-05 |
| cg05154454_BC21 | chr9 | 110337773 | exon_1;exc | TXNDC8;TX | -0.332229 | 3.67E-08 | 1.30E-05 |
| cg15175162_BC21 | chr4 | 15656034 | TSS1500;TS | FBXL5;FBXI | -0.328543 | 3.67E-08 | 1.30E-05 |
| cg12225766_BC21 | chr2 | 60731993 | | | 0.244728 | 3.68E-08 | 1.30E-05 |
| cg06193998_TC21 | chr13 | 20701600 | 5UTR;exon | IL17D;IL17I | 0.204883 | 3.68E-08 | 1.30E-05 |
| cg17938220_BC21 | chr6 | 157774126 | | | -0.134663 | 3.69E-08 | 1.31E-05 |
| cg12303102_BC21 | chr10 | 73415091 | TSS1500;TS | ANXA7;AN) | 0.273442 | 3.69E-08 | 1.31E-05 |
| cg11624078_BC21 | chr10 | 132736296 | | | 0.339027 | 3.70E-08 | 1.31E-05 |
| cg17085844_TC21 | chr1 | 23845184 | 3UTR;exon | FUCA1;FUC | 0.296583 | 3.70E-08 | 1.31E-05 |
| cg14076729_TC21 | chr1 | 37660956 | | | -0.174887 | 3.70E-08 | 1.31E-05 |
| cg18601186_TC21 | chr11 | 13145918 | | | -0.351554 | 3.71E-08 | 1.31E-05 |
| cg27655946_TC21 | chr20 | 62316403 | | | -0.18525 | 3.71E-08 | 1.31E-05 |
| cg23693998_TC21 | chr18 | 69043910 | | | -0.15266 | 3.71E-08 | 1.31E-05 |
| cg10577820_BC21 | chr7 | 2937994 | exon_9;exc | CARD11;CA | -0.225252 | 3.71E-08 | 1.31E-05 |
| cg27506810_TC21 | chr19 | 12295359 | TSS1500;TS | ZNF44;ZNF | -0.335781 | 3.72E-08 | 1.31E-05 |
| cg05182627_BC21 | chr12 | 49924238 | exon_3 | LINC02395 | -0.115791 | 3.72E-08 | 1.31E-05 |
| cg20403142_TC21 | chr4 | 56861143 | | | -0.335163 | 3.72E-08 | 1.31E-05 |
| cg07403971_BC21 | chr14 | 95194370 | | | 0.227937 | 3.73E-08 | 1.32E-05 |
| cg21820873_TC11 | chr11 | 64371247 | | | 0.206615 | 3.73E-08 | 1.32E-05 |
| cg23846851_TC21 | chr20 | 58298357 | | | 0.379325 | 3.74E-08 | 1.32E-05 |
| cg21530605_BC21 | chr7 | 105571380 | | | -0.181275 | 3.74E-08 | 1.32E-05 |
| cg01908666_TC21 | chr1 | 210569636 | | | 0.196708 | 3.74E-08 | 1.32E-05 |
| cg19566658_TC11 | chr7 | 100868619 | exon_4 | TRIP6 | -0.166466 | 3.75E-08 | 1.32E-05 |
| cg21366655_TC21 | chr14 | 103118202 | | | -0.237589 | 3.75E-08 | 1.32E-05 |
| cg03364381_BC21 | chr21 | 41679300 | TSS200 | LINC00111 | -0.497144 | 3.76E-08 | 1.32E-05 |
| cg19374905_TC21 | chr2 | 217978471 | TSS1500 | TNS1-AS1 | -0.332254 | 3.77E-08 | 1.32E-05 |
| cg19200827_BC21 | chr11 | 97943756 | | | 0.190968 | 3.77E-08 | 1.32E-05 |
| cg13840713_TC21 | chr16 | 57859636 | | | 0.166539 | 3.78E-08 | 1.33E-05 |
| cg14445396_BC21 | chr9 | 135555040 | | | -0.340966 | 3.78E-08 | 1.33E-05 |
| cg01496458_BC21 | chr1 | 158612254 | exon_2;3U | OR10Z1;OF | -0.240267 | 3.79E-08 | 1.33E-05 |
| cg23612492_TC21 | chr14 | 100424026 | | | -0.186113 | 3.79E-08 | 1.33E-05 |
| cg06220761_TC21 | chr10 | 7413556 | | | -0.217415 | 3.79E-08 | 1.33E-05 |
| cg13763000_BC21 | chr9 | 73986532 | | | -0.372995 | 3.79E-08 | 1.33E-05 |
| cg16298921_TC21 | chr9 | 25003841 | | | -0.271855 | 3.80E-08 | 1.33E-05 |

| | | | | | | | |
|---|---|---|---|---|---|---|---|
| cg24369388_TC21 | chr2 | 238382453 | | | 0.264896 | 3.81E-08 | 1.33E-05 |
| cg18522690_BC21 | chr22 | 36470464 | | | 0.197358 | 3.82E-08 | 1.33E-05 |
| cg05585734_TC21 | chr14 | 35332430 | | | -0.093212 | 3.83E-08 | 1.34E-05 |
| cg01571775_BC21 | chr7 | 4306544 | | | 0.239333 | 3.83E-08 | 1.34E-05 |
| cg25243771_TC21 | chr1 | 40782584 | TSS1500;TS | KCNQ4;KCI | -0.206726 | 3.84E-08 | 1.34E-05 |
| cg26301577_TC21 | chr6 | 46736125 | TSS1500;TS | PLA2G7;PL | 0.195619 | 3.84E-08 | 1.34E-05 |
| cg15342933_TC21 | chr11 | 18287916 | exon_16;e | HPS5;HPS5 | 0.299978 | 3.86E-08 | 1.35E-05 |
| cg25185001_TC11 | chr7 | 151149041 | exon_2 | GBX1 | -0.192832 | 3.87E-08 | 1.35E-05 |
| cg06988244_BC21 | chr4 | 116078217 | | | 0.410733 | 3.87E-08 | 1.35E-05 |
| cg19859781_TC21 | chr13 | 39600548 | | | 0.367288 | 3.87E-08 | 1.35E-05 |
| cg01908798_BC21 | chr1 | 210586877 | | | 0.179855 | 3.87E-08 | 1.35E-05 |
| cg01799353_BC21 | chr1 | 200254308 | | | -0.133192 | 3.88E-08 | 1.35E-05 |
| cg01476044_TC21 | chr7 | 141995493 | TSS1500;TS | MGAM;MG | 0.228149 | 3.89E-08 | 1.35E-05 |
| cg14761693_BC21 | chr8 | 125214482 | | | 0.258474 | 3.89E-08 | 1.35E-05 |
| cg20568322_BC11 | chr1 | 201946581 | TSS200 | LMOD1 | 0.15969 | 3.90E-08 | 1.36E-05 |
| cg22742114_BC21 | chr17 | 58332381 | TSS1500 | MIR142 | 0.31439 | 3.90E-08 | 1.36E-05 |
| cg05677014_BC21 | chr14 | 74913777 | | | -0.183804 | 3.90E-08 | 1.36E-05 |
| cg18716829_TC21 | chr4 | 70628558 | TSS200 | ENAM | -0.334995 | 3.91E-08 | 1.36E-05 |
| cg13263950_BC21 | chr1 | 35573795 | | | -0.173812 | 3.92E-08 | 1.36E-05 |
| cg05085383_TC21 | chr12 | 121275211 | TSS1500 | CAMKK2 | 0.394797 | 3.92E-08 | 1.36E-05 |
| cg19493223_TC21 | chr11 | 42522279 | | | -0.252028 | 3.92E-08 | 1.36E-05 |
| cg13555278_BC21 | chr1 | 26021993 | 5UTR;exon | EXTL1;EXTL | -0.1518 | 3.92E-08 | 1.36E-05 |
| cg05918890_TC21 | chr3 | 184460371 | | | 0.165541 | 3.93E-08 | 1.36E-05 |
| cg22895078_BC11 | chr17 | 71975642 | | | -0.301537 | 3.94E-08 | 1.37E-05 |
| cg14874907_TC21 | chr10 | 31693751 | | | -0.286911 | 3.94E-08 | 1.37E-05 |
| cg12112870_TC11 | chr8 | 85200696 | | | 0.226062 | 3.94E-08 | 1.37E-05 |
| cg03923641_TC21 | chr2 | 189540769 | | | -0.238779 | 3.94E-08 | 1.37E-05 |
| cg10549986_TC11 | chr2 | 6878022 | exon_1 | RSAD2 | 0.221271 | 3.94E-08 | 1.37E-05 |
| cg22637730_TC21 | chr3 | 160755766 | TSS1500 | PPM1L | -0.343169 | 3.95E-08 | 1.37E-05 |
| cg26819180_TC21 | chr6 | 168677153 | | | -0.184464 | 3.95E-08 | 1.37E-05 |
| cg17148613_BC21 | chr13 | 19820157 | | | 0.325942 | 3.96E-08 | 1.37E-05 |
| cg21325760_BC11 | chr15 | 23646255 | exon_1 | MAGEL2 | 0.177257 | 3.96E-08 | 1.37E-05 |
| cg15231222_BC21 | chr16 | 1675360 | exon_21;3 | CRAMP1;C | -0.206908 | 3.96E-08 | 1.37E-05 |
| cg12379775_BC21 | chr20 | 64038438 | exon_6;ex | C20orf204; | 0.197135 | 3.97E-08 | 1.37E-05 |
| cg23957912_TC21 | chr4 | 186375544 | | | -0.280017 | 3.97E-08 | 1.37E-05 |
| cg26535834_BC21 | chr2 | 238518346 | | | -0.312696 | 3.97E-08 | 1.37E-05 |
| cg09353378_BC21 | chr17 | 35947392 | | | 0.308196 | 3.98E-08 | 1.37E-05 |
| cg16615744_BC21 | chr14 | 22319248 | | | -0.310923 | 3.98E-08 | 1.37E-05 |
| cg06562836_TC21 | chr5 | 153492058 | TSS200;TSS | GRIA1;GRI | -0.254441 | 3.98E-08 | 1.37E-05 |
| cg03420871_BC21 | chr2 | 121568200 | | | -0.29064 | 3.99E-08 | 1.37E-05 |
| cg09611128_BC21 | chr6 | 39182103 | | | -0.208006 | 3.99E-08 | 1.37E-05 |
| cg03410711_BC21 | chr10 | 91634501 | TSS1500 | PPP1R3C | 0.196973 | 4.00E-08 | 1.38E-05 |
| cg17332897_TC21 | chr2 | 71276959 | | | 0.240567 | 4.00E-08 | 1.38E-05 |
| cg14006566_BC21 | chr14 | 31577247 | TSS1500 | NUBPL | -0.271808 | 4.00E-08 | 1.38E-05 |
| cg15142913_TC21 | chr8 | 141430150 | | | -0.322146 | 4.00E-08 | 1.38E-05 |
| cg17999686_TC11 | chr7 | 103990363 | TSS1500;TS | RELN;RELN | -0.33945 | 4.01E-08 | 1.38E-05 |
| cg20794541_BC21 | chr6 | 146879181 | | | -0.147203 | 4.01E-08 | 1.38E-05 |

| | | | | | | | |
|---|---|---|---|---|---|---|---|
| cg02346062_TC21 | chr6 | 24490940 | TSS1500 | GPLD1 | -0.181351 | 4.01E-08 | 1.38E-05 |
| cg07230440_BC21 | chr12 | 129872667 | | | -0.2911 | 4.02E-08 | 1.38E-05 |
| cg04124636_TC21 | chr16 | 983408 | | | -0.319243 | 4.02E-08 | 1.38E-05 |
| cg24662533_BC21 | chr6 | 53829472 | | | 0.171061 | 4.02E-08 | 1.38E-05 |
| cg15983280_BC21 | chr11 | 7677995 | | | -0.265233 | 4.02E-08 | 1.38E-05 |
| cg23503501_BC21 | chr22 | 45413912 | exon_1;TSS | RIBC2;SMC | -0.795807 | 4.02E-08 | 1.38E-05 |
| cg01476137_BC21 | chr14 | 101953659 | | | -0.224209 | 4.03E-08 | 1.38E-05 |
| cg07521808_BC21 | chr17 | 5032851 | 3UTR;exon | SLC52A1;SI | -0.193311 | 4.03E-08 | 1.38E-05 |
| cg05529886_BC21 | chr3 | 63343928 | | | -0.415933 | 4.03E-08 | 1.38E-05 |
| cg18023504_TC21 | chr7 | 115497061 | | | -0.152392 | 4.04E-08 | 1.38E-05 |
| cg13767755_BC21 | chr2 | 222298081 | TSS200 | CCDC140 | -0.285452 | 4.05E-08 | 1.38E-05 |
| cg05204898_BC21 | chr2 | 98577107 | exon_25;e | INPP4A;INI | -0.224751 | 4.05E-08 | 1.38E-05 |
| cg18640964_BC21 | chr8 | 54476816 | | | -0.150098 | 4.06E-08 | 1.39E-05 |
| cg26325867_BC21 | chr6 | 41430955 | | | -0.114111 | 4.07E-08 | 1.39E-05 |
| cg00098628_BC21 | chr1 | 43791541 | | | -0.433093 | 4.08E-08 | 1.39E-05 |
| cg04786693_BC21 | chr6 | 137172694 | | | -0.318373 | 4.08E-08 | 1.39E-05 |
| cg26493189_BC21 | chr1 | 166602396 | | | -0.398854 | 4.10E-08 | 1.40E-05 |
| cg20036294_BC21 | chr1 | 155397049 | | | 0.352174 | 4.10E-08 | 1.40E-05 |
| cg06373470_TC21 | chr10 | 86972118 | TSS1500;TS | ADIRF-AS1; | -0.200968 | 4.11E-08 | 1.40E-05 |
| cg14855089_BC21 | chr1 | 230133375 | | | -0.263704 | 4.11E-08 | 1.40E-05 |
| cg19127138_BC11 | chr7 | 65731914 | | | -0.272535 | 4.11E-08 | 1.40E-05 |
| cg25398905_TC21 | chr2 | 201694903 | | | -0.250767 | 4.11E-08 | 1.40E-05 |
| cg04097219_TC21 | chr5 | 143250184 | | | -0.266184 | 4.12E-08 | 1.40E-05 |
| cg12053462_TC21 | chr1 | 90704454 | | | -0.359538 | 4.12E-08 | 1.40E-05 |
| cg18012213_BC21 | chr3 | 66854027 | | | 0.323506 | 4.12E-08 | 1.40E-05 |
| cg06668495_TC21 | chr3 | 12609299 | exon_4;exo | RAF1;RAF1 | 0.257915 | 4.13E-08 | 1.41E-05 |
| cg13195486_TC21 | chr1 | 218284540 | TSS1500 | RRP15 | -0.192077 | 4.15E-08 | 1.41E-05 |
| cg07766402_BC21 | chr17 | 2240290 | | | 0.251688 | 4.15E-08 | 1.41E-05 |
| cg11334870_BC21 | chr5 | 172114823 | | | -0.29517 | 4.15E-08 | 1.41E-05 |
| cg10513594_TC21 | chr3 | 196411 | TSS1500;TS | CHL1;CHL1 | 0.259694 | 4.15E-08 | 1.41E-05 |
| cg02412256_BC21 | chr21 | 34036444 | | | -0.212714 | 4.15E-08 | 1.41E-05 |
| cg17451422_TC11 | chr2 | 61066118 | TSS200;TSS | SANBR;SAN | 0.217624 | 4.16E-08 | 1.41E-05 |
| cg12137682_TC21 | chr2 | 172829482 | | | -0.339178 | 4.17E-08 | 1.41E-05 |
| cg04953373_BC21 | chr6 | 166479274 | | | -0.305796 | 4.17E-08 | 1.41E-05 |
| cg24841310_BC11 | chr8 | 130304090 | | | -0.265699 | 4.17E-08 | 1.41E-05 |
| cg13931845_TC21 | chr9 | 87279950 | | | 0.208802 | 4.17E-08 | 1.41E-05 |
| cg06235560_BC21 | chr16 | 79505577 | | | -0.169564 | 4.17E-08 | 1.41E-05 |
| cg02636177_BC21 | chr4 | 71334614 | | | -0.133393 | 4.18E-08 | 1.41E-05 |
| cg15975724_BC21 | chr11 | 6632241 | exon_6 | DCHS1 | 0.171767 | 4.19E-08 | 1.42E-05 |
| cg15769876_BC21 | chr5 | 14773737 | | | -0.414219 | 4.19E-08 | 1.42E-05 |
| cg25662846_TC21 | chr21 | 15905562 | | | 0.242295 | 4.20E-08 | 1.42E-05 |
| cg20359323_TC21 | chr8 | 119965416 | | | -0.271527 | 4.20E-08 | 1.42E-05 |
| cg07721458_TC21 | chr19 | 55173924 | | | -0.39474 | 4.20E-08 | 1.42E-05 |
| cg14971320_BC11 | chr10 | 43203622 | | | -0.254521 | 4.21E-08 | 1.42E-05 |
| cg16509370_BC21 | chr8 | 26595522 | | | 0.288917 | 4.21E-08 | 1.42E-05 |
| cg00096603_BC21 | chr4 | 94650971 | | | 0.209611 | 4.22E-08 | 1.42E-05 |
| cg02558684_BC21 | chr9 | 94174544 | TSS1500 | MIRLET7A1 | -0.271251 | 4.23E-08 | 1.43E-05 |

| | | | | | | | |
|---|---|---|---|---|---|---|---|
| cg06991314_BC21 | chr6 | 120774268 | | | -0.218994 | 4.23E-08 | 1.43E-05 |
| cg11210138_TC21 | chr17 | 48156583 | TSS200 | MIR1203 | -0.158309 | 4.23E-08 | 1.43E-05 |
| cg06950392_BC11 | chr19 | 11102888 | | | 0.151292 | 4.24E-08 | 1.43E-05 |
| cg17786067_TC21 | chr4 | 122925101 | | | -0.243914 | 4.24E-08 | 1.43E-05 |
| cg14604706_BC21 | chr15 | 39247979 | | | -0.31048 | 4.25E-08 | 1.43E-05 |
| cg26081051_BC21 | chr12 | 31849413 | | | -0.145764 | 4.26E-08 | 1.43E-05 |
| cg09290397_BC21 | chr14 | 93232428 | | | -0.29079 | 4.26E-08 | 1.43E-05 |
| cg16466831_BC21 | chr16 | 1772345 | exon_3;exo | MRPS34;M | -0.246716 | 4.26E-08 | 1.43E-05 |
| cg13753443_BC21 | chr9 | 72474258 | | | -0.13935 | 4.27E-08 | 1.44E-05 |
| cg05449213_BC21 | chr6 | 130811283 | | | -0.297909 | 4.28E-08 | 1.44E-05 |
| cg09726646_BC11 | chr21 | 31344005 | | | 0.246099 | 4.28E-08 | 1.44E-05 |
| cg00125480_TC21 | chr21 | 35675013 | | | -0.21088 | 4.29E-08 | 1.44E-05 |
| cg01173733_BC21 | chr13 | 100580316 | | | 0.288126 | 4.30E-08 | 1.44E-05 |
| cg25963048_BC21 | chr17 | 42176249 | exon_5 | KCNH4 | -0.211887 | 4.31E-08 | 1.45E-05 |
| cg17477578_TC21 | chr8 | 128540668 | | | -0.095086 | 4.32E-08 | 1.45E-05 |
| cg08209375_TC21 | chr17 | 10119497 | | | 0.136275 | 4.32E-08 | 1.45E-05 |
| cg07950786_BC21 | chr1 | 109764950 | TSS1500;TS | EPS8L3;EPS | -0.219714 | 4.32E-08 | 1.45E-05 |
| cg27510960_BC21 | chr7 | 4857609 | | | -0.339978 | 4.32E-08 | 1.45E-05 |
| cg06272998_BC21 | chr4 | 6367573 | | | -0.201502 | 4.33E-08 | 1.45E-05 |
| cg01799623_BC21 | chr1 | 200279888 | | | -0.115714 | 4.33E-08 | 1.45E-05 |
| cg01257890_TC21 | chr16 | 20918987 | | | 0.193002 | 4.34E-08 | 1.45E-05 |
| cg07208649_TC21 | chr17 | 4788931 | TSS200;TSS | GLTPD2;GL | -0.175374 | 4.34E-08 | 1.45E-05 |
| cg09837799_TC21 | chr3 | 81974705 | | | -0.184717 | 4.35E-08 | 1.45E-05 |
| cg04110061_TC21 | chr16 | 81199111 | | | 0.295179 | 4.35E-08 | 1.45E-05 |
| cg04610178_TC21 | chr7 | 140240068 | | | 0.215647 | 4.35E-08 | 1.45E-05 |
| cg11445094_TC11 | chr6 | 28554060 | | | -0.250283 | 4.35E-08 | 1.45E-05 |
| cg15403900_TC21 | chr16 | 85634243 | | | -0.11098 | 4.36E-08 | 1.45E-05 |
| cg05338505_BC21 | chr7 | 5570043 | | | 0.217978 | 4.36E-08 | 1.45E-05 |
| cg24505241_TC21 | chr3 | 107218885 | | | -0.219443 | 4.36E-08 | 1.46E-05 |
| cg26160573_TC21 | chr2 | 130355442 | TSS1500;TS | PTPN18;PT | -0.170929 | 4.36E-08 | 1.46E-05 |
| cg01507634_BC21 | chr1 | 160117608 | | | -0.12202 | 4.37E-08 | 1.46E-05 |
| cg19614454_BC21 | chr6 | 138944438 | | | -0.310759 | 4.38E-08 | 1.46E-05 |
| cg21857204_BC21 | chr14 | 59819091 | | | -0.302949 | 4.39E-08 | 1.46E-05 |
| cg08886418_TC21 | chr5 | 172235893 | | | -0.250452 | 4.39E-08 | 1.46E-05 |
| cg03572011_TC21 | chr17 | 45151877 | exon_1;3U | HEXIM1;HE | -0.171822 | 4.39E-08 | 1.46E-05 |
| cg07856568_TC21 | chr6 | 34476984 | | | -0.115676 | 4.39E-08 | 1.46E-05 |
| cg18068949_TC21 | chr12 | 109727219 | | | -0.196212 | 4.40E-08 | 1.46E-05 |
| cg12189289_TC21 | chr9 | 83965725 | | | -0.220933 | 4.40E-08 | 1.46E-05 |
| cg20590643_BC21 | chr14 | 51299725 | | | -0.24912 | 4.41E-08 | 1.47E-05 |
| cg18921025_TC21 | chr6 | 30984287 | | | -0.188568 | 4.41E-08 | 1.47E-05 |
| cg09545384_TC21 | chr20 | 15699568 | | | -0.177808 | 4.42E-08 | 1.47E-05 |
| cg21197958_TC21 | chr17 | 38585133 | | | 0.090221 | 4.42E-08 | 1.47E-05 |
| cg21278129_BC21 | chr19 | 7732469 | TSS1500;TS | CLEC4G;CL | -0.200266 | 4.42E-08 | 1.47E-05 |
| cg00822607_TC11 | chr16 | 30571597 | exon_1 | ZNF688 | -0.167753 | 4.43E-08 | 1.47E-05 |
| cg00449934_BC21 | chr1 | 114976244 | | | -0.308418 | 4.43E-08 | 1.47E-05 |
| cg24718197_BC11 | chr17 | 81518683 | | | -0.165053 | 4.43E-08 | 1.47E-05 |
| cg18848287_BC21 | chr7 | 5072010 | TSS200 | RBAKDN | -0.391564 | 4.44E-08 | 1.47E-05 |

| | | | | | | | |
|---|---|---|---|---|---|---|---|
| cg03161804_TC21 | chr3 | 148279656 | TSS1500 | LINC02046 | -0.262501 | 4.44E-08 | 1.47E-05 |
| cg01264197_BC21 | chr3 | 127389343 | | | -0.627757 | 4.44E-08 | 1.47E-05 |
| cg21336877_BC21 | chr22 | 25726441 | exon_20;3U | GRK3;GRK3 | -0.14095 | 4.45E-08 | 1.47E-05 |
| cg11472320_BC21 | chr19 | 11237332 | | | -0.115646 | 4.46E-08 | 1.48E-05 |
| cg00341906_TC21 | chr9 | 2933229 | | | -0.182483 | 4.46E-08 | 1.48E-05 |
| cg06895843_BC21 | chr3 | 128384679 | | | -0.20427 | 4.46E-08 | 1.48E-05 |
| cg14112545_BC21 | chr9 | 111692012 | exon_23;ex | SHOC1;SHC | -0.173525 | 4.46E-08 | 1.48E-05 |
| cg22379697_BC21 | chr20 | 41475866 | | | 0.25216 | 4.47E-08 | 1.48E-05 |
| cg00894477_BC21 | chr3 | 59424869 | | | 0.17806 | 4.48E-08 | 1.48E-05 |
| cg09581888_TC21 | chr6 | 52724586 | | | -0.26844 | 4.49E-08 | 1.48E-05 |
| cg03229996_TC21 | chr16 | 88424611 | | | -0.156739 | 4.50E-08 | 1.48E-05 |
| cg18749832_BC21 | chr13 | 51919727 | | | -0.21277 | 4.50E-08 | 1.48E-05 |
| cg12776618_TC21 | chr8 | 80998379 | | | -0.147852 | 4.50E-08 | 1.48E-05 |
| cg03212678_BC21 | chr11 | 800670 | | | -0.169652 | 4.50E-08 | 1.48E-05 |
| cg15918906_TC21 | chr19 | 52068879 | | | 0.387869 | 4.50E-08 | 1.48E-05 |
| cg00350503_TC21 | chr19 | 13502800 | | | 0.280063 | 4.50E-08 | 1.48E-05 |
| cg23766254_TC11 | chr17 | 44354491 | exon_8 | FAM171A2 | -0.273822 | 4.50E-08 | 1.48E-05 |
| cg13598232_BC21 | chr6 | 33316971 | exon_2;5U | ZBTB22;ZB | -0.222153 | 4.51E-08 | 1.49E-05 |
| cg11349980_BC21 | chr10 | 970755 | | | -0.156683 | 4.53E-08 | 1.49E-05 |
| cg02790967_BC21 | chr7 | 7567391 | 5UTR;exon | MIOS;MIOS | -0.325806 | 4.53E-08 | 1.49E-05 |
| cg05660634_BC21 | chr15 | 38672908 | | | 0.223604 | 4.53E-08 | 1.49E-05 |
| cg25538012_BC21 | chr22 | 43254290 | | | -0.152146 | 4.54E-08 | 1.49E-05 |
| cg06497051_BC21 | chr1 | 156291409 | 5UTR;exon | SMG5;SMG | -0.155431 | 4.54E-08 | 1.49E-05 |
| cg17576261_TC21 | chr12 | 49046214 | | | 0.168829 | 4.54E-08 | 1.49E-05 |
| cg01675596_TC21 | chr8 | 144686512 | TSS1500;TS | ARHGAP39 | -0.383706 | 4.54E-08 | 1.49E-05 |
| cg03068051_TC21 | chr1 | 64469486 | TSS1500 | CACHD1 | -0.159176 | 4.54E-08 | 1.49E-05 |
| cg11625723_BC21 | chr10 | 7469380 | | | -0.173998 | 4.54E-08 | 1.49E-05 |
| cg24859029_TC21 | chr19 | 7910527 | exon_6;exo | MAP2K7;M | -0.254636 | 4.55E-08 | 1.49E-05 |
| cg02603875_BC21 | chr5 | 153491989 | TSS200;TSS | GRIA1;GRIA | -0.207438 | 4.55E-08 | 1.49E-05 |
| cg27110569_BC21 | chr1 | 178544903 | | | -0.465202 | 4.56E-08 | 1.49E-05 |
| cg02345255_BC21 | chr11 | 134583537 | | | 0.196128 | 4.57E-08 | 1.50E-05 |
| cg25484319_BC21 | chr14 | 67695587 | | | 0.312092 | 4.58E-08 | 1.50E-05 |
| cg14120049_BC11 | chr19 | 35755507 | 3UTR;exon | HSPB6;HSP | -0.342836 | 4.58E-08 | 1.50E-05 |
| cg24169178_TC21 | chr1 | 33093269 | 5UTR;exon | AZIN2;AZIN | -0.221284 | 4.59E-08 | 1.50E-05 |
| cg25091367_BC21 | chr4 | 7287780 | | | 0.189586 | 4.59E-08 | 1.50E-05 |
| cg13520102_BC21 | chr15 | 87672593 | | | -0.33614 | 4.60E-08 | 1.50E-05 |
| cg22885214_TC21 | chr11 | 8009017 | | | -0.202891 | 4.60E-08 | 1.50E-05 |
| cg26333986_TC21 | chr22 | 38755011 | TSS200;TSS | SUN2;SUN2 | 0.282691 | 4.60E-08 | 1.50E-05 |
| cg05499776_BC21 | chr22 | 36286233 | | | -0.139483 | 4.61E-08 | 1.50E-05 |
| cg13433390_BC21 | chr9 | 74799833 | | | -0.304689 | 4.61E-08 | 1.51E-05 |
| cg22064635_TC21 | chr3 | 111621725 | | | -0.168686 | 4.62E-08 | 1.51E-05 |
| cg15952487_TC21 | chr1 | 158330943 | exon_2 | CD1B | -0.184622 | 4.62E-08 | 1.51E-05 |
| cg12799537_BC21 | chr12 | 55814125 | | | -0.276796 | 4.62E-08 | 1.51E-05 |
| cg24590766_TC21 | chr1 | 44970695 | | | -0.205377 | 4.62E-08 | 1.51E-05 |
| cg04866691_BC21 | chr3 | 48586349 | exon_27 | COL7A1 | -0.1978 | 4.63E-08 | 1.51E-05 |
| cg20652404_TC21 | chr15 | 73926563 | 5UTR;exon | LOXL1;LOX | -0.172346 | 4.63E-08 | 1.51E-05 |
| cg04647162_TC21 | chr17 | 59409716 | | | -0.216565 | 4.63E-08 | 1.51E-05 |

| | | | | | | | |
|---|---|---|---|---|---|---|---|
| cg21092682_BC21 | chr4 | 74308162 | TSS1500;TS | EPGN;EPGN | 0.299226 | 4.64E-08 | 1.51E-05 |
| cg07886153_TC21 | chr12 | 80762975 | TSS200;TSS | LINC01490 | -0.270031 | 4.64E-08 | 1.51E-05 |
| cg09048665_TC11 | chr16 | 654976 | | | -0.179276 | 4.65E-08 | 1.51E-05 |
| cg26888312_BC21 | chr15 | 69309627 | | | -0.218238 | 4.66E-08 | 1.51E-05 |
| cg20012848_BC11 | chr12 | 52041636 | | | -0.196775 | 4.66E-08 | 1.51E-05 |
| cg11931123_TC21 | chr16 | 55508919 | TSS200 | LPCAT2 | -0.310772 | 4.68E-08 | 1.52E-05 |
| cg12000761_TC21 | chr19 | 49597266 | exon_4 | PRR12 | -0.284688 | 4.68E-08 | 1.52E-05 |
| cg22250571_BC21 | chr16 | 30973026 | | | -0.175789 | 4.69E-08 | 1.52E-05 |
| cg00734311_BC11 | chr6 | 139726406 | | | -0.313828 | 4.70E-08 | 1.52E-05 |
| cg23106378_BC11 | chr8 | 143549751 | | | -0.440076 | 4.70E-08 | 1.52E-05 |
| cg09484977_BC21 | chr6 | 41626107 | | | -0.362942 | 4.70E-08 | 1.52E-05 |
| cg27516506_BC11 | chr22 | 37199493 | TSS200 | C1QTNF6 | -0.303537 | 4.70E-08 | 1.52E-05 |
| cg16563569_TC21 | chr1 | 17193313 | | | -0.215737 | 4.71E-08 | 1.52E-05 |
| cg13653173_BC21 | chr6 | 32876892 | | | -0.286964 | 4.71E-08 | 1.53E-05 |
| cg14576839_TC21 | chr5 | 178940232 | TSS1500;TS | ZNF454;ZN | -0.16351 | 4.72E-08 | 1.53E-05 |
| cg21985441_TC21 | chr16 | 55036212 | | | -0.128341 | 4.72E-08 | 1.53E-05 |
| cg23752348_TC21 | chr6 | 30074360 | | | -0.188234 | 4.72E-08 | 1.53E-05 |
| cg08977179_BC21 | chr4 | 96352439 | | | -0.205021 | 4.72E-08 | 1.53E-05 |
| cg18813545_BC21 | chr3 | 9125501 | | | -0.152455 | 4.74E-08 | 1.53E-05 |
| cg27169539_BC21 | chr14 | 21476629 | 5UTR;exon | RAB2B;RAB | 0.365694 | 4.75E-08 | 1.53E-05 |
| cg00010692_TC11 | chr1 | 977689 | | | 0.508915 | 4.75E-08 | 1.53E-05 |
| cg21125559_BC21 | chr13 | 96140840 | | | -0.335652 | 4.75E-08 | 1.54E-05 |
| cg12260277_BC21 | chr9 | 37443032 | | | -0.306877 | 4.77E-08 | 1.54E-05 |
| cg05549350_BC21 | chr15 | 24859394 | | | -0.240291 | 4.78E-08 | 1.54E-05 |
| cg02032593_BC21 | chr3 | 127523558 | | | -0.225254 | 4.79E-08 | 1.54E-05 |
| cg04477101_BC21 | chr17 | 2371736 | | | 0.200945 | 4.79E-08 | 1.54E-05 |
| cg22859309_BC21 | chr14 | 96383666 | | | -0.336487 | 4.79E-08 | 1.54E-05 |
| cg16806046_TC21 | chr13 | 39306851 | | | -0.286381 | 4.80E-08 | 1.55E-05 |
| cg21103567_BC21 | chr19 | 13275849 | | | 0.136073 | 4.82E-08 | 1.55E-05 |
| cg06892123_TC21 | chr10 | 71656129 | | | -0.156497 | 4.84E-08 | 1.56E-05 |
| cg03950798_BC21 | chr4 | 34667817 | | | -0.307629 | 4.86E-08 | 1.56E-05 |
| cg11701421_BC21 | chr17 | 65678064 | | | -0.369295 | 4.86E-08 | 1.56E-05 |
| cg05167523_BC21 | chr13 | 94188990 | TSS1500 | GPC6-AS1 | 0.294451 | 4.87E-08 | 1.57E-05 |
| cg09947542_TC21 | chr1 | 158115648 | | | -0.105743 | 4.87E-08 | 1.57E-05 |
| cg25898882_BC21 | chr21 | 41973674 | | | -0.20599 | 4.87E-08 | 1.57E-05 |
| cg06993255_BC21 | chr8 | 144620849 | | | -0.210574 | 4.88E-08 | 1.57E-05 |
| cg07093570_BC21 | chr1 | 70487697 | | | -0.187294 | 4.89E-08 | 1.57E-05 |
| cg01049355_TC21 | chr11 | 18509072 | | | -0.388467 | 4.89E-08 | 1.57E-05 |
| cg03706479_BC21 | chr11 | 73788864 | | | -0.746665 | 4.89E-08 | 1.57E-05 |
| cg10627436_TC21 | chr14 | 101957817 | | | -0.220595 | 4.89E-08 | 1.57E-05 |
| cg07175786_BC21 | chr9 | 111178323 | | | 0.28964 | 4.89E-08 | 1.57E-05 |
| cg10954560_BC21 | chr6 | 46793107 | TSS1500 | MEP1A | -0.246886 | 4.90E-08 | 1.57E-05 |
| cg26154897_BC21 | chr22 | 43171884 | exon_11 | TTLL12 | -0.129139 | 4.90E-08 | 1.57E-05 |
| cg04034803_TC21 | chr2 | 203759110 | | | -0.158498 | 4.90E-08 | 1.57E-05 |
| cg15762048_TC21 | chr10 | 128098981 | 3UTR;exon | MKI67;MKI | -0.31577 | 4.90E-08 | 1.57E-05 |
| cg10295188_BC11 | chr6 | 154938331 | | | -0.280926 | 4.91E-08 | 1.57E-05 |
| cg01749549_TC21 | chr1 | 246817790 | | | -0.243698 | 4.93E-08 | 1.58E-05 |

| | | | | | | | |
|---|---|---|---|---|---|---|---|
| cg12740693_TC21 | chr8 | 75473007 | | | -0.398591 | 4.93E-08 | 1.58E-05 |
| cg19817820_BC11 | chr10 | 133200287 | | | -0.183833 | 4.93E-08 | 1.58E-05 |
| cg04422266_TC21 | chr12 | 110679933 | | | -0.22403 | 4.93E-08 | 1.58E-05 |
| cg06485774_TC21 | chr1 | 184890191 | exon_4 | NIBAN1 | -0.261008 | 4.93E-08 | 1.58E-05 |
| cg21149639_BC21 | chr4 | 8192916 | | | 0.14404 | 4.95E-08 | 1.58E-05 |
| cg04141974_TC21 | chr2 | 217881159 | | | -0.210177 | 4.95E-08 | 1.58E-05 |
| cg09864183_BC21 | chr5 | 137141962 | | | 0.269329 | 4.95E-08 | 1.58E-05 |
| cg05318071_BC21 | chr2 | 118087700 | TSS1500;TS | INSIG2;INSI | 0.174096 | 4.95E-08 | 1.58E-05 |
| cg00306641_BC21 | chr16 | 70538647 | | | -0.157932 | 4.97E-08 | 1.58E-05 |
| cg04368724_BC21 | chr6 | 31792816 | exon_4 | VARS1 | 0.275174 | 4.97E-08 | 1.58E-05 |
| cg18008675_TC21 | chr2 | 172968208 | | | -0.26845 | 4.97E-08 | 1.58E-05 |
| cg27606137_BC21 | chr4 | 37890195 | TSS1500;TS | TBC1D1;TB | -0.287594 | 4.97E-08 | 1.58E-05 |
| cg13074173_TC21 | chr8 | 143379837 | exon_9 | RHPN1 | 0.183797 | 4.98E-08 | 1.59E-05 |
| cg11845532_BC21 | chr12 | 3252830 | | | 0.196337 | 5.00E-08 | 1.59E-05 |
| cg02230941_BC21 | chr14 | 85482583 | | | -0.242025 | 5.00E-08 | 1.59E-05 |
| cg18692621_BC21 | chr17 | 55774965 | | | -0.337815 | 5.00E-08 | 1.59E-05 |
| cg20089532_BC21 | chr10 | 48252506 | TSS1500 | FRMPD2 | 0.168762 | 5.00E-08 | 1.59E-05 |
| cg02219447_BC21 | chr15 | 85627935 | | | -0.850586 | 5.00E-08 | 1.59E-05 |
| cg09493945_BC21 | chr6 | 42178003 | | | -0.146133 | 5.01E-08 | 1.59E-05 |
| cg16060369_TC11 | chr10 | 43203386 | exon_3;exo | RASGEF1A; | -0.200329 | 5.02E-08 | 1.60E-05 |
| cg07446376_BC21 | chr1 | 3347005 | | | -0.164149 | 5.03E-08 | 1.60E-05 |
| cg17120795_TC21 | chr7 | 94446184 | | | -0.230036 | 5.05E-08 | 1.60E-05 |
| cg15851696_BC21 | chr2 | 182318547 | | | -0.28123 | 5.05E-08 | 1.60E-05 |
| cg16812425_TC21 | chr4 | 176237681 | | | -0.350249 | 5.05E-08 | 1.60E-05 |
| cg05495155_TC21 | chr19 | 44800607 | 3UTR;exon | CBLC;CBLC; | 0.424433 | 5.05E-08 | 1.60E-05 |
| cg11061136_BC21 | chr5 | 176542422 | TSS200 | CDHR2 | -0.276337 | 5.07E-08 | 1.61E-05 |
| cg07020967_BC21 | chr12 | 44016695 | | | -0.092156 | 5.08E-08 | 1.61E-05 |
| cg11250268_BC21 | chr20 | 63666395 | | | -0.193371 | 5.08E-08 | 1.61E-05 |
| cg11659341_BC21 | chr15 | 52858215 | | | -0.248924 | 5.09E-08 | 1.61E-05 |
| cg07332724_TC21 | chr12 | 54379330 | | | -0.146833 | 5.09E-08 | 1.61E-05 |
| cg19642408_TC21 | chr2 | 9754206 | | | -0.292966 | 5.10E-08 | 1.61E-05 |
| cg16757449_TC21 | chr7 | 73849148 | | | -0.129094 | 5.10E-08 | 1.61E-05 |
| cg27557317_TC21 | chr1 | 26233877 | TSS1500;TS | CEP85;CEP | 0.20215 | 5.11E-08 | 1.61E-05 |
| cg03234419_BC21 | chr1 | 161231355 | exon_6;exo | NR1I3;NR1 | 0.225313 | 5.11E-08 | 1.61E-05 |
| cg26352827_BC21 | chr16 | 3304591 | TSS1500;TS | ZNF75A;ZN | -0.309917 | 5.11E-08 | 1.61E-05 |
| cg21263146_BC21 | chr11 | 36016842 | | | 0.257614 | 5.12E-08 | 1.62E-05 |
| cg06143361_TC11 | chr3 | 42803715 | | | -0.201149 | 5.12E-08 | 1.62E-05 |
| cg22440004_TC21 | chr17 | 34069017 | | | -0.187928 | 5.13E-08 | 1.62E-05 |
| cg07585749_TC21 | chr20 | 13325287 | | | -0.189028 | 5.13E-08 | 1.62E-05 |
| cg22527127_BC21 | chr17 | 40774695 | | | 0.306491 | 5.13E-08 | 1.62E-05 |
| cg11918751_TC21 | chr1 | 197266721 | | | -0.303681 | 5.13E-08 | 1.62E-05 |
| cg12005026_BC21 | chr10 | 87937017 | | | 0.11712 | 5.13E-08 | 1.62E-05 |
| cg25641330_BC21 | chr6 | 54805369 | | | -0.354809 | 5.14E-08 | 1.62E-05 |
| cg05368740_TC11 | chr19 | 35755981 | | | -0.346773 | 5.14E-08 | 1.62E-05 |
| cg26217974_TC21 | chr2 | 42623326 | | | 0.19145 | 5.14E-08 | 1.62E-05 |
| cg00213606_TC11 | chr15 | 22926080 | exon_14;e) | CYFIP1;CYF | 0.260199 | 5.15E-08 | 1.62E-05 |
| cg19093056_BC11 | chr8 | 100951651 | TSS1500;TS | YWHAZ;YW | -0.332398 | 5.16E-08 | 1.62E-05 |

| | | | | | | | |
|---|---|---|---|---|---|---|---|
| cg18564686_TC21 | chr2 | 10029548 | | | 0.28806 | 5.17E-08 | 1.62E-05 |
| cg25153233_BC21 | chr20 | 39225626 | TSS1500 | LINC01734 | -0.184297 | 5.17E-08 | 1.63E-05 |
| cg02469297_TC11 | chr16 | 23755204 | | | -0.264565 | 5.17E-08 | 1.63E-05 |
| cg01583021_TC21 | chr2 | 241199126 | | | -0.164942 | 5.18E-08 | 1.63E-05 |
| cg21142107_BC21 | chr17 | 19482611 | | | -0.299055 | 5.20E-08 | 1.63E-05 |
| cg16362201_BC21 | chr4 | 56652290 | | | -0.171631 | 5.20E-08 | 1.63E-05 |
| cg22828383_BC11 | chr12 | 124517645 | | | 0.1388 | 5.21E-08 | 1.64E-05 |
| cg27020036_TC21 | chr5 | 180216370 | | | -0.188986 | 5.22E-08 | 1.64E-05 |
| cg15921669_TC21 | chr12 | 27973597 | TSS1500 | PTHLH | 0.196755 | 5.22E-08 | 1.64E-05 |
| cg18144198_BC21 | chr12 | 115508075 | | | -0.211125 | 5.22E-08 | 1.64E-05 |
| cg06821107_BC21 | chr9 | 133376677 | TSS1500;TS | SURF4;SUR | 0.279605 | 5.22E-08 | 1.64E-05 |
| cg00989803_TC21 | chr1 | 81920679 | | | -0.232531 | 5.26E-08 | 1.65E-05 |
| cg00489772_BC21 | chr1 | 3858514 | TSS1500 | CEP104 | -0.447562 | 5.27E-08 | 1.65E-05 |
| cg01165781_BC11 | chr1 | 27349626 | | | -0.27896 | 5.27E-08 | 1.65E-05 |
| cg10091866_TC21 | chr1 | 185735417 | | | -0.328256 | 5.27E-08 | 1.65E-05 |
| cg15282819_BC21 | chr16 | 12903735 | | | -0.216279 | 5.28E-08 | 1.65E-05 |
| cg07970752_TC11 | chr16 | 767463 | exon_16;ex | MSLN;MSL | 0.497723 | 5.28E-08 | 1.65E-05 |
| cg27212089_BC21 | chr13 | 20869875 | | | -0.126637 | 5.28E-08 | 1.65E-05 |
| cg17435575_TC21 | chr6 | 71165127 | | | -0.224633 | 5.29E-08 | 1.65E-05 |
| cg24201941_TC21 | chr10 | 97937277 | | | -0.141968 | 5.30E-08 | 1.65E-05 |
| cg02416105_BC21 | chr2 | 174076095 | | | 0.319804 | 5.30E-08 | 1.65E-05 |
| cg14549249_TC21 | chr7 | 3043907 | TSS200;TSS | CARD11;CA | -0.247126 | 5.31E-08 | 1.66E-05 |
| cg19104757_TC11 | chr7 | 226563 | | | -0.137447 | 5.31E-08 | 1.66E-05 |
| cg09305680_TC21 | chr8 | 116765830 | TSS1500 | UTP23 | -0.22528 | 5.31E-08 | 1.66E-05 |
| cg15073016_BC21 | chr16 | 11023992 | | | 0.251824 | 5.33E-08 | 1.66E-05 |
| cg10454127_BC21 | chr16 | 86352485 | | | -0.107148 | 5.33E-08 | 1.66E-05 |
| cg02087795_TC21 | chr10 | 124264422 | | | 0.20899 | 5.33E-08 | 1.66E-05 |
| cg09749643_TC21 | chr3 | 20013486 | TSS1500;TS | PP2D1;PP2 | -0.301417 | 5.34E-08 | 1.66E-05 |
| cg07088771_TC21 | chr6 | 32090069 | | | -0.430959 | 5.34E-08 | 1.66E-05 |
| cg10438391_TC11 | chr8 | 143549745 | | | -0.504917 | 5.34E-08 | 1.66E-05 |
| cg20801491_TC21 | chr5 | 76952695 | TSS1500 | CRHBP | -0.149835 | 5.35E-08 | 1.66E-05 |
| cg11414276_TC21 | chr12 | 2057665 | | | -0.155057 | 5.36E-08 | 1.67E-05 |
| cg19954481_BC21 | chr2 | 95107249 | | | -0.127006 | 5.36E-08 | 1.67E-05 |
| cg23782616_TC11 | chr18 | 63949292 | TSS200 | HMSD | -0.254205 | 5.36E-08 | 1.67E-05 |
| cg13793262_TC21 | chr10 | 123688038 | exon_3 | GPR26 | -0.345172 | 5.37E-08 | 1.67E-05 |
| cg23080898_TC21 | chr17 | 81253440 | | | 0.238566 | 5.37E-08 | 1.67E-05 |
| cg08605991_TC21 | chr13 | 45852103 | TSS1500 | SIAH3 | 0.421465 | 5.38E-08 | 1.67E-05 |
| cg21717644_BC21 | chr16 | 17541129 | | | -0.266359 | 5.38E-08 | 1.67E-05 |
| cg16032894_TC21 | chr11 | 30583864 | | | -0.368648 | 5.38E-08 | 1.67E-05 |
| cg14140554_TC21 | chr6 | 157383007 | | | -0.438911 | 5.38E-08 | 1.67E-05 |
| cg00616135_TC21 | chr15 | 63121415 | TSS1500;TS | LACTB;LAC | -0.371699 | 5.38E-08 | 1.67E-05 |
| cg19014664_TC21 | chr5 | 876961 | | | 0.245182 | 5.38E-08 | 1.67E-05 |
| cg25938010_BC21 | chr6 | 30192303 | | | 0.155965 | 5.38E-08 | 1.67E-05 |
| cg00274888_TC21 | chr14 | 45274821 | | | -0.239013 | 5.39E-08 | 1.67E-05 |
| cg03935379_TC21 | chr5 | 1282588 | exon_3;exo | TERT;TERT; | -0.197861 | 5.39E-08 | 1.67E-05 |
| cg07068045_BC13 | chr11 | 69497434 | | | -0.313699 | 5.40E-08 | 1.67E-05 |
| cg04281994_TC21 | chr2 | 231388438 | | | 0.186104 | 5.40E-08 | 1.67E-05 |

| | | | | | | | |
|---|---|---|---|---|---|---|---|
| cg23195757_BC21 | chr11 | 5205049 | | | -0.14154 | 5.41E-08 | 1.67E-05 |
| cg02319094_BC21 | chr11 | 73269821 | TSS200 | P2RY6 | -0.110797 | 5.42E-08 | 1.67E-05 |
| cg00626167_TC21 | chr12 | 116697375 | | | -0.207677 | 5.42E-08 | 1.67E-05 |
| cg11557984_TC21 | chr3 | 177934706 | | | 0.250173 | 5.42E-08 | 1.67E-05 |
| cg12667716_BC21 | chr4 | 39700281 | | | 0.237901 | 5.42E-08 | 1.67E-05 |
| cg12910107_TC21 | chr2 | 27029646 | | | 0.272325 | 5.43E-08 | 1.68E-05 |
| cg23589976_TC21 | chr7 | 44701573 | exon_20;ex | OGDH;OGD | 0.297141 | 5.43E-08 | 1.68E-05 |
| cg24859092_BC21 | chr1 | 160654863 | | | -0.212482 | 5.43E-08 | 1.68E-05 |
| cg13748570_BC21 | chr17 | 64002718 | 3UTR;exon_ | ICAM2;ICAI | -0.348699 | 5.44E-08 | 1.68E-05 |
| cg03109701_BC21 | chr12 | 116736953 | TSS1500;TS | RNFT2;RNF | -0.148321 | 5.44E-08 | 1.68E-05 |
| cg00594278_BC21 | chr22 | 39900403 | TSS1500 | GRAP2 | 0.193298 | 5.46E-08 | 1.68E-05 |
| cg25478161_BC21 | chr15 | 44664060 | TSS1500;TS | SPG11;SPG | 0.163851 | 5.48E-08 | 1.69E-05 |
| cg09745989_BC21 | chr5 | 119471016 | | | -0.256467 | 5.48E-08 | 1.69E-05 |
| cg15677870_BC21 | chr13 | 103517125 | | | -0.137165 | 5.49E-08 | 1.69E-05 |
| cg06780673_BC21 | chr10 | 87607949 | | | 0.259358 | 5.50E-08 | 1.69E-05 |
| cg23317381_BC21 | chr8 | 144991904 | | | -0.408397 | 5.50E-08 | 1.69E-05 |
| cg19348178_TC21 | chr7 | 41028794 | | | -0.293651 | 5.51E-08 | 1.69E-05 |
| cg10888707_BC21 | chr6 | 90537289 | | | 0.272582 | 5.53E-08 | 1.70E-05 |
| cg02286761_TC21 | chr19 | 14204882 | | | -0.20744 | 5.53E-08 | 1.70E-05 |
| cg13983281_TC21 | chr9 | 96442139 | | | -0.120646 | 5.53E-08 | 1.70E-05 |
| cg07573942_TC21 | chr5 | 1253540 | 3UTR;exon_ | TERT;TERT; | -0.164488 | 5.53E-08 | 1.70E-05 |
| cg03733065_TC21 | chr2 | 162733282 | | | 0.219634 | 5.54E-08 | 1.70E-05 |
| cg07407218_BC21 | chr1 | 50330628 | | | 0.328213 | 5.56E-08 | 1.70E-05 |
| cg04484523_BC21 | chr1 | 209705044 | | | 0.291786 | 5.56E-08 | 1.71E-05 |
| cg13557129_BC21 | chr12 | 45065463 | TSS200 | RACGAP1P | -0.222903 | 5.57E-08 | 1.71E-05 |
| cg22947679_BC21 | chr1 | 117366553 | TSS1500 | MAN1A2 | -0.179319 | 5.57E-08 | 1.71E-05 |
| cg02331890_BC21 | chr2 | 2662700 | | | -0.125076 | 5.58E-08 | 1.71E-05 |
| cg13038847_TC21 | chr22 | 27678000 | | | 0.314425 | 5.58E-08 | 1.71E-05 |
| cg25933195_TC11 | chr19 | 12722742 | TSS1500;TS | TNPO2;TNI | 0.277083 | 5.58E-08 | 1.71E-05 |
| cg05295329_BC21 | chr2 | 38215456 | | | -0.158464 | 5.58E-08 | 1.71E-05 |
| cg16189627_BC21 | chr11 | 34052980 | | | 0.259108 | 5.59E-08 | 1.71E-05 |
| cg06598290_TC21 | chr8 | 20235645 | | | 0.258699 | 5.59E-08 | 1.71E-05 |
| cg26437267_BC21 | chr1 | 107136102 | | | 0.336872 | 5.59E-08 | 1.71E-05 |
| cg15516836_BC21 | chr20 | 33886534 | | | 0.220727 | 5.60E-08 | 1.71E-05 |
| cg14876571_BC21 | chr9 | 131950215 | | | -0.335101 | 5.61E-08 | 1.71E-05 |
| cg12751207_TC21 | chr15 | 72116725 | TSS200;TSS | SENP8;SEN | -0.244216 | 5.61E-08 | 1.71E-05 |
| cg00140798_TC21 | chr18 | 24429136 | | | -0.380803 | 5.61E-08 | 1.71E-05 |
| cg12793943_BC21 | chr12 | 125431342 | | | -0.215357 | 5.61E-08 | 1.71E-05 |
| cg24345118_TC21 | chr19 | 27775046 | | | 0.422095 | 5.61E-08 | 1.71E-05 |
| cg05265931_BC21 | chr5 | 31432423 | | | 0.259151 | 5.62E-08 | 1.71E-05 |
| cg19426625_TC21 | chr2 | 9378195 | | | 0.142125 | 5.62E-08 | 1.71E-05 |
| cg14476984_BC21 | chr5 | 58863645 | | | 0.28383 | 5.63E-08 | 1.72E-05 |
| cg19353326_TC21 | chr13 | 112160769 | TSS200 | LOC10050€ | 0.348201 | 5.63E-08 | 1.72E-05 |
| cg00467338_BC21 | chr2 | 86332496 | | | 0.354265 | 5.64E-08 | 1.72E-05 |
| cg04908088_BC21 | chr13 | 20350924 | | | 0.191195 | 5.65E-08 | 1.72E-05 |
| cg04336930_BC21 | chr10 | 30743177 | | | 0.226742 | 5.65E-08 | 1.72E-05 |
| cg03126633_TC21 | chr13 | 30143243 | | | 0.150645 | 5.66E-08 | 1.72E-05 |

| | | | | | | | |
|---|---|---|---|---|---|---|---|
| cg27633538_BC21 | chr12 | 57296565 | | | -0.259977 | 5.66E-08 | 1.72E-05 |
| cg16207991_BC21 | chr6 | 12751917 | | | -0.380952 | 5.66E-08 | 1.72E-05 |
| cg19376551_TC21 | chr19 | 46226244 | | | -0.289788 | 5.67E-08 | 1.72E-05 |
| cg23220343_BC21 | chr10 | 114286171 | exon_11;e | VWA2;VW | -0.197143 | 5.68E-08 | 1.73E-05 |
| cg07841463_BC21 | chr5 | 37370865 | exon_1;5U | NUP155;NU | -0.420276 | 5.68E-08 | 1.73E-05 |
| cg22683010_TC21 | chr2 | 2566511 | | | 0.208539 | 5.68E-08 | 1.73E-05 |
| cg17050275_BC21 | chr7 | 5387620 | | | -0.280338 | 5.71E-08 | 1.73E-05 |
| cg02509011_TC21 | chr4 | 183363101 | | | 0.157944 | 5.71E-08 | 1.73E-05 |
| cg13614604_BC21 | chr9 | 36793497 | | | -0.23516 | 5.71E-08 | 1.73E-05 |
| cg17811241_TC21 | chr1 | 187085407 | | | -0.191714 | 5.72E-08 | 1.73E-05 |
| cg15639378_BC21 | chr19 | 34125007 | | | -0.228068 | 5.73E-08 | 1.73E-05 |
| cg16820616_TC21 | chr11 | 30584034 | TSS200 | MPPED2 | -0.280762 | 5.73E-08 | 1.73E-05 |
| cg17710087_BC21 | chr12 | 60704610 | | | -0.246586 | 5.73E-08 | 1.73E-05 |
| cg27077677_BC21 | chr14 | 68328680 | | | -0.243931 | 5.73E-08 | 1.73E-05 |
| cg01537972_TC21 | chr1 | 163259732 | | | -0.426352 | 5.73E-08 | 1.73E-05 |
| cg13990539_BC21 | chr10 | 101013650 | | | 0.235618 | 5.73E-08 | 1.73E-05 |

**Supplementary Table S4.3. Significant Genome-wide CpGs in Perinatal SI v NC cohort at 8w post-partum.**

| Probe ID | Chr | Position | Gene Featu | Gene Anno | Log FC | *p* -value | FDR |
|---|---|---|---|---|---|---|---|
| cg16386488_TC21 | chr15 | 62886494 | | | 0.470317 | 4.08E-13 | 3.55E-07 |
| cg16269049_BC21 | chr16 | 3713855 | | | 0.194501 | 2.77E-12 | 8.27E-07 |
| cg05360109_BC21 | chr1 | 6737442 | | | 0.293567 | 3.08E-12 | 8.27E-07 |
| cg03764767_TC21 | chr1 | 2406771 | exon_4;exo | PEX10;PEX | 0.245695 | 3.83E-12 | 8.27E-07 |
| cg12223243_BC21 | chr2 | 60416338 | | | 0.676922 | 4.75E-12 | 8.27E-07 |
| cg16661769_TC21 | chr14 | 84252969 | | | 0.376415 | 7.19E-12 | 1.01E-06 |
| cg07976887_TC21 | chr22 | 22685727 | | | 0.303785 | 8.61E-12 | 1.01E-06 |
| cg25382573_TC11 | chr12 | 92703068 | 5UTR;exon | PLEKHG7;P | 0.211448 | 9.25E-12 | 1.01E-06 |
| cg15498667_TC11 | chr8 | 92877703 | | | 0.223561 | 1.22E-11 | 1.18E-06 |
| cg14601891_BC21 | chr9 | 105655279 | | | 0.357974 | 2.44E-11 | 1.92E-06 |
| cg22854517_BC21 | chr5 | 162096768 | | | 0.373409 | 2.76E-11 | 1.92E-06 |
| cg09548578_BC21 | chr1 | 22839217 | | | 0.436067 | 3.07E-11 | 1.92E-06 |
| cg19879170_BC21 | chr14 | 93745794 | | | 0.231588 | 3.24E-11 | 1.92E-06 |
| cg09407650_BC21 | chr16 | 85286550 | | | 0.396024 | 3.31E-11 | 1.92E-06 |
| cg02134663_TC21 | chr1 | 234167266 | | | 0.242364 | 3.50E-11 | 1.92E-06 |
| cg05058115_TC11 | chr16 | 15650152 | TSS200 | NDE1 | -0.3397 | 3.54E-11 | 1.92E-06 |
| cg21251209_TC21 | chr16 | 21194799 | | | 0.186402 | 3.98E-11 | 2.04E-06 |
| cg26560367_BC21 | chr19 | 12295432 | TSS1500;TS | ZNF44;ZNF | 0.268438 | 4.78E-11 | 2.26E-06 |
| cg02349066_BC21 | chr1 | 182588411 | | | 0.244454 | 4.94E-11 | 2.26E-06 |
| cg09798466_TC21 | chr11 | 6458211 | exon_3;exo | TRIM3;TRIN | 0.208272 | 5.39E-11 | 2.34E-06 |
| cg04984171_BC21 | chr3 | 58260497 | | | 0.223409 | 7.27E-11 | 3.01E-06 |
| cg02363500_TC21 | chr17 | 19750953 | | | 0.231287 | 9.72E-11 | 3.49E-06 |
| cg18229466_TC21 | chr5 | 43297578 | | | 0.366996 | 9.82E-11 | 3.49E-06 |
| cg01828078_TC21 | chr1 | 202631369 | | | 0.419177 | 1.02E-10 | 3.49E-06 |
| cg07793819_BC21 | chr4 | 176555077 | | | 0.354377 | 1.02E-10 | 3.49E-06 |
| cg11917968_TC21 | chr6 | 168829646 | | | 0.318229 | 1.04E-10 | 3.49E-06 |
| cg17722404_TC11 | chr12 | 62739007 | | | 0.156861 | 1.12E-10 | 3.60E-06 |
| cg03766541_BC21 | chr7 | 4805863 | | | 0.246852 | 1.17E-10 | 3.60E-06 |
| cg07744392_BC21 | chr1 | 151792013 | TSS1500;TS | TDRKH;TDF | 0.40218 | 1.20E-10 | 3.60E-06 |
| cg20988620_BC21 | chr1 | 10232116 | | | 0.289103 | 1.27E-10 | 3.69E-06 |
| cg20447079_TC11 | chr7 | 50590684 | 3UTR;exon | GRB10;GRE | 0.248068 | 1.37E-10 | 3.74E-06 |
| cg02244698_BC21 | chr6 | 168170599 | | | -0.38798 | 1.42E-10 | 3.74E-06 |
| cg22302512_TC21 | chr19 | 18495001 | | | 0.388846 | 1.42E-10 | 3.74E-06 |
| cg26942031_BC21 | chr6 | 163491364 | | | 0.355792 | 1.47E-10 | 3.77E-06 |
| cg14621698_BC21 | chr1 | 109393088 | 5UTR;exon | SORT1;SOR | 0.325692 | 1.65E-10 | 4.08E-06 |
| cg08012287_BC21 | chr11 | 66547827 | | | 0.170931 | 1.69E-10 | 4.08E-06 |
| cg01305328_BC11 | chr10 | 103967490 | 5UTR;exon | SLK;SLK;SLI | 0.320221 | 1.81E-10 | 4.19E-06 |
| cg22295444_TC21 | chr17 | 19707753 | exon_8;exo | SLC47A2;SI | 0.138588 | 1.83E-10 | 4.19E-06 |
| cg00602502_TC21 | chr15 | 25843933 | | | 0.378878 | 1.93E-10 | 4.26E-06 |
| cg17747948_TC21 | chr11 | 73665312 | | | 0.184355 | 1.98E-10 | 4.26E-06 |
| cg26133767_TC21 | chr8 | 47600196 | | | 0.251958 | 2.05E-10 | 4.26E-06 |
| cg25569590_BC21 | chr7 | 51289406 | | | 0.220866 | 2.06E-10 | 4.26E-06 |
| cg16149927_TC11 | chr21 | 43778711 | | | 0.243721 | 2.14E-10 | 4.28E-06 |

| | | | | | | | |
|---|---|---|---|---|---|---|---|
| cg22111970_TC21 | chr15 | 68342590 | | | 0.177718 | 2.18E-10 | 4.28E-06 |
| cg01631738_TC21 | chr12 | 52369914 | | | 0.26468 | 2.21E-10 | 4.28E-06 |
| cg04806392_TC21 | chr18 | 24732142 | | | 0.196719 | 2.29E-10 | 4.32E-06 |
| cg13803234_BC21 | chr14 | 68364096 | | | 0.177964 | 2.35E-10 | 4.35E-06 |
| cg18335991_BC21 | chr15 | 74432221 | | | 0.175994 | 2.54E-10 | 4.60E-06 |
| cg10395806_BC11 | chr11 | 70434495 | | | 0.262384 | 2.78E-10 | 4.94E-06 |
| cg06101841_TC21 | chr3 | 48720615 | | | 0.439224 | 3.01E-10 | 5.15E-06 |
| cg14764790_BC21 | chr3 | 62415754 | | | 0.283328 | 3.02E-10 | 5.15E-06 |
| cg16500852_BC21 | chr5 | 177608557 | exon_4 | B4GALT7 | 0.257426 | 3.41E-10 | 5.71E-06 |
| cg16134741_TC21 | chr12 | 1841780 | | | 0.154755 | 3.53E-10 | 5.80E-06 |
| cg09929056_TC21 | chr8 | 138425452 | | | 0.336694 | 3.70E-10 | 5.96E-06 |
| cg26114881_TC21 | chr22 | 23118490 | | | 0.211667 | 3.83E-10 | 6.06E-06 |
| cg15742758_TC21 | chr2 | 236217592 | | | 0.331285 | 4.15E-10 | 6.44E-06 |
| cg00001610_TC21 | chr12 | 30691298 | | | 0.361076 | 4.93E-10 | 7.46E-06 |
| cg01559644_BC21 | chr1 | 178188511 | | | 0.285319 | 4.97E-10 | 7.46E-06 |
| cg23330006_TC21 | chr12 | 106113935 | | | 0.249211 | 5.41E-10 | 7.60E-06 |
| cg24058386_BC21 | chr1 | 156424125 | | | 0.235824 | 5.45E-10 | 7.60E-06 |
| cg00909706_BC21 | chr1 | 9151777 | exon_1;exo | MIR34A;M | 0.187785 | 5.46E-10 | 7.60E-06 |
| cg09977446_TC11 | chr4 | 141133622 | TSS200 | RNF150 | -0.2663 | 5.47E-10 | 7.60E-06 |
| cg22832301_BC21 | chr15 | 88836458 | | | 0.184415 | 5.51E-10 | 7.60E-06 |
| cg19050484_BC11 | chr5 | 466906 | exon_13;3U | EXOC3;EXC | 0.174282 | 6.22E-10 | 8.46E-06 |
| cg17842157_BC11 | chr17 | 44123620 | 5UTR;exon | HDAC5;HD | -0.64346 | 6.41E-10 | 8.57E-06 |
| cg10741238_BC21 | chr7 | 20447296 | | | 0.387386 | 6.62E-10 | 8.73E-06 |
| cg26068305_BC21 | chr3 | 136949796 | exon_3;3U | NCK1;NCK1 | 0.513509 | 7.08E-10 | 9.03E-06 |
| cg17476499_TC21 | chr12 | 6904932 | 5UTR;exon | LRRC23;LR | -0.51922 | 7.19E-10 | 9.03E-06 |
| cg19036041_BC21 | chr1 | 155237161 | | | 0.207738 | 7.26E-10 | 9.03E-06 |
| cg01797899_TC21 | chr14 | 22509211 | | | 0.176017 | 7.26E-10 | 9.03E-06 |
| cg00342761_TC21 | chr6 | 152380490 | TSS200 | SYNE1-AS1 | 0.263178 | 7.59E-10 | 9.30E-06 |
| cg14191082_BC21 | chr20 | 41357929 | exon_17;ex | LPIN3;LPIN | 0.210442 | 7.80E-10 | 9.42E-06 |
| cg07132585_BC21 | chr7 | 151852945 | | | 0.230745 | 8.06E-10 | 9.61E-06 |
| cg12581446_BC21 | chr10 | 6103087 | | | -0.43532 | 8.19E-10 | 9.63E-06 |
| cg21314058_TC21 | chr14 | 93762358 | | | 0.225724 | 8.43E-10 | 9.78E-06 |
| cg18283910_BC21 | chr12 | 124737545 | | | 0.309106 | 8.63E-10 | 9.88E-06 |
| cg14843888_TC21 | chr3 | 53496220 | | | 0.246679 | 8.74E-10 | 9.88E-06 |
| cg21321814_TC21 | chr3 | 146252591 | TSS1500;TS | PLSCR4;PLS | 0.516254 | 8.98E-10 | 1.00E-05 |
| cg08730319_TC21 | chr5 | 153497128 | | | 0.287604 | 9.09E-10 | 1.00E-05 |
| cg00470817_BC21 | chr12 | 6632716 | TSS1500 | LPAR5 | 0.220594 | 9.26E-10 | 1.00E-05 |
| cg17990180_TC21 | chr1 | 155776627 | | | 0.249136 | 9.32E-10 | 1.00E-05 |
| cg19814100_TC21 | chr11 | 111451284 | | | 0.157222 | 9.80E-10 | 1.02E-05 |
| cg04571584_BC21 | chr1 | 112505323 | | | 0.126163 | 9.80E-10 | 1.02E-05 |
| cg00817475_BC21 | chr7 | 22823884 | TSS1500;TS | TOMM7;TO | 0.34494 | 1.00E-09 | 1.02E-05 |
| cg01572884_TC21 | chr1 | 110067644 | | | 0.12288 | 1.02E-09 | 1.02E-05 |
| cg26177616_BC21 | chr4 | 153429328 | | | 0.238297 | 1.03E-09 | 1.02E-05 |
| cg08980461_BC21 | chr9 | 121651234 | TSS1500 | DAB2IP | 0.231336 | 1.03E-09 | 1.02E-05 |
| cg08075765_BC21 | chr5 | 66898065 | | | 0.268401 | 1.03E-09 | 1.02E-05 |
| cg13851454_BC21 | chr22 | 27243668 | | | 0.352688 | 1.05E-09 | 1.02E-05 |
| cg16318442_BC21 | chr2 | 230661369 | | | 0.489641 | 1.09E-09 | 1.05E-05 |

| | | | | | | | |
|---|---|---|---|---|---|---|---|
| cg06700133_TC21 | chr6 | 84847750 | | | 0.257146 | 1.22E-09 | 1.17E-05 |
| cg04227094_BC21 | chr2 | 226995906 | exon_9;3U | RHBDD1;RI | 0.228086 | 1.27E-09 | 1.18E-05 |
| cg06957256_BC21 | chr1 | 167216022 | | | 0.403754 | 1.27E-09 | 1.18E-05 |
| cg10502150_BC21 | chr17 | 8338806 | TSS1500;TS | ODF4;ODF4 | 0.319334 | 1.28E-09 | 1.18E-05 |
| cg04193787_TC21 | chr7 | 132206696 | | | 0.248451 | 1.29E-09 | 1.18E-05 |
| cg00652502_BC21 | chr1 | 41394969 | | | 0.185841 | 1.34E-09 | 1.21E-05 |
| cg25846273_TC11 | chr21 | 37368059 | | | -0.22436 | 1.43E-09 | 1.29E-05 |
| cg08563925_BC21 | chr5 | 135547230 | | | 0.256287 | 1.49E-09 | 1.32E-05 |
| cg09011861_TC21 | chr4 | 3243827 | exon_67;3I | HTT;HTT;H | 0.246445 | 1.50E-09 | 1.32E-05 |
| cg14021168_TC21 | chr17 | 8338859 | TSS1500;TS | ODF4;ODF4 | 0.322463 | 1.56E-09 | 1.36E-05 |
| cg04158627_BC21 | chr2 | 219039379 | | | 0.329416 | 1.67E-09 | 1.42E-05 |
| cg14240629_BC21 | chr9 | 124948148 | 3UTR;exon | SCAI;SCAI;S | 0.45399 | 1.67E-09 | 1.42E-05 |
| cg16824370_BC21 | chr11 | 102176913 | | | 0.23427 | 1.71E-09 | 1.43E-05 |
| cg06674436_BC11 | chr5 | 1278749 | exon_6;exo | TERT;TERT; | -0.28421 | 1.71E-09 | 1.43E-05 |
| cg22166173_BC21 | chr6 | 70396022 | | | -0.26698 | 1.74E-09 | 1.43E-05 |
| cg14074251_TC21 | chr2 | 219434394 | TSS1500 | SPEG | 0.305127 | 1.75E-09 | 1.43E-05 |
| cg16900671_BC21 | chr15 | 31159811 | | | 0.238298 | 1.85E-09 | 1.50E-05 |
| cg15914497_BC21 | chr8 | 29371256 | | | 0.22898 | 1.94E-09 | 1.56E-05 |
| cg04281159_TC21 | chr1 | 25027875 | | | 0.179054 | 1.96E-09 | 1.56E-05 |
| cg06574575_TC21 | chr19 | 18215024 | | | 0.127385 | 2.01E-09 | 1.59E-05 |
| cg03266904_TC11 | chr7 | 135509937 | 5UTR;exon | CNOT4;CN( | -0.33122 | 2.12E-09 | 1.65E-05 |
| cg27522658_TC21 | chr9 | 123851911 | | | 0.257295 | 2.15E-09 | 1.65E-05 |
| cg16279982_TC21 | chr11 | 45848786 | | | 0.319695 | 2.16E-09 | 1.65E-05 |
| cg14484149_TC21 | chr8 | 65490337 | | | 0.396799 | 2.17E-09 | 1.65E-05 |
| cg08223003_TC11 | chr8 | 133273077 | | | 0.134287 | 2.20E-09 | 1.66E-05 |
| cg25768270_BC21 | chr10 | 76624772 | | | 0.13639 | 2.22E-09 | 1.66E-05 |
| cg26860750_TC21 | chr10 | 97196053 | | | 0.256496 | 2.23E-09 | 1.66E-05 |
| cg25725899_BC21 | chr12 | 117141696 | | | 0.249105 | 2.26E-09 | 1.66E-05 |
| cg21509568_BC21 | chr4 | 151227773 | 5UTR;exon | SH3D19;SH | 0.179433 | 2.29E-09 | 1.67E-05 |
| cg07367113_TC21 | chr5 | 891092 | | | 0.404237 | 2.32E-09 | 1.68E-05 |
| cg17945048_TC21 | chr1 | 109723454 | | | 0.290915 | 2.40E-09 | 1.69E-05 |
| cg02858402_BC21 | chr1 | 204214391 | TSS1500 | GOLT1A | 0.310258 | 2.41E-09 | 1.69E-05 |
| cg22516914_BC21 | chr21 | 42836922 | | | 0.273546 | 2.42E-09 | 1.69E-05 |
| cg04473763_TC21 | chr6 | 88170046 | | | 0.248405 | 2.43E-09 | 1.69E-05 |
| cg20289312_TC21 | chr1 | 33219981 | | | 0.17734 | 2.43E-09 | 1.69E-05 |
| cg18390071_BC21 | chr12 | 132274111 | TSS1500 | LOC10013( | 0.315426 | 2.50E-09 | 1.72E-05 |
| cg13860280_BC21 | chr1 | 230663151 | exon_4;exo | COG2;COG | 0.287944 | 2.52E-09 | 1.72E-05 |
| cg07018857_TC11 | chr2 | 238912746 | | | 0.196483 | 2.58E-09 | 1.76E-05 |
| cg12827842_TC21 | chr16 | 86120668 | | | 0.359815 | 2.67E-09 | 1.80E-05 |
| cg23993315_BC21 | chr19 | 5114944 | | | -0.21395 | 2.69E-09 | 1.80E-05 |
| cg12821203_BC21 | chr22 | 41753455 | | | 0.261004 | 2.71E-09 | 1.80E-05 |
| cg05727666_BC21 | chr1 | 66863226 | | | 0.42491 | 2.77E-09 | 1.83E-05 |
| cg15640112_TC21 | chr5 | 52710238 | | | 0.300202 | 2.82E-09 | 1.84E-05 |
| cg19874685_BC21 | chr15 | 77493148 | | | 0.27001 | 2.90E-09 | 1.88E-05 |
| cg11994358_BC21 | chr3 | 49861031 | | | 0.337939 | 2.96E-09 | 1.91E-05 |
| cg00876532_BC21 | chr7 | 130441969 | TSS1500;TS | CEP41;CEP | 0.320915 | 3.09E-09 | 1.97E-05 |
| cg20853155_TC21 | chr15 | 96296429 | | | 0.432859 | 3.10E-09 | 1.97E-05 |

| | | | | | | | |
|---|---|---|---|---|---|---|---|
| cg09289463_BC21 | chr19 | 5787855 | | | 0.206171 | 3.17E-09 | 2.00E-05 |
| cg17382065_BC21 | chr3 | 123149800 | | | 0.2325 | 3.24E-09 | 2.03E-05 |
| cg09782171_BC21 | chr10 | 125626396 | | | 0.264603 | 3.27E-09 | 2.03E-05 |
| cg14154655_BC21 | chr8 | 29459388 | | | 0.188003 | 3.28E-09 | 2.03E-05 |
| cg22362864_BC21 | chr1 | 241984543 | | | 0.186841 | 3.39E-09 | 2.07E-05 |
| cg23085846_TC21 | chr20 | 63891165 | exon_7;3U | TPD52L2;TI | 0.12289 | 3.40E-09 | 2.07E-05 |
| cg22166284_BC21 | chr11 | 88103463 | | | 0.314175 | 3.55E-09 | 2.14E-05 |
| cg16281806_BC21 | chr19 | 19636925 | exon_12;ex | GMIP;GMII | 0.327611 | 4.02E-09 | 2.41E-05 |
| cg17724082_BC21 | chr21 | 34204898 | | | 0.15247 | 4.04E-09 | 2.41E-05 |
| cg18070189_BC21 | chr14 | 99579273 | | | 0.2146 | 4.17E-09 | 2.45E-05 |
| cg16405520_BC21 | chr11 | 61599293 | | | 0.155053 | 4.17E-09 | 2.45E-05 |
| cg05182365_BC21 | chr14 | 106211716 | | | 0.23026 | 4.25E-09 | 2.46E-05 |
| cg11680826_TC21 | chr19 | 37219192 | 5UTR;exon | ZNF383;ZN | 0.329109 | 4.25E-09 | 2.46E-05 |
| cg06821966_BC21 | chr4 | 175748394 | | | -0.24892 | 4.26E-09 | 2.46E-05 |
| cg12315735_TC21 | chr11 | 124617420 | exon_3 | PANX3 | 0.219791 | 4.31E-09 | 2.47E-05 |
| cg01913046_BC21 | chr20 | 11001812 | | | 0.276264 | 4.38E-09 | 2.48E-05 |
| cg07456278_BC21 | chr3 | 53566636 | | | 0.178666 | 4.43E-09 | 2.48E-05 |
| cg24286230_BC21 | chr10 | 45425744 | | | 0.157135 | 4.45E-09 | 2.48E-05 |
| cg17149814_BC21 | chr12 | 310688 | | | 0.23113 | 4.46E-09 | 2.48E-05 |
| cg01437356_BC21 | chr2 | 1778630 | | | 0.366449 | 4.49E-09 | 2.49E-05 |
| cg18089086_TC21 | chr15 | 73767974 | TSS1500;TS | INSYN1-AS | 0.162309 | 4.65E-09 | 2.54E-05 |
| cg11366948_TC21 | chr21 | 39095430 | | | 0.272978 | 4.67E-09 | 2.54E-05 |
| cg06935846_BC21 | chr2 | 178108857 | TSS1500 | PDE11A | 0.26532 | 4.67E-09 | 2.54E-05 |
| cg25377656_TC21 | chr20 | 50294859 | | | 0.095706 | 4.72E-09 | 2.55E-05 |
| cg11557932_TC11 | chr6 | 4806071 | | | 0.266997 | 4.95E-09 | 2.64E-05 |
| cg25429578_BC21 | chr5 | 177511406 | TSS1500;TS | DOK3;DOK | 0.184905 | 4.95E-09 | 2.64E-05 |
| cg05384127_TC21 | chr22 | 26671443 | TSS1500;ex | MIATNB;M | 0.23817 | 4.98E-09 | 2.64E-05 |
| cg12450012_TC21 | chr11 | 70436381 | exon_19;ex | CTTN;CTTN | 0.273907 | 5.02E-09 | 2.64E-05 |
| cg10754659_TC21 | chr7 | 38070549 | | | 0.304702 | 5.09E-09 | 2.67E-05 |
| cg00794607_TC21 | chr20 | 31555348 | | | 0.18533 | 5.12E-09 | 2.67E-05 |
| cg22480687_TC21 | chr17 | 37687095 | 3UTR;exon | HNF1B;HNI | 0.315198 | 5.33E-09 | 2.76E-05 |
| cg10413089_TC21 | chr15 | 22951641 | | | 0.240249 | 5.57E-09 | 2.86E-05 |
| cg21320221_TC21 | chr17 | 81835121 | TSS200 | PPP1R27 | 0.263985 | 5.59E-09 | 2.86E-05 |
| cg23112817_BC21 | chr17 | 82267803 | | | 0.159193 | 5.68E-09 | 2.89E-05 |
| cg11829392_TC21 | chr7 | 143409817 | TSS1500 | EPHA1 | 0.252641 | 5.71E-09 | 2.89E-05 |
| cg25677709_BC21 | chr5 | 150520897 | 5UTR;exon | NDST1;NDS | 0.171433 | 5.82E-09 | 2.92E-05 |
| cg09448665_TC21 | chr6 | 37692036 | | | 0.257539 | 5.86E-09 | 2.92E-05 |
| cg08851943_BC21 | chr6 | 39115036 | 5UTR;exon | SAYSD1;SA | -0.27711 | 5.88E-09 | 2.92E-05 |
| cg19057304_BC21 | chr2 | 207820085 | | | 0.31651 | 6.08E-09 | 3.01E-05 |
| cg01472464_TC21 | chr2 | 3379307 | TSS1500;TS | TRAPPC12; | 0.441437 | 6.17E-09 | 3.03E-05 |
| cg06450130_TC21 | chr4 | 37483331 | | | 0.208581 | 6.22E-09 | 3.03E-05 |
| cg24016574_BC21 | chr17 | 77034796 | | | 0.271331 | 6.29E-09 | 3.03E-05 |
| cg05856951_BC21 | chr16 | 4495327 | TSS1500;TS | HMOX2;HN | 0.371855 | 6.30E-09 | 3.03E-05 |
| cg17164090_BC21 | chr12 | 114620703 | | | -0.37498 | 6.30E-09 | 3.03E-05 |
| cg25906360_TC21 | chr10 | 125773775 | | | -0.39015 | 6.42E-09 | 3.06E-05 |
| cg27044180_TC21 | chr7 | 55189035 | TSS200 | EGFR-AS1 | 0.299495 | 6.44E-09 | 3.06E-05 |
| cg06672272_TC21 | chr4 | 187063258 | | | -0.27654 | 6.50E-09 | 3.08E-05 |

| | | | | | | | |
|---|---|---|---|---|---|---|---|
| cg24981310_TC21 | chr14 | 57493813 | exon_1;5U | CCDC198;C | 0.287508 | 6.59E-09 | 3.10E-05 |
| cg10081660_BC21 | chr1 | 205566081 | | | 0.25795 | 6.65E-09 | 3.10E-05 |
| cg08535918_TC21 | chr16 | 56222836 | | | 0.194587 | 6.66E-09 | 3.10E-05 |
| cg00116468_BC21 | chr1 | 5090789 | | | 0.23595 | 6.93E-09 | 3.20E-05 |
| cg11317625_BC21 | chr8 | 8618821 | | | -0.24929 | 6.95E-09 | 3.20E-05 |
| cg25229964_TC21 | chr1 | 26177132 | TSS1500;TS | CNKSR1;CN | 0.178503 | 7.14E-09 | 3.26E-05 |
| cg16659071_BC21 | chr2 | 207256880 | | | 0.226091 | 7.15E-09 | 3.26E-05 |
| cg25617157_BC21 | chr2 | 6241583 | | | -0.25507 | 7.31E-09 | 3.31E-05 |
| cg19719042_BC21 | chr10 | 63720872 | | | 0.264773 | 7.60E-09 | 3.43E-05 |
| cg13951597_BC21 | chr17 | 15943096 | | | 0.316895 | 7.66E-09 | 3.44E-05 |
| cg01755929_BC21 | chr1 | 193126852 | | | 0.369146 | 7.71E-09 | 3.44E-05 |
| cg22883321_BC21 | chr10 | 92181499 | | | 0.298509 | 7.74E-09 | 3.44E-05 |
| cg09640065_BC21 | chr2 | 142643079 | | | 0.458922 | 8.03E-09 | 3.54E-05 |
| cg18212999_BC21 | chr12 | 120998899 | | | 0.24671 | 8.13E-09 | 3.56E-05 |
| cg06825478_TC21 | chr6 | 31578290 | exon_4;3U | TNF;TNF | 0.263134 | 8.14E-09 | 3.56E-05 |
| cg23659062_TC21 | chr1 | 167119421 | exon_5 | STYXL2 | 0.222383 | 8.38E-09 | 3.64E-05 |
| cg05785686_BC11 | chr2 | 19901745 | exon_1 | TTC32 | 0.301299 | 8.40E-09 | 3.64E-05 |
| cg26053498_BC21 | chr7 | 2301215 | | | 0.307498 | 8.47E-09 | 3.65E-05 |
| cg23999081_TC21 | chr1 | 178058898 | | | 0.275685 | 8.58E-09 | 3.66E-05 |
| cg18599162_TC21 | chr15 | 30925214 | exon_9 | FAN1 | -0.36445 | 8.58E-09 | 3.66E-05 |
| cg01043895_BC21 | chr1 | 89763973 | | | 0.30176 | 8.66E-09 | 3.67E-05 |
| cg04826516_BC11 | chr16 | 1351897 | TSS200;TSS | GNPTG;TSF | 0.351002 | 8.70E-09 | 3.68E-05 |
| cg24707640_BC21 | chr19 | 33810226 | | | 0.295368 | 8.82E-09 | 3.71E-05 |
| cg05748265_TC21 | chr5 | 41671055 | | | 0.244061 | 9.18E-09 | 3.84E-05 |
| cg13930739_BC21 | chr20 | 19751408 | | | 0.377343 | 9.24E-09 | 3.85E-05 |
| cg27378337_BC21 | chr8 | 140465024 | | | -0.39075 | 9.35E-09 | 3.86E-05 |
| cg14265434_TC21 | chr6 | 153002746 | TSS200;TSS | MTRF1L;M | -0.27361 | 9.48E-09 | 3.86E-05 |
| cg10583856_BC21 | chr3 | 84881783 | TSS1500 | LINC02025 | 0.262434 | 9.48E-09 | 3.86E-05 |
| cg08529105_BC21 | chr5 | 132777703 | 5UTR;exon | SEPTIN8;SE | -0.28881 | 9.54E-09 | 3.86E-05 |
| cg20060396_BC21 | chr6 | 25652587 | | | 0.493001 | 9.56E-09 | 3.86E-05 |
| cg00637875_BC21 | chr1 | 227192621 | | | 0.196572 | 9.59E-09 | 3.86E-05 |
| cg16416322_BC21 | chr9 | 37603184 | TSS200;TSS | FRMPD1;FI | 0.233596 | 9.59E-09 | 3.86E-05 |
| cg05786516_TC21 | chr15 | 44633083 | | | 0.25614 | 9.63E-09 | 3.86E-05 |
| cg10308445_BC21 | chr2 | 196840538 | 3UTR;exon | PGAP1;PG/ | 0.299253 | 9.67E-09 | 3.86E-05 |
| cg26793846_TC21 | chr22 | 42142987 | | | -0.5771 | 9.82E-09 | 3.90E-05 |
| cg11259183_BC11 | chr19 | 39320718 | | | 0.390295 | 9.88E-09 | 3.91E-05 |
| cg25043967_BC21 | chr20 | 19993665 | | | 0.253166 | 9.93E-09 | 3.91E-05 |
| cg09795027_TC21 | chr17 | 76010359 | exon_22;e | EVPL;EVPL | 0.184795 | 9.97E-09 | 3.91E-05 |
| cg01847389_TC21 | chr20 | 54070725 | TSS200;TSS | BCAS1;BCA | 0.286687 | 1.04E-08 | 4.01E-05 |
| cg24408057_TC21 | chr19 | 4277336 | | | 0.223189 | 1.04E-08 | 4.01E-05 |
| cg01932519_TC21 | chr3 | 16414858 | | | 0.205216 | 1.04E-08 | 4.01E-05 |
| cg09759637_BC21 | chr2 | 11496075 | | | -0.27876 | 1.05E-08 | 4.02E-05 |
| cg13988652_BC21 | chr5 | 180807518 | | | 0.368982 | 1.05E-08 | 4.02E-05 |
| cg23828300_TC21 | chr19 | 12984987 | | | 0.201011 | 1.05E-08 | 4.02E-05 |
| cg01147014_TC21 | chr1 | 104463455 | | | -0.21449 | 1.06E-08 | 4.04E-05 |
| cg07677174_BC21 | chr6 | 105553284 | | | 0.168175 | 1.08E-08 | 4.10E-05 |
| cg10042026_BC21 | chr6 | 122723015 | | | 0.162751 | 1.11E-08 | 4.17E-05 |

| | | | | | | | |
|---|---|---|---|---|---|---|---|
| cg21569094_BC21 | chr6 | 3252784 | | | 0.418737 | 1.13E-08 | 4.22E-05 |
| cg13051944_BC21 | chr8 | 121170567 | | | 0.221999 | 1.13E-08 | 4.22E-05 |
| cg13845059_BC21 | chr9 | 84258378 | | | 0.143667 | 1.16E-08 | 4.32E-05 |
| cg22696982_TC11 | chr2 | 176638177 | TSS1500 | LINC01116 | 0.18442 | 1.18E-08 | 4.36E-05 |
| cg17725140_TC21 | chr20 | 41348567 | | | 0.247871 | 1.18E-08 | 4.36E-05 |
| cg16349774_BC21 | chr1 | 981869 | TSS1500 | PERM1 | 0.159464 | 1.20E-08 | 4.42E-05 |
| cg03120836_BC21 | chr9 | 104992255 | | | 0.302825 | 1.23E-08 | 4.48E-05 |
| cg15196314_TC11 | chr19 | 51108897 | TSS1500 | CTU1 | -0.3598 | 1.26E-08 | 4.57E-05 |
| cg02660161_TC21 | chr2 | 62050789 | | | 0.315474 | 1.27E-08 | 4.57E-05 |
| cg12043230_BC21 | chr12 | 2296146 | | | 0.124959 | 1.27E-08 | 4.57E-05 |
| cg01773788_TC21 | chr1 | 196270748 | | | 0.37214 | 1.28E-08 | 4.57E-05 |
| cg25063189_BC21 | chr10 | 132533749 | | | 0.149976 | 1.29E-08 | 4.57E-05 |
| cg11890641_BC21 | chr7 | 849727 | | | 0.287489 | 1.29E-08 | 4.57E-05 |
| cg17011686_BC21 | chr1 | 173825948 | TSS1500;TS | CENPL;CEN | 0.408614 | 1.29E-08 | 4.57E-05 |
| cg18491926_BC21 | chr2 | 203781268 | | | 0.25677 | 1.29E-08 | 4.57E-05 |
| cg02733432_BC21 | chr10 | 124691076 | | | 0.265872 | 1.32E-08 | 4.65E-05 |
| cg11614621_BC21 | chr22 | 46610445 | | | 0.232423 | 1.33E-08 | 4.67E-05 |
| cg12106818_TC21 | chr3 | 49123896 | exon_24 | LAMB2 | 0.254773 | 1.34E-08 | 4.69E-05 |
| cg08289346_TC21 | chr6 | 34121573 | | | 0.1541 | 1.36E-08 | 4.72E-05 |
| cg22110158_TC21 | chr11 | 130166647 | | | 0.244822 | 1.36E-08 | 4.72E-05 |
| cg10748688_BC21 | chr16 | 58503726 | 5UTR;exon_ | NDRG4;ND | 0.197703 | 1.39E-08 | 4.81E-05 |
| cg25513073_BC21 | chr11 | 2460411 | TSS1500 | KCNQ1 | 0.204107 | 1.40E-08 | 4.81E-05 |
| cg21371458_TC21 | chr1 | 147517888 | TSS200 | LINC00624 | 0.958694 | 1.42E-08 | 4.85E-05 |
| cg23668222_TC21 | chr8 | 41684574 | exon_37;e | ANK1;ANK1 | -1.21567 | 1.42E-08 | 4.86E-05 |
| cg07450552_BC21 | chr5 | 139439730 | TSS1500 | DNAJC18 | -0.33322 | 1.44E-08 | 4.88E-05 |
| cg03074172_BC21 | chr2 | 84022851 | | | 0.333935 | 1.45E-08 | 4.90E-05 |
| cg10157276_BC21 | chr1 | 9419098 | | | 0.223341 | 1.47E-08 | 4.94E-05 |
| cg02128159_BC21 | chr1 | 233395262 | | | 0.335034 | 1.48E-08 | 4.94E-05 |
| cg24998002_BC21 | chr6 | 135021783 | | | 0.338627 | 1.48E-08 | 4.94E-05 |
| cg06795233_BC21 | chr2 | 218872670 | | | 0.238701 | 1.48E-08 | 4.94E-05 |
| cg09466770_BC21 | chr2 | 114460450 | TSS1500;TS | DPP10;DPF | 0.35466 | 1.50E-08 | 4.97E-05 |
| cg00169026_TC21 | chr9 | 75201348 | | | 0.226452 | 1.50E-08 | 4.97E-05 |
| cg11848876_BC21 | chr16 | 4258819 | | | -0.30028 | 1.51E-08 | 4.98E-05 |
| cg00955967_BC11 | chr4 | 147158565 | | | 0.202141 | 1.54E-08 | 5.05E-05 |
| cg19766988_TC21 | chr19 | 10121035 | TSS1500 | EIF3G | -0.34832 | 1.57E-08 | 5.14E-05 |
| cg22807251_TC21 | chr15 | 98519038 | | | 0.298821 | 1.58E-08 | 5.14E-05 |
| cg17172417_TC21 | chr12 | 1924683 | | | 0.309708 | 1.59E-08 | 5.14E-05 |
| cg11514097_BC21 | chr1 | 2256323 | | | 0.254798 | 1.59E-08 | 5.14E-05 |
| cg22941061_TC21 | chr17 | 74971678 | | | 0.112462 | 1.61E-08 | 5.20E-05 |
| cg21813747_TC21 | chr1 | 19643437 | 5UTR;exon_ | NBL1;NBL1 | 0.256869 | 1.64E-08 | 5.26E-05 |
| cg18220816_TC21 | chr14 | 101945328 | | | 0.303548 | 1.65E-08 | 5.26E-05 |
| cg04170298_TC21 | chr15 | 86262332 | | | 0.351426 | 1.67E-08 | 5.29E-05 |
| cg25870748_TC21 | chr21 | 39666845 | exon_3;3U | B3GALT5;B | 0.330156 | 1.67E-08 | 5.29E-05 |
| cg21947280_BC21 | chr7 | 95096993 | | | 0.243597 | 1.67E-08 | 5.29E-05 |
| cg12878830_BC11 | chr10 | 73656663 | TSS1500 | SYNPO2L | 0.135038 | 1.68E-08 | 5.29E-05 |
| cg01080907_TC21 | chr20 | 57310790 | | | 0.200252 | 1.70E-08 | 5.29E-05 |
| cg13454365_TC21 | chr9 | 16681937 | | | 0.27076 | 1.70E-08 | 5.29E-05 |

| | | | | | | | |
|---|---|---|---|---|---|---|---|
| cg19933015_BC11 | chr9 | 83708283 | exon_1;TSS | UBQLN1-AS | -0.33351 | 1.70E-08 | 5.29E-05 |
| cg05070392_BC21 | chr3 | 193879204 | | | 0.19621 | 1.71E-08 | 5.29E-05 |
| cg13604623_TC21 | chr11 | 113875000 | | | -0.87162 | 1.72E-08 | 5.29E-05 |
| cg02477677_TC21 | chr17 | 40356057 | exon_9;exo | RARA;RARA | 0.241663 | 1.72E-08 | 5.29E-05 |
| cg18637476_BC21 | chr19 | 17042112 | | | 0.255014 | 1.72E-08 | 5.29E-05 |
| cg11346426_BC21 | chr7 | 86914825 | exon_13;ex | ELAPOR2;E | 0.23881 | 1.73E-08 | 5.30E-05 |
| cg02592586_TC21 | chr2 | 60603122 | | | 0.188035 | 1.74E-08 | 5.32E-05 |
| cg14436056_TC21 | chr8 | 53759332 | | | 0.24533 | 1.77E-08 | 5.37E-05 |
| cg11751876_BC21 | chr5 | 180140375 | | | 0.117765 | 1.77E-08 | 5.37E-05 |
| cg10772263_BC21 | chr10 | 71986898 | | | 0.284644 | 1.79E-08 | 5.42E-05 |
| cg18672919_TC21 | chr13 | 42883221 | | | 0.26004 | 1.80E-08 | 5.42E-05 |
| cg22706980_TC21 | chr19 | 8430892 | | | 0.273108 | 1.81E-08 | 5.42E-05 |
| cg17574699_TC21 | chr12 | 48970770 | | | 0.180267 | 1.82E-08 | 5.42E-05 |
| cg21030597_BC21 | chr1 | 37656099 | | | 0.20113 | 1.82E-08 | 5.42E-05 |
| cg22762455_BC21 | chr7 | 105906541 | | | 0.202621 | 1.83E-08 | 5.43E-05 |
| cg10773014_TC21 | chr5 | 135533994 | | | 0.265548 | 1.83E-08 | 5.43E-05 |
| cg25859224_BC21 | chr1 | 248037754 | | | 0.330807 | 1.85E-08 | 5.45E-05 |
| cg06432884_BC21 | chr2 | 74266560 | | | 0.490299 | 1.86E-08 | 5.46E-05 |
| cg12401762_TC21 | chr22 | 37123547 | | | -0.2345 | 1.86E-08 | 5.46E-05 |
| cg24260931_TC21 | chr10 | 3106134 | | | -0.24272 | 1.89E-08 | 5.51E-05 |
| cg20719304_TC21 | chr12 | 108410032 | | | 0.413072 | 1.91E-08 | 5.55E-05 |
| cg12012114_TC21 | chr21 | 42464378 | | | 0.132257 | 1.92E-08 | 5.55E-05 |
| cg04654562_BC21 | chr3 | 23751443 | | | 0.115792 | 1.92E-08 | 5.55E-05 |
| cg19717352_TC21 | chr14 | 75727223 | | | 0.426147 | 1.93E-08 | 5.56E-05 |
| cg23628760_TC21 | chr11 | 1359125 | | | 0.255786 | 1.95E-08 | 5.59E-05 |
| cg10837116_BC21 | chr12 | 52676149 | | | 0.137733 | 1.98E-08 | 5.64E-05 |
| cg05304178_TC21 | chr13 | 114059476 | | | 0.168858 | 1.98E-08 | 5.64E-05 |
| cg20203628_BC21 | chr1 | 202631352 | | | 0.437933 | 2.01E-08 | 5.72E-05 |
| cg06510002_TC21 | chr6 | 43254772 | | | 0.162987 | 2.03E-08 | 5.76E-05 |
| cg18587092_BC21 | chr12 | 122602045 | | | 0.471605 | 2.05E-08 | 5.79E-05 |
| cg09513758_BC21 | chr1 | 109983746 | TSS1500;TS | AHCYL1;AH | 0.221416 | 2.07E-08 | 5.82E-05 |
| cg14960030_BC21 | chr11 | 74071501 | | | 0.299478 | 2.08E-08 | 5.84E-05 |
| cg00551146_TC21 | chr10 | 13972579 | TSS1500 | FRMD4A | 0.291664 | 2.10E-08 | 5.88E-05 |
| cg10779242_BC21 | chr7 | 152077801 | | | 0.213313 | 2.11E-08 | 5.88E-05 |
| cg04458869_TC11 | chr2 | 168456177 | TSS200;TSS | CERS6;CER | 0.346414 | 2.12E-08 | 5.89E-05 |
| cg06271128_TC21 | chr10 | 34666597 | | | 0.226667 | 2.13E-08 | 5.90E-05 |
| cg22398192_BC21 | chr6 | 10515000 | | | 0.319708 | 2.16E-08 | 5.97E-05 |
| cg01357997_BC21 | chr9 | 126293785 | | | 0.130461 | 2.20E-08 | 6.06E-05 |
| cg25911248_TC21 | chr3 | 12408655 | exon_5;3U | PPARG;PPA | 0.388336 | 2.21E-08 | 6.06E-05 |
| cg13063369_TC21 | chr11 | 119755084 | | | 0.218907 | 2.24E-08 | 6.09E-05 |
| cg10914859_BC21 | chr9 | 88990221 | TSS1500;TS | S1PR3;S1PI | 0.145815 | 2.24E-08 | 6.09E-05 |
| cg11333664_TC21 | chr1 | 73749822 | | | 0.354611 | 2.24E-08 | 6.09E-05 |
| cg17844017_BC21 | chr11 | 250601 | | | 0.286187 | 2.26E-08 | 6.12E-05 |
| cg07083327_TC21 | chr4 | 76808708 | | | 0.252371 | 2.26E-08 | 6.12E-05 |
| cg01023590_TC21 | chr10 | 69313346 | | | 0.241953 | 2.33E-08 | 6.27E-05 |
| cg01064150_BC21 | chr3 | 119377185 | | | 0.182886 | 2.38E-08 | 6.34E-05 |
| cg10376517_TC21 | chr1 | 17826564 | 3UTR;exon | ACTL8;ACT | 0.328311 | 2.38E-08 | 6.34E-05 |

| | | | | | | | |
|---|---|---|---|---|---|---|---|
| cg01807508_BC21 | chr1 | 200979644 | exon_30;e› | KIF21B;KIF. | 0.186943 | 2.38E-08 | 6.34E-05 |
| cg17534192_TC21 | chr15 | 72756801 | | | 0.189107 | 2.39E-08 | 6.34E-05 |
| cg00014830_BC21 | chr17 | 79791934 | | | 0.189785 | 2.39E-08 | 6.34E-05 |
| cg03023900_TC21 | chr17 | 64007876 | TSS1500 | ICAM2 | 0.268728 | 2.40E-08 | 6.34E-05 |
| cg15122081_BC21 | chr1 | 53591026 | exon_5 | GLIS1 | 0.122624 | 2.41E-08 | 6.35E-05 |
| cg10431512_TC21 | chr17 | 1478470 | exon_11;e› | MYO1C;MY | 0.191446 | 2.44E-08 | 6.40E-05 |
| cg24444530_TC21 | chr19 | 35476368 | | | 0.163496 | 2.51E-08 | 6.54E-05 |
| cg17258765_TC21 | chr1 | 24876126 | | | 0.231979 | 2.53E-08 | 6.54E-05 |
| cg20873526_TC21 | chr15 | 98512641 | | | 0.24502 | 2.53E-08 | 6.54E-05 |
| cg00380266_TC21 | chr8 | 121635655 | | | 0.441109 | 2.54E-08 | 6.54E-05 |
| cg08881278_TC21 | chr5 | 171858551 | | | 0.196904 | 2.54E-08 | 6.54E-05 |
| cg14173476_BC11 | chr17 | 19287163 | | | 0.161456 | 2.54E-08 | 6.54E-05 |
| cg14261176_BC21 | chr9 | 84239654 | | | 0.231476 | 2.54E-08 | 6.54E-05 |
| cg22295812_BC21 | chr16 | 66731145 | | | 0.242512 | 2.56E-08 | 6.55E-05 |
| cg22059580_TC21 | chr10 | 101972813 | | | 0.182923 | 2.56E-08 | 6.55E-05 |
| cg21281293_TC21 | chr16 | 23826277 | | | 0.160923 | 2.59E-08 | 6.60E-05 |
| cg06866011_TC21 | chr1 | 11067918 | | | 0.283633 | 2.61E-08 | 6.63E-05 |
| cg13601615_BC21 | chr8 | 140451142 | exon_2;exc | TRAPPC9;T | -0.3345 | 2.62E-08 | 6.64E-05 |
| cg09108754_BC21 | chr22 | 24133125 | | | 0.170893 | 2.64E-08 | 6.69E-05 |
| cg02115610_BC21 | chr11 | 108938900 | | | 0.348476 | 2.66E-08 | 6.71E-05 |
| cg03666588_BC21 | chr3 | 119660082 | exon_1;exc | POPDC2;P( | 0.225044 | 2.67E-08 | 6.71E-05 |
| cg02411879_TC11 | chr17 | 40819144 | exon_7;exc | KRT10;KRT | -0.2562 | 2.71E-08 | 6.79E-05 |
| cg13708995_BC21 | chr5 | 135545205 | | | 0.24386 | 2.75E-08 | 6.84E-05 |
| cg16040564_TC21 | chr1 | 107967909 | | | 0.655744 | 2.76E-08 | 6.84E-05 |
| cg17494750_BC21 | chr11 | 116645048 | | | 0.158139 | 2.76E-08 | 6.84E-05 |
| cg18771854_BC21 | chr4 | 6732847 | | | 0.168782 | 2.77E-08 | 6.84E-05 |
| cg27278382_BC11 | chr3 | 48609289 | exon_2 | UQCRC1 | -0.25751 | 2.77E-08 | 6.84E-05 |
| cg16509547_BC21 | chr1 | 221488341 | | | 0.190893 | 2.77E-08 | 6.84E-05 |
| cg23092048_BC21 | chr12 | 14701011 | | | 0.172489 | 2.79E-08 | 6.86E-05 |
| cg22656199_TC21 | chr9 | 122194398 | | | 0.241272 | 2.81E-08 | 6.87E-05 |
| cg05355381_BC21 | chr10 | 61785887 | | | 0.214759 | 2.82E-08 | 6.87E-05 |
| cg00711090_TC21 | chr19 | 56507245 | TSS1500;TS | ZNF471;ZN | 0.332584 | 2.82E-08 | 6.87E-05 |
| cg26865909_BC21 | chr2 | 152048417 | | | 0.277351 | 2.84E-08 | 6.90E-05 |
| cg15936790_BC21 | chr16 | 1451678 | exon_15;e› | CLCN7;CLC | 0.210499 | 2.85E-08 | 6.91E-05 |
| cg09615453_BC11 | chr17 | 50274382 | TSS200 | TMEM92 | 0.241923 | 2.87E-08 | 6.94E-05 |
| cg15038926_BC21 | chr10 | 104038388 | | | 0.173016 | 2.90E-08 | 6.98E-05 |
| cg09411369_BC21 | chr3 | 184303896 | | | 0.179066 | 2.97E-08 | 7.14E-05 |
| cg21075784_BC21 | chr19 | 54133649 | | | -0.38834 | 2.99E-08 | 7.17E-05 |
| cg01893450_BC21 | chr18 | 54558008 | | | 0.196902 | 3.01E-08 | 7.18E-05 |
| cg10465989_BC21 | chr18 | 26460191 | | | 0.347173 | 3.06E-08 | 7.28E-05 |
| cg18570947_TC21 | chr19 | 11435293 | TSS1500;TS | PRKCSH;PR | 0.407902 | 3.09E-08 | 7.34E-05 |
| cg00818106_BC21 | chr10 | 132407999 | | | 0.352994 | 3.10E-08 | 7.34E-05 |
| cg17126234_TC21 | chr15 | 53717064 | | | 0.334622 | 3.10E-08 | 7.34E-05 |
| cg06745656_TC21 | chr17 | 39149475 | | | 0.303816 | 3.15E-08 | 7.44E-05 |
| cg12773787_BC21 | chr1 | 38226761 | | | 0.303001 | 3.18E-08 | 7.46E-05 |
| cg21417545_TC21 | chr10 | 96992647 | exon_8;3U | LCOR;LCOF | 0.256369 | 3.18E-08 | 7.46E-05 |
| cg23141629_BC21 | chr13 | 45218447 | | | 0.474158 | 3.19E-08 | 7.46E-05 |

| | | | | | | | |
|---|---|---|---|---|---|---|---|
| cg19555906_TC21 | chr17 | 76871336 | TSS1500 | MGAT5B | 0.168008 | 3.22E-08 | 7.50E-05 |
| cg09510108_BC21 | chr3 | 50473644 | | | 0.24503 | 3.24E-08 | 7.50E-05 |
| cg11094568_TC21 | chr2 | 3377113 | | | 0.370269 | 3.25E-08 | 7.50E-05 |
| cg19707031_TC21 | chr21 | 46346051 | | | 0.207884 | 3.25E-08 | 7.50E-05 |
| cg18141902_TC21 | chr1 | 2864710 | | | 0.114347 | 3.26E-08 | 7.50E-05 |
| cg07232711_TC21 | chr11 | 86148836 | | | 0.357128 | 3.26E-08 | 7.50E-05 |
| cg00100491_TC21 | chr2 | 233981614 | | | 0.330487 | 3.28E-08 | 7.50E-05 |
| cg02845413_BC21 | chr1 | 230084119 | | | 0.343182 | 3.28E-08 | 7.50E-05 |
| cg03653236_BC21 | chr11 | 36266778 | | | 0.124786 | 3.29E-08 | 7.50E-05 |
| cg17493253_TC21 | chr2 | 23501361 | | | 0.297868 | 3.34E-08 | 7.60E-05 |
| cg24555935_TC21 | chr20 | 33182169 | | | 0.201879 | 3.35E-08 | 7.61E-05 |
| cg14266730_TC21 | chr19 | 35977908 | | | -0.23779 | 3.36E-08 | 7.61E-05 |
| cg07367459_TC21 | chr9 | 135506636 | TSS200 | LOC10192[illegible] | 0.200107 | 3.39E-08 | 7.65E-05 |
| cg09373597_TC21 | chr3 | 138948933 | | | 0.194396 | 3.39E-08 | 7.65E-05 |
| cg24749602_BC21 | chr1 | 171511015 | | | 0.203217 | 3.41E-08 | 7.66E-05 |
| cg02136975_BC21 | chr13 | 111588223 | | | 0.307929 | 3.44E-08 | 7.72E-05 |
| cg03942086_TC21 | chr1 | 3164917 | | | 0.223377 | 3.47E-08 | 7.73E-05 |
| cg06052478_BC21 | chr19 | 39473247 | exon_24;e[illegible] | SUPT5H;SU[illegible] | 0.218436 | 3.47E-08 | 7.73E-05 |
| cg20014585_BC21 | chr14 | 73904909 | exon_8;ex[illegible] | ZNF410;ZN | 0.172827 | 3.48E-08 | 7.73E-05 |
| cg16106179_BC21 | chr20 | 58688474 | TSS1500;T[illegible] | NPEPL1;NP | 0.254121 | 3.51E-08 | 7.73E-05 |
| cg06547490_BC11 | chr10 | 132408549 | | | 0.321681 | 3.52E-08 | 7.73E-05 |
| cg17044643_BC21 | chr1 | 24335327 | | | 0.224852 | 3.52E-08 | 7.73E-05 |
| cg20140101_TC21 | chr15 | 25903617 | | | 0.15287 | 3.52E-08 | 7.73E-05 |
| cg09689385_TC21 | chr8 | 103375811 | exon_2;ex[illegible] | CTHRC1;CT | 0.354405 | 3.53E-08 | 7.73E-05 |
| cg11894388_BC21 | chr1 | 25568983 | | | 0.234354 | 3.53E-08 | 7.73E-05 |
| cg05560934_TC21 | chr3 | 136078738 | | | 0.343967 | 3.56E-08 | 7.79E-05 |
| cg25490986_BC21 | chr20 | 59338228 | | | 0.225866 | 3.61E-08 | 7.86E-05 |
| cg24998848_BC21 | chr15 | 25346540 | | | 0.437027 | 3.62E-08 | 7.88E-05 |
| cg00714597_BC21 | chr10 | 97505479 | | | 0.275293 | 3.68E-08 | 7.97E-05 |
| cg23014713_BC11 | chr15 | 90227192 | | | 0.122201 | 3.70E-08 | 7.97E-05 |
| cg17316947_BC21 | chr1 | 47332797 | TSS1500;T[illegible] | CMPK1;CM[illegible] | 0.261139 | 3.70E-08 | 7.97E-05 |
| cg12507091_BC21 | chr5 | 146621295 | | | 0.189003 | 3.70E-08 | 7.97E-05 |
| cg01344914_TC21 | chr6 | 30195877 | | | 0.227794 | 3.72E-08 | 7.99E-05 |
| cg24656269_BC11 | chr11 | 132401687 | | | 0.226179 | 3.73E-08 | 8.00E-05 |
| cg25569760_TC21 | chr8 | 6660763 | | | 0.308858 | 3.76E-08 | 8.03E-05 |
| cg00582562_BC21 | chr17 | 48311103 | | | 0.429808 | 3.79E-08 | 8.06E-05 |
| cg25976755_TC21 | chr11 | 2460265 | TSS1500 | KCNQ1 | 0.209032 | 3.80E-08 | 8.06E-05 |
| cg14606328_TC21 | chr21 | 43757758 | exon_10;3[illegible] | PDXK;PDX[illegible] | 0.193157 | 3.80E-08 | 8.06E-05 |
| cg17598937_TC21 | chr1 | 36582201 | | | 0.335845 | 3.85E-08 | 8.15E-05 |
| cg09390448_BC22 | chr6 | 33608000 | | | 0.121866 | 3.86E-08 | 8.16E-05 |
| cg02862897_TC21 | chr17 | 81835212 | TSS200 | PPP1R27 | 0.167937 | 3.90E-08 | 8.22E-05 |
| cg02511172_BC21 | chr1 | 232949463 | TSS1500;T[illegible] | NTPCR;NT[illegible] | 0.298862 | 3.92E-08 | 8.22E-05 |
| cg10226103_TC21 | chr12 | 113775067 | TSS1500;T[illegible] | LINC01234[illegible] | 0.31228 | 3.92E-08 | 8.22E-05 |
| cg24038782_BC21 | chr13 | 77584282 | | | 0.224119 | 3.93E-08 | 8.22E-05 |
| cg16949533_BC21 | chr2 | 86562113 | | | 0.28898 | 3.95E-08 | 8.23E-05 |
| cg04825027_TC21 | chr3 | 45227962 | | | 0.166196 | 3.98E-08 | 8.23E-05 |
| cg06435639_TC21 | chr8 | 37899424 | exon_1;ex[illegible] | RAB11FIP1 | -0.2418 | 3.98E-08 | 8.23E-05 |

| | | | | | | | |
|---|---|---|---|---|---|---|---|
| cg26348253_BC21 | chr6 | 2576614 | | | 0.235065 | 3.99E-08 | 8.23E-05 |
| cg14104626_TC21 | chr5 | 157561392 | | | 0.186757 | 3.99E-08 | 8.23E-05 |
| cg22239855_BC21 | chr17 | 16107406 | | | -0.44485 | 4.00E-08 | 8.23E-05 |
| cg21366688_TC11 | chr6 | 134170283 | exon_14;e› | SGK1;SGK1 | 0.18828 | 4.00E-08 | 8.23E-05 |
| cg06816718_TC11 | chr14 | 92685103 | exon_8;exc | RIN3;RIN3 | 0.178662 | 4.02E-08 | 8.26E-05 |
| cg22074272_BC21 | chr17 | 4197527 | exon_8;exc | ANKFY1;AN | 0.417285 | 4.07E-08 | 8.33E-05 |
| cg10198174_TC21 | chr2 | 24108959 | | | -0.51551 | 4.20E-08 | 8.59E-05 |
| cg22197320_TC21 | chr15 | 88480156 | exon_6;3U | MRPS11;M | 0.159344 | 4.23E-08 | 8.62E-05 |
| cg25779645_BC21 | chr20 | 44336385 | TSS1500 | R3HDML | 0.196719 | 4.25E-08 | 8.63E-05 |
| cg21688941_BC21 | chr6 | 13709810 | | | 0.389987 | 4.27E-08 | 8.66E-05 |
| cg06017461_BC21 | chr18 | 48154484 | | | 0.251072 | 4.28E-08 | 8.66E-05 |
| cg06627827_TC21 | chr1 | 72432985 | | | 0.273774 | 4.29E-08 | 8.66E-05 |
| cg23590049_TC21 | chr4 | 89309163 | TSS1500 | GPRIN3 | 0.324921 | 4.31E-08 | 8.68E-05 |
| cg18525432_BC21 | chr20 | 35559594 | exon_41 | FER1L4 | -0.2263 | 4.35E-08 | 8.74E-05 |
| cg23840491_TC21 | chr19 | 535548 | | | 0.153087 | 4.40E-08 | 8.79E-05 |
| cg22873218_BC21 | chr17 | 68972214 | | | 0.19087 | 4.40E-08 | 8.79E-05 |
| cg12298582_BC21 | chr3 | 194132370 | | | 0.223976 | 4.40E-08 | 8.79E-05 |
| cg08846394_TC21 | chr19 | 17772536 | exon_10;e› | FCHO1;FCH | 0.197653 | 4.42E-08 | 8.80E-05 |
| cg10853432_TC21 | chr16 | 28507570 | TSS1500 | IL27 | 0.197922 | 4.43E-08 | 8.80E-05 |
| cg15921824_BC21 | chr7 | 530189 | | | 0.148964 | 4.44E-08 | 8.80E-05 |
| cg17619724_TC21 | chr12 | 52147526 | | | 0.19549 | 4.45E-08 | 8.81E-05 |
| cg23731781_TC21 | chr1 | 156908178 | exon_9;exc | PEAR1;PEA | 0.275562 | 4.47E-08 | 8.82E-05 |
| cg16520327_TC21 | chr11 | 67652148 | TSS1500 | ACY3 | 0.247578 | 4.49E-08 | 8.84E-05 |
| cg24652786_TC21 | chr1 | 160645450 | | | 0.181748 | 4.51E-08 | 8.85E-05 |
| cg17079125_BC21 | chr11 | 129433513 | | | 0.148369 | 4.54E-08 | 8.89E-05 |
| cg07478690_BC21 | chr6 | 118351568 | | | 0.221629 | 4.58E-08 | 8.96E-05 |
| cg20831871_TC21 | chr13 | 68257802 | | | 0.197795 | 4.60E-08 | 8.96E-05 |
| cg07331956_TC21 | chr18 | 37362183 | | | 0.223695 | 4.60E-08 | 8.96E-05 |
| cg00004723_BC21 | chr19 | 19444598 | | | 0.182513 | 4.63E-08 | 8.99E-05 |
| cg14604386_TC11 | chr6 | 119349717 | 5UTR;exon | MAN1A1;M | 0.263751 | 4.67E-08 | 9.03E-05 |
| cg17648210_TC11 | chr21 | 44932902 | | | 0.317397 | 4.67E-08 | 9.03E-05 |
| cg21261158_BC11 | chr13 | 114109937 | TSS200 | RASA3-IT1 | 0.237107 | 4.69E-08 | 9.04E-05 |
| cg26038461_TC21 | chr2 | 24885493 | | | 0.220118 | 4.71E-08 | 9.04E-05 |
| cg05174899_BC11 | chr17 | 36601740 | 5UTR;exon | MRM1;MR | -0.33368 | 4.74E-08 | 9.04E-05 |
| cg16725562_BC21 | chr14 | 101945399 | | | 0.375065 | 4.75E-08 | 9.04E-05 |
| cg14060518_TC21 | chr5 | 135545277 | | | 0.391185 | 4.75E-08 | 9.04E-05 |
| cg11029367_TC21 | chr3 | 124986301 | | | 0.241759 | 4.78E-08 | 9.04E-05 |
| cg13284285_BC21 | chr5 | 16787444 | | | 0.271818 | 4.78E-08 | 9.04E-05 |
| cg25172728_TC21 | chr20 | 33653647 | | | 0.162594 | 4.78E-08 | 9.04E-05 |
| cg02384338_BC11 | chr16 | 19872647 | exon_2;exc | GPRC5B;GI | 0.204173 | 4.79E-08 | 9.04E-05 |
| cg17036014_TC21 | chr16 | 46483216 | | | 0.240452 | 4.80E-08 | 9.04E-05 |
| cg23029013_BC21 | chr17 | 79167544 | | | 0.19358 | 4.80E-08 | 9.04E-05 |
| cg22722760_BC21 | chr4 | 41749609 | TSS1500 | PHOX2B | 0.157281 | 4.81E-08 | 9.04E-05 |
| cg14799536_TC21 | chr1 | 201531265 | | | 0.248503 | 4.81E-08 | 9.04E-05 |
| cg24032619_TC21 | chr11 | 67675315 | TSS1500;TS | ALDH3B2;A | 0.177474 | 4.83E-08 | 9.05E-05 |
| cg21805788_TC21 | chr1 | 108591750 | | | 0.11402 | 4.85E-08 | 9.08E-05 |
| cg01245530_BC21 | chr17 | 44147779 | | | 0.29232 | 4.88E-08 | 9.08E-05 |

| | | | | | | | |
|---|---|---|---|---|---|---|---|
| cg02993987_BC21 | chr3 | 66139788 | | | 0.383227 | 4.88E-08 | 9.08E-05 |
| cg23002268_BC11 | chr5 | 172772315 | TSS1500 | DUSP1 | -0.25772 | 4.89E-08 | 9.08E-05 |
| cg14094975_TC21 | chr9 | 109633060 | | | 0.256542 | 4.90E-08 | 9.08E-05 |
| cg24873258_TC21 | chr14 | 104591063 | 3UTR;exon | TMEM179; | 0.190724 | 4.91E-08 | 9.08E-05 |
| cg04134859_BC21 | chr7 | 73683362 | exon_1;TSS | DNAJC30;B | 0.237383 | 4.95E-08 | 9.14E-05 |
| cg05436346_BC21 | chr3 | 123637567 | | | 0.260403 | 4.97E-08 | 9.14E-05 |
| cg14982164_TC21 | chr13 | 78355903 | | | 0.176277 | 5.00E-08 | 9.14E-05 |
| cg23313924_TC21 | chr14 | 22481840 | | | 0.292134 | 5.00E-08 | 9.14E-05 |
| cg16669927_TC21 | chr11 | 78495560 | | | -0.33249 | 5.02E-08 | 9.14E-05 |
| cg20360474_TC21 | chr15 | 48202132 | exon_2;3U | CTXN2;CTX | 0.270663 | 5.03E-08 | 9.14E-05 |
| cg06513368_BC21 | chr16 | 86930196 | | | 0.218141 | 5.03E-08 | 9.14E-05 |
| cg17315417_BC21 | chr8 | 138083967 | TSS1500 | LOC401478 | 0.374953 | 5.04E-08 | 9.14E-05 |
| cg16750732_BC21 | chr9 | 133714027 | | | 0.152119 | 5.05E-08 | 9.14E-05 |
| cg08847071_BC21 | chr2 | 41414725 | | | 0.338942 | 5.05E-08 | 9.14E-05 |
| cg23875707_BC21 | chr19 | 1361623 | | | 0.219301 | 5.06E-08 | 9.14E-05 |
| cg08177925_TC21 | chr15 | 65623070 | TSS1500;TS | SLC24A1;SI | 0.342242 | 5.07E-08 | 9.14E-05 |
| cg27149150_BC21 | chr1 | 7389791 | TSS200;TSS | CAMTA1-A | 0.169699 | 5.10E-08 | 9.18E-05 |
| cg06728690_BC21 | chr7 | 139334985 | | | -0.2883 | 5.14E-08 | 9.24E-05 |
| cg17762975_TC21 | chr8 | 10524408 | TSS1500;TS | PRSS55;PRS | 0.154862 | 5.19E-08 | 9.28E-05 |
| cg06034578_BC21 | chr3 | 195839815 | | | 0.149222 | 5.19E-08 | 9.28E-05 |
| cg05814752_TC21 | chr7 | 123636103 | | | 0.541428 | 5.19E-08 | 9.28E-05 |
| cg13974692_BC21 | chr17 | 64506445 | TSS1500;TS | CEP95;CEP | -0.46092 | 5.23E-08 | 9.31E-05 |
| cg08852005_BC21 | chr15 | 99252604 | TSS1500;TS | TTC23;TTC2 | 0.160874 | 5.24E-08 | 9.31E-05 |
| cg20768117_BC21 | chr15 | 24764270 | | | -0.26312 | 5.25E-08 | 9.32E-05 |
| cg17353900_TC21 | chr17 | 10197693 | | | 0.17648 | 5.30E-08 | 9.39E-05 |
| cg20473642_BC21 | chr17 | 63973127 | TSS1500 | SCN4A | 0.178697 | 5.31E-08 | 9.39E-05 |
| cg01905270_TC21 | chr7 | 30970844 | | | 0.200194 | 5.33E-08 | 9.41E-05 |
| cg07529461_BC21 | chr1 | 15983465 | | | 0.187199 | 5.36E-08 | 9.44E-05 |
| cg09304165_BC21 | chr3 | 52016379 | | | 0.18512 | 5.38E-08 | 9.45E-05 |
| cg12948116_TC11 | chr12 | 14365992 | | | -0.36194 | 5.39E-08 | 9.45E-05 |
| cg12746554_BC21 | chr20 | 62823643 | | | 0.130484 | 5.41E-08 | 9.46E-05 |
| cg03088955_TC21 | chr22 | 38686197 | 3UTR;exon | JOSD1;JOSI | 0.230136 | 5.44E-08 | 9.51E-05 |
| cg00800365_TC21 | chr15 | 89775541 | TSS1500 | MESP2 | -0.2508 | 5.47E-08 | 9.54E-05 |
| cg03114157_TC21 | chr9 | 93618354 | | | 0.199351 | 5.48E-08 | 9.54E-05 |
| cg23070393_TC21 | chr17 | 80923714 | | | 0.178694 | 5.52E-08 | 9.59E-05 |
| cg16567310_TC21 | chr11 | 70179242 | | | 0.140652 | 5.53E-08 | 9.59E-05 |
| cg21186722_TC21 | chr18 | 2905300 | | | -0.32816 | 5.55E-08 | 9.60E-05 |
| cg25376452_BC21 | chr20 | 50224923 | | | 0.146693 | 5.56E-08 | 9.60E-05 |
| cg14228484_TC11 | chr2 | 96761708 | exon_1 | CNNM4 | 0.171024 | 5.61E-08 | 9.66E-05 |
| cg17212019_BC21 | chr8 | 25067286 | | | 0.165591 | 5.67E-08 | 9.74E-05 |
| cg19560241_BC21 | chr14 | 59626760 | | | 0.273348 | 5.68E-08 | 9.75E-05 |
| cg02318080_TC21 | chr7 | 157089021 | | | 0.185444 | 5.70E-08 | 9.75E-05 |
| cg26126690_TC21 | chr3 | 134338131 | | | 0.224459 | 5.74E-08 | 9.79E-05 |
| cg09249615_BC21 | chr6 | 20117519 | | | 0.265008 | 5.74E-08 | 9.79E-05 |
