## Supplementary Table S5 for "Early Epigenetic Biomarkers for Perinatal Suicidal Ideation: DNA Methylation Signatures Across the Peripartum Period"

**Supplementary Table S5.1. Significant Differentially methylated regions between perinatal SI v NC at 17w gestation ranked by Fisher's p-value.**

| Chr | Start | End | Width | *n* CpGs | Minimum smoothed FDR | Stouffer | HMFDR | Fisher | Mean differential | Overlapping genes |
|---|---|---|---|---|---|---|---|---|---|---|
| chr6 | 31722219 | 31724983 | 2765 | 38 | 6.06E-65 | 4.31E-68 | 3.05E-05 | 6.35E-68 | -0.013769828 | MPIG6B |
| chr20 | 37518640 | 37521733 | 3094 | 46 | 2.32E-46 | 3.74E-60 | 0.00014 | 3.48E-58 | -0.015294028 | NNAT, BLCAP |
| chr20 | 64056201 | 64058007 | 1807 | 14 | 4.27E-37 | 2.43E-34 | 9.29E-06 | 1.12E-34 | -0.015770116 | TCEA2, AL355803.1 |
| chr11 | 57498577 | 57500794 | 2218 | 9 | 2.71E-33 | 1.26E-30 | 2.86E-06 | 5.62E-32 | -0.020640805 | SLC43A1 |
| chr5 | 141412285 | 141413903 | 1619 | 13 | 1.95E-32 | 4.72E-33 | 1.11E-05 | 5.62E-32 | 0.015827182 | PCDHGA1, PCDHGA2, PCDHGA3, PCDHGB1, PCDHGA4, PCDHGB2, PCDHGA5, PCDHGB3, PCDHGA6, PCDHGA7, PCDHGB4, PCDHGA8, PCDHGB5, PCDHGA9, PCDHGB6, PCDHGA10 |
| chr1 | 74732527 | 74734410 | 1884 | 18 | 9.89E-32 | 4.46E-35 | 0.00013 | 5.62E-32 | 0.013320046 | TYW3, CRYZ |
| chr6 | 113903970 | 113904718 | 749 | 5 | 2.85E-29 | 1.27E-24 | 2.76E-07 | 8.09E-26 | -0.033650726 | LINC02880 |
| chr17 | 35752646 | 35753882 | 1237 | 8 | 2.12E-28 | 2.73E-24 | 2.28E-06 | 1.10E-25 | -0.018915259 | TAF15, GAS2L2 |
| chr7 | 2606038 | 2609461 | 3424 | 20 | 2.57E-28 | 3.66E-17 | 1.02E-06 | 9.05E-25 | -0.009213124 | IQCE |
| chr7 | 27142816 | 27148072 | 5257 | 66 | 1.27E-27 | 1.92E-59 | 0.00056 | 2.80E-51 | -0.020810195 | HOXA-AS3, HOXA3, AC004080.6, HOXA5, HOXA6, AC004080.5 |
| chr15 | 42079313 | 42080422 | 1110 | 9 | 5.05E-26 | 1.72E-19 | 2.55E-07 | 1.88E-23 | 0.019596411 | PLA2G4D |
| chr16 | 67963393 | 67965141 | 1749 | 12 | 7.11E-25 | 2.14E-17 | 1.82E-05 | 8.36E-24 | -0.02129601 | SLC12A4 |
| chr11 | 124751933 | 124752919 | 987 | 9 | 1.10E-24 | 2.62E-19 | 2.39E-05 | 1.46E-22 | -0.019248257 | VSIG2, ESAM |
| chr7 | 44962166 | 44963381 | 1216 | 9 | 5.03E-24 | 4.69E-19 | 1.57E-05 | 9.04E-21 | -0.028403539 | MYO1G |
| chr19 | 37333908 | 37335106 | 1199 | 10 | 5.42E-24 | 1.10E-24 | 7.79E-05 | 1.31E-23 | -0.0465997 | ZNF875 |
| chr6 | 31680873 | 31681951 | 1079 | 25 | 1.34E-23 | 6.18E-09 | 2.44E-06 | 7.09E-15 | -0.004133643 | LY6G5C |
| chr22 | 19663525 | 19664296 | 772 | 6 | 3.17E-23 | 1.01E-18 | 8.08E-07 | 1.95E-20 | -0.017179178 | NA |
| chr1 | 1627365 | 1630551 | 3187 | 15 | 1.07E-22 | 1.77E-25 | 4.82E-05 | 8.08E-27 | -0.020170251 | MIB2 |
| chr15 | 74201299 | 74203699 | 2401 | 18 | 1.30E-22 | 8.77E-23 | 3.41E-06 | 5.51E-24 | -0.012867903 | CCDC33, STRA6 |
| chr4 | 121932309 | 121933975 | 1667 | 14 | 3.12E-22 | 6.53E-14 | 2.62E-05 | 2.75E-19 | -0.019543469 | TRPC3 |
| chr17 | 40014070 | 40015444 | 1375 | 11 | 4.43E-22 | 1.55E-14 | 5.51E-06 | 4.98E-19 | -0.011104406 | CSF3, AC090844.3 |
| chr19 | 13053491 | 13056109 | 2619 | 12 | 8.62E-22 | 1.01E-21 | 4.62E-06 | 9.05E-25 | -0.012762606 | NFIX |
| chr3 | 49132567 | 49133910 | 1344 | 11 | 1.22E-21 | 1.34E-22 | 0.00013 | 1.29E-21 | -0.028589208 | LAMB2 |
| chr7 | 2760562 | 2763808 | 3247 | 19 | 1.37E-21 | 2.95E-23 | 3.83E-05 | 2.02E-27 | -0.016873387 | AMZ1, GNA12 |
| chr6 | 31765657 | 31767666 | 2010 | 21 | 2.02E-21 | 2.80E-25 | 0.00068 | 2.98E-23 | -0.014299286 | VWA7 |
| chr7 | 56092716 | 56093327 | 612 | 10 | 2.35E-21 | 3.52E-14 | 2.44E-06 | 4.31E-20 | -0.014381323 | PHKG1 |
| chr20 | 19886099 | 19886779 | 681 | 9 | 4.72E-21 | 3.56E-18 | 3.45E-05 | 1.21E-19 | -0.008979311 | RIN2 |
| chr22 | 50577957 | 50579294 | 1338 | 16 | 4.87E-21 | 7.85E-27 | 0.00038 | 5.17E-23 | -0.024297365 | CPT1B, CHKB-CPT1B, CHKB |
| chr2 | 27078327 | 27079075 | 749 | 9 | 1.27E-20 | 4.38E-23 | 7.72E-05 | 6.63E-21 | -0.0241416 | EMILIN1 |
| chr19 | 16065487 | 16067760 | 2274 | 14 | 1.97E-20 | 5.82E-19 | 2.70E-05 | 1.63E-21 | -0.009179019 | LINC01855, TPM4 |
| chr1 | 174447908 | 174449077 | 1170 | 6 | 4.89E-20 | 7.91E-15 | 6.61E-07 | 2.05E-16 | -0.022718404 | RABGAP1L, GPR52 |
| chr6 | 31538791 | 31541859 | 3069 | 34 | 6.34E-20 | 9.78E-13 | 0.00018 | 7.95E-18 | -0.005893964 | DDX39B, ATP6V1G2-DDX39B, SNORD84 |
| chr6 | 35017059 | 35018106 | 1048 | 5 | 9.85E-20 | 8.98E-16 | 1.55E-06 | 5.19E-18 | -0.023099034 | ANKS1A |
| chr1 | 8197859 | 8198286 | 428 | 3 | 1.14E-19 | 5.25E-19 | 1.83E-07 | 1.44E-18 | -0.027663935 | NA |
| chr19 | 45233753 | 45235599 | 1847 | 12 | 1.62E-19 | 4.19E-16 | 0.00015 | 1.21E-18 | -0.018404684 | MARK4, EXOC3L2 |
| chr19 | 2783383 | 2785893 | 2511 | 20 | 1.71E-19 | 5.21E-07 | 1.11E-05 | 7.25E-14 | -0.006257134 | THOP1 |
| chr3 | 42935464 | 42937398 | 1935 | 16 | 3.34E-19 | 1.03E-19 | 0.00034 | 4.68E-19 | -0.018850805 | AC092042.3, KRBOX1, KRBOX1-AS1 |
| chr17 | 73261841 | 73263142 | 1302 | 10 | 3.79E-19 | 3.76E-13 | 9.89E-06 | 4.69E-16 | -0.010923366 | CPSF4L |
| chr10 | 79159485 | 79160914 | 1430 | 5 | 4.76E-19 | 2.54E-19 | 1.07E-07 | 1.25E-21 | -0.020011995 | ZMIZ1 |
| chr12 | 47752684 | 47753692 | 1009 | 4 | 5.14E-19 | 6.31E-14 | 4.48E-07 | 2.14E-16 | -0.017584758 | RAPGEF3 |
| chr21 | 33401797 | 33403153 | 1357 | 11 | 9.34E-19 | 1.24E-15 | 5.34E-05 | 5.99E-17 | -0.007629782 | NA |
| chr19 | 40241454 | 40242656 | 1203 | 7 | 1.18E-18 | 2.16E-07 | 8.50E-08 | 4.43E-13 | -0.006711749 | AKT2 |
| chr19 | 40279431 | 40281005 | 1575 | 6 | 1.25E-18 | 1.67E-21 | 1.28E-06 | 1.08E-20 | -0.016796867 | AKT2 |
| chr1 | 41503891 | 41504306 | 416 | 3 | 1.31E-18 | 6.22E-13 | 6.49E-08 | 1.81E-15 | -0.024173132 | NA |
| chr6 | 32837621 | 32842481 | 4861 | 61 | 1.51E-18 | 2.25E-10 | 5.11E-05 | 5.19E-18 | -0.001353367 | AL669918.1, TAP2, PSMB8 |
| chr11 | 66334390 | 66337677 | 3288 | 23 | 4.37E-18 | 1.05E-11 | 1.56E-05 | 1.44E-18 | -0.007385709 | AP001107.6, RIN1, BRMS1 |
| chr13 | 114108813 | 114110586 | 1774 | 8 | 4.67E-18 | 1.24E-17 | 7.03E-05 | 5.90E-17 | -0.01389855 | RASA3, RASA3-IT1 |
| chr22 | 29030844 | 29032120 | 1277 | 5 | 5.47E-18 | 6.53E-14 | 1.81E-05 | 1.81E-15 | -0.02232325 | ZNRF3, ZNRF3-AS1 |
| chr22 | 38317106 | 38318766 | 1661 | 14 | 6.88E-18 | 2.23E-21 | 0.00025 | 7.63E-19 | -0.009672929 | CSNK1E, TPTEP2-CSNK1E |
| chr6 | 111759899 | 111760335 | 437 | 6 | 1.07E-17 | 2.62E-19 | 2.93E-05 | 5.85E-18 | -0.019824566 | FYN |
| chr20 | 63733449 | 63737212 | 3764 | 18 | 1.28E-17 | 2.85E-27 | 4.23E-06 | 4.53E-26 | -0.013421706 | ZGPAT, AL121845.3, LIME1 |
| chr11 | 1297647 | 1299983 | 2337 | 12 | 1.46E-17 | 5.81E-17 | 2.19E-06 | 4.49E-19 | -0.005465907 | TOLLIP |
| chr20 | 63685496 | 63687235 | 1740 | 11 | 1.47E-17 | 2.53E-14 | 1.96E-06 | 5.52E-15 | -0.009400249 | RTEL1, RTEL1-TNFRSF6B |
| chr1 | 159899373 | 159901125 | 1753 | 10 | 2.01E-17 | 3.12E-14 | 8.19E-05 | 1.81E-15 | -0.014934259 | AL590560.5, CFAP45, MIR4259 |
| chr9 | 128732813 | 128733711 | 899 | 20 | 2.49E-17 | 1.72E-09 | 0.00011 | 3.02E-14 | -0.006846826 | AL441992.1, ZER1 |
| chr6 | 30687919 | 30689692 | 1774 | 19 | 2.95E-17 | 4.47E-13 | 0.00013 | 4.28E-14 | -0.003732241 | NRM |
| chr20 | 62666308 | 62668456 | 2149 | 11 | 3.04E-17 | 1.80E-14 | 4.09E-06 | 1.25E-15 | -0.00613381 | SLCO4A1, SLCO4A1-AS1 |
| chr9 | 134880436 | 134881040 | 605 | 6 | 3.29E-17 | 3.71E-15 | 5.86E-07 | 7.85E-16 | -0.015602108 | FCN2 |
| chr12 | 4808909 | 4810425 | 1517 | 12 | 4.90E-17 | 7.92E-11 | 0.00024 | 2.28E-14 | 0.016333695 | GALNT8, KCNA6, AC005833.2 |
| chr17 | 17792064 | 17793078 | 1015 | 7 | 4.90E-17 | 2.22E-10 | 1.94E-05 | 1.49E-14 | -0.007513512 | RAI1 |
| chr17 | 74767468 | 74768846 | 1379 | 5 | 5.07E-17 | 1.34E-16 | 4.68E-06 | 1.21E-16 | -0.016792344 | SLC9A3R1 |
| chr10 | 133377084 | 133380489 | 3406 | 38 | 6.09E-17 | 1.56E-15 | 4.20E-05 | 4.56E-19 | -0.003545915 | PAOX, AL360181.3 |
| chr8 | 41660616 | 41661881 | 1266 | 8 | 7.23E-17 | 3.68E-10 | 2.74E-05 | 1.01E-13 | -0.006958739 | AC113133.1, ANK1 |
| chr13 | 112978330 | 112981915 | 3586 | 15 | 7.73E-17 | 1.02E-16 | 7.04E-06 | 3.95E-23 | -0.020573488 | MCF2L |
| chr5 | 172904730 | 172905682 | 953 | 6 | 8.73E-17 | 2.29E-14 | 1.28E-06 | 7.27E-16 | -0.010364707 | ERGIC1 |
| chr7 | 29145929 | 29147403 | 1475 | 16 | 8.77E-17 | 3.31E-07 | 7.54E-05 | 6.16E-12 | -0.009212009 | AC004593.2, CHN2, CPVL |
| chr20 | 62677728 | 62678765 | 1038 | 5 | 1.23E-16 | 5.17E-15 | 1.51E-05 | 4.27E-16 | -0.028902297 | SLCO4A1 |
| chr6 | 32196366 | 32198140 | 1775 | 31 | 1.84E-16 | 1.41E-12 | 0.0009 | 6.27E-14 | -0.004261666 | NOTCH4 |
| chr7 | 92608062 | 92609050 | 989 | 8 | 1.84E-16 | 1.00E-15 | 6.11E-05 | 5.74E-16 | -0.017319541 | CDK6 |
| chr9 | 133978737 | 133979646 | 910 | 5 | 1.86E-16 | 1.10E-16 | 2.88E-05 | 5.74E-16 | -0.02655985 | VAV2 |
| chr11 | 67439292 | 67440591 | 1300 | 8 | 2.00E-16 | 1.87E-18 | 0.00011 | 4.42E-17 | -0.013432387 | CORO1B |
| chr1 | 112359593 | 112360607 | 1015 | 4 | 2.37E-16 | 3.19E-14 | 1.57E-06 | 4.27E-15 | -0.014938404 | LINC02884 |
| chr12 | 52009727 | 52011096 | 1370 | 13 | 2.60E-16 | 3.11E-09 | 2.62E-05 | 1.27E-13 | -0.008197798 | TAMALIN |
| chr7 | 55156513 | 55158516 | 2004 | 13 | 3.49E-16 | 6.36E-14 | 0.00015 | 2.11E-15 | 0.01105283 | EGFR |
| chr12 | 117032736 | 117033310 | 575 | 6 | 3.52E-16 | 8.05E-18 | 3.01E-05 | 2.08E-16 | -0.023541058 | NA |
| chr20 | 31637522 | 31638674 | 1153 | 10 | 3.82E-16 | 6.29E-14 | 2.71E-05 | 3.27E-15 | -0.012672605 | COX4I2 |
| chr6 | 33426009 | 33431089 | 5081 | 45 | 4.16E-16 | 3.07E-16 | 5.16E-05 | 6.02E-22 | -0.002165228 | SYNGAP1 |
| chr1 | 27528003 | 27529826 | 1824 | 7 | 4.54E-16 | 3.92E-10 | 4.09E-06 | 4.88E-13 | -0.010619365 | BX293535.1 |
| chr16 | 81500967 | 81501476 | 510 | 4 | 4.76E-16 | 1.28E-09 | 1.42E-07 | 6.91E-13 | -0.016628889 | CMIP |
| chr12 | 50103254 | 50104810 | 1557 | 8 | 4.81E-16 | 2.14E-09 | 3.56E-05 | 1.57E-12 | -0.01057363 | GPD1 |
| chr7 | 128910357 | 128911703 | 1347 | 12 | 5.27E-16 | 3.47E-12 | 6.21E-05 | 1.10E-14 | -0.007670153 | KCP |
| chr8 | 143950469 | 143952445 | 1977 | 15 | 5.46E-16 | 3.27E-17 | 7.56E-07 | 5.04E-21 | -0.006434066 | PLEC |
| chr22 | 24425610 | 24428394 | 2785 | 15 | 6.70E-16 | 6.18E-17 | 7.09E-05 | 1.48E-17 | -0.004861694 | SPECC1L-ADORA2A, ADORA2A |
| chr6 | 6587856 | 6588842 | 987 | 8 | 9.99E-16 | 5.48E-16 | 6.50E-05 | 1.44E-15 | -0.017062786 | LY86, LY86-AS1 |
| chr12 | 56993035 | 56994400 | 1366 | 8 | 1.20E-15 | 6.70E-10 | 2.66E-09 | 1.28E-13 | -0.00867051 | NA |
| chr17 | 82442321 | 82445856 | 3536 | 15 | 1.39E-15 | 4.25E-19 | 1.29E-05 | 1.36E-19 | -0.007346533 | HEXD, CYBC1 |
| chr7 | 882208 | 884217 | 2010 | 11 | 1.62E-15 | 1.28E-12 | 3.01E-05 | 1.37E-15 | -0.007999264 | SUN1, GET4, AC073957.3 |
| chr11 | 1024148 | 1025969 | 1822 | 9 | 1.72E-15 | 1.42E-12 | 1.83E-06 | 1.73E-15 | -0.009074146 | MUC6 |
| chr16 | 70687793 | 70689698 | 1906 | 6 | 3.12E-15 | 7.94E-09 | 6.80E-08 | 7.48E-12 | -0.008296818 | VAC14 |
| chr22 | 46043867 | 46044589 | 723 | 4 | 4.33E-15 | 5.84E-09 | 4.82E-06 | 3.05E-11 | -0.013969179 | LINC00899 |
| chr16 | 53504580 | 53504748 | 169 | 2 | 4.87E-15 | 3.64E-12 | 3.96E-07 | 6.01E-12 | -0.029884868 | NA |
| chr6 | 31715274 | 31715575 | 302 | 6 | 5.32E-15 | 2.36E-16 | 8.36E-05 | 5.37E-15 | -0.016271212 | LY6G6F-LY6G6D, LY6G6D |
| chr19 | 2727824 | 2729363 | 1540 | 6 | 7.22E-15 | 2.03E-06 | 1.07E-07 | 6.59E-11 | -0.005800748 | AC006538.2, AC006538.1 |
| chr9 | 127950104 | 127951481 | 1378 | 10 | 7.24E-15 | 1.22E-12 | 1.28E-05 | 3.24E-14 | -0.006068581 | FAM102A |
| chr16 | 57632681 | 57633138 | 458 | 3 | 8.60E-15 | 1.56E-14 | 2.53E-07 | 1.87E-14 | -0.022883051 | ADGRG1 |
| chr9 | 131421275 | 131423519 | 2245 | 8 | 9.91E-15 | 6.45E-15 | 3.81E-06 | 1.77E-18 | -0.012265014 | PRRC2B |
| chr12 | 124517461 | 124517928 | 468 | 4 | 1.10E-14 | 3.71E-15 | 2.76E-05 | 3.82E-14 | -0.020561383 | NCOR2 |
| chr11 | 13961369 | 13963049 | 1681 | 14 | 1.37E-14 | 1.27E-11 | 7.91E-05 | 1.83E-13 | -0.016830438 | SPON1 |
| chr6 | 166691113 | 166692076 | 964 | 7 | 2.23E-14 | 1.55E-14 | 3.93E-05 | 7.10E-16 | -0.011585051 | RPS6KA2 |
| chr11 | 47247407 | 47248983 | 1577 | 12 | 2.32E-14 | 7.05E-09 | 4.94E-05 | 9.01E-11 | -0.0024795 | NR1H3, ACP2 |
| chr13 | 51845129 | 51845592 | 464 | 9 | 2.44E-14 | 8.98E-11 | 7.24E-06 | 4.70E-14 | -0.012278016 | AL162377.3, TMEM272 |
| chr6 | 116060446 | 116061717 | 1272 | 8 | 2.75E-14 | 3.19E-14 | 0.00013 | 1.10E-14 | -0.01471283 | FRK |
| chr19 | 728176 | 728385 | 210 | 2 | 2.76E-14 | 5.41E-14 | 4.79E-07 | 1.35E-13 | -0.049426896 | PALM |
| chr3 | 184319342 | 184320304 | 963 | 9 | 2.80E-14 | 2.40E-07 | 2.51E-05 | 8.33E-11 | -0.007788912 | EIF4G1 |
| chr1 | 1432917 | 1434086 | 1170 | 4 | 3.38E-14 | 2.64E-08 | 1.77E-07 | 5.50E-10 | -0.0098697 | LINC01770 |
| chr11 | 1830414 | 1831344 | 931 | 6 | 3.53E-14 | 3.09E-11 | 3.99E-06 | 1.84E-12 | -0.013708562 | SYT8 |
| chr15 | 90081844 | 90083416 | 1573 | 7 | 3.75E-14 | 5.01E-11 | 1.17E-05 | 5.86E-13 | -0.009098471 | ZNF710, ZNF710-AS1, IDH2 |
| chr19 | 49887807 | 49888628 | 822 | 5 | 3.79E-14 | 1.57E-15 | 4.69E-05 | 1.39E-14 | -0.014798468 | TBC1D17, MIR4750 |
| chr5 | 175680094 | 175681426 | 1333 | 9 | 3.84E-14 | 9.44E-13 | 2.88E-05 | 5.89E-14 | -0.006304348 | HRH2 |
| chr19 | 813735 | 815275 | 1541 | 7 | 4.64E-14 | 2.58E-10 | 2.45E-06 | 3.66E-12 | -0.009482223 | PLPPR3 |
| chr1 | 2025634 | 2026366 | 733 | 5 | 5.06E-14 | 3.19E-14 | 4.68E-06 | 3.11E-14 | -0.02094731 | GABRD |
| chr11 | 65541746 | 65542153 | 408 | 3 | 5.40E-14 | 9.59E-09 | 3.83E-09 | 2.28E-11 | -0.00334116 | LTBP3 |
| chr10 | 104274714 | 104275410 | 697 | 11 | 5.73E-14 | 1.78E-08 | 2.99E-05 | 1.18E-11 | -0.011112646 | GSTO2 |
| chr20 | 63907630 | 63908522 | 893 | 5 | 5.77E-14 | 1.88E-12 | 1.88E-05 | 4.63E-13 | -0.01650601 | DNAJC5 |
| chr1 | 64171709 | 64172090 | 382 | 3 | 5.77E-14 | 1.53E-14 | 3.16E-06 | 6.07E-14 | 0.021293298 | ROR1 |
| chr7 | 908609 | 910197 | 1589 | 6 | 5.84E-14 | 2.11E-11 | 1.36E-05 | 5.13E-13 | -0.043782148 | ADAP1, COX19 |
| chr13 | 75759864 | 75760730 | 867 | 5 | 6.12E-14 | 1.04E-07 | 9.86E-06 | 1.10E-11 | -0.01769162 | AL137782.1, LMO7 |
| chr2 | 96858194 | 96859272 | 1079 | 5 | 6.59E-14 | 2.14E-12 | 2.03E-05 | 2.04E-12 | -0.012449722 | NA |
| chr22 | 46660313 | 46661492 | 1180 | 5 | 7.12E-14 | 2.94E-09 | 2.75E-05 | 9.21E-12 | -0.008288649 | GRAMD4 |
| chr2 | 137963733 | 137964756 | 1024 | 9 | 7.67E-14 | 1.35E-12 | 8.10E-06 | 3.34E-15 | -0.016589025 | HNMT |
| chr6 | 33320032 | 33322631 | 2600 | 45 | 7.84E-14 | 7.77E-10 | 0.00541 | 1.49E-13 | -0.001957864 | DAXX, SMIM40 |
| chr5 | 79689602 | 79690337 | 736 | 9 | 8.32E-14 | 1.27E-16 | 0.00065 | 6.42E-15 | -0.02168153 | CMYA5 |
| chr14 | 104700403 | 104702302 | 1900 | 11 | 9.58E-14 | 9.02E-12 | 0.00024 | 2.93E-14 | -0.008940992 | INF2 |
| chr7 | 111727617 | 111728791 | 1175 | 6 | 9.59E-14 | 9.75E-14 | 3.10E-05 | 2.59E-13 | -0.018801813 | DOCK4 |
| chr12 | 7060009 | 7061241 | 1233 | 11 | 1.01E-13 | 4.45E-11 | 7.47E-05 | 3.66E-12 | -0.008501047 | C1S |
| chr6 | 3324167 | 3324933 | 767 | 6 | 1.01E-13 | 1.14E-10 | 5.98E-05 | 4.10E-12 | -0.013241051 | SLC22A23 |
| chr3 | 46923615 | 46924042 | 428 | 6 | 1.02E-13 | 5.23E-13 | 3.99E-06 | 1.04E-13 | -0.010079049 | CCDC12 |
| chr6 | 32149788 | 32154384 | 4597 | 73 | 1.16E-13 | 5.49E-22 | 0.00457 | 1.07E-21 | -0.005798052 | PPT2, PPT2-EGFL8, PRRT1, AL662884.4, AL662884.3 |
| chr7 | 995011 | 997017 | 2007 | 7 | 1.24E-13 | 5.53E-14 | 3.60E-06 | 5.45E-15 | -0.010732594 | C7orf50 |
| chr18 | 11150044 | 11151284 | 1241 | 3 | 1.29E-13 | 1.71E-05 | 5.76E-08 | 1.25E-08 | -0.007403434 | NA |
| chr16 | 89096446 | 89097392 | 947 | 6 | 1.43E-13 | 7.13E-12 | 1.87E-05 | 3.14E-12 | -0.01813856 | ACSF3 |
| chr18 | 10453627 | 10455466 | 1840 | 11 | 1.50E-13 | 1.92E-14 | 3.38E-05 | 9.14E-16 | -0.006230467 | APCDD1 |
| chr11 | 111918086 | 111918969 | 884 | 10 | 1.61E-13 | 2.07E-10 | 0.00012 | 6.09E-12 | -0.005517828 | HSPB2-C11orf52, C11orf52, CRYAB |
| chr2 | 130987925 | 130988231 | 307 | 3 | 1.61E-13 | 1.70E-11 | 5.14E-08 | 1.26E-12 | -0.02054082 | ARHGEF4 |
| chr16 | 1457264 | 1459205 | 1942 | 8 | 1.63E-13 | 6.53E-14 | 2.83E-05 | 9.98E-16 | -0.032472717 | CLCN7 |
| chr2 | 74440473 | 74442260 | 1788 | 11 | 1.70E-13 | 2.27E-08 | 1.88E-05 | 6.00E-10 | -0.01073277 | RTKN |
| chr11 | 94767446 | 94769713 | 2268 | 21 | 1.83E-13 | 5.56E-07 | 4.21E-06 | 2.10E-14 | -0.004720995 | AMOTL1 |
| chr21 | 43732302 | 43734889 | 2588 | 9 | 1.91E-13 | 3.74E-17 | 3.01E-05 | 3.22E-16 | -0.018869316 | PDXK |
| chr12 | 130336908 | 130338273 | 1366 | 13 | 2.08E-13 | 2.81E-10 | 0.0006 | 7.72E-12 | -0.038109832 | PIWIL1 |
| chr6 | 29486846 | 29487755 | 910 | 11 | 2.23E-13 | 4.84E-09 | 6.40E-05 | 1.88E-12 | -0.0074852 | MAS1L |
| chr17 | 8309877 | 8311381 | 1505 | 7 | 2.63E-13 | 3.29E-08 | 6.36E-05 | 1.57E-11 | -0.01455803 | ARHGEF15 |
| chr10 | 5534229 | 5535203 | 975 | 5 | 2.79E-13 | 6.84E-11 | 5.14E-08 | 1.96E-12 | -0.009221977 | NA |
| chr7 | 35037986 | 35039499 | 1514 | 14 | 3.04E-13 | 6.56E-07 | 0.00025 | 5.76E-10 | -0.013584594 | AC005400.1, DPY19L1 |
| chr5 | 141320332 | 141321109 | 778 | 15 | 3.09E-13 | 2.64E-10 | 1.31E-05 | 1.18E-12 | 0.004114407 | AC005618.4, TAF7 |
| chr12 | 113333063 | 113333403 | 341 | 4 | 3.37E-13 | 1.15E-13 | 1.12E-05 | 3.67E-13 | -0.01983262 | SLC8B1 |
| chr12 | 121535520 | 121536400 | 881 | 4 | 3.46E-13 | 1.98E-09 | 3.65E-06 | 4.72E-11 | -0.029082336 | KDM2B |
| chr19 | 11142751 | 11143369 | 619 | 4 | 3.50E-13 | 2.87E-14 | 1.26E-05 | 1.73E-13 | -0.014669288 | SPC24 |
| chr2 | 168796295 | 168796874 | 580 | 4 | 3.76E-13 | 8.66E-12 | 4.56E-06 | 1.99E-12 | -0.015479804 | NOSTRIN |
| chr6 | 31791635 | 31793278 | 1644 | 22 | 4.15E-13 | 1.26E-13 | 3.59E-05 | 1.52E-14 | -0.006486815 | VARS1 |
| chr15 | 93028758 | 93030805 | 2048 | 7 | 4.22E-13 | 8.85E-09 | 1.79E-05 | 9.03E-12 | -0.009299095 | NA |
| chr20 | 2535610 | 2536967 | 1358 | 11 | 4.76E-13 | 1.08E-11 | 7.67E-05 | 3.48E-13 | -0.008991492 | TMC2 |
| chr15 | 100544302 | 100545183 | 882 | 8 | 5.40E-13 | 4.04E-07 | 5.03E-05 | 7.57E-10 | 0.03303792 | CERS3 |
| chr1 | 3230765 | 3232246 | 1482 | 8 | 6.72E-13 | 4.12E-10 | 1.09E-05 | 9.15E-12 | -0.00232557 | PRDM16 |
| chr1 | 31575091 | 31577024 | 1934 | 16 | 7.80E-13 | 5.84E-09 | 7.41E-05 | 3.14E-12 | -0.006590278 | LINC01226, TINAGL1, AC114488.1 |
| chr2 | 104754951 | 104756331 | 1381 | 9 | 8.14E-13 | 4.89E-08 | 2.57E-05 | 1.32E-10 | -0.010127174 | LINC01114 |
| chr14 | 101518634 | 101519029 | 396 | 3 | 8.49E-13 | 5.01E-14 | 1.69E-05 | 3.04E-13 | -0.025280459 | NA |
| chr1 | 31181219 | 31182398 | 1180 | 9 | 1.01E-12 | 1.48E-12 | 0.00037 | 3.30E-12 | -0.019485468 | NKAIN1 |
| chr11 | 64122370 | 64123909 | 1540 | 4 | 1.05E-12 | 2.40E-06 | 5.85E-06 | 9.18E-10 | -0.018163209 | MACROD1 |
| chr1 | 110780387 | 110781120 | 734 | 6 | 1.20E-12 | 5.32E-08 | 6.05E-06 | 1.72E-11 | -0.012282878 | NA |

| | | | | | | | | | | |
|---|---|---|---|---|---|---|---|---|---|---|
| chr10 | 133277061 | 133278760 | 1700 | 8 | 1.20E-12 | 2.94E-08 | 3.25E-06 | 6.37E-10 | -0.004158822 | TUBGCP2 |
| chr12 | 11706826 | 11707718 | 893 | 5 | 1.43E-12 | 1.65E-06 | 5.14E-08 | 4.22E-11 | -0.012077919 | ETV6 |
| chr1 | 223143292 | 223144508 | 1217 | 11 | 1.46E-12 | 4.95E-08 | 7.89E-05 | 5.56E-11 | -0.010087234 | AL359979.2 |
| chr3 | 119780902 | 119782340 | 1439 | 8 | 1.54E-12 | 6.79E-14 | 3.10E-05 | 7.14E-14 | -0.018519913 | AC069444.2, NR1I2 |
| chr11 | 70281474 | 70282011 | 538 | 3 | 1.55E-12 | 3.12E-08 | 5.76E-08 | 2.96E-10 | -0.006254194 | PPFIA1 |
| chr15 | 72197075 | 72198949 | 1875 | 11 | 1.76E-12 | 1.10E-09 | 1.20E-05 | 9.71E-14 | -0.006016702 | GRAMD2A |
| chr2 | 69437185 | 69438826 | 1642 | 15 | 1.78E-12 | 3.30E-09 | 7.34E-05 | 2.63E-11 | -0.002800257 | NFU1 |
| chr16 | 88743987 | 88744752 | 766 | 5 | 2.00E-12 | 8.00E-09 | 1.09E-05 | 3.14E-12 | -0.01664607 | AC138028.1, PIEZO1 |
| chr12 | 113241751 | 113242690 | 940 | 5 | 2.11E-12 | 2.52E-14 | 5.73E-05 | 2.37E-13 | -0.016599165 | TPCN1 |
| chr9 | 128148716 | 128149513 | 798 | 7 | 2.16E-12 | 7.27E-11 | 9.97E-05 | 3.20E-11 | -0.007582188 | LCN2 |
| chr1 | 1217200 | 1219473 | 2274 | 8 | 2.25E-12 | 3.42E-07 | 8.25E-06 | 1.96E-12 | 0.000634463 | SDF4 |
| chr22 | 42644440 | 42647751 | 3312 | 15 | 2.35E-12 | 5.03E-10 | 4.34E-05 | 9.91E-15 | -0.008404429 | CYB5R3 |
| chr17 | 82602138 | 82604352 | 2215 | 15 | 2.59E-12 | 1.65E-10 | 6.94E-06 | 2.16E-13 | -0.006003243 | FOXK2, AC124283.1 |
| chr2 | 84290197 | 84290753 | 557 | 7 | 2.91E-12 | 9.85E-13 | 5.86E-05 | 1.38E-12 | -0.007400082 | FUNDC2P2 |
| chr13 | 52739394 | 52740678 | 1285 | 9 | 3.41E-12 | 1.68E-08 | 7.87E-06 | 4.22E-10 | -0.007055675 | CNMD |
| chr1 | 205449150 | 205450619 | 1470 | 9 | 3.52E-12 | 1.22E-12 | 9.70E-05 | 3.04E-12 | -0.01006542 | LEMD1, BLACAT1 |
| chr22 | 23306067 | 23306575 | 509 | 2 | 3.65E-12 | 1.48E-06 | 1.01E-08 | 9.67E-10 | -0.005228409 | BCR |
| chr1 | 27349626 | 27350749 | 1124 | 7 | 3.89E-12 | 1.00E-10 | 0.00023 | 4.99E-12 | -0.025738931 | SYTL1 |
| chr9 | 137302596 | 137304299 | 1704 | 8 | 4.34E-12 | 5.20E-07 | 9.27E-05 | 1.30E-09 | -0.011126421 | NA |
| chr10 | 102450726 | 102452656 | 1931 | 12 | 4.36E-12 | 2.55E-09 | 1.21E-05 | 8.75E-12 | -0.005057732 | RPARP-AS1, C10orf95 |
| chr16 | 22092028 | 22092830 | 803 | 7 | 4.38E-12 | 1.89E-08 | 7.70E-06 | 4.72E-11 | -0.009550721 | VWA3A, AC009019.1 |
| chr4 | 41537290 | 41538212 | 923 | 5 | 4.54E-12 | 9.17E-08 | 1.64E-06 | 5.92E-10 | -0.013359978 | LIMCH1 |
| chr1 | 202965274 | 202966994 | 1721 | 7 | 4.58E-12 | 3.27E-05 | 7.29E-07 | 9.18E-08 | -0.002813745 | CYB5R1 |
| chr2 | 3656398 | 3658091 | 1694 | 10 | 4.62E-12 | 3.21E-13 | 0.00024 | 1.11E-12 | -0.02745536 | NA |
| chr6 | 13014259 | 13014989 | 731 | 7 | 4.62E-12 | 5.39E-12 | 3.75E-05 | 4.63E-12 | -0.009339495 | PHACTR1 |
| chr19 | 42278181 | 42278959 | 779 | 4 | 4.82E-12 | 3.67E-11 | 4.03E-07 | 4.81E-12 | -0.010535833 | CIC |
| chr2 | 127643267 | 127644840 | 1574 | 7 | 4.92E-12 | 9.88E-09 | 2.49E-05 | 1.26E-11 | -0.006737573 | LIMS2 |
| chr17 | 75633002 | 75635705 | 2704 | 14 | 4.94E-12 | 6.79E-14 | 3.25E-05 | 1.38E-16 | -0.007078371 | SMIM5, RECQL5 |
| chr19 | 6375391 | 6376565 | 1175 | 8 | 5.20E-12 | 1.70E-09 | 8.72E-06 | 8.60E-12 | -0.007168447 | PSPN |
| chr6 | 32879170 | 32880068 | 899 | 14 | 5.52E-12 | 1.26E-12 | 0.00149 | 2.13E-12 | -0.018489691 | PPP1R2P1 |
| chr2 | 64405502 | 64406070 | 569 | 2 | 5.73E-12 | 9.37E-05 | 2.33E-07 | 1.11E-07 | -0.014789131 | LGALSL-DT |
| chr1 | 2213434 | 2214905 | 1472 | 8 | 5.78E-12 | 1.72E-11 | 8.56E-05 | 4.83E-13 | -0.012645825 | AL590822.1 |
| chr17 | 64006047 | 64008172 | 2126 | 11 | 5.83E-12 | 4.58E-19 | 0.00012 | 3.91E-18 | -0.009782617 | ICAM2 |
| chr2 | 132584713 | 132586084 | 1372 | 10 | 5.98E-12 | 3.26E-14 | 0.0006 | 4.19E-13 | -0.012040277 | GPR39 |
| chr16 | 87847605 | 87850046 | 2442 | 9 | 5.98E-12 | 9.63E-15 | 9.09E-07 | 3.02E-16 | -0.006546098 | SLC7A5 |
| chr6 | 34514034 | 34514902 | 869 | 10 | 5.99E-12 | 1.17E-14 | 0.00025 | 9.06E-14 | -0.029570509 | PACSIN1 |
| chr11 | 64130203 | 64132024 | 1822 | 5 | 5.99E-12 | 8.05E-12 | 7.47E-06 | 8.73E-13 | -0.016453203 | MACROD1 |
| chr22 | 38213364 | 38215362 | 1999 | 9 | 6.34E-12 | 5.92E-13 | 0.00014 | 8.82E-13 | -0.011664558 | MAFF, PLA2G6 |
| chr11 | 85854432 | 85855698 | 1267 | 14 | 6.39E-12 | 2.03E-10 | 3.24E-05 | 6.72E-12 | -0.001155941 | CCDC83, AP000974.1 |
| chr6 | 31877333 | 31878472 | 1140 | 11 | 6.53E-12 | 5.70E-11 | 2.80E-05 | 1.66E-11 | -0.004578495 | EHMT2-AS1, SLC44A4 |
| chr17 | 7179243 | 7180200 | 958 | 10 | 6.96E-12 | 3.55E-11 | 3.04E-05 | 1.62E-12 | -0.007649675 | ASGR1 |
| chr9 | 68714833 | 68715492 | 660 | 2 | 6.96E-12 | 2.69E-05 | 5.76E-08 | 1.41E-08 | -0.014572783 | PIP5K1B |
| chr12 | 14773383 | 14774870 | 1488 | 15 | 7.94E-12 | 3.57E-07 | 9.54E-05 | 3.68E-10 | -0.010930904 | H2AJ |
| chr1 | 162631280 | 162633391 | 2112 | 11 | 7.97E-12 | 6.30E-10 | 5.59E-05 | 5.04E-12 | -0.007104827 | DDR2 |
| chr3 | 184144711 | 184145406 | 696 | 3 | 7.99E-12 | 1.41E-08 | 4.63E-07 | 1.71E-09 | -0.010758771 | EIF2B5 |
| chr6 | 32068753 | 32071181 | 2429 | 38 | 8.36E-12 | 9.80E-09 | 6.67E-05 | 1.35E-13 | -0.002316718 | TNXB |
| chr5 | 141507735 | 141509267 | 1533 | 5 | 8.69E-12 | 1.56E-13 | 2.62E-05 | 1.03E-12 | -0.021093674 | PCDHGA1, PCDHGA2, PCDHGA3, PCDHGB1, PCDHGA4, PCDHGB2, PCDHGA5, PCDHGB3, PCDHGA6, PCDHGA7, PCDHGB4, PCDHGA8, PCDHGB5, PCDHGA9, PCDHGB6, PCDHGA10, PCDHGB7, PCDHGA11, PCDHGA12, PCDHGC3, PCDHGC4, PCDHGC5 |
| chr11 | 8810564 | 8812355 | 1792 | 15 | 9.74E-12 | 2.07E-07 | 0.00028 | 7.51E-10 | -0.006731858 | DENND2B |
| chr7 | 74480243 | 74480779 | 537 | 5 | 1.05E-11 | 9.65E-13 | 1.70E-05 | 4.56E-12 | -0.019140997 | GTF2IRD1 |
| chr11 | 57903589 | 57904728 | 1140 | 5 | 1.08E-11 | 3.25E-12 | 8.36E-05 | 3.42E-12 | -0.034304579 | NA |
| chr1 | 61076539 | 61077339 | 801 | 15 | 1.11E-11 | 0.001627 | 5.53E-07 | 1.90E-07 | -0.002573131 | NFIA |
| chr17 | 75723972 | 75724760 | 789 | 9 | 1.13E-11 | 1.67E-08 | 4.42E-05 | 1.26E-10 | -0.007422437 | ITGB4 |
| chr6 | 27162243 | 27162429 | 187 | 2 | 1.22E-11 | 1.34E-11 | 1.56E-07 | 1.21E-11 | -0.035324301 | NA |
| chr13 | 48585017 | 48587172 | 2156 | 6 | 1.22E-11 | 6.29E-14 | 2.98E-05 | 1.12E-13 | -0.016371058 | RB1 |
| chr6 | 149630240 | 149630828 | 589 | 2 | 1.26E-11 | 6.43E-06 | 6.80E-08 | 1.37E-08 | -0.005375689 | KATNA1 |
| chr7 | 114415019 | 114415364 | 346 | 7 | 1.26E-11 | 7.76E-13 | 0.00035 | 1.10E-11 | -0.017879192 | FOXP2, AC073626.1 |
| chr11 | 118892150 | 118893782 | 1633 | 11 | 1.36E-11 | 1.21E-09 | 6.22E-05 | 8.29E-12 | 0.005136672 | CXCR5 |
| chr2 | 238099145 | 238100861 | 1717 | 14 | 1.37E-11 | 1.15E-13 | 7.78E-05 | 4.70E-14 | 0.019252483 | UBE2F-SCLY, SCLY, ESPNL |
| chr12 | 57847417 | 57848532 | 1116 | 6 | 1.40E-11 | 6.28E-08 | 1.03E-05 | 6.86E-09 | -0.00612364 | NA |
| chr19 | 5913843 | 5915026 | 1184 | 8 | 1.41E-11 | 4.47E-09 | 1.38E-05 | 2.32E-11 | -0.004367334 | AC104532.1, CAPS, AC104532.2 |
| chr14 | 21022886 | 21024577 | 1692 | 14 | 1.47E-11 | 6.65E-08 | 0.00039 | 1.35E-09 | -0.004828006 | TPPP2, AL161668.4, NDRG2, MIR6717 |
| chr14 | 104680452 | 104682057 | 1606 | 8 | 1.52E-11 | 1.96E-05 | 1.76E-06 | 1.48E-09 | -0.004733028 | INF2 |
| chr12 | 8038773 | 8039050 | 278 | 2 | 1.55E-11 | 3.32E-10 | 5.14E-08 | 5.60E-11 | -0.034037836 | FOXJ2 |
| chr1 | 156076553 | 156077718 | 1166 | 10 | 1.56E-11 | 9.79E-10 | 0.00035 | 1.10E-10 | -0.010329533 | MEX3A |
| chr1 | 46569643 | 46570103 | 461 | 3 | 1.57E-11 | 3.16E-11 | 1.83E-06 | 3.11E-11 | -0.007232549 | MKNK1-AS1, MKNK1 |
| chr6 | 31968024 | 31971545 | 3522 | 59 | 1.63E-11 | 2.79E-11 | 0.0009 | 1.73E-15 | -0.000305957 | SKIV2L, STK19, DXO |
| chr11 | 10693628 | 10694220 | 593 | 12 | 1.63E-11 | 9.15E-10 | 0.00061 | 1.37E-10 | -0.007393269 | IRAG1 |
| chr10 | 48465127 | 48466208 | 1082 | 4 | 1.66E-11 | 1.56E-08 | 4.77E-06 | 1.72E-09 | -0.012874179 | ARHGAP22 |
| chr12 | 110743051 | 110744288 | 1238 | 9 | 1.77E-11 | 5.23E-06 | 2.13E-06 | 4.99E-09 | -0.006353018 | NA |
| chr4 | 40750134 | 40751230 | 1097 | 6 | 1.78E-11 | 6.43E-07 | 4.28E-05 | 1.05E-09 | -0.012523689 | NSUN7, AC131953.2 |
| chr7 | 27179725 | 27181364 | 1640 | 11 | 1.83E-11 | 2.10E-07 | 9.13E-06 | 1.61E-09 | -0.004382291 | AC004080.3, HOXA10, HOXA11 |
| chr6 | 33162919 | 33164541 | 1623 | 18 | 1.84E-11 | 1.64E-07 | 6.82E-05 | 3.08E-10 | -0.00181973 | COL11A2 |
| chr9 | 123372890 | 123374654 | 1765 | 5 | 1.86E-11 | 8.36E-10 | 8.81E-06 | 7.87E-12 | 0.027123244 | CRB2 |
| chr6 | 41656058 | 41656669 | 612 | 4 | 1.90E-11 | 1.12E-08 | 1.12E-05 | 8.62E-11 | -0.012548499 | NA |
| chr16 | 29820859 | 29822399 | 1541 | 8 | 1.97E-11 | 1.44E-11 | 8.40E-05 | 1.66E-11 | -0.022814993 | PAGR1, AC120114.4, MVP, AC009133.1 |
| chr17 | 39671416 | 39672761 | 1346 | 4 | 2.16E-11 | 1.50E-05 | 4.62E-06 | 3.96E-08 | -0.022664174 | PGAP3 |
| chr6 | 31161552 | 31162739 | 1188 | 20 | 2.16E-11 | 1.40E-06 | 4.99E-05 | 3.08E-10 | -0.002964209 | TCF19 |
| chr14 | 72670265 | 72671298 | 1034 | 6 | 2.17E-11 | 1.59E-13 | 3.34E-06 | 2.54E-13 | -0.016393032 | DPF3 |
| chr10 | 5550814 | 5551986 | 1173 | 6 | 2.37E-11 | 1.19E-15 | 2.33E-05 | 5.45E-15 | -0.013142958 | AL732437.3 |
| chr3 | 196211177 | 196211565 | 389 | 6 | 2.44E-11 | 2.54E-11 | 2.51E-05 | 9.62E-12 | -0.009275638 | SLC51A, ZDHHC19 |
| chr1 | 43979530 | 43980529 | 1000 | 10 | 2.47E-11 | 2.15E-10 | 2.60E-05 | 2.09E-11 | -0.001734155 | B4GALT2 |
| chr12 | 132172421 | 132173334 | 914 | 7 | 2.52E-11 | 1.55E-10 | 0.00023 | 3.55E-11 | -0.013670902 | NA |
| chr7 | 150851677 | 150852384 | 708 | 8 | 2.57E-11 | 6.18E-12 | 0.00029 | 6.99E-12 | -0.01323776 | AOC1 |
| chr6 | 52575009 | 52576270 | 1262 | 5 | 2.60E-11 | 5.40E-08 | 1.38E-05 | 5.22E-09 | -0.011071975 | TRAM2 |
| chr15 | 63121232 | 63122290 | 1059 | 9 | 2.88E-11 | 1.10E-09 | 8.17E-05 | 1.67E-10 | -0.003790384 | LACTB |
| chr10 | 7475253 | 7475871 | 619 | 5 | 3.00E-11 | 1.15E-13 | 0.00024 | 2.44E-12 | -0.024620783 | NA |
| chr9 | 107486104 | 107488508 | 2405 | 8 | 3.11E-11 | 1.57E-15 | 0.00013 | 9.23E-15 | -0.016454993 | KLF4 |
| chr1 | 207822407 | 207823986 | 1580 | 7 | 3.16E-11 | 9.93E-11 | 0.00013 | 3.53E-11 | -0.014172767 | MIR29B2CHG |
| chr2 | 218333573 | 218334523 | 951 | 4 | 3.23E-11 | 1.89E-09 | 1.74E-05 | 4.12E-10 | -0.02215386 | PNKD, CATIP-AS2 |
| chr17 | 82235576 | 82239484 | 3909 | 19 | 3.31E-11 | 2.48E-16 | 0.00027 | 1.67E-17 | -0.006785855 | SLC16A3, MIR6787, CSNK1D |
| chr10 | 113665274 | 113666140 | 867 | 5 | 3.41E-11 | 2.16E-10 | 5.25E-06 | 1.08E-10 | -0.007222483 | NA |
| chr7 | 70775798 | 70777230 | 1433 | 4 | 3.42E-11 | 7.02E-14 | 4.95E-05 | 7.18E-13 | -0.029903661 | AUTS2 |
| chr16 | 8956401 | 8957428 | 1028 | 5 | 3.78E-11 | 3.50E-08 | 4.56E-06 | 2.46E-11 | -0.010585265 | USP7 |
| chr11 | 74008638 | 74009361 | 724 | 5 | 3.81E-11 | 6.96E-12 | 5.80E-05 | 2.66E-11 | -0.004657555 | UCP3 |
| chr6 | 168377545 | 168377632 | 88 | 3 | 3.88E-11 | 2.49E-12 | 2.19E-05 | 1.03E-11 | -0.03516164 | NA |
| chr3 | 47421991 | 47423009 | 1019 | 5 | 4.03E-11 | 6.44E-10 | 5.53E-05 | 1.22E-10 | -0.009382738 | SCAP |
| chr11 | 14971240 | 14973990 | 2751 | 41 | 4.12E-11 | 1.91E-14 | 0.00259 | 1.51E-13 | -0.003402784 | CALCB, CALCA |
| chr13 | 51895806 | 51896252 | 447 | 2 | 4.25E-11 | 7.78E-12 | 1.95E-06 | 1.67E-11 | -0.027055602 | NA |
| chr22 | 23249978 | 23251574 | 1597 | 7 | 4.81E-11 | 1.14E-08 | 8.10E-06 | 6.77E-11 | -0.008903513 | BCR |
| chr11 | 75263046 | 75264126 | 1081 | 4 | 4.82E-11 | 7.07E-13 | 2.61E-05 | 4.18E-12 | -0.010019435 | ARRB1 |
| chr20 | 25039789 | 25040220 | 432 | 2 | 4.83E-11 | 6.12E-08 | 1.04E-07 | 2.63E-09 | -0.015638608 | ACSS1 |
| chr9 | 87509819 | 87510533 | 715 | 2 | 4.85E-11 | 3.31E-09 | 1.56E-07 | 6.89E-10 | -0.0250651 | DAPK1 |
| chr1 | 6599145 | 6600343 | 1199 | 7 | 5.07E-11 | 1.52E-10 | 8.10E-05 | 2.56E-11 | -0.007393867 | KLHL21 |
| chr20 | 64106465 | 64108300 | 1836 | 15 | 5.08E-11 | 3.03E-05 | 0.00029 | 3.27E-09 | -0.005321446 | MYT1, NPBWR2 |
| chr3 | 165837315 | 165838144 | 830 | 5 | 5.47E-11 | 1.11E-10 | 2.40E-05 | 6.56E-11 | -0.021698222 | LINC01322, BCHE |
| chr3 | 12758486 | 12759634 | 1149 | 8 | 5.57E-11 | 6.85E-09 | 1.38E-05 | 1.79E-10 | -0.007157518 | TMEM40 |
| chr12 | 113246186 | 113246824 | 639 | 4 | 5.65E-11 | 1.35E-12 | 5.95E-05 | 8.02E-12 | -0.015760962 | TPCN1 |
| chr9 | 75962028 | 75962909 | 882 | 5 | 5.71E-11 | 1.07E-07 | 1.83E-06 | 1.36E-09 | -0.006038533 | PCSK5 |
| chr6 | 30343539 | 30345041 | 1503 | 8 | 5.74E-11 | 2.17E-06 | 9.60E-07 | 2.13E-08 | -0.001366124 | TRIM39, TRIM39-RPP21 |
| chr2 | 189878936 | 189879659 | 724 | 5 | 5.86E-11 | 1.08E-08 | 9.41E-06 | 3.32E-10 | -0.0162469 | C2orf88 |
| chr7 | 373616 | 376953 | 3338 | 26 | 6.08E-11 | 2.13E-10 | 0.0016 | 4.42E-15 | -0.004623173 | NA |
| chr2 | 168801953 | 168802672 | 720 | 8 | 6.27E-11 | 2.11E-07 | 7.00E-05 | 2.59E-10 | -0.008659917 | NOSTRIN |
| chr10 | 93060029 | 93062990 | 2962 | 19 | 6.51E-11 | 3.26E-14 | 0.00071 | 2.50E-14 | -0.011693039 | CYP26C1, AL358613.1 |
| chr12 | 124543248 | 124544461 | 1214 | 7 | 6.95E-11 | 8.34E-09 | 0.00019 | 1.28E-10 | -0.011713695 | NCOR2 |
| chr12 | 54259281 | 54260306 | 1026 | 7 | 7.39E-11 | 1.07E-12 | 4.71E-05 | 2.93E-12 | -0.014590738 | CBX5 |
| chr3 | 132653421 | 132654554 | 1134 | 6 | 7.43E-11 | 5.32E-07 | 3.41E-06 | 1.78E-09 | -0.003853967 | UBA5, ACAD11, NPHP3-ACAD11 |
| chr15 | 100901131 | 100902608 | 1478 | 6 | 7.43E-11 | 4.52E-07 | 1.30E-06 | 3.37E-11 | -0.01061229 | ALDH1A3, AC015712.2 |
| chr4 | 56656629 | 56658604 | 1976 | 11 | 7.49E-11 | 2.29E-10 | 0.00015 | 5.01E-11 | -0.013790962 | HOPX |
| chr11 | 45919494 | 45920511 | 1018 | 4 | 7.54E-11 | 9.78E-13 | 4.35E-05 | 5.77E-12 | -0.014599062 | NA |
| chr17 | 76141688 | 76142657 | 970 | 5 | 7.76E-11 | 2.28E-07 | 1.50E-05 | 1.35E-08 | -0.008024794 | RNF157-AS1, RNF157 |
| chr14 | 91396469 | 91396563 | 95 | 4 | 7.96E-11 | 1.91E-11 | 1.05E-05 | 2.64E-11 | -0.019689733 | CCDC88C |
| chr9 | 137694764 | 137695687 | 924 | 4 | 8.38E-11 | 2.71E-10 | 3.98E-05 | 1.97E-10 | -0.011690302 | AL590627.2, EHMT1 |
| chr16 | 54062428 | 54063874 | 1447 | 6 | 8.43E-11 | 3.04E-12 | 6.40E-05 | 4.30E-12 | -0.011464773 | FTO |
| chr14 | 103724341 | 103725132 | 792 | 6 | 8.60E-11 | 1.05E-10 | 0.00013 | 2.81E-11 | -0.007513123 | ZFYVE21 |
| chr7 | 911907 | 912244 | 338 | 2 | 8.61E-11 | 5.07E-11 | 3.07E-06 | 1.06E-10 | -0.038233725 | ADAP1, COX19 |
| chr9 | 136078756 | 136079082 | 327 | 3 | 8.65E-11 | 1.76E-10 | 3.01E-06 | 1.49E-10 | -0.015257296 | NACC2 |
| chr2 | 88055539 | 88057421 | 1883 | 13 | 8.85E-11 | 5.47E-05 | 2.93E-06 | 4.33E-08 | -0.001613736 | KRCC1 |
| chr11 | 47267804 | 47269179 | 1376 | 8 | 9.07E-11 | 9.34E-08 | 0.00078 | 3.66E-09 | -0.00716141 | NR1H3, MADD |
| chr1 | 230713773 | 230715086 | 1314 | 10 | 9.11E-11 | 3.71E-12 | 0.00091 | 2.28E-11 | -0.016327833 | AGT, AL512328.1 |
| chr14 | 21722899 | 21723641 | 743 | 7 | 9.60E-11 | 1.49E-08 | 0.00028 | 3.60E-10 | -0.015170153 | NA |
| chr6 | 30890829 | 30893183 | 2355 | 21 | 9.70E-11 | 2.29E-09 | 0.00042 | 5.15E-12 | -0.006006355 | DDR1, MIR4640 |
| chr15 | 43266657 | 43267386 | 730 | 8 | 9.75E-11 | 5.48E-06 | 4.34E-05 | 8.67E-10 | -0.005830816 | TGM5 |
| chr19 | 2330964 | 2332720 | 1757 | 5 | 9.97E-11 | 3.19E-14 | 3.60E-05 | 2.71E-13 | -0.019568682 | AC005258.1, SPPL2B |
| chr1 | 232319202 | 232319258 | 57 | 2 | 9.97E-11 | 3.87E-11 | 3.99E-06 | 8.62E-11 | -0.022940103 | NA |
| chr16 | 1532319 | 1534515 | 2197 | 14 | 1.03E-10 | 1.92E-14 | 0.00403 | 1.78E-12 | -0.024539847 | TMEM204, AL031719.1, IFT140 |
| chr6 | 32761085 | 32763082 | 1998 | 26 | 1.03E-10 | 9.14E-12 | 0.00117 | 3.14E-12 | -0.010699606 | HLA-DQB2 |
| chr4 | 174522256 | 174523797 | 1542 | 13 | 1.11E-10 | 1.40E-09 | 0.00014 | 5.11E-10 | -0.008105346 | AC096751.2, HPGD, AC096751.1 |
| chr11 | 118180545 | 118180682 | 138 | 2 | 1.13E-10 | 2.18E-11 | 3.72E-06 | 4.88E-11 | -0.028711677 | NA |
| chr19 | 2041347 | 2042593 | 1247 | 8 | 1.13E-10 | 6.41E-09 | 7.89E-05 | 8.62E-11 | -0.008948876 | MKNK2 |
| chr7 | 151187244 | 151188567 | 1324 | 15 | 1.14E-10 | 3.46E-08 | 1.84E-06 | 1.49E-09 | -0.003115692 | ASB10 |
| chr7 | 130940714 | 130941800 | 1087 | 6 | 1.17E-10 | 8.77E-15 | 0.00019 | 1.82E-13 | -0.024407683 | AC016831.6, LINC-PINT |
| chr2 | 97723610 | 97725094 | 1485 | 11 | 1.17E-10 | 1.05E-12 | 0.00074 | 5.53E-12 | -0.010996859 | ZAP70 |
| chr11 | 69643827 | 69645117 | 1291 | 6 | 1.18E-10 | 9.06E-08 | 5.62E-05 | 6.70E-09 | -0.008033125 | CCND1 |
| chr8 | 144399604 | 144401049 | 1446 | 7 | 1.20E-10 | 3.25E-06 | 0.00011 | 2.07E-08 | -0.007525491 | CPSF1, MIR1234, MIR6849 |
| chr1 | 21254877 | 21255606 | 730 | 3 | 1.30E-10 | 3.09E-08 | 5.90E-07 | 2.07E-09 | -0.009149848 | ECE1 |
| chr22 | 45903476 | 45904145 | 670 | 3 | 1.33E-10 | 2.45E-11 | 1.52E-05 | 5.96E-11 | -0.01665191 | NA |
| chr16 | 1535643 | 1536304 | 662 | 5 | 1.39E-10 | 4.25E-11 | 0.00013 | 1.42E-10 | -0.029908854 | TMEM204, IFT140 |
| chr17 | 82374135 | 82374402 | 268 | 6 | 1.46E-10 | 2.16E-10 | 7.95E-05 | 1.17E-10 | -0.007890236 | UTS2R |
| chr1 | 118184908 | 118185603 | 696 | 12 | 1.54E-10 | 0.000122 | 6.75E-05 | 2.49E-09 | -0.002849161 | AL391557.1, SPAG17 |
| chr11 | 63872053 | 63873568 | 1516 | 5 | 1.56E-10 | 2.08E-08 | 1.30E-05 | 2.88E-09 | -0.007307372 | MARK2 |
| chr19 | 58000792 | 58003678 | 2887 | 15 | 1.60E-10 | 4.85E-11 | 0.00022 | 2.26E-14 | -0.009030941 | AC008969.1, ZNF606 |
| chr17 | 4901379 | 4903770 | 2392 | 16 | 1.64E-10 | 3.71E-15 | 0.00208 | 1.04E-13 | -0.016214386 | C17orf107, CHRNE |
| chr16 | 67966546 | 67968141 | 1596 | 10 | 1.68E-10 | 2.25E-07 | 0.00015 | 1.33E-08 | -0.00619456 | SLC12A4 |
| chr7 | 102299567 | 102301272 | 1706 | 9 | 1.71E-10 | 2.66E-11 | 0.00082 | 7.15E-11 | -0.010895925 | SH2B2 |
| chr5 | 140639059 | 140639960 | 902 | 14 | 1.78E-10 | 9.16E-09 | 0.00024 | 1.22E-11 | -0.005870689 | TMCO6, NDUFA2 |
| chr1 | 58813847 | 58815287 | 1441 | 8 | 1.80E-10 | 2.49E-08 | 0.00024 | 1.81E-09 | -0.010847089 | LINC01135 |
| chr2 | 73269559 | 73271355 | 1797 | 10 | 1.80E-10 | 1.10E-08 | 8.77E-06 | 1.32E-09 | -0.008154898 | FBXO41 |
| chr6 | 167290103 | 167292262 | 2160 | 10 | 1.99E-10 | 6.77E-12 | 0.00019 | 1.34E-12 | -0.012043787 | UNC93A |
| chr7 | 157667785 | 157668618 | 834 | 6 | 2.00E-10 | 1.74E-05 | 3.98E-05 | 8.41E-09 | -0.009736504 | PTPRN2 |
| chr1 | 3206490 | 3207969 | 1480 | 7 | 2.02E-10 | 1.51E-06 | 4.23E-05 | 5.22E-08 | -0.005912062 | PRDM16, AL512383.1 |
| chr15 | 101188029 | 101189746 | 1718 | 11 | 2.10E-10 | 4.38E-10 | 0.00056 | 1.45E-10 | -0.007235626 | CHSY1 |
| chr17 | 80779473 | 80780572 | 1100 | 8 | 2.13E-10 | 2.47E-11 | 4.51E-05 | 1.24E-11 | -0.019652518 | RPTOR |

| | | | | | | | | | | |
|---|---|---|---|---|---|---|---|---|---|---|
| chr10 | 99731841 | 99733710 | 1870 | 20 | 2.20E-10 | 6.67E-07 | 2.13E-06 | 7.14E-11 | -0.000179524 | CUTC, AL133353.2, COX15 |
| chr11 | 73645271 | 73646696 | 1426 | 13 | 2.21E-10 | 7.05E-11 | 0.00038 | 2.94E-11 | -0.009115411 | PLEKHB1 |
| chr17 | 68539751 | 68540489 | 739 | 4 | 2.21E-10 | 6.63E-11 | 7.53E-05 | 1.75E-10 | -0.010031298 | PRKAR1A, FAM20A |
| chr5 | 72442952 | 72443665 | 714 | 3 | 2.27E-10 | 5.78E-05 | 4.95E-08 | 1.36E-08 | -0.005291259 | ZNF366 |
| chr1 | 162366192 | 162367585 | 1394 | 10 | 2.35E-10 | 6.62E-08 | 2.18E-05 | 4.69E-09 | -0.006279985 | NOS1AP, AL512785.2 |
| chr22 | 23845947 | 23846639 | 693 | 4 | 2.52E-10 | 6.41E-09 | 1.31E-05 | 1.15E-09 | -0.009518513 | NA |
| chr7 | 39130897 | 39131513 | 617 | 6 | 2.71E-10 | 1.05E-12 | 0.00053 | 1.97E-11 | -0.065721821 | POU6F2 |
| chr14 | 100549176 | 100550052 | 877 | 4 | 2.75E-10 | 8.97E-09 | 6.54E-05 | 1.15E-09 | -0.015931848 | BEGAIN |
| chr17 | 13543821 | 13544536 | 716 | 5 | 2.78E-10 | 3.87E-11 | 4.11E-05 | 1.08E-10 | -0.00782659 | HS3ST3A1 |
| chr10 | 75282768 | 75283850 | 1083 | 7 | 2.81E-10 | 3.09E-08 | 2.95E-05 | 3.16E-10 | -0.011009683 | ZNF503-AS1, AC010997.3 |
| chr6 | 29626704 | 29627884 | 1181 | 12 | 2.88E-10 | 1.68E-11 | 0.00253 | 1.52E-10 | -0.009537556 | GABBR1 |
| chr22 | 30546650 | 30547507 | 858 | 4 | 2.91E-10 | 1.70E-08 | 1.65E-05 | 7.78E-10 | -0.008613456 | SEC14L6 |
| chr8 | 17001536 | 17002612 | 1077 | 14 | 2.93E-10 | 1.35E-10 | 0.00155 | 1.23E-10 | -0.012298048 | FGF20 |
| chr17 | 50114883 | 50116348 | 1466 | 4 | 2.93E-10 | 1.45E-07 | 4.32E-07 | 2.49E-09 | -0.009440544 | SAMD14 |
| chr18 | 23357240 | 23358245 | 1006 | 3 | 2.95E-10 | 5.52E-10 | 2.18E-05 | 5.64E-10 | -0.01501082 | TMEM241 |
| chr19 | 14265016 | 14266148 | 1133 | 5 | 2.95E-10 | 2.21E-11 | 8.27E-05 | 9.92E-11 | -0.022536192 | AC011509.2 |
| chr5 | 14440383 | 14440965 | 583 | 6 | 2.98E-10 | 7.68E-09 | 8.70E-06 | 3.67E-10 | -0.008723248 | TRIO |
| chr1 | 26862779 | 26864148 | 1370 | 11 | 3.08E-10 | 3.22E-12 | 0.00092 | 2.63E-11 | -0.007902547 | SFN |
| chr10 | 71805683 | 71806209 | 527 | 5 | 3.12E-10 | 9.93E-11 | 7.47E-05 | 2.11E-10 | -0.016196069 | CDH23 |
| chr4 | 3447573 | 3449043 | 1471 | 6 | 3.14E-10 | 2.01E-06 | 4.62E-07 | 1.92E-09 | -0.004545052 | HGFAC |
| chr3 | 52064606 | 52065545 | 940 | 6 | 3.14E-10 | 1.67E-11 | 6.69E-05 | 3.17E-11 | -0.009026535 | NA |
| chr7 | 105648499 | 105649771 | 1273 | 6 | 3.18E-10 | 5.60E-12 | 0.00014 | 3.12E-11 | -0.012229455 | ATXN7L1 |
| chr9 | 129619382 | 129620876 | 1495 | 10 | 3.33E-10 | 6.38E-07 | 7.66E-05 | 2.13E-08 | -0.000772707 | NTMT1, C9orf50 |
| chr13 | 99085110 | 99085531 | 422 | 4 | 3.42E-10 | 3.00E-09 | 9.80E-06 | 1.48E-09 | -0.020295217 | DOCK9 |
| chr12 | 50241149 | 50241760 | 612 | 3 | 3.56E-10 | 9.42E-05 | 1.88E-07 | 8.70E-09 | -0.013941597 | LIMA1 |
| chr7 | 2248509 | 2250040 | 1532 | 7 | 3.59E-10 | 7.78E-11 | 0.0002 | 2.32E-10 | -0.01616404 | NUDT1 |
| chr17 | 75645595 | 75647297 | 1703 | 14 | 3.85E-10 | 1.60E-09 | 0.00012 | 2.63E-11 | -0.005613014 | SMIM6, RECQL5 |
| chr6 | 30642815 | 30644553 | 1739 | 11 | 3.85E-10 | 7.40E-12 | 2.61E-05 | 5.33E-13 | -0.003941428 | ATAT1 |
| chr16 | 85484673 | 85487285 | 2613 | 13 | 4.02E-10 | 8.26E-19 | 0.00027 | 2.19E-16 | -0.011530028 | GSE1 |
| chr1 | 6359021 | 6361186 | 2166 | 12 | 4.35E-10 | 8.79E-08 | 5.68E-05 | 8.29E-11 | -0.007206716 | ACOT7 |
| chr15 | 99507445 | 99508295 | 851 | 7 | 4.56E-10 | 6.77E-13 | 0.00018 | 4.41E-12 | -0.020725225 | NA |
| chr13 | 114100469 | 114102226 | 1758 | 7 | 4.67E-10 | 4.34E-12 | 4.50E-06 | 2.77E-12 | -0.006793339 | RASA3 |
| chr2 | 241900515 | 241901670 | 1156 | 6 | 4.77E-10 | 0.001 | 8.08E-07 | 1.70E-06 | -0.000380631 | LINC01237, FAM240C |
| chr16 | 3443133 | 3444155 | 1023 | 15 | 5.01E-10 | 2.42E-13 | 0.00416 | 1.58E-11 | -0.016020129 | AC025283.2, NAA60, ZNF597 |
| chr1 | 3198256 | 3199080 | 825 | 7 | 5.22E-10 | 0.001919 | 2.28E-07 | 3.40E-07 | -0.001182997 | PRDM16 |
| chr2 | 131039865 | 131041495 | 1631 | 8 | 5.28E-10 | 3.03E-10 | 0.00057 | 2.28E-11 | -0.02568295 | ARHGEF4 |
| chr22 | 29342192 | 29344379 | 2188 | 4 | 5.28E-10 | 3.54E-14 | 2.51E-06 | 1.19E-13 | -0.017629411 | AP1B1 |
| chr13 | 98106038 | 98106241 | 204 | 2 | 5.31E-10 | 9.90E-11 | 3.99E-06 | 2.06E-10 | -0.058280488 | NA |
| chr7 | 985037 | 986727 | 1691 | 11 | 5.35E-10 | 6.45E-07 | 0.00016 | 9.21E-10 | -0.008940894 | CYP2W1 |
| chr19 | 46300655 | 46301271 | 617 | 4 | 5.35E-10 | 1.67E-11 | 7.44E-05 | 1.11E-10 | -0.016287218 | HIF3A |
| chr9 | 136052328 | 136053980 | 1653 | 10 | 5.38E-10 | 1.86E-07 | 0.00021 | 5.77E-09 | -0.005935503 | NACC2 |
| chr6 | 6758849 | 6759907 | 1059 | 7 | 5.44E-10 | 1.05E-09 | 8.70E-06 | 2.26E-10 | -0.007256457 | AL136361.1 |
| chr17 | 4560456 | 4562091 | 1636 | 12 | 5.51E-10 | 5.99E-08 | 8.21E-05 | 9.83E-10 | -0.006073646 | GGT6 |
| chr16 | 85444974 | 85446346 | 1373 | 10 | 5.65E-10 | 2.59E-09 | 4.85E-05 | 9.57E-10 | -0.006481336 | GSE1 |
| chr7 | 99011060 | 99013487 | 2428 | 16 | 5.65E-10 | 4.55E-12 | 0.00065 | 3.59E-13 | -0.001616471 | TRRAP |
| chr5 | 176352526 | 176354051 | 1526 | 5 | 6.03E-10 | 5.44E-10 | 3.75E-05 | 5.75E-10 | -0.000447224 | AC138956.1, KIAA1191 |
| chr2 | 12313440 | 12314259 | 820 | 4 | 6.04E-10 | 6.45E-10 | 4.22E-05 | 8.85E-10 | -0.015629126 | MIR3681HG |
| chr9 | 83219180 | 83219799 | 620 | 4 | 6.20E-10 | 8.40E-11 | 9.46E-05 | 2.64E-10 | -0.020462629 | AL137847.1 |
| chr11 | 67279429 | 67283242 | 3814 | 13 | 6.25E-10 | 3.19E-13 | 3.96E-05 | 2.70E-14 | -0.006004937 | GRK2 |
| chr16 | 80541826 | 80541875 | 50 | 2 | 6.25E-10 | 1.43E-07 | 4.89E-06 | 8.74E-08 | 0.005609072 | DYNLRB2, AC105411.1 |
| chr8 | 125578159 | 125579780 | 1622 | 6 | 6.29E-10 | 2.90E-11 | 7.42E-05 | 4.55E-12 | -0.014282853 | NA |
| chr15 | 65368384 | 65368742 | 359 | 2 | 6.49E-10 | 2.44E-10 | 2.06E-06 | 3.84E-10 | -0.020285189 | IGDCC3 |
| chr17 | 81251509 | 81252174 | 666 | 6 | 6.98E-10 | 2.09E-09 | 1.47E-05 | 5.77E-10 | -0.018647081 | AC027601.3, SLC38A10 |
| chr10 | 21507223 | 21509769 | 2547 | 12 | 7.20E-10 | 7.43E-12 | 8.35E-05 | 7.18E-13 | -0.006145003 | NA |
| chr22 | 46085219 | 46086729 | 1511 | 14 | 7.27E-10 | 2.23E-05 | 0.00049 | 1.52E-08 | -0.002397453 | MIRLET7BHG |
| chr3 | 62875035 | 62876250 | 1216 | 14 | 7.39E-10 | 2.93E-06 | 8.42E-07 | 2.69E-08 | -0.000370495 | CADPS |
| chr2 | 128090235 | 128091635 | 1401 | 11 | 7.44E-10 | 4.91E-08 | 0.00012 | 6.59E-10 | 0.003424201 | UGGT1 |
| chr3 | 46700002 | 46701588 | 1587 | 9 | 7.46E-10 | 5.84E-08 | 4.09E-05 | 5.50E-10 | -0.004220537 | TMIE |
| chr8 | 134754150 | 134754607 | 458 | 2 | 7.72E-10 | 1.63E-09 | 8.08E-07 | 1.13E-09 | -0.006685306 | NA |
| chr5 | 120615832 | 120616132 | 301 | 2 | 7.90E-10 | 3.63E-06 | 2.42E-07 | 4.71E-08 | -0.011028442 | PRR16 |
| chr16 | 88523510 | 88524001 | 492 | 5 | 7.97E-10 | 3.91E-09 | 2.40E-05 | 8.28E-10 | -0.01642062 | ZFPM1, AC116552.1 |
| chr9 | 20356461 | 20357210 | 750 | 3 | 8.11E-10 | 2.42E-06 | 2.21E-06 | 1.11E-08 | -0.014736287 | MLLT3 |
| chr22 | 43168724 | 43168807 | 84 | 2 | 8.17E-10 | 1.16E-10 | 9.01E-06 | 2.76E-10 | -0.027055675 | TTLL12 |
| chr5 | 112861549 | 112862325 | 777 | 5 | 8.66E-10 | 8.33E-08 | 7.84E-05 | 1.76E-08 | -0.013687684 | AC008575.1, SRP19 |
| chr2 | 64679842 | 64680574 | 733 | 3 | 8.67E-10 | 1.17E-06 | 2.53E-07 | 2.06E-08 | -0.002201844 | SERTAD2 |
| chr12 | 26271763 | 26272278 | 516 | 6 | 8.85E-10 | 1.46E-08 | 8.40E-05 | 4.03E-10 | -0.01134797 | SSPN, AC022509.2, AC022509.1 |
| chr16 | 57809609 | 57810660 | 1052 | 10 | 9.08E-10 | 0.000109 | 3.25E-06 | 2.91E-07 | -0.003849388 | AC092118.1, KIFC3 |
| chr16 | 87173159 | 87173723 | 565 | 2 | 9.22E-10 | 7.28E-08 | 1.43E-06 | 2.29E-08 | -0.011100706 | C16orf95 |
| chr20 | 63953970 | 63954926 | 957 | 3 | 9.25E-10 | 1.72E-10 | 1.99E-05 | 4.12E-10 | -0.014384772 | UCKL1-AS1, UCKL1 |
| chr3 | 195114927 | 195116118 | 1192 | 4 | 9.40E-10 | 3.25E-06 | 1.59E-06 | 5.78E-08 | -0.006029477 | XXYLT1 |
| chr20 | 63283460 | 63284927 | 1468 | 12 | 9.46E-10 | 2.66E-11 | 9.29E-05 | 1.15E-10 | -0.010023592 | ARFGAP1 |
| chr22 | 24431969 | 24433085 | 1117 | 8 | 9.66E-10 | 1.42E-05 | 2.68E-05 | 3.46E-08 | -0.004465163 | SPECC1L-ADORA2A, ADORA2A, ADORA2A-AS1 |
| chr8 | 28775806 | 28776961 | 1156 | 5 | 9.75E-10 | 7.78E-11 | 2.54E-05 | 3.69E-11 | -0.020819886 | INTS9 |
| chr1 | 44800860 | 44801673 | 814 | 3 | 9.80E-10 | 1.14E-06 | 3.82E-05 | 7.63E-08 | -0.012133595 | PLK3 |
| chr3 | 194270718 | 194271567 | 850 | 4 | 9.87E-10 | 6.25E-08 | 2.29E-05 | 1.36E-08 | -0.010509876 | NA |
| chr7 | 151199327 | 151199922 | 596 | 3 | 1.02E-09 | 5.90E-10 | 3.04E-05 | 9.61E-10 | -0.006503689 | IQCA1L |
| chr10 | 4279574 | 4280152 | 579 | 3 | 1.05E-09 | 0.000467 | 3.84E-07 | 5.18E-07 | 0.00529373 | NA |
| chr6 | 31729503 | 31734031 | 4529 | 38 | 1.07E-09 | 5.05E-17 | 0.0004 | 5.72E-20 | -0.002924766 | DDAH2, CLIC1 |
| chr11 | 64660528 | 64662244 | 1717 | 10 | 1.10E-09 | 1.59E-07 | 0.00075 | 4.65E-09 | -0.008746988 | NRXN2 |
| chr2 | 189178811 | 189180257 | 1447 | 8 | 1.10E-09 | 4.10E-12 | 0.00018 | 8.32E-12 | -0.014199795 | COL5A2 |
| chr1 | 184426172 | 184427170 | 999 | 2 | 1.12E-09 | 3.23E-12 | 2.58E-07 | 4.78E-12 | -0.016581745 | C1orf21 |
| chr2 | 26562592 | 26563178 | 587 | 6 | 1.13E-09 | 0.000363 | 7.36E-05 | 2.67E-07 | -0.008253139 | FAM166C |
| chr5 | 149140209 | 149142557 | 2349 | 18 | 1.13E-09 | 2.78E-09 | 0.0016 | 8.73E-12 | -0.008773638 | ABLIM3 |
| chr6 | 33287103 | 33288007 | 905 | 18 | 1.15E-09 | 7.32E-11 | 0.00145 | 6.89E-10 | -0.004067853 | WDR46, MIR6873 |
| chr15 | 69593949 | 69594714 | 766 | 5 | 1.15E-09 | 2.02E-10 | 9.49E-05 | 2.14E-10 | -0.010537531 | DRAIC |
| chr3 | 53243379 | 53243733 | 355 | 3 | 1.15E-09 | 1.30E-10 | 5.69E-05 | 5.67E-10 | -0.019919599 | TKT |
| chr16 | 31213913 | 31215439 | 1527 | 7 | 1.16E-09 | 2.82E-10 | 0.00019 | 4.86E-10 | -0.009185724 | TRIM72 |
| chr6 | 291124 | 293285 | 2162 | 11 | 1.18E-09 | 1.47E-13 | 0.00308 | 8.15E-12 | -0.075359296 | DUSP22, AL365272.1 |
| chr20 | 33792025 | 33794372 | 2348 | 8 | 1.25E-09 | 4.36E-10 | 0.00014 | 5.63E-12 | -0.007452742 | ZNF341, ZNF341-AS1 |
| chr11 | 73648317 | 73648942 | 626 | 4 | 1.25E-09 | 6.16E-09 | 5.68E-05 | 3.91E-09 | -0.008787757 | PLEKHB1 |
| chr3 | 12816770 | 12818373 | 1604 | 10 | 1.28E-09 | 7.62E-08 | 0.00026 | 1.13E-09 | -0.008219369 | CAND2 |
| chr19 | 15507655 | 15508544 | 890 | 9 | 1.36E-09 | 2.46E-07 | 0.00042 | 3.27E-09 | -0.002910594 | CYP4F22 |
| chr3 | 123621071 | 123621967 | 897 | 5 | 1.37E-09 | 5.47E-10 | 0.00012 | 1.35E-09 | -0.00557856 | MYLK-AS1, MYLK |
| chr6 | 31571762 | 31572682 | 921 | 18 | 1.38E-09 | 4.79E-08 | 0.00279 | 1.31E-09 | -0.00741481 | LTA |
| chr11 | 128451213 | 128451854 | 642 | 3 | 1.39E-09 | 2.54E-06 | 1.97E-06 | 4.05E-08 | -0.0141529 | NA |
| chr16 | 4239416 | 4239923 | 508 | 5 | 1.40E-09 | 4.36E-10 | 0.00011 | 1.94E-09 | -0.012349286 | SRL |
| chr11 | 95088455 | 95090439 | 1985 | 9 | 1.40E-09 | 3.98E-06 | 2.23E-05 | 5.50E-10 | -0.005749314 | ENDOD1 |
| chr12 | 116318089 | 116319364 | 1276 | 8 | 1.44E-09 | 3.78E-11 | 0.00022 | 7.31E-11 | -0.008078796 | NA |
| chr15 | 77875142 | 77875457 | 316 | 2 | 1.46E-09 | 1.96E-10 | 8.51E-06 | 4.43E-10 | -0.045563521 | NA |
| chr7 | 101941100 | 101942379 | 1280 | 4 | 1.47E-09 | 3.75E-08 | 3.84E-05 | 1.08E-09 | -0.012450943 | CUX1 |
| chr22 | 36286194 | 36286974 | 781 | 8 | 1.58E-09 | 3.83E-08 | 0.00015 | 7.47E-10 | -0.006348724 | MYH9, MIR6819 |
| chr3 | 49929340 | 49930813 | 1474 | 15 | 1.62E-09 | 2.64E-09 | 0.00232 | 5.65E-10 | -0.004657639 | MON1A |
| chr20 | 38259270 | 38260388 | 1119 | 5 | 1.66E-09 | 4.02E-05 | 7.98E-06 | 2.90E-07 | -0.011398942 | AL359555.3, AL359555.4, KIAA1755 |
| chr3 | 101778631 | 101780877 | 2247 | 19 | 1.66E-09 | 3.59E-09 | 2.47E-05 | 2.63E-11 | -0.003693863 | NXPE3 |
| chr1 | 34885120 | 34886026 | 907 | 8 | 1.69E-09 | 6.93E-07 | 3.42E-05 | 1.01E-08 | -0.009458245 | DLGAP3 |
| chr17 | 39648712 | 39649336 | 625 | 2 | 1.69E-09 | 2.04E-06 | 1.39E-06 | 1.68E-07 | -0.014969285 | STARD3 |
| chr9 | 134652926 | 134653425 | 500 | 6 | 1.69E-09 | 4.98E-08 | 1.83E-06 | 1.05E-09 | -0.005926624 | COL5A1, COL5A1-AS1 |
| chr16 | 88473453 | 88474584 | 1132 | 7 | 1.71E-09 | 1.19E-07 | 2.94E-05 | 5.43E-09 | -0.00734882 | ZFPM1 |
| chr7 | 2277001 | 2277940 | 940 | 10 | 1.72E-09 | 2.09E-07 | 9.58E-05 | 2.13E-09 | -0.011537416 | SNX8 |
| chr21 | 43762708 | 43763031 | 324 | 2 | 1.73E-09 | 2.99E-06 | 3.66E-07 | 6.60E-08 | -0.005014729 | NA |
| chr17 | 7419396 | 7420774 | 1379 | 12 | 1.78E-09 | 1.93E-06 | 3.43E-05 | 1.02E-08 | -0.004707025 | NLGN2, AC113189.2, SPEM1 |
| chr13 | 113483704 | 113484401 | 698 | 7 | 1.79E-09 | 1.80E-06 | 3.62E-05 | 1.42E-08 | -0.002351654 | DCUN1D2 |
| chr9 | 134441389 | 134442757 | 1369 | 3 | 1.83E-09 | 3.83E-06 | 3.66E-07 | 2.53E-08 | -0.005747138 | NA |
| chr10 | 103658465 | 103659784 | 1320 | 6 | 1.85E-09 | 8.29E-12 | 7.36E-05 | 5.04E-11 | -0.005646668 | SH3PXD2A |
| chr18 | 62437098 | 62437960 | 863 | 4 | 1.90E-09 | 4.38E-07 | 9.45E-05 | 8.99E-09 | -0.018497603 | NA |
| chr17 | 59381937 | 59382159 | 223 | 2 | 1.90E-09 | 1.58E-07 | 8.83E-08 | 3.80E-09 | -0.012475846 | YPEL2 |
| chr19 | 48716845 | 48717228 | 384 | 6 | 1.98E-09 | 6.53E-10 | 0.00022 | 1.08E-09 | -0.010914216 | MAMSTR |
| chr13 | 112204631 | 112206142 | 1512 | 8 | 1.99E-09 | 1.21E-09 | 7.70E-05 | 9.18E-11 | 0.010964601 | NA |
| chr19 | 801569 | 802198 | 630 | 2 | 2.00E-09 | 3.11E-05 | 1.47E-06 | 5.82E-07 | -0.014128254 | PTBP1 |
| chr17 | 19747877 | 19749222 | 1346 | 13 | 2.01E-09 | 9.94E-10 | 0.0009 | 8.32E-10 | -0.006437723 | ALDH3A1 |
| chr16 | 85376354 | 85377868 | 1515 | 5 | 2.11E-09 | 1.70E-08 | 6.02E-06 | 2.72E-09 | -0.008217438 | GSE1 |
| chr3 | 53110040 | 53110600 | 561 | 8 | 2.14E-09 | 1.58E-07 | 0.0001 | 2.21E-09 | -0.009756011 | RFT1 |
| chr3 | 188037725 | 188038230 | 506 | 2 | 2.14E-09 | 0.000118 | 2.03E-06 | 1.27E-06 | -0.008261924 | NA |
| chr16 | 85371048 | 85371538 | 491 | 2 | 2.18E-09 | 4.47E-08 | 1.67E-06 | 1.85E-08 | -0.021754799 | GSE1 |
| chr1 | 40317041 | 40318651 | 1611 | 12 | 2.19E-09 | 6.41E-10 | 0.00367 | 1.20E-09 | -0.008582986 | COL9A2 |
| chr16 | 1537308 | 1537841 | 534 | 5 | 2.21E-09 | 2.16E-08 | 0.00013 | 1.31E-08 | -0.011872713 | TMEM204, IFT140 |
| chr14 | 103801726 | 103802784 | 1059 | 3 | 2.21E-09 | 1.11E-05 | 3.70E-06 | 2.07E-09 | -0.011307946 | PPP1R13B |
| chr7 | 151237219 | 151238684 | 1466 | 11 | 2.22E-09 | 2.56E-06 | 0.00013 | 3.15E-09 | -0.004681237 | CHPF2, MIR671 |
| chr5 | 54985370 | 54986631 | 1262 | 12 | 2.30E-09 | 3.09E-11 | 0.00042 | 8.22E-11 | -0.012582648 | ESM1 |
| chr12 | 6912823 | 6914742 | 1920 | 14 | 2.30E-09 | 7.85E-08 | 0.00025 | 1.11E-10 | -0.006024745 | LRRC23, ENO2 |
| chr14 | 100307262 | 100307950 | 689 | 3 | 2.37E-09 | 9.29E-08 | 5.38E-05 | 4.05E-08 | -0.012135402 | MIR345 |
| chr16 | 82062826 | 82063870 | 1045 | 5 | 2.40E-09 | 1.80E-11 | 0.0002 | 1.39E-10 | -0.007763306 | HSD17B2, AC092142.1 |
| chr7 | 100865584 | 100866956 | 1373 | 9 | 2.42E-09 | 7.51E-10 | 0.00103 | 9.88E-10 | -0.010525078 | SLC12A9 |
| chr21 | 42108095 | 42109593 | 1499 | 7 | 2.48E-09 | 3.42E-12 | 0.00082 | 8.62E-11 | -0.020325519 | UMODL1, UMODL1-AS1 |
| chr12 | 124393051 | 124393293 | 243 | 2 | 2.50E-09 | 7.42E-09 | 2.32E-06 | 6.30E-09 | -0.019124556 | NCOR2 |
| chr15 | 74726333 | 74727861 | 1529 | 16 | 2.54E-09 | 7.34E-08 | 0.00386 | 8.16E-09 | -0.012496914 | NA |
| chr1 | 9842381 | 9843703 | 1323 | 3 | 2.56E-09 | 1.42E-06 | 1.84E-06 | 1.02E-07 | -0.007584692 | NA |
| chr5 | 139461958 | 139463191 | 1234 | 6 | 2.65E-09 | 3.62E-08 | 0.00018 | 8.20E-09 | -0.007746174 | AC142391.1, ECSCR |
| chr3 | 129059717 | 129059895 | 179 | 6 | 2.70E-09 | 6.56E-09 | 0.00023 | 4.58E-09 | -0.010621422 | NA |
| chr5 | 173824379 | 173825201 | 823 | 4 | 2.71E-09 | 1.21E-06 | 2.48E-06 | 5.78E-08 | -0.002942515 | NA |
| chr14 | 97206693 | 97207728 | 1036 | 5 | 2.71E-09 | 2.74E-07 | 9.19E-06 | 1.65E-08 | -0.0033514 | AL158800.1 |
| chr1 | 2534836 | 2536369 | 1534 | 5 | 2.78E-09 | 7.90E-08 | 0.00017 | 1.35E-08 | -0.0146651 | AL139246.5 |
| chr17 | 44389831 | 44390763 | 933 | 6 | 2.88E-09 | 1.09E-08 | 0.00021 | 6.17E-09 | -0.018338624 | NA |
| chr1 | 33429904 | 33430289 | 386 | 2 | 2.96E-09 | 2.54E-07 | 3.25E-06 | 9.82E-08 | -0.010631376 | PHC2 |
| chr1 | 35572517 | 35574054 | 1538 | 11 | 2.98E-09 | 7.64E-11 | 0.00262 | 4.82E-10 | -0.011251881 | TFAP2E, TFAP2E-AS1 |
| chr12 | 1936193 | 1937802 | 1610 | 8 | 3.02E-09 | 2.78E-08 | 0.00011 | 2.46E-09 | -0.006934565 | LINC00940 |
| chr2 | 199910282 | 199911562 | 1281 | 11 | 3.05E-09 | 5.26E-05 | 4.79E-07 | 9.60E-08 | 0.011520009 | C2orf69, AC097717.1 |
| chr7 | 1087125 | 1088131 | 1007 | 6 | 3.08E-09 | 1.31E-07 | 5.20E-05 | 1.37E-08 | -0.005354322 | GPER1, C7orf50 |
| chr3 | 63863019 | 63864091 | 1073 | 9 | 3.17E-09 | 0.000473 | 3.17E-06 | 1.60E-06 | -0.000475094 | ATXN7, THOC7 |
| chr15 | 64761397 | 64763106 | 1710 | 9 | 3.24E-09 | 1.58E-09 | 1.90E-05 | 4.83E-13 | -0.006118251 | RBPMS2, MIR1272 |
| chr9 | 134783534 | 134784500 | 967 | 3 | 3.30E-09 | 6.88E-11 | 4.50E-05 | 3.01E-10 | -0.01808359 | COL5A1 |
| chr17 | 40501058 | 40502887 | 1830 | 12 | 3.30E-09 | 3.19E-09 | 0.00021 | 1.15E-10 | -0.0044442 | TNS4 |
| chr3 | 126343107 | 126343805 | 699 | 4 | 3.33E-09 | 8.06E-11 | 0.00023 | 6.30E-10 | -0.021849925 | KLF15 |
| chr3 | 125990115 | 125991383 | 1269 | 6 | 3.34E-09 | 9.18E-08 | 9.39E-06 | 2.32E-10 | 0.046336714 | NA |
| chr21 | 44294818 | 44295645 | 828 | 4 | 3.36E-09 | 2.06E-07 | 4.79E-05 | 2.21E-08 | -0.012270349 | AIRE |
| chr1 | 2084535 | 2085350 | 816 | 2 | 3.37E-09 | 9.53E-05 | 2.53E-06 | 1.54E-06 | -0.003389721 | PRKCZ |
| chr1 | 153633239 | 153634342 | 1104 | 13 | 3.38E-09 | 1.30E-08 | 0.00196 | 1.46E-08 | -0.006228461 | CHTOP, AL162258.1, AL162258.2 |
| chr1 | 94690001 | 94690438 | 438 | 2 | 3.47E-09 | 6.79E-06 | 1.83E-07 | 4.50E-08 | -0.006243687 | SLC44A3-AS1 |
| chr17 | 8397658 | 8399161 | 1504 | 9 | 3.48E-09 | 4.12E-12 | 0.00055 | 1.49E-11 | -0.012303813 | RNF222 |
| chr1 | 44787407 | 44788947 | 1541 | 10 | 3.52E-09 | 1.60E-08 | 0.00052 | 8.16E-10 | -0.004142054 | BEST4 |
| chr3 | 50155616 | 50158049 | 2434 | 11 | 3.55E-09 | 7.21E-08 | 1.15E-05 | 1.57E-09 | -0.008808229 | SEMA3F, SEMA3F-AS1 |
| chr3 | 53845710 | 53847243 | 1534 | 12 | 3.64E-09 | 0.000321 | 0.0002 | 7.80E-08 | -0.004812913 | IL17RB, CHDH |

| | | | | | | | | | | |
|---|---|---|---|---|---|---|---|---|---|---|
| chr12 | 19546570 | 19546898 | 329 | 3 | 3.73E-09 | 4.47E-10 | 1.15E-05 | 9.51E-10 | -0.016141558 | AEBP2 |
| chr2 | 108942139 | 108943278 | 1140 | 7 | 3.79E-09 | 4.16E-06 | 5.41E-05 | 5.05E-08 | -0.003141109 | EDAR |
| chr12 | 53048538 | 53050178 | 1641 | 10 | 3.87E-09 | 1.07E-12 | 0.00014 | 8.39E-13 | -0.008008212 | TNS2, AC068888.1 |
| chr5 | 1404955 | 1406250 | 1296 | 7 | 3.89E-09 | 4.45E-08 | 5.29E-05 | 9.28E-09 | -0.008088938 | SLC6A3 |
| chr5 | 144536124 | 144536984 | 861 | 3 | 3.92E-09 | 2.54E-05 | 2.28E-07 | 8.42E-08 | -0.00448699 | NA |
| chr21 | 34886126 | 34887500 | 1375 | 15 | 3.94E-09 | 3.11E-13 | 0.01168 | 1.11E-10 | -0.031434686 | RUNX1 |
| chr2 | 3101244 | 3101574 | 331 | 3 | 3.97E-09 | 1.63E-08 | 1.22E-05 | 1.39E-08 | -0.013132563 | LINC01250 |
| chr1 | 24909371 | 24909954 | 584 | 4 | 4.02E-09 | 4.00E-09 | 5.25E-06 | 2.50E-09 | -0.011534049 | RUNX3 |
| chr1 | 161198661 | 161202501 | 3841 | 27 | 4.04E-09 | 9.92E-12 | 0.00034 | 4.74E-14 | -0.004887796 | NDUFS2, ADAMTS4 |
| chr17 | 48563412 | 48564742 | 1331 | 6 | 4.04E-09 | 1.63E-11 | 0.00071 | 3.00E-10 | -0.019203302 | HOXB-AS3, HOXB3 |
| chr15 | 41892724 | 41892733 | 10 | 2 | 4.12E-09 | 3.56E-07 | 4.86E-07 | 2.97E-08 | -0.013049035 | SPTBN5 |
| chr11 | 77196438 | 77197113 | 676 | 2 | 4.21E-09 | 8.19E-08 | 5.12E-06 | 5.93E-08 | -0.002461727 | MYO7A |
| chr7 | 151010225 | 151010742 | 518 | 2 | 4.27E-09 | 1.53E-09 | 1.31E-05 | 3.11E-09 | -0.0176689 | NOS3 |
| chr20 | 41318302 | 41318810 | 509 | 3 | 4.28E-09 | 1.46E-07 | 1.35E-05 | 2.60E-09 | -0.012695583 | NA |
| chr16 | 70400629 | 70401308 | 680 | 7 | 4.29E-09 | 9.44E-06 | 6.79E-06 | 1.08E-09 | -0.003922764 | ST3GAL2 |
| chr17 | 16485979 | 16486797 | 819 | 3 | 4.31E-09 | 5.10E-11 | 5.97E-05 | 2.41E-10 | -0.039287524 | LRRC75A |
| chr14 | 23146942 | 23147590 | 649 | 6 | 4.40E-09 | 0.000959 | 1.56E-07 | 2.27E-07 | -0.003497365 | SLC7A8 |
| chr9 | 121299032 | 121300025 | 994 | 11 | 4.45E-09 | 3.46E-08 | 0.00064 | 2.12E-08 | -0.005238521 | GSN |
| chr1 | 18784240 | 18784595 | 356 | 4 | 4.50E-09 | 1.35E-09 | 0.00011 | 1.86E-09 | -0.08903495 | NA |
| chr15 | 48336990 | 48337369 | 380 | 2 | 4.60E-09 | 4.39E-10 | 1.30E-05 | 9.82E-10 | -0.006257154 | DUT |
| chr9 | 114348052 | 114349083 | 1032 | 4 | 4.67E-09 | 2.84E-06 | 3.06E-06 | 5.26E-08 | -0.010037369 | AKNA |
| chr16 | 30090359 | 30092721 | 2363 | 18 | 4.70E-09 | 1.85E-06 | 3.21E-05 | 1.34E-08 | -0.002359734 | TBX6, YPEL3 |
| chr3 | 23819821 | 23820325 | 505 | 2 | 4.71E-09 | 3.15E-07 | 4.98E-07 | 2.81E-08 | -0.014917443 | UBE2E1 |
| chr20 | 59007732 | 59008654 | 923 | 18 | 4.73E-09 | 6.37E-08 | 0.00242 | 1.70E-08 | -0.004052589 | NA |
| chr14 | 99174134 | 99174814 | 681 | 3 | 4.85E-09 | 4.43E-08 | 3.05E-05 | 1.76E-08 | -0.035637334 | BCL11B |
| chr17 | 81405650 | 81406906 | 1257 | 7 | 4.90E-09 | 3.23E-09 | 0.00038 | 1.67E-10 | -0.021969141 | BAHCC1 |
| chr19 | 35115630 | 35117266 | 1637 | 11 | 4.95E-09 | 3.13E-06 | 1.65E-05 | 7.36E-09 | 0.005598321 | AC020907.6, FXYD3 |
| chr12 | 51026755 | 51027442 | 688 | 2 | 5.03E-09 | 0.000104 | 3.88E-05 | 2.16E-05 | -0.00703249 | SLC11A2 |
| chr6 | 167146363 | 167147313 | 951 | 7 | 5.09E-09 | 3.25E-10 | 0.00056 | 7.47E-10 | -0.029610579 | TCP10L2 |
| chr11 | 62610462 | 62611729 | 1268 | 8 | 5.17E-09 | 1.35E-06 | 3.83E-05 | 8.58E-08 | -0.00503678 | ROM1, EML3 |
| chr12 | 54472715 | 54474539 | 1825 | 11 | 5.18E-09 | 2.08E-11 | 0.00284 | 4.86E-10 | -0.00849303 | AC079313.2, GTSF1 |
| chr21 | 46136884 | 46138300 | 1417 | 3 | 5.20E-09 | 2.83E-08 | 3.59E-06 | 4.69E-09 | -0.004810055 | FTCD |
| chr17 | 44905664 | 44905899 | 236 | 2 | 5.22E-09 | 1.87E-06 | 1.83E-06 | 2.10E-07 | -0.01235137 | GFAP |
| chr11 | 36036901 | 36037175 | 275 | 2 | 5.29E-09 | 9.93E-09 | 8.08E-07 | 4.36E-09 | -0.016009359 | LDLRAD3 |
| chr10 | 102775263 | 102776364 | 1102 | 13 | 5.35E-09 | 3.19E-10 | 0.00148 | 8.32E-10 | -0.005977324 | WBP1L |
| chr19 | 11673431 | 11674433 | 1003 | 12 | 5.38E-09 | 2.71E-11 | 0.00106 | 1.73E-10 | 0.014899835 | AC008543.1 |
| chr22 | 46112114 | 46113735 | 1622 | 11 | 5.41E-09 | 4.77E-12 | 0.00028 | 1.61E-11 | -0.005665459 | MIRLET7BHG, MIRLET7A3, MIR4763, MIRLET7B |
| chr5 | 68225313 | 68226704 | 1392 | 8 | 5.41E-09 | 2.26E-08 | 0.00015 | 2.32E-08 | -0.006159552 | PIK3R1 |
| chr6 | 30206348 | 30208015 | 1668 | 12 | 5.43E-09 | 1.11E-07 | 0.0024 | 1.83E-08 | -0.006216512 | TRIM26 |
| chr1 | 39268138 | 39269470 | 1333 | 6 | 5.45E-09 | 9.49E-09 | 7.93E-05 | 1.45E-09 | -0.010676041 | MACF1 |
| chr20 | 3042346 | 3044197 | 1852 | 9 | 5.48E-09 | 3.13E-08 | 4.47E-05 | 1.14E-10 | -0.005398553 | GNRH2 |
| chr12 | 128381627 | 128382190 | 564 | 5 | 5.54E-09 | 4.07E-09 | 0.00014 | 4.99E-09 | 0.024899027 | TMEM132C |
| chr5 | 136057836 | 136059173 | 1338 | 7 | 5.82E-09 | 1.69E-11 | 0.00032 | 6.74E-11 | -0.011632703 | TGFBI |
| chr4 | 158208862 | 158209439 | 578 | 2 | 5.83E-09 | 3.97E-06 | 1.45E-06 | 2.47E-07 | -0.009515495 | TMEM144 |
| chr4 | 16573700 | 16574491 | 792 | 3 | 5.85E-09 | 2.37E-07 | 5.12E-06 | 1.61E-08 | -0.017058415 | LDB2 |
| chr2 | 120239477 | 120240049 | 573 | 5 | 5.86E-09 | 1.06E-07 | 7.28E-05 | 1.76E-08 | -0.012185002 | NA |
| chr20 | 62553282 | 62554692 | 1411 | 12 | 5.87E-09 | 1.47E-10 | 0.00103 | 6.37E-10 | -0.005964506 | MIR1-1HG, MIR1-1 |
| chr1 | 27360583 | 27361971 | 1389 | 7 | 6.01E-09 | 1.04E-09 | 3.65E-05 | 8.96E-10 | -0.011625152 | MAP3K6 |
| chr11 | 124777348 | 124778097 | 750 | 4 | 6.06E-09 | 2.08E-07 | 5.30E-05 | 1.38E-08 | -0.011601773 | MSANTD2 |
| chr22 | 37103411 | 37104481 | 1071 | 7 | 6.12E-09 | 1.01E-07 | 0.00043 | 3.67E-09 | -0.008933211 | TMPRSS6 |
| chr5 | 149857064 | 149857507 | 444 | 3 | 6.15E-09 | 4.72E-09 | 1.00E-05 | 5.24E-09 | -0.011804146 | NA |
| chr2 | 222655692 | 222656964 | 1273 | 15 | 6.31E-09 | 2.98E-07 | 3.84E-05 | 3.83E-09 | 0.003432476 | FARSB |
| chr1 | 56818601 | 56820592 | 1992 | 11 | 6.35E-09 | 1.55E-07 | 1.56E-05 | 5.83E-12 | -0.007962348 | FYB2 |
| chr10 | 44910557 | 44911399 | 843 | 8 | 6.37E-09 | 4.47E-08 | 7.82E-05 | 3.96E-09 | -0.020891081 | TMEM72, TMEM72-AS1, AL353801.2 |
| chr8 | 23003371 | 23004881 | 1511 | 5 | 6.57E-09 | 4.92E-08 | 1.41E-05 | 4.99E-09 | -0.007082039 | RHOBTB2, AC107959.1 |
| chr16 | 28069945 | 28070959 | 1015 | 6 | 6.60E-09 | 4.02E-08 | 5.80E-05 | 6.28E-09 | -0.001373766 | NA |
| chr1 | 10530424 | 10530615 | 192 | 2 | 6.65E-09 | 8.30E-09 | 1.13E-06 | 4.56E-09 | -0.0310112 | PEX14 |
| chr17 | 81858628 | 81859757 | 1130 | 7 | 6.67E-09 | 1.26E-08 | 0.00021 | 1.20E-09 | -0.00992422 | P4HB |
| chr5 | 90390487 | 90391380 | 894 | 3 | 6.81E-09 | 1.21E-06 | 1.63E-05 | 3.29E-08 | -0.008667297 | NA |
| chr1 | 6235308 | 6237068 | 1761 | 15 | 6.82E-09 | 3.67E-05 | 1.36E-05 | 3.08E-08 | -0.00380657 | LINC00337, ICMT |
| chr9 | 136671759 | 136672295 | 537 | 4 | 6.95E-09 | 1.31E-06 | 2.01E-05 | 2.30E-07 | -0.005682877 | EGFL7 |
| chr9 | 128097927 | 128099140 | 1214 | 9 | 6.98E-09 | 6.54E-07 | 5.69E-05 | 3.11E-08 | -0.008592055 | SLC25A25 |
| chr3 | 129313983 | 129314761 | 779 | 3 | 6.99E-09 | 1.68E-08 | 4.65E-05 | 1.62E-08 | -0.00835414 | NA |
| chr9 | 136885048 | 136885965 | 918 | 4 | 7.00E-09 | 8.02E-08 | 0.00018 | 4.27E-08 | -0.014391282 | TRAF2 |
| chr8 | 54134519 | 54135123 | 605 | 7 | 7.03E-09 | 7.16E-08 | 0.00011 | 3.71E-08 | -0.007342969 | NA |
| chr10 | 30027503 | 30028998 | 1496 | 4 | 7.13E-09 | 2.37E-07 | 6.61E-06 | 2.27E-08 | -0.00916958 | JCAD |
| chr17 | 6433903 | 6436146 | 2244 | 14 | 7.29E-09 | 3.51E-07 | 1.57E-06 | 1.44E-09 | -0.002483105 | AIPL1 |
| chr1 | 236393865 | 236395165 | 1301 | 11 | 7.47E-09 | 7.78E-07 | 0.00019 | 4.99E-08 | -0.009490727 | EDARADD |
| chr8 | 57278827 | 57280687 | 1861 | 13 | 7.72E-09 | 4.01E-06 | 9.41E-05 | 2.47E-09 | -0.01156684 | LINC00588 |
| chr17 | 81082261 | 81084030 | 1770 | 9 | 7.73E-09 | 1.11E-06 | 0.00011 | 3.06E-09 | -0.002844334 | BAIAP2 |
| chr1 | 56414901 | 56416386 | 1486 | 5 | 7.80E-09 | 1.42E-06 | 2.31E-05 | 1.54E-07 | -0.008675262 | LINC01767, AC119674.1, AC119674.2 |
| chr7 | 4416488 | 4416961 | 474 | 8 | 7.98E-09 | 5.41E-10 | 7.57E-05 | 9.75E-10 | 0.019931474 | NA |
| chr5 | 115297096 | 115297771 | 676 | 4 | 8.04E-09 | 0.033615 | 2.77E-06 | 6.15E-06 | 0.000303141 | NA |
| chr15 | 74888759 | 74890545 | 1787 | 14 | 8.30E-09 | 7.02E-06 | 0.00035 | 4.79E-08 | -0.001042849 | MPI |
| chr1 | 41484566 | 41485146 | 581 | 5 | 8.44E-09 | 5.48E-08 | 0.00016 | 1.15E-08 | 0.019055091 | EDN2 |
| chr8 | 141431315 | 141431488 | 174 | 3 | 8.60E-09 | 2.38E-09 | 8.72E-06 | 2.86E-09 | -0.011822838 | PTP4A3 |
| chr12 | 132070462 | 132071090 | 629 | 2 | 8.68E-09 | 5.93E-08 | 1.57E-06 | 2.13E-08 | -0.010673459 | EP400 |
| chr5 | 157563655 | 157564502 | 848 | 3 | 8.70E-09 | 0.000191 | 4.32E-07 | 4.50E-07 | -0.013167997 | ADAM19 |
| chr5 | 169945278 | 169945815 | 538 | 3 | 8.70E-09 | 0.000107 | 7.29E-07 | 1.40E-07 | -0.003045542 | DOCK2, INSYN2B |
| chr15 | 83067608 | 83068208 | 601 | 4 | 8.75E-09 | 2.43E-06 | 7.21E-06 | 1.88E-07 | -0.00873122 | AC022558.2, AC024270.1 |
| chr2 | 104851994 | 104853369 | 1376 | 10 | 8.82E-09 | 1.23E-05 | 1.57E-05 | 2.46E-07 | 0.001835692 | AC018730.1, POU3F3, PANTR1 |
| chr1 | 3365885 | 3366924 | 1040 | 4 | 8.85E-09 | 9.74E-11 | 0.00011 | 5.38E-10 | -0.017741016 | PRDM16 |
| chr21 | 45490645 | 45491564 | 920 | 3 | 8.85E-09 | 9.39E-08 | 2.02E-05 | 7.31E-08 | -0.010413518 | COL18A1 |
| chr18 | 31447522 | 31447922 | 401 | 6 | 8.86E-09 | 1.03E-07 | 4.29E-05 | 7.44E-09 | -0.010249895 | DSG3 |
| chr11 | 66316692 | 66317982 | 1291 | 10 | 8.89E-09 | 1.09E-05 | 4.96E-05 | 1.09E-07 | -0.005929846 | AP001107.3, CD248 |
| chr9 | 134323518 | 134324364 | 847 | 3 | 9.00E-09 | 5.54E-05 | 1.01E-06 | 4.18E-07 | -0.012003794 | RXRA |
| chr19 | 1826374 | 1826958 | 585 | 4 | 9.03E-09 | 3.48E-09 | 7.58E-05 | 8.79E-09 | -0.008729 | REXO1 |
| chr2 | 79280114 | 79280142 | 29 | 3 | 9.12E-09 | 5.19E-07 | 2.59E-07 | 8.16E-09 | -0.01424853 | CTNNA2, AC011754.2 |
| chr22 | 17266863 | 17266927 | 65 | 3 | 9.32E-09 | 1.39E-09 | 9.71E-05 | 5.51E-09 | -0.020777622 | NA |
| chr7 | 32070913 | 32072302 | 1390 | 11 | 9.41E-09 | 7.24E-07 | 0.00047 | 1.21E-07 | -0.008874573 | PDE1C |
| chr19 | 44140696 | 44141915 | 1220 | 11 | 9.56E-09 | 1.02E-07 | 0.00028 | 2.22E-08 | -0.008325643 | ZNF234 |
| chr6 | 134540103 | 134540546 | 444 | 2 | 9.56E-09 | 1.65E-06 | 8.08E-07 | 1.01E-07 | -0.00751662 | AL078590.2 |
| chr14 | 103879608 | 103881421 | 1814 | 9 | 9.69E-09 | 2.57E-11 | 0.00108 | 5.10E-10 | -0.007701932 | AL132712.2, AL132712.1 |
| chr13 | 108214101 | 108214806 | 706 | 4 | 9.79E-09 | 2.57E-06 | 2.58E-05 | 5.43E-07 | -0.021113699 | LIG4 |
| chr9 | 136995736 | 136998079 | 2344 | 14 | 9.80E-09 | 6.17E-10 | 0.00083 | 8.72E-11 | -0.007194948 | CLIC3 |
| chr13 | 51644865 | 51645471 | 607 | 2 | 9.97E-09 | 1.18E-05 | 1.84E-06 | 5.14E-07 | -0.004867963 | WDFY2 |
| chr14 | 50659400 | 50659579 | 180 | 2 | 1.03E-08 | 4.66E-09 | 1.02E-05 | 7.80E-09 | -0.020942918 | SAV1 |
| chr6 | 33589881 | 33591826 | 1946 | 6 | 1.06E-08 | 1.12E-09 | 1.01E-05 | 1.38E-10 | -0.007012335 | LINC00336 |
| chr10 | 133237118 | 133238500 | 1383 | 18 | 1.06E-08 | 2.45E-09 | 0.00025 | 9.44E-10 | -0.008545494 | VENTX |
| chr18 | 69956835 | 69958187 | 1353 | 8 | 1.07E-08 | 4.16E-10 | 0.00079 | 1.09E-09 | -0.011606976 | CD226 |
| chr22 | 22558737 | 22559831 | 1095 | 13 | 1.09E-08 | 7.79E-07 | 0.00016 | 5.93E-08 | -0.006045453 | LL22NC03-63E9.3, PRAME |
| chr12 | 128117712 | 128118857 | 1146 | 10 | 1.12E-08 | 8.71E-07 | 0.00012 | 1.79E-09 | -0.004636158 | LINC02368, LINC02369 |
| chr17 | 39663297 | 39664887 | 1591 | 7 | 1.12E-08 | 1.48E-07 | 0.00016 | 2.04E-09 | -0.008184281 | STARD3 |
| chr19 | 7507110 | 7508080 | 971 | 5 | 1.13E-08 | 8.68E-08 | 5.35E-05 | 1.37E-08 | -0.012393483 | TEX45, AC008878.1 |
| chr22 | 36818820 | 36820023 | 1204 | 8 | 1.14E-08 | 5.40E-09 | 0.0005 | 1.81E-09 | -0.010367977 | Z82185.1, PVALB |
| chr19 | 5692611 | 5694005 | 1395 | 4 | 1.16E-08 | 1.43E-08 | 1.33E-05 | 1.15E-09 | -0.007437156 | LONP1 |
| chr3 | 128074540 | 128075988 | 1449 | 8 | 1.16E-08 | 7.58E-05 | 9.49E-06 | 9.29E-08 | -0.007751482 | RUVBL1-AS1, RUVBL1, RNU2-37P |
| chr1 | 153789725 | 153790513 | 789 | 8 | 1.17E-08 | 2.74E-11 | 0.00238 | 5.37E-10 | -0.014273118 | GATAD2B |
| chr12 | 3220238 | 3220395 | 158 | 4 | 1.17E-08 | 2.51E-06 | 1.91E-06 | 1.07E-08 | -0.001580776 | TSPAN9 |
| chr1 | 31986777 | 31987329 | 553 | 2 | 1.17E-08 | 1.18E-07 | 1.45E-06 | 3.25E-08 | -0.01151744 | AL136115.3 |
| chr17 | 31564472 | 31565468 | 997 | 6 | 1.17E-08 | 0.00211 | 1.94E-06 | 4.59E-07 | -0.005436902 | NA |
| chr9 | 95770693 | 95772691 | 1999 | 8 | 1.17E-08 | 9.34E-14 | 0.0016 | 6.91E-12 | -0.030714082 | AL354861.3, LINC00476 |
| chr15 | 66994276 | 66994565 | 290 | 4 | 1.18E-08 | 6.12E-07 | 1.91E-05 | 8.92E-09 | -0.014441039 | AC087482.1 |
| chr12 | 122132309 | 122134168 | 1860 | 14 | 1.19E-08 | 7.77E-09 | 0.00189 | 8.84E-10 | -0.003961065 | MLXIP |
| chr1 | 27366850 | 27367899 | 1050 | 11 | 1.19E-08 | 2.63E-07 | 0.00036 | 8.26E-08 | -0.003785935 | MAP3K6 |
| chr3 | 172709211 | 172710636 | 1426 | 5 | 1.22E-08 | 1.92E-07 | 8.34E-05 | 6.09E-08 | -0.015348421 | NCEH1 |
| chr1 | 75796114 | 75797509 | 1396 | 12 | 1.24E-08 | 5.98E-07 | 0.00118 | 4.45E-09 | 0.006250103 | MSH4 |
| chr22 | 44638885 | 44639639 | 755 | 3 | 1.24E-08 | 1.13E-06 | 2.55E-05 | 9.10E-08 | -0.018886131 | NA |
| chr21 | 32484879 | 32485130 | 252 | 4 | 1.28E-08 | 5.66E-05 | 5.58E-07 | 3.36E-08 | -0.01010581 | EVA1C, CFAP298-TCP10L |
| chr7 | 130008678 | 130009356 | 679 | 4 | 1.28E-08 | 1.02E-06 | 8.07E-05 | 1.66E-07 | -0.008411683 | AC073320.1 |
| chr1 | 207811449 | 207812875 | 1427 | 4 | 1.28E-08 | 0.000568 | 6.59E-07 | 9.31E-07 | -0.00910518 | MIR29B2CHG |
| chr14 | 96311277 | 96311609 | 333 | 4 | 1.30E-08 | 0.000111 | 7.24E-06 | 4.63E-08 | -0.008037098 | ATG2B |
| chr11 | 379296 | 380593 | 1298 | 5 | 1.30E-08 | 4.45E-10 | 4.42E-05 | 1.23E-09 | -0.012531408 | B4GALNT4 |
| chr1 | 27153615 | 27155901 | 2287 | 14 | 1.31E-08 | 7.97E-06 | 1.90E-05 | 1.72E-09 | -0.000201955 | SLC9A1 |
| chr9 | 136975842 | 136977046 | 1205 | 6 | 1.32E-08 | 1.08E-06 | 0.00015 | 1.49E-07 | -0.010212483 | PTGDS |
| chr5 | 176836532 | 176836985 | 454 | 2 | 1.34E-08 | 3.01E-08 | 4.15E-06 | 2.48E-08 | 0.034279925 | UNC5A |
| chr20 | 3408253 | 3408888 | 636 | 3 | 1.34E-08 | 0.000203 | 1.62E-06 | 4.20E-07 | 0.003680098 | AL117334.1 |
| chr16 | 1492732 | 1496778 | 4047 | 22 | 1.35E-08 | 4.71E-09 | 2.57E-05 | 8.22E-13 | -0.002841665 | TELO2 |
| chr6 | 31861734 | 31866401 | 4668 | 70 | 1.36E-08 | 2.14E-12 | 0.00111 | 5.85E-18 | -0.001232487 | NEU1, SLC44A4 |
| chr7 | 152138555 | 152139713 | 1159 | 7 | 1.36E-08 | 1.61E-05 | 1.83E-06 | 1.45E-07 | -0.002129186 | KMT2C |
| chr17 | 7379857 | 7381343 | 1487 | 14 | 1.40E-08 | 2.10E-08 | 0.00167 | 1.63E-08 | -0.008264386 | TNK1 |
| chr3 | 61560546 | 61561754 | 1209 | 14 | 1.41E-08 | 3.37E-05 | 3.58E-06 | 1.38E-07 | 0.001051884 | PTPRG |
| chr22 | 44675460 | 44676004 | 545 | 3 | 1.44E-08 | 1.11E-06 | 5.73E-05 | 3.77E-07 | -0.005946911 | PRR5 |
| chr14 | 102461625 | 102462100 | 476 | 2 | 1.44E-08 | 8.35E-10 | 1.64E-05 | 1.81E-09 | -0.023056663 | TECPR2 |
| chr7 | 2525941 | 2527277 | 1337 | 11 | 1.45E-08 | 0.014662 | 8.25E-06 | 9.52E-07 | -0.00311991 | LFNG, MIR4648 |
| chr19 | 7848983 | 7849638 | 656 | 4 | 1.46E-08 | 0.000725 | 1.83E-07 | 2.99E-07 | -0.003517337 | EVI5L |
| chr18 | 78543815 | 78544675 | 861 | 6 | 1.46E-08 | 1.55E-05 | 3.01E-05 | 2.22E-08 | 0.00291126 | Y_RNA |
| chr19 | 1411455 | 1413125 | 1671 | 5 | 1.46E-08 | 6.42E-11 | 0.00011 | 2.20E-10 | -0.010275041 | DAZAP1 |
| chr5 | 132656533 | 132657138 | 606 | 4 | 1.47E-08 | 2.51E-07 | 0.00017 | 3.29E-07 | -0.009713723 | IL13, TH2LCRR |
| chr9 | 101486644 | 101487654 | 1011 | 10 | 1.48E-08 | 1.14E-06 | 0.00013 | 2.11E-08 | 0.009331052 | PGAP4 |
| chr7 | 149777179 | 149777786 | 608 | 2 | 1.51E-08 | 4.10E-05 | 1.38E-06 | 5.92E-07 | -0.01150909 | SSPOP |
| chr12 | 104999191 | 104999247 | 57 | 2 | 1.56E-08 | 1.94E-09 | 1.15E-05 | 3.77E-09 | -0.011908655 | C12orf45 |
| chr20 | 44930205 | 44930921 | 717 | 3 | 1.57E-08 | 8.08E-05 | 2.13E-06 | 4.07E-07 | -0.002678128 | PABPC1L |
| chr19 | 18761875 | 18762458 | 584 | 4 | 1.60E-08 | 1.65E-05 | 3.12E-05 | 6.43E-08 | -0.013000952 | CRTC1 |
| chr7 | 81367172 | 81367587 | 416 | 2 | 1.61E-08 | 2.56E-07 | 8.42E-07 | 3.76E-08 | -0.025298579 | NA |
| chr1 | 9262606 | 9263122 | 517 | 2 | 1.62E-08 | 1.58E-06 | 4.71E-06 | 4.17E-07 | 0.003723065 | H6PD |
| chr15 | 75722958 | 75723994 | 1037 | 8 | 1.63E-08 | 5.33E-09 | 6.71E-05 | 1.89E-09 | -0.008320292 | NA |
| chr17 | 48570425 | 48571220 | 796 | 5 | 1.65E-08 | 4.06E-07 | 3.78E-05 | 1.88E-07 | -0.009582389 | HOXB-AS3, HOXB3 |
| chr19 | 1957362 | 1958360 | 999 | 4 | 1.65E-08 | 1.51E-08 | 1.03E-05 | 7.38E-09 | -0.010952403 | CSNK1G2 |
| chr2 | 74383400 | 74385685 | 2286 | 9 | 1.66E-08 | 4.75E-10 | 7.66E-05 | 2.53E-12 | -0.008021604 | DCTN1-AS1, DCTN1 |
| chr15 | 65101695 | 65102535 | 841 | 5 | 1.67E-08 | 2.22E-08 | 0.00022 | 1.71E-08 | -0.0094745 | UBAP1L |
| chr4 | 183852795 | 183853390 | 596 | 5 | 1.68E-08 | 0.001476 | 8.29E-06 | 1.10E-06 | -0.003751268 | STOX2 |
| chr16 | 721506 | 723253 | 1748 | 7 | 1.71E-08 | 2.15E-09 | 6.79E-05 | 3.56E-10 | -0.008346306 | ANTKMT, CCDC78 |
| chr11 | 417189 | 418251 | 1063 | 9 | 1.73E-08 | 3.53E-08 | 2.57E-05 | 9.59E-09 | -0.006305349 | SIGIRR, ANO9 |
| chr14 | 59919757 | 59920961 | 1205 | 7 | 1.74E-08 | 9.42E-05 | 0.00026 | 5.92E-07 | -0.005783895 | LRRC9, AL133299.1 |
| chr22 | 30290356 | 30291712 | 1357 | 5 | 1.74E-08 | 3.41E-07 | 2.68E-05 | 1.42E-07 | -0.00095068 | AC004997.1 |
| chr2 | 120820590 | 120821222 | 633 | 2 | 1.76E-08 | 5.60E-06 | 2.44E-06 | 4.86E-07 | -0.016855186 | GLI2 |
| chr8 | 29905341 | 29905618 | 278 | 3 | 1.78E-08 | 1.70E-05 | 1.42E-07 | 4.62E-08 | -0.002550573 | NA |
| chr17 | 80781579 | 80782041 | 463 | 4 | 1.80E-08 | 1.14E-08 | 0.00031 | 2.62E-08 | -0.02194533 | RPTOR |
| chr3 | 196338235 | 196338698 | 464 | 10 | 1.83E-08 | 8.88E-06 | 5.11E-05 | 2.21E-08 | -0.004832365 | TM4SF19-TCTEX1D2, TM4SF19 |
| chr19 | 39130543 | 39131320 | 778 | 4 | 1.84E-08 | 5.24E-07 | 2.54E-05 | 5.05E-08 | -0.00921927 | PAK4 |
| chr8 | 143825270 | 143826052 | 783 | 2 | 1.84E-08 | 7.74E-06 | 2.63E-06 | 6.10E-07 | -0.008889208 | PUF60 |

| | | | | | | | | | | |
|---|---|---|---|---|---|---|---|---|---|---|
| chr2 | 112784514 | 112784816 | 303 | 7 | 1.85E-08 | 1.45E-10 | 0.00134 | 1.88E-09 | -0.015987469 | NA |
| chr9 | 127965733 | 127966215 | 483 | 3 | 1.86E-08 | 2.18E-05 | 2.71E-07 | 1.16E-07 | -0.005541925 | FAM102A |
| chr1 | 212750241 | 212750438 | 198 | 2 | 1.86E-08 | 2.32E-09 | 1.47E-05 | 4.66E-09 | -0.015092259 | NSL1 |
| chr16 | 66737687 | 66738116 | 430 | 5 | 1.87E-08 | 4.14E-08 | 7.87E-06 | 1.35E-08 | -0.008317014 | AC018557.3, DYNC1LI2 |
| chr10 | 35398101 | 35399010 | 910 | 3 | 1.90E-08 | 1.81E-07 | 3.96E-06 | 5.12E-08 | -0.005755688 | CCNY |
| chr4 | 133148884 | 133149664 | 781 | 10 | 1.92E-08 | 7.82E-07 | 4.50E-06 | 3.29E-08 | 2.85E-05 | PCDH10, AC105383.1 |
| chr6 | 128639896 | 128641080 | 1185 | 4 | 1.93E-08 | 1.80E-05 | 1.89E-06 | 3.79E-07 | -0.007570728 | NA |
| chr21 | 45283582 | 45285912 | 2331 | 7 | 1.93E-08 | 1.60E-10 | 7.31E-05 | 5.96E-12 | -0.002916837 | POFUT2 |
| chr16 | 89219664 | 89221017 | 1354 | 6 | 1.97E-08 | 6.42E-07 | 2.10E-05 | 1.68E-07 | -0.003101249 | ZNF778 |
| chr5 | 170782120 | 170783436 | 1317 | 7 | 2.02E-08 | 5.46E-07 | 3.16E-05 | 2.21E-08 | -0.007582997 | GABRP, AC008514.1 |
| chr6 | 37696206 | 37697275 | 1070 | 6 | 2.04E-08 | 1.71E-06 | 0.00022 | 7.91E-08 | 0.000961718 | MDGA1 |
| chr1 | 203764786 | 203765431 | 646 | 10 | 2.06E-08 | 6.66E-10 | 0.00339 | 3.67E-09 | -0.010000688 | LAX1 |
| chr5 | 39760698 | 39761135 | 438 | 2 | 2.09E-08 | 2.91E-06 | 5.86E-07 | 1.01E-07 | -0.025065571 | NA |
| chr13 | 40562834 | 40563408 | 575 | 6 | 2.11E-08 | 3.06E-05 | 5.86E-05 | 1.75E-07 | -0.005761756 | AL133318.1, FOXO1 |
| chr13 | 97275465 | 97276247 | 783 | 6 | 2.12E-08 | 8.71E-07 | 1.54E-05 | 3.74E-08 | -0.001924494 | MBNL2 |
| chr19 | 55038046 | 55038980 | 935 | 11 | 2.13E-08 | 5.12E-09 | 0.00576 | 5.77E-09 | -0.018543756 | AC011476.3, GP6, AC011476.2 |
| chr8 | 142784755 | 142785888 | 1134 | 6 | 2.13E-08 | 3.05E-05 | 1.36E-05 | 6.75E-07 | -0.006149067 | AC083841.2, LY6D |
| chr11 | 124898345 | 124898949 | 605 | 4 | 2.15E-08 | 1.46E-05 | 6.77E-05 | 4.85E-07 | -0.007246543 | ROBO4 |
| chr8 | 123022168 | 123022506 | 339 | 3 | 2.18E-08 | 2.90E-06 | 2.76E-07 | 4.05E-08 | -0.007778517 | AC104316.1, DERL1 |
| chr14 | 104084633 | 104085216 | 584 | 4 | 2.22E-08 | 2.08E-06 | 2.71E-05 | 7.22E-07 | 0.000132277 | NA |
| chr2 | 199954053 | 199956078 | 2026 | 19 | 2.23E-08 | 1.40E-05 | 1.12E-05 | 7.52E-09 | -5.14E-06 | C2orf69, MAIP1, TYW5 |
| chr7 | 1022546 | 1023721 | 1176 | 12 | 2.24E-08 | 1.24E-05 | 0.00077 | 4.15E-08 | -0.003604298 | C7orf50, MIR339 |
| chr20 | 63654573 | 63654792 | 220 | 2 | 2.25E-08 | 4.86E-08 | 5.95E-05 | 9.44E-08 | -0.012099232 | STMN3 |
| chr4 | 16585498 | 16586293 | 796 | 2 | 2.28E-08 | 5.84E-09 | 2.31E-06 | 5.28E-09 | -0.008394742 | LDB2 |
| chr16 | 87688613 | 87689707 | 1095 | 3 | 2.30E-08 | 1.56E-06 | 3.62E-05 | 1.06E-07 | -0.004660358 | JPH3 |
| chr20 | 44714288 | 44715356 | 1069 | 13 | 2.31E-08 | 4.47E-09 | 0.00066 | 1.58E-09 | -0.007053613 | CCN5, KCNK15-AS1 |
| chr12 | 59811561 | 59812150 | 590 | 4 | 2.33E-08 | 4.66E-06 | 7.10E-05 | 2.58E-07 | -0.001550099 | AC079905.1 |
| chr1 | 236516769 | 236517461 | 693 | 3 | 2.34E-08 | 1.99E-06 | 9.18E-07 | 5.37E-08 | -0.011007214 | NA |
| chr3 | 53666114 | 53666236 | 123 | 3 | 2.36E-08 | 7.64E-10 | 0.00013 | 3.37E-09 | -0.045594375 | CACNA1D |
| chr19 | 57708712 | 57709933 | 1222 | 12 | 2.38E-08 | 2.48E-10 | 0.00406 | 1.08E-09 | -0.010211533 | ZNF551, AC003006.1, ZNF154 |
| chr17 | 48572948 | 48576349 | 3402 | 19 | 2.39E-08 | 5.32E-13 | 0.00035 | 5.04E-14 | -0.009764736 | HOXB-AS3, HOXB3, HOXB4 |
| chr14 | 75831633 | 75832767 | 1135 | 5 | 2.44E-08 | 2.83E-07 | 0.00015 | 5.71E-08 | -0.010156613 | TTLL5, AF107885.2 |
| chr21 | 46318196 | 46319225 | 1030 | 8 | 2.44E-08 | 8.41E-08 | 7.08E-05 | 1.22E-08 | -0.004405352 | C21orf58 |
| chr20 | 57974559 | 57975429 | 871 | 2 | 2.45E-08 | 3.56E-07 | 1.68E-06 | 7.41E-08 | -0.005730947 | NA |
| chr12 | 117188972 | 117189673 | 702 | 10 | 2.45E-08 | 0.000798 | 4.68E-06 | 1.86E-06 | -0.003528227 | FBXO21 |
| chr6 | 3848400 | 3849584 | 1185 | 27 | 2.52E-08 | 2.35E-07 | 0.00736 | 2.41E-08 | 0.008224333 | AL391422.3, AL391422.2, FAM50B |
| chr2 | 98822440 | 98823534 | 1095 | 5 | 2.54E-08 | 3.12E-08 | 6.05E-05 | 2.65E-09 | -0.011506865 | CRACDL |
| chr7 | 129387453 | 129388800 | 1348 | 7 | 2.55E-08 | 2.05E-05 | 1.59E-06 | 1.32E-07 | -0.001069043 | AHCYL2 |
| chr13 | 99655367 | 99655932 | 566 | 4 | 2.56E-08 | 1.14E-05 | 1.13E-06 | 2.00E-07 | -0.006083866 | CLYBL |
| chr12 | 1510194 | 1510440 | 247 | 2 | 2.58E-08 | 3.56E-09 | 1.30E-05 | 6.72E-09 | -0.019708365 | NA |
| chr17 | 4003346 | 4005303 | 1958 | 10 | 2.63E-08 | 1.97E-10 | 0.00011 | 1.18E-11 | -0.006787254 | ZZEF1 |
| chr22 | 43996485 | 43997048 | 564 | 5 | 2.68E-08 | 2.46E-07 | 0.00035 | 2.55E-08 | -0.010495656 | SAMM50 |
| chr16 | 2232956 | 2233717 | 762 | 3 | 2.68E-08 | 4.20E-05 | 4.56E-06 | 1.64E-06 | -0.003182998 | E4F1 |
| chr1 | 19981413 | 19981604 | 192 | 4 | 2.70E-08 | 1.57E-06 | 2.68E-05 | 4.65E-08 | -0.010499957 | NA |
| chr11 | 61917054 | 61918276 | 1223 | 6 | 2.75E-08 | 1.10E-05 | 9.52E-05 | 1.38E-07 | -0.000708577 | RAB3IL1 |
| chr1 | 155186928 | 155188465 | 1538 | 4 | 2.77E-08 | 1.86E-08 | 5.38E-05 | 1.55E-08 | -0.013795031 | AL713999.2, MUC1 |
| chr16 | 57815234 | 57816736 | 1503 | 3 | 2.81E-08 | 1.83E-08 | 3.89E-06 | 4.43E-09 | -0.008485406 | AC092118.1, KIFC3 |
| chr1 | 222543762 | 222544201 | 440 | 4 | 2.81E-08 | 1.02E-07 | 2.65E-06 | 1.89E-08 | -0.006207687 | HHIPL2 |
| chr2 | 127107005 | 127107855 | 851 | 12 | 2.83E-08 | 9.26E-05 | 4.52E-05 | 3.84E-07 | -0.002479689 | BIN1 |
| chr2 | 109318272 | 109318404 | 133 | 3 | 2.83E-08 | 3.11E-09 | 4.36E-05 | 5.43E-09 | -0.008441359 | SH3RF3 |
| chr22 | 36510592 | 36511656 | 1065 | 5 | 2.88E-08 | 2.48E-08 | 1.61E-05 | 1.24E-08 | -0.006604953 | EIF3D |
| chr9 | 134051735 | 134054549 | 2815 | 12 | 2.89E-08 | 9.65E-13 | 7.22E-05 | 3.77E-14 | -0.00700554 | AL445931.1, BRD3 |
| chr1 | 15736627 | 15738843 | 2217 | 9 | 2.90E-08 | 2.03E-10 | 1.62E-05 | 4.54E-11 | -0.004583639 | SLC25A34, AL450998.2 |
| chr17 | 58529167 | 58530238 | 1072 | 6 | 2.98E-08 | 0.001313 | 4.09E-06 | 3.70E-06 | -0.002821133 | SEPTIN4-AS1, SEPTIN4 |
| chr5 | 64768022 | 64769627 | 1606 | 15 | 3.02E-08 | 0.000107 | 1.93E-05 | 5.93E-08 | -8.20E-05 | CWC27, SREK1IP1 |
| chr7 | 47350576 | 47351544 | 969 | 3 | 3.02E-08 | 5.43E-06 | 3.51E-06 | 4.63E-07 | -0.00730731 | TNS3 |
| chr12 | 131848475 | 131851862 | 3388 | 14 | 3.03E-08 | 3.50E-12 | 1.83E-05 | 6.90E-16 | -0.018311917 | MMP17 |
| chr7 | 959071 | 960700 | 1630 | 9 | 3.08E-08 | 1.42E-11 | 0.00035 | 1.45E-11 | -0.009082164 | COX19 |
| chr20 | 33674211 | 33675685 | 1475 | 11 | 3.10E-08 | 4.22E-07 | 0.00065 | 2.21E-08 | -0.003756648 | AL121906.1, NECAB3, E2F1 |
| chr19 | 32675217 | 32676546 | 1330 | 10 | 3.10E-08 | 0.000283 | 1.56E-05 | 9.76E-06 | 0.002371878 | RGS9BP, ANKRD27 |
| chr17 | 82316900 | 82318258 | 1359 | 8 | 3.11E-08 | 1.94E-06 | 0.00017 | 6.20E-08 | -0.006704839 | CD7 |
| chr11 | 34570363 | 34571308 | 946 | 6 | 3.16E-08 | 1.24E-08 | 0.00022 | 1.62E-08 | -0.009514521 | LINC02707 |
| chr1 | 169460460 | 169461226 | 767 | 8 | 3.18E-08 | 0.000439 | 4.78E-05 | 1.05E-06 | -0.01058612 | CCDC181 |
| chr1 | 6385572 | 6386174 | 603 | 11 | 3.22E-08 | 4.25E-07 | 0.00018 | 2.97E-08 | -0.006678479 | ACOT7 |
| chr1 | 2104844 | 2106037 | 1194 | 6 | 3.28E-08 | 3.25E-08 | 0.00021 | 1.69E-08 | -0.009138845 | PRKCZ |
| chr1 | 19715572 | 19715585 | 14 | 2 | 3.29E-08 | 3.31E-09 | 2.49E-05 | 7.04E-09 | -0.019308163 | TMCO4 |
| chr11 | 118918469 | 118919100 | 632 | 3 | 3.31E-08 | 8.46E-06 | 6.28E-05 | 3.10E-07 | -0.011552956 | BCL9L |
| chr15 | 98899170 | 98900437 | 1268 | 8 | 3.40E-08 | 1.35E-08 | 0.00064 | 3.29E-08 | -0.008550091 | IGF1R |
| chr20 | 62707925 | 62708187 | 263 | 2 | 3.41E-08 | 6.76E-07 | 1.28E-05 | 4.60E-07 | -0.015240506 | NA |
| chr5 | 133924500 | 133924777 | 278 | 2 | 3.44E-08 | 4.84E-08 | 8.87E-06 | 5.21E-08 | -0.017452756 | NA |
| chr16 | 67661064 | 67662278 | 1215 | 19 | 3.45E-08 | 1.21E-06 | 0.00193 | 1.34E-08 | -0.002313949 | PARD6A |
| chr10 | 8053997 | 8055391 | 1395 | 27 | 3.49E-08 | 6.06E-05 | 0.0017 | 5.63E-07 | 0.000181215 | GATA3 |
| chr16 | 8866976 | 8868057 | 1082 | 9 | 3.49E-08 | 6.39E-08 | 0.00243 | 7.47E-08 | -0.016165276 | CARHSP1 |
| chr7 | 116432986 | 116433208 | 223 | 3 | 3.51E-08 | 3.68E-07 | 9.93E-07 | 2.99E-08 | -0.004169232 | CAV2, AC002066.1 |
| chr15 | 58864679 | 58865658 | 980 | 6 | 3.52E-08 | 9.20E-09 | 0.00027 | 1.25E-09 | -0.016236342 | ZNF444P1, RNF111, AC090515.4 |
| chr6 | 7505281 | 7505400 | 120 | 3 | 3.52E-08 | 1.17E-09 | 0.00012 | 4.99E-09 | -0.046937094 | NA |
| chr1 | 153533175 | 153534101 | 927 | 4 | 3.55E-08 | 0.000646 | 1.18E-06 | 1.86E-07 | -0.006021025 | BX470102.1 |
| chr4 | 139303836 | 139304432 | 597 | 2 | 3.56E-08 | 6.38E-06 | 3.03E-05 | 3.65E-06 | -0.006846476 | NAA15 |
| chr11 | 118338623 | 118339302 | 680 | 3 | 3.60E-08 | 1.10E-09 | 6.54E-05 | 3.88E-09 | -0.016095688 | CD3D |
| chr1 | 43932814 | 43933397 | 584 | 5 | 3.60E-08 | 7.38E-06 | 3.90E-05 | 1.47E-06 | -0.005717712 | ARTN |
| chr10 | 127906163 | 127907521 | 1359 | 9 | 3.62E-08 | 6.71E-08 | 0.00121 | 4.96E-09 | 0.013212832 | PTPRE |
| chr3 | 37992279 | 37993647 | 1369 | 8 | 3.65E-08 | 1.12E-06 | 4.66E-06 | 1.36E-08 | -0.00338911 | VILL |
| chr4 | 48944587 | 48944782 | 196 | 3 | 3.65E-08 | 2.68E-09 | 8.45E-05 | 7.49E-09 | -0.051990476 | NA |
| chr13 | 50133291 | 50134284 | 994 | 6 | 3.66E-08 | 0.000241 | 2.60E-05 | 1.62E-07 | -0.005442354 | DLEU1 |
| chr16 | 1354913 | 1356321 | 1409 | 3 | 3.66E-08 | 1.59E-06 | 1.62E-05 | 6.32E-07 | -0.004866704 | GNPTG |
| chr6 | 142146920 | 142147519 | 600 | 12 | 3.71E-08 | 7.20E-09 | 0.0008 | 5.33E-09 | 0.001833773 | VTA1 |
| chr16 | 10859887 | 10860011 | 125 | 2 | 3.72E-08 | 5.59E-09 | 9.61E-06 | 8.99E-09 | -0.016228576 | NA |
| chr1 | 16217189 | 16217725 | 537 | 9 | 3.75E-08 | 5.07E-05 | 9.78E-05 | 2.35E-07 | -0.001265854 | ANO7L1 |
| chr14 | 103533403 | 103534752 | 1350 | 8 | 3.75E-08 | 4.85E-05 | 1.04E-05 | 4.02E-08 | -0.002960177 | TRMT61A |
| chr9 | 134566376 | 134566407 | 32 | 2 | 3.81E-08 | 8.98E-09 | 4.68E-06 | 1.03E-08 | -0.003774604 | NA |
| chr15 | 50547129 | 50548020 | 892 | 5 | 3.82E-08 | 0.000238 | 8.39E-06 | 2.67E-07 | 0.010012992 | NA |
| chr5 | 134845015 | 134846459 | 1445 | 17 | 3.86E-08 | 8.95E-07 | 3.94E-05 | 7.57E-08 | -0.00149418 | DDX46, C5orf24 |
| chr20 | 62710665 | 62711283 | 619 | 3 | 3.98E-08 | 3.15E-05 | 5.85E-06 | 1.31E-07 | -0.018718059 | NTSR1 |
| chr5 | 174617516 | 174618597 | 1082 | 8 | 4.02E-08 | 1.17E-05 | 2.72E-06 | 1.44E-07 | -0.004014945 | AC108112.1 |
| chr9 | 69820617 | 69821141 | 525 | 9 | 4.05E-08 | 7.17E-05 | 0.00113 | 2.51E-07 | -0.015550114 | C9orf135, C9orf135-DT |
| chr3 | 58049305 | 58049934 | 630 | 3 | 4.06E-08 | 1.15E-09 | 7.34E-05 | 4.33E-09 | -0.015267173 | FLNB |
| chr4 | 77075091 | 77075687 | 597 | 3 | 4.07E-08 | 8.65E-07 | 4.16E-05 | 6.58E-07 | -3.67E-05 | CCNI |
| chr16 | 10938539 | 10939813 | 1275 | 5 | 4.07E-08 | 6.78E-06 | 6.92E-06 | 7.70E-07 | -0.004869943 | CIITA, DEXI |
| chr10 | 13345198 | 13346567 | 1370 | 8 | 4.11E-08 | 5.34E-10 | 0.00103 | 3.04E-09 | -0.015921743 | SEPHS1 |
| chr6 | 106077618 | 106078221 | 604 | 2 | 4.12E-08 | 2.72E-07 | 2.45E-06 | 8.60E-08 | -0.00569429 | PRDM1, ATG5 |
| chr19 | 1244376 | 1246542 | 2167 | 8 | 4.13E-08 | 5.11E-09 | 3.16E-05 | 2.25E-11 | -0.009109556 | ATP5F1D |
| chr22 | 44025688 | 44026902 | 1215 | 10 | 4.17E-08 | 8.53E-06 | 0.00057 | 8.82E-08 | -0.007822159 | PARVB |
| chr7 | 1843361 | 1844756 | 1396 | 8 | 4.34E-08 | 6.41E-11 | 0.00404 | 1.91E-09 | -0.024379068 | AC104129.1, MAD1L1, AC069288.1, MIR4655 |
| chr1 | 36124343 | 36124880 | 538 | 2 | 4.34E-08 | 1.79E-07 | 7.87E-06 | 1.34E-07 | -0.011875873 | COL8A2 |
| chr12 | 113243861 | 113244510 | 650 | 4 | 4.44E-08 | 7.79E-06 | 4.75E-06 | 3.61E-07 | -0.006948056 | TPCN1 |
| chr1 | 64343317 | 64343852 | 536 | 2 | 4.49E-08 | 3.53E-07 | 2.27E-06 | 9.56E-08 | -0.018504759 | NA |
| chr16 | 67525538 | 67526653 | 1116 | 3 | 4.50E-08 | 6.18E-09 | 2.51E-06 | 2.04E-09 | -0.006470769 | RIPOR1, AC027682.6 |
| chr22 | 23566988 | 23567032 | 45 | 3 | 4.50E-08 | 6.94E-09 | 6.13E-05 | 1.91E-08 | -0.020367012 | NA |
| chr17 | 40541804 | 40542757 | 954 | 5 | 4.51E-08 | 6.57E-06 | 0.00011 | 2.75E-07 | -0.008670012 | NA |
| chr17 | 29166186 | 29167277 | 1092 | 6 | 4.52E-08 | 1.36E-05 | 8.69E-06 | 6.09E-08 | -0.002392787 | MYO18A |
| chr9 | 135504839 | 135505629 | 791 | 3 | 4.61E-08 | 2.47E-06 | 8.03E-05 | 4.26E-07 | -0.006468729 | AL161452.1 |
| chr20 | 50916375 | 50917194 | 820 | 3 | 4.61E-08 | 2.52E-07 | 1.21E-06 | 1.10E-08 | -0.009382581 | ADNP |
| chr17 | 7589483 | 7590833 | 1351 | 12 | 4.64E-08 | 3.85E-06 | 0.00061 | 1.63E-08 | -0.004009245 | MPDU1, SOX15 |
| chr10 | 92045141 | 92046113 | 973 | 8 | 4.65E-08 | 1.67E-10 | 0.00234 | 3.04E-09 | -0.01080296 | NA |
| chr6 | 37648634 | 37650738 | 2105 | 14 | 4.65E-08 | 3.26E-14 | 0.00266 | 3.29E-12 | 0.041187638 | MDGA1 |
| chr22 | 42940043 | 42940428 | 386 | 3 | 4.76E-08 | 1.16E-07 | 4.15E-06 | 4.09E-08 | -0.003916998 | PACSIN2 |
| chr16 | 2175169 | 2176394 | 1226 | 13 | 4.86E-08 | 1.54E-06 | 0.00035 | 4.93E-08 | -0.001165903 | TRAF7 |
| chr5 | 81232611 | 81233521 | 911 | 12 | 4.90E-08 | 1.11E-10 | 0.0037 | 3.00E-09 | -0.007439484 | CKMT2, CKMT2-AS1 |
| chr16 | 57797833 | 57798848 | 1016 | 11 | 4.90E-08 | 4.27E-09 | 0.00319 | 9.87E-09 | -0.010762046 | AC092118.1, KIFC3 |
| chr12 | 117967860 | 117968235 | 376 | 4 | 4.92E-08 | 1.77E-06 | 1.33E-05 | 2.92E-07 | 0.011243144 | KSR2 |
| chr6 | 43020707 | 43021539 | 833 | 6 | 4.92E-08 | 7.66E-05 | 0.00011 | 1.63E-05 | -0.002347422 | KLHDC3 |
| chr14 | 103704153 | 103706962 | 2810 | 11 | 4.95E-08 | 8.11E-08 | 4.13E-05 | 9.10E-13 | -0.008330796 | KLC1, XRCC3 |
| chr13 | 36214325 | 36215544 | 1220 | 10 | 4.99E-08 | 1.54E-05 | 4.42E-05 | 2.57E-07 | -0.002587896 | SOHLH2, CCDC169-SOHLH2 |
| chr3 | 120450936 | 120452229 | 1294 | 8 | 5.11E-08 | 1.67E-07 | 0.00018 | 3.35E-08 | -0.006139417 | AC126182.3, FSTL1 |
| chr14 | 63203993 | 63205019 | 1027 | 13 | 5.12E-08 | 6.51E-08 | 0.00027 | 1.58E-08 | 0.019841561 | RHOJ |
| chr13 | 87673989 | 87674663 | 675 | 4 | 5.23E-08 | 7.72E-06 | 0.0001 | 5.44E-07 | 0.005761452 | SLITRK5, MIR4500HG |
| chr17 | 50266033 | 50267003 | 971 | 4 | 5.27E-08 | 5.80E-06 | 4.55E-06 | 3.05E-07 | -0.008616814 | NA |
| chr22 | 49823171 | 49824705 | 1535 | 11 | 5.28E-08 | 3.08E-11 | 0.00317 | 3.67E-10 | -0.005448361 | BRD1 |
| chr17 | 38572446 | 38573775 | 1330 | 6 | 5.33E-08 | 0.000106 | 5.12E-06 | 7.82E-07 | -0.006443837 | SRCIN1 |
| chr12 | 120266915 | 120268212 | 1298 | 3 | 5.33E-08 | 1.33E-07 | 1.35E-05 | 7.78E-08 | -0.012796036 | NA |
| chr8 | 10525411 | 10526253 | 843 | 5 | 5.40E-08 | 1.89E-05 | 1.83E-05 | 2.16E-07 | -0.00357248 | PRSS55, PRSS51 |
| chr5 | 33891788 | 33892600 | 813 | 10 | 5.41E-08 | 0.000414 | 5.16E-06 | 3.49E-07 | -0.002339899 | ADAMTS12 |
| chr1 | 156579741 | 156581115 | 1375 | 4 | 5.41E-08 | 7.28E-06 | 2.18E-05 | 2.18E-06 | -0.002212414 | TTC24 |
| chr1 | 46131646 | 46132784 | 1139 | 11 | 5.45E-08 | 4.06E-07 | 0.00047 | 9.10E-08 | 0.00305687 | PIK3R3, P3R3URF-PIK3R3 |
| chr10 | 133166799 | 133167482 | 684 | 7 | 5.49E-08 | 2.61E-07 | 0.00025 | 1.27E-08 | -0.00389373 | KNDC1 |
| chr5 | 1768612 | 1769916 | 1305 | 5 | 5.49E-08 | 0.000304 | 2.08E-05 | 1.01E-06 | -0.003758991 | NA |
| chr21 | 34890990 | 34891511 | 522 | 4 | 5.51E-08 | 5.10E-09 | 0.0003 | 2.53E-08 | -0.028682142 | RUNX1 |
| chr2 | 46697687 | 46698242 | 556 | 5 | 5.64E-08 | 0.000225 | 3.61E-06 | 5.88E-07 | -0.004077062 | NA |
| chr2 | 241509128 | 241510350 | 1223 | 12 | 5.67E-08 | 1.13E-05 | 0.00016 | 2.44E-07 | -0.004545104 | STK25 |
| chr9 | 31335755 | 31335770 | 16 | 2 | 5.73E-08 | 5.84E-09 | 2.35E-05 | 1.16E-08 | -0.032471303 | NA |
| chr11 | 1006985 | 1012101 | 5117 | 28 | 5.74E-08 | 2.96E-12 | 0.00045 | 3.27E-15 | -0.003324597 | AP2A2 |
| chr17 | 6474378 | 6475319 | 942 | 5 | 5.77E-08 | 3.97E-05 | 3.50E-05 | 2.03E-07 | 0.000309853 | PITPNM3 |
| chr12 | 131946906 | 131947482 | 577 | 2 | 5.95E-08 | 1.60E-06 | 2.06E-06 | 2.18E-07 | -0.009904953 | NA |
| chr13 | 111370713 | 111371353 | 641 | 2 | 5.95E-08 | 1.52E-06 | 2.19E-06 | 2.24E-07 | -0.022540299 | NA |
| chr5 | 151568745 | 151568945 | 201 | 2 | 5.98E-08 | 1.89E-07 | 3.75E-05 | 2.76E-07 | -0.006582001 | FAT2 |
| chr19 | 19489347 | 19490656 | 1310 | 6 | 6.00E-08 | 4.59E-06 | 3.62E-05 | 4.59E-07 | -0.005900578 | GATAD2A |
| chr1 | 2318457 | 2319421 | 965 | 6 | 6.04E-08 | 6.90E-07 | 2.33E-05 | 1.01E-07 | -0.005494212 | AL590822.3 |
| chr2 | 130354946 | 130355813 | 868 | 5 | 6.11E-08 | 2.55E-05 | 9.59E-05 | 1.54E-07 | -0.009318344 | NA |
| chr2 | 174012629 | 174013385 | 757 | 5 | 6.12E-08 | 3.25E-08 | 6.75E-05 | 6.30E-08 | -0.013318767 | AC016737.2 |
| chr1 | 116167148 | 116168322 | 1175 | 6 | 6.25E-08 | 8.49E-07 | 0.00059 | 2.16E-07 | -0.013209214 | NA |
| chr6 | 15243692 | 15244647 | 956 | 5 | 6.29E-08 | 2.00E-07 | 4.00E-05 | 1.77E-07 | -0.009723853 | AL136162.1 |
| chr2 | 10140493 | 10142435 | 1943 | 9 | 6.40E-08 | 6.43E-07 | 1.47E-05 | 5.99E-09 | -0.003089134 | RRM2 |
| chr3 | 129560586 | 129561297 | 712 | 4 | 6.40E-08 | 3.60E-06 | 5.84E-05 | 1.63E-07 | -0.011842437 | PLXND1 |
| chr17 | 7613542 | 7614379 | 838 | 13 | 6.44E-08 | 0.000149 | 0.00036 | 5.60E-07 | -0.0051896 | SHBG, FXR2 |
| chr22 | 49958736 | 49959709 | 974 | 4 | 6.48E-08 | 2.50E-05 | 2.41E-05 | 9.84E-07 | -0.004336843 | NA |
| chr13 | 113022624 | 113024710 | 2087 | 12 | 6.52E-08 | 4.41E-11 | 0.00081 | 1.34E-10 | 0.004250281 | MCF2L |
| chr17 | 9072573 | 9073203 | 631 | 3 | 6.57E-08 | 3.07E-07 | 5.42E-05 | 1.57E-07 | -0.008081078 | NTN1 |
| chr7 | 73978367 | 73979495 | 1129 | 7 | 6.58E-08 | 3.76E-06 | 0.00015 | 1.33E-07 | -0.006592024 | NA |
| chr11 | 976950 | 977562 | 613 | 5 | 6.61E-08 | 0.000254 | 4.79E-05 | 5.91E-07 | -0.007236887 | AP2A2 |
| chr2 | 95053237 | 95053647 | 411 | 5 | 6.63E-08 | 1.39E-08 | 0.00047 | 1.83E-08 | -0.01133567 | MAL |
| chr22 | 24583286 | 24583996 | 711 | 11 | 6.69E-08 | 7.70E-08 | 0.00246 | 2.04E-08 | -0.004134917 | AP000356.5 |
| chr8 | 106617905 | 106618031 | 127 | 3 | 6.76E-08 | 3.43E-09 | 0.00014 | 1.27E-08 | -0.040461682 | OXR1, AC090579.1 |

| | | | | | | | | | | |
|---|---|---|---|---|---|---|---|---|---|---|
| chr9 | 127877460 | 127878507 | 1048 | 11 | 6.77E-08 | 2.23E-07 | 0.00201 | 7.12E-08 | -0.004339493 | AK1, AL157935.2 |
| chr16 | 30673716 | 30674631 | 916 | 3 | 6.86E-08 | 2.16E-06 | 5.60E-06 | 2.44E-07 | -0.009992513 | NA |
| chr11 | 66677912 | 66678473 | 562 | 10 | 6.90E-08 | 1.45E-06 | 7.61E-05 | 2.23E-07 | 0.001417464 | NA |
| chr1 | 154867337 | 154869504 | 2168 | 7 | 6.98E-08 | 1.40E-10 | 0.00021 | 5.68E-10 | -0.03780539 | KCNN3 |
| chr12 | 121229052 | 121229777 | 726 | 5 | 6.98E-08 | 4.37E-05 | 1.61E-05 | 1.53E-07 | -0.002824844 | P2RX4 |
| chr3 | 10028406 | 10028611 | 206 | 2 | 7.08E-08 | 8.28E-06 | 4.43E-07 | 1.19E-07 | -0.007905518 | FANCD2 |
| chr9 | 75088703 | 75089621 | 919 | 5 | 7.11E-08 | 2.63E-06 | 5.89E-05 | 6.01E-08 | 0.004873532 | OSTF1 |
| chr17 | 19534378 | 19534940 | 563 | 6 | 7.11E-08 | 1.71E-05 | 8.07E-05 | 1.55E-07 | -0.003995723 | SLC47A1 |
| chr1 | 47687535 | 47688450 | 916 | 5 | 7.26E-08 | 5.73E-08 | 0.00015 | 4.21E-08 | -0.011976369 | NA |
| chr7 | 56078409 | 56079564 | 1156 | 3 | 7.28E-08 | 3.40E-09 | 2.52E-05 | 5.42E-09 | -0.012400042 | SUMF2 |
| chr11 | 2383154 | 2384605 | 1452 | 5 | 7.32E-08 | 4.73E-08 | 2.67E-05 | 3.14E-08 | -0.007692918 | CD81 |
| chr14 | 68363583 | 68364381 | 799 | 5 | 7.35E-08 | 1.90E-07 | 0.0004 | 1.25E-07 | -0.014936886 | RAD51B |
| chr3 | 50450228 | 50450855 | 628 | 5 | 7.47E-08 | 1.02E-07 | 7.37E-05 | 1.47E-07 | -0.02057985 | CACNA2D2 |
| chr1 | 47190189 | 47191181 | 993 | 6 | 7.49E-08 | 2.22E-08 | 0.00064 | 3.29E-08 | -0.00917893 | PDZK1IP1 |
| chr20 | 63877587 | 63877811 | 225 | 2 | 7.54E-08 | 3.19E-08 | 7.11E-06 | 3.44E-08 | -0.009942562 | TPD52L2 |
| chr11 | 836653 | 838487 | 1835 | 7 | 7.55E-08 | 4.21E-08 | 0.00026 | 2.90E-08 | -0.006726608 | CD151, POLR2L |
| chr6 | 28526419 | 28527107 | 689 | 6 | 7.55E-08 | 6.71E-06 | 0.00019 | 2.09E-07 | -0.009208001 | GPX5, GPX6 |
| chr14 | 105091752 | 105092828 | 1077 | 5 | 7.55E-08 | 4.71E-06 | 1.95E-05 | 2.04E-08 | -0.009556502 | NA |
| chr20 | 3084697 | 3085368 | 672 | 10 | 7.60E-08 | 2.68E-09 | 0.0017 | 1.17E-08 | -0.010779093 | AVP |
| chr12 | 121452360 | 121453545 | 1186 | 8 | 7.60E-08 | 9.43E-11 | 0.00114 | 1.50E-09 | -0.007472892 | KDM2B |
| chr1 | 197775386 | 197776165 | 780 | 13 | 7.78E-08 | 0.000129 | 1.80E-05 | 5.25E-06 | -0.00255394 | DENND1B |
| chr2 | 42857289 | 42857305 | 17 | 2 | 7.81E-08 | 1.13E-08 | 4.00E-05 | 2.29E-08 | -0.018526274 | NA |
| chr2 | 37323881 | 37324748 | 868 | 8 | 7.86E-08 | 2.35E-05 | 0.00015 | 5.57E-07 | 0.000893102 | PRKD3 |
| chr2 | 238644242 | 238644765 | 524 | 3 | 7.94E-08 | 7.32E-08 | 6.84E-05 | 5.22E-08 | -0.084723121 | LINC01937 |
| chr15 | 44676457 | 44677341 | 885 | 7 | 8.04E-08 | 7.55E-08 | 2.57E-05 | 5.42E-08 | -0.00578028 | PATL2 |
| chr2 | 9472993 | 9474615 | 1623 | 15 | 8.05E-08 | 2.57E-09 | 0.00748 | 7.18E-09 | -0.007784831 | CPSF3, IAH1 |
| chr6 | 31945498 | 31946488 | 991 | 9 | 8.15E-08 | 1.91E-05 | 0.00085 | 3.79E-07 | -0.007129714 | C2, AL645922.1, CFB |
| chr5 | 71718582 | 71719605 | 1024 | 15 | 8.24E-08 | 2.54E-07 | 0.00077 | 1.34E-08 | -0.000445568 | CARTPT |
| chr17 | 19364810 | 19365156 | 347 | 3 | 8.36E-08 | 6.12E-05 | 1.67E-06 | 2.01E-07 | 0.0049124 | AC124066.1, B9D1 |
| chr1 | 2968520 | 2968679 | 160 | 4 | 8.48E-08 | 4.40E-09 | 0.0002 | 1.16E-08 | -0.023516639 | AL589702.1 |
| chr16 | 47004386 | 47004908 | 523 | 2 | 8.55E-08 | 0.000396 | 2.38E-06 | 1.84E-06 | -0.003590411 | AC018845.1 |
| chr14 | 104688162 | 104688990 | 829 | 8 | 8.61E-08 | 4.38E-07 | 0.00114 | 1.12E-07 | -0.007566627 | INF2 |
| chr2 | 176149739 | 176151134 | 1396 | 16 | 8.82E-08 | 7.10E-07 | 0.00051 | 1.45E-07 | -0.009750465 | HOXD3, MIR10B |
| chr2 | 27496677 | 27497573 | 897 | 4 | 8.83E-08 | 1.71E-07 | 0.00024 | 9.17E-08 | -0.009674582 | GCKR |
| chr1 | 24930538 | 24932188 | 1651 | 22 | 8.92E-08 | 1.38E-06 | 0.00153 | 9.44E-09 | -0.002545702 | RUNX3 |
| chr3 | 48595051 | 48596008 | 958 | 12 | 9.02E-08 | 9.74E-06 | 0.00168 | 1.04E-07 | -0.006731693 | COL7A1 |
| chr4 | 17097267 | 17097640 | 374 | 2 | 9.04E-08 | 0.000127 | 6.68E-07 | 3.89E-07 | -0.008864015 | NA |
| chr19 | 39205161 | 39205494 | 334 | 2 | 9.04E-08 | 0.000137 | 3.65E-06 | 2.45E-06 | -0.011562705 | NA |
| chr1 | 170662190 | 170663282 | 1093 | 6 | 9.05E-08 | 0.201305 | 1.58E-06 | 1.60E-05 | 0.005234856 | PRRX1 |
| chr3 | 53882617 | 53883516 | 900 | 5 | 9.05E-08 | 0.000205 | 3.65E-06 | 5.46E-07 | -0.004804232 | NA |
| chr16 | 29825406 | 29825946 | 541 | 3 | 9.10E-08 | 1.92E-05 | 7.21E-05 | 3.87E-07 | -0.017849803 | AC120114.4, MVP |
| chr6 | 41967027 | 41967748 | 722 | 2 | 9.36E-08 | 8.12E-08 | 6.71E-06 | 6.71E-08 | -0.015273606 | CCND3 |
| chr1 | 151164798 | 151165773 | 976 | 6 | 9.40E-08 | 2.18E-07 | 0.00011 | 3.10E-07 | -0.010826086 | SCNM1, LYSMD1 |
| chr19 | 6271949 | 6272603 | 655 | 5 | 9.41E-08 | 4.69E-07 | 7.79E-05 | 2.54E-07 | -0.009356642 | MLLT1 |
| chr19 | 37251579 | 37252054 | 476 | 7 | 9.46E-08 | 1.12E-09 | 0.00183 | 8.99E-09 | -0.010740752 | LINC01535, AC016590.1 |
| chr9 | 120843841 | 120844750 | 910 | 5 | 9.57E-08 | 1.90E-05 | 1.09E-05 | 1.21E-06 | -0.012474363 | CUTALP |
| chr1 | 34866048 | 34866884 | 837 | 6 | 9.60E-08 | 4.02E-08 | 5.25E-05 | 4.37E-08 | -0.001320558 | DLGAP3 |
| chr4 | 80196493 | 80198319 | 1827 | 12 | 9.64E-08 | 2.72E-12 | 0.00904 | 3.42E-10 | -0.027126807 | PRDM8, AC021127.1 |
| chr8 | 20972535 | 20973775 | 1241 | 6 | 9.67E-08 | 3.68E-08 | 2.22E-05 | 4.06E-10 | -0.005968241 | LINC02153, AC021613.1 |
| chr17 | 39636485 | 39637067 | 583 | 10 | 9.83E-08 | 0.002239 | 0.00023 | 1.85E-05 | -0.003189127 | PPP1R1B |
| chr7 | 141664619 | 141665510 | 892 | 3 | 9.83E-08 | 5.40E-05 | 1.90E-06 | 1.02E-06 | -0.004570879 | DENND11 |
| chr11 | 65779517 | 65781970 | 2454 | 16 | 9.90E-08 | 9.77E-08 | 4.42E-05 | 4.27E-10 | -0.002744962 | AP5B1 |
| chr13 | 77740330 | 77743027 | 2698 | 14 | 1.01E-07 | 9.90E-10 | 0.00017 | 3.62E-11 | -0.009271917 | SLAIN1 |
| chr14 | 68963552 | 68964912 | 1361 | 3 | 1.01E-07 | 2.42E-06 | 7.78E-06 | 2.25E-07 | -0.008782082 | ACTN1 |
| chr1 | 11847929 | 11848889 | 961 | 9 | 1.02E-07 | 2.46E-06 | 0.00121 | 7.37E-08 | -0.012676219 | CLCN6, NPPA |
| chr19 | 49889660 | 49889997 | 338 | 2 | 1.03E-07 | 5.06E-06 | 1.33E-05 | 1.82E-06 | -0.022195278 | IL4I1 |
| chr20 | 63750985 | 63751204 | 220 | 2 | 1.03E-07 | 1.01E-07 | 8.60E-05 | 1.99E-07 | -0.031089054 | ZBTB46 |
| chr5 | 141149859 | 141150886 | 1028 | 13 | 1.04E-07 | 3.12E-07 | 0.0006 | 7.10E-08 | 0.016081625 | PCDHB6, AC244517.2, AC244517.11 |
| chr3 | 193852467 | 193852621 | 155 | 3 | 1.04E-07 | 7.77E-09 | 7.54E-05 | 1.99E-08 | -0.011708336 | NA |
| chr14 | 91867421 | 91867969 | 549 | 4 | 1.05E-07 | 1.52E-06 | 2.86E-06 | 1.29E-07 | -0.00254904 | TC2N |
| chr2 | 162768556 | 162768824 | 269 | 4 | 1.05E-07 | 7.53E-09 | 0.00013 | 1.58E-08 | -0.016903299 | KCNH7 |
| chr1 | 19390011 | 19391451 | 1441 | 6 | 1.06E-07 | 9.67E-07 | 0.0001 | 2.61E-07 | -0.002105752 | CAPZB |
| chr7 | 1473099 | 1474987 | 1889 | 10 | 1.07E-07 | 2.10E-09 | 0.00101 | 5.86E-09 | -0.005396239 | INTS1 |
| chr10 | 101060570 | 101063116 | 2547 | 21 | 1.07E-07 | 6.59E-08 | 0.00095 | 3.36E-10 | -0.003049408 | KAZALD1, AL133215.2 |
| chr3 | 48503946 | 48505219 | 1274 | 12 | 1.09E-07 | 6.27E-09 | 0.00046 | 1.06E-08 | -0.004493244 | SHISA5 |
| chr16 | 69106891 | 69107575 | 685 | 11 | 1.10E-07 | 7.62E-06 | 0.00074 | 2.54E-07 | -0.005316719 | HAS3 |
| chr8 | 123001115 | 123002238 | 1124 | 3 | 1.11E-07 | 1.42E-06 | 6.29E-05 | 2.99E-07 | -0.015993989 | NA |
| chr18 | 46656884 | 46658400 | 1517 | 12 | 1.11E-07 | 2.57E-10 | 0.00114 | 7.57E-10 | -0.006262636 | LOXHD1 |
| chr8 | 22563516 | 22563860 | 345 | 2 | 1.12E-07 | 0.000471 | 2.93E-06 | 2.37E-06 | -0.005714413 | SORBS3 |
| chr16 | 48365168 | 48366652 | 1485 | 12 | 1.12E-07 | 5.59E-06 | 1.55E-05 | 7.99E-08 | -0.000431593 | SIAH1 |
| chr2 | 7031355 | 7032166 | 812 | 7 | 1.12E-07 | 1.37E-06 | 8.70E-06 | 2.49E-07 | -0.007381231 | RNF144A |
| chr7 | 90211510 | 90212729 | 1220 | 14 | 1.13E-07 | 2.98E-07 | 0.00024 | 2.49E-07 | -0.00239042 | STEAP2, STEAP2-AS1 |
| chr18 | 51136810 | 51138061 | 1252 | 4 | 1.14E-07 | 1.05E-06 | 1.31E-05 | 3.55E-07 | -0.000160123 | NA |
| chr19 | 37931978 | 37932329 | 352 | 5 | 1.14E-07 | 3.20E-09 | 0.00054 | 1.80E-08 | -0.011808625 | SIPA1L3 |
| chr11 | 62508132 | 62508434 | 303 | 2 | 1.15E-07 | 4.93E-07 | 1.83E-06 | 9.56E-08 | -0.021530472 | AHNAK |
| chr16 | 56404413 | 56404424 | 12 | 3 | 1.16E-07 | 0.002028 | 2.42E-07 | 4.58E-07 | -0.003163551 | AMFR |
| chr20 | 10637057 | 10637205 | 149 | 2 | 1.17E-07 | 1.09E-08 | 3.76E-05 | 2.20E-08 | -0.018985667 | NA |
| chr1 | 41918229 | 41919489 | 1261 | 13 | 1.17E-07 | 1.94E-07 | 0.00776 | 1.34E-07 | -0.019452913 | HIVEP3 |
| chr11 | 47233916 | 47234905 | 990 | 4 | 1.19E-07 | 3.97E-08 | 7.75E-05 | 2.80E-08 | -0.008112673 | DDB2 |
| chr17 | 51164157 | 51165808 | 1652 | 11 | 1.19E-07 | 2.63E-07 | 0.0011 | 2.69E-08 | -0.005234151 | NME1-NME2, NME2 |
| chr17 | 40316709 | 40317537 | 829 | 6 | 1.20E-07 | 2.94E-08 | 0.00075 | 4.64E-08 | -0.009595001 | RARA |
| chr1 | 7441560 | 7442166 | 607 | 4 | 1.21E-07 | 1.21E-07 | 0.00016 | 1.22E-07 | -0.012360247 | CAMTA1 |
| chr2 | 218368723 | 218368955 | 233 | 4 | 1.21E-07 | 2.22E-08 | 0.0003 | 6.64E-08 | -0.011833596 | NA |
| chr5 | 36960533 | 36961457 | 925 | 3 | 1.21E-07 | 9.74E-08 | 5.67E-06 | 3.85E-08 | 0.000108365 | NIPBL |
| chr9 | 137435043 | 137436234 | 1192 | 7 | 1.21E-07 | 1.10E-08 | 0.0003 | 2.23E-08 | -0.008275989 | ENTPD8 |
| chr3 | 52403630 | 52404252 | 623 | 4 | 1.23E-07 | 3.77E-06 | 0.00014 | 7.11E-07 | -0.002936772 | BAP1 |
| chr6 | 31622149 | 31624009 | 1861 | 24 | 1.26E-07 | 1.01E-07 | 0.0005 | 8.38E-09 | -0.002741653 | PRRC2A, SNORA38 |
| chr13 | 113364809 | 113365484 | 676 | 4 | 1.27E-07 | 7.00E-05 | 3.99E-06 | 1.48E-06 | -0.000109898 | NA |
| chr7 | 22444879 | 22445263 | 385 | 2 | 1.27E-07 | 1.41E-06 | 5.26E-06 | 4.24E-07 | -0.021568164 | STEAP1B |
| chr1 | 52920946 | 52921531 | 586 | 2 | 1.28E-07 | 1.28E-05 | 5.79E-05 | 8.62E-06 | -0.012378828 | ECHDC2 |
| chr6 | 53544570 | 53546057 | 1488 | 13 | 1.28E-07 | 3.32E-07 | 0.00027 | 2.69E-08 | 0.00027929 | GCLC |
| chr3 | 42907134 | 42908598 | 1465 | 5 | 1.29E-07 | 3.44E-05 | 1.22E-05 | 1.01E-06 | -0.005670204 | AC099329.2, AC092042.3, KRBOX1, ZNF662 |
| chr1 | 45508269 | 45509054 | 786 | 3 | 1.29E-07 | 2.33E-08 | 5.98E-05 | 5.42E-08 | -0.007018257 | MMACHC |
| chr17 | 32943336 | 32943997 | 662 | 3 | 1.30E-07 | 5.63E-08 | 4.58E-05 | 6.09E-08 | -0.00675416 | TMEM98 |
| chr6 | 111551958 | 111553257 | 1300 | 4 | 1.30E-07 | 2.44E-07 | 0.00015 | 1.53E-07 | -0.011186462 | TRAF3IP2-AS1 |
| chr12 | 6384355 | 6385644 | 1290 | 8 | 1.30E-07 | 1.42E-06 | 0.00014 | 1.50E-07 | -0.007364656 | LTBR |
| chr2 | 85693751 | 85695354 | 1604 | 8 | 1.31E-07 | 1.15E-06 | 7.99E-05 | 2.81E-09 | -0.007601356 | GNLY |
| chr19 | 17304168 | 17304414 | 247 | 4 | 1.32E-07 | 3.56E-07 | 0.00071 | 3.85E-07 | -0.009527761 | MRPL34, ABHD8 |
| chr12 | 6199488 | 6200125 | 638 | 6 | 1.32E-07 | 0.000107 | 8.23E-05 | 1.70E-06 | -0.00907964 | CD9 |
| chr7 | 2701702 | 2702418 | 717 | 4 | 1.32E-07 | 1.42E-06 | 3.56E-05 | 9.23E-07 | -0.006863407 | AMZ1 |
| chr1 | 33048128 | 33049257 | 1130 | 3 | 1.33E-07 | 0.00017 | 8.87E-06 | 4.64E-06 | -0.008572299 | AK2 |
| chr12 | 6332502 | 6334116 | 1615 | 12 | 1.34E-07 | 1.84E-08 | 0.00283 | 7.77E-08 | 0.010764897 | TNFRSF1A |
| chr16 | 57637241 | 57640081 | 2841 | 15 | 1.34E-07 | 1.20E-12 | 0.00032 | 1.50E-12 | -0.008907643 | ADGRG1 |
| chr16 | 67189830 | 67190438 | 609 | 4 | 1.34E-07 | 3.28E-06 | 4.45E-05 | 1.03E-06 | -0.00796208 | EXOC3L1 |
| chr19 | 799621 | 800532 | 912 | 4 | 1.34E-07 | 6.29E-08 | 6.19E-05 | 9.24E-08 | -0.014291748 | PTBP1 |
| chr10 | 14478612 | 14478742 | 131 | 2 | 1.35E-07 | 2.31E-07 | 1.57E-06 | 5.26E-08 | -0.020256201 | NA |
| chr11 | 69088355 | 69089483 | 1129 | 7 | 1.35E-07 | 2.82E-07 | 0.00018 | 2.86E-08 | -0.007761305 | TPCN2, AP003071.5 |
| chr17 | 3956683 | 3958340 | 1658 | 7 | 1.35E-07 | 1.04E-06 | 3.61E-05 | 1.01E-07 | -0.005749052 | ATP2A3 |
| chr17 | 57518357 | 57519427 | 1071 | 6 | 1.35E-07 | 4.83E-05 | 5.12E-05 | 1.50E-06 | -0.005752195 | MSI2 |
| chr6 | 167705375 | 167705646 | 272 | 4 | 1.35E-07 | 8.29E-09 | 0.00029 | 2.40E-08 | -0.031863894 | NA |
| chr14 | 35385769 | 35386796 | 1028 | 5 | 1.36E-07 | 1.97E-06 | 2.94E-05 | 2.45E-07 | -0.006265907 | NA |
| chr10 | 62818709 | 62819886 | 1178 | 9 | 1.37E-07 | 1.94E-08 | 0.00066 | 1.63E-08 | -0.01196305 | EGR2 |
| chr3 | 46405507 | 46407211 | 1705 | 10 | 1.40E-07 | 2.15E-08 | 0.00024 | 9.46E-09 | -0.007242808 | CCRL2, CCR5AS |
| chr6 | 31884649 | 31887932 | 3284 | 44 | 1.40E-07 | 1.21E-08 | 0.00042 | 1.99E-12 | -0.001550966 | EHMT2 |
| chr9 | 98086992 | 98087418 | 427 | 2 | 1.41E-07 | 2.89E-07 | 0.00013 | 5.42E-07 | -0.010924008 | TRIM14 |
| chr19 | 39248575 | 39249861 | 1287 | 8 | 1.42E-07 | 0.000573 | 6.50E-05 | 2.70E-07 | -0.004688415 | IFNL4 |
| chr3 | 14508928 | 14509506 | 579 | 18 | 1.43E-07 | 2.12E-07 | 0.00385 | 1.28E-08 | -0.004910652 | GRIP2 |
| chr3 | 195155720 | 195156640 | 921 | 4 | 1.43E-07 | 0.000186 | 6.00E-06 | 9.03E-07 | 0.001394615 | XXYLT1 |
| chr20 | 62988905 | 62989623 | 719 | 3 | 1.43E-07 | 1.30E-07 | 2.58E-05 | 9.82E-08 | -0.018713088 | NA |
| chr12 | 122226425 | 122227834 | 1410 | 15 | 1.45E-07 | 5.41E-08 | 0.00211 | 3.11E-09 | -0.004447553 | AC048338.2, DIABLO, AC048338.1 |
| chr22 | 26391500 | 26392041 | 542 | 2 | 1.46E-07 | 1.38E-05 | 7.23E-06 | 1.95E-06 | -0.001446014 | NA |
| chr1 | 41496396 | 41497107 | 712 | 5 | 1.47E-07 | 0.000158 | 2.55E-06 | 2.26E-06 | -0.005188102 | NA |
| chr1 | 153967006 | 153969428 | 2423 | 22 | 1.48E-07 | 5.89E-07 | 0.00091 | 1.97E-10 | -0.000724619 | CREB3L4, SLC39A1, AL358472.7 |
| chr21 | 42836922 | 42837246 | 325 | 4 | 1.50E-07 | 3.93E-07 | 8.40E-05 | 1.11E-07 | -0.007606452 | NA |
| chr6 | 41426418 | 41427492 | 1075 | 6 | 1.51E-07 | 1.28E-07 | 0.00054 | 9.56E-08 | -0.009344982 | NA |
| chr16 | 1030144 | 1030808 | 665 | 5 | 1.53E-07 | 2.27E-08 | 7.89E-05 | 4.84E-08 | -0.011895095 | NA |
| chr2 | 171893453 | 171894735 | 1283 | 13 | 1.54E-07 | 0.000116 | 8.46E-05 | 7.54E-07 | -0.00177354 | SLC25A12 |
| chr12 | 49636826 | 49637783 | 958 | 3 | 1.54E-07 | 2.32E-08 | 6.83E-05 | 3.74E-08 | -0.012352669 | PRPF40B, FMNL3 |
| chr6 | 30882430 | 30883976 | 1547 | 18 | 1.54E-07 | 5.06E-09 | 0.00171 | 1.06E-08 | -0.006168723 | DDR1 |
| chr1 | 234863022 | 234863640 | 619 | 2 | 1.54E-07 | 0.000108 | 3.01E-06 | 1.87E-06 | -0.012552402 | AL391832.4 |
| chr2 | 54662829 | 54663765 | 937 | 6 | 1.55E-07 | 1.47E-08 | 0.00036 | 4.22E-08 | -0.003029972 | SPTBN1, AC093110.1 |
| chr14 | 93041065 | 93041537 | 473 | 3 | 1.56E-07 | 7.06E-07 | 5.39E-05 | 4.11E-07 | -0.012124465 | ITPK1 |
| chr2 | 217978471 | 217979479 | 1009 | 7 | 1.57E-07 | 6.81E-08 | 0.00166 | 1.43E-08 | -0.018368345 | TNS1-AS1, TNS1 |
| chr2 | 60495178 | 60495667 | 490 | 4 | 1.58E-07 | 6.67E-05 | 1.00E-05 | 5.79E-07 | -0.007775364 | BCL11A |
| chr14 | 51359909 | 51360700 | 792 | 5 | 1.58E-07 | 0.000142 | 1.28E-05 | 6.53E-08 | -0.005734948 | LINC00640, AL358332.1 |
| chr22 | 17872278 | 17872737 | 460 | 4 | 1.59E-07 | 4.63E-07 | 1.32E-05 | 1.51E-07 | -0.00857679 | MICAL3 |
| chr1 | 101847052 | 101847740 | 689 | 4 | 1.60E-07 | 3.55E-07 | 0.00012 | 2.54E-07 | 0.026321157 | OLFM3 |
| chr1 | 159825472 | 159826868 | 1397 | 10 | 1.60E-07 | 1.56E-09 | 0.00093 | 2.35E-09 | -0.009291376 | SLAMF8 |
| chr2 | 88170654 | 88172155 | 1502 | 7 | 1.61E-07 | 4.14E-08 | 0.0004 | 3.27E-08 | -0.01068149 | THNSL2 |
| chr14 | 92952040 | 92953209 | 1170 | 8 | 1.61E-07 | 4.82E-07 | 2.54E-05 | 4.79E-08 | -0.006068607 | ITPK1 |
| chr7 | 157851468 | 157851973 | 506 | 5 | 1.62E-07 | 3.39E-05 | 4.24E-06 | 6.76E-07 | 0.008264413 | PTPRN2 |
| chr7 | 90402035 | 90402349 | 315 | 3 | 1.67E-07 | 0.000822 | 4.56E-05 | 1.86E-06 | -0.012119618 | CLDN12 |
| chr10 | 58511745 | 58512458 | 714 | 4 | 1.67E-07 | 7.20E-06 | 1.47E-05 | 3.46E-07 | -0.014501486 | NA |
| chr20 | 34544523 | 34545461 | 939 | 4 | 1.69E-07 | 2.89E-05 | 2.60E-05 | 5.23E-06 | -0.009955368 | NA |
| chr21 | 38491526 | 38491847 | 322 | 3 | 1.71E-07 | 1.22E-07 | 4.63E-05 | 6.18E-08 | -0.020842711 | ERG |
| chr19 | 4531626 | 4532028 | 403 | 4 | 1.71E-07 | 5.43E-09 | 0.00017 | 2.41E-08 | -0.014737195 | PLIN5, AC011498.4 |
| chr6 | 14116886 | 14117672 | 787 | 7 | 1.72E-07 | 5.21E-07 | 0.00072 | 1.78E-07 | -0.010785056 | CD83 |
| chr2 | 88857429 | 88858416 | 988 | 9 | 1.73E-07 | 2.74E-07 | 0.00047 | 6.98E-08 | -0.005138385 | AC244205.1, IGKC |
| chr16 | 56519449 | 56520802 | 1354 | 15 | 1.74E-07 | 7.16E-08 | 0.00112 | 3.94E-08 | -0.007812387 | BBS2 |
| chr4 | 146242015 | 146242461 | 447 | 2 | 1.75E-07 | 0.00019 | 8.29E-06 | 6.30E-06 | -0.024131819 | AC097372.1 |
| chr1 | 159945779 | 159946682 | 904 | 7 | 1.75E-07 | 0.000112 | 5.93E-05 | 8.57E-07 | -0.006646852 | NA |
| chr7 | 16882075 | 16882797 | 723 | 3 | 1.76E-07 | 8.44E-06 | 1.25E-05 | 7.46E-07 | -0.005753262 | NA |
| chr18 | 76348641 | 76349712 | 1072 | 4 | 1.76E-07 | 7.63E-05 | 7.11E-06 | 6.94E-07 | 0.00451215 | NA |
| chr19 | 42287074 | 42288206 | 1133 | 7 | 1.76E-07 | 4.05E-06 | 0.00013 | 3.54E-07 | -0.003542954 | CIC |
| chr10 | 84244525 | 84245956 | 1432 | 9 | 1.77E-07 | 2.19E-07 | 0.00011 | 4.29E-08 | -0.003841651 | RGR |
| chr18 | 46559196 | 46560082 | 887 | 7 | 1.77E-07 | 4.82E-05 | 2.94E-05 | 1.03E-06 | -0.00431434 | LOXHD1 |
| chr20 | 62692919 | 62693309 | 391 | 2 | 1.77E-07 | 9.29E-06 | 2.28E-06 | 5.83E-07 | -0.026522093 | NA |
| chr19 | 58304854 | 58305356 | 503 | 6 | 1.77E-07 | 9.46E-06 | 3.25E-05 | 8.59E-08 | -0.009237184 | AC020915.4, ZNF8-ERVK3-1, ERVK3-1 |
| chr17 | 75827688 | 75829146 | 1459 | 7 | 1.78E-07 | 4.66E-10 | 0.00086 | 3.92E-09 | -0.017343528 | UNC13D |
| chr13 | 99087128 | 99088134 | 1007 | 6 | 1.80E-07 | 1.38E-06 | 1.66E-05 | 1.41E-07 | 0.001053428 | DOCK9-DT |
| chr1 | 44723354 | 44724847 | 1494 | 11 | 1.80E-07 | 2.43E-07 | 0.00176 | 1.00E-07 | -0.005461373 | ARMH1 |
| chr13 | 113086176 | 113087291 | 1116 | 7 | 1.81E-07 | 7.30E-06 | 2.93E-05 | 7.27E-07 | -0.002572322 | MCF2L |

| | | | | | | | | | | |
|---|---|---|---|---|---|---|---|---|---|---|
| chr11 | 614521 | 615519 | 999 | 5 | 1.82E-07 | 1.41E-06 | 0.00064 | 4.25E-07 | -0.010104782 | IRF7 |
| chr2 | 74148001 | 74148974 | 974 | 9 | 1.87E-07 | 1.52E-05 | 0.00028 | 1.86E-07 | -0.002741083 | BOLA3-AS1 |
| chr12 | 3188532 | 3188637 | 106 | 2 | 1.89E-07 | 2.41E-08 | 1.65E-05 | 3.81E-08 | -0.01177948 | TSPAN9 |
| chr2 | 114661960 | 114662869 | 910 | 8 | 1.90E-07 | 2.39E-07 | 0.00171 | 4.27E-08 | 0.017260353 | DPP10 |
| chr10 | 61661600 | 61663477 | 1878 | 14 | 1.90E-07 | 4.92E-09 | 0.00029 | 6.28E-09 | -0.00598076 | CABCOCO1 |
| chr19 | 18588015 | 18588821 | 807 | 11 | 1.90E-07 | 2.38E-09 | 0.00164 | 1.74E-08 | 0.016903431 | REX1BD, CRLF1 |
| chr1 | 31742099 | 31743102 | 1004 | 6 | 1.92E-07 | 7.63E-08 | 0.00028 | 6.62E-08 | -0.00094469 | ADGRB2 |
| chr11 | 123559867 | 123560454 | 588 | 8 | 1.93E-07 | 7.88E-07 | 0.00137 | 1.16E-07 | -0.012537913 | GRAMD1B |
| chr1 | 180953493 | 180954961 | 1469 | 11 | 1.94E-07 | 6.08E-11 | 0.00628 | 2.64E-09 | -0.019452156 | AL162431.1, KIAA1614-AS1 |
| chr1 | 109273579 | 109274695 | 1117 | 5 | 1.98E-07 | 9.46E-08 | 8.94E-05 | 1.46E-07 | -0.012092425 | CELSR2 |
| chr7 | 101231544 | 101232275 | 732 | 3 | 2.00E-07 | 5.72E-06 | 6.55E-06 | 6.09E-07 | -0.005500999 | CLDN15 |
| chr18 | 47035347 | 47036547 | 1201 | 11 | 2.02E-07 | 3.01E-07 | 0.00127 | 9.44E-08 | 0.007894575 | KATNAL2, ELOA2 |
| chr11 | 65612040 | 65613625 | 1586 | 7 | 2.02E-07 | 1.55E-07 | 0.00259 | 1.73E-07 | -0.009146091 | MAP3K11 |
| chr4 | 47464324 | 47465235 | 912 | 4 | 2.03E-07 | 0.000215 | 8.39E-06 | 7.54E-06 | -0.005319447 | AC107398.4 |
| chr1 | 16158173 | 16158316 | 144 | 2 | 2.04E-07 | 6.30E-07 | 7.67E-05 | 9.52E-07 | -0.011520929 | NA |
| chr17 | 49610626 | 49611316 | 691 | 2 | 2.04E-07 | 1.63E-05 | 1.08E-05 | 2.95E-06 | -0.015763742 | SPOP |
| chr1 | 32361242 | 32363246 | 2005 | 14 | 2.05E-07 | 1.43E-07 | 7.41E-05 | 1.16E-08 | -0.002331096 | TSSK3, FAM229A |
| chr16 | 30488926 | 30489159 | 234 | 2 | 2.05E-07 | 7.54E-08 | 2.76E-05 | 1.17E-07 | -0.019298138 | ITGAL, AC116348.3 |
| chr21 | 44425168 | 44425612 | 445 | 4 | 2.07E-07 | 2.01E-06 | 0.00019 | 3.47E-07 | -0.010653127 | TRPM2, TRPM2-AS |
| chr9 | 127825272 | 127826401 | 1130 | 4 | 2.09E-07 | 6.02E-06 | 3.72E-05 | 2.65E-07 | -0.007918125 | ENG |
| chr3 | 50191438 | 50192593 | 1156 | 4 | 2.10E-07 | 5.78E-07 | 5.40E-05 | 4.98E-08 | -0.007794981 | GNAT1 |
| chr2 | 186486308 | 186487256 | 949 | 7 | 2.10E-07 | 6.70E-05 | 5.35E-05 | 9.12E-07 | 0.002725749 | ZC3H15 |
| chr7 | 2103872 | 2104944 | 1073 | 5 | 2.11E-07 | 3.33E-07 | 0.00069 | 4.10E-08 | -0.015652715 | MAD1L1, AC069288.1 |
| chr5 | 139680087 | 139680891 | 805 | 5 | 2.11E-07 | 0.000314 | 2.44E-05 | 1.10E-05 | -0.003153496 | CXXC5 |
| chr12 | 1628747 | 1631234 | 2488 | 16 | 2.12E-07 | 4.57E-07 | 1.21E-05 | 5.62E-10 | -0.004751217 | WNT5B |
| chr15 | 27714327 | 27714849 | 523 | 3 | 2.13E-07 | 2.35E-07 | 6.50E-06 | 8.74E-08 | -0.006728203 | NA |
| chr8 | 38429807 | 38430634 | 828 | 4 | 2.14E-07 | 4.29E-09 | 0.00026 | 1.57E-08 | -0.01727032 | FGFR1 |
| chr16 | 89915323 | 89916210 | 888 | 6 | 2.18E-07 | 2.68E-09 | 0.00063 | 2.33E-08 | -0.006577741 | AC092143.3, MC1R |
| chr11 | 1838151 | 1839565 | 1415 | 13 | 2.19E-07 | 3.79E-05 | 0.00015 | 4.54E-07 | -0.005497817 | TNNI2 |
| chr3 | 64214123 | 64214856 | 734 | 3 | 2.19E-07 | 1.61E-07 | 5.20E-05 | 1.27E-07 | -0.016322192 | PRICKLE2 |
| chr8 | 96159784 | 96161107 | 1324 | 8 | 2.20E-07 | 1.16E-05 | 7.78E-05 | 5.14E-07 | 0.001394193 | GDF6 |
| chr3 | 133745619 | 133748460 | 2842 | 15 | 2.20E-07 | 1.19E-09 | 0.00072 | 2.36E-11 | -0.006560171 | INHCAP, TF |
| chr6 | 169577298 | 169578291 | 994 | 5 | 2.22E-07 | 3.14E-08 | 0.00063 | 6.26E-08 | -0.018676129 | AL031315.1, WDR27 |
| chr20 | 31812084 | 31812619 | 536 | 3 | 2.23E-07 | 4.19E-05 | 2.40E-05 | 4.14E-07 | -0.016581698 | NA |
| chr7 | 2221266 | 2221871 | 606 | 5 | 2.23E-07 | 0.000317 | 2.86E-05 | 7.66E-07 | -0.00605088 | MAD1L1, AC069288.1 |
| chr17 | 1936696 | 1937914 | 1219 | 4 | 2.23E-07 | 1.27E-08 | 0.00023 | 5.07E-08 | -0.012361976 | RTN4RL1 |
| chr11 | 66559296 | 66560898 | 1603 | 7 | 2.23E-07 | 6.39E-08 | 0.00029 | 6.89E-09 | -0.012048887 | ACTN3, AP002748.2 |
| chr11 | 27486097 | 27486550 | 454 | 3 | 2.23E-07 | 1.07E-05 | 6.54E-05 | 2.79E-07 | -0.011501688 | NA |
| chr1 | 155940732 | 155941570 | 839 | 6 | 2.23E-07 | 2.00E-06 | 3.27E-05 | 1.26E-07 | -0.006980541 | NA |
| chr1 | 227318812 | 227319595 | 784 | 6 | 2.24E-07 | 0.000171 | 3.35E-05 | 3.24E-06 | -0.01539111 | NA |
| chr5 | 171842130 | 171842573 | 444 | 3 | 2.25E-07 | 2.82E-06 | 5.84E-05 | 3.79E-07 | -0.010256607 | NA |
| chr10 | 71773650 | 71774581 | 932 | 9 | 2.25E-07 | 7.51E-07 | 0.00072 | 3.36E-07 | -0.005852196 | CDH23 |
| chr14 | 23366003 | 23366838 | 836 | 5 | 2.26E-07 | 3.51E-06 | 0.00056 | 1.35E-06 | -0.009294406 | NA |
| chr1 | 222711570 | 222712065 | 496 | 3 | 2.28E-07 | 0.000173 | 1.31E-05 | 6.25E-06 | -0.001124769 | AIDA |
| chr7 | 1497852 | 1498970 | 1119 | 3 | 2.28E-07 | 4.25E-08 | 8.15E-05 | 1.00E-07 | -0.004413657 | INTS1 |
| chr13 | 42781378 | 42781980 | 603 | 4 | 2.28E-07 | 0.00064 | 5.12E-05 | 1.20E-06 | -0.013310277 | FAM216B |
| chr1 | 9438095 | 9439946 | 1852 | 5 | 2.29E-07 | 7.78E-11 | 2.53E-05 | 1.52E-10 | -0.009949325 | LINC02606 |
| chr11 | 11569763 | 11570189 | 427 | 4 | 2.30E-07 | 7.38E-06 | 0.00021 | 1.52E-07 | -0.013631083 | AC104031.1, GALNT18 |
| chr12 | 57521593 | 57522628 | 1036 | 8 | 2.30E-07 | 2.01E-06 | 0.00209 | 9.45E-07 | -0.005457252 | MBD6, DDIT3 |
| chr6 | 30041968 | 30043130 | 1163 | 6 | 2.30E-07 | 2.77E-08 | 0.00017 | 7.61E-08 | -0.00704132 | ZNRD1ASP |
| chr11 | 6320119 | 6321280 | 1162 | 15 | 2.31E-07 | 1.71E-07 | 0.00133 | 9.63E-08 | -0.005628907 | AC068733.3, CAVIN3 |
| chr16 | 84091116 | 84091788 | 673 | 3 | 2.32E-07 | 0.000199 | 1.46E-06 | 1.01E-06 | -0.005728274 | MBTPS1 |
| chr9 | 137220319 | 137221772 | 1454 | 12 | 2.33E-07 | 1.71E-05 | 0.00034 | 3.61E-08 | -0.007288541 | RNF208 |
| chr22 | 29553834 | 29555306 | 1473 | 9 | 2.37E-07 | 5.25E-07 | 1.76E-05 | 3.79E-08 | -0.002212035 | THOC5, NIPSNAP1 |
| chr16 | 24845470 | 24846280 | 811 | 10 | 2.37E-07 | 1.35E-06 | 0.00061 | 2.32E-07 | -0.007527315 | SLC5A11 |
| chr6 | 30189958 | 30190923 | 966 | 13 | 2.38E-07 | 0.115169 | 7.53E-07 | 7.83E-06 | -0.001685713 | TRIM26 |
| chr9 | 136858953 | 136860633 | 1681 | 9 | 2.38E-07 | 4.66E-06 | 6.95E-05 | 1.02E-07 | -0.005115428 | MAMDC4 |
| chr6 | 32190376 | 32191750 | 1375 | 13 | 2.39E-07 | 4.89E-05 | 0.00025 | 2.70E-06 | -0.002006079 | GPSM3 |
| chr21 | 39451353 | 39452531 | 1179 | 8 | 2.40E-07 | 1.42E-09 | 0.00475 | 2.19E-08 | -0.010181796 | GET1-SH3BGR, SH3BGR |
| chr10 | 102393943 | 102395054 | 1112 | 17 | 2.43E-07 | 0.000282 | 4.93E-05 | 2.19E-06 | 0.00040011 | NFKB2 |
| chr10 | 35231678 | 35232591 | 914 | 3 | 2.43E-07 | 6.58E-07 | 9.96E-06 | 2.91E-09 | -0.003773744 | NA |
| chr2 | 16854463 | 16854881 | 419 | 3 | 2.44E-07 | 0.000108 | 3.82E-06 | 1.30E-06 | -0.004767793 | AC008069.1 |
| chr1 | 19077999 | 19078550 | 552 | 2 | 2.44E-07 | 2.12E-05 | 2.86E-06 | 1.02E-06 | -0.00208001 | UBR4 |
| chr15 | 70420739 | 70421253 | 515 | 2 | 2.44E-07 | 0.000233 | 2.45E-06 | 1.77E-06 | -0.010228742 | NA |
| chr3 | 124351590 | 124352074 | 485 | 2 | 2.45E-07 | 0.001196 | 3.01E-06 | 2.70E-06 | 0.018601634 | KALRN |
| chr7 | 30920094 | 30920610 | 517 | 4 | 2.47E-07 | 2.23E-05 | 5.76E-05 | 6.91E-07 | -0.011202593 | AC004691.2, AQP1 |
| chr9 | 113500011 | 113500440 | 430 | 3 | 2.47E-07 | 3.11E-06 | 1.46E-05 | 4.80E-07 | -0.009163907 | RGS3 |
| chr3 | 11582052 | 11583059 | 1008 | 9 | 2.49E-07 | 3.29E-06 | 0.00022 | 2.84E-07 | -0.005699441 | VGLL4 |
| chr2 | 118185062 | 118187472 | 2411 | 14 | 2.49E-07 | 2.15E-08 | 0.00042 | 5.96E-11 | -0.004970206 | THORLNC |
| chr15 | 70195730 | 70196714 | 985 | 3 | 2.50E-07 | 1.60E-10 | 3.11E-05 | 5.75E-10 | -0.013929098 | AC026583.1 |
| chr16 | 1444475 | 1445121 | 647 | 8 | 2.51E-07 | 4.39E-07 | 0.00012 | 1.40E-07 | -0.005876671 | CCDC154, CLCN7 |
| chr11 | 65316503 | 65317142 | 640 | 5 | 2.51E-07 | 1.90E-07 | 8.19E-05 | 1.32E-07 | -0.00544761 | AP000944.5, CDC42EP2 |
| chr11 | 45803101 | 45804345 | 1245 | 11 | 2.54E-07 | 1.11E-06 | 0.00052 | 3.59E-07 | -0.00683769 | SLC35C1 |
| chr19 | 651367 | 652016 | 650 | 2 | 2.55E-07 | 0.000451 | 6.87E-06 | 6.08E-06 | -0.004596574 | RNF126 |
| chr4 | 1740574 | 1741724 | 1151 | 5 | 2.59E-07 | 3.31E-09 | 0.0002 | 1.34E-08 | -0.010623558 | TACC3 |
| chr17 | 49013254 | 49014910 | 1657 | 11 | 2.60E-07 | 1.12E-09 | 0.00023 | 6.02E-09 | -0.023035987 | IGF2BP1, AC091133.4 |
| chr18 | 25327355 | 25328258 | 904 | 5 | 2.61E-07 | 9.64E-06 | 6.16E-05 | 3.97E-07 | -0.013792079 | ZNF521 |
| chr4 | 73620363 | 73620958 | 596 | 13 | 2.63E-07 | 2.72E-05 | 0.00011 | 8.97E-07 | 0.000955429 | RASSF6 |
| chr8 | 143726795 | 143727864 | 1070 | 4 | 2.63E-07 | 2.77E-08 | 0.00027 | 4.42E-08 | -0.025322544 | FAM83H |
| chr12 | 51440110 | 51440664 | 555 | 4 | 2.63E-07 | 0.000349 | 2.21E-06 | 2.11E-06 | -0.004891844 | SLC4A8 |
| chr6 | 90218320 | 90219564 | 1245 | 4 | 2.63E-07 | 2.44E-09 | 4.53E-05 | 6.02E-09 | -0.010594744 | BACH2 |
| chr13 | 36430399 | 36432474 | 2076 | 32 | 2.64E-07 | 6.52E-06 | 0.00232 | 9.46E-09 | 0.0024583 | CCNA1 |
| chr22 | 28986614 | 28987108 | 495 | 5 | 2.64E-07 | 0.008354 | 1.44E-06 | 2.56E-06 | -0.003091124 | ZNRF3 |
| chr13 | 45372333 | 45373336 | 1004 | 4 | 2.64E-07 | 4.70E-06 | 8.94E-05 | 1.67E-07 | -0.012977479 | TPT1-AS1 |
| chr4 | 98142951 | 98144291 | 1341 | 11 | 2.66E-07 | 1.38E-08 | 0.00679 | 5.23E-08 | -0.009444828 | STPG2 |
| chr4 | 907997 | 908286 | 290 | 4 | 2.69E-07 | 6.56E-08 | 0.00029 | 8.68E-08 | -0.01206308 | GAK |
| chr6 | 33278213 | 33280937 | 2725 | 32 | 2.69E-07 | 4.16E-10 | 0.00028 | 5.86E-14 | -0.004289363 | B3GALT4, WDR46 |
| chr2 | 239249403 | 239250052 | 650 | 3 | 2.70E-07 | 1.80E-06 | 1.70E-05 | 7.71E-07 | -0.009971805 | HDAC4 |
| chr16 | 89413814 | 89414366 | 553 | 3 | 2.71E-07 | 9.89E-06 | 4.32E-06 | 8.32E-07 | -0.005352529 | ANKRD11 |
| chr12 | 52904599 | 52905526 | 928 | 8 | 2.71E-07 | 1.06E-06 | 0.00109 | 2.43E-07 | -0.006173118 | KRT8 |
| chr17 | 50123765 | 50125824 | 2060 | 7 | 2.71E-07 | 4.81E-10 | 0.00018 | 2.05E-10 | -0.007038014 | SAMD14 |
| chr3 | 57143154 | 57143875 | 722 | 3 | 2.71E-07 | 1.25E-06 | 4.17E-05 | 6.37E-07 | -0.009661411 | IL17RD |
| chr19 | 15170296 | 15170796 | 501 | 6 | 2.71E-07 | 1.43E-07 | 3.04E-05 | 5.50E-08 | -0.003728441 | NOTCH3 |
| chr19 | 3521012 | 3521452 | 441 | 5 | 2.71E-07 | 1.21E-07 | 7.45E-05 | 1.28E-07 | -0.007161232 | FZR1, AC005787.1 |
| chr8 | 100474207 | 100474864 | 658 | 3 | 2.71E-07 | 2.10E-06 | 2.46E-05 | 5.81E-07 | -0.008906244 | NA |
| chr5 | 179265690 | 179265805 | 116 | 2 | 2.71E-07 | 2.92E-08 | 2.47E-05 | 5.05E-08 | 0.046053437 | ADAMTS2 |
| chr3 | 119579236 | 119579964 | 729 | 12 | 2.72E-07 | 0.000558 | 0.00012 | 7.50E-07 | -0.004399938 | ADPRH, AC073352.2 |
| chr13 | 20118386 | 20119277 | 892 | 7 | 2.72E-07 | 2.09E-07 | 0.00037 | 1.75E-07 | 0.009618181 | NA |
| chr3 | 23804630 | 23806003 | 1374 | 12 | 2.72E-07 | 2.34E-06 | 7.33E-05 | 1.37E-07 | -0.002490284 | UBE2E1, UBE2E1-AS1 |
| chr8 | 79610089 | 79611085 | 997 | 6 | 2.78E-07 | 6.08E-05 | 1.70E-05 | 2.15E-06 | -0.011535116 | NA |
| chr16 | 67652929 | 67654793 | 1865 | 8 | 2.81E-07 | 3.85E-11 | 0.00163 | 5.44E-10 | -0.020880056 | CARMIL2 |
| chr14 | 93927151 | 93927445 | 295 | 7 | 2.82E-07 | 2.28E-06 | 0.00067 | 1.16E-07 | -0.004286726 | FAM181A |
| chr9 | 131278339 | 131278813 | 475 | 4 | 2.82E-07 | 2.32E-06 | 0.00024 | 2.03E-07 | -0.010302761 | NA |
| chr10 | 99331592 | 99331724 | 133 | 2 | 2.82E-07 | 3.23E-06 | 3.27E-06 | 4.68E-07 | 0.000324471 | CNNM1 |
| chr20 | 44450102 | 44450455 | 354 | 4 | 2.83E-07 | 4.92E-07 | 4.27E-05 | 1.34E-07 | -0.009470656 | LINC01620 |
| chr19 | 14560048 | 14560700 | 653 | 4 | 2.83E-07 | 1.57E-05 | 1.38E-05 | 5.44E-07 | -0.008108219 | TECR, DNAJB1 |
| chr20 | 26209692 | 26209718 | 27 | 3 | 2.84E-07 | 2.17E-08 | 0.00015 | 5.68E-08 | 0.015608505 | MIR663AHG |
| chr8 | 143277490 | 143279145 | 1656 | 8 | 2.84E-07 | 1.07E-06 | 9.24E-05 | 5.21E-08 | -0.015403054 | NA |
| chr10 | 70782635 | 70783514 | 880 | 5 | 2.84E-07 | 3.06E-06 | 0.00015 | 2.50E-07 | -0.017766354 | TBATA |
| chr11 | 64246365 | 64248102 | 1738 | 13 | 2.84E-07 | 6.48E-11 | 0.00265 | 2.17E-10 | -0.000660656 | PPP1R14B-AS1, PPP1R14B, AP001453.2 |
| chr7 | 55964133 | 55965274 | 1142 | 14 | 2.87E-07 | 5.17E-06 | 0.00012 | 3.69E-07 | -0.000689955 | NIPSNAP2 |
| chr12 | 989497 | 991432 | 1936 | 17 | 2.88E-07 | 1.17E-06 | 0.00037 | 1.15E-08 | -0.001779239 | ERC1, RAD52, AC004803.1 |
| chr7 | 28958462 | 28959608 | 1147 | 9 | 2.88E-07 | 8.02E-05 | 0.00028 | 8.01E-06 | -0.005301372 | AC005013.1 |
| chr17 | 39174208 | 39175804 | 1597 | 8 | 2.89E-07 | 1.24E-09 | 0.00111 | 3.64E-09 | -0.009878949 | AC004408.1, CACNB1 |
| chr5 | 141181486 | 141184586 | 3101 | 18 | 2.91E-07 | 2.15E-08 | 0.00022 | 1.97E-11 | 0.012071411 | AC244517.8, PCDHB16, AC244517.6, AC244517.11, AC244517.4 |
| chr19 | 4970765 | 4971476 | 712 | 2 | 2.94E-07 | 1.15E-07 | 2.19E-05 | 1.54E-07 | -0.040058687 | KDM4B |
| chr18 | 6929477 | 6930453 | 977 | 5 | 2.94E-07 | 9.76E-08 | 0.0002 | 1.72E-08 | -0.001625077 | LINC00668 |
| chr20 | 5190322 | 5190732 | 411 | 2 | 2.95E-07 | 1.51E-07 | 1.05E-05 | 1.37E-07 | -0.008655688 | CDS2 |
| chr2 | 151973675 | 151974700 | 1026 | 9 | 2.95E-07 | 7.26E-09 | 0.00221 | 1.27E-08 | -0.010872612 | AC068547.1, CACNB4 |
| chr3 | 36993239 | 36994134 | 896 | 25 | 2.95E-07 | 5.71E-05 | 0.00048 | 4.65E-06 | 0.000597237 | MLH1 |
| chr1 | 32885967 | 32886988 | 1022 | 13 | 2.97E-07 | 2.69E-05 | 0.00044 | 2.93E-07 | 0.00326413 | HPCA |
| chr14 | 76918857 | 76920158 | 1302 | 5 | 2.97E-07 | 2.04E-08 | 9.98E-05 | 1.83E-08 | -0.012213787 | NA |
| chr11 | 118921088 | 118921359 | 272 | 3 | 2.98E-07 | 3.83E-08 | 0.00011 | 9.40E-08 | -0.013823996 | BCL9L |
| chr2 | 126983110 | 126983556 | 447 | 3 | 3.00E-07 | 0.027772 | 1.91E-06 | 5.41E-06 | -0.003934953 | NA |
| chr1 | 11478044 | 11478808 | 765 | 6 | 3.00E-07 | 4.61E-06 | 8.36E-05 | 2.76E-06 | -0.00485124 | NA |
| chr20 | 34558044 | 34559939 | 1896 | 13 | 3.01E-07 | 2.95E-07 | 0.00192 | 1.99E-08 | -0.000825858 | MAP1LC3A |
| chr14 | 105862948 | 105864863 | 1916 | 13 | 3.03E-07 | 1.05E-09 | 0.00499 | 7.80E-09 | 0.020464008 | IGHJ6, IGHJ3P, IGHJ5, IGHJ4, IGHJ3, IGHJ2P |
| chr10 | 14008306 | 14009839 | 1534 | 12 | 3.03E-07 | 4.35E-11 | 0.00386 | 6.98E-10 | -0.015033383 | FRMD4A |
| chr22 | 36463947 | 36464784 | 838 | 4 | 3.05E-07 | 7.78E-08 | 0.00017 | 1.10E-07 | -0.003453477 | NA |
| chr1 | 108878875 | 108879562 | 688 | 2 | 3.06E-07 | 3.80E-08 | 3.96E-05 | 7.04E-08 | -0.01874211 | GPSM2, AKNAD1 |
| chr9 | 136074738 | 136076026 | 1289 | 8 | 3.07E-07 | 9.12E-09 | 0.00129 | 3.71E-08 | -0.00971544 | NACC2 |
| chr14 | 103712195 | 103712758 | 564 | 5 | 3.08E-07 | 5.67E-07 | 0.00114 | 2.56E-07 | -0.008566293 | KLC1, XRCC3 |
| chr19 | 33131657 | 33132629 | 973 | 12 | 3.08E-07 | 2.26E-06 | 0.00014 | 3.39E-08 | -0.006210897 | WDR88 |
| chr1 | 16119617 | 16121071 | 1455 | 3 | 3.09E-07 | 1.24E-08 | 7.11E-06 | 1.06E-08 | -0.009329747 | NA |
| chr18 | 79680800 | 79681810 | 1011 | 8 | 3.10E-07 | 7.22E-09 | 0.00234 | 5.69E-08 | -0.009851977 | CTDP1 |
| chr17 | 38657163 | 38657545 | 383 | 3 | 3.10E-07 | 5.56E-08 | 6.09E-05 | 1.08E-07 | -0.014551075 | NA |
| chr7 | 100168753 | 100169511 | 759 | 9 | 3.10E-07 | 4.89E-07 | 0.00059 | 5.58E-07 | -0.006616593 | NA |
| chr1 | 89820631 | 89822540 | 1910 | 15 | 3.10E-07 | 0.000103 | 3.51E-06 | 6.33E-07 | -0.000128914 | AC093423.3, LRRC8D, AC099568.2 |
| chr16 | 84240391 | 84240956 | 566 | 4 | 3.11E-07 | 0.01157 | 5.04E-06 | 1.48E-05 | -0.006784974 | NA |
| chr18 | 58628862 | 58629375 | 514 | 10 | 3.13E-07 | 1.99E-07 | 0.00227 | 8.12E-08 | -0.006422403 | ALPK2 |
| chr16 | 85351168 | 85351241 | 74 | 2 | 3.14E-07 | 1.14E-07 | 1.54E-05 | 1.34E-07 | -0.013009019 | GSE1, AC092377.1 |
| chr7 | 151081035 | 151082623 | 1589 | 23 | 3.14E-07 | 3.11E-09 | 0.00175 | 2.28E-09 | -0.002108862 | TMUB1 |
| chr17 | 7235067 | 7235755 | 689 | 3 | 3.15E-07 | 4.75E-06 | 0.00018 | 4.81E-06 | -0.009752941 | PHF23 |
| chr2 | 20353326 | 20353717 | 392 | 2 | 3.19E-07 | 0.000337 | 2.55E-06 | 1.97E-06 | -0.00450068 | NA |
| chr1 | 6034111 | 6035888 | 1778 | 9 | 3.20E-07 | 1.81E-09 | 0.00119 | 5.35E-09 | -0.006237189 | KCNAB2 |
| chr11 | 133810534 | 133811681 | 1148 | 5 | 3.20E-07 | 3.97E-06 | 3.23E-05 | 1.08E-06 | -0.000555546 | NA |
| chr5 | 134568542 | 134568984 | 443 | 3 | 3.20E-07 | 0.000248 | 4.32E-06 | 4.52E-06 | -0.007430984 | JADE2 |
| chr22 | 30356135 | 30357153 | 1019 | 12 | 3.21E-07 | 4.92E-05 | 0.00012 | 1.24E-06 | 0.000963416 | CCDC157, SF3A1 |
| chr12 | 6123651 | 6125624 | 1974 | 8 | 3.21E-07 | 2.15E-05 | 2.14E-05 | 7.81E-10 | -0.006541653 | VWF |
| chr17 | 48598530 | 48599611 | 1082 | 5 | 3.23E-07 | 1.25E-08 | 0.00054 | 5.09E-08 | -0.01664966 | HOXB-AS3, HOXB3, HOXB6 |
| chr21 | 44284903 | 44286218 | 1316 | 13 | 3.24E-07 | 1.01E-07 | 0.00543 | 6.98E-08 | -0.00944284 | AIRE |
| chr2 | 27129086 | 27129935 | 850 | 5 | 3.24E-07 | 0.001961 | 7.76E-07 | 2.92E-06 | -0.005190419 | ABHD1 |
| chr8 | 133244375 | 133244882 | 508 | 4 | 3.25E-07 | 1.60E-07 | 1.82E-05 | 1.12E-07 | -0.010393065 | NDRG1 |
| chr15 | 31338533 | 31339887 | 1355 | 8 | 3.27E-07 | 1.10E-09 | 0.00023 | 7.64E-10 | -0.007834666 | KLF13 |
| chr7 | 106009596 | 106010104 | 509 | 4 | 3.27E-07 | 2.08E-06 | 3.92E-05 | 4.63E-07 | -0.008152354 | CDHR3 |
| chr17 | 48591751 | 48592667 | 917 | 6 | 3.28E-07 | 3.51E-07 | 0.00065 | 2.47E-07 | -0.009414299 | HOXB-AS3, HOXB3, HOXB5 |
| chr14 | 21476783 | 21477699 | 917 | 13 | 3.29E-07 | 0.000171 | 0.00013 | 3.99E-06 | 0.000913218 | TOX4, RAB2B |
| chr2 | 216080873 | 216082426 | 1554 | 15 | 3.30E-07 | 8.09E-06 | 0.00034 | 1.47E-07 | 0.000818399 | TMEM169, PECR |
| chr14 | 103728964 | 103729666 | 703 | 4 | 3.32E-07 | 1.91E-06 | 0.00023 | 9.73E-07 | -0.011385337 | ZFYVE21 |
| chr7 | 5481659 | 5482139 | 481 | 4 | 3.36E-07 | 1.31E-06 | 7.23E-06 | 1.86E-07 | -0.005535187 | FBXL18 |
| chr22 | 41689597 | 41690542 | 946 | 3 | 3.38E-07 | 0.019601 | 1.05E-05 | 2.67E-05 | -0.009231325 | C22orf46, SNU13 |
| chr8 | 143874147 | 143874613 | 467 | 3 | 3.38E-07 | 7.17E-05 | 2.89E-05 | 2.04E-06 | -0.012832374 | EPPK1 |

| | | | | | | | | | | |
|---|---|---|---|---|---|---|---|---|---|---|
| chr11 | 128621349 | 128622280 | 932 | 7 | 3.40E-07 | 6.69E-06 | 3.38E-05 | 3.61E-07 | -0.006556931 | AP001122.1 |
| chr17 | 43389035 | 43389844 | 810 | 6 | 3.40E-07 | 4.75E-05 | 7.05E-05 | 1.28E-06 | 0.000568249 | LINC00910 |
| chr19 | 33430149 | 33430877 | 729 | 4 | 3.40E-07 | 1.89E-05 | 5.11E-05 | 7.16E-07 | -0.011183856 | PEPD |
| chr12 | 6039924 | 6041001 | 1078 | 4 | 3.40E-07 | 3.97E-06 | 6.70E-05 | 5.69E-07 | -0.01291344 | VWF |
| chr3 | 52411080 | 52411706 | 627 | 9 | 3.40E-07 | 5.38E-06 | 0.00244 | 2.34E-06 | -0.00480581 | PHF7 |
| chr8 | 20274218 | 20274921 | 704 | 4 | 3.40E-07 | 0.000112 | 1.21E-06 | 9.45E-07 | -0.0033435 | LZTS1 |
| chr6 | 33267727 | 33268576 | 850 | 10 | 3.40E-07 | 7.79E-06 | 0.00018 | 2.67E-07 | -0.003148666 | VPS52 |
| chr12 | 25976732 | 25977247 | 516 | 2 | 3.42E-07 | 8.36E-06 | 3.61E-06 | 8.32E-07 | -0.007353959 | RASSF8 |
| chr2 | 27274887 | 27276329 | 1443 | 15 | 3.42E-07 | 1.13E-06 | 0.00023 | 4.09E-09 | -0.002427196 | DNAJC5G, SLC30A3 |
| chr19 | 35755050 | 35757565 | 2516 | 20 | 3.43E-07 | 2.18E-08 | 0.00491 | 1.13E-09 | -0.009171794 | AC002398.2, HSPB6 |
| chr20 | 17873591 | 17874929 | 1339 | 5 | 3.43E-07 | 6.81E-07 | 0.00021 | 5.11E-07 | -0.01150635 | NA |
| chr16 | 87407650 | 87408053 | 404 | 4 | 3.45E-07 | 1.21E-08 | 0.00062 | 6.13E-08 | -0.022373847 | ZCCHC14 |
| chr9 | 134411584 | 134412573 | 990 | 4 | 3.46E-07 | 2.10E-09 | 0.0002 | 1.03E-08 | -0.011898519 | RXRA |
| chr19 | 17376967 | 17377776 | 810 | 10 | 3.49E-07 | 1.65E-05 | 4.39E-05 | 8.14E-07 | -0.006216612 | PLVAP |
| chr12 | 65947939 | 65948588 | 650 | 3 | 3.51E-07 | 1.21E-06 | 0.00014 | 6.27E-07 | -0.015667375 | HMGA2, AC090673.1 |
| chr1 | 5882574 | 5883115 | 542 | 5 | 3.51E-07 | 7.33E-05 | 2.01E-06 | 6.22E-07 | -0.00510435 | NPHP4 |
| chr4 | 40516007 | 40517077 | 1071 | 9 | 3.53E-07 | 8.93E-10 | 0.00057 | 5.59E-09 | -0.007568671 | RBM47 |
| chr16 | 66801417 | 66802317 | 901 | 13 | 3.53E-07 | 7.07E-06 | 0.00239 | 5.99E-07 | -0.00195774 | TERB1 |
| chr22 | 37984970 | 37986154 | 1185 | 6 | 3.53E-07 | 1.68E-08 | 0.00013 | 5.95E-09 | -0.00499372 | POLR2F, SOX10 |
| chr1 | 39080474 | 39081406 | 933 | 7 | 3.57E-07 | 1.94E-05 | 9.17E-05 | 1.20E-06 | -0.00829988 | MACF1 |
| chr2 | 236971908 | 236972147 | 240 | 2 | 3.58E-07 | 7.54E-08 | 4.38E-05 | 1.33E-07 | -0.024714745 | AC012063.1 |
| chr1 | 37689448 | 37691685 | 2238 | 16 | 3.59E-07 | 1.45E-05 | 0.0017 | 6.29E-07 | -0.014225335 | C1orf109 |
| chr5 | 16758113 | 16758414 | 302 | 3 | 3.60E-07 | 5.17E-06 | 1.95E-06 | 2.65E-07 | -0.013591548 | MYO10 |
| chr5 | 150759095 | 150760150 | 1056 | 9 | 3.61E-07 | 2.62E-06 | 0.00061 | 1.45E-06 | -0.003510789 | DCTN4 |
| chr17 | 8223747 | 8224829 | 1083 | 6 | 3.62E-07 | 4.98E-07 | 0.00069 | 4.16E-07 | -0.009287211 | LINC00324, CTC1 |
| chr10 | 70131125 | 70132337 | 1213 | 5 | 3.63E-07 | 1.34E-06 | 1.47E-05 | 1.53E-07 | -0.008605734 | AIFM2 |
| chr9 | 33448228 | 33448384 | 157 | 3 | 3.64E-07 | 1.37E-07 | 0.00016 | 2.79E-07 | -0.01125585 | NA |
| chr10 | 119680333 | 119681054 | 722 | 4 | 3.64E-07 | 6.92E-06 | 0.00011 | 3.32E-07 | -0.013780054 | NA |
| chr8 | 98826090 | 98826433 | 344 | 5 | 3.68E-07 | 1.47E-05 | 2.45E-05 | 5.38E-07 | -0.008107857 | STK3 |
| chr4 | 184168559 | 184168890 | 332 | 4 | 3.70E-07 | 5.75E-07 | 6.48E-05 | 6.89E-08 | 0.001247184 | ENPP6 |
| chr2 | 106064882 | 106065753 | 872 | 10 | 3.70E-07 | 7.37E-05 | 0.0018 | 6.22E-07 | -0.006214336 | ECRG4 |
| chr2 | 46844940 | 46845768 | 829 | 5 | 3.70E-07 | 2.78E-08 | 2.89E-05 | 6.86E-09 | -0.008069447 | LINC01119 |
| chr21 | 31347258 | 31347715 | 458 | 2 | 3.71E-07 | 1.57E-07 | 1.28E-05 | 1.55E-07 | -0.006653891 | TIAM1 |
| chr17 | 82872756 | 82873123 | 368 | 4 | 3.73E-07 | 3.34E-05 | 7.17E-06 | 8.25E-07 | -0.005319221 | TBCD |
| chr13 | 40987319 | 40987702 | 384 | 2 | 3.74E-07 | 0.001414 | 2.28E-06 | 1.98E-06 | -0.008930111 | ELF1 |
| chr7 | 1982740 | 1983296 | 557 | 3 | 3.76E-07 | 6.34E-05 | 4.21E-06 | 1.43E-06 | -0.004014506 | MAD1L1, AC069288.1 |
| chr10 | 17230765 | 17231188 | 424 | 2 | 3.77E-07 | 0.000797 | 5.56E-06 | 5.30E-06 | 0.009798023 | VIM |
| chr14 | 50872354 | 50872498 | 145 | 4 | 3.78E-07 | 2.78E-08 | 0.00069 | 1.32E-07 | -0.013380403 | ABHD12B, PYGL |
| chr12 | 6215882 | 6216552 | 671 | 3 | 3.79E-07 | 1.88E-05 | 1.17E-05 | 1.98E-06 | -0.010795122 | CD9 |
| chr1 | 6519524 | 6520465 | 942 | 9 | 3.80E-07 | 2.11E-07 | 0.00075 | 1.17E-07 | -0.005408238 | PLEKHG5 |
| chr1 | 1437297 | 1437936 | 640 | 3 | 3.81E-07 | 1.36E-06 | 2.40E-05 | 7.82E-07 | -0.010640605 | VWA1 |
| chr3 | 111673644 | 111674757 | 1114 | 16 | 3.83E-07 | 5.66E-05 | 4.78E-05 | 5.68E-07 | 0.001187123 | PLCXD2 |
| chr11 | 46380501 | 46382067 | 1567 | 17 | 3.83E-07 | 1.49E-05 | 0.0009 | 1.05E-07 | 0.00012411 | DGKZ, MDK |
| chr17 | 50662147 | 50663020 | 874 | 2 | 3.84E-07 | 3.09E-07 | 2.48E-05 | 3.54E-07 | -0.013075151 | ABCC3 |
| chr11 | 128942567 | 128943793 | 1227 | 18 | 3.87E-07 | 1.20E-05 | 0.00235 | 2.54E-07 | -0.001952158 | TP53AIP1 |
| chr17 | 76527342 | 76528964 | 1623 | 10 | 3.94E-07 | 2.79E-09 | 0.0003 | 5.16E-10 | -0.009943448 | PRCD, CYGB |
| chr11 | 1219856 | 1222070 | 2215 | 13 | 3.94E-07 | 3.79E-11 | 0.0002 | 6.75E-11 | -0.006859439 | AC061979.1 |
| chr19 | 12111666 | 12111871 | 206 | 2 | 3.96E-07 | 2.92E-05 | 4.91E-06 | 1.98E-06 | -0.005714559 | AC022415.2, ZNF788P, ZNF20 |
| chr11 | 117327015 | 117328282 | 1268 | 16 | 3.97E-07 | 1.27E-06 | 0.00108 | 1.18E-07 | 5.30E-05 | CEP164, AP000892.1 |
| chr4 | 742540 | 744411 | 1872 | 13 | 3.98E-07 | 2.51E-10 | 0.00017 | 1.18E-10 | -0.005743754 | PCGF3 |
| chr22 | 26463988 | 26464859 | 872 | 6 | 4.00E-07 | 8.83E-09 | 0.00144 | 5.65E-08 | -0.008224914 | HPS4 |
| chr14 | 100290916 | 100291540 | 625 | 6 | 4.04E-07 | 6.25E-05 | 5.76E-05 | 1.55E-06 | -0.004046839 | AL157871.1, AL157871.6, SLC25A29 |
| chr16 | 85752137 | 85753570 | 1434 | 5 | 4.04E-07 | 6.78E-10 | 0.00045 | 5.65E-09 | -0.012271322 | NA |
| chr10 | 133309242 | 133310365 | 1124 | 12 | 4.05E-07 | 1.94E-05 | 0.00012 | 9.95E-07 | -0.002113147 | ZNF511, ZNF511-PRAP1, TUBGCP2 |
| chr10 | 73109026 | 73109786 | 761 | 3 | 4.05E-07 | 2.90E-06 | 1.11E-05 | 7.91E-07 | -0.012312501 | NA |
| chr9 | 136693949 | 136694961 | 1013 | 17 | 4.05E-07 | 2.74E-08 | 0.00918 | 1.16E-07 | -0.006174177 | NA |
| chr11 | 130897483 | 130897865 | 383 | 2 | 4.08E-07 | 9.37E-08 | 1.77E-05 | 1.20E-07 | -0.018729459 | SNX19 |
| chr3 | 113532334 | 113533793 | 1460 | 10 | 4.09E-07 | 1.48E-07 | 0.00016 | 6.59E-08 | 0.001451782 | SIDT1 |
| chr22 | 27757333 | 27757753 | 421 | 3 | 4.10E-07 | 0.000224 | 2.69E-06 | 1.82E-06 | -0.001611359 | MN1 |
| chr3 | 186108341 | 186110551 | 2211 | 19 | 4.11E-07 | 2.59E-07 | 0.00116 | 8.69E-09 | -0.000675806 | ETV5, DGKG |
| chr1 | 203272820 | 203273510 | 691 | 7 | 4.12E-07 | 1.04E-05 | 0.00069 | 6.30E-07 | -0.007896643 | LINC01353, CHIT1 |
| chr19 | 50715543 | 50717526 | 1984 | 14 | 4.14E-07 | 4.65E-10 | 0.00078 | 3.83E-10 | -0.004856787 | SHANK1 |
| chr17 | 73506875 | 73507722 | 848 | 4 | 4.16E-07 | 7.85E-08 | 3.71E-05 | 7.80E-08 | -0.008686321 | SDK2 |
| chr17 | 351884 | 353296 | 1413 | 12 | 4.16E-07 | 1.77E-05 | 7.47E-06 | 1.72E-08 | -0.010176119 | AC129507.2, RPH3AL |
| chr7 | 726222 | 726747 | 526 | 6 | 4.17E-07 | 7.99E-06 | 0.0005 | 1.43E-06 | 0.034801369 | DNAAF5, PRKAR1B |
| chr12 | 6974317 | 6974585 | 269 | 4 | 4.17E-07 | 1.58E-06 | 8.09E-05 | 1.40E-07 | -0.004680703 | EMG1 |
| chr11 | 72013631 | 72014544 | 914 | 6 | 4.22E-07 | 3.80E-07 | 0.00126 | 6.49E-07 | -0.005589768 | AP002490.1, NUMA1 |
| chr1 | 2466765 | 2466961 | 197 | 2 | 4.22E-07 | 5.86E-07 | 2.52E-05 | 5.89E-07 | -0.025250256 | PLCH2 |
| chr12 | 121797074 | 121798458 | 1385 | 6 | 4.22E-07 | 3.32E-10 | 0.0015 | 4.72E-09 | -0.009607586 | AC084018.1, RHOF, LINC01089 |
| chr1 | 153774161 | 153775644 | 1484 | 13 | 4.22E-07 | 1.30E-06 | 0.0016 | 1.01E-07 | -0.006855531 | INTS3, SLC27A3 |
| chr11 | 65916842 | 65918403 | 1562 | 7 | 4.25E-07 | 4.91E-07 | 4.69E-05 | 4.09E-08 | -0.003842214 | C11orf68 |
| chr5 | 138283129 | 138283579 | 451 | 2 | 4.25E-07 | 0.001917 | 2.23E-06 | 1.97E-06 | -0.003769965 | NA |
| chr19 | 1624495 | 1627486 | 2992 | 10 | 4.31E-07 | 5.34E-09 | 0.00011 | 6.10E-11 | -0.006626534 | TCF3 |
| chr1 | 41360356 | 41361184 | 829 | 6 | 4.31E-07 | 2.76E-08 | 0.00145 | 1.04E-07 | -0.009224178 | NA |
| chr11 | 111255261 | 111256065 | 805 | 6 | 4.31E-07 | 8.78E-07 | 7.44E-05 | 2.21E-07 | -0.003015718 | C11orf53 |
| chr19 | 58037873 | 58038538 | 666 | 4 | 4.32E-07 | 1.25E-08 | 0.00037 | 4.98E-08 | -0.034577635 | ZSCAN1 |
| chr19 | 1355911 | 1356445 | 535 | 10 | 4.33E-07 | 1.58E-07 | 0.00212 | 9.71E-08 | -0.009697515 | PWWP3A |
| chr17 | 18268004 | 18268566 | 563 | 4 | 4.33E-07 | 2.82E-06 | 5.79E-05 | 5.20E-07 | -0.010238914 | AC127537.1 |
| chr20 | 45124230 | 45125011 | 782 | 9 | 4.34E-07 | 2.25E-07 | 0.00086 | 6.98E-08 | -0.007503129 | WFDC12 |
| chr3 | 121431868 | 121432339 | 472 | 2 | 4.34E-07 | 0.002359 | 2.45E-06 | 2.23E-06 | -0.014077574 | POLQ |
| chr1 | 202295785 | 202296022 | 238 | 2 | 4.34E-07 | 0.000833 | 8.83E-07 | 6.46E-07 | -0.007890309 | LGR6 |
| chr11 | 45180190 | 45181753 | 1564 | 6 | 4.34E-07 | 2.68E-08 | 8.26E-05 | 2.32E-08 | -0.00861173 | PRDM11 |
| chr10 | 80413779 | 80413866 | 88 | 4 | 4.34E-07 | 3.53E-05 | 1.53E-05 | 6.91E-07 | -0.009427633 | PRXL2A |
| chr20 | 63235608 | 63236717 | 1110 | 9 | 4.35E-07 | 3.11E-08 | 0.0007 | 6.49E-08 | -0.009563035 | BIRC7 |
| chr1 | 37947662 | 37948385 | 724 | 4 | 4.43E-07 | 0.004645 | 3.50E-06 | 1.18E-05 | -0.004913032 | NA |
| chr3 | 12898787 | 12900023 | 1237 | 9 | 4.44E-07 | 5.98E-07 | 4.27E-05 | 1.08E-07 | -0.002622751 | IQSEC1 |
| chr19 | 39154294 | 39155534 | 1241 | 5 | 4.44E-07 | 4.80E-06 | 0.00026 | 7.73E-07 | -0.006397596 | PAK4 |
| chr1 | 226821425 | 226821544 | 120 | 3 | 4.45E-07 | 1.30E-08 | 0.00032 | 5.12E-08 | -0.010670845 | NA |
| chr7 | 53035001 | 53036308 | 1308 | 9 | 4.48E-07 | 0.000182 | 0.00049 | 2.31E-06 | -0.0015936 | POM121L12 |
| chr12 | 6329151 | 6330282 | 1132 | 7 | 4.49E-07 | 1.02E-06 | 0.00017 | 2.70E-07 | -0.003637622 | TNFRSF1A |
| chr8 | 60997707 | 60998390 | 684 | 2 | 4.49E-07 | 6.38E-08 | 1.55E-05 | 8.30E-08 | -0.024445483 | NA |
| chr11 | 62189388 | 62190223 | 836 | 5 | 4.50E-07 | 3.20E-05 | 7.87E-06 | 2.05E-07 | -0.004235266 | SCGB1D1 |
| chr17 | 17506610 | 17507102 | 493 | 3 | 4.50E-07 | 6.60E-06 | 3.94E-05 | 4.17E-07 | -0.011129376 | AC020558.6, PEMT |
| chr20 | 45114660 | 45116237 | 1578 | 11 | 4.52E-07 | 1.02E-07 | 0.0004 | 3.10E-08 | -0.005680056 | WFDC5 |
| chr17 | 17281126 | 17281746 | 621 | 11 | 4.54E-07 | 2.54E-07 | 0.00145 | 2.77E-07 | 0.003035403 | COPS3 |
| chr12 | 109096302 | 109096665 | 364 | 3 | 4.57E-07 | 6.88E-05 | 4.07E-06 | 2.25E-06 | -0.001895829 | NA |
| chr2 | 44932006 | 44933722 | 1717 | 17 | 4.57E-07 | 4.53E-07 | 0.00031 | 8.51E-09 | -0.005123034 | AC012354.2, LINC01833 |
| chr19 | 1650225 | 1650453 | 229 | 4 | 4.57E-07 | 1.51E-06 | 1.05E-05 | 3.04E-07 | -0.004312324 | TCF3 |
| chr12 | 69699570 | 69700520 | 951 | 4 | 4.57E-07 | 5.81E-06 | 6.00E-05 | 6.38E-07 | -0.000962967 | NA |
| chr22 | 20392964 | 20394381 | 1418 | 12 | 4.59E-07 | 1.28E-06 | 9.69E-05 | 1.45E-07 | -0.000916934 | ZNF74 |
| chr16 | 66483837 | 66485271 | 1435 | 5 | 4.61E-07 | 9.56E-07 | 7.69E-06 | 1.31E-07 | -0.003105879 | BEAN1, AC010542.3 |
| chr13 | 30409358 | 30409965 | 608 | 2 | 4.61E-07 | 1.36E-05 | 2.04E-05 | 4.42E-06 | -0.013612285 | NA |
| chr12 | 12890195 | 12891228 | 1034 | 12 | 4.62E-07 | 1.38E-05 | 9.37E-05 | 2.55E-07 | -0.004987383 | NA |
| chr17 | 5019831 | 5020954 | 1124 | 7 | 4.76E-07 | 0.000473 | 0.00019 | 1.42E-06 | -0.003883461 | KIF1C, KIF1C-AS1 |
| chr16 | 3730897 | 3731829 | 933 | 7 | 4.78E-07 | 2.37E-05 | 7.50E-05 | 1.13E-07 | -0.002737006 | CREBBP |
| chr17 | 36885190 | 36885733 | 544 | 6 | 4.80E-07 | 7.59E-06 | 2.64E-05 | 5.57E-07 | -0.010004029 | LHX1-DT |
| chr12 | 130813942 | 130814171 | 230 | 3 | 4.81E-07 | 3.91E-06 | 1.76E-05 | 3.49E-07 | -0.004612875 | STX2 |
| chr19 | 19501025 | 19501497 | 473 | 5 | 4.84E-07 | 2.82E-07 | 0.00035 | 3.04E-07 | -0.002912908 | GATAD2A |
| chr15 | 41774624 | 41775864 | 1241 | 6 | 4.85E-07 | 7.91E-07 | 0.00154 | 6.99E-07 | -0.012565062 | MAPKBP1 |
| chr3 | 197959447 | 197960857 | 1411 | 15 | 4.85E-07 | 7.10E-05 | 6.68E-05 | 1.01E-06 | 0.001724752 | LMLN, IQCG |
| chr1 | 16212518 | 16212846 | 329 | 4 | 4.86E-07 | 1.68E-08 | 0.00095 | 8.85E-08 | -0.011804254 | ARHGEF19 |
| chr20 | 49675412 | 49675994 | 583 | 3 | 4.86E-07 | 4.94E-07 | 0.00012 | 4.79E-07 | -0.007386708 | B4GALT5 |
| chr12 | 31728533 | 31729346 | 814 | 9 | 4.87E-07 | 1.76E-06 | 0.00108 | 2.32E-06 | 0.001703954 | AC023157.2, AMN1 |
| chr9 | 95316847 | 95318340 | 1494 | 17 | 4.87E-07 | 6.20E-07 | 5.80E-05 | 4.62E-08 | -2.78E-05 | FANCC |
| chr1 | 154337322 | 154337587 | 266 | 4 | 4.94E-07 | 1.43E-07 | 0.0001 | 9.17E-08 | -0.012351321 | ATP8B2 |
| chr3 | 45554498 | 45555335 | 838 | 4 | 4.94E-07 | 8.00E-07 | 2.35E-05 | 4.60E-07 | -0.008412454 | LARS2 |
| chr11 | 845621 | 847072 | 1452 | 8 | 4.95E-07 | 5.77E-07 | 0.00052 | 1.04E-07 | -0.006132959 | TSPAN4 |
| chr15 | 69482846 | 69483353 | 508 | 2 | 4.96E-07 | 7.20E-06 | 7.22E-06 | 1.39E-06 | -0.003294204 | DRAIC |
| chr13 | 113861082 | 113862844 | 1763 | 17 | 4.96E-07 | 1.26E-07 | 0.00452 | 1.02E-07 | -0.002285065 | GAS6 |
| chr11 | 102606710 | 102607313 | 604 | 5 | 4.96E-07 | 9.08E-05 | 4.56E-06 | 1.50E-06 | -0.002409844 | AP000851.1, MMP20 |
| chr6 | 31780110 | 31782755 | 2646 | 16 | 4.99E-07 | 2.52E-06 | 5.39E-05 | 4.04E-09 | -0.001835478 | VARS1 |
| chr6 | 137491168 | 137491467 | 300 | 2 | 5.00E-07 | 2.39E-06 | 8.67E-06 | 8.32E-07 | -0.019006035 | NA |
| chr1 | 53956737 | 53957529 | 793 | 2 | 5.00E-07 | 4.47E-09 | 1.72E-05 | 8.68E-09 | 0.001219853 | LRRC42 |
| chr12 | 66061909 | 66061969 | 61 | 2 | 5.00E-07 | 4.89E-08 | 3.72E-05 | 8.71E-08 | -0.028517558 | NA |
| chr15 | 90213546 | 90213595 | 50 | 2 | 5.02E-07 | 1.66E-06 | 9.20E-06 | 6.92E-07 | -0.012668571 | SEMA4B |
| chr8 | 141131356 | 141131873 | 518 | 3 | 5.03E-07 | 0.004962 | 2.17E-06 | 4.71E-06 | -0.001142809 | DENND3 |
| chr1 | 155255973 | 155256403 | 431 | 5 | 5.05E-07 | 2.67E-07 | 0.00019 | 5.53E-07 | -0.004941761 | SCAMP3 |
| chr17 | 75821619 | 75822484 | 866 | 5 | 5.05E-07 | 9.44E-06 | 0.00119 | 7.96E-07 | -0.00830523 | UNK |
| chr4 | 5894872 | 5896164 | 1293 | 5 | 5.06E-07 | 8.86E-10 | 0.00037 | 6.01E-09 | -0.006772892 | NA |
| chr17 | 40019136 | 40020432 | 1297 | 4 | 5.10E-07 | 4.80E-08 | 0.00016 | 1.50E-07 | -0.010107305 | MED24 |
| chr17 | 78568881 | 78569591 | 711 | 8 | 5.10E-07 | 3.46E-07 | 6.28E-05 | 1.28E-07 | -0.004474255 | DNAH17 |
| chr12 | 6042078 | 6042536 | 459 | 4 | 5.14E-07 | 1.31E-07 | 6.37E-05 | 9.03E-08 | -0.011897796 | VWF |
| chr20 | 36093841 | 36094328 | 488 | 2 | 5.15E-07 | 2.52E-06 | 2.81E-05 | 1.84E-06 | 0.008089267 | EPB41L1 |
| chr2 | 110731812 | 110732918 | 1107 | 9 | 5.19E-07 | 0.000106 | 1.15E-05 | 1.98E-07 | -0.000515337 | ACOXL |
| chr1 | 169629761 | 169631466 | 1706 | 8 | 5.20E-07 | 6.54E-06 | 0.00014 | 1.50E-08 | -0.00719558 | SELP |
| chr20 | 58692120 | 58692624 | 505 | 7 | 5.23E-07 | 3.18E-08 | 0.00116 | 7.78E-08 | -0.011633435 | STX16-NPEPL1, NPEPL1 |
| chr2 | 203327485 | 203328655 | 1171 | 13 | 5.24E-07 | 5.32E-06 | 0.00244 | 2.32E-07 | -0.000546895 | ABI2, AC080075.1 |
| chr12 | 57726454 | 57727767 | 1314 | 4 | 5.25E-07 | 2.67E-06 | 9.60E-05 | 2.88E-06 | -0.005191582 | AGAP2-AS1, AGAP2 |
| chr15 | 75903110 | 75903714 | 605 | 4 | 5.26E-07 | 2.57E-05 | 0.00022 | 3.30E-06 | 0.007448832 | NA |
| chr2 | 101308205 | 101309692 | 1488 | 10 | 5.28E-07 | 1.27E-05 | 9.92E-05 | 3.32E-07 | -0.003119503 | MIR5696, RNF149 |
| chr19 | 1110401 | 1112328 | 1928 | 7 | 5.28E-07 | 1.69E-06 | 0.00016 | 2.56E-07 | -0.004791832 | SBNO2 |
| chr1 | 240596810 | 240597377 | 568 | 3 | 5.30E-07 | 6.94E-05 | 4.50E-05 | 1.01E-06 | -0.01825676 | GREM2 |
| chr12 | 42240436 | 42241005 | 570 | 2 | 5.31E-07 | 2.29E-05 | 4.76E-06 | 1.75E-06 | -0.008838053 | PPHLN1 |
| chr5 | 32613160 | 32613203 | 44 | 2 | 5.31E-07 | 1.20E-07 | 4.71E-05 | 2.04E-07 | -0.004184191 | NA |
| chr10 | 104314523 | 104316199 | 1677 | 8 | 5.34E-07 | 2.03E-07 | 7.93E-05 | 9.83E-10 | -0.003326003 | ITPRIP |
| chr16 | 85313820 | 85314275 | 456 | 3 | 5.34E-07 | 1.32E-06 | 2.17E-05 | 4.97E-07 | -0.01195062 | GSE1 |
| chr14 | 51094844 | 51096716 | 1873 | 20 | 5.34E-07 | 9.73E-08 | 0.00431 | 1.10E-08 | -0.001836811 | TRIM9 |
| chr6 | 111258691 | 111259736 | 1046 | 17 | 5.35E-07 | 2.38E-07 | 0.00197 | 2.26E-07 | -0.001447987 | MFSD4B, AL360227.1 |
| chr15 | 43184476 | 43185198 | 723 | 6 | 5.37E-07 | 7.54E-05 | 0.00044 | 6.53E-06 | -0.003333824 | TMEM62, CCNDBP1, AC068724.3, AC068724.4, AC068724.1 |
| chr9 | 127763585 | 127764040 | 456 | 4 | 5.37E-07 | 1.83E-06 | 3.31E-05 | 6.90E-07 | -0.00749522 | SH2D3C |
| chr19 | 41998579 | 41999260 | 682 | 5 | 5.40E-07 | 6.33E-09 | 0.00098 | 5.15E-08 | -0.015223488 | GRIK5 |
| chr17 | 45270224 | 45271156 | 933 | 2 | 5.40E-07 | 6.83E-09 | 7.78E-06 | 9.97E-09 | -0.015626695 | MAP3K14 |
| chr2 | 74548302 | 74549283 | 982 | 10 | 5.42E-07 | 8.73E-06 | 0.00021 | 1.14E-06 | -0.003729893 | DOK1, LOXL3 |
| chr1 | 1625069 | 1625514 | 446 | 4 | 5.42E-07 | 3.16E-06 | 5.53E-05 | 1.01E-07 | -0.005762555 | MIB2 |
| chr1 | 109914738 | 109915561 | 824 | 5 | 5.42E-07 | 3.43E-05 | 4.43E-05 | 1.16E-06 | -0.005445002 | CSF1 |
| chr1 | 232293043 | 232293122 | 80 | 2 | 5.43E-07 | 6.11E-08 | 2.71E-05 | 9.63E-08 | -0.021588579 | NA |
| chr13 | 112992379 | 112992585 | 207 | 3 | 5.43E-07 | 7.45E-08 | 0.00036 | 2.34E-07 | -0.018718016 | MCF2L |
| chr7 | 72336002 | 72337199 | 1198 | 5 | 5.43E-07 | 3.20E-07 | 2.68E-05 | 3.29E-08 | 0.00541959 | CALN1 |
| chr7 | 99002999 | 99003176 | 178 | 3 | 5.45E-07 | 7.10E-07 | 5.89E-05 | 1.98E-07 | -0.007807764 | TRRAP |
| chr20 | 45887336 | 45888462 | 1127 | 6 | 5.47E-07 | 0.000286 | 1.89E-05 | 6.35E-06 | -0.001373358 | SPATA25 |
| chr11 | 45699612 | 45701259 | 1648 | 5 | 5.48E-07 | 8.27E-07 | 6.82E-05 | 1.60E-07 | -0.006131913 | NA |
| chr5 | 34804641 | 34805178 | 538 | 2 | 5.49E-07 | 2.34E-07 | 1.74E-05 | 2.46E-07 | -0.013413494 | RAI14 |

| | | | | | | | | | | |
|---|---|---|---|---|---|---|---|---|---|---|
| chr21 | 44994612 | 44995489 | 878 | 6 | 5.50E-07 | 1.22E-07 | 0.00015 | 4.62E-08 | -0.007510752 | LINC00165 |
| chr22 | 38865084 | 38865717 | 634 | 2 | 5.51E-07 | 0.000758 | 1.15E-05 | 1.16E-05 | -0.008260174 | CBX6 |
| chr4 | 173499963 | 173502486 | 2524 | 11 | 5.52E-07 | 2.22E-11 | 0.0016 | 2.37E-11 | -0.015361236 | NA |
| chr12 | 12695582 | 12696654 | 1073 | 11 | 5.54E-07 | 1.18E-08 | 0.0014 | 5.78E-08 | -0.005912027 | GPR19 |
| chr1 | 246900735 | 246900840 | 106 | 3 | 5.58E-07 | 8.63E-05 | 5.90E-07 | 4.28E-07 | -0.002384481 | AHCTF1 |
| chr10 | 89281669 | 89282770 | 1102 | 3 | 5.58E-07 | 2.20E-05 | 1.32E-05 | 2.72E-06 | -0.002562782 | LIPA |
| chr17 | 39659878 | 39661617 | 1740 | 6 | 5.58E-07 | 1.37E-06 | 2.97E-05 | 4.28E-08 | -0.008035101 | STARD3 |
| chr19 | 58118534 | 58119245 | 712 | 8 | 5.60E-07 | 3.64E-05 | 0.00012 | 6.38E-07 | 0.001213875 | NA |
| chr20 | 4949750 | 4950024 | 275 | 3 | 5.62E-07 | 7.86E-08 | 5.51E-05 | 1.42E-07 | -0.016892768 | SLC23A2 |
| chr20 | 24745764 | 24746431 | 668 | 3 | 5.62E-07 | 2.92E-08 | 7.57E-05 | 5.71E-08 | -0.016805822 | NA |
| chr20 | 58849466 | 58850460 | 995 | 3 | 5.63E-07 | 6.68E-07 | 0.00058 | 1.71E-06 | -0.013900198 | GNAS, GNAS-AS1 |
| chr10 | 5619604 | 5620061 | 458 | 3 | 5.63E-07 | 1.42E-07 | 0.00045 | 4.15E-07 | -0.013289151 | LASTR |
| chr19 | 817794 | 817832 | 39 | 2 | 5.63E-07 | 6.42E-07 | 2.56E-06 | 1.53E-07 | -0.010547836 | PLPPR3 |
| chr19 | 29817228 | 29817512 | 285 | 3 | 5.64E-07 | 2.88E-08 | 0.00033 | 1.02E-07 | -0.014703304 | CCNE1 |
| chr2 | 102718463 | 102719580 | 1118 | 6 | 5.65E-07 | 1.88E-05 | 9.57E-05 | 1.65E-06 | -0.00458068 | MFSD9 |
| chr10 | 104304961 | 104305978 | 1018 | 5 | 5.65E-07 | 6.25E-08 | 0.00037 | 4.84E-08 | -0.008829919 | NA |
| chr13 | 98301017 | 98301310 | 294 | 3 | 5.65E-07 | 0.014895 | 3.28E-06 | 3.63E-06 | 0.003752426 | FARP1 |
| chr17 | 75763528 | 75765025 | 1498 | 7 | 5.66E-07 | 3.65E-06 | 0.00226 | 1.61E-06 | -0.007311795 | GALK1 |
| chr6 | 158167731 | 158168825 | 1095 | 16 | 5.66E-07 | 1.30E-06 | 2.75E-05 | 2.60E-07 | -0.000113522 | GTF2H5, SERAC1 |
| chr16 | 30833868 | 30834829 | 962 | 6 | 5.67E-07 | 7.02E-05 | 7.13E-05 | 9.23E-07 | 0.000440181 | BCL7C |
| chr17 | 81128632 | 81129169 | 538 | 7 | 5.68E-07 | 2.84E-06 | 1.41E-05 | 3.22E-07 | -0.003217434 | AATK |
| chr19 | 1970993 | 1971282 | 290 | 2 | 5.73E-07 | 0.000225 | 7.73E-06 | 6.06E-06 | -0.003925018 | CSNK1G2 |
| chr22 | 39908867 | 39910112 | 1246 | 3 | 5.74E-07 | 6.45E-07 | 2.55E-05 | 4.31E-07 | -0.011845397 | GRAP2 |
| chr10 | 113704260 | 113704951 | 692 | 3 | 5.77E-07 | 1.58E-07 | 5.70E-05 | 2.32E-07 | -0.008173375 | CASP7 |
| chr2 | 74374304 | 74375463 | 1160 | 8 | 5.77E-07 | 6.29E-06 | 0.00161 | 6.49E-07 | -0.004594906 | DCTN1 |
| chr9 | 121566489 | 121567235 | 747 | 7 | 5.83E-07 | 3.77E-06 | 0.00029 | 6.46E-07 | -0.003917033 | DAB2IP |
| chr12 | 104546833 | 104547367 | 535 | 3 | 5.85E-07 | 5.89E-07 | 5.96E-05 | 3.45E-07 | -0.021342425 | CHST11 |
| chr17 | 81821273 | 81822652 | 1380 | 26 | 5.86E-07 | 9.67E-07 | 0.00615 | 2.61E-07 | -0.006512543 | MCRIP1 |
| chr7 | 99081991 | 99082603 | 613 | 3 | 5.94E-07 | 3.03E-07 | 1.10E-05 | 1.43E-07 | 0.000120297 | SMURF1 |
| chr15 | 90645809 | 90646198 | 390 | 2 | 5.94E-07 | 1.55E-05 | 3.25E-06 | 1.01E-06 | -0.005061769 | CRTC3-AS1 |
| chr17 | 2904299 | 2905461 | 1163 | 4 | 6.02E-07 | 4.88E-07 | 3.57E-05 | 8.60E-08 | -0.011428322 | RAP1GAP2 |
| chr7 | 102293052 | 102293262 | 211 | 2 | 6.04E-07 | 1.19E-07 | 2.23E-05 | 1.59E-07 | -0.035459432 | SH2B2, MIR4285 |
| chr6 | 108689405 | 108690085 | 681 | 5 | 6.05E-07 | 2.46E-06 | 1.36E-05 | 4.20E-07 | -0.009190734 | NA |
| chr1 | 159828998 | 159829272 | 275 | 3 | 6.06E-07 | 5.61E-06 | 0.00011 | 1.04E-06 | -0.007671275 | SLAMF8 |
| chr20 | 32727792 | 32728473 | 682 | 5 | 6.09E-07 | 6.87E-07 | 9.32E-05 | 2.70E-08 | -0.011415342 | FO393400.1, COMMD7 |
| chr3 | 33382369 | 33382956 | 588 | 2 | 6.11E-07 | 0.002524 | 4.16E-06 | 4.24E-06 | -0.003071163 | FBXL2 |
| chr17 | 42562683 | 42563263 | 581 | 8 | 6.11E-07 | 9.09E-06 | 0.00078 | 1.25E-06 | -0.006539988 | COASY |
| chr17 | 40552147 | 40552557 | 411 | 5 | 6.12E-07 | 3.32E-06 | 0.00021 | 1.09E-07 | -0.005293567 | NA |
| chr2 | 241773994 | 241775607 | 1614 | 6 | 6.20E-07 | 1.61E-06 | 4.13E-05 | 1.37E-07 | -0.011509848 | NA |
| chr20 | 63665872 | 63666395 | 524 | 3 | 6.21E-07 | 3.37E-07 | 8.76E-05 | 2.78E-07 | -0.012113856 | RTEL1, RTEL1-TNFRSF6B |
| chr11 | 57321892 | 57322801 | 910 | 7 | 6.24E-07 | 1.67E-06 | 0.00012 | 7.00E-07 | -0.007815651 | TNKS1BP1 |
| chr6 | 110813803 | 110814651 | 849 | 10 | 6.25E-07 | 1.50E-05 | 0.00036 | 5.10E-07 | -0.00213711 | CDK19 |
| chr15 | 93312521 | 93313066 | 546 | 4 | 6.27E-07 | 3.25E-06 | 4.85E-05 | 5.16E-07 | -0.001318692 | AC091078.1 |
| chr5 | 128377827 | 128378364 | 538 | 4 | 6.32E-07 | 1.03E-05 | 2.44E-06 | 4.79E-07 | -0.003797825 | FBN2 |
| chr3 | 169812239 | 169813132 | 894 | 16 | 6.35E-07 | 6.68E-07 | 0.00922 | 4.39E-07 | -0.008915594 | LRRC34 |
| chr5 | 87412495 | 87413309 | 815 | 13 | 6.35E-07 | 1.37E-06 | 0.00092 | 7.29E-07 | 0.001834313 | AC018754.1, CCNH |
| chr14 | 34131133 | 34131701 | 569 | 6 | 6.41E-07 | 1.19E-05 | 0.00021 | 1.83E-06 | -0.005870844 | EGLN3 |
| chr19 | 8259867 | 8259890 | 24 | 2 | 6.41E-07 | 6.28E-07 | 3.03E-06 | 1.68E-07 | -0.00853795 | CERS4 |
| chr1 | 29767272 | 29767418 | 147 | 2 | 6.42E-07 | 5.19E-08 | 6.71E-05 | 1.02E-07 | -0.040895514 | AL645944.2 |
| chr17 | 50164994 | 50166346 | 1353 | 10 | 6.43E-07 | 1.52E-06 | 0.00114 | 1.77E-07 | -0.004159684 | SGCA, AC015909.3 |
| chr16 | 68370894 | 68371503 | 610 | 5 | 6.47E-07 | 1.46E-06 | 0.00038 | 6.94E-07 | -0.007323758 | SMPD3 |
| chr3 | 58536962 | 58537843 | 882 | 5 | 6.52E-07 | 9.26E-08 | 0.00102 | 2.79E-07 | -0.010875226 | ACOX2 |
| chr1 | 12616755 | 12617258 | 504 | 4 | 6.53E-07 | 7.35E-07 | 0.00173 | 2.05E-06 | -0.005373665 | DHRS3 |
| chr10 | 131726053 | 131726635 | 583 | 5 | 6.53E-07 | 1.09E-08 | 0.0013 | 6.15E-08 | 0.023757504 | NA |
| chr12 | 121520457 | 121521029 | 573 | 4 | 6.59E-07 | 7.11E-08 | 0.00014 | 6.01E-08 | -0.006062528 | KDM2B |
| chr1 | 156291127 | 156292480 | 1354 | 10 | 6.60E-07 | 2.37E-08 | 0.00568 | 1.01E-07 | -0.009879286 | TMEM79, GLMP |
| chr10 | 70376607 | 70377264 | 658 | 6 | 6.60E-07 | 1.81E-05 | 0.00046 | 7.40E-07 | -0.006627556 | LRRC20 |
| chr11 | 76719471 | 76720168 | 698 | 6 | 6.60E-07 | 4.67E-06 | 0.00018 | 2.79E-07 | -0.003463969 | GUCY2EP |
| chr2 | 109006025 | 109006110 | 86 | 3 | 6.64E-07 | 1.85E-05 | 5.39E-05 | 2.19E-07 | -0.009287391 | NA |
| chr3 | 15809458 | 15810031 | 574 | 2 | 6.70E-07 | 2.09E-05 | 5.24E-06 | 1.82E-06 | -0.002705777 | ANKRD28 |
| chr10 | 125005578 | 125006184 | 607 | 7 | 6.71E-07 | 3.25E-06 | 0.00018 | 4.58E-07 | -0.001947954 | CTBP2 |
| chr14 | 75453372 | 75453495 | 124 | 3 | 6.71E-07 | 1.19E-07 | 0.00023 | 2.93E-07 | -0.013894495 | JDP2 |
| chr18 | 5891603 | 5892428 | 826 | 7 | 6.73E-07 | 3.50E-05 | 0.00018 | 2.45E-06 | -0.01670723 | AP001021.1, TMEM200C |
| chr1 | 30778047 | 30778460 | 414 | 2 | 6.80E-07 | 2.64E-07 | 2.12E-05 | 2.92E-07 | -0.012619299 | NA |
| chr3 | 47848470 | 47849986 | 1517 | 12 | 6.81E-07 | 2.26E-08 | 0.00142 | 1.68E-08 | -0.003559845 | DHX30, MIR1226 |
| chr12 | 120546204 | 120546660 | 457 | 4 | 6.82E-07 | 1.16E-05 | 7.16E-06 | 7.28E-07 | -0.010604173 | RNF10 |
| chr16 | 67942461 | 67944547 | 2087 | 18 | 6.84E-07 | 4.40E-09 | 0.00075 | 3.62E-10 | -0.003196136 | LCAT, SLC12A4 |
| chr2 | 134719486 | 134719855 | 370 | 2 | 6.85E-07 | 0.000151 | 7.61E-06 | 5.45E-06 | -0.00375458 | AC110620.2 |
| chr7 | 105655418 | 105655579 | 162 | 2 | 6.86E-07 | 6.64E-08 | 4.74E-05 | 1.20E-07 | -0.01681836 | ATXN7L1 |
| chr16 | 88759402 | 88760021 | 620 | 3 | 6.94E-07 | 4.33E-06 | 0.00013 | 1.79E-06 | -0.023069303 | PIEZO1 |
| chr20 | 60308164 | 60308405 | 242 | 5 | 6.95E-07 | 1.26E-08 | 0.00107 | 6.49E-08 | -0.017163907 | MIR646HG |
| chr1 | 204094304 | 204095018 | 715 | 2 | 6.96E-07 | 5.67E-07 | 1.54E-05 | 4.48E-07 | -0.017972241 | SOX13 |
| chr19 | 40425790 | 40426369 | 580 | 11 | 6.96E-07 | 2.27E-05 | 0.00015 | 4.65E-07 | 0.002236059 | AC010271.2, SERTAD1 |
| chr8 | 60526271 | 60526306 | 36 | 2 | 6.97E-07 | 9.40E-07 | 8.87E-06 | 4.59E-07 | -0.004524941 | RAB2A |
| chr15 | 74407312 | 74407440 | 129 | 2 | 6.97E-07 | 9.69E-08 | 2.05E-05 | 1.31E-07 | -0.015420042 | NA |
| chr17 | 41822265 | 41822601 | 337 | 2 | 6.98E-07 | 0.00023 | 3.16E-06 | 2.31E-06 | -0.008165733 | FKBP10 |
| chr12 | 122096880 | 122098522 | 1643 | 9 | 6.98E-07 | 7.14E-11 | 0.00043 | 9.73E-11 | -0.007092606 | MLXIP |
| chr11 | 47979746 | 47980388 | 643 | 5 | 6.99E-07 | 0.000103 | 0.0001 | 4.35E-06 | -0.006946911 | NA |
| chr6 | 32405843 | 32407895 | 2053 | 9 | 7.00E-07 | 7.69E-08 | 0.00024 | 7.49E-09 | -0.01042098 | TSBP1-AS1, BTNL2 |
| chr17 | 7294056 | 7294768 | 713 | 10 | 7.05E-07 | 0.000735 | 0.00012 | 2.78E-05 | -0.000676888 | YBX2 |
| chr7 | 129210535 | 129212089 | 1555 | 18 | 7.07E-07 | 4.06E-07 | 0.00277 | 2.30E-07 | -0.002950861 | SMO, AC011005.4 |
| chr5 | 134958336 | 134958786 | 451 | 2 | 7.11E-07 | 4.22E-06 | 5.50E-06 | 8.32E-07 | 0.00093955 | PCBD2 |
| chr4 | 154465126 | 154465195 | 70 | 2 | 7.17E-07 | 5.93E-08 | 6.40E-05 | 1.13E-07 | -0.02389268 | DCHS2 |
| chr2 | 96760537 | 96761708 | 1172 | 11 | 7.21E-07 | 0.000203 | 9.99E-05 | 9.73E-07 | -0.000208848 | CNNM4 |
| chr19 | 48497145 | 48499825 | 2681 | 7 | 7.22E-07 | 5.50E-15 | 0.00058 | 2.41E-13 | -0.03491411 | LMTK3 |
| chr14 | 99262602 | 99262944 | 343 | 6 | 7.22E-07 | 4.35E-06 | 0.00095 | 3.16E-07 | -0.015409224 | BCL11B |
| chr5 | 142685974 | 142686680 | 707 | 10 | 7.24E-07 | 6.78E-06 | 0.00019 | 5.27E-07 | -0.003744698 | FGF1 |
| chr9 | 34379716 | 34382301 | 2586 | 22 | 7.26E-07 | 1.09E-11 | 0.00107 | 4.17E-12 | -0.00411201 | C9orf24 |
| chr15 | 75234040 | 75234883 | 844 | 4 | 7.26E-07 | 4.37E-07 | 0.00011 | 5.23E-07 | -0.008213541 | AC113208.3 |
| chr1 | 8871615 | 8873059 | 1445 | 8 | 7.29E-07 | 6.85E-08 | 0.00014 | 1.36E-07 | -0.008827661 | ENO1 |
| chr16 | 85978009 | 85979234 | 1226 | 7 | 7.29E-07 | 9.46E-08 | 0.00073 | 1.30E-07 | -0.007664986 | AC092723.4 |
| chr4 | 183904291 | 183905326 | 1036 | 9 | 7.33E-07 | 0.000875 | 2.95E-05 | 8.94E-06 | -0.002193409 | STOX2 |
| chr7 | 982577 | 983520 | 944 | 12 | 7.36E-07 | 2.55E-05 | 0.0011 | 1.24E-06 | -0.008137065 | CYP2W1 |
| chr1 | 200872518 | 200873762 | 1245 | 10 | 7.37E-07 | 5.47E-05 | 0.00104 | 2.95E-06 | -0.005228539 | GPR25 |
| chr3 | 42148243 | 42149115 | 873 | 6 | 7.38E-07 | 4.11E-07 | 0.00045 | 5.49E-07 | -0.014214781 | TRAK1 |
| chr13 | 113853813 | 113855779 | 1967 | 8 | 7.38E-07 | 3.44E-07 | 0.00059 | 1.51E-08 | -0.010587119 | GAS6 |
| chr1 | 14959060 | 14959285 | 226 | 2 | 7.38E-07 | 5.84E-07 | 1.15E-05 | 3.89E-07 | -0.018288089 | KAZN |
| chr16 | 85261877 | 85262614 | 738 | 4 | 7.38E-07 | 3.10E-05 | 8.23E-05 | 1.24E-06 | -0.000642343 | GSE1, AC092275.1 |
| chr17 | 39199708 | 39200814 | 1107 | 11 | 7.40E-07 | 0.000169 | 9.39E-06 | 1.97E-06 | 0.000754171 | RPL19 |
| chr6 | 31761871 | 31763523 | 1653 | 8 | 7.40E-07 | 2.74E-09 | 0.00129 | 7.27E-09 | -0.003809354 | MSH5, MSH5-SAPCD1, SAPCD1 |
| chr17 | 80836052 | 80836073 | 22 | 2 | 7.40E-07 | 1.02E-07 | 3.92E-05 | 1.69E-07 | -0.037425338 | RPTOR |
| chr7 | 151709087 | 151709722 | 636 | 3 | 7.42E-07 | 3.89E-06 | 0.00011 | 1.01E-06 | -0.006378162 | PRKAG2 |
| chr2 | 233307404 | 233307916 | 513 | 10 | 7.44E-07 | 2.09E-05 | 0.00016 | 3.47E-07 | -0.005212497 | SAG |
| chr9 | 133814536 | 133814863 | 328 | 2 | 7.48E-07 | 1.21E-07 | 7.59E-05 | 2.31E-07 | -0.011494963 | VAV2 |
| chr8 | 144007972 | 144009294 | 1323 | 4 | 7.50E-07 | 1.04E-06 | 2.89E-05 | 2.12E-08 | -0.009570028 | PARP10 |
| chr12 | 94177416 | 94177809 | 394 | 2 | 7.50E-07 | 0.000132 | 3.25E-06 | 2.13E-06 | -0.008896719 | PLXNC1, AC123567.2 |
| chr12 | 122279871 | 122280493 | 623 | 3 | 7.51E-07 | 4.82E-06 | 0.00019 | 2.02E-06 | -0.01866654 | CLIP1 |
| chr10 | 50374276 | 50374955 | 680 | 2 | 7.52E-07 | 2.35E-07 | 2.34E-05 | 2.77E-07 | -0.012368096 | SGMS1 |
| chr17 | 48629976 | 48630419 | 444 | 2 | 7.53E-07 | 2.05E-06 | 1.21E-05 | 9.48E-07 | -0.000351351 | HOXB-AS4, HOXB7 |
| chr2 | 240424505 | 240425004 | 500 | 4 | 7.56E-07 | 0.000367 | 6.82E-06 | 1.80E-06 | -0.005382978 | NA |
| chr20 | 37711222 | 37711986 | 765 | 2 | 7.62E-07 | 1.77E-06 | 8.87E-06 | 7.01E-07 | -0.015183434 | CTNNBL1 |
| chr19 | 50334315 | 50334731 | 417 | 3 | 7.62E-07 | 0.000114 | 8.87E-06 | 5.51E-06 | -0.005060219 | NR1H2, NAPSB |
| chr16 | 2035817 | 2037376 | 1560 | 5 | 7.67E-07 | 3.86E-05 | 1.28E-05 | 2.02E-06 | -0.001368366 | SLC9A3R2 |
| chr10 | 132402948 | 132403184 | 237 | 2 | 7.67E-07 | 3.66E-07 | 0.00014 | 6.78E-07 | -0.006744323 | PWWP2B |
| chr9 | 128940172 | 128940785 | 614 | 3 | 7.68E-07 | 9.20E-09 | 0.00029 | 3.67E-08 | -0.010139316 | PHYHD1 |
| chr19 | 38894127 | 38894899 | 773 | 4 | 7.71E-07 | 2.37E-05 | 6.16E-05 | 1.77E-06 | 0.004117098 | SIRT2 |
| chr16 | 87830825 | 87831456 | 632 | 5 | 7.71E-07 | 1.77E-06 | 9.50E-05 | 6.56E-07 | -0.00300357 | SLC7A5 |
| chr2 | 234171164 | 234171950 | 787 | 6 | 7.72E-07 | 9.50E-06 | 9.73E-05 | 7.57E-07 | -0.003686011 | NA |
| chr18 | 6284001 | 6284571 | 571 | 5 | 7.72E-07 | 7.37E-05 | 0.00014 | 1.02E-06 | -0.001993893 | L3MBTL4 |
| chr19 | 5036846 | 5038163 | 1318 | 6 | 7.75E-07 | 1.28E-06 | 2.55E-05 | 5.37E-08 | -0.005703874 | KDM4B |
| chr3 | 186664627 | 186666085 | 1459 | 8 | 7.78E-07 | 4.39E-10 | 0.00031 | 3.62E-10 | -0.010457921 | HRG, AC068631.1 |
| chr10 | 70888339 | 70889681 | 1343 | 11 | 7.85E-07 | 3.46E-05 | 0.00025 | 6.42E-07 | 0.004116897 | PCBD1 |
| chr12 | 9624305 | 9625291 | 987 | 3 | 7.90E-07 | 2.67E-05 | 1.53E-05 | 2.29E-06 | -0.006047884 | AC092821.3 |
| chr17 | 30956852 | 30957030 | 179 | 2 | 7.96E-07 | 2.76E-07 | 5.82E-05 | 4.42E-07 | -0.017665865 | ADAP2, AC138207.1 |
| chr17 | 3794699 | 3795041 | 343 | 7 | 8.01E-07 | 0.002216 | 1.04E-06 | 1.52E-06 | -0.003294715 | ITGAE |
| chr1 | 2028003 | 2028463 | 461 | 5 | 8.06E-07 | 1.30E-06 | 0.00042 | 6.42E-07 | -0.007314277 | GABRD |
| chr17 | 81200396 | 81201332 | 937 | 4 | 8.11E-07 | 1.50E-05 | 2.74E-05 | 3.94E-06 | -0.006039038 | CEP131 |
| chr16 | 4623917 | 4625386 | 1470 | 14 | 8.11E-07 | 1.29E-08 | 0.00247 | 6.37E-09 | -0.001332132 | MGRN1 |
| chr1 | 52574716 | 52574819 | 104 | 2 | 8.12E-07 | 2.90E-07 | 7.21E-06 | 1.81E-07 | -0.007882734 | NA |
| chr5 | 10521628 | 10522680 | 1053 | 5 | 8.14E-07 | 3.54E-06 | 5.93E-05 | 1.29E-06 | -0.007014074 | LINC02213 |
| chr6 | 43181303 | 43182465 | 1163 | 9 | 8.16E-07 | 4.38E-07 | 0.00064 | 2.64E-07 | -0.004750641 | SRF, CUL9 |
| chr10 | 80289216 | 80290102 | 887 | 6 | 8.20E-07 | 1.27E-06 | 0.00134 | 5.63E-07 | -0.01523773 | MAT1A |
| chr3 | 43285294 | 43286537 | 1244 | 10 | 8.22E-07 | 0.000134 | 6.52E-05 | 3.19E-06 | 0.000455752 | SNRK |
| chr2 | 96115668 | 96116704 | 1037 | 7 | 8.25E-07 | 2.72E-06 | 0.00086 | 1.23E-06 | -0.004331391 | ADRA2B |
| chr17 | 44751458 | 44751925 | 468 | 2 | 8.26E-07 | 6.54E-08 | 6.60E-05 | 1.26E-07 | -6.63E-06 | DBF4B |
| chr12 | 57039616 | 57040282 | 667 | 3 | 8.29E-07 | 1.08E-05 | 0.00013 | 2.04E-06 | -0.010710971 | MYO1A |
| chr17 | 58755190 | 58756064 | 875 | 14 | 8.33E-07 | 0.000174 | 0.0004 | 2.44E-06 | -6.83E-05 | PPM1E |
| chr17 | 58525012 | 58525390 | 379 | 3 | 8.33E-07 | 1.78E-06 | 9.59E-06 | 5.04E-07 | -0.0081917 | SEPTIN4-AS1, SEPTIN4 |
| chr1 | 158253723 | 158254144 | 422 | 4 | 8.41E-07 | 1.33E-06 | 4.51E-05 | 3.35E-07 | -0.012064696 | NA |
| chr17 | 41634797 | 41635969 | 1173 | 5 | 8.45E-07 | 8.64E-06 | 0.00012 | 5.16E-07 | -0.003416323 | KRT42P |
| chr7 | 76358895 | 76359988 | 1094 | 13 | 8.48E-07 | 1.28E-06 | 0.00092 | 5.17E-07 | -0.001589213 | YWHAG |
| chr3 | 152834195 | 152836266 | 2072 | 15 | 8.48E-07 | 4.35E-07 | 0.00113 | 2.14E-08 | -0.001160049 | P2RY1 |
| chr6 | 31127499 | 31128136 | 638 | 6 | 8.48E-07 | 1.44E-05 | 4.85E-05 | 9.26E-07 | -0.008253151 | PSORS1C1 |
| chr4 | 80335339 | 80336674 | 1336 | 14 | 8.50E-07 | 0.000559 | 6.62E-05 | 4.05E-06 | 0.000931514 | FGF5, CFAP299 |
| chr11 | 85719027 | 85720136 | 1110 | 5 | 8.50E-07 | 0.000351 | 3.98E-06 | 1.47E-06 | -0.006800398 | SYTL2 |
| chr5 | 154466388 | 154466992 | 605 | 3 | 8.51E-07 | 0.000145 | 4.31E-06 | 2.21E-06 | 0.000147979 | NA |
| chr20 | 841856 | 842539 | 684 | 3 | 8.53E-07 | 3.66E-07 | 0.00017 | 5.92E-07 | -0.011583587 | FAM110A |
| chr7 | 99372738 | 99375110 | 2373 | 14 | 8.53E-07 | 1.72E-08 | 0.00093 | 1.20E-09 | -0.002970924 | AC004922.1, ARPC1B |
| chr6 | 3427887 | 3427999 | 113 | 2 | 8.55E-07 | 9.84E-07 | 3.58E-06 | 2.51E-07 | -0.021198748 | SLC22A23 |
| chr6 | 31629136 | 31630054 | 919 | 12 | 8.56E-07 | 5.12E-05 | 9.85E-05 | 2.90E-06 | -0.002069207 | PRRC2A |
| chr13 | 102394580 | 102395410 | 831 | 7 | 8.56E-07 | 1.43E-06 | 0.00013 | 2.99E-07 | 0.001413081 | FGF14-AS2, FGF14 |
| chr4 | 150722914 | 150723075 | 162 | 2 | 8.56E-07 | 6.86E-08 | 7.36E-05 | 1.34E-07 | -0.027640221 | LRBA |
| chr12 | 53753977 | 53755019 | 1043 | 6 | 8.59E-07 | 3.21E-06 | 0.00064 | 1.20E-06 | 0.021966098 | AC076968.2, AC076968.1, CISTR |
| chr9 | 94800030 | 94800058 | 29 | 2 | 8.64E-07 | 2.07E-06 | 1.01E-05 | 8.44E-07 | -0.018573805 | AOPEP |
| chr7 | 65954198 | 65954301 | 104 | 6 | 8.67E-07 | 2.00E-06 | 7.64E-05 | 3.30E-07 | -0.00829827 | VKORC1L1 |
| chr1 | 54327955 | 54328692 | 738 | 2 | 8.81E-07 | 3.65E-08 | 3.72E-05 | 6.71E-08 | -0.005913434 | SSBP3 |
| chr6 | 3350667 | 3351159 | 493 | 2 | 8.81E-07 | 0.000383 | 7.87E-06 | 6.87E-06 | -0.007938676 | SLC22A23 |
| chr16 | 68357141 | 68357352 | 212 | 2 | 8.82E-07 | 6.94E-08 | 7.50E-05 | 1.36E-07 | -0.012798465 | PRMT7 |
| chr17 | 17202543 | 17203525 | 983 | 9 | 8.85E-07 | 1.07E-07 | 0.00011 | 1.70E-08 | 3.47E-05 | MPRIP, PLD6, AC055811.2 |
| chr18 | 50281322 | 50282705 | 1384 | 15 | 8.86E-07 | 7.36E-05 | 0.00036 | 1.52E-06 | -0.000409801 | MBD1, CXXC1 |
| chr6 | 152636986 | 152638520 | 1535 | 16 | 8.92E-07 | 5.75E-09 | 0.00353 | 4.70E-08 | -0.006005219 | SYNE1 |
| chr19 | 14561916 | 14562241 | 326 | 3 | 8.92E-07 | 5.85E-07 | 3.65E-05 | 4.14E-07 | -0.017318193 | TECR |

| | | | | | | | | | | |
|---|---|---|---|---|---|---|---|---|---|---|
| chr14 | 105485267 | 105487125 | 1859 | 17 | 8.95E-07 | 3.29E-10 | 0.0066 | 5.54E-09 | -0.004142949 | CRIP1 |
| chr9 | 133441805 | 133442109 | 305 | 2 | 8.95E-07 | 1.62E-06 | 5.25E-06 | 4.61E-07 | -0.008126109 | ADAMTS13 |
| chr6 | 169794433 | 169794483 | 51 | 2 | 8.97E-07 | 3.26E-07 | 1.67E-05 | 3.08E-07 | -0.014054742 | LINC00574, LINC00242 |
| chr17 | 393721 | 394577 | 857 | 3 | 9.02E-07 | 1.91E-08 | 0.00027 | 6.97E-08 | -0.030896106 | NA |
| chr19 | 40266013 | 40266359 | 347 | 3 | 9.09E-07 | 0.006924 | 3.44E-06 | 8.63E-06 | -0.008557637 | AKT2 |
| chr8 | 25479425 | 25479998 | 574 | 5 | 9.11E-07 | 4.99E-07 | 0.00017 | 2.66E-07 | -0.003353313 | CDCA2 |
| chr7 | 73880889 | 73881600 | 712 | 2 | 9.12E-07 | 2.99E-06 | 1.94E-05 | 1.66E-06 | -0.010334173 | NA |
| chr5 | 75053819 | 75054990 | 1172 | 8 | 9.16E-07 | 7.31E-08 | 0.00208 | 1.31E-07 | -0.01321042 | NA |
| chr22 | 29689896 | 29690963 | 1068 | 4 | 9.18E-07 | 2.83E-06 | 4.46E-05 | 1.65E-06 | -0.005469081 | NF2 |
| chr8 | 1988969 | 1989757 | 789 | 5 | 9.19E-07 | 6.64E-08 | 0.00086 | 1.63E-07 | -0.015997455 | KBTBD11 |
| chr3 | 13064178 | 13064936 | 759 | 2 | 9.19E-07 | 1.58E-07 | 5.95E-05 | 2.71E-07 | -0.007189354 | IQSEC1 |
| chr7 | 129755323 | 129756238 | 916 | 5 | 9.22E-07 | 9.12E-08 | 0.00018 | 1.40E-07 | -0.002899076 | NRF1 |
| chr15 | 41881887 | 41883132 | 1246 | 5 | 9.23E-07 | 8.55E-05 | 0.00031 | 7.97E-06 | -0.007100916 | SPTBN5 |
| chr12 | 128365651 | 128366358 | 708 | 7 | 9.25E-07 | 5.78E-05 | 3.15E-05 | 6.89E-07 | 7.67E-05 | TMEM132C |
| chr19 | 42368857 | 42369796 | 940 | 8 | 9.26E-07 | 2.64E-08 | 0.00262 | 1.06E-07 | -0.00089861 | MEGF8 |
| chr16 | 610965 | 611482 | 518 | 4 | 9.27E-07 | 1.08E-07 | 0.00033 | 2.53E-07 | -0.010223978 | RAB40C |
| chr16 | 2183797 | 2184570 | 774 | 6 | 9.38E-07 | 2.86E-05 | 1.15E-05 | 1.24E-06 | -0.004077141 | CASKIN1 |
| chr11 | 77147602 | 77148341 | 740 | 5 | 9.44E-07 | 1.25E-08 | 0.002 | 9.40E-08 | -0.013497323 | MYO7A |
| chr16 | 67327251 | 67328065 | 815 | 3 | 9.45E-07 | 0.011835 | 1.74E-05 | 4.70E-05 | -0.007215885 | LRRC36 |
| chr15 | 36886554 | 36887578 | 1025 | 7 | 9.51E-07 | 3.55E-07 | 0.00024 | 5.99E-07 | -0.001914668 | AC018563.1 |
| chr6 | 109235711 | 109236601 | 891 | 5 | 9.52E-07 | 1.29E-07 | 5.84E-05 | 1.77E-07 | -0.022397468 | CCDC162P |
| chr12 | 52908016 | 52908426 | 411 | 3 | 9.53E-07 | 1.85E-06 | 1.38E-05 | 5.96E-07 | -0.012945166 | KRT8 |
| chr4 | 74117148 | 74117520 | 373 | 2 | 9.54E-07 | 2.61E-07 | 2.01E-05 | 2.82E-07 | -0.011601605 | MTHFD2L |
| chr9 | 113461756 | 113462376 | 621 | 2 | 9.54E-07 | 6.23E-08 | 4.57E-05 | 1.12E-07 | -0.007829583 | RGS3 |
| chr20 | 53012803 | 53013730 | 928 | 3 | 9.57E-07 | 9.80E-08 | 0.00017 | 1.82E-07 | -0.011403401 | TSHZ2 |
| chr17 | 56153058 | 56153635 | 578 | 10 | 9.59E-07 | 2.91E-06 | 0.00057 | 3.16E-07 | -0.001724964 | ANKFN1 |
| chr18 | 79506058 | 79507148 | 1091 | 6 | 9.59E-07 | 1.97E-08 | 0.00021 | 5.50E-08 | -0.007692064 | NFATC1 |
| chr11 | 72081160 | 72082051 | 892 | 5 | 9.60E-07 | 2.08E-05 | 0.00024 | 1.35E-05 | -0.004668054 | AP000812.4, LRTOMT |
| chr16 | 30895021 | 30896569 | 1549 | 9 | 9.70E-07 | 8.31E-06 | 0.00129 | 1.23E-06 | -0.007104094 | MIR762HG, AC135048.2 |
| chr8 | 23353704 | 23355224 | 1521 | 11 | 9.77E-07 | 1.81E-05 | 0.00021 | 2.95E-07 | -0.004968224 | AC090197.1, LOXL2 |
| chr7 | 4742073 | 4743012 | 940 | 5 | 9.82E-07 | 1.58E-09 | 0.00149 | 1.67E-08 | -0.005549035 | FOXK1 |
| chr6 | 13413473 | 13414033 | 561 | 3 | 9.88E-07 | 2.01E-07 | 2.86E-05 | 1.67E-07 | -0.009438736 | GFOD1 |
| chr2 | 78993384 | 78994140 | 757 | 4 | 9.89E-07 | 3.36E-05 | 3.59E-05 | 6.09E-06 | 0.010234961 | NA |
| chr1 | 224928892 | 224930115 | 1224 | 12 | 9.92E-07 | 3.52E-05 | 0.00011 | 1.74E-06 | -0.001676964 | DNAH14 |
| chr16 | 8961636 | 8962759 | 1124 | 11 | 9.93E-07 | 5.73E-09 | 0.00646 | 6.93E-08 | -0.005086486 | AC022167.3, USP7 |
| chr3 | 133805316 | 133806354 | 1039 | 10 | 9.95E-07 | 0.000126 | 0.00012 | 6.02E-07 | -0.000850679 | SRPRB, AC080128.2 |
| chr1 | 228106700 | 228108427 | 1728 | 8 | 1.00E-06 | 5.68E-06 | 0.0005 | 1.81E-07 | -0.005177238 | MRPL55 |
| chr10 | 44829205 | 44829551 | 347 | 2 | 1.01E-06 | 8.12E-08 | 6.64E-05 | 1.54E-07 | -0.007017094 | TMEM72-AS1 |
| chr5 | 169980435 | 169980937 | 503 | 6 | 1.01E-06 | 2.61E-07 | 0.0006 | 1.96E-07 | -0.014237716 | DOCK2, INSYN2B |
| chr8 | 145002187 | 145003534 | 1348 | 18 | 1.01E-06 | 0.000375 | 1.70E-05 | 2.26E-06 | 0.001406351 | ZNF252P-AS1, ZNF252P |
| chr20 | 37413982 | 37414364 | 383 | 2 | 1.02E-06 | 9.38E-06 | 7.01E-06 | 1.55E-06 | -0.006376228 | NA |
| chr10 | 101066240 | 101066868 | 629 | 3 | 1.02E-06 | 7.84E-06 | 8.17E-05 | 3.32E-06 | -0.004364001 | KAZALD1 |
| chr17 | 81962256 | 81963255 | 1000 | 7 | 1.02E-06 | 0.081531 | 5.25E-06 | 1.42E-05 | -0.003725343 | NA |
| chr19 | 3189498 | 3189959 | 462 | 3 | 1.02E-06 | 1.64E-07 | 6.45E-05 | 2.66E-07 | -0.007888066 | NCLN |
| chr2 | 226215933 | 226215972 | 40 | 2 | 1.02E-06 | 1.16E-07 | 3.93E-05 | 1.89E-07 | -0.015569341 | NA |
| chr4 | 134201255 | 134202481 | 1227 | 9 | 1.03E-06 | 1.02E-07 | 0.00688 | 4.36E-07 | -0.007318699 | AC015631.1, PABPC4L |
| chr17 | 3801177 | 3801813 | 637 | 6 | 1.03E-06 | 3.99E-08 | 0.00137 | 1.64E-07 | -0.010577909 | ITGAE |
| chr17 | 74203227 | 74204421 | 1195 | 14 | 1.04E-06 | 6.56E-06 | 0.00051 | 1.72E-06 | 0.001334664 | RPL38 |
| chr21 | 42234747 | 42235809 | 1063 | 5 | 1.05E-06 | 5.77E-07 | 0.0005 | 4.20E-07 | -0.007542888 | ABCG1 |
| chr3 | 45884624 | 45885177 | 554 | 3 | 1.05E-06 | 2.26E-06 | 2.49E-05 | 1.17E-06 | -0.008565736 | LZTFL1 |
| chr12 | 90808755 | 90809228 | 474 | 3 | 1.05E-06 | 0.00324 | 1.17E-05 | 2.76E-06 | -0.011764414 | LINC02822, AC112481.1 |
| chr13 | 23324225 | 23324890 | 666 | 5 | 1.05E-06 | 1.04E-07 | 0.00074 | 2.67E-07 | -0.012806701 | SGCG |
| chr16 | 1344642 | 1345841 | 1200 | 5 | 1.05E-06 | 1.09E-08 | 0.00158 | 8.86E-08 | -0.009153694 | BAIAP3 |
| chr11 | 34160419 | 34161023 | 605 | 2 | 1.05E-06 | 5.07E-08 | 5.73E-05 | 9.82E-08 | -0.007810646 | ABTB2 |
| chr11 | 64117052 | 64117307 | 256 | 2 | 1.05E-06 | 0.004252 | 3.25E-06 | 3.19E-06 | -0.002914553 | FLRT1, MACROD1 |
| chr22 | 30308487 | 30309956 | 1470 | 6 | 1.06E-06 | 1.23E-06 | 0.00054 | 2.00E-07 | -0.006985216 | TBC1D10A |
| chr14 | 104772306 | 104773591 | 1286 | 15 | 1.06E-06 | 1.85E-06 | 0.00071 | 2.11E-07 | -0.00246387 | AKT1 |
| chr22 | 47392866 | 47393417 | 552 | 2 | 1.06E-06 | 1.11E-05 | 7.87E-06 | 1.87E-06 | -0.010561603 | NA |
| chr17 | 43671151 | 43671310 | 160 | 2 | 1.07E-06 | 3.11E-06 | 4.09E-06 | 5.57E-07 | -0.013162758 | NA |
| chr1 | 26157758 | 26158738 | 981 | 4 | 1.07E-06 | 0.000166 | 9.83E-05 | 1.62E-05 | -0.005992649 | NA |
| chr15 | 74802441 | 74803168 | 728 | 9 | 1.07E-06 | 4.30E-05 | 3.67E-05 | 1.66E-06 | -0.000247571 | CSK |
| chr10 | 84337033 | 84337305 | 273 | 2 | 1.08E-06 | 6.66E-07 | 8.87E-06 | 3.63E-07 | -0.010363077 | CCSER2 |
| chr11 | 57600570 | 57600619 | 50 | 2 | 1.08E-06 | 5.53E-05 | 2.03E-06 | 1.02E-06 | -0.005974735 | SERPING1 |
| chr11 | 64007754 | 64008925 | 1172 | 6 | 1.08E-06 | 2.71E-09 | 0.00132 | 1.26E-08 | 0.005144702 | MACROD1 |
| chr6 | 155945621 | 155945744 | 124 | 2 | 1.09E-06 | 0.001379 | 1.79E-06 | 1.41E-06 | -0.0105642 | AL589693.1 |
| chr11 | 993372 | 996018 | 2647 | 11 | 1.09E-06 | 1.23E-06 | 1.41E-05 | 4.99E-09 | -0.001300755 | AP2A2 |
| chr1 | 205014926 | 205015466 | 541 | 2 | 1.09E-06 | 2.05E-05 | 1.10E-05 | 3.40E-06 | -0.013594371 | NFASC |
| chr14 | 60734376 | 60735259 | 884 | 11 | 1.10E-06 | 4.72E-06 | 3.93E-05 | 6.49E-07 | 0.000457031 | MNAT1 |
| chr12 | 111773213 | 111773876 | 664 | 3 | 1.10E-06 | 2.03E-05 | 4.44E-06 | 1.33E-06 | -0.005947119 | AC002996.1, ALDH2 |
| chr15 | 71189097 | 71189382 | 286 | 3 | 1.10E-06 | 0.000257 | 1.96E-06 | 1.57E-06 | -0.01008981 | THSD4 |
| chr2 | 178108857 | 178109585 | 729 | 5 | 1.11E-06 | 6.43E-05 | 2.08E-05 | 6.48E-06 | -0.006983474 | NA |
| chr6 | 76861185 | 76861645 | 461 | 2 | 1.11E-06 | 8.45E-08 | 5.69E-05 | 1.55E-07 | -0.0210314 | AL355612.1 |
| chr15 | 67039575 | 67039874 | 300 | 2 | 1.11E-06 | 5.58E-07 | 1.13E-05 | 3.69E-07 | -0.010571233 | AC087482.1 |
| chr2 | 85705946 | 85706728 | 783 | 3 | 1.11E-06 | 1.22E-05 | 0.00016 | 1.92E-06 | -0.020090838 | NA |
| chr10 | 113369368 | 113369947 | 580 | 3 | 1.11E-06 | 4.64E-06 | 2.42E-05 | 1.56E-06 | -0.015251658 | NA |
| chr5 | 42811850 | 42812742 | 893 | 7 | 1.11E-06 | 1.03E-08 | 0.004 | 1.06E-07 | -0.009860864 | SELENOP |
| chr9 | 131189827 | 131190154 | 328 | 4 | 1.12E-06 | 3.31E-06 | 1.57E-05 | 6.17E-07 | -0.005953916 | NUP214, AL157938.3 |
| chr10 | 5547777 | 5548307 | 531 | 2 | 1.12E-06 | 4.06E-05 | 2.36E-05 | 9.08E-06 | -0.012749714 | NA |
| chr6 | 30684870 | 30686022 | 1153 | 16 | 1.12E-06 | 6.72E-08 | 0.01455 | 3.09E-07 | 0.006151335 | PPP1R18 |
| chr1 | 156491806 | 156492053 | 248 | 3 | 1.12E-06 | 1.27E-05 | 6.94E-06 | 1.38E-06 | -0.008345585 | MEF2D |
| chr15 | 61211209 | 61211818 | 610 | 3 | 1.12E-06 | 0.012335 | 3.04E-05 | 8.74E-06 | -0.010109209 | RORA |
| chr6 | 7051070 | 7051954 | 885 | 5 | 1.13E-06 | 5.55E-06 | 0.0003 | 2.99E-07 | -0.015480095 | NA |
| chr3 | 100708545 | 100709562 | 1018 | 15 | 1.13E-06 | 6.65E-06 | 0.00169 | 1.28E-06 | 0.00280317 | TFG |
| chr8 | 102728525 | 102728595 | 71 | 2 | 1.14E-06 | 1.40E-06 | 5.87E-06 | 4.55E-07 | -0.013988868 | NA |
| chr2 | 11374750 | 11375122 | 373 | 4 | 1.14E-06 | 1.57E-06 | 6.58E-05 | 3.50E-07 | -0.006631631 | LINC00570 |
| chr1 | 35558291 | 35558872 | 582 | 5 | 1.14E-06 | 5.71E-05 | 0.00019 | 1.26E-05 | -0.007703624 | NCDN |
| chr10 | 30182235 | 30182244 | 10 | 2 | 1.14E-06 | 3.76E-07 | 7.14E-05 | 6.11E-07 | -0.017086882 | NA |
| chr11 | 47257280 | 47258035 | 756 | 7 | 1.15E-06 | 0.000174 | 0.00033 | 2.01E-06 | -0.00433755 | NR1H3 |
| chr11 | 119095420 | 119096612 | 1193 | 10 | 1.15E-06 | 6.31E-06 | 5.39E-05 | 1.12E-07 | 0.002216749 | H2AX, DPAGT1 |
| chr9 | 124734286 | 124734792 | 507 | 2 | 1.15E-06 | 2.03E-06 | 1.11E-05 | 8.93E-07 | -0.015164798 | NR6A1 |
| chr7 | 152156004 | 152156411 | 408 | 4 | 1.15E-06 | 1.00E-05 | 4.09E-05 | 8.28E-07 | -0.002806127 | KMT2C |
| chr15 | 68700412 | 68700734 | 323 | 2 | 1.15E-06 | 1.25E-05 | 3.51E-06 | 9.67E-07 | -0.017598867 | CORO2B |
| chr8 | 97259026 | 97259721 | 696 | 4 | 1.15E-06 | 6.02E-08 | 0.00047 | 1.52E-07 | -0.02735034 | NA |
| chr1 | 228734355 | 228735564 | 1210 | 14 | 1.15E-06 | 7.68E-05 | 0.00019 | 1.37E-06 | -0.001824442 | RHOU |
| chr1 | 226309539 | 226311234 | 1696 | 16 | 1.16E-06 | 0.000202 | 0.00027 | 2.00E-06 | -0.003912469 | LIN9 |
| chr11 | 119692767 | 119693728 | 962 | 5 | 1.16E-06 | 3.15E-08 | 0.0002 | 9.82E-08 | -0.011171049 | NECTIN1 |
| chr7 | 17297718 | 17298754 | 1037 | 9 | 1.16E-06 | 2.22E-07 | 0.00051 | 4.22E-07 | -0.002220117 | AHR, AC073332.1 |
| chr3 | 19933991 | 19934770 | 780 | 5 | 1.17E-06 | 5.00E-05 | 0.00014 | 8.51E-07 | -0.013513222 | EFHB |
| chr14 | 37610843 | 37611699 | 857 | 6 | 1.17E-06 | 1.51E-08 | 0.00149 | 7.78E-08 | 0.00171995 | TTC6 |
| chr19 | 44071394 | 44072721 | 1328 | 14 | 1.17E-06 | 1.27E-05 | 0.00454 | 8.02E-07 | -0.002441683 | AC067968.1, ZNF284 |
| chr10 | 13729107 | 13729465 | 359 | 5 | 1.17E-06 | 5.99E-07 | 0.00011 | 2.54E-07 | -0.003251183 | AL157392.2, FRMD4A |
| chr16 | 75062213 | 75063139 | 927 | 4 | 1.18E-06 | 1.06E-06 | 7.62E-05 | 7.49E-07 | -0.00675313 | ZNRF1 |
| chr17 | 57662624 | 57663103 | 480 | 2 | 1.18E-06 | 1.02E-06 | 2.99E-05 | 9.59E-07 | -0.014389694 | MSI2 |
| chr15 | 100914114 | 100914608 | 495 | 2 | 1.18E-06 | 6.72E-08 | 6.56E-05 | 1.30E-07 | -0.000965718 | ALDH1A3, AC015712.2 |
| chr17 | 81459987 | 81462703 | 2717 | 13 | 1.19E-06 | 9.44E-11 | 0.00081 | 1.19E-10 | -0.007321194 | BAHCC1, AC110285.4 |
| chr12 | 106954861 | 106955730 | 870 | 10 | 1.19E-06 | 3.65E-06 | 0.00499 | 8.40E-07 | -0.008938486 | TMEM263, AC007541.1 |
| chr19 | 6238545 | 6239530 | 986 | 5 | 1.19E-06 | 4.01E-08 | 0.0013 | 1.80E-07 | -0.010213459 | MLLT1 |
| chr14 | 100285921 | 100286439 | 519 | 4 | 1.19E-06 | 0.000127 | 1.48E-05 | 6.97E-06 | -0.006165961 | AL157871.6 |
| chr7 | 50732281 | 50733266 | 986 | 8 | 1.20E-06 | 0.000116 | 0.00044 | 3.66E-06 | -0.001152001 | GRB10 |
| chr3 | 49902693 | 49902812 | 120 | 2 | 1.20E-06 | 4.93E-05 | 1.49E-05 | 6.84E-06 | -0.003212755 | AC105935.2, MST1R |
| chr15 | 66618702 | 66619484 | 783 | 7 | 1.20E-06 | 1.79E-06 | 0.00047 | 2.74E-07 | 0.000776231 | LINC01169 |
| chr19 | 55657394 | 55658079 | 686 | 4 | 1.20E-06 | 7.58E-08 | 0.00041 | 2.07E-07 | -0.016573088 | U2AF2 |
| chr11 | 60039691 | 60040576 | 886 | 7 | 1.21E-06 | 0.000126 | 7.35E-05 | 5.06E-06 | -0.005522101 | OOSP2 |
| chr3 | 197750009 | 197751588 | 1580 | 10 | 1.21E-06 | 6.84E-06 | 0.0009 | 7.19E-07 | -0.001573774 | FYTTD1 |
| chr12 | 18752440 | 18752655 | 216 | 2 | 1.21E-06 | 2.03E-06 | 4.32E-06 | 4.59E-07 | -0.00576997 | NA |
| chr4 | 88697260 | 88698661 | 1402 | 18 | 1.22E-06 | 1.81E-06 | 0.00369 | 2.02E-06 | -0.004244499 | AC098582.1, HERC3, NAP1L5 |
| chr8 | 1257291 | 1257710 | 420 | 3 | 1.22E-06 | 1.06E-05 | 1.14E-05 | 1.36E-06 | 0.000583272 | DLGAP2 |
| chr3 | 52286816 | 52288090 | 1275 | 15 | 1.23E-06 | 1.01E-06 | 0.00098 | 1.71E-07 | -0.001204281 | GLYCTK, WDR82 |
| chr3 | 197519966 | 197520998 | 1033 | 6 | 1.23E-06 | 6.65E-06 | 2.92E-05 | 2.50E-07 | -0.002607078 | BDH1 |
| chr3 | 9703224 | 9703776 | 553 | 8 | 1.23E-06 | 8.64E-05 | 0.00033 | 1.80E-06 | -0.006115953 | AC022382.2 |
| chr9 | 134473457 | 134473913 | 457 | 2 | 1.23E-06 | 2.83E-05 | 7.47E-06 | 2.86E-06 | -0.003470384 | NA |
| chr6 | 33206391 | 33207911 | 1521 | 10 | 1.23E-06 | 8.98E-05 | 0.00105 | 1.07E-05 | -0.004676013 | HSD17B8, MIR219A1 |
| chr18 | 62324525 | 62326166 | 1642 | 9 | 1.23E-06 | 2.06E-05 | 0.00022 | 2.59E-07 | -0.00146268 | TNFRSF11A |
| chr1 | 35559888 | 35560593 | 706 | 2 | 1.24E-06 | 2.61E-06 | 0.00015 | 3.90E-06 | -0.007551033 | NCDN |
| chr4 | 89836969 | 89838467 | 1499 | 15 | 1.24E-06 | 1.14E-08 | 0.00046 | 2.87E-08 | 8.67E-05 | SNCA-AS1, SNCA |
| chr6 | 43516889 | 43517687 | 799 | 13 | 1.24E-06 | 0.000116 | 0.00015 | 5.18E-06 | 0.001553467 | POLR1C, YIPF3 |
| chr15 | 38564814 | 38566433 | 1620 | 15 | 1.25E-06 | 7.20E-05 | 0.00065 | 2.09E-06 | -0.001720142 | RASGRP1 |
| chr12 | 132847635 | 132848750 | 1116 | 11 | 1.25E-06 | 2.22E-08 | 0.00069 | 6.09E-08 | -0.004360482 | CHFR |
| chr16 | 56354996 | 56355117 | 122 | 5 | 1.25E-06 | 9.19E-08 | 0.00061 | 1.87E-07 | -0.010012493 | GNAO1 |
| chr17 | 1560190 | 1560754 | 565 | 3 | 1.25E-06 | 2.51E-06 | 4.57E-05 | 1.85E-06 | -0.008578744 | PITPNA |
| chr1 | 24322361 | 24323004 | 644 | 6 | 1.25E-06 | 1.16E-06 | 0.00318 | 7.55E-07 | -0.008671355 | GRHL3 |
| chr15 | 77646283 | 77646330 | 48 | 3 | 1.26E-06 | 5.33E-05 | 8.75E-05 | 9.77E-07 | -0.008205037 | LINGO1-AS1, LINGO1 |
| chr6 | 127517748 | 127518899 | 1152 | 4 | 1.26E-06 | 1.61E-05 | 7.24E-05 | 6.53E-07 | -0.013550108 | AL096711.2, SOGA3 |
| chr12 | 125064989 | 125065804 | 816 | 12 | 1.27E-06 | 1.23E-05 | 0.00017 | 3.93E-07 | -0.002423832 | AACS |
| chr1 | 112707320 | 112708155 | 836 | 10 | 1.28E-06 | 5.79E-06 | 0.00551 | 9.21E-07 | -0.003190491 | RHOC, AL603832.3 |
| chr1 | 161209211 | 161209945 | 735 | 6 | 1.28E-06 | 1.78E-06 | 9.32E-05 | 6.08E-07 | -0.005079662 | NDUFS2 |
| chr10 | 23094824 | 23095757 | 934 | 6 | 1.29E-06 | 0.003005 | 2.14E-05 | 1.06E-05 | -0.008840016 | MSRB2, AL139815.1 |
| chr12 | 89349109 | 89349552 | 444 | 3 | 1.29E-06 | 9.83E-05 | 7.29E-05 | 1.88E-06 | 0.017437656 | DUSP6 |
| chr1 | 84847396 | 84847831 | 436 | 2 | 1.29E-06 | 5.10E-07 | 1.96E-05 | 4.69E-07 | -0.013693289 | LPAR3 |
| chr16 | 10580800 | 10581173 | 374 | 4 | 1.30E-06 | 7.33E-07 | 0.00072 | 7.26E-07 | -0.011811952 | NA |
| chr13 | 110567559 | 110567974 | 416 | 3 | 1.30E-06 | 6.58E-08 | 0.0003 | 1.87E-07 | -0.006902137 | NA |
| chr13 | 40480765 | 40481451 | 687 | 4 | 1.30E-06 | 8.68E-08 | 0.00051 | 3.10E-07 | -0.012553739 | LINC00598, AL133318.1 |
| chr6 | 13295329 | 13296165 | 837 | 10 | 1.31E-06 | 7.85E-06 | 0.00052 | 8.59E-07 | -0.003485256 | AL008729.1, TBC1D7 |
| chr20 | 18587207 | 18587793 | 587 | 6 | 1.33E-06 | 3.43E-06 | 0.00031 | 1.45E-06 | 0.010448276 | AL121900.1 |
| chr19 | 39254375 | 39255097 | 723 | 6 | 1.34E-06 | 1.56E-07 | 0.0013 | 2.39E-07 | -0.011861868 | MSRB1P1 |
| chr12 | 113403812 | 113403863 | 52 | 2 | 1.35E-06 | 2.16E-07 | 4.59E-05 | 3.30E-07 | -0.026143301 | SDS |
| chr1 | 110165739 | 110166431 | 693 | 3 | 1.35E-06 | 1.22E-07 | 0.00013 | 2.05E-07 | -0.006890161 | SLC6A17, SLC6A17-AS1 |
| chr8 | 19312921 | 19313960 | 1040 | 11 | 1.35E-06 | 0.005557 | 0.00018 | 8.21E-06 | -0.003179526 | SH2D4A |
| chr6 | 125834627 | 125834994 | 368 | 2 | 1.36E-06 | 8.85E-08 | 8.09E-05 | 1.72E-07 | -0.018434885 | NCOA7 |
| chr7 | 74234127 | 74234574 | 448 | 3 | 1.36E-06 | 1.09E-05 | 8.90E-05 | 5.69E-07 | -0.008209426 | RFC2 |
| chr14 | 99247093 | 99247694 | 602 | 3 | 1.36E-06 | 0.000167 | 5.16E-06 | 4.52E-06 | -0.006399862 | BCL11B |
| chr1 | 19640477 | 19640934 | 458 | 2 | 1.36E-06 | 5.74E-07 | 0.00014 | 1.01E-06 | -0.010000989 | NBL1, MICOS10-NBL1 |
| chr9 | 6689890 | 6690431 | 542 | 2 | 1.36E-06 | 9.52E-06 | 1.92E-05 | 3.43E-06 | -0.011035385 | NA |
| chr20 | 43945270 | 43945984 | 715 | 6 | 1.36E-06 | 3.57E-06 | 5.62E-05 | 6.57E-07 | -0.005073474 | TOX2 |
| chr7 | 151060768 | 151063035 | 2268 | 17 | 1.37E-06 | 5.94E-08 | 0.00086 | 1.06E-08 | -0.003986095 | SLC4A2, AC010973.3 |
| chr11 | 118910441 | 118912414 | 1974 | 14 | 1.37E-06 | 7.74E-11 | 0.00542 | 1.72E-09 | -0.008053566 | MIR4492, BCL9L |
| chr7 | 6653399 | 6655805 | 2407 | 9 | 1.37E-06 | 4.28E-08 | 0.00039 | 6.27E-08 | -0.00147951 | ZNF316 |
| chr7 | 1571058 | 1571511 | 454 | 5 | 1.37E-06 | 1.29E-07 | 0.00053 | 3.32E-07 | -0.013595641 | PSMG3-AS1 |
| chr2 | 241372430 | 241373619 | 1190 | 5 | 1.38E-06 | 4.30E-08 | 0.00018 | 3.09E-08 | -0.004578653 | FARP2, AC005104.2 |
| chr2 | 199851452 | 199852015 | 564 | 2 | 1.38E-06 | 3.09E-05 | 5.29E-05 | 1.45E-05 | -0.009932489 | NA |
| chr10 | 125094527 | 125094640 | 114 | 2 | 1.38E-06 | 8.04E-06 | 9.71E-06 | 1.85E-06 | -0.006778499 | CTBP2 |
| chr2 | 43838120 | 43839859 | 1740 | 21 | 1.38E-06 | 3.21E-06 | 0.00311 | 5.37E-07 | -0.001424738 | ABCG8, ABCG5 |

| | | | | | | | | | | |
|---|---|---|---|---|---|---|---|---|---|---|
| chr14 | 93956765 | 93958393 | 1629 | 12 | 1.39E-06 | 7.31E-07 | 0.00184 | 4.48E-07 | -0.003726129 | ASB2 |
| chr2 | 162244421 | 162244983 | 563 | 3 | 1.39E-06 | 8.03E-06 | 3.38E-05 | 3.50E-06 | -0.008408988 | FAP |
| chr2 | 9770354 | 9771756 | 1403 | 6 | 1.39E-06 | 3.12E-08 | 0.00063 | 5.85E-08 | -0.008209341 | NA |
| chr2 | 143161158 | 143162225 | 1068 | 4 | 1.39E-06 | 7.77E-09 | 4.58E-05 | 7.01E-09 | -0.006296424 | ARHGAP15, AC013437.1 |
| chr4 | 11367725 | 11368391 | 667 | 3 | 1.39E-06 | 7.79E-06 | 1.39E-05 | 1.29E-06 | -0.009686131 | NA |
| chr11 | 126339453 | 126339602 | 150 | 2 | 1.39E-06 | 2.95E-07 | 8.24E-05 | 5.13E-07 | -0.01101734 | DCPS |
| chr6 | 158060370 | 158061140 | 771 | 3 | 1.39E-06 | 5.12E-07 | 2.85E-05 | 4.27E-07 | -0.013355795 | SYNJ2 |
| chr10 | 45594537 | 45595359 | 823 | 12 | 1.40E-06 | 3.07E-07 | 0.00837 | 3.65E-07 | -0.001804863 | MARCHF8 |
| chr7 | 5387200 | 5388383 | 1184 | 6 | 1.41E-06 | 4.01E-05 | 8.35E-05 | 5.83E-07 | -0.003076023 | TNRC18 |
| chr1 | 94246509 | 94246850 | 342 | 3 | 1.41E-06 | 4.46E-07 | 0.00036 | 1.01E-06 | -0.015572942 | ARHGAP29 |
| chr12 | 560986 | 561861 | 876 | 5 | 1.41E-06 | 2.19E-06 | 8.36E-05 | 5.43E-07 | -0.004292818 | B4GALNT3 |
| chr2 | 9315686 | 9315862 | 177 | 2 | 1.41E-06 | 1.14E-06 | 5.92E-06 | 4.01E-07 | -0.005837695 | ASAP2 |
| chr22 | 44067354 | 44067901 | 548 | 3 | 1.41E-06 | 4.70E-06 | 6.40E-05 | 3.55E-06 | -0.004439769 | PARVB |
| chr12 | 6310854 | 6311477 | 624 | 7 | 1.41E-06 | 5.84E-05 | 0.00154 | 1.41E-05 | -0.002386387 | PLEKHG6 |
| chr15 | 31049261 | 31049345 | 85 | 2 | 1.42E-06 | 2.65E-07 | 2.31E-05 | 3.04E-07 | -0.026602817 | TRPM1 |
| chr3 | 66463723 | 66464684 | 962 | 5 | 1.42E-06 | 0.000264 | 5.68E-05 | 2.87E-06 | -0.005480524 | LRIG1 |
| chr12 | 57752390 | 57753642 | 1253 | 17 | 1.42E-06 | 3.43E-05 | 0.00024 | 1.21E-06 | -0.000375883 | CDK4 |
| chr7 | 159097286 | 159097672 | 387 | 4 | 1.43E-06 | 4.38E-06 | 6.60E-05 | 9.12E-07 | -0.007113382 | VIPR2 |
| chr5 | 134110627 | 134110766 | 140 | 2 | 1.44E-06 | 1.64E-07 | 5.15E-05 | 2.71E-07 | -0.017594771 | NA |
| chr6 | 30953815 | 30954266 | 452 | 4 | 1.45E-06 | 1.94E-05 | 3.95E-05 | 4.76E-06 | -0.006676527 | MUCL3, SFTA2, HCG21 |
| chr16 | 8958632 | 8958905 | 274 | 2 | 1.46E-06 | 0.000337 | 3.01E-06 | 2.32E-06 | -0.007415393 | USP7 |
| chr13 | 39810311 | 39810606 | 296 | 2 | 1.46E-06 | 1.57E-07 | 8.54E-05 | 2.89E-07 | -0.02668868 | NA |
| chr9 | 129401505 | 129402386 | 882 | 3 | 1.46E-06 | 2.35E-07 | 5.52E-05 | 2.39E-07 | -0.013084408 | NA |
| chr2 | 234989006 | 234989207 | 202 | 2 | 1.46E-06 | 9.89E-06 | 2.72E-06 | 7.09E-07 | -0.01744818 | SH3BP4 |
| chr17 | 42015959 | 42018604 | 2646 | 25 | 1.47E-06 | 0.000175 | 0.00055 | 4.62E-08 | 0.00020183 | NKIRAS2, DNAJC7 |
| chr17 | 74213017 | 74214204 | 1188 | 13 | 1.47E-06 | 1.76E-06 | 0.00117 | 4.84E-07 | 0.000854844 | TTYH2, MGC16275 |
| chr1 | 6131810 | 6132358 | 549 | 4 | 1.47E-06 | 2.88E-05 | 0.00013 | 9.41E-07 | -0.005723154 | CHD5 |
| chr9 | 34651065 | 34652041 | 977 | 8 | 1.48E-06 | 7.35E-07 | 0.00154 | 5.78E-08 | -0.008685256 | AL162231.3, IL11RA |
| chr2 | 227717357 | 227717772 | 416 | 4 | 1.48E-06 | 0.00013 | 0.00011 | 7.97E-06 | -0.002963913 | SLC19A3 |
| chr19 | 40217325 | 40218908 | 1584 | 9 | 1.48E-06 | 5.61E-06 | 9.54E-05 | 1.01E-07 | 0.002077934 | TTC9B |
| chr16 | 85395916 | 85396489 | 574 | 3 | 1.49E-06 | 1.19E-06 | 0.00054 | 2.10E-06 | -0.009782934 | GSE1 |
| chr6 | 117538683 | 117539282 | 600 | 2 | 1.50E-06 | 0.003356 | 2.10E-05 | 2.71E-05 | -0.007869757 | DCBLD1, AL132671.2 |
| chr9 | 123438498 | 123438934 | 437 | 2 | 1.50E-06 | 9.38E-06 | 1.15E-05 | 2.32E-06 | -0.010408681 | DENND1A |
| chr11 | 113778298 | 113779957 | 1660 | 7 | 1.50E-06 | 1.70E-09 | 0.00046 | 1.25E-09 | -0.009866261 | CLDN25 |
| chr1 | 3246358 | 3248078 | 1721 | 12 | 1.51E-06 | 2.08E-07 | 0.00569 | 2.32E-07 | -0.006502453 | PRDM16 |
| chr20 | 32761448 | 32763365 | 1918 | 12 | 1.51E-06 | 5.32E-07 | 0.0019 | 1.36E-08 | -0.004079157 | DNMT3B |
| chr16 | 57175786 | 57176395 | 610 | 5 | 1.52E-06 | 5.70E-05 | 1.66E-05 | 8.40E-07 | -0.004791189 | PSME3IP1 |
| chr3 | 121645071 | 121645313 | 243 | 2 | 1.52E-06 | 8.83E-06 | 3.68E-05 | 5.09E-06 | -0.001429452 | HCLS1 |
| chr19 | 12996327 | 12997403 | 1077 | 5 | 1.52E-06 | 0.000613 | 4.70E-05 | 3.02E-05 | -0.006622746 | NFIX |
| chr9 | 27124552 | 27125097 | 546 | 2 | 1.52E-06 | 3.57E-05 | 7.47E-06 | 3.16E-06 | -0.017643071 | TEK |
| chr3 | 12610595 | 12610731 | 137 | 3 | 1.53E-06 | 2.97E-07 | 6.47E-05 | 3.39E-07 | -0.003044763 | RAF1 |
| chr6 | 105362709 | 105363615 | 907 | 2 | 1.53E-06 | 2.74E-07 | 1.13E-05 | 2.24E-07 | -0.009701816 | PREP |
| chr22 | 29309055 | 29309474 | 420 | 3 | 1.53E-06 | 8.43E-06 | 3.95E-05 | 1.22E-06 | -0.007712278 | GAS2L1 |
| chr7 | 5382217 | 5383252 | 1036 | 3 | 1.53E-06 | 8.25E-08 | 0.00023 | 2.08E-07 | -0.010998899 | TNRC18 |
| chr19 | 17300857 | 17301496 | 640 | 5 | 1.54E-06 | 0.002463 | 0.00013 | 5.11E-07 | -0.002061598 | MRPL34, ABHD8 |
| chr14 | 23041684 | 23042097 | 414 | 7 | 1.54E-06 | 0.000163 | 3.66E-06 | 3.32E-06 | -0.00270206 | NA |
| chr17 | 1010758 | 1011526 | 769 | 3 | 1.54E-06 | 3.86E-07 | 3.98E-05 | 4.11E-07 | -0.003483684 | ABR |
| chr11 | 443853 | 444548 | 696 | 3 | 1.55E-06 | 5.22E-06 | 9.44E-05 | 1.34E-06 | -0.011147156 | NA |
| chr7 | 77415556 | 77415983 | 428 | 5 | 1.55E-06 | 0.000451 | 2.19E-05 | 3.46E-06 | -0.000652506 | GSAP |
| chr9 | 21512332 | 21512412 | 81 | 2 | 1.56E-06 | 2.22E-06 | 4.91E-06 | 5.34E-07 | -0.010325892 | MIR31HG |
| chr2 | 218872670 | 218874010 | 1341 | 6 | 1.56E-06 | 2.14E-09 | 0.00015 | 1.22E-09 | -0.022774274 | WNT6 |
| chr17 | 68378561 | 68379054 | 494 | 3 | 1.56E-06 | 1.65E-07 | 0.00015 | 3.86E-07 | -0.016918251 | ARSG |
| chr12 | 130249465 | 130249613 | 149 | 3 | 1.57E-06 | 2.67E-05 | 5.18E-06 | 8.97E-07 | -0.010319012 | NA |
| chr16 | 1988930 | 1990022 | 1093 | 11 | 1.58E-06 | 4.10E-06 | 0.00501 | 2.77E-06 | -0.008477671 | SYNGR3, AC005606.2 |
| chr19 | 50927693 | 50928219 | 527 | 4 | 1.58E-06 | 1.51E-07 | 0.00032 | 2.76E-07 | 1.70E-05 | NA |
| chr8 | 17808038 | 17808652 | 615 | 2 | 1.58E-06 | 2.22E-06 | 2.75E-05 | 1.67E-06 | 0.009838325 | AC027117.2, AC027117.1 |
| chr15 | 49319655 | 49319657 | 3 | 2 | 1.58E-06 | 3.56E-07 | 1.22E-05 | 2.81E-07 | 0.002708209 | GALK2 |
| chr1 | 13700095 | 13701086 | 992 | 10 | 1.59E-06 | 1.34E-05 | 0.00229 | 1.81E-06 | 0.00627599 | PRDM2 |
| chr1 | 1757277 | 1759543 | 2267 | 13 | 1.60E-06 | 3.04E-08 | 0.00242 | 1.54E-08 | -0.008130575 | NADK |
| chr1 | 205504094 | 205504685 | 592 | 11 | 1.60E-06 | 5.63E-05 | 7.16E-05 | 3.24E-06 | -0.006046174 | CDK18 |
| chr3 | 138714665 | 138715303 | 639 | 4 | 1.60E-06 | 1.70E-06 | 5.75E-05 | 1.23E-06 | 0.001013021 | PIK3CB |
| chr5 | 141418528 | 141421019 | 2492 | 11 | 1.60E-06 | 1.03E-07 | 0.0012 | 7.49E-09 | 0.015128815 | PCDHGA1, PCDHGA2, PCDHGA3, PCDHGB1, PCDHGA4, PCDHGB2, PCDHGA5, PCDHGB3, PCDHGA6, PCDHGA7, PCDHGB4, PCDHGA8, PCDHGB5, PCDHGA9, PCDHGB6, PCDHGA10, PCDHGB7 |
| chr9 | 127927919 | 127929130 | 1212 | 6 | 1.60E-06 | 3.03E-07 | 0.00044 | 9.15E-07 | -0.007141737 | PIP5KL1 |
| chr1 | 74197295 | 74197996 | 702 | 5 | 1.61E-06 | 0.000792 | 0.00011 | 6.17E-05 | -0.00425813 | LRRIQ3 |
| chr20 | 54070468 | 54071385 | 918 | 6 | 1.61E-06 | 7.32E-08 | 0.00291 | 4.37E-07 | -0.016223078 | BCAS1 |
| chr7 | 99390943 | 99391106 | 164 | 3 | 1.61E-06 | 1.64E-06 | 9.96E-06 | 4.58E-07 | -0.004397122 | AC004922.1, ARPC1B |
| chr6 | 12751381 | 12752176 | 796 | 3 | 1.62E-06 | 9.72E-08 | 0.00014 | 2.46E-07 | -0.006139649 | PHACTR1 |
| chr7 | 114413559 | 114414008 | 450 | 5 | 1.62E-06 | 9.87E-06 | 0.00273 | 7.51E-06 | -0.008980422 | FOXP2 |
| chr11 | 105045032 | 105046046 | 1015 | 4 | 1.62E-06 | 2.92E-05 | 0.00012 | 4.87E-07 | 0.000494829 | CARD16 |
| chr22 | 27980653 | 27980940 | 288 | 3 | 1.62E-06 | 1.48E-07 | 0.00015 | 2.70E-07 | -0.003954627 | TTC28-AS1, TTC28 |
| chr19 | 1271020 | 1273550 | 2531 | 10 | 1.62E-06 | 2.51E-10 | 0.00186 | 5.10E-10 | -0.010976423 | CIRBP |
| chr19 | 35605081 | 35605425 | 345 | 2 | 1.63E-06 | 0.001349 | 3.08E-06 | 2.80E-06 | -0.004166019 | NA |
| chr19 | 44623511 | 44623879 | 369 | 2 | 1.64E-06 | 6.52E-06 | 6.53E-06 | 1.20E-06 | -0.006132258 | IGSF23 |
| chr20 | 6123206 | 6124239 | 1034 | 10 | 1.64E-06 | 5.33E-08 | 0.00557 | 1.96E-07 | -0.0097177 | NA |
| chr4 | 48830588 | 48831570 | 983 | 19 | 1.65E-06 | 7.56E-05 | 0.00272 | 1.21E-06 | 0.001065917 | OCIAD1 |
| chr19 | 35123419 | 35124734 | 1316 | 7 | 1.65E-06 | 8.38E-09 | 0.00333 | 8.84E-08 | -0.008800642 | AC020907.6, FXYD3, LGI4 |
| chr2 | 223837550 | 223838713 | 1164 | 8 | 1.65E-06 | 6.68E-07 | 0.00052 | 6.92E-08 | -0.00241911 | AP1S3, AC093884.1 |
| chr8 | 143246719 | 143247705 | 987 | 16 | 1.66E-06 | 4.58E-05 | 0.00021 | 3.19E-06 | 0.001346837 | ZFP41, AC138696.1 |
| chr10 | 79088252 | 79088386 | 135 | 2 | 1.66E-06 | 1.29E-07 | 8.36E-05 | 2.47E-07 | -0.028170717 | ZMIZ1 |
| chr13 | 28483579 | 28483689 | 111 | 2 | 1.67E-06 | 2.90E-05 | 2.49E-06 | 1.02E-06 | -0.006023606 | FLT1 |
| chr16 | 2841861 | 2842911 | 1051 | 7 | 1.67E-06 | 9.17E-08 | 0.00063 | 3.08E-07 | -0.014805435 | PRSS30P |
| chr6 | 36026984 | 36027048 | 65 | 2 | 1.67E-06 | 0.001246 | 6.23E-06 | 6.34E-06 | -0.005052807 | NA |
| chr20 | 2860109 | 2860971 | 863 | 6 | 1.67E-06 | 3.53E-06 | 5.30E-05 | 1.52E-06 | -0.003062698 | VPS16 |
| chr21 | 46246070 | 46246929 | 860 | 7 | 1.68E-06 | 7.49E-06 | 3.33E-05 | 2.69E-07 | -0.003853748 | MCM3AP-AS1, AP001469.2, MCM3AP |
| chr22 | 41941218 | 41941653 | 436 | 3 | 1.68E-06 | 0.000403 | 3.51E-06 | 4.24E-06 | -0.005614481 | CENPM |
| chr12 | 6229238 | 6230370 | 1133 | 4 | 1.69E-06 | 1.13E-06 | 0.00035 | 2.63E-07 | -0.010203808 | CD9 |
| chr8 | 123537313 | 123538418 | 1106 | 3 | 1.69E-06 | 5.70E-05 | 1.41E-05 | 5.67E-06 | -0.004851963 | FBXO32 |
| chr19 | 3480365 | 3480942 | 578 | 8 | 1.69E-06 | 3.59E-09 | 0.01036 | 9.86E-08 | -0.024130016 | SMIM24 |
| chr7 | 128829687 | 128830365 | 679 | 8 | 1.69E-06 | 0.000644 | 2.80E-05 | 5.22E-06 | 0.00523464 | NA |
| chr6 | 136544587 | 136545181 | 595 | 4 | 1.70E-06 | 2.71E-06 | 0.00012 | 3.54E-07 | -0.00221039 | MAP7 |
| chr13 | 112696631 | 112697356 | 726 | 5 | 1.70E-06 | 0.000497 | 6.76E-05 | 9.65E-06 | -0.002638582 | ATP11A |
| chr11 | 18411953 | 18412807 | 855 | 9 | 1.70E-06 | 1.04E-07 | 0.00298 | 3.05E-07 | -0.034744491 | LDHC |
| chr10 | 60882855 | 60883480 | 626 | 3 | 1.71E-06 | 2.49E-07 | 7.74E-05 | 2.61E-07 | -0.005670638 | RHOBTB1 |
| chr6 | 33670536 | 33671145 | 610 | 5 | 1.71E-06 | 4.84E-05 | 7.56E-05 | 2.89E-06 | -0.004705537 | ITPR3 |
| chr16 | 89368835 | 89369377 | 543 | 2 | 1.71E-06 | 1.09E-05 | 1.36E-05 | 2.89E-06 | -0.008076271 | ANKRD11 |
| chr14 | 24369787 | 24370020 | 234 | 2 | 1.71E-06 | 1.38E-05 | 8.86E-05 | 1.17E-05 | -0.002795763 | NFATC4 |
| chr16 | 85309675 | 85310330 | 656 | 6 | 1.71E-06 | 9.19E-07 | 0.00076 | 5.80E-07 | -0.004033569 | GSE1 |
| chr2 | 68773954 | 68775272 | 1319 | 20 | 1.73E-06 | 9.40E-07 | 0.00217 | 5.43E-07 | 0.000257454 | ARHGAP25 |
| chr16 | 84013936 | 84014463 | 528 | 2 | 1.73E-06 | 1.37E-06 | 1.77E-05 | 9.12E-07 | -0.013310809 | SLC38A8 |
| chr12 | 54052235 | 54053160 | 926 | 11 | 1.75E-06 | 1.51E-08 | 0.00491 | 1.72E-07 | 0.015906078 | HOXC4 |
| chr16 | 87724942 | 87725794 | 853 | 4 | 1.75E-06 | 8.64E-06 | 0.0001 | 2.16E-06 | -0.007415719 | KLHDC4 |
| chr10 | 96359096 | 96359947 | 852 | 6 | 1.75E-06 | 0.005115 | 0.00015 | 3.80E-06 | -0.004988846 | OPALIN |
| chr1 | 202631352 | 202631369 | 18 | 2 | 1.76E-06 | 1.72E-07 | 4.17E-05 | 2.66E-07 | -0.033572081 | SYT2 |
| chr5 | 889761 | 890324 | 564 | 5 | 1.77E-06 | 3.97E-06 | 0.00123 | 6.70E-06 | 0.00042229 | BRD9 |
| chr14 | 33566512 | 33566978 | 467 | 2 | 1.77E-06 | 5.87E-05 | 6.05E-06 | 3.17E-06 | -0.01056717 | NPAS3 |
| chr1 | 21620073 | 21620944 | 872 | 6 | 1.78E-06 | 6.45E-08 | 0.00099 | 2.61E-07 | -0.015178606 | RAP1GAP |
| chr20 | 63934433 | 63934855 | 423 | 2 | 1.78E-06 | 1.83E-06 | 2.68E-05 | 1.43E-06 | -0.005986026 | DNAJC5 |
| chr16 | 87461106 | 87461742 | 637 | 3 | 1.78E-06 | 4.14E-07 | 0.00019 | 5.05E-07 | -0.012291939 | ZCCHC14 |
| chr19 | 18645087 | 18645259 | 173 | 2 | 1.78E-06 | 2.51E-05 | 3.72E-06 | 1.43E-06 | -0.011643483 | KLHL26 |
| chr16 | 70646201 | 70646914 | 714 | 8 | 1.78E-06 | 6.79E-06 | 0.00107 | 6.00E-07 | -0.003748275 | IL34 |
| chr3 | 107521628 | 107521798 | 171 | 2 | 1.79E-06 | 1.01E-05 | 3.35E-05 | 5.27E-06 | -0.009993663 | NA |
| chr11 | 46332072 | 46333733 | 1662 | 20 | 1.80E-06 | 2.80E-06 | 0.00144 | 4.68E-08 | -0.002183321 | DGKZ |
| chr16 | 30760655 | 30763816 | 3162 | 31 | 1.81E-06 | 1.84E-07 | 0.00064 | 9.59E-10 | -0.002026024 | PHKG2, RNF40, CCDC189 |
| chr15 | 74443138 | 74443757 | 620 | 3 | 1.81E-06 | 2.83E-06 | 1.16E-05 | 8.10E-07 | -0.008818644 | NA |
| chr1 | 44783294 | 44784227 | 934 | 4 | 1.81E-06 | 0.060065 | 1.26E-05 | 3.24E-05 | -0.003105568 | BEST4 |
| chr2 | 233014004 | 233014439 | 436 | 4 | 1.82E-06 | 7.21E-05 | 0.00024 | 3.14E-06 | -0.006394484 | AC106876.1 |
| chr17 | 8383201 | 8383928 | 728 | 8 | 1.82E-06 | 7.63E-08 | 0.00107 | 1.44E-07 | 0.000551065 | RPL26 |
| chr4 | 87821433 | 87821708 | 276 | 2 | 1.83E-06 | 0.012925 | 4.91E-06 | 5.54E-06 | -0.004257785 | MEPE |
| chr1 | 19084244 | 19084612 | 369 | 3 | 1.83E-06 | 0.000511 | 4.21E-06 | 5.63E-06 | 0.018435208 | UBR4 |
| chr7 | 152646280 | 152646880 | 601 | 2 | 1.83E-06 | 5.31E-07 | 2.58E-05 | 5.51E-07 | -0.008253098 | XRCC2 |
| chr21 | 33197756 | 33197834 | 79 | 2 | 1.83E-06 | 9.99E-07 | 8.33E-05 | 1.45E-06 | -0.01089293 | NA |
| chr10 | 121974726 | 121975828 | 1103 | 15 | 1.84E-06 | 8.03E-07 | 0.002 | 4.11E-07 | 0.005556776 | NSMCE4A |
| chr16 | 67395614 | 67396551 | 938 | 4 | 1.84E-06 | 2.00E-07 | 0.00046 | 4.28E-07 | -0.011261305 | ZDHHC1 |
| chr7 | 368660 | 370710 | 2051 | 16 | 1.86E-06 | 7.92E-05 | 0.00017 | 1.31E-07 | -0.004216631 | NA |
| chr16 | 82056341 | 82056742 | 402 | 2 | 1.86E-06 | 1.45E-05 | 1.06E-05 | 2.73E-06 | -0.014811325 | HSD17B2, AC092142.1 |
| chr7 | 30860444 | 30860988 | 545 | 2 | 1.86E-06 | 3.13E-06 | 3.93E-05 | 2.59E-06 | -0.006238541 | INMT-MINDY4, MINDY4, AC004691.2 |
| chr3 | 57544146 | 57545577 | 1432 | 9 | 1.86E-06 | 1.74E-07 | 0.00011 | 2.75E-08 | -0.006670846 | DNAH12 |
| chr1 | 160354238 | 160354753 | 516 | 2 | 1.87E-06 | 4.38E-05 | 2.05E-05 | 8.45E-06 | 0.003163674 | NCSTN |
| chr16 | 25215100 | 25215781 | 682 | 6 | 1.87E-06 | 1.61E-06 | 0.00126 | 8.21E-07 | -0.008678688 | AQP8 |
| chr11 | 691247 | 692353 | 1107 | 6 | 1.87E-06 | 5.34E-07 | 0.00031 | 2.40E-07 | -0.005583757 | DEAF1 |
| chr2 | 99451542 | 99452050 | 509 | 4 | 1.88E-06 | 4.61E-06 | 1.15E-05 | 9.85E-07 | -0.000745485 | REV1 |
| chr2 | 10716846 | 10717754 | 909 | 5 | 1.88E-06 | 1.34E-06 | 7.01E-05 | 6.70E-07 | -0.005175627 | NA |
| chr17 | 75011348 | 75013736 | 2389 | 15 | 1.88E-06 | 3.66E-05 | 0.00035 | 5.78E-08 | -0.00172976 | MRPL58 |
| chr5 | 177607316 | 177608575 | 1260 | 7 | 1.88E-06 | 9.61E-07 | 0.00233 | 3.37E-07 | -0.006107919 | B4GALT7 |
| chr2 | 20327323 | 20327916 | 594 | 3 | 1.89E-06 | 8.43E-06 | 8.10E-05 | 6.44E-06 | -0.006054417 | PUM2 |
| chr16 | 55670847 | 55671782 | 936 | 4 | 1.90E-06 | 3.95E-05 | 3.43E-05 | 6.43E-06 | -0.004093347 | SLC6A2 |
| chr19 | 35513331 | 35514493 | 1163 | 7 | 1.90E-06 | 1.20E-07 | 0.00037 | 2.96E-07 | -0.011208772 | DMKN |
| chr14 | 90618078 | 90618479 | 402 | 2 | 1.90E-06 | 5.13E-06 | 1.56E-05 | 2.03E-06 | -0.0093831 | TTC7B |
| chr19 | 48628853 | 48628994 | 142 | 2 | 1.90E-06 | 0.001346 | 5.74E-06 | 5.82E-06 | -0.006581149 | SPHK2 |
| chr5 | 95285225 | 95285813 | 589 | 4 | 1.90E-06 | 4.76E-06 | 1.78E-05 | 8.51E-07 | -0.00211906 | NA |
| chr11 | 120659184 | 120661324 | 2141 | 8 | 1.90E-06 | 2.42E-06 | 0.00023 | 1.32E-07 | -0.008790979 | GRIK4 |
| chr15 | 52569174 | 52570209 | 1036 | 14 | 1.91E-06 | 2.62E-06 | 0.00288 | 8.21E-07 | -0.004222306 | ARPP19 |
| chr10 | 72309073 | 72309906 | 834 | 4 | 1.91E-06 | 9.09E-06 | 0.00015 | 2.88E-07 | -0.011302876 | NA |
| chr12 | 132318861 | 132320157 | 1297 | 8 | 1.91E-06 | 7.37E-07 | 0.00484 | 7.11E-07 | -0.007358913 | GALNT9 |
| chr8 | 101154698 | 101155466 | 769 | 3 | 1.91E-06 | 5.12E-07 | 0.00011 | 8.48E-07 | -0.01253712 | NA |
| chr17 | 47848238 | 47849056 | 819 | 2 | 1.92E-06 | 2.56E-08 | 4.69E-05 | 5.13E-08 | -0.032918383 | SP6 |
| chr14 | 97977814 | 97979085 | 1272 | 9 | 1.93E-06 | 1.47E-07 | 2.60E-05 | 8.36E-09 | -0.008266945 | LINC01550 |
| chr17 | 82437098 | 82437529 | 432 | 3 | 1.93E-06 | 3.13E-06 | 5.15E-05 | 7.96E-07 | -0.003327199 | HEXD |
| chr16 | 86087243 | 86087787 | 545 | 4 | 1.93E-06 | 2.01E-05 | 6.33E-05 | 4.17E-06 | -0.003693778 | AC135012.1 |
| chr19 | 7845810 | 7846665 | 856 | 9 | 1.94E-06 | 8.80E-07 | 0.00019 | 8.93E-07 | -0.004550005 | EVI5L |
| chr2 | 100877610 | 100878418 | 809 | 5 | 1.95E-06 | 5.98E-06 | 0.00079 | 1.36E-06 | -0.001643469 | NPAS2 |
| chr17 | 40025969 | 40026259 | 291 | 3 | 1.95E-06 | 3.75E-05 | 5.54E-05 | 3.69E-06 | -0.012480918 | MED24 |
| chr3 | 184633614 | 184634279 | 666 | 2 | 1.96E-06 | 2.32E-06 | 3.04E-05 | 1.82E-06 | -0.012491731 | NA |
| chr16 | 73172095 | 73173314 | 1220 | 6 | 1.96E-06 | 0.014232 | 5.60E-06 | 3.15E-05 | -0.000749219 | ZFHX3 |
| chr2 | 118988924 | 118989438 | 515 | 2 | 1.97E-06 | 4.25E-06 | 1.55E-05 | 1.81E-06 | -0.012894299 | MARCO |
| chr1 | 7502244 | 7502502 | 259 | 3 | 1.97E-06 | 8.70E-08 | 0.00027 | 2.47E-07 | -0.019473757 | CAMTA1 |
| chr4 | 104901431 | 104901476 | 46 | 2 | 1.97E-06 | 3.32E-07 | 1.84E-05 | 3.29E-07 | -0.017412591 | AC096577.1, AC004053.1 |
| chr1 | 156368360 | 156369495 | 1136 | 11 | 1.97E-06 | 9.97E-05 | 0.00013 | 4.17E-06 | -0.00298787 | RHBG |
| chr12 | 34327112 | 34327434 | 323 | 4 | 1.97E-06 | 3.16E-07 | 0.00021 | 2.70E-07 | -0.004761241 | NA |
| chr22 | 31754132 | 31754948 | 817 | 5 | 1.98E-06 | 1.21E-05 | 0.00185 | 5.82E-06 | -0.009018756 | DEPDC5, Z82190.2 |
| chr15 | 45586807 | 45587821 | 1015 | 23 | 1.98E-06 | 2.02E-05 | 0.00379 | 8.32E-07 | 0.00039581 | BLOC1S6, AC090527.2 |
| chr16 | 57372067 | 57373043 | 977 | 9 | 1.99E-06 | 1.20E-05 | 0.00049 | 4.03E-07 | -0.007097986 | CX3CL1 |

| | | | | | | | | | | |
|---|---|---|---|---|---|---|---|---|---|---|
| chr22 | 44245599 | 44246096 | 498 | 4 | 1.99E-06 | 0.000291 | 1.53E-05 | 7.85E-06 | -0.004642455 | SHISAL1 |
| chr17 | 47872311 | 47872936 | 626 | 7 | 1.99E-06 | 5.32E-07 | 0.00263 | 6.47E-07 | -0.013511366 | NA |
| chr8 | 1970174 | 1971694 | 1521 | 5 | 1.99E-06 | 1.77E-06 | 2.08E-05 | 3.11E-08 | -0.00894221 | KBTBD11-OT1, AC019257.1 |
| chr12 | 6963113 | 6964101 | 989 | 12 | 2.00E-06 | 1.41E-06 | 0.00419 | 8.25E-07 | -0.005412638 | MIR200CHG, MIR200C, MIR141 |
| chr20 | 50713432 | 50713916 | 485 | 4 | 2.00E-06 | 5.78E-07 | 9.92E-05 | 3.66E-07 | -0.008591367 | NA |
| chr1 | 52354825 | 52355082 | 258 | 3 | 2.00E-06 | 6.30E-07 | 3.34E-05 | 5.13E-07 | -0.011088172 | CC2D1B |
| chr10 | 603079 | 603618 | 540 | 2 | 2.00E-06 | 2.45E-05 | 1.24E-05 | 4.15E-06 | -0.003567518 | DIP2C |
| chr15 | 31433819 | 31434152 | 334 | 3 | 2.02E-06 | 8.87E-06 | 6.93E-05 | 8.24E-07 | -0.007601594 | KLF13 |
| chr11 | 120562068 | 120563287 | 1220 | 4 | 2.03E-06 | 6.71E-07 | 0.00027 | 5.94E-07 | -0.008012327 | GRIK4 |
| chr2 | 95316036 | 95316922 | 887 | 3 | 2.03E-06 | 1.24E-07 | 0.00017 | 2.68E-07 | -0.012953299 | KCNIP3 |
| chr8 | 56016251 | 56017316 | 1066 | 3 | 2.04E-06 | 6.11E-07 | 2.60E-05 | 1.21E-07 | -0.009891944 | NA |
| chr13 | 28100468 | 28101373 | 906 | 7 | 2.04E-06 | 0.001926 | 3.37E-05 | 1.37E-05 | -0.004160611 | FLT3 |
| chr1 | 18917297 | 18918200 | 904 | 3 | 2.05E-06 | 2.87E-07 | 0.00014 | 4.17E-07 | -0.005628995 | IFFO2 |
| chr22 | 39472178 | 39472751 | 574 | 3 | 2.05E-06 | 9.42E-06 | 5.16E-05 | 1.37E-06 | -0.008859388 | MGAT3 |
| chr2 | 101330644 | 101330966 | 323 | 4 | 2.06E-06 | 1.03E-07 | 0.00049 | 2.83E-07 | -0.008223393 | NA |
| chr5 | 73418513 | 73418917 | 405 | 2 | 2.06E-06 | 2.67E-06 | 0.00012 | 3.75E-06 | 0.019465356 | NA |
| chr10 | 70602536 | 70604066 | 1531 | 13 | 2.06E-06 | 5.05E-07 | 8.30E-05 | 1.47E-07 | 0.002018596 | PRF1 |
| chr5 | 172958448 | 172959665 | 1218 | 20 | 2.07E-06 | 1.75E-06 | 0.00071 | 7.19E-08 | -0.000736537 | RPL26L1, AC008429.1 |
| chr7 | 38317128 | 38318083 | 956 | 6 | 2.08E-06 | 2.78E-08 | 0.00145 | 1.53E-07 | -0.014505586 | TRGV9 |
| chr1 | 205680661 | 205681346 | 686 | 10 | 2.08E-06 | 3.44E-05 | 0.0013 | 3.97E-06 | -0.002056928 | NA |
| chr3 | 129893383 | 129894892 | 1510 | 10 | 2.08E-06 | 4.12E-07 | 0.00026 | 2.67E-07 | -0.000783618 | TMCC1-AS1, TMCC1 |
| chr1 | 27871863 | 27872745 | 883 | 11 | 2.08E-06 | 8.40E-06 | 0.00266 | 9.13E-07 | -0.003095059 | THEMIS2 |
| chr16 | 86680930 | 86681804 | 875 | 7 | 2.08E-06 | 1.56E-07 | 0.00062 | 3.34E-07 | -0.012334496 | NA |
| chr17 | 28717667 | 28718876 | 1210 | 18 | 2.09E-06 | 2.58E-06 | 0.00666 | 6.42E-07 | -0.004140675 | RAB34 |
| chr9 | 88281211 | 88281296 | 86 | 3 | 2.09E-06 | 1.15E-07 | 0.0002 | 2.49E-07 | -0.020712768 | NA |
| chr8 | 29336927 | 29337406 | 480 | 5 | 2.09E-06 | 2.11E-07 | 0.00062 | 3.61E-07 | -0.002963645 | DUSP4 |
| chr17 | 8475438 | 8475907 | 470 | 5 | 2.10E-06 | 9.96E-05 | 0.00034 | 4.92E-06 | -0.007830225 | NDEL1, MYH10 |
| chr17 | 80065297 | 80065729 | 433 | 3 | 2.10E-06 | 4.32E-05 | 1.49E-05 | 5.63E-06 | -0.005186264 | CCDC40 |
| chr17 | 16491304 | 16492336 | 1033 | 7 | 2.10E-06 | 0.000219 | 0.00046 | 9.40E-07 | -0.001744416 | LRRC75A |
| chr14 | 64503947 | 64505241 | 1295 | 14 | 2.11E-06 | 2.95E-06 | 0.00081 | 3.87E-07 | 0.000700989 | ZBTB1, ZBTB25 |
| chr11 | 117316676 | 117317510 | 835 | 3 | 2.11E-06 | 4.41E-06 | 0.00011 | 4.39E-06 | -0.006351876 | CEP164, AP000892.1 |
| chr11 | 821283 | 822402 | 1120 | 6 | 2.12E-06 | 4.67E-05 | 4.30E-05 | 1.33E-06 | -0.006904992 | PNPLA2 |
| chr7 | 99598032 | 99598845 | 814 | 6 | 2.12E-06 | 2.32E-06 | 0.0011 | 7.21E-07 | -0.013045026 | AC005020.2, TMEM225B |
| chr6 | 33197627 | 33198388 | 762 | 10 | 2.12E-06 | 2.70E-05 | 0.00017 | 3.13E-06 | -0.003771417 | RXRB |
| chr7 | 158855490 | 158857005 | 1516 | 11 | 2.12E-06 | 2.35E-06 | 0.00031 | 4.15E-07 | -0.000927638 | DYNC2I1 |
| chr17 | 7018904 | 7020207 | 1304 | 6 | 2.13E-06 | 3.70E-05 | 6.44E-05 | 2.50E-06 | -0.006081656 | AC040977.2, MIR497HG |
| chr3 | 186241807 | 186242651 | 845 | 4 | 2.14E-06 | 2.47E-07 | 0.00049 | 5.32E-07 | -0.024146584 | DGKG |
| chr1 | 41381011 | 41382764 | 1754 | 8 | 2.15E-06 | 1.84E-07 | 0.00024 | 5.98E-09 | -0.003216999 | FOXO6 |
| chr14 | 37172667 | 37173185 | 519 | 9 | 2.15E-06 | 4.31E-05 | 1.28E-05 | 2.73E-06 | -0.00448885 | SLC25A21-AS1 |
| chr6 | 31700055 | 31701708 | 1654 | 6 | 2.15E-06 | 6.05E-05 | 1.83E-05 | 1.18E-06 | -0.003030306 | ABHD16A, AL662899.1, MIR4646 |
| chr3 | 124636827 | 124636854 | 28 | 2 | 2.16E-06 | 1.54E-06 | 5.73E-05 | 1.80E-06 | 0.036847821 | KALRN |
| chr7 | 155412160 | 155412709 | 550 | 3 | 2.16E-06 | 2.85E-05 | 7.51E-06 | 2.26E-06 | 4.37E-05 | AC008060.5 |
| chr1 | 2304744 | 2305761 | 1018 | 6 | 2.16E-06 | 0.000362 | 1.12E-05 | 1.17E-05 | -0.004042772 | SKI |
| chr3 | 45688997 | 45689913 | 917 | 19 | 2.16E-06 | 0.001032 | 8.17E-05 | 4.86E-06 | -0.000187683 | SACM1L, LIMD1-AS1 |
| chr3 | 126101336 | 126102421 | 1086 | 11 | 2.17E-06 | 9.92E-06 | 1.80E-05 | 8.41E-07 | -0.003226552 | SLC41A3 |
| chr19 | 34780003 | 34780019 | 17 | 2 | 2.17E-06 | 2.59E-07 | 3.03E-05 | 3.34E-07 | -0.021182852 | AC020910.3 |
| chr10 | 97713091 | 97715184 | 2094 | 14 | 2.18E-06 | 7.77E-09 | 0.00063 | 7.88E-10 | -0.003577574 | MARVELD1 |
| chr14 | 105370933 | 105371144 | 212 | 2 | 2.19E-06 | 2.58E-06 | 9.78E-05 | 3.39E-06 | -0.012815319 | PACS2, AL928654.4 |
| chr19 | 58009091 | 58009905 | 815 | 5 | 2.19E-06 | 2.18E-08 | 0.00151 | 1.22E-07 | -0.010488844 | AC008969.1 |
| chr17 | 78644828 | 78646013 | 1186 | 12 | 2.20E-06 | 2.72E-06 | 0.00012 | 5.13E-07 | -0.003949408 | NA |
| chr10 | 112373477 | 112374142 | 666 | 7 | 2.20E-06 | 9.09E-07 | 0.00138 | 9.24E-07 | -0.007655018 | ACSL5 |
| chr4 | 74795739 | 74796151 | 413 | 3 | 2.20E-06 | 1.44E-06 | 0.0003 | 2.03E-06 | -0.011443189 | NA |
| chr12 | 54183951 | 54184384 | 434 | 2 | 2.20E-06 | 3.64E-05 | 3.90E-05 | 1.27E-05 | -0.003733833 | SMUG1 |
| chr7 | 134778952 | 134779544 | 593 | 6 | 2.21E-06 | 9.47E-07 | 0.0006 | 1.03E-06 | -0.012556536 | CALD1 |
| chr16 | 1776039 | 1778258 | 2220 | 10 | 2.21E-06 | 4.23E-09 | 0.00036 | 5.98E-09 | -0.005142266 | EME2, SPSB3 |
| chr14 | 52791449 | 52792377 | 929 | 9 | 2.22E-06 | 8.36E-06 | 0.00068 | 1.71E-06 | -0.006848089 | AL139317.5, GNPNAT1 |
| chr22 | 31944708 | 31946158 | 1451 | 16 | 2.22E-06 | 1.71E-05 | 0.00078 | 7.05E-07 | 0.000257951 | Z82190.2, YWHAH, C22orf24 |
| chr19 | 1854024 | 1854820 | 797 | 6 | 2.22E-06 | 4.90E-08 | 0.00167 | 2.71E-07 | -0.008023173 | KLF16 |
| chr1 | 158024940 | 158025522 | 583 | 4 | 2.23E-06 | 3.72E-05 | 1.28E-05 | 3.28E-06 | -0.00820223 | KIRREL1 |
| chr17 | 49677614 | 49678472 | 859 | 10 | 2.23E-06 | 1.24E-07 | 0.00069 | 3.01E-07 | 0.002230776 | SPOP |
| chr17 | 41549633 | 41550176 | 544 | 2 | 2.24E-06 | 0.000266 | 2.19E-05 | 1.78E-05 | -0.00851028 | LINC00974 |
| chr17 | 50539741 | 50540276 | 536 | 2 | 2.24E-06 | 8.34E-06 | 3.42E-05 | 4.68E-06 | -0.004607567 | EPN3 |
| chr1 | 6460134 | 6461430 | 1297 | 9 | 2.25E-06 | 2.17E-08 | 0.00489 | 2.03E-07 | -0.005016328 | ESPN, TNFRSF25 |
| chr8 | 22058745 | 22059504 | 760 | 7 | 2.27E-06 | 2.76E-08 | 0.0032 | 2.61E-07 | -0.009181974 | DMTN |
| chr8 | 926825 | 927456 | 632 | 5 | 2.28E-06 | 2.17E-05 | 0.00065 | 3.67E-06 | -0.005034084 | DLGAP2 |
| chr11 | 68942923 | 68943772 | 850 | 5 | 2.29E-06 | 2.61E-06 | 3.73E-05 | 8.61E-07 | -0.005835927 | NA |
| chr2 | 64312433 | 64312508 | 76 | 2 | 2.30E-06 | 6.50E-07 | 6.19E-05 | 9.22E-07 | -0.017209717 | NA |
| chr2 | 25197998 | 25198136 | 139 | 2 | 2.30E-06 | 4.24E-07 | 7.62E-05 | 6.88E-07 | -0.017698939 | NA |
| chr19 | 1618152 | 1618612 | 461 | 2 | 2.30E-06 | 1.15E-06 | 0.00017 | 1.93E-06 | -0.0072366 | TCF3 |
| chr2 | 239139112 | 239139779 | 668 | 5 | 2.31E-06 | 6.09E-05 | 0.00068 | 1.03E-05 | -0.004952903 | HDAC4 |
| chr8 | 101051560 | 101052010 | 451 | 6 | 2.31E-06 | 1.78E-07 | 0.00072 | 2.45E-07 | -0.010537959 | FLJ42969 |
| chr18 | 50317816 | 50318282 | 467 | 2 | 2.32E-06 | 6.75E-06 | 1.48E-05 | 2.32E-06 | -0.001931943 | NA |
| chr3 | 57150596 | 57151164 | 569 | 2 | 2.33E-06 | 2.47E-07 | 6.49E-05 | 4.11E-07 | -0.016102188 | IL17RD |
| chr6 | 167993250 | 167993518 | 269 | 4 | 2.33E-06 | 2.46E-07 | 0.00014 | 2.71E-07 | -0.02223339 | KIF25-AS1 |
| chr8 | 144290588 | 144291105 | 518 | 6 | 2.34E-06 | 4.22E-06 | 0.00088 | 1.38E-06 | -0.010068081 | BOP1 |
| chr22 | 19940538 | 19942654 | 2117 | 18 | 2.34E-06 | 4.76E-05 | 0.00141 | 1.70E-07 | -0.003529666 | COMT, TXNRD2 |
| chr17 | 4594928 | 4595437 | 510 | 5 | 2.34E-06 | 4.25E-05 | 0.00014 | 2.45E-06 | -0.006543942 | SMTNL2 |
| chr16 | 68235580 | 68236979 | 1400 | 14 | 2.34E-06 | 1.21E-09 | 0.00801 | 1.04E-08 | -0.006791427 | ESRP2, AC020978.4 |
| chr1 | 148328900 | 148329798 | 899 | 6 | 2.34E-06 | 1.11E-06 | 0.001 | 6.04E-07 | -0.026519059 | AC245100.1, LINC01138 |
| chr7 | 143477120 | 143477360 | 241 | 2 | 2.35E-06 | 1.10E-06 | 0.00022 | 1.91E-06 | -0.007526351 | EPHA1-AS1 |
| chr16 | 56657310 | 56658165 | 856 | 8 | 2.35E-06 | 2.41E-06 | 0.0002 | 6.32E-07 | 1.83E-06 | MT1F |
| chr1 | 15343956 | 15345026 | 1071 | 7 | 2.35E-06 | 0.000509 | 1.38E-05 | 3.50E-07 | -0.007085327 | FHAD1 |
| chr14 | 100304759 | 100305585 | 827 | 3 | 2.35E-06 | 2.11E-06 | 0.00014 | 1.62E-06 | -0.015768328 | SLC25A29 |
| chr12 | 131087697 | 131088852 | 1156 | 6 | 2.36E-06 | 1.13E-06 | 9.84E-05 | 6.94E-07 | -0.004216661 | ADGRD1 |
| chr6 | 135218627 | 135218969 | 343 | 2 | 2.37E-06 | 3.59E-07 | 6.07E-05 | 5.62E-07 | -0.018669899 | MYB |
| chr10 | 102597278 | 102597648 | 371 | 3 | 2.37E-06 | 2.70E-07 | 0.00011 | 5.14E-07 | -0.012288356 | SUFU |
| chr2 | 73877258 | 73877758 | 501 | 2 | 2.37E-06 | 8.40E-07 | 2.74E-05 | 8.04E-07 | -0.013072225 | AC073046.2 |
| chr4 | 69994967 | 69995143 | 177 | 3 | 2.37E-06 | 2.23E-07 | 0.00077 | 6.94E-07 | -0.013401485 | NA |
| chr10 | 102115740 | 102116212 | 473 | 3 | 2.38E-06 | 1.87E-06 | 0.00021 | 3.02E-06 | -0.00700029 | LDB1 |
| chr9 | 129109192 | 129109739 | 548 | 4 | 2.39E-06 | 0.000735 | 6.56E-05 | 2.17E-05 | -0.006328015 | CRAT |
| chr2 | 85772375 | 85772750 | 376 | 3 | 2.39E-06 | 1.73E-07 | 0.00015 | 3.79E-07 | -0.020959916 | ATOH8 |
| chr1 | 15151710 | 15152599 | 890 | 10 | 2.40E-06 | 0.000107 | 0.00013 | 2.68E-06 | -0.003056407 | TMEM51, TMEM51-AS1 |
| chr6 | 109446629 | 109447431 | 803 | 4 | 2.40E-06 | 5.46E-07 | 0.00015 | 2.54E-07 | -0.016575895 | MICAL1 |
| chr4 | 151226238 | 151227058 | 821 | 4 | 2.40E-06 | 5.77E-07 | 0.00025 | 1.07E-06 | -0.008574501 | SH3D19 |
| chr17 | 79949804 | 79951648 | 1845 | 16 | 2.40E-06 | 2.72E-05 | 0.0002 | 8.81E-08 | -0.007923015 | TBC1D16 |
| chr3 | 195859925 | 195860669 | 745 | 5 | 2.41E-06 | 2.01E-06 | 0.00011 | 7.50E-07 | -0.009663097 | LINC01983 |
| chr5 | 345735 | 346872 | 1138 | 6 | 2.41E-06 | 3.10E-09 | 0.00137 | 3.60E-08 | -0.01089811 | PDCD6-AHRR, AHRR |
| chr12 | 53197265 | 53197982 | 718 | 6 | 2.41E-06 | 2.74E-08 | 0.00243 | 2.22E-07 | -0.011119615 | ITGB7 |
| chr2 | 38484004 | 38484559 | 556 | 3 | 2.41E-06 | 1.78E-06 | 0.00013 | 1.39E-06 | -0.008002597 | LINC02613 |
| chr6 | 168302879 | 168303474 | 596 | 2 | 2.41E-06 | 0.000117 | 1.64E-05 | 1.06E-05 | -0.006584319 | DACT2 |
| chr9 | 111631698 | 111632043 | 346 | 3 | 2.42E-06 | 0.000148 | 1.16E-05 | 8.77E-06 | 0.001292309 | DNAJC25, DNAJC25-GNG10 |
| chr3 | 46208533 | 46209724 | 1192 | 5 | 2.43E-06 | 1.23E-06 | 0.00042 | 2.79E-07 | -0.013663538 | CCR3 |
| chr14 | 102217275 | 102219616 | 2342 | 11 | 2.43E-06 | 5.43E-06 | 1.46E-05 | 7.38E-09 | -0.000410108 | WDR20 |
| chr12 | 94675196 | 94675924 | 729 | 2 | 2.44E-06 | 3.01E-08 | 5.39E-05 | 6.04E-08 | -0.017324987 | NA |
| chr10 | 119178237 | 119179002 | 766 | 12 | 2.44E-06 | 4.67E-05 | 0.00021 | 2.14E-06 | 9.18E-05 | PRDX3 |
| chr1 | 3323148 | 3323663 | 516 | 3 | 2.44E-06 | 1.62E-05 | 2.57E-05 | 2.80E-06 | -0.011917065 | PRDM16, AL354743.2 |
| chr11 | 117918851 | 117919132 | 282 | 2 | 2.44E-06 | 9.54E-06 | 6.74E-06 | 1.51E-06 | -0.006253506 | TMPRSS13 |
| chr7 | 27168666 | 27170279 | 1614 | 21 | 2.45E-06 | 8.09E-07 | 0.00688 | 2.61E-07 | -0.001191109 | HOXA10-AS, HOXA9, AC004080.3, MIR196B |
| chr3 | 46233381 | 46233822 | 442 | 2 | 2.47E-06 | 2.74E-07 | 5.63E-05 | 4.35E-07 | -0.014429147 | CCR3 |
| chr5 | 149129249 | 149129952 | 704 | 2 | 2.47E-06 | 3.54E-07 | 4.16E-05 | 4.89E-07 | -0.010095877 | SH3TC2-DT |
| chr13 | 105410789 | 105410823 | 35 | 3 | 2.47E-06 | 7.74E-08 | 0.00047 | 2.57E-07 | -0.032286642 | AL138954.1 |
| chr1 | 28505270 | 28507179 | 1910 | 21 | 2.47E-06 | 1.65E-06 | 0.00011 | 1.67E-07 | 0.001185549 | RCC1, SNHG3 |
| chr6 | 35490428 | 35490639 | 212 | 2 | 2.47E-06 | 3.15E-07 | 0.00012 | 5.81E-07 | -0.025878885 | TEAD3 |
| chr17 | 39737385 | 39738160 | 776 | 8 | 2.47E-06 | 7.54E-07 | 0.0016 | 3.40E-07 | -0.004669859 | GRB7 |
| chr3 | 152299017 | 152300030 | 1014 | 5 | 2.49E-06 | 0.001204 | 1.78E-05 | 2.98E-05 | -0.003450938 | MBNL1 |
| chr2 | 109241604 | 109242968 | 1365 | 3 | 2.49E-06 | 0.000101 | 1.72E-05 | 1.11E-06 | -0.005251986 | SH3RF3 |
| chr12 | 56449503 | 56450555 | 1053 | 9 | 2.52E-06 | 2.51E-05 | 0.00063 | 1.45E-06 | 0.00026232 | MIP, AC097104.1 |
| chr19 | 46496017 | 46496583 | 567 | 9 | 2.52E-06 | 2.51E-07 | 0.00137 | 8.06E-07 | -0.009638086 | AC011484.1, PPP5D1 |
| chr18 | 74315341 | 74316140 | 800 | 9 | 2.53E-06 | 0.000123 | 6.65E-05 | 9.56E-06 | -0.001600221 | C18orf63 |
| chr20 | 61260777 | 61261360 | 584 | 3 | 2.54E-06 | 7.44E-07 | 0.00017 | 8.00E-07 | -0.010393965 | CDH4 |
| chr11 | 794764 | 795582 | 819 | 7 | 2.54E-06 | 6.55E-06 | 0.00275 | 1.33E-06 | -0.006051918 | SLC25A22 |
| chr8 | 69831884 | 69835471 | 3588 | 34 | 2.54E-06 | 4.92E-08 | 0.00021 | 9.30E-11 | -0.001013756 | AC079089.1, SLCO5A1 |
| chr19 | 33499555 | 33499611 | 57 | 2 | 2.56E-06 | 7.41E-06 | 3.30E-06 | 7.25E-07 | -0.012551867 | PEPD |
| chr4 | 1365350 | 1366013 | 664 | 3 | 2.56E-06 | 1.33E-07 | 0.00037 | 3.58E-07 | -0.009822562 | UVSSA |
| chr17 | 82387345 | 82388163 | 819 | 4 | 2.57E-06 | 5.81E-07 | 0.00013 | 7.09E-07 | -0.010080698 | NA |
| chr13 | 112874061 | 112874687 | 627 | 3 | 2.57E-06 | 2.00E-06 | 0.0002 | 1.59E-06 | -0.012731577 | ATP11A |
| chr10 | 119670219 | 119670236 | 18 | 2 | 2.57E-06 | 5.98E-07 | 0.00017 | 1.07E-06 | -0.008180094 | BAG3 |
| chr13 | 98002461 | 98002909 | 449 | 3 | 2.58E-06 | 4.33E-07 | 0.00012 | 6.41E-07 | -0.006503983 | IPO5 |
| chr6 | 44727611 | 44728402 | 792 | 6 | 2.58E-06 | 2.43E-05 | 0.00096 | 2.59E-06 | -0.002763713 | AL133334.1 |
| chr2 | 173221257 | 173221711 | 455 | 2 | 2.58E-06 | 0.00059 | 6.20E-06 | 5.72E-06 | -0.004920526 | MAP3K20, MAP3K20-AS1 |
| chr14 | 103628282 | 103628737 | 456 | 4 | 2.58E-06 | 2.26E-06 | 0.00017 | 3.12E-06 | -0.002765079 | KLC1, AL139300.1 |
| chr19 | 47512364 | 47513603 | 1240 | 4 | 2.58E-06 | 1.78E-08 | 0.00021 | 5.82E-08 | -0.009896819 | NAPA |
| chr10 | 95168319 | 95168900 | 582 | 3 | 2.59E-06 | 1.42E-05 | 7.36E-05 | 2.59E-06 | -0.011912365 | AL157834.4 |
| chr19 | 35744208 | 35745943 | 1736 | 18 | 2.60E-06 | 1.63E-05 | 0.002 | 5.33E-08 | -0.00147882 | PSENEN, AD000671.1, AD000671.2, U2AF1L4 |
| chr9 | 38068130 | 38068764 | 635 | 3 | 2.60E-06 | 8.57E-05 | 0.00024 | 1.44E-05 | 0.003688243 | AL138752.2, SHB |
| chr5 | 178730517 | 178731085 | 569 | 10 | 2.61E-06 | 1.55E-05 | 0.00141 | 1.60E-06 | -0.006147636 | ZNF354A |
| chr16 | 86722970 | 86723168 | 199 | 2 | 2.62E-06 | 1.39E-05 | 1.57E-05 | 3.71E-06 | -0.012247051 | LINC02188 |
| chr9 | 131125106 | 131126140 | 1035 | 12 | 2.65E-06 | 6.65E-06 | 0.00045 | 6.02E-07 | 0.000980663 | NUP214 |
| chr16 | 5065303 | 5066141 | 839 | 6 | 2.65E-06 | 3.50E-07 | 0.00138 | 3.81E-07 | -0.006227468 | ALG1, C16orf89 |
| chr6 | 30738693 | 30739305 | 613 | 9 | 2.66E-06 | 5.25E-05 | 2.37E-05 | 2.13E-06 | -0.003081277 | FLOT1 |
| chr7 | 99029373 | 99030217 | 845 | 4 | 2.66E-06 | 3.65E-06 | 0.00017 | 9.07E-07 | -0.018794183 | TRRAP, SMURF1 |
| chr7 | 83648625 | 83649278 | 654 | 11 | 2.66E-06 | 1.50E-05 | 0.00273 | 5.14E-07 | 0.00297752 | SEMA3E |
| chr4 | 85827140 | 85827887 | 748 | 8 | 2.66E-06 | 9.05E-07 | 0.00183 | 1.54E-06 | -0.008333736 | ARHGAP24 |
| chr20 | 4170394 | 4171656 | 1263 | 5 | 2.67E-06 | 9.76E-08 | 4.43E-05 | 3.11E-08 | -0.007753776 | SMOX |
| chr8 | 122781927 | 122783665 | 1739 | 6 | 2.67E-06 | 3.50E-07 | 0.00015 | 2.41E-07 | -0.002178673 | ZHX2 |
| chr17 | 28572194 | 28572668 | 475 | 3 | 2.67E-06 | 2.12E-05 | 9.06E-05 | 1.34E-05 | -0.00809281 | AC005726.1 |
| chr19 | 38676487 | 38677156 | 670 | 3 | 2.69E-06 | 1.56E-05 | 3.13E-05 | 3.38E-06 | -0.010476683 | ACTN4 |
| chr2 | 238287578 | 238288306 | 729 | 3 | 2.70E-06 | 3.36E-05 | 0.00012 | 1.24E-05 | -0.006116912 | PER2 |
| chr7 | 100488476 | 100488822 | 347 | 4 | 2.71E-06 | 1.64E-06 | 0.00034 | 5.87E-07 | -0.0033974 | NYAP1 |
| chr3 | 128993156 | 128994788 | 1633 | 13 | 2.72E-06 | 1.49E-07 | 0.00052 | 2.88E-08 | -0.002402499 | CFAP92 |
| chr19 | 39533369 | 39534309 | 941 | 4 | 2.74E-06 | 5.96E-06 | 9.65E-05 | 1.23E-06 | -0.007638656 | AC011500.3 |
| chr5 | 110737572 | 110738155 | 584 | 6 | 2.74E-06 | 0.028497 | 9.59E-06 | 3.03E-05 | -0.002386801 | SLC25A46, TMEM232 |
| chr7 | 159007856 | 159008405 | 550 | 6 | 2.75E-06 | 0.000161 | 4.26E-05 | 2.00E-06 | -0.001529316 | LINC00689 |
| chr11 | 46743263 | 46744067 | 805 | 3 | 2.76E-06 | 0.000108 | 9.19E-06 | 5.68E-06 | -0.001693391 | CKAP5 |
| chr2 | 235614259 | 235615227 | 969 | 4 | 2.76E-06 | 7.27E-06 | 0.00033 | 2.03E-06 | -0.005372521 | AGAP1 |
| chr5 | 141155309 | 141156229 | 921 | 7 | 2.77E-06 | 3.82E-06 | 0.00041 | 1.28E-06 | 0.010229121 | PCDHB17P, AC244517.2, AC244517.11 |
| chr12 | 104577581 | 104578204 | 624 | 3 | 2.77E-06 | 0.000251 | 0.00035 | 1.25E-05 | -0.005550928 | CHST11 |
| chr1 | 198858262 | 198859373 | 1112 | 8 | 2.78E-06 | 0.004571 | 2.49E-06 | 3.59E-06 | -0.002050316 | MIR181A1HG, MIR181B1, MIR181A1 |
| chr2 | 46914158 | 46914798 | 641 | 3 | 2.78E-06 | 0.000445 | 1.33E-05 | 1.42E-05 | -0.007999186 | MCFD2 |
| chr7 | 920122 | 922205 | 2084 | 17 | 2.79E-06 | 1.75E-08 | 0.00048 | 4.07E-09 | 0.006000604 | ADAP1, COX19 |
| chr1 | 234592033 | 234592517 | 485 | 2 | 2.80E-06 | 8.53E-06 | 1.81E-05 | 3.09E-06 | -0.007142628 | NA |
| chr1 | 92485798 | 92486976 | 1179 | 20 | 2.80E-06 | 1.54E-05 | 0.00533 | 4.69E-06 | 0.000557313 | GFI1 |

| | | | | | | | | | | |
|---|---|---|---|---|---|---|---|---|---|---|
| chr1 | 25683855 | 25684049 | 195 | 2 | 2.82E-06 | 2.82E-07 | 0.00012 | 5.22E-07 | -0.018776487 | MAN1C1 |
| chr15 | 75579385 | 75579933 | 549 | 13 | 2.82E-06 | 7.20E-06 | 0.00328 | 2.12E-06 | -0.001712014 | AC105036.3 |
| chr16 | 56915029 | 56915052 | 24 | 2 | 2.83E-06 | 5.79E-06 | 1.18E-05 | 1.79E-06 | -0.003849184 | SLC12A3 |
| chr16 | 15605025 | 15605322 | 298 | 2 | 2.83E-06 | 1.13E-06 | 0.00011 | 1.76E-06 | -0.015998595 | BMERB1, MARF1 |
| chr12 | 48969965 | 48971340 | 1376 | 8 | 2.84E-06 | 2.48E-05 | 0.00013 | 2.51E-06 | -0.005018663 | WNT10B |
| chr22 | 31246874 | 31248572 | 1699 | 11 | 2.86E-06 | 1.80E-08 | 9.51E-05 | 1.20E-09 | -0.004811161 | LIMK2 |
| chr11 | 112073696 | 112075123 | 1428 | 15 | 2.86E-06 | 5.77E-07 | 0.009 | 2.54E-07 | 0.002485493 | NKAPD1, PIH1D2 |
| chr5 | 172684366 | 172685208 | 843 | 3 | 2.87E-06 | 0.000664 | 1.32E-05 | 1.65E-05 | -0.006223662 | NEURL1B |
| chr6 | 166556764 | 166557239 | 476 | 3 | 2.87E-06 | 1.89E-07 | 0.00032 | 4.68E-07 | -0.017157776 | RPS6KA2 |
| chr13 | 29924474 | 29925051 | 578 | 3 | 2.88E-06 | 0.000127 | 8.40E-05 | 5.23E-06 | -0.006696706 | LINC00572 |
| chr16 | 89963075 | 89964430 | 1356 | 4 | 2.89E-06 | 2.37E-08 | 0.00031 | 6.25E-08 | -0.004285583 | DEF8 |
| chr1 | 54152982 | 54154030 | 1049 | 10 | 2.89E-06 | 1.65E-06 | 0.00136 | 2.06E-07 | -0.005696895 | AL357673.1, CDCP2 |
| chr1 | 1778158 | 1779746 | 1589 | 12 | 2.91E-06 | 1.60E-07 | 0.00309 | 5.57E-07 | 0.00026563 | NADK |
| chr10 | 15325638 | 15325982 | 345 | 3 | 2.91E-06 | 3.95E-07 | 8.09E-05 | 5.50E-07 | 0.014643554 | FAM171A1 |
| chr5 | 66209574 | 66210480 | 907 | 3 | 2.92E-06 | 2.22E-06 | 0.00012 | 2.85E-06 | -0.010129665 | LINC02065 |
| chr7 | 38177907 | 38178939 | 1033 | 13 | 2.94E-06 | 3.20E-05 | 1.36E-05 | 1.96E-06 | 0.000687272 | STARD3NL |
| chr8 | 80121362 | 80121508 | 147 | 2 | 2.96E-06 | 4.83E-07 | 0.0001 | 8.15E-07 | -0.021946524 | TPD52 |
| chr19 | 3249445 | 3249790 | 346 | 2 | 2.96E-06 | 2.84E-07 | 7.14E-05 | 4.79E-07 | -0.011485222 | CELF5 |
| chr11 | 69493276 | 69494000 | 725 | 4 | 2.97E-06 | 2.33E-06 | 0.00067 | 1.78E-06 | -0.004105003 | LINC01488 |
| chr17 | 78278059 | 78279052 | 994 | 5 | 2.97E-06 | 1.21E-06 | 0.00047 | 6.14E-07 | -0.015356138 | LINC01993 |
| chr1 | 31059682 | 31060139 | 458 | 2 | 2.98E-06 | 6.60E-07 | 8.26E-05 | 1.02E-06 | 0.004461632 | PUM1 |
| chr11 | 86906219 | 86906540 | 322 | 4 | 3.00E-06 | 2.01E-07 | 0.00018 | 4.14E-07 | -0.012535714 | PRSS23, AP001528.1 |
| chr18 | 10726520 | 10726668 | 149 | 4 | 3.00E-06 | 9.77E-08 | 0.0009 | 3.56E-07 | -0.021743438 | KIAA0895LP1, PIEZO2 |
| chr5 | 148637235 | 148637861 | 627 | 3 | 3.00E-06 | 0.000762 | 4.75E-06 | 4.33E-06 | -0.006134318 | HTR4 |
| chr6 | 85677825 | 85679315 | 1491 | 13 | 3.00E-06 | 1.92E-07 | 0.00124 | 3.35E-07 | 0.002198302 | SNHG5 |
| chr15 | 39140083 | 39140699 | 617 | 3 | 3.00E-06 | 7.04E-08 | 0.00023 | 1.92E-07 | -0.010347803 | LINC02694, AC013652.1 |
| chr19 | 1815426 | 1816628 | 1203 | 14 | 3.00E-06 | 1.66E-05 | 0.00723 | 1.47E-06 | -0.002199622 | AC012615.2, REXO1, MIR1909 |
| chr1 | 209207978 | 209208432 | 455 | 3 | 3.00E-06 | 0.007391 | 4.83E-06 | 1.19E-05 | -0.0066933 | AC092810.2 |
| chr14 | 102096443 | 102096758 | 316 | 3 | 3.01E-06 | 4.93E-07 | 0.00015 | 5.43E-07 | -0.008272224 | HSP90AA1 |
| chr5 | 172733296 | 172734261 | 966 | 3 | 3.02E-06 | 8.57E-05 | 1.65E-05 | 8.28E-06 | -0.002763347 | NA |
| chr2 | 96642680 | 96642843 | 164 | 4 | 3.03E-06 | 2.18E-05 | 7.63E-05 | 1.07E-06 | -0.00791694 | FER1L5, KANSL3 |
| chr3 | 189630885 | 189631534 | 650 | 9 | 3.03E-06 | 1.52E-05 | 0.00324 | 1.29E-06 | -0.011911084 | TP63 |
| chr9 | 136116996 | 136117429 | 434 | 3 | 3.03E-06 | 7.97E-06 | 0.0002 | 3.36E-06 | -0.00604177 | TMEM250 |
| chr10 | 89837450 | 89838266 | 817 | 5 | 3.03E-06 | 1.11E-06 | 0.00027 | 1.97E-06 | -0.017882704 | LINC00865, MARK2P16 |
| chr2 | 100561785 | 100562318 | 534 | 4 | 3.04E-06 | 1.03E-06 | 0.00109 | 2.24E-06 | -0.008764654 | NA |
| chr2 | 158795047 | 158795507 | 461 | 5 | 3.04E-06 | 8.12E-08 | 0.00334 | 5.14E-07 | -0.019650236 | DAPL1 |
| chr7 | 87344649 | 87345928 | 1280 | 15 | 3.05E-06 | 0.002206 | 5.74E-05 | 1.06E-05 | 8.02E-05 | AC005045.1, AC005045.2, CROT, TP53TG1 |
| chr4 | 95838863 | 95839878 | 1016 | 4 | 3.06E-06 | 0.001541 | 8.93E-05 | 2.29E-05 | -0.005718164 | NA |
| chr10 | 46107800 | 46108276 | 477 | 6 | 3.06E-06 | 2.92E-07 | 0.00089 | 4.27E-07 | -0.010047944 | NA |
| chr12 | 49868811 | 49869203 | 393 | 2 | 3.06E-06 | 6.24E-07 | 5.29E-05 | 8.41E-07 | -0.011805328 | FAIM2 |
| chr4 | 111323570 | 111323974 | 405 | 2 | 3.07E-06 | 6.01E-07 | 3.93E-05 | 7.30E-07 | -0.00968256 | NA |
| chr3 | 68931796 | 68932947 | 1152 | 14 | 3.08E-06 | 5.79E-07 | 0.00343 | 3.55E-07 | 0.000871646 | TAFA4 |
| chr16 | 85638569 | 85639993 | 1425 | 4 | 3.08E-06 | 1.56E-06 | 0.00068 | 1.80E-06 | -0.003486849 | GSE1 |
| chr22 | 37831817 | 37832194 | 378 | 2 | 3.08E-06 | 0.002412 | 7.13E-06 | 7.82E-06 | -0.002380683 | ANKRD54 |
| chr8 | 93741796 | 93742010 | 215 | 2 | 3.08E-06 | 4.04E-05 | 0.00044 | 4.69E-05 | -0.014468199 | AC010834.2 |
| chr1 | 4161328 | 4162022 | 695 | 6 | 3.09E-06 | 9.78E-06 | 4.15E-05 | 1.41E-06 | -0.000438528 | NA |
| chr1 | 165826026 | 165826077 | 52 | 2 | 3.09E-06 | 0.011248 | 1.09E-05 | 1.38E-05 | -0.003291328 | TMCO1 |
| chr4 | 109803678 | 109804547 | 870 | 2 | 3.09E-06 | 1.31E-07 | 2.49E-05 | 1.79E-07 | -0.013384014 | NA |
| chr2 | 74465048 | 74465569 | 522 | 9 | 3.09E-06 | 0.003646 | 1.95E-06 | 9.15E-06 | -0.000480206 | AC005041.5, MOGS |
| chr2 | 27211259 | 27212373 | 1115 | 16 | 3.10E-06 | 7.23E-05 | 0.0011 | 2.55E-06 | -0.000958837 | ATRAID, SLC5A6 |
| chr3 | 43364132 | 43364819 | 688 | 3 | 3.12E-06 | 0.00024 | 1.30E-05 | 8.90E-06 | -0.003397348 | SNRK, ANO10 |
| chr2 | 13022152 | 13022451 | 300 | 4 | 3.13E-06 | 4.59E-06 | 6.65E-05 | 7.17E-07 | -0.005557493 | AC093912.1 |
| chr22 | 23142121 | 23142769 | 649 | 6 | 3.13E-06 | 0.000458 | 8.82E-05 | 3.56E-05 | 0.000268757 | RSPH14 |
| chr7 | 94004718 | 94005673 | 956 | 4 | 3.13E-06 | 0.000126 | 0.0008 | 1.26E-05 | -0.013350649 | BET1-AS1 |
| chr17 | 7868282 | 7868334 | 53 | 3 | 3.14E-06 | 9.46E-08 | 0.00058 | 3.16E-07 | -0.008531345 | NAA38 |
| chr17 | 27380402 | 27381182 | 781 | 6 | 3.16E-06 | 0.000377 | 5.73E-05 | 1.25E-05 | -0.008367224 | NA |
| chr7 | 99557972 | 99558846 | 875 | 18 | 3.17E-06 | 0.000182 | 0.00076 | 2.64E-06 | 0.001019213 | ZNF655, AC005020.2, FAM200A |
| chr17 | 37066516 | 37068394 | 1879 | 7 | 3.17E-06 | 1.65E-08 | 9.89E-05 | 1.12E-08 | -0.012328534 | NA |
| chr11 | 92981635 | 92981727 | 93 | 2 | 3.20E-06 | 1.51E-06 | 1.84E-05 | 9.95E-07 | 0.001639191 | MTNR1B |
| chr8 | 143212611 | 143213264 | 654 | 5 | 3.20E-06 | 7.79E-07 | 7.20E-05 | 8.35E-07 | -0.008909297 | GPIHBP1 |
| chr22 | 36456027 | 36456594 | 568 | 2 | 3.20E-06 | 7.66E-07 | 8.46E-05 | 1.17E-06 | -0.019001822 | NA |
| chr13 | 53028366 | 53029151 | 786 | 4 | 3.21E-06 | 0.000177 | 3.94E-05 | 1.26E-05 | -0.009260859 | OLFM4 |
| chr7 | 21426633 | 21427445 | 813 | 4 | 3.21E-06 | 1.06E-05 | 0.00053 | 7.19E-06 | -0.005429414 | NA |
| chr20 | 38428539 | 38428723 | 185 | 2 | 3.22E-06 | 7.10E-06 | 0.0001 | 7.60E-06 | -0.004812934 | SNHG17 |
| chr11 | 2929173 | 2930383 | 1211 | 20 | 3.23E-06 | 5.00E-06 | 0.02393 | 5.57E-06 | 0.001232275 | PHLDA2 |
| chr12 | 19515471 | 19515682 | 212 | 2 | 3.23E-06 | 5.74E-06 | 6.38E-06 | 1.10E-06 | -0.001254681 | AEBP2 |
| chr12 | 53486541 | 53486880 | 340 | 2 | 3.25E-06 | 1.24E-06 | 0.00014 | 2.00E-06 | -0.008088803 | MAP3K12 |
| chr6 | 24489443 | 24489993 | 551 | 6 | 3.26E-06 | 6.37E-07 | 0.00016 | 7.21E-07 | -0.006207006 | GPLD1 |
| chr9 | 120874227 | 120874710 | 484 | 2 | 3.27E-06 | 2.56E-07 | 9.00E-05 | 4.58E-07 | -0.000144097 | PHF19 |
| chr5 | 80952984 | 80953188 | 205 | 3 | 3.27E-06 | 0.002171 | 2.18E-06 | 4.32E-06 | -0.001519777 | RASGRF2-AS1 |
| chr10 | 131939843 | 131940498 | 656 | 5 | 3.28E-06 | 1.49E-06 | 7.74E-05 | 6.49E-07 | -0.009479842 | PPP2R2D |
| chr7 | 128121307 | 128121699 | 393 | 2 | 3.28E-06 | 7.56E-05 | 1.18E-05 | 6.65E-06 | -0.002459042 | NA |
| chr11 | 26330942 | 26332083 | 1142 | 9 | 3.28E-06 | 0.001107 | 0.00316 | 6.04E-05 | -0.007524943 | ANO3, ANO3-AS1 |
| chr16 | 680898 | 682232 | 1335 | 15 | 3.28E-06 | 2.18E-05 | 0.00207 | 1.86E-06 | -0.002819292 | STUB1, JMJD8 |
| chr11 | 64137801 | 64138632 | 832 | 5 | 3.28E-06 | 0.000386 | 3.25E-06 | 5.46E-06 | -0.003255874 | MACROD1 |
| chr10 | 114239084 | 114240157 | 1074 | 9 | 3.29E-06 | 8.53E-07 | 0.00603 | 9.23E-07 | -0.003717052 | VWA2 |
| chr4 | 34668153 | 34668596 | 444 | 3 | 3.29E-06 | 0.000868 | 4.34E-06 | 6.35E-06 | -0.005921858 | AC093689.1 |
| chr1 | 234721912 | 234722344 | 433 | 2 | 3.30E-06 | 1.64E-05 | 2.55E-05 | 5.81E-06 | -0.014291333 | NA |
| chr13 | 50801284 | 50801709 | 426 | 2 | 3.30E-06 | 7.74E-07 | 9.41E-05 | 1.21E-06 | -0.009539269 | DLEU1, DLEU7 |
| chr22 | 50013986 | 50014001 | 16 | 2 | 3.31E-06 | 2.30E-06 | 0.00011 | 3.17E-06 | -0.00928258 | NA |
| chr8 | 142985426 | 142985433 | 8 | 2 | 3.31E-06 | 4.95E-07 | 6.53E-05 | 7.45E-07 | -0.004204471 | LY6E-DT |
| chr1 | 15329268 | 15330140 | 873 | 5 | 3.31E-06 | 1.18E-05 | 0.00013 | 2.13E-06 | -0.006516445 | FHAD1, FHAD1-AS1 |
| chr20 | 63536589 | 63537110 | 522 | 3 | 3.32E-06 | 0.000415 | 0.0002 | 5.56E-05 | -0.005812236 | PTK6 |
| chr12 | 12979543 | 12980197 | 655 | 8 | 3.33E-06 | 2.72E-05 | 0.00013 | 3.22E-06 | -0.003956215 | GPRC5D-AS1, HEBP1 |
| chr4 | 174831545 | 174831630 | 86 | 2 | 3.33E-06 | 1.08E-06 | 1.28E-05 | 6.32E-07 | -0.008062168 | ADAM29 |
| chr17 | 35981520 | 35982541 | 1022 | 8 | 3.33E-06 | 8.70E-06 | 0.00095 | 3.97E-06 | -0.007811857 | NA |
| chr3 | 179261061 | 179261554 | 494 | 4 | 3.34E-06 | 3.02E-05 | 0.00024 | 2.31E-06 | 0.017257559 | KCNMB3, LRRFIP1P1 |
| chr5 | 122126711 | 122127249 | 539 | 3 | 3.34E-06 | 6.05E-07 | 0.00014 | 9.45E-07 | -0.016837316 | AC010255.1 |
| chr11 | 4585358 | 4585471 | 114 | 3 | 3.34E-06 | 6.73E-07 | 9.36E-05 | 9.15E-07 | -0.007209515 | OR52I2 |
| chr4 | 1569922 | 1570437 | 516 | 2 | 3.34E-06 | 9.01E-07 | 6.67E-05 | 1.24E-06 | -0.011970837 | NA |
| chr14 | 104724781 | 104725932 | 1152 | 5 | 3.35E-06 | 6.17E-05 | 0.00068 | 1.13E-05 | -0.011642706 | ADSS1 |
| chr2 | 70871810 | 70872132 | 323 | 3 | 3.35E-06 | 6.56E-07 | 9.47E-05 | 7.82E-07 | -0.010667812 | NA |
| chr10 | 79163130 | 79163133 | 4 | 2 | 3.35E-06 | 7.42E-07 | 0.00011 | 1.21E-06 | -0.011799574 | ZMIZ1 |
| chr10 | 73167540 | 73168124 | 585 | 12 | 3.39E-06 | 0.001893 | 0.00013 | 1.29E-05 | 0.001222325 | FAM149B1, ECD |
| chr19 | 53537520 | 53538602 | 1083 | 11 | 3.39E-06 | 4.92E-05 | 7.03E-05 | 1.66E-07 | -0.009645998 | ZNF331 |
| chr13 | 76456002 | 76456639 | 638 | 3 | 3.39E-06 | 0.000257 | 4.71E-06 | 4.78E-06 | -0.00754171 | NA |
| chr1 | 156924742 | 156925199 | 458 | 2 | 3.39E-06 | 5.59E-06 | 3.62E-05 | 3.72E-06 | -0.010285791 | LRRC71 |
| chr7 | 157279832 | 157280742 | 911 | 5 | 3.39E-06 | 3.03E-05 | 0.00017 | 2.38E-06 | -0.007014233 | NA |
| chr17 | 17172607 | 17173421 | 815 | 3 | 3.39E-06 | 1.38E-06 | 8.85E-05 | 4.65E-07 | -0.006789213 | MPRIP, AC055811.1 |
| chr22 | 38724579 | 38726166 | 1588 | 6 | 3.41E-06 | 4.86E-07 | 0.0002 | 9.26E-08 | -0.005269542 | GTPBP1 |
| chr12 | 106066356 | 106066790 | 435 | 2 | 3.45E-06 | 1.03E-06 | 0.00014 | 1.69E-06 | -0.005023933 | NUAK1 |
| chr6 | 144007915 | 144009208 | 1294 | 20 | 3.46E-06 | 1.46E-06 | 0.00396 | 4.88E-07 | -0.002729997 | PLAGL1, HYMAI |
| chr14 | 105194116 | 105194539 | 424 | 2 | 3.46E-06 | 4.06E-07 | 0.00014 | 7.44E-07 | -0.022650394 | NA |
| chr17 | 74379639 | 74380652 | 1014 | 4 | 3.46E-06 | 4.67E-07 | 0.00049 | 9.45E-07 | -0.008780781 | NA |
| chr7 | 100675598 | 100676738 | 1141 | 6 | 3.46E-06 | 6.97E-05 | 0.00055 | 5.27E-06 | -0.000942843 | GNB2 |
| chr5 | 140647018 | 140648364 | 1347 | 18 | 3.47E-06 | 2.87E-06 | 0.00358 | 2.96E-07 | 0.002304639 | IK, MIR3655, NDUFA2 |
| chr7 | 151380181 | 151381172 | 992 | 5 | 3.49E-06 | 4.25E-06 | 0.00055 | 3.29E-06 | -0.006787106 | WDR86 |
| chr6 | 87266085 | 87266677 | 593 | 2 | 3.49E-06 | 3.76E-07 | 4.90E-05 | 5.47E-07 | -0.005605462 | NA |
| chr8 | 144308995 | 144310346 | 1352 | 7 | 3.49E-06 | 5.21E-06 | 3.45E-05 | 9.99E-07 | -0.002992663 | HSF1 |
| chr19 | 40358686 | 40359576 | 891 | 8 | 3.51E-06 | 3.05E-06 | 0.00159 | 8.67E-07 | -0.00462448 | PLD3, AC118344.2 |
| chr17 | 77734019 | 77735278 | 1260 | 4 | 3.51E-06 | 1.56E-06 | 0.00019 | 1.64E-06 | -0.01251789 | NA |
| chr1 | 222814156 | 222815097 | 942 | 9 | 3.52E-06 | 1.12E-06 | 0.00693 | 1.31E-06 | -0.00629484 | DISP1 |
| chr3 | 40477867 | 40478135 | 269 | 3 | 3.52E-06 | 1.99E-05 | 0.00014 | 4.16E-06 | -0.009612042 | ZNF619, ZNF620 |
| chr2 | 76985355 | 76985962 | 608 | 5 | 3.52E-06 | 6.43E-06 | 9.36E-05 | 7.59E-07 | -0.019378236 | LRRTM4 |
| chr15 | 74010702 | 74011463 | 762 | 3 | 3.52E-06 | 1.72E-05 | 1.13E-05 | 2.58E-06 | -0.006358629 | PML |
| chr1 | 117643153 | 117643343 | 191 | 3 | 3.53E-06 | 3.45E-05 | 1.68E-05 | 1.29E-06 | -0.006983663 | NA |
| chr7 | 23298224 | 23299473 | 1250 | 14 | 3.54E-06 | 5.67E-07 | 0.00067 | 8.83E-07 | -0.00058955 | MALSU1 |
| chr6 | 31158596 | 31160086 | 1491 | 23 | 3.54E-06 | 3.35E-05 | 0.02851 | 8.18E-06 | -0.005643726 | TCF19 |
| chr5 | 389240 | 389648 | 409 | 3 | 3.54E-06 | 0.003558 | 1.08E-05 | 8.64E-06 | -0.004750686 | PDCD6-AHRR, AHRR |
| chr11 | 12285903 | 12286478 | 576 | 3 | 3.56E-06 | 2.08E-05 | 0.00014 | 3.15E-06 | -0.01525991 | MICAL2 |
| chr22 | 36814913 | 36815221 | 309 | 3 | 3.57E-06 | 3.60E-05 | 0.00015 | 2.08E-06 | 0.015297514 | PVALB |
| chr9 | 95044115 | 95044498 | 384 | 2 | 3.57E-06 | 1.41E-05 | 1.98E-05 | 4.45E-06 | -0.006135441 | AOPEP |
| chr7 | 116499126 | 116500539 | 1414 | 23 | 3.57E-06 | 1.91E-06 | 0.00013 | 5.89E-08 | -0.003461656 | CAV2, AC002066.1 |
| chr1 | 15944345 | 15945547 | 1203 | 8 | 3.57E-06 | 0.00388 | 0.0001 | 2.51E-05 | -0.002376061 | ZBTB17 |
| chr16 | 86806861 | 86807047 | 187 | 2 | 3.57E-06 | 3.33E-07 | 6.69E-05 | 5.42E-07 | 0.035960128 | NA |
| chr2 | 104946023 | 104946357 | 335 | 2 | 3.57E-06 | 2.23E-06 | 1.76E-05 | 1.29E-06 | -0.020912667 | MRPS9-AS2 |
| chr13 | 48000993 | 48001791 | 799 | 13 | 3.57E-06 | 1.79E-05 | 0.00104 | 1.69E-06 | -0.005436385 | SUCLA2-AS1, SUCLA2 |
| chr17 | 60390193 | 60390939 | 747 | 4 | 3.57E-06 | 0.001906 | 7.16E-06 | 1.04E-05 | 0.011095423 | USP32 |
| chr8 | 140047188 | 140047989 | 802 | 6 | 3.58E-06 | 1.03E-06 | 0.00275 | 9.30E-07 | -0.01496139 | TRAPPC9 |
| chr19 | 11418720 | 11419462 | 743 | 9 | 3.58E-06 | 8.84E-05 | 4.64E-05 | 3.05E-06 | -0.002140801 | RGL3 |
| chr18 | 11856430 | 11857785 | 1356 | 8 | 3.58E-06 | 1.72E-08 | 0.00216 | 5.79E-08 | -0.004312249 | GNAL, AP005137.2 |
| chr6 | 30101832 | 30103215 | 1384 | 21 | 3.61E-06 | 1.97E-05 | 0.00773 | 1.63E-06 | -0.002257538 | TRIM31 |
| chr7 | 1647128 | 1648720 | 1593 | 8 | 3.61E-06 | 9.70E-07 | 0.00036 | 1.54E-07 | -0.002144349 | NA |
| chr9 | 136670275 | 136670702 | 428 | 5 | 3.61E-06 | 5.32E-06 | 0.00013 | 2.84E-06 | -0.001583581 | EGFL7, MIR126 |
| chr7 | 22109434 | 22109914 | 481 | 2 | 3.62E-06 | 5.85E-05 | 3.49E-05 | 1.52E-05 | -0.010221925 | NA |
| chr1 | 202282833 | 202283427 | 595 | 3 | 3.62E-06 | 3.54E-05 | 3.00E-05 | 6.88E-06 | -0.008080263 | LGR6 |
| chr3 | 177934186 | 177935207 | 1022 | 5 | 3.63E-06 | 4.73E-05 | 3.27E-05 | 6.52E-06 | -0.007441345 | AC007953.1 |
| chr13 | 47932732 | 47933305 | 574 | 2 | 3.64E-06 | 3.58E-06 | 3.10E-05 | 2.50E-06 | -0.007391227 | SUCLA2 |
| chr4 | 8188850 | 8189262 | 413 | 2 | 3.65E-06 | 1.18E-06 | 4.71E-05 | 1.35E-06 | -0.017194638 | SH3TC1 |
| chr5 | 141373509 | 141374088 | 580 | 11 | 3.67E-06 | 3.73E-05 | 0.00055 | 1.56E-06 | 0.008386486 | PCDHGA1, PCDHGA2, PCDHGA3, PCDHGB1, PCDHGA4, PCDHGB2, PCDHGA5, PCDHGB3, PCDHGA6 |
| chr3 | 188899845 | 188900591 | 747 | 8 | 3.68E-06 | 3.40E-05 | 2.05E-05 | 2.63E-06 | -0.003561326 | NA |
| chr8 | 129679110 | 129679310 | 201 | 2 | 3.68E-06 | 5.56E-07 | 8.05E-05 | 8.70E-07 | -0.019527471 | CCDC26 |
| chr17 | 4533285 | 4534254 | 970 | 5 | 3.68E-06 | 0.000182 | 0.0003 | 5.48E-06 | -0.006526494 | SPNS2 |
| chr1 | 225580293 | 225580399 | 107 | 2 | 3.68E-06 | 3.79E-05 | 9.19E-06 | 3.95E-06 | -0.011723365 | ENAH |
| chr9 | 97981242 | 97982015 | 774 | 3 | 3.69E-06 | 2.28E-06 | 0.0002 | 2.72E-06 | -0.011675653 | NA |
| chr6 | 31774787 | 31777185 | 2399 | 25 | 3.70E-06 | 1.75E-08 | 0.00034 | 1.38E-09 | -0.002093963 | VWA7 |
| chr1 | 221335725 | 221336118 | 394 | 3 | 3.70E-06 | 5.17E-06 | 0.00014 | 4.01E-06 | -0.020301223 | LINC02817 |
| chr1 | 12006503 | 12006836 | 334 | 2 | 3.70E-06 | 5.46E-07 | 0.00011 | 9.21E-07 | -0.007243086 | MFN2 |
| chr1 | 228145315 | 228145967 | 653 | 5 | 3.71E-06 | 7.75E-05 | 0.00014 | 9.74E-06 | -0.002846787 | GUK1 |
| chr2 | 156331903 | 156334235 | 2333 | 18 | 3.71E-06 | 1.26E-07 | 9.59E-05 | 1.74E-08 | 0.002001132 | NR4A2 |
| chr17 | 50397018 | 50398399 | 1382 | 14 | 3.72E-06 | 4.25E-06 | 0.00165 | 5.18E-08 | -0.006769184 | AC004707.1, LRRC59 |
| chr1 | 7616794 | 7617168 | 375 | 3 | 3.72E-06 | 0.000111 | 5.00E-06 | 3.74E-06 | -0.010857535 | CAMTA1 |
| chr3 | 196867824 | 196868573 | 750 | 5 | 3.73E-06 | 0.003704 | 0.00037 | 0.00016 | -0.000118721 | SENP5 |
| chr4 | 16075701 | 16076362 | 662 | 7 | 3.73E-06 | 5.84E-06 | 0.0007 | 2.45E-06 | -0.004237691 | PROM1 |
| chr19 | 54106066 | 54106775 | 710 | 3 | 3.74E-06 | 3.86E-06 | 0.00031 | 3.45E-06 | -0.004550068 | NDUFA3 |
| chr3 | 52295070 | 52295494 | 425 | 3 | 3.74E-06 | 2.75E-06 | 0.00018 | 2.44E-06 | -0.003240218 | GLYCTK, GLYCTK-AS1 |
| chr16 | 11742169 | 11743235 | 1067 | 14 | 3.74E-06 | 5.88E-06 | 0.00173 | 2.30E-06 | -0.000578972 | AC007613.1, TXNDC11 |
| chr10 | 72243882 | 72244313 | 432 | 2 | 3.75E-06 | 2.29E-06 | 4.01E-05 | 2.07E-06 | -0.00899587 | NA |
| chr6 | 31880251 | 31881495 | 1245 | 6 | 3.75E-06 | 0.002161 | 1.30E-05 | 8.41E-06 | -0.001180995 | EHMT2-AS1, EHMT2 |
| chr2 | 174771771 | 174772836 | 1066 | 5 | 3.76E-06 | 4.96E-06 | 0.00012 | 3.41E-06 | -0.009107747 | AC010894.2, CHRNA1 |
| chr8 | 1948271 | 1948869 | 599 | 5 | 3.77E-06 | 5.43E-05 | 0.00053 | 1.17E-05 | -0.006979198 | AC019257.8, ARHGEF10 |
| chr11 | 122882391 | 122883381 | 991 | 14 | 3.77E-06 | 5.01E-05 | 0.00296 | 4.68E-06 | -0.00058825 | JHY |

| | | | | | | | | | | |
|---|---|---|---|---|---|---|---|---|---|---|
| chr22 | 31139686 | 31140599 | 914 | 7 | 3.78E-06 | 2.08E-05 | 4.31E-05 | 1.84E-06 | -0.007780322 | PLA2G3 |
| chr19 | 18149520 | 18150324 | 805 | 5 | 3.79E-06 | 1.38E-08 | 0.00289 | 1.28E-07 | -0.008458109 | MAST3, AC007192.2 |
| chr14 | 20468712 | 20470061 | 1350 | 15 | 3.80E-06 | 0.000259 | 0.00015 | 5.54E-06 | 0.000710032 | PNP |
| chr15 | 66706660 | 66707707 | 1048 | 6 | 3.80E-06 | 1.49E-05 | 0.00205 | 5.85E-06 | -0.00371437 | SMAD6 |
| chr6 | 134175731 | 134176733 | 1003 | 10 | 3.83E-06 | 6.86E-06 | 0.00262 | 2.90E-06 | -0.004303265 | SGK1 |
| chr1 | 51793756 | 51794000 | 245 | 4 | 3.84E-06 | 3.00E-06 | 9.74E-05 | 1.01E-06 | -0.008807297 | OSBPL9, AL050343.2, NRDC |
| chr7 | 266995 | 267639 | 645 | 18 | 3.85E-06 | 5.67E-05 | 0.00352 | 4.49E-06 | -0.005425799 | NA |
| chr1 | 53725209 | 53725604 | 396 | 2 | 3.85E-06 | 3.76E-05 | 8.18E-06 | 3.54E-06 | -0.016894015 | GLIS1 |
| chr7 | 2814878 | 2815450 | 573 | 7 | 3.85E-06 | 0.016683 | 0.00011 | 7.89E-06 | -0.001940641 | GNA12 |
| chr8 | 121262805 | 121263302 | 498 | 2 | 3.85E-06 | 7.02E-05 | 1.28E-05 | 6.90E-06 | -0.000954267 | NA |
| chr7 | 28356133 | 28356412 | 280 | 2 | 3.86E-06 | 0.069438 | 6.00E-06 | 7.04E-06 | -0.003165632 | CREB5 |
| chr5 | 42840412 | 42840596 | 185 | 4 | 3.89E-06 | 3.91E-06 | 3.98E-05 | 7.71E-07 | -0.011029762 | AC008945.2, SELENOP |
| chr22 | 19758633 | 19759503 | 871 | 6 | 3.89E-06 | 7.12E-08 | 0.00124 | 3.77E-07 | -0.010975233 | TBX1 |
| chr17 | 79834592 | 79836291 | 1700 | 5 | 3.89E-06 | 2.88E-06 | 3.72E-05 | 2.99E-07 | -0.004099298 | CBX4 |
| chr1 | 206946677 | 206947283 | 607 | 4 | 3.89E-06 | 1.65E-06 | 0.00016 | 1.69E-06 | -0.006511548 | NA |
| chr3 | 3102491 | 3102890 | 400 | 2 | 3.90E-06 | 0.000105 | 1.05E-05 | 6.66E-06 | 0.002937851 | IL5RA |
| chr16 | 89003527 | 89004239 | 713 | 4 | 3.90E-06 | 5.02E-06 | 0.00123 | 5.00E-06 | -0.017942469 | NA |
| chr12 | 51905740 | 51907189 | 1450 | 7 | 3.93E-06 | 5.90E-08 | 0.00068 | 9.03E-08 | -0.006333765 | ACVRL1 |
| chr5 | 6737665 | 6738294 | 630 | 3 | 3.94E-06 | 1.21E-06 | 0.00011 | 1.64E-06 | -0.007560389 | TENT4A |
| chr1 | 6024589 | 6025450 | 862 | 5 | 3.95E-06 | 0.00012 | 2.57E-05 | 1.09E-05 | -0.005557103 | KCNAB2 |
| chr14 | 55085506 | 55085965 | 460 | 2 | 3.95E-06 | 0.00011 | 8.91E-06 | 5.81E-06 | -0.00263115 | NA |
| chr18 | 70261311 | 70261723 | 413 | 2 | 3.95E-06 | 2.47E-07 | 0.00011 | 4.58E-07 | 0.003359757 | NA |
| chr1 | 2592170 | 2593451 | 1282 | 5 | 3.96E-06 | 5.14E-06 | 0.0004 | 2.76E-06 | -0.014841227 | MMEL1 |
| chr14 | 70637971 | 70638405 | 435 | 2 | 3.96E-06 | 1.23E-06 | 3.29E-05 | 1.18E-06 | -0.009456208 | AL357153.3, AL357153.1 |
| chr7 | 2195934 | 2196770 | 837 | 3 | 3.96E-06 | 3.63E-05 | 6.40E-05 | 6.89E-06 | -0.005775673 | MAD1L1, AC069288.1 |
| chr11 | 74748272 | 74749512 | 1241 | 9 | 3.96E-06 | 4.87E-06 | 8.47E-05 | 4.14E-07 | 0.002390569 | RNF169 |
| chr16 | 49491816 | 49492863 | 1048 | 15 | 3.97E-06 | 2.26E-07 | 0.00407 | 1.57E-07 | -0.004306904 | ZNF423 |
| chr1 | 19650629 | 19650951 | 323 | 2 | 3.97E-06 | 4.32E-07 | 8.66E-05 | 7.19E-07 | -0.010064444 | NBL1, MICOS10-NBL1 |
| chr1 | 156141127 | 156141580 | 454 | 2 | 3.98E-06 | 2.86E-05 | 2.12E-05 | 6.96E-06 | -0.005843381 | NA |
| chr1 | 33161148 | 33161554 | 407 | 3 | 3.98E-06 | 1.89E-07 | 0.00055 | 5.63E-07 | -0.013604748 | TRIM62 |
| chr11 | 27701175 | 27702242 | 1068 | 15 | 3.98E-06 | 3.13E-05 | 0.00283 | 1.90E-06 | 0.002498487 | BDNF, AC103796.1 |
| chr16 | 88939068 | 88940047 | 980 | 11 | 3.98E-06 | 7.26E-06 | 0.00081 | 2.42E-06 | -0.006385964 | AC092384.1, CBFA2T3, AC092384.3 |
| chr18 | 48974460 | 48974832 | 373 | 2 | 4.00E-06 | 0.000545 | 2.51E-05 | 2.40E-05 | -0.004624749 | NA |
| chr11 | 45929982 | 45930686 | 705 | 3 | 4.03E-06 | 4.13E-07 | 0.00059 | 1.15E-06 | -0.020630683 | PHF21A |
| chr4 | 10093113 | 10093559 | 447 | 4 | 4.03E-06 | 4.42E-05 | 0.0002 | 4.01E-06 | -0.010600721 | WDR1 |
| chr6 | 125952790 | 125953919 | 1130 | 4 | 4.03E-06 | 7.08E-07 | 0.00015 | 1.15E-06 | -0.011155054 | NA |
| chr2 | 74106554 | 74107427 | 874 | 3 | 4.04E-06 | 3.41E-07 | 0.00015 | 5.53E-07 | -0.004219442 | TET3 |
| chr14 | 105044977 | 105045969 | 993 | 6 | 4.04E-06 | 2.51E-05 | 0.00023 | 2.95E-07 | -0.004512178 | NA |
| chr19 | 53037520 | 53038034 | 515 | 4 | 4.05E-06 | 6.59E-08 | 0.00145 | 3.35E-07 | -0.026060979 | ZNF702P |
| chr22 | 24145443 | 24145842 | 400 | 2 | 4.07E-06 | 0.000674 | 8.61E-06 | 8.26E-06 | -0.00848827 | CABIN1 |
| chr10 | 46024210 | 46024459 | 250 | 3 | 4.07E-06 | 0.001587 | 3.40E-05 | 1.44E-05 | -0.010094537 | NCOA4 |
| chr10 | 29687960 | 29688406 | 447 | 3 | 4.08E-06 | 0.000194 | 0.0002 | 7.04E-06 | -0.008398821 | SVIL |
| chr17 | 77253649 | 77255005 | 1357 | 5 | 4.10E-06 | 2.88E-07 | 0.00011 | 4.88E-07 | -0.001728687 | NA |
| chr3 | 107986518 | 107987398 | 881 | 2 | 4.11E-06 | 1.55E-07 | 4.34E-05 | 2.47E-07 | -0.021235839 | NA |
| chr21 | 44329802 | 44330229 | 428 | 6 | 4.11E-06 | 1.52E-05 | 0.0012 | 1.25E-06 | -0.007310103 | AP001062.3, CFAP410 |
| chr12 | 114407375 | 114409238 | 1864 | 18 | 4.12E-06 | 1.30E-07 | 0.01452 | 1.45E-07 | -0.001199635 | TBX5-AS1, TBX5 |
| chr3 | 172126956 | 172128093 | 1138 | 3 | 4.13E-06 | 5.32E-08 | 0.00032 | 1.66E-07 | -0.012356018 | FNDC3B |
| chr7 | 98126222 | 98126480 | 259 | 6 | 4.13E-06 | 6.30E-05 | 0.00033 | 1.65E-06 | -0.009156621 | LMTK2 |
| chr1 | 41890733 | 41890996 | 264 | 2 | 4.13E-06 | 1.41E-06 | 5.00E-05 | 1.59E-06 | -0.007617166 | HIVEP3 |
| chr2 | 234454611 | 234454975 | 365 | 4 | 4.14E-06 | 3.12E-07 | 0.00061 | 6.93E-07 | 0.039513097 | AC097713.1 |
| chr11 | 112085923 | 112087148 | 1226 | 28 | 4.17E-06 | 4.83E-05 | 0.00285 | 3.31E-06 | -0.000399312 | SDHD, AP002884.2, TIMM8B |
| chr2 | 3126075 | 3126577 | 503 | 4 | 4.18E-06 | 0.000139 | 8.07E-05 | 4.06E-06 | -0.001419781 | NA |
| chr14 | 64737837 | 64737913 | 77 | 3 | 4.19E-06 | 2.77E-07 | 0.00053 | 7.74E-07 | -0.012714084 | PLEKHG3 |
| chr17 | 81301509 | 81301682 | 174 | 2 | 4.19E-06 | 2.14E-05 | 5.10E-06 | 1.79E-06 | -0.010134832 | NA |
| chr19 | 46125207 | 46125833 | 627 | 2 | 4.20E-06 | 2.05E-05 | 6.75E-05 | 1.31E-05 | -0.017053196 | AC007785.1 |
| chr7 | 778672 | 780760 | 2089 | 10 | 4.22E-06 | 8.86E-08 | 6.56E-05 | 6.94E-09 | -0.003079677 | DNAAF5 |
| chr3 | 172315355 | 172315695 | 341 | 3 | 4.22E-06 | 4.19E-05 | 9.75E-06 | 2.02E-06 | -0.005728047 | FNDC3B |
| chr1 | 94236269 | 94236879 | 611 | 2 | 4.23E-06 | 3.73E-06 | 9.47E-05 | 4.50E-06 | 0.00840235 | ARHGAP29 |
| chr6 | 134949918 | 134950663 | 746 | 6 | 4.24E-06 | 5.21E-05 | 2.33E-05 | 3.66E-06 | -0.003327151 | ALDH8A1 |
| chr4 | 80185618 | 80186031 | 414 | 3 | 4.25E-06 | 2.32E-06 | 0.00063 | 4.82E-06 | -0.015948364 | PRDM8, AC021127.1 |
| chr6 | 52307175 | 52307207 | 33 | 2 | 4.25E-06 | 3.49E-06 | 1.15E-05 | 1.29E-06 | -0.025827072 | NA |
| chr8 | 145007647 | 145007953 | 307 | 3 | 4.26E-06 | 4.60E-07 | 7.89E-05 | 6.71E-07 | -0.013934557 | NA |
| chr1 | 43367560 | 43368768 | 1209 | 16 | 4.27E-06 | 0.000305 | 0.00113 | 4.01E-06 | 0.000632873 | ELOVL1 |
| chr2 | 23030201 | 23031543 | 1343 | 5 | 4.30E-06 | 2.31E-07 | 5.49E-05 | 5.19E-08 | -0.002903479 | AC018467.1 |
| chr12 | 130017458 | 130018041 | 584 | 8 | 4.32E-06 | 5.51E-07 | 0.00068 | 8.51E-07 | 0.012876559 | NA |
| chr17 | 41913934 | 41914438 | 505 | 4 | 4.33E-06 | 0.002291 | 5.07E-05 | 1.23E-05 | -0.003533605 | ACLY |
| chr2 | 8282064 | 8282252 | 189 | 5 | 4.34E-06 | 1.13E-05 | 1.69E-05 | 1.59E-06 | -0.006723599 | LINC00299 |
| chr14 | 89701507 | 89702458 | 952 | 8 | 4.37E-06 | 3.70E-05 | 0.00184 | 3.56E-06 | -0.00573956 | AL137230.2 |
| chr12 | 107845693 | 107846108 | 416 | 3 | 4.37E-06 | 7.13E-07 | 0.00013 | 1.19E-06 | -0.033035096 | AC126177.3 |
| chr7 | 159230645 | 159231137 | 493 | 3 | 4.37E-06 | 4.55E-05 | 1.72E-05 | 1.54E-06 | -0.004426471 | NA |
| chr8 | 30812332 | 30813230 | 899 | 6 | 4.39E-06 | 3.74E-05 | 0.00091 | 2.70E-06 | 0.001123827 | PPP2CB |
| chr11 | 125920497 | 125920996 | 500 | 3 | 4.39E-06 | 4.42E-05 | 1.33E-05 | 5.15E-06 | -6.06E-05 | DDX25, AP000842.2 |
| chr12 | 49045674 | 49046705 | 1032 | 8 | 4.43E-06 | 6.79E-06 | 0.00125 | 9.45E-07 | -0.003059264 | KMT2D |
| chr12 | 88580360 | 88580888 | 529 | 11 | 4.43E-06 | 0.019003 | 2.88E-05 | 4.02E-05 | 0.001872005 | AC024941.2, KITLG |
| chr22 | 46107245 | 46108740 | 1496 | 11 | 4.43E-06 | 8.86E-05 | 0.00124 | 4.88E-06 | -0.00243363 | MIRLET7BHG |
| chr7 | 150736505 | 150736989 | 485 | 4 | 4.43E-06 | 0.01433 | 2.02E-05 | 8.18E-05 | -0.004731734 | GIMAP1-GIMAP5, GIMAP5 |
| chr1 | 10303573 | 10303967 | 395 | 3 | 4.43E-06 | 1.99E-06 | 2.75E-05 | 1.15E-06 | -0.011501737 | KIF1B |
| chr19 | 710823 | 711646 | 824 | 4 | 4.44E-06 | 1.03E-07 | 0.00042 | 2.58E-07 | -0.006613493 | PALM |
| chr10 | 86668056 | 86668672 | 617 | 8 | 4.45E-06 | 2.16E-06 | 0.00197 | 1.52E-06 | -0.006101423 | LDB3 |
| chr9 | 134214847 | 134215316 | 470 | 2 | 4.45E-06 | 5.94E-06 | 4.42E-05 | 4.36E-06 | -0.010668827 | NA |
| chr1 | 16043335 | 16043770 | 436 | 5 | 4.46E-06 | 3.31E-05 | 3.41E-05 | 1.19E-06 | -0.005654609 | CLCNKB |
| chr15 | 99447117 | 99447750 | 634 | 4 | 4.49E-06 | 6.58E-07 | 0.00048 | 9.85E-07 | -0.019831064 | NA |
| chr1 | 109624887 | 109625956 | 1070 | 9 | 4.49E-06 | 7.23E-05 | 0.00241 | 4.41E-06 | -0.002535288 | AMPD2 |
| chr1 | 63023430 | 63024060 | 631 | 3 | 4.49E-06 | 6.83E-05 | 2.57E-05 | 4.37E-07 | -0.003117394 | NA |
| chr3 | 39150724 | 39150964 | 241 | 3 | 4.49E-06 | 2.17E-06 | 3.62E-05 | 1.25E-06 | -0.008485139 | AC092053.3, CSRNP1 |
| chr5 | 132822654 | 132822933 | 280 | 2 | 4.51E-06 | 8.61E-05 | 1.29E-05 | 7.51E-06 | -0.007048141 | SHROOM1 |
| chr11 | 118522735 | 118523189 | 455 | 2 | 4.52E-06 | 0.001931 | 2.69E-05 | 3.29E-05 | -0.010275533 | KMT2A, AP001267.3 |
| chr17 | 57420054 | 57420862 | 809 | 4 | 4.52E-06 | 1.87E-05 | 7.89E-05 | 9.39E-06 | -0.006585853 | MSI2 |
| chr19 | 3961088 | 3962196 | 1109 | 13 | 4.53E-06 | 9.09E-06 | 0.00135 | 1.68E-06 | -0.002988038 | DAPK3, MIR637 |
| chr9 | 137454376 | 137455517 | 1142 | 11 | 4.53E-06 | 2.24E-05 | 0.00179 | 1.88E-06 | -0.006115521 | NSMF |
| chr7 | 2111549 | 2112453 | 905 | 7 | 4.53E-06 | 9.04E-05 | 0.00011 | 8.41E-07 | -0.006453375 | MAD1L1, AC069288.1 |
| chr6 | 107709447 | 107709535 | 89 | 3 | 4.54E-06 | 7.05E-07 | 4.30E-05 | 6.33E-07 | -0.009223665 | SCML4 |
| chr18 | 7011218 | 7011464 | 247 | 4 | 4.54E-06 | 1.16E-07 | 0.00141 | 5.16E-07 | -0.027698595 | LAMA1 |
| chr17 | 48500743 | 48501162 | 420 | 3 | 4.55E-06 | 0.000129 | 9.14E-05 | 3.11E-06 | -0.012221995 | NA |
| chr16 | 56607461 | 56608571 | 1111 | 11 | 4.56E-06 | 5.41E-05 | 0.00025 | 4.17E-06 | -0.004539882 | NA |
| chr1 | 50048077 | 50048788 | 712 | 4 | 4.56E-06 | 0.00012 | 0.00114 | 5.92E-05 | -0.00449623 | ELAVL4 |
| chr8 | 142304821 | 142305137 | 317 | 4 | 4.57E-06 | 7.83E-06 | 3.54E-05 | 3.09E-06 | -0.004037671 | TSNARE1 |
| chr4 | 173537668 | 173538592 | 925 | 4 | 4.58E-06 | 0.000392 | 2.40E-05 | 1.87E-05 | 0.004648047 | HAND2-AS1 |
| chr11 | 118519485 | 118520132 | 648 | 5 | 4.61E-06 | 5.55E-06 | 0.00079 | 1.47E-06 | -0.008538945 | KMT2A, AP001267.3 |
| chr2 | 144519749 | 144522003 | 2255 | 23 | 4.61E-06 | 1.24E-08 | 0.00059 | 2.25E-09 | -0.004743417 | ZEB2-AS1, AC009951.1, AC009951.3, ZEB2 |
| chr8 | 141300592 | 141300973 | 382 | 5 | 4.61E-06 | 1.60E-05 | 6.98E-05 | 2.34E-06 | -0.001621451 | SLC45A4 |
| chr16 | 2071746 | 2072884 | 1139 | 6 | 4.62E-06 | 9.29E-06 | 0.00131 | 5.33E-06 | -0.003299152 | TSC2 |
| chr2 | 190342516 | 190343509 | 994 | 10 | 4.63E-06 | 3.20E-05 | 0.00441 | 4.63E-06 | -0.003917131 | HIBCH |
| chr22 | 43343569 | 43344224 | 656 | 7 | 4.63E-06 | 0.000205 | 0.00013 | 4.82E-06 | -0.007471969 | NA |
| chr22 | 38981242 | 38982377 | 1136 | 4 | 4.64E-06 | 4.71E-07 | 0.00044 | 3.21E-07 | -0.019845485 | APOBEC3A, APOBEC3B |
| chr8 | 129726577 | 129726695 | 119 | 2 | 4.64E-06 | 6.83E-05 | 3.30E-06 | 1.81E-06 | -0.010641482 | NA |
| chr13 | 106530191 | 106530744 | 554 | 4 | 4.64E-06 | 9.99E-06 | 8.40E-05 | 7.04E-06 | -0.007304883 | EFNB2 |
| chr8 | 96333264 | 96333845 | 582 | 8 | 4.65E-06 | 1.57E-05 | 0.00248 | 2.56E-06 | -0.001861687 | PTDSS1 |
| chr17 | 560345 | 560898 | 554 | 3 | 4.68E-06 | 6.33E-07 | 0.00021 | 8.26E-07 | -0.027537968 | VPS53 |
| chr11 | 709627 | 710392 | 766 | 4 | 4.70E-06 | 5.84E-07 | 0.00239 | 2.00E-06 | -0.010789194 | EPS8L2, AP006621.4 |
| chr16 | 89769950 | 89770659 | 710 | 6 | 4.70E-06 | 2.55E-06 | 0.00192 | 1.61E-06 | -0.006460759 | FANCA |
| chr12 | 49577533 | 49578346 | 814 | 4 | 4.71E-06 | 1.40E-06 | 7.45E-05 | 1.36E-06 | -0.007045692 | PRPF40B |
| chr12 | 95997134 | 95997474 | 341 | 4 | 4.71E-06 | 0.000225 | 1.16E-05 | 1.03E-05 | -0.005347373 | AC007298.2 |
| chr19 | 8585639 | 8586202 | 564 | 5 | 4.72E-06 | 2.60E-06 | 0.00025 | 1.60E-06 | -0.005070792 | ADAMTS10 |
| chr2 | 239311348 | 239311545 | 198 | 3 | 4.72E-06 | 1.07E-05 | 2.28E-05 | 1.49E-06 | -0.003034182 | HDAC4 |
| chr9 | 120477767 | 120478427 | 661 | 2 | 4.72E-06 | 2.18E-06 | 4.04E-05 | 2.00E-06 | -0.016081859 | CDK5RAP2 |
| chr6 | 137539184 | 137539702 | 519 | 3 | 4.74E-06 | 1.39E-06 | 0.00023 | 1.47E-06 | -0.011047592 | NA |
| chr3 | 129440735 | 129441769 | 1035 | 6 | 4.75E-06 | 2.30E-07 | 0.00556 | 1.28E-06 | -0.006402202 | IFT122 |
| chr3 | 25457614 | 25458236 | 623 | 4 | 4.77E-06 | 1.70E-05 | 0.00013 | 5.22E-06 | -0.005642868 | RARB |
| chr8 | 54023045 | 54023629 | 585 | 4 | 4.78E-06 | 4.98E-05 | 0.00024 | 6.51E-06 | -0.011311084 | NA |
| chr7 | 134549045 | 134549077 | 33 | 2 | 4.79E-06 | 1.88E-06 | 4.67E-05 | 1.92E-06 | -0.025427953 | NA |
| chr7 | 39519150 | 39519606 | 457 | 2 | 4.80E-06 | 6.79E-06 | 2.84E-05 | 3.65E-06 | -0.015554572 | NA |
| chr21 | 38659371 | 38659429 | 59 | 2 | 4.81E-06 | 1.14E-06 | 0.00011 | 1.77E-06 | -0.009780285 | ERG |
| chr10 | 2934025 | 2934267 | 243 | 2 | 4.83E-06 | 4.35E-07 | 0.0001 | 7.48E-07 | -0.023035639 | NA |
| chr7 | 48454559 | 48455736 | 1178 | 8 | 4.85E-06 | 1.58E-07 | 0.00299 | 4.02E-07 | -0.006799645 | ABCA13 |
| chr6 | 13470420 | 13470955 | 536 | 2 | 4.87E-06 | 4.20E-06 | 3.89E-05 | 3.16E-06 | -0.010677881 | GFOD1 |
| chr21 | 42865863 | 42866390 | 528 | 3 | 4.87E-06 | 5.29E-05 | 6.10E-05 | 1.82E-05 | -0.008062736 | WDR4 |
| chr9 | 129809902 | 129810421 | 520 | 3 | 4.87E-06 | 1.78E-05 | 3.75E-05 | 6.05E-06 | -0.001728998 | TOR1B |
| chr9 | 95095242 | 95095626 | 385 | 3 | 4.87E-06 | 2.61E-07 | 0.00077 | 8.06E-07 | -0.024751778 | NA |
| chr3 | 131027627 | 131028317 | 691 | 6 | 4.87E-06 | 7.79E-06 | 0.00181 | 4.29E-06 | -0.0030993 | NEK11, ASTE1 |
| chr1 | 16967448 | 16968277 | 830 | 4 | 4.90E-06 | 1.89E-07 | 0.00011 | 9.86E-08 | -0.012557454 | CROCC |
| chr9 | 79687302 | 79687817 | 516 | 2 | 4.96E-06 | 7.14E-06 | 2.54E-05 | 3.44E-06 | -0.001599732 | TLE4 |
| chr6 | 32166152 | 32168026 | 1875 | 42 | 4.98E-06 | 6.38E-07 | 0.00055 | 7.15E-09 | -0.001729102 | PPT2-EGFL8, EGFL8 |
| chr19 | 3786483 | 3787091 | 609 | 8 | 4.98E-06 | 0.000945 | 2.07E-05 | 1.62E-05 | -0.003105448 | MATK |
| chr22 | 30972579 | 30972982 | 404 | 3 | 4.99E-06 | 1.66E-06 | 6.76E-05 | 1.50E-06 | -0.003489243 | TUG1, AC004542.6 |
| chr16 | 8861694 | 8862589 | 896 | 8 | 4.99E-06 | 6.56E-07 | 0.00173 | 1.12E-06 | -0.00357112 | CARHSP1 |
| chr12 | 132405150 | 132406191 | 1042 | 8 | 5.00E-06 | 2.31E-05 | 0.00091 | 2.62E-06 | -0.00835796 | NA |
| chr3 | 146544387 | 146545360 | 974 | 12 | 5.01E-06 | 8.64E-08 | 0.01247 | 5.24E-07 | 0.005459567 | PLSCR1 |
| chr1 | 2879628 | 2880126 | 499 | 4 | 5.02E-06 | 0.00031 | 1.80E-05 | 4.95E-06 | -0.00519618 | NA |
| chr20 | 3916989 | 3917489 | 501 | 5 | 5.03E-06 | 2.14E-05 | 8.66E-05 | 2.14E-06 | -0.003379459 | PANK2 |
| chr14 | 64221295 | 64221546 | 252 | 2 | 5.03E-06 | 9.16E-07 | 7.67E-05 | 1.32E-06 | -0.015520673 | SYNE2, ESR2 |
| chr9 | 68540362 | 68541562 | 1201 | 3 | 5.03E-06 | 4.11E-08 | 0.00018 | 1.20E-07 | -0.015067143 | TMEM252-DT, TMEM252 |
| chr2 | 86195171 | 86196151 | 981 | 13 | 5.03E-06 | 4.32E-05 | 0.00022 | 3.98E-06 | 0.003396516 | AC009309.1, IMMT |
| chr2 | 127205754 | 127206077 | 324 | 9 | 5.03E-06 | 1.60E-06 | 0.00695 | 1.10E-06 | -0.008350098 | CYP27C1 |
| chr5 | 138055289 | 138055494 | 206 | 3 | 5.03E-06 | 1.91E-07 | 0.00046 | 5.47E-07 | -0.018446701 | NA |
| chr4 | 54539697 | 54539721 | 25 | 2 | 5.04E-06 | 3.59E-07 | 0.00013 | 6.57E-07 | 0.027730296 | NA |
| chr19 | 5341584 | 5342090 | 507 | 2 | 5.06E-06 | 7.47E-05 | 5.71E-05 | 2.51E-05 | 0.013370762 | NA |
| chr9 | 34990118 | 34990401 | 284 | 5 | 5.08E-06 | 6.87E-05 | 0.00025 | 2.61E-06 | 0.005719738 | DNAJB5 |
| chr6 | 29357340 | 29357500 | 161 | 3 | 5.08E-06 | 2.30E-06 | 6.36E-05 | 1.49E-06 | -0.011408496 | OR5V1 |
| chr2 | 28319953 | 28320431 | 479 | 2 | 5.08E-06 | 5.94E-05 | 1.34E-05 | 6.80E-06 | -0.003481164 | BABAM2 |
| chr8 | 92017871 | 92019342 | 1472 | 7 | 5.08E-06 | 3.11E-07 | 0.00021 | 8.30E-08 | -0.005949288 | RUNX1T1 |
| chr21 | 33268612 | 33269202 | 591 | 3 | 5.10E-06 | 5.23E-06 | 0.00011 | 4.58E-06 | -0.009067678 | AP000295.1, IL10RB |
| chr3 | 195762837 | 195763438 | 602 | 8 | 5.12E-06 | 1.81E-08 | 0.0116 | 3.53E-07 | 0.045311187 | MUC4 |
| chr20 | 62938761 | 62939525 | 765 | 5 | 5.17E-06 | 0.001089 | 8.27E-05 | 1.57E-05 | 0.000218653 | GID8 |
| chr6 | 28589433 | 28590336 | 904 | 23 | 5.17E-06 | 4.01E-06 | 0.00902 | 2.83E-06 | -0.003842997 | AL049543.1, ZBED9 |
| chr14 | 35347123 | 35347760 | 638 | 3 | 5.19E-06 | 1.58E-07 | 0.00056 | 4.92E-07 | -0.021000474 | NA |
| chr14 | 94095711 | 94096172 | 462 | 2 | 5.19E-06 | 9.38E-05 | 3.62E-05 | 1.94E-05 | 0.001439787 | IFI27L1 |
| chr22 | 24592053 | 24592301 | 249 | 2 | 5.19E-06 | 1.95E-05 | 0.00016 | 1.97E-05 | -0.002898111 | AP000356.5, LRRC75B |
| chr9 | 137431584 | 137431905 | 322 | 2 | 5.20E-06 | 6.10E-05 | 2.05E-05 | 9.91E-06 | -0.006206919 | NOXA1 |
| chr7 | 108526753 | 108528322 | 1570 | 8 | 5.21E-06 | 4.47E-06 | 0.00014 | 7.91E-07 | -0.004610493 | PNPLA8 |
| chr4 | 3763459 | 3765442 | 1984 | 7 | 5.23E-06 | 8.50E-07 | 0.00045 | 2.37E-07 | -0.009518727 | NA |
| chr6 | 32177369 | 32179002 | 1634 | 34 | 5.24E-06 | 0.000193 | 0.00393 | 9.96E-07 | 0.001333414 | RNF5, AGPAT1 |

| | | | | | | | | | | |
|---|---|---|---|---|---|---|---|---|---|---|
| chr6 | 28574624 | 28575916 | 1293 | 9 | 5.25E-06 | 2.23E-07 | 0.00265 | 3.22E-07 | -0.004114353 | ZBED9 |
| chr15 | 28191760 | 28192013 | 254 | 4 | 5.25E-06 | 6.89E-05 | 0.00014 | 2.92E-06 | -0.006345305 | HERC2 |
| chr2 | 24793027 | 24793713 | 687 | 18 | 5.26E-06 | 1.95E-05 | 0.00548 | 2.02E-06 | 0.000410065 | CENPO, PTRHD1 |
| chr2 | 237600186 | 237600519 | 334 | 2 | 5.28E-06 | 5.85E-07 | 0.00016 | 1.04E-06 | -0.013156347 | RAB17 |
| chr12 | 132178883 | 132179338 | 456 | 8 | 5.29E-06 | 0.00013 | 0.0007 | 1.52E-06 | 0.013476923 | NA |
| chr17 | 64083746 | 64084817 | 1072 | 3 | 5.29E-06 | 1.56E-07 | 4.46E-05 | 1.52E-07 | -0.010287522 | ERN1 |
| chr9 | 124262486 | 124263003 | 518 | 4 | 5.30E-06 | 7.01E-07 | 0.00109 | 1.84E-06 | -0.010480566 | NEK6, AL162724.1 |
| chr3 | 113948542 | 113948962 | 421 | 3 | 5.30E-06 | 3.10E-05 | 0.00047 | 1.02E-05 | -0.012952914 | ZDHHC23 |
| chr1 | 67054509 | 67055804 | 1296 | 7 | 5.31E-06 | 2.37E-05 | 0.0003 | 8.87E-07 | -0.00741968 | NA |
| chr11 | 1760275 | 1761660 | 1386 | 8 | 5.33E-06 | 3.20E-09 | 0.00098 | 9.44E-09 | -0.006068874 | AC068580.3, AC068580.4, CTSD |
| chr1 | 154154986 | 154155967 | 982 | 9 | 5.34E-06 | 2.98E-07 | 0.00419 | 1.45E-06 | -0.01342317 | NUP210L, TPM3 |
| chr9 | 138052150 | 138053085 | 936 | 3 | 5.34E-06 | 4.69E-06 | 8.42E-05 | 3.30E-06 | -0.006975629 | CACNA1B |
| chr4 | 140078569 | 140078993 | 425 | 2 | 5.36E-06 | 1.20E-05 | 1.53E-05 | 3.32E-06 | -0.003197931 | MAML3 |
| chr6 | 159239774 | 159240636 | 863 | 4 | 5.37E-06 | 2.33E-05 | 0.00022 | 1.43E-05 | -0.007323636 | FNDC1 |
| chr9 | 128091293 | 128092434 | 1142 | 6 | 5.38E-06 | 0.002445 | 0.00028 | 9.11E-05 | -0.007834669 | SLC25A25, AL360268.1 |
| chr14 | 75430639 | 75432243 | 1605 | 8 | 5.40E-06 | 4.00E-09 | 0.00067 | 1.19E-08 | -0.00686587 | JDP2 |
| chr9 | 137279504 | 137281784 | 2281 | 9 | 5.40E-06 | 1.12E-11 | 0.00394 | 6.31E-10 | -0.006514713 | TOR4A |
| chr5 | 149777103 | 149777653 | 551 | 2 | 5.40E-06 | 8.55E-07 | 9.47E-05 | 1.32E-06 | -0.011801824 | PPARGC1B |
| chr1 | 41582628 | 41582940 | 313 | 2 | 5.40E-06 | 6.42E-07 | 6.58E-05 | 9.29E-07 | -0.009892556 | HIVEP3 |
| chr1 | 230332422 | 230333584 | 1163 | 7 | 5.41E-06 | 5.81E-07 | 0.00193 | 1.19E-06 | 0.020207666 | PGBD5 |
| chr10 | 84173544 | 84174042 | 499 | 3 | 5.43E-06 | 9.88E-05 | 2.28E-05 | 1.26E-05 | -0.006318012 | C10orf99 |
| chr19 | 17247936 | 17248431 | 496 | 2 | 5.46E-06 | 7.18E-06 | 5.37E-05 | 5.56E-06 | -0.007200548 | AC010646.1 |
| chr11 | 44728533 | 44728914 | 382 | 2 | 5.47E-06 | 1.78E-06 | 0.0001 | 2.53E-06 | -0.018313112 | TSPAN18, TSPAN18-AS1 |
| chr15 | 71748425 | 71748454 | 30 | 2 | 5.48E-06 | 0.00037 | 7.07E-06 | 6.06E-06 | -0.00243511 | THSD4 |
| chr19 | 1057920 | 1058263 | 344 | 2 | 5.51E-06 | 7.24E-06 | 0.00018 | 9.46E-06 | 0.003226052 | ABCA7 |
| chr3 | 127722516 | 127722924 | 409 | 3 | 5.53E-06 | 2.23E-07 | 0.00045 | 6.28E-07 | -0.007140179 | MGLL |
| chr3 | 51395997 | 51396069 | 73 | 2 | 5.53E-06 | 1.94E-05 | 5.29E-06 | 1.79E-06 | -0.01332964 | RBM15B, DCAF1 |
| chr10 | 12388860 | 12389271 | 412 | 4 | 5.55E-06 | 8.39E-07 | 0.00022 | 1.02E-06 | -0.011001669 | CAMK1D |
| chr17 | 78351896 | 78351977 | 82 | 2 | 5.56E-06 | 1.96E-06 | 0.00012 | 2.88E-06 | -0.012508447 | NA |
| chr11 | 128290388 | 128290742 | 355 | 3 | 5.56E-06 | 7.10E-07 | 0.00021 | 9.53E-07 | -0.012362989 | NA |
| chr19 | 18915436 | 18915885 | 450 | 4 | 5.57E-06 | 3.66E-05 | 1.32E-05 | 3.28E-06 | -0.005567487 | COPE, AC002985.1 |
| chr16 | 396077 | 400051 | 3975 | 20 | 5.57E-06 | 1.20E-08 | 0.00049 | 2.11E-12 | -0.002569592 | NME4 |
| chr8 | 141219662 | 141220322 | 661 | 6 | 5.60E-06 | 2.38E-06 | 0.00055 | 1.53E-06 | -0.002838064 | SLC45A4 |
| chr3 | 32570949 | 32571591 | 643 | 10 | 5.60E-06 | 0.000473 | 5.54E-05 | 1.87E-05 | 0.003160184 | NA |
| chr21 | 31871921 | 31872801 | 881 | 5 | 5.61E-06 | 3.95E-05 | 0.00039 | 8.12E-06 | 0.007882081 | NA |
| chr19 | 35351950 | 35352379 | 430 | 3 | 5.61E-06 | 0.000146 | 0.00046 | 2.73E-05 | -0.004771824 | FFAR1 |
| chr3 | 33219240 | 33219712 | 473 | 8 | 5.63E-06 | 0.000123 | 0.00122 | 7.05E-06 | -0.008202817 | NA |
| chr8 | 142543308 | 142544236 | 929 | 3 | 5.66E-06 | 2.51E-05 | 7.65E-05 | 8.10E-06 | -0.00460945 | ADGRB1 |
| chr2 | 239308757 | 239309360 | 604 | 6 | 5.67E-06 | 8.93E-07 | 0.00034 | 1.02E-06 | -0.009627618 | HDAC4 |
| chr14 | 92539055 | 92540813 | 1759 | 5 | 5.67E-06 | 8.36E-10 | 0.00061 | 7.49E-09 | -0.020737943 | RIN3 |
| chr8 | 1764484 | 1764846 | 363 | 7 | 5.68E-06 | 5.00E-05 | 0.00018 | 5.16E-06 | -0.010637458 | CLN8, AC019257.8, AC100810.1 |
| chr1 | 39025268 | 39026423 | 1156 | 8 | 5.68E-06 | 1.13E-06 | 0.00115 | 2.34E-07 | -0.002328973 | NDUFS5 |
| chr17 | 8466699 | 8467230 | 532 | 4 | 5.70E-06 | 0.00043 | 0.00019 | 1.94E-05 | -0.001202463 | NDEL1 |
| chr17 | 1773628 | 1773935 | 308 | 2 | 5.70E-06 | 1.60E-06 | 0.00014 | 2.50E-06 | -0.014680898 | SERPINF1 |
| chr6 | 152701877 | 152702144 | 268 | 2 | 5.70E-06 | 2.09E-06 | 2.19E-05 | 1.40E-06 | -0.019537601 | MYCT1 |
| chr10 | 107165109 | 107165944 | 836 | 5 | 5.71E-06 | 3.69E-05 | 7.13E-05 | 1.01E-05 | 0.009595969 | NA |
| chr5 | 173689432 | 173690378 | 947 | 3 | 5.71E-06 | 1.10E-06 | 0.00017 | 1.84E-06 | -0.019458623 | LINC01942 |
| chr1 | 15975737 | 15976804 | 1068 | 12 | 5.71E-06 | 0.001771 | 8.73E-05 | 1.97E-06 | -0.001767471 | ZBTB17 |
| chr4 | 100790874 | 100791283 | 410 | 3 | 5.72E-06 | 7.51E-05 | 2.81E-05 | 5.93E-06 | 0.00377589 | EMCN, LINC01217 |
| chr14 | 92512760 | 92515322 | 2563 | 15 | 5.72E-06 | 1.08E-05 | 0.00011 | 9.35E-08 | -0.005231493 | RIN3 |
| chr10 | 125816335 | 125817493 | 1159 | 4 | 5.75E-06 | 0.00031 | 1.47E-05 | 9.40E-06 | -0.002881301 | UROS |
| chr20 | 59674177 | 59674226 | 50 | 2 | 5.75E-06 | 9.11E-07 | 0.0002 | 1.60E-06 | -0.024868941 | PHACTR3 |
| chr20 | 35560146 | 35560673 | 528 | 3 | 5.76E-06 | 2.32E-05 | 3.21E-05 | 5.64E-06 | -0.009810216 | FER1L4 |
| chr2 | 241354431 | 241355067 | 637 | 3 | 5.77E-06 | 3.47E-05 | 7.58E-05 | 1.56E-05 | -0.010026759 | NA |
| chr2 | 176124211 | 176124858 | 648 | 4 | 5.77E-06 | 8.08E-05 | 0.00093 | 1.51E-05 | -0.012084862 | HOXD9, HOXD-AS2 |
| chr9 | 35563315 | 35564163 | 849 | 9 | 5.78E-06 | 1.67E-07 | 0.00367 | 5.67E-07 | -0.012193446 | FAM166B |
| chr12 | 57737228 | 57737898 | 671 | 3 | 5.78E-06 | 2.60E-05 | 0.00152 | 4.48E-05 | 0.000572791 | AGAP2 |
| chr1 | 20685195 | 20685793 | 599 | 5 | 5.78E-06 | 2.77E-06 | 8.07E-05 | 2.09E-06 | -0.006976024 | KIF17 |
| chr4 | 119519072 | 119519687 | 616 | 2 | 5.79E-06 | 2.06E-06 | 3.62E-05 | 1.82E-06 | -0.005058915 | AC093752.1, PDE5A |
| chr8 | 28374597 | 28374659 | 63 | 2 | 5.81E-06 | 4.76E-06 | 0.00015 | 6.48E-06 | -0.011411528 | ZNF395, FBXO16 |
| chr1 | 202809315 | 202810369 | 1055 | 8 | 5.82E-06 | 4.38E-07 | 0.0005 | 8.51E-07 | 0.016324762 | AC098934.2 |
| chr10 | 71439916 | 71440235 | 320 | 2 | 5.85E-06 | 0.000407 | 6.13E-06 | 5.35E-06 | -0.003570904 | CDH23 |
| chr5 | 132665029 | 132665994 | 966 | 5 | 5.86E-06 | 1.95E-06 | 0.00187 | 2.52E-06 | -0.003246086 | NA |
| chr20 | 50242492 | 50243672 | 1181 | 5 | 5.87E-06 | 0.000181 | 0.00068 | 8.79E-06 | -0.004864896 | NA |
| chr4 | 6986917 | 6987857 | 941 | 15 | 5.88E-06 | 3.69E-06 | 0.00317 | 1.25E-06 | 0.000186536 | TBC1D14, AC097382.3 |
| chr2 | 43464693 | 43464702 | 10 | 2 | 5.88E-06 | 1.05E-06 | 8.34E-05 | 1.51E-06 | -0.003658303 | THADA |
| chr14 | 102318909 | 102319813 | 905 | 11 | 5.89E-06 | 0.000641 | 0.00013 | 8.08E-07 | -0.000543757 | ZNF839 |
| chr1 | 42172750 | 42173908 | 1159 | 4 | 5.90E-06 | 2.47E-05 | 2.13E-05 | 7.17E-07 | -0.007322291 | NA |
| chr12 | 108846273 | 108846757 | 485 | 3 | 5.90E-06 | 1.08E-05 | 2.42E-05 | 3.18E-06 | -0.004487851 | SSH1 |
| chr9 | 35790474 | 35791801 | 1328 | 8 | 5.91E-06 | 2.96E-06 | 0.00193 | 1.96E-06 | -0.013798231 | NPR2 |
| chr6 | 1684281 | 1684333 | 53 | 2 | 5.91E-06 | 4.96E-07 | 8.47E-05 | 8.04E-07 | -0.010940624 | GMDS |
| chr18 | 76351900 | 76352352 | 453 | 3 | 5.93E-06 | 2.79E-05 | 0.00015 | 4.60E-06 | -0.020380896 | NA |
| chr14 | 90331719 | 90332714 | 996 | 18 | 5.94E-06 | 3.27E-05 | 0.00039 | 1.95E-06 | -0.000181966 | NRDE2 |
| chr18 | 54827303 | 54828310 | 1008 | 5 | 5.94E-06 | 0.002192 | 0.00099 | 4.70E-05 | 0.000681867 | RAB27B |
| chr12 | 51970915 | 51971438 | 524 | 4 | 5.94E-06 | 2.03E-05 | 5.95E-05 | 6.83E-06 | -0.010514513 | ACVR1B |
| chr1 | 117339863 | 117340216 | 354 | 2 | 5.98E-06 | 3.97E-06 | 2.07E-05 | 2.07E-06 | -0.014376774 | NA |
| chr1 | 158353692 | 158354681 | 990 | 4 | 5.98E-06 | 6.62E-05 | 1.81E-05 | 3.72E-06 | -0.006797484 | CD1E |
| chr11 | 2412238 | 2412988 | 751 | 4 | 6.00E-06 | 1.23E-06 | 0.00022 | 1.57E-06 | -0.008380965 | TRPM5 |
| chr14 | 103519890 | 103520130 | 241 | 4 | 6.00E-06 | 0.000197 | 1.02E-05 | 5.65E-06 | -0.004626507 | CKB |
| chr2 | 181457541 | 181458171 | 631 | 8 | 6.02E-06 | 1.21E-05 | 7.05E-05 | 3.23E-06 | 0.00105301 | ITGA4 |
| chr16 | 838938 | 839432 | 495 | 5 | 6.03E-06 | 0.000691 | 7.65E-05 | 4.71E-06 | -0.002274335 | AL023882.1 |
| chr9 | 136883478 | 136883971 | 494 | 3 | 6.06E-06 | 3.06E-05 | 0.00022 | 1.66E-05 | -0.000577813 | TRAF2 |
| chr21 | 42261792 | 42261805 | 14 | 2 | 6.08E-06 | 7.79E-07 | 8.35E-05 | 1.18E-06 | -0.016818378 | ABCG1 |
| chr15 | 85778471 | 85778577 | 107 | 2 | 6.08E-06 | 1.04E-06 | 0.00015 | 1.73E-06 | -0.013482325 | KLHL25 |
| chr1 | 19678921 | 19679251 | 331 | 5 | 6.10E-06 | 3.74E-06 | 0.00015 | 1.47E-06 | -0.004562864 | HTR6 |
| chr2 | 165294937 | 165295308 | 372 | 3 | 6.11E-06 | 4.03E-05 | 0.00014 | 6.13E-06 | -0.016409144 | SCN2A |
| chr17 | 10026784 | 10027563 | 780 | 3 | 6.11E-06 | 4.91E-06 | 0.00033 | 7.89E-06 | -0.006179073 | GAS7 |
| chr7 | 1557116 | 1559138 | 2023 | 17 | 6.13E-06 | 1.26E-08 | 0.00254 | 1.69E-08 | -0.006551596 | TMEM184A |
| chr14 | 92644212 | 92644505 | 294 | 2 | 6.13E-06 | 0.003084 | 5.57E-06 | 6.01E-06 | -0.004385979 | RIN3 |
| chr1 | 156591133 | 156593152 | 2020 | 14 | 6.21E-06 | 8.70E-08 | 0.00014 | 2.09E-08 | -0.004653046 | NAXE |
| chr5 | 34042871 | 34043619 | 749 | 8 | 6.22E-06 | 1.77E-05 | 0.00078 | 4.20E-06 | -0.005791384 | C1QTNF3-AMACR, C1QTNF3 |
| chr21 | 43302195 | 43302978 | 784 | 2 | 6.23E-06 | 2.18E-06 | 0.00011 | 3.06E-06 | -0.006539166 | NA |
| chr7 | 26191138 | 26191781 | 644 | 2 | 6.24E-06 | 2.00E-06 | 3.95E-05 | 1.86E-06 | -0.01270356 | HNRNPA2B1 |
| chr1 | 6321842 | 6322190 | 349 | 3 | 6.25E-06 | 2.21E-06 | 5.22E-05 | 1.33E-06 | -0.002508369 | ACOT7 |
| chr5 | 132812974 | 132815355 | 2382 | 16 | 6.27E-06 | 1.12E-05 | 2.08E-05 | 6.09E-09 | -0.000370861 | SOWAHA |
| chr10 | 102132552 | 102133076 | 525 | 11 | 6.27E-06 | 6.58E-07 | 0.0041 | 8.61E-07 | 0.001307741 | PPRC1 |
| chr17 | 21285942 | 21286547 | 606 | 4 | 6.28E-06 | 1.44E-05 | 0.00025 | 9.53E-06 | 0.004601171 | MAP2K3 |
| chr8 | 144414423 | 144414938 | 516 | 3 | 6.29E-06 | 8.67E-06 | 0.00013 | 3.23E-06 | -0.006053399 | SLC39A4 |
| chr5 | 5572437 | 5573080 | 644 | 3 | 6.31E-06 | 1.28E-06 | 9.40E-05 | 1.52E-06 | -0.019846075 | NA |
| chr18 | 77016882 | 77017595 | 714 | 14 | 6.32E-06 | 2.07E-06 | 0.00114 | 1.17E-06 | -0.000537271 | MBP |
| chr5 | 141647583 | 141648090 | 508 | 4 | 6.32E-06 | 0.000135 | 5.75E-05 | 9.26E-06 | -0.004444241 | FCHSD1 |
| chr16 | 66999511 | 67000670 | 1160 | 10 | 6.35E-06 | 1.66E-06 | 0.0015 | 1.47E-06 | -0.006140804 | CES4A |
| chr5 | 174995551 | 174996457 | 907 | 5 | 6.37E-06 | 1.21E-06 | 0.00023 | 1.47E-06 | -0.009912828 | NA |
| chr7 | 106050366 | 106050772 | 407 | 2 | 6.37E-06 | 0.000151 | 9.61E-06 | 6.84E-06 | -0.010196165 | NA |
| chr8 | 107331740 | 107332354 | 615 | 3 | 6.38E-06 | 3.86E-06 | 0.00026 | 2.20E-06 | -0.011760877 | ANGPT1 |
| chr1 | 234769939 | 234771157 | 1219 | 4 | 6.40E-06 | 1.97E-07 | 0.00101 | 7.39E-07 | -0.005759192 | NA |
| chr19 | 2749082 | 2749321 | 240 | 4 | 6.41E-06 | 6.31E-06 | 0.00029 | 1.24E-06 | 0.012982332 | NA |
| chr7 | 2256363 | 2257418 | 1056 | 4 | 6.43E-06 | 1.77E-07 | 0.00154 | 6.51E-07 | -0.004552094 | SNX8 |
| chr3 | 155807187 | 155807194 | 8 | 2 | 6.44E-06 | 0.000133 | 5.89E-05 | 3.39E-05 | -0.011052999 | AC104472.3 |
| chr1 | 33375565 | 33376330 | 766 | 5 | 6.46E-06 | 7.08E-07 | 0.00042 | 4.54E-07 | -0.000106012 | PHC2 |
| chr7 | 5494781 | 5496117 | 1337 | 15 | 6.48E-06 | 5.12E-06 | 0.00069 | 1.91E-07 | -0.004154625 | FBXL18, MIR589 |
| chr1 | 149936420 | 149937540 | 1121 | 11 | 6.48E-06 | 1.60E-07 | 0.01012 | 8.53E-07 | -0.008837589 | MTMR11 |
| chr16 | 4681297 | 4682433 | 1137 | 7 | 6.48E-06 | 1.93E-07 | 0.00258 | 7.65E-07 | -0.008677979 | MGRN1 |
| chr1 | 9823971 | 9825459 | 1489 | 14 | 6.48E-06 | 1.07E-06 | 0.00233 | 1.72E-07 | -0.004020557 | CLSTN1 |
| chr1 | 155134238 | 155135309 | 1072 | 8 | 6.48E-06 | 0.000312 | 8.45E-05 | 2.47E-05 | -0.002755073 | EFNA1 |
| chr7 | 44380810 | 44381836 | 1027 | 3 | 6.50E-06 | 1.78E-06 | 4.55E-05 | 1.68E-07 | -0.005858607 | NUDCD3 |
| chr13 | 20567183 | 20567625 | 443 | 7 | 6.50E-06 | 3.65E-05 | 0.00013 | 7.64E-06 | -0.002898034 | IFT88 |
| chr19 | 47509964 | 47510683 | 720 | 4 | 6.52E-06 | 8.03E-05 | 2.94E-05 | 1.03E-05 | -0.005415082 | NAPA |
| chr22 | 46079660 | 46080790 | 1131 | 6 | 6.53E-06 | 4.71E-08 | 0.00203 | 2.29E-07 | -0.004923896 | MIRLET7BHG |
| chr7 | 100156149 | 100156609 | 461 | 4 | 6.53E-06 | 0.000939 | 0.00041 | 3.42E-05 | -0.005532686 | MAP11, MIR4658 |
| chr2 | 201806435 | 201806860 | 426 | 5 | 6.54E-06 | 3.66E-06 | 0.00017 | 1.51E-06 | -0.002099731 | CDK15 |
| chr6 | 157236782 | 157236962 | 181 | 2 | 6.54E-06 | 1.11E-06 | 0.00023 | 1.92E-06 | -0.016438722 | NA |
| chr15 | 73925876 | 73926966 | 1091 | 12 | 6.55E-06 | 1.48E-07 | 0.00723 | 9.13E-07 | -0.010483881 | LOXL1, LOXL1-AS1 |
| chr15 | 31329426 | 31330244 | 819 | 4 | 6.56E-06 | 9.09E-08 | 0.00217 | 4.80E-07 | -0.014570005 | KLF13 |
| chr2 | 159202547 | 159202916 | 370 | 3 | 6.56E-06 | 3.33E-06 | 2.40E-05 | 1.49E-06 | -0.003494624 | TANC1 |
| chr2 | 127418315 | 127418878 | 564 | 4 | 6.57E-06 | 9.64E-06 | 8.76E-05 | 2.33E-06 | -0.003686402 | PROC |
| chr2 | 230994590 | 230995231 | 642 | 4 | 6.57E-06 | 1.61E-05 | 0.00163 | 1.27E-05 | -0.004191216 | SPATA3, SPATA3-AS1 |
| chr19 | 35118628 | 35119207 | 580 | 4 | 6.57E-06 | 9.09E-06 | 0.00027 | 3.38E-06 | -0.007826699 | AC020907.6, FXYD3 |
| chr6 | 28895767 | 28896937 | 1171 | 20 | 6.57E-06 | 3.73E-05 | 0.0114 | 2.14E-05 | 0.000434762 | HCG14 |
| chr13 | 30107790 | 30109060 | 1271 | 4 | 6.57E-06 | 6.07E-09 | 0.00072 | 3.46E-08 | -0.006595318 | LINC00365 |
| chr1 | 173123184 | 173123543 | 360 | 2 | 6.58E-06 | 3.01E-05 | 1.09E-05 | 4.11E-06 | -0.019386625 | NA |
| chr13 | 98840388 | 98840580 | 193 | 3 | 6.58E-06 | 4.38E-06 | 3.45E-05 | 1.17E-06 | -0.009375171 | DOCK9 |
| chr19 | 45973307 | 45974833 | 1527 | 10 | 6.59E-06 | 4.24E-06 | 0.00012 | 1.73E-07 | -0.000289224 | NOVA2 |
| chr1 | 180926732 | 180928010 | 1279 | 5 | 6.59E-06 | 1.85E-06 | 0.00011 | 1.73E-06 | -0.006591997 | KIAA1614 |
| chr2 | 173991713 | 173992021 | 309 | 2 | 6.60E-06 | 1.15E-06 | 5.33E-05 | 1.40E-06 | -0.011249625 | NA |
| chr2 | 235024413 | 235024560 | 148 | 3 | 6.62E-06 | 6.98E-05 | 9.95E-05 | 2.73E-06 | -0.012874625 | SH3BP4 |
| chr6 | 32741693 | 32742132 | 440 | 3 | 6.62E-06 | 0.000624 | 1.82E-05 | 2.09E-05 | -0.008955991 | HLA-DQA2 |
| chr1 | 40160465 | 40162025 | 1561 | 12 | 6.62E-06 | 9.89E-06 | 0.00056 | 3.07E-07 | -0.001904687 | RLF |
| chr1 | 166988496 | 166989852 | 1357 | 18 | 6.64E-06 | 5.29E-06 | 6.22E-05 | 7.34E-07 | -0.002927845 | MAEL |
| chr17 | 17714025 | 17714688 | 664 | 5 | 6.65E-06 | 5.59E-07 | 0.00074 | 1.04E-06 | -0.009576069 | RAI1 |
| chr11 | 70804351 | 70804416 | 66 | 2 | 6.65E-06 | 9.11E-07 | 0.00017 | 1.58E-06 | -0.018698704 | SHANK2 |
| chr9 | 83622519 | 83623229 | 711 | 8 | 6.66E-06 | 3.72E-06 | 0.00231 | 2.68E-06 | -0.004286547 | IDNK |
| chr1 | 155983168 | 155983536 | 369 | 2 | 6.67E-06 | 4.98E-07 | 0.00012 | 8.66E-07 | -0.011516727 | ARHGEF2 |
| chr1 | 19437986 | 19438327 | 342 | 3 | 6.68E-06 | 2.78E-05 | 2.42E-05 | 5.01E-06 | -0.006771719 | CAPZB |
| chr19 | 52269782 | 52270143 | 362 | 2 | 6.69E-06 | 8.15E-06 | 0.00034 | 1.21E-05 | 0.000728752 | ZNF766 |
| chr9 | 136514631 | 136515273 | 643 | 3 | 6.69E-06 | 2.94E-06 | 0.00018 | 3.85E-06 | -0.005150009 | NOTCH1 |
| chr22 | 36921755 | 36923719 | 1965 | 6 | 6.69E-06 | 2.24E-05 | 0.00014 | 4.85E-07 | -0.005881129 | CSF2RB |
| chr4 | 8475835 | 8476188 | 354 | 3 | 6.70E-06 | 6.47E-06 | 3.89E-05 | 2.93E-06 | -0.008269155 | TRMT44 |
| chr17 | 82897964 | 82898362 | 399 | 2 | 6.70E-06 | 0.000956 | 3.74E-05 | 4.05E-05 | -0.010178052 | TBCD |
| chr1 | 32246636 | 32247179 | 544 | 4 | 6.71E-06 | 1.79E-05 | 2.56E-05 | 3.96E-06 | -0.006814727 | NA |
| chr10 | 117140490 | 117140953 | 464 | 4 | 6.74E-06 | 2.10E-05 | 5.53E-05 | 7.51E-06 | 0.003921309 | NA |
| chr13 | 111092825 | 111093514 | 690 | 4 | 6.74E-06 | 3.72E-06 | 0.00017 | 2.97E-06 | -0.008716359 | NA |
| chr19 | 45732963 | 45733817 | 855 | 7 | 6.75E-06 | 1.18E-05 | 0.00095 | 1.18E-06 | -0.005706358 | BHMG1 |
| chr6 | 39322383 | 39323101 | 719 | 7 | 6.75E-06 | 0.017042 | 7.07E-06 | 1.59E-05 | -0.002620009 | KCNK16 |
| chr1 | 201058273 | 201058942 | 670 | 2 | 6.76E-06 | 5.53E-06 | 5.47E-05 | 4.66E-06 | -0.008178648 | CACNA1S |
| chr18 | 79616639 | 79617119 | 481 | 3 | 6.76E-06 | 1.57E-05 | 0.00035 | 7.82E-06 | -0.034136138 | NA |
| chr17 | 29069943 | 29070782 | 840 | 5 | 6.78E-06 | 1.14E-08 | 0.00165 | 8.50E-08 | -0.013462534 | NA |
| chr2 | 241749671 | 241750421 | 751 | 4 | 6.79E-06 | 1.33E-05 | 0.00017 | 5.34E-06 | -0.009945691 | D2HGDH |
| chr19 | 40282289 | 40282880 | 592 | 4 | 6.79E-06 | 6.50E-06 | 0.00016 | 5.82E-06 | -0.004414216 | AKT2, MIR641 |
| chr20 | 4685777 | 4686270 | 494 | 8 | 6.80E-06 | 0.000102 | 0.0003 | 2.70E-06 | 0.001631058 | NA |
| chr16 | 27267519 | 27268566 | 1048 | 7 | 6.81E-06 | 1.87E-05 | 0.00315 | 1.90E-06 | 0.015252473 | NSMCE1-DT, NSMCE1 |
| chr6 | 158643960 | 158644370 | 411 | 5 | 6.82E-06 | 2.31E-05 | 0.00094 | 8.48E-06 | 0.001219136 | DYNLT1 |

| | | | | | | | | | | |
|---|---|---|---|---|---|---|---|---|---|---|
| chr7 | 30135839 | 30136648 | 810 | 4 | 6.82E-06 | 1.84E-06 | 0.00062 | 2.66E-06 | -0.007730116 | MTURN |
| chr7 | 1746330 | 1747735 | 1406 | 6 | 6.83E-06 | 1.34E-06 | 0.00138 | 1.16E-06 | 0.001095676 | ELFN1 |
| chr9 | 93575353 | 93575700 | 348 | 4 | 6.84E-06 | 1.18E-05 | 0.00033 | 2.24E-06 | -0.012888557 | NA |
| chr17 | 62693854 | 62694011 | 158 | 3 | 6.86E-06 | 1.12E-06 | 0.00017 | 1.92E-06 | -0.009163337 | NA |
| chr11 | 119686855 | 119687487 | 633 | 3 | 6.92E-06 | 1.09E-06 | 0.00024 | 2.10E-06 | -0.010409924 | NECTIN1 |
| chr16 | 19698081 | 19699292 | 1212 | 5 | 6.93E-06 | 1.35E-06 | 0.00022 | 9.73E-07 | -0.004511622 | VPS35L |
| chr17 | 2391280 | 2392131 | 852 | 5 | 6.93E-06 | 0.000458 | 0.00049 | 6.92E-06 | -0.012749394 | MNT |
| chr7 | 45968478 | 45969072 | 595 | 3 | 6.93E-06 | 1.85E-06 | 0.00076 | 4.01E-06 | -0.006386748 | AC073115.2 |
| chr4 | 15655209 | 15656260 | 1052 | 7 | 6.94E-06 | 9.91E-05 | 0.00016 | 1.69E-06 | -0.009386085 | FBXL5 |
| chr6 | 109436443 | 109437075 | 633 | 2 | 6.95E-06 | 1.74E-06 | 4.69E-05 | 1.82E-06 | -0.010672925 | PPIL6 |
| chr15 | 39247836 | 39248383 | 548 | 3 | 6.96E-06 | 1.23E-06 | 0.00014 | 1.92E-06 | -0.007501618 | AC013652.1 |
| chr10 | 128069666 | 128070143 | 478 | 5 | 6.97E-06 | 4.70E-06 | 7.47E-05 | 2.23E-06 | -0.00686811 | PTPRE |
| chr2 | 121710289 | 121710587 | 299 | 2 | 6.97E-06 | 0.000156 | 6.86E-06 | 4.86E-06 | -0.007631742 | NIFK-AS1 |
| chr8 | 123158875 | 123159181 | 307 | 3 | 6.97E-06 | 1.87E-05 | 5.69E-05 | 8.80E-06 | -0.015377702 | AC068228.3 |
| chr16 | 833622 | 833846 | 225 | 3 | 6.98E-06 | 1.54E-06 | 0.00052 | 2.64E-06 | -0.011233163 | AL023882.1 |
| chr15 | 74798279 | 74798741 | 463 | 3 | 6.98E-06 | 1.96E-06 | 7.59E-05 | 1.51E-06 | -0.007469986 | CSK |
| chr6 | 159967892 | 159968685 | 794 | 9 | 7.02E-06 | 0.000694 | 9.74E-05 | 5.19E-05 | -0.002172545 | NA |
| chr10 | 99316068 | 99316481 | 414 | 2 | 7.03E-06 | 4.47E-05 | 1.27E-05 | 5.64E-06 | -0.008670715 | NA |
| chr6 | 31652395 | 31654662 | 2268 | 43 | 7.04E-06 | 5.02E-06 | 0.00067 | 5.13E-08 | 0.000131044 | APOM, BAG6 |
| chr1 | 202858152 | 202860611 | 2460 | 6 | 7.06E-06 | 2.41E-06 | 0.00026 | 2.44E-07 | -0.005885433 | TUBA5P |
| chr19 | 55162412 | 55164218 | 1807 | 5 | 7.07E-06 | 1.16E-07 | 0.00018 | 5.46E-08 | -0.009575617 | AC010327.4, DNAAF3 |
| chr9 | 127903885 | 127904778 | 894 | 3 | 7.07E-06 | 1.65E-06 | 0.00037 | 2.74E-06 | -0.011313495 | AL157935.2, ST6GALNAC6 |
| chr16 | 11105286 | 11105335 | 50 | 2 | 7.07E-06 | 1.09E-06 | 3.09E-05 | 1.03E-06 | -0.007441524 | CLEC16A |
| chr5 | 132011022 | 132011649 | 628 | 7 | 7.08E-06 | 4.58E-05 | 0.0002 | 1.43E-05 | -0.005053911 | AC034228.1, AC034228.3, ACSL6 |
| chr11 | 62393322 | 62393621 | 300 | 4 | 7.09E-06 | 2.47E-07 | 0.00153 | 9.38E-07 | -0.042250835 | ASRGL1, AP003064.1 |
| chr20 | 17505166 | 17505177 | 12 | 2 | 7.09E-06 | 1.41E-06 | 5.07E-05 | 1.60E-06 | 0.003583399 | BFSP1 |
| chr14 | 95460894 | 95461138 | 245 | 3 | 7.10E-06 | 0.000111 | 7.37E-06 | 3.24E-06 | -0.004460063 | SYNE3 |
| chr17 | 29073613 | 29074594 | 982 | 6 | 7.13E-06 | 0.000288 | 5.29E-05 | 7.99E-06 | -0.006929883 | MYO18A, TIAF1 |
| chr11 | 790142 | 791124 | 983 | 9 | 7.13E-06 | 0.001214 | 0.00271 | 2.49E-05 | -0.005644863 | SLC25A22 |
| chr6 | 151445244 | 151445787 | 544 | 3 | 7.14E-06 | 4.62E-07 | 0.0003 | 7.63E-07 | -0.012392706 | RMND1 |
| chr2 | 96239652 | 96240483 | 832 | 5 | 7.15E-06 | 4.86E-05 | 0.00026 | 3.66E-06 | -0.001764806 | STARD7-AS1 |
| chr6 | 89886341 | 89887202 | 862 | 7 | 7.15E-06 | 8.26E-07 | 0.00174 | 8.67E-07 | -0.001353846 | AL353692.1 |
| chr22 | 37803752 | 37804214 | 463 | 4 | 7.19E-06 | 4.98E-07 | 0.00132 | 1.53E-06 | -0.008594939 | NA |
| chr1 | 227558410 | 227559181 | 772 | 5 | 7.19E-06 | 2.92E-07 | 0.00223 | 1.08E-06 | -0.023004554 | NA |
| chr8 | 92966440 | 92967276 | 837 | 4 | 7.20E-06 | 2.78E-05 | 9.64E-06 | 1.06E-06 | -0.006004435 | TRIQK |
| chr19 | 15237701 | 15238565 | 865 | 4 | 7.20E-06 | 0.007158 | 0.00019 | 0.00024 | -0.002102149 | BRD4 |
| chr6 | 170159060 | 170161250 | 2191 | 8 | 7.21E-06 | 2.17E-06 | 0.00044 | 1.39E-08 | -0.002917922 | AL596442.1, AL596442.3 |
| chr14 | 92939592 | 92939737 | 146 | 3 | 7.21E-06 | 2.95E-07 | 0.0003 | 7.46E-07 | -0.030807172 | ITPK1 |
| chr6 | 118652146 | 118652565 | 420 | 8 | 7.21E-06 | 4.54E-06 | 0.00406 | 2.33E-06 | -0.010874304 | CEP85L |
| chr11 | 72821510 | 72822619 | 1110 | 7 | 7.24E-06 | 3.01E-08 | 0.00801 | 3.28E-07 | -0.019883032 | ATG16L2 |
| chr14 | 80210597 | 80211816 | 1220 | 8 | 7.24E-06 | 1.60E-05 | 0.00051 | 5.65E-06 | -0.00858406 | DIO2-AS1, DIO2 |
| chr14 | 100297827 | 100298140 | 314 | 2 | 7.24E-06 | 3.43E-06 | 0.00034 | 5.68E-06 | -0.011263482 | SLC25A29 |
| chr1 | 205057373 | 205058510 | 1138 | 6 | 7.28E-06 | 7.28E-05 | 0.00031 | 5.04E-06 | -0.004141172 | CNTN2 |
| chr1 | 153460004 | 153460584 | 581 | 3 | 7.30E-06 | 7.86E-05 | 1.30E-05 | 3.55E-06 | -0.007086625 | S100A7 |
| chr2 | 173074515 | 173075027 | 513 | 3 | 7.32E-06 | 6.02E-05 | 0.00067 | 2.48E-05 | -0.01110432 | NA |
| chr6 | 27757409 | 27757629 | 221 | 3 | 7.33E-06 | 3.22E-07 | 0.00052 | 8.17E-07 | -0.014914111 | NA |
| chr22 | 21937278 | 21938087 | 810 | 8 | 7.33E-06 | 4.29E-07 | 0.00672 | 1.35E-06 | -0.005184577 | PPM1F |
| chr6 | 1624151 | 1625521 | 1371 | 9 | 7.35E-06 | 8.53E-07 | 0.0008 | 2.54E-07 | -0.00531895 | GMDS |
| chr1 | 9128183 | 9128629 | 447 | 2 | 7.35E-06 | 2.62E-05 | 9.93E-05 | 1.95E-05 | -0.010527833 | GPR157 |
| chr9 | 35675390 | 35676059 | 670 | 5 | 7.36E-06 | 1.71E-05 | 0.00181 | 8.16E-06 | -0.009540252 | CA9, ARHGEF39 |
| chr12 | 57603215 | 57604119 | 905 | 3 | 7.37E-06 | 4.70E-06 | 0.00029 | 2.73E-06 | -0.012297266 | PIP4K2C |
| chr14 | 105456750 | 105457328 | 579 | 3 | 7.37E-06 | 2.61E-06 | 0.00017 | 2.78E-06 | -0.007007048 | MTA1 |
| chr15 | 78004995 | 78005481 | 487 | 3 | 7.37E-06 | 9.38E-05 | 4.37E-05 | 9.43E-06 | -0.00961443 | TBC1D2B |
| chr5 | 140965876 | 140966818 | 943 | 13 | 7.38E-06 | 4.56E-05 | 0.00047 | 3.99E-06 | -0.001525888 | PCDHA1, PCDHA2, PCDHA3, PCDHA4, PCDHA5, PCDHA6, PCDHA7, PCDHA8, PCDHA9, PCDHA10, PCDHA11, PCDHA12, PCDHA13, PCDHAC1, PCDHAC2, AC010223.1 |
| chr7 | 149071007 | 149072128 | 1122 | 4 | 7.38E-06 | 1.03E-09 | 0.00031 | 6.45E-09 | -0.016837678 | ZNF786 |
| chr7 | 739826 | 740099 | 274 | 2 | 7.39E-06 | 4.53E-06 | 0.00031 | 7.16E-06 | -0.00673283 | DNAAF5 |
| chr17 | 73361138 | 73361313 | 176 | 3 | 7.39E-06 | 3.16E-06 | 0.00061 | 4.38E-06 | -0.008929174 | SDK2 |
| chr11 | 2825451 | 2826028 | 578 | 5 | 7.41E-06 | 3.32E-05 | 0.00119 | 6.66E-06 | -0.010585051 | KCNQ1 |
| chr11 | 45907437 | 45908269 | 833 | 5 | 7.48E-06 | 2.31E-05 | 0.00015 | 1.86E-06 | -0.00395564 | NA |
| chr16 | 70390906 | 70391294 | 389 | 3 | 7.48E-06 | 4.67E-05 | 0.0002 | 3.94E-06 | -0.003609192 | AC012184.1, ST3GAL2 |
| chr4 | 69758919 | 69759585 | 667 | 3 | 7.49E-06 | 3.49E-07 | 0.0006 | 9.26E-07 | 0.001563649 | SULT1B1 |
| chr6 | 32814628 | 32815570 | 943 | 6 | 7.51E-06 | 4.12E-05 | 0.00054 | 7.04E-06 | -0.007813105 | HLA-DOB, AL669918.1 |
| chr6 | 130020875 | 130021485 | 611 | 8 | 7.52E-06 | 6.31E-06 | 0.00231 | 1.38E-06 | 0.002015162 | L3MBTL3 |
| chr15 | 95825980 | 95825984 | 5 | 2 | 7.53E-06 | 1.74E-06 | 4.43E-05 | 1.77E-06 | -0.01716181 | AC015574.1 |
| chr19 | 37403079 | 37404070 | 992 | 6 | 7.53E-06 | 0.000234 | 5.42E-05 | 8.06E-06 | -0.018371877 | AC008806.1 |
| chr5 | 177085646 | 177086310 | 665 | 4 | 7.55E-06 | 3.01E-07 | 0.00126 | 1.00E-06 | -0.006602517 | NA |
| chr10 | 117424937 | 117425186 | 250 | 4 | 7.55E-06 | 2.32E-07 | 0.00116 | 7.56E-07 | -0.01334774 | NA |
| chr16 | 22373394 | 22373628 | 235 | 2 | 7.55E-06 | 9.47E-06 | 4.34E-05 | 5.92E-06 | -0.015593019 | CDR2 |
| chr4 | 69057349 | 69057667 | 319 | 3 | 7.60E-06 | 5.71E-05 | 2.29E-05 | 7.14E-06 | 0.010287866 | UGT2B7 |
| chr12 | 107319963 | 107321049 | 1087 | 6 | 7.60E-06 | 1.63E-05 | 0.0004 | 1.81E-06 | 0.001869098 | BTBD11 |
| chr22 | 43277737 | 43278273 | 537 | 3 | 7.62E-06 | 0.021954 | 6.74E-06 | 1.58E-05 | -0.003442463 | SCUBE1-AS1, SCUBE1 |
| chr1 | 11983455 | 11983598 | 144 | 2 | 7.62E-06 | 0.004371 | 6.84E-06 | 7.64E-06 | -0.001806784 | MFN2 |
| chr12 | 7075706 | 7076569 | 864 | 3 | 7.62E-06 | 1.51E-05 | 4.83E-05 | 5.14E-06 | -0.007989561 | NA |
| chr9 | 83920657 | 83921128 | 472 | 4 | 7.63E-06 | 0.025413 | 1.32E-05 | 6.77E-05 | 0.003389648 | KIF27 |
| chr14 | 22836339 | 22837300 | 962 | 7 | 7.63E-06 | 8.42E-07 | 0.00089 | 1.53E-06 | -0.008877918 | MMP14 |
| chr19 | 10011566 | 10012100 | 535 | 5 | 7.63E-06 | 7.54E-06 | 4.91E-05 | 2.70E-06 | -0.01294007 | NA |
| chr16 | 87414094 | 87414508 | 415 | 2 | 7.68E-06 | 3.27E-05 | 9.82E-05 | 2.23E-05 | -0.005688877 | ZCCHC14 |
| chr8 | 143878761 | 143879849 | 1089 | 6 | 7.70E-06 | 2.86E-06 | 0.00071 | 3.28E-06 | -0.011999528 | NA |
| chr11 | 47416232 | 47416517 | 286 | 2 | 7.70E-06 | 8.71E-07 | 0.00018 | 1.52E-06 | -0.013848844 | SLC39A13 |
| chr5 | 20576284 | 20577142 | 859 | 3 | 7.71E-06 | 0.000139 | 1.38E-05 | 6.96E-06 | -0.004690415 | NA |
| chr7 | 149415227 | 149416105 | 879 | 7 | 7.72E-06 | 7.26E-08 | 0.00635 | 5.57E-07 | -0.015890121 | NA |
| chr15 | 23973895 | 23974164 | 270 | 2 | 7.73E-06 | 0.001441 | 1.09E-05 | 1.19E-05 | -0.015078994 | PWRN4 |
| chr6 | 169231044 | 169231665 | 622 | 4 | 7.73E-06 | 6.10E-05 | 2.21E-05 | 7.59E-06 | -0.003304814 | BX322234.1, THBS2 |
| chr1 | 229507761 | 229508530 | 770 | 11 | 7.76E-06 | 0.000467 | 0.00251 | 1.34E-05 | 0.001489399 | AL121990.1, NUP133 |
| chr19 | 11237763 | 11239951 | 2189 | 14 | 7.77E-06 | 1.16E-08 | 0.00048 | 1.70E-08 | -0.004335155 | ANGPTL8, DOCK6 |
| chr18 | 76486818 | 76487277 | 460 | 3 | 7.77E-06 | 6.75E-05 | 1.89E-05 | 5.45E-06 | -0.010815132 | ZNF516 |
| chr16 | 27723617 | 27724012 | 396 | 2 | 7.79E-06 | 0.000423 | 2.05E-05 | 1.88E-05 | -0.010543573 | KATNIP |
| chr11 | 66480895 | 66481150 | 256 | 2 | 7.80E-06 | 0.003186 | 3.26E-05 | 4.36E-05 | -0.002449717 | DPP3 |
| chr16 | 89828240 | 89828900 | 661 | 4 | 7.81E-06 | 0.000954 | 1.62E-05 | 2.00E-05 | -0.00097189 | SPIRE2 |
| chr18 | 79950649 | 79951645 | 997 | 8 | 7.81E-06 | 7.82E-06 | 0.00045 | 5.87E-06 | -0.00081311 | SLC66A2 |
| chr7 | 2523550 | 2524622 | 1073 | 7 | 7.82E-06 | 2.75E-05 | 0.00124 | 2.16E-06 | -0.007878368 | LFNG |
| chr3 | 69948081 | 69948609 | 529 | 2 | 7.83E-06 | 1.02E-06 | 6.88E-05 | 1.38E-06 | 0.001046074 | MITF |
| chr7 | 47344399 | 47344739 | 341 | 5 | 7.84E-06 | 0.000341 | 9.59E-06 | 6.06E-06 | -0.001867459 | TNS3 |
| chr13 | 107451212 | 107451495 | 284 | 2 | 7.88E-06 | 5.32E-07 | 0.00016 | 9.50E-07 | -0.026148745 | FAM155A |
| chr3 | 157540422 | 157540711 | 290 | 2 | 7.92E-06 | 1.89E-06 | 3.62E-05 | 1.70E-06 | -0.010831092 | NA |
| chr10 | 73910341 | 73911704 | 1364 | 14 | 7.95E-06 | 1.71E-05 | 0.00124 | 6.28E-07 | -0.002582618 | PLAU, C10orf55 |
| chr15 | 68812959 | 68813752 | 794 | 2 | 7.96E-06 | 0.013662 | 8.74E-05 | 0.00017 | -0.00547558 | ANP32A |
| chr13 | 113457549 | 113458274 | 726 | 7 | 7.99E-06 | 0.000529 | 0.0009 | 1.17E-05 | -0.002839625 | DCUN1D2 |
| chr19 | 2703117 | 2703894 | 778 | 2 | 8.00E-06 | 0.000209 | 0.00076 | 0.0002 | -0.005371913 | NA |
| chr2 | 177694172 | 177694222 | 51 | 2 | 8.01E-06 | 7.59E-06 | 1.69E-05 | 2.73E-06 | -0.015586028 | PDE11A-AS1, PDE11A, PDE11A |
| chr15 | 45401488 | 45402258 | 771 | 11 | 8.01E-06 | 0.000179 | 0.0074 | 1.59E-05 | -0.005128925 | GATM |
| chr2 | 62504891 | 62505757 | 867 | 4 | 8.04E-06 | 0.004343 | 5.50E-05 | 1.99E-05 | -0.006522574 | TMEM17 |
| chr11 | 18635321 | 18635728 | 408 | 2 | 8.06E-06 | 1.27E-05 | 0.00045 | 1.85E-05 | -0.005486401 | NA |
| chr2 | 102049983 | 102050216 | 234 | 2 | 8.06E-06 | 5.32E-06 | 1.78E-05 | 2.27E-06 | -0.010920399 | AC007271.1 |
| chr15 | 69389222 | 69389693 | 472 | 2 | 8.10E-06 | 3.01E-05 | 4.35E-05 | 1.24E-05 | -0.003901286 | PAQR5 |
| chr2 | 47155148 | 47156581 | 1434 | 14 | 8.12E-06 | 1.67E-05 | 0.00018 | 4.60E-07 | -0.002081287 | STPG4, AC073283.3 |
| chr19 | 1173052 | 1173636 | 585 | 5 | 8.12E-06 | 5.70E-06 | 0.00065 | 4.01E-06 | -0.009005958 | SBNO2 |
| chr6 | 167825243 | 167825896 | 654 | 5 | 8.12E-06 | 1.50E-05 | 0.00168 | 1.54E-05 | -0.003319038 | AFDN-DT |
| chr12 | 132505959 | 132507007 | 1049 | 6 | 8.12E-06 | 7.10E-07 | 0.00037 | 7.22E-07 | -0.01026006 | FBRSL1 |
| chr9 | 134838359 | 134840034 | 1676 | 6 | 8.16E-06 | 3.30E-09 | 0.00023 | 7.44E-09 | -0.001066344 | COL5A1 |
| chr16 | 27225801 | 27226398 | 598 | 5 | 8.18E-06 | 0.006593 | 0.0003 | 2.50E-05 | -0.008739307 | NSMCE1 |
| chr19 | 48468267 | 48468939 | 673 | 5 | 8.21E-06 | 0.055837 | 0.00075 | 0.0018 | -0.003174072 | AC008403.1, AC008403.2 |
| chr18 | 45836995 | 45837576 | 582 | 4 | 8.21E-06 | 1.13E-06 | 0.0024 | 3.70E-06 | -0.014201712 | SIGLEC15 |
| chr1 | 26702216 | 26702799 | 584 | 3 | 8.22E-06 | 3.61E-05 | 0.00011 | 7.00E-06 | -0.008540643 | ARID1A |
| chr7 | 668091 | 668159 | 69 | 2 | 8.24E-06 | 7.43E-07 | 7.10E-05 | 1.08E-06 | -0.014244515 | PRKAR1B |
| chr1 | 225942666 | 225942857 | 192 | 2 | 8.24E-06 | 3.76E-06 | 0.00013 | 5.08E-06 | -0.012964332 | NA |
| chr14 | 105086238 | 105087238 | 1001 | 5 | 8.29E-06 | 2.69E-07 | 0.00096 | 6.40E-07 | 0.002528841 | NA |
| chr16 | 1299974 | 1300102 | 129 | 2 | 8.29E-06 | 7.91E-05 | 2.94E-05 | 1.53E-05 | -0.001185295 | NA |
| chr2 | 108287496 | 108288047 | 552 | 3 | 8.30E-06 | 2.88E-06 | 0.00029 | 3.43E-06 | -0.011387231 | NA |
| chr10 | 114178753 | 114179820 | 1068 | 12 | 8.32E-06 | 7.39E-05 | 0.00495 | 5.12E-06 | -0.003414811 | TDRD1 |
| chr4 | 98929100 | 98930130 | 1031 | 16 | 8.32E-06 | 1.11E-06 | 0.00638 | 1.65E-06 | -0.005322392 | AC019131.1, EIF4E |
| chr1 | 230112975 | 230114609 | 1635 | 6 | 8.32E-06 | 1.47E-06 | 5.47E-05 | 4.63E-08 | -0.003857901 | GALNT2 |
| chr17 | 55444143 | 55444314 | 172 | 2 | 8.32E-06 | 2.00E-06 | 0.00011 | 2.81E-06 | -0.001643148 | NA |
| chr8 | 28361152 | 28361734 | 583 | 3 | 8.33E-06 | 8.37E-05 | 0.00024 | 6.44E-06 | -0.010426768 | ZNF395, FBXO16 |
| chr5 | 177995781 | 177996339 | 559 | 4 | 8.33E-06 | 0.001755 | 1.84E-05 | 2.78E-05 | -0.003370647 | PROP1 |
| chr17 | 42843789 | 42844561 | 773 | 4 | 8.35E-06 | 2.00E-05 | 0.00016 | 3.51E-06 | -0.006645593 | NA |
| chr17 | 17783925 | 17784520 | 596 | 4 | 8.36E-06 | 1.83E-06 | 0.00103 | 3.66E-06 | -0.006541965 | RAI1 |
| chr9 | 83063935 | 83064085 | 151 | 2 | 8.38E-06 | 0.002974 | 6.06E-05 | 8.46E-05 | -0.01499145 | AL499602.1 |
| chr6 | 28336146 | 28336985 | 840 | 11 | 8.38E-06 | 4.89E-06 | 0.00475 | 1.20E-06 | -0.001803307 | ZSCAN31 |
| chr17 | 3473940 | 3475510 | 1571 | 9 | 8.39E-06 | 1.35E-05 | 5.39E-05 | 1.08E-06 | -0.005301924 | ASPA, SPATA22 |
| chr6 | 138545728 | 138545988 | 261 | 7 | 8.40E-06 | 0.000315 | 1.20E-05 | 5.06E-06 | -0.003539376 | NHSL1 |
| chr6 | 149059740 | 149060468 | 729 | 2 | 8.42E-06 | 2.11E-06 | 0.00019 | 3.40E-06 | -0.028761713 | UST |
| chr20 | 31587655 | 31587822 | 168 | 2 | 8.43E-06 | 5.12E-05 | 1.19E-05 | 5.69E-06 | -0.002322251 | NA |
| chr22 | 35478309 | 35478958 | 650 | 3 | 8.45E-06 | 1.73E-06 | 0.00042 | 2.54E-06 | -0.011662233 | NA |
| chr17 | 63476577 | 63477568 | 992 | 11 | 8.46E-06 | 0.000908 | 6.15E-05 | 1.81E-05 | 0.000330694 | ACE |
| chr1 | 232578784 | 232579236 | 453 | 3 | 8.46E-06 | 0.000731 | 5.60E-06 | 8.15E-06 | -0.008296841 | SIPA1L2 |
| chr2 | 105398080 | 105398622 | 543 | 3 | 8.51E-06 | 0.000613 | 4.71E-05 | 5.17E-05 | -0.008266029 | FHL2 |
| chr22 | 44442875 | 44443242 | 368 | 4 | 8.51E-06 | 3.87E-05 | 8.47E-05 | 2.97E-06 | -0.007748378 | NA |
| chr1 | 205321204 | 205322087 | 884 | 8 | 8.52E-06 | 5.86E-06 | 0.00029 | 1.83E-06 | 0.002669141 | NUAK2 |
| chr10 | 119608241 | 119608724 | 484 | 3 | 8.52E-06 | 1.77E-05 | 3.02E-05 | 4.16E-06 | -0.010061721 | NA |
| chr5 | 6793052 | 6793311 | 260 | 3 | 8.53E-06 | 5.48E-06 | 2.47E-05 | 2.14E-06 | -0.011212618 | LINC02236 |
| chr3 | 64023541 | 64024070 | 530 | 7 | 8.53E-06 | 2.59E-05 | 0.00089 | 9.28E-06 | -0.004569775 | PSMD6 |
| chr15 | 82850457 | 82851146 | 690 | 4 | 8.56E-06 | 2.73E-06 | 0.0006 | 3.53E-06 | -0.006293315 | HOMER2 |
| chr3 | 42978232 | 42979907 | 1676 | 16 | 8.56E-06 | 6.55E-07 | 0.00065 | 9.18E-08 | -0.00449001 | AC092042.3, GASK1A |
| chr9 | 27108917 | 27109494 | 578 | 7 | 8.56E-06 | 3.55E-05 | 0.00056 | 1.72E-06 | -0.013151602 | TEK |
| chr2 | 159758882 | 159759161 | 280 | 2 | 8.62E-06 | 0.001658 | 1.13E-05 | 1.27E-05 | -0.005727116 | MARCHF7 |
| chr19 | 7380820 | 7380964 | 145 | 5 | 8.63E-06 | 3.71E-07 | 0.00093 | 1.02E-06 | -0.010716288 | AC119396.1 |
| chr4 | 1211168 | 1212362 | 1195 | 7 | 8.64E-06 | 2.47E-06 | 0.00154 | 1.09E-06 | -0.004586436 | CTBP1-AS, CTBP1 |
| chr6 | 32224247 | 32225458 | 1212 | 5 | 8.64E-06 | 9.06E-07 | 0.00105 | 1.84E-06 | -0.009052637 | NA |
| chr9 | 127969148 | 127969780 | 633 | 3 | 8.65E-06 | 4.70E-07 | 0.00054 | 1.26E-06 | -0.010911053 | FAM102A |
| chr7 | 2007186 | 2007328 | 143 | 4 | 8.65E-06 | 0.015214 | 2.10E-06 | 1.02E-05 | -0.001481941 | MAD1L1, AC069288.1 |
| chr16 | 10876343 | 10877393 | 1051 | 10 | 8.68E-06 | 3.47E-06 | 0.00039 | 1.65E-06 | 0.003628739 | CIITA, AC133065.1 |
| chr2 | 26971447 | 26971568 | 122 | 2 | 8.70E-06 | 3.41E-06 | 0.0002 | 5.22E-06 | -0.012634793 | MAPRE3, AC013472.1 |
| chr15 | 34795572 | 34796206 | 635 | 6 | 8.71E-06 | 7.23E-05 | 0.00075 | 5.84E-06 | -0.006443689 | AC087457.1 |
| chr8 | 37939940 | 37940638 | 699 | 8 | 8.72E-06 | 0.000118 | 0.00033 | 4.14E-06 | -0.001662879 | GOT1L1, AC144573.1 |
| chr8 | 109596070 | 109597103 | 1034 | 3 | 8.73E-06 | 1.81E-06 | 0.00022 | 2.89E-06 | -0.006162395 | SYBU |
| chr20 | 4614041 | 4614491 | 451 | 2 | 8.75E-06 | 4.75E-07 | 0.00015 | 8.53E-07 | -0.025664362 | NA |
| chr5 | 78758130 | 78758825 | 696 | 2 | 8.76E-06 | 3.20E-06 | 3.95E-05 | 2.63E-06 | -0.006724169 | LHFPL2 |
| chr4 | 1414769 | 1415769 | 1001 | 6 | 8.79E-06 | 6.94E-06 | 0.00108 | 2.05E-06 | -0.012228117 | NA |
| chr17 | 1562986 | 1563789 | 804 | 7 | 8.79E-06 | 0.000607 | 0.00047 | 1.01E-05 | -0.007414096 | NA |
| chr11 | 44304410 | 44305465 | 1056 | 5 | 8.81E-06 | 1.86E-05 | 5.23E-05 | 5.30E-06 | 0.003536224 | ALX4 |
| chr3 | 44886553 | 44887031 | 479 | 2 | 8.83E-06 | 1.54E-05 | 3.98E-05 | 7.72E-06 | -0.007019083 | TGM4 |

| | | | | | | | | | | |
|---|---|---|---|---|---|---|---|---|---|---|
| chr8 | 28386304 | 28387252 | 949 | 13 | 8.83E-06 | 1.22E-06 | 0.01369 | 1.31E-06 | -0.000699292 | ZNF395, FBXO16 |
| chr2 | 3177031 | 3177593 | 563 | 6 | 8.87E-06 | 2.61E-05 | 0.0006 | 2.17E-06 | -0.002281467 | NA |
| chr11 | 82790830 | 82791523 | 694 | 3 | 8.88E-06 | 2.30E-06 | 0.00027 | 3.62E-06 | -0.012976269 | LINC02734 |
| chr17 | 82356575 | 82357063 | 489 | 3 | 8.88E-06 | 3.40E-05 | 0.0001 | 1.01E-05 | -0.005746912 | NA |
| chr1 | 157127108 | 157127116 | 9 | 2 | 8.88E-06 | 4.25E-06 | 1.82E-05 | 2.00E-06 | -0.003117788 | ETV3 |
| chr14 | 105059078 | 105059264 | 187 | 2 | 8.88E-06 | 6.79E-06 | 1.31E-05 | 2.11E-06 | -0.009807858 | GPR132 |
| chr17 | 1480876 | 1482265 | 1390 | 8 | 8.88E-06 | 2.33E-06 | 0.00024 | 4.43E-07 | -0.005482964 | MYO1C |
| chr7 | 2127681 | 2127868 | 188 | 3 | 8.90E-06 | 0.002981 | 5.16E-06 | 4.19E-06 | -0.002146184 | MAD1L1, AC069288.1 |
| chr19 | 37942000 | 37942807 | 808 | 3 | 8.92E-06 | 0.001073 | 1.51E-05 | 2.02E-05 | -0.004658184 | SIPA1L3 |
| chr6 | 73353799 | 73354871 | 1073 | 7 | 8.92E-06 | 3.09E-07 | 0.00116 | 6.77E-07 | -0.019841546 | DPPA5 |
| chr7 | 4731312 | 4731351 | 40 | 2 | 8.94E-06 | 1.08E-05 | 0.00058 | 1.68E-05 | -0.015660707 | FOXK1 |
| chr8 | 133552269 | 133553239 | 971 | 3 | 8.94E-06 | 1.50E-08 | 8.70E-05 | 2.66E-08 | -0.010750032 | ST3GAL1 |
| chr17 | 3896972 | 3897100 | 129 | 2 | 8.95E-06 | 8.86E-07 | 9.05E-05 | 1.35E-06 | -0.00948185 | P2RX1 |
| chr14 | 105471677 | 105473335 | 1659 | 8 | 8.99E-06 | 2.70E-05 | 9.90E-05 | 9.23E-07 | -0.004226461 | CRIP2 |
| chr22 | 46045919 | 46046730 | 812 | 3 | 9.05E-06 | 8.68E-07 | 0.00042 | 1.75E-06 | -0.010796034 | NA |
| chr20 | 653858 | 654206 | 349 | 4 | 9.05E-06 | 2.01E-05 | 4.73E-05 | 7.29E-06 | -0.012139708 | AL121758.1 |
| chr9 | 134010914 | 134011244 | 331 | 2 | 9.05E-06 | 5.32E-06 | 0.00057 | 8.95E-06 | -0.010423211 | NA |
| chr1 | 151846124 | 151846446 | 323 | 3 | 9.06E-06 | 3.84E-07 | 0.00034 | 9.50E-07 | -0.010680411 | C2CD4D-AS1 |
| chr1 | 47180257 | 47180771 | 515 | 3 | 9.08E-06 | 4.06E-05 | 5.63E-05 | 5.40E-06 | -0.012515838 | LINC00853 |
| chr1 | 1072350 | 1073262 | 913 | 5 | 9.12E-06 | 0.000426 | 4.71E-05 | 8.93E-06 | -0.004312568 | RNF223 |
| chr15 | 88631220 | 88631876 | 657 | 5 | 9.12E-06 | 1.02E-05 | 0.00027 | 2.64E-06 | -0.002821014 | AEN |
| chr8 | 130514394 | 130514942 | 549 | 2 | 9.14E-06 | 1.03E-06 | 7.59E-05 | 1.45E-06 | -0.018206555 | NA |
| chr2 | 240713735 | 240713884 | 150 | 2 | 9.20E-06 | 6.43E-06 | 3.01E-05 | 3.64E-06 | -0.010982047 | KIF1A |
| chr2 | 148876091 | 148876131 | 41 | 2 | 9.20E-06 | 3.11E-05 | 3.70E-05 | 1.12E-05 | 0.001969144 | KIF5C, KIF5C-AS1 |
| chr15 | 84908734 | 84908753 | 20 | 2 | 9.22E-06 | 6.33E-07 | 0.00016 | 1.12E-06 | -0.012180972 | SLC28A1 |
| chr10 | 69961493 | 69962294 | 802 | 3 | 9.23E-06 | 9.74E-06 | 4.31E-05 | 4.74E-06 | -0.009168832 | COL13A1 |
| chr1 | 161074302 | 161075190 | 889 | 5 | 9.25E-06 | 0.000199 | 0.00013 | 8.86E-06 | -0.00070404 | NECTIN4 |
| chr9 | 123302784 | 123303431 | 648 | 4 | 9.27E-06 | 3.16E-06 | 0.00041 | 5.42E-06 | -0.007539134 | NA |
| chr9 | 124941033 | 124942435 | 1403 | 10 | 9.28E-06 | 4.35E-05 | 0.00022 | 1.36E-06 | -0.001494673 | GOLGA1 |
| chr7 | 101825429 | 101825866 | 438 | 3 | 9.29E-06 | 0.000108 | 2.58E-05 | 1.31E-05 | -0.00736437 | CUX1 |
| chr4 | 3774747 | 3775939 | 1193 | 4 | 9.29E-06 | 4.40E-06 | 0.00068 | 4.11E-06 | 0.009094334 | NA |
| chr6 | 30676458 | 30677055 | 598 | 7 | 9.30E-06 | 1.43E-05 | 0.00054 | 1.23E-06 | -0.002870903 | PPP1R18 |
| chr8 | 39106778 | 39107973 | 1196 | 15 | 9.30E-06 | 0.000287 | 0.00012 | 4.97E-07 | -0.005341976 | ADAM32 |
| chr17 | 80924389 | 80924569 | 181 | 3 | 9.31E-06 | 2.02E-05 | 3.78E-05 | 6.95E-06 | -0.000185084 | RPTOR |
| chr4 | 144645048 | 144645823 | 776 | 6 | 9.32E-06 | 2.58E-05 | 0.00107 | 1.10E-05 | -0.014566598 | AC098588.3, AC098588.2, HHIP-AS1 |
| chr10 | 131774090 | 131774697 | 608 | 4 | 9.36E-06 | 7.82E-05 | 0.00014 | 4.66E-06 | -0.006561319 | LINC01164 |
| chr4 | 95547811 | 95548483 | 673 | 9 | 9.36E-06 | 9.71E-07 | 0.00212 | 1.52E-06 | -0.006581661 | UNC5C |
| chr12 | 6942902 | 6943368 | 467 | 3 | 9.40E-06 | 2.01E-06 | 0.00078 | 4.22E-06 | -0.012287047 | C12orf57 |
| chr17 | 31468046 | 31468451 | 406 | 3 | 9.41E-06 | 2.25E-05 | 4.59E-05 | 4.60E-06 | -0.004412388 | RAB11FIP4 |
| chr1 | 165941059 | 165941522 | 464 | 4 | 9.43E-06 | 0.000109 | 9.04E-06 | 5.15E-06 | -0.000242112 | AL606495.1 |
| chr8 | 92062293 | 92062701 | 409 | 2 | 9.45E-06 | 0.00331 | 1.76E-05 | 2.23E-05 | -0.005832595 | RUNX1T1 |
| chr2 | 86542578 | 86542932 | 355 | 2 | 9.48E-06 | 3.57E-05 | 1.31E-05 | 5.26E-06 | 0.00766701 | CHMP3, RNF103-CHMP3 |
| chr12 | 123151223 | 123152170 | 948 | 8 | 9.53E-06 | 0.000305 | 0.00152 | 4.89E-06 | -0.003420275 | NA |
| chr15 | 76863087 | 76863294 | 208 | 3 | 9.53E-06 | 9.86E-05 | 1.14E-05 | 6.16E-06 | -0.011004207 | SCAPER |
| chr3 | 10290776 | 10291971 | 1196 | 8 | 9.54E-06 | 1.35E-05 | 0.00421 | 7.17E-06 | -0.00424838 | GHRLOS, GHRL |
| chr1 | 217076899 | 217078137 | 1239 | 6 | 9.54E-06 | 5.34E-07 | 0.00101 | 4.07E-07 | -0.016766798 | ESRRG |
| chr5 | 1551756 | 1552144 | 389 | 6 | 9.55E-06 | 2.76E-06 | 0.00184 | 2.70E-06 | -0.008448306 | NA |
| chr8 | 60847594 | 60848379 | 786 | 3 | 9.56E-06 | 2.84E-06 | 0.00012 | 3.32E-06 | -0.01258434 | CHD7 |
| chr11 | 35042194 | 35043334 | 1141 | 3 | 9.56E-06 | 2.35E-07 | 5.51E-05 | 2.73E-07 | -0.007055751 | NA |
| chr10 | 29534987 | 29535637 | 651 | 3 | 9.57E-06 | 1.87E-05 | 3.01E-05 | 5.16E-06 | -0.006710009 | SVIL |
| chr20 | 32074696 | 32075073 | 378 | 2 | 9.57E-06 | 7.66E-05 | 1.97E-05 | 1.06E-05 | -0.006070758 | HCK |
| chr5 | 42745266 | 42745527 | 262 | 2 | 9.58E-06 | 0.00031 | 6.94E-06 | 5.75E-06 | -0.017284271 | NA |
| chr8 | 143367746 | 143368796 | 1051 | 9 | 9.59E-06 | 2.45E-07 | 0.00205 | 2.87E-07 | -0.000997823 | RHPN1-AS1 |
| chr19 | 7522040 | 7523114 | 1075 | 11 | 9.60E-06 | 1.66E-07 | 0.00097 | 6.68E-07 | 0.001669362 | MCOLN1 |
| chr5 | 116626390 | 116626888 | 499 | 3 | 9.62E-06 | 7.87E-07 | 0.00044 | 1.47E-06 | -0.017375106 | NA |
| chr8 | 142542134 | 142542300 | 167 | 3 | 9.62E-06 | 8.00E-07 | 0.0003 | 1.64E-06 | -0.005118242 | ADGRB1 |
| chr16 | 85234451 | 85235254 | 804 | 3 | 9.64E-06 | 0.002002 | 1.36E-05 | 2.10E-05 | -0.012844849 | GSE1, AC092275.1 |
| chr4 | 38455511 | 38456016 | 506 | 3 | 9.64E-06 | 0.001015 | 1.65E-05 | 6.52E-06 | -0.005567826 | LINC01258 |
| chr21 | 34836278 | 34836916 | 639 | 4 | 9.67E-06 | 6.47E-06 | 0.00034 | 3.08E-06 | -0.005593298 | AP000331.1, RUNX1 |
| chr9 | 69204100 | 69204646 | 547 | 3 | 9.68E-06 | 0.003224 | 4.74E-05 | 7.28E-05 | -0.003637797 | AL358113.1, TJP2 |
| chr17 | 7348954 | 7350401 | 1448 | 9 | 9.68E-06 | 1.68E-08 | 0.00022 | 4.50E-08 | -0.024428244 | ACAP1 |
| chr6 | 70302688 | 70303686 | 999 | 7 | 9.68E-06 | 4.53E-06 | 8.74E-05 | 3.78E-06 | -0.00727908 | COL9A1 |
| chr11 | 76142998 | 76143700 | 703 | 3 | 9.68E-06 | 3.49E-06 | 6.55E-05 | 2.71E-06 | -0.005003683 | UVRAG |
| chr7 | 107602825 | 107603344 | 520 | 3 | 9.70E-06 | 1.66E-05 | 9.20E-05 | 5.65E-06 | -0.002485998 | DUS4L-BCAP29, BCAP29 |
| chr3 | 40386636 | 40387523 | 888 | 13 | 9.71E-06 | 4.01E-06 | 0.00713 | 3.93E-06 | -0.006746755 | ENTPD3, ENTPD3-AS1 |
| chr4 | 54656957 | 54657657 | 701 | 5 | 9.72E-06 | 1.58E-05 | 0.00102 | 1.02E-05 | -0.009881 | NA |
| chr17 | 63819327 | 63819552 | 226 | 2 | 9.75E-06 | 2.86E-05 | 1.41E-05 | 5.09E-06 | -0.015587479 | FTSJ3 |
| chr8 | 11808508 | 11809534 | 1027 | 10 | 9.78E-06 | 6.22E-05 | 0.00365 | 1.47E-05 | -0.014719664 | FDFT1 |
| chr8 | 55961179 | 55961504 | 326 | 2 | 9.79E-06 | 1.63E-06 | 0.00022 | 2.73E-06 | -0.016969281 | LYN |
| chr5 | 140874110 | 140875484 | 1375 | 10 | 9.81E-06 | 0.00015 | 0.00122 | 6.45E-06 | -0.00161217 | PCDHA1, PCDHA2, PCDHA3, PCDHA4, PCDHA5, PCDHA6, PCDHA7, PCDHA8, PCDHA9, PCDHA10, PCDHA11, PCDHA12, AC005609.3 |
| chr14 | 52266401 | 52267094 | 694 | 4 | 9.83E-06 | 2.58E-06 | 0.00226 | 5.75E-06 | -0.016343807 | NA |
| chr6 | 33407580 | 33408285 | 706 | 5 | 9.83E-06 | 5.49E-06 | 0.00289 | 2.16E-06 | -0.005792981 | KIFC1 |
| chr19 | 45091748 | 45092829 | 1082 | 4 | 9.84E-06 | 1.15E-05 | 0.00179 | 9.29E-06 | 0.01203749 | MARK4, PPP1R37, GEMIN7-AS1 |
| chr19 | 281291 | 281467 | 177 | 4 | 9.85E-06 | 9.05E-07 | 0.00049 | 1.21E-06 | -0.012504162 | PLPP2 |
| chr22 | 31122721 | 31122977 | 257 | 5 | 9.86E-06 | 1.84E-06 | 0.00014 | 1.62E-06 | -0.010834523 | INPP5J |
| chr17 | 49971251 | 49971911 | 661 | 5 | 9.87E-06 | 2.22E-06 | 0.0011 | 3.71E-06 | -0.004794636 | DLX4 |
| chr15 | 62267143 | 62267553 | 411 | 2 | 9.88E-06 | 2.84E-06 | 5.93E-05 | 2.96E-06 | -0.006893699 | GOLGA2P11 |
| chr2 | 31139292 | 31140426 | 1135 | 6 | 9.88E-06 | 4.85E-07 | 0.00053 | 8.95E-07 | -0.00595666 | GALNT14 |
| chr1 | 22056705 | 22058274 | 1570 | 4 | 9.88E-06 | 3.15E-09 | 0.00025 | 1.68E-08 | -0.010162147 | CDC42 |
| chr12 | 53756832 | 53757159 | 328 | 2 | 9.89E-06 | 8.28E-06 | 4.17E-05 | 5.27E-06 | -0.01173558 | AC076968.2, AC076968.1, CISTR |
| chr7 | 2998420 | 2999414 | 995 | 5 | 9.91E-06 | 6.43E-07 | 0.00141 | 1.12E-06 | -0.000244109 | RN7SKP130, CARD11 |
| chr10 | 79315310 | 79315735 | 426 | 7 | 9.93E-06 | 1.65E-05 | 7.03E-05 | 4.11E-06 | -0.002196694 | ZMIZ1 |
| chr11 | 44937339 | 44937573 | 235 | 2 | 9.94E-06 | 3.15E-05 | 8.00E-05 | 1.94E-05 | -0.006599892 | TP53I11 |
| chr17 | 14205511 | 14206354 | 844 | 3 | 9.97E-06 | 0.00725 | 4.57E-05 | 4.54E-05 | -0.004398304 | COX10 |
| chr10 | 129768463 | 129769483 | 1021 | 8 | 1.00E-05 | 5.38E-05 | 0.00197 | 1.72E-05 | -0.004715154 | MGMT, AL157832.1 |
| chr3 | 128495617 | 128497279 | 1663 | 4 | 1.01E-05 | 3.27E-08 | 0.00048 | 1.08E-07 | -0.021147406 | GATA2-AS1 |
| chr9 | 137012788 | 137013239 | 452 | 3 | 1.01E-05 | 0.000197 | 0.00033 | 1.02E-05 | -0.002253313 | ABCA2 |
| chr8 | 129786761 | 129787357 | 597 | 3 | 1.01E-05 | 8.28E-05 | 0.00051 | 3.09E-05 | -0.009985373 | NA |
| chr22 | 30286807 | 30287436 | 630 | 5 | 1.01E-05 | 2.70E-05 | 0.00016 | 4.37E-06 | -0.006250904 | CASTOR1, AC004997.1 |
| chr15 | 70199939 | 70200556 | 618 | 2 | 1.01E-05 | 5.56E-07 | 0.00015 | 9.85E-07 | -0.016167843 | NA |
| chr2 | 74003183 | 74003596 | 414 | 2 | 1.02E-05 | 2.10E-05 | 5.39E-05 | 1.16E-05 | -0.014102861 | TET3 |
| chr1 | 4600286 | 4600505 | 220 | 2 | 1.02E-05 | 2.42E-06 | 7.46E-05 | 2.90E-06 | -0.023819161 | NA |
| chr17 | 67070207 | 67070610 | 404 | 2 | 1.02E-05 | 0.002584 | 1.47E-05 | 1.81E-05 | -0.003697204 | HELZ |
| chr17 | 78224527 | 78224874 | 348 | 4 | 1.02E-05 | 5.30E-05 | 5.61E-05 | 3.76E-06 | -0.002471485 | BIRC5 |
| chr17 | 7857276 | 7858479 | 1204 | 18 | 1.02E-05 | 3.35E-05 | 0.001 | 1.18E-06 | -0.001040215 | CYB5D1, NAA38 |
| chr7 | 80700224 | 80700371 | 148 | 2 | 1.02E-05 | 2.68E-05 | 1.16E-05 | 4.12E-06 | -0.009557097 | NA |
| chr17 | 81158143 | 81158283 | 141 | 2 | 1.02E-05 | 0.000855 | 2.23E-05 | 2.36E-05 | -0.000964461 | AATK |
| chr6 | 53063819 | 53064685 | 867 | 4 | 1.03E-05 | 0.001232 | 0.00206 | 0.00011 | -0.005689757 | FBXO9 |
| chr6 | 109465875 | 109466732 | 858 | 7 | 1.03E-05 | 1.67E-05 | 0.00068 | 2.00E-06 | -0.003274956 | MICAL1, ZBTB24 |
| chr10 | 885703 | 886569 | 867 | 3 | 1.03E-05 | 2.77E-07 | 8.03E-05 | 2.44E-07 | -0.00677571 | LARP4B |
| chr5 | 142216089 | 142216131 | 43 | 2 | 1.03E-05 | 7.72E-07 | 0.00012 | 1.27E-06 | -0.024031947 | NA |
| chr6 | 31639563 | 31639857 | 295 | 4 | 1.03E-05 | 4.53E-06 | 0.00027 | 3.16E-06 | -0.00885241 | BAG6 |
| chr11 | 1559462 | 1560243 | 782 | 4 | 1.03E-05 | 5.89E-07 | 0.00153 | 1.81E-06 | -0.011012894 | DUSP8 |
| chr7 | 111393587 | 111393647 | 61 | 2 | 1.04E-05 | 7.31E-07 | 0.00015 | 1.27E-06 | -0.014919279 | IMMP2L |
| chr4 | 95051315 | 95051639 | 325 | 8 | 1.05E-05 | 0.000207 | 0.00054 | 2.95E-06 | -0.00496625 | BMPR1B |
| chr4 | 87219109 | 87219284 | 176 | 2 | 1.05E-05 | 0.000186 | 2.26E-05 | 1.66E-05 | -0.014436068 | KLHL8 |
| chr20 | 57392447 | 57394630 | 2184 | 10 | 1.05E-05 | 3.19E-10 | 0.00271 | 9.53E-10 | -0.01023381 | RBM38, RBM38-AS1 |
| chr4 | 145480595 | 145482362 | 1768 | 10 | 1.05E-05 | 5.38E-05 | 0.00068 | 3.59E-07 | -0.003040197 | SMAD1 |
| chr6 | 13923681 | 13924703 | 1023 | 8 | 1.05E-05 | 2.09E-06 | 0.00067 | 1.67E-06 | -0.007392833 | RNF182 |
| chr17 | 969522 | 970212 | 691 | 5 | 1.05E-05 | 5.73E-05 | 0.00029 | 3.07E-05 | -0.008597263 | NXN |
| chr3 | 129121414 | 129122604 | 1191 | 17 | 1.05E-05 | 2.78E-05 | 0.0025 | 3.38E-06 | -0.001828498 | RAB43, ISY1-RAB43 |
| chr9 | 32565611 | 32566589 | 979 | 3 | 1.06E-05 | 6.44E-06 | 0.00013 | 6.70E-06 | -0.010841714 | SMIM27, NDUFB6, DFFBP1 |
| chr3 | 156120243 | 156120836 | 594 | 7 | 1.06E-05 | 0.002821 | 8.04E-05 | 2.16E-05 | -0.003737115 | KCNAB1 |
| chr19 | 43727261 | 43727337 | 77 | 2 | 1.06E-05 | 1.12E-06 | 8.55E-05 | 1.62E-06 | -0.014752679 | NA |
| chr5 | 58455398 | 58456295 | 898 | 5 | 1.06E-05 | 3.60E-05 | 8.07E-05 | 8.11E-06 | -0.003266799 | PLK2 |
| chr2 | 169733298 | 169733980 | 683 | 9 | 1.06E-05 | 2.99E-07 | 0.00307 | 1.25E-06 | -0.002708312 | KLHL23 |
| chr6 | 14228855 | 14228907 | 53 | 2 | 1.06E-05 | 7.10E-07 | 0.00018 | 1.28E-06 | -0.019993765 | NA |
| chr22 | 50176197 | 50179031 | 2835 | 9 | 1.06E-05 | 2.54E-11 | 0.00015 | 1.32E-10 | -0.014032861 | PANX2 |
| chr19 | 11495529 | 11496469 | 941 | 5 | 1.07E-05 | 4.03E-06 | 0.00097 | 8.79E-07 | -0.005140223 | AC008481.3, ZNF653, MIR7974 |
| chr14 | 21432661 | 21432912 | 252 | 3 | 1.07E-05 | 0.000127 | 1.33E-05 | 9.27E-06 | -0.002560502 | CHD8 |
| chr2 | 129337182 | 129337486 | 305 | 2 | 1.07E-05 | 9.23E-07 | 0.00012 | 1.49E-06 | -0.010695532 | NA |
| chr16 | 57627387 | 57627970 | 584 | 5 | 1.07E-05 | 9.80E-05 | 0.00013 | 4.61E-05 | -0.004230102 | ADGRG1, AC018552.3 |
| chr16 | 85717137 | 85718172 | 1036 | 4 | 1.07E-05 | 1.95E-07 | 0.00029 | 3.53E-07 | -0.009840145 | C16orf74 |
| chr9 | 135743350 | 135743410 | 61 | 2 | 1.07E-05 | 1.48E-05 | 9.69E-06 | 2.59E-06 | -0.011536712 | KCNT1 |
| chr2 | 135741145 | 135741947 | 803 | 10 | 1.07E-05 | 4.92E-05 | 0.00025 | 4.02E-06 | -0.000962214 | UBXN4 |
| chr14 | 95134033 | 95134902 | 870 | 5 | 1.07E-05 | 0.000229 | 2.27E-05 | 1.94E-05 | -0.004837284 | DICER1 |
| chr21 | 41442855 | 41443739 | 885 | 4 | 1.07E-05 | 3.91E-05 | 6.86E-05 | 1.03E-05 | -0.002756403 | MX1, AP001610.2 |
| chr11 | 45470433 | 45471121 | 689 | 3 | 1.08E-05 | 3.49E-07 | 0.00058 | 9.53E-07 | -0.021729849 | AC103855.3 |
| chr6 | 117602109 | 117603117 | 1009 | 13 | 1.08E-05 | 0.002879 | 0.00043 | 1.80E-05 | 0.001509971 | GOPC |
| chr11 | 66367798 | 66368156 | 359 | 5 | 1.08E-05 | 3.00E-06 | 0.0012 | 2.06E-06 | -0.004986247 | SLC29A2 |
| chr22 | 50268995 | 50269426 | 432 | 3 | 1.09E-05 | 7.06E-05 | 0.00011 | 1.08E-05 | -0.010965039 | MAPK11 |
| chr2 | 105961875 | 105962293 | 419 | 4 | 1.09E-05 | 0.000207 | 2.60E-05 | 5.95E-06 | 0.000170645 | AC009505.1 |
| chr7 | 2436691 | 2437352 | 662 | 2 | 1.09E-05 | 4.04E-06 | 7.19E-05 | 4.27E-06 | -0.012860138 | CHST12 |
| chr12 | 14283834 | 14284728 | 895 | 4 | 1.09E-05 | 2.84E-06 | 3.93E-05 | 5.43E-07 | -0.004940836 | NA |
| chr17 | 78979862 | 78980975 | 1114 | 10 | 1.09E-05 | 2.84E-05 | 0.00287 | 6.72E-06 | -0.007783264 | LGALS3BP |
| chr7 | 140054271 | 140054951 | 681 | 3 | 1.10E-05 | 1.19E-05 | 2.24E-05 | 2.88E-06 | -0.005532576 | PARP12 |
| chr2 | 73981475 | 73982033 | 559 | 6 | 1.10E-05 | 2.47E-07 | 0.001 | 1.01E-06 | -0.00754039 | NA |
| chr7 | 66378654 | 66378697 | 44 | 2 | 1.10E-05 | 8.72E-05 | 7.76E-06 | 4.66E-06 | 0.002323493 | TPST1, LINC00174 |
| chr10 | 44005043 | 44005881 | 839 | 7 | 1.10E-05 | 1.56E-07 | 0.00386 | 7.27E-07 | -0.00599263 | NA |
| chr6 | 36139573 | 36139875 | 303 | 2 | 1.11E-05 | 5.73E-06 | 7.87E-05 | 5.78E-06 | -0.015912739 | MAPK13 |
| chr1 | 1939723 | 1940594 | 872 | 6 | 1.11E-05 | 6.57E-06 | 0.00042 | 1.29E-06 | -0.003970559 | CFAP74 |
| chr3 | 49172596 | 49172788 | 193 | 2 | 1.11E-05 | 1.33E-05 | 6.96E-05 | 9.94E-06 | -0.012690354 | KLHDC8B |
| chr1 | 11745708 | 11746207 | 500 | 3 | 1.11E-05 | 6.37E-05 | 2.96E-05 | 7.56E-06 | -0.003746553 | AGTRAP |
| chr2 | 32839827 | 32840434 | 608 | 3 | 1.11E-05 | 0.153638 | 1.45E-05 | 5.77E-05 | -0.001940986 | LINC00486 |
| chr4 | 89438495 | 89438602 | 108 | 3 | 1.12E-05 | 4.06E-07 | 0.00068 | 1.16E-06 | -0.011859605 | AC093866.1 |
| chr21 | 42376655 | 42376657 | 3 | 2 | 1.12E-05 | 1.11E-06 | 0.00019 | 1.90E-06 | -0.024733991 | TMPRSS3 |
| chr7 | 22853700 | 22854305 | 606 | 4 | 1.12E-05 | 4.12E-05 | 7.62E-05 | 5.25E-06 | -0.006660087 | SNHG26 |
| chr4 | 42392285 | 42392746 | 462 | 3 | 1.12E-05 | 9.37E-06 | 6.36E-05 | 2.10E-06 | -0.007601742 | NA |
| chr12 | 53335662 | 53336622 | 961 | 11 | 1.12E-05 | 7.32E-06 | 0.00116 | 7.29E-07 | -0.001988865 | SP7 |
| chr3 | 172358059 | 172358629 | 571 | 3 | 1.12E-05 | 5.66E-05 | 3.76E-05 | 7.38E-06 | -0.004419369 | FNDC3B |
| chr17 | 82021297 | 82022225 | 929 | 5 | 1.12E-05 | 1.71E-06 | 0.00914 | 8.52E-06 | -0.005642579 | CENPX |
| chr11 | 124535969 | 124536798 | 830 | 3 | 1.12E-05 | 9.40E-07 | 9.27E-05 | 1.28E-06 | -0.020603973 | NA |
| chr1 | 167126356 | 167126510 | 155 | 2 | 1.12E-05 | 0.003058 | 1.11E-05 | 1.32E-05 | -0.003465421 | STYXL2, GPA33 |
| chr3 | 184037719 | 184038056 | 338 | 2 | 1.12E-05 | 1.72E-05 | 2.49E-05 | 5.88E-06 | -0.012954249 | HTR3D |
| chr12 | 14554163 | 14554315 | 153 | 3 | 1.13E-05 | 0.004586 | 5.60E-06 | 6.62E-06 | -0.003282095 | PLBD1 |
| chr12 | 52814135 | 52814978 | 844 | 7 | 1.13E-05 | 7.57E-05 | 0.00011 | 9.80E-07 | -0.003126497 | NA |
| chr13 | 28494963 | 28495919 | 957 | 6 | 1.14E-05 | 6.04E-08 | 0.00262 | 3.15E-07 | -0.005457945 | AL139005.1, FLT1 |
| chr11 | 119298293 | 119298465 | 173 | 2 | 1.14E-05 | 6.74E-05 | 5.05E-05 | 2.17E-05 | -0.002437629 | CBL |
| chr18 | 77087294 | 77088124 | 831 | 7 | 1.14E-05 | 3.68E-07 | 0.00421 | 1.36E-06 | -0.012531285 | MBP |
| chr2 | 42100861 | 42101270 | 410 | 2 | 1.14E-05 | 3.12E-06 | 0.00017 | 4.65E-06 | 0.007637098 | NA |
| chr16 | 25118904 | 25119300 | 397 | 2 | 1.15E-05 | 0.000131 | 1.57E-05 | 1.07E-05 | -0.008473131 | LCMT1 |
| chr17 | 39708184 | 39710176 | 1993 | 11 | 1.15E-05 | 9.77E-08 | 0.00121 | 2.90E-08 | -0.005630918 | ERBB2 |

| | | | | | | | | | | |
|---|---|---|---|---|---|---|---|---|---|---|
| chr3 | 52232039 | 52232416 | 378 | 4 | 1.15E-05 | 8.88E-06 | 0.00015 | 6.84E-06 | -0.005726882 | TWF2 |
| chr11 | 67285521 | 67286458 | 938 | 3 | 1.15E-05 | 0.001838 | 0.00012 | 2.43E-05 | -0.007705673 | GRK2 |
| chr2 | 100105560 | 100106967 | 1408 | 11 | 1.16E-05 | 0.000364 | 0.00101 | 2.71E-06 | 0.000274295 | AC092667.1, AFF3 |
| chr21 | 38661859 | 38662498 | 640 | 4 | 1.16E-05 | 8.54E-07 | 0.00063 | 1.37E-06 | -0.008441935 | NA |
| chr1 | 15941962 | 15943100 | 1139 | 6 | 1.16E-05 | 4.46E-07 | 0.00168 | 1.84E-06 | -0.004489003 | ZBTB17 |
| chr3 | 22370633 | 22371741 | 1109 | 5 | 1.16E-05 | 1.62E-07 | 0.00328 | 8.53E-07 | -0.035113491 | ZNF385D |
| chr17 | 79137695 | 79138077 | 383 | 5 | 1.16E-05 | 2.19E-07 | 0.00255 | 8.99E-07 | -0.016331296 | RBFOX3 |
| chr2 | 64859332 | 64859688 | 357 | 3 | 1.17E-05 | 2.91E-06 | 0.00117 | 6.15E-06 | -0.012193394 | LINC01800 |
| chr16 | 75550064 | 75550628 | 565 | 4 | 1.17E-05 | 0.000421 | 8.09E-05 | 2.94E-06 | -0.01030264 | TMEM231, AC025287.1 |
| chr10 | 119873692 | 119874142 | 451 | 4 | 1.17E-05 | 5.89E-06 | 0.00051 | 4.59E-06 | -0.011381975 | MCMBP |
| chr1 | 43359625 | 43360233 | 609 | 3 | 1.18E-05 | 2.09E-05 | 0.00015 | 1.44E-05 | -0.007282788 | CDC20 |
| chr16 | 58391213 | 58392370 | 1158 | 10 | 1.18E-05 | 6.37E-05 | 0.00161 | 3.57E-06 | -0.001030356 | GINS3, AC009118.2 |
| chr12 | 88142122 | 88142798 | 677 | 12 | 1.18E-05 | 0.000233 | 0.0013 | 7.04E-06 | 0.000746714 | TMTC3 |
| chr12 | 51917276 | 51917980 | 705 | 3 | 1.18E-05 | 9.28E-06 | 2.72E-05 | 2.50E-06 | -0.007453633 | ACVRL1 |
| chr7 | 35988345 | 35989125 | 781 | 2 | 1.18E-05 | 2.71E-07 | 6.56E-05 | 4.48E-07 | -0.004047531 | AC083864.5 |
| chr13 | 77917847 | 77918171 | 325 | 3 | 1.18E-05 | 0.000153 | 6.15E-05 | 2.96E-05 | -0.002026033 | EDNRB |
| chr22 | 23245051 | 23245340 | 290 | 2 | 1.18E-05 | 9.06E-07 | 0.00014 | 1.52E-06 | -0.014463851 | BCR |
| chr18 | 11750099 | 11750588 | 490 | 5 | 1.18E-05 | 2.57E-06 | 0.00342 | 8.12E-06 | -0.00738446 | GNAL |
| chr9 | 3732719 | 3733226 | 508 | 4 | 1.18E-05 | 0.000164 | 0.00011 | 2.77E-06 | -0.001634452 | NA |
| chr12 | 14843209 | 14844282 | 1074 | 12 | 1.19E-05 | 4.33E-05 | 0.00047 | 5.22E-06 | -0.003996687 | C12orf60, AC007655.1, ART4 |
| chr19 | 57578176 | 57578517 | 342 | 2 | 1.19E-05 | 5.63E-05 | 9.99E-05 | 3.19E-05 | -0.006549248 | ZIK1, ZNF416 |
| chr2 | 232014263 | 232015588 | 1326 | 6 | 1.19E-05 | 3.20E-07 | 0.00057 | 6.38E-07 | -0.004285499 | DIS3L2, AC019130.1 |
| chr8 | 135456242 | 135457537 | 1296 | 9 | 1.19E-05 | 4.27E-06 | 0.00013 | 1.37E-06 | -0.000274664 | KHDRBS3, AC040914.1 |
| chr1 | 153536611 | 153537159 | 549 | 5 | 1.19E-05 | 0.00023 | 0.00057 | 3.08E-05 | -0.004703188 | S100A5 |
| chr10 | 101889231 | 101890404 | 1174 | 5 | 1.19E-05 | 9.64E-06 | 0.00023 | 1.05E-05 | -0.005403344 | ARMH3 |
| chr1 | 56967536 | 56968431 | 896 | 6 | 1.19E-05 | 6.76E-05 | 0.00015 | 1.00E-05 | -0.004030976 | AL161740.1 |
| chr14 | 68248745 | 68249132 | 388 | 2 | 1.20E-05 | 0.000159 | 2.56E-05 | 1.76E-05 | -0.005268203 | RAD51B |
| chr2 | 102185201 | 102187525 | 2325 | 13 | 1.20E-05 | 5.80E-07 | 0.0005 | 1.36E-08 | -0.008984829 | IL1RL2 |
| chr12 | 111363908 | 111364443 | 536 | 3 | 1.20E-05 | 3.92E-05 | 0.00054 | 2.08E-05 | -0.00844021 | PHETA1 |
| chr6 | 7540601 | 7541635 | 1035 | 11 | 1.20E-05 | 1.79E-06 | 0.00276 | 4.17E-06 | -0.001146681 | DSP, AL031058.1 |
| chr2 | 120711244 | 120712127 | 884 | 4 | 1.20E-05 | 2.33E-06 | 0.00052 | 3.86E-06 | 0.002074193 | NA |
| chr1 | 226408721 | 226408967 | 247 | 2 | 1.21E-05 | 7.21E-05 | 7.00E-05 | 2.88E-05 | -0.005894335 | NA |
| chr15 | 49064000 | 49064466 | 467 | 2 | 1.21E-05 | 0.000119 | 2.32E-05 | 1.47E-05 | -0.013150149 | NA |
| chr1 | 165543743 | 165544124 | 382 | 6 | 1.21E-05 | 5.97E-06 | 0.00034 | 3.04E-06 | -0.009392619 | LRRC52, LRRC52-AS1 |
| chr3 | 39502024 | 39502835 | 812 | 8 | 1.21E-05 | 5.67E-07 | 0.01146 | 2.32E-06 | -0.027673407 | MOBP, AC092058.1 |
| chr12 | 6851257 | 6852782 | 1526 | 18 | 1.21E-05 | 0.000418 | 0.00285 | 1.80E-05 | 0.000745283 | USP5, CDCA3 |
| chr12 | 3951615 | 3951859 | 245 | 3 | 1.21E-05 | 9.64E-06 | 2.19E-05 | 2.74E-06 | -0.006325352 | NA |
| chr17 | 55276506 | 55276543 | 38 | 2 | 1.21E-05 | 1.45E-06 | 4.93E-05 | 1.62E-06 | -0.013648548 | HLF |
| chr2 | 143720347 | 143720415 | 69 | 3 | 1.21E-05 | 2.76E-06 | 6.00E-05 | 1.74E-06 | -0.010633035 | ARHGAP15, AC092652.1 |
| chr11 | 67395351 | 67396046 | 696 | 7 | 1.21E-05 | 1.26E-06 | 0.00087 | 2.56E-06 | -0.004113072 | RAD9A |
| chr19 | 50512447 | 50512573 | 127 | 3 | 1.22E-05 | 0.002459 | 6.86E-06 | 1.38E-05 | -0.002071151 | ASPDH |
| chr17 | 80941855 | 80942912 | 1058 | 6 | 1.22E-05 | 7.82E-07 | 0.00034 | 6.09E-07 | -0.006253614 | RPTOR, AC127496.3 |
| chr14 | 94113607 | 94114787 | 1181 | 4 | 1.22E-05 | 7.13E-06 | 0.00077 | 6.55E-06 | -0.002003172 | IFI27 |
| chr4 | 183960915 | 183961058 | 144 | 2 | 1.22E-05 | 2.61E-06 | 0.00021 | 4.17E-06 | -0.019429924 | STOX2 |
| chr17 | 4537521 | 4537695 | 175 | 2 | 1.22E-05 | 2.95E-06 | 0.00017 | 4.48E-06 | -0.012385832 | SPNS2 |
| chr21 | 42063921 | 42064170 | 250 | 2 | 1.22E-05 | 0.001083 | 2.17E-05 | 2.39E-05 | -0.002000103 | UMODL1 |
| chr3 | 127676344 | 127677171 | 828 | 4 | 1.22E-05 | 5.53E-06 | 0.00126 | 9.62E-06 | -0.004710255 | ABTB1 |
| chr17 | 7675729 | 7677592 | 1864 | 10 | 1.22E-05 | 2.00E-07 | 0.00081 | 7.11E-08 | -0.003220645 | TP53 |
| chr13 | 49792054 | 49793204 | 1151 | 13 | 1.23E-05 | 0.003376 | 0.00023 | 3.82E-05 | 0.00106869 | AL136301.1, KPNA3 |
| chr12 | 111099391 | 111100038 | 648 | 5 | 1.23E-05 | 2.88E-05 | 0.00097 | 1.05E-05 | 0.001158224 | CUX2 |
| chr15 | 91856364 | 91856667 | 304 | 2 | 1.23E-05 | 0.004624 | 2.13E-05 | 2.83E-05 | -0.008530134 | SLCO3A1 |
| chr12 | 110224827 | 110225166 | 340 | 3 | 1.23E-05 | 0.003289 | 1.28E-05 | 1.84E-05 | -0.009000344 | NA |
| chr18 | 12493161 | 12493167 | 7 | 2 | 1.23E-05 | 6.06E-05 | 1.66E-05 | 8.21E-06 | -0.00658968 | SPIRE1 |
| chr17 | 5486142 | 5486981 | 840 | 15 | 1.24E-05 | 0.000815 | 0.0009 | 4.63E-06 | 0.001162246 | MIS12, DERL2 |
| chr20 | 36856497 | 36857015 | 519 | 3 | 1.24E-05 | 4.98E-05 | 5.71E-05 | 7.04E-06 | -0.010493093 | SOGA1 |
| chr1 | 7572687 | 7572980 | 294 | 2 | 1.25E-05 | 2.34E-06 | 5.10E-05 | 2.38E-06 | -0.020098389 | CAMTA1 |
| chr17 | 48093321 | 48093666 | 346 | 2 | 1.25E-05 | 3.97E-05 | 1.50E-05 | 6.22E-06 | -0.005014608 | CBX1 |
| chr19 | 12064292 | 12065207 | 916 | 11 | 1.25E-05 | 0.000633 | 0.00065 | 5.84E-06 | -0.00122745 | AC008770.4, ZNF844 |
| chr1 | 9443990 | 9444370 | 381 | 3 | 1.25E-05 | 3.12E-06 | 0.00014 | 2.77E-06 | -0.012210586 | NA |
| chr19 | 29874534 | 29875016 | 483 | 2 | 1.26E-05 | 8.52E-05 | 2.35E-05 | 1.29E-05 | -0.003285906 | NA |
| chr3 | 124406736 | 124407103 | 368 | 2 | 1.26E-05 | 8.56E-06 | 7.10E-05 | 7.33E-06 | -0.009375139 | KALRN |
| chr1 | 36397406 | 36398337 | 932 | 11 | 1.26E-05 | 6.29E-06 | 0.00103 | 3.78E-06 | 0.000670987 | LSM10 |
| chr18 | 12702186 | 12703073 | 888 | 17 | 1.27E-05 | 5.55E-06 | 0.01098 | 1.37E-06 | 0.001137816 | PSMG2, CEP76 |
| chr6 | 33896912 | 33897289 | 378 | 5 | 1.27E-05 | 3.15E-07 | 0.00091 | 1.14E-06 | -0.012664876 | AL138889.1, LINC01016 |
| chr19 | 36711494 | 36712085 | 592 | 3 | 1.27E-05 | 4.96E-05 | 0.00012 | 7.69E-06 | -0.000394353 | ZNF567 |
| chr3 | 52237315 | 52237708 | 394 | 2 | 1.27E-05 | 4.96E-05 | 2.28E-05 | 9.77E-06 | -0.004306188 | TWF2 |
| chr16 | 88822865 | 88823450 | 586 | 3 | 1.27E-05 | 3.17E-06 | 7.41E-05 | 2.92E-06 | -0.004343701 | GALNS |
| chr13 | 113105457 | 113105972 | 516 | 7 | 1.27E-05 | 3.90E-05 | 0.00382 | 3.50E-06 | -0.01095169 | F7 |
| chr7 | 4834654 | 4835345 | 692 | 5 | 1.27E-05 | 0.000476 | 0.00017 | 8.02E-06 | -0.005486423 | RADIL |
| chr16 | 3642021 | 3642928 | 908 | 5 | 1.27E-05 | 1.49E-06 | 0.00059 | 1.32E-06 | -0.004360545 | DNASE1 |
| chr1 | 156676273 | 156676954 | 682 | 6 | 1.27E-05 | 6.00E-05 | 0.0041 | 7.23E-06 | -0.011064943 | NES |
| chr17 | 76688422 | 76688952 | 531 | 3 | 1.28E-05 | 2.05E-06 | 0.00019 | 3.15E-06 | -0.004957528 | MXRA7 |
| chr12 | 55743999 | 55744569 | 571 | 2 | 1.28E-05 | 1.72E-05 | 0.00013 | 1.66E-05 | -0.008540727 | GDF11 |
| chr8 | 78798249 | 78798868 | 620 | 5 | 1.28E-05 | 8.15E-05 | 0.00068 | 5.48E-06 | -0.000191299 | IL7 |
| chr9 | 93592615 | 93592809 | 195 | 2 | 1.28E-05 | 1.21E-06 | 9.55E-05 | 1.78E-06 | -0.009318131 | PHF2 |
| chr7 | 711372 | 712325 | 954 | 6 | 1.28E-05 | 4.48E-06 | 0.0013 | 4.06E-06 | -0.010142752 | PRKAR1B |
| chr19 | 40623976 | 40624136 | 161 | 4 | 1.29E-05 | 3.20E-05 | 0.00015 | 2.64E-06 | -0.003020109 | LTBP4 |
| chr17 | 79977908 | 79979305 | 1398 | 11 | 1.29E-05 | 1.25E-07 | 0.00596 | 2.59E-07 | -0.002377616 | TBC1D16 |
| chr11 | 70124422 | 70124827 | 406 | 4 | 1.29E-05 | 0.000104 | 0.00014 | 7.69E-06 | -0.01270802 | ANO1 |
| chr5 | 142868918 | 142869808 | 891 | 6 | 1.30E-05 | 6.44E-06 | 0.00022 | 3.72E-06 | 0.00230104 | ARHGAP26, ARHGAP26-AS1 |
| chr12 | 6232167 | 6233391 | 1225 | 4 | 1.30E-05 | 3.53E-08 | 0.00094 | 1.77E-07 | -0.010411601 | CD9 |
| chr1 | 230572564 | 230572727 | 164 | 2 | 1.30E-05 | 4.47E-05 | 8.25E-06 | 3.86E-06 | -0.016406751 | NA |
| chr16 | 11280866 | 11281596 | 731 | 8 | 1.30E-05 | 1.76E-05 | 4.99E-05 | 4.53E-06 | -0.001485267 | RMI2, PRM1 |
| chr2 | 219571908 | 219572596 | 689 | 7 | 1.31E-05 | 7.35E-07 | 0.00362 | 1.87E-06 | -0.009798312 | INHA |
| chr4 | 1576205 | 1576876 | 672 | 7 | 1.31E-05 | 1.11E-05 | 0.00043 | 3.12E-06 | -0.002702536 | AC147067.1 |
| chr1 | 248837717 | 248838850 | 1134 | 12 | 1.31E-05 | 3.82E-07 | 0.00602 | 3.89E-07 | 0.00155019 | ZNF672 |
| chr20 | 53711780 | 53712077 | 298 | 2 | 1.31E-05 | 0.000274 | 1.83E-05 | 1.52E-05 | -0.008000399 | NA |
| chr11 | 73964967 | 73966035 | 1069 | 5 | 1.31E-05 | 3.04E-05 | 0.00013 | 2.12E-05 | 0.004894193 | DNAJB13, AP003717.1 |
| chr15 | 40055846 | 40056102 | 257 | 2 | 1.32E-05 | 3.42E-06 | 0.00018 | 5.09E-06 | -0.013601525 | SRP14-AS1 |
| chr11 | 82898882 | 82899281 | 400 | 2 | 1.32E-05 | 1.21E-05 | 4.89E-05 | 7.51E-06 | -0.01188145 | PRCP |
| chr1 | 64173662 | 64173742 | 81 | 2 | 1.32E-05 | 2.63E-05 | 9.39E-06 | 3.36E-06 | -0.005968632 | ROR1 |
| chr12 | 112937597 | 112938488 | 892 | 8 | 1.32E-05 | 1.50E-06 | 0.0034 | 3.01E-06 | -0.004307678 | OAS3, AC004551.1 |
| chr8 | 124280304 | 124280769 | 466 | 4 | 1.32E-05 | 1.87E-07 | 0.00259 | 9.11E-07 | -0.009727177 | NA |
| chr7 | 156124208 | 156124465 | 258 | 3 | 1.32E-05 | 2.52E-05 | 9.93E-05 | 4.52E-06 | -0.00725135 | NA |
| chr14 | 105269159 | 105269560 | 402 | 4 | 1.32E-05 | 5.71E-05 | 9.45E-05 | 5.03E-06 | -0.006305956 | BRF1 |
| chr9 | 37437438 | 37437869 | 432 | 2 | 1.32E-05 | 4.75E-06 | 4.58E-05 | 3.82E-06 | -0.003689993 | NA |
| chr8 | 143633776 | 143633854 | 79 | 3 | 1.33E-05 | 2.96E-05 | 2.93E-05 | 6.79E-06 | -0.00479627 | NA |
| chr14 | 89027217 | 89028423 | 1207 | 4 | 1.33E-05 | 1.77E-06 | 0.0004 | 1.56E-06 | -0.004995456 | NA |
| chr19 | 11033341 | 11034152 | 812 | 8 | 1.33E-05 | 2.76E-05 | 0.00068 | 3.70E-06 | -0.001463315 | SMARCA4, AC006127.1 |
| chr20 | 45911894 | 45912514 | 621 | 6 | 1.33E-05 | 0.000292 | 0.00052 | 1.38E-05 | -0.005071687 | PLTP |
| chr3 | 64088280 | 64088731 | 452 | 2 | 1.34E-05 | 0.001081 | 4.22E-05 | 4.70E-05 | -0.013842888 | PRICKLE2-AS1 |
| chr9 | 70862962 | 70863474 | 513 | 4 | 1.34E-05 | 1.11E-05 | 0.00028 | 2.63E-06 | -0.004793162 | TRPM3 |
| chr17 | 29590873 | 29592614 | 1742 | 9 | 1.34E-05 | 1.72E-06 | 0.00031 | 3.47E-07 | -0.007061986 | ABHD15-AS1, ANKRD13B, GIT1, AC104564.4 |
| chr1 | 6634271 | 6634766 | 496 | 4 | 1.34E-05 | 4.22E-06 | 5.49E-05 | 2.76E-06 | -0.006294694 | THAP3, DNAJC11 |
| chr15 | 90062386 | 90063262 | 877 | 3 | 1.35E-05 | 4.20E-05 | 3.21E-05 | 4.78E-06 | -0.008398759 | ZNF710 |
| chr2 | 85594684 | 85595603 | 920 | 7 | 1.35E-05 | 0.002386 | 0.00099 | 0.00038 | -0.004842921 | NA |
| chr3 | 52830643 | 52830891 | 249 | 5 | 1.35E-05 | 1.96E-06 | 0.00032 | 2.32E-06 | -0.00197426 | ITIH4, AC006254.1 |
| chr7 | 8436468 | 8436891 | 424 | 4 | 1.35E-05 | 4.94E-06 | 0.00042 | 7.01E-06 | 0.011193342 | NXPH1 |
| chr20 | 35539775 | 35540197 | 423 | 3 | 1.35E-05 | 2.61E-06 | 0.00034 | 3.29E-06 | 0.012193532 | NA |
| chr6 | 41784938 | 41785493 | 556 | 3 | 1.35E-05 | 3.05E-06 | 0.00174 | 7.62E-06 | -0.008444514 | AL365205.1, PRICKLE4, FRS3 |
| chr2 | 211534819 | 211535512 | 694 | 4 | 1.36E-05 | 6.79E-07 | 0.00016 | 9.62E-07 | -0.012651254 | ERBB4 |
| chr5 | 69502493 | 69503513 | 1021 | 6 | 1.36E-05 | 0.000392 | 4.41E-05 | 8.15E-06 | -0.00493922 | OCLN |
| chr18 | 78500568 | 78500974 | 407 | 2 | 1.36E-05 | 9.89E-05 | 6.41E-05 | 3.18E-05 | 0.000229638 | NA |
| chr18 | 13544055 | 13544069 | 15 | 2 | 1.36E-05 | 5.72E-06 | 2.60E-05 | 3.06E-06 | -0.00960604 | LDLRAD4 |
| chr3 | 141230230 | 141230873 | 644 | 2 | 1.36E-05 | 0.005112 | 0.00021 | 0.00041 | -0.002614008 | PXYLP1 |
| chr10 | 125060778 | 125061243 | 466 | 2 | 1.36E-05 | 1.35E-06 | 0.00023 | 2.32E-06 | -0.010543841 | CTBP2 |
| chr10 | 102493258 | 102493708 | 451 | 2 | 1.36E-05 | 6.95E-07 | 0.00018 | 1.25E-06 | -0.012285628 | ACTR1A |
| chr12 | 106684540 | 106685123 | 584 | 8 | 1.36E-05 | 4.29E-05 | 7.57E-05 | 8.16E-06 | -0.003044541 | RFX4, AC079385.1, AC079385.2 |
| chr19 | 45645334 | 45646252 | 919 | 7 | 1.36E-05 | 0.003609 | 0.00133 | 2.80E-05 | -0.002228448 | EML2 |
| chr19 | 48695643 | 48695977 | 335 | 5 | 1.36E-05 | 6.49E-05 | 6.88E-05 | 1.35E-05 | -0.00611458 | FUT2 |
| chr1 | 156703503 | 156703966 | 464 | 2 | 1.37E-05 | 3.23E-05 | 3.84E-05 | 1.17E-05 | -0.007933191 | AL590666.4, CRABP2 |
| chr1 | 26772746 | 26773759 | 1014 | 6 | 1.37E-05 | 0.000134 | 5.69E-05 | 4.82E-06 | -0.002380389 | ARID1A |
| chr14 | 104708169 | 104708305 | 137 | 2 | 1.37E-05 | 5.84E-06 | 0.00055 | 9.64E-06 | -0.011936558 | INF2 |
| chr17 | 77318748 | 77320301 | 1554 | 14 | 1.37E-05 | 3.57E-06 | 0.01396 | 1.36E-06 | -0.007190155 | SEPTIN9 |
| chr10 | 87792812 | 87792927 | 116 | 2 | 1.37E-05 | 1.70E-06 | 0.00027 | 2.93E-06 | -0.013417246 | ATAD1 |
| chr1 | 183205102 | 183205725 | 624 | 4 | 1.37E-05 | 0.002845 | 0.00015 | 1.35E-05 | -0.010747267 | LAMC2 |
| chr22 | 29004205 | 29005096 | 892 | 2 | 1.38E-05 | 4.97E-08 | 5.51E-05 | 9.56E-08 | -0.017301033 | ZNRF3, ZNRF3-IT1 |
| chr22 | 23278999 | 23279262 | 264 | 3 | 1.38E-05 | 0.000907 | 7.48E-06 | 9.54E-06 | -0.01010101 | BCR |
| chr7 | 6501701 | 6501890 | 190 | 2 | 1.38E-05 | 1.69E-05 | 5.19E-05 | 9.74E-06 | -0.002882538 | GRID2IP |
| chr11 | 68773090 | 68774395 | 1306 | 6 | 1.38E-05 | 1.80E-06 | 5.42E-05 | 1.56E-07 | -0.003970953 | CPT1A |
| chr17 | 7888312 | 7890096 | 1785 | 9 | 1.38E-05 | 0.000279 | 0.00021 | 1.59E-06 | -0.004559003 | CHD3 |
| chr22 | 21772415 | 21774451 | 2037 | 7 | 1.38E-05 | 2.10E-08 | 0.00148 | 5.85E-08 | -0.004565357 | MAPK1 |
| chr11 | 2805721 | 2806526 | 806 | 4 | 1.39E-05 | 3.90E-07 | 0.00109 | 1.34E-06 | -0.002671539 | KCNQ1 |
| chr19 | 3383485 | 3383728 | 244 | 3 | 1.39E-05 | 4.69E-06 | 6.95E-05 | 3.28E-06 | -0.005120571 | NFIC |
| chr1 | 36516823 | 36517856 | 1034 | 4 | 1.39E-05 | 0.000508 | 0.00015 | 5.08E-05 | -0.002643813 | NA |
| chr22 | 50248042 | 50249153 | 1112 | 4 | 1.40E-05 | 1.07E-06 | 0.00027 | 1.15E-06 | -0.010407524 | HDAC10, MAPK12 |
| chr20 | 441906 | 442398 | 493 | 5 | 1.40E-05 | 0.007227 | 2.57E-05 | 1.26E-05 | -0.002843124 | TBC1D20 |
| chr11 | 8932659 | 8933590 | 932 | 7 | 1.40E-05 | 3.28E-06 | 0.00169 | 4.39E-06 | -0.005668154 | C11orf16 |
| chr14 | 91285379 | 91286083 | 705 | 6 | 1.40E-05 | 3.93E-06 | 0.00017 | 1.75E-06 | 0.006714552 | CCDC88C |
| chr1 | 203798326 | 203799389 | 1064 | 5 | 1.40E-05 | 2.76E-06 | 8.51E-05 | 3.29E-07 | -0.003680316 | ZC3H11A, ZBED6 |
| chr19 | 47495678 | 47495764 | 87 | 2 | 1.41E-05 | 3.79E-06 | 7.33E-05 | 4.10E-06 | -0.013673168 | NAPA-AS1, NAPA |
| chr5 | 20305562 | 20305826 | 265 | 4 | 1.41E-05 | 6.70E-06 | 0.00024 | 3.31E-06 | -0.010115634 | CDH18-AS1, CDH18 |
| chr3 | 66544732 | 66544940 | 209 | 2 | 1.41E-05 | 1.43E-06 | 8.21E-05 | 1.95E-06 | 0.00118672 | NA |
| chr2 | 42763817 | 42764765 | 949 | 10 | 1.41E-05 | 5.97E-06 | 0.00503 | 6.55E-06 | -0.005340456 | OXER1 |
| chr15 | 66564795 | 66566257 | 1463 | 10 | 1.41E-05 | 2.05E-06 | 0.00115 | 6.22E-07 | -0.00479875 | LCTL |
| chr19 | 4676190 | 4678030 | 1841 | 8 | 1.41E-05 | 4.98E-07 | 0.00055 | 3.06E-07 | -0.004751193 | DPP9 |
| chr13 | 50527504 | 50529237 | 1734 | 5 | 1.41E-05 | 1.13E-06 | 2.69E-05 | 5.79E-08 | -0.010813562 | DLEU1, DLEU7 |
| chr15 | 40941503 | 40941799 | 297 | 3 | 1.41E-05 | 1.09E-06 | 0.00163 | 3.13E-06 | -0.00373133 | NA |
| chr2 | 191151000 | 191152660 | 1661 | 13 | 1.41E-05 | 2.05E-05 | 0.00051 | 6.91E-06 | -0.000865847 | STAT4 |
| chr19 | 18501208 | 18502200 | 993 | 4 | 1.41E-05 | 2.74E-07 | 0.00106 | 8.93E-07 | -0.009182425 | ELL |
| chr7 | 151410551 | 151411725 | 1175 | 11 | 1.41E-05 | 2.29E-07 | 0.00974 | 1.76E-06 | -0.007181719 | WDR86-AS1, WDR86 |
| chr17 | 1031777 | 1033696 | 1920 | 11 | 1.41E-05 | 5.82E-07 | 0.00086 | 5.50E-08 | -0.013239676 | ABR |
| chr9 | 78288391 | 78288482 | 92 | 2 | 1.42E-05 | 2.19E-06 | 3.93E-05 | 1.98E-06 | -0.017225499 | NA |
| chr18 | 63751843 | 63753100 | 1258 | 6 | 1.42E-05 | 3.09E-05 | 0.00034 | 2.71E-06 | -0.012146318 | SERPINB7 |
| chr5 | 158958377 | 158958435 | 59 | 2 | 1.42E-05 | 1.06E-06 | 0.00012 | 1.69E-06 | -0.018386457 | EBF1 |
| chr1 | 22257163 | 22257344 | 182 | 2 | 1.42E-05 | 4.95E-06 | 0.00013 | 6.33E-06 | -0.010173733 | NA |
| chr2 | 11347829 | 11348642 | 814 | 6 | 1.42E-05 | 6.01E-06 | 0.00023 | 2.46E-06 | -0.006221625 | ROCK2 |
| chr1 | 31771822 | 31771856 | 35 | 2 | 1.42E-05 | 8.57E-06 | 5.90E-05 | 6.64E-06 | -0.009950971 | NA |
| chr3 | 187519208 | 187519514 | 307 | 3 | 1.42E-05 | 6.91E-07 | 0.00068 | 1.65E-06 | -0.010955247 | NA |
| chr7 | 4191253 | 4191422 | 170 | 3 | 1.42E-05 | 3.98E-06 | 0.00022 | 2.63E-06 | -0.004399676 | SDK1 |
| chr1 | 234534079 | 234534940 | 862 | 2 | 1.43E-05 | 1.04E-06 | 6.14E-05 | 1.35E-06 | -0.016321256 | NA |
| chr17 | 60158930 | 60159440 | 511 | 3 | 1.43E-05 | 1.86E-06 | 0.0004 | 3.70E-06 | -0.007131334 | CA4 |

| | | | | | | | | | | |
|---|---|---|---|---|---|---|---|---|---|---|
| chr7 | 100610910 | 100611327 | 418 | 3 | 1.43E-05 | 6.77E-05 | 6.17E-05 | 2.16E-05 | -0.007204206 | NA |
| chr6 | 122995452 | 122996730 | 1279 | 17 | 1.43E-05 | 0.00017 | 0.00015 | 6.54E-06 | 0.001980384 | CLVS2, AL591428.1 |
| chr11 | 65625827 | 65626312 | 486 | 3 | 1.43E-05 | 0.001031 | 9.32E-05 | 1.16E-05 | -0.004749916 | PCNX3 |
| chr1 | 38559037 | 38559797 | 761 | 5 | 1.44E-05 | 3.98E-05 | 0.0004 | 6.15E-06 | -0.006184431 | NA |
| chr10 | 31944309 | 31944586 | 278 | 2 | 1.44E-05 | 0.000111 | 1.09E-05 | 6.95E-06 | -0.011474638 | NA |
| chr20 | 3908250 | 3908538 | 289 | 2 | 1.44E-05 | 1.34E-05 | 2.43E-05 | 5.00E-06 | -0.007805489 | PANK2 |
| chr1 | 228554986 | 228555678 | 693 | 6 | 1.44E-05 | 6.75E-07 | 0.00167 | 2.14E-06 | -0.032004076 | NA |
| chr19 | 5851111 | 5851751 | 641 | 9 | 1.44E-05 | 1.36E-05 | 0.00018 | 5.04E-06 | -0.005654991 | AC024592.2, FUT3 |
| chr11 | 118609861 | 118610310 | 450 | 2 | 1.44E-05 | 5.89E-05 | 6.75E-05 | 2.52E-05 | -0.006597492 | PHLDB1 |
| chr6 | 163820761 | 163821209 | 449 | 2 | 1.44E-05 | 1.26E-05 | 0.00011 | 1.19E-05 | -0.015505872 | AL078602.1 |
| chr19 | 13057196 | 13057282 | 87 | 2 | 1.44E-05 | 6.06E-06 | 0.0002 | 8.34E-06 | -0.010775744 | NFIX |
| chr5 | 141369791 | 141370593 | 803 | 9 | 1.45E-05 | 7.12E-05 | 0.00046 | 5.00E-06 | 0.00743117 | PCDHGA1, PCDHGA2, PCDHGA3, PCDHGB1, PCDHGA4, PCDHGB2, PCDHGA5, PCDHGB3 |
| chr1 | 31855223 | 31855546 | 324 | 2 | 1.46E-05 | 6.13E-05 | 2.25E-05 | 1.07E-05 | -0.00673364 | AL354919.1, AL136115.2 |
| chr5 | 59038849 | 59039353 | 505 | 4 | 1.46E-05 | 4.21E-06 | 0.00033 | 5.87E-06 | 0.001764507 | PDE4D |
| chr21 | 42782247 | 42783234 | 988 | 5 | 1.46E-05 | 6.97E-06 | 6.36E-05 | 6.02E-07 | -0.006181467 | NA |
| chr17 | 50559833 | 50560829 | 997 | 22 | 1.46E-05 | 1.99E-05 | 0.00786 | 4.02E-06 | 0.001822146 | CACNA1G, CACNA1G-AS1 |
| chr17 | 48482965 | 48483645 | 681 | 4 | 1.47E-05 | 0.000122 | 3.92E-05 | 1.02E-05 | -0.007674364 | NA |
| chr19 | 19109901 | 19110765 | 865 | 4 | 1.47E-05 | 2.70E-07 | 0.00112 | 8.24E-07 | -0.015374192 | SLC25A42 |
| chr3 | 195178470 | 195179558 | 1089 | 3 | 1.48E-05 | 9.54E-06 | 4.32E-05 | 8.26E-07 | -0.012661306 | XXYLT1 |
| chr16 | 3590453 | 3591319 | 867 | 3 | 1.48E-05 | 3.52E-06 | 0.00011 | 4.01E-06 | -0.012211683 | SLX4 |
| chr11 | 2395991 | 2396927 | 937 | 6 | 1.48E-05 | 2.95E-07 | 0.00201 | 9.22E-07 | -0.00156546 | CD81 |
| chr16 | 11123616 | 11123759 | 144 | 2 | 1.48E-05 | 1.64E-05 | 1.89E-05 | 4.69E-06 | -0.001985962 | CLEC16A |
| chr3 | 31562731 | 31563182 | 452 | 3 | 1.48E-05 | 4.04E-06 | 0.00013 | 3.91E-06 | -0.007620548 | STT3B |
| chr18 | 24112448 | 24112657 | 210 | 2 | 1.49E-05 | 6.61E-06 | 4.34E-05 | 4.63E-06 | -0.002647602 | TTC39C |
| chr12 | 111754789 | 111754959 | 171 | 3 | 1.49E-05 | 2.94E-05 | 3.52E-05 | 5.64E-06 | -0.005366227 | ACAD10, AC002996.1 |
| chr14 | 102585103 | 102585683 | 581 | 3 | 1.49E-05 | 1.19E-05 | 4.72E-05 | 5.80E-06 | -0.00866384 | NA |
| chr6 | 135187694 | 135188381 | 688 | 3 | 1.49E-05 | 3.51E-06 | 0.00013 | 3.40E-06 | -0.011715464 | MYB |
| chr14 | 104170296 | 104170724 | 429 | 4 | 1.49E-05 | 0.266816 | 7.24E-06 | 5.82E-05 | -0.003804236 | KIF26A |
| chr16 | 676786 | 677212 | 427 | 2 | 1.49E-05 | 6.44E-05 | 0.0005 | 6.97E-05 | -0.01649775 | RHBDL1 |
| chr15 | 79064695 | 79064969 | 275 | 5 | 1.49E-05 | 6.81E-06 | 0.00042 | 1.96E-06 | -0.005382594 | RASGRF1 |
| chr14 | 105466045 | 105466946 | 902 | 6 | 1.49E-05 | 0.00056 | 0.00016 | 9.57E-06 | -0.002407987 | MTA1 |
| chr15 | 22786088 | 22787484 | 1397 | 16 | 1.49E-05 | 6.78E-06 | 0.0003 | 1.65E-07 | -0.003131357 | NIPA1 |
| chr11 | 86174162 | 86174660 | 499 | 2 | 1.49E-05 | 3.72E-05 | 2.92E-05 | 1.03E-05 | -0.004065837 | NA |
| chr2 | 207624346 | 207625656 | 1311 | 18 | 1.50E-05 | 8.92E-05 | 0.00076 | 2.68E-06 | 0.000791061 | METTL21A |
| chr1 | 247517800 | 247518629 | 830 | 11 | 1.50E-05 | 3.75E-07 | 0.01557 | 2.49E-06 | 0.017750829 | GCSAML |
| chr1 | 117563674 | 117564426 | 753 | 3 | 1.50E-05 | 9.29E-06 | 2.86E-05 | 1.73E-06 | -0.004308576 | NA |
| chr7 | 2114036 | 2114788 | 753 | 6 | 1.50E-05 | 0.011452 | 0.0002 | 2.61E-05 | -0.003121738 | MAD1L1, AC069288.1 |
| chr6 | 46734946 | 46735514 | 569 | 7 | 1.50E-05 | 2.96E-05 | 0.00138 | 8.52E-06 | 0.000621532 | PLA2G7 |
| chr2 | 25332694 | 25332862 | 169 | 2 | 1.50E-05 | 0.000845 | 8.73E-06 | 8.68E-06 | -0.005683622 | DNMT3A |
| chr16 | 88633880 | 88635164 | 1285 | 6 | 1.51E-05 | 3.09E-07 | 0.00213 | 9.12E-07 | -0.005965172 | NA |
| chr16 | 83952777 | 83953860 | 1084 | 7 | 1.51E-05 | 1.64E-06 | 0.00026 | 4.84E-07 | -0.015997739 | OSGIN1 |
| chr2 | 102250419 | 102251225 | 807 | 5 | 1.51E-05 | 0.000121 | 0.00233 | 1.51E-05 | -0.011099904 | NA |
| chr11 | 1469508 | 1470202 | 695 | 4 | 1.51E-05 | 4.81E-07 | 0.00196 | 1.76E-06 | -0.000199117 | MOB2 |
| chr15 | 63535679 | 63535983 | 305 | 2 | 1.51E-05 | 9.22E-07 | 0.00021 | 1.64E-06 | -0.0069805 | USP3 |
| chr10 | 117348518 | 117348575 | 58 | 2 | 1.51E-05 | 2.33E-06 | 4.53E-05 | 2.24E-06 | -0.006941799 | PDZD8 |
| chr1 | 2505444 | 2506219 | 776 | 5 | 1.51E-05 | 1.21E-05 | 0.00159 | 8.04E-06 | -0.006678097 | PLCH2 |
| chr3 | 46744467 | 46745164 | 698 | 5 | 1.52E-05 | 0.005497 | 0.00037 | 2.61E-05 | -0.005597782 | AC109583.1, PRSS45P, AC109583.4 |
| chr3 | 45333470 | 45333676 | 207 | 2 | 1.52E-05 | 1.27E-06 | 0.00012 | 1.95E-06 | -0.014488262 | NA |
| chr6 | 4610462 | 4610946 | 485 | 4 | 1.53E-05 | 1.03E-05 | 0.00029 | 6.54E-06 | -0.010897942 | KU-MEL-3 |
| chr10 | 126598229 | 126598697 | 469 | 3 | 1.53E-05 | 1.81E-06 | 0.00094 | 4.39E-06 | -0.013155512 | C10orf90 |
| chr3 | 123296596 | 123296855 | 260 | 2 | 1.53E-05 | 2.58E-06 | 5.36E-05 | 2.63E-06 | -0.008471432 | ADCY5 |
| chr16 | 81364170 | 81365290 | 1121 | 4 | 1.53E-05 | 2.99E-05 | 0.00034 | 1.41E-06 | -0.00353579 | GAN |
| chr11 | 65606792 | 65607732 | 941 | 10 | 1.53E-05 | 4.14E-06 | 0.00069 | 4.30E-06 | -0.00213546 | MAP3K11 |
| chr7 | 98619540 | 98619791 | 252 | 5 | 1.53E-05 | 1.68E-06 | 0.00118 | 1.95E-06 | -0.011384671 | NPTX2 |
| chr20 | 35280058 | 35280724 | 667 | 3 | 1.53E-05 | 2.67E-06 | 0.00074 | 4.46E-06 | -0.007102514 | EIF6 |
| chr8 | 143290672 | 143291766 | 1095 | 14 | 1.54E-05 | 4.05E-05 | 0.00094 | 1.55E-06 | -0.001373779 | ZNF696 |
| chr14 | 95620447 | 95620868 | 422 | 4 | 1.54E-05 | 1.27E-06 | 0.0015 | 3.04E-06 | -0.013867139 | NA |
| chr16 | 85634243 | 85635194 | 952 | 2 | 1.54E-05 | 8.60E-07 | 0.00011 | 1.37E-06 | -0.011479134 | GSE1 |
| chr1 | 25922063 | 25922894 | 832 | 5 | 1.54E-05 | 6.15E-06 | 0.00066 | 8.24E-06 | -0.006094772 | NA |
| chr1 | 110675457 | 110675932 | 476 | 5 | 1.54E-05 | 1.37E-06 | 0.00329 | 2.01E-06 | -0.017385082 | NA |
| chr2 | 106342749 | 106343069 | 321 | 5 | 1.54E-05 | 0.000305 | 5.30E-05 | 7.42E-06 | -0.002403044 | NA |
| chr7 | 100093504 | 100094319 | 816 | 14 | 1.55E-05 | 4.51E-05 | 0.0033 | 4.23E-06 | -0.003008647 | MCM7, MIR25, MIR93, MIR106B |
| chr5 | 1911987 | 1912053 | 67 | 2 | 1.56E-05 | 8.88E-06 | 1.65E-05 | 2.94E-06 | -0.007211747 | NA |
| chr3 | 123066711 | 123067244 | 534 | 10 | 1.56E-05 | 0.000103 | 0.00095 | 4.17E-06 | -0.004006848 | PDIA5 |
| chr2 | 43170979 | 43171200 | 222 | 4 | 1.56E-05 | 2.20E-05 | 0.00014 | 4.66E-06 | -0.009696768 | NA |
| chr17 | 38554542 | 38555062 | 521 | 2 | 1.56E-05 | 3.61E-06 | 0.0002 | 5.45E-06 | -0.014022699 | SRCIN1 |
| chr13 | 87677945 | 87678359 | 415 | 3 | 1.56E-05 | 2.33E-05 | 0.00021 | 6.70E-06 | 0.004404011 | SLITRK5 |
| chr17 | 45277371 | 45277722 | 352 | 2 | 1.56E-05 | 0.012355 | 1.36E-05 | 1.83E-05 | -0.020616285 | MAP3K14 |
| chr20 | 47228215 | 47228518 | 304 | 2 | 1.56E-05 | 3.51E-06 | 4.79E-05 | 3.12E-06 | -0.004490881 | ZMYND8 |
| chr4 | 948554 | 948917 | 364 | 4 | 1.57E-05 | 9.53E-05 | 0.00083 | 2.62E-05 | -0.00492111 | TMEM175 |
| chr8 | 95267789 | 95268967 | 1179 | 7 | 1.57E-05 | 0.000113 | 0.00131 | 2.15E-05 | -0.002885262 | C8orf37-AS1, C8orf37 |
| chr5 | 135190434 | 135191062 | 629 | 4 | 1.57E-05 | 6.75E-05 | 0.00034 | 1.03E-05 | -0.011269341 | C5orf66 |
| chr12 | 124845183 | 124845345 | 163 | 4 | 1.57E-05 | 3.19E-05 | 0.0007 | 2.41E-06 | -0.003626941 | SCARB1 |
| chr1 | 152982604 | 152983710 | 1107 | 4 | 1.57E-05 | 2.36E-05 | 0.00016 | 1.67E-05 | -0.012727569 | NA |
| chr2 | 10084074 | 10084625 | 552 | 3 | 1.57E-05 | 5.69E-05 | 0.00051 | 1.63E-05 | -0.014057924 | AC104794.2 |
| chr2 | 176763349 | 176764024 | 676 | 3 | 1.57E-05 | 0.000266 | 8.72E-05 | 2.32E-05 | -0.00383686 | LINC01117, AC092162.2 |
| chr17 | 78375325 | 78375798 | 474 | 3 | 1.57E-05 | 9.03E-06 | 7.65E-05 | 5.96E-06 | -0.005964291 | NA |
| chr15 | 89655948 | 89656887 | 940 | 4 | 1.58E-05 | 9.47E-06 | 0.00072 | 3.96E-06 | -0.009216312 | NA |
| chr16 | 88337708 | 88337811 | 104 | 3 | 1.58E-05 | 2.22E-06 | 0.00064 | 4.63E-06 | 0.002908144 | NA |
| chr5 | 1950098 | 1950728 | 631 | 6 | 1.58E-05 | 6.24E-06 | 0.0009 | 2.08E-06 | -0.001654663 | AC124852.1 |
| chr14 | 24130316 | 24132599 | 2284 | 14 | 1.58E-05 | 2.27E-07 | 0.00229 | 1.12E-08 | -0.005994691 | AL136295.3, FITM1 |
| chr12 | 121648617 | 121649452 | 836 | 2 | 1.58E-05 | 2.74E-07 | 6.31E-05 | 4.48E-07 | -0.009416023 | MORN3 |
| chr8 | 22747380 | 22748320 | 941 | 5 | 1.58E-05 | 0.000159 | 0.00031 | 1.87E-05 | -0.005609111 | AC055854.1, PEBP4 |
| chr17 | 55283353 | 55283697 | 345 | 2 | 1.58E-05 | 7.32E-05 | 2.56E-05 | 1.29E-05 | -0.005691481 | HLF |
| chr3 | 46677237 | 46678070 | 834 | 5 | 1.59E-05 | 4.06E-06 | 0.00046 | 1.79E-06 | -0.006975699 | ALS2CL |
| chr14 | 85530151 | 85530529 | 379 | 4 | 1.59E-05 | 0.001198 | 8.76E-05 | 0.00011 | -0.002752939 | FLRT2, AL049775.1 |
| chr11 | 86672388 | 86672898 | 511 | 11 | 1.60E-05 | 1.74E-05 | 0.01076 | 1.27E-05 | -0.005141648 | ME3 |
| chr13 | 113001056 | 113002110 | 1055 | 15 | 1.60E-05 | 4.47E-06 | 0.00763 | 2.02E-06 | -0.006454289 | MCF2L |
| chr12 | 57627512 | 57628246 | 735 | 6 | 1.60E-05 | 1.13E-06 | 0.00494 | 2.13E-06 | -0.004512791 | B4GALNT1 |
| chr3 | 9946460 | 9947871 | 1412 | 8 | 1.61E-05 | 4.06E-07 | 0.01409 | 2.80E-06 | -0.0119108 | PRRT3-AS1, PRRT3 |
| chr16 | 8642363 | 8642576 | 214 | 4 | 1.61E-05 | 1.36E-05 | 5.05E-05 | 4.42E-06 | -0.007968048 | METTL22 |
| chr12 | 6594559 | 6594945 | 387 | 2 | 1.61E-05 | 1.94E-06 | 9.36E-05 | 2.63E-06 | -0.002957283 | AC006064.6, CHD4 |
| chr7 | 101822425 | 101822479 | 55 | 2 | 1.61E-05 | 2.50E-05 | 1.60E-05 | 5.21E-06 | -0.005915292 | CUX1, AC005096.1 |
| chr10 | 125124071 | 125124418 | 348 | 3 | 1.61E-05 | 2.15E-05 | 0.00012 | 7.16E-06 | -0.006264994 | CTBP2 |
| chr22 | 23838305 | 23839478 | 1174 | 11 | 1.61E-05 | 3.07E-06 | 0.0003 | 4.41E-06 | 0.000490251 | DERL3 |
| chr5 | 15500161 | 15500724 | 564 | 3 | 1.61E-05 | 0.000573 | 5.01E-05 | 2.44E-05 | 0.001911406 | FBXL7 |
| chr16 | 87554953 | 87555713 | 761 | 4 | 1.62E-05 | 1.53E-06 | 0.00152 | 2.74E-06 | -0.012017937 | NA |
| chr17 | 6712329 | 6712527 | 199 | 2 | 1.62E-05 | 1.38E-05 | 0.0003 | 1.83E-05 | -0.007355651 | SLC13A5 |
| chr12 | 113149482 | 113149885 | 404 | 6 | 1.63E-05 | 8.92E-06 | 0.00144 | 2.77E-06 | -0.008242726 | CFAP73 |
| chr1 | 93179756 | 93180613 | 858 | 15 | 1.63E-05 | 0.00157 | 0.00054 | 7.75E-05 | 0.000923321 | CCDC18, TMED5 |
| chr5 | 55159736 | 55160228 | 493 | 7 | 1.63E-05 | 5.54E-06 | 0.0012 | 4.36E-06 | -0.010322708 | GPX8, CDC20B |
| chr15 | 41543840 | 41545202 | 1363 | 13 | 1.63E-05 | 8.38E-05 | 0.00281 | 1.19E-05 | -0.002439362 | RPAP1 |
| chr4 | 4404117 | 4404473 | 357 | 6 | 1.64E-05 | 0.000654 | 4.22E-05 | 1.25E-05 | -0.003615368 | NSG1 |
| chr5 | 180389678 | 180390084 | 407 | 5 | 1.64E-05 | 1.80E-05 | 5.03E-05 | 7.29E-06 | -0.009977325 | NA |
| chr16 | 67999956 | 68000529 | 574 | 5 | 1.64E-05 | 2.65E-05 | 0.0015 | 5.53E-06 | -0.008191081 | DUS2, DPEP2 |
| chr17 | 38885714 | 38885948 | 235 | 2 | 1.64E-05 | 1.06E-05 | 6.94E-05 | 8.45E-06 | -0.006332812 | LASP1 |
| chr9 | 131265734 | 131266861 | 1128 | 3 | 1.64E-05 | 7.60E-09 | 7.41E-05 | 1.94E-08 | -0.011568292 | FAM78A |
| chr11 | 1017797 | 1017827 | 31 | 2 | 1.64E-05 | 1.44E-05 | 2.95E-05 | 6.05E-06 | -0.006045788 | MUC6 |
| chr7 | 42911260 | 42911586 | 327 | 3 | 1.64E-05 | 0.000166 | 9.25E-05 | 7.21E-06 | -0.00596411 | C7orf25, AC010132.3 |
| chr9 | 137293956 | 137294573 | 618 | 2 | 1.64E-05 | 5.24E-07 | 0.00014 | 9.24E-07 | -0.012391638 | BX255925.1 |
| chr16 | 67237069 | 67237519 | 451 | 4 | 1.64E-05 | 0.000176 | 6.35E-05 | 9.03E-06 | -0.003088426 | FHOD1 |
| chr12 | 132867775 | 132868186 | 412 | 2 | 1.64E-05 | 0.000239 | 1.89E-05 | 1.52E-05 | -0.007557515 | CHFR, AC127070.4 |
| chr14 | 74552177 | 74552478 | 302 | 4 | 1.64E-05 | 1.13E-05 | 0.00041 | 3.91E-06 | -0.003481721 | AC013451.1, LTBP2 |
| chr2 | 85601097 | 85602015 | 919 | 4 | 1.65E-05 | 5.25E-07 | 0.0026 | 2.08E-06 | -0.007524944 | TMEM150A |
| chr2 | 130809049 | 130809154 | 106 | 2 | 1.65E-05 | 2.63E-06 | 5.94E-05 | 2.80E-06 | -0.01007594 | NA |
| chr20 | 37525077 | 37525986 | 910 | 7 | 1.65E-05 | 0.000444 | 0.0004 | 2.60E-05 | -0.004745462 | BLCAP |
| chr8 | 143273712 | 143274846 | 1135 | 5 | 1.65E-05 | 2.31E-06 | 0.00076 | 1.40E-06 | -0.007105691 | AC138696.1, GLI4 |
| chr10 | 27153888 | 27154475 | 588 | 9 | 1.65E-05 | 1.00E-05 | 0.00238 | 6.54E-06 | 0.001283597 | YME1L1 |
| chr8 | 104466455 | 104466987 | 533 | 3 | 1.66E-05 | 0.009293 | 0.00025 | 0.00035 | -0.005053393 | DPYS |
| chr1 | 114002193 | 114002293 | 101 | 2 | 1.66E-05 | 7.98E-06 | 4.14E-05 | 5.12E-06 | -0.002454202 | OLFML3, AL591742.1 |
| chr7 | 2616790 | 2617713 | 924 | 3 | 1.66E-05 | 2.48E-07 | 0.00019 | 5.63E-07 | -0.008657525 | NA |
| chr22 | 37559920 | 37560422 | 503 | 4 | 1.66E-05 | 1.71E-05 | 0.00011 | 5.83E-06 | -0.010822348 | NA |
| chr7 | 100469424 | 100469906 | 483 | 2 | 1.66E-05 | 2.71E-05 | 3.10E-05 | 9.12E-06 | -0.007375886 | TSC22D4 |
| chr1 | 228276348 | 228277095 | 748 | 5 | 1.66E-05 | 2.80E-06 | 0.00281 | 4.14E-06 | -0.023893077 | OBSCN |
| chr22 | 46509049 | 46509441 | 393 | 3 | 1.66E-05 | 2.46E-06 | 0.00011 | 2.99E-06 | -0.004352455 | CELSR1 |
| chr11 | 73323414 | 73323496 | 83 | 2 | 1.66E-05 | 2.26E-06 | 5.71E-05 | 2.45E-06 | -0.014185096 | ARHGEF17 |
| chr18 | 60010579 | 60011280 | 702 | 4 | 1.67E-05 | 0.001578 | 2.40E-05 | 1.50E-05 | -0.000720548 | AC090377.1, ENTR1P1 |
| chr17 | 42520065 | 42520582 | 518 | 2 | 1.67E-05 | 2.88E-05 | 0.00047 | 3.65E-05 | -0.011803967 | ATP6V0A1 |
| chr4 | 2135127 | 2135787 | 661 | 4 | 1.67E-05 | 8.05E-07 | 0.00141 | 2.13E-06 | -0.008762567 | POLN, HAUS3 |
| chr11 | 126366739 | 126367126 | 388 | 3 | 1.67E-05 | 6.91E-06 | 0.00039 | 5.30E-06 | -0.013847174 | ST3GAL4 |
| chr3 | 49939673 | 49940555 | 883 | 15 | 1.67E-05 | 0.000232 | 0.00029 | 8.53E-06 | 7.84E-05 | RBM6 |
| chr4 | 153452361 | 153452684 | 324 | 2 | 1.68E-05 | 1.33E-06 | 0.00015 | 2.14E-06 | -0.032883595 | NA |
| chr15 | 67522467 | 67522542 | 76 | 2 | 1.68E-05 | 0.00423 | 2.75E-05 | 3.72E-05 | -0.011514337 | C15orf61 |
| chr17 | 8177244 | 8177335 | 92 | 3 | 1.68E-05 | 0.000364 | 3.84E-05 | 3.15E-05 | -0.002923367 | AC129492.4 |
| chr17 | 48156352 | 48157163 | 812 | 6 | 1.68E-05 | 3.66E-07 | 0.0036 | 1.69E-06 | -0.010744655 | SKAP1, MIR1203 |
| chr11 | 134234419 | 134234988 | 570 | 6 | 1.68E-05 | 0.003693 | 2.16E-05 | 1.72E-05 | -0.000764756 | VPS26B |
| chr11 | 108221670 | 108222953 | 1284 | 36 | 1.68E-05 | 3.58E-05 | 0.0009 | 2.17E-06 | 4.42E-05 | ATM, NPAT |
| chr9 | 83956989 | 83957351 | 363 | 5 | 1.68E-05 | 0.03501 | 8.10E-06 | 4.71E-05 | -0.002324382 | NA |
| chr18 | 39454030 | 39454755 | 726 | 2 | 1.68E-05 | 5.77E-07 | 8.76E-05 | 9.22E-07 | 0.001698312 | MIR924HG |
| chr8 | 143613523 | 143614186 | 664 | 5 | 1.68E-05 | 1.87E-05 | 0.00578 | 4.02E-05 | -0.004976407 | GFUS |
| chr2 | 96933542 | 96934222 | 681 | 3 | 1.68E-05 | 1.44E-05 | 3.81E-05 | 4.39E-06 | -0.002341909 | FAM178B |
| chr10 | 132776335 | 132776940 | 606 | 5 | 1.69E-05 | 2.50E-05 | 0.00014 | 9.36E-06 | -0.001637621 | INPP5A |
| chr16 | 89248274 | 89248912 | 639 | 5 | 1.69E-05 | 0.00157 | 3.24E-05 | 1.91E-05 | 0.001953971 | NA |
| chr17 | 4484690 | 4485184 | 495 | 4 | 1.69E-05 | 0.000533 | 0.00044 | 1.03E-05 | -0.00475463 | SPNS3, AC127521.1 |
| chr22 | 37496896 | 37497403 | 508 | 2 | 1.69E-05 | 2.28E-06 | 9.41E-05 | 3.03E-06 | -0.004749346 | CARD10 |
| chr6 | 13395793 | 13395959 | 167 | 3 | 1.70E-05 | 0.000115 | 5.93E-05 | 4.29E-06 | -0.009622594 | GFOD1 |
| chr1 | 161177188 | 161178569 | 1382 | 17 | 1.70E-05 | 1.56E-07 | 0.00946 | 5.30E-07 | -0.001803433 | PPOX, B4GALT3 |
| chr12 | 12716845 | 12718602 | 1758 | 19 | 1.70E-05 | 0.001154 | 4.70E-05 | 7.00E-06 | 0.001905371 | CDKN1B |
| chr1 | 1302341 | 1303879 | 1539 | 7 | 1.71E-05 | 3.42E-06 | 0.00031 | 1.39E-07 | -0.005361116 | ACAP3 |
| chr8 | 132963836 | 132964468 | 633 | 3 | 1.71E-05 | 6.25E-05 | 0.0002 | 4.35E-05 | -0.00528494 | TG |
| chr4 | 3121241 | 3122440 | 1200 | 5 | 1.72E-05 | 5.61E-09 | 0.00179 | 4.95E-08 | -0.010452112 | HTT |
| chr15 | 88905039 | 88905618 | 580 | 2 | 1.72E-05 | 8.95E-07 | 0.0002 | 1.58E-06 | -0.007168802 | MFGE8 |
| chr19 | 11338481 | 11338488 | 8 | 2 | 1.73E-05 | 1.32E-05 | 6.19E-05 | 9.20E-06 | -0.011156761 | RAB3D |
| chr22 | 46103076 | 46103216 | 141 | 2 | 1.73E-05 | 0.000308 | 2.65E-05 | 2.22E-05 | -0.006404428 | MIRLET7BHG |
| chr7 | 7968309 | 7969318 | 1010 | 11 | 1.73E-05 | 2.96E-05 | 0.00167 | 4.29E-06 | 0.000252338 | AC007161.3, GLCCI1, GLCCI1-DT |
| chr11 | 64856282 | 64856464 | 183 | 2 | 1.73E-05 | 4.60E-06 | 0.00039 | 7.49E-06 | -0.012049259 | EHD1 |
| chr16 | 48421805 | 48421916 | 112 | 2 | 1.73E-05 | 2.14E-05 | 5.38E-05 | 1.17E-05 | -0.005553633 | SIAH1 |
| chr1 | 15799719 | 15800235 | 517 | 3 | 1.73E-05 | 0.001793 | 6.76E-05 | 1.86E-05 | -0.004567317 | NA |
| chr18 | 31937266 | 31937268 | 3 | 2 | 1.73E-05 | 4.06E-06 | 2.68E-05 | 2.50E-06 | -0.00114963 | TRAPPC8 |
| chr11 | 65427462 | 65427568 | 107 | 4 | 1.74E-05 | 7.79E-07 | 0.00146 | 2.72E-06 | -0.015763377 | NEAT1 |
| chr9 | 135817898 | 135818481 | 584 | 4 | 1.74E-05 | 0.073763 | 0.00013 | 0.00157 | -0.003522201 | CAMSAP1 |
| chr19 | 751098 | 752097 | 1000 | 6 | 1.75E-05 | 0.000179 | 0.0012 | 8.21E-06 | -0.007264452 | MISP |

| | | | | | | | | | | |
|---|---|---|---|---|---|---|---|---|---|---|
| chr6 | 33193330 | 33194072 | 743 | 13 | 1.75E-05 | 8.00E-05 | 0.00037 | 2.63E-05 | -0.005457871 | RXRB |
| chr9 | 12814398 | 12814644 | 247 | 3 | 1.75E-05 | 0.000175 | 7.87E-06 | 6.78E-06 | -0.007657674 | LURAP1L |
| chr15 | 74433321 | 74434565 | 1245 | 17 | 1.75E-05 | 8.70E-06 | 0.00177 | 1.11E-06 | -0.001287576 | SEMA7A |
| chr11 | 93624919 | 93625111 | 193 | 2 | 1.75E-05 | 1.11E-06 | 0.00023 | 1.95E-06 | -0.018406978 | NA |
| chr15 | 51971400 | 51972561 | 1162 | 12 | 1.75E-05 | 1.64E-06 | 0.00191 | 1.11E-06 | -0.001110559 | MAPK6, LEO1 |
| chr18 | 23137447 | 23138337 | 891 | 5 | 1.75E-05 | 6.10E-05 | 0.00022 | 6.33E-06 | -0.008626118 | CABLES1 |
| chr20 | 43238523 | 43238953 | 431 | 3 | 1.75E-05 | 6.91E-07 | 0.00089 | 1.82E-06 | -0.019313343 | NA |
| chr14 | 50092192 | 50093184 | 993 | 4 | 1.76E-05 | 5.41E-05 | 8.85E-05 | 5.03E-06 | -0.003834713 | LINC01588 |
| chr3 | 47009313 | 47010056 | 744 | 5 | 1.76E-05 | 1.26E-06 | 0.00171 | 1.02E-06 | -0.014179138 | NBEAL2 |
| chr19 | 33509153 | 33509171 | 19 | 2 | 1.76E-05 | 1.59E-06 | 0.00026 | 2.73E-06 | -0.032782383 | PEPD |
| chr8 | 73419433 | 73419877 | 445 | 3 | 1.76E-05 | 0.00011 | 0.00024 | 6.66E-05 | -0.006332274 | NA |
| chr21 | 43024605 | 43024723 | 119 | 3 | 1.77E-05 | 8.58E-06 | 3.75E-05 | 3.85E-06 | -0.002996169 | PKNOX1 |
| chr12 | 107580956 | 107581522 | 567 | 3 | 1.77E-05 | 0.000262 | 0.00179 | 0.00031 | -0.007882449 | BTBD11 |
| chr12 | 129337643 | 129337714 | 72 | 2 | 1.77E-05 | 2.71E-05 | 1.04E-05 | 3.72E-06 | -0.00336457 | TMEM132D |
| chr13 | 112592963 | 112592994 | 32 | 2 | 1.77E-05 | 1.24E-06 | 0.00015 | 2.02E-06 | -0.010261319 | NA |
| chr15 | 51737804 | 51738549 | 746 | 9 | 1.78E-05 | 2.29E-05 | 0.00264 | 4.07E-06 | -0.002804371 | LYSMD2 |
| chr10 | 118046949 | 118047461 | 513 | 6 | 1.78E-05 | 0.000153 | 0.00018 | 1.02E-05 | -0.00443465 | CASC2 |
| chr4 | 88706740 | 88706990 | 251 | 2 | 1.78E-05 | 4.06E-06 | 4.29E-05 | 3.28E-06 | -0.007202534 | AC098582.1, HERC3 |
| chr8 | 143126895 | 143127268 | 374 | 4 | 1.78E-05 | 8.84E-05 | 1.50E-05 | 5.90E-06 | -0.001587891 | NA |
| chr2 | 5697797 | 5698126 | 330 | 3 | 1.78E-05 | 1.90E-05 | 0.00035 | 8.28E-06 | 0.015855616 | SOX11, AC010729.2 |
| chr7 | 44847679 | 44848359 | 681 | 10 | 1.78E-05 | 0.000346 | 0.00236 | 2.85E-05 | -0.000160067 | H2AZ2 |
| chr6 | 149094400 | 149094467 | 68 | 2 | 1.78E-05 | 3.11E-06 | 0.00014 | 4.41E-06 | -0.011385551 | NA |
| chr19 | 45123775 | 45124275 | 501 | 2 | 1.78E-05 | 1.51E-05 | 4.11E-05 | 7.76E-06 | -0.009299827 | MARK4, PPP1R37 |
| chr20 | 48650636 | 48651025 | 390 | 3 | 1.78E-05 | 0.004256 | 1.08E-05 | 2.35E-05 | -0.002527637 | PREX1 |
| chr13 | 95301687 | 95302280 | 594 | 4 | 1.79E-05 | 6.13E-05 | 0.00091 | 2.43E-05 | -0.021570662 | NA |
| chr16 | 35573489 | 35574108 | 620 | 2 | 1.79E-05 | 1.06E-05 | 9.69E-05 | 1.00E-05 | -0.008178528 | NA |
| chr7 | 114286864 | 114287474 | 611 | 2 | 1.79E-05 | 8.38E-07 | 0.00013 | 1.39E-06 | -0.000675926 | FOXP2 |
| chr8 | 141174880 | 141175348 | 469 | 4 | 1.79E-05 | 0.003018 | 1.78E-05 | 2.69E-05 | -0.001856107 | DENND3 |
| chr19 | 49580322 | 49580660 | 339 | 7 | 1.79E-05 | 2.64E-05 | 0.00119 | 7.80E-06 | -0.00134286 | PRRG2, NOSIP |
| chr19 | 17821123 | 17821607 | 485 | 8 | 1.79E-05 | 1.53E-06 | 0.00434 | 3.89E-06 | -0.009430174 | INSL3 |
| chr16 | 28837377 | 28838407 | 1031 | 4 | 1.79E-05 | 4.69E-07 | 0.00053 | 1.28E-06 | -0.004540897 | AC009093.11 |
| chr12 | 52192851 | 52193325 | 475 | 2 | 1.80E-05 | 1.24E-05 | 0.00085 | 1.99E-05 | -0.004862784 | LINC00592 |
| chr2 | 172455165 | 172455369 | 205 | 3 | 1.80E-05 | 6.21E-07 | 0.00092 | 1.71E-06 | -0.017131245 | ITGA6 |
| chr2 | 177392240 | 177394726 | 2487 | 16 | 1.80E-05 | 2.21E-06 | 0.00011 | 3.93E-08 | -0.00232363 | AGPS, NFE2L2, AC019080.1 |
| chr17 | 10357995 | 10358238 | 244 | 2 | 1.80E-05 | 5.35E-06 | 3.50E-05 | 3.53E-06 | -0.008289765 | MYH13 |
| chr18 | 44679279 | 44680169 | 891 | 11 | 1.81E-05 | 0.009244 | 7.78E-05 | 0.00012 | -0.001449135 | SETBP1-DT |
| chr5 | 663524 | 663780 | 257 | 3 | 1.82E-05 | 0.000722 | 1.52E-05 | 2.01E-05 | -0.002809277 | CEP72, TPPP |
| chr17 | 50362010 | 50362329 | 320 | 2 | 1.82E-05 | 9.16E-06 | 7.26E-05 | 7.80E-06 | -0.012252809 | XYLT2 |
| chr17 | 8019152 | 8019856 | 705 | 3 | 1.82E-05 | 2.08E-07 | 0.00061 | 6.35E-07 | -0.010364531 | GUCY2D |
| chr1 | 245895837 | 245896313 | 477 | 2 | 1.82E-05 | 6.45E-06 | 0.00015 | 8.14E-06 | -0.01123673 | SMYD3 |
| chr1 | 17247683 | 17247835 | 153 | 2 | 1.82E-05 | 0.002198 | 1.28E-05 | 1.51E-05 | -0.004259691 | NA |
| chr12 | 107685607 | 107686173 | 567 | 11 | 1.82E-05 | 0.000373 | 0.00548 | 2.31E-05 | 0.000290709 | PWP1 |
| chr16 | 2238664 | 2239309 | 646 | 4 | 1.83E-05 | 0.004201 | 3.22E-05 | 1.39E-05 | -0.004241463 | DNASE1L2 |
| chr6 | 12289630 | 12290546 | 917 | 12 | 1.83E-05 | 6.26E-05 | 0.00315 | 8.59E-06 | 0.001782961 | EDN1 |
| chr9 | 131259061 | 131259621 | 561 | 3 | 1.83E-05 | 3.76E-06 | 0.00044 | 3.85E-06 | -0.007951834 | FAM78A |
| chr22 | 45865335 | 45866556 | 1222 | 3 | 1.83E-05 | 7.19E-06 | 4.47E-05 | 1.22E-06 | -0.005833043 | NA |
| chr1 | 2911479 | 2911728 | 250 | 5 | 1.83E-05 | 7.79E-06 | 0.0003 | 2.27E-06 | -0.004058353 | NA |
| chr20 | 56711261 | 56711276 | 16 | 2 | 1.83E-05 | 2.43E-06 | 0.0001 | 3.29E-06 | -0.023509785 | NA |
| chr16 | 1435400 | 1436535 | 1136 | 6 | 1.84E-05 | 2.79E-05 | 4.63E-05 | 7.72E-06 | -0.002239635 | CCDC154 |
| chr15 | 41001333 | 41001656 | 324 | 2 | 1.84E-05 | 0.000174 | 1.36E-05 | 1.02E-05 | -0.004298687 | AC020661.3, INO80 |
| chr20 | 18141176 | 18142250 | 1075 | 7 | 1.84E-05 | 1.49E-07 | 0.00047 | 1.73E-07 | -0.003356338 | KAT14, PET117 |
| chr8 | 17664028 | 17664443 | 416 | 3 | 1.84E-05 | 0.021327 | 1.09E-05 | 3.09E-05 | -0.002474569 | MTUS1 |
| chr16 | 2087579 | 2089113 | 1535 | 11 | 1.84E-05 | 0.000114 | 0.00056 | 2.63E-06 | -0.00250978 | TSC2, PKD1 |
| chr6 | 34277491 | 34277564 | 74 | 2 | 1.85E-05 | 6.78E-06 | 6.64E-05 | 6.00E-06 | -0.011475083 | AL354740.1 |
| chr3 | 137509389 | 137509794 | 406 | 3 | 1.85E-05 | 3.45E-05 | 8.46E-05 | 5.51E-06 | -0.023611229 | NA |
| chr9 | 38023008 | 38023881 | 874 | 5 | 1.86E-05 | 5.46E-07 | 0.0026 | 2.45E-06 | -0.006068955 | AL138752.2, SHB |
| chr19 | 11505033 | 11505217 | 185 | 2 | 1.86E-05 | 0.000196 | 5.25E-05 | 3.56E-05 | 0.003297094 | AC008481.3, ZNF653 |
| chr1 | 65174851 | 65175251 | 401 | 2 | 1.86E-05 | 0.000747 | 2.57E-05 | 2.64E-05 | -0.002295824 | AK4 |
| chr6 | 165776528 | 165776646 | 119 | 3 | 1.87E-05 | 1.06E-06 | 0.00056 | 2.22E-06 | -0.010411064 | PDE10A |
| chr12 | 95790802 | 95791862 | 1061 | 10 | 1.87E-05 | 7.23E-05 | 0.0006 | 1.19E-05 | 0.002645157 | NTN4, AC090001.1 |
| chr12 | 21819141 | 21819531 | 391 | 3 | 1.88E-05 | 3.60E-05 | 4.35E-05 | 3.65E-06 | -0.013307063 | ABCC9 |
| chr12 | 43834803 | 43835908 | 1106 | 5 | 1.88E-05 | 0.000148 | 0.00018 | 2.17E-05 | 0.000790642 | NA |
| chr6 | 43075372 | 43076093 | 722 | 7 | 1.88E-05 | 0.000236 | 6.75E-05 | 2.02E-05 | -0.003322255 | NA |
| chr15 | 73364907 | 73365460 | 554 | 3 | 1.88E-05 | 1.72E-05 | 4.85E-05 | 7.51E-06 | -0.010339522 | HCN4 |
| chr5 | 129905717 | 129906607 | 891 | 9 | 1.88E-05 | 0.00012 | 0.00147 | 4.51E-06 | -0.007267081 | CHSY3, AC008591.1 |
| chr8 | 17647309 | 17647737 | 429 | 2 | 1.88E-05 | 3.34E-06 | 9.40E-05 | 4.12E-06 | -0.000269427 | MTUS1 |
| chr16 | 75238408 | 75239130 | 723 | 4 | 1.88E-05 | 3.83E-05 | 0.0007 | 1.64E-05 | -0.00754982 | BCAR1 |
| chr10 | 49418592 | 49419308 | 717 | 6 | 1.90E-05 | 2.12E-07 | 0.00359 | 9.05E-07 | -0.006562673 | AL138760.1 |
| chr22 | 24514219 | 24515465 | 1247 | 6 | 1.90E-05 | 8.88E-06 | 0.00043 | 1.57E-06 | -0.004363379 | UPB1 |
| chr12 | 53719678 | 53719800 | 123 | 2 | 1.91E-05 | 2.22E-06 | 6.34E-05 | 2.53E-06 | -0.004311797 | CALCOCO1 |
| chr2 | 27122980 | 27123897 | 918 | 11 | 1.91E-05 | 0.000419 | 0.00129 | 1.95E-05 | -0.00174945 | ABHD1 |
| chr5 | 121851650 | 121852533 | 884 | 10 | 1.91E-05 | 0.000211 | 0.00089 | 2.61E-06 | -0.002114773 | FTMT |
| chr5 | 141100616 | 141101683 | 1068 | 10 | 1.91E-05 | 0.000173 | 0.00244 | 7.19E-06 | 0.014338348 | PCDHB3, AC244517.2 |
| chr12 | 92540576 | 92540785 | 210 | 2 | 1.91E-05 | 2.04E-06 | 8.71E-05 | 2.70E-06 | -0.013541694 | NA |
| chr7 | 5073011 | 5073408 | 398 | 3 | 1.91E-05 | 1.00E-05 | 0.00069 | 1.72E-05 | -0.017529074 | RBAK-RBAKDN, RBAKDN |
| chr16 | 83807435 | 83808652 | 1218 | 15 | 1.91E-05 | 1.15E-06 | 0.00227 | 2.97E-06 | -0.000693699 | HSBP1, AC009119.1 |
| chr11 | 9530196 | 9530557 | 362 | 3 | 1.92E-05 | 7.20E-06 | 0.00014 | 7.66E-06 | -0.006213967 | NA |
| chr10 | 3785008 | 3786142 | 1135 | 13 | 1.92E-05 | 0.001048 | 0.00203 | 1.76E-05 | 0.000125554 | KLF6 |
| chr17 | 72819097 | 72819581 | 485 | 2 | 1.92E-05 | 0.000132 | 2.51E-05 | 1.62E-05 | -0.00676729 | SLC39A11 |
| chr17 | 64975474 | 64976240 | 767 | 9 | 1.92E-05 | 4.69E-06 | 0.00153 | 3.91E-06 | 0.001816775 | AMZ2P1 |
| chr1 | 22600653 | 22601636 | 984 | 5 | 1.92E-05 | 2.58E-06 | 0.00068 | 2.63E-06 | -0.005670328 | EPHA8 |
| chr16 | 89078081 | 89078403 | 323 | 3 | 1.92E-05 | 1.20E-05 | 0.00023 | 9.26E-06 | -0.004891539 | NA |
| chr17 | 40797033 | 40797260 | 228 | 2 | 1.93E-05 | 2.30E-06 | 0.00018 | 3.64E-06 | -0.022783109 | KRT28 |
| chr16 | 27516395 | 27516457 | 63 | 2 | 1.93E-05 | 6.21E-06 | 2.57E-05 | 3.19E-06 | -0.003549622 | GTF3C1 |
| chr6 | 16064692 | 16064913 | 222 | 3 | 1.93E-05 | 3.74E-06 | 0.00015 | 2.80E-06 | -0.007133395 | NA |
| chr8 | 142291787 | 142292374 | 588 | 5 | 1.93E-05 | 3.59E-05 | 0.00105 | 1.52E-05 | -0.003159087 | TSNARE1 |
| chr1 | 218344975 | 218345788 | 814 | 12 | 1.93E-05 | 0.000265 | 0.00056 | 1.78E-05 | -0.002467586 | TGFB2, TGFB2-AS1 |
| chr4 | 150579040 | 150580587 | 1548 | 8 | 1.93E-05 | 6.83E-06 | 0.00015 | 1.35E-07 | -0.002211951 | AC110813.1, LRBA |
| chr12 | 119176039 | 119176565 | 527 | 5 | 1.94E-05 | 3.59E-05 | 0.00025 | 5.21E-06 | -0.011242538 | HSPB8, AC084880.4 |
| chr7 | 27189008 | 27189544 | 537 | 3 | 1.94E-05 | 2.71E-06 | 0.00074 | 5.69E-06 | -0.011101818 | HOXA11-AS, AC004080.2 |
| chr22 | 29663522 | 29663737 | 216 | 2 | 1.94E-05 | 4.58E-06 | 4.42E-05 | 3.63E-06 | -0.01394908 | NF2 |
| chr10 | 45454164 | 45454472 | 309 | 3 | 1.94E-05 | 6.80E-06 | 0.00125 | 1.36E-05 | -0.012875468 | NA |
| chr1 | 9720634 | 9721592 | 959 | 10 | 1.95E-05 | 1.83E-05 | 0.00573 | 7.03E-06 | -0.00166124 | PIK3CD |
| chr16 | 11642286 | 11642858 | 573 | 2 | 1.95E-05 | 4.66E-06 | 0.00048 | 7.79E-06 | -0.01910774 | NA |
| chr10 | 5445619 | 5446665 | 1047 | 11 | 1.95E-05 | 6.93E-06 | 0.00186 | 1.47E-06 | -0.006047118 | NET1 |
| chr13 | 24740244 | 24740626 | 383 | 2 | 1.95E-05 | 3.51E-05 | 2.95E-05 | 1.02E-05 | -0.003872055 | NA |
| chr10 | 99436404 | 99436666 | 263 | 2 | 1.95E-05 | 4.51E-05 | 1.50E-05 | 6.62E-06 | -0.006552165 | AL391684.1 |
| chr3 | 38495467 | 38496089 | 623 | 4 | 1.96E-05 | 0.000437 | 0.00016 | 5.13E-05 | -0.008447631 | NA |
| chr12 | 122942860 | 122943415 | 556 | 2 | 1.96E-05 | 1.67E-05 | 4.68E-05 | 9.05E-06 | -0.005995875 | ABCB9 |
| chr1 | 119711554 | 119712222 | 669 | 12 | 1.96E-05 | 0.000124 | 0.00023 | 1.69E-05 | 0.001372374 | PHGDH |
| chr9 | 138075494 | 138075950 | 457 | 6 | 1.96E-05 | 4.07E-05 | 3.86E-05 | 5.32E-06 | -0.005067254 | CACNA1B |
| chr6 | 136064701 | 136065273 | 573 | 6 | 1.97E-05 | 0.000301 | 6.45E-05 | 6.53E-06 | -0.003944785 | PDE7B, AL138828.1 |
| chr7 | 23022106 | 23022518 | 413 | 3 | 1.97E-05 | 7.59E-06 | 0.00017 | 5.62E-06 | 0.001855444 | NA |
| chr2 | 219029836 | 219030695 | 860 | 5 | 1.97E-05 | 2.53E-05 | 4.93E-05 | 9.45E-06 | 0.000740834 | CFAP65 |
| chr7 | 114087557 | 114088451 | 895 | 5 | 1.97E-05 | 3.30E-05 | 0.00094 | 5.73E-06 | -0.008950152 | FOXP2 |
| chr10 | 126905055 | 126905848 | 794 | 10 | 1.97E-05 | 5.87E-05 | 0.0022 | 7.55E-06 | -0.000827153 | DOCK1 |
| chr16 | 89286121 | 89286488 | 368 | 4 | 1.97E-05 | 1.40E-05 | 0.00076 | 3.86E-06 | -0.000920252 | ANKRD11 |
| chr2 | 445069 | 445430 | 362 | 3 | 1.97E-05 | 1.80E-06 | 0.00063 | 4.12E-06 | -0.01096555 | NA |
| chr17 | 41965117 | 41965560 | 444 | 2 | 1.97E-05 | 0.004442 | 5.41E-05 | 7.94E-05 | -0.005830992 | TTC25 |
| chr11 | 88098725 | 88099814 | 1090 | 4 | 1.97E-05 | 0.00102 | 3.62E-05 | 3.59E-05 | -0.004965343 | AP005436.2 |
| chr19 | 3547752 | 3548075 | 324 | 3 | 1.97E-05 | 0.000131 | 8.52E-05 | 1.53E-05 | -0.002630877 | AC005786.3, MFSD12 |
| chr16 | 533676 | 534723 | 1048 | 4 | 1.97E-05 | 8.15E-06 | 3.93E-05 | 5.15E-07 | -0.002100862 | CAPN15 |
| chr16 | 31148237 | 31148599 | 363 | 5 | 1.98E-05 | 1.45E-06 | 0.00121 | 3.03E-06 | -0.016363392 | PRSS36 |
| chr2 | 27379708 | 27381019 | 1312 | 14 | 1.98E-05 | 0.001588 | 0.00071 | 3.95E-05 | -0.00072988 | ZNF513 |
| chr5 | 43302031 | 43302379 | 349 | 2 | 1.98E-05 | 1.14E-06 | 0.00024 | 2.00E-06 | -0.012617894 | HMGCS1 |
| chr18 | 13654050 | 13654468 | 419 | 3 | 1.98E-05 | 4.11E-05 | 7.65E-05 | 9.64E-06 | -0.009701519 | NA |
| chr1 | 112606073 | 112606133 | 61 | 2 | 1.98E-05 | 0.002014 | 7.87E-06 | 8.60E-06 | 0.006688426 | ST7L |
| chr3 | 197028817 | 197029102 | 286 | 2 | 1.99E-05 | 0.000597 | 6.52E-05 | 6.25E-05 | -0.010086281 | MELTF |
| chr19 | 46678157 | 46678635 | 479 | 5 | 2.00E-05 | 8.89E-05 | 0.00011 | 7.82E-06 | -0.003178184 | PRKD2 |
| chr12 | 128815724 | 128816063 | 340 | 2 | 2.00E-05 | 4.66E-06 | 0.00025 | 7.04E-06 | 0.009271552 | SLC15A4 |
| chr16 | 88576364 | 88577122 | 759 | 6 | 2.00E-05 | 0.000502 | 5.33E-05 | 6.85E-06 | -0.003344729 | ZC3H18 |
| chr11 | 62613792 | 62614259 | 468 | 3 | 2.00E-05 | 2.61E-05 | 0.00012 | 1.05E-05 | -0.003774617 | ROM1 |
| chr11 | 117225386 | 117225598 | 213 | 2 | 2.00E-05 | 1.66E-05 | 8.51E-05 | 1.31E-05 | -0.004683139 | PCSK7 |
| chr22 | 37055685 | 37055786 | 102 | 2 | 2.00E-05 | 0.000572 | 1.12E-05 | 1.07E-05 | -0.007107281 | KCTD17 |
| chr16 | 81220087 | 81221114 | 1028 | 9 | 2.00E-05 | 9.50E-06 | 0.00335 | 5.49E-06 | 0.016755611 | PKD1L2 |
| chr16 | 1314964 | 1316379 | 1416 | 6 | 2.00E-05 | 1.45E-05 | 0.00018 | 1.23E-06 | -0.003519052 | UBE2I |
| chr1 | 113978950 | 113979563 | 614 | 7 | 2.01E-05 | 1.33E-06 | 0.0021 | 3.87E-06 | -0.006919676 | OLFML3 |
| chr11 | 65966156 | 65967144 | 989 | 4 | 2.01E-05 | 0.004496 | 4.67E-05 | 1.94E-05 | -0.002314209 | SART1 |
| chr5 | 67166056 | 67167160 | 1105 | 5 | 2.01E-05 | 8.30E-08 | 0.0011 | 4.43E-07 | -0.014948235 | MAST4 |
| chr10 | 5651368 | 5651621 | 254 | 3 | 2.01E-05 | 1.38E-06 | 0.00071 | 2.97E-06 | -0.005277318 | ASB13 |
| chr10 | 132248574 | 132249110 | 537 | 5 | 2.02E-05 | 4.04E-06 | 0.00379 | 8.73E-06 | 0.02002483 | STK32C |
| chr3 | 195892691 | 195893276 | 586 | 5 | 2.02E-05 | 5.67E-07 | 0.00074 | 1.86E-06 | -0.012310958 | TNK2 |
| chr12 | 98931064 | 98931620 | 557 | 3 | 2.02E-05 | 3.85E-05 | 0.00016 | 9.27E-06 | -0.00998353 | ANKS1B |
| chr19 | 4768560 | 4769228 | 669 | 7 | 2.02E-05 | 7.56E-05 | 0.0063 | 3.94E-05 | -0.007079282 | MIR7-3HG |
| chr14 | 99197798 | 99198873 | 1076 | 6 | 2.03E-05 | 2.39E-05 | 0.00011 | 4.59E-06 | -0.006416421 | BCL11B |
| chr11 | 2600043 | 2600135 | 93 | 2 | 2.03E-05 | 1.92E-05 | 1.35E-05 | 3.96E-06 | -0.005910659 | KCNQ1 |
| chr16 | 65601740 | 65602224 | 485 | 4 | 2.03E-05 | 5.10E-07 | 0.00387 | 2.32E-06 | -0.017382162 | AC092138.2 |
| chr1 | 89787829 | 89787911 | 83 | 2 | 2.03E-05 | 1.73E-06 | 0.00029 | 3.00E-06 | -0.007507883 | AC093423.3 |
| chr12 | 132496866 | 132497045 | 180 | 3 | 2.03E-05 | 9.54E-05 | 0.00027 | 7.98E-06 | -0.009382062 | FBRSL1 |
| chr11 | 65618440 | 65618657 | 218 | 3 | 2.03E-05 | 1.69E-06 | 0.00069 | 3.12E-06 | -0.002606946 | PCNX3 |
| chr20 | 4181835 | 4182515 | 681 | 4 | 2.03E-05 | 3.76E-05 | 0.00025 | 9.60E-06 | -0.001103974 | SMOX |
| chr4 | 6268953 | 6269430 | 478 | 6 | 2.03E-05 | 1.07E-05 | 0.00034 | 6.88E-06 | -0.004857827 | NA |
| chr11 | 68754981 | 68755017 | 37 | 2 | 2.03E-05 | 9.86E-06 | 1.91E-05 | 3.50E-06 | -0.001217725 | CPT1A |
| chr6 | 45421576 | 45422547 | 972 | 14 | 2.03E-05 | 3.94E-05 | 0.00031 | 2.51E-06 | 0.000713995 | RUNX2, AL096865.1 |
| chr17 | 39864819 | 39865243 | 425 | 2 | 2.04E-05 | 3.15E-05 | 0.00013 | 2.52E-05 | -0.017002379 | NA |
| chr20 | 31548526 | 31549087 | 562 | 4 | 2.04E-05 | 0.000632 | 6.43E-05 | 1.99E-05 | -0.003903526 | HM13 |
| chr12 | 57897067 | 57897620 | 554 | 4 | 2.04E-05 | 1.43E-05 | 0.00021 | 7.52E-06 | -0.007612776 | NA |
| chr15 | 89630095 | 89630574 | 480 | 2 | 2.04E-05 | 4.17E-05 | 4.71E-05 | 1.60E-05 | -0.004826095 | TICRR, KIF7 |
| chr12 | 49030355 | 49030919 | 565 | 5 | 2.04E-05 | 0.000142 | 5.24E-05 | 1.13E-05 | 0.003024215 | KMT2D |
| chr20 | 62977711 | 62978255 | 545 | 4 | 2.04E-05 | 0.000153 | 5.36E-05 | 1.09E-05 | -0.001284715 | NA |
| chr1 | 228172830 | 228173097 | 268 | 2 | 2.04E-05 | 5.68E-05 | 2.57E-05 | 1.16E-05 | -0.003322127 | IBA57 |
| chr9 | 128636905 | 128637037 | 133 | 4 | 2.04E-05 | 4.94E-05 | 0.0012 | 4.68E-06 | -0.00395898 | DYNC2I2 |
| chr14 | 37709198 | 37709649 | 452 | 2 | 2.04E-05 | 8.49E-06 | 5.21E-05 | 6.13E-06 | -0.009512365 | TTC6 |
| chr5 | 15615915 | 15616273 | 359 | 3 | 2.05E-05 | 4.54E-06 | 0.00033 | 6.54E-06 | -0.005779671 | FBXL7, CTD-2350J17.1 |
| chr10 | 58665755 | 58665973 | 219 | 2 | 2.05E-05 | 2.77E-06 | 6.56E-05 | 3.06E-06 | -0.025333744 | BICC1 |
| chr10 | 31603100 | 31603711 | 612 | 5 | 2.05E-05 | 3.39E-06 | 0.00026 | 2.61E-06 | -0.00344595 | AL161935.1 |
| chr2 | 128442050 | 128442535 | 486 | 2 | 2.05E-05 | 1.71E-06 | 0.00029 | 2.96E-06 | -0.011504362 | NA |
| chr19 | 3500753 | 3501212 | 460 | 11 | 2.06E-05 | 7.02E-05 | 0.00148 | 1.06E-05 | -0.001196934 | NA |
| chr6 | 111721108 | 111721883 | 776 | 4 | 2.06E-05 | 1.35E-05 | 0.00191 | 6.87E-06 | -0.006488891 | FYN |
| chr9 | 18238549 | 18238709 | 161 | 2 | 2.06E-05 | 3.48E-06 | 4.65E-05 | 3.05E-06 | -0.008966073 | ADAMTSL1 |
| chr1 | 225811028 | 225811871 | 844 | 4 | 2.06E-05 | 2.83E-06 | 0.00079 | 5.54E-06 | -0.013038267 | EPHX1 |
| chr17 | 17813692 | 17814653 | 962 | 11 | 2.06E-05 | 2.52E-06 | 0.00376 | 2.62E-06 | -0.003053234 | SREBF1, MIR33B |

| | | | | | | | | | | |
|---|---|---|---|---|---|---|---|---|---|---|
| chr16 | 17277894 | 17278165 | 272 | 2 | 2.07E-05 | 5.01E-06 | 4.57E-05 | 3.96E-06 | -0.010240858 | XYLT1 |
| chr15 | 75455409 | 75456338 | 930 | 11 | 2.07E-05 | 0.092112 | 2.51E-06 | 9.64E-05 | 0.000563904 | SIN3A |
| chr7 | 1529433 | 1529967 | 535 | 4 | 2.07E-05 | 2.13E-06 | 0.00264 | 7.16E-06 | -0.006397179 | NA |
| chr16 | 21157202 | 21158417 | 1216 | 9 | 2.09E-05 | 0.001651 | 0.0001 | 2.45E-06 | -0.008443033 | TMEM159, DNAH3 |
| chr11 | 131760570 | 131760754 | 185 | 3 | 2.09E-05 | 1.09E-06 | 0.00068 | 2.26E-06 | -0.038105827 | NTM |
| chr1 | 159853897 | 159854319 | 423 | 3 | 2.10E-05 | 5.00E-05 | 0.00013 | 2.27E-05 | -0.003757734 | AL590560.2, SNHG28, VSIG8 |
| chr5 | 1886195 | 1887469 | 1275 | 16 | 2.10E-05 | 0.007087 | 0.00102 | 2.81E-05 | -0.000603764 | CTD-2194D22.4, IRX4 |
| chr17 | 2303446 | 2304201 | 756 | 9 | 2.10E-05 | 2.19E-05 | 0.00096 | 5.78E-06 | 0.002560474 | SRR, SMG6 |
| chr14 | 77301301 | 77301457 | 157 | 3 | 2.10E-05 | 2.78E-05 | 0.00019 | 3.96E-06 | -0.009465053 | POMT2 |
| chr19 | 8395106 | 8395115 | 10 | 2 | 2.10E-05 | 8.43E-06 | 4.90E-05 | 5.90E-06 | -0.010677954 | RAB11B |
| chr12 | 67647538 | 67647958 | 421 | 4 | 2.10E-05 | 3.81E-05 | 0.00017 | 8.17E-06 | -0.007090248 | NA |
| chr18 | 63132213 | 63133190 | 978 | 4 | 2.11E-05 | 1.43E-07 | 0.00027 | 2.32E-07 | -0.006166362 | BCL2 |
| chr22 | 30889510 | 30890884 | 1375 | 6 | 2.12E-05 | 8.07E-09 | 0.00044 | 2.62E-08 | -0.007397156 | OSBP2 |
| chr7 | 47536546 | 47537359 | 814 | 4 | 2.12E-05 | 3.08E-05 | 0.00051 | 6.34E-06 | -0.010776857 | TNS3 |
| chr16 | 74695567 | 74696301 | 735 | 4 | 2.13E-05 | 7.32E-06 | 0.00118 | 9.46E-06 | -0.017919154 | MLKL |
| chr3 | 195443851 | 195444202 | 352 | 2 | 2.13E-05 | 0.000331 | 4.53E-05 | 3.73E-05 | -0.006514256 | NA |
| chr1 | 1069720 | 1070018 | 299 | 3 | 2.13E-05 | 4.10E-06 | 0.00054 | 7.71E-06 | -0.011130912 | NA |
| chr1 | 41510401 | 41510922 | 522 | 4 | 2.13E-05 | 2.81E-06 | 0.00025 | 3.28E-06 | -0.004726495 | HIVEP3 |
| chr15 | 81374052 | 81374506 | 455 | 6 | 2.13E-05 | 4.49E-05 | 0.00072 | 5.63E-06 | -0.001673874 | TMC3-AS1, TMC3 |
| chr20 | 4192145 | 4193558 | 1414 | 6 | 2.13E-05 | 2.44E-06 | 0.00015 | 5.59E-07 | -0.006552802 | LINC01433 |
| chr19 | 2248303 | 2249331 | 1029 | 7 | 2.14E-05 | 0.001919 | 0.00012 | 1.52E-05 | -0.001640328 | SF3A2, AMH |
| chr20 | 45823882 | 45824162 | 281 | 3 | 2.14E-05 | 1.47E-05 | 0.00047 | 1.77E-05 | -0.010651464 | TNNC2 |
| chr7 | 142749433 | 142749448 | 16 | 2 | 2.14E-05 | 2.50E-05 | 1.20E-05 | 4.10E-06 | -0.008674903 | NA |
| chr12 | 104741927 | 104742561 | 635 | 2 | 2.15E-05 | 4.61E-06 | 0.00016 | 6.36E-06 | -0.014995097 | CHST11 |
| chr12 | 10030573 | 10031251 | 679 | 7 | 2.15E-05 | 0.000113 | 6.90E-05 | 8.95E-06 | -0.012033844 | CLEC9A, AC024224.2 |
| chr6 | 169641324 | 169641863 | 540 | 4 | 2.15E-05 | 8.23E-05 | 0.00066 | 1.77E-05 | -0.003800073 | AL031315.1, WDR27 |
| chr6 | 18154870 | 18155362 | 493 | 8 | 2.15E-05 | 0.000132 | 6.36E-05 | 8.08E-06 | 0.00027726 | KDM1B, TPMT |
| chr19 | 10289632 | 10290075 | 444 | 6 | 2.15E-05 | 0.000417 | 3.68E-05 | 2.36E-05 | 0.001857142 | ICAM5 |
| chr17 | 29699002 | 29699473 | 472 | 2 | 2.16E-05 | 6.69E-05 | 8.35E-05 | 3.15E-05 | -0.007538846 | ABHD15-AS1, SSH2 |
| chr12 | 57815509 | 57816130 | 622 | 5 | 2.17E-05 | 1.19E-05 | 0.00041 | 5.79E-06 | -0.005889842 | AVIL |
| chr12 | 117046501 | 117046781 | 281 | 4 | 2.17E-05 | 0.001369 | 2.10E-05 | 1.71E-05 | -0.003367425 | TESC |
| chr3 | 47824301 | 47825327 | 1027 | 8 | 2.17E-05 | 3.89E-05 | 0.00016 | 1.49E-05 | -0.00280399 | DHX30 |
| chr10 | 63976587 | 63977288 | 702 | 4 | 2.17E-05 | 0.002038 | 2.58E-05 | 3.28E-05 | -0.00387153 | AC013287.1 |
| chr16 | 1026540 | 1027624 | 1085 | 6 | 2.18E-05 | 3.71E-05 | 0.00079 | 6.86E-06 | -0.004039347 | NA |
| chr1 | 51329029 | 51329668 | 640 | 6 | 2.18E-05 | 0.000629 | 0.00011 | 2.28E-05 | -0.00425444 | TTC39A-AS1, TTC39A |
| chr10 | 103800810 | 103801310 | 501 | 2 | 2.18E-05 | 2.02E-05 | 4.30E-05 | 9.62E-06 | -0.016130069 | SH3PXD2A |
| chr6 | 157592484 | 157593190 | 707 | 7 | 2.19E-05 | 2.19E-06 | 0.00455 | 2.97E-06 | 0.0103027 | ZDHHC14 |
| chr17 | 42108344 | 42109366 | 1023 | 4 | 2.19E-05 | 3.36E-06 | 1.00E-04 | 1.27E-06 | -0.004518172 | DHX58 |
| chr15 | 90100534 | 90100894 | 361 | 7 | 2.19E-05 | 2.03E-06 | 0.00089 | 4.55E-06 | -0.008930476 | IDH2 |
| chr6 | 37576769 | 37577272 | 504 | 2 | 2.19E-05 | 1.03E-05 | 5.47E-05 | 7.21E-06 | -0.014327936 | NA |
| chr8 | 11013681 | 11013861 | 181 | 2 | 2.19E-05 | 3.01E-05 | 2.15E-05 | 7.20E-06 | -0.013566547 | XKR6 |
| chr16 | 84964925 | 84965476 | 552 | 3 | 2.19E-05 | 2.17E-06 | 0.00028 | 3.94E-06 | -0.000256082 | NA |
| chr19 | 3006113 | 3006722 | 610 | 5 | 2.19E-05 | 5.72E-07 | 0.00411 | 2.60E-06 | -0.015927523 | TLE2 |
| chr12 | 52249955 | 52250480 | 526 | 2 | 2.19E-05 | 5.83E-06 | 0.00012 | 6.99E-06 | -0.027994489 | KRT7, KRT86, KRT87P |
| chr5 | 168232360 | 168233609 | 1250 | 4 | 2.20E-05 | 2.31E-06 | 0.00028 | 2.04E-07 | -0.006608162 | TENM2, CTB-178M22.2 |
| chr4 | 849530 | 850040 | 511 | 4 | 2.20E-05 | 4.19E-05 | 4.09E-05 | 7.91E-06 | 0.000106326 | GAK |
| chr1 | 2207318 | 2208649 | 1332 | 12 | 2.20E-05 | 2.53E-06 | 0.0016 | 6.02E-07 | -0.003568646 | FAAP20 |
| chr12 | 103672432 | 103672879 | 448 | 4 | 2.20E-05 | 3.83E-06 | 0.00034 | 4.14E-06 | -0.006821643 | STAB2 |
| chr1 | 182614933 | 182615385 | 453 | 5 | 2.21E-05 | 0.00034 | 2.41E-05 | 1.94E-05 | 0.001202274 | LINC01686 |
| chr15 | 62569756 | 62570224 | 469 | 2 | 2.21E-05 | 2.40E-05 | 6.14E-05 | 1.37E-05 | -0.01729007 | TLN2 |
| chr19 | 2424795 | 2425754 | 960 | 4 | 2.21E-05 | 1.32E-06 | 0.00258 | 4.54E-06 | -0.014238901 | TMPRSS9, TIMM13 |
| chr7 | 122886354 | 122887297 | 944 | 13 | 2.21E-05 | 5.16E-05 | 0.00019 | 8.45E-06 | 0.002214167 | CADPS2 |
| chr15 | 68651608 | 68652176 | 569 | 2 | 2.21E-05 | 8.45E-06 | 0.00025 | 1.16E-05 | -0.01145482 | CORO2B |
| chr10 | 7817660 | 7818165 | 506 | 4 | 2.21E-05 | 2.76E-05 | 0.0026 | 3.71E-05 | -0.01816821 | NA |
| chr4 | 1359079 | 1360560 | 1482 | 7 | 2.21E-05 | 9.08E-05 | 8.03E-05 | 4.76E-07 | -0.003172516 | UVSSA, AC078852.1 |
| chr8 | 142317015 | 142317452 | 438 | 4 | 2.21E-05 | 0.039078 | 7.37E-06 | 2.87E-05 | -0.001751034 | TSNARE1 |
| chr19 | 5962076 | 5962725 | 650 | 3 | 2.22E-05 | 0.000512 | 0.00078 | 0.00035 | -0.001263172 | RANBP3 |
| chr16 | 85041357 | 85041643 | 287 | 4 | 2.22E-05 | 1.54E-06 | 0.00114 | 2.83E-06 | -0.012441061 | KIAA0513 |
| chr1 | 116382572 | 116383167 | 596 | 5 | 2.22E-05 | 0.000186 | 0.00015 | 1.51E-05 | -0.00757992 | ATP1A1, ATP1A1-AS1 |
| chr10 | 92204542 | 92205173 | 632 | 2 | 2.22E-05 | 3.56E-05 | 0.00015 | 2.92E-05 | -0.005117322 | CPEB3 |
| chr7 | 113919867 | 113920197 | 331 | 3 | 2.23E-05 | 0.000191 | 1.80E-05 | 1.18E-05 | -0.005897667 | PPP1R3A |
| chr22 | 41240937 | 41241463 | 527 | 5 | 2.23E-05 | 7.10E-05 | 0.00094 | 1.47E-05 | -0.004081762 | NA |
| chr1 | 27725191 | 27725260 | 70 | 2 | 2.23E-05 | 9.11E-06 | 0.00071 | 1.49E-05 | -0.012147864 | AL020997.5 |
| chr8 | 142378825 | 142379407 | 583 | 4 | 2.23E-05 | 0.000192 | 7.49E-05 | 3.46E-05 | -0.00355299 | TSNARE1 |
| chr16 | 15874826 | 15875007 | 182 | 2 | 2.23E-05 | 0.000711 | 9.00E-06 | 8.82E-06 | -0.009554356 | CEP20 |
| chr7 | 134443715 | 134444357 | 643 | 3 | 2.23E-05 | 1.50E-06 | 0.00026 | 2.76E-06 | -0.007018855 | AKR1B1 |
| chr12 | 19203994 | 19204305 | 312 | 3 | 2.24E-05 | 7.63E-05 | 1.00E-04 | 1.55E-05 | -0.009597181 | PLEKHA5 |
| chr6 | 27865091 | 27865777 | 687 | 10 | 2.25E-05 | 0.000309 | 0.00088 | 9.20E-06 | -0.001157631 | H2BC15, H2AC16 |
| chr3 | 193551771 | 193551807 | 37 | 2 | 2.25E-05 | 0.001621 | 2.14E-05 | 2.52E-05 | -0.003134308 | ATP13A4 |
| chr19 | 42290957 | 42291867 | 911 | 6 | 2.25E-05 | 2.20E-05 | 8.19E-05 | 7.79E-06 | -0.005823613 | CIC |
| chr2 | 107985905 | 107986737 | 833 | 12 | 2.25E-05 | 9.81E-06 | 0.00114 | 3.24E-06 | -0.005634391 | SLC5A7 |
| chr9 | 136334280 | 136334455 | 176 | 4 | 2.25E-05 | 3.61E-05 | 0.00025 | 5.63E-06 | -0.00421888 | GPSM1 |
| chr7 | 1966833 | 1966883 | 51 | 2 | 2.25E-05 | 3.66E-05 | 4.67E-05 | 1.47E-05 | -0.001522567 | MAD1L1, AC069288.1 |
| chr19 | 48454959 | 48455786 | 828 | 9 | 2.26E-05 | 5.10E-05 | 0.00117 | 7.85E-06 | -0.002415887 | GRWD1, KCNJ14 |
| chr1 | 234962914 | 234963375 | 462 | 5 | 2.26E-05 | 1.11E-06 | 0.00585 | 5.10E-06 | 0.007046643 | AL391832.2 |
| chr17 | 17147019 | 17147136 | 118 | 3 | 2.26E-05 | 4.77E-06 | 0.00034 | 3.39E-06 | -0.002356715 | MPRIP |
| chr20 | 57407259 | 57407922 | 664 | 5 | 2.26E-05 | 0.000134 | 4.65E-05 | 1.49E-05 | -0.004270322 | RBM38 |
| chr11 | 70303075 | 70303199 | 125 | 2 | 2.26E-05 | 2.85E-06 | 0.00016 | 4.31E-06 | -0.018337211 | PPFIA1, AP002336.2 |
| chr3 | 37573315 | 37573580 | 266 | 3 | 2.27E-05 | 7.99E-06 | 6.60E-05 | 5.44E-06 | -0.008007555 | ITGA9 |
| chr17 | 7094740 | 7095665 | 926 | 4 | 2.27E-05 | 1.14E-07 | 0.00057 | 3.42E-07 | -0.019490349 | NA |
| chr19 | 45439519 | 45440607 | 1089 | 6 | 2.27E-05 | 4.11E-07 | 0.007 | 2.48E-06 | -0.00384756 | ERCC1 |
| chr14 | 51828048 | 51828650 | 603 | 3 | 2.27E-05 | 9.35E-06 | 4.50E-05 | 4.22E-06 | -0.008870933 | GNG2 |
| chr10 | 63711568 | 63712363 | 796 | 2 | 2.28E-05 | 9.67E-07 | 7.49E-05 | 1.36E-06 | -0.012362467 | AC013287.1 |
| chr9 | 5510497 | 5510595 | 99 | 2 | 2.29E-05 | 3.51E-06 | 0.00033 | 5.78E-06 | -0.030427933 | PDCD1LG2, AL162253.2 |
| chr7 | 100433592 | 100434255 | 664 | 3 | 2.29E-05 | 3.81E-06 | 0.00036 | 6.60E-06 | -0.009639529 | MEPCE |
| chr17 | 42552978 | 42553570 | 593 | 4 | 2.29E-05 | 5.88E-05 | 0.0001 | 1.17E-05 | -0.000684956 | AC067852.1, HSD17B1, AC067852.2 |
| chr14 | 99970950 | 99971614 | 665 | 6 | 2.29E-05 | 2.46E-05 | 0.00033 | 1.34E-05 | 2.64E-05 | EVL |
| chr6 | 33771629 | 33771876 | 248 | 4 | 2.29E-05 | 1.32E-06 | 0.00063 | 2.53E-06 | -0.015279678 | LEMD2 |
| chr17 | 75831300 | 75831606 | 307 | 2 | 2.30E-05 | 0.000709 | 4.90E-05 | 4.95E-05 | -0.005128727 | UNC13D |
| chr11 | 2708323 | 2708899 | 577 | 4 | 2.30E-05 | 0.000135 | 3.74E-05 | 2.08E-05 | -0.006413499 | KCNQ1 |
| chr7 | 752698 | 753081 | 384 | 3 | 2.30E-05 | 9.38E-06 | 0.00034 | 1.32E-05 | -0.000861198 | DNAAF5 |
| chr15 | 23036126 | 23036248 | 123 | 2 | 2.30E-05 | 3.47E-06 | 0.00011 | 4.51E-06 | -0.013677177 | TUBGCP5 |
| chr2 | 68364783 | 68365605 | 823 | 6 | 2.30E-05 | 5.77E-07 | 0.00219 | 2.59E-06 | -0.004208993 | PLEK, AC015969.1 |
| chr11 | 69470865 | 69471584 | 720 | 10 | 2.31E-05 | 2.82E-07 | 0.00707 | 2.09E-06 | -0.010155892 | NA |
| chr10 | 96396275 | 96396363 | 89 | 3 | 2.31E-05 | 7.17E-05 | 0.00034 | 8.38E-06 | -0.014117236 | TLL2 |
| chr14 | 64756023 | 64756242 | 220 | 2 | 2.31E-05 | 1.55E-06 | 0.00023 | 2.63E-06 | -0.021323509 | SPTB |
| chr12 | 123966748 | 123967238 | 491 | 2 | 2.31E-05 | 7.56E-05 | 3.10E-05 | 1.57E-05 | -0.00977876 | CCDC92 |
| chr1 | 52365448 | 52366648 | 1201 | 10 | 2.31E-05 | 0.002494 | 0.00027 | 1.09E-05 | 0.001587433 | AL513218.2, CC2D1B |
| chr1 | 26354099 | 26354676 | 578 | 8 | 2.32E-05 | 5.98E-06 | 0.00032 | 4.13E-06 | 0.000836273 | CRYBG2 |
| chr16 | 84451993 | 84452120 | 128 | 2 | 2.32E-05 | 8.02E-05 | 1.38E-05 | 7.91E-06 | -0.003625152 | ATP2C2 |
| chr20 | 46899490 | 46899877 | 388 | 2 | 2.32E-05 | 0.000258 | 4.23E-05 | 3.25E-05 | -0.008029083 | EYA2 |
| chr12 | 8032080 | 8032481 | 402 | 5 | 2.32E-05 | 0.000194 | 0.00031 | 5.77E-05 | -0.005004019 | NA |
| chr19 | 36032503 | 36033552 | 1050 | 7 | 2.33E-05 | 1.37E-05 | 0.00096 | 3.41E-06 | -0.006550663 | AC002116.2, CLIP3 |
| chr7 | 155296612 | 155296744 | 133 | 3 | 2.33E-05 | 5.45E-06 | 0.00022 | 5.95E-06 | -0.008593161 | AC144652.1 |
| chr5 | 173671109 | 173671432 | 324 | 2 | 2.33E-05 | 2.63E-06 | 0.00035 | 4.53E-06 | -0.017535177 | NA |
| chr6 | 27838242 | 27839064 | 823 | 11 | 2.33E-05 | 0.000314 | 0.0004 | 4.45E-06 | 0.000759544 | H2BC15, H2AC15 |
| chr20 | 38572737 | 38573151 | 415 | 2 | 2.34E-05 | 4.85E-05 | 0.0003 | 4.76E-05 | -0.013503976 | RALGAPB |
| chr11 | 62536271 | 62536600 | 330 | 4 | 2.34E-05 | 0.000129 | 0.0001 | 4.87E-06 | -0.003719733 | AHNAK |
| chr1 | 26170616 | 26171780 | 1165 | 6 | 2.35E-05 | 0.022065 | 0.00017 | 0.00068 | -0.003930828 | ZNF593, AL391650.1 |
| chr1 | 193104736 | 193106061 | 1326 | 12 | 2.35E-05 | 3.19E-05 | 0.00022 | 8.57E-06 | 0.001370443 | GLRX2 |
| chr7 | 50641854 | 50641928 | 75 | 3 | 2.35E-05 | 1.91E-06 | 0.00028 | 2.43E-06 | -0.003079521 | AC005153.1, GRB10 |
| chr7 | 45921249 | 45923425 | 2177 | 34 | 2.35E-05 | 0.0002 | 0.00271 | 2.37E-06 | 0.000855407 | IGFBP3 |
| chr14 | 99351043 | 99351793 | 751 | 2 | 2.36E-05 | 1.38E-06 | 8.19E-05 | 1.88E-06 | -0.012144239 | NA |
| chr9 | 137310648 | 137311710 | 1063 | 4 | 2.36E-05 | 1.31E-05 | 0.00056 | 1.57E-06 | -0.008279026 | EXD3 |
| chr20 | 51003099 | 51004142 | 1044 | 4 | 2.36E-05 | 2.21E-06 | 0.00025 | 7.31E-07 | -0.004109507 | AL050404.1, KCNG1 |
| chr20 | 63973733 | 63975146 | 1414 | 6 | 2.37E-05 | 1.57E-06 | 0.00016 | 2.67E-07 | -0.005157246 | SAMD10 |
| chr19 | 4968093 | 4969076 | 984 | 5 | 2.37E-05 | 3.38E-07 | 0.00104 | 7.22E-07 | -0.003753388 | NA |
| chr9 | 37011475 | 37011601 | 127 | 2 | 2.37E-05 | 2.38E-06 | 9.32E-05 | 3.12E-06 | -0.023682651 | PAX5 |
| chr1 | 18106466 | 18106698 | 233 | 3 | 2.37E-05 | 3.54E-06 | 0.00249 | 9.39E-06 | 0.019416594 | NA |
| chr16 | 3595434 | 3596053 | 620 | 4 | 2.38E-05 | 6.14E-06 | 0.00117 | 7.58E-06 | -0.005947071 | SLX4 |
| chr2 | 70509286 | 70509730 | 445 | 2 | 2.38E-05 | 2.74E-06 | 0.00011 | 3.76E-06 | -0.006451331 | TGFA |
| chr19 | 51531716 | 51532033 | 318 | 4 | 2.39E-05 | 7.48E-06 | 0.00051 | 6.55E-06 | -0.001853372 | SIGLEC6 |
| chr3 | 53083535 | 53084151 | 617 | 2 | 2.39E-05 | 8.05E-06 | 0.00034 | 1.19E-05 | -0.014018928 | AC096887.1 |
| chr7 | 151448149 | 151449316 | 1168 | 6 | 2.39E-05 | 3.00E-06 | 0.00047 | 5.57E-07 | -0.014751104 | NA |
| chr15 | 41772797 | 41773496 | 700 | 5 | 2.39E-05 | 3.00E-05 | 0.00258 | 1.47E-05 | -0.008959931 | MGA |
| chr14 | 88562057 | 88563373 | 1317 | 17 | 2.39E-05 | 1.02E-05 | 0.00779 | 1.62E-06 | -0.001465517 | ZC3H14 |
| chr19 | 35281754 | 35282815 | 1062 | 12 | 2.39E-05 | 5.13E-06 | 0.00887 | 2.17E-06 | -0.006756343 | HAMP |
| chr6 | 154932803 | 154933206 | 404 | 2 | 2.39E-05 | 0.001622 | 2.01E-05 | 2.36E-05 | -0.00633269 | TIAM2 |
| chr2 | 47402519 | 47403648 | 1130 | 15 | 2.39E-05 | 0.000409 | 0.00159 | 8.24E-06 | 0.00171169 | MSH2 |
| chr8 | 133033493 | 133033955 | 463 | 2 | 2.40E-05 | 0.002832 | 2.23E-05 | 2.83E-05 | -0.003917215 | TG |
| chr3 | 100260075 | 100260747 | 673 | 11 | 2.40E-05 | 0.000102 | 0.00907 | 2.49E-05 | -0.00505157 | AC129803.1 |
| chr11 | 67367656 | 67368426 | 771 | 5 | 2.40E-05 | 3.13E-06 | 0.00172 | 5.08E-06 | -0.009560995 | RAD9A, AP003419.1, CLCF1 |
| chr1 | 2203919 | 2204778 | 860 | 4 | 2.40E-05 | 1.49E-06 | 0.00126 | 4.50E-06 | 3.98E-05 | FAAP20 |
| chr21 | 44348225 | 44348437 | 213 | 4 | 2.41E-05 | 5.39E-07 | 0.00187 | 1.97E-06 | -0.009648148 | AP001063.1 |
| chr2 | 119695056 | 119695581 | 526 | 2 | 2.41E-05 | 2.18E-06 | 0.00013 | 3.19E-06 | -0.014821602 | NA |
| chr2 | 238569724 | 238569873 | 150 | 3 | 2.41E-05 | 8.86E-07 | 0.00085 | 2.21E-06 | -0.014449932 | LINC01937 |
| chr20 | 36540576 | 36541483 | 908 | 8 | 2.41E-05 | 6.43E-06 | 0.00401 | 1.34E-05 | -0.010229935 | DLGAP4-AS1 |
| chr11 | 66693248 | 66693314 | 67 | 2 | 2.41E-05 | 3.63E-06 | 0.00043 | 6.15E-06 | -0.007527687 | SPTBN2 |
| chr17 | 3532502 | 3532930 | 429 | 4 | 2.42E-05 | 9.81E-05 | 0.00012 | 9.74E-06 | -0.003561769 | TRPV3 |
| chr1 | 10056658 | 10056722 | 65 | 2 | 2.42E-05 | 1.25E-05 | 3.02E-05 | 5.64E-06 | -0.005529498 | UBE4B |
| chr1 | 43143760 | 43143978 | 219 | 3 | 2.42E-05 | 1.20E-05 | 0.00044 | 7.17E-06 | -0.018040376 | NA |
| chr3 | 172045423 | 172045451 | 29 | 2 | 2.42E-05 | 3.61E-06 | 8.35E-05 | 4.17E-06 | -0.011653626 | FNDC3B |
| chr5 | 32394604 | 32395396 | 793 | 4 | 2.42E-05 | 1.72E-06 | 0.00056 | 1.51E-06 | 0.011398735 | ZFR |
| chr2 | 149587022 | 149588320 | 1299 | 16 | 2.42E-05 | 7.18E-05 | 0.00336 | 6.64E-06 | -0.002783457 | MMADHC-DT, MMADHC |
| chr16 | 85171080 | 85173047 | 1968 | 6 | 2.43E-05 | 1.10E-06 | 0.00057 | 8.50E-08 | -0.010058547 | GSE1 |
| chr8 | 124710625 | 124711031 | 407 | 4 | 2.43E-05 | 1.04E-06 | 0.00102 | 2.51E-06 | -0.005800796 | MTSS1 |
| chr10 | 71625746 | 71625809 | 64 | 2 | 2.43E-05 | 2.10E-06 | 0.00014 | 3.19E-06 | -0.027532913 | CDH23 |
| chr12 | 51028658 | 51029044 | 387 | 6 | 2.44E-05 | 3.73E-05 | 0.00013 | 6.61E-06 | -0.004735691 | NA |
| chr7 | 949564 | 950514 | 951 | 4 | 2.46E-05 | 1.75E-06 | 0.00047 | 1.34E-06 | -0.00912196 | ADAP1, COX19 |
| chr1 | 41505458 | 41505509 | 52 | 2 | 2.46E-05 | 0.000976 | 3.18E-05 | 3.50E-05 | -0.008487278 | NA |
| chr2 | 53859717 | 53860604 | 888 | 13 | 2.46E-05 | 4.98E-08 | 0.00899 | 6.42E-07 | -0.009056242 | ASB3, GPR75 |
| chr19 | 5993434 | 5994346 | 913 | 4 | 2.47E-05 | 4.81E-07 | 0.0016 | 1.47E-06 | -0.007879136 | AC011444.1, RFX2 |
| chr11 | 62817243 | 62817345 | 103 | 2 | 2.47E-05 | 5.32E-05 | 1.70E-05 | 7.89E-06 | -0.008007618 | STX5 |
| chr16 | 88637822 | 88639308 | 1487 | 10 | 2.47E-05 | 7.66E-06 | 0.0031 | 2.14E-06 | 0.007824779 | IL17C |
| chr2 | 238257821 | 238258282 | 462 | 2 | 2.47E-05 | 4.71E-05 | 3.94E-05 | 1.49E-05 | -0.001887406 | PER2 |
| chr6 | 167720854 | 167721927 | 1074 | 6 | 2.48E-05 | 0.00024 | 0.00459 | 2.48E-05 | -0.00279067 | NA |
| chr7 | 116861258 | 116861504 | 247 | 2 | 2.48E-05 | 0.000101 | 6.16E-05 | 3.12E-05 | -0.00479209 | CAPZA2 |
| chr6 | 30164044 | 30164938 | 895 | 8 | 2.48E-05 | 6.52E-05 | 0.00441 | 2.85E-05 | -0.004562799 | TRIM15 |
| chr8 | 130356187 | 130357350 | 1164 | 4 | 2.48E-05 | 0.042818 | 2.42E-05 | 5.72E-05 | -0.003274813 | ASAP1 |
| chr1 | 229403483 | 229403924 | 442 | 3 | 2.48E-05 | 7.32E-06 | 5.59E-05 | 4.32E-06 | -0.006612231 | NA |
| chr6 | 31689690 | 31690399 | 710 | 5 | 2.48E-05 | 0.000686 | 0.00052 | 1.67E-05 | -0.004081089 | ABHD16A, AL662899.1 |
| chr16 | 89098545 | 89099197 | 653 | 5 | 2.48E-05 | 1.81E-06 | 0.00315 | 5.21E-06 | -0.011170162 | ACSF3 |

| | | | | | | | | | | |
|---|---|---|---|---|---|---|---|---|---|---|
| chr2 | 73737017 | 73737707 | 691 | 10 | 2.49E-05 | 2.08E-06 | 0.00849 | 4.06E-06 | -0.001940813 | TPRKB |
| chr12 | 57764510 | 57764754 | 245 | 2 | 2.49E-05 | 0.0001 | 2.62E-05 | 1.52E-05 | -0.001167136 | CYP27B1 |
| chr1 | 204126459 | 204127287 | 829 | 3 | 2.49E-05 | 0.000104 | 8.20E-05 | 2.62E-06 | -0.007036238 | SOX13 |
| chr21 | 33324228 | 33324731 | 504 | 7 | 2.50E-05 | 0.000129 | 0.00036 | 5.33E-06 | 0.000935187 | IFNAR1 |
| chr8 | 27489091 | 27490958 | 1868 | 11 | 2.50E-05 | 3.25E-05 | 0.00017 | 3.62E-08 | -0.008140988 | EPHX2 |
| chr11 | 65592850 | 65593842 | 993 | 6 | 2.50E-05 | 1.11E-06 | 0.00125 | 3.51E-06 | -0.014739002 | KCNK7 |
| chr5 | 1933063 | 1933488 | 426 | 3 | 2.50E-05 | 0.0001 | 0.00057 | 3.17E-05 | -0.00500016 | AC124852.1, AC126768.3 |
| chr16 | 89994364 | 89994760 | 397 | 5 | 2.50E-05 | 0.000736 | 7.64E-05 | 1.66E-05 | -0.007285819 | AFG3L1P |
| chr8 | 1435393 | 1435631 | 239 | 4 | 2.50E-05 | 1.18E-06 | 0.00118 | 2.25E-06 | -0.003383869 | DLGAP2 |
| chr3 | 126654979 | 126655995 | 1017 | 9 | 2.51E-05 | 2.12E-05 | 0.00139 | 3.98E-06 | -0.002261351 | TXNRD3 |
| chr15 | 92586577 | 92586937 | 361 | 4 | 2.51E-05 | 1.45E-06 | 0.00109 | 3.98E-06 | -0.007358056 | AC091544.2 |
| chr16 | 21952109 | 21953183 | 1075 | 11 | 2.51E-05 | 0.006209 | 0.00077 | 4.15E-05 | 0.00125193 | NA |
| chr15 | 94080667 | 94081230 | 564 | 4 | 2.51E-05 | 1.96E-06 | 0.00035 | 2.37E-06 | 0.000143218 | LINC01581 |
| chr14 | 22547261 | 22547385 | 125 | 4 | 2.51E-05 | 6.85E-06 | 0.0015 | 4.36E-06 | -0.002135446 | NA |
| chr11 | 1310267 | 1311463 | 1197 | 6 | 2.52E-05 | 1.63E-07 | 0.00533 | 1.15E-06 | 0.0108143 | TOLLIP-AS1 |
| chr4 | 102826348 | 102827340 | 993 | 9 | 2.53E-05 | 2.22E-06 | 0.00513 | 1.77E-06 | 0.000644342 | UBE2D3 |
| chr3 | 64267858 | 64268436 | 579 | 7 | 2.53E-05 | 2.09E-06 | 0.00222 | 3.19E-06 | -0.007596932 | PRICKLE2 |
| chr12 | 52079230 | 52080263 | 1034 | 7 | 2.53E-05 | 0.000245 | 0.00207 | 1.77E-05 | -0.004865851 | SMIM41, AC025259.1 |
| chr19 | 15377029 | 15377180 | 152 | 2 | 2.53E-05 | 8.09E-06 | 0.00024 | 1.12E-05 | -0.011335517 | AKAP8 |
| chr22 | 37601496 | 37601805 | 310 | 2 | 2.53E-05 | 6.16E-06 | 5.48E-05 | 5.05E-06 | -0.012814022 | NA |
| chr1 | 89768544 | 89769081 | 538 | 4 | 2.54E-05 | 4.71E-06 | 0.00015 | 1.97E-06 | -0.003458967 | LRRC8C, AC093423.3 |
| chr11 | 1033157 | 1033345 | 189 | 3 | 2.54E-05 | 0.029176 | 1.02E-05 | 3.36E-05 | -0.002698267 | MUC6 |
| chr1 | 1540295 | 1541281 | 987 | 5 | 2.54E-05 | 1.20E-05 | 0.00613 | 3.41E-05 | -0.004029887 | TMEM240 |
| chr5 | 175656978 | 175657247 | 270 | 3 | 2.55E-05 | 1.65E-06 | 0.00152 | 4.36E-06 | -0.006134953 | NA |
| chr5 | 501748 | 502176 | 429 | 5 | 2.55E-05 | 4.41E-05 | 0.00286 | 8.01E-06 | -0.005605277 | SLC9A3 |
| chr12 | 107660596 | 107661065 | 470 | 3 | 2.55E-05 | 0.000206 | 0.00045 | 1.51E-05 | -0.009846234 | NA |
| chr19 | 2334566 | 2335086 | 521 | 5 | 2.55E-05 | 1.54E-06 | 0.00237 | 2.70E-06 | -0.006670896 | AC005258.1, SPPL2B |
| chr8 | 141440131 | 141440315 | 185 | 2 | 2.55E-05 | 4.05E-06 | 0.00014 | 5.60E-06 | -0.008750143 | MROH5 |
| chr21 | 41957468 | 41958336 | 869 | 3 | 2.55E-05 | 1.11E-06 | 0.00081 | 2.65E-06 | -0.022952148 | NA |
| chr7 | 150955697 | 150956361 | 665 | 11 | 2.56E-05 | 3.29E-05 | 0.00144 | 1.10E-05 | -0.005057935 | KCNH2 |
| chr3 | 197458246 | 197459285 | 1040 | 7 | 2.57E-05 | 1.54E-06 | 0.00619 | 1.10E-06 | -0.016904272 | AC128709.3, AC128709.2 |
| chr20 | 56403469 | 56403798 | 330 | 4 | 2.57E-05 | 0.000742 | 1.77E-05 | 1.57E-05 | -0.001770959 | CSTF1 |
| chr3 | 186611753 | 186612278 | 526 | 5 | 2.57E-05 | 0.018635 | 1.52E-05 | 4.20E-05 | -0.001224941 | AC068631.1 |
| chr9 | 17134571 | 17134972 | 402 | 6 | 2.57E-05 | 3.43E-05 | 0.00012 | 8.72E-06 | -0.005696483 | NA |
| chr11 | 72553941 | 72554353 | 413 | 2 | 2.57E-05 | 1.94E-06 | 0.00018 | 3.12E-06 | -0.019438766 | NA |
| chr19 | 18931911 | 18932530 | 620 | 5 | 2.57E-05 | 0.001004 | 5.61E-05 | 9.17E-06 | 0.000806205 | AC002985.1, HOMER3 |
| chr4 | 2792047 | 2792910 | 864 | 9 | 2.57E-05 | 0.001261 | 6.28E-05 | 2.71E-05 | -0.00330867 | NA |
| chr1 | 150009705 | 150010453 | 749 | 5 | 2.58E-05 | 1.34E-05 | 0.0006 | 3.19E-06 | 0.004234163 | OTUD7B |
| chr1 | 53894105 | 53894885 | 781 | 6 | 2.58E-05 | 0.000163 | 4.54E-05 | 2.15E-05 | -0.003050367 | DIO1 |
| chr14 | 50823418 | 50823451 | 34 | 2 | 2.59E-05 | 5.88E-06 | 0.00027 | 8.70E-06 | -0.01563126 | AL133485.2, NIN |
| chr10 | 97444970 | 97446545 | 1576 | 17 | 2.59E-05 | 3.36E-06 | 0.00267 | 4.02E-07 | 0.000526697 | ZDHHC16, EXOSC1 |
| chr7 | 127058102 | 127058775 | 674 | 4 | 2.59E-05 | 0.000373 | 0.00017 | 1.47E-05 | -4.91E-05 | GRM8, MIR592 |
| chr1 | 1061889 | 1062967 | 1079 | 4 | 2.60E-05 | 1.56E-06 | 0.00043 | 3.13E-06 | -0.009449219 | AL390719.1, AL645608.8 |
| chr1 | 44806965 | 44810865 | 3901 | 21 | 2.60E-05 | 1.30E-10 | 0.00055 | 4.52E-11 | -0.005286338 | BTBD19, TCTEX1D4 |
| chr5 | 142982264 | 142982558 | 295 | 2 | 2.60E-05 | 1.14E-05 | 0.00075 | 1.82E-05 | -0.01249773 | ARHGAP26 |
| chr22 | 46830280 | 46830606 | 327 | 3 | 2.61E-05 | 3.07E-06 | 0.00014 | 3.96E-06 | -0.007441817 | TBC1D22A |
| chr20 | 49936169 | 49936996 | 828 | 7 | 2.62E-05 | 0.000106 | 0.00015 | 7.89E-06 | 0.0013032 | RNF114 |
| chr1 | 9151777 | 9152462 | 686 | 4 | 2.62E-05 | 0.001233 | 6.60E-05 | 5.60E-05 | -0.00359488 | MIR34AHG, MIR34A |
| chr8 | 29054109 | 29054276 | 168 | 2 | 2.62E-05 | 1.02E-05 | 0.00016 | 1.21E-05 | -0.010201694 | HMBOX1 |
| chr3 | 119463906 | 119464998 | 1093 | 5 | 2.62E-05 | 3.03E-05 | 0.00024 | 6.33E-06 | -0.007228081 | TMEM39A |
| chr11 | 69658377 | 69658948 | 572 | 5 | 2.62E-05 | 2.65E-05 | 0.00062 | 8.93E-06 | -0.007050138 | LTO1 |
| chr5 | 146878090 | 146879440 | 1351 | 19 | 2.63E-05 | 2.62E-06 | 0.01005 | 6.28E-07 | -0.002173319 | PPP2R2B |
| chr12 | 57128084 | 57129527 | 1444 | 17 | 2.63E-05 | 0.000102 | 0.00663 | 4.82E-06 | -0.001640405 | LRP1, STAT6 |
| chr4 | 186708078 | 186709148 | 1071 | 11 | 2.63E-05 | 1.98E-05 | 0.00109 | 1.37E-06 | -0.000864951 | FAT1 |
| chr1 | 16243064 | 16243394 | 331 | 3 | 2.63E-05 | 1.02E-05 | 0.00024 | 3.39E-06 | -0.005360915 | NA |
| chr8 | 66178674 | 66179675 | 1002 | 8 | 2.63E-05 | 2.01E-06 | 0.00786 | 6.64E-06 | -0.006066169 | NA |
| chr14 | 24302998 | 24303029 | 32 | 2 | 2.64E-05 | 4.79E-06 | 0.00034 | 7.64E-06 | -0.003863917 | NOP9 |
| chr12 | 111777524 | 111777978 | 455 | 2 | 2.64E-05 | 1.28E-06 | 0.00025 | 2.22E-06 | -0.017576459 | AC002996.1, ALDH2 |
| chr10 | 17229007 | 17229520 | 514 | 9 | 2.64E-05 | 0.000395 | 0.00112 | 5.42E-05 | -0.00269766 | VIM, VIM-AS1 |
| chr9 | 133787750 | 133788215 | 466 | 3 | 2.64E-05 | 0.000288 | 0.00043 | 1.44E-05 | -0.023450196 | VAV2 |
| chr12 | 64882629 | 64882861 | 233 | 2 | 2.64E-05 | 9.21E-05 | 4.17E-05 | 2.17E-05 | -0.005774559 | AC078815.1 |
| chr11 | 6405270 | 6406134 | 865 | 7 | 2.64E-05 | 0.008679 | 1.98E-05 | 0.00015 | -0.002191731 | APBB1 |
| chr17 | 82588485 | 82589248 | 764 | 6 | 2.64E-05 | 2.67E-06 | 0.00387 | 7.21E-06 | -0.000617959 | FOXK2 |
| chr1 | 236182988 | 236183224 | 237 | 2 | 2.65E-05 | 0.001554 | 2.04E-05 | 2.39E-05 | -0.003845925 | GPR137B |
| chr20 | 62952185 | 62952896 | 712 | 11 | 2.65E-05 | 1.69E-05 | 0.00286 | 2.80E-06 | 0.006042577 | SLC17A9 |
| chr17 | 8697435 | 8697785 | 351 | 4 | 2.66E-05 | 3.36E-06 | 0.00027 | 4.43E-06 | -0.007089043 | NA |
| chr18 | 11850908 | 11851990 | 1083 | 12 | 2.66E-05 | 0.006705 | 7.44E-05 | 3.17E-05 | -0.000398647 | GNAL, CHMP1B, CHMP1B-AS1 |
| chr3 | 30894578 | 30895065 | 488 | 10 | 2.66E-05 | 2.36E-06 | 0.00195 | 4.27E-06 | 0.00312249 | GADL1 |
| chr8 | 81720739 | 81721855 | 1117 | 9 | 2.67E-05 | 6.63E-06 | 0.00214 | 1.91E-06 | -0.000134054 | ZFAND1 |
| chr8 | 666458 | 667669 | 1212 | 3 | 2.68E-05 | 4.69E-07 | 0.00106 | 1.41E-06 | -0.004640606 | ERICH1 |
| chr6 | 17281571 | 17282469 | 899 | 9 | 2.68E-05 | 0.001242 | 0.00048 | 8.49E-06 | -0.001127492 | RBM24 |
| chr21 | 32545638 | 32545699 | 62 | 2 | 2.68E-05 | 9.06E-06 | 2.57E-05 | 4.07E-06 | -0.003672962 | CFAP298-TCP10L, TCP10L |
| chr19 | 42095337 | 42096126 | 790 | 6 | 2.68E-05 | 0.000132 | 0.00276 | 6.94E-06 | -0.005065834 | POU2F2 |
| chr3 | 9932619 | 9933849 | 1231 | 12 | 2.69E-05 | 1.61E-05 | 0.00277 | 1.61E-06 | 0.000254503 | IL17RC, AC018809.3, CRELD1 |
| chr20 | 25032139 | 25032887 | 749 | 8 | 2.70E-05 | 1.14E-05 | 0.00121 | 8.13E-06 | -0.002185688 | ACSS1 |
| chr6 | 33270991 | 33272438 | 1448 | 33 | 2.70E-05 | 4.12E-05 | 0.0135 | 8.16E-06 | -0.000131872 | RPS18, VPS52 |
| chr9 | 122228353 | 122229377 | 1025 | 16 | 2.71E-05 | 0.001483 | 0.0005 | 3.42E-05 | -0.002087037 | LHX6 |
| chr16 | 85935313 | 85936044 | 732 | 4 | 2.71E-05 | 1.26E-05 | 0.00174 | 9.86E-06 | -0.007999264 | AC092723.5, LINC02132 |
| chr11 | 61498897 | 61499283 | 387 | 2 | 2.71E-05 | 2.41E-06 | 0.00015 | 3.66E-06 | -0.008729001 | AP003108.1 |
| chr11 | 3166041 | 3167336 | 1296 | 22 | 2.72E-05 | 0.000216 | 0.00022 | 1.55E-05 | -0.002393815 | OSBPL5 |
| chr11 | 63984805 | 63985756 | 952 | 7 | 2.72E-05 | 6.23E-05 | 0.00032 | 4.67E-06 | -0.007169254 | AP000721.1 |
| chr12 | 3454102 | 3454920 | 819 | 5 | 2.72E-05 | 6.79E-07 | 0.00654 | 3.71E-06 | -0.027285416 | PRMT8 |
| chr7 | 149496136 | 149496433 | 298 | 2 | 2.73E-05 | 6.52E-06 | 0.00017 | 8.54E-06 | -0.020314935 | ZNF746 |
| chr7 | 1479063 | 1479807 | 745 | 3 | 2.73E-05 | 2.09E-05 | 0.00069 | 2.04E-05 | -0.006421341 | INTS1 |
| chr9 | 91817391 | 91817738 | 348 | 3 | 2.73E-05 | 1.61E-05 | 4.80E-05 | 6.35E-06 | -0.007218491 | ROR2 |
| chr3 | 38305034 | 38305950 | 917 | 6 | 2.73E-05 | 5.89E-07 | 0.0039 | 2.70E-06 | -0.001967947 | SLC22A14 |
| chr12 | 6646650 | 6646922 | 273 | 2 | 2.74E-05 | 5.17E-06 | 0.00052 | 8.63E-06 | -0.020779875 | ACRBP |
| chr1 | 109075816 | 109076267 | 452 | 8 | 2.74E-05 | 0.000523 | 0.0006 | 1.23E-05 | 0.001793717 | TAF13 |
| chr7 | 2558345 | 2558874 | 530 | 5 | 2.74E-05 | 0.00024 | 0.00039 | 3.15E-05 | -0.006249656 | NA |
| chr19 | 10566815 | 10567339 | 525 | 5 | 2.75E-05 | 1.52E-05 | 0.00055 | 2.07E-05 | -0.007176627 | CDKN2D |
| chr9 | 114503092 | 114503749 | 658 | 6 | 2.76E-05 | 1.58E-05 | 0.00132 | 1.40E-05 | -0.007520902 | WHRN |
| chr11 | 81388893 | 81388953 | 61 | 2 | 2.77E-05 | 0.002185 | 8.73E-06 | 9.72E-06 | -0.003406237 | AP003464.1 |
| chr5 | 135336364 | 135336606 | 243 | 2 | 2.77E-05 | 2.55E-05 | 4.42E-05 | 1.14E-05 | -0.006415631 | C5orf66, MACROH2A1 |
| chr17 | 43648746 | 43649101 | 356 | 2 | 2.77E-05 | 6.81E-06 | 0.00018 | 8.95E-06 | -0.006766039 | MEOX1 |
| chr2 | 95273919 | 95274957 | 1039 | 8 | 2.77E-05 | 8.08E-05 | 0.00118 | 1.10E-05 | -0.003648758 | PROM2 |
| chr2 | 234450921 | 234451019 | 99 | 2 | 2.77E-05 | 0.000248 | 1.28E-05 | 1.03E-05 | -0.010501784 | AC097713.1, LINC01891 |
| chr11 | 64935449 | 64936165 | 717 | 9 | 2.78E-05 | 1.81E-06 | 0.0045 | 3.70E-06 | -0.00465242 | GPHA2 |
| chr5 | 31825182 | 31825846 | 665 | 4 | 2.78E-05 | 4.23E-05 | 8.17E-05 | 6.63E-06 | -0.003404876 | PDZD2 |
| chr19 | 40222348 | 40223250 | 903 | 3 | 2.78E-05 | 1.54E-05 | 0.00011 | 1.16E-05 | -0.014484019 | CCNP |
| chr10 | 88952868 | 88953657 | 790 | 9 | 2.78E-05 | 0.001966 | 0.00027 | 1.49E-05 | 0.000118934 | STAMBPL1, ACTA2 |
| chr3 | 127849284 | 127849680 | 397 | 2 | 2.79E-05 | 0.007689 | 2.79E-05 | 4.01E-05 | -0.002292808 | MGLL |
| chr8 | 27396492 | 27396513 | 22 | 2 | 2.80E-05 | 6.93E-06 | 7.43E-05 | 6.49E-06 | -0.00946337 | PTK2B |
| chr8 | 1018906 | 1019381 | 476 | 5 | 2.80E-05 | 0.001888 | 0.00025 | 2.88E-05 | -0.015272698 | DLGAP2, AC129915.3 |
| chr3 | 141456458 | 141457194 | 737 | 2 | 2.80E-05 | 8.66E-06 | 0.00017 | 1.07E-05 | -0.011841031 | NA |
| chr19 | 795310 | 795467 | 158 | 3 | 2.80E-05 | 3.80E-06 | 0.00108 | 8.54E-06 | -0.003458777 | NA |
| chr21 | 45592731 | 45593289 | 559 | 3 | 2.80E-05 | 1.37E-05 | 0.00123 | 2.34E-05 | -0.003676739 | NA |
| chr7 | 1541581 | 1542047 | 467 | 5 | 2.80E-05 | 6.75E-06 | 0.00118 | 1.55E-05 | -0.008001439 | MAFK |
| chr22 | 41366298 | 41366495 | 198 | 3 | 2.81E-05 | 1.35E-05 | 0.00298 | 2.80E-05 | -0.00484891 | NA |
| chr3 | 52685125 | 52686506 | 1382 | 14 | 2.81E-05 | 1.46E-05 | 0.00051 | 1.62E-06 | 0.002273099 | GNL3, PBRM1 |
| chr3 | 138608316 | 138608886 | 571 | 11 | 2.81E-05 | 4.55E-05 | 0.00597 | 1.35E-05 | -0.003889432 | FAIM |
| chr3 | 195093825 | 195094843 | 1019 | 6 | 2.81E-05 | 2.54E-06 | 0.00052 | 5.36E-07 | -0.00420867 | XXYLT1-AS1, XXYLT1 |
| chr15 | 64387134 | 64388096 | 963 | 12 | 2.82E-05 | 0.003164 | 0.00036 | 2.41E-05 | 8.52E-05 | TRIP4, PCLAF |
| chr15 | 68354618 | 68354999 | 382 | 2 | 2.82E-05 | 0.001162 | 2.54E-05 | 2.81E-05 | -0.00472596 | ITGA11 |
| chr10 | 77866392 | 77867145 | 754 | 6 | 2.82E-05 | 0.02265 | 1.30E-05 | 5.33E-05 | -0.001470513 | AL391421.1, DLG5 |
| chr11 | 112806878 | 112807184 | 307 | 2 | 2.83E-05 | 2.61E-06 | 0.00027 | 4.36E-06 | -0.030951429 | NA |
| chr17 | 42379254 | 42379466 | 213 | 2 | 2.84E-05 | 1.86E-05 | 3.56E-05 | 7.97E-06 | -0.00854828 | STAT3 |
| chr1 | 15729144 | 15729723 | 580 | 6 | 2.84E-05 | 0.000858 | 0.00044 | 2.79E-05 | -0.000965441 | PLEKHM2, AL450998.2 |
| chr2 | 10908804 | 10909729 | 926 | 2 | 2.84E-05 | 8.70E-07 | 9.41E-05 | 1.34E-06 | -0.003562305 | NA |
| chr17 | 44847759 | 44848364 | 606 | 4 | 2.84E-05 | 9.94E-06 | 0.00023 | 9.48E-06 | -0.003706784 | HIGD1B |
| chr7 | 130734833 | 130735171 | 339 | 6 | 2.84E-05 | 0.010936 | 6.55E-05 | 3.99E-05 | 0.007886355 | NA |
| chr1 | 155261975 | 155262588 | 614 | 13 | 2.85E-05 | 0.000426 | 0.00016 | 1.47E-05 | -1.32E-05 | SCAMP3 |
| chr8 | 135709769 | 135710290 | 522 | 2 | 2.85E-05 | 6.05E-06 | 8.52E-05 | 6.27E-06 | -0.009940439 | NA |
| chr2 | 197676931 | 197677326 | 396 | 5 | 2.85E-05 | 0.001697 | 0.00015 | 2.11E-05 | -0.004689772 | NA |
| chr8 | 101042002 | 101042336 | 335 | 2 | 2.86E-05 | 5.59E-06 | 7.04E-05 | 5.39E-06 | -0.005355292 | FLJ42969 |
| chr17 | 56888605 | 56888840 | 236 | 4 | 2.86E-05 | 3.66E-06 | 0.00045 | 4.93E-06 | -0.004300709 | TRIM25 |
| chr8 | 85466029 | 85466086 | 58 | 2 | 2.86E-05 | 2.27E-05 | 3.91E-05 | 9.64E-06 | 0.00633014 | CA2 |
| chr6 | 73461214 | 73462019 | 806 | 12 | 2.86E-05 | 3.43E-06 | 0.00215 | 4.71E-06 | 0.00028279 | MTO1 |
| chr6 | 69867167 | 69867714 | 548 | 7 | 2.86E-05 | 0.00647 | 1.15E-05 | 5.46E-05 | 0.001813138 | COL19A1, LMBRD1 |
| chr19 | 38293141 | 38293482 | 342 | 2 | 2.88E-05 | 9.69E-06 | 0.00061 | 1.54E-05 | -0.007320407 | AC011479.1 |
| chr11 | 26993926 | 26994444 | 519 | 8 | 2.88E-05 | 4.32E-07 | 0.00422 | 2.05E-06 | -0.016149528 | FIBIN |
| chr12 | 93887345 | 93887760 | 416 | 3 | 2.88E-05 | 1.74E-05 | 0.00034 | 8.41E-06 | -0.019676563 | CRADD |
| chr4 | 654963 | 656112 | 1150 | 9 | 2.88E-05 | 1.76E-05 | 0.00062 | 1.20E-05 | -0.004788262 | PDE6B, PDE6B-AS1 |
| chr6 | 157669883 | 157669917 | 35 | 2 | 2.89E-05 | 3.16E-06 | 0.00012 | 4.22E-06 | -0.022541042 | ZDHHC14 |
| chr11 | 2399419 | 2400061 | 643 | 3 | 2.89E-05 | 0.000223 | 0.00109 | 6.85E-05 | -0.009839296 | NA |
| chr19 | 45712173 | 45712921 | 749 | 6 | 2.89E-05 | 3.19E-06 | 0.00183 | 4.17E-06 | -0.002983041 | FBXO46 |
| chr12 | 116149067 | 116149463 | 397 | 3 | 2.90E-05 | 0.000139 | 0.00013 | 1.26E-05 | -0.010121426 | MED13L |
| chr2 | 111139849 | 111140005 | 157 | 2 | 2.90E-05 | 5.90E-06 | 4.42E-05 | 4.33E-06 | -0.006815793 | BCL2L11, MIR4435-2HG |
| chr11 | 119311425 | 119311866 | 442 | 2 | 2.90E-05 | 6.34E-06 | 0.0002 | 8.69E-06 | -0.005657312 | CBL, MCAM |
| chr10 | 110530095 | 110530573 | 479 | 7 | 2.90E-05 | 1.56E-05 | 0.0013 | 6.13E-06 | -0.008812357 | NA |
| chr2 | 10303506 | 10304324 | 819 | 7 | 2.90E-05 | 2.26E-06 | 0.00031 | 1.79E-06 | -0.000677241 | HPCAL1 |
| chr19 | 45039228 | 45039452 | 225 | 3 | 2.91E-05 | 0.000125 | 2.93E-05 | 1.76E-05 | 0.008260104 | CLASRP |
| chr1 | 2276420 | 2276910 | 491 | 4 | 2.91E-05 | 4.85E-07 | 0.00329 | 2.14E-06 | -0.015661335 | SKI |
| chr1 | 205002705 | 205003690 | 986 | 4 | 2.91E-05 | 3.82E-07 | 0.00197 | 1.59E-06 | -0.013654768 | NFASC |
| chr4 | 845740 | 845910 | 171 | 3 | 2.92E-05 | 8.85E-07 | 0.00132 | 2.52E-06 | -0.009850485 | AC139887.5 |
| chr1 | 40708899 | 40709527 | 629 | 8 | 2.92E-05 | 3.60E-06 | 0.00511 | 4.69E-06 | -0.009071327 | NFYC |
| chr6 | 31120411 | 31121380 | 970 | 7 | 2.92E-05 | 9.83E-08 | 0.00758 | 7.50E-07 | 0.018822479 | PSORS1C1, CDSN |
| chr2 | 16227956 | 16228367 | 412 | 4 | 2.92E-05 | 5.17E-05 | 0.00014 | 2.88E-05 | -0.011656177 | AC010745.2, AC010745.3 |
| chr3 | 194135654 | 194136881 | 1228 | 12 | 2.93E-05 | 0.000611 | 0.00043 | 5.87E-05 | 0.001636498 | HES1 |
| chr11 | 69080002 | 69080809 | 808 | 5 | 2.94E-05 | 6.30E-07 | 0.00041 | 1.60E-06 | -0.007058894 | TPCN2, AP003071.5 |
| chr3 | 12354055 | 12354415 | 361 | 2 | 2.94E-05 | 0.001363 | 1.83E-05 | 2.09E-05 | -0.01434329 | PPARG |
| chr5 | 160007763 | 160008426 | 664 | 4 | 2.94E-05 | 0.000251 | 0.00116 | 5.76E-05 | -0.006269437 | NA |
| chr1 | 54764027 | 54764284 | 258 | 2 | 2.95E-05 | 0.060025 | 4.65E-05 | 7.96E-05 | 0.002626607 | PARS2 |
| chr1 | 3120352 | 3121148 | 797 | 6 | 2.95E-05 | 5.12E-05 | 0.00105 | 8.66E-06 | -0.004526004 | PRDM16 |
| chr4 | 52995775 | 52996354 | 580 | 2 | 2.95E-05 | 1.39E-06 | 0.00018 | 2.30E-06 | -0.016614842 | SCFD2 |
| chr16 | 27242822 | 27243071 | 250 | 2 | 2.95E-05 | 1.18E-05 | 5.02E-05 | 7.48E-06 | -0.020152808 | NSMCE1 |
| chr5 | 172044487 | 172045072 | 586 | 3 | 2.95E-05 | 1.55E-06 | 0.00087 | 3.66E-06 | -0.007598442 | STK10 |
| chr7 | 50142961 | 50143473 | 513 | 3 | 2.95E-05 | 1.96E-05 | 0.00016 | 1.41E-05 | -0.005192481 | SPATA48 |
| chr1 | 109213041 | 109213792 | 752 | 7 | 2.95E-05 | 1.64E-05 | 0.00055 | 1.30E-05 | -0.002944941 | NA |
| chr6 | 151805045 | 151805803 | 759 | 8 | 2.96E-05 | 1.63E-06 | 0.01047 | 9.40E-06 | 0.010215757 | ESR1 |
| chr20 | 63819679 | 63820046 | 368 | 3 | 2.96E-05 | 1.16E-05 | 0.00032 | 5.65E-06 | -0.016554078 | ZBTB46 |
| chr14 | 22002157 | 22003035 | 879 | 3 | 2.96E-05 | 3.80E-07 | 0.00066 | 1.03E-06 | -0.012997154 | NA |
| chr11 | 65890175 | 65890988 | 814 | 10 | 2.96E-05 | 0.071639 | 2.04E-05 | 3.20E-05 | 0.001063071 | CCDC85B |

| | | | | | | | | | | |
|---|---|---|---|---|---|---|---|---|---|---|
| chr15 | 75713520 | 75713802 | 283 | 3 | 2.96E-05 | 2.29E-06 | 0.00104 | 5.49E-06 | -0.01409592 | NA |
| chr1 | 204071874 | 204072721 | 848 | 5 | 2.97E-05 | 0.00017 | 8.95E-05 | 2.01E-05 | -0.001359253 | AL592146.2 |
| chr12 | 122127236 | 122127691 | 456 | 3 | 2.97E-05 | 4.24E-05 | 0.00011 | 4.63E-06 | -0.002503405 | MLXIP |
| chr9 | 136823486 | 136823895 | 410 | 4 | 2.97E-05 | 0.000103 | 0.00034 | 1.11E-05 | -0.002098421 | AL355987.2, RABL6 |
| chr7 | 129776128 | 129776154 | 27 | 2 | 2.97E-05 | 3.32E-05 | 0.0005 | 4.15E-05 | -0.015609537 | NA |
| chr16 | 5033738 | 5034242 | 505 | 8 | 2.97E-05 | 2.75E-05 | 0.00114 | 6.95E-06 | 0.001012128 | NAGPA-AS1, ALG1, NAGPA |
| chr10 | 86957607 | 86958636 | 1030 | 10 | 2.97E-05 | 1.64E-06 | 0.00674 | 3.22E-06 | -0.009912268 | SNCG, MMRN2 |
| chr6 | 11778669 | 11779708 | 1040 | 6 | 2.98E-05 | 2.76E-08 | 0.00244 | 2.20E-07 | -0.009251627 | ADTRP |
| chr8 | 130802807 | 130803176 | 370 | 3 | 2.98E-05 | 0.000342 | 3.11E-05 | 1.41E-05 | -0.012860151 | ADCY8 |
| chr12 | 125105745 | 125107094 | 1350 | 5 | 2.99E-05 | 6.03E-05 | 0.00013 | 2.04E-06 | -0.002909158 | AACS |
| chr14 | 92650168 | 92650657 | 490 | 3 | 3.00E-05 | 1.28E-05 | 0.00054 | 9.53E-06 | -0.008693173 | RIN3 |
| chr7 | 156178260 | 156178704 | 445 | 6 | 3.00E-05 | 2.00E-05 | 0.00186 | 9.01E-06 | -0.008331867 | NA |
| chr7 | 100572987 | 100573994 | 1008 | 5 | 3.00E-05 | 0.000162 | 0.00032 | 7.26E-06 | -0.004490704 | SAP25, AC069281.2 |
| chr12 | 123571416 | 123571829 | 414 | 3 | 3.01E-05 | 6.88E-05 | 9.32E-05 | 4.32E-06 | -0.005716197 | NA |
| chr2 | 11656660 | 11656891 | 232 | 4 | 3.01E-05 | 9.09E-07 | 0.0025 | 3.11E-06 | -0.018146979 | NA |
| chr15 | 97652578 | 97653103 | 526 | 6 | 3.01E-05 | 3.61E-07 | 0.00887 | 2.61E-06 | -0.028307635 | LINC00923 |
| chr10 | 117375470 | 117376471 | 1002 | 8 | 3.01E-05 | 1.57E-05 | 0.00124 | 3.31E-06 | -0.002623768 | NA |
| chr17 | 82596094 | 82597142 | 1049 | 10 | 3.02E-05 | 3.19E-05 | 0.00076 | 1.65E-06 | -0.005377014 | FOXK2 |
| chr4 | 957672 | 958578 | 907 | 5 | 3.02E-05 | 9.09E-07 | 0.00188 | 2.37E-06 | -0.004531362 | TMEM175 |
| chr4 | 152115951 | 152116018 | 68 | 2 | 3.02E-05 | 2.61E-06 | 0.0001 | 3.48E-06 | -0.014847482 | AC079340.3 |
| chr17 | 27349175 | 27349895 | 721 | 5 | 3.02E-05 | 7.63E-05 | 0.00029 | 7.04E-06 | -0.004871256 | AC026254.2 |
| chr10 | 104333901 | 104334408 | 508 | 8 | 3.02E-05 | 3.32E-06 | 0.00233 | 3.33E-06 | -0.002209967 | ITPRIP |
| chr12 | 96989360 | 96990069 | 710 | 2 | 3.03E-05 | 1.40E-06 | 6.28E-05 | 1.73E-06 | -0.014923963 | AC007656.2 |
| chr12 | 85279930 | 85280916 | 987 | 13 | 3.03E-05 | 1.87E-05 | 0.00126 | 3.96E-06 | 7.79E-05 | ALX1 |
| chr12 | 113135125 | 113136004 | 880 | 12 | 3.03E-05 | 0.047158 | 2.96E-05 | 6.50E-05 | -0.000167862 | RASAL1 |
| chr11 | 1460206 | 1460549 | 344 | 3 | 3.03E-05 | 6.30E-05 | 6.64E-05 | 2.22E-05 | -0.008838766 | BRSK2 |
| chr19 | 55578441 | 55579320 | 880 | 3 | 3.03E-05 | 8.88E-06 | 0.00108 | 1.27E-05 | -0.001829435 | ZNF579 |
| chr11 | 110713156 | 110714004 | 849 | 10 | 3.04E-05 | 7.80E-06 | 0.00053 | 2.80E-06 | -0.001784347 | ARHGAP20 |
| chr19 | 1359968 | 1360279 | 312 | 5 | 3.04E-05 | 3.71E-05 | 0.00013 | 6.55E-06 | -0.002954634 | PWWP3A |
| chr12 | 121831278 | 121831769 | 492 | 3 | 3.04E-05 | 9.08E-05 | 0.00061 | 3.87E-05 | -0.006864969 | SETD1B |
| chr22 | 45413438 | 45414162 | 725 | 13 | 3.05E-05 | 3.75E-07 | 0.00325 | 2.17E-06 | -0.013081258 | RIBC2, SMC1B |
| chr1 | 154707739 | 154708918 | 1180 | 7 | 3.05E-05 | 0.001491 | 0.00035 | 4.18E-05 | -0.002237931 | KCNN3 |
| chr13 | 98576701 | 98577750 | 1050 | 4 | 3.05E-05 | 5.29E-05 | 0.00102 | 2.71E-05 | -0.003726593 | STK24-AS1, STK24 |
| chr17 | 40175807 | 40176805 | 999 | 5 | 3.05E-05 | 2.75E-05 | 0.00084 | 2.09E-05 | -0.003432442 | NA |
| chr14 | 103536036 | 103536317 | 282 | 2 | 3.05E-05 | 2.75E-05 | 6.45E-05 | 1.54E-05 | -0.005495166 | TRMT61A |
| chr11 | 73787068 | 73787903 | 836 | 8 | 3.06E-05 | 1.15E-06 | 0.00421 | 2.80E-06 | -0.006560676 | MRPL48 |
| chr17 | 12706303 | 12706639 | 337 | 3 | 3.06E-05 | 0.000312 | 0.00016 | 1.55E-05 | -0.014931227 | MYOCD |
| chr1 | 27113972 | 27114655 | 684 | 5 | 3.06E-05 | 0.00046 | 0.00112 | 1.15E-05 | -0.00596575 | SLC9A1 |
| chr11 | 75835245 | 75835831 | 587 | 3 | 3.07E-05 | 3.17E-05 | 5.76E-05 | 1.27E-05 | -0.005060365 | UVRAG, Y RNA |
| chr12 | 114693984 | 114694210 | 227 | 3 | 3.07E-05 | 1.65E-05 | 0.00088 | 1.54E-05 | -0.002777438 | AC026765.3 |
| chr19 | 7224309 | 7224702 | 394 | 5 | 3.07E-05 | 0.000405 | 1.81E-05 | 1.03E-05 | -0.004468678 | INSR |
| chr7 | 120858520 | 120859614 | 1095 | 5 | 3.09E-05 | 3.34E-07 | 0.00016 | 1.70E-07 | -0.011717397 | NA |
| chr19 | 19335821 | 19336018 | 198 | 2 | 3.09E-05 | 1.03E-05 | 4.18E-05 | 6.13E-06 | -0.005250405 | MAU2 |
| chr1 | 212272014 | 212272524 | 511 | 3 | 3.09E-05 | 5.89E-06 | 0.00057 | 6.57E-06 | -0.010508873 | NA |
| chr14 | 104175066 | 104176154 | 1089 | 5 | 3.09E-05 | 0.00074 | 0.00026 | 1.39E-05 | -0.003691854 | KIF26A |
| chr10 | 89260710 | 89261072 | 363 | 4 | 3.10E-05 | 0.000497 | 0.00025 | 9.45E-06 | -0.004276437 | LIPA |
| chr3 | 52704865 | 52705530 | 666 | 9 | 3.10E-05 | 2.01E-06 | 0.01788 | 8.53E-06 | -0.004375244 | SPCS1, GLT8D1 |
| chr17 | 31857978 | 31858350 | 373 | 2 | 3.10E-05 | 8.30E-06 | 0.0001 | 8.63E-06 | -0.018048436 | COPRS |
| chr16 | 4601731 | 4602236 | 506 | 2 | 3.11E-05 | 1.88E-06 | 0.0002 | 3.10E-06 | -0.014444268 | NA |
| chr5 | 179878602 | 179879210 | 609 | 2 | 3.11E-05 | 0.026665 | 0.00016 | 0.00043 | -0.002574333 | TBC1D9B |
| chr11 | 78079750 | 78080471 | 722 | 12 | 3.11E-05 | 9.08E-05 | 0.0015 | 7.66E-06 | -0.002002044 | NDUFC2-KCTD14, NDUFC2 |
| chr9 | 136148440 | 136148892 | 453 | 2 | 3.11E-05 | 2.04E-06 | 0.00023 | 3.40E-06 | 0.006484337 | NA |
| chr7 | 156944887 | 156945476 | 590 | 2 | 3.12E-05 | 7.83E-06 | 0.00011 | 8.35E-06 | -0.017667148 | AC006357.1 |
| chr14 | 100778269 | 100778521 | 253 | 2 | 3.12E-05 | 0.011412 | 2.19E-05 | 3.09E-05 | -0.002287442 | NA |
| chr22 | 19950494 | 19951018 | 525 | 7 | 3.12E-05 | 0.000242 | 0.00167 | 2.65E-05 | -0.005255527 | COMT |
| chr12 | 111470845 | 111471341 | 497 | 3 | 3.12E-05 | 0.000482 | 0.00027 | 2.26E-05 | -0.005565843 | ATXN2 |
| chr19 | 21750584 | 21750943 | 360 | 9 | 3.13E-05 | 0.000473 | 0.00036 | 1.04E-05 | -0.001267096 | CCNYL6, ZNF100 |
| chr3 | 50319740 | 50320245 | 506 | 7 | 3.13E-05 | 1.82E-05 | 0.00086 | 5.83E-06 | -0.004240165 | HYAL2, TUSC2 |
| chr19 | 49339903 | 49340665 | 763 | 5 | 3.14E-05 | 6.80E-05 | 0.00446 | 1.33E-05 | -0.012890851 | CD37, AC011450.1, TEAD2 |
| chr2 | 3463409 | 3463642 | 234 | 5 | 3.14E-05 | 0.000339 | 2.08E-05 | 1.86E-05 | -0.005556347 | TRAPPC12 |
| chr1 | 8424357 | 8425316 | 960 | 6 | 3.15E-05 | 4.04E-05 | 0.00593 | 1.91E-05 | 0.020656673 | RERE-AS1, RERE |
| chr17 | 16981424 | 16982398 | 975 | 6 | 3.15E-05 | 7.79E-07 | 0.00381 | 9.23E-07 | -0.002725707 | NA |
| chr12 | 56256449 | 56257182 | 734 | 2 | 3.15E-05 | 5.02E-06 | 0.00051 | 8.38E-06 | -0.00764581 | ANKRD52 |
| chr1 | 1011495 | 1011963 | 469 | 6 | 3.16E-05 | 0.000466 | 0.00013 | 1.69E-05 | -0.009813057 | ISG15 |
| chr22 | 48535968 | 48536512 | 545 | 3 | 3.16E-05 | 9.51E-06 | 0.00021 | 9.47E-06 | -0.002117335 | TAFA5 |
| chr11 | 92882766 | 92883952 | 1187 | 7 | 3.17E-05 | 2.97E-07 | 0.00824 | 2.25E-06 | -0.010213599 | FAT3 |
| chr7 | 156578625 | 156578919 | 295 | 3 | 3.17E-05 | 0.000358 | 3.99E-05 | 1.02E-05 | -0.009324007 | LINC01006 |
| chr2 | 134679718 | 134680140 | 423 | 3 | 3.17E-05 | 1.09E-06 | 0.00104 | 2.87E-06 | -0.024391179 | TMEM163 |
| chr7 | 4783176 | 4784324 | 1149 | 6 | 3.18E-05 | 0.000123 | 0.00074 | 1.83E-05 | -0.000304078 | AP5Z1 |
| chr6 | 35317207 | 35317519 | 313 | 3 | 3.19E-05 | 0.001796 | 6.93E-05 | 2.92E-05 | -0.003146548 | DEF6 |
| chr20 | 63803038 | 63803963 | 926 | 3 | 3.19E-05 | 1.05E-05 | 0.00071 | 1.59E-05 | -0.014173517 | ZBTB46 |
| chr16 | 804168 | 804640 | 473 | 4 | 3.19E-05 | 9.78E-05 | 0.0004 | 3.11E-05 | -0.007679556 | NA |
| chr17 | 10659085 | 10660073 | 989 | 5 | 3.20E-05 | 3.09E-05 | 0.00043 | 1.54E-06 | -0.006010289 | NA |
| chr15 | 90903534 | 90904519 | 986 | 7 | 3.20E-05 | 0.000109 | 0.00227 | 3.53E-05 | -0.006107514 | MAN2A2 |
| chr14 | 105326973 | 105327848 | 876 | 6 | 3.20E-05 | 2.77E-05 | 0.00016 | 9.23E-06 | -0.002020225 | PACS2 |
| chr10 | 3636626 | 3636893 | 268 | 4 | 3.21E-05 | 8.78E-05 | 0.00014 | 1.05E-05 | -0.003142751 | NA |
| chr4 | 41359202 | 41360098 | 897 | 10 | 3.21E-05 | 0.000586 | 0.00057 | 9.64E-05 | -0.002672953 | LIMCH1 |
| chr16 | 50210163 | 50210801 | 639 | 2 | 3.22E-05 | 7.92E-06 | 9.81E-05 | 8.10E-06 | -0.012451512 | TENT4B |
| chr1 | 3352688 | 3352914 | 227 | 3 | 3.23E-05 | 1.05E-06 | 0.00128 | 2.89E-06 | -0.023072223 | PRDM16 |
| chr7 | 29190009 | 29190101 | 93 | 2 | 3.23E-05 | 0.000217 | 1.08E-05 | 8.31E-06 | -0.007497295 | AC004593.2, CHN2, CPVL |
| chr11 | 18455606 | 18456133 | 528 | 6 | 3.23E-05 | 2.31E-07 | 0.00745 | 1.85E-06 | -0.030324473 | LDHAL6A |
| chr20 | 10673299 | 10674183 | 885 | 8 | 3.23E-05 | 0.000144 | 0.002 | 5.26E-06 | 0.001503806 | AL050403.2, JAG1 |
| chr17 | 17078144 | 17078582 | 439 | 3 | 3.23E-05 | 5.10E-06 | 0.00034 | 7.64E-06 | -0.004299959 | MPRIP |
| chr17 | 1571309 | 1571585 | 277 | 5 | 3.23E-05 | 8.11E-07 | 0.00186 | 2.68E-06 | -0.00780758 | SLC43A2 |
| chr1 | 9847014 | 9847217 | 204 | 2 | 3.24E-05 | 9.74E-06 | 0.00066 | 1.56E-05 | -0.005663044 | NA |
| chr3 | 142723692 | 142725091 | 1400 | 14 | 3.24E-05 | 4.90E-05 | 0.00415 | 1.02E-05 | 0.000222529 | TRPC1, AC072028.1 |
| chr3 | 133491400 | 133491764 | 365 | 2 | 3.24E-05 | 0.000218 | 3.70E-05 | 2.72E-05 | -0.004713287 | AC022296.2 |
| chr11 | 86151564 | 86151780 | 217 | 3 | 3.24E-05 | 1.10E-05 | 0.00011 | 7.20E-06 | 0.008833156 | NA |
| chr13 | 52848246 | 52848339 | 94 | 2 | 3.24E-05 | 0.000173 | 7.87E-06 | 5.85E-06 | 0.014155637 | PCDH8 |
| chr22 | 50251951 | 50252417 | 467 | 3 | 3.24E-05 | 6.69E-06 | 0.00095 | 1.32E-05 | -0.010026887 | MAPK12 |
| chr1 | 30878321 | 30879407 | 1087 | 5 | 3.24E-05 | 4.46E-05 | 0.00015 | 1.57E-06 | -0.007223187 | SDC3 |
| chr17 | 10731032 | 10731215 | 184 | 3 | 3.24E-05 | 3.58E-06 | 0.00021 | 5.08E-06 | -0.020473899 | TMEM220-AS1, AC015908.7 |
| chr11 | 77191947 | 77192601 | 655 | 6 | 3.24E-05 | 1.78E-06 | 0.00238 | 4.36E-06 | 0.014401331 | MYO7A |
| chr13 | 51454282 | 51454870 | 589 | 2 | 3.24E-05 | 5.40E-05 | 0.00056 | 6.30E-05 | -0.014168359 | INTS6-AS1, RPS4XP16 |
| chr7 | 2478554 | 2479190 | 637 | 4 | 3.24E-05 | 1.96E-06 | 0.00175 | 5.82E-06 | -0.008443007 | NA |
| chr8 | 41652360 | 41652680 | 321 | 2 | 3.25E-05 | 7.71E-06 | 0.00069 | 1.27E-05 | -0.006885665 | NA |
| chr16 | 68083557 | 68084538 | 982 | 6 | 3.25E-05 | 0.000184 | 0.00209 | 4.02E-05 | -0.00881375 | AC130462.2 |
| chr2 | 239078066 | 239078614 | 549 | 3 | 3.26E-05 | 0.001222 | 0.00075 | 7.22E-05 | -0.00427597 | HDAC4 |
| chr17 | 1223407 | 1224448 | 1042 | 4 | 3.26E-05 | 3.56E-06 | 0.00044 | 1.65E-06 | -0.006246823 | ABR |
| chr2 | 96711955 | 96712293 | 339 | 2 | 3.26E-05 | 4.70E-06 | 0.00031 | 7.38E-06 | -0.005732629 | LMAN2L |
| chr6 | 30614043 | 30614961 | 919 | 7 | 3.27E-05 | 1.31E-05 | 0.00113 | 1.40E-05 | -0.006632918 | PPP1R10 |
| chr1 | 2157945 | 2158116 | 172 | 2 | 3.27E-05 | 1.55E-05 | 0.00095 | 2.47E-05 | -0.004053896 | PRKCZ |
| chr7 | 4227732 | 4227780 | 49 | 3 | 3.28E-05 | 8.38E-06 | 0.00053 | 6.59E-06 | -0.004481373 | SDK1 |
| chr2 | 196259719 | 196260101 | 383 | 3 | 3.28E-05 | 1.55E-05 | 8.56E-05 | 5.52E-06 | -0.015257855 | HECW2-AS1, HECW2 |
| chr3 | 48189358 | 48189768 | 411 | 2 | 3.29E-05 | 8.65E-06 | 0.00076 | 1.43E-05 | 0.001467657 | NA |
| chr9 | 116355540 | 116355808 | 269 | 3 | 3.29E-05 | 1.95E-06 | 0.00074 | 3.69E-06 | -0.01562263 | PAPPA |
| chr9 | 131402499 | 131403005 | 507 | 3 | 3.29E-05 | 1.68E-06 | 0.00101 | 3.82E-06 | -0.010016385 | PRRC2B |
| chr4 | 2270755 | 2270843 | 89 | 2 | 3.29E-05 | 0.002445 | 1.30E-05 | 1.55E-05 | -0.002860405 | ZFYVE28 |
| chr22 | 42959932 | 42960666 | 735 | 6 | 3.29E-05 | 5.17E-05 | 0.00111 | 2.87E-05 | -0.007683888 | PACSIN2 |
| chr14 | 74963699 | 74964261 | 563 | 4 | 3.29E-05 | 1.43E-06 | 0.00173 | 3.84E-06 | -0.014947393 | NA |
| chr12 | 132786326 | 132786755 | 430 | 4 | 3.30E-05 | 0.000206 | 0.00015 | 1.68E-05 | -0.008122275 | GOLGA3 |
| chr12 | 123479036 | 123479619 | 584 | 2 | 3.30E-05 | 2.54E-06 | 0.00028 | 4.27E-06 | -0.004291738 | RILPL1 |
| chr17 | 29019714 | 29020242 | 529 | 6 | 3.30E-05 | 2.01E-06 | 0.00131 | 2.44E-06 | -0.010565262 | PIPOX |
| chr10 | 5412221 | 5413114 | 894 | 8 | 3.30E-05 | 0.000374 | 0.00155 | 5.12E-06 | 0.002135749 | NET1 |
| chr1 | 6603604 | 6604208 | 605 | 7 | 3.31E-05 | 1.53E-05 | 0.00208 | 1.11E-05 | -0.006392778 | KLHL21 |
| chr19 | 35152982 | 35153667 | 686 | 2 | 3.31E-05 | 2.98E-06 | 0.00036 | 5.09E-06 | -0.018758442 | FXYD7 |
| chr17 | 2723697 | 2724034 | 338 | 2 | 3.31E-05 | 1.37E-05 | 0.00023 | 1.70E-05 | -0.009780775 | AC005696.4 |
| chr14 | 100728975 | 100730661 | 1687 | 4 | 3.31E-05 | 4.87E-08 | 0.00018 | 9.18E-08 | -0.013062285 | DLK1 |
| chr10 | 28281368 | 28282393 | 1026 | 5 | 3.32E-05 | 4.73E-06 | 0.00347 | 8.69E-06 | -0.009967516 | MPP7 |
| chr14 | 100128095 | 100128306 | 212 | 2 | 3.32E-05 | 7.20E-05 | 4.71E-05 | 2.13E-05 | -0.008423723 | EVL |
| chr14 | 24299567 | 24300548 | 982 | 16 | 3.32E-05 | 3.90E-05 | 0.00962 | 1.82E-05 | -0.001259553 | NOP9, DHRS1 |
| chr15 | 89627855 | 89628965 | 1111 | 9 | 3.32E-05 | 1.53E-05 | 0.0004 | 4.17E-06 | -0.002835032 | TICRR, KIF7 |
| chr6 | 166627398 | 166628452 | 1055 | 9 | 3.33E-05 | 9.79E-07 | 0.01152 | 4.34E-06 | 0.002906113 | RPS6KA2 |
| chr15 | 72305945 | 72307231 | 1287 | 9 | 3.33E-05 | 1.61E-06 | 0.00524 | 4.78E-07 | -0.00696386 | AC009690.2, AC009690.3, CELF6, AC009690.1 |
| chr22 | 21958585 | 21958717 | 133 | 3 | 3.33E-05 | 0.001247 | 1.17E-05 | 1.85E-05 | -0.00199457 | PPM1F-AS1, TOP3B |
| chr1 | 6645702 | 6645830 | 129 | 2 | 3.34E-05 | 3.28E-05 | 0.00019 | 3.07E-05 | -0.001669729 | DNAJC11 |
| chr13 | 35696669 | 35697772 | 1104 | 9 | 3.34E-05 | 5.63E-06 | 0.00338 | 9.24E-07 | -0.005719475 | LINC00445 |
| chr17 | 49566885 | 49567315 | 431 | 2 | 3.34E-05 | 7.03E-06 | 0.00047 | 1.12E-05 | -0.000114784 | AC006487.1, AC006487.2 |
| chr9 | 128994233 | 128995082 | 850 | 2 | 3.35E-05 | 2.86E-07 | 0.00013 | 5.36E-07 | -0.000240177 | NUP188 |
| chr19 | 43623957 | 43624178 | 222 | 2 | 3.35E-05 | 7.05E-06 | 0.00014 | 8.56E-06 | -0.017047149 | CADM4 |
| chr7 | 5374008 | 5374543 | 536 | 4 | 3.35E-05 | 5.67E-05 | 0.00255 | 1.63E-05 | -0.001957184 | TNRC18 |
| chr18 | 62959070 | 62959707 | 638 | 3 | 3.36E-05 | 7.88E-05 | 0.00042 | 6.98E-05 | 0.001783338 | PHLPP1 |
| chr22 | 31082663 | 31082758 | 96 | 3 | 3.36E-05 | 4.75E-05 | 0.0002 | 2.65E-05 | -0.007071785 | SMTN, AC005005.3 |
| chr11 | 17352977 | 17353566 | 590 | 5 | 3.37E-05 | 3.48E-05 | 0.00092 | 9.04E-06 | 0.002451301 | NCR3LG1 |
| chr9 | 4666266 | 4667099 | 834 | 12 | 3.38E-05 | 1.21E-05 | 0.00362 | 3.99E-06 | 0.000710607 | SPATA6L |
| chr11 | 4094523 | 4095843 | 1321 | 12 | 3.39E-05 | 7.29E-05 | 0.00028 | 1.34E-05 | 0.000920064 | RRM1 |
| chr16 | 72062003 | 72062709 | 707 | 5 | 3.39E-05 | 1.62E-06 | 0.00163 | 3.43E-06 | -0.003886267 | TXNL4B |
| chr16 | 275026 | 275184 | 159 | 3 | 3.39E-05 | 8.38E-05 | 0.0003 | 4.48E-05 | -0.007967141 | Z69667.1, RGS11 |
| chr2 | 181915996 | 181916572 | 577 | 3 | 3.40E-05 | 1.79E-05 | 0.00021 | 1.83E-05 | -0.003195588 | ITPRID2 |
| chr2 | 233711817 | 233711834 | 18 | 2 | 3.41E-05 | 2.93E-05 | 3.62E-05 | 1.06E-05 | -0.005362425 | UGT1A8, UGT1A10, UGT1A9, UGT1A7, UGT1A6 |
| chr2 | 10844417 | 10844505 | 89 | 2 | 3.41E-05 | 2.17E-06 | 0.00026 | 3.67E-06 | -0.023145888 | LINC01954 |
| chr6 | 33433703 | 33433765 | 63 | 5 | 3.41E-05 | 1.81E-06 | 0.0053 | 6.70E-06 | -0.014153663 | SYNGAP1 |
| chr1 | 53890233 | 53890895 | 663 | 4 | 3.41E-05 | 1.34E-05 | 0.00138 | 2.53E-05 | -0.013459296 | NA |
| chr22 | 50018779 | 50019157 | 379 | 2 | 3.42E-05 | 2.30E-05 | 4.36E-05 | 1.06E-05 | -0.001121263 | TTLL8 |
| chr17 | 50689573 | 50690693 | 1121 | 6 | 3.42E-05 | 6.47E-06 | 0.00121 | 7.27E-07 | -0.004911756 | ABCC3 |
| chr6 | 30156463 | 30157165 | 703 | 10 | 3.43E-05 | 0.000811 | 0.00188 | 1.93E-05 | -0.00366276 | TRIM10 |
| chr7 | 50705433 | 50706520 | 1088 | 6 | 3.43E-05 | 3.06E-06 | 0.00301 | 6.54E-06 | -0.006673385 | GRB10 |
| chr10 | 100806486 | 100806719 | 234 | 2 | 3.43E-05 | 9.46E-06 | 4.53E-05 | 6.09E-06 | -0.009410636 | PAX2 |
| chr16 | 67155726 | 67156741 | 1016 | 7 | 3.43E-05 | 4.31E-06 | 0.00183 | 1.88E-06 | -0.009104678 | TRADD |
| chr22 | 37581418 | 37581817 | 400 | 4 | 3.43E-05 | 0.000149 | 0.00013 | 1.82E-05 | -0.008681636 | LGALS2 |
| chr1 | 60970179 | 60970378 | 200 | 3 | 3.44E-05 | 1.79E-06 | 0.00084 | 4.18E-06 | -0.017918313 | NFIA, NFIA-AS2 |
| chr6 | 156827604 | 156828343 | 740 | 2 | 3.44E-05 | 4.40E-06 | 0.00021 | 6.55E-06 | -0.013319922 | ARID1B |
| chr8 | 115419831 | 115419930 | 100 | 3 | 3.45E-05 | 0.00013 | 0.00021 | 6.37E-06 | -0.012183972 | TRPS1 |
| chr1 | 228512185 | 228512882 | 698 | 8 | 3.45E-05 | 1.38E-06 | 0.00515 | 4.00E-06 | -0.002780822 | BTNL10 |
| chr13 | 31005150 | 31005263 | 114 | 3 | 3.46E-05 | 2.72E-05 | 2.28E-05 | 5.94E-06 | -0.000927935 | NA |
| chr9 | 131481564 | 131481751 | 188 | 2 | 3.46E-05 | 9.10E-06 | 6.40E-05 | 7.21E-06 | -0.01425963 | PRRC2B |
| chr8 | 126840767 | 126840818 | 52 | 2 | 3.46E-05 | 4.29E-06 | 6.69E-05 | 4.32E-06 | -0.004643898 | PCAT1, AC024382.1 |
| chr7 | 152180505 | 152181090 | 586 | 2 | 3.47E-05 | 8.86E-06 | 0.00014 | 1.03E-05 | -0.004119086 | KMT2C |
| chr11 | 134098534 | 134098592 | 59 | 2 | 3.47E-05 | 0.001903 | 1.05E-05 | 1.18E-05 | -0.003987564 | JAM3 |
| chr22 | 39456276 | 39456857 | 582 | 5 | 3.47E-05 | 0.000104 | 0.0004 | 5.01E-06 | -0.007506237 | NA |
| chr20 | 62926624 | 62927953 | 1330 | 11 | 3.48E-05 | 5.03E-06 | 0.00459 | 3.71E-07 | -0.001701713 | DIDO1 |
| chr20 | 36406527 | 36406707 | 181 | 2 | 3.49E-05 | 4.31E-06 | 0.00044 | 7.21E-06 | -0.004887526 | DLGAP4 |

| | | | | | | | | | | |
|---|---|---|---|---|---|---|---|---|---|---|
| chr1 | 151046658 | 151047251 | 594 | 3 | 3.49E-05 | 1.19E-05 | 0.00106 | 2.15E-05 | -0.00333364 | BNIPL |
| chr16 | 78098933 | 78099395 | 463 | 6 | 3.49E-05 | 0.009543 | 0.00038 | 0.00046 | -0.001169571 | NA |
| chr10 | 52771503 | 52771949 | 447 | 5 | 3.50E-05 | 1.08E-05 | 0.00072 | 4.96E-06 | -0.004727919 | MBL2 |
| chr1 | 53837934 | 53839179 | 1246 | 14 | 3.51E-05 | 3.06E-05 | 0.00061 | 1.71E-05 | 0.001141604 | NDC1 |
| chr11 | 18588151 | 18588915 | 765 | 10 | 3.51E-05 | 0.002467 | 0.00027 | 4.02E-05 | -1.13E-06 | SPTY2D1OS, UEVLD |
| chr10 | 3763060 | 3763249 | 190 | 4 | 3.51E-05 | 3.19E-06 | 0.00013 | 3.80E-06 | -0.006992041 | AL450322.2 |
| chr9 | 133594026 | 133594137 | 112 | 2 | 3.52E-05 | 4.71E-06 | 0.00021 | 6.90E-06 | -0.010588484 | FAM163B |
| chr20 | 63157310 | 63157631 | 322 | 2 | 3.52E-05 | 0.000142 | 5.40E-05 | 3.25E-05 | -0.006057096 | NA |
| chr2 | 98508407 | 98508829 | 423 | 4 | 3.52E-05 | 4.98E-05 | 0.0002 | 8.70E-06 | -0.008865186 | INPP4A |
| chr11 | 78198075 | 78198378 | 304 | 2 | 3.53E-05 | 9.20E-05 | 0.0001 | 4.28E-05 | 0.000162065 | USP35 |
| chr11 | 22362351 | 22362749 | 399 | 4 | 3.54E-05 | 1.26E-05 | 0.00053 | 7.53E-06 | -0.010051688 | SLC17A6 |
| chr6 | 12715629 | 12715905 | 277 | 2 | 3.54E-05 | 0.000416 | 7.65E-05 | 6.49E-05 | -0.014038996 | NA |
| chr11 | 32088641 | 32088777 | 137 | 2 | 3.54E-05 | 5.88E-06 | 0.00041 | 9.35E-06 | -0.012419464 | AL035078.4 |
| chr7 | 92179284 | 92179803 | 520 | 8 | 3.54E-05 | 1.21E-06 | 0.00295 | 4.33E-06 | -0.010238764 | CYP51A1-AS1, AC000120.2, AC000120.3, LRRD1 |
| chr12 | 15712952 | 15713265 | 314 | 5 | 3.54E-05 | 0.000469 | 8.43E-05 | 1.09E-05 | -0.003657978 | EPS8 |
| chr13 | 113074176 | 113075063 | 888 | 7 | 3.54E-05 | 0.000107 | 0.00021 | 9.87E-06 | -0.002961891 | MCF2L |
| chr7 | 114921438 | 114922261 | 824 | 9 | 3.55E-05 | 5.60E-05 | 0.00253 | 2.10E-05 | -0.0059792 | NA |
| chr16 | 2081717 | 2083768 | 2052 | 12 | 3.55E-05 | 6.42E-07 | 0.00015 | 2.00E-07 | -0.00432026 | TSC2 |
| chr2 | 157597077 | 157597889 | 813 | 13 | 3.55E-05 | 3.57E-05 | 0.00095 | 4.64E-06 | 0.001369808 | ACVR1C |
| chr14 | 105248688 | 105249589 | 902 | 6 | 3.55E-05 | 0.001572 | 0.00901 | 9.61E-05 | -0.005825575 | BTBD6, BRF1 |
| chr20 | 35296001 | 35296294 | 294 | 2 | 3.55E-05 | 2.30E-06 | 0.00028 | 3.92E-06 | -0.010618169 | NA |
| chr7 | 157983987 | 157984341 | 355 | 3 | 3.55E-05 | 3.68E-06 | 0.00047 | 6.66E-06 | -0.008028619 | PTPRN2 |
| chr5 | 134873252 | 134874334 | 1083 | 11 | 3.56E-05 | 7.98E-05 | 0.00279 | 5.67E-06 | -0.004404001 | NA |
| chr8 | 124538420 | 124539451 | 1032 | 15 | 3.57E-05 | 3.17E-05 | 0.00466 | 9.06E-06 | 0.00177402 | NDUFB9, TATDN1 |
| chr20 | 32520744 | 32520860 | 117 | 2 | 3.57E-05 | 1.50E-05 | 3.68E-05 | 7.16E-06 | -0.004010473 | NOL4L |
| chr5 | 74866398 | 74867898 | 1501 | 12 | 3.58E-05 | 1.44E-05 | 0.00031 | 9.33E-07 | -0.001494352 | AC010501.2, FAM169A |
| chr21 | 45287866 | 45288263 | 398 | 10 | 3.58E-05 | 0.061461 | 5.85E-06 | 5.40E-05 | 0.000256337 | LINC00205, POFUT2 |
| chr14 | 104116406 | 104116908 | 503 | 5 | 3.58E-05 | 3.93E-05 | 0.00011 | 1.42E-05 | -0.002658025 | NA |
| chr14 | 22956561 | 22957383 | 823 | 8 | 3.58E-05 | 7.77E-05 | 0.00031 | 1.42E-05 | 0.00015635 | HAUS4, AL132780.3, MIR4707 |
| chr16 | 30984306 | 30985501 | 1196 | 11 | 3.58E-05 | 1.91E-06 | 0.00431 | 1.87E-06 | -0.005312499 | SETD1A, HSD3B7, AC135048.4 |
| chr7 | 108613296 | 108613727 | 432 | 2 | 3.58E-05 | 1.78E-06 | 0.0003 | 3.10E-06 | 0.004310664 | AC005487.1 |
| chr6 | 6610805 | 6611069 | 265 | 2 | 3.58E-05 | 7.27E-05 | 6.53E-05 | 2.75E-05 | -0.007980845 | LY86, LY86-AS1 |
| chr7 | 151815935 | 151816242 | 308 | 3 | 3.58E-05 | 0.000469 | 5.15E-05 | 3.67E-05 | -0.001254472 | PRKAG2 |
| chr10 | 87861983 | 87862964 | 982 | 32 | 3.58E-05 | 2.72E-05 | 0.03158 | 2.19E-05 | 9.48E-05 | KLLN |
| chr17 | 34637027 | 34637827 | 801 | 5 | 3.59E-05 | 1.81E-05 | 0.00013 | 6.83E-06 | -0.003654316 | TMEM132E |
| chr16 | 2857516 | 2857858 | 343 | 4 | 3.59E-05 | 1.91E-06 | 0.00234 | 6.06E-06 | -0.027999833 | PRSS22 |
| chr5 | 94111499 | 94112647 | 1149 | 12 | 3.59E-05 | 3.60E-06 | 0.00721 | 1.93E-06 | -0.001774449 | AC117528.1, FAM172A |
| chr14 | 100896714 | 100896855 | 142 | 2 | 3.59E-05 | 4.06E-06 | 0.00042 | 6.83E-06 | -0.023659408 | AL117190.1, MEG8, RTL1 |
| chr12 | 32106404 | 32107898 | 1495 | 13 | 3.59E-05 | 3.33E-06 | 0.00544 | 1.67E-06 | 0.002138944 | BICD1, AC048344.1 |
| chr22 | 46536936 | 46537772 | 837 | 8 | 3.60E-05 | 0.000226 | 0.00053 | 2.02E-05 | 0.002077787 | CELSR1 |
| chr17 | 45143526 | 45144561 | 1036 | 7 | 3.60E-05 | 1.01E-06 | 0.00351 | 1.96E-06 | -0.010006112 | ACBD4 |
| chr1 | 93345269 | 93346562 | 1294 | 14 | 3.60E-05 | 3.63E-06 | 0.00724 | 1.98E-06 | 0.000658707 | DR1, CCDC18-AS1 |
| chr8 | 144756560 | 144756674 | 115 | 2 | 3.60E-05 | 0.027029 | 6.03E-05 | 0.00011 | -0.00334358 | NA |
| chr19 | 46373352 | 46374453 | 1102 | 3 | 3.61E-05 | 6.02E-07 | 0.00016 | 6.71E-07 | -0.011324139 | PPP5C |
| chr19 | 41262098 | 41262597 | 500 | 11 | 3.61E-05 | 0.001508 | 0.00024 | 2.15E-05 | 0.000366762 | HNRNPUL1 |
| chr14 | 104752214 | 104753280 | 1067 | 11 | 3.62E-05 | 0.003352 | 0.00442 | 3.82E-05 | 0.00026811 | SIVA1 |
| chr4 | 109912298 | 109912788 | 491 | 4 | 3.62E-05 | 0.001948 | 4.85E-05 | 3.62E-05 | -0.00517411 | NA |
| chr6 | 52236807 | 52237138 | 332 | 2 | 3.63E-05 | 0.002759 | 1.85E-05 | 2.32E-05 | -0.007631896 | IL17F |
| chr19 | 14380355 | 14381265 | 911 | 4 | 3.63E-05 | 1.90E-05 | 0.00031 | 9.62E-06 | -0.015496232 | ADGRE5 |
| chr7 | 53811510 | 53812096 | 587 | 7 | 3.63E-05 | 1.73E-06 | 0.00572 | 4.57E-06 | -0.007776893 | LINC01446 |
| chr10 | 133126960 | 133127325 | 366 | 4 | 3.64E-05 | 0.003385 | 3.95E-05 | 1.67E-05 | -0.000448817 | ADGRA1 |
| chr11 | 36030496 | 36030631 | 136 | 2 | 3.65E-05 | 3.05E-06 | 0.00013 | 4.25E-06 | -0.0084874 | LDLRAD3 |
| chr15 | 40330384 | 40330512 | 129 | 2 | 3.66E-05 | 0.001494 | 1.69E-05 | 1.94E-05 | -0.010429414 | NA |
| chr4 | 183765548 | 183765620 | 73 | 2 | 3.66E-05 | 9.14E-06 | 3.75E-05 | 5.26E-06 | -0.024222562 | AC108477.1 |
| chr2 | 157246544 | 157246823 | 280 | 2 | 3.66E-05 | 1.44E-05 | 6.49E-05 | 1.00E-05 | 0.008983498 | NA |
| chr17 | 81656573 | 81656988 | 416 | 5 | 3.66E-05 | 0.015486 | 0.00029 | 4.88E-05 | -0.003066413 | PDE6G |
| chr13 | 30445536 | 30446331 | 796 | 5 | 3.66E-05 | 6.67E-07 | 0.00223 | 1.56E-06 | -0.009009234 | NA |
| chr16 | 87391689 | 87392565 | 877 | 12 | 3.67E-05 | 0.000473 | 0.00365 | 1.21E-05 | 0.000852593 | MAP1LC3B, FBXO31 |
| chr17 | 3647181 | 3647618 | 438 | 4 | 3.67E-05 | 0.000179 | 0.0004 | 2.06E-05 | -0.003161055 | CTNS |
| chr11 | 61076120 | 61076173 | 54 | 2 | 3.67E-05 | 2.39E-06 | 0.00023 | 3.94E-06 | -0.011267979 | NA |
| chr3 | 134312171 | 134313106 | 936 | 5 | 3.67E-05 | 5.68E-07 | 0.00648 | 3.21E-06 | -0.009935798 | NA |
| chr16 | 89593861 | 89594291 | 431 | 5 | 3.67E-05 | 1.22E-05 | 0.0012 | 6.28E-06 | -0.011495935 | CPNE7 |
| chr7 | 100687424 | 100688643 | 1220 | 9 | 3.68E-05 | 9.66E-07 | 0.01029 | 2.56E-06 | -0.004340832 | GIGYF1 |
| chr3 | 157089113 | 157089902 | 790 | 7 | 3.68E-05 | 2.17E-05 | 0.00363 | 1.41E-05 | -0.006495938 | LINC00881, LINC00880 |
| chr7 | 353732 | 354230 | 499 | 7 | 3.68E-05 | 1.71E-06 | 0.00556 | 3.29E-06 | -0.003925163 | NA |
| chr9 | 35771182 | 35771227 | 46 | 2 | 3.68E-05 | 3.49E-06 | 0.00043 | 5.93E-06 | -0.020319409 | NA |
| chr22 | 41899663 | 41899987 | 325 | 2 | 3.68E-05 | 0.000273 | 6.58E-05 | 4.89E-05 | -0.002344971 | SREBF2 |
| chr16 | 89100610 | 89101073 | 464 | 4 | 3.69E-05 | 4.91E-06 | 0.00115 | 8.60E-06 | -0.012082889 | ACSF3 |
| chr1 | 36471960 | 36472801 | 842 | 7 | 3.69E-05 | 1.31E-05 | 0.01027 | 4.42E-06 | -0.004951144 | CSF3R |
| chr4 | 75035421 | 75036169 | 749 | 4 | 3.69E-05 | 2.80E-05 | 0.00128 | 1.74E-05 | -0.002944674 | PARM1 |
| chr17 | 50602790 | 50603193 | 404 | 4 | 3.69E-05 | 1.55E-05 | 0.00056 | 6.98E-06 | -0.001362884 | CACNA1G |
| chr6 | 30940525 | 30940996 | 472 | 5 | 3.70E-05 | 1.52E-06 | 0.00182 | 4.41E-06 | -0.009182109 | MUCL3, SFTA2 |
| chr12 | 122944480 | 122944828 | 349 | 3 | 3.70E-05 | 0.011427 | 1.31E-05 | 3.07E-05 | -0.005158104 | ABCB9 |
| chr12 | 124607825 | 124608658 | 834 | 4 | 3.71E-05 | 4.07E-06 | 0.00049 | 2.99E-06 | -0.005274068 | NA |
| chr3 | 186456228 | 186456505 | 278 | 2 | 3.72E-05 | 0.000754 | 3.23E-05 | 3.36E-05 | -0.005740544 | LINC02020, LINC02052 |
| chr6 | 166318389 | 166318480 | 92 | 2 | 3.72E-05 | 1.45E-05 | 7.69E-05 | 1.12E-05 | -0.017740349 | NA |
| chr11 | 83071439 | 83071961 | 523 | 10 | 3.72E-05 | 8.86E-06 | 0.00329 | 8.64E-06 | 0.00126865 | RAB30 |
| chr17 | 8371557 | 8372159 | 603 | 3 | 3.72E-05 | 0.000454 | 0.00014 | 2.59E-05 | -0.002796964 | AC135178.4, KRBA2, AC135178.3 |
| chr17 | 81285583 | 81286163 | 581 | 4 | 3.72E-05 | 9.14E-06 | 0.00123 | 1.04E-05 | -0.004458421 | SLC38A10 |
| chr12 | 109246311 | 109246720 | 410 | 2 | 3.73E-05 | 0.00043 | 2.94E-05 | 2.69E-05 | -0.002886983 | ACACB |
| chr10 | 122020509 | 122020856 | 348 | 2 | 3.73E-05 | 6.70E-05 | 0.00013 | 4.09E-05 | -0.005583236 | TACC2 |
| chr15 | 78540181 | 78541163 | 983 | 14 | 3.73E-05 | 1.40E-05 | 0.01063 | 8.02E-06 | 0.001204123 | PSMA4 |
| chr1 | 204191256 | 204191813 | 558 | 4 | 3.73E-05 | 1.97E-06 | 0.00154 | 5.61E-06 | -0.008888942 | KISS1 |
| chr21 | 33542326 | 33543286 | 961 | 19 | 3.74E-05 | 5.36E-05 | 0.0013 | 8.95E-06 | -0.000281471 | SON, GART |
| chr12 | 49752480 | 49752978 | 499 | 4 | 3.74E-05 | 2.93E-05 | 0.00015 | 8.30E-06 | -0.006355477 | TMBIM6 |
| chr3 | 71244543 | 71245229 | 687 | 5 | 3.74E-05 | 3.35E-05 | 0.00345 | 3.43E-06 | -0.010550163 | FOXP1, AC097634.4 |
| chr6 | 81752725 | 81753306 | 582 | 9 | 3.75E-05 | 3.68E-06 | 0.00786 | 4.29E-06 | -0.000231552 | TENT5A |
| chr17 | 82786957 | 82787359 | 403 | 3 | 3.75E-05 | 1.15E-05 | 0.00011 | 7.42E-06 | -0.006040544 | TBCD |
| chr1 | 160363984 | 160364321 | 338 | 2 | 3.75E-05 | 6.02E-05 | 4.17E-05 | 1.78E-05 | -0.006092215 | NA |
| chr22 | 23198093 | 23198274 | 182 | 2 | 3.76E-05 | 2.84E-06 | 0.00017 | 4.34E-06 | -0.010864047 | BCR |
| chr12 | 51270442 | 51270871 | 430 | 9 | 3.76E-05 | 9.94E-06 | 0.00101 | 4.79E-06 | 0.00063052 | DAZAP2 |
| chr16 | 58196347 | 58196993 | 647 | 4 | 3.77E-05 | 0.000167 | 0.00062 | 1.22E-05 | -0.005079425 | CSNK2A2 |
| chr22 | 50299549 | 50300461 | 913 | 4 | 3.78E-05 | 1.21E-06 | 0.00244 | 4.26E-06 | -0.021713753 | PLXNB2 |
| chr1 | 45550451 | 45551190 | 740 | 15 | 3.79E-05 | 1.25E-06 | 0.01149 | 4.10E-06 | -0.001425558 | AKR1A1 |
| chr2 | 46977595 | 46977953 | 359 | 2 | 3.79E-05 | 0.000111 | 3.73E-05 | 2.15E-05 | -0.004804536 | TTC7A |
| chr2 | 3378665 | 3378803 | 139 | 3 | 3.79E-05 | 0.005925 | 0.03247 | 0.00976 | -0.002523836 | NA |
| chr16 | 1378015 | 1379264 | 1250 | 6 | 3.79E-05 | 1.54E-05 | 0.00688 | 1.72E-05 | -0.001633254 | UNKL |
| chr1 | 16227676 | 16227907 | 232 | 2 | 3.79E-05 | 9.94E-05 | 0.00201 | 0.00015 | 0.008100608 | ANO7L1 |
| chr5 | 131258671 | 131258737 | 67 | 2 | 3.80E-05 | 6.05E-06 | 6.36E-05 | 5.40E-06 | 0.02700407 | CDC42SE2 |
| chr19 | 38674874 | 38674889 | 16 | 2 | 3.80E-05 | 1.33E-05 | 8.12E-05 | 1.08E-05 | -0.008976606 | ACTN4 |
| chr9 | 6715738 | 6716878 | 1141 | 5 | 3.80E-05 | 1.72E-05 | 0.00035 | 2.87E-06 | 0.005133198 | AL354707.1 |
| chr12 | 57296495 | 57296805 | 311 | 4 | 3.80E-05 | 7.65E-06 | 0.00069 | 7.71E-06 | 0.0027293 | R3HDM2 |
| chr22 | 20974103 | 20974452 | 350 | 3 | 3.80E-05 | 2.45E-05 | 8.97E-05 | 1.11E-05 | -0.007576226 | AIFM3 |
| chr10 | 71318821 | 71319344 | 524 | 7 | 3.80E-05 | 2.54E-05 | 0.00287 | 5.71E-06 | -0.001697655 | SLC29A3 |
| chr15 | 81189754 | 81189778 | 25 | 2 | 3.81E-05 | 4.73E-06 | 0.00025 | 7.17E-06 | -0.016327909 | IL16 |
| chr6 | 170269043 | 170269504 | 462 | 2 | 3.81E-05 | 0.002699 | 9.84E-05 | 0.00015 | -0.003297594 | AL109910.2 |
| chr1 | 50104940 | 50104959 | 20 | 2 | 3.83E-05 | 0.003362 | 3.04E-05 | 4.07E-05 | -0.008578852 | ELAVL4 |
| chr2 | 62195849 | 62196581 | 733 | 8 | 3.83E-05 | 3.67E-05 | 0.00022 | 4.50E-06 | 0.002084305 | B3GNT2 |
| chr11 | 76781631 | 76782832 | 1202 | 6 | 3.83E-05 | 0.005894 | 0.00266 | 0.00013 | -0.005835386 | TSKU, AP003119.2 |
| chr4 | 56316251 | 56316473 | 223 | 2 | 3.83E-05 | 1.80E-05 | 9.14E-05 | 1.44E-05 | -0.005614619 | CRACD |
| chr2 | 26612485 | 26612698 | 214 | 2 | 3.84E-05 | 7.99E-06 | 6.29E-05 | 6.55E-06 | -0.031469346 | CIB4 |
| chr1 | 21610191 | 21611234 | 1044 | 5 | 3.84E-05 | 2.17E-06 | 0.00023 | 7.00E-07 | -0.004703272 | RAP1GAP |
| chr14 | 105437017 | 105437196 | 180 | 2 | 3.84E-05 | 3.39E-05 | 9.00E-05 | 2.17E-05 | -0.003971717 | TEX22, MTA1 |
| chr1 | 27960929 | 27961271 | 343 | 2 | 3.84E-05 | 6.38E-06 | 0.00048 | 1.03E-05 | -0.008532467 | XKR8 |
| chr15 | 28940107 | 28940639 | 533 | 2 | 3.84E-05 | 5.21E-06 | 0.00023 | 7.64E-06 | -0.009885429 | APBA2 |
| chr3 | 58938290 | 58938630 | 341 | 2 | 3.85E-05 | 2.79E-05 | 8.95E-05 | 1.91E-05 | -0.004610372 | CFAP20DC-AS1, CFAP20DC |
| chr10 | 24721809 | 24721950 | 142 | 2 | 3.85E-05 | 8.64E-06 | 0.00073 | 1.42E-05 | -0.031929197 | ARHGAP21 |
| chr11 | 47524050 | 47524753 | 704 | 8 | 3.86E-05 | 3.97E-06 | 0.00466 | 4.83E-06 | -0.007205354 | CELF1 |
| chr1 | 198540539 | 198541932 | 1394 | 7 | 3.86E-05 | 1.44E-05 | 0.00095 | 5.03E-07 | -0.003628316 | ATP6V1G3 |
| chr6 | 27019796 | 27020510 | 715 | 10 | 3.87E-05 | 6.91E-07 | 0.00771 | 2.16E-06 | -0.006469773 | LINC00240, AL590062.1 |
| chr19 | 6746173 | 6746527 | 355 | 3 | 3.88E-05 | 9.76E-05 | 7.24E-05 | 2.17E-05 | -0.003207265 | TRIP10 |
| chr2 | 170928100 | 170929053 | 954 | 15 | 3.88E-05 | 8.76E-05 | 0.00027 | 2.34E-06 | -0.003427439 | GORASP2 |
| chr14 | 68971550 | 68971641 | 92 | 2 | 3.88E-05 | 3.47E-05 | 1.90E-05 | 7.05E-06 | -0.010782365 | ACTN1 |
| chr17 | 28368118 | 28368417 | 300 | 2 | 3.88E-05 | 6.95E-05 | 5.02E-05 | 2.19E-05 | -0.003228658 | AC002094.1, SARM1, VTN |
| chr2 | 197311256 | 197311709 | 454 | 4 | 3.89E-05 | 3.49E-06 | 0.00223 | 7.26E-06 | -0.010233496 | AC010746.2 |
| chr1 | 26220568 | 26220886 | 319 | 3 | 3.89E-05 | 0.002005 | 2.29E-05 | 3.68E-05 | -0.007773074 | ZPLD2P |
| chr17 | 44004226 | 44005024 | 799 | 11 | 3.89E-05 | 3.71E-07 | 0.01519 | 2.75E-06 | -0.006267891 | NAGS, PYY |
| chr16 | 1527915 | 1528806 | 892 | 9 | 3.89E-05 | 1.85E-05 | 0.00224 | 3.60E-06 | -0.008007431 | TMEM204, IFT140 |
| chr6 | 108251475 | 108252257 | 783 | 6 | 3.90E-05 | 3.26E-05 | 0.00021 | 1.35E-05 | -0.00396171 | Z98742.2, SNX3 |
| chr15 | 67107542 | 67108235 | 694 | 4 | 3.90E-05 | 4.62E-06 | 0.00087 | 7.05E-06 | -0.007618006 | SMAD3 |
| chr12 | 6621143 | 6621433 | 291 | 2 | 3.91E-05 | 1.37E-05 | 0.00012 | 1.34E-05 | -0.014435063 | LPAR5 |
| chr9 | 137025621 | 137027519 | 1899 | 8 | 3.91E-05 | 1.66E-08 | 0.00057 | 1.82E-08 | -0.007450324 | C9orf139, ABCA2 |
| chr12 | 50400305 | 50401192 | 888 | 12 | 3.91E-05 | 8.49E-05 | 0.0036 | 9.57E-06 | 0.002128257 | LARP4 |
| chr17 | 9054347 | 9054773 | 427 | 4 | 3.92E-05 | 0.000998 | 9.01E-05 | 1.59E-05 | -0.00319268 | NTN1 |
| chr14 | 50531762 | 50532684 | 923 | 8 | 3.92E-05 | 8.96E-05 | 0.00342 | 2.99E-05 | -3.85E-05 | ATL1, MAP4K5 |
| chr11 | 118622244 | 118622681 | 438 | 3 | 3.92E-05 | 0.004214 | 6.68E-05 | 5.77E-05 | -0.006918506 | PHLDB1 |
| chr1 | 109622416 | 109622619 | 204 | 4 | 3.93E-05 | 0.000153 | 4.85E-05 | 1.92E-05 | -0.004313809 | AMPD2 |
| chr7 | 149831742 | 149832544 | 803 | 2 | 3.94E-05 | 5.16E-07 | 0.00016 | 9.30E-07 | -0.013633803 | SSPOP |
| chr5 | 132181144 | 132181504 | 361 | 5 | 3.94E-05 | 2.64E-06 | 0.00152 | 3.81E-06 | -0.004204287 | NA |
| chr5 | 141095402 | 141096027 | 626 | 4 | 3.94E-05 | 0.001391 | 3.25E-05 | 3.50E-05 | 0.008191408 | PCDHB2, AC244517.5 |
| chr21 | 33524244 | 33524889 | 646 | 3 | 3.94E-05 | 1.66E-06 | 0.00046 | 3.53E-06 | -0.004997118 | GART |
| chr4 | 55793977 | 55794294 | 318 | 4 | 3.95E-05 | 2.10E-05 | 0.00214 | 9.26E-06 | -0.013351369 | NA |
| chr12 | 6067522 | 6067675 | 154 | 2 | 3.95E-05 | 1.51E-05 | 3.33E-05 | 6.79E-06 | -0.008381092 | VWF |
| chr5 | 81393977 | 81394639 | 663 | 6 | 3.95E-05 | 4.92E-05 | 0.00118 | 1.17E-05 | 0.006450321 | ACOT12 |
| chr6 | 31559916 | 31561138 | 1223 | 8 | 3.96E-05 | 7.25E-06 | 0.00105 | 1.87E-06 | 0.002243075 | NA |
| chr18 | 24560441 | 24561002 | 562 | 3 | 3.97E-05 | 0.000305 | 3.56E-05 | 1.03E-05 | -0.001070926 | LINC01915 |
| chr12 | 132784084 | 132784254 | 171 | 4 | 3.97E-05 | 1.71E-05 | 0.00015 | 5.25E-06 | -0.005448181 | GOLGA3 |
| chr20 | 3785615 | 3786314 | 700 | 5 | 3.97E-05 | 5.73E-05 | 0.00266 | 2.12E-05 | -0.009246021 | CENPB |
| chr14 | 71586177 | 71586644 | 468 | 6 | 3.97E-05 | 0.001367 | 0.00011 | 8.43E-05 | -0.009314316 | SIPA1L1, AC004974.1 |
| chr3 | 138835319 | 138835674 | 356 | 2 | 3.98E-05 | 0.000147 | 9.52E-05 | 5.17E-05 | -0.009644775 | NA |
| chr16 | 30004704 | 30005553 | 850 | 4 | 3.98E-05 | 9.40E-07 | 0.00213 | 2.80E-06 | -0.010860732 | INO80E, DOC2A |
| chr12 | 77230094 | 77230173 | 80 | 2 | 3.98E-05 | 3.91E-06 | 0.00012 | 5.09E-06 | -0.014618623 | LINC02464 |
| chr6 | 28533367 | 28534139 | 773 | 7 | 3.98E-05 | 0.001592 | 0.0003 | 1.35E-05 | -0.005045426 | GPX5 |
| chr17 | 1269207 | 1269772 | 566 | 4 | 3.99E-05 | 0.001169 | 4.13E-05 | 6.21E-05 | -0.004097242 | NA |
| chr1 | 1421062 | 1421981 | 920 | 10 | 3.99E-05 | 6.78E-05 | 0.00387 | 2.07E-05 | -0.003878435 | AL391244.1, ANKRD65 |
| chr10 | 79443111 | 79444813 | 1703 | 7 | 4.00E-05 | 2.03E-07 | 9.28E-05 | 5.68E-08 | -0.004743147 | ZCCHC24 |
| chr9 | 137256896 | 137257254 | 359 | 2 | 4.00E-05 | 0.000991 | 3.05E-05 | 3.36E-05 | -0.001362368 | NELFB |
| chr10 | 102621765 | 102621894 | 130 | 2 | 4.02E-05 | 3.43E-06 | 0.00035 | 5.72E-06 | -0.015486688 | SUFU |
| chr17 | 75552860 | 75553010 | 151 | 2 | 4.02E-05 | 3.15E-06 | 0.00016 | 4.64E-06 | -0.019502555 | LLGL2 |
| chr6 | 168842386 | 168843757 | 1372 | 6 | 4.02E-05 | 9.37E-07 | 0.00056 | 3.53E-07 | -0.005488549 | NA |
| chr10 | 80539850 | 80540395 | 546 | 5 | 4.02E-05 | 7.57E-05 | 0.00139 | 4.23E-05 | -0.004223961 | SH2D4B |
| chr20 | 64001224 | 64001770 | 547 | 3 | 4.03E-05 | 6.82E-05 | 0.00045 | 3.58E-05 | -0.005570676 | PRPF6 |

| | | | | | | | | | | |
|---|---|---|---|---|---|---|---|---|---|---|
| chr2 | 104856882 | 104857927 | 1046 | 4 | 4.03E-05 | 2.20E-05 | 0.00014 | 6.62E-06 | -0.001498907 | AC018730.1, POU3F3 |
| chr10 | 6191639 | 6191811 | 173 | 4 | 4.04E-05 | 0.000437 | 0.00017 | 1.25E-05 | -0.006894512 | PFKFB3 |
| chr11 | 118980312 | 118980319 | 8 | 2 | 4.04E-05 | 6.80E-06 | 9.27E-05 | 7.05E-06 | -0.012130994 | FOXR1 |
| chr10 | 18400106 | 18401019 | 914 | 11 | 4.04E-05 | 6.58E-07 | 0.00628 | 2.95E-06 | 0.014095447 | CACNB2 |
| chr1 | 63198942 | 63199197 | 256 | 2 | 4.06E-05 | 1.17E-05 | 5.00E-05 | 7.42E-06 | -0.02076399 | LINC00466 |
| chr2 | 120314145 | 120315473 | 1329 | 4 | 4.06E-05 | 1.58E-06 | 0.00125 | 2.99E-06 | 0.003641626 | NA |
| chr5 | 135131125 | 135131310 | 186 | 2 | 4.06E-05 | 5.75E-05 | 5.16E-05 | 2.03E-05 | -0.003681453 | C5orf66, AC008406.2, AC008406.1 |
| chr7 | 48175071 | 48175700 | 630 | 2 | 4.06E-05 | 6.37E-06 | 0.00011 | 7.06E-06 | 0.015998265 | ABCA13 |
| chr9 | 135699131 | 135700008 | 878 | 8 | 4.07E-05 | 0.000764 | 0.00024 | 1.99E-05 | -0.005319065 | SOHLH1 |
| chr3 | 184244081 | 184244296 | 216 | 4 | 4.07E-05 | 0.000629 | 0.0003 | 1.39E-05 | -0.006549297 | ALG3 |
| chr3 | 128971195 | 128971351 | 157 | 2 | 4.08E-05 | 1.63E-05 | 0.00014 | 1.66E-05 | -0.007101352 | CFAP92 |
| chr14 | 55286386 | 55286656 | 271 | 2 | 4.08E-05 | 7.00E-06 | 7.54E-05 | 6.56E-06 | -0.012807286 | FBXO34 |
| chr2 | 218975787 | 218976752 | 966 | 5 | 4.08E-05 | 6.43E-06 | 0.00424 | 1.49E-05 | -0.008329845 | LINC00608 |
| chr2 | 157837523 | 157838298 | 776 | 4 | 4.09E-05 | 0.000158 | 0.00173 | 4.31E-05 | -0.009847733 | ACVR1 |
| chr9 | 129289247 | 129289285 | 39 | 2 | 4.09E-05 | 4.30E-06 | 0.00029 | 6.80E-06 | -0.009724628 | AL161785.1 |
| chr19 | 45157028 | 45157097 | 70 | 2 | 4.10E-05 | 0.002046 | 1.81E-05 | 2.18E-05 | -0.005782126 | MARK4, NKPD1 |
| chr21 | 43319486 | 43319506 | 21 | 2 | 4.10E-05 | 2.61E-06 | 0.00025 | 4.31E-06 | -0.009258332 | NA |
| chr16 | 68364416 | 68365802 | 1387 | 5 | 4.10E-05 | 1.91E-06 | 0.00013 | 2.54E-07 | -0.009481595 | SMPD3 |
| chr7 | 45080499 | 45080687 | 189 | 2 | 4.10E-05 | 2.36E-05 | 6.74E-05 | 1.44E-05 | -0.008733153 | NACAD |
| chr1 | 9183541 | 9183620 | 80 | 2 | 4.10E-05 | 2.83E-05 | 0.00105 | 4.20E-05 | -0.015041288 | LNCTAM34A, MIR34AHG |
| chr12 | 124964496 | 124964569 | 74 | 2 | 4.11E-05 | 2.15E-05 | 3.05E-05 | 7.88E-06 | -0.004769662 | DHX37 |
| chr12 | 59736673 | 59737064 | 392 | 2 | 4.11E-05 | 2.22E-06 | 0.00033 | 3.87E-06 | -0.019322552 | SLC16A7 |
| chr15 | 99100268 | 99100855 | 588 | 6 | 4.11E-05 | 0.019381 | 1.17E-05 | 2.57E-05 | -0.000626121 | SYNM, AC036108.1 |
| chr22 | 40239585 | 40239773 | 189 | 2 | 4.12E-05 | 0.002972 | 1.33E-05 | 1.65E-05 | -0.005206494 | TNRC6B |
| chr17 | 50189232 | 50190876 | 1645 | 7 | 4.14E-05 | 4.59E-06 | 0.00018 | 7.11E-07 | -0.001224166 | COL1A1 |
| chr7 | 158782942 | 158783243 | 302 | 5 | 4.14E-05 | 6.49E-06 | 0.00023 | 5.25E-06 | -0.00591303 | ESYT2 |
| chr2 | 117837046 | 117837739 | 694 | 3 | 4.14E-05 | 6.34E-05 | 0.00045 | 1.72E-05 | -0.005511657 | AC009404.1 |
| chr20 | 64064781 | 64065438 | 658 | 4 | 4.14E-05 | 6.70E-06 | 0.00117 | 1.38E-05 | -0.009282878 | TCEA2 |
| chr7 | 149003195 | 149004312 | 1118 | 5 | 4.15E-05 | 3.22E-05 | 8.36E-05 | 5.06E-06 | -0.003737927 | PDIA4 |
| chr4 | 3384445 | 3384502 | 58 | 2 | 4.15E-05 | 5.62E-06 | 0.00029 | 8.52E-06 | -0.011432181 | RGS12 |
| chr8 | 129989132 | 129989477 | 346 | 3 | 4.15E-05 | 1.27E-06 | 0.00116 | 3.26E-06 | -0.009838138 | CYRIB |
| chr17 | 40069707 | 40070197 | 491 | 3 | 4.15E-05 | 1.10E-05 | 0.00047 | 1.63E-05 | -0.010561367 | THRA |
| chr3 | 124520782 | 124521183 | 402 | 2 | 4.16E-05 | 0.00058 | 2.70E-05 | 2.65E-05 | -0.002461956 | KALRN |
| chr4 | 2926155 | 2927038 | 884 | 6 | 4.17E-05 | 2.63E-06 | 0.0015 | 2.19E-06 | -0.003448 | ADD1 |
| chr6 | 29630214 | 29630918 | 705 | 6 | 4.17E-05 | 0.000503 | 0.00052 | 5.69E-05 | 0.001235433 | GABBR1 |
| chr2 | 230821321 | 230821836 | 516 | 2 | 4.17E-05 | 7.37E-06 | 0.00011 | 8.09E-06 | -0.011480434 | NA |
| chr13 | 110915924 | 110916059 | 136 | 3 | 4.18E-05 | 0.000342 | 0.00106 | 5.35E-05 | 0.002131811 | AL442128.2 |
| chr11 | 57567578 | 57568257 | 680 | 10 | 4.19E-05 | 2.82E-05 | 0.00605 | 1.37E-05 | 0.000131986 | UBE2L6 |
| chr17 | 1516778 | 1517255 | 478 | 7 | 4.20E-05 | 5.89E-05 | 0.00037 | 8.94E-06 | -0.000556677 | PITPNA-AS1 |
| chr10 | 122475392 | 122475935 | 544 | 2 | 4.20E-05 | 3.70E-06 | 0.00019 | 5.56E-06 | -0.011393472 | HTRA1 |
| chr17 | 27494008 | 27494623 | 616 | 3 | 4.20E-05 | 0.000749 | 3.26E-05 | 3.96E-05 | -0.002815916 | KSR1 |
| chr15 | 28974333 | 28974375 | 43 | 3 | 4.21E-05 | 7.10E-06 | 9.62E-05 | 4.91E-06 | -0.007921409 | APBA2, AC127522.1 |
| chr19 | 39133720 | 39134172 | 453 | 3 | 4.22E-05 | 3.09E-05 | 0.00012 | 1.28E-05 | -0.010885689 | PAK4 |
| chr19 | 53787483 | 53788724 | 1242 | 14 | 4.22E-05 | 1.02E-06 | 0.00299 | 9.32E-07 | -0.002797172 | MIR371A, MIR372, MIR373, AC008753.3, MIR371B |
| chr1 | 161039673 | 161040281 | 609 | 6 | 4.22E-05 | 0.002981 | 0.00013 | 0.00037 | -0.000805122 | USF1 |
| chr17 | 17221065 | 17221969 | 905 | 9 | 4.22E-05 | 0.000113 | 0.00215 | 1.42E-05 | 0.000521079 | AC055811.2, FLCN |
| chr16 | 914974 | 915857 | 884 | 4 | 4.22E-05 | 2.87E-06 | 0.00308 | 8.94E-06 | -0.000224665 | LMF1 |
| chr12 | 52949730 | 52950575 | 846 | 4 | 4.23E-05 | 9.08E-05 | 0.00453 | 6.81E-05 | -0.013087128 | KRT18, KRT8 |
| chr19 | 46625713 | 46626380 | 668 | 5 | 4.24E-05 | 2.19E-06 | 0.00282 | 5.47E-06 | -0.013304464 | NA |
| chr17 | 66578903 | 66580761 | 1859 | 6 | 4.24E-05 | 2.26E-07 | 0.00056 | 1.53E-07 | -0.009975459 | PRKCA |
| chr12 | 68745610 | 68746049 | 440 | 9 | 4.25E-05 | 0.000544 | 0.00157 | 3.40E-05 | -0.006136162 | NUP107 |
| chr7 | 151071231 | 151071634 | 404 | 4 | 4.25E-05 | 7.46E-05 | 0.00013 | 2.31E-05 | -0.002363641 | SLC4A2 |
| chr1 | 59043910 | 59044015 | 106 | 2 | 4.25E-05 | 7.57E-05 | 3.15E-05 | 1.59E-05 | -0.012780144 | LINC01358 |
| chr3 | 113441224 | 113441974 | 751 | 12 | 4.26E-05 | 1.57E-06 | 0.01038 | 4.55E-06 | -0.010584784 | CFAP44, AC112128.1 |
| chr19 | 7533136 | 7533907 | 772 | 6 | 4.26E-05 | 1.50E-05 | 0.00212 | 2.04E-05 | -0.00622309 | MCOLN1, AC008878.2 |
| chr20 | 38986098 | 38986928 | 831 | 3 | 4.27E-05 | 1.31E-06 | 0.00035 | 1.81E-06 | -0.023515566 | DHX35 |
| chr6 | 34145793 | 34147050 | 1258 | 11 | 4.28E-05 | 1.91E-06 | 0.01857 | 7.03E-06 | -0.007997017 | GRM4 |
| chr16 | 87411765 | 87412474 | 710 | 5 | 4.28E-05 | 0.000997 | 0.00057 | 4.42E-05 | -0.003058756 | ZCCHC14 |
| chr11 | 60852151 | 60852870 | 720 | 4 | 4.29E-05 | 2.28E-05 | 0.00125 | 1.23E-05 | -0.003560447 | PTGDR2 |
| chr15 | 78319296 | 78319318 | 23 | 2 | 4.29E-05 | 1.50E-05 | 2.94E-05 | 6.16E-06 | -0.027436264 | NA |
| chr7 | 135232124 | 135232392 | 269 | 2 | 4.30E-05 | 4.80E-05 | 0.0016 | 7.14E-05 | -0.00790369 | STRA8 |
| chr13 | 113453239 | 113454398 | 1160 | 13 | 4.30E-05 | 5.75E-05 | 0.00437 | 6.57E-06 | -0.002551344 | ADPRHL1 |
| chr8 | 98601842 | 98602258 | 417 | 2 | 4.30E-05 | 6.01E-05 | 4.49E-05 | 1.87E-05 | -0.008376455 | STK3 |
| chr10 | 14879551 | 14880383 | 833 | 3 | 4.31E-05 | 2.39E-06 | 0.00056 | 3.43E-06 | -0.001986307 | SUV39H2 |
| chr15 | 40339957 | 40341615 | 1659 | 12 | 4.31E-05 | 2.41E-07 | 0.00226 | 1.09E-07 | -0.004290287 | CCDC9B |
| chr12 | 69639257 | 69639684 | 428 | 4 | 4.33E-05 | 7.04E-05 | 0.00081 | 1.30E-05 | -0.005663173 | NA |
| chr7 | 43444822 | 43445116 | 295 | 3 | 4.33E-05 | 3.28E-06 | 0.00039 | 5.96E-06 | -0.007239709 | HECW1 |
| chr6 | 133240918 | 133242244 | 1327 | 23 | 4.33E-05 | 0.000768 | 0.00019 | 5.89E-05 | -0.000209046 | EYA4 |
| chr1 | 12506006 | 12507142 | 1137 | 6 | 4.33E-05 | 7.22E-06 | 0.00079 | 2.78E-06 | -0.005466153 | VPS13D |
| chr1 | 1131719 | 1132000 | 282 | 4 | 4.33E-05 | 5.22E-06 | 0.00052 | 6.10E-06 | -0.009545768 | NA |
| chr18 | 8720330 | 8720397 | 68 | 2 | 4.33E-05 | 6.16E-06 | 0.00031 | 9.28E-06 | -0.009068265 | MTCL1 |
| chr19 | 55483493 | 55484277 | 785 | 4 | 4.34E-05 | 1.88E-05 | 0.0008 | 2.29E-05 | -0.003009453 | ZNF628 |
| chr17 | 41967829 | 41968478 | 650 | 7 | 4.34E-05 | 0.000174 | 0.00085 | 1.48E-05 | -0.003323161 | CNP |
| chr14 | 68167428 | 68167666 | 239 | 2 | 4.34E-05 | 6.49E-06 | 0.00012 | 7.43E-06 | -0.012308489 | RAD51B |
| chr2 | 9318796 | 9319467 | 672 | 8 | 4.34E-05 | 0.000589 | 1.73E-05 | 2.01E-05 | -0.001811353 | ASAP2 |
| chr10 | 110685037 | 110685094 | 58 | 2 | 4.34E-05 | 0.001339 | 1.44E-05 | 1.63E-05 | -0.003276922 | RBM20 |
| chr11 | 78903194 | 78904017 | 824 | 5 | 4.35E-05 | 1.03E-06 | 0.00484 | 4.15E-06 | -0.015657497 | TENM4 |
| chr2 | 30155341 | 30156716 | 1376 | 4 | 4.35E-05 | 2.41E-07 | 0.00072 | 6.66E-07 | -0.000338618 | YPEL5 |
| chr20 | 1329807 | 1330299 | 493 | 4 | 4.35E-05 | 0.012213 | 0.0004 | 4.92E-05 | -0.005897128 | SDCBP2-AS1, AL136531.2 |
| chr20 | 48690900 | 48691477 | 578 | 3 | 4.35E-05 | 3.28E-06 | 0.00113 | 6.54E-06 | -0.018696333 | PREX1 |
| chr22 | 17257797 | 17258244 | 448 | 2 | 4.35E-05 | 9.00E-06 | 9.72E-05 | 8.85E-06 | -0.006585963 | ADA2, CECR3 |
| chr1 | 119923460 | 119923808 | 349 | 2 | 4.36E-05 | 3.69E-05 | 6.00E-05 | 1.77E-05 | -0.000985001 | NOTCH2 |
| chr1 | 6585034 | 6585403 | 370 | 5 | 4.36E-05 | 0.048529 | 4.41E-05 | 0.00017 | -0.004287333 | ZBTB48 |
| chr16 | 27209203 | 27209909 | 707 | 2 | 4.37E-05 | 0.000113 | 0.00112 | 0.00014 | -0.00111965 | KDM8 |
| chr17 | 78997312 | 78998480 | 1169 | 6 | 4.38E-05 | 0.00099 | 0.00011 | 1.69E-06 | -0.0028573 | CANT1 |
| chr17 | 81691025 | 81692211 | 1187 | 6 | 4.38E-05 | 0.000839 | 0.00053 | 8.76E-06 | -0.002724925 | HGS |
| chr15 | 69986620 | 69986872 | 253 | 3 | 4.38E-05 | 1.17E-05 | 0.0001 | 7.59E-06 | -0.020325098 | NA |
| chr9 | 21392294 | 21393117 | 824 | 3 | 4.38E-05 | 0.000169 | 5.42E-05 | 1.04E-05 | -0.008107307 | NA |
| chr7 | 151248521 | 151249357 | 837 | 13 | 4.38E-05 | 0.000347 | 0.00958 | 7.89E-06 | -0.000186846 | AC005486.1, SMARCD3 |
| chr11 | 10449436 | 10450140 | 705 | 5 | 4.39E-05 | 0.000436 | 0.00147 | 9.31E-05 | 0.015997661 | AMPD3 |
| chr11 | 12110913 | 12111786 | 874 | 6 | 4.39E-05 | 9.73E-06 | 0.00111 | 9.98E-06 | -0.004687944 | MICAL2 |
| chr17 | 8142297 | 8142345 | 49 | 3 | 4.39E-05 | 0.000387 | 0.00013 | 1.29E-05 | -0.004621326 | PER1 |
| chr5 | 56058400 | 56058959 | 560 | 6 | 4.39E-05 | 7.61E-05 | 0.00174 | 1.42E-05 | -0.007934164 | AC008892.1 |
| chr6 | 165752798 | 165753343 | 546 | 3 | 4.39E-05 | 1.29E-05 | 0.00056 | 1.60E-05 | -0.005656451 | PDE10A |
| chr2 | 7420694 | 7421076 | 383 | 3 | 4.40E-05 | 0.000696 | 2.60E-05 | 3.24E-05 | -0.008028631 | AC013460.1 |
| chr10 | 119827473 | 119827914 | 442 | 2 | 4.40E-05 | 5.38E-06 | 0.00017 | 7.33E-06 | -0.005003648 | INPP5F |
| chr9 | 34521335 | 34522066 | 732 | 4 | 4.40E-05 | 6.45E-06 | 0.00476 | 1.60E-05 | -0.000506669 | AL160270.1, ENHO |
| chr6 | 16237689 | 16237925 | 237 | 2 | 4.41E-05 | 2.23E-05 | 0.00095 | 3.37E-05 | -0.00931334 | NA |
| chr16 | 85187141 | 85188162 | 1022 | 4 | 4.41E-05 | 1.77E-06 | 0.00042 | 2.07E-06 | -0.001576103 | GSE1 |
| chr1 | 113811659 | 113812442 | 784 | 8 | 4.42E-05 | 4.02E-05 | 0.00454 | 2.17E-05 | -0.001500851 | AL137856.1, RSBN1 |
| chr17 | 44328451 | 44328816 | 366 | 2 | 4.42E-05 | 3.44E-05 | 4.52E-05 | 1.38E-05 | -0.00733438 | AC003043.1 |
| chr1 | 243931466 | 243932113 | 648 | 6 | 4.42E-05 | 1.40E-06 | 0.0027 | 4.47E-06 | -0.009909181 | LINC02774 |
| chr1 | 980678 | 981118 | 441 | 3 | 4.43E-05 | 0.001021 | 9.11E-05 | 6.90E-05 | 0.001544911 | PERM1 |
| chr2 | 99305915 | 99306316 | 402 | 2 | 4.43E-05 | 4.26E-05 | 5.57E-05 | 1.83E-05 | 0.000551543 | AC079447.1 |
| chr5 | 159208367 | 159208665 | 299 | 3 | 4.43E-05 | 0.024647 | 0.00014 | 0.00047 | 0.002238626 | RNF145 |
| chr8 | 21623842 | 21623952 | 111 | 2 | 4.43E-05 | 3.42E-05 | 2.20E-05 | 7.87E-06 | 0.001880428 | NA |
| chr15 | 101138140 | 101138856 | 717 | 3 | 4.44E-05 | 1.87E-06 | 0.00071 | 4.22E-06 | -0.009831496 | NA |
| chr6 | 34022067 | 34022404 | 338 | 2 | 4.44E-05 | 8.89E-05 | 9.57E-05 | 4.02E-05 | -0.012523599 | GRM4 |
| chr5 | 141478080 | 141478540 | 461 | 7 | 4.44E-05 | 2.39E-05 | 0.00513 | 5.49E-06 | 0.015852331 | PCDHGA1, PCDHGA2, PCDHGA3, PCDHGB1, PCDHGA4, PCDHGB2, PCDHGA5, PCDHGB3, PCDHGA6, PCDHGA7, PCDHGB4, PCDHGA8, PCDHGB5, PCDHGA9, PCDHGB6, PCDHGA10, PCDHGB7, PCDHGA11, PCDHGA12, PCDHGC3 |
| chr1 | 154500949 | 154501868 | 920 | 4 | 4.44E-05 | 8.89E-06 | 0.00168 | 1.78E-05 | -0.019140201 | SHE |
| chr12 | 11863796 | 11863947 | 152 | 2 | 4.44E-05 | 8.74E-05 | 2.56E-05 | 1.40E-05 | -0.007178741 | ETV6 |
| chr6 | 17599717 | 17600763 | 1047 | 10 | 4.44E-05 | 0.00082 | 7.94E-05 | 2.76E-05 | 0.000925137 | FAM8A1, AL138824.1 |
| chr22 | 40405775 | 40406387 | 613 | 3 | 4.44E-05 | 0.002278 | 2.35E-05 | 4.20E-05 | -0.002263922 | AL022238.3, SGSM3 |
| chr19 | 45780148 | 45780303 | 156 | 2 | 4.45E-05 | 0.000203 | 0.00548 | 0.00036 | -0.013221285 | DMPK, AC011530.1 |
| chr9 | 69171313 | 69171355 | 43 | 2 | 4.45E-05 | 1.94E-05 | 9.58E-05 | 1.55E-05 | -0.011485854 | AL358113.1, TJP2 |
| chr17 | 34811784 | 34811788 | 5 | 2 | 4.46E-05 | 4.74E-06 | 8.19E-05 | 5.12E-06 | -0.013671579 | NA |
| chr4 | 185021375 | 185021877 | 503 | 5 | 4.46E-05 | 9.63E-06 | 0.0014 | 1.11E-05 | 0.004072689 | NA |
| chr11 | 4577803 | 4578407 | 605 | 2 | 4.46E-05 | 0.000186 | 0.00015 | 7.93E-05 | 0.002731338 | C11orf40 |
| chr10 | 125048419 | 125048636 | 218 | 3 | 4.46E-05 | 2.27E-06 | 0.00044 | 4.52E-06 | -0.003941813 | CTBP2 |
| chr22 | 35258651 | 35258673 | 23 | 2 | 4.46E-05 | 4.16E-06 | 0.00026 | 6.50E-06 | -0.007933631 | HMGXB4 |
| chr14 | 101025657 | 101026069 | 413 | 6 | 4.47E-05 | 0.000432 | 7.05E-05 | 1.41E-05 | -0.00819692 | MEG8, MIR323A, MIR758 |
| chr19 | 10304073 | 10304881 | 809 | 2 | 4.48E-05 | 1.71E-06 | 0.00029 | 2.95E-06 | -0.018909922 | ZGLP1 |
| chr8 | 123042269 | 123042906 | 638 | 12 | 4.48E-05 | 4.46E-05 | 0.00419 | 1.10E-05 | 0.001954499 | TBC1D31, DERL1 |
| chr18 | 58340781 | 58341063 | 283 | 3 | 4.49E-05 | 0.000154 | 2.97E-05 | 1.34E-05 | -0.001312828 | NEDD4L |
| chr14 | 91352153 | 91352324 | 172 | 3 | 4.49E-05 | 6.50E-06 | 0.00014 | 7.09E-06 | -0.009319502 | CCDC88C |
| chr11 | 334251 | 334561 | 311 | 5 | 4.49E-05 | 1.08E-05 | 0.0019 | 4.64E-06 | -0.009449979 | NA |
| chr16 | 2044176 | 2044877 | 702 | 6 | 4.49E-05 | 0.001582 | 0.00061 | 4.62E-05 | -0.006040931 | NTHL1 |
| chr16 | 50062919 | 50062956 | 38 | 2 | 4.49E-05 | 0.000168 | 1.33E-05 | 9.71E-06 | -0.007045342 | AC007610.1 |
| chr7 | 29989615 | 29990522 | 908 | 13 | 4.49E-05 | 3.87E-05 | 0.01991 | 6.04E-06 | -0.003952955 | FKBP14-AS1, SCRN1 |
| chr7 | 22356353 | 22357025 | 673 | 7 | 4.50E-05 | 2.05E-05 | 0.00118 | 1.27E-05 | -0.002904102 | RAPGEF5 |
| chr16 | 3138214 | 3138603 | 390 | 4 | 4.50E-05 | 1.45E-05 | 0.00036 | 1.87E-05 | -0.005111751 | ZNF213, ZNF213-AS1 |
| chr2 | 24359784 | 24360462 | 679 | 4 | 4.50E-05 | 0.00199 | 0.00503 | 0.00074 | -0.000297257 | ITSN2 |
| chr6 | 17392498 | 17393594 | 1097 | 11 | 4.50E-05 | 1.03E-05 | 0.00877 | 6.48E-06 | -0.005150954 | CAP2 |
| chr12 | 131755403 | 131755710 | 308 | 2 | 4.50E-05 | 0.007025 | 2.65E-05 | 3.72E-05 | -0.002482959 | SFSWAP |
| chr19 | 39372278 | 39372445 | 168 | 2 | 4.52E-05 | 1.10E-05 | 4.67E-05 | 6.84E-06 | -0.003522978 | SAMD4B |
| chr12 | 29784652 | 29784815 | 164 | 3 | 4.52E-05 | 0.000118 | 0.00064 | 0.00011 | -0.010067929 | TMTC1 |
| chr16 | 85626495 | 85626928 | 434 | 3 | 4.53E-05 | 2.28E-06 | 0.00139 | 5.86E-06 | -0.021025016 | GSE1 |
| chr13 | 112489397 | 112490016 | 620 | 2 | 4.54E-05 | 4.20E-06 | 0.0002 | 6.19E-06 | -0.004731424 | TUBGCP3 |
| chr17 | 40071387 | 40072556 | 1170 | 4 | 4.54E-05 | 1.64E-06 | 0.00059 | 1.55E-06 | -0.011377817 | THRA |
| chr16 | 769064 | 769526 | 463 | 3 | 4.54E-05 | 7.83E-06 | 0.00347 | 1.94E-05 | -0.029324288 | AL031258.1, MSLNL |
| chr12 | 90938393 | 90938702 | 310 | 4 | 4.55E-05 | 0.007235 | 1.54E-05 | 3.14E-05 | -0.006872737 | LINC00615, CCER1 |
| chr19 | 10308066 | 10308175 | 110 | 2 | 4.56E-05 | 7.66E-06 | 8.46E-05 | 7.41E-06 | -0.005765171 | ZGLP1, AC011511.1 |
| chr1 | 180519946 | 180520017 | 72 | 2 | 4.57E-05 | 4.35E-05 | 1.95E-05 | 8.09E-06 | -0.014465345 | OVAAL |
| chr6 | 33455039 | 33456325 | 1287 | 21 | 4.57E-05 | 0.000172 | 0.0072 | 1.09E-05 | 0.004496661 | ZBTB9 |
| chr4 | 163344593 | 163345024 | 432 | 3 | 4.57E-05 | 7.99E-05 | 0.00021 | 1.82E-05 | -0.008810896 | NPY5R, NPY1R |
| chr12 | 109154090 | 109155182 | 1093 | 10 | 4.57E-05 | 3.10E-06 | 0.00242 | 9.24E-07 | 0.003085621 | ACACB |
| chr13 | 39765797 | 39765886 | 90 | 2 | 4.57E-05 | 0.000117 | 1.50E-05 | 9.77E-06 | -0.012120396 | COG6 |
| chr8 | 133101882 | 133103417 | 1536 | 11 | 4.58E-05 | 0.000565 | 0.00125 | 4.65E-06 | -0.00312027 | TG, SLA |
| chr1 | 92217846 | 92218264 | 419 | 8 | 4.58E-05 | 0.000154 | 0.00304 | 1.28E-05 | 0.000505234 | C1orf146 |
| chr19 | 44757021 | 44757747 | 727 | 7 | 4.59E-05 | 2.35E-05 | 0.00133 | 4.05E-06 | -0.002573983 | BCL3 |
| chr17 | 80790456 | 80790690 | 235 | 2 | 4.59E-05 | 0.001701 | 2.62E-05 | 3.14E-05 | -0.001645167 | RPTOR |
| chr8 | 1899833 | 1900241 | 409 | 5 | 4.59E-05 | 4.79E-06 | 0.00202 | 6.25E-06 | -0.017095607 | AC019257.8, ARHGEF10 |
| chr17 | 21126383 | 21127057 | 675 | 9 | 4.60E-05 | 1.04E-05 | 0.00086 | 8.50E-06 | 0.002170745 | DHRS7B |
| chr10 | 6173540 | 6174471 | 932 | 3 | 4.61E-05 | 2.92E-06 | 0.00014 | 2.40E-06 | -0.004992727 | PFKFB3 |
| chr5 | 57216447 | 57216866 | 420 | 2 | 4.61E-05 | 0.001447 | 5.29E-05 | 6.31E-05 | -0.012065705 | GPBP1 |
| chr19 | 12834484 | 12834732 | 249 | 6 | 4.61E-05 | 0.000557 | 0.00067 | 2.08E-05 | -0.002130774 | MAST1, HOOK2, RTBDN |
| chr8 | 123181444 | 123182121 | 678 | 8 | 4.61E-05 | 0.006488 | 0.00128 | 9.66E-05 | -0.002093713 | FAM83A, AC068228.1 |
| chr6 | 11805785 | 11806458 | 674 | 3 | 4.62E-05 | 1.80E-05 | 0.00095 | 2.90E-05 | -0.006126776 | ADTRP |
| chr13 | 85584976 | 85585750 | 775 | 3 | 4.62E-05 | 4.17E-06 | 0.00045 | 7.04E-06 | -0.005595562 | NA |
| chr8 | 71827727 | 71828094 | 368 | 3 | 4.62E-05 | 1.77E-05 | 0.00013 | 1.44E-05 | 0.000326039 | NA |
| chr20 | 3470645 | 3471633 | 989 | 12 | 4.64E-05 | 4.01E-05 | 0.00079 | 7.34E-06 | 0.001631437 | ATRN |
| chr3 | 196949104 | 196949357 | 254 | 2 | 4.65E-05 | 0.002961 | 4.15E-05 | 5.56E-05 | -0.002288215 | PIGZ |

| | | | | | | | | | | |
|---|---|---|---|---|---|---|---|---|---|---|
| chr1 | 46594973 | 46595584 | 612 | 4 | 4.65E-05 | 2.16E-06 | 0.00228 | 4.56E-06 | -0.00803849 | MKNK1 |
| chr1 | 239387278 | 239387949 | 672 | 4 | 4.65E-05 | 0.000134 | 8.28E-05 | 2.74E-05 | 0.002999652 | CHRM3 |
| chr11 | 66637584 | 66638347 | 764 | 4 | 4.65E-05 | 8.38E-06 | 0.00124 | 1.82E-05 | -0.005850069 | RBM14-RBM4 |
| chr17 | 1227082 | 1227163 | 82 | 2 | 4.65E-05 | 1.48E-05 | 6.35E-05 | 1.01E-05 | -0.006832745 | ABR |
| chr12 | 52652009 | 52652321 | 313 | 3 | 4.66E-05 | 0.000195 | 0.00018 | 3.45E-05 | -0.00430422 | KRT2 |
| chr6 | 32186670 | 32187364 | 695 | 5 | 4.66E-05 | 1.03E-05 | 0.00015 | 6.51E-06 | -0.00708542 | PBX2 |
| chr17 | 522943 | 523296 | 354 | 5 | 4.67E-05 | 0.001554 | 3.43E-05 | 3.22E-05 | -0.000900182 | AC015853.3, VPS53 |
| chr10 | 15096606 | 15097208 | 603 | 11 | 4.67E-05 | 0.000135 | 0.00133 | 1.25E-05 | 0.001605341 | RPP38, RPP38-DT |
| chr4 | 1718084 | 1718712 | 629 | 5 | 4.68E-05 | 1.02E-05 | 0.001 | 9.03E-06 | -0.006737441 | TACC3, TMEM129 |
| chr2 | 232231581 | 232231743 | 163 | 2 | 4.68E-05 | 0.000216 | 2.62E-05 | 1.99E-05 | -0.004777462 | DIS3L2 |
| chr2 | 201909705 | 201910045 | 341 | 2 | 4.68E-05 | 2.05E-05 | 5.31E-05 | 1.12E-05 | 0.002982882 | NA |
| chr1 | 210374553 | 210374612 | 60 | 2 | 4.68E-05 | 0.01678 | 1.72E-05 | 2.40E-05 | -0.011111174 | HHAT |
| chr3 | 134029467 | 134029968 | 502 | 5 | 4.68E-05 | 1.24E-05 | 0.00127 | 9.93E-06 | -0.000493496 | SLCO2A1 |
| chr9 | 137674402 | 137674440 | 39 | 2 | 4.68E-05 | 1.42E-05 | 0.00014 | 1.48E-05 | -0.004480742 | AL590627.2, EHMT1 |
| chr13 | 112694077 | 112694408 | 332 | 2 | 4.68E-05 | 0.008307 | 2.78E-05 | 4.02E-05 | -0.007339976 | ATP11A |
| chr9 | 82404951 | 82405135 | 185 | 3 | 4.69E-05 | 1.52E-05 | 0.00015 | 8.09E-06 | -0.014393313 | AL162726.3 |
| chr2 | 3194125 | 3194435 | 311 | 4 | 4.69E-05 | 0.00288 | 0.0004 | 1.72E-05 | -0.001156675 | EIPR1 |
| chr1 | 116123863 | 116124010 | 148 | 2 | 4.69E-05 | 0.000608 | 4.67E-05 | 4.55E-05 | -0.000719192 | MAB21L3 |
| chr8 | 96327310 | 96328392 | 1083 | 7 | 4.69E-05 | 4.32E-07 | 0.00249 | 1.82E-06 | -0.006859421 | PTDSS1 |
| chr2 | 69520270 | 69520872 | 603 | 3 | 4.70E-05 | 5.73E-05 | 0.00053 | 4.88E-05 | -0.001618774 | AAK1 |
| chr20 | 37251194 | 37251766 | 573 | 3 | 4.71E-05 | 0.000568 | 9.19E-05 | 2.35E-05 | -0.007683514 | GHRH |
| chr14 | 68824907 | 68825139 | 233 | 3 | 4.71E-05 | 4.99E-05 | 0.00017 | 8.50E-06 | -0.009600381 | NA |
| chr9 | 128240071 | 128240764 | 694 | 4 | 4.72E-05 | 3.25E-05 | 0.00059 | 1.32E-05 | -0.002147774 | DNM1 |
| chr2 | 74537898 | 74538789 | 892 | 3 | 4.72E-05 | 1.15E-06 | 0.00012 | 7.50E-07 | -0.007503702 | LOXL3 |
| chr6 | 10419451 | 10420179 | 729 | 10 | 4.72E-05 | 0.000243 | 5.42E-05 | 2.17E-05 | -0.002098899 | TFAP2A |
| chr6 | 33414780 | 33415635 | 856 | 5 | 4.73E-05 | 0.000168 | 0.00138 | 5.77E-05 | -0.003761433 | PHF1 |
| chr10 | 119377245 | 119377342 | 98 | 2 | 4.73E-05 | 7.62E-05 | 6.62E-05 | 2.85E-05 | 0.00153767 | GRK5 |
| chr6 | 18491376 | 18491722 | 347 | 2 | 4.73E-05 | 8.62E-06 | 9.07E-05 | 8.34E-06 | 0.003020931 | NA |
| chr11 | 66850231 | 66851670 | 1440 | 8 | 4.73E-05 | 1.96E-06 | 0.00307 | 1.24E-06 | -0.004417566 | PC |
| chr20 | 62143214 | 62143585 | 372 | 7 | 4.74E-05 | 0.000201 | 9.72E-05 | 2.08E-05 | 0.002030295 | PSMA7 |
| chr6 | 7237747 | 7237806 | 60 | 2 | 4.75E-05 | 1.20E-05 | 7.58E-05 | 9.65E-06 | -0.004725977 | RREB1 |
| chr12 | 94078894 | 94079447 | 554 | 3 | 4.76E-05 | 4.64E-05 | 0.0003 | 1.84E-05 | -0.004599297 | NA |
| chr13 | 112807186 | 112807731 | 546 | 7 | 4.76E-05 | 7.85E-06 | 0.00772 | 9.08E-06 | -0.001296311 | ATP11A |
| chr16 | 265565 | 265914 | 350 | 3 | 4.76E-05 | 5.18E-05 | 0.00063 | 4.03E-05 | -0.007996534 | FAM234A |
| chr22 | 30204700 | 30204718 | 19 | 2 | 4.77E-05 | 5.61E-06 | 0.00012 | 6.77E-06 | -0.00896244 | AC002378.1 |
| chr22 | 26589588 | 26590460 | 873 | 7 | 4.77E-05 | 1.40E-05 | 0.00129 | 9.53E-06 | -1.43E-05 | TPST2 |
| chr6 | 26285130 | 26285942 | 813 | 9 | 4.77E-05 | 3.91E-06 | 0.01143 | 5.93E-06 | 0.002300269 | H4C8 |
| chr2 | 219178002 | 219178808 | 807 | 11 | 4.78E-05 | 5.68E-05 | 0.00253 | 9.03E-06 | -0.000458705 | RETREG2, CNPPD1 |
| chr14 | 101069122 | 101069833 | 712 | 4 | 4.79E-05 | 1.72E-05 | 0.00035 | 1.95E-05 | -0.004848077 | MEG9 |
| chr7 | 155456979 | 155458205 | 1227 | 9 | 4.79E-05 | 0.005164 | 0.00108 | 0.00029 | 0.001220383 | EN2, AC008060.3 |
| chr6 | 24356768 | 24357757 | 990 | 7 | 4.80E-05 | 3.50E-07 | 0.00849 | 2.47E-06 | -0.009914074 | KAAG1, DCDC2 |
| chr3 | 13622980 | 13623522 | 543 | 3 | 4.80E-05 | 5.40E-05 | 0.00013 | 2.38E-05 | -0.006852748 | FBLN2 |
| chr19 | 4902564 | 4902887 | 324 | 6 | 4.80E-05 | 0.000482 | 0.00099 | 1.81E-05 | -0.004993229 | ARRDC5 |
| chr17 | 39612530 | 39612551 | 22 | 2 | 4.80E-05 | 1.77E-05 | 6.37E-05 | 1.15E-05 | -0.010034036 | NA |
| chr6 | 11042875 | 11043135 | 261 | 3 | 4.80E-05 | 9.74E-06 | 0.00029 | 6.86E-06 | -0.020938591 | ELOVL2 |
| chr11 | 45805709 | 45807481 | 1773 | 9 | 4.81E-05 | 3.76E-05 | 0.00059 | 1.02E-06 | -0.005779626 | SLC35C1 |
| chr11 | 35281552 | 35282294 | 743 | 3 | 4.82E-05 | 4.68E-05 | 5.69E-05 | 2.00E-06 | -0.00749624 | SLC1A2-AS1, SLC1A2 |
| chr9 | 130462314 | 130462578 | 265 | 3 | 4.83E-05 | 1.32E-05 | 0.00016 | 7.76E-06 | -0.010593896 | ASS1 |
| chr2 | 204438272 | 204438377 | 106 | 2 | 4.83E-05 | 3.27E-06 | 0.00023 | 5.15E-06 | -0.004364085 | NA |
| chr12 | 11879394 | 11879561 | 168 | 2 | 4.83E-05 | 1.42E-05 | 0.00083 | 2.22E-05 | -0.009269228 | ETV6 |
| chr12 | 6182253 | 6182775 | 523 | 2 | 4.84E-05 | 1.93E-05 | 8.62E-05 | 1.47E-05 | -0.017146907 | NA |
| chr19 | 49361521 | 49362085 | 565 | 5 | 4.84E-05 | 0.00056 | 9.17E-05 | 4.47E-05 | -0.005128545 | DKKL1, TEAD2 |
| chr20 | 46998807 | 46998906 | 100 | 2 | 4.84E-05 | 0.000621 | 1.32E-05 | 1.31E-05 | 0.018171612 | EYA2 |
| chr6 | 31580330 | 31581382 | 1053 | 19 | 4.85E-05 | 0.000728 | 0.00026 | 5.99E-05 | -0.001290463 | LTB |
| chr11 | 8092758 | 8092849 | 92 | 2 | 4.85E-05 | 4.24E-06 | 0.00026 | 6.60E-06 | -0.015417343 | TUB |
| chr2 | 25169687 | 25169716 | 30 | 2 | 4.85E-05 | 0.000255 | 0.00483 | 0.00043 | -0.007558305 | NA |
| chr7 | 12571063 | 12571982 | 920 | 8 | 4.85E-05 | 2.18E-06 | 0.01276 | 6.96E-06 | -0.008001888 | SCIN, AC005281.1 |
| chr6 | 4482649 | 4482756 | 108 | 2 | 4.86E-05 | 1.40E-05 | 6.88E-05 | 1.02E-05 | -0.003879852 | AL162718.1 |
| chr5 | 66827874 | 66828760 | 887 | 10 | 4.86E-05 | 8.84E-05 | 0.00248 | 1.73E-05 | -0.004016474 | MAST4 |
| chr9 | 96116780 | 96117140 | 361 | 2 | 4.86E-05 | 0.00048 | 0.00012 | 0.0001 | 0.004405146 | ERCC6L2 |
| chr20 | 31260401 | 31260702 | 302 | 2 | 4.88E-05 | 3.59E-05 | 7.52E-05 | 2.01E-05 | -0.022666342 | NA |
| chr10 | 129606325 | 129606505 | 181 | 3 | 4.88E-05 | 2.00E-06 | 0.00102 | 4.59E-06 | -0.029683076 | MGMT |
| chr1 | 98045967 | 98046638 | 672 | 8 | 4.88E-05 | 0.000405 | 5.40E-05 | 3.84E-05 | -0.00091736 | MIR137HG, MIR137 |
| chr6 | 133837140 | 133837161 | 22 | 2 | 4.88E-05 | 0.000191 | 0.00015 | 8.28E-05 | -0.000810042 | LINC01312, TARID |
| chr16 | 85492236 | 85492330 | 95 | 2 | 4.89E-05 | 3.03E-06 | 0.00036 | 5.15E-06 | -0.019047686 | GSE1 |
| chr17 | 75036518 | 75036914 | 397 | 2 | 4.89E-05 | 5.40E-06 | 0.00015 | 7.13E-06 | -0.034619747 | KCTD2 |
| chr8 | 934647 | 935128 | 482 | 4 | 4.89E-05 | 4.63E-06 | 0.00078 | 8.62E-06 | -0.005892572 | DLGAP2 |
| chr15 | 58552160 | 58552384 | 225 | 3 | 4.89E-05 | 8.53E-06 | 1.00E-04 | 6.05E-06 | -0.015445941 | LIPC |
| chr1 | 157841513 | 157842115 | 603 | 5 | 4.90E-05 | 6.43E-05 | 0.00014 | 1.32E-05 | -0.003502874 | CD5L |
| chr7 | 47979608 | 47980116 | 509 | 7 | 4.90E-05 | 2.90E-05 | 0.00358 | 1.48E-05 | -0.012222183 | HUS1 |
| chr5 | 10559738 | 10560028 | 291 | 2 | 4.90E-05 | 0.001718 | 1.99E-05 | 2.36E-05 | -0.01012553 | NA |
| chr2 | 101559426 | 101559834 | 409 | 2 | 4.90E-05 | 9.08E-05 | 3.98E-05 | 2.08E-05 | -0.006932559 | AC093894.2 |
| chr10 | 61453346 | 61454026 | 681 | 10 | 4.90E-05 | 0.000187 | 0.00413 | 4.35E-05 | -0.00484933 | TMEM26-AS1, TMEM26 |
| chr10 | 80158216 | 80158877 | 662 | 2 | 4.91E-05 | 4.32E-05 | 0.00027 | 4.25E-05 | -0.020405459 | ANXA11 |
| chr13 | 46386873 | 46387812 | 940 | 17 | 4.91E-05 | 5.14E-06 | 0.01464 | 3.73E-06 | 0.001287031 | RUBCNL |
| chr19 | 55836481 | 55837523 | 1043 | 8 | 4.91E-05 | 0.000116 | 0.00187 | 5.25E-05 | 0.001035995 | NLRP4, NLRP11 |
| chr8 | 139711341 | 139711795 | 455 | 3 | 4.91E-05 | 2.49E-05 | 7.78E-05 | 1.27E-05 | -0.006330762 | NA |
| chr3 | 151283226 | 151283647 | 422 | 2 | 4.91E-05 | 0.00081 | 3.10E-05 | 3.29E-05 | -0.006822692 | MED12L |
| chr16 | 11032321 | 11033180 | 860 | 2 | 4.92E-05 | 1.30E-06 | 9.58E-05 | 1.87E-06 | -0.009910155 | CLEC16A |
| chr16 | 31179798 | 31180538 | 741 | 6 | 4.93E-05 | 0.040628 | 4.30E-05 | 0.00029 | 0.00131666 | FUS |
| chr7 | 38632043 | 38632597 | 555 | 5 | 4.93E-05 | 0.00034 | 9.92E-05 | 1.50E-05 | -0.007780867 | NA |
| chr15 | 47961434 | 47961982 | 549 | 3 | 4.93E-05 | 1.44E-06 | 0.00179 | 4.11E-06 | -0.009067208 | AC092078.2, AC092078.3 |
| chr19 | 9538200 | 9539564 | 1365 | 13 | 4.93E-05 | 2.49E-05 | 0.00388 | 8.63E-06 | -0.000201011 | ZNF426-DT, ZNF426 |
| chr3 | 126064125 | 126064499 | 375 | 2 | 4.94E-05 | 5.45E-06 | 0.00052 | 9.02E-06 | 0.001167635 | SLC41A3 |
| chr11 | 70934318 | 70935176 | 859 | 5 | 4.94E-05 | 2.11E-05 | 0.00077 | 1.05E-05 | -2.07E-05 | SHANK2 |
| chr3 | 184229837 | 184230534 | 698 | 10 | 4.94E-05 | 3.05E-05 | 0.00087 | 1.80E-05 | 0.002434427 | VWA5B2 |
| chr10 | 70785297 | 70785717 | 421 | 4 | 4.95E-05 | 2.41E-05 | 0.00069 | 1.11E-05 | -0.008571307 | TBATA |
| chr7 | 1077782 | 1078280 | 499 | 6 | 4.95E-05 | 1.12E-05 | 0.00301 | 8.16E-06 | -0.004075773 | AC091729.1, C7orf50 |
| chr16 | 4160850 | 4161239 | 390 | 2 | 4.95E-05 | 6.53E-06 | 0.00013 | 7.83E-06 | -0.03242577 | NA |
| chr11 | 77124848 | 77125414 | 567 | 4 | 4.96E-05 | 9.55E-06 | 0.0022 | 9.50E-06 | -0.006309966 | CAPN5 |
| chr19 | 46377759 | 46378176 | 418 | 2 | 4.96E-05 | 0.000128 | 3.85E-05 | 2.34E-05 | -0.017163844 | PPP5C |
| chr9 | 133374149 | 133374994 | 846 | 2 | 4.96E-05 | 1.87E-06 | 0.00021 | 3.09E-06 | -0.01194849 | SURF4 |
| chr22 | 39959038 | 39959407 | 370 | 3 | 4.96E-05 | 0.000279 | 7.95E-05 | 2.19E-05 | -0.004767937 | GRAP2 |
| chr13 | 110475158 | 110475254 | 97 | 2 | 4.97E-05 | 1.16E-05 | 6.77E-05 | 8.85E-06 | -0.007613787 | COL4A2 |
| chr19 | 40209279 | 40209713 | 435 | 2 | 4.97E-05 | 1.48E-05 | 0.00011 | 1.37E-05 | -0.010087572 | MAP3K10 |
| chr11 | 66080629 | 66080676 | 48 | 2 | 4.98E-05 | 4.15E-06 | 0.00033 | 6.74E-06 | -0.004643324 | PACS1 |
| chr19 | 17943551 | 17943976 | 426 | 3 | 4.98E-05 | 8.64E-06 | 0.00104 | 1.23E-05 | -0.003236946 | CCDC124 |
| chr6 | 33187053 | 33187358 | 306 | 3 | 4.99E-05 | 0.000136 | 0.00023 | 2.37E-05 | -0.004088916 | COL11A2 |
| chr1 | 85611898 | 85612254 | 357 | 3 | 4.99E-05 | 2.06E-05 | 0.00013 | 1.34E-05 | -0.003040464 | NA |
| chr5 | 139658899 | 139659441 | 543 | 2 | 4.99E-05 | 1.25E-05 | 0.00012 | 1.27E-05 | -0.010614316 | CXXC5 |
| chr2 | 134218207 | 134218584 | 378 | 2 | 4.99E-05 | 0.000294 | 3.21E-05 | 2.64E-05 | 0.005853045 | MGAT5 |
| chr22 | 20966965 | 20967775 | 811 | 6 | 5.00E-05 | 7.16E-05 | 0.00288 | 2.52E-05 | -0.005576248 | AIFM3 |
| chr4 | 67724029 | 67724133 | 105 | 3 | 5.00E-05 | 3.08E-05 | 5.94E-05 | 7.59E-06 | -0.004572417 | UBA6-AS1, AC079880.1 |
| chr17 | 74309328 | 74310076 | 749 | 5 | 5.00E-05 | 1.56E-05 | 0.00132 | 1.13E-05 | -0.008015727 | DNAI2, AC103809.1 |
| chr6 | 36442491 | 36443439 | 949 | 14 | 5.00E-05 | 5.34E-05 | 0.00558 | 1.52E-05 | 0.000407807 | KCTD20, PXT1 |
| chr7 | 5394822 | 5395435 | 614 | 2 | 5.00E-05 | 2.10E-06 | 0.00031 | 3.66E-06 | -0.011583249 | TNRC18 |
| chr14 | 98891061 | 98891164 | 104 | 3 | 5.00E-05 | 0.000862 | 1.02E-05 | 1.51E-05 | 0.014233255 | NA |
| chr19 | 11537811 | 11538647 | 837 | 4 | 5.00E-05 | 2.57E-05 | 0.00019 | 1.05E-05 | -0.002133355 | NA |
| chr20 | 37095591 | 37096497 | 907 | 14 | 5.01E-05 | 0.000252 | 0.00034 | 2.70E-05 | -1.64E-05 | AL136172.1, RBL1 |
| chr11 | 95697506 | 95697972 | 467 | 3 | 5.02E-05 | 0.001546 | 0.0005 | 1.99E-05 | -0.008314587 | AP000820.2 |
| chr8 | 26513036 | 26513518 | 483 | 3 | 5.02E-05 | 0.119359 | 7.74E-05 | 0.0005 | 0.003223783 | PNMA2 |
| chr11 | 47441069 | 47441516 | 448 | 3 | 5.03E-05 | 0.003444 | 0.00029 | 0.00014 | -0.005201101 | RAPSN |
| chr9 | 4904699 | 4905203 | 505 | 2 | 5.04E-05 | 3.07E-06 | 0.00025 | 4.98E-06 | 0.016532378 | NA |
| chr22 | 46547028 | 46547090 | 63 | 2 | 5.04E-05 | 5.55E-06 | 0.00058 | 9.31E-06 | -0.012585971 | AL021392.1 |
| chr8 | 1866579 | 1866987 | 409 | 6 | 5.04E-05 | 0.000513 | 0.00406 | 2.64E-05 | -0.005943507 | AC019257.8, ARHGEF10 |
| chr14 | 102891254 | 102891899 | 646 | 2 | 5.04E-05 | 3.97E-06 | 0.00019 | 5.90E-06 | -0.003006988 | TRAF3 |
| chr17 | 41812520 | 41813145 | 626 | 14 | 5.04E-05 | 7.16E-05 | 0.0323 | 3.93E-05 | -0.004287607 | FKBP10, P3H4 |
| chr3 | 159844835 | 159844995 | 161 | 2 | 5.04E-05 | 3.76E-06 | 0.00019 | 5.63E-06 | -0.022829448 | IQCJ-SCHIP1, SCHIP1 |
| chr16 | 87067855 | 87069085 | 1231 | 11 | 5.04E-05 | 1.94E-07 | 0.0029 | 9.50E-07 | -0.004581337 | LINC02181, AC106745.1 |
| chr1 | 25871364 | 25872230 | 867 | 4 | 5.05E-05 | 1.18E-05 | 0.00112 | 3.64E-06 | -0.007506661 | PAQR7 |
| chr20 | 49381610 | 49381792 | 183 | 2 | 5.06E-05 | 1.30E-05 | 4.85E-05 | 7.83E-06 | -0.009948393 | AL035685.1, KCNB1 |
| chr16 | 2236286 | 2236785 | 500 | 10 | 5.06E-05 | 9.39E-06 | 0.02377 | 1.96E-05 | -0.006993256 | DNASE1L2, AC009065.1 |
| chr20 | 47058242 | 47058336 | 95 | 2 | 5.06E-05 | 0.000287 | 2.40E-05 | 1.98E-05 | -0.007418514 | EYA2 |
| chr15 | 93048535 | 93049192 | 658 | 4 | 5.07E-05 | 9.07E-06 | 0.00173 | 1.51E-05 | -0.019519038 | RGMA |
| chr19 | 53671352 | 53672165 | 814 | 5 | 5.07E-05 | 6.27E-06 | 0.00154 | 8.63E-06 | -0.009324795 | MIR1323 |
| chr10 | 31928353 | 31929040 | 688 | 9 | 5.07E-05 | 5.61E-05 | 0.00104 | 1.52E-05 | 0.000848024 | ARHGAP12 |
| chr19 | 47407292 | 47407323 | 32 | 2 | 5.07E-05 | 6.92E-06 | 9.05E-05 | 7.08E-06 | 0.00663175 | MEIS3 |
| chr13 | 83883593 | 83883747 | 155 | 2 | 5.07E-05 | 5.33E-06 | 0.00051 | 8.85E-06 | 0.020292684 | AL355481.1 |
| chr21 | 33569239 | 33569609 | 371 | 2 | 5.07E-05 | 0.001344 | 2.68E-05 | 3.09E-05 | -0.004415535 | SON, DONSON |
| chr4 | 186610140 | 186610362 | 223 | 2 | 5.09E-05 | 7.15E-06 | 0.00021 | 9.74E-06 | -0.010447957 | FAT1 |
| chr1 | 36323787 | 36324459 | 673 | 10 | 5.09E-05 | 2.21E-05 | 0.00331 | 9.68E-06 | -0.002955957 | SH3D21, EVA1B |
| chr11 | 1280204 | 1280938 | 735 | 3 | 5.10E-05 | 5.64E-05 | 0.00014 | 3.36E-05 | -0.00277439 | TOLLIP |
| chr1 | 229318162 | 229318341 | 180 | 3 | 5.11E-05 | 5.10E-06 | 0.00191 | 1.17E-05 | -0.004922781 | NA |
| chr2 | 173355472 | 173356977 | 1506 | 6 | 5.11E-05 | 2.44E-05 | 0.00184 | 1.13E-05 | -0.00552032 | CDCA7, AC092573.1 |
| chr20 | 62831888 | 62832816 | 929 | 5 | 5.11E-05 | 1.84E-06 | 0.00166 | 2.18E-06 | 0.001129415 | COL9A3 |
| chr8 | 30656191 | 30657297 | 1107 | 3 | 5.13E-05 | 0.000117 | 8.60E-05 | 6.81E-06 | 0.002658403 | GTF2E2 |
| chr10 | 58326978 | 58327537 | 560 | 7 | 5.13E-05 | 2.23E-05 | 0.00084 | 1.24E-05 | -0.00195804 | AC016396.2 |
| chr13 | 99565670 | 99566298 | 629 | 5 | 5.14E-05 | 2.31E-06 | 0.00159 | 6.68E-06 | -0.02793626 | NA |
| chr20 | 62796573 | 62797285 | 713 | 6 | 5.14E-05 | 1.14E-06 | 0.00066 | 1.85E-06 | -0.000719826 | MRGBP |
| chr7 | 12653746 | 12654040 | 295 | 2 | 5.14E-05 | 0.004866 | 2.17E-05 | 2.89E-05 | -0.008722384 | SCIN |
| chr1 | 224329642 | 224330428 | 787 | 11 | 5.14E-05 | 0.00035 | 0.00171 | 2.56E-05 | 0.000425508 | NVL |
| chr22 | 22892536 | 22893685 | 1150 | 5 | 5.15E-05 | 3.81E-06 | 0.00107 | 1.56E-06 | -0.003614994 | IGLL5 |
| chr3 | 159764624 | 159765088 | 465 | 4 | 5.15E-05 | 0.000758 | 8.42E-05 | 4.40E-05 | 0.000733084 | IQCJ-SCHIP1 |
| chr20 | 1226095 | 1226739 | 645 | 6 | 5.15E-05 | 5.63E-05 | 0.00224 | 2.85E-05 | 0.001307099 | RAD21L1 |
| chr3 | 194628308 | 194628418 | 111 | 2 | 5.15E-05 | 6.31E-05 | 2.01E-05 | 9.87E-06 | -0.003461018 | TMEM44 |
| chr14 | 76825959 | 76826390 | 432 | 5 | 5.15E-05 | 7.64E-05 | 0.0002 | 1.58E-05 | -0.004038313 | LRRC74A, ANGEL1 |
| chr9 | 124280405 | 124280581 | 177 | 2 | 5.15E-05 | 3.28E-06 | 0.00042 | 5.64E-06 | -0.011429916 | NEK6 |
| chr2 | 200735186 | 200735913 | 728 | 3 | 5.16E-05 | 0.000114 | 7.93E-05 | 2.26E-05 | -0.025786901 | AOX3P-AOX2P, LINC01792 |
| chr1 | 160982861 | 160982913 | 53 | 2 | 5.16E-05 | 0.006337 | 2.28E-05 | 3.12E-05 | -0.003343082 | NA |
| chr6 | 41653213 | 41653494 | 282 | 4 | 5.16E-05 | 0.000111 | 4.31E-05 | 1.28E-05 | -0.004257475 | MDFI |
| chr10 | 126476471 | 126476590 | 120 | 2 | 5.16E-05 | 4.46E-06 | 0.00013 | 5.83E-06 | -0.026034162 | C10orf90 |
| chr20 | 63345869 | 63346718 | 850 | 3 | 5.16E-05 | 6.45E-07 | 0.00122 | 1.90E-06 | -0.013952694 | CHRNA4 |
| chr2 | 26693024 | 26693707 | 684 | 4 | 5.17E-05 | 2.69E-05 | 0.00021 | 8.61E-06 | 0.007337646 | KCNK3 |
| chr6 | 31950722 | 31951253 | 532 | 2 | 5.17E-05 | 0.000131 | 9.61E-05 | 4.91E-05 | -0.005530203 | AL645922.1, CFB |
| chr5 | 151770779 | 151771349 | 571 | 5 | 5.17E-05 | 1.24E-05 | 0.00391 | 1.84E-05 | 0.001669559 | G3BP1, ATOX1, AC091982.3 |
| chr17 | 64320733 | 64321407 | 675 | 5 | 5.17E-05 | 1.51E-05 | 0.00169 | 4.32E-06 | -0.008344734 | PECAM1 |
| chr2 | 10449572 | 10449947 | 376 | 3 | 5.17E-05 | 1.40E-05 | 0.00468 | 3.32E-05 | -0.008839305 | ODC1-DT |
| chr16 | 921556 | 922115 | 560 | 4 | 5.17E-05 | 0.000354 | 0.00011 | 3.59E-05 | 0.001536454 | LMF1-AS1, LMF1 |
| chr22 | 29337784 | 29337985 | 202 | 2 | 5.17E-05 | 0.005566 | 5.19E-05 | 7.77E-05 | -0.004125594 | AP1B1 |
| chr1 | 204079995 | 204080458 | 464 | 2 | 5.17E-05 | 1.76E-05 | 0.00016 | 1.83E-05 | -0.011535555 | SOX13 |
| chr18 | 23155573 | 23156658 | 1086 | 6 | 5.19E-05 | 2.72E-06 | 0.00127 | 2.77E-06 | -0.005262381 | CABLES1 |
| chr16 | 11136431 | 11136865 | 435 | 4 | 5.20E-05 | 6.12E-05 | 0.00069 | 1.40E-05 | -0.013103592 | CLEC16A |

| | | | | | | | | | | |
|---|---|---|---|---|---|---|---|---|---|---|
| chr19 | 2052182 | 2052565 | 384 | 2 | 5.20E-05 | 2.18E-05 | 0.00076 | 3.21E-05 | -0.0167635 | NA |
| chr6 | 166508272 | 166508402 | 131 | 3 | 5.22E-05 | 2.86E-05 | 0.00067 | 1.83E-05 | -0.005879837 | RPS6KA2 |
| chr8 | 143842047 | 143842721 | 675 | 4 | 5.22E-05 | 0.004995 | 0.00026 | 2.94E-05 | -0.001192573 | NA |
| chr5 | 168261097 | 168261612 | 516 | 2 | 5.22E-05 | 1.28E-05 | 0.00045 | 1.85E-05 | 0.000598517 | TENM2, CTB-178M22.2 |
| chr17 | 44197755 | 44198183 | 429 | 4 | 5.22E-05 | 5.85E-05 | 0.00068 | 2.27E-05 | -0.006485364 | ATXN7L3 |
| chr11 | 69669665 | 69670467 | 803 | 6 | 5.22E-05 | 3.94E-07 | 0.00688 | 2.63E-06 | -0.004504001 | LTO1 |
| chr18 | 5544209 | 5544662 | 454 | 6 | 5.23E-05 | 0.001876 | 0.00017 | 0.00028 | 0.002189478 | EPB41L3 |
| chr6 | 31661311 | 31663146 | 1836 | 11 | 5.24E-05 | 0.000396 | 0.00013 | 7.63E-07 | -0.003705353 | GPANK1 |
| chr14 | 50766553 | 50766880 | 328 | 2 | 5.24E-05 | 4.07E-05 | 4.18E-05 | 1.44E-05 | -0.0055003 | NIN |
| chr10 | 120436405 | 120436608 | 204 | 2 | 5.25E-05 | 5.70E-06 | 0.00011 | 6.68E-06 | -0.00580147 | AC073587.1 |
| chr14 | 93017653 | 93017858 | 206 | 4 | 5.25E-05 | 3.56E-06 | 0.0006 | 6.53E-06 | -0.004656077 | ITPK1 |
| chr1 | 64214215 | 64214621 | 407 | 2 | 5.25E-05 | 6.32E-06 | 0.00015 | 7.94E-06 | -0.009268088 | UBE2U |
| chr15 | 92332357 | 92332529 | 173 | 2 | 5.25E-05 | 0.00032 | 1.89E-05 | 1.63E-05 | -0.004200071 | NA |
| chr17 | 3864983 | 3865305 | 323 | 2 | 5.26E-05 | 7.03E-05 | 5.77E-05 | 2.46E-05 | -0.012527464 | CAMKK1 |
| chr7 | 1054218 | 1055172 | 955 | 8 | 5.26E-05 | 4.28E-06 | 0.00674 | 1.17E-05 | -0.004728609 | GPR146, C7orf50 |
| chr3 | 48429736 | 48430586 | 851 | 10 | 5.27E-05 | 1.02E-06 | 0.01278 | 5.12E-06 | -0.00478853 | PLXNB1 |
| chr1 | 3111666 | 3112013 | 348 | 5 | 5.28E-05 | 0.000157 | 0.0013 | 5.14E-05 | -0.003798292 | PRDM16 |
| chr16 | 1870814 | 1871074 | 261 | 3 | 5.28E-05 | 0.000941 | 3.93E-05 | 2.46E-05 | -0.012704535 | MEIOB |
| chr4 | 153790658 | 153791428 | 771 | 5 | 5.28E-05 | 0.000149 | 0.003 | 0.00013 | -0.002408733 | NA |
| chr1 | 61005552 | 61006174 | 623 | 4 | 5.28E-05 | 1.44E-06 | 0.00322 | 4.79E-06 | -0.012612091 | NFIA, NFIA-AS2 |
| chr6 | 16681274 | 16682030 | 757 | 4 | 5.29E-05 | 2.78E-06 | 0.0002 | 2.49E-06 | -0.007860082 | ATXN1 |
| chr1 | 40039925 | 40040850 | 926 | 14 | 5.29E-05 | 7.26E-05 | 0.00021 | 2.62E-05 | 0.000875837 | CAP1 |
| chr13 | 111998124 | 111998352 | 229 | 3 | 5.29E-05 | 7.31E-06 | 0.00038 | 6.88E-06 | 0.003760628 | NA |
| chr18 | 23141817 | 23141882 | 66 | 2 | 5.30E-05 | 3.69E-06 | 0.00021 | 5.63E-06 | -0.013220063 | CABLES1 |
| chr12 | 12338477 | 12339197 | 721 | 5 | 5.31E-05 | 1.55E-05 | 0.00063 | 8.38E-06 | -0.00153486 | RPL23AP66, MANSC1 |
| chr17 | 46808986 | 46809481 | 496 | 3 | 5.31E-05 | 0.000591 | 0.00054 | 2.31E-05 | -0.003661688 | WNT3 |
| chr7 | 128870130 | 128870264 | 135 | 2 | 5.32E-05 | 4.80E-06 | 0.00042 | 7.89E-06 | -0.015006478 | ATP6V1FNB, KCP |
| chr11 | 108122571 | 108122740 | 170 | 2 | 5.33E-05 | 0.000225 | 0.00015 | 8.64E-05 | -0.004786767 | ACAT1 |
| chr12 | 13002228 | 13003197 | 970 | 2 | 5.33E-05 | 6.87E-07 | 9.83E-05 | 1.11E-06 | -0.00550854 | HTR7P1, AC023790.2 |
| chr2 | 3667346 | 3667449 | 104 | 2 | 5.33E-05 | 1.49E-05 | 0.00017 | 1.66E-05 | -0.006014216 | ALLC |
| chr4 | 163475181 | 163475231 | 51 | 2 | 5.33E-05 | 3.58E-05 | 0.00012 | 2.61E-05 | 0.001038212 | NA |
| chr11 | 114784442 | 114784558 | 117 | 2 | 5.33E-05 | 8.11E-06 | 6.57E-05 | 6.79E-06 | -0.009457658 | NA |
| chr22 | 37007937 | 37008987 | 1051 | 6 | 5.33E-05 | 2.38E-06 | 0.00566 | 7.33E-06 | -0.006698846 | NA |
| chr2 | 28337136 | 28337415 | 280 | 2 | 5.33E-05 | 0.000262 | 2.40E-05 | 1.93E-05 | -0.003520022 | BABAM2 |
| chr16 | 57486075 | 57487139 | 1065 | 10 | 5.33E-05 | 0.000158 | 0.0034 | 1.72E-05 | 0.000589236 | DOK4 |
| chr3 | 167620557 | 167621050 | 494 | 5 | 5.33E-05 | 0.000285 | 5.16E-05 | 1.55E-05 | -0.006210222 | WDR49 |
| chr13 | 99015722 | 99016179 | 458 | 5 | 5.33E-05 | 0.000291 | 0.00044 | 9.14E-06 | -0.006259236 | DOCK9 |
| chr1 | 58391714 | 58392566 | 853 | 4 | 5.34E-05 | 2.54E-06 | 0.00186 | 6.40E-06 | -0.020200188 | DAB1 |
| chr3 | 52529617 | 52530110 | 494 | 2 | 5.34E-05 | 1.85E-05 | 0.00059 | 2.67E-05 | -0.00841708 | NT5DC2 |
| chr19 | 35736805 | 35737163 | 359 | 2 | 5.35E-05 | 2.06E-05 | 0.00029 | 2.49E-05 | -0.000985495 | KMT2B |
| chr2 | 26251584 | 26252023 | 440 | 3 | 5.35E-05 | 2.96E-05 | 0.00032 | 1.41E-05 | -0.005583497 | HADHB |
| chr12 | 3821757 | 3822324 | 568 | 4 | 5.36E-05 | 1.51E-06 | 0.00178 | 4.82E-06 | -0.002759955 | PARP11 |
| chr14 | 92263707 | 92264471 | 765 | 7 | 5.36E-05 | 1.76E-07 | 0.01165 | 1.65E-06 | -0.009718959 | NA |
| chr2 | 8500932 | 8501341 | 410 | 4 | 5.37E-05 | 7.74E-06 | 0.00225 | 8.29E-06 | -0.006052236 | AC092617.1, LINC01814 |
| chr2 | 240574501 | 240575351 | 851 | 7 | 5.37E-05 | 5.10E-05 | 0.00354 | 3.27E-05 | -0.001488562 | RNPEPL1 |
| chr20 | 57692731 | 57693389 | 659 | 4 | 5.38E-05 | 6.54E-06 | 0.00148 | 8.67E-06 | -0.008566371 | PMEPA1 |
| chr19 | 3721261 | 3721834 | 574 | 10 | 5.38E-05 | 2.50E-05 | 0.00112 | 1.03E-05 | 0.006142065 | TJP3 |
| chr13 | 113847829 | 113848427 | 599 | 6 | 5.39E-05 | 2.06E-05 | 0.00084 | 2.67E-05 | -0.004175433 | GAS6 |
| chr2 | 26483285 | 26483562 | 278 | 2 | 5.40E-05 | 0.002161 | 3.99E-05 | 5.07E-05 | -0.002512511 | OTOF |
| chr10 | 23693850 | 23695061 | 1212 | 12 | 5.40E-05 | 0.000138 | 0.00643 | 1.71E-05 | 5.87E-05 | KIAA1217 |
| chr10 | 102612925 | 102613264 | 340 | 2 | 5.40E-05 | 0.003924 | 3.11E-05 | 4.27E-05 | -0.010048607 | SUFU |
| chr17 | 7854615 | 7855822 | 1208 | 16 | 5.40E-05 | 1.09E-06 | 0.00786 | 1.60E-06 | -0.003826885 | KDM6B, TMEM88 |
| chr6 | 96649620 | 96650170 | 551 | 4 | 5.41E-05 | 2.47E-06 | 0.00121 | 6.54E-06 | -0.010260272 | NA |
| chr4 | 56169782 | 56170237 | 456 | 3 | 5.42E-05 | 0.001166 | 3.11E-05 | 4.56E-05 | 0.004185338 | CRACD |
| chr14 | 38207505 | 38208571 | 1067 | 12 | 5.43E-05 | 6.18E-05 | 0.00391 | 1.27E-05 | -0.002886627 | SSTR1 |
| chr22 | 39364031 | 39364919 | 889 | 5 | 5.44E-05 | 0.002051 | 0.00201 | 0.00061 | -0.002274074 | SYNGR1 |
| chr19 | 18157094 | 18157913 | 820 | 2 | 5.45E-05 | 1.79E-06 | 0.00025 | 3.06E-06 | -0.013562984 | AC007192.1, PIK3R2 |
| chr9 | 78440976 | 78441420 | 445 | 4 | 5.45E-05 | 0.000138 | 0.00016 | 3.48E-05 | -0.001630774 | NA |
| chr17 | 17626185 | 17626525 | 341 | 2 | 5.47E-05 | 0.001539 | 2.55E-05 | 2.96E-05 | -0.006711993 | NA |
| chr10 | 79194762 | 79194912 | 151 | 2 | 5.47E-05 | 8.40E-06 | 0.00073 | 1.38E-05 | -0.009653913 | ZMIZ1 |
| chr20 | 50267334 | 50268623 | 1290 | 6 | 5.47E-05 | 1.17E-06 | 0.00198 | 1.31E-06 | -0.009691051 | PELATON |
| chr1 | 26190132 | 26191095 | 964 | 7 | 5.48E-05 | 1.04E-06 | 0.00534 | 2.65E-06 | -0.007139645 | CATSPER4 |
| chr19 | 35121330 | 35121354 | 25 | 2 | 5.49E-05 | 5.94E-06 | 0.00013 | 7.38E-06 | -0.008511689 | AC020907.6, FXYD3 |
| chr16 | 21188318 | 21188606 | 289 | 3 | 5.49E-05 | 3.16E-06 | 0.00051 | 5.16E-06 | -0.011075938 | NA |
| chr15 | 22963667 | 22963866 | 200 | 2 | 5.49E-05 | 1.06E-05 | 0.00017 | 1.27E-05 | -0.005269737 | CYFIP1 |
| chr16 | 861759 | 861857 | 99 | 3 | 5.50E-05 | 0.001572 | 1.75E-05 | 1.70E-05 | -0.002272451 | AL023882.1, LMF1 |
| chr2 | 219626708 | 219627608 | 901 | 9 | 5.50E-05 | 9.33E-06 | 0.00129 | 5.96E-06 | -0.00512351 | SLC4A3 |
| chr6 | 111615326 | 111615955 | 630 | 3 | 5.51E-05 | 2.85E-06 | 0.00078 | 6.25E-06 | -0.016899531 | NA |
| chr9 | 33103946 | 33104140 | 195 | 2 | 5.51E-05 | 1.17E-05 | 0.00027 | 1.54E-05 | -0.002942226 | B4GALT1 |
| chr5 | 863179 | 863634 | 456 | 2 | 5.53E-05 | 1.87E-05 | 0.00011 | 1.63E-05 | -0.008236686 | BRD9 |
| chr21 | 42498913 | 42499561 | 649 | 3 | 5.53E-05 | 0.000392 | 0.00013 | 2.35E-05 | -0.008320472 | SLC37A1 |
| chr14 | 22529358 | 22529420 | 63 | 3 | 5.53E-05 | 2.79E-05 | 0.00045 | 2.30E-05 | -0.008429552 | NA |
| chr9 | 93207316 | 93207782 | 467 | 2 | 5.53E-05 | 3.96E-06 | 0.00047 | 6.75E-06 | -0.009434228 | WNK2 |
| chr14 | 94052890 | 94053117 | 228 | 2 | 5.54E-05 | 0.000235 | 3.45E-05 | 2.62E-05 | -0.00954916 | DDX24 |
| chr6 | 46011278 | 46011612 | 335 | 3 | 5.55E-05 | 1.51E-05 | 0.00165 | 2.23E-05 | 0.001769743 | CLIC5 |
| chr9 | 133359159 | 133359422 | 264 | 2 | 5.56E-05 | 7.17E-06 | 0.00012 | 8.05E-06 | -0.008692936 | SURF2 |
| chr11 | 64759768 | 64761097 | 1330 | 10 | 5.56E-05 | 9.46E-06 | 0.00289 | 2.13E-06 | -0.005763928 | PYGM |
| chr5 | 126370989 | 126371425 | 437 | 4 | 5.56E-05 | 1.83E-06 | 0.00127 | 4.82E-06 | -0.006098752 | GRAMD2B |
| chr11 | 1273621 | 1273842 | 222 | 3 | 5.58E-05 | 0.002115 | 5.45E-05 | 2.41E-05 | -0.003497892 | NA |
| chr3 | 97972540 | 97973406 | 867 | 6 | 5.60E-05 | 0.000541 | 0.00276 | 2.60E-05 | -0.003915791 | AC026100.1 |
| chr1 | 2239773 | 2240440 | 668 | 4 | 5.60E-05 | 8.93E-05 | 0.00374 | 3.71E-05 | -0.005587368 | SKI |
| chr11 | 65647327 | 65647789 | 463 | 3 | 5.60E-05 | 1.80E-05 | 0.00092 | 1.86E-05 | 0.000711317 | SIPA1 |
| chr1 | 54376532 | 54376731 | 200 | 2 | 5.63E-05 | 0.033656 | 2.31E-05 | 3.40E-05 | -0.007072248 | SSBP3 |
| chr19 | 49057963 | 49058742 | 780 | 4 | 5.63E-05 | 0.010496 | 5.97E-05 | 7.36E-05 | -0.002783089 | CGB7, AC008687.8, AC008687.7 |
| chr21 | 10417729 | 10417768 | 40 | 2 | 5.63E-05 | 9.98E-06 | 5.39E-05 | 6.99E-06 | -0.011285224 | BAGE2 |
| chr17 | 3048802 | 3049473 | 672 | 7 | 5.64E-05 | 5.17E-06 | 0.00291 | 1.24E-05 | 0.001236933 | NA |
| chr2 | 68837228 | 68837479 | 252 | 4 | 5.64E-05 | 2.14E-05 | 0.00066 | 9.53E-06 | -0.0142164 | LINC01888, LINC01890 |
| chr6 | 68633943 | 68634810 | 868 | 9 | 5.66E-05 | 0.000118 | 0.0106 | 1.43E-05 | 0.000577296 | AL391807.1 |
| chr11 | 8593909 | 8595006 | 1098 | 15 | 5.67E-05 | 0.002049 | 0.00412 | 0.00019 | 0.001776544 | STK33 |
| chr12 | 49777802 | 49778141 | 340 | 2 | 5.67E-05 | 7.93E-06 | 0.00014 | 9.45E-06 | -0.009248623 | NA |
| chr13 | 37599203 | 37599715 | 513 | 4 | 5.68E-05 | 0.001113 | 8.36E-05 | 2.48E-05 | -0.00582609 | NA |
| chr1 | 54850808 | 54851633 | 826 | 6 | 5.68E-05 | 1.63E-05 | 0.00308 | 9.98E-06 | -0.002944221 | DHCR24 |
| chr3 | 48465512 | 48466219 | 708 | 10 | 5.69E-05 | 0.030777 | 0.00296 | 8.66E-05 | 0.004262218 | ATRIP, TREX1 |
| chr5 | 88877937 | 88878027 | 91 | 2 | 5.69E-05 | 0.002686 | 1.76E-05 | 2.18E-05 | -0.012891286 | MEF2C |
| chr19 | 18461568 | 18461890 | 323 | 5 | 5.69E-05 | 0.000141 | 5.09E-05 | 1.36E-05 | 0.000441888 | ELL |
| chr5 | 1088455 | 1089145 | 691 | 6 | 5.70E-05 | 1.47E-05 | 0.00388 | 1.71E-05 | -0.0012938 | SLC12A7 |
| chr6 | 31833906 | 31835312 | 1407 | 24 | 5.71E-05 | 3.70E-06 | 0.00786 | 8.02E-07 | -0.000956976 | AL671762.1, SNHG32, SNORD48 |
| chr6 | 43629027 | 43629923 | 897 | 15 | 5.71E-05 | 0.010539 | 4.73E-05 | 0.00011 | 0.000468863 | MAD2L1BP, GTPBP2 |
| chr1 | 32014355 | 32014638 | 284 | 3 | 5.71E-05 | 0.000207 | 9.76E-05 | 5.31E-05 | -0.007363651 | KHDRBS1 |
| chr15 | 72344064 | 72344138 | 75 | 2 | 5.71E-05 | 4.81E-06 | 0.00016 | 6.58E-06 | -0.009154421 | AC009690.2, AC009690.1, HEXA |
| chr15 | 85703526 | 85703579 | 54 | 3 | 5.72E-05 | 4.98E-06 | 0.00168 | 1.18E-05 | -0.017599567 | AKAP13 |
| chr11 | 133162677 | 133162755 | 79 | 2 | 5.72E-05 | 7.34E-06 | 0.00016 | 9.30E-06 | -0.021293744 | OPCML |
| chr1 | 1047880 | 1048349 | 470 | 5 | 5.73E-05 | 7.20E-06 | 0.00383 | 2.07E-05 | -0.001562906 | AGRN |
| chr11 | 123061648 | 123062833 | 1186 | 13 | 5.74E-05 | 0.000131 | 0.00013 | 4.17E-06 | 0.001781993 | AP000926.2, HSPA8 |
| chr3 | 51711930 | 51713082 | 1153 | 6 | 5.74E-05 | 4.88E-05 | 0.0002 | 1.22E-05 | -0.003549006 | GRM2 |
| chr3 | 19542719 | 19543565 | 847 | 2 | 5.74E-05 | 5.78E-07 | 0.00013 | 9.93E-07 | -0.020325576 | AC061958.1 |
| chr6 | 1554370 | 1555337 | 968 | 7 | 5.74E-05 | 9.87E-07 | 0.00408 | 5.30E-06 | -0.017846127 | NA |
| chr2 | 39138271 | 39138394 | 124 | 2 | 5.75E-05 | 4.85E-05 | 4.15E-05 | 1.57E-05 | -0.004131152 | NA |
| chr7 | 98228980 | 98229712 | 733 | 2 | 5.75E-05 | 0.002163 | 0.00087 | 0.00091 | -0.001967478 | TECPR1 |
| chr9 | 133421545 | 133422289 | 745 | 10 | 5.75E-05 | 9.07E-06 | 0.00535 | 6.09E-06 | -0.004046946 | ADAMTS13 |
| chr5 | 66239000 | 66239170 | 171 | 2 | 5.76E-05 | 7.74E-06 | 9.32E-05 | 7.79E-06 | -0.011082445 | NA |
| chr8 | 1902297 | 1902759 | 463 | 3 | 5.76E-05 | 1.05E-05 | 0.00115 | 1.76E-05 | -0.004100575 | AC019257.8, ARHGEF10 |
| chr17 | 42110482 | 42110511 | 30 | 2 | 5.77E-05 | 1.43E-05 | 8.37E-05 | 1.16E-05 | -0.008994486 | DHX58 |
| chr1 | 42339135 | 42339432 | 298 | 3 | 5.77E-05 | 7.86E-05 | 4.23E-05 | 1.38E-05 | -0.001147817 | NA |
| chr13 | 113552890 | 113553606 | 717 | 2 | 5.77E-05 | 1.91E-06 | 0.00022 | 3.17E-06 | -0.014114214 | TMCO3 |
| chr6 | 168044496 | 168044934 | 439 | 4 | 5.78E-05 | 0.176497 | 4.42E-05 | 0.00016 | -0.000174003 | KIF25 |
| chr11 | 34977729 | 34977799 | 71 | 2 | 5.78E-05 | 0.000387 | 1.97E-05 | 1.78E-05 | 0.011770121 | PDHX |
| chr19 | 17842664 | 17842965 | 302 | 2 | 5.81E-05 | 2.51E-05 | 0.00029 | 2.89E-05 | -0.01019442 | JAK3 |
| chr12 | 121580992 | 121581855 | 864 | 9 | 5.81E-05 | 1.51E-05 | 0.02246 | 4.34E-05 | -0.009239799 | KDM2B-DT, KDM2B |
| chr17 | 80182261 | 80182753 | 493 | 6 | 5.81E-05 | 1.01E-05 | 0.00053 | 9.49E-06 | -0.003333499 | CARD14 |
| chr17 | 81228172 | 81228635 | 464 | 5 | 5.81E-05 | 1.73E-05 | 0.00119 | 2.65E-05 | 0.008661296 | TEPSIN |
| chr12 | 54292120 | 54292722 | 603 | 4 | 5.82E-05 | 8.41E-06 | 0.00028 | 8.24E-06 | -0.006244779 | AC078778.1, NFE2 |
| chr2 | 99179855 | 99179859 | 5 | 2 | 5.82E-05 | 0.000124 | 8.25E-05 | 4.32E-05 | -0.007192783 | AC079447.1, AC092587.1, MITD1 |
| chr15 | 88913008 | 88914010 | 1003 | 12 | 5.83E-05 | 0.000187 | 0.00123 | 1.40E-05 | 0.001651243 | MFGE8 |
| chr12 | 131609191 | 131609555 | 365 | 3 | 5.83E-05 | 0.000173 | 4.88E-05 | 2.01E-05 | 0.002798735 | NA |
| chr2 | 230778950 | 230778962 | 13 | 2 | 5.83E-05 | 8.80E-05 | 3.01E-05 | 1.64E-05 | -0.00874037 | CAB39 |
| chr4 | 166527453 | 166527644 | 192 | 3 | 5.84E-05 | 0.000726 | 1.60E-05 | 1.74E-05 | 0.011034102 | NA |
| chr12 | 132167914 | 132167975 | 62 | 3 | 5.84E-05 | 8.93E-06 | 0.00387 | 2.21E-05 | -0.014337598 | NA |
| chr5 | 141392456 | 141393555 | 1100 | 8 | 5.85E-05 | 1.64E-05 | 0.00098 | 7.05E-06 | 0.010842024 | PCDHGA1, PCDHGA2, PCDHGA3, PCDHGB1, PCDHGA4, PCDHGB2, PCDHGA5, PCDHGB3, PCDHGA6, PCDHGA7, PCDHGB4, PCDHGA8 |
| chr5 | 172877856 | 172878239 | 384 | 2 | 5.87E-05 | 0.000346 | 0.00019 | 0.00013 | -0.008356776 | ERGIC1 |
| chr2 | 168773913 | 168774150 | 238 | 2 | 5.88E-05 | 0.002922 | 1.89E-05 | 2.39E-05 | -0.007033684 | CERS6, CERS6-AS1 |
| chr2 | 28913653 | 28913695 | 43 | 2 | 5.88E-05 | 4.16E-05 | 5.87E-05 | 1.86E-05 | 0.001352739 | WDR43, SNORD92 |
| chr6 | 107653306 | 107653400 | 95 | 3 | 5.88E-05 | 5.29E-06 | 0.00024 | 5.97E-06 | -0.003373847 | SOBP |
| chr8 | 142198131 | 142198521 | 391 | 4 | 5.88E-05 | 5.01E-05 | 0.0003 | 1.68E-05 | -0.011486944 | LINC00051 |
| chr14 | 92107488 | 92107643 | 156 | 3 | 5.88E-05 | 0.00088 | 0.00039 | 9.00E-05 | -0.003392721 | NA |
| chr8 | 105908555 | 105908750 | 196 | 2 | 5.89E-05 | 1.10E-05 | 6.68E-05 | 8.46E-06 | -0.016763577 | ZFPM2-AS1 |
| chr19 | 19745157 | 19745421 | 265 | 2 | 5.89E-05 | 3.36E-06 | 0.00044 | 5.77E-06 | -0.006761041 | AC011477.6 |
| chr17 | 79770125 | 79770285 | 161 | 2 | 5.91E-05 | 1.78E-05 | 6.07E-05 | 1.12E-05 | -0.010546312 | NA |
| chr2 | 16097035 | 16097097 | 63 | 2 | 5.91E-05 | 3.63E-06 | 0.00036 | 6.01E-06 | -0.015935405 | AC130710.1 |
| chr1 | 36150244 | 36150429 | 186 | 4 | 5.92E-05 | 1.75E-05 | 0.00173 | 3.25E-05 | -0.007176666 | TRAPPC3 |
| chr17 | 75240285 | 75241251 | 967 | 4 | 5.93E-05 | 1.50E-05 | 0.00034 | 1.28E-05 | -0.004075845 | GGA3 |
| chr2 | 237497465 | 237497835 | 371 | 3 | 5.93E-05 | 2.53E-05 | 9.61E-05 | 1.24E-05 | -0.011266853 | MLPH |
| chr3 | 171334610 | 171335204 | 595 | 3 | 5.94E-05 | 5.02E-06 | 0.00041 | 8.54E-06 | -0.002329217 | TNIK |
| chr17 | 43013159 | 43013326 | 168 | 2 | 5.94E-05 | 0.00373 | 1.99E-05 | 2.59E-05 | -0.002922326 | IFI35 |
| chr11 | 9363276 | 9364433 | 1158 | 5 | 5.95E-05 | 3.37E-05 | 0.00015 | 1.36E-06 | -0.000188861 | NA |
| chr7 | 157373762 | 157373765 | 4 | 2 | 5.96E-05 | 2.25E-05 | 4.52E-05 | 1.07E-05 | -0.004632902 | DNAJB6 |
| chr20 | 34255472 | 34255970 | 499 | 4 | 5.97E-05 | 0.00016 | 0.00013 | 2.56E-05 | -0.004707442 | ASIP, AL035458.2 |
| chr17 | 19281680 | 19281794 | 115 | 2 | 5.97E-05 | 0.001143 | 2.55E-05 | 2.83E-05 | -0.003516095 | EPN2 |
| chr1 | 179088691 | 179088759 | 69 | 3 | 5.97E-05 | 5.93E-06 | 0.00019 | 7.32E-06 | -0.007515345 | TOR3A |
| chr11 | 68929619 | 68929966 | 348 | 2 | 5.97E-05 | 0.000383 | 7.40E-05 | 6.10E-05 | -0.008431019 | IGHMBP2 |
| chr22 | 26656609 | 26657970 | 1362 | 18 | 5.97E-05 | 5.89E-06 | 0.01107 | 2.40E-06 | -0.000907828 | MIAT, Z99774.5 |
| chr18 | 78841647 | 78841952 | 306 | 3 | 5.97E-05 | 1.64E-05 | 7.62E-05 | 1.00E-05 | -0.007235433 | NA |
| chr16 | 87183958 | 87184034 | 77 | 2 | 5.98E-05 | 5.12E-06 | 0.00018 | 7.13E-06 | -0.003773018 | C16orf95 |
| chr11 | 70434985 | 70435946 | 962 | 7 | 5.98E-05 | 9.17E-05 | 0.00294 | 2.34E-05 | -0.002942964 | CTTN |
| chr12 | 102480508 | 102481194 | 687 | 6 | 5.99E-05 | 1.19E-05 | 0.0005 | 1.06E-05 | -0.001381166 | IGF1 |
| chr11 | 69671718 | 69672295 | 578 | 4 | 5.99E-05 | 1.45E-06 | 0.00248 | 5.03E-06 | -0.003610735 | LTO1 |
| chr4 | 185470542 | 185472390 | 1849 | 18 | 5.99E-05 | 1.65E-06 | 0.00325 | 1.94E-07 | -0.001993598 | AC106897.1, CCDC110 |
| chr7 | 2133897 | 2133961 | 65 | 2 | 5.99E-05 | 8.39E-05 | 3.82E-05 | 1.93E-05 | -0.006279807 | MAD1L1, AC069288.1 |
| chr15 | 57016940 | 57017343 | 404 | 2 | 6.00E-05 | 3.84E-05 | 0.00012 | 2.74E-05 | -0.005052698 | TCF12 |
| chr16 | 2075674 | 2076532 | 859 | 9 | 6.01E-05 | 0.001408 | 0.00053 | 5.10E-05 | -0.000901556 | TSC2 |
| chr11 | 46291212 | 46291500 | 289 | 2 | 6.02E-05 | 3.32E-05 | 8.52E-05 | 2.08E-05 | -0.004374455 | CREB3L1 |
| chr15 | 89802768 | 89802863 | 96 | 3 | 6.04E-05 | 4.70E-06 | 0.00138 | 1.06E-05 | -0.034373753 | ANPEP |

| | | | | | | | | | | |
|---|---|---|---|---|---|---|---|---|---|---|
| chr3 | 10108119 | 10108540 | 422 | 10 | 6.04E-05 | 0.130912 | 1.92E-05 | 0.00013 | -0.002924095 | FANCD2OS |
| chr17 | 82917863 | 82918521 | 659 | 7 | 6.05E-05 | 0.00045 | 0.0006 | 3.08E-05 | -0.007181369 | TBCD, AC087222.1 |
| chr10 | 131259906 | 131260725 | 820 | 7 | 6.05E-05 | 0.005137 | 8.76E-05 | 9.03E-06 | -0.004144067 | TCERG1L |
| chr5 | 160506512 | 160507405 | 894 | 4 | 6.05E-05 | 4.82E-06 | 0.00157 | 1.13E-05 | -0.010747281 | NA |
| chr5 | 32174362 | 32174940 | 579 | 9 | 6.06E-05 | 0.001709 | 6.20E-05 | 1.33E-05 | 0.001651241 | AC025181.2 |
| chr11 | 33256200 | 33257170 | 971 | 10 | 6.06E-05 | 0.00093 | 0.0023 | 4.49E-05 | -0.004470421 | HIPK3 |
| chr4 | 2790225 | 2790855 | 631 | 2 | 6.07E-05 | 5.69E-06 | 0.00025 | 8.38E-06 | -0.012709158 | NA |
| chr8 | 143940821 | 143943352 | 2532 | 14 | 6.07E-05 | 1.42E-06 | 0.00058 | 3.48E-08 | -0.002194515 | PLEC |
| chr4 | 40189581 | 40189614 | 34 | 2 | 6.07E-05 | 0.000195 | 3.82E-05 | 2.70E-05 | -0.004012321 | AC095057.4 |
| chr2 | 197447256 | 197447310 | 55 | 2 | 6.08E-05 | 7.79E-05 | 2.01E-05 | 1.09E-05 | -0.010794981 | NA |
| chr1 | 31240086 | 31240481 | 396 | 5 | 6.09E-05 | 5.28E-05 | 0.00306 | 1.78E-05 | -0.010549628 | NA |
| chr12 | 6318332 | 6318608 | 277 | 3 | 6.09E-05 | 5.02E-06 | 0.00026 | 7.41E-06 | -0.013844047 | PLEKHG6 |
| chr18 | 46172733 | 46174083 | 1351 | 15 | 6.09E-05 | 3.20E-06 | 0.00487 | 1.59E-06 | -0.002242714 | C18orf25 |
| chr19 | 35776537 | 35776608 | 72 | 3 | 6.09E-05 | 1.02E-05 | 0.00141 | 1.59E-05 | -0.005743709 | ARHGAP33 |
| chr15 | 39801104 | 39801697 | 594 | 4 | 6.09E-05 | 2.09E-05 | 0.0006 | 1.36E-05 | -0.000711076 | AC023908.2, GPR176 |
| chr5 | 142108033 | 142108344 | 312 | 2 | 6.10E-05 | 1.33E-05 | 0.0005 | 1.95E-05 | -0.006444803 | NA |
| chr3 | 128361975 | 128362754 | 780 | 6 | 6.10E-05 | 5.17E-06 | 0.00087 | 7.12E-06 | -0.002464062 | EEFSEC |
| chr13 | 51975402 | 51975451 | 50 | 3 | 6.11E-05 | 5.10E-06 | 0.00089 | 7.64E-06 | -0.005835961 | ATP7B |
| chr22 | 32371107 | 32371175 | 69 | 3 | 6.11E-05 | 4.66E-06 | 0.00037 | 6.70E-06 | 0.000920664 | RFPL3S |
| chr8 | 51809097 | 51810190 | 1094 | 10 | 6.12E-05 | 0.000165 | 0.00117 | 4.86E-06 | -0.002585883 | PXDNL, AC090186.1 |
| chr10 | 122301318 | 122301353 | 36 | 2 | 6.13E-05 | 3.99E-06 | 0.00026 | 6.27E-06 | -0.018221001 | BTBD16 |
| chr22 | 37048359 | 37048945 | 587 | 2 | 6.13E-05 | 1.08E-05 | 0.00014 | 1.20E-05 | -0.008739696 | NA |
| chr16 | 46556846 | 46556995 | 150 | 2 | 6.13E-05 | 6.48E-06 | 0.00018 | 8.56E-06 | -0.021691671 | ANKRD26P1 |
| chr17 | 1025413 | 1025611 | 199 | 2 | 6.13E-05 | 6.49E-06 | 0.00064 | 1.08E-05 | -0.006771289 | ABR |
| chr3 | 137767371 | 137768344 | 974 | 6 | 6.13E-05 | 2.72E-05 | 0.00043 | 2.51E-05 | 0.001678469 | NA |
| chr10 | 29634991 | 29635924 | 934 | 7 | 6.14E-05 | 4.86E-06 | 0.004 | 7.50E-06 | -0.006465468 | SVIL |
| chr4 | 127632514 | 127633172 | 659 | 12 | 6.14E-05 | 0.001753 | 0.00041 | 2.65E-05 | -0.000464197 | INTU |
| chr21 | 46393110 | 46393940 | 831 | 6 | 6.14E-05 | 0.000115 | 0.0006 | 2.03E-05 | -0.003989687 | PCNT |
| chr22 | 40678678 | 40679424 | 747 | 8 | 6.14E-05 | 0.000203 | 0.00074 | 2.55E-05 | -0.003231583 | MCHR1 |
| chr1 | 205919896 | 205920305 | 410 | 2 | 6.15E-05 | 2.75E-05 | 9.93E-05 | 2.01E-05 | -0.010448787 | SLC26A9 |
| chr15 | 42208144 | 42208645 | 502 | 12 | 6.15E-05 | 0.017537 | 3.59E-05 | 7.10E-05 | 0.004865529 | AC036103.1, VPS39 |
| chr19 | 53158502 | 53159470 | 969 | 10 | 6.15E-05 | 0.000262 | 0.00021 | 3.63E-05 | 0.001312165 | ZNF347, ZNF665 |
| chr3 | 21750942 | 21751917 | 976 | 10 | 6.16E-05 | 5.75E-06 | 0.00495 | 1.18E-05 | 0.009757379 | ZNF385D |
| chr2 | 10588380 | 10588446 | 67 | 2 | 6.16E-05 | 0.017014 | 4.96E-05 | 8.23E-05 | -0.002622093 | NOL10 |
| chr21 | 43756188 | 43756566 | 379 | 2 | 6.18E-05 | 2.15E-05 | 0.00024 | 2.45E-05 | -0.003341293 | PDXK |
| chr19 | 50702899 | 50702901 | 3 | 2 | 6.18E-05 | 8.12E-06 | 0.0006 | 1.31E-05 | -0.019078238 | SHANK1 |
| chr9 | 137416137 | 137417994 | 1858 | 7 | 6.18E-05 | 1.01E-06 | 0.00024 | 1.63E-07 | -0.015905718 | EXD3 |
| chr5 | 176641457 | 176641609 | 153 | 4 | 6.19E-05 | 0.001664 | 1.89E-05 | 2.43E-05 | -0.002134582 | EIF4E1B |
| chr5 | 180897512 | 180897804 | 293 | 2 | 6.20E-05 | 6.53E-06 | 0.00063 | 1.08E-05 | 0.002406602 | NA |
| chr22 | 30471686 | 30472276 | 591 | 7 | 6.20E-05 | 3.76E-06 | 0.00977 | 5.63E-06 | -0.009634335 | SEC14L3 |
| chr12 | 109572531 | 109574263 | 1733 | 24 | 6.20E-05 | 2.00E-06 | 0.0103 | 8.16E-07 | 0.000657991 | MVK, MMAB |
| chr7 | 157860684 | 157861478 | 795 | 5 | 6.21E-05 | 2.54E-06 | 0.00629 | 1.10E-05 | -0.004486806 | PTPRN2-AS1, PTPRN2 |
| chr2 | 109613330 | 109613564 | 235 | 3 | 6.21E-05 | 0.017437 | 0.00012 | 0.00036 | 0.000930672 | SEPTIN10 |
| chr8 | 27772825 | 27773631 | 807 | 6 | 6.21E-05 | 9.57E-05 | 0.00337 | 4.37E-05 | -0.013062493 | ESCO2 |
| chr3 | 175150837 | 175151331 | 495 | 3 | 6.21E-05 | 0.000111 | 0.00048 | 2.39E-05 | -0.013233051 | NAALADL2 |
| chr6 | 35424469 | 35424959 | 491 | 5 | 6.22E-05 | 2.39E-06 | 0.00284 | 6.52E-06 | -0.001921091 | PPARD |
| chr6 | 33451092 | 33452450 | 1359 | 18 | 6.22E-05 | 0.000435 | 0.0002 | 2.12E-05 | -0.00245657 | SYNGAP1, SYNGAP1-AS1 |
| chr2 | 231892270 | 231892704 | 435 | 5 | 6.23E-05 | 0.000101 | 0.00017 | 1.14E-05 | -0.005097556 | MIR1471 |
| chr1 | 236762577 | 236762665 | 89 | 3 | 6.23E-05 | 2.74E-06 | 0.00054 | 5.65E-06 | -0.023218679 | ACTN2 |
| chr6 | 134437352 | 134437666 | 315 | 2 | 6.23E-05 | 0.015586 | 3.12E-05 | 4.78E-05 | -0.008677705 | LINC01010, CT69 |
| chr5 | 1462705 | 1463735 | 1031 | 6 | 6.23E-05 | 2.64E-05 | 0.00201 | 1.81E-05 | -0.003494476 | LPCAT1 |
| chr12 | 123436166 | 123437310 | 1145 | 13 | 6.24E-05 | 1.41E-06 | 0.00329 | 2.43E-06 | -0.002604217 | RILPL2 |
| chr1 | 11151217 | 11151628 | 412 | 3 | 6.25E-05 | 3.22E-05 | 0.00019 | 2.27E-05 | -0.003569827 | MTOR |
| chr16 | 57939184 | 57939985 | 802 | 2 | 6.25E-05 | 7.25E-07 | 0.00013 | 1.24E-06 | -0.00225566 | CNGB1 |
| chr2 | 233463823 | 233464246 | 424 | 4 | 6.25E-05 | 5.15E-05 | 0.00043 | 1.58E-05 | -0.002444919 | DGKD |
| chr12 | 94924981 | 94925315 | 335 | 2 | 6.26E-05 | 1.61E-05 | 7.74E-05 | 1.21E-05 | -0.003759208 | NDUFA12 |
| chr9 | 136746950 | 136746956 | 7 | 2 | 6.27E-05 | 0.049851 | 2.95E-05 | 4.60E-05 | -0.003240968 | AL355987.1, LCN6, MIR6722 |
| chr17 | 63621986 | 63622485 | 500 | 9 | 6.27E-05 | 3.07E-05 | 0.00145 | 1.21E-05 | -0.001287713 | MAP3K3 |
| chr1 | 20652289 | 20652692 | 404 | 3 | 6.27E-05 | 2.23E-05 | 0.00044 | 2.82E-05 | -0.005830135 | DDOST |
| chr5 | 109997250 | 109997524 | 275 | 2 | 6.28E-05 | 0.006232 | 2.19E-05 | 2.97E-05 | -0.007050589 | NA |
| chr12 | 50763240 | 50764176 | 937 | 12 | 6.28E-05 | 5.79E-05 | 0.00033 | 1.18E-05 | 0.000961503 | ATF1 |
| chr10 | 104253776 | 104254807 | 1032 | 13 | 6.28E-05 | 0.019795 | 4.98E-05 | 0.00013 | -0.000669552 | GSTO1 |
| chr15 | 67519942 | 67520807 | 866 | 5 | 6.29E-05 | 0.000733 | 0.00176 | 8.56E-05 | -0.000948188 | IQCH-AS1 |
| chr10 | 126459273 | 126459435 | 163 | 2 | 6.29E-05 | 0.000287 | 3.08E-05 | 2.53E-05 | 0.001176981 | AL589787.1, C10orf90 |
| chr19 | 35666499 | 35666923 | 425 | 6 | 6.29E-05 | 1.13E-05 | 0.00175 | 1.26E-05 | -0.004577632 | UPK1A |
| chr20 | 35955500 | 35955544 | 45 | 2 | 6.29E-05 | 0.000388 | 5.85E-05 | 4.94E-05 | -0.005168251 | CNBD2, SCAND1 |
| chr2 | 241225894 | 241226103 | 210 | 3 | 6.29E-05 | 3.93E-05 | 9.95E-05 | 1.22E-05 | -0.003599784 | NA |
| chr16 | 8935907 | 8936236 | 330 | 4 | 6.30E-05 | 0.001262 | 0.00087 | 3.67E-05 | -0.006572763 | USP7 |
| chr1 | 3133817 | 3134566 | 750 | 6 | 6.30E-05 | 1.07E-06 | 0.00216 | 2.74E-06 | -0.004550542 | PRDM16 |
| chr3 | 52711646 | 52711859 | 214 | 2 | 6.30E-05 | 5.31E-06 | 0.00018 | 7.33E-06 | -0.004284388 | NEK4 |
| chr8 | 2098668 | 2099180 | 513 | 4 | 6.30E-05 | 0.000143 | 3.55E-05 | 1.81E-05 | -0.006246707 | MYOM2 |
| chr1 | 184974759 | 184975156 | 398 | 6 | 6.31E-05 | 1.63E-05 | 0.0026 | 7.95E-06 | -0.007143917 | NA |
| chr8 | 104329110 | 104329268 | 159 | 2 | 6.32E-05 | 0.012425 | 4.90E-05 | 7.94E-05 | -0.005717207 | NA |
| chr1 | 201766361 | 201766671 | 311 | 2 | 6.32E-05 | 3.66E-06 | 0.00043 | 6.23E-06 | -0.012176758 | NAV1, IPO9-AS1 |
| chr5 | 9380185 | 9380390 | 206 | 2 | 6.32E-05 | 9.16E-06 | 0.00023 | 1.23E-05 | -0.009731216 | AC091906.1, SEMA5A |
| chr17 | 9575901 | 9576648 | 748 | 15 | 6.33E-05 | 0.000113 | 0.00068 | 9.08E-06 | 0.002392637 | CFAP52, STX8 |
| chr14 | 55795104 | 55795496 | 393 | 2 | 6.34E-05 | 0.00028 | 5.29E-05 | 4.06E-05 | -0.006181134 | LINC00520 |
| chr14 | 64823010 | 64823949 | 940 | 11 | 6.35E-05 | 3.31E-05 | 0.00513 | 3.38E-06 | -0.000378631 | SPTB |
| chr2 | 24563337 | 24563395 | 59 | 3 | 6.36E-05 | 9.94E-06 | 0.00013 | 7.01E-06 | 0.001705414 | NCOA1 |
| chr2 | 119747016 | 119747223 | 208 | 2 | 6.36E-05 | 3.73E-06 | 0.00046 | 6.38E-06 | 0.000103941 | NA |
| chr2 | 42809658 | 42809851 | 194 | 2 | 6.37E-05 | 0.013292 | 4.70E-05 | 7.60E-05 | 0.002300203 | NA |
| chr17 | 21200050 | 21200415 | 366 | 2 | 6.38E-05 | 3.05E-05 | 8.36E-05 | 1.95E-05 | -0.010476423 | TMEM11 |
| chr1 | 24294586 | 24295183 | 598 | 4 | 6.39E-05 | 6.28E-05 | 0.00246 | 1.67E-05 | -0.018862415 | NA |
| chr17 | 69327360 | 69327835 | 476 | 6 | 6.39E-05 | 0.00172 | 0.00031 | 2.30E-05 | 0.001717376 | NA |
| chr11 | 59615936 | 59616822 | 887 | 7 | 6.40E-05 | 0.000358 | 0.00305 | 3.27E-05 | 0.003247905 | AP000442.1 |
| chr11 | 78023160 | 78024006 | 847 | 8 | 6.41E-05 | 5.78E-06 | 0.00521 | 3.11E-06 | -0.007593176 | AP003032.1, KCTD14, NDUFC2-KCTD14 |
| chr5 | 68533600 | 68534411 | 812 | 4 | 6.41E-05 | 1.21E-05 | 0.00071 | 6.09E-06 | -0.008285523 | AC010280.1 |
| chr3 | 81761848 | 81762297 | 450 | 7 | 6.41E-05 | 9.48E-05 | 0.00027 | 3.24E-05 | -0.001858938 | NA |
| chr6 | 137866345 | 137866735 | 391 | 5 | 6.41E-05 | 0.021811 | 4.55E-05 | 0.00033 | 0.001661824 | WAKMAR2 |
| chr18 | 63949156 | 63949628 | 473 | 8 | 6.42E-05 | 0.00019 | 0.00254 | 1.32E-05 | -0.005992682 | HMSD |
| chr14 | 77316956 | 77317104 | 149 | 2 | 6.42E-05 | 7.23E-06 | 0.00012 | 8.35E-06 | -0.014193626 | POMT2 |
| chr4 | 86950963 | 86951281 | 319 | 3 | 6.42E-05 | 0.000546 | 6.36E-05 | 4.76E-05 | -0.009748628 | AFF1 |
| chr21 | 39011351 | 39011888 | 538 | 4 | 6.43E-05 | 0.000956 | 8.35E-05 | 2.45E-05 | -0.000221332 | AF064858.1 |
| chr5 | 132490919 | 132491592 | 674 | 5 | 6.44E-05 | 0.001043 | 0.00057 | 2.08E-05 | -0.001520365 | AC116366.2 |
| chr9 | 3954809 | 3954988 | 180 | 2 | 6.44E-05 | 7.25E-05 | 5.36E-05 | 2.36E-05 | 0.0143367 | GLIS3 |
| chr11 | 35236793 | 35237057 | 265 | 2 | 6.46E-05 | 0.000535 | 3.18E-05 | 3.08E-05 | -0.010541629 | NA |
| chr10 | 11164809 | 11165489 | 681 | 10 | 6.46E-05 | 0.100104 | 0.00738 | 0.00067 | 0.00143882 | CELF2 |
| chr8 | 22552061 | 22552730 | 670 | 4 | 6.47E-05 | 0.00025 | 0.00015 | 6.46E-05 | -0.006876075 | SORBS3, AC087854.1 |
| chr8 | 47997664 | 47998218 | 555 | 2 | 6.48E-05 | 2.24E-06 | 0.00034 | 3.91E-06 | -0.019628974 | NA |
| chr1 | 40313460 | 40315233 | 1774 | 4 | 6.49E-05 | 1.44E-08 | 9.96E-05 | 2.07E-08 | -0.01068243 | COL9A2 |
| chr3 | 58316523 | 58316778 | 256 | 2 | 6.50E-05 | 0.004645 | 2.14E-05 | 2.84E-05 | -0.003417729 | RPP14, HTD2 |
| chr12 | 108523616 | 108524494 | 879 | 5 | 6.51E-05 | 9.36E-06 | 0.00071 | 4.78E-06 | -0.002987025 | FICD, SART3 |
| chr19 | 31925218 | 31925234 | 17 | 2 | 6.52E-05 | 1.52E-05 | 4.55E-05 | 8.34E-06 | 0.004261341 | LINC01837 |
| chr16 | 81936208 | 81936387 | 180 | 3 | 6.53E-05 | 0.041215 | 1.69E-05 | 2.36E-05 | -0.003494078 | PLCG2 |
| chr6 | 11111661 | 11111884 | 224 | 3 | 6.55E-05 | 2.34E-05 | 9.65E-05 | 1.05E-05 | -0.015181727 | SMIM13, ERVFRD-1 |
| chr8 | 29350609 | 29351513 | 905 | 10 | 6.56E-05 | 4.05E-05 | 0.00232 | 5.64E-06 | -1.94E-05 | AC084262.2, DUSP4 |
| chr5 | 78492837 | 78493210 | 374 | 2 | 6.57E-05 | 2.88E-05 | 8.91E-05 | 1.95E-05 | -0.009589572 | LHFPL2 |
| chr17 | 59892800 | 59893624 | 825 | 13 | 6.58E-05 | 0.028231 | 3.04E-05 | 0.00014 | 0.000320174 | RPS6KB1, TUBD1 |
| chr6 | 30988173 | 30988826 | 654 | 8 | 6.59E-05 | 0.000868 | 0.0003 | 0.00014 | -0.004939037 | MUC21 |
| chr8 | 16567650 | 16567670 | 21 | 2 | 6.59E-05 | 3.84E-06 | 0.00047 | 6.57E-06 | -0.012338279 | NA |
| chr19 | 53631539 | 53632043 | 505 | 6 | 6.59E-05 | 0.000918 | 9.17E-05 | 1.84E-05 | -0.003042843 | NA |
| chr20 | 561481 | 561816 | 336 | 2 | 6.61E-05 | 1.13E-05 | 0.00015 | 1.26E-05 | -0.00772101 | NA |
| chr14 | 68975185 | 68975980 | 796 | 3 | 6.62E-05 | 2.69E-06 | 0.00071 | 5.66E-06 | -0.010955571 | ACTN1 |
| chr20 | 45578649 | 45578904 | 256 | 2 | 6.62E-05 | 0.00054 | 2.51E-05 | 2.40E-05 | -0.007160275 | WFDC8 |
| chr11 | 120532576 | 120532923 | 348 | 2 | 6.62E-05 | 3.77E-05 | 5.52E-05 | 1.69E-05 | -0.00514583 | GRIK4 |
| chr5 | 40678916 | 40679761 | 846 | 12 | 6.63E-05 | 0.00113 | 0.00013 | 3.31E-05 | 0.001094405 | TTC33 |
| chr3 | 49412860 | 49413472 | 613 | 5 | 6.63E-05 | 0.000751 | 0.00036 | 4.80E-05 | -0.00444555 | TCTA, RHOA |
| chr6 | 32047306 | 32047936 | 631 | 10 | 6.64E-05 | 0.010732 | 0.00139 | 0.00018 | -0.002403887 | TNXB |
| chr17 | 864140 | 865008 | 869 | 7 | 6.64E-05 | 9.84E-07 | 0.00214 | 3.16E-06 | -0.008882147 | NXN |
| chr8 | 22142463 | 22142805 | 343 | 4 | 6.64E-05 | 0.000895 | 6.35E-05 | 6.84E-05 | -0.006515498 | NA |
| chr1 | 205679059 | 205679208 | 150 | 2 | 6.65E-05 | 1.76E-05 | 0.00056 | 2.53E-05 | -0.017556048 | SLC45A3 |
| chr8 | 21022365 | 21023064 | 700 | 3 | 6.65E-05 | 5.32E-06 | 0.00264 | 1.34E-05 | -0.007087543 | AC021613.1 |
| chr8 | 142834945 | 142835835 | 891 | 4 | 6.66E-05 | 0.00011 | 0.00249 | 0.00012 | -0.002291423 | GML |
| chr11 | 5624094 | 5624473 | 380 | 4 | 6.66E-05 | 3.16E-05 | 0.0004 | 2.29E-05 | -0.002772582 | TRIM6-TRIM34, TRIM34, AC104389.5 |
| chr3 | 128573471 | 128574475 | 1005 | 5 | 6.67E-05 | 7.05E-07 | 0.00266 | 2.07E-06 | -0.008859749 | LINC01565 |
| chr10 | 77843691 | 77844013 | 323 | 2 | 6.67E-05 | 0.000945 | 3.04E-05 | 3.31E-05 | -0.004933452 | DLG5 |
| chr6 | 119148411 | 119149224 | 814 | 3 | 6.68E-05 | 3.66E-06 | 0.00072 | 5.33E-06 | -0.009156748 | AL137009.1, FAM184A |
| chr6 | 170022523 | 170022991 | 469 | 4 | 6.69E-05 | 6.42E-06 | 0.00718 | 2.09E-05 | -0.005427234 | NA |
| chr17 | 42754792 | 42754971 | 180 | 2 | 6.69E-05 | 0.001302 | 1.83E-05 | 2.07E-05 | -0.00445335 | RAMP2-AS1 |
| chr19 | 49168246 | 49168566 | 321 | 2 | 6.69E-05 | 0.001606 | 2.68E-05 | 3.19E-05 | -0.004292119 | TRPM4 |
| chr17 | 57862026 | 57862229 | 204 | 3 | 6.69E-05 | 1.67E-05 | 0.00156 | 2.72E-05 | -0.012715205 | CUEDC1 |
| chr11 | 134031783 | 134032615 | 833 | 6 | 6.69E-05 | 5.37E-06 | 0.00463 | 1.03E-05 | -0.008861926 | LINC02731 |
| chr2 | 170363467 | 170364170 | 704 | 3 | 6.69E-05 | 1.30E-06 | 0.00092 | 3.19E-06 | -0.069010539 | MYO3B, MYO3B-AS1 |
| chr2 | 48567682 | 48567928 | 247 | 2 | 6.69E-05 | 8.39E-05 | 6.22E-05 | 2.85E-05 | 0.010629127 | STON1, STON1-GTF2A1L |
| chr9 | 26975799 | 26975927 | 129 | 2 | 6.70E-05 | 4.37E-06 | 0.00032 | 6.97E-06 | -0.010746907 | IFT74 |
| chr22 | 24406921 | 24407280 | 360 | 2 | 6.70E-05 | 1.38E-05 | 0.00078 | 2.15E-05 | -0.0132807 | SPECC1L, SPECC1L-ADORA2A |
| chr5 | 127031592 | 127031765 | 174 | 3 | 6.70E-05 | 2.73E-05 | 0.00068 | 2.96E-05 | -0.012305476 | NA |
| chr13 | 110344013 | 110344380 | 368 | 2 | 6.71E-05 | 0.000142 | 8.34E-05 | 4.61E-05 | -0.002037358 | COL4A2 |
| chr22 | 27639099 | 27639559 | 461 | 5 | 6.71E-05 | 0.001721 | 0.00029 | 2.46E-05 | -0.002924975 | NA |
| chr9 | 135993028 | 135993322 | 295 | 2 | 6.72E-05 | 0.001399 | 3.40E-05 | 3.99E-05 | -0.008473266 | NA |
| chr19 | 45097931 | 45098665 | 735 | 4 | 6.72E-05 | 1.17E-05 | 0.00065 | 1.39E-05 | -0.00742984 | MARK4, PPP1R37 |
| chr19 | 7916483 | 7916820 | 338 | 5 | 6.73E-05 | 0.002955 | 0.00036 | 6.71E-05 | 1.25E-05 | TGFBR3L |
| chr16 | 31134330 | 31134372 | 43 | 2 | 6.73E-05 | 3.21E-05 | 9.31E-05 | 2.14E-05 | -0.009580331 | PRSS8 |
| chr13 | 20722326 | 20722838 | 513 | 4 | 6.73E-05 | 0.000453 | 7.67E-05 | 3.28E-05 | -0.001197235 | IL17D |
| chr5 | 54224074 | 54224428 | 355 | 3 | 6.75E-05 | 2.36E-06 | 0.00118 | 5.88E-06 | -0.010886438 | ARL15 |
| chr15 | 39582212 | 39582575 | 364 | 2 | 6.76E-05 | 2.55E-05 | 0.00065 | 3.54E-05 | -0.004300601 | THBS1 |
| chr10 | 129569237 | 129569359 | 123 | 3 | 6.76E-05 | 0.000147 | 4.23E-05 | 1.29E-05 | -0.006455164 | MGMT |
| chr20 | 45580388 | 45580603 | 216 | 2 | 6.76E-05 | 6.78E-05 | 3.16E-05 | 1.52E-05 | -0.009070778 | NA |
| chr18 | 50561686 | 50561992 | 307 | 2 | 6.77E-05 | 1.82E-05 | 0.00023 | 2.11E-05 | 0.026292821 | MAPK4 |
| chr6 | 30655470 | 30656258 | 789 | 6 | 6.78E-05 | 0.000211 | 0.00012 | 3.91E-05 | 0.001083151 | DHX16 |
| chr11 | 47449335 | 47449901 | 567 | 6 | 6.78E-05 | 0.003558 | 0.00239 | 0.00029 | -0.005650882 | NA |
| chr20 | 9344229 | 9344955 | 727 | 4 | 6.78E-05 | 2.37E-06 | 0.00226 | 4.98E-06 | -0.021282658 | PLCB4 |
| chr12 | 85836026 | 85837047 | 1022 | 9 | 6.78E-05 | 1.83E-06 | 0.00676 | 6.35E-06 | -0.017637664 | RASSF9 |
| chr17 | 40074946 | 40075096 | 151 | 2 | 6.78E-05 | 3.46E-05 | 7.62E-05 | 1.99E-05 | -0.008642293 | THRA |
| chr20 | 21302550 | 21303722 | 1173 | 13 | 6.79E-05 | 0.000459 | 0.01164 | 4.33E-05 | 0.000571722 | XRN2, AL117332.1 |
| chr2 | 186693785 | 186694601 | 817 | 7 | 6.80E-05 | 0.003702 | 0.00038 | 5.42E-05 | 0.000107334 | FAM171B, AC017101.1 |
| chr16 | 85652126 | 85652388 | 263 | 2 | 6.80E-05 | 2.23E-05 | 0.00136 | 3.55E-05 | -0.015649667 | GSE1 |
| chr16 | 56651410 | 56651990 | 581 | 6 | 6.80E-05 | 0.000116 | 0.00192 | 2.10E-05 | -0.003696645 | AC026461.2, MT1B |
| chr7 | 112931190 | 112931421 | 232 | 2 | 6.80E-05 | 2.32E-05 | 5.19E-05 | 1.19E-05 | -0.011762657 | BMT2 |
| chr17 | 77185172 | 77186088 | 917 | 4 | 6.81E-05 | 0.004564 | 0.00014 | 0.00032 | -0.001213019 | SEC14L1 |
| chr1 | 45698030 | 45698374 | 345 | 2 | 6.81E-05 | 7.33E-05 | 4.42E-05 | 2.05E-05 | -0.005061354 | IPP |
| chr19 | 8507575 | 8508109 | 535 | 2 | 6.82E-05 | 1.81E-05 | 0.00015 | 1.82E-05 | -0.005593105 | NA |

| | | | | | | | | | | |
|---|---|---|---|---|---|---|---|---|---|---|
| chr4 | 40467586 | 40467619 | 34 | 2 | 6.82E-05 | 1.06E-05 | 0.00079 | 1.72E-05 | -0.003982488 | RBM47 |
| chr7 | 157568348 | 157569402 | 1055 | 9 | 6.82E-05 | 1.91E-06 | 0.00744 | 5.74E-06 | -0.003272892 | PTPRN2 |
| chr10 | 129032565 | 129033009 | 445 | 6 | 6.83E-05 | 0.001403 | 0.00011 | 2.37E-05 | -0.002604683 | NA |
| chr10 | 91390159 | 91390261 | 103 | 2 | 6.83E-05 | 6.73E-06 | 0.00012 | 7.66E-06 | -0.014859871 | NA |
| chr1 | 6681130 | 6681714 | 585 | 8 | 6.83E-05 | 1.90E-05 | 0.00839 | 1.22E-05 | -0.004815139 | DNAJC11 |
| chr16 | 569455 | 570796 | 1342 | 11 | 6.85E-05 | 2.43E-05 | 0.00025 | 5.75E-06 | 0.000373824 | PIGQ, NHLRC4, Z98883.1 |
| chr6 | 138405570 | 138406150 | 581 | 2 | 6.85E-05 | 1.33E-05 | 0.00077 | 2.08E-05 | -0.016792836 | HEBP2 |
| chr7 | 45106362 | 45106891 | 530 | 2 | 6.85E-05 | 4.76E-05 | 0.001 | 6.51E-05 | -0.005273802 | TBRG4 |
| chr4 | 1173199 | 1173926 | 728 | 6 | 6.86E-05 | 0.000133 | 0.01284 | 0.00012 | -0.005689383 | SPON2 |
| chr1 | 155173819 | 155175681 | 1863 | 10 | 6.86E-05 | 0.000121 | 0.00245 | 9.02E-06 | -0.001392009 | TRIM46, AL713999.2 |
| chr13 | 40439227 | 40439598 | 372 | 3 | 6.86E-05 | 9.96E-06 | 0.00013 | 8.69E-06 | -0.008548273 | LINC00598 |
| chr22 | 41515641 | 41515731 | 91 | 3 | 6.86E-05 | 1.00E-05 | 0.00037 | 1.42E-05 | -0.005536912 | ACO2 |
| chr22 | 50196204 | 50196316 | 113 | 2 | 6.87E-05 | 0.000169 | 2.08E-05 | 1.50E-05 | -0.004768424 | TRABD |
| chr5 | 149062661 | 149063436 | 776 | 8 | 6.89E-05 | 5.88E-05 | 0.00094 | 1.23E-05 | -0.003798704 | SH3TC2-DT, SH3TC2 |
| chr4 | 186889068 | 186889629 | 562 | 4 | 6.89E-05 | 2.97E-05 | 0.00011 | 1.70E-05 | 0.000330995 | NA |
| chr2 | 28750282 | 28750711 | 430 | 3 | 6.89E-05 | 0.000118 | 0.00138 | 6.26E-05 | -0.008452897 | AC092164.1 |
| chr22 | 47164823 | 47164935 | 113 | 3 | 6.90E-05 | 0.000456 | 3.16E-05 | 2.34E-05 | -0.003943741 | TBC1D22A |
| chr16 | 72371430 | 72371464 | 35 | 2 | 6.90E-05 | 1.28E-05 | 8.07E-05 | 1.05E-05 | -0.013547329 | LINC01572 |
| chr5 | 177336724 | 177337454 | 731 | 5 | 6.90E-05 | 7.94E-05 | 0.00102 | 9.15E-06 | -0.002520371 | LMAN2 |
| chr12 | 53972976 | 53974038 | 1063 | 10 | 6.90E-05 | 0.000563 | 0.00065 | 1.30E-05 | 0.001781492 | HOXC11, HOTAIR |
| chr17 | 50202896 | 50203071 | 176 | 4 | 6.90E-05 | 4.05E-05 | 0.0002 | 2.09E-05 | -0.006083058 | AC015909.1 |
| chr16 | 58110599 | 58111421 | 823 | 3 | 6.91E-05 | 3.87E-06 | 0.00032 | 5.04E-06 | -0.012024104 | NA |
| chr2 | 16336158 | 16336330 | 173 | 2 | 6.92E-05 | 0.000106 | 2.52E-05 | 1.50E-05 | 0.0093307 | NA |
| chr4 | 169175302 | 169175370 | 69 | 2 | 6.92E-05 | 7.79E-06 | 9.31E-05 | 7.83E-06 | -0.007752763 | SH3RF1 |
| chr3 | 61564705 | 61564961 | 257 | 3 | 6.92E-05 | 1.58E-05 | 0.00016 | 1.01E-05 | 0.009883808 | PTPRG |
| chr5 | 35229686 | 35230690 | 1005 | 11 | 6.93E-05 | 6.93E-06 | 0.00429 | 8.41E-06 | -0.006151713 | PRLR |
| chr10 | 114684481 | 114684704 | 224 | 4 | 6.94E-05 | 3.39E-05 | 0.00015 | 1.67E-05 | -0.009330343 | ABLIM1 |
| chr12 | 51817813 | 51818004 | 192 | 2 | 6.94E-05 | 0.000129 | 3.22E-05 | 2.03E-05 | -0.006713493 | AC068987.2, AC068987.3, FIGNL2 |
| chr20 | 34280825 | 34281223 | 399 | 4 | 6.94E-05 | 0.001163 | 3.05E-05 | 2.92E-05 | -0.001799035 | AL035458.2, AHCY |
| chr7 | 117791865 | 117792082 | 218 | 2 | 6.94E-05 | 7.53E-06 | 0.00024 | 1.05E-05 | -0.015294608 | CTTNBP2 |
| chr4 | 145882703 | 145882858 | 156 | 2 | 6.95E-05 | 1.27E-05 | 0.00013 | 1.31E-05 | -0.010333167 | ZNF827 |
| chr3 | 158961597 | 158962086 | 490 | 4 | 6.96E-05 | 3.10E-06 | 0.00334 | 7.47E-06 | -0.002577983 | NA |
| chr20 | 57054483 | 57054692 | 210 | 4 | 6.97E-05 | 0.000311 | 4.41E-05 | 1.50E-05 | 0.01199944 | NA |
| chr2 | 235053777 | 235054284 | 508 | 3 | 6.97E-05 | 0.000127 | 0.00023 | 2.14E-05 | -0.000690896 | SH3BP4 |
| chr10 | 117240572 | 117240761 | 190 | 4 | 6.98E-05 | 2.03E-05 | 0.00051 | 2.17E-05 | -0.010263495 | AL731557.1 |
| chr13 | 114319808 | 114320054 | 247 | 3 | 6.99E-05 | 0.000279 | 3.98E-05 | 1.76E-05 | -0.005107715 | CHAMP1 |
| chr11 | 131929393 | 131929807 | 415 | 4 | 6.99E-05 | 0.001138 | 2.77E-05 | 3.48E-05 | 0.002330355 | NTM |
| chr14 | 72938081 | 72938172 | 92 | 2 | 6.99E-05 | 9.33E-06 | 0.00019 | 1.18E-05 | -0.003428788 | DCAF4 |
| chr2 | 40453147 | 40453471 | 325 | 2 | 7.00E-05 | 0.00078 | 4.88E-05 | 5.06E-05 | -0.003010666 | SLC8A1 |
| chr22 | 29062549 | 29062735 | 187 | 5 | 7.01E-05 | 2.84E-06 | 0.00094 | 5.85E-06 | -0.008041072 | NA |
| chr10 | 3104357 | 3105282 | 926 | 7 | 7.01E-05 | 9.09E-06 | 0.00296 | 1.42E-05 | -0.000740654 | PFKP |
| chr1 | 2980023 | 2980270 | 248 | 2 | 7.02E-05 | 8.45E-05 | 5.41E-05 | 2.57E-05 | -0.00497837 | NA |
| chr11 | 126403069 | 126403364 | 296 | 8 | 7.02E-05 | 0.000227 | 0.00098 | 1.83E-05 | -0.00406886 | ST3GAL4 |
| chr16 | 30922616 | 30923406 | 791 | 12 | 7.03E-05 | 0.001167 | 0.00015 | 4.45E-05 | 0.000697964 | FBXL19, FBXL19-AS1 |
| chr20 | 63980505 | 63980584 | 80 | 2 | 7.04E-05 | 7.28E-06 | 0.00063 | 1.20E-05 | -0.013578683 | NA |
| chr16 | 47143569 | 47144448 | 880 | 9 | 7.04E-05 | 0.000192 | 0.00161 | 1.58E-05 | -0.000749085 | ITFG1-AS1, NETO2 |
| chr2 | 98869340 | 98869380 | 41 | 2 | 7.04E-05 | 2.17E-05 | 0.00012 | 1.89E-05 | -0.01265348 | CRACDL |
| chr16 | 67198277 | 67198654 | 378 | 3 | 7.05E-05 | 1.75E-05 | 0.00021 | 1.77E-05 | -0.009490349 | E2F4 |
| chr3 | 122614117 | 122614210 | 94 | 2 | 7.06E-05 | 9.14E-05 | 5.33E-05 | 2.63E-05 | -0.011947182 | PARP15 |
| chr20 | 33030403 | 33030843 | 441 | 3 | 7.07E-05 | 1.39E-05 | 0.00029 | 1.65E-05 | -0.001030413 | NA |
| chr22 | 38334804 | 38335292 | 489 | 2 | 7.07E-05 | 3.24E-05 | 0.00013 | 2.57E-05 | -0.002832757 | TPTEP2-CSNK1E |
| chr3 | 194632703 | 194633723 | 1021 | 14 | 7.08E-05 | 3.28E-05 | 0.00638 | 4.19E-05 | 0.001666053 | AC046143.1, TMEM44 |
| chr12 | 123115590 | 123115866 | 277 | 2 | 7.09E-05 | 8.45E-06 | 0.00022 | 1.13E-05 | -0.013184336 | PITPNM2 |
| chr15 | 81003267 | 81003643 | 377 | 2 | 7.10E-05 | 2.72E-05 | 0.00026 | 2.97E-05 | -0.007631416 | TLNRD1 |
| chr12 | 57619675 | 57620926 | 1252 | 12 | 7.12E-05 | 1.39E-05 | 0.00454 | 2.56E-06 | 0.001575807 | AC025165.6, SLC26A10 |
| chr6 | 108589268 | 108589507 | 240 | 2 | 7.12E-05 | 0.005713 | 3.62E-05 | 5.18E-05 | -0.00656024 | FOXO3 |
| chr14 | 100949554 | 100949791 | 238 | 3 | 7.12E-05 | 1.50E-05 | 0.00034 | 1.71E-05 | -0.010853467 | MEG8 |
| chr6 | 29044492 | 29045082 | 591 | 4 | 7.13E-05 | 2.52E-06 | 0.00464 | 9.15E-06 | -0.011583381 | AL662791.2, OR2W1 |
| chr2 | 23347082 | 23347185 | 104 | 2 | 7.14E-05 | 4.43E-05 | 0.00012 | 3.12E-05 | -0.006048476 | NA |
| chr6 | 43114182 | 43114558 | 377 | 3 | 7.16E-05 | 0.000115 | 0.00026 | 1.51E-05 | -0.005879522 | PTK7 |
| chr11 | 1694188 | 1694763 | 576 | 6 | 7.18E-05 | 0.00345 | 2.57E-05 | 6.39E-05 | -0.001722966 | NA |
| chr12 | 6226704 | 6226791 | 88 | 2 | 7.19E-05 | 9.30E-06 | 0.00029 | 1.30E-05 | -0.012025119 | CD9 |
| chr16 | 70589996 | 70590527 | 532 | 4 | 7.19E-05 | 7.73E-05 | 0.00098 | 2.81E-05 | -0.0093359 | IL34 |
| chr6 | 160257904 | 160258359 | 456 | 3 | 7.20E-05 | 2.78E-05 | 0.00133 | 4.31E-05 | -0.005765672 | SLC22A2 |
| chr3 | 84943674 | 84943716 | 43 | 2 | 7.20E-05 | 4.80E-06 | 0.00025 | 7.27E-06 | -0.016372292 | AC132660.2 |
| chr11 | 1088424 | 1088927 | 504 | 2 | 7.20E-05 | 9.95E-05 | 7.88E-05 | 3.72E-05 | -0.003475159 | MUC2 |
| chr7 | 157503237 | 157503465 | 229 | 2 | 7.21E-05 | 5.22E-06 | 0.00049 | 8.63E-06 | -0.023565998 | AC006372.1 |
| chr7 | 108001045 | 108001932 | 888 | 4 | 7.21E-05 | 0.000235 | 0.00137 | 0.00024 | -0.006651939 | LAMB1 |
| chr12 | 80935787 | 80936633 | 847 | 6 | 7.21E-05 | 3.68E-05 | 0.00016 | 1.98E-05 | -0.005397945 | ACSS3, LIN7A, MIR618 |
| chr10 | 94227729 | 94228102 | 374 | 2 | 7.21E-05 | 3.70E-05 | 6.45E-05 | 1.86E-05 | -0.011467318 | PLCE1 |
| chr2 | 201257140 | 201257586 | 447 | 3 | 7.22E-05 | 0.008982 | 0.00021 | 0.00071 | -0.001766368 | CASP8 |
| chr6 | 43016553 | 43016957 | 405 | 4 | 7.22E-05 | 7.49E-06 | 0.00321 | 1.56E-05 | -0.005669143 | KLHDC3, MEA1 |
| chr22 | 24166648 | 24167224 | 577 | 6 | 7.22E-05 | 0.002725 | 0.00118 | 4.35E-05 | -0.004266981 | CABIN1 |
| chr4 | 99510575 | 99511109 | 535 | 4 | 7.23E-05 | 1.01E-05 | 0.00029 | 1.03E-05 | -0.00670118 | C4orf17 |
| chr6 | 132814353 | 132815208 | 856 | 13 | 7.23E-05 | 0.000119 | 0.00043 | 1.14E-05 | 0.001602752 | RPS12 |
| chr11 | 66700714 | 66701103 | 390 | 4 | 7.24E-05 | 1.50E-05 | 0.00098 | 1.10E-05 | -0.000437844 | SPTBN2 |
| chr14 | 23376661 | 23377270 | 610 | 6 | 7.24E-05 | 8.14E-05 | 0.00165 | 1.20E-05 | -0.00688798 | CMTM5 |
| chr1 | 26938693 | 26938824 | 132 | 2 | 7.24E-05 | 4.71E-06 | 0.00031 | 7.41E-06 | -0.005583839 | NUDC |
| chr16 | 66605150 | 66605878 | 729 | 5 | 7.24E-05 | 9.63E-06 | 0.00716 | 2.37E-05 | 0.001551393 | CMTM3 |
| chr11 | 76674599 | 76675016 | 418 | 3 | 7.25E-05 | 7.87E-06 | 0.00138 | 1.60E-05 | -0.002163485 | NA |
| chr5 | 122967166 | 122967670 | 505 | 2 | 7.26E-05 | 1.53E-05 | 0.00024 | 1.87E-05 | -0.009428119 | SNX24 |
| chr17 | 1039701 | 1040495 | 795 | 7 | 7.26E-05 | 3.18E-06 | 0.00665 | 4.92E-06 | -0.008158287 | ABR |
| chr2 | 112568561 | 112568589 | 29 | 2 | 7.27E-05 | 0.000354 | 3.30E-05 | 2.85E-05 | -0.001517547 | POLR1B |
| chr19 | 46786153 | 46786354 | 202 | 3 | 7.28E-05 | 9.04E-06 | 0.00109 | 1.76E-05 | -0.00920332 | SLC1A5 |
| chr6 | 108968594 | 108968876 | 283 | 2 | 7.29E-05 | 0.004568 | 2.97E-05 | 4.11E-05 | -0.010297343 | ARMC2 |
| chr20 | 4926315 | 4927211 | 897 | 3 | 7.30E-05 | 1.68E-06 | 0.00019 | 2.41E-06 | -0.003958463 | SLC23A2 |
| chr19 | 1998687 | 1999323 | 637 | 4 | 7.30E-05 | 1.64E-06 | 0.00281 | 5.31E-06 | -0.014679927 | BTBD2 |
| chr3 | 55616144 | 55617015 | 872 | 2 | 7.30E-05 | 3.33E-07 | 0.00011 | 5.99E-07 | -0.021582894 | ERC2 |
| chr9 | 72527011 | 72527395 | 385 | 4 | 7.31E-05 | 6.81E-06 | 0.00068 | 9.94E-06 | -0.008464033 | TMC1 |
| chr2 | 178531158 | 178531487 | 330 | 4 | 7.31E-05 | 0.000138 | 0.00474 | 5.54E-05 | -0.009357726 | TTN-AS1, TTN |
| chr20 | 57694519 | 57694923 | 405 | 2 | 7.32E-05 | 1.33E-05 | 0.0006 | 2.01E-05 | -0.001839828 | PMEPA1 |
| chr4 | 896120 | 896421 | 302 | 3 | 7.32E-05 | 1.39E-05 | 0.00082 | 1.58E-05 | -0.004373241 | GAK |
| chr17 | 37519014 | 37519207 | 194 | 2 | 7.33E-05 | 0.002155 | 1.97E-05 | 2.41E-05 | -0.002553394 | SYNRG |
| chr6 | 13573802 | 13574139 | 338 | 6 | 7.33E-05 | 0.000433 | 0.00034 | 3.66E-05 | 0.001199267 | NA |
| chr4 | 40856948 | 40857327 | 380 | 8 | 7.34E-05 | 0.000659 | 0.00093 | 1.72E-05 | 0.000717827 | APBB2 |
| chr4 | 1898937 | 1900738 | 1802 | 8 | 7.34E-05 | 6.37E-06 | 0.00174 | 7.14E-07 | -0.003722004 | NSD2 |
| chr20 | 23086396 | 23087172 | 777 | 9 | 7.35E-05 | 3.48E-05 | 0.00054 | 1.63E-05 | 0.005015822 | NA |
| chr7 | 151915028 | 151915409 | 382 | 2 | 7.35E-05 | 2.34E-05 | 0.00021 | 2.50E-05 | -0.00028205 | NA |
| chr16 | 282515 | 283155 | 641 | 9 | 7.36E-05 | 9.08E-05 | 0.00225 | 9.21E-06 | -0.004756638 | ARHGDIG |
| chr7 | 129774727 | 129775054 | 328 | 11 | 7.36E-05 | 0.000148 | 0.00346 | 2.15E-05 | -0.001925906 | MIR96, MIR183 |
| chr4 | 139622785 | 139623923 | 1139 | 4 | 7.36E-05 | 0.000599 | 0.00028 | 3.27E-05 | 0.001641321 | AC112236.1 |
| chr19 | 47698206 | 47699006 | 801 | 5 | 7.37E-05 | 3.10E-06 | 0.00356 | 9.94E-06 | -0.004741567 | AC010519.1, BICRA |
| chr1 | 13760828 | 13761387 | 560 | 3 | 7.37E-05 | 4.11E-05 | 0.00071 | 2.29E-05 | -0.008497407 | PRDM2 |
| chr20 | 1336956 | 1337602 | 647 | 5 | 7.37E-05 | 0.000343 | 0.005 | 1.58E-05 | -0.012774562 | SDCBP2-AS1, AL136531.2 |
| chr8 | 94271977 | 94272343 | 367 | 2 | 7.37E-05 | 1.82E-05 | 0.0003 | 2.28E-05 | -0.007417447 | NA |
| chr6 | 89840593 | 89840778 | 186 | 3 | 7.38E-05 | 0.000375 | 0.00015 | 1.25E-05 | 0.002358126 | CASP8AP2 |
| chr17 | 81991387 | 81991841 | 455 | 2 | 7.39E-05 | 0.000139 | 0.00117 | 0.00017 | -0.01730215 | ASPSCR1 |
| chr20 | 64049617 | 64050668 | 1052 | 8 | 7.39E-05 | 4.21E-06 | 0.00518 | 5.32E-06 | -0.005890331 | TCEA2, SOX18 |
| chr10 | 128157393 | 128157682 | 290 | 2 | 7.40E-05 | 3.70E-05 | 0.00016 | 3.16E-05 | -0.008185581 | NA |
| chr3 | 38109619 | 38109634 | 16 | 2 | 7.40E-05 | 1.66E-05 | 0.00015 | 1.73E-05 | -0.007461162 | DLEC1, ACAA1 |
| chr8 | 140238889 | 140239628 | 740 | 5 | 7.41E-05 | 1.42E-05 | 0.00056 | 9.35E-06 | -0.002309054 | TRAPPC9 |
| chr18 | 34223103 | 34224036 | 934 | 11 | 7.41E-05 | 0.000509 | 0.00784 | 3.24E-05 | 0.000653488 | AC104985.1, NOL4 |
| chr9 | 37340214 | 37340563 | 350 | 2 | 7.41E-05 | 0.002736 | 3.11E-05 | 4.07E-05 | -0.009492859 | ZCCHC7 |
| chr11 | 45157813 | 45158303 | 491 | 2 | 7.42E-05 | 4.75E-06 | 0.00038 | 7.72E-06 | -0.010697901 | PRDM11 |
| chr18 | 62714824 | 62715531 | 708 | 14 | 7.43E-05 | 3.39E-06 | 0.01544 | 9.49E-06 | -0.000831793 | NA |
| chr2 | 183123830 | 183124339 | 510 | 9 | 7.43E-05 | 0.001505 | 0.00194 | 2.78E-05 | 0.003417071 | NUP35 |
| chr12 | 116598898 | 116599415 | 518 | 7 | 7.43E-05 | 0.000156 | 0.00047 | 1.49E-05 | -0.000679935 | AC125603.3 |
| chr2 | 174125259 | 174125430 | 172 | 2 | 7.44E-05 | 0.000297 | 2.88E-05 | 2.38E-05 | -0.010724318 | OLA1 |
| chr1 | 32882074 | 32882514 | 441 | 3 | 7.46E-05 | 2.53E-06 | 0.00058 | 5.39E-06 | -0.012470496 | NA |
| chr13 | 99384885 | 99386290 | 1406 | 6 | 7.46E-05 | 1.92E-05 | 0.00011 | 2.29E-06 | -0.003044451 | UBAC2 |
| chr1 | 226945516 | 226946100 | 585 | 2 | 7.48E-05 | 8.55E-06 | 0.00017 | 1.07E-05 | -0.015341004 | AL391628.1, COQ8A |
| chr1 | 23188692 | 23189058 | 367 | 2 | 7.48E-05 | 1.12E-05 | 0.00013 | 1.20E-05 | -0.010159839 | NA |
| chr17 | 80331684 | 80332096 | 413 | 2 | 7.48E-05 | 0.000152 | 4.85E-05 | 3.06E-05 | -0.002638124 | RNF213 |
| chr1 | 27673481 | 27673565 | 85 | 2 | 7.48E-05 | 0.000123 | 4.84E-05 | 2.80E-05 | -0.002881542 | AL445490.1 |
| chr8 | 127890664 | 127891582 | 919 | 5 | 7.48E-05 | 0.001265 | 0.00011 | 7.49E-05 | -0.004788819 | PVT1, AC084123.1 |
| chr17 | 82649004 | 82650106 | 1103 | 6 | 7.48E-05 | 1.65E-06 | 0.00506 | 3.65E-06 | -0.012042233 | AC124283.5 |
| chr13 | 52924961 | 52925448 | 488 | 2 | 7.48E-05 | 4.75E-06 | 0.00049 | 7.93E-06 | -0.009448972 | NA |
| chr10 | 110496260 | 110496387 | 128 | 2 | 7.49E-05 | 4.02E-05 | 0.00056 | 4.95E-05 | -0.01307856 | AL355512.1 |
| chr18 | 23330892 | 23331538 | 647 | 6 | 7.49E-05 | 1.11E-05 | 0.00971 | 2.48E-05 | -0.00232287 | TMEM241 |
| chr11 | 28109193 | 28109469 | 277 | 3 | 7.49E-05 | 0.000521 | 0.00012 | 7.71E-05 | -0.009488321 | METTL15 |
| chr2 | 205765267 | 205765504 | 238 | 2 | 7.50E-05 | 0.053606 | 4.26E-05 | 7.14E-05 | -0.004128828 | NRP2 |
| chr6 | 166803371 | 166803733 | 363 | 5 | 7.51E-05 | 0.006302 | 0.00029 | 3.09E-05 | -0.003222444 | RPS6KA2 |
| chr1 | 156848800 | 156849601 | 802 | 6 | 7.52E-05 | 8.00E-07 | 0.00784 | 4.96E-06 | -0.009593228 | NTRK1, INSRR |
| chr6 | 683859 | 684123 | 265 | 3 | 7.52E-05 | 4.24E-06 | 0.0009 | 8.95E-06 | -0.036830256 | EXOC2 |
| chr2 | 47488717 | 47488958 | 242 | 2 | 7.53E-05 | 1.51E-05 | 0.00083 | 2.37E-05 | -0.011796195 | MSH2 |
| chr8 | 22927744 | 22928199 | 456 | 5 | 7.53E-05 | 3.79E-05 | 0.00015 | 1.53E-05 | 0.000249193 | PEBP4 |
| chr1 | 87211541 | 87212244 | 704 | 4 | 7.54E-05 | 0.004178 | 0.00041 | 5.14E-05 | 0.005547365 | NA |
| chr1 | 202010350 | 202010810 | 461 | 7 | 7.54E-05 | 0.003928 | 7.24E-05 | 0.00016 | 0.000625078 | ELF3, ELF3-AS1 |
| chr2 | 227887016 | 227887090 | 75 | 2 | 7.55E-05 | 0.000402 | 2.08E-05 | 1.88E-05 | -0.009280245 | DAW1 |
| chr12 | 108561226 | 108562076 | 851 | 11 | 7.56E-05 | 0.000686 | 0.00013 | 6.93E-05 | 0.000449701 | SART3 |
| chr12 | 57832968 | 57833632 | 665 | 2 | 7.57E-05 | 1.07E-05 | 0.00085 | 1.75E-05 | -0.004070414 | CTDSP2 |
| chr16 | 81497507 | 81497995 | 489 | 3 | 7.57E-05 | 3.08E-06 | 0.00115 | 6.33E-06 | 0.002545528 | CMIP |
| chr11 | 124841952 | 124842396 | 445 | 3 | 7.58E-05 | 3.26E-05 | 0.00075 | 3.90E-05 | -0.014489012 | NA |
| chr11 | 1053344 | 1053393 | 50 | 2 | 7.58E-05 | 1.44E-05 | 0.001 | 2.31E-05 | -0.010166345 | LINC02688 |
| chr20 | 31818779 | 31819585 | 807 | 11 | 7.58E-05 | 1.48E-05 | 0.00199 | 6.43E-06 | -0.004061832 | MYLK2 |
| chr19 | 1163486 | 1164146 | 661 | 3 | 7.58E-05 | 8.69E-06 | 0.00044 | 1.13E-05 | -0.007155616 | SBNO2 |
| chr6 | 31869662 | 31871552 | 1891 | 15 | 7.58E-05 | 0.000203 | 0.00038 | 7.02E-07 | -0.002342906 | SLC44A4 |
| chr14 | 19875763 | 19876235 | 473 | 3 | 7.58E-05 | 0.000881 | 0.00014 | 1.86E-05 | -0.002484985 | OR4K2 |
| chr11 | 2422639 | 2424212 | 1574 | 18 | 7.58E-05 | 6.04E-05 | 0.00075 | 1.82E-06 | -0.002005985 | TRPM5 |
| chr19 | 5894199 | 5895045 | 847 | 3 | 7.58E-05 | 4.30E-05 | 0.00308 | 6.98E-05 | -0.008608862 | AC024592.3, NDUFA11 |
| chr1 | 203285834 | 203285875 | 42 | 2 | 7.60E-05 | 3.21E-05 | 0.00014 | 2.65E-05 | -0.012501535 | LINC01353 |
| chr2 | 1207762 | 1209193 | 1432 | 7 | 7.61E-05 | 5.56E-07 | 0.00048 | 6.71E-07 | -0.000714883 | SNTG2 |
| chr3 | 142947266 | 142947634 | 369 | 4 | 7.61E-05 | 6.93E-06 | 0.00138 | 1.02E-05 | -0.021181982 | NA |
| chr5 | 150904886 | 150905615 | 730 | 11 | 7.62E-05 | 9.01E-05 | 0.00284 | 8.80E-06 | -0.007018261 | ZNF300 |
| chr15 | 69145422 | 69145998 | 577 | 2 | 7.62E-05 | 5.66E-06 | 0.00024 | 8.24E-06 | -0.013341251 | NA |
| chr17 | 7014559 | 7015521 | 963 | 12 | 7.62E-05 | 0.021657 | 0.00037 | 0.00025 | -2.58E-05 | RNASEK-C17orf49, C17orf49, AC040977.2 |
| chr11 | 66876030 | 66876604 | 575 | 4 | 7.63E-05 | 7.67E-06 | 0.00304 | 1.37E-05 | -0.004604928 | PC |
| chr6 | 89315086 | 89316303 | 1218 | 7 | 7.64E-05 | 2.05E-05 | 0.00149 | 2.14E-05 | -0.002670997 | GABRR2 |
| chr20 | 50138063 | 50138862 | 800 | 3 | 7.64E-05 | 7.75E-06 | 0.00011 | 1.70E-06 | -0.004995101 | PEDS1-UBE2V1, PEDS1 |
| chr8 | 143738221 | 143738656 | 436 | 5 | 7.65E-05 | 2.66E-05 | 0.00053 | 1.28E-05 | -0.003556762 | IQANK1, FAM83H |
| chr21 | 32412755 | 32413680 | 926 | 4 | 7.65E-05 | 0.001079 | 0.00064 | 0.00025 | -0.003645422 | EVA1C, CFAP298-TCP10L |
| chr17 | 6556032 | 6556256 | 225 | 3 | 7.65E-05 | 1.50E-05 | 0.00092 | 2.56E-05 | -0.001378508 | PITPNM3 |

| | | | | | | | | | | |
|---|---|---|---|---|---|---|---|---|---|---|
| chr12 | 116592349 | 116592666 | 318 | 2 | 7.66E-05 | 5.43E-06 | 0.00034 | 8.49E-06 | -0.015465673 | NA |
| chr1 | 6784459 | 6785159 | 701 | 6 | 7.69E-05 | 4.73E-05 | 0.00781 | 2.85E-05 | 0.000472198 | CAMTA1-DT |
| chr2 | 70887116 | 70888240 | 1125 | 7 | 7.70E-05 | 9.77E-06 | 0.00108 | 9.74E-06 | 0.003948187 | LINC01143 |
| chr17 | 79024956 | 79025634 | 679 | 4 | 7.72E-05 | 0.000404 | 0.00088 | 3.12E-05 | -0.001035647 | C1QTNF1, C1QTNF1-AS1 |
| chr9 | 133563019 | 133563584 | 566 | 2 | 7.73E-05 | 0.001331 | 0.00023 | 0.00027 | -0.011785604 | ADAMTSL2 |
| chr11 | 111512190 | 111512943 | 754 | 8 | 7.73E-05 | 0.001554 | 0.00222 | 1.71E-05 | -0.001837207 | AP002008.4, MIR34B, BTG4 |
| chr12 | 119676798 | 119677205 | 408 | 2 | 7.74E-05 | 6.70E-05 | 8.95E-05 | 3.29E-05 | -0.003900216 | PRKAB1 |
| chr15 | 81331733 | 81331878 | 146 | 2 | 7.74E-05 | 0.000227 | 8.15E-05 | 5.52E-05 | -0.005983648 | TMC3-AS1, TMC3 |
| chr11 | 1499401 | 1500168 | 768 | 6 | 7.74E-05 | 3.13E-05 | 0.00076 | 8.71E-06 | -0.001841394 | MOB2 |
| chr11 | 118142716 | 118143060 | 345 | 5 | 7.75E-05 | 2.50E-05 | 0.00238 | 2.00E-05 | -0.014193801 | SCN4B |
| chr10 | 21069956 | 21070773 | 818 | 7 | 7.75E-05 | 6.36E-06 | 0.00046 | 9.01E-06 | -0.006921008 | NEBL |
| chr11 | 75559024 | 75559169 | 146 | 2 | 7.75E-05 | 5.03E-05 | 5.68E-05 | 2.02E-05 | -0.007572917 | NA |
| chr17 | 51397863 | 51398208 | 346 | 3 | 7.76E-05 | 1.60E-05 | 0.0002 | 1.35E-05 | -0.014405807 | LINC02073 |
| chr3 | 143118722 | 143120180 | 1459 | 9 | 7.76E-05 | 3.66E-06 | 0.00737 | 1.05E-05 | -0.000160918 | CHST2 |
| chr22 | 36371275 | 36371539 | 265 | 2 | 7.77E-05 | 2.07E-05 | 0.00075 | 3.06E-05 | -0.012022134 | MYH9 |
| chr9 | 123033004 | 123033656 | 653 | 6 | 7.77E-05 | 0.000376 | 3.72E-05 | 4.37E-05 | -0.00606336 | RABGAP1 |
| chr1 | 15022227 | 15022605 | 379 | 2 | 7.79E-05 | 0.000294 | 6.84E-05 | 5.21E-05 | -0.00743041 | KAZN |
| chr1 | 16167648 | 16168004 | 357 | 2 | 7.79E-05 | 0.005559 | 7.08E-05 | 0.00011 | 0.00279644 | NA |
| chr6 | 132537299 | 132538346 | 1048 | 6 | 7.80E-05 | 2.09E-06 | 0.00126 | 5.53E-06 | -0.009186589 | TAAR9 |
| chr19 | 6501767 | 6502546 | 780 | 7 | 7.80E-05 | 0.000446 | 0.00044 | 1.12E-05 | 0.000665353 | TUBB4A |
| chr19 | 53013775 | 53013932 | 158 | 4 | 7.81E-05 | 4.53E-05 | 0.00015 | 1.24E-05 | -0.005330398 | ERVV-1, ZNF702P |
| chr9 | 87888589 | 87889177 | 589 | 3 | 7.82E-05 | 9.92E-05 | 0.00011 | 1.46E-05 | -0.006937332 | SPATA31E1 |
| chr13 | 98482826 | 98483732 | 907 | 9 | 7.84E-05 | 0.00017 | 0.00185 | 2.42E-05 | -0.0009435 | STK24 |
| chr18 | 55587045 | 55587832 | 788 | 7 | 7.84E-05 | 0.000238 | 0.00021 | 6.17E-05 | 0.002664255 | TCF4 |
| chr11 | 7538077 | 7538175 | 99 | 2 | 7.84E-05 | 1.38E-05 | 0.00011 | 1.30E-05 | -0.016531791 | PPFIBP2 |
| chr13 | 36879292 | 36879789 | 498 | 6 | 7.85E-05 | 0.000168 | 0.00288 | 2.81E-05 | 0.010137323 | SMAD9 |
| chr10 | 14089328 | 14089834 | 507 | 5 | 7.85E-05 | 0.001591 | 0.00028 | 4.52E-05 | -0.004532683 | FRMD4A |
| chr16 | 50275260 | 50276035 | 776 | 2 | 7.85E-05 | 9.46E-06 | 0.00013 | 1.06E-05 | -0.008390448 | ADCY7 |
| chr14 | 23551377 | 23552268 | 892 | 12 | 7.85E-05 | 5.94E-05 | 0.00127 | 1.81E-05 | -0.000251814 | ZFHX2-AS1, ZFHX2 |
| chr6 | 157148116 | 157149077 | 962 | 7 | 7.85E-05 | 0.000497 | 0.00035 | 2.83E-05 | -0.003897387 | ARID1B |
| chr14 | 104780120 | 104780371 | 252 | 3 | 7.86E-05 | 2.37E-05 | 0.00016 | 1.78E-05 | -0.007860738 | AKT1 |
| chr11 | 69666349 | 69666966 | 618 | 3 | 7.87E-05 | 0.000464 | 0.0001 | 7.73E-05 | 0.003949441 | LTO1 |
| chr7 | 158829637 | 158830480 | 844 | 11 | 7.87E-05 | 5.05E-05 | 0.00026 | 1.34E-05 | 0.000820883 | ESYT2 |
| chr13 | 35476651 | 35477612 | 962 | 10 | 7.90E-05 | 2.32E-05 | 0.00318 | 1.47E-05 | -0.00319082 | NBEA, MAB21L1 |
| chr15 | 88625212 | 88626031 | 820 | 3 | 7.90E-05 | 0.000513 | 0.00052 | 2.78E-05 | -0.009775487 | AEN |
| chr3 | 187735400 | 187736651 | 1252 | 8 | 7.90E-05 | 0.009969 | 0.00319 | 0.00047 | -0.004862885 | BCL6 |
| chr16 | 85208624 | 85209343 | 720 | 4 | 7.94E-05 | 0.000104 | 0.0007 | 4.11E-05 | -0.005581318 | GSE1 |
| chr10 | 102225903 | 102226511 | 609 | 11 | 7.94E-05 | 5.88E-05 | 0.00065 | 1.40E-05 | -0.002181921 | ELOVL3 |
| chr17 | 29150139 | 29150626 | 488 | 3 | 7.95E-05 | 2.22E-05 | 0.00123 | 2.68E-05 | -0.008774503 | AC024619.3, MYO18A |
| chr2 | 231781196 | 231781976 | 781 | 15 | 7.97E-05 | 2.57E-05 | 0.00174 | 1.21E-05 | 0.001093423 | COPS7B, PDE6D |
| chr2 | 27011004 | 27011226 | 223 | 2 | 7.97E-05 | 0.0006 | 2.93E-05 | 2.88E-05 | -0.004486838 | MAPRE3, MAPRE3-AS1 |
| chr17 | 21171081 | 21171766 | 686 | 4 | 7.97E-05 | 0.002459 | 0.00032 | 0.00047 | -0.003870877 | DHRS7B |
| chr17 | 27559767 | 27560725 | 959 | 6 | 7.98E-05 | 6.18E-07 | 0.00259 | 1.84E-06 | -0.00720291 | KSR1 |
| chr22 | 39446457 | 39446498 | 42 | 2 | 8.00E-05 | 6.23E-06 | 0.00017 | 8.24E-06 | -0.010093624 | NA |
| chr2 | 162177672 | 162177884 | 213 | 2 | 8.00E-05 | 0.000315 | 3.19E-05 | 2.69E-05 | -0.011343034 | FAP |
| chr5 | 134086538 | 134087035 | 498 | 2 | 8.01E-05 | 2.80E-05 | 0.0001 | 2.07E-05 | -0.010514952 | NA |
| chr11 | 66618857 | 66619178 | 322 | 3 | 8.02E-05 | 5.25E-06 | 0.00132 | 1.09E-05 | -0.009233953 | RBM14, RBM14-RBM4 |
| chr19 | 11018959 | 11019661 | 703 | 3 | 8.02E-05 | 8.31E-06 | 0.00068 | 1.33E-05 | -0.00612416 | SMARCA4 |
| chr17 | 50686804 | 50687605 | 802 | 5 | 8.02E-05 | 3.87E-06 | 0.00244 | 8.72E-06 | -0.005787906 | ABCC3 |
| chr7 | 4819494 | 4819817 | 324 | 3 | 8.03E-05 | 0.000122 | 0.00029 | 7.02E-05 | -0.01462782 | RADIL |
| chr5 | 123813409 | 123814022 | 614 | 2 | 8.03E-05 | 2.05E-06 | 0.00033 | 3.59E-06 | -0.027559152 | NA |
| chr20 | 36573207 | 36574074 | 868 | 7 | 8.04E-05 | 0.000173 | 0.00022 | 1.87E-05 | 0.000954615 | TGIF2, DLGAP4-AS1 |
| chr5 | 141417325 | 141417525 | 201 | 2 | 8.04E-05 | 0.000115 | 0.00031 | 8.86E-05 | 0.020354308 | PCDHGA1, PCDHGA2, PCDHGA3, PCDHGB1, PCDHGA4, PCDHGB2, PCDHGA5, PCDHGB3, PCDHGA6, PCDHGA7, PCDHGB4, PCDHGA8, PCDHGB5, PCDHGA9, PCDHGB6, PCDHGA10 |
| chr4 | 38563673 | 38564049 | 377 | 4 | 8.05E-05 | 5.74E-05 | 0.00037 | 1.66E-05 | -0.002587948 | LINC02278 |
| chr19 | 38663799 | 38664192 | 394 | 3 | 8.06E-05 | 1.75E-05 | 0.00037 | 1.95E-05 | -0.006287474 | ACTN4 |
| chr19 | 2858856 | 2859155 | 300 | 4 | 8.06E-05 | 3.71E-06 | 0.00083 | 7.97E-06 | -0.016258164 | ZNF555 |
| chr19 | 9826752 | 9827971 | 1220 | 11 | 8.07E-05 | 2.13E-05 | 0.00087 | 4.33E-06 | 0.00095751 | UBL5, FBXL12 |
| chr4 | 2251076 | 2251135 | 60 | 2 | 8.08E-05 | 3.91E-05 | 0.00131 | 5.77E-05 | -0.002696447 | MXD4 |
| chr12 | 14257911 | 14258601 | 691 | 4 | 8.08E-05 | 1.09E-05 | 0.00023 | 1.09E-05 | -0.005943202 | NA |
| chr5 | 92660925 | 92661005 | 81 | 2 | 8.09E-05 | 4.25E-05 | 0.00015 | 3.40E-05 | -0.001803471 | AC026780.1 |
| chr5 | 169302550 | 169302843 | 294 | 3 | 8.09E-05 | 7.02E-05 | 0.00056 | 1.90E-05 | -0.009953983 | NA |
| chr14 | 64114295 | 64114320 | 26 | 2 | 8.09E-05 | 1.02E-05 | 0.00078 | 1.66E-05 | -0.012866383 | SYNE2, ESR2 |
| chr3 | 141425180 | 141427453 | 2274 | 7 | 8.09E-05 | 9.83E-08 | 0.00234 | 4.51E-08 | -0.010571535 | ZBTB38 |
| chr1 | 203860353 | 203860762 | 410 | 3 | 8.09E-05 | 7.11E-05 | 0.00033 | 1.95E-05 | -0.006477619 | NA |
| chr17 | 2511925 | 2512913 | 989 | 9 | 8.10E-05 | 3.16E-05 | 0.00257 | 8.15E-06 | -0.0029841 | NA |
| chr5 | 172833461 | 172834512 | 1052 | 11 | 8.11E-05 | 2.80E-06 | 0.00859 | 4.17E-06 | 0.005184871 | ERGIC1 |
| chr10 | 133246971 | 133247982 | 1012 | 11 | 8.11E-05 | 9.10E-06 | 0.00015 | 2.80E-06 | -0.006726115 | MIR202HG, MIR202 |
| chr20 | 38580111 | 38580752 | 642 | 5 | 8.11E-05 | 4.39E-06 | 0.00447 | 1.56E-05 | -0.009096527 | NA |
| chr14 | 22839085 | 22839840 | 756 | 4 | 8.11E-05 | 2.20E-05 | 0.00095 | 1.10E-05 | -0.007835955 | MMP14 |
| chr16 | 726037 | 726501 | 465 | 4 | 8.12E-05 | 1.25E-05 | 0.00148 | 1.43E-05 | -0.009414439 | CCDC78 |
| chr16 | 50644993 | 50645738 | 746 | 4 | 8.13E-05 | 1.32E-05 | 0.00177 | 1.66E-05 | -0.006424294 | NKD1 |
| chr21 | 41801220 | 41801681 | 462 | 6 | 8.14E-05 | 1.41E-06 | 0.01001 | 7.20E-06 | -0.009137281 | PRDM15 |
| chr5 | 61736705 | 61737111 | 407 | 3 | 8.14E-05 | 8.46E-05 | 0.00015 | 1.87E-05 | -0.003629219 | C5orf64 |
| chr11 | 58507737 | 58508145 | 409 | 2 | 8.15E-05 | 0.006954 | 4.16E-05 | 6.21E-05 | -0.001487805 | OR5B21 |
| chr14 | 74922577 | 74923360 | 784 | 7 | 8.15E-05 | 0.00137 | 0.01176 | 0.00023 | -0.005855759 | RPS6KL1 |
| chr13 | 95553041 | 95553850 | 810 | 5 | 8.16E-05 | 8.32E-06 | 0.00224 | 1.52E-05 | 0.012770265 | CLDN10 |
| chr1 | 15390870 | 15391101 | 232 | 2 | 8.16E-05 | 1.67E-05 | 7.65E-05 | 1.24E-05 | -0.001565776 | FHAD1 |
| chr9 | 135253482 | 135253885 | 404 | 2 | 8.17E-05 | 4.32E-05 | 0.00033 | 4.53E-05 | 0.024033084 | NA |
| chr8 | 47968386 | 47968915 | 530 | 4 | 8.17E-05 | 3.61E-05 | 0.00146 | 1.67E-05 | -0.002801111 | MCM4 |
| chr5 | 40688357 | 40688422 | 66 | 2 | 8.18E-05 | 2.58E-05 | 7.24E-05 | 1.60E-05 | 0.002945494 | PTGER4, TTC33 |
| chr10 | 60911378 | 60911470 | 93 | 3 | 8.19E-05 | 0.000183 | 0.00047 | 2.13E-05 | -0.006058083 | RHOBTB1 |
| chr1 | 16176089 | 16176398 | 310 | 2 | 8.19E-05 | 7.98E-06 | 0.00047 | 1.25E-05 | -0.011979136 | NA |
| chr1 | 66361752 | 66361852 | 101 | 2 | 8.19E-05 | 6.26E-05 | 4.08E-05 | 1.78E-05 | -0.004711101 | PDE4B |
| chr5 | 126600144 | 126600861 | 718 | 10 | 8.20E-05 | 2.85E-05 | 0.00707 | 9.66E-06 | -0.002408733 | NA |
| chr9 | 117897364 | 117897811 | 448 | 3 | 8.20E-05 | 4.54E-05 | 6.67E-05 | 1.84E-05 | 0.002799278 | AL160272.2 |
| chr15 | 80946097 | 80946780 | 684 | 2 | 8.21E-05 | 2.22E-06 | 0.00026 | 3.76E-06 | -0.003209965 | CEMIP, AC027808.1, MESD |
| chr2 | 127220818 | 127221003 | 186 | 3 | 8.22E-05 | 3.44E-05 | 0.00056 | 1.37E-05 | -0.007251864 | NA |
| chr18 | 63317252 | 63318036 | 785 | 3 | 8.24E-05 | 0.061428 | 0.00012 | 0.00072 | -0.008119196 | BCL2 |
| chr3 | 9361662 | 9361840 | 179 | 2 | 8.24E-05 | 6.51E-05 | 0.00015 | 4.46E-05 | -0.004037149 | SRGAP3, AC026191.1, THUMPD3-AS1 |
| chr12 | 124779275 | 124779572 | 298 | 3 | 8.24E-05 | 8.64E-06 | 0.00043 | 1.07E-05 | -0.005334231 | SCARB1 |
| chr6 | 28537131 | 28537652 | 522 | 5 | 8.24E-05 | 2.83E-06 | 0.00325 | 7.72E-06 | -0.011350082 | NA |
| chr17 | 55783818 | 55784356 | 539 | 4 | 8.25E-05 | 5.07E-06 | 0.0003 | 5.85E-06 | -0.012021337 | PCTP |
| chr1 | 205947127 | 205947512 | 386 | 2 | 8.26E-05 | 0.000297 | 5.12E-05 | 4.02E-05 | -0.00906479 | NA |
| chr12 | 12017966 | 12018320 | 355 | 2 | 8.26E-05 | 1.14E-05 | 0.00013 | 1.20E-05 | -0.01420398 | NA |
| chr2 | 240782505 | 240782864 | 360 | 4 | 8.27E-05 | 3.00E-05 | 0.00062 | 1.54E-05 | -0.00348978 | KIF1A |
| chr6 | 30203706 | 30204365 | 660 | 4 | 8.27E-05 | 0.003298 | 0.00012 | 1.70E-05 | -0.001545216 | TRIM26 |
| chr4 | 148054254 | 148054789 | 536 | 2 | 8.27E-05 | 1.70E-05 | 0.00017 | 1.84E-05 | -0.002848121 | ARHGAP10 |
| chr3 | 9216349 | 9216889 | 541 | 5 | 8.27E-05 | 0.002439 | 0.00023 | 2.86E-05 | -0.002896606 | SRGAP3 |
| chr8 | 143527910 | 143528384 | 475 | 3 | 8.27E-05 | 0.000106 | 0.00064 | 2.15E-05 | -0.004313877 | ZC3H3 |
| chr7 | 128407387 | 128407398 | 12 | 2 | 8.27E-05 | 1.94E-05 | 0.00085 | 2.94E-05 | -0.009967003 | IMPDH1 |
| chr7 | 1909183 | 1909611 | 429 | 4 | 8.28E-05 | 3.95E-05 | 0.00032 | 3.40E-05 | -0.002379312 | MAD1L1, AC069288.1 |
| chr16 | 28847021 | 28847121 | 101 | 2 | 8.28E-05 | 0.000112 | 0.00059 | 0.00011 | -0.009752055 | AC009093.11, SH2B1 |
| chr5 | 146378816 | 146379318 | 503 | 6 | 8.29E-05 | 9.11E-07 | 0.01032 | 5.44E-06 | -0.012860586 | NA |
| chr14 | 100992917 | 100993202 | 286 | 2 | 8.29E-05 | 7.34E-05 | 5.19E-05 | 2.31E-05 | 0.01020084 | MEG8 |
| chr3 | 187701502 | 187702233 | 732 | 6 | 8.30E-05 | 4.19E-05 | 0.00034 | 7.58E-06 | -0.003185054 | RTP2 |
| chr13 | 51994875 | 51995790 | 916 | 2 | 8.30E-05 | 4.54E-07 | 0.00012 | 8.04E-07 | -0.031724007 | ATP7B |
| chr14 | 101932899 | 101933435 | 537 | 5 | 8.31E-05 | 1.56E-05 | 0.00023 | 1.37E-05 | 0.000417174 | NA |
| chr22 | 19713992 | 19714348 | 357 | 5 | 8.31E-05 | 0.000436 | 0.00033 | 7.69E-05 | -0.004088799 | NA |
| chr16 | 90031237 | 90031353 | 117 | 4 | 8.31E-05 | 3.43E-06 | 0.00292 | 9.26E-06 | -0.003207506 | GAS8 |
| chr16 | 81531539 | 81532831 | 1293 | 6 | 8.33E-05 | 1.01E-05 | 0.00026 | 1.10E-05 | -0.003447036 | CMIP |
| chr19 | 35224349 | 35224721 | 373 | 2 | 8.35E-05 | 6.75E-06 | 0.00057 | 1.10E-05 | -0.006580435 | NA |
| chr3 | 150216431 | 150216655 | 225 | 2 | 8.36E-05 | 1.48E-05 | 8.44E-05 | 1.20E-05 | -0.009751125 | AC117386.1 |
| chr10 | 73867085 | 73867862 | 778 | 3 | 8.36E-05 | 2.78E-05 | 0.00019 | 1.45E-05 | -0.008100773 | CAMK2G |
| chr3 | 15498630 | 15499158 | 529 | 7 | 8.38E-05 | 5.09E-05 | 0.00027 | 1.78E-05 | -0.008142176 | COLQ |
| chr14 | 75276981 | 75277307 | 327 | 2 | 8.40E-05 | 2.35E-05 | 9.70E-05 | 1.78E-05 | -0.001065814 | NA |
| chr5 | 28809441 | 28810295 | 855 | 6 | 8.40E-05 | 8.50E-05 | 0.00112 | 4.47E-05 | -0.004209469 | LINC02109 |
| chr17 | 74763143 | 74763790 | 648 | 3 | 8.41E-05 | 0.03369 | 0.00044 | 0.00061 | -0.006925991 | SLC9A3R1 |
| chr16 | 89154108 | 89154601 | 494 | 3 | 8.43E-05 | 9.79E-06 | 0.00112 | 1.38E-05 | -0.007729731 | ACSF3 |
| chr12 | 6607391 | 6607604 | 214 | 6 | 8.43E-05 | 2.12E-05 | 0.0069 | 2.02E-05 | 0.002645189 | CHD4 |
| chr16 | 85321834 | 85322293 | 460 | 2 | 8.43E-05 | 5.24E-06 | 0.00043 | 8.55E-06 | -0.011926458 | GSE1 |
| chr6 | 168570094 | 168570455 | 362 | 5 | 8.44E-05 | 2.91E-05 | 0.00188 | 1.31E-05 | -0.006246379 | SMOC2 |
| chr8 | 23532275 | 23532717 | 443 | 2 | 8.44E-05 | 3.71E-05 | 0.00011 | 2.54E-05 | -0.010332448 | SLC25A37 |
| chr6 | 45480198 | 45480496 | 299 | 2 | 8.44E-05 | 3.59E-05 | 6.37E-05 | 1.81E-05 | 0.01980137 | RUNX2 |
| chr22 | 50527353 | 50527694 | 342 | 4 | 8.45E-05 | 2.37E-05 | 0.00155 | 2.51E-05 | -0.013820523 | TYMP |
| chr11 | 69205439 | 69206174 | 736 | 6 | 8.45E-05 | 4.95E-06 | 0.00456 | 5.85E-06 | -0.003174618 | NA |
| chr6 | 26045227 | 26045417 | 191 | 5 | 8.46E-05 | 1.28E-05 | 0.00013 | 1.10E-05 | 0.004456676 | H3C3 |
| chr1 | 198465777 | 198466321 | 545 | 2 | 8.49E-05 | 5.23E-06 | 0.0003 | 8.09E-06 | 0.003730195 | AL450352.1 |
| chr11 | 1073496 | 1074009 | 514 | 4 | 8.49E-05 | 0.009646 | 6.96E-05 | 0.00015 | -0.005684374 | NA |
| chr12 | 58919473 | 58920102 | 630 | 9 | 8.50E-05 | 0.003873 | 5.39E-05 | 0.0001 | 0.000384427 | LRIG3 |
| chr1 | 153932913 | 153933424 | 512 | 6 | 8.50E-05 | 4.48E-05 | 0.00536 | 2.71E-05 | -0.004915351 | AL358472.5, DENND4B |
| chr1 | 117540733 | 117541157 | 425 | 3 | 8.50E-05 | 5.35E-05 | 0.00022 | 2.36E-05 | -0.006059084 | NA |
| chr3 | 43532254 | 43532269 | 16 | 2 | 8.52E-05 | 5.17E-06 | 0.00039 | 8.31E-06 | -0.004232937 | ANO10 |
| chr16 | 3180725 | 3180846 | 122 | 2 | 8.52E-05 | 1.43E-05 | 0.00014 | 1.50E-05 | -0.050995288 | NA |
| chr8 | 142988130 | 142988280 | 151 | 2 | 8.52E-05 | 0.001343 | 2.56E-05 | 2.92E-05 | -0.00850569 | LY6E-DT |
| chr5 | 36071400 | 36071762 | 363 | 2 | 8.53E-05 | 0.000159 | 0.00012 | 6.17E-05 | 0.035402681 | NA |
| chr5 | 500087 | 500450 | 364 | 5 | 8.54E-05 | 0.000267 | 6.16E-05 | 2.85E-05 | -0.003163124 | SLC9A3 |
| chr2 | 191245005 | 191245932 | 928 | 9 | 8.55E-05 | 1.57E-05 | 0.00199 | 2.96E-05 | -0.000299948 | MYO1B, AC092614.1 |
| chr15 | 82852662 | 82853605 | 944 | 5 | 8.56E-05 | 7.23E-07 | 0.00319 | 2.81E-06 | -0.014580723 | HOMER2 |
| chr3 | 75965201 | 75965466 | 266 | 3 | 8.56E-05 | 9.26E-05 | 6.36E-05 | 1.54E-05 | -0.016012506 | ROBO2 |
| chr7 | 12370400 | 12370802 | 403 | 3 | 8.57E-05 | 0.000143 | 0.00053 | 1.76E-05 | -0.004766635 | VWDE |
| chr7 | 154912864 | 154913347 | 484 | 3 | 8.57E-05 | 1.76E-05 | 0.00549 | 4.20E-05 | 0.001020295 | NA |
| chr12 | 57464649 | 57465232 | 584 | 5 | 8.59E-05 | 2.89E-05 | 0.00501 | 3.53E-05 | -0.00302627 | GLI1 |
| chr11 | 1084585 | 1085487 | 903 | 7 | 8.60E-05 | 0.000346 | 0.00052 | 1.89E-05 | -0.005732957 | MUC2 |
| chr4 | 105141121 | 105141243 | 123 | 2 | 8.60E-05 | 9.93E-05 | 3.62E-05 | 1.99E-05 | -0.005144193 | NA |
| chr1 | 5833522 | 5833847 | 326 | 3 | 8.60E-05 | 2.05E-05 | 0.00045 | 1.26E-05 | -0.000596294 | NA |
| chr12 | 69739512 | 69739888 | 377 | 4 | 8.60E-05 | 0.014211 | 9.58E-05 | 0.0004 | 0.002828105 | RAB3IP |
| chr7 | 18770589 | 18771247 | 659 | 2 | 8.60E-05 | 2.51E-06 | 0.00029 | 4.26E-06 | -0.01352156 | HDAC9 |
| chr14 | 104399947 | 104400299 | 353 | 2 | 8.60E-05 | 3.96E-05 | 7.22E-05 | 2.09E-05 | -0.013319746 | NA |
| chr17 | 8056589 | 8057088 | 500 | 3 | 8.61E-05 | 2.05E-05 | 0.00023 | 2.02E-05 | -0.005524112 | AC129492.2 |
| chr3 | 194350155 | 194351041 | 887 | 3 | 8.65E-05 | 6.07E-06 | 0.00335 | 1.57E-05 | -0.006562845 | CPN2 |
| chr2 | 44361295 | 44362140 | 846 | 17 | 8.65E-05 | 0.000489 | 0.01238 | 2.37E-05 | 0.000890249 | CAMKMT, PREPL |
| chr2 | 126655116 | 126657251 | 2136 | 17 | 8.66E-05 | 5.22E-06 | 0.00978 | 4.25E-06 | -0.003152039 | GYPC |
| chr7 | 151078976 | 151079387 | 412 | 3 | 8.67E-05 | 4.74E-06 | 0.0017 | 1.08E-05 | -0.004878454 | FASTK |
| chr8 | 48473317 | 48473701 | 385 | 2 | 8.69E-05 | 0.000375 | 4.35E-05 | 3.72E-05 | -0.007777525 | NA |
| chr20 | 44609091 | 44610051 | 961 | 4 | 8.70E-05 | 0.004147 | 5.33E-05 | 1.82E-05 | -0.004989687 | PKIG |
| chr19 | 58516468 | 58517911 | 1444 | 6 | 8.70E-05 | 2.26E-07 | 0.00099 | 2.42E-07 | -0.00372722 | ZBTB45 |
| chr6 | 33304787 | 33305351 | 565 | 6 | 8.70E-05 | 0.000216 | 0.00225 | 7.91E-05 | -0.003048063 | TAPBP |
| chr11 | 4607804 | 4608682 | 879 | 12 | 8.71E-05 | 1.02E-06 | 0.01703 | 7.02E-06 | 0.004154208 | TRIM68 |
| chr7 | 151850884 | 151851163 | 280 | 4 | 8.72E-05 | 3.41E-05 | 0.00035 | 1.46E-05 | -0.010429444 | PRKAG2 |
| chr16 | 81616526 | 81617072 | 547 | 2 | 8.72E-05 | 5.06E-06 | 0.00032 | 7.91E-06 | -0.012116091 | CMIP, AC092135.2 |
| chr12 | 2874260 | 2874489 | 230 | 3 | 8.73E-05 | 0.000243 | 6.26E-05 | 2.50E-05 | -0.002923515 | FOXM1 |
| chr7 | 116524054 | 116524896 | 843 | 16 | 8.73E-05 | 0.000436 | 0.00069 | 4.87E-05 | -0.00105447 | NA |

| | | | | | | | | | | |
|---|---|---|---|---|---|---|---|---|---|---|
| chr9 | 104927366 | 104928489 | 1124 | 10 | 8.74E-05 | 9.88E-05 | 0.00647 | 4.46E-05 | -0.000748622 | AL359182.1, ABCA1 |
| chr14 | 49621542 | 49621880 | 339 | 3 | 8.75E-05 | 0.000136 | 0.00047 | 9.86E-05 | -0.010597354 | MGAT2, AL139099.1 |
| chr11 | 101956246 | 101956373 | 128 | 2 | 8.75E-05 | 0.001314 | 6.88E-05 | 8.15E-05 | -0.009367806 | CEP126 |
| chr7 | 157406175 | 157407334 | 1160 | 6 | 8.76E-05 | 0.00019 | 0.00017 | 6.77E-06 | -0.00395178 | DNAJB6 |
| chr16 | 56789509 | 56789604 | 96 | 2 | 8.78E-05 | 5.22E-06 | 0.00046 | 8.57E-06 | -0.023865128 | NUP93 |
| chr1 | 11042840 | 11043386 | 547 | 3 | 8.78E-05 | 0.000272 | 4.00E-05 | 2.79E-05 | -0.005855276 | MASP2 |
| chr18 | 72542790 | 72543259 | 470 | 4 | 8.78E-05 | 4.58E-05 | 0.00262 | 3.36E-05 | 0.004710838 | CBLN2 |
| chr3 | 66582912 | 66583357 | 446 | 4 | 8.79E-05 | 2.05E-05 | 0.00033 | 1.52E-05 | -0.009367546 | NA |
| chr2 | 96832599 | 96833005 | 407 | 2 | 8.79E-05 | 0.022298 | 5.16E-05 | 8.73E-05 | -0.002044852 | CNNM3, ANKRD23 |
| chr10 | 50619286 | 50619707 | 422 | 2 | 8.79E-05 | 5.17E-05 | 0.00027 | 4.86E-05 | -0.006119074 | SGMS1 |
| chr15 | 64503755 | 64504041 | 287 | 2 | 8.79E-05 | 0.001866 | 3.07E-05 | 3.78E-05 | -0.007117409 | ZNF609 |
| chr19 | 38229986 | 38230212 | 227 | 4 | 8.80E-05 | 0.000222 | 3.59E-05 | 2.77E-05 | -0.007369405 | NA |
| chr12 | 121552779 | 121552889 | 111 | 2 | 8.80E-05 | 7.64E-06 | 0.00056 | 1.23E-05 | -0.011561894 | KDM2B |
| chr5 | 159049735 | 159049945 | 211 | 2 | 8.80E-05 | 3.91E-05 | 0.00011 | 2.62E-05 | -0.014296431 | EBF1 |
| chr8 | 23321473 | 23322258 | 786 | 3 | 8.80E-05 | 3.84E-06 | 0.0013 | 8.65E-06 | -0.056786756 | LOXL2 |
| chr15 | 33968232 | 33968971 | 740 | 8 | 8.81E-05 | 2.47E-05 | 0.0018 | 1.14E-05 | 0.000122062 | CHRM5, AVEN |
| chr5 | 138869745 | 138869976 | 232 | 2 | 8.82E-05 | 8.27E-06 | 9.47E-05 | 8.24E-06 | 0.002111061 | CTNNA1, LRRTM2 |
| chr3 | 87226479 | 87227593 | 1115 | 13 | 8.82E-05 | 0.000486 | 0.00179 | 2.33E-05 | 0.001288372 | CHMP2B |
| chr6 | 33297239 | 33298312 | 1074 | 14 | 8.82E-05 | 0.000137 | 0.00893 | 4.03E-05 | -0.001378462 | PFDN6, RGL2 |
| chr12 | 49647584 | 49648327 | 744 | 2 | 8.82E-05 | 1.74E-06 | 0.00023 | 2.93E-06 | -0.004636456 | FMNL3 |
| chr10 | 13712779 | 13713042 | 264 | 2 | 8.82E-05 | 0.012057 | 2.62E-05 | 3.81E-05 | -0.009555369 | FRMD4A |
| chr2 | 11073691 | 11073935 | 245 | 2 | 8.84E-05 | 4.01E-05 | 5.19E-05 | 1.67E-05 | -0.012007221 | NA |
| chr3 | 11509185 | 11509867 | 683 | 2 | 8.84E-05 | 1.72E-06 | 0.00028 | 2.97E-06 | -0.005448501 | ATG7 |
| chr9 | 137688741 | 137689305 | 565 | 2 | 8.85E-05 | 7.58E-05 | 0.00013 | 4.38E-05 | -0.010213577 | AL590627.2, EHMT1 |
| chr2 | 127060372 | 127060650 | 279 | 7 | 8.85E-05 | 0.000859 | 9.67E-05 | 2.82E-05 | -0.002457437 | BIN1 |
| chr13 | 98890365 | 98890462 | 98 | 3 | 8.85E-05 | 5.71E-05 | 4.57E-05 | 1.44E-05 | -0.006757605 | DOCK9 |
| chr2 | 3585743 | 3586481 | 739 | 6 | 8.85E-05 | 7.22E-05 | 0.00174 | 1.25E-05 | -0.001518628 | NA |
| chr10 | 35604696 | 35606456 | 1761 | 9 | 8.86E-05 | 1.03E-07 | 0.00331 | 7.55E-07 | -0.009954076 | GJD4, AL121749.1 |
| chr12 | 52068636 | 52069584 | 949 | 8 | 8.86E-05 | 0.000588 | 0.01116 | 0.00014 | 0.011574146 | ATG101 |
| chr1 | 151825956 | 151826790 | 835 | 5 | 8.86E-05 | 0.000551 | 0.00047 | 4.54E-05 | -0.006768637 | RORC |
| chr18 | 50375265 | 50375649 | 385 | 3 | 8.87E-05 | 0.000968 | 0.00157 | 0.00049 | 0.009777382 | SKA1 |
| chr6 | 7165414 | 7166039 | 626 | 2 | 8.87E-05 | 4.60E-06 | 0.00033 | 7.29E-06 | -0.009263694 | RREB1 |
| chr4 | 7471164 | 7471570 | 407 | 3 | 8.87E-05 | 5.11E-05 | 9.65E-05 | 1.68E-05 | -0.000941137 | SORCS2 |
| chr17 | 80804432 | 80804753 | 322 | 2 | 8.87E-05 | 5.74E-05 | 0.00065 | 6.88E-05 | -0.007525708 | RPTOR, AC016245.1 |
| chr16 | 68643461 | 68644066 | 606 | 2 | 8.88E-05 | 0.078509 | 0.07315 | 0.08242 | -0.006861655 | CDH3 |
| chr2 | 96507976 | 96509078 | 1103 | 14 | 8.88E-05 | 1.81E-05 | 0.00794 | 9.07E-06 | -0.00166126 | NEURL3 |
| chr2 | 136990412 | 136991263 | 852 | 6 | 8.89E-05 | 0.000519 | 0.00241 | 3.36E-05 | -0.00215961 | THSD7B |
| chr22 | 46056357 | 46056460 | 104 | 2 | 8.89E-05 | 0.000122 | 8.12E-05 | 4.23E-05 | -0.006397964 | PRR34-AS1, MIRLET7BHG, AL121672.1 |
| chr5 | 141223279 | 141223698 | 420 | 7 | 8.90E-05 | 6.54E-06 | 0.00371 | 9.87E-06 | 0.011090764 | PCDHB14, AC244517.10, AC244517.11 |
| chr11 | 92915518 | 92916449 | 932 | 6 | 8.90E-05 | 0.001866 | 0.00039 | 5.99E-05 | -0.002831054 | AP003171.2, LINC02746 |
| chr11 | 47714580 | 47715389 | 810 | 8 | 8.91E-05 | 0.000131 | 0.00021 | 2.05E-05 | -0.002328234 | AGBL2 |
| chr7 | 151123257 | 151124085 | 829 | 4 | 8.91E-05 | 0.001085 | 0.0005 | 0.00013 | -0.005012306 | AGAP3 |
| chr7 | 99031838 | 99033165 | 1328 | 7 | 8.92E-05 | 1.81E-06 | 0.00046 | 1.05E-06 | -0.003589352 | TRRAP, AC004893.1, SMURF1 |
| chr5 | 157266564 | 157266804 | 241 | 4 | 8.92E-05 | 1.50E-05 | 0.00139 | 2.59E-05 | 0.002300447 | CYFIP2, AC009185.1 |
| chr6 | 30913335 | 30914031 | 697 | 22 | 8.93E-05 | 1.80E-05 | 0.00322 | 2.26E-05 | -0.005140908 | GTF2H4, AL669830.1 |
| chr16 | 84045355 | 84045517 | 163 | 2 | 8.93E-05 | 6.53E-05 | 7.79E-05 | 2.95E-05 | -0.013618064 | NA |
| chr2 | 49974275 | 49974367 | 93 | 3 | 8.94E-05 | 2.89E-06 | 0.00166 | 7.19E-06 | 0.035364467 | NRXN1 |
| chr15 | 57706386 | 57707075 | 690 | 11 | 8.94E-05 | 9.49E-05 | 0.01131 | 1.89E-05 | 0.001617199 | GCOM1, POLR2M |
| chr16 | 89936059 | 89936484 | 426 | 4 | 8.95E-05 | 1.20E-05 | 0.0002 | 1.17E-05 | -0.003330709 | AC092143.1, TUBB3 |
| chr16 | 69190509 | 69190979 | 471 | 3 | 8.95E-05 | 0.000101 | 0.00087 | 3.00E-05 | 0.009185789 | UTP4, SNTB2 |
| chr1 | 965044 | 965170 | 127 | 2 | 8.97E-05 | 0.000232 | 0.00371 | 0.00037 | -0.005637383 | KLHL17 |
| chr11 | 5634746 | 5634829 | 84 | 2 | 8.98E-05 | 1.51E-05 | 0.00014 | 1.53E-05 | -0.004307185 | TRIM6-TRIM34, TRIM34, AC104389.5 |
| chr15 | 89751240 | 89751545 | 306 | 9 | 8.99E-05 | 0.000114 | 0.00135 | 2.30E-05 | -0.00395998 | MESP1 |
| chr12 | 132151009 | 132151757 | 749 | 7 | 8.99E-05 | 3.23E-05 | 0.00243 | 8.48E-06 | -0.003565756 | NOC4L |
| chr16 | 88454340 | 88454504 | 165 | 2 | 9.00E-05 | 3.00E-05 | 0.00016 | 2.72E-05 | 0.004411754 | ZFPM1 |
| chr2 | 28979946 | 28980478 | 533 | 5 | 9.00E-05 | 2.72E-05 | 0.00286 | 2.88E-05 | -0.001781491 | TOGARAM2 |
| chr10 | 124946489 | 124947091 | 603 | 3 | 9.01E-05 | 0.000528 | 6.19E-05 | 3.56E-05 | -0.01015353 | ZRANB1 |
| chr5 | 139383123 | 139383781 | 659 | 6 | 9.01E-05 | 6.06E-06 | 0.00124 | 1.06E-05 | -0.006955718 | SLC23A1 |
| chr16 | 67441493 | 67441793 | 301 | 3 | 9.01E-05 | 0.000284 | 2.57E-05 | 2.27E-05 | -0.009052208 | ATP6V0D1 |
| chr1 | 154993592 | 154993842 | 251 | 3 | 9.03E-05 | 6.50E-05 | 0.00094 | 7.04E-05 | -0.000157462 | LENEP |
| chr2 | 115439376 | 115439603 | 228 | 2 | 9.03E-05 | 0.00017 | 3.15E-05 | 2.19E-05 | -0.004634075 | DPP10 |
| chr13 | 27450672 | 27451457 | 786 | 10 | 9.04E-05 | 0.000133 | 0.00936 | 4.97E-05 | -0.002154336 | NA |
| chr12 | 131834472 | 131834547 | 76 | 2 | 9.05E-05 | 0.083284 | 6.07E-05 | 0.00011 | -0.009350974 | MMP17 |
| chr2 | 111123117 | 111123498 | 382 | 3 | 9.05E-05 | 0.001672 | 7.41E-05 | 4.40E-05 | -0.006905126 | BCL2L11, MIR4435-2HG |
| chr12 | 113240027 | 113240601 | 575 | 2 | 9.06E-05 | 1.00E-05 | 0.00056 | 1.56E-05 | -0.012734516 | TPCN1 |
| chr1 | 247833925 | 247834032 | 108 | 2 | 9.06E-05 | 1.15E-05 | 9.48E-05 | 1.05E-05 | -0.000414338 | OR6R1P |
| chr10 | 62075041 | 62075477 | 437 | 2 | 9.07E-05 | 9.00E-05 | 0.0006 | 9.45E-05 | 0.006795636 | ARID5B |
| chr9 | 18791474 | 18791826 | 353 | 3 | 9.07E-05 | 0.000367 | 3.26E-05 | 3.08E-05 | -0.001185064 | ADAMTSL1 |
| chr17 | 1004403 | 1005450 | 1048 | 5 | 9.08E-05 | 5.08E-06 | 0.00508 | 1.14E-05 | -0.00729048 | ABR |
| chr7 | 138750579 | 138751041 | 463 | 4 | 9.09E-05 | 3.99E-05 | 0.00067 | 3.90E-05 | -0.004612947 | ATP6V0A4 |
| chr17 | 48026241 | 48026625 | 385 | 3 | 9.10E-05 | 3.78E-05 | 0.00016 | 2.64E-05 | -0.005411251 | COPZ2 |
| chr17 | 81648522 | 81648831 | 310 | 3 | 9.10E-05 | 9.66E-05 | 0.00035 | 2.24E-05 | 0.018205714 | TSPAN10 |
| chr1 | 210253013 | 210253214 | 202 | 2 | 9.10E-05 | 1.54E-05 | 0.00107 | 2.48E-05 | -0.007822718 | NA |
| chr20 | 45338300 | 45338833 | 534 | 4 | 9.11E-05 | 0.00095 | 0.00062 | 9.25E-05 | -0.000839997 | SDC4 |
| chr12 | 22624475 | 22624531 | 57 | 3 | 9.12E-05 | 0.000408 | 0.0018 | 0.00011 | -0.004942499 | AC087241.2 |
| chr7 | 158509344 | 158509587 | 244 | 2 | 9.13E-05 | 0.000273 | 4.42E-05 | 3.44E-05 | -0.006533135 | PTPRN2 |
| chr9 | 83980236 | 83981017 | 782 | 14 | 9.13E-05 | 1.74E-06 | 0.01168 | 5.49E-06 | -2.90E-05 | RMI1, HNRNPK |
| chr7 | 130485130 | 130485915 | 786 | 7 | 9.14E-05 | 0.000238 | 0.00039 | 4.46E-05 | -0.001021042 | NA |
| chr13 | 110707188 | 110707372 | 185 | 2 | 9.14E-05 | 0.000301 | 7.24E-05 | 5.53E-05 | 0.004677635 | CARS2 |
| chr1 | 171742121 | 171742720 | 600 | 13 | 9.15E-05 | 0.0012 | 0.00203 | 2.85E-05 | -2.28E-06 | NA |
| chr16 | 57645823 | 57646828 | 1006 | 5 | 9.16E-05 | 1.27E-06 | 0.0057 | 5.45E-06 | -0.008204442 | ADGRG1 |
| chr11 | 1036307 | 1036938 | 632 | 7 | 9.17E-05 | 3.34E-06 | 0.01438 | 1.57E-05 | 0.009734315 | MUC6 |
| chr9 | 126833194 | 126833301 | 108 | 4 | 9.17E-05 | 9.27E-06 | 0.00221 | 8.45E-06 | -0.002416517 | ZBTB43 |
| chr5 | 66143991 | 66145303 | 1313 | 18 | 9.18E-05 | 1.97E-05 | 0.00804 | 9.75E-06 | -3.90E-05 | SREK1, AC025442.1 |
| chr15 | 41904462 | 41905473 | 1012 | 5 | 9.19E-05 | 2.38E-06 | 0.00349 | 6.16E-06 | -0.006508585 | EHD4 |
| chr6 | 31682958 | 31683153 | 196 | 6 | 9.19E-05 | 0.000381 | 0.06038 | 0.0022 | 0.025351141 | LY6G5C |
| chr1 | 150260405 | 150260738 | 334 | 3 | 9.20E-05 | 0.000168 | 6.53E-05 | 3.71E-05 | -0.001350637 | CA14 |
| chr9 | 132903464 | 132904081 | 618 | 3 | 9.20E-05 | 7.98E-06 | 0.00097 | 1.54E-05 | -0.005069497 | TSC1 |
| chr5 | 69485967 | 69485975 | 9 | 2 | 9.21E-05 | 6.79E-06 | 0.00054 | 1.10E-05 | -0.012443465 | AC145146.1 |
| chr6 | 169725877 | 169726260 | 384 | 2 | 9.21E-05 | 3.04E-05 | 0.00044 | 3.74E-05 | -0.003937122 | AL354892.1 |
| chr16 | 88327952 | 88328469 | 518 | 4 | 9.21E-05 | 0.000134 | 0.00189 | 6.66E-05 | -0.00292051 | NA |
| chr1 | 154219686 | 154220171 | 486 | 8 | 9.21E-05 | 7.70E-05 | 0.00112 | 1.78E-05 | -0.004480027 | C1orf43 |
| chr10 | 127441302 | 127441595 | 294 | 3 | 9.21E-05 | 3.33E-05 | 7.62E-05 | 1.65E-05 | -0.00512906 | DOCK1 |
| chr7 | 30801494 | 30801767 | 274 | 2 | 9.22E-05 | 4.99E-05 | 0.00025 | 4.60E-05 | -0.004652815 | INMT-MINDY4, MINDY4, AC004691.1 |
| chr17 | 81056739 | 81057411 | 673 | 4 | 9.23E-05 | 0.000573 | 0.00012 | 0.00011 | -0.006384077 | BAIAP2 |
| chr3 | 141345970 | 141346177 | 208 | 2 | 9.23E-05 | 5.44E-05 | 0.00015 | 4.00E-05 | -0.0086799 | PXYLP1, ZBTB38, AC117383.1 |
| chr4 | 1644039 | 1644452 | 414 | 4 | 9.23E-05 | 0.000391 | 0.00022 | 7.96E-05 | -0.001570251 | FAM53A |
| chr3 | 12286585 | 12288246 | 1662 | 17 | 9.24E-05 | 1.81E-05 | 0.00295 | 3.77E-06 | -0.004073848 | PPARG |
| chr12 | 65603426 | 65603949 | 524 | 3 | 9.25E-05 | 0.000244 | 0.00041 | 0.00013 | -0.011843899 | LINC02454, AC025419.1 |
| chr20 | 44651821 | 44652443 | 623 | 7 | 9.27E-05 | 0.001798 | 0.00026 | 2.85E-05 | 0.001599043 | ADA |
| chr17 | 13776370 | 13776507 | 138 | 3 | 9.27E-05 | 0.001001 | 4.83E-05 | 2.02E-05 | -0.004191477 | COX10-AS1 |
| chr16 | 50100020 | 50100161 | 142 | 2 | 9.28E-05 | 0.000716 | 2.22E-05 | 2.27E-05 | -0.004310772 | HEATR3 |
| chr3 | 160566215 | 160566594 | 380 | 3 | 9.28E-05 | 0.000203 | 0.0031 | 0.00018 | -0.02201691 | KRT8P12 |
| chr11 | 71116373 | 71116411 | 39 | 2 | 9.28E-05 | 7.97E-06 | 0.00015 | 9.64E-06 | -0.02103721 | SHANK2 |
| chr20 | 50345757 | 50346084 | 328 | 2 | 9.29E-05 | 4.89E-05 | 0.00012 | 3.21E-05 | -0.006560151 | NA |
| chr7 | 101363292 | 101363682 | 391 | 2 | 9.30E-05 | 0.00478 | 0.00021 | 0.0004 | 0.002187938 | COL26A1 |
| chr3 | 5121935 | 5122841 | 907 | 9 | 9.30E-05 | 0.000201 | 0.00039 | 3.09E-05 | -0.000983523 | ARL8B |
| chr10 | 11747384 | 11747811 | 428 | 2 | 9.31E-05 | 1.16E-05 | 0.00019 | 1.40E-05 | 0.007275774 | ECHDC3 |
| chr15 | 93422722 | 93422796 | 75 | 2 | 9.33E-05 | 0.000126 | 4.83E-05 | 2.83E-05 | -0.005591998 | AC091078.1 |
| chr8 | 108081804 | 108082652 | 849 | 7 | 9.33E-05 | 0.000138 | 0.00095 | 3.21E-05 | 0.000768502 | RSPO2 |
| chr1 | 185316714 | 185317590 | 877 | 10 | 9.35E-05 | 0.000349 | 0.00119 | 5.25E-05 | 0.000596807 | IVNS1ABP |
| chr12 | 47772252 | 47772933 | 682 | 3 | 9.36E-05 | 0.010305 | 0.00104 | 0.00157 | -0.000745383 | SLC48A1 |
| chr12 | 6866605 | 6867957 | 1353 | 17 | 9.37E-05 | 2.90E-05 | 0.00633 | 1.08E-05 | -0.00120967 | USP5, TPI1 |
| chr5 | 122915894 | 122916113 | 220 | 2 | 9.37E-05 | 1.08E-05 | 0.00013 | 1.17E-05 | -0.000515316 | SNX24 |
| chr2 | 238891079 | 238891627 | 549 | 5 | 9.38E-05 | 0.001258 | 0.00149 | 5.21E-05 | -0.00242901 | TWIST2 |
| chr2 | 152558513 | 152558815 | 303 | 2 | 9.39E-05 | 1.37E-05 | 0.00018 | 1.57E-05 | -0.019426664 | FMNL2 |
| chr2 | 594380 | 594509 | 130 | 2 | 9.40E-05 | 1.94E-05 | 0.00047 | 2.65E-05 | -0.007385472 | NA |
| chr5 | 1728328 | 1729141 | 814 | 4 | 9.42E-05 | 0.000857 | 7.68E-05 | 8.72E-05 | -0.001522847 | NA |
| chr4 | 864860 | 865441 | 582 | 3 | 9.42E-05 | 6.02E-05 | 0.00116 | 7.22E-05 | -0.001608767 | GAK |
| chr11 | 66491725 | 66492280 | 556 | 5 | 9.43E-05 | 1.84E-05 | 0.00046 | 1.14E-05 | -0.002285099 | DPP3 |
| chr10 | 78408889 | 78409115 | 227 | 3 | 9.43E-05 | 5.38E-06 | 0.00056 | 9.29E-06 | -0.024881202 | AC012560.1, LINC00595 |
| chr15 | 29113823 | 29113933 | 111 | 3 | 9.43E-05 | 0.000437 | 2.69E-05 | 2.82E-05 | -0.001578502 | APBA2 |
| chr1 | 220992251 | 220992617 | 367 | 3 | 9.43E-05 | 6.97E-06 | 0.0009 | 1.05E-05 | -0.008214138 | NA |
| chr16 | 9090242 | 9090628 | 387 | 3 | 9.43E-05 | 0.000352 | 0.00108 | 6.40E-05 | -0.008294493 | NA |
| chr18 | 58864657 | 58864727 | 71 | 2 | 9.44E-05 | 0.000187 | 0.00036 | 0.00013 | -0.016819183 | ZNF532 |
| chr11 | 132186662 | 132186992 | 331 | 2 | 9.44E-05 | 0.000523 | 4.69E-05 | 4.39E-05 | -0.014495676 | NTM |
| chr5 | 611896 | 612418 | 523 | 8 | 9.45E-05 | 0.000147 | 0.00134 | 1.67E-05 | 0.00076518 | CEP72, AC106772.1 |
| chr14 | 65412289 | 65412678 | 390 | 4 | 9.46E-05 | 0.000193 | 0.00043 | 3.30E-05 | -0.000729947 | FUT8, FUT8-AS1 |
| chr12 | 1798581 | 1799137 | 557 | 3 | 9.47E-05 | 0.000118 | 8.25E-05 | 3.82E-05 | -0.002038814 | CACNA2D4 |
| chr1 | 46450575 | 46450747 | 173 | 3 | 9.48E-05 | 0.000259 | 0.00011 | 2.65E-05 | -0.005440141 | FAAHP1, LINC01398 |
| chr2 | 95384978 | 95385695 | 718 | 3 | 9.52E-05 | 2.61E-06 | 0.00094 | 6.05E-06 | -0.006023025 | KCNIP3 |
| chr7 | 116210701 | 116210996 | 296 | 2 | 9.53E-05 | 7.11E-06 | 0.00023 | 9.93E-06 | 0.004056339 | TES, AC073130.3 |
| chr6 | 111703013 | 111703312 | 300 | 2 | 9.53E-05 | 5.39E-05 | 7.39E-05 | 2.55E-05 | -0.00416598 | FYN |
| chr14 | 105490324 | 105491285 | 962 | 11 | 9.55E-05 | 0.001261 | 0.00246 | 0.00029 | -0.000555273 | AL928654.3, TEDC1 |
| chr6 | 3054482 | 3054586 | 105 | 2 | 9.56E-05 | 5.20E-05 | 0.00224 | 8.08E-05 | -0.021533116 | NA |
| chr1 | 9910199 | 9911057 | 859 | 15 | 9.60E-05 | 6.02E-05 | 0.00474 | 1.31E-05 | -3.74E-06 | CTNNBIP1 |
| chr1 | 92792731 | 92792762 | 32 | 3 | 9.62E-05 | 3.51E-05 | 6.98E-05 | 1.49E-05 | -0.0073791 | NA |
| chr10 | 78247466 | 78248724 | 1259 | 6 | 9.62E-05 | 7.61E-05 | 3.82E-05 | 1.62E-06 | -0.004122622 | AC012560.1, LINC00595 |
| chr13 | 20128783 | 20129250 | 468 | 5 | 9.63E-05 | 0.000183 | 0.00126 | 3.20E-05 | 0.010439699 | NA |
| chr18 | 11179066 | 11179361 | 296 | 2 | 9.63E-05 | 1.02E-05 | 0.00018 | 1.25E-05 | -0.010346648 | NA |
| chr3 | 127598731 | 127599301 | 571 | 3 | 9.64E-05 | 0.008119 | 0.00014 | 9.89E-05 | -0.006015736 | MCM2 |
| chr9 | 105354843 | 105355211 | 369 | 2 | 9.64E-05 | 5.56E-06 | 0.00039 | 8.84E-06 | -0.008824886 | SLC44A1 |
| chr4 | 105057495 | 105057863 | 369 | 2 | 9.64E-05 | 1.06E-05 | 0.0002 | 1.33E-05 | 0.014963531 | AC096577.1 |
| chr3 | 58149168 | 58149904 | 737 | 3 | 9.65E-05 | 0.009551 | 5.72E-05 | 0.00012 | -0.001319297 | FLNB |
| chr16 | 672515 | 673047 | 533 | 4 | 9.65E-05 | 7.70E-05 | 0.00015 | 3.29E-05 | -0.002650853 | RHOT2 |
| chr13 | 29023310 | 29023961 | 652 | 5 | 9.65E-05 | 8.26E-06 | 0.00206 | 1.14E-05 | -0.007504431 | MTUS2 |
| chr5 | 1325626 | 1326256 | 631 | 4 | 9.67E-05 | 0.001352 | 4.36E-05 | 7.46E-05 | -0.002862499 | CLPTM1L |
| chr19 | 30668666 | 30668829 | 164 | 2 | 9.67E-05 | 0.000107 | 9.61E-05 | 4.46E-05 | -0.011918894 | ZNF536 |
| chr15 | 37396696 | 37397044 | 349 | 3 | 9.67E-05 | 9.17E-05 | 0.00063 | 2.00E-05 | 0.001709714 | AC068875.1 |
| chr1 | 41022268 | 41023158 | 891 | 5 | 9.67E-05 | 1.17E-05 | 0.01416 | 4.27E-05 | -0.003267912 | SLFNL1-AS1, SLFNL1 |
| chr11 | 409149 | 410307 | 1159 | 7 | 9.68E-05 | 1.54E-05 | 0.00091 | 2.40E-06 | -0.00578888 | SIGIRR |
| chr11 | 61892193 | 61892718 | 526 | 5 | 9.70E-05 | 2.45E-05 | 0.0014 | 1.43E-05 | -0.007287773 | NA |
| chr7 | 73842030 | 73842252 | 223 | 4 | 9.71E-05 | 0.00052 | 0.0006 | 7.59E-05 | -0.013064707 | METTL27 |
| chr11 | 70422398 | 70422881 | 484 | 3 | 9.72E-05 | 1.55E-05 | 0.00053 | 2.17E-05 | -0.012205748 | CTTN |
| chr12 | 5086285 | 5086385 | 101 | 2 | 9.73E-05 | 0.000657 | 1.99E-05 | 2.01E-05 | -0.003925594 | NA |
| chr17 | 79787865 | 79788660 | 796 | 2 | 9.73E-05 | 3.80E-06 | 0.00032 | 6.18E-06 | -0.016038335 | CBX2 |
| chr5 | 179619113 | 179619120 | 8 | 2 | 9.73E-05 | 1.06E-05 | 0.00058 | 1.65E-05 | -0.021623049 | HNRNPH1 |
| chr20 | 32457172 | 32457523 | 352 | 2 | 9.75E-05 | 1.86E-05 | 0.00088 | 2.85E-05 | -0.023806778 | NOL4L |
| chr12 | 20369302 | 20370583 | 1282 | 9 | 9.77E-05 | 6.81E-05 | 0.00333 | 3.14E-05 | -0.00418943 | PDE3A, AC129102.1 |
| chr22 | 25199385 | 25200343 | 959 | 9 | 9.77E-05 | 1.89E-05 | 0.00155 | 1.21E-05 | -0.006497131 | CRYBB3 |
| chr19 | 41844264 | 41844737 | 474 | 9 | 9.77E-05 | 0.001648 | 0.00018 | 7.45E-05 | -0.002704462 | LYPD4 |
| chr3 | 196988374 | 196988761 | 388 | 3 | 9.82E-05 | 9.32E-05 | 0.00061 | 1.96E-05 | -0.003376089 | MELTF |

| | | | | | | | | | | |
|---|---|---|---|---|---|---|---|---|---|---|
| chr4 | 3441088 | 3441898 | 811 | 7 | 9.82E-05 | 0.001532 | 0.00057 | 0.00014 | -0.006607311 | NA |
| chr7 | 121313795 | 121314495 | 701 | 7 | 9.83E-05 | 6.86E-06 | 0.0095 | 8.30E-06 | -0.013800011 | NA |
| chr6 | 111631863 | 111632453 | 591 | 2 | 9.84E-05 | 2.58E-06 | 0.00038 | 4.49E-06 | -0.029132707 | NA |
| chr1 | 32272662 | 32274151 | 1490 | 5 | 9.84E-05 | 2.48E-05 | 0.00205 | 7.18E-06 | -0.007418844 | LCK |
| chr13 | 32117385 | 32117888 | 504 | 2 | 9.85E-05 | 1.68E-05 | 0.00016 | 1.79E-05 | -0.012388832 | FRY |
| chr22 | 46408629 | 46409063 | 435 | 3 | 9.87E-05 | 0.00055 | 5.12E-05 | 3.32E-05 | -0.00327155 | CELSR1 |
| chr10 | 112447163 | 112447731 | 569 | 5 | 9.87E-05 | 2.81E-05 | 0.00592 | 5.79E-05 | 0.001737754 | VTI1A |
| chr14 | 69994033 | 69994203 | 171 | 2 | 9.89E-05 | 1.31E-05 | 0.00017 | 1.49E-05 | -0.026165247 | SMOC1 |
| chr5 | 140785353 | 140785476 | 124 | 3 | 9.92E-05 | 0.005245 | 0.00013 | 7.01E-05 | 0.009341736 | NA |
| chr3 | 32780145 | 32780438 | 294 | 3 | 9.92E-05 | 0.001942 | 0.00029 | 7.85E-05 | -0.002095559 | NA |
| chr11 | 72849081 | 72849279 | 199 | 2 | 9.92E-05 | 0.001026 | 2.57E-05 | 2.80E-05 | -0.007088277 | FCHSD2 |
| chr3 | 14305834 | 14306122 | 289 | 3 | 9.92E-05 | 2.30E-05 | 0.00018 | 1.84E-05 | -0.013476904 | NA |
| chr3 | 195104281 | 195104586 | 306 | 2 | 9.92E-05 | 0.034984 | 5.54E-05 | 9.82E-05 | -0.012282682 | XXYLT1 |
| chr7 | 105680645 | 105681008 | 364 | 2 | 9.92E-05 | 2.70E-05 | 0.00015 | 2.42E-05 | -0.007333741 | ATXN7L1 |
| chr11 | 1485706 | 1486117 | 412 | 5 | 9.94E-05 | 7.00E-05 | 0.00015 | 2.50E-05 | -0.005254703 | MOB2 |
| chr12 | 57615924 | 57616929 | 1006 | 4 | 9.94E-05 | 0.001323 | 9.89E-05 | 3.56E-05 | -0.004085915 | ARHGEF25, AC025165.6, AC025165.1 |
| chr16 | 9092200 | 9093165 | 966 | 5 | 9.96E-05 | 1.09E-05 | 0.00634 | 3.03E-05 | 0.001385049 | C16orf72 |
| chr6 | 25925822 | 25926205 | 384 | 3 | 9.97E-05 | 6.65E-06 | 0.00086 | 8.75E-06 | -0.007467194 | SLC17A2 |
| chr10 | 17201827 | 17202394 | 568 | 5 | 9.97E-05 | 4.77E-05 | 0.00049 | 1.78E-05 | -0.004374598 | TRDMT1 |
| chr2 | 172021445 | 172022232 | 788 | 4 | 9.98E-05 | 1.27E-05 | 0.00097 | 5.81E-06 | -0.008351117 | METAP1D |
| chr16 | 87959377 | 87959942 | 566 | 6 | 9.98E-05 | 4.62E-05 | 0.01375 | 3.96E-05 | -0.013445225 | BANP |
| chr3 | 188946843 | 188947677 | 835 | 11 | 1.00E-04 | 9.24E-05 | 0.00031 | 2.84E-05 | -0.004580195 | TPRG1, TPRG1-AS1 |
| chr7 | 1545816 | 1546135 | 320 | 3 | 0.000100445 | 0.000102 | 0.00367 | 0.00013 | -0.009089579 | TMEM184A |
| chr7 | 530005 | 531160 | 1156 | 6 | 0.00010052 | 5.57E-05 | 0.0018 | 1.84E-05 | -0.008652594 | NA |
| chr8 | 119873120 | 119874210 | 1091 | 12 | 0.00010054 | 0.000348 | 0.00152 | 1.58E-05 | -0.000647092 | DEPTOR, AP005717.1 |
| chr9 | 35073749 | 35074100 | 352 | 4 | 0.000100574 | 0.002027 | 0.0001 | 7.97E-05 | -0.004465291 | VCP, FANCG |
| chr16 | 53510911 | 53511743 | 833 | 3 | 0.000100731 | 2.34E-06 | 0.00104 | 5.44E-06 | -0.000554582 | NA |
| chr10 | 102242189 | 102242238 | 50 | 2 | 0.000100885 | 7.40E-05 | 0.00028 | 6.32E-05 | -0.009113108 | NA |
| chr17 | 72394632 | 72394734 | 103 | 2 | 0.000101026 | 8.88E-06 | 0.00073 | 1.46E-05 | -0.021234271 | LINC00511 |
| chr4 | 145870115 | 145870550 | 436 | 2 | 0.000101116 | 2.27E-05 | 0.00065 | 3.22E-05 | -0.017027151 | ZNF827 |
| chr5 | 141431159 | 141432075 | 917 | 9 | 0.000101242 | 0.000357 | 0.00077 | 0.00013 | 0.010693976 | PCDHGA1, PCDHGA2, PCDHGA3, PCDHGB1, PCDHGA4, PCDHGB2, PCDHGA5, PCDHGB3, PCDHGA6, PCDHGA7, PCDHGB4, PCDHGA8, PCDHGB5, PCDHGA9, PCDHGB6, PCDHGA10, PCDHGB7, PCDHGA11, PCDHGA12 |
| chr4 | 76087028 | 76087367 | 340 | 2 | 0.000101306 | 6.82E-05 | 6.10E-05 | 2.52E-05 | -0.020097879 | ART3 |
| chr21 | 42309547 | 42309649 | 103 | 2 | 0.000101316 | 1.79E-05 | 0.0006 | 2.61E-05 | -0.011075344 | NA |
| chr1 | 3708748 | 3708860 | 113 | 3 | 0.00010155 | 4.06E-06 | 0.00266 | 1.08E-05 | 0.019729506 | TP73 |
| chr17 | 65074693 | 65074928 | 236 | 2 | 0.000101605 | 3.72E-05 | 5.95E-05 | 1.77E-05 | -0.014615161 | NA |
| chr5 | 1473474 | 1473959 | 486 | 4 | 0.000101621 | 6.04E-05 | 0.0018 | 7.60E-05 | 0.000303632 | LPCAT1 |
| chr15 | 68256840 | 68257394 | 555 | 6 | 0.000101632 | 3.70E-05 | 0.00341 | 1.87E-05 | -0.004215664 | CLN6 |
| chr2 | 191835991 | 191836788 | 798 | 4 | 0.000101694 | 3.74E-06 | 0.00149 | 6.31E-06 | -0.003719099 | CAVIN2 |
| chr10 | 73248263 | 73248593 | 331 | 4 | 0.000101694 | 4.28E-05 | 0.00305 | 6.37E-05 | -0.002694716 | DNAJC9-AS1, AC016394.2 |
| chr17 | 63440805 | 63441601 | 797 | 8 | 0.000102106 | 0.002317 | 0.00021 | 2.65E-05 | -0.002116628 | CYB561 |
| chr1 | 7975245 | 7975522 | 278 | 2 | 0.000102126 | 0.000748 | 3.25E-05 | 3.38E-05 | -0.004346481 | PARK7 |
| chr6 | 53293725 | 53294180 | 456 | 2 | 0.000102152 | 2.35E-05 | 0.00013 | 2.07E-05 | -0.008922754 | ELOVL5 |
| chr16 | 66469102 | 66469781 | 680 | 5 | 0.00010218 | 0.000255 | 0.0014 | 3.29E-05 | -0.001987477 | BEAN1 |
| chr15 | 74208976 | 74209430 | 455 | 6 | 0.000102294 | 0.000138 | 0.00103 | 2.32E-05 | -0.008055957 | CCDC33, STRA6 |
| chr1 | 42152370 | 42152835 | 466 | 5 | 0.000102337 | 0.000263 | 0.00165 | 7.58E-05 | -0.003791799 | NA |
| chr10 | 93756653 | 93757706 | 1054 | 7 | 0.000102365 | 1.80E-05 | 0.00273 | 2.36E-05 | -0.003351634 | NA |
| chr7 | 42931744 | 42932391 | 648 | 12 | 0.000102401 | 0.00014 | 0.00966 | 1.95E-05 | 0.000347913 | MRPL32, AC010132.3, PSMA2 |
| chr1 | 89994441 | 89995390 | 950 | 10 | 0.000102414 | 0.001941 | 0.00013 | 6.25E-05 | 0.000639624 | ZNF326 |
| chr8 | 116852267 | 116852560 | 294 | 2 | 0.000102527 | 6.91E-05 | 0.00015 | 4.58E-05 | -0.007455881 | RAD21 |
| chr1 | 919538 | 920269 | 732 | 9 | 0.000102579 | 0.000126 | 0.00298 | 7.10E-05 | -0.003448875 | LINC02593 |
| chr15 | 78265777 | 78266117 | 341 | 4 | 0.000102651 | 0.000582 | 9.45E-05 | 5.16E-05 | -0.010466032 | DNAJA4 |
| chr16 | 30398587 | 30399108 | 522 | 3 | 0.000102667 | 6.34E-06 | 0.00182 | 1.44E-05 | -0.0093798 | ZNF48 |
| chr11 | 774038 | 774070 | 33 | 2 | 0.000102791 | 0.000213 | 4.23E-05 | 3.06E-05 | -0.006181363 | GATD1 |
| chr8 | 24441117 | 24441258 | 142 | 2 | 0.00010286 | 0.000668 | 4.96E-05 | 4.93E-05 | -0.005887167 | ADAM7, AC120193.1 |
| chr4 | 165209623 | 165209967 | 345 | 4 | 0.000102931 | 0.007747 | 0.00074 | 8.39E-05 | -0.003924032 | KLHL2 |
| chr19 | 16373703 | 16374238 | 536 | 3 | 0.000103084 | 0.002147 | 3.76E-05 | 5.38E-05 | -0.001507649 | EPS15L1 |
| chr7 | 158739003 | 158739061 | 59 | 3 | 0.000103153 | 0.000941 | 5.74E-05 | 4.12E-05 | -0.010083001 | ESYT2 |
| chr16 | 49859211 | 49859620 | 410 | 2 | 0.00010318 | 0.000368 | 0.00021 | 0.00014 | -0.007186459 | NA |
| chr2 | 201262787 | 201262819 | 33 | 2 | 0.000103205 | 5.43E-05 | 6.96E-05 | 2.46E-05 | 0.00587831 | CASP8 |
| chr1 | 24730553 | 24730924 | 372 | 3 | 0.000103401 | 3.93E-06 | 0.00149 | 8.48E-06 | -0.018566827 | NA |
| chr6 | 38595514 | 38595879 | 366 | 3 | 0.000103479 | 0.000105 | 5.69E-05 | 2.72E-05 | -0.001588763 | BTBD9 |
| chr1 | 97796788 | 97796852 | 65 | 2 | 0.00010352 | 3.18E-05 | 4.78E-05 | 1.38E-05 | -0.007677723 | DPYD |
| chr16 | 3654318 | 3655228 | 911 | 6 | 0.000103663 | 4.75E-05 | 0.00068 | 2.72E-05 | -0.005007983 | DNASE1, TRAP1 |
| chr9 | 133840276 | 133840819 | 544 | 2 | 0.000103663 | 1.80E-05 | 0.00029 | 2.23E-05 | -0.013645934 | VAV2 |
| chr1 | 225926711 | 225926865 | 155 | 2 | 0.000103727 | 0.000487 | 2.68E-05 | 2.51E-05 | -0.006508055 | NA |
| chr1 | 17440421 | 17440786 | 366 | 3 | 0.000103838 | 3.05E-05 | 0.00264 | 5.06E-05 | -0.012203017 | NA |
| chr17 | 58082089 | 58082474 | 386 | 3 | 0.000103853 | 0.004392 | 2.95E-05 | 5.76E-05 | -0.003197714 | AC015813.3 |
| chr3 | 49869981 | 49870399 | 419 | 8 | 0.000103992 | 6.33E-05 | 0.00808 | 5.82E-05 | -0.005158475 | NA |
| chr3 | 57959550 | 57959968 | 419 | 3 | 0.000104029 | 5.04E-06 | 0.00104 | 1.05E-05 | 0.031754615 | NA |
| chr17 | 2361672 | 2363703 | 2032 | 10 | 0.000104042 | 3.42E-05 | 5.51E-05 | 5.99E-07 | -0.003364925 | SGSM2 |
| chr11 | 2591551 | 2591646 | 96 | 4 | 0.000104057 | 3.41E-05 | 0.00069 | 1.19E-05 | -0.004156632 | KCNQ1 |
| chr1 | 3184864 | 3186542 | 1679 | 6 | 0.000104071 | 6.62E-08 | 0.00117 | 1.99E-07 | -0.003703282 | PRDM16 |
| chr7 | 157500844 | 157500912 | 69 | 3 | 0.000104071 | 3.40E-05 | 0.00114 | 2.78E-05 | -0.001680599 | NA |
| chr2 | 708656 | 708791 | 136 | 2 | 0.000104197 | 1.20E-05 | 0.00071 | 1.88E-05 | -0.007972103 | NA |
| chr9 | 137315420 | 137315646 | 227 | 2 | 0.000104322 | 9.20E-05 | 0.00164 | 0.00013 | -0.007362651 | EXD3 |
| chr4 | 184097325 | 184097963 | 639 | 3 | 0.000104383 | 0.000521 | 0.00055 | 0.00027 | -0.012832855 | ENPP6 |
| chr9 | 95126989 | 95127785 | 797 | 7 | 0.000104477 | 9.68E-05 | 0.00027 | 1.65E-05 | -0.008946188 | FANCC |
| chr2 | 127608749 | 127609019 | 271 | 3 | 0.000104574 | 9.59E-06 | 0.00084 | 1.58E-05 | -0.013389035 | MYO7B |
| chr1 | 201829038 | 201829331 | 294 | 7 | 0.000104623 | 0.000584 | 0.00048 | 3.09E-05 | -2.25E-05 | IPO9, IPO9-AS1 |
| chr7 | 95485368 | 95486042 | 675 | 10 | 0.000104674 | 7.69E-05 | 0.00268 | 1.03E-05 | -0.002828271 | ASB4 |
| chr2 | 43229348 | 43229700 | 353 | 3 | 0.000104675 | 0.000558 | 7.37E-05 | 6.77E-05 | -0.005773367 | AC010883.1 |
| chr11 | 111976592 | 111977440 | 849 | 8 | 0.000104677 | 0.003544 | 0.00157 | 0.0002 | -0.004405369 | DIXDC1 |
| chr16 | 31488560 | 31489338 | 779 | 6 | 0.000104694 | 3.90E-05 | 0.00852 | 2.51E-05 | -0.008569475 | SLC5A2 |
| chr3 | 127604807 | 127605122 | 316 | 3 | 0.000104777 | 0.000818 | 6.03E-05 | 7.18E-05 | -0.005686942 | MCM2 |
| chr5 | 1752553 | 1752749 | 197 | 2 | 0.000104845 | 0.004293 | 0.0001 | 0.00018 | -0.005317247 | NA |
| chr15 | 31023992 | 31024657 | 666 | 4 | 0.000104881 | 4.63E-06 | 0.00185 | 7.64E-06 | -0.007242909 | AC009562.1, TRPM1 |
| chr2 | 204546801 | 204547063 | 263 | 2 | 0.000104903 | 3.20E-05 | 0.00172 | 5.00E-05 | -0.008690497 | PARD3B |
| chr6 | 7312607 | 7313949 | 1343 | 9 | 0.000105027 | 0.001214 | 0.00061 | 6.59E-06 | -9.81E-05 | SSR1 |
| chr9 | 34457131 | 34457900 | 770 | 5 | 0.000105031 | 5.50E-05 | 0.00267 | 2.61E-05 | -0.005501244 | DNAI1, FAM219A |
| chr20 | 41037825 | 41038125 | 301 | 2 | 0.000105162 | 1.07E-05 | 0.00021 | 1.35E-05 | -0.005086969 | TOP1 |
| chr4 | 2486706 | 2486854 | 149 | 2 | 0.000105183 | 2.10E-05 | 7.20E-05 | 1.40E-05 | -0.01430024 | RNF4 |
| chr21 | 36384099 | 36384807 | 709 | 4 | 0.000105196 | 0.000118 | 0.00013 | 3.81E-05 | 0.000173142 | MORC3 |
| chr7 | 73683932 | 73684288 | 357 | 4 | 0.000105234 | 4.45E-05 | 0.00206 | 4.21E-05 | 0.005891813 | BUD23 |
| chr10 | 125104031 | 125104184 | 154 | 4 | 0.000105276 | 1.26E-05 | 0.00143 | 1.02E-05 | -0.0054978 | CTBP2 |
| chr2 | 177212241 | 177212596 | 356 | 4 | 0.000105391 | 0.000155 | 0.00094 | 4.23E-05 | 0.001569749 | HNRNPA3, AC079305.2 |
| chr14 | 104774735 | 104775855 | 1121 | 7 | 0.000105535 | 0.000147 | 0.00046 | 1.51E-05 | -0.003406825 | AKT1 |
| chr17 | 81839162 | 81839497 | 336 | 4 | 0.000105535 | 1.03E-05 | 0.00052 | 1.19E-05 | -0.007718497 | NA |
| chr15 | 90878913 | 90878939 | 27 | 2 | 0.000105548 | 0.002272 | 5.80E-05 | 7.68E-05 | -0.003286814 | FURIN |
| chr6 | 91816521 | 91816598 | 78 | 3 | 0.00010558 | 2.31E-05 | 7.63E-05 | 1.25E-05 | -0.004928216 | NA |
| chr11 | 57481911 | 57482731 | 821 | 4 | 0.000105607 | 0.000107 | 0.0002 | 3.72E-05 | -0.007614596 | NA |
| chr19 | 45788217 | 45788494 | 278 | 2 | 0.000105607 | 0.000144 | 9.99E-05 | 5.30E-05 | -0.003266693 | DMWD |
| chr12 | 50842276 | 50842985 | 710 | 10 | 0.000105649 | 0.00024 | 0.00141 | 4.09E-05 | -0.006298545 | TMPRSS12 |
| chr17 | 36575208 | 36575210 | 3 | 2 | 0.000105801 | 0.000994 | 2.83E-05 | 3.11E-05 | -0.008617014 | GGNBP2, AC243732.1 |
| chr3 | 9750456 | 9750986 | 531 | 3 | 0.00010588 | 0.000194 | 0.00509 | 0.00037 | -0.003528911 | OGG1 |
| chr8 | 123166496 | 123166503 | 8 | 2 | 0.000105933 | 1.16E-05 | 0.0008 | 1.85E-05 | -0.026819268 | NA |
| chr20 | 19798663 | 19799513 | 851 | 4 | 0.00010604 | 1.38E-05 | 0.00092 | 1.09E-05 | -0.004493321 | RIN2 |
| chr2 | 216774483 | 216774809 | 327 | 2 | 0.000106141 | 9.55E-05 | 5.48E-05 | 2.76E-05 | -0.009268105 | AC007563.2 |
| chr14 | 20723595 | 20724067 | 473 | 3 | 0.000106278 | 1.16E-05 | 0.00032 | 1.52E-05 | -0.024908557 | NA |
| chr16 | 2512449 | 2513035 | 587 | 2 | 0.00010635 | 1.84E-05 | 0.00088 | 2.84E-05 | -0.010272457 | AC093525.2 |
| chr11 | 72088229 | 72088569 | 341 | 3 | 0.000106408 | 4.66E-06 | 0.00252 | 1.18E-05 | -0.007769152 | AP000812.4, LRTOMT, LAMTOR1 |
| chr5 | 160312694 | 160313434 | 741 | 7 | 0.000106408 | 8.58E-05 | 0.0018 | 3.66E-05 | 0.002793828 | CCNJL |
| chr3 | 38003186 | 38003628 | 443 | 2 | 0.000106408 | 2.84E-05 | 0.0005 | 3.66E-05 | -0.008231019 | VILL |
| chr10 | 122220615 | 122220993 | 379 | 2 | 0.000106408 | 3.75E-05 | 9.51E-05 | 2.40E-05 | -0.011618761 | TACC2 |
| chr7 | 36303223 | 36303242 | 20 | 2 | 0.000106532 | 8.27E-06 | 0.00055 | 1.32E-05 | -0.02308136 | NA |
| chr16 | 69924395 | 69925521 | 1127 | 7 | 0.000106654 | 3.48E-06 | 0.00107 | 7.80E-06 | -0.003925966 | WWP2 |
| chr19 | 43465498 | 43465896 | 399 | 6 | 0.000106705 | 9.33E-05 | 0.00019 | 2.87E-05 | -0.0092055 | LYPD3 |
| chr21 | 36330926 | 36331119 | 194 | 2 | 0.000106765 | 0.000808 | 2.69E-05 | 2.84E-05 | -0.00751319 | MORC3 |
| chr2 | 56182639 | 56183541 | 903 | 12 | 0.000106796 | 0.00606 | 0.00341 | 0.00016 | -0.001980159 | AC007744.1, AC007743.1 |
| chr4 | 3371188 | 3372144 | 957 | 7 | 0.000106887 | 0.000228 | 0.00152 | 3.25E-05 | -0.006204106 | RGS12 |
| chr17 | 81330388 | 81331001 | 614 | 14 | 0.000106944 | 1.87E-05 | 0.00027 | 1.49E-05 | -0.002870122 | TMEM105 |
| chr10 | 129873109 | 129873496 | 388 | 4 | 0.000107006 | 1.98E-05 | 0.00191 | 2.44E-05 | -0.007121024 | EBF3 |
| chr2 | 239080360 | 239081478 | 1119 | 6 | 0.000107009 | 0.00024 | 0.00059 | 6.72E-05 | -0.00260989 | HDAC4 |
| chr16 | 22321943 | 22321994 | 52 | 2 | 0.000107011 | 5.07E-05 | 4.34E-05 | 1.67E-05 | -0.004978799 | POLR3E |
| chr20 | 63374037 | 63374159 | 123 | 3 | 0.000107011 | 0.000112 | 0.00104 | 3.95E-05 | 0.004159623 | CHRNA4 |
| chr2 | 24076306 | 24076776 | 471 | 8 | 0.000107078 | 4.34E-05 | 0.0026 | 2.29E-05 | -0.000551424 | FAM228B, SF3B6 |
| chr10 | 110007083 | 110007121 | 39 | 2 | 0.000107085 | 0.000212 | 0.00278 | 0.00032 | -0.016948147 | ADD3, ADD3-AS1 |
| chr14 | 59483998 | 59484911 | 914 | 14 | 0.000107111 | 8.42E-06 | 0.01183 | 7.59E-06 | 0.00086591 | JKAMP, L3HYPDH, AL121694.1 |
| chr11 | 112322239 | 112322795 | 557 | 3 | 0.000107226 | 3.68E-06 | 0.00232 | 9.62E-06 | -0.002015803 | LINC02762 |
| chr12 | 4813798 | 4814363 | 566 | 6 | 0.000107269 | 0.001168 | 0.0001 | 4.31E-05 | -0.003643244 | GALNT8, AC005833.2 |
| chr2 | 206785753 | 206786113 | 361 | 2 | 0.000107269 | 0.001814 | 4.11E-05 | 5.05E-05 | -0.010916646 | FASTKD2 |
| chr1 | 116133482 | 116133851 | 370 | 2 | 0.000107383 | 5.73E-05 | 9.48E-05 | 3.12E-05 | -0.008028076 | MAB21L3 |
| chr20 | 51432731 | 51432861 | 131 | 2 | 0.000107476 | 1.89E-05 | 0.00028 | 2.29E-05 | -0.004647872 | NFATC2 |
| chr1 | 207454147 | 207455038 | 892 | 11 | 0.000107494 | 0.006425 | 0.00037 | 0.00021 | 0.001124574 | CR2 |
| chr4 | 94253065 | 94253424 | 360 | 2 | 0.000107595 | 1.68E-05 | 0.00037 | 2.23E-05 | -0.00093065 | SMARCAD1 |
| chr7 | 44115567 | 44115965 | 399 | 2 | 0.000107595 | 2.14E-05 | 0.0002 | 2.28E-05 | -0.007624361 | POLD2 |
| chr5 | 134436592 | 134437451 | 860 | 5 | 0.000107637 | 9.77E-06 | 0.00081 | 5.49E-06 | -0.008864208 | AC005355.1, AC109454.3 |
| chr1 | 162548292 | 162548368 | 77 | 2 | 0.000107674 | 0.000432 | 2.37E-05 | 2.16E-05 | -0.009056413 | NA |
| chr2 | 118839839 | 118840010 | 172 | 2 | 0.000108108 | 3.67E-05 | 6.36E-05 | 1.83E-05 | -0.021328019 | NA |
| chr3 | 119279306 | 119280063 | 758 | 3 | 0.00010811 | 1.55E-05 | 0.00011 | 7.19E-06 | -0.007499254 | B4GALT4-AS1 |
| chr5 | 6714424 | 6714974 | 551 | 8 | 0.000108112 | 0.026968 | 1.65E-05 | 0.00017 | -0.001633209 | TENT4A |
| chr16 | 660674 | 661611 | 938 | 5 | 0.00010819 | 0.000103 | 0.00173 | 0.00012 | 0.003866578 | WDR90 |
| chr7 | 1692592 | 1692642 | 51 | 3 | 0.00010819 | 0.005217 | 1.97E-05 | 4.50E-05 | -0.001441138 | ELFN1 |
| chr1 | 156858410 | 156859216 | 807 | 9 | 0.000108253 | 5.22E-06 | 0.00922 | 1.43E-05 | -0.007595666 | NTRK1, INSRR |
| chr11 | 46027112 | 46027422 | 311 | 2 | 0.000108277 | 0.002111 | 3.55E-05 | 4.42E-05 | -0.006547519 | PHF21A |
| chr6 | 101393282 | 101394028 | 747 | 4 | 0.0001083 | 0.000265 | 0.00168 | 7.72E-05 | -0.001965179 | GRIK2 |
| chr13 | 99605911 | 99606293 | 383 | 3 | 0.000108308 | 0.001234 | 0.00127 | 0.00083 | -0.010074873 | NA |
| chr20 | 6736335 | 6736636 | 302 | 2 | 0.000108323 | 0.000934 | 7.19E-05 | 7.80E-05 | 0.009698455 | NA |
| chr1 | 181190538 | 181191005 | 468 | 3 | 0.000108343 | 8.62E-05 | 0.00011 | 3.77E-05 | -0.009008438 | AL358393.1 |
| chr1 | 19436272 | 19436864 | 593 | 3 | 0.000108348 | 0.000508 | 5.84E-05 | 4.63E-05 | -0.006490107 | CAPZB |
| chr3 | 114178703 | 114178899 | 197 | 3 | 0.000108471 | 0.000136 | 6.44E-05 | 3.04E-05 | 0.005376913 | DRD3 |
| chr7 | 157937470 | 157937572 | 103 | 2 | 0.000108608 | 7.79E-06 | 0.00026 | 1.10E-05 | -0.004667825 | PTPRN2 |
| chr1 | 228306569 | 228307410 | 842 | 5 | 0.000108609 | 1.02E-05 | 0.00109 | 7.84E-06 | -0.008085289 | OBSCN |
| chr17 | 63930372 | 63930581 | 210 | 3 | 0.000108731 | 2.86E-05 | 0.00023 | 2.54E-05 | -0.006355456 | AC127029.3, CD79B |
| chr12 | 64639171 | 64639556 | 386 | 3 | 0.000108739 | 0.000168 | 5.69E-05 | 3.30E-05 | -0.000452638 | RASSF3 |
| chr11 | 68215075 | 68215236 | 162 | 2 | 0.000108844 | 1.63E-05 | 0.00113 | 2.63E-05 | -0.014285187 | NA |
| chr2 | 208186637 | 208187354 | 718 | 2 | 0.000108881 | 5.82E-06 | 0.00056 | 9.64E-06 | -0.011779567 | C2orf80 |
| chr19 | 16495667 | 16496285 | 619 | 10 | 0.000108894 | 3.74E-05 | 0.00789 | 2.93E-05 | -0.005984937 | CALR3, AC008764.1 |
| chr1 | 201896868 | 201897225 | 358 | 3 | 0.000108944 | 0.000182 | 0.00076 | 4.49E-05 | -0.003711489 | AL513217.1, LMOD1 |
| chr1 | 206469611 | 206470034 | 424 | 4 | 0.000109053 | 0.001285 | 0.001 | 7.23E-05 | -0.007264481 | NA |
| chr12 | 12356373 | 12358289 | 1917 | 19 | 0.000109073 | 3.38E-05 | 0.00036 | 3.41E-06 | -0.001398497 | BORCS5, LOH12CR2 |
| chr2 | 240461109 | 240461357 | 249 | 2 | 0.000109287 | 0.003343 | 5.72E-05 | 8.08E-05 | -0.005337038 | GPC1 |
| chr19 | 43634698 | 43635171 | 474 | 2 | 0.00010932 | 1.49E-05 | 0.0002 | 1.73E-05 | -0.01299796 | CADM4 |

| | | | | | | | | | | |
|---|---|---|---|---|---|---|---|---|---|---|
| chr11 | 117478633 | 117478963 | 331 | 2 | 0.000109382 | 2.70E-05 | 0.0001 | 2.00E-05 | -0.00759352 | DSCAML1 |
| chr4 | 139013683 | 139014034 | 352 | 2 | 0.000109453 | 9.03E-05 | 6.93E-05 | 3.22E-05 | -0.006727495 | NA |
| chr14 | 101628593 | 101628802 | 210 | 3 | 0.000109502 | 1.48E-05 | 0.00089 | 2.40E-05 | -0.004595402 | NA |
| chr9 | 114524630 | 114525434 | 805 | 3 | 0.000109734 | 8.01E-05 | 0.0001 | 6.43E-06 | -0.011189173 | NA |
| chr6 | 73299098 | 73299318 | 221 | 2 | 0.000109737 | 4.53E-05 | 0.00144 | 6.67E-05 | -0.013829873 | AC019205.1, KHDC1 |
| chr21 | 41819839 | 41820203 | 365 | 4 | 0.000109784 | 0.000955 | 0.00014 | 5.24E-05 | -0.002381696 | PRDM15 |
| chr9 | 96047946 | 96048249 | 304 | 2 | 0.000109833 | 1.00E-05 | 0.00045 | 1.52E-05 | -0.014328274 | ERCC6L2 |
| chr9 | 35739294 | 35740121 | 828 | 3 | 0.000109903 | 3.39E-06 | 0.00075 | 6.54E-06 | -0.00083603 | GBA2 |
| chr15 | 78088000 | 78088742 | 743 | 2 | 0.000109911 | 3.19E-06 | 0.00024 | 5.09E-06 | -0.006339222 | SH2D7 |
| chr2 | 73079510 | 73079681 | 172 | 2 | 0.000109938 | 1.64E-05 | 0.00024 | 1.99E-05 | -0.006657719 | RAB11FIP5 |
| chr19 | 1420405 | 1420631 | 227 | 2 | 0.000109961 | 1.69E-05 | 0.00106 | 2.69E-05 | -0.00416238 | DAZAP1 |
| chr15 | 60651337 | 60652811 | 1475 | 4 | 0.000110197 | 1.14E-07 | 0.00023 | 2.00E-07 | -0.005391794 | AC009560.4, RORA |
| chr20 | 1423591 | 1424068 | 478 | 3 | 0.00011026 | 2.42E-05 | 0.00014 | 1.54E-05 | -0.005596878 | NA |
| chr11 | 65040609 | 65041166 | 558 | 10 | 0.00011036 | 0.000124 | 0.00191 | 2.50E-05 | -0.001618101 | SAC3D1 |
| chr19 | 37551516 | 37551883 | 368 | 4 | 0.000110449 | 7.32E-05 | 0.00031 | 3.57E-05 | 0.008343099 | ZNF571-AS1, ZNF540 |
| chr16 | 89737653 | 89737867 | 215 | 2 | 0.000110585 | 2.81E-05 | 0.00014 | 2.42E-05 | -0.009418422 | ZNF276, FANCA |
| chr9 | 137368597 | 137369163 | 567 | 4 | 0.000110745 | 0.00159 | 0.0001 | 6.28E-05 | -0.003658213 | EXD3 |
| chr3 | 127287444 | 127287762 | 319 | 3 | 0.000110863 | 0.000584 | 9.35E-05 | 6.51E-05 | -0.002595668 | PRR20G |
| chr2 | 96987513 | 96987702 | 190 | 3 | 0.000110922 | 0.000304 | 6.76E-05 | 3.69E-05 | -0.005260018 | NA |
| chr9 | 135352018 | 135353618 | 1601 | 5 | 0.000110935 | 4.83E-07 | 0.00059 | 2.86E-07 | -0.004379107 | AL353615.1 |
| chr17 | 80743084 | 80743332 | 249 | 3 | 0.000110977 | 1.89E-05 | 0.00029 | 1.37E-05 | -0.013530983 | RPTOR |
| chr5 | 125003062 | 125003952 | 891 | 2 | 0.000111044 | 8.30E-07 | 0.00016 | 1.44E-06 | -0.017250915 | LINC02240 |
| chr11 | 597295 | 598325 | 1031 | 5 | 0.000111143 | 0.000911 | 0.00018 | 9.04E-05 | -0.00270029 | PHRF1 |
| chr12 | 94602900 | 94603522 | 623 | 2 | 0.000111149 | 6.13E-05 | 0.00013 | 3.95E-05 | -0.014279529 | TMCC3 |
| chr18 | 80107989 | 80108256 | 268 | 3 | 0.000111349 | 0.000123 | 0.00022 | 2.38E-05 | -0.007307855 | NA |
| chr14 | 102038514 | 102039283 | 770 | 5 | 0.000111376 | 5.94E-05 | 0.00033 | 2.64E-05 | -0.002113187 | DYNC1H1, AL118558.1 |
| chr20 | 58905189 | 58905443 | 255 | 3 | 0.000111477 | 9.74E-05 | 0.00066 | 4.35E-05 | 0.000114857 | GNAS |
| chr5 | 171308182 | 171308969 | 788 | 6 | 0.000111499 | 5.12E-05 | 0.00308 | 3.93E-05 | 0.005499378 | AC091980.2 |
| chr8 | 132106144 | 132106608 | 465 | 3 | 0.000111507 | 1.10E-05 | 0.00121 | 2.08E-05 | 0.003806768 | HHLA1 |
| chr21 | 28992574 | 28993426 | 853 | 10 | 0.000111538 | 5.92E-05 | 0.00225 | 3.70E-05 | -0.002009701 | LTN1 |
| chr6 | 230908 | 230959 | 52 | 3 | 0.000111596 | 0.000454 | 5.39E-05 | 3.68E-05 | -0.002060304 | NA |
| chr2 | 11634016 | 11634348 | 333 | 5 | 0.000111633 | 7.91E-05 | 0.00018 | 1.74E-05 | -0.003932399 | GREB1 |
| chr13 | 51817059 | 51818015 | 957 | 4 | 0.000111733 | 1.20E-05 | 0.00043 | 1.56E-05 | -0.005279363 | AL162377.3, TMEM272 |
| chr17 | 58219357 | 58220154 | 798 | 14 | 0.000111779 | 0.000202 | 0.01007 | 4.03E-05 | -0.004887851 | LPO, AC005962.2, MKS1 |
| chr5 | 39365288 | 39365631 | 344 | 3 | 0.000111884 | 1.51E-05 | 0.00088 | 1.69E-05 | -0.009626599 | C9 |
| chr14 | 92659257 | 92659672 | 416 | 2 | 0.000111919 | 4.17E-05 | 0.00015 | 3.27E-05 | -0.003900258 | RIN3 |
| chr17 | 80213837 | 80214105 | 269 | 2 | 0.000111919 | 1.90E-05 | 0.0012 | 3.03E-05 | -0.005135702 | SGSH |
| chr8 | 76705245 | 76705906 | 662 | 4 | 0.000111924 | 0.013375 | 0.00039 | 6.84E-05 | -0.00507393 | ZFHX4 |
| chr22 | 32457984 | 32458374 | 391 | 4 | 0.000111943 | 0.000172 | 0.00015 | 3.59E-05 | -0.010457168 | BPIFC |
| chr7 | 29193852 | 29194259 | 408 | 4 | 0.000111992 | 7.25E-06 | 0.00409 | 1.72E-05 | -0.008487661 | AC004593.2, CHN2, CPVL, AC004593.1 |
| chr4 | 588148 | 588845 | 698 | 3 | 0.000111992 | 5.08E-06 | 0.00267 | 1.30E-05 | -0.016340071 | AC116565.1 |
| chr19 | 3607271 | 3607610 | 340 | 2 | 0.000111992 | 0.000742 | 0.00066 | 0.00042 | -0.011800672 | CACTIN-AS1 |
| chr6 | 31693084 | 31693401 | 318 | 4 | 0.000112172 | 9.89E-05 | 0.0001 | 2.88E-05 | -0.003865099 | ABHD16A, AL662899.1 |
| chr21 | 38472429 | 38472614 | 186 | 2 | 0.000112172 | 1.29E-05 | 0.00081 | 2.04E-05 | -0.013964327 | ERG |
| chr8 | 42317167 | 42317741 | 575 | 3 | 0.000112274 | 1.76E-05 | 0.00072 | 2.23E-05 | -0.005659684 | IKBKB |
| chr7 | 1013143 | 1014715 | 1573 | 6 | 0.000112446 | 1.25E-06 | 0.00379 | 1.99E-06 | -0.007845821 | C7orf50 |
| chr17 | 34961402 | 34962322 | 921 | 12 | 0.000112463 | 7.50E-05 | 0.01763 | 8.18E-05 | 0.000205922 | ZNF830, CCT6B |
| chr11 | 3020342 | 3020768 | 427 | 2 | 0.000112463 | 0.000214 | 7.36E-05 | 4.92E-05 | -0.002356596 | CARS1 |
| chr8 | 29487050 | 29487082 | 33 | 3 | 0.000112635 | 0.051002 | 1.74E-05 | 3.25E-05 | -0.003007744 | NA |
| chr8 | 141331211 | 141332200 | 990 | 4 | 0.000112656 | 7.83E-06 | 0.00111 | 9.86E-06 | -0.007621768 | NA |
| chr5 | 82588493 | 82588520 | 28 | 2 | 0.00011289 | 0.000403 | 4.20E-05 | 3.68E-05 | -0.003501409 | NA |
| chr14 | 99568639 | 99569251 | 613 | 3 | 0.000112969 | 5.14E-06 | 0.00182 | 1.13E-05 | -0.011914756 | CCDC85C |
| chr12 | 51216560 | 51216904 | 345 | 2 | 0.000112979 | 0.005194 | 0.0002 | 0.00039 | -0.002142376 | POU6F1 |
| chr6 | 113564619 | 113565138 | 520 | 3 | 0.000113044 | 3.63E-06 | 0.00201 | 8.95E-06 | -0.015916091 | NA |
| chr3 | 182740852 | 182740987 | 136 | 3 | 0.000113078 | 5.94E-06 | 0.00154 | 1.31E-05 | -0.016269577 | NA |
| chr12 | 122946132 | 122946600 | 469 | 3 | 0.000113078 | 6.72E-05 | 0.00026 | 5.09E-05 | -0.006784206 | ABCB9 |
| chr7 | 129761368 | 129761411 | 44 | 2 | 0.000113118 | 3.07E-05 | 7.95E-05 | 1.90E-05 | -0.000958237 | NA |
| chr11 | 69466546 | 69466909 | 364 | 2 | 0.000113154 | 0.000111 | 0.00172 | 0.00016 | -0.003243174 | NA |
| chr22 | 41367941 | 41368138 | 198 | 2 | 0.000113367 | 4.79E-05 | 0.00122 | 6.79E-05 | -0.005214884 | TEF |
| chr16 | 23640947 | 23641853 | 907 | 10 | 0.000113396 | 0.021017 | 2.08E-05 | 0.00018 | 0.001026591 | DCTN5, PALB2 |
| chr19 | 43498838 | 43499468 | 631 | 3 | 0.0001134 | 0.030769 | 6.68E-05 | 0.00031 | -0.002374442 | PHLDB3 |
| chr5 | 5886161 | 5886955 | 795 | 4 | 0.000113523 | 4.96E-05 | 0.00064 | 1.93E-05 | -0.000749874 | NA |
| chr7 | 452150 | 452404 | 255 | 5 | 0.000113721 | 1.34E-06 | 0.0084 | 6.70E-06 | -0.018901435 | NA |
| chr7 | 1080124 | 1081224 | 1101 | 8 | 0.000113816 | 0.000399 | 0.00052 | 3.39E-05 | -0.00115953 | AC091729.2, C7orf50 |
| chr20 | 63884721 | 63885719 | 999 | 3 | 0.000113816 | 2.01E-05 | 7.07E-05 | 2.89E-06 | -0.006203499 | TPD52L2 |
| chr5 | 138561537 | 138562271 | 735 | 5 | 0.000113917 | 9.17E-05 | 0.00029 | 2.24E-05 | -0.003089048 | HSPA9 |
| chr7 | 2085339 | 2085888 | 550 | 5 | 0.000113918 | 2.80E-05 | 0.0016 | 8.02E-06 | -0.005159935 | MAD1L1, AC069288.1 |
| chr2 | 75654411 | 75654804 | 394 | 5 | 0.000114013 | 0.002344 | 3.74E-05 | 4.66E-05 | -0.002707113 | MRPL19, GCFC2 |
| chr17 | 80904730 | 80905044 | 315 | 4 | 0.000114023 | 2.08E-05 | 0.00092 | 1.80E-05 | -0.004279057 | RPTOR |
| chr3 | 111712520 | 111712655 | 136 | 3 | 0.000114084 | 6.11E-06 | 0.00164 | 1.24E-05 | -0.015951405 | PLCXD2 |
| chr16 | 3857665 | 3857754 | 90 | 3 | 0.000114113 | 6.50E-06 | 0.00075 | 1.23E-05 | -0.009981833 | CREBBP |
| chr2 | 44948742 | 44949412 | 671 | 3 | 0.000114193 | 0.001356 | 0.00017 | 0.00012 | -0.004345485 | NA |
| chr1 | 54604050 | 54604088 | 39 | 2 | 0.000114193 | 1.21E-05 | 0.00033 | 1.68E-05 | -0.012042206 | ACOT11 |
| chr17 | 38443442 | 38443767 | 326 | 2 | 0.000114419 | 0.000804 | 0.0016 | 0.00072 | -0.00848401 | ARHGAP23 |
| chr8 | 140098508 | 140099632 | 1125 | 6 | 0.000114457 | 3.73E-05 | 0.00124 | 8.80E-06 | -0.004580824 | TRAPPC9, PEG13 |
| chr2 | 187570239 | 187570385 | 147 | 2 | 0.000114577 | 1.67E-05 | 0.0002 | 1.88E-05 | -0.01519056 | NA |
| chr21 | 45302274 | 45302862 | 589 | 3 | 0.000114663 | 4.32E-05 | 0.0002 | 3.30E-05 | -0.010754671 | BX322557.2, LINC00315 |
| chr10 | 129962497 | 129962514 | 18 | 2 | 0.000114704 | 9.99E-06 | 0.00082 | 1.64E-05 | 0.009245021 | EBF3 |
| chr6 | 6851134 | 6851214 | 81 | 2 | 0.00011474 | 0.024032 | 2.42E-05 | 3.56E-05 | -0.006757017 | NA |
| chr1 | 2493734 | 2494596 | 863 | 6 | 0.000114776 | 0.000207 | 0.00024 | 3.96E-05 | -0.004179599 | PLCH2, AL139246.4 |
| chr17 | 42608624 | 42609800 | 1177 | 21 | 0.000114868 | 1.57E-05 | 0.00619 | 8.79E-06 | 0.000819716 | TUBG1, RETREG3 |
| chr11 | 1473478 | 1473898 | 421 | 4 | 0.000114879 | 0.00018 | 7.36E-05 | 2.93E-05 | -0.003299048 | MOB2 |
| chr12 | 98894695 | 98895517 | 823 | 10 | 0.000114916 | 5.85E-05 | 0.00249 | 2.30E-05 | -0.000221132 | ANKS1B |
| chr1 | 109112581 | 109112921 | 341 | 5 | 0.000114916 | 2.19E-05 | 0.00062 | 2.84E-05 | -0.001778725 | C1orf194 |
| chr9 | 125467514 | 125467823 | 310 | 2 | 0.000114939 | 0.000431 | 4.15E-05 | 3.69E-05 | -0.003241168 | MAPKAP1 |
| chr17 | 81919438 | 81919631 | 194 | 3 | 0.000114942 | 9.64E-06 | 0.00136 | 1.95E-05 | -0.010357084 | SIRT7, MAFG |
| chr6 | 37589601 | 37589918 | 318 | 2 | 0.000115045 | 1.09E-05 | 0.00033 | 1.54E-05 | -0.002927834 | NA |
| chr1 | 109465008 | 109465130 | 123 | 3 | 0.000115053 | 0.000123 | 6.09E-05 | 3.08E-05 | -0.00935287 | NA |
| chr1 | 3454156 | 3454770 | 615 | 8 | 0.000115118 | 0.000254 | 0.0008 | 2.50E-05 | -6.26E-06 | ARHGEF16 |
| chr18 | 74453380 | 74453692 | 313 | 2 | 0.000115467 | 3.38E-05 | 0.00012 | 2.54E-05 | -0.004236931 | DIPK1C |
| chr11 | 74005120 | 74005322 | 203 | 2 | 0.00011555 | 1.27E-05 | 0.00024 | 1.60E-05 | -0.011025884 | UCP3 |
| chr5 | 160399815 | 160400194 | 380 | 7 | 0.000115551 | 1.20E-05 | 0.00143 | 1.12E-05 | 0.000514517 | ZBED8 |
| chr16 | 67892414 | 67893448 | 1035 | 11 | 0.000115601 | 7.96E-05 | 0.00212 | 1.38E-05 | 0.001098612 | PSKH1 |
| chr1 | 16237926 | 16238194 | 269 | 3 | 0.000115641 | 4.85E-05 | 0.0055 | 0.0001 | -5.71E-06 | NA |
| chr10 | 27413845 | 27414618 | 774 | 6 | 0.000115708 | 2.06E-05 | 0.00976 | 1.66E-05 | -0.019558314 | PTCHD3 |
| chr19 | 18248695 | 18248762 | 68 | 5 | 0.000115708 | 0.006185 | 0.00186 | 3.31E-05 | -0.004158485 | AC008397.2 |
| chr11 | 63762627 | 63763525 | 899 | 3 | 0.000115708 | 1.10E-05 | 0.00223 | 2.14E-05 | -0.012877041 | C11orf95 |
| chr16 | 88902488 | 88903219 | 732 | 5 | 0.000115793 | 1.29E-05 | 0.00533 | 2.38E-05 | -0.004618561 | CBFA2T3 |
| chr8 | 143982622 | 143983006 | 385 | 4 | 0.000115843 | 2.56E-05 | 0.00146 | 2.42E-05 | -0.005435184 | PARP10 |
| chr8 | 139799688 | 139799980 | 293 | 2 | 0.000115843 | 0.000116 | 5.18E-05 | 2.88E-05 | -0.004371605 | TRAPPC9 |
| chr9 | 137170088 | 137172669 | 2582 | 23 | 0.000115906 | 1.21E-09 | 0.00951 | 2.19E-08 | -0.006215143 | AL929554.1, TMEM210 |
| chr2 | 3287415 | 3287630 | 216 | 3 | 0.000115955 | 0.000124 | 0.00013 | 3.50E-05 | -0.011166219 | EIPR1 |
| chr9 | 130397582 | 130397847 | 266 | 2 | 0.000116144 | 2.57E-05 | 0.00036 | 3.12E-05 | -0.009401509 | HMCN2 |
| chr16 | 88405808 | 88406133 | 326 | 3 | 0.000116273 | 1.76E-05 | 0.00056 | 2.36E-05 | -0.012988667 | ZNF469 |
| chr17 | 69828245 | 69828359 | 115 | 2 | 0.000116421 | 8.02E-06 | 0.0003 | 1.16E-05 | -0.02029299 | LINC01483 |
| chr2 | 134268539 | 134268554 | 16 | 2 | 0.000116551 | 6.60E-06 | 0.00065 | 1.10E-05 | -0.008427114 | MGAT5 |
| chr8 | 38421772 | 38422351 | 580 | 6 | 0.000116798 | 4.89E-05 | 0.00302 | 2.59E-05 | 0.00184129 | AC087623.1, FGFR1 |
| chr10 | 116255818 | 116256049 | 232 | 3 | 0.000116878 | 9.13E-05 | 0.00019 | 2.77E-05 | -0.013438323 | GFRA1 |
| chr12 | 124294592 | 124294986 | 395 | 4 | 0.000116966 | 0.000976 | 0.0004 | 0.00022 | -0.006622895 | RFLNA |
| chr3 | 47443419 | 47443754 | 336 | 3 | 0.000116971 | 0.000782 | 0.00012 | 6.53E-05 | -0.00952979 | SCAP |
| chr13 | 110646177 | 110646339 | 163 | 2 | 0.000117148 | 8.46E-05 | 5.89E-05 | 2.75E-05 | -0.01092228 | CARS2 |
| chr19 | 56838924 | 56839135 | 212 | 3 | 0.000117238 | 1.12E-05 | 0.00243 | 2.41E-05 | -0.019301618 | ZIM2, PEG3 |
| chr7 | 4706806 | 4707585 | 780 | 7 | 0.000117252 | 0.000238 | 0.00165 | 3.60E-05 | -0.006147759 | FOXK1 |
| chr17 | 49035699 | 49036264 | 566 | 7 | 0.000117338 | 7.47E-05 | 0.00058 | 1.33E-05 | -0.004912964 | IGF2BP1 |
| chr7 | 50308578 | 50309274 | 697 | 4 | 0.000117412 | 0.00063 | 0.00011 | 4.21E-05 | -0.005302927 | IKZF1 |
| chr4 | 1578405 | 1578650 | 246 | 3 | 0.000117456 | 0.000106 | 0.00348 | 0.00012 | -0.028811032 | AC147067.1 |
| chr3 | 186596675 | 186596927 | 253 | 4 | 0.000117608 | 1.90E-05 | 0.00022 | 1.25E-05 | -0.006571652 | AC068631.2, AC068631.1 |
| chr11 | 117535572 | 117536052 | 481 | 2 | 0.0001178 | 2.19E-05 | 0.0002 | 2.34E-05 | -0.012831923 | DSCAML1 |
| chr11 | 116789355 | 116790672 | 1318 | 6 | 0.000117809 | 2.55E-05 | 0.00237 | 3.95E-06 | -0.003767464 | APOA5 |
| chr9 | 134338937 | 134339393 | 457 | 3 | 0.00011794 | 4.21E-05 | 0.00037 | 3.25E-05 | -0.010435584 | RXRA |
| chr5 | 74684502 | 74685665 | 1164 | 8 | 0.000118019 | 0.001405 | 0.00028 | 4.06E-05 | -0.001317887 | HEXB |
| chr10 | 58651112 | 58651367 | 256 | 2 | 0.000118052 | 7.20E-06 | 0.00068 | 1.20E-05 | -0.062436354 | BICC1 |
| chr18 | 23238385 | 23238388 | 4 | 2 | 0.000118053 | 1.60E-05 | 0.00016 | 1.71E-05 | -0.014224754 | CABLES1, TMEM241 |
| chr7 | 4137719 | 4137769 | 51 | 3 | 0.000118115 | 0.026309 | 3.59E-05 | 5.25E-05 | -0.00254372 | SDK1 |
| chr7 | 151601947 | 151602082 | 136 | 2 | 0.000118207 | 0.003059 | 5.69E-05 | 7.91E-05 | -0.001149282 | PRKAG2 |
| chr2 | 224882559 | 224882677 | 119 | 2 | 0.000118241 | 1.92E-05 | 0.00018 | 2.06E-05 | 0.017873279 | DOCK10 |
| chr20 | 58990642 | 58991274 | 633 | 3 | 0.000118313 | 1.71E-05 | 0.00124 | 2.44E-05 | -0.003691006 | NELFCD |
| chr4 | 143513805 | 143514585 | 781 | 6 | 0.000118355 | 0.000772 | 0.0014 | 6.22E-05 | -0.000589413 | SMARCA5, SMARCA5-AS1 |
| chr1 | 229433183 | 229433354 | 172 | 4 | 0.000118387 | 1.95E-05 | 0.01253 | 6.20E-05 | 0.019806462 | ACTA1 |
| chr17 | 78315787 | 78316374 | 588 | 3 | 0.000118453 | 8.10E-06 | 0.00115 | 1.62E-05 | -0.009840035 | AC087645.2 |
| chr2 | 176508682 | 176509488 | 807 | 4 | 0.000118466 | 0.000413 | 0.00011 | 7.07E-05 | -0.00780047 | LINC01117 |
| chr6 | 43308740 | 43309518 | 779 | 12 | 0.000118511 | 4.38E-06 | 0.01212 | 9.62E-06 | -0.002786243 | CRIP3, ZNF318 |
| chr17 | 82870377 | 82870710 | 334 | 2 | 0.000118578 | 0.000152 | 0.00026 | 9.62E-05 | -0.003212805 | TBCD |
| chr14 | 106803322 | 106803611 | 290 | 4 | 0.000118622 | 6.01E-05 | 0.00073 | 2.56E-05 | -0.01035365 | NA |
| chr16 | 17368087 | 17368279 | 193 | 2 | 0.000118705 | 0.000317 | 4.41E-05 | 3.58E-05 | 0.024256001 | XYLT1 |
| chr2 | 174486921 | 174487699 | 779 | 10 | 0.000118792 | 0.00016 | 0.00139 | 1.76E-05 | 0.002376117 | AC010894.1, GPR155 |
| chr10 | 44316868 | 44317094 | 227 | 2 | 0.000118854 | 5.05E-05 | 5.79E-05 | 2.07E-05 | -0.015125895 | NA |
| chr12 | 56424825 | 56425276 | 452 | 3 | 0.000119049 | 6.23E-06 | 0.00125 | 1.31E-05 | -0.009694655 | TIMELESS |
| chr1 | 204686019 | 204686250 | 232 | 3 | 0.000119115 | 1.53E-05 | 0.00051 | 1.26E-05 | -0.007231185 | NA |
| chr22 | 39297532 | 39297873 | 342 | 2 | 0.000119119 | 1.04E-05 | 0.00069 | 1.66E-05 | -0.016251013 | NA |
| chr22 | 45923864 | 45924146 | 283 | 2 | 0.000119143 | 0.000108 | 0.00023 | 7.47E-05 | -0.013643639 | WNT7B |
| chr9 | 89557026 | 89557043 | 18 | 2 | 0.000119401 | 6.94E-06 | 0.00055 | 1.13E-05 | 0.022704754 | NA |
| chr1 | 153958643 | 153959646 | 1004 | 10 | 0.000119438 | 1.60E-05 | 0.01413 | 4.01E-05 | -0.000570642 | SLC39A1, AL358472.7 |
| chr8 | 27543900 | 27544525 | 626 | 2 | 0.000119438 | 8.67E-06 | 0.00066 | 1.41E-05 | 0.000479315 | EPHX2 |
| chr13 | 112588341 | 112588965 | 625 | 8 | 0.000119486 | 2.75E-05 | 0.01539 | 3.95E-05 | -0.013558975 | NA |
| chr2 | 105743739 | 105744252 | 514 | 2 | 0.000119549 | 0.000107 | 0.00259 | 0.00017 | -0.011039468 | NA |
| chr8 | 37067132 | 37067192 | 61 | 2 | 0.000119595 | 7.65E-06 | 0.00037 | 1.16E-05 | 0.005517952 | NA |
| chr8 | 143380332 | 143381729 | 1398 | 7 | 0.000119652 | 8.57E-08 | 0.00081 | 2.81E-07 | -0.003035751 | RHPN1 |
| chr14 | 92885199 | 92885628 | 430 | 2 | 0.000119783 | 2.78E-05 | 0.00056 | 3.69E-05 | -0.01025332 | NA |
| chr8 | 2010764 | 2010955 | 192 | 2 | 0.000119791 | 0.000156 | 5.12E-05 | 3.21E-05 | -0.003515576 | NA |
| chr8 | 71832739 | 71832962 | 224 | 2 | 0.000119826 | 0.000148 | 6.35E-05 | 3.75E-05 | -0.008146112 | MSC-AS1 |
| chr1 | 90719471 | 90719884 | 414 | 6 | 0.000119921 | 0.000466 | 0.00134 | 9.99E-05 | 0.000374398 | AC092805.1 |
| chr14 | 100726383 | 100727095 | 713 | 9 | 0.00012015 | 2.52E-05 | 0.00061 | 1.60E-05 | 0.00373421 | DLK1 |
| chr14 | 50944452 | 50945374 | 923 | 13 | 0.000120371 | 2.08E-05 | 0.00876 | 2.58E-05 | -0.003809321 | PYGL |
| chr1 | 53292971 | 53293288 | 318 | 2 | 0.000120461 | 5.86E-05 | 9.85E-05 | 3.24E-05 | -0.003249746 | LRP8 |
| chr2 | 69386975 | 69387986 | 1012 | 10 | 0.000120581 | 5.69E-05 | 0.00071 | 4.87E-06 | -0.002254068 | GFPT1 |
| chr16 | 14186326 | 14186852 | 527 | 8 | 0.000120583 | 8.22E-05 | 0.00295 | 1.91E-05 | -0.00602405 | MRTFB |
| chr19 | 10713003 | 10713426 | 424 | 6 | 0.000120583 | 2.59E-05 | 0.00373 | 2.88E-05 | -0.00403422 | QTRT1 |
| chr3 | 44975176 | 44975408 | 233 | 4 | 0.000120625 | 0.000119 | 0.00011 | 2.38E-05 | -0.00166889 | EXOSC7, ZDHHC3 |
| chr1 | 160189435 | 160190976 | 1542 | 14 | 0.000120734 | 3.00E-06 | 0.0018 | 1.00E-06 | -0.004964557 | CASQ1 |
| chr10 | 104232367 | 104233741 | 1375 | 8 | 0.000120782 | 0.000414 | 0.00029 | 1.17E-05 | -0.000407269 | NA |
| chr18 | 11821777 | 11822137 | 361 | 5 | 0.000120871 | 0.038612 | 2.34E-05 | 8.07E-05 | -0.001293054 | GNAL |

| | | | | | | | | | | |
|---|---|---|---|---|---|---|---|---|---|---|
| chr14 | 21092324 | 21092416 | 93 | 2 | 0.000121204 | 1.05E-05 | 0.0003 | 1.46E-05 | -0.006904513 | ZNF219 |
| chr6 | 127462435 | 127463151 | 717 | 2 | 0.000121338 | 4.75E-06 | 0.00043 | 7.84E-06 | -0.012534917 | AL096711.2 |
| chr15 | 75346772 | 75347543 | 772 | 13 | 0.00012142 | 3.32E-06 | 0.00422 | 1.11E-05 | -0.004221386 | NEIL1, AC068338.3 |
| chr3 | 179852895 | 179853378 | 484 | 2 | 0.000121451 | 1.29E-05 | 0.0003 | 1.72E-05 | -0.002947072 | PEX5L |
| chr7 | 141554579 | 141555005 | 427 | 2 | 0.000121451 | 7.87E-06 | 0.00061 | 1.28E-05 | -0.010405991 | AGK |
| chr8 | 109691181 | 109691231 | 51 | 2 | 0.000121516 | 2.32E-05 | 0.0008 | 3.39E-05 | 0.022169458 | SYBU |
| chr1 | 1260151 | 1261367 | 1217 | 5 | 0.000121534 | 2.74E-07 | 0.00251 | 1.11E-06 | -0.014829467 | UBE2J2 |
| chr6 | 166235878 | 166236148 | 271 | 2 | 0.000121666 | 4.78E-05 | 0.00042 | 5.26E-05 | -0.010116448 | NA |
| chr15 | 39594940 | 39595892 | 953 | 5 | 0.000121758 | 0.000352 | 0.00026 | 2.15E-05 | -0.004283475 | THBS1, FSIP1 |
| chr14 | 99273179 | 99273754 | 576 | 4 | 0.000121773 | 4.06E-05 | 0.00154 | 2.22E-05 | 0.001067077 | NA |
| chr11 | 73970236 | 73970292 | 57 | 2 | 0.000121946 | 4.02E-05 | 7.38E-05 | 2.14E-05 | -0.011910784 | DNAJB13, AP003717.1 |
| chr4 | 138924933 | 138924942 | 10 | 2 | 0.000122025 | 0.000244 | 3.75E-05 | 2.85E-05 | -0.004515211 | AC109927.1 |
| chr2 | 9738687 | 9739382 | 696 | 4 | 0.000122042 | 7.07E-06 | 0.00457 | 2.14E-05 | -0.028382176 | NA |
| chr20 | 2706214 | 2706717 | 504 | 3 | 0.000122135 | 0.004034 | 0.00015 | 0.00019 | -0.001607474 | EBF4 |
| chr1 | 10672006 | 10672687 | 682 | 8 | 0.000122161 | 4.76E-05 | 0.00182 | 1.48E-05 | -0.003696987 | CASZ1 |
| chr15 | 71744079 | 71744397 | 319 | 2 | 0.000122183 | 6.61E-06 | 0.00064 | 1.10E-05 | 0.012090394 | THSD4 |
| chr1 | 12570187 | 12570253 | 67 | 3 | 0.000122282 | 1.64E-05 | 0.00115 | 1.83E-05 | -0.003775414 | DHRS3 |
| chr1 | 51989476 | 51991246 | 1771 | 15 | 0.000122368 | 5.77E-07 | 0.01038 | 8.95E-07 | -0.002103796 | RAB3B |
| chr1 | 161721224 | 161721873 | 650 | 3 | 0.000122368 | 5.37E-05 | 0.00184 | 5.52E-05 | -0.012788493 | FCRLB |
| chr14 | 68212075 | 68212312 | 238 | 2 | 0.000122391 | 0.000116 | 7.57E-05 | 3.90E-05 | -0.004018776 | RAD51B |
| chr10 | 93068848 | 93069333 | 486 | 3 | 0.000122454 | 4.44E-06 | 0.00311 | 1.19E-05 | -0.010421558 | CYP26C1, AL358613.3 |
| chr7 | 20365348 | 20365824 | 477 | 2 | 0.000122496 | 1.75E-05 | 0.0002 | 1.96E-05 | -0.000586706 | ITGB8 |
| chr9 | 22118631 | 22118864 | 234 | 2 | 0.000122635 | 0.005332 | 4.05E-05 | 5.84E-05 | -0.008380117 | CDKN2B-AS1 |
| chr9 | 128867642 | 128867711 | 70 | 2 | 0.000122825 | 1.13E-05 | 0.00015 | 1.28E-05 | -0.010899049 | AL441992.2, KYAT1 |
| chr17 | 39927101 | 39927866 | 766 | 8 | 0.000122899 | 0.000437 | 0.0012 | 7.58E-05 | 0.000222039 | AC090844.2, ORMDL3 |
| chr7 | 140696970 | 140697580 | 611 | 10 | 0.000123038 | 0.000474 | 0.00384 | 1.82E-05 | -0.000320778 | NDUFB2, NDUFB2-AS1 |
| chr12 | 7189868 | 7190738 | 871 | 11 | 0.000123088 | 0.000853 | 0.00092 | 6.58E-05 | -0.005663403 | PEX5 |
| chr7 | 2556636 | 2557091 | 456 | 4 | 0.000123207 | 2.07E-05 | 0.00096 | 2.88E-05 | -0.010524273 | NA |
| chr19 | 1315505 | 1315963 | 459 | 5 | 0.000123231 | 7.21E-05 | 0.00342 | 2.22E-05 | 0.000856088 | NA |
| chr18 | 58599586 | 58599914 | 329 | 2 | 0.000123441 | 0.000228 | 7.08E-05 | 4.88E-05 | -0.00357603 | ALPK2 |
| chr12 | 51820974 | 51823474 | 2501 | 11 | 0.000123457 | 3.22E-07 | 0.00101 | 1.57E-07 | -0.005992021 | AC068987.2, FIGNL2 |
| chr13 | 75838838 | 75839543 | 706 | 2 | 0.00012364 | 2.83E-06 | 0.00034 | 4.82E-06 | -0.019714861 | LMO7 |
| chr16 | 2720210 | 2720900 | 691 | 8 | 0.000123642 | 0.000189 | 0.00106 | 2.26E-05 | -0.003062707 | PRSS27 |
| chr2 | 31276096 | 31276805 | 710 | 3 | 0.000123761 | 3.83E-06 | 0.00071 | 7.64E-06 | -0.000270476 | NA |
| chr9 | 134536391 | 134536399 | 9 | 2 | 0.000123783 | 1.55E-05 | 0.00109 | 2.50E-05 | -0.006527575 | AL669970.3 |
| chr11 | 76787657 | 76788291 | 635 | 3 | 0.00012379 | 3.47E-05 | 0.00011 | 1.44E-05 | -0.006678356 | TSKU |
| chr10 | 113045106 | 113045247 | 142 | 2 | 0.000123914 | 8.02E-06 | 0.00041 | 1.23E-05 | -0.004999083 | TCF7L2 |
| chr21 | 46219724 | 46220307 | 584 | 6 | 0.000123933 | 0.002559 | 4.20E-05 | 6.63E-05 | -0.003321534 | AP001469.1, LSS |
| chr19 | 13938280 | 13939010 | 731 | 7 | 0.000123996 | 9.46E-06 | 0.00652 | 2.82E-05 | -0.008929713 | PODNL1 |
| chr8 | 143249108 | 143249761 | 654 | 2 | 0.00012407 | 6.64E-05 | 0.00282 | 0.0001 | -0.003985195 | ZFP41, AC138696.1 |
| chr22 | 20956439 | 20957382 | 944 | 7 | 0.000124088 | 6.48E-05 | 0.00013 | 1.26E-05 | -0.006185704 | LINC01637 |
| chr19 | 15116408 | 15116880 | 473 | 2 | 0.000124171 | 1.94E-05 | 0.00029 | 2.37E-05 | -0.007612087 | ILVBL |
| chr2 | 113062137 | 113062447 | 311 | 4 | 0.000124233 | 0.020636 | 1.30E-05 | 3.43E-05 | -0.002205355 | IL36RN |
| chr11 | 6562752 | 6563656 | 905 | 4 | 0.000124328 | 0.000195 | 0.00172 | 0.00018 | 0.000884655 | DNHD1 |
| chr10 | 101050382 | 101050849 | 468 | 3 | 0.000124495 | 0.01825 | 0.00066 | 0.00138 | -0.002931743 | NA |
| chr1 | 160629449 | 160629715 | 267 | 2 | 0.000124628 | 4.44E-05 | 0.00068 | 5.65E-05 | -0.011083047 | SLAMF1 |
| chr22 | 23768436 | 23769150 | 715 | 6 | 0.000124709 | 0.00934 | 3.46E-05 | 7.91E-05 | -0.00326832 | MMP11 |
| chr17 | 11881371 | 11882406 | 1036 | 5 | 0.000124716 | 0.000396 | 7.64E-05 | 1.38E-05 | -0.005623326 | DNAH9, AC005209.1 |
| chr2 | 3280226 | 3280390 | 165 | 2 | 0.00012477 | 2.38E-05 | 0.00132 | 3.72E-05 | 0.004427268 | EIPR1 |
| chr16 | 85453380 | 85453418 | 39 | 2 | 0.00012477 | 8.27E-06 | 0.00063 | 1.34E-05 | -0.022082479 | GSE1 |
| chr15 | 76139507 | 76139656 | 150 | 2 | 0.000124797 | 1.72E-05 | 0.00045 | 2.38E-05 | -0.009777681 | TMEM266 |
| chr11 | 70670453 | 70670940 | 488 | 5 | 0.000124828 | 0.00051 | 0.0002 | 3.49E-05 | 0.011287241 | SHANK2 |
| chr13 | 113637131 | 113637738 | 608 | 5 | 0.000125007 | 0.000995 | 0.00418 | 7.43E-05 | -9.18E-05 | TFDP1 |
| chr15 | 28099127 | 28099584 | 458 | 11 | 0.000125074 | 4.27E-05 | 0.00151 | 2.02E-05 | 0.001317838 | OCA2 |
| chr14 | 24292652 | 24292853 | 202 | 4 | 0.000125193 | 0.000133 | 0.00053 | 2.16E-05 | -0.002754337 | AL096870.12, DHRS1 |
| chr11 | 66370978 | 66371871 | 894 | 10 | 0.000125234 | 5.81E-05 | 0.00437 | 1.87E-05 | 0.003227518 | SLC29A2 |
| chr5 | 150364605 | 150364718 | 114 | 3 | 0.000125241 | 0.000126 | 3.71E-05 | 2.10E-05 | -0.001927304 | TCOF1 |
| chr22 | 31089131 | 31089557 | 427 | 3 | 0.000125347 | 1.65E-05 | 0.00055 | 2.35E-05 | -0.008466868 | SMTN |
| chr1 | 957568 | 958034 | 467 | 2 | 0.000125347 | 0.000124 | 0.00277 | 0.0002 | -0.010248311 | NOC2L |
| chr1 | 203889328 | 203889703 | 376 | 2 | 0.000125347 | 1.81E-05 | 0.00017 | 1.92E-05 | 0.000832669 | NA |
| chr18 | 58862523 | 58863557 | 1035 | 12 | 0.000125392 | 7.58E-05 | 0.00629 | 4.07E-05 | -0.002852532 | ZNF532 |
| chr12 | 100472788 | 100473024 | 237 | 2 | 0.000125401 | 0.008213 | 0.00055 | 0.00112 | 0.005929137 | NA |
| chr11 | 2418802 | 2419156 | 355 | 3 | 0.000125445 | 0.000105 | 0.00096 | 3.73E-05 | -0.007622276 | TRPM5 |
| chr15 | 73997946 | 73998392 | 447 | 4 | 0.000125473 | 0.000968 | 0.00062 | 4.28E-05 | -0.004858674 | PML |
| chr16 | 21170340 | 21171150 | 811 | 3 | 0.000125476 | 0.000328 | 0.00015 | 1.02E-05 | -0.002912349 | TMEM159 |
| chr10 | 34110027 | 34110730 | 704 | 6 | 0.00012561 | 0.000194 | 8.52E-05 | 2.63E-05 | -0.001875446 | PARD3 |
| chr4 | 118684456 | 118684712 | 257 | 2 | 0.000125825 | 0.003541 | 0.00094 | 0.00123 | -0.002565543 | METTL14-DT |
| chr5 | 68899327 | 68899679 | 353 | 2 | 0.000125882 | 4.55E-05 | 8.94E-05 | 2.62E-05 | -0.001711364 | AC093523.1 |
| chr12 | 47777753 | 47778307 | 555 | 4 | 0.000125949 | 3.39E-05 | 0.00026 | 1.37E-05 | -0.006692124 | SLC48A1 |
| chr1 | 1292909 | 1294766 | 1858 | 9 | 0.000126117 | 5.06E-08 | 0.00447 | 2.83E-07 | -0.004364226 | ACAP3 |
| chr6 | 109381526 | 109382735 | 1210 | 16 | 0.000126268 | 9.86E-05 | 0.01096 | 5.40E-06 | 0.000336713 | CD164 |
| chr1 | 42189915 | 42190433 | 519 | 5 | 0.000126615 | 0.00044 | 0.00068 | 1.83E-05 | -0.00180919 | FOXJ3 |
| chr12 | 13255728 | 13256540 | 813 | 3 | 0.000126672 | 7.30E-06 | 0.00106 | 1.47E-05 | -0.010381557 | AC022276.1 |
| chr18 | 57545362 | 57545572 | 211 | 3 | 0.000126987 | 4.48E-05 | 0.0007 | 1.86E-05 | -0.012845945 | FECH |
| chr5 | 1385517 | 1386035 | 519 | 5 | 0.000127117 | 0.000236 | 0.0003 | 4.46E-05 | 0.001904849 | AC026748.3 |
| chr3 | 159839704 | 159840465 | 762 | 9 | 0.000127152 | 3.20E-07 | 0.01509 | 4.39E-06 | -0.02012334 | IQCJ-SCHIP1, SCHIP1 |
| chr1 | 35769373 | 35769888 | 516 | 5 | 0.000127167 | 0.029809 | 0.00237 | 0.0007 | -0.000506463 | CLSPN |
| chr14 | 20873414 | 20873712 | 299 | 4 | 0.000127171 | 0.001024 | 8.19E-05 | 4.52E-05 | -0.006959204 | AL133371.3 |
| chr8 | 6806153 | 6807001 | 849 | 6 | 0.00012718 | 0.002007 | 0.00472 | 0.0001 | -0.003357855 | NA |
| chr19 | 5968200 | 5968688 | 489 | 3 | 0.000127601 | 1.24E-05 | 0.00099 | 1.96E-05 | -0.00793991 | RANBP3 |
| chr1 | 13779011 | 13779710 | 700 | 6 | 0.000127703 | 0.001725 | 0.00028 | 5.32E-05 | -0.002772144 | PRDM2 |
| chr16 | 88742592 | 88742744 | 153 | 2 | 0.000127845 | 2.47E-05 | 0.00112 | 3.77E-05 | 0.005794801 | PIEZO1 |
| chr4 | 87733987 | 87734322 | 336 | 2 | 0.000127868 | 2.42E-05 | 0.00024 | 2.68E-05 | -0.012597521 | NA |
| chr1 | 157700920 | 157701596 | 677 | 6 | 0.000128028 | 0.000109 | 0.00363 | 6.32E-05 | -0.006565796 | NA |
| chr12 | 133036151 | 133036847 | 697 | 3 | 0.000128223 | 0.002294 | 0.00077 | 0.00036 | -0.006625143 | ZNF84-DT |
| chr18 | 46077296 | 46077722 | 427 | 2 | 0.000128326 | 0.0002 | 5.35E-05 | 3.66E-05 | -0.007168631 | NA |
| chr3 | 25665333 | 25666018 | 686 | 3 | 0.000128361 | 8.06E-05 | 0.00034 | 8.95E-06 | 0.006448645 | NA |
| chr13 | 27380265 | 27380637 | 373 | 2 | 0.00012843 | 5.86E-05 | 8.53E-05 | 2.96E-05 | -0.013441318 | NA |
| chr1 | 116110246 | 116110940 | 695 | 5 | 0.000128455 | 0.000118 | 0.00203 | 3.36E-05 | -0.005194746 | NA |
| chr16 | 29662863 | 29663297 | 435 | 6 | 0.000128474 | 0.002021 | 2.58E-05 | 5.17E-05 | 0.001641775 | SPN, QPRT |
| chr1 | 209652327 | 209652533 | 207 | 5 | 0.000128517 | 2.42E-05 | 0.00088 | 2.19E-05 | -0.001681619 | LAMB3 |
| chr17 | 16352246 | 16352570 | 325 | 4 | 0.000128517 | 1.44E-05 | 0.00055 | 2.06E-05 | -0.005526861 | CENPV |
| chr7 | 2938116 | 2939087 | 972 | 3 | 0.000128573 | 0.000775 | 0.00039 | 9.07E-05 | -3.53E-05 | CARD11 |
| chr9 | 127898120 | 127898896 | 777 | 7 | 0.000128619 | 2.77E-05 | 0.00365 | 2.72E-05 | -0.004299682 | AL157935.2, ST6GALNAC6 |
| chr12 | 2890258 | 2890960 | 703 | 11 | 0.000128724 | 0.001789 | 0.00077 | 3.78E-05 | -0.000577071 | TULP3 |
| chr19 | 289762 | 290403 | 642 | 3 | 0.000128727 | 0.000374 | 0.00025 | 3.09E-05 | -0.007947823 | PLPP2 |
| chr12 | 6925662 | 6926023 | 362 | 2 | 0.000128727 | 3.49E-05 | 0.00077 | 4.74E-05 | -0.007944883 | ATN1 |
| chr17 | 81259593 | 81259687 | 95 | 2 | 0.000128743 | 5.46E-05 | 0.00011 | 3.38E-05 | -0.008462645 | SLC38A10 |
| chr2 | 136126176 | 136126546 | 371 | 2 | 0.00012884 | 1.24E-05 | 0.00027 | 1.63E-05 | -0.011728514 | NA |
| chr2 | 119159110 | 119159809 | 700 | 4 | 0.000128849 | 1.32E-05 | 0.00016 | 9.43E-06 | -0.000319961 | NA |
| chr6 | 16409913 | 16410075 | 163 | 3 | 0.00012885 | 7.61E-06 | 0.00041 | 1.18E-05 | -0.012747647 | ATXN1 |
| chr3 | 53232318 | 53232625 | 308 | 4 | 0.000129559 | 5.68E-06 | 0.00463 | 1.42E-05 | -0.006521114 | TKT |
| chr1 | 39410115 | 39410387 | 273 | 3 | 0.000129565 | 4.91E-05 | 0.00774 | 0.00011 | -0.008317523 | MACF1 |
| chr5 | 177132333 | 177132880 | 548 | 4 | 0.000129573 | 9.39E-06 | 0.00504 | 2.56E-05 | -0.018827642 | NA |
| chr22 | 49303470 | 49303782 | 313 | 2 | 0.000129573 | 0.000161 | 5.53E-05 | 3.48E-05 | -0.004302566 | NA |
| chr10 | 79081741 | 79081980 | 240 | 3 | 0.000129603 | 3.49E-05 | 0.0008 | 1.91E-05 | -0.008721244 | ZMIZ1 |
| chr17 | 75891127 | 75891570 | 444 | 5 | 0.000129604 | 4.53E-05 | 0.00012 | 2.65E-05 | -0.000548736 | TRIM65 |
| chr20 | 57713727 | 57714024 | 298 | 2 | 0.000129649 | 0.000149 | 0.00073 | 0.00015 | -0.011962819 | NA |
| chr19 | 48391437 | 48391773 | 337 | 9 | 0.000129834 | 0.00682 | 0.0001 | 6.66E-05 | -0.000917976 | KDELR1 |
| chr20 | 44355087 | 44355939 | 853 | 11 | 0.000129933 | 0.004837 | 0.00012 | 5.84E-05 | -0.002754464 | HNF4A, AL117382.1 |
| chr3 | 45928982 | 45929360 | 379 | 2 | 0.00012997 | 8.88E-06 | 0.00066 | 1.43E-05 | -0.015166649 | FYCO1 |
| chr1 | 225907585 | 225907638 | 54 | 2 | 0.000129972 | 2.78E-05 | 0.00013 | 2.33E-05 | -0.00428894 | LEFTY1, AL117348.2 |
| chr2 | 237158303 | 237159536 | 1234 | 3 | 0.000129993 | 2.49E-06 | 0.00012 | 7.39E-07 | -0.006729933 | NA |
| chr13 | 111956485 | 111957231 | 747 | 3 | 0.000129993 | 0.000113 | 0.00223 | 8.80E-05 | -0.002410572 | NA |
| chr12 | 50072727 | 50073271 | 545 | 3 | 0.000130035 | 9.28E-06 | 0.00166 | 1.84E-05 | -0.009496948 | ASIC1 |
| chr19 | 16598621 | 16598623 | 3 | 2 | 0.0001301 | 1.58E-05 | 0.00019 | 1.80E-05 | -0.006945189 | AC008764.1, MED26, AC008764.4 |
| chr16 | 2740807 | 2741086 | 280 | 4 | 0.000130101 | 0.000367 | 0.00027 | 2.98E-05 | -0.005652184 | SRRM2-AS1 |
| chr13 | 112441804 | 112441889 | 86 | 2 | 0.000130101 | 0.000107 | 6.07E-05 | 3.17E-05 | -0.007824932 | NA |
| chr1 | 1313722 | 1313987 | 266 | 3 | 0.000130114 | 0.000287 | 0.00024 | 9.97E-05 | -0.003769626 | INTS11 |
| chr3 | 126988260 | 126989637 | 1378 | 7 | 0.000130224 | 4.03E-06 | 0.00101 | 1.01E-06 | -0.001207923 | PLXNA1 |
| chr14 | 76424084 | 76424610 | 527 | 3 | 0.000130315 | 2.95E-05 | 0.00024 | 1.36E-05 | -0.007566918 | ESRRB |
| chr7 | 1493414 | 1493866 | 453 | 2 | 0.000130435 | 1.03E-05 | 0.00073 | 1.66E-05 | -0.005787374 | INTS1 |
| chr5 | 53913037 | 53913068 | 32 | 2 | 0.00013055 | 0.000129 | 0.00011 | 5.24E-05 | -0.006198458 | ARL15 |
| chr19 | 40413338 | 40413853 | 516 | 4 | 0.00013057 | 9.10E-05 | 0.00103 | 3.56E-05 | -0.015160894 | PRX |
| chr16 | 85615143 | 85615875 | 733 | 3 | 0.000130639 | 3.41E-05 | 0.0035 | 6.81E-05 | -0.008504123 | GSE1 |
| chr17 | 50767648 | 50768397 | 750 | 5 | 0.000130701 | 1.04E-05 | 0.00247 | 1.66E-05 | -0.003751526 | NA |
| chr15 | 79844870 | 79845346 | 477 | 3 | 0.000130788 | 0.000919 | 0.00054 | 3.13E-05 | -0.003327261 | MTHFS, ST20-MTHFS |
| chr8 | 10839061 | 10839179 | 119 | 3 | 0.000130906 | 0.002718 | 0.00033 | 0.00013 | -0.001413073 | AC105001.2, PINX1 |
| chr19 | 54257959 | 54257964 | 6 | 2 | 0.000131002 | 0.01473 | 7.86E-05 | 0.00015 | -0.004254071 | NA |
| chr12 | 130953257 | 130953995 | 739 | 8 | 0.000131069 | 0.001148 | 0.00055 | 0.00011 | -0.003913845 | ADGRD1 |
| chr16 | 16101762 | 16101769 | 8 | 2 | 0.000131195 | 1.76E-05 | 0.00032 | 2.24E-05 | -0.021135683 | ABCC1 |
| chr6 | 30885478 | 30886234 | 757 | 8 | 0.000131246 | 1.95E-05 | 0.01092 | 3.88E-05 | -0.009736377 | DDR1 |
| chr11 | 44615941 | 44616096 | 156 | 3 | 0.000132037 | 0.000287 | 0.00014 | 2.40E-05 | 0.000135208 | CD82 |
| chr17 | 58324262 | 58325308 | 1047 | 5 | 0.000132112 | 2.89E-05 | 0.00035 | 9.16E-06 | -0.009905357 | TSPOAP1-AS1, TSPOAP1 |
| chr5 | 34930139 | 34930513 | 375 | 3 | 0.000132112 | 0.000256 | 0.00035 | 0.00014 | 0.002539883 | DNAJC21 |
| chr17 | 38706875 | 38706963 | 89 | 2 | 0.000132143 | 1.60E-05 | 0.00075 | 2.45E-05 | -0.016542554 | MLLT6 |
| chr1 | 64133023 | 64133655 | 633 | 3 | 0.000132322 | 0.002801 | 4.13E-05 | 7.47E-05 | -0.008458673 | ROR1, ROR1-AS1 |
| chr5 | 157730985 | 157731874 | 890 | 10 | 0.000132342 | 5.56E-05 | 0.00299 | 5.56E-06 | 0.001217642 | THG1L |
| chr15 | 51884032 | 51884492 | 461 | 3 | 0.000132427 | 0.000331 | 0.00019 | 4.91E-05 | -0.000725389 | TMOD3 |
| chr17 | 81309938 | 81310159 | 222 | 3 | 0.000132446 | 0.000937 | 0.00031 | 0.00028 | -0.012886943 | NA |
| chr11 | 124891761 | 124893115 | 1355 | 3 | 0.000132464 | 1.51E-06 | 0.00151 | 4.07E-06 | -0.00817716 | AP003501.2, ROBO4 |
| chr12 | 9950006 | 9950221 | 216 | 2 | 0.000132537 | 1.05E-05 | 0.00039 | 1.53E-05 | 0.009255538 | CLEC12A-AS1 |
| chr11 | 68043438 | 68043727 | 290 | 3 | 0.000132587 | 5.19E-05 | 0.00022 | 3.23E-05 | -0.005562117 | TCIRG1 |
| chr12 | 106302477 | 106302895 | 419 | 6 | 0.000132589 | 0.003226 | 0.00275 | 7.72E-05 | 0.002377988 | TCP11L2, CKAP4 |
| chr10 | 81116572 | 81116583 | 12 | 2 | 0.000132597 | 3.31E-05 | 5.89E-05 | 1.63E-05 | -0.012458248 | NA |
| chr10 | 35598413 | 35598669 | 257 | 4 | 0.000132749 | 0.003494 | 0.00012 | 3.22E-05 | -0.010212498 | NA |
| chr9 | 92765527 | 92766217 | 691 | 4 | 0.000132861 | 0.000174 | 0.00032 | 1.58E-05 | -0.002968564 | NA |
| chr3 | 149780320 | 149780548 | 229 | 3 | 0.000132979 | 2.36E-05 | 0.00018 | 1.73E-05 | 0.001830376 | ANKUB1 |
| chr6 | 44660823 | 44661666 | 844 | 4 | 0.000132989 | 4.70E-06 | 0.00213 | 1.24E-05 | -0.002826578 | NA |
| chr16 | 4701266 | 4701806 | 541 | 4 | 0.000133116 | 0.000169 | 7.14E-05 | 2.94E-05 | -0.00408107 | ANKS3 |
| chr1 | 180586624 | 180586783 | 160 | 2 | 0.000133637 | 9.94E-06 | 0.00082 | 1.64E-05 | -0.005831213 | NA |
| chr8 | 143150236 | 143150684 | 449 | 2 | 0.000133663 | 1.59E-05 | 0.00026 | 1.98E-05 | -0.004031965 | NA |
| chr16 | 2284255 | 2284963 | 709 | 5 | 0.000133886 | 1.54E-05 | 0.00183 | 2.98E-05 | -0.002550051 | ABCA3 |
| chr11 | 33869371 | 33870253 | 883 | 7 | 0.000133909 | 2.56E-05 | 0.01093 | 6.19E-05 | -0.000244434 | LMO2 |
| chr6 | 30909018 | 30909469 | 452 | 4 | 0.000134083 | 0.000385 | 0.00086 | 0.00026 | 0.0056319 | GTF2H4, AL669830.1 |
| chr8 | 95679465 | 95679958 | 494 | 3 | 0.000134236 | 9.93E-06 | 0.00035 | 1.24E-05 | -0.004246227 | C8orf37-AS1 |
| chr10 | 3069226 | 3069260 | 35 | 2 | 0.000134261 | 0.000101 | 0.00071 | 0.00011 | 0.000426655 | PFKP |
| chr16 | 69319855 | 69321109 | 1255 | 7 | 0.000134347 | 8.35E-07 | 0.00247 | 2.59E-06 | -0.002100685 | AC026464.4, VPS4A, COG8 |
| chr6 | 168381687 | 168382302 | 616 | 6 | 0.000134377 | 0.004457 | 0.00034 | 0.00029 | -0.001973482 | NA |
| chr5 | 14462979 | 14464903 | 1925 | 6 | 0.000134401 | 1.57E-07 | 0.00014 | 1.35E-07 | -0.009806806 | TRIO |
| chr6 | 564561 | 564609 | 49 | 3 | 0.000134414 | 1.76E-05 | 0.00162 | 3.27E-05 | -0.006482127 | EXOC2 |
| chr3 | 12708631 | 12709191 | 561 | 2 | 0.000134423 | 1.37E-05 | 0.00027 | 1.76E-05 | -0.006550965 | NA |

| | | | | | | | | | | |
|---|---|---|---|---|---|---|---|---|---|---|
| chr19 | 19225341 | 19225612 | 272 | 5 | 0.000134476 | 0.000709 | 3.27E-05 | 3.74E-05 | -0.011788569 | NCAN |
| chr19 | 1254067 | 1254436 | 370 | 3 | 0.000134484 | 0.000124 | 0.00047 | 9.82E-05 | -0.005261242 | MIDN |
| chr11 | 20160341 | 20161149 | 809 | 11 | 0.000134553 | 4.92E-05 | 0.00096 | 1.01E-05 | -0.002485511 | DBX1 |
| chr22 | 37905403 | 37905570 | 168 | 2 | 0.000134612 | 0.000252 | 0.00013 | 8.34E-05 | -0.013827446 | NA |
| chr14 | 23307273 | 23307772 | 500 | 3 | 0.000134645 | 2.36E-05 | 0.00063 | 2.91E-05 | -0.007806843 | BCL2L2, BCL2L2-PABPN1 |
| chr10 | 43456374 | 43456978 | 605 | 5 | 0.0001348 | 3.69E-06 | 0.0039 | 9.45E-06 | -0.003587829 | ZNF487 |
| chr19 | 54114231 | 54114551 | 321 | 6 | 0.000134801 | 6.42E-05 | 0.00042 | 4.42E-05 | -6.34E-06 | TFPT |
| chr1 | 212167205 | 212167282 | 78 | 3 | 0.000134971 | 1.62E-05 | 0.00248 | 3.16E-05 | -0.008711864 | NA |
| chr6 | 107069218 | 107070385 | 1168 | 3 | 0.000134975 | 9.35E-07 | 0.00039 | 1.51E-06 | -0.001568491 | BEND3 |
| chr11 | 62151890 | 62152526 | 637 | 6 | 0.00013516 | 1.67E-05 | 0.0004 | 1.98E-05 | -0.002755867 | INCENP |
| chr8 | 42950107 | 42950233 | 127 | 3 | 0.00013516 | 2.58E-05 | 0.00069 | 1.51E-05 | -0.004759092 | HOOK3 |
| chr19 | 19528565 | 19528787 | 223 | 6 | 0.000135574 | 0.000274 | 0.00028 | 8.61E-05 | -0.005951286 | NDUFA13, AC011448.1 |
| chr8 | 17632912 | 17633043 | 132 | 2 | 0.000135625 | 2.61E-05 | 0.00077 | 3.72E-05 | -0.006343992 | PDGFRL |
| chr2 | 394115 | 394572 | 458 | 5 | 0.000135629 | 2.23E-06 | 0.00455 | 8.54E-06 | -0.013301775 | AC105393.2 |
| chr1 | 12476435 | 12476477 | 43 | 3 | 0.000135681 | 8.53E-05 | 0.00011 | 3.07E-05 | -0.001228666 | VPS13D |
| chr2 | 27306920 | 27308016 | 1097 | 7 | 0.000135802 | 0.00027 | 0.03251 | 0.0002 | -0.007715897 | TRIM54, UCN |
| chr4 | 110475720 | 110475999 | 280 | 3 | 0.000135838 | 0.000594 | 0.00081 | 4.47E-05 | -0.013836051 | ENPEP |
| chr18 | 35713727 | 35713796 | 70 | 2 | 0.000135925 | 9.39E-06 | 0.00059 | 1.49E-05 | -0.013377149 | NA |
| chr9 | 137777180 | 137778169 | 990 | 3 | 0.000135956 | 1.19E-05 | 8.97E-05 | 3.70E-06 | -0.003136619 | EHMT1, AL590627.1 |
| chr1 | 28258884 | 28259594 | 711 | 9 | 0.000135973 | 4.75E-06 | 0.0117 | 8.95E-06 | -0.002411238 | SESN2 |
| chr20 | 18793591 | 18794643 | 1053 | 12 | 0.000135993 | 8.92E-05 | 0.00379 | 2.18E-05 | 0.003041429 | LINC00653, LINC00652 |
| chr20 | 19688528 | 19688538 | 11 | 2 | 0.000136044 | 5.16E-05 | 9.97E-05 | 3.02E-05 | -0.010486578 | SLC24A3 |
| chr1 | 26994000 | 26994654 | 655 | 5 | 0.000136058 | 0.00152 | 0.00425 | 4.92E-05 | 0.00178681 | TRNP1 |
| chr8 | 63208151 | 63208548 | 398 | 2 | 0.000136137 | 0.004306 | 0.0001 | 0.00017 | -0.003973037 | YTHDF3 |
| chr14 | 91947153 | 91948558 | 1406 | 14 | 0.000136168 | 0.000523 | 0.00058 | 5.43E-06 | -0.00400325 | FBLN5 |
| chr10 | 132758857 | 132759523 | 667 | 4 | 0.000136168 | 5.80E-06 | 0.00316 | 1.48E-05 | -0.003882216 | INPP5A |
| chr15 | 52179384 | 52179520 | 137 | 2 | 0.000136273 | 8.92E-05 | 0.00011 | 4.35E-05 | -0.00166214 | GNB5 |
| chr15 | 93073714 | 93074604 | 891 | 12 | 0.000136499 | 0.000355 | 0.0062 | 2.62E-05 | -0.008833407 | RGMA |
| chr1 | 247315930 | 247316244 | 315 | 2 | 0.000136556 | 5.59E-05 | 0.00126 | 7.81E-05 | -0.003644549 | ZNF496 |
| chr11 | 35814163 | 35814269 | 107 | 3 | 0.000136559 | 1.71E-05 | 0.0007 | 1.86E-05 | -0.01298032 | TRIM44 |
| chr14 | 35292625 | 35293087 | 463 | 3 | 0.000136575 | 0.001166 | 0.0068 | 0.00082 | 0.000356181 | AL121594.1, PSMA6 |
| chr5 | 191012 | 191988 | 977 | 13 | 0.000136718 | 1.23E-06 | 0.02371 | 1.00E-05 | -0.013221828 | LRRC14B |
| chr3 | 133036995 | 133037280 | 286 | 5 | 0.000136722 | 0.001068 | 0.0004 | 0.00017 | -0.002079821 | AC079942.1 |
| chr21 | 30349110 | 30349759 | 650 | 3 | 0.000136756 | 1.22E-05 | 0.00038 | 1.03E-05 | -0.005219846 | NA |
| chr1 | 111749052 | 111749254 | 203 | 2 | 0.000136969 | 0.000177 | 4.10E-05 | 2.78E-05 | -0.006945288 | INKA2, AL049557.1 |
| chr12 | 132017324 | 132018091 | 768 | 6 | 0.000137056 | 2.26E-05 | 0.00068 | 1.88E-05 | -0.001454149 | EP400 |
| chr12 | 48957158 | 48958082 | 925 | 9 | 0.000137154 | 0.000466 | 0.00367 | 1.72E-05 | -0.001571111 | AC073610.2, ARF3 |
| chr8 | 27430522 | 27430888 | 367 | 7 | 0.000137173 | 0.100856 | 7.47E-06 | 0.00011 | -0.000655594 | PTK2B |
| chr6 | 85448173 | 85448640 | 468 | 3 | 0.000137318 | 0.001738 | 0.00012 | 9.23E-05 | -0.001435253 | NA |
| chr13 | 39148796 | 39148922 | 127 | 2 | 0.000137334 | 0.00068 | 2.87E-05 | 2.91E-05 | -0.018559294 | AL354809.1 |
| chr6 | 7728443 | 7729561 | 1119 | 5 | 0.000137406 | 5.78E-07 | 0.00576 | 3.15E-06 | -0.00597715 | BMP6 |
| chr12 | 130426131 | 130426186 | 56 | 2 | 0.000137527 | 2.68E-05 | 0.00034 | 3.19E-05 | -0.008000756 | AC063926.3, RIMBP2 |
| chr11 | 1456481 | 1456882 | 402 | 2 | 0.000137751 | 2.28E-05 | 0.00017 | 2.27E-05 | -0.001967307 | BRSK2 |
| chr17 | 46875331 | 46875339 | 9 | 2 | 0.000137968 | 1.85E-05 | 0.00021 | 2.09E-05 | -0.011217513 | WNT9B |
| chr17 | 49132184 | 49132804 | 621 | 10 | 0.00013809 | 0.000231 | 0.01369 | 0.00012 | 0.011193963 | B4GALNT2 |
| chr13 | 89553762 | 89554478 | 717 | 3 | 0.000138255 | 0.000524 | 0.00025 | 7.78E-06 | -0.005747172 | LINC02336, LINC00353 |
| chr7 | 142783759 | 142784406 | 648 | 2 | 0.000138355 | 2.10E-05 | 0.00071 | 3.07E-05 | -0.019938495 | NA |
| chr4 | 719759 | 720552 | 794 | 7 | 0.000138358 | 0.003645 | 0.00619 | 0.00028 | -0.002353763 | PCGF3 |
| chr14 | 104841254 | 104841559 | 306 | 2 | 0.000138398 | 5.01E-05 | 8.95E-05 | 2.78E-05 | -0.010453941 | NA |
| chr11 | 66956957 | 66957687 | 731 | 2 | 0.000138458 | 1.54E-05 | 0.00076 | 2.38E-05 | -0.009954893 | PC |
| chr9 | 124453390 | 124453768 | 379 | 5 | 0.000138562 | 0.001546 | 0.00014 | 4.52E-05 | 0.004634206 | ADGRD2 |
| chr16 | 85463979 | 85464436 | 458 | 4 | 0.000138646 | 0.002847 | 0.00027 | 5.12E-05 | -0.003824943 | GSE1 |
| chr6 | 105329659 | 105330331 | 673 | 2 | 0.000138723 | 3.98E-06 | 0.00041 | 6.69E-06 | -0.006847678 | PREP |
| chr16 | 2530832 | 2531832 | 1001 | 10 | 0.000138863 | 0.000108 | 0.008 | 3.27E-05 | -0.003973397 | AMDHD2, CEMP1 |
| chr1 | 165910845 | 165911187 | 343 | 3 | 0.000138941 | 6.34E-06 | 0.00203 | 1.42E-05 | -0.007058246 | UCK2 |
| chr18 | 48943177 | 48943569 | 393 | 3 | 0.000139036 | 0.000623 | 0.0008 | 0.00017 | -0.003508576 | SMAD7 |
| chr22 | 38947294 | 38947508 | 215 | 3 | 0.000139256 | 2.49E-05 | 0.00077 | 2.30E-05 | -0.010683959 | NA |
| chr1 | 111909285 | 111909588 | 304 | 5 | 0.000139435 | 0.002577 | 0.00022 | 3.51E-05 | -0.002514681 | KCND3-AS1, KCND3 |
| chr2 | 1163154 | 1163547 | 394 | 2 | 0.000139554 | 7.76E-05 | 0.0005 | 8.00E-05 | -0.009428789 | SNTG2 |
| chr20 | 50189953 | 50190391 | 439 | 3 | 0.000139569 | 0.026178 | 0.0113 | 0.00563 | -0.001055267 | CEBPB-AS1 |
| chr17 | 81994266 | 81994652 | 387 | 4 | 0.000139695 | 0.000353 | 0.00031 | 4.75E-05 | -0.001593428 | ASPSCR1 |
| chr17 | 81455504 | 81456113 | 610 | 2 | 0.000139754 | 0.000127 | 0.00053 | 0.00012 | -0.011361114 | BAHCC1 |
| chr3 | 129975174 | 129975822 | 649 | 5 | 0.000139902 | 0.000149 | 0.00399 | 5.57E-05 | -0.000542044 | TRH |
| chr20 | 62137153 | 62137220 | 68 | 2 | 0.00014001 | 9.33E-06 | 0.00048 | 1.44E-05 | -0.026010351 | PSMA7 |
| chr2 | 172944273 | 172944516 | 244 | 2 | 0.000140036 | 8.12E-06 | 0.00065 | 1.33E-05 | -0.069115408 | RAPGEF4 |
| chr11 | 13339399 | 13339798 | 400 | 2 | 0.000140039 | 1.62E-05 | 0.00082 | 2.51E-05 | -0.009863346 | ARNTL |
| chr8 | 17676989 | 17677003 | 15 | 2 | 0.000140082 | 3.89E-05 | 0.00176 | 6.00E-05 | -0.017282168 | MTUS1 |
| chr1 | 41160777 | 41161296 | 520 | 4 | 0.000140301 | 1.67E-05 | 0.0015 | 2.48E-05 | -0.010304675 | SCMH1 |
| chr1 | 6929972 | 6930284 | 313 | 2 | 0.000140481 | 7.74E-06 | 0.00068 | 1.27E-05 | -0.004583232 | CAMTA1 |
| chr11 | 13314012 | 13315127 | 1116 | 3 | 0.000140859 | 2.96E-05 | 0.00017 | 4.05E-06 | -0.004159332 | ARNTL |
| chr3 | 119468499 | 119468931 | 433 | 6 | 0.000140947 | 0.001285 | 0.00318 | 5.06E-05 | -0.012273817 | TMEM39A |
| chr22 | 40408644 | 40409007 | 364 | 3 | 0.000140995 | 0.005135 | 0.00016 | 0.00041 | -0.001664049 | AL022238.3, SGSM3 |
| chr8 | 17576116 | 17577579 | 1464 | 17 | 0.000141003 | 1.15E-05 | 0.00092 | 2.22E-06 | 0.005407411 | PDGFRL |
| chr5 | 149405094 | 149406253 | 1160 | 4 | 0.000141107 | 1.95E-06 | 0.00264 | 5.93E-06 | -0.010758414 | NA |
| chr17 | 22216815 | 22217520 | 706 | 4 | 0.000141258 | 0.001085 | 4.16E-05 | 6.57E-05 | -0.016919891 | NA |
| chr11 | 108486816 | 108487027 | 212 | 2 | 0.000141343 | 0.001229 | 3.62E-05 | 4.13E-05 | -0.002921479 | POGLUT3 |
| chr20 | 62394087 | 62394222 | 136 | 2 | 0.000141388 | 0.000393 | 0.00689 | 0.00069 | -0.003011562 | CABLES2 |
| chr4 | 2761826 | 2762678 | 853 | 2 | 0.000141489 | 4.70E-06 | 0.00039 | 7.65E-06 | -0.011153076 | NA |
| chr3 | 106404943 | 106405072 | 130 | 2 | 0.000141592 | 3.27E-05 | 0.0004 | 3.87E-05 | 0.00061442 | AC079382.2 |
| chr5 | 29396013 | 29396046 | 34 | 2 | 0.000141786 | 7.91E-06 | 0.00072 | 1.31E-05 | -0.00918171 | LINC02064 |
| chr10 | 102869054 | 102869202 | 149 | 4 | 0.000141909 | 0.000138 | 3.38E-05 | 9.88E-06 | -0.011954894 | BORCS7-ASMT |
| chr16 | 28985720 | 28986138 | 419 | 3 | 0.000141914 | 9.66E-05 | 0.00633 | 0.00019 | -0.007974011 | AC009093.11, AC109460.3, LAT |
| chr10 | 86346487 | 86346594 | 108 | 2 | 0.000141914 | 0.001728 | 3.61E-05 | 4.36E-05 | -0.007062946 | GRID1 |
| chr12 | 49942881 | 49943248 | 368 | 3 | 0.000142107 | 6.47E-05 | 0.00087 | 2.63E-05 | -0.016865304 | NA |
| chr15 | 85592267 | 85592433 | 167 | 2 | 0.000142107 | 0.000125 | 5.06E-05 | 2.91E-05 | -0.013409799 | AKAP13 |
| chr3 | 48287768 | 48288012 | 245 | 2 | 0.000142216 | 2.01E-05 | 0.00013 | 1.87E-05 | -0.011507566 | ZNF589 |
| chr3 | 15601061 | 15602023 | 963 | 21 | 0.000142247 | 1.99E-05 | 0.03517 | 4.31E-05 | 0.000699676 | BTD, HACL1 |
| chr7 | 4727775 | 4728616 | 842 | 4 | 0.00014243 | 0.002774 | 0.00044 | 5.80E-05 | -0.000587853 | FOXK1, AC092428.1 |
| chr16 | 3035981 | 3036537 | 557 | 4 | 0.000142453 | 0.001187 | 0.00623 | 0.00082 | -0.002363216 | AC108134.1, BICDL2 |
| chr2 | 208124524 | 208124928 | 405 | 6 | 0.000142497 | 0.000491 | 6.19E-05 | 2.17E-05 | -0.002603725 | CRYGD |
| chr6 | 26242893 | 26243407 | 515 | 2 | 0.000142497 | 8.16E-06 | 0.00047 | 1.27E-05 | -0.006974275 | NA |
| chr20 | 45827512 | 45827845 | 334 | 5 | 0.000142583 | 0.000164 | 0.00061 | 2.80E-05 | -0.003709769 | TNNC2 |
| chr22 | 37063471 | 37063473 | 3 | 2 | 0.000142631 | 2.01E-05 | 0.00016 | 2.01E-05 | -0.016618113 | NA |
| chr7 | 41096971 | 41097711 | 741 | 4 | 0.00014273 | 0.002053 | 0.00012 | 2.76E-05 | -0.004675673 | LINC01449 |
| chr1 | 3697626 | 3699198 | 1573 | 8 | 0.000142758 | 7.52E-05 | 0.00015 | 1.69E-06 | -0.003059784 | TP73 |
| chr2 | 25252936 | 25253251 | 316 | 3 | 0.000143155 | 0.000572 | 7.42E-05 | 7.19E-05 | -0.007429242 | DNMT3A |
| chr10 | 119442042 | 119442469 | 428 | 2 | 0.00014316 | 0.003634 | 0.00013 | 0.00023 | -0.008933626 | GRK5 |
| chr3 | 156290981 | 156291362 | 382 | 5 | 0.00014334 | 0.000133 | 0.00372 | 7.65E-05 | -0.006326946 | KCNAB1, AC091607.2 |
| chr7 | 108004630 | 108004895 | 266 | 2 | 0.000143935 | 0.00197 | 8.93E-05 | 0.00012 | -0.012842987 | NA |
| chr5 | 150649653 | 150650156 | 504 | 3 | 0.000143956 | 3.17E-05 | 0.00013 | 1.38E-05 | -0.005694539 | SYNPO |
| chr16 | 2039865 | 2039896 | 32 | 2 | 0.000144022 | 3.13E-05 | 0.00037 | 3.67E-05 | -0.00604635 | NTHL1 |
| chr5 | 32585633 | 32585789 | 157 | 3 | 0.000144146 | 5.25E-05 | 0.00014 | 3.09E-05 | -0.000787996 | SUB1 |
| chr12 | 109534942 | 109535474 | 533 | 5 | 0.000144166 | 0.00011 | 0.00012 | 3.25E-05 | -0.001343152 | UBE3B |
| chr18 | 79782921 | 79783269 | 349 | 3 | 0.000144166 | 3.09E-05 | 0.00014 | 2.22E-05 | -0.008503783 | NA |
| chr13 | 74290220 | 74290645 | 426 | 3 | 0.000144387 | 5.03E-06 | 0.00256 | 1.28E-05 | -0.029512938 | AL353660.1 |
| chr2 | 74552672 | 74553340 | 669 | 8 | 0.000144544 | 1.24E-05 | 0.00718 | 1.63E-05 | -0.008324899 | DOK1, LOXL3 |
| chr17 | 6650857 | 6651651 | 795 | 6 | 0.000144544 | 2.57E-05 | 0.00085 | 1.22E-05 | 0.003047317 | MED31 |
| chr3 | 112409238 | 112409293 | 56 | 2 | 0.000144653 | 0.001756 | 0.00014 | 0.00019 | -0.00633306 | NA |
| chr10 | 4825581 | 4826320 | 740 | 13 | 0.000144669 | 0.00017 | 0.0008 | 2.53E-05 | -0.004889629 | AKR1E2 |
| chr2 | 20246552 | 20246594 | 43 | 4 | 0.000144747 | 9.99E-06 | 0.00128 | 1.39E-05 | -0.004056251 | NA |
| chr15 | 31236051 | 31236792 | 742 | 4 | 0.000144821 | 9.54E-06 | 0.00105 | 8.55E-06 | -0.009017052 | NA |
| chr17 | 17686157 | 17686568 | 412 | 3 | 0.000144937 | 3.84E-05 | 0.0017 | 3.95E-05 | -0.013953478 | RAI1 |
| chr4 | 173169407 | 173169851 | 445 | 4 | 0.000145062 | 2.87E-05 | 0.00108 | 2.62E-05 | -0.003043653 | GALNT7, AC105285.1 |
| chr11 | 118929808 | 118930460 | 653 | 4 | 0.000145188 | 0.000114 | 0.002 | 8.78E-05 | -0.000956488 | UPK2 |
| chr14 | 32724899 | 32725125 | 227 | 2 | 0.000145253 | 1.22E-05 | 0.00026 | 1.59E-05 | -0.021875222 | AKAP6 |
| chr11 | 61744338 | 61744637 | 300 | 3 | 0.000145389 | 2.38E-05 | 0.00182 | 4.01E-05 | -0.004960616 | DAGLA |
| chr22 | 38069498 | 38069808 | 311 | 4 | 0.000145431 | 4.56E-05 | 0.0014 | 3.64E-05 | -0.005210291 | PICK1, AL031587.2 |
| chr8 | 81633191 | 81633453 | 263 | 2 | 0.000145482 | 1.46E-05 | 0.00024 | 1.81E-05 | -0.011354181 | NA |
| chr6 | 29011433 | 29011721 | 289 | 11 | 0.00014563 | 0.000163 | 0.0034 | 1.85E-05 | -8.59E-05 | NA |
| chr15 | 74614661 | 74615405 | 745 | 9 | 0.000145651 | 0.005906 | 0.00256 | 0.00067 | -8.31E-05 | CLK3, AC100835.1 |
| chr6 | 30730807 | 30731586 | 780 | 13 | 0.000145863 | 4.34E-05 | 0.02414 | 1.98E-05 | -0.005266045 | FLOT1 |
| chr6 | 1575051 | 1575751 | 701 | 3 | 0.000145878 | 8.64E-06 | 0.00132 | 1.76E-05 | -0.008191221 | NA |
| chr2 | 113071678 | 113071891 | 214 | 3 | 0.000145904 | 9.17E-05 | 0.00097 | 6.47E-05 | -0.003247283 | IL1F10 |
| chr16 | 2498127 | 2498613 | 487 | 4 | 0.000146202 | 0.00011 | 0.00333 | 4.93E-05 | -0.004093584 | TBC1D24, AC093525.2 |
| chr17 | 7316000 | 7316652 | 653 | 3 | 0.000146309 | 8.14E-05 | 0.00455 | 0.00016 | -0.00120338 | AC026954.2, NEURL4 |
| chr7 | 27185777 | 27186710 | 934 | 13 | 0.000146346 | 0.000246 | 0.02896 | 0.00039 | -0.008050568 | HOXA11-AS, AC004080.14, AC004080.8, AC004080.2 |
| chr6 | 88142219 | 88142251 | 33 | 2 | 0.000146346 | 1.38E-05 | 0.00015 | 1.49E-05 | -0.006167734 | CNR1 |
| chr3 | 195063676 | 195063991 | 316 | 2 | 0.000146739 | 4.00E-05 | 0.00028 | 4.06E-05 | -0.033247809 | NA |
| chr16 | 13935400 | 13935964 | 565 | 8 | 0.000146871 | 1.40E-05 | 0.00355 | 1.17E-05 | -0.002267727 | ERCC4, AC010401.1 |
| chr10 | 100967172 | 100967482 | 311 | 2 | 0.000146871 | 1.03E-05 | 0.00079 | 1.68E-05 | -0.00523144 | NA |
| chr2 | 60530505 | 60530767 | 263 | 2 | 0.000147063 | 0.003977 | 4.46E-05 | 6.31E-05 | -0.008815716 | BCL11A |
| chr19 | 33177002 | 33178151 | 1150 | 3 | 0.000147114 | 1.11E-06 | 0.00103 | 2.78E-06 | -0.002035075 | LRP3, AC008738.1 |
| chr12 | 79538725 | 79538766 | 42 | 2 | 0.000147347 | 0.00047 | 6.07E-05 | 5.44E-05 | -0.008774322 | NA |
| chr10 | 69629457 | 69629823 | 367 | 2 | 0.000147389 | 1.58E-05 | 1.60E-05 | 4.07E-06 | -0.017647985 | NA |
| chr14 | 51459073 | 51459562 | 490 | 2 | 0.000147492 | 1.31E-05 | 0.00033 | 1.78E-05 | -0.000953041 | FRMD6-AS2 |
| chr11 | 94171470 | 94171742 | 273 | 2 | 0.000147544 | 4.58E-05 | 0.00018 | 3.81E-05 | -0.005594114 | PANX1 |
| chr7 | 79454319 | 79454880 | 562 | 11 | 0.000147549 | 0.000197 | 0.00336 | 4.62E-05 | -0.003952197 | MAGI2-AS3 |
| chr11 | 118606319 | 118606691 | 373 | 4 | 0.000147658 | 4.14E-05 | 0.0003 | 2.85E-05 | -0.002208131 | PHLDB1 |
| chr2 | 102027436 | 102027785 | 350 | 3 | 0.00014771 | 1.58E-05 | 0.00043 | 1.28E-05 | -0.008203763 | IL1R2 |
| chr6 | 33006702 | 33007583 | 882 | 13 | 0.000147877 | 1.85E-05 | 0.0103 | 1.35E-05 | -0.002906139 | HLA-DOA |
| chr1 | 155034690 | 155034778 | 89 | 2 | 0.000148016 | 0.000621 | 6.98E-05 | 6.77E-05 | -0.007762586 | DCST1 |
| chr6 | 33312936 | 33313545 | 610 | 8 | 0.000148074 | 0.002343 | 0.03732 | 0.00395 | 0.001032269 | TAPBP |
| chr2 | 26401360 | 26402790 | 1431 | 14 | 0.000148171 | 8.48E-07 | 0.00541 | 2.13E-06 | -0.004428732 | DRC1 |
| chr16 | 89305126 | 89305574 | 449 | 5 | 0.000148278 | 9.85E-05 | 0.00328 | 2.09E-05 | -0.001884487 | ANKRD11 |
| chr16 | 27631918 | 27632371 | 454 | 4 | 0.0001484 | 0.000323 | 8.69E-05 | 4.34E-05 | -0.00115756 | KATNIP |
| chr2 | 127675036 | 127676685 | 1650 | 10 | 0.000148504 | 0.000759 | 0.00846 | 0.00013 | 0.00212267 | LIMS2 |
| chr16 | 1677430 | 1678947 | 1518 | 9 | 0.000148574 | 0.000666 | 0.00538 | 3.09E-05 | 0.000175252 | CRAMP1, AL031708.1, JPT2 |
| chr1 | 31296400 | 31297358 | 959 | 14 | 0.000148631 | 0.000205 | 0.00174 | 4.14E-05 | 0.000921064 | ZCCHC17, SNRNP40 |
| chr14 | 67564443 | 67564496 | 54 | 2 | 0.000148641 | 6.12E-05 | 5.70E-05 | 2.26E-05 | -0.012618655 | PLEKHH1 |
| chr7 | 158428164 | 158429084 | 921 | 7 | 0.000148806 | 0.000235 | 0.00029 | 1.42E-05 | -0.004067786 | PTPRN2 |
| chr12 | 111685874 | 111686396 | 523 | 12 | 0.000149222 | 0.000191 | 0.00472 | 2.40E-05 | -0.000180032 | ACAD10, BRAP |
| chr10 | 52561662 | 52562030 | 369 | 2 | 0.000149239 | 0.007338 | 4.38E-05 | 6.62E-05 | -0.016428376 | LNCAROD |
| chr20 | 63490565 | 63491019 | 455 | 4 | 0.000149269 | 0.00012 | 8.97E-05 | 3.19E-05 | -0.007883001 | EEF1A2 |
| chr2 | 230224324 | 230224926 | 603 | 2 | 0.000149531 | 0.000896 | 0.00014 | 0.00015 | -0.006214104 | SP140, SP110 |
| chr16 | 4883715 | 4884049 | 335 | 2 | 0.000149689 | 0.000228 | 0.00018 | 9.86E-05 | -0.000889564 | PPL |
| chr5 | 58583026 | 58583616 | 591 | 10 | 0.000149751 | 0.000408 | 0.0045 | 5.34E-05 | 0.001961241 | RAB3C |
| chr10 | 125823105 | 125823719 | 615 | 12 | 0.000149877 | 0.00029 | 0.00204 | 2.71E-05 | -0.000489784 | BCCIP, UROS |
| chr5 | 173054026 | 173054328 | 303 | 3 | 0.000149982 | 2.73E-05 | 0.00033 | 2.96E-05 | -0.017183291 | NA |
| chr19 | 38315771 | 38316553 | 783 | 10 | 0.000150005 | 0.000805 | 0.00013 | 3.55E-05 | 0.000571298 | YIF1B |
| chr2 | 186696516 | 186696594 | 79 | 2 | 0.000150034 | 0.000191 | 4.65E-05 | 3.20E-05 | -0.002412358 | FAM171B |
| chr8 | 144512633 | 144513642 | 1010 | 11 | 0.000150088 | 0.000427 | 0.01882 | 5.25E-05 | -0.001813617 | AC084125.4, RECQL4 |
| chr2 | 1680176 | 1680498 | 323 | 2 | 0.000150276 | 3.44E-05 | 0.00013 | 2.74E-05 | -0.008754789 | PXDN |

| | | | | | | | | | | |
|---|---|---|---|---|---|---|---|---|---|---|
| chr20 | 37295190 | 37295782 | 593 | 4 | 0.00015042 | 4.75E-06 | 0.00656 | 1.67E-05 | -0.005286974 | MANBAL |
| chr19 | 18834344 | 18834402 | 59 | 2 | 0.000150427 | 3.28E-05 | 0.00022 | 3.26E-05 | -0.017798319 | UPF1 |
| chr14 | 100348927 | 100348935 | 9 | 2 | 0.000150476 | 2.92E-05 | 7.33E-05 | 1.74E-05 | -0.006193039 | AL157871.4, WARS1 |
| chr1 | 27489789 | 27490735 | 947 | 14 | 0.000150506 | 0.000149 | 0.01019 | 1.80E-05 | 0.001202966 | WASF2 |
| chr17 | 48582389 | 48582735 | 347 | 7 | 0.000150513 | 0.00013 | 0.00539 | 5.03E-05 | -0.00586664 | HOXB-AS3, HOXB3 |
| chr2 | 241242666 | 241242902 | 237 | 2 | 0.000150653 | 0.001172 | 5.80E-05 | 6.66E-05 | -0.007015617 | HDLBP |
| chr12 | 91178365 | 91178499 | 135 | 3 | 0.000150813 | 4.43E-05 | 0.00279 | 6.14E-05 | -0.0109362 | DCN |
| chr9 | 22374348 | 22374623 | 276 | 3 | 0.000150836 | 1.23E-05 | 0.00058 | 1.92E-05 | -0.009808653 | NA |
| chr19 | 45900773 | 45901452 | 680 | 3 | 0.000150897 | 6.57E-06 | 0.00324 | 1.67E-05 | -0.010012392 | MYPOP |
| chr12 | 105106864 | 105107584 | 721 | 12 | 0.000150965 | 0.000306 | 0.00457 | 6.27E-05 | 0.000887723 | WASHC4, ALDH1L2 |
| chr10 | 76112200 | 76112700 | 501 | 4 | 0.000151038 | 1.02E-05 | 0.0034 | 2.47E-05 | -0.010670233 | LRMDA |
| chr14 | 64736612 | 64736633 | 22 | 2 | 0.000151114 | 0.003108 | 4.75E-05 | 6.54E-05 | -0.004490962 | PLEKHG3 |
| chr2 | 196276595 | 196276855 | 261 | 2 | 0.000151202 | 0.001462 | 3.95E-05 | 4.66E-05 | -0.012955151 | HECW2 |
| chr13 | 111623286 | 111623999 | 714 | 6 | 0.000151393 | 0.002801 | 0.00069 | 0.00014 | -0.002082043 | AL359649.1, LINC02337 |
| chr2 | 151289356 | 151290430 | 1075 | 11 | 0.000151413 | 1.34E-05 | 0.00427 | 4.38E-06 | 0.002076036 | NMI |
| chr1 | 203182778 | 203183167 | 390 | 3 | 0.000151559 | 3.75E-05 | 0.00021 | 2.97E-05 | 0.000793378 | CHI3L1 |
| chr20 | 10651483 | 10651524 | 42 | 2 | 0.000151602 | 1.09E-05 | 0.00086 | 1.78E-05 | -0.01757703 | JAG1 |
| chr16 | 88718155 | 88718524 | 370 | 4 | 0.000151629 | 0.000326 | 0.00017 | 7.10E-05 | -0.005266553 | PIEZO1 |
| chr1 | 84141752 | 84141870 | 119 | 2 | 0.00015175 | 2.39E-05 | 9.95E-05 | 1.83E-05 | -0.015935927 | PRKACB |
| chr19 | 49113395 | 49113744 | 350 | 2 | 0.000151857 | 9.89E-05 | 0.00178 | 0.00014 | -0.025859494 | NA |
| chr22 | 17980634 | 17980880 | 247 | 2 | 0.000151972 | 2.50E-05 | 0.00145 | 3.96E-05 | -0.022733813 | MICAL3, MIR648 |
| chr20 | 23049003 | 23049465 | 463 | 3 | 0.000151992 | 0.007889 | 0.00025 | 0.00071 | 0.002502769 | THBD |
| chr11 | 50268028 | 50268305 | 278 | 3 | 0.000152238 | 6.70E-06 | 0.0007 | 9.39E-06 | 0.000237888 | AC109635.4 |
| chr12 | 72029903 | 72030540 | 638 | 3 | 0.000152395 | 1.79E-05 | 0.00089 | 2.78E-05 | -0.00547505 | TPH2 |
| chr1 | 114581586 | 114582562 | 977 | 11 | 0.000152436 | 0.000156 | 0.00912 | 1.97E-05 | -0.000322976 | BCAS2 |
| chr2 | 230848007 | 230848266 | 260 | 3 | 0.000152436 | 0.000151 | 8.31E-05 | 3.41E-05 | -0.01103311 | NA |
| chr17 | 29594329 | 29594647 | 319 | 3 | 0.000152879 | 0.002297 | 6.73E-05 | 0.00012 | -0.004583643 | ABHD15-AS1, ANKRD13B |
| chr2 | 37343257 | 37343461 | 205 | 2 | 0.000152887 | 0.003702 | 7.01E-05 | 0.0001 | -0.01163008 | QPCT, AC007391.2 |
| chr4 | 27100998 | 27101305 | 308 | 4 | 0.000152925 | 0.000121 | 0.0014 | 2.47E-05 | -0.006948303 | NA |
| chr16 | 1350672 | 1350860 | 189 | 5 | 0.000153024 | 0.000376 | 0.00013 | 2.43E-05 | -0.006634922 | TSR3 |
| chr1 | 6221823 | 6222146 | 324 | 2 | 0.000153035 | 0.075784 | 4.83E-05 | 8.40E-05 | -0.00281714 | ICMT |
| chr3 | 50592277 | 50592634 | 358 | 2 | 0.000153056 | 5.46E-05 | 0.00013 | 3.67E-05 | -0.01029877 | HEMK1 |
| chr7 | 1218945 | 1219735 | 791 | 2 | 0.000153056 | 3.37E-05 | 0.00043 | 4.04E-05 | -0.01300792 | NA |
| chr1 | 145927476 | 145927977 | 502 | 11 | 0.000153281 | 3.53E-05 | 0.0049 | 2.97E-05 | 0.001777838 | LIX1L-AS1, AC243547.3, RBM8A |
| chr20 | 57519469 | 57519736 | 268 | 4 | 0.000153662 | 5.37E-05 | 0.00017 | 2.88E-05 | -0.004096297 | CTCFL |
| chr7 | 26363044 | 26363211 | 168 | 3 | 0.000153816 | 0.001317 | 0.00068 | 3.27E-05 | -0.01146088 | SNX10 |
| chr1 | 54274009 | 54274487 | 479 | 2 | 0.000153842 | 1.90E-05 | 0.00072 | 2.82E-05 | -0.006222094 | SSBP3 |
| chr8 | 68436651 | 68437090 | 440 | 2 | 0.000153856 | 1.34E-05 | 0.00068 | 2.06E-05 | -0.006653591 | C8orf34 |
| chr19 | 14529230 | 14530227 | 998 | 21 | 0.000154007 | 0.000113 | 0.01019 | 1.87E-05 | 0.000724963 | TECR, MIR639, DNAJB1 |
| chr20 | 58103788 | 58104173 | 386 | 3 | 0.000154075 | 0.005369 | 3.19E-05 | 6.77E-05 | -0.003262228 | NA |
| chr19 | 14136329 | 14137278 | 950 | 11 | 0.000154229 | 0.001011 | 0.00864 | 6.03E-05 | -2.91E-05 | AC022098.1, ASF1B |
| chr6 | 136550571 | 136551414 | 844 | 14 | 0.000154423 | 4.47E-06 | 0.01031 | 1.36E-05 | -0.004379797 | AL024508.1, MAP7 |
| chr5 | 140664397 | 140664867 | 471 | 10 | 0.000154479 | 0.002045 | 0.0008 | 3.30E-05 | -0.000104949 | WDR55 |
| chr12 | 57242959 | 57243197 | 239 | 2 | 0.000154505 | 0.000135 | 0.00052 | 0.00012 | -0.000542328 | NA |
| chr19 | 12806272 | 12806954 | 683 | 13 | 0.000154727 | 5.83E-05 | 0.00391 | 2.46E-05 | -0.000287553 | THSD8, RNASEH2A, HOOK2 |
| chr6 | 30341768 | 30342538 | 771 | 17 | 0.000154869 | 0.000147 | 0.00147 | 2.81E-05 | -5.45E-05 | TRIM39, TRIM39-RPP21 |
| chr8 | 1957114 | 1958340 | 1227 | 7 | 0.000155037 | 3.51E-06 | 0.00044 | 3.29E-07 | -0.004384149 | AC019257.8, ARHGEF10, AC019257.9 |
| chr4 | 47981436 | 47981646 | 211 | 4 | 0.000155151 | 0.000645 | 8.09E-05 | 6.17E-05 | -0.011070165 | NIPAL1, CNGA1 |
| chr4 | 140947598 | 140947839 | 242 | 2 | 0.000155201 | 9.12E-06 | 0.00066 | 1.47E-05 | -0.006910841 | RNF150 |
| chr17 | 48111736 | 48111751 | 16 | 2 | 0.000155213 | 1.20E-05 | 0.00025 | 1.55E-05 | 0.011102903 | SNX11 |
| chr22 | 23792567 | 23792636 | 70 | 2 | 0.000155321 | 3.70E-05 | 0.00182 | 5.76E-05 | -0.010798283 | SMARCB1 |
| chr17 | 47850527 | 47851418 | 892 | 9 | 0.000155387 | 0.002612 | 0.00129 | 7.35E-05 | -0.003975218 | SP6 |
| chr15 | 67254253 | 67254880 | 628 | 8 | 0.000155424 | 0.002025 | 0.00015 | 4.20E-05 | 0.000532573 | IQCH, AAGAB |
| chr15 | 40806638 | 40807073 | 436 | 6 | 0.000155622 | 0.000238 | 0.0016 | 4.38E-05 | -0.003186163 | DNAJC17 |
| chr12 | 94074983 | 94075451 | 469 | 2 | 0.000155671 | 8.53E-06 | 0.00073 | 1.40E-05 | -0.021523804 | NA |
| chr9 | 98263658 | 98263972 | 315 | 4 | 0.000156111 | 2.87E-05 | 0.00036 | 2.39E-05 | 0.000184752 | NA |
| chr11 | 36566667 | 36567263 | 597 | 4 | 0.0001562 | 9.97E-05 | 0.00046 | 9.26E-06 | -0.005304883 | RAG1 |
| chr17 | 43224565 | 43225402 | 838 | 4 | 0.000156268 | 2.51E-06 | 0.00233 | 5.64E-06 | -0.009702631 | CCDC200 |
| chr11 | 1480743 | 1481635 | 893 | 7 | 0.000156329 | 9.38E-06 | 0.00029 | 3.74E-06 | -0.002360484 | MOB2 |
| chr6 | 32852252 | 32854501 | 2250 | 50 | 0.000156556 | 0.000138 | 0.00674 | 1.73E-06 | 0.001134114 | PSMB9, TAP1 |
| chr15 | 99438781 | 99439141 | 361 | 3 | 0.000156692 | 0.001853 | 0.00072 | 3.73E-05 | -0.009598596 | NA |
| chr12 | 109773516 | 109774506 | 991 | 9 | 0.000156723 | 3.25E-06 | 0.00373 | 2.65E-06 | 0.009340731 | NA |
| chr4 | 168585610 | 168585969 | 360 | 3 | 0.000156734 | 1.26E-05 | 0.00094 | 2.01E-05 | -0.012809371 | PALLD |
| chr16 | 4696989 | 4697167 | 179 | 2 | 0.000156969 | 0.004185 | 3.25E-05 | 4.48E-05 | -0.002094025 | ANKS3 |
| chr10 | 13160710 | 13161051 | 342 | 2 | 0.000157288 | 0.000129 | 0.00012 | 5.69E-05 | -0.001607486 | NA |
| chr1 | 155933563 | 155933847 | 285 | 2 | 0.00015739 | 0.000269 | 0.00309 | 0.0004 | -0.005594926 | KHDC4 |
| chr1 | 27391717 | 27391991 | 275 | 4 | 0.00015749 | 5.51E-05 | 0.00055 | 3.90E-05 | -0.007690096 | NA |
| chr7 | 29482427 | 29482674 | 248 | 2 | 0.000157641 | 0.009374 | 6.39E-05 | 0.00011 | -0.002268859 | AC004593.2, CHN2 |
| chr9 | 123401804 | 123403190 | 1387 | 7 | 0.000157687 | 7.61E-05 | 0.0054 | 3.10E-05 | -0.003855118 | DENND1A, MIR601 |
| chr14 | 59630406 | 59631054 | 649 | 11 | 0.000157702 | 0.00024 | 0.00082 | 3.70E-05 | 0.001638341 | RTN1 |
| chr17 | 46171101 | 46172500 | 1400 | 9 | 0.000157765 | 0.000138 | 0.00046 | 1.34E-05 | -0.005879651 | KANSL1 |
| chr8 | 60741417 | 60741726 | 310 | 2 | 0.000157824 | 3.61E-05 | 0.00035 | 4.02E-05 | -0.004182513 | CHD7 |
| chr7 | 149249920 | 149250272 | 353 | 2 | 0.000157862 | 0.001729 | 4.30E-05 | 5.27E-05 | -0.002368312 | ZNF212 |
| chr17 | 76370243 | 76370497 | 255 | 2 | 0.000157862 | 6.01E-05 | 0.00011 | 3.41E-05 | -0.001103265 | PRPSAP1 |
| chr14 | 61995251 | 61995802 | 552 | 4 | 0.000157864 | 5.26E-06 | 0.00397 | 1.47E-05 | -0.003489124 | SYT16 |
| chr11 | 67415017 | 67416145 | 1129 | 13 | 0.00015822 | 1.23E-05 | 0.00495 | 5.12E-06 | -0.003059726 | CARNS1, PPP1CA |
| chr1 | 11321878 | 11322262 | 385 | 2 | 0.000158326 | 6.13E-05 | 0.00011 | 3.50E-05 | -0.01107825 | NA |
| chr1 | 63878205 | 63878553 | 349 | 2 | 0.000158331 | 0.000289 | 6.15E-05 | 4.68E-05 | -0.00197179 | ROR1 |
| chr11 | 67398170 | 67398236 | 67 | 2 | 0.00015846 | 2.06E-05 | 0.00122 | 3.26E-05 | -0.007488592 | RAD9A, PPP1CA |
| chr5 | 122844082 | 122844355 | 274 | 4 | 0.000158593 | 4.25E-05 | 0.00152 | 5.95E-05 | -0.006795311 | SNX24 |
| chr1 | 46667622 | 46668281 | 660 | 5 | 0.000158715 | 0.000141 | 0.02325 | 0.00052 | 0.001538565 | ATPAF1 |
| chr17 | 19702400 | 19702622 | 223 | 2 | 0.000158717 | 0.003463 | 5.30E-05 | 7.45E-05 | 0.01041668 | SLC47A2 |
| chr1 | 109100562 | 109100857 | 296 | 4 | 0.000158744 | 0.000324 | 9.90E-05 | 4.72E-05 | 0.001200163 | SCARNA2, AL356488.2 |
| chr22 | 21628498 | 21628962 | 465 | 3 | 0.000158781 | 0.000297 | 0.00052 | 0.00011 | -0.007810081 | YDJC |
| chr11 | 22626063 | 22626561 | 499 | 6 | 0.000158863 | 0.000255 | 0.0129 | 0.00014 | -0.00385779 | GAS2 |
| chr19 | 55673994 | 55674616 | 623 | 5 | 0.000158908 | 0.018556 | 3.92E-05 | 0.0002 | -0.002830141 | U2AF2, AC008735.2 |
| chr6 | 33175159 | 33175499 | 341 | 4 | 0.000158908 | 0.0003 | 0.00026 | 2.63E-05 | -0.004511128 | COL11A2 |
| chr11 | 70603114 | 70603211 | 98 | 3 | 0.000158977 | 1.00E-05 | 0.00075 | 1.76E-05 | -0.011617584 | SHANK2 |
| chr3 | 56356005 | 56356338 | 334 | 2 | 0.000158977 | 1.54E-05 | 0.00027 | 1.95E-05 | -0.009846332 | ERC2 |
| chr19 | 55621317 | 55621850 | 534 | 3 | 0.000159181 | 6.69E-06 | 0.00167 | 1.44E-05 | -0.006547902 | ZNF784 |
| chr1 | 43299946 | 43300535 | 590 | 5 | 0.000159361 | 8.53E-05 | 0.00396 | 5.07E-05 | -0.005787894 | NA |
| chr8 | 41933350 | 41933716 | 367 | 2 | 0.000159421 | 1.40E-05 | 0.00034 | 1.89E-05 | -0.00653163 | KAT6A |
| chr11 | 73876333 | 73877385 | 1053 | 15 | 0.000159593 | 0.002526 | 0.0068 | 0.00022 | 0.00016914 | PAAF1, COA4 |
| chr11 | 75339055 | 75339103 | 49 | 2 | 0.00015985 | 7.06E-05 | 9.31E-05 | 3.47E-05 | -0.009081923 | ARRB1 |
| chr9 | 26946621 | 26947401 | 781 | 12 | 0.000160105 | 3.37E-05 | 0.00659 | 2.43E-05 | -0.000512244 | IFT74, PLAA |
| chr2 | 96895666 | 96895992 | 327 | 3 | 0.000160206 | 0.000104 | 0.00089 | 7.10E-05 | -0.005712646 | FAM178B |
| chr19 | 50832910 | 50833296 | 387 | 3 | 0.000160206 | 0.005059 | 0.00058 | 0.00023 | -0.00757317 | AC011523.1, KLK15 |
| chr7 | 155491636 | 155492064 | 429 | 3 | 0.00016028 | 0.000238 | 0.00075 | 5.49E-05 | 0.026362636 | CNPY1, AC009403.2 |
| chr7 | 157540124 | 157541255 | 1132 | 10 | 0.000160476 | 1.39E-06 | 0.00143 | 1.82E-06 | -0.002631174 | PTPRN2 |
| chr22 | 43155302 | 43155745 | 444 | 3 | 0.000160497 | 6.55E-06 | 0.00131 | 1.13E-05 | -0.012508984 | TSPO |
| chr17 | 3004601 | 3005075 | 475 | 5 | 0.000160528 | 2.52E-05 | 0.00149 | 2.88E-05 | -0.008984748 | RAP1GAP2 |
| chr2 | 148936941 | 148937188 | 248 | 3 | 0.000160591 | 1.81E-05 | 0.00044 | 2.28E-05 | -0.01032853 | KIF5C |
| chr12 | 121549519 | 121549673 | 155 | 2 | 0.000160799 | 1.06E-05 | 0.00071 | 1.70E-05 | -0.003801503 | KDM2B |
| chr17 | 44250792 | 44251319 | 528 | 4 | 0.000160802 | 1.46E-05 | 0.00291 | 2.53E-05 | -0.002482602 | SLC4A1 |
| chr20 | 64034646 | 64036164 | 1519 | 5 | 0.000160815 | 3.84E-06 | 0.00223 | 6.06E-06 | -0.008253396 | C20orf204 |
| chr7 | 47304981 | 47305152 | 172 | 4 | 0.000160815 | 1.24E-05 | 0.00114 | 1.61E-05 | -0.006150153 | TNS3 |
| chr7 | 152074773 | 152074835 | 63 | 2 | 0.000161004 | 1.39E-05 | 0.00026 | 1.77E-05 | -0.010753497 | GALNT11 |
| chr19 | 5953395 | 5954681 | 1287 | 5 | 0.00016105 | 6.38E-07 | 0.00381 | 2.74E-06 | -0.009954732 | RANBP3 |
| chr9 | 129103146 | 129103462 | 317 | 3 | 0.000161153 | 2.13E-05 | 0.00099 | 2.18E-05 | -0.013223216 | CRAT |
| chr22 | 48549363 | 48549584 | 222 | 2 | 0.000161266 | 0.000223 | 0.0001 | 6.54E-05 | -0.003057037 | TAFA5 |
| chr16 | 974642 | 974838 | 197 | 2 | 0.000161383 | 1.73E-05 | 0.00115 | 2.77E-05 | -0.006848834 | LMF1 |
| chr10 | 7103703 | 7103721 | 19 | 2 | 0.000161632 | 0.000376 | 3.11E-05 | 2.76E-05 | -0.003898409 | AL392086.3 |
| chr20 | 2380698 | 2381395 | 698 | 8 | 0.000161765 | 9.78E-05 | 0.00421 | 3.58E-05 | -0.003615749 | TGM6 |
| chr3 | 195863125 | 195863185 | 61 | 3 | 0.000161834 | 2.19E-05 | 0.00059 | 2.69E-05 | -0.014252214 | NA |
| chr12 | 30795778 | 30796480 | 703 | 6 | 0.000161845 | 0.000271 | 0.00713 | 4.76E-05 | 0.002676008 | LINC00941, AC010198.2 |
| chr5 | 141398775 | 141398889 | 115 | 4 | 0.000161921 | 0.000107 | 0.00098 | 2.74E-05 | 0.018202502 | PCDHGA1, PCDHGA2, PCDHGA3, PCDHGB1, PCDHGA4, PCDHGB2, PCDHGA5, PCDHGB3, PCDHGA6, PCDHGA7, PCDHGB4, PCDHGA8, PCDHGB5 |
| chr8 | 144444804 | 144445243 | 440 | 4 | 0.000161989 | 0.001197 | 0.00035 | 4.46E-05 | 0.000612017 | NA |
| chr2 | 189441350 | 189441765 | 416 | 8 | 0.000162114 | 0.000445 | 0.00612 | 9.37E-05 | 0.002056755 | WDR75 |
| chr2 | 70258013 | 70258624 | 612 | 16 | 0.00016213 | 0.0001 | 0.0045 | 1.74E-05 | -0.000800576 | PCYOX1 |
| chr20 | 23629123 | 23629293 | 171 | 2 | 0.00016213 | 1.87E-05 | 0.00015 | 1.86E-05 | -0.013832681 | CST3 |
| chr1 | 169367744 | 169368622 | 879 | 13 | 0.000162159 | 0.006909 | 0.00026 | 3.88E-05 | 0.001536548 | BLZF1, NME7 |
| chr10 | 6195322 | 6196117 | 796 | 3 | 0.000162398 | 3.63E-06 | 0.00045 | 4.07E-06 | -0.010241149 | PFKFB3 |
| chr1 | 171136642 | 171137725 | 1084 | 5 | 0.000162403 | 5.02E-06 | 0.00047 | 1.76E-06 | -0.009002176 | BX284613.2 |
| chr17 | 17001562 | 17002098 | 537 | 4 | 0.00016243 | 0.000466 | 0.00027 | 4.17E-05 | 0.00349125 | NA |
| chr11 | 67606168 | 67607197 | 1030 | 13 | 0.000162439 | 0.00724 | 0.0003 | 4.50E-05 | 0.000523307 | AP003385.3, NDUFV1, C11orf72 |
| chr18 | 31203590 | 31203990 | 401 | 3 | 0.000162457 | 5.28E-06 | 0.0017 | 1.23E-05 | -0.011805589 | NA |
| chr20 | 49307121 | 49307253 | 133 | 2 | 0.000162457 | 2.33E-05 | 0.00011 | 1.88E-05 | -0.01215147 | KCNB1 |
| chr5 | 111092568 | 111092974 | 407 | 4 | 0.000162474 | 0.0006 | 0.00024 | 0.0002 | 0.001524208 | WDR36 |
| chr13 | 51624587 | 51625818 | 1232 | 3 | 0.000162525 | 2.44E-07 | 0.00027 | 5.59E-07 | 0.002337849 | WDFY2 |
| chr22 | 23222292 | 23222680 | 389 | 4 | 0.000162653 | 2.01E-05 | 0.00243 | 3.39E-05 | -0.004717027 | BCR |
| chr5 | 160419536 | 160420136 | 601 | 5 | 0.000162657 | 0.001025 | 0.00096 | 0.0002 | 0.006697094 | SLU7 |
| chr7 | 2647584 | 2648297 | 714 | 7 | 0.000162705 | 3.14E-05 | 0.00886 | 3.56E-05 | -0.00451951 | TTYH3 |
| chr1 | 153978367 | 153978803 | 437 | 4 | 0.000162714 | 0.000187 | 0.00086 | 0.00011 | -0.003652204 | AL358472.2, AL358472.6 |
| chr6 | 33085815 | 33086014 | 200 | 2 | 0.000162822 | 9.62E-05 | 5.94E-05 | 2.95E-05 | -0.003891963 | HLA-DPB1 |
| chr11 | 12101974 | 12103804 | 1831 | 3 | 0.000163007 | 2.20E-08 | 0.00033 | 8.19E-08 | -0.002294332 | MICAL2 |
| chr3 | 167734935 | 167735288 | 354 | 7 | 0.00016309 | 0.00078 | 2.68E-05 | 2.88E-05 | 0.001261605 | SERPINI1, PDCD10 |
| chr17 | 50213098 | 50213486 | 389 | 4 | 0.000163109 | 7.05E-06 | 0.00091 | 1.37E-05 | -0.011974723 | AC015909.1, AC015909.4 |
| chr12 | 122857118 | 122857188 | 71 | 3 | 0.000163215 | 1.95E-05 | 0.0017 | 2.79E-05 | -0.002666943 | HIP1R |
| chr22 | 29300648 | 29300954 | 307 | 2 | 0.000163259 | 9.44E-05 | 0.00011 | 4.47E-05 | -0.006914285 | NA |
| chr11 | 68405772 | 68405930 | 159 | 2 | 0.000163401 | 1.44E-05 | 0.00054 | 2.12E-05 | -0.005048823 | LRP5 |
| chr11 | 77112872 | 77113479 | 608 | 4 | 0.000163417 | 5.53E-05 | 0.0004 | 2.98E-05 | -0.005241962 | CAPN5 |
| chr1 | 62318598 | 62319131 | 534 | 5 | 0.000163581 | 1.52E-05 | 0.00412 | 2.63E-05 | -0.003182242 | KANK4 |
| chr19 | 1422485 | 1423968 | 1484 | 6 | 0.000163623 | 4.70E-08 | 0.00227 | 2.21E-07 | -0.006504702 | DAZAP1 |
| chr6 | 144215933 | 144216469 | 537 | 4 | 0.000163739 | 9.57E-05 | 0.00051 | 4.13E-05 | 0.003065001 | NA |
| chr10 | 124654327 | 124654601 | 275 | 2 | 0.00016374 | 0.000246 | 8.07E-05 | 5.63E-05 | 0.004536783 | AC068896.1, FAM53B |
| chr9 | 97913448 | 97913809 | 362 | 3 | 0.000163867 | 0.002419 | 5.11E-05 | 4.13E-05 | -0.009401533 | TRMO |
| chr19 | 12783178 | 12783498 | 321 | 4 | 0.000164004 | 0.000521 | 0.00011 | 5.87E-05 | 0.002665319 | HOOK2 |
| chr19 | 41348913 | 41350831 | 1919 | 6 | 0.000164043 | 1.11E-06 | 0.00094 | 1.18E-06 | 0.000879681 | TGFB1 |
| chr14 | 105204532 | 105205677 | 1146 | 4 | 0.000164181 | 1.34E-07 | 0.00056 | 3.79E-07 | -0.013636143 | NA |
| chr5 | 173578322 | 173578654 | 333 | 3 | 0.000164333 | 0.000199 | 7.65E-05 | 3.68E-05 | -0.002567035 | AC008663.2 |
| chr17 | 50131112 | 50131116 | 5 | 2 | 0.00016443 | 3.91E-05 | 0.00044 | 4.54E-05 | -0.010219162 | NA |
| chr9 | 129257088 | 129257782 | 695 | 2 | 0.000164443 | 1.48E-05 | 0.00063 | 2.22E-05 | -0.008661409 | AL158151.3, AL158151.1 |
| chr11 | 63919682 | 63920465 | 784 | 5 | 0.000164647 | 1.27E-05 | 0.00054 | 1.44E-05 | -0.011729277 | NA |
| chr14 | 22535286 | 22535535 | 250 | 2 | 0.000164854 | 0.045871 | 4.90E-05 | 8.47E-05 | -0.004631199 | NA |
| chr7 | 157467732 | 157468123 | 392 | 2 | 0.000164948 | 0.001379 | 0.00117 | 0.00086 | -0.00474953 | AC006372.3 |
| chr20 | 64261481 | 64261489 | 9 | 2 | 0.000165026 | 9.18E-06 | 0.00076 | 1.51E-05 | -0.013069129 | PCMTD2 |
| chr6 | 157275390 | 157275548 | 159 | 4 | 0.000165052 | 2.14E-05 | 0.00482 | 4.02E-05 | -0.008435215 | NA |
| chr1 | 39883023 | 39883657 | 635 | 8 | 0.00016524 | 0.001142 | 0.0033 | 7.27E-05 | 0.000594476 | TRIT1 |
| chr5 | 133428275 | 133428707 | 433 | 2 | 0.00016526 | 0.001611 | 7.14E-05 | 8.91E-05 | -0.004355778 | FSTL4 |
| chr19 | 15934898 | 15935430 | 533 | 3 | 0.000165302 | 1.68E-05 | 0.00279 | 3.57E-05 | -0.021179868 | NA |
| chr2 | 234953071 | 234953757 | 687 | 2 | 0.000165381 | 4.25E-06 | 0.00033 | 6.88E-06 | -0.031076512 | SH3BP4 |
| chr10 | 74897281 | 74897533 | 253 | 2 | 0.000165389 | 3.97E-05 | 0.00018 | 3.43E-05 | -0.010389914 | KAT6B |

| | | | | | | | | | | |
|---|---|---|---|---|---|---|---|---|---|---|
| chr20 | 63295851 | 63296129 | 279 | 3 | 0.000165502 | 0.000104 | 0.00013 | 4.44E-05 | 0.012718105 | COL20A1 |
| chr16 | 56972228 | 56972580 | 353 | 2 | 0.000165553 | 2.21E-05 | 0.00066 | 3.17E-05 | -0.007460467 | CETP |
| chr19 | 35501889 | 35502269 | 381 | 6 | 0.000165671 | 0.046507 | 1.38E-05 | 0.00014 | -0.000726428 | DMKN |
| chr7 | 195195 | 196130 | 936 | 7 | 0.000165937 | 1.09E-05 | 0.00239 | 2.40E-05 | -0.003410845 | FAM20C |
| chr7 | 45460517 | 45460785 | 269 | 3 | 0.00016604 | 0.000207 | 0.00044 | 3.80E-05 | -0.006794048 | AC073325.1 |
| chr17 | 32265014 | 32265227 | 214 | 2 | 0.000166088 | 2.03E-05 | 0.0005 | 2.78E-05 | -0.015892435 | AC026620.2 |
| chr14 | 93016382 | 93016506 | 125 | 3 | 0.00016624 | 7.23E-05 | 0.00012 | 3.50E-05 | -0.001780628 | ITPK1 |
| chr11 | 67428150 | 67429035 | 886 | 10 | 0.000166733 | 1.51E-05 | 0.00185 | 1.53E-05 | 0.00115281 | RPS6KB2 |
| chr14 | 60649788 | 60650444 | 657 | 6 | 0.000166836 | 0.000961 | 0.00069 | 3.13E-05 | 0.00239576 | SIX1 |
| chr11 | 31988458 | 31988982 | 525 | 2 | 0.000166836 | 1.20E-05 | 0.00075 | 1.90E-05 | 0.000634442 | PAUPAR, AL035078.4 |
| chr12 | 8058214 | 8058795 | 582 | 4 | 0.000167148 | 1.42E-05 | 0.0038 | 2.05E-05 | -0.024759076 | C3AR1 |
| chr8 | 9375060 | 9375402 | 343 | 3 | 0.000167293 | 9.25E-06 | 0.00096 | 1.73E-05 | -0.001624321 | AC022784.1, AC021242.2 |
| chr11 | 47586208 | 47586758 | 551 | 7 | 0.000167786 | 1.08E-05 | 0.00487 | 2.13E-05 | -0.003903158 | FAM180B |
| chr5 | 175773398 | 175773451 | 54 | 2 | 0.000167805 | 0.000573 | 3.28E-05 | 3.22E-05 | 0.007335164 | NA |
| chr10 | 119818617 | 119819495 | 879 | 9 | 0.000167828 | 7.40E-06 | 0.02153 | 1.39E-05 | -0.008778015 | INPP5F, AL133461.1 |
| chr17 | 48586032 | 48587285 | 1254 | 5 | 0.000167838 | 3.99E-06 | 0.00168 | 5.65E-06 | -0.00629266 | HOXB-AS3, HOXB3 |
| chr8 | 1828029 | 1828181 | 153 | 2 | 0.000167838 | 0.0107 | 5.20E-05 | 8.39E-05 | -0.006650873 | AC019257.8, ARHGEF10 |
| chr16 | 4242046 | 4242676 | 631 | 9 | 0.000167857 | 0.000212 | 0.00983 | 1.14E-05 | -0.003878285 | SRL |
| chr16 | 87457981 | 87458465 | 485 | 2 | 0.000168076 | 6.96E-05 | 0.00047 | 7.23E-05 | 0.003430494 | ZCCHC14 |
| chr3 | 21829582 | 21829594 | 13 | 2 | 0.000168192 | 4.81E-05 | 0.00206 | 7.43E-05 | 0.008340236 | ZNF385D |
| chr11 | 60777793 | 60777853 | 61 | 2 | 0.000168409 | 9.47E-06 | 0.00074 | 1.54E-05 | -0.011994 | NA |
| chr10 | 47406809 | 47407071 | 263 | 2 | 0.000168485 | 8.00E-05 | 7.15E-05 | 3.10E-05 | -0.011448888 | NA |
| chr1 | 9291926 | 9292816 | 891 | 8 | 0.000168488 | 0.000326 | 0.00034 | 2.44E-05 | -0.005740653 | NA |
| chr9 | 105447424 | 105447503 | 80 | 2 | 0.000168541 | 2.79E-05 | 0.00047 | 3.55E-05 | -0.008151753 | NA |
| chr19 | 9435353 | 9436076 | 724 | 12 | 0.000168561 | 0.000939 | 0.00071 | 3.53E-05 | -0.000924775 | ZNF266 |
| chr7 | 107165025 | 107165378 | 354 | 2 | 0.000168629 | 3.45E-05 | 0.00164 | 5.32E-05 | -0.008253809 | NA |
| chr5 | 181201689 | 181201855 | 167 | 2 | 0.000168629 | 0.000455 | 0.00013 | 0.00011 | 0.006695006 | TRIM7-AS1, TRIM7 |
| chr21 | 44586174 | 44586533 | 360 | 2 | 0.000168703 | 6.94E-05 | 9.84E-05 | 3.56E-05 | -0.010067464 | KRTAP10-4, TSPEAR |
| chr7 | 1549659 | 1550220 | 562 | 6 | 0.000168717 | 7.57E-06 | 0.00635 | 2.05E-05 | -0.001608296 | TMEM184A |
| chr10 | 119165209 | 119165581 | 373 | 3 | 0.000168742 | 0.017113 | 0.00114 | 0.00233 | 0.001472973 | SFXN4 |
| chr18 | 58590891 | 58590938 | 48 | 2 | 0.000168799 | 5.10E-05 | 5.94E-05 | 2.11E-05 | -0.004389848 | ALPK2 |
| chr2 | 208359501 | 208359952 | 452 | 7 | 0.000168861 | 5.53E-05 | 0.00198 | 3.71E-05 | -0.010024267 | PTH2R |
| chr19 | 42424087 | 42424915 | 829 | 5 | 0.000169084 | 6.24E-06 | 0.0016 | 1.48E-05 | -0.007042254 | LIPE-AS1, LIPE, AC011497.1 |
| chr18 | 11861792 | 11862071 | 280 | 2 | 0.000169118 | 7.47E-05 | 0.00011 | 3.90E-05 | -0.005926619 | GNAL |
| chr2 | 57886146 | 57886391 | 246 | 2 | 0.000169237 | 1.14E-05 | 0.00049 | 1.70E-05 | -0.008077707 | NA |
| chr6 | 41287080 | 41287352 | 273 | 5 | 0.000169437 | 0.000369 | 0.00123 | 3.57E-05 | 0.003371745 | NA |
| chr20 | 63741397 | 63741980 | 584 | 3 | 0.000169484 | 0.000447 | 0.00273 | 0.00039 | -0.005043016 | SLC2A4RG |
| chr3 | 127304076 | 127304279 | 204 | 3 | 0.000169549 | 1.14E-05 | 0.00039 | 1.60E-05 | 0.022129071 | NA |
| chr9 | 33170588 | 33170851 | 264 | 2 | 0.00016978 | 0.000945 | 6.57E-05 | 7.16E-05 | -0.005583664 | B4GALT1-AS1 |
| chr11 | 1227127 | 1227778 | 652 | 6 | 0.000169886 | 2.63E-06 | 0.01124 | 1.34E-05 | -0.007276145 | MUC5B |
| chr1 | 180196801 | 180197539 | 739 | 3 | 0.000170128 | 0.000267 | 0.00125 | 0.00028 | -0.002738039 | QSOX1 |
| chr8 | 143148803 | 143149213 | 411 | 3 | 0.00017016 | 9.73E-05 | 8.98E-05 | 3.09E-05 | -0.004489311 | NA |
| chr11 | 96335761 | 96335818 | 58 | 2 | 0.0001702 | 1.06E-05 | 0.00045 | 1.59E-05 | -0.011149803 | MAML2 |
| chr11 | 84274027 | 84274316 | 290 | 2 | 0.000170264 | 0.00033 | 0.00011 | 8.31E-05 | -0.006350403 | DLG2 |
| chr12 | 65962577 | 65963303 | 727 | 3 | 0.000170296 | 5.91E-06 | 0.00057 | 8.41E-06 | -0.013004728 | HMGA2 |
| chr10 | 11344632 | 11345625 | 994 | 3 | 0.000170303 | 0.003188 | 0.00012 | 4.17E-05 | -0.0102785 | NA |
| chr11 | 67403057 | 67403139 | 83 | 2 | 0.000170409 | 0.000159 | 0.00182 | 0.00022 | -0.011397609 | PPP1CA |
| chr2 | 128346566 | 128346890 | 325 | 2 | 0.000170753 | 0.000232 | 0.00014 | 8.26E-05 | -0.006162217 | NA |
| chr12 | 47707022 | 47707059 | 38 | 2 | 0.000170876 | 0.000231 | 7.56E-05 | 5.17E-05 | -0.015423805 | AC004241.1 |
| chr9 | 135657885 | 135658231 | 347 | 2 | 0.000171242 | 3.90E-05 | 0.0002 | 3.56E-05 | -9.24E-05 | NA |
| chr4 | 56314221 | 56314876 | 656 | 3 | 0.000171411 | 2.09E-05 | 0.00079 | 2.23E-05 | -0.005774901 | CRACD |
| chr19 | 47741062 | 47741262 | 201 | 3 | 0.000171525 | 0.000371 | 3.75E-05 | 3.42E-05 | -0.002698345 | EHD2 |
| chr2 | 29064057 | 29064469 | 413 | 3 | 0.000171569 | 0.000728 | 0.00101 | 9.94E-05 | -0.002692589 | PCARE |
| chr11 | 17388635 | 17389160 | 526 | 6 | 0.000171581 | 0.001267 | 0.00081 | 0.00015 | 0.003542672 | KCNJ11 |
| chr20 | 47361723 | 47361735 | 13 | 2 | 0.000171585 | 1.52E-05 | 0.00038 | 2.08E-05 | -0.012087322 | NA |
| chr5 | 66958646 | 66959192 | 547 | 5 | 0.000171673 | 0.000205 | 0.00086 | 0.00017 | -0.010816308 | MAST4 |
| chr3 | 48634829 | 48635196 | 368 | 4 | 0.000171777 | 0.069335 | 4.11E-05 | 0.00035 | 0.001555653 | SLC26A6 |
| chr7 | 45974224 | 45974525 | 302 | 2 | 0.00017186 | 1.26E-05 | 0.00043 | 1.83E-05 | -0.021513002 | AC073115.2 |
| chr13 | 72781771 | 72782069 | 299 | 6 | 0.000171976 | 1.08E-05 | 0.00172 | 1.37E-05 | -0.00028696 | DIS3 |
| chr3 | 119145782 | 119146631 | 850 | 12 | 0.000172012 | 0.000224 | 0.00678 | 2.23E-05 | -0.002250215 | TEX55, IGSF11 |
| chr1 | 10467838 | 10468036 | 199 | 2 | 0.000172357 | 3.75E-05 | 9.14E-05 | 2.35E-05 | -0.015720031 | DFFA |
| chr3 | 32371614 | 32372346 | 733 | 5 | 0.000172359 | 7.77E-05 | 0.00229 | 9.09E-05 | -0.009315748 | NA |
| chr12 | 124406418 | 124406668 | 251 | 2 | 0.000172372 | 0.000426 | 8.86E-05 | 7.46E-05 | -0.002244366 | NCOR2 |
| chr9 | 108565538 | 108565792 | 255 | 2 | 0.000172433 | 0.000276 | 5.10E-05 | 3.91E-05 | -0.009519135 | NA |
| chr18 | 51196508 | 51196813 | 306 | 3 | 0.000172475 | 0.009512 | 7.75E-05 | 0.00011 | 0.001536387 | MEX3C |
| chr2 | 219381830 | 219382081 | 252 | 4 | 0.00017249 | 0.000625 | 0.00022 | 4.15E-05 | -0.003549686 | DNPEP |
| chr11 | 88169663 | 88169768 | 106 | 2 | 0.00017249 | 1.71E-05 | 0.00016 | 1.80E-05 | 0.033224925 | RAB38 |
| chr22 | 49964604 | 49964681 | 78 | 2 | 0.000172672 | 1.69E-05 | 0.00063 | 2.49E-05 | -0.011649884 | NA |
| chr5 | 143811629 | 143812206 | 578 | 6 | 0.000172677 | 1.57E-05 | 0.00439 | 2.59E-05 | -0.008582091 | AC008696.2, HMHB1 |
| chr4 | 1250061 | 1250572 | 512 | 9 | 0.000172879 | 3.10E-06 | 0.01963 | 1.39E-05 | -0.018755604 | CTBP1-DT |
| chr12 | 120047040 | 120047430 | 391 | 2 | 0.000172879 | 1.33E-05 | 0.00087 | 2.12E-05 | -0.006544685 | BICDL1 |
| chr21 | 42395576 | 42396246 | 671 | 7 | 0.000172956 | 0.000475 | 0.0002 | 4.31E-05 | -0.001674244 | TMPRSS3 |
| chr11 | 68342637 | 68343122 | 486 | 3 | 0.000173032 | 2.79E-05 | 0.00162 | 3.93E-05 | -0.00470704 | LRP5 |
| chr1 | 161307292 | 161307358 | 67 | 2 | 0.000173249 | 3.68E-05 | 0.00017 | 3.23E-05 | 0.004897323 | MPZ |
| chr6 | 149450671 | 149451210 | 540 | 4 | 0.000173305 | 0.000862 | 0.00289 | 6.52E-05 | -0.010573981 | ZC3H12D |
| chr3 | 183636015 | 183636388 | 374 | 3 | 0.000173402 | 0.001179 | 0.00014 | 0.0001 | 0.002536083 | KLHL24 |
| chr14 | 91257544 | 91258252 | 709 | 7 | 0.000173446 | 0.002007 | 0.0014 | 1.34E-05 | -0.004985423 | NA |
| chr4 | 2817125 | 2817666 | 542 | 2 | 0.00017356 | 0.000469 | 0.0016 | 0.0005 | -0.01144212 | SH3BP2 |
| chr14 | 69770787 | 69771366 | 580 | 4 | 0.000173637 | 0.002337 | 0.00011 | 4.81E-05 | -0.002200516 | SRSF5 |
| chr9 | 131429415 | 131430245 | 831 | 7 | 0.000173665 | 0.003828 | 0.00122 | 7.51E-05 | -0.003213775 | PRRC2B |
| chr4 | 637790 | 637945 | 156 | 3 | 0.000173761 | 4.07E-05 | 0.00022 | 2.09E-05 | 0.001759989 | PDE6B |
| chr1 | 190474765 | 190474851 | 87 | 2 | 0.000173764 | 2.22E-05 | 0.00106 | 3.41E-05 | -0.013695826 | BRINP3 |
| chr7 | 30322539 | 30322563 | 25 | 2 | 0.000173783 | 4.22E-05 | 0.00032 | 4.40E-05 | -0.013428121 | ZNRF2 |
| chr7 | 29563440 | 29563925 | 486 | 10 | 0.000173805 | 0.000798 | 0.00115 | 4.65E-05 | -0.00064368 | PRR15, AC007255.1 |
| chr10 | 79113846 | 79114040 | 195 | 2 | 0.000173811 | 9.03E-05 | 7.91E-05 | 3.55E-05 | -0.010978956 | ZMIZ1 |
| chr17 | 2666735 | 2667064 | 330 | 2 | 0.000173811 | 0.000128 | 0.0002 | 7.66E-05 | -0.004671072 | PAFAH1B1 |
| chr20 | 46028476 | 46028585 | 110 | 3 | 0.000173841 | 1.43E-05 | 0.00141 | 2.19E-05 | -0.019422765 | SLC12A5 |
| chr1 | 168280529 | 168280881 | 353 | 4 | 0.000173855 | 2.92E-05 | 0.00113 | 2.07E-05 | -0.003523669 | TBX19 |
| chr3 | 133832180 | 133832412 | 233 | 2 | 0.00017392 | 2.07E-05 | 0.00027 | 2.45E-05 | -0.003935206 | RAB6B |
| chr19 | 1013775 | 1014482 | 708 | 3 | 0.000173963 | 3.56E-06 | 0.0025 | 9.39E-06 | -0.005196445 | TMEM259 |
| chr8 | 118105036 | 118105064 | 29 | 2 | 0.000173963 | 3.09E-05 | 8.29E-05 | 1.96E-05 | -0.004072606 | EXT1 |
| chr14 | 101587621 | 101588213 | 593 | 5 | 0.000174027 | 0.003083 | 0.00178 | 9.19E-05 | -0.000621364 | NA |
| chr15 | 70775523 | 70775803 | 281 | 2 | 0.00017403 | 0.02353 | 4.26E-05 | 6.96E-05 | -0.004929883 | AC009269.3 |
| chr14 | 59465315 | 59465603 | 289 | 9 | 0.000174153 | 0.028342 | 0.0001 | 9.51E-05 | -0.002743683 | GPR135, L3HYPDH |
| chr3 | 46749401 | 46750972 | 1572 | 7 | 0.000174178 | 4.35E-07 | 0.00066 | 3.85E-07 | -0.010371678 | AC109583.1, PRSS45P, AC109583.4 |
| chr21 | 33479974 | 33480550 | 577 | 8 | 0.000174275 | 0.002582 | 0.00023 | 0.00012 | -0.000823807 | TMEM50B |
| chr18 | 75428830 | 75429086 | 257 | 2 | 0.000174288 | 5.57E-05 | 0.00126 | 7.80E-05 | -0.005600408 | NA |
| chr7 | 76999310 | 76999356 | 47 | 2 | 0.000174474 | 1.04E-05 | 0.00055 | 1.61E-05 | -0.006182311 | DTX2P1-UPK3BP1-PMS2P11, DTX2P1 |
| chr1 | 202274570 | 202274763 | 194 | 2 | 0.00017449 | 3.79E-05 | 0.0016 | 5.78E-05 | -0.011181207 | LGR6 |
| chr6 | 36758683 | 36758937 | 255 | 3 | 0.000174611 | 0.013205 | 7.58E-05 | 4.57E-05 | -0.005619001 | CPNE5 |
| chr7 | 44581561 | 44582605 | 1045 | 11 | 0.000174682 | 0.000839 | 0.00407 | 0.00011 | 0.000376831 | AC004938.2, TMED4 |
| chr1 | 226364289 | 226364453 | 165 | 2 | 0.000174682 | 0.009278 | 3.75E-05 | 5.64E-05 | -0.003219817 | PARP1 |
| chr17 | 973235 | 973323 | 89 | 2 | 0.00017489 | 0.002431 | 2.97E-05 | 3.80E-05 | 0.004568741 | NXN |
| chr11 | 70362906 | 70363410 | 505 | 3 | 0.000174998 | 9.14E-06 | 0.00106 | 1.37E-05 | -0.007906986 | PPFIA1, AP002336.2 |
| chr17 | 9021884 | 9022435 | 552 | 6 | 0.000175197 | 0.000108 | 0.00026 | 3.95E-05 | -6.36E-05 | NTN1 |
| chr11 | 3617026 | 3617228 | 203 | 4 | 0.00017531 | 0.000183 | 8.72E-05 | 4.05E-05 | -0.001411083 | TRPC2 |
| chr19 | 45160929 | 45160940 | 12 | 2 | 0.000175339 | 0.000298 | 0.00707 | 0.00054 | -0.019658671 | MARK4 |
| chr4 | 152064204 | 152064246 | 43 | 2 | 0.00017577 | 0.000132 | 4.23E-05 | 2.57E-05 | -0.016073795 | NA |
| chr3 | 189829501 | 189829989 | 489 | 2 | 0.000175775 | 0.000125 | 0.00025 | 8.46E-05 | -0.000803243 | TP63, MIR944 |
| chr16 | 81294331 | 81294484 | 154 | 3 | 0.000175778 | 1.25E-05 | 0.00101 | 1.65E-05 | -0.014376924 | NA |
| chr1 | 9734796 | 9735164 | 369 | 3 | 0.000175912 | 1.96E-05 | 0.00275 | 4.04E-05 | -0.003018942 | CLSTN1 |
| chr2 | 165127723 | 165127956 | 234 | 2 | 0.000176094 | 5.68E-05 | 7.99E-05 | 2.78E-05 | -0.005352958 | AC019197.1, SCN3A |
| chr13 | 39038136 | 39038966 | 831 | 13 | 0.000176095 | 0.000961 | 0.00962 | 4.46E-05 | 0.000489553 | NHLRC3 |
| chr15 | 74022367 | 74023198 | 832 | 8 | 0.000176262 | 0.001237 | 0.0001 | 5.27E-05 | -0.002078949 | PML |
| chr22 | 29767002 | 29767719 | 718 | 10 | 0.000176274 | 0.000384 | 0.00059 | 1.27E-05 | 0.001355294 | UQCR10, ZMAT5, AC004882.3 |
| chr1 | 153616451 | 153617052 | 602 | 4 | 0.000176453 | 6.86E-05 | 0.00043 | 1.14E-05 | -0.004596378 | S100A14 |
| chr17 | 77477262 | 77477887 | 626 | 6 | 0.000176559 | 5.51E-05 | 0.00202 | 3.62E-05 | -0.004585312 | SEPTIN9 |
| chr12 | 108515279 | 108515782 | 504 | 4 | 0.000176562 | 0.006346 | 0.00139 | 0.00057 | 0.001829644 | FICD |
| chr5 | 93573723 | 93573728 | 6 | 2 | 0.000176572 | 4.10E-05 | 0.00041 | 4.64E-05 | 0.001460864 | NR2F1-AS1 |
| chr17 | 13011519 | 13011710 | 192 | 2 | 0.000176615 | 5.36E-05 | 7.50E-05 | 2.57E-05 | -0.003317518 | ELAC2 |
| chr17 | 45169516 | 45169923 | 408 | 3 | 0.000176667 | 0.003084 | 0.00021 | 0.00035 | 0.001696474 | HEXIM2, AC138150.2 |
| chr18 | 79688054 | 79688162 | 109 | 3 | 0.000176755 | 7.40E-05 | 0.00025 | 3.04E-05 | 0.002737641 | CTDP1 |
| chr15 | 69093494 | 69093604 | 111 | 2 | 0.00017693 | 2.10E-05 | 0.00017 | 2.13E-05 | -0.014391253 | EWSAT1, AC027088.2 |
| chr8 | 23084341 | 23084519 | 179 | 4 | 0.000176981 | 1.76E-05 | 0.00269 | 2.12E-05 | 0.04321416 | AC107959.4, AC107959.5 |
| chr4 | 113761548 | 113762219 | 672 | 8 | 0.000177171 | 0.000379 | 0.00121 | 1.11E-05 | 0.000968724 | CAMK2D |
| chr17 | 28842698 | 28843025 | 328 | 7 | 0.000177319 | 0.000128 | 0.00141 | 2.71E-05 | -0.000201282 | FAM222B |
| chr8 | 144011562 | 144011786 | 225 | 3 | 0.00017734 | 2.92E-05 | 0.0008 | 2.41E-05 | -0.003790242 | PARP10 |
| chr12 | 9115783 | 9116197 | 415 | 5 | 0.000177351 | 2.31E-05 | 0.00389 | 4.49E-05 | -0.005972933 | A2M |
| chr22 | 18028414 | 18029315 | 902 | 4 | 0.000177351 | 3.70E-05 | 0.00043 | 6.61E-06 | -0.011625172 | NA |
| chr17 | 41717598 | 41718016 | 419 | 3 | 0.000177429 | 3.06E-05 | 0.00024 | 2.62E-05 | -0.008450758 | HAP1 |
| chr20 | 15196435 | 15196863 | 429 | 3 | 0.000177667 | 1.44E-05 | 0.00179 | 2.50E-05 | -0.037504664 | MACROD2 |
| chr5 | 149357534 | 149358197 | 664 | 11 | 0.000177698 | 6.65E-05 | 0.01977 | 2.56E-05 | -0.00021534 | PCYOX1L, GRPEL2-AS1 |
| chr11 | 85810796 | 85811051 | 256 | 4 | 0.000177897 | 4.98E-05 | 0.00807 | 0.00012 | -0.00046317 | SYTL2 |
| chr11 | 57597368 | 57597615 | 248 | 7 | 0.000178005 | 0.000514 | 0.00268 | 5.21E-05 | -0.004226647 | SERPING1 |
| chr4 | 112516074 | 112516753 | 680 | 6 | 0.00017804 | 5.06E-05 | 0.00121 | 1.50E-05 | 0.001619487 | NEUROG2-AS1, NEUROG2 |
| chr6 | 163355027 | 163355750 | 724 | 3 | 0.00017804 | 3.47E-05 | 0.00296 | 6.66E-05 | -0.015744819 | NA |
| chr10 | 49298703 | 49299242 | 540 | 9 | 0.000178068 | 0.00032 | 0.0099 | 5.04E-05 | 0.003266851 | C10orf71, C10orf71-AS1 |
| chr4 | 2824437 | 2824693 | 257 | 5 | 0.000178088 | 0.001783 | 2.68E-05 | 6.93E-05 | -0.001907761 | SH3BP2 |
| chr10 | 94687707 | 94688098 | 392 | 4 | 0.000178364 | 0.000164 | 0.00029 | 3.58E-05 | -0.006566339 | CYP2C18 |
| chr20 | 31721663 | 31723055 | 1393 | 14 | 0.000178452 | 1.75E-05 | 0.00869 | 8.86E-06 | 0.003106685 | ABALON, BCL2L1 |
| chr8 | 80051488 | 80051800 | 313 | 3 | 0.000178456 | 1.30E-05 | 0.00139 | 2.14E-05 | -0.007881138 | AC036214.3, TPD52 |
| chr11 | 64803676 | 64804713 | 1038 | 5 | 0.00017847 | 8.93E-05 | 0.00317 | 3.03E-05 | -0.002723752 | MEN1 |
| chr17 | 42980046 | 42980645 | 600 | 16 | 0.000178562 | 1.63E-05 | 0.0029 | 2.01E-05 | 0.000718298 | RUNDC1, PTGES3L-AARSD1, PTGES3L |
| chr17 | 39714158 | 39714204 | 47 | 2 | 0.000178576 | 0.000503 | 0.00016 | 0.00013 | -0.00994685 | ERBB2 |
| chr1 | 35451754 | 35452192 | 439 | 2 | 0.000178671 | 1.56E-05 | 0.00101 | 2.50E-05 | -0.011405381 | KIAA0319L |
| chr7 | 105389038 | 105389448 | 411 | 5 | 0.000179215 | 3.66E-05 | 0.0031 | 1.13E-05 | -0.000954819 | SRPK2 |
| chr12 | 132202367 | 132203184 | 818 | 6 | 0.000179222 | 4.03E-06 | 0.00676 | 1.25E-05 | -0.005524484 | GALNT9 |
| chr7 | 100582955 | 100583468 | 514 | 3 | 0.000179283 | 9.33E-05 | 0.00304 | 0.00016 | -0.002976506 | LRCH4 |
| chr5 | 55714040 | 55714961 | 922 | 3 | 0.000179548 | 2.50E-05 | 0.0007 | 3.30E-05 | -0.005257746 | SLC38A9 |
| chr22 | 20531549 | 20531749 | 201 | 4 | 0.000179639 | 1.84E-05 | 0.01136 | 5.74E-05 | -0.011081585 | MED15 |
| chr10 | 129790877 | 129790925 | 49 | 3 | 0.000179776 | 6.69E-06 | 0.00191 | 1.40E-05 | -0.012377354 | LINC02666 |
| chr5 | 154340269 | 154340446 | 178 | 2 | 0.000179853 | 8.60E-05 | 0.00013 | 4.72E-05 | -0.003839933 | GALNT10, SAP30L-AS1 |
| chr12 | 51895009 | 51895434 | 426 | 2 | 0.000179931 | 3.16E-05 | 0.00019 | 3.02E-05 | -0.008691874 | NA |
| chr3 | 101677513 | 101678173 | 661 | 7 | 0.000180075 | 0.000872 | 0.00296 | 6.13E-05 | 0.002426678 | ZBTB11-AS1 |
| chr9 | 130635846 | 130636110 | 265 | 2 | 0.000180166 | 1.61E-05 | 0.00029 | 2.05E-05 | -0.003813211 | FUBP3 |
| chr2 | 27426276 | 27426614 | 339 | 3 | 0.000180508 | 0.000201 | 7.49E-05 | 4.62E-05 | -0.000657385 | NA |
| chr20 | 31996656 | 31996728 | 73 | 2 | 0.000180522 | 0.020519 | 3.92E-05 | 6.23E-05 | -0.003682438 | XKR7 |
| chr12 | 49035137 | 49035236 | 100 | 2 | 0.000180528 | 9.77E-05 | 0.0001 | 4.37E-05 | -0.002375807 | KMT2D |
| chr1 | 200409835 | 200410236 | 402 | 6 | 0.000180536 | 0.000195 | 0.00599 | 8.95E-05 | -0.000662583 | AC104461.1, ZNF281 |
| chr10 | 101595808 | 101596040 | 233 | 2 | 0.000180539 | 1.77E-05 | 0.00078 | 2.69E-05 | -0.00623385 | DPCD |
| chr1 | 30900305 | 30900329 | 25 | 2 | 0.000180644 | 2.11E-05 | 0.00015 | 2.05E-05 | -0.008006363 | SDC3 |
| chr2 | 176817098 | 176817180 | 83 | 2 | 0.000180662 | 1.22E-05 | 0.00083 | 1.96E-05 | -0.010693684 | LINC01117, AC092162.2 |

| | | | | | | | | | | |
|---|---|---|---|---|---|---|---|---|---|---|
| chr17 | 29046384 | 29046487 | 104 | 2 | 0.000180824 | 3.18E-05 | 0.00015 | 2.72E-05 | -0.011618392 | PIPOX |
| chr20 | 31900685 | 31901234 | 550 | 4 | 0.000180835 | 2.08E-05 | 0.00052 | 1.82E-05 | -0.003817447 | TTLL9 |
| chr13 | 24149126 | 24149777 | 652 | 3 | 0.00018088 | 0.001443 | 7.08E-05 | 7.05E-05 | -0.003613631 | AL359736.1, SPATA13 |
| chr3 | 49429755 | 49429937 | 183 | 3 | 0.00018117 | 2.11E-05 | 0.00235 | 3.72E-05 | -0.003512174 | NA |
| chr1 | 59316496 | 59316843 | 348 | 2 | 0.000181312 | 1.72E-05 | 0.00109 | 2.74E-05 | 0.000568878 | FGGY |
| chr2 | 60463207 | 60463342 | 136 | 2 | 0.000181443 | 1.81E-05 | 0.00044 | 2.47E-05 | -0.008736914 | BCL11A |
| chr15 | 25204771 | 25204984 | 214 | 3 | 0.000181687 | 1.24E-05 | 0.00134 | 1.73E-05 | -0.004895814 | SNHG14 |
| chr11 | 79054692 | 79054988 | 297 | 2 | 0.00018172 | 1.68E-05 | 0.0003 | 2.13E-05 | -0.001111036 | TENM4 |
| chr3 | 195097375 | 195097399 | 25 | 2 | 0.000181904 | 0.000294 | 4.41E-05 | 3.50E-05 | -0.013745081 | XXYLT1 |
| chr14 | 22472631 | 22473189 | 559 | 2 | 0.000181934 | 0.001076 | 0.0004 | 0.00037 | -0.003329306 | AC244502.1 |
| chr18 | 63411473 | 63411549 | 77 | 2 | 0.000182059 | 1.24E-05 | 0.00035 | 1.72E-05 | -0.008637244 | VPS4B |
| chr17 | 18258303 | 18258951 | 649 | 13 | 0.000182189 | 0.000134 | 0.0062 | 3.72E-05 | 0.001768737 | FLII |
| chr9 | 128382444 | 128383076 | 633 | 3 | 0.000182446 | 1.39E-05 | 0.00303 | 3.09E-05 | -0.004744254 | URM1 |
| chr15 | 51281000 | 51281389 | 390 | 2 | 0.000182503 | 5.34E-05 | 0.00047 | 5.92E-05 | -0.005647288 | MIR4713HG, CYP19A1 |
| chr7 | 99785601 | 99785627 | 27 | 2 | 0.000182952 | 3.27E-05 | 8.41E-05 | 2.05E-05 | -0.004266703 | NA |
| chr5 | 150616141 | 150616489 | 349 | 3 | 0.000182958 | 1.30E-05 | 0.00411 | 3.09E-05 | -0.015240625 | SYNPO |
| chr19 | 40380991 | 40381061 | 71 | 2 | 0.000183017 | 2.03E-05 | 0.00014 | 1.91E-05 | -8.20E-05 | HIPK4 |
| chr9 | 126804135 | 126804764 | 630 | 3 | 0.000183034 | 0.000346 | 0.00032 | 0.00012 | -0.005701398 | NA |
| chr4 | 68349587 | 68350385 | 799 | 15 | 0.000183097 | 0.000114 | 0.01491 | 2.09E-05 | -0.000386963 | YTHDC1 |
| chr18 | 80197145 | 80197171 | 27 | 2 | 0.000183103 | 1.76E-05 | 0.00035 | 2.30E-05 | -0.019276892 | AC139100.1, PARD6G |
| chr1 | 111743357 | 111743953 | 597 | 2 | 0.000183169 | 1.43E-05 | 0.00039 | 1.98E-05 | -0.009564287 | INKA2-AS1, INKA2 |
| chr12 | 50025455 | 50026250 | 796 | 14 | 0.000183694 | 0.000128 | 0.00163 | 2.33E-05 | -5.39E-05 | RACGAP1 |
| chr4 | 11624542 | 11625012 | 471 | 2 | 0.000183793 | 1.10E-05 | 0.00046 | 1.65E-05 | -0.010045539 | NA |
| chr7 | 926933 | 928941 | 2009 | 12 | 0.000183807 | 1.78E-06 | 0.00454 | 2.81E-07 | -0.008154655 | ADAP1, COX19 |
| chr4 | 15905273 | 15905866 | 594 | 2 | 0.000184059 | 1.52E-05 | 0.00049 | 2.18E-05 | 0.000697508 | NA |
| chr6 | 29587165 | 29587537 | 373 | 3 | 0.00018406 | 6.56E-05 | 0.00027 | 3.00E-05 | -0.005705799 | OR2H2, GABBR1 |
| chr7 | 141695896 | 141696101 | 206 | 3 | 0.000184129 | 0.000786 | 0.00045 | 3.33E-05 | -0.003835669 | DENND11 |
| chr7 | 155831094 | 155831614 | 521 | 4 | 0.000184358 | 3.17E-05 | 0.00049 | 3.30E-05 | 0.000627979 | NA |
| chr1 | 1590042 | 1590108 | 67 | 3 | 0.000184389 | 0.004016 | 0.00013 | 3.13E-05 | -0.003205182 | NA |
| chr6 | 142900812 | 142901052 | 241 | 2 | 0.00018443 | 2.50E-05 | 0.00023 | 2.68E-05 | -0.011181695 | HIVEP2 |
| chr8 | 42502625 | 42503369 | 745 | 2 | 0.000184727 | 6.89E-05 | 0.00263 | 0.00011 | -0.009877796 | SLC20A2 |
| chr5 | 141488563 | 141489119 | 557 | 5 | 0.000184761 | 0.001104 | 0.00052 | 8.43E-05 | 0.009814109 | PCDHGA1, PCDHGA2, PCDHGA3, PCDHGB1, PCDHGA4, PCDHGB2, PCDHGA5, PCDHGB3, PCDHGA6, PCDHGA7, PCDHGB4, PCDHGA8, PCDHGB5, PCDHGA9, PCDHGB6, PCDHGA10, PCDHGB7, PCDHGA11, PCDHGA12, PCDHGC3, PCDHGC4, PCDHGC5 |
| chr1 | 182791008 | 182791295 | 288 | 2 | 0.00018483 | 0.005292 | 5.69E-05 | 8.61E-05 | -0.006123162 | NPL |
| chr15 | 40116331 | 40116998 | 668 | 2 | 0.000184888 | 3.97E-06 | 0.00042 | 6.70E-06 | -0.007333729 | NA |
| chr15 | 59286255 | 59286700 | 446 | 2 | 0.000185009 | 1.16E-05 | 0.00058 | 1.78E-05 | -0.00689386 | MYO1E |
| chr10 | 44386486 | 44387151 | 666 | 6 | 0.000185058 | 0.00041 | 0.00018 | 2.62E-05 | -0.007108621 | CXCL12 |
| chr8 | 121639922 | 121640391 | 470 | 3 | 0.000185104 | 0.089278 | 0.00019 | 0.00143 | 0.001207345 | HAS2-AS1, HAS2 |
| chr1 | 154553262 | 154553686 | 425 | 4 | 0.000185173 | 0.000213 | 0.00016 | 8.03E-05 | -0.001372453 | UBE2Q1-AS1, UBE2Q1 |
| chr19 | 12668243 | 12668559 | 317 | 4 | 0.000185534 | 0.000505 | 0.0005 | 0.00019 | -0.002390626 | WDR83, AC010422.5, AC010422.8, WDR83OS |
| chr7 | 1946610 | 1947005 | 396 | 5 | 0.000185788 | 4.16E-05 | 0.00024 | 3.52E-05 | -0.011499427 | MAD1L1, AC069288.1 |
| chr4 | 1214359 | 1214522 | 164 | 3 | 0.000185855 | 7.96E-05 | 0.0015 | 0.00011 | -0.001626773 | CTBP1-AS, CTBP1 |
| chr16 | 664968 | 665591 | 624 | 5 | 0.000185916 | 0.001711 | 0.00018 | 0.00012 | 0.002297121 | WDR90 |
| chr11 | 75336067 | 75337062 | 996 | 3 | 0.000185931 | 3.99E-05 | 0.00012 | 4.17E-06 | -0.005138322 | ARRB1 |
| chr20 | 50998739 | 50998751 | 13 | 2 | 0.000185931 | 3.03E-05 | 0.00092 | 4.35E-05 | -0.003754239 | NA |
| chr3 | 111541355 | 111541936 | 582 | 4 | 0.000186019 | 1.28E-05 | 0.00106 | 1.48E-05 | 0.00767724 | CD96 |
| chr6 | 4079108 | 4079820 | 713 | 11 | 0.000186118 | 0.026821 | 0.00166 | 6.45E-05 | 0.000164958 | C6orf201, FAM217A |
| chr5 | 174793901 | 174794186 | 286 | 4 | 0.00018619 | 0.000479 | 3.89E-05 | 3.13E-05 | 0.011947238 | AC113346.2 |
| chr5 | 95788436 | 95788692 | 257 | 2 | 0.000186249 | 0.000155 | 6.91E-05 | 4.13E-05 | -0.003741273 | RHOBTB3, GLRX |
| chr1 | 239405317 | 239405484 | 168 | 3 | 0.000186292 | 0.002134 | 3.25E-05 | 4.21E-05 | -0.009879082 | CHRM3 |
| chr6 | 28838515 | 28838893 | 379 | 10 | 0.000186516 | 0.000982 | 0.00356 | 6.56E-05 | 0.000926811 | AL662890.1 |
| chr7 | 111108812 | 111108963 | 152 | 2 | 0.000186516 | 9.38E-05 | 0.00016 | 5.66E-05 | -0.010496353 | LRRN3, IMMP2L |
| chr12 | 52367314 | 52367632 | 319 | 5 | 0.00018654 | 9.60E-05 | 0.00256 | 4.79E-05 | -0.009809695 | KRT85 |
| chr15 | 70069378 | 70069904 | 527 | 2 | 0.000186559 | 0.011071 | 0.00015 | 0.00035 | -0.002411522 | TLE3 |
| chr17 | 79993730 | 79994014 | 285 | 3 | 0.000186688 | 1.43E-05 | 0.0018 | 2.37E-05 | -0.008614374 | TBC1D16 |
| chr14 | 104589919 | 104590023 | 105 | 2 | 0.000186739 | 3.06E-05 | 0.00096 | 4.42E-05 | -0.007141648 | C14orf180, TMEM179 |
| chr5 | 68188384 | 68188751 | 368 | 2 | 0.000186798 | 0.000489 | 0.00132 | 0.00047 | -0.004599786 | NA |
| chr6 | 166463002 | 166463550 | 549 | 7 | 0.000186973 | 1.33E-05 | 0.0085 | 2.07E-05 | -0.012748385 | RPS6KA2, RPS6KA2-IT1 |
| chr7 | 75073706 | 75074062 | 357 | 9 | 0.000187209 | 4.39E-05 | 0.00709 | 3.65E-05 | 0.000884803 | RCC1L |
| chr17 | 80152657 | 80152983 | 327 | 2 | 0.000187447 | 2.09E-05 | 0.0004 | 2.73E-05 | -0.009129278 | NA |
| chr19 | 5967062 | 5967152 | 91 | 2 | 0.000187786 | 0.015612 | 0.00031 | 0.00079 | -0.004703258 | RANBP3 |
| chr19 | 10110235 | 10110470 | 236 | 4 | 0.000187789 | 0.000678 | 0.00274 | 0.0003 | -0.002404431 | PPAN-P2RY11, PPAN |
| chr14 | 55440404 | 55441185 | 782 | 9 | 0.000187944 | 4.58E-05 | 0.00068 | 2.56E-05 | 0.011329615 | TBPL2 |
| chr8 | 48620797 | 48621122 | 326 | 5 | 0.000187979 | 0.004796 | 0.00052 | 0.00013 | -0.006105986 | AC022915.2, AC022915.1, LINC02847 |
| chr8 | 116471950 | 116472577 | 628 | 3 | 0.000188346 | 4.75E-06 | 0.00294 | 1.24E-05 | -0.019615395 | NA |
| chr4 | 88467147 | 88467596 | 450 | 3 | 0.000188377 | 0.001243 | 0.00011 | 7.34E-05 | -0.003857306 | HERC5 |
| chr22 | 20999775 | 21000043 | 269 | 3 | 0.000188811 | 0.001375 | 0.00034 | 0.00037 | -0.001092482 | THAP7 |
| chr7 | 87414306 | 87414920 | 615 | 2 | 0.000188982 | 1.41E-05 | 0.00036 | 1.93E-05 | -0.010624074 | ABCB4 |
| chr11 | 14259196 | 14259611 | 416 | 6 | 0.000189143 | 4.96E-06 | 0.00712 | 1.65E-05 | 0.022846704 | SPON1 |
| chr18 | 13380488 | 13380561 | 74 | 2 | 0.000189145 | 1.51E-05 | 0.00047 | 2.15E-05 | -0.018773752 | LDLRAD4 |
| chr7 | 129993451 | 129993792 | 342 | 2 | 0.000189303 | 0.000143 | 8.82E-05 | 4.82E-05 | -0.002067906 | AC073320.1 |
| chr20 | 34525011 | 34525671 | 661 | 3 | 0.000189318 | 4.44E-05 | 0.00045 | 4.23E-05 | -0.004632735 | ITCH, DYNLRB1 |
| chr1 | 31200176 | 31200248 | 73 | 2 | 0.000189446 | 0.000285 | 5.39E-05 | 4.15E-05 | -0.018601384 | NKAIN1 |
| chr14 | 24135502 | 24136573 | 1072 | 14 | 0.000189529 | 4.49E-05 | 0.00805 | 5.71E-06 | -0.000790641 | PSME1 |
| chr3 | 105938388 | 105938718 | 331 | 3 | 0.000189544 | 3.57E-05 | 0.00094 | 2.62E-05 | -0.013638291 | NA |
| chr7 | 90228834 | 90229180 | 347 | 2 | 0.000189602 | 5.74E-05 | 0.00014 | 4.02E-05 | -0.006007578 | STEAP2 |
| chr4 | 38804404 | 38805112 | 709 | 5 | 0.000189893 | 3.10E-05 | 0.00363 | 1.85E-05 | -0.005950742 | TLR1 |
| chr8 | 85245135 | 85245706 | 572 | 13 | 0.000189961 | 0.000129 | 0.01818 | 1.71E-05 | -0.001646559 | CA13, AC011773.3 |
| chr1 | 162613383 | 162613891 | 509 | 4 | 0.000190018 | 8.87E-06 | 0.00172 | 2.08E-05 | -0.010075245 | NA |
| chr11 | 64770743 | 64771091 | 349 | 2 | 0.000190092 | 6.80E-05 | 0.0008 | 8.30E-05 | -0.003342017 | SF1 |
| chr10 | 91141104 | 91141474 | 371 | 3 | 0.000190315 | 8.49E-06 | 0.00321 | 2.04E-05 | -0.012992752 | NA |
| chr2 | 231126278 | 231126403 | 126 | 3 | 0.000190639 | 6.07E-05 | 0.00013 | 2.64E-05 | -0.008196963 | PSMD1 |
| chr7 | 151640621 | 151640823 | 203 | 3 | 0.000190639 | 3.02E-05 | 0.00056 | 1.85E-05 | -0.007139265 | PRKAG2 |
| chr20 | 50909704 | 50910395 | 692 | 8 | 0.000190648 | 0.001624 | 0.00312 | 5.84E-05 | -0.000927132 | ADNP |
| chr11 | 72870077 | 72870148 | 72 | 2 | 0.000190789 | 1.16E-05 | 0.00067 | 1.82E-05 | 0.003321137 | FCHSD2 |
| chr5 | 96874527 | 96874691 | 165 | 3 | 0.000190841 | 0.000287 | 0.00011 | 4.66E-05 | -0.004247315 | AC009126.1 |
| chr20 | 33282969 | 33283546 | 578 | 6 | 0.000190892 | 4.25E-06 | 0.01125 | 2.17E-05 | -0.00552042 | BPIFB1 |
| chr6 | 47785980 | 47786269 | 290 | 2 | 0.000190991 | 5.46E-05 | 9.66E-05 | 3.07E-05 | -0.006779738 | OPN5 |
| chr9 | 137230222 | 137230861 | 640 | 8 | 0.00019102 | 1.76E-05 | 0.01142 | 2.20E-05 | -0.006781749 | SLC34A3 |
| chr1 | 178495196 | 178495588 | 393 | 2 | 0.000191021 | 6.09E-05 | 0.00013 | 3.94E-05 | 0.001674677 | CLEC20A |
| chr9 | 136448882 | 136449110 | 229 | 3 | 0.000191226 | 0.006168 | 4.12E-05 | 9.19E-05 | -0.001280009 | SEC16A |
| chr16 | 27314351 | 27314733 | 383 | 2 | 0.00019125 | 0.000342 | 0.00613 | 0.00059 | 0.000811249 | IL4R |
| chr10 | 522600 | 522613 | 14 | 2 | 0.00019138 | 4.11E-05 | 0.00023 | 3.87E-05 | -0.004234471 | DIP2C |
| chr1 | 43619294 | 43619391 | 98 | 2 | 0.000191774 | 1.45E-05 | 0.00065 | 2.19E-05 | -0.004080783 | PTPRF |
| chr5 | 150052439 | 150053208 | 770 | 2 | 0.000191774 | 4.02E-06 | 0.00047 | 6.85E-06 | -0.010315661 | HMGXB3 |
| chr16 | 90039842 | 90040201 | 360 | 2 | 0.000191774 | 1.05E-05 | 0.00077 | 1.70E-05 | -0.003235969 | GAS8, URAHP |
| chr1 | 1442145 | 1442276 | 132 | 2 | 0.000191816 | 2.15E-05 | 0.00106 | 3.32E-05 | -0.005514115 | VWA1 |
| chr22 | 38179922 | 38180080 | 159 | 2 | 0.000191929 | 0.000448 | 0.00025 | 0.00017 | -0.012282822 | PLA2G6 |
| chr1 | 39047654 | 39048042 | 389 | 2 | 0.000192279 | 1.24E-05 | 0.00051 | 1.85E-05 | -0.035800827 | NA |
| chr12 | 113794897 | 113795132 | 236 | 4 | 0.000192298 | 4.41E-06 | 0.00488 | 1.49E-05 | -0.020075809 | NA |
| chr8 | 43073386 | 43073625 | 240 | 2 | 0.000192298 | 1.44E-05 | 0.00088 | 2.28E-05 | -0.014903226 | AC110275.1, FNTA, RNU1-124P |
| chr15 | 72784294 | 72785101 | 808 | 9 | 0.000192434 | 3.84E-05 | 0.00265 | 4.60E-05 | -0.001650117 | ADPGK-AS1, ADPGK |
| chr15 | 101121324 | 101121629 | 306 | 5 | 0.000192556 | 3.81E-05 | 0.00077 | 1.79E-05 | -0.01049846 | NA |
| chr22 | 46455268 | 46455746 | 479 | 2 | 0.000192979 | 6.57E-06 | 0.00057 | 1.08E-05 | -0.011011751 | CELSR1 |
| chr17 | 39153296 | 39153563 | 268 | 3 | 0.000193367 | 4.03E-05 | 0.00088 | 5.43E-05 | 0.004338285 | PLXDC1 |
| chr2 | 208025510 | 208026078 | 569 | 11 | 0.00019339 | 1.76E-05 | 0.00585 | 1.31E-05 | 0.007235928 | PLEKHM3 |
| chr11 | 132307711 | 132307799 | 89 | 3 | 0.000193632 | 0.003704 | 2.81E-05 | 4.47E-05 | -0.001414142 | NTM |
| chr16 | 27068396 | 27068597 | 202 | 2 | 0.000193775 | 5.30E-05 | 4.71E-05 | 1.83E-05 | -0.011479868 | C16orf82 |
| chr11 | 57527537 | 57527849 | 313 | 2 | 0.000193995 | 3.72E-05 | 0.00021 | 3.48E-05 | -0.012107603 | NA |
| chr16 | 89885294 | 89885541 | 248 | 3 | 0.000194078 | 0.003626 | 7.47E-05 | 7.69E-05 | -0.003271538 | TCF25 |
| chr22 | 20456472 | 20456636 | 165 | 2 | 0.00019419 | 2.01E-05 | 0.00027 | 2.41E-05 | -0.004586319 | KLHL22 |
| chr14 | 30621674 | 30622466 | 793 | 12 | 0.000194244 | 0.000121 | 0.0017 | 2.17E-05 | 0.000789334 | SCFD1 |
| chr6 | 43522342 | 43523008 | 667 | 2 | 0.000194246 | 7.59E-06 | 0.00046 | 1.19E-05 | -0.007677114 | POLR1C, XPO5 |
| chr10 | 119158206 | 119158287 | 82 | 3 | 0.000194318 | 2.81E-05 | 0.00018 | 2.36E-05 | -0.00032709 | SFXN4 |
| chr2 | 25702125 | 25702181 | 57 | 2 | 0.000194443 | 0.000172 | 4.30E-05 | 2.88E-05 | -0.003226579 | NA |
| chr1 | 42035873 | 42036086 | 214 | 4 | 0.000194493 | 1.40E-05 | 0.00319 | 2.06E-05 | 0.000415381 | HIVEP3 |
| chr3 | 48631430 | 48631753 | 324 | 2 | 0.000194702 | 0.016675 | 0.00028 | 0.00073 | -0.004415178 | SLC26A6 |
| chr1 | 145997501 | 145997772 | 272 | 2 | 0.000194721 | 0.004751 | 0.00074 | 0.00117 | 0.003354024 | NA |
| chr6 | 10555620 | 10556357 | 738 | 8 | 0.0001949 | 6.18E-06 | 0.00534 | 1.74E-05 | -0.007847452 | GCNT2 |
| chr6 | 44342028 | 44342190 | 163 | 2 | 0.000194954 | 0.000313 | 0.00012 | 8.47E-05 | -0.012928336 | AL353588.1 |
| chr1 | 26173856 | 26174105 | 250 | 2 | 0.00019504 | 4.75E-05 | 0.00012 | 3.19E-05 | -0.00115458 | NA |
| chr11 | 65978390 | 65979283 | 894 | 4 | 0.000195144 | 9.07E-06 | 0.00309 | 2.18E-05 | -0.003267366 | SART1 |
| chr18 | 55589788 | 55590648 | 861 | 11 | 0.000195249 | 0.006798 | 0.00165 | 0.00011 | -0.001933433 | AC022031.2, TCF4 |
| chr16 | 2963744 | 2965091 | 1348 | 14 | 0.000195515 | 3.06E-05 | 0.00325 | 2.93E-06 | -0.000116743 | KREMEN2 |
| chr17 | 81086395 | 81087878 | 1484 | 4 | 0.000195575 | 5.00E-06 | 0.00062 | 4.53E-06 | -0.005050044 | BAIAP2 |
| chr6 | 156847466 | 156847700 | 235 | 2 | 0.000195684 | 1.99E-05 | 0.00065 | 2.88E-05 | -0.007319325 | ARID1B |
| chr7 | 1846641 | 1847281 | 641 | 5 | 0.000195831 | 9.22E-06 | 0.01425 | 3.74E-05 | -0.026140783 | AC104129.1, MAD1L1, AC069288.1 |
| chr10 | 118668929 | 118669693 | 765 | 4 | 0.000196057 | 2.61E-06 | 0.00384 | 8.01E-06 | -0.015127822 | NA |
| chr15 | 22887486 | 22887975 | 490 | 3 | 0.000196165 | 9.38E-06 | 0.00095 | 1.60E-05 | 0.000996434 | CYFIP1 |
| chr2 | 233854455 | 233854932 | 478 | 13 | 0.000196402 | 0.004659 | 0.00096 | 6.31E-05 | 0.000922747 | HJURP |
| chr8 | 22877598 | 22878481 | 884 | 5 | 0.000196515 | 7.12E-06 | 0.00557 | 2.17E-05 | -0.010558111 | AC037441.1, PEBP4 |
| chr13 | 52159999 | 52160297 | 299 | 3 | 0.00019653 | 0.022231 | 0.02039 | 0.02274 | 3.17E-05 | NA |
| chr15 | 101304854 | 101305319 | 466 | 4 | 0.000196575 | 9.87E-05 | 0.00109 | 3.41E-05 | -0.003652082 | AC023024.1, PCSK6 |
| chr5 | 108747061 | 108747258 | 198 | 3 | 0.000196592 | 0.000121 | 0.00029 | 7.38E-05 | -0.009162969 | NA |
| chr20 | 44911561 | 44911668 | 108 | 3 | 0.000196738 | 1.94E-05 | 0.00085 | 2.47E-05 | -0.020718634 | PABPC1L |
| chr8 | 80078168 | 80078224 | 57 | 2 | 0.0001971 | 4.19E-05 | 7.84E-05 | 2.29E-05 | -0.005241033 | AC036214.3, TPD52 |
| chr1 | 167428298 | 167428671 | 374 | 2 | 0.000197131 | 1.50E-05 | 0.00051 | 2.17E-05 | -0.012805204 | NA |
| chr14 | 22870644 | 22870660 | 17 | 2 | 0.000197131 | 0.000154 | 0.0009 | 0.00017 | -0.010591118 | NA |
| chr2 | 104659143 | 104659695 | 553 | 6 | 0.000197166 | 2.20E-05 | 0.00415 | 2.95E-05 | 0.017666969 | AC013402.1, AC068057.3 |
| chr4 | 55079227 | 55079673 | 447 | 3 | 0.000197166 | 1.87E-05 | 0.00036 | 2.26E-05 | -0.012657858 | AC111194.2, KDR |
| chr12 | 27752804 | 27752961 | 158 | 3 | 0.000197508 | 1.43E-05 | 0.00072 | 1.84E-05 | -0.005261491 | MRPS35, Y RNA |
| chr16 | 19210830 | 19211154 | 325 | 5 | 0.000197632 | 3.44E-05 | 0.00232 | 2.17E-05 | -0.006479721 | SYT17 |
| chr10 | 133024160 | 133025152 | 993 | 8 | 0.000197702 | 7.95E-05 | 0.00065 | 7.26E-06 | 0.003400102 | NA |
| chr2 | 218884037 | 218884483 | 447 | 3 | 0.000197851 | 7.75E-06 | 0.00164 | 1.62E-05 | -0.008263255 | WNT10A |
| chr1 | 204565813 | 204565872 | 60 | 2 | 0.000198153 | 1.31E-05 | 0.00039 | 1.84E-05 | -0.021658772 | NA |
| chr12 | 1930586 | 1930712 | 127 | 2 | 0.00019822 | 4.66E-05 | 0.00017 | 3.79E-05 | -0.012900817 | LINC00940 |
| chr16 | 593447 | 593706 | 260 | 3 | 0.000198229 | 0.000331 | 0.00189 | 9.80E-05 | -0.006046911 | RAB40C |
| chr10 | 58819157 | 58819719 | 563 | 2 | 0.000198331 | 1.24E-05 | 0.00086 | 1.99E-05 | -0.000266919 | BICC1 |
| chr4 | 6993363 | 6994031 | 669 | 3 | 0.000198387 | 4.40E-06 | 0.00188 | 1.08E-05 | 0.004959398 | TBC1D14 |
| chr2 | 48107461 | 48107824 | 364 | 3 | 0.000198389 | 7.20E-05 | 0.00021 | 3.59E-05 | -0.003318823 | AC092650.1 |
| chr12 | 123268639 | 123268725 | 87 | 2 | 0.000198497 | 0.001415 | 9.93E-05 | 0.00012 | -0.007965676 | CDK2AP1 |
| chr7 | 18494671 | 18494993 | 323 | 3 | 0.000198542 | 1.02E-05 | 0.00074 | 1.72E-05 | -0.006072979 | HDAC9 |
| chr9 | 136249846 | 136250203 | 358 | 3 | 0.000198637 | 3.66E-05 | 0.0012 | 3.65E-05 | -0.004840649 | CR392000.1, CCDC187 |
| chr2 | 235044085 | 235044660 | 576 | 3 | 0.000198679 | 3.23E-05 | 0.0013 | 4.87E-05 | -0.014961318 | SH3BP4 |
| chr16 | 1559907 | 1561051 | 1145 | 6 | 0.000198828 | 7.09E-06 | 0.00163 | 1.26E-05 | -0.004656931 | IFT140 |
| chr6 | 25726230 | 25726728 | 499 | 7 | 0.000198856 | 0.000193 | 0.00166 | 4.02E-05 | -0.001912035 | H2AC1 |
| chr21 | 43720950 | 43721050 | 101 | 2 | 0.000198951 | 3.04E-05 | 0.00011 | 2.30E-05 | -0.008841519 | PDXK |
| chr9 | 86343409 | 86343422 | 14 | 2 | 0.00019928 | 1.48E-05 | 0.00105 | 2.39E-05 | 0.000169141 | TUT7 |
| chr1 | 24857668 | 24857672 | 5 | 2 | 0.000199305 | 4.75E-05 | 7.21E-05 | 2.33E-05 | -0.007247376 | NA |
| chr11 | 13366802 | 13367291 | 490 | 2 | 0.000199313 | 4.10E-05 | 0.0019 | 6.39E-05 | -0.005538314 | ARNTL |
| chr17 | 48579328 | 48580895 | 1568 | 12 | 0.000199357 | 1.04E-06 | 0.01115 | 2.53E-06 | -0.009797744 | HOXB-AS3, HOXB3, AC103702.1, MIR10A |
| chr6 | 31803067 | 31803505 | 439 | 4 | 0.00019936 | 0.001363 | 0.0002 | 3.34E-05 | -0.001826259 | LSM2 |
| chr16 | 2271167 | 2271186 | 20 | 2 | 0.000199386 | 1.92E-05 | 9.89E-05 | 1.57E-05 | -0.004614602 | AC009065.4 |

| | | | | | | | | | | |
|---|---|---|---|---|---|---|---|---|---|---|
| chr11 | 94152657 | 94153101 | 445 | 3 | 0.000199582 | 2.21E-05 | 0.00372 | 4.77E-05 | -0.049759414 | PANX1 |
| chr16 | 1705944 | 1706770 | 827 | 10 | 0.00019989 | 6.21E-05 | 0.0068 | 2.38E-05 | 0.000446117 | MAPK8IP3 |
| chr1 | 177914802 | 177914987 | 186 | 2 | 0.000199891 | 1.82E-05 | 0.00115 | 2.90E-05 | 0.002283713 | AL136114.1 |
| chr15 | 84868154 | 84868340 | 187 | 2 | 0.000200474 | 1.77E-05 | 0.00119 | 2.85E-05 | -0.003549265 | ALPK3 |
| chr3 | 58236347 | 58237800 | 1454 | 12 | 0.000200481 | 9.45E-05 | 0.00048 | 3.42E-06 | -0.000263702 | ABHD6 |
| chr19 | 9768641 | 9769019 | 379 | 9 | 0.000200536 | 0.000991 | 0.00056 | 4.56E-05 | 0.000899101 | ZNF846 |
| chr1 | 169893368 | 169893947 | 580 | 6 | 0.000200698 | 0.000247 | 0.00261 | 8.84E-05 | 0.001811875 | SCYL3 |
| chr7 | 141763369 | 141763771 | 403 | 3 | 0.000200808 | 1.20E-05 | 0.0007 | 1.14E-05 | -0.003239968 | SSBP1 |
| chr3 | 10816710 | 10817267 | 558 | 4 | 0.000200837 | 0.001024 | 0.00117 | 7.72E-05 | -0.011341765 | SLC6A11 |
| chr7 | 66088828 | 66089070 | 243 | 4 | 0.000200858 | 0.001143 | 5.96E-05 | 4.42E-05 | -0.002191315 | ASL, AC068533.4 |
| chr3 | 169579297 | 169579777 | 481 | 3 | 0.0002009 | 1.51E-05 | 0.00149 | 2.14E-05 | -0.01658338 | MECOM |
| chr8 | 133051706 | 133051979 | 274 | 3 | 0.0002009 | 0.000125 | 0.00026 | 4.35E-05 | -0.005209354 | TG, SLA |
| chr5 | 160370393 | 160370944 | 552 | 12 | 0.000200952 | 9.25E-06 | 0.00468 | 1.34E-05 | -0.002393593 | C1QTNF2 |
| chr15 | 40892676 | 40893605 | 930 | 4 | 0.000201013 | 2.61E-05 | 0.00482 | 5.54E-05 | -0.006871058 | NA |
| chr5 | 149732041 | 149732707 | 667 | 5 | 0.000201111 | 3.72E-05 | 0.00061 | 2.88E-05 | -0.007104437 | PPARGC1B |
| chr17 | 50527508 | 50527548 | 41 | 3 | 0.000201208 | 6.31E-06 | 0.00281 | 1.55E-05 | -0.01300314 | MYCBPAP, AC021491.2 |
| chr8 | 90939339 | 90939525 | 187 | 3 | 0.000201265 | 0.000955 | 0.00062 | 3.89E-05 | -0.004362561 | NECAB1 |
| chr2 | 8623773 | 8624003 | 231 | 2 | 0.000201605 | 8.08E-05 | 7.81E-05 | 3.32E-05 | 0.00283906 | NA |
| chr1 | 210327044 | 210328622 | 1579 | 12 | 0.000201717 | 9.17E-05 | 0.00075 | 4.87E-06 | -0.001289963 | HHAT |
| chr13 | 45682682 | 45682765 | 84 | 2 | 0.000201717 | 8.95E-05 | 6.22E-05 | 2.94E-05 | -0.011183359 | LINC01055 |
| chr6 | 75151971 | 75152377 | 407 | 2 | 0.000201763 | 1.29E-05 | 0.00062 | 1.96E-05 | 0.000961093 | COL12A1 |
| chr6 | 149217017 | 149217389 | 373 | 2 | 0.000201983 | 0.019889 | 0.0001 | 0.00022 | -0.002383735 | NA |
| chr14 | 21570572 | 21571073 | 502 | 4 | 0.000202081 | 0.001013 | 0.00019 | 8.91E-05 | -0.007018414 | OR10G3 |
| chr4 | 163473534 | 163474135 | 602 | 10 | 0.000202218 | 4.47E-05 | 0.00749 | 3.45E-05 | -0.004473775 | TKTL2 |
| chr9 | 135770269 | 135770916 | 648 | 6 | 0.000202288 | 3.00E-06 | 0.01324 | 1.49E-05 | -0.008512413 | KCNT1 |
| chr19 | 805124 | 805238 | 115 | 2 | 0.000202359 | 0.002672 | 4.42E-05 | 5.89E-05 | -0.003075474 | PTBP1 |
| chr7 | 4739088 | 4739711 | 624 | 8 | 0.000202657 | 1.15E-05 | 0.0031 | 1.98E-05 | -0.007194078 | FOXK1 |
| chr2 | 159904733 | 159905235 | 503 | 11 | 0.000203014 | 7.04E-05 | 0.00736 | 2.14E-05 | -6.10E-06 | LY75 |
| chr12 | 52380046 | 52380363 | 318 | 3 | 0.000203014 | 1.64E-05 | 0.00149 | 2.89E-05 | -0.043648948 | KRT84 |
| chr3 | 172594733 | 172594809 | 77 | 3 | 0.000203223 | 7.94E-06 | 0.00288 | 1.91E-05 | -0.045401914 | LINC02068 |
| chr5 | 176969682 | 176969996 | 315 | 2 | 0.000203251 | 0.001243 | 0.00016 | 0.0002 | -0.002821664 | UIMC1 |
| chr3 | 46110954 | 46111273 | 320 | 2 | 0.000203302 | 0.000293 | 6.76E-05 | 5.15E-05 | -0.015102645 | NA |
| chr2 | 151711003 | 151711235 | 233 | 2 | 0.00020338 | 0.007682 | 0.00011 | 0.00021 | -0.014299981 | NEB |
| chr16 | 85610148 | 85610777 | 630 | 5 | 0.000203463 | 0.001141 | 0.00187 | 0.00021 | -0.003638224 | GSE1 |
| chr17 | 79097324 | 79097784 | 461 | 5 | 0.000203484 | 5.66E-05 | 0.00103 | 2.02E-05 | -0.009067627 | RBFOX3 |
| chr19 | 18339991 | 18340763 | 773 | 14 | 0.000203606 | 2.36E-05 | 0.00873 | 2.29E-05 | 0.003306306 | PGPEP1 |
| chr17 | 44212303 | 44213051 | 749 | 6 | 0.000203935 | 0.00282 | 0.0013 | 3.95E-05 | -0.004622961 | AC004596.1, UBTF |
| chr19 | 42320048 | 42320428 | 381 | 3 | 0.000204047 | 5.06E-05 | 0.00107 | 3.34E-05 | -0.006161933 | TMEM145 |
| chr1 | 212485629 | 212485807 | 179 | 2 | 0.000204059 | 0.017687 | 7.91E-05 | 0.00016 | -0.002386497 | NA |
| chr16 | 87696912 | 87697298 | 387 | 4 | 0.000204126 | 0.000654 | 0.00075 | 6.91E-05 | -0.003101616 | JPH3, KLHDC4 |
| chr2 | 3181359 | 3181641 | 283 | 2 | 0.000204148 | 0.003624 | 4.93E-05 | 6.96E-05 | -0.00506902 | NA |
| chr1 | 4711925 | 4712203 | 279 | 3 | 0.000204156 | 0.000456 | 5.22E-05 | 4.93E-05 | 0.001820005 | AJAP1 |
| chr11 | 60996799 | 60996830 | 32 | 2 | 0.000204247 | 2.30E-05 | 0.00014 | 2.09E-05 | -0.009938141 | CD6 |
| chr12 | 52407089 | 52407286 | 198 | 2 | 0.000204534 | 0.002464 | 0.00011 | 0.00016 | -0.006211379 | NA |
| chr14 | 105493779 | 105494044 | 266 | 2 | 0.000204535 | 4.89E-05 | 0.00069 | 6.13E-05 | -0.004027877 | TEDC1 |
| chr8 | 27610266 | 27610654 | 389 | 3 | 0.000204635 | 4.57E-05 | 0.0017 | 5.72E-05 | -0.009606194 | CLU, MIR6843 |
| chr12 | 56138509 | 56138771 | 263 | 2 | 0.000204635 | 9.13E-05 | 0.00014 | 5.31E-05 | -0.006120042 | ESYT1 |
| chr12 | 79787011 | 79787473 | 463 | 2 | 0.000205011 | 1.48E-05 | 0.00057 | 2.18E-05 | -0.013380379 | PPP1R12A |
| chr10 | 96612195 | 96612670 | 476 | 2 | 0.000205158 | 1.19E-05 | 0.00063 | 1.84E-05 | -0.004087197 | PIK3AP1 |
| chr1 | 200866537 | 200866760 | 224 | 2 | 0.000205321 | 0.000212 | 5.93E-05 | 4.09E-05 | -0.009790398 | NA |
| chr17 | 36601048 | 36601180 | 133 | 3 | 0.000205385 | 5.37E-05 | 7.45E-05 | 2.10E-05 | -0.002029554 | NA |
| chr14 | 54622755 | 54622761 | 7 | 2 | 0.000205757 | 1.44E-05 | 0.00033 | 1.91E-05 | 0.007146817 | SAMD4A |
| chr2 | 237414037 | 237414784 | 748 | 8 | 0.000205882 | 3.67E-05 | 0.0047 | 2.27E-05 | -0.005471381 | COL6A3 |
| chr1 | 161214515 | 161214738 | 224 | 3 | 0.000206094 | 0.000489 | 0.00197 | 0.00013 | -0.006585775 | NDUFS2 |
| chr14 | 104988889 | 104989065 | 177 | 2 | 0.000206099 | 0.000797 | 0.00767 | 0.0013 | -0.008992411 | CLBA1 |
| chr19 | 2947574 | 2947907 | 334 | 2 | 0.000206149 | 0.000139 | 9.87E-05 | 5.16E-05 | 0.003108816 | NA |
| chr20 | 34042452 | 34042567 | 116 | 2 | 0.000206166 | 1.43E-05 | 0.00103 | 2.30E-05 | -0.011658721 | RALY |
| chr1 | 110631837 | 110632543 | 707 | 3 | 0.000206212 | 2.54E-06 | 0.00082 | 5.53E-06 | -0.009077662 | NA |
| chr5 | 115962156 | 115963023 | 868 | 10 | 0.000206336 | 0.000333 | 0.00493 | 8.12E-05 | 0.003090558 | LVRN |
| chr1 | 168236020 | 168236081 | 62 | 2 | 0.000206572 | 3.20E-05 | 0.00159 | 4.95E-05 | -0.003297923 | SFT2D2 |
| chr7 | 1534462 | 1534777 | 316 | 4 | 0.000206662 | 9.09E-05 | 0.00019 | 4.36E-05 | -0.003648946 | MAFK |
| chr3 | 136983379 | 136983387 | 9 | 2 | 0.000206781 | 0.000722 | 5.16E-05 | 5.22E-05 | -0.001436883 | IL20RB |
| chr19 | 10929336 | 10929608 | 273 | 3 | 0.000206805 | 7.00E-05 | 0.00216 | 0.00011 | -0.005690983 | TIMM29 |
| chr7 | 80646108 | 80646568 | 461 | 6 | 0.000206997 | 0.001509 | 0.00065 | 9.82E-05 | -0.006954504 | CD36 |
| chr10 | 48671659 | 48671869 | 211 | 3 | 0.000207254 | 0.00062 | 5.90E-05 | 5.91E-05 | -0.001999373 | AC068898.1 |
| chr4 | 7287996 | 7288167 | 172 | 4 | 0.000207283 | 0.000561 | 0.00022 | 4.63E-05 | -0.012210715 | SORCS2 |
| chr11 | 118322847 | 118322904 | 58 | 2 | 0.000207426 | 2.96E-05 | 0.00011 | 2.22E-05 | -0.002659964 | AP001582.1 |
| chr8 | 144705079 | 144705467 | 389 | 2 | 0.000207933 | 0.000106 | 0.00023 | 7.40E-05 | -0.007428063 | AF186192.2 |
| chr17 | 39618604 | 39618681 | 78 | 3 | 0.000208191 | 9.37E-05 | 0.00142 | 0.00011 | -0.001508631 | NA |
| chr13 | 112898068 | 112898458 | 391 | 4 | 0.000208413 | 0.000153 | 0.00139 | 3.33E-05 | 0.002505088 | MCF2L |
| chr13 | 51324375 | 51324812 | 438 | 3 | 0.000208659 | 0.000301 | 0.00029 | 4.66E-05 | -0.00341955 | NA |
| chr6 | 160132744 | 160133642 | 899 | 5 | 0.000208703 | 6.05E-06 | 0.00328 | 1.57E-05 | -0.0158416 | SLC22A1 |
| chr1 | 54172464 | 54172817 | 354 | 4 | 0.000208819 | 0.010096 | 0.00036 | 0.00019 | -0.000649781 | AL357673.1, CYB5RL |
| chr3 | 45818318 | 45818680 | 363 | 4 | 0.000208882 | 0.000608 | 5.56E-05 | 4.66E-05 | 0.001572855 | NA |
| chr6 | 168032511 | 168032756 | 246 | 2 | 0.000208932 | 9.14E-05 | 0.00032 | 7.63E-05 | -0.019195087 | KIF25 |
| chr4 | 123008223 | 123008691 | 469 | 3 | 0.000208958 | 0.000162 | 7.81E-05 | 4.33E-05 | -0.003636582 | SPATA5 |
| chr4 | 1544738 | 1544904 | 167 | 2 | 0.000209093 | 1.23E-05 | 0.0007 | 1.93E-05 | -0.017384569 | NA |
| chr13 | 51912447 | 51912886 | 440 | 2 | 0.000209093 | 0.000306 | 0.00681 | 0.00055 | -0.023862646 | NA |
| chr1 | 202007789 | 202008262 | 474 | 4 | 0.000209111 | 0.000451 | 0.00027 | 0.00012 | -0.00478348 | ELF3, ELF3-AS1 |
| chr7 | 80512179 | 80512191 | 13 | 2 | 0.000209382 | 1.72E-05 | 0.00114 | 2.76E-05 | 0.006451921 | CD36 |
| chr8 | 70071113 | 70071742 | 630 | 9 | 0.000209406 | 7.18E-05 | 0.00722 | 2.22E-05 | 0.000678308 | PRDM14 |
| chr19 | 47514943 | 47515622 | 680 | 11 | 0.000209496 | 0.002249 | 0.00359 | 6.42E-05 | -0.000767495 | NAPA |
| chr1 | 47532965 | 47533491 | 527 | 5 | 0.000209571 | 3.77E-05 | 0.00149 | 1.96E-05 | -0.003934717 | NA |
| chr20 | 35246759 | 35246892 | 134 | 2 | 0.000209586 | 2.63E-05 | 0.00014 | 2.30E-05 | -0.004242562 | MMP24, MMP24OS |
| chr17 | 3280188 | 3280254 | 67 | 3 | 0.000209721 | 0.002045 | 0.00021 | 3.30E-05 | -0.022369203 | OR3A2 |
| chr11 | 72380499 | 72380927 | 429 | 2 | 0.000210046 | 6.07E-05 | 0.00048 | 6.57E-05 | 9.58E-05 | CLPB |
| chr2 | 235812039 | 235812609 | 571 | 4 | 0.000210189 | 1.17E-05 | 0.00333 | 1.86E-05 | -0.001263887 | AGAP1 |
| chr3 | 183278721 | 183278911 | 191 | 2 | 0.000210249 | 0.002189 | 4.86E-05 | 6.31E-05 | -0.011074541 | B3GNT5, MCF2L2 |
| chr14 | 45135100 | 45136150 | 1051 | 9 | 0.000210299 | 0.001337 | 0.01332 | 0.00019 | 0.002168703 | FANCM, FKBP3 |
| chr6 | 44157736 | 44158833 | 1098 | 5 | 0.000210494 | 3.79E-06 | 0.00063 | 2.17E-06 | 0.002944799 | CAPN11 |
| chr4 | 87848777 | 87849071 | 295 | 3 | 0.000210541 | 0.007726 | 0.00015 | 6.43E-05 | 0.013872441 | NA |
| chr17 | 16654571 | 16654639 | 69 | 3 | 0.000210643 | 6.19E-05 | 0.00154 | 4.88E-05 | -0.015933502 | AC098850.2 |
| chr1 | 110192002 | 110192552 | 551 | 4 | 0.000210796 | 8.92E-05 | 0.00148 | 3.27E-05 | -0.003295943 | SLC6A17 |
| chr14 | 20324562 | 20324609 | 48 | 2 | 0.000210878 | 2.51E-05 | 0.00023 | 2.71E-05 | -0.009004564 | CCNB1IP1 |
| chr19 | 55536708 | 55537226 | 519 | 6 | 0.000210937 | 1.99E-05 | 0.01042 | 4.56E-05 | -0.011025489 | SBK2 |
| chr10 | 128568432 | 128568631 | 200 | 4 | 0.000210976 | 7.10E-06 | 0.00167 | 1.56E-05 | 0.005498533 | NA |
| chr6 | 31962546 | 31963098 | 553 | 7 | 0.000211373 | 0.010665 | 0.00017 | 0.00017 | -0.001930546 | SKIV2L |
| chr10 | 3494149 | 3494581 | 433 | 4 | 0.000211389 | 0.00052 | 0.00053 | 4.06E-05 | -0.003380508 | LINC02669 |
| chr19 | 38004854 | 38005386 | 533 | 4 | 0.000211483 | 5.97E-06 | 0.00625 | 2.01E-05 | -0.013269249 | SIPA1L3 |
| chr14 | 45110358 | 45110743 | 386 | 3 | 0.000211802 | 0.000437 | 0.0017 | 0.0001 | -0.004630179 | PRPF39 |
| chr4 | 8747531 | 8747869 | 339 | 5 | 0.00021183 | 0.000834 | 0.00024 | 5.85E-05 | -0.004885588 | AC209005.1 |
| chr22 | 42565773 | 42566246 | 474 | 2 | 0.000211845 | 1.39E-05 | 0.00057 | 2.07E-05 | -0.00554322 | SERHL2, RRP7BP |
| chr1 | 103108531 | 103109402 | 872 | 10 | 0.000211845 | 0.000341 | 0.01065 | 8.10E-05 | -0.00686064 | COL11A1 |
| chr8 | 140555985 | 140556299 | 315 | 3 | 0.000211845 | 3.54E-05 | 0.00091 | 4.78E-05 | -0.004630741 | AGO2 |
| chr12 | 126406135 | 126406416 | 282 | 3 | 0.00021206 | 0.000333 | 6.38E-05 | 4.46E-05 | 0.003754766 | AC007368.1 |
| chr9 | 128551464 | 128552263 | 800 | 5 | 0.000212214 | 0.000644 | 0.00226 | 5.60E-05 | -0.004365647 | AL356481.1 |
| chr9 | 137074174 | 137074295 | 122 | 2 | 0.000212347 | 1.47E-05 | 0.00079 | 2.28E-05 | -0.010183236 | NA |
| chr2 | 97737760 | 97737829 | 70 | 3 | 0.00021235 | 0.000307 | 0.00029 | 3.34E-05 | -0.002178791 | ZAP70 |
| chr7 | 101818543 | 101818835 | 293 | 3 | 0.000212547 | 1.11E-05 | 0.0042 | 2.68E-05 | -0.012346018 | CUX1 |
| chr19 | 54952730 | 54952762 | 33 | 3 | 0.000212605 | 0.000496 | 0.00011 | 8.61E-05 | -0.002339422 | NLRP7 |
| chr12 | 6863830 | 6864102 | 273 | 2 | 0.000212661 | 0.001506 | 4.68E-05 | 5.63E-05 | -0.00242437 | USP5 |
| chr19 | 49511730 | 49512523 | 794 | 8 | 0.000212664 | 0.005131 | 0.003 | 0.00051 | -0.00406851 | FCGRT |
| chr19 | 40226611 | 40226995 | 385 | 9 | 0.000212969 | 0.000142 | 0.00133 | 9.62E-05 | -0.006693208 | CCNP |
| chr1 | 161259424 | 161259619 | 196 | 2 | 0.000213368 | 0.000141 | 0.00017 | 7.59E-05 | -0.013102208 | PCP4L1 |
| chr2 | 241210260 | 241210337 | 78 | 2 | 0.000213411 | 2.76E-05 | 0.00143 | 4.30E-05 | 0.002848544 | ANO7 |
| chr4 | 39527169 | 39527894 | 726 | 13 | 0.000213874 | 0.000516 | 0.00095 | 4.69E-05 | 0.000899027 | UGDH |
| chr19 | 40256679 | 40257186 | 508 | 4 | 0.000213931 | 9.52E-05 | 0.00017 | 3.31E-05 | -0.004127491 | AKT2 |
| chr1 | 6273736 | 6274258 | 523 | 4 | 0.000213932 | 1.29E-05 | 0.00168 | 1.63E-05 | -0.004870089 | ACOT7 |
| chr3 | 49999414 | 49999462 | 49 | 2 | 0.000213947 | 2.05E-05 | 0.00036 | 2.62E-05 | -0.006243379 | RBM6 |
| chr6 | 151764112 | 151765412 | 1301 | 7 | 0.000214095 | 3.95E-05 | 0.00027 | 2.79E-06 | -0.009988822 | ESR1 |
| chr7 | 151876828 | 151877458 | 631 | 13 | 0.000214178 | 0.001229 | 0.00023 | 2.30E-05 | 0.000820813 | PRKAG2-AS1, PRKAG2 |
| chr6 | 149445454 | 149445909 | 456 | 2 | 0.000214313 | 3.49E-05 | 0.00109 | 5.06E-05 | -0.0056907 | NA |
| chr4 | 48932954 | 48933639 | 686 | 4 | 0.000214353 | 3.12E-06 | 0.00314 | 8.93E-06 | 0.005243275 | NA |
| chr1 | 7650659 | 7651078 | 420 | 4 | 0.000214609 | 0.008115 | 0.00016 | 6.62E-05 | -0.000388659 | CAMTA1 |
| chr9 | 124282214 | 124282712 | 499 | 2 | 0.000214906 | 5.29E-06 | 0.00045 | 8.64E-06 | -0.01088086 | NEK6 |
| chr1 | 8194371 | 8194593 | 223 | 2 | 0.000215158 | 0.000528 | 4.57E-05 | 4.31E-05 | -0.01176781 | NA |
| chr22 | 19785835 | 19785892 | 58 | 2 | 0.000215541 | 1.63E-05 | 0.00057 | 2.38E-05 | -0.008048949 | GNB1L |
| chr17 | 76446256 | 76446285 | 30 | 3 | 0.000215602 | 1.10E-05 | 0.00119 | 2.04E-05 | -0.007838584 | UBE2O |
| chr3 | 101849659 | 101850201 | 543 | 3 | 0.000215612 | 0.000362 | 0.00238 | 0.00025 | 0.006420293 | NFKBIZ |
| chr11 | 119764316 | 119764624 | 309 | 2 | 0.00021593 | 0.000273 | 6.96E-05 | 5.15E-05 | -0.005112135 | NA |
| chr15 | 43302045 | 43302419 | 375 | 5 | 0.000215981 | 0.000283 | 0.00044 | 6.55E-05 | 0.004193124 | AC009852.1, TGM7 |
| chr1 | 75397443 | 75397700 | 258 | 2 | 0.000216098 | 1.44E-05 | 0.00086 | 2.26E-05 | -0.006829728 | SLC44A5 |
| chr14 | 99478957 | 99479670 | 714 | 4 | 0.000216295 | 0.00056 | 0.00106 | 0.00026 | -0.005387563 | SETD3 |
| chr2 | 28682995 | 28683326 | 332 | 2 | 0.00021633 | 9.89E-05 | 0.00047 | 9.33E-05 | -0.003477215 | NA |
| chr19 | 53647646 | 53648345 | 700 | 6 | 0.00021638 | 0.000139 | 0.00011 | 2.09E-05 | -0.002544598 | NA |
| chr5 | 83802919 | 83803442 | 524 | 3 | 0.000216609 | 0.000114 | 0.00094 | 3.95E-05 | -0.006689437 | NA |
| chr7 | 27896736 | 27897024 | 289 | 2 | 0.000216809 | 3.41E-05 | 0.00022 | 3.33E-05 | -0.000415137 | JAZF1 |
| chr19 | 48606941 | 48607191 | 251 | 4 | 0.000216893 | 7.05E-05 | 0.0005 | 6.86E-05 | -0.004795269 | SPACA4, FAM83E |
| chr1 | 230336310 | 230336414 | 105 | 2 | 0.000216962 | 7.68E-05 | 0.00031 | 6.76E-05 | -0.013688306 | PGBD5 |
| chr1 | 211346226 | 211346791 | 566 | 4 | 0.000217022 | 0.003966 | 4.47E-05 | 8.37E-05 | -0.004485509 | TRAF5 |
| chr8 | 97774730 | 97775123 | 394 | 5 | 0.000217218 | 0.002183 | 0.00013 | 9.07E-05 | -0.000737158 | LAPTM4B |
| chr10 | 102409844 | 102410465 | 622 | 5 | 0.000217545 | 2.94E-05 | 0.00478 | 3.95E-05 | -0.006706126 | PSD |
| chr10 | 97318945 | 97319326 | 382 | 9 | 0.000217612 | 0.00065 | 0.00598 | 3.98E-05 | -0.00082098 | FRAT1 |
| chr12 | 6093546 | 6094103 | 558 | 2 | 0.000217615 | 1.18E-05 | 0.00015 | 1.33E-05 | 0.012502505 | VWF |
| chr14 | 91332778 | 91333013 | 236 | 2 | 0.000217732 | 0.000173 | 8.76E-05 | 5.23E-05 | -0.00754862 | CCDC88C |
| chr8 | 649525 | 650940 | 1416 | 8 | 0.000217787 | 9.03E-06 | 0.00258 | 5.16E-06 | 0.034170662 | ERICH1 |
| chr5 | 3393758 | 3394384 | 627 | 6 | 0.000217832 | 1.74E-05 | 0.00055 | 2.68E-05 | -0.014410319 | LINC01019 |
| chr6 | 31010463 | 31010469 | 7 | 2 | 0.000217938 | 0.000141 | 0.00017 | 7.46E-05 | -0.007285585 | NA |
| chr14 | 63659805 | 63659996 | 192 | 2 | 0.000217968 | 2.77E-05 | 0.00015 | 2.52E-05 | 0.005138256 | AL136038.4 |
| chr16 | 31371301 | 31371815 | 515 | 6 | 0.000217989 | 2.25E-05 | 0.00401 | 3.24E-05 | -0.002994958 | ITGAX |
| chr9 | 129436543 | 129436721 | 179 | 2 | 0.000218083 | 3.09E-05 | 0.0009 | 4.41E-05 | -0.020215358 | AL353803.2 |
| chr15 | 50181657 | 50182035 | 379 | 7 | 0.000218088 | 2.23E-05 | 0.00149 | 2.35E-05 | -0.009129567 | ATP8B4 |
| chr2 | 138087607 | 138087722 | 116 | 2 | 0.000218164 | 2.43E-05 | 0.00015 | 2.30E-05 | -0.013760149 | NA |
| chr10 | 133195884 | 133196009 | 126 | 2 | 0.00021817 | 0.000958 | 0.00051 | 0.00041 | -0.001281967 | KNDC1 |
| chr9 | 116688292 | 116688734 | 443 | 3 | 0.00021857 | 0.000123 | 0.00032 | 5.77E-05 | -0.005044986 | TRIM32, ASTN2 |
| chr5 | 36067239 | 36067589 | 351 | 4 | 0.000218651 | 0.002904 | 0.00125 | 8.72E-05 | -0.003630578 | UGT3A2 |
| chr11 | 2521154 | 2521821 | 668 | 5 | 0.000218677 | 0.000989 | 0.0004 | 4.38E-05 | -0.001261106 | KCNQ1 |
| chr5 | 181126022 | 181126165 | 144 | 2 | 0.000218703 | 6.58E-05 | 0.00094 | 8.41E-05 | -0.004928294 | OR2V1 |
| chr19 | 5661414 | 5661771 | 358 | 4 | 0.000218951 | 0.000139 | 0.00099 | 4.44E-05 | -0.000785714 | SAFB |
| chr2 | 241665947 | 241666707 | 761 | 5 | 0.000219245 | 0.000585 | 0.03867 | 0.00198 | -0.003070305 | ATG4B |
| chr9 | 128877545 | 128877868 | 324 | 2 | 0.000219245 | 1.92E-05 | 0.00041 | 2.56E-05 | -0.005791205 | AL441992.2, KYAT1 |
| chr4 | 4542180 | 4543004 | 825 | 11 | 0.000219246 | 0.000543 | 0.00601 | 7.81E-05 | -0.000441052 | STX18-AS1, STX18 |

| | | | | | | | | | | |
|---|---|---|---|---|---|---|---|---|---|---|
| chr6 | 33851292 | 33852010 | 719 | 3 | 0.000219254 | 4.43E-06 | 0.00269 | 1.16E-05 | -0.011068686 | NA |
| chr6 | 119117096 | 119117417 | 322 | 2 | 0.000219299 | 2.28E-05 | 0.00029 | 2.68E-05 | -0.006166682 | AL137009.1, FAM184A |
| chr9 | 14910218 | 14911009 | 792 | 4 | 0.000219613 | 0.000356 | 0.01103 | 0.00031 | -0.015580686 | FREM1 |
| chr7 | 157736643 | 157737013 | 371 | 2 | 0.000219672 | 6.49E-05 | 0.00169 | 9.39E-05 | -0.008980642 | PTPRN2 |
| chr5 | 173123641 | 173124194 | 554 | 2 | 0.000219674 | 9.58E-06 | 0.00066 | 1.54E-05 | -0.013312477 | CREBRF |
| chr7 | 2750694 | 2750925 | 232 | 3 | 0.000219677 | 9.04E-05 | 8.49E-05 | 2.90E-05 | -0.004300663 | AMZ1, GNA12 |
| chr19 | 8209969 | 8210199 | 231 | 3 | 0.000219677 | 2.82E-05 | 0.00363 | 5.29E-05 | -0.015109537 | CERS4 |
| chr7 | 38304490 | 38304854 | 365 | 2 | 0.000219852 | 1.96E-05 | 0.00125 | 3.13E-05 | -0.020504627 | NA |
| chr5 | 337733 | 338078 | 346 | 2 | 0.000219891 | 1.34E-05 | 0.00075 | 2.09E-05 | -0.001345244 | PDCD6-AHRR, AHRR |
| chr5 | 115179644 | 115180298 | 655 | 7 | 0.000220155 | 9.96E-05 | 0.00459 | 8.12E-05 | 0.001024411 | TRIM36 |
| chr10 | 102831487 | 102831806 | 320 | 3 | 0.000220222 | 0.000373 | 6.86E-05 | 5.65E-05 | -0.004400054 | WBP1L, CYP17A1 |
| chr2 | 10344130 | 10344454 | 325 | 3 | 0.000220667 | 3.70E-05 | 0.00021 | 3.05E-05 | -0.012670943 | HPCAL1 |
| chr12 | 125123123 | 125123582 | 460 | 2 | 0.000221062 | 1.01E-05 | 0.00081 | 1.65E-05 | 0.000346567 | AACS |
| chr16 | 67939885 | 67940173 | 289 | 3 | 0.000221117 | 1.61E-05 | 0.00071 | 2.11E-05 | -0.008393449 | LCAT |
| chr16 | 81888650 | 81889238 | 589 | 7 | 0.000221178 | 1.40E-06 | 0.0174 | 1.00E-05 | -0.010198629 | PLCG2 |
| chr6 | 30198355 | 30199129 | 775 | 20 | 0.000221192 | 2.43E-05 | 0.00542 | 3.09E-05 | -0.001276602 | TRIM26 |
| chr11 | 130197966 | 130198553 | 588 | 4 | 0.000221541 | 0.000131 | 7.67E-05 | 3.88E-05 | -0.005476576 | ST14 |
| chr8 | 80029811 | 80030585 | 775 | 12 | 0.00022169 | 6.62E-05 | 0.01383 | 2.17E-05 | 0.001590868 | MRPS28, AC036214.3 |
| chr5 | 92328920 | 92329074 | 155 | 2 | 0.00022169 | 0.001371 | 6.69E-05 | 8.01E-05 | -0.006856364 | AC114316.1 |
| chr7 | 140348752 | 140349150 | 399 | 2 | 0.000221697 | 5.00E-05 | 0.00017 | 4.00E-05 | -0.00351402 | SLC37A3 |
| chr11 | 10807862 | 10808456 | 595 | 7 | 0.000221746 | 0.01694 | 0.00203 | 0.0007 | -0.001244332 | EIF4G2 |
| chr2 | 97913194 | 97913822 | 629 | 4 | 0.000221843 | 0.01936 | 6.95E-05 | 0.00019 | -0.002075831 | TMEM131 |
| chr10 | 44908274 | 44908280 | 7 | 2 | 0.000222202 | 1.21E-05 | 0.0009 | 1.96E-05 | -0.023038235 | TMEM72-AS1, AL353801.2 |
| chr8 | 139957310 | 139957317 | 8 | 2 | 0.000222203 | 1.44E-05 | 0.00084 | 2.27E-05 | -0.00544593 | TRAPPC9 |
| chr6 | 33410233 | 33410739 | 507 | 9 | 0.000222205 | 9.48E-06 | 0.01395 | 4.14E-05 | -0.000933422 | PHF1 |
| chr16 | 4692731 | 4693514 | 784 | 6 | 0.000222421 | 2.35E-05 | 0.00841 | 3.47E-05 | -0.002765347 | NA |
| chr22 | 31086936 | 31087921 | 986 | 3 | 0.000222565 | 4.33E-05 | 0.00012 | 3.39E-06 | -0.004160399 | SMTN |
| chr10 | 116590865 | 116591096 | 232 | 4 | 0.00022269 | 0.000134 | 0.0007 | 2.85E-05 | -0.002029297 | PNLIPRP1 |
| chr10 | 132141894 | 132142030 | 137 | 4 | 0.000223035 | 0.001073 | 5.61E-05 | 4.05E-05 | -0.005067915 | JAKMIP3 |
| chr5 | 32098444 | 32098754 | 311 | 2 | 0.000223035 | 0.001288 | 0.00038 | 0.00039 | -0.004563111 | PDZD2 |
| chr1 | 11099202 | 11099976 | 775 | 11 | 0.000223188 | 0.005388 | 0.00013 | 7.06E-05 | -0.000575578 | EXOSC10-AS1, EXOSC10 |
| chr17 | 76135468 | 76135671 | 204 | 3 | 0.000223202 | 7.00E-06 | 0.00341 | 1.77E-05 | -0.012947261 | NA |
| chr1 | 183635375 | 183636355 | 981 | 16 | 0.000223252 | 1.92E-05 | 0.00393 | 1.58E-05 | 0.001707814 | RGL1, ARPC5 |
| chr17 | 48627240 | 48627656 | 417 | 3 | 0.000223296 | 0.000454 | 0.00162 | 0.00028 | -0.006265953 | HOXB7 |
| chr19 | 44793530 | 44794073 | 544 | 2 | 0.000223317 | 2.21E-05 | 0.00038 | 2.81E-05 | -0.007563648 | CBLC |
| chr12 | 1915811 | 1916273 | 463 | 2 | 0.000223563 | 4.05E-05 | 0.00071 | 5.30E-05 | -0.010774364 | CACNA2D4 |
| chr16 | 29926354 | 29927129 | 776 | 9 | 0.000223635 | 0.000107 | 0.016 | 4.36E-05 | -0.002051182 | AC120114.1 |
| chr20 | 20894850 | 20894858 | 9 | 2 | 0.000223925 | 1.81E-05 | 0.00064 | 2.65E-05 | -0.015332646 | NA |
| chr11 | 1943910 | 1944227 | 318 | 3 | 0.000224027 | 7.65E-06 | 0.0028 | 1.83E-05 | -0.016035083 | NA |
| chr9 | 127771278 | 127771971 | 694 | 8 | 0.000224053 | 1.46E-05 | 0.01326 | 1.46E-05 | -0.002235548 | SH2D3C |
| chr6 | 34723456 | 34723901 | 446 | 2 | 0.000224678 | 1.63E-05 | 0.00054 | 2.36E-05 | -0.010760955 | NA |
| chr9 | 20805002 | 20805200 | 199 | 3 | 0.000224742 | 1.03E-05 | 0.00131 | 2.04E-05 | -0.026488484 | FOCAD |
| chr21 | 45073254 | 45073521 | 268 | 4 | 0.000224787 | 0.000467 | 0.00188 | 4.66E-05 | -0.010789193 | SSR4P1 |
| chr11 | 117428405 | 117428478 | 74 | 2 | 0.00022516 | 0.000165 | 3.62E-05 | 2.43E-05 | -0.006076346 | DSCAML1 |
| chr12 | 116066802 | 116067030 | 229 | 2 | 0.000225388 | 0.000152 | 7.73E-05 | 4.46E-05 | -0.008127304 | MED13L |
| chr1 | 206828848 | 206829316 | 469 | 2 | 0.000225578 | 1.11E-05 | 0.00079 | 1.78E-05 | -0.013213456 | IL19 |
| chr2 | 96860876 | 96861337 | 462 | 5 | 0.000225582 | 0.000913 | 0.00031 | 5.22E-05 | -0.000856701 | SEMA4C |
| chr19 | 2319506 | 2319508 | 3 | 2 | 0.000225655 | 3.57E-05 | 0.00158 | 5.46E-05 | -0.008069526 | AC005258.1 |
| chr2 | 207505015 | 207505308 | 294 | 3 | 0.000225789 | 1.02E-05 | 0.00114 | 1.92E-05 | -0.02354717 | AC009226.1 |
| chr15 | 41337641 | 41337890 | 250 | 2 | 0.000225789 | 0.000407 | 8.70E-05 | 7.25E-05 | -0.00457125 | AC087721.2, NUSAP1 |
| chr19 | 35008371 | 35008417 | 47 | 2 | 0.000225817 | 0.003261 | 0.00021 | 0.00036 | -0.002816415 | GRAMD1A |
| chr12 | 42904815 | 42904936 | 122 | 2 | 0.000225928 | 2.03E-05 | 0.00024 | 2.34E-05 | -0.012243281 | NA |
| chr10 | 9239048 | 9239485 | 438 | 3 | 0.000226273 | 1.35E-05 | 0.00078 | 2.24E-05 | -0.015681636 | NA |
| chr11 | 96155494 | 96155863 | 370 | 2 | 0.000226407 | 9.02E-05 | 0.001 | 0.00011 | -0.009596313 | MAML2 |
| chr11 | 1897046 | 1897220 | 175 | 2 | 0.000226462 | 7.81E-05 | 0.00016 | 5.21E-05 | -0.018294419 | LINC01150 |
| chr11 | 104164985 | 104165033 | 49 | 2 | 0.000226722 | 0.000174 | 0.00298 | 0.00028 | -0.00028275 | NA |
| chr1 | 32871656 | 32871982 | 327 | 3 | 0.000227081 | 8.27E-06 | 0.0033 | 2.03E-05 | -0.010299939 | FNDC5 |
| chr15 | 90955723 | 90957663 | 1941 | 12 | 0.000227255 | 3.11E-06 | 0.00225 | 2.25E-06 | -0.0070546 | RCCD1 |
| chr3 | 194296692 | 194297382 | 691 | 7 | 0.000227284 | 7.38E-06 | 0.00551 | 1.74E-05 | -0.016760502 | LINC00887 |
| chr12 | 124500726 | 124501109 | 384 | 3 | 0.000227337 | 4.32E-05 | 0.00338 | 7.40E-05 | -0.012039731 | NCOR2 |
| chr19 | 17969986 | 17970811 | 826 | 3 | 0.000227465 | 0.000686 | 0.00059 | 2.53E-05 | -0.004053448 | KCNN1 |
| chr9 | 124260259 | 124260842 | 584 | 2 | 0.000227983 | 1.84E-05 | 0.00041 | 2.47E-05 | -0.01927656 | NEK6, AL162724.2 |
| chr8 | 1195761 | 1195806 | 46 | 2 | 0.000228027 | 2.44E-05 | 0.00031 | 2.88E-05 | -0.008791934 | DLGAP2 |
| chr2 | 70978954 | 70979398 | 445 | 3 | 0.000228042 | 0.000431 | 0.00112 | 9.57E-05 | -0.009735247 | ANKRD53, AC007040.2 |
| chr1 | 175505301 | 175505397 | 97 | 3 | 0.000228115 | 8.88E-06 | 0.00144 | 1.77E-05 | -0.021817027 | TNR |
| chr12 | 6540183 | 6540828 | 646 | 5 | 0.000228133 | 3.31E-05 | 0.0073 | 6.31E-05 | -0.008831022 | IFFO1 |
| chr12 | 50112451 | 50112606 | 156 | 2 | 0.000228175 | 0.000191 | 0.00546 | 0.00034 | 0.002673726 | COX14, AC074032.1 |
| chr12 | 49351198 | 49351623 | 426 | 5 | 0.000228191 | 8.04E-05 | 0.00259 | 6.01E-05 | -0.006206969 | DNAJC22 |
| chr5 | 39100917 | 39101278 | 362 | 2 | 0.00022884 | 2.61E-05 | 0.00029 | 2.97E-05 | -0.004568151 | AC008964.1 |
| chr22 | 39291061 | 39292930 | 1870 | 8 | 0.000229118 | 1.52E-05 | 0.00012 | 6.97E-07 | -0.004694792 | NA |
| chr4 | 184756395 | 184756431 | 37 | 2 | 0.000229129 | 0.000119 | 6.27E-05 | 3.39E-05 | -0.002521274 | ACSL1 |
| chr2 | 55133708 | 55134174 | 467 | 3 | 0.000229187 | 1.50E-05 | 0.00331 | 3.17E-05 | 0.000567457 | NA |
| chr3 | 141368277 | 141368521 | 245 | 5 | 0.000229196 | 4.33E-06 | 0.012 | 2.02E-05 | -0.017355683 | ZBTB38 |
| chr6 | 41894252 | 41894466 | 215 | 3 | 0.000229275 | 0.000251 | 0.0007 | 3.37E-05 | -0.007899244 | USP49 |
| chr4 | 104491310 | 104491727 | 418 | 6 | 0.000229347 | 0.000191 | 0.00271 | 7.60E-05 | -0.001136628 | CXXC4-AS1, CXXC4 |
| chr17 | 73312055 | 73312667 | 613 | 10 | 0.000229696 | 4.64E-05 | 0.00896 | 2.27E-05 | -0.002157528 | NA |
| chr15 | 65355765 | 65356152 | 388 | 4 | 0.000229696 | 6.55E-05 | 0.0005 | 4.72E-05 | -0.000792886 | IGDCC3 |
| chr7 | 96321817 | 96322496 | 680 | 13 | 0.000229889 | 0.005977 | 0.00038 | 7.01E-05 | -1.91E-05 | SLC25A13 |
| chr12 | 5043173 | 5043380 | 208 | 3 | 0.000229993 | 0.000221 | 0.00016 | 8.53E-05 | 0.011784553 | NA |
| chr2 | 124025666 | 124026121 | 456 | 3 | 0.000230008 | 1.98E-05 | 0.00126 | 2.18E-05 | 0.010975965 | CNTNAP5 |
| chr1 | 46613080 | 46613294 | 215 | 3 | 0.000230112 | 0.000123 | 0.0008 | 3.79E-05 | -0.003074281 | MKNK1, MOB3C |
| chr1 | 151024657 | 151024734 | 78 | 3 | 0.000230138 | 0.000264 | 6.58E-05 | 3.96E-05 | -0.000763868 | PRUNE1 |
| chr16 | 2303937 | 2304019 | 83 | 2 | 0.0002302 | 4.11E-05 | 9.78E-05 | 2.59E-05 | -0.001471391 | ABCA3 |
| chr12 | 6493465 | 6494262 | 798 | 11 | 0.000230381 | 2.54E-06 | 0.01406 | 1.24E-05 | 0.000345512 | NCAPD2, MRPL51 |
| chr2 | 164560354 | 164560663 | 310 | 2 | 0.000230429 | 2.17E-05 | 0.00033 | 2.68E-05 | -0.004681523 | GRB14 |
| chr13 | 77001844 | 77001943 | 100 | 2 | 0.000230506 | 1.92E-05 | 0.00123 | 3.07E-05 | -0.01408537 | CLN5, AC001226.2, FBXL3 |
| chr6 | 30331932 | 30332236 | 305 | 3 | 0.000230508 | 0.000152 | 0.0006 | 4.33E-05 | -0.002079014 | TRIM39, TRIM39-RPP21 |
| chr12 | 62602632 | 62602708 | 77 | 3 | 0.000230514 | 0.000456 | 0.00017 | 0.0001 | 0.001252251 | LINC01465 |
| chr11 | 65497210 | 65497926 | 717 | 9 | 0.000230652 | 8.03E-06 | 0.00557 | 1.87E-05 | 0.00191853 | MALAT1 |
| chr10 | 132371019 | 132371043 | 25 | 2 | 0.000230706 | 5.00E-05 | 0.00091 | 6.66E-05 | -0.004274386 | LRRC27 |
| chr16 | 979745 | 980166 | 422 | 2 | 0.000230885 | 0.002062 | 0.00484 | 0.00231 | -0.008974681 | LMF1, CEROX1 |
| chr5 | 135233003 | 135233148 | 146 | 2 | 0.000230906 | 2.92E-05 | 0.00017 | 2.69E-05 | -0.003721206 | C5orf66 |
| chr3 | 170037807 | 170038162 | 356 | 6 | 0.000230974 | 0.006003 | 0.00038 | 0.00011 | 0.000714947 | GPR160 |
| chr8 | 144425986 | 144426416 | 431 | 2 | 0.000231424 | 3.10E-05 | 0.00094 | 4.46E-05 | -0.009280519 | VPS28 |
| chr10 | 21739514 | 21739609 | 96 | 2 | 0.000231424 | 3.18E-05 | 0.00024 | 3.26E-05 | -0.008247725 | MLLT10 |
| chr2 | 10041998 | 10042108 | 111 | 2 | 0.000231669 | 0.001303 | 9.47E-05 | 0.00011 | -0.002332951 | NA |
| chr6 | 30552165 | 30552628 | 464 | 8 | 0.000231764 | 0.000702 | 0.00495 | 0.00012 | -0.001616925 | GNL1 |
| chr7 | 66628728 | 66629109 | 382 | 7 | 0.000231869 | 8.17E-05 | 0.00223 | 1.99E-05 | 0.000882001 | KCTD7, AC027644.4 |
| chr2 | 241553747 | 241554391 | 645 | 2 | 0.000231869 | 1.17E-05 | 0.00077 | 1.86E-05 | 0.006347912 | BOK, BOK-AS1 |
| chr5 | 122311045 | 122311624 | 580 | 6 | 0.000232062 | 0.000195 | 0.00136 | 2.87E-05 | 0.007474225 | SNCAIP, AC113349.2 |
| chr17 | 49708777 | 49709064 | 288 | 3 | 0.000232323 | 6.08E-05 | 0.00254 | 8.40E-05 | -0.006555631 | AC015795.1, SLC35B1 |
| chr21 | 32727276 | 32728484 | 1209 | 13 | 0.000232428 | 0.000131 | 0.00778 | 7.34E-05 | 0.00017389 | PAXBP1-AS1, SYNJ1 |
| chr12 | 14771098 | 14771521 | 424 | 8 | 0.000232474 | 0.006551 | 7.09E-05 | 8.40E-05 | 0.002357549 | H4-16 |
| chr6 | 48099294 | 48099650 | 357 | 2 | 0.000232513 | 2.46E-05 | 0.00031 | 2.90E-05 | -0.006345739 | AL353138.1 |
| chr14 | 54349084 | 54349454 | 371 | 3 | 0.000232733 | 2.12E-05 | 0.00234 | 3.74E-05 | -0.026971703 | NA |
| chr18 | 725626 | 725885 | 260 | 3 | 0.000232835 | 0.00013 | 0.00027 | 4.15E-05 | -0.011753055 | YES1 |
| chr20 | 63952202 | 63952593 | 392 | 5 | 0.000233053 | 0.005395 | 0.00065 | 8.73E-05 | -0.003438955 | UCKL1 |
| chr2 | 109028475 | 109028908 | 434 | 2 | 0.000233083 | 1.77E-05 | 0.00076 | 2.68E-05 | -0.008595832 | NA |
| chr19 | 1228889 | 1229185 | 297 | 2 | 0.000233083 | 0.000162 | 0.00012 | 6.38E-05 | -0.003434362 | CBARP |
| chr11 | 66466481 | 66467403 | 923 | 9 | 0.000233188 | 0.003435 | 0.00275 | 0.00037 | 0.001520371 | PELI3, MRPL11 |
| chr9 | 124274687 | 124274793 | 107 | 2 | 0.000233282 | 2.83E-05 | 0.00061 | 3.80E-05 | -0.003412292 | NEK6 |
| chr11 | 64879814 | 64880452 | 639 | 7 | 0.000233451 | 9.68E-05 | 0.00109 | 2.50E-05 | -0.004108473 | EHD1 |
| chr1 | 3334049 | 3334327 | 279 | 2 | 0.000233675 | 0.000216 | 0.001 | 0.00023 | -0.006789031 | PRDM16 |
| chr17 | 42853147 | 42854035 | 889 | 5 | 0.000233888 | 0.063384 | 0.00014 | 0.00054 | -0.003214441 | AOC3 |
| chr19 | 44701088 | 44701256 | 169 | 4 | 0.000233988 | 8.58E-05 | 8.61E-05 | 1.73E-05 | -0.004667918 | CEACAM16, AC243964.3 |
| chr12 | 14972417 | 14973086 | 670 | 5 | 0.00023401 | 4.07E-05 | 0.00669 | 2.37E-05 | -0.005324479 | PDE6H |
| chr10 | 79153805 | 79153940 | 136 | 2 | 0.000234165 | 0.0002 | 5.75E-05 | 3.90E-05 | -0.008375873 | ZMIZ1 |
| chr2 | 189775136 | 189775294 | 159 | 2 | 0.000234238 | 0.007814 | 4.98E-05 | 7.73E-05 | -0.003338094 | ORMDL1 |
| chr9 | 93566119 | 93566247 | 129 | 2 | 0.00023427 | 5.44E-05 | 0.00041 | 5.77E-05 | 0.004710208 | NA |
| chr7 | 151032731 | 151032989 | 259 | 2 | 0.000234279 | 5.00E-05 | 0.00113 | 6.97E-05 | -0.004960633 | ABCB8 |
| chr7 | 94657199 | 94657930 | 732 | 17 | 0.000234364 | 0.003333 | 0.0142 | 0.00049 | -0.003274937 | PEG10 |
| chr11 | 66666350 | 66666522 | 173 | 3 | 0.000234395 | 3.57E-05 | 0.00182 | 5.77E-05 | -0.005597277 | RBM4, RBM4B |
| chr19 | 7395441 | 7395785 | 345 | 2 | 0.000234395 | 0.017561 | 0.00275 | 0.0054 | -0.011086306 | AC008878.3, ARHGEF18 |
| chr15 | 77072177 | 77072459 | 283 | 4 | 0.000234565 | 2.45E-05 | 0.0025 | 4.08E-05 | -0.003665207 | TSPAN3 |
| chr5 | 172559062 | 172559767 | 706 | 6 | 0.000234654 | 1.05E-05 | 0.00072 | 1.67E-05 | -0.002855993 | LINC01944 |
| chr16 | 29694830 | 29695060 | 231 | 4 | 0.000234675 | 4.05E-06 | 0.00728 | 1.52E-05 | -0.00243478 | QPRT |
| chr12 | 56236577 | 56236690 | 114 | 2 | 0.000234813 | 0.000766 | 0.00013 | 0.00013 | -0.002106932 | SLC39A5 |
| chr1 | 43558479 | 43558561 | 83 | 3 | 0.000234923 | 1.00E-05 | 0.0017 | 2.11E-05 | -0.015063901 | PTPRF |
| chr11 | 62727577 | 62727995 | 419 | 5 | 0.000234953 | 0.047478 | 0.00169 | 0.00059 | 0.001432356 | NA |
| chr4 | 2341480 | 2341664 | 185 | 3 | 0.000235159 | 0.015018 | 6.38E-05 | 9.19E-05 | -0.000837314 | ZFYVE28 |
| chr17 | 68457253 | 68458170 | 918 | 11 | 0.000235245 | 4.46E-05 | 0.00271 | 2.87E-05 | 0.000408579 | AC007780.1, WIPI1 |
| chr9 | 76458639 | 76459294 | 656 | 10 | 0.00023543 | 2.67E-05 | 0.00297 | 2.06E-05 | 0.000707553 | GCNT1 |
| chr4 | 169619458 | 169619793 | 336 | 3 | 0.000235434 | 0.002394 | 0.00058 | 8.35E-05 | -0.005957457 | CLCN3 |
| chr12 | 56146896 | 56147155 | 260 | 2 | 0.000235434 | 1.33E-05 | 0.00088 | 2.11E-05 | -0.012082948 | NA |
| chr12 | 77030448 | 77030494 | 47 | 3 | 0.000235495 | 4.30E-05 | 0.00014 | 2.46E-05 | -0.004296857 | E2F7 |
| chr8 | 77000090 | 77000596 | 507 | 9 | 0.000235557 | 0.000221 | 0.00626 | 3.34E-05 | -0.000183455 | PEX2 |
| chr2 | 149141318 | 149141320 | 3 | 2 | 0.000235634 | 0.000146 | 0.00097 | 0.00017 | 0.00189132 | LYPD6B |
| chr9 | 129483345 | 129483643 | 299 | 4 | 0.00023581 | 2.07E-05 | 0.00129 | 1.96E-05 | -0.007633135 | LINC00963 |
| chr2 | 241726107 | 241726651 | 545 | 5 | 0.000235823 | 0.000108 | 0.00172 | 3.07E-05 | -0.004476259 | ING5 |
| chr16 | 78199598 | 78199670 | 73 | 2 | 0.000236103 | 0.000497 | 4.99E-05 | 4.57E-05 | -0.003193579 | WWOX |
| chr9 | 124271733 | 124271971 | 239 | 3 | 0.000236234 | 0.002944 | 0.00165 | 0.00017 | -0.004602687 | NEK6 |
| chr12 | 107929615 | 107929716 | 102 | 3 | 0.000236521 | 1.73E-05 | 0.00033 | 2.10E-05 | -0.022998139 | NA |
| chr2 | 143519200 | 143519705 | 506 | 3 | 0.000236525 | 8.94E-06 | 0.00182 | 1.80E-05 | -0.004797939 | ARHGAP15, AC079793.1 |
| chr13 | 49585526 | 49586214 | 689 | 11 | 0.000236758 | 3.17E-05 | 0.01884 | 3.24E-05 | -0.002774105 | RCBTB1 |
| chr3 | 169147301 | 169147425 | 125 | 3 | 0.000237072 | 0.016988 | 0.0022 | 0.00371 | 0.000904191 | MECOM |
| chr7 | 87876146 | 87877535 | 1390 | 12 | 0.000237158 | 1.99E-06 | 0.00272 | 1.92E-06 | 0.001238796 | DBF4, SLC25A40 |
| chr16 | 81238332 | 81238676 | 345 | 5 | 0.000237272 | 8.58E-06 | 0.00449 | 1.83E-05 | -0.01182609 | NA |
| chr9 | 33167496 | 33168525 | 1030 | 7 | 0.000237553 | 7.33E-05 | 0.00141 | 1.41E-05 | -0.003636294 | B4GALT1-AS1 |
| chr17 | 43779487 | 43779687 | 201 | 2 | 0.000237707 | 0.001942 | 0.00028 | 0.00037 | 0.002131596 | NA |
| chr17 | 45150938 | 45151204 | 267 | 3 | 0.000237806 | 6.53E-05 | 0.00098 | 7.20E-05 | -0.013652493 | HEXIM1, AC138150.1 |
| chr6 | 165942925 | 165943540 | 616 | 3 | 0.000237822 | 5.91E-05 | 0.00072 | 6.78E-05 | -0.00200962 | PDE10A |
| chr8 | 125341182 | 125341730 | 549 | 2 | 0.000237897 | 1.34E-05 | 0.00072 | 2.08E-05 | -0.004610125 | NSMCE2 |
| chr10 | 124642611 | 124642856 | 246 | 4 | 0.000237931 | 0.012145 | 0.00021 | 0.00062 | -0.003117623 | AC068896.1, FAM53B |
| chr16 | 16009554 | 16009711 | 158 | 3 | 0.00023808 | 3.56E-05 | 0.00019 | 2.47E-05 | -0.003864351 | ABCC1 |
| chr5 | 34687014 | 34687460 | 447 | 6 | 0.000238156 | 0.000286 | 0.00098 | 5.86E-05 | -0.005325252 | RAI14 |
| chr12 | 263540 | 263789 | 250 | 3 | 0.000238216 | 9.21E-05 | 0.00132 | 0.00011 | -0.00836415 | NA |
| chr16 | 50065985 | 50066622 | 638 | 3 | 0.000238513 | 0.000565 | 0.00442 | 0.00044 | 0.002423816 | HEATR3, AC007610.1 |
| chr1 | 223712736 | 223713120 | 385 | 4 | 0.000238569 | 0.006532 | 0.03006 | 0.00433 | -0.001045227 | CAPN2 |
| chr1 | 40303997 | 40304517 | 521 | 4 | 0.000238627 | 1.04E-05 | 0.00417 | 2.59E-05 | -0.001499228 | COL9A2 |
| chr3 | 39809638 | 39810150 | 513 | 3 | 0.000238686 | 0.008914 | 0.00249 | 0.00096 | 0.001350641 | MYRIP |
| chr10 | 118241754 | 118241856 | 103 | 3 | 0.000238686 | 4.51E-06 | 0.0001 | 4.34E-06 | -0.028140753 | AL354863.1 |
| chr11 | 117098577 | 117099286 | 710 | 7 | 0.000238714 | 0.000117 | 0.01101 | 5.32E-05 | -0.008992895 | AP000936.1 |

| | | | | | | | | | | |
|---|---|---|---|---|---|---|---|---|---|---|
| chr2 | 233348834 | 233349137 | 304 | 2 | 0.000238729 | 1.81E-05 | 0.00063 | 2.65E-05 | -0.011340894 | NA |
| chr2 | 38602679 | 38603477 | 799 | 8 | 0.000238829 | 0.000111 | 0.00709 | 0.0001 | 0.00048821 | HNRNPLL |
| chr8 | 76853837 | 76854030 | 194 | 2 | 0.000239086 | 0.004555 | 5.77E-05 | 8.59E-05 | -0.002358599 | ZFHX4 |
| chr15 | 65424072 | 65424092 | 21 | 2 | 0.000239266 | 0.000546 | 9.95E-05 | 8.95E-05 | -0.007834021 | NA |
| chr10 | 60905647 | 60905856 | 210 | 2 | 0.000239295 | 2.11E-05 | 0.00029 | 2.53E-05 | -0.006078422 | RHOBTB1 |
| chr6 | 110903077 | 110903408 | 332 | 2 | 0.000239409 | 2.92E-05 | 0.0005 | 3.74E-05 | -0.004505167 | NA |
| chr17 | 28042513 | 28042914 | 402 | 3 | 0.00023957 | 0.001071 | 0.00046 | 0.00021 | -0.00309796 | NLK |
| chr8 | 140534274 | 140534391 | 118 | 2 | 0.000239667 | 9.94E-05 | 0.00014 | 5.41E-05 | -0.003339579 | AGO2 |
| chr6 | 154218963 | 154219371 | 409 | 4 | 0.000239976 | 0.002098 | 0.00439 | 5.37E-05 | -0.00746018 | OPRM1, IPCEF1, AL357075.4 |
| chr1 | 91950757 | 91951184 | 428 | 5 | 0.000240126 | 0.000272 | 0.00161 | 3.58E-05 | 0.002427419 | BRDT |
| chr9 | 214612 | 215128 | 517 | 4 | 0.000240263 | 4.65E-05 | 0.0026 | 5.18E-05 | -0.006789972 | DOCK8, DOCK8-AS1 |
| chr2 | 3388084 | 3388184 | 101 | 2 | 0.000240263 | 1.69E-05 | 0.0002 | 1.92E-05 | -0.00321716 | TRAPPC12 |
| chr1 | 1033662 | 1033877 | 216 | 3 | 0.000240539 | 0.000417 | 0.00017 | 4.03E-05 | 0.005827229 | AGRN |
| chr13 | 23374010 | 23374024 | 15 | 2 | 0.000240599 | 4.09E-05 | 0.00101 | 5.74E-05 | -0.009077863 | RPLP1P13, SACS |
| chr12 | 39337606 | 39337851 | 246 | 2 | 0.000240861 | 0.000538 | 7.62E-05 | 7.02E-05 | -0.014886517 | KIF21A |
| chr8 | 29080916 | 29081091 | 176 | 3 | 0.000240892 | 0.000235 | 0.00019 | 3.39E-05 | -0.003952364 | KIF13B |
| chr3 | 52841359 | 52842242 | 884 | 4 | 0.000241069 | 3.67E-05 | 0.00132 | 3.98E-05 | -0.00384509 | STIMATE-MUSTN1, STIMATE |
| chr2 | 190699284 | 190699585 | 302 | 3 | 0.000241261 | 2.09E-05 | 0.00049 | 2.68E-05 | -0.009198118 | AC006460.1 |
| chr16 | 52608040 | 52608645 | 606 | 4 | 0.000241869 | 0.001375 | 0.00118 | 0.0002 | -0.003558355 | AC026462.3, CASC16, AC026462.2 |
| chr9 | 127118923 | 127119126 | 204 | 3 | 0.000241892 | 6.03E-05 | 0.00069 | 3.01E-05 | -0.007625653 | RALGPS1, ANGPTL2 |
| chr4 | 87530194 | 87531107 | 914 | 5 | 0.000242146 | 0.00029 | 0.00022 | 2.63E-05 | -0.004033775 | SPARCL1 |
| chr7 | 100485461 | 100485505 | 45 | 2 | 0.000242146 | 8.50E-05 | 0.00024 | 6.47E-05 | -0.007292217 | NYAP1 |
| chr14 | 89544547 | 89544836 | 290 | 2 | 0.00024238 | 9.89E-05 | 9.74E-05 | 4.31E-05 | -0.008696389 | FOXN3, AL137230.2 |
| chr7 | 116574003 | 116574070 | 68 | 2 | 0.000242483 | 0.000106 | 6.25E-05 | 3.21E-05 | 0.000776952 | COMETT |
| chr6 | 33315832 | 33316765 | 934 | 19 | 0.000242496 | 0.000456 | 0.0182 | 6.11E-05 | -0.000827917 | ZBTB22 |
| chr1 | 54807964 | 54808512 | 549 | 3 | 0.000242573 | 3.74E-05 | 0.00072 | 4.79E-05 | -0.00646026 | LEXM |
| chr7 | 55526994 | 55527569 | 576 | 5 | 0.000242704 | 0.004032 | 0.00011 | 5.89E-05 | -0.005564879 | VOPP1 |
| chr22 | 35716855 | 35717465 | 611 | 3 | 0.000242753 | 1.50E-05 | 0.00217 | 3.09E-05 | -0.010741673 | NA |
| chr7 | 27174274 | 27175296 | 1023 | 16 | 0.000242842 | 0.000737 | 0.00012 | 1.92E-05 | -0.00063425 | HOXA9, AC004080.3, HOXA10 |
| chr3 | 69739833 | 69740048 | 216 | 3 | 0.00024307 | 0.02711 | 6.81E-05 | 0.0001 | -0.002207377 | MITF |
| chr6 | 33124466 | 33125025 | 560 | 4 | 0.000243257 | 0.000412 | 0.00667 | 0.00044 | -0.000828898 | HLA-DPB2 |
| chr6 | 11771045 | 11771607 | 563 | 4 | 0.000243338 | 7.41E-05 | 0.00029 | 4.64E-05 | -0.011098491 | ADTRP |
| chr7 | 102396151 | 102396900 | 750 | 11 | 0.000243534 | 5.51E-05 | 0.00502 | 2.31E-05 | 0.000455081 | PRKRIP1 |
| chr8 | 93166763 | 93166952 | 190 | 2 | 0.000243592 | 2.15E-05 | 0.00017 | 2.17E-05 | -0.011682711 | C8orf87 |
| chr9 | 136586655 | 136587293 | 639 | 2 | 0.000243811 | 0.000114 | 0.00064 | 0.00012 | -0.008034339 | NA |
| chr10 | 131916201 | 131916222 | 22 | 2 | 0.000243813 | 5.82E-05 | 0.00034 | 5.72E-05 | -0.003182568 | PPP2R2D |
| chr16 | 58167791 | 58168121 | 331 | 2 | 0.000244092 | 8.45E-05 | 0.00038 | 7.76E-05 | -0.000877314 | CSNK2A2 |
| chr5 | 177049467 | 177049532 | 66 | 3 | 0.000244187 | 3.34E-05 | 0.0002 | 2.80E-05 | -0.003094083 | ZNF346 |
| chr16 | 88322182 | 88322647 | 466 | 4 | 0.00024429 | 0.000182 | 0.00069 | 9.26E-05 | 0.001394448 | NA |
| chr22 | 45518596 | 45518791 | 196 | 4 | 0.000244538 | 0.000334 | 8.44E-05 | 5.27E-05 | -0.004386838 | FBLN1 |
| chr1 | 203126619 | 203126860 | 242 | 2 | 0.000244597 | 9.88E-05 | 0.00016 | 5.94E-05 | 0.016081177 | ADORA1, AC105940.2 |
| chr20 | 36460423 | 36460771 | 349 | 4 | 0.000244734 | 0.000176 | 0.00013 | 2.96E-05 | -0.005001141 | DLGAP4 |
| chr12 | 31326804 | 31327366 | 563 | 4 | 0.00024483 | 0.000113 | 0.00606 | 0.0001 | -0.008344484 | SINHCAF |
| chr11 | 72676020 | 72676170 | 151 | 2 | 0.00024483 | 0.001111 | 0.00023 | 0.00025 | -0.018091422 | NA |
| chr12 | 89472681 | 89472893 | 213 | 2 | 0.00024483 | 0.000358 | 5.76E-05 | 4.74E-05 | -0.006674278 | POC1B |
| chr13 | 50140897 | 50141568 | 672 | 5 | 0.000244878 | 0.000148 | 0.00019 | 4.61E-05 | -0.001557989 | DLEU1 |
| chr4 | 55117120 | 55117367 | 248 | 2 | 0.00024488 | 8.72E-05 | 0.00011 | 4.32E-05 | -0.015663236 | KDR |
| chr15 | 29003922 | 29004466 | 545 | 3 | 0.000244988 | 1.71E-05 | 0.00082 | 1.67E-05 | -0.004481899 | APBA2 |
| chr1 | 1019703 | 1019783 | 81 | 3 | 0.000245258 | 0.010077 | 1.85E-05 | 5.15E-05 | -0.003052457 | NA |
| chr5 | 9545864 | 9546643 | 780 | 8 | 0.000245303 | 0.004271 | 0.00023 | 8.50E-05 | 0.002604099 | SNHG18, SEMA5A |
| chr15 | 55897264 | 55897376 | 113 | 2 | 0.000245358 | 0.069944 | 0.00012 | 0.00032 | -0.010970757 | NEDD4 |
| chr19 | 46016799 | 46016879 | 81 | 2 | 0.00024544 | 0.000406 | 3.42E-05 | 3.05E-05 | 0.001938207 | CCDC61 |
| chr12 | 2794232 | 2794397 | 166 | 3 | 0.000245566 | 0.00018 | 0.00475 | 0.00028 | -0.007496118 | ITFG2-AS1, CBX3P4 |
| chr15 | 72289865 | 72290607 | 743 | 3 | 0.000246 | 4.72E-05 | 0.00066 | 2.91E-05 | -0.003006528 | AC009690.2, AC009690.3, CELF6, AC009690.1 |
| chr3 | 49013392 | 49014076 | 685 | 3 | 0.00024618 | 4.88E-05 | 0.00028 | 3.66E-05 | -0.004606385 | WDR6 |
| chr22 | 35617513 | 35618309 | 797 | 9 | 0.000246464 | 0.275659 | 0.00174 | 0.00145 | -0.001198754 | MB |
| chr8 | 101138604 | 101139151 | 548 | 3 | 0.000246532 | 6.06E-05 | 0.00104 | 3.20E-05 | 0.004133111 | NA |
| chr18 | 79356151 | 79356310 | 160 | 3 | 0.000246667 | 2.06E-05 | 0.00041 | 2.31E-05 | -0.004040451 | ATP9B |
| chr10 | 1056669 | 1056977 | 309 | 7 | 0.000246673 | 1.36E-05 | 0.00526 | 2.50E-05 | 0.000811178 | WDR37 |
| chr8 | 124549018 | 124549994 | 977 | 4 | 0.000246875 | 3.39E-06 | 0.0012 | 4.60E-06 | -6.88E-05 | NDUFB9 |
| chr2 | 216003171 | 216003386 | 216 | 3 | 0.00024692 | 1.98E-05 | 0.00046 | 2.49E-05 | -0.007796881 | MREG, PECR |
| chr1 | 35078869 | 35079485 | 617 | 9 | 0.00024701 | 3.60E-05 | 0.0148 | 4.15E-05 | -0.0014302 | ZMYM1 |
| chr22 | 41597353 | 41597718 | 366 | 3 | 0.000247042 | 0.000107 | 0.00133 | 5.02E-05 | -0.008053522 | NA |
| chr5 | 141853898 | 141854328 | 431 | 3 | 0.000247062 | 6.75E-06 | 0.00357 | 1.74E-05 | -0.006205481 | PCDH1 |
| chr9 | 122346767 | 122346841 | 75 | 2 | 0.000247079 | 2.13E-05 | 0.00062 | 3.03E-05 | -0.017812354 | NA |
| chr13 | 99124821 | 99125331 | 511 | 3 | 0.000247258 | 9.93E-06 | 0.00103 | 1.83E-05 | 0.013971466 | NA |
| chr11 | 126185229 | 126185562 | 334 | 3 | 0.00024734 | 2.39E-05 | 0.00161 | 2.92E-05 | -0.000870421 | NA |
| chr14 | 76538128 | 76538530 | 403 | 3 | 0.000247584 | 0.003153 | 0.00012 | 7.14E-05 | -0.003893978 | AC008050.1 |
| chr9 | 130444512 | 130444907 | 396 | 4 | 0.000247707 | 0.00059 | 0.00049 | 9.35E-05 | 0.00148785 | NA |
| chr10 | 45181550 | 45181593 | 44 | 2 | 0.000248087 | 0.000484 | 5.76E-05 | 5.21E-05 | -0.003388731 | NA |
| chr8 | 144416498 | 144417072 | 575 | 10 | 0.000248183 | 5.70E-05 | 0.00139 | 2.81E-05 | -0.001796043 | SLC39A4 |
| chr9 | 93316887 | 93318044 | 1158 | 3 | 0.000248221 | 1.31E-06 | 0.00077 | 3.16E-06 | -0.018547766 | WNK2 |
| chr13 | 114152981 | 114153227 | 247 | 3 | 0.000248233 | 0.000617 | 0.00114 | 6.34E-05 | -0.013091283 | NA |
| chr16 | 52076173 | 52076176 | 4 | 2 | 0.000248271 | 7.53E-05 | 0.0006 | 8.28E-05 | -0.007129191 | C16orf97 |
| chr15 | 81000087 | 81000585 | 499 | 3 | 0.000248573 | 0.001103 | 0.00237 | 0.00032 | 0.000515772 | NA |
| chr20 | 43563911 | 43564196 | 286 | 2 | 0.000248638 | 0.000136 | 0.00039 | 0.00011 | -0.005942872 | Z98752.3, SGK2 |
| chr21 | 46378889 | 46379348 | 460 | 5 | 0.000248946 | 0.000323 | 0.00711 | 7.49E-05 | -0.001779571 | PCNT |
| chr15 | 40695170 | 40695592 | 423 | 7 | 0.000249181 | 0.000114 | 0.00666 | 0.00012 | 0.000432997 | RAD51 |
| chr16 | 88781899 | 88782564 | 666 | 3 | 0.000249456 | 0.001655 | 0.00014 | 2.88E-05 | -0.005086122 | PIEZO1 |
| chr6 | 15505221 | 15505854 | 634 | 4 | 0.000249695 | 0.000295 | 0.00344 | 0.00012 | -0.008208584 | JARID2 |
| chr8 | 22597960 | 22598473 | 514 | 2 | 0.000250228 | 2.42E-05 | 0.00144 | 3.81E-05 | -0.01448935 | PDLIM2, AC037459.1 |
| chr7 | 115788674 | 115789145 | 472 | 2 | 0.000250402 | 0.004145 | 0.00023 | 0.00043 | -0.009199785 | NA |
| chr22 | 30305166 | 30305681 | 516 | 2 | 0.000250647 | 2.26E-05 | 0.00068 | 3.23E-05 | -0.005207192 | TBC1D10A |
| chr16 | 35207416 | 35207797 | 382 | 4 | 0.000250849 | 2.07E-05 | 0.00223 | 1.20E-05 | -0.018895439 | NA |
| chr6 | 142949254 | 142949650 | 397 | 2 | 0.00025101 | 4.78E-05 | 0.00073 | 6.09E-05 | -0.008639488 | AL355304.1, HIVEP2 |
| chr20 | 62149610 | 62150114 | 505 | 5 | 0.000251028 | 0.000944 | 0.00012 | 5.25E-05 | -0.001241658 | SS18L1 |
| chr6 | 168414797 | 168415358 | 562 | 3 | 0.000251154 | 0.007754 | 0.00584 | 0.00308 | -0.004712752 | NA |
| chr2 | 98935646 | 98935798 | 153 | 2 | 0.000251197 | 9.15E-05 | 0.00032 | 7.66E-05 | -0.006072665 | CRACDL |
| chr17 | 78358540 | 78359065 | 526 | 4 | 0.000251324 | 1.02E-05 | 0.00266 | 1.89E-05 | -0.020751205 | SOCS3 |
| chr19 | 15379404 | 15380021 | 618 | 10 | 0.000251681 | 0.000296 | 0.00053 | 4.49E-05 | 0.001124472 | AC005785.1, AKAP8 |
| chr8 | 30638284 | 30638611 | 328 | 2 | 0.000252119 | 3.41E-05 | 0.00023 | 3.38E-05 | -0.004899592 | SMIM18, GTF2E2 |
| chr13 | 100588400 | 100588971 | 572 | 7 | 0.000252191 | 7.18E-05 | 0.00351 | 1.10E-05 | -0.005135245 | GGACT |
| chr8 | 52009080 | 52009691 | 612 | 3 | 0.000252207 | 0.000158 | 0.00102 | 7.01E-05 | -0.009677031 | AC064807.2 |
| chr2 | 9251441 | 9251767 | 327 | 2 | 0.000252565 | 1.35E-05 | 0.00095 | 2.17E-05 | -0.018267967 | ASAP2 |
| chr6 | 150882280 | 150882322 | 43 | 2 | 0.000252742 | 0.000132 | 5.76E-05 | 3.33E-05 | -0.002437524 | MTHFD1L |
| chr11 | 119309412 | 119309430 | 19 | 2 | 0.000252837 | 0.002574 | 4.27E-05 | 5.62E-05 | -0.00269115 | CBL, MCAM |
| chr3 | 191212351 | 191212497 | 147 | 2 | 0.000252865 | 1.44E-05 | 0.00031 | 1.90E-05 | -0.017646511 | OSTN |
| chr1 | 9249830 | 9250198 | 369 | 3 | 0.000252913 | 0.082872 | 3.69E-05 | 0.00013 | -0.005884923 | H6PD |
| chr20 | 32581762 | 32581837 | 76 | 2 | 0.000253343 | 2.77E-05 | 0.00079 | 3.93E-05 | -0.010166837 | NOL4L-DT, NOL4L |
| chr3 | 47323931 | 47324421 | 491 | 2 | 0.000253478 | 1.76E-05 | 0.00058 | 2.55E-05 | -0.01254907 | KLHL18 |
| chr6 | 30172548 | 30172936 | 389 | 4 | 0.000253509 | 0.005227 | 3.90E-05 | 3.43E-05 | -0.004469125 | TRIM15 |
| chr7 | 44749647 | 44749869 | 223 | 2 | 0.000253611 | 0.000875 | 0.00019 | 0.0002 | -0.007310916 | ZMIZ2 |
| chr1 | 109249217 | 109249926 | 710 | 8 | 0.000253657 | 0.017167 | 8.26E-05 | 0.00022 | 0.000923843 | CELSR2 |
| chr20 | 17968557 | 17969219 | 663 | 17 | 0.000253825 | 2.31E-05 | 0.02225 | 3.19E-05 | -7.54E-05 | MGME1, SNX5, OVOL2 |
| chr12 | 131600108 | 131600439 | 332 | 2 | 0.0002544 | 4.72E-05 | 0.00027 | 4.49E-05 | -0.005726579 | NA |
| chr6 | 15513156 | 15513446 | 291 | 3 | 0.000254664 | 7.94E-05 | 0.00113 | 9.56E-05 | -0.006550326 | JARID2 |
| chr4 | 184733889 | 184734549 | 661 | 10 | 0.000254759 | 0.000265 | 0.00407 | 6.79E-05 | -0.00017685 | CENPU |
| chr21 | 46289750 | 46290495 | 746 | 2 | 0.000254832 | 3.70E-06 | 0.00045 | 6.33E-06 | -0.004362589 | YBEY |
| chr16 | 1361747 | 1362265 | 519 | 4 | 0.000255091 | 1.56E-05 | 0.00201 | 2.92E-05 | -0.002262958 | GNPTG |
| chr12 | 14938461 | 14938972 | 512 | 5 | 0.000255103 | 5.36E-06 | 0.00625 | 1.48E-05 | -0.002969294 | ERP27 |
| chr2 | 29114731 | 29115770 | 1040 | 13 | 0.000255201 | 5.71E-05 | 0.00221 | 1.83E-05 | -0.000847864 | CLIP4 |
| chr20 | 24027434 | 24027686 | 253 | 2 | 0.00025533 | 4.26E-05 | 0.00015 | 3.36E-05 | -0.007511891 | NA |
| chr19 | 10627606 | 10627776 | 171 | 2 | 0.000255353 | 0.000119 | 7.13E-05 | 3.76E-05 | 0.002604207 | SLC44A2 |
| chr4 | 112301139 | 112301840 | 702 | 2 | 0.000255525 | 8.48E-06 | 0.0006 | 1.36E-05 | -0.009602384 | ALPK1 |
| chr6 | 18122488 | 18123074 | 587 | 12 | 0.000255693 | 4.36E-05 | 0.00397 | 2.52E-05 | -0.003467138 | NHLRC1 |
| chr4 | 38109459 | 38110165 | 707 | 7 | 0.000255727 | 0.000141 | 0.01586 | 9.16E-05 | -0.002302591 | TBC1D1 |
| chr15 | 31325256 | 31325561 | 306 | 5 | 0.000255819 | 1.95E-05 | 0.00566 | 3.96E-05 | -0.005509705 | NA |
| chr16 | 67483938 | 67484276 | 339 | 5 | 0.00025595 | 3.67E-05 | 0.00245 | 4.17E-05 | -0.005466545 | AC009061.2 |
| chr21 | 36699023 | 36699519 | 497 | 5 | 0.00025595 | 0.006932 | 0.00211 | 0.00037 | 0.000428653 | SIM2, AP000697.1 |
| chr5 | 14357934 | 14358044 | 111 | 2 | 0.000256133 | 1.80E-05 | 0.00059 | 2.61E-05 | -0.014187371 | TRIO, AC016549.1 |
| chr1 | 156123764 | 156124157 | 394 | 3 | 0.000256249 | 0.00211 | 0.00024 | 0.00021 | -0.008094884 | LMNA |
| chr15 | 78860282 | 78860578 | 297 | 2 | 0.00025632 | 1.14E-05 | 0.00084 | 1.84E-05 | 0.008871295 | MORF4L1 |
| chr3 | 183263844 | 183264454 | 611 | 3 | 0.000256391 | 4.48E-05 | 0.00038 | 2.07E-05 | -0.009140469 | B3GNT5, MCF2L2 |
| chr4 | 41256773 | 41257266 | 494 | 6 | 0.000256391 | 8.34E-05 | 0.00161 | 3.40E-05 | -0.002294422 | UCHL1 |
| chr7 | 149873842 | 149874167 | 326 | 10 | 0.000256723 | 5.17E-05 | 0.02085 | 6.76E-05 | -0.000163158 | ATP6V0E2, ATP6V0E2-AS1 |
| chr14 | 75200775 | 75201386 | 612 | 3 | 0.000257168 | 2.28E-05 | 0.00055 | 1.55E-05 | -0.004612583 | NA |
| chr2 | 226801645 | 226802011 | 367 | 2 | 0.000257279 | 3.03E-05 | 0.0014 | 4.63E-05 | -0.019261751 | AC010735.2 |
| chr11 | 46542279 | 46543175 | 897 | 6 | 0.000257336 | 3.58E-05 | 0.00129 | 8.48E-06 | -0.006824095 | AMBRA1 |
| chr2 | 233868161 | 233868439 | 279 | 4 | 0.000257336 | 4.49E-05 | 0.00131 | 4.90E-05 | 0.012221872 | MSL3P1 |
| chr20 | 50817851 | 50818537 | 687 | 2 | 0.000257358 | 3.92E-06 | 0.00023 | 6.05E-06 | -0.010815139 | BCAS4 |
| chr14 | 75560730 | 75561064 | 335 | 4 | 0.000257474 | 0.007835 | 7.97E-05 | 9.01E-05 | -0.006572022 | NA |
| chr3 | 129586880 | 129587206 | 327 | 2 | 0.000257474 | 2.35E-05 | 0.00083 | 3.46E-05 | -0.01161378 | PLXND1 |
| chr19 | 17716794 | 17717183 | 390 | 3 | 0.000257579 | 1.42E-05 | 0.00111 | 2.52E-05 | -0.009353207 | NA |
| chr7 | 158532332 | 158532426 | 95 | 2 | 0.000257773 | 0.000121 | 0.00034 | 9.48E-05 | 0.015070937 | PTPRN2 |
| chr5 | 14445392 | 14445554 | 163 | 2 | 0.000257907 | 8.37E-05 | 0.00084 | 9.94E-05 | -0.003674281 | TRIO |
| chr22 | 50282384 | 50282691 | 308 | 2 | 0.000258082 | 0.000113 | 0.00196 | 0.00017 | -0.007427186 | PLXNB2 |
| chr7 | 111852945 | 111852971 | 27 | 2 | 0.000258332 | 0.000102 | 0.00101 | 0.00013 | -0.002921795 | DOCK4 |
| chr1 | 1348300 | 1348352 | 53 | 2 | 0.000258349 | 0.000177 | 0.00061 | 0.00016 | -0.007543403 | DVL1 |
| chr5 | 132060445 | 132060664 | 220 | 6 | 0.000258417 | 0.005451 | 0.00021 | 7.65E-05 | -0.007597386 | IL3 |
| chr12 | 114624854 | 114625206 | 353 | 2 | 0.00025846 | 0.000145 | 7.77E-05 | 4.39E-05 | -0.007519782 | NA |
| chr3 | 160448960 | 160449089 | 130 | 3 | 0.000258472 | 0.003476 | 0.00422 | 0.0006 | 0.008420792 | AC079594.1, TRIM59 |
| chr19 | 11354635 | 11354883 | 249 | 3 | 0.000258472 | 0.022737 | 0.0004 | 0.00012 | -0.003279839 | CCDC159 |
| chr1 | 23162840 | 23162865 | 26 | 2 | 0.000258875 | 0.000294 | 0.00012 | 8.52E-05 | -0.000608393 | LUZP1 |
| chr20 | 31562525 | 31563115 | 591 | 4 | 0.000259327 | 8.72E-05 | 0.00147 | 3.25E-05 | -0.009904823 | HM13 |
| chr2 | 187264007 | 187264082 | 76 | 2 | 0.000259565 | 1.44E-05 | 0.00087 | 2.27E-05 | -0.032734466 | AC007319.1 |
| chr13 | 51801640 | 51801973 | 334 | 2 | 0.000259578 | 5.28E-05 | 0.00024 | 4.70E-05 | -0.011015262 | AL162377.3, DHRS12 |
| chr18 | 10403968 | 10404190 | 223 | 3 | 0.000259589 | 5.72E-05 | 0.00055 | 3.45E-05 | -0.009288777 | NA |
| chr3 | 151130339 | 151130618 | 280 | 3 | 0.000259634 | 0.009767 | 3.55E-05 | 9.85E-05 | -0.006334287 | MED12L |
| chr1 | 64471662 | 64471845 | 184 | 3 | 0.000259651 | 0.001067 | 0.00011 | 0.00013 | -0.007359456 | CACHD1 |
| chr9 | 21994753 | 21995736 | 984 | 9 | 0.000259745 | 0.000808 | 0.00027 | 1.76E-05 | -0.00065758 | AL359922.1, CDKN2B-AS1, AL449423.1, CDKN2A |
| chr8 | 123206837 | 123207485 | 649 | 5 | 0.000259767 | 3.49E-05 | 0.00577 | 3.67E-05 | -0.021029652 | FAM83A |
| chr10 | 102502626 | 102502857 | 232 | 6 | 0.000259768 | 2.32E-05 | 0.00553 | 3.72E-05 | 0.001407546 | ACTR1A |
| chr7 | 83589021 | 83589091 | 71 | 2 | 0.00025984 | 2.14E-05 | 0.00017 | 2.17E-05 | -0.007795361 | SEMA3E |
| chr2 | 40512375 | 40512923 | 549 | 6 | 0.000260437 | 0.000858 | 0.00212 | 8.12E-05 | -0.000587705 | SLC8A1 |
| chr16 | 71844805 | 71844971 | 167 | 3 | 0.000260639 | 9.34E-05 | 0.00085 | 6.63E-05 | -0.003548129 | NA |
| chr15 | 52529181 | 52529720 | 540 | 8 | 0.000260646 | 0.011406 | 0.0003 | 0.00011 | 0.000550339 | NA |
| chr4 | 4856158 | 4857087 | 930 | 7 | 0.000260965 | 0.00019 | 0.0028 | 7.73E-05 | 0.001401091 | NA |
| chr12 | 120100859 | 120100988 | 130 | 2 | 0.000261016 | 5.47E-05 | 0.00013 | 3.69E-05 | -0.001218082 | RAB35 |
| chr15 | 75048947 | 75049661 | 715 | 5 | 0.000261099 | 0.000283 | 0.003 | 0.00011 | -0.006262691 | PPCDC |
| chr10 | 133268504 | 133268775 | 272 | 4 | 0.000261139 | 0.00188 | 0.0001 | 5.27E-05 | -0.001598545 | ADAM8 |
| chr12 | 6582411 | 6582639 | 229 | 3 | 0.000261228 | 0.002076 | 0.00014 | 0.00013 | -0.004904176 | AC006064.2, AC006064.6, CHD4 |
| chr8 | 141427798 | 141428207 | 410 | 4 | 0.000261376 | 1.45E-05 | 0.00805 | 4.19E-05 | 6.40E-05 | PTP4A3 |

| | | | | | | | | | | |
|---|---|---|---|---|---|---|---|---|---|---|
| chr19 | 48681300 | 48681371 | 72 | 2 | 0.000261396 | 2.61E-05 | 0.0012 | 4.00E-05 | -0.012335657 | SEC1P |
| chr12 | 133236126 | 133236433 | 308 | 2 | 0.000261509 | 7.87E-05 | 0.00071 | 8.99E-05 | -0.027204423 | NA |
| chr11 | 67650487 | 67650934 | 448 | 11 | 0.000261652 | 7.25E-07 | 0.0327 | 1.06E-05 | -0.020366103 | ACY3 |
| chr12 | 93826260 | 93826328 | 69 | 2 | 0.000261654 | 0.000527 | 4.47E-05 | 4.21E-05 | -0.005063719 | CRADD |
| chr14 | 36773166 | 36773730 | 565 | 2 | 0.000261784 | 1.92E-05 | 0.0005 | 2.65E-05 | -0.022036157 | SLC25A21 |
| chr3 | 42536188 | 42536392 | 205 | 5 | 0.00026192 | 0.004595 | 5.96E-05 | 5.83E-05 | -0.001781191 | VIPR1 |
| chr1 | 38050986 | 38051215 | 230 | 3 | 0.00026192 | 1.22E-05 | 0.00147 | 1.85E-05 | -0.003550865 | MIR3659HG |
| chr11 | 12069203 | 12069292 | 90 | 3 | 0.000262075 | 1.50E-05 | 0.00089 | 2.30E-05 | -0.008441543 | AC124276.1 |
| chr6 | 168034956 | 168035419 | 464 | 5 | 0.000262201 | 3.57E-05 | 0.00272 | 3.10E-05 | 0.02125994 | KIF25 |
| chr19 | 16382872 | 16383207 | 336 | 2 | 0.000262324 | 0.000105 | 0.00039 | 9.10E-05 | -0.008825282 | EPS15L1 |
| chr17 | 8121778 | 8122157 | 380 | 3 | 0.000262338 | 0.000262 | 0.00055 | 0.00014 | 0.00224771 | HES7 |
| chr6 | 1686005 | 1686512 | 508 | 2 | 0.000262387 | 1.31E-05 | 0.00084 | 2.07E-05 | -0.011996628 | GMDS |
| chr1 | 202341696 | 202342364 | 669 | 12 | 0.000262847 | 8.03E-06 | 0.02145 | 2.37E-05 | 0.001578512 | UBE2T |
| chr8 | 141973778 | 141974213 | 436 | 3 | 0.000262913 | 1.23E-05 | 0.00081 | 2.10E-05 | -0.000528505 | NA |
| chr19 | 53982067 | 53982476 | 410 | 7 | 0.00026296 | 0.01102 | 0.00053 | 0.00019 | -0.004690343 | CACNG8, MIR935 |
| chr19 | 20052031 | 20052218 | 188 | 3 | 0.00026296 | 0.000355 | 5.41E-05 | 3.16E-05 | -0.000772448 | NA |
| chr2 | 181909909 | 181909957 | 49 | 2 | 0.000263142 | 0.000749 | 4.84E-05 | 4.96E-05 | -0.005182849 | ITPRID2 |
| chr17 | 27498903 | 27499458 | 556 | 2 | 0.000263161 | 2.14E-05 | 0.00062 | 3.05E-05 | -0.005216707 | KSR1 |
| chr12 | 57552300 | 57552474 | 175 | 2 | 0.000263758 | 2.47E-05 | 0.00127 | 3.84E-05 | -0.006356525 | KIF5A |
| chr20 | 37411846 | 37412500 | 655 | 2 | 0.00026388 | 1.64E-05 | 0.00023 | 1.96E-05 | -0.006064104 | NA |
| chr1 | 1984905 | 1985279 | 375 | 3 | 0.000264018 | 4.38E-05 | 0.00026 | 3.75E-05 | -0.00995485 | CFAP74 |
| chr7 | 73438361 | 73438688 | 328 | 2 | 0.0002641 | 6.43E-05 | 0.00015 | 4.38E-05 | -0.007515089 | NA |
| chr1 | 109630336 | 109630490 | 155 | 2 | 0.000264113 | 0.000911 | 8.43E-05 | 9.11E-05 | -0.003517321 | AMPD2 |
| chr4 | 1374644 | 1374827 | 184 | 3 | 0.000264354 | 3.75E-05 | 0.00196 | 4.81E-05 | -0.004876029 | UVSSA |
| chr13 | 36920607 | 36920857 | 251 | 4 | 0.000264477 | 0.000279 | 0.00218 | 0.00017 | -0.00741359 | SMAD9 |
| chr11 | 130208341 | 130208754 | 414 | 2 | 0.000264632 | 0.000151 | 0.00143 | 0.0002 | -0.003355237 | ST14 |
| chr1 | 66142646 | 66143036 | 391 | 2 | 0.000264703 | 8.45E-05 | 0.00011 | 4.33E-05 | -0.009535722 | PDE4B |
| chr19 | 14162864 | 14162901 | 38 | 2 | 0.000264703 | 2.34E-05 | 0.00132 | 3.67E-05 | -0.005310412 | AC022098.1, ADGRL1 |
| chr12 | 75327029 | 75327062 | 34 | 2 | 0.000264822 | 4.94E-05 | 9.75E-05 | 2.89E-05 | -0.009118252 | CAPS2 |
| chr5 | 38967177 | 38967621 | 445 | 4 | 0.000264834 | 0.001359 | 0.00035 | 6.79E-05 | -0.003509815 | RICTOR |
| chr11 | 73307957 | 73309684 | 1728 | 15 | 0.000264971 | 4.09E-05 | 0.00085 | 1.66E-06 | -0.002049286 | ARHGEF17, AP002761.3 |
| chr7 | 5500849 | 5501129 | 281 | 4 | 0.000264989 | 8.61E-05 | 0.00134 | 5.22E-05 | -0.002618633 | FBXL18 |
| chr4 | 7481670 | 7481850 | 181 | 2 | 0.000265126 | 4.06E-05 | 0.00014 | 3.12E-05 | -0.006645214 | SORCS2 |
| chr7 | 100602976 | 100603537 | 562 | 5 | 0.000265128 | 0.000191 | 0.0054 | 0.00035 | -0.005593994 | PCOLCE, PCOLCE-AS1 |
| chr4 | 83618416 | 83618740 | 325 | 2 | 0.00026518 | 1.36E-05 | 0.001 | 2.18E-05 | -0.002797995 | NA |
| chr10 | 21932445 | 21932759 | 315 | 2 | 0.000265214 | 2.41E-05 | 0.0006 | 3.32E-05 | -0.001009897 | DNAJC1 |
| chr1 | 220045821 | 220046088 | 268 | 3 | 0.000265668 | 0.000182 | 0.00018 | 7.38E-05 | 0.000722463 | EPRS1 |
| chr13 | 112833976 | 112835097 | 1122 | 6 | 0.000266473 | 6.94E-07 | 0.00238 | 2.70E-06 | -0.003202127 | ATP11A |
| chr10 | 89958127 | 89958584 | 458 | 2 | 0.0002665 | 1.46E-05 | 0.00103 | 2.36E-05 | -0.015291074 | LINC01374 |
| chr1 | 56963846 | 56963998 | 153 | 2 | 0.000266681 | 0.000596 | 6.07E-05 | 5.82E-05 | -0.019696007 | AL161740.1, C8B |
| chr5 | 175677954 | 175678347 | 394 | 2 | 0.000266967 | 0.000595 | 0.0017 | 0.0006 | -0.009397136 | HRH2 |
| chr15 | 40369674 | 40370140 | 467 | 4 | 0.000267087 | 0.000706 | 0.00084 | 9.72E-05 | -0.001978445 | DISP2 |
| chr18 | 46072890 | 46073568 | 679 | 2 | 0.00026738 | 4.42E-05 | 0.00123 | 6.38E-05 | -0.009206665 | NA |
| chr10 | 96129323 | 96130040 | 718 | 14 | 0.000267729 | 0.001585 | 0.00154 | 8.95E-05 | -0.002967336 | ZNF518A, AC021037.1 |
| chr6 | 106787375 | 106787748 | 374 | 2 | 0.000267789 | 0.000101 | 0.00018 | 6.48E-05 | -0.007628126 | LINC02532 |
| chr1 | 33080381 | 33081624 | 1244 | 16 | 0.000267846 | 9.86E-06 | 0.00356 | 4.11E-06 | 0.003369083 | AZIN2, AK2 |
| chr11 | 45686320 | 45686483 | 164 | 3 | 0.000267872 | 8.68E-05 | 0.00011 | 3.56E-05 | 0.002039485 | NA |
| chr7 | 130668653 | 130669233 | 581 | 14 | 0.000267908 | 4.18E-06 | 0.0219 | 1.66E-05 | 0.000301358 | AC234644.2, COPG2, TSGA13 |
| chr3 | 16349164 | 16349190 | 27 | 2 | 0.000267914 | 0.001608 | 3.98E-05 | 4.79E-05 | -0.00270002 | OXNAD1, RFTN1 |
| chr7 | 144835840 | 144836378 | 539 | 10 | 0.00026794 | 0.00083 | 0.00262 | 0.00012 | 0.002057563 | TPK1 |
| chr10 | 69416745 | 69416983 | 239 | 3 | 0.000268042 | 0.003292 | 0.00047 | 0.00061 | -0.004751285 | TACR2 |
| chr15 | 26891068 | 26891934 | 867 | 18 | 0.000268219 | 2.88E-05 | 0.01711 | 1.96E-05 | 0.001103736 | GABRA5, GABRB3 |
| chr13 | 110438086 | 110438322 | 237 | 4 | 0.000268769 | 0.000791 | 0.00011 | 5.46E-05 | -3.96E-05 | COL4A2 |
| chr21 | 43703780 | 43703794 | 15 | 2 | 0.000268978 | 3.49E-05 | 0.00079 | 4.77E-05 | -0.005914936 | NA |
| chr16 | 69798291 | 69798928 | 638 | 5 | 0.000269019 | 0.000697 | 0.00276 | 0.00011 | -0.002188508 | WWP2 |
| chr14 | 53093420 | 53093753 | 334 | 2 | 0.000269131 | 2.72E-05 | 0.00034 | 3.22E-05 | -0.010650749 | DDHD1 |
| chr4 | 6587066 | 6587184 | 119 | 2 | 0.00026917 | 7.29E-05 | 7.62E-05 | 3.09E-05 | -0.004940865 | MAN2B2 |
| chr1 | 19054240 | 19054932 | 693 | 3 | 0.000269186 | 0.000336 | 0.00018 | 9.30E-05 | -0.009532203 | NA |
| chr18 | 74597773 | 74598427 | 655 | 10 | 0.000269305 | 0.033645 | 5.78E-05 | 0.0005 | 0.000949903 | ZNF407, LINC00909 |
| chr2 | 60881867 | 60882020 | 154 | 2 | 0.000269583 | 0.06953 | 0.00068 | 0.0022 | 0.003604241 | REL |
| chr15 | 88877641 | 88878116 | 476 | 4 | 0.000269775 | 6.83E-05 | 0.00079 | 3.88E-05 | -0.003019312 | HAPLN3 |
| chr10 | 102140780 | 102140795 | 16 | 3 | 0.00026996 | 0.000118 | 0.00015 | 3.72E-05 | -0.001328363 | PPRC1 |
| chr19 | 35257572 | 35257886 | 315 | 2 | 0.000269967 | 1.38E-05 | 0.00101 | 2.22E-05 | -0.015130576 | LSR |
| chr1 | 109185692 | 109186544 | 853 | 6 | 0.000270187 | 1.54E-05 | 0.00247 | 2.39E-05 | -0.006145077 | ELAPOR1 |
| chr9 | 134139365 | 134139810 | 446 | 3 | 0.000270289 | 0.000274 | 0.0007 | 0.00014 | -0.003634995 | WDR5 |
| chr19 | 49503225 | 49503542 | 318 | 3 | 0.00027062 | 3.73E-05 | 0.00068 | 3.52E-05 | -0.007173694 | NA |
| chr16 | 1768698 | 1769059 | 362 | 3 | 0.000270676 | 0.000191 | 0.00083 | 4.38E-05 | -0.005332861 | MAPK8IP3 |
| chr16 | 78045672 | 78046171 | 500 | 5 | 0.000270914 | 1.71E-05 | 0.00564 | 2.18E-05 | 0.005270945 | CLEC3A, AC079414.1 |
| chr16 | 30676428 | 30676792 | 365 | 3 | 0.000271052 | 4.30E-05 | 0.00101 | 5.37E-05 | -0.000932625 | NA |
| chr19 | 48636900 | 48637081 | 182 | 2 | 0.000271388 | 3.13E-05 | 0.00061 | 4.14E-05 | 0.002520462 | DBP |
| chr2 | 466712 | 467135 | 424 | 8 | 0.000271638 | 0.000373 | 0.00263 | 4.81E-05 | -0.001042949 | NA |
| chr6 | 31636097 | 31636513 | 417 | 3 | 0.000272189 | 0.001211 | 0.00038 | 0.00023 | -0.004738393 | PRRC2A |
| chr6 | 33294963 | 33295698 | 736 | 11 | 0.000272199 | 6.74E-05 | 0.02081 | 5.48E-05 | -0.000230071 | PFDN6, RGL2 |
| chr7 | 42961101 | 42961718 | 618 | 2 | 0.00027222 | 1.32E-05 | 0.00059 | 1.98E-05 | -0.001408101 | AC005537.1 |
| chr2 | 127681373 | 127681824 | 452 | 4 | 0.000272291 | 0.002319 | 0.00264 | 0.00022 | -0.01244748 | LIMS2 |
| chr20 | 25213365 | 25214042 | 678 | 2 | 0.000272583 | 0.020224 | 0.00045 | 0.00119 | -0.002710784 | ENTPD6 |
| chr11 | 61793551 | 61793747 | 197 | 2 | 0.000272684 | 7.31E-05 | 0.00118 | 9.69E-05 | -0.00691149 | FEN1, FADS2 |
| chr13 | 72755918 | 72755992 | 75 | 2 | 0.000272895 | 0.012976 | 5.41E-05 | 8.95E-05 | -0.008127137 | BORA, DIS3 |
| chr3 | 114651749 | 114652253 | 505 | 3 | 0.00027307 | 0.001609 | 0.00013 | 4.69E-05 | -0.0033553 | ZBTB20 |
| chr22 | 47859710 | 47860325 | 616 | 3 | 0.000273264 | 0.000726 | 0.00023 | 2.60E-05 | 0.0014626 | AL117329.1 |
| chr1 | 241957928 | 241958525 | 598 | 3 | 0.000273432 | 0.000126 | 0.00195 | 0.00012 | -0.006374135 | BECN2 |
| chr8 | 27465950 | 27466149 | 200 | 2 | 0.000273662 | 0.000234 | 6.93E-05 | 4.83E-05 | -0.001364486 | CHRNA2 |
| chr1 | 153509948 | 153509971 | 24 | 2 | 0.000273665 | 4.79E-05 | 0.00115 | 6.73E-05 | -0.012728529 | NA |
| chr1 | 45566598 | 45566824 | 227 | 2 | 0.00027374 | 4.09E-05 | 0.0003 | 4.25E-05 | -0.003101911 | AKR1A1 |
| chr10 | 102066723 | 102066948 | 226 | 2 | 0.000274014 | 4.25E-05 | 0.00183 | 6.56E-05 | -0.005717528 | HPS6 |
| chr11 | 58576318 | 58577056 | 739 | 4 | 0.000274018 | 7.92E-05 | 0.00413 | 9.21E-05 | -0.002332585 | LPXN |
| chr7 | 139722344 | 139722697 | 354 | 3 | 0.000274046 | 2.57E-05 | 0.0017 | 3.13E-05 | -0.010521984 | HIPK2 |
| chr14 | 26595548 | 26596540 | 993 | 5 | 0.000274092 | 4.82E-05 | 0.00371 | 4.23E-05 | -0.009859353 | NOVA1 |
| chr2 | 222305246 | 222305651 | 406 | 2 | 0.000274092 | 0.000158 | 0.00017 | 8.03E-05 | -0.004079795 | NA |
| chr1 | 11858750 | 11859497 | 748 | 10 | 0.000274178 | 0.000103 | 0.00307 | 6.70E-05 | -0.007036755 | NPPB |
| chr20 | 3406456 | 3406720 | 265 | 2 | 0.000274213 | 0.000383 | 0.00209 | 0.00047 | -0.009682031 | AL117334.1, C20orf194 |
| chr7 | 76264634 | 76265358 | 725 | 4 | 0.000274332 | 9.09E-06 | 0.00209 | 1.66E-05 | -0.00526899 | SRRM3 |
| chr7 | 964841 | 965453 | 613 | 5 | 0.000274657 | 6.64E-05 | 0.00377 | 3.34E-05 | -0.018594072 | COX19 |
| chr1 | 205285954 | 205286491 | 538 | 3 | 0.000275305 | 1.18E-05 | 0.00264 | 2.47E-05 | -0.005678185 | NA |
| chr18 | 74481165 | 74481175 | 11 | 2 | 0.000275712 | 7.55E-05 | 0.00015 | 4.80E-05 | -0.013008569 | NA |
| chr11 | 124248939 | 124249117 | 179 | 3 | 0.000275951 | 0.000107 | 0.00013 | 3.65E-05 | -0.000124424 | OR8G1 |
| chr19 | 16862784 | 16862999 | 216 | 2 | 0.000276341 | 2.46E-05 | 0.0006 | 3.38E-05 | -0.009322964 | SIN3B |
| chr1 | 53438323 | 53439063 | 741 | 7 | 0.000276413 | 0.000311 | 0.00054 | 0.00011 | -0.001803506 | SLC25A3P1 |
| chr17 | 63675597 | 63675742 | 146 | 2 | 0.000276525 | 2.42E-05 | 0.00039 | 3.03E-05 | -0.00657769 | MAP3K3 |
| chr6 | 29429219 | 29429525 | 307 | 3 | 0.000276545 | 0.000135 | 0.00044 | 7.82E-05 | -0.003180683 | OR5V1, OR11A1 |
| chr2 | 3485358 | 3485464 | 107 | 2 | 0.000276599 | 4.00E-05 | 0.00165 | 6.10E-05 | -0.006911273 | NA |
| chr6 | 144660000 | 144660067 | 68 | 4 | 0.000276726 | 2.25E-05 | 0.00204 | 2.75E-05 | -0.00949849 | UTRN |
| chr17 | 33648402 | 33648599 | 198 | 2 | 0.000276757 | 1.86E-05 | 0.00054 | 2.63E-05 | -0.000180023 | ASIC2 |
| chr22 | 42437465 | 42437530 | 66 | 2 | 0.000277019 | 1.32E-05 | 0.00086 | 2.10E-05 | -0.004459588 | NA |
| chr17 | 82005163 | 82005815 | 653 | 4 | 0.000277232 | 3.57E-05 | 0.00298 | 4.36E-05 | -0.007410616 | ASPSCR1 |
| chr1 | 95072819 | 95073281 | 463 | 11 | 0.000277265 | 0.004752 | 0.00086 | 5.28E-05 | -0.000715154 | ALG14 |
| chr2 | 45323359 | 45323368 | 10 | 2 | 0.000277415 | 0.002036 | 4.41E-05 | 5.58E-05 | -0.004715211 | LINC01121 |
| chr14 | 92917120 | 92917645 | 526 | 2 | 0.000277482 | 1.55E-05 | 0.00076 | 2.39E-05 | -0.01533794 | NA |
| chr20 | 57779626 | 57779980 | 355 | 2 | 0.000277625 | 2.12E-05 | 0.00095 | 3.25E-05 | -0.00592799 | NA |
| chr16 | 48538784 | 48539130 | 347 | 2 | 0.000277691 | 0.000117 | 0.00014 | 6.07E-05 | -0.006083369 | N4BP1 |
| chr16 | 89571981 | 89572111 | 131 | 2 | 0.000277709 | 1.67E-05 | 0.00071 | 2.52E-05 | -0.012001555 | NA |
| chr13 | 43785279 | 43785860 | 582 | 4 | 0.000277992 | 0.001438 | 9.92E-05 | 0.00012 | -0.004133757 | ENOX1 |
| chr17 | 4480027 | 4480587 | 561 | 3 | 0.000278002 | 0.000546 | 0.00012 | 9.80E-05 | -0.006476543 | SPNS3 |
| chr2 | 215335816 | 215335900 | 85 | 2 | 0.000278301 | 0.009282 | 4.00E-05 | 6.10E-05 | -0.00287866 | ATIC |
| chr17 | 44186678 | 44187503 | 826 | 15 | 0.000278675 | 4.20E-05 | 0.00054 | 4.83E-05 | 0.00095991 | TMUB2, ASB16-AS1 |
| chr19 | 50786303 | 50786395 | 93 | 4 | 0.00027895 | 0.000889 | 3.01E-05 | 4.45E-05 | 0.00395116 | AC010325.1 |
| chr16 | 85390796 | 85390947 | 152 | 2 | 0.000279236 | 2.70E-05 | 0.00028 | 3.04E-05 | -0.011545572 | GSE1 |
| chr11 | 61749117 | 61749926 | 810 | 3 | 0.000279342 | 1.14E-05 | 0.00145 | 2.26E-05 | -0.009196532 | MYRF-AS1 |
| chr5 | 39506250 | 39506533 | 284 | 3 | 0.000279658 | 8.50E-05 | 0.00051 | 4.52E-05 | -0.001064602 | NA |
| chr19 | 47331745 | 47332218 | 474 | 7 | 0.000279727 | 0.000133 | 0.00399 | 3.79E-05 | -0.005381681 | C5AR2 |
| chr14 | 22588984 | 22589206 | 223 | 3 | 0.00028023 | 0.000237 | 0.00045 | 0.00011 | 0.001669194 | DAD1 |
| chr5 | 135489776 | 135490205 | 430 | 3 | 0.000280414 | 0.001064 | 0.00174 | 0.00021 | 0.000138404 | NA |
| chr7 | 6531343 | 6531485 | 143 | 4 | 0.000280475 | 5.64E-05 | 0.00208 | 7.81E-05 | 0.001232443 | GRID2IP |
| chr1 | 87154588 | 87154800 | 213 | 2 | 0.000280836 | 0.004967 | 7.57E-05 | 0.00012 | -0.007612923 | AC093155.3, LINC01140 |
| chr19 | 45382542 | 45382820 | 279 | 4 | 0.000280944 | 6.32E-05 | 0.0008 | 3.78E-05 | -0.014018798 | PPP1R13L |
| chr15 | 78034938 | 78035098 | 161 | 2 | 0.00028118 | 2.53E-05 | 0.00033 | 3.03E-05 | -0.007184989 | TBC1D2B |
| chr6 | 36349632 | 36349799 | 168 | 2 | 0.000281204 | 0.00031 | 0.00011 | 7.86E-05 | -0.006733171 | NA |
| chr7 | 4307868 | 4308476 | 609 | 5 | 0.000281226 | 6.94E-05 | 0.00405 | 3.26E-05 | -0.004438909 | NA |
| chr4 | 39447355 | 39447982 | 628 | 4 | 0.000281315 | 3.73E-05 | 0.00358 | 7.28E-05 | 0.013066311 | KLB |
| chr5 | 172754316 | 172754355 | 40 | 2 | 0.000281657 | 4.00E-05 | 0.00174 | 6.15E-05 | -0.012855167 | NA |
| chr1 | 151189681 | 151191299 | 1619 | 10 | 0.000281717 | 1.81E-05 | 0.00048 | 2.75E-06 | -6.57E-05 | VPS72 |
| chr6 | 169864762 | 169865056 | 295 | 2 | 0.000281903 | 0.000569 | 0.00012 | 0.00011 | -0.007974402 | NA |
| chr19 | 49876701 | 49876765 | 65 | 2 | 0.000281903 | 0.162059 | 0.03781 | 0.0759 | 0.000880069 | PNKP, AKT1S1 |
| chr8 | 27471190 | 27471667 | 478 | 3 | 0.000282271 | 0.000937 | 0.00097 | 4.95E-05 | -0.005772842 | CHRNA2 |
| chr5 | 132369627 | 132370050 | 424 | 6 | 0.000282399 | 0.00056 | 0.0007 | 8.58E-05 | 0.000209005 | SLC22A5, MIR3936HG |
| chr15 | 65057268 | 65057921 | 654 | 3 | 0.000282429 | 0.000516 | 0.00024 | 0.00015 | -0.003016643 | RASL12 |
| chr15 | 74461444 | 74461671 | 228 | 6 | 0.000282622 | 0.007206 | 0.00085 | 0.0011 | 0.001341692 | UBL7-AS1 |
| chr8 | 102388904 | 102389345 | 442 | 3 | 0.000282848 | 0.000588 | 0.00052 | 0.00026 | -0.004539919 | UBR5 |
| chr3 | 136721617 | 136721627 | 11 | 2 | 0.000282848 | 0.000267 | 7.62E-05 | 5.52E-05 | -0.000252399 | STAG1 |
| chr14 | 22280950 | 22281075 | 126 | 3 | 0.000283085 | 0.001683 | 9.91E-05 | 7.71E-05 | -0.004651059 | AC244502.1 |
| chr20 | 47633621 | 47634090 | 470 | 3 | 0.000283389 | 0.000389 | 9.32E-05 | 5.61E-05 | -0.006184962 | NCOA3 |
| chr14 | 65282852 | 65282966 | 115 | 2 | 0.000283446 | 3.67E-05 | 0.00141 | 5.51E-05 | -0.015873537 | AL355076.2 |
| chr10 | 131954452 | 131954823 | 372 | 5 | 0.000283676 | 0.000369 | 0.00018 | 0.00013 | -0.002732131 | PPP2R2D |
| chr12 | 94046704 | 94047291 | 588 | 3 | 0.000283852 | 2.31E-05 | 0.00086 | 3.12E-05 | -0.010722345 | NA |
| chr6 | 10414117 | 10414423 | 307 | 8 | 0.000283928 | 0.00019 | 0.00607 | 7.94E-05 | -0.002356133 | TFAP2A-AS1, TFAP2A |
| chr11 | 495007 | 495198 | 192 | 4 | 0.000284062 | 0.000736 | 0.00079 | 5.60E-05 | -0.002743446 | RNH1 |
| chr2 | 197378672 | 197378843 | 172 | 2 | 0.000284115 | 2.09E-05 | 0.00132 | 3.33E-05 | -0.011349051 | NA |
| chr1 | 207103673 | 207104236 | 564 | 10 | 0.000284268 | 1.57E-05 | 0.0026 | 3.05E-05 | -0.003160248 | C4BPA |
| chr17 | 2095135 | 2095405 | 271 | 3 | 0.000284433 | 0.000232 | 0.0001 | 5.61E-05 | -0.007649533 | AC090617.4, SMG6 |
| chr4 | 168318569 | 168318930 | 362 | 8 | 0.000284491 | 2.36E-05 | 0.00604 | 4.96E-05 | 0.002044799 | DDX60 |
| chr6 | 41736134 | 41736470 | 337 | 4 | 0.000284728 | 0.001809 | 0.0008 | 0.00033 | -0.007009028 | TFEB |
| chr8 | 38734847 | 38735116 | 270 | 2 | 0.000285065 | 1.97E-05 | 0.00097 | 3.04E-05 | -0.014519539 | TACC1 |
| chr2 | 64064284 | 64064666 | 383 | 2 | 0.000285218 | 2.47E-05 | 0.00066 | 3.46E-05 | -0.015485267 | NA |
| chr11 | 100720017 | 100720163 | 147 | 2 | 0.000285299 | 0.009523 | 4.41E-05 | 6.84E-05 | -0.015639381 | ARHGAP42 |
| chr10 | 129415313 | 129415726 | 414 | 3 | 0.000285596 | 5.04E-05 | 5.51E-05 | 1.57E-05 | 0.000221269 | NA |
| chr20 | 41603995 | 41604140 | 146 | 2 | 0.000285633 | 2.12E-05 | 0.00128 | 3.37E-05 | 0.003050304 | CHD6 |
| chr14 | 75717650 | 75718155 | 506 | 3 | 0.000285744 | 0.014484 | 6.86E-05 | 0.00017 | -0.002861344 | TTLL5 |
| chr20 | 37314785 | 37315177 | 393 | 3 | 0.000285831 | 0.005414 | 0.00011 | 0.00027 | -0.005479597 | MANBAL |
| chr5 | 168187835 | 168188402 | 568 | 6 | 0.000286299 | 0.000291 | 0.00232 | 3.68E-05 | -0.003115731 | TENM2 |
| chr5 | 1800492 | 1801028 | 537 | 6 | 0.000286326 | 3.40E-05 | 0.00064 | 2.53E-05 | 0.003253144 | MRPL36 |
| chr15 | 65094583 | 65095144 | 562 | 2 | 0.000286809 | 1.60E-05 | 0.00068 | 2.42E-05 | -0.009707595 | UBAP1L |
| chr10 | 102120070 | 102120902 | 833 | 14 | 0.000286975 | 0.018545 | 0.00018 | 0.00047 | 0.00022577 | LDB1 |

| | | | | | | | | | | |
|---|---|---|---|---|---|---|---|---|---|---|
| chr1 | 151534133 | 151534201 | 69 | 2 | 0.000287174 | 3.57E-05 | 0.00015 | 2.96E-05 | -0.004071382 | CGN |
| chr17 | 75616697 | 75616951 | 255 | 4 | 0.000287204 | 0.000977 | 0.00019 | 4.71E-05 | -0.003750895 | MYO15B |
| chr17 | 8189170 | 8189606 | 437 | 3 | 0.000287204 | 0.004919 | 0.00997 | 0.00285 | -0.006511679 | AC129492.6, BORCS6 |
| chr2 | 60989477 | 60989575 | 99 | 2 | 0.000287216 | 0.001984 | 4.35E-05 | 5.49E-05 | -0.007543374 | PUS10 |
| chr4 | 83197676 | 83198020 | 345 | 3 | 0.000287267 | 0.001023 | 0.00101 | 6.24E-05 | -0.007393408 | NA |
| chr9 | 6008588 | 6008675 | 88 | 4 | 0.000287318 | 1.48E-05 | 0.00325 | 2.84E-05 | -0.001233915 | NA |
| chr4 | 139849422 | 139849442 | 21 | 2 | 0.000287539 | 0.000105 | 0.00018 | 6.57E-05 | -0.013026492 | MAML3 |
| chr7 | 142934109 | 142934169 | 61 | 4 | 0.000287706 | 0.004074 | 3.70E-05 | 6.29E-05 | -0.002965547 | NA |
| chr4 | 134327467 | 134327791 | 325 | 3 | 0.000287706 | 2.71E-05 | 0.00145 | 3.49E-05 | -0.006567195 | PES1P1, AC015631.1 |
| chr1 | 207801837 | 207802354 | 518 | 8 | 0.00028791 | 0.000116 | 0.00085 | 3.21E-05 | -0.004064728 | MIR29B2CHG, MIR29C |
| chr2 | 7037541 | 7037722 | 182 | 2 | 0.000287971 | 3.61E-05 | 0.00038 | 4.11E-05 | -0.008198444 | RNF144A |
| chr13 | 44219135 | 44219148 | 14 | 2 | 0.000288079 | 5.89E-05 | 0.00013 | 3.82E-05 | -0.004313372 | SMIM2-AS1 |
| chr16 | 545486 | 546899 | 1414 | 11 | 0.000288404 | 6.74E-05 | 0.00096 | 2.60E-06 | -0.005247131 | CAPN15 |
| chr11 | 113162442 | 113163673 | 1232 | 3 | 0.000288749 | 1.95E-06 | 0.0011 | 4.64E-06 | -0.010012667 | NCAM1 |
| chr8 | 57993113 | 57993415 | 303 | 3 | 0.000288818 | 0.00034 | 0.00018 | 8.55E-05 | -0.007955908 | NA |
| chr2 | 199471710 | 199471957 | 248 | 3 | 0.000289218 | 0.000675 | 0.00248 | 0.00065 | 0.005742775 | SATB2-AS1 |
| chr19 | 4277325 | 4277659 | 335 | 4 | 0.000289552 | 9.27E-06 | 0.00615 | 2.66E-05 | -0.01381844 | NA |
| chr22 | 28772147 | 28772804 | 658 | 11 | 0.000289815 | 0.005327 | 0.00053 | 8.95E-05 | 0.000168726 | CCDC117 |
| chr15 | 99249417 | 99250206 | 790 | 10 | 0.000290019 | 4.77E-06 | 0.02417 | 1.28E-05 | -0.018774601 | TTC23 |
| chr18 | 2571395 | 2571748 | 354 | 8 | 0.000290129 | 0.000645 | 7.96E-05 | 3.38E-05 | 0.001098697 | NDC80, METTL4 |
| chr10 | 101001213 | 101002644 | 1432 | 6 | 0.000290396 | 0.000135 | 0.00022 | 5.22E-06 | -0.004965188 | LZTS2 |
| chr11 | 76408732 | 76409069 | 338 | 3 | 0.000290711 | 1.10E-05 | 0.00158 | 2.12E-05 | -0.006573397 | GVQW3 |
| chr3 | 40457724 | 40458277 | 554 | 5 | 0.000290775 | 0.001946 | 0.00179 | 0.00052 | 0.004518819 | RPL14 |
| chr17 | 1190735 | 1191261 | 527 | 7 | 0.000291092 | 7.01E-05 | 0.00034 | 4.36E-05 | -0.005068494 | ABR |
| chr16 | 30418723 | 30418827 | 105 | 3 | 0.000291305 | 8.12E-05 | 0.00018 | 4.79E-05 | -0.003362735 | ZNF771 |
| chr19 | 13024817 | 13024994 | 178 | 3 | 0.000291305 | 2.96E-06 | 0.00201 | 7.80E-06 | -0.02540396 | NFIX |
| chr2 | 100974636 | 100974720 | 85 | 2 | 0.00029147 | 7.57E-05 | 8.82E-05 | 3.49E-05 | -0.005365812 | NPAS2, NPAS2-AS1 |
| chr17 | 16569485 | 16570154 | 670 | 3 | 0.000291624 | 0.001482 | 0.00991 | 0.00191 | -0.011299598 | NA |
| chr8 | 142612238 | 142612649 | 412 | 2 | 0.000292311 | 8.13E-05 | 0.0024 | 0.00013 | -0.004602065 | ARC |
| chr2 | 173184313 | 173184504 | 192 | 2 | 0.000292773 | 1.55E-05 | 0.00109 | 2.50E-05 | -0.012533913 | MAP3K20, MAP3K20-AS1 |
| chr12 | 130933398 | 130933512 | 115 | 2 | 0.000292905 | 5.67E-05 | 0.00036 | 5.73E-05 | -0.007803133 | NA |
| chr12 | 52871139 | 52871306 | 168 | 3 | 0.000292923 | 9.51E-06 | 0.00257 | 2.13E-05 | -0.016830344 | NA |
| chr14 | 22977843 | 22978583 | 741 | 5 | 0.000293468 | 0.000247 | 0.00268 | 0.00011 | -0.002490846 | AL132780.3, AJUBA |
| chr12 | 123601671 | 123602356 | 686 | 11 | 0.00029352 | 0.000739 | 0.00053 | 4.92E-05 | -0.000161395 | DDX55 |
| chr11 | 72039495 | 72039875 | 381 | 3 | 0.000293617 | 2.64E-05 | 0.00232 | 5.02E-05 | -0.010553781 | NUMA1 |
| chr3 | 197509497 | 197510283 | 787 | 6 | 0.000293697 | 5.10E-05 | 0.00715 | 2.60E-05 | 0.005693291 | BDH1 |
| chr2 | 112541419 | 112542206 | 788 | 12 | 0.000293866 | 0.000146 | 0.01908 | 0.00024 | 0.000297191 | TTL, POLR1B |
| chr19 | 47729525 | 47729731 | 207 | 3 | 0.000293869 | 3.32E-05 | 0.00377 | 5.92E-05 | -0.001581731 | AC010519.1, EHD2 |
| chr7 | 123624646 | 123625019 | 374 | 5 | 0.000293957 | 0.000141 | 0.00114 | 4.49E-05 | -0.014362096 | ASB15, ASB15-AS1 |
| chr2 | 108614145 | 108614173 | 29 | 2 | 0.000294683 | 0.00089 | 9.96E-05 | 0.00011 | -0.002092988 | LIMS1 |
| chr17 | 45145920 | 45145949 | 30 | 2 | 0.000294692 | 0.005016 | 0.00012 | 0.00021 | 0.013298026 | NA |
| chr14 | 91392921 | 91393063 | 143 | 2 | 0.000294818 | 4.22E-05 | 0.00014 | 3.28E-05 | -0.011481154 | CCDC88C |
| chr15 | 77641346 | 77641650 | 305 | 5 | 0.000294846 | 3.89E-05 | 0.0007 | 2.99E-05 | -0.005162569 | LINGO1 |
| chr12 | 6935624 | 6935659 | 36 | 2 | 0.000294982 | 4.91E-05 | 0.00015 | 3.74E-05 | -0.013418409 | ATN1 |
| chr1 | 36115484 | 36116150 | 667 | 2 | 0.000295059 | 1.24E-05 | 0.00057 | 1.88E-05 | -0.012252623 | COL8A2 |
| chr1 | 2233103 | 2233163 | 61 | 2 | 0.00029517 | 5.89E-05 | 0.00248 | 9.18E-05 | -0.00946851 | SKI |
| chr3 | 49555992 | 49556471 | 480 | 2 | 0.000295181 | 2.38E-05 | 0.00136 | 3.73E-05 | 0.002198409 | BSN |
| chr4 | 7541206 | 7541589 | 384 | 2 | 0.0002955 | 3.21E-05 | 0.00167 | 5.00E-05 | -0.007371859 | SORCS2 |
| chr10 | 80642697 | 80642906 | 210 | 2 | 0.000295575 | 2.73E-05 | 0.00037 | 3.30E-05 | -0.004915653 | SH2D4B |
| chr1 | 95233271 | 95233757 | 487 | 7 | 0.000295682 | 7.05E-06 | 0.01784 | 3.20E-05 | -0.016585806 | TLCD4-RWDD3, AC092802.1 |
| chr12 | 109116242 | 109116829 | 588 | 5 | 0.000295761 | 0.000103 | 0.00413 | 0.00014 | -0.009997428 | AC007637.1, ACACB |
| chr7 | 141660169 | 141660423 | 255 | 2 | 0.000296069 | 0.00323 | 5.79E-05 | 8.16E-05 | -0.006093993 | DENND11 |
| chr2 | 144332589 | 144333289 | 701 | 10 | 0.000296422 | 9.21E-05 | 0.00102 | 2.31E-05 | -0.001809629 | NA |
| chr19 | 40383941 | 40384207 | 267 | 4 | 0.000296422 | 0.05294 | 2.75E-05 | 0.00011 | -0.003331197 | HIPK4 |
| chr10 | 93612283 | 93612705 | 423 | 7 | 0.000296437 | 1.50E-05 | 0.00257 | 2.62E-05 | -0.001875863 | PDE6C |
| chr20 | 33523577 | 33524274 | 698 | 2 | 0.000296547 | 8.09E-06 | 0.0006 | 1.31E-05 | -0.017678528 | CBFA2T2 |
| chr19 | 50720482 | 50720725 | 244 | 3 | 0.000296593 | 2.96E-05 | 0.00194 | 4.89E-05 | -0.010336448 | NA |
| chr8 | 122985564 | 122986065 | 502 | 3 | 0.00029669 | 1.15E-05 | 0.00207 | 2.41E-05 | -0.011135973 | AC104316.2 |
| chr1 | 15445659 | 15446604 | 946 | 2 | 0.000296798 | 3.22E-06 | 0.00025 | 5.18E-06 | -0.016629308 | CTRC |
| chr1 | 25805402 | 25805692 | 291 | 2 | 0.000297328 | 0.000255 | 0.00037 | 0.00016 | -0.005401836 | SELENON |
| chr3 | 49166475 | 49167115 | 641 | 7 | 0.000297349 | 1.76E-05 | 0.00504 | 2.73E-05 | -0.005013377 | NA |
| chr17 | 63573366 | 63573450 | 85 | 2 | 0.000297349 | 3.80E-05 | 0.0011 | 5.48E-05 | -0.004978741 | DCAF7 |
| chr10 | 45374772 | 45374931 | 160 | 2 | 0.000297372 | 4.30E-05 | 0.00145 | 6.40E-05 | -0.030521123 | ALOX5 |
| chr20 | 10173338 | 10173396 | 59 | 3 | 0.000297509 | 9.90E-05 | 0.00016 | 4.89E-05 | -0.001141169 | AL354824.1, AL354824.2, SNAP25-AS1 |
| chr19 | 12401150 | 12401293 | 144 | 6 | 0.000297598 | 8.80E-05 | 0.00018 | 3.08E-05 | 0.00139944 | AC008758.4, ZNF799, AC008758.5 |
| chr4 | 48681368 | 48681370 | 3 | 2 | 0.000297671 | 1.85E-05 | 0.00054 | 2.62E-05 | -0.006320182 | FRYL |
| chr17 | 74843350 | 74843611 | 262 | 2 | 0.000297874 | 4.89E-05 | 0.00141 | 7.11E-05 | -0.013528221 | GRIN2C |
| chr5 | 65722119 | 65722773 | 655 | 8 | 0.000297987 | 0.061935 | 9.52E-05 | 0.00026 | 6.71E-05 | NLN, SGTB |
| chr21 | 34668879 | 34669035 | 157 | 2 | 0.000298146 | 0.000104 | 0.00305 | 0.00017 | -0.018306948 | CLIC6 |
| chr16 | 49827140 | 49827769 | 630 | 4 | 0.000298558 | 0.000394 | 0.00145 | 0.00018 | -0.004955559 | ZNF423 |
| chr11 | 131186177 | 131186440 | 264 | 2 | 0.000298568 | 0.001064 | 0.00084 | 0.0006 | -0.010121858 | NA |
| chr15 | 74317853 | 74318342 | 490 | 4 | 0.000298587 | 7.53E-05 | 0.00024 | 4.74E-05 | -0.007884403 | CCDC33, AC023300.2 |
| chr6 | 63319965 | 63320323 | 359 | 4 | 0.000298631 | 0.002103 | 2.92E-05 | 6.20E-05 | -0.005618743 | LGSN |
| chr17 | 72187589 | 72187685 | 97 | 2 | 0.000298788 | 0.000479 | 8.35E-05 | 7.39E-05 | -0.012147887 | AC007461.1, SOX9-AS1 |
| chr10 | 11741795 | 11742059 | 265 | 5 | 0.000298855 | 0.000429 | 0.00024 | 0.0001 | -0.008676334 | NA |
| chr5 | 83077853 | 83078125 | 273 | 4 | 0.000299265 | 0.000267 | 0.00128 | 0.00028 | 0.007523451 | XRCC4, TMEM167A |
| chr4 | 1509841 | 1511532 | 1692 | 8 | 0.000299354 | 6.58E-08 | 0.00671 | 7.14E-07 | -0.012794136 | NA |
| chr17 | 81133283 | 81133459 | 177 | 5 | 0.000299354 | 0.000221 | 0.00092 | 7.00E-05 | -0.002409772 | AATK, MIR1250 |
| chr7 | 100014906 | 100015150 | 245 | 3 | 0.000299527 | 0.000438 | 0.00291 | 0.00019 | -0.007228152 | NA |
| chr17 | 78891794 | 78891907 | 114 | 2 | 0.000299663 | 0.000133 | 0.00176 | 0.00019 | -0.007291529 | TIMP2, CEP295NL |
| chr4 | 79491886 | 79492335 | 450 | 2 | 0.000301316 | 2.79E-05 | 0.00048 | 3.57E-05 | -0.008513822 | LINC00989 |
| chr14 | 73251322 | 73251471 | 150 | 2 | 0.000301329 | 0.001552 | 6.39E-05 | 7.83E-05 | -0.004603591 | PAPLN |
| chr2 | 219224769 | 219225081 | 313 | 2 | 0.000301455 | 0.000113 | 0.00077 | 0.00013 | -0.003387873 | ATG9A |
| chr21 | 46266298 | 46267156 | 859 | 3 | 0.000301508 | 2.31E-06 | 0.00154 | 6.03E-06 | -0.003534913 | MCM3AP |
| chr17 | 47969294 | 47969702 | 409 | 2 | 0.000301708 | 7.06E-05 | 0.00186 | 0.0001 | 0.002397776 | AC018521.1, CDK5RAP3 |
| chr9 | 131609675 | 131609804 | 130 | 2 | 0.000301712 | 3.92E-05 | 0.00196 | 6.14E-05 | -0.00910995 | RAPGEF1 |
| chr11 | 61150954 | 61151114 | 161 | 2 | 0.00030196 | 3.05E-05 | 0.00158 | 4.75E-05 | -0.007886707 | VPS37C |
| chr17 | 81911530 | 81912121 | 592 | 13 | 0.000302027 | 0.000125 | 0.00698 | 8.79E-05 | -0.002919116 | SIRT7 |
| chr1 | 1134549 | 1134660 | 112 | 2 | 0.00030244 | 2.85E-05 | 0.00017 | 2.65E-05 | -0.008458808 | NA |
| chr9 | 91898289 | 91899052 | 764 | 2 | 0.000302532 | 1.34E-05 | 0.00098 | 2.15E-05 | -0.010685829 | ROR2 |
| chr13 | 27922276 | 27922675 | 400 | 3 | 0.00030321 | 0.001614 | 0.00725 | 0.00142 | -0.000320047 | PDX1 |
| chr4 | 760115 | 760150 | 36 | 2 | 0.00030345 | 0.000432 | 0.00014 | 0.00011 | -0.010140718 | PCGF3 |
| chr20 | 32359532 | 32359921 | 390 | 7 | 0.000303513 | 3.54E-05 | 0.00638 | 4.42E-05 | -0.001600351 | ASXL1 |
| chr6 | 169775060 | 169775350 | 291 | 2 | 0.000303787 | 0.002134 | 5.14E-05 | 6.63E-05 | -0.004841249 | ERMARD |
| chr15 | 75359480 | 75360169 | 690 | 4 | 0.000303841 | 1.07E-05 | 0.00089 | 1.41E-05 | -0.0008763 | MAN2C1 |
| chr17 | 35940069 | 35940107 | 39 | 2 | 0.000303857 | 0.00068 | 0.00013 | 0.00012 | -0.005477029 | LYZL6 |
| chr20 | 49630945 | 49631199 | 255 | 2 | 0.000303857 | 4.95E-05 | 0.00017 | 3.92E-05 | -0.012504376 | NA |
| chr9 | 131522912 | 131523307 | 396 | 2 | 0.000304309 | 2.63E-05 | 0.00054 | 3.50E-05 | -0.000912442 | POMT1 |
| chr13 | 113152085 | 113152448 | 364 | 2 | 0.000304342 | 0.000109 | 0.0004 | 9.50E-05 | -0.006767622 | AL137002.2 |
| chr1 | 99645831 | 99646393 | 563 | 6 | 0.000304675 | 0.000571 | 0.00223 | 0.00012 | 0.011467206 | PALMD |
| chr6 | 43129632 | 43129936 | 305 | 3 | 0.000304797 | 4.38E-05 | 0.00837 | 0.0001 | -0.008530475 | PTK7 |
| chr8 | 55142361 | 55142392 | 32 | 3 | 0.000304806 | 1.17E-05 | 0.00254 | 2.57E-05 | -0.012933134 | XKR4 |
| chr11 | 47409225 | 47409513 | 289 | 5 | 0.000305033 | 4.95E-05 | 0.01036 | 7.65E-05 | -0.004020794 | SLC39A13, AC090559.1 |
| chr18 | 13278008 | 13278119 | 112 | 2 | 0.000305066 | 2.06E-05 | 0.00068 | 2.99E-05 | 0.000666477 | LDLRAD4 |
| chr19 | 42200554 | 42200678 | 125 | 2 | 0.000305096 | 0.000254 | 0.00011 | 7.13E-05 | -0.008189054 | DEDD2 |
| chr7 | 101915854 | 101916328 | 475 | 8 | 0.00030517 | 0.156255 | 6.27E-05 | 0.00169 | -0.001931954 | CUX1 |
| chr19 | 2038365 | 2038601 | 237 | 3 | 0.000305421 | 0.006196 | 4.16E-05 | 0.0001 | -0.000848369 | MKNK2 |
| chr5 | 40735549 | 40736530 | 982 | 2 | 0.000305558 | 3.71E-06 | 0.00043 | 6.30E-06 | 0.0067505 | TTC33 |
| chr16 | 566212 | 566251 | 40 | 3 | 0.00030592 | 0.000301 | 0.00064 | 0.00017 | -0.004942289 | NA |
| chr3 | 12908235 | 12908421 | 187 | 4 | 0.000305951 | 0.000247 | 0.00012 | 6.77E-05 | -0.000617251 | IQSEC1 |
| chr11 | 67616074 | 67616569 | 496 | 7 | 0.000305977 | 1.18E-05 | 0.02442 | 5.86E-05 | 0.027514806 | DOC2GP |
| chr16 | 70417124 | 70418020 | 897 | 2 | 0.000306191 | 2.93E-06 | 0.00037 | 5.04E-06 | -0.012232564 | ST3GAL2 |
| chr3 | 14267771 | 14267824 | 54 | 2 | 0.000306199 | 3.11E-05 | 0.00023 | 3.19E-05 | -0.000146711 | NA |
| chr6 | 32459974 | 32460091 | 118 | 2 | 0.000306326 | 5.75E-05 | 0.00093 | 7.52E-05 | -0.064258337 | HLA-DRB9 |
| chr1 | 223141263 | 223141446 | 184 | 2 | 0.000306347 | 0.000193 | 9.44E-05 | 5.77E-05 | -0.006863428 | TLR5 |
| chr2 | 12006251 | 12006565 | 315 | 2 | 0.000306347 | 0.00245 | 0.00209 | 0.00169 | -0.006728924 | MIR3681HG |
| chr17 | 17809295 | 17809753 | 459 | 3 | 0.000306542 | 1.48E-05 | 0.00229 | 3.09E-05 | -0.005670055 | RAI1 |
| chr20 | 48372801 | 48372966 | 166 | 2 | 0.000306544 | 0.005435 | 5.11E-05 | 7.63E-05 | -0.00201357 | NA |
| chr10 | 3149078 | 3149414 | 337 | 2 | 0.000306724 | 9.37E-05 | 0.00034 | 7.99E-05 | -0.000813947 | PITRM1-AS1, PITRM1 |
| chr11 | 64317494 | 64318548 | 1055 | 20 | 0.00030685 | 0.000122 | 0.00678 | 9.01E-06 | 0.000277031 | PRDX5, TRMT112 |
| chr16 | 27341946 | 27342093 | 148 | 2 | 0.00030708 | 0.000214 | 0.00013 | 7.70E-05 | -0.006354552 | IL4R |
| chr2 | 222670365 | 222670436 | 72 | 2 | 0.000307159 | 0.000174 | 0.00023 | 9.88E-05 | -0.00607984 | NA |
| chr20 | 20216262 | 20216266 | 5 | 2 | 0.000307471 | 6.06E-05 | 0.0021 | 9.18E-05 | -0.022058458 | CFAP61 |
| chr3 | 179266811 | 179267070 | 260 | 4 | 0.000307593 | 3.16E-05 | 0.00349 | 6.66E-05 | -0.013305161 | KCNMB3 |
| chr7 | 50528583 | 50528839 | 257 | 2 | 0.000307593 | 1.98E-05 | 0.00071 | 2.91E-05 | -0.004836438 | FIGNL1, DDC |
| chr9 | 99182618 | 99183343 | 726 | 5 | 0.000307728 | 2.50E-05 | 0.00548 | 2.62E-05 | -0.004653994 | NA |
| chr7 | 2521342 | 2521482 | 141 | 3 | 0.000307947 | 1.64E-05 | 0.00292 | 3.49E-05 | -0.014484035 | LFNG |
| chr1 | 58718209 | 58718989 | 781 | 2 | 0.000307988 | 4.63E-06 | 0.00047 | 7.71E-06 | -0.014627115 | AL136985.3 |
| chr6 | 34234377 | 34234791 | 415 | 2 | 0.000308385 | 2.18E-05 | 0.00114 | 3.39E-05 | -0.009124618 | NA |
| chr8 | 133571902 | 133572449 | 548 | 7 | 0.00030859 | 8.48E-05 | 0.00471 | 7.00E-05 | 0.000565165 | ST3GAL1 |
| chr1 | 2251173 | 2251999 | 827 | 3 | 0.000308903 | 0.00244 | 0.00188 | 0.00036 | -0.005451349 | SKI |
| chr10 | 116324189 | 116325075 | 887 | 12 | 0.00030899 | 0.00038 | 0.00642 | 5.48E-05 | -0.005463977 | CCDC172 |
| chr17 | 17738902 | 17739385 | 484 | 3 | 0.000309167 | 0.000126 | 0.00128 | 0.00016 | -0.006273781 | RAI1 |
| chr14 | 20116103 | 20116740 | 638 | 3 | 0.000309375 | 7.04E-06 | 0.0014 | 1.51E-05 | -0.026807268 | OR4K17 |
| chr1 | 89631937 | 89632422 | 486 | 6 | 0.000309458 | 0.001068 | 0.00053 | 6.20E-05 | -0.005285621 | AC093423.2, LRRC8C-DT |
| chr15 | 80563116 | 80563243 | 128 | 5 | 0.000309659 | 8.67E-06 | 0.0034 | 2.33E-05 | -0.007161503 | ARNT2 |
| chr16 | 47723639 | 47723810 | 172 | 3 | 0.000309728 | 0.000136 | 8.35E-05 | 4.17E-05 | -0.005679618 | NA |
| chr2 | 128503985 | 128504226 | 242 | 2 | 0.000309818 | 0.000404 | 0.00013 | 9.85E-05 | -0.011421018 | NA |
| chr17 | 40307910 | 40308427 | 518 | 2 | 0.000309818 | 8.03E-05 | 0.00017 | 5.41E-05 | -0.015894003 | NA |
| chr2 | 47066246 | 47066631 | 386 | 3 | 0.00030983 | 6.74E-05 | 0.00147 | 5.35E-05 | -0.009204608 | TTC7A, STPG4, AC073283.3 |
| chr4 | 76421905 | 76421918 | 14 | 2 | 0.000310189 | 5.94E-05 | 0.00069 | 7.17E-05 | -0.006036262 | NA |
| chr19 | 50666403 | 50667099 | 697 | 5 | 0.000310584 | 0.000899 | 0.00088 | 0.00012 | 0.002221154 | SHANK1 |
| chr1 | 78044693 | 78044884 | 192 | 2 | 0.000310824 | 0.005343 | 0.00024 | 0.00047 | -0.003088081 | GIPC2 |
| chr10 | 100790294 | 100790910 | 617 | 2 | 0.000310948 | 1.57E-05 | 0.00064 | 2.36E-05 | -0.001993649 | PAX2 |
| chr10 | 61015216 | 61015300 | 85 | 2 | 0.000310995 | 0.000808 | 6.34E-05 | 6.60E-05 | -0.006487178 | NA |
| chr15 | 34336530 | 34336999 | 470 | 5 | 0.000311243 | 0.001469 | 0.00121 | 0.00014 | -0.006223515 | SLC12A6 |
| chr2 | 101272957 | 101273292 | 336 | 8 | 0.000311308 | 1.06E-05 | 0.00382 | 2.17E-05 | -0.002805588 | RNF149, SNORD89 |
| chr1 | 15352180 | 15352348 | 169 | 2 | 0.000311489 | 0.003092 | 5.44E-05 | 7.55E-05 | -0.004853938 | FHAD1 |
| chr12 | 80186630 | 80186635 | 6 | 2 | 0.000311576 | 0.000139 | 0.00014 | 6.77E-05 | -0.00656958 | OTOGL |
| chr9 | 127955002 | 127955139 | 138 | 2 | 0.000311695 | 8.98E-05 | 0.00267 | 0.00014 | -0.007546681 | FAM102A |
| chr12 | 108302785 | 108303165 | 381 | 3 | 0.000311979 | 0.000134 | 0.00048 | 3.26E-05 | -0.006756495 | CMKLR1 |
| chr4 | 73998879 | 73998992 | 114 | 3 | 0.000312054 | 0.002511 | 7.95E-05 | 3.86E-05 | -0.013717309 | AC097709.1 |
| chr9 | 130331578 | 130331895 | 318 | 2 | 0.000312054 | 4.45E-05 | 0.00024 | 4.15E-05 | -0.011408205 | HMCN2 |
| chr4 | 15132956 | 15132959 | 4 | 2 | 0.000312683 | 1.87E-05 | 0.00093 | 2.88E-05 | -0.025980046 | C1QTNF7-AS1 |
| chr18 | 28037568 | 28037977 | 410 | 2 | 0.000312692 | 0.017941 | 0.00018 | 0.00046 | -0.010061521 | CDH2 |
| chr3 | 141737452 | 141737463 | 12 | 2 | 0.000312861 | 2.44E-05 | 0.00083 | 3.56E-05 | 0.018531521 | NA |
| chr6 | 13364183 | 13364322 | 140 | 2 | 0.000312861 | 2.79E-05 | 0.00138 | 4.31E-05 | -0.005106143 | GFOD1 |
| chr10 | 68338663 | 68338814 | 152 | 2 | 0.000312904 | 2.94E-05 | 0.0003 | 3.30E-05 | -0.005425719 | HNRNPH3 |
| chr9 | 136722990 | 136723300 | 311 | 3 | 0.000312911 | 2.38E-05 | 0.00529 | 5.35E-05 | -0.007332605 | DIPK1B, SNHG7 |
| chr11 | 68796899 | 68797009 | 111 | 2 | 0.000312911 | 0.002591 | 4.95E-05 | 6.61E-05 | -0.007075001 | CPT1A |
| chr17 | 81901933 | 81902769 | 837 | 10 | 0.000313021 | 0.004933 | 0.00072 | 9.95E-05 | -0.003738693 | NPB, PCYT2 |
| chr17 | 73743200 | 73743387 | 188 | 3 | 0.000313024 | 1.76E-05 | 0.0015 | 2.75E-05 | -0.029978325 | AC125421.1 |

| | | | | | | | | | | |
|---|---|---|---|---|---|---|---|---|---|---|
| chr8 | 10828511 | 10828560 | 50 | 2 | 0.000313277 | 0.010256 | 4.22E-05 | 6.55E-05 | -0.005716859 | AC105001.2, PINX1 |
| chr8 | 8892409 | 8893007 | 599 | 4 | 0.000313454 | 5.50E-05 | 0.00233 | 9.62E-05 | -0.005732205 | MFHAS1 |
| chr4 | 39044196 | 39044572 | 377 | 5 | 0.000313534 | 0.000251 | 0.00012 | 4.51E-05 | 0.001807654 | NA |
| chr16 | 2026056 | 2027208 | 1153 | 9 | 0.000314357 | 5.70E-05 | 0.00788 | 5.98E-05 | 0.000326036 | SLC9A3R2 |
| chr7 | 5607701 | 5608289 | 589 | 5 | 0.000314525 | 5.25E-05 | 0.00749 | 0.00012 | -0.003107766 | NA |
| chr19 | 41440696 | 41441281 | 586 | 4 | 0.000314544 | 8.10E-05 | 0.00274 | 0.00011 | -0.006105415 | DMAC2 |
| chr17 | 75897343 | 75897936 | 594 | 5 | 0.000314716 | 0.004948 | 0.00043 | 0.00067 | -0.004321375 | AC087289.2 |
| chr10 | 48915158 | 48915299 | 142 | 2 | 0.000314788 | 0.003448 | 4.13E-05 | 5.64E-05 | 0.003706007 | WDFY4, AC060234.3 |
| chr15 | 68230328 | 68230737 | 410 | 4 | 0.000314993 | 0.00014 | 0.00196 | 0.00016 | -0.008195598 | CLN6 |
| chr14 | 72559580 | 72559779 | 200 | 2 | 0.000315076 | 5.07E-05 | 0.00062 | 6.16E-05 | -0.009547639 | RGS6, AC004828.1 |
| chr5 | 141870110 | 141870255 | 146 | 2 | 0.000315238 | 0.000323 | 0.00078 | 0.00027 | -0.008400987 | PCDH1 |
| chr17 | 77884187 | 77884460 | 274 | 3 | 0.000315881 | 8.29E-05 | 0.00086 | 9.53E-05 | -0.006680205 | NA |
| chr10 | 29878163 | 29878273 | 111 | 2 | 0.000316075 | 4.86E-05 | 0.00012 | 3.28E-05 | -0.007204885 | NA |
| chr11 | 71179564 | 71179832 | 269 | 2 | 0.000316105 | 7.28E-05 | 0.00014 | 4.53E-05 | -0.002440159 | SHANK2 |
| chr13 | 98044576 | 98044606 | 31 | 2 | 0.000316171 | 1.76E-05 | 0.00087 | 2.72E-05 | -0.014612836 | NA |
| chr19 | 3669202 | 3670248 | 1047 | 4 | 0.000316199 | 6.47E-05 | 0.00018 | 8.48E-06 | -0.008031607 | PIP5K1C |
| chr10 | 11170947 | 11171433 | 487 | 3 | 0.000316479 | 2.11E-05 | 0.00166 | 3.26E-05 | 0.002799717 | CELF2, AL136320.1 |
| chr11 | 35585394 | 35585550 | 157 | 3 | 0.000316803 | 8.12E-06 | 0.00181 | 1.76E-05 | -0.016612094 | NA |
| chr1 | 160801906 | 160802208 | 303 | 3 | 0.000316911 | 1.69E-05 | 0.0017 | 3.24E-05 | -0.001065008 | LY9 |
| chr17 | 57432562 | 57432681 | 120 | 2 | 0.000316932 | 4.22E-05 | 0.00025 | 4.09E-05 | -0.008587966 | MSI2 |
| chr1 | 84690041 | 84690883 | 843 | 10 | 0.000317058 | 0.012122 | 9.09E-05 | 2.55E-05 | 0.000135569 | SSX2IP |
| chr20 | 19210501 | 19210882 | 382 | 2 | 0.000317058 | 0.000127 | 0.00169 | 0.00018 | -0.010633472 | AL136090.2 |
| chr17 | 27293361 | 27293924 | 564 | 6 | 0.000317181 | 0.003208 | 0.00076 | 0.00024 | -0.003355032 | MIR4522 |
| chr1 | 8944656 | 8945415 | 760 | 4 | 0.000317188 | 0.001816 | 0.00117 | 0.00083 | -0.001875218 | NA |
| chr2 | 7004455 | 7004809 | 355 | 3 | 0.000317274 | 0.000686 | 0.00031 | 7.28E-05 | -0.030537148 | RNF144A |
| chr10 | 129841051 | 129841742 | 692 | 3 | 0.000317373 | 0.086063 | 0.15506 | 0.13438 | -0.000666994 | EBF3 |
| chr2 | 69893804 | 69894556 | 753 | 12 | 0.000317622 | 0.000103 | 0.00668 | 3.34E-05 | 0.003263723 | SNRNP27 |
| chr3 | 150520421 | 150520676 | 256 | 3 | 0.000317695 | 3.31E-05 | 0.00164 | 4.02E-05 | -0.010700769 | NA |
| chr14 | 69797094 | 69797326 | 233 | 3 | 0.000317746 | 4.33E-05 | 0.00065 | 4.53E-05 | -0.006282813 | SLC10A1 |
| chr3 | 45011038 | 45011397 | 360 | 2 | 0.000318119 | 3.35E-05 | 0.00136 | 5.04E-05 | -0.010786993 | EXOSC7, CLEC3B |
| chr12 | 132493191 | 132493496 | 306 | 2 | 0.000318148 | 7.48E-05 | 0.00225 | 0.00011 | -0.004108624 | FBRSL1 |
| chr13 | 90119911 | 90120151 | 241 | 2 | 0.000318403 | 0.000202 | 8.49E-05 | 5.46E-05 | -0.018329722 | NA |
| chr1 | 19013883 | 19014151 | 269 | 2 | 0.000318866 | 0.000371 | 8.55E-05 | 6.91E-05 | -0.004345801 | NA |
| chr13 | 91398229 | 91398900 | 672 | 11 | 0.000318942 | 0.002951 | 0.00011 | 0.00015 | 0.002466888 | GPC5 |
| chr6 | 167734080 | 167734487 | 408 | 3 | 0.000319059 | 0.001498 | 0.00012 | 5.61E-05 | -0.012637932 | NA |
| chr17 | 82983596 | 82983682 | 87 | 3 | 0.000319136 | 1.53E-05 | 0.00101 | 2.54E-05 | -0.007000718 | B3GNTL1 |
| chr19 | 1019200 | 1019363 | 164 | 2 | 0.00031915 | 1.97E-05 | 0.00126 | 3.15E-05 | -0.010041823 | TMEM259 |
| chr19 | 41429248 | 41429562 | 315 | 3 | 0.000319413 | 0.000107 | 0.00081 | 3.67E-05 | -0.001082711 | NA |
| chr4 | 8198534 | 8198822 | 289 | 3 | 0.00031943 | 3.06E-05 | 0.00278 | 5.75E-05 | -0.008583709 | SH3TC1 |
| chr9 | 128684925 | 128685362 | 438 | 4 | 0.000319587 | 5.33E-05 | 0.00732 | 0.00011 | -0.00517899 | SET |
| chr6 | 166870347 | 166870865 | 519 | 2 | 0.000319984 | 0.01143 | 0.00739 | 0.00836 | -0.001368085 | RPS6KA2, AL159163.1 |
| chr8 | 69466280 | 69466525 | 246 | 5 | 0.000320059 | 0.000266 | 0.00021 | 2.49E-05 | 0.007642997 | NA |
| chr6 | 158233843 | 158233972 | 130 | 2 | 0.000320147 | 3.63E-05 | 0.00145 | 5.48E-05 | -0.010001017 | TULP4 |
| chr1 | 61052135 | 61052204 | 70 | 2 | 0.000320315 | 3.68E-05 | 0.00184 | 5.74E-05 | -0.03160229 | NFIA, NFIA-AS2 |
| chr3 | 76419195 | 76419353 | 159 | 2 | 0.000320706 | 4.47E-05 | 0.0017 | 6.78E-05 | -0.007810223 | ROBO2 |
| chr19 | 42374595 | 42374661 | 67 | 2 | 0.000320875 | 2.88E-05 | 0.00161 | 4.53E-05 | -0.004624288 | MEGF8 |
| chr1 | 151459373 | 151459910 | 538 | 5 | 0.000320953 | 0.001636 | 0.00145 | 5.79E-05 | -0.000509829 | POGZ |
| chr19 | 14487529 | 14487616 | 88 | 3 | 0.000321099 | 0.000412 | 0.00059 | 4.65E-05 | 0.004286198 | GIPC1 |
| chr20 | 3734614 | 3734663 | 50 | 2 | 0.000321104 | 4.52E-05 | 0.00099 | 6.22E-05 | -0.005169777 | HSPA12B |
| chr10 | 104301091 | 104301293 | 203 | 2 | 0.000321484 | 3.77E-05 | 0.00192 | 5.90E-05 | 0.001031748 | GSTO2 |
| chr10 | 102658220 | 102658550 | 331 | 3 | 0.000321554 | 0.000294 | 0.00014 | 7.80E-05 | -0.004657146 | TRIM8 |
| chr19 | 49661216 | 49661231 | 16 | 2 | 0.000321756 | 2.86E-05 | 0.00055 | 3.76E-05 | 0.002107139 | IRF3 |
| chr1 | 234697279 | 234697566 | 288 | 2 | 0.000322071 | 2.26E-05 | 0.0008 | 3.32E-05 | -0.000941726 | NA |
| chr3 | 114384220 | 114384498 | 279 | 3 | 0.000322492 | 0.000867 | 0.00062 | 0.00043 | -0.013105157 | ZBTB20-AS1, ZBTB20 |
| chr11 | 119380386 | 119380573 | 188 | 3 | 0.000322963 | 0.006606 | 8.60E-05 | 0.00015 | -0.005829607 | USP2-AS1, AP003396.3, USP2 |
| chr10 | 48508903 | 48509747 | 845 | 3 | 0.000323025 | 1.94E-06 | 0.00141 | 4.95E-06 | -0.009891899 | ARHGAP22 |
| chr22 | 31272364 | 31273221 | 858 | 5 | 0.000323044 | 0.00081 | 0.00016 | 8.78E-05 | -0.004601255 | LIMK2 |
| chr9 | 882695 | 882740 | 46 | 2 | 0.000323129 | 5.64E-05 | 0.00011 | 3.44E-05 | -0.015930447 | DMRT1 |
| chr2 | 232876929 | 232877210 | 282 | 3 | 0.000323259 | 0.000124 | 0.00051 | 8.05E-05 | -0.009387563 | SNORC |
| chr1 | 25564047 | 25564105 | 59 | 2 | 0.000323259 | 0.000446 | 0.00174 | 0.00049 | -0.007088005 | LDLRAP1 |
| chr10 | 14557024 | 14557291 | 268 | 2 | 0.000323329 | 0.02291 | 0.00025 | 0.0007 | -0.00555231 | FAM107B |
| chr11 | 123483723 | 123483938 | 216 | 2 | 0.000323343 | 1.71E-05 | 0.00115 | 2.74E-05 | -0.022266864 | GRAMD1B |
| chr1 | 56831617 | 56832037 | 421 | 3 | 0.000323671 | 2.32E-05 | 0.00126 | 3.55E-05 | -0.007591206 | NA |
| chr3 | 12978149 | 12978424 | 276 | 2 | 0.000323884 | 8.10E-05 | 0.00014 | 4.79E-05 | -0.003183805 | IQSEC1 |
| chr19 | 43414716 | 43415384 | 669 | 8 | 0.00032395 | 7.01E-06 | 0.01629 | 2.95E-05 | -0.002002972 | TEX101 |
| chr1 | 8327097 | 8327436 | 340 | 2 | 0.000324064 | 8.22E-05 | 0.00099 | 0.0001 | -0.006175375 | SLC45A1 |
| chr17 | 28405818 | 28406050 | 233 | 4 | 0.000324383 | 0.389419 | 0.00018 | 0.00439 | 0.001317333 | AC015917.2, SLC46A1 |
| chr3 | 23916999 | 23917583 | 585 | 13 | 0.000324839 | 0.000176 | 0.00532 | 3.37E-05 | 0.000373441 | RPL15, NKIRAS1 |
| chr15 | 100530357 | 100530454 | 98 | 2 | 0.000325292 | 0.006163 | 7.73E-05 | 0.00013 | -0.006677504 | CERS3 |
| chr16 | 83948831 | 83949040 | 210 | 2 | 0.0003253 | 6.88E-05 | 0.00105 | 8.95E-05 | 0.000219966 | MLYCD, OSGIN1 |
| chr4 | 186726043 | 186726536 | 494 | 6 | 0.000325478 | 4.48E-05 | 0.00305 | 5.54E-05 | 0.00077983 | FAT1 |
| chr2 | 172821320 | 172821949 | 630 | 5 | 0.000325493 | 0.000793 | 0.00013 | 0.00011 | -0.003352216 | RAPGEF4 |
| chr15 | 58310403 | 58310738 | 336 | 3 | 0.000325493 | 0.000167 | 0.00143 | 5.98E-05 | -0.008780831 | ALDH1A2 |
| chr11 | 64185927 | 64186215 | 289 | 8 | 0.000325662 | 0.012903 | 8.07E-05 | 0.00055 | -0.001524183 | STIP1 |
| chr17 | 15256902 | 15257088 | 187 | 3 | 0.000325831 | 0.000303 | 0.0004 | 0.00011 | -0.011232689 | PMP22 |
| chr7 | 44045019 | 44045287 | 269 | 2 | 0.000325969 | 0.013687 | 0.00058 | 0.00139 | 0.002275866 | DBNL |
| chr20 | 42980773 | 42981090 | 318 | 2 | 0.000326057 | 0.00018 | 0.00193 | 0.00025 | -0.006705567 | PTPRT |
| chr5 | 140345508 | 140345665 | 158 | 2 | 0.000326202 | 0.000375 | 0.00011 | 8.73E-05 | 0.000140091 | HBEGF |
| chr4 | 15938239 | 15938715 | 477 | 6 | 0.000326285 | 6.51E-05 | 0.01301 | 4.27E-05 | -0.008631037 | FGFBP1 |
| chr3 | 184142466 | 184142642 | 177 | 2 | 0.0003265 | 7.49E-05 | 0.00027 | 6.24E-05 | -0.003194607 | EIF2B5 |
| chr1 | 11805585 | 11806359 | 775 | 15 | 0.00032662 | 2.76E-05 | 0.00245 | 4.38E-05 | 0.00085707 | CLCN6, MTHFR |
| chr12 | 108827631 | 108827854 | 224 | 6 | 0.00032662 | 1.63E-05 | 0.00833 | 3.82E-05 | -0.003199555 | SSH1 |
| chr2 | 37648508 | 37648940 | 433 | 4 | 0.000326847 | 1.65E-05 | 0.00154 | 2.51E-05 | 0.003021281 | CDC42EP3 |
| chr6 | 90537940 | 90538654 | 715 | 2 | 0.000326847 | 6.13E-05 | 0.00098 | 8.01E-05 | -0.013331348 | MAP3K7 |
| chr19 | 17027241 | 17028048 | 808 | 5 | 0.000327004 | 3.50E-06 | 0.0054 | 7.71E-06 | -0.00730137 | NA |
| chr3 | 169668195 | 169668441 | 247 | 3 | 0.000327089 | 0.00012 | 0.00021 | 6.43E-05 | 0.002014436 | NA |
| chr10 | 11831963 | 11832285 | 323 | 2 | 0.000327207 | 2.97E-05 | 0.00043 | 3.66E-05 | -0.015079067 | PROSER2 |
| chr10 | 62057086 | 62057151 | 66 | 2 | 0.000327349 | 0.001376 | 4.85E-05 | 5.75E-05 | -0.003915696 | ARID5B |
| chr7 | 826465 | 826818 | 354 | 4 | 0.000327431 | 6.60E-05 | 0.00064 | 3.49E-05 | -0.005278234 | SUN1 |
| chr13 | 50387292 | 50387535 | 244 | 2 | 0.000327474 | 0.00289 | 7.36E-05 | 0.00011 | -0.008147397 | DLEU1 |
| chr11 | 36156149 | 36156352 | 204 | 2 | 0.000327754 | 0.004786 | 5.46E-05 | 8.10E-05 | -0.006711291 | LDLRAD3 |
| chr3 | 184275783 | 184276285 | 503 | 8 | 0.000327861 | 5.25E-05 | 0.00112 | 2.03E-05 | -0.004523193 | EEF1AKMT4-ECE2, ECE2 |
| chr7 | 100463988 | 100464440 | 453 | 3 | 0.000327951 | 1.29E-05 | 0.00341 | 2.92E-05 | -0.004924287 | C7orf61, TSC22D4 |
| chr5 | 61394919 | 61394929 | 11 | 2 | 0.000328067 | 0.000326 | 6.28E-05 | 4.95E-05 | 0.002224894 | ZSWIM6 |
| chr10 | 7578583 | 7578741 | 159 | 2 | 0.000328073 | 3.67E-05 | 0.00035 | 4.07E-05 | -0.008832765 | ITIH5 |
| chr13 | 111548294 | 111548689 | 396 | 6 | 0.000328222 | 0.004663 | 5.48E-05 | 0.00012 | -0.002400075 | NA |
| chr11 | 119340730 | 119341537 | 808 | 7 | 0.000328594 | 0.01515 | 0.00165 | 0.00024 | -0.004867318 | C1QTNF5, MFRP |
| chr20 | 19885026 | 19885088 | 63 | 2 | 0.00032864 | 0.000196 | 0.00264 | 0.0003 | -0.026120593 | RIN2 |
| chr10 | 60817576 | 60817870 | 295 | 2 | 0.000328665 | 0.000157 | 0.00015 | 7.29E-05 | -0.004533821 | NA |
| chr12 | 101788656 | 101788922 | 267 | 2 | 0.000328722 | 0.000706 | 9.95E-05 | 9.78E-05 | 0.003639807 | GNPTAB |
| chr8 | 81153499 | 81153549 | 51 | 2 | 0.000328763 | 0.009118 | 3.72E-05 | 5.57E-05 | -0.005659811 | AC079209.1 |
| chr14 | 100873690 | 100875274 | 1585 | 11 | 0.000328929 | 1.84E-05 | 0.00141 | 1.62E-06 | -0.001048679 | AL117190.1, MIR337, MIR665 |
| chr19 | 10718252 | 10719127 | 876 | 14 | 0.00032896 | 0.003516 | 0.00087 | 0.00022 | -0.00056661 | DNM2, MIR638 |
| chr15 | 45630725 | 45630925 | 201 | 2 | 0.000329597 | 4.39E-05 | 0.00057 | 5.36E-05 | -0.007483685 | AC090527.2 |
| chr3 | 143314443 | 143314736 | 294 | 4 | 0.000329953 | 0.000338 | 0.00029 | 5.11E-05 | -0.004999498 | SLC9A9 |
| chr1 | 21176820 | 21177628 | 809 | 12 | 0.000330077 | 0.000251 | 0.00255 | 3.60E-05 | -0.000881866 | AL031005.2, EIF4G3 |
| chr22 | 38569406 | 38569929 | 524 | 6 | 0.000330077 | 3.05E-05 | 0.00241 | 5.75E-05 | -0.00946312 | DMC1 |
| chr1 | 42817448 | 42817760 | 313 | 6 | 0.000330205 | 0.000192 | 0.00163 | 4.41E-05 | -0.00062037 | ERMAP |
| chr2 | 85710786 | 85711157 | 372 | 2 | 0.000330381 | 4.67E-05 | 0.0003 | 4.64E-05 | -0.018921502 | NA |
| chr22 | 40388321 | 40388941 | 621 | 5 | 0.000330627 | 0.029662 | 0.00204 | 0.00062 | -0.006565807 | ADSL, AL022238.3, SGSM3 |
| chr19 | 6522627 | 6522722 | 96 | 2 | 0.000331005 | 1.82E-05 | 0.00097 | 2.84E-05 | -0.010592634 | NA |
| chr5 | 151023905 | 151023914 | 10 | 2 | 0.000331522 | 1.66E-05 | 0.00095 | 2.61E-05 | -0.011067873 | GPX3 |
| chr1 | 11658118 | 11658402 | 285 | 2 | 0.000331642 | 3.68E-05 | 0.0003 | 3.89E-05 | -0.006886401 | FBXO44 |
| chr12 | 132314967 | 132315120 | 154 | 2 | 0.000331733 | 2.31E-05 | 0.00132 | 3.64E-05 | -0.008980822 | GALNT9 |
| chr10 | 133281799 | 133281978 | 180 | 2 | 0.000332132 | 0.01781 | 0.00084 | 0.00208 | -0.00169003 | TUBGCP2 |
| chr11 | 64150235 | 64150626 | 392 | 2 | 0.000332346 | 3.80E-05 | 0.00048 | 4.55E-05 | -0.026915994 | MACROD1 |
| chr16 | 68698201 | 68698671 | 471 | 4 | 0.000332688 | 0.000125 | 0.00434 | 4.61E-05 | -0.002772565 | CDH3 |
| chr2 | 241196704 | 241196769 | 66 | 2 | 0.000333402 | 2.87E-05 | 0.0014 | 4.44E-05 | -0.007244933 | ANO7 |
| chr16 | 1180935 | 1181723 | 789 | 4 | 0.00033398 | 0.000636 | 0.00282 | 0.00027 | -0.001180838 | CACNA1H |
| chr11 | 68761599 | 68761629 | 31 | 2 | 0.00033398 | 3.37E-05 | 0.0004 | 3.96E-05 | -0.003401364 | CPT1A |
| chr12 | 47819822 | 47820030 | 209 | 4 | 0.000334596 | 0.000169 | 0.00042 | 5.11E-05 | 0.002644693 | HDAC7 |
| chr19 | 42074753 | 42074922 | 170 | 4 | 0.00033474 | 5.90E-05 | 0.00023 | 4.24E-05 | -0.002533767 | ZNF574 |
| chr11 | 5872508 | 5872897 | 390 | 2 | 0.000334999 | 0.00015 | 0.0004 | 0.00012 | -0.014422988 | TRIM5 |
| chr4 | 1797978 | 1798721 | 744 | 5 | 0.000335701 | 0.07509 | 0.00057 | 0.00187 | -0.001738689 | FGFR3 |
| chr3 | 194071476 | 194072296 | 821 | 6 | 0.000335706 | 0.001024 | 0.00053 | 5.62E-05 | -0.007638364 | AC024559.2 |
| chr18 | 22559479 | 22559679 | 201 | 2 | 0.000335781 | 0.000103 | 0.00037 | 8.85E-05 | 0.003519804 | NA |
| chr19 | 6712346 | 6712582 | 237 | 6 | 0.000336063 | 0.000932 | 0.00038 | 5.17E-05 | -0.002408099 | C3 |
| chr5 | 6765315 | 6765873 | 559 | 5 | 0.000336111 | 1.64E-05 | 0.00295 | 3.87E-05 | -0.006519177 | NA |
| chr9 | 129822471 | 129822664 | 194 | 2 | 0.00033677 | 0.000248 | 8.19E-05 | 5.73E-05 | -0.007947589 | TOR1A |
| chr1 | 204141631 | 204142125 | 495 | 5 | 0.00033678 | 0.001721 | 0.00035 | 9.70E-05 | -0.00163503 | ERLNC1, ETNK2 |
| chr5 | 1438260 | 1438416 | 157 | 3 | 0.0003368 | 0.00027 | 0.00056 | 0.00016 | -0.005475335 | SLC6A3 |
| chr20 | 36091743 | 36091940 | 198 | 3 | 0.000336828 | 0.000149 | 0.00021 | 6.11E-05 | -0.00558375 | EPB41L1 |
| chr6 | 155154426 | 155154519 | 94 | 3 | 0.000336977 | 8.34E-05 | 0.00031 | 5.97E-05 | -0.012604431 | TIAM2 |
| chr10 | 807213 | 807385 | 173 | 3 | 0.000337302 | 4.21E-05 | 0.00651 | 9.34E-05 | -0.001418101 | LARP4B |
| chr13 | 94443683 | 94443846 | 164 | 3 | 0.000337489 | 1.50E-05 | 0.00141 | 2.70E-05 | -0.005670837 | DCT |
| chr8 | 23458581 | 23458945 | 365 | 3 | 0.00033749 | 0.000233 | 0.00043 | 0.00011 | -0.006674872 | AC104561.1 |
| chr1 | 93448264 | 93448495 | 232 | 2 | 0.000337562 | 9.02E-05 | 0.00153 | 0.00013 | -0.000254421 | FNBP1L |
| chr20 | 31484110 | 31484315 | 206 | 2 | 0.000337621 | 4.04E-05 | 0.00174 | 6.20E-05 | -0.016536189 | REM1 |
| chr16 | 424271 | 424528 | 258 | 4 | 0.000337838 | 0.000139 | 0.00238 | 0.00014 | -0.01476298 | AL023881.1 |
| chr20 | 38164920 | 38165926 | 1007 | 12 | 0.000338055 | 0.000212 | 0.00115 | 4.33E-05 | -0.002887754 | TGM2 |
| chr10 | 73415091 | 73415378 | 288 | 2 | 0.000338065 | 0.017318 | 0.00038 | 0.00098 | -0.003677517 | NA |
| chr10 | 1468829 | 1469082 | 254 | 4 | 0.000338144 | 9.44E-05 | 0.00019 | 4.35E-05 | -0.015700887 | ADARB2 |
| chr2 | 101617245 | 101617956 | 712 | 2 | 0.000338333 | 8.76E-06 | 0.00061 | 1.41E-05 | -0.013186666 | NA |
| chr12 | 68334497 | 68335265 | 769 | 2 | 0.000338475 | 9.22E-06 | 0.00078 | 1.52E-05 | -0.012516435 | LINC02384 |
| chr13 | 111961429 | 111961458 | 30 | 2 | 0.000338477 | 3.99E-05 | 0.00048 | 4.75E-05 | -6.77E-05 | NA |
| chr2 | 190562154 | 190562345 | 192 | 2 | 0.000338855 | 3.23E-05 | 0.00052 | 4.09E-05 | -0.008700404 | AC108047.1 |
| chr8 | 144082852 | 144083177 | 326 | 5 | 0.000339517 | 0.000754 | 0.00063 | 0.00015 | 0.002271657 | GPAA1 |
| chr1 | 17745337 | 17745347 | 11 | 2 | 0.00033958 | 2.10E-05 | 0.00059 | 2.96E-05 | -0.02056424 | LINC02810 |
| chr7 | 97013482 | 97013848 | 367 | 2 | 0.000339646 | 0.000276 | 0.00011 | 7.39E-05 | 0.017415212 | DLX6-AS1 |
| chr16 | 72198203 | 72198745 | 543 | 3 | 0.000339653 | 1.59E-05 | 0.00152 | 2.97E-05 | -0.002543841 | NA |
| chr2 | 47218131 | 47218494 | 364 | 2 | 0.000339779 | 1.85E-05 | 0.00104 | 2.90E-05 | -0.013345105 | EPCAM-DT |
| chr4 | 4694531 | 4694590 | 60 | 2 | 0.000339937 | 0.007398 | 0.00022 | 0.00047 | -0.004250172 | STX18-AS1 |
| chr5 | 111757148 | 111757589 | 442 | 6 | 0.000339952 | 2.97E-05 | 0.01968 | 0.00011 | 0.001291435 | NREP |
| chr1 | 84960879 | 84960903 | 25 | 2 | 0.000340377 | 2.63E-05 | 0.00032 | 3.09E-05 | -0.005236249 | MCOLN2 |
| chr20 | 59251400 | 59251411 | 12 | 2 | 0.000340484 | 0.001758 | 6.09E-05 | 7.66E-05 | -0.003233787 | ZNF831 |
| chr1 | 109620536 | 109620908 | 373 | 8 | 0.000340664 | 1.37E-05 | 0.0056 | 2.58E-05 | -0.002857993 | AMPD2 |
| chr17 | 81036658 | 81037168 | 511 | 4 | 0.000340758 | 0.000187 | 0.00146 | 9.42E-05 | -0.009756922 | BAIAP2 |
| chr19 | 1065185 | 1066350 | 1166 | 10 | 0.000340764 | 7.25E-05 | 0.00526 | 4.90E-05 | -0.00459836 | ABCA7, ARHGAP45 |
| chr21 | 30634257 | 30634452 | 196 | 4 | 0.000340946 | 0.003656 | 0.0004 | 8.01E-05 | 0.005864505 | NA |
| chr6 | 43702782 | 43703175 | 394 | 3 | 0.000341024 | 2.18E-05 | 0.00305 | 4.32E-05 | -0.01060237 | NA |
| chr1 | 156381191 | 156381942 | 752 | 4 | 0.000341142 | 6.52E-06 | 0.00101 | 1.07E-05 | -0.005162049 | RHBG |
| chr8 | 94266464 | 94266501 | 38 | 2 | 0.000341192 | 8.81E-05 | 0.00014 | 5.08E-05 | -0.010753992 | NA |

| | | | | | | | | | | |
|---|---|---|---|---|---|---|---|---|---|---|
| chr1 | 60973133 | 60973669 | 537 | 3 | 0.00034133 | 1.50E-05 | 0.0022 | 3.09E-05 | -0.011987643 | NFIA, NFIA-AS2 |
| chr8 | 1892628 | 1892899 | 272 | 2 | 0.000341431 | 0.000136 | 0.00011 | 5.66E-05 | -0.004917696 | AC019257.8, ARHGEF10 |
| chr6 | 30213804 | 30214311 | 508 | 16 | 0.00034148 | 9.84E-06 | 0.01288 | 5.53E-06 | 0.000708071 | NA |
| chr6 | 79538061 | 79538731 | 671 | 3 | 0.000341542 | 0.001453 | 0.00597 | 0.002 | 0.0144916 | AL391840.3 |
| chr11 | 61022919 | 61022946 | 28 | 2 | 0.000341878 | 0.000129 | 0.00321 | 0.00021 | -0.011148567 | NA |
| chr1 | 84505328 | 84505830 | 503 | 6 | 0.000342204 | 0.001747 | 0.0008 | 0.00043 | 0.00093371 | GNG5, AL359762.3 |
| chr10 | 112183639 | 112184356 | 718 | 11 | 0.000342263 | 0.000141 | 0.00134 | 4.42E-05 | -0.001634585 | GPAM |
| chr5 | 139482247 | 139482571 | 325 | 7 | 0.000342399 | 0.000117 | 0.00774 | 7.72E-05 | 0.00155427 | STING1 |
| chr2 | 23630314 | 23630362 | 49 | 2 | 0.000342437 | 2.73E-05 | 0.00138 | 4.24E-05 | -0.004083046 | KLHL29 |
| chr9 | 90642955 | 90643530 | 576 | 6 | 0.000342726 | 0.000135 | 0.00064 | 1.07E-05 | 0.00391523 | NA |
| chr2 | 101830807 | 101831044 | 238 | 3 | 0.000342789 | 5.08E-05 | 0.00135 | 4.21E-05 | -0.002875789 | MAP4K4 |
| chr18 | 73235536 | 73235613 | 78 | 3 | 0.00034299 | 0.000452 | 7.78E-05 | 5.85E-05 | -0.006005432 | LINC02864 |
| chr6 | 44228469 | 44228675 | 207 | 2 | 0.000343099 | 0.001234 | 0.00055 | 0.0005 | -0.00333137 | SLC29A1 |
| chr1 | 10429905 | 10430179 | 275 | 7 | 0.000343432 | 0.0005 | 0.02551 | 0.00031 | -0.007322543 | CENPS-CORT, AL139424.1 |
| chr16 | 68264601 | 68265075 | 475 | 3 | 0.000343489 | 0.003883 | 0.00036 | 0.00068 | -0.000189067 | SLC7A6 |
| chr11 | 45722049 | 45722386 | 338 | 7 | 0.000343604 | 0.000417 | 7.66E-05 | 8.48E-05 | -0.005920687 | AC044839.1 |
| chr11 | 104059770 | 104060116 | 347 | 2 | 0.000343724 | 2.99E-05 | 0.0005 | 3.79E-05 | -0.025728255 | PDGFD |
| chr7 | 193229 | 193732 | 504 | 5 | 0.000344772 | 0.010076 | 0.00015 | 0.0001 | 0.001375543 | FAM20C, AC093627.3 |
| chr19 | 45273184 | 45273318 | 135 | 2 | 0.000345 | 0.000117 | 0.00013 | 5.79E-05 | -0.009059225 | MARK4 |
| chr17 | 77602168 | 77602283 | 116 | 2 | 0.000345047 | 3.17E-05 | 0.00068 | 4.27E-05 | -0.01053511 | AC021683.5 |
| chr2 | 241632927 | 241633529 | 603 | 6 | 0.000345199 | 0.000131 | 0.00384 | 3.69E-05 | -0.002020455 | THAP4 |
| chr1 | 230980125 | 230980251 | 127 | 2 | 0.00034545 | 0.000629 | 0.00077 | 0.00042 | -0.010881449 | ARV1 |
| chr5 | 178984786 | 178984971 | 186 | 2 | 0.00034552 | 0.000132 | 0.00049 | 0.00012 | -0.004151469 | AC104117.3, GRM6 |
| chr19 | 48394606 | 48394903 | 298 | 6 | 0.000345568 | 0.000614 | 0.00111 | 8.95E-05 | -0.000213561 | GRIN2D |
| chr5 | 150137529 | 150138205 | 677 | 2 | 0.000345636 | 1.14E-05 | 0.00066 | 1.79E-05 | -0.010869609 | PDGFRB |
| chr19 | 35874612 | 35874898 | 287 | 4 | 0.00034588 | 0.000438 | 0.0031 | 5.04E-05 | -0.003749875 | APLP1 |
| chr2 | 239246587 | 239246717 | 131 | 2 | 0.00034635 | 0.000329 | 0.00195 | 0.00041 | -0.013163342 | HDAC4 |
| chr20 | 48807773 | 48807796 | 24 | 2 | 0.000346979 | 2.50E-05 | 0.00085 | 3.65E-05 | -0.003408547 | PREX1 |
| chr22 | 37661178 | 37661566 | 389 | 2 | 0.00034708 | 2.78E-05 | 0.00075 | 3.90E-05 | -0.026268757 | Z83844.2, PDXP |
| chr1 | 86422260 | 86422946 | 687 | 4 | 0.000347506 | 8.73E-06 | 0.00234 | 2.18E-05 | -0.014764303 | NA |
| chr18 | 70381585 | 70381644 | 60 | 2 | 0.000348244 | 0.000667 | 8.11E-05 | 7.98E-05 | -0.015275525 | LINC01910 |
| chr6 | 15017604 | 15017652 | 49 | 2 | 0.000348256 | 2.86E-05 | 0.00067 | 3.93E-05 | -0.02414847 | AL138720.1 |
| chr22 | 24231326 | 24231819 | 494 | 4 | 0.000348305 | 0.000197 | 0.002 | 5.28E-05 | -0.007689211 | GGT5 |
| chr1 | 15337686 | 15337889 | 204 | 3 | 0.000348439 | 0.000664 | 0.00031 | 6.04E-05 | -0.004497041 | FHAD1, FHAD1-AS1 |
| chr11 | 47189497 | 47189549 | 53 | 2 | 0.000348515 | 1.87E-05 | 0.00115 | 2.97E-05 | -0.009495543 | NA |
| chr19 | 42170963 | 42171624 | 662 | 2 | 0.000348782 | 2.53E-05 | 0.00039 | 3.16E-05 | -0.006132188 | POU2F2 |
| chr3 | 126482054 | 126482227 | 174 | 2 | 0.000349031 | 5.41E-05 | 0.00015 | 3.98E-05 | 0.006187113 | UROC1 |
| chr3 | 184317720 | 184317739 | 20 | 3 | 0.000349411 | 0.000189 | 0.00013 | 4.46E-05 | -0.006505652 | EIF4G1 |
| chr15 | 89242957 | 89243391 | 435 | 3 | 0.000349411 | 0.000323 | 0.00473 | 0.00052 | -0.010705856 | NA |
| chr8 | 941456 | 941513 | 58 | 2 | 0.000349442 | 0.000266 | 0.00012 | 8.09E-05 | -0.009934906 | DLGAP2 |
| chr1 | 53689078 | 53689797 | 720 | 2 | 0.000349606 | 0.001342 | 0.00149 | 0.00097 | -0.006021044 | GLIS1 |
| chr21 | 42338271 | 42338734 | 464 | 3 | 0.000349673 | 0.000149 | 0.00038 | 0.0001 | -0.005681119 | NA |
| chr21 | 31181306 | 31181535 | 230 | 2 | 0.00034984 | 0.000166 | 0.00203 | 0.00024 | -0.020145252 | TIAM1 |
| chr2 | 32265520 | 32265766 | 247 | 6 | 0.000350177 | 0.000587 | 0.00013 | 7.32E-05 | -0.002612067 | NLRC4 |
| chr9 | 95025265 | 95025965 | 701 | 2 | 0.000350235 | 7.77E-05 | 0.00075 | 9.06E-05 | -0.008148147 | AOPEP |
| chr12 | 131066193 | 131066323 | 131 | 2 | 0.000350392 | 2.59E-05 | 0.0011 | 3.93E-05 | 0.000212294 | ADGRD1 |
| chr15 | 32671954 | 32672558 | 605 | 6 | 0.000350433 | 1.61E-05 | 0.00288 | 2.35E-05 | -0.002733722 | SCG5, SCG5-AS1 |
| chr9 | 69387658 | 69387945 | 288 | 3 | 0.000350433 | 0.000464 | 0.00031 | 4.33E-05 | -0.001318065 | FAM189A2 |
| chr4 | 36399976 | 36400264 | 289 | 2 | 0.000351002 | 0.000123 | 0.00013 | 5.76E-05 | -0.00637533 | NA |
| chr9 | 92735341 | 92735740 | 400 | 3 | 0.000351005 | 3.70E-05 | 0.00071 | 4.53E-05 | -0.004273558 | BICD2 |
| chr21 | 38255349 | 38255460 | 112 | 2 | 0.000351644 | 0.000226 | 0.00257 | 0.00033 | 0.010082696 | KCNJ15 |
| chr17 | 73231410 | 73231613 | 204 | 3 | 0.000351704 | 0.004408 | 0.00065 | 0.00077 | -0.000809017 | FAM104A |
| chr11 | 69648889 | 69649244 | 356 | 2 | 0.000352066 | 0.002659 | 0.0001 | 0.00016 | 0.002155358 | CCND1 |
| chr3 | 119599564 | 119600086 | 523 | 4 | 0.000352245 | 0.002864 | 6.22E-05 | 0.00012 | -0.0023726 | PLA1A |
| chr9 | 135639443 | 135639666 | 224 | 4 | 0.000352428 | 0.000613 | 0.00283 | 4.77E-05 | 0.001010711 | GLT6D1 |
| chr17 | 44558410 | 44558421 | 12 | 2 | 0.000353308 | 0.000144 | 0.0013 | 0.00018 | -0.013183326 | FZD2 |
| chr14 | 38290521 | 38290977 | 457 | 5 | 0.000353392 | 2.58E-05 | 0.00503 | 2.98E-05 | -0.017563515 | AL355835.1 |
| chr22 | 47783898 | 47784162 | 265 | 2 | 0.000353456 | 9.99E-05 | 0.00013 | 5.26E-05 | -0.014415746 | AL117329.1 |
| chr15 | 45429391 | 45429394 | 4 | 2 | 0.00035383 | 0.000514 | 0.00021 | 0.00016 | -0.001461728 | NA |
| chr2 | 174326646 | 174327028 | 383 | 2 | 0.000354075 | 0.000172 | 0.00013 | 7.14E-05 | 0.00355778 | LINC01305 |
| chr10 | 6115073 | 6115488 | 416 | 3 | 0.000354188 | 2.40E-05 | 0.00213 | 3.87E-05 | 0.001399238 | RBM17 |
| chr5 | 95777600 | 95777614 | 15 | 2 | 0.000354714 | 0.000145 | 0.0001 | 5.49E-05 | -0.015406015 | RHOBTB3, GLRX |
| chr7 | 129532170 | 129532426 | 257 | 3 | 0.000354718 | 9.75E-05 | 0.00023 | 4.13E-05 | -0.003209323 | NA |
| chr8 | 142473274 | 142473989 | 716 | 4 | 0.000354819 | 0.028358 | 0.00011 | 0.00011 | -0.004484128 | ADGRB1 |
| chr2 | 113059055 | 113059466 | 412 | 4 | 0.000355024 | 4.05E-05 | 0.0008 | 3.65E-05 | -0.005250043 | IL36RN |
| chr1 | 205671962 | 205672414 | 453 | 2 | 0.000355308 | 2.79E-05 | 0.00063 | 3.78E-05 | -0.011845123 | SLC45A3 |
| chr1 | 19406292 | 19406373 | 82 | 2 | 0.00035534 | 5.63E-05 | 0.00018 | 4.34E-05 | -0.014470218 | CAPZB |
| chr11 | 66052147 | 66052595 | 449 | 7 | 0.000355366 | 5.83E-05 | 0.0023 | 4.21E-05 | 0.000624603 | SF3B2 |
| chr16 | 70429262 | 70430108 | 847 | 3 | 0.000355572 | 1.09E-05 | 0.00044 | 1.10E-05 | -0.007222095 | ST3GAL2 |
| chr8 | 6787583 | 6788140 | 558 | 2 | 0.00035563 | 1.30E-05 | 0.00093 | 2.09E-05 | -0.010029952 | AF233439.2 |
| chr22 | 18110209 | 18110843 | 635 | 9 | 0.000355715 | 0.000293 | 0.00161 | 5.68E-05 | 0.001069162 | PEX26, TUBA8, AC008079.1 |
| chr17 | 41777524 | 41777832 | 309 | 2 | 0.000355899 | 0.039395 | 0.00025 | 0.00073 | -0.005431223 | JUP |
| chr6 | 26224697 | 26225071 | 375 | 6 | 0.000356041 | 0.00011 | 0.00456 | 0.00012 | -0.016656144 | H3C6 |
| chr8 | 1876738 | 1876773 | 36 | 2 | 0.000356159 | 5.32E-05 | 0.00021 | 4.51E-05 | -0.009862386 | AC019257.8, ARHGEF10 |
| chr11 | 8080246 | 8080479 | 234 | 4 | 0.000356346 | 8.64E-05 | 0.00262 | 4.88E-05 | -0.011928777 | TUB |
| chr12 | 51047660 | 51047751 | 92 | 2 | 0.000356365 | 9.17E-05 | 0.0018 | 0.00013 | -0.001489477 | NA |
| chr3 | 179071811 | 179072549 | 739 | 12 | 0.000356814 | 4.51E-05 | 0.0067 | 4.45E-05 | 0.001207655 | ZMAT3 |
| chr15 | 41838452 | 41838794 | 343 | 6 | 0.00035682 | 0.008383 | 0.00252 | 0.00015 | -0.004163731 | JMJD7-PLA2G4B, PLA2G4B |
| chr7 | 114423046 | 114423169 | 124 | 2 | 0.000356839 | 7.25E-05 | 0.00012 | 4.08E-05 | -0.004028582 | FOXP2 |
| chr7 | 154984915 | 154985121 | 207 | 2 | 0.000357033 | 0.000146 | 0.00017 | 7.71E-05 | -0.003861999 | PAXIP1 |
| chr19 | 43826513 | 43827175 | 663 | 5 | 0.000357458 | 2.02E-05 | 0.00432 | 1.92E-05 | -0.011447073 | LYPD5 |
| chr17 | 76888386 | 76888431 | 46 | 3 | 0.000358121 | 0.060707 | 4.67E-05 | 0.0002 | -0.001884759 | MGAT5B |
| chr5 | 177493020 | 177493515 | 496 | 4 | 0.000358273 | 8.08E-05 | 0.00329 | 7.65E-05 | -0.005449634 | PDLIM7 |
| chr7 | 953580 | 953781 | 202 | 2 | 0.000358284 | 6.54E-05 | 0.00064 | 7.58E-05 | -0.010179711 | ADAP1, COX19 |
| chr1 | 34918719 | 34919068 | 350 | 2 | 0.000358479 | 2.07E-05 | 0.00099 | 3.19E-05 | -0.01473071 | DLGAP3 |
| chr20 | 53713784 | 53713847 | 64 | 2 | 0.000358569 | 0.002678 | 6.28E-05 | 8.62E-05 | -0.006082956 | NA |
| chr21 | 29130410 | 29131154 | 745 | 8 | 0.0003592 | 0.038627 | 0.00084 | 9.97E-05 | -0.002728144 | MAP3K7CL |
| chr11 | 35112661 | 35112685 | 25 | 2 | 0.000359436 | 2.86E-05 | 0.00107 | 4.27E-05 | -0.008636096 | NA |
| chr11 | 64833512 | 64833519 | 8 | 2 | 0.000359582 | 0.032602 | 0.00022 | 0.00064 | -0.004606131 | CDC42BPG |
| chr4 | 41612380 | 41612825 | 446 | 5 | 0.000359595 | 2.63E-05 | 0.00112 | 3.23E-05 | -0.007674272 | LIMCH1, AC108050.1 |
| chr11 | 62394768 | 62395173 | 406 | 3 | 0.000359656 | 5.29E-05 | 0.0011 | 5.52E-05 | -0.010115494 | NA |
| chr17 | 17476895 | 17477321 | 427 | 7 | 0.000359741 | 0.000508 | 0.00086 | 4.98E-05 | -0.001762525 | MED9 |
| chr22 | 23690239 | 23690714 | 476 | 4 | 0.000360946 | 0.000106 | 0.00054 | 4.86E-05 | -0.002057843 | RGL4, GUSBP11, AP000347.1 |
| chr15 | 90912119 | 90912177 | 59 | 3 | 0.000360971 | 2.02E-05 | 0.00159 | 3.67E-05 | -0.009198005 | MAN2A2 |
| chr17 | 79130784 | 79131344 | 561 | 3 | 0.000361013 | 0.000102 | 0.00269 | 0.0001 | -0.005633465 | RBFOX3 |
| chr7 | 92806455 | 92806658 | 204 | 2 | 0.000361053 | 0.000446 | 7.68E-05 | 6.66E-05 | -0.015788856 | CDK6 |
| chr13 | 33015864 | 33016355 | 492 | 9 | 0.000361152 | 0.000466 | 0.00323 | 4.11E-05 | -0.001243022 | NA |
| chr7 | 70787964 | 70787967 | 4 | 2 | 0.000361178 | 1.38E-05 | 0.00099 | 2.21E-05 | -0.000858604 | AUTS2 |
| chr16 | 87316947 | 87317501 | 555 | 8 | 0.000361328 | 3.95E-05 | 0.00017 | 2.84E-05 | -0.000552932 | C16orf95, AC010531.1 |
| chr17 | 5394767 | 5395150 | 384 | 2 | 0.00036143 | 5.02E-05 | 0.00174 | 7.54E-05 | -0.007618629 | NUP88 |
| chr18 | 54269342 | 54269644 | 303 | 10 | 0.000361489 | 6.28E-05 | 0.00054 | 2.25E-05 | 8.43E-05 | POLI, AC093462.1 |
| chr1 | 107252095 | 107252381 | 287 | 2 | 0.000362178 | 3.20E-05 | 0.00151 | 4.91E-05 | -0.011281646 | NTNG1 |
| chr9 | 123112670 | 123112990 | 321 | 2 | 0.000362297 | 4.19E-05 | 0.00205 | 6.57E-05 | -0.007926659 | MIR600HG, STRBP |
| chr21 | 43787067 | 43787245 | 179 | 2 | 0.000362445 | 2.39E-05 | 0.001 | 3.60E-05 | 0.012320155 | NA |
| chr16 | 1375081 | 1375255 | 175 | 2 | 0.000362859 | 0.000508 | 0.00059 | 0.00031 | -0.004260424 | UNKL |
| chr17 | 32487496 | 32488016 | 521 | 7 | 0.000363067 | 0.000121 | 0.00186 | 5.49E-05 | -0.005103773 | CDK5R1 |
| chr1 | 180085258 | 180085448 | 191 | 2 | 0.000363118 | 0.002828 | 6.28E-05 | 8.70E-05 | -0.001363146 | CEP350 |
| chr3 | 41900208 | 41900234 | 27 | 2 | 0.000363143 | 2.44E-05 | 0.00146 | 3.86E-05 | 0.000255844 | ULK4 |
| chr5 | 40990170 | 40990261 | 92 | 2 | 0.000363386 | 0.000117 | 8.72E-05 | 4.38E-05 | -0.006177418 | NA |
| chr1 | 3607861 | 3607895 | 35 | 2 | 0.000364108 | 0.000126 | 0.00027 | 8.79E-05 | 0.003459999 | MEGF6 |
| chr12 | 97159441 | 97159741 | 301 | 2 | 0.000364371 | 5.89E-05 | 0.00022 | 4.87E-05 | -0.012351942 | AC007656.2, AC007656.4 |
| chr1 | 1307350 | 1307515 | 166 | 4 | 0.000364732 | 0.027305 | 0.00029 | 5.95E-05 | 0.000468682 | ACAP3 |
| chr4 | 1792912 | 1793003 | 92 | 2 | 0.000364746 | 0.000824 | 0.00913 | 0.0014 | -0.000969134 | NA |
| chr9 | 123742007 | 123742085 | 79 | 2 | 0.000364923 | 0.000106 | 9.07E-05 | 4.27E-05 | -0.003734793 | AL390774.2, DENND1A |
| chr16 | 30947994 | 30948341 | 348 | 6 | 0.000364946 | 0.007646 | 0.00051 | 0.00043 | -0.004197023 | FBXL19 |
| chr4 | 8372834 | 8372970 | 137 | 2 | 0.000365157 | 0.000197 | 0.00265 | 0.0003 | 0.0006735 | ACOX3 |
| chr7 | 27114325 | 27114686 | 362 | 4 | 0.000365269 | 0.036758 | 0.00022 | 0.00057 | -0.003641561 | HOXA-AS2, HOXA3 |
| chr22 | 50562771 | 50563271 | 501 | 8 | 0.000365431 | 0.001958 | 0.0001 | 0.00023 | 0.002037014 | SYCE3 |
| chr22 | 39388459 | 39388791 | 333 | 5 | 0.000365739 | 0.000721 | 0.00086 | 0.00025 | 0.012192331 | NA |
| chr1 | 27287541 | 27288028 | 488 | 5 | 0.000366103 | 2.37E-05 | 0.0048 | 4.29E-05 | -0.003173713 | WDTC1 |
| chr4 | 140135588 | 140135888 | 301 | 2 | 0.000366223 | 2.47E-05 | 0.00073 | 3.52E-05 | -0.012912413 | MAML3 |
| chr1 | 22830253 | 22830362 | 110 | 2 | 0.000366623 | 3.51E-05 | 0.00032 | 3.83E-05 | -0.005920044 | EPHB2 |
| chr2 | 100142552 | 100142697 | 146 | 3 | 0.000366653 | 4.76E-05 | 0.00445 | 9.44E-05 | 0.006845244 | AFF3 |
| chr19 | 18280519 | 18281073 | 555 | 3 | 0.000366798 | 0.000791 | 0.00806 | 0.00131 | 0.002901357 | JUND |
| chr19 | 29145466 | 29145896 | 431 | 2 | 0.000367016 | 0.000141 | 0.0046 | 0.00025 | 0.016975203 | NA |
| chr20 | 24987963 | 24988213 | 251 | 2 | 0.00036715 | 0.000116 | 0.00063 | 0.00012 | -0.008624836 | APMAP |
| chr21 | 26841244 | 26841909 | 666 | 2 | 0.00036745 | 0.007933 | 0.00087 | 0.00163 | -0.004031263 | ADAMTS1 |
| chr12 | 51920457 | 51920826 | 370 | 2 | 0.000367524 | 5.04E-05 | 0.00108 | 6.96E-05 | -0.003326817 | ACVRL1 |
| chr20 | 44020608 | 44020732 | 125 | 2 | 0.000367679 | 3.38E-05 | 0.00027 | 3.56E-05 | -0.012760031 | TOX2 |
| chr7 | 148783938 | 148784815 | 878 | 3 | 0.000367906 | 8.28E-06 | 0.00145 | 1.47E-05 | 7.34E-05 | CUL1 |
| chr16 | 75224575 | 75224882 | 308 | 2 | 0.000368227 | 4.84E-05 | 0.0003 | 4.77E-05 | -0.011950638 | CTRB1 |
| chr5 | 181243531 | 181244165 | 635 | 12 | 0.000368531 | 0.00127 | 0.00019 | 0.00027 | 0.000582127 | RACK1 |
| chr1 | 11653483 | 11654212 | 730 | 9 | 0.000368958 | 3.01E-05 | 0.00794 | 7.85E-05 | -0.006172581 | FBXO2 |
| chr2 | 66437213 | 66437677 | 465 | 8 | 0.000369375 | 0.000825 | 0.00169 | 5.69E-05 | -0.000532764 | MEIS1 |
| chr4 | 8290210 | 8291074 | 865 | 4 | 0.000369542 | 5.13E-06 | 0.00886 | 1.93E-05 | 0.01815868 | HTRA3 |
| chr13 | 110248151 | 110248346 | 196 | 3 | 0.000369587 | 0.001257 | 0.00109 | 6.72E-05 | -0.002620197 | COL4A1 |
| chr13 | 98729552 | 98729566 | 15 | 2 | 0.000369762 | 0.000106 | 0.00013 | 5.57E-05 | -0.033331331 | SLC15A1 |
| chr6 | 13710748 | 13710862 | 115 | 2 | 0.000369785 | 0.016735 | 0.00015 | 0.00036 | -0.011159871 | RANBP9 |
| chr9 | 98973811 | 98973870 | 60 | 2 | 0.000370116 | 3.57E-05 | 0.00024 | 3.56E-05 | 0.005185955 | COL15A1 |
| chr19 | 3707946 | 3708207 | 262 | 7 | 0.000370211 | 0.000796 | 0.00059 | 7.00E-05 | -0.005380001 | NA |
| chr3 | 50246705 | 50246970 | 266 | 5 | 0.000370343 | 0.003084 | 0.00362 | 0.00036 | -0.003532033 | GNAI2 |
| chr10 | 123775370 | 123775923 | 554 | 3 | 0.000370899 | 0.000263 | 0.0011 | 5.60E-05 | -0.005295574 | CPXM2 |
| chr5 | 142645050 | 142645274 | 225 | 4 | 0.000371056 | 0.000201 | 0.00349 | 8.74E-05 | -0.006552391 | SPRY4-AS1, FGF1 |
| chr2 | 240245720 | 240245879 | 160 | 4 | 0.000371562 | 0.002345 | 0.0002 | 7.29E-05 | -0.001464486 | AC124861.1 |
| chr4 | 36281562 | 36281653 | 92 | 3 | 0.000371562 | 0.000134 | 0.01565 | 0.00038 | 0.003244545 | DTHD1 |
| chr15 | 44458659 | 44458671 | 13 | 3 | 0.000371562 | 0.000263 | 0.00022 | 4.46E-05 | -0.00988391 | CTDSPL2 |
| chr8 | 51647198 | 51647447 | 250 | 2 | 0.00037188 | 2.00E-05 | 0.00117 | 3.16E-05 | 0.012312567 | PXDNL |
| chr17 | 1087251 | 1087668 | 418 | 2 | 0.000372632 | 3.32E-05 | 0.00055 | 4.24E-05 | -0.020985467 | ABR |
| chr12 | 66679531 | 66679851 | 321 | 3 | 0.000372728 | 0.003339 | 0.00138 | 0.00044 | -0.006953177 | GRIP1 |
| chr17 | 49512817 | 49512964 | 148 | 4 | 0.000372796 | 3.83E-05 | 0.00045 | 3.69E-05 | -0.004519605 | NGFR, AC006487.1 |
| chr5 | 142235804 | 142236500 | 697 | 2 | 0.000374177 | 6.56E-06 | 0.00064 | 1.09E-05 | 0.006120753 | NA |
| chr6 | 165846450 | 165847084 | 635 | 6 | 0.000374315 | 1.03E-05 | 0.01076 | 2.97E-05 | 0.000399984 | PDE10A |
| chr4 | 22388083 | 22388123 | 41 | 2 | 0.000374413 | 0.00013 | 9.74E-05 | 4.94E-05 | -0.002626813 | ADGRA3 |
| chr6 | 112531029 | 112531154 | 126 | 3 | 0.000374569 | 1.71E-05 | 0.00174 | 3.09E-05 | -0.009845552 | NA |
| chr16 | 83137463 | 83137694 | 232 | 3 | 0.000374632 | 1.35E-05 | 0.00341 | 3.03E-05 | -0.038703015 | CDH13 |
| chr7 | 121872341 | 121872473 | 133 | 2 | 0.000374637 | 0.000139 | 0.00219 | 0.00021 | 0.000348728 | NA |
| chr5 | 173764645 | 173764992 | 348 | 2 | 0.000375931 | 7.77E-05 | 0.00033 | 6.95E-05 | -0.018070808 | AC008674.1 |
| chr10 | 113679108 | 113679547 | 440 | 9 | 0.000375962 | 7.91E-05 | 0.03829 | 0.00027 | 0.002559574 | CASP7 |
| chr10 | 3264361 | 3264395 | 35 | 2 | 0.000376202 | 7.43E-05 | 9.52E-05 | 3.63E-05 | -0.011370302 | AL139280.1 |
| chr1 | 233115354 | 233115371 | 18 | 2 | 0.000376234 | 4.42E-05 | 0.00024 | 4.18E-05 | -0.011289473 | PCNX2 |
| chr4 | 26859639 | 26860435 | 797 | 5 | 0.000376334 | 0.018168 | 0.00186 | 0.0006 | -0.002319816 | STIM2, STIM2-AS1 |
| chr7 | 8443619 | 8443939 | 321 | 3 | 0.000376352 | 0.000245 | 0.00011 | 5.28E-05 | 0.001511575 | NXPH1 |

| | | | | | | | | | | |
|---|---|---|---|---|---|---|---|---|---|---|
| chr3 | 33113965 | 33114578 | 614 | 4 | 0.000376599 | 0.00452 | 0.00338 | 0.00354 | 0.00029897 | CRTAP |
| chr3 | 49294413 | 49294830 | 418 | 4 | 0.000376759 | 1.43E-05 | 0.00373 | 3.30E-05 | -0.005306874 | USP4 |
| chr11 | 17013244 | 17013297 | 54 | 2 | 0.000377175 | 5.18E-05 | 0.00203 | 7.94E-05 | -0.017205671 | PLEKHA7 |
| chr2 | 28361135 | 28361137 | 3 | 2 | 0.000377508 | 3.11E-05 | 0.00148 | 4.78E-05 | -0.011942384 | NA |
| chr2 | 218748884 | 218748887 | 4 | 2 | 0.000377932 | 0.000134 | 8.15E-05 | 4.42E-05 | -0.006930247 | TTLL4 |
| chr19 | 18428303 | 18429031 | 729 | 4 | 0.000377946 | 0.000232 | 0.00051 | 2.80E-05 | -0.004394518 | SSBP4 |
| chr2 | 64627139 | 64627271 | 133 | 2 | 0.000378053 | 0.000508 | 0.0004 | 0.00025 | -0.007847724 | NA |
| chr6 | 42781864 | 42782062 | 199 | 4 | 0.000378395 | 0.004345 | 0.00013 | 0.00026 | 0.000440378 | BICRAL |
| chr2 | 207770881 | 207771639 | 759 | 9 | 0.00037872 | 1.27E-05 | 0.01076 | 2.27E-05 | -0.003796873 | NA |
| chr2 | 48715065 | 48715070 | 6 | 2 | 0.000378907 | 1.97E-05 | 0.00128 | 3.15E-05 | -0.011782499 | STON1-GTF2A1L, GTF2A1L, AC073082.1, LHCGR |
| chr2 | 365090 | 365559 | 470 | 5 | 0.000379086 | 0.000293 | 0.00025 | 6.27E-05 | -0.00753427 | NA |
| chr12 | 16353312 | 16353407 | 96 | 3 | 0.000379098 | 7.58E-05 | 0.00082 | 4.71E-05 | -0.01281438 | MGST1 |
| chr4 | 122378492 | 122379503 | 1012 | 17 | 0.00037952 | 0.001055 | 0.01226 | 9.31E-05 | -0.000500436 | ADAD1 |
| chr20 | 11301552 | 11301735 | 184 | 4 | 0.000379908 | 0.001442 | 4.52E-05 | 6.12E-05 | 0.00494316 | NA |
| chr16 | 89103450 | 89103799 | 350 | 2 | 0.00038007 | 0.000383 | 0.00043 | 0.00022 | -0.003759099 | ACSF3 |
| chr17 | 7080491 | 7081161 | 671 | 5 | 0.000380169 | 0.00097 | 0.00579 | 0.00013 | -0.006066907 | NA |
| chr13 | 77026318 | 77026440 | 123 | 3 | 0.000380345 | 0.001753 | 0.00088 | 0.00057 | 0.001614805 | AC001226.2, FBXL3 |
| chr2 | 85341713 | 85341795 | 83 | 2 | 0.000380648 | 0.00641 | 5.17E-05 | 7.89E-05 | -0.002178486 | PEBP1P2 |
| chr1 | 153945810 | 153946240 | 431 | 4 | 0.000380657 | 0.000276 | 0.00059 | 7.46E-05 | 0.000320145 | DENND4B |
| chr7 | 113144620 | 113144862 | 243 | 3 | 0.000381451 | 0.001463 | 0.00014 | 0.00014 | 0.006057863 | AC073346.1 |
| chr7 | 156516486 | 156516664 | 179 | 3 | 0.000381459 | 1.21E-05 | 0.00355 | 2.78E-05 | -0.008080522 | LINC01006 |
| chr12 | 132416369 | 132416564 | 196 | 2 | 0.000381459 | 6.07E-05 | 0.00023 | 5.12E-05 | -0.002446957 | NA |
| chr3 | 33799178 | 33799433 | 256 | 2 | 0.000381466 | 0.022837 | 7.62E-05 | 0.00015 | 0.001221374 | PDCD6IP |
| chr11 | 67358238 | 67358258 | 21 | 2 | 0.000381616 | 0.000204 | 8.61E-05 | 5.55E-05 | 0.002174863 | RAD9A, AP003419.1 |
| chr19 | 42218564 | 42218758 | 195 | 3 | 0.000381696 | 0.013072 | 0.00217 | 0.00042 | -0.001058578 | DEDD2, AC006486.3 |
| chr18 | 46756436 | 46757136 | 701 | 2 | 0.000382313 | 0.004566 | 0.00028 | 0.00052 | 0.005195744 | AC090241.2, ST8SIA5 |
| chr20 | 23638888 | 23639206 | 319 | 4 | 0.000382317 | 9.35E-05 | 0.0018 | 0.00011 | -0.006223123 | NA |
| chr19 | 649039 | 649464 | 426 | 3 | 0.000382347 | 4.33E-05 | 0.001 | 5.74E-05 | -0.002745753 | RNF126 |
| chr19 | 18434252 | 18434758 | 507 | 3 | 0.000382437 | 0.000731 | 0.00032 | 0.00015 | -0.007629719 | SSBP4, ISYNA1 |
| chr17 | 43943380 | 43943478 | 99 | 2 | 0.000382726 | 0.000359 | 0.00783 | 0.00065 | -0.004220279 | NA |
| chr11 | 316088 | 316456 | 369 | 4 | 0.000382816 | 1.56E-05 | 0.01117 | 4.88E-05 | -0.020970049 | NA |
| chr16 | 57357743 | 57358286 | 544 | 5 | 0.000383147 | 1.30E-05 | 0.00814 | 2.58E-05 | -0.00839494 | NA |
| chr17 | 46912839 | 46913119 | 281 | 2 | 0.000385073 | 0.000173 | 0.00182 | 0.00024 | -0.006144367 | NA |
| chr10 | 25821011 | 25821221 | 211 | 2 | 0.000386107 | 6.34E-05 | 0.00162 | 9.14E-05 | -0.016220766 | NA |
| chr19 | 10869324 | 10869405 | 82 | 2 | 0.000386148 | 0.023582 | 5.51E-05 | 9.56E-05 | -0.003287698 | C19orf38 |
| chr9 | 135251162 | 135251316 | 155 | 2 | 0.000386159 | 2.70E-05 | 0.00053 | 3.54E-05 | -0.013139231 | AL390778.2 |
| chr4 | 3430110 | 3430137 | 28 | 2 | 0.000386259 | 0.000105 | 0.00024 | 7.56E-05 | -0.004343243 | RGS12 |
| chr6 | 168024138 | 168024599 | 462 | 3 | 0.000386296 | 0.000224 | 0.00038 | 0.00012 | -0.000541674 | KIF25 |
| chr14 | 104883118 | 104883439 | 322 | 5 | 0.000386834 | 0.0001 | 0.00321 | 6.74E-05 | -0.002839718 | CEP170B |
| chr17 | 12788836 | 12789455 | 620 | 10 | 0.000387164 | 3.84E-05 | 0.01394 | 2.86E-05 | -0.004884931 | AC005358.1 |
| chr9 | 127485898 | 127485928 | 31 | 2 | 0.000387363 | 2.10E-05 | 0.00103 | 3.24E-05 | -0.005643786 | LRSAM1 |
| chr6 | 71289359 | 71289405 | 47 | 3 | 0.00038754 | 0.00019 | 0.00037 | 9.16E-05 | 0.001154386 | OGFRL1, AL589935.1 |
| chr14 | 27730965 | 27731014 | 50 | 2 | 0.000387893 | 7.52E-05 | 0.00029 | 6.47E-05 | -0.000256131 | AL390334.1 |
| chr15 | 85501333 | 85501396 | 64 | 2 | 0.000388191 | 0.000174 | 7.86E-05 | 4.79E-05 | -0.006564299 | AKAP13 |
| chr22 | 26565271 | 26565653 | 383 | 4 | 0.000388256 | 0.000146 | 0.0025 | 7.41E-05 | -0.003134246 | TPST2 |
| chr12 | 6786608 | 6786939 | 332 | 3 | 0.00038876 | 5.04E-05 | 0.00078 | 3.89E-05 | -0.010671368 | CD4 |
| chr20 | 63588494 | 63588615 | 122 | 2 | 0.000388966 | 0.000946 | 0.00022 | 0.00023 | -0.004242929 | GMEB2 |
| chr22 | 23178820 | 23179173 | 354 | 3 | 0.000388974 | 0.015338 | 0.00033 | 0.00115 | -0.004025145 | NA |
| chr12 | 3241700 | 3241709 | 10 | 2 | 0.000389291 | 0.040593 | 5.65E-05 | 0.0001 | -0.003414432 | TSPAN9 |
| chr1 | 1543447 | 1543858 | 412 | 3 | 0.000389751 | 0.000433 | 0.00121 | 0.00036 | -0.006951211 | SSU72 |
| chr11 | 11941360 | 11941626 | 267 | 3 | 0.000389793 | 0.000572 | 8.76E-05 | 8.17E-05 | -0.004802728 | USP47 |
| chr14 | 58396379 | 58396644 | 266 | 2 | 0.000389793 | 0.000371 | 0.0002 | 0.00014 | -0.010364304 | TOMM20L |
| chr10 | 49188380 | 49189027 | 648 | 7 | 0.000389992 | 0.000188 | 0.00117 | 5.77E-05 | -0.00368351 | TMEM273 |
| chr19 | 1955396 | 1955420 | 25 | 2 | 0.000390051 | 0.000226 | 0.00181 | 0.0003 | -0.005987362 | CSNK1G2 |
| chr14 | 104977065 | 104977295 | 231 | 2 | 0.00039046 | 0.001544 | 0.00101 | 0.00084 | -0.006842091 | AHNAK2 |
| chr2 | 176601853 | 176602191 | 339 | 3 | 0.000391013 | 5.40E-05 | 0.00075 | 6.59E-05 | 0.004492391 | LINC01117 |
| chr1 | 214609212 | 214609311 | 100 | 2 | 0.000391478 | 0.007897 | 5.16E-05 | 8.05E-05 | -0.002473846 | CENPF |
| chr8 | 141643555 | 141644291 | 737 | 7 | 0.000391668 | 7.26E-05 | 0.00044 | 5.77E-05 | -0.006132754 | AC025839.1 |
| chr3 | 174191497 | 174191703 | 207 | 2 | 0.00039209 | 7.90E-05 | 0.00014 | 4.87E-05 | -0.01768442 | NLGN1 |
| chr19 | 19063454 | 19064385 | 932 | 11 | 0.000392362 | 0.001592 | 0.00399 | 0.00023 | -0.000165493 | SLC25A42 |
| chr3 | 128051480 | 128051985 | 506 | 4 | 0.000392522 | 0.000228 | 0.00294 | 7.51E-05 | 0.002623257 | SEC61A1, MGLL |
| chr14 | 103738734 | 103739322 | 589 | 4 | 0.000393151 | 0.02418 | 6.89E-05 | 0.00043 | 0.001758287 | PPP1R13B |
| chr16 | 89343788 | 89343914 | 127 | 2 | 0.000393151 | 7.71E-05 | 7.58E-05 | 3.16E-05 | -0.002051039 | ANKRD11 |
| chr4 | 1294047 | 1294100 | 54 | 2 | 0.000393795 | 3.46E-05 | 0.00112 | 5.04E-05 | -0.005720528 | MAEA |
| chr20 | 43508030 | 43508744 | 715 | 4 | 0.000394188 | 0.000803 | 0.00134 | 0.00059 | -0.00391524 | AL031681.2, L3MBTL1 |
| chr12 | 56265498 | 56265924 | 427 | 4 | 0.000394353 | 0.000426 | 0.00112 | 0.00014 | -0.002836211 | NA |
| chr20 | 5030626 | 5030736 | 111 | 2 | 0.000394447 | 5.03E-05 | 0.0003 | 4.90E-05 | -0.008577671 | NA |
| chr10 | 98463209 | 98463297 | 89 | 2 | 0.000394608 | 0.001016 | 6.28E-05 | 6.95E-05 | -0.001408403 | HPSE2 |
| chr11 | 18209073 | 18209414 | 342 | 6 | 0.000395184 | 4.09E-06 | 0.00656 | 1.45E-05 | -0.018362021 | SLC25A51P4 |
| chr9 | 27102421 | 27102899 | 479 | 4 | 0.000395455 | 0.000269 | 0.00232 | 4.72E-05 | -0.012567471 | AL355432.1 |
| chr6 | 31947678 | 31948351 | 674 | 2 | 0.000395631 | 0.001928 | 0.01617 | 0.00331 | -0.00699701 | AL645922.1, CFB |
| chr4 | 53899302 | 53899676 | 375 | 3 | 0.000395676 | 1.49E-05 | 0.00309 | 3.27E-05 | -0.010105205 | AC058822.1 |
| chr8 | 95154648 | 95154773 | 126 | 2 | 0.000395809 | 2.59E-05 | 0.00025 | 2.84E-05 | -0.007607287 | PLEKHF2 |
| chr4 | 8205331 | 8205598 | 268 | 4 | 0.000396286 | 2.66E-05 | 0.00234 | 3.91E-05 | -0.015741132 | SH3TC1 |
| chr16 | 29995857 | 29996644 | 788 | 13 | 0.000396442 | 4.14E-05 | 0.03022 | 6.55E-05 | 0.001210152 | INO80E, HIRIP3 |
| chr8 | 122316287 | 122316508 | 222 | 2 | 0.0003968 | 0.004385 | 6.45E-05 | 9.70E-05 | -0.005033828 | NA |
| chr20 | 41097634 | 41098300 | 667 | 2 | 0.000397013 | 4.84E-05 | 0.00166 | 7.25E-05 | -0.000679608 | TOP1, PLCG1-AS1 |
| chr3 | 39154408 | 39154668 | 261 | 4 | 0.00039722 | 0.009598 | 0.00023 | 0.00034 | -0.002471253 | AC092053.3, AC092053.2, CSRNP1 |
| chr6 | 52498899 | 52498985 | 87 | 3 | 0.000397517 | 7.99E-05 | 0.00062 | 4.30E-05 | -0.003263149 | EFHC1, TRAM2 |
| chr2 | 65350635 | 65350802 | 168 | 2 | 0.000397592 | 0.000163 | 9.47E-05 | 5.39E-05 | -0.009713773 | SPRED2 |
| chr1 | 26531175 | 26531283 | 109 | 3 | 0.000397618 | 7.77E-05 | 0.00073 | 7.84E-05 | -0.009819234 | RPS6KA1 |
| chr11 | 66721167 | 66721551 | 385 | 7 | 0.000397769 | 0.001211 | 0.00032 | 0.00014 | -0.002530519 | SPTBN2 |
| chr13 | 110489483 | 110489718 | 236 | 6 | 0.000398024 | 0.004307 | 0.00018 | 7.53E-05 | -0.003304619 | COL4A2 |
| chr2 | 237858583 | 237858859 | 277 | 4 | 0.000398053 | 4.86E-05 | 0.00934 | 0.00011 | -0.021806289 | NA |
| chr12 | 120761680 | 120761734 | 55 | 3 | 0.000398739 | 0.000153 | 0.0003 | 7.57E-05 | -0.00608859 | NA |
| chr14 | 21040069 | 21040602 | 534 | 3 | 0.000398739 | 0.000848 | 0.00091 | 6.10E-05 | -0.00844935 | NDRG2 |
| chr5 | 75336537 | 75337428 | 892 | 10 | 0.000398916 | 5.88E-05 | 0.00923 | 1.60E-05 | 0.000434272 | HMGCR, AC008897.2 |
| chr16 | 83830567 | 83830704 | 138 | 3 | 0.000399089 | 9.06E-05 | 0.00033 | 3.89E-05 | -0.00460329 | NA |
| chr5 | 159330448 | 159330651 | 204 | 2 | 0.000399089 | 0.005071 | 0.00014 | 0.00027 | 0.010440779 | AC008691.1, IL12B |
| chr17 | 27384285 | 27384453 | 169 | 2 | 0.000399089 | 3.17E-05 | 0.0002 | 3.07E-05 | -0.013439502 | NA |
| chr21 | 43740903 | 43741534 | 632 | 4 | 0.00039941 | 5.21E-05 | 0.01991 | 0.00018 | -0.041518891 | PDXK |
| chr19 | 3572518 | 3572959 | 442 | 8 | 0.000399656 | 0.000101 | 0.00491 | 0.0002 | 0.001213851 | HMG20B, MFSD12 |
| chr9 | 34957257 | 34958003 | 747 | 6 | 0.000399943 | 0.000477 | 0.02227 | 0.00056 | -0.00144314 | PHF24 |
| chr16 | 57838482 | 57838523 | 42 | 2 | 0.000400132 | 2.07E-05 | 0.00132 | 3.30E-05 | -0.00613479 | KIFC3 |
| chr8 | 100105674 | 100105855 | 182 | 6 | 0.00040051 | 0.000477 | 0.0018 | 9.97E-05 | -0.002699555 | RGS22 |
| chr22 | 24894583 | 24895247 | 665 | 3 | 0.00040051 | 6.18E-06 | 0.00213 | 1.38E-05 | -0.019379118 | SGSM1 |
| chr2 | 71601068 | 71601163 | 96 | 2 | 0.00040051 | 0.003794 | 5.94E-05 | 8.63E-05 | -0.008073098 | DYSF |
| chr7 | 100436315 | 100436831 | 517 | 10 | 0.00040065 | 0.000863 | 0.00052 | 4.36E-05 | 0.00126822 | AC092849.1, PPP1R35-AS1, PPP1R35 |
| chr1 | 110480030 | 110480209 | 180 | 3 | 0.00040068 | 0.000877 | 0.00043 | 0.00026 | -0.003216747 | AL358215.3 |
| chr7 | 135336032 | 135336398 | 367 | 2 | 0.000400869 | 2.29E-05 | 0.00105 | 3.51E-05 | -0.012742059 | NA |
| chr11 | 116792762 | 116793088 | 327 | 3 | 0.000401029 | 0.000449 | 0.00204 | 0.00035 | -0.006061039 | NA |
| chr17 | 82064171 | 82064740 | 570 | 3 | 0.000402088 | 8.55E-05 | 0.01082 | 0.00021 | -0.006140781 | DUS1L |
| chr3 | 19946036 | 19946323 | 288 | 5 | 0.000402153 | 0.001946 | 0.00082 | 9.28E-05 | -0.006097872 | EFHB |
| chr5 | 151758895 | 151759236 | 342 | 4 | 0.00040315 | 0.035218 | 0.00255 | 0.00896 | -0.002871507 | AC091982.1, ATOX1 |
| chr9 | 113564372 | 113564998 | 627 | 7 | 0.000403422 | 7.83E-06 | 0.0114 | 2.00E-05 | -0.009134169 | RGS3 |
| chr11 | 125111613 | 125112262 | 650 | 13 | 0.000403611 | 0.00085 | 0.00101 | 0.00014 | 0.000493359 | TMEM218 |
| chr7 | 158501900 | 158501974 | 75 | 3 | 0.000404049 | 0.046962 | 6.16E-05 | 0.00029 | -0.003047411 | PTPRN2 |
| chr2 | 237432964 | 237433150 | 187 | 2 | 0.000404123 | 2.39E-05 | 0.00089 | 3.54E-05 | -0.014334088 | AC112721.2 |
| chr1 | 228207199 | 228207444 | 246 | 2 | 0.000405136 | 0.426724 | 0.42507 | 0.50741 | 0.001813341 | OBSCN-AS1 |
| chr6 | 43008931 | 43009456 | 526 | 3 | 0.000405238 | 6.10E-05 | 0.00244 | 6.98E-05 | 0.001650182 | PPP2R5D |
| chr7 | 50382568 | 50382676 | 109 | 2 | 0.000405319 | 0.001914 | 9.27E-05 | 0.00012 | 0.004844165 | IKZF1 |
| chr1 | 20488407 | 20488420 | 14 | 2 | 0.000405839 | 0.000286 | 9.27E-05 | 6.68E-05 | -0.022479602 | NA |
| chr3 | 13496481 | 13496704 | 224 | 4 | 0.000406197 | 0.011001 | 0.00088 | 8.29E-05 | -0.008336946 | HDAC11 |
| chr3 | 12484251 | 12484813 | 563 | 10 | 0.000406541 | 0.000167 | 0.01285 | 6.21E-05 | -2.20E-05 | TSEN2 |
| chr11 | 84923082 | 84923691 | 610 | 9 | 0.000406823 | 4.06E-05 | 0.02315 | 6.78E-05 | 0.011204704 | DLG2 |
| chr6 | 169158801 | 169159301 | 501 | 5 | 0.000406844 | 3.90E-05 | 0.02309 | 0.00015 | 0.001962344 | LINC01615 |
| chr18 | 28175677 | 28175978 | 302 | 3 | 0.000407061 | 0.01374 | 0.00015 | 0.00062 | 0.000324865 | CDH2 |
| chr12 | 12824328 | 12824582 | 255 | 2 | 0.000407211 | 2.52E-05 | 0.00101 | 3.78E-05 | -0.003653108 | APOLD1, DDX47 |
| chr6 | 44176292 | 44176476 | 185 | 3 | 0.000407318 | 5.86E-05 | 0.00023 | 3.38E-05 | -0.006019177 | CAPN11 |
| chr17 | 37742749 | 37743298 | 550 | 5 | 0.000407547 | 0.015472 | 0.00088 | 0.0002 | 0.001833648 | HNF1B |
| chr17 | 80855485 | 80855901 | 417 | 3 | 0.000407549 | 3.47E-05 | 0.0024 | 6.32E-05 | -0.007132563 | RPTOR |
| chr4 | 713545 | 713667 | 123 | 3 | 0.000407671 | 5.01E-05 | 0.00056 | 5.47E-05 | -0.00586665 | PCGF3 |
| chr1 | 112699217 | 112699544 | 328 | 3 | 0.000407772 | 0.000303 | 0.00011 | 6.20E-05 | -0.001407127 | MOV10 |
| chr1 | 2860648 | 2860940 | 293 | 3 | 0.000407947 | 0.006409 | 0.00168 | 0.00198 | -0.004449279 | NA |
| chr11 | 123939972 | 123940403 | 432 | 2 | 0.000408081 | 3.32E-05 | 0.00096 | 4.74E-05 | -0.003612324 | OR4D5 |
| chr7 | 101843741 | 101844654 | 914 | 2 | 0.000408131 | 1.79E-06 | 0.0003 | 3.12E-06 | 0.014017343 | CUX1 |
| chr14 | 102033961 | 102034135 | 175 | 4 | 0.000408217 | 4.33E-05 | 0.00369 | 4.95E-05 | -0.002405425 | DYNC1H1 |
| chr16 | 55326270 | 55326421 | 152 | 2 | 0.000408381 | 0.0001 | 0.00038 | 8.73E-05 | -0.015159954 | IRX6, AC109462.1 |
| chr2 | 229715809 | 229715858 | 50 | 2 | 0.000408605 | 0.000105 | 0.00054 | 0.0001 | -0.005709937 | NA |
| chr11 | 118532942 | 118533132 | 191 | 2 | 0.000409062 | 0.000109 | 0.00023 | 7.57E-05 | -0.006005552 | TMEM25 |
| chr12 | 55926917 | 55927424 | 508 | 11 | 0.000409309 | 0.001048 | 0.00184 | 0.00015 | -0.000299587 | DGKA, PYM1 |
| chr11 | 123166228 | 123166434 | 207 | 3 | 0.000409492 | 0.000149 | 9.52E-05 | 4.21E-05 | -0.008287802 | CLMP |
| chr4 | 75513930 | 75514243 | 314 | 5 | 0.000409904 | 0.00028 | 0.00562 | 0.00029 | -0.004108242 | THAP6, RCHY1 |
| chr11 | 60699100 | 60699590 | 491 | 4 | 0.000410102 | 6.27E-05 | 0.00084 | 3.56E-05 | -0.00279254 | MS4A8 |
| chr10 | 697821 | 698033 | 213 | 3 | 0.000410102 | 0.046328 | 0.00099 | 0.00046 | 0.00260392 | NA |
| chr10 | 78528987 | 78529018 | 32 | 2 | 0.000410643 | 4.50E-05 | 0.00019 | 3.90E-05 | -0.001176255 | AC012560.1, LINC00595 |
| chr17 | 37406956 | 37407495 | 540 | 12 | 0.000410732 | 0.000119 | 0.0023 | 6.36E-05 | 0.001318121 | TADA2A |
| chr3 | 66139788 | 66139797 | 10 | 2 | 0.000411399 | 0.002806 | 5.89E-05 | 8.12E-05 | -0.008316008 | SLC25A26 |
| chr19 | 40750688 | 40751212 | 525 | 9 | 0.0004114 | 0.001281 | 8.09E-05 | 5.46E-05 | 0.000887486 | SNRPA, C19orf54 |
| chr8 | 67065109 | 67065212 | 104 | 7 | 0.000411806 | 0.000833 | 0.0044 | 0.00017 | -0.000862805 | CSPP1, COPS5 |
| chr12 | 56646415 | 56646702 | 288 | 4 | 0.000411946 | 0.000264 | 0.00548 | 0.00037 | -0.00284919 | NA |
| chr4 | 40514623 | 40514874 | 252 | 4 | 0.000412308 | 2.01E-05 | 0.01021 | 5.69E-05 | -0.013401057 | RBM47 |
| chr2 | 100110504 | 100110514 | 11 | 2 | 0.000412702 | 7.99E-05 | 0.00054 | 8.40E-05 | -0.006213016 | AFF3 |
| chr12 | 66130801 | 66131245 | 445 | 9 | 0.000412975 | 1.43E-05 | 0.01478 | 2.21E-05 | -0.001468351 | LLPH-DT, AC078927.1 |
| chr4 | 83013614 | 83013803 | 190 | 3 | 0.00041308 | 1.77E-05 | 0.00584 | 4.28E-05 | -0.033458034 | NA |
| chr16 | 15502130 | 15502566 | 437 | 8 | 0.000413547 | 0.000397 | 0.00048 | 3.98E-05 | -0.002312504 | AC140504.1, BMERB1 |
| chr8 | 54381976 | 54382322 | 347 | 6 | 0.00041373 | 5.64E-05 | 0.00241 | 4.23E-05 | -0.003688948 | AC027250.2 |
| chr6 | 169765589 | 169765809 | 221 | 2 | 0.000414036 | 0.000168 | 0.00039 | 0.00013 | -0.011958381 | ERMARD |
| chr10 | 80183107 | 80183261 | 155 | 2 | 0.000414456 | 0.002055 | 7.34E-05 | 9.69E-05 | -0.005537882 | ANXA11 |
| chr7 | 157743989 | 157744035 | 47 | 2 | 0.000414493 | 6.71E-05 | 0.00011 | 3.70E-05 | -0.008232428 | PTPRN2 |
| chr19 | 2231869 | 2232171 | 303 | 2 | 0.000414535 | 0.000172 | 0.00014 | 7.34E-05 | -0.004253364 | DOT1L, PLEKHJ1 |
| chr11 | 11949937 | 11950029 | 93 | 2 | 0.000414658 | 0.000123 | 0.00019 | 7.38E-05 | -0.004979866 | USP47 |
| chr7 | 151868029 | 151868510 | 482 | 2 | 0.000415208 | 2.58E-05 | 0.0015 | 4.09E-05 | -0.006231375 | PRKAG2 |
| chr16 | 69482210 | 69482349 | 140 | 3 | 0.00041549 | 1.39E-05 | 0.00457 | 3.30E-05 | -0.020719013 | NA |
| chr9 | 5269207 | 5269568 | 362 | 2 | 0.000415606 | 2.54E-05 | 0.00133 | 3.96E-05 | 0.011980015 | NA |
| chr1 | 2296799 | 2296955 | 157 | 3 | 0.000415678 | 0.012967 | 8.05E-05 | 0.00016 | -0.004161458 | SKI |
| chr10 | 133189214 | 133189472 | 259 | 3 | 0.00041611 | 0.000206 | 0.00099 | 7.41E-05 | -0.002357782 | KNDC1 |
| chr7 | 44057808 | 44058036 | 229 | 2 | 0.000417004 | 0.000113 | 0.00117 | 0.00014 | 0.000786307 | DBNL |
| chr21 | 41166957 | 41166967 | 11 | 2 | 0.000417004 | 0.00095 | 0.0056 | 0.00137 | -0.019780356 | NA |
| chr16 | 3017625 | 3018127 | 503 | 5 | 0.000417214 | 0.205181 | 0.00095 | 0.01458 | -0.000436222 | CLDN6 |
| chr22 | 35431143 | 35431210 | 68 | 2 | 0.000417222 | 0.000269 | 0.00023 | 0.00013 | -0.006974161 | NA |
| chr12 | 4587291 | 4587490 | 200 | 3 | 0.000417379 | 4.08E-05 | 0.00051 | 4.19E-05 | -0.003018635 | DYRK4 |

| | | | | | | | | | | |
|---|---|---|---|---|---|---|---|---|---|---|
| chr12 | 2052964 | 2053527 | 564 | 11 | 0.000417587 | 0.011251 | 0.00325 | 0.00017 | 0.000810578 | CACNA1C |
| chr15 | 71214629 | 71214961 | 333 | 3 | 0.000417777 | 0.004776 | 0.0006 | 5.75E-05 | -0.011408794 | THSD4 |
| chr2 | 239104951 | 239105013 | 63 | 2 | 0.000417852 | 0.0001 | 0.00011 | 4.81E-05 | -0.002258986 | HDAC4 |
| chr22 | 42431635 | 42431748 | 114 | 2 | 0.000418386 | 0.000234 | 0.00113 | 0.00026 | -0.008490089 | NFAM1 |
| chr17 | 2712114 | 2712426 | 313 | 5 | 0.000418468 | 2.06E-05 | 0.0051 | 2.31E-05 | 0.012505977 | AC005696.1, CLUH |
| chr6 | 154448538 | 154448647 | 110 | 2 | 0.000418468 | 2.21E-05 | 0.00123 | 3.47E-05 | -0.011995456 | AL357075.4, CNKSR3 |
| chr17 | 304429 | 304880 | 452 | 5 | 0.000418907 | 3.71E-05 | 0.005 | 3.72E-05 | 0.034403127 | RPH3AL |
| chr6 | 73695156 | 73695325 | 170 | 2 | 0.000419541 | 0.005112 | 0.03116 | 0.00878 | -0.00676944 | AL590428.1 |
| chr6 | 30555536 | 30557335 | 1800 | 51 | 0.000419977 | 0.032994 | 0.00122 | 4.76E-05 | 0.00067879 | PRR3, GNL1 |
| chr17 | 39994349 | 39994720 | 372 | 3 | 0.000420591 | 0.002507 | 0.00535 | 0.00087 | -0.002295189 | PSMD3 |
| chr10 | 973086 | 973221 | 136 | 2 | 0.000420611 | 0.000196 | 0.00015 | 8.38E-05 | 0.00666334 | AL359878.1 |
| chr6 | 162444096 | 162444226 | 131 | 3 | 0.000420625 | 0.002877 | 0.00013 | 0.0001 | -0.004323339 | PRKN |
| chr9 | 131518744 | 131518884 | 141 | 2 | 0.000420963 | 4.66E-05 | 0.00039 | 5.02E-05 | -0.002627751 | POMT1, AL358781.2 |
| chr19 | 16052353 | 16052569 | 217 | 2 | 0.000421345 | 0.000104 | 0.00014 | 5.53E-05 | -0.005276286 | OR1AB1P |
| chr6 | 166252410 | 166252799 | 390 | 4 | 0.000421713 | 0.000239 | 0.00094 | 4.56E-05 | -0.004061629 | AL121956.1 |
| chr15 | 58409487 | 58410345 | 859 | 2 | 0.000422543 | 4.77E-06 | 0.00045 | 7.90E-06 | -0.0213598 | ALDH1A2 |
| chr3 | 138707734 | 138708196 | 463 | 4 | 0.000422776 | 4.20E-05 | 0.00149 | 6.76E-05 | -0.006631173 | PIK3CB |
| chr16 | 89704387 | 89705296 | 910 | 2 | 0.000422926 | 3.29E-06 | 0.00038 | 5.59E-06 | -0.008272552 | NA |
| chr20 | 50850716 | 50850879 | 164 | 2 | 0.000424646 | 0.0001 | 0.00014 | 5.56E-05 | -0.003914824 | BCAS4 |
| chr15 | 40900688 | 40900845 | 158 | 4 | 0.00042497 | 6.27E-05 | 0.00088 | 3.63E-05 | -0.000282131 | VPS18 |
| chr9 | 137243430 | 137243746 | 317 | 3 | 0.000425081 | 0.000546 | 0.00062 | 0.00018 | -0.004458565 | TUBB4B, FAM166A |
| chr1 | 147891994 | 147892179 | 186 | 2 | 0.000425352 | 0.000158 | 0.00011 | 5.80E-05 | -0.005221077 | NA |
| chr7 | 50492826 | 50492988 | 163 | 2 | 0.000425848 | 5.74E-05 | 7.89E-05 | 2.77E-05 | -0.002770993 | FIGNL1, DDC |
| chr12 | 131753087 | 131753244 | 158 | 3 | 0.000426076 | 0.000478 | 0.0001 | 7.54E-05 | -0.00012846 | SFSWAP |
| chr11 | 882319 | 882636 | 318 | 2 | 0.000426259 | 8.35E-05 | 0.00245 | 0.00013 | -0.010447346 | CHID1 |
| chr19 | 17079449 | 17079587 | 139 | 2 | 0.000426543 | 5.31E-05 | 0.00044 | 5.77E-05 | -0.006413488 | MYO9B |
| chr19 | 42256531 | 42256590 | 60 | 2 | 0.000426754 | 3.41E-05 | 0.00077 | 4.65E-05 | -0.008788963 | NA |
| chr1 | 243470804 | 243471297 | 494 | 2 | 0.000427006 | 4.20E-05 | 0.00114 | 6.01E-05 | -0.009206609 | SDCCAG8 |
| chr9 | 74834165 | 74834382 | 218 | 2 | 0.000427148 | 0.000108 | 0.00014 | 5.64E-05 | -0.001269254 | TRPM6 |
| chr12 | 103929589 | 103929912 | 324 | 4 | 0.000427552 | 0.000155 | 0.00491 | 0.00019 | 0.001847021 | TTC41P |
| chr21 | 46648491 | 46648836 | 346 | 5 | 0.000427553 | 0.006894 | 0.00669 | 0.00023 | -0.001697157 | PRMT2 |
| chr10 | 49524251 | 49524450 | 200 | 2 | 0.000427586 | 0.000191 | 0.00058 | 0.00017 | -0.007145606 | ERCC6 |
| chr11 | 68870100 | 68870632 | 533 | 2 | 0.000427629 | 2.62E-05 | 0.00097 | 3.89E-05 | -0.038317304 | NA |
| chr5 | 959565 | 960089 | 525 | 2 | 0.000427715 | 2.47E-05 | 0.0013 | 3.85E-05 | -0.013198681 | NA |
| chr7 | 1950919 | 1951433 | 515 | 2 | 0.00042786 | 0.000137 | 0.0017 | 0.00019 | -0.015159895 | MAD1L1, AC069288.1 |
| chr10 | 6221469 | 6221643 | 175 | 2 | 0.000428491 | 0.000191 | 0.00073 | 0.00018 | -0.005753056 | PFKFB3 |
| chr1 | 15081261 | 15081439 | 179 | 2 | 0.000428901 | 2.57E-05 | 0.00085 | 3.74E-05 | -0.046262137 | KAZN |
| chr10 | 99537751 | 99537885 | 135 | 2 | 0.000429036 | 6.49E-05 | 0.00185 | 9.56E-05 | -0.01994372 | NA |
| chr20 | 59410364 | 59411117 | 754 | 2 | 0.00042929 | 7.22E-06 | 0.00064 | 1.19E-05 | -0.006459554 | NA |
| chr6 | 170403620 | 170404120 | 501 | 5 | 0.000429896 | 0.001155 | 0.00012 | 9.82E-05 | -0.002105597 | FAM120B |
| chr10 | 121500841 | 121501037 | 197 | 2 | 0.000429939 | 2.86E-05 | 0.00067 | 3.92E-05 | -0.007706171 | FGFR2 |
| chr7 | 99416150 | 99416226 | 77 | 2 | 0.000430323 | 0.000908 | 7.20E-05 | 7.76E-05 | -0.005451371 | BUD31 |
| chr19 | 5929831 | 5929892 | 62 | 2 | 0.000430368 | 3.51E-05 | 0.00033 | 3.87E-05 | -0.005697793 | RANBP3 |
| chr7 | 28481560 | 28481722 | 163 | 3 | 0.000430668 | 9.92E-05 | 0.00021 | 4.70E-05 | 0.003492532 | CREB5 |
| chr2 | 200812433 | 200812855 | 423 | 4 | 0.000431028 | 0.00113 | 7.78E-05 | 0.00011 | -0.004118469 | BZW1 |
| chr8 | 139789934 | 139790374 | 441 | 6 | 0.000431333 | 9.89E-05 | 0.00613 | 4.15E-05 | -0.000366845 | TRAPPC9 |
| chr15 | 75451412 | 75451921 | 510 | 9 | 0.000432105 | 0.010513 | 0.00243 | 0.00047 | 0.001192434 | SIN3A |
| chr21 | 44592541 | 44592639 | 99 | 4 | 0.000432114 | 1.66E-05 | 0.00943 | 4.86E-05 | -0.017220494 | KRTAP10-4, TSPEAR |
| chr17 | 8392496 | 8393001 | 506 | 4 | 0.000432364 | 0.00034 | 0.00042 | 0.00012 | -0.003508096 | RNF222 |
| chr1 | 10475330 | 10475528 | 199 | 2 | 0.000432783 | 0.00032 | 0.00017 | 0.00011 | 0.002585569 | PEX14 |
| chr7 | 13989329 | 13989889 | 561 | 11 | 0.000433067 | 5.53E-05 | 0.00829 | 4.14E-05 | -0.000385816 | ETV1 |
| chr13 | 44877038 | 44877135 | 98 | 3 | 0.000433266 | 0.000623 | 0.00029 | 5.60E-05 | -0.00432034 | NA |
| chr3 | 46565860 | 46566109 | 250 | 2 | 0.000433266 | 0.000332 | 0.00563 | 0.00057 | -0.005604315 | LRRC2 |
| chr19 | 48490649 | 48490786 | 138 | 2 | 0.000433266 | 2.10E-05 | 0.00132 | 3.34E-05 | -0.017999605 | LMTK3 |
| chr7 | 144050329 | 144050503 | 175 | 3 | 0.000433343 | 0.037267 | 3.38E-05 | 0.00013 | -0.004872392 | OR2A5 |
| chr10 | 31827126 | 31827529 | 404 | 3 | 0.000433343 | 0.000203 | 0.00015 | 6.22E-05 | -0.002674215 | ARHGAP12 |
| chr8 | 13116235 | 13116393 | 159 | 3 | 0.000433444 | 0.000117 | 0.00364 | 0.00021 | -0.008109459 | DLC1 |
| chr7 | 2138425 | 2138630 | 206 | 2 | 0.000433776 | 6.20E-05 | 0.00245 | 9.60E-05 | -0.016968617 | MAD1L1, AC069288.1 |
| chr3 | 49029543 | 49030075 | 533 | 7 | 0.000433874 | 0.016847 | 0.00065 | 0.00053 | -0.000102281 | AC137630.3, QRICH1 |
| chr19 | 43619675 | 43620146 | 472 | 11 | 0.000434551 | 0.000236 | 0.02044 | 0.00018 | 0.001417508 | NA |
| chr21 | 32842076 | 32842119 | 44 | 2 | 0.000434798 | 0.001275 | 0.00012 | 0.00014 | -0.005034355 | C21orf62-AS1 |
| chr14 | 29354499 | 29354781 | 283 | 3 | 0.000435098 | 3.23E-05 | 0.00053 | 3.84E-05 | -0.002065862 | AL135878.1, AL133166.1 |
| chr12 | 49131429 | 49131708 | 280 | 8 | 0.000435401 | 0.005443 | 0.00213 | 0.00032 | 0.00163668 | AC011603.2 |
| chr2 | 199324741 | 199324971 | 231 | 2 | 0.000435466 | 0.000102 | 0.00044 | 9.33E-05 | -0.005466913 | SATB2 |
| chr3 | 124787114 | 124787232 | 119 | 2 | 0.000435853 | 0.000156 | 9.67E-05 | 5.37E-05 | -0.007747083 | ITGB5-AS1, ITGB5 |
| chr16 | 50309434 | 50310412 | 979 | 3 | 0.0004362 | 1.78E-06 | 0.00159 | 4.76E-06 | -0.006481979 | ADCY7 |
| chr16 | 4262875 | 4263397 | 523 | 2 | 0.000436335 | 0.002185 | 0.00086 | 0.00091 | -0.002555761 | TFAP4 |
| chr12 | 89351100 | 89351415 | 316 | 5 | 0.00043648 | 0.002726 | 0.00099 | 0.00014 | -6.91E-05 | AC024909.1, DUSP6 |
| chr6 | 30672518 | 30673238 | 721 | 27 | 0.000436664 | 0.029396 | 0.00469 | 0.00033 | 0.000500954 | DHX16 |
| chr16 | 85591901 | 85592065 | 165 | 3 | 0.000436901 | 0.031947 | 8.12E-05 | 0.00034 | -0.004048794 | GSE1 |
| chr21 | 26167788 | 26167902 | 115 | 3 | 0.000436943 | 1.62E-05 | 0.0033 | 3.40E-05 | -0.014700942 | AP001439.1, APP |
| chr3 | 179562858 | 179563377 | 520 | 5 | 0.000437015 | 0.001272 | 0.00022 | 0.00033 | 0.001501247 | ACTL6A |
| chr12 | 75305084 | 75305098 | 15 | 2 | 0.000437222 | 3.69E-05 | 0.00173 | 5.70E-05 | -0.014571081 | CAPS2 |
| chr17 | 7010202 | 7010627 | 426 | 2 | 0.000437868 | 3.05E-05 | 0.00167 | 4.78E-05 | -0.011601847 | ALOX12, ALOX12-AS1, AC040977.2 |
| chr9 | 36036454 | 36036738 | 285 | 4 | 0.000437912 | 0.006101 | 0.00546 | 0.00046 | -0.020062473 | NA |
| chr9 | 110408622 | 110408901 | 280 | 4 | 0.000438103 | 0.000523 | 0.00011 | 7.57E-05 | -0.002405758 | SVEP1 |
| chr8 | 127959813 | 127959871 | 59 | 2 | 0.000438681 | 0.0002 | 5.74E-05 | 3.89E-05 | -0.006520099 | PVT1 |
| chr20 | 51768739 | 51769221 | 483 | 5 | 0.000438717 | 0.006362 | 0.00011 | 0.00011 | -0.008395735 | NA |
| chr19 | 6772359 | 6772467 | 109 | 3 | 0.000438949 | 0.00431 | 0.00068 | 0.00011 | 0.002757891 | NA |
| chr15 | 89411329 | 89411760 | 432 | 5 | 0.000439041 | 2.72E-05 | 0.00207 | 4.50E-05 | 0.001453695 | NA |
| chr4 | 2426434 | 2427241 | 808 | 3 | 0.000439871 | 9.09E-06 | 0.0003 | 7.72E-06 | -0.009525952 | CFAP99 |
| chr1 | 99790645 | 99790853 | 209 | 2 | 0.000440018 | 0.002069 | 0.0008 | 0.00085 | -0.005865553 | NA |
| chr15 | 30903354 | 30903872 | 519 | 11 | 0.000440041 | 0.019099 | 0.00162 | 0.00012 | 0.001059452 | FAN1 |
| chr2 | 98604970 | 98605232 | 263 | 2 | 0.000440557 | 8.18E-05 | 0.00034 | 7.29E-05 | -0.011880493 | COA5 |
| chr19 | 6371611 | 6371638 | 28 | 2 | 0.000440715 | 2.88E-05 | 0.00021 | 2.92E-05 | 0.000612084 | NA |
| chr16 | 28294212 | 28294500 | 289 | 2 | 0.000440765 | 0.002899 | 0.00015 | 0.00025 | -0.007796976 | SBK1 |
| chr19 | 48746675 | 48747195 | 521 | 6 | 0.000440925 | 0.00019 | 0.0049 | 8.39E-05 | -0.005382434 | IZUMO1 |
| chr15 | 65211469 | 65211491 | 23 | 2 | 0.000441055 | 9.14E-05 | 0.00344 | 0.00015 | -0.009673647 | CILP |
| chr1 | 23368008 | 23368456 | 449 | 5 | 0.000441147 | 0.020073 | 0.00703 | 0.0045 | 0.001373177 | ZNF436 |
| chr8 | 17735400 | 17735503 | 104 | 2 | 0.0004419 | 0.000181 | 0.00016 | 8.09E-05 | -0.0059236 | MTUS1 |
| chr19 | 1088783 | 1088988 | 206 | 2 | 0.000442097 | 0.001538 | 9.86E-05 | 0.00013 | -0.004048417 | POLR2E |
| chr15 | 42920648 | 42921159 | 512 | 10 | 0.000442257 | 0.006152 | 0.0015 | 0.00036 | -0.000376394 | TTBK2 |
| chr20 | 32051088 | 32051090 | 3 | 2 | 0.000442704 | 0.000107 | 0.00254 | 0.00017 | -0.01021712 | NA |
| chr21 | 42105933 | 42106677 | 745 | 3 | 0.000442743 | 7.63E-06 | 0.00354 | 1.91E-05 | -0.005176559 | UMODL1, UMODL1-AS1 |
| chr12 | 64451671 | 64452430 | 760 | 11 | 0.000442752 | 0.000549 | 0.01023 | 0.00019 | 0.000873405 | TBK1, AC135279.3 |
| chr8 | 94217370 | 94217644 | 275 | 4 | 0.000442906 | 0.007984 | 0.05248 | 0.01528 | -0.00330936 | NA |
| chr12 | 68452005 | 68452155 | 151 | 3 | 0.000443039 | 5.53E-05 | 0.0009 | 4.27E-05 | -0.039425299 | NA |
| chr16 | 26594854 | 26595139 | 286 | 3 | 0.000443039 | 0.006236 | 0.00011 | 9.39E-05 | -0.001480673 | AC002331.1 |
| chr10 | 60389201 | 60389571 | 371 | 2 | 0.000443382 | 0.035562 | 0.00122 | 0.00344 | -0.002314199 | ANK3 |
| chr5 | 80210680 | 80210900 | 221 | 2 | 0.0004434 | 5.83E-05 | 0.00062 | 6.86E-05 | -0.005678609 | SERINC5 |
| chr7 | 12719382 | 12719626 | 245 | 2 | 0.000443429 | 5.83E-05 | 0.00032 | 5.59E-05 | -0.028140612 | NA |
| chr5 | 80257328 | 80257787 | 460 | 3 | 0.000444293 | 2.47E-05 | 0.0042 | 5.02E-05 | -0.010637059 | NA |
| chr3 | 167902559 | 167902692 | 134 | 2 | 0.000444523 | 3.05E-05 | 0.00123 | 4.58E-05 | -0.003110925 | LRRC77P, AC026353.1 |
| chr9 | 129362204 | 129362378 | 175 | 2 | 0.000445014 | 3.70E-05 | 0.00057 | 4.65E-05 | -0.005741084 | NA |
| chr14 | 96264736 | 96265028 | 293 | 2 | 0.000445251 | 0.004884 | 0.00138 | 0.00188 | -0.005659461 | BDKRB1, AL355102.4 |
| chr3 | 49459320 | 49459600 | 281 | 2 | 0.000445731 | 0.000233 | 0.00014 | 8.37E-05 | -0.006648242 | NA |
| chr21 | 44089658 | 44089849 | 192 | 2 | 0.000445731 | 0.000184 | 0.00019 | 9.11E-05 | -0.006652445 | TRAPPC10 |
| chr16 | 31229435 | 31229674 | 240 | 2 | 0.000445889 | 0.000551 | 8.94E-05 | 8.20E-05 | -0.006886049 | TRIM72 |
| chr6 | 132824227 | 132824497 | 271 | 2 | 0.000446242 | 3.43E-05 | 0.00181 | 5.36E-05 | -0.012636589 | NA |
| chr16 | 85316038 | 85316370 | 333 | 2 | 0.000447056 | 6.79E-05 | 0.00055 | 7.48E-05 | -0.007989374 | GSE1 |
| chr1 | 19613336 | 19613646 | 311 | 2 | 0.000447281 | 5.20E-05 | 0.00035 | 5.31E-05 | -0.01537634 | MICOS10, NBL1, MICOS10-NBL1 |
| chr19 | 46042227 | 46042347 | 121 | 2 | 0.000448377 | 0.000266 | 0.00025 | 0.00013 | -0.01142684 | IGFL4 |
| chr14 | 36659173 | 36660397 | 1225 | 6 | 0.000448496 | 0.000202 | 0.00108 | 1.45E-05 | 0.006702402 | PAX9 |
| chr17 | 80965786 | 80966179 | 394 | 3 | 0.000448835 | 0.000726 | 0.0143 | 0.0013 | -0.003470518 | RPTOR |
| chr11 | 67374797 | 67375091 | 295 | 2 | 0.000449126 | 0.001359 | 0.0032 | 0.00144 | -0.008580419 | RAD9A, AP003419.3 |
| chr2 | 160416906 | 160417008 | 103 | 3 | 0.000449152 | 0.008266 | 6.47E-05 | 0.00016 | -0.003591977 | RBMS1 |
| chr8 | 19816959 | 19817385 | 427 | 11 | 0.000449305 | 0.002358 | 0.00991 | 6.96E-05 | 0.000124857 | NA |
| chr2 | 241742412 | 241742689 | 278 | 2 | 0.000449493 | 5.54E-05 | 0.00046 | 6.04E-05 | -0.009239899 | D2HGDH |
| chr14 | 105220064 | 105220245 | 182 | 3 | 0.000449527 | 0.017703 | 0.0001 | 0.00039 | -0.001903022 | BRF1 |
| chr8 | 144163045 | 144163285 | 241 | 3 | 0.000449827 | 0.001034 | 0.00072 | 0.0003 | -0.003043907 | MROH1 |
| chr4 | 52050685 | 52051709 | 1025 | 13 | 0.000449893 | 0.000336 | 0.00373 | 8.68E-05 | 0.002905645 | SPATA18 |
| chr12 | 68344230 | 68344563 | 334 | 3 | 0.000450292 | 3.40E-05 | 0.00366 | 5.93E-05 | -0.001328032 | LINC02384 |
| chr1 | 108830482 | 108830876 | 395 | 3 | 0.000450447 | 1.26E-05 | 0.00378 | 2.89E-05 | -0.015590626 | AKNAD1 |
| chr22 | 30396685 | 30396935 | 251 | 4 | 0.000450532 | 0.01356 | 0.00439 | 0.00321 | 0.003482325 | RNF215 |
| chr1 | 152110449 | 152110553 | 105 | 2 | 0.000450573 | 1.51E-05 | 0.00092 | 2.39E-05 | 0.010413577 | TCHH |
| chr17 | 38588378 | 38588816 | 439 | 2 | 0.000450626 | 2.89E-05 | 0.00136 | 4.44E-05 | 0.003210237 | SRCIN1 |
| chr1 | 220809854 | 220809951 | 98 | 3 | 0.000450632 | 0.005296 | 0.00068 | 0.00046 | -0.000477303 | MTARC1, AL445423.3 |
| chr3 | 172312264 | 172312283 | 20 | 2 | 0.0004508 | 0.000118 | 0.00015 | 6.29E-05 | -0.005176165 | FNDC3B |
| chr7 | 48663367 | 48663457 | 91 | 3 | 0.000451281 | 0.000682 | 0.00012 | 6.63E-05 | -0.007382652 | AC091770.1 |
| chr17 | 32008086 | 32008106 | 21 | 2 | 0.000451414 | 0.00094 | 0.00015 | 0.00017 | 0.001147719 | LRRC37B |
| chr19 | 17488902 | 17489313 | 412 | 3 | 0.000451486 | 1.20E-05 | 0.00485 | 2.97E-05 | -0.045085824 | SLC27A1, AC010618.3 |
| chr16 | 70419282 | 70419524 | 243 | 2 | 0.000451501 | 0.000131 | 0.00016 | 6.89E-05 | -0.010122758 | ST3GAL2 |
| chr13 | 31162374 | 31162917 | 544 | 9 | 0.000453225 | 0.002231 | 0.00377 | 0.00257 | 0.001239115 | HSPH1 |
| chr9 | 137463224 | 137463254 | 31 | 2 | 0.000453323 | 0.002028 | 8.09E-05 | 0.00011 | -0.00386627 | PNPLA7 |
| chr16 | 3067861 | 3068214 | 354 | 3 | 0.000453495 | 0.000381 | 0.0003 | 0.00013 | -0.005134764 | IL32 |
| chr20 | 50027884 | 50027996 | 113 | 2 | 0.000453588 | 3.11E-05 | 0.00053 | 3.99E-05 | -0.012246885 | NA |
| chr10 | 26216140 | 26216316 | 177 | 3 | 0.000453778 | 2.99E-05 | 0.00416 | 5.82E-05 | 0.004399725 | NA |
| chr1 | 154956979 | 154957539 | 561 | 6 | 0.000453905 | 0.00114 | 0.0056 | 8.58E-05 | -0.00324121 | PYGO2 |
| chr17 | 35808884 | 35809423 | 540 | 10 | 0.00045421 | 3.21E-06 | 0.01635 | 9.63E-06 | -0.000779696 | TAF15, AC015849.2 |
| chr19 | 12092383 | 12092829 | 447 | 4 | 0.000454224 | 0.00016 | 0.00366 | 0.0003 | 0.001781803 | AC022415.2, ZNF788P |
| chr17 | 40549508 | 40549643 | 136 | 2 | 0.000454598 | 0.000151 | 0.00059 | 0.00014 | -0.009090099 | NA |
| chr9 | 120928308 | 120928393 | 86 | 3 | 0.000455106 | 3.08E-05 | 0.00149 | 4.74E-05 | -0.013453458 | TRAF1 |
| chr17 | 32323693 | 32324412 | 720 | 2 | 0.000455188 | 1.06E-05 | 0.00074 | 1.70E-05 | -0.01533454 | RHBDL3 |
| chr7 | 128937539 | 128938122 | 584 | 11 | 0.000455215 | 0.001516 | 0.00073 | 8.29E-05 | 0.001736514 | IRF5 |
| chr4 | 177444695 | 177444859 | 165 | 4 | 0.000455355 | 2.91E-05 | 0.00215 | 3.70E-05 | 0.007628553 | AC078881.1 |
| chr8 | 60852832 | 60852909 | 78 | 3 | 0.000455798 | 6.05E-05 | 0.00201 | 7.11E-05 | -0.00362735 | CHD7 |
| chr3 | 24148999 | 24149312 | 314 | 2 | 0.000455829 | 3.92E-05 | 0.00054 | 4.81E-05 | -0.010432343 | THRB |
| chr3 | 71168050 | 71168126 | 77 | 2 | 0.000455829 | 5.53E-05 | 0.00218 | 8.51E-05 | -0.010050065 | FOXP1, AC097634.4 |
| chr20 | 23656482 | 23656587 | 106 | 2 | 0.00045583 | 2.77E-05 | 0.00033 | 3.24E-05 | -0.012213331 | AL121894.3 |
| chr11 | 92352265 | 92352373 | 109 | 3 | 0.000456016 | 0.001136 | 5.49E-05 | 7.20E-05 | -0.005100335 | FAT3 |
| chr1 | 2300486 | 2301042 | 557 | 5 | 0.000456032 | 0.004173 | 0.00119 | 3.78E-05 | 0.001990786 | SKI |
| chr1 | 99850369 | 99850830 | 462 | 8 | 0.000456069 | 0.001379 | 0.01398 | 0.00025 | 0.002603182 | AGL |
| chr19 | 11453627 | 11453724 | 98 | 4 | 0.000456088 | 0.000139 | 0.00227 | 0.00015 | -0.005073425 | ELAVL3 |
| chr14 | 103806848 | 103807252 | 405 | 2 | 0.000456115 | 2.79E-05 | 0.00101 | 4.13E-05 | -0.025776725 | PPP1R13B |
| chr8 | 98883589 | 98883911 | 323 | 2 | 0.000456381 | 3.54E-05 | 0.00065 | 4.62E-05 | -0.000630251 | STK3 |
| chr10 | 488912 | 488982 | 71 | 2 | 0.000456627 | 0.000192 | 0.00011 | 6.42E-05 | -0.004031105 | DIP2C |
| chr12 | 96571412 | 96571434 | 23 | 2 | 0.000456627 | 0.060394 | 5.37E-05 | 9.52E-05 | -0.007496397 | CFAP54 |
| chr6 | 31740449 | 31740735 | 287 | 6 | 0.000456639 | 0.006184 | 0.02236 | 0.00492 | 5.81E-05 | MSH5, MSH5-SAPCD1 |
| chr11 | 69047981 | 69048214 | 234 | 3 | 0.000456856 | 0.000935 | 0.00331 | 0.00057 | -0.007645711 | NA |
| chr17 | 75502046 | 75502461 | 416 | 4 | 0.000456875 | 0.001613 | 0.00081 | 0.00013 | -0.001100454 | CASKIN2 |
| chr18 | 76619430 | 76619510 | 81 | 2 | 0.000456875 | 0.001108 | 6.46E-05 | 7.34E-05 | -0.007085643 | LINC00683 |
| chr17 | 45261673 | 45262148 | 476 | 6 | 0.000457342 | 0.000113 | 0.00747 | 4.77E-05 | 0.00181515 | MAP3K14-AS1, SPATA32 |

| | | | | | | | | | | |
|---|---|---|---|---|---|---|---|---|---|---|
| chr13 | 55653072 | 55653405 | 334 | 2 | 0.000457384 | 7.29E-05 | 0.00014 | 4.50E-05 | -0.006395213 | NA |
| chr21 | 37417665 | 37417809 | 145 | 4 | 0.000457593 | 1.53E-05 | 0.01094 | 4.81E-05 | -0.001152737 | DYRK1A |
| chr19 | 39315882 | 39316125 | 244 | 2 | 0.000457597 | 0.046711 | 0.00203 | 0.00575 | -0.001542615 | LRFN1 |
| chr15 | 28112301 | 28112332 | 32 | 2 | 0.000457718 | 0.003849 | 8.08E-05 | 0.00013 | 0.00218289 | HERC2 |
| chr20 | 62401931 | 62401952 | 22 | 2 | 0.000457838 | 6.64E-05 | 0.0028 | 0.0001 | -0.005831697 | CABLES2 |
| chr17 | 38344354 | 38344664 | 311 | 2 | 0.000457964 | 0.000493 | 0.00787 | 0.00087 | 0.001170075 | NA |
| chr7 | 74254607 | 74254900 | 294 | 6 | 0.000458048 | 0.023618 | 0.00148 | 0.00097 | 0.000998966 | NA |
| chr1 | 204434478 | 204434569 | 92 | 3 | 0.000458182 | 0.004997 | 0.00075 | 0.00025 | -0.006335019 | AL606489.1, PIK3C2B |
| chr11 | 63710548 | 63710737 | 190 | 2 | 0.000458379 | 3.08E-05 | 0.00066 | 4.15E-05 | -0.012298383 | RTN3 |
| chr15 | 52749799 | 52749813 | 15 | 2 | 0.000458379 | 0.000951 | 6.20E-05 | 6.76E-05 | -0.003232694 | NA |
| chr12 | 54054945 | 54054985 | 41 | 2 | 0.000458486 | 0.000278 | 0.00196 | 0.00036 | 0.023549819 | HOXC4 |
| chr20 | 45771525 | 45771561 | 37 | 2 | 0.000458737 | 0.051196 | 5.41E-05 | 9.59E-05 | -0.004317071 | WFDC3 |
| chr19 | 8595874 | 8596380 | 507 | 3 | 0.000459428 | 2.38E-05 | 0.00411 | 5.15E-05 | -0.005846593 | ADAMTS10 |
| chr21 | 42495815 | 42495910 | 96 | 2 | 0.000459484 | 0.002023 | 0.00065 | 0.00072 | 0.007628769 | RSPH1 |
| chr1 | 110472677 | 110472891 | 215 | 2 | 0.00045967 | 0.000631 | 8.48E-05 | 8.21E-05 | -0.003617986 | AL358215.2 |
| chr8 | 141228858 | 141229224 | 367 | 3 | 0.000459856 | 0.000132 | 0.01177 | 0.00033 | -0.002204084 | SLC45A4 |
| chr17 | 81845822 | 81846237 | 416 | 2 | 0.000460171 | 0.001364 | 0.01271 | 0.00233 | -0.003917274 | P4HB |
| chr15 | 87940673 | 87940836 | 164 | 2 | 0.000460464 | 4.51E-05 | 0.00064 | 5.62E-05 | -0.009495039 | NTRK3 |
| chr12 | 119804526 | 119804709 | 184 | 4 | 0.000460795 | 0.000568 | 0.00048 | 6.62E-05 | -0.004676271 | CIT |
| chr15 | 66918473 | 66918658 | 186 | 2 | 0.000460817 | 8.23E-05 | 0.00086 | 9.88E-05 | -0.002651895 | NA |
| chr6 | 106539578 | 106539798 | 221 | 3 | 0.000460988 | 5.68E-05 | 0.0013 | 4.76E-05 | -0.005706333 | CRYBG1 |
| chr6 | 159983419 | 159983659 | 241 | 2 | 0.000461001 | 9.87E-05 | 0.0003 | 7.82E-05 | 0.007715244 | IGF2R |
| chr2 | 98556528 | 98556853 | 326 | 2 | 0.000461211 | 3.94E-05 | 0.00056 | 4.89E-05 | -0.006294899 | INPP4A |
| chr12 | 48716644 | 48717070 | 427 | 7 | 0.00046183 | 0.069471 | 5.80E-05 | 0.00013 | -0.000383426 | CCNT1 |
| chr1 | 16154254 | 16154308 | 55 | 2 | 0.000462083 | 0.000542 | 0.00042 | 0.00027 | -0.011005174 | EPHA2 |
| chr5 | 887073 | 887304 | 232 | 4 | 0.000462609 | 7.29E-05 | 0.00068 | 4.88E-05 | -0.002473736 | BRD9 |
| chr6 | 37320723 | 37320862 | 140 | 2 | 0.000462813 | 3.16E-05 | 0.00066 | 4.23E-05 | -0.012310572 | TBC1D22B |
| chr2 | 219332324 | 219332552 | 229 | 2 | 0.000463666 | 0.001501 | 0.00111 | 0.00087 | -0.011973224 | RESP18 |
| chr15 | 100866753 | 100866794 | 42 | 2 | 0.000463666 | 0.000285 | 0.00013 | 8.47E-05 | -0.008018735 | AC015712.6, AC015712.4, AC015712.5 |
| chr2 | 70995395 | 70995525 | 131 | 3 | 0.000463884 | 0.002687 | 0.00873 | 0.00227 | 0.002211461 | AC007040.1 |
| chr21 | 42637790 | 42637945 | 156 | 2 | 0.000464065 | 2.46E-05 | 0.0011 | 3.74E-05 | -0.007818323 | NA |
| chr4 | 1192681 | 1192823 | 143 | 2 | 0.000464098 | 4.29E-05 | 0.00119 | 6.17E-05 | -0.005123641 | SPON2 |
| chr1 | 235093182 | 235093605 | 424 | 5 | 0.000464434 | 1.17E-05 | 0.00674 | 3.52E-05 | -0.009277887 | NA |
| chr2 | 227327376 | 227328065 | 690 | 6 | 0.000465132 | 1.95E-05 | 0.00369 | 3.35E-05 | -0.003788154 | MFF |
| chr17 | 33905554 | 33906105 | 552 | 2 | 0.00046542 | 1.62E-05 | 0.00111 | 2.61E-05 | -0.009072407 | ASIC2, AC024610.2 |
| chr17 | 35735376 | 35735554 | 179 | 2 | 0.00046542 | 2.77E-05 | 0.00029 | 3.13E-05 | -0.005046148 | TAF15, RASL10B |
| chr16 | 89321803 | 89321988 | 186 | 3 | 0.000465519 | 0.001078 | 0.00239 | 0.00028 | -0.003691943 | AC137932.2, ANKRD11 |
| chr7 | 129188924 | 129189524 | 601 | 4 | 0.000465835 | 0.005959 | 0.00204 | 0.00253 | 0.000175022 | SMO |
| chr2 | 231530558 | 231530752 | 195 | 4 | 0.000465924 | 0.005369 | 0.00917 | 0.00211 | -0.006790767 | NA |
| chr9 | 117708275 | 117708616 | 342 | 3 | 0.000466009 | 0.00061 | 0.00128 | 7.60E-05 | -0.004682818 | AL160272.2, TLR4 |
| chr12 | 130070185 | 130070650 | 466 | 7 | 0.000466129 | 0.000363 | 0.00224 | 6.12E-05 | -0.010749701 | LINC02419 |
| chr10 | 133029074 | 133029184 | 111 | 2 | 0.000466911 | 0.007632 | 5.29E-05 | 8.25E-05 | -0.007478014 | NA |
| chr7 | 2903689 | 2903851 | 163 | 2 | 0.000466932 | 2.15E-05 | 0.00087 | 3.24E-05 | -0.010124496 | NA |
| chr16 | 684766 | 685282 | 517 | 4 | 0.000466966 | 0.041711 | 0.00995 | 0.00711 | -0.004000401 | WDR24 |
| chr2 | 20177886 | 20178123 | 238 | 2 | 0.000467199 | 0.000862 | 8.93E-05 | 9.43E-05 | -0.004290273 | NA |
| chr1 | 156053045 | 156053579 | 535 | 5 | 0.000467282 | 0.018168 | 0.00053 | 0.00157 | -2.12E-05 | UBQLN4 |
| chr15 | 90086216 | 90087176 | 961 | 5 | 0.000467885 | 0.001121 | 0.00628 | 0.00017 | -0.003534986 | IDH2 |
| chr12 | 97557552 | 97557934 | 383 | 2 | 0.00046816 | 3.93E-05 | 0.00064 | 5.04E-05 | -0.015634216 | RMST |
| chr17 | 47664984 | 47665072 | 89 | 2 | 0.000468636 | 0.000149 | 0.00071 | 0.00015 | -0.005446478 | KPNB1 |
| chr20 | 59032984 | 59033036 | 53 | 2 | 0.00046923 | 0.001888 | 0.0013 | 0.0011 | -0.011361841 | NA |
| chr6 | 3806597 | 3806746 | 150 | 2 | 0.000469326 | 2.74E-05 | 0.00098 | 4.04E-05 | -0.006779578 | NA |
| chr15 | 25117041 | 25117273 | 233 | 2 | 0.000469425 | 7.07E-05 | 0.00023 | 5.66E-05 | -0.020905503 | SNHG14 |
| chr2 | 169360697 | 169360891 | 195 | 2 | 0.000469652 | 8.44E-05 | 0.00138 | 0.00011 | -0.008966572 | LRP2 |
| chr2 | 202072885 | 202073480 | 596 | 5 | 0.000469772 | 0.001705 | 0.00116 | 8.75E-05 | 0.000222026 | KIAA2012 |
| chr11 | 2328280 | 2328523 | 244 | 3 | 0.000469924 | 0.000161 | 0.00138 | 5.66E-05 | 0.000537983 | NA |
| chr3 | 188725192 | 188725321 | 130 | 2 | 0.000470489 | 9.74E-05 | 0.00013 | 5.27E-05 | -0.003769172 | LPP |
| chr2 | 46898527 | 46898934 | 408 | 2 | 0.00047062 | 7.66E-05 | 2.93E-05 | 1.50E-05 | -0.01016577 | NA |
| chr15 | 90193012 | 90193234 | 223 | 2 | 0.000470659 | 5.41E-05 | 0.00112 | 7.45E-05 | -0.016942828 | SEMA4B |
| chr12 | 124463196 | 124463522 | 327 | 2 | 0.000470712 | 4.18E-05 | 0.00067 | 5.35E-05 | 0.00084243 | NCOR2 |
| chr10 | 122846389 | 122846447 | 59 | 3 | 0.000470767 | 0.000106 | 0.00108 | 9.32E-05 | 5.35E-05 | AC073585.1 |
| chr17 | 17847541 | 17847596 | 56 | 2 | 0.000471257 | 2.47E-05 | 0.00137 | 3.88E-05 | -0.016638937 | TOM1L2 |
| chr20 | 45306721 | 45306888 | 168 | 5 | 0.000471823 | 7.49E-06 | 0.00328 | 1.67E-05 | -0.013056693 | RBPJL, MATN4 |
| chr21 | 38989685 | 38990303 | 619 | 5 | 0.000472665 | 6.91E-06 | 0.01223 | 2.79E-05 | -0.046156574 | AF064858.1 |
| chr5 | 10519793 | 10519971 | 179 | 2 | 0.000472665 | 0.001303 | 7.44E-05 | 8.80E-05 | -0.004157634 | LINC02213 |
| chr17 | 48554548 | 48554792 | 245 | 3 | 0.000472676 | 0.000642 | 0.00023 | 0.00014 | -0.004485627 | HOXB-AS3, HOXB3 |
| chr9 | 21778791 | 21778814 | 24 | 2 | 0.00047279 | 2.42E-05 | 0.00145 | 3.83E-05 | -0.022095309 | NA |
| chr17 | 75436094 | 75436166 | 73 | 2 | 0.00047279 | 2.42E-05 | 0.00146 | 3.83E-05 | -0.021046844 | NA |
| chr14 | 51599924 | 51599963 | 40 | 3 | 0.000473499 | 2.21E-05 | 0.00136 | 3.24E-05 | -0.004310697 | FRMD6, FRMD6-AS2 |
| chr1 | 50417657 | 50417722 | 66 | 2 | 0.000473575 | 0.000301 | 0.00161 | 0.00036 | 0.000562001 | DMRTA2 |
| chr9 | 78295972 | 78296169 | 198 | 3 | 0.000473642 | 5.72E-05 | 0.00354 | 9.19E-05 | -0.013359656 | NA |
| chr9 | 127983191 | 127983338 | 148 | 2 | 0.00047398 | 7.60E-05 | 0.00054 | 8.08E-05 | -0.007370829 | NA |
| chr1 | 119808750 | 119808824 | 75 | 2 | 0.000474042 | 0.013421 | 9.04E-05 | 0.00018 | -0.011486558 | REG4 |
| chr4 | 48102541 | 48102631 | 91 | 2 | 0.000474078 | 4.73E-05 | 0.00186 | 7.21E-05 | -0.008942084 | TXK |
| chr18 | 23872302 | 23872931 | 630 | 8 | 0.000474143 | 0.000102 | 0.00056 | 5.67E-05 | -0.003981747 | LAMA3 |
| chr6 | 26017711 | 26019157 | 1447 | 13 | 0.000474532 | 2.85E-06 | 0.00147 | 2.58E-06 | -0.007220562 | H1-1 |
| chr1 | 44031559 | 44032022 | 464 | 5 | 0.000474635 | 1.10E-05 | 0.007 | 3.05E-05 | 0.000229495 | AL139220.2 |
| chr3 | 150226310 | 150226625 | 316 | 2 | 0.000475189 | 4.54E-05 | 0.00167 | 6.85E-05 | -0.007924217 | NA |
| chr19 | 3570832 | 3571003 | 172 | 2 | 0.000475543 | 3.03E-05 | 0.00095 | 4.37E-05 | -0.008104357 | MFSD12 |
| chr11 | 67260914 | 67261680 | 767 | 3 | 0.000475554 | 0.002414 | 0.00026 | 0.0002 | -0.005380583 | AP001885.3 |
| chr17 | 39064635 | 39065215 | 581 | 2 | 0.000476688 | 1.83E-05 | 0.00107 | 2.89E-05 | -0.004787322 | RDM1P5, PLXDC1 |
| chr17 | 81172503 | 81173056 | 554 | 5 | 0.000476963 | 0.027668 | 0.00283 | 0.00023 | -0.002516399 | PVALEF |
| chr16 | 88497839 | 88498339 | 501 | 3 | 0.000477446 | 0.000243 | 0.00095 | 5.35E-05 | -0.002941115 | ZFPM1 |
| chr6 | 170083805 | 170084086 | 282 | 3 | 0.000477895 | 9.19E-05 | 0.0041 | 0.00015 | -0.001939142 | NA |
| chr5 | 56606802 | 56606934 | 133 | 2 | 0.000477976 | 6.43E-05 | 0.00264 | 0.0001 | 0.003688471 | NA |
| chr6 | 157036383 | 157036579 | 197 | 2 | 0.000478428 | 4.19E-05 | 0.00071 | 5.46E-05 | -0.011043655 | ARID1B |
| chr6 | 10432905 | 10433223 | 319 | 2 | 0.000478472 | 6.63E-05 | 0.00083 | 8.21E-05 | 0.000252166 | LINC00518 |
| chr16 | 88579036 | 88579124 | 89 | 2 | 0.000480022 | 5.32E-05 | 0.00033 | 5.28E-05 | -0.008065125 | ZC3H18 |
| chr18 | 45063325 | 45063971 | 647 | 6 | 0.000481023 | 0.000382 | 0.00018 | 8.24E-05 | 0.000423073 | SETBP1 |
| chr2 | 64908765 | 64908880 | 116 | 2 | 0.000481353 | 0.003465 | 4.01E-05 | 5.49E-05 | -0.011993271 | LINC02245 |
| chr3 | 64025109 | 64025228 | 120 | 2 | 0.000481353 | 0.000152 | 0.00083 | 0.00016 | -0.005478293 | NA |
| chr14 | 105468869 | 105469398 | 530 | 3 | 0.000481368 | 2.21E-05 | 0.00465 | 4.88E-05 | -0.00793004 | MTA1, AL928654.2 |
| chr1 | 246932723 | 246932815 | 93 | 3 | 0.00048147 | 0.001043 | 0.00058 | 0.00039 | -0.016122953 | NA |
| chr3 | 49422624 | 49423133 | 510 | 5 | 0.000481618 | 0.000236 | 0.00032 | 0.00013 | -0.005658658 | AC104452.1, AMT, NICN1 |
| chr16 | 24919802 | 24919982 | 181 | 2 | 0.0004821 | 3.52E-05 | 0.00064 | 4.57E-05 | 0.000148407 | ARHGAP17 |
| chr11 | 117176598 | 117176602 | 5 | 2 | 0.000482465 | 7.18E-05 | 0.0029 | 0.00011 | 0.000822984 | PAFAH1B2 |
| chr17 | 9240924 | 9240964 | 41 | 2 | 0.000482494 | 4.20E-05 | 0.00106 | 5.92E-05 | -0.010765625 | NTN1 |
| chr12 | 123458032 | 123458147 | 116 | 4 | 0.000482978 | 0.000767 | 0.00022 | 0.00015 | 0.001962021 | SNRNP35 |
| chr11 | 1302794 | 1303395 | 602 | 3 | 0.000483097 | 0.001644 | 0.0002 | 9.94E-05 | -0.004911367 | TOLLIP |
| chr1 | 153357305 | 153357406 | 102 | 2 | 0.00048389 | 0.007205 | 0.01224 | 0.00812 | -0.006165259 | NA |
| chr1 | 1393454 | 1393664 | 211 | 3 | 0.000484278 | 3.63E-05 | 0.00353 | 7.14E-05 | -0.002839976 | CCNL2 |
| chr12 | 54188017 | 54188095 | 79 | 3 | 0.000484883 | 0.078877 | 0.00017 | 0.00074 | -0.002616748 | SMUG1 |
| chr1 | 23978076 | 23978247 | 172 | 2 | 0.000484894 | 0.000129 | 0.00012 | 5.65E-05 | -0.009660122 | SRSF10 |
| chr4 | 71231256 | 71231281 | 26 | 2 | 0.000485201 | 6.44E-05 | 0.00232 | 9.85E-05 | -0.006615235 | SLC4A4 |
| chr17 | 4795024 | 4795104 | 81 | 2 | 0.000485227 | 0.000105 | 0.0036 | 0.00018 | -0.005761165 | NA |
| chr6 | 154085529 | 154085706 | 178 | 2 | 0.000485501 | 0.000544 | 0.00065 | 0.00035 | -0.000935094 | OPRM1 |
| chr14 | 102910577 | 102910816 | 240 | 3 | 0.000485656 | 0.002374 | 0.00015 | 0.00011 | -0.003042983 | TRAF3 |
| chr13 | 112116494 | 112116583 | 90 | 2 | 0.000485656 | 0.000684 | 0.00018 | 0.00017 | -0.005850227 | NA |
| chr1 | 168225992 | 168226320 | 329 | 4 | 0.000485894 | 0.000159 | 0.00291 | 0.00011 | 0.000366462 | SFT2D2 |
| chr15 | 63218253 | 63218743 | 491 | 3 | 0.000486126 | 0.000834 | 0.00023 | 6.64E-05 | -0.007461017 | RAB8B |
| chr19 | 18161049 | 18161274 | 226 | 3 | 0.000486318 | 0.001148 | 0.00021 | 0.00025 | -0.00794943 | AC007192.1, PIK3R2 |
| chr16 | 69132247 | 69132443 | 197 | 4 | 0.000486868 | 0.001229 | 0.02084 | 0.00297 | -0.003143854 | UTP4, CHTF8 |
| chr7 | 48088328 | 48088554 | 227 | 7 | 0.000487043 | 0.00041 | 0.00868 | 0.00061 | 0.000547458 | NA |
| chr3 | 47250675 | 47251185 | 511 | 3 | 0.000487648 | 2.17E-05 | 0.00348 | 4.29E-05 | -0.005334138 | KIF9 |
| chr10 | 122506596 | 122506870 | 275 | 3 | 0.000487896 | 0.000119 | 0.00192 | 8.00E-05 | -0.003386434 | HTRA1 |
| chr14 | 24190598 | 24190767 | 170 | 3 | 0.000488194 | 0.00015 | 0.00039 | 3.70E-05 | -0.002739445 | AL136295.22, TM9SF1, AL136295.1 |
| chr20 | 36942072 | 36942141 | 70 | 3 | 0.000488389 | 0.002645 | 0.00104 | 6.42E-05 | 0.001570625 | SAMHD1 |
| chr11 | 78209046 | 78209683 | 638 | 4 | 0.000488596 | 0.00329 | 0.01629 | 0.00402 | -0.000825866 | USP35 |
| chr10 | 75210598 | 75210915 | 318 | 7 | 0.000488688 | 0.001942 | 0.00026 | 0.0001 | 0.001393759 | VDAC2 |
| chr22 | 37925825 | 37926032 | 208 | 2 | 0.000488824 | 0.014966 | 0.00084 | 0.00198 | -0.002010375 | MICALL1 |
| chr2 | 239390934 | 239391357 | 424 | 5 | 0.000488831 | 0.000291 | 0.00575 | 0.00012 | 0.001265202 | HDAC4 |
| chr11 | 65116439 | 65116757 | 319 | 2 | 0.000488913 | 0.002165 | 0.00041 | 0.00053 | -0.000378376 | ZNHIT2 |
| chr22 | 38221464 | 38222575 | 1112 | 6 | 0.000489814 | 1.10E-05 | 0.00105 | 8.91E-06 | -0.00292126 | TMEM184B |
| chr5 | 61332025 | 61332549 | 525 | 7 | 0.000489901 | 0.013823 | 0.0017 | 0.00012 | -0.000101986 | ZSWIM6 |
| chr1 | 151372647 | 151372863 | 217 | 5 | 0.000490049 | 3.84E-05 | 0.0141 | 7.23E-05 | -0.004096691 | SELENBP1 |
| chr21 | 41812086 | 41812106 | 21 | 2 | 0.000490068 | 2.73E-05 | 0.0007 | 3.79E-05 | -0.001958523 | PRDM15 |
| chr21 | 46391284 | 46391435 | 152 | 2 | 0.000490183 | 0.000377 | 0.00011 | 8.78E-05 | 0.002346176 | PCNT |
| chr1 | 21534485 | 21534836 | 352 | 3 | 0.000490346 | 0.000102 | 0.00024 | 6.45E-05 | 0.002654826 | ALPL |
| chr2 | 233481686 | 233481899 | 214 | 2 | 0.000490543 | 0.00072 | 0.00015 | 0.00015 | -0.003707414 | USP40 |
| chr12 | 11729960 | 11730072 | 113 | 2 | 0.000490816 | 0.000152 | 0.00024 | 9.19E-05 | -0.007927102 | ETV6 |
| chr14 | 22497359 | 22497648 | 290 | 2 | 0.000491516 | 8.64E-05 | 0.0011 | 0.00011 | -0.011549651 | NA |
| chr4 | 1113414 | 1113479 | 66 | 3 | 0.00049154 | 1.51E-05 | 0.00054 | 1.09E-05 | -0.016530596 | RNF212 |
| chr10 | 133358962 | 133358987 | 26 | 2 | 0.000492571 | 0.000125 | 0.00343 | 0.00021 | -0.009203116 | NA |
| chr17 | 67467925 | 67468181 | 257 | 2 | 0.000492771 | 4.23E-05 | 0.00055 | 5.15E-05 | -0.008166455 | PITPNC1 |
| chr12 | 119988814 | 119988980 | 167 | 2 | 0.000493098 | 0.000131 | 0.00013 | 6.11E-05 | 0.00327815 | NA |
| chr3 | 128069614 | 128069730 | 117 | 3 | 0.000493154 | 0.00098 | 0.00039 | 7.14E-05 | -0.000907596 | SEC61A1, RUVBL1 |
| chr7 | 823053 | 823194 | 142 | 3 | 0.000493413 | 2.50E-05 | 0.00138 | 3.26E-05 | -0.005596698 | SUN1 |
| chr15 | 92820334 | 92820534 | 201 | 3 | 0.000493628 | 0.001282 | 0.00012 | 0.00013 | 0.002690416 | CHASERR |
| chr2 | 241776692 | 241776973 | 282 | 6 | 0.000493725 | 3.05E-05 | 0.00182 | 3.55E-05 | -0.005701997 | GAL3ST2 |
| chr19 | 48453008 | 48453632 | 625 | 3 | 0.000493744 | 0.009961 | 0.00017 | 0.00061 | -0.001190359 | GRWD1 |
| chr10 | 77857385 | 77857442 | 58 | 2 | 0.000493842 | 0.000301 | 0.00012 | 8.43E-05 | -0.007012711 | DLG5 |
| chr4 | 140047681 | 140047885 | 205 | 2 | 0.000494034 | 0.000542 | 0.00012 | 0.0001 | -0.008885124 | MAML3 |
| chr13 | 46641786 | 46641988 | 203 | 2 | 0.000494157 | 3.41E-05 | 0.00112 | 4.97E-05 | -0.023391185 | LRCH1 |
| chr10 | 89328294 | 89328393 | 100 | 2 | 0.000494181 | 0.007129 | 0.00012 | 0.00023 | -0.003031503 | IFIT3, LIPA |
| chr7 | 128860209 | 128860420 | 212 | 3 | 0.000494348 | 0.000178 | 0.00038 | 6.44E-05 | -0.00944919 | FLNC-AS1 |
| chr17 | 39114903 | 39115122 | 220 | 2 | 0.000494592 | 2.50E-05 | 0.00149 | 3.96E-05 | -0.009626194 | PLXDC1 |
| chr12 | 55827694 | 55827863 | 170 | 2 | 0.000494804 | 0.006756 | 0.00021 | 0.00045 | -0.003850266 | DNAJC14 |
| chr2 | 26943505 | 26943682 | 178 | 2 | 0.000494813 | 0.000184 | 0.00468 | 0.00032 | 0.011969291 | DPYSL5 |
| chr14 | 75400314 | 75400528 | 215 | 2 | 0.000494814 | 4.33E-05 | 0.00097 | 5.99E-05 | -0.009593361 | NA |
| chr10 | 27745863 | 27746279 | 417 | 10 | 0.000495456 | 0.009243 | 0.00385 | 0.00017 | 0.001495536 | MKX-AS1, MKX |
| chr3 | 100333796 | 100334815 | 1020 | 12 | 0.000495458 | 7.29E-05 | 0.00126 | 1.95E-05 | -0.001792708 | NIT2 |
| chr2 | 20674534 | 20674842 | 309 | 2 | 0.000495668 | 7.25E-05 | 0.00025 | 5.95E-05 | -0.000980588 | GDF7 |
| chr1 | 99727922 | 99728281 | 360 | 2 | 0.000495825 | 0.000929 | 0.00054 | 0.00042 | 0.016400442 | FRRS1 |
| chr5 | 177093023 | 177093268 | 246 | 4 | 0.000495994 | 0.043895 | 0.00015 | 0.00118 | -0.001833189 | FGFR4 |
| chr16 | 77713629 | 77713662 | 34 | 2 | 0.000496321 | 0.063755 | 5.71E-05 | 0.0001 | -0.002899925 | NA |
| chr10 | 86960091 | 86960459 | 369 | 4 | 0.00049641 | 0.000514 | 0.00058 | 0.00019 | -0.006065677 | SNCG, MMRN2 |
| chr11 | 78419190 | 78419411 | 222 | 2 | 0.000496482 | 6.86E-05 | 0.00207 | 0.0001 | -0.008605497 | AP003086.1 |
| chr10 | 101005470 | 101005647 | 178 | 3 | 0.000496511 | 0.000108 | 0.00152 | 6.79E-05 | -0.002927742 | LZTS2 |
| chr12 | 124413323 | 124413429 | 107 | 2 | 0.000496715 | 5.74E-05 | 0.00024 | 4.97E-05 | -0.003717244 | NCOR2 |
| chr16 | 56838973 | 56839033 | 61 | 2 | 0.000496715 | 0.005473 | 5.96E-05 | 9.19E-05 | -0.007237431 | NUP93 |
| chr19 | 49154975 | 49155355 | 381 | 4 | 0.000497672 | 0.002991 | 5.78E-05 | 0.00015 | -0.004885926 | HRC |
| chr16 | 27460926 | 27461199 | 274 | 4 | 0.000497993 | 0.000185 | 0.00047 | 6.18E-05 | -0.006900829 | GTF3C1 |
| chr6 | 103898043 | 103898064 | 22 | 2 | 0.000498396 | 4.44E-05 | 0.00045 | 5.06E-05 | -0.007816015 | NA |

| | | | | | | | | | | |
|---|---|---|---|---|---|---|---|---|---|---|
| chr5 | 157049297 | 157049865 | 569 | 2 | 0.000498861 | 0.000106 | 0.0034 | 0.00018 | 0.008067165 | HAVCR1 |
| chr2 | 190636561 | 190637423 | 863 | 2 | 0.000499402 | 4.16E-06 | 0.00048 | 7.05E-06 | -0.017729229 | AC006460.2 |
| chr6 | 169214299 | 169214354 | 56 | 2 | 0.000499441 | 6.33E-05 | 0.00114 | 8.51E-05 | -0.003455218 | BX322234.1 |
| chr13 | 20717034 | 20717214 | 181 | 2 | 0.000499478 | 0.000299 | 0.00292 | 0.00044 | -1.26E-06 | IL17D |
| chr9 | 19050016 | 19050208 | 193 | 2 | 0.000499715 | 0.000369 | 0.00034 | 0.00019 | 0.001361904 | RRAGA |
| chr15 | 41115887 | 41116118 | 232 | 2 | 0.000500737 | 9.64E-06 | 5.56E-05 | 6.96E-06 | -0.002990449 | INO80 |
| chr19 | 45076283 | 45076856 | 574 | 12 | 0.000500915 | 5.80E-06 | 0.02729 | 3.25E-05 | -0.00324128 | ZNF296, GEMIN7-AS1 |
| chr19 | 15272783 | 15272832 | 50 | 2 | 0.000500915 | 5.78E-05 | 0.00147 | 8.31E-05 | -0.005859728 | BRD4 |
| chr11 | 4044975 | 4045392 | 418 | 2 | 0.000501372 | 0.005551 | 0.00088 | 0.00144 | -0.004466693 | STIM1 |
| chr5 | 359462 | 359820 | 359 | 2 | 0.000501789 | 4.15E-05 | 0.00066 | 5.30E-05 | -0.005800553 | PDCD6-AHRR, AHRR |
| chr11 | 76860414 | 76860566 | 153 | 5 | 0.000501825 | 3.86E-05 | 0.00105 | 5.44E-05 | -0.009171229 | NA |
| chr3 | 49829956 | 49830140 | 185 | 2 | 0.000502209 | 0.000403 | 0.00091 | 0.00034 | -0.003382543 | TRAIP |
| chr3 | 57128004 | 57128011 | 8 | 2 | 0.000502248 | 0.000163 | 0.0001 | 5.77E-05 | -0.004266476 | IL17RD |
| chr16 | 67423760 | 67423772 | 13 | 2 | 0.000502846 | 2.92E-05 | 0.00083 | 4.16E-05 | -0.016387034 | NA |
| chr12 | 53676510 | 53676826 | 317 | 8 | 0.000502847 | 0.064323 | 0.00054 | 0.00041 | 0.002895912 | ATP5MC2 |
| chr3 | 72887550 | 72887565 | 16 | 2 | 0.0005029 | 0.000209 | 0.0004 | 0.00015 | 0.004502259 | NA |
| chr8 | 51898814 | 51899387 | 574 | 10 | 0.000503038 | 0.020086 | 0.00278 | 0.00107 | -0.000420674 | AC064807.1, PCMTD1 |
| chr6 | 33859236 | 33859382 | 147 | 2 | 0.000503038 | 4.94E-05 | 0.0016 | 7.34E-05 | -0.004556371 | NA |
| chr20 | 40962320 | 40962785 | 466 | 2 | 0.0005031 | 9.43E-06 | 0.00064 | 1.51E-05 | -0.01103501 | NA |
| chr4 | 1502754 | 1502815 | 62 | 2 | 0.000503173 | 9.53E-05 | 0.00031 | 7.82E-05 | -0.005420445 | NA |
| chr14 | 77922385 | 77922393 | 9 | 2 | 0.000503238 | 9.51E-05 | 0.00013 | 5.15E-05 | -0.003631086 | ADCK1 |
| chr14 | 77000739 | 77001048 | 310 | 3 | 0.000503267 | 2.43E-05 | 0.00155 | 4.27E-05 | -0.008795007 | NA |
| chr6 | 47697487 | 47697556 | 70 | 2 | 0.000503387 | 0.000957 | 0.00011 | 0.00012 | -0.012446306 | ADGRF2, ADGRF4 |
| chr11 | 35141692 | 35141703 | 12 | 2 | 0.000503778 | 3.39E-05 | 0.00108 | 4.91E-05 | 0.004418499 | CD44 |
| chr20 | 25196391 | 25196704 | 314 | 2 | 0.000503972 | 0.00019 | 0.00538 | 0.00034 | -0.010904048 | ENTPD6 |
| chr4 | 183444894 | 183445068 | 175 | 3 | 0.000504117 | 0.005537 | 5.49E-05 | 0.00013 | 0.002695242 | CDKN2AIP |
| chr12 | 43806417 | 43806888 | 472 | 10 | 0.000504412 | 0.00017 | 0.00282 | 5.60E-05 | 7.41E-05 | NA |
| chr1 | 111755348 | 111755868 | 521 | 8 | 0.000504448 | 0.000138 | 0.00092 | 3.59E-05 | 0.001669342 | DDX20, INKA2, AL049557.1 |
| chr2 | 151828558 | 151828808 | 251 | 9 | 0.000504848 | 1.27E-05 | 0.00833 | 3.03E-05 | 0.001832906 | AC068547.1 |
| chr6 | 166333438 | 166333445 | 8 | 2 | 0.000505427 | 4.06E-05 | 0.00036 | 4.42E-05 | -0.02187574 | SFT2D1 |
| chr20 | 43569004 | 43569426 | 423 | 4 | 0.000505825 | 0.003557 | 0.00332 | 0.00025 | -0.005146155 | Z98752.3, SGK2 |
| chr19 | 42254728 | 42255079 | 352 | 6 | 0.000505912 | 0.000457 | 0.00201 | 6.55E-05 | -0.000583304 | AC006486.1, ERF |
| chr2 | 18910843 | 18910975 | 133 | 2 | 0.000505929 | 0.003654 | 6.57E-05 | 9.63E-05 | -0.007031264 | NA |
| chr6 | 157510759 | 157511148 | 390 | 6 | 0.000506159 | 4.95E-06 | 0.01586 | 2.42E-05 | -0.015746468 | ZDHHC14 |
| chr11 | 76211133 | 76211279 | 147 | 3 | 0.000506186 | 1.50E-05 | 0.00331 | 3.15E-05 | -0.022214663 | LINC02761 |
| chr11 | 2376025 | 2376120 | 96 | 4 | 0.000506257 | 0.000141 | 0.00423 | 0.00021 | -0.003526555 | CD81-AS1 |
| chr19 | 4325444 | 4326126 | 683 | 3 | 0.000506431 | 1.33E-05 | 0.0045 | 3.19E-05 | -0.005804931 | STAP2 |
| chr9 | 94666585 | 94666699 | 115 | 3 | 0.000506644 | 0.004943 | 0.00011 | 8.47E-05 | -0.006637216 | NA |
| chr2 | 109130279 | 109130298 | 20 | 3 | 0.000506709 | 0.000273 | 0.00991 | 0.00058 | -0.022729809 | SH3RF3 |
| chr7 | 10300837 | 10300894 | 58 | 2 | 0.000507859 | 2.79E-05 | 0.00157 | 4.38E-05 | -0.016724236 | NA |
| chr19 | 58384377 | 58384447 | 71 | 2 | 0.000508037 | 0.000188 | 0.00015 | 7.94E-05 | -0.005751119 | NA |
| chr19 | 17831216 | 17831412 | 197 | 2 | 0.000508081 | 0.000154 | 0.00148 | 0.0002 | -0.004994556 | JAK3 |
| chr3 | 73404389 | 73404639 | 251 | 2 | 0.000508572 | 0.000124 | 0.00028 | 8.84E-05 | 0.001956303 | PDZRN3 |
| chr5 | 96935226 | 96936118 | 893 | 13 | 0.000508717 | 0.064034 | 0.00427 | 0.00168 | 0.000515662 | LNPEP, AC009126.1 |
| chr1 | 39560473 | 39560848 | 376 | 4 | 0.000508814 | 0.002749 | 0.00208 | 0.00022 | -0.005887024 | PABPC4 |
| chr2 | 74481261 | 74481640 | 380 | 2 | 0.000508992 | 4.70E-05 | 0.00045 | 5.29E-05 | 0.015098732 | CCDC142 |
| chr11 | 74467417 | 74467838 | 422 | 8 | 0.000509162 | 2.31E-05 | 0.01806 | 4.35E-05 | -0.002693866 | AP001372.1, KCNE3 |
| chr2 | 118973724 | 118974239 | 516 | 3 | 0.000509651 | 0.074513 | 6.09E-05 | 0.0003 | -0.012145423 | MARCO |
| chr14 | 99035039 | 99035414 | 376 | 3 | 0.000509761 | 5.04E-05 | 0.0038 | 8.59E-05 | -0.022317855 | NA |
| chr5 | 132943886 | 132943978 | 93 | 2 | 0.000510016 | 3.18E-05 | 0.00151 | 4.88E-05 | -0.009977471 | AC010240.2, AFF4 |
| chr7 | 157970561 | 157970624 | 64 | 2 | 0.00051051 | 0.000105 | 0.00013 | 5.40E-05 | -0.006069558 | PTPRN2 |
| chr2 | 215311992 | 215312580 | 589 | 9 | 0.000510752 | 0.000626 | 0.01676 | 0.00064 | 0.00354687 | ATIC |
| chr1 | 178093551 | 178094207 | 657 | 12 | 0.000510907 | 0.255483 | 0.00046 | 0.00034 | 0.001456913 | RASAL2, RASAL2-AS1 |
| chr20 | 10309309 | 10309604 | 296 | 3 | 0.000511092 | 0.000835 | 0.00011 | 8.20E-05 | -0.002243334 | SNAP25-AS1 |
| chr1 | 151007610 | 151007738 | 129 | 2 | 0.000511474 | 0.002532 | 0.00412 | 0.00247 | -0.003295474 | MINDY1 |
| chr15 | 26893885 | 26894049 | 165 | 3 | 0.000511485 | 3.39E-05 | 0.00759 | 7.86E-05 | -0.030300641 | GABRA5, GABRB3 |
| chr11 | 1757294 | 1757397 | 104 | 3 | 0.000511517 | 2.88E-05 | 0.00395 | 5.49E-05 | -0.009880054 | AC068580.4, CTSD |
| chr14 | 55199314 | 55199489 | 176 | 2 | 0.000511675 | 0.000372 | 0.00012 | 9.38E-05 | -0.003598905 | NA |
| chr6 | 39102403 | 39102527 | 125 | 2 | 0.00051212 | 2.70E-05 | 0.00137 | 4.19E-05 | -0.02239695 | NA |
| chr11 | 77117651 | 77117849 | 199 | 2 | 0.000512913 | 2.92E-05 | 0.00135 | 4.47E-05 | -0.005465892 | CAPN5 |
| chr14 | 22541101 | 22541540 | 440 | 2 | 0.000513147 | 7.20E-05 | 0.00285 | 0.00011 | -0.011906528 | NA |
| chr17 | 49016836 | 49016890 | 55 | 2 | 0.000513245 | 3.67E-05 | 0.00109 | 5.28E-05 | -0.021447567 | IGF2BP1 |
| chr19 | 19051949 | 19052002 | 54 | 2 | 0.000513463 | 0.00055 | 7.52E-05 | 6.96E-05 | -0.003486083 | ARMC6 |
| chr7 | 151308985 | 151309259 | 275 | 2 | 0.000513745 | 5.37E-05 | 0.00078 | 6.81E-05 | -0.015598121 | NA |
| chr18 | 12407131 | 12407140 | 10 | 2 | 0.000514029 | 0.000433 | 0.00856 | 0.00079 | 0.013291438 | NA |
| chr11 | 117929347 | 117929561 | 215 | 3 | 0.000514173 | 0.014349 | 7.66E-05 | 7.82E-05 | -0.007371097 | TMPRSS13 |
| chr8 | 37637545 | 37637761 | 217 | 2 | 0.000514352 | 2.59E-05 | 0.00152 | 4.10E-05 | -0.003852486 | AC137579.1 |
| chr22 | 19876785 | 19876933 | 149 | 2 | 0.000514353 | 3.50E-05 | 0.00062 | 4.53E-05 | -0.01472854 | TXNRD2 |
| chr17 | 7573478 | 7573740 | 263 | 2 | 0.000514439 | 0.000213 | 0.00147 | 0.00026 | 0.000711752 | SENP3-EIF4A1, EIF4A1, AC016876.2 |
| chr17 | 82418996 | 82419008 | 13 | 2 | 0.000514548 | 0.001164 | 0.00682 | 0.00171 | 0.002562243 | HEXD |
| chr20 | 62716331 | 62716390 | 60 | 2 | 0.00051468 | 2.70E-05 | 0.00139 | 4.19E-05 | -0.015484179 | NTSR1 |
| chr17 | 45284353 | 45284571 | 219 | 3 | 0.000514691 | 0.000794 | 0.00165 | 9.69E-05 | 0.000928933 | MAP3K14 |
| chr2 | 233381140 | 233381507 | 368 | 2 | 0.000514798 | 4.79E-05 | 0.00227 | 7.49E-05 | 0.010275637 | DGKD |
| chr3 | 157504043 | 157504241 | 199 | 3 | 0.000514852 | 0.000672 | 0.00077 | 0.00011 | -0.003827734 | VEPH1 |
| chr20 | 63287834 | 63288098 | 265 | 2 | 0.000514921 | 0.000806 | 0.00267 | 0.00091 | 0.000191974 | ARFGAP1 |
| chr19 | 56106306 | 56106691 | 386 | 3 | 0.00051496 | 3.64E-05 | 0.00179 | 6.17E-05 | -0.009182159 | ZNF787 |
| chr12 | 50086840 | 50087108 | 269 | 3 | 0.000515175 | 3.90E-05 | 0.0021 | 6.80E-05 | -0.0048009 | SMARCD1 |
| chr5 | 37249772 | 37250140 | 369 | 4 | 0.000515625 | 8.08E-05 | 0.00028 | 5.61E-05 | -0.004363809 | AC025449.2 |
| chr12 | 123850947 | 123850988 | 42 | 2 | 0.000515632 | 0.003025 | 6.16E-05 | 8.64E-05 | -0.002893523 | DNAH10 |
| chr16 | 73482596 | 73483230 | 635 | 4 | 0.000515952 | 4.28E-05 | 0.00068 | 2.73E-05 | -0.015704419 | LINC01568, ZFHX3 |
| chr13 | 32539651 | 32539868 | 218 | 2 | 0.00051642 | 0.000579 | 0.00113 | 0.00049 | -0.001734403 | NA |
| chr17 | 82968138 | 82968236 | 99 | 3 | 0.000517183 | 0.594091 | 5.09E-05 | 0.00034 | -0.00245324 | B3GNTL1 |
| chr17 | 63972908 | 63973359 | 452 | 6 | 0.000517587 | 0.002573 | 0.00038 | 8.95E-05 | 7.07E-05 | SCN4A, AC127029.2 |
| chr1 | 245352836 | 245353346 | 511 | 2 | 0.000517674 | 0.006621 | 0.00362 | 0.0041 | -0.011741177 | KIF26B |
| chr9 | 34194961 | 34195074 | 114 | 2 | 0.000517742 | 2.37E-05 | 0.00109 | 3.62E-05 | -0.012552969 | UBAP1 |
| chr1 | 53195557 | 53195776 | 220 | 3 | 0.000517882 | 0.006648 | 0.00032 | 0.00045 | -0.000959124 | NA |
| chr2 | 42566733 | 42567259 | 527 | 3 | 0.00051835 | 0.001849 | 0.00293 | 0.00095 | -0.005623746 | MTA3 |
| chr10 | 93567548 | 93568127 | 580 | 5 | 0.000518412 | 0.001463 | 0.00059 | 0.0001 | 0.004403739 | FFAR4 |
| chr20 | 62892523 | 62892845 | 323 | 2 | 0.000518428 | 3.13E-05 | 0.00145 | 4.79E-05 | -0.008493652 | DIDO1 |
| chr6 | 24799353 | 24799529 | 177 | 3 | 0.000518533 | 2.72E-05 | 0.00591 | 6.20E-05 | -0.004625189 | AL512428.1 |
| chr19 | 18264657 | 18265163 | 507 | 3 | 0.000518896 | 0.005578 | 0.00021 | 7.94E-05 | -0.002250674 | IQCN |
| chr12 | 108768614 | 108768865 | 252 | 5 | 0.000519208 | 7.19E-06 | 0.0056 | 2.30E-05 | -0.007111161 | NA |
| chr5 | 178151569 | 178151697 | 129 | 2 | 0.000519268 | 7.85E-05 | 0.0002 | 5.65E-05 | -0.006241203 | NHP2 |
| chr15 | 69413979 | 69414200 | 222 | 7 | 0.000519318 | 0.000438 | 0.00859 | 0.00022 | -0.000633829 | AC027237.3 |
| chr10 | 69801372 | 69801455 | 84 | 2 | 0.000519424 | 0.000163 | 0.00014 | 7.26E-05 | 0.004392858 | NA |
| chr4 | 183796893 | 183797062 | 170 | 2 | 0.000519553 | 6.63E-05 | 0.00069 | 7.81E-05 | -0.009911588 | NA |
| chr3 | 111890168 | 111890322 | 155 | 2 | 0.000520311 | 2.64E-05 | 0.00154 | 4.17E-05 | -0.006478574 | PHLDB2 |
| chr7 | 107580523 | 107580710 | 188 | 6 | 0.000520565 | 0.00031 | 0.00182 | 9.94E-05 | -0.001457636 | DUS4L-BCAP29, BCAP29 |
| chr5 | 1483318 | 1484199 | 882 | 5 | 0.000520565 | 3.92E-06 | 0.00662 | 1.47E-05 | -0.001736704 | LPCAT1 |
| chr3 | 42773301 | 42773456 | 156 | 6 | 0.000520826 | 0.001751 | 0.00166 | 4.75E-05 | -0.001074096 | LINC02158, AC006059.2 |
| chr12 | 67441873 | 67442661 | 789 | 5 | 0.000520826 | 1.73E-06 | 0.011 | 9.57E-06 | -0.017963889 | LINC02420 |
| chr2 | 53971201 | 53971523 | 323 | 3 | 0.000520826 | 0.002594 | 0.001 | 0.00033 | 0.002395431 | ACYP2 |
| chr16 | 1747203 | 1747382 | 180 | 2 | 0.000520828 | 0.000474 | 0.00043 | 0.00025 | -0.001825294 | MAPK8IP3 |
| chr17 | 74952254 | 74952411 | 158 | 2 | 0.000521137 | 0.000138 | 0.00017 | 7.38E-05 | -0.015069321 | HID1 |
| chr20 | 52893102 | 52893116 | 15 | 2 | 0.000521137 | 0.000191 | 9.85E-05 | 5.93E-05 | -0.011788549 | NA |
| chr1 | 19643437 | 19644027 | 591 | 13 | 0.000521184 | 0.000625 | 0.00393 | 0.00027 | -0.002752188 | NBL1, MICOS10-NBL1 |
| chr14 | 76697155 | 76697208 | 54 | 2 | 0.000521342 | 2.75E-05 | 0.00055 | 3.63E-05 | -0.016149546 | AC008050.1 |
| chr19 | 4400882 | 4401421 | 540 | 3 | 0.00052142 | 0.001015 | 0.00146 | 0.00069 | -0.005561962 | NA |
| chr10 | 74103283 | 74103473 | 191 | 2 | 0.000521448 | 5.12E-05 | 0.00032 | 5.09E-05 | -0.004340073 | VCL |
| chr6 | 2798390 | 2798678 | 289 | 2 | 0.000521459 | 3.97E-05 | 0.00071 | 5.21E-05 | -0.00231923 | NA |
| chr16 | 74729755 | 74730380 | 626 | 2 | 0.000522324 | 0.000104 | 0.00263 | 0.00016 | -0.01091122 | FA2H |
| chr8 | 41723316 | 41723414 | 99 | 2 | 0.000522539 | 0.001476 | 9.09E-05 | 0.00011 | -0.005371468 | ANK1 |
| chr18 | 49813258 | 49813864 | 607 | 5 | 0.00052261 | 0.106387 | 0.0188 | 0.02455 | 0.001430877 | ACAA2, AC090227.2 |
| chr11 | 8016496 | 8016536 | 41 | 2 | 0.000522612 | 0.000203 | 0.00343 | 0.00033 | -0.016471105 | CASC23 |
| chr4 | 1804795 | 1805062 | 268 | 4 | 0.000522701 | 6.18E-05 | 0.01026 | 0.00012 | -0.003698743 | FGFR3 |
| chr12 | 76032813 | 76033055 | 243 | 3 | 0.000523006 | 0.000163 | 0.00075 | 6.60E-05 | -0.013707489 | AC011611.4, PHLDA1 |
| chr17 | 41696495 | 41696659 | 165 | 2 | 0.000523187 | 2.84E-05 | 0.00126 | 4.33E-05 | -0.016206603 | NA |
| chr1 | 228103167 | 228103476 | 310 | 5 | 0.000523384 | 0.000798 | 0.0005 | 0.00017 | 0.001990453 | C1orf35 |
| chr12 | 29086754 | 29087014 | 261 | 2 | 0.000523735 | 4.46E-05 | 0.00052 | 5.29E-05 | -0.007865038 | NA |
| chr14 | 68981467 | 68981670 | 204 | 2 | 0.000524303 | 3.58E-05 | 0.00084 | 4.92E-05 | -0.007192582 | ACTN1-AS1 |
| chr2 | 96739311 | 96740142 | 832 | 11 | 0.000524614 | 0.00011 | 0.00585 | 2.96E-05 | 0.002336404 | LMAN2L |
| chr12 | 49202171 | 49202219 | 49 | 2 | 0.000524622 | 0.000129 | 0.00015 | 6.69E-05 | -0.012866788 | TUBA1C |
| chr2 | 102616936 | 102617274 | 339 | 3 | 0.00052463 | 6.04E-05 | 0.00082 | 7.13E-05 | -0.003491516 | NA |
| chr1 | 173603042 | 173603231 | 190 | 3 | 0.000526231 | 0.00074 | 0.00162 | 0.0001 | -0.011881822 | AL139142.1, SLC9C2, AL139142.2 |
| chr9 | 83304894 | 83304915 | 22 | 2 | 0.000526302 | 4.11E-05 | 0.00039 | 4.57E-05 | -0.00587604 | FRMD3 |
| chr19 | 48444748 | 48444948 | 201 | 2 | 0.000526921 | 0.000747 | 0.00543 | 0.00112 | -0.017064932 | GRIN2D |
| chr1 | 46455481 | 46456074 | 594 | 5 | 0.000526941 | 0.005344 | 0.00018 | 0.00041 | -0.004015755 | FAAHP1 |
| chr16 | 4409618 | 4409693 | 76 | 2 | 0.000527329 | 2.00E-05 | 0.00053 | 2.78E-05 | -0.005475112 | CORO7-PAM16, CORO7 |
| chr1 | 22051919 | 22052557 | 639 | 7 | 0.000527997 | 6.83E-05 | 0.00933 | 6.12E-05 | 0.000204528 | CDC42, CDC42-AS1 |
| chr5 | 1268661 | 1268951 | 291 | 4 | 0.000528056 | 5.68E-05 | 0.00793 | 0.00013 | -0.005885379 | TERT |
| chr15 | 98650237 | 98650943 | 707 | 5 | 0.000528326 | 0.000109 | 0.00408 | 9.58E-05 | 0.002009333 | IGF1R |
| chr4 | 2243308 | 2243590 | 283 | 3 | 0.000528556 | 5.83E-05 | 0.00088 | 4.90E-05 | 0.003817975 | NA |
| chr3 | 179322077 | 179322799 | 723 | 4 | 0.000528592 | 0.00029 | 0.00563 | 0.00024 | -0.00318558 | NA |
| chr20 | 48976011 | 48976088 | 78 | 2 | 0.000528688 | 4.44E-05 | 0.00035 | 4.72E-05 | -0.003177093 | ARFGEF2 |
| chr16 | 30956695 | 30956884 | 190 | 4 | 0.000529187 | 0.00929 | 0.00527 | 0.00128 | -0.004138759 | AC135048.3 |
| chr5 | 116480068 | 116480456 | 389 | 3 | 0.000529276 | 0.010178 | 0.0005 | 7.89E-05 | -0.011076644 | SEMA6A-AS1, SEMA6A |
| chr17 | 34156541 | 34156621 | 81 | 2 | 0.000529766 | 0.000152 | 0.00161 | 0.00021 | -0.013947373 | AC004147.2, ASIC2 |
| chr10 | 113043248 | 113043331 | 84 | 2 | 0.000529804 | 3.14E-05 | 0.00089 | 4.45E-05 | -0.014598708 | TCF7L2 |
| chr11 | 5929568 | 5930203 | 636 | 6 | 0.00053008 | 0.000169 | 0.0003 | 6.59E-05 | -0.001782768 | TRIM5 |
| chr16 | 75594285 | 75594328 | 44 | 2 | 0.000530321 | 2.75E-05 | 0.00141 | 4.28E-05 | -0.014704935 | AC025287.1 |
| chr16 | 89574888 | 89574952 | 65 | 2 | 0.000530508 | 0.015984 | 0.00011 | 0.00026 | 0.003785164 | NA |
| chr20 | 62427539 | 62427810 | 272 | 7 | 0.000530756 | 4.35E-05 | 0.00189 | 4.49E-05 | -0.008288002 | RBBP8NL |
| chr21 | 46254290 | 46254320 | 31 | 2 | 0.000531341 | 4.89E-05 | 0.0023 | 7.65E-05 | -0.010910044 | MCM3AP-AS1, MCM3AP |
| chr5 | 139298685 | 139298818 | 134 | 2 | 0.000531395 | 8.78E-05 | 0.00022 | 6.40E-05 | -0.012120801 | MATR3, MATR3 |
| chr16 | 553568 | 553743 | 176 | 2 | 0.000531401 | 0.001496 | 7.78E-05 | 9.59E-05 | -0.006294142 | CAPN15, Z97986.1 |
| chr19 | 29666404 | 29666789 | 386 | 2 | 0.000531401 | 0.000113 | 0.00258 | 0.00018 | -0.016448513 | PLEKHF1 |
| chr11 | 95230827 | 95231653 | 827 | 11 | 0.000531883 | 1.40E-05 | 0.0233 | 3.49E-05 | 0.000767474 | AP000787.1, SESN3 |
| chr5 | 96768922 | 96769004 | 83 | 2 | 0.000532218 | 0.000219 | 9.97E-05 | 6.37E-05 | -0.00207489 | CAST, ERAP1 |
| chr17 | 79090268 | 79090306 | 39 | 2 | 0.000532426 | 0.04627 | 0.00029 | 0.00088 | -0.003343924 | RBFOX3 |
| chr12 | 26196536 | 26196670 | 135 | 2 | 0.000532482 | 0.000106 | 0.00357 | 0.00018 | -0.009414863 | SSPN |
| chr21 | 43361304 | 43361370 | 67 | 2 | 0.000532687 | 0.0004 | 0.00025 | 0.00016 | 0.014446948 | LINC01679 |
| chr5 | 115031836 | 115031965 | 130 | 2 | 0.000532748 | 0.000169 | 0.00012 | 6.61E-05 | 0.007344236 | AC094104.2 |
| chr19 | 5090951 | 5091280 | 330 | 2 | 0.000532917 | 0.001748 | 0.00109 | 0.00094 | -0.002979401 | KDM4B |
| chr17 | 80339848 | 80339863 | 16 | 2 | 0.000533123 | 7.61E-05 | 2.78E-05 | 1.43E-05 | -0.002719897 | RNF213 |
| chr6 | 31471162 | 31471306 | 145 | 3 | 0.000533306 | 0.000424 | 0.00015 | 0.00011 | 0.003137587 | HCP5 |
| chr14 | 68855471 | 68855531 | 61 | 3 | 0.000533372 | 0.000247 | 0.00019 | 6.07E-05 | -0.002224988 | NA |
| chr16 | 828130 | 828441 | 312 | 3 | 0.000533823 | 0.000433 | 0.00211 | 0.00033 | -0.005350473 | AL023882.1 |
| chr12 | 132837708 | 132837985 | 278 | 4 | 0.000534564 | 3.07E-05 | 0.00274 | 5.06E-05 | -0.007747587 | CHFR |
| chr15 | 65088791 | 65088808 | 18 | 2 | 0.000534687 | 7.09E-05 | 0.00296 | 0.00011 | -0.001179267 | NA |
| chr1 | 3738897 | 3739049 | 153 | 2 | 0.000534833 | 8.96E-05 | 0.00264 | 0.00014 | -0.005555471 | TP73-AS1 |
| chr1 | 23959455 | 23959792 | 338 | 8 | 0.000535027 | 0.006882 | 0.00846 | 0.003 | 0.001283487 | PNRC2 |
| chr20 | 63573555 | 63574207 | 653 | 3 | 0.000535297 | 0.108826 | 0.00451 | 0.01509 | -0.004542943 | HELZ2 |

| | | | | | | | | | | |
|---|---|---|---|---|---|---|---|---|---|---|
| chr13 | 24085660 | 24085785 | 126 | 2 | 0.000535367 | 2.09E-05 | 0.00053 | 2.89E-05 | -0.022210311 | AL359736.1, SPATA13 |
| chr3 | 195911459 | 195912065 | 607 | 5 | 0.00053649 | 0.00058 | 0.0007 | 7.39E-05 | -0.006415002 | TNK2-AS1, TNK2 |
| chr19 | 14027827 | 14028325 | 499 | 12 | 0.00053674 | 3.51E-05 | 0.01686 | 3.52E-05 | -0.003758652 | RLN3 |
| chr10 | 6465191 | 6465667 | 477 | 3 | 0.00053674 | 6.57E-05 | 0.00114 | 4.60E-05 | -0.005526601 | PRKCQ |
| chr15 | 67165246 | 67165741 | 496 | 5 | 0.000537244 | 0.000157 | 0.01372 | 0.0003 | -0.003018296 | SMAD3 |
| chr19 | 43718822 | 43718920 | 99 | 2 | 0.000537281 | 0.006625 | 0.00018 | 0.00037 | -0.00219541 | IRGC |
| chr6 | 35085308 | 35085493 | 186 | 2 | 0.000537329 | 4.34E-05 | 0.00048 | 5.06E-05 | -0.007597868 | ANKS1A |
| chr1 | 64598353 | 64598483 | 131 | 2 | 0.000537707 | 3.24E-05 | 0.00158 | 5.01E-05 | -0.018863057 | CACHD1 |
| chr16 | 24495627 | 24495658 | 32 | 2 | 0.000538135 | 2.87E-05 | 0.00023 | 3.00E-05 | -0.019688457 | NA |
| chr20 | 60137997 | 60138359 | 363 | 3 | 0.000538512 | 4.91E-05 | 0.00282 | 8.87E-05 | 0.008202582 | MIR646HG |
| chr7 | 931391 | 931494 | 104 | 2 | 0.000539394 | 3.58E-05 | 0.00132 | 5.34E-05 | -0.014469245 | ADAP1, COX19 |
| chr1 | 113226290 | 113226646 | 357 | 2 | 0.000539718 | 2.55E-05 | 0.0015 | 4.04E-05 | 0.002133637 | NA |
| chr12 | 57632684 | 57632922 | 239 | 4 | 0.000540066 | 0.030562 | 0.00412 | 0.0109 | 0.000992423 | B4GALNT1 |
| chr5 | 16616696 | 16616725 | 30 | 2 | 0.000541062 | 0.021254 | 0.00142 | 0.00343 | 0.001936925 | AC024588.1, RETREG1 |
| chr1 | 17610127 | 17610147 | 21 | 2 | 0.000541667 | 0.000102 | 0.00192 | 0.00015 | -0.010017827 | ARHGEF10L |
| chr17 | 82461998 | 82462369 | 372 | 4 | 0.000541693 | 1.37E-05 | 0.00452 | 3.67E-05 | -0.003715853 | NARF |
| chr8 | 144503743 | 144504154 | 412 | 5 | 0.000542399 | 0.003119 | 0.00179 | 0.0003 | -0.004063835 | GPT, AC084125.2 |
| chr5 | 132830207 | 132830638 | 432 | 3 | 0.000542788 | 0.000464 | 0.00055 | 0.00026 | 0.002336619 | SHROOM1 |
| chr16 | 8848745 | 8848910 | 166 | 3 | 0.000542976 | 4.33E-05 | 0.0019 | 7.27E-05 | -0.003458277 | PMM2, AC022167.2 |
| chr5 | 180786189 | 180786302 | 114 | 2 | 0.00054313 | 5.73E-05 | 0.00043 | 6.08E-05 | -0.013030688 | MGAT1 |
| chr12 | 51846584 | 51846761 | 178 | 2 | 0.000543278 | 0.000293 | 0.00046 | 0.0002 | -0.009143962 | FIGNL2 |
| chr6 | 117906716 | 117907219 | 504 | 4 | 0.000543497 | 0.015088 | 0.00473 | 0.00149 | -0.004871399 | NA |
| chr12 | 131799967 | 131800139 | 173 | 4 | 0.000543679 | 8.15E-05 | 0.00814 | 0.00016 | -0.003203502 | NA |
| chr4 | 105930722 | 105931017 | 296 | 2 | 0.000543832 | 0.000114 | 0.00028 | 8.47E-05 | -0.003321126 | NPNT, AC109361.2 |
| chr21 | 29073653 | 29074129 | 477 | 11 | 0.000544246 | 0.000291 | 0.02352 | 0.00012 | -0.001938731 | CCT8 |
| chr10 | 1210399 | 1210429 | 31 | 2 | 0.000545427 | 3.44E-05 | 0.00058 | 4.42E-05 | 0.003144114 | LINC00200, ADARB2 |
| chr1 | 92229986 | 92230007 | 22 | 2 | 0.000545892 | 0.00016 | 0.00015 | 7.41E-05 | -0.001084095 | C1orf146 |
| chr10 | 68693392 | 68693571 | 180 | 2 | 0.000545926 | 0.00254 | 0.00019 | 0.0003 | -0.00387279 | TET1 |
| chr15 | 58474676 | 58474686 | 11 | 2 | 0.000546201 | 5.62E-05 | 0.00071 | 6.90E-05 | -0.002693353 | LIPC, ALDH1A2, LIPC-AS1 |
| chr1 | 1331166 | 1331401 | 236 | 6 | 0.000546295 | 7.63E-06 | 0.01133 | 2.80E-05 | -0.008540549 | TAS1R3 |
| chr14 | 88479224 | 88479697 | 474 | 2 | 0.000546386 | 0.000326 | 0.00702 | 0.00058 | 0.007780289 | PTPN21 |
| chr6 | 89639346 | 89639611 | 266 | 3 | 0.000546509 | 0.0002 | 0.00516 | 0.00039 | -0.013730593 | NA |
| chr13 | 67229763 | 67229779 | 17 | 2 | 0.000546542 | 0.000269 | 0.00013 | 8.28E-05 | 0.003699503 | PCDH9 |
| chr2 | 207067641 | 207067658 | 18 | 2 | 0.000546585 | 9.40E-05 | 0.00026 | 7.16E-05 | -0.011238108 | NA |
| chr4 | 2942415 | 2942867 | 453 | 4 | 0.000547448 | 1.21E-05 | 0.00751 | 3.36E-05 | -0.007051961 | NOP14-AS1, NOP14 |
| chr11 | 2965275 | 2965311 | 37 | 3 | 0.000547448 | 7.55E-05 | 0.00088 | 7.94E-05 | -0.011440734 | NAP1L4 |
| chr4 | 2625287 | 2625519 | 233 | 6 | 0.000547957 | 3.02E-05 | 0.00863 | 7.49E-05 | -0.000277076 | RNF4, FAM193A |
| chr17 | 7905336 | 7905511 | 176 | 2 | 0.000548205 | 0.001164 | 7.58E-05 | 8.71E-05 | 0.004608314 | CHD3 |
| chr11 | 118572901 | 118573151 | 251 | 4 | 0.000548696 | 0.000298 | 0.00104 | 0.00015 | -0.000156292 | ARCN1, IFT46 |
| chr1 | 31756229 | 31756550 | 322 | 4 | 0.000548997 | 0.005526 | 0.01238 | 0.001 | -0.005337054 | ADGRB2 |
| chr1 | 936600 | 936855 | 256 | 2 | 0.000549061 | 5.27E-05 | 0.00067 | 6.47E-05 | -0.000726356 | SAMD11 |
| chr6 | 168050221 | 168050231 | 11 | 2 | 0.000549061 | 0.009947 | 0.00026 | 0.00061 | -0.003415316 | NA |
| chr7 | 1807931 | 1808126 | 196 | 2 | 0.00054912 | 6.70E-05 | 0.00027 | 5.81E-05 | 0.003970869 | NA |
| chr9 | 95434235 | 95434443 | 209 | 2 | 0.000549148 | 4.95E-05 | 0.00127 | 7.06E-05 | -0.032705581 | NA |
| chr12 | 110936365 | 110936559 | 195 | 4 | 0.000549696 | 0.000266 | 0.00209 | 9.23E-05 | 0.009114147 | LINC01405 |
| chr20 | 13639373 | 13639470 | 98 | 4 | 0.00054976 | 1.85E-05 | 0.0006 | 2.62E-05 | 0.01595031 | NA |
| chr8 | 55978325 | 55978776 | 452 | 2 | 0.000549801 | 2.62E-05 | 0.00138 | 4.08E-05 | -0.008012068 | LYN |
| chr20 | 33662229 | 33662320 | 92 | 4 | 0.000549888 | 0.017743 | 5.95E-05 | 0.00027 | -0.001595723 | NECAB3 |
| chr2 | 166887908 | 166887956 | 49 | 2 | 0.000550144 | 0.00034 | 0.00287 | 0.00048 | -0.010118583 | NA |
| chr20 | 31555151 | 31555348 | 198 | 2 | 0.000550325 | 3.66E-05 | 0.00189 | 5.72E-05 | 0.00132618 | HM13 |
| chr6 | 12718176 | 12718298 | 123 | 2 | 0.000550466 | 5.51E-05 | 0.00039 | 5.74E-05 | 0.000899415 | PHACTR1 |
| chr1 | 205121961 | 205122587 | 627 | 11 | 0.000550496 | 0.000109 | 0.00395 | 5.80E-05 | 2.59E-05 | RBBP5 |
| chr1 | 162377992 | 162378260 | 269 | 3 | 0.000550496 | 0.000162 | 0.00023 | 5.29E-05 | -0.00561177 | AL512785.2 |
| chr17 | 28743717 | 28744101 | 385 | 7 | 0.000550923 | 0.001123 | 0.00019 | 9.97E-05 | 0.000165828 | TRAF4 |
| chr1 | 12594887 | 12595227 | 341 | 2 | 0.000551478 | 0.000214 | 0.00042 | 0.00015 | -0.004695166 | DHRS3 |
| chr7 | 999304 | 999323 | 20 | 2 | 0.000551592 | 0.006035 | 8.36E-05 | 0.00014 | -0.001592236 | C7orf50 |
| chr22 | 49788908 | 49789092 | 185 | 2 | 0.000551704 | 2.39E-05 | 0.00123 | 3.71E-05 | -0.010913809 | BRD1 |
| chr19 | 34926798 | 34927312 | 515 | 6 | 0.000552714 | 0.001364 | 0.00646 | 0.00014 | 0.000690705 | ZNF30, ZNF30-AS1 |
| chr2 | 4656577 | 4656928 | 352 | 3 | 0.000552819 | 7.83E-05 | 0.00031 | 6.10E-05 | -0.004826548 | NA |
| chr11 | 126416274 | 126416598 | 325 | 4 | 0.000552893 | 0.000144 | 0.00724 | 0.00024 | -0.006325136 | ST3GAL4 |
| chr19 | 2061153 | 2061850 | 698 | 5 | 0.000552985 | 0.007564 | 0.00054 | 0.00025 | 0.001408628 | NA |
| chr22 | 20240165 | 20240714 | 550 | 3 | 0.00055368 | 5.17E-05 | 0.00396 | 8.35E-05 | -0.008223943 | NA |
| chr4 | 44726508 | 44726942 | 435 | 10 | 0.000554319 | 8.56E-05 | 0.01179 | 5.95E-05 | -0.000608013 | GNPDA2 |
| chr2 | 47274005 | 47274231 | 227 | 3 | 0.000554481 | 0.000431 | 0.00113 | 8.58E-05 | 0.001003521 | EPCAM-DT |
| chr5 | 111069995 | 111070351 | 357 | 2 | 0.000555282 | 6.60E-05 | 0.00064 | 7.62E-05 | -0.015848251 | TSLP |
| chr8 | 27375749 | 27375904 | 156 | 2 | 0.000555648 | 6.34E-05 | 0.00042 | 6.54E-05 | -0.006747282 | PTK2B |
| chr11 | 77994621 | 77995136 | 516 | 6 | 0.000555878 | 0.000673 | 9.02E-05 | 0.00014 | 0.000403828 | INTS4 |
| chr2 | 169768969 | 169769002 | 34 | 2 | 0.000556281 | 0.000133 | 0.00053 | 0.00012 | 0.022566503 | KLHL23 |
| chr18 | 23133795 | 23134368 | 574 | 5 | 0.000556385 | 2.06E-05 | 0.00758 | 6.19E-05 | -0.007069788 | NA |
| chr1 | 20413266 | 20413707 | 442 | 5 | 0.000556459 | 0.000539 | 0.00199 | 5.06E-05 | -0.004657819 | LINC01141 |
| chr7 | 159156681 | 159156989 | 309 | 2 | 0.000557623 | 0.018635 | 0.0005 | 0.0013 | -0.00274968 | NA |
| chr12 | 123445570 | 123445577 | 8 | 2 | 0.000557623 | 0.02596 | 7.95E-05 | 0.00016 | -0.006490156 | NA |
| chr17 | 7403888 | 7404253 | 366 | 9 | 0.00055789 | 0.001013 | 0.00979 | 7.52E-05 | 0.000293007 | TMEM256-PLSCR3, TMEM256 |
| chr2 | 169695038 | 169695235 | 198 | 3 | 0.000558038 | 0.000338 | 0.02277 | 0.00097 | -0.010190898 | PHOSPHO2, KLHL23 |
| chr4 | 112146133 | 112146243 | 111 | 3 | 0.000558118 | 0.00156 | 8.86E-05 | 0.00012 | 0.001679654 | FAM241A |
| chr1 | 155202007 | 155202149 | 143 | 3 | 0.000558602 | 0.000803 | 0.00073 | 6.84E-05 | -0.011967053 | THBS3-AS1, THBS3 |
| chr8 | 17722604 | 17722627 | 24 | 3 | 0.000558875 | 0.000198 | 0.00036 | 7.12E-05 | -0.001243547 | MTUS1 |
| chr5 | 1033403 | 1033961 | 559 | 4 | 0.000559088 | 0.00013 | 0.00274 | 0.00015 | -0.003572843 | NKD2 |
| chr6 | 8102498 | 8102686 | 189 | 6 | 0.000559105 | 4.43E-05 | 0.00071 | 2.46E-05 | 0.001724269 | EEF1E1-BLOC1S5, EEF1E1 |
| chr6 | 33172619 | 33172682 | 64 | 2 | 0.000559652 | 0.000276 | 0.00317 | 0.00042 | -0.010678047 | COL11A2 |
| chr14 | 61269807 | 61269862 | 56 | 3 | 0.000559893 | 0.00032 | 0.00031 | 6.37E-05 | 0.003772782 | PRKCH |
| chr16 | 27888165 | 27888317 | 153 | 3 | 0.000560109 | 0.000304 | 0.00056 | 0.00012 | 0.003551239 | GSG1L |
| chr12 | 6568243 | 6568671 | 429 | 6 | 0.000560388 | 0.000627 | 0.00491 | 8.43E-05 | 0.001018125 | NOP2, AC006064.6 |
| chr13 | 31827923 | 31827929 | 7 | 2 | 0.000560603 | 3.16E-05 | 0.00098 | 4.55E-05 | -0.017553572 | NA |
| chr4 | 1305354 | 1305367 | 14 | 2 | 0.000560966 | 0.001934 | 4.97E-05 | 6.28E-05 | -0.001715245 | MAEA |
| chr1 | 115842384 | 115842508 | 125 | 3 | 0.000561081 | 0.003342 | 0.00123 | 9.59E-05 | -0.013856196 | NHLH2 |
| chr11 | 70711340 | 70711414 | 75 | 3 | 0.000561142 | 8.27E-06 | 0.00313 | 1.98E-05 | -0.005755756 | SHANK2 |
| chr17 | 80139089 | 80139347 | 259 | 4 | 0.000561204 | 0.000239 | 0.00046 | 9.84E-05 | 0.001295079 | EIF4A3 |
| chr3 | 187016060 | 187016270 | 211 | 2 | 0.000561204 | 6.55E-05 | 0.00069 | 7.74E-05 | -0.013015291 | ST6GAL1 |
| chr12 | 47750520 | 47750584 | 65 | 3 | 0.000561323 | 0.00011 | 0.00033 | 5.73E-05 | -0.008527612 | RAPGEF3 |
| chr3 | 184363304 | 184363612 | 309 | 7 | 0.000561605 | 2.58E-05 | 0.0091 | 4.65E-05 | 0.00192785 | POLR2H |
| chr11 | 124717619 | 124717730 | 112 | 2 | 0.000562077 | 6.48E-05 | 0.0024 | 9.95E-05 | -0.018475806 | NA |
| chr2 | 10837750 | 10838019 | 270 | 2 | 0.000562792 | 9.09E-05 | 0.0024 | 0.00014 | -0.008343076 | PDIA6 |
| chr1 | 8077367 | 8077504 | 138 | 3 | 0.000562959 | 0.0001 | 0.00031 | 6.66E-05 | -0.001340363 | AL034417.2 |
| chr16 | 991053 | 991181 | 129 | 3 | 0.000562959 | 2.82E-05 | 0.00403 | 5.48E-05 | -0.004510532 | AC009041.1 |
| chr6 | 143965620 | 143965668 | 49 | 2 | 0.000563066 | 0.000492 | 0.00018 | 0.00015 | -0.013653699 | PLAGL1 |
| chr4 | 8135125 | 8135394 | 270 | 5 | 0.00056433 | 0.076436 | 0.00019 | 0.00088 | 0.001567122 | ABLIM2 |
| chr4 | 2008974 | 2009354 | 381 | 7 | 0.000564401 | 5.33E-05 | 0.02088 | 0.0001 | 0.000828543 | NELFA |
| chr17 | 7719077 | 7719501 | 425 | 2 | 0.000564568 | 0.000338 | 0.00148 | 0.00038 | -0.008463226 | DNAH2 |
| chr16 | 1193447 | 1193622 | 176 | 3 | 0.000564656 | 0.000106 | 0.00225 | 0.00015 | 0.000771915 | CACNA1H |
| chr19 | 55301205 | 55301313 | 109 | 2 | 0.00056538 | 4.94E-05 | 0.00114 | 6.91E-05 | -0.009519864 | BRSK1 |
| chr5 | 141402710 | 141402943 | 234 | 7 | 0.000565806 | 0.002141 | 0.00088 | 0.00014 | 0.012377418 | PCDHGA1, PCDHGA2, PCDHGA3, PCDHGB1, PCDHGA4, PCDHGB2, PCDHGA5, PCDHGB3, PCDHGA6, PCDHGA7, PCDHGB4, PCDHGA8, PCDHGB5, PCDHGA9 |
| chr1 | 180276387 | 180276416 | 30 | 2 | 0.000566821 | 0.000512 | 8.98E-05 | 8.04E-05 | -0.007424414 | LHX4, ACBD6 |
| chr6 | 42083398 | 42083557 | 160 | 2 | 0.000566943 | 0.008442 | 0.0001 | 0.0002 | -0.005685335 | TAF8 |
| chr14 | 104581112 | 104581516 | 405 | 3 | 0.000566988 | 3.89E-05 | 0.00718 | 8.70E-05 | -0.01055581 | C14orf180, TMEM179 |
| chr3 | 161373010 | 161373387 | 378 | 3 | 0.000567 | 5.36E-05 | 0.00325 | 7.93E-05 | -0.007838945 | NA |
| chr7 | 2543627 | 2543907 | 281 | 3 | 0.000567013 | 0.001289 | 0.0005 | 9.95E-05 | -0.002348198 | BRAT1 |
| chr9 | 117744901 | 117745153 | 253 | 4 | 0.000568079 | 6.25E-05 | 0.00623 | 8.09E-05 | 0.004636982 | AL160272.2 |
| chr1 | 3312402 | 3312511 | 110 | 2 | 0.000568139 | 6.80E-05 | 0.00227 | 0.0001 | -0.007096877 | PRDM16 |
| chr3 | 124780806 | 124781146 | 341 | 3 | 0.000568495 | 0.000258 | 0.00091 | 0.00023 | -0.007164375 | ITGB5 |
| chr9 | 134328318 | 134328520 | 203 | 4 | 0.000568702 | 0.000345 | 0.00196 | 6.66E-05 | -0.003808611 | RXRA |
| chr21 | 32297678 | 32297884 | 207 | 2 | 0.000568899 | 0.021559 | 0.00015 | 0.0004 | -0.005966873 | MRAP |
| chr16 | 17293879 | 17294086 | 208 | 2 | 0.000569456 | 5.18E-05 | 0.00043 | 5.62E-05 | -0.003781207 | XYLT1 |
| chr7 | 149862281 | 149862382 | 102 | 3 | 0.000569563 | 5.34E-05 | 0.00205 | 6.19E-05 | -0.003626126 | ZNF862, AC004877.1 |
| chr6 | 32162785 | 32163477 | 693 | 8 | 0.000570144 | 0.000674 | 0.01165 | 0.00029 | -0.002048455 | PPT2, PPT2-EGFL8 |
| chr20 | 58447999 | 58448017 | 19 | 2 | 0.000570684 | 4.65E-05 | 0.00072 | 5.92E-05 | -0.01053894 | VAPB |
| chr16 | 17483777 | 17483903 | 127 | 2 | 0.000570697 | 5.91E-05 | 0.00029 | 5.46E-05 | -0.000342386 | NA |
| chr10 | 32347269 | 32347479 | 211 | 4 | 0.000570745 | 2.49E-05 | 0.00084 | 1.10E-05 | 0.00177195 | AL391839.1, EPC1 |
| chr11 | 67468756 | 67468794 | 39 | 2 | 0.000571691 | 0.013346 | 0.00076 | 0.00175 | -0.002686398 | TMEM134 |
| chr4 | 152936971 | 152937057 | 87 | 2 | 0.000571763 | 0.035571 | 0.00282 | 0.00703 | -0.001565258 | FHDC1 |
| chr16 | 82152596 | 82152712 | 117 | 2 | 0.000571763 | 3.04E-05 | 0.00142 | 4.65E-05 | -0.055846346 | MPHOSPH6 |
| chr18 | 63970065 | 63970207 | 143 | 3 | 0.000572046 | 6.08E-05 | 0.00307 | 8.99E-05 | 0.000239443 | HMSD, SERPINB8, AC009802.1 |
| chr7 | 36291493 | 36291724 | 232 | 2 | 0.000572403 | 5.34E-05 | 0.0023 | 8.30E-05 | -0.009775427 | EEPD1 |
| chr10 | 48289763 | 48289809 | 47 | 2 | 0.000572512 | 0.000177 | 0.00036 | 0.00013 | -0.015593373 | NA |
| chr4 | 54277102 | 54277390 | 289 | 2 | 0.000573 | 0.000184 | 0.00231 | 0.00027 | -0.025092629 | AC058822.1, PDGFRA |
| chr9 | 137227002 | 137227017 | 16 | 2 | 0.000573227 | 2.48E-05 | 0.00069 | 3.50E-05 | -0.014408335 | NA |
| chr5 | 9712505 | 9712680 | 176 | 2 | 0.000573289 | 0.000205 | 0.00017 | 9.10E-05 | -0.00159603 | LINC02112 |
| chr16 | 88087253 | 88087419 | 167 | 5 | 0.000573349 | 2.14E-05 | 0.00203 | 3.00E-05 | -0.006186638 | AC134312.3, AC134312.4 |
| chr6 | 168079172 | 168079333 | 162 | 2 | 0.000573395 | 0.00179 | 8.19E-05 | 0.00011 | -0.005171482 | FRMD1 |
| chr5 | 135136127 | 135136284 | 158 | 2 | 0.000573567 | 6.04E-05 | 0.00241 | 9.33E-05 | -0.017474017 | C5orf66, AC008406.2 |
| chr7 | 158387643 | 158387804 | 162 | 3 | 0.00057374 | 3.39E-05 | 0.00152 | 5.08E-05 | -0.015693642 | PTPRN2 |
| chr3 | 49199367 | 49199894 | 528 | 6 | 0.000574007 | 1.44E-05 | 0.00665 | 3.73E-05 | -0.013266359 | IHO1 |
| chr17 | 74784788 | 74785109 | 322 | 2 | 0.000574084 | 0.000154 | 0.00046 | 0.00013 | -0.005108489 | TMEM104 |
| chr8 | 26053363 | 26053542 | 180 | 4 | 0.000574356 | 0.000254 | 0.00205 | 5.96E-05 | 0.007136996 | NA |
| chr11 | 33658674 | 33658786 | 113 | 4 | 0.00057457 | 0.003026 | 0.00029 | 0.0001 | -0.003930595 | KIAA1549L |
| chr8 | 100949368 | 100949614 | 247 | 2 | 0.000574789 | 5.00E-05 | 0.00172 | 7.50E-05 | -0.014030449 | YWHAZ |
| chr17 | 3891279 | 3891534 | 256 | 3 | 0.000574804 | 0.000121 | 0.00071 | 9.11E-05 | -0.007062104 | CAMKK1 |
| chr7 | 98211530 | 98211689 | 160 | 2 | 0.000574921 | 0.0002 | 0.00093 | 0.00021 | 0.012189638 | BHLHA15 |
| chr11 | 2185862 | 2185928 | 67 | 2 | 0.000575179 | 2.61E-05 | 0.00117 | 3.98E-05 | -0.016110089 | NA |
| chr7 | 105061876 | 105062359 | 484 | 3 | 0.000575535 | 0.012613 | 7.65E-05 | 0.00021 | -0.011140685 | KMT2E |
| chr19 | 50210657 | 50210815 | 159 | 3 | 0.000575624 | 3.28E-05 | 0.00107 | 4.91E-05 | -0.007322136 | MYH14 |
| chr12 | 52520263 | 52520625 | 363 | 6 | 0.000576674 | 0.000432 | 0.00102 | 5.92E-05 | 0.006934491 | KRT5 |
| chr12 | 123075541 | 123075932 | 392 | 3 | 0.00057683 | 2.75E-05 | 0.00444 | 5.89E-05 | -0.009252786 | PITPNM2 |
| chr16 | 73067170 | 73067365 | 196 | 2 | 0.000577048 | 6.09E-05 | 0.0024 | 9.41E-05 | -0.016291279 | ZFHX3 |
| chr11 | 119314874 | 119314961 | 88 | 2 | 0.000577504 | 0.000332 | 0.005 | 0.00055 | -0.003656369 | MCAM |
| chr4 | 2844240 | 2844428 | 189 | 2 | 0.000577513 | 0.048948 | 0.00083 | 0.00257 | 0.002651063 | ADD1 |
| chr7 | 38331075 | 38331273 | 199 | 2 | 0.00057767 | 6.07E-05 | 0.00225 | 9.30E-05 | -0.010326333 | NA |
| chr19 | 18358138 | 18358150 | 13 | 2 | 0.000577675 | 8.70E-05 | 0.00022 | 6.30E-05 | -0.003137398 | PGPEP1 |
| chr4 | 732736 | 732908 | 173 | 2 | 0.000577922 | 4.91E-05 | 0.00227 | 7.66E-05 | -0.025355789 | PCGF3 |
| chr19 | 6111467 | 6111530 | 64 | 2 | 0.000578122 | 0.000151 | 0.00057 | 0.00014 | -0.0165574 | AC011444.3, RFX2 |
| chr17 | 81684033 | 81684329 | 297 | 6 | 0.000578176 | 0.0024 | 0.0003 | 9.42E-05 | 0.002076182 | HGS |
| chr16 | 2949020 | 2949033 | 14 | 2 | 0.000578342 | 3.08E-05 | 0.00146 | 4.74E-05 | -0.00453307 | FLYWCH1, AC004034.1 |
| chr10 | 98407029 | 98407128 | 100 | 2 | 0.000578838 | 9.21E-05 | 0.00046 | 8.79E-05 | -0.005013845 | PYROXD2 |
| chr12 | 27332751 | 27333142 | 392 | 5 | 0.000579208 | 5.30E-05 | 0.0007 | 4.40E-05 | 0.001734228 | ARNTL2 |
| chr1 | 2418114 | 2418758 | 645 | 5 | 0.000579261 | 8.02E-06 | 0.01113 | 2.73E-05 | -0.008180869 | NA |
| chr18 | 3451278 | 3451752 | 475 | 12 | 0.000579656 | 0.004018 | 0.00013 | 0.0002 | 0.00035738 | TGIF1 |
| chr18 | 35950379 | 35950597 | 219 | 3 | 0.000579741 | 0.000753 | 0.00063 | 0.00021 | 0.005239163 | NA |
| chr2 | 182715467 | 182715967 | 501 | 6 | 0.000579792 | 0.000374 | 0.00441 | 0.00011 | 0.001756468 | NA |
| chr9 | 131095748 | 131096161 | 414 | 3 | 0.000579818 | 0.049643 | 0.0004 | 0.00121 | -0.002953916 | NA |
| chr16 | 3012295 | 3012794 | 500 | 8 | 0.000579931 | 3.68E-05 | 0.04407 | 0.00022 | -0.014431766 | NA |
| chr16 | 9108546 | 9108549 | 4 | 2 | 0.000580459 | 6.96E-05 | 0.00061 | 7.83E-05 | -0.007463552 | C16orf72, AC087190.3 |
| chr19 | 2754565 | 2754749 | 185 | 2 | 0.000580974 | 0.000353 | 0.00025 | 0.00015 | 0.01138994 | SGTA |

| | | | | | | | | | | |
|---|---|---|---|---|---|---|---|---|---|---|
| chr1 | 3276716 | 3277223 | 508 | 2 | 0.00058098 | 0.000259 | 0.00379 | 0.00041 | -0.004195674 | PRDM16 |
| chr14 | 101065610 | 101066015 | 406 | 6 | 0.000582239 | 0.003282 | 0.01162 | 0.00114 | -0.002545209 | MIR369, MIR410 |
| chr16 | 72928000 | 72928483 | 484 | 3 | 0.000582302 | 2.01E-05 | 0.00442 | 4.40E-05 | -0.008611562 | ZFHX3 |
| chr3 | 73625505 | 73625664 | 160 | 4 | 0.00058234 | 0.001225 | 0.00541 | 0.00151 | -0.012275917 | PDZRN3-AS1, AC117489.1 |
| chr17 | 82082095 | 82082231 | 137 | 3 | 0.000582752 | 0.000419 | 0.00034 | 0.00012 | -0.003557441 | FASN |
| chr17 | 43394370 | 43394429 | 60 | 2 | 0.000582864 | 0.000161 | 0.00096 | 0.00018 | -0.009268706 | NA |
| chr9 | 5833864 | 5834034 | 171 | 2 | 0.00058292 | 0.000437 | 0.00011 | 8.90E-05 | -0.004348282 | NA |
| chr1 | 1281723 | 1281750 | 28 | 2 | 0.000583737 | 0.000977 | 0.01124 | 0.00171 | -0.006693473 | SCNN1D |
| chr7 | 106661945 | 106662102 | 158 | 3 | 0.000584437 | 0.000147 | 0.00732 | 0.00033 | -0.000726835 | AC004917.1 |
| chr10 | 110844141 | 110844175 | 35 | 2 | 0.000584437 | 4.66E-05 | 0.0004 | 5.09E-05 | -0.01111088 | NA |
| chr20 | 61873720 | 61873827 | 108 | 2 | 0.000584523 | 0.00086 | 5.84E-05 | 6.20E-05 | -0.003427576 | CDH4 |
| chr11 | 62707263 | 62707272 | 10 | 2 | 0.000584675 | 0.024358 | 0.0062 | 0.01091 | -0.001687745 | BSCL2, HNRNPUL2-BSCL2 |
| chr6 | 37705932 | 37706464 | 533 | 4 | 0.000585555 | 1.69E-05 | 0.00199 | 2.30E-05 | -0.008861587 | NA |
| chr22 | 19130652 | 19131026 | 375 | 3 | 0.000585626 | 0.000156 | 0.00383 | 0.00026 | -0.004769226 | ESS2 |
| chr2 | 1766420 | 1766615 | 196 | 2 | 0.000585932 | 8.93E-05 | 0.00027 | 7.07E-05 | -0.005508189 | NA |
| chr10 | 119412347 | 119412571 | 225 | 2 | 0.000586478 | 0.000128 | 0.00366 | 0.00022 | -0.013286571 | GRK5 |
| chr16 | 22660076 | 22660080 | 5 | 2 | 0.000586478 | 3.43E-05 | 0.00087 | 4.78E-05 | -0.006919895 | NA |
| chr21 | 45923350 | 45923802 | 453 | 2 | 0.000586641 | 0.000162 | 0.00062 | 0.00015 | -0.009541963 | PCBP3 |
| chr17 | 81299960 | 81300040 | 81 | 2 | 0.000586707 | 0.000184 | 0.0003 | 0.00012 | -0.007199459 | NA |
| chr13 | 52949850 | 52949890 | 41 | 2 | 0.000586959 | 3.00E-05 | 0.00159 | 4.67E-05 | -0.010176173 | NA |
| chr1 | 17121855 | 17122113 | 259 | 2 | 0.000587642 | 6.40E-05 | 0.0005 | 6.92E-05 | -0.010095008 | NA |
| chr5 | 95783447 | 95783690 | 244 | 2 | 0.000587688 | 5.66E-05 | 0.00065 | 6.79E-05 | -0.005291917 | RHOBTB3, GLRX |
| chr1 | 4019135 | 4019226 | 92 | 2 | 0.000587962 | 1.63E-05 | 0.00109 | 2.62E-05 | -0.009925925 | AL805961.1 |
| chr2 | 239144704 | 239145225 | 522 | 3 | 0.000588008 | 0.000276 | 0.00676 | 0.00038 | 0.005190227 | HDAC4 |
| chr2 | 3650831 | 3651272 | 442 | 4 | 0.000588389 | 0.002434 | 0.00164 | 0.00095 | 0.001591641 | NA |
| chr2 | 98347973 | 98348228 | 256 | 2 | 0.000588627 | 7.71E-05 | 0.00296 | 0.00012 | -0.013715004 | CNGA3, AC092675.2, AC092675.1 |
| chr12 | 47504858 | 47505533 | 676 | 4 | 0.000588656 | 5.70E-05 | 0.00202 | 2.15E-05 | -0.005693495 | NA |
| chr17 | 66302875 | 66303444 | 570 | 2 | 0.000589321 | 3.00E-05 | 0.00075 | 4.16E-05 | -0.000874086 | PRKCA |
| chr3 | 38634385 | 38634544 | 160 | 2 | 0.00058934 | 7.03E-05 | 0.00047 | 7.32E-05 | -0.010687669 | SCN5A |
| chr1 | 156661443 | 156661714 | 272 | 4 | 0.00058984 | 3.24E-05 | 0.00218 | 5.19E-05 | -0.004657768 | NA |
| chr12 | 53299538 | 53300041 | 504 | 11 | 0.000589938 | 0.005471 | 0.01963 | 0.00069 | -0.002317981 | AC073611.1, MYG1, MYG1-AS1 |
| chr12 | 103906138 | 103906241 | 104 | 2 | 0.000590175 | 4.21E-05 | 0.0017 | 6.44E-05 | -0.013187792 | TTC41P |
| chr1 | 25775264 | 25775535 | 272 | 2 | 0.000590851 | 3.38E-05 | 0.00128 | 5.04E-05 | -0.012931007 | MAN1C1 |
| chr2 | 85614950 | 85615570 | 621 | 4 | 0.000591858 | 0.00386 | 0.01004 | 0.00373 | -0.001418349 | USP39 |
| chr12 | 110468245 | 110468683 | 439 | 9 | 0.000591858 | 0.002106 | 0.00017 | 0.00052 | -0.000194207 | FAM216A, GPN3 |
| chr6 | 44226768 | 44227030 | 263 | 3 | 0.000592073 | 3.61E-05 | 0.00504 | 7.62E-05 | -0.003366953 | SLC29A1 |
| chr10 | 419856 | 420035 | 180 | 3 | 0.000592303 | 0.000179 | 0.01557 | 0.00049 | -0.003159489 | DIP2C |
| chr9 | 126397364 | 126397484 | 121 | 2 | 0.000592305 | 9.20E-05 | 0.00117 | 0.00012 | 0.037130295 | MVB12B |
| chr19 | 48111455 | 48111516 | 62 | 3 | 0.000592362 | 0.000159 | 0.00072 | 0.00013 | -0.010348297 | NA |
| chr6 | 122063973 | 122063982 | 10 | 2 | 0.000592459 | 4.51E-05 | 0.00044 | 5.08E-05 | -0.008204507 | NA |
| chr17 | 2883701 | 2883745 | 45 | 2 | 0.000592833 | 4.46E-05 | 0.00072 | 5.75E-05 | -0.011777219 | RAP1GAP2 |
| chr7 | 4681574 | 4681777 | 204 | 2 | 0.000593154 | 0.000102 | 0.00311 | 0.00017 | -0.006927589 | NA |
| chr6 | 166634354 | 166634389 | 36 | 2 | 0.000593619 | 7.29E-05 | 0.00211 | 0.00011 | -0.015850342 | RPS6KA2 |
| chr9 | 127754416 | 127754537 | 122 | 2 | 0.000593718 | 0.015564 | 0.00023 | 0.0006 | -0.001887584 | AL162586.2, SH2D3C |
| chr17 | 80990971 | 80991657 | 687 | 9 | 0.000594026 | 2.21E-05 | 0.01532 | 5.06E-05 | -0.001893526 | CHMP6 |
| chr11 | 5516101 | 5516800 | 700 | 2 | 0.000594436 | 0.036428 | 0.0017 | 0.00464 | -0.002388167 | AC104389.5, UBQLNL |
| chr2 | 109501507 | 109501646 | 140 | 3 | 0.000594679 | 0.001096 | 0.00541 | 0.00097 | -0.001843724 | SH3RF3 |
| chr10 | 133202083 | 133202171 | 89 | 2 | 0.000594681 | 6.88E-05 | 0.00269 | 0.00011 | -0.004237966 | KNDC1 |
| chr17 | 48107184 | 48107267 | 84 | 5 | 0.000594891 | 0.002555 | 0.00017 | 8.20E-05 | 0.001825752 | SNX11 |
| chr5 | 77972507 | 77972800 | 294 | 3 | 0.000594961 | 7.20E-05 | 0.00021 | 4.85E-05 | 0.001727557 | NA |
| chr11 | 57752979 | 57753516 | 538 | 7 | 0.000594995 | 0.01267 | 6.34E-05 | 0.00028 | 0.000339491 | AP001931.2, AP001931.1, CTNND1 |
| chr12 | 112167905 | 112167960 | 56 | 3 | 0.000595733 | 0.000145 | 0.00014 | 6.16E-05 | -0.004795624 | HECTD4 |
| chr18 | 34976140 | 34976144 | 5 | 2 | 0.000595895 | 2.44E-05 | 0.00113 | 3.72E-05 | -0.013433364 | NA |
| chr18 | 47749750 | 47749805 | 56 | 2 | 0.000596068 | 5.55E-05 | 0.00035 | 5.60E-05 | -0.003483029 | NA |
| chr15 | 48402841 | 48402956 | 116 | 2 | 0.000596753 | 0.001001 | 8.10E-05 | 8.99E-05 | -0.005275582 | NA |
| chr12 | 109518270 | 109518325 | 56 | 2 | 0.000597131 | 0.000455 | 0.00158 | 0.00048 | -0.000978011 | UBE3B |
| chr12 | 23845987 | 23846111 | 125 | 4 | 0.000597903 | 0.000704 | 0.00021 | 8.15E-05 | -0.003140112 | SOX5 |
| chr1 | 234091880 | 234091908 | 29 | 2 | 0.000598008 | 0.000704 | 0.00017 | 0.00017 | -0.004999295 | SLC35F3 |
| chr7 | 106057732 | 106057903 | 172 | 2 | 0.000598617 | 0.020459 | 0.00018 | 0.00047 | -0.008274954 | NA |
| chr18 | 46104066 | 46104239 | 174 | 8 | 0.000598656 | 2.05E-05 | 0.00823 | 4.22E-05 | 3.81E-05 | ATP5F1A |
| chr17 | 39626049 | 39626432 | 384 | 3 | 0.000598662 | 0.000315 | 0.01232 | 0.00073 | -0.015300996 | NA |
| chr1 | 6378290 | 6378308 | 19 | 2 | 0.000599166 | 3.03E-05 | 0.00167 | 4.75E-05 | -0.004732455 | ACOT7 |
| chr2 | 74674773 | 74674806 | 34 | 3 | 0.000599478 | 0.000162 | 0.00101 | 0.00017 | -0.002317449 | SEMA4F |
| chr22 | 39209868 | 39209880 | 13 | 2 | 0.000599595 | 3.06E-05 | 0.00044 | 3.75E-05 | -0.004107704 | NA |
| chr19 | 10996131 | 10996541 | 411 | 4 | 0.000599648 | 0.000461 | 0.00018 | 0.0001 | -0.002253336 | SMARCA4 |
| chr2 | 98663522 | 98663873 | 352 | 6 | 0.000599761 | 0.002144 | 0.00051 | 0.00027 | -0.006525348 | MGAT4A |
| chr16 | 71484715 | 71484931 | 217 | 2 | 0.000599761 | 0.000929 | 0.0019 | 0.00087 | 0.004431748 | AC010547.4, ZNF19 |
| chr12 | 6765209 | 6765543 | 335 | 2 | 0.000599986 | 0.000723 | 0.0063 | 0.00114 | -0.002268171 | PTMS, MLF2 |
| chr20 | 38805515 | 38805909 | 395 | 7 | 0.000600114 | 5.08E-05 | 0.00935 | 9.31E-05 | -0.000231906 | PPP1R16B |
| chr1 | 84164260 | 84164367 | 108 | 3 | 0.000600412 | 0.039819 | 0.0006 | 0.00324 | -0.007055234 | PRKACB |
| chr12 | 13104447 | 13104778 | 332 | 3 | 0.000600676 | 0.016685 | 0.00056 | 0.00138 | -0.004658404 | FAM234B |
| chr9 | 136506147 | 136506571 | 425 | 2 | 0.000600954 | 0.000323 | 0.00675 | 0.00057 | -0.012176469 | NOTCH1 |
| chr17 | 44948195 | 44948287 | 93 | 4 | 0.000601288 | 0.007984 | 0.02328 | 0.00425 | -0.00878152 | AC015936.1 |
| chr20 | 1312916 | 1313038 | 123 | 2 | 0.000601993 | 0.012031 | 0.00057 | 0.0013 | -0.003174438 | SDCBP2 |
| chr17 | 4881922 | 4882166 | 245 | 2 | 0.000602274 | 3.45E-05 | 0.00125 | 5.12E-05 | -0.008897374 | MINK1 |
| chr11 | 62541020 | 62541338 | 319 | 3 | 0.000602491 | 5.88E-05 | 0.00421 | 0.00011 | -0.013274169 | AP001363.1, AHNAK |
| chr10 | 30001728 | 30001886 | 159 | 2 | 0.000602553 | 4.80E-05 | 0.00205 | 7.42E-05 | -0.004408919 | NA |
| chr10 | 43124323 | 43124445 | 123 | 2 | 0.000602928 | 0.000108 | 0.00034 | 8.79E-05 | -0.00585581 | RET |
| chr9 | 134056791 | 134056813 | 23 | 2 | 0.000603156 | 0.000272 | 0.00035 | 0.00016 | -0.010467338 | AL445931.1, BRD3 |
| chr14 | 68780620 | 68781441 | 822 | 3 | 0.000603229 | 7.20E-05 | 0.00357 | 0.00013 | -0.006516564 | NA |
| chr5 | 115602848 | 115602943 | 96 | 3 | 0.000603378 | 0.026565 | 0.00182 | 0.00347 | -0.005073968 | AC010226.1, TMED7-TICAM2 |
| chr11 | 76824520 | 76825025 | 506 | 2 | 0.000603782 | 3.10E-05 | 0.00133 | 4.70E-05 | -0.013398422 | NA |
| chr12 | 49973508 | 49974327 | 820 | 2 | 0.00060407 | 1.76E-05 | 0.00077 | 2.67E-05 | -0.011542873 | AQP6 |
| chr22 | 23193949 | 23194041 | 93 | 2 | 0.000604183 | 0.004256 | 0.00016 | 0.00029 | -0.002880469 | BCR |
| chr11 | 61920464 | 61920661 | 198 | 4 | 0.000604293 | 0.000115 | 0.00337 | 0.00011 | -0.005107246 | NA |
| chr9 | 132487714 | 132488229 | 516 | 2 | 0.000604342 | 0.00441 | 0.00632 | 0.00434 | -0.017682816 | CFAP77 |
| chr14 | 100145995 | 100146005 | 11 | 2 | 0.000604496 | 0.000277 | 0.00086 | 0.00026 | -0.00639 | DEGS2 |
| chr3 | 184777579 | 184777895 | 317 | 4 | 0.000604625 | 2.43E-05 | 0.00284 | 4.62E-05 | 0.002400452 | NA |
| chr20 | 25221682 | 25222050 | 369 | 3 | 0.000604625 | 0.000818 | 0.00083 | 7.26E-05 | -0.004171673 | ENTPD6 |
| chr12 | 113435425 | 113435632 | 208 | 3 | 0.000604648 | 2.78E-05 | 0.00259 | 4.31E-05 | -0.004232877 | SDSL |
| chr2 | 207613999 | 207614077 | 79 | 2 | 0.000604648 | 0.004285 | 0.00031 | 0.00055 | -0.002514492 | METTL21A |
| chr1 | 92483780 | 92484051 | 272 | 7 | 0.000604792 | 0.014021 | 0.00016 | 0.00019 | 0.000118987 | GFI1 |
| chr6 | 42748572 | 42748965 | 394 | 2 | 0.000605024 | 8.37E-05 | 0.00066 | 9.21E-05 | -0.00862459 | BICRAL |
| chr1 | 2108221 | 2108232 | 12 | 2 | 0.000605039 | 7.96E-05 | 0.00198 | 0.00012 | -0.005765535 | PRKCZ |
| chr6 | 158066509 | 158066576 | 68 | 3 | 0.000605133 | 9.26E-05 | 0.00033 | 6.65E-05 | -0.002944549 | SYNJ2 |
| chr12 | 6929560 | 6929658 | 99 | 2 | 0.000605178 | 0.000496 | 0.00014 | 0.00012 | -0.007947266 | ATN1 |
| chr16 | 14482647 | 14482794 | 148 | 2 | 0.000605391 | 7.61E-05 | 0.00025 | 6.12E-05 | -0.003195475 | PARN |
| chr21 | 36499916 | 36500195 | 280 | 4 | 0.000605553 | 9.97E-05 | 0.00032 | 5.95E-05 | -0.004184772 | AP000695.2, CLDN14 |
| chr16 | 11125164 | 11125848 | 685 | 3 | 0.000605716 | 0.000674 | 0.00117 | 8.48E-05 | 0.001715388 | CLEC16A |
| chr2 | 85578498 | 85578508 | 11 | 2 | 0.000606065 | 0.000552 | 0.00077 | 0.00039 | 0.00443592 | VAMP8 |
| chr10 | 129758893 | 129759205 | 313 | 4 | 0.000607005 | 5.33E-05 | 0.00256 | 6.44E-05 | -0.005799942 | MGMT |
| chr6 | 157822160 | 157822575 | 416 | 3 | 0.000607611 | 0.003042 | 0.00095 | 0.00036 | -0.011343637 | SNX9 |
| chr2 | 236264153 | 236264478 | 326 | 6 | 0.000607917 | 0.000185 | 0.00363 | 9.57E-05 | 0.00193988 | AC019068.1, ASB18 |
| chr4 | 88378581 | 88379004 | 424 | 9 | 0.000608059 | 1.54E-05 | 0.0227 | 2.85E-05 | -0.004045656 | HERC6 |
| chr3 | 77096767 | 77096987 | 221 | 2 | 0.000608203 | 2.51E-05 | 0.00028 | 2.86E-05 | -0.009467999 | ROBO2 |
| chr10 | 1725529 | 1725721 | 193 | 3 | 0.000608324 | 0.026188 | 0.00012 | 0.00063 | -0.002653512 | ADARB2 |
| chr8 | 144614642 | 144614782 | 141 | 3 | 0.000608456 | 0.000211 | 0.00018 | 8.46E-05 | -0.001289088 | NA |
| chr6 | 126643389 | 126643480 | 92 | 2 | 0.000608856 | 0.000538 | 0.00015 | 0.00013 | -0.006857898 | AL356534.1 |
| chr14 | 102213850 | 102214092 | 243 | 4 | 0.000609032 | 5.82E-05 | 0.00384 | 8.22E-05 | -0.003345696 | WDR20 |
| chr4 | 140153240 | 140153246 | 7 | 2 | 0.000609334 | 0.001964 | 0.00011 | 0.00015 | 0.002411807 | MAML3 |
| chr10 | 132023238 | 132023726 | 489 | 4 | 0.000610403 | 0.000173 | 0.00142 | 8.95E-05 | 0.009719329 | NA |
| chr3 | 191328591 | 191328786 | 196 | 3 | 0.000610798 | 0.008333 | 0.00733 | 0.00142 | -0.010322554 | UTS2B |
| chr2 | 96547825 | 96547905 | 81 | 2 | 0.000612614 | 4.46E-05 | 0.00091 | 6.05E-05 | -0.003596582 | ARID5A |
| chr20 | 38891525 | 38891940 | 416 | 2 | 0.000612915 | 0.000176 | 0.00058 | 0.00016 | -0.003153646 | PPP1R16B |
| chr10 | 61999410 | 62000113 | 704 | 2 | 0.000613275 | 1.49E-05 | 0.00089 | 2.36E-05 | 0.002496413 | ARID5B |
| chr12 | 82686900 | 82687610 | 711 | 7 | 0.000613524 | 0.000157 | 0.00253 | 4.94E-05 | 0.001584034 | TMTC2 |
| chr1 | 2933676 | 2933947 | 272 | 2 | 0.000614222 | 0.000416 | 5.81E-05 | 5.02E-05 | 0.000261577 | NA |
| chr4 | 686325 | 686549 | 225 | 3 | 0.000614496 | 0.005485 | 0.00065 | 0.0008 | -0.004216729 | SLC49A3 |
| chr2 | 47782591 | 47782727 | 137 | 3 | 0.000614501 | 0.000385 | 0.0008 | 0.00028 | -0.005215807 | MSH6 |
| chr22 | 48049156 | 48049450 | 295 | 2 | 0.000615848 | 0.009756 | 0.00076 | 0.00158 | -0.004737734 | NA |
| chr15 | 89003871 | 89004342 | 472 | 2 | 0.000615925 | 2.87E-05 | 0.00148 | 4.46E-05 | -0.006168736 | NA |
| chr19 | 2799623 | 2800624 | 1002 | 5 | 0.000615965 | 4.64E-05 | 0.00162 | 5.60E-05 | -0.003439633 | THOP1 |
| chr7 | 140672772 | 140672955 | 184 | 4 | 0.000616179 | 0.001423 | 0.00089 | 6.18E-05 | -0.000300279 | ADCK2, DENND2A |
| chr4 | 122151739 | 122151868 | 130 | 2 | 0.000616814 | 0.004565 | 6.22E-05 | 9.35E-05 | -0.012240237 | NA |
| chr7 | 38299937 | 38300080 | 144 | 2 | 0.000617426 | 6.94E-05 | 0.00044 | 7.04E-05 | -0.014784201 | TRGV10 |
| chr11 | 118971703 | 118972290 | 588 | 5 | 0.000618944 | 0.000126 | 0.00525 | 6.67E-05 | -0.010397164 | FOXR1 |
| chr6 | 2515063 | 2515103 | 41 | 2 | 0.000618944 | 1.99E-05 | 0.00065 | 2.88E-05 | -0.007361189 | GMDS-DT, AL359852.2 |
| chr12 | 96254922 | 96255325 | 404 | 3 | 0.000619236 | 2.47E-05 | 0.00034 | 2.71E-05 | -0.013112698 | ELK3 |
| chr22 | 50085945 | 50086112 | 168 | 3 | 0.000619613 | 0.515534 | 0.34488 | 0.53566 | -0.001687652 | NA |
| chr10 | 72356083 | 72356137 | 55 | 2 | 0.000619619 | 0.000294 | 0.00041 | 0.00018 | -0.00652348 | NA |
| chr1 | 152515223 | 152515380 | 158 | 3 | 0.000620088 | 0.000834 | 0.01862 | 0.00181 | -0.007748587 | CRCT1 |
| chr17 | 7767882 | 7768012 | 131 | 2 | 0.000620088 | 0.009642 | 9.07E-05 | 0.00017 | -0.005134104 | DNAH2 |
| chr22 | 42079016 | 42079446 | 431 | 4 | 0.0006202 | 0.004452 | 0.02695 | 0.00344 | -0.005384619 | PHETA2 |
| chr2 | 216014344 | 216014996 | 653 | 4 | 0.000620679 | 0.000396 | 0.00688 | 0.00031 | -0.00519007 | MREG, PECR |
| chr11 | 9139216 | 9139707 | 492 | 3 | 0.000620779 | 3.91E-05 | 0.00233 | 6.44E-05 | -0.004156346 | DENND5A |
| chr11 | 74003091 | 74003755 | 665 | 3 | 0.000620786 | 1.92E-05 | 0.00523 | 4.42E-05 | -0.004438086 | UCP3 |
| chr1 | 20212832 | 20213116 | 285 | 3 | 0.000622094 | 0.000476 | 0.00082 | 0.00025 | 0.002259304 | NA |
| chr11 | 26135224 | 26135481 | 258 | 2 | 0.000622131 | 4.35E-05 | 0.00081 | 5.78E-05 | -0.021544183 | NA |
| chr9 | 132945266 | 132945511 | 246 | 4 | 0.0006222 | 0.000109 | 0.00837 | 0.00022 | -0.004524236 | GFI1B, TSC1 |
| chr16 | 951653 | 951743 | 91 | 2 | 0.000622663 | 4.86E-05 | 0.00085 | 6.41E-05 | -0.003350376 | LMF1 |
| chr9 | 105694442 | 105694700 | 259 | 3 | 0.000623175 | 0.000136 | 0.00136 | 0.00017 | 0.003249448 | TMEM38B |
| chr7 | 21169304 | 21169719 | 416 | 4 | 0.000623692 | 0.000292 | 0.00841 | 0.00048 | -0.01103632 | NA |
| chr7 | 157577203 | 157577268 | 66 | 3 | 0.000624539 | 3.72E-05 | 0.00624 | 8.16E-05 | -0.015653505 | PTPRN2 |
| chr20 | 50788113 | 50788206 | 94 | 2 | 0.000625094 | 3.23E-05 | 0.00155 | 4.97E-05 | -0.01410833 | NA |
| chr1 | 204257429 | 204257699 | 271 | 6 | 0.000625249 | 0.000197 | 0.00017 | 4.23E-05 | -0.00272094 | PLEKHA6 |
| chr4 | 44653084 | 44653098 | 15 | 2 | 0.000625291 | 4.98E-05 | 0.00119 | 7.00E-05 | 0.001000522 | YIPF7 |
| chr15 | 49434088 | 49434135 | 48 | 2 | 0.000625291 | 0.001258 | 9.32E-05 | 0.00011 | -0.001446361 | FGF7, FAM227B |
| chr16 | 4316838 | 4316967 | 130 | 2 | 0.000625372 | 0.004588 | 0.00012 | 0.00021 | -0.009173363 | GLIS2 |
| chr2 | 127025892 | 127025930 | 39 | 3 | 0.000625724 | 0.000222 | 0.00667 | 0.00037 | 0.002067778 | AC012508.3, AC012508.1 |
| chr4 | 182826080 | 182826193 | 114 | 2 | 0.000626312 | 0.000228 | 0.00115 | 0.00025 | -0.017133052 | NA |
| chr6 | 168692591 | 168692913 | 323 | 2 | 0.000626312 | 0.033156 | 0.0039 | 0.00884 | -0.002880588 | NA |
| chr3 | 184137705 | 184137974 | 270 | 3 | 0.000626844 | 0.000121 | 0.00096 | 9.78E-05 | -0.005003642 | EIF2B5 |
| chr1 | 43590987 | 43591428 | 442 | 5 | 0.000627264 | 0.000482 | 0.00118 | 8.68E-05 | -0.001888387 | PTPRF |
| chr20 | 13008079 | 13008405 | 327 | 3 | 0.0006278 | 0.000694 | 0.00015 | 0.00013 | -0.0054147 | LINC01723 |
| chr16 | 85229102 | 85229314 | 213 | 3 | 0.000627824 | 0.00016 | 0.00018 | 7.55E-05 | -0.000667457 | GSE1, AC092275.1 |
| chr15 | 72293458 | 72293596 | 139 | 3 | 0.000628297 | 2.46E-05 | 0.0048 | 5.46E-05 | -0.002513064 | AC009690.2, AC009690.3, CELF6, AC009690.1 |
| chr6 | 32953744 | 32954028 | 285 | 3 | 0.000628348 | 9.58E-05 | 0.00803 | 0.00021 | 0.007481555 | HLA-DMA |
| chr14 | 74975171 | 74975218 | 48 | 2 | 0.000629505 | 0.000129 | 0.00027 | 8.88E-05 | -0.008637564 | NA |
| chr1 | 227728218 | 227728364 | 147 | 5 | 0.000630062 | 0.00019 | 0.00164 | 0.00012 | 0.00130793 | NA |
| chr14 | 105415342 | 105415631 | 290 | 2 | 0.000630062 | 8.02E-05 | 0.00166 | 0.00011 | 0.004192488 | TEX22 |
| chr2 | 226800195 | 226800237 | 43 | 2 | 0.000630293 | 9.66E-05 | 0.00253 | 0.00015 | -0.00165027 | AC010735.2 |
| chr10 | 117968729 | 117968803 | 75 | 2 | 0.000630398 | 4.05E-05 | 0.0007 | 5.27E-05 | -0.010460694 | NA |

| | | | | | | | | | | |
|---|---|---|---|---|---|---|---|---|---|---|
| chr15 | 63899027 | 63899452 | 426 | 3 | 0.000630606 | 2.77E-05 | 0.00199 | 5.03E-05 | -0.014868823 | NA |
| chr16 | 11273257 | 11273445 | 189 | 2 | 0.000630962 | 0.000729 | 0.00371 | 0.00096 | -0.006402036 | RMI2, PRM3 |
| chr17 | 50461568 | 50461860 | 293 | 2 | 0.000631291 | 6.78E-05 | 0.00287 | 0.00011 | -0.007469925 | ACSF2 |
| chr9 | 128371591 | 128371987 | 397 | 3 | 0.00063152 | 0.002992 | 5.53E-05 | 0.0001 | 0.000999259 | URM1 |
| chr15 | 88257336 | 88257773 | 438 | 4 | 0.00063186 | 1.66E-05 | 0.00162 | 2.02E-05 | 0.001895891 | NTRK3-AS1 |
| chr7 | 158974135 | 158974444 | 310 | 3 | 0.000632628 | 2.87E-05 | 0.00588 | 6.38E-05 | -0.00576575 | NA |
| chr10 | 3750730 | 3751055 | 326 | 4 | 0.000632885 | 0.000256 | 0.00644 | 0.00012 | -0.005929662 | AL450322.2 |
| chr2 | 85354752 | 85355235 | 484 | 9 | 0.0006339 | 9.88E-05 | 0.01084 | 8.11E-05 | -0.001453481 | ELMOD3 |
| chr16 | 67151437 | 67151624 | 188 | 2 | 0.000634227 | 0.165711 | 0.1221 | 0.16029 | -0.004008139 | NA |
| chr9 | 133121034 | 133121544 | 511 | 2 | 0.00063437 | 6.07E-05 | 0.00209 | 9.19E-05 | 0.003959248 | RALGDS, AL162417.1 |
| chr16 | 1471616 | 1472108 | 493 | 4 | 0.000634398 | 2.10E-05 | 0.00596 | 4.72E-05 | -0.013660054 | CLCN7, AL031600.2 |
| chr19 | 50164479 | 50164544 | 66 | 2 | 0.000634565 | 0.002595 | 1.00E-04 | 0.00015 | -0.01129348 | NA |
| chr2 | 102142147 | 102142149 | 3 | 2 | 0.000634744 | 3.87E-05 | 0.0015 | 5.83E-05 | -0.02332729 | IL1R1 |
| chr2 | 233976232 | 233976655 | 424 | 2 | 0.000634976 | 3.93E-05 | 0.00183 | 6.09E-05 | -0.012899188 | TRPM8 |
| chr11 | 70570164 | 70570204 | 41 | 2 | 0.000635229 | 6.74E-05 | 0.00267 | 0.00011 | -0.005102132 | SHANK2 |
| chr17 | 14299439 | 14299599 | 161 | 2 | 0.000636437 | 9.00E-05 | 0.00201 | 0.00013 | 0.002699371 | NA |
| chr1 | 6302413 | 6302832 | 420 | 2 | 0.000636591 | 5.42E-05 | 0.0013 | 7.66E-05 | -0.006773811 | ACOT7 |
| chr14 | 24337531 | 24337575 | 45 | 2 | 0.000637731 | 0.016632 | 0.0011 | 0.00257 | 0.002988087 | RIPK3 |
| chr2 | 218417691 | 218417973 | 283 | 3 | 0.000637747 | 9.29E-05 | 0.00241 | 8.09E-05 | 0.002438552 | NA |
| chr14 | 77041118 | 77041191 | 74 | 2 | 0.000637822 | 0.000569 | 0.00773 | 0.00098 | -0.011402476 | LINC02288 |
| chr1 | 174799518 | 174799837 | 320 | 4 | 0.000637857 | 0.00026 | 0.01704 | 0.00067 | -0.015391915 | RABGAP1L |
| chr13 | 112207185 | 112207204 | 20 | 3 | 0.000638031 | 0.000246 | 0.00462 | 0.00043 | 0.018896881 | NA |
| chr22 | 24460626 | 24460909 | 284 | 2 | 0.000638094 | 4.30E-05 | 0.00182 | 6.62E-05 | -0.01067854 | ADORA2A-AS1 |
| chr1 | 35882615 | 35883028 | 414 | 6 | 0.000638235 | 0.003483 | 0.00186 | 0.00021 | -0.002043357 | AGO1 |
| chr12 | 56221748 | 56222276 | 529 | 15 | 0.000638419 | 2.30E-05 | 0.02298 | 4.63E-05 | 0.00040276 | NABP2, RNF41 |
| chr8 | 1702935 | 1703031 | 97 | 2 | 0.000639449 | 0.0002 | 0.00039 | 0.00014 | 0.001404985 | DLGAP2 |
| chr10 | 44925373 | 44925548 | 176 | 2 | 0.000640193 | 0.000241 | 0.00013 | 8.26E-05 | -0.012435387 | TMEM72, TMEM72-AS1 |
| chr4 | 3389448 | 3389768 | 321 | 4 | 0.000640538 | 4.43E-05 | 0.00206 | 7.10E-05 | -0.020079737 | RGS12 |
| chr15 | 88033040 | 88033089 | 50 | 3 | 0.000640659 | 0.002715 | 0.00201 | 0.00015 | -0.004834045 | NTRK3 |
| chr22 | 46625327 | 46626270 | 944 | 6 | 0.000640812 | 0.007897 | 0.00085 | 0.00166 | -0.00411618 | GRAMD4 |
| chr2 | 69643760 | 69643764 | 5 | 2 | 0.000641521 | 2.95E-05 | 0.00159 | 4.61E-05 | 0.004367644 | AAK1 |
| chr6 | 109793850 | 109794259 | 410 | 2 | 0.000641577 | 3.88E-05 | 0.00194 | 6.06E-05 | -0.003953848 | FIG4 |
| chr1 | 110449908 | 110450263 | 356 | 2 | 0.000641882 | 0.000844 | 0.00092 | 0.00056 | -0.01212798 | NA |
| chr12 | 101697441 | 101697995 | 555 | 7 | 0.000642295 | 0.000168 | 0.00556 | 0.00012 | 0.000169001 | CHPT1 |
| chr19 | 2540909 | 2541106 | 198 | 2 | 0.000642299 | 4.23E-05 | 0.00209 | 6.64E-05 | -0.004453023 | GNG7 |
| chr7 | 1953717 | 1953925 | 209 | 3 | 0.000642709 | 5.74E-05 | 0.00111 | 6.63E-05 | -0.002682497 | MAD1L1, AC069288.1 |
| chr22 | 40819820 | 40819935 | 116 | 2 | 0.000642854 | 0.001029 | 0.00117 | 0.00072 | -0.008589211 | NA |
| chr4 | 140196013 | 140196018 | 6 | 2 | 0.000642855 | 4.71E-05 | 0.0005 | 5.45E-05 | -0.011983918 | NA |
| chr12 | 49069723 | 49070024 | 302 | 4 | 0.000643297 | 0.046084 | 0.00013 | 0.00122 | -0.000408622 | RHEBL1 |
| chr4 | 54447292 | 54447988 | 697 | 3 | 0.000644199 | 9.70E-06 | 0.00382 | 2.37E-05 | 0.003596146 | NA |
| chr6 | 27205795 | 27206212 | 418 | 5 | 0.000645159 | 0.00018 | 0.00079 | 6.33E-05 | 0.004468497 | NA |
| chr17 | 28612280 | 28612686 | 407 | 3 | 0.000645234 | 8.31E-05 | 0.00212 | 0.00011 | -0.005481449 | SPAG5-AS1, AC005726.1, RSKR |
| chr5 | 181164594 | 181164704 | 111 | 2 | 0.000645297 | 0.000239 | 0.00012 | 7.64E-05 | -0.015486899 | NA |
| chr11 | 14644701 | 14645127 | 427 | 4 | 0.00064537 | 0.000121 | 0.00421 | 9.88E-05 | 0.001907551 | PDE3B |
| chr10 | 103677033 | 103677047 | 15 | 2 | 0.000645425 | 0.000257 | 0.00173 | 0.00032 | -0.007970432 | SH3PXD2A |
| chr10 | 117343013 | 117343085 | 73 | 2 | 0.00064599 | 0.000125 | 0.00017 | 6.96E-05 | -0.009709741 | PDZD8 |
| chr13 | 25172335 | 25172506 | 172 | 6 | 0.000646593 | 2.50E-05 | 0.00698 | 4.69E-05 | 0.014779747 | LINC01053 |
| chr12 | 14835084 | 14835189 | 106 | 2 | 0.000646828 | 3.40E-05 | 0.00148 | 5.18E-05 | -0.011824669 | C12orf60, ART4 |
| chr8 | 118952201 | 118952397 | 197 | 6 | 0.000647628 | 0.300519 | 0.00204 | 0.00144 | 0.000845679 | NA |
| chr5 | 123208881 | 123208909 | 29 | 2 | 0.000647701 | 2.68E-05 | 0.00155 | 4.22E-05 | 0.000949514 | NA |
| chr3 | 186057519 | 186057681 | 163 | 2 | 0.000648304 | 2.99E-05 | 0.00124 | 4.50E-05 | -0.011952672 | ETV5 |
| chr2 | 201260365 | 201260587 | 223 | 5 | 0.000648755 | 0.000118 | 0.00361 | 7.46E-05 | 0.000119894 | CASP8 |
| chr17 | 75783954 | 75784490 | 537 | 6 | 0.000649385 | 0.00248 | 0.00169 | 0.00146 | -0.000913838 | H3-3B |
| chr19 | 16828473 | 16828893 | 421 | 2 | 0.000649414 | 0.000156 | 0.001 | 0.00018 | 0.010160028 | NA |
| chr19 | 17334203 | 17334264 | 62 | 2 | 0.000649807 | 0.000388 | 0.00254 | 0.00051 | -0.009983488 | ANO8 |
| chr10 | 33156180 | 33156402 | 223 | 2 | 0.000649905 | 7.45E-05 | 0.00054 | 7.93E-05 | -0.011504031 | NA |
| chr1 | 111346069 | 111346537 | 469 | 10 | 0.000650044 | 0.000254 | 0.00267 | 0.00019 | 0.000840211 | NA |
| chr16 | 88316118 | 88316176 | 59 | 2 | 0.000650849 | 0.001034 | 0.0001 | 0.00012 | -0.008102781 | NA |
| chr5 | 141923915 | 141924057 | 143 | 2 | 0.000651006 | 8.91E-05 | 0.00331 | 0.00015 | -0.000412987 | DELE1 |
| chr7 | 2355491 | 2355512 | 22 | 3 | 0.00065106 | 0.00487 | 0.00233 | 0.00136 | -0.002400913 | EIF3B |
| chr15 | 63842214 | 63842256 | 43 | 2 | 0.000651177 | 3.38E-05 | 0.00163 | 5.22E-05 | -0.010937987 | NA |
| chr17 | 3935030 | 3935149 | 120 | 3 | 0.000651427 | 0.000191 | 0.0005 | 7.73E-05 | -0.007703731 | ATP2A3 |
| chr14 | 21392550 | 21392790 | 241 | 4 | 0.000651526 | 0.094087 | 3.38E-05 | 0.0003 | -0.001531663 | CHD8 |
| chr12 | 18714504 | 18714506 | 3 | 2 | 0.000651734 | 0.000754 | 5.24E-05 | 5.37E-05 | -0.005212483 | PLCZ1 |
| chr3 | 5002494 | 5003259 | 766 | 3 | 0.00065181 | 0.001503 | 0.00102 | 0.0003 | -0.004410705 | NA |
| chr19 | 4058386 | 4058393 | 8 | 2 | 0.000651839 | 9.27E-05 | 0.00143 | 0.00013 | 0.015667879 | ZBTB7A |
| chr22 | 44133879 | 44134494 | 616 | 2 | 0.000652047 | 2.40E-05 | 0.00128 | 3.73E-05 | -0.056771978 | PARVB |
| chr6 | 158649927 | 158650074 | 148 | 5 | 0.000652052 | 0.000552 | 0.00196 | 0.00022 | 0.003155944 | SYTL3 |
| chr7 | 34657881 | 34658403 | 523 | 4 | 0.000652214 | 0.006116 | 0.00069 | 0.00149 | -0.004362358 | NPSR1, NPSR1-AS1 |
| chr16 | 30650916 | 30651041 | 126 | 5 | 0.000653138 | 0.103744 | 0.0001 | 0.00083 | -9.30E-05 | PRR14 |
| chr5 | 1001766 | 1002116 | 351 | 2 | 0.000653331 | 3.71E-05 | 0.00176 | 5.75E-05 | -0.01046185 | NA |
| chr22 | 25219951 | 25220238 | 288 | 2 | 0.000654009 | 0.000408 | 0.00287 | 0.00056 | -0.015245961 | CRYBB2 |
| chr19 | 36053781 | 36054367 | 587 | 5 | 0.000655039 | 0.308885 | 0.00039 | 0.00644 | 0.000812421 | THAP8 |
| chr11 | 36593483 | 36593801 | 319 | 2 | 0.000655306 | 0.000212 | 0.00576 | 0.00038 | 0.000957567 | RAG2 |
| chr1 | 59309583 | 59309701 | 119 | 3 | 0.000655331 | 0.00465 | 0.00035 | 0.00053 | -0.003526417 | FGGY |
| chr8 | 120537240 | 120537500 | 261 | 3 | 0.00065647 | 4.09E-05 | 0.00187 | 6.28E-05 | -0.006009763 | MTBP, SNTB1 |
| chr12 | 118161136 | 118161188 | 53 | 2 | 0.000656515 | 0.000378 | 0.0003 | 0.00018 | -0.009245303 | TAOK3 |
| chr10 | 244370 | 244408 | 39 | 3 | 0.00065669 | 0.004486 | 0.00123 | 8.40E-05 | -0.005658108 | ZMYND11 |
| chr15 | 58490486 | 58490489 | 4 | 2 | 0.000657589 | 0.045389 | 4.52E-05 | 7.68E-05 | 0.002586989 | LIPC, ALDH1A2, LIPC-AS1 |
| chr7 | 73313430 | 73313506 | 77 | 2 | 0.000658608 | 0.000108 | 0.00281 | 0.00017 | -0.003896882 | TRIM50 |
| chr8 | 124474629 | 124474816 | 188 | 4 | 0.000658928 | 0.004799 | 0.00458 | 0.00367 | 0.000948798 | NA |
| chr10 | 11814183 | 11814194 | 12 | 2 | 0.000658975 | 2.64E-05 | 0.00084 | 3.81E-05 | -0.019756706 | NA |
| chr15 | 77787995 | 77788003 | 9 | 2 | 0.000659364 | 0.000107 | 0.00126 | 0.00014 | -0.006278328 | AC105133.1, LINGO1 |
| chr1 | 156153395 | 156153709 | 315 | 6 | 0.000659989 | 0.003994 | 0.00044 | 0.00032 | -0.002471707 | SEMA4A |
| chr6 | 41887427 | 41887717 | 291 | 3 | 0.000660313 | 2.61E-05 | 0.00399 | 5.10E-05 | -0.007916848 | USP49 |
| chr2 | 238289653 | 238289796 | 144 | 2 | 0.000660568 | 0.000237 | 0.00059 | 0.00019 | -0.010985382 | AC012485.3, PER2 |
| chr14 | 94110601 | 94110872 | 272 | 4 | 0.000660976 | 0.000299 | 0.00037 | 0.00014 | 8.60E-06 | IFI27 |
| chr4 | 6889475 | 6889824 | 350 | 3 | 0.000661199 | 2.10E-05 | 0.00537 | 4.81E-05 | -0.028836327 | NA |
| chr11 | 68302787 | 68302792 | 6 | 2 | 0.00066141 | 0.00522 | 7.10E-05 | 0.00011 | -0.004732036 | NA |
| chr15 | 73796527 | 73796608 | 82 | 2 | 0.000661531 | 5.49E-05 | 0.00122 | 7.66E-05 | -0.003579764 | NA |
| chr10 | 59175817 | 59175903 | 87 | 2 | 0.000662404 | 0.000141 | 0.00302 | 0.00023 | -0.024244678 | NA |
| chr12 | 48681755 | 48682043 | 289 | 4 | 0.000662522 | 0.004617 | 8.31E-05 | 0.00016 | 0.000977275 | KANSL2 |
| chr12 | 55842750 | 55843257 | 508 | 5 | 0.000662524 | 0.000895 | 0.00168 | 0.0001 | -0.005381702 | MMP19 |
| chr6 | 146028502 | 146029078 | 577 | 6 | 0.000662725 | 0.01198 | 0.02538 | 0.00656 | 0.000960753 | GRM1 |
| chr20 | 7936565 | 7936670 | 106 | 2 | 0.000662967 | 0.057544 | 8.25E-05 | 0.00018 | -0.002904658 | HAO1 |
| chr12 | 32679035 | 32679140 | 106 | 5 | 0.000663897 | 2.49E-05 | 0.01455 | 8.74E-05 | 0.000746619 | NA |
| chr10 | 209739 | 209932 | 194 | 4 | 0.000663897 | 0.02854 | 0.00012 | 0.00034 | -0.002181704 | ZMYND11 |
| chr17 | 82315366 | 82315446 | 81 | 3 | 0.000663978 | 2.13E-05 | 0.00287 | 4.28E-05 | -0.011996126 | CD7 |
| chr6 | 30076147 | 30076482 | 336 | 5 | 0.000664113 | 0.002185 | 0.0116 | 0.00067 | -0.010161507 | NA |
| chr13 | 78595579 | 78595737 | 159 | 4 | 0.000664383 | 0.000313 | 0.00021 | 9.95E-05 | -0.001491388 | OBI1-AS1 |
| chr7 | 53087273 | 53087414 | 142 | 2 | 0.000665134 | 1.76E-05 | 0.00093 | 2.75E-05 | -0.004902415 | NA |
| chr1 | 155071085 | 155071491 | 407 | 5 | 0.00066521 | 0.001329 | 0.00102 | 0.00034 | -0.005866955 | AL691442.1 |
| chr9 | 114377635 | 114377955 | 321 | 5 | 0.000665256 | 0.000524 | 0.00518 | 0.00028 | -0.002319375 | AKNA |
| chr17 | 58489601 | 58489760 | 160 | 2 | 0.000665456 | 0.000805 | 0.00024 | 0.00023 | -0.005016797 | MTMR4 |
| chr13 | 113597357 | 113597407 | 51 | 3 | 0.000666117 | 6.76E-05 | 0.00608 | 0.00013 | -0.003177938 | TFDP1 |
| chr20 | 62182515 | 62182863 | 349 | 6 | 0.000666216 | 0.052862 | 0.00265 | 0.00014 | -0.000763429 | NA |
| chr16 | 1336074 | 1336289 | 216 | 4 | 0.000666403 | 0.003529 | 0.00267 | 0.00014 | -0.006679503 | BAIAP3 |
| chr11 | 2313502 | 2313823 | 322 | 4 | 0.000666462 | 2.15E-05 | 0.00568 | 4.58E-05 | -0.007635314 | TSPAN32 |
| chr8 | 139914983 | 139915082 | 100 | 2 | 0.000666462 | 0.000789 | 0.00327 | 0.00098 | 0.000911382 | AC021744.1, TRAPPC9 |
| chr10 | 68472431 | 68472826 | 396 | 2 | 0.000666852 | 0.064991 | 0.12525 | 0.09392 | -0.003112984 | NA |
| chr12 | 116881459 | 116881772 | 314 | 4 | 0.000667258 | 0.000543 | 0.00065 | 0.00015 | 0.00089493 | NA |
| chr20 | 62352866 | 62352950 | 85 | 3 | 0.000667693 | 0.001404 | 0.00014 | 0.00018 | -0.005628624 | LAMA5 |
| chr20 | 49826678 | 49826853 | 176 | 2 | 0.000667719 | 5.77E-05 | 0.00225 | 8.88E-05 | -0.006997021 | SLC9A8 |
| chr3 | 138348211 | 138348281 | 71 | 3 | 0.000668788 | 0.000208 | 0.00185 | 0.00021 | 0.00249627 | MRAS |
| chr6 | 109094513 | 109094870 | 358 | 6 | 0.000669181 | 0.001378 | 0.00639 | 0.00013 | 0.001058211 | SESN1 |
| chr2 | 238253403 | 238253481 | 79 | 2 | 0.000669847 | 0.032197 | 0.00078 | 0.00224 | -0.002028452 | AC012485.1, PER2 |
| chr14 | 39174466 | 39174906 | 441 | 3 | 0.000669998 | 0.052567 | 0.00473 | 0.00077 | 8.85E-05 | AL132639.3 |
| chr1 | 2342296 | 2342519 | 224 | 3 | 0.000670194 | 0.000122 | 0.00027 | 7.50E-05 | -0.003700846 | MORN1 |
| chr1 | 109613217 | 109613488 | 272 | 2 | 0.000670362 | 0.03917 | 0.00172 | 0.00478 | -0.002134941 | GNAT2 |
| chr6 | 155084986 | 155085164 | 179 | 2 | 0.000671398 | 0.001303 | 0.00053 | 0.0005 | -0.00411335 | TIAM2 |
| chr10 | 14548663 | 14549087 | 425 | 5 | 0.000671802 | 0.001413 | 0.00225 | 0.00025 | -0.003830045 | FAM107B |
| chr16 | 64950600 | 64951074 | 475 | 6 | 0.000671836 | 0.00016 | 0.01912 | 0.00014 | -0.000253754 | CDH11 |
| chr6 | 168444018 | 168444075 | 58 | 2 | 0.000672144 | 0.001294 | 0.0003 | 0.00033 | -0.007330626 | SMOC2 |
| chr2 | 45675547 | 45675603 | 57 | 3 | 0.000672306 | 2.28E-05 | 0.00367 | 4.58E-05 | -0.011055584 | PRKCE |
| chr13 | 49933705 | 49933747 | 43 | 2 | 0.000672499 | 3.70E-05 | 0.00118 | 5.40E-05 | -0.005230969 | SPRYD7 |
| chr16 | 75433525 | 75433874 | 350 | 4 | 0.000672544 | 0.000451 | 0.00429 | 0.00029 | 0.002013853 | AC009163.7, AC009163.5 |
| chr6 | 36777103 | 36777277 | 175 | 2 | 0.000673506 | 0.000187 | 0.00015 | 8.17E-05 | -0.014705162 | CPNE5 |
| chr10 | 49783843 | 49783856 | 14 | 2 | 0.000674066 | 3.55E-05 | 0.00141 | 5.34E-05 | -0.018335733 | NA |
| chr8 | 105669352 | 105669366 | 15 | 2 | 0.000674195 | 0.001144 | 8.14E-05 | 9.37E-05 | -0.005432518 | ZFPM2, ZFPM2-AS1, AC103853.1 |
| chr10 | 101830612 | 101830830 | 219 | 2 | 0.000674834 | 0.000689 | 0.01024 | 0.00123 | 0.003581593 | KCNIP2 |
| chr8 | 41796886 | 41797100 | 215 | 4 | 0.000674891 | 0.0038 | 0.00084 | 0.00021 | 0.001496948 | ANK1 |
| chr12 | 122775204 | 122775371 | 168 | 3 | 0.000676005 | 0.000962 | 0.00139 | 0.00016 | 0.001442641 | CCDC62 |
| chr22 | 46903456 | 46903492 | 37 | 2 | 0.000676005 | 4.03E-05 | 0.00087 | 5.51E-05 | -0.00325552 | TBC1D22A |
| chr8 | 48521757 | 48521772 | 16 | 2 | 0.000676249 | 6.63E-05 | 0.00031 | 6.03E-05 | -0.014823221 | NA |
| chr16 | 2786680 | 2786815 | 136 | 3 | 0.000676631 | 0.051158 | 0.00012 | 0.00071 | -0.005084794 | PRSS33 |
| chr3 | 12814378 | 12814482 | 105 | 2 | 0.00067684 | 0.001477 | 0.00074 | 0.00067 | -0.005492124 | CAND2 |
| chr6 | 117359785 | 117360363 | 579 | 4 | 0.000677028 | 2.74E-05 | 0.00243 | 5.15E-05 | -0.012222674 | ROS1, AL132671.2 |
| chr2 | 200586580 | 200587353 | 774 | 2 | 0.000677028 | 0.000167 | 0.00465 | 0.00029 | -0.011330877 | AOX1 |
| chr11 | 125883156 | 125883215 | 60 | 2 | 0.000677062 | 3.44E-05 | 0.00104 | 4.94E-05 | -0.007605425 | NA |
| chr11 | 117696718 | 117697060 | 343 | 2 | 0.000677206 | 3.78E-05 | 0.00145 | 5.70E-05 | -0.00109136 | DSCAML1 |
| chr8 | 143325911 | 143325956 | 46 | 2 | 0.000677457 | 0.000141 | 0.00099 | 0.00016 | -0.000898254 | TOP1MT |
| chr6 | 7141791 | 7141833 | 43 | 2 | 0.000677652 | 0.000438 | 0.00427 | 0.00067 | -0.010346993 | RREB1 |
| chr5 | 57482859 | 57483293 | 435 | 3 | 0.000677985 | 5.73E-05 | 0.00069 | 4.73E-05 | -0.004871115 | AC025470.2 |
| chr1 | 910430 | 910815 | 386 | 3 | 0.000678007 | 3.45E-05 | 0.00204 | 5.66E-05 | -0.018722936 | AL645608.6 |
| chr6 | 33289724 | 33289979 | 256 | 8 | 0.000678009 | 6.31E-05 | 0.04627 | 0.00027 | 0.000808501 | PFDN6 |
| chr10 | 101036343 | 101036533 | 191 | 2 | 0.000678298 | 7.88E-05 | 0.00083 | 9.42E-05 | -0.008307965 | SFXN3 |
| chr7 | 102304414 | 102304662 | 249 | 2 | 0.000678856 | 3.54E-05 | 0.00167 | 5.46E-05 | -0.006984433 | SH2B2 |
| chr14 | 101031869 | 101032006 | 138 | 5 | 0.000678924 | 0.003124 | 0.00187 | 0.00012 | -0.001773596 | MEG8, MIR543 |
| chr10 | 80078564 | 80078717 | 154 | 2 | 0.000679358 | 0.07027 | 0.01494 | 0.03021 | 0.001218358 | TMEM254, TMEM254-AS1 |
| chr10 | 13666083 | 13666265 | 183 | 8 | 0.000679551 | 0.000244 | 0.00643 | 6.25E-05 | -0.002555989 | AL157392.3, FRMD4A |
| chr15 | 98911173 | 98911325 | 153 | 2 | 0.000679675 | 0.000266 | 0.00015 | 9.44E-05 | -0.007499086 | IGF1R |
| chr2 | 231483162 | 231483623 | 462 | 3 | 0.00067978 | 0.000557 | 0.0283 | 0.0016 | -0.014071674 | NCL |
| chr20 | 62547675 | 62548322 | 648 | 2 | 0.000680012 | 4.22E-05 | 0.00108 | 5.96E-05 | -0.012638253 | MIR1-1HG-AS1 |
| chr16 | 88770310 | 88770777 | 468 | 2 | 0.00068085 | 0.000388 | 0.00125 | 0.00039 | -0.009840494 | PIEZO1 |
| chr22 | 38733291 | 38733365 | 75 | 2 | 0.000682476 | 0.000802 | 0.00462 | 0.00112 | -0.008379963 | GTPBP1 |
| chr20 | 44475155 | 44475387 | 233 | 5 | 0.000682477 | 0.000238 | 0.00307 | 9.50E-05 | -0.004216212 | NA |
| chr4 | 188032523 | 188032815 | 293 | 3 | 0.00068254 | 0.000103 | 0.00033 | 6.74E-05 | 0.006868677 | NA |
| chr15 | 67806713 | 67806806 | 94 | 2 | 0.00068274 | 0.003986 | 0.00024 | 0.00042 | -0.001468136 | MAP2K5 |
| chr7 | 139915009 | 139915025 | 17 | 2 | 0.000683194 | 0.000107 | 0.00018 | 6.62E-05 | -0.003941393 | TBXAS1 |
| chr17 | 81872258 | 81872320 | 63 | 2 | 0.000683206 | 0.000487 | 0.00028 | 0.00019 | 0.006006208 | NA |
| chr21 | 26889127 | 26889343 | 217 | 2 | 0.000685289 | 0.00129 | 0.00013 | 0.00016 | -0.005877571 | NA |
| chr8 | 144178636 | 144179035 | 400 | 3 | 0.000685719 | 0.000789 | 0.00058 | 9.47E-05 | -0.004727876 | MROH1 |

| | | | | | | | | | | |
|---|---|---|---|---|---|---|---|---|---|---|
| chr7 | 42227670 | 42227784 | 115 | 3 | 0.000685889 | 1.08E-05 | 0.00119 | 2.04E-05 | -0.019241875 | GLI3 |
| chr15 | 43207392 | 43207711 | 320 | 3 | 0.000686099 | 0.000146 | 0.00742 | 0.00028 | -0.001047632 | EPB42 |
| chr6 | 157324350 | 157324533 | 184 | 2 | 0.00068627 | 0.000728 | 0.00044 | 0.00033 | 0.003662833 | AL390955.2 |
| chr20 | 62015281 | 62015816 | 536 | 3 | 0.000686915 | 0.004134 | 0.00269 | 0.00266 | -0.001296041 | TAF4 |
| chr4 | 6070443 | 6070507 | 65 | 3 | 0.000687159 | 2.05E-05 | 0.00507 | 4.64E-05 | -0.024531583 | C4orf50, JAKMIP1 |
| chr6 | 167747195 | 167747338 | 144 | 3 | 0.000687255 | 0.001443 | 0.00246 | 0.00084 | -0.003878239 | NA |
| chr10 | 3015759 | 3015983 | 225 | 4 | 0.000687375 | 9.69E-05 | 0.00104 | 9.69E-05 | -0.002290158 | NA |
| chr1 | 1426096 | 1426286 | 191 | 3 | 0.000688258 | 0.000147 | 0.00247 | 0.00022 | -0.014925932 | TMEM88B |
| chr3 | 54120779 | 54121213 | 435 | 4 | 0.000688277 | 0.010138 | 0.01716 | 0.01174 | -0.001204017 | AC115282.1 |
| chr17 | 76948693 | 76949137 | 445 | 5 | 0.000688635 | 0.000254 | 0.00072 | 8.99E-05 | -0.005165508 | MGAT5B |
| chr1 | 1537761 | 1539019 | 1259 | 13 | 0.000688848 | 0.000134 | 0.00239 | 2.10E-05 | -0.001430898 | TMEM240 |
| chr10 | 75029456 | 75029553 | 98 | 2 | 0.00068915 | 0.001829 | 5.14E-05 | 6.43E-05 | 0.002795947 | KAT6B |
| chr15 | 48205454 | 48205456 | 3 | 2 | 0.00068965 | 0.000338 | 0.00095 | 0.00031 | -0.004520369 | SLC12A1 |
| chr7 | 140326477 | 140326575 | 99 | 2 | 0.000689867 | 4.53E-05 | 0.00071 | 5.80E-05 | -0.015743755 | SLC37A3 |
| chr21 | 42127576 | 42127930 | 355 | 4 | 0.00069031 | 0.000258 | 0.01156 | 0.00066 | -0.001042104 | UMODL1 |
| chr12 | 120723641 | 120723675 | 35 | 2 | 0.000691075 | 0.001498 | 0.00013 | 0.00017 | -0.007901754 | NA |
| chr22 | 17832006 | 17832250 | 245 | 4 | 0.00069143 | 5.14E-05 | 0.00693 | 9.26E-05 | -0.005644389 | MICAL3 |
| chr22 | 50599950 | 50600294 | 345 | 4 | 0.00069164 | 0.000947 | 0.00043 | 0.00011 | -0.01191151 | CHKB |
| chr19 | 5977955 | 5978270 | 316 | 3 | 0.00069173 | 0.014177 | 0.01756 | 0.00616 | 0.000899158 | RANBP3 |
| chr2 | 11669555 | 11669732 | 178 | 2 | 0.000691772 | 0.006877 | 0.01173 | 0.00773 | -0.0100455 | NTSR2 |
| chr4 | 168882813 | 168882887 | 75 | 2 | 0.000692752 | 0.007488 | 8.98E-05 | 0.00016 | -0.004462196 | PALLD, CBR4 |
| chr12 | 51197942 | 51198297 | 356 | 5 | 0.000692888 | 0.000101 | 0.00348 | 4.31E-05 | -0.003044755 | POU6F1 |
| chr5 | 154690584 | 154690825 | 242 | 3 | 0.000693407 | 0.000383 | 0.00049 | 0.00019 | -0.005134843 | NA |
| chr1 | 38990623 | 38991119 | 497 | 5 | 0.000693458 | 0.006984 | 0.00654 | 0.00434 | -0.004049258 | NA |
| chr1 | 40344619 | 40344742 | 124 | 2 | 0.000693941 | 0.000117 | 0.00106 | 0.00014 | -0.003096618 | NA |
| chr10 | 376041 | 376247 | 207 | 3 | 0.000694087 | 2.42E-05 | 0.0018 | 4.38E-05 | -0.006727757 | DIP2C |
| chr1 | 54542032 | 54542063 | 32 | 3 | 0.000694206 | 0.007364 | 0.00134 | 0.0022 | 0.001848995 | NA |
| chr7 | 123534142 | 123534565 | 424 | 7 | 0.000695151 | 0.000367 | 0.01034 | 9.99E-05 | -0.003185862 | IQUB |
| chr11 | 133951110 | 133951271 | 162 | 4 | 0.000695289 | 0.000258 | 0.00118 | 5.29E-05 | 0.014941137 | IGSF9B |
| chr14 | 100156427 | 100156935 | 509 | 4 | 0.000695289 | 0.00032 | 0.00345 | 0.00025 | -0.005349174 | DEGS2 |
| chr17 | 58192406 | 58192457 | 52 | 3 | 0.000696907 | 0.000407 | 0.00488 | 0.00039 | -0.003923275 | NA |
| chr8 | 1864448 | 1864543 | 96 | 2 | 0.000697037 | 0.026471 | 0.00032 | 0.0009 | -0.002199133 | AC019257.8, ARHGEF10 |
| chr12 | 57590867 | 57591038 | 172 | 6 | 0.000697172 | 0.003165 | 0.004 | 0.00048 | 0.001837694 | NA |
| chr20 | 35092996 | 35093420 | 425 | 6 | 0.000697286 | 0.004086 | 0.01173 | 0.00199 | -0.002286591 | NA |
| chr12 | 48845598 | 48845694 | 97 | 2 | 0.00069734 | 0.009693 | 7.67E-05 | 0.00014 | 0.002481129 | DDX23 |
| chr7 | 30027325 | 30027388 | 64 | 2 | 0.000697476 | 0.004571 | 0.01428 | 0.00628 | 0.003832213 | FKBP14-AS1 |
| chr2 | 6764920 | 6765330 | 411 | 2 | 0.000697623 | 3.71E-05 | 0.00152 | 5.64E-05 | 0.001333008 | LINC00487 |
| chr7 | 55588206 | 55588305 | 100 | 2 | 0.000697925 | 3.73E-05 | 0.00139 | 5.60E-05 | -0.00496346 | AC099681.1 |
| chr5 | 14760586 | 14760778 | 193 | 3 | 0.00069812 | 0.001203 | 8.11E-05 | 0.00011 | -0.000877968 | ANKH |
| chr6 | 108294845 | 108295084 | 240 | 3 | 0.000698161 | 0.00717 | 0.01316 | 0.0044 | -0.002764665 | AFG1L |
| chr12 | 93448866 | 93449055 | 190 | 2 | 0.000698161 | 8.58E-05 | 0.00017 | 5.61E-05 | -0.006904877 | NA |
| chr17 | 17691405 | 17691963 | 559 | 2 | 0.000698393 | 2.70E-05 | 0.00155 | 4.24E-05 | -0.008244016 | RAI1 |
| chr2 | 236177120 | 236177165 | 46 | 2 | 0.000698419 | 0.000237 | 0.00027 | 0.00013 | -0.023166026 | AC019068.1 |
| chr15 | 51367222 | 51367414 | 193 | 4 | 0.000698522 | 0.000476 | 0.00063 | 0.00028 | -0.007659953 | GLDN, AC020891.1 |
| chr18 | 32471668 | 32471840 | 173 | 3 | 0.000698522 | 0.007949 | 0.00491 | 0.0057 | -0.006498184 | AC025887.2 |
| chr3 | 57536611 | 57536774 | 164 | 2 | 0.000698606 | 0.000837 | 0.00014 | 0.00015 | -0.006725492 | DNAH12 |
| chr2 | 219229497 | 219229795 | 299 | 7 | 0.000698789 | 0.000103 | 0.00053 | 1.24E-05 | 0.00239403 | ANKZF1, ATG9A |
| chr9 | 70168135 | 70168175 | 41 | 2 | 0.000698976 | 0.015338 | 7.55E-05 | 0.00014 | -0.010557638 | MAMDC2, MAMDC2-AS1 |
| chr12 | 13043886 | 13044321 | 436 | 8 | 0.000699001 | 1.14E-05 | 0.00666 | 4.71E-05 | -0.01093907 | NA |
| chr2 | 30225663 | 30226440 | 778 | 2 | 0.000699734 | 6.92E-06 | 0.00059 | 1.14E-05 | -0.009433851 | NA |
| chr14 | 24316641 | 24316727 | 87 | 2 | 0.000700874 | 5.42E-05 | 0.00067 | 6.63E-05 | -0.00435639 | LTB4R |
| chr11 | 46617715 | 46617929 | 215 | 3 | 0.00070088 | 0.024068 | 0.01417 | 0.01778 | 0.000429226 | ATG13, HARBI1 |
| chr22 | 37745636 | 37745827 | 192 | 3 | 0.000700937 | 0.001254 | 0.00144 | 0.00026 | 0.004877488 | TRIOBP |
| chr12 | 128266426 | 128266503 | 78 | 3 | 0.000701128 | 0.000157 | 0.00203 | 8.67E-05 | -0.00828859 | NA |
| chr12 | 49323183 | 49323497 | 315 | 5 | 0.000701482 | 0.002626 | 0.03996 | 0.00531 | -0.00060344 | TROAP, AC125611.4 |
| chr4 | 21948230 | 21948550 | 321 | 3 | 0.000701509 | 0.000478 | 0.00114 | 0.00023 | -0.005814613 | KCNIP4 |
| chr2 | 27583200 | 27583559 | 360 | 4 | 0.000701661 | 0.001571 | 0.02654 | 0.00392 | 0.002422563 | ZNF512, AC074091.2 |
| chr1 | 3510843 | 3511034 | 192 | 3 | 0.000702293 | 0.000352 | 0.00037 | 9.55E-05 | -0.007293468 | MEGF6 |
| chr11 | 126359871 | 126359896 | 26 | 2 | 0.000702428 | 0.000103 | 0.00147 | 0.00014 | -0.008334979 | ST3GAL4 |
| chr8 | 144994542 | 144994772 | 231 | 3 | 0.000702569 | 2.33E-05 | 0.0044 | 4.89E-05 | -0.001327896 | AF235103.1, ZNF252P |
| chr8 | 22703409 | 22703932 | 524 | 6 | 0.000702623 | 0.000303 | 0.01425 | 0.00014 | -0.010802122 | AC055854.1, AC105046.2 |
| chr2 | 233775638 | 233775724 | 87 | 2 | 0.000702623 | 0.000113 | 0.00052 | 0.00011 | -0.005819524 | MROH2A |
| chr13 | 106491632 | 106492005 | 374 | 4 | 0.000703693 | 0.00159 | 0.00011 | 0.00016 | 0.008014346 | AL138689.2, EFNB2 |
| chr17 | 47732361 | 47732439 | 79 | 2 | 0.000704438 | 0.005601 | 0.0077 | 0.00557 | -0.006169835 | NA |
| chr8 | 13437603 | 13438674 | 1072 | 3 | 0.000704467 | 5.02E-05 | 0.00018 | 5.18E-06 | 0.011019365 | DLC1 |
| chr12 | 102095590 | 102095849 | 260 | 2 | 0.000704856 | 3.97E-05 | 0.00187 | 6.18E-05 | -0.005795836 | NUP37 |
| chr17 | 796814 | 797432 | 619 | 4 | 0.000704987 | 0.000109 | 0.00295 | 5.13E-05 | -0.006413415 | NA |
| chr11 | 68119759 | 68119768 | 10 | 2 | 0.000705288 | 0.008272 | 0.0002 | 0.00045 | -0.002904675 | CHKA |
| chr10 | 72328267 | 72328431 | 165 | 2 | 0.000705622 | 0.00033 | 0.00013 | 9.51E-05 | -0.007983362 | NA |
| chr18 | 9743560 | 9743591 | 32 | 2 | 0.000706138 | 0.00107 | 0.01397 | 0.00194 | -0.009270897 | RAB31 |
| chr13 | 113089250 | 113089549 | 300 | 2 | 0.000706408 | 0.010224 | 0.00175 | 0.00311 | 0.000746925 | MCF2L |
| chr7 | 76400471 | 76400564 | 94 | 2 | 0.000706888 | 1.16E-05 | 0.00083 | 1.86E-05 | -0.003693788 | ZP3, SSC4D |
| chr20 | 51507391 | 51507595 | 205 | 2 | 0.00070704 | 4.17E-05 | 0.00193 | 6.48E-05 | -0.008298269 | NFATC2 |
| chr3 | 46665543 | 46665551 | 9 | 2 | 0.000707364 | 4.98E-05 | 0.00026 | 4.62E-05 | -0.001429475 | NA |
| chr16 | 31002920 | 31002983 | 64 | 2 | 0.000707364 | 0.026167 | 0.00396 | 0.00822 | -0.001532203 | STX1B |
| chr2 | 2872192 | 2872410 | 219 | 2 | 0.000707787 | 0.000923 | 0.00252 | 0.00098 | -0.001075086 | NA |
| chr12 | 6989593 | 6989761 | 169 | 2 | 0.000708032 | 0.001113 | 0.00014 | 0.00017 | 0.00339624 | EMG1, C1S, LPCAT3 |
| chr12 | 98592955 | 98593109 | 155 | 3 | 0.000708454 | 0.021454 | 0.01483 | 0.01077 | -0.006254784 | NA |
| chr14 | 54776351 | 54776434 | 84 | 2 | 0.000709036 | 7.43E-05 | 0.00058 | 8.11E-05 | -0.001555951 | SAMD4A |
| chr20 | 50224631 | 50224923 | 293 | 2 | 0.000709427 | 9.86E-05 | 0.00361 | 0.00016 | -0.002638525 | NA |
| chr22 | 38655812 | 38655936 | 125 | 3 | 0.000710086 | 0.00612 | 0.00462 | 0.00066 | -0.007429651 | FAM227A |
| chr11 | 1437184 | 1437208 | 25 | 2 | 0.000710574 | 0.00035 | 0.00026 | 0.00016 | -0.015482133 | BRSK2 |
| chr6 | 24934003 | 24934041 | 39 | 2 | 0.00071066 | 0.006135 | 7.64E-05 | 0.00013 | -0.007824489 | RIPOR2 |
| chr8 | 81496559 | 81497053 | 495 | 2 | 0.000711069 | 2.68E-05 | 0.00155 | 4.23E-05 | 1.78E-05 | AC018616.1 |
| chr1 | 117173770 | 117173789 | 20 | 2 | 0.000711379 | 0.001309 | 0.0001 | 0.00013 | -0.005226649 | VTCN1 |
| chr12 | 56127600 | 56128169 | 570 | 10 | 0.000711581 | 0.007301 | 0.00038 | 0.00034 | 0.001176323 | ESYT1, AC034102.6 |
| chr5 | 61229682 | 61229832 | 151 | 2 | 0.000711774 | 6.92E-05 | 0.00038 | 6.74E-05 | -0.012677718 | SMIM15-AS1, LINC02057 |
| chr17 | 40064742 | 40065309 | 568 | 2 | 0.000711805 | 0.001391 | 0.00133 | 0.00093 | -0.01004893 | THRA |
| chr11 | 2414951 | 2415021 | 71 | 2 | 0.000712029 | 7.30E-05 | 0.00036 | 6.83E-05 | -0.00706197 | TRPM5 |
| chr18 | 58195444 | 58195640 | 197 | 4 | 0.000712064 | 0.004034 | 0.00205 | 0.00021 | 0.000114073 | NEDD4L, AC107896.1 |
| chr5 | 946162 | 946222 | 61 | 3 | 0.000712477 | 0.000119 | 0.01135 | 0.0003 | -0.012857572 | NA |
| chr7 | 43907030 | 43907370 | 341 | 4 | 0.000712669 | 0.002238 | 0.00859 | 0.00103 | -0.00921297 | URGCP |
| chr21 | 45509703 | 45510045 | 343 | 4 | 0.00071277 | 6.49E-05 | 0.00304 | 8.09E-05 | -0.003702957 | COL18A1, SLC19A1 |
| chr12 | 2384313 | 2384401 | 89 | 2 | 0.000713554 | 0.02271 | 9.14E-05 | 0.00019 | 0.002351073 | CACNA1C |
| chr17 | 42998137 | 42998470 | 334 | 9 | 0.000713929 | 7.44E-05 | 0.01046 | 0.00015 | -0.000648042 | RPL27 |
| chr10 | 117134017 | 117134091 | 75 | 2 | 0.000714504 | 0.011024 | 0.05075 | 0.0186 | 0.006033894 | VAX1 |
| chr4 | 4482280 | 4482354 | 75 | 2 | 0.000714682 | 0.008087 | 0.00011 | 0.00021 | -0.002076518 | STX18 |
| chr19 | 56404153 | 56404836 | 684 | 8 | 0.000714827 | 0.003093 | 0.00166 | 0.00055 | -0.001733681 | ZNF583 |
| chr7 | 50781588 | 50782026 | 439 | 3 | 0.000714926 | 2.61E-05 | 0.00178 | 3.26E-05 | -0.014594642 | GRB10 |
| chr17 | 61483119 | 61483238 | 120 | 2 | 0.000715938 | 5.13E-05 | 0.00038 | 5.42E-05 | -0.004948359 | TBX4 |
| chr3 | 51941881 | 51942411 | 531 | 12 | 0.00071595 | 0.000618 | 0.0255 | 0.00011 | 0.000267552 | PARP3, RRP9 |
| chr9 | 127715886 | 127716099 | 214 | 5 | 0.000717267 | 6.97E-05 | 0.00288 | 7.24E-05 | -0.000330891 | CFAP157, TTC16, PTRH1 |
| chr15 | 55587891 | 55588026 | 136 | 2 | 0.000717549 | 0.415006 | 0.00573 | 0.01887 | 0.001264479 | PYGO1 |
| chr2 | 11847794 | 11847965 | 172 | 2 | 0.000717881 | 0.000171 | 0.00015 | 7.82E-05 | -0.003430712 | MIR3681HG |
| chr2 | 100125431 | 100125467 | 37 | 2 | 0.000717955 | 8.37E-05 | 0.00036 | 7.52E-05 | -0.006915383 | AFF3 |
| chr12 | 93127534 | 93127554 | 21 | 2 | 0.000718081 | 0.000265 | 0.0002 | 0.00011 | -0.003926319 | AC138123.1 |
| chr4 | 70241839 | 70241865 | 27 | 2 | 0.000718178 | 0.002072 | 0.00011 | 0.00015 | -0.005751634 | NA |
| chr17 | 81025364 | 81025562 | 199 | 3 | 0.000718436 | 0.000302 | 0.0133 | 0.00074 | 0.003372137 | AC127496.5, AC127496.2 |
| chr17 | 44041692 | 44041949 | 258 | 2 | 0.000719215 | 3.43E-05 | 0.00031 | 3.74E-05 | -0.011805853 | LSM12 |
| chr3 | 32503857 | 32503981 | 125 | 3 | 0.000719979 | 0.018383 | 0.00033 | 0.00128 | -0.001790094 | NA |
| chr1 | 239908423 | 239908507 | 85 | 2 | 0.000720501 | 0.000933 | 0.00156 | 0.00079 | 0.003488776 | CHRM3 |
| chr8 | 60517268 | 60517598 | 331 | 2 | 0.000720923 | 0.000272 | 0.0004 | 0.00017 | 0.00283691 | RAB2A |
| chr15 | 65303660 | 65303991 | 332 | 3 | 0.000721102 | 0.026346 | 0.00927 | 0.00758 | -0.004876276 | NA |
| chr4 | 176195488 | 176195858 | 371 | 9 | 0.000721286 | 7.80E-05 | 0.00124 | 4.32E-05 | 0.000295298 | SPATA4 |
| chr3 | 50476646 | 50476776 | 131 | 2 | 0.000722169 | 0.000794 | 0.0001 | 0.0001 | -0.003732772 | CACNA2D2 |
| chr1 | 34792257 | 34792460 | 204 | 3 | 0.000722556 | 0.002249 | 0.00094 | 0.00032 | -0.007455495 | SMIM12 |
| chr1 | 20138516 | 20138793 | 278 | 4 | 0.000722953 | 0.00019 | 0.00234 | 0.00016 | -0.003949167 | NA |
| chr11 | 13495937 | 13497129 | 1193 | 4 | 0.000723177 | 1.08E-05 | 0.00021 | 2.21E-06 | -0.003143636 | PTH |
| chr2 | 25010146 | 25010294 | 149 | 2 | 0.000723198 | 0.00041 | 8.35E-05 | 7.00E-05 | -0.015363033 | DNAJC27-AS1, AC013267.1 |
| chr8 | 11739663 | 11739742 | 80 | 2 | 0.000723376 | 4.19E-05 | 0.00103 | 5.87E-05 | -0.028945197 | GATA4 |
| chr6 | 36874431 | 36874475 | 45 | 2 | 0.00072365 | 0.000189 | 0.00542 | 0.00034 | 0.003678378 | C6orf89, PPIL1 |
| chr16 | 89198933 | 89198956 | 24 | 3 | 0.000724059 | 0.001899 | 0.00321 | 0.00034 | -0.003360767 | SLC22A31 |
| chr20 | 47358118 | 47358302 | 185 | 3 | 0.00072482 | 4.74E-05 | 0.00747 | 0.00011 | -0.008879975 | NA |
| chr3 | 83329825 | 83329856 | 32 | 2 | 0.00072492 | 4.76E-05 | 0.00226 | 7.45E-05 | -0.004249827 | NA |
| chr3 | 9878851 | 9879014 | 164 | 4 | 0.000725274 | 0.000213 | 0.00341 | 5.94E-05 | -0.001585706 | CIDEC |
| chr1 | 22637112 | 22637121 | 10 | 2 | 0.000725495 | 0.001257 | 0.00077 | 0.00063 | -0.009750111 | C1QA |
| chr7 | 130393347 | 130393406 | 60 | 2 | 0.000726877 | 0.000116 | 0.0005 | 0.00011 | -0.0125521 | NA |
| chr11 | 129364482 | 129364965 | 484 | 2 | 0.000727071 | 3.86E-05 | 0.0001 | 2.54E-05 | 0.008725205 | NA |
| chr1 | 164558568 | 164558675 | 108 | 2 | 0.0007276 | 0.130182 | 0.01044 | 0.02776 | 0.005071911 | PBX1 |
| chr12 | 1932415 | 1932541 | 127 | 2 | 0.000727839 | 6.89E-05 | 0.00094 | 8.72E-05 | -0.013039381 | LINC00940 |
| chr1 | 205084332 | 205084592 | 261 | 2 | 0.000727932 | 0.000219 | 0.00013 | 7.76E-05 | 0.002406835 | TMEM81 |
| chr6 | 29850302 | 29850523 | 222 | 6 | 0.000728232 | 2.56E-05 | 0.00805 | 5.10E-05 | -0.018818734 | AL645929.2 |
| chr9 | 127961147 | 127961165 | 19 | 2 | 0.000728365 | 4.02E-05 | 0.00198 | 6.29E-05 | -0.007299086 | FAM102A |
| chr17 | 13024401 | 13024657 | 257 | 2 | 0.000728454 | 6.69E-05 | 0.00016 | 4.66E-05 | -0.006878004 | NA |
| chr6 | 52147533 | 52147592 | 60 | 2 | 0.000728719 | 3.33E-05 | 0.00172 | 5.19E-05 | -0.005398993 | LINCMD1 |
| chr10 | 85558578 | 85558647 | 70 | 3 | 0.000729121 | 0.02751 | 5.33E-05 | 0.00022 | -0.010357974 | NA |
| chr9 | 130428862 | 130428986 | 125 | 2 | 0.000729246 | 0.024961 | 9.29E-05 | 0.0002 | -0.003022197 | HMCN2 |
| chr3 | 3109995 | 3110056 | 62 | 2 | 0.00073009 | 3.73E-05 | 0.00046 | 4.45E-05 | -0.016184095 | IL5RA |
| chr4 | 182708279 | 182708319 | 41 | 2 | 0.000730354 | 3.28E-05 | 0.00066 | 4.36E-05 | 0.010000346 | TENM3 |
| chr3 | 114351810 | 114351893 | 84 | 4 | 0.000730375 | 9.26E-05 | 0.00642 | 0.00016 | -0.002325751 | ZBTB20-AS1, ZBTB20 |
| chr19 | 3808584 | 3808864 | 281 | 3 | 0.000730447 | 0.000166 | 0.00087 | 7.60E-05 | -0.003731701 | ZFR2 |
| chr9 | 86353565 | 86354387 | 823 | 4 | 0.000731034 | 0.003808 | 0.00516 | 0.00115 | -0.002278196 | TUT7 |
| chr8 | 142347329 | 142347957 | 629 | 2 | 0.000731825 | 0.002294 | 0.01174 | 0.00348 | -0.008377118 | TSNARE1 |
| chr5 | 93582362 | 93582366 | 5 | 2 | 0.000731978 | 0.001491 | 0.00016 | 0.00021 | 0.008724998 | NR2F1-AS1 |
| chr17 | 19870215 | 19870308 | 94 | 2 | 0.000732093 | 0.000524 | 0.00012 | 0.0001 | -0.005849734 | NA |
| chr1 | 110677585 | 110677674 | 90 | 2 | 0.000732157 | 0.000618 | 0.0001 | 9.65E-05 | -0.010532497 | NA |
| chr9 | 77954371 | 77954404 | 34 | 2 | 0.000732543 | 7.61E-05 | 0.0003 | 6.61E-05 | -0.013243737 | GNAQ |
| chr8 | 66540487 | 66540495 | 9 | 2 | 0.00073341 | 0.000144 | 0.00084 | 0.00016 | -0.018488531 | NA |
| chr9 | 131406631 | 131406666 | 36 | 2 | 0.000734011 | 0.002709 | 0.00012 | 0.00018 | -0.009816304 | PRRC2B |
| chr14 | 92609500 | 92609663 | 164 | 2 | 0.000734193 | 4.81E-05 | 0.00092 | 6.46E-05 | -0.014887371 | RIN3 |
| chr19 | 44954633 | 44955159 | 527 | 9 | 0.000734605 | 0.003775 | 0.01959 | 0.00419 | 0.000706836 | CLPTM1 |
| chr15 | 73442403 | 73442846 | 444 | 3 | 0.000734645 | 0.00011 | 0.00038 | 8.17E-05 | -0.002287124 | NA |
| chr15 | 101053488 | 101053780 | 293 | 2 | 0.00073502 | 0.000443 | 0.00106 | 0.00039 | -0.001602371 | LRRK1, AC090907.2 |
| chr12 | 55965426 | 55965666 | 241 | 3 | 0.000735086 | 5.90E-05 | 0.00603 | 0.00013 | -0.007871908 | PMEL |
| chr1 | 117077192 | 117077217 | 26 | 2 | 0.000735205 | 0.00034 | 0.00017 | 0.00012 | -0.00548106 | TTF2 |
| chr1 | 150925380 | 150925916 | 537 | 2 | 0.000735534 | 0.001139 | 0.0118 | 0.00197 | 0.029167322 | NA |
| chr3 | 120682211 | 120682318 | 108 | 4 | 0.000737113 | 0.000101 | 0.00338 | 0.00012 | -0.00827582 | HGD |
| chr17 | 2388340 | 2388499 | 160 | 2 | 0.000737256 | 0.000424 | 6.56E-05 | 5.66E-05 | -0.00527075 | MNT |
| chr10 | 80155257 | 80155411 | 155 | 2 | 0.000737475 | 8.35E-05 | 0.00242 | 0.00013 | -0.002041478 | ANXA11 |
| chr19 | 4685703 | 4685803 | 101 | 2 | 0.000737632 | 8.68E-05 | 0.00212 | 0.00013 | -0.002435647 | DPP9-AS1, DPP9 |
| chr9 | 35908120 | 35908431 | 312 | 2 | 0.000738874 | 8.00E-05 | 0.0032 | 0.00013 | 0.016592605 | NA |
| chr11 | 65505561 | 65505609 | 49 | 2 | 0.000739123 | 0.002912 | 0.00051 | 0.00071 | -0.012732966 | MALAT1 |

| | | | | | | | | | | |
|---|---|---|---|---|---|---|---|---|---|---|
| chr16 | 70753519 | 70753596 | 78 | 2 | 0.000739371 | 0.000172 | 0.00019 | 8.81E-05 | -0.005600376 | VAC14 |
| chr22 | 40418962 | 40419097 | 136 | 2 | 0.000739716 | 0.000211 | 0.00042 | 0.00015 | -0.005836401 | MRTFA |
| chr10 | 61780266 | 61780565 | 300 | 2 | 0.000740717 | 4.38E-05 | 0.00135 | 6.42E-05 | -0.016203103 | NA |
| chr17 | 42095093 | 42095143 | 51 | 2 | 0.00074089 | 5.39E-05 | 0.00247 | 8.46E-05 | -0.009266853 | ZNF385C |
| chr7 | 73570731 | 73570824 | 94 | 4 | 0.000741371 | 0.00017 | 0.00091 | 7.34E-05 | -0.002650624 | TBL2 |
| chr14 | 64704586 | 64704590 | 5 | 2 | 0.000741382 | 0.012355 | 5.96E-05 | 0.0001 | -0.002636858 | PLEKHG3 |
| chr16 | 66961200 | 66961558 | 359 | 4 | 0.000741413 | 5.94E-05 | 0.00176 | 8.49E-05 | -0.00813248 | AC009084.1, CES3 |
| chr3 | 31703703 | 31704138 | 436 | 5 | 0.000742609 | 0.001516 | 0.00076 | 0.00032 | -0.003818625 | OSBPL10-AS1, OSBPL10 |
| chr17 | 79798851 | 79798917 | 67 | 2 | 0.000742656 | 0.018249 | 4.48E-05 | 7.34E-05 | 0.001714519 | CBX8 |
| chr3 | 107240970 | 107241367 | 398 | 4 | 0.000743467 | 0.023883 | 0.0039 | 0.00277 | -0.001431053 | DUBR |
| chr16 | 81450852 | 81450987 | 136 | 3 | 0.000744205 | 0.007753 | 0.00073 | 0.00011 | 0.000635252 | CMIP |
| chr19 | 33683759 | 33684130 | 372 | 7 | 0.000744425 | 0.098794 | 0.00299 | 0.00322 | 0.002847011 | CHST8 |
| chr11 | 118635574 | 118635634 | 61 | 2 | 0.000744964 | 0.015968 | 0.00072 | 0.00175 | -0.00741629 | PHLDB1 |
| chr7 | 1709045 | 1709214 | 170 | 3 | 0.0007452 | 0.000847 | 0.0081 | 0.00135 | -0.005388709 | ELFN1 |
| chr11 | 8031699 | 8031873 | 175 | 2 | 0.000746157 | 8.71E-05 | 0.0029 | 0.00014 | 0.001928641 | TUB |
| chr12 | 47803699 | 47803708 | 10 | 2 | 0.000746202 | 3.69E-05 | 0.0016 | 5.64E-05 | -0.012605617 | HDAC7 |
| chr2 | 241735632 | 241736040 | 409 | 4 | 0.000746674 | 0.000828 | 0.00064 | 0.00028 | -0.002556802 | D2HGDH |
| chr15 | 50220803 | 50220855 | 53 | 2 | 0.000747189 | 0.002095 | 5.56E-05 | 7.23E-05 | 0.006740644 | SLC27A2 |
| chr11 | 76670643 | 76670893 | 251 | 5 | 0.000747223 | 0.000257 | 0.00159 | 1.83E-05 | 0.00105594 | LRRC32 |
| chr2 | 240434994 | 240435306 | 313 | 3 | 0.000747475 | 0.000369 | 0.00182 | 0.00027 | 0.000440748 | NA |
| chr18 | 76379735 | 76379831 | 97 | 2 | 0.000748013 | 7.16E-05 | 0.00186 | 0.0001 | -0.006477699 | ZNF516 |
| chr22 | 19881196 | 19881388 | 193 | 4 | 0.000748437 | 0.000438 | 0.00022 | 9.58E-05 | -0.001690779 | TXNRD2 |
| chr9 | 136410691 | 136410774 | 84 | 6 | 0.00074936 | 0.013629 | 0.00248 | 0.00017 | 0.000310911 | PMPCA |
| chr12 | 109814872 | 109815225 | 354 | 6 | 0.000749362 | 0.00036 | 0.00111 | 5.54E-05 | -0.006982389 | TRPV4 |
| chr11 | 133065067 | 133065225 | 159 | 2 | 0.000749523 | 0.043601 | 0.04482 | 0.04602 | -0.006784194 | AP004782.1, OPCML |
| chr10 | 49362827 | 49363124 | 298 | 2 | 0.000749822 | 6.86E-05 | 0.00059 | 7.68E-05 | -0.001056598 | NA |
| chr8 | 22693207 | 22693318 | 112 | 3 | 0.000750613 | 0.483072 | 0.08204 | 0.24593 | 0.001447204 | AC055854.1, EGR3 |
| chr11 | 915377 | 915440 | 64 | 2 | 0.000750992 | 0.005825 | 0.00275 | 0.00325 | -0.002436284 | NA |
| chr10 | 774827 | 774895 | 69 | 2 | 0.000751434 | 0.374638 | 0.00561 | 0.01848 | -0.002777454 | NA |
| chr6 | 31670469 | 31670567 | 99 | 2 | 0.000752098 | 0.69685 | 0.64159 | 0.7771 | -0.000265775 | AL662899.2, LY6G5B |
| chr18 | 79625172 | 79625290 | 119 | 2 | 0.000754084 | 0.000107 | 0.00033 | 8.66E-05 | 0.004755713 | NA |
| chr11 | 108538180 | 108538254 | 75 | 4 | 0.000754311 | 3.20E-05 | 0.01164 | 8.85E-05 | -0.013460917 | EXPH5 |
| chr3 | 132721830 | 132722015 | 186 | 3 | 0.000754311 | 0.048609 | 0.00084 | 0.00446 | 0.000751723 | NPHP3-AS1, NPHP3-ACAD11, NPHP3 |
| chr10 | 47321962 | 47322040 | 79 | 3 | 0.000754416 | 0.011063 | 0.003 | 0.00106 | -0.003209423 | NA |
| chr19 | 1166721 | 1166872 | 152 | 2 | 0.00075597 | 6.97E-05 | 0.00214 | 0.00011 | -0.005755379 | SBNO2 |
| chr9 | 130845889 | 130845979 | 91 | 2 | 0.000756068 | 0.010954 | 0.00018 | 0.00043 | -0.008864292 | ABL1 |
| chr22 | 46421464 | 46421538 | 75 | 2 | 0.000756446 | 0.000567 | 9.25E-05 | 8.52E-05 | -0.007036577 | CELSR1 |
| chr11 | 99555301 | 99555332 | 32 | 3 | 0.000756612 | 0.001297 | 0.00027 | 0.0003 | 0.002139665 | CNTN5 |
| chr10 | 70599157 | 70599185 | 29 | 2 | 0.000756943 | 3.58E-05 | 0.00114 | 5.20E-05 | -0.008056252 | PRF1 |
| chr3 | 126069418 | 126069478 | 61 | 2 | 0.000757198 | 0.00017 | 0.00021 | 9.23E-05 | -0.003646119 | SLC41A3 |
| chr6 | 35663219 | 35663232 | 14 | 2 | 0.000757769 | 0.000137 | 0.00025 | 8.85E-05 | -0.009891535 | FKBP5 |
| chr21 | 44733166 | 44733223 | 58 | 2 | 0.00075889 | 0.000148 | 0.00022 | 8.81E-05 | -0.010652132 | NA |
| chr10 | 133077188 | 133077295 | 108 | 2 | 0.000759296 | 0.034275 | 0.00144 | 0.00394 | -0.006703455 | ADGRA1 |
| chr6 | 27177790 | 27177907 | 118 | 2 | 0.000759696 | 0.003074 | 0.01537 | 0.00476 | 0.002490064 | NA |
| chr22 | 46658170 | 46658376 | 207 | 3 | 0.000759937 | 4.02E-05 | 0.00324 | 6.23E-05 | -0.002376426 | GRAMD4 |
| chr16 | 12661979 | 12662039 | 61 | 2 | 0.000760678 | 5.57E-05 | 0.00238 | 8.65E-05 | -0.022214448 | CPPED1 |
| chr8 | 56993607 | 56993804 | 198 | 2 | 0.000760709 | 0.004103 | 0.00042 | 0.0007 | 0.001467477 | BPNT2 |
| chr1 | 11766703 | 11766721 | 19 | 2 | 0.000762067 | 0.005374 | 0.00015 | 0.0003 | -0.003407164 | C1orf167 |
| chr19 | 56836084 | 56836130 | 47 | 2 | 0.000762067 | 5.93E-05 | 0.0026 | 9.30E-05 | -0.000772384 | ZIM2, PEG3 |
| chr13 | 110715346 | 110715568 | 223 | 6 | 0.000762364 | 0.008673 | 6.47E-05 | 0.0001 | 0.000576604 | ING1 |
| chr15 | 99251291 | 99251725 | 435 | 6 | 0.000763208 | 0.000367 | 0.00468 | 0.0001 | -5.21E-06 | LRRC28 |
| chr4 | 103198646 | 103198894 | 249 | 5 | 0.000763771 | 0.01287 | 0.00579 | 0.00264 | 0.002494506 | NA |
| chr11 | 17474717 | 17474926 | 210 | 2 | 0.000764435 | 8.53E-05 | 0.00018 | 5.77E-05 | -0.006217617 | ABCC8 |
| chr12 | 57546040 | 57546356 | 317 | 2 | 0.000764883 | 0.00012 | 0.00229 | 0.00018 | -0.007256939 | KIF5A, DCTN2 |
| chr1 | 235182250 | 235182454 | 205 | 2 | 0.000764899 | 9.96E-05 | 0.00243 | 0.00015 | -0.010183808 | ARID4B |
| chr4 | 8473070 | 8473134 | 65 | 2 | 0.000764968 | 0.000386 | 0.00506 | 0.00063 | -0.006928734 | TRMT44 |
| chr5 | 850986 | 851343 | 358 | 7 | 0.000765463 | 3.60E-05 | 0.01929 | 0.00011 | -0.001220972 | ZDHHC11, BRD9 |
| chr11 | 3232351 | 3232666 | 316 | 8 | 0.000765665 | 5.33E-05 | 0.00565 | 0.00018 | -0.008642366 | AC109309.1, MRGPRE |
| chr7 | 105760536 | 105760550 | 15 | 2 | 0.000765701 | 0.000108 | 0.00032 | 8.65E-05 | -0.005410377 | ATXN7L1 |
| chr3 | 148863913 | 148864014 | 102 | 2 | 0.000766533 | 4.25E-05 | 0.00139 | 6.29E-05 | -0.012266254 | AC092979.1 |
| chr13 | 113460189 | 113460268 | 80 | 2 | 0.00076725 | 0.000107 | 0.00385 | 0.00018 | -0.010875551 | DCUN1D2 |
| chr20 | 23051521 | 23051529 | 9 | 2 | 0.000767749 | 0.000187 | 0.00435 | 0.00032 | -0.012506578 | NA |
| chr10 | 125149604 | 125149652 | 49 | 2 | 0.000768119 | 3.96E-05 | 0.00175 | 6.11E-05 | -0.015074128 | CTBP2 |
| chr11 | 71478339 | 71478923 | 585 | 5 | 0.000768611 | 0.000338 | 0.00249 | 6.41E-05 | -0.007277344 | NADSYN1 |
| chr6 | 31664543 | 31664764 | 222 | 4 | 0.000769189 | 0.000936 | 0.00019 | 0.00012 | 0.001916046 | GPANK1 |
| chr17 | 7548470 | 7548574 | 105 | 3 | 0.000769926 | 0.000766 | 0.00098 | 0.00046 | -0.000144558 | NA |
| chr1 | 223180874 | 223180968 | 95 | 3 | 0.000770591 | 0.000232 | 0.00283 | 0.00025 | 0.001990357 | NA |
| chr19 | 52397629 | 52397735 | 107 | 8 | 0.000771197 | 0.000254 | 0.00655 | 7.96E-05 | 0.004549046 | ZNF528-AS1 |
| chr7 | 157875243 | 157875359 | 117 | 2 | 0.000771197 | 0.002143 | 0.00751 | 0.00283 | -0.01659197 | PTPRN2 |
| chr1 | 38153305 | 38153538 | 234 | 2 | 0.000771221 | 0.000131 | 0.0043 | 0.00023 | 0.019306547 | AL390839.1 |
| chr22 | 50461265 | 50461580 | 316 | 2 | 0.000771241 | 3.98E-05 | 0.00182 | 6.17E-05 | -0.002895896 | SBF1 |
| chr7 | 129785310 | 129785490 | 181 | 4 | 0.000771661 | 8.26E-05 | 0.00384 | 6.45E-05 | 0.004607218 | NA |
| chr9 | 136943207 | 136943305 | 99 | 2 | 0.000772166 | 0.001329 | 0.01572 | 0.0024 | 0.000502946 | FBXW5 |
| chr12 | 122984737 | 122985122 | 386 | 3 | 0.000772272 | 4.55E-05 | 0.00114 | 6.27E-05 | -0.009863815 | PITPNM2 |
| chr1 | 19815169 | 19815288 | 120 | 3 | 0.000772548 | 0.001026 | 0.0003 | 0.0003 | -0.000952165 | AL391883.1, RNF186 |
| chr8 | 144374640 | 144374830 | 191 | 3 | 0.000772654 | 0.003647 | 0.0005 | 0.00041 | -0.00106772 | ADCK5 |
| chr2 | 46366084 | 46366132 | 49 | 3 | 0.000772809 | 4.83E-05 | 0.00358 | 8.58E-05 | -0.003938274 | EPAS1 |
| chr3 | 98036757 | 98036783 | 27 | 2 | 0.000772863 | 0.000239 | 0.00025 | 0.00012 | 0.010197246 | NA |
| chr12 | 95740040 | 95740042 | 3 | 2 | 0.000773378 | 7.49E-05 | 0.0008 | 8.93E-05 | -0.009576258 | NTN4 |
| chr9 | 136839758 | 136840558 | 801 | 2 | 0.000774846 | 0.000191 | 0.00086 | 0.0002 | -0.000628048 | RABL6 |
| chr3 | 194686200 | 194686316 | 117 | 2 | 0.000775784 | 0.252259 | 0.00877 | 0.02664 | 0.001165484 | FAM43A |
| chr10 | 1609007 | 1609098 | 92 | 2 | 0.000776445 | 0.001948 | 0.00013 | 0.00019 | -0.005686112 | ADARB2 |
| chr10 | 127196870 | 127196928 | 59 | 2 | 0.0007768 | 4.50E-05 | 0.00044 | 5.08E-05 | 0.008964521 | DOCK1 |
| chr1 | 167630482 | 167630593 | 112 | 2 | 0.000776933 | 0.002981 | 0.00328 | 0.00244 | 0.004233462 | RCSD1, AL031733.2 |
| chr5 | 176361869 | 176362235 | 367 | 10 | 0.000776951 | 0.002478 | 0.00524 | 0.00056 | -0.00030511 | NA |
| chr5 | 139517458 | 139517882 | 425 | 3 | 0.000776974 | 8.19E-05 | 0.00909 | 0.0002 | -0.007699301 | NA |
| chr11 | 130262434 | 130262449 | 16 | 2 | 0.000778302 | 0.000129 | 0.00042 | 0.00011 | 0.000670176 | ZBTB44 |
| chr19 | 1466498 | 1466648 | 151 | 2 | 0.000779402 | 0.007566 | 0.00551 | 0.00563 | 0.018153631 | APC2, C19orf25 |
| chr4 | 10685495 | 10685851 | 357 | 3 | 0.000779548 | 0.001019 | 0.00073 | 0.00021 | 0.002200512 | AC084048.1 |
| chr17 | 16378200 | 16378600 | 401 | 2 | 0.000779711 | 0.000316 | 0.00212 | 0.00041 | -0.001132368 | NA |
| chr4 | 1713045 | 1713277 | 233 | 4 | 0.000779755 | 0.002424 | 0.00038 | 0.00057 | 0.001856159 | AC016773.1, TACC3 |
| chr13 | 100674870 | 100675063 | 194 | 7 | 0.000781025 | 0.000294 | 0.00113 | 0.00014 | 0.000498663 | TMTC4 |
| chr12 | 49059855 | 49060207 | 353 | 3 | 0.000782503 | 0.008886 | 9.27E-05 | 0.00025 | 0.001579292 | KMT2D |
| chr6 | 31631836 | 31632178 | 343 | 7 | 0.000782557 | 0.008478 | 0.00164 | 0.00083 | -0.000880148 | PRRC2A |
| chr5 | 177327986 | 177328198 | 213 | 2 | 0.000782817 | 0.002826 | 0.02423 | 0.00505 | -0.017114338 | NA |
| chr7 | 156843426 | 156843583 | 158 | 2 | 0.000783222 | 4.11E-05 | 0.00166 | 6.26E-05 | 0.001282631 | LMBR1 |
| chr4 | 55592203 | 55592455 | 253 | 5 | 0.000785142 | 5.65E-05 | 0.0021 | 2.72E-05 | -0.004266866 | PDCL2 |
| chr6 | 83065768 | 83065835 | 68 | 2 | 0.000785241 | 0.062759 | 0.08546 | 0.07785 | 0.00192051 | UBE3D |
| chr20 | 9069047 | 9069172 | 126 | 2 | 0.00078526 | 0.003834 | 0.02234 | 0.00635 | 0.002604111 | PLCB4 |
| chr10 | 110501437 | 110501859 | 423 | 2 | 0.000786446 | 0.001261 | 0.00051 | 0.00048 | -0.000960149 | DUSP5 |
| chr8 | 108084071 | 108084178 | 108 | 3 | 0.000786833 | 0.001108 | 0.00885 | 0.001 | -0.013698884 | NA |
| chr1 | 57860362 | 57860538 | 177 | 2 | 0.000788192 | 9.88E-05 | 0.00083 | 0.00011 | -0.009038705 | DAB1-AS1, DAB1 |
| chr11 | 70824775 | 70824960 | 186 | 3 | 0.000788464 | 3.10E-05 | 0.00503 | 6.41E-05 | -0.023619461 | SHANK2 |
| chr9 | 130387555 | 130387671 | 117 | 2 | 0.000788852 | 0.000665 | 0.00011 | 0.00011 | -0.008884992 | HMCN2 |
| chr17 | 43831743 | 43832348 | 606 | 5 | 0.000790065 | 0.001493 | 0.00034 | 0.0002 | -0.004125501 | MPP3 |
| chr4 | 83109701 | 83109977 | 277 | 4 | 0.000790377 | 0.004319 | 9.94E-05 | 0.00027 | 0.001287044 | PLAC8 |
| chr4 | 979366 | 979389 | 24 | 2 | 0.000791139 | 0.001076 | 9.69E-05 | 0.00011 | -0.00666962 | DGKQ, SLC26A1 |
| chr2 | 5932007 | 5932021 | 15 | 2 | 0.000791911 | 0.000678 | 0.00032 | 0.00026 | -0.006622333 | NA |
| chr17 | 17586322 | 17586340 | 19 | 2 | 0.000792352 | 0.003014 | 8.37E-05 | 0.00013 | -0.010532077 | PEMT |
| chr9 | 121822182 | 121822195 | 14 | 2 | 0.000792506 | 5.06E-05 | 0.00202 | 7.77E-05 | -0.011186336 | TTLL11 |
| chr3 | 48919307 | 48919616 | 310 | 5 | 0.000792733 | 9.77E-05 | 0.00344 | 7.32E-05 | 0.00233622 | ARIH2 |
| chr10 | 102917150 | 102917163 | 14 | 2 | 0.000792733 | 0.005285 | 0.03192 | 0.00908 | 0.015954748 | AL356608.3 |
| chr19 | 3697924 | 3698225 | 302 | 3 | 0.000792814 | 0.001947 | 0.01331 | 0.00213 | -0.006297088 | PIP5K1C |
| chr12 | 7155757 | 7156257 | 501 | 2 | 0.000794631 | 0.000129 | 0.00415 | 0.00022 | -0.006237685 | CLSTN3 |
| chr2 | 240875640 | 240875777 | 138 | 2 | 0.000794739 | 0.024043 | 0.00014 | 0.00037 | -0.002540863 | AGXT |
| chr2 | 81466998 | 81467322 | 325 | 6 | 0.000794745 | 0.061516 | 0.00022 | 0.00015 | -0.001165398 | LINC01815 |
| chr13 | 111144737 | 111144785 | 49 | 3 | 0.000797322 | 0.000855 | 0.00022 | 0.00014 | -0.004427735 | ARHGEF7 |
| chr22 | 46289831 | 46289879 | 49 | 3 | 0.000797859 | 0.00012 | 7.58E-05 | 3.58E-05 | 0.007016796 | TTC38 |
| chr1 | 212558579 | 212558742 | 164 | 2 | 0.000797869 | 0.01221 | 0.00049 | 0.00115 | 0.002215239 | AC092803.1 |
| chr7 | 25179924 | 25179991 | 68 | 2 | 0.000798402 | 0.309901 | 0.19221 | 0.2762 | 0.000732407 | C7orf31 |
| chr19 | 57614018 | 57614026 | 9 | 2 | 0.000798842 | 4.24E-06 | 0.00047 | 7.17E-06 | -0.009020873 | NA |
| chr19 | 49961456 | 49961524 | 69 | 2 | 0.00079905 | 0.0058 | 0.00016 | 0.00032 | -0.0157064 | NA |
| chr2 | 172429083 | 172429365 | 283 | 2 | 0.000799956 | 0.002102 | 0.0055 | 0.00247 | -0.009059877 | ITGA6 |
| chr17 | 55732203 | 55732960 | 758 | 2 | 0.000800239 | 6.03E-05 | 0.00028 | 5.48E-05 | -0.013687246 | NA |
| chr3 | 71131264 | 71131302 | 39 | 2 | 0.000800302 | 0.002036 | 0.00031 | 0.00042 | -0.005525994 | FOXP1, AC097634.4 |
| chr10 | 133179642 | 133179888 | 247 | 3 | 0.000800462 | 6.70E-05 | 0.01092 | 0.00017 | -0.007076542 | KNDC1 |
| chr5 | 126718446 | 126718459 | 14 | 2 | 0.000800482 | 4.56E-05 | 0.00109 | 6.40E-05 | -0.003867245 | NA |
| chr7 | 128343601 | 128343707 | 107 | 2 | 0.000801523 | 0.16247 | 0.00691 | 0.02067 | 0.00040072 | RBM28 |
| chr2 | 234981323 | 234981436 | 114 | 2 | 0.000801625 | 8.15E-05 | 0.00322 | 0.00013 | -0.014661657 | SH3BP4 |
| chr17 | 76354861 | 76354961 | 101 | 3 | 0.000801762 | 0.000224 | 0.01673 | 0.00061 | -0.011314455 | PRPSAP1 |
| chr5 | 1794900 | 1795025 | 126 | 2 | 0.000801968 | 0.000227 | 0.00021 | 0.00011 | 0.008894446 | NA |
| chr2 | 232378364 | 232378688 | 325 | 3 | 0.000801999 | 0.003719 | 0.00018 | 0.00029 | -0.004242677 | NA |
| chr8 | 140210078 | 140210221 | 144 | 2 | 0.000802068 | 6.16E-05 | 0.0006 | 7.11E-05 | -0.007051995 | TRAPPC9 |
| chr9 | 37118451 | 37119068 | 618 | 2 | 0.000802094 | 3.76E-05 | 0.00039 | 4.27E-05 | 0.005435661 | AL512604.2 |
| chr11 | 560396 | 560868 | 473 | 5 | 0.000802497 | 0.08211 | 0.21302 | 0.17974 | 0.000452014 | RASSF7, LMNTD2 |
| chr2 | 3184026 | 3184166 | 141 | 3 | 0.000803405 | 2.21E-05 | 0.00649 | 5.28E-05 | -0.002248401 | NA |
| chr10 | 75099625 | 75099683 | 59 | 2 | 0.000803615 | 0.530789 | 0.27028 | 0.41231 | -0.001588994 | SAMD8, DUSP13 |
| chr3 | 58497699 | 58497741 | 43 | 2 | 0.000803663 | 0.000667 | 0.00022 | 0.0002 | -0.005854879 | KCTD6 |
| chr10 | 132522495 | 132522529 | 35 | 2 | 0.00080456 | 8.64E-05 | 7.82E-05 | 3.44E-05 | -0.009306886 | NA |
| chr2 | 205682420 | 205682513 | 94 | 5 | 0.000804661 | 0.000491 | 0.00157 | 0.00034 | 0.000646772 | NRP2 |
| chr11 | 3095635 | 3095653 | 19 | 2 | 0.000805058 | 0.015557 | 0.00021 | 0.00053 | -0.004417915 | OSBPL5 |
| chr4 | 1981375 | 1981557 | 183 | 3 | 0.000805567 | 1.51E-05 | 0.00364 | 3.28E-05 | -0.007313823 | NSD2 |
| chr13 | 19344385 | 19344446 | 62 | 2 | 0.000805615 | 0.000189 | 0.00104 | 0.00021 | -0.00123366 | ANKRD26P3 |
| chr20 | 31513982 | 31514020 | 39 | 2 | 0.000806719 | 0.012218 | 0.00091 | 0.00198 | 0.002182413 | NA |
| chr17 | 79938051 | 79938222 | 172 | 2 | 0.000806793 | 4.83E-05 | 0.0019 | 7.38E-05 | -0.005740066 | TBC1D16 |
| chr16 | 87701210 | 87701271 | 62 | 2 | 0.000808263 | 0.000397 | 0.00039 | 0.00021 | -0.008583103 | KLHDC4 |
| chr22 | 19906219 | 19906245 | 27 | 2 | 0.000808812 | 0.000145 | 0.00082 | 0.00016 | -0.004890868 | TXNRD2 |
| chr22 | 38313548 | 38313631 | 84 | 2 | 0.000808858 | 5.10E-05 | 0.00137 | 7.37E-05 | -0.010550842 | CSNK1E, TPTEP2-CSNK1E |
| chr8 | 96644844 | 96644869 | 26 | 2 | 0.000809732 | 2.09E-05 | 0.00104 | 3.24E-05 | -0.017523367 | NA |
| chr1 | 3706538 | 3706825 | 288 | 3 | 0.000810485 | 0.006596 | 0.00026 | 7.34E-05 | -0.009334403 | TP73 |
| chr16 | 23420545 | 23420551 | 7 | 2 | 0.000811588 | 5.86E-05 | 0.00013 | 3.90E-05 | -0.002046101 | COG7 |
| chr11 | 66871837 | 66872107 | 271 | 7 | 0.000812407 | 8.18E-06 | 0.02037 | 3.95E-05 | -0.00176663 | PC |
| chr16 | 2171490 | 2171576 | 87 | 2 | 0.000812748 | 0.009795 | 0.01507 | 0.0109 | -0.002578009 | TRAF7 |
| chr10 | 102715530 | 102715583 | 54 | 3 | 0.000813238 | 6.83E-05 | 0.01131 | 0.00017 | -0.014444421 | SFXN2 |
| chr19 | 58499537 | 58499643 | 107 | 2 | 0.000813697 | 0.000409 | 0.00077 | 0.00032 | 0.000655181 | SLC27A5 |
| chr7 | 105013071 | 105013336 | 266 | 4 | 0.000813981 | 0.041254 | 0.00034 | 0.00273 | 0.006315429 | NA |
| chr17 | 18950160 | 18950206 | 47 | 2 | 0.000814511 | 0.000362 | 0.00148 | 0.0004 | -0.00091159 | NA |
| chr21 | 42159791 | 42159852 | 62 | 2 | 0.000814627 | 0.000104 | 0.00134 | 0.00014 | 0.000739472 | NA |
| chr6 | 122009976 | 122009992 | 17 | 2 | 0.00081549 | 0.000534 | 0.00018 | 0.00015 | -0.004828129 | NA |
| chr5 | 141338394 | 141338772 | 379 | 6 | 0.000816422 | 0.000412 | 0.00386 | 0.00015 | 0.005409173 | PCDHGA1, PCDHGA2 |
| chr12 | 132415122 | 132415180 | 59 | 2 | 0.000816832 | 0.002361 | 0.00036 | 0.00049 | -0.002248544 | NA |
| chr12 | 110345884 | 110346182 | 299 | 4 | 0.000817476 | 0.000799 | 0.00381 | 0.00039 | -0.004847626 | ATP2A2 |

| | | | | | | | | | | |
|---|---|---|---|---|---|---|---|---|---|---|
| chr21 | 45474873 | 45475269 | 397 | 2 | 0.000817848 | 0.000189 | 0.00078 | 0.00019 | -0.003491053 | COL18A1 |
| chr15 | 35546313 | 35546532 | 220 | 6 | 0.000819029 | 0.010114 | 0.02495 | 0.00818 | -0.000675818 | DPH6-DT |
| chr12 | 2652329 | 2652383 | 55 | 2 | 0.000819389 | 0.000158 | 7.80E-05 | 4.58E-05 | -0.002621384 | CACNA1C |
| chr11 | 72793812 | 72794081 | 270 | 6 | 0.000819699 | 8.45E-05 | 0.0169 | 0.00021 | 0.000425602 | AP002381.2 |
| chr6 | 31117257 | 31117458 | 202 | 2 | 0.000820321 | 4.26E-05 | 0.00181 | 6.57E-05 | -0.015607014 | PSORS1C1, CDSN |
| chr12 | 51683755 | 51683876 | 122 | 2 | 0.000820583 | 0.000124 | 0.0002 | 7.66E-05 | -0.005901861 | SCN8A |
| chr17 | 42679331 | 42679347 | 17 | 2 | 0.000820641 | 0.001528 | 0.00034 | 0.0004 | 0.012354243 | CCR10 |
| chr19 | 17114530 | 17114575 | 46 | 2 | 0.00082158 | 8.26E-05 | 0.003 | 0.00013 | -0.005538506 | MYO9B |
| chr16 | 4347872 | 4347894 | 23 | 2 | 0.000822147 | 4.64E-05 | 0.00127 | 6.67E-05 | -0.007418804 | PAM16, CORO7-PAM16, AC012676.5 |
| chr5 | 6745654 | 6745859 | 206 | 4 | 0.000825262 | 8.55E-05 | 0.00092 | 4.38E-05 | 0.016038882 | TENT4A |
| chr12 | 116987974 | 116988063 | 90 | 2 | 0.000825356 | 0.015367 | 0.00039 | 0.00098 | 0.002157824 | FBXW8 |
| chr17 | 73642392 | 73642890 | 499 | 2 | 0.000825462 | 2.45E-05 | 0.00145 | 3.87E-05 | -0.01896492 | AC032019.1, SDK2 |
| chr8 | 22024487 | 22024928 | 442 | 8 | 0.000826186 | 0.001025 | 0.00309 | 0.00075 | -0.001139914 | NPM2 |
| chr11 | 133983043 | 133983098 | 56 | 2 | 0.000826186 | 0.000347 | 0.00028 | 0.00016 | -0.003342838 | NA |
| chr8 | 22365718 | 22366004 | 287 | 3 | 0.000826387 | 0.000265 | 0.00062 | 0.00013 | -0.002687448 | NA |
| chr22 | 31954574 | 31954609 | 36 | 2 | 0.000826681 | 6.04E-05 | 0.00112 | 8.15E-05 | -0.000624302 | Z82190.2, YWHAH |
| chr10 | 2973469 | 2973546 | 78 | 2 | 0.000827685 | 7.00E-05 | 0.00046 | 7.24E-05 | -0.014759025 | NA |
| chr17 | 1742511 | 1742902 | 392 | 4 | 0.000827932 | 6.13E-05 | 0.00153 | 6.55E-05 | -0.002209286 | SERPINF2 |
| chr11 | 65976665 | 65976688 | 24 | 3 | 0.000828124 | 0.026061 | 0.00126 | 0.00013 | -0.000747483 | SART1 |
| chr17 | 59619348 | 59619632 | 285 | 8 | 0.000828847 | 0.00014 | 0.01453 | 0.00038 | 0.000752971 | AC091271.1 |
| chr9 | 121859010 | 121859059 | 50 | 2 | 0.000829708 | 6.43E-05 | 0.00053 | 7.07E-05 | -0.009223614 | TTLL11 |
| chr3 | 129073180 | 129073260 | 81 | 2 | 0.000830439 | 0.078899 | 0.01778 | 0.03547 | 0.001370632 | NA |
| chr11 | 85664807 | 85665189 | 383 | 7 | 0.000831071 | 0.001692 | 0.00014 | 0.00018 | 0.000127637 | CREBZF |
| chr13 | 23569196 | 23569551 | 356 | 3 | 0.000831388 | 0.000216 | 0.0004 | 0.00013 | -0.006239855 | NA |
| chr8 | 142615103 | 142615170 | 68 | 3 | 0.000831462 | 0.001264 | 0.03403 | 0.00331 | -0.007184878 | NA |
| chr3 | 49094625 | 49094646 | 22 | 2 | 0.000832809 | 1.43E-05 | 0.00043 | 2.01E-05 | 0.000412028 | NA |
| chr19 | 3382106 | 3382129 | 24 | 2 | 0.000832809 | 0.001978 | 0.00153 | 0.00124 | -0.002025948 | NFIC |
| chr2 | 25328706 | 25328791 | 86 | 3 | 0.000834184 | 0.001085 | 0.00016 | 0.00019 | -0.001060627 | DNMT3A, MIR1301 |
| chr10 | 27242302 | 27242433 | 132 | 4 | 0.000836587 | 0.000154 | 0.00217 | 8.53E-05 | 9.31E-05 | ACBD5 |
| chr13 | 88345411 | 88345666 | 256 | 2 | 0.000836632 | 4.47E-05 | 0.00174 | 6.79E-05 | 0.004957099 | NA |
| chr19 | 52171601 | 52171978 | 378 | 6 | 0.000837574 | 2.47E-05 | 0.00655 | 4.82E-05 | -0.000829234 | AC010320.1, ZNF836 |
| chr15 | 76059677 | 76059706 | 30 | 2 | 0.00083777 | 0.000159 | 0.00438 | 0.00028 | 0.003583489 | NRG4 |
| chr7 | 158431799 | 158432391 | 593 | 2 | 0.000838135 | 1.84E-05 | 0.0007 | 2.74E-05 | -0.00848557 | PTPRN2 |
| chr6 | 151689968 | 151690065 | 98 | 3 | 0.000838323 | 0.149603 | 0.06928 | 0.06478 | -0.007693204 | ESR1 |
| chr7 | 106108812 | 106108882 | 71 | 2 | 0.000838323 | 0.000179 | 0.00018 | 8.64E-05 | -0.011119799 | SYPL1 |
| chr14 | 101774575 | 101774650 | 76 | 2 | 0.000838689 | 5.90E-05 | 0.00087 | 7.57E-05 | -0.006085459 | PPP2R5C |
| chr22 | 45891584 | 45891726 | 143 | 2 | 0.000839743 | 0.03632 | 0.00026 | 0.00078 | -0.010060896 | NA |
| chr1 | 202166643 | 202166769 | 127 | 2 | 0.000840029 | 4.65E-05 | 0.00212 | 7.24E-05 | -0.010045925 | NA |
| chr4 | 70151521 | 70151539 | 19 | 2 | 0.000840846 | 0.000854 | 0.0001 | 0.00011 | -0.010832546 | PRR27 |
| chr10 | 72009369 | 72009416 | 48 | 2 | 0.000840846 | 0.000491 | 0.00031 | 0.00021 | -0.009978336 | CHST3 |
| chr3 | 21274671 | 21274676 | 6 | 2 | 0.000841117 | 0.000542 | 0.00011 | 0.0001 | 0.001861186 | NA |
| chr20 | 35467425 | 35467586 | 162 | 2 | 0.000841507 | 5.08E-05 | 0.00221 | 7.90E-05 | 0.001024727 | CEP250 |
| chr17 | 36807736 | 36807768 | 33 | 2 | 0.000841689 | 0.000101 | 0.0027 | 0.00016 | -0.014581082 | NA |
| chr5 | 157778244 | 157778251 | 8 | 2 | 0.000842717 | 5.59E-05 | 0.00072 | 6.91E-05 | -0.009846273 | NA |
| chr1 | 35180714 | 35180768 | 55 | 2 | 0.000842893 | 0.000238 | 0.00448 | 0.0004 | -0.014662773 | SFPQ |
| chr9 | 122913281 | 122913397 | 117 | 5 | 0.00084348 | 0.022717 | 0.00062 | 0.0002 | -0.000310268 | ZBTB6 |
| chr19 | 10179553 | 10179717 | 165 | 3 | 0.00084348 | 0.000846 | 0.00026 | 0.00011 | -0.000943763 | DNMT1 |
| chr3 | 185552310 | 185552572 | 263 | 3 | 0.000843764 | 0.060755 | 0.00406 | 0.00104 | 0.004933581 | LIPH |
| chr6 | 166306945 | 166307261 | 317 | 2 | 0.000845554 | 0.022776 | 0.06305 | 0.03463 | -0.008923273 | PRR18 |
| chr14 | 49820397 | 49820446 | 50 | 2 | 0.000845697 | 5.48E-05 | 0.00078 | 6.94E-05 | -0.004230717 | NEMF |
| chr11 | 63766136 | 63766622 | 487 | 3 | 0.00084652 | 8.28E-05 | 0.00171 | 8.23E-05 | -0.00562806 | C11orf95 |
| chr6 | 41072653 | 41072920 | 268 | 9 | 0.000847528 | 0.022506 | 0.00045 | 0.00038 | 0.001248396 | OARD1 |
| chr17 | 22552862 | 22552864 | 3 | 2 | 0.000847672 | 9.06E-05 | 0.00032 | 7.59E-05 | -0.022832505 | NA |
| chr17 | 40106837 | 40106845 | 9 | 2 | 0.000847672 | 0.000239 | 0.00243 | 0.00034 | -0.007377228 | NA |
| chr1 | 1310317 | 1310532 | 216 | 3 | 0.000847717 | 0.000179 | 0.00383 | 0.00021 | -0.004180654 | PUSL1 |
| chr6 | 1636727 | 1636732 | 6 | 2 | 0.000847972 | 8.52E-05 | 0.00141 | 0.00012 | 0.000919767 | GMDS |
| chr17 | 41963106 | 41963135 | 30 | 2 | 0.00084823 | 0.004947 | 0.0017 | 0.00219 | -0.002900541 | TTC25 |
| chr1 | 27033265 | 27033401 | 137 | 2 | 0.000848804 | 4.42E-05 | 0.00197 | 6.85E-05 | -0.007533778 | NA |
| chr10 | 99521130 | 99521446 | 317 | 2 | 0.000849404 | 0.110484 | 0.11011 | 0.12283 | -0.007845968 | NA |
| chr2 | 127707013 | 127707365 | 353 | 2 | 0.000853099 | 4.19E-05 | 0.00206 | 6.58E-05 | -0.010693871 | WDR33 |
| chr5 | 75512378 | 75512468 | 91 | 2 | 0.000853312 | 0.993006 | 0.89656 | 0.98105 | -8.12E-05 | POLK |
| chr7 | 128849398 | 128849502 | 105 | 3 | 0.000853542 | 0.000357 | 0.00154 | 0.00037 | 0.000882688 | FLNC |
| chr8 | 41817984 | 41818050 | 67 | 2 | 0.000854009 | 5.12E-05 | 0.00107 | 7.04E-05 | 0.00372216 | ANK1 |
| chr7 | 82950764 | 82950935 | 172 | 3 | 0.000854166 | 9.57E-05 | 0.00073 | 9.03E-05 | -0.009796531 | PCLO |
| chr1 | 10032241 | 10032571 | 331 | 2 | 0.000855719 | 0.002749 | 0.00932 | 0.00368 | 0.005253163 | NA |
| chr11 | 10294207 | 10294385 | 179 | 6 | 0.000855818 | 0.000111 | 0.00189 | 4.78E-05 | -0.00028501 | SBF2 |
| chr1 | 42851428 | 42851520 | 93 | 3 | 0.000855902 | 0.056958 | 0.00167 | 0.00056 | -0.002732357 | ZNF691 |
| chr17 | 2232578 | 2232663 | 86 | 2 | 0.000856406 | 0.000212 | 3.04E-05 | 2.28E-05 | -0.006666598 | SMG6 |
| chr3 | 197364312 | 197364324 | 13 | 2 | 0.000857298 | 4.04E-05 | 0.00023 | 3.86E-05 | -0.007123488 | NA |
| chr12 | 116698235 | 116698253 | 19 | 2 | 0.000857378 | 0.012376 | 0.00014 | 0.00033 | -0.004044931 | NA |
| chr15 | 73369301 | 73369428 | 128 | 4 | 0.000857604 | 0.000313 | 6.31E-05 | 5.25E-05 | -0.001784198 | NA |
| chr11 | 6728444 | 6728453 | 10 | 2 | 0.000860531 | 0.000124 | 0.00042 | 0.00011 | -0.024908027 | GVINP1 |
| chr8 | 66432066 | 66432485 | 420 | 8 | 0.000860689 | 0.068213 | 0.0007 | 0.00383 | -0.00324142 | AC009879.2, AC009879.4 |
| chr3 | 43179206 | 43179287 | 82 | 2 | 0.000861737 | 5.44E-05 | 0.00245 | 8.53E-05 | 0.001949572 | NA |
| chr17 | 79981368 | 79981757 | 390 | 6 | 0.000862548 | 0.019977 | 0.0013 | 0.00032 | -0.002705219 | TBC1D16 |
| chr1 | 33321248 | 33321260 | 13 | 2 | 0.000862824 | 0.015775 | 0.06365 | 0.02634 | -0.004871347 | AL513327.1 |
| chr4 | 74157732 | 74158029 | 298 | 7 | 0.000862829 | 0.000577 | 0.00237 | 0.00075 | 0.00163137 | MTHFD2L, AC093677.2 |
| chr16 | 23453254 | 23453490 | 237 | 8 | 0.00086354 | 0.000579 | 0.00018 | 5.94E-05 | -0.002659249 | AC008915.2 |
| chr19 | 19618907 | 19618951 | 45 | 3 | 0.000863603 | 0.015226 | 0.01835 | 0.01544 | 0.001086649 | NA |
| chr22 | 41347604 | 41347628 | 25 | 2 | 0.000863752 | 6.94E-05 | 0.00154 | 9.81E-05 | -0.004014833 | ZC3H7B |
| chr16 | 2520165 | 2520186 | 22 | 2 | 0.000863839 | 2.76E-05 | 0.00098 | 4.07E-05 | 0.003163935 | AC093525.2, ATP6V0C, AC093525.1 |
| chr16 | 27748130 | 27748196 | 67 | 2 | 0.000864547 | 0.00014 | 0.00038 | 0.00011 | 0.003776099 | KATNIP |
| chr17 | 80010408 | 80010682 | 275 | 3 | 0.00086578 | 3.74E-05 | 0.00259 | 6.22E-05 | -0.003489534 | TBC1D16 |
| chr19 | 48613481 | 48613582 | 102 | 2 | 0.000865807 | 0.000223 | 0.00324 | 0.00035 | 0.000142004 | FAM83E |
| chr5 | 149041573 | 149041588 | 16 | 2 | 0.000866656 | 0.001321 | 8.15E-05 | 9.77E-05 | -0.00512576 | SH3TC2 |
| chr1 | 156908399 | 156908527 | 129 | 2 | 0.00086667 | 0.45727 | 0.43249 | 0.528 | -0.001207826 | PEAR1 |
| chr17 | 12429289 | 12429317 | 29 | 2 | 0.000867281 | 0.001875 | 9.47E-05 | 0.00013 | -0.003892871 | NA |
| chr17 | 39715492 | 39715792 | 301 | 2 | 0.000868079 | 0.018875 | 0.00211 | 0.00453 | -0.002259916 | ERBB2 |
| chr14 | 99391966 | 99392022 | 57 | 2 | 0.000869018 | 0.00293 | 0.00011 | 0.00017 | -0.011004858 | NA |
| chr2 | 156434950 | 156435314 | 365 | 4 | 0.000870305 | 0.00155 | 0.02535 | 0.00266 | -0.006265403 | GPD2 |
| chr16 | 55656218 | 55656336 | 119 | 4 | 0.00087047 | 0.001159 | 0.00086 | 0.00063 | -0.001090111 | SLC6A2 |
| chr4 | 6918285 | 6918386 | 102 | 2 | 0.000870629 | 0.000145 | 0.00014 | 6.79E-05 | -0.014315141 | TBC1D14 |
| chr10 | 43667333 | 43667619 | 287 | 3 | 0.000870694 | 3.42E-05 | 0.00058 | 4.16E-05 | 0.011780387 | ZNF32-AS3 |
| chr3 | 117997375 | 117997614 | 240 | 3 | 0.000871008 | 0.038481 | 0.05337 | 0.03685 | 0.001951367 | AC092691.1 |
| chr6 | 32055606 | 32055632 | 27 | 2 | 0.000872354 | 5.00E-05 | 0.00072 | 6.32E-05 | -0.006740398 | TNXB |
| chr3 | 100210846 | 100210906 | 61 | 2 | 0.000873498 | 0.000121 | 0.00025 | 8.34E-05 | -0.01424804 | TMEM30CP, AC129803.1 |
| chr2 | 33706618 | 33706656 | 39 | 2 | 0.000875044 | 3.37E-05 | 0.00178 | 5.27E-05 | -0.00740723 | NA |
| chr6 | 43049583 | 43049629 | 47 | 2 | 0.000875344 | 0.002853 | 0.00012 | 0.00018 | -0.002148472 | KLC4, CUL7 |
| chr4 | 76959202 | 76959215 | 14 | 3 | 0.000875427 | 0.00297 | 0.0032 | 0.00033 | -0.007200858 | SEPTIN11 |
| chr17 | 82153543 | 82153598 | 56 | 2 | 0.000875549 | 0.000399 | 0.00588 | 0.00067 | -0.00342172 | AC129510.1, CCDC57 |
| chr16 | 65177056 | 65177096 | 41 | 2 | 0.000876383 | 0.000947 | 0.00012 | 0.00013 | -0.005027943 | NA |
| chr17 | 909447 | 909494 | 48 | 2 | 0.000876488 | 8.45E-05 | 0.00076 | 9.71E-05 | 0.042614905 | NXN |
| chr14 | 105461947 | 105462053 | 107 | 2 | 0.000877554 | 0.000352 | 0.00725 | 0.00063 | -0.002981672 | MTA1 |
| chr3 | 184386327 | 184386592 | 266 | 4 | 0.000878493 | 0.000154 | 0.03077 | 0.00062 | -0.011990855 | CHRD |
| chr8 | 134799673 | 134799683 | 11 | 2 | 0.000879609 | 0.000432 | 0.00024 | 0.00017 | -0.006998117 | NA |
| chr17 | 76053656 | 76053893 | 238 | 5 | 0.00088038 | 0.000429 | 0.00145 | 0.00031 | -0.003477266 | SRP68 |
| chr12 | 132579924 | 132579979 | 56 | 3 | 0.000880628 | 5.56E-05 | 0.00096 | 6.79E-05 | -0.000712563 | FBRSL1 |
| chr19 | 39151434 | 39151530 | 97 | 2 | 0.000880883 | 5.17E-05 | 0.00239 | 8.11E-05 | -0.010258086 | PAK4 |
| chr14 | 60684322 | 60684375 | 54 | 2 | 0.000883223 | 0.011582 | 8.98E-05 | 0.00018 | -0.00865392 | NA |
| chr1 | 180533633 | 180533652 | 20 | 2 | 0.000883502 | 9.37E-05 | 0.00047 | 8.96E-05 | -0.015641002 | OVAAL |
| chr17 | 68351659 | 68351808 | 150 | 2 | 0.000883904 | 4.72E-05 | 0.00165 | 7.06E-05 | 0.000509357 | ARSG |
| chr18 | 75161752 | 75161806 | 55 | 2 | 0.000884001 | 0.000554 | 0.00392 | 0.00079 | -0.011777399 | NA |
| chr6 | 166678595 | 166678669 | 75 | 2 | 0.000884732 | 0.000151 | 0.00022 | 8.78E-05 | -0.014937609 | RPS6KA2 |
| chr19 | 1513380 | 1513514 | 135 | 3 | 0.000885783 | 0.002777 | 0.00027 | 0.00032 | -0.005864667 | PLK5, ADAMTSL5 |
| chr16 | 66513163 | 66513246 | 84 | 2 | 0.000886446 | 6.25E-05 | 0.00196 | 9.34E-05 | -0.006605259 | AC010542.3, TK2 |
| chr8 | 81641616 | 81641653 | 38 | 2 | 0.000888114 | 0.031807 | 6.52E-05 | 0.00012 | 0.002282708 | NA |
| chr9 | 109477730 | 109477831 | 102 | 2 | 0.000888851 | 7.78E-05 | 0.00314 | 0.00013 | -0.013355662 | PTPN3 |
| chr8 | 37667667 | 37667721 | 55 | 2 | 0.000890049 | 4.75E-05 | 0.0015 | 7.00E-05 | 0.000443107 | AC137579.1 |
| chr11 | 14891743 | 14891805 | 63 | 3 | 0.0008905 | 0.006665 | 0.00718 | 0.00515 | 0.000979992 | CYP2R1 |
| chr8 | 80404644 | 80404673 | 30 | 2 | 0.000893171 | 0.000253 | 0.00016 | 9.42E-05 | -0.006833579 | AC034114.2 |
| chr15 | 25093865 | 25093921 | 57 | 2 | 0.000893844 | 0.001078 | 8.60E-05 | 9.78E-05 | -0.009483119 | SNHG14, AC124312.5 |
| chr2 | 120424737 | 120424751 | 15 | 2 | 0.000893905 | 0.000101 | 0.00031 | 8.08E-05 | -0.008742401 | NA |
| chr22 | 30246991 | 30247074 | 84 | 5 | 0.000894053 | 0.017407 | 8.33E-05 | 0.00051 | 0.001468494 | AC004264.1 |
| chr5 | 113294980 | 113295114 | 135 | 4 | 0.000895227 | 0.03152 | 5.72E-05 | 0.00039 | 0.001513263 | MCC |
| chr11 | 125002504 | 125002651 | 148 | 2 | 0.0008954 | 5.65E-05 | 0.00099 | 7.51E-05 | -0.008959706 | CCDC15 |
| chr1 | 924464 | 924642 | 179 | 3 | 0.000896399 | 0.002731 | 0.00077 | 0.00059 | 0.002443029 | SAMD11 |
| chr17 | 56596851 | 56596977 | 127 | 2 | 0.000896455 | 0.000563 | 0.00166 | 0.00058 | -0.001538454 | NA |
| chr1 | 2882670 | 2882789 | 120 | 2 | 0.000896636 | 0.00055 | 0.00957 | 0.001 | -0.005945749 | NA |
| chr9 | 137235684 | 137235816 | 133 | 3 | 0.00089729 | 4.54E-05 | 0.00468 | 8.61E-05 | -0.013040547 | SLC34A3 |
| chr5 | 170389751 | 170389872 | 122 | 3 | 0.000897942 | 9.11E-05 | 0.00071 | 7.46E-05 | -0.010460766 | KCNIP1, KCNIP1-OT1 |
| chr11 | 829035 | 829050 | 16 | 2 | 0.000900454 | 0.020715 | 0.04381 | 0.02824 | -0.005942465 | CRACR2B, AP006621.1 |
| chr1 | 203007798 | 203007975 | 178 | 3 | 0.000900846 | 0.009899 | 0.01023 | 0.00787 | -0.00110286 | TMEM183A |
| chr11 | 47261417 | 47261448 | 32 | 2 | 0.000903534 | 0.000258 | 0.00637 | 0.00046 | -0.01203064 | NR1H3 |
| chr4 | 57065657 | 57065662 | 6 | 2 | 0.000903624 | 0.000999 | 0.00013 | 0.00015 | -0.004754874 | IGFBP7 |
| chr11 | 67443829 | 67443908 | 80 | 5 | 0.000904964 | 0.007179 | 0.00084 | 0.00016 | 0.004803619 | NA |
| chr11 | 71452336 | 71452382 | 47 | 2 | 0.000905372 | 0.190517 | 0.19619 | 0.22271 | -0.001270558 | DHCR7 |
| chr11 | 9838474 | 9838960 | 487 | 2 | 0.000905522 | 0.00016 | 0.00474 | 0.00028 | -0.004372984 | SBF2 |
| chr19 | 56148686 | 56148851 | 166 | 4 | 0.000906603 | 3.59E-05 | 0.00539 | 6.82E-05 | -0.004346774 | ZNF444 |
| chr10 | 128540870 | 128540922 | 53 | 3 | 0.000906637 | 0.223152 | 0.01023 | 0.04598 | 0.001898045 | NA |
| chr10 | 110919670 | 110919722 | 53 | 2 | 0.000908529 | 0.039227 | 0.00306 | 0.00772 | 0.001532999 | SHOC2 |
| chr1 | 151059825 | 151059875 | 51 | 5 | 0.000908657 | 0.00393 | 0.03905 | 0.00429 | 0.000566104 | CDC42SE1 |
| chr2 | 218880375 | 218880529 | 155 | 4 | 0.000908657 | 9.56E-05 | 0.00025 | 4.65E-05 | 0.00144529 | NA |
| chr19 | 13575012 | 13575121 | 110 | 2 | 0.000908848 | 4.89E-05 | 0.00154 | 7.23E-05 | 0.025198549 | CACNA1A |
| chr8 | 141147005 | 141147041 | 37 | 2 | 0.000908908 | 0.011805 | 0.00054 | 0.00125 | -0.002067985 | DENND3 |
| chr17 | 1934470 | 1934554 | 85 | 3 | 0.000909039 | 0.031332 | 0.04638 | 0.02414 | -0.001972451 | NA |
| chr11 | 44605395 | 44605399 | 5 | 2 | 0.000909396 | 0.000448 | 0.00026 | 0.00018 | -0.001301298 | CD82 |
| chr9 | 98456082 | 98456108 | 27 | 2 | 0.000909951 | 0.000204 | 0.00482 | 0.00035 | -0.00795416 | GABBR2 |
| chr1 | 54331144 | 54331153 | 10 | 2 | 0.000911368 | 7.76E-05 | 0.00086 | 9.39E-05 | -0.006604425 | SSBP3 |
| chr3 | 72255931 | 72256040 | 110 | 2 | 0.000912302 | 5.20E-05 | 0.00125 | 7.37E-05 | -0.012441572 | LINC00877 |
| chr9 | 123369030 | 123369578 | 549 | 2 | 0.000912707 | 0.011441 | 0.00702 | 0.00813 | 0.00807696 | CRB2 |
| chr12 | 1918827 | 1918990 | 164 | 2 | 0.000913929 | 0.001173 | 0.00824 | 0.00183 | -0.012140991 | AC005342.2 |
| chr2 | 61471272 | 61471278 | 7 | 2 | 0.000914941 | 0.001524 | 0.0002 | 0.00025 | 0.002517773 | AC016727.1 |
| chr9 | 33452951 | 33453245 | 295 | 2 | 0.000915455 | 4.46E-05 | 0.00213 | 6.98E-05 | -0.012450519 | NA |
| chr12 | 126357489 | 126357576 | 88 | 2 | 0.000915565 | 7.56E-05 | 0.00094 | 9.41E-05 | -0.0130506 | AC007368.1 |
| chr4 | 14889383 | 14889424 | 42 | 2 | 0.000915986 | 0.000141 | 0.00038 | 0.00011 | -0.00568139 | NA |
| chr14 | 103848865 | 103848886 | 22 | 2 | 0.000916089 | 0.001256 | 0.01016 | 0.00205 | -0.005246767 | LINC00637 |
| chr4 | 38663973 | 38663992 | 20 | 5 | 0.000916217 | 5.94E-05 | 0.00766 | 0.00011 | 0.0012836 | KLF3-AS1 |
| chr11 | 2211333 | 2211360 | 28 | 3 | 0.000917469 | 4.56E-05 | 0.00669 | 9.69E-05 | -0.003222063 | NA |
| chr10 | 101901514 | 101901519 | 6 | 2 | 0.000918917 | 9.52E-05 | 0.00224 | 0.00014 | -0.006589272 | ARMH3 |
| chr10 | 122688412 | 122688516 | 105 | 2 | 0.000919192 | 4.83E-05 | 0.00169 | 7.25E-05 | -0.004385166 | NA |
| chr1 | 1399411 | 1399434 | 24 | 5 | 0.000920504 | 0.000777 | 0.0054 | 0.00042 | 0.00210296 | NA |
| chr12 | 107773725 | 107773953 | 229 | 2 | 0.000920754 | 0.008885 | 0.00045 | 0.00097 | 0.005506329 | NA |

| | | | | | | | | | | |
|---|---|---|---|---|---|---|---|---|---|---|
| chr18 | 58047026 | 58047161 | 136 | 4 | 0.00092085 | 0.000649 | 0.00128 | 0.00017 | -0.002988374 | NEDD4L |
| chr11 | 62671382 | 62671845 | 464 | 15 | 0.00092085 | 0.001944 | 0.01701 | 0.00024 | -0.000532521 | UQCC3, LBHD1 |
| chr10 | 49505957 | 49505974 | 18 | 2 | 0.000921435 | 5.78E-05 | 0.00191 | 8.68E-05 | -0.005113638 | ERCC6 |
| chr10 | 101783415 | 101783502 | 88 | 5 | 0.000921473 | 0.116194 | 0.00019 | 0.00168 | 0.001520473 | NA |
| chr19 | 488516 | 488565 | 50 | 2 | 0.000922494 | 0.000738 | 0.00021 | 0.00019 | 0.003146982 | NA |
| chr12 | 124847448 | 124847637 | 190 | 3 | 0.000924202 | 0.002059 | 0.00022 | 0.00022 | -0.008487576 | SCARB1 |
| chr22 | 27686660 | 27686797 | 138 | 2 | 0.0009275 | 0.01977 | 5.22E-05 | 8.79E-05 | -0.002971071 | AL121885.2 |
| chr2 | 97995868 | 97995896 | 29 | 2 | 0.000927532 | 0.011769 | 0.00045 | 0.00105 | 0.000888181 | TMEM131 |
| chr17 | 80187696 | 80187773 | 78 | 2 | 0.000927532 | 0.001768 | 0.01115 | 0.00278 | -0.006346913 | CARD14 |
| chr16 | 54177613 | 54177676 | 64 | 3 | 0.000927534 | 0.000624 | 0.00134 | 0.00013 | -0.000364352 | NA |
| chr5 | 93348431 | 93348458 | 28 | 2 | 0.000928454 | 7.90E-05 | 0.00192 | 0.00012 | -0.006175551 | NA |
| chr1 | 231161770 | 231161871 | 102 | 2 | 0.000929165 | 0.000156 | 0.00474 | 0.00027 | 0.008059756 | NA |
| chr14 | 103334791 | 103334806 | 16 | 4 | 0.000929558 | 0.021928 | 0.01893 | 0.00754 | 0.001965591 | EIF5 |
| chr1 | 69432303 | 69432317 | 15 | 2 | 0.000931603 | 0.000241 | 9.95E-05 | 6.61E-05 | -0.001364498 | NA |
| chr12 | 6961154 | 6961666 | 513 | 6 | 0.000932239 | 0.005212 | 0.00041 | 0.00064 | -0.002542416 | PTPN6 |
| chr5 | 180899142 | 180899186 | 45 | 3 | 0.000934536 | 9.13E-05 | 0.00283 | 0.00012 | -0.003166112 | BTNL8 |
| chr1 | 1160227 | 1160357 | 131 | 2 | 0.000936168 | 8.46E-05 | 0.00332 | 0.00014 | -0.004315097 | NA |
| chr20 | 37396139 | 37396266 | 128 | 5 | 0.000936276 | 6.12E-05 | 0.00168 | 7.50E-05 | -0.00470684 | SRC |
| chr13 | 96091329 | 96091500 | 172 | 3 | 0.000936734 | 0.352428 | 0.04706 | 0.14748 | -0.001373619 | HS6ST3 |
| chr11 | 57791505 | 57791594 | 90 | 3 | 0.000937022 | 0.002594 | 6.95E-05 | 0.00012 | -0.001784408 | AP001931.2, AP001931.1, CTNND1 |
| chr12 | 65883081 | 65883194 | 114 | 2 | 0.000939647 | 0.048104 | 0.00072 | 0.00223 | -0.00622109 | HMGA2 |
| chr7 | 45104475 | 45104553 | 79 | 4 | 0.000940476 | 0.003488 | 0.00033 | 0.00025 | -0.003114547 | TBRG4, SNORA5A |
| chr6 | 27290947 | 27290988 | 42 | 2 | 0.000940685 | 0.000151 | 6.20E-05 | 3.72E-05 | -0.011628053 | POM121L2 |
| chr6 | 36722473 | 36722483 | 11 | 2 | 0.000940685 | 6.84E-05 | 0.00032 | 6.25E-05 | -0.009248282 | RAB44 |
| chr15 | 55742603 | 55742619 | 17 | 2 | 0.000941822 | 0.001376 | 0.01079 | 0.00225 | -0.000741387 | PRTG |
| chr6 | 165819472 | 165819653 | 182 | 3 | 0.000942021 | 3.18E-05 | 0.00464 | 6.36E-05 | 0.02164831 | PDE10A |
| chr6 | 151808253 | 151808265 | 13 | 2 | 0.000942047 | 0.035326 | 0.06713 | 0.04828 | 0.001684075 | ESR1 |
| chr11 | 68799277 | 68799344 | 68 | 3 | 0.000942319 | 0.00046 | 0.00029 | 0.00017 | -0.008146106 | CPT1A |
| chr12 | 52611232 | 52611250 | 19 | 2 | 0.000947192 | 0.000168 | 0.00075 | 0.00017 | -0.003999336 | KRT73-AS1, KRT73 |
| chr8 | 93699890 | 93700167 | 278 | 4 | 0.000950618 | 0.006208 | 0.00433 | 0.00156 | -0.000829525 | CIBAR1, LINC00535 |
| chr2 | 100417808 | 100417833 | 26 | 4 | 0.000951205 | 5.12E-05 | 0.00282 | 9.53E-05 | 0.005962909 | NA |
| chr10 | 104121069 | 104121167 | 99 | 3 | 0.000952665 | 0.000617 | 0.02336 | 0.00164 | 0.000251545 | NA |
| chr3 | 137998738 | 137998765 | 28 | 3 | 0.000952686 | 0.051768 | 0.06209 | 0.05862 | -0.001978115 | CLDN18 |
| chr11 | 68980085 | 68980167 | 83 | 2 | 0.000952827 | 0.000105 | 0.0038 | 0.00018 | -0.009445618 | MRGPRD |
| chr13 | 98880082 | 98880118 | 37 | 2 | 0.000954436 | 5.75E-05 | 0.00227 | 8.86E-05 | -0.010532149 | DOCK9 |
| chr17 | 82941430 | 82941577 | 148 | 2 | 0.00095532 | 0.002598 | 0.00013 | 0.00021 | -0.001120931 | TBCD |
| chr13 | 45852103 | 45852128 | 26 | 2 | 0.000957576 | 0.101187 | 0.16218 | 0.14012 | -0.007113222 | NA |
| chr3 | 45594201 | 45594210 | 10 | 2 | 0.000959517 | 0.007614 | 0.01217 | 0.0084 | -0.002254583 | LIMD1 |
| chr7 | 4689322 | 4690289 | 968 | 7 | 0.000961031 | 0.002936 | 0.00102 | 3.82E-05 | -0.003081164 | FOXK1 |
| chr16 | 86892580 | 86892644 | 65 | 2 | 0.000961239 | 4.93E-05 | 0.00227 | 7.69E-05 | -0.023398639 | NA |
| chr1 | 181737284 | 181737313 | 30 | 2 | 0.000965196 | 0.00047 | 0.00014 | 0.00011 | -0.016607355 | CACNA1E |
| chr8 | 99893650 | 99893787 | 138 | 3 | 0.000967564 | 0.147635 | 0.16913 | 0.19885 | -8.73E-05 | AC105328.1, COX6C |
| chr16 | 2223297 | 2223331 | 35 | 3 | 0.00096791 | 0.125592 | 0.19561 | 0.19345 | 0.000474218 | NA |
| chr4 | 888809 | 888851 | 43 | 2 | 0.0009762 | 6.32E-05 | 0.00024 | 5.35E-05 | -0.000369028 | GAK |
| chr7 | 136868341 | 136868423 | 83 | 3 | 0.000979834 | 0.059277 | 0.00081 | 0.00445 | -0.011081784 | AC009264.1 |
| chr6 | 17619396 | 17619408 | 13 | 2 | 0.000983109 | 0.00077 | 0.00036 | 0.0003 | -0.010232571 | NUP153 |
| chr1 | 14223724 | 14223953 | 230 | 2 | 0.000983917 | 8.64E-05 | 0.00057 | 9.02E-05 | -0.003188827 | KAZN, AL357873.1 |
| chr17 | 59107021 | 59107051 | 31 | 2 | 0.000985885 | 0.754523 | 0.57538 | 0.74286 | 4.35E-06 | AC099850.1 |
| chr9 | 113303184 | 113303352 | 169 | 2 | 0.000988178 | 0.008852 | 0.00165 | 0.0028 | -0.000473265 | RNF183 |

**Supplementary Table S5.2. Significant Differentially methylated regions between perinatal SI v NC at 38w gestation ranked by Fisher's p-value.**

| Chr | Start | End | Width | *n* CpGs | Minimum smoothed FDR | Stouffer | HMFDR | Fisher | Mean differential | Overlapping genes |
|---|---|---|---|---|---|---|---|---|---|---|
| chr6 | 30127170 | 30128025 | 856 | 25 | 1.89E-99 | 4.24E-69 | 1.55E-07 | 1.15E-81 | -0.02303068 | NA |
| chr6 | 30070227 | 30072024 | 1798 | 36 | 6.83E-81 | 3.55E-67 | 1.81E-06 | 2.96E-71 | -0.02953086 | PPP1R11, RNF39 |
| chr5 | 177399261 | 177403142 | 3882 | 16 | 1.46E-66 | 2.29E-50 | 3.01E-09 | 1.30E-64 | -0.0242388 | PFN3, F12 |
| chr6 | 30909962 | 30918384 | 8423 | 94 | 5.06E-40 | 1.42E-42 | 7.01E-06 | 1.06E-63 | -0.00558348 | GTF2H4, AL669830.1, VARS2 |
| chr6 | 33313541 | 33324252 | 10712 | 181 | 8.39E-27 | 4.39E-43 | 2.65E-05 | 9.25E-57 | -0.00209096 | TAPBP, ZBTB22, DAXX, SMIM40, SMIM40 |
| chr1 | 43348221 | 43349364 | 1144 | 10 | 9.86E-73 | 3.17E-46 | 9.87E-09 | 1.14E-56 | -0.06378684 | MPL, AL139289.2 |
| chr21 | 34884019 | 34888434 | 4416 | 21 | 7.72E-47 | 1.18E-51 | 9.81E-06 | 1.03E-53 | -0.03820896 | AP000331.1, RUNX1 |
| chr22 | 45412782 | 45415067 | 2286 | 16 | 1.16E-61 | 4.71E-42 | 1.13E-07 | 8.77E-53 | -0.02833576 | RIBC2, SMC1B |
| chr6 | 32146548 | 32155257 | 8710 | 125 | 4.51E-21 | 2.50E-35 | 0.000133 | 1.74E-48 | -0.00342033 | PPT2, PPT2-EGFL8, PRRT1, AL662884.4, AL662884.3 |
| chr4 | 2817125 | 2818752 | 1628 | 10 | 1.12E-48 | 9.37E-46 | 3.80E-07 | 1.47E-47 | -0.03118683 | SH3BP2 |
| chr5 | 1849461 | 1852725 | 3265 | 13 | 1.80E-48 | 3.22E-40 | 6.62E-08 | 1.05E-45 | 0.029011987 | AC025183.1 |
| chr6 | 28584805 | 28590336 | 5532 | 63 | 1.28E-22 | 2.08E-32 | 3.96E-07 | 1.36E-43 | -0.00337876 | AL049543.1, ZBED9 |
| chr17 | 48601357 | 48606047 | 4691 | 28 | 5.48E-36 | 8.28E-39 | 4.03E-05 | 1.74E-43 | -0.0232499 | HOXB-AS3, HOXB3, HOXB6 |
| chr6 | 32879170 | 32880068 | 899 | 14 | 4.26E-46 | 4.98E-44 | 4.63E-07 | 4.40E-42 | -0.02536592 | PPP1R2P1 |
| chr19 | 35754424 | 35759262 | 4839 | 34 | 1.69E-42 | 1.17E-27 | 6.42E-07 | 2.56E-41 | -0.00913777 | AD000671.1, LIN37, AC002398.2, PROSER3, HSPB6 |
| chr17 | 4785274 | 4789965 | 4692 | 23 | 1.74E-21 | 2.47E-37 | 7.91E-07 | 2.83E-41 | -0.01344295 | AC233723.2, GLTPD2, VMO1 |
| chr6 | 32192297 | 32199062 | 6766 | 83 | 4.30E-24 | 7.88E-28 | 5.38E-05 | 3.62E-40 | -0.00187462 | GPSM3, NOTCH4 |
| chr17 | 41519724 | 41525408 | 5685 | 23 | 3.79E-24 | 3.17E-25 | 8.12E-08 | 1.31E-39 | -0.00410088 | KRT15, KRT19 |
| chr9 | 128732813 | 128733711 | 899 | 20 | 3.90E-48 | 1.07E-33 | 1.84E-06 | 3.17E-39 | -0.00913653 | AL441992.1, ZER1 |
| chr6 | 33161048 | 33169156 | 8109 | 79 | 5.30E-18 | 7.51E-31 | 4.94E-05 | 1.43E-38 | -0.00330427 | COL11A2 |
| chr19 | 48740170 | 48741708 | 1539 | 10 | 1.42E-46 | 4.61E-33 | 1.98E-09 | 8.75E-38 | -0.02529941 | RASIP1, IZUMO1 |
| chr9 | 114098157 | 114099773 | 1617 | 12 | 2.18E-46 | 6.13E-30 | 5.55E-07 | 2.85E-36 | -0.02003394 | KIF12 |
| chr6 | 37648634 | 37650738 | 2105 | 14 | 8.42E-24 | 7.70E-40 | 2.28E-06 | 4.70E-36 | -0.04742967 | MDGA1 |
| chr11 | 70826260 | 70828140 | 1881 | 10 | 7.16E-35 | 4.09E-39 | 1.26E-06 | 4.97E-36 | 0.033014479 | SHANK2 |
| chr1 | 227558410 | 227561018 | 2609 | 12 | 1.12E-21 | 7.14E-38 | 2.31E-06 | 2.46E-35 | -0.02759749 | NA |
| chr17 | 80890288 | 80893047 | 2760 | 15 | 5.33E-29 | 3.71E-38 | 1.32E-05 | 5.55E-35 | -0.02485667 | RPTOR |
| chr10 | 43202560 | 43204773 | 2214 | 9 | 1.20E-42 | 4.15E-30 | 3.52E-07 | 3.20E-34 | -0.02660492 | RASGEF1A |
| chr6 | 31538946 | 31542630 | 3685 | 47 | 2.94E-29 | 2.49E-28 | 4.12E-05 | 6.36E-34 | -0.00366623 | DDX39B-AS1, DDX39B, ATP6V1G2-DDX39B, SNORD84 |
| chr8 | 43276117 | 43277364 | 1248 | 8 | 9.49E-37 | 4.42E-34 | 4.58E-08 | 9.66E-34 | -0.04867978 | AC022616.1 |
| chr1 | 217076899 | 217079171 | 2273 | 8 | 5.57E-29 | 1.58E-28 | 4.76E-08 | 9.66E-34 | 0.026410906 | ESRRG |
| chr17 | 81821273 | 81823119 | 1847 | 27 | 1.21E-38 | 8.29E-30 | 3.48E-05 | 9.71E-34 | -0.00953522 | MCRIP1 |
| chr2 | 231392270 | 231396296 | 4027 | 19 | 1.05E-19 | 3.63E-27 | 1.83E-06 | 9.02E-32 | 0.010386135 | AC017104.1, B3GNT7 |
| chr10 | 86967302 | 86972118 | 4817 | 27 | 8.46E-27 | 1.90E-19 | 1.38E-06 | 9.07E-32 | -0.00870098 | ADIRF, AGAP11, MMRN2, ADIRF-AS1, AL136982.7 |
| chr16 | 1430029 | 1433191 | 3163 | 15 | 1.54E-28 | 8.80E-28 | 1.08E-06 | 5.63E-31 | 0.014888781 | PERCC1 |
| chr6 | 36960796 | 36962950 | 2155 | 7 | 6.98E-34 | 2.84E-30 | 2.38E-07 | 6.71E-31 | -0.04119583 | PI16 |
| chr8 | 143549354 | 143550305 | 952 | 8 | 4.40E-33 | 2.26E-31 | 2.66E-07 | 6.71E-31 | -0.03053917 | AC067930.3 |
| chr11 | 30582920 | 30586810 | 3891 | 25 | 1.31E-33 | 1.78E-21 | 5.05E-06 | 9.88E-31 | -0.00806767 | MPPED2-AS1, MPPED2 |
| chr6 | 32851615 | 32856164 | 4550 | 63 | 1.75E-22 | 8.78E-22 | 8.51E-06 | 2.55E-30 | -0.00241573 | PSMB9, TAP1 |
| chr1 | 153627003 | 153629180 | 2178 | 23 | 2.60E-28 | 1.62E-32 | 0.000647 | 3.39E-30 | -0.03552153 | S100A1, S100A13, AL162258.1 |
| chr17 | 17699305 | 17700870 | 1566 | 7 | 5.77E-34 | 1.31E-27 | 4.72E-08 | 6.67E-30 | -0.02570483 | RAI1 |
| chr15 | 73925876 | 73927607 | 1732 | 15 | 1.03E-36 | 1.00E-23 | 5.26E-06 | 7.36E-30 | -0.01276055 | LOXL1, LOXL1-AS1 |
| chr16 | 981944 | 985774 | 3831 | 13 | 1.51E-26 | 2.17E-26 | 3.92E-06 | 4.11E-29 | -0.01553924 | SOX8 |
| chr4 | 39445817 | 39447982 | 2166 | 11 | 4.87E-32 | 7.84E-24 | 5.08E-06 | 7.12E-29 | -0.02137864 | KLB |
| chr7 | 73742887 | 73743687 | 801 | 5 | 8.91E-34 | 2.47E-29 | 2.75E-08 | 7.71E-29 | -0.04262947 | NA |
| chr6 | 30552165 | 30558248 | 6084 | 87 | 6.29E-17 | 2.39E-18 | 9.50E-06 | 3.68E-28 | -0.00032691 | PRR3, GNL1 |
| chr11 | 94767446 | 94769713 | 2268 | 21 | 8.39E-27 | 5.66E-20 | 4.79E-08 | 4.81E-28 | -0.00572049 | AMOTL1 |
| chr2 | 231387343 | 231388908 | 1566 | 7 | 2.43E-30 | 6.54E-26 | 4.82E-07 | 7.17E-28 | 0.013022433 | NA |
| chr6 | 30073304 | 30077291 | 3988 | 45 | 1.46E-33 | 2.51E-19 | 6.12E-06 | 8.48E-28 | -0.00397845 | RNF39 |
| chr2 | 48617056 | 48617929 | 874 | 9 | 9.41E-31 | 5.50E-29 | 5.14E-06 | 1.71E-27 | -0.03190263 | STON1-GTF2A1L, GTF2A1L |
| chr17 | 42783980 | 42785890 | 1911 | 13 | 1.25E-19 | 6.00E-29 | 1.65E-05 | 1.77E-27 | -0.01664412 | WNK4 |
| chr20 | 37518640 | 37523281 | 4642 | 51 | 2.21E-21 | 7.84E-24 | 1.42E-07 | 1.79E-27 | -0.01059521 | NNAT, BLCAP |
| chr11 | 2884701 | 2887241 | 2541 | 32 | 1.34E-26 | 1.07E-21 | 3.65E-05 | 2.16E-27 | -0.00332742 | CDKN1C |
| chr12 | 4269101 | 4274604 | 5504 | 55 | 6.39E-17 | 3.62E-20 | 2.77E-05 | 3.74E-27 | 0.000372198 | CCND2, AC008012.1, CCND2-AS1 |
| chr8 | 126556083 | 126559068 | 2986 | 28 | 4.41E-27 | 1.32E-22 | 5.30E-05 | 4.27E-27 | -0.00543525 | PCAT1, LRATD2 |
| chr6 | 34145516 | 34147050 | 1535 | 13 | 8.42E-34 | 2.73E-23 | 4.72E-08 | 4.54E-27 | -0.01422736 | GRM4 |
| chr22 | 20112508 | 20116552 | 4045 | 18 | 4.44E-17 | 7.11E-22 | 1.82E-06 | 4.67E-27 | -0.00311114 | RANBP1, TRMT2A, MIR6816 |
| chr12 | 14567166 | 14569286 | 2121 | 17 | 1.37E-36 | 8.62E-18 | 3.02E-06 | 6.47E-27 | -0.00963594 | PLBD1-AS1, PLBD1 |
| chr6 | 30134398 | 30137421 | 3024 | 16 | 2.58E-23 | 2.40E-24 | 7.42E-06 | 6.98E-27 | -0.00422346 | TRIM40 |
| chr7 | 27184411 | 27186710 | 2300 | 38 | 3.21E-26 | 1.89E-18 | 5.76E-07 | 1.48E-26 | 0.001494986 | HOXA11-AS, AC004080.7, AC004080.14, AC004080.8, HOXA11, AC004080.2 |
| chr1 | 236522712 | 236525264 | 2553 | 25 | 1.24E-26 | 5.85E-23 | 1.57E-05 | 1.55E-26 | -0.00928773 | LGALS8, LGALS8-AS1 |
| chr1 | 26861503 | 26863839 | 2337 | 13 | 7.26E-26 | 4.43E-22 | 6.24E-08 | 3.97E-26 | -0.00681363 | SFN |
| chr7 | 2723340 | 2725776 | 2437 | 13 | 1.20E-26 | 4.72E-26 | 1.96E-07 | 4.63E-26 | 0.01155621 | AMZ1 |
| chr10 | 7408150 | 7413752 | 5603 | 32 | 8.71E-26 | 6.59E-15 | 3.13E-07 | 4.63E-26 | -0.00261428 | AL139125.1, SFMBT2 |
| chr14 | 73245548 | 73247023 | 1476 | 11 | 6.52E-32 | 1.28E-21 | 1.44E-07 | 4.65E-26 | -0.0106476 | PAPLN, AC004846.1, RNU6-419P |
| chr6 | 32166152 | 32171062 | 4911 | 67 | 7.96E-15 | 2.72E-20 | 3.60E-06 | 4.65E-26 | -0.00124646 | PPT2-EGFL8, EGFL8, AGPAT1, MIR6721 |
| chr6 | 30152254 | 30157165 | 4912 | 34 | 7.76E-14 | 1.19E-18 | 2.57E-05 | 5.51E-26 | 0.009004815 | TRIM10 |
| chr16 | 57371163 | 57373043 | 1881 | 10 | 7.82E-26 | 4.42E-23 | 4.34E-07 | 6.12E-26 | -0.01103348 | CX3CL1 |
| chr6 | 170244045 | 170246188 | 2144 | 8 | 9.11E-21 | 2.52E-28 | 1.21E-05 | 6.26E-26 | 0.036250807 | NA |
| chr6 | 10880853 | 10884658 | 3806 | 25 | 1.51E-15 | 1.74E-21 | 9.14E-05 | 7.03E-26 | -0.00851963 | AL024498.2, AL357497.1, GCM2 |
| chr2 | 73267949 | 73270946 | 2998 | 12 | 1.17E-27 | 1.15E-24 | 1.36E-06 | 7.23E-26 | -0.01597707 | FBXO41 |
| chr17 | 8798781 | 8800023 | 1243 | 10 | 4.16E-31 | 1.02E-22 | 1.47E-06 | 8.63E-26 | -0.03119398 | MFSD6L |
| chr2 | 236566825 | 236570372 | 3548 | 16 | 3.57E-27 | 4.85E-20 | 2.01E-07 | 1.05E-25 | -0.00988492 | ACKR3 |
| chr15 | 29674623 | 29677516 | 2894 | 15 | 1.50E-28 | 1.99E-21 | 3.65E-07 | 1.53E-25 | -0.0202156 | AC022613.1 |
| chr6 | 31791709 | 31793278 | 1570 | 21 | 9.21E-33 | 3.19E-19 | 6.27E-06 | 2.32E-25 | 0.007755391 | VARS1 |
| chr1 | 27349193 | 27350749 | 1557 | 8 | 1.95E-29 | 1.14E-19 | 2.68E-06 | 2.35E-25 | -0.03227847 | SYTL1 |
| chr15 | 72155183 | 72156547 | 1365 | 8 | 1.48E-30 | 4.21E-22 | 7.31E-07 | 2.53E-25 | -0.02274269 | AC020779.2 |
| chr22 | 38317106 | 38318766 | 1661 | 14 | 1.84E-27 | 1.49E-25 | 3.24E-06 | 3.31E-25 | -0.00940958 | CSNK1E, TPTEP2-CSNK1E |
| chr10 | 86957607 | 86960459 | 2853 | 18 | 9.55E-19 | 1.42E-26 | 8.32E-05 | 3.60E-25 | -0.01077102 | SNCG, MMRN2 |
| chr12 | 100355663 | 100357366 | 1704 | 12 | 3.35E-26 | 7.56E-26 | 2.73E-06 | 3.73E-25 | -0.01431984 | SLC17A8 |
| chr13 | 67226773 | 67230933 | 4161 | 22 | 2.48E-20 | 3.48E-20 | 2.77E-07 | 4.59E-25 | 0.007525361 | PCDH9 |
| chr18 | 79962271 | 79964692 | 2422 | 16 | 4.72E-30 | 4.14E-18 | 7.34E-08 | 4.69E-25 | -0.01000288 | HSBP1L1, AC114341.2 |
| chr6 | 30686645 | 30691915 | 5271 | 69 | 1.51E-11 | 2.02E-16 | 4.16E-05 | 5.01E-25 | -0.00111937 | PPP1R18, NRM |
| chr20 | 47351460 | 47353857 | 2398 | 12 | 1.07E-19 | 2.72E-20 | 1.17E-05 | 7.37E-25 | 0.014770422 | AL031666.1, ZMYND8 |
| chr22 | 35298164 | 35301653 | 3490 | 19 | 2.72E-19 | 3.92E-15 | 6.70E-08 | 1.00E-24 | -0.00437585 | TOM1, AL008635.1 |
| chr3 | 194296692 | 194297382 | 691 | 7 | 1.24E-26 | 1.49E-25 | 5.15E-06 | 1.10E-24 | -0.03233615 | LINC00887 |
| chr15 | 25862235 | 25865359 | 3125 | 36 | 4.11E-19 | 3.37E-23 | 0.000167 | 1.46E-24 | -0.00235468 | AC016266.1, ATP10A |
| chr17 | 6994996 | 6997480 | 2485 | 25 | 1.44E-19 | 7.58E-27 | 0.001208 | 1.55E-24 | -0.01865526 | ALOX12, ALOX12-AS1, AC040977.2, AC040977.1 |
| chr5 | 141338365 | 141339793 | 1429 | 16 | 1.10E-28 | 3.66E-22 | 3.27E-06 | 1.84E-24 | -0.01944888 | PCDHGA1, PCDHGA2 |
| chr1 | 2228810 | 2232370 | 3561 | 13 | 1.66E-15 | 3.43E-19 | 4.34E-07 | 2.41E-24 | -0.01110471 | SKI |
| chr10 | 100518882 | 100520794 | 1913 | 17 | 1.18E-28 | 1.44E-21 | 8.67E-07 | 2.85E-24 | -0.01271479 | SEC31B, AL133352.1 |
| chr11 | 65591629 | 65593842 | 2214 | 9 | 1.70E-26 | 1.08E-23 | 9.16E-07 | 3.10E-24 | -0.02171791 | EHBP1L1, KCNK7 |
| chr1 | 3856748 | 3858770 | 2023 | 18 | 2.75E-29 | 3.67E-12 | 3.01E-06 | 4.56E-24 | -0.01135509 | DFFB, CEP104 |
| chr22 | 43109886 | 43111659 | 1774 | 30 | 3.74E-26 | 4.07E-17 | 9.26E-05 | 5.50E-24 | -0.00407064 | BIK |
| chr3 | 49132567 | 49133910 | 1344 | 11 | 7.25E-25 | 7.84E-24 | 1.86E-05 | 7.49E-24 | -0.03343937 | LAMB2 |
| chr17 | 7434641 | 7440702 | 6062 | 36 | 1.13E-13 | 1.96E-17 | 1.23E-05 | 7.94E-24 | -0.00354773 | AC113189.2, AC113189.12, TMEM102, FGF11, AC113189.4, AC113189.1, AC113189.3 |
| chr14 | 105475199 | 105480056 | 4858 | 19 | 2.50E-14 | 5.15E-24 | 9.80E-05 | 8.48E-24 | -0.02143566 | CRIP2 |
| chr6 | 30325804 | 30330164 | 4361 | 48 | 6.87E-14 | 8.53E-21 | 0.002344 | 9.58E-24 | -0.00243872 | TRIM39, TRIM39-RPP21, HCG17, HCG18 |
| chr2 | 222297052 | 222300601 | 3550 | 33 | 7.19E-23 | 5.22E-15 | 2.25E-07 | 1.54E-23 | -0.00371582 | CCDC140, PAX3 |
| chr1 | 92481775 | 92485279 | 3505 | 32 | 7.00E-20 | 5.94E-19 | 1.39E-05 | 1.73E-23 | -0.00152529 | GFI1 |
| chr20 | 62815610 | 62817182 | 1573 | 35 | 3.00E-29 | 4.86E-16 | 4.19E-05 | 3.06E-23 | -0.00597404 | COL9A3 |
| chr20 | 25147493 | 25150233 | 2741 | 18 | 2.66E-18 | 3.34E-17 | 2.96E-05 | 3.38E-23 | -0.00533435 | AL035252.2 |
| chr19 | 45590772 | 45592304 | 1533 | 8 | 3.25E-25 | 1.22E-19 | 6.97E-08 | 5.64E-23 | -0.01918076 | OPA3, GPR4 |
| chr10 | 45672178 | 45673735 | 1558 | 19 | 4.35E-25 | 2.52E-18 | 2.24E-06 | 6.42E-23 | -0.00442329 | ZFAND4 |
| chr5 | 169298175 | 169302843 | 4669 | 24 | 5.96E-19 | 1.55E-18 | 8.39E-06 | 6.81E-23 | -0.00472815 | SLIT3 |
| chr12 | 6765209 | 6768574 | 3366 | 16 | 1.01E-25 | 1.25E-15 | 8.58E-08 | 6.99E-23 | -0.0078742 | PTMS, MLF2 |
| chr5 | 153489449 | 153492243 | 2795 | 20 | 3.14E-20 | 1.19E-15 | 4.01E-06 | 1.18E-22 | -0.00980806 | GRIA1 |
| chr4 | 76420098 | 76421918 | 1821 | 9 | 6.03E-24 | 5.03E-21 | 1.03E-07 | 1.28E-22 | -0.02194449 | CCDC158 |
| chr17 | 49013254 | 49014910 | 1657 | 11 | 9.06E-23 | 2.35E-22 | 8.28E-05 | 1.64E-22 | 0.038234633 | IGF2BP1, AC091133.4 |
| chr11 | 61391146 | 61393018 | 1873 | 16 | 1.04E-36 | 3.77E-10 | 1.62E-08 | 1.74E-22 | -0.0073183 | TMEM216 |
| chr9 | 114929484 | 114931572 | 2089 | 11 | 3.45E-22 | 4.92E-18 | 4.93E-07 | 1.80E-22 | 0.010095176 | DELEC1, TNFSF8 |
| chr2 | 240144337 | 240145680 | 1344 | 9 | 6.86E-22 | 6.46E-22 | 1.05E-05 | 1.82E-22 | -0.02028701 | OTOS |
| chr2 | 144517307 | 144521241 | 3935 | 40 | 1.33E-14 | 3.44E-15 | 6.11E-06 | 2.61E-22 | -0.00080946 | ZEB2-AS1, AC009951.2, AC009951.1, AC009951.3, ZEB2 |
| chr11 | 637038 | 641042 | 4005 | 16 | 2.03E-16 | 2.17E-22 | 5.37E-05 | 3.56E-22 | -0.0119903 | DRD4 |
| chr10 | 62048161 | 62049579 | 1419 | 15 | 1.40E-24 | 1.26E-19 | 2.60E-06 | 4.48E-22 | -0.0113075 | ARID5B |
| chr7 | 11831909 | 11833119 | 1211 | 12 | 2.52E-28 | 5.78E-17 | 1.74E-06 | 4.85E-22 | -0.02263961 | THSD7A |
| chr10 | 133199071 | 133202171 | 3101 | 8 | 3.81E-15 | 1.53E-20 | 3.12E-06 | 6.02E-22 | -0.01343698 | KNDC1 |
| chr2 | 185738037 | 185739405 | 1369 | 18 | 1.26E-26 | 4.11E-17 | 3.16E-06 | 6.24E-22 | -0.00657002 | FSIP2, FSIP2-AS1, FSIP2-AS2 |
| chr2 | 86807677 | 86809959 | 2283 | 17 | 1.97E-21 | 2.65E-14 | 1.58E-06 | 8.02E-22 | -0.00981971 | CD8A |
| chr4 | 173526696 | 173531810 | 5115 | 41 | 2.93E-13 | 3.21E-16 | 3.04E-07 | 8.56E-22 | -0.00096744 | HAND2-AS1, HAND2 |
| chr12 | 6619852 | 6621433 | 1582 | 6 | 5.87E-19 | 8.59E-21 | 6.50E-07 | 1.47E-21 | -0.01546368 | LPAR5 |
| chr17 | 9958395 | 9960740 | 2346 | 15 | 1.62E-21 | 8.55E-21 | 4.58E-05 | 1.57E-21 | -0.00987093 | GAS7 |
| chr15 | 26628325 | 26629938 | 1614 | 14 | 1.71E-24 | 1.63E-20 | 2.95E-05 | 1.57E-21 | -0.02556102 | GABRB3 |
| chr1 | 78044693 | 78046445 | 1753 | 16 | 8.11E-26 | 5.66E-20 | 3.19E-05 | 1.59E-21 | -0.01028952 | GIPC2 |
| chr18 | 77248673 | 77250413 | 1741 | 31 | 1.29E-20 | 1.38E-16 | 1.04E-07 | 1.59E-21 | 0.002469846 | GALR1 |
| chr6 | 41100270 | 41101309 | 1040 | 10 | 7.06E-24 | 1.94E-21 | 7.30E-05 | 2.12E-21 | 0.028375006 | ADCY10P1, AL031778.1 |
| chr6 | 31968024 | 31973272 | 5249 | 105 | 3.27E-10 | 1.96E-15 | 0.003739 | 3.08E-21 | -0.00172472 | SKIV2L, STK19, DXO |
| chr2 | 15939565 | 15942909 | 3345 | 28 | 2.58E-18 | 2.30E-17 | 0.000202 | 3.11E-21 | 0.001686635 | MYCN, MYCNOS |
| chr8 | 100950473 | 100953511 | 3039 | 33 | 3.20E-15 | 2.56E-13 | 8.88E-06 | 3.28E-21 | 0.000485529 | YWHAZ |
| chr11 | 457243 | 458052 | 810 | 6 | 9.68E-28 | 2.11E-18 | 4.91E-08 | 3.83E-21 | 0.011424617 | PTDSS2 |
| chr19 | 39031591 | 39032986 | 1396 | 11 | 3.94E-25 | 4.00E-18 | 1.09E-05 | 4.10E-21 | -0.016644 | FBXO27 |
| chr12 | 70719287 | 70720207 | 921 | 5 | 3.63E-25 | 3.83E-21 | 5.60E-08 | 4.17E-21 | -0.02795304 | PTPRR |
| chr17 | 45143526 | 45147862 | 4337 | 27 | 8.10E-16 | 4.14E-14 | 3.18E-05 | 4.25E-21 | 0.002731462 | ACBD4, AC142472.1 |
| chr2 | 120738822 | 120742134 | 3313 | 10 | 2.73E-15 | 8.97E-20 | 2.59E-05 | 4.60E-21 | 0.033098281 | GLI2 |
| chr2 | 219433825 | 219435520 | 1696 | 14 | 2.61E-27 | 1.55E-18 | 2.38E-07 | 4.67E-21 | -0.00864064 | SPEG |
| chr12 | 57725760 | 57728558 | 2799 | 13 | 4.16E-16 | 8.50E-17 | 5.75E-07 | 4.67E-21 | -0.00843406 | AGAP2-AS1, AGAP2 |
| chr9 | 133199499 | 133200672 | 1174 | 6 | 3.46E-24 | 7.71E-20 | 5.79E-08 | 5.26E-21 | -0.03049544 | NA |
| chr1 | 154154612 | 154155967 | 1356 | 11 | 8.56E-24 | 1.63E-20 | 1.50E-05 | 5.64E-21 | 0.023178717 | NUP210L, TPM3 |
| chr19 | 55535322 | 55538101 | 2780 | 14 | 1.81E-22 | 3.61E-18 | 3.04E-06 | 7.60E-21 | -0.01013652 | SBK2 |
| chr11 | 65354189 | 65357298 | 3110 | 17 | 1.47E-15 | 1.11E-18 | 8.04E-05 | 7.60E-21 | -0.00456165 | DPF2, TIGD3 |
| chr4 | 3369261 | 3371279 | 2019 | 22 | 1.20E-26 | 1.36E-15 | 3.20E-06 | 8.06E-21 | 0.005847728 | RGS12 |
| chr17 | 82233761 | 82236182 | 2422 | 17 | 1.28E-20 | 1.47E-17 | 6.69E-07 | 9.15E-21 | -0.0062195 | SLC16A3 |
| chr12 | 131035338 | 131036642 | 1305 | 9 | 3.28E-18 | 4.19E-22 | 1.63E-05 | 9.30E-21 | 0.010802992 | ADGRD1, AC078925.1 |
| chr6 | 163352282 | 163356474 | 4193 | 11 | 1.70E-14 | 8.18E-20 | 3.04E-05 | 9.51E-21 | -0.01752365 | AL031121.3 |
| chr2 | 218291819 | 218292952 | 1134 | 12 | 9.54E-27 | 4.65E-13 | 4.74E-07 | 9.66E-21 | -0.00732829 | PNKD, TMBIM1 |
| chr1 | 25905974 | 25907987 | 2014 | 26 | 3.07E-29 | 1.30E-14 | 1.72E-05 | 1.00E-20 | -0.0072732 | STMN1, MIR3917 |
| chr11 | 66545281 | 66548399 | 3119 | 21 | 2.35E-16 | 2.00E-18 | 0.00012 | 1.02E-20 | -0.00350041 | ACTN3, ZDHHC24 |
| chr6 | 31136528 | 31140801 | 4274 | 28 | 3.78E-12 | 1.12E-18 | 4.30E-05 | 1.44E-20 | -0.00021173 | PSORS1C1, POLR2LP1, PSORS1C2 |
| chr11 | 2699580 | 2701483 | 1904 | 38 | 8.03E-21 | 7.42E-17 | 0.00032 | 1.66E-20 | -0.00331345 | KCNQ1, KCNQ1OT1 |
| chr19 | 11177713 | 11178700 | 988 | 9 | 1.80E-24 | 1.83E-15 | 8.97E-08 | 1.71E-20 | -0.01003019 | KANK2 |
| chr16 | 85484673 | 85487434 | 2762 | 14 | 2.18E-16 | 1.97E-21 | 0.000244 | 1.94E-20 | -0.01156497 | GSE1 |
| chr19 | 3006113 | 3007746 | 1634 | 8 | 9.19E-25 | 9.26E-16 | 6.75E-07 | 2.25E-20 | -0.01965332 | TLE2 |
| chr6 | 30100879 | 30104607 | 3729 | 57 | 4.72E-22 | 2.06E-13 | 0.000102 | 2.28E-20 | -3.89E-05 | TRIM31 |
| chr17 | 7929161 | 7933627 | 4467 | 35 | 1.28E-12 | 9.86E-20 | 0.002766 | 2.88E-20 | -0.00524548 | CNTROB, KCNAB3, AC104581.2, TRAPPC1 |

| | | | | | | | | | | |
|---|---|---|---|---|---|---|---|---|---|---|
| chr4 | 98142951 | 98144291 | 1341 | 11 | 4.22E-20 | 1.93E-19 | 3.74E-05 | 3.51E-20 | -0.01749689 | STPG2 |
| chr8 | 49909344 | 49912189 | 2846 | 25 | 2.67E-18 | 5.23E-15 | 1.70E-05 | 3.76E-20 | 0.002753785 | SNTG1 |
| chr8 | 22365718 | 22367521 | 1804 | 16 | 3.24E-29 | 4.61E-13 | 6.45E-06 | 4.37E-20 | -0.01012273 | SLC39A14 |
| chr8 | 124997239 | 125000192 | 2954 | 18 | 7.21E-17 | 1.92E-13 | 4.25E-06 | 4.45E-20 | -0.00526342 | SQLE, AC009908.1 |
| chr2 | 217977429 | 217979479 | 2051 | 9 | 3.14E-21 | 3.84E-20 | 3.45E-06 | 5.49E-20 | -0.02470001 | TNS1-AS1, TNS1 |
| chr12 | 52040896 | 52044434 | 3539 | 12 | 2.46E-11 | 1.12E-18 | 4.70E-06 | 5.68E-20 | -0.00409654 | NR4A1 |
| chr19 | 57708712 | 57710254 | 1543 | 13 | 1.85E-20 | 2.37E-20 | 0.000101 | 5.75E-20 | -0.00995787 | ZNF551, AC003006.1, ZNF154 |
| chr11 | 2898459 | 2899946 | 1488 | 18 | 8.54E-22 | 1.65E-17 | 1.36E-06 | 6.10E-20 | 0.008952567 | SLC22A18, SLC22A18AS |
| chr7 | 1557765 | 1559686 | 1922 | 21 | 3.21E-26 | 9.08E-15 | 1.03E-05 | 6.74E-20 | -0.00801635 | TMEM184A |
| chr6 | 32402860 | 32407309 | 4450 | 21 | 2.18E-08 | 6.51E-16 | 6.07E-05 | 6.78E-20 | -0.00784108 | TSBP1-AS1, BTNL2 |
| chr6 | 106509124 | 106510599 | 1476 | 10 | 9.28E-28 | 1.48E-14 | 1.10E-07 | 6.99E-20 | -0.01406709 | CRYBG1 |
| chr17 | 44353741 | 44355673 | 1933 | 8 | 6.32E-23 | 5.74E-19 | 3.07E-06 | 7.65E-20 | -0.02407873 | FAM171A2 |
| chr9 | 128191264 | 128193778 | 2515 | 19 | 6.97E-18 | 1.13E-15 | 0.000238 | 7.70E-20 | -0.01172934 | DNM1, CIZ1 |
| chr11 | 64637582 | 64638579 | 998 | 8 | 3.85E-24 | 1.30E-11 | 1.75E-06 | 1.04E-19 | -0.01501733 | NRXN2 |
| chr7 | 1053321 | 1059440 | 6120 | 36 | 7.24E-08 | 2.69E-19 | 0.000184 | 1.14E-19 | -0.00532439 | GPR146, C7orf50, AC073957.2 |
| chr16 | 87957087 | 87959942 | 2856 | 17 | 4.76E-16 | 2.39E-18 | 0.000363 | 1.20E-19 | 0.01075398 | BANP |
| chr1 | 67830496 | 67833979 | 3484 | 19 | 1.35E-16 | 4.93E-15 | 1.00E-05 | 1.23E-19 | -0.00504664 | GNG12-AS1, GNG12 |
| chr2 | 231900464 | 231901044 | 581 | 5 | 4.92E-23 | 1.50E-17 | 1.83E-07 | 1.23E-19 | -0.0152379 | NA |
| chr4 | 53591478 | 53592842 | 1365 | 14 | 8.83E-18 | 2.15E-16 | 0.000159 | 1.59E-19 | -0.01468386 | AC058822.1, LNX1 |
| chr6 | 33199834 | 33202400 | 2567 | 46 | 2.31E-23 | 6.05E-13 | 8.05E-07 | 1.77E-19 | -0.00131767 | SLC39A7, RXRB |
| chr18 | 78725100 | 78727352 | 2253 | 6 | 2.84E-15 | 1.34E-20 | 4.67E-06 | 1.85E-19 | -0.01051454 | NA |
| chr1 | 43338192 | 43340047 | 1856 | 7 | 4.74E-25 | 6.03E-13 | 1.60E-08 | 2.02E-19 | 0.00748587 | MPL |
| chr12 | 51887191 | 51888420 | 1230 | 12 | 2.84E-22 | 6.54E-16 | 3.21E-05 | 2.03E-19 | -0.01741805 | ANKRD33 |
| chr10 | 133163793 | 133168297 | 4505 | 20 | 4.87E-12 | 7.98E-16 | 1.33E-05 | 2.03E-19 | -0.00542773 | KNDC1 |
| chr13 | 44159809 | 44162096 | 2288 | 8 | 7.06E-19 | 3.19E-13 | 4.72E-08 | 2.30E-19 | 0.00072287 | SMIM2-AS1, SMIM2 |
| chr16 | 24844783 | 24846280 | 1498 | 12 | 5.85E-18 | 1.65E-18 | 1.05E-05 | 2.52E-19 | -0.01310788 | SLC5A11 |
| chr8 | 123181444 | 123183195 | 1752 | 19 | 9.72E-19 | 3.06E-16 | 3.04E-05 | 2.75E-19 | -0.01195002 | FAM83A, AC068228.1 |
| chr2 | 127810622 | 127811939 | 1318 | 11 | 4.45E-24 | 5.16E-16 | 2.95E-07 | 2.98E-19 | -0.01096832 | WDR33 |
| chr6 | 43769070 | 43770703 | 1634 | 13 | 1.94E-23 | 1.87E-16 | 2.21E-06 | 3.03E-19 | -0.00180127 | VEGFA, AL136131.3 |
| chr19 | 50357475 | 50359032 | 1558 | 9 | 1.72E-21 | 3.34E-17 | 7.88E-06 | 3.33E-19 | -0.02433451 | NR1H2, NAPSA |
| chr2 | 128900667 | 128902372 | 1706 | 6 | 1.11E-19 | 8.54E-20 | 3.76E-06 | 3.44E-19 | -0.02977356 | NA |
| chr17 | 29004767 | 29007146 | 2380 | 22 | 1.49E-18 | 8.54E-20 | 4.94E-05 | 3.53E-19 | 0.000148613 | PIPOX, SEZ6 |
| chr19 | 3480365 | 3480942 | 578 | 8 | 1.06E-19 | 2.78E-21 | 0.000103 | 3.64E-19 | -0.03798393 | SMIM24 |
| chr15 | 85627824 | 85628490 | 667 | 3 | 6.21E-21 | 7.71E-20 | 5.60E-08 | 3.75E-19 | -0.03292711 | AKAP13, AC087286.2 |
| chr13 | 98977716 | 98979277 | 1562 | 9 | 5.96E-18 | 1.19E-15 | 3.07E-06 | 3.78E-19 | -0.00468792 | DOCK9 |
| chr16 | 31107465 | 31109328 | 1864 | 15 | 3.29E-18 | 6.77E-14 | 1.80E-05 | 3.90E-19 | -0.00502825 | BCKDK |
| chr2 | 68837228 | 68838309 | 1082 | 5 | 3.92E-21 | 9.85E-19 | 4.29E-07 | 3.94E-19 | -0.02317471 | LINC01888, LINC01890 |
| chr16 | 2170245 | 2175995 | 5751 | 28 | 7.91E-10 | 1.83E-14 | 2.16E-05 | 4.66E-19 | -0.00257147 | TRAF7 |
| chr2 | 176149281 | 176152902 | 3622 | 32 | 1.88E-15 | 2.60E-13 | 0.000276 | 5.18E-19 | 0.007697865 | HOXD3, MIR10B, HOXD4 |
| chr6 | 155216461 | 155217850 | 1390 | 9 | 1.17E-21 | 5.23E-15 | 1.68E-07 | 5.23E-19 | -0.013803 | TIAM2 |
| chr16 | 10180207 | 10183160 | 2954 | 26 | 5.39E-14 | 1.64E-16 | 0.000468 | 5.62E-19 | 0.000902873 | GRIN2A |
| chr16 | 2209922 | 2213060 | 3139 | 22 | 1.94E-12 | 3.59E-19 | 0.000309 | 6.06E-19 | -0.0037853 | AC009065.7, BRICD5, PGP |
| chr3 | 66220292 | 66221930 | 1639 | 17 | 6.89E-19 | 4.14E-18 | 3.38E-05 | 6.15E-19 | -0.00817763 | SLC25A26 |
| chr5 | 41869271 | 41871235 | 1965 | 18 | 1.78E-16 | 5.00E-20 | 0.001163 | 6.18E-19 | -0.01056281 | AC034222.2, OXCT1-AS1, OXCT1 |
| chr1 | 35571982 | 35574284 | 2303 | 15 | 4.53E-20 | 6.37E-15 | 4.80E-06 | 6.34E-19 | -0.00959886 | TFAP2E, TFAP2E-AS1 |
| chr6 | 170248014 | 170248975 | 962 | 5 | 3.65E-21 | 1.21E-19 | 3.37E-07 | 6.57E-19 | 0.040978605 | NA |
| chr12 | 54378904 | 54379752 | 849 | 4 | 8.99E-22 | 4.28E-19 | 1.18E-07 | 7.51E-19 | -0.02774918 | AC079313.2, AC079313.1, ZNF385A |
| chr20 | 64047902 | 64050075 | 2174 | 14 | 3.76E-21 | 8.77E-15 | 1.10E-05 | 7.51E-19 | -0.00921705 | TCEA2, SOX18 |
| chr1 | 95232822 | 95234309 | 1488 | 17 | 1.93E-25 | 1.83E-15 | 2.53E-05 | 8.25E-19 | -0.01372498 | TLCD4-RWDD3, RWDD3, AC092802.1 |
| chr17 | 40013825 | 40015444 | 1620 | 12 | 2.38E-19 | 1.70E-18 | 2.52E-05 | 8.25E-19 | -0.00694241 | CSF3, AC090844.3 |
| chr1 | 1354415 | 1355684 | 1270 | 10 | 2.03E-18 | 8.71E-21 | 0.000125 | 8.49E-19 | -0.02808399 | MXRA8 |
| chr10 | 21496632 | 21501073 | 4442 | 23 | 1.29E-10 | 9.40E-13 | 4.04E-06 | 8.49E-19 | -0.00089662 | MIR1915HG, MIR1915 |
| chr12 | 113148946 | 113150458 | 1513 | 8 | 1.90E-19 | 2.11E-18 | 9.23E-06 | 8.75E-19 | -0.01064967 | CFAP73 |
| chr4 | 52861264 | 52862756 | 1493 | 7 | 1.18E-22 | 3.70E-16 | 4.07E-07 | 8.91E-19 | -0.02480238 | LINC01618, RASL11B |
| chr6 | 36959052 | 36959472 | 421 | 4 | 4.61E-23 | 1.50E-17 | 3.58E-08 | 9.06E-19 | -0.01368653 | PI16 |
| chr10 | 117541272 | 117547148 | 5877 | 37 | 7.77E-10 | 1.27E-15 | 6.65E-05 | 9.62E-19 | -0.00088518 | EMX2, EMX2OS |
| chr17 | 55722091 | 55723329 | 1239 | 8 | 5.56E-22 | 8.74E-16 | 1.32E-05 | 9.69E-19 | -0.01780728 | TMEM100 |
| chr19 | 11236683 | 11239951 | 3269 | 18 | 5.32E-15 | 6.22E-16 | 6.05E-06 | 1.03E-18 | -0.00515428 | ANGPTL8, DOCK6 |
| chr7 | 108001045 | 108004125 | 3081 | 22 | 1.92E-18 | 2.16E-15 | 7.11E-07 | 1.12E-18 | -0.00171078 | LAMB1 |
| chr1 | 26094617 | 26094858 | 242 | 2 | 4.56E-22 | 4.97E-19 | 5.44E-09 | 1.20E-18 | -0.03310727 | NA |
| chr6 | 33188387 | 33191576 | 3190 | 23 | 2.98E-14 | 2.16E-17 | 0.00019 | 1.26E-18 | -0.00238395 | COL11A2 |
| chr17 | 82917617 | 82918521 | 905 | 8 | 1.81E-19 | 2.07E-19 | 6.51E-06 | 1.31E-18 | -0.01251489 | TBCD, AC087222.1 |
| chr21 | 33641521 | 33644567 | 3047 | 23 | 3.12E-12 | 9.11E-14 | 2.24E-06 | 1.40E-18 | 0.001726962 | ITSN1, AP000311.1, CRYZL1 |
| chr12 | 113056226 | 113058363 | 2138 | 12 | 1.81E-16 | 1.25E-15 | 1.31E-05 | 1.42E-18 | -0.00550479 | DTX1 |
| chr16 | 67395547 | 67396551 | 1005 | 5 | 1.46E-26 | 1.46E-13 | 3.82E-09 | 1.53E-18 | -0.01778531 | ZDHHC1 |
| chr11 | 44309812 | 44311642 | 1831 | 41 | 1.32E-17 | 3.84E-14 | 0.000158 | 1.59E-18 | -0.00024661 | ALX4 |
| chr10 | 1240639 | 1242418 | 1780 | 10 | 4.51E-15 | 1.10E-18 | 3.48E-05 | 1.70E-18 | 0.004611555 | LINC00200, ADARB2 |
| chr5 | 142587196 | 142587485 | 290 | 2 | 5.99E-23 | 2.11E-18 | 3.03E-10 | 1.72E-18 | -0.04086833 | SPRY4-AS1 |
| chr21 | 44284625 | 44286830 | 2206 | 15 | 4.72E-17 | 8.26E-20 | 0.001069 | 1.96E-18 | -0.01131179 | AIRE |
| chr10 | 132064866 | 132067111 | 2246 | 11 | 5.82E-26 | 1.19E-11 | 3.22E-08 | 1.96E-18 | 0.007707552 | JAKMIP3 |
| chr14 | 100725535 | 100729793 | 4259 | 23 | 4.85E-11 | 2.06E-14 | 2.71E-05 | 2.03E-18 | 0.006412149 | DLK1 |
| chr3 | 46676879 | 46678716 | 1838 | 9 | 2.72E-20 | 1.21E-12 | 4.67E-08 | 2.14E-18 | -0.00632458 | ALS2CL |
| chr17 | 7853892 | 7859098 | 5207 | 45 | 3.76E-10 | 8.04E-13 | 2.95E-05 | 2.22E-18 | -0.00030724 | KDM6B, TMEM88, CYB5D1, AC104581.4, NAA38 |
| chr6 | 32128202 | 32130476 | 2275 | 49 | 4.74E-22 | 2.59E-10 | 3.20E-06 | 2.24E-18 | -0.00060756 | ATF6B, FKBPL |
| chr6 | 106910661 | 106911426 | 766 | 2 | 4.27E-18 | 1.02E-18 | 4.22E-09 | 2.24E-18 | 0.029361247 | NA |
| chr4 | 1172482 | 1173926 | 1445 | 17 | 9.06E-23 | 2.97E-13 | 9.43E-06 | 2.24E-18 | -0.00596208 | SPON2 |
| chr6 | 30716563 | 30722067 | 5505 | 81 | 3.25E-09 | 9.57E-12 | 0.000399 | 2.30E-18 | -0.00101891 | TUBB, MDC1 |
| chr11 | 68683150 | 68685470 | 2321 | 14 | 3.31E-21 | 4.19E-13 | 1.06E-06 | 2.37E-18 | -0.0046914 | GAL |
| chr3 | 27719917 | 27724891 | 4975 | 37 | 6.43E-10 | 2.48E-16 | 0.00041 | 2.41E-18 | -0.00303354 | EOMES |
| chr2 | 85611353 | 85612599 | 1247 | 18 | 1.60E-24 | 2.86E-11 | 2.76E-07 | 2.56E-18 | -0.00393619 | USP39, C2orf68 |
| chr6 | 31571762 | 31575909 | 4148 | 37 | 1.94E-12 | 1.13E-13 | 0.000161 | 2.76E-18 | -0.00176657 | LTA, TNF |
| chr6 | 32086784 | 32090619 | 3836 | 45 | 4.84E-15 | 8.04E-13 | 1.09E-05 | 2.93E-18 | -0.00294864 | TNXB |
| chr9 | 113563741 | 113565462 | 1722 | 10 | 1.89E-17 | 2.02E-19 | 3.57E-05 | 3.13E-18 | -0.01299076 | RGS3 |
| chr1 | 236393865 | 236396375 | 2511 | 21 | 1.67E-12 | 6.92E-10 | 2.41E-05 | 3.36E-18 | -0.00781995 | EDARADD |
| chr18 | 22417093 | 22419206 | 2114 | 12 | 4.53E-21 | 2.12E-13 | 4.79E-08 | 3.37E-18 | -0.00762943 | CTAGE1, ATP7BP1 |
| chr16 | 3443133 | 3444155 | 1023 | 15 | 6.73E-18 | 1.47E-16 | 2.74E-05 | 3.95E-18 | -0.01056095 | AC025283.2, NAA60, ZNF597 |
| chr12 | 124543248 | 124544461 | 1214 | 7 | 6.41E-23 | 9.65E-13 | 1.75E-06 | 4.22E-18 | 0.014601345 | NCOR2 |
| chr3 | 39105640 | 39108753 | 3114 | 21 | 1.13E-14 | 2.79E-13 | 1.54E-06 | 4.32E-18 | 0.000247836 | TTC21A, GORASP1 |
| chr14 | 89700897 | 89702458 | 1562 | 11 | 1.49E-19 | 1.30E-14 | 5.41E-05 | 4.61E-18 | 0.001778706 | AL137230.2 |
| chr11 | 3232088 | 3233274 | 1187 | 15 | 3.81E-18 | 1.34E-17 | 0.000319 | 4.70E-18 | -0.01020399 | AC109309.1, MRGPRE |
| chr15 | 67063159 | 67065023 | 1865 | 7 | 1.06E-17 | 9.10E-15 | 4.89E-05 | 4.83E-18 | -0.07568435 | SMAD3, AC087482.1 |
| chr10 | 124741572 | 124744165 | 2594 | 12 | 5.25E-16 | 5.73E-14 | 6.01E-06 | 5.26E-18 | -0.00201512 | AC068896.1, FAM53B |
| chr17 | 8867591 | 8869102 | 1512 | 11 | 3.01E-17 | 5.23E-15 | 1.69E-06 | 5.78E-18 | 0.007618368 | PIK3R6 |
| chr1 | 65791228 | 65793002 | 1775 | 12 | 3.59E-20 | 9.55E-17 | 2.51E-05 | 6.04E-18 | -0.00889765 | PDE4B |
| chr13 | 111066590 | 111067684 | 1095 | 8 | 2.35E-22 | 9.38E-16 | 1.74E-06 | 6.23E-18 | -0.01057705 | NA |
| chr13 | 51845129 | 51845592 | 464 | 9 | 6.25E-19 | 1.58E-18 | 1.66E-05 | 6.52E-18 | -0.01181527 | AL162377.3, TMEM272 |
| chr11 | 117876315 | 117878709 | 2395 | 24 | 7.26E-16 | 5.36E-15 | 5.80E-05 | 6.87E-18 | -0.00591833 | FXYD6-FXYD2, FXYD6 |
| chr6 | 31157860 | 31160086 | 2227 | 40 | 3.02E-23 | 8.20E-11 | 0.000177 | 6.96E-18 | -0.00358956 | TCF19, CCHCR1 |
| chr10 | 132627581 | 132629157 | 1577 | 4 | 2.32E-20 | 8.16E-17 | 3.31E-09 | 7.44E-18 | -0.02039362 | INPP5A |
| chr6 | 29748759 | 29753238 | 4480 | 42 | 1.14E-12 | 1.52E-13 | 3.05E-05 | 7.78E-18 | -0.00231269 | Y_RNA, AL645939.5, HLA-F-AS1, MICE, IFITM4P, AL645939.4 |
| chr7 | 2277001 | 2277940 | 940 | 10 | 6.10E-23 | 2.68E-12 | 3.04E-07 | 7.79E-18 | -0.01980954 | SNX8 |
| chr10 | 131979938 | 131982119 | 2182 | 13 | 3.47E-17 | 1.39E-14 | 1.37E-06 | 7.79E-18 | -0.01056135 | AL162274.1, BNIP3 |
| chr15 | 40357133 | 40358929 | 1797 | 12 | 3.65E-15 | 1.68E-15 | 2.68E-05 | 7.79E-18 | -0.00139419 | DISP2 |
| chr17 | 74736293 | 74737739 | 1447 | 18 | 2.96E-18 | 4.07E-17 | 4.34E-06 | 8.29E-18 | -2.95E-07 | RAB37 |
| chr2 | 112541078 | 112543464 | 2387 | 23 | 1.61E-16 | 5.90E-13 | 1.60E-06 | 9.27E-18 | 0.001869708 | TTL, POLR1B |
| chr12 | 50102423 | 50105622 | 3200 | 15 | 2.61E-14 | 9.02E-13 | 1.04E-07 | 9.37E-18 | -0.00592655 | GPD1 |
| chr10 | 131997955 | 131998108 | 154 | 3 | 1.04E-20 | 2.16E-17 | 3.59E-08 | 9.53E-18 | -0.02710303 | AL162274.3 |
| chr9 | 128880078 | 128884149 | 4072 | 20 | 1.37E-09 | 1.03E-11 | 4.03E-06 | 1.03E-17 | -0.00214186 | LRRC8A, AL441992.2, KYAT1 |
| chr15 | 48330601 | 48334980 | 4380 | 24 | 2.73E-11 | 2.18E-14 | 0.000468 | 1.13E-17 | -0.00238518 | DUT, AC023355.1 |
| chr17 | 38967385 | 38968448 | 1064 | 13 | 5.70E-19 | 4.23E-18 | 0.000367 | 1.20E-17 | -0.01723141 | FBXO47 |
| chr2 | 1744340 | 1745304 | 965 | 8 | 6.44E-22 | 2.37E-15 | 3.08E-07 | 1.23E-17 | -0.01085664 | PXDN |
| chr15 | 70096223 | 70099330 | 3108 | 30 | 4.04E-15 | 1.91E-11 | 1.12E-06 | 1.31E-17 | 0.001833293 | TLE3 |
| chr9 | 33238802 | 33240962 | 2161 | 7 | 1.85E-18 | 7.81E-16 | 1.08E-06 | 1.34E-17 | -0.00880636 | SPINK4 |
| chr19 | 49063755 | 49065705 | 1951 | 10 | 1.68E-17 | 3.65E-14 | 1.17E-05 | 1.34E-17 | -0.01379668 | AC008687.3, AC008687.8, NTF4 |
| chr18 | 36127954 | 36130820 | 2867 | 25 | 1.18E-17 | 6.10E-13 | 5.95E-06 | 1.38E-17 | 0.000185627 | ELP2, SLC39A6 |
| chr11 | 27718280 | 27723010 | 4731 | 31 | 1.06E-09 | 1.17E-14 | 9.17E-05 | 1.53E-17 | 0.001869978 | BDNF, AC103796.1 |
| chr1 | 150256748 | 150258387 | 1640 | 11 | 3.97E-17 | 2.32E-16 | 0.000127 | 1.56E-17 | -0.01059175 | CA14, AC242988.1 |
| chr10 | 99521846 | 99524090 | 2245 | 15 | 1.20E-14 | 1.19E-17 | 5.13E-05 | 1.75E-17 | -0.0172145 | NA |
| chr2 | 202033626 | 202036747 | 3122 | 16 | 2.44E-11 | 1.00E-14 | 6.83E-05 | 1.84E-17 | -0.01708181 | FZD7 |
| chr12 | 29380524 | 29382049 | 1526 | 14 | 7.58E-16 | 4.89E-11 | 2.69E-07 | 1.90E-17 | -0.00702126 | ERGIC2 |
| chr11 | 18633339 | 18635728 | 2390 | 14 | 1.41E-17 | 2.18E-12 | 1.42E-07 | 1.98E-17 | -0.00462684 | SPTY2D1 |
| chr10 | 129961203 | 129966155 | 4953 | 27 | 1.49E-08 | 5.09E-15 | 9.09E-05 | 2.07E-17 | -0.00101366 | EBF3 |
| chr11 | 70824300 | 70824960 | 661 | 4 | 9.61E-21 | 1.07E-16 | 5.44E-07 | 2.14E-17 | 0.028677639 | SHANK2 |
| chr17 | 82331561 | 82334727 | 3167 | 17 | 1.81E-18 | 3.41E-12 | 2.75E-08 | 2.22E-17 | -0.005301 | SECTM1 |
| chr7 | 45920768 | 45922977 | 2210 | 35 | 8.83E-18 | 3.61E-15 | 6.75E-05 | 2.65E-17 | 0.001948855 | IGFBP3 |
| chr6 | 149864052 | 149865352 | 1301 | 15 | 5.10E-23 | 1.75E-14 | 1.27E-07 | 2.66E-17 | -0.00264877 | RAET1E-AS1, AL355312.5, LRP11 |
| chr6 | 82362579 | 82365470 | 2892 | 23 | 4.13E-13 | 1.30E-15 | 0.000337 | 2.70E-17 | -0.00457492 | TPBG, AL121977.2 |
| chr4 | 8072010 | 8072805 | 796 | 11 | 4.32E-19 | 8.90E-14 | 2.87E-05 | 2.72E-17 | 0.009127852 | ABLIM2 |
| chr10 | 122162796 | 122164003 | 1208 | 13 | 3.60E-26 | 1.19E-10 | 4.20E-07 | 2.75E-17 | -0.00982705 | TACC2 |
| chr3 | 57757131 | 57757872 | 742 | 10 | 1.51E-19 | 3.17E-15 | 2.42E-05 | 2.91E-17 | -0.01146743 | SLMAP |
| chr5 | 126599667 | 126601449 | 1783 | 13 | 6.09E-17 | 7.71E-18 | 0.000313 | 3.07E-17 | -0.00644162 | PHAX |
| chr2 | 207770881 | 207771639 | 759 | 9 | 2.31E-22 | 9.17E-11 | 3.93E-07 | 3.14E-17 | -0.00583705 | NA |
| chr10 | 624847 | 625688 | 842 | 7 | 1.49E-19 | 5.44E-15 | 9.47E-06 | 3.20E-17 | 0.020459014 | DIP2C |
| chr18 | 46656884 | 46658400 | 1517 | 12 | 1.36E-19 | 3.47E-16 | 8.21E-06 | 3.33E-17 | -0.0058837 | LOXHD1 |
| chr8 | 41306534 | 41310962 | 4429 | 27 | 4.90E-15 | 5.80E-11 | 3.39E-07 | 3.56E-17 | -0.00324902 | SFRP1 |
| chr9 | 114503718 | 114506112 | 2395 | 17 | 4.83E-17 | 8.90E-16 | 2.47E-06 | 3.68E-17 | -0.00408542 | WHRN |
| chr7 | 2607955 | 2610498 | 2544 | 17 | 3.15E-11 | 4.77E-17 | 0.000179 | 3.68E-17 | 0.002138556 | IQCE |
| chr9 | 126479497 | 126481631 | 2135 | 8 | 8.53E-20 | 1.07E-14 | 3.75E-07 | 3.88E-17 | -0.01200915 | MVB12B |
| chr22 | 45162728 | 45165040 | 2313 | 12 | 2.43E-13 | 1.21E-13 | 6.33E-07 | 3.97E-17 | -0.00090124 | NUP50, NUP50-DT |
| chr3 | 42905280 | 42907590 | 2311 | 12 | 1.69E-18 | 1.32E-13 | 3.39E-06 | 4.04E-17 | -0.00896881 | AC099329.2, AC092042.3, KRBOX1, ZNF662 |
| chr2 | 20676460 | 20677795 | 1336 | 5 | 2.34E-16 | 1.30E-15 | 3.51E-07 | 4.11E-17 | -0.01316205 | GDF7 |
| chr10 | 130188928 | 130191585 | 2658 | 14 | 6.85E-13 | 1.58E-15 | 8.54E-05 | 4.19E-17 | -0.01263348 | NA |
| chr19 | 45900773 | 45903583 | 2811 | 17 | 4.66E-21 | 2.03E-10 | 3.60E-07 | 4.56E-17 | -0.00268365 | MYPOP |
| chr7 | 4631629 | 4633808 | 2180 | 7 | 6.22E-14 | 1.42E-17 | 5.15E-05 | 5.03E-17 | -0.01332472 | NA |
| chr1 | 204869749 | 204870747 | 999 | 6 | 9.03E-21 | 3.83E-12 | 8.14E-08 | 5.09E-17 | -0.0122345 | NFASC |
| chr8 | 143378960 | 143381729 | 2770 | 15 | 8.61E-14 | 1.86E-15 | 5.71E-06 | 5.32E-17 | -9.83E-05 | RHPN1 |
| chr1 | 112743318 | 112744616 | 1299 | 7 | 3.71E-19 | 4.18E-16 | 1.38E-05 | 5.56E-17 | -0.0151383 | NA |
| chr2 | 235384227 | 235384966 | 740 | 5 | 9.03E-20 | 7.64E-16 | 1.44E-07 | 5.59E-17 | -0.00890985 | NA |
| chr1 | 27321210 | 27325108 | 3899 | 18 | 1.00E-10 | 3.62E-17 | 0.000273 | 5.87E-17 | -0.00236998 | TMEM222 |
| chr11 | 62705516 | 62710152 | 4637 | 31 | 7.61E-10 | 1.20E-13 | 1.97E-05 | 5.95E-17 | -0.00063736 | GNG3, BSCL2, HNRNPUL2-BSCL2 |
| chr2 | 239318494 | 239319523 | 1030 | 4 | 3.28E-22 | 1.25E-15 | 9.86E-07 | 6.08E-17 | -0.09846288 | HDAC4 |
| chr2 | 9472993 | 9474615 | 1623 | 15 | 4.67E-21 | 9.74E-14 | 0.000199 | 6.08E-17 | -0.0110219 | CPSF3, IAH1 |
| chr5 | 149140209 | 149142060 | 1852 | 17 | 1.65E-22 | 3.17E-13 | 8.12E-06 | 6.67E-17 | -0.00771568 | ABLIM3 |
| chr12 | 50842276 | 50843923 | 1648 | 11 | 3.25E-17 | 2.77E-14 | 6.10E-06 | 6.73E-17 | -0.01240577 | TMPRSS12 |
| chr6 | 170162451 | 170163230 | 780 | 9 | 1.76E-19 | 1.17E-14 | 5.53E-08 | 7.02E-17 | 0.010769876 | AL596442.1 |
| chr6 | 31727775 | 31731122 | 3348 | 59 | 1.17E-10 | 2.12E-12 | 0.00048 | 7.02E-17 | -0.00241911 | DDAH2, CLIC1 |
| chr16 | 4112202 | 4115885 | 3684 | 16 | 3.23E-09 | 3.13E-15 | 1.52E-05 | 7.02E-17 | -0.0048356 | ADCY9 |
| chr2 | 172092540 | 172096265 | 3726 | 14 | 2.01E-11 | 6.16E-12 | 2.81E-05 | 7.38E-17 | -0.0045903 | NA |

| | | | | | | | | | | |
|---|---|---|---|---|---|---|---|---|---|---|
| chr20 | 345821 | 348372 | 2552 | 15 | 3.38E-16 | 1.25E-15 | 0.000266 | 7.64E-17 | -0.00199657 | NRSN2, NRSN2-AS1 |
| chr5 | 172769150 | 172772664 | 3515 | 33 | 2.89E-13 | 2.80E-10 | 3.41E-05 | 7.82E-17 | 0.000607928 | AC022217.3, DUSP1 |
| chr8 | 66428513 | 66433532 | 5020 | 29 | 1.65E-11 | 1.57E-13 | 5.72E-06 | 7.82E-17 | -0.00319085 | RRS1, AC009879.2, AC009879.3, ADHFE1, AC009879.4, RRS1-AS1 |
| chr5 | 150946020 | 150947300 | 1281 | 9 | 6.60E-17 | 1.28E-17 | 7.55E-05 | 8.70E-17 | -0.01552615 | ZNF300P1 |
| chr10 | 121973878 | 121975828 | 1951 | 17 | 1.14E-14 | 7.33E-15 | 9.53E-05 | 8.78E-17 | -0.00527752 | NSMCE4A |
| chr4 | 1368431 | 1370725 | 2295 | 13 | 8.71E-14 | 5.20E-13 | 4.13E-06 | 9.06E-17 | -0.0032947 | UVSSA |
| chr7 | 94655958 | 94657930 | 1973 | 62 | 4.84E-18 | 2.30E-13 | 0.000271 | 9.26E-17 | -0.00289039 | PEG10, SGCE |
| chr15 | 38672882 | 38673218 | 337 | 4 | 3.83E-19 | 2.21E-17 | 1.85E-06 | 9.35E-17 | 0.026702071 | LINC02694 |
| chr15 | 101188029 | 101189746 | 1718 | 11 | 1.17E-17 | 2.53E-14 | 7.10E-06 | 9.35E-17 | -0.00862537 | CHSY1 |
| chr4 | 83010560 | 83013803 | 3244 | 21 | 6.15E-14 | 3.66E-14 | 2.45E-05 | 1.01E-16 | 0.003864307 | LIN54 |
| chr14 | 21089479 | 21091465 | 1987 | 9 | 2.30E-15 | 1.13E-14 | 1.35E-06 | 1.02E-16 | -0.00455939 | ARHGEF40, ZNF219 |
| chr5 | 157264745 | 157268541 | 3797 | 22 | 4.19E-10 | 4.33E-15 | 0.000102 | 1.06E-16 | 0.004515747 | CYFIP2, AC009185.1 |
| chr6 | 30741651 | 30745665 | 4015 | 70 | 2.77E-14 | 1.90E-11 | 0.001911 | 1.08E-16 | -0.00040603 | IER3-AS1, HCG20, FLOT1, IER3 |
| chr16 | 50287479 | 50288245 | 767 | 7 | 2.37E-20 | 5.07E-14 | 1.71E-06 | 1.09E-16 | 0.011528705 | ADCY7 |
| chr5 | 1168117 | 1170269 | 2153 | 9 | 3.11E-16 | 1.22E-12 | 9.64E-06 | 1.09E-16 | -0.00615515 | NA |
| chr6 | 34514034 | 34514902 | 869 | 10 | 4.59E-16 | 4.13E-18 | 0.000304 | 1.09E-16 | -0.03327315 | PACSIN1 |
| chr1 | 20184228 | 20188029 | 3802 | 17 | 3.96E-11 | 4.41E-09 | 6.37E-07 | 1.09E-16 | -0.00317002 | UBXN10, PLA2G2C |
| chr15 | 73333545 | 73335271 | 1727 | 7 | 1.83E-13 | 2.37E-16 | 9.10E-06 | 1.09E-16 | -0.01420334 | AC068397.2, HCN4 |
| chr1 | 54885986 | 54888538 | 2553 | 14 | 1.82E-12 | 1.02E-15 | 8.17E-05 | 1.11E-16 | -0.00392454 | AC096536.2, DHCR24-DT, DHCR24 |
| chr5 | 168291119 | 168292737 | 1619 | 16 | 1.77E-16 | 3.44E-16 | 9.56E-05 | 1.29E-16 | -0.00034326 | WWC1 |
| chr19 | 29872109 | 29874048 | 1940 | 9 | 2.60E-17 | 7.10E-12 | 4.71E-06 | 1.35E-16 | -0.01633933 | NA |
| chr7 | 126377778 | 126378732 | 955 | 4 | 2.94E-19 | 2.16E-15 | 1.41E-07 | 1.39E-16 | -0.0114492 | NA |
| chr3 | 10108119 | 10108803 | 685 | 11 | 2.75E-17 | 2.15E-17 | 4.30E-05 | 1.40E-16 | -0.00789537 | FANCD2OS |
| chr8 | 144500019 | 144501305 | 1287 | 6 | 4.34E-18 | 6.88E-17 | 9.83E-06 | 1.42E-16 | 0.022546646 | PPP1R16A, AC084125.2 |
| chr1 | 8423330 | 8425481 | 2152 | 17 | 3.03E-20 | 1.57E-10 | 3.21E-05 | 1.51E-16 | -0.01228107 | RERE-AS1, RERE |
| chr5 | 45694421 | 45697197 | 2777 | 13 | 3.82E-11 | 3.26E-14 | 1.17E-05 | 1.62E-16 | -0.01055502 | HCN1 |
| chr2 | 238644242 | 238644765 | 524 | 3 | 4.03E-20 | 1.19E-15 | 6.25E-08 | 1.74E-16 | -0.12660972 | LINC01937 |
| chr12 | 117097437 | 117100522 | 3086 | 12 | 2.84E-10 | 3.18E-13 | 0.000104 | 1.74E-16 | -0.00462603 | TESC-AS1, TESC |
| chr19 | 38975181 | 38976795 | 1615 | 15 | 3.58E-20 | 1.66E-12 | 9.73E-05 | 1.85E-16 | -0.00978653 | AC011455.8, FBXO17 |
| chr2 | 105037185 | 105038912 | 1728 | 19 | 1.16E-14 | 3.58E-14 | 0.000243 | 1.85E-16 | -0.00197849 | MRPS9, MRPS9-AS2 |
| chr20 | 31485406 | 31485773 | 368 | 6 | 7.86E-19 | 1.97E-17 | 1.25E-05 | 1.92E-16 | -0.0299224 | NA |
| chr8 | 101223699 | 101224603 | 905 | 8 | 4.33E-17 | 8.03E-18 | 0.000184 | 2.07E-16 | -0.03764757 | AP001330.2 |
| chr1 | 3471283 | 3473745 | 2463 | 11 | 3.31E-12 | 4.47E-12 | 5.58E-07 | 2.31E-16 | -0.00920571 | ARHGEF16 |
| chr11 | 119422667 | 119425047 | 2381 | 19 | 2.05E-14 | 1.69E-15 | 0.000148 | 2.32E-16 | -0.0026026 | USP2-AS1, THY1 |
| chr17 | 81961702 | 81964796 | 3095 | 16 | 1.50E-12 | 2.04E-12 | 8.88E-05 | 2.42E-16 | -0.00341445 | NOTUM |
| chr20 | 62549734 | 62552219 | 2486 | 24 | 2.83E-16 | 1.41E-13 | 2.81E-05 | 2.48E-16 | 0.002901185 | MIR1-1HG, MIR1-1HG-AS1 |
| chr11 | 8766345 | 8767790 | 1446 | 6 | 3.20E-13 | 2.16E-16 | 4.07E-07 | 2.54E-16 | 0.000382693 | DENND2B |
| chr12 | 124516188 | 124517928 | 1741 | 7 | 5.88E-13 | 6.77E-17 | 3.66E-06 | 2.63E-16 | 0.008572841 | AC073592.1, NCOR2 |
| chr3 | 50192593 | 50195026 | 2434 | 11 | 3.98E-13 | 1.83E-15 | 1.85E-05 | 2.64E-16 | -0.016899 | GNAT1 |
| chr1 | 50023568 | 50025281 | 1714 | 16 | 8.42E-15 | 3.58E-12 | 1.35E-05 | 2.66E-16 | -0.00633476 | ELAVL4, AGBL4 |
| chr11 | 8932659 | 8933590 | 932 | 7 | 1.76E-19 | 1.77E-14 | 1.39E-06 | 2.66E-16 | -0.00807412 | C11orf16 |
| chr6 | 137044302 | 137046100 | 1799 | 17 | 4.98E-20 | 9.71E-14 | 3.96E-06 | 2.77E-16 | 0.000261418 | IL20RA |
| chr12 | 16347029 | 16349126 | 2098 | 10 | 1.69E-10 | 2.22E-15 | 8.53E-05 | 2.77E-16 | -0.00833721 | MGST1 |
| chr13 | 66994341 | 66995995 | 1655 | 4 | 1.27E-12 | 3.07E-17 | 7.05E-06 | 2.90E-16 | -0.01078047 | PCDH9-AS4, PCDH9 |
| chr2 | 171574162 | 171574306 | 145 | 4 | 8.49E-18 | 3.78E-16 | 1.30E-07 | 3.12E-16 | -0.01617977 | NA |
| chr8 | 22703409 | 22704723 | 1315 | 9 | 1.64E-17 | 6.02E-15 | 4.44E-05 | 3.12E-16 | -0.01385445 | AC055854.1, AC105046.2 |
| chr16 | 29961837 | 29963381 | 1545 | 14 | 6.21E-20 | 1.11E-11 | 1.16E-05 | 3.20E-16 | -0.00346944 | TMEM219 |
| chr5 | 133113411 | 133114802 | 1392 | 5 | 1.92E-18 | 1.79E-15 | 6.42E-08 | 3.21E-16 | -0.00831767 | AC113410.2 |
| chr5 | 121850784 | 121852533 | 1750 | 11 | 5.91E-18 | 3.32E-14 | 6.74E-07 | 3.21E-16 | -0.00525694 | FTMT |
| chr6 | 32838922 | 32842481 | 3560 | 37 | 1.23E-12 | 8.39E-13 | 0.000455 | 3.21E-16 | 0.00064811 | PSMB8 |
| chr6 | 34154725 | 34156106 | 1382 | 10 | 6.40E-23 | 7.70E-09 | 1.48E-07 | 3.34E-16 | -0.00424145 | GRM4 |
| chr19 | 50722591 | 50724145 | 1555 | 8 | 6.15E-17 | 7.50E-17 | 4.04E-05 | 3.48E-16 | -0.0184795 | CLEC11A |
| chr12 | 132283022 | 132285524 | 2503 | 12 | 6.47E-13 | 1.94E-16 | 0.000129 | 3.49E-16 | 0.021353108 | AC148477.4, GALNT9 |
| chr10 | 43406464 | 43410067 | 3604 | 28 | 2.49E-10 | 7.63E-11 | 0.000203 | 3.59E-16 | -0.00170483 | HNRNPF |
| chr6 | 28920618 | 28924302 | 3685 | 59 | 2.02E-13 | 1.60E-12 | 2.98E-05 | 3.61E-16 | 0.000341351 | TRIM27 |
| chr20 | 62666212 | 62668456 | 2245 | 12 | 7.16E-15 | 5.26E-15 | 1.97E-05 | 3.86E-16 | -0.00396588 | SLCO4A1, SLCO4A1-AS1 |
| chr11 | 66267210 | 66269461 | 2252 | 47 | 2.14E-16 | 3.06E-11 | 1.84E-06 | 3.91E-16 | 0.000165372 | KLC2, RAB1B, AP001107.7 |
| chr19 | 43465498 | 43467330 | 1833 | 10 | 1.75E-15 | 2.06E-13 | 6.81E-05 | 3.92E-16 | -0.01292262 | LYPD3 |
| chr8 | 100558883 | 100560579 | 1697 | 14 | 2.25E-16 | 7.20E-14 | 1.80E-05 | 4.00E-16 | -0.00096824 | ANKRD46 |
| chr16 | 67963393 | 67965141 | 1749 | 12 | 8.12E-15 | 9.61E-17 | 5.95E-05 | 4.17E-16 | -0.02061533 | SLC12A4 |
| chr8 | 66436653 | 66436857 | 205 | 2 | 1.90E-19 | 3.30E-16 | 4.86E-09 | 4.46E-16 | -0.01680425 | AC009879.2, AC009879.3, ADHFE1 |
| chr21 | 34890329 | 34891511 | 1183 | 5 | 3.44E-19 | 1.40E-11 | 6.27E-06 | 4.46E-16 | -0.03180376 | RUNX1 |
| chr17 | 19744660 | 19746756 | 2097 | 14 | 1.39E-12 | 2.19E-15 | 7.05E-05 | 4.46E-16 | -0.00094391 | ALDH3A1 |
| chr1 | 100538078 | 100539565 | 1488 | 7 | 2.38E-17 | 1.14E-12 | 2.51E-05 | 4.85E-16 | -0.06526571 | GPR88 |
| chr16 | 30445004 | 30446576 | 1573 | 19 | 6.69E-19 | 6.98E-13 | 6.25E-08 | 5.01E-16 | 0.005023163 | SEPHS2 |
| chr21 | 41677813 | 41679300 | 1488 | 9 | 5.50E-17 | 3.69E-15 | 3.47E-06 | 5.78E-16 | -0.02243314 | LINC00111 |
| chr17 | 83048945 | 83051475 | 2531 | 6 | 6.97E-17 | 1.38E-11 | 1.61E-07 | 5.91E-16 | -0.01606121 | B3GNTL1 |
| chr22 | 38843597 | 38845472 | 1876 | 10 | 3.72E-21 | 4.74E-12 | 2.36E-09 | 6.21E-16 | -0.00504582 | NPTXR |
| chr9 | 127846644 | 127848664 | 2021 | 10 | 1.69E-15 | 2.97E-15 | 2.31E-05 | 6.32E-16 | -0.00197068 | ENG |
| chr19 | 16495094 | 16496672 | 1579 | 15 | 6.49E-23 | 1.74E-12 | 3.85E-07 | 6.33E-16 | -0.00847073 | C19orf44, CALR3, AC008764.1 |
| chr1 | 246788746 | 246790582 | 1837 | 12 | 2.68E-18 | 1.06E-13 | 2.47E-06 | 6.33E-16 | 0.003795492 | LINC01341, KIF28P |
| chr17 | 30969437 | 30971334 | 1898 | 18 | 5.57E-21 | 4.28E-10 | 2.60E-05 | 6.96E-16 | -0.00642895 | AC138207.9, RNF135 |
| chr20 | 58850102 | 58852118 | 2017 | 35 | 1.61E-16 | 6.28E-12 | 0.000658 | 7.14E-16 | -0.00697459 | GNAS, AL132655.4, AL132655.5, GNAS-AS1 |
| chr10 | 1187245 | 1189288 | 2044 | 9 | 2.46E-15 | 9.56E-16 | 2.58E-05 | 7.23E-16 | -0.01229197 | LINC00200, ADARB2 |
| chr2 | 205762797 | 205764590 | 1794 | 13 | 1.66E-15 | 3.08E-17 | 0.001603 | 7.33E-16 | -0.02891946 | NRP2, AC007362.1 |
| chr10 | 11004306 | 11005623 | 1318 | 12 | 1.53E-13 | 2.34E-12 | 1.84E-07 | 7.72E-16 | -0.004742 | CELF2 |
| chr11 | 113386756 | 113388186 | 1431 | 11 | 3.05E-17 | 6.26E-15 | 8.84E-06 | 8.06E-16 | -0.00954645 | ANKK1 |
| chr6 | 30600755 | 30602386 | 1632 | 18 | 9.54E-13 | 1.68E-13 | 0.00023 | 8.07E-16 | -0.00308957 | PPP1R10 |
| chr17 | 4745271 | 4746643 | 1373 | 8 | 2.76E-15 | 2.05E-14 | 6.70E-07 | 8.17E-16 | -0.01720994 | ZMYND15 |
| chr1 | 204213829 | 204214527 | 699 | 13 | 4.11E-19 | 6.54E-13 | 3.38E-07 | 8.59E-16 | -0.00533144 | GOLT1A |
| chr20 | 62677728 | 62678765 | 1038 | 5 | 2.11E-17 | 7.10E-16 | 8.97E-06 | 8.60E-16 | -0.02558966 | SLCO4A1 |
| chr3 | 132720834 | 132722529 | 1696 | 14 | 1.07E-24 | 2.77E-10 | 2.58E-06 | 8.64E-16 | -0.00545885 | NPHP3-AS1, NPHP3-ACAD11, NPHP3 |
| chr2 | 43676088 | 43676872 | 785 | 9 | 6.19E-16 | 5.81E-16 | 9.78E-05 | 8.73E-16 | -0.02279546 | PLEKHH2, C1GALT1C1L |
| chr10 | 74824229 | 74826892 | 2664 | 16 | 8.23E-12 | 1.38E-12 | 3.77E-05 | 9.13E-16 | 0.001060116 | KAT6B |
| chr10 | 73771638 | 73773673 | 2036 | 18 | 1.54E-13 | 9.02E-16 | 0.000367 | 9.14E-16 | -0.00315232 | SEC24C, FUT11, AC022400.7 |
| chr1 | 161079357 | 161079887 | 531 | 6 | 8.19E-17 | 1.26E-16 | 4.64E-05 | 9.76E-16 | -0.02055452 | NECTIN4 |
| chr10 | 100346323 | 100348000 | 1678 | 15 | 3.53E-16 | 5.82E-13 | 1.92E-05 | 1.02E-15 | -0.00340505 | SCD, AL139819.1 |
| chr6 | 29721967 | 29724622 | 2656 | 30 | 4.74E-12 | 7.71E-14 | 3.61E-05 | 1.05E-15 | 0.005787377 | HLA-F, AL645939.1 |
| chr16 | 84406178 | 84408376 | 2199 | 9 | 2.66E-12 | 7.60E-15 | 6.09E-06 | 1.11E-15 | 0.004173632 | ATP2C2 |
| chr2 | 27306285 | 27309099 | 2815 | 21 | 1.17E-12 | 7.60E-15 | 0.000303 | 1.14E-15 | -0.00294125 | TRIM54, UCN |
| chr9 | 91159687 | 91160396 | 710 | 5 | 5.73E-16 | 2.62E-16 | 1.88E-05 | 1.14E-15 | 0.007544521 | LINC00484, LINC00484, AL158071.3, AL158071.2 |
| chr7 | 5071857 | 5073408 | 1552 | 15 | 2.67E-15 | 3.80E-13 | 5.38E-06 | 1.14E-15 | -0.01685531 | RBAK-RBAKDN, RBAKDN |
| chr1 | 933323 | 933966 | 644 | 3 | 1.07E-18 | 2.37E-15 | 1.23E-08 | 1.16E-15 | -0.0487171 | SAMD11 |
| chr5 | 101782629 | 101784062 | 1434 | 6 | 1.02E-13 | 7.96E-17 | 3.38E-05 | 1.18E-15 | -0.04348163 | NA |
| chr8 | 76680522 | 76683221 | 2700 | 20 | 3.49E-11 | 2.03E-12 | 1.73E-05 | 1.18E-15 | -0.00243289 | ZFHX4, ZFHX4-AS1 |
| chr11 | 10807453 | 10809425 | 1973 | 24 | 2.08E-17 | 1.81E-10 | 3.47E-05 | 1.22E-15 | 0.000595924 | AC116535.1, EIF4G2 |
| chr8 | 23705405 | 23707661 | 2257 | 21 | 6.92E-13 | 2.33E-11 | 0.000184 | 1.31E-15 | -0.00419178 | AC012574.1, NKX2-6 |
| chr15 | 78618757 | 78622025 | 3269 | 20 | 1.20E-13 | 6.37E-13 | 4.84E-05 | 1.32E-15 | 0.002881424 | CHRNA3 |
| chr2 | 172086773 | 172088902 | 2130 | 15 | 4.81E-19 | 2.17E-08 | 1.84E-06 | 1.41E-15 | -0.00240877 | DLX1 |
| chr1 | 114757100 | 114759349 | 2250 | 16 | 8.64E-15 | 2.57E-12 | 1.07E-05 | 1.42E-15 | 0.001337657 | CSDE1 |
| chr19 | 1062179 | 1064939 | 2761 | 7 | 1.34E-13 | 1.44E-11 | 1.32E-05 | 1.46E-15 | -0.01365958 | ABCA7 |
| chr3 | 42935464 | 42936912 | 1449 | 15 | 1.27E-16 | 2.30E-15 | 0.000111 | 1.47E-15 | -0.01647966 | AC092042.3, KRBOX1, KRBOX1-AS1 |
| chr16 | 4271639 | 4274279 | 2641 | 14 | 2.55E-12 | 3.22E-13 | 3.37E-05 | 1.52E-15 | 0.002259117 | TFAP4 |
| chr20 | 1630879 | 1631065 | 187 | 4 | 3.51E-17 | 2.37E-15 | 3.74E-06 | 1.56E-15 | -0.00496802 | SIRPG |
| chr10 | 1410835 | 1413132 | 2298 | 8 | 1.14E-10 | 6.41E-13 | 3.00E-06 | 1.56E-15 | 0.000950056 | ADARB2 |
| chr17 | 1231612 | 1231617 | 6 | 3 | 3.43E-17 | 3.22E-16 | 1.99E-06 | 1.62E-15 | -0.03104531 | NA |
| chr17 | 41047369 | 41047757 | 389 | 3 | 6.38E-21 | 1.46E-12 | 1.83E-08 | 1.63E-15 | -0.03067844 | NA |
| chr6 | 127342055 | 127344736 | 2682 | 20 | 2.70E-16 | 1.29E-10 | 1.83E-05 | 1.72E-15 | -0.00330717 | ECHDC1 |
| chr11 | 64290299 | 64292122 | 1824 | 16 | 5.34E-17 | 2.88E-13 | 4.52E-05 | 1.80E-15 | -0.00492136 | KCNK4, KCNK4-TEX40 |
| chr1 | 180187002 | 180187017 | 16 | 2 | 5.10E-18 | 9.26E-16 | 4.16E-08 | 1.86E-15 | -0.06531989 | QSOX1 |
| chr4 | 98928502 | 98930130 | 1629 | 20 | 7.47E-16 | 1.17E-14 | 0.000159 | 1.86E-15 | -0.00448501 | AC019131.1, EIF4E |
| chr17 | 4899237 | 4902691 | 3455 | 23 | 3.18E-12 | 6.90E-13 | 0.000124 | 1.87E-15 | -0.01230107 | C17orf107, CHRNE |
| chr12 | 12465201 | 12467589 | 2389 | 6 | 7.57E-14 | 1.92E-13 | 1.93E-07 | 1.88E-15 | 0.009847804 | BORCS5 |
| chr17 | 81165073 | 81166872 | 1800 | 12 | 5.11E-12 | 1.61E-13 | 3.07E-05 | 1.89E-15 | -0.00870051 | PVALEF, AATK |
| chr1 | 1898336 | 1899254 | 919 | 5 | 4.10E-22 | 2.25E-11 | 1.47E-08 | 2.06E-15 | 0.01266695 | NA |
| chr7 | 158585426 | 158589442 | 4017 | 17 | 6.97E-11 | 6.65E-11 | 0.000103 | 2.18E-15 | -0.00110755 | PTPRN2 |
| chr20 | 21827072 | 21828168 | 1097 | 8 | 2.11E-15 | 1.66E-14 | 1.12E-05 | 2.19E-15 | -0.0163451 | NA |
| chr13 | 112342032 | 112345718 | 3687 | 13 | 3.95E-09 | 6.89E-11 | 4.80E-06 | 2.26E-15 | 0.010804355 | NA |
| chr17 | 8397658 | 8399161 | 1504 | 9 | 1.33E-14 | 3.81E-15 | 1.03E-05 | 2.40E-15 | 0.012424339 | RNF222 |
| chr16 | 68447910 | 68449651 | 1742 | 15 | 1.14E-20 | 7.75E-11 | 2.44E-06 | 2.42E-15 | -0.00652707 | AC099521.3, SMPD3 |
| chr22 | 50525396 | 50528507 | 3112 | 18 | 4.62E-12 | 8.78E-10 | 0.000117 | 2.48E-15 | 0.004814269 | SCO2, TYMP |
| chr20 | 64164305 | 64167678 | 3374 | 14 | 1.26E-11 | 2.21E-11 | 1.35E-05 | 2.54E-15 | -0.00830885 | MYT1 |
| chr10 | 67883442 | 67885254 | 1813 | 14 | 2.01E-20 | 2.26E-10 | 7.46E-08 | 2.58E-15 | -0.0085554 | SIRT1 |
| chr16 | 30029595 | 30031929 | 2335 | 14 | 4.96E-14 | 2.39E-11 | 1.22E-06 | 2.65E-15 | 0.004188532 | TLCD3B |
| chr9 | 133346180 | 133349299 | 3120 | 21 | 6.08E-14 | 4.90E-12 | 1.27E-06 | 2.75E-15 | -0.0010244 | RPL7A, MED22 |
| chr1 | 155199749 | 155200767 | 1019 | 7 | 8.04E-23 | 1.80E-08 | 2.83E-08 | 2.84E-15 | -0.00727591 | THBS3-AS1, THBS3 |
| chr11 | 2399738 | 2402617 | 2880 | 40 | 1.14E-13 | 2.20E-11 | 0.000597 | 3.01E-15 | 0.000124964 | TSSC4 |
| chr22 | 20999520 | 21002815 | 3296 | 21 | 8.07E-12 | 1.70E-11 | 2.08E-05 | 3.12E-15 | -0.00010171 | THAP7-AS1, THAP7 |
| chr11 | 14903920 | 14905458 | 1539 | 8 | 3.12E-17 | 1.21E-12 | 1.25E-06 | 3.12E-15 | -0.00226706 | CALCB |
| chr18 | 63948230 | 63949628 | 1399 | 10 | 2.60E-16 | 3.39E-10 | 4.58E-06 | 3.19E-15 | -0.00767397 | HMSD |
| chr19 | 35712930 | 35714672 | 1743 | 5 | 2.94E-18 | 2.18E-12 | 8.93E-07 | 3.25E-15 | -0.01158865 | ZBTB32 |
| chr8 | 21782588 | 21782792 | 205 | 3 | 1.74E-16 | 6.56E-16 | 2.86E-06 | 3.35E-15 | -0.0203406 | GFRA2 |
| chr20 | 43187986 | 43191296 | 3311 | 17 | 1.05E-09 | 7.09E-12 | 8.21E-06 | 3.45E-15 | -8.28E-06 | AL021395.1, PTPRT |
| chr2 | 118844970 | 118849521 | 4552 | 31 | 6.89E-08 | 3.79E-14 | 0.001844 | 3.45E-15 | -0.00133032 | EN1 |
| chr17 | 79791193 | 79793686 | 2494 | 12 | 2.16E-12 | 4.53E-12 | 7.22E-05 | 3.55E-15 | -0.0114381 | CBX8 |
| chr12 | 12890195 | 12891622 | 1428 | 16 | 2.46E-16 | 1.72E-13 | 2.90E-05 | 3.65E-15 | -0.00512272 | GPRC5A |
| chr12 | 94262159 | 94263632 | 1474 | 7 | 8.09E-15 | 2.26E-14 | 2.85E-05 | 3.69E-15 | -0.00843036 | PLXNC1, AC073655.3 |
| chr4 | 48985007 | 48986784 | 1778 | 15 | 3.80E-18 | 1.20E-12 | 1.88E-05 | 3.70E-15 | -0.00684109 | CWH43 |
| chr12 | 121888023 | 121889230 | 1208 | 15 | 1.35E-15 | 2.81E-14 | 0.00023 | 3.71E-15 | -0.00112715 | PSMD9, AC069503.2, AC069503.5 |
| chr6 | 166440323 | 166440395 | 73 | 2 | 3.66E-17 | 1.88E-15 | 5.18E-08 | 3.89E-15 | 0.060959926 | RPS6KA2 |
| chr5 | 14900903 | 14901858 | 956 | 4 | 1.21E-17 | 8.08E-11 | 6.74E-07 | 4.12E-15 | -0.02472486 | NA |
| chr2 | 118087256 | 118088713 | 1458 | 14 | 1.50E-20 | 6.11E-10 | 7.73E-07 | 4.25E-15 | 0.003829027 | INSIG2 |
| chr1 | 160115043 | 160116560 | 1518 | 7 | 4.57E-18 | 9.35E-13 | 3.96E-07 | 4.27E-15 | -0.01261877 | ATP1A2 |
| chr6 | 24356768 | 24358587 | 1820 | 15 | 4.35E-20 | 4.28E-10 | 4.70E-05 | 4.29E-15 | -0.00763729 | KAAG1, DCDC2 |
| chr6 | 28633492 | 28633742 | 251 | 12 | 4.92E-16 | 3.80E-17 | 0.000666 | 4.29E-15 | -0.03194731 | AL121932.1 |
| chr8 | 143611790 | 143614701 | 2912 | 12 | 6.11E-09 | 1.99E-11 | 2.43E-05 | 4.36E-15 | 0.000578107 | GFUS |
| chr11 | 65791358 | 65794234 | 2877 | 11 | 5.41E-11 | 1.02E-12 | 4.30E-05 | 4.37E-15 | -0.00452079 | OVOL1 |
| chr14 | 20195862 | 20197929 | 2068 | 11 | 3.91E-14 | 1.30E-15 | 5.90E-05 | 4.40E-15 | -0.01138 | OR11G2 |
| chr19 | 44212281 | 44213056 | 776 | 14 | 1.39E-19 | 1.69E-10 | 1.39E-05 | 4.52E-15 | 0.0018518 | ZNF227 |
| chr20 | 62427539 | 62429495 | 1957 | 12 | 8.44E-14 | 4.30E-15 | 2.97E-05 | 4.67E-15 | -0.00803782 | AL121832.1, RBBP8NL |
| chr14 | 91253264 | 91254474 | 1211 | 12 | 2.50E-18 | 1.17E-11 | 1.36E-06 | 4.70E-15 | -0.00718322 | GPR68 |
| chr12 | 89524291 | 89526873 | 2583 | 29 | 3.46E-16 | 1.81E-10 | 4.52E-08 | 4.70E-15 | -0.00033305 | POC1B-AS1, POC1B, POC1B-GALNT4, GALNT4 |
| chr12 | 124293882 | 124295759 | 1878 | 12 | 1.01E-18 | 8.96E-10 | 5.74E-07 | 4.71E-15 | -0.00425839 | RFLNA |
| chr5 | 77639121 | 77641686 | 2566 | 11 | 5.98E-19 | 9.58E-11 | 1.47E-08 | 4.79E-15 | -0.00525116 | NA |
| chr12 | 48296666 | 48297253 | 588 | 6 | 3.97E-17 | 8.19E-14 | 9.65E-06 | 4.80E-15 | -0.01593581 | NA |
| chr3 | 129120765 | 129123207 | 2443 | 23 | 5.27E-15 | 8.19E-10 | 2.53E-06 | 4.80E-15 | -0.00310327 | RAB43, ISY1-RAB43 |
| chr3 | 14508928 | 14509506 | 579 | 18 | 7.91E-17 | 1.42E-10 | 7.65E-05 | 4.84E-15 | -0.00399226 | GRIP2 |
| chr8 | 38104400 | 38106402 | 2003 | 22 | 1.03E-16 | 3.35E-09 | 2.81E-06 | 4.85E-15 | -0.00202987 | ASH2L |

| | | | | | | | | | | |
|---|---|---|---|---|---|---|---|---|---|---|
| chr6 | 83859038 | 83860669 | 1632 | 11 | 1.79E-15 | 1.07E-11 | 3.09E-05 | 4.88E-15 | -0.00257453 | CYB5R4 |
| chr6 | 30343849 | 30346763 | 2915 | 43 | 1.23E-14 | 5.55E-12 | 0.000707 | 4.95E-15 | -0.00041617 | TRIM39-RPP21, RPP21 |
| chr10 | 84200759 | 84201891 | 1133 | 7 | 4.03E-15 | 2.23E-15 | 2.56E-05 | 4.95E-15 | -0.00805224 | CDHR1 |
| chr1 | 46302299 | 46303726 | 1428 | 13 | 2.36E-17 | 2.68E-09 | 1.15E-06 | 5.05E-15 | 0.00081726 | UQCRH, LRRC41 |
| chr8 | 718053 | 719656 | 1604 | 10 | 5.86E-17 | 1.54E-12 | 3.11E-07 | 5.13E-15 | -0.00135068 | ERICH1 |
| chr17 | 39736588 | 39738691 | 2104 | 11 | 7.14E-16 | 3.15E-14 | 7.20E-05 | 5.14E-15 | -0.00941244 | GRB7 |
| chr6 | 30189115 | 30192862 | 3748 | 29 | 3.10E-14 | 3.72E-10 | 8.13E-06 | 5.16E-15 | 1.61E-05 | TRIM26 |
| chr10 | 89224915 | 89225305 | 391 | 4 | 4.05E-19 | 9.49E-15 | 1.44E-07 | 5.39E-15 | -0.03784807 | LIPA |
| chr7 | 95324341 | 95325143 | 803 | 10 | 4.94E-15 | 3.30E-16 | 0.000166 | 5.40E-15 | -0.03141507 | PON1 |
| chr16 | 876082 | 879333 | 3252 | 13 | 1.91E-09 | 3.94E-14 | 0.000119 | 5.50E-15 | -0.00696843 | LMF1 |
| chr15 | 98420965 | 98422246 | 1282 | 9 | 4.67E-18 | 1.45E-13 | 1.29E-06 | 6.30E-15 | -0.01351541 | LINC02351 |
| chr5 | 567142 | 567723 | 582 | 3 | 3.33E-16 | 1.09E-13 | 1.97E-08 | 6.30E-15 | -0.00867779 | NA |
| chr10 | 132785404 | 132787986 | 2583 | 34 | 9.22E-14 | 9.67E-11 | 0.000423 | 6.39E-15 | 0.00095685 | NKX6-2 |
| chr9 | 116685983 | 116689586 | 3604 | 18 | 4.70E-11 | 1.10E-13 | 7.40E-05 | 6.42E-15 | -0.00069369 | TRIM32, ASTN2 |
| chr5 | 50968628 | 50970879 | 2252 | 20 | 2.70E-17 | 7.39E-09 | 1.45E-06 | 6.43E-15 | -0.00396626 | AC008808.1, LINC02106 |
| chr3 | 9946460 | 9947982 | 1523 | 9 | 5.17E-17 | 6.12E-14 | 1.47E-06 | 6.43E-15 | -0.01760429 | PRRT3-AS1, PRRT3 |
| chr4 | 151673604 | 151675093 | 1490 | 8 | 8.41E-17 | 1.88E-13 | 1.19E-06 | 6.64E-15 | -0.0082771 | AC092611.1, GATB |
| chr19 | 19537090 | 19538731 | 1642 | 14 | 1.37E-14 | 1.35E-15 | 0.000548 | 6.67E-15 | -0.00968958 | YJEFN3, CILP2 |
| chr16 | 3017356 | 3019455 | 2100 | 19 | 2.60E-15 | 1.26E-11 | 1.57E-05 | 6.74E-15 | -0.00469922 | TNFRSF12A, CLDN6 |
| chr1 | 118983437 | 118987521 | 4085 | 21 | 1.12E-10 | 3.51E-12 | 9.16E-05 | 6.78E-15 | -0.00440487 | TBX15 |
| chr12 | 106580447 | 106583611 | 3165 | 25 | 2.32E-11 | 1.05E-11 | 6.10E-05 | 6.79E-15 | 0.000466558 | RFX4, AC079385.1 |
| chr19 | 10140733 | 10144663 | 3931 | 13 | 7.74E-10 | 1.31E-13 | 7.98E-05 | 7.01E-15 | -0.00101791 | DNMT1 |
| chr11 | 36294836 | 36296887 | 2052 | 7 | 1.10E-15 | 1.67E-12 | 4.72E-08 | 7.10E-15 | 2.49E-05 | PRR5L |
| chr22 | 38081795 | 38085158 | 3364 | 20 | 8.54E-10 | 7.58E-12 | 8.10E-06 | 7.10E-15 | -0.00045008 | SLC16A8, BAIAP2L2 |
| chr5 | 23506629 | 23507547 | 919 | 7 | 3.26E-15 | 4.60E-16 | 9.58E-05 | 7.16E-15 | -0.0281061 | PRDM9 |
| chr3 | 128462479 | 128464350 | 1872 | 9 | 1.37E-17 | 4.70E-12 | 3.19E-07 | 7.17E-15 | 0.002202127 | DNAJB8-AS1, DNAJB8 |
| chr20 | 58887743 | 58891082 | 3340 | 61 | 8.16E-10 | 2.75E-11 | 0.000452 | 7.31E-15 | -0.00131445 | GNAS, AL121917.1 |
| chr19 | 7533136 | 7536121 | 2986 | 29 | 2.27E-13 | 1.01E-12 | 2.99E-05 | 7.31E-15 | -0.00155141 | MCOLN1, AC008878.2, PNPLA6 |
| chr7 | 44107615 | 44111573 | 3959 | 16 | 5.21E-08 | 9.15E-12 | 6.80E-05 | 7.40E-15 | -0.00399376 | AEBP1, MIR4649 |
| chr20 | 62062892 | 62067208 | 4317 | 16 | 5.66E-07 | 1.44E-11 | 0.00045 | 7.52E-15 | -0.00367099 | MIR3195, AL137077.2, TAF4 |
| chr17 | 45149366 | 45151877 | 2512 | 8 | 3.72E-11 | 3.21E-14 | 3.51E-06 | 7.74E-15 | -9.08E-05 | HEXIM1, AC138150.1 |
| chr6 | 33007306 | 33010998 | 3693 | 48 | 2.24E-10 | 1.17E-11 | 0.000953 | 7.82E-15 | 0.001887869 | HLA-DOA |
| chr3 | 181695031 | 181697060 | 2030 | 13 | 1.42E-13 | 6.16E-12 | 7.14E-06 | 7.92E-15 | -0.00360993 | SOX2-OT, AC125613.1 |
| chr6 | 30655470 | 30656992 | 1523 | 15 | 1.29E-12 | 1.35E-12 | 0.000399 | 7.92E-15 | -0.00343414 | DHX16 |
| chr8 | 70070020 | 70072682 | 2663 | 19 | 1.77E-11 | 4.13E-10 | 1.72E-05 | 8.12E-15 | -0.00275926 | PRDM14 |
| chr19 | 50049765 | 50051253 | 1489 | 13 | 1.33E-16 | 3.86E-13 | 3.71E-05 | 8.66E-15 | -0.00690166 | ZNF473, AC010624.1, AC010624.3 |
| chr12 | 85036215 | 85037219 | 1005 | 15 | 4.04E-17 | 1.12E-08 | 5.77E-07 | 8.83E-15 | 0.002353779 | LRRIQ1, TSPAN19 |
| chr11 | 1639283 | 1640389 | 1107 | 7 | 7.64E-16 | 2.07E-14 | 4.18E-07 | 9.23E-15 | -0.00319888 | NA |
| chr11 | 14971963 | 14974224 | 2262 | 39 | 9.64E-17 | 1.25E-09 | 7.34E-06 | 9.36E-15 | -0.00116077 | CALCB, CALCA |
| chr8 | 143941796 | 143946213 | 4418 | 29 | 1.55E-08 | 9.79E-14 | 0.001434 | 9.38E-15 | -0.00036512 | PLEC, MIR661 |
| chr13 | 36879292 | 36880276 | 985 | 8 | 4.15E-16 | 6.02E-14 | 0.000118 | 9.49E-15 | -0.01597389 | SMAD9 |
| chr13 | 44648370 | 44651408 | 3039 | 7 | 3.94E-13 | 4.55E-12 | 9.11E-08 | 9.53E-15 | -0.00860277 | NA |
| chr1 | 40837684 | 40838763 | 1080 | 6 | 4.97E-14 | 1.38E-15 | 3.21E-05 | 9.60E-15 | -0.01685392 | KCNQ4 |
| chr2 | 66461010 | 66462675 | 1666 | 4 | 6.36E-17 | 2.75E-13 | 5.54E-09 | 1.02E-14 | 0.000340762 | MEIS1 |
| chr3 | 126393718 | 126395863 | 2146 | 16 | 8.26E-18 | 9.91E-10 | 1.59E-06 | 1.03E-14 | -0.00607594 | CFAP100, CCDC37-DT |
| chr16 | 3022206 | 3024536 | 2331 | 20 | 3.80E-12 | 6.74E-12 | 1.86E-06 | 1.03E-14 | -0.00025784 | TNFRSF12A, THOC6, HCFC1R1 |
| chr1 | 156291127 | 156292480 | 1354 | 10 | 3.84E-17 | 6.70E-11 | 4.21E-06 | 1.07E-14 | -0.0110873 | TMEM79, GLMP |
| chr20 | 31721153 | 31724160 | 3008 | 23 | 4.04E-10 | 7.10E-13 | 7.33E-05 | 1.08E-14 | -0.00118528 | ABALON, BCL2L1 |
| chr3 | 53195475 | 53197162 | 1688 | 6 | 4.11E-16 | 1.22E-10 | 5.46E-06 | 1.10E-14 | -0.00050748 | AC097015.1 |
| chr3 | 45688315 | 45689913 | 1599 | 24 | 1.68E-15 | 1.65E-10 | 6.87E-06 | 1.13E-14 | 0.000595029 | SACM1L, LIMD1-AS1 |
| chr10 | 101013036 | 101015715 | 2680 | 8 | 1.45E-13 | 3.48E-11 | 4.20E-06 | 1.13E-14 | -0.01118898 | PDZD7 |
| chr10 | 23280382 | 23280735 | 354 | 2 | 2.31E-16 | 5.63E-15 | 8.71E-08 | 1.17E-14 | -0.03708726 | C10orf67 |
| chr11 | 125624979 | 125626857 | 1879 | 17 | 1.58E-15 | 2.55E-11 | 4.95E-05 | 1.20E-14 | -0.00012838 | STT3A, CHEK1 |
| chr4 | 157075030 | 157077049 | 2020 | 18 | 1.67E-17 | 3.95E-11 | 4.12E-05 | 1.20E-14 | -0.00265111 | GLRB |
| chr6 | 29457011 | 29460138 | 3128 | 13 | 8.21E-11 | 2.11E-12 | 1.58E-05 | 1.25E-14 | 0.001635422 | OR2H1, OR11A1 |
| chr1 | 44806008 | 44808928 | 2921 | 20 | 9.31E-12 | 6.70E-11 | 1.85E-05 | 1.27E-14 | -0.00413058 | BTBD19, TCTEX1D4 |
| chr6 | 88047179 | 88048326 | 1148 | 9 | 3.84E-15 | 8.05E-16 | 3.14E-05 | 1.31E-14 | -0.0184535 | SPACA1, AL136096.1 |
| chr17 | 29589840 | 29591743 | 1904 | 11 | 4.75E-18 | 1.86E-12 | 1.10E-05 | 1.36E-14 | -0.0091581 | ABHD15-AS1, ANKRD13B, GIT1, AC104564.4 |
| chr10 | 1021432 | 1022823 | 1392 | 8 | 2.22E-11 | 1.33E-15 | 0.000136 | 1.39E-14 | -0.00801785 | IDI2-AS1, IDI2 |
| chr1 | 31575686 | 31578018 | 2333 | 21 | 5.32E-15 | 4.78E-11 | 3.07E-06 | 1.39E-14 | -0.00537132 | LINC01226, TINAGL1, AC114488.1 |
| chr11 | 3156392 | 3157794 | 1403 | 7 | 2.71E-13 | 4.33E-15 | 1.64E-05 | 1.39E-14 | -0.01617422 | OSBPL5 |
| chr6 | 30450731 | 30451774 | 1044 | 20 | 1.98E-20 | 1.07E-10 | 5.73E-05 | 1.49E-14 | -0.00806945 | AL662873.1 |
| chr1 | 153687832 | 153690607 | 2776 | 8 | 7.14E-09 | 7.95E-15 | 1.25E-05 | 1.49E-14 | -0.00561485 | NPR1, MIR8083 |
| chr1 | 2237209 | 2241220 | 4012 | 12 | 1.02E-08 | 4.98E-10 | 1.39E-05 | 1.49E-14 | -0.0035364 | SKI |
| chr5 | 56952033 | 56953533 | 1501 | 6 | 1.70E-18 | 9.88E-11 | 1.36E-07 | 1.50E-14 | -0.00558543 | MIER3 |
| chr7 | 92608062 | 92609050 | 989 | 8 | 9.79E-16 | 1.05E-13 | 1.68E-07 | 1.53E-14 | 0.008979653 | CDK6 |
| chr9 | 135253165 | 135253885 | 721 | 4 | 1.18E-15 | 2.06E-13 | 1.41E-07 | 1.54E-14 | -0.00532528 | NA |
| chr15 | 72116232 | 72120233 | 4002 | 26 | 3.30E-12 | 3.25E-08 | 8.85E-06 | 1.56E-14 | -0.00284061 | SENP8, MYO9A |
| chr1 | 217088993 | 217090635 | 1643 | 14 | 7.14E-13 | 2.76E-13 | 0.000688 | 1.58E-14 | 0.00774945 | ESRRG |
| chr10 | 113819664 | 113820834 | 1171 | 7 | 2.22E-19 | 2.89E-10 | 1.61E-07 | 1.62E-14 | -0.00539572 | AL592546.1 |
| chr10 | 419168 | 422909 | 3742 | 22 | 1.58E-08 | 6.76E-10 | 3.86E-05 | 1.66E-14 | -0.00176661 | DIP2C |
| chr1 | 11700489 | 11701709 | 1221 | 9 | 2.10E-18 | 2.64E-11 | 2.57E-07 | 1.70E-14 | 0.014763188 | DRAXIN |
| chr9 | 87497171 | 87498678 | 1508 | 17 | 1.65E-14 | 7.16E-13 | 2.74E-05 | 1.70E-14 | -0.00118623 | DAPK1 |
| chr15 | 24674453 | 24676036 | 1584 | 12 | 2.89E-15 | 1.10E-12 | 1.87E-06 | 1.72E-14 | 0.00277844 | PWRN1, AC090983.2, NPAP1 |
| chr17 | 8015704 | 8016834 | 1131 | 7 | 7.28E-14 | 2.21E-13 | 1.07E-06 | 1.76E-14 | -0.0065789 | GUCY2D, AC104581.3 |
| chr20 | 62641960 | 62644782 | 2823 | 14 | 1.28E-11 | 6.41E-13 | 0.000642 | 1.79E-14 | -0.0038313 | SLCO4A1, AL357033.1 |
| chr19 | 12294362 | 12295608 | 1247 | 9 | 4.95E-17 | 1.27E-11 | 3.43E-06 | 1.81E-14 | -0.00776437 | ZNF44 |
| chr17 | 50467363 | 50469757 | 2395 | 18 | 1.57E-13 | 3.62E-12 | 8.44E-05 | 1.81E-14 | -0.006246 | ACSF2, CHAD |
| chr22 | 46798577 | 46799198 | 622 | 4 | 1.72E-15 | 5.75E-15 | 1.08E-06 | 1.81E-14 | -0.01928482 | TBC1D22A |
| chr19 | 54162643 | 54164522 | 1880 | 10 | 2.98E-18 | 5.60E-12 | 1.31E-06 | 1.84E-14 | -0.01264417 | TMC4 |
| chr3 | 157119801 | 157121332 | 1532 | 8 | 2.80E-15 | 3.27E-15 | 0.000209 | 1.86E-14 | -0.01583291 | LINC00881, LINC00880 |
| chr6 | 31631836 | 31636331 | 4496 | 41 | 5.02E-09 | 4.73E-11 | 0.000721 | 1.87E-14 | -0.00117937 | PRRC2A, MIR6832 |
| chr9 | 89603379 | 89606461 | 3083 | 18 | 5.04E-11 | 7.53E-11 | 0.000254 | 1.88E-14 | -0.00129488 | GADD45G, AL161910.1 |
| chr4 | 108166156 | 108169424 | 3269 | 24 | 1.04E-13 | 7.07E-12 | 1.86E-05 | 1.93E-14 | -0.00224954 | LEF1-AS1, LEF1 |
| chr14 | 24188107 | 24190767 | 2661 | 23 | 6.28E-10 | 5.73E-11 | 5.24E-05 | 1.93E-14 | -0.00046894 | AL136295.22, IPO4, AL136295.4, TM9SF1, AL136295.1 |
| chr16 | 89296639 | 89297817 | 1179 | 10 | 8.97E-16 | 1.69E-14 | 8.58E-06 | 1.93E-14 | -0.00696746 | AC137932.1, ANKRD11 |
| chr1 | 3981457 | 3982431 | 975 | 3 | 2.55E-14 | 1.34E-14 | 1.02E-07 | 1.98E-14 | 0.012886598 | LINC02780 |
| chr16 | 89922502 | 89925728 | 3227 | 14 | 6.48E-11 | 3.06E-12 | 4.23E-05 | 2.08E-14 | -1.16E-05 | AC092143.1, TUBB3, AC092143.2 |
| chr1 | 91404058 | 91406191 | 2134 | 14 | 5.03E-12 | 5.75E-10 | 5.77E-05 | 2.15E-14 | 0.003853757 | HFM1 |
| chr12 | 110845722 | 110847175 | 1454 | 14 | 8.15E-16 | 1.46E-11 | 4.80E-07 | 2.16E-14 | -0.00351684 | AC002375.1, CCDC63 |
| chr9 | 137220798 | 137224013 | 3216 | 18 | 2.31E-11 | 2.46E-10 | 1.74E-06 | 2.19E-14 | -0.00165469 | RNF208 |
| chr14 | 103941798 | 103943300 | 1503 | 11 | 1.32E-17 | 9.20E-08 | 2.96E-07 | 2.21E-14 | -0.00394447 | TDRD9, RD3L |
| chr11 | 69470865 | 69472374 | 1510 | 11 | 7.65E-16 | 1.18E-12 | 3.49E-05 | 2.24E-14 | -0.01217474 | NA |
| chr9 | 137020598 | 137022984 | 2387 | 6 | 6.66E-12 | 1.60E-14 | 8.92E-06 | 2.27E-14 | -0.00580567 | ABCA2 |
| chr1 | 1279916 | 1281750 | 1835 | 7 | 9.20E-21 | 1.76E-11 | 5.61E-09 | 2.31E-14 | -0.00499965 | LINC01786, SCNN1D |
| chr6 | 64733925 | 64734757 | 833 | 3 | 1.15E-20 | 2.30E-08 | 7.72E-10 | 2.39E-14 | -0.01916964 | EYS |
| chr7 | 92831992 | 92836548 | 4557 | 31 | 5.52E-08 | 5.24E-12 | 0.000826 | 2.40E-14 | -0.00101713 | CDK6-AS1, CDK6 |
| chr12 | 1795336 | 1798202 | 2867 | 13 | 1.59E-13 | 1.39E-11 | 1.27E-05 | 2.44E-14 | -0.00593636 | CACNA2D4 |
| chr3 | 177933441 | 177935413 | 1973 | 7 | 4.13E-12 | 1.85E-12 | 3.59E-06 | 2.45E-14 | 0.010808556 | AC007953.1 |
| chr13 | 48093630 | 48096552 | 2923 | 14 | 2.38E-11 | 2.38E-12 | 0.000371 | 2.48E-14 | -0.00159978 | MED4 |
| chr17 | 81858628 | 81861714 | 3087 | 22 | 1.53E-09 | 3.76E-12 | 3.26E-05 | 2.48E-14 | -0.00160398 | P4HB |
| chr17 | 49131153 | 49133460 | 2308 | 19 | 2.99E-14 | 2.96E-11 | 3.89E-06 | 2.51E-14 | -0.00506605 | B4GALNT2 |
| chr10 | 99367335 | 99368265 | 931 | 4 | 6.19E-14 | 5.53E-15 | 6.78E-06 | 2.52E-14 | -0.00494151 | CNNM1 |
| chr17 | 1604349 | 1605953 | 1605 | 11 | 3.42E-15 | 3.52E-14 | 0.000938 | 2.53E-14 | -0.04311186 | SLC43A2, RN7SL105P |
| chr6 | 38639469 | 38641592 | 2124 | 16 | 4.33E-15 | 1.41E-09 | 2.44E-05 | 2.58E-14 | -0.00115171 | BTBD9 |
| chr22 | 42292542 | 42293372 | 831 | 4 | 6.30E-16 | 1.55E-14 | 4.44E-07 | 2.58E-14 | 0.001408179 | TCF20 |
| chr3 | 66404963 | 66408546 | 3584 | 11 | 1.79E-10 | 5.47E-09 | 4.78E-07 | 2.62E-14 | -0.00169093 | LRIG1 |
| chr12 | 109832796 | 109833919 | 1124 | 12 | 2.29E-13 | 1.28E-10 | 1.18E-05 | 2.63E-14 | -0.00934881 | MIR4497, TRPV4 |
| chr11 | 67373608 | 67375591 | 1984 | 15 | 1.70E-11 | 9.40E-13 | 8.48E-06 | 2.68E-14 | -0.00319377 | RAD9A, AP003419.3, CLCF1 |
| chr1 | 2103648 | 2106037 | 2390 | 10 | 3.00E-12 | 1.43E-14 | 2.19E-05 | 2.69E-14 | -0.00632142 | PRKCZ |
| chr9 | 127494062 | 127495380 | 1319 | 7 | 9.42E-17 | 3.39E-13 | 1.28E-07 | 2.70E-14 | -0.00406435 | LRSAM1 |
| chr12 | 122020699 | 122022567 | 1869 | 8 | 2.47E-15 | 2.57E-11 | 1.20E-05 | 2.77E-14 | -0.00588619 | BCL7A |
| chr17 | 75632450 | 75633430 | 981 | 10 | 6.26E-14 | 4.82E-13 | 0.000171 | 2.79E-14 | -0.00412742 | RECQL5 |
| chr16 | 915780 | 919468 | 3689 | 18 | 1.09E-09 | 9.29E-11 | 0.000183 | 2.81E-14 | -0.0027638 | LMF1 |
| chr3 | 169148737 | 169149686 | 950 | 10 | 6.32E-20 | 1.00E-09 | 6.83E-07 | 2.82E-14 | -0.00223535 | MECOM |
| chr17 | 54898795 | 54900099 | 1305 | 10 | 2.44E-17 | 3.03E-12 | 2.80E-05 | 2.92E-14 | 0.008316967 | TOM1L1 |
| chr2 | 74130400 | 74131096 | 697 | 4 | 3.98E-15 | 2.99E-15 | 2.75E-05 | 2.92E-14 | -0.07687201 | AC073263.1 |
| chr14 | 38209332 | 38211650 | 2319 | 10 | 6.46E-11 | 3.43E-11 | 8.01E-06 | 2.92E-14 | -0.0078771 | SSTR1 |
| chr5 | 142324381 | 142326648 | 2268 | 20 | 5.47E-11 | 9.30E-13 | 0.000239 | 2.93E-14 | -0.00168368 | SPRY4-AS1, SPRY4 |
| chr3 | 50503580 | 50505535 | 1956 | 15 | 9.69E-16 | 2.73E-12 | 5.90E-05 | 2.96E-14 | -0.00254491 | CACNA2D2 |
| chr1 | 149898874 | 149900612 | 1739 | 18 | 2.58E-13 | 2.81E-11 | 1.94E-05 | 3.05E-14 | -0.00119093 | BOLA1 |
| chr14 | 74347149 | 74349883 | 2735 | 12 | 1.83E-12 | 2.39E-12 | 2.17E-05 | 3.08E-14 | -0.00654356 | VRTN |
| chr17 | 76353893 | 76354961 | 1069 | 11 | 6.63E-25 | 6.76E-07 | 1.64E-06 | 3.08E-14 | -0.00639365 | PRPSAP1 |
| chr11 | 65546137 | 65547442 | 1306 | 8 | 5.63E-17 | 3.39E-13 | 2.82E-05 | 3.11E-14 | -0.01292092 | LTBP3 |
| chr16 | 12901141 | 12903827 | 2687 | 20 | 1.87E-11 | 9.71E-11 | 7.18E-06 | 3.17E-14 | -0.00100285 | SHISA9 |
| chr1 | 36099580 | 36100932 | 1353 | 8 | 6.54E-18 | 5.94E-11 | 3.55E-07 | 3.18E-14 | -0.01453328 | COL8A2 |
| chr11 | 645870 | 646580 | 711 | 4 | 7.36E-18 | 4.26E-09 | 2.82E-08 | 3.18E-14 | 0.026596523 | DEAF1 |
| chr19 | 33130739 | 33132629 | 1891 | 13 | 1.37E-13 | 1.45E-11 | 0.000111 | 3.25E-14 | -0.0043847 | WDR88 |
| chr3 | 142888438 | 142890056 | 1619 | 17 | 2.48E-15 | 1.97E-10 | 4.71E-05 | 3.26E-14 | -0.00826769 | PCOLCE2 |
| chr4 | 55553943 | 55554949 | 1007 | 3 | 1.77E-14 | 2.49E-14 | 1.02E-07 | 3.29E-14 | -0.02454861 | NA |
| chr13 | 70961041 | 70961065 | 25 | 2 | 6.70E-16 | 2.14E-14 | 3.61E-08 | 3.29E-14 | 0.064279731 | NA |
| chr5 | 140699508 | 140700990 | 1483 | 15 | 5.18E-14 | 3.89E-12 | 3.72E-06 | 3.36E-14 | -0.00472056 | ZMAT2 |
| chr3 | 50313317 | 50313333 | 17 | 3 | 2.33E-16 | 1.44E-11 | 5.39E-08 | 3.42E-14 | -0.00866908 | NA |
| chr7 | 6526260 | 6529667 | 3408 | 8 | 6.37E-10 | 1.31E-08 | 2.57E-06 | 3.42E-14 | -0.0120228 | GRID2IP |
| chr1 | 44723354 | 44725438 | 2085 | 13 | 5.68E-14 | 2.16E-11 | 3.81E-05 | 3.44E-14 | -0.00554308 | ARMH1 |
| chr12 | 56993035 | 56994657 | 1623 | 11 | 1.52E-12 | 3.87E-13 | 1.27E-05 | 3.53E-14 | 0.005675454 | GPR182 |
| chr17 | 80880166 | 80880860 | 695 | 7 | 6.89E-15 | 1.16E-14 | 6.69E-05 | 3.54E-14 | -0.01329703 | RPTOR |
| chr5 | 16615815 | 16617952 | 2138 | 17 | 9.05E-15 | 6.98E-09 | 5.10E-05 | 3.56E-14 | 0.006680262 | AC024588.1, AC022113.1, RETREG1 |
| chr5 | 33937077 | 33938804 | 1728 | 6 | 3.85E-13 | 5.84E-14 | 1.06E-05 | 3.62E-14 | -0.0341107 | RXFP3 |
| chr5 | 157142380 | 157144034 | 1655 | 15 | 4.89E-15 | 1.73E-10 | 2.45E-06 | 3.66E-14 | -0.00059594 | ITK, HAVCR2, MED7 |
| chr11 | 65857302 | 65861767 | 4466 | 38 | 7.08E-10 | 2.67E-10 | 0.000186 | 3.73E-14 | 0.000585033 | MUS81, CFL1 |
| chr1 | 50967639 | 50970458 | 2820 | 21 | 1.25E-09 | 1.41E-13 | 0.000185 | 3.73E-14 | -0.00063344 | CDKN2C |
| chr2 | 200305080 | 200308031 | 2952 | 23 | 2.59E-10 | 2.90E-10 | 0.000197 | 3.86E-14 | -0.00013647 | SPATS2L |
| chr17 | 80443989 | 80444626 | 638 | 6 | 1.32E-14 | 3.15E-14 | 1.97E-05 | 3.97E-14 | -0.02758188 | NA |
| chr1 | 204152311 | 204153280 | 970 | 9 | 1.65E-18 | 1.17E-11 | 3.02E-08 | 3.98E-14 | -0.00626256 | NA |
| chr13 | 106374185 | 106376670 | 2486 | 12 | 9.23E-17 | 1.47E-11 | 2.38E-07 | 3.99E-14 | -0.00288343 | LINC00460 |
| chr10 | 126302011 | 126302907 | 897 | 3 | 1.84E-17 | 8.84E-13 | 6.46E-08 | 4.11E-14 | -0.02442195 | ADAM12 |
| chr2 | 234289059 | 234290764 | 1706 | 5 | 2.76E-11 | 1.68E-13 | 6.47E-07 | 4.11E-14 | -0.00645939 | NA |
| chr16 | 67942461 | 67945226 | 2766 | 21 | 3.27E-12 | 5.65E-10 | 2.12E-05 | 4.19E-14 | -0.00107794 | LCAT, SLC12A4 |
| chr19 | 35992832 | 35995070 | 2239 | 10 | 3.02E-15 | 4.17E-13 | 0.000163 | 4.22E-14 | -0.01733287 | NA |
| chr12 | 52246326 | 52249955 | 3630 | 12 | 2.83E-09 | 5.44E-13 | 8.51E-05 | 4.27E-14 | -0.00589677 | KRT7, KRT86, KRT7-AS |
| chr3 | 57163392 | 57166464 | 3073 | 15 | 1.62E-12 | 7.43E-11 | 7.89E-06 | 4.27E-14 | 0.002264529 | IL17RD |
| chr12 | 132506516 | 132507007 | 492 | 5 | 6.54E-16 | 1.70E-13 | 1.23E-05 | 4.40E-14 | 0.016083619 | FBRSL1 |
| chr7 | 100602141 | 100604890 | 2750 | 18 | 9.91E-11 | 1.82E-10 | 1.91E-05 | 4.40E-14 | -0.00338889 | PCOLCE, PCOLCE-AS1 |
| chr6 | 30570402 | 30574416 | 4015 | 38 | 9.54E-11 | 1.32E-09 | 4.94E-05 | 4.41E-14 | -0.00119182 | ABCF1 |
| chr1 | 34780091 | 34781895 | 1805 | 11 | 1.40E-17 | 3.06E-11 | 3.52E-06 | 4.53E-14 | -0.00805461 | GJB3, SMIM12, AL121988.1 |
| chr13 | 50132447 | 50133907 | 1461 | 12 | 1.04E-16 | 5.62E-10 | 7.52E-06 | 4.54E-14 | -0.00642436 | DLEU1 |
| chr20 | 59190260 | 59191841 | 1582 | 13 | 1.06E-19 | 2.72E-10 | 5.54E-09 | 4.64E-14 | -0.00627487 | ZNF831 |

| | | | | | | | | | | |
|---|---|---|---|---|---|---|---|---|---|---|
| chr17 | 28717151 | 28718876 | 1726 | 21 | 6.88E-13 | 1.37E-12 | 5.01E-05 | 4.64E-14 | -0.00402515 | RAB34 |
| chr6 | 26195260 | 26196843 | 1584 | 7 | 3.16E-14 | 9.42E-14 | 1.91E-05 | 4.68E-14 | 0.016386168 | H1-12P, AL031777.2, H3C4 |
| chr16 | 3364173 | 3366070 | 1898 | 10 | 1.20E-14 | 2.67E-12 | 6.68E-07 | 4.68E-14 | -0.01394561 | AC025283.2 |
| chr3 | 46462979 | 46465756 | 2778 | 19 | 4.12E-14 | 4.52E-12 | 3.91E-05 | 4.68E-14 | -0.00446678 | LTF |
| chr12 | 95857770 | 95859541 | 1772 | 19 | 3.20E-17 | 1.24E-08 | 1.11E-05 | 4.83E-14 | -0.00452113 | SNRPF, AC090001.1 |
| chr1 | 20138516 | 20139921 | 1406 | 7 | 7.94E-14 | 3.17E-09 | 3.59E-08 | 4.99E-14 | -0.00631624 | PLA2G2F |
| chr11 | 118358921 | 118360007 | 1087 | 13 | 1.16E-16 | 7.90E-11 | 7.85E-09 | 5.01E-14 | -0.00436235 | UBE4A |
| chr18 | 79432641 | 79434663 | 2023 | 7 | 1.51E-11 | 4.89E-13 | 1.23E-05 | 5.03E-14 | 0.002208838 | NFATC1 |
| chr1 | 63011751 | 63012943 | 1193 | 7 | 7.16E-17 | 3.64E-11 | 4.75E-06 | 5.03E-14 | -0.00829439 | AL162400.3, LINC01739 |
| chr6 | 33898332 | 33900661 | 2330 | 19 | 3.07E-09 | 1.02E-12 | 0.000877 | 5.04E-14 | -0.0020915 | AL138889.1, MIR7159 |
| chr3 | 113210825 | 113211935 | 1111 | 7 | 2.53E-20 | 5.26E-11 | 1.04E-07 | 5.06E-14 | -0.00759021 | BOC |
| chr7 | 149872375 | 149873842 | 1468 | 14 | 4.12E-16 | 6.40E-12 | 3.78E-05 | 5.32E-14 | -0.00519159 | ATP6V0E2, ATP6V0E2-AS1 |
| chr19 | 50047027 | 50047235 | 209 | 4 | 5.77E-15 | 6.50E-14 | 6.29E-06 | 5.35E-14 | -0.01774098 | ZNF473, AC010624.3 |
| chr20 | 3795836 | 3800004 | 4169 | 18 | 8.70E-14 | 5.85E-09 | 9.17E-07 | 5.35E-14 | -0.00337606 | CDC25B |
| chr6 | 32843718 | 32847314 | 3597 | 56 | 3.30E-08 | 1.20E-11 | 0.001831 | 5.39E-14 | 0.000505288 | PSMB8-AS1, PSMB9, PSMB8, TAP1 |
| chr10 | 133161373 | 133162537 | 1165 | 6 | 2.72E-16 | 3.65E-12 | 1.09E-06 | 5.43E-14 | -0.01009575 | KNDC1 |
| chr19 | 8208621 | 8210006 | 1386 | 16 | 1.02E-14 | 1.60E-12 | 0.000137 | 5.43E-14 | -0.00369756 | CERS4 |
| chr9 | 136531130 | 136532719 | 1590 | 5 | 2.01E-10 | 7.27E-15 | 2.56E-05 | 5.43E-14 | -0.02950597 | NOTCH1 |
| chr22 | 41799059 | 41801034 | 1976 | 15 | 7.46E-16 | 1.99E-09 | 1.76E-05 | 5.48E-14 | 0.00632047 | MEI1, CCDC134 |
| chr3 | 52703562 | 52706970 | 3409 | 29 | 5.14E-10 | 1.93E-10 | 0.000485 | 5.50E-14 | 0.000509058 | SPCS1, GLT8D1 |
| chr15 | 88610436 | 88610965 | 530 | 4 | 6.18E-15 | 1.12E-14 | 1.50E-05 | 5.56E-14 | -0.02558722 | NA |
| chr11 | 111298320 | 111300605 | 2286 | 27 | 9.27E-15 | 5.95E-11 | 9.30E-05 | 5.57E-14 | -0.00033423 | COLCA2, COLCA1 |
| chr8 | 33812839 | 33813101 | 263 | 2 | 1.74E-15 | 2.85E-14 | 1.79E-07 | 5.61E-14 | -0.06248435 | AF279873.3 |
| chr4 | 179465442 | 179466714 | 1273 | 6 | 2.82E-15 | 2.10E-11 | 6.74E-07 | 5.61E-14 | -0.00393341 | NA |
| chr16 | 66999511 | 67001485 | 1975 | 13 | 3.81E-14 | 7.03E-12 | 7.43E-05 | 5.73E-14 | -0.005025 | CES4A |
| chr14 | 23305262 | 23307772 | 2511 | 11 | 1.89E-14 | 8.16E-11 | 1.04E-07 | 5.84E-14 | -0.00221662 | BCL2L2, BCL2L2-PABPN1 |
| chr9 | 110427704 | 110428509 | 806 | 3 | 2.32E-13 | 1.87E-14 | 1.05E-06 | 5.89E-14 | -0.02299196 | SVEP1 |
| chr6 | 30672377 | 30673443 | 1067 | 35 | 1.17E-14 | 2.00E-10 | 1.09E-05 | 5.90E-14 | -0.00073471 | DHX16 |
| chr12 | 132570439 | 132572434 | 1996 | 9 | 9.51E-12 | 1.41E-13 | 0.000158 | 5.90E-14 | -0.0016809 | FBRSL1 |
| chr11 | 74310778 | 74312174 | 1397 | 15 | 2.90E-16 | 1.42E-10 | 7.64E-05 | 5.99E-14 | -0.0072537 | P4HA3-AS1, P4HA3 |
| chr6 | 33390634 | 33393325 | 2692 | 46 | 3.10E-12 | 2.51E-10 | 0.00017 | 5.99E-14 | -0.00063543 | KIFC1 |
| chr8 | 6561457 | 6563702 | 2246 | 15 | 6.32E-09 | 6.52E-15 | 0.000586 | 5.99E-14 | -0.01764319 | MCPH1, ANGPT2 |
| chr19 | 17264135 | 17266203 | 2069 | 10 | 4.23E-14 | 2.32E-12 | 4.01E-05 | 6.25E-14 | -0.00795636 | USHBP1 |
| chr5 | 172452606 | 172455236 | 2631 | 13 | 2.22E-09 | 1.61E-12 | 7.37E-05 | 6.30E-14 | -0.00271207 | SH3PXD2B |
| chr2 | 85668878 | 85669882 | 1005 | 5 | 1.46E-15 | 6.50E-14 | 2.17E-06 | 6.37E-14 | 0.008394027 | NA |
| chr2 | 98311901 | 98312435 | 535 | 5 | 1.54E-14 | 7.56E-15 | 1.41E-05 | 6.41E-14 | -0.03223278 | VWA3B |
| chr4 | 1244207 | 1246476 | 2270 | 7 | 1.30E-09 | 4.61E-14 | 4.35E-05 | 6.46E-14 | 0.007016222 | CTBP1 |
| chr9 | 95459306 | 95460889 | 1584 | 6 | 2.54E-13 | 5.45E-14 | 4.12E-05 | 6.55E-14 | 0.018791072 | PTCH1 |
| chr2 | 74548302 | 74550282 | 1981 | 21 | 2.45E-13 | 7.88E-11 | 1.00E-05 | 6.59E-14 | -0.0034687 | DOK1, LOXL3 |
| chr1 | 6496188 | 6498053 | 1866 | 14 | 2.34E-12 | 6.35E-12 | 0.000316 | 6.59E-14 | -0.00301309 | PLEKHG5 |
| chr19 | 51390550 | 51391645 | 1096 | 8 | 4.43E-12 | 1.05E-13 | 8.08E-06 | 6.59E-14 | -0.00488812 | C19orf84 |
| chr4 | 1347043 | 1349912 | 2870 | 22 | 4.66E-14 | 4.29E-11 | 0.000145 | 6.59E-14 | -0.00244456 | UVSSA |
| chr7 | 100872762 | 100875433 | 2672 | 15 | 4.04E-16 | 1.74E-08 | 8.06E-06 | 6.66E-14 | -0.01428362 | TRIP6, SRRT |
| chr12 | 53750701 | 53753103 | 2403 | 13 | 2.74E-09 | 1.45E-11 | 7.05E-06 | 6.82E-14 | 0.002619655 | AC076968.2, CISTR |
| chr16 | 11254576 | 11257058 | 2483 | 22 | 2.55E-10 | 5.04E-15 | 0.000792 | 6.89E-14 | -0.00307184 | RMI2, SOCS1 |
| chr8 | 142785313 | 142787815 | 2503 | 19 | 5.58E-12 | 6.80E-11 | 7.09E-05 | 7.03E-14 | -0.00540184 | AC083841.2, LY6D |
| chr1 | 7561678 | 7563027 | 1350 | 6 | 5.88E-14 | 1.04E-12 | 4.49E-07 | 7.04E-14 | -0.00385369 | CAMTA1 |
| chr6 | 41784484 | 41787348 | 2865 | 15 | 1.86E-13 | 1.61E-09 | 2.42E-05 | 7.04E-14 | -0.00527051 | AL365205.1, PRICKLE4, FRS3 |
| chr6 | 169789683 | 169790986 | 1304 | 15 | 2.63E-13 | 6.12E-09 | 4.38E-05 | 7.06E-14 | 0.003499611 | LINC00574, LINC00242 |
| chr12 | 49292836 | 49294533 | 1698 | 10 | 2.62E-13 | 2.24E-13 | 7.22E-06 | 7.30E-14 | -0.00624208 | PRPH, AC125611.4 |
| chr16 | 67246064 | 67249335 | 3272 | 25 | 3.11E-09 | 1.88E-13 | 0.001346 | 7.30E-14 | 0.000350853 | SLC9A5, FHOD1 |
| chr12 | 54369297 | 54370587 | 1291 | 6 | 1.71E-11 | 4.01E-14 | 3.48E-05 | 7.33E-14 | -0.02831291 | AC079313.2, AC079313.1, ZNF385A |
| chr12 | 92143649 | 92147000 | 3352 | 23 | 7.43E-08 | 9.14E-13 | 6.93E-05 | 7.35E-14 | 0.003505078 | AC025164.1, BTG1 |
| chr21 | 28745038 | 28746339 | 1302 | 6 | 2.05E-14 | 2.27E-14 | 6.59E-05 | 7.63E-14 | 0.008763784 | NA |
| chr15 | 90929829 | 90930890 | 1062 | 12 | 1.38E-14 | 6.25E-14 | 0.000946 | 7.84E-14 | -0.00932638 | UNC45A, HDDC3 |
| chr7 | 368660 | 371153 | 2494 | 17 | 1.45E-12 | 2.84E-12 | 1.43E-05 | 8.12E-14 | 0.000320393 | NA |
| chr8 | 141253368 | 141255147 | 1780 | 16 | 1.35E-12 | 6.86E-11 | 8.28E-05 | 8.15E-14 | -0.00025869 | AC011676.2, SLC45A4 |
| chr1 | 6205011 | 6206693 | 1683 | 14 | 4.37E-17 | 3.46E-08 | 8.99E-06 | 8.20E-14 | -0.00083453 | RNF207, RPL22, RNF207-AS1 |
| chr5 | 54985370 | 54986506 | 1137 | 11 | 2.55E-15 | 3.22E-13 | 8.91E-05 | 8.20E-14 | -0.01541356 | ESM1 |
| chr7 | 150339955 | 150342390 | 2436 | 9 | 6.41E-09 | 2.26E-12 | 1.42E-05 | 8.20E-14 | 0.014224206 | AC005586.1, RARRES2 |
| chr2 | 25167555 | 25170072 | 2518 | 20 | 1.89E-11 | 1.42E-08 | 5.03E-05 | 8.38E-14 | -0.00355642 | POMC |
| chr9 | 127937606 | 127939233 | 1628 | 14 | 6.38E-16 | 1.20E-09 | 2.53E-05 | 8.47E-14 | -0.00572196 | AL157935.1, DPM2 |
| chr16 | 2963226 | 2965091 | 1866 | 17 | 2.07E-13 | 2.89E-14 | 0.001303 | 8.56E-14 | -0.00050854 | KREMEN2 |
| chr16 | 68235138 | 68236595 | 1458 | 13 | 3.31E-19 | 8.65E-11 | 1.78E-05 | 8.65E-14 | -0.00613772 | ESRP2 |
| chr10 | 71214008 | 71214937 | 930 | 4 | 2.37E-16 | 9.65E-13 | 9.13E-08 | 8.67E-14 | -0.01240967 | UNC5B |
| chr18 | 62329863 | 62330774 | 912 | 6 | 5.91E-15 | 2.25E-13 | 0.000103 | 8.69E-14 | 0.024551797 | TNFRSF11A |
| chr19 | 44071394 | 44072721 | 1328 | 14 | 1.05E-14 | 1.72E-10 | 3.44E-06 | 8.91E-14 | -0.00391209 | AC067968.1, ZNF284 |
| chr4 | 175812258 | 175814309 | 2052 | 13 | 7.56E-16 | 3.51E-10 | 7.75E-06 | 8.92E-14 | -0.01146152 | GPM6A |
| chr4 | 1247939 | 1250716 | 2778 | 18 | 2.64E-12 | 9.74E-14 | 0.000916 | 9.10E-14 | -0.0086303 | CTBP1-DT, CTBP1 |
| chr7 | 27241295 | 27243994 | 2700 | 35 | 1.40E-11 | 8.84E-11 | 0.000451 | 9.10E-14 | -0.00173201 | EVX1, EVX1-AS |
| chr7 | 99010522 | 99012217 | 1696 | 11 | 9.09E-11 | 1.81E-11 | 9.69E-05 | 9.37E-14 | 0.000504293 | TRRAP |
| chr3 | 186193419 | 186194697 | 1279 | 11 | 1.35E-15 | 3.10E-12 | 6.11E-06 | 9.48E-14 | 0.010529043 | DGKG |
| chr16 | 88921766 | 88924338 | 2573 | 12 | 5.84E-13 | 1.78E-11 | 4.60E-05 | 9.60E-14 | -0.00324024 | CBFA2T3 |
| chr6 | 31196922 | 31198056 | 1135 | 13 | 1.98E-15 | 3.87E-11 | 0.000159 | 9.70E-14 | -0.00755565 | HCG27, AL662844.4 |
| chr22 | 41832720 | 41834895 | 2176 | 16 | 3.41E-13 | 3.65E-08 | 4.70E-06 | 9.79E-14 | -0.00476684 | SREBF2, SREBF2-AS1 |
| chr3 | 128993332 | 128995301 | 1970 | 15 | 1.45E-11 | 5.95E-11 | 3.46E-05 | 9.79E-14 | -0.00252973 | CFAP92 |
| chr12 | 122895900 | 122897443 | 1544 | 14 | 1.25E-16 | 3.30E-07 | 6.56E-07 | 9.83E-14 | -0.00534749 | VPS37B |
| chr7 | 74082405 | 74084448 | 2044 | 16 | 4.51E-16 | 1.24E-10 | 0.000175 | 9.91E-14 | -0.00211101 | LIMK1 |
| chr1 | 1923034 | 1924634 | 1601 | 5 | 1.65E-12 | 1.02E-12 | 7.79E-06 | 1.02E-13 | 0.01108816 | CFAP74 |
| chr10 | 87862227 | 87863603 | 1377 | 45 | 6.92E-19 | 5.26E-08 | 4.66E-05 | 1.02E-13 | 0.0005171 | KLLN |
| chr19 | 55703738 | 55704576 | 839 | 5 | 5.91E-14 | 1.39E-14 | 1.14E-05 | 1.07E-13 | -0.0265557 | EPN1 |
| chr1 | 162411991 | 162413210 | 1220 | 5 | 2.22E-14 | 6.39E-12 | 5.14E-06 | 1.08E-13 | -0.06853906 | SH2D1B |
| chr7 | 13987945 | 13990057 | 2113 | 21 | 8.67E-18 | 7.72E-10 | 9.53E-06 | 1.10E-13 | -0.00405305 | ETV1 |
| chr20 | 25081618 | 25084571 | 2954 | 21 | 8.80E-10 | 2.11E-12 | 0.000382 | 1.10E-13 | 0.002845647 | VSX1 |
| chr11 | 18411953 | 18412807 | 855 | 9 | 1.87E-13 | 1.55E-13 | 0.000445 | 1.11E-13 | -0.06845987 | LDHC |
| chr1 | 111270523 | 111271586 | 1064 | 9 | 1.91E-16 | 3.40E-10 | 2.40E-07 | 1.11E-13 | -0.01007804 | NA |
| chr16 | 49698313 | 49699466 | 1154 | 6 | 1.18E-15 | 7.62E-11 | 4.95E-05 | 1.12E-13 | -0.03031833 | ZNF423 |
| chr2 | 106065375 | 106066953 | 1579 | 10 | 2.92E-17 | 1.46E-11 | 4.70E-05 | 1.12E-13 | -0.00978717 | ECRG4 |
| chr11 | 6603179 | 6604515 | 1337 | 18 | 1.03E-12 | 7.07E-12 | 1.66E-05 | 1.14E-13 | -0.00070588 | ILK, RRP8, AC091564.2 |
| chr3 | 27712148 | 27715170 | 3023 | 19 | 1.67E-11 | 4.31E-07 | 1.96E-07 | 1.16E-13 | 4.97E-05 | LINC02084 |
| chr1 | 23018695 | 23020919 | 2225 | 17 | 1.44E-15 | 1.18E-09 | 1.37E-06 | 1.17E-13 | -0.00069699 | KDM1A, AL031428.1 |
| chr6 | 31942839 | 31946528 | 3690 | 19 | 6.31E-09 | 3.13E-10 | 4.86E-05 | 1.19E-13 | -0.00290611 | C2, AL645922.1, CFB |
| chr17 | 81402736 | 81405865 | 3130 | 11 | 2.74E-09 | 4.11E-15 | 0.000559 | 1.20E-13 | -0.00812994 | BAHCC1 |
| chr11 | 116832254 | 116832872 | 619 | 4 | 3.55E-15 | 7.50E-13 | 1.59E-07 | 1.21E-13 | -0.005557 | APOC3 |
| chr9 | 109329407 | 109329964 | 558 | 4 | 1.32E-16 | 3.69E-12 | 2.05E-07 | 1.22E-13 | -0.01274801 | NA |
| chr12 | 119333979 | 119335037 | 1059 | 11 | 7.66E-16 | 1.11E-10 | 3.47E-06 | 1.24E-13 | -0.0163252 | CCDC60 |
| chr1 | 20494061 | 20496623 | 2563 | 18 | 8.82E-11 | 1.32E-10 | 6.15E-05 | 1.24E-13 | -0.00317391 | NA |
| chr1 | 2389925 | 2392957 | 3033 | 18 | 2.33E-07 | 2.46E-15 | 0.000475 | 1.24E-13 | 0.001139454 | RER1, MORN1 |
| chr6 | 28258316 | 28259704 | 1389 | 14 | 1.00E-13 | 1.87E-14 | 0.000625 | 1.25E-13 | -0.01318544 | NKAPL |
| chr19 | 49909559 | 49909826 | 268 | 7 | 4.81E-16 | 2.32E-10 | 2.97E-08 | 1.25E-13 | -0.00311616 | IL4I1, NUP62, AC011452.1 |
| chr13 | 42039308 | 42041855 | 2548 | 15 | 1.80E-11 | 2.14E-10 | 5.39E-05 | 1.25E-13 | 0.006151301 | DGKH |
| chr16 | 87789024 | 87790733 | 1710 | 6 | 4.30E-12 | 1.84E-12 | 1.60E-05 | 1.26E-13 | -0.00301737 | AC126696.1 |
| chr17 | 49994321 | 49996394 | 2074 | 16 | 1.85E-15 | 8.00E-09 | 2.56E-06 | 1.28E-13 | -0.0033966 | DLX3 |
| chr1 | 1939056 | 1940594 | 1539 | 7 | 7.46E-17 | 5.59E-11 | 6.60E-07 | 1.31E-13 | -0.00450688 | CFAP74 |
| chr20 | 46200963 | 46201801 | 839 | 4 | 1.69E-13 | 1.61E-14 | 2.50E-05 | 1.33E-13 | -0.02568981 | CDH22 |
| chr3 | 156606163 | 156606944 | 782 | 3 | 1.57E-12 | 3.21E-14 | 5.79E-06 | 1.33E-13 | -0.02747722 | NA |
| chr12 | 85836026 | 85837299 | 1274 | 10 | 6.95E-15 | 6.01E-12 | 0.000146 | 1.34E-13 | -0.01722859 | RASSF9 |
| chr12 | 53049273 | 53050178 | 906 | 8 | 1.26E-16 | 1.17E-11 | 2.34E-05 | 1.35E-13 | -0.01021516 | TNS2, AC068888.1 |
| chr1 | 156675764 | 156678314 | 2551 | 15 | 1.32E-09 | 4.37E-13 | 0.000639 | 1.36E-13 | -0.00707151 | NES |
| chr4 | 105144698 | 105146875 | 2178 | 20 | 9.57E-13 | 3.59E-09 | 8.00E-05 | 1.40E-13 | 0.001678845 | TET2 |
| chr6 | 126339099 | 126340482 | 1384 | 13 | 2.27E-15 | 2.18E-11 | 1.18E-05 | 1.43E-13 | -0.00339502 | CENPW, AL356534.1 |
| chr9 | 129487635 | 129489619 | 1985 | 12 | 2.20E-12 | 1.25E-12 | 6.01E-05 | 1.43E-13 | -0.0027296 | LINC00963 |
| chr8 | 144412141 | 144414496 | 2356 | 11 | 4.27E-12 | 6.15E-14 | 7.46E-05 | 1.44E-13 | -0.01369119 | SLC39A4 |
| chr17 | 82583673 | 82584843 | 1171 | 10 | 1.27E-15 | 1.59E-12 | 2.09E-05 | 1.46E-13 | 0.011057736 | FOXK2 |
| chr5 | 1727289 | 1730028 | 2740 | 9 | 1.05E-08 | 1.88E-13 | 3.74E-05 | 1.46E-13 | -0.00165466 | AC112176.1 |
| chr17 | 4489137 | 4490129 | 993 | 4 | 2.78E-15 | 2.29E-13 | 1.12E-05 | 1.48E-13 | -0.05156437 | NA |
| chr20 | 2655370 | 2656857 | 1488 | 14 | 3.41E-11 | 1.02E-12 | 0.000154 | 1.49E-13 | -0.00061603 | NOP56, SNORD86, SNORD56 |
| chr11 | 46295128 | 46296976 | 1849 | 8 | 1.80E-13 | 2.56E-11 | 1.05E-05 | 1.49E-13 | -0.01046921 | CREB3L1 |
| chr5 | 43037021 | 43037980 | 960 | 9 | 2.07E-14 | 7.24E-14 | 0.00017 | 1.49E-13 | -0.01924664 | AC025171.1 |
| chr11 | 69012085 | 69013889 | 1805 | 15 | 9.05E-15 | 9.94E-13 | 0.000453 | 1.49E-13 | -0.00512728 | MRGPRF-AS1, MRGPRF |
| chr5 | 138257276 | 138257894 | 619 | 6 | 1.08E-15 | 4.72E-11 | 2.31E-07 | 1.51E-13 | -0.00368418 | GFRA3 |
| chr9 | 19047932 | 19050397 | 2466 | 17 | 9.88E-13 | 3.38E-08 | 4.58E-06 | 1.56E-13 | -0.00218026 | RRAGA, SAXO1 |
| chr22 | 30564870 | 30566339 | 1470 | 8 | 1.29E-11 | 3.25E-11 | 2.08E-06 | 1.56E-13 | -0.0062343 | GAL3ST1 |
| chr2 | 66702803 | 66704643 | 1841 | 7 | 1.84E-17 | 1.17E-08 | 8.24E-07 | 1.57E-13 | -0.00921487 | LINC01798, LINC01797 |
| chr13 | 113842669 | 113844288 | 1620 | 11 | 2.09E-15 | 2.74E-11 | 5.95E-06 | 1.57E-13 | 0.005224564 | GAS6-AS1, GAS6 |
| chr19 | 29617061 | 29618180 | 1120 | 5 | 1.78E-16 | 1.24E-09 | 3.15E-07 | 1.57E-13 | -0.00705353 | POP4 |
| chr2 | 27078327 | 27079075 | 749 | 9 | 2.49E-13 | 6.57E-15 | 0.000597 | 1.57E-13 | -0.01788058 | EMILIN1 |
| chr10 | 104313648 | 104315310 | 1663 | 7 | 5.50E-16 | 3.61E-13 | 5.21E-06 | 1.58E-13 | -0.00516582 | AL162742.1, ITPRIP |
| chr4 | 74374716 | 74376001 | 1286 | 8 | 3.68E-16 | 9.19E-10 | 9.63E-07 | 1.58E-13 | -0.00078388 | EREG |
| chr20 | 1993823 | 1994800 | 978 | 12 | 1.76E-17 | 7.28E-08 | 5.38E-08 | 1.59E-13 | -0.00860209 | PDYN-AS1, PDYN |
| chr16 | 11500701 | 11502061 | 1361 | 5 | 6.05E-14 | 5.44E-12 | 2.25E-05 | 1.61E-13 | 0.019966396 | AC099489.1 |
| chr4 | 681348 | 683849 | 2502 | 7 | 2.83E-10 | 6.53E-14 | 3.50E-05 | 1.63E-13 | 0.013941298 | MYL5, SLC49A3 |
| chr17 | 18145878 | 18147775 | 1898 | 5 | 4.65E-14 | 1.90E-12 | 7.45E-07 | 1.65E-13 | -0.00619621 | MYO15A |
| chr12 | 48334626 | 48335028 | 403 | 3 | 1.05E-15 | 1.30E-12 | 4.19E-07 | 1.69E-13 | -0.02295049 | NA |
| chr16 | 50740748 | 50742191 | 1444 | 13 | 2.49E-16 | 1.38E-10 | 2.31E-06 | 1.70E-13 | -0.00361532 | CYLD, AC007728.2 |
| chr6 | 122778389 | 122780292 | 1904 | 10 | 4.39E-14 | 4.97E-14 | 0.000369 | 1.71E-13 | -0.01307831 | FABP7 |
| chr19 | 12056304 | 12057106 | 803 | 5 | 1.90E-16 | 1.93E-09 | 1.89E-07 | 1.78E-13 | -0.01524928 | AC008770.4 |
| chr16 | 69564022 | 69567395 | 3374 | 21 | 9.87E-09 | 1.53E-12 | 0.000624 | 1.78E-13 | 0.002194322 | NFAT5, MIR1538 |
| chr17 | 80844641 | 80849123 | 4483 | 20 | 7.60E-07 | 2.79E-12 | 0.000604 | 1.79E-13 | -0.00413667 | RPTOR |
| chr11 | 62689467 | 62691293 | 1827 | 12 | 2.75E-13 | 4.24E-11 | 5.97E-05 | 1.80E-13 | -0.00123766 | LRRN4CL, BSCL2, HNRNPUL2-BSCL2 |
| chr19 | 11577838 | 11579182 | 1345 | 14 | 2.02E-14 | 6.43E-13 | 5.83E-05 | 1.81E-13 | 0.003446886 | ZNF627, ACP5 |
| chr3 | 38305034 | 38305950 | 917 | 6 | 8.77E-15 | 1.02E-11 | 9.99E-06 | 1.82E-13 | -0.00509706 | SLC22A14 |
| chr12 | 49972044 | 49973508 | 1465 | 10 | 4.52E-15 | 1.41E-12 | 1.89E-05 | 1.84E-13 | -0.0039952 | AQP6 |
| chr15 | 71808335 | 71812068 | 3734 | 18 | 9.63E-12 | 8.00E-09 | 3.57E-05 | 1.84E-13 | -0.00196993 | NR2E3 |
| chr1 | 1574453 | 1575611 | 1159 | 13 | 1.32E-12 | 1.02E-13 | 0.000216 | 1.85E-13 | -0.0036971 | AL645728.1, SSU72 |
| chr20 | 49567136 | 49569637 | 2502 | 12 | 6.84E-11 | 1.13E-12 | 5.56E-06 | 1.86E-13 | -0.00583038 | PTGIS |
| chr2 | 63054248 | 63058356 | 4109 | 22 | 2.42E-06 | 3.06E-13 | 0.002615 | 1.87E-13 | -0.00105512 | OTX1 |
| chr6 | 29552921 | 29554004 | 1084 | 30 | 1.67E-17 | 1.56E-09 | 0.000117 | 1.88E-13 | 0.006950867 | OR2I1P |
| chr17 | 44707429 | 44709654 | 2226 | 15 | 1.79E-12 | 1.77E-13 | 9.10E-05 | 1.89E-13 | 0.002343716 | DBF4B |
| chr11 | 65116757 | 65118396 | 1640 | 13 | 8.18E-14 | 2.39E-12 | 5.23E-05 | 1.90E-13 | -0.00187415 | AP003068.3, AP003068.4, ZNHIT2 |
| chr14 | 67411425 | 67412668 | 1244 | 15 | 8.69E-17 | 1.03E-10 | 4.50E-05 | 1.90E-13 | -0.00412493 | PLEK2 |
| chr17 | 64129290 | 64131014 | 1725 | 13 | 7.08E-15 | 2.08E-09 | 2.37E-06 | 1.95E-13 | -0.00239019 | ERN1 |
| chr12 | 53102945 | 53103522 | 578 | 8 | 2.47E-15 | 2.68E-10 | 1.04E-06 | 1.96E-13 | 0.0076557 | SOAT2 |
| chr22 | 35840227 | 35840713 | 487 | 8 | 6.48E-14 | 1.48E-12 | 7.61E-06 | 1.96E-13 | -0.00833571 | RBFOX2 |
| chr8 | 85463677 | 85465438 | 1762 | 12 | 4.22E-11 | 1.10E-09 | 6.78E-05 | 1.96E-13 | -0.00467749 | CA2, CA3-AS1 |
| chr14 | 81219249 | 81222140 | 2892 | 27 | 2.40E-09 | 1.38E-11 | 0.001314 | 1.99E-13 | 0.000214168 | AL136040.1, GTF2A1 |
| chr8 | 142304821 | 142307203 | 2383 | 9 | 1.00E-08 | 2.91E-11 | 2.14E-05 | 1.99E-13 | -0.00223209 | TSNARE1 |
| chr1 | 1166896 | 1170138 | 3243 | 21 | 9.39E-08 | 5.95E-11 | 0.000177 | 2.02E-13 | -0.0005596 | MIR200B, MIR200A, MIR429, AL390719.2 |

| | | | | | | | | | | |
|---|---|---|---|---|---|---|---|---|---|---|
| chr8 | 143953573 | 143954718 | 1146 | 12 | 8.42E-15 | 3.93E-12 | 0.000123 | 2.03E-13 | -0.00982596 | PLEC |
| chr16 | 1379014 | 1380366 | 1353 | 11 | 8.48E-16 | 3.01E-10 | 3.58E-06 | 2.08E-13 | -0.00840154 | UNKL |
| chr17 | 39721093 | 39723404 | 2312 | 7 | 1.20E-14 | 2.68E-09 | 1.05E-05 | 2.11E-13 | -0.00364076 | ERBB2 |
| chr1 | 26317743 | 26318822 | 1080 | 12 | 3.48E-13 | 2.82E-15 | 0.001306 | 2.11E-13 | 0.014146304 | CD52, UBXN11 |
| chr13 | 77697247 | 77699029 | 1783 | 20 | 1.21E-11 | 3.83E-12 | 0.000262 | 2.13E-13 | -0.00021194 | SLAIN1, MIR3665 |
| chr9 | 108854781 | 108856149 | 1369 | 9 | 1.12E-15 | 1.11E-09 | 2.14E-06 | 2.16E-13 | -0.00040844 | ACTL7B |
| chr12 | 262750 | 263789 | 1040 | 11 | 1.43E-15 | 7.09E-12 | 1.54E-06 | 2.17E-13 | -0.00581349 | SLC6A13 |
| chr7 | 90210551 | 90212729 | 2179 | 20 | 4.59E-11 | 1.12E-09 | 0.000153 | 2.25E-13 | 0.005564771 | STEAP2, STEAP2-AS1 |
| chr19 | 18437194 | 18439372 | 2179 | 19 | 8.85E-15 | 1.55E-08 | 1.35E-05 | 2.26E-13 | -0.00500666 | ISYNA1 |
| chr2 | 219001350 | 219002991 | 1642 | 14 | 9.59E-21 | 1.42E-06 | 8.28E-07 | 2.26E-13 | 0.000954047 | AC097468.1, MIR375, CFAP65 |
| chr6 | 87321387 | 87323130 | 1744 | 12 | 1.02E-16 | 3.92E-08 | 2.22E-06 | 2.28E-13 | -0.00650857 | SMIM8, GJB7 |
| chr4 | 75939737 | 75941759 | 2023 | 14 | 1.54E-14 | 2.73E-12 | 2.03E-05 | 2.30E-13 | -0.00071192 | NAAA, SDAD1 |
| chr13 | 21667981 | 21671591 | 3611 | 25 | 5.23E-09 | 1.69E-10 | 0.000319 | 2.30E-13 | 0.000499574 | FGF9 |
| chr17 | 2053325 | 2055557 | 2233 | 47 | 2.95E-17 | 1.56E-09 | 3.53E-05 | 2.35E-13 | -0.00041923 | HIC1 |
| chr11 | 66464891 | 66466877 | 1987 | 17 | 3.84E-17 | 2.14E-09 | 0.000103 | 2.39E-13 | -0.00264342 | PELI3, MRPL11 |
| chr1 | 52141690 | 52142795 | 1106 | 13 | 3.64E-15 | 3.95E-11 | 1.94E-05 | 2.39E-13 | -0.00201013 | ZFYVE9 |
| chr3 | 186929142 | 186931376 | 2235 | 14 | 3.57E-09 | 1.78E-11 | 0.000157 | 2.39E-13 | -0.00242206 | ST6GAL1 |
| chr6 | 30110303 | 30111885 | 1583 | 19 | 1.00E-14 | 2.99E-11 | 0.000426 | 2.39E-13 | -0.02023874 | TRIM31-AS1, TRIM31 |
| chr6 | 33195008 | 33197021 | 2014 | 17 | 2.57E-17 | 1.09E-10 | 7.18E-08 | 2.42E-13 | 0.000510495 | RXRB |
| chr4 | 2961537 | 2964114 | 2578 | 18 | 1.62E-14 | 4.45E-08 | 1.91E-05 | 2.42E-13 | -0.00205439 | NOP14-AS1, GRK4, NOP14 |
| chr2 | 1676477 | 1678640 | 2164 | 14 | 1.59E-09 | 5.05E-12 | 7.10E-05 | 2.42E-13 | -0.0037907 | PXDN |
| chr14 | 104723063 | 104724781 | 1719 | 10 | 5.45E-15 | 1.04E-09 | 6.65E-07 | 2.45E-13 | -0.00122722 | ADSS1 |
| chr15 | 43799203 | 43801231 | 2029 | 17 | 2.11E-16 | 2.70E-07 | 2.50E-07 | 2.47E-13 | -0.00284083 | SERF2, HYPK, SERINC4 |
| chr8 | 28347516 | 28348761 | 1246 | 4 | 2.27E-13 | 3.76E-14 | 2.43E-05 | 2.47E-13 | -0.02323093 | ZNF395, FBXO16 |
| chr15 | 69288440 | 69289252 | 813 | 5 | 1.41E-14 | 5.46E-10 | 4.59E-08 | 2.53E-13 | -0.00440023 | AC026992.1, AC026992.2 |
| chr22 | 42214864 | 42217559 | 2696 | 8 | 1.59E-08 | 2.11E-12 | 4.49E-06 | 2.55E-13 | 0.004501731 | TCF20 |
| chr21 | 10521044 | 10521566 | 523 | 5 | 1.68E-13 | 9.47E-14 | 4.46E-06 | 2.58E-13 | -0.02015459 | AF254983.1, TPTE |
| chr6 | 137494511 | 137498130 | 3620 | 17 | 9.13E-09 | 2.89E-12 | 8.75E-05 | 2.60E-13 | -0.0030671 | NA |
| chr1 | 39921995 | 39922723 | 729 | 4 | 2.18E-16 | 3.47E-10 | 3.52E-07 | 2.64E-13 | -0.01515172 | NA |
| chr22 | 31139686 | 31141083 | 1398 | 10 | 3.26E-16 | 8.22E-11 | 4.51E-05 | 2.71E-13 | -0.01018607 | PLA2G3 |
| chr6 | 98842408 | 98845193 | 2786 | 19 | 7.02E-12 | 5.45E-09 | 0.000103 | 2.71E-13 | -0.00257649 | NA |
| chr6 | 46687569 | 46689444 | 1876 | 16 | 3.57E-12 | 1.38E-12 | 0.001053 | 2.72E-13 | -0.00431491 | TDRD6, AL591242.1 |
| chr16 | 89827690 | 89828900 | 1211 | 8 | 9.19E-15 | 7.49E-11 | 2.59E-05 | 2.73E-13 | -0.01555254 | SPIRE2 |
| chr13 | 21326253 | 21327510 | 1258 | 6 | 3.24E-14 | 1.62E-13 | 6.98E-05 | 2.73E-13 | -0.04044382 | LINC00539 |
| chr2 | 68642939 | 68644722 | 1784 | 12 | 7.41E-12 | 7.30E-14 | 0.000671 | 2.73E-13 | -0.0027248 | APLF, PROKR1 |
| chr4 | 1804435 | 1806859 | 2425 | 24 | 3.45E-15 | 1.12E-05 | 1.14E-06 | 2.80E-13 | 0.000229712 | FGFR3 |
| chr22 | 35914533 | 35915760 | 1228 | 5 | 5.20E-11 | 1.44E-10 | 3.72E-06 | 2.80E-13 | -0.01273215 | RBFOX2 |
| chr7 | 32298538 | 32299885 | 1348 | 14 | 4.48E-14 | 2.24E-11 | 0.000561 | 2.83E-13 | -0.00918805 | PDE1C |
| chr19 | 4908669 | 4911463 | 2795 | 26 | 4.94E-13 | 1.28E-09 | 6.24E-05 | 2.84E-13 | -0.00044358 | UHRF1 |
| chr16 | 85088580 | 85089948 | 1369 | 5 | 1.11E-17 | 1.05E-10 | 7.71E-09 | 2.89E-13 | -0.0064265 | KIAA0513 |
| chr2 | 172555171 | 172557116 | 1946 | 23 | 7.10E-10 | 1.72E-12 | 0.00307 | 2.95E-13 | -0.00309654 | PDK1, AC078883.1 |
| chr14 | 104275111 | 104276437 | 1327 | 7 | 3.55E-15 | 2.91E-12 | 2.49E-05 | 2.98E-13 | -0.01364994 | LINC02691 |
| chr1 | 55038521 | 55040207 | 1687 | 16 | 7.54E-14 | 1.05E-08 | 3.09E-05 | 2.98E-13 | -0.0050782 | PCSK9 |
| chr10 | 76976714 | 76978045 | 1332 | 5 | 1.44E-16 | 4.29E-11 | 4.45E-07 | 3.01E-13 | -0.00604747 | KCNMA1-AS1, KCNMA1 |
| chr2 | 190533634 | 190535884 | 2251 | 14 | 8.72E-13 | 2.19E-12 | 6.44E-06 | 3.02E-13 | 0.009114067 | AC108047.1, NEMP2 |
| chr1 | 32273892 | 32276018 | 2127 | 8 | 4.17E-10 | 3.60E-13 | 9.47E-06 | 3.02E-13 | -0.00805383 | LCK |
| chr2 | 241815165 | 241816803 | 1639 | 7 | 5.67E-12 | 2.38E-12 | 1.30E-05 | 3.03E-13 | -0.00469616 | NEU4 |
| chr11 | 111514299 | 111515544 | 1246 | 10 | 7.82E-15 | 1.49E-11 | 3.47E-06 | 3.04E-13 | -0.00438407 | HOATZ, AP002008.1 |
| chr20 | 31483923 | 31484315 | 393 | 3 | 6.78E-18 | 1.32E-09 | 2.38E-07 | 3.06E-13 | -0.02238029 | REM1 |
| chr4 | 153788226 | 153791428 | 3203 | 45 | 6.75E-09 | 7.24E-12 | 0.005547 | 3.07E-13 | -0.00073173 | SFRP2 |
| chr6 | 133240230 | 133241638 | 1409 | 33 | 3.38E-12 | 7.46E-11 | 0.000237 | 3.07E-13 | 0.001014668 | EYA4 |
| chr20 | 63036385 | 63037117 | 733 | 7 | 3.05E-14 | 9.85E-14 | 0.000155 | 3.12E-13 | 0.027250539 | LINC01749, LINC00029 |
| chr16 | 8904549 | 8905866 | 1318 | 4 | 4.09E-16 | 5.55E-12 | 1.50E-07 | 3.18E-13 | -0.00807841 | USP7 |
| chr19 | 7943073 | 7945089 | 2017 | 16 | 2.41E-11 | 2.56E-11 | 7.01E-05 | 3.24E-13 | -0.00025506 | AC010336.7, TIMM44 |
| chr19 | 49153743 | 49156042 | 2300 | 11 | 2.18E-11 | 3.89E-12 | 0.000271 | 3.25E-13 | -0.00988041 | HRC |
| chr6 | 31778430 | 31781939 | 3510 | 29 | 1.45E-09 | 6.40E-12 | 0.002103 | 3.25E-13 | -0.00066448 | VARS1, Y RNA |
| chr3 | 11846053 | 11847889 | 1837 | 15 | 2.77E-14 | 7.13E-10 | 4.19E-05 | 3.29E-13 | -0.00066968 | TAMM41 |
| chr2 | 27494438 | 27496677 | 2240 | 12 | 2.42E-13 | 2.76E-12 | 2.48E-05 | 3.32E-13 | -0.00230136 | FNDC4 |
| chr12 | 51026117 | 51028416 | 2300 | 20 | 5.22E-11 | 4.15E-10 | 6.52E-05 | 3.32E-13 | -0.00227652 | SLC11A2 |
| chr17 | 81122782 | 81126324 | 3543 | 18 | 7.33E-10 | 1.10E-11 | 0.001106 | 3.33E-13 | -0.00278386 | MIR3065, AATK, MIR657, MIR338 |
| chr10 | 101134168 | 101137112 | 2945 | 16 | 2.04E-10 | 3.33E-13 | 0.000865 | 3.35E-13 | -0.00520745 | TLX1 |
| chr6 | 32068753 | 32070685 | 1933 | 33 | 1.11E-12 | 5.96E-10 | 0.000188 | 3.36E-13 | -0.00101142 | TNXB |
| chr2 | 127218153 | 127219983 | 1831 | 8 | 2.27E-11 | 8.45E-10 | 3.36E-05 | 3.36E-13 | -0.00138446 | CYP27C1 |
| chr14 | 39113206 | 39114626 | 1421 | 12 | 1.46E-14 | 5.80E-10 | 1.60E-05 | 3.38E-13 | -0.00309113 | GEMIN2 |
| chr11 | 58605516 | 58606005 | 490 | 4 | 4.39E-14 | 1.25E-13 | 7.36E-06 | 3.38E-13 | 0.002310593 | ZFP91, ZFP91-CNTF |
| chr19 | 38613936 | 38614403 | 468 | 2 | 2.58E-17 | 6.35E-12 | 1.15E-09 | 3.42E-13 | 0.005986232 | MAP4K1 |
| chr22 | 49673405 | 49674788 | 1384 | 3 | 3.22E-12 | 5.60E-13 | 9.44E-08 | 3.43E-13 | -0.00498003 | NA |
| chr6 | 168691734 | 168692913 | 1180 | 8 | 7.60E-15 | 7.58E-12 | 1.81E-05 | 3.47E-13 | -0.00338272 | NA |
| chr1 | 33625064 | 33627499 | 2436 | 12 | 5.67E-09 | 4.77E-13 | 0.000909 | 3.47E-13 | -0.00629309 | CSMD2 |
| chr16 | 1135381 | 1136286 | 906 | 8 | 1.71E-13 | 1.11E-11 | 2.86E-05 | 3.52E-13 | 0.004699069 | NA |
| chr17 | 47855275 | 47857315 | 2041 | 6 | 4.02E-15 | 1.38E-10 | 1.94E-06 | 3.52E-13 | -0.00973021 | SP6 |
| chr19 | 57190652 | 57191933 | 1282 | 24 | 4.26E-16 | 1.22E-10 | 0.000362 | 3.59E-13 | -0.00487469 | ZNF264 |
| chr1 | 118999210 | 119001706 | 2497 | 19 | 7.58E-11 | 1.51E-10 | 9.51E-05 | 3.65E-13 | -0.00237998 | AL139420.1, AL139420.2 |
| chr2 | 239137666 | 239141212 | 3547 | 15 | 3.12E-11 | 2.50E-10 | 9.16E-06 | 3.69E-13 | -0.00127921 | HDAC4 |
| chr11 | 46385566 | 46387951 | 2386 | 8 | 3.74E-09 | 2.37E-11 | 8.86E-06 | 3.79E-13 | -0.00443805 | CHRM4 |
| chr12 | 132423442 | 132424244 | 803 | 8 | 2.72E-12 | 5.64E-14 | 0.000108 | 3.83E-13 | 0.019547899 | NA |
| chr19 | 40279431 | 40281005 | 1575 | 6 | 1.66E-15 | 1.18E-09 | 2.82E-08 | 3.83E-13 | 0.010285629 | AKT2 |
| chr6 | 33416042 | 33418779 | 2738 | 60 | 2.54E-13 | 8.32E-08 | 0.000436 | 3.83E-13 | 9.05E-05 | PHF1, CUTA |
| chr11 | 114398779 | 114400949 | 2171 | 20 | 2.13E-13 | 1.04E-12 | 0.000658 | 3.85E-13 | 0.000313167 | RBM7, AP002373.1, C11orf71 |
| chr6 | 116875832 | 116877059 | 1228 | 5 | 3.23E-18 | 7.68E-10 | 1.50E-07 | 3.89E-13 | -0.01739989 | NA |
| chr15 | 99997099 | 99999994 | 2896 | 9 | 4.73E-12 | 4.88E-09 | 1.01E-06 | 3.93E-13 | 0.003160006 | ADAMTS17 |
| chr7 | 88218927 | 88221074 | 2148 | 22 | 6.13E-16 | 6.34E-08 | 1.37E-05 | 3.94E-13 | -0.00044391 | SRI-AS1, AC003991.1, SRI |
| chr12 | 131894449 | 131897439 | 2991 | 19 | 1.41E-10 | 3.83E-09 | 0.000518 | 4.02E-13 | -0.00352679 | ULK1, AC131009.4 |
| chr9 | 16058432 | 16058438 | 7 | 2 | 1.78E-14 | 3.73E-13 | 4.92E-08 | 4.04E-13 | -0.02144902 | CCDC171 |
| chr9 | 114907037 | 114908324 | 1288 | 4 | 3.42E-14 | 1.02E-12 | 6.83E-06 | 4.06E-13 | -0.0169969 | DELEC1, TNFSF8 |
| chr14 | 100881021 | 100884689 | 3669 | 25 | 2.07E-10 | 5.10E-11 | 1.73E-05 | 4.06E-13 | 0.000294557 | AL117190.1, MIR431, MIR433, MIR127, MIR432, RTL1 |
| chr8 | 19602161 | 19603557 | 1397 | 11 | 5.53E-14 | 9.89E-11 | 6.20E-05 | 4.09E-13 | -0.00898325 | CSGALNACT1 |
| chr21 | 45285681 | 45288591 | 2911 | 18 | 1.84E-11 | 4.80E-11 | 2.59E-06 | 4.12E-13 | -0.00076352 | LINC00205, POFUT2 |
| chr1 | 40978906 | 40982108 | 3203 | 23 | 3.80E-08 | 1.40E-09 | 0.000132 | 4.19E-13 | -0.00343447 | CTPS1 |
| chr17 | 40316709 | 40318489 | 1781 | 15 | 1.81E-14 | 4.60E-11 | 5.34E-05 | 4.26E-13 | -0.00432109 | RARA, AC080112.4 |
| chr4 | 10456425 | 10458212 | 1788 | 14 | 3.26E-15 | 1.75E-07 | 1.68E-07 | 4.26E-13 | -0.0058105 | AC005599.1, ZNF518B |
| chr9 | 132669615 | 132671642 | 2028 | 19 | 1.12E-12 | 7.92E-11 | 4.02E-06 | 4.30E-13 | -0.00011293 | GTF3C4, DDX31 |
| chr22 | 18048262 | 18048825 | 564 | 5 | 1.41E-15 | 5.05E-12 | 2.17E-07 | 4.37E-13 | 0.018709118 | NA |
| chr14 | 93231428 | 93232908 | 1481 | 9 | 1.96E-15 | 1.86E-10 | 3.76E-06 | 4.43E-13 | -0.02094479 | NA |
| chr1 | 116167148 | 116168718 | 1571 | 8 | 2.55E-17 | 3.47E-08 | 7.29E-06 | 4.47E-13 | -0.01345168 | NA |
| chr14 | 21023484 | 21026388 | 2905 | 30 | 1.79E-09 | 3.03E-11 | 0.001755 | 4.49E-13 | -0.00361946 | TPPP2, AL161668.4, NDRG2 |
| chr8 | 56073867 | 56075439 | 1573 | 13 | 6.54E-17 | 6.02E-08 | 2.63E-08 | 4.52E-13 | 0.001757873 | CERNA3, RPS20, SNORD54 |
| chr11 | 830883 | 833811 | 2929 | 15 | 1.70E-11 | 9.38E-11 | 3.08E-05 | 4.58E-13 | -0.00597055 | CRACR2B, CD151, AP006621.1 |
| chr3 | 193688210 | 193688978 | 769 | 2 | 1.29E-12 | 3.03E-13 | 1.46E-07 | 4.62E-13 | 0.016932623 | OPA1 |
| chr6 | 44290050 | 44290961 | 912 | 3 | 3.60E-16 | 1.98E-10 | 7.72E-10 | 4.64E-13 | -0.01979302 | TMEM151B, AL353588.1, TCTE1 |
| chr11 | 62545465 | 62547848 | 2384 | 12 | 8.71E-12 | 3.65E-13 | 3.45E-05 | 4.78E-13 | -0.00381101 | AP001363.2, AHNAK |
| chr15 | 99249572 | 99251939 | 2368 | 22 | 1.54E-10 | 6.79E-12 | 4.04E-05 | 4.86E-13 | -0.00881874 | LRRC28, TTC23 |
| chr1 | 155940732 | 155942778 | 2047 | 9 | 3.86E-11 | 6.96E-09 | 4.57E-05 | 4.94E-13 | -0.00723306 | RXFP4 |
| chr16 | 31148237 | 31149072 | 836 | 6 | 6.22E-13 | 2.04E-13 | 9.05E-06 | 4.94E-13 | -0.01933 | PRSS36 |
| chr11 | 914329 | 915440 | 1112 | 11 | 1.08E-12 | 5.36E-13 | 8.47E-05 | 4.96E-13 | -0.00473758 | CHID1 |
| chr15 | 40470208 | 40471564 | 1357 | 13 | 7.32E-17 | 2.41E-06 | 2.09E-05 | 4.97E-13 | -0.00389949 | CHST14 |
| chr1 | 228406132 | 228407979 | 1848 | 14 | 3.41E-13 | 7.25E-10 | 3.82E-05 | 4.97E-13 | -0.00393869 | AL139288.1, TRIM11, TRIM17 |
| chr19 | 14495088 | 14497280 | 2193 | 14 | 6.58E-11 | 1.43E-08 | 3.06E-05 | 4.97E-13 | -0.003349 | GIPC1 |
| chr6 | 29964966 | 29966194 | 1229 | 6 | 2.23E-14 | 2.34E-12 | 1.52E-05 | 5.00E-13 | -0.01066142 | NA |
| chr9 | 130185257 | 130186170 | 914 | 4 | 5.64E-13 | 4.89E-13 | 1.74E-06 | 5.04E-13 | -0.01257005 | NCS1 |
| chr11 | 63762048 | 63763525 | 1478 | 5 | 2.33E-12 | 9.39E-14 | 5.40E-05 | 5.16E-13 | -0.01960501 | C11orf95 |
| chr19 | 49057963 | 49059522 | 1560 | 6 | 7.41E-16 | 1.03E-11 | 2.80E-07 | 5.23E-13 | -0.02370721 | CGB7, AC008687.8, AC008687.7 |
| chr12 | 54032102 | 54033852 | 1751 | 17 | 8.54E-16 | 1.56E-08 | 2.87E-05 | 5.27E-13 | -0.00119685 | AC012531.3, HOXC4, AC012531.2, HOXC5 |
| chr4 | 3247290 | 3249659 | 2370 | 17 | 5.22E-11 | 3.87E-09 | 1.47E-06 | 5.34E-13 | -0.00166681 | MSANTD1 |
| chr10 | 11532279 | 11533103 | 825 | 6 | 3.11E-14 | 1.35E-11 | 5.19E-06 | 5.48E-13 | -0.0029177 | USP6NL |
| chr11 | 8681034 | 8684254 | 3221 | 23 | 8.89E-10 | 1.30E-10 | 2.19E-05 | 5.50E-13 | 0.001779251 | RPL27A, SNORA3A, TRIM66 |
| chr6 | 31619693 | 31623093 | 3401 | 50 | 2.57E-09 | 1.37E-10 | 0.000124 | 5.50E-13 | -0.00132365 | PRRC2A, SNORA38 |
| chr5 | 174327529 | 174329769 | 2241 | 5 | 6.62E-12 | 3.92E-09 | 1.48E-08 | 5.58E-13 | -0.00823158 | AC025752.1 |
| chr3 | 46756438 | 46757520 | 1083 | 3 | 5.91E-14 | 1.27E-12 | 1.02E-07 | 5.65E-13 | -0.01513332 | AC109583.1, AC109583.4, PRSS43P |
| chr11 | 69653569 | 69655052 | 1484 | 15 | 3.07E-12 | 1.91E-10 | 0.000152 | 5.65E-13 | -0.00353512 | CCND1, LTO1 |
| chr13 | 112992379 | 112994578 | 2200 | 12 | 4.45E-08 | 3.89E-11 | 4.82E-05 | 5.69E-13 | 0.003275334 | MCF2L |
| chr10 | 80077628 | 80079425 | 1798 | 19 | 2.75E-10 | 3.69E-11 | 0.000262 | 5.74E-13 | -0.00066993 | TMEM254, TMEM254-AS1 |
| chr8 | 144441645 | 144445243 | 3599 | 18 | 4.87E-08 | 3.58E-10 | 0.000125 | 5.77E-13 | 0.000100418 | TONSL |
| chr12 | 132523747 | 132526527 | 2781 | 15 | 6.03E-08 | 2.22E-12 | 0.000103 | 5.81E-13 | -0.00217659 | FBRSL1 |
| chr7 | 111727617 | 111728791 | 1175 | 6 | 4.24E-14 | 2.52E-11 | 2.63E-05 | 5.83E-13 | -0.02534169 | DOCK4 |
| chr18 | 3622400 | 3624899 | 2500 | 8 | 1.78E-09 | 5.73E-14 | 0.000132 | 5.86E-13 | -0.01397479 | DLGAP1 |
| chr18 | 46172733 | 46174083 | 1351 | 15 | 1.39E-14 | 1.68E-08 | 2.51E-05 | 5.96E-13 | 0.000811402 | C18orf25 |
| chr5 | 1672203 | 1673395 | 1193 | 7 | 2.06E-14 | 3.84E-12 | 5.67E-06 | 5.96E-13 | 0.020959493 | NA |
| chr10 | 97425507 | 97427364 | 1858 | 11 | 5.88E-11 | 2.09E-10 | 7.45E-06 | 6.10E-13 | -0.00899627 | PGAM1, RRP12 |
| chr4 | 73981383 | 73982299 | 917 | 9 | 2.71E-14 | 4.79E-14 | 2.80E-05 | 6.20E-13 | -0.03278735 | PF4 |
| chr22 | 41543977 | 41545490 | 1514 | 12 | 3.49E-12 | 5.24E-09 | 1.71E-06 | 6.26E-13 | 0.001156166 | POLR3H |
| chr2 | 230908374 | 230908815 | 442 | 4 | 8.33E-15 | 1.25E-11 | 3.96E-06 | 6.31E-13 | -0.01087865 | GPR55 |
| chr14 | 100547811 | 100550052 | 2242 | 8 | 3.21E-10 | 2.85E-11 | 0.000111 | 6.31E-13 | -0.00707625 | BEGAIN |
| chr11 | 73645271 | 73646696 | 1426 | 13 | 1.47E-12 | 1.25E-12 | 0.000504 | 6.31E-13 | -0.00982663 | PLEKHB1 |
| chr13 | 109783764 | 109785941 | 2178 | 8 | 3.49E-13 | 3.87E-13 | 5.35E-05 | 6.38E-13 | -0.01008801 | AL162497.1, IRS2 |
| chr5 | 88144304 | 88146152 | 1849 | 11 | 7.61E-10 | 2.40E-12 | 0.00062 | 6.52E-13 | -0.00932517 | AC114971.1 |
| chr2 | 219041394 | 219042861 | 1468 | 15 | 3.73E-19 | 3.66E-06 | 5.39E-08 | 6.53E-13 | 0.003268065 | CFAP65 |
| chr5 | 69368082 | 69371026 | 2945 | 33 | 1.45E-11 | 1.60E-09 | 7.73E-05 | 6.65E-13 | 0.000375795 | RAD17, AK6, TAF9 |
| chr17 | 67375784 | 67378482 | 2699 | 20 | 3.82E-11 | 1.41E-09 | 0.000104 | 6.65E-13 | 0.000906804 | PITPNC1 |
| chr10 | 11270191 | 11270895 | 705 | 4 | 2.77E-16 | 7.59E-10 | 8.50E-09 | 6.67E-13 | -0.01271252 | CELF2 |
| chr9 | 93182919 | 93185777 | 2859 | 13 | 7.91E-09 | 4.16E-10 | 2.19E-05 | 6.70E-13 | -0.00455739 | WNK2 |
| chr6 | 166721664 | 166722944 | 1281 | 6 | 1.75E-10 | 3.76E-14 | 0.000153 | 6.77E-13 | -0.00237188 | RPS6KA2 |
| chr11 | 2171804 | 2173144 | 1341 | 9 | 3.93E-14 | 2.54E-09 | 1.48E-05 | 6.89E-13 | -0.00334529 | MIR4686, TH |
| chr8 | 143140598 | 143141957 | 1360 | 8 | 2.88E-12 | 6.90E-13 | 7.08E-05 | 6.97E-13 | -0.01743997 | NA |
| chr8 | 10525411 | 10526253 | 843 | 5 | 1.44E-18 | 2.22E-07 | 1.10E-09 | 7.12E-13 | -0.00911471 | PRSS55, PRSS51 |
| chr11 | 77301461 | 77302894 | 1434 | 7 | 2.45E-12 | 4.22E-13 | 2.81E-05 | 7.13E-13 | -0.00795098 | GDPD4 |
| chr16 | 22162263 | 22164644 | 2382 | 9 | 3.72E-08 | 8.69E-13 | 2.85E-05 | 7.13E-13 | -0.00765827 | NA |
| chr19 | 48390717 | 48391773 | 1057 | 11 | 2.29E-13 | 1.02E-11 | 0.000122 | 7.16E-13 | 0.004649194 | KDELR1 |
| chr17 | 75729643 | 75731980 | 2338 | 15 | 3.44E-10 | 8.38E-10 | 0.000126 | 7.16E-13 | -0.0021834 | ITGB4 |
| chr15 | 63121232 | 63122290 | 1059 | 9 | 1.37E-16 | 3.08E-10 | 4.07E-06 | 7.21E-13 | -0.0057711 | LACTB |
| chr10 | 1159282 | 1160750 | 1469 | 11 | 3.65E-12 | 5.45E-14 | 0.000493 | 7.26E-13 | -0.01220563 | LINC00200 |
| chr6 | 30013627 | 30013918 | 292 | 4 | 8.74E-14 | 1.70E-12 | 6.30E-06 | 7.38E-13 | -0.00594721 | ZNRD1ASP |
| chr16 | 2970388 | 2971801 | 1414 | 7 | 1.05E-12 | 7.51E-13 | 3.01E-05 | 7.65E-13 | -0.00398389 | PAQR4, PKMYT1 |
| chr1 | 23958016 | 23960188 | 2173 | 19 | 5.22E-15 | 2.54E-08 | 0.000352 | 7.70E-13 | 0.001033949 | PNRC2 |
| chr18 | 74457425 | 74458722 | 1298 | 10 | 4.10E-13 | 3.05E-11 | 5.12E-05 | 7.78E-13 | -0.01370748 | DIPK1C |
| chr16 | 13927809 | 13928943 | 1135 | 3 | 3.92E-17 | 7.81E-11 | 1.98E-09 | 7.83E-13 | -0.02303458 | ERCC4 |
| chr1 | 39080474 | 39082329 | 1856 | 12 | 1.22E-09 | 5.85E-10 | 6.58E-05 | 7.90E-13 | -0.00640938 | MACF1 |
| chr4 | 742977 | 744411 | 1435 | 11 | 3.19E-13 | 5.60E-12 | 2.15E-05 | 7.91E-13 | -0.00827201 | PCGF3 |

| | | | | | | | | | | |
|---|---|---|---|---|---|---|---|---|---|---|
| chr16 | 85642686 | 85643255 | 570 | 5 | 2.20E-13 | 1.39E-13 | 7.47E-05 | 7.95E-13 | -0.01136793 | GSE1 |
| chr6 | 32175904 | 32178771 | 2868 | 42 | 3.75E-11 | 2.49E-08 | 0.000459 | 7.95E-13 | -0.0017297 | RNF5, AGPAT1 |
| chr8 | 143929485 | 143931143 | 1659 | 5 | 2.16E-11 | 9.66E-13 | 1.28E-06 | 7.97E-13 | -0.02386616 | PLEC |
| chr11 | 65180591 | 65184581 | 3991 | 29 | 2.49E-10 | 1.37E-07 | 5.36E-06 | 8.21E-13 | 0.000209439 | CAPN1, AP003068.2 |
| chr6 | 100466047 | 100467929 | 1883 | 12 | 1.37E-13 | 3.97E-11 | 2.58E-05 | 8.27E-13 | 0.000198347 | NA |
| chr1 | 27356854 | 27358003 | 1150 | 10 | 4.61E-16 | 1.45E-10 | 5.51E-07 | 8.37E-13 | 0.008587598 | MAP3K6 |
| chr3 | 128360793 | 128362754 | 1962 | 9 | 3.18E-12 | 4.43E-09 | 1.03E-05 | 8.45E-13 | 0.000576104 | EEFSEC |
| chr2 | 95159288 | 95160447 | 1160 | 11 | 1.39E-15 | 8.23E-13 | 1.70E-05 | 8.50E-13 | -0.00252028 | ZNF514 |
| chr15 | 22979070 | 22980532 | 1463 | 10 | 1.01E-13 | 1.77E-12 | 0.000168 | 8.65E-13 | 0.002233117 | CYFIP1 |
| chr11 | 43577422 | 43580886 | 3465 | 19 | 1.22E-09 | 2.79E-10 | 0.000134 | 8.68E-13 | 0.000791767 | AC068205.1, AC068205.2 |
| chr2 | 100606607 | 100607158 | 552 | 7 | 1.09E-13 | 5.11E-11 | 1.17E-05 | 8.70E-13 | 0.015515776 | LINC01849 |
| chr3 | 61688334 | 61689310 | 977 | 3 | 4.92E-16 | 1.02E-10 | 1.04E-07 | 8.70E-13 | -0.0218641 | PTPRG |
| chr18 | 78977834 | 78980962 | 3129 | 24 | 3.09E-12 | 2.27E-08 | 6.19E-07 | 8.71E-13 | -0.00070854 | SALL3, LINC01896 |
| chr16 | 67652929 | 67654450 | 1522 | 7 | 8.39E-12 | 1.25E-13 | 0.000202 | 8.71E-13 | -0.03842882 | CARMIL2 |
| chr3 | 156672429 | 156675807 | 3379 | 33 | 5.74E-10 | 3.32E-07 | 0.000139 | 8.91E-13 | -0.00034659 | TIPARP, TIPARP-AS1 |
| chr11 | 78574250 | 78575212 | 963 | 12 | 2.34E-14 | 2.36E-09 | 3.92E-07 | 8.95E-13 | -0.0006304 | NARS2 |
| chr16 | 721506 | 723253 | 1748 | 7 | 2.06E-14 | 2.03E-08 | 1.92E-07 | 8.97E-13 | -0.00788864 | ANTKMT, CCDC78 |
| chr7 | 80133640 | 80136078 | 2439 | 18 | 7.86E-09 | 2.91E-11 | 0.000567 | 9.10E-13 | 0.001964767 | GNAI1 |
| chr17 | 1647683 | 1651012 | 3330 | 17 | 1.85E-09 | 8.12E-10 | 0.000291 | 9.49E-13 | -0.00125594 | RILP, PRPF8 |
| chr22 | 45007612 | 45010664 | 3053 | 26 | 2.98E-11 | 6.62E-08 | 0.001292 | 9.53E-13 | 0.001488533 | PHF21B |
| chr17 | 58692026 | 58693063 | 1038 | 19 | 9.96E-13 | 9.26E-11 | 0.000381 | 9.62E-13 | -0.00439488 | RAD51C, TEX14 |
| chr1 | 44354401 | 44356140 | 1740 | 18 | 7.85E-15 | 3.82E-10 | 0.000288 | 9.69E-13 | -0.00205069 | ERI3 |
| chr10 | 133464626 | 133465643 | 1018 | 7 | 1.10E-13 | 1.98E-11 | 9.19E-06 | 9.80E-13 | -0.02740443 | SCART1 |
| chr2 | 1685057 | 1688411 | 3355 | 12 | 8.01E-08 | 1.02E-13 | 0.001169 | 9.83E-13 | -0.00036315 | PXDN |
| chr11 | 90222710 | 90224308 | 1599 | 17 | 1.23E-12 | 8.69E-13 | 0.00059 | 9.84E-13 | -0.00719066 | AP002364.1, CHORDC1 |
| chr16 | 22205925 | 22207162 | 1238 | 12 | 1.24E-12 | 1.08E-12 | 8.70E-05 | 9.90E-13 | 0.000221583 | EEF2K |
| chr8 | 22602860 | 22605678 | 2819 | 21 | 8.02E-09 | 7.05E-11 | 0.000257 | 1.00E-12 | -0.00013207 | C8orf58, CCAR2 |
| chr18 | 2906034 | 2907474 | 1441 | 8 | 8.31E-14 | 7.48E-11 | 9.80E-07 | 1.00E-12 | -0.00549139 | EMILIN2 |
| chr11 | 44950042 | 44952099 | 2058 | 21 | 1.34E-09 | 1.35E-11 | 0.000675 | 1.00E-12 | -0.00240164 | TP53I11 |
| chr6 | 33294545 | 33296757 | 2213 | 33 | 1.65E-11 | 2.63E-11 | 0.000458 | 1.01E-12 | -0.00046814 | PFDN6, RGL2 |
| chr2 | 240432128 | 240433708 | 1581 | 5 | 1.30E-13 | 2.35E-12 | 4.10E-06 | 1.02E-12 | 0.007659476 | NA |
| chr9 | 132877698 | 132878825 | 1128 | 11 | 1.35E-13 | 2.60E-13 | 0.000335 | 1.03E-12 | -0.00463371 | SPACA9, AK8 |
| chr1 | 155126351 | 155128035 | 1685 | 15 | 2.93E-13 | 5.82E-11 | 0.000116 | 1.04E-12 | 0.001210889 | EFNA1 |
| chr5 | 176446936 | 176448950 | 2015 | 16 | 4.63E-12 | 2.84E-11 | 0.000953 | 1.04E-12 | 0.000724612 | FAF2 |
| chr2 | 110204457 | 110205460 | 1004 | 11 | 9.48E-15 | 6.60E-11 | 0.000194 | 1.04E-12 | -0.00262468 | NPHP1 |
| chr22 | 37983211 | 37986707 | 3497 | 12 | 1.08E-08 | 1.93E-09 | 7.22E-05 | 1.05E-12 | -0.00212321 | POLR2F, SOX10 |
| chr6 | 29792381 | 29793511 | 1131 | 13 | 1.19E-14 | 6.73E-10 | 6.15E-06 | 1.05E-12 | 0.000752995 | HLA-V, HCG4 |
| chr10 | 35639680 | 35642307 | 2628 | 12 | 4.45E-12 | 1.34E-11 | 1.79E-05 | 1.05E-12 | -0.00043435 | AL121749.2, FZD8, MIR4683 |
| chr6 | 31960118 | 31963098 | 2981 | 21 | 2.72E-09 | 8.96E-11 | 0.001051 | 1.07E-12 | 0.000403704 | SKIV2L |
| chr5 | 173323447 | 173325165 | 1719 | 10 | 1.87E-14 | 1.06E-12 | 0.000122 | 1.07E-12 | -0.01567684 | STC2 |
| chr17 | 48156352 | 48157163 | 812 | 6 | 1.37E-14 | 2.77E-11 | 2.92E-06 | 1.07E-12 | -0.01186769 | SKAP1, MIR1203 |
| chr20 | 63292665 | 63296129 | 3465 | 14 | 1.01E-06 | 3.97E-11 | 6.45E-05 | 1.07E-12 | -0.00228044 | COL20A1 |
| chr18 | 58764422 | 58765259 | 838 | 3 | 5.38E-15 | 2.23E-11 | 5.54E-08 | 1.07E-12 | 0.011578836 | NA |
| chr22 | 45226078 | 45228598 | 2521 | 11 | 1.64E-09 | 1.07E-10 | 7.41E-05 | 1.10E-12 | -0.00732383 | KIAA0930 |
| chr11 | 66567613 | 66569240 | 1628 | 13 | 1.92E-14 | 5.21E-11 | 9.35E-05 | 1.10E-12 | -0.00616764 | CTSF |
| chr16 | 88870808 | 88870900 | 93 | 3 | 7.72E-14 | 2.39E-12 | 1.11E-07 | 1.11E-12 | 0.035640648 | NA |
| chr18 | 28175347 | 28178470 | 3124 | 18 | 5.86E-07 | 8.22E-13 | 0.000918 | 1.11E-12 | 0.002233451 | CDH2 |
| chr6 | 30197875 | 30199626 | 1752 | 26 | 3.89E-12 | 4.69E-09 | 0.000284 | 1.15E-12 | -0.00065278 | TRIM26 |
| chr15 | 64989108 | 64990790 | 1683 | 13 | 3.56E-12 | 2.12E-11 | 3.78E-05 | 1.16E-12 | 0.000321888 | SPG21 |
| chr17 | 42681364 | 42684107 | 2744 | 15 | 5.55E-08 | 4.52E-12 | 0.00026 | 1.16E-12 | -0.0051713 | AC100793.2, CNTNAP1, CCR10, AC100793.3 |
| chr1 | 40132722 | 40133518 | 797 | 5 | 4.77E-14 | 1.04E-11 | 6.54E-06 | 1.17E-12 | -0.03134516 | OAZ1P1 |
| chr2 | 3784921 | 3786226 | 1306 | 5 | 1.98E-11 | 3.90E-13 | 2.80E-05 | 1.20E-12 | -0.0097774 | DCDC2C |
| chr6 | 32046197 | 32048338 | 2142 | 24 | 4.14E-11 | 4.08E-09 | 0.000298 | 1.20E-12 | -0.00360412 | TNXB |
| chr10 | 35604509 | 35606456 | 1948 | 11 | 3.49E-11 | 4.24E-14 | 0.001675 | 1.21E-12 | -0.01063735 | GJD4, AL121749.1 |
| chr17 | 50208813 | 50211177 | 2365 | 6 | 1.14E-11 | 1.26E-11 | 6.65E-06 | 1.22E-12 | -0.01275266 | AC015909.1 |
| chr2 | 74441545 | 74442934 | 1390 | 10 | 1.21E-13 | 1.03E-10 | 0.000307 | 1.22E-12 | -0.01270953 | RTKN |
| chr12 | 65120693 | 65123220 | 2528 | 16 | 1.68E-09 | 9.47E-09 | 9.72E-06 | 1.22E-12 | 0.002903864 | WIF1 |
| chr2 | 237133025 | 237133708 | 684 | 2 | 1.34E-14 | 2.57E-12 | 1.62E-08 | 1.23E-12 | -0.02843633 | NA |
| chr2 | 68156804 | 68158494 | 1691 | 16 | 7.61E-14 | 2.79E-10 | 8.23E-05 | 1.26E-12 | 0.002858083 | PNO1, WDR92, AC017083.3 |
| chr11 | 45803749 | 45806529 | 2781 | 23 | 7.41E-11 | 9.52E-10 | 0.000607 | 1.26E-12 | -0.00422661 | SLC35C1 |
| chr2 | 61144745 | 61145893 | 1149 | 10 | 4.85E-14 | 3.98E-09 | 1.60E-05 | 1.26E-12 | -0.00314653 | AC016747.4, C2orf74, AC016747.1 |
| chr5 | 1049117 | 1052108 | 2992 | 11 | 9.53E-08 | 1.48E-12 | 0.000462 | 1.27E-12 | -0.00382749 | SLC12A7 |
| chr7 | 103989171 | 103990682 | 1512 | 11 | 2.20E-15 | 5.19E-10 | 4.05E-06 | 1.27E-12 | -0.00112377 | RELN |
| chr10 | 112373477 | 112375334 | 1858 | 12 | 2.12E-10 | 1.91E-10 | 2.32E-05 | 1.27E-12 | 0.006654292 | ACSL5 |
| chr18 | 50193227 | 50196436 | 3210 | 10 | 8.05E-09 | 2.61E-12 | 0.000182 | 1.28E-12 | 0.004492966 | MYO5B |
| chr13 | 48411988 | 48413029 | 1042 | 4 | 3.86E-13 | 6.42E-13 | 5.30E-06 | 1.29E-12 | -0.02942861 | RB1, LPAR6 |
| chr19 | 14431797 | 14433071 | 1275 | 7 | 7.97E-16 | 3.18E-11 | 2.05E-05 | 1.29E-12 | -0.0056579 | PKN1 |
| chr2 | 120222421 | 120223787 | 1367 | 10 | 1.16E-17 | 6.45E-09 | 3.83E-07 | 1.30E-12 | -0.00397657 | TMEM185B |
| chr7 | 100367422 | 100368389 | 968 | 3 | 1.08E-13 | 9.89E-13 | 5.02E-07 | 1.30E-12 | 0.00499729 | STAG3L5P-PVRIG2P-PILRB, PILRB, PILRA |
| chr22 | 37210293 | 37211997 | 1705 | 9 | 4.10E-13 | 3.98E-09 | 3.37E-07 | 1.31E-12 | -0.00033439 | SSTR3 |
| chr8 | 144285389 | 144286772 | 1384 | 8 | 1.24E-12 | 4.27E-10 | 4.88E-07 | 1.34E-12 | 0.000557127 | BOP1 |
| chr2 | 201618981 | 201619860 | 880 | 12 | 2.27E-13 | 1.98E-12 | 8.29E-05 | 1.34E-12 | -0.00910832 | C2CD6 |
| chr17 | 48726440 | 48729573 | 3134 | 24 | 1.42E-08 | 5.80E-09 | 8.29E-05 | 1.36E-12 | 0.001836241 | HOXB13 |
| chr1 | 228157623 | 228158646 | 1024 | 7 | 1.79E-12 | 1.12E-11 | 8.03E-06 | 1.37E-12 | 0.003845382 | GJC2 |
| chr1 | 33279733 | 33281131 | 1399 | 8 | 2.41E-12 | 2.11E-10 | 7.65E-05 | 1.38E-12 | 0.000335771 | ZNF362 |
| chr6 | 30212911 | 30214786 | 1876 | 49 | 4.87E-12 | 1.81E-09 | 0.000163 | 1.38E-12 | 0.000348531 | TRIM26 |
| chr10 | 117554560 | 117556273 | 1714 | 5 | 4.48E-19 | 1.16E-08 | 3.96E-08 | 1.40E-12 | 0.019922307 | NA |
| chr2 | 138501170 | 138502729 | 1560 | 15 | 1.38E-12 | 1.73E-11 | 4.16E-05 | 1.40E-12 | -0.00339081 | SPOPL, AC114763.1 |
| chr12 | 56228274 | 56230158 | 1885 | 9 | 3.50E-10 | 4.06E-12 | 2.15E-05 | 1.45E-12 | -0.00278266 | NABP2, SLC39A5 |
| chr8 | 1772938 | 1773558 | 621 | 5 | 2.77E-12 | 4.67E-13 | 3.62E-05 | 1.47E-12 | 0.018186139 | CLN8, AC019257.8 |
| chr1 | 46673060 | 46674298 | 1239 | 6 | 2.60E-13 | 5.12E-13 | 5.57E-05 | 1.48E-12 | 0.00678515 | TEX38, EFCAB14-AS1, ATPAF1 |
| chr1 | 201421078 | 201422974 | 1897 | 12 | 4.31E-10 | 1.14E-12 | 5.71E-05 | 1.51E-12 | -0.00614128 | TNNI1 |
| chr7 | 65731795 | 65731914 | 120 | 2 | 1.49E-13 | 8.28E-13 | 7.81E-07 | 1.51E-12 | -0.03838499 | AC073107.1 |
| chr2 | 2565712 | 2566546 | 835 | 5 | 9.03E-13 | 8.13E-10 | 3.10E-06 | 1.52E-12 | -0.00018568 | NA |
| chr17 | 76943330 | 76944863 | 1534 | 8 | 1.84E-10 | 8.18E-09 | 5.69E-06 | 1.52E-12 | -0.01192047 | MGAT5B |
| chr22 | 50300121 | 50303245 | 3125 | 9 | 5.97E-08 | 7.52E-13 | 0.000903 | 1.54E-12 | 0.000771772 | PLXNB2 |
| chr6 | 33287695 | 33290471 | 2777 | 50 | 5.44E-14 | 7.13E-10 | 0.000412 | 1.58E-12 | -6.69E-05 | PFDN6, MIR6834, WDR46 |
| chr8 | 142537259 | 142538222 | 964 | 2 | 5.98E-12 | 8.85E-13 | 6.22E-07 | 1.59E-12 | -0.01063642 | ADGRB1 |
| chr1 | 18557880 | 18558014 | 135 | 3 | 2.21E-13 | 2.40E-12 | 2.05E-06 | 1.60E-12 | -0.01426439 | NA |
| chr8 | 71843823 | 71846226 | 2404 | 20 | 2.02E-12 | 5.92E-09 | 2.64E-05 | 1.60E-12 | -0.0012226 | MSC-AS1, MSC |
| chr3 | 126101336 | 126103648 | 2313 | 15 | 3.69E-09 | 7.67E-12 | 0.000135 | 1.60E-12 | -0.00405512 | ALDH1L1-AS1, SLC41A3, ALDH1L1 |
| chr10 | 132625722 | 132626299 | 578 | 4 | 4.29E-14 | 2.72E-11 | 1.12E-07 | 1.62E-12 | -0.01497858 | INPP5A |
| chr13 | 112967724 | 112969530 | 1807 | 13 | 1.30E-11 | 1.17E-11 | 8.68E-05 | 1.62E-12 | 0.002960905 | MCF2L, MCF2L-AS1 |
| chr13 | 26221631 | 26223020 | 1390 | 23 | 1.42E-11 | 5.65E-13 | 9.89E-05 | 1.63E-12 | -0.00124219 | AL138966.2, RNF6 |
| chr2 | 235811594 | 235813182 | 1589 | 12 | 1.10E-14 | 5.70E-09 | 4.11E-06 | 1.64E-12 | -0.003486 | AGAP1 |
| chr4 | 3510523 | 3513073 | 2551 | 12 | 7.85E-08 | 2.87E-13 | 0.000618 | 1.64E-12 | -0.00031142 | LRPAP1 |
| chr1 | 26161100 | 26162497 | 1398 | 7 | 4.12E-12 | 1.10E-11 | 0.000102 | 1.67E-12 | -0.01331767 | FAM110D |
| chr6 | 70412156 | 70413974 | 1819 | 12 | 1.88E-14 | 1.38E-10 | 6.48E-06 | 1.68E-12 | -0.00388148 | FAM135A, FAM135A-AS1 |
| chr1 | 228211861 | 228213762 | 1902 | 10 | 9.95E-16 | 3.78E-07 | 8.37E-06 | 1.68E-12 | -0.00840421 | OBSCN, OBSCN-AS1 |
| chr10 | 87503821 | 87505843 | 2023 | 16 | 3.02E-12 | 3.89E-13 | 0.002535 | 1.73E-12 | -0.00339441 | MIR4678, MINPP1, AL355334.2 |
| chr2 | 38075444 | 38077662 | 2219 | 31 | 2.22E-12 | 8.06E-08 | 4.40E-05 | 1.73E-12 | -0.00260568 | CYP1B1-AS1, CYP1B1 |
| chr11 | 72012929 | 72014544 | 1616 | 12 | 2.89E-11 | 1.44E-11 | 0.000379 | 1.74E-12 | -0.00464311 | AP002490.1, NUMA1 |
| chr7 | 158956513 | 158958493 | 1981 | 11 | 1.72E-11 | 5.32E-11 | 1.52E-06 | 1.75E-12 | -0.02287004 | DYNC2I1, AC004863.1 |
| chr2 | 165571994 | 165572746 | 753 | 5 | 3.73E-22 | 2.00E-06 | 1.49E-10 | 1.76E-12 | -0.01398064 | CSRNP3 |
| chr11 | 73786643 | 73788864 | 2222 | 12 | 1.08E-10 | 1.31E-09 | 4.96E-06 | 1.76E-12 | -0.00974416 | MRPL48 |
| chr3 | 147407327 | 147409406 | 2080 | 29 | 6.90E-12 | 6.04E-10 | 0.000635 | 1.77E-12 | -0.00300465 | ZIC1 |
| chr19 | 36245370 | 36246667 | 1298 | 9 | 6.34E-15 | 4.81E-09 | 2.49E-08 | 1.79E-12 | 0.008009454 | ZNF565 |
| chr7 | 142796464 | 142798162 | 1699 | 18 | 5.64E-14 | 9.05E-10 | 3.05E-05 | 1.79E-12 | 0.010511697 | TRBJ2-2, TRBJ2-2P, TRBJ2-3, TRBJ2-4, TRBJ2-5, TRBJ2-6, TRBJ2-7 |
| chr3 | 121749180 | 121750321 | 1142 | 13 | 1.40E-13 | 1.31E-10 | 1.97E-05 | 1.79E-12 | 0.001161335 | GOLGB1 |
| chr22 | 50577957 | 50579294 | 1338 | 16 | 4.56E-11 | 1.60E-14 | 0.006739 | 1.80E-12 | -0.02079225 | CPT1B, CHKB-CPT1B, CHKB |
| chr11 | 66859723 | 66862680 | 2958 | 16 | 5.11E-09 | 2.69E-08 | 0.00015 | 1.81E-12 | -0.00286661 | LRFN4, PC |
| chr1 | 115314464 | 115314917 | 454 | 4 | 3.11E-14 | 8.14E-11 | 4.68E-07 | 1.81E-12 | -0.01384922 | NGF-AS1, NGF |
| chr17 | 7583013 | 7584535 | 1523 | 24 | 8.48E-14 | 1.22E-10 | 1.18E-05 | 1.82E-12 | -0.00273538 | MPDU1, AC016876.1 |
| chr6 | 33426547 | 33429345 | 2799 | 27 | 6.58E-11 | 6.00E-10 | 0.000218 | 1.82E-12 | 0.000360587 | SYNGAP1 |
| chr11 | 126366431 | 126367126 | 696 | 4 | 5.72E-13 | 8.52E-13 | 1.73E-05 | 1.87E-12 | 0.0187226 | ST3GAL4 |
| chr16 | 2061074 | 2065528 | 4455 | 22 | 1.96E-06 | 4.70E-10 | 0.000565 | 1.89E-12 | -0.00088033 | TSC2 |
| chr14 | 104578647 | 104579910 | 1264 | 10 | 6.35E-11 | 9.20E-11 | 2.87E-05 | 1.90E-12 | -0.00490521 | C14orf180, TMEM179 |
| chr16 | 87066351 | 87069085 | 2735 | 18 | 2.76E-08 | 9.21E-11 | 3.16E-05 | 1.94E-12 | 0.003914458 | LINC02181, AC106745.1 |
| chr5 | 154629709 | 154631560 | 1852 | 6 | 8.15E-16 | 5.40E-10 | 8.78E-07 | 1.96E-12 | -0.00306476 | NA |
| chr11 | 83192826 | 83194503 | 1678 | 20 | 3.76E-17 | 2.72E-08 | 3.60E-06 | 1.99E-12 | 0.002661655 | ANKRD42, AP000873.2 |
| chr5 | 91279324 | 91279642 | 319 | 5 | 5.62E-13 | 1.41E-13 | 0.000223 | 2.01E-12 | -0.03161016 | LUCAT1 |
| chr2 | 238060079 | 238061550 | 1472 | 14 | 6.19E-10 | 3.62E-10 | 2.67E-05 | 2.01E-12 | 0.000562763 | UBE2F-SCLY, SCLY |
| chr7 | 116524054 | 116526116 | 2063 | 25 | 1.08E-12 | 3.91E-07 | 2.45E-06 | 2.02E-12 | 0.001912265 | CAV1 |
| chr21 | 44329802 | 44332577 | 2776 | 14 | 4.28E-11 | 1.75E-08 | 6.81E-06 | 2.02E-12 | -0.00333538 | AP001062.1, AP001062.3, CFAP410 |
| chr12 | 681752 | 683111 | 1360 | 3 | 1.59E-11 | 8.10E-13 | 2.95E-06 | 2.04E-12 | -0.01362703 | LINC02455 |
| chr6 | 89886341 | 89887872 | 1532 | 11 | 9.29E-18 | 1.27E-08 | 2.74E-07 | 2.06E-12 | 0.001656539 | AL353692.1 |
| chr4 | 55101969 | 55102505 | 537 | 3 | 3.59E-13 | 4.39E-11 | 1.09E-07 | 2.06E-12 | -0.01009139 | KDR |
| chr15 | 75348393 | 75349188 | 796 | 6 | 4.73E-15 | 8.80E-11 | 1.48E-05 | 2.11E-12 | -0.01175232 | NEIL1 |
| chr13 | 99983476 | 99986192 | 2717 | 19 | 2.05E-10 | 4.53E-10 | 9.76E-05 | 2.11E-12 | -0.00097814 | ZIC2 |
| chr17 | 319066 | 321643 | 2578 | 17 | 3.68E-10 | 7.02E-11 | 0.001618 | 2.11E-12 | -0.00952304 | AC129507.4, RPH3AL |
| chr1 | 207644658 | 207645548 | 891 | 9 | 2.21E-13 | 1.37E-06 | 1.22E-05 | 2.14E-12 | -0.01482002 | CR1L, CD46P1 |
| chr4 | 80206665 | 80207818 | 1154 | 7 | 1.83E-13 | 8.43E-13 | 0.00013 | 2.15E-12 | -0.01782129 | NA |
| chr1 | 28886313 | 28888280 | 1968 | 15 | 4.85E-12 | 1.96E-09 | 1.10E-05 | 2.17E-12 | -0.00253228 | EPB41 |
| chr22 | 30635799 | 30637765 | 1967 | 10 | 2.24E-15 | 2.75E-11 | 1.07E-05 | 2.19E-12 | -0.0128924 | SLC35E4 |
| chr2 | 128026818 | 128028672 | 1855 | 17 | 5.70E-13 | 2.24E-06 | 6.53E-06 | 2.19E-12 | 0.001683442 | SAP130 |
| chr10 | 42951844 | 42952602 | 759 | 3 | 6.10E-14 | 2.44E-12 | 3.33E-07 | 2.21E-12 | -0.01987782 | NA |
| chr10 | 114648623 | 114648808 | 186 | 2 | 1.30E-13 | 1.97E-12 | 1.00E-07 | 2.21E-12 | 0.027563818 | ABLIM1 |
| chr5 | 34655232 | 34657881 | 2650 | 17 | 1.15E-09 | 1.07E-09 | 5.12E-05 | 2.25E-12 | -0.00060885 | RAI14, AC025754.1 |
| chr9 | 137204977 | 137207465 | 2489 | 15 | 3.27E-10 | 2.23E-07 | 3.14E-05 | 2.30E-12 | 0.001311928 | NDOR1, TMEM203 |
| chr13 | 50127365 | 50129453 | 2089 | 15 | 3.52E-12 | 1.97E-08 | 0.000103 | 2.31E-12 | -0.00229291 | DLEU1, AL137060.3 |
| chr6 | 137865521 | 137867893 | 2373 | 23 | 1.01E-12 | 4.45E-08 | 4.36E-05 | 2.35E-12 | -0.00019911 | TNFAIP3, WAKMAR2 |
| chr8 | 54456202 | 54458453 | 2252 | 15 | 6.10E-09 | 2.78E-10 | 0.000101 | 2.35E-12 | 0.001035974 | SOX17 |
| chr16 | 281821 | 283737 | 1917 | 12 | 6.27E-12 | 6.86E-12 | 0.000114 | 2.35E-12 | -0.00332032 | Z69667.1, ARHGDIG, PDIA2 |
| chr5 | 179378557 | 179379746 | 1190 | 7 | 1.60E-13 | 9.18E-12 | 3.62E-05 | 2.36E-12 | -0.00518231 | AC109479.3, AC109479.1 |
| chr16 | 4690693 | 4691494 | 802 | 3 | 5.18E-11 | 8.28E-13 | 8.00E-06 | 2.39E-12 | -0.01718434 | MGRN1 |
| chr2 | 174394841 | 174396918 | 2078 | 23 | 6.09E-12 | 3.47E-10 | 0.001037 | 2.40E-12 | -0.00089809 | SCRN3, CIR1 |
| chr9 | 136482173 | 136484721 | 2549 | 16 | 1.04E-09 | 4.69E-10 | 2.11E-05 | 2.40E-12 | -0.00032823 | C9orf163, SEC16A |
| chr15 | 70851859 | 70853989 | 2131 | 14 | 1.05E-11 | 1.90E-10 | 0.000185 | 2.42E-12 | -0.00203948 | LRRC49, LARP6 |
| chr4 | 185470542 | 185472390 | 1849 | 18 | 1.81E-10 | 8.37E-11 | 0.000781 | 2.42E-12 | 0.008886382 | AC106897.1, CCDC110 |
| chr12 | 257320 | 258405 | 1086 | 6 | 1.83E-13 | 2.23E-11 | 8.27E-06 | 2.43E-12 | -0.01292549 | SLC6A13 |
| chr17 | 75978455 | 75980702 | 2248 | 17 | 7.13E-12 | 6.75E-11 | 1.00E-05 | 2.43E-12 | 0.001083262 | TEN1-CDK3, TEN1, ACOX1 |
| chr5 | 38445118 | 38446090 | 973 | 12 | 4.68E-13 | 2.31E-10 | 0.000384 | 2.45E-12 | 0.012449912 | EGFLAM |
| chr16 | 2750867 | 2752710 | 1844 | 21 | 1.97E-13 | 1.06E-09 | 0.000385 | 2.46E-12 | -0.00222587 | SRRM2, SRRM2-AS1 |
| chr13 | 77918081 | 77920327 | 2247 | 48 | 3.13E-13 | 1.17E-07 | 8.19E-05 | 2.46E-12 | -0.00017303 | OBI1-AS1, EDNRB |
| chr3 | 35678049 | 35680266 | 2218 | 15 | 1.08E-10 | 4.16E-12 | 0.002233 | 2.46E-12 | -0.00167689 | ARPP21 |
| chr13 | 94701767 | 94703560 | 1794 | 9 | 1.81E-11 | 7.53E-11 | 4.14E-05 | 2.50E-12 | -0.00559513 | SOX21-AS1, LINC00391 |
| chr12 | 7107121 | 7109717 | 2597 | 25 | 1.04E-11 | 4.33E-11 | 0.001423 | 2.54E-12 | 0.002003094 | C1RL-AS1, C1RL |
| chr10 | 132335680 | 132337256 | 1577 | 13 | 9.53E-11 | 2.45E-10 | 0.000525 | 2.56E-12 | -0.01379522 | LRRC27 |

| | | | | | | | | | | |
|---|---|---|---|---|---|---|---|---|---|---|
| chr10 | 14837194 | 14839097 | 1904 | 16 | 1.17E-10 | 2.14E-11 | 0.000126 | 2.57E-12 | 0.00142057 | AC069544.2, HSPA14, CDNF |
| chr17 | 39224444 | 39226446 | 2003 | 15 | 3.86E-09 | 2.63E-10 | 2.66E-05 | 2.58E-12 | -0.00028277 | STAC2 |
| chr9 | 94638475 | 94640847 | 2373 | 14 | 7.53E-12 | 2.20E-09 | 0.000571 | 2.64E-12 | -0.00365993 | FBP1 |
| chr13 | 24159543 | 24161398 | 1856 | 13 | 5.38E-13 | 2.87E-10 | 3.41E-05 | 2.64E-12 | -0.00221204 | AL359736.1, SPATA13 |
| chr17 | 3048372 | 3049473 | 1102 | 12 | 3.47E-10 | 1.32E-09 | 3.30E-06 | 2.64E-12 | 0.009581546 | NA |
| chr12 | 93570370 | 93573935 | 3566 | 37 | 3.69E-10 | 1.97E-07 | 0.000133 | 2.71E-12 | -0.00078181 | SOCS2, SOCS2-AS1 |
| chr4 | 39457749 | 39459535 | 1787 | 17 | 8.75E-15 | 1.88E-07 | 2.76E-06 | 2.71E-12 | 0.001681069 | LIAS, RPL9 |
| chr20 | 61965332 | 61965691 | 360 | 3 | 7.58E-13 | 7.93E-13 | 1.08E-05 | 2.74E-12 | -0.02536803 | TAF4 |
| chr10 | 80453842 | 80455900 | 2059 | 10 | 5.52E-10 | 3.95E-11 | 5.82E-05 | 2.74E-12 | -0.00187371 | TSPAN14 |
| chr9 | 33473447 | 33475073 | 1627 | 14 | 8.92E-13 | 2.87E-10 | 9.84E-05 | 2.76E-12 | 0.000398514 | NOL6 |
| chr20 | 63696181 | 63697358 | 1178 | 11 | 2.65E-15 | 1.74E-10 | 4.28E-05 | 2.76E-12 | -0.02263732 | RTEL1, RTEL1-TNFRSF6B, TNFRSF6B |
| chr19 | 51000978 | 51003017 | 2040 | 8 | 1.14E-12 | 4.92E-12 | 1.27E-05 | 2.76E-12 | -0.02549935 | AC011483.1, KLK8, AC011473.4, KLK9 |
| chr1 | 1425119 | 1426286 | 1168 | 9 | 9.06E-11 | 1.41E-12 | 0.000561 | 2.84E-12 | -0.01067018 | TMEM88B |
| chr20 | 35292186 | 35293132 | 947 | 10 | 1.35E-14 | 1.98E-08 | 3.40E-06 | 2.89E-12 | -0.00628367 | FAM83C |
| chr19 | 1007844 | 1009173 | 1330 | 7 | 2.97E-13 | 5.04E-10 | 4.45E-06 | 2.90E-12 | -0.00655512 | GRIN3B |
| chr3 | 123375205 | 123375803 | 599 | 2 | 1.68E-12 | 1.62E-12 | 7.88E-07 | 2.92E-12 | -0.01806898 | ADCY5 |
| chr3 | 141385549 | 141387747 | 2199 | 10 | 5.99E-11 | 2.88E-09 | 2.13E-06 | 2.95E-12 | -0.01175979 | ZBTB38 |
| chr20 | 45812185 | 45813380 | 1196 | 18 | 1.38E-15 | 7.72E-10 | 3.41E-05 | 3.00E-12 | -0.00233468 | UBE2C |
| chr9 | 95509167 | 95511067 | 1901 | 9 | 4.43E-14 | 6.53E-10 | 9.35E-07 | 3.01E-12 | -0.00013478 | PTCH1 |
| chr3 | 101676076 | 101678173 | 2098 | 23 | 1.23E-09 | 5.79E-07 | 3.19E-05 | 3.01E-12 | -0.0012255 | ZBTB11-AS1, ZBTB11 |
| chr17 | 2040267 | 2042660 | 2394 | 15 | 1.15E-10 | 3.91E-08 | 5.54E-05 | 3.03E-12 | 0.007637143 | DPH1, OVCA2, AC090617.10 |
| chr2 | 71330786 | 71332465 | 1680 | 10 | 3.24E-11 | 6.97E-10 | 0.000114 | 3.08E-12 | 0.003412523 | ZNF638 |
| chr21 | 16587624 | 16589804 | 2181 | 8 | 4.93E-14 | 4.68E-10 | 1.48E-05 | 3.09E-12 | -0.00721007 | MIR99AHG |
| chr6 | 1593799 | 1595887 | 2089 | 11 | 2.07E-10 | 1.33E-08 | 1.52E-06 | 3.14E-12 | 0.012709356 | NA |
| chr1 | 152908643 | 152909944 | 1302 | 4 | 2.89E-13 | 2.48E-12 | 9.44E-06 | 3.16E-12 | -0.01318965 | IVL |
| chr14 | 52791050 | 52792377 | 1328 | 11 | 1.25E-12 | 1.47E-09 | 1.36E-05 | 3.18E-12 | -0.00513187 | AL139317.5, GNPNAT1 |
| chr9 | 136995736 | 136997419 | 1684 | 11 | 2.32E-12 | 1.83E-12 | 0.000184 | 3.21E-12 | 0.004841699 | CLIC3 |
| chr10 | 133236487 | 133238500 | 2014 | 27 | 4.78E-10 | 2.45E-12 | 0.009874 | 3.22E-12 | -0.00413406 | VENTX |
| chr1 | 1263657 | 1265478 | 1822 | 6 | 7.00E-10 | 2.29E-13 | 0.000246 | 3.24E-12 | -0.00087775 | UBE2J2 |
| chr17 | 2049521 | 2052187 | 2667 | 16 | 1.12E-08 | 7.06E-08 | 5.39E-05 | 3.26E-12 | -0.00207724 | MIR132, MIR212 |
| chr2 | 162072808 | 162075785 | 2978 | 18 | 6.59E-11 | 2.87E-11 | 8.33E-06 | 3.30E-12 | -0.00179062 | DPP4-DT, DPP4 |
| chr4 | 1046907 | 1048437 | 1531 | 6 | 1.37E-13 | 7.82E-12 | 2.09E-05 | 3.30E-12 | -0.01461875 | NA |
| chr9 | 95434235 | 95435224 | 990 | 3 | 4.72E-11 | 8.28E-13 | 1.68E-05 | 3.30E-12 | -0.03613332 | NA |
| chr8 | 60903184 | 60904339 | 1156 | 4 | 2.47E-12 | 3.87E-11 | 4.43E-06 | 3.30E-12 | -0.01608064 | AC113143.2 |
| chr11 | 66850385 | 66853023 | 2639 | 9 | 3.91E-12 | 1.25E-11 | 1.24E-05 | 3.32E-12 | 0.003674161 | PC |
| chr2 | 41854026 | 41854516 | 491 | 2 | 1.16E-12 | 1.88E-12 | 9.04E-07 | 3.44E-12 | -0.01393121 | NA |
| chr20 | 59159747 | 59160470 | 724 | 3 | 1.13E-15 | 1.78E-10 | 3.37E-08 | 3.50E-12 | 0.021075138 | ZNF831 |
| chr11 | 69003571 | 69006123 | 2553 | 11 | 1.05E-08 | 7.02E-11 | 0.000128 | 3.50E-12 | -0.00614076 | AP003071.3, MRGPRF |
| chr3 | 187669022 | 187670949 | 1928 | 16 | 1.24E-14 | 1.19E-08 | 3.88E-05 | 3.54E-12 | 0.00798215 | SST |
| chr8 | 100870861 | 100871952 | 1092 | 3 | 8.70E-11 | 2.18E-12 | 1.21E-06 | 3.54E-12 | -0.01338341 | NA |
| chr1 | 3745298 | 3748116 | 2819 | 18 | 1.37E-08 | 1.52E-09 | 5.09E-05 | 3.55E-12 | -0.00329188 | CCDC27, TP73-AS1 |
| chr18 | 32470375 | 32471840 | 1466 | 26 | 6.02E-14 | 1.58E-09 | 1.62E-05 | 3.61E-12 | 0.000156374 | AC025887.2, GAREM1 |
| chr3 | 55485560 | 55488807 | 3248 | 23 | 1.16E-11 | 2.35E-09 | 4.12E-06 | 3.61E-12 | 0.000411411 | WNT5A-AS1, WNT5A |
| chr7 | 100560090 | 100560720 | 631 | 3 | 1.69E-14 | 7.34E-12 | 1.38E-07 | 3.62E-12 | -0.00559899 | AGFG2 |
| chr12 | 57844449 | 57847762 | 3314 | 21 | 6.77E-09 | 1.42E-10 | 0.000628 | 3.62E-12 | -0.00089014 | CTDSP2 |
| chr20 | 1224967 | 1226739 | 1773 | 13 | 9.58E-13 | 6.60E-11 | 9.78E-05 | 3.67E-12 | -0.00036043 | RAD21L1 |
| chr15 | 86316986 | 86318521 | 1536 | 6 | 2.51E-09 | 2.33E-12 | 4.80E-06 | 3.67E-12 | -0.01396676 | AGBL1, AGBL1-AS1 |
| chr11 | 66691313 | 66691610 | 298 | 4 | 3.29E-13 | 7.59E-12 | 4.28E-06 | 3.68E-12 | -0.00678726 | SPTBN2 |
| chr2 | 154697618 | 154700278 | 2661 | 16 | 5.92E-09 | 8.31E-10 | 8.11E-05 | 3.68E-12 | -0.00159409 | KCNJ3, AC061961.1 |
| chr10 | 101235888 | 101239005 | 3118 | 22 | 1.65E-07 | 1.86E-07 | 4.20E-05 | 3.68E-12 | -0.0024539 | LBX1-AS1 |
| chr21 | 37454813 | 37455560 | 748 | 2 | 7.25E-12 | 2.47E-12 | 2.91E-07 | 3.73E-12 | -0.00643065 | DYRK1A |
| chr5 | 42756000 | 42757713 | 1714 | 11 | 2.79E-13 | 1.31E-10 | 0.000148 | 3.75E-12 | -0.01546383 | CCDC152 |
| chr1 | 224182453 | 224183906 | 1454 | 15 | 7.91E-12 | 4.98E-07 | 7.17E-06 | 3.77E-12 | 0.00059383 | DEGS1 |
| chr1 | 44813032 | 44814012 | 981 | 7 | 4.55E-14 | 3.86E-11 | 6.21E-05 | 3.77E-12 | -0.0129585 | BTBD19 |
| chr1 | 3019713 | 3022899 | 3187 | 15 | 4.29E-08 | 1.05E-10 | 0.000895 | 3.78E-12 | -0.00293758 | ACTRT2 |
| chr5 | 1799995 | 1801790 | 1796 | 15 | 2.22E-15 | 1.20E-08 | 0.000111 | 3.82E-12 | -0.00221789 | NDUFS6, MRPL36 |
| chr21 | 46424098 | 46426321 | 2224 | 9 | 5.83E-08 | 1.60E-11 | 9.71E-05 | 3.84E-12 | 0.010868467 | PCNT |
| chr1 | 3727976 | 3728557 | 582 | 3 | 2.06E-15 | 7.44E-11 | 1.47E-08 | 3.88E-12 | -0.01980701 | TP73 |
| chr16 | 78240456 | 78241740 | 1285 | 6 | 2.72E-13 | 3.79E-10 | 6.17E-06 | 3.92E-12 | -0.00719355 | WWOX, WWOX-AS1 |
| chr10 | 22334892 | 22336945 | 2054 | 14 | 7.47E-11 | 3.65E-12 | 0.000516 | 3.97E-12 | -0.00204984 | NA |
| chr6 | 28140830 | 28142218 | 1389 | 16 | 5.20E-12 | 1.30E-08 | 0.000147 | 4.08E-12 | -0.00046359 | ZKSCAN8 |
| chr16 | 67841362 | 67843510 | 2149 | 18 | 1.92E-10 | 2.75E-10 | 5.96E-05 | 4.08E-12 | -0.0016084 | THAP11, CENPT |
| chr3 | 30605886 | 30607525 | 1640 | 20 | 1.68E-10 | 1.99E-10 | 0.000121 | 4.10E-12 | -0.00208467 | TGFBR2 |
| chr2 | 197514353 | 197516776 | 2424 | 29 | 2.12E-10 | 2.34E-10 | 0.001376 | 4.12E-12 | 0.000634886 | HSPE1-MOB4, MOB4, HSPD1 |
| chr2 | 41870409 | 41871224 | 816 | 5 | 1.32E-13 | 5.79E-10 | 1.71E-07 | 4.16E-12 | -0.01484612 | LINC01913 |
| chr16 | 58462638 | 58464914 | 2277 | 24 | 2.19E-08 | 9.88E-10 | 0.000308 | 4.16E-12 | -0.00126933 | NDRG4 |
| chr11 | 111918086 | 111918969 | 884 | 10 | 1.49E-12 | 9.22E-11 | 0.000221 | 4.21E-12 | -0.00795582 | HSPB2-C11orf52, C11orf52, CRYAB |
| chr11 | 6611298 | 6612870 | 1573 | 18 | 7.39E-11 | 1.25E-09 | 1.20E-05 | 4.31E-12 | 0.000424355 | TAF10, AC091564.7, TPP1 |
| chr21 | 32277335 | 32279278 | 1944 | 11 | 3.14E-11 | 1.03E-07 | 1.85E-06 | 4.35E-12 | -0.00206359 | MIS18A-AS1, MIS18A |
| chr4 | 99088835 | 99090059 | 1225 | 10 | 8.98E-18 | 2.73E-09 | 2.58E-08 | 4.37E-12 | 0.000304952 | AP002026.1 |
| chr12 | 103586239 | 103587335 | 1097 | 7 | 1.74E-14 | 1.92E-08 | 3.73E-07 | 4.40E-12 | -0.0131503 | STAB2 |
| chr7 | 52120136 | 52121015 | 880 | 2 | 2.99E-10 | 2.45E-12 | 7.53E-07 | 4.40E-12 | -0.0528165 | NA |
| chr11 | 70712714 | 70714106 | 1393 | 8 | 4.32E-13 | 2.12E-10 | 3.07E-05 | 4.40E-12 | -0.01218533 | SHANK2 |
| chr14 | 99361260 | 99363495 | 2236 | 10 | 1.37E-10 | 1.95E-09 | 4.19E-05 | 4.44E-12 | -0.00539039 | NA |
| chr8 | 129983171 | 129984468 | 1298 | 6 | 3.21E-13 | 7.54E-12 | 3.12E-05 | 4.44E-12 | -0.00528107 | CYRIB |
| chr17 | 78129289 | 78130805 | 1517 | 8 | 9.22E-15 | 6.12E-12 | 3.24E-05 | 4.48E-12 | 0.001125015 | TMC8, TMC6 |
| chr20 | 62142102 | 62144674 | 2573 | 20 | 9.78E-11 | 1.95E-07 | 8.76E-05 | 4.52E-12 | 0.001470429 | SS18L1, PSMA7 |
| chr1 | 62783526 | 62785053 | 1528 | 14 | 2.69E-10 | 4.50E-12 | 0.00168 | 4.54E-12 | -0.00799877 | ATG4C |
| chr9 | 94174544 | 94176038 | 1495 | 5 | 1.18E-09 | 1.27E-10 | 2.47E-06 | 4.54E-12 | 0.006390547 | MIRLET7A1HG, MIRLET7A1 |
| chr7 | 139635533 | 139636218 | 686 | 5 | 1.23E-15 | 1.68E-08 | 1.99E-06 | 4.54E-12 | 0.010167123 | HIPK2 |
| chr10 | 102132552 | 102134080 | 1529 | 13 | 1.85E-11 | 6.30E-07 | 2.46E-05 | 4.54E-12 | 0.001302971 | PPRC1 |
| chr5 | 82277487 | 82279380 | 1894 | 17 | 8.65E-11 | 1.66E-09 | 0.000167 | 4.55E-12 | -0.00265891 | RPS23 |
| chr10 | 99658726 | 99659870 | 1145 | 14 | 3.94E-12 | 2.37E-09 | 0.000118 | 4.55E-12 | -0.00252784 | ENTPD7, AL133353.2, AL133353.1 |
| chr14 | 101023078 | 101025795 | 2718 | 28 | 1.57E-08 | 2.21E-10 | 0.000566 | 4.56E-12 | -0.00231693 | MEG8, MIR411, MIR299, MIR380, MIR1197, MIR323A |
| chr12 | 124425534 | 124427636 | 2103 | 10 | 2.04E-09 | 3.97E-12 | 6.26E-05 | 4.60E-12 | -0.00224518 | NCOR2 |
| chr8 | 141275046 | 141275272 | 227 | 4 | 1.14E-12 | 6.19E-12 | 8.76E-06 | 4.67E-12 | -0.01954116 | SLC45A4 |
| chr5 | 33522276 | 33523691 | 1416 | 4 | 1.25E-10 | 8.65E-11 | 1.02E-05 | 4.75E-12 | -0.00844579 | ADAMTS12 |
| chr4 | 573748 | 575385 | 1638 | 6 | 5.17E-14 | 2.66E-10 | 7.54E-06 | 4.76E-12 | 0.008346704 | TMEM271 |
| chr4 | 105707958 | 105710850 | 2893 | 32 | 6.44E-08 | 2.91E-11 | 0.005264 | 4.81E-12 | -0.00183905 | ARHGEF38, GSTCD, INTS12 |
| chr3 | 44623634 | 44625518 | 1885 | 15 | 1.46E-15 | 2.16E-10 | 3.62E-06 | 4.89E-12 | -0.00343077 | ZNF197, ZKSCAN7-AS1, ZNF197-AS1 |
| chr14 | 39174064 | 39176528 | 2465 | 20 | 6.68E-11 | 2.27E-07 | 3.09E-07 | 4.91E-12 | -0.00101698 | PNN, AL132639.3 |
| chr4 | 8029400 | 8031501 | 2102 | 5 | 6.86E-13 | 1.21E-10 | 7.87E-07 | 4.92E-12 | -0.01120734 | ABLIM2 |
| chr20 | 25281098 | 25282552 | 1455 | 6 | 1.12E-10 | 2.32E-10 | 1.84E-06 | 4.95E-12 | -0.01886014 | PYGB |
| chr7 | 139648090 | 139649123 | 1034 | 5 | 7.20E-16 | 3.05E-09 | 1.39E-07 | 4.96E-12 | -0.00622349 | HIPK2 |
| chr13 | 32538338 | 32539868 | 1531 | 16 | 1.13E-15 | 5.73E-08 | 9.51E-06 | 5.00E-12 | -0.00328773 | N4BP2L2 |
| chr1 | 114853414 | 114855452 | 2039 | 13 | 7.13E-10 | 2.07E-11 | 0.000134 | 5.01E-12 | -0.00869738 | SYCP1 |
| chr19 | 2044541 | 2046775 | 2235 | 11 | 3.14E-13 | 2.35E-08 | 6.20E-08 | 5.12E-12 | -9.74E-05 | MKNK2 |
| chr8 | 11807849 | 11809534 | 1686 | 19 | 1.05E-12 | 2.13E-12 | 0.001135 | 5.12E-12 | 0.00696401 | FDFT1 |
| chr13 | 99990717 | 99992403 | 1687 | 13 | 5.01E-10 | 2.50E-09 | 3.81E-05 | 5.14E-12 | -0.00235646 | NA |
| chr12 | 103964913 | 103966534 | 1622 | 14 | 2.34E-14 | 4.90E-09 | 4.13E-06 | 5.15E-12 | 0.006599599 | TDG, C12orf73 |
| chr17 | 41811839 | 41813145 | 1307 | 26 | 8.13E-12 | 1.97E-09 | 0.001004 | 5.15E-12 | -0.0015536 | FKBP10, P3H4 |
| chr7 | 101598103 | 101598738 | 636 | 3 | 1.01E-14 | 1.32E-10 | 2.97E-08 | 5.18E-12 | -0.00621522 | NA |
| chr7 | 27190633 | 27192531 | 1899 | 7 | 3.74E-11 | 6.19E-12 | 4.78E-05 | 5.23E-12 | 0.024037933 | AC004080.2 |
| chr6 | 31679057 | 31681951 | 2895 | 38 | 1.90E-09 | 1.08E-09 | 0.00024 | 5.24E-12 | -0.00110288 | LY6G5C |
| chr7 | 3245963 | 3246821 | 859 | 2 | 3.02E-12 | 6.72E-12 | 6.47E-08 | 5.25E-12 | -0.02034903 | NA |
| chr14 | 105445294 | 105446403 | 1110 | 7 | 1.76E-13 | 9.16E-11 | 6.75E-06 | 5.25E-12 | 0.001232412 | TEX22, MTA1 |
| chr1 | 159713557 | 159715894 | 2338 | 11 | 6.47E-12 | 1.10E-08 | 9.52E-07 | 5.53E-12 | -0.00538758 | CRP |
| chr6 | 3283762 | 3284700 | 939 | 6 | 3.01E-14 | 1.37E-07 | 4.01E-06 | 5.55E-12 | -0.00094129 | PSMG4, SLC22A23 |
| chr1 | 177164400 | 177165318 | 919 | 16 | 1.57E-12 | 1.78E-08 | 9.55E-05 | 5.57E-12 | -0.00024057 | ASTN1 |
| chr15 | 69297916 | 69299414 | 1499 | 13 | 7.66E-15 | 7.01E-08 | 4.25E-05 | 5.59E-12 | -0.00289977 | PAQR5, AC026992.2 |
| chr22 | 30889058 | 30890884 | 1827 | 7 | 1.33E-08 | 4.59E-10 | 7.42E-06 | 5.63E-12 | -0.0059533 | OSBP2 |
| chr6 | 28634766 | 28635660 | 895 | 28 | 4.55E-13 | 5.98E-10 | 0.000165 | 5.66E-12 | -0.00036817 | AL121932.2 |
| chr17 | 48617830 | 48621236 | 3407 | 17 | 2.53E-08 | 1.00E-09 | 0.000348 | 5.87E-12 | 4.22E-05 | HOXB7, HOXB9 |
| chr12 | 114734697 | 114736968 | 2272 | 11 | 5.34E-08 | 1.85E-11 | 9.86E-06 | 5.89E-12 | -0.00747343 | AC026765.3, AC026765.4 |
| chr22 | 39501575 | 39502841 | 1267 | 15 | 5.16E-14 | 1.16E-08 | 0.000317 | 5.96E-12 | -0.00439013 | MIEF1 |
| chr17 | 75783271 | 75785226 | 1956 | 20 | 1.21E-13 | 2.81E-08 | 1.56E-05 | 5.97E-12 | -0.00098668 | UNK, H3-3B, MIR4738 |
| chr6 | 127517748 | 127519600 | 1853 | 18 | 2.81E-10 | 9.85E-11 | 0.000153 | 5.97E-12 | -0.00301081 | C6orf58, AL096711.2, SOGA3 |
| chr6 | 656504 | 658301 | 1798 | 11 | 1.77E-11 | 2.21E-11 | 0.000645 | 5.98E-12 | 0.005611017 | EXOC2, HUS1B |
| chr7 | 156543923 | 156544515 | 593 | 5 | 8.51E-13 | 1.51E-10 | 1.73E-05 | 5.98E-12 | -0.00454212 | LINC01006 |
| chr14 | 22765066 | 22767516 | 2451 | 21 | 9.67E-09 | 1.65E-07 | 3.79E-05 | 5.98E-12 | -0.00146411 | OXA1L, AL160314.2 |
| chr12 | 641955 | 644625 | 2671 | 14 | 5.61E-09 | 4.01E-12 | 0.00048 | 6.03E-12 | -0.00421905 | NINJ2-AS1, NINJ2 |
| chr19 | 4401411 | 4403596 | 2186 | 18 | 7.96E-09 | 2.15E-08 | 1.18E-05 | 6.03E-12 | 8.55E-05 | CHAF1A |
| chr2 | 226794690 | 226796575 | 1886 | 9 | 7.90E-15 | 5.53E-09 | 4.93E-06 | 6.04E-12 | -0.00513885 | IRS1 |
| chr1 | 21176086 | 21177628 | 1543 | 13 | 7.70E-13 | 4.14E-10 | 2.90E-05 | 6.08E-12 | 0.000495669 | AL031005.2, EIF4G3 |
| chr11 | 1867735 | 1870469 | 2735 | 16 | 1.66E-08 | 1.73E-09 | 4.08E-05 | 6.08E-12 | -0.00272587 | LSP1 |
| chr14 | 101632042 | 101633261 | 1220 | 7 | 3.69E-16 | 6.21E-09 | 6.18E-09 | 6.14E-12 | -0.00026353 | AL049836.1 |
| chr9 | 134163226 | 134165604 | 2379 | 12 | 1.36E-09 | 2.51E-09 | 2.70E-05 | 6.19E-12 | -0.00358628 | RNU6ATAC |
| chr3 | 122912876 | 122914006 | 1131 | 4 | 9.51E-14 | 2.78E-12 | 1.74E-05 | 6.24E-12 | -0.02672151 | SEMA5B |
| chr12 | 44874380 | 44877284 | 2905 | 35 | 4.36E-10 | 1.44E-08 | 1.64E-05 | 6.25E-12 | -0.00034119 | NELL2 |
| chr19 | 3771685 | 3773247 | 1563 | 9 | 8.95E-13 | 1.18E-06 | 1.33E-06 | 6.35E-12 | -0.00662672 | RAX2 |
| chr3 | 48656164 | 48657632 | 1469 | 8 | 3.68E-13 | 3.76E-11 | 3.43E-05 | 6.38E-12 | -0.01276899 | CELSR3 |
| chr14 | 91277293 | 91278101 | 809 | 5 | 1.81E-16 | 7.62E-09 | 3.57E-09 | 6.43E-12 | -0.00323165 | CCDC88C |
| chr7 | 156127528 | 156127814 | 287 | 4 | 1.33E-13 | 1.17E-09 | 1.03E-07 | 6.50E-12 | -0.00981946 | NA |
| chr15 | 51341184 | 51342386 | 1203 | 7 | 1.07E-11 | 1.68E-09 | 4.69E-05 | 6.53E-12 | -0.01684991 | GLDN |
| chr11 | 57600570 | 57600619 | 50 | 2 | 3.31E-14 | 7.63E-11 | 5.44E-09 | 6.57E-12 | 0.013106416 | SERPING1 |
| chr2 | 127196936 | 127197993 | 1058 | 5 | 1.43E-12 | 3.29E-12 | 3.57E-06 | 6.59E-12 | 0.008477882 | CYP27C1 |
| chr8 | 56445226 | 56448054 | 2829 | 25 | 2.44E-09 | 2.72E-11 | 0.000136 | 6.60E-12 | -0.00442041 | AC012349.1, PENK |
| chr10 | 72060733 | 72063048 | 2316 | 8 | 1.50E-08 | 3.41E-12 | 0.000211 | 6.73E-12 | -0.00187053 | SPOCK2 |
| chr8 | 133248764 | 133249858 | 1095 | 5 | 2.08E-11 | 1.01E-12 | 1.00E-04 | 6.73E-12 | -0.01489443 | NDRG1 |
| chr9 | 2933121 | 2933464 | 344 | 4 | 1.23E-12 | 9.10E-12 | 2.31E-06 | 6.79E-12 | -0.00253583 | CARM1P1 |
| chr2 | 218417691 | 218418548 | 858 | 4 | 3.29E-14 | 2.60E-10 | 5.22E-06 | 6.89E-12 | -0.01280453 | NA |
| chr15 | 22916353 | 22918780 | 2428 | 18 | 1.23E-12 | 7.39E-08 | 1.32E-05 | 6.93E-12 | -0.00066955 | CYFIP1 |
| chr2 | 3779020 | 3780321 | 1302 | 7 | 2.71E-11 | 4.89E-13 | 0.000294 | 6.93E-12 | 0.028793091 | DCDC2C |
| chr7 | 151187244 | 151188879 | 1636 | 16 | 2.92E-11 | 3.97E-13 | 0.00129 | 6.96E-12 | -0.00493359 | ASB10 |
| chr9 | 113340024 | 113341214 | 1191 | 14 | 5.88E-15 | 9.24E-08 | 1.99E-08 | 6.99E-12 | -0.00114297 | WDR31 |
| chr16 | 29805146 | 29809061 | 3916 | 24 | 8.07E-09 | 1.09E-07 | 8.87E-05 | 6.99E-12 | -0.00056504 | KIF22, MAZ, AC009133.5, AC009133.2, AC009133.1 |
| chr3 | 48659976 | 48661683 | 1708 | 11 | 1.44E-16 | 1.03E-10 | 5.71E-07 | 7.01E-12 | -0.00601819 | CELSR3 |
| chr18 | 79719176 | 79719525 | 350 | 3 | 6.75E-13 | 1.15E-11 | 2.79E-07 | 7.13E-12 | -0.00813046 | CTDP1 |
| chr16 | 81005720 | 81007793 | 2074 | 22 | 1.78E-12 | 6.52E-08 | 8.29E-06 | 7.21E-12 | 0.000659569 | CENPN, AC009070.1, CMC2 |
| chr8 | 123042126 | 123043945 | 1820 | 16 | 1.89E-16 | 6.66E-08 | 1.96E-05 | 7.30E-12 | -0.00210002 | TBC1D31, DERL1 |
| chr15 | 64892702 | 64894246 | 1545 | 6 | 5.19E-14 | 1.92E-10 | 2.05E-06 | 7.31E-12 | -0.00684236 | AC069368.1 |
| chr5 | 180619901 | 180621893 | 1993 | 6 | 6.20E-08 | 1.86E-12 | 2.51E-05 | 7.33E-12 | -0.00181975 | FLT4 |
| chr6 | 118651858 | 118653674 | 1817 | 14 | 1.92E-16 | 3.83E-07 | 6.69E-06 | 7.34E-12 | -0.00968281 | CEP85L |
| chr15 | 98899170 | 98900437 | 1268 | 8 | 2.47E-15 | 1.29E-08 | 8.58E-07 | 7.34E-12 | -0.00764094 | IGF1R |
| chr1 | 17984862 | 17984909 | 48 | 2 | 4.27E-13 | 1.72E-11 | 2.98E-08 | 7.34E-12 | 0.031988151 | NA |
| chr6 | 30626409 | 30627841 | 1433 | 19 | 6.08E-13 | 8.11E-10 | 0.000105 | 7.35E-12 | -0.00059814 | ATAT1 |
| chr3 | 129305063 | 129305869 | 807 | 5 | 7.81E-12 | 1.03E-10 | 2.09E-07 | 7.37E-12 | -0.01049218 | HMCES |
| chr6 | 31759932 | 31763133 | 3202 | 12 | 6.83E-11 | 3.98E-09 | 1.78E-05 | 7.39E-12 | -0.00385973 | MSH5, MSH5-SAPCD1, SAPCD1 |

| | | | | | | | | | | |
|---|---|---|---|---|---|---|---|---|---|---|
| chr3 | 15331808 | 15333540 | 1733 | 14 | 4.18E-12 | 9.07E-10 | 9.41E-07 | 7.40E-12 | -0.00016215 | SH3BP5 |
| chr9 | 34665326 | 34667182 | 1857 | 12 | 6.38E-12 | 1.14E-10 | 8.69E-05 | 7.43E-12 | -0.0044944 | AL162231.2, AL162231.4, AL162231.1 |
| chr6 | 16305473 | 16306626 | 1154 | 6 | 1.78E-14 | 1.33E-06 | 4.63E-07 | 7.52E-12 | -0.00570234 | ATXN1 |
| chr5 | 159104124 | 159106934 | 2811 | 15 | 1.56E-09 | 8.84E-08 | 1.27E-05 | 7.52E-12 | -0.00597753 | LINC02202, AC134043.3 |
| chr11 | 132792560 | 132793068 | 509 | 4 | 5.18E-12 | 1.24E-12 | 5.53E-05 | 7.60E-12 | -0.05379996 | OPCML |
| chr1 | 21289370 | 21291557 | 2188 | 14 | 3.39E-11 | 6.78E-09 | 1.88E-05 | 7.66E-12 | -0.00080905 | ECE1 |
| chr15 | 24877329 | 24879066 | 1738 | 7 | 6.83E-10 | 3.70E-09 | 1.17E-05 | 7.81E-12 | -0.02289735 | AC090983.2, SNRPN |
| chr3 | 13019415 | 13021665 | 2251 | 6 | 2.28E-09 | 5.09E-10 | 8.99E-06 | 7.83E-12 | -0.00565647 | IQSEC1 |
| chr22 | 46363641 | 46364781 | 1141 | 6 | 3.07E-15 | 2.56E-08 | 5.60E-08 | 7.83E-12 | -0.00542693 | CELSR1 |
| chr17 | 82320403 | 82321799 | 1397 | 10 | 3.69E-13 | 2.35E-10 | 1.05E-06 | 7.86E-12 | -0.005505 | SECTM1 |
| chr8 | 1061674 | 1062465 | 792 | 5 | 1.75E-15 | 1.13E-09 | 2.22E-07 | 7.91E-12 | -0.00658695 | DLGAP2 |
| chr18 | 3297178 | 3297367 | 190 | 3 | 2.69E-12 | 2.88E-12 | 5.73E-06 | 7.98E-12 | -0.02994216 | AP001025.1 |
| chr4 | 164977514 | 164978244 | 731 | 11 | 6.38E-13 | 5.90E-12 | 0.000742 | 8.06E-12 | -0.02213573 | TRIM61 |
| chr17 | 76193850 | 76194437 | 588 | 2 | 7.78E-13 | 6.28E-12 | 2.38E-07 | 8.06E-12 | -0.01125557 | RNF157 |
| chr10 | 114404363 | 114406098 | 1736 | 13 | 2.60E-11 | 3.47E-11 | 3.34E-05 | 8.08E-12 | -0.00562557 | AFAP1L2 |
| chr10 | 133389018 | 133391117 | 2100 | 9 | 2.15E-07 | 7.53E-12 | 8.42E-05 | 8.11E-12 | 0.018773797 | PAOX, AL360181.3 |
| chr1 | 155321433 | 155325476 | 4044 | 23 | 1.88E-06 | 4.49E-08 | 6.46E-05 | 8.24E-12 | -0.0010326 | RUSC1, RUSC1-AS1 |
| chr19 | 44642946 | 44644140 | 1195 | 11 | 2.35E-13 | 1.01E-08 | 3.31E-05 | 8.25E-12 | -0.00263123 | PVR, AC243964.3 |
| chr16 | 54935794 | 54939667 | 3874 | 22 | 1.07E-07 | 1.23E-10 | 0.001536 | 8.26E-12 | -0.00092363 | AC106738.1, AC106738.2 |
| chr9 | 136901841 | 136902759 | 919 | 5 | 2.18E-13 | 4.89E-11 | 1.32E-06 | 8.29E-12 | -0.01347644 | TRAF2 |
| chr3 | 113019095 | 113020676 | 1582 | 16 | 2.43E-11 | 5.15E-11 | 0.000394 | 8.29E-12 | -0.00188342 | AC078785.1, NEPRO |
| chr9 | 124449940 | 124451559 | 1620 | 9 | 4.11E-10 | 1.67E-12 | 0.000561 | 8.32E-12 | -0.00591949 | ADGRD2 |
| chr7 | 4784024 | 4786256 | 2233 | 10 | 5.63E-10 | 2.51E-10 | 0.00011 | 8.33E-12 | -0.00247028 | AP5Z1 |
| chr5 | 1243599 | 1244810 | 1212 | 8 | 9.31E-13 | 1.91E-10 | 1.56E-05 | 8.36E-12 | 0.005214508 | SLC6A18 |
| chr12 | 6913902 | 6915814 | 1913 | 15 | 2.48E-11 | 6.07E-11 | 0.000105 | 8.37E-12 | 0.0048147 | LRRC23, ENO2 |
| chr19 | 41924786 | 41925689 | 904 | 3 | 3.25E-11 | 2.75E-12 | 1.03E-05 | 8.38E-12 | -0.02798094 | ARHGEF1, ERFL |
| chr16 | 4649880 | 4651292 | 1413 | 3 | 1.43E-14 | 5.15E-11 | 7.18E-08 | 8.38E-12 | -0.00727196 | MGRN1 |
| chr7 | 27102808 | 27104683 | 1876 | 21 | 8.09E-14 | 1.42E-11 | 0.003161 | 8.56E-12 | -0.01314102 | NA |
| chr11 | 76209813 | 76211279 | 1467 | 6 | 4.58E-10 | 1.66E-12 | 0.000184 | 8.67E-12 | -0.01346459 | WNT11, LINC02761 |
| chr2 | 601927 | 602673 | 747 | 3 | 7.29E-13 | 2.87E-12 | 1.10E-05 | 8.80E-12 | -0.00572984 | NA |
| chr8 | 113436769 | 113438908 | 2140 | 17 | 7.32E-13 | 1.32E-10 | 0.000349 | 8.82E-12 | -0.0017492 | AC055788.1, CSMD3 |
| chr2 | 25341533 | 25343359 | 1827 | 16 | 7.46E-16 | 2.51E-06 | 1.58E-06 | 8.83E-12 | 0.001226129 | DNMT3A |
| chr5 | 177484178 | 177485609 | 1432 | 8 | 8.09E-14 | 1.51E-09 | 9.93E-06 | 9.08E-12 | -0.00652094 | PDLIM7 |
| chr10 | 100866419 | 100866895 | 477 | 3 | 1.05E-14 | 5.46E-08 | 1.68E-07 | 9.09E-12 | -0.00988023 | NA |
| chr7 | 139339467 | 139341638 | 2172 | 24 | 1.16E-09 | 7.30E-09 | 0.000274 | 9.09E-12 | 0.000630318 | FMC1, LUC7L2, FMC1-LUC7L2 |
| chr4 | 46992731 | 46995256 | 2526 | 16 | 2.65E-09 | 4.34E-10 | 0.000224 | 9.30E-12 | -0.00065543 | GABRB1, GABRA4 |
| chr14 | 103715042 | 103715838 | 797 | 11 | 1.62E-11 | 1.07E-11 | 0.000604 | 9.35E-12 | 7.36E-05 | ZFYVE21, XRCC3 |
| chr2 | 112645038 | 112647548 | 2511 | 15 | 1.11E-11 | 2.58E-07 | 3.19E-06 | 9.35E-12 | -0.00035099 | SLC20A1, AC079922.2 |
| chr12 | 120639787 | 120641293 | 1507 | 12 | 1.28E-11 | 4.84E-10 | 0.000211 | 9.35E-12 | 2.42E-05 | CABP1, AC125616.1 |
| chr8 | 38650622 | 38651262 | 641 | 5 | 1.65E-12 | 3.73E-11 | 8.95E-06 | 9.39E-12 | -0.01613697 | NA |
| chr22 | 26656609 | 26657970 | 1362 | 18 | 3.85E-14 | 1.37E-09 | 3.14E-05 | 9.43E-12 | -0.00073242 | MIAT, Z99774.5 |
| chr20 | 64254543 | 64257589 | 3047 | 15 | 2.41E-11 | 2.89E-06 | 2.05E-06 | 9.54E-12 | -0.00089648 | PCMTD2 |
| chr17 | 48794220 | 48795509 | 1290 | 8 | 9.14E-11 | 1.56E-10 | 3.89E-05 | 9.57E-12 | 0.000821419 | TTLL6 |
| chr14 | 104773120 | 104775855 | 2736 | 17 | 2.64E-08 | 6.11E-09 | 9.12E-05 | 9.59E-12 | -0.00090719 | AKT1 |
| chr13 | 44050497 | 44050959 | 463 | 2 | 4.27E-12 | 5.27E-12 | 1.57E-06 | 9.72E-12 | -0.02302334 | NA |
| chr9 | 128392068 | 128393540 | 1473 | 6 | 3.38E-12 | 3.98E-12 | 6.40E-05 | 9.72E-12 | -0.00839272 | MIR219B, AL359091.1, MIR219A2 |
| chr7 | 100868003 | 100868619 | 617 | 4 | 3.19E-14 | 1.68E-11 | 2.07E-06 | 9.80E-12 | -0.01743404 | TRIP6, MIR6875 |
| chr14 | 31576698 | 31577971 | 1274 | 4 | 5.72E-14 | 4.47E-10 | 2.06E-06 | 9.87E-12 | -0.01016787 | NUBPL |
| chr19 | 51019638 | 51021507 | 1870 | 14 | 1.15E-10 | 4.42E-09 | 0.000204 | 9.92E-12 | -0.00077788 | KLK10 |
| chr7 | 27146935 | 27149151 | 2217 | 17 | 6.78E-10 | 8.64E-08 | 0.000563 | 9.92E-12 | -0.00999778 | HOXA-AS3, HOXA3, AC004080.6, HOXA6, AC004080.5 |
| chr10 | 70440856 | 70442959 | 2104 | 16 | 9.39E-13 | 4.58E-09 | 0.000282 | 9.93E-12 | -0.00641349 | NODAL |
| chr7 | 1713760 | 1714273 | 514 | 6 | 1.22E-12 | 2.30E-11 | 1.04E-05 | 9.93E-12 | -0.01650567 | ELFN1 |
| chr19 | 35509407 | 35511151 | 1745 | 17 | 1.77E-14 | 3.70E-06 | 2.17E-06 | 9.96E-12 | 0.000797978 | DMKN |
| chr17 | 44898511 | 44900618 | 2108 | 20 | 2.11E-10 | 3.44E-11 | 0.000347 | 1.01E-11 | -0.00281783 | CCDC103, FAM187A, EFTUD2 |
| chr3 | 39406400 | 39407461 | 1062 | 12 | 1.22E-15 | 1.96E-06 | 5.54E-06 | 1.01E-11 | -0.00075686 | RPSA |
| chr6 | 29427617 | 29429525 | 1909 | 12 | 2.67E-10 | 4.28E-11 | 0.000875 | 1.01E-11 | -0.00528733 | OR5V1, OR11A1 |
| chr3 | 112990135 | 112991496 | 1362 | 16 | 6.20E-11 | 4.23E-12 | 0.001224 | 1.01E-11 | -0.00420826 | GTPBP8, AC074044.1 |
| chr1 | 180247479 | 180249073 | 1595 | 3 | 3.96E-12 | 2.20E-11 | 6.61E-07 | 1.01E-11 | -0.01747027 | LHX4 |
| chr2 | 31580148 | 31582405 | 2258 | 15 | 1.02E-10 | 9.58E-12 | 3.64E-05 | 1.03E-11 | 0.005448534 | SRD5A2 |
| chr3 | 179036512 | 179037409 | 898 | 5 | 5.72E-11 | 2.06E-10 | 1.50E-05 | 1.04E-11 | -0.01246011 | ZMAT3 |
| chr3 | 119779133 | 119782055 | 2923 | 9 | 3.65E-09 | 3.83E-11 | 9.09E-05 | 1.04E-11 | -0.01476981 | AC069444.2, NR1I2 |
| chr16 | 3962935 | 3964294 | 1360 | 8 | 3.32E-12 | 5.88E-10 | 1.01E-05 | 1.04E-11 | 0.009291728 | ADCY9 |
| chr17 | 83182349 | 83184327 | 1979 | 7 | 2.41E-10 | 2.39E-10 | 9.38E-06 | 1.05E-11 | -0.0045643 | AC139099.2 |
| chr18 | 79616639 | 79617589 | 951 | 5 | 3.64E-13 | 2.33E-11 | 1.18E-05 | 1.05E-11 | -0.050452 | NA |
| chr1 | 24318435 | 24319902 | 1468 | 18 | 1.36E-13 | 6.77E-07 | 1.92E-05 | 1.06E-11 | 0.002678194 | GRHL3, GRHL3-AS1 |
| chr17 | 7241048 | 7243438 | 2391 | 24 | 5.00E-09 | 1.24E-08 | 0.000126 | 1.08E-11 | 0.000121491 | AC120057.3, GABARAP, AC120057.2 |
| chr9 | 132212516 | 132214304 | 1789 | 6 | 5.10E-11 | 1.30E-08 | 1.15E-05 | 1.08E-11 | -0.00969587 | NTNG2 |
| chr7 | 100100355 | 100102078 | 1724 | 27 | 3.31E-10 | 5.30E-08 | 6.24E-05 | 1.09E-11 | 0.000981382 | AP4M1, MCM7 |
| chr10 | 75400072 | 75402166 | 2095 | 16 | 3.00E-12 | 1.39E-09 | 2.08E-05 | 1.09E-11 | 0.000339218 | ZNF503-AS2, AC010997.3, ZNF503 |
| chr14 | 103126268 | 103127658 | 1391 | 11 | 1.50E-14 | 2.12E-07 | 3.04E-05 | 1.10E-11 | -0.00932372 | TNFAIP2 |
| chr19 | 50475900 | 50476908 | 1009 | 13 | 9.44E-12 | 9.64E-09 | 0.00013 | 1.12E-11 | -0.00063532 | EMC10, FAM71E1 |
| chr19 | 33418844 | 33419722 | 879 | 2 | 1.57E-09 | 6.18E-12 | 1.59E-06 | 1.13E-11 | -0.00102616 | PEPD |
| chr1 | 211829157 | 211832068 | 2912 | 20 | 2.31E-09 | 9.72E-10 | 3.48E-05 | 1.13E-11 | -0.00307223 | LPGAT1-AS1, LPGAT1 |
| chr9 | 127877460 | 127878507 | 1048 | 11 | 4.00E-14 | 6.88E-08 | 1.86E-05 | 1.15E-11 | -0.00575881 | AK1, AL157935.2 |
| chr2 | 234545041 | 234545249 | 209 | 3 | 5.59E-12 | 3.41E-12 | 1.12E-05 | 1.15E-11 | -0.01653663 | NA |
| chr1 | 54405252 | 54408317 | 3066 | 13 | 8.17E-09 | 2.03E-09 | 1.87E-05 | 1.17E-11 | -0.00037746 | SSBP3 |
| chr2 | 27408263 | 27410368 | 2106 | 17 | 3.75E-09 | 8.01E-08 | 5.67E-05 | 1.17E-11 | -0.00262314 | PPM1G |
| chr21 | 36964681 | 36966158 | 1478 | 6 | 7.65E-13 | 4.47E-10 | 8.13E-06 | 1.18E-11 | 0.000689856 | HLCS |
| chr5 | 135029830 | 135032529 | 2700 | 14 | 3.32E-11 | 8.13E-10 | 1.33E-05 | 1.18E-11 | 0.007869354 | PITX1 |
| chr7 | 1994212 | 1994971 | 760 | 2 | 1.61E-17 | 2.29E-08 | 7.72E-10 | 1.19E-11 | 0.002846184 | MAD1L1, AC069288.1 |
| chr11 | 70063982 | 70065687 | 1706 | 10 | 1.57E-09 | 2.68E-09 | 3.11E-05 | 1.19E-11 | -0.00879406 | LINC02753 |
| chr17 | 19409711 | 19411305 | 1595 | 9 | 1.15E-10 | 9.09E-12 | 0.000195 | 1.20E-11 | -0.00967862 | RNF112 |
| chr19 | 48618337 | 48620299 | 1963 | 24 | 1.70E-15 | 2.77E-08 | 1.84E-05 | 1.21E-11 | 0.00097263 | SPHK2, RPL18, AC022154.1 |
| chr4 | 8010443 | 8010831 | 389 | 4 | 1.53E-13 | 1.07E-08 | 1.30E-07 | 1.21E-11 | -0.01218857 | ABLIM2 |
| chr19 | 43503993 | 43506722 | 2730 | 16 | 1.32E-10 | 6.78E-08 | 3.95E-05 | 1.22E-11 | -0.00020729 | PHLDB3, ETHE1 |
| chr5 | 181114779 | 181115623 | 845 | 9 | 2.45E-13 | 6.16E-10 | 7.51E-05 | 1.23E-11 | -0.00966079 | AC008620.2 |
| chr5 | 179731568 | 179733711 | 2144 | 16 | 9.84E-09 | 1.72E-10 | 8.80E-05 | 1.24E-11 | -0.00080035 | CANX, MAML1 |
| chr2 | 213284132 | 213285011 | 880 | 13 | 2.81E-18 | 5.14E-09 | 1.50E-06 | 1.24E-11 | -0.00386449 | SPAG16, SPAG16-DT |
| chr19 | 43607298 | 43607975 | 678 | 3 | 2.99E-15 | 1.02E-08 | 4.03E-07 | 1.25E-11 | 0.023777046 | SRRM5, ZNF428 |
| chr9 | 4835493 | 4836601 | 1109 | 4 | 4.72E-15 | 6.81E-10 | 1.30E-07 | 1.26E-11 | -0.01456677 | RCL1 |
| chr20 | 16747588 | 16749279 | 1692 | 8 | 7.63E-11 | 1.31E-10 | 2.09E-05 | 1.26E-11 | -0.0086818 | OTOR |
| chr7 | 101915854 | 101917686 | 1833 | 14 | 2.03E-18 | 4.89E-07 | 1.96E-08 | 1.26E-11 | -0.00100558 | CUX1 |
| chr19 | 47479578 | 47480800 | 1223 | 4 | 4.40E-13 | 1.16E-08 | 1.56E-07 | 1.26E-11 | -0.01074 | KPTN |
| chr1 | 37889333 | 37890624 | 1292 | 6 | 3.72E-12 | 2.26E-10 | 8.45E-07 | 1.27E-11 | 0.005622017 | INPP5B |
| chr5 | 139389275 | 139391152 | 1878 | 11 | 4.19E-15 | 8.92E-09 | 3.20E-07 | 1.27E-11 | 0.006430865 | MZB1, PROB1 |
| chr17 | 82646871 | 82649788 | 2918 | 10 | 1.75E-10 | 6.07E-11 | 7.79E-06 | 1.27E-11 | 0.008108981 | AC124283.5, WDR45B |
| chr9 | 123425517 | 123426307 | 791 | 2 | 7.20E-12 | 1.28E-11 | 1.54E-07 | 1.29E-11 | 0.000914178 | DENND1A |
| chr2 | 225399549 | 225401083 | 1535 | 14 | 4.68E-11 | 4.29E-11 | 0.000146 | 1.29E-11 | -0.00658212 | NYAP2 |
| chr2 | 11541279 | 11542738 | 1460 | 7 | 1.19E-13 | 3.59E-09 | 5.64E-08 | 1.31E-11 | -0.00870428 | GREB1 |
| chr9 | 95504361 | 95506829 | 2469 | 9 | 4.99E-12 | 3.30E-08 | 1.31E-05 | 1.31E-11 | 0.008955265 | AL161729.1, PTCH1 |
| chr12 | 20368701 | 20371114 | 2414 | 16 | 4.88E-13 | 7.47E-09 | 1.34E-05 | 1.33E-11 | -0.00215716 | PDE3A, AC129102.1 |
| chr16 | 28877604 | 28879830 | 2227 | 9 | 6.53E-12 | 2.24E-08 | 4.63E-05 | 1.33E-11 | -0.00325295 | AC009093.11, ATP2A1, ATP2A1-AS1 |
| chr14 | 100825265 | 100828093 | 2829 | 24 | 2.33E-11 | 9.14E-09 | 9.17E-07 | 1.34E-11 | -0.00035768 | MEG3, AL117190.3 |
| chr6 | 41370607 | 41372320 | 1714 | 9 | 5.21E-14 | 7.73E-10 | 5.30E-06 | 1.34E-11 | -0.00632707 | NA |
| chr6 | 28863125 | 28864895 | 1771 | 36 | 1.60E-12 | 2.67E-07 | 1.34E-05 | 1.35E-11 | 0.00041366 | LINC01623 |
| chr1 | 31762307 | 31765418 | 3112 | 14 | 4.47E-11 | 4.27E-07 | 6.73E-06 | 1.35E-11 | -0.00239757 | ADGRB2 |
| chr9 | 94082756 | 94084442 | 1687 | 8 | 1.68E-11 | 4.86E-10 | 1.14E-05 | 1.35E-11 | -0.00010808 | PTPDC1 |
| chr20 | 767789 | 768976 | 1188 | 11 | 1.17E-12 | 3.05E-11 | 0.000771 | 1.36E-11 | -0.01564508 | SLC52A3 |
| chr2 | 3600555 | 3601997 | 1443 | 13 | 6.30E-10 | 1.15E-08 | 7.58E-05 | 1.36E-11 | 0.001862908 | COLEC11 |
| chr11 | 7019849 | 7020790 | 942 | 17 | 4.22E-12 | 3.54E-10 | 0.000612 | 1.37E-11 | 0.004220342 | NLRP14, ZNF214 |
| chr17 | 27879818 | 27880020 | 203 | 2 | 1.56E-12 | 1.08E-11 | 3.12E-07 | 1.39E-11 | -0.01475873 | AC005697.2, AC005697.1, LYRM9 |
| chr22 | 40678153 | 40679988 | 1836 | 13 | 1.22E-09 | 1.19E-09 | 0.000165 | 1.39E-11 | 0.003092327 | MCHR1 |
| chr4 | 13656766 | 13657493 | 728 | 3 | 3.61E-13 | 8.21E-11 | 8.92E-07 | 1.40E-11 | -0.02396855 | LINC01182 |
| chr16 | 77434380 | 77436324 | 1945 | 16 | 7.74E-10 | 6.50E-09 | 8.90E-05 | 1.40E-11 | -0.00479874 | AC025284.1, ADAMTS18 |
| chr12 | 48350078 | 48351911 | 1834 | 16 | 5.34E-10 | 5.02E-10 | 0.000819 | 1.41E-11 | 0.004261885 | AC090115.1, ZNF641 |
| chr11 | 5938428 | 5938983 | 556 | 7 | 3.09E-11 | 5.20E-13 | 0.001121 | 1.41E-11 | -0.04102797 | OR56A3, TRIM5 |
| chr17 | 7613542 | 7614810 | 1269 | 14 | 7.20E-13 | 2.31E-09 | 0.000315 | 1.43E-11 | -0.00395531 | SHBG, FXR2 |
| chr11 | 72079233 | 72081634 | 2402 | 22 | 3.62E-11 | 1.42E-09 | 0.000151 | 1.44E-11 | -0.00090037 | AP000812.4, LRTOMT, NUMA1 |
| chr6 | 33123790 | 33125332 | 1543 | 12 | 9.12E-10 | 6.81E-08 | 0.000259 | 1.44E-11 | -0.0126551 | HLA-DPB2 |
| chr18 | 79485369 | 79486369 | 1001 | 4 | 9.69E-13 | 2.22E-11 | 4.87E-06 | 1.45E-11 | -0.01513501 | NFATC1 |
| chr16 | 931766 | 934767 | 3002 | 10 | 2.27E-07 | 3.66E-09 | 1.85E-05 | 1.45E-11 | -0.00317171 | LMF1-AS1, LMF1 |
| chr10 | 77637317 | 77639249 | 1933 | 18 | 2.77E-15 | 8.80E-07 | 3.08E-06 | 1.46E-11 | -0.00309151 | KCNMA1 |
| chr6 | 168823730 | 168825460 | 1731 | 7 | 4.83E-12 | 3.92E-11 | 5.45E-06 | 1.47E-11 | -0.01234932 | NA |
| chr10 | 71240748 | 71241656 | 909 | 2 | 1.72E-11 | 1.82E-11 | 1.05E-07 | 1.47E-11 | 0.030970878 | UNC5B |
| chr17 | 14307867 | 14309908 | 2042 | 6 | 7.81E-10 | 4.38E-10 | 2.81E-05 | 1.48E-11 | 0.000295906 | HS3ST3B1 |
| chr22 | 44025023 | 44027105 | 2083 | 12 | 8.11E-12 | 3.31E-09 | 0.000201 | 1.49E-11 | -0.00716036 | PARVB |
| chr12 | 3752401 | 3753644 | 1244 | 15 | 1.90E-12 | 5.11E-11 | 9.03E-06 | 1.49E-11 | 0.003588163 | AC006207.1, CRACR2A |
| chr9 | 37033753 | 37036497 | 2745 | 19 | 1.87E-11 | 2.52E-09 | 0.000328 | 1.50E-11 | -0.00321573 | PAX5 |
| chr15 | 89242957 | 89244493 | 1537 | 15 | 2.93E-10 | 1.04E-11 | 0.001386 | 1.50E-11 | -0.00208117 | FANCI |
| chr1 | 236762577 | 236762665 | 89 | 3 | 3.66E-12 | 1.56E-11 | 1.40E-06 | 1.50E-11 | -0.03829343 | ACTN2 |
| chr5 | 2755765 | 2757974 | 2210 | 14 | 6.99E-09 | 2.46E-10 | 1.78E-05 | 1.53E-11 | 1.37E-05 | NA |
| chr10 | 132036937 | 132037912 | 976 | 7 | 1.14E-15 | 2.09E-08 | 1.81E-08 | 1.53E-11 | -0.00307302 | JAKMIP3 |
| chr18 | 63318036 | 63320922 | 2887 | 35 | 6.26E-07 | 5.71E-08 | 0.001332 | 1.55E-11 | -0.00153855 | BCL2 |
| chr3 | 15600848 | 15602396 | 1549 | 28 | 1.04E-10 | 8.92E-11 | 0.001854 | 1.58E-11 | 0.000515295 | BTD, HACL1 |
| chr2 | 70257395 | 70258624 | 1230 | 19 | 1.07E-10 | 1.25E-09 | 0.000137 | 1.61E-11 | -0.00210923 | PCYOX1 |
| chr1 | 31181219 | 31182398 | 1180 | 9 | 1.88E-10 | 2.07E-12 | 0.001028 | 1.61E-11 | -0.01010543 | NKAIN1 |
| chr17 | 80220813 | 80221719 | 907 | 8 | 3.85E-13 | 3.41E-11 | 4.06E-05 | 1.61E-11 | -0.00558551 | SLC26A11, SGSH |
| chr7 | 100641127 | 100642747 | 1621 | 11 | 2.46E-11 | 4.57E-10 | 0.000925 | 1.61E-11 | 0.002381552 | TFR2 |
| chr1 | 151372330 | 151374201 | 1872 | 12 | 2.10E-09 | 2.45E-11 | 0.000122 | 1.61E-11 | -0.00302226 | SELENBP1 |
| chr17 | 80760080 | 80761823 | 1744 | 8 | 1.69E-08 | 7.15E-11 | 1.99E-05 | 1.61E-11 | 0.003839857 | RPTOR |
| chr19 | 2542839 | 2544445 | 1607 | 8 | 2.00E-10 | 1.22E-12 | 0.00042 | 1.63E-11 | -0.0112693 | GNG7 |
| chr4 | 41537290 | 41538212 | 923 | 5 | 1.05E-11 | 1.38E-09 | 9.68E-06 | 1.63E-11 | -0.00638284 | LIMCH1 |
| chr13 | 52848246 | 52849914 | 1669 | 14 | 4.55E-14 | 3.37E-09 | 8.24E-06 | 1.63E-11 | -0.00193239 | PCDH8 |
| chr15 | 83282911 | 83286097 | 3187 | 35 | 3.88E-08 | 2.05E-09 | 0.001246 | 1.64E-11 | -0.00340119 | AC103876.1, BNC1 |
| chr9 | 132578810 | 132581389 | 2580 | 7 | 4.37E-08 | 4.94E-10 | 3.21E-05 | 1.65E-11 | 0.010107314 | NA |
| chr11 | 126101889 | 126102829 | 941 | 6 | 9.52E-12 | 4.34E-10 | 2.61E-06 | 1.65E-11 | -0.01931349 | AP000821.1 |
| chr12 | 14802798 | 14804586 | 1789 | 17 | 5.04E-11 | 8.88E-10 | 0.000217 | 1.66E-11 | 0.000421382 | C12orf60, WBP11 |
| chr6 | 42745667 | 42746810 | 1144 | 9 | 5.00E-12 | 4.29E-11 | 0.000184 | 1.67E-11 | -0.00029336 | TBCC |
| chr5 | 141244315 | 141246226 | 1912 | 15 | 1.20E-11 | 4.88E-09 | 0.000414 | 1.68E-11 | -0.00460435 | AC244517.10, PCDHB15, AC244517.11 |
| chr12 | 109711220 | 109713318 | 2099 | 10 | 7.55E-11 | 2.48E-09 | 4.62E-05 | 1.68E-11 | -0.00514317 | NA |
| chr10 | 133182815 | 133183967 | 1153 | 7 | 3.64E-13 | 2.28E-10 | 2.01E-05 | 1.69E-11 | -0.00636927 | KNDC1 |
| chr9 | 137422437 | 137423587 | 1151 | 8 | 1.55E-10 | 3.06E-11 | 0.000426 | 1.69E-11 | 0.00058384 | NOXA1, EXD3 |
| chr13 | 113914538 | 113915932 | 1395 | 6 | 5.85E-13 | 3.19E-09 | 3.92E-06 | 1.70E-11 | 0.008227971 | LINC00452 |
| chr11 | 111378934 | 111380019 | 1086 | 15 | 5.39E-12 | 2.26E-10 | 1.03E-05 | 1.70E-11 | -0.0117498 | POU2AF1 |
| chr15 | 101104477 | 101105759 | 1283 | 3 | 1.58E-11 | 3.92E-09 | 1.01E-06 | 1.70E-11 | -0.00482344 | NA |
| chr11 | 7087676 | 7088917 | 1242 | 9 | 6.44E-12 | 1.44E-10 | 0.000226 | 1.70E-11 | -0.02383111 | NA |
| chr21 | 34668472 | 34669871 | 1400 | 11 | 2.09E-12 | 9.20E-12 | 0.000592 | 1.73E-11 | -0.00412585 | CLIC6 |
| chr8 | 23083446 | 23084519 | 1074 | 5 | 4.56E-11 | 5.44E-12 | 6.29E-05 | 1.74E-11 | -0.0449819 | AC107959.2, AC107959.4, AC107959.5 |

| | | | | | | | | | | |
|---|---|---|---|---|---|---|---|---|---|---|
| chr1 | 9727157 | 9729924 | 2768 | 10 | 2.30E-08 | 1.43E-10 | 0.000148 | 1.74E-11 | -0.00624667 | PIK3CD, CLSTN1 |
| chr11 | 49433662 | 49434649 | 988 | 4 | 1.23E-13 | 8.22E-09 | 7.54E-08 | 1.74E-11 | -0.01207231 | AC135977.1 |
| chr22 | 41204974 | 41205577 | 604 | 12 | 6.64E-15 | 2.77E-06 | 1.14E-07 | 1.75E-11 | 0.002069963 | L3MBTL2 |
| chr16 | 769064 | 770155 | 1092 | 6 | 5.33E-12 | 7.07E-11 | 1.35E-05 | 1.75E-11 | 0.003854586 | AL031258.1, MSLNL |
| chr14 | 104642384 | 104644092 | 1709 | 4 | 1.96E-10 | 1.77E-10 | 1.75E-06 | 1.76E-11 | 0.002316791 | NA |
| chr5 | 149771134 | 149772124 | 991 | 7 | 1.26E-13 | 4.68E-07 | 4.91E-06 | 1.76E-11 | 0.001056469 | PPARGC1B |
| chr22 | 30920197 | 30922559 | 2363 | 14 | 8.34E-10 | 4.34E-10 | 9.64E-06 | 1.76E-11 | 0.008705967 | MORC2-AS1 |
| chr7 | 5387200 | 5388383 | 1184 | 6 | 2.81E-13 | 1.64E-10 | 3.48E-06 | 1.76E-11 | 0.001351417 | TNRC18 |
| chr3 | 120450936 | 120452229 | 1294 | 8 | 2.23E-13 | 2.26E-10 | 0.000125 | 1.76E-11 | -0.00723072 | AC126182.3, FSTL1 |
| chr3 | 50340324 | 50343249 | 2926 | 23 | 7.19E-09 | 1.22E-07 | 1.83E-05 | 1.77E-11 | -0.00150898 | ZMYND10-AS1, RASSF1, ZMYND10 |
| chr5 | 134114547 | 134116497 | 1951 | 15 | 1.10E-12 | 5.81E-09 | 1.18E-05 | 1.78E-11 | 0.001109968 | TCF7 |
| chr11 | 69639655 | 69641933 | 2279 | 33 | 1.55E-10 | 9.21E-08 | 7.70E-05 | 1.80E-11 | -0.00150482 | CCND1 |
| chr17 | 1178931 | 1181359 | 2429 | 14 | 1.54E-10 | 2.96E-09 | 0.000194 | 1.83E-11 | 0.000189269 | ABR |
| chr10 | 66615375 | 66616686 | 1312 | 4 | 2.86E-11 | 6.16E-11 | 3.51E-06 | 1.85E-11 | -0.00898708 | CTNNA3 |
| chr1 | 44799637 | 44801673 | 2037 | 17 | 7.58E-11 | 4.19E-09 | 1.28E-05 | 1.85E-11 | -0.00354092 | PLK3 |
| chr15 | 89774798 | 89777378 | 2581 | 14 | 3.20E-09 | 5.53E-09 | 0.000107 | 1.90E-11 | -0.00247182 | MESP2 |
| chr22 | 42614435 | 42616553 | 2119 | 15 | 4.10E-09 | 4.50E-08 | 2.60E-06 | 1.92E-11 | -0.0034193 | RNU12, Z93241.1, POLDIP3 |
| chr3 | 112974050 | 112975690 | 1641 | 9 | 3.24E-14 | 2.61E-07 | 2.25E-07 | 1.95E-11 | -0.00386973 | CD200R1 |
| chr12 | 113092651 | 113093660 | 1010 | 6 | 4.78E-11 | 1.46E-09 | 5.70E-06 | 1.96E-11 | -0.01279939 | DTX1 |
| chr9 | 83219180 | 83219799 | 620 | 4 | 6.77E-12 | 4.52E-12 | 3.64E-05 | 1.98E-11 | -0.02023679 | AL137847.1 |
| chr6 | 43170442 | 43173190 | 2749 | 22 | 1.58E-08 | 1.43E-08 | 0.000385 | 2.00E-11 | 0.00015627 | SRF |
| chr15 | 42919094 | 42921860 | 2767 | 20 | 1.92E-08 | 3.98E-09 | 0.00151 | 2.01E-11 | -0.00246836 | TTBK2 |
| chr6 | 42924128 | 42926385 | 2258 | 10 | 2.49E-11 | 1.71E-10 | 2.04E-05 | 2.04E-11 | -0.0017005 | PTCRA |
| chr2 | 240350865 | 240351653 | 789 | 8 | 2.48E-13 | 8.92E-11 | 6.40E-05 | 2.04E-11 | 0.014816357 | NA |
| chr7 | 85041808 | 85042028 | 221 | 4 | 1.98E-11 | 3.44E-12 | 4.52E-05 | 2.04E-11 | 4.15E-06 | SEMA3D |
| chr3 | 138114652 | 138116169 | 1518 | 17 | 9.09E-11 | 1.43E-08 | 0.000338 | 2.04E-11 | -0.01074053 | DZIP1L |
| chr22 | 43150776 | 43152882 | 2107 | 18 | 5.32E-09 | 4.23E-09 | 7.40E-05 | 2.05E-11 | -0.00136339 | TSPO |
| chr19 | 10361589 | 10364236 | 2648 | 9 | 3.29E-10 | 2.11E-10 | 6.76E-07 | 2.06E-11 | -0.00351938 | TYK2 |
| chr2 | 9386542 | 9388531 | 1990 | 8 | 1.35E-08 | 2.11E-10 | 4.30E-05 | 2.06E-11 | -0.00120398 | ASAP2 |
| chr15 | 71115741 | 71119003 | 3263 | 13 | 2.01E-07 | 2.05E-10 | 0.000637 | 2.08E-11 | 0.001478971 | THSD4 |
| chr7 | 29239429 | 29240041 | 613 | 2 | 5.74E-14 | 1.11E-10 | 1.77E-08 | 2.10E-11 | 0.00096366 | AC004593.2, CHN2 |
| chr13 | 44436526 | 44438749 | 2224 | 22 | 2.10E-09 | 5.63E-11 | 0.001788 | 2.12E-11 | -0.00196138 | AL139184.1, TSC22D1 |
| chr6 | 36947722 | 36950126 | 2405 | 9 | 7.73E-10 | 1.02E-09 | 3.26E-05 | 2.13E-11 | -0.00925865 | PI16 |
| chr8 | 126433316 | 126434575 | 1260 | 7 | 8.86E-13 | 6.83E-11 | 4.18E-05 | 2.15E-11 | -0.00184533 | NA |
| chr8 | 124654287 | 124655544 | 1258 | 5 | 5.21E-11 | 9.64E-11 | 6.92E-06 | 2.16E-11 | 0.011584282 | MTSS1 |
| chr14 | 64502656 | 64505271 | 2616 | 21 | 5.54E-09 | 2.97E-11 | 0.005768 | 2.16E-11 | -1.34E-05 | ZBTB1, ZBTB25 |
| chr14 | 68789091 | 68790260 | 1170 | 5 | 1.62E-10 | 2.39E-12 | 0.000157 | 2.17E-11 | -0.01257474 | ZFP36L1 |
| chr1 | 109269235 | 109270759 | 1525 | 7 | 2.24E-11 | 2.60E-10 | 1.38E-06 | 2.18E-11 | -0.00597516 | CELSR2 |
| chr4 | 4858045 | 4860513 | 2469 | 30 | 1.03E-09 | 8.13E-10 | 0.002015 | 2.18E-11 | -0.00057604 | MSX1 |
| chr21 | 44293055 | 44294824 | 1770 | 6 | 8.48E-11 | 7.60E-12 | 0.000195 | 2.19E-11 | -0.00268166 | AIRE |
| chr4 | 188659177 | 188659721 | 545 | 7 | 2.59E-13 | 1.93E-08 | 2.27E-05 | 2.22E-11 | -0.01146325 | LINC01060 |
| chr15 | 31069815 | 31071701 | 1887 | 9 | 7.44E-10 | 3.06E-11 | 0.000275 | 2.24E-11 | 0.000923716 | TRPM1 |
| chr16 | 57637731 | 57640081 | 2351 | 13 | 2.80E-08 | 1.86E-12 | 0.000494 | 2.24E-11 | 0.006077007 | ADGRG1 |
| chr12 | 52146994 | 52148083 | 1090 | 5 | 2.88E-13 | 3.30E-11 | 5.48E-06 | 2.28E-11 | -0.00618183 | NA |
| chr4 | 24965787 | 24966350 | 564 | 2 | 1.09E-12 | 2.75E-11 | 1.36E-07 | 2.29E-11 | -0.00477257 | CCDC149 |
| chr12 | 54015701 | 54017689 | 1989 | 27 | 3.19E-14 | 5.75E-07 | 1.72E-06 | 2.29E-11 | -0.00086849 | AC012531.3, HOXC6, HOXC4, AC012531.2 |
| chr11 | 443853 | 444548 | 696 | 3 | 3.81E-12 | 9.56E-12 | 5.28E-06 | 2.29E-11 | -0.00776687 | NA |
| chr7 | 1541581 | 1543493 | 1913 | 13 | 6.07E-09 | 5.14E-10 | 0.000416 | 2.29E-11 | 0.001781605 | MAFK, TMEM184A |
| chr17 | 49889503 | 49890989 | 1487 | 6 | 1.84E-09 | 6.83E-11 | 4.83E-05 | 2.32E-11 | -0.01477623 | AC027801.2 |
| chr6 | 147202523 | 147205231 | 2709 | 15 | 5.99E-08 | 1.35E-11 | 3.51E-05 | 2.32E-11 | 1.05E-05 | STXBP5, STXBP5-AS1 |
| chr6 | 32974048 | 32976718 | 2671 | 16 | 4.17E-10 | 9.82E-10 | 0.000288 | 2.34E-11 | -0.00349857 | BRD2 |
| chr7 | 26857634 | 26858201 | 568 | 8 | 2.51E-11 | 2.71E-12 | 0.000195 | 2.34E-11 | -0.01294503 | SKAP2 |
| chr15 | 74135842 | 74137276 | 1435 | 8 | 1.09E-10 | 3.32E-08 | 8.30E-06 | 2.35E-11 | -0.00198906 | ISLR2 |
| chr8 | 28889557 | 28891582 | 2026 | 26 | 2.41E-10 | 4.06E-10 | 0.002044 | 2.36E-11 | 8.47E-05 | HMBOX1, INTS9 |
| chr17 | 39729357 | 39731527 | 2171 | 16 | 7.74E-10 | 7.12E-07 | 1.22E-05 | 2.38E-11 | -0.00069233 | ERBB2, MIEN1 |
| chr5 | 115179407 | 115181726 | 2320 | 20 | 1.62E-11 | 1.68E-07 | 1.99E-05 | 2.38E-11 | -0.00281151 | TRIM36 |
| chr9 | 121299032 | 121300025 | 994 | 11 | 2.14E-14 | 5.43E-07 | 8.28E-06 | 2.38E-11 | -0.00567114 | GSN |
| chr5 | 38845027 | 38846641 | 1615 | 12 | 5.81E-14 | 2.22E-07 | 3.18E-05 | 2.38E-11 | -0.00174734 | OSMR, OSMR-AS1 |
| chr10 | 110496260 | 110497863 | 1604 | 11 | 5.35E-13 | 1.06E-11 | 0.000283 | 2.39E-11 | -0.00790288 | AL355512.1 |
| chr3 | 108757098 | 108758031 | 934 | 7 | 2.10E-11 | 3.10E-10 | 2.43E-05 | 2.41E-11 | 0.008998727 | RETNLB |
| chr7 | 128454656 | 128455676 | 1021 | 6 | 1.04E-15 | 6.32E-10 | 7.02E-06 | 2.44E-11 | -0.00999163 | NA |
| chr12 | 23949873 | 23951423 | 1551 | 12 | 1.42E-12 | 3.70E-07 | 2.54E-05 | 2.45E-11 | -0.00227417 | SOX5 |
| chr7 | 4203137 | 4204740 | 1604 | 10 | 1.36E-09 | 1.56E-07 | 4.78E-06 | 2.46E-11 | -0.0180291 | SDK1 |
| chr2 | 9754206 | 9754409 | 204 | 2 | 2.47E-14 | 1.41E-11 | 1.53E-06 | 2.46E-11 | -0.02903966 | NA |
| chr9 | 100097838 | 100099067 | 1230 | 7 | 1.23E-13 | 3.13E-10 | 5.87E-05 | 2.46E-11 | -0.00364227 | ERP44 |
| chr8 | 10727884 | 10729795 | 1912 | 10 | 4.59E-10 | 7.27E-11 | 0.000151 | 2.49E-11 | -0.00521634 | AC105001.1, SOX7, AC105001.2 |
| chr22 | 32353813 | 32354894 | 1082 | 8 | 8.63E-10 | 3.69E-10 | 0.000119 | 2.50E-11 | -0.00286742 | RFPL3 |
| chr6 | 33116212 | 33117693 | 1482 | 20 | 2.62E-11 | 2.25E-11 | 0.000843 | 2.50E-11 | -0.00956755 | HLA-DPB2 |
| chr1 | 11735352 | 11737336 | 1985 | 14 | 4.44E-14 | 5.64E-07 | 3.15E-05 | 2.51E-11 | 0.000539193 | AGTRAP |
| chr12 | 120649133 | 120650605 | 1473 | 9 | 7.57E-11 | 1.35E-11 | 0.000342 | 2.51E-11 | -0.01153465 | CABP1 |
| chr16 | 4935484 | 4937531 | 2048 | 12 | 1.39E-09 | 5.80E-11 | 0.000141 | 2.51E-11 | -0.00206401 | PPL |
| chr8 | 42206501 | 42208223 | 1723 | 10 | 1.67E-09 | 6.74E-12 | 0.000146 | 2.53E-11 | -0.00728038 | PLAT |
| chr18 | 75203571 | 75205495 | 1925 | 10 | 6.04E-12 | 7.43E-11 | 0.000372 | 2.56E-11 | 0.028599435 | ZADH2 |
| chr11 | 2160636 | 2162190 | 1555 | 12 | 1.20E-11 | 2.58E-10 | 2.23E-05 | 2.61E-11 | -0.00877768 | INS-IGF2, INS |
| chr12 | 57200822 | 57201822 | 1001 | 8 | 8.00E-12 | 1.72E-11 | 0.000101 | 2.62E-11 | -0.00754923 | LRP1 |
| chr10 | 129466333 | 129467830 | 1498 | 36 | 1.67E-12 | 1.02E-07 | 0.003718 | 2.63E-11 | 0.002661239 | MGMT |
| chr17 | 60092162 | 60092800 | 639 | 2 | 9.55E-12 | 2.02E-11 | 4.33E-07 | 2.66E-11 | -0.03277571 | NA |
| chr2 | 1604957 | 1606060 | 1104 | 9 | 5.80E-12 | 9.24E-11 | 3.88E-05 | 2.66E-11 | -0.01605309 | AC141930.1 |
| chr20 | 37294593 | 37297188 | 2596 | 11 | 1.21E-10 | 1.47E-07 | 5.26E-07 | 2.67E-11 | 0.002485878 | MANBAL |
| chr1 | 16043335 | 16043770 | 436 | 5 | 8.76E-13 | 2.78E-10 | 3.43E-06 | 2.67E-11 | -0.0077822 | CLCNKB |
| chr17 | 7280897 | 7282232 | 1336 | 17 | 1.02E-11 | 3.63E-10 | 0.000652 | 2.68E-11 | 0.00209141 | SLC2A4 |
| chr17 | 81097561 | 81098695 | 1135 | 6 | 3.41E-11 | 3.80E-12 | 0.000122 | 2.68E-11 | -0.01425535 | BAIAP2 |
| chr12 | 133180765 | 133182013 | 1249 | 13 | 6.02E-13 | 2.47E-09 | 0.000432 | 2.68E-11 | -0.00522789 | AC026786.1, ZNF268 |
| chr13 | 45851244 | 45852128 | 885 | 12 | 1.51E-13 | 8.32E-10 | 5.28E-06 | 2.69E-11 | 0.007930002 | SIAH3 |
| chr1 | 42359133 | 42359141 | 9 | 2 | 5.81E-12 | 1.50E-11 | 1.84E-06 | 2.70E-11 | -0.03213548 | NA |
| chr7 | 93043170 | 93044607 | 1438 | 5 | 1.47E-12 | 2.85E-11 | 1.04E-05 | 2.71E-11 | -0.02096754 | NA |
| chr14 | 93009629 | 93009726 | 98 | 2 | 2.38E-13 | 2.21E-11 | 3.56E-07 | 2.71E-11 | -0.0537818 | ITPK1 |
| chr1 | 231929445 | 231930043 | 599 | 5 | 3.33E-11 | 1.07E-10 | 3.59E-05 | 2.71E-11 | -0.0120627 | DISC1, DISC1-IT1 |
| chr4 | 1948220 | 1949151 | 932 | 5 | 4.28E-14 | 9.45E-09 | 5.82E-07 | 2.72E-11 | -0.00118028 | NSD2 |
| chr3 | 125119313 | 125121452 | 2140 | 10 | 2.72E-09 | 4.23E-10 | 0.000152 | 2.73E-11 | -0.00660138 | SLC12A8 |
| chr4 | 26320251 | 26322686 | 2436 | 18 | 3.57E-10 | 1.00E-06 | 6.31E-06 | 2.73E-11 | -0.00152777 | RBPJ |
| chr4 | 119627182 | 119629624 | 2443 | 18 | 7.80E-09 | 1.10E-08 | 0.000654 | 2.73E-11 | 0.001493347 | PDE5A |
| chr2 | 10911602 | 10913811 | 2210 | 14 | 7.48E-11 | 7.88E-09 | 1.51E-06 | 2.75E-11 | -0.00084827 | KCNF1 |
| chr2 | 120252279 | 120254447 | 2169 | 14 | 4.10E-10 | 1.16E-08 | 2.33E-05 | 2.76E-11 | -0.00429962 | RALB |
| chr20 | 53208777 | 53210020 | 1244 | 5 | 1.58E-09 | 1.80E-09 | 1.05E-05 | 2.77E-11 | -0.01029989 | TSHZ2 |
| chr7 | 4106158 | 4107946 | 1789 | 8 | 1.15E-10 | 3.10E-10 | 5.04E-05 | 2.78E-11 | -0.00513798 | SDK1 |
| chr5 | 60842934 | 60845073 | 2140 | 14 | 2.20E-10 | 5.96E-10 | 0.000192 | 2.78E-11 | -0.00073244 | ELOVL7 |
| chr14 | 55440404 | 55441647 | 1244 | 10 | 2.50E-10 | 4.85E-08 | 4.43E-05 | 2.78E-11 | -0.00838273 | TBPL2 |
| chr11 | 128942567 | 128943793 | 1227 | 18 | 5.24E-11 | 8.86E-11 | 0.002654 | 2.82E-11 | -0.00152392 | TP53AIP1 |
| chr20 | 64188788 | 64189601 | 814 | 3 | 7.83E-12 | 1.71E-11 | 2.38E-06 | 2.83E-11 | -0.01887243 | MYT1 |
| chr7 | 21945034 | 21947054 | 2021 | 13 | 4.24E-11 | 3.88E-08 | 7.66E-05 | 2.83E-11 | 0.001928705 | CDCA7L |
| chr2 | 38665094 | 38667056 | 1963 | 13 | 1.02E-09 | 2.74E-09 | 0.000661 | 2.84E-11 | -0.00689965 | GALM |
| chr1 | 65262792 | 65265749 | 2958 | 13 | 2.54E-08 | 2.63E-10 | 0.000217 | 2.84E-11 | 0.00071996 | DNAJC6 |
| chr10 | 102419109 | 102420722 | 1614 | 23 | 2.99E-12 | 1.65E-09 | 0.000314 | 2.85E-11 | 0.000459542 | FBXL15, PSD |
| chr6 | 134052347 | 134054048 | 1702 | 10 | 7.58E-16 | 8.94E-07 | 7.52E-06 | 2.86E-11 | -0.00041266 | SLC2A12 |
| chr11 | 393606 | 394857 | 1252 | 9 | 6.26E-14 | 8.48E-09 | 4.58E-06 | 2.87E-11 | -0.0032335 | PKP3 |
| chr1 | 22635539 | 22638108 | 2570 | 9 | 5.04E-07 | 1.14E-10 | 0.000207 | 2.87E-11 | -0.00983354 | C1QA |
| chr18 | 55401406 | 55402886 | 1481 | 9 | 2.46E-11 | 6.91E-10 | 0.000264 | 2.87E-11 | -0.01513385 | TCF4 |
| chr7 | 151011880 | 151014361 | 2482 | 10 | 1.45E-08 | 2.65E-08 | 2.65E-05 | 2.89E-11 | -0.00721358 | NOS3, ATG9B |
| chr16 | 86710157 | 86710715 | 559 | 4 | 1.15E-12 | 3.69E-10 | 1.03E-05 | 2.89E-11 | -0.01085622 | LINC02188 |
| chr8 | 123274871 | 123276251 | 1381 | 7 | 3.07E-14 | 2.08E-08 | 2.32E-08 | 2.91E-11 | 0.003115889 | ZHX1 |
| chr19 | 54114231 | 54115786 | 1556 | 19 | 4.34E-11 | 4.91E-08 | 0.000143 | 2.98E-11 | -0.00131977 | PRPF31, TFPT |
| chr10 | 1199003 | 1200396 | 1394 | 9 | 3.35E-12 | 2.82E-08 | 9.20E-05 | 2.98E-11 | -0.01495141 | LINC00200, ADARB2 |
| chr2 | 61536234 | 61538955 | 2722 | 22 | 6.18E-08 | 4.71E-07 | 0.001214 | 2.99E-11 | -0.00062111 | XPO1 |
| chr6 | 32967577 | 32970133 | 2557 | 44 | 7.28E-09 | 8.20E-08 | 6.18E-05 | 3.01E-11 | 8.82E-06 | BRD2, HLA-DMA |
| chr19 | 18315963 | 18316772 | 810 | 3 | 3.30E-12 | 2.68E-11 | 7.05E-06 | 3.01E-11 | 0.031040871 | LSM4 |
| chr16 | 31711804 | 31714596 | 2793 | 19 | 4.21E-10 | 2.45E-07 | 3.09E-05 | 3.01E-11 | -0.00010766 | CLUHP3, ZNF720, AC074050.3 |
| chr16 | 87436477 | 87437472 | 996 | 4 | 3.62E-16 | 3.40E-09 | 2.17E-08 | 3.02E-11 | -0.01475783 | ZCCHC14 |
| chr1 | 9996779 | 9997912 | 1134 | 11 | 7.13E-12 | 3.09E-09 | 3.79E-05 | 3.04E-11 | -0.00675272 | RBP7 |
| chr11 | 1750325 | 1752175 | 1851 | 12 | 5.07E-10 | 8.86E-11 | 0.00026 | 3.07E-11 | -0.00196478 | IFITM10, AC068580.4 |
| chr4 | 52656654 | 52658717 | 2064 | 10 | 1.56E-08 | 2.36E-11 | 0.001628 | 3.13E-11 | -0.00071288 | USP46 |
| chr3 | 32818975 | 32821947 | 2973 | 12 | 1.50E-08 | 1.04E-11 | 0.001069 | 3.14E-11 | 0.001001461 | TRIM71 |
| chr20 | 35604396 | 35606159 | 1764 | 6 | 8.36E-11 | 1.12E-06 | 4.77E-07 | 3.14E-11 | -0.00450045 | FER1L4 |
| chr22 | 46389325 | 46391521 | 2197 | 6 | 5.72E-09 | 1.60E-09 | 8.73E-06 | 3.17E-11 | -0.0061179 | CELSR1 |
| chr1 | 212613001 | 212615258 | 2258 | 12 | 7.95E-08 | 1.08E-11 | 0.000818 | 3.18E-11 | -0.00266855 | ATF3 |
| chr1 | 81799252 | 81801156 | 1905 | 10 | 7.09E-14 | 1.08E-06 | 3.56E-05 | 3.20E-11 | -0.00585422 | ADGRL2 |
| chr5 | 177365894 | 177367404 | 1511 | 7 | 1.32E-13 | 4.23E-10 | 1.51E-07 | 3.20E-11 | -0.00749906 | RGS14 |
| chr14 | 24130316 | 24132599 | 2284 | 14 | 2.03E-07 | 6.42E-10 | 0.000192 | 3.21E-11 | 0.00495545 | AL136295.3, FITM1 |
| chr12 | 31729150 | 31730181 | 1032 | 11 | 1.76E-16 | 6.02E-09 | 8.12E-08 | 3.25E-11 | 0.004725704 | AC023157.2 |
| chr6 | 54845601 | 54846678 | 1078 | 10 | 2.06E-12 | 1.30E-10 | 8.13E-05 | 3.26E-11 | -0.00453982 | NA |
| chr22 | 17418362 | 17419239 | 878 | 4 | 3.56E-11 | 6.12E-12 | 4.35E-05 | 3.27E-11 | -0.00896782 | CECR2, FO681548.1 |
| chr14 | 24366988 | 24369196 | 2209 | 24 | 1.59E-09 | 1.87E-10 | 0.000363 | 3.27E-11 | -0.00355312 | NFATC4 |
| chr7 | 1856267 | 1857191 | 925 | 6 | 1.53E-11 | 3.59E-12 | 0.000397 | 3.28E-11 | -0.02574852 | MAD1L1, AC069288.1 |
| chr6 | 166861907 | 166863079 | 1173 | 14 | 5.41E-14 | 1.25E-07 | 2.75E-06 | 3.29E-11 | 0.001190383 | RPS6KA2, AL159163.1 |
| chr5 | 66958488 | 66960885 | 2398 | 10 | 8.54E-09 | 3.20E-10 | 6.01E-05 | 3.33E-11 | 0.003658671 | MAST4 |
| chr21 | 43732453 | 43734889 | 2437 | 8 | 1.84E-09 | 1.13E-10 | 2.61E-05 | 3.35E-11 | 0.023608633 | PDXK |
| chr11 | 133366227 | 133366349 | 123 | 5 | 9.66E-12 | 1.17E-10 | 1.30E-05 | 3.36E-11 | -0.01797145 | OPCML |
| chr1 | 11858330 | 11859818 | 1489 | 14 | 6.28E-11 | 1.99E-09 | 1.69E-05 | 3.36E-11 | -0.00627994 | NPPB |
| chr8 | 1963422 | 1965258 | 1837 | 8 | 2.48E-10 | 2.79E-07 | 6.16E-07 | 3.36E-11 | 0.007689402 | NA |
| chr6 | 167666208 | 167666985 | 778 | 4 | 1.23E-15 | 3.03E-08 | 3.36E-08 | 3.37E-11 | 0.004002162 | LINC02538 |
| chr8 | 11847911 | 11849290 | 1380 | 6 | 1.13E-11 | 1.44E-11 | 3.10E-05 | 3.37E-11 | -0.01303999 | CTSB |
| chr1 | 21814276 | 21815066 | 791 | 8 | 1.17E-12 | 3.30E-10 | 1.36E-06 | 3.37E-11 | -0.00601387 | LDLRAD2 |
| chr17 | 41538703 | 41541034 | 2332 | 5 | 1.72E-12 | 2.41E-07 | 9.44E-08 | 3.38E-11 | -0.0087669 | NA |
| chr1 | 206603202 | 206603719 | 518 | 2 | 7.32E-16 | 9.96E-09 | 1.98E-09 | 3.38E-11 | -0.01450801 | EIF2D |
| chr4 | 139656828 | 139657915 | 1088 | 4 | 2.13E-13 | 2.76E-08 | 1.32E-06 | 3.41E-11 | -0.0294965 | NA |
| chr17 | 72475467 | 72476466 | 1000 | 3 | 1.93E-12 | 1.93E-10 | 2.84E-06 | 3.42E-11 | -0.01493772 | LINC00511 |
| chr20 | 47626082 | 47627951 | 1870 | 9 | 1.71E-08 | 2.15E-10 | 0.000159 | 3.43E-11 | -0.00277273 | NCOA3 |
| chr16 | 2018941 | 2020860 | 1920 | 11 | 3.72E-09 | 2.03E-09 | 4.03E-05 | 3.44E-11 | -0.00485638 | NPW |
| chr17 | 80543779 | 80545282 | 1504 | 11 | 1.29E-14 | 4.38E-08 | 1.88E-06 | 3.44E-11 | -0.00535666 | RPTOR |
| chr1 | 161765334 | 161766223 | 890 | 8 | 1.18E-15 | 2.54E-09 | 2.81E-06 | 3.45E-11 | -0.01482379 | AL359541.1 |
| chr17 | 7674651 | 7677592 | 2942 | 17 | 8.17E-09 | 4.22E-07 | 4.06E-05 | 3.46E-11 | -0.0009626 | TP53 |
| chr4 | 4216400 | 4217451 | 1052 | 8 | 2.17E-13 | 9.89E-09 | 1.87E-05 | 3.46E-11 | -0.0031946 | OTOP1 |
| chr5 | 38467146 | 38469015 | 1870 | 11 | 4.42E-10 | 3.83E-07 | 1.18E-05 | 3.47E-11 | -0.00175249 | AC010457.1 |
| chr19 | 55301205 | 55302850 | 1646 | 9 | 3.14E-12 | 8.63E-09 | 5.53E-06 | 3.49E-11 | -0.00038424 | BRSK1 |
| chr6 | 31886980 | 31889323 | 2344 | 41 | 1.92E-09 | 6.95E-10 | 0.000257 | 3.49E-11 | 0.000234182 | EHMT2 |
| chr11 | 70688356 | 70688613 | 258 | 2 | 3.63E-15 | 3.99E-08 | 1.23E-09 | 3.51E-11 | -0.01599425 | SHANK2 |
| chr11 | 67522571 | 67524060 | 1490 | 13 | 8.92E-12 | 1.81E-09 | 3.08E-05 | 3.54E-11 | -0.00158208 | CABP2 |
| chr15 | 90529525 | 90530917 | 1393 | 11 | 2.83E-10 | 1.17E-08 | 5.70E-05 | 3.54E-11 | -0.00152436 | CRTC3 |

| | | | | | | | | | | |
|---|---|---|---|---|---|---|---|---|---|---|
| chr1 | 182838439 | 182839856 | 1418 | 15 | 3.44E-12 | 3.31E-10 | 0.001332 | 3.56E-11 | 0.000427113 | DHX9, AL355999.1 |
| chr19 | 11293778 | 11295499 | 1722 | 10 | 6.39E-09 | 2.35E-10 | 3.52E-05 | 3.58E-11 | -0.00083582 | NA |
| chr9 | 14530861 | 14532007 | 1147 | 3 | 6.16E-12 | 1.26E-11 | 9.93E-06 | 3.59E-11 | -0.00961204 | AL159169.1 |
| chr15 | 39471977 | 39472643 | 667 | 3 | 5.51E-12 | 5.84E-11 | 1.54E-06 | 3.59E-11 | -0.00591642 | NA |
| chr6 | 90537289 | 90537940 | 652 | 4 | 6.57E-13 | 4.89E-10 | 2.64E-06 | 3.61E-11 | 0.007235232 | MAP3K7 |
| chr3 | 40386636 | 40387926 | 1291 | 15 | 1.08E-11 | 1.54E-08 | 0.000272 | 3.67E-11 | 0.008826934 | ENTPD3, ENTPD3-AS1 |
| chr10 | 79443111 | 79444813 | 1703 | 7 | 2.29E-11 | 1.59E-09 | 9.93E-05 | 3.67E-11 | 0.003123435 | ZCCHC24 |
| chr10 | 92572745 | 92575079 | 2335 | 16 | 1.27E-09 | 9.09E-08 | 2.74E-05 | 3.68E-11 | -0.00209415 | KIF11, IDE |
| chr8 | 17001786 | 17003367 | 1582 | 14 | 2.23E-11 | 3.72E-11 | 0.002728 | 3.69E-11 | -0.01081187 | FGF20 |
| chr20 | 13106879 | 13107477 | 599 | 2 | 1.78E-13 | 1.07E-10 | 4.74E-08 | 3.71E-11 | -0.00730452 | SPTLC3 |
| chr4 | 143512693 | 143515191 | 2499 | 25 | 1.57E-08 | 7.67E-07 | 4.00E-06 | 3.71E-11 | 6.95E-05 | SMARCA5, SMARCA5-AS1 |
| chr21 | 43361304 | 43362617 | 1314 | 10 | 3.37E-19 | 6.81E-07 | 3.57E-07 | 3.75E-11 | 0.007682857 | LINC01679 |
| chr9 | 136820871 | 136822147 | 1277 | 9 | 4.41E-12 | 1.08E-09 | 1.28E-05 | 3.76E-11 | -0.01085169 | AL355987.2, RABL6 |
| chr16 | 75266092 | 75268483 | 2392 | 21 | 7.46E-09 | 8.07E-08 | 3.24E-05 | 3.78E-11 | -0.00092568 | BCAR1 |
| chr19 | 42254728 | 42256590 | 1863 | 11 | 1.28E-09 | 8.48E-08 | 4.94E-05 | 3.79E-11 | -0.00292649 | AC006486.1, ERF |
| chr11 | 85854432 | 85855698 | 1267 | 14 | 6.29E-14 | 1.14E-07 | 3.46E-05 | 3.80E-11 | -0.00280855 | CCDC83, AP000974.1 |
| chr20 | 33282363 | 33283546 | 1184 | 11 | 3.85E-13 | 1.83E-07 | 3.45E-05 | 3.82E-11 | -0.00348529 | BPIFB1 |
| chr11 | 2270770 | 2272435 | 1666 | 41 | 2.16E-12 | 1.53E-09 | 3.19E-05 | 3.83E-11 | -0.00574588 | ASCL2 |
| chr12 | 119365110 | 119365812 | 703 | 6 | 4.03E-16 | 3.88E-06 | 4.59E-07 | 3.84E-11 | 0.004255589 | CCDC60, AC002070.1 |
| chr4 | 70627534 | 70628615 | 1082 | 4 | 5.64E-09 | 2.87E-10 | 2.12E-06 | 3.87E-11 | -0.01174511 | NA |
| chr4 | 74932320 | 74935136 | 2817 | 16 | 2.23E-07 | 1.30E-10 | 0.001199 | 3.88E-11 | -0.00248181 | PARM1 |
| chr2 | 121902463 | 121902545 | 83 | 3 | 2.15E-11 | 1.10E-11 | 2.30E-05 | 3.88E-11 | -0.03473411 | AC011246.1 |
| chr17 | 48597971 | 48599611 | 1641 | 6 | 1.45E-12 | 1.21E-10 | 0.000142 | 3.99E-11 | -0.01733396 | HOXB-AS3, HOXB3, HOXB6 |
| chr21 | 25734129 | 25736220 | 2092 | 28 | 1.15E-10 | 4.13E-09 | 0.000153 | 4.03E-11 | 0.000356736 | GABPA, ATP5PF |
| chr22 | 35312654 | 35313640 | 987 | 3 | 2.57E-10 | 1.36E-11 | 1.32E-05 | 4.10E-11 | -0.03903128 | TOM1 |
| chr12 | 132153189 | 132155630 | 2442 | 12 | 2.45E-08 | 1.34E-08 | 6.66E-05 | 4.10E-11 | 0.002592822 | NA |
| chr14 | 30558433 | 30559857 | 1425 | 14 | 1.72E-11 | 1.53E-09 | 0.000152 | 4.10E-11 | -0.00218658 | G2E3, G2E3-AS1 |
| chr13 | 49495353 | 49496630 | 1278 | 14 | 1.09E-12 | 7.46E-08 | 1.41E-05 | 4.11E-11 | 0.001813066 | PHF11 |
| chr17 | 1977322 | 1978039 | 718 | 8 | 2.41E-11 | 1.46E-11 | 0.000213 | 4.11E-11 | -0.00690973 | RTN4RL1 |
| chr3 | 50154511 | 50157185 | 2675 | 17 | 5.38E-08 | 3.62E-10 | 0.002561 | 4.16E-11 | -0.00200778 | SEMA3F, SEMA3F-AS1 |
| chr5 | 122077007 | 122078939 | 1933 | 24 | 1.86E-10 | 6.05E-08 | 0.000342 | 4.18E-11 | 0.001507089 | LOX |
| chr1 | 161158927 | 161161205 | 2279 | 20 | 1.23E-09 | 1.37E-06 | 1.63E-05 | 4.18E-11 | -0.00103013 | USP21, AL590714.1 |
| chr4 | 40437805 | 40438887 | 1083 | 4 | 6.91E-14 | 1.23E-08 | 1.96E-07 | 4.23E-11 | -0.00235459 | RBM47 |
| chr4 | 76434322 | 76435354 | 1033 | 9 | 5.13E-13 | 1.68E-09 | 7.84E-07 | 4.25E-11 | -0.01488731 | SHROOM3 |
| chr12 | 6450996 | 6452381 | 1386 | 13 | 1.21E-10 | 7.88E-11 | 0.000204 | 4.25E-11 | 0.001979111 | CD27, TAPBPL, CD27-AS1 |
| chr3 | 5024779 | 5027046 | 2268 | 6 | 6.29E-14 | 6.66E-06 | 2.80E-08 | 4.26E-11 | -0.0051881 | NA |
| chr17 | 81700515 | 81702811 | 2297 | 10 | 3.30E-18 | 1.38E-06 | 2.14E-07 | 4.26E-11 | -0.00391051 | HGS, AC139530.1 |
| chr2 | 219297960 | 219299668 | 1709 | 7 | 1.15E-11 | 2.74E-09 | 4.56E-05 | 4.26E-11 | -0.00108128 | AC114803.1, PTPRN |
| chr19 | 45250122 | 45251129 | 1008 | 6 | 1.33E-12 | 3.76E-09 | 1.01E-06 | 4.28E-11 | -0.00644934 | MARK4 |
| chr16 | 3304260 | 3306149 | 1890 | 20 | 1.57E-15 | 7.15E-09 | 7.08E-06 | 4.29E-11 | -0.00785922 | ZNF75A, TIGD7 |
| chr4 | 173500226 | 173501757 | 1532 | 9 | 5.57E-10 | 4.70E-11 | 0.000911 | 4.29E-11 | -0.01534101 | NA |
| chr20 | 3732148 | 3734663 | 2516 | 11 | 1.63E-08 | 4.58E-07 | 3.46E-05 | 4.33E-11 | -0.00295198 | HSPA12B |
| chr11 | 38646688 | 38648656 | 1969 | 7 | 4.07E-09 | 3.42E-07 | 1.48E-06 | 4.33E-11 | -0.00300461 | LINC01493 |
| chr7 | 154748615 | 154748870 | 256 | 2 | 4.76E-13 | 1.31E-10 | 4.72E-08 | 4.34E-11 | -0.02673053 | DPP6 |
| chr19 | 7848983 | 7850631 | 1649 | 7 | 2.26E-09 | 2.41E-07 | 3.56E-05 | 4.35E-11 | -0.00555593 | EVI5L |
| chr2 | 218781088 | 218782637 | 1550 | 11 | 1.24E-11 | 1.08E-10 | 0.000444 | 4.36E-11 | -0.00071157 | CYP27A1 |
| chr16 | 15434083 | 15436016 | 1934 | 15 | 1.33E-08 | 3.78E-10 | 0.000569 | 4.37E-11 | 0.001592605 | AC140504.1, BMERB1 |
| chr17 | 81869293 | 81872258 | 2966 | 16 | 1.06E-08 | 1.10E-07 | 8.22E-05 | 4.42E-11 | -6.07E-05 | ARHGDIA |
| chr11 | 35942810 | 35944772 | 1963 | 6 | 9.89E-12 | 4.28E-10 | 6.71E-07 | 4.45E-11 | 0.003557336 | LDLRAD3 |
| chr17 | 41526972 | 41529660 | 2689 | 15 | 7.45E-07 | 1.22E-10 | 0.002327 | 4.45E-11 | -0.0028596 | KRT19 |
| chr10 | 30743177 | 30744316 | 1140 | 4 | 7.24E-11 | 8.22E-11 | 2.69E-06 | 4.48E-11 | 0.001592389 | NA |
| chr4 | 112408829 | 112409592 | 764 | 3 | 1.38E-10 | 1.47E-11 | 1.62E-05 | 4.48E-11 | -6.38E-05 | ALPK1 |
| chr11 | 2443592 | 2444807 | 1216 | 17 | 1.51E-11 | 2.19E-11 | 0.00267 | 4.50E-11 | -0.00779678 | KCNQ1 |
| chr1 | 145885633 | 145887548 | 1916 | 13 | 1.44E-10 | 9.17E-07 | 1.87E-06 | 4.51E-11 | -0.00402047 | ANKRD35 |
| chr19 | 48574726 | 48575772 | 1047 | 4 | 1.84E-09 | 1.52E-11 | 4.94E-05 | 4.52E-11 | -0.02120813 | SULT2B1, AC008403.3 |
| chr3 | 52678591 | 52680479 | 1889 | 5 | 2.32E-11 | 5.80E-10 | 4.94E-06 | 4.52E-11 | 0.006669897 | PBRM1 |
| chr2 | 27889906 | 27891482 | 1577 | 22 | 2.63E-13 | 9.17E-08 | 1.79E-05 | 4.53E-11 | 0.000141111 | MRPL33, BABAM2, RBKS |
| chr11 | 22829320 | 22830359 | 1040 | 15 | 4.44E-12 | 1.45E-09 | 0.000217 | 4.53E-11 | -0.00328154 | AC006299.1, SVIP |
| chr12 | 121085974 | 121086305 | 332 | 2 | 2.33E-12 | 6.70E-11 | 1.32E-07 | 4.53E-11 | -0.01116439 | NA |
| chr5 | 176541647 | 176543362 | 1716 | 12 | 1.21E-15 | 1.06E-07 | 5.01E-06 | 4.56E-11 | 0.001735286 | CDHR2 |
| chr6 | 166097394 | 166098426 | 1033 | 6 | 1.33E-12 | 3.71E-08 | 3.44E-08 | 4.56E-11 | -0.00226143 | AL627443.3 |
| chr8 | 22578215 | 22580113 | 1899 | 15 | 5.29E-11 | 3.24E-07 | 4.43E-05 | 4.56E-11 | -0.00122233 | PDLIM2 |
| chr19 | 42368857 | 42370736 | 1880 | 9 | 1.19E-08 | 1.74E-10 | 2.05E-05 | 4.56E-11 | -0.00539617 | MEGF8 |
| chr2 | 26345367 | 26347006 | 1640 | 18 | 6.68E-13 | 5.52E-06 | 2.91E-06 | 4.58E-11 | 0.001100444 | SELENOI, ADGRF3 |
| chr7 | 135169454 | 135171022 | 1569 | 21 | 2.14E-10 | 4.84E-08 | 0.000284 | 4.58E-11 | -0.00243021 | AC083862.1, AC083862.2, CYREN |
| chr1 | 93447245 | 93449308 | 2064 | 14 | 4.12E-12 | 5.62E-08 | 8.74E-06 | 4.59E-11 | -1.69E-06 | FNBP1L |
| chr8 | 118073892 | 118074523 | 632 | 4 | 2.29E-13 | 2.67E-07 | 1.49E-06 | 4.59E-11 | -0.02204534 | EXT1 |
| chr13 | 99199406 | 99201668 | 2263 | 22 | 2.93E-10 | 2.36E-08 | 1.23E-06 | 4.59E-11 | 0.00107507 | UBAC2, UBAC2-AS1 |
| chr10 | 79314020 | 79315735 | 1716 | 9 | 2.26E-13 | 6.62E-09 | 7.56E-06 | 4.60E-11 | -0.00170187 | ZMIZ1 |
| chr8 | 37694158 | 37696553 | 2396 | 16 | 7.71E-10 | 1.97E-07 | 8.04E-05 | 4.60E-11 | -0.00075577 | ZNF703 |
| chr9 | 100427505 | 100429476 | 1972 | 10 | 2.74E-11 | 1.68E-07 | 3.27E-05 | 4.62E-11 | 0.000323754 | MSANTD3 |
| chr6 | 56806569 | 56808050 | 1482 | 3 | 4.70E-10 | 2.25E-10 | 5.93E-07 | 4.62E-11 | 0.002667193 | DST |
| chr11 | 440206 | 442753 | 2548 | 14 | 8.74E-09 | 2.39E-08 | 2.43E-05 | 4.66E-11 | -0.00374158 | ANO9, RN7SL838P |
| chr1 | 7953914 | 7954590 | 677 | 7 | 3.24E-14 | 5.59E-08 | 5.53E-08 | 4.66E-11 | -0.00432207 | PARK7 |
| chr12 | 132985777 | 132986866 | 1090 | 16 | 5.27E-13 | 3.86E-08 | 1.66E-05 | 4.68E-11 | -0.0005629 | ZNF26 |
| chr6 | 28907577 | 28908995 | 1419 | 9 | 3.26E-11 | 8.67E-09 | 1.29E-05 | 4.70E-11 | 0.000721096 | TRIM27 |
| chr19 | 35665162 | 35666499 | 1338 | 6 | 8.34E-16 | 1.72E-07 | 1.28E-07 | 4.71E-11 | -0.00407907 | NA |
| chr3 | 196941627 | 196943732 | 2106 | 22 | 1.94E-09 | 5.66E-10 | 0.002229 | 4.72E-11 | -0.00113008 | NCBP2-AS1, NCBP2AS2, NCBP2 |
| chr5 | 1106983 | 1108916 | 1934 | 14 | 5.79E-08 | 1.64E-10 | 0.000785 | 4.72E-11 | -0.01237331 | SLC12A7 |
| chr15 | 60643931 | 60644877 | 947 | 2 | 2.04E-08 | 2.67E-11 | 2.93E-06 | 4.75E-11 | -0.01344051 | AC009560.4, RORA |
| chr3 | 49198393 | 49200067 | 1675 | 11 | 4.27E-11 | 1.88E-10 | 0.000679 | 4.75E-11 | -0.01259821 | IHO1 |
| chr14 | 104801249 | 104803465 | 2217 | 9 | 8.61E-10 | 1.47E-08 | 1.54E-05 | 4.81E-11 | -0.00178017 | ZBTB42 |
| chr12 | 114696396 | 114698503 | 2108 | 45 | 1.62E-08 | 4.83E-09 | 0.004177 | 4.82E-11 | -0.0002334 | AC026765.3 |
| chr12 | 132618038 | 132619861 | 1824 | 13 | 6.29E-13 | 7.17E-08 | 7.65E-05 | 4.85E-11 | -0.0023518 | P2RX2 |
| chr16 | 909749 | 911353 | 1605 | 10 | 1.27E-09 | 4.80E-08 | 0.000176 | 4.85E-11 | -0.00154176 | LMF1 |
| chr12 | 57487267 | 57488743 | 1477 | 10 | 2.15E-11 | 6.35E-10 | 7.32E-06 | 4.86E-11 | -0.00058676 | MARS1, ARHGAP9 |
| chr18 | 657219 | 658937 | 1719 | 18 | 4.17E-10 | 6.84E-09 | 0.000373 | 4.90E-11 | -0.00087574 | TYMS, TYMSOS, AP001178.1 |
| chr17 | 44004226 | 44005556 | 1331 | 13 | 8.18E-11 | 3.93E-10 | 0.000157 | 4.92E-11 | -0.00529964 | NAGS, PYY |
| chr17 | 42658541 | 42659562 | 1022 | 8 | 5.82E-16 | 7.75E-07 | 4.46E-06 | 5.01E-11 | 0.001579129 | TUBG2 |
| chr9 | 70043058 | 70045166 | 2109 | 10 | 2.98E-07 | 1.25E-11 | 7.52E-05 | 5.01E-11 | -0.00263017 | MAMDC2, MAMDC2-AS1 |
| chr3 | 43105903 | 43106510 | 608 | 13 | 3.62E-14 | 7.39E-08 | 3.38E-07 | 5.03E-11 | -0.00629755 | POMGNT2 |
| chr2 | 178479998 | 178481020 | 1023 | 11 | 1.31E-11 | 2.70E-09 | 8.31E-05 | 5.03E-11 | 8.70E-05 | PLEKHA3 |
| chr9 | 35649909 | 35651291 | 1383 | 7 | 1.40E-11 | 3.77E-09 | 1.17E-05 | 5.04E-11 | -0.00472906 | SIT1 |
| chr18 | 46518863 | 46520904 | 2042 | 11 | 3.41E-11 | 1.15E-07 | 8.22E-05 | 5.10E-11 | -0.00344275 | LOXHD1 |
| chr12 | 109727219 | 109727647 | 429 | 3 | 1.61E-11 | 3.74E-11 | 1.90E-06 | 5.10E-11 | -0.01399853 | FAM222A |
| chr17 | 39709347 | 39711533 | 2187 | 12 | 1.83E-10 | 7.15E-10 | 2.21E-05 | 5.14E-11 | -0.0021802 | ERBB2 |
| chr17 | 61399429 | 61402200 | 2772 | 23 | 1.17E-07 | 1.08E-10 | 0.000438 | 5.15E-11 | -0.0005135 | TBX2, AC005746.2, TBX2-AS1 |
| chr12 | 54336272 | 54337302 | 1031 | 4 | 8.84E-14 | 5.40E-08 | 3.05E-06 | 5.18E-11 | -0.00752986 | AC078778.1, COPZ1, MIR148B |
| chr18 | 34222448 | 34224601 | 2154 | 19 | 5.27E-11 | 7.93E-11 | 0.000327 | 5.19E-11 | -0.00015839 | AC104985.1, NOL4 |
| chr15 | 70024683 | 70024785 | 103 | 3 | 1.09E-12 | 1.73E-09 | 3.31E-08 | 5.20E-11 | -0.0204533 | NA |
| chr11 | 119094939 | 119096828 | 1890 | 12 | 1.33E-10 | 2.87E-10 | 0.000393 | 5.25E-11 | -0.00137872 | H2AX, DPAGT1 |
| chr1 | 156858410 | 156859970 | 1561 | 11 | 2.14E-12 | 2.99E-09 | 0.000469 | 5.27E-11 | -0.00821075 | NTRK1, INSRR |
| chr13 | 49792054 | 49793446 | 1393 | 16 | 2.40E-12 | 3.97E-07 | 3.45E-05 | 5.35E-11 | 0.001577912 | AL136301.1, KPNA3 |
| chr6 | 31161552 | 31162739 | 1188 | 20 | 4.15E-12 | 2.71E-07 | 1.77E-05 | 5.35E-11 | -0.00395579 | TCF19 |
| chr2 | 25250660 | 25253251 | 2592 | 14 | 1.07E-07 | 1.86E-10 | 9.98E-05 | 5.37E-11 | 0.003004238 | DNMT3A |
| chr13 | 112060455 | 112063393 | 2939 | 12 | 1.24E-07 | 9.82E-09 | 0.000135 | 5.42E-11 | -0.00288424 | SOX1-OT |
| chr4 | 6889250 | 6889824 | 575 | 4 | 5.70E-11 | 7.79E-12 | 0.000162 | 5.45E-11 | 0.02724839 | NA |
| chr7 | 151340605 | 151342482 | 1878 | 11 | 1.08E-09 | 8.63E-09 | 0.00063 | 5.46E-11 | -0.00292311 | NUB1 |
| chr2 | 164840050 | 164842673 | 2624 | 21 | 1.60E-08 | 3.59E-08 | 0.000153 | 5.46E-11 | -0.00139121 | AC019197.1, COBLL1 |
| chr7 | 151172787 | 151174745 | 1959 | 9 | 1.18E-10 | 9.58E-09 | 0.000303 | 5.52E-11 | 0.000534871 | GBX1 |
| chr17 | 47649050 | 47650424 | 1375 | 15 | 6.26E-12 | 8.20E-08 | 3.43E-05 | 5.56E-11 | 0.001312243 | KPNB1, AC025682.1 |
| chr4 | 77156759 | 77158393 | 1635 | 15 | 1.34E-10 | 2.58E-07 | 0.000423 | 5.61E-11 | 0.000846027 | CCNG2 |
| chr11 | 12263356 | 12263919 | 564 | 4 | 2.69E-12 | 3.47E-08 | 4.17E-06 | 5.61E-11 | -0.00343284 | MICAL2 |
| chr20 | 63685496 | 63687164 | 1669 | 10 | 7.00E-12 | 1.70E-09 | 0.000503 | 5.62E-11 | -0.0010744 | RTEL1, RTEL1-TNFRSF6B |
| chr4 | 188681007 | 188681031 | 25 | 2 | 1.49E-11 | 3.19E-11 | 2.63E-06 | 5.65E-11 | -0.01094299 | LINC01060 |
| chr5 | 175867904 | 175868610 | 707 | 3 | 1.36E-12 | 1.20E-10 | 4.64E-06 | 5.67E-11 | -0.00295895 | CPLX2 |
| chr17 | 39699566 | 39701581 | 2016 | 15 | 5.04E-09 | 3.13E-09 | 3.13E-05 | 5.67E-11 | -0.00457053 | ERBB2 |
| chr17 | 73164278 | 73166254 | 1977 | 16 | 1.09E-08 | 7.89E-08 | 0.000122 | 5.69E-11 | 0.002111965 | AC097641.1, SSTR2 |
| chr10 | 100228266 | 100231030 | 2765 | 15 | 1.65E-09 | 2.73E-06 | 6.78E-06 | 5.71E-11 | -0.00462632 | AL138921.1, CHUK |
| chr3 | 157542187 | 157543484 | 1298 | 14 | 9.25E-15 | 6.83E-07 | 6.61E-05 | 5.74E-11 | -0.00790547 | SLC66A1L |
| chr1 | 65146349 | 65149598 | 3250 | 19 | 1.31E-06 | 4.04E-10 | 0.000453 | 5.76E-11 | -0.00158626 | AK4 |
| chr5 | 167599258 | 167600102 | 845 | 3 | 2.38E-13 | 8.82E-11 | 4.97E-07 | 5.78E-11 | 0.001388856 | TENM2 |
| chr10 | 133310565 | 133312355 | 1791 | 5 | 2.61E-10 | 7.06E-12 | 0.000173 | 5.79E-11 | -0.00576038 | ZNF511, ZNF511-PRAP1, TUBGCP2 |
| chr9 | 68809413 | 68810341 | 929 | 2 | 1.93E-10 | 6.18E-11 | 2.68E-07 | 5.79E-11 | -0.00512597 | PIP5K1B |
| chr1 | 177970090 | 177970760 | 671 | 9 | 2.22E-12 | 4.15E-10 | 2.49E-05 | 5.83E-11 | -0.007462 | SEC16B, CRYZL2P-SEC16B |
| chr16 | 22166250 | 22166942 | 693 | 4 | 9.11E-11 | 1.22E-10 | 3.24E-06 | 5.86E-11 | 0.003325335 | SDR42E2 |
| chr22 | 17562850 | 17564346 | 1497 | 9 | 2.74E-10 | 7.10E-10 | 1.37E-05 | 5.86E-11 | -0.0081704 | SLC25A18, AC007666.2 |
| chr17 | 79835200 | 79837113 | 1914 | 10 | 1.84E-08 | 6.40E-12 | 4.86E-05 | 5.89E-11 | -0.00186484 | CBX4 |
| chr6 | 41545115 | 41546694 | 1580 | 10 | 8.24E-12 | 1.24E-10 | 0.000291 | 5.90E-11 | -0.00406466 | FOXP4, FOXP4-AS1 |
| chr7 | 130440585 | 130441969 | 1385 | 18 | 3.76E-12 | 4.23E-07 | 8.32E-05 | 5.90E-11 | -0.00211232 | CEP41 |
| chr16 | 67278010 | 67279708 | 1699 | 14 | 8.11E-11 | 6.65E-09 | 0.0006 | 5.90E-11 | -0.00266942 | PLEKHG4 |
| chr2 | 9374610 | 9376094 | 1485 | 6 | 2.71E-08 | 1.30E-09 | 8.44E-06 | 5.93E-11 | -0.0047413 | ASAP2 |
| chr19 | 7141690 | 7143336 | 1647 | 9 | 1.21E-09 | 3.43E-07 | 4.98E-06 | 5.95E-11 | -0.00301566 | INSR |
| chr3 | 47409228 | 47409536 | 309 | 6 | 1.14E-11 | 1.03E-07 | 7.35E-06 | 6.02E-11 | -0.00327708 | PTPN23 |
| chr17 | 19716097 | 19717455 | 1359 | 7 | 4.12E-15 | 3.32E-06 | 4.82E-07 | 6.02E-11 | -0.0065185 | SLC47A2 |
| chr16 | 85181070 | 85182563 | 1494 | 6 | 1.25E-10 | 1.03E-10 | 4.99E-05 | 6.03E-11 | -0.01334042 | GSE1 |
| chr1 | 234585831 | 234585833 | 3 | 2 | 6.53E-12 | 4.08E-11 | 1.01E-06 | 6.05E-11 | -0.00554169 | NA |
| chr6 | 16352090 | 16353011 | 922 | 3 | 5.09E-10 | 7.28E-08 | 1.32E-06 | 6.06E-11 | -0.00489318 | ATXN1 |
| chr5 | 37370778 | 37371829 | 1052 | 17 | 1.10E-11 | 7.46E-09 | 6.84E-06 | 6.06E-11 | -0.00153854 | NUP155 |
| chr16 | 11349176 | 11350248 | 1073 | 6 | 1.18E-11 | 1.47E-09 | 5.82E-06 | 6.07E-11 | -0.00982147 | RMI2, AC009121.2 |
| chr3 | 64443931 | 64444991 | 1061 | 5 | 4.03E-13 | 1.31E-10 | 1.62E-05 | 6.10E-11 | 0.018098997 | PRICKLE2 |
| chr19 | 53723978 | 53724022 | 45 | 2 | 1.01E-12 | 6.90E-09 | 3.82E-09 | 6.16E-11 | -0.01389289 | NA |
| chr17 | 28613908 | 28615197 | 1290 | 10 | 1.60E-10 | 1.74E-10 | 0.000202 | 6.16E-11 | 0.005138377 | SPAG5-AS1, AC005726.1, RSKR, KIAA0100 |
| chr7 | 74042556 | 74042750 | 195 | 2 | 1.47E-12 | 4.19E-11 | 1.02E-06 | 6.19E-11 | 0.044324438 | ELN |
| chr8 | 144755148 | 144756674 | 1527 | 13 | 1.13E-11 | 1.43E-08 | 3.59E-05 | 6.25E-11 | 0.000269754 | ZNF251 |
| chr18 | 31102171 | 31103831 | 1661 | 9 | 1.26E-10 | 7.16E-09 | 3.17E-06 | 6.26E-11 | -0.00442821 | DSCAS, DSC2 |
| chr8 | 104466455 | 104467634 | 1180 | 12 | 1.79E-11 | 9.90E-10 | 3.38E-05 | 6.26E-11 | -0.00624539 | DPYS |
| chr19 | 37333908 | 37335106 | 1199 | 10 | 7.91E-10 | 9.65E-13 | 0.004534 | 6.28E-11 | -0.0284274 | ZNF875 |
| chr3 | 122947001 | 122948539 | 1539 | 3 | 6.11E-09 | 1.69E-11 | 3.69E-05 | 6.29E-11 | -0.00757384 | SEMA5B |
| chr17 | 39652086 | 39654667 | 2582 | 9 | 3.40E-07 | 1.37E-08 | 7.01E-05 | 6.31E-11 | -0.00539732 | STARD3 |
| chr21 | 33915076 | 33916695 | 1620 | 13 | 6.25E-11 | 1.45E-11 | 0.000476 | 6.32E-11 | -8.39E-05 | LINC00649, AP000311.1, ATP5PO |
| chr19 | 42410349 | 42411220 | 872 | 5 | 1.90E-13 | 7.05E-10 | 9.89E-07 | 6.34E-11 | -0.00446101 | LIPE-AS1, LIPE |
| chr21 | 36576411 | 36577745 | 1335 | 12 | 7.83E-11 | 3.59E-09 | 1.61E-05 | 6.38E-11 | -0.00586384 | CLDN14 |
| chr15 | 74410713 | 74412082 | 1370 | 8 | 2.97E-09 | 1.74E-09 | 0.000129 | 6.38E-11 | -0.00228802 | SEMA7A, MIR6881 |
| chr8 | 104665273 | 104665673 | 401 | 2 | 2.36E-12 | 1.08E-10 | 1.21E-07 | 6.43E-11 | 0.044036407 | ZFPM2 |
| chr4 | 183904291 | 183905933 | 1643 | 12 | 7.20E-12 | 6.35E-10 | 7.91E-06 | 6.51E-11 | -0.00060289 | STOX2 |
| chr22 | 44496443 | 44498489 | 2047 | 14 | 7.27E-11 | 1.06E-06 | 1.96E-05 | 6.52E-11 | -0.00157727 | RTL6 |
| chr15 | 92470912 | 92472419 | 1508 | 7 | 1.73E-08 | 2.14E-11 | 6.06E-05 | 6.59E-11 | -0.00378285 | C15orf32, AC090985.2 |

| | | | | | | | | | | |
|---|---|---|---|---|---|---|---|---|---|---|
| chr6 | 168625216 | 168626538 | 1323 | 10 | 2.11E-14 | 6.62E-09 | 1.51E-07 | 6.59E-11 | -0.00729908 | SMOC2 |
| chr19 | 40348113 | 40349595 | 1483 | 18 | 2.06E-11 | 4.89E-09 | 3.25E-05 | 6.61E-11 | -0.003617 | PLD3, C19orf47 |
| chr13 | 113044094 | 113045212 | 1119 | 15 | 2.74E-13 | 5.55E-07 | 2.39E-06 | 6.63E-11 | -0.0002561 | MCF2L |
| chr3 | 183098598 | 183099838 | 1241 | 13 | 5.00E-11 | 4.25E-10 | 0.000549 | 6.65E-11 | -0.00348496 | MCCC1 |
| chr6 | 138943963 | 138944438 | 476 | 3 | 3.17E-12 | 5.93E-11 | 1.89E-06 | 6.65E-11 | -0.00641934 | REPS1 |
| chr10 | 126215903 | 126216622 | 720 | 2 | 1.02E-10 | 4.74E-11 | 8.73E-07 | 6.66E-11 | -0.03109038 | ADAM12 |
| chr5 | 78985059 | 78986970 | 1912 | 12 | 1.24E-10 | 2.52E-09 | 1.17E-05 | 6.68E-11 | 0.000197229 | ARSB |
| chr19 | 15418152 | 15420059 | 1908 | 23 | 2.80E-11 | 1.17E-07 | 3.05E-05 | 6.69E-11 | -0.00022153 | AKAP8L, WIZ |
| chr4 | 56319901 | 56320570 | 670 | 3 | 7.97E-13 | 1.16E-09 | 3.96E-08 | 6.76E-11 | 0.003414971 | CRACD |
| chr1 | 174874616 | 174876024 | 1409 | 9 | 1.14E-13 | 8.69E-10 | 8.41E-05 | 6.77E-11 | 0.011085475 | RABGAP1L |
| chr12 | 1082783 | 1083407 | 625 | 4 | 1.14E-14 | 1.19E-07 | 1.52E-06 | 6.79E-11 | -0.01498797 | ERC1 |
| chr2 | 218857813 | 218859434 | 1622 | 7 | 6.51E-14 | 5.87E-09 | 6.08E-07 | 6.83E-11 | 0.003362934 | NA |
| chr11 | 799221 | 800865 | 1645 | 7 | 4.48E-11 | 2.26E-09 | 3.34E-06 | 6.83E-11 | -0.00167731 | PIDD1 |
| chr11 | 120299990 | 120300538 | 549 | 5 | 1.23E-10 | 2.57E-11 | 3.43E-05 | 6.89E-11 | -0.01368414 | POU2F3 |
| chr1 | 16155935 | 16157199 | 1265 | 13 | 9.63E-17 | 8.91E-07 | 5.45E-07 | 6.92E-11 | -0.00389232 | AL451042.2, EPHA2 |
| chr6 | 30170985 | 30172936 | 1952 | 22 | 6.67E-10 | 1.74E-07 | 0.000456 | 6.92E-11 | -0.00429484 | TRIM15 |
| chr2 | 70948409 | 70949406 | 998 | 7 | 5.17E-15 | 2.01E-08 | 8.43E-07 | 6.94E-11 | -0.00669385 | VAX2, ATP6V1B1, AC007040.2, ATP6V1B1-AS1 |
| chr11 | 22624809 | 22626104 | 1296 | 21 | 3.63E-11 | 3.55E-07 | 6.89E-05 | 6.96E-11 | 0.000474638 | GAS2, FANCF |
| chr19 | 39833547 | 39834765 | 1219 | 17 | 4.21E-12 | 2.12E-08 | 2.19E-06 | 7.00E-11 | 0.000694622 | DYRK1B, FBL |
| chr13 | 23722879 | 23724053 | 1175 | 3 | 1.58E-13 | 7.57E-09 | 4.18E-08 | 7.00E-11 | -0.01201779 | NA |
| chr17 | 59153840 | 59156267 | 2428 | 23 | 1.55E-07 | 4.83E-09 | 8.81E-05 | 7.03E-11 | 0.001745187 | PRR11, SKA2 |
| chr1 | 976221 | 976615 | 395 | 2 | 3.03E-12 | 7.42E-11 | 3.11E-07 | 7.09E-11 | 0.066080425 | PERM1 |
| chr14 | 57060826 | 57060908 | 83 | 2 | 1.03E-11 | 1.08E-10 | 1.47E-07 | 7.11E-11 | -0.01928178 | OTX2-AS1 |
| chr19 | 54430919 | 54432467 | 1549 | 4 | 2.04E-18 | 9.64E-08 | 1.48E-08 | 7.17E-11 | -0.01717711 | TTYH1, AC245884.1 |
| chr4 | 137532162 | 137533804 | 1643 | 12 | 1.13E-09 | 2.37E-09 | 0.00029 | 7.21E-11 | 0.002760956 | PCDH18 |
| chr2 | 98086645 | 98087472 | 828 | 13 | 2.73E-14 | 1.56E-09 | 4.14E-05 | 7.26E-11 | -0.00520292 | VWA3B |
| chr13 | 30931839 | 30934234 | 2396 | 16 | 4.89E-10 | 2.76E-09 | 1.81E-06 | 7.32E-11 | -0.00912428 | TEX26, TEX26-AS1 |
| chr20 | 59351156 | 59351306 | 151 | 2 | 8.22E-12 | 1.20E-10 | 1.34E-07 | 7.33E-11 | -0.01485016 | NA |
| chr16 | 1435969 | 1437179 | 1211 | 6 | 1.11E-13 | 1.47E-09 | 2.31E-07 | 7.34E-11 | 0.00407864 | AL032819.2, CCDC154 |
| chr12 | 114403866 | 114407418 | 3553 | 23 | 5.94E-07 | 9.13E-08 | 0.000459 | 7.39E-11 | -0.00023028 | TBX5 |
| chr17 | 28370031 | 28372532 | 2502 | 12 | 1.95E-10 | 1.09E-07 | 3.58E-06 | 7.45E-11 | -0.00268871 | AC002094.1, SARM1, VTN |
| chr5 | 96584046 | 96584415 | 370 | 4 | 3.69E-12 | 8.38E-10 | 9.09E-07 | 7.47E-11 | 0.002153358 | AC104123.1, CAST |
| chr16 | 72956368 | 72957445 | 1078 | 5 | 1.91E-11 | 1.73E-07 | 3.72E-06 | 7.58E-11 | -0.0043522 | ZFHX3 |
| chr1 | 183804741 | 183805629 | 889 | 9 | 1.55E-11 | 2.24E-07 | 1.46E-06 | 7.59E-11 | 0.000373753 | RGL1 |
| chr9 | 114155901 | 114156989 | 1089 | 5 | 8.34E-13 | 6.05E-08 | 7.53E-06 | 7.75E-11 | -0.01169508 | COL27A1 |
| chr14 | 95193949 | 95195728 | 1780 | 12 | 4.84E-11 | 2.55E-07 | 4.18E-06 | 7.75E-11 | -0.00117662 | CLMN |
| chr3 | 59424869 | 59426258 | 1390 | 3 | 1.11E-09 | 3.31E-09 | 1.83E-06 | 7.75E-11 | 0.005241564 | AC126121.3 |
| chr6 | 44660823 | 44663357 | 2535 | 7 | 1.66E-08 | 2.85E-09 | 5.97E-06 | 7.75E-11 | -0.00810149 | NA |
| chr1 | 15407276 | 15407789 | 514 | 3 | 6.31E-13 | 1.52E-09 | 6.42E-08 | 7.76E-11 | -0.01164073 | AL031283.1 |
| chr11 | 61361100 | 61362889 | 1790 | 21 | 4.29E-10 | 1.98E-11 | 0.000512 | 7.76E-11 | -0.00081517 | TMEM138, CYB561A3 |
| chr14 | 103522439 | 103523746 | 1308 | 13 | 8.55E-13 | 8.70E-08 | 0.000157 | 7.89E-11 | -0.00652402 | CKB |
| chr1 | 43284969 | 43286514 | 1546 | 12 | 6.33E-12 | 1.86E-08 | 0.000593 | 7.89E-11 | -0.007302 | C1orf210 |
| chr15 | 41772797 | 41775306 | 2510 | 18 | 7.20E-08 | 4.87E-10 | 0.000635 | 7.92E-11 | -0.00421531 | MGA, MAPKBP1 |
| chr15 | 82646864 | 82648775 | 1912 | 21 | 7.89E-11 | 1.59E-08 | 2.59E-05 | 8.00E-11 | -0.00310102 | CPEB1-AS1, CPEB1 |
| chr12 | 97275630 | 97276490 | 861 | 5 | 2.93E-10 | 3.80E-11 | 2.37E-05 | 8.02E-11 | -0.01273175 | LINC02409 |
| chr3 | 48187336 | 48189768 | 2433 | 15 | 2.86E-10 | 2.21E-07 | 7.01E-06 | 8.04E-11 | -0.00217301 | CDC25A |
| chr11 | 122653235 | 122654936 | 1702 | 4 | 2.06E-09 | 2.94E-10 | 1.71E-06 | 8.04E-11 | -0.02081588 | NA |
| chr5 | 112825864 | 112827386 | 1523 | 5 | 8.34E-09 | 4.59E-10 | 4.87E-06 | 8.09E-11 | -0.00568674 | APC, AC008575.1 |
| chr13 | 98752252 | 98753015 | 764 | 8 | 3.25E-14 | 1.15E-05 | 3.89E-08 | 8.16E-11 | 0.002898011 | SLC15A1 |
| chr4 | 55096744 | 55098787 | 2044 | 7 | 2.44E-08 | 1.50E-07 | 5.10E-06 | 8.22E-11 | -0.00394196 | KDR |
| chr1 | 51989476 | 51991575 | 2100 | 17 | 1.29E-08 | 4.64E-10 | 0.001246 | 8.22E-11 | -0.00382096 | RAB3B |
| chr5 | 141343511 | 141345153 | 1643 | 14 | 3.80E-12 | 8.77E-10 | 5.81E-05 | 8.31E-11 | -0.00871852 | PCDHGA1, PCDHGA2, PCDHGA3 |
| chr22 | 43409959 | 43411423 | 1465 | 8 | 9.88E-10 | 1.98E-09 | 0.000442 | 8.36E-11 | -0.00944292 | MPPED1 |
| chr11 | 66688068 | 66688991 | 924 | 3 | 5.28E-16 | 2.94E-07 | 3.31E-09 | 8.36E-11 | 0.00122076 | SPTBN2 |
| chr14 | 34714391 | 34716231 | 1841 | 17 | 2.42E-13 | 1.47E-09 | 0.000495 | 8.36E-11 | 0.006438215 | CFL2 |
| chr1 | 3244462 | 3246483 | 2022 | 10 | 6.02E-10 | 4.33E-09 | 3.49E-05 | 8.44E-11 | -0.00166836 | PRDM16 |
| chr18 | 11856392 | 11857785 | 1394 | 9 | 7.66E-10 | 1.48E-11 | 0.001179 | 8.46E-11 | -0.00763912 | GNAL, AP005137.2 |
| chr1 | 218344436 | 218346234 | 1799 | 18 | 1.28E-10 | 2.37E-07 | 5.07E-05 | 8.60E-11 | 0.001189376 | TGFB2, TGFB2-AS1 |
| chr7 | 158450677 | 158452126 | 1450 | 5 | 1.33E-09 | 5.60E-11 | 6.41E-05 | 8.73E-11 | 0.010455428 | PTPRN2 |
| chr17 | 21275164 | 21276465 | 1302 | 7 | 4.00E-12 | 1.08E-10 | 6.46E-06 | 8.74E-11 | 0.003106032 | NA |
| chr14 | 101406963 | 101409739 | 2777 | 10 | 6.02E-08 | 1.53E-09 | 0.00013 | 8.74E-11 | -0.0022845 | LINC00524 |
| chr8 | 134779489 | 134780231 | 743 | 3 | 2.46E-12 | 2.27E-10 | 1.23E-06 | 8.78E-11 | 0.025959183 | NA |
| chr19 | 43982977 | 43984731 | 1755 | 14 | 5.32E-12 | 9.89E-09 | 7.54E-06 | 8.78E-11 | 0.001381839 | ZNF155 |
| chr15 | 41230474 | 41232052 | 1579 | 21 | 2.75E-09 | 1.77E-09 | 0.005121 | 8.78E-11 | -0.00236829 | CHP1, EXD1 |
| chr7 | 73734841 | 73735638 | 798 | 5 | 7.96E-12 | 3.21E-11 | 4.22E-05 | 8.78E-11 | 0.020472351 | ABHD11-AS1 |
| chr20 | 31818249 | 31819585 | 1337 | 13 | 3.41E-11 | 5.98E-08 | 1.55E-05 | 8.94E-11 | -0.00417253 | MYLK2 |
| chr17 | 10197693 | 10200111 | 2419 | 8 | 4.36E-07 | 1.04E-09 | 4.15E-05 | 8.94E-11 | 0.000616934 | GAS7 |
| chr1 | 217136226 | 217138718 | 2493 | 19 | 9.35E-09 | 1.72E-06 | 2.09E-05 | 8.95E-11 | 0.000906239 | ESRRG |
| chr12 | 110612682 | 110614462 | 1781 | 14 | 8.19E-14 | 3.25E-08 | 5.34E-07 | 9.06E-11 | -0.00025562 | TCTN1 |
| chr14 | 21475689 | 21477539 | 1851 | 22 | 7.05E-11 | 2.11E-07 | 7.18E-06 | 9.06E-11 | -0.00090274 | TOX4, RAB2B |
| chr1 | 114625344 | 114625965 | 622 | 5 | 4.84E-11 | 7.20E-11 | 2.54E-05 | 9.12E-11 | -0.01507494 | DENND2C |
| chr11 | 12008917 | 12010602 | 1686 | 16 | 3.11E-14 | 8.67E-08 | 1.15E-05 | 9.22E-11 | -0.00601389 | DKK3 |
| chr1 | 2014569 | 2016027 | 1459 | 9 | 2.11E-09 | 3.32E-10 | 0.000505 | 9.27E-11 | -0.00588873 | AL391845.2 |
| chr9 | 133030026 | 133031133 | 1108 | 10 | 1.26E-11 | 3.92E-10 | 0.000375 | 9.30E-11 | 0.000975625 | GTF3C5 |
| chr11 | 132099835 | 132100678 | 844 | 3 | 2.78E-14 | 2.32E-06 | 8.31E-09 | 9.36E-11 | -0.02211301 | NTM |
| chr11 | 113272458 | 113274452 | 1995 | 11 | 3.63E-11 | 1.58E-06 | 6.03E-06 | 9.38E-11 | -0.00095221 | NCAM1, NCAM1-AS1 |
| chr2 | 20106778 | 20107972 | 1195 | 3 | 1.78E-08 | 2.87E-11 | 3.06E-05 | 9.40E-11 | -0.03110808 | AC098828.1 |
| chr7 | 100851371 | 100853225 | 1855 | 10 | 6.90E-10 | 2.37E-08 | 0.000158 | 9.40E-11 | -0.00499763 | SLC12A9, SLC12A9-AS1 |
| chr17 | 75405192 | 75406686 | 1495 | 16 | 1.04E-10 | 8.84E-08 | 0.000723 | 9.54E-11 | -0.00149058 | MIR3678, GRB2 |
| chr19 | 38849073 | 38850809 | 1737 | 9 | 8.22E-13 | 1.11E-06 | 2.30E-06 | 9.57E-11 | -0.00113946 | HNRNPL |
| chr2 | 120465958 | 120467103 | 1146 | 10 | 1.19E-10 | 8.73E-10 | 0.000637 | 9.57E-11 | -0.01583466 | LINC01101 |
| chr12 | 56128167 | 56129486 | 1320 | 11 | 6.67E-14 | 5.29E-09 | 7.22E-05 | 9.60E-11 | 0.0025879 | ESYT1, AC034102.6 |
| chr2 | 234496042 | 234498509 | 2468 | 16 | 5.79E-08 | 7.22E-09 | 0.000872 | 9.61E-11 | -0.00250366 | ARL4C |
| chr9 | 91873686 | 91874589 | 904 | 4 | 1.80E-09 | 2.79E-11 | 2.36E-05 | 9.64E-11 | 0.005262596 | ROR2 |
| chr14 | 101054032 | 101056094 | 2063 | 18 | 4.22E-10 | 1.14E-10 | 0.003104 | 9.66E-11 | -3.38E-05 | MIR382, MIR134, MIR668, MIR485 |
| chr6 | 33270991 | 33273249 | 2259 | 38 | 3.98E-09 | 5.08E-09 | 0.00854 | 9.68E-11 | 0.001158872 | RPS18, VPS52 |
| chr14 | 103684458 | 103686619 | 2162 | 11 | 1.20E-12 | 2.54E-07 | 8.99E-07 | 9.71E-11 | -0.00046961 | KLC1, AL139300.1 |
| chr5 | 159099100 | 159101032 | 1933 | 16 | 3.63E-08 | 4.88E-10 | 0.000145 | 9.71E-11 | 0.000975374 | LINC02202, EBF1 |
| chr8 | 143933942 | 143935522 | 1581 | 9 | 8.82E-11 | 1.93E-10 | 0.000178 | 9.73E-11 | 0.02187532 | PLEC |
| chr5 | 151721314 | 151721553 | 240 | 2 | 2.94E-11 | 5.59E-11 | 3.37E-06 | 9.80E-11 | -0.01863362 | CLMAT3 |
| chr5 | 141181450 | 141183825 | 2376 | 18 | 2.37E-08 | 2.90E-07 | 5.50E-05 | 9.82E-11 | -0.0131753 | AC244517.8, PCDHB16, AC244517.6, AC244517.11, AC244517.4 |
| chr13 | 78606886 | 78608487 | 1602 | 9 | 3.70E-12 | 8.59E-08 | 1.40E-06 | 9.84E-11 | -0.00619328 | OBI1-AS1 |
| chr1 | 216805084 | 216806610 | 1527 | 7 | 9.00E-09 | 2.99E-11 | 3.39E-05 | 9.84E-11 | -0.00474732 | ESRRG |
| chr11 | 133913400 | 133913704 | 305 | 2 | 3.73E-13 | 1.38E-10 | 1.92E-07 | 9.88E-11 | -0.01167385 | IGSF9B |
| chr17 | 29084579 | 29084911 | 333 | 4 | 2.86E-12 | 1.58E-10 | 1.34E-06 | 9.91E-11 | 0.004421762 | MYO18A |
| chr3 | 101646023 | 101646449 | 427 | 2 | 3.46E-14 | 2.39E-08 | 3.57E-09 | 9.97E-11 | 0.008858838 | NA |
| chr17 | 40054128 | 40055428 | 1301 | 15 | 1.07E-12 | 2.09E-07 | 1.91E-05 | 9.98E-11 | -0.00445503 | MED24 |
| chr2 | 2330994 | 2333471 | 2478 | 12 | 1.39E-07 | 1.54E-07 | 1.06E-05 | 9.99E-11 | -0.00990913 | MYT1L |
| chr8 | 31032193 | 31034257 | 2065 | 34 | 2.88E-08 | 2.27E-08 | 9.93E-05 | 1.00E-10 | -0.00025851 | WRN, PURG |
| chr1 | 212484890 | 212486686 | 1797 | 7 | 1.02E-10 | 8.36E-09 | 1.93E-05 | 1.00E-10 | -0.00498004 | NA |
| chr15 | 55196215 | 55198284 | 2070 | 16 | 1.00E-13 | 6.34E-06 | 1.24E-07 | 1.01E-10 | 0.000484294 | RSL24D1 |
| chr1 | 162366192 | 162367585 | 1394 | 10 | 3.58E-14 | 4.06E-09 | 3.98E-05 | 1.02E-10 | -0.00786756 | NOS1AP, AL512785.2 |
| chr9 | 106620669 | 106622502 | 1834 | 19 | 2.15E-11 | 4.60E-08 | 0.00021 | 1.02E-10 | -0.0006296 | LINC01505 |
| chr1 | 220093518 | 220094753 | 1236 | 7 | 3.68E-11 | 1.55E-08 | 4.05E-06 | 1.02E-10 | 0.003959641 | IARS2 |
| chr7 | 50590684 | 50593155 | 2472 | 10 | 3.88E-08 | 1.94E-08 | 2.86E-05 | 1.03E-10 | 0.002333724 | GRB10 |
| chr5 | 71586328 | 71587620 | 1293 | 12 | 1.17E-11 | 4.91E-09 | 3.60E-05 | 1.03E-10 | -0.00018965 | MCCC2 |
| chr5 | 161846833 | 161848995 | 2163 | 26 | 2.55E-09 | 2.59E-07 | 0.000739 | 1.03E-10 | 0.001159877 | GABRA1 |
| chr17 | 1186829 | 1188121 | 1293 | 14 | 5.33E-09 | 1.20E-10 | 0.000707 | 1.03E-10 | -0.0017591 | ABR |
| chr5 | 178940114 | 178941619 | 1506 | 13 | 6.37E-11 | 2.06E-10 | 5.15E-06 | 1.04E-10 | -0.00308079 | ZNF454 |
| chr19 | 16325860 | 16326551 | 692 | 3 | 8.65E-12 | 1.04E-09 | 4.28E-07 | 1.04E-10 | -0.01539232 | KLF2 |
| chr8 | 19755667 | 19756896 | 1230 | 4 | 3.34E-11 | 1.42E-09 | 1.21E-05 | 1.04E-10 | -0.03675352 | CSGALNACT1 |
| chr7 | 44621945 | 44624455 | 2511 | 9 | 1.64E-07 | 4.74E-11 | 7.51E-05 | 1.04E-10 | -0.00323536 | OGDH |
| chr4 | 3363172 | 3363715 | 544 | 5 | 3.92E-12 | 1.97E-08 | 6.46E-06 | 1.05E-10 | 0.011031978 | RGS12 |
| chr11 | 10303708 | 10306323 | 2616 | 26 | 3.13E-08 | 1.60E-09 | 0.008896 | 1.05E-10 | 0.000827828 | ADM |
| chr11 | 18011433 | 18013803 | 2371 | 16 | 9.10E-09 | 7.84E-11 | 0.000608 | 1.05E-10 | -0.00121887 | SERGEF, AC055860.1 |
| chr3 | 50276262 | 50278953 | 2692 | 12 | 5.18E-10 | 5.89E-09 | 5.04E-06 | 1.05E-10 | -0.00474233 | SEMA3B |
| chr7 | 99503058 | 99506198 | 3141 | 22 | 2.11E-09 | 3.94E-07 | 1.00E-05 | 1.06E-10 | -0.00037238 | ZNF789, ZKSCAN5, ZNF394 |
| chr11 | 61302591 | 61303426 | 836 | 6 | 3.24E-11 | 3.15E-11 | 0.000533 | 1.07E-10 | 0.00250548 | DDB1 |
| chr8 | 143867311 | 143869641 | 2331 | 10 | 2.63E-07 | 6.56E-09 | 0.000127 | 1.08E-10 | -0.00295357 | EPPK1 |
| chr16 | 260538 | 261150 | 613 | 4 | 2.18E-12 | 2.43E-09 | 4.07E-06 | 1.08E-10 | -0.00023414 | FAM234A |
| chr1 | 163470018 | 163470030 | 13 | 2 | 2.79E-11 | 8.26E-11 | 8.05E-07 | 1.08E-10 | -0.02858001 | NA |
| chr12 | 213084 | 215581 | 2498 | 22 | 2.61E-09 | 1.95E-09 | 3.45E-05 | 1.08E-10 | -0.00450212 | SLC6A12 |
| chr17 | 7888312 | 7890658 | 2347 | 10 | 1.06E-09 | 1.92E-06 | 1.92E-05 | 1.08E-10 | -0.00336431 | CHD3 |
| chr8 | 104329110 | 104330905 | 1796 | 9 | 5.96E-08 | 9.98E-11 | 0.000172 | 1.09E-10 | -0.04859811 | DPYS |
| chr6 | 31821384 | 31822917 | 1534 | 10 | 1.01E-08 | 8.65E-11 | 0.000475 | 1.10E-10 | -0.00087588 | NA |
| chr17 | 83073373 | 83075508 | 2136 | 7 | 9.10E-12 | 1.32E-06 | 1.64E-05 | 1.10E-10 | 0.007631502 | NA |
| chr9 | 96501839 | 96501845 | 7 | 2 | 2.47E-12 | 6.48E-10 | 3.56E-08 | 1.10E-10 | 0.004906829 | CDC14B |
| chr10 | 31320418 | 31321825 | 1408 | 14 | 6.17E-13 | 2.02E-08 | 2.87E-05 | 1.10E-10 | 0.001406356 | ZEB1, ZEB1-AS1 |
| chr8 | 143668520 | 143669268 | 749 | 2 | 4.95E-11 | 1.15E-10 | 3.56E-07 | 1.11E-10 | -0.01582063 | NA |
| chr12 | 131808784 | 131809157 | 374 | 5 | 2.13E-10 | 1.26E-11 | 0.000382 | 1.11E-10 | -0.0365979 | NA |
| chr10 | 35174312 | 35175701 | 1390 | 8 | 2.32E-10 | 3.52E-08 | 9.00E-07 | 1.11E-10 | -0.0026645 | CREM |
| chr3 | 52826179 | 52827558 | 1380 | 3 | 7.00E-11 | 8.78E-08 | 2.50E-07 | 1.11E-10 | -0.01135754 | ITIH4, AC006254.1 |
| chr4 | 70914630 | 70915070 | 441 | 2 | 5.50E-14 | 1.37E-08 | 4.86E-09 | 1.11E-10 | 0.00270859 | MOB1B |
| chr4 | 15655251 | 15656684 | 1434 | 7 | 1.08E-13 | 9.66E-08 | 2.92E-06 | 1.11E-10 | -0.01544912 | FBXL5 |
| chr12 | 52368101 | 52369914 | 1814 | 5 | 1.94E-08 | 2.61E-09 | 1.51E-05 | 1.12E-10 | -0.00947376 | NA |
| chr10 | 133158230 | 133160101 | 1872 | 8 | 4.78E-10 | 5.46E-08 | 1.54E-05 | 1.12E-10 | -0.00801826 | NA |
| chr3 | 133673950 | 133674812 | 863 | 6 | 4.77E-13 | 2.89E-08 | 1.76E-06 | 1.12E-10 | -0.01771629 | INHCAP |
| chr3 | 13479185 | 13481240 | 2056 | 34 | 2.46E-08 | 3.81E-08 | 0.00032 | 1.13E-10 | -5.66E-05 | HDAC11, HDAC11-AS1 |
| chr22 | 20964475 | 20966061 | 1587 | 12 | 3.69E-12 | 6.32E-10 | 0.000268 | 1.15E-10 | 0.000537372 | LINC01637, AIFM3 |
| chr5 | 133611810 | 133613451 | 1642 | 10 | 2.46E-11 | 5.96E-08 | 2.59E-06 | 1.16E-10 | -9.83E-05 | FSTL4 |
| chr7 | 151439962 | 151441209 | 1248 | 11 | 1.16E-13 | 2.49E-07 | 9.54E-06 | 1.16E-10 | -0.00908824 | CRYGN |
| chr1 | 167517363 | 167519922 | 2560 | 10 | 1.19E-07 | 1.01E-07 | 7.78E-05 | 1.16E-10 | -0.00056971 | AKR1D1P1, CD247 |
| chr5 | 149405845 | 149406955 | 1111 | 7 | 3.26E-11 | 2.80E-07 | 2.90E-06 | 1.16E-10 | -0.00414628 | CARMN |
| chr6 | 33171752 | 33173375 | 1624 | 12 | 1.65E-10 | 5.93E-09 | 0.000145 | 1.17E-10 | -0.00613957 | COL11A2 |
| chr19 | 48745866 | 48747431 | 1566 | 9 | 2.84E-14 | 1.41E-07 | 4.76E-06 | 1.17E-10 | -0.01047835 | IZUMO1 |
| chr16 | 30956172 | 30957814 | 1643 | 15 | 1.85E-10 | 7.56E-09 | 7.59E-06 | 1.17E-10 | 0.00110195 | AC135048.1, ORAI3, SETD1A, AC135048.3 |
| chr14 | 74955252 | 74956699 | 1448 | 14 | 2.45E-11 | 6.54E-10 | 0.000626 | 1.18E-10 | -0.00423225 | PGF |
| chr3 | 109100465 | 109101952 | 1488 | 6 | 5.71E-09 | 5.56E-11 | 0.000131 | 1.19E-10 | -0.00830788 | MORC1-AS1, MORC1 |
| chr17 | 40417524 | 40418910 | 1387 | 18 | 1.17E-10 | 2.86E-08 | 0.001069 | 1.20E-10 | 0.000438167 | TOP2A |
| chr1 | 935411 | 937357 | 1947 | 11 | 2.64E-08 | 2.98E-10 | 2.01E-05 | 1.20E-10 | -0.02123137 | SAMD11 |
| chr6 | 43628015 | 43630357 | 2343 | 23 | 2.71E-09 | 1.41E-08 | 0.000301 | 1.22E-10 | -0.00208273 | MAD2L1BP, GTPBP2 |
| chr8 | 142699979 | 142701555 | 1577 | 11 | 1.83E-13 | 3.41E-09 | 0.00017 | 1.22E-10 | -0.00780291 | LY6K |
| chr4 | 39526649 | 39528604 | 1956 | 23 | 4.84E-09 | 1.34E-07 | 0.000435 | 1.22E-10 | -0.00078524 | UGDH-AS1, UGDH |
| chr1 | 23368008 | 23370251 | 2244 | 25 | 7.29E-10 | 6.68E-07 | 7.92E-05 | 1.22E-10 | 0.001085027 | ZNF436-AS1, ZNF436 |
| chr1 | 21279558 | 21280274 | 717 | 7 | 1.45E-11 | 1.06E-09 | 8.27E-05 | 1.23E-10 | -0.00144992 | ECE1 |
| chr3 | 42159774 | 42160406 | 633 | 7 | 2.17E-11 | 4.55E-10 | 0.000158 | 1.23E-10 | -0.01889575 | TRAK1 |
| chr13 | 61415279 | 61417531 | 2253 | 17 | 1.99E-07 | 8.82E-10 | 4.18E-05 | 1.23E-10 | -0.00254193 | PCDH20, AL592490.1 |
| chr17 | 39979559 | 39981236 | 1678 | 18 | 4.95E-12 | 1.44E-05 | 1.57E-07 | 1.23E-10 | 0.000866874 | PSMD3 |

| | | | | | | | | | | |
|---|---|---|---|---|---|---|---|---|---|---|
| chr2 | 10914868 | 10916299 | 1432 | 4 | 9.61E-10 | 3.30E-10 | 1.36E-05 | 1.24E-10 | -0.00273307 | NA |
| chr11 | 118887226 | 118888692 | 1467 | 8 | 3.96E-10 | 8.36E-11 | 0.000502 | 1.24E-10 | -0.00668093 | CXCR5, AP004609.1 |
| chr12 | 127133779 | 127134163 | 385 | 3 | 7.34E-11 | 8.03E-11 | 8.84E-06 | 1.25E-10 | -0.02968777 | AC079949.3 |
| chr10 | 17129696 | 17131063 | 1368 | 6 | 1.46E-11 | 1.26E-06 | 9.01E-06 | 1.25E-10 | -0.00248019 | CUBN |
| chr11 | 9573657 | 9576030 | 2374 | 16 | 5.93E-09 | 1.38E-08 | 9.54E-06 | 1.25E-10 | -0.00209246 | WEE1 |
| chr12 | 117140788 | 117141696 | 909 | 4 | 3.53E-12 | 2.87E-10 | 1.25E-05 | 1.25E-10 | -0.01252341 | TESC-AS1 |
| chr7 | 97022603 | 97025476 | 2874 | 27 | 1.82E-10 | 1.95E-08 | 0.001046 | 1.25E-10 | -0.00097016 | DLX5 |
| chr9 | 136806569 | 136807842 | 1274 | 9 | 2.42E-11 | 1.10E-09 | 6.97E-05 | 1.25E-10 | -0.00459474 | CCDC183, AL355987.4, AL355987.2, CCDC183-AS1 |
| chr16 | 71670 | 74113 | 2444 | 11 | 5.20E-09 | 1.04E-09 | 0.00053 | 1.26E-10 | -0.00343588 | RHBDF1 |
| chr17 | 3595871 | 3598490 | 2620 | 12 | 8.21E-06 | 1.82E-10 | 0.000615 | 1.26E-10 | -0.00209063 | TRPV1, AC027796.3 |
| chr14 | 61994665 | 61996213 | 1549 | 8 | 1.43E-11 | 7.44E-08 | 6.84E-06 | 1.27E-10 | -0.00350385 | SYT16 |
| chr1 | 110111892 | 110113886 | 1995 | 11 | 1.01E-09 | 1.83E-06 | 2.70E-05 | 1.28E-10 | -0.00221988 | UBL4B |
| chr13 | 74133008 | 74135242 | 2235 | 15 | 7.09E-09 | 9.93E-09 | 0.000628 | 1.28E-10 | -6.18E-05 | NA |
| chr1 | 44774728 | 44776684 | 1957 | 19 | 5.30E-11 | 3.70E-08 | 4.56E-05 | 1.28E-10 | -0.00142396 | RPS8, SNORD55, SNORD46, AL592166.1 |
| chr1 | 205121713 | 205122792 | 1080 | 13 | 1.63E-10 | 3.30E-11 | 0.001415 | 1.29E-10 | 0.000108364 | RBBP5 |
| chr5 | 150053973 | 150055598 | 1626 | 4 | 8.69E-14 | 2.22E-08 | 8.86E-07 | 1.30E-10 | -0.00263542 | CSF1R |
| chr1 | 93878579 | 93880740 | 2162 | 25 | 7.54E-10 | 4.51E-07 | 0.001301 | 1.30E-10 | -0.0020161 | DNTTIP2 |
| chr10 | 17643266 | 17644701 | 1436 | 13 | 7.59E-10 | 6.34E-10 | 0.000335 | 1.30E-10 | 0.002412123 | STAM, STAM-AS1 |
| chr2 | 241270091 | 241273007 | 2917 | 9 | 9.19E-08 | 7.90E-09 | 1.88E-05 | 1.30E-10 | 0.002139927 | HDLBP |
| chr3 | 9878851 | 9880076 | 1226 | 7 | 3.50E-12 | 7.29E-08 | 6.07E-05 | 1.32E-10 | -0.00558297 | CIDEC |
| chr11 | 19777368 | 19778087 | 720 | 8 | 1.50E-11 | 2.18E-10 | 0.000404 | 1.32E-10 | -0.0040438 | NAV2 |
| chr5 | 176618745 | 176620484 | 1740 | 9 | 9.48E-09 | 1.47E-11 | 0.002573 | 1.32E-10 | -0.0111389 | SNCB |
| chr5 | 95645631 | 95647069 | 1439 | 14 | 5.88E-12 | 4.96E-09 | 3.13E-05 | 1.33E-10 | 0.0005029 | RFESD |
| chr8 | 18084243 | 18086481 | 2239 | 21 | 1.55E-11 | 7.63E-09 | 0.000665 | 1.33E-10 | 0.00404816 | AC124242.1, ASAH1 |
| chr11 | 19712068 | 19715102 | 3035 | 23 | 4.57E-12 | 5.11E-07 | 1.33E-05 | 1.33E-10 | -0.00034107 | NAV2, AC009549.1 |
| chr19 | 49495965 | 49497200 | 1236 | 14 | 1.48E-09 | 2.40E-10 | 0.000323 | 1.34E-10 | 0.001591944 | RPS11 |
| chr3 | 167379776 | 167381081 | 1306 | 12 | 1.70E-13 | 2.12E-07 | 7.00E-08 | 1.34E-10 | -0.00161453 | ZBBX |
| chr3 | 48594525 | 48596008 | 1484 | 13 | 6.24E-10 | 9.19E-10 | 0.000589 | 1.34E-10 | -0.00803783 | COL7A1 |
| chr11 | 59795445 | 59795740 | 296 | 3 | 8.49E-12 | 4.69E-10 | 2.38E-06 | 1.34E-10 | 0.00194349 | STX3 |
| chr17 | 48722278 | 48723676 | 1399 | 19 | 1.87E-12 | 4.77E-10 | 0.000587 | 1.35E-10 | 0.000216819 | PRAC2, AC091179.3, PRAC1 |
| chr10 | 97936224 | 97937277 | 1054 | 5 | 6.69E-11 | 3.01E-08 | 3.00E-06 | 1.35E-10 | -0.00292377 | CRTAC1 |
| chr16 | 57809609 | 57810660 | 1052 | 10 | 8.00E-10 | 3.42E-08 | 1.50E-05 | 1.35E-10 | -0.00138054 | AC092118.1, KIFC3 |
| chr19 | 2255420 | 2257397 | 1978 | 18 | 4.33E-08 | 5.54E-09 | 0.000156 | 1.35E-10 | -0.00230995 | JSRP1 |
| chr17 | 82587144 | 82588568 | 1425 | 14 | 4.74E-11 | 2.34E-09 | 0.001329 | 1.35E-10 | -0.00434863 | FOXK2, AC124283.2 |
| chr13 | 20700817 | 20702706 | 1890 | 7 | 1.01E-13 | 1.08E-09 | 2.94E-06 | 1.35E-10 | 0.001136377 | IL17D, AL161772.1 |
| chr12 | 126689708 | 126691155 | 1448 | 7 | 2.36E-10 | 2.17E-11 | 0.000439 | 1.35E-10 | -0.00509238 | LINC02824, AC006065.4, AC006065.3 |
| chr10 | 8053158 | 8055837 | 2680 | 43 | 1.43E-08 | 3.69E-07 | 0.000356 | 1.35E-10 | 3.15E-06 | GATA3, GATA3-AS1 |
| chr17 | 5013724 | 5015071 | 1348 | 5 | 1.92E-09 | 7.15E-11 | 5.18E-05 | 1.36E-10 | -0.00246978 | KIF1C |
| chr2 | 230219996 | 230221523 | 1528 | 9 | 2.26E-13 | 3.48E-05 | 4.97E-07 | 1.36E-10 | 0.003787862 | SP140, SP110 |
| chr6 | 34696438 | 34697379 | 942 | 9 | 1.81E-10 | 8.88E-09 | 6.43E-05 | 1.37E-10 | 0.004579656 | AL451165.2, ILRUN |
| chr11 | 65026374 | 65027977 | 1604 | 12 | 1.76E-11 | 2.79E-10 | 0.000646 | 1.37E-10 | -0.00445068 | ARL2-SNX15, SNX15, AP000436.1 |
| chr2 | 6826267 | 6826616 | 350 | 3 | 9.87E-12 | 1.55E-10 | 1.33E-06 | 1.37E-10 | -0.01208669 | NRIR |
| chr10 | 24207667 | 24209166 | 1500 | 11 | 1.60E-11 | 1.43E-06 | 2.05E-07 | 1.37E-10 | 0.002281605 | KIAA1217 |
| chr17 | 17714025 | 17714688 | 664 | 5 | 4.99E-11 | 1.27E-09 | 2.07E-05 | 1.37E-10 | 0.010430051 | RAI1 |
| chr16 | 4987460 | 4989187 | 1728 | 4 | 3.47E-07 | 4.20E-10 | 1.39E-05 | 1.37E-10 | -0.00665073 | SEC14L5 |
| chr8 | 58659321 | 58661330 | 2010 | 19 | 2.88E-08 | 8.24E-08 | 0.000314 | 1.39E-10 | 0.000567691 | NSMAF |
| chr7 | 97398635 | 97399503 | 869 | 2 | 1.42E-09 | 9.68E-11 | 1.30E-06 | 1.39E-10 | -0.01380717 | NA |
| chr19 | 53864250 | 53866427 | 2178 | 17 | 1.03E-11 | 1.49E-06 | 5.51E-07 | 1.40E-10 | 0.002603252 | MYADM, MYADM-AS1 |
| chr9 | 35675390 | 35676368 | 979 | 9 | 3.57E-12 | 2.32E-08 | 0.000192 | 1.40E-10 | -0.00865392 | CA9, ARHGEF39 |
| chr19 | 3868874 | 3870880 | 2007 | 15 | 1.34E-10 | 9.56E-06 | 2.40E-06 | 1.40E-10 | 0.008723326 | ZFR2 |
| chr16 | 67661559 | 67663680 | 2122 | 15 | 5.76E-10 | 2.02E-08 | 0.000183 | 1.41E-10 | -0.00146957 | PARD6A, ENKD1 |
| chr1 | 155026400 | 155026803 | 404 | 6 | 2.24E-12 | 5.22E-06 | 1.58E-07 | 1.42E-10 | -0.00183644 | DCST2 |
| chr2 | 31137729 | 31139292 | 1564 | 17 | 2.90E-12 | 3.12E-07 | 2.15E-05 | 1.43E-10 | 0.00132782 | GALNT14 |
| chr3 | 128073848 | 128076728 | 2881 | 12 | 6.08E-07 | 2.97E-10 | 3.55E-05 | 1.43E-10 | -0.00429177 | RUVBL1-AS1, RUVBL1, RNU2-37P |
| chr15 | 42010144 | 42011094 | 951 | 5 | 2.86E-11 | 2.21E-08 | 1.67E-05 | 1.44E-10 | -0.00648251 | PLA2G4E |
| chr2 | 227164330 | 227165208 | 879 | 11 | 2.73E-12 | 3.92E-10 | 0.000574 | 1.44E-10 | -0.00325648 | COL4A3, COL4A4 |
| chr11 | 36273417 | 36275079 | 1663 | 8 | 1.01E-10 | 6.99E-08 | 7.27E-05 | 1.44E-10 | -0.00210409 | COMMD9 |
| chr14 | 93007721 | 93008609 | 889 | 4 | 8.49E-12 | 1.20E-10 | 4.85E-06 | 1.45E-10 | -0.01039629 | ITPK1 |
| chr17 | 47970268 | 47971580 | 1313 | 21 | 7.93E-10 | 4.39E-11 | 0.003612 | 1.46E-10 | -0.00026981 | AC018521.1, CDK5RAP3 |
| chr4 | 3441235 | 3442420 | 1186 | 10 | 6.66E-15 | 7.17E-06 | 1.56E-05 | 1.47E-10 | -0.00072651 | HGFAC |
| chr3 | 10357695 | 10358864 | 1170 | 6 | 4.42E-08 | 2.39E-10 | 1.08E-05 | 1.48E-10 | -0.00534342 | ATP2B2 |
| chr2 | 195656573 | 195658306 | 1734 | 20 | 3.64E-09 | 2.49E-09 | 0.000108 | 1.49E-10 | -0.0005141 | SLC39A10 |
| chr17 | 1934294 | 1935145 | 852 | 9 | 1.06E-12 | 7.48E-09 | 7.67E-06 | 1.49E-10 | -0.00383859 | RTN4RL1 |
| chr3 | 46692903 | 46694372 | 1470 | 13 | 1.65E-10 | 1.15E-08 | 0.000691 | 1.49E-10 | -0.0030717 | ALS2CL |
| chr3 | 38002684 | 38004435 | 1752 | 5 | 4.39E-10 | 8.73E-10 | 4.12E-06 | 1.49E-10 | -0.01172777 | VILL |
| chr2 | 152617955 | 152619545 | 1591 | 7 | 1.06E-07 | 7.85E-09 | 0.00011 | 1.49E-10 | 0.001584414 | FMNL2 |
| chr17 | 81976978 | 81978685 | 1708 | 14 | 3.41E-08 | 5.14E-08 | 1.10E-05 | 1.50E-10 | 0.001246536 | ASPSCR1 |
| chr22 | 35256958 | 35258106 | 1149 | 10 | 3.38E-12 | 1.82E-06 | 1.61E-06 | 1.50E-10 | 0.003278303 | HMGXB4 |
| chr5 | 179906851 | 179908704 | 1854 | 9 | 1.87E-07 | 3.68E-11 | 0.000954 | 1.52E-10 | -0.00462108 | TBC1D9B |
| chr1 | 8925086 | 8926014 | 929 | 3 | 2.87E-10 | 3.56E-10 | 7.30E-07 | 1.52E-10 | -0.01318698 | NA |
| chr5 | 75717360 | 75718759 | 1400 | 10 | 3.12E-11 | 1.18E-09 | 3.53E-05 | 1.53E-10 | -0.00369778 | POC5 |
| chr3 | 23809763 | 23811524 | 1762 | 19 | 2.20E-10 | 2.34E-07 | 6.91E-05 | 1.53E-10 | 0.003010623 | UBE2E1 |
| chr5 | 979820 | 981074 | 1255 | 7 | 2.05E-10 | 4.08E-10 | 6.27E-05 | 1.54E-10 | -0.00898178 | AC122719.2 |
| chr17 | 77184678 | 77186088 | 1411 | 6 | 2.25E-11 | 1.16E-08 | 9.83E-06 | 1.54E-10 | -0.00141737 | SEC14L1 |
| chr17 | 47732361 | 47733873 | 1513 | 14 | 2.41E-11 | 1.03E-08 | 4.38E-05 | 1.55E-10 | -0.00333052 | TBX21 |
| chr15 | 47961434 | 47961982 | 549 | 3 | 3.12E-12 | 1.12E-08 | 9.62E-08 | 1.56E-10 | 0.024613383 | AC092078.2, AC092078.3 |
| chr17 | 82940405 | 82941577 | 1173 | 8 | 1.15E-10 | 1.70E-10 | 4.16E-05 | 1.56E-10 | -0.00265186 | TBCD |
| chr18 | 58193947 | 58195640 | 1694 | 12 | 1.02E-10 | 8.35E-09 | 9.56E-05 | 1.57E-10 | -0.00155755 | NEDD4L, AC107896.1 |
| chr1 | 63321423 | 63322886 | 1464 | 12 | 1.60E-11 | 1.24E-08 | 2.53E-05 | 1.58E-10 | 0.000673246 | FOXD3, FOXD3-AS1 |
| chr19 | 4151821 | 4153716 | 1896 | 12 | 1.21E-09 | 1.98E-08 | 9.35E-06 | 1.58E-10 | -0.00138826 | CREB3L3 |
| chr10 | 96384649 | 96386221 | 1573 | 5 | 3.85E-08 | 1.70E-08 | 1.23E-05 | 1.59E-10 | -0.00981358 | TLL2 |
| chr9 | 133701542 | 133702617 | 1076 | 9 | 8.17E-11 | 1.31E-10 | 0.00086 | 1.60E-10 | -0.02522641 | SARDH |
| chr14 | 49635184 | 49636506 | 1323 | 12 | 1.04E-13 | 8.40E-05 | 1.16E-05 | 1.60E-10 | -0.0041789 | DNAAF2 |
| chr18 | 56638056 | 56639206 | 1151 | 14 | 6.66E-11 | 1.31E-10 | 0.000215 | 1.61E-10 | 0.005697774 | TXNL1 |
| chr6 | 33593176 | 33593672 | 497 | 9 | 2.14E-10 | 2.87E-11 | 0.00021 | 1.62E-10 | -0.01207836 | LINC00336 |
| chr17 | 57542051 | 57542784 | 734 | 3 | 4.76E-13 | 5.36E-08 | 7.94E-07 | 1.62E-10 | 0.0297102 | MSI2 |
| chr19 | 3610338 | 3611498 | 1161 | 6 | 7.92E-13 | 3.04E-08 | 5.26E-07 | 1.63E-10 | 0.000827404 | CACTIN-AS1, CACTIN |
| chr1 | 222589147 | 222590726 | 1580 | 15 | 2.58E-09 | 5.41E-11 | 0.003504 | 1.63E-10 | -0.00107874 | TAF1A-AS1, TAF1A |
| chr1 | 27604129 | 27604992 | 864 | 9 | 5.90E-13 | 5.45E-09 | 1.31E-06 | 1.64E-10 | -0.00458671 | AHDC1 |
| chr1 | 90840098 | 90840120 | 23 | 2 | 2.13E-11 | 1.47E-10 | 6.08E-07 | 1.64E-10 | 0.027560769 | LINC02788, LINC02609 |
| chr11 | 34357428 | 34359658 | 2231 | 13 | 1.28E-08 | 1.22E-09 | 0.000358 | 1.64E-10 | -0.00251179 | ABTB2 |
| chr14 | 99522209 | 99523552 | 1344 | 10 | 6.37E-11 | 1.04E-09 | 7.72E-05 | 1.64E-10 | -0.00666469 | CCNK, CCDC85C |
| chr10 | 103492963 | 103494362 | 1400 | 10 | 6.52E-14 | 3.85E-07 | 1.78E-05 | 1.66E-10 | -0.00415408 | NEURL1, NEURL1-AS1 |
| chr19 | 7732145 | 7732469 | 325 | 6 | 1.37E-11 | 6.52E-08 | 2.99E-06 | 1.66E-10 | -0.01236624 | CLEC4G, AC008763.4 |
| chr4 | 186162276 | 186163368 | 1093 | 3 | 1.30E-11 | 1.74E-10 | 5.67E-06 | 1.67E-10 | -0.01842651 | FAM149A |
| chr3 | 150972464 | 150973165 | 702 | 10 | 2.21E-11 | 3.79E-09 | 2.92E-06 | 1.68E-10 | -0.00456887 | CLRN1-AS1, AC020636.1, CLRN1 |
| chr19 | 35291112 | 35293185 | 2074 | 10 | 2.90E-11 | 9.49E-10 | 4.09E-05 | 1.69E-10 | -0.00382204 | MAG |
| chr8 | 117863813 | 117864082 | 270 | 2 | 9.04E-13 | 1.66E-08 | 6.32E-09 | 1.70E-10 | -0.05098684 | EXT1 |
| chr14 | 105172626 | 105174898 | 2273 | 10 | 6.16E-08 | 3.62E-10 | 0.000393 | 1.70E-10 | -0.00425574 | AL512356.1, NUDT14 |
| chr17 | 15782995 | 15783159 | 165 | 3 | 2.51E-11 | 5.52E-10 | 6.74E-06 | 1.70E-10 | 0.022639495 | NA |
| chr1 | 153310314 | 153312114 | 1801 | 8 | 3.96E-10 | 2.99E-07 | 1.58E-05 | 1.70E-10 | -0.00600126 | PGLYRP3 |
| chr9 | 137454376 | 137456052 | 1677 | 12 | 2.89E-14 | 2.76E-08 | 2.90E-06 | 1.70E-10 | -0.0011379 | NSMF |
| chr6 | 122006323 | 122008980 | 2658 | 8 | 9.17E-08 | 2.43E-09 | 2.65E-05 | 1.71E-10 | -0.01112036 | NA |
| chr16 | 11023168 | 11024847 | 1680 | 5 | 3.56E-13 | 3.08E-09 | 2.99E-06 | 1.71E-10 | 0.003696704 | CLEC16A |
| chr20 | 64084407 | 64085253 | 847 | 10 | 1.73E-10 | 9.38E-11 | 0.000723 | 1.72E-10 | 0.003556863 | OPRL1 |
| chr1 | 58249827 | 58251435 | 1609 | 16 | 1.43E-10 | 2.72E-08 | 9.38E-05 | 1.72E-10 | -0.00661179 | DAB1 |
| chr2 | 216081276 | 216083020 | 1745 | 17 | 2.14E-09 | 8.88E-07 | 0.000632 | 1.72E-10 | -0.00056336 | TMEM169, PECR |
| chr21 | 42315172 | 42316626 | 1455 | 10 | 4.58E-12 | 5.47E-09 | 1.21E-06 | 1.74E-10 | -0.00140604 | TFF3 |
| chr8 | 48735020 | 48735960 | 941 | 13 | 5.93E-12 | 5.02E-07 | 1.91E-05 | 1.74E-10 | 0.005412822 | EFCAB1 |
| chr10 | 133218650 | 133219000 | 351 | 4 | 6.59E-12 | 2.91E-09 | 8.80E-07 | 1.74E-10 | -0.01098887 | KNDC1 |
| chr9 | 127223267 | 127224515 | 1249 | 6 | 1.62E-15 | 5.56E-07 | 2.78E-07 | 1.75E-10 | -0.00946066 | GARNL3 |
| chr19 | 44746906 | 44747923 | 1018 | 7 | 4.60E-17 | 1.57E-06 | 7.28E-07 | 1.75E-10 | -0.00358556 | BCL3 |
| chr4 | 73998522 | 74000683 | 2162 | 7 | 4.20E-10 | 2.36E-07 | 8.00E-06 | 1.75E-10 | -0.00315542 | AC097709.1, CXCL5 |
| chr3 | 170036443 | 170039030 | 2588 | 14 | 2.83E-08 | 2.73E-07 | 0.000156 | 1.75E-10 | -5.73E-05 | GPR160 |
| chr17 | 28896965 | 28898752 | 1788 | 16 | 1.52E-09 | 1.04E-09 | 7.93E-05 | 1.76E-10 | -0.00131069 | AC024267.6, FLOT2, DHRS13 |
| chr7 | 149238970 | 149240096 | 1127 | 12 | 2.92E-12 | 1.93E-08 | 8.86E-05 | 1.76E-10 | 0.002213703 | ZNF212 |
| chr1 | 13623258 | 13625420 | 2163 | 8 | 1.96E-07 | 6.56E-07 | 1.66E-05 | 1.76E-10 | -0.00258697 | RNA5SP41 |
| chr19 | 4303820 | 4305242 | 1423 | 11 | 2.67E-09 | 1.39E-09 | 5.95E-05 | 1.77E-10 | -0.00607749 | FSD1 |
| chr6 | 39792165 | 39793819 | 1655 | 14 | 9.76E-16 | 4.16E-06 | 0.000102 | 1.77E-10 | 0.007528945 | DAAM2 |
| chr1 | 47022584 | 47023871 | 1288 | 11 | 1.23E-12 | 1.20E-07 | 2.35E-05 | 1.77E-10 | -0.00320022 | CYP4X1 |
| chr2 | 10080328 | 10081219 | 892 | 10 | 3.97E-12 | 5.53E-08 | 1.53E-06 | 1.77E-10 | 0.000354508 | CYS1 |
| chr6 | 143677497 | 143678578 | 1082 | 17 | 3.20E-12 | 1.22E-07 | 1.97E-05 | 1.77E-10 | -0.00428584 | PHACTR2 |
| chr19 | 57389221 | 57390377 | 1157 | 8 | 5.88E-10 | 1.40E-07 | 0.000294 | 1.79E-10 | 0.000441114 | AC003002.1, ZNF548, AC003002.3 |
| chr13 | 24270011 | 24271896 | 1886 | 14 | 9.26E-11 | 9.09E-08 | 2.09E-05 | 1.81E-10 | 0.003011503 | AL359736.1, SPATA13 |
| chr7 | 150946827 | 150948004 | 1178 | 7 | 6.24E-11 | 6.89E-11 | 3.03E-05 | 1.81E-10 | -0.01094508 | KCNH2 |
| chr19 | 3720614 | 3721834 | 1221 | 11 | 3.31E-12 | 1.08E-07 | 0.000143 | 1.81E-10 | 0.008996003 | TJP3 |
| chr18 | 5890839 | 5892754 | 1916 | 10 | 2.37E-10 | 7.62E-09 | 0.000592 | 1.82E-10 | -0.01415642 | AP001021.1, TMEM200C |
| chr17 | 76708076 | 76708151 | 76 | 2 | 1.32E-11 | 1.13E-10 | 2.38E-06 | 1.83E-10 | -0.02303966 | MXRA7 |
| chr19 | 58554133 | 58555333 | 1201 | 13 | 4.41E-12 | 1.64E-08 | 0.000432 | 1.83E-10 | -0.00223145 | CHMP2A |
| chr17 | 13598098 | 13598987 | 890 | 3 | 6.25E-09 | 1.07E-07 | 1.39E-06 | 1.85E-10 | 0.006806644 | HS3ST3A1 |
| chr6 | 139114675 | 139114995 | 321 | 2 | 7.76E-11 | 1.04E-10 | 4.85E-06 | 1.85E-10 | 0.004416703 | NA |
| chr12 | 5043173 | 5044982 | 1810 | 11 | 3.27E-10 | 1.07E-07 | 7.13E-06 | 1.85E-10 | -0.00850313 | KCNA5 |
| chr19 | 7550305 | 7551661 | 1357 | 7 | 9.69E-10 | 1.82E-10 | 0.000144 | 1.86E-10 | -0.01449175 | PNPLA6 |
| chr15 | 49046054 | 49047196 | 1143 | 13 | 3.00E-13 | 1.90E-06 | 1.67E-06 | 1.86E-10 | 0.002012779 | SECISBP2L |
| chr1 | 7178885 | 7178938 | 54 | 2 | 1.55E-11 | 1.17E-10 | 2.19E-06 | 1.86E-10 | -0.02502112 | CAMTA1 |
| chr3 | 16264273 | 16266850 | 2578 | 15 | 3.96E-08 | 8.74E-09 | 0.000146 | 1.87E-10 | -0.00114406 | OXNAD1, DPH3 |
| chr16 | 54929312 | 54932559 | 3248 | 21 | 8.49E-08 | 1.92E-07 | 0.000796 | 1.87E-10 | -0.00099377 | IRX5 |
| chr13 | 48318415 | 48319038 | 624 | 2 | 6.56E-12 | 1.10E-10 | 3.48E-06 | 1.88E-10 | -0.02829314 | RB1, PPP1R26P1 |
| chr1 | 37995278 | 37996224 | 947 | 6 | 2.93E-10 | 3.15E-11 | 0.000454 | 1.88E-10 | -0.03579693 | NA |
| chr8 | 103499548 | 103501890 | 2343 | 17 | 2.35E-07 | 7.13E-09 | 0.001056 | 1.89E-10 | 0.000208169 | RIMS2, AC012213.5, AC012213.1 |
| chr11 | 61273414 | 61274522 | 1109 | 6 | 7.69E-13 | 8.99E-08 | 6.70E-07 | 1.90E-10 | -0.00633734 | VWCE |
| chr8 | 26577173 | 26579202 | 2030 | 16 | 3.25E-08 | 6.03E-11 | 0.002922 | 1.90E-10 | -0.00053808 | DPYSL2 |
| chr15 | 66291880 | 66294155 | 2276 | 14 | 1.67E-07 | 8.93E-09 | 0.000118 | 1.90E-10 | 0.001440343 | DIS3L, AC055855.2 |
| chr16 | 68221680 | 68223974 | 2295 | 6 | 1.19E-08 | 5.75E-07 | 2.13E-06 | 1.90E-10 | -3.49E-05 | NFATC3 |
| chr17 | 50570701 | 50573301 | 2601 | 7 | 9.07E-07 | 1.44E-10 | 0.000369 | 1.90E-10 | -0.00292087 | CACNA1G |
| chr2 | 11391480 | 11391883 | 404 | 3 | 1.17E-13 | 1.30E-07 | 1.14E-06 | 1.91E-10 | -0.01395091 | LINC00570 |
| chr16 | 19717455 | 19718976 | 1522 | 16 | 1.78E-10 | 1.81E-09 | 5.41E-05 | 1.92E-10 | -0.00038921 | IQCK, KNOP1 |
| chr6 | 31579927 | 31582464 | 2538 | 32 | 1.43E-09 | 1.85E-07 | 0.001642 | 1.92E-10 | -0.00010133 | LTB |
| chr16 | 736393 | 738268 | 1876 | 13 | 1.04E-08 | 2.82E-09 | 0.000307 | 1.93E-10 | -0.01744735 | CIAO3 |
| chr3 | 61807387 | 61807949 | 563 | 2 | 2.00E-13 | 4.58E-09 | 1.59E-08 | 1.93E-10 | -0.00675513 | PTPRG |
| chr12 | 46266966 | 46269080 | 2115 | 19 | 3.90E-10 | 6.26E-08 | 0.001895 | 1.93E-10 | -9.59E-05 | SLC38A1 |
| chr6 | 4774075 | 4775462 | 1388 | 11 | 2.08E-10 | 3.66E-09 | 3.66E-05 | 1.93E-10 | -0.0013605 | CDYL, CDYL-AS1 |
| chr12 | 55927328 | 55930032 | 2705 | 21 | 2.29E-08 | 7.02E-08 | 0.000389 | 1.93E-10 | 0.001518011 | DGKA, AC025162.2, PYM1 |
| chr7 | 138917189 | 138918916 | 1728 | 12 | 7.66E-10 | 3.38E-06 | 1.04E-06 | 1.94E-10 | -0.00179956 | KIAA1549 |
| chr20 | 21125191 | 21126046 | 856 | 9 | 4.00E-14 | 6.63E-08 | 1.84E-05 | 1.94E-10 | -0.003451 | KIZ |
| chr12 | 119667263 | 119668487 | 1225 | 16 | 1.47E-10 | 4.75E-09 | 0.001551 | 1.95E-10 | -8.42E-05 | PRKAB1, AC002070.1 |
| chr22 | 38569406 | 38570986 | 1581 | 12 | 2.38E-08 | 3.62E-10 | 0.000744 | 1.96E-10 | -0.00481239 | DMC1 |
| chr6 | 31651736 | 31653988 | 2253 | 58 | 7.88E-08 | 1.29E-08 | 0.011931 | 1.97E-10 | -0.00117443 | APOM, BAG6 |

| | | | | | | | | | | |
|---|---|---|---|---|---|---|---|---|---|---|
| chr1 | 68384108 | 68384464 | 357 | 2 | 1.24E-11 | 3.27E-10 | 1.98E-07 | 1.97E-10 | -0.01553804 | NA |
| chr6 | 83708214 | 83710108 | 1895 | 26 | 3.38E-09 | 2.33E-10 | 0.005927 | 1.98E-10 | -0.0035783 | SNAP91 |
| chr17 | 205040 | 206284 | 1245 | 4 | 6.50E-10 | 3.81E-10 | 2.41E-05 | 2.00E-10 | -0.01669152 | NA |
| chr7 | 2998035 | 2999414 | 1380 | 7 | 7.15E-11 | 1.28E-09 | 2.56E-05 | 2.00E-10 | -0.00020821 | RN7SKP130, CARD11 |
| chr19 | 8525683 | 8526892 | 1210 | 5 | 5.95E-09 | 4.78E-11 | 0.000299 | 2.00E-10 | -0.03359079 | MYO1F |
| chr11 | 65612040 | 65613886 | 1847 | 8 | 6.32E-10 | 1.06E-08 | 0.000145 | 2.00E-10 | -0.0067657 | MAP3K11 |
| chr1 | 155134238 | 155136902 | 2665 | 21 | 1.05E-07 | 6.03E-07 | 0.000392 | 2.01E-10 | -0.00219866 | EFNA1, SLC50A1 |
| chr1 | 45499348 | 45501253 | 1906 | 15 | 4.14E-11 | 1.70E-07 | 1.28E-05 | 2.01E-10 | 0.000580665 | MMACHC, CCDC163 |
| chr11 | 134609463 | 134610171 | 709 | 3 | 1.94E-10 | 3.15E-10 | 7.04E-06 | 2.01E-10 | 0.001496028 | NA |
| chr1 | 221458200 | 221458614 | 415 | 2 | 1.54E-11 | 2.54E-10 | 3.19E-07 | 2.02E-10 | 0.006707917 | NA |
| chr6 | 61680790 | 61682032 | 1243 | 5 | 4.50E-10 | 1.89E-08 | 5.01E-05 | 2.03E-10 | 0.004504374 | KHDRBS2-OT1, KHDRBS2 |
| chr19 | 7293259 | 7295605 | 2347 | 12 | 4.20E-07 | 7.34E-09 | 6.95E-05 | 2.03E-10 | -0.00375534 | INSR |
| chr12 | 25250746 | 25252383 | 1638 | 23 | 2.28E-10 | 1.21E-07 | 3.05E-05 | 2.03E-10 | -0.00090776 | KRAS |
| chr14 | 69547831 | 69548756 | 926 | 6 | 1.79E-11 | 3.03E-09 | 4.59E-06 | 2.05E-10 | -0.00585557 | NA |
| chr5 | 80034273 | 80036027 | 1755 | 14 | 1.94E-08 | 2.86E-08 | 9.04E-05 | 2.07E-10 | -0.00438301 | THBS4 |
| chr16 | 29862858 | 29864478 | 1621 | 17 | 3.10E-12 | 3.76E-08 | 0.000335 | 2.07E-10 | -0.00262083 | AC120114.2, CDIPTOSP, CDIPT |
| chr9 | 136361280 | 136363122 | 1843 | 7 | 6.13E-10 | 5.29E-08 | 0.000178 | 2.08E-10 | -0.0064933 | DNLZ, CARD9 |
| chr22 | 49657958 | 49658952 | 995 | 5 | 1.99E-11 | 1.63E-09 | 4.14E-05 | 2.09E-10 | -0.02441341 | NA |
| chr5 | 69517842 | 69518631 | 790 | 2 | 6.41E-11 | 2.87E-10 | 2.74E-07 | 2.09E-10 | -0.01198796 | OCLN |
| chr3 | 194346751 | 194347411 | 661 | 3 | 1.52E-10 | 2.17E-10 | 7.98E-06 | 2.09E-10 | -0.00051392 | CPN2 |
| chr1 | 172658880 | 172659123 | 244 | 3 | 4.55E-11 | 2.07E-10 | 2.84E-06 | 2.10E-10 | -0.01314964 | FASLG |
| chr3 | 123113718 | 123114314 | 597 | 3 | 1.82E-10 | 7.83E-11 | 1.51E-05 | 2.10E-10 | 0.045865281 | PDIA5 |
| chr2 | 45008880 | 45011303 | 2424 | 16 | 1.67E-08 | 2.68E-07 | 2.47E-06 | 2.10E-10 | 0.000214223 | SIX2 |
| chr1 | 42766550 | 42767852 | 1303 | 15 | 1.11E-09 | 3.66E-09 | 0.000637 | 2.12E-10 | 0.000492766 | C1orf50, AC098484.3, P3H1 |
| chr1 | 37555761 | 37557393 | 1633 | 14 | 8.87E-12 | 1.36E-08 | 3.74E-05 | 2.14E-10 | -0.0001572 | AL034379.1, DNALI1 |
| chr4 | 15681299 | 15682232 | 934 | 14 | 1.14E-10 | 1.24E-09 | 0.000278 | 2.14E-10 | 0.001002376 | FAM200B, AC114744.2, FBXL5 |
| chr6 | 27295078 | 27296492 | 1415 | 4 | 4.28E-12 | 7.29E-09 | 1.80E-06 | 2.15E-10 | -0.0117301 | POM121L2 |
| chr11 | 1758122 | 1759094 | 973 | 7 | 2.58E-10 | 3.09E-09 | 1.24E-05 | 2.15E-10 | -0.00597192 | AC068580.4, CTSD |
| chr3 | 160755224 | 160757548 | 2325 | 17 | 7.63E-08 | 8.19E-08 | 5.38E-06 | 2.15E-10 | -0.00363276 | PPM1L |
| chr16 | 86720723 | 86722574 | 1852 | 6 | 8.47E-10 | 5.92E-09 | 2.31E-05 | 2.15E-10 | -0.00492035 | LINC02188, LINC02189 |
| chr7 | 120857505 | 120858675 | 1171 | 10 | 9.07E-13 | 4.81E-07 | 2.03E-06 | 2.15E-10 | -0.0067061 | TSPAN12 |
| chr4 | 184800336 | 184800430 | 95 | 3 | 9.00E-11 | 1.89E-10 | 8.39E-06 | 2.15E-10 | -0.01470849 | ACSL1 |
| chr2 | 88729046 | 88729273 | 228 | 3 | 5.23E-11 | 1.09E-10 | 8.00E-06 | 2.16E-10 | -0.02028472 | RPIA |
| chr19 | 35811875 | 35813177 | 1303 | 6 | 9.78E-15 | 2.47E-08 | 1.55E-07 | 2.16E-10 | -0.01387843 | PRODH2 |
| chr7 | 78039926 | 78040874 | 949 | 7 | 2.05E-11 | 5.61E-08 | 3.07E-07 | 2.17E-10 | -0.00329767 | MAGI2 |
| chr16 | 2043360 | 2044877 | 1518 | 8 | 1.44E-11 | 7.37E-09 | 1.67E-05 | 2.17E-10 | -0.00738488 | NTHL1 |
| chr5 | 159050699 | 159053027 | 2329 | 9 | 1.50E-07 | 2.84E-10 | 0.000101 | 2.17E-10 | 0.00092025 | EBF1 |
| chr16 | 30010375 | 30012459 | 2085 | 19 | 6.47E-08 | 4.26E-08 | 0.000149 | 2.17E-10 | 0.005305759 | DOC2A |
| chr12 | 52650714 | 52651029 | 316 | 2 | 3.65E-12 | 2.14E-10 | 5.61E-07 | 2.20E-10 | -0.015069 | KRT2 |
| chr11 | 119120339 | 119122383 | 2045 | 16 | 2.13E-09 | 9.71E-09 | 0.000316 | 2.20E-10 | -0.00040561 | HINFP |
| chr12 | 108668185 | 108668300 | 116 | 5 | 1.22E-11 | 1.77E-08 | 2.63E-07 | 2.21E-10 | 0.001695301 | CORO1C |
| chr4 | 119519072 | 119519687 | 616 | 2 | 2.33E-13 | 1.74E-09 | 3.84E-08 | 2.21E-10 | 0.005836875 | AC093752.1, PDE5A |
| chr1 | 158753467 | 158754952 | 1486 | 7 | 1.14E-10 | 6.37E-08 | 2.05E-05 | 2.22E-10 | -5.18E-05 | OR6K6, OR6N1 |
| chr5 | 125317295 | 125317554 | 260 | 2 | 5.68E-12 | 2.33E-09 | 3.07E-08 | 2.22E-10 | -0.01134073 | LINC02240 |
| chr21 | 40846625 | 40847927 | 1303 | 12 | 3.39E-13 | 6.33E-06 | 1.17E-06 | 2.24E-10 | -0.00700001 | DSCAM |
| chr14 | 67694934 | 67696866 | 1933 | 14 | 4.45E-09 | 1.23E-06 | 5.25E-06 | 2.25E-10 | 0.00110098 | RDH11 |
| chr9 | 129026040 | 129026910 | 871 | 4 | 1.07E-10 | 1.33E-10 | 1.25E-05 | 2.25E-10 | -0.00377162 | SH3GLB2 |
| chr1 | 155397049 | 155398176 | 1128 | 3 | 3.15E-10 | 8.45E-10 | 1.81E-06 | 2.27E-10 | -0.00699096 | ASH1L |
| chr13 | 113220457 | 113221155 | 699 | 3 | 4.62E-12 | 1.99E-09 | 4.28E-07 | 2.27E-10 | 0.006173147 | CUL4A |
| chr14 | 76827921 | 76829112 | 1192 | 7 | 7.46E-10 | 1.55E-11 | 0.001101 | 2.27E-10 | -0.01055876 | LRRC74A |
| chr12 | 7128316 | 7130618 | 2303 | 26 | 8.80E-09 | 8.67E-07 | 0.000597 | 2.27E-10 | 0.001092417 | CLSTN3, RBP5, AC018653.3 |
| chr7 | 155450556 | 155452032 | 1477 | 8 | 4.39E-11 | 7.87E-09 | 1.43E-05 | 2.28E-10 | 0.004066825 | AC008060.3 |
| chr3 | 194369403 | 194370814 | 1412 | 10 | 3.28E-10 | 1.97E-08 | 0.000345 | 2.28E-10 | -0.01153362 | LRRC15 |
| chr13 | 36430399 | 36432611 | 2213 | 33 | 6.26E-09 | 4.20E-07 | 0.000645 | 2.31E-10 | 0.001440874 | CCNA1 |
| chr10 | 21533161 | 21535518 | 2358 | 20 | 2.09E-07 | 1.04E-08 | 0.001145 | 2.31E-10 | 0.0019336 | MLLT10 |
| chr10 | 58384364 | 58385692 | 1329 | 17 | 1.08E-09 | 1.99E-10 | 0.002724 | 2.31E-10 | -0.00098789 | TFAM |
| chr2 | 75645830 | 75647327 | 1498 | 13 | 4.10E-11 | 4.85E-07 | 6.76E-05 | 2.33E-10 | -0.00100448 | MRPL19 |
| chr5 | 1253540 | 1255501 | 1962 | 11 | 4.50E-08 | 1.13E-07 | 5.12E-06 | 2.33E-10 | -0.00498248 | AC114291.1, TERT |
| chr6 | 31629070 | 31630736 | 1667 | 20 | 2.73E-13 | 2.14E-08 | 8.89E-05 | 2.34E-10 | 0.00151641 | PRRC2A |
| chr4 | 7068324 | 7069434 | 1111 | 9 | 6.07E-13 | 1.91E-08 | 1.03E-05 | 2.34E-10 | -0.0039058 | NA |
| chr20 | 35171345 | 35172214 | 870 | 7 | 4.35E-11 | 1.86E-08 | 4.75E-06 | 2.34E-10 | -0.00040219 | PROCR |
| chr12 | 110742283 | 110744014 | 1732 | 11 | 1.61E-09 | 1.13E-08 | 0.000728 | 2.34E-10 | -0.00343789 | PPP1CC |
| chr11 | 10568263 | 10570080 | 1818 | 11 | 2.82E-10 | 5.82E-10 | 3.87E-05 | 2.34E-10 | -0.00297752 | IRAG1-AS1, LYVE1 |
| chr19 | 19262757 | 19264599 | 1843 | 12 | 7.83E-10 | 2.98E-07 | 5.13E-05 | 2.36E-10 | -0.00317973 | AC138430.1, HAPLN4, TM6SF2 |
| chr18 | 3411489 | 3412784 | 1296 | 14 | 9.85E-10 | 4.98E-09 | 0.001963 | 2.37E-10 | -0.00410111 | TGIF1 |
| chr11 | 67443341 | 67444851 | 1511 | 13 | 4.76E-10 | 1.51E-08 | 2.40E-05 | 2.37E-10 | 0.000896471 | CORO1B |
| chr2 | 675921 | 678866 | 2946 | 21 | 4.36E-07 | 1.42E-09 | 0.00248 | 2.38E-10 | -0.00574409 | TMEM18-DT, TMEM18 |
| chr3 | 196537816 | 196537908 | 93 | 2 | 3.41E-11 | 6.53E-10 | 1.12E-07 | 2.39E-10 | -0.03502355 | NA |
| chr1 | 75130234 | 75132202 | 1969 | 10 | 4.42E-10 | 7.24E-09 | 4.78E-05 | 2.41E-10 | -0.00185964 | LHX8, AC099786.1, AC099786.2 |
| chr19 | 53808925 | 53809750 | 826 | 5 | 2.06E-14 | 6.82E-05 | 3.99E-09 | 2.43E-10 | -0.00667202 | NLRP12 |
| chr1 | 114153728 | 114155826 | 2099 | 19 | 2.27E-09 | 5.42E-08 | 3.89E-05 | 2.43E-10 | 0.001636643 | SYT6 |
| chr6 | 30616819 | 30618691 | 1873 | 37 | 5.12E-11 | 5.99E-08 | 0.003071 | 2.43E-10 | -7.98E-05 | MRPS18B, PPP1R10 |
| chr16 | 1079666 | 1081907 | 2242 | 12 | 1.56E-09 | 7.29E-08 | 0.00025 | 2.43E-10 | 0.005820591 | SSTR5 |
| chr7 | 27220483 | 27221370 | 888 | 7 | 2.43E-10 | 8.28E-11 | 0.000175 | 2.44E-10 | 0.010757863 | NA |
| chr10 | 508771 | 509605 | 835 | 3 | 1.32E-14 | 2.81E-08 | 2.10E-07 | 2.44E-10 | 0.003631997 | DIP2C |
| chr16 | 4943226 | 4943542 | 317 | 3 | 1.63E-10 | 8.44E-11 | 2.94E-05 | 2.45E-10 | -0.02743659 | PPL |
| chr2 | 151423000 | 151423585 | 586 | 2 | 1.67E-10 | 1.62E-10 | 2.07E-06 | 2.45E-10 | -0.01080695 | RIF1 |
| chr19 | 47349047 | 47349752 | 706 | 11 | 1.22E-11 | 1.06E-06 | 3.76E-05 | 2.48E-10 | 0.001419567 | DHX34 |
| chr13 | 94704788 | 94706670 | 1883 | 13 | 1.44E-10 | 7.99E-08 | 0.000496 | 2.48E-10 | -0.00459236 | SOX21-AS1, LINC00391 |
| chr9 | 108562243 | 108562393 | 151 | 2 | 9.07E-11 | 1.44E-10 | 4.14E-06 | 2.48E-10 | -0.00099737 | NA |
| chr6 | 170293823 | 170295915 | 2093 | 13 | 2.85E-08 | 1.60E-08 | 0.000112 | 2.50E-10 | 0.00227435 | FAM120B, DLL1 |
| chr17 | 66301464 | 66302875 | 1412 | 11 | 1.04E-09 | 9.79E-09 | 2.09E-05 | 2.50E-10 | -0.00403531 | PRKCA |
| chr17 | 2029101 | 2030626 | 1526 | 12 | 4.99E-12 | 2.21E-08 | 4.05E-06 | 2.50E-10 | -0.00456858 | DPH1 |
| chr9 | 36168598 | 36169206 | 609 | 4 | 2.38E-10 | 4.17E-11 | 0.000171 | 2.50E-10 | -0.00464359 | NA |
| chr5 | 66238654 | 66239170 | 517 | 3 | 1.76E-11 | 6.70E-10 | 4.10E-07 | 2.51E-10 | -0.01190543 | NA |
| chr6 | 99424910 | 99426067 | 1158 | 19 | 1.06E-10 | 6.07E-08 | 0.000684 | 2.51E-10 | 7.02E-05 | AL513550.1, PNISR |
| chr18 | 31317085 | 31318944 | 1860 | 9 | 9.33E-08 | 8.20E-11 | 0.000601 | 2.51E-10 | -0.00914788 | DSG1 |
| chr6 | 33210820 | 33213368 | 2549 | 28 | 1.12E-11 | 1.59E-06 | 2.87E-05 | 2.52E-10 | -0.00025518 | RING1 |
| chr17 | 48944337 | 48945249 | 913 | 14 | 1.64E-11 | 1.37E-07 | 7.87E-06 | 2.53E-10 | 0.003184338 | SNF8 |
| chr1 | 39407405 | 39410115 | 2711 | 15 | 3.32E-08 | 7.54E-09 | 0.000914 | 2.53E-10 | -0.00047429 | MACF1 |
| chr17 | 50129385 | 50130298 | 914 | 6 | 1.30E-12 | 1.99E-08 | 8.87E-07 | 2.53E-10 | 0.004854534 | SAMD14 |
| chr1 | 156874062 | 156876544 | 2483 | 11 | 7.80E-08 | 8.13E-10 | 0.000427 | 2.53E-10 | -0.0065947 | NTRK1 |
| chr17 | 75046697 | 75048896 | 2200 | 14 | 1.13E-08 | 1.63E-08 | 0.000337 | 2.54E-10 | 0.002278017 | KCTD2, ATP5PD |
| chr1 | 25819925 | 25821388 | 1464 | 12 | 1.59E-09 | 1.98E-08 | 0.000226 | 2.55E-10 | -0.00161063 | AL020996.2, MTFR1L, AL020996.1 |
| chr12 | 113135013 | 113136399 | 1387 | 18 | 5.02E-12 | 7.10E-09 | 0.00027 | 2.55E-10 | 1.28E-06 | RASAL1 |
| chr7 | 27142816 | 27144697 | 1882 | 40 | 1.50E-12 | 9.98E-09 | 0.017308 | 2.56E-10 | -0.01759347 | HOXA-AS3, HOXA3, AC004080.6, HOXA5 |
| chr16 | 70685374 | 70687793 | 2420 | 13 | 4.61E-12 | 2.64E-06 | 0.000154 | 2.56E-10 | -0.00428176 | MTSS2, VAC14 |
| chr2 | 218731748 | 218731814 | 67 | 2 | 2.91E-11 | 1.56E-10 | 3.18E-06 | 2.57E-10 | -0.03776548 | TTLL4 |
| chr11 | 134178264 | 134179061 | 798 | 6 | 1.03E-10 | 4.63E-10 | 2.36E-05 | 2.59E-10 | 0.000156772 | AP001775.2, NCAPD3 |
| chr8 | 139629268 | 139630992 | 1725 | 7 | 6.59E-11 | 2.41E-09 | 3.91E-05 | 2.62E-10 | 0.003042339 | KCNK9 |
| chr19 | 11453627 | 11454868 | 1242 | 7 | 1.70E-10 | 2.62E-09 | 8.51E-06 | 2.63E-10 | -0.0056794 | ELAVL3 |
| chr11 | 62269462 | 62270092 | 631 | 5 | 1.69E-11 | 4.64E-09 | 9.50E-07 | 2.64E-10 | -0.01045823 | NA |
| chr10 | 3485804 | 3486533 | 730 | 3 | 4.02E-10 | 8.52E-10 | 7.84E-06 | 2.64E-10 | -0.01526299 | LINC02669 |
| chr19 | 52638367 | 52638893 | 527 | 5 | 3.25E-12 | 4.88E-09 | 1.48E-06 | 2.65E-10 | -0.02044406 | ZNF83 |
| chr16 | 3111759 | 3113227 | 1469 | 11 | 3.99E-11 | 9.87E-11 | 0.001073 | 2.65E-10 | -0.01739089 | ZNF205, ZNF213-AS1 |
| chr4 | 56681181 | 56682124 | 944 | 12 | 4.28E-11 | 1.20E-09 | 0.002034 | 2.65E-10 | 0.024129256 | HOPX |
| chr16 | 27400815 | 27402960 | 2146 | 12 | 1.39E-08 | 4.20E-09 | 0.000118 | 2.66E-10 | 0.005583758 | IL21R |
| chr4 | 7800465 | 7801286 | 822 | 7 | 3.67E-11 | 3.09E-09 | 5.95E-05 | 2.69E-10 | -0.00623468 | AC112254.1, AFAP1 |
| chr8 | 37330917 | 37332621 | 1705 | 6 | 1.90E-09 | 4.38E-10 | 6.33E-06 | 2.69E-10 | -0.00299098 | AC091182.1 |
| chr11 | 126354801 | 126356259 | 1459 | 13 | 3.39E-12 | 1.63E-06 | 0.000323 | 2.69E-10 | 0.000748977 | ST3GAL4, GSEC |
| chr17 | 57420856 | 57421279 | 424 | 4 | 1.96E-11 | 7.74E-10 | 1.55E-06 | 2.70E-10 | 0.003945171 | MSI2 |
| chr17 | 19376984 | 19379841 | 2858 | 22 | 2.15E-15 | 1.64E-07 | 3.46E-07 | 2.70E-10 | -0.00354051 | MAPK7, B9D1 |
| chr3 | 63544005 | 63545413 | 1409 | 4 | 1.83E-11 | 7.33E-09 | 1.51E-06 | 2.71E-10 | -0.0129262 | SYNPR, SYNPR-AS1 |
| chr4 | 184983861 | 184984186 | 326 | 6 | 4.55E-12 | 9.80E-07 | 1.70E-06 | 2.71E-10 | -0.00119629 | NA |
| chr12 | 6074857 | 6076114 | 1258 | 6 | 9.27E-10 | 1.78E-10 | 0.000112 | 2.72E-10 | -0.00283863 | VWF |
| chr2 | 127016523 | 127017304 | 782 | 5 | 2.38E-13 | 1.15E-06 | 1.33E-06 | 2.73E-10 | -0.00579135 | NA |
| chr1 | 210366879 | 210367079 | 201 | 2 | 1.86E-11 | 1.32E-09 | 6.46E-08 | 2.74E-10 | -0.01080105 | HHAT, AL034351.4 |
| chr20 | 33993010 | 33994619 | 1610 | 12 | 1.20E-09 | 8.94E-10 | 0.00026 | 2.75E-10 | 0.001259673 | RALY, RALY-AS1 |
| chr8 | 37774781 | 37775872 | 1092 | 5 | 1.32E-10 | 1.38E-09 | 1.26E-06 | 2.76E-10 | -0.00718092 | PLPBP |
| chr1 | 196608880 | 196609733 | 854 | 6 | 4.95E-12 | 8.01E-09 | 0.000201 | 2.77E-10 | -0.00906198 | KCNT2 |
| chr17 | 58323359 | 58324877 | 1519 | 6 | 4.11E-11 | 1.63E-06 | 8.02E-07 | 2.77E-10 | 0.000566237 | TSPOAP1-AS1, TSPOAP1 |
| chr16 | 81715663 | 81716705 | 1043 | 3 | 1.70E-10 | 1.61E-10 | 1.18E-05 | 2.78E-10 | -0.02755898 | NA |
| chr6 | 35296833 | 35298412 | 1580 | 12 | 2.40E-10 | 3.18E-08 | 1.96E-06 | 2.79E-10 | -0.00140206 | DEF6 |
| chr1 | 2330793 | 2331484 | 692 | 4 | 9.54E-14 | 3.00E-05 | 8.93E-07 | 2.79E-10 | -0.00710318 | MORN1 |
| chr5 | 123422566 | 123424217 | 1652 | 14 | 1.72E-09 | 1.34E-08 | 0.000578 | 2.81E-10 | -0.00206087 | CEP120 |
| chr12 | 53499664 | 53502033 | 2370 | 26 | 6.12E-08 | 1.85E-09 | 0.004527 | 2.81E-10 | -0.00028887 | TARBP2, MAP3K12, AC023509.3 |
| chr6 | 33298636 | 33300294 | 1659 | 41 | 1.91E-10 | 1.02E-09 | 0.001112 | 2.82E-10 | 0.000381233 | RGL2, TAPBP |
| chr2 | 27355808 | 27357892 | 2085 | 17 | 2.64E-08 | 2.94E-07 | 9.07E-05 | 2.83E-10 | -0.00088687 | AC074117.1, GTF3C2 |
| chr17 | 50905259 | 50905856 | 598 | 4 | 5.80E-13 | 1.38E-07 | 5.75E-06 | 2.84E-10 | -0.00718146 | TOB1-AS1 |
| chr11 | 57499628 | 57500794 | 1167 | 7 | 6.28E-10 | 2.43E-11 | 0.000724 | 2.85E-10 | -0.01135289 | SLC43A1 |
| chr16 | 27213537 | 27215149 | 1613 | 12 | 2.52E-09 | 8.50E-11 | 0.000523 | 2.86E-10 | -0.00205655 | KDM8, AC109449.1 |
| chr13 | 110615050 | 110616367 | 1318 | 18 | 3.72E-09 | 1.31E-06 | 2.23E-05 | 2.86E-10 | 0.002048351 | NAXD, AL139385.1 |
| chr11 | 123183688 | 123184858 | 1171 | 3 | 4.48E-08 | 1.19E-10 | 2.57E-05 | 2.86E-10 | -0.01359055 | CLMP |
| chr4 | 305520 | 305581 | 62 | 3 | 5.62E-11 | 1.83E-09 | 1.75E-07 | 2.87E-10 | -0.02190918 | NA |
| chr4 | 784548 | 786092 | 1545 | 9 | 6.71E-11 | 1.38E-09 | 0.00049 | 2.88E-10 | -0.01222 | CPLX1 |
| chr2 | 217847972 | 217848924 | 953 | 6 | 7.61E-11 | 3.95E-09 | 4.68E-05 | 2.89E-10 | -0.00250445 | TNS1 |
| chr20 | 64098532 | 64099410 | 879 | 4 | 9.88E-10 | 6.46E-09 | 6.52E-06 | 2.92E-10 | -0.00443246 | OPRL1, AL121581.3 |
| chr12 | 130838415 | 130838877 | 463 | 4 | 2.68E-12 | 1.43E-09 | 9.95E-06 | 2.93E-10 | -0.01616177 | STX2 |
| chr11 | 67420721 | 67421650 | 930 | 10 | 1.52E-11 | 1.07E-08 | 6.70E-05 | 2.93E-10 | -0.00403197 | CARNS1, PPP1CA |
| chr8 | 60921814 | 60923289 | 1476 | 9 | 1.09E-10 | 9.94E-10 | 3.15E-05 | 2.93E-10 | -0.01072916 | AC022182.1 |
| chr12 | 107092451 | 107094096 | 1646 | 14 | 5.88E-14 | 2.03E-07 | 4.11E-06 | 2.95E-10 | -0.0004212 | AC078929.1, CRY1 |
| chr11 | 69144208 | 69144595 | 388 | 3 | 2.46E-11 | 2.46E-10 | 1.54E-05 | 2.96E-10 | -0.01327892 | AP003071.5 |
| chr9 | 111794787 | 111795534 | 748 | 7 | 4.34E-11 | 1.24E-09 | 5.26E-06 | 2.96E-10 | -0.0122328 | SHOC1 |
| chr15 | 49619978 | 49622067 | 2090 | 14 | 2.95E-08 | 2.99E-09 | 0.001705 | 2.96E-10 | 0.000159603 | DTWD1, FAM227B |
| chr1 | 52055518 | 52057658 | 2141 | 18 | 1.38E-09 | 7.02E-06 | 0.000152 | 2.97E-10 | -0.00054288 | BTF3L4 |
| chr2 | 3377574 | 3378803 | 1230 | 16 | 1.18E-10 | 1.62E-09 | 0.000114 | 2.97E-10 | -0.00108938 | EIPR1 |
| chr16 | 1770948 | 1772345 | 1398 | 15 | 2.98E-10 | 6.66E-06 | 5.26E-06 | 2.98E-10 | -0.0004602 | NME3, MRPS34 |
| chr9 | 101397863 | 101399734 | 1872 | 16 | 7.95E-09 | 6.11E-09 | 0.000243 | 2.98E-10 | -0.00150868 | ZNF189, MRPL50 |
| chr2 | 222311448 | 222313023 | 1576 | 13 | 2.76E-10 | 7.80E-10 | 0.000999 | 3.04E-10 | -0.00321162 | NA |
| chr3 | 61560546 | 61562270 | 1725 | 16 | 3.55E-11 | 6.88E-08 | 0.000432 | 3.05E-10 | 0.001547795 | PTPRG |
| chr6 | 106782530 | 106784428 | 1899 | 6 | 2.08E-08 | 2.41E-06 | 5.18E-06 | 3.06E-10 | -0.00654947 | MIR587, LINC02532 |
| chr9 | 37786127 | 37786920 | 794 | 5 | 9.18E-12 | 1.83E-10 | 2.58E-05 | 3.07E-10 | 0.003893917 | AL138752.2, EXOSC3 |
| chr2 | 203936132 | 203936854 | 723 | 7 | 2.17E-11 | 9.95E-09 | 1.48E-06 | 3.07E-10 | -0.01579447 | ICOS |
| chr2 | 172102484 | 172104153 | 1670 | 14 | 1.19E-11 | 1.07E-05 | 6.42E-06 | 3.07E-10 | -0.00110878 | DLX2-DT, DLX2 |
| chr5 | 1087469 | 1089685 | 2217 | 18 | 1.51E-08 | 1.52E-07 | 7.36E-05 | 3.09E-10 | -0.00013689 | SLC12A7 |
| chr16 | 66832195 | 66833217 | 1023 | 3 | 3.39E-12 | 1.96E-06 | 8.12E-08 | 3.12E-10 | -0.00640098 | NAE1 |
| chr2 | 84515778 | 84516811 | 1034 | 14 | 2.04E-11 | 2.19E-07 | 9.42E-05 | 3.16E-10 | -0.0036788 | DNAH6 |
| chr7 | 157853913 | 157855314 | 1402 | 10 | 4.06E-11 | 1.10E-08 | 3.57E-06 | 3.16E-10 | -0.01096103 | PTPRN2-AS1, PTPRN2 |

| | | | | | | | | | | |
|---|---|---|---|---|---|---|---|---|---|---|
| chr15 | 26883132 | 26883787 | 656 | 4 | 1.55E-11 | 1.10E-07 | 2.94E-05 | 3.16E-10 | -0.00606121 | GABRA5, GABRB3 |
| chr4 | 176001405 | 176002408 | 1004 | 9 | 3.45E-13 | 2.62E-07 | 1.91E-05 | 3.17E-10 | -0.00177201 | GPM6A |
| chr8 | 116765126 | 116767046 | 1921 | 17 | 7.54E-13 | 4.45E-08 | 6.56E-06 | 3.18E-10 | 0.000509526 | UTP23, EIF3H |
| chr1 | 2605903 | 2606335 | 433 | 6 | 4.33E-12 | 3.58E-07 | 4.19E-06 | 3.18E-10 | -0.00189346 | MMEL1 |
| chr2 | 189878736 | 189879659 | 924 | 6 | 3.10E-10 | 1.43E-10 | 7.11E-05 | 3.19E-10 | -0.01594956 | C2orf88 |
| chr16 | 66548689 | 66550594 | 1906 | 18 | 6.90E-08 | 2.17E-09 | 0.003313 | 3.19E-10 | -0.00574849 | AC010542.4, Y RNA, TK2 |
| chr1 | 221772255 | 221772472 | 218 | 2 | 2.86E-12 | 5.81E-08 | 6.70E-09 | 3.19E-10 | -0.00389585 | NA |
| chr1 | 161962203 | 161962422 | 220 | 2 | 5.29E-11 | 4.15E-10 | 3.77E-07 | 3.21E-10 | 0.049781132 | ATF6 |
| chr19 | 8423443 | 8424898 | 1456 | 4 | 6.21E-14 | 2.73E-07 | 2.16E-07 | 3.21E-10 | -0.01429616 | MARCHF2, AC136469.1 |
| chr5 | 87412495 | 87413786 | 1292 | 15 | 6.08E-10 | 2.63E-09 | 0.000508 | 3.22E-10 | 0.000372611 | AC018754.1, CCNH |
| chr7 | 143027287 | 143027894 | 608 | 4 | 4.28E-15 | 8.73E-05 | 2.32E-09 | 3.22E-10 | 0.005635248 | NA |
| chr6 | 31926778 | 31927821 | 1044 | 10 | 1.47E-10 | 2.62E-09 | 0.000166 | 3.22E-10 | -0.01060322 | C2, AL645922.1 |
| chr11 | 125111218 | 125112828 | 1611 | 16 | 6.83E-10 | 2.08E-08 | 0.003438 | 3.23E-10 | 0.001658023 | TMEM218 |
| chr16 | 81198934 | 81199111 | 178 | 2 | 9.12E-11 | 2.41E-10 | 1.46E-06 | 3.28E-10 | 0.010961307 | PKD1L2 |
| chr1 | 54996453 | 54998894 | 2442 | 12 | 1.27E-06 | 1.63E-10 | 0.002442 | 3.28E-10 | 0.000937577 | NA |
| chr7 | 156604958 | 156606693 | 1736 | 12 | 2.07E-10 | 2.58E-07 | 6.57E-05 | 3.29E-10 | -0.00478555 | LINC01006 |
| chr8 | 57260012 | 57261042 | 1031 | 4 | 5.16E-10 | 6.70E-09 | 1.15E-06 | 3.29E-10 | 0.016506374 | NA |
| chr5 | 150904323 | 150905615 | 1293 | 12 | 3.25E-11 | 2.96E-09 | 0.000487 | 3.29E-10 | -0.00902698 | ZNF300 |
| chr4 | 34667817 | 34668596 | 780 | 4 | 2.41E-10 | 3.27E-08 | 2.38E-06 | 3.29E-10 | -0.01524218 | AC093689.1 |
| chr13 | 113860070 | 113861093 | 1024 | 11 | 1.42E-10 | 4.30E-09 | 0.000651 | 3.29E-10 | -0.00069257 | GAS6 |
| chr6 | 10885897 | 10887896 | 2000 | 14 | 1.76E-09 | 1.28E-08 | 0.000213 | 3.30E-10 | -0.0036713 | AL024498.2, SYCP2L |
| chr20 | 63656647 | 63659023 | 2377 | 17 | 1.28E-06 | 1.51E-10 | 0.000942 | 3.30E-10 | -0.00466881 | RTEL1, STMN3 |
| chr17 | 48100634 | 48101247 | 614 | 5 | 6.19E-13 | 4.08E-09 | 1.72E-06 | 3.31E-10 | -0.00297757 | CBX1 |
| chr12 | 122225743 | 122228378 | 2636 | 24 | 1.56E-07 | 4.01E-07 | 0.00016 | 3.31E-10 | -0.00098369 | AC048338.2, DIABLO, AC048338.1 |
| chr1 | 201377519 | 201379978 | 2460 | 13 | 6.11E-07 | 8.77E-09 | 0.000762 | 3.31E-10 | -0.00105785 | TNNT2, AC119427.2 |
| chr9 | 113220769 | 113222199 | 1431 | 10 | 2.05E-12 | 5.77E-08 | 1.37E-05 | 3.32E-10 | 0.000664553 | SLC31A1, FKBP15 |
| chr4 | 3469670 | 3472010 | 2341 | 8 | 4.08E-07 | 5.63E-09 | 0.000124 | 3.33E-10 | -0.00130052 | DOK7 |
| chr12 | 45291462 | 45292992 | 1531 | 7 | 5.91E-09 | 2.02E-07 | 1.03E-05 | 3.33E-10 | -0.00012254 | ANO6 |
| chr4 | 167233624 | 167235975 | 2352 | 24 | 4.03E-08 | 1.18E-06 | 1.31E-05 | 3.34E-10 | -0.00063642 | SPOCK3 |
| chr11 | 134383715 | 134385401 | 1687 | 12 | 1.73E-11 | 2.46E-09 | 6.84E-05 | 3.34E-10 | -0.00514209 | B3GAT1 |
| chr2 | 27369786 | 27371337 | 1552 | 19 | 2.26E-11 | 4.12E-07 | 7.65E-06 | 3.34E-10 | -0.00017069 | SNX17, EIF2B4 |
| chr15 | 25437928 | 25440221 | 2294 | 22 | 1.78E-09 | 1.32E-07 | 0.00032 | 3.35E-10 | -0.00038164 | UBE3A |
| chr17 | 74669355 | 74669443 | 89 | 2 | 8.08E-13 | 5.63E-10 | 2.58E-07 | 3.37E-10 | -0.00883189 | NA |
| chr16 | 67159112 | 67160436 | 1325 | 14 | 5.86E-11 | 4.63E-08 | 8.22E-05 | 3.38E-10 | 0.000151541 | FBXL8, TRADD |
| chr4 | 7797464 | 7798488 | 1025 | 5 | 8.07E-12 | 4.01E-08 | 1.12E-06 | 3.38E-10 | -0.00141606 | AFAP1 |
| chr1 | 162677020 | 162678577 | 1558 | 4 | 4.72E-09 | 9.74E-11 | 3.32E-05 | 3.41E-10 | -0.01055484 | DDR2 |
| chr1 | 230916670 | 230917958 | 1289 | 4 | 5.58E-11 | 2.91E-07 | 3.76E-05 | 3.43E-10 | -0.0070775 | TTC13 |
| chr11 | 57349188 | 57350192 | 1005 | 5 | 2.62E-11 | 1.95E-09 | 9.58E-05 | 3.44E-10 | -0.01036086 | P2RX3 |
| chr2 | 233352340 | 233354605 | 2266 | 13 | 4.43E-09 | 7.93E-07 | 1.42E-05 | 3.44E-10 | 7.48E-05 | DGKD, AC013726.1 |
| chr7 | 4832906 | 4835350 | 2445 | 11 | 7.03E-13 | 2.69E-06 | 3.27E-07 | 3.44E-10 | -0.00093168 | RADIL |
| chr2 | 56182639 | 56183937 | 1299 | 15 | 6.23E-10 | 9.14E-11 | 0.000266 | 3.45E-10 | 1.45E-05 | AC007744.1, AC007743.1 |
| chr7 | 27133012 | 27135533 | 2522 | 7 | 3.93E-07 | 3.63E-08 | 1.05E-05 | 3.46E-10 | -0.00352183 | HOXA-AS2, HOXA-AS3, HOXA3, AC004080.6 |
| chr20 | 62710665 | 62712020 | 1356 | 5 | 4.67E-11 | 3.36E-08 | 7.15E-06 | 3.47E-10 | -0.00896484 | NTSR1 |
| chr20 | 62413946 | 62414302 | 357 | 3 | 1.09E-10 | 2.22E-10 | 6.97E-06 | 3.48E-10 | -0.00549541 | RBBP8NL |
| chr20 | 59299542 | 59301146 | 1605 | 14 | 1.19E-09 | 6.81E-10 | 0.000383 | 3.48E-10 | 0.003336758 | EDN3 |
| chr1 | 153989544 | 153991117 | 1574 | 14 | 2.83E-12 | 3.53E-08 | 0.000849 | 3.50E-10 | -0.00133441 | RPS27 |
| chr1 | 227219445 | 227220022 | 578 | 2 | 1.29E-11 | 7.16E-10 | 1.94E-07 | 3.50E-10 | 0.002528391 | CDC42BPA |
| chr1 | 151824460 | 151825956 | 1497 | 4 | 1.01E-10 | 1.68E-10 | 4.13E-05 | 3.51E-10 | 0.005682859 | RORC |
| chr14 | 36661007 | 36662793 | 1787 | 11 | 3.84E-10 | 5.96E-08 | 1.35E-05 | 3.51E-10 | 0.000738482 | PAX9 |
| chr10 | 93567044 | 93568127 | 1084 | 8 | 8.36E-11 | 5.96E-10 | 0.000135 | 3.51E-10 | -0.00774131 | FFAR4 |
| chr8 | 144397800 | 144400256 | 2457 | 12 | 7.37E-07 | 3.71E-09 | 0.001698 | 3.54E-10 | -0.00328571 | CPSF1, MIR1234 |
| chr5 | 77180333 | 77181506 | 1174 | 5 | 1.07E-10 | 7.15E-09 | 1.92E-05 | 3.54E-10 | 0.000181238 | AC022414.1 |
| chr7 | 4767124 | 4767925 | 802 | 7 | 5.01E-10 | 4.95E-09 | 0.000437 | 3.54E-10 | -0.00701225 | FOXK1 |
| chr16 | 4891219 | 4892392 | 1174 | 3 | 2.18E-11 | 4.06E-09 | 4.23E-06 | 3.54E-10 | 0.017097764 | PPL |
| chr18 | 11750099 | 11752593 | 2495 | 21 | 2.82E-07 | 2.11E-07 | 0.000483 | 3.55E-10 | -0.00253891 | GNAL |
| chr10 | 132735639 | 132737476 | 1838 | 9 | 4.96E-10 | 1.85E-07 | 3.44E-06 | 3.57E-10 | 0.003583693 | INPP5A |
| chr22 | 36286194 | 36286974 | 781 | 8 | 3.30E-12 | 5.01E-09 | 3.69E-06 | 3.62E-10 | -0.00720172 | MYH9, MIR6819 |
| chr19 | 17648430 | 17649562 | 1133 | 6 | 5.09E-09 | 2.60E-10 | 3.25E-05 | 3.62E-10 | -0.00740527 | UNC13A |
| chr12 | 56092808 | 56094528 | 1721 | 9 | 3.17E-10 | 7.95E-10 | 2.54E-05 | 3.62E-10 | 0.000921598 | ERBB3 |
| chr6 | 24933687 | 24934041 | 355 | 4 | 2.17E-12 | 1.26E-07 | 1.05E-06 | 3.64E-10 | -0.01193514 | RIPOR2 |
| chr7 | 157866070 | 157866180 | 111 | 2 | 1.60E-11 | 2.28E-10 | 3.01E-06 | 3.65E-10 | -0.01074152 | PTPRN2 |
| chr16 | 13350289 | 13351281 | 993 | 5 | 4.25E-12 | 6.95E-09 | 6.98E-06 | 3.67E-10 | 0.001111508 | U91319.1 |
| chr17 | 38751916 | 38753353 | 1438 | 17 | 2.55E-09 | 7.56E-08 | 4.95E-05 | 3.67E-10 | 0.001595772 | PSMB3 |
| chr16 | 35207416 | 35208749 | 1334 | 10 | 2.35E-09 | 6.66E-09 | 0.000559 | 3.68E-10 | -0.01541034 | AC023824.3, TP53TG3GP |
| chr4 | 41611544 | 41612941 | 1398 | 10 | 6.19E-11 | 9.14E-09 | 0.000354 | 3.70E-10 | -0.00977593 | LIMCH1, AC108050.1 |
| chr4 | 74157502 | 74158932 | 1431 | 13 | 1.51E-10 | 2.66E-09 | 0.000117 | 3.70E-10 | 0.00038035 | MTHFD2L, AC093677.2 |
| chr8 | 9902210 | 9904395 | 2186 | 11 | 8.09E-13 | 8.54E-09 | 4.44E-05 | 3.70E-10 | 0.002437869 | MIR124-1HG, MIR124-1 |
| chr8 | 109539214 | 109540667 | 1454 | 21 | 5.96E-10 | 1.36E-08 | 0.000458 | 3.72E-10 | 0.001436291 | EBAG9 |
| chr3 | 126558850 | 126559387 | 538 | 5 | 6.55E-11 | 3.34E-09 | 4.17E-05 | 3.72E-10 | -0.00489928 | C3orf22 |
| chr6 | 81751596 | 81754194 | 2599 | 18 | 5.15E-10 | 4.76E-07 | 4.98E-06 | 3.74E-10 | 0.000589405 | TENT5A |
| chr5 | 169263814 | 169264467 | 654 | 2 | 4.35E-12 | 1.31E-09 | 1.04E-07 | 3.74E-10 | -0.01305196 | SLIT3 |
| chr6 | 41038606 | 41039953 | 1348 | 7 | 2.54E-11 | 1.03E-09 | 5.00E-06 | 3.75E-10 | -0.0070516 | UNC5CL, OARD1 |
| chr11 | 285974 | 287216 | 1243 | 7 | 1.67E-10 | 7.94E-08 | 0.000148 | 3.75E-10 | -0.00357636 | NA |
| chr19 | 35114117 | 35116317 | 2201 | 15 | 1.76E-08 | 3.74E-07 | 4.29E-05 | 3.76E-10 | 0.000988011 | AC020907.6, FXYD3 |
| chr1 | 46455481 | 46456172 | 692 | 6 | 4.81E-11 | 4.91E-09 | 3.28E-05 | 3.78E-10 | -0.00679956 | FAAHP1 |
| chr17 | 78132825 | 78134224 | 1400 | 9 | 1.16E-13 | 1.93E-08 | 1.69E-06 | 3.78E-10 | -0.00434124 | TMC8 |
| chr17 | 30824258 | 30825950 | 1693 | 10 | 1.55E-09 | 4.12E-07 | 9.30E-06 | 3.79E-10 | -0.00296968 | CRLF3 |
| chr1 | 212949487 | 212951788 | 2302 | 15 | 3.22E-08 | 6.34E-09 | 0.000348 | 3.79E-10 | -0.00242978 | VASH2 |
| chr9 | 134698163 | 134698842 | 680 | 5 | 6.96E-13 | 6.29E-09 | 1.85E-07 | 3.79E-10 | -0.00516108 | COL5A1 |
| chr1 | 152509386 | 152511199 | 1814 | 9 | 5.80E-13 | 1.89E-06 | 1.43E-07 | 3.80E-10 | -0.00712939 | LCE5A |
| chr6 | 168048437 | 168049528 | 1092 | 7 | 2.42E-11 | 2.17E-07 | 8.00E-07 | 3.83E-10 | -0.00418315 | NA |
| chr22 | 33724486 | 33726490 | 2005 | 6 | 8.55E-09 | 1.31E-07 | 5.86E-07 | 3.83E-10 | -0.00636267 | LARGE-AS1, LARGE1, LARGE-IT1 |
| chr1 | 31644477 | 31645944 | 1468 | 18 | 1.55E-09 | 1.58E-08 | 0.00076 | 3.86E-10 | 0.002628457 | PEF1-AS1, AC114488.2, PEF1 |
| chr2 | 120564124 | 120564597 | 474 | 3 | 8.15E-11 | 3.40E-10 | 3.24E-06 | 3.87E-10 | -0.01477068 | NA |
| chr15 | 59208771 | 59209209 | 439 | 3 | 6.90E-13 | 1.35E-06 | 6.51E-08 | 3.88E-10 | 0.00348427 | MYO1E |
| chr5 | 77086556 | 77087959 | 1404 | 15 | 4.96E-09 | 1.93E-07 | 0.000496 | 3.88E-10 | 0.003178246 | AC008581.2, ZBED3-AS1, AC022414.1, ZBED3 |
| chr17 | 79977908 | 79979305 | 1398 | 11 | 3.50E-12 | 3.19E-08 | 0.000175 | 3.92E-10 | -0.00373677 | TBC1D16 |
| chr1 | 35560593 | 35561010 | 418 | 3 | 1.95E-13 | 3.32E-06 | 1.36E-08 | 3.94E-10 | -0.00602422 | NCDN |
| chr22 | 42364641 | 42365598 | 958 | 3 | 1.62E-09 | 1.13E-10 | 5.67E-05 | 3.94E-10 | -0.02876365 | LINC01315 |
| chr11 | 506354 | 507970 | 1617 | 18 | 8.82E-12 | 3.48E-06 | 8.38E-05 | 3.94E-10 | -0.00115123 | RNH1 |
| chr11 | 16737975 | 16739986 | 2012 | 21 | 5.92E-07 | 8.18E-09 | 0.00173 | 3.95E-10 | 0.001570747 | C11orf58, SOX6 |
| chr1 | 34976133 | 34977895 | 1763 | 9 | 1.52E-09 | 2.71E-08 | 6.01E-06 | 3.95E-10 | -0.00221491 | AC114490.3, AC114490.1 |
| chr2 | 197786156 | 197787625 | 1470 | 13 | 7.41E-11 | 7.66E-10 | 6.65E-05 | 3.96E-10 | 0.005365998 | BOLL |
| chr19 | 4889308 | 4891949 | 2642 | 13 | 7.78E-08 | 0.0001331 | 6.17E-06 | 3.96E-10 | -0.00342008 | ARRDC5 |
| chr9 | 34457442 | 34459062 | 1621 | 12 | 7.18E-08 | 5.14E-09 | 0.000178 | 3.97E-10 | 0.000438664 | DNAI1, FAM219A |
| chr7 | 4208236 | 4210548 | 2313 | 6 | 3.75E-07 | 2.73E-10 | 0.000173 | 3.98E-10 | -0.0069985 | SDK1 |
| chr7 | 55157050 | 55158516 | 1467 | 10 | 1.11E-14 | 5.08E-06 | 1.63E-06 | 3.99E-10 | -0.00456605 | EGFR |
| chr19 | 17309230 | 17310121 | 892 | 11 | 3.12E-11 | 1.79E-06 | 3.15E-05 | 3.99E-10 | -0.00013235 | DDA1, ABHD8 |
| chr1 | 211325620 | 211327568 | 1949 | 16 | 1.85E-09 | 8.13E-10 | 0.000568 | 4.00E-10 | 0.00161351 | TRAF5 |
| chr6 | 144007284 | 144009208 | 1925 | 22 | 6.73E-12 | 1.09E-08 | 0.000151 | 4.04E-10 | -0.00231567 | AL109755.2, PLAGL1, HYMAI |
| chr1 | 24864287 | 24865340 | 1054 | 3 | 7.29E-11 | 1.01E-09 | 3.48E-06 | 4.06E-10 | -0.01552936 | NA |
| chr3 | 184372842 | 184374552 | 1711 | 7 | 2.33E-09 | 1.07E-09 | 3.77E-05 | 4.07E-10 | -0.00159968 | THPO |
| chr12 | 57219187 | 57220399 | 1213 | 8 | 1.44E-08 | 3.75E-09 | 0.000145 | 4.08E-10 | -0.01188117 | NXPH4 |
| chr22 | 45284548 | 45285818 | 1271 | 10 | 1.22E-11 | 6.61E-08 | 5.63E-06 | 4.08E-10 | -0.00400074 | UPK3A |
| chr9 | 132480529 | 132481417 | 889 | 2 | 4.12E-15 | 3.86E-07 | 4.85E-09 | 4.08E-10 | -0.00661389 | CFAP77 |
| chr15 | 64386956 | 64388096 | 1141 | 14 | 3.21E-09 | 2.00E-10 | 0.001629 | 4.08E-10 | -0.00085534 | TRIP4, PCLAF |
| chr1 | 230377531 | 230378224 | 694 | 6 | 4.84E-15 | 0.0006488 | 7.72E-10 | 4.14E-10 | 0.001686223 | PGBD5 |
| chr2 | 3194125 | 3194435 | 311 | 4 | 1.76E-10 | 1.93E-10 | 0.000106 | 4.15E-10 | 0.003162736 | EIPR1 |
| chr16 | 3256524 | 3257814 | 1291 | 6 | 2.41E-09 | 2.64E-10 | 0.000392 | 4.16E-10 | -0.01127152 | MEFV |
| chr20 | 32456252 | 32457523 | 1272 | 4 | 1.13E-09 | 3.35E-09 | 1.66E-05 | 4.18E-10 | -0.01421567 | NOL4L |
| chr6 | 168066879 | 168068236 | 1358 | 8 | 1.86E-13 | 6.26E-06 | 3.40E-07 | 4.18E-10 | 0.001333847 | FRMD1 |
| chr19 | 7473198 | 7473537 | 340 | 3 | 1.08E-11 | 3.71E-07 | 2.45E-06 | 4.18E-10 | -0.04803948 | NA |
| chr1 | 116908921 | 116910888 | 1968 | 16 | 7.98E-11 | 1.38E-07 | 1.73E-05 | 4.19E-10 | -0.0027987 | PTGFRN, AL157904.1 |
| chr17 | 51164910 | 51167713 | 2804 | 26 | 1.89E-07 | 3.20E-09 | 0.000559 | 4.20E-10 | -0.00104003 | NME1-NME2, NME2 |
| chr11 | 33672320 | 33672605 | 286 | 2 | 5.00E-12 | 7.70E-09 | 2.80E-08 | 4.22E-10 | -0.01350356 | KIAA1549L, AL049629.1 |
| chr2 | 119431336 | 119432721 | 1386 | 7 | 2.34E-07 | 1.60E-10 | 0.000636 | 4.22E-10 | -0.00783829 | TMEM37 |
| chr17 | 42832262 | 42833904 | 1643 | 18 | 4.23E-09 | 8.16E-11 | 0.006229 | 4.22E-10 | 0.002533059 | PSME3, BECN1 |
| chr6 | 33454125 | 33457098 | 2974 | 51 | 3.17E-07 | 1.25E-06 | 0.000616 | 4.22E-10 | 0.000139057 | ZBTB9, SYNGAP1-AS1 |
| chr3 | 37174726 | 37177586 | 2861 | 14 | 1.86E-06 | 3.39E-07 | 0.0011 | 4.24E-10 | 0.002947365 | LRRFIP2 |
| chr10 | 67852743 | 67852851 | 109 | 2 | 3.95E-12 | 1.42E-08 | 1.83E-08 | 4.24E-10 | 0.050431399 | NA |
| chr20 | 1735624 | 1735986 | 363 | 4 | 1.20E-10 | 5.77E-10 | 2.14E-05 | 4.30E-10 | -0.01016199 | AL109809.4 |
| chr17 | 80068780 | 80069258 | 479 | 3 | 1.17E-12 | 2.85E-09 | 4.88E-07 | 4.31E-10 | 0.008497929 | CCDC40 |
| chr6 | 167985961 | 167987708 | 1748 | 10 | 7.61E-07 | 8.28E-09 | 3.40E-05 | 4.32E-10 | -0.00071462 | NA |
| chr5 | 159094539 | 159095891 | 1353 | 6 | 1.20E-11 | 9.20E-08 | 1.13E-05 | 4.32E-10 | -0.00044437 | EBF1 |
| chr16 | 62595613 | 62596368 | 756 | 5 | 1.23E-10 | 2.84E-07 | 4.75E-06 | 4.34E-10 | -0.0161394 | NA |
| chr19 | 50968108 | 50970567 | 2460 | 11 | 3.37E-07 | 1.48E-08 | 2.45E-05 | 4.37E-10 | -0.00633225 | AC011483.1, KLK6 |
| chr11 | 67451770 | 67453333 | 1564 | 11 | 1.05E-10 | 1.07E-07 | 0.000212 | 4.37E-10 | -0.00408155 | CABP4, GPR152 |
| chr4 | 41747192 | 41748989 | 1798 | 15 | 8.08E-14 | 4.27E-07 | 6.97E-05 | 4.39E-10 | -0.00429921 | PHOX2B-AS1, PHOX2B |
| chr1 | 112512440 | 112513768 | 1329 | 5 | 2.70E-10 | 1.68E-08 | 3.77E-07 | 4.40E-10 | -0.00569673 | WNT2B |
| chr3 | 142595651 | 142597240 | 1590 | 14 | 2.37E-08 | 5.91E-08 | 0.002871 | 4.40E-10 | 0.010443597 | PLS1 |
| chr17 | 76725669 | 76728129 | 2461 | 23 | 1.58E-07 | 2.45E-08 | 0.00059 | 4.43E-10 | -0.00150109 | METTL23, JMJD6 |
| chr11 | 47578499 | 47579630 | 1132 | 18 | 4.31E-11 | 5.67E-07 | 0.00024 | 4.44E-10 | -0.00056484 | NDUFS3, KBTBD4 |
| chr1 | 154404615 | 154406195 | 1581 | 14 | 1.71E-10 | 1.34E-07 | 0.000101 | 4.44E-10 | 0.001488247 | IL6R, IL6R-AS1 |
| chr17 | 79397847 | 79399864 | 2018 | 8 | 7.54E-07 | 1.38E-08 | 2.39E-05 | 4.44E-10 | 0.002409416 | RBFOX3 |
| chr17 | 41059642 | 41060435 | 794 | 7 | 4.75E-10 | 4.81E-09 | 5.43E-05 | 4.45E-10 | -0.0119928 | KRTAP2-3 |
| chr1 | 54152982 | 54154030 | 1049 | 10 | 8.10E-10 | 2.47E-09 | 0.00021 | 4.45E-10 | -0.00714333 | AL357673.1, CDCP2 |
| chr11 | 47425812 | 47426983 | 1172 | 14 | 1.12E-09 | 5.28E-06 | 5.12E-05 | 4.46E-10 | -0.00236972 | PSMC3 |
| chr2 | 98577090 | 98578291 | 1202 | 6 | 1.27E-09 | 1.60E-07 | 2.83E-06 | 4.46E-10 | -0.00016851 | INPP4A |
| chr3 | 42732404 | 42733567 | 1164 | 5 | 2.43E-09 | 5.13E-09 | 1.62E-05 | 4.47E-10 | -0.0088615 | CCDC13-AS1, AC006059.3, CCDC13, AC006059.2 |
| chr5 | 141099131 | 141101186 | 2056 | 15 | 1.97E-07 | 1.69E-10 | 0.003157 | 4.51E-10 | -0.0085487 | PCDHB3, AC244517.2 |
| chr19 | 1580401 | 1581002 | 602 | 5 | 8.80E-12 | 1.34E-06 | 1.53E-05 | 4.51E-10 | -0.00608998 | MBD3, AC005943.1 |
| chr10 | 100622124 | 100623065 | 942 | 3 | 2.28E-09 | 4.47E-10 | 3.48E-06 | 4.53E-10 | -0.01908733 | NA |
| chr15 | 39933045 | 39934704 | 1660 | 14 | 1.31E-11 | 3.88E-07 | 9.41E-05 | 4.54E-10 | -0.0005563 | EIF2AK4 |
| chr1 | 230133375 | 230133813 | 439 | 2 | 2.20E-11 | 3.59E-10 | 1.40E-06 | 4.55E-10 | -0.00278033 | GALNT2 |
| chr14 | 59254455 | 59255384 | 930 | 4 | 4.80E-11 | 6.03E-10 | 3.12E-06 | 4.55E-10 | -0.01037082 | DAAM1 |
| chr3 | 9280097 | 9280230 | 134 | 2 | 2.02E-10 | 2.56E-10 | 6.79E-06 | 4.55E-10 | -0.02086036 | SRGAP3 |
| chr14 | 54489819 | 54490683 | 865 | 3 | 1.04E-17 | 6.30E-06 | 2.08E-09 | 4.60E-10 | 0.010822767 | NA |
| chr7 | 159064320 | 159065939 | 1620 | 7 | 4.48E-08 | 3.19E-07 | 5.52E-05 | 4.60E-10 | 0.003116207 | VIPR2 |
| chr15 | 41115662 | 41117092 | 1431 | 14 | 1.90E-09 | 3.01E-09 | 9.20E-05 | 4.60E-10 | 0.000910237 | INO80 |
| chr6 | 130364340 | 130366250 | 1911 | 12 | 5.42E-10 | 8.04E-10 | 3.78E-05 | 4.60E-10 | -0.00541897 | TMEM200A, SAMD3 |
| chr14 | 39169827 | 39171450 | 1624 | 15 | 3.38E-10 | 1.96E-07 | 0.000114 | 4.61E-10 | -0.00048194 | TRAPPC6B |
| chr22 | 44664489 | 44665562 | 1074 | 5 | 1.04E-08 | 1.14E-09 | 3.90E-05 | 4.61E-10 | -0.00117 | NA |
| chr5 | 142685533 | 142687203 | 1671 | 13 | 2.63E-11 | 5.15E-08 | 6.89E-06 | 4.63E-10 | -0.00596027 | FGF1 |
| chr14 | 73026713 | 73027711 | 999 | 15 | 6.58E-13 | 7.57E-07 | 3.77E-05 | 4.64E-10 | 0.001022375 | ZFYVE1 |
| chr11 | 107770910 | 107771383 | 474 | 3 | 4.12E-12 | 1.32E-08 | 8.34E-08 | 4.64E-10 | 0.024741586 | NA |
| chr12 | 109020927 | 109022148 | 1222 | 9 | 6.41E-11 | 2.37E-07 | 1.45E-05 | 4.64E-10 | 0.008769985 | SVOP, AC190387.1 |
| chr16 | 2235251 | 2236936 | 1686 | 18 | 5.97E-14 | 6.76E-07 | 6.38E-07 | 4.65E-10 | -0.0036249 | E4F1, DNASE1L2, AC009065.1 |
| chr19 | 56367667 | 56368625 | 959 | 14 | 6.30E-11 | 1.02E-08 | 0.000889 | 4.65E-10 | -0.00172237 | ZNF542P, ZSCAN5A, AC006116.8 |

| | | | | | | | | | | |
|---|---|---|---|---|---|---|---|---|---|---|
| chr11 | 116788124 | 116790962 | 2839 | 14 | 1.68E-08 | 6.76E-08 | 7.35E-05 | 4.65E-10 | -0.00278288 | APOA5 |
| chr6 | 36386455 | 36388385 | 1931 | 18 | 4.73E-08 | 1.01E-09 | 0.000671 | 4.66E-10 | 0.001942228 | ETV7-AS1, ETV7 |
| chr1 | 151327306 | 151328428 | 1123 | 16 | 2.70E-13 | 3.26E-07 | 0.000154 | 4.66E-10 | -0.00339345 | AL391069.2, PI4KB |
| chr22 | 37135898 | 37137697 | 1800 | 7 | 8.50E-09 | 1.69E-09 | 0.000161 | 4.66E-10 | -0.00728947 | IL2RB |
| chr17 | 43378775 | 43379201 | 427 | 5 | 3.83E-10 | 1.00E-09 | 7.41E-05 | 4.67E-10 | 5.16E-05 | LINC00910 |
| chr17 | 78278059 | 78279052 | 994 | 5 | 1.49E-09 | 4.84E-11 | 0.000643 | 4.71E-10 | -0.01855829 | LINC01993 |
| chr6 | 30982889 | 30984398 | 1510 | 12 | 4.80E-10 | 3.39E-07 | 4.69E-06 | 4.75E-10 | -0.00234874 | MUC21 |
| chr1 | 16212518 | 16212846 | 329 | 4 | 3.46E-10 | 2.00E-10 | 3.26E-05 | 4.79E-10 | -0.01273554 | ARHGEF19 |
| chr14 | 100439401 | 100440352 | 952 | 3 | 3.69E-09 | 1.53E-10 | 4.89E-05 | 4.79E-10 | -0.0207304 | WDR25 |
| chr4 | 128683346 | 128684088 | 743 | 2 | 4.85E-10 | 4.15E-10 | 1.11E-06 | 4.80E-10 | -0.02658853 | NA |
| chr17 | 41918403 | 41919682 | 1280 | 14 | 5.83E-11 | 7.22E-09 | 0.000547 | 4.81E-10 | -0.00225893 | ACLY |
| chr8 | 143648021 | 143648392 | 372 | 4 | 8.61E-11 | 2.09E-10 | 3.64E-05 | 4.81E-10 | -0.00687135 | ZNF623 |
| chr11 | 61566913 | 61568128 | 1216 | 7 | 3.87E-10 | 2.57E-08 | 4.19E-05 | 4.82E-10 | 0.008940511 | SYT7 |
| chr10 | 96227509 | 96227805 | 297 | 4 | 9.40E-11 | 4.22E-09 | 1.42E-05 | 4.89E-10 | -0.00335551 | BLNK |
| chr20 | 63688908 | 63690364 | 1457 | 7 | 8.41E-10 | 6.95E-09 | 1.15E-05 | 4.91E-10 | -0.00083873 | RTEL1, RTEL1-TNFRSF6B |
| chr16 | 30673716 | 30674631 | 916 | 3 | 6.92E-14 | 1.15E-07 | 4.36E-06 | 4.94E-10 | -0.00761695 | NA |
| chr2 | 11784799 | 11787123 | 2325 | 12 | 9.28E-07 | 2.00E-09 | 3.50E-05 | 4.94E-10 | -0.00247022 | LPIN1 |
| chr11 | 3056692 | 3057809 | 1118 | 10 | 1.52E-12 | 2.09E-08 | 5.19E-06 | 4.96E-10 | 0.005679511 | AC108448.3, CARS1 |
| chr17 | 1478265 | 1479645 | 1381 | 8 | 8.80E-09 | 1.07E-09 | 7.81E-05 | 4.97E-10 | -0.00354389 | MYO1C |
| chr7 | 138773123 | 138775017 | 1895 | 7 | 1.33E-11 | 1.00E-08 | 1.41E-05 | 4.98E-10 | -0.00660609 | ATP6V0A4 |
| chr20 | 37527523 | 37529362 | 1840 | 48 | 3.92E-08 | 3.66E-09 | 0.008887 | 4.99E-10 | -0.00040838 | BLCAP |
| chr3 | 100491738 | 100492778 | 1041 | 10 | 7.44E-12 | 2.55E-08 | 3.28E-05 | 4.99E-10 | -0.00587069 | TMEM45A |
| chr6 | 44257365 | 44258783 | 1419 | 18 | 8.98E-11 | 1.70E-08 | 0.000357 | 5.02E-10 | -0.00322087 | SLC35B2, NFKBIE |
| chr20 | 10432776 | 10435810 | 3035 | 22 | 3.40E-06 | 1.03E-06 | 0.000375 | 5.04E-10 | -0.0022579 | SLX4IP, MKKS |
| chr1 | 3657943 | 3659110 | 1168 | 5 | 1.06E-09 | 2.14E-10 | 8.49E-05 | 5.07E-10 | -0.02152733 | TP73, TP73-AS3 |
| chr9 | 127950104 | 127952602 | 2499 | 12 | 1.42E-07 | 6.66E-09 | 1.81E-05 | 5.12E-10 | -0.00311277 | FAM102A |
| chr7 | 881455 | 883879 | 2425 | 12 | 3.81E-07 | 6.36E-10 | 0.000331 | 5.15E-10 | -0.00266007 | SUN1, GET4, AC073957.3 |
| chr17 | 40019136 | 40020432 | 1297 | 4 | 4.73E-12 | 2.67E-08 | 3.43E-06 | 5.16E-10 | -0.01070077 | MED24 |
| chr14 | 91835026 | 91836919 | 1894 | 10 | 3.79E-08 | 1.09E-08 | 0.000483 | 5.18E-10 | 0.003069364 | TC2N |
| chr9 | 123401804 | 123403285 | 1482 | 8 | 3.42E-10 | 4.35E-08 | 6.47E-05 | 5.24E-10 | -0.00643604 | DENND1A, MIR601 |
| chr11 | 59793058 | 59794278 | 1221 | 5 | 1.73E-10 | 2.42E-09 | 3.11E-05 | 5.24E-10 | -0.00457776 | STX3 |
| chr4 | 7643807 | 7645132 | 1326 | 3 | 1.87E-08 | 1.05E-09 | 5.80E-06 | 5.27E-10 | 0.007786497 | SORCS2 |
| chr5 | 119268047 | 119269138 | 1092 | 15 | 2.96E-11 | 8.47E-07 | 0.000516 | 5.27E-10 | -0.00207823 | TNFAIP8 |
| chr12 | 109131254 | 109132002 | 749 | 7 | 1.29E-10 | 8.63E-10 | 4.92E-05 | 5.28E-10 | -0.01548357 | ACACB |
| chr2 | 263991 | 266641 | 2651 | 24 | 5.06E-07 | 1.01E-08 | 0.002693 | 5.31E-10 | -6.32E-05 | ACP1, SH3YL1 |
| chr22 | 35410337 | 35411239 | 903 | 8 | 9.09E-10 | 1.02E-10 | 0.000538 | 5.31E-10 | 0.005817904 | MCM5 |
| chr16 | 85901950 | 85903890 | 1941 | 11 | 7.30E-08 | 2.61E-09 | 3.03E-05 | 5.34E-10 | -0.00180126 | IRF8 |
| chr1 | 87211541 | 87212729 | 1189 | 6 | 5.00E-12 | 8.33E-07 | 1.55E-07 | 5.35E-10 | -0.00991016 | LINC02801 |
| chr13 | 29023310 | 29024571 | 1262 | 6 | 9.00E-15 | 2.33E-05 | 1.32E-09 | 5.36E-10 | -0.00210064 | MTUS2 |
| chr7 | 116511867 | 116512186 | 320 | 4 | 2.73E-10 | 2.92E-10 | 1.53E-05 | 5.36E-10 | -0.01476864 | NA |
| chr11 | 73268749 | 73269954 | 1206 | 6 | 2.87E-10 | 4.38E-09 | 3.43E-06 | 5.36E-10 | -0.00455985 | P2RY6 |
| chr8 | 122780095 | 122782604 | 2510 | 19 | 1.11E-08 | 6.56E-09 | 2.10E-05 | 5.38E-10 | 0.000746432 | AC016405.2, ZHX2, AC016405.3 |
| chr2 | 181790286 | 181791364 | 1079 | 3 | 5.24E-11 | 1.89E-08 | 4.75E-06 | 5.39E-10 | 0.021134393 | NA |
| chr13 | 113454685 | 113456592 | 1908 | 9 | 1.06E-08 | 1.01E-08 | 0.000122 | 5.47E-10 | -0.0004984 | DCUN1D2 |
| chr17 | 41571969 | 41572710 | 742 | 8 | 3.87E-11 | 1.96E-08 | 4.79E-07 | 5.50E-10 | -0.01006086 | KRT9 |
| chr16 | 56452351 | 56452424 | 74 | 3 | 2.90E-12 | 1.51E-10 | 7.94E-05 | 5.51E-10 | -0.01880885 | OGFOD1 |
| chr14 | 76267984 | 76268262 | 279 | 5 | 1.01E-09 | 7.04E-11 | 0.000271 | 5.52E-10 | 0.036511589 | NA |
| chr9 | 133657079 | 133657625 | 547 | 5 | 3.48E-12 | 2.42E-06 | 1.21E-06 | 5.54E-10 | -0.01053861 | DBH, DBH-AS1 |
| chr6 | 30060473 | 30062233 | 1761 | 40 | 1.55E-09 | 6.00E-09 | 0.006942 | 5.57E-10 | -9.96E-05 | POLR1H, AL669914.4, ZNRD1ASP |
| chr5 | 77972091 | 77973518 | 1428 | 9 | 1.13E-08 | 1.68E-08 | 6.17E-06 | 5.57E-10 | 0.000227395 | NA |
| chr14 | 67619374 | 67620622 | 1249 | 14 | 1.50E-11 | 1.53E-06 | 0.000205 | 5.58E-10 | -0.00197286 | ARG2 |
| chr12 | 94185889 | 94187603 | 1715 | 8 | 8.01E-07 | 1.13E-10 | 0.000118 | 5.59E-10 | 0.000865739 | PLXNC1, AC123567.2 |
| chr11 | 119316681 | 119317954 | 1274 | 13 | 7.18E-11 | 2.87E-07 | 1.12E-06 | 5.60E-10 | 0.000464526 | MCAM |
| chr19 | 39195295 | 39196928 | 1634 | 7 | 6.61E-08 | 9.10E-08 | 0.000102 | 5.61E-10 | -0.00993899 | NA |
| chr6 | 30259097 | 30260654 | 1558 | 21 | 1.93E-10 | 3.61E-06 | 0.000114 | 5.62E-10 | -0.0029457 | HLA-L, HCG17 |
| chr1 | 109764187 | 109765290 | 1104 | 6 | 1.94E-10 | 4.20E-09 | 2.95E-06 | 5.62E-10 | -0.00484431 | GSTM5, AL158847.1 |
| chr8 | 33566158 | 33567302 | 1145 | 8 | 4.26E-09 | 1.93E-10 | 6.93E-05 | 5.62E-10 | 0.000954435 | RNF122 |
| chr1 | 18105906 | 18107300 | 1395 | 7 | 5.78E-11 | 4.85E-09 | 9.41E-05 | 5.65E-10 | -0.00198773 | NA |
| chr11 | 1832685 | 1835077 | 2393 | 17 | 2.62E-06 | 7.69E-11 | 0.002786 | 5.66E-10 | -0.00445819 | SYT8 |
| chr14 | 100964719 | 100966977 | 2259 | 7 | 1.22E-09 | 7.48E-08 | 1.63E-06 | 5.66E-10 | -0.00918842 | MEG8, SNORD114-9 |
| chr4 | 53376771 | 53378873 | 2103 | 17 | 2.31E-07 | 1.70E-08 | 0.000883 | 5.66E-10 | -0.0031709 | FIP1L1, AC058822.1 |
| chr5 | 36151072 | 36152485 | 1414 | 19 | 2.34E-12 | 1.86E-06 | 1.24E-05 | 5.67E-10 | 0.000860074 | SKP2, LMBRD2 |
| chr12 | 56266542 | 56267927 | 1386 | 19 | 3.83E-10 | 2.31E-09 | 0.000331 | 5.71E-10 | 0.000172415 | COQ10A, AC073896.5 |
| chr16 | 654976 | 656648 | 1673 | 11 | 6.22E-10 | 1.45E-05 | 4.34E-06 | 5.75E-10 | -0.00279884 | WDR90, AL022341.2 |
| chr10 | 27743028 | 27745318 | 2291 | 17 | 4.84E-08 | 1.16E-08 | 0.000895 | 5.75E-10 | -0.00028046 | MKX-AS1, MKX |
| chr19 | 52269052 | 52269782 | 731 | 10 | 2.41E-11 | 3.09E-08 | 2.59E-05 | 5.76E-10 | -0.00354618 | ZNF766 |
| chr2 | 23527165 | 23528680 | 1516 | 4 | 3.30E-09 | 2.39E-09 | 1.66E-05 | 5.78E-10 | -0.00211768 | KLHL29 |
| chr10 | 48944654 | 48945554 | 901 | 2 | 4.26E-12 | 5.46E-09 | 5.24E-08 | 5.78E-10 | -0.00952444 | WDFY4 |
| chr2 | 231463735 | 231465893 | 2159 | 17 | 3.97E-10 | 1.03E-05 | 8.40E-05 | 5.79E-10 | 0.000719031 | NCL |
| chr18 | 48085176 | 48086091 | 916 | 3 | 2.74E-09 | 1.38E-09 | 1.60E-06 | 5.79E-10 | -0.00999588 | ZBTB7C |
| chr4 | 17584178 | 17586161 | 1984 | 6 | 7.24E-08 | 7.03E-08 | 9.11E-06 | 5.79E-10 | 0.001306831 | LAP3 |
| chr7 | 100893313 | 100896295 | 2983 | 20 | 3.27E-06 | 2.57E-08 | 0.004097 | 5.79E-10 | -0.00178672 | ACHE |
| chr2 | 63048043 | 63051076 | 3034 | 46 | 4.70E-07 | 1.70E-08 | 0.002571 | 5.81E-10 | -0.00094136 | OTX1, AC007098.1 |
| chr8 | 47179801 | 47180548 | 748 | 5 | 1.20E-12 | 2.70E-06 | 3.72E-08 | 5.84E-10 | -0.01210954 | AC120036.1 |
| chr17 | 28505566 | 28506741 | 1176 | 5 | 5.91E-11 | 2.09E-08 | 5.55E-07 | 5.84E-10 | -0.00404172 | FOXN1, AC005726.1 |
| chr5 | 110737572 | 110739477 | 1906 | 19 | 4.64E-11 | 9.23E-08 | 0.000147 | 5.85E-10 | -0.00040982 | SLC25A46, TMEM232 |
| chr20 | 13092164 | 13092451 | 288 | 2 | 8.41E-11 | 3.40E-10 | 5.70E-06 | 5.85E-10 | 0.026440908 | SPTLC3 |
| chr16 | 56108277 | 56109465 | 1189 | 4 | 2.23E-08 | 1.64E-10 | 6.75E-05 | 5.85E-10 | -0.0105599 | AC040168.2, AC040168.1 |
| chr6 | 33203988 | 33206391 | 2404 | 30 | 3.56E-09 | 9.50E-08 | 4.55E-05 | 5.88E-10 | -0.00163555 | SLC39A7, HSD17B8 |
| chr17 | 42760297 | 42761684 | 1388 | 13 | 2.53E-09 | 6.64E-08 | 0.001327 | 5.89E-10 | -0.00250657 | RAMP2, RAMP2-AS1 |
| chr16 | 1492732 | 1494647 | 1916 | 17 | 4.60E-08 | 1.02E-07 | 8.49E-05 | 5.89E-10 | -0.00170208 | TELO2 |
| chr16 | 11463768 | 11465516 | 1749 | 6 | 7.79E-07 | 1.68E-10 | 0.000343 | 5.89E-10 | -0.00378838 | AC099489.3, AC099489.1 |
| chr14 | 101559749 | 101561460 | 1712 | 23 | 5.20E-09 | 5.80E-07 | 0.000662 | 5.91E-10 | -0.00091536 | DIO3, DIO3OS, MIR1247 |
| chr8 | 143741908 | 143743677 | 1770 | 8 | 9.43E-09 | 6.69E-09 | 0.000598 | 5.96E-10 | -0.00799442 | IQANK1 |
| chr12 | 76877675 | 76879689 | 2015 | 16 | 1.59E-08 | 3.32E-10 | 0.001105 | 5.97E-10 | -0.00100212 | AC124784.1, CSRP2 |
| chr5 | 126463756 | 126465086 | 1331 | 6 | 2.21E-09 | 4.35E-08 | 1.11E-05 | 5.98E-10 | -0.00233057 | GRAMD2B, AC093535.1 |
| chr14 | 77270391 | 77271737 | 1347 | 11 | 9.34E-09 | 1.41E-09 | 0.000231 | 6.00E-10 | -0.00715746 | NGB |
| chr21 | 36059741 | 36061248 | 1508 | 18 | 1.07E-11 | 4.46E-08 | 3.77E-06 | 6.01E-10 | 0.000310405 | SETD4-AS1, SETD4 |
| chr2 | 202871143 | 202872472 | 1330 | 13 | 6.73E-11 | 6.72E-08 | 2.19E-05 | 6.07E-10 | -0.00145718 | ICA1L |
| chr21 | 31248828 | 31249762 | 935 | 2 | 1.19E-10 | 1.71E-09 | 1.64E-07 | 6.07E-10 | -0.00562235 | TIAM1 |
| chr7 | 1065685 | 1066397 | 713 | 6 | 2.73E-14 | 2.14E-06 | 3.36E-08 | 6.13E-10 | 0.007505986 | C7orf50 |
| chr11 | 13667704 | 13668641 | 938 | 10 | 1.89E-10 | 4.03E-08 | 0.00038 | 6.13E-10 | -0.00485953 | NA |
| chr11 | 12044122 | 12044941 | 820 | 4 | 3.51E-09 | 7.45E-09 | 1.38E-05 | 6.13E-10 | -0.00845301 | LINC02547 |
| chr1 | 7960672 | 7961902 | 1231 | 13 | 3.38E-10 | 9.63E-09 | 0.000184 | 6.20E-10 | -0.00042579 | PARK7 |
| chr5 | 66054012 | 66054922 | 911 | 5 | 1.00E-16 | 8.63E-05 | 1.72E-08 | 6.22E-10 | -0.00508204 | ERBIN |
| chr12 | 48118693 | 48119384 | 692 | 4 | 3.21E-11 | 1.52E-09 | 1.94E-05 | 6.22E-10 | 0.0069844 | PFKM |
| chr8 | 37493463 | 37495320 | 1858 | 7 | 2.44E-07 | 2.60E-10 | 0.000568 | 6.22E-10 | -0.00794266 | LINC01605 |
| chr9 | 72443980 | 72445235 | 1256 | 4 | 2.38E-09 | 4.18E-08 | 5.64E-06 | 6.22E-10 | 0.00816034 | NA |
| chr5 | 69093122 | 69094530 | 1409 | 14 | 3.59E-11 | 3.38E-08 | 0.000142 | 6.23E-10 | 0.001297209 | SLC30A5 |
| chr10 | 100654669 | 100656958 | 2290 | 16 | 1.58E-09 | 3.26E-06 | 4.84E-06 | 6.26E-10 | 0.001130417 | NA |
| chr5 | 177097385 | 177098354 | 970 | 5 | 1.91E-13 | 4.65E-08 | 1.10E-07 | 6.27E-10 | -0.00592141 | FGFR4 |
| chr2 | 23198559 | 23199593 | 1035 | 6 | 3.33E-09 | 5.77E-09 | 6.09E-05 | 6.28E-10 | 0.005205582 | AC018467.1 |
| chr3 | 42978232 | 42979907 | 1676 | 16 | 1.78E-10 | 3.18E-06 | 0.000346 | 6.29E-10 | -0.00301183 | AC092042.3, GASK1A |
| chr3 | 45795678 | 45797178 | 1501 | 14 | 9.99E-12 | 5.89E-07 | 1.62E-05 | 6.30E-10 | -0.0048775 | SLC6A20 |
| chr1 | 2529839 | 2530925 | 1087 | 11 | 8.88E-10 | 5.66E-09 | 9.80E-05 | 6.31E-10 | 0.00158482 | AL139246.5, HES5 |
| chr10 | 129569237 | 129569359 | 123 | 3 | 4.62E-10 | 3.30E-10 | 9.03E-06 | 6.38E-10 | 0.010711974 | MGMT |
| chr1 | 111448032 | 111449770 | 1739 | 20 | 1.48E-11 | 3.77E-08 | 0.00094 | 6.38E-10 | -0.00135164 | ATP5PB, WDR77 |
| chr12 | 106599768 | 106601223 | 1456 | 7 | 2.08E-08 | 3.26E-08 | 6.78E-05 | 6.38E-10 | -0.00701183 | RFX4, AC079385.1 |
| chr5 | 141121341 | 141122542 | 1202 | 16 | 9.94E-11 | 3.15E-08 | 0.000134 | 6.39E-10 | -0.00249949 | PCDHB4, AC244517.2 |
| chr11 | 73403119 | 73404388 | 1270 | 4 | 4.24E-12 | 1.68E-08 | 9.72E-07 | 6.41E-10 | -0.002197 | FAM168A |
| chr11 | 31369223 | 31370756 | 1534 | 20 | 9.12E-10 | 1.28E-09 | 0.000207 | 6.42E-10 | -0.000158 | DNAJC24, DCDC1 |
| chr3 | 52830643 | 52831549 | 907 | 6 | 6.07E-11 | 2.78E-08 | 3.54E-06 | 6.42E-10 | -0.01377949 | ITIH4, AC006254.1 |
| chr12 | 105083748 | 105084963 | 1216 | 14 | 5.05E-11 | 2.90E-07 | 0.000175 | 6.45E-10 | -0.0020106 | ALDH1L2 |
| chr17 | 78231018 | 78232545 | 1528 | 12 | 1.67E-11 | 7.51E-09 | 0.000211 | 6.46E-10 | -0.00132338 | TMEM235 |
| chr8 | 89984374 | 89985070 | 697 | 11 | 1.96E-10 | 6.98E-08 | 0.000442 | 6.53E-10 | -0.00411579 | NBN |
| chr8 | 142753121 | 142754154 | 1034 | 6 | 2.57E-10 | 1.02E-07 | 5.87E-05 | 6.53E-10 | -0.00709555 | NA |
| chr22 | 31334252 | 31335310 | 1059 | 3 | 1.71E-09 | 2.47E-10 | 3.79E-05 | 6.55E-10 | 0.047432992 | PIK3IP1-DT, PATZ1 |
| chr6 | 33250905 | 33252264 | 1360 | 12 | 2.68E-12 | 1.70E-08 | 7.33E-05 | 6.56E-10 | -0.00251508 | HCG25, VPS52 |
| chr10 | 95422139 | 95423731 | 1593 | 4 | 1.47E-08 | 7.26E-09 | 1.54E-05 | 6.59E-10 | -0.00365342 | SORBS1 |
| chr2 | 112275084 | 112276140 | 1057 | 12 | 3.54E-10 | 1.35E-09 | 0.000246 | 6.59E-10 | -0.0010307 | ZC3H6 |
| chr5 | 1200918 | 1201863 | 946 | 8 | 1.33E-10 | 5.17E-09 | 0.000148 | 6.60E-10 | 0.011030693 | SLC6A19 |
| chr6 | 135054313 | 135055779 | 1467 | 15 | 2.99E-10 | 3.19E-08 | 0.001174 | 6.60E-10 | 0.000379833 | AL353596.1, HBS1L |
| chr6 | 143536181 | 143537679 | 1499 | 9 | 5.81E-09 | 9.27E-08 | 0.000199 | 6.60E-10 | 0.003504761 | PHACTR2 |
| chr5 | 32709753 | 32712131 | 2379 | 23 | 1.21E-08 | 6.65E-08 | 0.001512 | 6.64E-10 | 0.001080141 | NPR3 |
| chr2 | 172079610 | 172081204 | 1595 | 9 | 7.02E-10 | 8.58E-07 | 2.85E-06 | 6.64E-10 | -0.00700672 | METAP1D |
| chr8 | 27992658 | 27993800 | 1143 | 11 | 4.00E-12 | 2.04E-06 | 5.78E-06 | 6.64E-10 | -0.00605102 | SCARA5 |
| chr7 | 141430399 | 141431601 | 1203 | 4 | 2.84E-09 | 2.25E-10 | 5.07E-05 | 6.66E-10 | -0.01701622 | TMEM178B, AC005692.2 |
| chr15 | 43368923 | 43371384 | 2462 | 17 | 3.85E-08 | 1.97E-09 | 0.002526 | 6.69E-10 | -0.00266724 | TUBGCP4, ZSCAN29 |
| chr18 | 57145799 | 57147072 | 1274 | 9 | 5.31E-10 | 9.29E-08 | 0.000199 | 6.70E-10 | -0.00612932 | AC100775.1 |
| chr3 | 190941839 | 190942111 | 273 | 2 | 1.71E-11 | 1.10E-08 | 3.75E-08 | 6.74E-10 | -0.01601852 | NA |
| chr10 | 92591619 | 92593574 | 1956 | 13 | 6.78E-07 | 3.42E-09 | 0.000637 | 6.77E-10 | -2.72E-05 | KIF11 |
| chr10 | 110918164 | 110919891 | 1728 | 19 | 1.05E-09 | 5.45E-09 | 0.000706 | 6.78E-10 | -0.00124138 | SHOC2, BBIP1 |
| chr7 | 151061447 | 151063035 | 1589 | 14 | 1.45E-10 | 1.59E-10 | 0.000537 | 6.78E-10 | -0.00308001 | SLC4A2, AC010973.3 |
| chr5 | 96431846 | 96433601 | 1756 | 19 | 4.38E-09 | 3.79E-07 | 0.001305 | 6.78E-10 | -0.0015177 | AC104123.1, PCSK1 |
| chr20 | 59221186 | 59222983 | 1798 | 7 | 3.38E-08 | 2.88E-09 | 2.34E-05 | 6.83E-10 | -0.00031829 | ZNF831 |
| chr11 | 130143462 | 130144161 | 700 | 9 | 5.16E-11 | 2.05E-06 | 2.48E-05 | 6.83E-10 | -0.00378016 | APLP2 |
| chr22 | 42649203 | 42649984 | 782 | 10 | 4.27E-12 | 3.69E-09 | 9.85E-05 | 6.87E-10 | -0.01413134 | CYB5R3 |
| chr18 | 58670632 | 58672429 | 1798 | 15 | 7.34E-09 | 3.39E-05 | 2.75E-05 | 6.95E-10 | 0.000864719 | MALT1, AC104971.1 |
| chr1 | 119139931 | 119141453 | 1523 | 16 | 2.49E-10 | 3.44E-10 | 0.002486 | 7.03E-10 | 0.000799649 | WARS2-AS1, WARS2 |
| chr3 | 51941355 | 51943369 | 2015 | 18 | 4.11E-10 | 2.42E-05 | 2.01E-05 | 7.03E-10 | 0.002225377 | PARP3, RRP9 |
| chr8 | 1702935 | 1703898 | 964 | 3 | 5.82E-10 | 4.11E-08 | 4.96E-07 | 7.03E-10 | -0.00258172 | DLGAP2 |
| chr7 | 94393370 | 94394942 | 1573 | 13 | 7.51E-10 | 5.45E-09 | 0.000547 | 7.08E-10 | -0.01327928 | COL1A2, AC002074.1 |
| chr3 | 40476356 | 40477254 | 899 | 7 | 3.47E-13 | 1.93E-07 | 3.29E-07 | 7.11E-10 | -0.00807336 | ZNF619, ZNF620 |
| chr1 | 27615183 | 27615675 | 493 | 4 | 3.42E-11 | 2.58E-07 | 4.22E-07 | 7.11E-10 | -0.00495129 | FGR |
| chr3 | 127722516 | 127722924 | 409 | 3 | 1.42E-10 | 6.82E-10 | 1.33E-05 | 7.13E-10 | -0.00932022 | MGLL |
| chr10 | 77821619 | 77822788 | 1170 | 5 | 1.75E-09 | 7.90E-09 | 8.24E-06 | 7.16E-10 | -0.00726059 | DLG5 |
| chr6 | 33276395 | 33277582 | 1188 | 15 | 4.95E-12 | 1.61E-07 | 0.000434 | 7.18E-10 | -0.00208993 | RPS18, B3GALT4 |
| chr19 | 18896502 | 18897245 | 744 | 8 | 5.49E-11 | 5.78E-09 | 7.63E-06 | 7.19E-10 | -0.00923007 | CERS1 |
| chr7 | 1225999 | 1227592 | 1594 | 6 | 6.79E-09 | 1.25E-09 | 0.000109 | 7.22E-10 | 0.027561263 | NA |
| chr3 | 123066711 | 123068073 | 1363 | 13 | 3.19E-08 | 4.56E-09 | 0.001868 | 7.22E-10 | -0.00165015 | PDIA5 |
| chr5 | 33891978 | 33892992 | 1015 | 11 | 1.20E-13 | 1.66E-06 | 1.17E-05 | 7.27E-10 | -0.00679343 | ADAMTS12 |
| chr19 | 9139970 | 9140847 | 878 | 13 | 8.92E-12 | 8.92E-06 | 1.02E-06 | 7.28E-10 | -0.00245313 | ZNF317 |
| chr12 | 109458690 | 109460487 | 1798 | 9 | 4.79E-08 | 4.88E-10 | 0.001502 | 7.29E-10 | -0.00162462 | KCTD10 |
| chr12 | 13043886 | 13044893 | 1008 | 12 | 6.53E-10 | 8.57E-09 | 0.002223 | 7.30E-10 | 0.006184776 | FAM234B |
| chr19 | 2307593 | 2309262 | 1670 | 12 | 6.93E-10 | 2.49E-08 | 4.09E-05 | 7.30E-10 | 0.002354288 | AC005258.1 |
| chr6 | 31863563 | 31866189 | 2627 | 40 | 2.81E-08 | 1.03E-06 | 0.000812 | 7.33E-10 | 0.000157413 | SLC44A4 |
| chr4 | 54228332 | 54230063 | 1732 | 10 | 1.66E-12 | 3.52E-06 | 1.79E-06 | 7.33E-10 | -0.00534699 | AC058822.1, PDGFRA |
| chr18 | 35939339 | 35939617 | 279 | 3 | 6.67E-11 | 6.08E-09 | 2.38E-07 | 7.33E-10 | -0.01317554 | NA |

| | | | | | | | | | | |
|---|---|---|---|---|---|---|---|---|---|---|
| chr11 | 107565267 | 107567119 | 1853 | 13 | 1.79E-10 | 4.97E-07 | 0.000141 | 7.38E-10 | -5.63E-05 | ALKBH8 |
| chr20 | 63453196 | 63454498 | 1303 | 5 | 3.46E-08 | 1.38E-10 | 0.000299 | 7.38E-10 | 0.020763008 | KCNQ2 |
| chr2 | 120847986 | 120848242 | 257 | 3 | 2.09E-12 | 2.80E-06 | 4.86E-09 | 7.39E-10 | 0.003551021 | GLI2 |
| chr7 | 29955300 | 29956387 | 1088 | 4 | 9.63E-10 | 6.17E-09 | 1.08E-06 | 7.40E-10 | -0.00366896 | SCRN1 |
| chr16 | 89315496 | 89316571 | 1076 | 7 | 9.83E-12 | 3.81E-06 | 1.41E-05 | 7.45E-10 | 0.007421732 | ANKRD11 |
| chr3 | 154120831 | 154122929 | 2099 | 22 | 8.29E-09 | 7.64E-08 | 0.000813 | 7.45E-10 | -0.0015255 | ARHGEF26, ARHGEF26-AS1 |
| chr1 | 230067294 | 230068779 | 1486 | 11 | 2.55E-10 | 9.61E-09 | 6.07E-05 | 7.46E-10 | 0.000968142 | GALNT2 |
| chr22 | 39495849 | 39496493 | 645 | 4 | 4.86E-10 | 6.21E-08 | 1.57E-05 | 7.46E-10 | -0.00884732 | NA |
| chr6 | 3848009 | 3849584 | 1576 | 29 | 5.80E-09 | 4.14E-09 | 0.012003 | 7.47E-10 | -0.00273953 | AL391422.3, AL391422.2, FAM50B |
| chr22 | 24950224 | 24950787 | 564 | 3 | 8.16E-10 | 4.04E-10 | 2.84E-05 | 7.49E-10 | -0.01254072 | NA |
| chr11 | 17476261 | 17478385 | 2125 | 15 | 4.54E-09 | 5.60E-08 | 1.52E-05 | 7.52E-10 | -0.0006347 | AC124798.3, ABCC8 |
| chr8 | 64378784 | 64381177 | 2394 | 15 | 2.19E-08 | 1.75E-05 | 6.06E-05 | 7.53E-10 | -0.00641663 | MIR124-2HG, MIR124-2 |
| chr7 | 77797497 | 77799532 | 2036 | 30 | 9.78E-10 | 3.63E-07 | 2.96E-05 | 7.53E-10 | -0.00045649 | PHTF2, TMEM60 |
| chr10 | 133082728 | 133084227 | 1500 | 8 | 2.20E-08 | 1.38E-09 | 0.000451 | 7.53E-10 | -0.00717062 | ADGRA1, ADGRA1-AS1 |
| chr10 | 96830817 | 96833080 | 2264 | 8 | 4.29E-07 | 2.22E-08 | 1.41E-05 | 7.56E-10 | -0.00529123 | LCOR |
| chr1 | 235648067 | 235649540 | 1474 | 9 | 2.74E-11 | 3.87E-08 | 0.000252 | 7.57E-10 | 0.011426688 | GNG4 |
| chr1 | 181190538 | 181192167 | 1630 | 6 | 1.95E-12 | 2.20E-08 | 1.92E-07 | 7.59E-10 | -0.0046882 | AL358393.1 |
| chr1 | 160083899 | 160085820 | 1922 | 7 | 1.06E-10 | 4.94E-08 | 1.25E-05 | 7.67E-10 | -0.00882794 | KCNJ9 |
| chr7 | 35253285 | 35254819 | 1535 | 13 | 1.24E-09 | 6.34E-08 | 0.000266 | 7.69E-10 | -0.00072027 | TBX20 |
| chr7 | 158996812 | 158997799 | 988 | 6 | 3.64E-10 | 1.00E-09 | 0.000336 | 7.70E-10 | 0.038390946 | NA |
| chr13 | 110265298 | 110266731 | 1434 | 9 | 1.23E-12 | 7.75E-07 | 7.67E-06 | 7.77E-10 | 0.000490224 | COL4A1, AL161773.1 |
| chr7 | 74252660 | 74254900 | 2241 | 15 | 4.20E-08 | 7.67E-07 | 1.91E-05 | 7.77E-10 | -0.00157195 | RFC2 |
| chr1 | 98660995 | 98662231 | 1237 | 14 | 4.12E-12 | 2.39E-06 | 5.93E-05 | 7.77E-10 | -0.00253409 | SNX7, AC095031.1 |
| chr16 | 58195619 | 58196993 | 1375 | 7 | 6.37E-10 | 2.27E-07 | 3.00E-05 | 7.80E-10 | -0.00367656 | CSNK2A2 |
| chr3 | 122183595 | 122184797 | 1203 | 14 | 1.96E-10 | 3.64E-09 | 0.000139 | 7.80E-10 | -0.00239625 | CASR |
| chr15 | 67818486 | 67819944 | 1459 | 10 | 5.37E-11 | 1.49E-07 | 1.36E-05 | 7.83E-10 | 0.000741138 | SKOR1 |
| chr18 | 5295081 | 5297826 | 2746 | 26 | 1.37E-08 | 9.22E-07 | 0.000333 | 7.83E-10 | 0.001523019 | ZBTB14 |
| chr10 | 80205682 | 80208062 | 2381 | 21 | 6.03E-07 | 1.02E-09 | 0.006797 | 7.88E-10 | -0.00774349 | LINC00857 |
| chr2 | 167292891 | 167294541 | 1651 | 9 | 3.79E-07 | 4.03E-09 | 0.000203 | 7.90E-10 | -0.00244425 | AC073050.1 |
| chr15 | 101329288 | 101330083 | 796 | 5 | 3.61E-09 | 4.45E-10 | 0.000111 | 7.96E-10 | -0.00603781 | PCSK6 |
| chr1 | 2029504 | 2030643 | 1140 | 8 | 4.09E-11 | 7.81E-08 | 2.80E-05 | 7.97E-10 | -0.00173578 | GABRD |
| chr1 | 109909511 | 109911743 | 2233 | 16 | 6.66E-09 | 1.51E-06 | 0.000135 | 7.98E-10 | -0.00088626 | CSF1 |
| chr3 | 64010930 | 64011096 | 167 | 2 | 2.47E-11 | 4.77E-10 | 5.40E-06 | 7.98E-10 | -0.01015858 | PSMD6-AS2, PSMD6 |
| chr1 | 115286058 | 115288178 | 2121 | 10 | 2.15E-10 | 5.41E-07 | 1.59E-05 | 7.98E-10 | -0.00200497 | NGF-AS1, NGF |
| chr12 | 120367347 | 120368814 | 1468 | 5 | 5.90E-09 | 3.63E-10 | 0.000113 | 8.00E-10 | 0.002102254 | MSI1 |
| chr12 | 27021373 | 27023275 | 1903 | 13 | 4.80E-11 | 1.14E-06 | 3.95E-05 | 8.03E-10 | -0.00518393 | MED21 |
| chr9 | 6719411 | 6720040 | 630 | 2 | 9.10E-10 | 5.34E-10 | 3.34E-06 | 8.06E-10 | -0.00903466 | AL354707.1 |
| chr9 | 135555040 | 135555869 | 830 | 3 | 3.29E-13 | 3.57E-06 | 1.71E-06 | 8.10E-10 | -0.01108034 | NA |
| chr8 | 144791705 | 144793219 | 1515 | 15 | 8.58E-10 | 8.71E-10 | 0.000952 | 8.11E-10 | 0.001477205 | RPL8, MIR6850 |
| chr12 | 309285 | 311009 | 1725 | 4 | 2.21E-08 | 1.77E-10 | 0.000105 | 8.14E-10 | -0.00793354 | KDM5A |
| chr1 | 116667019 | 116668477 | 1459 | 15 | 7.00E-09 | 1.36E-08 | 0.000103 | 8.14E-10 | -0.00072873 | IGSF3 |
| chr2 | 27716811 | 27717205 | 395 | 3 | 1.00E-10 | 9.27E-09 | 4.27E-06 | 8.18E-10 | -0.01731895 | NA |
| chr11 | 66475606 | 66475903 | 298 | 4 | 3.36E-11 | 5.05E-07 | 5.10E-07 | 8.20E-10 | -0.00637261 | PELI3, AP002748.4 |
| chr1 | 26020597 | 26021993 | 1397 | 5 | 9.27E-11 | 2.75E-07 | 2.46E-06 | 8.20E-10 | -0.00470025 | EXTL1 |
| chr21 | 45867103 | 45868807 | 1705 | 7 | 4.77E-07 | 3.96E-07 | 1.79E-05 | 8.21E-10 | -0.00111277 | PCBP3, AJ011931.2 |
| chr20 | 25226611 | 25227807 | 1197 | 4 | 6.45E-13 | 3.79E-06 | 1.79E-06 | 8.25E-10 | -0.02699411 | ENTPD6, AL035252.4 |
| chr8 | 144318456 | 144320326 | 1871 | 6 | 2.72E-09 | 6.53E-08 | 6.07E-05 | 8.25E-10 | -0.00799803 | DGAT1 |
| chr1 | 8373638 | 8375035 | 1398 | 4 | 6.20E-09 | 3.54E-10 | 9.37E-05 | 8.25E-10 | -0.00747667 | RERE |
| chr11 | 32434523 | 32436332 | 1810 | 18 | 1.73E-08 | 1.10E-06 | 9.22E-05 | 8.25E-10 | 0.000638045 | WT1-AS, AL049692.6, AL049692.1, WT1 |
| chr13 | 26760325 | 26761405 | 1081 | 12 | 1.03E-11 | 3.25E-09 | 0.000174 | 8.25E-10 | -0.00166802 | GPR12 |
| chr20 | 63940738 | 63942433 | 1696 | 9 | 1.35E-10 | 1.81E-08 | 0.00029 | 8.25E-10 | 0.01291938 | UCKL1, MIR1914 |
| chr14 | 91364391 | 91365392 | 1002 | 6 | 8.38E-12 | 4.13E-07 | 6.73E-07 | 8.27E-10 | 0.00253973 | CCDC88C |
| chr14 | 97977814 | 97978670 | 857 | 6 | 3.34E-11 | 1.24E-07 | 7.46E-05 | 8.32E-10 | 0.015146701 | LINC01550 |
| chr14 | 64740480 | 64742099 | 1620 | 6 | 3.10E-09 | 1.61E-07 | 6.13E-06 | 8.32E-10 | -0.00417242 | PLEKHG3 |
| chr5 | 81971128 | 81972547 | 1420 | 9 | 2.39E-10 | 1.89E-08 | 0.000326 | 8.36E-10 | -0.00115774 | ATG10 |
| chr15 | 75578913 | 75579933 | 1021 | 15 | 1.27E-09 | 1.15E-07 | 0.000659 | 8.37E-10 | -0.00250227 | PTPN9, AC105036.3 |
| chr14 | 103225024 | 103225767 | 744 | 10 | 2.73E-11 | 1.22E-07 | 0.000103 | 8.38E-10 | -0.00578179 | NA |
| chr8 | 96260559 | 96262338 | 1780 | 13 | 1.96E-15 | 2.47E-06 | 9.89E-07 | 8.41E-10 | -0.00060434 | PTDSS1, MTERF3 |
| chr1 | 39453742 | 39455035 | 1294 | 5 | 1.17E-12 | 8.04E-06 | 9.43E-07 | 8.42E-10 | -0.00048117 | MACF1 |
| chr15 | 78565506 | 78566247 | 742 | 5 | 6.32E-14 | 1.89E-07 | 8.30E-07 | 8.42E-10 | -0.00237491 | CHRNA5 |
| chr13 | 30306690 | 30308136 | 1447 | 16 | 3.09E-08 | 4.84E-09 | 0.003469 | 8.43E-10 | -0.00225217 | KATNAL1 |
| chr5 | 127517209 | 127518262 | 1054 | 16 | 1.41E-10 | 1.66E-09 | 0.000866 | 8.52E-10 | 0.002424058 | PRRC1 |
| chr1 | 35506231 | 35506928 | 698 | 3 | 1.71E-11 | 7.66E-08 | 1.19E-06 | 8.56E-10 | -0.01231425 | KIAA0319L |
| chr8 | 143695739 | 143697206 | 1468 | 9 | 5.79E-11 | 6.03E-08 | 0.000162 | 8.63E-10 | -0.00171911 | ZNF707, LINC02878 |
| chr19 | 41998579 | 41999260 | 682 | 5 | 3.84E-10 | 2.49E-10 | 0.00028 | 8.69E-10 | -0.01557977 | GRIK5 |
| chr5 | 100902282 | 100904519 | 2238 | 17 | 5.70E-08 | 3.70E-08 | 9.57E-05 | 8.70E-10 | 0.000451732 | ST8SIA4 |
| chr7 | 155456324 | 155457604 | 1281 | 13 | 2.77E-12 | 8.50E-08 | 1.11E-06 | 8.73E-10 | -0.00053253 | AC008060.3 |
| chr19 | 58395670 | 58397083 | 1414 | 10 | 2.08E-10 | 2.33E-06 | 7.13E-07 | 8.79E-10 | -0.00013635 | RNF225 |
| chr15 | 73442056 | 73443353 | 1298 | 15 | 1.75E-08 | 7.03E-08 | 0.00027 | 8.79E-10 | -0.00254971 | REC114 |
| chr4 | 39697696 | 39698689 | 994 | 10 | 2.54E-14 | 2.29E-08 | 8.28E-07 | 8.81E-10 | 0.001652076 | UBE2K |
| chr1 | 165543161 | 165544479 | 1319 | 11 | 6.53E-12 | 6.21E-07 | 9.01E-07 | 8.81E-10 | -0.00431747 | LRRC52, LRRC52-AS1 |
| chr6 | 43260911 | 43263325 | 2415 | 9 | 6.91E-08 | 5.13E-07 | 3.31E-05 | 8.81E-10 | -0.0038794 | TTBK1 |
| chr14 | 103102029 | 103103060 | 1032 | 5 | 2.78E-11 | 3.28E-07 | 2.50E-05 | 8.81E-10 | -0.01731973 | EXOC3L4 |
| chr16 | 86993716 | 86995321 | 1606 | 6 | 5.00E-09 | 1.86E-08 | 4.11E-06 | 8.84E-10 | -0.00864907 | NA |
| chr13 | 26742359 | 26743244 | 886 | 4 | 8.86E-11 | 4.67E-09 | 5.20E-05 | 8.93E-10 | -0.04923719 | NA |
| chr12 | 131917926 | 131919804 | 1879 | 9 | 2.85E-09 | 1.09E-07 | 3.27E-05 | 8.93E-10 | -0.00126358 | ULK1 |
| chr18 | 3602088 | 3604909 | 2822 | 12 | 2.37E-07 | 1.11E-06 | 0.000392 | 8.96E-10 | 0.001529233 | DLGAP1-AS2, DLGAP1 |
| chr4 | 52017201 | 52018265 | 1065 | 4 | 5.55E-13 | 1.43E-07 | 1.91E-07 | 8.99E-10 | -0.01154651 | LRRC66 |
| chr1 | 43649516 | 43651422 | 1907 | 15 | 1.81E-07 | 3.95E-08 | 0.001023 | 8.99E-10 | 0.001925701 | KDM4A, AL451062.2 |
| chr10 | 26696868 | 26698191 | 1324 | 7 | 1.16E-10 | 1.16E-07 | 4.53E-06 | 8.99E-10 | -0.00356021 | PDSS1, SELENOOLP, AL390961.3 |
| chr13 | 112756035 | 112757663 | 1629 | 7 | 2.96E-08 | 8.36E-09 | 0.000442 | 9.00E-10 | -0.00444803 | ATP11A |
| chr16 | 28924345 | 28926155 | 1811 | 14 | 3.55E-11 | 3.98E-08 | 2.66E-06 | 9.05E-10 | 0.000964042 | AC009093.11, RABEP2 |
| chr1 | 246689115 | 246690312 | 1198 | 5 | 7.02E-12 | 4.52E-07 | 1.83E-06 | 9.08E-10 | -0.01345225 | AL591848.1 |
| chr19 | 12064292 | 12065207 | 916 | 11 | 6.70E-11 | 2.94E-07 | 0.00053 | 9.08E-10 | -0.00223444 | AC008770.4, ZNF844 |
| chr22 | 17476634 | 17477599 | 966 | 9 | 4.01E-11 | 5.26E-09 | 8.74E-05 | 9.09E-10 | 0.010087288 | CECR2 |
| chr15 | 39918728 | 39920324 | 1597 | 10 | 1.01E-08 | 7.07E-08 | 1.24E-05 | 9.12E-10 | -0.00550438 | GPR176 |
| chr8 | 1200340 | 1201175 | 836 | 5 | 2.25E-13 | 9.75E-06 | 1.62E-08 | 9.14E-10 | -0.00674688 | DLGAP2 |
| chr12 | 40105460 | 40107696 | 2237 | 12 | 4.77E-07 | 2.09E-09 | 0.000916 | 9.19E-10 | -0.00069059 | SLC2A13 |
| chr16 | 77431794 | 77433298 | 1505 | 5 | 1.78E-16 | 1.56E-06 | 1.02E-07 | 9.19E-10 | 0.007875577 | ADAMTS18 |
| chr9 | 19408002 | 19408798 | 797 | 7 | 7.90E-12 | 3.16E-07 | 4.43E-06 | 9.21E-10 | -0.00833591 | AL391834.3 |
| chr19 | 45381642 | 45382820 | 1179 | 5 | 2.67E-09 | 1.51E-05 | 4.08E-06 | 9.21E-10 | 0.002005698 | PPP1R13L |
| chr12 | 56338006 | 56339268 | 1263 | 6 | 8.34E-09 | 3.01E-08 | 0.000108 | 9.21E-10 | 0.004116106 | IL23A |
| chr22 | 19823860 | 19823966 | 107 | 2 | 2.04E-11 | 3.79E-07 | 9.03E-09 | 9.23E-10 | 0.017312505 | GNB1L |
| chr7 | 151080784 | 151084141 | 3358 | 37 | 7.96E-06 | 7.06E-07 | 0.006202 | 9.23E-10 | -0.00035493 | FASTK, TMUB1 |
| chr10 | 23343600 | 23344589 | 990 | 10 | 2.81E-11 | 3.60E-08 | 0.000412 | 9.23E-10 | -0.00776901 | AL606469.1, C10orf67 |
| chr2 | 102185962 | 102187525 | 1564 | 13 | 1.12E-08 | 2.70E-10 | 0.000678 | 9.24E-10 | -0.00775951 | IL1RL2 |
| chr15 | 34977820 | 34978206 | 387 | 3 | 1.13E-11 | 4.76E-08 | 6.24E-08 | 9.26E-10 | -0.00835838 | NA |
| chr5 | 16740948 | 16742245 | 1298 | 6 | 3.71E-11 | 3.04E-08 | 2.47E-06 | 9.29E-10 | -0.00828169 | MYO10 |
| chr22 | 30821243 | 30823177 | 1935 | 14 | 2.50E-10 | 8.51E-09 | 0.000106 | 9.30E-10 | 0.000831748 | OSBP2 |
| chr16 | 70579429 | 70580041 | 613 | 6 | 7.47E-10 | 8.87E-09 | 4.33E-05 | 9.30E-10 | -0.00700767 | IL34 |
| chr7 | 44113397 | 44114268 | 872 | 6 | 3.69E-12 | 3.06E-07 | 3.85E-07 | 9.32E-10 | -0.00174518 | AEBP1 |
| chr14 | 76538128 | 76538530 | 403 | 3 | 2.61E-11 | 2.56E-07 | 5.45E-06 | 9.32E-10 | 0.004778098 | AC008050.1 |
| chr11 | 73664759 | 73666046 | 1288 | 4 | 4.22E-10 | 1.33E-08 | 1.35E-06 | 9.35E-10 | -0.00505746 | NA |
| chr3 | 64224763 | 64226318 | 1556 | 10 | 7.10E-10 | 1.04E-09 | 2.59E-05 | 9.47E-10 | 0.00724641 | PRICKLE2 |
| chr8 | 143212045 | 143213264 | 1220 | 7 | 3.71E-08 | 6.81E-10 | 0.000207 | 9.48E-10 | -0.00378311 | GPIHBP1 |
| chr12 | 49923060 | 49924883 | 1824 | 11 | 7.29E-09 | 4.97E-06 | 3.99E-06 | 9.50E-10 | -0.00192805 | LINC02395 |
| chr12 | 49044262 | 49046705 | 2444 | 13 | 2.49E-07 | 1.29E-06 | 5.03E-06 | 9.52E-10 | 0.00021162 | KMT2D |
| chr3 | 122659717 | 122660839 | 1123 | 4 | 4.06E-10 | 5.51E-10 | 3.96E-05 | 9.60E-10 | 0.00343767 | EIF4BP8 |
| chr9 | 34620094 | 34621671 | 1578 | 12 | 4.56E-14 | 9.30E-06 | 1.58E-07 | 9.62E-10 | -0.00476714 | DCTN3, ARID3C |
| chr17 | 81238214 | 81239881 | 1668 | 14 | 6.72E-12 | 1.51E-06 | 9.46E-05 | 9.64E-10 | 0.000762135 | NDUFAF8, TEPSIN |
| chr8 | 67063639 | 67065212 | 1574 | 18 | 4.09E-10 | 6.99E-08 | 8.00E-06 | 9.64E-10 | -0.00036274 | CSPP1, COPS5 |
| chr5 | 149344463 | 149346031 | 1569 | 15 | 1.28E-09 | 6.68E-07 | 7.75E-05 | 9.64E-10 | -0.00099185 | GRPEL2, AC131025.3 |
| chr1 | 209703492 | 209705044 | 1553 | 11 | 3.67E-09 | 2.30E-05 | 5.07E-06 | 9.64E-10 | -0.00125541 | HSD11B1, HSD11B1-AS1 |
| chr19 | 35747404 | 35748982 | 1579 | 13 | 4.88E-09 | 7.45E-11 | 0.000796 | 9.64E-10 | 0.000316235 | PSENEN, AD000671.1, LIN37, AD000671.3 |
| chr9 | 92654059 | 92656580 | 2522 | 5 | 9.82E-07 | 8.17E-10 | 8.11E-05 | 9.66E-10 | -0.002239 | IPPK |
| chr1 | 24648844 | 24649795 | 952 | 2 | 1.75E-08 | 5.69E-10 | 7.09E-06 | 9.68E-10 | 0.006882824 | SRRM1 |
| chr17 | 28523304 | 28525514 | 2211 | 10 | 1.24E-11 | 2.69E-06 | 7.36E-06 | 9.69E-10 | -0.00178793 | FOXN1, AC005726.1 |
| chr2 | 112583637 | 112585161 | 1525 | 16 | 2.46E-10 | 3.19E-10 | 0.001675 | 9.70E-10 | -0.00310595 | CHCHD5 |
| chr17 | 44628393 | 44628716 | 324 | 2 | 1.79E-10 | 1.07E-09 | 8.87E-07 | 9.72E-10 | -0.04555058 | NA |
| chr4 | 165326375 | 165328087 | 1713 | 16 | 4.26E-10 | 6.49E-08 | 0.000376 | 9.75E-10 | 4.98E-07 | MSMO1 |
| chr1 | 9128183 | 9129615 | 1433 | 12 | 7.07E-11 | 3.36E-07 | 3.14E-05 | 9.79E-10 | -0.00066573 | GPR157 |
| chr21 | 41303365 | 41304855 | 1491 | 9 | 9.79E-10 | 1.14E-06 | 9.28E-06 | 9.84E-10 | -0.00525156 | FAM3B |
| chr12 | 56103289 | 56105976 | 2688 | 14 | 1.67E-08 | 5.25E-08 | 1.35E-05 | 9.90E-10 | -0.0008243 | ERBB3, AC034102.2, PA2G4, AC034102.4 |
| chr16 | 71489241 | 71490339 | 1099 | 12 | 2.53E-09 | 3.42E-10 | 0.003303 | 9.91E-10 | -0.00752644 | AC010547.4, ZNF19 |
| chr12 | 130204555 | 130204775 | 221 | 2 | 2.96E-10 | 8.07E-10 | 1.88E-06 | 9.92E-10 | -0.04871198 | NA |
| chr2 | 239274382 | 239275435 | 1054 | 8 | 9.89E-11 | 2.10E-09 | 1.30E-05 | 9.93E-10 | -0.00802485 | HDAC4 |
| chr7 | 1218813 | 1219735 | 923 | 3 | 5.86E-10 | 1.25E-09 | 2.27E-06 | 9.93E-10 | -0.01553275 | NA |
| chr8 | 95129665 | 95130443 | 779 | 2 | 1.22E-08 | 5.80E-10 | 7.84E-06 | 9.98E-10 | 0.008152485 | NA |
| chr4 | 1960361 | 1961115 | 755 | 6 | 9.07E-10 | 7.10E-10 | 0.000248 | 1.00E-09 | -0.00383832 | NSD2 |
| chr17 | 21283477 | 21285156 | 1680 | 18 | 2.70E-09 | 2.05E-07 | 0.000314 | 1.00E-09 | -0.0036855 | MAP2K3 |
| chr10 | 14604456 | 14605754 | 1299 | 6 | 7.86E-12 | 3.97E-08 | 3.27E-05 | 1.00E-09 | -0.01148411 | FAM107B |
| chr16 | 18800472 | 18802197 | 1726 | 19 | 2.19E-08 | 1.67E-07 | 0.000511 | 1.01E-09 | 0.001311856 | AC138811.2, ARL6IP1 |
| chr3 | 69052071 | 69052978 | 908 | 12 | 1.93E-12 | 1.19E-05 | 1.40E-05 | 1.01E-09 | -0.0039301 | AC109587.1, TMF1 |
| chr11 | 57640369 | 57641278 | 910 | 4 | 1.98E-12 | 9.93E-07 | 4.43E-07 | 1.01E-09 | 0.010486312 | AP000662.1, MIR130A |
| chr8 | 16914647 | 16915515 | 869 | 4 | 6.41E-12 | 4.04E-09 | 1.46E-06 | 1.01E-09 | 0.002258236 | AC011586.2 |
| chr2 | 96289363 | 96290504 | 1142 | 3 | 4.97E-11 | 7.02E-08 | 2.01E-07 | 1.01E-09 | 0.001926956 | SNRNP200 |
| chr16 | 25031486 | 25032697 | 1212 | 6 | 3.55E-09 | 1.30E-06 | 2.86E-05 | 1.01E-09 | -0.00437589 | AC133552.2 |
| chr17 | 48718498 | 48720936 | 2439 | 11 | 3.75E-07 | 1.02E-07 | 7.46E-05 | 1.01E-09 | -0.00538757 | NA |
| chr11 | 102955361 | 102956627 | 1267 | 11 | 2.30E-11 | 6.25E-08 | 6.54E-05 | 1.02E-09 | -0.0062569 | MMP13 |
| chr8 | 142247211 | 142249141 | 1931 | 5 | 6.30E-07 | 5.37E-09 | 4.68E-05 | 1.02E-09 | -0.00685308 | TSNARE1, RN7SL260P |
| chr6 | 28673845 | 28674617 | 773 | 12 | 8.43E-12 | 3.93E-06 | 4.96E-07 | 1.02E-09 | 0.001709209 | NA |
| chr2 | 159711049 | 159712646 | 1598 | 12 | 1.94E-08 | 4.60E-08 | 3.09E-05 | 1.03E-09 | 0.001530672 | MARCHF7, AC009961.1 |
| chr6 | 169659318 | 169660832 | 1515 | 5 | 7.00E-08 | 6.95E-09 | 4.91E-05 | 1.03E-09 | -0.00100398 | AL031315.1, WDR27 |
| chr7 | 27863872 | 27865027 | 1156 | 7 | 6.94E-09 | 9.95E-09 | 0.000131 | 1.03E-09 | -0.00329137 | JAZF1 |
| chr6 | 33409335 | 33411273 | 1939 | 23 | 7.52E-10 | 3.34E-08 | 0.001221 | 1.03E-09 | 0.000562223 | KIFC1, PHF1 |
| chr7 | 100427275 | 100429848 | 2574 | 20 | 3.99E-07 | 3.71E-08 | 0.000135 | 1.03E-09 | -0.00130228 | MEPCE, ZCWPW1 |
| chr12 | 92224946 | 92225148 | 203 | 2 | 1.08E-11 | 6.21E-07 | 8.57E-09 | 1.04E-09 | -0.00620639 | AC025164.1 |
| chr12 | 43758309 | 43759414 | 1106 | 12 | 2.37E-09 | 1.82E-10 | 0.000561 | 1.05E-09 | 0.002478017 | IRAK4, PUS7L |
| chr19 | 8584960 | 8586202 | 1243 | 8 | 1.14E-09 | 2.58E-07 | 1.38E-05 | 1.06E-09 | -0.00312291 | ADAMTS10 |
| chr1 | 182400586 | 182401879 | 1294 | 6 | 2.07E-09 | 9.97E-09 | 9.04E-05 | 1.06E-09 | -0.00370432 | TEDDM1 |
| chr11 | 67390973 | 67392299 | 1327 | 13 | 1.37E-13 | 1.28E-06 | 3.95E-05 | 1.07E-09 | -0.0005022 | RAD9A |
| chr11 | 20600652 | 20601713 | 1062 | 8 | 1.05E-10 | 6.41E-09 | 9.06E-05 | 1.07E-09 | 0.001391761 | SLC6A5 |
| chr6 | 10414395 | 10416854 | 2460 | 24 | 1.71E-09 | 2.54E-08 | 6.69E-05 | 1.07E-09 | -0.00127099 | TFAP2A-AS1, TFAP2A |
| chr6 | 143539963 | 143540046 | 84 | 2 | 4.10E-10 | 1.03E-09 | 1.23E-06 | 1.07E-09 | 0.014786731 | PHACTR2 |
| chr8 | 101764104 | 101764980 | 877 | 2 | 6.40E-11 | 4.88E-09 | 1.32E-07 | 1.07E-09 | -0.0069157 | NCALD |
| chr16 | 30377465 | 30380067 | 2603 | 15 | 8.42E-07 | 4.25E-06 | 0.00017 | 1.07E-09 | -0.00011196 | MYLPF, ZNF48, SEPTIN1, AC116348.4 |
| chr2 | 70705149 | 70705590 | 442 | 2 | 5.65E-13 | 5.25E-09 | 1.25E-07 | 1.08E-09 | -0.01293702 | ADD2 |
| chr15 | 88894897 | 88896583 | 1687 | 12 | 2.58E-09 | 4.69E-07 | 1.49E-05 | 1.08E-09 | -0.00145694 | HAPLN3 |

| | | | | | | | | | | |
|---|---|---|---|---|---|---|---|---|---|---|
| chr13 | 27373213 | 27374914 | 1702 | 6 | 5.55E-07 | 1.42E-08 | 7.16E-05 | 1.08E-09 | -0.01549497 | LINC01079 |
| chr22 | 24100593 | 24101667 | 1075 | 6 | 6.80E-10 | 1.58E-08 | 3.62E-06 | 1.09E-09 | -0.00120656 | CABIN1 |
| chr10 | 8046348 | 8048199 | 1852 | 5 | 7.06E-09 | 1.94E-10 | 0.000267 | 1.09E-09 | 0.016956649 | GATA3 |
| chr11 | 60783802 | 60784793 | 992 | 5 | 2.12E-13 | 3.44E-07 | 4.26E-07 | 1.09E-09 | -0.0027456 | NA |
| chr8 | 141787724 | 141788646 | 923 | 7 | 1.57E-08 | 1.05E-09 | 0.000697 | 1.09E-09 | -0.00544978 | NA |
| chr5 | 140362698 | 140363718 | 1021 | 3 | 5.74E-09 | 2.98E-10 | 9.78E-05 | 1.09E-09 | -0.01906443 | SLC4A9 |
| chr8 | 106271273 | 106271994 | 722 | 3 | 1.14E-09 | 3.42E-10 | 5.54E-05 | 1.09E-09 | 0.001755912 | AC027031.2 |
| chr1 | 33471832 | 33473303 | 1472 | 12 | 7.90E-12 | 3.01E-07 | 1.04E-06 | 1.10E-09 | -0.0012752 | ZSCAN20 |
| chr7 | 151075797 | 151077542 | 1746 | 10 | 3.72E-10 | 8.13E-08 | 0.000286 | 1.10E-09 | -0.00153054 | SLC4A2, AC010973.2, FASTK |
| chr7 | 140026203 | 140026284 | 82 | 2 | 6.82E-11 | 2.57E-09 | 2.85E-07 | 1.10E-09 | -0.00224323 | PARP12 |
| chr12 | 132352275 | 132354826 | 2552 | 10 | 3.12E-06 | 1.64E-06 | 0.000114 | 1.11E-09 | 0.0070372 | AC148477.8 |
| chr16 | 50065206 | 50066904 | 1699 | 10 | 9.56E-08 | 1.29E-07 | 1.53E-05 | 1.12E-09 | -0.00305472 | HEATR3, AC007610.1 |
| chr9 | 110337773 | 110338453 | 681 | 2 | 5.20E-10 | 1.06E-09 | 1.30E-06 | 1.12E-09 | 0.001762949 | TXNDC8 |
| chr12 | 123584621 | 123585952 | 1332 | 11 | 5.24E-12 | 1.16E-08 | 0.000181 | 1.12E-09 | -0.00475111 | TMED2, TMED2-DT |
| chr10 | 62814901 | 62816754 | 1854 | 15 | 1.57E-11 | 2.19E-07 | 4.06E-06 | 1.12E-09 | 0.000942614 | EGR2 |
| chr16 | 19065899 | 19068124 | 2226 | 18 | 2.09E-10 | 1.37E-05 | 1.03E-06 | 1.12E-09 | 0.000849379 | AC099518.2, COQ7, AC099518.4 |
| chr14 | 59630406 | 59632416 | 2011 | 16 | 2.91E-07 | 1.29E-06 | 0.000794 | 1.12E-09 | 0.000719624 | RTN1 |
| chr10 | 70052171 | 70054156 | 1986 | 19 | 1.43E-08 | 3.04E-08 | 0.001634 | 1.12E-09 | -0.00032415 | MACROH2A2 |
| chr16 | 1344642 | 1345841 | 1200 | 5 | 3.55E-11 | 5.52E-08 | 6.10E-06 | 1.12E-09 | -0.00992312 | BAIAP3 |
| chr7 | 2631344 | 2633355 | 2012 | 13 | 8.05E-10 | 4.03E-08 | 0.000208 | 1.12E-09 | 0.000318408 | TTYH3 |
| chr1 | 64743785 | 64744737 | 953 | 6 | 5.37E-12 | 2.43E-06 | 5.18E-06 | 1.13E-09 | -0.00864216 | NA |
| chr1 | 12616755 | 12618755 | 2001 | 18 | 1.34E-09 | 4.76E-05 | 1.97E-05 | 1.13E-09 | 0.001768645 | DHRS3 |
| chr5 | 72307761 | 72308848 | 1088 | 6 | 9.97E-09 | 7.44E-11 | 0.001779 | 1.13E-09 | -0.00442339 | MRPS27 |
| chr12 | 40691484 | 40693718 | 2235 | 13 | 6.05E-07 | 4.39E-11 | 0.003951 | 1.13E-09 | -0.00328454 | CNTN1 |
| chr16 | 712760 | 713037 | 278 | 2 | 1.25E-11 | 4.60E-09 | 1.51E-07 | 1.13E-09 | 0.011286953 | NA |
| chr19 | 7915773 | 7916820 | 1048 | 11 | 4.62E-09 | 5.70E-08 | 9.22E-05 | 1.14E-09 | 0.003062717 | TGFBR3L |
| chr19 | 45601310 | 45602246 | 937 | 6 | 1.55E-17 | 1.00E-05 | 2.18E-07 | 1.14E-09 | -0.00525059 | OPA3, GPR4 |
| chr14 | 91460292 | 91460304 | 13 | 3 | 4.79E-10 | 1.47E-09 | 1.15E-05 | 1.14E-09 | -0.00319759 | PPP4R3A |
| chr12 | 14769625 | 14771521 | 1897 | 14 | 8.38E-10 | 3.58E-09 | 0.001221 | 1.14E-09 | -1.92E-05 | H4-16 |
| chr20 | 38164920 | 38165926 | 1007 | 13 | 1.50E-09 | 3.49E-08 | 0.000782 | 1.15E-09 | 5.69E-05 | TGM2 |
| chr9 | 120934675 | 120936687 | 2013 | 4 | 2.15E-06 | 4.45E-08 | 3.22E-05 | 1.15E-09 | -0.00629703 | NA |
| chr22 | 21866766 | 21868307 | 1542 | 14 | 2.28E-10 | 1.67E-08 | 4.45E-05 | 1.15E-09 | -0.00076457 | MAPK1 |
| chr16 | 67847059 | 67848172 | 1114 | 9 | 4.39E-12 | 1.12E-06 | 3.65E-05 | 1.16E-09 | -0.00091172 | NUTF2, CENPT |
| chr21 | 43928751 | 43929968 | 1218 | 6 | 1.68E-10 | 1.37E-09 | 1.02E-05 | 1.16E-09 | 0.002287327 | AGPAT3 |
| chr11 | 18455606 | 18456588 | 983 | 7 | 1.56E-09 | 1.82E-10 | 0.001551 | 1.17E-09 | -0.02790332 | LDHAL6A |
| chr6 | 30018983 | 30020445 | 1463 | 6 | 5.40E-08 | 6.56E-09 | 6.81E-05 | 1.17E-09 | -4.81E-05 | ZNRD1ASP |
| chr2 | 10550824 | 10552194 | 1371 | 5 | 3.75E-08 | 9.37E-09 | 4.82E-05 | 1.17E-09 | -0.00291844 | NA |
| chr16 | 2084078 | 2085838 | 1761 | 13 | 3.89E-07 | 2.03E-08 | 0.000268 | 1.17E-09 | -0.00088443 | TSC2 |
| chr1 | 93591690 | 93593082 | 1393 | 8 | 1.18E-08 | 4.78E-11 | 0.003336 | 1.17E-09 | 0.015817493 | BCAR3-AS1, BCAR3 |
| chr15 | 79058010 | 79058417 | 408 | 2 | 3.25E-11 | 5.45E-09 | 1.36E-07 | 1.17E-09 | -0.01753481 | RASGRF1 |
| chr2 | 46465 | 47889 | 1425 | 15 | 1.76E-09 | 3.91E-09 | 0.000125 | 1.18E-09 | 0.005118612 | FAM110C |
| chr15 | 44287945 | 44289777 | 1833 | 17 | 3.50E-08 | 6.92E-07 | 0.000254 | 1.18E-09 | -0.00197738 | GOLM2 |
| chr11 | 10750466 | 10751743 | 1278 | 12 | 4.86E-09 | 8.00E-09 | 0.000883 | 1.19E-09 | -0.00204978 | CTR9 |
| chr5 | 32089248 | 32090767 | 1520 | 7 | 4.03E-08 | 3.28E-08 | 7.91E-05 | 1.19E-09 | -0.00306505 | PDZD2 |
| chr10 | 100561979 | 100563712 | 1734 | 7 | 2.78E-10 | 1.41E-09 | 3.05E-05 | 1.19E-09 | -0.01446669 | NA |
| chr16 | 1070876 | 1072688 | 1813 | 9 | 1.35E-08 | 2.71E-08 | 5.95E-05 | 1.19E-09 | -0.01251244 | SSTR5-AS1 |
| chr2 | 234454611 | 234454975 | 365 | 4 | 1.67E-09 | 2.32E-10 | 0.000278 | 1.20E-09 | -0.04616631 | AC097713.1 |
| chr15 | 39247836 | 39248383 | 548 | 3 | 3.27E-10 | 1.79E-09 | 1.85E-06 | 1.20E-09 | -0.01213847 | AC013652.1 |
| chr11 | 70655498 | 70656572 | 1075 | 4 | 7.51E-12 | 4.44E-08 | 1.24E-07 | 1.20E-09 | -0.01693621 | SHANK2 |
| chr1 | 214383399 | 214384812 | 1414 | 11 | 4.69E-10 | 2.51E-07 | 0.000242 | 1.20E-09 | -0.0021152 | PTPN14 |
| chr5 | 55159349 | 55160228 | 880 | 8 | 4.23E-09 | 1.66E-10 | 0.000678 | 1.20E-09 | -0.00683004 | GPX8, CDC20B |
| chr19 | 39934509 | 39935740 | 1232 | 7 | 5.96E-08 | 5.99E-08 | 6.28E-05 | 1.20E-09 | -0.0006827 | FCGBP |
| chr20 | 46342463 | 46342695 | 233 | 2 | 1.28E-10 | 4.45E-09 | 1.72E-07 | 1.21E-09 | -0.01648372 | NA |
| chr19 | 18017619 | 18017957 | 339 | 3 | 1.29E-10 | 9.36E-09 | 4.60E-07 | 1.21E-09 | -0.00246022 | NA |
| chr12 | 121038223 | 121040332 | 2110 | 13 | 3.72E-08 | 7.77E-07 | 3.91E-06 | 1.21E-09 | -0.00111416 | OASL |
| chr1 | 226127182 | 226128156 | 975 | 4 | 2.80E-08 | 4.17E-10 | 8.61E-05 | 1.22E-09 | -0.01020069 | AL512343.1 |
| chr2 | 51032037 | 51033601 | 1565 | 17 | 2.57E-09 | 6.96E-08 | 0.001744 | 1.22E-09 | 0.002146977 | AC007402.1, NRXN1 |
| chr19 | 57477492 | 57478606 | 1115 | 13 | 6.55E-12 | 1.10E-06 | 2.23E-07 | 1.23E-09 | -0.00033784 | AC003005.2, AC004076.1, ZNF772 |
| chr1 | 151765265 | 151767203 | 1939 | 7 | 4.66E-09 | 1.99E-08 | 2.75E-05 | 1.23E-09 | -0.0027095 | OAZ3, AL589765.2, AL589765.4, AL589765.5 |
| chr9 | 34646240 | 34646686 | 447 | 7 | 1.33E-09 | 4.57E-10 | 0.000133 | 1.23E-09 | -0.00505967 | GALT, AL162231.3 |
| chr3 | 195931 | 197353 | 1423 | 19 | 3.37E-12 | 6.73E-06 | 6.02E-06 | 1.23E-09 | 0.003348302 | CHL1, CHL1-AS2 |
| chr2 | 208124524 | 208125398 | 875 | 9 | 4.48E-11 | 2.09E-08 | 1.03E-05 | 1.23E-09 | -0.005869 | CRYGD |
| chr1 | 116570996 | 116572685 | 1690 | 8 | 6.22E-12 | 4.31E-08 | 6.75E-06 | 1.23E-09 | -0.00188349 | CD58 |
| chr1 | 85307876 | 85308380 | 505 | 2 | 4.38E-11 | 3.39E-09 | 2.45E-07 | 1.24E-09 | 0.00379842 | AL078459.1 |
| chr2 | 222301716 | 222304008 | 2293 | 13 | 7.73E-08 | 2.70E-08 | 0.000121 | 1.24E-09 | -0.00398864 | CCDC140 |
| chr4 | 165461509 | 165461962 | 454 | 2 | 1.28E-09 | 7.08E-10 | 1.04E-05 | 1.24E-09 | -0.01538027 | CPE |
| chr11 | 119417664 | 119418280 | 617 | 4 | 7.38E-12 | 2.06E-07 | 7.74E-08 | 1.25E-09 | -4.56E-05 | USP2-AS1, THY1-AS1, THY1 |
| chr14 | 24440788 | 24443191 | 2404 | 22 | 2.89E-10 | 7.01E-08 | 0.000928 | 1.25E-09 | -0.00066819 | KHNYN, AL132800.1, SDR39U1 |
| chr22 | 45309384 | 45311395 | 2012 | 13 | 6.07E-07 | 5.87E-10 | 0.004062 | 1.25E-09 | 0.000224512 | FAM118A |
| chr2 | 29748163 | 29749093 | 931 | 2 | 7.59E-11 | 2.48E-09 | 3.77E-07 | 1.26E-09 | -0.0179951 | ALK |
| chr1 | 3213015 | 3213986 | 972 | 6 | 3.94E-10 | 2.76E-08 | 2.64E-05 | 1.26E-09 | -0.00323125 | PRDM16 |
| chr11 | 2992079 | 2993707 | 1629 | 24 | 1.30E-11 | 7.72E-06 | 7.66E-05 | 1.27E-09 | -4.24E-05 | NAP1L4 |
| chr11 | 7677418 | 7678099 | 682 | 3 | 5.60E-10 | 2.70E-09 | 1.79E-06 | 1.27E-09 | -0.01327507 | NA |
| chr4 | 55110329 | 55111404 | 1076 | 5 | 2.43E-09 | 2.61E-08 | 2.43E-05 | 1.27E-09 | -0.00913087 | KDR |
| chr16 | 30394093 | 30395968 | 1876 | 20 | 1.12E-12 | 2.17E-06 | 2.20E-06 | 1.28E-09 | -0.00089064 | ZNF48, SEPTIN1 |
| chr6 | 149933306 | 149934622 | 1317 | 5 | 3.13E-09 | 4.56E-09 | 1.63E-05 | 1.28E-09 | -0.00057319 | AL583835.2 |
| chr1 | 184407526 | 184408391 | 866 | 5 | 3.44E-13 | 7.24E-07 | 6.56E-08 | 1.28E-09 | 0.009018634 | C1orf21, AL078645.1 |
| chr8 | 80265144 | 80266159 | 1016 | 3 | 3.14E-08 | 1.88E-09 | 1.06E-05 | 1.28E-09 | -0.00868554 | AC034114.2, AC104212.3 |
| chr13 | 52450021 | 52452103 | 2083 | 13 | 2.48E-08 | 3.03E-08 | 0.000173 | 1.29E-09 | -0.00221641 | VPS36 |
| chr14 | 69766882 | 69769071 | 2190 | 19 | 1.38E-07 | 7.07E-09 | 0.002184 | 1.29E-09 | -0.00086714 | SRSF5 |
| chr15 | 26096105 | 26096415 | 311 | 2 | 1.54E-10 | 2.50E-09 | 3.93E-07 | 1.30E-09 | -0.02454452 | NA |
| chr1 | 59997670 | 59998231 | 562 | 2 | 1.53E-09 | 7.92E-10 | 6.57E-06 | 1.30E-09 | -0.01301777 | C1orf87 |
| chr10 | 122560275 | 122561289 | 1015 | 5 | 4.50E-11 | 1.48E-06 | 5.82E-07 | 1.30E-09 | -0.00599696 | DMBT1 |
| chr6 | 42177801 | 42178405 | 605 | 6 | 1.94E-10 | 5.43E-07 | 3.23E-06 | 1.31E-09 | -0.00992174 | AL096814.2, GUCA1A |
| chr6 | 107689589 | 107690240 | 652 | 2 | 1.16E-14 | 6.01E-07 | 1.05E-08 | 1.31E-09 | -0.01012141 | NA |
| chr7 | 23245795 | 23247941 | 2147 | 11 | 2.03E-06 | 9.20E-09 | 0.000315 | 1.31E-09 | -0.00125554 | GPNMB |
| chr8 | 101051560 | 101052010 | 451 | 6 | 1.34E-09 | 4.34E-10 | 0.000374 | 1.31E-09 | 0.01121828 | FLJ42969 |
| chr17 | 81118152 | 81119081 | 930 | 4 | 3.97E-11 | 1.72E-08 | 6.84E-06 | 1.32E-09 | 0.00510249 | AATK |
| chr14 | 23468998 | 23469948 | 951 | 12 | 4.38E-11 | 5.56E-08 | 8.03E-05 | 1.32E-09 | -0.00184441 | NGDN |
| chr17 | 81308374 | 81310115 | 1742 | 14 | 3.60E-09 | 6.39E-07 | 0.000849 | 1.33E-09 | -0.00960838 | LINC00482 |
| chr11 | 20597466 | 20599368 | 1903 | 13 | 6.14E-12 | 1.23E-06 | 3.75E-06 | 1.33E-09 | 0.002812707 | NA |
| chr1 | 51616266 | 51617622 | 1357 | 11 | 2.98E-09 | 8.62E-10 | 0.000485 | 1.33E-09 | -0.00107117 | OSBPL9 |
| chr4 | 10018736 | 10019731 | 996 | 5 | 1.12E-09 | 2.39E-10 | 0.000366 | 1.34E-09 | -0.02210166 | SLC2A9 |
| chr1 | 231186569 | 231189052 | 2484 | 15 | 4.03E-08 | 1.05E-07 | 0.000126 | 1.34E-09 | -0.00723341 | TRIM67, TRIM67-AS1 |
| chr1 | 50959604 | 50960742 | 1139 | 18 | 6.42E-10 | 4.20E-08 | 8.13E-05 | 1.34E-09 | -0.00023607 | FAF1 |
| chr10 | 34110027 | 34110916 | 890 | 7 | 3.04E-09 | 6.71E-09 | 8.75E-05 | 1.35E-09 | 0.002327165 | PARD3 |
| chr2 | 17877836 | 17880059 | 2224 | 15 | 1.22E-10 | 3.70E-07 | 7.26E-05 | 1.35E-09 | -0.00051432 | KCNS3 |
| chr19 | 15507655 | 15509203 | 1549 | 10 | 1.09E-09 | 8.47E-08 | 0.000131 | 1.35E-09 | -0.00740135 | CYP4F22 |
| chr19 | 1356151 | 1357565 | 1415 | 10 | 8.99E-09 | 7.45E-09 | 4.40E-05 | 1.36E-09 | -0.00022876 | PWWP3A |
| chr5 | 172837361 | 172837978 | 618 | 4 | 6.76E-10 | 2.48E-08 | 1.60E-05 | 1.37E-09 | 0.003845563 | ERGIC1 |
| chr6 | 42890462 | 42891285 | 824 | 14 | 4.40E-11 | 1.03E-07 | 5.16E-05 | 1.37E-09 | -0.00325346 | C6orf226 |
| chr3 | 52404252 | 52407064 | 2813 | 12 | 1.71E-09 | 1.20E-06 | 0.000101 | 1.37E-09 | -0.00044488 | BAP1 |
| chr9 | 134219973 | 134220561 | 589 | 3 | 1.09E-10 | 8.80E-10 | 9.73E-06 | 1.37E-09 | -0.0063685 | NA |
| chr10 | 103395972 | 103396933 | 962 | 21 | 9.06E-10 | 7.56E-08 | 0.002308 | 1.37E-09 | 0.00057472 | PDCD11, ATP5MD |
| chr11 | 3165319 | 3167336 | 2018 | 30 | 3.47E-09 | 2.22E-08 | 0.000516 | 1.38E-09 | -0.00153422 | OSBPL5 |
| chr16 | 85594042 | 85594830 | 789 | 4 | 6.09E-11 | 1.41E-06 | 4.89E-07 | 1.38E-09 | 0.003373628 | GSE1, AC092127.1 |
| chr7 | 155532902 | 155534550 | 1649 | 14 | 4.49E-08 | 5.09E-08 | 2.95E-05 | 1.39E-09 | 0.003339113 | CNPY1, AC009403.2 |
| chr19 | 18271777 | 18272509 | 733 | 2 | 3.22E-09 | 9.85E-10 | 3.36E-06 | 1.39E-09 | -0.0193463 | IQCN |
| chr2 | 206253529 | 206253564 | 36 | 2 | 7.02E-10 | 9.47E-10 | 3.90E-06 | 1.39E-09 | -0.0151345 | GPR1-AS |
| chr16 | 28063063 | 28064927 | 1865 | 11 | 9.76E-11 | 7.04E-07 | 3.58E-05 | 1.39E-09 | -0.00270271 | GSG1L |
| chr1 | 111199893 | 111201486 | 1594 | 12 | 1.23E-10 | 6.57E-09 | 0.000695 | 1.39E-09 | -0.00763376 | CHI3L2, DENND2D |
| chr21 | 42467222 | 42467580 | 359 | 3 | 1.23E-09 | 4.94E-10 | 3.61E-05 | 1.40E-09 | -0.02036607 | NA |
| chr4 | 3407290 | 3408200 | 911 | 6 | 5.95E-10 | 1.43E-09 | 0.000298 | 1.40E-09 | -0.00236235 | RGS12 |
| chr11 | 32438829 | 32439666 | 838 | 5 | 3.63E-11 | 4.36E-08 | 1.49E-05 | 1.41E-09 | 0.005468794 | WT1-AS, AL049692.3 |
| chr17 | 37066105 | 37066759 | 655 | 6 | 8.94E-10 | 3.56E-10 | 0.000378 | 1.41E-09 | -0.0156122 | NA |
| chr7 | 4268187 | 4269403 | 1217 | 8 | 1.05E-10 | 6.13E-09 | 0.000117 | 1.42E-09 | 0.006380683 | SDK1 |
| chr3 | 56775609 | 56776775 | 1167 | 6 | 2.39E-13 | 1.32E-05 | 5.18E-08 | 1.42E-09 | 0.001528604 | ARHGEF3 |
| chr12 | 6229238 | 6230370 | 1133 | 4 | 3.26E-10 | 2.55E-09 | 6.57E-06 | 1.42E-09 | -0.00880464 | CD9 |
| chr16 | 86293984 | 86294180 | 197 | 3 | 8.44E-10 | 4.57E-10 | 5.21E-05 | 1.42E-09 | -0.01156462 | LINC01081 |
| chr2 | 12716296 | 12718047 | 1752 | 14 | 7.47E-09 | 3.98E-09 | 0.001587 | 1.43E-09 | 0.001570485 | TRIB2, AC009486.2 |
| chr22 | 41261332 | 41262107 | 776 | 2 | 6.00E-12 | 6.64E-09 | 1.46E-07 | 1.43E-09 | 0.002154025 | RANGAP1 |
| chr12 | 52947902 | 52948986 | 1085 | 11 | 5.25E-10 | 1.15E-07 | 0.000266 | 1.44E-09 | -0.00412591 | KRT18, KRT8 |
| chr22 | 20186460 | 20186752 | 293 | 2 | 4.87E-10 | 1.10E-09 | 2.59E-06 | 1.44E-09 | -0.00921168 | NA |
| chr16 | 87541087 | 87542049 | 963 | 6 | 1.27E-10 | 2.88E-09 | 6.48E-05 | 1.45E-09 | -0.00805393 | NA |
| chr15 | 26117260 | 26118694 | 1435 | 6 | 1.52E-07 | 3.94E-08 | 6.63E-05 | 1.45E-09 | -0.00668171 | LINC00929 |
| chr1 | 1092140 | 1093498 | 1359 | 8 | 1.51E-10 | 0.0006213 | 9.46E-08 | 1.45E-09 | -0.00193763 | C1orf159 |
| chr1 | 29123333 | 29124959 | 1627 | 14 | 3.38E-10 | 3.66E-08 | 0.000293 | 1.45E-09 | -0.00070084 | TMEM200B |
| chr17 | 1834543 | 1834661 | 119 | 3 | 4.21E-10 | 1.91E-08 | 2.62E-06 | 1.45E-09 | 0.001757791 | RPA1 |
| chr19 | 36527532 | 36529270 | 1739 | 16 | 6.19E-09 | 2.74E-06 | 5.61E-05 | 1.45E-09 | 0.00147287 | AC092295.1, ZNF260 |
| chr12 | 6688497 | 6690454 | 1958 | 18 | 9.02E-08 | 1.20E-07 | 0.001798 | 1.45E-09 | 0.00206083 | ZNF384 |
| chr1 | 16145506 | 16146984 | 1479 | 4 | 4.84E-09 | 2.01E-08 | 1.52E-05 | 1.45E-09 | -0.0152943 | EPHA2 |
| chr7 | 156397331 | 156397401 | 71 | 3 | 1.25E-10 | 1.85E-06 | 1.55E-07 | 1.46E-09 | -0.00850699 | LINC01006 |
| chr1 | 19305249 | 19305476 | 228 | 3 | 3.27E-10 | 2.44E-08 | 5.37E-07 | 1.46E-09 | 0.000429516 | AKR7A2 |
| chr3 | 12554353 | 12557551 | 3199 | 15 | 1.22E-09 | 5.30E-05 | 9.79E-05 | 1.46E-09 | -0.00210423 | MKRN2, MKRN2OS |
| chr19 | 17468530 | 17470763 | 2234 | 17 | 7.27E-07 | 2.67E-07 | 0.000129 | 1.46E-09 | -0.00306639 | SLC27A1 |
| chr1 | 202807746 | 202808490 | 745 | 5 | 2.41E-12 | 2.14E-07 | 2.08E-06 | 1.46E-09 | -0.00037205 | KDM5B |
| chr6 | 17985177 | 17985936 | 760 | 3 | 2.13E-09 | 4.93E-10 | 4.69E-05 | 1.47E-09 | 0.020167459 | KIF13A |
| chr4 | 168479694 | 168481147 | 1454 | 12 | 1.28E-09 | 3.16E-09 | 0.000173 | 1.47E-09 | 0.003897217 | DDX60L |
| chr14 | 99619800 | 99620088 | 289 | 2 | 9.23E-10 | 8.39E-10 | 1.11E-05 | 1.47E-09 | -0.01793325 | AL160313.1 |
| chr2 | 68251196 | 68252137 | 942 | 7 | 8.83E-11 | 2.77E-06 | 8.22E-06 | 1.47E-09 | 0.003843786 | AC017083.3, PPP3R1 |
| chr3 | 71574012 | 71575170 | 1159 | 6 | 2.21E-09 | 3.34E-09 | 7.73E-05 | 1.47E-09 | -0.00231063 | FOXP1, AC097634.4, FOXP1-IT1 |
| chr17 | 80779473 | 80780572 | 1100 | 8 | 1.54E-08 | 3.83E-09 | 0.000218 | 1.47E-09 | -0.01381346 | RPTOR |
| chr2 | 84458631 | 84460112 | 1482 | 13 | 4.31E-10 | 1.97E-07 | 0.000587 | 1.49E-09 | 0.001334329 | SUCLG1 |
| chr10 | 27242278 | 27243316 | 1039 | 8 | 1.82E-11 | 3.35E-06 | 6.01E-05 | 1.49E-09 | 0.019978559 | AL160291.1, ACBD5 |
| chr3 | 37974690 | 37975697 | 1008 | 4 | 7.85E-11 | 8.74E-09 | 1.82E-06 | 1.49E-09 | -0.00867487 | CTDSPL |
| chr9 | 121071794 | 121072339 | 546 | 4 | 2.38E-10 | 1.07E-09 | 2.08E-05 | 1.50E-09 | -0.00988369 | NA |
| chr5 | 368279 | 368973 | 695 | 5 | 1.31E-09 | 8.13E-10 | 7.41E-05 | 1.51E-09 | 0.012278973 | PDCD6-AHRR, AHRR |
| chr6 | 6320091 | 6322145 | 2055 | 13 | 9.40E-07 | 3.68E-08 | 8.04E-05 | 1.52E-09 | -0.00258447 | F13A1 |
| chr16 | 2511248 | 2514004 | 2757 | 17 | 6.93E-07 | 5.99E-08 | 0.000166 | 1.52E-09 | -0.00191052 | AC093525.2, ATP6V0C, AC093525.1 |
| chr3 | 127028761 | 127029938 | 1178 | 4 | 2.26E-12 | 4.32E-07 | 2.93E-07 | 1.53E-09 | -0.00404418 | PLXNA1 |
| chr3 | 157160723 | 157161294 | 572 | 11 | 6.94E-12 | 4.08E-07 | 3.23E-05 | 1.53E-09 | -0.00656516 | CCNL1 |
| chr10 | 46019369 | 46020229 | 861 | 7 | 1.69E-16 | 0.0002168 | 1.48E-08 | 1.53E-09 | -0.0073456 | NCOA4 |
| chr1 | 7379003 | 7379631 | 629 | 2 | 2.70E-09 | 9.52E-10 | 6.15E-06 | 1.53E-09 | -0.01335071 | CAMTA1 |
| chr2 | 176122210 | 176123777 | 1568 | 15 | 1.01E-08 | 4.61E-10 | 0.000422 | 1.53E-09 | -0.00225455 | HOXD9, HOXD-AS2 |
| chr11 | 70319066 | 70319916 | 851 | 6 | 7.95E-09 | 1.15E-07 | 7.09E-05 | 1.54E-09 | -0.00096203 | PPFIA1, AP002336.2 |
| chr11 | 69647892 | 69649504 | 1613 | 11 | 1.02E-08 | 2.25E-07 | 0.000148 | 1.55E-09 | -0.00634422 | CCND1 |
| chr5 | 1102560 | 1103795 | 1236 | 3 | 4.46E-10 | 9.91E-10 | 8.22E-06 | 1.56E-09 | -0.01978774 | SLC12A7 |
| chr18 | 79688054 | 79689259 | 1206 | 5 | 4.91E-08 | 8.84E-09 | 2.70E-05 | 1.56E-09 | -0.02387008 | CTDP1 |

| | | | | | | | | | | |
|---|---|---|---|---|---|---|---|---|---|---|
| chr1 | 232803635 | 232805309 | 1675 | 9 | 6.74E-10 | 8.80E-05 | 1.18E-05 | 1.57E-09 | -0.00411919 | MAP10 |
| chr2 | 25876964 | 25878694 | 1731 | 13 | 1.05E-08 | 9.74E-08 | 0.000446 | 1.57E-09 | -0.00059118 | ASXL2 |
| chr14 | 52313440 | 52314594 | 1155 | 12 | 4.91E-10 | 4.99E-08 | 0.000826 | 1.57E-09 | 0.00290387 | PTGER2 |
| chr16 | 179984 | 181390 | 1407 | 10 | 5.97E-09 | 8.32E-09 | 0.000934 | 1.57E-09 | 0.002236744 | HBQ1 |
| chr20 | 5119927 | 5122655 | 2729 | 12 | 1.83E-09 | 1.92E-07 | 1.66E-06 | 1.57E-09 | 0.00378736 | PCNA, AL121924.1 |
| chr19 | 3785575 | 3787091 | 1517 | 11 | 5.91E-12 | 3.73E-06 | 6.78E-07 | 1.57E-09 | -0.00355472 | MATK |
| chr9 | 135776665 | 135779262 | 2598 | 9 | 3.15E-06 | 1.66E-08 | 0.000212 | 1.57E-09 | -0.00412069 | KCNT1 |
| chr3 | 19348961 | 19349464 | 504 | 2 | 4.48E-12 | 1.52E-08 | 8.14E-08 | 1.58E-09 | -0.02057985 | KCNH8 |
| chr11 | 65640958 | 65642655 | 1698 | 10 | 4.47E-08 | 2.40E-09 | 0.000995 | 1.58E-09 | -0.00734229 | SIPA1 |
| chr2 | 152717369 | 152719203 | 1835 | 18 | 3.92E-08 | 1.23E-05 | 2.84E-06 | 1.59E-09 | 0.000613191 | ARL6IP6, PRPF40A |
| chr2 | 238097870 | 238100605 | 2736 | 16 | 2.75E-08 | 8.71E-07 | 0.000113 | 1.59E-09 | 0.011161115 | UBE2F-SCLY, SCLY, ESPNL |
| chr6 | 35181950 | 35182491 | 542 | 4 | 3.05E-09 | 8.23E-10 | 0.000109 | 1.59E-09 | -0.0156572 | NA |
| chr12 | 88141652 | 88142798 | 1147 | 17 | 4.21E-09 | 4.71E-09 | 0.001237 | 1.59E-09 | 0.001335149 | TMTC3, CEP290 |
| chr2 | 96550111 | 96550915 | 805 | 3 | 9.01E-14 | 4.55E-08 | 1.19E-07 | 1.59E-09 | -0.00313964 | ARID5A |
| chr7 | 139034697 | 139037094 | 2398 | 14 | 1.81E-07 | 5.97E-07 | 6.30E-05 | 1.59E-09 | -0.00283049 | ZC3HAV1L |
| chr8 | 85377990 | 85379411 | 1422 | 7 | 1.63E-09 | 2.57E-07 | 6.45E-05 | 1.59E-09 | -0.0041475 | CA3, CA1 |
| chr17 | 4948513 | 4950422 | 1910 | 11 | 3.81E-08 | 7.33E-09 | 0.000398 | 1.60E-09 | 0.00058684 | ENO3, PFN1 |
| chr7 | 142307647 | 142308504 | 858 | 4 | 7.10E-10 | 6.18E-09 | 1.11E-06 | 1.60E-09 | 0.005872438 | NA |
| chr19 | 41859248 | 41860597 | 1350 | 13 | 8.53E-10 | 4.16E-09 | 0.000551 | 1.60E-09 | -0.00192201 | RPS19 |
| chr6 | 166849243 | 166850860 | 1618 | 8 | 7.01E-07 | 3.85E-08 | 0.000145 | 1.60E-09 | -0.00755201 | RPS6KA2 |
| chr4 | 98604116 | 98605003 | 888 | 3 | 3.74E-11 | 8.17E-08 | 7.71E-06 | 1.60E-09 | -0.00015997 | TSPAN5 |
| chr3 | 25782987 | 25784734 | 1748 | 16 | 1.01E-08 | 4.53E-08 | 6.22E-05 | 1.61E-09 | 0.002878272 | OXSM, NGLY1 |
| chr11 | 57426457 | 57428067 | 1611 | 14 | 1.15E-10 | 3.58E-07 | 2.42E-05 | 1.62E-09 | -0.00401431 | SLC43A3 |
| chr17 | 64262293 | 64264083 | 1791 | 14 | 8.16E-09 | 2.65E-08 | 2.06E-05 | 1.62E-09 | 0.00293979 | TEX2 |
| chr6 | 73353799 | 73354871 | 1073 | 7 | 1.90E-10 | 6.99E-10 | 9.18E-05 | 1.62E-09 | -0.02281864 | DPPA5 |
| chr17 | 7012275 | 7014584 | 2310 | 23 | 1.87E-07 | 2.36E-09 | 0.013285 | 1.62E-09 | -0.00136196 | RNASEK, RNASEK-C17orf49, C17orf49, ALOX12-AS1, AC040977.2 |
| chr18 | 72867532 | 72869572 | 2041 | 7 | 1.73E-06 | 2.01E-10 | 0.001493 | 1.62E-09 | 0.000499026 | AC091138.1, NETO1 |
| chr17 | 49901574 | 49902366 | 793 | 5 | 1.18E-14 | 3.22E-05 | 1.83E-06 | 1.62E-09 | -0.01308536 | NA |
| chr11 | 70805448 | 70807117 | 1670 | 6 | 3.28E-08 | 4.10E-06 | 8.35E-06 | 1.63E-09 | -0.00355105 | SHANK2 |
| chr17 | 67043985 | 67045922 | 1938 | 11 | 2.77E-09 | 6.57E-09 | 0.001208 | 1.63E-09 | -0.00192115 | CACNG1 |
| chr1 | 45010446 | 45012901 | 2456 | 21 | 9.07E-08 | 1.09E-06 | 0.000677 | 1.63E-09 | -0.00172582 | UROD, HECTD3 |
| chr5 | 1555676 | 1556588 | 913 | 5 | 2.60E-11 | 2.38E-07 | 2.12E-05 | 1.63E-09 | -0.01079789 | NA |
| chr20 | 33666410 | 33667685 | 1276 | 5 | 1.31E-11 | 2.66E-09 | 8.97E-06 | 1.64E-09 | -0.01753506 | ACTL10, ACTL10, NECAB3 |
| chr1 | 210231709 | 210234135 | 2427 | 14 | 1.96E-06 | 1.35E-07 | 0.000296 | 1.64E-09 | 0.000155079 | SERTAD4, SERTAD4-AS1 |
| chr13 | 78994542 | 78995355 | 814 | 2 | 3.49E-10 | 3.27E-09 | 4.22E-07 | 1.64E-09 | 0.021522388 | NA |
| chr8 | 124271024 | 124271802 | 779 | 5 | 2.54E-10 | 3.03E-09 | 4.85E-06 | 1.64E-09 | -0.00097735 | AC090192.2 |
| chr17 | 77321864 | 77322986 | 1123 | 7 | 2.96E-09 | 2.05E-10 | 0.001069 | 1.65E-09 | -0.00087734 | SEPTIN9 |
| chr18 | 48345409 | 48345687 | 279 | 2 | 8.90E-11 | 1.24E-08 | 1.02E-07 | 1.66E-09 | -0.00860687 | ZBTB7C |
| chr19 | 12838034 | 12839337 | 1304 | 10 | 7.81E-10 | 2.58E-09 | 0.000212 | 1.66E-09 | -0.00338892 | MAST1, HOOK2 |
| chr7 | 124790699 | 124791628 | 930 | 7 | 5.64E-10 | 7.63E-08 | 0.000148 | 1.66E-09 | 0.013738726 | NA |
| chr6 | 12751381 | 12752176 | 796 | 3 | 2.73E-10 | 2.46E-09 | 2.24E-06 | 1.67E-09 | -0.01172216 | PHACTR1 |
| chr1 | 64966026 | 64967677 | 1652 | 8 | 8.13E-09 | 6.71E-06 | 2.90E-06 | 1.67E-09 | 0.00397797 | JAK1 |
| chr2 | 18719966 | 18721573 | 1608 | 4 | 1.50E-07 | 2.17E-09 | 5.11E-05 | 1.68E-09 | -0.0023379 | NA |
| chr2 | 239379809 | 239381709 | 1901 | 9 | 1.23E-08 | 3.27E-09 | 0.001978 | 1.68E-09 | 0.006851977 | HDAC4 |
| chr11 | 1271488 | 1273842 | 2355 | 8 | 1.30E-05 | 8.66E-10 | 0.001343 | 1.68E-09 | -0.00255201 | NA |
| chr15 | 101724109 | 101725600 | 1492 | 12 | 4.72E-12 | 7.82E-08 | 2.87E-05 | 1.69E-09 | -0.00678027 | TARS3 |
| chr1 | 20717538 | 20718930 | 1393 | 15 | 7.59E-11 | 1.15E-07 | 0.000364 | 1.69E-09 | -0.00196206 | KIF17 |
| chr16 | 7332186 | 7332863 | 678 | 8 | 6.57E-10 | 1.98E-08 | 6.35E-05 | 1.70E-09 | -0.00030332 | RBFOX1 |
| chr19 | 40749985 | 40751328 | 1344 | 19 | 2.42E-08 | 1.03E-06 | 0.000526 | 1.70E-09 | -0.00163219 | SNRPA, C19orf54 |
| chr7 | 74355045 | 74356016 | 972 | 4 | 1.50E-09 | 8.43E-09 | 6.83E-06 | 1.71E-09 | -0.01062425 | CLIP2 |
| chr6 | 35148679 | 35150705 | 2027 | 10 | 7.42E-08 | 1.06E-06 | 1.31E-05 | 1.71E-09 | -0.00507962 | NA |
| chr1 | 109667399 | 109668303 | 905 | 8 | 1.70E-10 | 7.99E-08 | 6.26E-05 | 1.71E-09 | -0.00566399 | GSTM4, GSTM2 |
| chr19 | 8307629 | 8308724 | 1096 | 13 | 3.55E-10 | 2.40E-07 | 4.00E-06 | 1.71E-09 | -0.00190305 | CD320, AC010323.1 |
| chr12 | 53980318 | 53982457 | 2140 | 8 | 1.70E-08 | 1.06E-09 | 0.00028 | 1.72E-09 | 0.010809263 | HOXC-AS3 |
| chr1 | 84297655 | 84298692 | 1038 | 7 | 3.52E-10 | 4.07E-08 | 0.000399 | 1.72E-09 | -0.00108658 | SAMD13 |
| chr2 | 238943517 | 238943530 | 14 | 2 | 8.14E-10 | 1.49E-09 | 2.00E-06 | 1.72E-09 | -0.02763922 | NA |
| chr2 | 38438614 | 38439128 | 515 | 2 | 2.25E-11 | 1.09E-08 | 1.22E-07 | 1.73E-09 | 0.01401148 | LINC02613 |
| chr16 | 52547069 | 52548552 | 1484 | 13 | 1.88E-10 | 2.30E-08 | 0.000166 | 1.73E-09 | -0.00319179 | TOX3 |
| chr16 | 30429243 | 30430732 | 1490 | 17 | 5.64E-10 | 7.24E-07 | 8.73E-06 | 1.73E-09 | -0.0012922 | ZNF771, DCTPP1 |
| chr6 | 90586416 | 90588499 | 2084 | 14 | 2.97E-09 | 4.08E-05 | 1.59E-05 | 1.73E-09 | -0.00331658 | MAP3K7 |
| chr2 | 218983536 | 218986550 | 3015 | 16 | 1.24E-06 | 3.78E-05 | 2.89E-05 | 1.73E-09 | -0.0021681 | LINC00608, FEV |
| chr16 | 67226758 | 67228663 | 1906 | 20 | 1.34E-07 | 6.94E-07 | 0.000531 | 1.73E-09 | -0.00046294 | TMEM208, LRRC29 |
| chr15 | 58270269 | 58270277 | 9 | 2 | 8.89E-10 | 1.29E-09 | 3.11E-06 | 1.73E-09 | -0.05016239 | ALDH1A2 |
| chr2 | 38097842 | 38098854 | 1013 | 5 | 1.37E-10 | 3.65E-08 | 1.67E-05 | 1.74E-09 | -0.0147336 | CYP1B1-AS1, CYP1B1 |
| chr5 | 115961281 | 115963391 | 2111 | 19 | 4.17E-09 | 3.82E-06 | 3.56E-05 | 1.74E-09 | -0.00427083 | LVRN |
| chr16 | 588109 | 589964 | 1856 | 17 | 1.53E-08 | 5.40E-07 | 0.00053 | 1.74E-09 | -0.00147866 | RAB40C |
| chr8 | 1454884 | 1456972 | 2089 | 10 | 3.64E-07 | 5.94E-08 | 2.85E-05 | 1.74E-09 | -0.00397638 | DLGAP2 |
| chr5 | 76082965 | 76084478 | 1514 | 14 | 1.97E-09 | 1.39E-07 | 4.67E-05 | 1.74E-09 | 0.000195297 | SV2C, AC026774.1 |
| chr5 | 113488059 | 113489569 | 1511 | 13 | 1.20E-08 | 1.26E-08 | 0.00069 | 1.75E-09 | -0.00550309 | MCC |
| chr8 | 126374960 | 126375726 | 767 | 2 | 3.27E-09 | 1.41E-09 | 2.45E-06 | 1.75E-09 | 0.004560943 | NA |
| chr19 | 45711716 | 45713369 | 1654 | 10 | 1.04E-07 | 5.62E-08 | 0.000252 | 1.75E-09 | -0.00254405 | FBXO46 |
| chr6 | 112346506 | 112347369 | 864 | 11 | 4.46E-11 | 4.20E-06 | 5.16E-05 | 1.76E-09 | -0.00618783 | AL365214.2, RFPL4B |
| chr16 | 409567 | 410746 | 1180 | 6 | 6.54E-11 | 1.49E-05 | 5.17E-06 | 1.76E-09 | 0.000366374 | NME4, DECR2 |
| chr10 | 80536435 | 80538312 | 1878 | 8 | 1.60E-07 | 1.30E-09 | 0.000529 | 1.76E-09 | -0.00484534 | SH2D4B |
| chr14 | 41606351 | 41607538 | 1188 | 9 | 1.89E-09 | 1.39E-09 | 0.001119 | 1.77E-09 | -0.00014135 | NA |
| chr6 | 15501038 | 15502157 | 1120 | 9 | 2.24E-09 | 3.91E-09 | 0.00078 | 1.77E-09 | -0.00713033 | JARID2 |
| chr15 | 59274475 | 59276513 | 2039 | 6 | 3.99E-06 | 1.14E-09 | 0.00011 | 1.77E-09 | -0.00466069 | MYO1E |
| chr11 | 89490813 | 89492215 | 1403 | 19 | 4.31E-09 | 1.18E-07 | 0.001204 | 1.77E-09 | -0.00436055 | NOX4 |
| chr17 | 80821222 | 80823129 | 1908 | 11 | 5.33E-09 | 3.33E-07 | 2.53E-05 | 1.78E-09 | -0.01244314 | RPTOR |
| chr17 | 79841367 | 79842340 | 974 | 6 | 3.36E-11 | 3.90E-07 | 6.65E-07 | 1.79E-09 | -0.00020741 | NA |
| chr10 | 110354422 | 110355307 | 886 | 4 | 3.05E-08 | 9.88E-10 | 7.18E-05 | 1.79E-09 | -0.01269252 | NA |
| chr9 | 123072787 | 123073666 | 880 | 3 | 1.11E-08 | 6.16E-10 | 5.49E-05 | 1.80E-09 | -0.00986352 | RABGAP1 |
| chr10 | 104300167 | 104300191 | 25 | 2 | 7.07E-11 | 4.91E-09 | 2.99E-07 | 1.81E-09 | -0.00998125 | GSTO2 |
| chr2 | 178807394 | 178807964 | 571 | 5 | 1.45E-09 | 4.83E-09 | 3.96E-06 | 1.81E-09 | -0.00156451 | TTN |
| chr9 | 69671472 | 69672905 | 1434 | 11 | 1.59E-09 | 7.78E-09 | 0.00037 | 1.81E-09 | -0.00197099 | AL162412.1, APBA1 |
| chr8 | 98041402 | 98041595 | 194 | 5 | 1.24E-09 | 7.23E-09 | 4.18E-05 | 1.81E-09 | -0.01199524 | RPL30 |
| chr10 | 74223390 | 74224235 | 846 | 3 | 7.53E-12 | 3.84E-08 | 3.81E-07 | 1.82E-09 | -0.01061008 | ADK |
| chr6 | 42048648 | 42050697 | 2050 | 20 | 3.22E-10 | 1.06E-07 | 0.000745 | 1.82E-09 | 0.000279415 | TAF8, CCND3 |
| chr20 | 49320721 | 49321070 | 350 | 3 | 6.50E-10 | 1.63E-09 | 5.27E-06 | 1.82E-09 | -0.0103295 | AL035685.1, KCNB1 |
| chr11 | 67650487 | 67650934 | 448 | 11 | 3.65E-09 | 1.74E-10 | 0.001749 | 1.83E-09 | 0.021081834 | ACY3 |
| chr3 | 194376079 | 194376406 | 328 | 3 | 6.13E-10 | 7.69E-10 | 3.57E-05 | 1.84E-09 | -0.01674992 | NA |
| chr3 | 155703946 | 155705379 | 1434 | 11 | 1.59E-10 | 1.52E-07 | 0.0001 | 1.84E-09 | -0.00170799 | PLCH1 |
| chr2 | 158734497 | 158735432 | 936 | 8 | 8.11E-11 | 1.26E-06 | 1.71E-06 | 1.85E-09 | -0.0043209 | AC005042.1, PKP4-AS1 |
| chr5 | 93591442 | 93592909 | 1468 | 5 | 1.03E-08 | 8.15E-09 | 7.43E-05 | 1.85E-09 | 0.018195332 | NR2F1 |
| chr22 | 41620203 | 41621976 | 1774 | 24 | 4.46E-09 | 1.66E-05 | 9.25E-05 | 1.85E-09 | 1.36E-05 | XRCC6, DESI1 |
| chr17 | 29013887 | 29014748 | 862 | 3 | 1.21E-09 | 1.19E-09 | 1.39E-05 | 1.85E-09 | -0.00582056 | PIPOX, AC024619.1 |
| chr19 | 46021548 | 46023492 | 1945 | 15 | 4.37E-09 | 3.61E-08 | 0.002669 | 1.85E-09 | -0.00659676 | PGLYRP1 |
| chr14 | 75126382 | 75128072 | 1691 | 18 | 3.11E-09 | 2.11E-07 | 0.002099 | 1.86E-09 | -0.00131551 | AL049780.2, NEK9 |
| chr8 | 102550744 | 102551470 | 727 | 7 | 4.52E-10 | 3.30E-09 | 0.001013 | 1.86E-09 | -0.00224792 | NA |
| chr9 | 4077672 | 4077749 | 78 | 2 | 1.07E-09 | 1.11E-09 | 8.22E-06 | 1.86E-09 | -0.04356677 | GLIS3 |
| chr11 | 2264572 | 2265911 | 1340 | 8 | 4.21E-09 | 1.10E-08 | 3.24E-05 | 1.86E-09 | -0.00182769 | NA |
| chr2 | 241665798 | 241668095 | 2298 | 11 | 1.66E-08 | 6.03E-09 | 3.81E-05 | 1.86E-09 | 0.001512182 | ATG4B |
| chr7 | 29609269 | 29609872 | 604 | 3 | 1.51E-11 | 5.31E-07 | 1.33E-06 | 1.86E-09 | -0.02265843 | ZNRF2P2 |
| chr6 | 149484460 | 149485596 | 1137 | 13 | 1.02E-10 | 1.75E-08 | 0.001406 | 1.87E-09 | 0.010326975 | ZC3H12D |
| chr6 | 32162878 | 32163945 | 1068 | 9 | 3.08E-10 | 1.22E-09 | 7.35E-05 | 1.87E-09 | -0.0015119 | PPT2, PPT2-EGFL8 |
| chr6 | 31805653 | 31808116 | 2464 | 18 | 1.85E-07 | 1.88E-09 | 0.002291 | 1.87E-09 | 0.001220135 | LSM2 |
| chr2 | 19989506 | 19990750 | 1245 | 16 | 9.60E-11 | 1.29E-07 | 0.000762 | 1.87E-09 | -0.0019186 | AC079145.1, WDR35 |
| chr20 | 46977992 | 46980227 | 2236 | 10 | 9.91E-08 | 3.29E-07 | 0.000305 | 1.87E-09 | -0.00712321 | EYA2 |
| chr11 | 34104976 | 34106759 | 1784 | 14 | 1.03E-07 | 5.73E-09 | 0.000646 | 1.87E-09 | -0.00148866 | NAT10 |
| chr1 | 20787150 | 20788411 | 1262 | 4 | 7.53E-12 | 6.82E-06 | 1.57E-07 | 1.87E-09 | -0.0090156 | HP1BP3 |
| chr15 | 42490322 | 42491551 | 1230 | 12 | 1.19E-11 | 0.0013811 | 3.29E-07 | 1.88E-09 | 0.00098752 | SNAP23, ZNF106 |
| chr8 | 18804496 | 18804875 | 380 | 2 | 6.23E-11 | 6.34E-09 | 2.37E-07 | 1.88E-09 | -0.01162061 | PSD3 |
| chr1 | 1242811 | 1244681 | 1871 | 8 | 4.63E-08 | 1.68E-10 | 0.001444 | 1.89E-09 | -0.00122084 | C1QTNF12 |
| chr6 | 30145297 | 30146262 | 966 | 8 | 2.09E-10 | 3.55E-09 | 0.000229 | 1.89E-09 | 0.005195297 | TRIM40 |
| chr12 | 121801629 | 121804700 | 3072 | 26 | 5.46E-06 | 1.68E-08 | 0.002516 | 1.89E-09 | 0.001028026 | AC084018.1, AC084018.2, SETD1B, RHOF, LINC01089 |
| chr14 | 91257544 | 91258475 | 932 | 9 | 2.82E-10 | 6.46E-08 | 1.94E-06 | 1.90E-09 | -0.00080344 | AL135818.2 |
| chr4 | 80196493 | 80198319 | 1827 | 12 | 3.87E-07 | 2.20E-10 | 0.005017 | 1.90E-09 | -0.01965141 | PRDM8, AC021127.1 |
| chr19 | 1102571 | 1105381 | 2811 | 19 | 1.51E-06 | 3.65E-09 | 0.00258 | 1.90E-09 | -0.00320265 | GPX4 |
| chr16 | 30571266 | 30573640 | 2375 | 27 | 8.14E-07 | 1.26E-08 | 0.006286 | 1.90E-09 | -0.00022527 | AC002310.2, ZNF688 |
| chr2 | 9692972 | 9693293 | 322 | 2 | 3.63E-10 | 2.42E-09 | 9.52E-07 | 1.90E-09 | 0.027936335 | AC082651.1, AC082651.4 |
| chr9 | 84075484 | 84075652 | 169 | 2 | 1.47E-11 | 2.96E-05 | 6.83E-09 | 1.91E-09 | -0.00835504 | AL390838.1 |
| chr12 | 11543225 | 11543960 | 736 | 3 | 1.46E-10 | 5.33E-09 | 1.03E-06 | 1.92E-09 | -0.00104836 | AC007450.1 |
| chr20 | 41999337 | 41999430 | 94 | 2 | 1.64E-10 | 2.60E-09 | 8.62E-07 | 1.92E-09 | 0.026037081 | AL049812.2, AL049812.1 |
| chr6 | 152982274 | 152984298 | 2025 | 17 | 2.94E-07 | 1.93E-09 | 0.003899 | 1.92E-09 | -0.00038679 | AL080276.2, FBXO5 |
| chr9 | 127705953 | 127707123 | 1171 | 6 | 1.67E-07 | 4.38E-10 | 0.000403 | 1.92E-09 | -0.00556775 | CFAP157, PTRH1 |
| chr8 | 124596288 | 124597355 | 1068 | 3 | 5.31E-13 | 2.05E-07 | 6.94E-08 | 1.93E-09 | -0.00094457 | MTSS1 |
| chr17 | 39029242 | 39029732 | 491 | 3 | 7.72E-12 | 1.82E-05 | 7.18E-08 | 1.93E-09 | 0.003435194 | LRRC37A11P |
| chr5 | 140711099 | 140711517 | 419 | 6 | 2.65E-09 | 1.40E-09 | 8.75E-05 | 1.93E-09 | -0.00354895 | VTRNA1-1 |
| chr12 | 110467599 | 110469804 | 2206 | 24 | 5.42E-07 | 4.72E-06 | 0.000278 | 1.93E-09 | 0.001460125 | FAM216A, GPN3 |
| chr17 | 72683722 | 72684634 | 913 | 4 | 2.19E-09 | 3.36E-09 | 8.11E-05 | 1.94E-09 | -0.00168724 | SLC39A11 |
| chr1 | 32885967 | 32887240 | 1274 | 14 | 5.47E-11 | 1.08E-07 | 6.73E-05 | 1.94E-09 | -0.00140315 | HPCA |
| chr14 | 104094200 | 104095348 | 1149 | 4 | 1.50E-09 | 2.71E-06 | 1.75E-06 | 1.94E-09 | -0.00070459 | ASPG |
| chr2 | 127807801 | 127807810 | 10 | 2 | 1.18E-09 | 1.11E-09 | 1.24E-05 | 1.95E-09 | 0.022839121 | WDR33 |
| chr5 | 31531838 | 31533151 | 1314 | 16 | 2.25E-09 | 7.94E-08 | 0.001138 | 1.95E-09 | 0.000969866 | C5orf22, DROSHA |
| chr18 | 63243814 | 63244931 | 1118 | 5 | 1.49E-09 | 3.87E-08 | 4.10E-05 | 1.95E-09 | 0.00164974 | BCL2 |
| chr19 | 34171731 | 34173628 | 1898 | 15 | 3.16E-10 | 9.74E-07 | 2.72E-06 | 1.96E-09 | -0.00068684 | LSM14A |
| chr11 | 705692 | 708371 | 2680 | 15 | 2.58E-06 | 1.48E-09 | 0.003246 | 1.96E-09 | -0.00101387 | EPS8L2, DEAF1 |
| chr5 | 171336475 | 171337148 | 674 | 9 | 1.14E-09 | 1.24E-09 | 0.000243 | 1.97E-09 | -0.00553052 | NA |
| chr21 | 26000072 | 26000146 | 75 | 4 | 3.42E-09 | 4.08E-10 | 0.000193 | 1.97E-09 | -0.02204956 | APP |
| chr16 | 21283626 | 21284244 | 619 | 5 | 7.60E-11 | 2.17E-07 | 7.15E-06 | 1.98E-09 | 0.002200301 | CRYM |
| chr11 | 65095679 | 65096810 | 1132 | 12 | 1.06E-09 | 2.31E-06 | 9.44E-05 | 1.98E-09 | -0.00232961 | VPS51 |
| chr7 | 2655563 | 2656585 | 1023 | 9 | 5.98E-11 | 1.12E-06 | 5.45E-07 | 1.98E-09 | -0.00205004 | TTYH3 |
| chr1 | 32179043 | 32180531 | 1489 | 20 | 7.60E-08 | 3.98E-07 | 0.001973 | 1.98E-09 | -0.00040964 | TXLNA |
| chr22 | 24986152 | 24986212 | 61 | 2 | 1.05E-09 | 1.44E-09 | 3.68E-06 | 1.99E-09 | 0.032863254 | KIAA1671 |
| chr7 | 30770017 | 30771612 | 1596 | 12 | 5.03E-11 | 3.62E-06 | 2.51E-06 | 1.99E-09 | -0.00322949 | INMT-MINDY4, MINDY4 |
| chr6 | 151111545 | 151112250 | 706 | 2 | 1.31E-11 | 2.10E-08 | 8.66E-08 | 2.00E-09 | -0.01012032 | AL138733.1 |
| chr2 | 47569642 | 47571117 | 1476 | 9 | 1.21E-11 | 1.10E-06 | 5.04E-06 | 2.00E-09 | 0.001776589 | MSH2, KCNK12 |
| chr1 | 198983067 | 198983525 | 459 | 8 | 1.63E-10 | 6.76E-07 | 1.12E-05 | 2.01E-09 | 0.001002657 | AC096631.1 |
| chr21 | 42879066 | 42880184 | 1119 | 16 | 6.47E-10 | 4.83E-08 | 0.00179 | 2.01E-09 | 0.002316795 | NDUFV3, WDR4 |
| chr12 | 52834032 | 52835758 | 1727 | 6 | 2.81E-10 | 5.61E-05 | 2.93E-07 | 2.03E-09 | -0.00463904 | KRT79 |
| chr7 | 31528387 | 31529051 | 665 | 4 | 5.45E-10 | 4.78E-09 | 5.59E-06 | 2.03E-09 | -0.01934915 | ITPRID1 |
| chr17 | 79776586 | 79778271 | 1686 | 10 | 1.40E-10 | 2.30E-08 | 0.000473 | 2.03E-09 | -0.00355975 | CBX2 |
| chr12 | 6279510 | 6279889 | 380 | 5 | 4.57E-12 | 1.19E-06 | 2.63E-08 | 2.03E-09 | 0.003146113 | NA |
| chr14 | 55270640 | 55272350 | 1711 | 19 | 2.05E-09 | 4.55E-08 | 0.000121 | 2.03E-09 | -0.00160829 | FBXO34, AL158801.2 |
| chr19 | 11088288 | 11090307 | 2020 | 12 | 1.29E-12 | 3.57E-06 | 1.17E-06 | 2.03E-09 | -0.0032252 | LDLR |
| chr17 | 43661525 | 43662908 | 1384 | 10 | 1.82E-08 | 8.24E-08 | 0.000975 | 2.04E-09 | -0.01175756 | MEOX1 |
| chr17 | 83077586 | 83078902 | 1317 | 5 | 6.42E-11 | 8.98E-09 | 4.82E-06 | 2.04E-09 | -0.00339532 | NA |
| chr3 | 94937889 | 94938204 | 316 | 6 | 8.91E-10 | 6.12E-08 | 5.82E-05 | 2.04E-09 | -0.00309029 | LINC00879, WDR82P1 |

| | | | | | | | | | | |
|---|---|---|---|---|---|---|---|---|---|---|
| chr9 | 135769700 | 135770916 | 1217 | 7 | 4.06E-15 | 6.05E-06 | 6.05E-06 | 2.04E-09 | 0.008094792 | KCNT1 |
| chr14 | 56310358 | 56311583 | 1226 | 8 | 1.22E-08 | 1.15E-09 | 0.00072 | 2.04E-09 | 0.013829525 | LINC02284, AL355073.1 |
| chr12 | 105562203 | 105562700 | 498 | 5 | 1.41E-09 | 6.74E-08 | 1.09E-05 | 2.05E-09 | -0.003426 | NA |
| chr12 | 1546589 | 1547391 | 803 | 6 | 1.44E-08 | 7.54E-10 | 0.000652 | 2.05E-09 | -0.00275841 | WNT5B |
| chr22 | 43254290 | 43254635 | 346 | 2 | 4.65E-10 | 2.12E-09 | 1.51E-06 | 2.06E-09 | -0.00405017 | SCUBE1 |
| chr16 | 681417 | 683842 | 2426 | 16 | 1.57E-07 | 3.96E-06 | 1.86E-05 | 2.06E-09 | -0.00027533 | STUB1, JMJD8 |
| chr2 | 133265318 | 133267046 | 1729 | 11 | 5.80E-07 | 3.92E-10 | 0.001368 | 2.06E-09 | -0.00474917 | NCKAP5-AS2, NCKAP5 |
| chr16 | 67506949 | 67507480 | 532 | 2 | 3.04E-11 | 1.20E-08 | 1.44E-07 | 2.07E-09 | 0.011313464 | NA |
| chr16 | 1096277 | 1097625 | 1349 | 9 | 2.03E-10 | 3.97E-07 | 4.92E-05 | 2.07E-09 | -0.0041966 | NA |
| chr20 | 571954 | 572323 | 370 | 3 | 9.68E-10 | 1.95E-09 | 4.71E-06 | 2.08E-09 | -0.01904018 | NA |
| chr12 | 48197751 | 48199035 | 1285 | 9 | 2.02E-07 | 2.64E-08 | 3.66E-05 | 2.08E-09 | -0.00806019 | AC024257.3 |
| chr3 | 184360683 | 184361785 | 1103 | 6 | 1.98E-12 | 2.60E-07 | 2.15E-07 | 2.09E-09 | -0.0052332 | POLR2H, CLCN2 |
| chr1 | 60970179 | 60971122 | 944 | 5 | 5.14E-11 | 1.55E-08 | 3.71E-05 | 2.10E-09 | -0.01452948 | NFIA, NFIA-AS2 |
| chr16 | 1557326 | 1559528 | 2203 | 12 | 1.25E-10 | 8.12E-07 | 2.74E-07 | 2.10E-09 | -0.00194739 | IFT140 |
| chr5 | 80217450 | 80218758 | 1309 | 3 | 3.17E-07 | 1.64E-09 | 2.26E-05 | 2.11E-09 | 0.034138783 | SERINC5 |
| chr15 | 63596321 | 63597699 | 1379 | 11 | 4.19E-09 | 2.27E-06 | 0.000102 | 2.11E-09 | -0.00458351 | FBXL22, USP3-AS1 |
| chr17 | 35432493 | 35433952 | 1460 | 16 | 7.05E-10 | 1.34E-09 | 0.007322 | 2.11E-09 | -0.01526806 | SLFN12 |
| chr2 | 190699284 | 190699585 | 302 | 3 | 6.46E-10 | 6.23E-09 | 1.14E-06 | 2.12E-09 | -0.00979589 | AC006460.1 |
| chr8 | 144356405 | 144358123 | 1719 | 11 | 6.25E-09 | 1.95E-08 | 0.000756 | 2.12E-09 | -0.00281208 | SLC52A2, FBXL6 |
| chr18 | 2946805 | 2948263 | 1459 | 7 | 2.56E-08 | 3.83E-07 | 0.000202 | 2.12E-09 | -0.00023033 | AP000919.1, LPIN2, CHORDC1P4 |
| chr12 | 119713722 | 119714322 | 601 | 6 | 3.30E-10 | 2.57E-08 | 5.81E-06 | 2.12E-09 | -0.00320389 | CIT, MIR1178 |
| chr4 | 15701889 | 15703221 | 1333 | 10 | 1.93E-08 | 4.65E-07 | 0.000303 | 2.12E-09 | -0.00191504 | AC114744.2, BST1 |
| chr18 | 48921367 | 48923019 | 1653 | 6 | 5.88E-15 | 2.15E-06 | 1.00E-06 | 2.14E-09 | -0.00384847 | SMAD7 |
| chr16 | 55655234 | 55657190 | 1957 | 20 | 7.02E-09 | 6.13E-07 | 5.29E-05 | 2.14E-09 | -0.00466631 | SLC6A2 |
| chr1 | 159465215 | 159466225 | 1011 | 4 | 2.17E-07 | 5.88E-10 | 0.000152 | 2.15E-09 | -0.00896624 | AL513323.1 |
| chr1 | 107057390 | 107058140 | 751 | 6 | 7.61E-11 | 2.14E-08 | 0.000119 | 2.15E-09 | -0.01009089 | PRMT6 |
| chr1 | 67190065 | 67190345 | 281 | 2 | 7.47E-10 | 1.81E-09 | 2.47E-06 | 2.16E-09 | -0.00733819 | IL23R, C1orf141 |
| chr15 | 34343026 | 34343675 | 650 | 12 | 1.30E-09 | 3.55E-07 | 6.30E-05 | 2.16E-09 | -0.00267404 | NUTM1, NOP10 |
| chr4 | 145178255 | 145179063 | 809 | 5 | 1.12E-10 | 4.36E-09 | 6.17E-05 | 2.16E-09 | -0.00843964 | OTUD4 |
| chr17 | 45893783 | 45895207 | 1425 | 12 | 8.84E-10 | 2.00E-08 | 0.000351 | 2.17E-09 | -0.00036693 | MAPT, MAPT-AS1 |
| chr4 | 151687110 | 151688096 | 987 | 6 | 2.05E-11 | 1.06E-08 | 6.03E-05 | 2.17E-09 | -0.00322687 | GATB |
| chr1 | 23905913 | 23906258 | 346 | 2 | 9.09E-11 | 4.75E-09 | 4.28E-07 | 2.17E-09 | 0.001537729 | CNR2 |
| chr2 | 207238023 | 207238672 | 650 | 3 | 1.26E-10 | 6.22E-08 | 1.04E-06 | 2.17E-09 | 0.007217788 | MYOSLID |
| chr16 | 77773 | 79563 | 1791 | 9 | 8.03E-08 | 5.28E-09 | 6.35E-05 | 2.17E-09 | -0.0037393 | MPG |
| chr2 | 160062008 | 160063319 | 1312 | 15 | 3.70E-11 | 3.48E-05 | 9.11E-06 | 2.18E-09 | -0.00196922 | PLA2R1 |
| chr5 | 38550049 | 38551243 | 1195 | 3 | 7.36E-10 | 9.13E-07 | 5.32E-07 | 2.18E-09 | -0.01804059 | LIFR |
| chr17 | 10119034 | 10119884 | 851 | 3 | 5.09E-11 | 6.06E-09 | 1.89E-06 | 2.19E-09 | 0.013071131 | GAS7 |
| chr14 | 21383346 | 21384878 | 1533 | 14 | 2.60E-11 | 8.65E-06 | 3.19E-06 | 2.19E-09 | 0.001492481 | AL135744.1, SUPT16H |
| chr12 | 64824948 | 64826372 | 1425 | 6 | 3.66E-09 | 1.83E-08 | 1.69E-05 | 2.20E-09 | -0.00597005 | TBC1D30, AC078815.1, AC025262.3 |
| chr5 | 1967135 | 1968173 | 1039 | 5 | 3.11E-10 | 4.07E-09 | 2.68E-05 | 2.21E-09 | 0.027863945 | AC124852.1, AC126768.2, AC126768.1 |
| chr6 | 31639095 | 31639857 | 763 | 5 | 2.43E-09 | 6.94E-10 | 0.000113 | 2.21E-09 | -0.00816875 | BAG6 |
| chr15 | 53759042 | 53759704 | 663 | 8 | 5.96E-12 | 5.74E-08 | 1.53E-05 | 2.21E-09 | -0.00655235 | WDR72 |
| chr17 | 58755190 | 58756494 | 1305 | 15 | 7.70E-11 | 5.06E-08 | 0.001484 | 2.21E-09 | 0.000756813 | PPM1E |
| chr7 | 42236405 | 42237811 | 1407 | 17 | 2.83E-11 | 2.94E-08 | 7.31E-05 | 2.22E-09 | 0.000431214 | GLI3 |
| chr16 | 1148408 | 1149524 | 1117 | 8 | 2.59E-11 | 1.45E-06 | 8.45E-07 | 2.22E-09 | -0.00402729 | AC120498.8 |
| chr10 | 78039139 | 78040283 | 1145 | 4 | 1.05E-09 | 3.32E-08 | 5.93E-06 | 2.22E-09 | -0.00720043 | RPS24 |
| chr15 | 40944777 | 40944997 | 221 | 2 | 1.50E-10 | 8.48E-09 | 2.22E-07 | 2.23E-09 | -0.00270139 | NA |
| chr10 | 133045954 | 133047680 | 1727 | 10 | 1.35E-08 | 8.21E-08 | 0.000608 | 2.23E-09 | -0.0016752 | NA |
| chr5 | 123087168 | 123089011 | 1844 | 11 | 4.20E-07 | 8.94E-08 | 0.000125 | 2.23E-09 | -0.00351476 | AC106786.1 |
| chr1 | 156704893 | 156706789 | 1897 | 17 | 1.09E-08 | 1.54E-07 | 0.000926 | 2.24E-09 | -0.00035922 | CRABP2 |
| chr3 | 157122445 | 157123469 | 1025 | 4 | 4.39E-11 | 1.77E-07 | 5.42E-06 | 2.25E-09 | -0.00361861 | LINC00881, LINC00880 |
| chr6 | 31588478 | 31588705 | 228 | 2 | 1.35E-10 | 1.30E-09 | 1.31E-05 | 2.26E-09 | -0.02251071 | LST1 |
| chr3 | 128336533 | 128337640 | 1108 | 5 | 1.17E-10 | 8.14E-08 | 6.83E-05 | 2.26E-09 | -0.00362801 | EEFSEC |
| chr16 | 68244520 | 68245710 | 1191 | 13 | 6.23E-09 | 1.52E-06 | 7.95E-06 | 2.27E-09 | -0.00115499 | PLA2G15 |
| chr19 | 11381163 | 11381969 | 807 | 5 | 5.79E-11 | 5.80E-09 | 5.20E-05 | 2.27E-09 | -0.01154867 | EPOR |
| chr11 | 57310635 | 57311888 | 1254 | 6 | 1.23E-08 | 1.44E-07 | 2.87E-05 | 2.29E-09 | -0.00121642 | TNKS1BP1 |
| chr6 | 28090409 | 28091430 | 1022 | 10 | 5.92E-09 | 2.39E-10 | 0.005475 | 2.29E-09 | -0.02905548 | ZSCAN12P1, ZSCAN16-AS1 |
| chr4 | 25312085 | 25313027 | 943 | 13 | 9.02E-10 | 4.12E-09 | 0.000302 | 2.29E-09 | -0.00298777 | ZCCHC4 |
| chr3 | 179652334 | 179654666 | 2333 | 18 | 2.48E-07 | 7.12E-07 | 0.000137 | 2.29E-09 | -9.50E-05 | USP13 |
| chr2 | 201858979 | 201860632 | 1654 | 7 | 1.24E-08 | 8.89E-07 | 8.52E-06 | 2.30E-09 | -0.00717956 | CDK15 |
| chr2 | 226690694 | 226691181 | 488 | 2 | 7.28E-10 | 1.93E-09 | 2.58E-06 | 2.31E-09 | -0.02954897 | NA |
| chr4 | 39366079 | 39367242 | 1164 | 13 | 3.76E-09 | 1.09E-07 | 0.000258 | 2.31E-09 | -0.00067875 | RFC1 |
| chr8 | 90645062 | 90646539 | 1478 | 8 | 1.72E-08 | 1.40E-06 | 8.60E-06 | 2.32E-09 | -0.00051787 | LINC00534, AC106038.1, TMEM64 |
| chr8 | 74983762 | 74985236 | 1475 | 15 | 1.42E-08 | 1.09E-05 | 0.000133 | 2.32E-09 | -0.00090499 | CRISPLD1 |
| chr3 | 48240027 | 48241415 | 1389 | 12 | 6.16E-10 | 4.92E-08 | 0.000167 | 2.32E-09 | -0.00251936 | ZNF589 |
| chr7 | 141014551 | 141016043 | 1493 | 19 | 1.03E-09 | 1.19E-07 | 0.000103 | 2.32E-09 | 0.003175878 | MRPS33 |
| chr1 | 99003767 | 99005814 | 2048 | 15 | 2.58E-07 | 3.08E-09 | 0.001019 | 2.32E-09 | -0.00789424 | AL445433.2, PLPPR5 |
| chr7 | 137820860 | 137821215 | 356 | 2 | 8.74E-10 | 1.73E-09 | 3.62E-06 | 2.33E-09 | -0.02409074 | DGKI |
| chr1 | 3230765 | 3232246 | 1482 | 8 | 2.45E-10 | 2.43E-09 | 0.00013 | 2.33E-09 | -0.00158229 | PRDM16 |
| chr4 | 112515076 | 112516829 | 1754 | 12 | 5.36E-09 | 5.36E-07 | 0.001126 | 2.33E-09 | 0.000549052 | NEUROG2-AS1, NEUROG2 |
| chr2 | 174771771 | 174772836 | 1066 | 5 | 4.05E-10 | 8.88E-08 | 1.76E-05 | 2.34E-09 | -0.01090941 | AC010894.2, CHRNA1 |
| chr16 | 31471956 | 31473401 | 1446 | 17 | 4.96E-10 | 1.16E-06 | 0.000476 | 2.34E-09 | -0.00358572 | TGFB1I1, AC026471.6 |
| chr10 | 133378294 | 133380489 | 2196 | 26 | 1.25E-06 | 6.71E-07 | 0.000758 | 2.34E-09 | -0.00097262 | PAOX, AL360181.3 |
| chr17 | 67390495 | 67391335 | 841 | 4 | 4.95E-10 | 3.97E-09 | 2.46E-05 | 2.35E-09 | -0.03449374 | PITPNC1 |
| chr19 | 12681188 | 12682937 | 1750 | 15 | 4.01E-08 | 1.65E-08 | 0.00114 | 2.35E-09 | -0.0001029 | AC010422.8, DHPS, AC010422.2 |
| chr14 | 101038793 | 101040601 | 1809 | 14 | 4.14E-09 | 2.94E-07 | 0.000496 | 2.35E-09 | -0.0061607 | MEG8, MIR376C, MIR376A2, MIR654, MIR376B |
| chr12 | 86028189 | 86029559 | 1371 | 5 | 1.15E-07 | 1.54E-09 | 6.66E-05 | 2.36E-09 | -0.00295381 | MGAT4C |
| chr14 | 50018442 | 50018932 | 491 | 2 | 1.09E-09 | 1.72E-09 | 4.03E-06 | 2.37E-09 | 0.001919594 | LINC01588 |
| chr11 | 10899263 | 10900691 | 1429 | 7 | 4.84E-09 | 2.64E-07 | 1.16E-05 | 2.37E-09 | 0.006035906 | ZBED5-AS1, AC069360.1 |
| chr1 | 180913140 | 180913545 | 406 | 4 | 2.32E-10 | 6.26E-09 | 2.47E-05 | 2.37E-09 | -0.01136941 | KIAA1614 |
| chr8 | 53593997 | 53594972 | 976 | 6 | 1.19E-09 | 4.00E-08 | 9.71E-05 | 2.37E-09 | -0.00216621 | NA |
| chr16 | 23148022 | 23149585 | 1564 | 6 | 1.82E-08 | 6.21E-09 | 8.72E-05 | 2.37E-09 | 0.000269992 | USP31 |
| chr11 | 68903278 | 68904739 | 1462 | 16 | 7.31E-08 | 8.67E-06 | 1.44E-05 | 2.37E-09 | -0.00213859 | IGHMBP2, MRPL21 |
| chr11 | 5595672 | 5597178 | 1507 | 11 | 1.73E-09 | 1.25E-09 | 0.007028 | 2.37E-09 | -0.01906619 | TRIM6, TRIM6-TRIM34, AC104389.5 |
| chr16 | 4787888 | 4788905 | 1018 | 10 | 3.01E-10 | 3.08E-08 | 0.000212 | 2.39E-09 | -0.00460663 | SMIM22, SEPTIN12, AC020663.4 |
| chr10 | 1363742 | 1364313 | 572 | 5 | 8.03E-10 | 6.90E-09 | 9.39E-05 | 2.40E-09 | -0.02336701 | ADARB2 |
| chr7 | 107563690 | 107565668 | 1979 | 22 | 5.84E-08 | 1.23E-06 | 0.002777 | 2.40E-09 | -0.00019551 | DUS4L, DUS4L-BCAP29, COG5 |
| chr3 | 123447714 | 123449539 | 1826 | 8 | 5.05E-10 | 9.41E-06 | 1.35E-06 | 2.41E-09 | -0.00199815 | ADCY5 |
| chr5 | 537234 | 539765 | 2532 | 11 | 5.41E-08 | 8.98E-07 | 8.24E-06 | 2.41E-09 | -0.00155687 | NA |
| chr15 | 23564798 | 23566328 | 1531 | 11 | 5.64E-08 | 7.76E-10 | 0.003278 | 2.42E-09 | 0.007571688 | MKRN3 |
| chr9 | 23820981 | 23823127 | 2147 | 17 | 1.89E-08 | 1.56E-08 | 0.005478 | 2.42E-09 | 0.000397234 | ELAVL2 |
| chr3 | 48472697 | 48473936 | 1240 | 10 | 1.06E-09 | 3.31E-09 | 0.000166 | 2.43E-09 | 0.000243271 | SHISA5 |
| chr1 | 153356585 | 153358300 | 1716 | 13 | 2.15E-08 | 1.15E-07 | 0.000445 | 2.43E-09 | -0.00050868 | S100A9 |
| chr14 | 21526228 | 21527593 | 1366 | 11 | 4.16E-10 | 1.96E-08 | 1.38E-05 | 2.45E-09 | -0.00156553 | SALL2 |
| chr6 | 36954535 | 36956154 | 1620 | 5 | 9.04E-11 | 2.15E-08 | 4.04E-06 | 2.45E-09 | 0.003524248 | PI16 |
| chr10 | 129958223 | 129959537 | 1315 | 8 | 6.81E-10 | 2.19E-07 | 1.50E-05 | 2.45E-09 | -0.00454451 | EBF3 |
| chr16 | 67966240 | 67967919 | 1680 | 10 | 7.00E-10 | 2.53E-09 | 6.01E-05 | 2.47E-09 | -0.00568371 | SLC12A4 |
| chr7 | 151754291 | 151755674 | 1384 | 3 | 3.56E-08 | 1.94E-09 | 6.52E-06 | 2.47E-09 | -0.01181264 | PRKAG2 |
| chr2 | 70984810 | 70985374 | 565 | 5 | 7.38E-10 | 2.80E-09 | 7.36E-05 | 2.48E-09 | -0.01107048 | ANKRD53, AC007040.2 |
| chr3 | 170181159 | 170182685 | 1527 | 12 | 8.87E-09 | 1.08E-10 | 0.003757 | 2.49E-09 | -0.00252314 | PHC3 |
| chr16 | 88698663 | 88700660 | 1998 | 10 | 8.29E-08 | 3.23E-09 | 0.001154 | 2.49E-09 | -0.01209364 | RNF166 |
| chr10 | 94323514 | 94324402 | 889 | 3 | 3.99E-11 | 1.24E-08 | 4.78E-06 | 2.51E-09 | 0.003459388 | PLCE1 |
| chr8 | 48912314 | 48913841 | 1528 | 7 | 1.06E-09 | 1.63E-07 | 4.20E-06 | 2.51E-09 | -0.0004383 | NA |
| chr9 | 15873831 | 15874431 | 601 | 3 | 3.70E-09 | 8.47E-10 | 6.64E-05 | 2.52E-09 | -0.01577568 | CCDC171 |
| chr2 | 218402475 | 218403292 | 818 | 7 | 2.93E-10 | 1.15E-07 | 9.65E-06 | 2.52E-09 | -0.00137426 | CTDSP1, MIR26B |
| chr18 | 712731 | 713205 | 475 | 9 | 5.07E-10 | 1.67E-09 | 0.001078 | 2.53E-09 | -0.02644939 | NA |
| chr10 | 25174421 | 25176574 | 2154 | 24 | 9.45E-08 | 1.39E-08 | 0.000464 | 2.55E-09 | 0.001225691 | GPR158, GPR158-AS1 |
| chr6 | 30184545 | 30186105 | 1561 | 10 | 6.13E-12 | 3.06E-05 | 6.92E-07 | 2.56E-09 | 0.001966576 | TRIM26 |
| chr2 | 21043628 | 21045280 | 1653 | 14 | 3.32E-08 | 2.12E-10 | 0.000473 | 2.56E-09 | -0.02150716 | APOB |
| chr19 | 11434594 | 11436305 | 1712 | 28 | 2.58E-09 | 2.84E-06 | 0.000621 | 2.57E-09 | -0.00045135 | PRKCSH, CCDC151 |
| chr2 | 69642830 | 69645055 | 2226 | 18 | 1.77E-07 | 3.10E-07 | 0.002436 | 2.57E-09 | -0.00085982 | ANXA4, AAK1 |
| chr19 | 10717470 | 10719127 | 1658 | 25 | 1.56E-10 | 5.45E-07 | 0.001199 | 2.58E-09 | -1.01E-05 | DNM2, MIR638 |
| chr1 | 176552057 | 176553369 | 1313 | 5 | 2.32E-09 | 7.33E-07 | 1.79E-06 | 2.58E-09 | 0.019116711 | PAPPA2 |
| chr15 | 52277223 | 52278415 | 1193 | 5 | 4.54E-09 | 3.76E-06 | 2.12E-06 | 2.58E-09 | -0.00193467 | MYO5C |
| chr5 | 135399242 | 135400908 | 1667 | 19 | 8.93E-09 | 1.08E-07 | 0.002967 | 2.59E-09 | -0.00468657 | AC026691.1, MACROH2A1 |
| chr1 | 5990567 | 5992612 | 2046 | 20 | 4.72E-07 | 5.12E-08 | 0.001564 | 2.60E-09 | 0.000769211 | KCNAB2, NPHP4 |
| chr4 | 154740697 | 154741942 | 1246 | 8 | 6.17E-13 | 2.90E-07 | 7.43E-06 | 2.61E-09 | -0.00791897 | LRAT |
| chr10 | 102916775 | 102917843 | 1069 | 6 | 4.09E-09 | 2.40E-08 | 0.000101 | 2.61E-09 | -0.01017878 | AL356608.3 |
| chr15 | 42548094 | 42549370 | 1277 | 16 | 5.71E-11 | 9.10E-07 | 0.000236 | 2.61E-09 | -0.00014529 | HAUS2, AC018362.3, LRRC57 |
| chr5 | 177307857 | 177308686 | 830 | 3 | 2.53E-12 | 7.68E-09 | 1.31E-06 | 2.62E-09 | 0.00930668 | MXD3 |
| chr2 | 70190281 | 70191740 | 1460 | 13 | 1.93E-11 | 7.48E-08 | 1.83E-06 | 2.62E-09 | -0.00190254 | C2orf42 |
| chr8 | 143872389 | 143874613 | 2225 | 10 | 5.25E-08 | 1.39E-06 | 8.60E-05 | 2.62E-09 | 0.002246157 | EPPK1 |
| chr6 | 12064204 | 12064742 | 539 | 2 | 1.23E-10 | 6.39E-09 | 4.03E-07 | 2.62E-09 | -0.01361081 | HIVEP1 |
| chr2 | 153870972 | 153871839 | 868 | 10 | 2.10E-10 | 2.37E-08 | 2.56E-05 | 2.63E-09 | -0.00322075 | NA |
| chr8 | 24905803 | 24906285 | 483 | 2 | 2.46E-10 | 4.40E-09 | 6.88E-07 | 2.63E-09 | -0.02580705 | AC120193.1 |
| chr9 | 15552208 | 15553453 | 1246 | 13 | 6.73E-10 | 9.64E-08 | 0.000174 | 2.64E-09 | 0.000335061 | CCDC171 |
| chr1 | 31798685 | 31798994 | 310 | 6 | 1.45E-09 | 2.72E-09 | 0.00028 | 2.64E-09 | -0.00881743 | SPOCD1 |
| chr7 | 151877407 | 151878592 | 1186 | 10 | 5.63E-18 | 0.0001177 | 2.36E-08 | 2.64E-09 | -0.00271776 | PRKAG2-AS1 |
| chr9 | 6413544 | 6414194 | 651 | 6 | 2.69E-11 | 3.41E-06 | 7.80E-06 | 2.64E-09 | -0.00214946 | UHRF2 |
| chr2 | 172829072 | 172829482 | 411 | 2 | 5.08E-10 | 2.99E-09 | 1.41E-06 | 2.65E-09 | -0.0127509 | RAPGEF4 |
| chr4 | 38118703 | 38118945 | 243 | 5 | 2.40E-09 | 1.83E-09 | 9.03E-05 | 2.65E-09 | 0.002645834 | TBC1D1 |
| chr8 | 6834865 | 6836805 | 1941 | 15 | 4.13E-09 | 5.45E-08 | 0.000587 | 2.66E-09 | -0.00093772 | GS1-24F4.2, XKR5 |
| chr7 | 128789638 | 128790615 | 978 | 3 | 7.30E-11 | 2.08E-08 | 4.89E-07 | 2.66E-09 | -0.00799386 | NA |
| chr11 | 126211033 | 126212105 | 1073 | 17 | 7.67E-10 | 3.39E-08 | 0.000345 | 2.67E-09 | 0.000635078 | FAM118B, RPUSD4 |
| chr6 | 137491168 | 137493219 | 2052 | 6 | 3.01E-07 | 9.18E-08 | 4.76E-05 | 2.67E-09 | -0.01064661 | OLIG3 |
| chr19 | 15113733 | 15114103 | 371 | 4 | 8.07E-11 | 7.68E-06 | 1.66E-06 | 2.67E-09 | 0.003814967 | SYDE1 |
| chr22 | 30762848 | 30764472 | 1625 | 10 | 2.98E-07 | 1.43E-07 | 0.0001 | 2.68E-09 | -0.00320429 | OSBP2 |
| chr11 | 48101293 | 48101364 | 72 | 2 | 5.03E-10 | 1.74E-08 | 1.46E-07 | 2.68E-09 | -0.00781137 | PTPRJ |
| chr4 | 658738 | 660583 | 1846 | 7 | 4.76E-07 | 7.95E-08 | 6.10E-05 | 2.68E-09 | -0.00362528 | PDE6B |
| chr6 | 110813803 | 110815285 | 1483 | 15 | 1.32E-09 | 3.28E-08 | 0.000137 | 2.68E-09 | -0.00113139 | CDK19 |
| chr6 | 43644168 | 43645658 | 1491 | 11 | 1.23E-10 | 2.18E-07 | 4.30E-05 | 2.68E-09 | -0.00244651 | RSPH9 |
| chr1 | 47423701 | 47424547 | 847 | 2 | 2.22E-09 | 3.60E-09 | 1.04E-06 | 2.71E-09 | 0.020511018 | NA |
| chr10 | 51699342 | 51700295 | 954 | 14 | 3.85E-09 | 2.89E-09 | 0.000768 | 2.71E-09 | -0.00105691 | PRKG1, CSTF2T |
| chr3 | 52238480 | 52240545 | 2066 | 15 | 7.06E-09 | 1.94E-08 | 0.000392 | 2.71E-09 | -0.00192612 | AC006252.1, TWF2 |
| chr7 | 1005389 | 1005952 | 564 | 5 | 6.20E-10 | 6.31E-08 | 3.60E-06 | 2.75E-09 | -0.00211787 | C7orf50 |
| chr15 | 42401250 | 42402755 | 1506 | 8 | 3.43E-12 | 5.18E-07 | 2.78E-06 | 2.75E-09 | 0.003292184 | AC012651.1, CAPN3 |
| chr6 | 128761928 | 128762179 | 252 | 4 | 1.48E-09 | 2.44E-09 | 1.24E-05 | 2.75E-09 | -0.00046426 | NA |
| chr7 | 130265528 | 130267359 | 1832 | 5 | 3.93E-08 | 1.55E-07 | 6.75E-06 | 2.75E-09 | -0.00460542 | CPA2 |
| chr19 | 10520199 | 10521097 | 899 | 4 | 8.40E-09 | 2.86E-09 | 0.000101 | 2.76E-09 | -0.01073438 | NA |
| chr16 | 25216762 | 25217388 | 627 | 8 | 1.24E-09 | 1.58E-07 | 0.000173 | 2.76E-09 | -0.00443228 | AQP8 |
| chr1 | 92486960 | 92488350 | 1391 | 13 | 7.73E-11 | 2.96E-08 | 2.56E-05 | 2.78E-09 | -0.00647031 | NA |
| chr2 | 53786051 | 53788095 | 2045 | 21 | 1.42E-09 | 1.87E-08 | 8.27E-05 | 2.78E-09 | 0.00082227 | ERLEC1, ASB3 |
| chr2 | 218994973 | 218996404 | 1432 | 7 | 1.35E-07 | 2.09E-09 | 0.000273 | 2.79E-09 | -0.00650956 | NA |
| chr18 | 36067507 | 36069981 | 2475 | 11 | 4.99E-06 | 8.65E-08 | 0.000589 | 2.79E-09 | -0.00266088 | RPRD1A |
| chr17 | 1376858 | 1377528 | 671 | 3 | 1.76E-10 | 6.64E-09 | 1.27E-06 | 2.79E-09 | -0.00397309 | YWHAE |
| chr1 | 99849534 | 99851081 | 1548 | 24 | 8.97E-09 | 1.64E-07 | 0.000544 | 2.79E-09 | 0.001017894 | AGL |
| chr10 | 93611744 | 93612705 | 962 | 9 | 1.73E-11 | 1.32E-06 | 6.54E-07 | 2.80E-09 | -0.00319576 | PDE6C |
| chr4 | 499051 | 500676 | 1626 | 14 | 1.36E-09 | 1.40E-07 | 0.000386 | 2.82E-09 | -0.00264239 | PIGG, ZNF721 |

| | | | | | | | | | | |
|---|---|---|---|---|---|---|---|---|---|---|
| chr7 | 7968309 | 7971064 | 2756 | 20 | 1.78E-07 | 5.94E-06 | 1.20E-05 | 2.82E-09 | 0.001734419 | AC007161.3, GLCCI1, GLCCI1-DT |
| chr3 | 78046579 | 78046957 | 379 | 4 | 2.53E-10 | 1.53E-08 | 8.75E-07 | 2.82E-09 | -0.00643636 | LINC02077 |
| chr2 | 241355571 | 241356584 | 1014 | 9 | 1.21E-10 | 1.30E-07 | 6.87E-06 | 2.83E-09 | -0.00214809 | FARP2 |
| chr10 | 24097682 | 24098215 | 534 | 2 | 6.25E-13 | 8.08E-06 | 1.12E-08 | 2.85E-09 | -0.02846887 | KIAA1217 |
| chr10 | 110006539 | 110008109 | 1571 | 13 | 1.41E-09 | 3.67E-08 | 4.62E-05 | 2.85E-09 | -0.00380816 | ADD3, ADD3-AS1 |
| chr16 | 11396333 | 11396582 | 250 | 3 | 3.47E-09 | 1.13E-09 | 5.08E-05 | 2.85E-09 | -0.01182257 | AC099489.1 |
| chr20 | 51022025 | 51023606 | 1582 | 8 | 1.00E-06 | 5.28E-07 | 6.31E-05 | 2.86E-09 | -0.00367934 | KCNG1 |
| chr8 | 17412129 | 17414026 | 1898 | 14 | 2.95E-08 | 9.58E-08 | 7.61E-05 | 2.86E-09 | -0.00541226 | MTMR7 |
| chr1 | 155196719 | 155198454 | 1736 | 12 | 7.96E-10 | 5.38E-07 | 5.70E-06 | 2.87E-09 | -0.00084389 | THBS3-AS1, THBS3 |
| chr7 | 35037986 | 35039499 | 1514 | 14 | 1.49E-14 | 2.61E-06 | 3.16E-07 | 2.87E-09 | -0.00879228 | AC005400.1, DPY19L1 |
| chr19 | 56837836 | 56838945 | 1110 | 9 | 9.88E-11 | 1.24E-08 | 0.000103 | 2.88E-09 | -0.00987731 | ZIM2, PEG3 |
| chr5 | 132830636 | 132831790 | 1155 | 9 | 8.68E-12 | 1.30E-06 | 6.38E-06 | 2.88E-09 | -0.0057201 | SHROOM1 |
| chr10 | 71377156 | 71377804 | 649 | 5 | 5.62E-10 | 2.04E-08 | 1.67E-05 | 2.88E-09 | -0.01903724 | SLC29A3 |
| chr6 | 109465875 | 109466732 | 858 | 7 | 1.86E-09 | 1.24E-07 | 6.23E-05 | 2.88E-09 | -0.00284218 | MICAL1, ZBTB24 |
| chr2 | 1841694 | 1843348 | 1655 | 9 | 1.72E-07 | 8.84E-06 | 1.98E-05 | 2.90E-09 | -0.00890546 | MYT1L |
| chr16 | 3579619 | 3581569 | 1951 | 8 | 2.31E-07 | 3.27E-07 | 0.000439 | 2.90E-09 | 0.006491917 | AC006111.1, SLX4 |
| chr15 | 48962921 | 48964061 | 1141 | 16 | 7.83E-10 | 1.58E-07 | 0.000152 | 2.90E-09 | -0.0002077 | SHC4 |
| chr1 | 61972288 | 61973349 | 1062 | 5 | 1.50E-10 | 1.13E-07 | 8.05E-07 | 2.94E-09 | -0.00245684 | PATJ |
| chr6 | 39229222 | 39230616 | 1395 | 9 | 5.79E-10 | 2.39E-07 | 2.72E-06 | 2.94E-09 | 0.000336999 | KCNK5 |
| chr6 | 29608552 | 29610536 | 1985 | 9 | 3.70E-08 | 1.53E-08 | 3.62E-05 | 2.95E-09 | -0.00102586 | GABBR1 |
| chr13 | 27922276 | 27923954 | 1679 | 7 | 1.16E-14 | 2.38E-06 | 1.39E-07 | 2.95E-09 | -0.00555971 | PDX1 |
| chr11 | 69370910 | 69371384 | 475 | 5 | 6.85E-10 | 2.38E-07 | 1.44E-05 | 2.95E-09 | -0.00591771 | NA |
| chr15 | 43266657 | 43267386 | 730 | 8 | 6.03E-10 | 1.10E-07 | 9.63E-05 | 2.96E-09 | -0.00492481 | TGM5 |
| chr15 | 97874042 | 97874984 | 943 | 6 | 1.59E-12 | 4.60E-06 | 4.57E-08 | 2.96E-09 | 0.0041472 | LINC00923 |
| chr11 | 75667227 | 75669528 | 2302 | 20 | 5.06E-07 | 9.54E-09 | 0.000791 | 2.97E-09 | -8.30E-05 | MAP6 |
| chr9 | 135559797 | 135561104 | 1308 | 5 | 8.65E-09 | 3.44E-08 | 1.97E-05 | 2.99E-09 | -0.01054114 | NA |
| chr10 | 129790877 | 129790925 | 49 | 3 | 3.11E-09 | 1.49E-09 | 2.09E-05 | 2.99E-09 | 0.016611859 | LINC02666 |
| chr11 | 3159884 | 3161539 | 1656 | 8 | 5.14E-09 | 7.68E-08 | 9.77E-05 | 2.99E-09 | -0.00782962 | OSBPL5 |
| chr10 | 21525609 | 21527338 | 1730 | 12 | 4.95E-12 | 1.52E-06 | 2.63E-05 | 2.99E-09 | 0.001216773 | MLLT10, AL358780.1, SKIDA1 |
| chr6 | 41430061 | 41430955 | 895 | 2 | 5.34E-09 | 3.48E-09 | 1.41E-06 | 3.00E-09 | -0.01747152 | NA |
| chr3 | 128425458 | 128427221 | 1764 | 10 | 2.92E-08 | 3.25E-08 | 0.000148 | 3.00E-09 | -0.00297187 | NA |
| chr7 | 975073 | 976094 | 1022 | 12 | 1.96E-10 | 1.18E-07 | 0.00106 | 3.01E-09 | -0.0043099 | COX19 |
| chr2 | 38215321 | 38215456 | 136 | 2 | 1.28E-09 | 3.18E-09 | 1.71E-06 | 3.01E-09 | -0.01805996 | CYP1B1-AS1, AC009229.1 |
| chr2 | 241795720 | 241796099 | 380 | 3 | 2.59E-10 | 3.42E-08 | 7.92E-07 | 3.01E-09 | -0.0174867 | GAL3ST2 |
| chr2 | 12006251 | 12007117 | 867 | 6 | 1.33E-08 | 2.08E-07 | 0.000435 | 3.01E-09 | 0.00981301 | MIR3681HG |
| chr3 | 129892672 | 129893726 | 1055 | 9 | 1.95E-13 | 1.17E-06 | 6.94E-08 | 3.02E-09 | -0.00057672 | TMCC1 |
| chr10 | 45180129 | 45180232 | 104 | 3 | 3.03E-09 | 9.92E-10 | 8.48E-05 | 3.02E-09 | 0.001984524 | AL358394.1 |
| chr4 | 3492655 | 3494573 | 1919 | 6 | 4.42E-09 | 3.09E-09 | 7.85E-05 | 3.02E-09 | -0.00258448 | DOK7 |
| chr9 | 14722244 | 14723606 | 1363 | 4 | 2.11E-07 | 7.88E-10 | 0.000203 | 3.02E-09 | -0.01499122 | CER1 |
| chr6 | 31127499 | 31128136 | 638 | 6 | 1.54E-09 | 4.56E-09 | 0.000112 | 3.02E-09 | -0.01108251 | PSORS1C1 |
| chr9 | 17627815 | 17627943 | 129 | 2 | 1.94E-09 | 1.77E-09 | 1.28E-05 | 3.02E-09 | -0.03981637 | SH3GL2 |
| chr17 | 80008541 | 80009375 | 835 | 8 | 1.07E-11 | 4.02E-07 | 2.04E-05 | 3.03E-09 | -0.00258029 | TBC1D16 |
| chr15 | 64567036 | 64567211 | 176 | 2 | 8.89E-11 | 4.49E-07 | 2.62E-08 | 3.03E-09 | -0.01072657 | ZNF609 |
| chr20 | 46688580 | 46690711 | 2132 | 17 | 2.27E-07 | 6.34E-08 | 0.000257 | 3.04E-09 | -0.00265244 | AL031055.1, TP53RK |
| chr7 | 98993291 | 98994788 | 1498 | 8 | 1.02E-07 | 1.05E-09 | 0.000371 | 3.04E-09 | -0.0011349 | TRRAP, AC004893.2 |
| chr15 | 93179270 | 93180021 | 752 | 3 | 2.47E-11 | 2.47E-08 | 4.93E-06 | 3.05E-09 | 0.001390498 | AC108457.1 |
| chr19 | 35513331 | 35514493 | 1163 | 7 | 4.54E-09 | 7.19E-10 | 0.000546 | 3.05E-09 | -0.00939056 | DMKN |
| chr3 | 196081384 | 196082640 | 1257 | 14 | 1.39E-09 | 5.63E-08 | 8.96E-06 | 3.06E-09 | -0.00010046 | TFRC |
| chr10 | 132057359 | 132058795 | 1437 | 6 | 6.34E-08 | 4.72E-09 | 0.000181 | 3.06E-09 | -0.00534995 | JAKMIP3 |
| chr22 | 41090831 | 41091757 | 927 | 7 | 6.87E-09 | 5.75E-08 | 7.81E-05 | 3.06E-09 | 0.005946167 | NA |
| chr12 | 111765991 | 111767341 | 1351 | 12 | 2.71E-10 | 1.45E-05 | 2.60E-06 | 3.07E-09 | 0.002235253 | AC002996.1, ALDH2 |
| chr9 | 131490166 | 131490467 | 302 | 3 | 4.10E-11 | 1.37E-05 | 3.80E-07 | 3.08E-09 | -0.00409382 | PRRC2B |
| chr17 | 77557517 | 77558576 | 1060 | 5 | 3.49E-09 | 1.45E-06 | 1.72E-05 | 3.09E-09 | -0.00325895 | AC021683.2 |
| chr1 | 3786986 | 3787995 | 1010 | 7 | 3.64E-09 | 6.75E-08 | 2.33E-05 | 3.09E-09 | -0.00808177 | LRRC47 |
| chr1 | 33081564 | 33082882 | 1319 | 8 | 1.97E-14 | 1.93E-05 | 2.34E-06 | 3.10E-09 | -0.00844729 | AZIN2 |
| chr1 | 17203210 | 17204520 | 1311 | 6 | 7.63E-10 | 4.34E-05 | 6.79E-06 | 3.10E-09 | -0.00297569 | NA |
| chr8 | 109580732 | 109582165 | 1434 | 5 | 1.83E-07 | 6.14E-09 | 0.000376 | 3.10E-09 | -0.00134628 | SYBU |
| chr1 | 224928892 | 224930115 | 1224 | 12 | 1.00E-08 | 6.25E-08 | 0.000109 | 3.10E-09 | -0.00047432 | DNAH14 |
| chr11 | 67487953 | 67489212 | 1260 | 4 | 3.15E-07 | 9.46E-09 | 2.76E-05 | 3.12E-09 | -0.00803053 | AIP |
| chr8 | 38046705 | 38047289 | 585 | 6 | 2.12E-10 | 3.28E-07 | 4.40E-05 | 3.13E-09 | -0.00517058 | EIF4EBP1 |
| chr11 | 47449081 | 47449901 | 821 | 11 | 5.48E-08 | 1.73E-10 | 0.00736 | 3.13E-09 | -0.00456147 | RAPSN |
| chr5 | 64689171 | 64690303 | 1133 | 7 | 6.61E-11 | 3.61E-07 | 2.83E-06 | 3.14E-09 | -0.00721356 | NA |
| chr2 | 9143011 | 9144763 | 1753 | 5 | 4.35E-11 | 1.03E-06 | 1.86E-06 | 3.14E-09 | 0.002346359 | AC093904.1 |
| chr1 | 178894326 | 178894534 | 209 | 2 | 1.85E-10 | 7.16E-08 | 6.46E-08 | 3.15E-09 | -0.00821546 | RALGPS2 |
| chr1 | 30816543 | 30817370 | 828 | 4 | 8.51E-14 | 8.52E-07 | 3.85E-08 | 3.15E-09 | -0.01604745 | NA |
| chr17 | 50173144 | 50174437 | 1294 | 5 | 4.21E-08 | 7.90E-08 | 5.17E-05 | 3.15E-09 | -0.00104079 | SGCA, H1-9P |
| chr17 | 1515986 | 1517255 | 1270 | 12 | 2.00E-09 | 4.02E-08 | 0.001035 | 3.16E-09 | 0.000982858 | PITPNA-AS1, INPP5K |
| chr12 | 54260004 | 54261844 | 1841 | 5 | 4.50E-09 | 4.42E-07 | 1.33E-05 | 3.16E-09 | 0.004888622 | CBX5 |
| chr7 | 94906305 | 94909096 | 2792 | 24 | 3.36E-08 | 1.12E-05 | 7.50E-05 | 3.16E-09 | 0.000183039 | PPP1R9A |
| chr13 | 94710575 | 94712421 | 1847 | 14 | 4.21E-09 | 4.45E-07 | 0.000425 | 3.16E-09 | -0.00150398 | SOX21-AS1, SOX21 |
| chr6 | 170064083 | 170064153 | 71 | 2 | 2.55E-10 | 1.45E-07 | 4.59E-08 | 3.17E-09 | -0.01216833 | NA |
| chr6 | 31891930 | 31892986 | 1057 | 16 | 1.53E-10 | 9.13E-06 | 7.47E-05 | 3.17E-09 | 0.00179959 | EHMT2 |
| chr15 | 25088970 | 25090598 | 1629 | 7 | 7.55E-10 | 1.29E-08 | 8.80E-06 | 3.17E-09 | -0.00752776 | SNHG14, SNORD116-22 |
| chr2 | 206641756 | 206642697 | 942 | 9 | 6.22E-11 | 4.43E-08 | 0.000296 | 3.18E-09 | -0.006279 | FAM237A |
| chr4 | 23888583 | 23889267 | 685 | 5 | 2.56E-13 | 9.45E-06 | 6.38E-07 | 3.19E-09 | -0.00687113 | PPARGC1A |
| chr3 | 58164732 | 58165902 | 1171 | 7 | 7.33E-10 | 1.68E-08 | 0.000107 | 3.20E-09 | -0.0028268 | FLNB, FLNB-AS1 |
| chr18 | 79576201 | 79577370 | 1170 | 7 | 4.74E-11 | 3.50E-06 | 2.80E-05 | 3.20E-09 | 0.000102481 | AC068473.3 |
| chr11 | 128823578 | 128825020 | 1443 | 11 | 7.70E-09 | 3.00E-08 | 6.14E-05 | 3.20E-09 | -0.01514928 | NA |
| chr5 | 132745651 | 132746402 | 752 | 2 | 1.01E-12 | 1.74E-07 | 4.20E-08 | 3.22E-09 | -0.0015825 | NA |
| chr6 | 31657924 | 31661158 | 3235 | 40 | 4.52E-07 | 1.78E-07 | 0.001363 | 3.22E-09 | -0.00052817 | APOM, C6orf47-AS1, C6orf47 |
| chr19 | 38510518 | 38511491 | 974 | 4 | 3.90E-09 | 3.13E-09 | 7.34E-06 | 3.23E-09 | -0.00528746 | RYR1 |
| chr19 | 55209812 | 55210453 | 642 | 5 | 9.57E-12 | 2.00E-06 | 1.45E-06 | 3.23E-09 | -0.01005331 | NA |
| chr3 | 86990000 | 86992292 | 2293 | 16 | 1.21E-07 | 5.21E-08 | 0.000261 | 3.24E-09 | -0.00275475 | VGLL3 |
| chr3 | 172749484 | 172751219 | 1736 | 13 | 5.57E-10 | 7.29E-08 | 0.000162 | 3.24E-09 | -0.00318032 | ECT2 |
| chr18 | 12947067 | 12948331 | 1265 | 14 | 7.54E-10 | 8.43E-08 | 8.93E-05 | 3.25E-09 | 0.001336797 | SEH1L |
| chr7 | 90243949 | 90245772 | 1824 | 16 | 7.84E-10 | 3.66E-06 | 0.000383 | 3.25E-09 | -0.004115 | CFAP69 |
| chr17 | 80720721 | 80721056 | 336 | 4 | 1.27E-09 | 6.14E-09 | 1.67E-05 | 3.25E-09 | -0.00875774 | RPTOR |
| chr9 | 35814226 | 35816326 | 2101 | 16 | 2.19E-09 | 1.93E-06 | 6.14E-05 | 3.25E-09 | 0.004056101 | TMEM8B, HINT2, AL133410.3 |
| chr2 | 3535980 | 3537184 | 1205 | 7 | 2.88E-09 | 9.42E-10 | 0.001321 | 3.26E-09 | -0.01637317 | AC108488.1, AC108488.2 |
| chr14 | 37546795 | 37546983 | 189 | 2 | 1.56E-10 | 2.60E-08 | 1.38E-07 | 3.26E-09 | -0.02150908 | MIPOL1 |
| chr20 | 62182183 | 62183616 | 1434 | 14 | 1.98E-09 | 2.68E-07 | 7.28E-05 | 3.26E-09 | 0.001139015 | SS18L1, MTG2 |
| chr4 | 2320230 | 2320980 | 751 | 6 | 8.47E-09 | 7.82E-09 | 0.000152 | 3.26E-09 | -0.00680459 | ZFYVE28 |
| chr2 | 211593195 | 211593348 | 154 | 2 | 1.99E-10 | 2.37E-09 | 4.98E-06 | 3.27E-09 | -0.01648495 | ERBB4 |
| chr2 | 232927180 | 232929211 | 2032 | 8 | 1.24E-08 | 7.38E-08 | 7.30E-05 | 3.27E-09 | -2.17E-05 | NGEF |
| chr19 | 7888331 | 7889099 | 769 | 10 | 4.93E-16 | 2.63E-05 | 3.75E-08 | 3.28E-09 | -0.00198345 | LRRC8E |
| chr19 | 56538320 | 56539465 | 1146 | 10 | 1.59E-10 | 1.68E-07 | 0.000691 | 3.29E-09 | 0.001029994 | ZFP28, AC005498.2 |
| chr1 | 31423092 | 31423615 | 524 | 4 | 2.60E-09 | 6.88E-09 | 2.10E-05 | 3.29E-09 | -0.01920542 | SERINC2 |
| chr6 | 157773509 | 157774126 | 618 | 3 | 5.64E-11 | 7.86E-07 | 1.71E-06 | 3.30E-09 | -0.00860521 | SNX9 |
| chr19 | 11617439 | 11618344 | 906 | 3 | 5.05E-09 | 9.90E-10 | 0.000118 | 3.30E-09 | -0.00473675 | ZNF627 |
| chr5 | 180135118 | 180136200 | 1083 | 6 | 1.87E-10 | 6.59E-08 | 1.45E-05 | 3.30E-09 | 0.005835244 | RASGEF1C |
| chr6 | 31814190 | 31815768 | 1579 | 26 | 3.62E-10 | 2.07E-07 | 4.32E-05 | 3.30E-09 | -0.00071652 | HSPA1A, HSPA1L |
| chr13 | 76886163 | 76887291 | 1129 | 15 | 6.05E-10 | 4.06E-07 | 2.94E-05 | 3.30E-09 | -0.00243214 | KCTD12 |
| chr19 | 35640778 | 35642010 | 1233 | 12 | 2.30E-09 | 9.41E-07 | 2.83E-05 | 3.31E-09 | -0.00240449 | ETV2 |
| chr11 | 61507775 | 61508975 | 1201 | 7 | 4.60E-08 | 1.08E-07 | 0.000185 | 3.31E-09 | -0.00312577 | MIR4488, LRRC10B |
| chr20 | 45406035 | 45407496 | 1462 | 17 | 3.37E-08 | 1.85E-07 | 0.000145 | 3.31E-09 | -0.00227201 | SYS1-DBNDD2, DBNDD2, TP53TG5 |
| chr20 | 18793591 | 18795501 | 1911 | 15 | 2.06E-11 | 8.67E-06 | 0.000135 | 3.31E-09 | 0.000160207 | LINC00653, LINC00652 |
| chr12 | 49961057 | 49962867 | 1811 | 8 | 1.82E-08 | 2.72E-10 | 0.001605 | 3.33E-09 | -0.01162635 | AQP5, AC025154.2 |
| chr13 | 32427614 | 32428683 | 1070 | 15 | 1.51E-09 | 6.04E-08 | 0.001433 | 3.35E-09 | 0.001347202 | N4BP2L1, AL137247.2 |
| chr5 | 58863645 | 58863661 | 17 | 2 | 5.36E-10 | 3.64E-09 | 1.72E-06 | 3.35E-09 | 0.020426339 | NA |
| chr17 | 45942466 | 45943412 | 947 | 3 | 2.50E-11 | 4.05E-06 | 2.53E-06 | 3.35E-09 | -0.01690372 | MAPT |
| chr3 | 9600784 | 9600946 | 163 | 3 | 3.84E-09 | 1.24E-09 | 5.85E-05 | 3.35E-09 | 0.023143888 | DUSP5P2 |
| chr22 | 35634613 | 35636470 | 1858 | 4 | 7.66E-06 | 3.28E-08 | 2.99E-05 | 3.35E-09 | -0.0008005 | MB, AL049747.1 |
| chr19 | 35449272 | 35449960 | 689 | 7 | 1.71E-10 | 2.22E-07 | 1.51E-05 | 3.36E-09 | -0.01005676 | FFAR2 |
| chr5 | 173214311 | 173215101 | 791 | 2 | 1.30E-08 | 2.33E-09 | 5.99E-06 | 3.36E-09 | -0.00695211 | NA |
| chr22 | 50485461 | 50486908 | 1448 | 11 | 6.99E-08 | 1.26E-06 | 3.41E-05 | 3.37E-09 | -0.0051277 | ADM2, MIOX |
| chr15 | 45169246 | 45169735 | 490 | 3 | 5.25E-10 | 6.56E-09 | 2.59E-06 | 3.38E-09 | -0.00382506 | SHF |
| chr1 | 154500949 | 154502324 | 1376 | 8 | 1.95E-08 | 7.10E-10 | 0.001307 | 3.39E-09 | -0.01358687 | TDRD10, SHE |
| chr1 | 197266721 | 197266971 | 251 | 4 | 7.23E-10 | 9.54E-09 | 2.51E-06 | 3.39E-09 | -0.00575516 | NA |
| chr12 | 57737562 | 57739628 | 2067 | 12 | 1.00E-06 | 6.96E-09 | 0.000578 | 3.39E-09 | -0.00778636 | TSPAN31, AGAP2 |
| chr8 | 56113103 | 56114793 | 1691 | 9 | 1.06E-06 | 7.66E-10 | 0.003209 | 3.39E-09 | -0.00391392 | MOS |
| chr16 | 89200183 | 89201632 | 1450 | 8 | 2.70E-07 | 6.69E-10 | 0.000985 | 3.40E-09 | -0.00182604 | SLC22A31 |
| chr2 | 20012010 | 20013070 | 1061 | 12 | 6.63E-13 | 2.21E-06 | 2.22E-05 | 3.41E-09 | -0.00355022 | MATN3 |
| chr1 | 24928512 | 24929889 | 1378 | 12 | 1.41E-10 | 7.21E-08 | 2.86E-05 | 3.41E-09 | 0.000628892 | RUNX3 |
| chr1 | 151761828 | 151763471 | 1644 | 9 | 3.74E-10 | 1.37E-05 | 5.61E-06 | 3.41E-09 | -0.00248901 | OAZ3, MRPL9, AL589765.4 |
| chr1 | 31815859 | 31817092 | 1234 | 8 | 3.70E-08 | 7.96E-07 | 0.000475 | 3.41E-09 | -0.00956055 | SPOCD1 |
| chr17 | 80105809 | 80107781 | 1973 | 7 | 2.01E-08 | 2.22E-08 | 0.000111 | 3.42E-09 | -0.00180746 | GAA |
| chr4 | 105551833 | 105553429 | 1597 | 9 | 1.41E-08 | 1.30E-06 | 4.60E-06 | 3.42E-09 | -6.28E-05 | ARHGEF38, AC004066.1 |
| chr10 | 100329924 | 100331164 | 1241 | 10 | 5.34E-10 | 2.30E-08 | 0.000195 | 3.42E-09 | -0.00827328 | PKD2L1 |
| chr8 | 934647 | 936038 | 1392 | 7 | 1.35E-08 | 2.57E-08 | 0.000117 | 3.43E-09 | -0.00084265 | DLGAP2 |
| chr6 | 25753504 | 25754320 | 817 | 4 | 3.23E-09 | 3.39E-09 | 5.34E-05 | 3.43E-09 | -0.00853091 | NA |
| chr8 | 10895637 | 10895727 | 91 | 3 | 4.39E-09 | 1.08E-09 | 0.000103 | 3.44E-09 | -0.00450833 | NA |
| chr11 | 79437326 | 79440144 | 2819 | 15 | 1.81E-06 | 2.09E-07 | 0.000429 | 3.44E-09 | -0.00100394 | TENM4 |
| chr11 | 69891517 | 69892616 | 1100 | 9 | 3.96E-10 | 1.31E-08 | 0.000211 | 3.45E-09 | -0.01341051 | NA |
| chr7 | 43112229 | 43113793 | 1565 | 16 | 4.71E-10 | 3.07E-07 | 7.10E-05 | 3.45E-09 | 0.001984862 | HECW1, AC005537.1 |
| chr11 | 10398492 | 10399429 | 938 | 2 | 3.95E-10 | 6.70E-09 | 6.35E-07 | 3.46E-09 | -0.01039464 | AMPD3, RNU6ATAC33P |
| chr16 | 67155231 | 67156741 | 1511 | 8 | 9.20E-10 | 5.87E-07 | 0.000154 | 3.46E-09 | -0.00862335 | TRADD |
| chr8 | 132479710 | 132481504 | 1795 | 14 | 1.90E-07 | 1.34E-08 | 0.001634 | 3.47E-09 | 0.001153267 | KCNQ3 |
| chr6 | 3749481 | 3751384 | 1904 | 7 | 4.15E-07 | 4.10E-08 | 5.25E-05 | 3.47E-09 | 0.003326088 | AL391422.4, PXDC1 |
| chr11 | 5401366 | 5402970 | 1605 | 4 | 5.07E-10 | 5.92E-08 | 8.58E-06 | 3.47E-09 | 0.003585648 | OR51J1, HBG2, AC104389.5, HBE1, OR51B5 |
| chr4 | 2843003 | 2844428 | 1426 | 16 | 3.45E-09 | 1.24E-06 | 3.62E-05 | 3.47E-09 | -0.00122525 | ADD1 |
| chr14 | 69580130 | 69580274 | 145 | 2 | 2.39E-09 | 2.03E-09 | 1.56E-05 | 3.48E-09 | -0.03586384 | NA |
| chr1 | 11398572 | 11399349 | 778 | 2 | 4.20E-11 | 1.07E-08 | 3.52E-07 | 3.49E-09 | 0.007335943 | NA |
| chr5 | 110086817 | 110087863 | 1047 | 3 | 5.98E-07 | 4.08E-09 | 2.84E-05 | 3.50E-09 | 0.00030118 | NA |
| chr10 | 28261272 | 28261643 | 372 | 3 | 1.98E-10 | 1.27E-08 | 3.53E-06 | 3.50E-09 | -0.00954016 | MPP7 |
| chr19 | 14380355 | 14381265 | 911 | 4 | 1.28E-10 | 2.95E-09 | 0.000104 | 3.50E-09 | 0.013302248 | ADGRE5 |
| chr16 | 66966120 | 66967822 | 1703 | 6 | 5.96E-07 | 3.40E-09 | 6.03E-05 | 3.51E-09 | -0.00333326 | CES3 |
| chr11 | 130102771 | 130103543 | 773 | 2 | 7.59E-13 | 2.79E-06 | 1.62E-08 | 3.51E-09 | 0.003166175 | APLP2 |
| chr1 | 205271114 | 205271782 | 669 | 4 | 2.62E-13 | 2.26E-06 | 7.31E-08 | 3.52E-09 | -0.00264712 | TMCC2 |
| chr8 | 53286972 | 53287159 | 188 | 2 | 2.32E-09 | 2.10E-09 | 1.33E-05 | 3.53E-09 | -0.01538744 | NA |
| chr17 | 55458602 | 55459187 | 586 | 3 | 7.48E-11 | 1.20E-06 | 8.66E-08 | 3.54E-09 | -0.00815984 | SMIM36 |
| chr16 | 29994535 | 29996928 | 2394 | 23 | 1.51E-08 | 1.57E-06 | 0.000282 | 3.56E-09 | -0.00041451 | INO80E, HIRIP3 |
| chr2 | 8176422 | 8177188 | 767 | 2 | 4.38E-08 | 2.15E-09 | 1.21E-05 | 3.57E-09 | 0.00454084 | LINC00298, LINC00299 |
| chr17 | 43021703 | 43022522 | 820 | 9 | 1.13E-09 | 5.15E-06 | 8.29E-05 | 3.57E-09 | -0.00061888 | VAT1 |
| chr12 | 89350373 | 89351896 | 1524 | 16 | 1.28E-09 | 4.39E-08 | 0.000409 | 3.57E-09 | -0.00163581 | AC024909.1, DUSP6 |
| chr16 | 20610704 | 20612673 | 1970 | 12 | 9.76E-09 | 8.03E-07 | 5.11E-05 | 3.59E-09 | -0.00295908 | ACSM3 |
| chr16 | 10743417 | 10744303 | 887 | 9 | 1.95E-11 | 1.55E-07 | 9.49E-06 | 3.62E-09 | -0.00421403 | NUBP1 |
| chr20 | 62378359 | 62379782 | 1424 | 5 | 1.51E-10 | 2.39E-06 | 5.81E-07 | 3.63E-09 | -0.00121955 | NA |
| chr13 | 113621246 | 113621355 | 110 | 2 | 1.64E-09 | 3.44E-09 | 2.38E-06 | 3.63E-09 | -0.01376938 | TFDP1 |

| | | | | | | | | | | |
|---|---|---|---|---|---|---|---|---|---|---|
| chr11 | 68041804 | 68043727 | 1924 | 14 | 3.83E-10 | 1.38E-05 | 1.03E-05 | 3.63E-09 | -0.00257473 | TCIRG1 |
| chr21 | 44657273 | 44657800 | 528 | 7 | 2.03E-09 | 5.16E-09 | 0.000415 | 3.63E-09 | -0.0201375 | TSPEAR |
| chr11 | 47267372 | 47269676 | 2305 | 17 | 1.02E-07 | 2.02E-06 | 9.72E-05 | 3.63E-09 | -0.00209783 | NR1H3, MADD |
| chr12 | 50110305 | 50112606 | 2302 | 20 | 3.13E-09 | 1.32E-05 | 2.61E-05 | 3.63E-09 | -0.0004821 | GPD1, COX14, AC074032.1 |
| chr7 | 1229545 | 1230201 | 657 | 4 | 6.04E-15 | 4.55E-06 | 1.68E-08 | 3.63E-09 | -0.00803308 | NA |
| chr19 | 43826513 | 43828159 | 1647 | 12 | 2.97E-08 | 7.00E-07 | 8.01E-05 | 3.64E-09 | -0.00724295 | ZNF283, LYPD5 |
| chr16 | 15855759 | 15858035 | 2277 | 10 | 1.08E-06 | 1.87E-06 | 8.10E-05 | 3.64E-09 | -0.00485417 | MYH11 |
| chr1 | 48221211 | 48223570 | 2360 | 8 | 6.17E-06 | 5.57E-09 | 0.00155 | 3.64E-09 | -0.00117097 | SLC5A9 |
| chr19 | 3028788 | 3030326 | 1539 | 12 | 4.18E-09 | 2.21E-07 | 1.05E-05 | 3.65E-09 | -0.0002354 | TLE2 |
| chr14 | 72925370 | 72927186 | 1817 | 15 | 6.70E-09 | 7.09E-09 | 0.006573 | 3.66E-09 | -0.00077776 | DCAF4 |
| chr5 | 65562725 | 65564074 | 1350 | 16 | 9.07E-11 | 1.82E-06 | 4.28E-05 | 3.67E-09 | -0.00189681 | PPWD1, CENPK |
| chr3 | 62372559 | 62374383 | 1825 | 15 | 9.87E-09 | 6.99E-08 | 0.000243 | 3.67E-09 | -0.00148507 | FEZF2 |
| chr13 | 112958641 | 112960001 | 1361 | 10 | 3.10E-09 | 1.61E-08 | 0.00072 | 3.68E-09 | -0.00642625 | MCF2L |
| chr10 | 119028854 | 119030443 | 1590 | 8 | 3.73E-08 | 8.10E-09 | 0.00079 | 3.69E-09 | -0.01202861 | NANOS1, AL157388.1 |
| chr1 | 32508286 | 32508816 | 531 | 2 | 2.35E-09 | 2.64E-09 | 5.51E-06 | 3.69E-09 | -0.01461011 | AL033529.1 |
| chr12 | 57011467 | 57013116 | 1650 | 3 | 5.15E-07 | 1.97E-09 | 1.71E-05 | 3.69E-09 | -0.008792 | TAC3 |
| chr12 | 6970313 | 6972016 | 1704 | 16 | 1.72E-08 | 8.85E-07 | 6.97E-05 | 3.71E-09 | -0.00121038 | EMG1, PHB2 |
| chr22 | 30968844 | 30971334 | 2491 | 13 | 1.30E-07 | 1.43E-09 | 0.000313 | 3.71E-09 | -0.00142251 | TUG1, AC004542.5, AC004542.4 |
| chr17 | 48577802 | 48579210 | 1409 | 18 | 5.94E-10 | 1.98E-05 | 5.22E-06 | 3.71E-09 | -0.00112664 | HOXB-AS3, HOXB3, HOXB4 |
| chr4 | 107930818 | 107932796 | 1979 | 15 | 3.14E-09 | 3.15E-06 | 6.65E-06 | 3.72E-09 | -0.00111786 | CYP2U1, CYP2U1-AS1 |
| chr17 | 81385322 | 81387942 | 2621 | 18 | 2.08E-05 | 1.44E-07 | 0.000267 | 3.72E-09 | -0.00276379 | AC110285.5, AC110285.1 |
| chr16 | 28984278 | 28986645 | 2368 | 22 | 1.36E-08 | 1.66E-06 | 2.12E-05 | 3.72E-09 | -0.00055237 | AC009093.11, SPNS1, AC109460.3, LAT |
| chr17 | 34504015 | 34504021 | 7 | 2 | 2.57E-09 | 2.20E-09 | 1.52E-05 | 3.73E-09 | -0.02224937 | NA |
| chr8 | 124673630 | 124673632 | 3 | 2 | 2.35E-09 | 2.65E-09 | 5.72E-06 | 3.73E-09 | -0.01303159 | MTSS1 |
| chr1 | 46179542 | 46180552 | 1011 | 6 | 1.89E-08 | 1.32E-08 | 3.10E-05 | 3.74E-09 | -0.00889678 | TSPAN1 |
| chr11 | 72224309 | 72225643 | 1335 | 12 | 1.26E-09 | 5.11E-07 | 0.000844 | 3.76E-09 | 0.001078608 | INPPL1 |
| chr13 | 113158340 | 113159823 | 1484 | 11 | 3.70E-10 | 1.36E-06 | 9.52E-05 | 3.76E-09 | -0.00208366 | PROZ |
| chr2 | 3105607 | 3105974 | 368 | 2 | 3.57E-10 | 9.44E-09 | 4.71E-07 | 3.77E-09 | -0.01532034 | LINC01250 |
| chr22 | 37822152 | 37823393 | 1242 | 9 | 1.84E-09 | 1.49E-08 | 8.40E-05 | 3.77E-09 | -0.00063188 | GALR3 |
| chr2 | 39095075 | 39095598 | 524 | 2 | 1.45E-10 | 1.21E-08 | 3.51E-07 | 3.79E-09 | 0.028983561 | SOS1 |
| chr16 | 81020449 | 81021543 | 1095 | 4 | 1.22E-07 | 2.42E-09 | 8.12E-05 | 3.79E-09 | -0.00090398 | CENPN, AC092718.2 |
| chr5 | 160198450 | 160199765 | 1316 | 10 | 9.94E-10 | 1.01E-09 | 0.002768 | 3.79E-09 | -0.00547909 | FABP6, FABP6-AS1 |
| chr17 | 21213538 | 21215264 | 1727 | 18 | 6.28E-08 | 2.34E-07 | 8.03E-05 | 3.80E-09 | -0.0013827 | AC087294.1, TMEM11 |
| chr8 | 128540558 | 128542457 | 1900 | 6 | 1.33E-07 | 2.74E-06 | 2.99E-06 | 3.80E-09 | -0.00493485 | LINC00824 |
| chr16 | 1533390 | 1534515 | 1126 | 11 | 1.77E-07 | 2.32E-10 | 0.00403 | 3.80E-09 | 0.023004981 | TMEM204, IFT140 |
| chr11 | 65646609 | 65647789 | 1181 | 7 | 6.38E-10 | 9.53E-07 | 6.37E-05 | 3.81E-09 | -0.0041293 | SIPA1 |
| chr1 | 2513917 | 2515242 | 1326 | 7 | 1.58E-09 | 3.50E-08 | 2.35E-05 | 3.83E-09 | -0.00323223 | PANK4 |
| chr17 | 4996961 | 4998289 | 1329 | 15 | 1.54E-08 | 1.34E-07 | 0.000203 | 3.83E-09 | -0.00143563 | KIF1C, INCA1 |
| chr2 | 19353476 | 19354189 | 714 | 5 | 9.56E-10 | 3.18E-08 | 1.03E-05 | 3.83E-09 | -0.00080135 | OSR1 |
| chr17 | 5281213 | 5282865 | 1653 | 14 | 4.37E-08 | 4.39E-07 | 0.0005 | 3.85E-09 | -0.00372598 | RABEP1 |
| chr22 | 42067255 | 42068231 | 977 | 4 | 8.03E-08 | 3.56E-09 | 2.49E-05 | 3.85E-09 | -0.00359863 | NAGA |
| chr20 | 37394872 | 37396266 | 1395 | 8 | 9.93E-11 | 1.22E-07 | 9.06E-06 | 3.86E-09 | -0.00491795 | SRC |
| chr19 | 5711299 | 5712161 | 863 | 6 | 8.52E-10 | 1.72E-07 | 5.41E-05 | 3.86E-09 | -0.0015172 | LONP1 |
| chr11 | 61627992 | 61628825 | 834 | 5 | 1.95E-10 | 5.42E-08 | 2.09E-05 | 3.86E-09 | -0.00796807 | RPLP0P2 |
| chr6 | 4426876 | 4427783 | 908 | 3 | 1.88E-11 | 1.61E-07 | 2.89E-07 | 3.86E-09 | -0.00067958 | AL162718.1 |
| chr5 | 64768022 | 64769627 | 1606 | 15 | 9.56E-11 | 1.25E-06 | 2.99E-05 | 3.86E-09 | -0.00034785 | CWC27, SREK1IP1 |
| chr11 | 121426848 | 121427445 | 598 | 5 | 9.34E-10 | 2.26E-08 | 1.42E-06 | 3.86E-09 | -0.0028437 | AP002365.1 |
| chr21 | 34525108 | 34527287 | 2180 | 16 | 1.35E-06 | 2.83E-07 | 0.001572 | 3.86E-09 | -0.00938519 | RCAN1 |
| chr9 | 119208842 | 119209219 | 378 | 4 | 9.62E-11 | 2.37E-07 | 7.84E-06 | 3.87E-09 | -0.00865458 | BRINP1 |
| chr14 | 102738249 | 102738449 | 201 | 2 | 9.17E-10 | 7.65E-09 | 6.51E-07 | 3.88E-09 | -0.00512871 | NA |
| chr8 | 81447332 | 81447564 | 233 | 4 | 5.01E-10 | 8.90E-10 | 0.000176 | 3.90E-09 | -0.01008096 | AC018616.1, PMP2 |
| chr14 | 81484030 | 81484781 | 752 | 4 | 8.54E-11 | 1.08E-06 | 1.27E-05 | 3.91E-09 | -0.00414121 | SEL1L |
| chr18 | 21120548 | 21120906 | 359 | 5 | 9.09E-10 | 2.46E-07 | 1.39E-05 | 3.91E-09 | -0.0069696 | NA |
| chr17 | 80191293 | 80192468 | 1176 | 7 | 3.40E-09 | 2.66E-08 | 4.92E-05 | 3.91E-09 | -0.00571408 | CARD14 |
| chr10 | 68231440 | 68233026 | 1587 | 11 | 5.77E-10 | 7.68E-07 | 5.68E-06 | 3.92E-09 | -0.00372436 | ATOH7 |
| chr6 | 26295764 | 26296433 | 670 | 3 | 6.41E-10 | 3.05E-09 | 2.27E-05 | 3.93E-09 | -0.02694808 | NA |
| chr16 | 1501665 | 1503695 | 2031 | 9 | 5.02E-07 | 3.36E-08 | 0.000421 | 3.93E-09 | -0.00026586 | TELO2 |
| chr10 | 23250818 | 23251705 | 888 | 5 | 3.23E-09 | 1.24E-08 | 2.29E-05 | 3.94E-09 | -0.009496 | C10orf67 |
| chr1 | 1006002 | 1008051 | 2050 | 12 | 6.21E-07 | 2.56E-07 | 0.000814 | 3.94E-09 | -0.00130917 | ISG15 |
| chr2 | 225040975 | 225043158 | 2184 | 11 | 8.80E-07 | 2.09E-06 | 0.00013 | 3.94E-09 | 0.001514642 | DOCK10 |
| chr8 | 92099612 | 92101005 | 1394 | 6 | 3.03E-13 | 7.56E-07 | 3.54E-07 | 3.96E-09 | -0.0143616 | RUNX1T1 |
| chr1 | 156727249 | 156729252 | 2004 | 22 | 6.09E-07 | 3.62E-08 | 0.003808 | 3.96E-09 | -0.00130246 | RRNAD1, ISG20L2 |
| chr5 | 12691345 | 12691565 | 221 | 2 | 2.21E-09 | 2.72E-09 | 6.79E-06 | 3.97E-09 | -0.02358495 | LINC01194 |
| chr10 | 131830936 | 131831765 | 830 | 2 | 6.69E-10 | 9.71E-09 | 4.92E-07 | 3.97E-09 | -0.01448972 | NA |
| chr9 | 96382913 | 96384492 | 1580 | 11 | 7.32E-10 | 1.03E-07 | 2.74E-06 | 3.99E-09 | 0.003415348 | SLC35D2 |
| chr12 | 49741050 | 49742322 | 1273 | 25 | 3.60E-09 | 3.09E-07 | 9.09E-05 | 3.99E-09 | -0.00105248 | TMBIM6 |
| chr2 | 33538764 | 33539701 | 938 | 4 | 6.33E-10 | 5.17E-09 | 1.60E-05 | 4.00E-09 | 0.003479364 | RASGRP3 |
| chr10 | 73358345 | 73360495 | 2151 | 18 | 3.14E-09 | 4.09E-07 | 0.002073 | 4.01E-09 | 0.000253396 | CFAP70 |
| chr15 | 85494718 | 85496416 | 1699 | 5 | 1.70E-07 | 8.82E-08 | 2.90E-05 | 4.02E-09 | 0.003409545 | AKAP13 |
| chr14 | 101809696 | 101811063 | 1368 | 13 | 3.13E-07 | 2.55E-08 | 0.001981 | 4.02E-09 | -0.00060155 | PPP2R5C, AL137779.2 |
| chr7 | 30993937 | 30994399 | 463 | 2 | 1.56E-10 | 1.49E-08 | 3.10E-07 | 4.02E-09 | 0.007534617 | NA |
| chr1 | 66929482 | 66930894 | 1413 | 13 | 5.73E-10 | 1.37E-07 | 6.06E-05 | 4.02E-09 | -0.00125929 | MIER1 |
| chr8 | 144353419 | 144355027 | 1609 | 11 | 2.19E-09 | 1.28E-07 | 0.001946 | 4.04E-09 | -0.01005389 | SLC52A2, TMEM249, AC233992.2 |
| chr11 | 3380770 | 3381544 | 775 | 2 | 1.16E-13 | 2.47E-05 | 1.33E-08 | 4.06E-09 | 0.008358676 | TSSC2 |
| chr6 | 168105993 | 168106250 | 258 | 2 | 2.95E-10 | 3.21E-09 | 4.18E-06 | 4.06E-09 | -0.02635361 | NA |
| chr4 | 3073757 | 3075367 | 1611 | 10 | 1.10E-08 | 2.79E-08 | 0.000585 | 4.10E-09 | 0.001433375 | HTT, HTT-AS |
| chr22 | 28829543 | 28830065 | 523 | 3 | 5.49E-09 | 1.09E-08 | 1.29E-05 | 4.10E-09 | -0.00054626 | Z93930.2 |
| chr10 | 14958622 | 14960095 | 1474 | 12 | 1.12E-09 | 2.92E-07 | 2.14E-05 | 4.12E-09 | 0.001469736 | MEIG1 |
| chr8 | 144333974 | 144335305 | 1332 | 7 | 8.44E-09 | 1.06E-07 | 8.32E-05 | 4.14E-09 | 0.011379854 | SLC52A2, SCRT1 |
| chr19 | 49596795 | 49597755 | 961 | 4 | 6.95E-11 | 1.61E-08 | 2.37E-06 | 4.15E-09 | -0.00102424 | PRR12 |
| chr12 | 49888156 | 49888953 | 798 | 3 | 6.81E-09 | 6.17E-09 | 2.50E-05 | 4.15E-09 | -0.00853746 | FAIM2 |
| chr2 | 106138507 | 106139854 | 1348 | 7 | 7.43E-08 | 5.62E-09 | 0.000215 | 4.15E-09 | 0.002216832 | UXS1 |
| chr8 | 20349372 | 20350686 | 1315 | 7 | 1.41E-10 | 3.46E-07 | 7.51E-05 | 4.18E-09 | -0.01353312 | AC023403.1 |
| chr17 | 42782380 | 42782889 | 510 | 3 | 1.46E-09 | 2.36E-09 | 3.79E-05 | 4.18E-09 | -0.0235103 | WNK4 |
| chr14 | 75984171 | 75985840 | 1670 | 13 | 2.92E-09 | 1.48E-07 | 4.66E-05 | 4.18E-09 | -0.00060647 | IFT43 |
| chr4 | 20421801 | 20422575 | 775 | 2 | 4.79E-08 | 2.58E-09 | 1.18E-05 | 4.18E-09 | -0.03089177 | SLIT2 |
| chr20 | 50304560 | 50305535 | 976 | 3 | 6.85E-08 | 7.57E-09 | 2.24E-05 | 4.19E-09 | 0.003327477 | LINC01270 |
| chr12 | 122525675 | 122527804 | 2130 | 19 | 2.52E-07 | 1.33E-08 | 0.003246 | 4.19E-09 | -0.00115702 | KNTC1, RSRC2 |
| chr7 | 47666320 | 47666876 | 557 | 2 | 3.34E-11 | 4.92E-08 | 1.17E-07 | 4.20E-09 | -0.00346456 | NA |
| chr19 | 14072206 | 14073164 | 959 | 13 | 4.22E-10 | 1.35E-08 | 5.26E-05 | 4.20E-09 | 0.000740417 | MISP3 |
| chr7 | 27739029 | 27740615 | 1587 | 21 | 2.40E-12 | 7.88E-05 | 2.31E-05 | 4.21E-09 | 0.000438656 | TAX1BP1, TAX1BP1-AS1 |
| chr1 | 17646516 | 17647062 | 547 | 3 | 4.93E-09 | 3.41E-09 | 1.59E-05 | 4.21E-09 | -0.00558457 | ARHGEF10L |
| chr14 | 23319935 | 23322005 | 2071 | 15 | 6.29E-08 | 6.66E-07 | 0.000386 | 4.22E-09 | 0.002147557 | BCL2L2-PABPN1, PABPN1 |
| chr17 | 43077091 | 43078026 | 936 | 3 | 6.47E-13 | 1.73E-06 | 3.09E-08 | 4.22E-09 | -0.00827863 | BRCA1 |
| chr5 | 1551756 | 1553019 | 1264 | 10 | 1.79E-07 | 4.69E-07 | 0.000137 | 4.24E-09 | -0.00407934 | NA |
| chr1 | 40092169 | 40093050 | 882 | 2 | 1.18E-09 | 1.00E-08 | 5.32E-07 | 4.26E-09 | 0.027021094 | PPT1 |
| chr1 | 200893598 | 200895095 | 1498 | 8 | 5.75E-07 | 3.17E-09 | 0.000839 | 4.26E-09 | -0.002207 | INAVA |
| chr6 | 29620798 | 29622366 | 1569 | 17 | 3.52E-11 | 6.86E-05 | 1.02E-05 | 4.28E-09 | -0.00023711 | GABBR1 |
| chr5 | 8457011 | 8457976 | 966 | 9 | 6.66E-09 | 6.23E-10 | 0.004095 | 4.31E-09 | 0.023358391 | MIR4458HG, LINC02226, AC091965.1 |
| chr7 | 2934360 | 2934545 | 186 | 2 | 1.88E-10 | 3.40E-08 | 1.57E-07 | 4.31E-09 | -0.00039066 | CARD11 |
| chr2 | 68239756 | 68240317 | 562 | 3 | 1.16E-09 | 2.85E-09 | 1.20E-05 | 4.31E-09 | -0.00499926 | AC017083.3, PPP3R1 |
| chr5 | 83720811 | 83721734 | 924 | 10 | 2.83E-10 | 5.80E-08 | 0.000326 | 4.33E-09 | -0.01321526 | HAPLN1 |
| chr17 | 4771548 | 4771997 | 450 | 5 | 5.98E-10 | 1.62E-08 | 8.80E-05 | 4.34E-09 | -0.00745508 | TM4SF5 |
| chr19 | 40690005 | 40691359 | 1355 | 16 | 7.76E-09 | 1.72E-08 | 0.000623 | 4.34E-09 | -0.00293063 | NUMBL |
| chr1 | 26959172 | 26961285 | 2114 | 12 | 2.15E-07 | 1.41E-07 | 8.93E-05 | 4.35E-09 | -0.00342312 | KDF1 |
| chr7 | 102432673 | 102433778 | 1106 | 6 | 3.49E-11 | 4.29E-07 | 4.44E-05 | 4.35E-09 | -0.00874462 | ORAI2, AC073517.1 |
| chr12 | 39322878 | 39323140 | 263 | 2 | 1.17E-09 | 6.05E-09 | 1.21E-06 | 4.35E-09 | 0.003156991 | KIF21A |
| chr14 | 23146125 | 23147422 | 1298 | 6 | 2.92E-10 | 1.22E-08 | 1.68E-05 | 4.36E-09 | -0.00596884 | SLC7A8 |
| chr15 | 57375132 | 57376981 | 1850 | 15 | 1.77E-07 | 1.40E-06 | 7.76E-05 | 4.36E-09 | -0.00256881 | CGNL1 |
| chr15 | 63676474 | 63677999 | 1526 | 4 | 3.69E-08 | 2.28E-08 | 7.23E-05 | 4.36E-09 | -0.00673302 | AC073167.1, HERC1 |
| chr22 | 42336383 | 42338689 | 2307 | 6 | 1.54E-06 | 1.11E-07 | 0.000352 | 4.37E-09 | -0.00569223 | TCF20 |
| chr3 | 71583526 | 71584911 | 1386 | 14 | 3.43E-09 | 1.04E-08 | 0.002547 | 4.38E-09 | 0.001007482 | AC097634.3, FOXP1, AC097634.4 |
| chr2 | 85851286 | 85851843 | 558 | 2 | 5.41E-09 | 2.62E-09 | 1.51E-05 | 4.38E-09 | 0.000123255 | ST3GAL5 |
| chr7 | 4727775 | 4728616 | 842 | 4 | 1.36E-13 | 3.52E-05 | 8.60E-08 | 4.39E-09 | -0.00117589 | FOXK1, AC092428.1 |
| chr12 | 124324446 | 124326362 | 1917 | 11 | 2.26E-06 | 1.68E-09 | 0.001426 | 4.39E-09 | -0.00307798 | NCOR2 |
| chr6 | 32216519 | 32217563 | 1045 | 12 | 1.57E-10 | 3.29E-07 | 2.76E-05 | 4.41E-09 | -0.00200704 | NOTCH4 |
| chr18 | 23155155 | 23156658 | 1504 | 7 | 4.93E-08 | 8.52E-10 | 0.00042 | 4.42E-09 | 0.002455483 | CABLES1 |
| chr11 | 68007258 | 68009106 | 1849 | 11 | 2.22E-07 | 4.79E-09 | 0.0011 | 4.42E-09 | 0.005822833 | ALDH3B1 |
| chr2 | 21269798 | 21270765 | 968 | 2 | 2.68E-09 | 9.81E-09 | 5.97E-07 | 4.45E-09 | 0.000812418 | AC018742.1 |
| chr19 | 2290844 | 2291873 | 1030 | 8 | 1.13E-09 | 1.48E-08 | 0.001284 | 4.45E-09 | -0.02011962 | AC005258.1, LINGO3 |
| chr22 | 50146732 | 50147109 | 378 | 9 | 1.55E-09 | 7.24E-09 | 0.00037 | 4.45E-09 | -0.01186321 | MOV10L1 |
| chr6 | 170209784 | 170211212 | 1429 | 4 | 4.49E-08 | 2.14E-08 | 6.97E-06 | 4.45E-09 | 0.002534457 | NA |
| chr2 | 120595083 | 120596501 | 1419 | 4 | 1.22E-06 | 6.50E-09 | 4.76E-05 | 4.47E-09 | -0.00576747 | NA |
| chr5 | 129905717 | 129907306 | 1590 | 10 | 2.03E-10 | 6.50E-09 | 0.000989 | 4.47E-09 | -0.00725851 | CHSY3, AC008591.1 |
| chr1 | 1944341 | 1944437 | 97 | 4 | 3.21E-09 | 8.28E-10 | 0.000449 | 4.48E-09 | 0.038716936 | CFAP74 |
| chr1 | 28735724 | 28737637 | 1914 | 22 | 8.99E-11 | 2.54E-05 | 5.50E-07 | 4.49E-09 | -0.00014024 | YTHDF2 |
| chr22 | 48644359 | 48645411 | 1053 | 3 | 4.84E-13 | 5.88E-06 | 2.58E-08 | 4.50E-09 | -0.0097673 | TAFA5 |
| chr19 | 45639316 | 45641044 | 1729 | 13 | 2.33E-09 | 2.02E-07 | 1.00E-05 | 4.50E-09 | -0.00182179 | EML2 |
| chr2 | 226906493 | 226907238 | 746 | 3 | 6.51E-09 | 3.66E-09 | 1.03E-05 | 4.51E-09 | 0.014752221 | RHBDD1 |
| chr1 | 6199085 | 6200300 | 1216 | 13 | 2.63E-10 | 5.63E-06 | 0.000102 | 4.51E-09 | -0.00175413 | RPL22 |
| chr1 | 42657545 | 42659161 | 1617 | 15 | 1.73E-08 | 1.69E-06 | 0.000138 | 4.51E-09 | 0.000943024 | PPIH, AC098484.4 |
| chr1 | 22024932 | 22025610 | 679 | 11 | 3.01E-12 | 1.04E-06 | 1.29E-07 | 4.51E-09 | 8.73E-06 | AL590556.3, LINC00339, CDC42, LINC01635 |
| chr14 | 54901625 | 54903637 | 2013 | 16 | 9.98E-08 | 3.43E-07 | 0.000772 | 4.51E-09 | 0.00129576 | GCH1 |
| chr6 | 63573751 | 63574436 | 686 | 3 | 2.19E-13 | 4.37E-07 | 6.27E-08 | 4.52E-09 | -0.00779958 | PTP4A1, AL135905.2 |
| chr11 | 236011 | 237407 | 1397 | 19 | 2.01E-08 | 6.82E-08 | 0.000433 | 4.52E-09 | 0.003273998 | PSMD13, SIRT3 |
| chr2 | 218245230 | 218245650 | 421 | 2 | 3.00E-12 | 0.0001716 | 1.25E-08 | 4.53E-09 | 0.004976511 | ARPC2 |
| chr12 | 6179096 | 6180681 | 1586 | 5 | 3.78E-07 | 1.87E-09 | 0.000186 | 4.53E-09 | -0.00597499 | NA |
| chr1 | 10297959 | 10299147 | 1189 | 7 | 1.19E-09 | 3.63E-06 | 2.77E-05 | 4.56E-09 | -0.00794348 | KIF1B, RNU6-37P |
| chr11 | 114447827 | 114448740 | 914 | 4 | 3.16E-09 | 3.42E-09 | 2.79E-05 | 4.56E-09 | -0.00461502 | REXO2 |
| chr5 | 93573723 | 93574828 | 1106 | 5 | 1.94E-11 | 9.68E-07 | 3.91E-06 | 4.57E-09 | -0.00347231 | NR2F1-AS1 |
| chr9 | 72384284 | 72385454 | 1171 | 3 | 1.19E-06 | 1.92E-09 | 5.43E-05 | 4.59E-09 | 0.005169884 | NA |
| chr7 | 158471292 | 158472869 | 1578 | 8 | 2.60E-10 | 1.37E-08 | 2.89E-05 | 4.59E-09 | -0.00600563 | PTPRN2 |
| chr14 | 105240971 | 105241918 | 948 | 5 | 5.73E-10 | 1.43E-08 | 1.50E-05 | 4.59E-09 | -0.00236495 | BRF1 |
| chr11 | 85809771 | 85811594 | 1824 | 14 | 5.12E-08 | 4.46E-07 | 5.64E-05 | 4.59E-09 | 0.000675718 | SYTL2 |
| chr14 | 92748123 | 92749838 | 1716 | 15 | 4.25E-12 | 0.0002682 | 9.01E-07 | 4.61E-09 | -0.00354826 | LGMN |
| chr1 | 239717142 | 239717553 | 412 | 3 | 2.78E-09 | 2.74E-09 | 3.43E-05 | 4.61E-09 | -0.01565914 | CHRM3, CHRM3-AS2 |
| chr12 | 93264938 | 93265903 | 966 | 4 | 4.74E-09 | 5.85E-09 | 5.61E-06 | 4.63E-09 | 0.002270695 | NA |
| chr10 | 297531 | 298629 | 1099 | 3 | 1.75E-08 | 6.94E-08 | 4.53E-06 | 4.64E-09 | -0.00448275 | DIP2C |
| chr1 | 54095907 | 54096870 | 964 | 6 | 1.34E-09 | 2.70E-08 | 0.000652 | 4.66E-09 | -0.00629512 | TCEANC2 |
| chr17 | 41377769 | 41377844 | 76 | 2 | 2.34E-09 | 5.25E-09 | 1.86E-06 | 4.66E-09 | -0.01538327 | KRT34 |
| chr1 | 20684306 | 20685793 | 1488 | 9 | 7.19E-09 | 1.35E-08 | 0.000294 | 4.66E-09 | -0.00695734 | KIF17 |
| chr13 | 112160342 | 112160832 | 491 | 4 | 2.26E-11 | 2.94E-07 | 2.68E-06 | 4.67E-09 | 0.007511946 | NA |
| chr16 | 55035236 | 55036212 | 977 | 2 | 1.41E-08 | 5.80E-09 | 1.54E-06 | 4.67E-09 | -0.01420451 | NA |
| chr19 | 39307110 | 39308739 | 1630 | 6 | 4.00E-08 | 1.05E-09 | 0.000815 | 4.67E-09 | -0.01259037 | LRFN1 |
| chr3 | 126517661 | 126519012 | 1352 | 10 | 1.39E-07 | 1.84E-09 | 0.005128 | 4.70E-09 | 0.000817533 | UROC1 |
| chr10 | 14908496 | 14909153 | 658 | 2 | 3.76E-12 | 5.50E-07 | 3.70E-08 | 4.70E-09 | -0.00206116 | DCLRE1C |
| chr11 | 102874732 | 102875942 | 1211 | 9 | 2.46E-10 | 2.15E-07 | 6.88E-05 | 4.70E-09 | 0.000581043 | MMP12 |
| chr6 | 10694312 | 10695604 | 1293 | 17 | 6.50E-10 | 7.84E-08 | 2.66E-05 | 4.71E-09 | 0.001697922 | PAK1IP1, C6orf52 |

| | | | | | | | | | | |
|---|---|---|---|---|---|---|---|---|---|---|
| chr13 | 110648970 | 110649427 | 458 | 6 | 1.63E-09 | 2.57E-06 | 0.000108 | 4.72E-09 | -0.00291704 | CARS2 |
| chr10 | 11017220 | 11018141 | 922 | 11 | 1.38E-09 | 5.87E-08 | 7.57E-05 | 4.72E-09 | -0.00075995 | CELF2 |
| chr16 | 6318209 | 6319406 | 1198 | 3 | 1.56E-07 | 5.47E-09 | 3.00E-05 | 4.73E-09 | -0.02050623 | RBFOX1 |
| chr2 | 120758572 | 120758957 | 386 | 4 | 3.68E-09 | 3.70E-09 | 2.60E-05 | 4.75E-09 | -0.01456833 | GLI2 |
| chr7 | 920122 | 921134 | 1013 | 12 | 3.24E-10 | 6.20E-08 | 6.36E-05 | 4.76E-09 | -0.00202933 | ADAP1, COX19 |
| chr20 | 1312916 | 1314214 | 1299 | 10 | 1.02E-08 | 1.92E-08 | 0.00084 | 4.76E-09 | -0.00500425 | SDCBP2 |
| chr6 | 168132827 | 168133358 | 532 | 4 | 1.49E-08 | 1.24E-09 | 0.000204 | 4.77E-09 | -0.04687567 | NA |
| chr8 | 81093750 | 81094182 | 433 | 2 | 9.78E-10 | 3.25E-09 | 7.65E-06 | 4.77E-09 | 0.043294672 | PAG1 |
| chr4 | 7371295 | 7371724 | 430 | 5 | 5.62E-09 | 1.15E-08 | 0.000444 | 4.79E-09 | 0.003267531 | SORCS2 |
| chr11 | 128694861 | 128696492 | 1632 | 5 | 1.98E-10 | 8.90E-08 | 1.08E-05 | 4.79E-09 | 0.002877494 | FLI1, SENCR |
| chr1 | 93681514 | 93682571 | 1058 | 13 | 1.78E-09 | 1.19E-08 | 0.001345 | 4.79E-09 | 0.000159859 | BCAR3 |
| chr19 | 38009567 | 38009754 | 188 | 4 | 9.66E-09 | 8.92E-10 | 0.000469 | 4.79E-09 | 0.000911738 | SIPA1L3 |
| chr14 | 92121370 | 92122474 | 1105 | 14 | 1.82E-10 | 8.11E-07 | 0.000837 | 4.80E-09 | 0.00328844 | CPSF2, NDUFB1 |
| chr4 | 5988863 | 5990746 | 1884 | 7 | 2.86E-06 | 2.87E-09 | 0.000745 | 4.80E-09 | 4.15E-06 | C4orf50 |
| chr20 | 1392949 | 1394209 | 1261 | 11 | 1.26E-08 | 3.33E-09 | 0.00097 | 4.81E-09 | -0.00273934 | AL136531.2, FKBP1A, MIR6869 |
| chr1 | 24236126 | 24236335 | 210 | 2 | 3.28E-09 | 2.94E-09 | 1.37E-05 | 4.81E-09 | -0.02091478 | NA |
| chr5 | 1222732 | 1223125 | 394 | 3 | 2.46E-09 | 2.13E-09 | 4.64E-05 | 4.84E-09 | 0.005313317 | SLC6A19 |
| chr5 | 180071114 | 180072824 | 1711 | 8 | 1.11E-07 | 2.30E-08 | 7.48E-05 | 4.84E-09 | 0.001634773 | RNF130 |
| chr22 | 24406563 | 24408059 | 1497 | 5 | 1.28E-09 | 7.14E-08 | 0.000136 | 4.84E-09 | -0.00981053 | SPECC1L, SPECC1L-ADORA2A |
| chr10 | 133188423 | 133189472 | 1050 | 4 | 1.78E-08 | 2.28E-09 | 3.80E-05 | 4.85E-09 | -0.00726382 | KNDC1 |
| chr17 | 44343982 | 44346015 | 2034 | 15 | 8.22E-08 | 1.16E-07 | 0.000108 | 4.86E-09 | -0.00094824 | GRN |
| chr15 | 88620697 | 88621815 | 1119 | 13 | 8.54E-10 | 1.69E-05 | 4.15E-06 | 4.87E-09 | 0.002105641 | AEN |
| chr11 | 118400176 | 118402214 | 2039 | 21 | 3.59E-14 | 1.85E-05 | 2.45E-07 | 4.89E-09 | 0.00074869 | AP001267.5, ATP5MG, AP001267.1 |
| chr2 | 39664201 | 39665472 | 1272 | 7 | 1.94E-11 | 2.05E-06 | 2.63E-06 | 4.92E-09 | -0.00594889 | MAP4K3-DT, TMEM178A |
| chr9 | 97852597 | 97853514 | 918 | 7 | 2.58E-09 | 2.19E-08 | 0.000111 | 4.92E-09 | -0.00148317 | FOXE1, PTCSC2 |
| chr10 | 5846388 | 5846656 | 269 | 2 | 1.34E-09 | 5.53E-09 | 1.92E-06 | 4.93E-09 | -0.02912049 | NA |
| chr20 | 2663560 | 2664788 | 1229 | 17 | 3.94E-10 | 3.43E-06 | 7.33E-08 | 4.96E-09 | -0.00255907 | AL049712.1, IDH3B |
| chr11 | 2139645 | 2141700 | 2056 | 47 | 3.08E-10 | 6.56E-06 | 0.000238 | 4.96E-09 | -0.00052319 | IGF2-AS, IGF2, AC132217.2, INS-IGF2 |
| chr12 | 86264490 | 86265230 | 741 | 4 | 1.48E-08 | 1.50E-08 | 1.19E-05 | 4.97E-09 | -0.01478371 | MGAT4C |
| chr8 | 119855466 | 119856511 | 1046 | 11 | 1.21E-12 | 5.68E-05 | 1.20E-07 | 4.97E-09 | -0.00302227 | AP005717.2, DSCC1 |
| chr1 | 202966193 | 202968182 | 1990 | 15 | 1.83E-08 | 1.56E-08 | 0.002336 | 4.97E-09 | -0.00066931 | CYB5R1 |
| chr17 | 80970606 | 80971513 | 908 | 7 | 3.86E-10 | 2.23E-08 | 3.24E-05 | 4.99E-09 | -0.01533949 | AC127496.1 |
| chr19 | 50667804 | 50669498 | 1695 | 9 | 1.00E-07 | 7.20E-09 | 0.001067 | 5.02E-09 | -0.00157047 | SHANK1 |
| chr14 | 101374100 | 101374595 | 496 | 4 | 5.71E-10 | 9.60E-07 | 8.39E-07 | 5.02E-09 | -0.00169833 | NA |
| chr1 | 151783563 | 151784498 | 936 | 3 | 2.65E-10 | 2.49E-08 | 1.09E-05 | 5.02E-09 | -0.01101731 | TDRKH |
| chr15 | 24847803 | 24848838 | 1036 | 6 | 8.57E-09 | 2.16E-07 | 6.31E-05 | 5.03E-09 | -0.00312546 | AC090983.2, SNRPN, AC090983.1 |
| chr7 | 101362551 | 101363292 | 742 | 12 | 8.19E-10 | 7.39E-08 | 1.89E-05 | 5.03E-09 | 0.006453248 | COL26A1 |
| chr2 | 33371198 | 33371480 | 283 | 2 | 2.26E-09 | 4.03E-09 | 4.44E-06 | 5.03E-09 | -0.01936362 | LTBP1 |
| chr2 | 65055329 | 65057128 | 1800 | 12 | 1.42E-08 | 8.75E-08 | 0.000585 | 5.03E-09 | 0.000169351 | CEP68, LINC02245 |
| chr10 | 48511259 | 48511879 | 621 | 4 | 4.69E-10 | 6.18E-08 | 9.24E-06 | 5.03E-09 | -0.0017225 | ARHGAP22, ARHGAP22-IT1 |
| chr14 | 24120768 | 24121283 | 516 | 3 | 6.05E-11 | 5.51E-09 | 8.19E-06 | 5.03E-09 | 0.000309665 | DCAF11, AL136295.3 |
| chr8 | 19938026 | 19939623 | 1598 | 14 | 2.78E-10 | 6.71E-08 | 0.000207 | 5.04E-09 | -0.00521326 | LPL |
| chr15 | 57705750 | 57707337 | 1588 | 18 | 2.48E-10 | 9.19E-06 | 8.34E-06 | 5.04E-09 | -0.00143312 | GCOM1, POLR2M |
| chr9 | 123111283 | 123112990 | 1708 | 6 | 1.39E-07 | 1.53E-09 | 0.000516 | 5.04E-09 | 0.010756122 | MIR600HG, STRBP, MIR600 |
| chr16 | 68287581 | 68287827 | 247 | 5 | 4.74E-09 | 1.04E-09 | 0.000498 | 5.05E-09 | 0.012318981 | SLC7A6, SLC7A6OS |
| chr5 | 140302762 | 140303647 | 886 | 12 | 7.03E-09 | 4.72E-08 | 0.002541 | 5.05E-09 | 0.001958392 | PFDN1 |
| chr19 | 58002158 | 58003678 | 1521 | 11 | 5.46E-08 | 1.64E-07 | 0.001996 | 5.06E-09 | -0.00633323 | AC008969.1, ZNF606 |
| chr9 | 87279950 | 87280695 | 746 | 2 | 1.97E-09 | 6.70E-09 | 1.42E-06 | 5.07E-09 | 0.009151515 | NA |
| chr12 | 120528453 | 120530133 | 1681 | 17 | 8.83E-09 | 3.88E-08 | 0.002248 | 5.11E-09 | -0.00219684 | COQ5 |
| chr15 | 39976220 | 39977013 | 794 | 6 | 1.01E-08 | 6.54E-10 | 0.001278 | 5.12E-09 | -0.02885654 | EIF2AK4 |
| chr11 | 74467223 | 74468504 | 1282 | 12 | 3.33E-11 | 5.74E-06 | 3.91E-06 | 5.12E-09 | 0.001062803 | AP001372.1, KCNE3 |
| chr8 | 80485366 | 80486858 | 1493 | 13 | 4.09E-08 | 2.90E-07 | 0.001464 | 5.12E-09 | 0.002378196 | ZBTB10, AC009812.1 |
| chr10 | 45373248 | 45374931 | 1684 | 13 | 6.16E-09 | 3.62E-05 | 0.000162 | 5.14E-09 | -0.00794675 | ALOX5 |
| chr6 | 169328568 | 169328838 | 271 | 2 | 4.03E-09 | 3.05E-09 | 1.81E-05 | 5.14E-09 | -0.01844321 | NA |
| chr10 | 8014699 | 8015043 | 345 | 4 | 4.90E-10 | 6.09E-07 | 5.19E-06 | 5.16E-09 | 0.000656752 | TAF3 |
| chr19 | 1921920 | 1923308 | 1389 | 5 | 1.00E-06 | 9.68E-10 | 0.000374 | 5.16E-09 | -0.00493069 | SCAMP4 |
| chr5 | 142108033 | 142109605 | 1573 | 16 | 8.59E-09 | 1.02E-07 | 0.000429 | 5.19E-09 | -0.00203343 | NDFIP1 |
| chr20 | 18224984 | 18225153 | 170 | 2 | 3.68E-10 | 1.97E-07 | 6.41E-08 | 5.20E-09 | -0.01646332 | NA |
| chr11 | 1879498 | 1880518 | 1021 | 6 | 2.14E-08 | 1.69E-08 | 0.00053 | 5.21E-09 | 0.007595872 | LSP1, MIR7847 |
| chr17 | 68292388 | 68294364 | 1977 | 6 | 1.85E-08 | 3.64E-07 | 8.01E-06 | 5.21E-09 | 0.002616501 | ARSG |
| chr14 | 20332593 | 20333519 | 927 | 10 | 3.75E-13 | 6.22E-07 | 4.42E-07 | 5.24E-09 | -0.00150153 | CCNB1IP1 |
| chr10 | 102394173 | 102395893 | 1721 | 20 | 4.16E-09 | 1.84E-07 | 0.000965 | 5.26E-09 | -0.00039127 | NFKB2 |
| chr4 | 6298106 | 6298672 | 567 | 4 | 6.32E-10 | 9.36E-09 | 1.35E-05 | 5.26E-09 | -0.00693151 | WFS1, AC116317.1 |
| chr12 | 68807697 | 68809062 | 1366 | 18 | 1.21E-10 | 4.39E-07 | 0.000155 | 5.27E-09 | 0.001150042 | MDM2 |
| chr21 | 42781363 | 42781438 | 76 | 2 | 1.02E-10 | 3.13E-09 | 1.84E-05 | 5.29E-09 | -0.03231576 | AP001628.2 |
| chr4 | 84496602 | 84499158 | 2557 | 20 | 2.58E-06 | 1.37E-07 | 0.001684 | 5.29E-09 | 0.000581004 | NKX6-1 |
| chr17 | 39573391 | 39574815 | 1425 | 7 | 3.16E-08 | 6.70E-10 | 0.001331 | 5.31E-09 | -0.00506468 | NA |
| chr7 | 373702 | 375595 | 1894 | 19 | 3.68E-07 | 1.18E-07 | 0.003452 | 5.31E-09 | -0.00238657 | NA |
| chr16 | 782110 | 783427 | 1318 | 12 | 7.44E-11 | 1.36E-07 | 6.00E-05 | 5.31E-09 | -0.00612842 | MSLNL |
| chr3 | 16932143 | 16933189 | 1047 | 5 | 1.33E-08 | 8.11E-10 | 0.000717 | 5.31E-09 | 0.025650739 | PLCL2 |
| chr4 | 102826715 | 102828428 | 1714 | 20 | 3.17E-07 | 4.02E-09 | 0.002446 | 5.32E-09 | 0.000570255 | UBE2D3-AS1, UBE2D3 |
| chr1 | 75785773 | 75786965 | 1193 | 12 | 7.09E-10 | 2.46E-08 | 3.36E-05 | 5.32E-09 | -0.00279916 | ACADM, RABGGTB |
| chr11 | 134562929 | 134563204 | 276 | 3 | 3.95E-11 | 3.73E-06 | 2.63E-08 | 5.32E-09 | -0.00721402 | NA |
| chr11 | 112960341 | 112962336 | 1996 | 16 | 3.20E-07 | 4.66E-08 | 0.000329 | 5.32E-09 | 0.003840834 | NCAM1, AP000802.1 |
| chr2 | 135984716 | 135986351 | 1636 | 20 | 2.60E-08 | 1.04E-06 | 8.65E-06 | 5.35E-09 | -0.00102506 | DARS-AS1, DARS1 |
| chr17 | 41554546 | 41555907 | 1362 | 6 | 4.46E-07 | 4.39E-09 | 0.000145 | 5.38E-09 | -0.00593249 | NA |
| chr6 | 118810570 | 118810725 | 156 | 3 | 3.22E-10 | 3.87E-07 | 3.42E-07 | 5.39E-09 | 0.007643767 | NA |
| chr9 | 133374994 | 133376844 | 1851 | 14 | 2.79E-08 | 1.31E-06 | 5.73E-06 | 5.41E-09 | 0.002399101 | STKLD1, SURF4 |
| chr1 | 78490074 | 78492094 | 2021 | 17 | 2.35E-09 | 5.19E-06 | 5.87E-05 | 5.42E-09 | -0.00654783 | PTGFR |
| chr12 | 114666469 | 114667714 | 1246 | 9 | 4.81E-08 | 7.10E-10 | 0.002016 | 5.42E-09 | 0.000284295 | NA |
| chr17 | 7483459 | 7484863 | 1405 | 17 | 1.87E-11 | 6.38E-07 | 0.000101 | 5.43E-09 | 0.000270513 | SLC35G6, POLR2A, ZBTB4 |
| chr19 | 1359367 | 1360766 | 1400 | 8 | 6.91E-07 | 2.73E-09 | 0.000658 | 5.43E-09 | -0.00322504 | PWWP3A |
| chr2 | 162733282 | 162733580 | 299 | 2 | 6.17E-10 | 6.62E-09 | 1.70E-06 | 5.43E-09 | 0.025589134 | KCNH7 |
| chr1 | 99969318 | 99971168 | 1851 | 23 | 2.20E-08 | 6.32E-05 | 8.33E-05 | 5.43E-09 | -0.00054123 | SLC35A3, AC118553.2, AC118553.1 |
| chr4 | 15478783 | 15479645 | 863 | 4 | 1.59E-09 | 2.17E-06 | 9.84E-05 | 5.44E-09 | -0.01547126 | CC2D2A |
| chr6 | 34063820 | 34064970 | 1151 | 4 | 2.27E-07 | 7.59E-09 | 3.39E-05 | 5.44E-09 | -0.00554921 | GRM4 |
| chr9 | 129203331 | 129204742 | 1412 | 5 | 2.10E-11 | 2.40E-08 | 5.99E-06 | 5.45E-09 | -0.00972832 | AL158151.3, AL158151.1 |
| chr19 | 48899167 | 48900837 | 1671 | 14 | 9.16E-11 | 2.85E-06 | 0.000412 | 5.46E-09 | -0.00028907 | NUCB1 |
| chr11 | 130158244 | 130160066 | 1823 | 12 | 4.13E-09 | 5.05E-07 | 5.64E-06 | 5.46E-09 | -0.0020213 | ST14 |
| chr1 | 245715480 | 245716311 | 832 | 2 | 7.42E-10 | 1.55E-08 | 4.82E-07 | 5.48E-09 | -0.01980868 | NA |
| chr15 | 37098920 | 37101388 | 2469 | 28 | 3.48E-08 | 3.30E-08 | 0.000102 | 5.48E-09 | -0.00143411 | AC078909.1, MEIS2 |
| chr2 | 70686040 | 70686044 | 5 | 2 | 1.16E-10 | 3.80E-08 | 2.00E-07 | 5.49E-09 | -0.01354603 | ADD2 |
| chr2 | 44291726 | 44291791 | 66 | 2 | 1.43E-09 | 2.69E-08 | 2.74E-07 | 5.50E-09 | -0.01389615 | AC013717.1, SLC3A1 |
| chr1 | 201468890 | 201470450 | 1561 | 10 | 2.50E-10 | 1.68E-08 | 2.75E-05 | 5.51E-09 | -0.00014809 | PHLDA3 |
| chr13 | 102872350 | 102873887 | 1538 | 5 | 5.02E-08 | 3.07E-06 | 4.14E-05 | 5.53E-09 | -0.00991799 | BIVM-ERCC5, ERCC5 |
| chr1 | 9687714 | 9689238 | 1525 | 7 | 3.99E-10 | 2.06E-06 | 4.45E-06 | 5.54E-09 | -0.00815834 | PIK3CD |
| chr16 | 28291027 | 28293342 | 2316 | 10 | 1.92E-08 | 7.25E-07 | 8.84E-05 | 5.54E-09 | 0.000223061 | AC138904.3, SBK1, AC138904.1 |
| chr12 | 94046704 | 94047291 | 588 | 3 | 1.33E-09 | 1.50E-08 | 5.23E-06 | 5.54E-09 | 0.014497799 | NA |
| chr16 | 3282899 | 3284273 | 1375 | 12 | 7.68E-11 | 2.11E-06 | 3.45E-06 | 5.57E-09 | -0.0022117 | ZNF263 |
| chr7 | 2525941 | 2527277 | 1337 | 11 | 8.21E-10 | 1.09E-07 | 9.16E-05 | 5.57E-09 | -0.0001066 | LFNG, MIR4648 |
| chr18 | 77162064 | 77162488 | 425 | 4 | 1.21E-08 | 9.79E-10 | 0.000572 | 5.57E-09 | 0.028752936 | NA |
| chr6 | 3421523 | 3422187 | 665 | 2 | 2.95E-08 | 3.34E-09 | 1.69E-05 | 5.58E-09 | -0.02529639 | SLC22A23 |
| chr11 | 70162026 | 70162931 | 906 | 3 | 4.48E-10 | 1.91E-08 | 3.14E-06 | 5.58E-09 | 0.003444017 | ANO1 |
| chr11 | 64878084 | 64880452 | 2369 | 19 | 1.92E-06 | 3.52E-08 | 0.002185 | 5.58E-09 | -0.00120409 | EHD1 |
| chr12 | 106170430 | 106171211 | 782 | 3 | 4.03E-12 | 3.46E-06 | 1.38E-07 | 5.62E-09 | -0.0046394 | AC010182.1 |
| chr19 | 21395648 | 21396168 | 521 | 5 | 2.09E-10 | 1.46E-09 | 0.000313 | 5.62E-09 | -0.002851 | NA |
| chr9 | 131842017 | 131842307 | 291 | 4 | 9.46E-10 | 3.46E-07 | 1.19E-05 | 5.63E-09 | -0.00397001 | NA |
| chr8 | 127793168 | 127794692 | 1525 | 12 | 1.42E-08 | 5.22E-08 | 0.00101 | 5.65E-09 | -0.0001055 | PVT1 |
| chr7 | 123557131 | 123558586 | 1456 | 20 | 3.80E-09 | 1.50E-05 | 0.000257 | 5.65E-09 | 0.000925842 | NDUFA5 |
| chr17 | 5418637 | 5420485 | 1849 | 27 | 8.13E-10 | 1.50E-06 | 0.000267 | 5.65E-09 | 0.000262231 | RPAIN, NUP88 |
| chr1 | 87327681 | 87329439 | 1759 | 15 | 3.77E-08 | 4.30E-06 | 8.08E-05 | 5.65E-09 | 0.002548657 | LMO4 |
| chr3 | 43621441 | 43623157 | 1717 | 14 | 3.33E-07 | 9.09E-08 | 0.000346 | 5.67E-09 | 0.00100715 | ANO10 |
| chr21 | 43757657 | 43758826 | 1170 | 6 | 6.09E-10 | 3.86E-06 | 5.56E-05 | 5.68E-09 | -0.00787591 | PDXK |
| chr18 | 44679595 | 44680829 | 1235 | 14 | 3.04E-11 | 2.08E-06 | 2.99E-05 | 5.69E-09 | 0.000345633 | SETBP1, SETBP1-DT |
| chr16 | 1010367 | 1011053 | 687 | 5 | 5.43E-09 | 3.17E-09 | 0.000189 | 5.70E-09 | -0.01404489 | NA |
| chr8 | 95117572 | 95117724 | 153 | 3 | 4.82E-09 | 4.90E-09 | 3.42E-05 | 5.71E-09 | 0.002010407 | NA |
| chr1 | 32250450 | 32251401 | 952 | 10 | 1.14E-08 | 3.46E-08 | 0.000706 | 5.72E-09 | 0.00151849 | LCK |
| chr14 | 105093217 | 105094291 | 1075 | 9 | 2.34E-11 | 2.22E-07 | 0.000231 | 5.75E-09 | -0.00308941 | LINC02298 |
| chr17 | 14303255 | 14304213 | 959 | 8 | 9.72E-09 | 8.28E-10 | 0.000883 | 5.76E-09 | -0.0145385 | HS3ST3B1, AC005224.3 |
| chr1 | 3543903 | 3545154 | 1252 | 3 | 2.31E-12 | 2.75E-07 | 1.45E-07 | 5.77E-09 | 0.000463852 | MEGF6 |
| chr1 | 227734540 | 227736414 | 1875 | 18 | 5.08E-08 | 7.99E-07 | 8.80E-05 | 5.79E-09 | -5.69E-05 | SNAP47, JMJD4 |
| chr11 | 102226543 | 102227536 | 994 | 6 | 3.60E-11 | 1.02E-07 | 1.67E-05 | 5.81E-09 | -0.00837193 | YAP1 |
| chr1 | 175757532 | 175757952 | 421 | 2 | 3.85E-09 | 3.77E-09 | 1.04E-05 | 5.81E-09 | -0.0122754 | NA |
| chr17 | 64001821 | 64002718 | 898 | 2 | 8.99E-09 | 7.23E-09 | 1.68E-06 | 5.81E-09 | 0.009102371 | PRR29, ICAM2 |
| chr15 | 41838452 | 41839881 | 1430 | 8 | 6.73E-08 | 7.73E-08 | 5.54E-05 | 5.81E-09 | -0.00177681 | JMJD7-PLA2G4B, PLA2G4B |
| chr10 | 27100147 | 27101463 | 1317 | 12 | 1.74E-09 | 4.82E-08 | 0.001115 | 5.81E-09 | -0.0068423 | ANKRD26 |
| chr16 | 64950600 | 64951611 | 1012 | 7 | 1.73E-09 | 2.93E-07 | 5.14E-06 | 5.82E-09 | -0.00274026 | CDH11 |
| chr7 | 2937403 | 2938821 | 1419 | 5 | 1.22E-10 | 4.90E-08 | 2.37E-06 | 5.82E-09 | -0.00366058 | CARD11 |
| chr12 | 130280515 | 130281698 | 1184 | 4 | 6.05E-10 | 1.34E-05 | 6.29E-06 | 5.82E-09 | 0.025631792 | NA |
| chr10 | 78164817 | 78165226 | 410 | 3 | 1.65E-09 | 9.68E-09 | 3.17E-06 | 5.83E-09 | 0.002852661 | NA |
| chr15 | 45521726 | 45523338 | 1613 | 15 | 2.13E-10 | 4.42E-07 | 1.84E-05 | 5.84E-09 | -0.00022273 | HMGN2P46, SLC30A4 |
| chr1 | 35642605 | 35642893 | 289 | 5 | 4.91E-10 | 5.64E-08 | 2.67E-05 | 5.85E-09 | 0.021176022 | NA |
| chr22 | 39133118 | 39133318 | 201 | 4 | 7.25E-10 | 6.06E-07 | 3.28E-07 | 5.85E-09 | -0.00548227 | AL031846.2, CBX7 |
| chr1 | 205679963 | 205681346 | 1384 | 14 | 1.43E-10 | 2.13E-05 | 1.52E-05 | 5.88E-09 | 8.70E-05 | SLC45A3 |
| chr6 | 47787589 | 47788274 | 686 | 6 | 1.26E-11 | 8.48E-07 | 4.17E-05 | 5.91E-09 | -0.00228192 | OPN5 |
| chr2 | 201182523 | 201183377 | 855 | 6 | 4.62E-09 | 2.08E-08 | 0.000528 | 5.91E-09 | -0.00236356 | CASP10 |
| chr2 | 235851610 | 235852906 | 1297 | 7 | 2.01E-07 | 4.55E-07 | 1.30E-05 | 5.92E-09 | -0.00383474 | AGAP1 |
| chr15 | 85380021 | 85381436 | 1416 | 9 | 1.32E-09 | 8.27E-09 | 0.000144 | 5.93E-09 | 0.000507046 | AKAP13, MIR7706 |
| chr11 | 118304070 | 118304960 | 891 | 6 | 1.17E-09 | 1.70E-08 | 0.000113 | 5.93E-09 | -0.00299247 | CD3E |
| chr4 | 704514 | 705905 | 1392 | 11 | 1.55E-08 | 4.56E-07 | 0.000138 | 5.94E-09 | -0.00452737 | PCGF3 |
| chr14 | 69398000 | 69399223 | 1224 | 22 | 1.39E-08 | 1.11E-07 | 0.001327 | 5.94E-09 | 0.000490498 | SLC39A9, ERH |
| chr16 | 80539819 | 80541875 | 2057 | 15 | 5.47E-08 | 2.27E-07 | 0.000109 | 5.97E-09 | 0.002763981 | DYNLRB2, AC105411.1 |
| chr12 | 6839074 | 6841108 | 2035 | 10 | 4.62E-07 | 6.01E-07 | 0.000566 | 6.02E-09 | -0.00288203 | P3H3, GNB3 |
| chr1 | 64186458 | 64186702 | 245 | 2 | 2.39E-09 | 6.14E-09 | 2.58E-06 | 6.04E-09 | 0.017056704 | NA |
| chr1 | 215566593 | 215568500 | 1908 | 10 | 2.32E-08 | 1.79E-05 | 1.28E-05 | 6.04E-09 | -0.00312223 | KCTD3 |
| chr7 | 102079927 | 102080667 | 741 | 7 | 1.45E-11 | 1.88E-05 | 4.64E-06 | 6.05E-09 | 0.001355005 | CUX1 |
| chr2 | 81194756 | 81196689 | 1934 | 4 | 7.07E-07 | 6.41E-09 | 6.32E-05 | 6.06E-09 | -0.00190783 | ANKRD11P1 |
| chr16 | 71440831 | 71441813 | 983 | 5 | 2.24E-09 | 4.04E-09 | 0.000309 | 6.07E-09 | -0.0227583 | TLE7 |
| chr10 | 2978941 | 2979499 | 559 | 2 | 4.56E-11 | 2.24E-08 | 3.79E-07 | 6.07E-09 | 0.002509204 | NA |
| chr5 | 41009238 | 41009382 | 145 | 2 | 4.85E-10 | 2.68E-07 | 6.42E-08 | 6.07E-09 | -0.00860405 | MROH2B |
| chr11 | 66479407 | 66480398 | 992 | 14 | 3.32E-10 | 1.39E-07 | 0.000757 | 6.08E-09 | -0.00258958 | DPP3, AP002748.4 |
| chr3 | 49716816 | 49718868 | 2053 | 8 | 2.07E-08 | 7.28E-07 | 6.95E-05 | 6.08E-09 | -0.00502373 | RNF123, AMIGO3, GMPPB |
| chr2 | 70301274 | 70302381 | 1108 | 11 | 2.52E-09 | 5.29E-07 | 3.02E-05 | 6.09E-09 | -0.00285228 | AC022201.2, FAM136A |
| chr8 | 10813493 | 10814393 | 901 | 3 | 3.13E-09 | 4.36E-09 | 4.53E-05 | 6.10E-09 | -0.02025901 | AC105001.2, PINX1 |
| chr9 | 137493353 | 137494152 | 800 | 4 | 5.19E-13 | 3.33E-07 | 2.41E-07 | 6.11E-09 | 0.007905955 | PNPLA7 |
| chr20 | 51470300 | 51470323 | 24 | 2 | 1.46E-09 | 4.38E-08 | 2.05E-07 | 6.11E-09 | -0.00616789 | NFATC2 |
| chr2 | 106108660 | 106108667 | 8 | 2 | 1.02E-09 | 3.78E-09 | 1.39E-05 | 6.11E-09 | -0.00085352 | UXS1 |
| chr10 | 96271504 | 96272682 | 1179 | 5 | 3.71E-09 | 5.12E-08 | 1.76E-05 | 6.12E-09 | -0.00948432 | BLNK |

| | | | | | | | | | | |
|---|---|---|---|---|---|---|---|---|---|---|
| chr7 | 55185519 | 55187378 | 1860 | 4 | 4.94E-07 | 3.87E-09 | 6.80E-05 | 6.12E-09 | -0.0082121 | EGFR, EGFR-AS1 |
| chr1 | 40257489 | 40258500 | 1012 | 19 | 1.11E-10 | 8.20E-06 | 1.85E-05 | 6.13E-09 | 0.000589944 | ZMPSTE24, AL050341.2 |
| chr5 | 68287770 | 68288826 | 1057 | 14 | 7.76E-10 | 4.75E-05 | 3.59E-05 | 6.14E-09 | 0.001036011 | PIK3R1 |
| chr13 | 113922822 | 113924375 | 1554 | 6 | 5.59E-08 | 1.06E-07 | 3.50E-05 | 6.14E-09 | -0.00559132 | LINC00452, LINC00565 |
| chr9 | 6703605 | 6704574 | 970 | 4 | 3.28E-09 | 5.29E-09 | 3.73E-05 | 6.14E-09 | 0.015147299 | LINC02851 |
| chr15 | 89455124 | 89455303 | 180 | 2 | 4.54E-09 | 3.69E-09 | 1.71E-05 | 6.14E-09 | -0.01845423 | NA |
| chr1 | 241627648 | 241628427 | 780 | 2 | 4.27E-10 | 2.09E-08 | 4.15E-07 | 6.15E-09 | 0.015484458 | OPN3 |
| chr8 | 125039498 | 125039765 | 268 | 2 | 2.46E-10 | 1.86E-07 | 7.84E-08 | 6.19E-09 | -0.00653016 | WASHC5 |
| chr10 | 48671659 | 48673011 | 1353 | 6 | 6.63E-11 | 4.80E-08 | 8.20E-06 | 6.20E-09 | -0.00168389 | AC068898.1 |
| chr11 | 65574731 | 65575870 | 1140 | 8 | 3.94E-10 | 1.22E-07 | 2.98E-05 | 6.21E-09 | 0.00458483 | NA |
| chr5 | 179631958 | 179632667 | 710 | 3 | 2.37E-10 | 1.23E-07 | 4.57E-06 | 6.22E-09 | -0.02177894 | HNRNPH1 |
| chr18 | 69957028 | 69958187 | 1160 | 7 | 7.92E-10 | 4.01E-08 | 5.16E-05 | 6.22E-09 | -0.00679204 | CD226 |
| chr3 | 195888431 | 195890167 | 1737 | 12 | 4.82E-08 | 2.28E-07 | 0.000212 | 6.23E-09 | -0.00080176 | TNK2 |
| chr7 | 175423 | 176739 | 1317 | 5 | 5.36E-11 | 3.20E-06 | 2.17E-07 | 6.25E-09 | -0.0041038 | AC093627.2 |
| chr6 | 216736 | 217658 | 923 | 3 | 1.98E-08 | 6.48E-08 | 1.54E-05 | 6.28E-09 | -0.00207114 | NA |
| chr10 | 132113413 | 132113912 | 500 | 3 | 4.40E-11 | 7.33E-08 | 7.94E-07 | 6.30E-09 | 0.003601072 | JAKMIP3 |
| chr22 | 42379293 | 42379388 | 96 | 2 | 1.04E-09 | 2.66E-08 | 3.37E-07 | 6.30E-09 | 0.002551633 | NA |
| chr3 | 186078635 | 186079927 | 1293 | 5 | 2.28E-08 | 4.53E-07 | 2.22E-05 | 6.30E-09 | -0.00299794 | ETV5-AS1, ETV5 |
| chr11 | 66720467 | 66722050 | 1584 | 9 | 1.15E-09 | 2.83E-06 | 0.000103 | 6.31E-09 | 0.000705742 | SPTBN2 |
| chr2 | 237699993 | 237701164 | 1172 | 4 | 7.53E-09 | 9.89E-09 | 6.29E-06 | 6.32E-09 | -0.00330072 | LRRFIP1 |
| chr15 | 98889996 | 98890622 | 627 | 3 | 9.83E-11 | 1.61E-08 | 1.64E-06 | 6.32E-09 | -0.00368807 | IGF1R, AC069029.1 |
| chr6 | 159128123 | 159129349 | 1227 | 6 | 3.94E-08 | 8.83E-10 | 0.00069 | 6.34E-09 | -0.00114692 | NA |
| chr1 | 166602058 | 166602454 | 397 | 3 | 3.16E-10 | 8.70E-06 | 1.81E-06 | 6.34E-09 | -0.008889 | NA |
| chr12 | 100141711 | 100143489 | 1779 | 13 | 1.05E-07 | 1.40E-07 | 0.000239 | 6.34E-09 | -0.00107437 | AC010203.2, UHRF1BP1L |
| chr18 | 21110830 | 21112377 | 1548 | 18 | 1.13E-07 | 2.23E-06 | 0.000628 | 6.34E-09 | 0.000765603 | ROCK1 |
| chr19 | 45346595 | 45346795 | 201 | 2 | 2.66E-09 | 4.14E-09 | 1.08E-05 | 6.36E-09 | 0.044026591 | KLC3 |
| chr6 | 30006245 | 30006744 | 500 | 4 | 3.60E-16 | 1.33E-06 | 5.69E-08 | 6.39E-09 | -0.01527317 | HLA-J, ZNRD1ASP, AL671277.2 |
| chr12 | 47963243 | 47964321 | 1079 | 14 | 1.13E-09 | 1.69E-06 | 2.02E-05 | 6.40E-09 | 0.00026835 | TMEM106C, AC004801.7 |
| chr12 | 69239040 | 69240237 | 1198 | 12 | 1.47E-08 | 3.86E-07 | 7.07E-05 | 6.45E-09 | 0.001101777 | CPSF6 |
| chr5 | 15927980 | 15928577 | 598 | 4 | 1.87E-11 | 8.58E-07 | 2.58E-07 | 6.45E-09 | -0.00394905 | FBXL7 |
| chr7 | 6046501 | 6047283 | 783 | 3 | 2.45E-08 | 7.88E-07 | 7.29E-06 | 6.46E-09 | 0.001453663 | EIF2AK1 |
| chr1 | 27346347 | 27347368 | 1022 | 4 | 4.11E-08 | 1.37E-08 | 0.000146 | 6.46E-09 | -0.00075148 | SYTL1 |
| chr1 | 2525975 | 2527437 | 1463 | 18 | 3.22E-08 | 1.42E-07 | 3.24E-05 | 6.50E-09 | -0.00030595 | PANK4 |
| chr11 | 68118927 | 68121206 | 2280 | 10 | 3.90E-07 | 3.60E-08 | 6.86E-05 | 6.50E-09 | -0.0016437 | CHKA |
| chr4 | 1311325 | 1312098 | 774 | 5 | 3.75E-11 | 1.15E-07 | 4.11E-05 | 6.51E-09 | -0.00248933 | MAEA |
| chr14 | 99737393 | 99738191 | 799 | 8 | 2.53E-11 | 4.52E-06 | 7.29E-06 | 6.51E-09 | 0.005510061 | EML1 |
| chr11 | 32072095 | 32072587 | 493 | 3 | 4.53E-09 | 1.41E-08 | 4.77E-06 | 6.51E-09 | -0.00131577 | AL035078.4, AL035078.2 |
| chr1 | 200374649 | 200375757 | 1109 | 4 | 1.70E-08 | 1.61E-08 | 5.88E-05 | 6.53E-09 | -0.00225987 | AC104461.1, LINC00862 |
| chr2 | 232455058 | 232456144 | 1087 | 6 | 2.74E-09 | 4.82E-09 | 2.67E-05 | 6.57E-09 | -0.00581676 | ALPI |
| chr12 | 52231702 | 52233792 | 2091 | 13 | 1.32E-07 | 6.51E-06 | 0.000553 | 6.57E-09 | 0.0010275 | KRT7 |
| chr1 | 90724809 | 90726605 | 1797 | 15 | 3.93E-08 | 1.64E-08 | 0.001047 | 6.57E-09 | -0.00156768 | AC092805.1 |
| chr17 | 44904807 | 44906400 | 1594 | 4 | 8.56E-14 | 1.35E-05 | 4.93E-07 | 6.58E-09 | -0.00736953 | CCDC103, FAM187A, GFAP |
| chr1 | 101081033 | 101081649 | 617 | 3 | 6.01E-12 | 6.04E-05 | 9.87E-09 | 6.58E-09 | 0.013953821 | AC093157.1 |
| chr16 | 55566009 | 55566732 | 724 | 4 | 1.82E-12 | 7.16E-06 | 9.46E-08 | 6.59E-09 | -0.0032027 | LPCAT2, CAPNS2 |
| chr10 | 13971216 | 13972651 | 1436 | 7 | 8.74E-08 | 2.96E-08 | 8.12E-05 | 6.59E-09 | 0.003623442 | FRMD4A |
| chr20 | 51412802 | 51414144 | 1343 | 6 | 2.94E-08 | 4.65E-08 | 0.000157 | 6.60E-09 | -0.01068738 | NFATC2 |
| chr5 | 158175657 | 158176829 | 1173 | 5 | 4.52E-10 | 7.63E-08 | 7.35E-06 | 6.61E-09 | -0.00898555 | AC025437.4, AC025437.5 |
| chr15 | 37094085 | 37095926 | 1842 | 8 | 2.02E-08 | 5.65E-07 | 0.000155 | 6.62E-09 | 0.002494294 | MEIS2 |
| chr1 | 3906846 | 3907883 | 1038 | 8 | 3.22E-10 | 8.60E-07 | 1.10E-06 | 6.62E-09 | -0.00692517 | LINC01134 |
| chr11 | 96343105 | 96343751 | 647 | 12 | 1.08E-09 | 7.03E-07 | 0.000148 | 6.62E-09 | 0.001591661 | MAML2 |
| chr6 | 50836107 | 50836461 | 355 | 6 | 2.28E-09 | 3.86E-07 | 5.37E-05 | 6.62E-09 | -0.00571213 | TFAP2B |
| chr5 | 2131410 | 2132764 | 1355 | 7 | 3.67E-08 | 2.49E-07 | 0.00013 | 6.62E-09 | -0.00060921 | NA |
| chr10 | 56360560 | 56361775 | 1216 | 14 | 7.07E-09 | 9.23E-07 | 0.000502 | 6.63E-09 | -0.00031225 | AC010996.1, ZWINT |
| chr11 | 65213348 | 65214125 | 778 | 9 | 4.04E-09 | 3.55E-09 | 0.000407 | 6.64E-09 | 0.010855887 | SLC22A20P |
| chr19 | 34772580 | 34774082 | 1503 | 17 | 2.60E-08 | 2.27E-05 | 0.000843 | 6.64E-09 | 0.000942509 | ZNF599 |
| chr2 | 73232714 | 73234400 | 1687 | 18 | 3.63E-10 | 1.59E-06 | 0.000216 | 6.64E-09 | 0.000246774 | CCT7, PRADC1 |
| chr6 | 168024599 | 168025984 | 1386 | 8 | 1.46E-09 | 4.90E-08 | 4.55E-05 | 6.65E-09 | -0.0065851 | KIF25 |
| chr20 | 63283460 | 63284450 | 991 | 9 | 5.06E-10 | 6.03E-08 | 4.44E-05 | 6.66E-09 | 0.006788177 | ARFGAP1 |
| chr14 | 105419194 | 105420628 | 1435 | 12 | 2.34E-08 | 1.03E-07 | 0.000146 | 6.66E-09 | -0.0010833 | TEX22, MTA1, AL928654.1 |
| chr9 | 33415280 | 33415869 | 590 | 5 | 3.01E-10 | 1.41E-08 | 1.03E-05 | 6.68E-09 | -0.00786166 | NA |
| chr19 | 17475804 | 17476624 | 821 | 2 | 2.17E-09 | 1.21E-08 | 9.72E-07 | 6.69E-09 | -0.01335707 | SLC27A1 |
| chr5 | 117415246 | 117415439 | 194 | 2 | 3.08E-10 | 3.06E-07 | 6.56E-08 | 6.69E-09 | 0.008799328 | NA |
| chr3 | 38953629 | 38954379 | 751 | 7 | 4.00E-10 | 6.73E-08 | 2.43E-05 | 6.70E-09 | -0.00183986 | SCN11A |
| chr16 | 27313020 | 27314733 | 1714 | 12 | 1.01E-08 | 2.74E-07 | 0.000133 | 6.70E-09 | 0.000237503 | IL4R, AC106739.1 |
| chr7 | 486047 | 486306 | 260 | 4 | 8.41E-10 | 8.64E-07 | 8.65E-06 | 6.70E-09 | -0.0034272 | AC147651.1 |
| chr11 | 8311453 | 8312275 | 823 | 2 | 1.60E-10 | 1.45E-08 | 7.66E-07 | 6.73E-09 | 0.00131725 | NA |
| chr1 | 28581122 | 28582493 | 1372 | 14 | 1.34E-08 | 2.17E-05 | 0.00041 | 6.74E-09 | -0.0002797 | SNHG12 |
| chr8 | 2071504 | 2072627 | 1124 | 6 | 4.64E-09 | 8.88E-09 | 7.20E-05 | 6.75E-09 | -0.00462789 | MYOM2 |
| chr15 | 100349478 | 100350791 | 1314 | 17 | 5.90E-09 | 9.40E-07 | 0.00038 | 6.78E-09 | -0.00017472 | SPATA41 |
| chr17 | 42851848 | 42853472 | 1625 | 7 | 4.48E-09 | 4.81E-09 | 3.12E-05 | 6.78E-09 | -0.0037781 | AOC3 |
| chr12 | 78863557 | 78864658 | 1102 | 14 | 2.06E-07 | 5.73E-09 | 0.000771 | 6.79E-09 | 0.001124067 | SYT1 |
| chr16 | 86387491 | 86388363 | 873 | 6 | 2.40E-08 | 4.18E-09 | 0.000638 | 6.79E-09 | 0.015356481 | NA |
| chr16 | 71894586 | 71896145 | 1560 | 14 | 1.06E-12 | 0.0002373 | 1.44E-07 | 6.79E-09 | 0.001669106 | IST1, ZNF821 |
| chr4 | 113978894 | 113980052 | 1159 | 13 | 4.72E-11 | 5.58E-06 | 4.04E-05 | 6.79E-09 | -0.00267871 | ARSJ |
| chr20 | 47361723 | 47361735 | 13 | 2 | 1.91E-09 | 4.21E-09 | 1.46E-05 | 6.79E-09 | 0.013967424 | NA |
| chr20 | 48364750 | 48365625 | 876 | 2 | 1.12E-07 | 4.56E-09 | 9.57E-06 | 6.79E-09 | -1.38E-05 | LINC00494, AL121888.1 |
| chr11 | 72112104 | 72113603 | 1500 | 18 | 1.57E-07 | 8.91E-08 | 0.005384 | 6.82E-09 | 0.000496928 | ANAPC15 |
| chr2 | 22281235 | 22281378 | 144 | 3 | 4.92E-09 | 7.72E-09 | 5.31E-06 | 6.84E-09 | -0.02067756 | AC096570.1 |
| chr17 | 10659085 | 10660073 | 989 | 5 | 8.10E-09 | 1.35E-09 | 0.000257 | 6.86E-09 | -0.00591806 | NA |
| chr15 | 90100534 | 90101446 | 913 | 8 | 2.19E-09 | 8.87E-09 | 0.000419 | 6.89E-09 | -0.00785712 | IDH2 |
| chr16 | 57283767 | 57285405 | 1639 | 14 | 9.25E-09 | 7.68E-06 | 0.000503 | 6.91E-09 | -0.00244993 | PLLP |
| chr10 | 71963406 | 71964162 | 757 | 4 | 1.06E-09 | 1.09E-08 | 4.44E-05 | 6.91E-09 | -0.00610082 | NA |
| chr20 | 56468415 | 56469621 | 1207 | 10 | 9.51E-08 | 3.47E-10 | 0.003752 | 6.91E-09 | 0.001822877 | RTF2 |
| chr22 | 18022945 | 18024248 | 1304 | 6 | 2.07E-10 | 1.25E-08 | 8.67E-05 | 6.93E-09 | -0.00300208 | MICAL3 |
| chr1 | 32107400 | 32108985 | 1586 | 19 | 7.49E-08 | 1.25E-05 | 2.18E-05 | 6.94E-09 | 0.000306486 | KPNA6 |
| chr5 | 27471922 | 27472435 | 514 | 4 | 7.52E-09 | 3.08E-09 | 0.000173 | 6.94E-09 | 0.005863674 | PURPL |
| chr8 | 118120497 | 118120939 | 443 | 2 | 2.19E-10 | 2.64E-08 | 3.93E-07 | 6.95E-09 | -0.01776718 | NA |
| chr17 | 41467061 | 41467836 | 776 | 5 | 9.84E-09 | 3.55E-09 | 0.000386 | 6.95E-09 | -0.01134092 | KRT32 |
| chr11 | 55935967 | 55937808 | 1842 | 6 | 3.15E-06 | 2.42E-07 | 6.48E-05 | 6.95E-09 | -0.01131888 | OR5I1 |
| chr13 | 43879047 | 43880519 | 1473 | 17 | 2.46E-09 | 6.24E-07 | 0.000113 | 6.96E-09 | 0.003001103 | LACC1, CCDC122 |
| chr12 | 6549779 | 6552578 | 2800 | 11 | 3.45E-07 | 1.05E-07 | 6.67E-05 | 6.99E-09 | -0.00320952 | IFFO1 |
| chr16 | 23803113 | 23803925 | 813 | 2 | 6.37E-11 | 1.10E-07 | 1.24E-07 | 7.00E-09 | -0.01524957 | NA |
| chr2 | 216412021 | 216413346 | 1326 | 17 | 3.57E-09 | 5.68E-08 | 0.002721 | 7.00E-09 | -0.00336574 | SMARCAL1, SMARCAL1-AS1 |
| chr22 | 25964978 | 25965350 | 373 | 3 | 3.18E-09 | 3.50E-09 | 2.88E-05 | 7.02E-09 | -0.00564493 | MYO18B, Z98949.2 |
| chr10 | 132883646 | 132885261 | 1616 | 7 | 9.05E-11 | 1.40E-07 | 0.000236 | 7.06E-09 | -0.00230804 | CFAP46 |
| chr11 | 62622027 | 62623612 | 1586 | 12 | 2.58E-06 | 1.26E-08 | 0.00123 | 7.07E-09 | -0.00012369 | B3GAT3 |
| chr1 | 86396329 | 86397641 | 1313 | 9 | 7.07E-09 | 2.71E-06 | 7.66E-05 | 7.08E-09 | 0.000338164 | ODF2L |
| chr1 | 59926027 | 59927261 | 1235 | 13 | 7.00E-10 | 3.51E-07 | 0.00016 | 7.08E-09 | -0.00199993 | CYP2J2 |
| chr12 | 78867615 | 78868101 | 487 | 2 | 9.22E-10 | 7.06E-09 | 2.91E-06 | 7.08E-09 | -0.03161405 | SYT1 |
| chr1 | 6148128 | 6148958 | 831 | 5 | 4.69E-09 | 3.31E-09 | 0.000126 | 7.08E-09 | -0.00457687 | CHD5 |
| chr2 | 219308869 | 219310397 | 1529 | 14 | 1.16E-10 | 0.0001697 | 1.74E-05 | 7.10E-09 | -0.00269661 | PTPRN |
| chr12 | 111761040 | 111761174 | 135 | 2 | 1.54E-10 | 1.57E-07 | 1.01E-07 | 7.11E-09 | 0.007006373 | AC002996.1 |
| chr4 | 16083059 | 16084669 | 1611 | 17 | 3.52E-08 | 9.13E-06 | 7.81E-05 | 7.11E-09 | -0.00127993 | PROM1 |
| chr2 | 74421179 | 74423324 | 2146 | 12 | 2.02E-07 | 2.09E-08 | 0.000371 | 7.11E-09 | 0.000568792 | WDR54, AC005041.1, C2orf81 |
| chr15 | 25134675 | 25136361 | 1687 | 5 | 3.66E-07 | 4.55E-09 | 0.000434 | 7.11E-09 | -0.0134636 | SNHG14, PWAR1 |
| chr22 | 30560967 | 30562802 | 1836 | 5 | 4.22E-07 | 4.70E-09 | 0.000181 | 7.11E-09 | -0.01180134 | GAL3ST1 |
| chr12 | 132202367 | 132203184 | 818 | 6 | 1.97E-10 | 7.02E-08 | 1.17E-05 | 7.13E-09 | -0.00161334 | GALNT9 |
| chr3 | 50268129 | 50269261 | 1133 | 8 | 1.53E-11 | 4.85E-07 | 1.79E-06 | 7.14E-09 | -0.00388256 | SEMA3B |
| chr16 | 83802391 | 83803037 | 647 | 2 | 1.02E-09 | 4.34E-09 | 1.73E-05 | 7.14E-09 | -0.00376108 | AC009119.1 |
| chr3 | 191235581 | 191235597 | 17 | 2 | 4.95E-09 | 5.51E-09 | 5.84E-06 | 7.14E-09 | 0.012336494 | OSTN |
| chr5 | 141349025 | 141350949 | 1925 | 15 | 1.33E-07 | 2.40E-06 | 0.0001 | 7.14E-09 | -0.00840545 | PCDHGA1, PCDHGA2, PCDHGA3, PCDHGB1, AC005618.3 |
| chr10 | 132441341 | 132442101 | 761 | 6 | 2.06E-09 | 3.03E-07 | 7.32E-06 | 7.16E-09 | 0.001104235 | AL451069.3 |
| chr11 | 60345440 | 60346254 | 815 | 2 | 7.54E-10 | 2.12E-08 | 5.28E-07 | 7.17E-09 | -0.00818253 | MS4A6E |
| chr4 | 102760939 | 102762226 | 1288 | 15 | 6.94E-09 | 3.37E-07 | 2.28E-05 | 7.17E-09 | -0.0017086 | MANBA |
| chr6 | 36844371 | 36844660 | 290 | 7 | 6.99E-10 | 7.37E-08 | 0.000153 | 7.18E-09 | -0.00728716 | Z85996.3 |
| chr17 | 79033436 | 79035074 | 1639 | 8 | 4.34E-10 | 3.55E-06 | 5.90E-07 | 7.19E-09 | -0.00385969 | C1QTNF1 |
| chr7 | 47582089 | 47583120 | 1032 | 11 | 4.14E-12 | 2.47E-05 | 0.000107 | 7.20E-09 | -0.00701937 | TNS3 |
| chr8 | 103414364 | 103415668 | 1305 | 22 | 1.58E-09 | 7.32E-07 | 0.001363 | 7.20E-09 | 0.00512584 | DCAF13, SLC25A32, AC012213.5 |
| chr17 | 39627626 | 39629399 | 1774 | 7 | 4.45E-07 | 3.77E-08 | 7.87E-05 | 7.21E-09 | -0.0073019 | PPP1R1B |
| chr12 | 57610232 | 57611975 | 1744 | 11 | 1.50E-12 | 1.61E-06 | 6.45E-05 | 7.21E-09 | -0.00216421 | ARHGEF25 |
| chr11 | 74747755 | 74748816 | 1062 | 8 | 5.61E-09 | 1.18E-06 | 0.000214 | 7.22E-09 | -0.00476946 | NA |
| chr10 | 71622179 | 71622238 | 60 | 2 | 4.42E-11 | 7.19E-06 | 2.58E-08 | 7.22E-09 | -0.01517481 | CDH23 |
| chr6 | 52419888 | 52421048 | 1161 | 13 | 2.51E-12 | 1.35E-07 | 2.30E-05 | 7.23E-09 | -0.00104321 | EFHC1 |
| chr19 | 2976791 | 2977845 | 1055 | 12 | 2.43E-11 | 1.46E-05 | 2.16E-05 | 7.23E-09 | -0.00162875 | TLE6 |
| chr7 | 2728932 | 2730776 | 1845 | 7 | 5.61E-09 | 4.82E-06 | 1.12E-05 | 7.23E-09 | 0.007121653 | AMZ1, GNA12 |
| chr18 | 50298634 | 50298993 | 360 | 5 | 1.55E-09 | 1.57E-07 | 2.59E-05 | 7.25E-09 | -0.01564151 | NA |
| chr19 | 41309944 | 41311124 | 1181 | 11 | 3.18E-09 | 3.44E-07 | 1.19E-05 | 7.25E-09 | -0.00226013 | CCDC97, TGFB1, AC011462.5 |
| chr5 | 442270 | 444113 | 1844 | 18 | 2.93E-08 | 2.80E-07 | 0.000258 | 7.25E-09 | 0.001719732 | EXOC3, EXOC3-AS1 |
| chr1 | 2919258 | 2920953 | 1696 | 5 | 3.07E-06 | 5.97E-09 | 0.000637 | 7.25E-09 | 8.96E-05 | NA |
| chr20 | 12762879 | 12762921 | 43 | 2 | 5.69E-09 | 4.41E-09 | 1.75E-05 | 7.25E-09 | 0.040354072 | NA |
| chr5 | 7849723 | 7850325 | 603 | 6 | 6.61E-09 | 4.76E-09 | 8.14E-05 | 7.26E-09 | -0.00204044 | C5orf49 |
| chr1 | 167935752 | 167938161 | 2410 | 21 | 1.25E-06 | 1.73E-06 | 0.000325 | 7.26E-09 | 0.000193082 | DCAF6, MPC2 |
| chr1 | 5876557 | 5877620 | 1064 | 6 | 5.17E-11 | 2.05E-06 | 1.55E-07 | 7.26E-09 | -0.00116019 | NPHP4 |
| chr13 | 102773039 | 102774822 | 1784 | 16 | 3.33E-08 | 8.01E-08 | 0.000148 | 7.28E-09 | -0.00182964 | TEX30 |
| chr14 | 24270078 | 24271430 | 1353 | 8 | 3.12E-11 | 2.30E-06 | 8.04E-06 | 7.28E-09 | 0.000476017 | AL096870.12, RABGGTA |
| chr14 | 69726243 | 69727264 | 1022 | 8 | 2.75E-10 | 3.02E-07 | 6.71E-05 | 7.30E-09 | -0.01236146 | SRSF5 |
| chr11 | 57248973 | 57249904 | 932 | 4 | 9.20E-09 | 1.61E-07 | 1.77E-05 | 7.31E-09 | -0.00869403 | NA |
| chr1 | 54979719 | 54981355 | 1637 | 13 | 1.36E-08 | 1.03E-08 | 0.001183 | 7.31E-09 | 0.001087072 | TMEM61, AL590440.2, AL590440.1 |
| chr15 | 31102082 | 31103018 | 937 | 4 | 6.11E-11 | 1.04E-07 | 4.16E-07 | 7.31E-09 | -0.00299086 | TRPM1 |
| chr3 | 115246132 | 115246880 | 749 | 2 | 9.34E-11 | 7.13E-08 | 1.78E-07 | 7.31E-09 | -0.00419046 | AC026341.1 |
| chr11 | 20386990 | 20388670 | 1681 | 17 | 6.60E-07 | 1.08E-06 | 0.000663 | 7.31E-09 | -0.00191772 | PRMT3 |
| chr17 | 82244469 | 82245401 | 933 | 6 | 3.18E-09 | 5.34E-08 | 0.000172 | 7.33E-09 | -0.00503861 | SLC16A3, AC132872.3, CSNK1D |
| chr11 | 70207148 | 70208000 | 853 | 4 | 1.81E-10 | 4.57E-08 | 1.01E-05 | 7.35E-09 | 0.004323789 | FADD, AP000879.1 |
| chr17 | 2088229 | 2089797 | 1569 | 9 | 1.50E-07 | 3.17E-08 | 1.40E-05 | 7.38E-09 | 0.001625375 | AC090617.4, SMG6 |
| chr9 | 84781510 | 84781865 | 356 | 2 | 4.12E-11 | 1.75E-07 | 9.94E-08 | 7.39E-09 | 0.004811322 | NTRK2 |
| chr3 | 157088696 | 157089902 | 1207 | 10 | 4.46E-08 | 4.01E-10 | 0.0051 | 7.40E-09 | -0.00554245 | LINC00881, LINC00880 |
| chr7 | 73700950 | 73702273 | 1324 | 5 | 2.73E-10 | 2.91E-06 | 3.30E-06 | 7.43E-09 | -0.0120792 | BUD23, STX1A |
| chr13 | 72780829 | 72782453 | 1625 | 17 | 2.17E-07 | 3.04E-07 | 0.000716 | 7.44E-09 | -0.00236561 | PIBF1, DIS3 |
| chr9 | 93095422 | 93096947 | 1526 | 15 | 1.11E-08 | 7.85E-06 | 7.50E-05 | 7.46E-09 | 0.003524119 | CARD19, AL451065.1 |
| chr1 | 220883894 | 220885463 | 1570 | 6 | 2.94E-07 | 4.67E-07 | 2.60E-05 | 7.46E-09 | -0.00326599 | AL445423.3, HLX |
| chr21 | 39321622 | 39322144 | 523 | 4 | 1.08E-09 | 1.55E-08 | 0.000127 | 7.46E-09 | -0.00764906 | BRWD1-AS1 |
| chr11 | 2132844 | 2134291 | 1448 | 10 | 2.18E-13 | 1.19E-05 | 6.57E-05 | 7.47E-09 | -0.00850253 | IGF2, AC132217.2, INS-IGF2, MIR483 |
| chr17 | 82302534 | 82303673 | 1140 | 4 | 3.21E-10 | 7.03E-08 | 5.81E-06 | 7.47E-09 | 0.010819224 | NA |
| chr3 | 25577300 | 25577775 | 476 | 2 | 5.91E-10 | 1.78E-08 | 7.09E-07 | 7.48E-09 | -0.0163383 | RARB |
| chr19 | 10701164 | 10702141 | 978 | 12 | 2.33E-09 | 4.69E-06 | 0.00021 | 7.49E-09 | -0.000482 | QTRT1 |
| chr9 | 137820055 | 137820774 | 720 | 5 | 2.74E-09 | 7.47E-08 | 4.85E-06 | 7.49E-09 | 0.003158953 | EHMT1 |
| chr2 | 10666 | 11618 | 953 | 3 | 4.11E-08 | 2.87E-08 | 1.20E-05 | 7.49E-09 | 0.004497226 | NA |
| chr7 | 129824448 | 129826161 | 1714 | 7 | 7.88E-09 | 6.14E-07 | 1.49E-05 | 7.51E-09 | -0.00152775 | NA |

| | | | | | | | | | | |
|---|---|---|---|---|---|---|---|---|---|---|
| chr12 | 130455381 | 130456761 | 1381 | 5 | 8.53E-07 | 4.05E-08 | 0.000129 | 7.52E-09 | -0.00676886 | RIMBP2 |
| chr15 | 52857918 | 52858215 | 298 | 3 | 1.54E-09 | 1.99E-08 | 2.09E-06 | 7.52E-09 | -0.01902173 | NA |
| chr8 | 92102819 | 92103872 | 1054 | 12 | 3.22E-09 | 6.50E-07 | 0.000106 | 7.52E-09 | -0.00313206 | RUNX1T1 |
| chr8 | 27614258 | 27615407 | 1150 | 15 | 5.77E-10 | 2.09E-07 | 3.39E-05 | 7.55E-09 | -0.00194663 | CLU |
| chr1 | 9384875 | 9385553 | 679 | 3 | 2.56E-08 | 1.46E-08 | 1.98E-05 | 7.57E-09 | -0.00455994 | NA |
| chr5 | 176416140 | 176417367 | 1228 | 9 | 1.86E-08 | 2.76E-08 | 0.000704 | 7.57E-09 | -0.0024371 | CLTB |
| chr3 | 139534095 | 139535692 | 1598 | 4 | 7.77E-07 | 2.98E-09 | 5.95E-05 | 7.57E-09 | -0.00732472 | AC046134.2, RBP1 |
| chr6 | 32116708 | 32118003 | 1296 | 15 | 9.70E-10 | 5.81E-07 | 0.000175 | 7.59E-09 | -0.00010707 | AL662884.2, ATF6B |
| chr3 | 116445395 | 116446096 | 702 | 8 | 2.28E-11 | 1.75E-06 | 6.91E-05 | 7.61E-09 | -0.01104584 | LSAMP |
| chr1 | 226363344 | 226364289 | 946 | 3 | 9.81E-11 | 2.74E-08 | 1.70E-06 | 7.62E-09 | 0.000134171 | PARP1 |
| chr20 | 57727401 | 57727576 | 176 | 4 | 3.85E-09 | 7.16E-07 | 2.90E-05 | 7.64E-09 | -0.00758195 | NA |
| chr13 | 28717238 | 28719485 | 2248 | 15 | 7.11E-08 | 1.40E-06 | 6.29E-05 | 7.64E-09 | 0.001328342 | SLC46A3 |
| chr5 | 132011022 | 132011649 | 628 | 7 | 3.55E-11 | 1.09E-07 | 0.000175 | 7.68E-09 | -0.00663109 | AC034228.1, AC034228.3, ACSL6 |
| chr4 | 76401436 | 76402001 | 566 | 3 | 1.25E-12 | 3.38E-06 | 1.51E-07 | 7.70E-09 | -0.01066417 | AC096743.2, CCDC158 |
| chr1 | 225740081 | 225740875 | 795 | 3 | 2.71E-12 | 6.82E-06 | 8.09E-08 | 7.72E-09 | 0.002428642 | AC099066.2 |
| chr6 | 93419171 | 93420152 | 982 | 13 | 2.78E-09 | 1.38E-07 | 2.69E-05 | 7.72E-09 | 0.00029862 | AL591519.1, EPHA7 |
| chr18 | 79950649 | 79952348 | 1700 | 14 | 8.62E-07 | 1.93E-06 | 0.000467 | 7.72E-09 | -0.00112015 | SLC66A2 |
| chr16 | 1175588 | 1177475 | 1888 | 8 | 1.92E-06 | 2.13E-08 | 0.000173 | 7.73E-09 | 0.000760885 | CACNA1H |
| chr2 | 218387810 | 218388095 | 286 | 4 | 1.32E-09 | 8.62E-07 | 1.57E-05 | 7.73E-09 | -0.01387708 | SLC11A1 |
| chr3 | 195894917 | 195896069 | 1153 | 7 | 2.77E-09 | 7.21E-08 | 7.64E-05 | 7.74E-09 | -0.00111974 | TNK2 |
| chr9 | 131839528 | 131840097 | 570 | 3 | 3.84E-09 | 1.89E-08 | 1.28E-05 | 7.74E-09 | -0.00464513 | NA |
| chr8 | 1914333 | 1916256 | 1924 | 6 | 8.04E-07 | 1.57E-07 | 0.000154 | 7.74E-09 | 0.001243043 | AC019257.8, ARHGEF10 |
| chr10 | 1395749 | 1395970 | 222 | 3 | 4.46E-09 | 1.27E-08 | 9.58E-06 | 7.75E-09 | -0.01003823 | ADARB2 |
| chr17 | 59847804 | 59848439 | 636 | 3 | 7.37E-11 | 3.16E-08 | 1.17E-06 | 7.77E-09 | -0.01364054 | NA |
| chr19 | 2342129 | 2342175 | 47 | 2 | 1.10E-09 | 1.52E-08 | 9.48E-07 | 7.77E-09 | 0.025056731 | SPPL2B |
| chr6 | 96468614 | 96469609 | 996 | 2 | 1.02E-07 | 6.58E-09 | 4.60E-06 | 7.77E-09 | -0.00575158 | UFL1-AS1 |
| chr10 | 31927103 | 31929040 | 1938 | 18 | 2.99E-07 | 1.37E-06 | 0.002077 | 7.77E-09 | -0.0036333 | ARHGAP12 |
| chr19 | 46296248 | 46297345 | 1098 | 8 | 6.20E-09 | 4.45E-08 | 0.00054 | 7.80E-09 | 0.003818853 | HIF3A |
| chr17 | 36728667 | 36728826 | 160 | 3 | 9.40E-09 | 4.86E-09 | 4.58E-05 | 7.84E-09 | 0.015471657 | NA |
| chr7 | 54203459 | 54203589 | 131 | 2 | 1.88E-09 | 4.87E-09 | 1.58E-05 | 7.84E-09 | -0.01824523 | AC092848.2 |
| chr7 | 66377450 | 66377500 | 51 | 2 | 1.33E-10 | 1.53E-07 | 1.14E-07 | 7.85E-09 | -0.00426944 | TPST1, LINC00174 |
| chr17 | 75824369 | 75825427 | 1059 | 4 | 2.55E-09 | 1.26E-07 | 1.24E-05 | 7.85E-09 | 0.005616266 | UNK |
| chr12 | 122266098 | 122267047 | 950 | 12 | 9.24E-09 | 2.21E-09 | 0.002108 | 7.89E-09 | 0.000672094 | AC048338.1, VPS33A |
| chr7 | 50400972 | 50402420 | 1449 | 6 | 2.13E-07 | 2.10E-09 | 0.000731 | 7.90E-09 | -0.00067597 | IKZF1 |
| chr4 | 2933965 | 2935654 | 1690 | 20 | 1.54E-08 | 6.67E-06 | 0.002039 | 7.90E-09 | 0.000403507 | NOP14-AS1, MFSD10 |
| chr6 | 43698623 | 43699000 | 378 | 2 | 3.15E-09 | 6.78E-09 | 4.48E-06 | 7.91E-09 | -0.02462738 | NA |
| chr16 | 56651142 | 56651990 | 849 | 8 | 2.54E-09 | 2.10E-08 | 2.48E-05 | 7.92E-09 | -0.00563349 | AC026461.2, MT1B |
| chr11 | 86671858 | 86673628 | 1771 | 18 | 1.04E-08 | 1.52E-05 | 3.64E-05 | 7.92E-09 | -0.00060779 | ME3 |
| chr17 | 1997433 | 1999728 | 2296 | 7 | 5.34E-06 | 3.43E-07 | 8.40E-05 | 7.92E-09 | -0.00553059 | AC099684.2, RTN4RL1 |
| chr10 | 555642 | 555876 | 235 | 4 | 8.10E-09 | 1.70E-08 | 5.86E-05 | 7.93E-09 | 0.029985171 | DIP2C |
| chr13 | 24679885 | 24681159 | 1275 | 13 | 1.63E-10 | 1.06E-05 | 1.45E-05 | 7.95E-09 | 0.001172272 | ATP12A |
| chr12 | 114693050 | 114694956 | 1907 | 11 | 3.84E-08 | 2.11E-07 | 0.000179 | 7.96E-09 | -0.00267621 | AC026765.3 |
| chr3 | 148741450 | 148742344 | 895 | 2 | 7.37E-12 | 8.34E-07 | 5.18E-08 | 7.96E-09 | -0.0056311 | AGTR1 |
| chr1 | 186126625 | 186127426 | 802 | 2 | 1.85E-10 | 6.22E-08 | 2.22E-07 | 7.99E-09 | -0.02093009 | HMCN1 |
| chr9 | 94807534 | 94809004 | 1471 | 3 | 2.36E-07 | 4.36E-09 | 5.54E-05 | 7.99E-09 | -0.00547968 | AOPEP |
| chr19 | 10919505 | 10921496 | 1992 | 6 | 4.50E-07 | 2.05E-09 | 0.000693 | 7.99E-09 | 0.000100196 | CARM1 |
| chr10 | 101030137 | 101032286 | 2150 | 19 | 2.43E-08 | 5.72E-07 | 0.002174 | 8.05E-09 | -0.00134853 | SFXN3, PDZD7 |
| chr3 | 50182462 | 50183483 | 1022 | 7 | 6.42E-10 | 2.26E-06 | 4.53E-06 | 8.05E-09 | -0.00026904 | SEMA3F |
| chr20 | 54115310 | 54116060 | 751 | 3 | 3.19E-09 | 7.23E-09 | 3.21E-05 | 8.06E-09 | -0.01980843 | NA |
| chr21 | 41859251 | 41860788 | 1538 | 4 | 8.81E-09 | 4.49E-09 | 0.000111 | 8.06E-09 | -0.00220564 | PRDM15 |
| chr17 | 28842098 | 28843909 | 1812 | 10 | 1.17E-07 | 9.59E-09 | 0.001769 | 8.07E-09 | -0.0036218 | FAM222B |
| chr7 | 138981950 | 138982782 | 833 | 5 | 3.20E-10 | 5.22E-08 | 3.88E-05 | 8.07E-09 | -0.00067977 | NA |
| chr11 | 72696643 | 72697467 | 825 | 5 | 2.81E-09 | 7.33E-09 | 0.000105 | 8.07E-09 | 0.000231824 | ARAP1 |
| chr17 | 83092752 | 83095254 | 2503 | 14 | 3.24E-07 | 4.58E-06 | 0.000116 | 8.08E-09 | -0.00085451 | METRNL |
| chr12 | 54548998 | 54550131 | 1134 | 14 | 1.76E-09 | 1.73E-06 | 2.90E-05 | 8.08E-09 | -0.00358129 | PDE1B |
| chr6 | 165846450 | 165847084 | 635 | 6 | 6.34E-10 | 1.68E-08 | 6.07E-05 | 8.09E-09 | 0.003826495 | PDE10A |
| chr17 | 4609228 | 4610413 | 1186 | 5 | 1.49E-07 | 1.29E-09 | 0.000426 | 8.09E-09 | -0.00435683 | NA |
| chr14 | 96362170 | 96364361 | 2192 | 17 | 3.77E-07 | 7.61E-07 | 0.000239 | 8.10E-09 | -0.00080501 | GSKIP, ATG2B |
| chr20 | 36830893 | 36832192 | 1300 | 3 | 1.24E-13 | 1.67E-06 | 7.56E-08 | 8.10E-09 | -0.00474892 | SOGA1 |
| chr8 | 52149971 | 52150753 | 783 | 4 | 4.82E-10 | 5.10E-08 | 1.11E-05 | 8.13E-09 | -0.01977678 | ST18 |
| chr14 | 103847259 | 103848886 | 1628 | 11 | 3.58E-07 | 6.23E-09 | 0.002581 | 8.13E-09 | -0.00213704 | LINC00637, PPP1R13B |
| chr6 | 30702765 | 30704104 | 1340 | 9 | 7.94E-12 | 1.31E-05 | 8.95E-07 | 8.13E-09 | -0.00338689 | MDC1-AS1, MDC1 |
| chr11 | 44754345 | 44754882 | 538 | 3 | 6.41E-09 | 1.76E-08 | 4.43E-06 | 8.15E-09 | -0.01167712 | TSPAN18 |
| chr1 | 109090430 | 109091626 | 1197 | 11 | 2.63E-08 | 4.30E-07 | 5.36E-05 | 8.15E-09 | -0.00062552 | TMEM167B, AL356488.3 |
| chr12 | 55842750 | 55844141 | 1392 | 8 | 2.72E-08 | 1.78E-07 | 0.000275 | 8.15E-09 | -0.00658294 | MMP19 |
| chr5 | 171772655 | 171773479 | 825 | 4 | 2.29E-08 | 1.23E-08 | 8.42E-05 | 8.16E-09 | -0.0037464 | NA |
| chr21 | 29024006 | 29025377 | 1372 | 15 | 1.00E-09 | 4.80E-07 | 0.000282 | 8.16E-09 | 0.000356034 | USP16, AF129075.2 |
| chr14 | 21722899 | 21723641 | 743 | 7 | 6.16E-09 | 7.04E-08 | 0.000242 | 8.16E-09 | -0.01188236 | NA |
| chr11 | 1346976 | 1347538 | 563 | 3 | 7.37E-09 | 1.05E-08 | 7.85E-06 | 8.16E-09 | 0.001265506 | NA |
| chr6 | 20497824 | 20498767 | 944 | 3 | 1.07E-09 | 8.24E-09 | 7.07E-06 | 8.16E-09 | 0.006682231 | NA |
| chr11 | 75581815 | 75583601 | 1787 | 9 | 8.13E-08 | 9.31E-08 | 0.000221 | 8.18E-09 | -0.00131401 | AP001922.5 |
| chr5 | 170388767 | 170390243 | 1477 | 9 | 3.90E-08 | 1.63E-08 | 2.36E-05 | 8.18E-09 | -0.00914615 | KCNIP1, KCNIP1-OT1, KCNMB1 |
| chr7 | 108885446 | 108885748 | 303 | 4 | 1.80E-09 | 2.62E-08 | 5.51E-06 | 8.21E-09 | -0.00786325 | NA |
| chr11 | 558659 | 559539 | 881 | 8 | 3.54E-10 | 3.80E-06 | 8.61E-05 | 8.21E-09 | -0.00419493 | LMNTD2-AS1, LMNTD2 |
| chr8 | 59119061 | 59120424 | 1364 | 16 | 1.87E-08 | 6.86E-07 | 0.000442 | 8.22E-09 | -0.00437621 | AC090152.1, TOX |
| chr3 | 153160574 | 153162312 | 1739 | 12 | 1.88E-07 | 1.32E-07 | 0.000405 | 8.25E-09 | 0.000665775 | RAP2B, AC117394.2 |
| chr20 | 44188740 | 44188962 | 223 | 4 | 7.75E-09 | 1.60E-09 | 0.000526 | 8.29E-09 | -0.00384131 | NA |
| chr16 | 30874643 | 30875907 | 1265 | 14 | 3.63E-08 | 4.15E-08 | 0.000203 | 8.30E-09 | -0.00168059 | MIR762HG, BCL7C, MIR4519, AC135048.2 |
| chr12 | 49187489 | 49189136 | 1648 | 10 | 1.35E-07 | 7.74E-10 | 0.002429 | 8.30E-09 | -0.00057127 | AC011603.2, TUBA1C, TUBA1A |
| chr17 | 18970752 | 18972208 | 1457 | 6 | 4.53E-09 | 4.67E-08 | 0.000128 | 8.31E-09 | -0.00284367 | SLC5A10, FAM83G |
| chr16 | 886480 | 888049 | 1570 | 6 | 6.57E-09 | 9.92E-09 | 0.000109 | 8.33E-09 | -0.00584889 | LMF1 |
| chr20 | 11219763 | 11219845 | 83 | 2 | 5.13E-11 | 4.75E-07 | 6.70E-08 | 8.38E-09 | -0.01010821 | NA |
| chr7 | 3972740 | 3972941 | 202 | 3 | 3.90E-09 | 1.41E-08 | 1.63E-05 | 8.42E-09 | -0.02006706 | SDK1 |
| chr17 | 17561539 | 17562368 | 830 | 9 | 1.54E-09 | 1.65E-06 | 0.000508 | 8.43E-09 | -0.01038314 | PEMT |
| chr4 | 110618196 | 110619836 | 1641 | 6 | 3.38E-10 | 8.77E-08 | 2.04E-05 | 8.46E-09 | -0.0073345 | PITX2 |
| chr8 | 74348987 | 74351364 | 2378 | 14 | 1.64E-07 | 0.0002342 | 1.35E-05 | 8.49E-09 | -0.00244634 | GDAP1, AC103952.1 |
| chr8 | 39838178 | 39839323 | 1146 | 9 | 1.44E-08 | 6.23E-06 | 2.31E-05 | 8.49E-09 | -0.00187851 | ADAM2 |
| chr3 | 49895598 | 49896370 | 773 | 5 | 3.15E-10 | 9.01E-07 | 3.12E-07 | 8.50E-09 | -0.0025141 | MST1R |
| chr12 | 111362190 | 111364443 | 2254 | 13 | 2.15E-07 | 5.54E-08 | 0.000163 | 8.50E-09 | -0.0028855 | PHETA1 |
| chr2 | 19360706 | 19361948 | 1243 | 6 | 4.27E-11 | 2.41E-07 | 2.37E-06 | 8.50E-09 | -0.00843609 | NA |
| chr7 | 39565930 | 39566681 | 752 | 16 | 3.68E-09 | 8.69E-06 | 0.000355 | 8.51E-09 | -0.00170643 | YAE1, AC011290.1 |
| chr14 | 67514798 | 67516089 | 1292 | 8 | 1.43E-10 | 8.64E-07 | 4.51E-07 | 8.54E-09 | -0.00024215 | TMEM229B |
| chr6 | 34531141 | 34532266 | 1126 | 6 | 8.95E-10 | 7.01E-08 | 6.43E-05 | 8.55E-09 | -0.01471698 | PACSIN1 |
| chr17 | 2371263 | 2372907 | 1645 | 11 | 5.53E-10 | 1.99E-06 | 4.67E-06 | 8.56E-09 | -0.0011983 | SGSM2 |
| chr11 | 92224282 | 92225633 | 1352 | 8 | 4.48E-08 | 9.76E-07 | 2.56E-05 | 8.59E-09 | 0.000404313 | NA |
| chr6 | 145686459 | 145687283 | 825 | 5 | 2.44E-10 | 9.63E-07 | 6.54E-06 | 8.60E-09 | -0.00140078 | EPM2A |
| chr5 | 140690179 | 140692090 | 1912 | 19 | 7.93E-09 | 2.96E-08 | 0.003156 | 8.62E-09 | -0.00213173 | HARS2, HARS1 |
| chr7 | 44761972 | 44762953 | 982 | 3 | 2.34E-08 | 1.36E-07 | 9.76E-07 | 8.63E-09 | 0.000139611 | ZMIZ2 |
| chr9 | 134781862 | 134782048 | 187 | 2 | 2.12E-09 | 5.16E-09 | 2.19E-05 | 8.63E-09 | -0.05268629 | COL5A1 |
| chr7 | 73842030 | 73842753 | 724 | 13 | 4.41E-11 | 8.91E-07 | 9.38E-06 | 8.63E-09 | -0.00581123 | METTL27 |
| chr1 | 225580293 | 225581332 | 1040 | 4 | 1.09E-10 | 1.87E-06 | 5.81E-07 | 8.66E-09 | -0.00462878 | ENAH |
| chr12 | 40554957 | 40555591 | 635 | 3 | 2.74E-09 | 2.14E-06 | 2.70E-06 | 8.66E-09 | -0.00320465 | MUC19 |
| chr15 | 65791247 | 65792960 | 1714 | 13 | 7.66E-08 | 5.00E-09 | 0.000805 | 8.69E-09 | -0.00014098 | RAB11A, DENND4A |
| chr11 | 110217431 | 110217724 | 294 | 2 | 3.45E-09 | 8.27E-09 | 3.57E-06 | 8.71E-09 | 0.007138137 | RDX |
| chr12 | 25193791 | 25195424 | 1634 | 14 | 7.89E-09 | 1.71E-06 | 0.001576 | 8.71E-09 | 0.003662436 | ETFRF1, CFAP94 |
| chr18 | 43396235 | 43396629 | 395 | 2 | 5.79E-11 | 1.14E-06 | 5.08E-08 | 8.72E-09 | -0.01009568 | NA |
| chr2 | 241944902 | 241947189 | 2288 | 10 | 2.19E-06 | 3.62E-09 | 0.000542 | 8.72E-09 | -0.0005549 | LINC01237 |
| chr17 | 28951596 | 28952382 | 787 | 5 | 1.10E-09 | 4.88E-08 | 8.43E-05 | 8.74E-09 | -0.00176715 | PIPOX, PHF12 |
| chr22 | 39291061 | 39291749 | 689 | 4 | 7.02E-12 | 5.12E-05 | 8.43E-08 | 8.76E-09 | -0.00596282 | NA |
| chr3 | 55512090 | 55513607 | 1518 | 5 | 2.98E-07 | 2.40E-09 | 0.000533 | 8.78E-09 | -0.00873831 | ERC2 |
| chr1 | 1068375 | 1070018 | 1644 | 9 | 1.53E-09 | 2.35E-05 | 2.29E-06 | 8.79E-09 | -0.00310844 | AL390719.1 |
| chr6 | 31170643 | 31171379 | 737 | 6 | 1.32E-09 | 1.28E-07 | 0.000121 | 8.81E-09 | -0.0102151 | POU5F1 |
| chr15 | 85840168 | 85840810 | 643 | 4 | 5.46E-10 | 4.64E-08 | 2.71E-05 | 8.84E-09 | -0.00506984 | NA |
| chr7 | 157124415 | 157125068 | 654 | 6 | 2.25E-09 | 2.09E-06 | 3.15E-05 | 8.87E-09 | -0.00388535 | NA |
| chr16 | 83903990 | 83904046 | 57 | 2 | 3.14E-09 | 3.00E-08 | 5.10E-07 | 8.88E-09 | 0.005710854 | MLYCD |
| chr8 | 27310586 | 27311739 | 1154 | 14 | 1.73E-09 | 6.18E-07 | 0.000332 | 8.91E-09 | -0.00052511 | PTK2B, TRIM35 |
| chr14 | 75147483 | 75148141 | 659 | 4 | 3.27E-09 | 1.02E-08 | 8.54E-06 | 8.93E-09 | -0.01769818 | TMED10 |
| chr7 | 158829637 | 158831164 | 1528 | 12 | 4.51E-08 | 9.68E-08 | 0.000135 | 8.93E-09 | 0.001692412 | ESYT2 |
| chr3 | 149010622 | 149011525 | 904 | 2 | 3.99E-07 | 5.53E-09 | 1.70E-05 | 8.93E-09 | -0.00199421 | GYG1 |
| chr1 | 234863022 | 234863640 | 619 | 2 | 1.60E-12 | 0.0001256 | 2.02E-08 | 8.96E-09 | -0.00366231 | AL391832.4 |
| chr22 | 30591495 | 30592488 | 994 | 13 | 1.01E-08 | 1.95E-08 | 0.000712 | 8.98E-09 | 0.001106789 | PES1 |
| chr1 | 22588441 | 22589160 | 720 | 3 | 3.84E-08 | 1.46E-08 | 1.17E-05 | 8.99E-09 | 0.003067996 | EPHA8 |
| chr1 | 6602908 | 6604208 | 1301 | 13 | 1.05E-08 | 3.25E-05 | 8.98E-05 | 9.02E-09 | -0.00151228 | KLHL21 |
| chr15 | 25189328 | 25190079 | 752 | 4 | 9.50E-08 | 7.57E-09 | 3.41E-05 | 9.02E-09 | -0.01357838 | SNHG14, SNORD115-11 |
| chr8 | 127266022 | 127266270 | 249 | 2 | 5.66E-09 | 6.37E-09 | 8.80E-06 | 9.02E-09 | -0.01370319 | PCAT1 |
| chr14 | 52928516 | 52929482 | 967 | 2 | 3.76E-07 | 5.76E-09 | 1.43E-05 | 9.03E-09 | -0.01677052 | AL139317.5, FERMT2 |
| chr16 | 28552479 | 28554106 | 1628 | 12 | 3.07E-09 | 5.16E-07 | 9.80E-05 | 9.06E-09 | -0.00401217 | AC009093.11, SGF29, AC020765.6, AC020765.2 |
| chr4 | 77818926 | 77820112 | 1187 | 8 | 7.74E-10 | 7.59E-08 | 0.00025 | 9.06E-09 | -0.00077585 | CNOT6L |
| chr22 | 46856791 | 46857950 | 1160 | 3 | 1.59E-13 | 0.0003193 | 2.08E-08 | 9.07E-09 | 0.006275385 | TBC1D22A |
| chr1 | 161739209 | 161740173 | 965 | 2 | 3.89E-08 | 9.70E-09 | 2.84E-06 | 9.07E-09 | -0.02338875 | NA |
| chr3 | 194728014 | 194729428 | 1415 | 3 | 1.98E-07 | 1.25E-08 | 1.49E-05 | 9.07E-09 | 0.002399355 | LINC01968 |
| chr1 | 172532086 | 172533282 | 1197 | 16 | 1.51E-08 | 1.09E-08 | 0.000823 | 9.09E-09 | -0.00035438 | SUCO |
| chr7 | 102239252 | 102240454 | 1203 | 6 | 2.43E-08 | 7.25E-09 | 0.00022 | 9.09E-09 | -0.00030825 | CUX1 |
| chr20 | 32207455 | 32209188 | 1734 | 14 | 2.62E-08 | 1.24E-07 | 0.000754 | 9.09E-09 | -0.00042337 | POFUT1, PLAGL2 |
| chr22 | 37044363 | 37045174 | 812 | 3 | 3.92E-09 | 1.97E-08 | 6.11E-06 | 9.10E-09 | -0.01953807 | NA |
| chr3 | 192957629 | 192958117 | 489 | 3 | 4.39E-09 | 6.46E-09 | 2.52E-05 | 9.10E-09 | -0.01795676 | NA |
| chr12 | 132591077 | 132593322 | 2246 | 10 | 2.71E-06 | 8.57E-08 | 0.001033 | 9.11E-09 | -0.00518425 | AC131212.1 |
| chr10 | 70146201 | 70147732 | 1532 | 14 | 1.37E-09 | 7.02E-06 | 2.48E-05 | 9.11E-09 | 0.003653348 | TYSND1, SAR1A |
| chr16 | 2028823 | 2029657 | 835 | 6 | 3.20E-09 | 1.49E-08 | 6.72E-05 | 9.13E-09 | -0.00202887 | SLC9A3R2 |
| chr5 | 122500631 | 122501101 | 471 | 2 | 3.79E-10 | 2.53E-08 | 6.57E-07 | 9.14E-09 | -0.01477292 | NA |
| chr10 | 123396953 | 123397697 | 745 | 2 | 4.00E-09 | 1.20E-08 | 1.94E-06 | 9.14E-09 | -0.01295474 | LINC02641 |
| chr12 | 57296495 | 57297687 | 1193 | 5 | 2.16E-08 | 3.74E-08 | 3.10E-06 | 9.14E-09 | 0.000800934 | R3HDM2 |
| chr16 | 86044305 | 86044692 | 388 | 2 | 1.41E-10 | 7.22E-08 | 2.36E-07 | 9.15E-09 | -0.0027373 | NA |
| chr5 | 43039765 | 43041387 | 1623 | 12 | 1.92E-10 | 1.37E-06 | 3.40E-05 | 9.17E-09 | -7.30E-05 | AC025171.1, ANXA2R |
| chr7 | 150323019 | 150323872 | 854 | 14 | 4.55E-09 | 2.33E-07 | 0.002284 | 9.18E-09 | -0.00335797 | ZBED6CL, LRRC61, ACTR3C |
| chr22 | 19890067 | 19891211 | 1145 | 3 | 9.98E-11 | 1.62E-08 | 4.20E-06 | 9.20E-09 | 0.00458319 | TXNRD2 |
| chr9 | 79145677 | 79146429 | 753 | 5 | 2.56E-09 | 7.16E-09 | 5.36E-05 | 9.20E-09 | 0.000564453 | AL512634.1 |
| chr16 | 86195022 | 86196585 | 1564 | 8 | 3.15E-08 | 9.69E-05 | 2.89E-05 | 9.20E-09 | -0.00721522 | LINC01082, AC135012.3 |
| chr1 | 61741478 | 61742973 | 1496 | 10 | 1.14E-07 | 8.16E-07 | 0.00074 | 9.20E-09 | 0.004526271 | PATJ, AC099791.2 |
| chr8 | 23224391 | 23225840 | 1450 | 15 | 4.74E-09 | 3.00E-07 | 0.000508 | 9.21E-09 | 0.001029241 | AC100861.1, TNFRSF10A, AC100861.2 |
| chr2 | 118713139 | 118714100 | 962 | 3 | 8.73E-08 | 3.41E-09 | 0.000111 | 9.22E-09 | -0.01062695 | NA |
| chr18 | 3450628 | 3451752 | 1125 | 16 | 7.24E-09 | 7.13E-09 | 0.002064 | 9.23E-09 | -0.00059616 | TGIF1 |
| chr17 | 76537157 | 76537771 | 615 | 5 | 1.84E-13 | 2.41E-06 | 2.08E-05 | 9.23E-09 | -0.00913168 | PRCD, CYGB |
| chr13 | 33642074 | 33643563 | 1490 | 5 | 3.78E-09 | 1.24E-08 | 5.52E-05 | 9.23E-09 | 0.003851098 | AL139383.1 |
| chr20 | 914870 | 916902 | 2033 | 8 | 1.57E-06 | 1.60E-05 | 0.000153 | 9.25E-09 | 0.00411655 | ANGPT4 |
| chr7 | 844651 | 845957 | 1307 | 6 | 5.39E-10 | 8.49E-07 | 2.17E-06 | 9.25E-09 | -0.00060906 | SUN1 |
| chr17 | 3660347 | 3661704 | 1358 | 6 | 2.47E-07 | 6.48E-09 | 0.000184 | 9.25E-09 | -0.00160347 | CTNS |
| chr17 | 48704655 | 48705241 | 587 | 4 | 2.21E-11 | 2.99E-07 | 2.38E-07 | 9.28E-09 | -0.01016008 | LINC02086 |

| | | | | | | | | | | |
|---|---|---|---|---|---|---|---|---|---|---|
| chr1 | 2205406 | 2206226 | 821 | 6 | 3.64E-09 | 1.43E-08 | 0.00018 | 9.28E-09 | -0.00592944 | FAAP20 |
| chr2 | 46385585 | 46385885 | 301 | 5 | 9.91E-09 | 1.07E-08 | 0.000179 | 9.28E-09 | -0.00775188 | EPAS1 |
| chr12 | 48969965 | 48972369 | 2405 | 17 | 2.58E-07 | 2.58E-07 | 0.002741 | 9.28E-09 | -0.00069613 | WNT10B |
| chr2 | 240566715 | 240568681 | 1967 | 10 | 7.56E-07 | 6.59E-08 | 0.000918 | 9.31E-09 | -0.0024122 | RNPEPL1, ANKMY1 |
| chr2 | 239715886 | 239716771 | 886 | 2 | 1.48E-08 | 7.78E-09 | 5.17E-06 | 9.33E-09 | -0.00125287 | NA |
| chr7 | 130732257 | 130733722 | 1466 | 6 | 2.27E-08 | 3.30E-07 | 2.69E-05 | 9.33E-09 | 0.002227854 | KLF14 |
| chr6 | 163148443 | 163149828 | 1386 | 7 | 1.46E-08 | 4.35E-09 | 4.51E-05 | 9.34E-09 | -0.01731518 | PACRG |
| chr16 | 30650213 | 30651931 | 1719 | 17 | 9.79E-07 | 6.05E-09 | 0.002003 | 9.34E-09 | -0.00148005 | PRR14 |
| chr2 | 43772387 | 43774113 | 1727 | 12 | 3.36E-07 | 4.20E-07 | 2.51E-05 | 9.35E-09 | -0.00052662 | DYNC2LI1 |
| chr1 | 211685841 | 211686435 | 595 | 5 | 1.42E-08 | 2.56E-08 | 0.000167 | 9.35E-09 | -0.00619014 | AC096637.2 |
| chr11 | 61587647 | 61588608 | 962 | 7 | 6.03E-09 | 4.68E-09 | 6.97E-05 | 9.35E-09 | -0.00223602 | AP002754.1 |
| chr2 | 27122980 | 27124070 | 1091 | 12 | 6.31E-08 | 6.26E-07 | 0.000201 | 9.35E-09 | -0.00155523 | ABHD1 |
| chr10 | 132135659 | 132136532 | 874 | 6 | 3.17E-11 | 1.24E-05 | 3.08E-07 | 9.38E-09 | -0.00571509 | JAKMIP3 |
| chr2 | 1207476 | 1208369 | 894 | 4 | 7.11E-09 | 5.11E-09 | 0.000114 | 9.38E-09 | -0.0035909 | SNTG2 |
| chr11 | 127000337 | 127001521 | 1185 | 14 | 3.05E-08 | 6.01E-06 | 0.000482 | 9.38E-09 | 0.00311787 | KIRREL3 |
| chr17 | 76946200 | 76947403 | 1204 | 4 | 6.96E-07 | 4.84E-09 | 0.000116 | 9.38E-09 | -0.00058091 | MGAT5B |
| chr2 | 69741470 | 69743507 | 2038 | 14 | 8.95E-07 | 4.33E-07 | 0.000255 | 9.38E-09 | -0.00016377 | ANXA4 |
| chr6 | 31664471 | 31666460 | 1990 | 51 | 3.36E-09 | 7.72E-07 | 0.004138 | 9.39E-09 | 0.000488606 | CSNK2B, AL662899.2, GPANK1 |
| chr9 | 84669016 | 84670832 | 1817 | 18 | 7.37E-08 | 5.29E-06 | 0.000223 | 9.41E-09 | 5.22E-05 | NTRK2 |
| chr8 | 144717821 | 144717878 | 58 | 2 | 5.33E-09 | 1.22E-08 | 2.06E-06 | 9.44E-09 | -0.01898321 | NA |
| chr10 | 124417856 | 124419850 | 1995 | 16 | 1.35E-06 | 1.74E-07 | 0.002016 | 9.45E-09 | -0.00400331 | OAT |
| chr6 | 32094383 | 32096481 | 2099 | 30 | 9.02E-09 | 6.34E-08 | 0.000209 | 9.45E-09 | -0.00903236 | TNXB |
| chr16 | 1050552 | 1050847 | 296 | 4 | 1.98E-10 | 1.13E-06 | 1.64E-07 | 9.51E-09 | -0.00749293 | NA |
| chr10 | 71378993 | 71379819 | 827 | 2 | 1.97E-07 | 5.71E-09 | 2.34E-05 | 9.54E-09 | -0.01331328 | SLC29A3 |
| chr2 | 88691215 | 88692087 | 873 | 14 | 1.32E-10 | 5.33E-06 | 2.49E-06 | 9.55E-09 | 0.001311224 | RPIA, AC062029.1 |
| chr7 | 95478252 | 95478926 | 675 | 4 | 2.01E-10 | 3.37E-08 | 5.59E-06 | 9.55E-09 | -0.01674322 | ASB4 |
| chr19 | 54464654 | 54465572 | 919 | 5 | 2.40E-10 | 3.55E-07 | 3.54E-05 | 9.56E-09 | -0.0068667 | CDC42EP5 |
| chr17 | 83087619 | 83087987 | 369 | 4 | 5.26E-10 | 1.56E-07 | 4.02E-07 | 9.57E-09 | -0.00275822 | METRNL |
| chr22 | 20506528 | 20508981 | 2454 | 17 | 8.39E-09 | 1.79E-06 | 2.64E-05 | 9.58E-09 | -0.00115017 | MED15 |
| chr20 | 53660221 | 53661033 | 813 | 4 | 7.12E-10 | 3.88E-07 | 4.29E-06 | 9.59E-09 | 0.005649811 | NA |
| chr8 | 134720053 | 134721171 | 1119 | 3 | 1.87E-08 | 1.41E-08 | 6.10E-06 | 9.60E-09 | 0.005503656 | NA |
| chr11 | 119726761 | 119727547 | 787 | 2 | 2.23E-13 | 2.10E-05 | 2.63E-08 | 9.61E-09 | -0.0116771 | NECTIN1 |
| chr8 | 142180752 | 142181211 | 460 | 4 | 5.25E-10 | 1.13E-07 | 9.00E-06 | 9.63E-09 | 0.009706031 | AC103758.1 |
| chr6 | 30209518 | 30210947 | 1430 | 9 | 5.26E-10 | 3.37E-06 | 0.000195 | 9.63E-09 | -0.00583634 | TRIM26 |
| chr16 | 55508418 | 55509254 | 837 | 10 | 5.20E-10 | 1.87E-06 | 4.30E-06 | 9.63E-09 | -0.00191579 | LPCAT2 |
| chr1 | 153921987 | 153923485 | 1499 | 15 | 1.79E-08 | 3.69E-07 | 0.000114 | 9.63E-09 | -0.00065764 | AL358472.5, GATAD2B |
| chr13 | 27237045 | 27238112 | 1068 | 3 | 3.35E-07 | 4.42E-09 | 4.98E-05 | 9.64E-09 | -0.00680951 | LINC02340, LINC00412 |
| chr1 | 204972011 | 204972178 | 168 | 4 | 3.35E-10 | 5.99E-05 | 8.66E-08 | 9.66E-09 | 0.005006111 | NFASC |
| chr2 | 100178268 | 100178916 | 649 | 2 | 1.26E-08 | 7.18E-09 | 7.60E-06 | 9.66E-09 | -0.01471658 | AFF3 |
| chr4 | 75629540 | 75631246 | 1707 | 12 | 1.85E-07 | 1.42E-07 | 0.000368 | 9.66E-09 | 0.005121447 | CDKL2 |
| chr8 | 115667900 | 115669656 | 1757 | 16 | 7.72E-07 | 2.89E-07 | 0.000716 | 9.66E-09 | 0.000801683 | TRPS1 |
| chr2 | 199464763 | 199466952 | 2190 | 16 | 2.51E-08 | 6.53E-08 | 0.001116 | 9.68E-09 | 0.000252587 | SATB2-AS1, SATB2 |
| chr2 | 64622921 | 64623042 | 122 | 2 | 1.55E-09 | 1.44E-07 | 1.51E-07 | 9.68E-09 | 0.007259069 | NA |
| chr8 | 47259691 | 47260723 | 1033 | 12 | 5.48E-12 | 8.59E-05 | 3.05E-05 | 9.71E-09 | -0.00270455 | NA |
| chr19 | 49325036 | 49325576 | 541 | 11 | 1.98E-08 | 1.07E-08 | 0.000577 | 9.72E-09 | -0.00403638 | SLC6A16 |
| chr5 | 141647583 | 141648368 | 786 | 6 | 6.13E-09 | 2.60E-08 | 6.43E-05 | 9.74E-09 | -0.00403778 | FCHSD1 |
| chr20 | 11890104 | 11891999 | 1896 | 16 | 5.11E-09 | 3.73E-07 | 0.000437 | 9.82E-09 | -0.00115354 | BTBD3 |
| chr1 | 230321885 | 230323074 | 1190 | 8 | 6.81E-09 | 3.36E-08 | 0.000536 | 9.92E-09 | -0.00334691 | PGBD5 |
| chr22 | 19431431 | 19432837 | 1407 | 11 | 8.60E-08 | 4.75E-09 | 0.002666 | 9.92E-09 | -0.00024327 | MRPL40, HIRA, C22orf39 |
| chr10 | 132653835 | 132655472 | 1638 | 4 | 4.03E-09 | 7.71E-09 | 6.98E-05 | 9.92E-09 | -0.0041663 | INPP5A |
| chr16 | 30055046 | 30055650 | 605 | 3 | 1.05E-08 | 1.03E-08 | 2.40E-05 | 9.94E-09 | -0.01703639 | AC093512.2 |
| chr1 | 64203139 | 64203916 | 778 | 11 | 1.10E-10 | 2.87E-06 | 3.64E-06 | 9.95E-09 | -0.00701306 | UBE2U |
| chr7 | 6164554 | 6165786 | 1233 | 8 | 1.91E-09 | 2.99E-05 | 0.00018 | 1.00E-08 | -0.00194936 | CYTH3 |
| chr2 | 95273591 | 95274957 | 1367 | 11 | 6.85E-13 | 3.74E-05 | 7.52E-08 | 1.00E-08 | 0.000724841 | PROM2 |
| chr3 | 52317171 | 52317947 | 777 | 6 | 1.36E-08 | 4.42E-09 | 0.000243 | 1.00E-08 | -0.01619485 | DNAH1 |
| chr20 | 43726302 | 43727706 | 1405 | 9 | 9.18E-13 | 6.28E-06 | 5.03E-07 | 1.01E-08 | -0.00597132 | GTSF1L |
| chr9 | 95004171 | 95005672 | 1502 | 7 | 1.02E-12 | 1.58E-05 | 5.85E-06 | 1.01E-08 | 0.000362272 | AOPEP |
| chr8 | 63768078 | 63768642 | 565 | 3 | 1.14E-12 | 1.75E-05 | 4.66E-08 | 1.01E-08 | -0.01934849 | LINC01414 |
| chr11 | 47272241 | 47273281 | 1041 | 4 | 3.20E-10 | 7.07E-07 | 1.62E-05 | 1.01E-08 | 0.000375013 | MADD |
| chr22 | 32206904 | 32207250 | 347 | 2 | 4.80E-09 | 6.02E-09 | 2.40E-05 | 1.01E-08 | 0.037551508 | SLC5A4-AS1 |
| chr13 | 35696669 | 35697772 | 1104 | 9 | 7.17E-10 | 5.96E-05 | 1.14E-06 | 1.01E-08 | 0.003043687 | LINC00445 |
| chr5 | 3980333 | 3980398 | 66 | 2 | 2.12E-09 | 1.18E-07 | 1.83E-07 | 1.01E-08 | -0.01780157 | NA |
| chr2 | 165954075 | 165955285 | 1211 | 5 | 2.64E-10 | 3.00E-07 | 3.10E-05 | 1.01E-08 | -0.00314703 | NA |
| chr7 | 157892224 | 157893004 | 781 | 2 | 4.08E-09 | 1.33E-08 | 2.02E-06 | 1.01E-08 | -0.01232094 | PTPRN2 |
| chr8 | 1950272 | 1952367 | 2096 | 13 | 6.67E-10 | 7.40E-07 | 0.001312 | 1.01E-08 | 0.00530273 | AC019257.8, ARHGEF10 |
| chr7 | 118901 | 119467 | 567 | 3 | 1.20E-09 | 6.79E-09 | 3.43E-05 | 1.01E-08 | 0.01102831 | AC093627.22 |
| chr3 | 49123896 | 49125020 | 1125 | 7 | 2.25E-10 | 3.99E-07 | 4.64E-06 | 1.01E-08 | -0.00024896 | LAMB2 |
| chr14 | 105588421 | 105588547 | 127 | 2 | 3.17E-09 | 3.46E-08 | 5.44E-07 | 1.01E-08 | -0.0070489 | NA |
| chr3 | 113890077 | 113890548 | 472 | 5 | 2.67E-10 | 2.05E-05 | 2.05E-07 | 1.01E-08 | -0.01130792 | GRAMD1C |
| chr15 | 92163721 | 92164823 | 1103 | 3 | 1.03E-09 | 2.63E-08 | 2.36E-06 | 1.01E-08 | -0.00361424 | SLCO3A1, AC116903.2, AC116903.1 |
| chr1 | 32753557 | 32754673 | 1117 | 8 | 6.93E-09 | 5.23E-06 | 7.76E-05 | 1.01E-08 | 0.003904208 | KIAA1522 |
| chr20 | 63789679 | 63791007 | 1329 | 8 | 2.77E-08 | 3.87E-06 | 6.41E-05 | 1.01E-08 | -0.00234494 | ZBTB46 |
| chr22 | 49703733 | 49705042 | 1310 | 7 | 3.90E-07 | 7.90E-10 | 0.002675 | 1.02E-08 | 0.014716783 | NA |
| chr7 | 1451644 | 1452983 | 1340 | 9 | 1.55E-10 | 8.14E-07 | 2.58E-06 | 1.02E-08 | -0.00294499 | MICALL2 |
| chr19 | 9945429 | 9945765 | 337 | 3 | 1.28E-11 | 0.0001258 | 1.48E-08 | 1.02E-08 | -0.01418112 | NA |
| chr1 | 47190189 | 47191181 | 993 | 6 | 2.79E-09 | 6.95E-08 | 1.47E-05 | 1.02E-08 | -0.00626187 | PDZK1IP1 |
| chr16 | 1098837 | 1101234 | 2398 | 6 | 2.70E-06 | 3.06E-07 | 0.000178 | 1.02E-08 | -0.00044803 | NA |
| chr12 | 21436674 | 21438461 | 1788 | 12 | 5.00E-07 | 1.83E-05 | 7.57E-05 | 1.02E-08 | -0.00097793 | PYROXD1 |
| chr4 | 127622877 | 127624038 | 1162 | 5 | 6.04E-09 | 5.50E-07 | 6.12E-06 | 1.02E-08 | -0.01063092 | INTU, AC097462.3 |
| chr17 | 82493027 | 82493723 | 697 | 5 | 2.94E-10 | 1.34E-07 | 5.82E-05 | 1.02E-08 | 0.001652273 | NA |
| chr5 | 88883193 | 88884329 | 1137 | 19 | 1.60E-10 | 1.41E-06 | 1.70E-05 | 1.02E-08 | 0.001615814 | MEF2C-AS1, MEF2C |
| chr1 | 8701684 | 8702251 | 568 | 2 | 1.04E-11 | 2.84E-07 | 1.09E-07 | 1.02E-08 | 0.024624719 | RERE |
| chr19 | 34480638 | 34481453 | 816 | 6 | 3.14E-10 | 9.41E-07 | 3.61E-05 | 1.02E-08 | -0.01225144 | NA |
| chr11 | 72690225 | 72690751 | 527 | 2 | 1.53E-10 | 4.26E-08 | 4.48E-07 | 1.02E-08 | 0.006774435 | ARAP1-AS1, AP003065.1, ARAP1 |
| chr11 | 35057992 | 35059321 | 1330 | 3 | 2.02E-09 | 1.24E-08 | 6.85E-06 | 1.02E-08 | 0.005878601 | NA |
| chr8 | 133381926 | 133382657 | 732 | 3 | 7.87E-08 | 3.25E-09 | 0.00018 | 1.03E-08 | -0.01220273 | NA |
| chr12 | 52976828 | 52977611 | 784 | 6 | 6.91E-10 | 5.38E-07 | 2.15E-06 | 1.03E-08 | -0.01465319 | NA |
| chr12 | 99259708 | 99260097 | 390 | 2 | 6.37E-09 | 7.22E-09 | 9.67E-06 | 1.03E-08 | -0.0534749 | ANKS1B |
| chr9 | 131097188 | 131098078 | 891 | 5 | 1.90E-09 | 8.04E-08 | 3.31E-05 | 1.03E-08 | -0.00382384 | AIF1L |
| chr20 | 52680529 | 52681434 | 906 | 4 | 9.61E-09 | 4.30E-09 | 0.000133 | 1.03E-08 | -0.00193719 | AL031674.1 |
| chr3 | 62376985 | 62378496 | 1512 | 11 | 1.68E-08 | 5.78E-09 | 0.000572 | 1.03E-08 | -0.00303448 | NA |
| chr4 | 75352689 | 75354028 | 1340 | 4 | 2.45E-07 | 4.02E-09 | 0.000292 | 1.03E-08 | 0.004840711 | AC025244.1, AC025244.2 |
| chr2 | 120791208 | 120792445 | 1238 | 6 | 1.12E-06 | 2.02E-08 | 0.00056 | 1.03E-08 | -0.00686469 | GLI2 |
| chr12 | 79914044 | 79915227 | 1184 | 5 | 2.75E-07 | 2.01E-08 | 4.04E-05 | 1.03E-08 | -0.01316014 | PPP1R12A |
| chr12 | 116736422 | 116737831 | 1410 | 5 | 8.82E-15 | 0.0001041 | 3.00E-06 | 1.04E-08 | -0.00633846 | SPRING1 |
| chr16 | 66960976 | 66961558 | 583 | 7 | 3.87E-10 | 3.97E-07 | 5.70E-06 | 1.04E-08 | -0.00632237 | AC009084.1, CES3 |
| chr14 | 74907576 | 74907705 | 130 | 2 | 4.97E-09 | 1.29E-08 | 2.34E-06 | 1.05E-08 | 0.007073111 | RPS6KL1 |
| chr15 | 40767429 | 40768624 | 1196 | 8 | 2.49E-09 | 1.51E-08 | 0.000711 | 1.05E-08 | -0.00123031 | GCHFR, DNAJC17 |
| chr11 | 66256133 | 66257856 | 1724 | 17 | 3.87E-10 | 8.37E-07 | 0.00094 | 1.05E-08 | -0.00112289 | KLC2 |
| chr7 | 92755562 | 92756076 | 515 | 2 | 5.98E-12 | 4.67E-05 | 2.58E-08 | 1.05E-08 | -0.00939465 | CDK6 |
| chr3 | 112085496 | 112086538 | 1043 | 14 | 2.62E-08 | 1.61E-08 | 0.001165 | 1.05E-08 | -0.00268193 | C3orf52 |
| chr20 | 63642618 | 63643763 | 1146 | 5 | 7.80E-08 | 3.57E-06 | 2.99E-05 | 1.05E-08 | 0.00619631 | STMN3 |
| chr12 | 121016330 | 121017903 | 1574 | 13 | 6.57E-08 | 3.79E-07 | 0.000184 | 1.05E-08 | -0.00086587 | C12orf43 |
| chr2 | 46812336 | 46812982 | 647 | 4 | 3.50E-09 | 5.42E-09 | 6.20E-05 | 1.06E-08 | -0.00859631 | LINC01118 |
| chr1 | 19761871 | 19762415 | 545 | 2 | 7.86E-10 | 2.58E-08 | 8.39E-07 | 1.06E-08 | -0.01234737 | TMCO4 |
| chr14 | 102210620 | 102210970 | 351 | 4 | 3.76E-09 | 1.78E-08 | 1.10E-05 | 1.06E-08 | -0.01217438 | WDR20 |
| chr16 | 49598650 | 49598992 | 343 | 2 | 1.53E-09 | 2.49E-08 | 8.81E-07 | 1.06E-08 | -0.00098803 | ZNF423 |
| chr7 | 28412447 | 28412670 | 224 | 5 | 7.19E-09 | 1.33E-08 | 0.000408 | 1.06E-08 | -0.02318406 | CREB5 |
| chr9 | 133715588 | 133716300 | 713 | 3 | 9.17E-10 | 4.96E-08 | 4.32E-06 | 1.06E-08 | -0.00945987 | SARDH |
| chr7 | 73741051 | 73741801 | 751 | 3 | 3.77E-10 | 9.71E-09 | 1.03E-05 | 1.06E-08 | -0.00972687 | NA |
| chr7 | 107168511 | 107170103 | 1593 | 15 | 4.59E-08 | 1.03E-06 | 0.000119 | 1.06E-08 | 0.001835248 | HBP1 |
| chr10 | 122460048 | 122461550 | 1503 | 15 | 5.79E-08 | 3.28E-06 | 0.000182 | 1.06E-08 | -0.00433558 | HTRA1, BX842242.1 |
| chr5 | 50751278 | 50751498 | 221 | 2 | 8.61E-10 | 2.40E-07 | 1.26E-07 | 1.06E-08 | 0.010771608 | PARP8 |
| chr4 | 1494091 | 1495496 | 1406 | 12 | 5.06E-10 | 3.75E-06 | 3.00E-05 | 1.07E-08 | -0.00758497 | NA |
| chr14 | 105299727 | 105301772 | 2046 | 21 | 5.80E-07 | 2.20E-08 | 0.000326 | 1.07E-08 | -0.0012802 | PACS2, BRF1 |
| chr15 | 55318376 | 55319683 | 1308 | 13 | 1.40E-10 | 5.33E-05 | 0.00017 | 1.07E-08 | -0.00123117 | PIGB, RAB27A, PIGBOS1 |
| chr6 | 35258854 | 35260542 | 1689 | 13 | 1.18E-10 | 1.96E-06 | 0.000101 | 1.07E-08 | -0.00043057 | ZNF76 |
| chr16 | 88908432 | 88909443 | 1012 | 5 | 5.60E-09 | 3.26E-07 | 3.90E-06 | 1.07E-08 | -0.0032408 | CBFA2T3 |
| chr12 | 6215882 | 6217162 | 1281 | 5 | 2.50E-12 | 0.0002979 | 6.24E-06 | 1.07E-08 | -0.0106026 | CD9 |
| chr11 | 61791969 | 61793747 | 1779 | 22 | 6.66E-09 | 0.0009519 | 2.27E-06 | 1.08E-08 | -0.00184554 | FEN1, FADS2, TMEM258, MIR611 |
| chr8 | 27389727 | 27391025 | 1299 | 4 | 1.76E-08 | 1.37E-08 | 5.15E-05 | 1.08E-08 | -0.02410535 | PTK2B |
| chr4 | 180827383 | 180828069 | 687 | 2 | 6.76E-10 | 1.45E-08 | 2.06E-06 | 1.09E-08 | -0.00924838 | NA |
| chr6 | 49815431 | 49816376 | 946 | 3 | 4.84E-09 | 3.88E-08 | 2.61E-05 | 1.09E-08 | 0.002482453 | AL121974.1 |
| chr1 | 11188980 | 11189704 | 725 | 6 | 1.95E-08 | 5.22E-09 | 0.00075 | 1.09E-08 | 0.000676907 | ANGPTL7, MTOR |
| chr18 | 49813841 | 49814937 | 1097 | 15 | 1.78E-08 | 7.67E-08 | 0.000108 | 1.09E-08 | -0.00231231 | SNHG22, SCARNA17, AC090227.1, ACAA2, AC090227.2 |
| chr12 | 108197793 | 108198345 | 553 | 2 | 2.38E-08 | 6.61E-09 | 2.25E-05 | 1.10E-08 | -0.02662063 | WSCD2 |
| chr11 | 83724155 | 83724987 | 833 | 4 | 2.53E-09 | 1.04E-08 | 1.87E-05 | 1.10E-08 | -0.0131505 | DLG2-AS2, DLG2 |
| chr7 | 90118924 | 90119784 | 861 | 7 | 8.33E-08 | 1.26E-06 | 0.000718 | 1.10E-08 | 0.004964589 | DPY19L2P4, STEAP2-AS1, AC004969.1 |
| chr1 | 207821974 | 207823114 | 1141 | 4 | 1.98E-11 | 1.58E-07 | 6.79E-06 | 1.10E-08 | 0.004752768 | MIR29B2CHG |
| chr20 | 36406527 | 36408321 | 1795 | 4 | 9.00E-06 | 3.11E-09 | 0.000183 | 1.11E-08 | -0.00509713 | DLGAP4 |
| chr19 | 1239607 | 1240653 | 1047 | 4 | 7.10E-14 | 0.0002169 | 6.51E-09 | 1.11E-08 | 0.004303386 | NA |
| chr17 | 41793006 | 41793312 | 307 | 2 | 2.48E-09 | 8.11E-09 | 8.34E-06 | 1.11E-08 | 0.016823272 | NA |
| chr16 | 10329691 | 10329760 | 70 | 2 | 8.98E-09 | 7.37E-09 | 1.24E-05 | 1.11E-08 | 0.012582404 | ATF7IP2 |
| chr19 | 3606465 | 3607610 | 1146 | 13 | 1.74E-07 | 2.37E-06 | 0.00013 | 1.11E-08 | 0.000395585 | CACTIN-AS1, TBXA2R |
| chr5 | 178130672 | 178131754 | 1083 | 8 | 7.81E-09 | 1.38E-08 | 0.000127 | 1.12E-08 | 0.000298792 | RMND5B |
| chr20 | 43797785 | 43798595 | 811 | 3 | 1.59E-09 | 2.39E-08 | 1.46E-05 | 1.12E-08 | 0.009754836 | NA |
| chr7 | 114084809 | 114086207 | 1399 | 5 | 6.53E-08 | 3.28E-09 | 0.000147 | 1.12E-08 | -0.00154869 | NA |
| chr6 | 30219374 | 30219747 | 374 | 2 | 9.37E-11 | 1.68E-06 | 5.45E-08 | 1.12E-08 | -0.00475118 | NA |
| chr16 | 86507874 | 86509486 | 1613 | 16 | 9.83E-08 | 1.22E-07 | 0.000553 | 1.12E-08 | -0.00149817 | FENDRR |
| chr17 | 82390306 | 82390426 | 121 | 3 | 1.32E-08 | 4.62E-09 | 6.16E-05 | 1.12E-08 | 0.004135705 | OGFOD3 |
| chr16 | 86951900 | 86952588 | 689 | 6 | 2.89E-08 | 4.89E-08 | 0.000375 | 1.13E-08 | -0.00427131 | AC093519.2 |
| chr3 | 36379339 | 36380601 | 1263 | 8 | 3.73E-09 | 7.29E-08 | 0.000134 | 1.13E-08 | -0.00470499 | STAC |
| chr1 | 152708026 | 152708687 | 662 | 5 | 2.48E-10 | 6.19E-06 | 3.81E-06 | 1.13E-08 | -0.0127318 | LCE4A |
| chr16 | 2686986 | 2687644 | 659 | 5 | 2.89E-08 | 2.48E-09 | 0.000699 | 1.13E-08 | 0.00758299 | KCTD5 |
| chr5 | 125409138 | 125410125 | 988 | 2 | 1.74E-06 | 6.79E-09 | 2.36E-05 | 1.13E-08 | -0.01490807 | LINC02240, AC116362.1 |
| chr3 | 32018598 | 32019059 | 462 | 2 | 1.03E-11 | 0.0001043 | 2.58E-08 | 1.14E-08 | -0.01414367 | OSBPL10 |
| chr7 | 16753337 | 16754453 | 1117 | 10 | 3.74E-09 | 5.26E-08 | 0.000428 | 1.14E-08 | 0.001452267 | TSPAN13 |
| chr6 | 167719876 | 167721574 | 1699 | 11 | 9.04E-08 | 3.91E-08 | 0.000333 | 1.14E-08 | 0.003462166 | NA |
| chr11 | 31810805 | 31812796 | 1992 | 25 | 8.90E-07 | 1.07E-08 | 0.008929 | 1.14E-08 | 0.000403437 | PAUPAR, AL035078.4, PAX6 |
| chr4 | 24245163 | 24245363 | 201 | 3 | 2.17E-09 | 2.44E-06 | 7.37E-06 | 1.14E-08 | -0.00185754 | NA |
| chr12 | 8081418 | 8082718 | 1301 | 12 | 3.98E-10 | 4.68E-07 | 5.85E-06 | 1.15E-08 | 0.001080128 | NECAP1, AC006511.5, AC006511.3 |
| chr15 | 73366707 | 73367598 | 892 | 5 | 6.36E-10 | 1.84E-07 | 1.16E-05 | 1.15E-08 | -0.00842867 | HCN4 |
| chr1 | 4710101 | 4712203 | 2103 | 10 | 2.16E-06 | 7.12E-08 | 0.001007 | 1.15E-08 | -0.00065865 | AJAP1 |
| chr5 | 154477725 | 154479407 | 1683 | 13 | 7.62E-08 | 2.18E-08 | 0.000345 | 1.15E-08 | 0.001315922 | HAND1 |
| chr9 | 97922487 | 97924114 | 1628 | 13 | 1.12E-06 | 1.20E-05 | 1.65E-05 | 1.16E-08 | 0.000210603 | TRMO |
| chr21 | 41360278 | 41360544 | 267 | 4 | 1.82E-09 | 1.37E-07 | 4.48E-06 | 1.16E-08 | 0.008713889 | NA |
| chr19 | 44703573 | 44704289 | 717 | 4 | 2.44E-09 | 9.14E-08 | 3.52E-06 | 1.16E-08 | -0.00351653 | CEACAM16, AC243964.3 |
| chr17 | 81566739 | 81567668 | 930 | 6 | 1.89E-09 | 5.25E-09 | 5.25E-05 | 1.16E-08 | 0.006360009 | NPLOC4 |

| | | | | | | | | | | |
|---|---|---|---|---|---|---|---|---|---|---|
| chr3 | 77096987 | 77098213 | 1227 | 9 | 7.37E-08 | 1.18E-06 | 0.001846 | 1.17E-08 | -0.00316536 | ROBO2 |
| chr7 | 64045336 | 64045883 | 548 | 7 | 2.90E-08 | 1.97E-09 | 0.000603 | 1.17E-08 | -0.01923136 | AC115220.1, ZNF727 |
| chr12 | 131003845 | 131004510 | 666 | 7 | 2.29E-08 | 1.60E-08 | 0.0004 | 1.17E-08 | 0.013941606 | ADGRD1 |
| chr8 | 108081222 | 108083160 | 1939 | 11 | 1.62E-08 | 2.35E-06 | 9.39E-05 | 1.17E-08 | -0.00032055 | RSPO2 |
| chr5 | 96279968 | 96280494 | 527 | 2 | 8.30E-09 | 8.51E-09 | 8.89E-06 | 1.17E-08 | -0.01450779 | AC104123.1 |
| chr5 | 96413004 | 96413294 | 291 | 3 | 9.87E-09 | 5.04E-09 | 6.90E-05 | 1.18E-08 | -0.01245411 | AC104123.1, PCSK1 |
| chr17 | 37359061 | 37360328 | 1268 | 9 | 2.48E-08 | 8.40E-08 | 0.00141 | 1.19E-08 | -0.00414582 | ACACA |
| chr9 | 125410496 | 125411111 | 616 | 3 | 2.76E-08 | 4.88E-09 | 8.40E-05 | 1.20E-08 | -0.03105717 | NA |
| chr9 | 98942879 | 98944335 | 1457 | 13 | 2.37E-09 | 2.28E-06 | 3.03E-05 | 1.20E-08 | 0.000285233 | COL15A1, AL136084.2 |
| chr2 | 6525180 | 6525189 | 10 | 2 | 2.84E-09 | 1.60E-08 | 2.12E-06 | 1.20E-08 | -0.02578919 | NA |
| chr1 | 220776666 | 220777163 | 498 | 2 | 1.57E-11 | 7.07E-06 | 3.89E-08 | 1.20E-08 | 0.004590133 | MTARC2 |
| chr8 | 55218958 | 55219064 | 107 | 2 | 1.06E-08 | 7.15E-09 | 2.54E-05 | 1.21E-08 | 0.000163107 | XKR4 |
| chr7 | 151392003 | 151392675 | 673 | 2 | 3.56E-10 | 3.28E-08 | 7.81E-07 | 1.21E-08 | -0.01043607 | WDR86 |
| chr11 | 132657052 | 132657543 | 492 | 3 | 7.97E-09 | 7.24E-09 | 6.40E-05 | 1.21E-08 | -0.01388302 | OPCML |
| chr14 | 100159602 | 100160675 | 1074 | 11 | 1.73E-09 | 5.58E-08 | 0.000812 | 1.21E-08 | -0.00020644 | DEGS2 |
| chr7 | 19106932 | 19108564 | 1633 | 6 | 3.84E-13 | 3.24E-05 | 5.71E-08 | 1.21E-08 | -0.00315477 | TWIST1 |
| chr8 | 69710890 | 69712765 | 1876 | 7 | 5.10E-07 | 2.63E-09 | 0.002155 | 1.21E-08 | -0.01567116 | SLCO5A1 |
| chr12 | 8822600 | 8824045 | 1446 | 7 | 2.09E-09 | 1.24E-06 | 9.50E-07 | 1.21E-08 | -0.00312322 | A2ML1, A2ML1-AS1 |
| chr2 | 129717461 | 129717859 | 399 | 2 | 7.16E-09 | 8.77E-09 | 9.05E-06 | 1.21E-08 | -0.01229208 | NA |
| chr17 | 38572209 | 38573274 | 1066 | 6 | 1.90E-09 | 3.37E-07 | 6.65E-05 | 1.22E-08 | -0.00025524 | SRCIN1 |
| chr11 | 75768780 | 75769681 | 902 | 7 | 6.67E-11 | 1.84E-06 | 2.31E-06 | 1.22E-08 | -0.00651469 | DGAT2 |
| chr19 | 43753437 | 43755419 | 1983 | 15 | 1.48E-09 | 1.02E-06 | 4.14E-05 | 1.22E-08 | -0.00156923 | SMG9 |
| chr11 | 34523830 | 34524237 | 408 | 2 | 9.97E-09 | 7.74E-09 | 1.58E-05 | 1.22E-08 | 0.005089015 | ELF5 |
| chr20 | 4685777 | 4687488 | 1712 | 12 | 1.31E-08 | 1.79E-06 | 8.91E-05 | 1.22E-08 | 0.000322834 | PRNP |
| chr1 | 2172136 | 2173447 | 1312 | 6 | 1.68E-07 | 5.56E-07 | 1.37E-05 | 1.22E-08 | -0.00129362 | PRKCZ |
| chr19 | 5046836 | 5047700 | 865 | 3 | 6.60E-08 | 9.73E-09 | 4.08E-05 | 1.22E-08 | 0.002885051 | KDM4B |
| chr13 | 114023830 | 114024636 | 807 | 4 | 5.91E-10 | 2.67E-06 | 7.88E-06 | 1.23E-08 | 0.004635334 | RASA3 |
| chr16 | 56370624 | 56370989 | 366 | 4 | 1.02E-09 | 4.13E-07 | 5.88E-07 | 1.23E-08 | -0.00037171 | AMFR |
| chr10 | 77197190 | 77197721 | 532 | 2 | 2.57E-10 | 8.92E-08 | 2.94E-07 | 1.23E-08 | -0.00611378 | KCNMA1 |
| chr11 | 69776615 | 69776787 | 173 | 3 | 4.30E-10 | 1.32E-08 | 4.62E-05 | 1.23E-08 | -0.01598691 | NA |
| chr12 | 53998950 | 54000756 | 1807 | 23 | 1.47E-08 | 4.21E-06 | 0.002626 | 1.23E-08 | 0.001112006 | AC012531.3, HOXC6, HOXC9, HOXC-AS1 |
| chr21 | 32727899 | 32729102 | 1204 | 14 | 8.92E-15 | 0.000935 | 2.02E-07 | 1.24E-08 | -0.00317301 | PAXBP1-AS1, SYNJ1 |
| chr13 | 52158776 | 52160297 | 1522 | 18 | 1.82E-09 | 3.15E-07 | 0.001638 | 1.24E-08 | -0.00081473 | NEK3 |
| chr12 | 110920384 | 110921158 | 775 | 12 | 5.92E-08 | 6.01E-09 | 0.00133 | 1.24E-08 | -0.00509943 | MYL2 |
| chr19 | 44499777 | 44501570 | 1794 | 12 | 3.81E-08 | 1.17E-06 | 4.32E-05 | 1.25E-08 | -0.00164916 | ZNF180 |
| chr4 | 186298076 | 186299308 | 1233 | 6 | 1.37E-08 | 4.86E-05 | 4.60E-06 | 1.25E-08 | -0.00789597 | F11-AS1 |
| chr1 | 119631366 | 119632818 | 1453 | 8 | 1.53E-07 | 1.29E-06 | 4.05E-05 | 1.25E-08 | 0.002333701 | ZNF697 |
| chr5 | 83076967 | 83078125 | 1159 | 20 | 1.23E-09 | 3.52E-07 | 0.000804 | 1.25E-08 | 0.000309155 | XRCC4, TMEM167A |
| chr20 | 57392622 | 57394630 | 2009 | 9 | 2.49E-06 | 3.55E-07 | 8.97E-05 | 1.25E-08 | 0.007881782 | RBM38, RBM38-AS1 |
| chr6 | 28574624 | 28575893 | 1270 | 8 | 1.49E-09 | 1.00E-07 | 9.33E-05 | 1.26E-08 | -0.00305765 | ZBED9 |
| chr19 | 45493114 | 45494943 | 1830 | 13 | 3.37E-07 | 3.03E-07 | 2.34E-05 | 1.26E-08 | -0.00087804 | PPM1N, RTN2 |
| chr4 | 168630602 | 168631881 | 1280 | 11 | 6.91E-13 | 0.0047958 | 1.49E-06 | 1.26E-08 | -0.00553822 | PALLD |
| chr15 | 41559766 | 41560440 | 675 | 5 | 1.54E-10 | 7.73E-08 | 3.49E-05 | 1.26E-08 | 0.016044122 | TYRO3 |
| chr16 | 61916828 | 61918272 | 1445 | 5 | 1.31E-07 | 7.90E-08 | 1.50E-05 | 1.26E-08 | 1.07E-06 | CDH8 |
| chr3 | 10246443 | 10248822 | 2380 | 15 | 4.26E-07 | 3.28E-07 | 0.000599 | 1.26E-08 | -0.00062408 | TATDN2 |
| chr11 | 130915641 | 130917184 | 1544 | 9 | 8.59E-09 | 3.77E-06 | 6.31E-06 | 1.26E-08 | -0.00823635 | SNX19 |
| chr1 | 156864002 | 156864786 | 785 | 4 | 5.49E-09 | 1.64E-08 | 6.36E-05 | 1.26E-08 | -0.00042131 | NTRK1 |
| chr22 | 43924293 | 43924624 | 332 | 2 | 6.13E-11 | 1.20E-08 | 4.29E-06 | 1.26E-08 | 0.013736943 | PNPLA3 |
| chr4 | 55271533 | 55272134 | 602 | 3 | 3.02E-12 | 0.000556 | 9.63E-09 | 1.26E-08 | -0.01720271 | NA |
| chr10 | 97771659 | 97772879 | 1221 | 11 | 7.81E-09 | 6.97E-08 | 0.000192 | 1.26E-08 | -0.00223708 | SFRP5 |
| chr17 | 7499045 | 7499354 | 310 | 7 | 2.45E-09 | 7.45E-06 | 8.38E-05 | 1.26E-08 | 0.002393647 | POLR2A |
| chr11 | 59556556 | 59558766 | 2211 | 11 | 4.82E-07 | 1.15E-07 | 0.000621 | 1.26E-08 | -0.00194401 | AP003778.1 |
| chr2 | 241652263 | 241653506 | 1244 | 6 | 1.17E-11 | 7.73E-06 | 1.18E-06 | 1.26E-08 | -0.00090164 | ATG4B |
| chr7 | 45071593 | 45072484 | 892 | 3 | 4.06E-09 | 1.39E-08 | 7.53E-06 | 1.26E-08 | 0.016880471 | CCM2 |
| chr15 | 66348206 | 66348760 | 555 | 2 | 7.98E-13 | 6.80E-07 | 8.66E-08 | 1.26E-08 | -0.01266043 | Y_RNA, TIPIN |
| chr20 | 31637522 | 31638674 | 1153 | 10 | 4.94E-08 | 1.03E-09 | 0.004669 | 1.27E-08 | -0.01005364 | COX4I2 |
| chr19 | 58278308 | 58279073 | 766 | 12 | 1.44E-09 | 1.09E-08 | 0.000167 | 1.27E-08 | -0.00014332 | AC020915.4, ZNF8, ZNF8-ERVK3-1, AC020915.2 |
| chr12 | 24901351 | 24902742 | 1392 | 13 | 1.72E-10 | 1.00E-05 | 3.80E-05 | 1.27E-08 | -0.00134215 | AC026310.2, BCAT1 |
| chr5 | 169980435 | 169980937 | 503 | 6 | 9.96E-09 | 4.33E-09 | 0.000155 | 1.27E-08 | -0.01326564 | DOCK2, INSYN2B |
| chr5 | 151214123 | 151215394 | 1272 | 6 | 3.25E-08 | 2.47E-09 | 0.000266 | 1.27E-08 | 0.006003265 | GM2A, CCDC69 |
| chr12 | 130473795 | 130474669 | 875 | 7 | 1.81E-08 | 4.84E-08 | 0.00019 | 1.27E-08 | -0.00130494 | RIMBP2 |
| chr4 | 907997 | 909787 | 1791 | 9 | 1.57E-06 | 1.70E-06 | 4.93E-06 | 1.29E-08 | -0.00031378 | GAK |
| chr1 | 109719744 | 109720147 | 404 | 2 | 1.96E-09 | 1.12E-08 | 5.38E-06 | 1.29E-08 | -0.01603562 | GSTM5 |
| chr22 | 35540590 | 35541484 | 895 | 11 | 1.04E-08 | 3.22E-07 | 0.000348 | 1.29E-08 | -0.0009134 | RASD2 |
| chr17 | 7251902 | 7252806 | 905 | 17 | 2.37E-08 | 3.63E-07 | 0.002436 | 1.29E-08 | 0.001040696 | ELP5, CTDNEP1, AC003688.1 |
| chr2 | 170712622 | 170714567 | 1946 | 10 | 5.81E-09 | 5.03E-06 | 3.83E-05 | 1.29E-08 | 0.00207662 | AC007405.1, LINC01124 |
| chr14 | 90811949 | 90812409 | 461 | 2 | 1.03E-09 | 3.62E-08 | 7.80E-07 | 1.29E-08 | -0.00515447 | TTC7B |
| chr11 | 14258787 | 14259611 | 825 | 7 | 7.74E-09 | 1.87E-08 | 0.000623 | 1.29E-08 | 0.027711573 | SPON1 |
| chr12 | 109679080 | 109680047 | 968 | 4 | 3.57E-08 | 1.22E-08 | 1.69E-05 | 1.29E-08 | -0.01352728 | NA |
| chr22 | 19149331 | 19151343 | 2013 | 10 | 2.89E-06 | 2.06E-06 | 0.000785 | 1.29E-08 | 0.003388601 | GSC2 |
| chr10 | 18700050 | 18700168 | 119 | 2 | 4.32E-10 | 3.95E-05 | 3.07E-08 | 1.30E-08 | -0.01972002 | NA |
| chr19 | 50786303 | 50786777 | 475 | 7 | 4.34E-10 | 1.61E-06 | 4.48E-05 | 1.30E-08 | 5.12E-05 | AC010325.1 |
| chr16 | 2712568 | 2712898 | 331 | 6 | 1.08E-09 | 5.87E-06 | 5.92E-06 | 1.30E-08 | -0.00433504 | PRSS27 |
| chr9 | 136109577 | 136110855 | 1279 | 7 | 2.13E-09 | 1.91E-08 | 0.000868 | 1.30E-08 | -0.00031356 | NA |
| chr19 | 19618239 | 19618951 | 713 | 9 | 2.04E-08 | 2.67E-07 | 0.000501 | 1.30E-08 | -0.00208306 | PBX4 |
| chr9 | 137271258 | 137273500 | 2243 | 8 | 2.40E-07 | 7.29E-06 | 7.29E-06 | 1.30E-08 | 0.003177272 | NELFB |
| chr12 | 101474116 | 101475589 | 1474 | 4 | 1.35E-07 | 4.65E-07 | 7.40E-06 | 1.30E-08 | -0.00127995 | SPIC |
| chr15 | 52568841 | 52570209 | 1369 | 16 | 2.84E-07 | 5.15E-06 | 0.001664 | 1.30E-08 | -0.00050711 | ARPP19 |
| chr8 | 134800643 | 134801643 | 1001 | 6 | 2.09E-08 | 5.01E-08 | 4.19E-05 | 1.30E-08 | 0.008517909 | NA |
| chr13 | 39857389 | 39857739 | 351 | 3 | 1.05E-09 | 3.44E-06 | 2.01E-07 | 1.31E-08 | -0.01228767 | NA |
| chr10 | 126557203 | 126557855 | 653 | 4 | 1.72E-08 | 2.93E-07 | 0.000111 | 1.31E-08 | -0.00032751 | C10orf90 |
| chr14 | 77023420 | 77023787 | 368 | 3 | 8.87E-10 | 1.59E-07 | 2.41E-06 | 1.31E-08 | -0.00447284 | NA |
| chr22 | 27677614 | 27678448 | 835 | 6 | 8.27E-10 | 4.89E-07 | 3.46E-06 | 1.31E-08 | 0.008733368 | AL121885.2 |
| chr5 | 177006244 | 177007752 | 1509 | 13 | 4.30E-10 | 1.37E-05 | 2.52E-05 | 1.31E-08 | -0.00109053 | UIMC1 |
| chr1 | 22260291 | 22260318 | 28 | 2 | 1.17E-08 | 7.78E-09 | 2.56E-05 | 1.31E-08 | -0.01852511 | NA |
| chr1 | 20484639 | 20485860 | 1222 | 8 | 4.31E-11 | 1.48E-06 | 8.40E-05 | 1.32E-08 | -0.00165761 | CAMK2N1 |
| chr7 | 100676738 | 100678464 | 1727 | 7 | 3.34E-06 | 8.70E-07 | 0.000146 | 1.32E-08 | -0.00260123 | GNB2 |
| chr17 | 9822086 | 9822611 | 526 | 4 | 5.75E-09 | 1.63E-08 | 6.65E-05 | 1.32E-08 | -0.00573387 | GLP2R |
| chr17 | 45404204 | 45406289 | 2086 | 10 | 8.64E-07 | 1.02E-07 | 5.33E-05 | 1.32E-08 | -0.00121071 | ARHGAP27 |
| chr10 | 69571772 | 69574211 | 2440 | 20 | 2.36E-06 | 3.32E-07 | 0.001777 | 1.32E-08 | 0.000692288 | NEUROG3 |
| chr12 | 12812491 | 12813530 | 1040 | 11 | 6.16E-09 | 2.01E-07 | 0.000948 | 1.32E-08 | -0.00157951 | APOLD1, DDX47 |
| chr17 | 68035332 | 68036759 | 1428 | 14 | 5.59E-08 | 1.72E-05 | 0.000102 | 1.32E-08 | -0.00010022 | KPNA2 |
| chr1 | 203493078 | 203493393 | 316 | 5 | 2.17E-10 | 1.25E-05 | 1.12E-07 | 1.32E-08 | -0.00586096 | NA |
| chr4 | 76801904 | 76802647 | 744 | 5 | 2.18E-09 | 2.65E-08 | 3.05E-05 | 1.32E-08 | -0.01867924 | SHROOM3-AS1, AC107072.1 |
| chr1 | 15760800 | 15761507 | 708 | 4 | 2.50E-09 | 1.56E-07 | 2.88E-05 | 1.32E-08 | -0.00472901 | FBLIM1 |
| chr14 | 61693940 | 61695769 | 1830 | 14 | 2.30E-08 | 3.33E-06 | 0.000174 | 1.32E-08 | 0.001446817 | HIF1A, HIF1A-AS1 |
| chr8 | 143922229 | 143923546 | 1318 | 5 | 2.91E-08 | 3.32E-08 | 5.47E-05 | 1.32E-08 | -0.00358906 | PLEC |
| chr19 | 45730797 | 45732418 | 1622 | 13 | 1.33E-08 | 3.77E-07 | 0.000211 | 1.33E-08 | -0.00042294 | FBXO46 |
| chr11 | 112481683 | 112483062 | 1380 | 6 | 2.75E-11 | 4.38E-06 | 1.79E-07 | 1.33E-08 | -0.00701168 | LINC02763 |
| chr11 | 6357147 | 6357461 | 315 | 2 | 2.51E-10 | 1.54E-06 | 6.48E-08 | 1.33E-08 | -0.01560692 | AC068733.3 |
| chr22 | 26321826 | 26322615 | 790 | 4 | 2.72E-09 | 2.00E-07 | 2.14E-05 | 1.33E-08 | -0.01048483 | SEZ6L |
| chr14 | 70223570 | 70223987 | 418 | 6 | 4.73E-08 | 1.02E-09 | 0.003297 | 1.33E-08 | 0.064002345 | AL160191.1 |
| chr2 | 170409028 | 170409239 | 212 | 2 | 3.07E-09 | 3.46E-08 | 8.72E-07 | 1.33E-08 | -0.02444981 | MYO3B |
| chr18 | 33577643 | 33579118 | 1476 | 13 | 3.52E-08 | 7.97E-07 | 0.000789 | 1.33E-08 | 0.000574818 | ASXL3, AC091198.1 |
| chr17 | 46817951 | 46819658 | 1708 | 14 | 2.94E-09 | 1.87E-05 | 9.56E-05 | 1.33E-08 | 0.000224508 | WNT3 |
| chr11 | 2902034 | 2904240 | 2207 | 21 | 9.76E-08 | 4.06E-08 | 0.002064 | 1.34E-08 | 0.003366333 | SLC22A18, SLC22A18AS |
| chr17 | 57256113 | 57257832 | 1720 | 16 | 2.12E-10 | 2.09E-06 | 0.000111 | 1.34E-08 | -0.0001944 | MSI2 |
| chr1 | 32765323 | 32766978 | 1656 | 10 | 3.19E-07 | 1.19E-07 | 0.000308 | 1.34E-08 | -0.00200565 | KIAA1522 |
| chr19 | 48636900 | 48638068 | 1169 | 18 | 1.81E-09 | 6.34E-08 | 0.000222 | 1.34E-08 | 0.000658588 | DBP, CA11 |
| chr14 | 35291410 | 35293087 | 1678 | 14 | 7.35E-08 | 1.61E-07 | 0.003435 | 1.34E-08 | -0.00130708 | AL121594.1, PSMA6 |
| chr2 | 204546186 | 204547063 | 878 | 3 | 2.74E-09 | 1.41E-07 | 2.45E-06 | 1.35E-08 | -5.19E-05 | PARD3B |
| chr1 | 44647014 | 44647916 | 903 | 2 | 8.81E-08 | 9.75E-09 | 9.55E-06 | 1.35E-08 | -0.0079162 | RNF220, TMEM53 |
| chr8 | 144685855 | 144686562 | 708 | 6 | 2.73E-12 | 2.61E-06 | 3.02E-06 | 1.35E-08 | -0.01741036 | NA |
| chr21 | 33542424 | 33543932 | 1509 | 19 | 4.53E-10 | 1.93E-06 | 0.000271 | 1.35E-08 | -0.00012772 | SON, GART |
| chr16 | 70267905 | 70268561 | 657 | 3 | 4.37E-11 | 8.62E-07 | 2.14E-07 | 1.35E-08 | -0.00438474 | AARS1 |
| chr14 | 85482583 | 85482601 | 19 | 2 | 7.49E-09 | 2.25E-08 | 1.59E-06 | 1.35E-08 | -0.00575955 | AL049775.2 |
| chr11 | 34051250 | 34053325 | 2076 | 14 | 7.18E-08 | 4.68E-05 | 5.98E-06 | 1.35E-08 | -0.0004678 | CAPRIN1, AC090469.1 |
| chr2 | 27211259 | 27213243 | 1985 | 25 | 1.95E-08 | 3.18E-06 | 0.000641 | 1.36E-08 | -0.00324904 | ATRAID, SLC5A6 |
| chr4 | 1208315 | 1209380 | 1066 | 18 | 3.72E-08 | 7.14E-08 | 0.007605 | 1.36E-08 | -0.00250955 | SPON2 |
| chr18 | 63775558 | 63777014 | 1457 | 4 | 1.36E-07 | 6.75E-08 | 3.07E-05 | 1.36E-08 | -0.01538525 | SERPINB7 |
| chr6 | 24741841 | 24742253 | 413 | 3 | 1.94E-09 | 3.50E-08 | 3.28E-06 | 1.36E-08 | -0.00153072 | LINC02828 |
| chr10 | 102450935 | 102452004 | 1070 | 10 | 7.83E-10 | 7.67E-07 | 0.000316 | 1.37E-08 | -0.00310528 | RPARP-AS1, C10orf95 |
| chr1 | 161153711 | 161155049 | 1339 | 12 | 5.44E-08 | 1.26E-06 | 8.29E-05 | 1.37E-08 | 0.000502691 | UFC1, AL590714.1 |
| chr11 | 18651055 | 18651249 | 195 | 2 | 3.60E-10 | 1.50E-05 | 3.72E-08 | 1.37E-08 | -0.0098794 | NA |
| chr19 | 4390017 | 4390307 | 291 | 3 | 2.37E-10 | 4.34E-06 | 2.64E-07 | 1.37E-08 | -0.00384016 | SH3GL1 |
| chr9 | 98173259 | 98174196 | 938 | 5 | 9.98E-11 | 9.28E-07 | 2.10E-06 | 1.37E-08 | 0.002329059 | CORO2A |
| chr4 | 6538336 | 6539389 | 1054 | 5 | 2.92E-09 | 1.02E-07 | 3.60E-05 | 1.37E-08 | 0.006711217 | PPP2R2C |
| chr1 | 44030804 | 44032850 | 2047 | 12 | 9.25E-08 | 5.85E-06 | 4.40E-05 | 1.38E-08 | -0.00010987 | AL139220.2, SLC6A9 |
| chr17 | 7462392 | 7463494 | 1103 | 6 | 1.29E-09 | 1.60E-06 | 0.000277 | 1.38E-08 | -0.00076825 | ZBTB4 |
| chr13 | 75870157 | 75871236 | 1080 | 11 | 2.44E-07 | 1.15E-08 | 0.002963 | 1.38E-08 | -0.01238431 | LMO7DN |
| chr2 | 176575102 | 176575800 | 699 | 2 | 7.24E-10 | 4.96E-08 | 6.02E-07 | 1.38E-08 | -0.00739206 | LINC01117 |
| chr6 | 170147245 | 170148173 | 929 | 7 | 5.65E-09 | 1.42E-07 | 5.19E-05 | 1.38E-08 | 0.007702481 | NA |
| chr7 | 1210402 | 1211702 | 1301 | 14 | 4.42E-08 | 1.00E-08 | 0.002628 | 1.38E-08 | 0.002286146 | NA |
| chr20 | 62831078 | 62831888 | 811 | 4 | 3.77E-11 | 8.10E-07 | 1.81E-07 | 1.38E-08 | -0.01657168 | COL9A3 |
| chr6 | 36678035 | 36679887 | 1853 | 15 | 9.90E-10 | 9.09E-08 | 6.36E-05 | 1.38E-08 | -0.0010045 | CDKN1A, DINOL |
| chr4 | 117085591 | 117086864 | 1274 | 10 | 1.10E-10 | 2.64E-08 | 2.71E-05 | 1.39E-08 | -0.00602307 | NA |
| chr8 | 22746080 | 22747914 | 1835 | 7 | 2.73E-06 | 4.55E-08 | 0.000369 | 1.39E-08 | -0.00556078 | AC055854.1, PEBP4 |
| chr3 | 157865165 | 157865590 | 426 | 2 | 3.44E-10 | 1.79E-07 | 2.01E-07 | 1.39E-08 | -0.0144606 | AC079943.2 |
| chr11 | 65382181 | 65384580 | 2400 | 14 | 2.72E-08 | 1.68E-07 | 7.06E-06 | 1.39E-08 | 0.000566058 | SLC25A45 |
| chr13 | 28137131 | 28139315 | 2185 | 17 | 6.25E-08 | 2.25E-07 | 1.61E-05 | 1.39E-08 | 0.000205546 | PAN3, PAN3-AS1 |
| chr15 | 78841989 | 78842365 | 377 | 2 | 1.18E-08 | 8.75E-09 | 1.80E-05 | 1.39E-08 | -0.00139505 | MORF4L1 |
| chr16 | 85138352 | 85139220 | 869 | 5 | 1.34E-08 | 1.37E-08 | 0.000218 | 1.39E-08 | -0.01237541 | LINC02139 |
| chr12 | 121924038 | 121925495 | 1458 | 3 | 1.54E-06 | 1.50E-08 | 3.52E-05 | 1.39E-08 | -0.01220788 | CFAP251 |
| chr12 | 15788358 | 15789257 | 900 | 7 | 3.72E-09 | 2.66E-08 | 0.000969 | 1.39E-08 | 0.002536446 | EPS8 |
| chr18 | 36643272 | 36644101 | 830 | 2 | 4.29E-10 | 8.75E-08 | 3.50E-07 | 1.39E-08 | -0.00983449 | FHOD3 |
| chr9 | 136107371 | 136107777 | 407 | 3 | 2.06E-09 | 4.38E-08 | 3.08E-05 | 1.39E-08 | 0.009142292 | NA |
| chr14 | 99273754 | 99275006 | 1253 | 5 | 1.76E-10 | 4.03E-06 | 1.37E-06 | 1.40E-08 | 0.003963895 | NA |
| chr19 | 36488931 | 36490073 | 1143 | 24 | 9.52E-10 | 4.07E-05 | 3.43E-05 | 1.40E-08 | 0.000778478 | AC092295.2, ZNF566 |
| chr12 | 459377 | 460567 | 1191 | 7 | 4.27E-09 | 2.60E-07 | 4.79E-05 | 1.40E-08 | 0.003810039 | B4GALNT3 |
| chr3 | 151459497 | 151461116 | 1620 | 7 | 1.94E-06 | 2.31E-07 | 0.002688 | 1.40E-08 | -0.02261043 | NA |
| chr6 | 33207836 | 33208896 | 1061 | 24 | 2.20E-08 | 4.86E-08 | 0.000203 | 1.40E-08 | 0.000552645 | MIR219A1, RING1 |
| chr6 | 41442244 | 41443390 | 1147 | 6 | 1.75E-11 | 1.06E-05 | 3.81E-05 | 1.40E-08 | -0.00917479 | NA |
| chr14 | 100956780 | 100956851 | 72 | 2 | 4.22E-09 | 8.80E-09 | 1.84E-05 | 1.40E-08 | -0.02623826 | MEG8 |
| chr6 | 29618995 | 29619710 | 716 | 4 | 2.46E-10 | 1.21E-07 | 5.29E-05 | 1.40E-08 | -0.00863819 | GABBR1 |
| chr20 | 41027958 | 41029446 | 1489 | 15 | 2.80E-08 | 2.83E-07 | 0.002135 | 1.40E-08 | -0.00079166 | TOP1 |
| chr8 | 142463664 | 142464588 | 925 | 8 | 3.12E-08 | 7.11E-08 | 0.000447 | 1.40E-08 | 0.002331655 | ADGRB1 |
| chr10 | 2498719 | 2500050 | 1332 | 6 | 3.32E-07 | 1.00E-08 | 0.000362 | 1.40E-08 | 6.66E-05 | AL713851.2, LINC02645 |
| chr18 | 62978677 | 62979438 | 762 | 3 | 3.96E-08 | 1.53E-08 | 4.13E-05 | 1.41E-08 | -0.0082798 | PHLPP1 |

| | | | | | | | | | | |
|---|---|---|---|---|---|---|---|---|---|---|
| chr4 | 1740574 | 1741223 | 650 | 4 | 4.41E-09 | 3.36E-07 | 6.41E-05 | 1.41E-08 | 0.013807851 | TACC3 |
| chr1 | 24730553 | 24730924 | 372 | 3 | 1.39E-08 | 6.66E-09 | 4.73E-05 | 1.41E-08 | -0.02271663 | NA |
| chr17 | 32793947 | 32794773 | 827 | 4 | 4.10E-07 | 5.79E-09 | 0.00011 | 1.41E-08 | -0.01344943 | MYO1D |
| chr22 | 46108496 | 46109673 | 1178 | 9 | 1.19E-07 | 5.21E-08 | 0.000275 | 1.41E-08 | -0.00194967 | MIRLET7BHG |
| chr12 | 38138133 | 38138911 | 779 | 4 | 4.44E-09 | 4.43E-08 | 5.85E-06 | 1.41E-08 | -0.01479008 | NA |
| chr19 | 4342592 | 4344005 | 1414 | 8 | 1.33E-08 | 2.92E-06 | 0.000145 | 1.41E-08 | 0.002946252 | AC104521.1, MPND, STAP2 |
| chr12 | 53984474 | 53985949 | 1476 | 18 | 6.13E-09 | 1.75E-07 | 0.000232 | 1.42E-08 | 0.000773041 | HOXC10, AC012531.3, HOXC-AS3 |
| chr5 | 139481225 | 139482902 | 1678 | 13 | 2.59E-08 | 7.86E-07 | 0.000287 | 1.42E-08 | -0.00086641 | STING1 |
| chr2 | 47368934 | 47370192 | 1259 | 10 | 6.91E-08 | 1.11E-09 | 0.003944 | 1.42E-08 | -0.00481408 | EPCAM |
| chr2 | 160493068 | 160494315 | 1248 | 14 | 1.69E-09 | 4.59E-06 | 9.67E-05 | 1.42E-08 | 0.000942732 | RBMS1 |
| chr8 | 123428965 | 123429961 | 997 | 3 | 1.02E-08 | 1.40E-08 | 4.10E-05 | 1.42E-08 | -0.00771934 | NTAQ1 |
| chr20 | 48522813 | 48523144 | 332 | 3 | 1.73E-08 | 7.09E-09 | 3.68E-05 | 1.42E-08 | 0.00383966 | NA |
| chr2 | 96949784 | 96950630 | 847 | 3 | 5.55E-09 | 2.86E-08 | 3.59E-05 | 1.42E-08 | -0.00285486 | FAM178B |
| chr12 | 89324531 | 89324571 | 41 | 2 | 1.86E-09 | 7.80E-07 | 9.26E-08 | 1.43E-08 | -0.00646943 | NA |
| chr6 | 113645492 | 113645785 | 294 | 2 | 7.36E-09 | 1.24E-08 | 5.81E-06 | 1.43E-08 | 0.00705433 | LINC02541 |
| chr19 | 39498594 | 39499415 | 822 | 12 | 1.64E-08 | 4.05E-08 | 0.001105 | 1.43E-08 | -0.00043137 | DLL3 |
| chr1 | 986446 | 986630 | 185 | 3 | 7.57E-09 | 5.15E-08 | 2.76E-05 | 1.44E-08 | -0.0103917 | NA |
| chr6 | 148342217 | 148343284 | 1068 | 14 | 1.80E-09 | 1.31E-05 | 0.000292 | 1.44E-08 | 0.001200932 | SASH1 |
| chr7 | 74571238 | 74571427 | 190 | 3 | 2.07E-09 | 3.93E-07 | 2.31E-07 | 1.44E-08 | -0.00919966 | GTF2IRD1 |
| chr11 | 3217988 | 3219229 | 1242 | 12 | 2.10E-09 | 4.33E-07 | 0.000336 | 1.44E-08 | -0.00560817 | MRGPRG-AS1, MRGPRG |
| chr14 | 105485834 | 105488102 | 2269 | 15 | 8.41E-08 | 6.21E-07 | 0.000302 | 1.44E-08 | -0.00186835 | CRIP1, AL928654.3 |
| chr12 | 100266676 | 100268000 | 1325 | 19 | 2.87E-09 | 2.40E-06 | 0.00104 | 1.44E-08 | 0.000122093 | SCYL2, DEPDC4 |
| chr6 | 41369228 | 41369500 | 273 | 4 | 1.64E-09 | 1.52E-07 | 2.04E-05 | 1.44E-08 | -0.01480968 | NA |
| chr5 | 171365204 | 171366038 | 835 | 4 | 3.71E-10 | 2.60E-06 | 1.10E-06 | 1.45E-08 | -0.00605596 | RPSAP71 |
| chr1 | 203475390 | 203475817 | 428 | 4 | 2.13E-09 | 2.12E-08 | 9.09E-06 | 1.45E-08 | -0.01059844 | PRELP |
| chr6 | 106129595 | 106129683 | 89 | 2 | 1.93E-09 | 6.13E-07 | 1.04E-07 | 1.45E-08 | 0.019052873 | ATG5 |
| chr11 | 133042310 | 133042414 | 105 | 2 | 1.10E-08 | 1.16E-08 | 7.39E-06 | 1.45E-08 | 0.012732163 | OPCML |
| chr8 | 1325369 | 1327144 | 1776 | 7 | 3.99E-08 | 2.86E-05 | 1.67E-06 | 1.46E-08 | -0.00450693 | DLGAP2 |
| chr4 | 6419738 | 6421644 | 1907 | 10 | 1.37E-06 | 2.31E-08 | 0.000857 | 1.46E-08 | -0.00280074 | PPP2R2C |
| chr19 | 1049134 | 1049288 | 155 | 2 | 4.22E-09 | 4.92E-08 | 6.73E-07 | 1.47E-08 | 0.003756159 | ABCA7 |
| chr11 | 65443129 | 65444238 | 1110 | 7 | 3.08E-10 | 3.03E-06 | 7.27E-06 | 1.47E-08 | 0.008841882 | NEAT1 |
| chr1 | 47986793 | 47987068 | 276 | 3 | 2.06E-08 | 6.28E-09 | 6.27E-05 | 1.47E-08 | -0.03096361 | TRABD2B |
| chr5 | 157574084 | 157575809 | 1726 | 9 | 1.55E-07 | 6.97E-08 | 0.000444 | 1.47E-08 | 0.000187342 | ADAM19 |
| chr3 | 10627298 | 10627827 | 530 | 3 | 4.50E-09 | 1.30E-08 | 1.12E-05 | 1.48E-08 | -0.0057303 | ATP2B2, ATP2B2-IT2 |
| chr14 | 92105929 | 92107643 | 1715 | 17 | 2.27E-09 | 2.65E-06 | 1.58E-06 | 1.48E-08 | 0.001424192 | ATXN3 |
| chr16 | 3026097 | 3028338 | 2242 | 7 | 1.25E-06 | 7.90E-09 | 0.000482 | 1.48E-08 | 0.00146469 | THOC6, BICDL2 |
| chr8 | 13159126 | 13159307 | 182 | 2 | 1.08E-08 | 1.09E-08 | 9.41E-06 | 1.49E-08 | -0.0105659 | AC106845.1, DLC1 |
| chr5 | 143403814 | 143405936 | 2123 | 24 | 6.45E-06 | 7.54E-08 | 0.001016 | 1.49E-08 | 0.000511683 | NR3C1 |
| chr7 | 130205083 | 130206521 | 1439 | 14 | 5.98E-09 | 5.66E-05 | 0.000374 | 1.49E-08 | 0.002286948 | AC087071.2, SSMEM1, TMEM209 |
| chr10 | 15461427 | 15461810 | 384 | 2 | 1.16E-08 | 9.79E-09 | 1.51E-05 | 1.49E-08 | -0.02559824 | NA |
| chr10 | 70602536 | 70604066 | 1531 | 13 | 2.10E-07 | 1.58E-07 | 0.000887 | 1.49E-08 | 0.001612809 | PRF1 |
| chr12 | 118649763 | 118650283 | 521 | 4 | 1.03E-10 | 2.44E-06 | 1.04E-06 | 1.49E-08 | -0.0089285 | NA |
| chr10 | 44999987 | 45001257 | 1271 | 16 | 1.57E-09 | 3.45E-07 | 0.002595 | 1.49E-08 | 0.004934429 | ZNF22, ZNF22-AS1 |
| chr17 | 81518503 | 81519144 | 642 | 6 | 1.01E-09 | 2.46E-07 | 3.00E-06 | 1.50E-08 | -0.00376411 | AC139149.1, ACTG1 |
| chr1 | 154193964 | 154194961 | 998 | 5 | 3.74E-09 | 2.03E-07 | 0.000287 | 1.50E-08 | 0.010031778 | TPM3 |
| chr2 | 208129649 | 208130366 | 718 | 8 | 4.17E-09 | 1.75E-07 | 1.42E-05 | 1.50E-08 | 0.000225441 | CRYGC |
| chr19 | 51224039 | 51225123 | 1085 | 5 | 4.95E-10 | 9.49E-07 | 3.46E-05 | 1.50E-08 | -0.00781681 | CD33 |
| chr8 | 33538939 | 33539891 | 953 | 4 | 1.44E-08 | 4.69E-06 | 1.78E-06 | 1.51E-08 | -0.0073158 | NA |
| chr3 | 52835272 | 52836527 | 1256 | 7 | 1.15E-08 | 5.23E-08 | 0.000276 | 1.51E-08 | -0.00205341 | AC006254.1, STIMATE-MUSTN1, STIMATE |
| chr9 | 14346425 | 14347754 | 1330 | 6 | 3.09E-09 | 1.57E-08 | 0.000113 | 1.51E-08 | -0.00482923 | AL136366.1, NFIB |
| chr10 | 133224140 | 133224942 | 803 | 4 | 2.21E-08 | 3.48E-08 | 0.000213 | 1.51E-08 | -0.00714942 | KNDC1 |
| chr19 | 1989529 | 1990897 | 1369 | 6 | 2.40E-08 | 6.43E-06 | 0.000102 | 1.51E-08 | -0.00151851 | AC005306.1, BTBD2 |
| chr13 | 34748316 | 34749223 | 908 | 3 | 8.03E-12 | 7.56E-06 | 1.88E-07 | 1.51E-08 | -0.02055903 | NA |
| chr16 | 3192737 | 3193552 | 816 | 7 | 1.48E-09 | 1.03E-07 | 5.94E-05 | 1.51E-08 | -0.0199728 | AJ003147.2 |
| chr8 | 132675098 | 132677132 | 2035 | 13 | 4.83E-06 | 2.00E-07 | 0.00185 | 1.51E-08 | -0.00305843 | LRRC6 |
| chr18 | 80107989 | 80109556 | 1568 | 10 | 4.63E-09 | 1.32E-06 | 0.000242 | 1.51E-08 | -0.00264887 | ADNP2 |
| chr12 | 113426087 | 113427604 | 1518 | 6 | 4.09E-07 | 3.49E-08 | 0.000179 | 1.51E-08 | -0.00705255 | SDSL, SDS |
| chr5 | 140525459 | 140526044 | 586 | 3 | 1.69E-08 | 6.27E-09 | 7.21E-05 | 1.51E-08 | -0.00794436 | ANKHD1, ANKHD1-EIF4EBP3 |
| chr17 | 909447 | 910149 | 703 | 3 | 1.23E-09 | 3.36E-08 | 5.03E-06 | 1.51E-08 | 0.034257224 | AC087392.1, NXN |
| chr8 | 101393194 | 101393713 | 520 | 5 | 4.64E-11 | 2.20E-05 | 1.28E-07 | 1.52E-08 | 0.002261947 | LINC02845 |
| chr18 | 59402163 | 59402854 | 692 | 4 | 2.22E-11 | 6.44E-06 | 5.82E-06 | 1.52E-08 | -0.00713337 | NA |
| chr8 | 141355540 | 141357695 | 2156 | 9 | 3.02E-08 | 6.50E-08 | 0.00083 | 1.52E-08 | 0.000167985 | GPR20 |
| chr17 | 40999863 | 41001214 | 1352 | 5 | 2.61E-07 | 9.71E-09 | 6.24E-05 | 1.52E-08 | -0.00851954 | KRTAP3-2 |
| chr1 | 44546484 | 44546621 | 138 | 3 | 6.35E-09 | 1.34E-07 | 2.40E-05 | 1.53E-08 | -0.00209141 | RNF220 |
| chr16 | 30406935 | 30408009 | 1075 | 16 | 1.08E-09 | 1.33E-06 | 0.000755 | 1.53E-08 | -0.00136185 | ZNF771 |
| chr20 | 36435793 | 36436406 | 614 | 2 | 2.25E-10 | 9.38E-08 | 3.77E-07 | 1.53E-08 | 0.003100647 | DLGAP4 |
| chr18 | 9912786 | 9914105 | 1320 | 15 | 4.22E-09 | 4.48E-07 | 9.95E-05 | 1.53E-08 | -0.000272 | VAPA, AC006238.1 |
| chr14 | 90674381 | 90675489 | 1109 | 6 | 2.58E-08 | 1.96E-06 | 2.71E-05 | 1.53E-08 | -0.00636566 | AL139193.1, TTC7B |
| chr12 | 122659574 | 122660305 | 732 | 3 | 6.10E-10 | 3.92E-07 | 1.99E-05 | 1.54E-08 | 0.026040518 | NA |
| chr4 | 7350297 | 7350508 | 212 | 2 | 8.26E-09 | 1.57E-08 | 4.03E-06 | 1.54E-08 | -0.02103998 | SORCS2 |
| chr6 | 99590477 | 99591600 | 1124 | 8 | 2.89E-08 | 6.68E-07 | 0.00017 | 1.54E-08 | -0.0008604 | NA |
| chr1 | 10471934 | 10472863 | 930 | 15 | 1.70E-09 | 1.21E-06 | 0.001418 | 1.54E-08 | 0.00051957 | PEX14, DFFA |
| chr5 | 5420250 | 5422083 | 1834 | 9 | 1.17E-06 | 5.88E-06 | 6.25E-05 | 1.54E-08 | -0.0019393 | ICE1, AC091978.1 |
| chr19 | 47471644 | 47473041 | 1398 | 9 | 1.73E-09 | 4.38E-06 | 0.000821 | 1.54E-08 | -0.0023032 | AC073548.2, SLC8A2 |
| chr3 | 113745969 | 113748080 | 2112 | 25 | 1.79E-06 | 2.69E-08 | 0.00094 | 1.55E-08 | 0.00206082 | ATP6V1A, NAA50, AC108693.2 |
| chr20 | 50320663 | 50321530 | 868 | 3 | 1.90E-12 | 5.01E-07 | 2.41E-07 | 1.55E-08 | -0.00413793 | LINC01271 |
| chr16 | 73498371 | 73499271 | 901 | 4 | 1.12E-07 | 4.00E-09 | 0.000384 | 1.55E-08 | -0.0019514 | LINC01568, ZFHX3 |
| chr1 | 200619882 | 200621346 | 1465 | 14 | 2.43E-07 | 3.21E-08 | 0.001967 | 1.55E-08 | -0.00059857 | KIF14 |
| chr19 | 5975702 | 5975860 | 159 | 2 | 4.98E-09 | 2.51E-08 | 1.80E-06 | 1.56E-08 | -0.00544947 | RANBP3 |
| chr10 | 32395211 | 32395672 | 462 | 2 | 1.86E-09 | 3.24E-08 | 1.25E-06 | 1.56E-08 | -0.00747182 | NA |
| chr17 | 81052249 | 81053317 | 1069 | 6 | 6.93E-10 | 4.32E-07 | 7.53E-06 | 1.56E-08 | -0.00549029 | BAIAP2 |
| chr11 | 69817349 | 69819206 | 1858 | 11 | 1.64E-08 | 5.26E-06 | 0.000662 | 1.56E-08 | 0.000971089 | FGF3 |
| chr11 | 1399883 | 1399979 | 97 | 3 | 5.48E-09 | 6.39E-09 | 8.69E-05 | 1.56E-08 | 0.012181579 | BRSK2 |
| chr17 | 75874414 | 75875451 | 1038 | 5 | 3.70E-09 | 1.24E-06 | 1.89E-05 | 1.56E-08 | 0.00170559 | TRIM47 |
| chr6 | 37536708 | 37537248 | 541 | 3 | 4.45E-12 | 7.49E-07 | 3.17E-07 | 1.56E-08 | -0.01166842 | NA |
| chr17 | 82005337 | 82006677 | 1341 | 5 | 3.30E-07 | 5.87E-09 | 0.000558 | 1.57E-08 | 0.010728526 | ASPSCR1 |
| chr17 | 49500360 | 49500492 | 133 | 2 | 2.28E-10 | 3.42E-05 | 3.68E-08 | 1.57E-08 | -0.01117757 | NGFR |
| chr5 | 178203998 | 178205258 | 1261 | 12 | 6.42E-09 | 7.39E-07 | 0.001047 | 1.57E-08 | 0.00068026 | HNRNPAB |
| chr16 | 86275582 | 86276214 | 633 | 5 | 6.34E-10 | 7.39E-06 | 1.62E-06 | 1.57E-08 | -0.01448055 | LINC01081 |
| chr16 | 90026272 | 90026347 | 76 | 2 | 9.18E-10 | 5.82E-06 | 5.18E-08 | 1.57E-08 | -0.00376925 | GAS8 |
| chr2 | 74529107 | 74530698 | 1592 | 22 | 3.29E-08 | 7.31E-07 | 0.002109 | 1.57E-08 | -9.49E-05 | HTRA2, AUP1 |
| chr22 | 32150060 | 32150991 | 932 | 4 | 2.61E-09 | 7.73E-08 | 9.55E-06 | 1.57E-08 | 0.002753146 | C22orf42 |
| chr2 | 106160115 | 106160759 | 645 | 6 | 2.87E-08 | 6.06E-09 | 0.001178 | 1.58E-08 | 0.00338626 | UXS1 |
| chr2 | 23642223 | 23643314 | 1092 | 6 | 1.19E-09 | 8.02E-08 | 6.54E-05 | 1.58E-08 | -0.00294168 | KLHL29 |
| chr5 | 74640228 | 74642340 | 2113 | 12 | 2.05E-07 | 1.18E-05 | 0.000174 | 1.59E-08 | 0.001305891 | HEXB, ENC1 |
| chr6 | 40599156 | 40599840 | 685 | 6 | 8.57E-09 | 6.92E-09 | 7.10E-05 | 1.59E-08 | -0.01887884 | NA |
| chr6 | 22035680 | 22036512 | 833 | 3 | 7.97E-09 | 1.95E-08 | 1.06E-05 | 1.59E-08 | -0.00904962 | CASC15 |
| chr20 | 13784435 | 13785664 | 1230 | 17 | 1.58E-09 | 3.39E-08 | 2.39E-05 | 1.59E-08 | 0.000636186 | NDUFAF5, ESF1 |
| chr2 | 138896777 | 138897274 | 498 | 2 | 1.11E-09 | 4.80E-08 | 8.02E-07 | 1.59E-08 | 0.000697603 | YY1P2 |
| chr21 | 44920602 | 44922003 | 1402 | 11 | 1.90E-10 | 1.33E-06 | 0.000857 | 1.59E-08 | -0.00582732 | ITGB2-AS1, ITGB2 |
| chr15 | 43329750 | 43331084 | 1335 | 19 | 1.31E-08 | 1.84E-06 | 0.003398 | 1.59E-08 | -0.00124807 | ADAL, LCMT2 |
| chr11 | 67437625 | 67439292 | 1668 | 13 | 8.19E-08 | 2.26E-07 | 0.000265 | 1.59E-08 | -0.00206088 | PTPRCAP, CORO1B |
| chr11 | 64370275 | 64371367 | 1093 | 5 | 7.39E-09 | 9.64E-07 | 2.37E-06 | 1.59E-08 | 0.003594572 | RPS6KA4 |
| chr9 | 96442139 | 96442225 | 87 | 2 | 8.60E-09 | 2.69E-08 | 1.71E-06 | 1.60E-08 | -0.00722808 | NA |
| chr11 | 9459828 | 9461895 | 2068 | 15 | 2.87E-07 | 7.66E-05 | 0.000114 | 1.60E-08 | -0.0010283 | ZNF143, AC132192.2 |
| chr17 | 41315162 | 41316300 | 1139 | 10 | 1.01E-08 | 2.38E-08 | 0.001153 | 1.60E-08 | -0.00487529 | KRTAP17-1 |
| chr13 | 20178285 | 20179266 | 982 | 2 | 6.10E-07 | 9.64E-09 | 2.76E-05 | 1.61E-08 | -0.00208952 | PPIAP28 |
| chr13 | 113446154 | 113446732 | 579 | 4 | 8.34E-09 | 2.01E-07 | 2.44E-05 | 1.61E-08 | 0.001070915 | ADPRHL1 |
| chr12 | 132319613 | 132320353 | 741 | 9 | 3.80E-09 | 5.84E-08 | 0.00024 | 1.61E-08 | 0.001840349 | GALNT9 |
| chr15 | 96223069 | 96223403 | 335 | 4 | 9.17E-09 | 1.89E-08 | 1.01E-05 | 1.62E-08 | -0.01209512 | AC012409.2, NR2F2-AS1 |
| chr15 | 51750924 | 51752874 | 1951 | 19 | 2.44E-07 | 3.74E-07 | 5.73E-05 | 1.62E-08 | -0.00069942 | TMOD2, AC026770.1, LYSMD2 |
| chr8 | 66961971 | 66963411 | 1441 | 11 | 5.32E-08 | 3.07E-09 | 0.002113 | 1.62E-08 | 0.000497432 | TCF24 |
| chr10 | 102493258 | 102493708 | 451 | 2 | 3.47E-10 | 2.17E-07 | 2.16E-07 | 1.62E-08 | -0.01554652 | ACTR1A |
| chr3 | 149376866 | 149377786 | 921 | 8 | 1.38E-09 | 1.36E-08 | 0.000289 | 1.62E-08 | 0.025152749 | TM4SF1-AS1, TM4SF1 |
| chr21 | 46312810 | 46313126 | 317 | 3 | 2.20E-09 | 1.50E-05 | 8.70E-07 | 1.63E-08 | -0.00142344 | C21orf58 |
| chr2 | 3214507 | 3215145 | 639 | 3 | 5.30E-09 | 7.49E-08 | 2.60E-05 | 1.63E-08 | -0.00084552 | EIPR1 |
| chr3 | 127610280 | 127610715 | 436 | 3 | 2.49E-08 | 5.64E-09 | 0.000138 | 1.63E-08 | -0.02218388 | MCM2 |
| chr5 | 173054026 | 173054328 | 303 | 3 | 2.34E-09 | 3.85E-08 | 3.42E-06 | 1.63E-08 | -0.0059701 | NA |
| chr11 | 72620481 | 72621545 | 1065 | 3 | 3.65E-07 | 6.56E-09 | 9.80E-05 | 1.63E-08 | -0.00376507 | PDE2A |
| chr12 | 119259023 | 119259609 | 587 | 4 | 1.80E-09 | 7.03E-06 | 9.81E-05 | 1.64E-08 | -0.02000314 | NA |
| chr1 | 186374926 | 186375866 | 941 | 16 | 3.44E-08 | 1.33E-07 | 0.000905 | 1.64E-08 | 0.001322049 | ODR4, TPR |
| chr1 | 175888392 | 175889365 | 974 | 4 | 4.01E-08 | 7.50E-08 | 5.91E-05 | 1.64E-08 | -0.01095426 | NA |
| chr6 | 109449430 | 109450631 | 1202 | 6 | 6.24E-10 | 3.50E-08 | 2.63E-05 | 1.64E-08 | 0.001387968 | MICAL1 |
| chr2 | 221569583 | 221570494 | 912 | 4 | 1.25E-09 | 2.57E-08 | 0.000147 | 1.64E-08 | -0.02781578 | EPHA4 |
| chr20 | 53800195 | 53801495 | 1301 | 4 | 2.20E-07 | 3.47E-07 | 2.90E-05 | 1.65E-08 | -0.00932424 | AC006076.1 |
| chr10 | 35231678 | 35232591 | 914 | 3 | 1.35E-08 | 9.19E-09 | 4.20E-05 | 1.65E-08 | 9.21E-05 | NA |
| chr10 | 88990163 | 88991512 | 1350 | 18 | 2.70E-11 | 1.50E-06 | 7.31E-06 | 1.65E-08 | 0.000803928 | FAS, ACTA2, AL157394.3 |
| chr12 | 97557552 | 97557934 | 383 | 2 | 2.64E-09 | 3.48E-08 | 1.27E-06 | 1.65E-08 | -0.00787724 | RMST |
| chr6 | 169250737 | 169251550 | 814 | 3 | 2.23E-08 | 4.61E-08 | 1.22E-05 | 1.65E-08 | -0.01636362 | THBS2 |
| chr2 | 207225479 | 207226941 | 1463 | 4 | 4.22E-07 | 9.63E-09 | 9.18E-05 | 1.65E-08 | 0.001480227 | MYOSLID, AC007879.3 |
| chr3 | 180989434 | 180991165 | 1732 | 15 | 1.76E-07 | 5.25E-08 | 0.000493 | 1.65E-08 | 0.003010649 | SOX2-OT, DNAJC19 |
| chr1 | 91948738 | 91949967 | 1230 | 13 | 1.86E-07 | 4.33E-07 | 0.000824 | 1.65E-08 | -0.0092302 | BRDT |
| chr8 | 17910002 | 17910606 | 605 | 7 | 8.49E-09 | 1.40E-07 | 3.85E-05 | 1.66E-08 | -0.00142278 | FGL1 |
| chr15 | 72320129 | 72321086 | 958 | 11 | 7.84E-11 | 2.22E-07 | 2.80E-05 | 1.66E-08 | -0.00459439 | AC009690.2, CELF6, AC009690.1 |
| chr10 | 132980409 | 132981249 | 841 | 3 | 3.52E-08 | 7.43E-09 | 9.13E-05 | 1.66E-08 | -0.01209856 | NA |
| chr8 | 33461756 | 33461885 | 130 | 2 | 1.18E-08 | 1.45E-08 | 6.05E-06 | 1.66E-08 | -0.01075383 | FUT10 |
| chr1 | 231335758 | 231337440 | 1683 | 10 | 1.09E-08 | 2.33E-07 | 2.01E-05 | 1.67E-08 | -0.0044271 | SPRTN, EXOC8 |
| chr11 | 3037974 | 3038141 | 168 | 3 | 2.09E-09 | 8.84E-08 | 2.37E-06 | 1.67E-08 | -0.00746174 | CARS1-AS1, CARS1 |
| chr17 | 65538222 | 65539105 | 884 | 7 | 4.06E-09 | 6.84E-08 | 0.000113 | 1.67E-08 | -0.00608707 | AXIN2, AC004805.1 |
| chr20 | 37763052 | 37763239 | 188 | 2 | 1.52E-08 | 9.94E-09 | 2.93E-05 | 1.67E-08 | -0.00190099 | CTNNBL1 |
| chr11 | 118348603 | 118349159 | 557 | 3 | 4.44E-10 | 3.60E-05 | 2.68E-07 | 1.67E-08 | -0.00803386 | CD3G |
| chr2 | 100561785 | 100562708 | 924 | 6 | 3.93E-09 | 5.00E-09 | 0.000721 | 1.68E-08 | 0.006355594 | NA |
| chr12 | 19439124 | 19440750 | 1627 | 12 | 6.46E-09 | 9.91E-06 | 9.95E-06 | 1.68E-08 | -0.00157479 | AEBP2 |
| chr3 | 127536709 | 127537923 | 1215 | 4 | 2.34E-08 | 2.41E-07 | 3.48E-05 | 1.68E-08 | -0.00313247 | LINC01471 |
| chr6 | 31811531 | 31812785 | 1255 | 8 | 1.88E-09 | 1.47E-07 | 8.27E-05 | 1.68E-08 | -0.00013088 | HSPA1L |
| chr1 | 40653603 | 40654676 | 1074 | 6 | 1.22E-07 | 9.97E-09 | 0.000695 | 1.68E-08 | -0.01179226 | RIMS3 |
| chr19 | 32599389 | 32599911 | 523 | 3 | 8.21E-11 | 2.38E-05 | 1.37E-07 | 1.69E-08 | -0.00688917 | ANKRD27 |
| chr19 | 863054 | 863226 | 173 | 3 | 1.73E-09 | 9.73E-08 | 9.89E-07 | 1.70E-08 | -0.02150109 | CFD |
| chr10 | 103478741 | 103480073 | 1333 | 8 | 1.10E-09 | 4.55E-06 | 3.99E-05 | 1.70E-08 | -0.00493067 | CALHM3, NEURL1-AS1 |
| chr22 | 26346964 | 26347743 | 780 | 6 | 2.20E-08 | 3.86E-08 | 0.000444 | 1.70E-08 | -0.0079643 | SEZ6L |
| chr5 | 179258056 | 179258786 | 731 | 2 | 5.44E-12 | 7.34E-07 | 1.14E-07 | 1.70E-08 | -0.01610856 | ADAMTS2 |
| chr16 | 8570594 | 8571186 | 593 | 4 | 2.37E-11 | 3.23E-07 | 1.30E-05 | 1.70E-08 | -0.01373194 | TMEM114 |
| chr2 | 88055249 | 88056265 | 1017 | 12 | 3.93E-08 | 1.64E-08 | 0.004112 | 1.70E-08 | 0.001808633 | KRCC1 |
| chr12 | 47884515 | 47885130 | 616 | 2 | 4.06E-09 | 2.18E-08 | 2.74E-06 | 1.70E-08 | -0.01021245 | AC121338.1, VDR |
| chr7 | 135246027 | 135246757 | 731 | 7 | 1.61E-09 | 2.75E-06 | 4.46E-05 | 1.70E-08 | -0.00257997 | STRA8, SLC23A4P |
| chr2 | 96632339 | 96632631 | 293 | 2 | 3.77E-10 | 2.22E-07 | 2.27E-07 | 1.72E-08 | -0.00611697 | KANSL3 |
| chr10 | 92690774 | 92691868 | 1095 | 12 | 1.23E-09 | 7.24E-07 | 0.00097 | 1.72E-08 | 0.002564314 | HHEX |
| chr12 | 121209325 | 121210290 | 966 | 14 | 5.92E-08 | 9.18E-06 | 0.000221 | 1.72E-08 | 0.000346713 | P2RX4 |

| | | | | | | | | | | |
|---|---|---|---|---|---|---|---|---|---|---|
| chr6 | 30932420 | 30933821 | 1402 | 8 | 8.60E-09 | 5.31E-07 | 5.81E-05 | 1.72E-08 | -0.00054702 | SFTA2, Y RNA |
| chr5 | 1483467 | 1484199 | 733 | 4 | 3.62E-11 | 1.09E-06 | 5.71E-06 | 1.72E-08 | -0.00099734 | LPCAT1 |
| chr11 | 8250714 | 8251374 | 661 | 2 | 8.93E-10 | 5.80E-08 | 7.37E-07 | 1.73E-08 | -0.01022558 | LMO1 |
| chr1 | 75357257 | 75358112 | 856 | 5 | 9.65E-11 | 2.42E-05 | 5.01E-07 | 1.73E-08 | -0.01246716 | SLC44A5 |
| chr6 | 111229718 | 111230461 | 744 | 3 | 1.02E-09 | 7.01E-08 | 1.63E-05 | 1.73E-08 | -0.00797968 | SLC16A10, AL360227.1 |
| chr11 | 615519 | 617778 | 2260 | 21 | 9.30E-06 | 8.76E-08 | 0.001423 | 1.74E-08 | -0.00091011 | IRF7, CDHR5 |
| chr16 | 88833962 | 88835339 | 1378 | 5 | 9.56E-07 | 5.51E-07 | 6.53E-05 | 1.74E-08 | 0.001342696 | GALNS |
| chr17 | 75694762 | 75695912 | 1151 | 3 | 5.60E-07 | 3.44E-06 | 1.47E-05 | 1.74E-08 | 0.006113226 | SAP30BP |
| chr7 | 156112957 | 156113783 | 827 | 2 | 1.06E-09 | 4.19E-08 | 1.10E-06 | 1.75E-08 | -0.00149612 | NA |
| chr17 | 79087574 | 79088536 | 963 | 4 | 1.59E-08 | 7.05E-09 | 9.42E-05 | 1.75E-08 | -0.01060761 | ENGASE |
| chr6 | 30041403 | 30043887 | 2485 | 8 | 1.41E-06 | 5.17E-06 | 0.001454 | 1.75E-08 | -0.0048259 | ZNRD1ASP |
| chr17 | 48012271 | 48012865 | 595 | 5 | 7.79E-09 | 9.48E-09 | 0.000154 | 1.76E-08 | -0.00794315 | NA |
| chr9 | 90960869 | 90961268 | 400 | 2 | 1.33E-09 | 7.64E-08 | 5.55E-07 | 1.76E-08 | 0.014110014 | AL355607.1 |
| chr1 | 3160792 | 3161960 | 1169 | 6 | 1.62E-10 | 6.29E-07 | 2.91E-05 | 1.76E-08 | -0.00485676 | PRDM16 |
| chr16 | 24538827 | 24540758 | 1932 | 19 | 3.44E-06 | 1.96E-06 | 0.000787 | 1.76E-08 | -0.00137114 | RBBP6 |
| chr5 | 33440415 | 33442410 | 1996 | 13 | 2.67E-09 | 1.02E-06 | 0.000281 | 1.76E-08 | -0.00133176 | TARS1, AC034231.1 |
| chr2 | 168912561 | 168913744 | 1184 | 6 | 5.58E-08 | 7.58E-09 | 0.000678 | 1.77E-08 | -0.01562684 | SPC25 |
| chr5 | 123034740 | 123036201 | 1462 | 6 | 2.71E-08 | 1.18E-05 | 3.55E-05 | 1.77E-08 | -0.00519469 | PPIC |
| chr3 | 44874426 | 44874981 | 556 | 5 | 1.44E-11 | 2.91E-06 | 5.15E-07 | 1.77E-08 | -0.00294259 | TGM4 |
| chr2 | 46844979 | 46845768 | 790 | 4 | 6.17E-09 | 1.05E-08 | 9.39E-05 | 1.77E-08 | -0.00893424 | LINC01119 |
| chr18 | 3446568 | 3448470 | 1903 | 14 | 8.38E-07 | 2.65E-07 | 0.001108 | 1.77E-08 | -0.00052408 | TGIF1 |
| chr14 | 77141183 | 77142921 | 1739 | 13 | 7.02E-08 | 3.22E-07 | 0.001181 | 1.78E-08 | -0.00543835 | AC007375.2, TMEM63C, AC007375.3, ZDHHC22 |
| chr20 | 2497390 | 2497495 | 106 | 2 | 1.51E-09 | 2.79E-06 | 6.94E-08 | 1.78E-08 | -0.00932456 | AL049650.1, ZNF343 |
| chr5 | 175468429 | 175468513 | 85 | 2 | 8.72E-09 | 3.85E-08 | 1.27E-06 | 1.78E-08 | -0.03049778 | NA |
| chr18 | 63489832 | 63490606 | 775 | 6 | 3.20E-09 | 1.62E-07 | 4.41E-05 | 1.78E-08 | -0.00790615 | SERPINB5 |
| chr8 | 22551049 | 22552171 | 1123 | 13 | 2.94E-07 | 6.82E-08 | 0.000502 | 1.78E-08 | -0.00288743 | SORBS3, AC087854.1 |
| chr1 | 94324418 | 94325371 | 954 | 3 | 1.99E-11 | 9.52E-06 | 3.92E-07 | 1.78E-08 | -0.01089931 | ARHGAP29-AS1, AC097059.1 |
| chr15 | 50908879 | 50909643 | 765 | 3 | 1.81E-12 | 9.32E-07 | 2.95E-06 | 1.78E-08 | -0.01171821 | AP4E1 |
| chr6 | 96014995 | 96015836 | 842 | 6 | 1.58E-09 | 3.54E-08 | 2.47E-05 | 1.79E-08 | -0.00674563 | AL034348.1 |
| chr17 | 56885167 | 56885451 | 285 | 3 | 6.29E-09 | 1.12E-07 | 1.43E-06 | 1.79E-08 | -0.00238739 | TRIM25 |
| chr5 | 52787024 | 52788579 | 1556 | 18 | 6.75E-09 | 3.59E-07 | 9.07E-05 | 1.79E-08 | -5.47E-05 | ITGA1, PELO, AC022126.1 |
| chr11 | 102466565 | 102467193 | 629 | 5 | 7.21E-09 | 1.47E-07 | 1.71E-05 | 1.80E-08 | 0.002933433 | TMEM123 |
| chr1 | 228321679 | 228322061 | 383 | 5 | 1.53E-08 | 1.46E-08 | 6.04E-05 | 1.80E-08 | -0.00395725 | OBSCN |
| chr22 | 37518852 | 37520279 | 1428 | 12 | 7.58E-09 | 1.61E-07 | 0.000695 | 1.80E-08 | -0.00253043 | CARD10 |
| chr17 | 81031050 | 81032867 | 1818 | 14 | 1.62E-06 | 3.11E-08 | 0.003518 | 1.80E-08 | -0.00477682 | BAIAP2-DT |
| chr5 | 66595217 | 66596050 | 834 | 4 | 2.29E-12 | 1.52E-05 | 9.28E-08 | 1.80E-08 | -0.00765403 | NA |
| chr14 | 56816597 | 56817810 | 1214 | 7 | 4.22E-07 | 1.54E-07 | 0.000192 | 1.80E-08 | -0.00668484 | OTX2-AS1, OTX2, AL161757.4 |
| chr6 | 28224679 | 28225505 | 827 | 13 | 1.61E-08 | 1.92E-07 | 5.72E-05 | 1.80E-08 | -0.00147102 | ZSCAN9 |
| chr14 | 103137927 | 103138970 | 1044 | 6 | 3.10E-09 | 1.61E-08 | 0.000492 | 1.81E-08 | -0.0114053 | NA |
| chr14 | 74018222 | 74019961 | 1740 | 13 | 1.38E-07 | 5.14E-08 | 0.002921 | 1.81E-08 | -9.11E-05 | BBOF1, ENTPD5 |
| chr11 | 69835463 | 69836221 | 759 | 2 | 8.20E-09 | 1.76E-08 | 4.88E-06 | 1.81E-08 | 0.009125512 | NA |
| chr11 | 118145532 | 118146783 | 1252 | 12 | 3.59E-10 | 6.04E-06 | 0.000111 | 1.82E-08 | 2.86E-05 | SCN4B |
| chr8 | 25479425 | 25479998 | 574 | 5 | 8.32E-10 | 4.62E-06 | 8.02E-05 | 1.82E-08 | -0.00129717 | CDCA2 |
| chr3 | 131381355 | 131381869 | 515 | 6 | 7.85E-09 | 4.13E-08 | 0.000459 | 1.83E-08 | 0.002523083 | NUDT16, AC010210.1 |
| chr17 | 17691405 | 17691963 | 559 | 2 | 7.69E-09 | 1.36E-08 | 1.02E-05 | 1.83E-08 | 0.003121188 | RAI1 |
| chr12 | 99131256 | 99132086 | 831 | 5 | 7.83E-10 | 1.00E-07 | 4.89E-05 | 1.83E-08 | -0.0092751 | ANKS1B |
| chr19 | 42528033 | 42529186 | 1154 | 5 | 1.12E-08 | 1.29E-08 | 0.000101 | 1.84E-08 | -0.00137296 | LIPE-AS1, CEACAM1 |
| chr17 | 78822878 | 78824050 | 1173 | 6 | 3.40E-09 | 2.63E-08 | 0.000456 | 1.84E-08 | -0.01010215 | USP36 |
| chr17 | 41254522 | 41254829 | 308 | 4 | 1.22E-08 | 2.64E-08 | 3.35E-05 | 1.84E-08 | -0.00867873 | NA |
| chr2 | 39704052 | 39704194 | 143 | 3 | 2.25E-08 | 1.34E-08 | 6.77E-05 | 1.84E-08 | 0.019713924 | TMEM178A |
| chr6 | 1311275 | 1313495 | 2221 | 13 | 1.29E-07 | 6.33E-08 | 2.01E-05 | 1.85E-08 | -8.71E-05 | FOXQ1 |
| chr19 | 57681594 | 57682730 | 1137 | 14 | 1.62E-08 | 8.14E-06 | 0.000306 | 1.86E-08 | -0.00426813 | ZNF551, AC003006.1 |
| chr18 | 56650635 | 56652119 | 1485 | 14 | 4.79E-09 | 9.08E-07 | 0.000572 | 1.86E-08 | -0.00133575 | WDR7, TXNL1 |
| chr1 | 109538939 | 109540302 | 1364 | 10 | 1.47E-07 | 7.82E-07 | 0.000346 | 1.86E-08 | -0.00340355 | GPR61, AL355310.3 |
| chr12 | 18562846 | 18563409 | 564 | 3 | 4.98E-08 | 6.48E-09 | 0.000148 | 1.86E-08 | -0.01053741 | PIK3C2G |
| chr17 | 14068301 | 14069702 | 1402 | 11 | 5.17E-08 | 5.64E-06 | 0.000212 | 1.86E-08 | 0.000927559 | COX10, COX10-AS1 |
| chr1 | 160117608 | 160118009 | 402 | 2 | 1.14E-09 | 3.69E-08 | 1.48E-06 | 1.87E-08 | -0.01648091 | ATP1A2 |
| chr15 | 79091043 | 79091638 | 596 | 15 | 1.76E-09 | 1.07E-07 | 0.00073 | 1.87E-08 | 0.001673248 | NA |
| chr21 | 23385676 | 23386110 | 435 | 3 | 2.51E-09 | 1.24E-07 | 1.16E-06 | 1.87E-08 | -0.01428477 | D21S2088E |
| chr20 | 44714288 | 44715356 | 1069 | 13 | 5.71E-08 | 6.08E-06 | 0.000289 | 1.87E-08 | -0.00439925 | CCN5, KCNK15-AS1 |
| chr15 | 74304492 | 74305238 | 747 | 4 | 1.17E-08 | 1.11E-08 | 0.00022 | 1.87E-08 | -0.01134784 | CCDC33, AC023300.3 |
| chr11 | 45884160 | 45886485 | 2326 | 16 | 5.24E-06 | 8.02E-08 | 0.000982 | 1.88E-08 | -0.00161271 | MAPK8IP1 |
| chr15 | 41886167 | 41886669 | 503 | 2 | 7.12E-09 | 1.82E-08 | 4.96E-06 | 1.88E-08 | -0.00596058 | SPTBN5 |
| chr19 | 57171796 | 57172517 | 722 | 5 | 1.21E-09 | 3.26E-07 | 7.72E-05 | 1.88E-08 | -0.00722877 | NA |
| chr2 | 236731948 | 236733325 | 1378 | 8 | 6.50E-14 | 3.81E-05 | 2.30E-07 | 1.88E-08 | -0.00882585 | AC084030.1 |
| chr4 | 92304313 | 92305775 | 1463 | 13 | 4.25E-07 | 1.13E-06 | 0.00164 | 1.88E-08 | 0.002678356 | GRID2 |
| chr2 | 232519779 | 232520538 | 760 | 7 | 1.59E-09 | 6.36E-07 | 1.32E-05 | 1.88E-08 | -0.01113724 | PRSS56 |
| chr7 | 15686541 | 15688222 | 1682 | 12 | 1.56E-07 | 3.24E-07 | 0.000272 | 1.89E-08 | 0.00117721 | MEOX2 |
| chr9 | 38363160 | 38363505 | 346 | 2 | 6.35E-10 | 4.95E-07 | 1.54E-07 | 1.89E-08 | -0.00814126 | AL135785.1 |
| chr2 | 109699326 | 109700397 | 1072 | 3 | 6.25E-10 | 1.17E-07 | 3.47E-06 | 1.89E-08 | -0.01704812 | NA |
| chr10 | 114285803 | 114286973 | 1171 | 5 | 1.81E-10 | 9.96E-07 | 3.11E-06 | 1.89E-08 | -0.00382234 | VWA2 |
| chr19 | 6737733 | 6739986 | 2254 | 20 | 8.75E-09 | 4.46E-06 | 9.38E-05 | 1.90E-08 | -0.00244271 | TRIP10 |
| chr8 | 9867039 | 9868389 | 1351 | 5 | 2.74E-09 | 5.64E-07 | 3.24E-06 | 1.90E-08 | -0.00595071 | NA |
| chr6 | 166423303 | 166424435 | 1133 | 4 | 1.02E-10 | 1.16E-07 | 1.31E-05 | 1.90E-08 | -0.00449564 | RPS6KA2 |
| chr19 | 1954452 | 1956254 | 1803 | 7 | 1.22E-05 | 4.02E-09 | 0.000375 | 1.91E-08 | -0.0025592 | CSNK1G2, CSNK1G2-AS1 |
| chr12 | 104485747 | 104485938 | 192 | 2 | 1.78E-08 | 1.16E-08 | 2.70E-05 | 1.91E-08 | 0.008713759 | CHST11 |
| chr19 | 11208700 | 11209095 | 396 | 6 | 8.43E-10 | 4.90E-07 | 4.01E-07 | 1.92E-08 | -0.00480741 | AC011472.1, AC011472.5, DOCK6 |
| chr2 | 199455732 | 199457021 | 1290 | 12 | 3.48E-08 | 5.21E-08 | 0.001948 | 1.92E-08 | 0.000278792 | SATB2 |
| chr2 | 3059343 | 3060330 | 988 | 11 | 2.03E-08 | 4.05E-07 | 0.000847 | 1.92E-08 | -0.00134081 | LINC01250 |
| chr16 | 68300153 | 68300716 | 564 | 3 | 1.06E-11 | 1.43E-06 | 1.96E-07 | 1.92E-08 | -0.0061834 | SLC7A6, SLC7A6OS |
| chr12 | 96401778 | 96401848 | 71 | 2 | 2.33E-10 | 5.89E-08 | 8.67E-07 | 1.92E-08 | -0.00823045 | NA |
| chr6 | 34028162 | 34028896 | 735 | 8 | 4.50E-09 | 1.56E-08 | 0.00107 | 1.92E-08 | -0.00410371 | GRM4 |
| chr11 | 124625631 | 124626465 | 835 | 2 | 9.22E-13 | 1.19E-05 | 5.18E-08 | 1.92E-08 | -0.00631741 | TBRG1 |
| chr11 | 112164078 | 112164956 | 879 | 8 | 1.44E-08 | 3.16E-07 | 2.90E-05 | 1.92E-08 | -0.00122615 | AP002884.2, IL18 |
| chr22 | 45335116 | 45337235 | 2120 | 6 | 1.03E-05 | 9.14E-09 | 0.000525 | 1.93E-08 | -0.0110579 | FAM118A |
| chr11 | 27472321 | 27473846 | 1526 | 15 | 2.56E-07 | 3.16E-07 | 0.000418 | 1.93E-08 | -0.00369491 | LGR4-AS1, LGR4 |
| chr1 | 224218156 | 224218582 | 427 | 3 | 1.41E-08 | 9.53E-09 | 5.04E-05 | 1.94E-08 | 0.015499134 | NA |
| chr14 | 23034488 | 23035712 | 1225 | 19 | 2.72E-10 | 1.13E-06 | 9.00E-05 | 1.94E-08 | -0.00041891 | PSMB5 |
| chr2 | 60756059 | 60757074 | 1016 | 8 | 1.97E-08 | 6.03E-08 | 0.000329 | 1.94E-08 | 0.001822325 | PAPOLG |
| chr11 | 14891134 | 14892466 | 1333 | 16 | 2.34E-08 | 3.73E-06 | 2.75E-05 | 1.94E-08 | -0.00061928 | CYP2R1 |
| chr4 | 54232891 | 54234274 | 1384 | 6 | 1.77E-10 | 1.91E-06 | 3.68E-06 | 1.95E-08 | 0.002431411 | AC058822.1, PDGFRA |
| chr1 | 26467343 | 26469045 | 1703 | 7 | 1.91E-07 | 2.09E-05 | 9.30E-05 | 1.95E-08 | -0.00310621 | DHDDS, DHDDS-AS1 |
| chr3 | 57597340 | 57598019 | 680 | 14 | 3.72E-09 | 5.25E-07 | 0.001054 | 1.95E-08 | 0.000626093 | ARF4-AS1, ARF4 |
| chr2 | 200888425 | 200889915 | 1491 | 16 | 4.38E-08 | 4.18E-07 | 0.000154 | 1.96E-08 | -5.18E-05 | NIF3L1, PPIL3 |
| chr11 | 70202301 | 70203362 | 1062 | 11 | 2.22E-10 | 2.46E-05 | 0.000182 | 1.96E-08 | 0.010234656 | FADD |
| chr7 | 129779190 | 129781073 | 1884 | 7 | 2.60E-07 | 3.54E-08 | 0.000695 | 1.96E-08 | -0.00085075 | NA |
| chr11 | 44905376 | 44906425 | 1050 | 7 | 8.79E-08 | 1.36E-07 | 0.000211 | 1.97E-08 | -0.00516459 | TSPAN18, TP53I11 |
| chr10 | 96672690 | 96673574 | 885 | 2 | 8.60E-08 | 1.65E-08 | 7.37E-06 | 1.98E-08 | 0.004228801 | PIK3AP1 |
| chr1 | 40784390 | 40785553 | 1164 | 5 | 6.42E-13 | 1.87E-06 | 4.29E-07 | 1.98E-08 | -0.00395313 | KCNQ4 |
| chr1 | 220527075 | 220528979 | 1905 | 13 | 1.45E-07 | 6.37E-07 | 0.001257 | 1.98E-08 | -0.00112531 | MARK1 |
| chr6 | 166478129 | 166479594 | 1466 | 8 | 2.02E-09 | 7.95E-06 | 3.45E-06 | 1.99E-08 | -0.00149323 | RPS6KA2 |
| chr16 | 55478985 | 55480558 | 1574 | 16 | 2.48E-10 | 3.16E-06 | 0.000385 | 2.00E-08 | 0.000434564 | MMP2 |
| chr2 | 174333552 | 174335100 | 1549 | 15 | 4.71E-10 | 1.21E-05 | 2.44E-06 | 2.00E-08 | -0.00230362 | SP9, AC018470.1 |
| chr6 | 35060713 | 35061372 | 660 | 4 | 2.28E-10 | 1.58E-06 | 7.07E-06 | 2.00E-08 | -0.00527414 | ANKS1A |
| chr12 | 121274535 | 121275899 | 1365 | 7 | 2.25E-11 | 1.15E-06 | 3.04E-06 | 2.00E-08 | 0.000247454 | CAMKK2 |
| chr3 | 127697248 | 127697924 | 677 | 3 | 1.19E-09 | 1.81E-07 | 7.04E-07 | 2.00E-08 | -0.00899767 | MGLL |
| chr10 | 132781483 | 132782759 | 1277 | 9 | 9.16E-08 | 1.39E-08 | 0.001096 | 2.00E-08 | -0.0017812 | INPP5A |
| chr1 | 77682577 | 77683881 | 1305 | 17 | 3.62E-07 | 0.0001487 | 0.000798 | 2.00E-08 | 0.000646213 | ZZZ3 |
| chr2 | 54483084 | 54483376 | 293 | 3 | 8.05E-09 | 5.99E-08 | 2.50E-05 | 2.01E-08 | -0.00700465 | SPTBN1 |
| chr1 | 20507321 | 20508842 | 1522 | 17 | 7.00E-07 | 7.37E-08 | 0.002912 | 2.01E-08 | 0.001147633 | MUL1 |
| chr3 | 50321880 | 50323259 | 1380 | 11 | 7.76E-09 | 9.90E-08 | 7.14E-05 | 2.01E-08 | -0.00560045 | HYAL2, TUSC2 |
| chr7 | 141995493 | 141995959 | 467 | 6 | 1.98E-10 | 5.40E-06 | 2.74E-06 | 2.02E-08 | -0.00093807 | MGAM |
| chr8 | 47831064 | 47831998 | 935 | 8 | 2.05E-10 | 1.51E-06 | 2.82E-05 | 2.02E-08 | -0.00146298 | PRKDC |
| chr6 | 230908 | 232111 | 1204 | 6 | 4.87E-09 | 3.76E-08 | 6.11E-05 | 2.02E-08 | -0.00576656 | NA |
| chr8 | 144713179 | 144714114 | 936 | 5 | 1.75E-08 | 2.55E-08 | 0.000193 | 2.02E-08 | -0.01225644 | AF186192.3 |
| chr14 | 64539233 | 64540247 | 1015 | 18 | 6.69E-10 | 9.07E-07 | 8.79E-05 | 2.03E-08 | -0.00453339 | HSPA2, AL049869.2 |
| chr10 | 128991117 | 128991228 | 112 | 3 | 3.96E-09 | 2.69E-06 | 5.86E-07 | 2.03E-08 | -0.01213761 | NA |
| chr16 | 47887300 | 47887689 | 390 | 4 | 2.30E-08 | 9.64E-09 | 9.64E-05 | 2.03E-08 | -0.00786057 | LINC02133 |
| chr11 | 67674596 | 67675660 | 1065 | 10 | 9.04E-10 | 7.82E-08 | 6.48E-05 | 2.03E-08 | 0.000701558 | ALDH3B2 |
| chr5 | 2005843 | 2007577 | 1735 | 10 | 3.07E-07 | 3.99E-06 | 4.01E-05 | 2.03E-08 | -0.00423687 | AC124852.1 |
| chr19 | 2731728 | 2733441 | 1714 | 5 | 8.31E-07 | 2.29E-08 | 3.34E-05 | 2.03E-08 | -0.00484575 | AC006538.2, SLC39A3 |
| chr16 | 89815812 | 89817580 | 1769 | 14 | 7.49E-09 | 7.95E-06 | 2.06E-06 | 2.04E-08 | -0.00141595 | FANCA |
| chr9 | 86354351 | 86355872 | 1522 | 12 | 2.35E-08 | 1.45E-06 | 0.000273 | 2.04E-08 | -0.00126402 | TUT7 |
| chr1 | 17371185 | 17372008 | 824 | 6 | 3.57E-08 | 1.04E-08 | 0.000229 | 2.04E-08 | -0.00196815 | NA |
| chr1 | 205347385 | 205348400 | 1016 | 3 | 7.87E-10 | 1.11E-07 | 2.47E-06 | 2.04E-08 | -0.00710496 | KLHDC8A |
| chr8 | 857799 | 858496 | 698 | 3 | 2.55E-08 | 8.24E-09 | 0.000131 | 2.05E-08 | 0.002410648 | DLGAP2 |
| chr7 | 45104475 | 45105735 | 1261 | 14 | 1.87E-07 | 4.39E-08 | 0.002225 | 2.05E-08 | -0.00044604 | TBRG4, SNORA5A, SNORA5C |
| chr2 | 190179755 | 190180583 | 829 | 7 | 1.66E-10 | 2.31E-07 | 7.28E-06 | 2.05E-08 | 0.005635575 | C2orf88 |
| chr18 | 75161752 | 75162379 | 628 | 4 | 3.19E-09 | 8.95E-08 | 0.000185 | 2.06E-08 | -0.01163544 | NA |
| chr16 | 84812816 | 84813330 | 515 | 2 | 2.13E-09 | 2.09E-08 | 4.72E-06 | 2.06E-08 | -1.62E-05 | NA |
| chr3 | 79239637 | 79240284 | 648 | 2 | 6.20E-09 | 1.94E-08 | 5.53E-06 | 2.06E-08 | -0.02353858 | ROBO1 |
| chr15 | 99733111 | 99734043 | 933 | 15 | 7.26E-11 | 1.09E-05 | 0.00038 | 2.07E-08 | -0.0027605 | LYSMD4 |
| chr5 | 2865815 | 2867249 | 1435 | 9 | 6.16E-09 | 1.52E-06 | 8.62E-05 | 2.07E-08 | -0.0044108 | NA |
| chr15 | 78641065 | 78642238 | 1174 | 8 | 6.65E-10 | 1.54E-07 | 0.000485 | 2.07E-08 | -0.00436287 | CHRNB4 |
| chr19 | 6475445 | 6476940 | 1496 | 6 | 5.95E-07 | 1.36E-07 | 5.67E-05 | 2.08E-08 | -0.00133881 | DENND1C |
| chr18 | 37243362 | 37244053 | 692 | 3 | 8.49E-10 | 3.00E-08 | 1.86E-05 | 2.08E-08 | -0.00132754 | AC015961.2, CELF4 |
| chr8 | 11995612 | 11997770 | 2159 | 9 | 1.30E-07 | 3.60E-05 | 7.00E-05 | 2.08E-08 | -0.0085337 | DEFB134 |
| chr7 | 30475670 | 30477126 | 1457 | 5 | 4.77E-07 | 5.25E-08 | 8.07E-05 | 2.08E-08 | 0.003850697 | NOD1 |
| chr3 | 115658160 | 115659539 | 1380 | 8 | 1.43E-08 | 4.78E-08 | 0.001112 | 2.08E-08 | 0.004232711 | GAP43, AC092468.1 |
| chr3 | 119172858 | 119173614 | 757 | 9 | 3.42E-10 | 6.65E-05 | 0.000112 | 2.08E-08 | 0.003914196 | AC083800.1, UPK1B |
| chr4 | 186724294 | 186726536 | 2243 | 15 | 5.60E-07 | 4.60E-08 | 0.003166 | 2.08E-08 | 0.000756129 | FAT1 |
| chr10 | 132405701 | 132406390 | 690 | 4 | 3.47E-08 | 2.40E-08 | 0.000161 | 2.08E-08 | -0.00164555 | PWWP2B |
| chr9 | 127714718 | 127716590 | 1873 | 17 | 1.34E-07 | 2.43E-06 | 0.000937 | 2.09E-08 | -0.00111487 | CFAP157, TTC16, PTRH1 |
| chr6 | 33422069 | 33423564 | 1496 | 12 | 3.42E-07 | 3.17E-06 | 0.000418 | 2.09E-08 | -0.00084326 | SYNGAP1 |
| chr13 | 112711760 | 112712606 | 847 | 2 | 2.98E-08 | 1.43E-08 | 1.48E-05 | 2.09E-08 | -0.01587688 | ATP11A |
| chr11 | 69152405 | 69153796 | 1392 | 6 | 6.44E-10 | 9.71E-08 | 0.000236 | 2.10E-08 | -0.01128745 | AP003071.5, AP003071.2 |
| chr8 | 142769823 | 142770667 | 845 | 7 | 3.38E-09 | 1.49E-06 | 0.000141 | 2.10E-08 | -0.00383097 | LYNX1-SLURP2, SLURP2 |
| chr22 | 24185823 | 24185887 | 65 | 2 | 1.12E-08 | 1.64E-08 | 9.24E-06 | 2.10E-08 | -0.00049252 | SUSD2 |
| chr20 | 50793743 | 50795684 | 1942 | 17 | 1.04E-07 | 1.80E-07 | 0.000427 | 2.10E-08 | 0.000582199 | BCAS4 |
| chr9 | 21967932 | 21969079 | 1148 | 16 | 3.64E-09 | 1.72E-07 | 0.004705 | 2.10E-08 | -0.00400861 | AL359922.1, CDKN2A |
| chr11 | 31799516 | 31800978 | 1463 | 10 | 1.69E-08 | 6.06E-07 | 7.33E-05 | 2.10E-08 | -0.00350134 | PAX6 |
| chr11 | 15615896 | 15616396 | 501 | 3 | 2.04E-11 | 2.15E-05 | 7.97E-08 | 2.10E-08 | -0.00329261 | AC087379.2, AC073172.1 |
| chr1 | 100894815 | 100896286 | 1472 | 18 | 4.01E-08 | 5.99E-07 | 0.000237 | 2.11E-08 | 0.001416704 | AC104506.1, SLC30A7, EXTL2 |
| chr8 | 141328399 | 141329135 | 737 | 3 | 1.79E-10 | 9.75E-07 | 4.29E-06 | 2.11E-08 | -0.00072661 | NA |
| chr5 | 142697964 | 142699152 | 1189 | 6 | 1.06E-09 | 6.20E-07 | 1.53E-05 | 2.12E-08 | -0.00645625 | FGF1 |
| chr17 | 8079192 | 8079936 | 745 | 9 | 1.39E-08 | 7.49E-08 | 0.00025 | 2.12E-08 | -0.00131687 | AC129492.1, ALOX12B |
| chr16 | 71300924 | 71301423 | 500 | 2 | 4.78E-08 | 1.26E-08 | 3.23E-05 | 2.12E-08 | -0.01931759 | AC106736.1 |
| chr1 | 226444858 | 226444900 | 43 | 2 | 2.24E-09 | 5.38E-08 | 1.12E-06 | 2.12E-08 | -0.01736679 | NA |

| | | | | | | | | | | |
|---|---|---|---|---|---|---|---|---|---|---|
| chr19 | 18110163 | 18111171 | 1009 | 6 | 1.86E-09 | 2.45E-08 | 0.000363 | 2.12E-08 | -7.37E-05 | MAST3 |
| chr22 | 43171045 | 43172577 | 1533 | 7 | 1.52E-09 | 1.94E-07 | 3.53E-06 | 2.13E-08 | -0.00588316 | TTLL12 |
| chr19 | 40284684 | 40286104 | 1421 | 13 | 7.16E-08 | 7.81E-09 | 0.000523 | 2.14E-08 | -0.00244571 | AKT2 |
| chr6 | 27138013 | 27139318 | 1306 | 7 | 1.81E-07 | 3.90E-07 | 0.000181 | 2.14E-08 | -0.0073538 | H4C9 |
| chr17 | 76736515 | 76738038 | 1524 | 22 | 1.18E-08 | 4.57E-08 | 0.005197 | 2.14E-08 | 0.000703102 | AC005837.2, MFSD11, SRSF2, MIR636 |
| chr7 | 92630669 | 92631574 | 906 | 4 | 1.36E-10 | 9.41E-07 | 4.82E-07 | 2.14E-08 | -0.00701412 | CDK6 |
| chr5 | 71718582 | 71719605 | 1024 | 15 | 5.79E-10 | 7.81E-06 | 1.25E-06 | 2.15E-08 | 0.00198408 | CARTPT |
| chr6 | 99993060 | 99994727 | 1668 | 16 | 9.52E-08 | 1.00E-06 | 0.001514 | 2.16E-08 | 0.001508079 | MCHR2-AS1, MCHR2 |
| chr7 | 27150616 | 27152568 | 1953 | 16 | 1.16E-06 | 1.17E-07 | 0.001071 | 2.16E-08 | 0.000826359 | HOXA-AS3, HOXA3, AC004080.6, AC004080.5 |
| chr12 | 109115554 | 109116962 | 1409 | 8 | 4.19E-08 | 3.22E-07 | 0.000193 | 2.16E-08 | -0.00884137 | AC007637.1, ACACB |
| chr1 | 6784254 | 6786100 | 1847 | 16 | 1.41E-07 | 6.06E-06 | 0.000158 | 2.16E-08 | 0.001229352 | CAMTA1, CAMTA1-DT |
| chr10 | 45199138 | 45199560 | 423 | 3 | 7.27E-09 | 1.32E-08 | 6.83E-05 | 2.16E-08 | -0.02023106 | NA |
| chr11 | 119134112 | 119134577 | 466 | 3 | 1.92E-09 | 1.58E-07 | 8.22E-06 | 2.17E-08 | -0.00101909 | HINFP |
| chr8 | 22057673 | 22059504 | 1832 | 11 | 1.63E-06 | 5.82E-08 | 0.001423 | 2.17E-08 | -0.00870998 | DMTN |
| chr14 | 24135502 | 24136573 | 1072 | 14 | 1.33E-07 | 6.82E-08 | 0.000458 | 2.18E-08 | 0.000384244 | PSME1 |
| chr9 | 15510455 | 15511669 | 1215 | 13 | 1.69E-07 | 1.37E-07 | 0.002179 | 2.18E-08 | 0.000391304 | PSIP1 |
| chr19 | 1415026 | 1415758 | 733 | 3 | 3.80E-08 | 1.80E-08 | 6.57E-05 | 2.19E-08 | -0.01566996 | DAZAP1 |
| chr14 | 55411517 | 55413019 | 1503 | 14 | 2.16E-07 | 1.80E-05 | 1.86E-05 | 2.19E-08 | -0.00145655 | ATG14 |
| chr16 | 84190235 | 84191474 | 1240 | 8 | 1.65E-09 | 5.68E-06 | 1.47E-05 | 2.19E-08 | -0.00043032 | ADAD2 |
| chr15 | 44663314 | 44664060 | 747 | 12 | 7.38E-10 | 1.76E-06 | 5.37E-06 | 2.19E-08 | 0.001639278 | SPG11 |
| chr14 | 77760684 | 77762035 | 1352 | 16 | 1.41E-09 | 3.42E-07 | 7.90E-05 | 2.19E-08 | 0.000167303 | SLIRP, C14orf178, SNW1 |
| chr3 | 19901674 | 19901679 | 6 | 2 | 1.30E-08 | 4.04E-08 | 1.75E-06 | 2.19E-08 | -0.01820355 | EFHB |
| chr11 | 67299988 | 67301546 | 1559 | 9 | 1.92E-08 | 8.01E-08 | 0.000911 | 2.19E-08 | -0.0041481 | ANKRD13D |
| chr17 | 10629772 | 10630098 | 327 | 4 | 6.75E-09 | 8.24E-06 | 2.00E-05 | 2.19E-08 | 0.000359351 | MYH3 |
| chr1 | 2451144 | 2452248 | 1105 | 7 | 4.98E-07 | 1.48E-07 | 0.000124 | 2.19E-08 | 0.004337957 | PLCH2 |
| chr12 | 10719619 | 10719725 | 107 | 2 | 1.61E-09 | 5.74E-06 | 6.70E-08 | 2.20E-08 | -0.01860481 | YBX3 |
| chr21 | 17612996 | 17613443 | 448 | 11 | 1.84E-08 | 1.38E-07 | 0.00035 | 2.20E-08 | -0.00365681 | BTG3-AS1 |
| chr1 | 192643281 | 192644522 | 1242 | 3 | 3.61E-08 | 1.14E-08 | 8.60E-05 | 2.20E-08 | 0.008104102 | RGS13, AL390957.1 |
| chr6 | 168016060 | 168017718 | 1659 | 9 | 7.56E-08 | 1.19E-07 | 0.001393 | 2.20E-08 | -0.0029734 | KIF25 |
| chr9 | 34178057 | 34179551 | 1495 | 10 | 2.17E-07 | 7.44E-08 | 0.000513 | 2.20E-08 | 0.000400772 | UBAP1 |
| chr7 | 149497992 | 149498774 | 783 | 6 | 5.09E-10 | 9.58E-08 | 1.28E-05 | 2.20E-08 | -0.00552745 | NA |
| chr7 | 1012756 | 1014813 | 2058 | 8 | 9.30E-07 | 1.47E-05 | 9.83E-05 | 2.20E-08 | 0.007864702 | C7orf50 |
| chr6 | 47309154 | 47310815 | 1662 | 13 | 4.61E-08 | 2.63E-06 | 3.71E-05 | 2.20E-08 | -0.00103098 | TNFRSF21 |
| chr14 | 104768293 | 104769190 | 898 | 6 | 4.37E-09 | 3.65E-08 | 1.69E-05 | 2.20E-08 | 0.006659088 | SIVA1 |
| chr6 | 71165008 | 71165127 | 120 | 2 | 1.11E-08 | 4.26E-08 | 1.66E-06 | 2.20E-08 | -0.00897031 | LYPLA1P3 |
| chr10 | 92290384 | 92292073 | 1690 | 19 | 1.99E-08 | 4.95E-07 | 0.001446 | 2.21E-08 | 5.21E-06 | MARCHF5, CPEB3 |
| chr7 | 157860684 | 157861478 | 795 | 5 | 1.97E-08 | 1.92E-08 | 0.000142 | 2.21E-08 | -0.00013679 | PTPRN2-AS1, PTPRN2 |
| chr15 | 55741408 | 55743002 | 1595 | 9 | 2.75E-06 | 1.50E-07 | 0.000318 | 2.21E-08 | -0.00431707 | PRTG |
| chr4 | 184009066 | 184010833 | 1768 | 16 | 5.16E-08 | 3.76E-07 | 0.000207 | 2.22E-08 | 0.001362149 | STOX2 |
| chr1 | 11059463 | 11061678 | 2216 | 13 | 3.76E-06 | 4.87E-08 | 0.002297 | 2.22E-08 | 3.09E-05 | SRM |
| chr1 | 155176025 | 155177590 | 1566 | 5 | 1.52E-06 | 6.72E-07 | 4.46E-05 | 2.22E-08 | 0.000417169 | TRIM46, AL713999.2 |
| chr10 | 122089399 | 122089783 | 385 | 2 | 3.79E-10 | 1.14E-06 | 1.24E-07 | 2.22E-08 | -0.00495246 | TACC2 |
| chr6 | 143960315 | 143961168 | 854 | 2 | 1.57E-09 | 5.39E-08 | 1.22E-06 | 2.22E-08 | -0.01342169 | PLAGL1 |
| chr5 | 80569989 | 80571418 | 1430 | 11 | 4.46E-09 | 4.72E-07 | 1.74E-05 | 2.22E-08 | -0.00455542 | ANKRD34B |
| chr3 | 184513150 | 184513925 | 776 | 4 | 1.09E-08 | 1.12E-06 | 1.94E-05 | 2.23E-08 | -0.01002594 | NA |
| chr8 | 85200696 | 85200711 | 16 | 2 | 1.22E-08 | 4.99E-08 | 1.36E-06 | 2.23E-08 | 0.023459171 | E2F5 |
| chr9 | 124861098 | 124862429 | 1332 | 13 | 1.41E-09 | 7.82E-05 | 5.32E-07 | 2.23E-08 | 0.00045502 | ARPC5L, RPL35 |
| chr18 | 75207106 | 75208242 | 1137 | 3 | 2.08E-08 | 8.27E-09 | 0.00011 | 2.23E-08 | -0.00887654 | ZADH2 |
| chr8 | 68331050 | 68332499 | 1450 | 12 | 1.79E-10 | 1.17E-07 | 0.000212 | 2.24E-08 | -0.00086179 | C8orf34, C8orf34-AS1 |
| chr19 | 57130478 | 57131630 | 1153 | 4 | 1.91E-08 | 6.79E-07 | 2.05E-05 | 2.24E-08 | 0.009670308 | USP29 |
| chr13 | 113016616 | 113017651 | 1036 | 5 | 1.93E-10 | 4.20E-05 | 9.17E-06 | 2.25E-08 | -0.00751383 | MCF2L |
| chr17 | 10838066 | 10838876 | 811 | 7 | 2.06E-08 | 5.91E-08 | 0.000154 | 2.25E-08 | -0.00442466 | AC015908.7, AC015908.6, PIRT |
| chr17 | 31337144 | 31338150 | 1007 | 6 | 5.18E-10 | 1.26E-05 | 6.63E-05 | 2.25E-08 | -0.002083 | NF1 |
| chr8 | 58272605 | 58273262 | 658 | 3 | 2.63E-08 | 3.19E-07 | 1.53E-05 | 2.26E-08 | -0.00450842 | NA |
| chr13 | 77326370 | 77327995 | 1626 | 13 | 2.84E-08 | 3.80E-06 | 0.000347 | 2.26E-08 | -0.00153449 | MYCBP2 |
| chr15 | 60591920 | 60593139 | 1220 | 12 | 6.55E-09 | 5.38E-07 | 0.00046 | 2.26E-08 | 0.00083895 | RORA-AS1, AC009560.1, RORA |
| chr14 | 73759724 | 73760796 | 1073 | 13 | 1.20E-08 | 3.31E-07 | 0.00013 | 2.26E-08 | -0.00100224 | MIDEAS |
| chr2 | 238890673 | 238891627 | 955 | 8 | 1.02E-09 | 3.04E-06 | 6.80E-05 | 2.26E-08 | -0.00384973 | TWIST2 |
| chr6 | 108294257 | 108295606 | 1350 | 14 | 5.08E-10 | 0.0006883 | 7.53E-05 | 2.27E-08 | -8.45E-05 | AFG1L |
| chr20 | 3162571 | 3162638 | 68 | 2 | 1.46E-09 | 2.76E-08 | 3.46E-06 | 2.27E-08 | -0.01843037 | LZTS3 |
| chr16 | 8297710 | 8298338 | 629 | 6 | 1.02E-08 | 3.28E-06 | 4.52E-05 | 2.27E-08 | -0.00507427 | NA |
| chr17 | 77741359 | 77741505 | 147 | 2 | 1.92E-08 | 1.52E-08 | 1.71E-05 | 2.27E-08 | -0.02758814 | NA |
| chr1 | 53142365 | 53143290 | 926 | 6 | 4.75E-08 | 3.65E-09 | 0.000951 | 2.27E-08 | -0.00631874 | SLC1A7 |
| chr2 | 63840387 | 63841571 | 1185 | 14 | 6.36E-08 | 4.56E-08 | 0.000191 | 2.27E-08 | 0.001412634 | UGP2 |
| chr6 | 125746892 | 125749572 | 2681 | 17 | 2.24E-07 | 1.72E-06 | 1.23E-05 | 2.27E-08 | 0.000267465 | HEY2, AL365259.1 |
| chr4 | 121950878 | 121952817 | 1940 | 11 | 1.05E-07 | 3.78E-07 | 0.000277 | 2.28E-08 | -0.00288924 | TRPC3 |
| chr2 | 237413631 | 237415397 | 1767 | 13 | 9.16E-08 | 5.71E-06 | 0.000188 | 2.28E-08 | -0.0030173 | COL6A3 |
| chr19 | 55149146 | 55150019 | 874 | 9 | 3.24E-08 | 3.43E-08 | 0.002597 | 2.29E-08 | -0.02278268 | TNNT1 |
| chr1 | 223666226 | 223667002 | 777 | 5 | 2.81E-11 | 1.39E-05 | 4.08E-06 | 2.29E-08 | -0.00540089 | NA |
| chr4 | 40192503 | 40193698 | 1196 | 7 | 2.64E-08 | 5.28E-09 | 0.001849 | 2.29E-08 | 0.005937341 | RHOH |
| chr16 | 1363158 | 1364717 | 1560 | 8 | 2.95E-08 | 1.62E-08 | 0.000435 | 2.30E-08 | -0.0023198 | GNPTG, UNKL |
| chr20 | 32858377 | 32859275 | 899 | 7 | 4.06E-09 | 8.47E-08 | 6.87E-05 | 2.31E-08 | 0.00205502 | AL035071.1, EFCAB8 |
| chr2 | 26478046 | 26479528 | 1483 | 5 | 1.13E-06 | 1.68E-08 | 4.63E-05 | 2.31E-08 | -0.00064703 | OTOF |
| chr14 | 50396523 | 50398476 | 1954 | 14 | 4.54E-08 | 9.49E-07 | 0.001386 | 2.31E-08 | -0.00239954 | AL359397.1, CDKL1 |
| chr10 | 72691615 | 72692904 | 1290 | 15 | 3.09E-08 | 8.39E-06 | 0.000279 | 2.31E-08 | -0.00025303 | MCU |
| chr20 | 44246203 | 44247451 | 1249 | 15 | 6.93E-10 | 1.49E-06 | 8.00E-05 | 2.32E-08 | -0.00489604 | GDAP1L1 |
| chr11 | 121655042 | 121656131 | 1090 | 7 | 2.24E-07 | 7.45E-09 | 0.000734 | 2.32E-08 | -0.00288696 | NA |
| chr9 | 136981129 | 136981832 | 704 | 4 | 6.52E-10 | 1.97E-08 | 0.000134 | 2.32E-08 | -0.02007562 | PTGDS, AL807752.7 |
| chr21 | 45072719 | 45073828 | 1110 | 6 | 1.75E-07 | 2.21E-08 | 0.001321 | 2.32E-08 | 0.008378518 | SSR4P1 |
| chr2 | 182522201 | 182523432 | 1232 | 9 | 1.55E-07 | 5.51E-09 | 0.001166 | 2.33E-08 | -0.0073509 | PDE1A |
| chr6 | 147142327 | 147142843 | 517 | 2 | 1.77E-09 | 6.81E-08 | 1.01E-06 | 2.34E-08 | 0.014772889 | STXBP5-AS1 |
| chr7 | 149734176 | 149734623 | 448 | 2 | 2.23E-10 | 2.50E-07 | 3.09E-07 | 2.34E-08 | -0.00407929 | KRBA1 |
| chr19 | 12665911 | 12667089 | 1179 | 21 | 4.76E-10 | 8.83E-06 | 1.54E-05 | 2.34E-08 | 7.48E-05 | WDR83, MAN2B1, AC010422.5 |
| chr11 | 61772978 | 61773956 | 979 | 6 | 1.01E-07 | 2.29E-07 | 6.07E-05 | 2.34E-08 | 0.000509768 | MYRF, TMEM258 |
| chr4 | 122620132 | 122621393 | 1262 | 7 | 8.42E-08 | 4.98E-07 | 7.32E-06 | 2.35E-08 | -0.00765041 | IL21-AS1, IL21 |
| chr1 | 175192390 | 175193417 | 1028 | 18 | 6.06E-11 | 9.92E-06 | 6.14E-05 | 2.35E-08 | -0.00211932 | KIAA0040 |
| chr3 | 133783201 | 133784593 | 1393 | 13 | 1.76E-08 | 4.48E-09 | 0.010419 | 2.35E-08 | -0.02341027 | TF, SRPRB |
| chr19 | 5804237 | 5806293 | 2057 | 8 | 5.98E-07 | 5.12E-07 | 0.000111 | 2.35E-08 | -0.00438817 | NA |
| chr14 | 88326593 | 88327397 | 805 | 13 | 6.91E-09 | 1.38E-07 | 0.00061 | 2.35E-08 | 0.003250842 | KCNK10 |
| chr21 | 42395576 | 42397140 | 1565 | 12 | 3.11E-06 | 3.93E-08 | 0.000698 | 2.36E-08 | -0.0007955 | TMPRSS3 |
| chr17 | 328906 | 330156 | 1251 | 4 | 1.03E-07 | 6.12E-07 | 7.62E-05 | 2.36E-08 | 0.008330222 | RPH3AL |
| chr2 | 74747604 | 74748109 | 506 | 2 | 8.61E-09 | 1.57E-08 | 1.79E-05 | 2.36E-08 | -0.01332297 | AC007387.2 |
| chr5 | 94111249 | 94112751 | 1503 | 14 | 1.76E-10 | 1.24E-06 | 4.11E-05 | 2.37E-08 | 0.004290762 | AC117528.1, FAM172A |
| chr19 | 23003219 | 23004081 | 863 | 4 | 1.51E-08 | 3.60E-07 | 3.30E-05 | 2.37E-08 | -0.01417841 | NA |
| chr1 | 234607366 | 234609032 | 1667 | 11 | 5.98E-07 | 1.58E-08 | 0.000706 | 2.37E-08 | -0.00050439 | AL160408.2, IRF2BP2 |
| chr7 | 142712124 | 142712475 | 352 | 2 | 1.93E-08 | 1.52E-08 | 2.18E-05 | 2.38E-08 | 0.035200446 | NA |
| chr6 | 7313227 | 7315273 | 2047 | 8 | 7.52E-08 | 0.0002099 | 2.61E-05 | 2.39E-08 | 0.00715229 | SSR1 |
| chr13 | 113650346 | 113651133 | 788 | 6 | 1.16E-10 | 3.56E-07 | 3.89E-05 | 2.39E-08 | -0.00324938 | ATP4B |
| chr19 | 41843872 | 41845409 | 1538 | 18 | 3.85E-09 | 8.24E-06 | 4.72E-05 | 2.39E-08 | -0.0008317 | DMRTC2, LYPD4 |
| chr14 | 64541923 | 64542989 | 1067 | 4 | 3.49E-09 | 8.66E-08 | 2.80E-05 | 2.40E-08 | -0.01167018 | HSPA2 |
| chr11 | 70102945 | 70104965 | 2021 | 11 | 3.80E-06 | 1.76E-08 | 0.000894 | 2.40E-08 | 0.001743224 | ANO1 |
| chr9 | 137229406 | 137230293 | 888 | 5 | 1.03E-10 | 3.36E-05 | 7.70E-05 | 2.40E-08 | 0.000457495 | RNF224 |
| chr20 | 48739629 | 48740677 | 1049 | 4 | 1.05E-07 | 1.09E-08 | 5.90E-05 | 2.40E-08 | 0.008954309 | PREX1 |
| chr17 | 39669910 | 39671416 | 1507 | 6 | 3.83E-07 | 1.12E-08 | 0.000325 | 2.40E-08 | 0.005914257 | PNMT, PGAP3 |
| chr12 | 2003953 | 2005116 | 1164 | 14 | 6.86E-09 | 3.05E-07 | 2.05E-05 | 2.40E-08 | -0.00234631 | CACNA1C, AC005342.1, DCP1B |
| chr1 | 53530997 | 53531580 | 584 | 3 | 5.88E-08 | 7.28E-08 | 3.40E-05 | 2.40E-08 | -0.00503761 | GLIS1 |
| chr3 | 128567811 | 128568517 | 707 | 3 | 1.04E-12 | 0.000292 | 1.97E-08 | 2.40E-08 | -0.00903967 | NA |
| chr10 | 25819603 | 25821221 | 1619 | 6 | 4.50E-08 | 3.37E-07 | 1.39E-05 | 2.40E-08 | -0.00417389 | NA |
| chr15 | 74201900 | 74203699 | 1800 | 17 | 2.01E-07 | 1.05E-06 | 0.000873 | 2.40E-08 | 0.004624209 | CCDC33, STRA6 |
| chr12 | 105676692 | 105676749 | 58 | 2 | 1.09E-08 | 1.52E-08 | 2.33E-05 | 2.41E-08 | -0.03554216 | NA |
| chr4 | 52751962 | 52752964 | 1003 | 4 | 6.94E-08 | 3.56E-07 | 4.43E-05 | 2.41E-08 | -0.00238731 | LINC01618 |
| chr2 | 731215 | 731561 | 347 | 5 | 5.91E-08 | 4.25E-09 | 0.00062 | 2.41E-08 | 0.050841741 | AC092159.1 |
| chr19 | 16566277 | 16567073 | 797 | 4 | 1.59E-07 | 8.43E-08 | 5.58E-05 | 2.41E-08 | -0.00792488 | AC008764.5, AC008764.1, SLC35E1 |
| chr5 | 134435318 | 134436697 | 1380 | 6 | 1.75E-08 | 6.50E-07 | 0.000125 | 2.41E-08 | -0.00705123 | AC109454.3 |
| chr11 | 73977517 | 73978397 | 881 | 4 | 2.04E-10 | 8.18E-07 | 3.33E-07 | 2.42E-08 | -0.00390963 | UCP2 |
| chr1 | 25812922 | 25814430 | 1509 | 7 | 1.49E-08 | 2.92E-06 | 6.51E-05 | 2.42E-08 | 0.000922403 | SELENON, AL020996.2 |
| chr10 | 963059 | 963478 | 420 | 3 | 8.93E-09 | 9.84E-08 | 5.70E-06 | 2.43E-08 | -0.00885185 | NA |
| chr3 | 52532585 | 52533617 | 1033 | 8 | 1.61E-09 | 1.18E-05 | 9.60E-06 | 2.43E-08 | -0.00551221 | NT5DC2 |
| chr17 | 47188380 | 47189773 | 1394 | 16 | 1.80E-11 | 0.0005204 | 6.80E-06 | 2.43E-08 | 0.000442123 | CDC27 |
| chr15 | 92711119 | 92711704 | 586 | 2 | 2.67E-09 | 5.15E-08 | 1.53E-06 | 2.43E-08 | -0.0069296 | FAM174B, AC091544.6 |
| chr18 | 10702002 | 10702427 | 426 | 5 | 2.34E-09 | 6.70E-06 | 2.29E-05 | 2.43E-08 | -0.0280458 | PIEZO2 |
| chr15 | 89003871 | 89004342 | 472 | 2 | 3.11E-10 | 4.13E-07 | 2.31E-07 | 2.45E-08 | -0.00667861 | NA |
| chr1 | 40708587 | 40709527 | 941 | 9 | 1.69E-08 | 2.66E-08 | 0.002545 | 2.45E-08 | 0.008023686 | NFYC |
| chr14 | 88562057 | 88563581 | 1525 | 18 | 2.57E-08 | 1.55E-07 | 0.005319 | 2.45E-08 | -0.00108554 | ZC3H14 |
| chr11 | 36744755 | 36745461 | 707 | 3 | 9.68E-09 | 4.55E-05 | 9.58E-07 | 2.45E-08 | 0.001905693 | NA |
| chr10 | 85448307 | 85449126 | 820 | 2 | 1.59E-08 | 3.02E-08 | 3.51E-06 | 2.46E-08 | -0.01187752 | AC025428.1 |
| chr4 | 2445173 | 2445874 | 702 | 2 | 4.33E-08 | 1.87E-08 | 1.08E-05 | 2.46E-08 | -0.00758542 | CFAP99 |
| chr9 | 121733556 | 121735702 | 2147 | 6 | 5.65E-07 | 2.40E-08 | 0.000342 | 2.46E-08 | -0.00207256 | DAB2IP |
| chr6 | 30619720 | 30620416 | 697 | 8 | 3.23E-09 | 2.84E-07 | 0.000614 | 2.47E-08 | -0.00199475 | MRPS18B |
| chr17 | 7266255 | 7266733 | 479 | 6 | 4.58E-08 | 5.71E-09 | 0.001309 | 2.47E-08 | -0.00897163 | NA |
| chr19 | 58115930 | 58116620 | 691 | 4 | 3.59E-09 | 6.57E-07 | 2.98E-06 | 2.47E-08 | 0.006992835 | ZSCAN18 |
| chr21 | 34807738 | 34808749 | 1012 | 3 | 2.28E-07 | 1.04E-08 | 9.42E-05 | 2.47E-08 | -0.02204499 | RUNX1 |
| chr8 | 41659666 | 41661201 | 1536 | 10 | 1.62E-07 | 1.91E-05 | 6.82E-06 | 2.47E-08 | -0.00071193 | MIR486-2, AC113133.1, ANK1, MIR486-1 |
| chr1 | 47710822 | 47711589 | 768 | 5 | 3.71E-10 | 3.06E-06 | 1.66E-05 | 2.48E-08 | -0.00855338 | NA |
| chr10 | 73839516 | 73841204 | 1689 | 7 | 2.56E-07 | 9.27E-07 | 0.000263 | 2.48E-08 | -0.00366312 | CAMK2G |
| chr5 | 194764 | 196047 | 1284 | 9 | 3.95E-10 | 7.39E-07 | 3.17E-05 | 2.48E-08 | -0.00039384 | LRRC14B, AC021087.4 |
| chr19 | 1167773 | 1168842 | 1070 | 5 | 1.11E-09 | 1.30E-07 | 2.56E-05 | 2.48E-08 | 0.001307317 | SBNO2 |
| chr10 | 22554678 | 22555009 | 332 | 3 | 6.02E-10 | 1.66E-05 | 6.47E-08 | 2.49E-08 | -0.00358728 | PIP4K2A |
| chr20 | 18507127 | 18508001 | 875 | 19 | 9.67E-10 | 5.58E-05 | 0.001372 | 2.49E-08 | 0.000301815 | SEC23B |
| chr2 | 200811349 | 200812855 | 1507 | 16 | 1.49E-09 | 1.69E-06 | 0.000303 | 2.49E-08 | 0.000358118 | BZW1, BZW1-AS1 |
| chr7 | 332436 | 333265 | 830 | 20 | 1.86E-09 | 1.30E-05 | 0.000516 | 2.49E-08 | -0.00356563 | AC188616.1 |
| chr5 | 174336052 | 174337097 | 1046 | 4 | 6.59E-10 | 4.43E-06 | 7.04E-07 | 2.49E-08 | -0.00852594 | LINC01411 |
| chr2 | 1540224 | 1541302 | 1079 | 8 | 3.49E-08 | 2.61E-08 | 0.001003 | 2.50E-08 | -0.00162027 | TPO, AC105450.1 |
| chr4 | 123049 | 123061 | 13 | 2 | 7.50E-09 | 1.99E-08 | 9.33E-06 | 2.50E-08 | -0.01559921 | NA |
| chr2 | 172109286 | 172110232 | 947 | 7 | 3.27E-08 | 2.87E-08 | 0.00216 | 2.50E-08 | 0.015384358 | DLX2-DT |
| chr8 | 143272698 | 143273712 | 1015 | 4 | 1.52E-08 | 8.23E-08 | 0.000126 | 2.50E-08 | -0.00120569 | AC138696.1, GLI4 |
| chr17 | 43029481 | 43029974 | 494 | 3 | 1.99E-08 | 1.62E-07 | 3.07E-05 | 2.51E-08 | 0.015690163 | RND2 |
| chr5 | 149296574 | 149297289 | 716 | 2 | 1.45E-11 | 1.64E-05 | 5.93E-08 | 2.51E-08 | 0.003063048 | AFAP1L1 |
| chr5 | 31193387 | 31194642 | 1256 | 12 | 1.31E-08 | 4.85E-06 | 0.000396 | 2.51E-08 | 0.001708234 | CDH6, AC113386.1 |
| chr4 | 94252129 | 94253065 | 937 | 5 | 1.69E-08 | 8.54E-08 | 3.63E-05 | 2.52E-08 | 0.001711127 | SMARCAD1 |
| chr2 | 165294343 | 165295308 | 966 | 4 | 3.38E-09 | 1.71E-07 | 2.81E-06 | 2.53E-08 | -0.01537636 | SCN2A |
| chr17 | 43483566 | 43484672 | 1107 | 15 | 1.19E-09 | 1.89E-05 | 1.25E-05 | 2.53E-08 | 0.001339262 | DHX8 |
| chr16 | 51113550 | 51114449 | 900 | 7 | 4.03E-08 | 2.49E-08 | 0.000356 | 2.53E-08 | -0.00157083 | NA |
| chr11 | 7673293 | 7674567 | 1275 | 12 | 1.20E-10 | 2.10E-05 | 1.47E-06 | 2.53E-08 | 0.001292893 | CYB5R2 |
| chr17 | 7178575 | 7180200 | 1626 | 11 | 1.36E-09 | 8.78E-08 | 0.000728 | 2.53E-08 | -0.00484329 | ASGR1 |
| chr4 | 4667402 | 4667909 | 508 | 3 | 2.32E-08 | 1.39E-08 | 9.69E-05 | 2.53E-08 | -0.00577156 | STX18-AS1 |
| chr7 | 27169266 | 27170655 | 1390 | 17 | 4.83E-08 | 4.94E-07 | 0.000967 | 2.54E-08 | -0.00014236 | HOXA10-AS, HOXA9, AC004080.3, MIR196B, HOXA10 |
| chr10 | 110501109 | 110501859 | 751 | 3 | 1.14E-09 | 5.18E-08 | 5.22E-06 | 2.55E-08 | -0.00553954 | DUSP5 |

| | | | | | | | | | | |
|---|---|---|---|---|---|---|---|---|---|---|
| chr20 | 4845146 | 4845354 | 209 | 2 | 5.39E-09 | 1.53E-07 | 5.02E-07 | 2.56E-08 | 0.038025809 | NA |
| chr15 | 47411752 | 47412313 | 562 | 5 | 6.32E-09 | 8.86E-07 | 4.53E-05 | 2.56E-08 | 9.53E-05 | SEMA6D |
| chr19 | 38432887 | 38433796 | 910 | 9 | 1.40E-07 | 9.37E-08 | 0.00105 | 2.57E-08 | 0.000603783 | RYR1 |
| chr22 | 21739214 | 21739694 | 481 | 2 | 1.23E-08 | 1.54E-08 | 3.35E-05 | 2.57E-08 | -0.02040292 | AP000553.8 |
| chr11 | 18321453 | 18322880 | 1428 | 17 | 5.67E-07 | 3.71E-07 | 0.000256 | 2.57E-08 | -0.00148147 | GTF2H1, HPS5 |
| chr5 | 88684481 | 88686334 | 1854 | 11 | 2.57E-08 | 2.85E-06 | 2.32E-05 | 2.57E-08 | -0.00355884 | MEF2C-AS2, LINC00461 |
| chr10 | 101061670 | 101063051 | 1382 | 13 | 2.18E-11 | 1.18E-05 | 0.00026 | 2.58E-08 | -0.00433729 | KAZALD1 |
| chr17 | 30377781 | 30378824 | 1044 | 7 | 1.51E-10 | 0.0003627 | 1.13E-05 | 2.58E-08 | 0.00818871 | NA |
| chr5 | 119470139 | 119471016 | 878 | 4 | 1.52E-09 | 1.61E-06 | 2.63E-06 | 2.58E-08 | -0.01074193 | HSD17B4 |
| chr5 | 177090235 | 177092454 | 2220 | 13 | 1.16E-06 | 1.28E-07 | 0.000754 | 2.58E-08 | -0.00400341 | FGFR4 |
| chr13 | 100588400 | 100589337 | 938 | 11 | 9.21E-09 | 4.99E-08 | 0.000412 | 2.59E-08 | 0.000883207 | GGACT |
| chr20 | 33673751 | 33675685 | 1935 | 13 | 1.70E-06 | 1.95E-07 | 0.000292 | 2.59E-08 | -0.00367312 | AL121906.1, NECAB3, E2F1 |
| chr15 | 77604864 | 77605841 | 978 | 3 | 1.89E-07 | 1.13E-08 | 7.02E-05 | 2.60E-08 | -0.01236775 | AC046168.2 |
| chr19 | 49426596 | 49427511 | 916 | 3 | 2.72E-09 | 1.66E-08 | 2.75E-05 | 2.60E-08 | -0.01397649 | GFY |
| chr8 | 48427936 | 48428462 | 527 | 4 | 6.02E-08 | 1.03E-08 | 0.00013 | 2.61E-08 | -0.00024357 | AC041040.1 |
| chr19 | 49961456 | 49961991 | 536 | 4 | 1.06E-09 | 1.57E-06 | 5.16E-07 | 2.61E-08 | -0.00994892 | NA |
| chr1 | 207817550 | 207818435 | 886 | 7 | 2.93E-11 | 1.45E-05 | 1.69E-05 | 2.62E-08 | 0.004704488 | MIR29B2CHG |
| chr9 | 37746065 | 37746779 | 715 | 3 | 3.04E-08 | 7.39E-08 | 3.69E-05 | 2.62E-08 | -0.00787793 | FRMPD1, AL138752.2 |
| chr16 | 68022472 | 68024008 | 1537 | 23 | 5.63E-10 | 1.47E-06 | 0.001103 | 2.62E-08 | 0.000661154 | DUS2, DDX28 |
| chr10 | 94544436 | 94545497 | 1062 | 6 | 7.15E-08 | 2.09E-06 | 0.000128 | 2.62E-08 | -0.00054136 | HELLS |
| chr20 | 59105656 | 59106630 | 975 | 3 | 2.74E-11 | 3.09E-05 | 5.39E-08 | 2.62E-08 | -0.00893426 | NA |
| chr1 | 246147109 | 246147844 | 736 | 3 | 2.26E-07 | 1.36E-08 | 0.000103 | 2.62E-08 | 0.002499089 | SMYD3 |
| chr16 | 979745 | 980619 | 875 | 10 | 5.88E-09 | 5.28E-09 | 0.001763 | 2.62E-08 | 0.005660207 | LMF1, CEROX1 |
| chr17 | 28721419 | 28722638 | 1220 | 7 | 1.67E-09 | 4.22E-08 | 0.00032 | 2.63E-08 | -0.00842316 | RPL23A, SNORD4A, AC010761.1 |
| chr1 | 227317020 | 227318320 | 1301 | 10 | 2.13E-07 | 3.67E-06 | 0.000293 | 2.63E-08 | -0.00319759 | CDC42BPA |
| chr3 | 72655023 | 72655986 | 964 | 8 | 2.42E-08 | 1.72E-07 | 0.000593 | 2.63E-08 | -0.01072989 | NA |
| chr7 | 44218112 | 44218455 | 344 | 2 | 1.01E-11 | 6.85E-07 | 1.87E-07 | 2.63E-08 | -0.01654358 | CAMK2B |
| chr2 | 88590998 | 88591020 | 23 | 2 | 2.67E-08 | 1.61E-08 | 2.97E-05 | 2.63E-08 | -0.0058082 | EIF2AK3 |
| chr22 | 19785835 | 19786375 | 541 | 3 | 6.24E-08 | 9.53E-09 | 0.000171 | 2.63E-08 | -0.01229863 | GNB1L |
| chr15 | 63276971 | 63277900 | 930 | 9 | 3.28E-09 | 4.33E-06 | 4.95E-05 | 2.64E-08 | 0.004559351 | APH1B |
| chr1 | 38871834 | 38873553 | 1720 | 11 | 6.07E-07 | 1.92E-06 | 0.000186 | 2.64E-08 | -0.00147508 | AL139260.1, AL139260.2, MYCBP, AL139260.3 |
| chr5 | 91311634 | 91313126 | 1493 | 5 | 5.67E-07 | 6.26E-08 | 5.73E-05 | 2.64E-08 | -0.00497951 | LUCAT1 |
| chr22 | 38769004 | 38769200 | 197 | 3 | 7.77E-09 | 8.40E-08 | 7.14E-06 | 2.64E-08 | -0.01402869 | SUN2 |
| chr9 | 86413870 | 86414225 | 356 | 4 | 3.23E-08 | 4.83E-08 | 9.21E-05 | 2.64E-08 | -0.00982827 | AL158828.1 |
| chr7 | 100634049 | 100634856 | 808 | 6 | 6.61E-11 | 1.41E-05 | 7.90E-07 | 2.64E-08 | -0.00227112 | TFR2 |
| chr1 | 111226215 | 111227669 | 1455 | 7 | 3.38E-10 | 6.34E-07 | 0.000339 | 2.64E-08 | -0.00944755 | CHI3L2 |
| chr8 | 93740578 | 93742010 | 1433 | 16 | 1.57E-08 | 1.07E-07 | 5.96E-05 | 2.64E-08 | -0.00097861 | RBM12B-AS1, AC010834.2, RBM12B |
| chr15 | 67109413 | 67110007 | 595 | 2 | 2.10E-08 | 1.62E-08 | 2.89E-05 | 2.64E-08 | 0.014231177 | SMAD3 |
| chr2 | 178522729 | 178523551 | 823 | 6 | 4.11E-08 | 1.15E-08 | 0.000331 | 2.64E-08 | -0.02146522 | TTN-AS1 |
| chr6 | 39901204 | 39902064 | 861 | 6 | 1.52E-07 | 8.25E-08 | 4.15E-05 | 2.64E-08 | -0.01065519 | DAAM2, MOCS1 |
| chr1 | 6624138 | 6625564 | 1427 | 10 | 1.69E-09 | 1.67E-05 | 1.12E-05 | 2.65E-08 | -0.00407613 | THAP3 |
| chr14 | 100569481 | 100570927 | 1447 | 11 | 4.00E-07 | 6.06E-07 | 0.000222 | 2.65E-08 | 0.012627613 | BEGAIN |
| chr11 | 65961255 | 65962164 | 910 | 9 | 2.01E-08 | 4.22E-06 | 7.21E-05 | 2.66E-08 | 0.000525555 | SART1 |
| chr17 | 64781180 | 64782295 | 1116 | 5 | 8.02E-09 | 3.45E-07 | 6.74E-06 | 2.66E-08 | -0.00404668 | ARHGAP27P1-BPTFP1-KPNA2P3, PLEKHM1P1 |
| chr20 | 36669644 | 36669664 | 21 | 3 | 1.57E-08 | 1.72E-07 | 2.78E-06 | 2.66E-08 | -0.00490991 | NDRG3 |
| chr1 | 32983290 | 32984532 | 1243 | 3 | 2.11E-06 | 2.34E-08 | 4.29E-05 | 2.66E-08 | -0.00225958 | NA |
| chr4 | 780605 | 781912 | 1308 | 6 | 4.40E-08 | 1.73E-06 | 0.000122 | 2.67E-08 | -0.00086162 | AC139887.2 |
| chr10 | 133179642 | 133180951 | 1310 | 9 | 5.08E-10 | 3.24E-06 | 7.32E-05 | 2.67E-08 | -0.00565449 | KNDC1 |
| chr2 | 112977800 | 112978467 | 668 | 6 | 8.20E-09 | 6.38E-08 | 7.53E-06 | 2.68E-08 | -0.00445429 | IL36G |
| chr9 | 34956994 | 34958144 | 1151 | 10 | 2.50E-10 | 0.0001042 | 3.31E-05 | 2.68E-08 | -0.00455812 | PHF24 |
| chr3 | 51526472 | 51527195 | 724 | 2 | 1.40E-11 | 4.32E-05 | 5.39E-08 | 2.68E-08 | -0.02063639 | NA |
| chr16 | 20740478 | 20742155 | 1678 | 10 | 3.80E-07 | 6.05E-06 | 1.78E-05 | 2.69E-08 | 0.001498549 | ACSM3, THUMPD1 |
| chr12 | 6752129 | 6753849 | 1721 | 16 | 1.66E-06 | 1.32E-07 | 0.002644 | 2.71E-08 | -0.00109042 | MLF2 |
| chr7 | 34041282 | 34042019 | 738 | 3 | 2.72E-10 | 9.83E-08 | 2.32E-06 | 2.71E-08 | 0.001308811 | BMPER |
| chr11 | 109122264 | 109122396 | 133 | 2 | 3.06E-09 | 5.93E-07 | 2.09E-07 | 2.72E-08 | 0.011571829 | NA |
| chr19 | 4677590 | 4679289 | 1700 | 11 | 4.16E-09 | 7.53E-08 | 0.00026 | 2.72E-08 | -0.00039974 | DPP9-AS1, DPP9 |
| chr19 | 12610095 | 12611856 | 1762 | 14 | 2.59E-08 | 8.30E-07 | 0.000437 | 2.72E-08 | 0.000533355 | ZNF791, ZNF490 |
| chr7 | 35538040 | 35538900 | 861 | 2 | 4.82E-12 | 1.48E-05 | 6.46E-08 | 2.72E-08 | -0.01170038 | NA |
| chr14 | 91888798 | 91889510 | 713 | 3 | 3.81E-10 | 7.29E-06 | 8.81E-07 | 2.72E-08 | -0.00417062 | FBLN5 |
| chr19 | 43745256 | 43745604 | 349 | 2 | 6.84E-09 | 4.73E-08 | 2.13E-06 | 2.72E-08 | -0.01540433 | SMG9 |
| chr13 | 35474755 | 35476936 | 2182 | 15 | 2.60E-06 | 2.09E-06 | 0.000479 | 2.72E-08 | -0.00606898 | NBEA, MAB21L1 |
| chr6 | 32894086 | 32895176 | 1091 | 28 | 4.56E-10 | 7.89E-06 | 0.000243 | 2.72E-08 | 9.64E-05 | NA |
| chr2 | 207978416 | 207979272 | 857 | 6 | 1.73E-07 | 8.73E-06 | 6.41E-05 | 2.73E-08 | -0.00308482 | PLEKHM3 |
| chr19 | 37655238 | 37656408 | 1171 | 12 | 5.44E-08 | 4.32E-07 | 0.000836 | 2.73E-08 | -8.49E-05 | ZFP30 |
| chr8 | 93705587 | 93706148 | 562 | 2 | 1.46E-10 | 1.78E-07 | 4.88E-07 | 2.74E-08 | 0.00093927 | CIBAR1 |
| chr1 | 58814618 | 58816250 | 1633 | 10 | 3.04E-08 | 5.32E-07 | 5.06E-05 | 2.74E-08 | -0.00235101 | LINC01135 |
| chr17 | 80781642 | 80782041 | 400 | 3 | 9.92E-10 | 7.38E-08 | 1.48E-05 | 2.74E-08 | -0.01953284 | RPTOR |
| chr6 | 36026984 | 36028982 | 1999 | 23 | 1.20E-08 | 8.00E-07 | 0.000121 | 2.75E-08 | 0.00129607 | MAPK14 |
| chr1 | 9234215 | 9234854 | 640 | 6 | 6.58E-09 | 1.47E-07 | 0.000257 | 2.76E-08 | -0.00528149 | H6PD |
| chr3 | 10570400 | 10571366 | 967 | 5 | 2.58E-07 | 2.10E-08 | 0.00053 | 2.76E-08 | -0.00654242 | ATP2B2 |
| chr19 | 56824439 | 56825165 | 727 | 5 | 8.74E-11 | 2.57E-05 | 3.27E-07 | 2.77E-08 | -0.0062949 | ZIM2, PEG3 |
| chr16 | 35752137 | 35754351 | 2215 | 12 | 2.65E-08 | 3.21E-05 | 0.000114 | 2.77E-08 | -0.00192632 | LINC02167, RNA5SP418, RNA5SP419, RNA5SP420 |
| chr14 | 45274821 | 45275177 | 357 | 3 | 4.66E-09 | 4.48E-07 | 2.16E-06 | 2.77E-08 | -0.00070984 | NA |
| chr4 | 82515846 | 82516259 | 414 | 2 | 1.49E-09 | 8.46E-08 | 1.03E-06 | 2.77E-08 | 0.014705603 | TMEM150C |
| chr8 | 98245295 | 98246230 | 936 | 2 | 3.29E-07 | 1.88E-08 | 1.74E-05 | 2.78E-08 | 0.005926362 | NIPAL2 |
| chr15 | 75043869 | 75044095 | 227 | 3 | 3.53E-10 | 1.58E-07 | 2.33E-05 | 2.78E-08 | 0.010993575 | PPCDC |
| chr16 | 67172831 | 67173837 | 1007 | 6 | 7.57E-10 | 1.20E-06 | 4.64E-05 | 2.78E-08 | -0.00666073 | NOL3 |
| chr1 | 100265126 | 100266734 | 1609 | 15 | 1.68E-07 | 3.39E-06 | 0.000683 | 2.78E-08 | -0.00086353 | RTCA, RTCA-AS1 |
| chr7 | 100611162 | 100612720 | 1559 | 16 | 7.39E-07 | 3.64E-09 | 0.00899 | 2.79E-08 | -0.00069383 | MOSPD3 |
| chr2 | 177332552 | 177333034 | 483 | 2 | 6.41E-11 | 2.04E-05 | 6.25E-08 | 2.79E-08 | -0.01435517 | NFE2L2, AC019080.1 |
| chr1 | 99790365 | 99791511 | 1147 | 4 | 6.80E-08 | 4.23E-06 | 7.70E-06 | 2.79E-08 | -0.00131525 | NA |
| chr10 | 526127 | 527393 | 1267 | 5 | 3.39E-09 | 8.81E-07 | 6.39E-05 | 2.79E-08 | 0.000253643 | DIP2C |
| chr10 | 125024146 | 125025444 | 1299 | 7 | 5.67E-09 | 5.39E-08 | 0.000356 | 2.79E-08 | -0.00160146 | CTBP2 |
| chr21 | 45257165 | 45257964 | 800 | 5 | 2.57E-08 | 1.56E-08 | 0.000227 | 2.79E-08 | -0.02601939 | LINC00334 |
| chr17 | 50560730 | 50562896 | 2167 | 14 | 3.22E-06 | 1.98E-08 | 0.001875 | 2.80E-08 | -0.00070276 | CACNA1G, CACNA1G-AS1 |
| chr9 | 129493724 | 129494613 | 890 | 3 | 8.05E-10 | 3.79E-07 | 8.15E-07 | 2.81E-08 | -0.00493026 | LINC00963 |
| chr13 | 83878859 | 83880360 | 1502 | 6 | 9.53E-08 | 1.28E-07 | 0.000726 | 2.81E-08 | -0.00838209 | AL355481.1, SLITRK1 |
| chr1 | 40782537 | 40783209 | 673 | 3 | 1.83E-10 | 3.61E-07 | 1.74E-06 | 2.82E-08 | -0.01365459 | NA |
| chr9 | 128503505 | 128505724 | 2220 | 14 | 9.43E-07 | 2.16E-06 | 0.000561 | 2.82E-08 | -0.00064298 | GLE1 |
| chr2 | 200774515 | 200775464 | 950 | 4 | 1.14E-08 | 2.52E-06 | 1.86E-05 | 2.84E-08 | -0.00515858 | AOX2P |
| chr15 | 74824692 | 74826373 | 1682 | 8 | 2.30E-08 | 9.80E-07 | 0.000131 | 2.84E-08 | -0.00267094 | LMAN1L, AC091230.1 |
| chr1 | 246900735 | 246900840 | 106 | 3 | 4.00E-08 | 1.57E-08 | 7.64E-05 | 2.84E-08 | -0.00135541 | AHCTF1 |
| chr16 | 65123760 | 65124545 | 786 | 6 | 4.44E-09 | 3.55E-07 | 1.14E-05 | 2.84E-08 | -0.01094377 | CDH11 |
| chr21 | 33971679 | 33971798 | 120 | 2 | 1.44E-09 | 4.47E-05 | 5.57E-08 | 2.84E-08 | -0.00960772 | LINC00649 |
| chr10 | 11868942 | 11869637 | 696 | 6 | 2.56E-08 | 3.11E-08 | 7.60E-05 | 2.84E-08 | 0.001640628 | PROSER2, PROSER2-AS1 |
| chr6 | 170096333 | 170096729 | 397 | 5 | 6.71E-08 | 7.70E-09 | 0.000525 | 2.84E-08 | 0.01141299 | AL603783.1 |
| chr6 | 69796136 | 69797338 | 1203 | 11 | 3.54E-08 | 5.65E-07 | 0.001442 | 2.85E-08 | 0.001088031 | LMBRD1 |
| chr22 | 46296053 | 46297451 | 1399 | 17 | 6.38E-09 | 1.87E-05 | 4.68E-05 | 2.86E-08 | 0.000260322 | GTSE1, GTSE1-DT |
| chr10 | 29687960 | 29689215 | 1256 | 4 | 6.41E-09 | 6.76E-06 | 3.11E-06 | 2.86E-08 | -0.00791117 | SVIL |
| chr15 | 65212186 | 65212810 | 625 | 5 | 8.20E-09 | 3.86E-07 | 3.57E-05 | 2.86E-08 | -0.00411922 | NA |
| chr1 | 230746857 | 230748331 | 1475 | 9 | 4.21E-07 | 4.90E-07 | 0.001276 | 2.87E-08 | -0.00356304 | CAPN9, AL512328.1 |
| chr3 | 108153389 | 108153582 | 194 | 2 | 1.59E-08 | 1.75E-08 | 3.08E-05 | 2.88E-08 | -0.01983091 | NA |
| chr2 | 25303827 | 25305461 | 1635 | 8 | 4.29E-08 | 7.37E-07 | 0.000322 | 2.88E-08 | -0.00574499 | DNMT3A |
| chr7 | 108103404 | 108104642 | 1239 | 3 | 1.95E-13 | 5.63E-05 | 6.42E-08 | 2.88E-08 | -0.00632392 | LAMB4 |
| chr6 | 40276244 | 40277295 | 1052 | 6 | 1.59E-09 | 7.28E-08 | 1.46E-05 | 2.89E-08 | -0.00589893 | NA |
| chr19 | 18152540 | 18154213 | 1674 | 15 | 1.36E-06 | 2.57E-07 | 0.000287 | 2.90E-08 | 0.001909059 | AC007192.1, PIK3R2 |
| chr5 | 128082411 | 128084527 | 2117 | 17 | 1.52E-06 | 1.16E-07 | 0.003447 | 2.90E-08 | -0.00147632 | SLC12A2, LINC01184 |
| chr17 | 38584357 | 38585133 | 777 | 2 | 2.00E-09 | 6.72E-08 | 1.50E-06 | 2.91E-08 | 0.001057578 | SRCIN1 |
| chr16 | 89101582 | 89102881 | 1300 | 5 | 1.37E-09 | 1.38E-07 | 9.24E-06 | 2.92E-08 | 0.011554309 | ACSF3 |
| chr5 | 35617585 | 35618281 | 697 | 9 | 3.68E-09 | 1.76E-07 | 0.000552 | 2.92E-08 | -0.0056742 | SPEF2 |
| chr11 | 134261784 | 134262920 | 1137 | 6 | 3.23E-08 | 5.39E-08 | 1.95E-05 | 2.92E-08 | -0.00280614 | ACAD8 |
| chr7 | 44188162 | 44189610 | 1449 | 7 | 6.11E-07 | 4.39E-08 | 0.000603 | 2.93E-08 | -0.00471408 | GCK |
| chr17 | 39193532 | 39195503 | 1972 | 8 | 7.09E-07 | 6.25E-06 | 1.62E-05 | 2.93E-08 | -0.00390217 | CACNB1 |
| chr20 | 31473019 | 31473831 | 813 | 6 | 9.85E-09 | 5.49E-07 | 7.36E-05 | 2.93E-08 | -0.00762552 | DEFB124 |
| chr1 | 61076539 | 61077978 | 1440 | 17 | 1.74E-08 | 2.54E-06 | 3.67E-05 | 2.94E-08 | -0.00098318 | NFIA |
| chr19 | 47273872 | 47276106 | 2235 | 15 | 5.07E-06 | 3.84E-06 | 0.000981 | 2.94E-08 | -3.90E-05 | INAFM1 |
| chr8 | 30044434 | 30044714 | 281 | 3 | 1.41E-08 | 5.27E-08 | 1.33E-05 | 2.94E-08 | -0.01323044 | NA |
| chr20 | 36951765 | 36953154 | 1390 | 13 | 5.87E-07 | 4.82E-08 | 0.000707 | 2.95E-08 | -0.00086506 | SAMHD1 |
| chr9 | 138052150 | 138053085 | 936 | 3 | 5.88E-08 | 9.48E-09 | 0.000272 | 2.95E-08 | -0.00885615 | CACNA1B |
| chr15 | 60626480 | 60628418 | 1939 | 5 | 2.63E-05 | 9.74E-09 | 0.000894 | 2.95E-08 | 0.000528924 | RORA-AS1, RORA |
| chr17 | 4599876 | 4600326 | 451 | 2 | 2.98E-11 | 4.72E-07 | 2.66E-07 | 2.95E-08 | -0.0122745 | SMTNL2 |
| chr1 | 180229393 | 180231454 | 2062 | 11 | 6.90E-10 | 5.00E-05 | 9.19E-05 | 2.96E-08 | 0.001405447 | LHX4 |
| chr3 | 194275544 | 194276931 | 1388 | 5 | 1.10E-06 | 5.80E-07 | 2.69E-05 | 2.96E-08 | -0.00513789 | LINC02048 |
| chr8 | 58552571 | 58553918 | 1348 | 18 | 9.42E-09 | 1.58E-05 | 0.001021 | 2.97E-08 | -0.00059842 | SDCBP |
| chr8 | 90363619 | 90363951 | 333 | 3 | 9.76E-10 | 3.94E-05 | 1.77E-05 | 2.97E-08 | -0.01100668 | LINC00534, AC004083.1 |
| chr1 | 202212741 | 202214151 | 1411 | 8 | 5.08E-07 | 1.99E-06 | 0.000165 | 2.98E-08 | -0.01013191 | LGR6 |
| chr20 | 22584309 | 22586532 | 2224 | 16 | 5.46E-06 | 8.24E-06 | 0.000152 | 2.98E-08 | 0.001138068 | FOXA2 |
| chr2 | 24792148 | 24793713 | 1566 | 22 | 6.38E-08 | 3.67E-06 | 0.000386 | 2.98E-08 | -0.0007912 | CENPO, PTRHD1 |
| chr19 | 1523061 | 1524039 | 979 | 9 | 7.44E-10 | 3.60E-06 | 1.70E-05 | 2.99E-08 | -0.00491502 | PLK5 |
| chr13 | 113637131 | 113638425 | 1295 | 11 | 1.65E-08 | 2.18E-07 | 0.00019 | 2.99E-08 | -0.00248214 | TFDP1 |
| chr10 | 132322816 | 132323402 | 587 | 3 | 3.75E-09 | 3.71E-08 | 8.68E-06 | 2.99E-08 | 0.001311362 | STK32C |
| chr1 | 109426027 | 109427311 | 1285 | 17 | 2.44E-09 | 5.17E-07 | 0.002138 | 2.99E-08 | 0.000497752 | PSMA5 |
| chr3 | 123314240 | 123315021 | 782 | 2 | 1.44E-08 | 4.41E-08 | 2.88E-06 | 2.99E-08 | -0.01636913 | ADCY5 |
| chr1 | 206506247 | 206507415 | 1169 | 9 | 2.96E-10 | 5.82E-07 | 2.28E-05 | 3.00E-08 | -0.00163454 | NA |
| chr12 | 2811891 | 2813767 | 1877 | 17 | 9.92E-07 | 8.28E-07 | 0.000641 | 3.01E-08 | 4.32E-05 | ITFG2, ITFG2-AS1, ITFG2-AS1 |
| chr1 | 85263482 | 85264269 | 788 | 2 | 4.60E-08 | 2.94E-08 | 6.19E-06 | 3.01E-08 | -0.00241152 | NA |
| chr20 | 58793698 | 58794294 | 597 | 3 | 8.12E-10 | 6.68E-06 | 1.42E-05 | 3.02E-08 | -0.00786564 | NA |
| chr5 | 140793373 | 140795164 | 1792 | 10 | 2.10E-06 | 2.30E-07 | 0.00032 | 3.02E-08 | -0.00770655 | PCDHA1, PCDHA2, AC005609.5 |
| chr8 | 132127878 | 132127953 | 76 | 2 | 1.88E-09 | 2.89E-06 | 1.12E-07 | 3.02E-08 | -0.01539541 | KCNQ3 |
| chr2 | 1839088 | 1839664 | 577 | 4 | 6.70E-08 | 1.65E-08 | 0.000125 | 3.02E-08 | -0.04490756 | MYT1L |
| chr16 | 362168 | 362523 | 356 | 3 | 2.91E-09 | 4.40E-08 | 4.01E-05 | 3.02E-08 | -0.0231697 | NA |
| chr19 | 5130629 | 5130840 | 212 | 2 | 8.47E-09 | 2.24E-08 | 1.30E-05 | 3.02E-08 | 0.043680961 | KDM4B |
| chr8 | 100156803 | 100158693 | 1891 | 18 | 2.39E-07 | 3.44E-06 | 0.000137 | 3.02E-08 | 0.000149183 | SPAG1 |
| chr1 | 193058371 | 193059462 | 1092 | 12 | 1.17E-09 | 3.10E-05 | 7.02E-06 | 3.03E-08 | -0.00177725 | RO60, UCHL5 |
| chr21 | 46378364 | 46379348 | 985 | 7 | 2.55E-09 | 4.43E-06 | 4.41E-05 | 3.03E-08 | -0.00392767 | PCNT |
| chr7 | 100687340 | 100688643 | 1304 | 11 | 1.24E-07 | 8.78E-07 | 0.000374 | 3.03E-08 | 0.002075841 | GIGYF1 |
| chr16 | 1323607 | 1324748 | 1142 | 5 | 2.97E-10 | 8.20E-06 | 2.71E-07 | 3.04E-08 | 0.00551218 | UBE2I |
| chr12 | 56333520 | 56334709 | 1190 | 17 | 2.67E-08 | 1.51E-08 | 0.011427 | 3.04E-08 | -0.00034832 | IL23A, PAN2 |
| chr7 | 130667396 | 130669233 | 1838 | 20 | 3.48E-07 | 1.95E-06 | 0.000793 | 3.04E-08 | 0.000580764 | AC234644.2, COPG2, TSGA13 |
| chr13 | 21038742 | 21039277 | 536 | 5 | 1.46E-07 | 1.65E-08 | 0.000125 | 3.04E-08 | -0.00492925 | LATS2 |
| chr6 | 87151543 | 87152763 | 1221 | 6 | 4.87E-06 | 2.61E-08 | 0.000797 | 3.05E-08 | 0.011736742 | AL139274.2 |
| chr8 | 143288974 | 143289881 | 908 | 6 | 7.21E-09 | 1.02E-07 | 0.000949 | 3.05E-08 | -0.03113092 | ZNF696 |
| chr11 | 64121712 | 64123214 | 1503 | 5 | 6.25E-07 | 4.03E-07 | 1.43E-05 | 3.06E-08 | 0.002705442 | MACROD1 |
| chr3 | 160398811 | 160401078 | 2268 | 29 | 3.82E-06 | 5.32E-06 | 0.00148 | 3.06E-08 | 0.000285195 | SMC4, AC079594.1, IFT80 |
| chr20 | 57519469 | 57520332 | 864 | 5 | 2.83E-08 | 2.15E-08 | 0.000154 | 3.07E-08 | -0.00686243 | CTCFL |
| chr4 | 89837906 | 89838467 | 562 | 5 | 4.74E-12 | 5.27E-07 | 6.29E-06 | 3.07E-08 | -0.00897278 | SNCA-AS1, SNCA |
| chr8 | 101756454 | 101757129 | 676 | 2 | 1.58E-09 | 1.20E-07 | 8.39E-07 | 3.09E-08 | -0.01374106 | NCALD |
| chr20 | 51594624 | 51595256 | 633 | 4 | 1.73E-08 | 3.53E-08 | 5.65E-05 | 3.09E-08 | -0.00376573 | NA |

| | | | | | | | | | | |
|---|---|---|---|---|---|---|---|---|---|---|
| chr6 | 143570378 | 143570838 | 461 | 2 | 5.35E-11 | 8.31E-06 | 8.22E-08 | 3.09E-08 | 0.00067656 | PHACTR2 |
| chr10 | 122849900 | 122851484 | 1585 | 8 | 4.41E-07 | 5.59E-05 | 5.68E-05 | 3.09E-08 | -0.00080703 | AC073585.1, FAM24B |
| chr13 | 113109444 | 113111083 | 1640 | 6 | 5.65E-07 | 1.05E-08 | 0.000351 | 3.09E-08 | 3.70E-05 | F7 |
| chr16 | 86578383 | 86580125 | 1743 | 7 | 2.34E-06 | 9.62E-08 | 0.000603 | 3.09E-08 | -0.00563688 | FOXL1 |
| chr12 | 121849713 | 121850104 | 392 | 6 | 4.11E-08 | 5.19E-08 | 0.000111 | 3.09E-08 | -0.01470229 | HPD |
| chr9 | 133635112 | 133636298 | 1187 | 8 | 2.45E-08 | 8.40E-08 | 0.000616 | 3.09E-08 | -0.00451551 | NA |
| chr10 | 101817836 | 101819554 | 1719 | 12 | 3.51E-08 | 3.50E-06 | 0.000177 | 3.10E-08 | 0.000510334 | KCNIP2-AS1, OGA |
| chr6 | 31669136 | 31670239 | 1104 | 9 | 3.58E-10 | 2.45E-06 | 3.62E-05 | 3.10E-08 | -0.0023726 | CSNK2B, AL662899.2, LY6G5B |
| chr4 | 114597639 | 114598437 | 799 | 11 | 1.96E-09 | 3.34E-07 | 0.000366 | 3.10E-08 | 0.007222522 | NA |
| chr3 | 47802873 | 47804182 | 1310 | 8 | 1.78E-08 | 4.34E-07 | 0.001777 | 3.10E-08 | 0.001388382 | DHX30 |
| chr11 | 62601922 | 62602398 | 477 | 6 | 5.70E-10 | 2.79E-05 | 0.000125 | 3.11E-08 | -0.01514247 | EML3 |
| chr7 | 55498086 | 55498928 | 843 | 3 | 3.02E-09 | 1.94E-07 | 2.94E-05 | 3.11E-08 | -0.01169386 | VOPP1 |
| chr18 | 76379437 | 76380959 | 1523 | 6 | 1.58E-08 | 2.19E-07 | 0.000255 | 3.12E-08 | -0.0009788 | ZNF516 |
| chr2 | 235502556 | 235504268 | 1713 | 4 | 2.20E-05 | 4.60E-08 | 0.000116 | 3.13E-08 | 0.021677894 | AGAP1 |
| chr1 | 42754125 | 42755004 | 880 | 5 | 1.20E-07 | 5.41E-06 | 1.83E-05 | 3.13E-08 | -0.00279313 | P3H1 |
| chr9 | 91978506 | 91979495 | 990 | 11 | 1.63E-08 | 1.61E-05 | 9.60E-05 | 3.13E-08 | -0.00472599 | NA |
| chr6 | 163635900 | 163637007 | 1108 | 4 | 9.91E-07 | 3.98E-08 | 4.82E-05 | 3.14E-08 | -0.00877386 | NA |
| chr19 | 50288592 | 50289279 | 688 | 2 | 1.81E-09 | 9.64E-08 | 1.09E-06 | 3.14E-08 | -0.01451575 | MYH14 |
| chr2 | 216498227 | 216499752 | 1526 | 11 | 7.15E-08 | 9.84E-06 | 0.000242 | 3.14E-08 | -0.00312821 | RPL37A, RPL37A-DT |
| chr3 | 195097375 | 195097399 | 25 | 2 | 6.49E-09 | 5.00E-08 | 2.59E-06 | 3.15E-08 | -0.00604605 | XXYLT1 |
| chr14 | 74909798 | 74910688 | 891 | 2 | 1.33E-07 | 2.05E-08 | 2.23E-05 | 3.15E-08 | -0.00775485 | RPS6KL1 |
| chr6 | 97669013 | 97669665 | 653 | 3 | 1.86E-10 | 3.95E-06 | 2.06E-07 | 3.16E-08 | 0.021896549 | AL589740.1 |
| chr2 | 136327332 | 136327496 | 165 | 2 | 3.36E-08 | 1.89E-08 | 3.76E-05 | 3.16E-08 | 0.026157163 | NA |
| chr11 | 94088856 | 94089574 | 719 | 5 | 1.84E-09 | 1.26E-05 | 1.19E-05 | 3.16E-08 | -0.01668694 | HEPHL1 |
| chr15 | 67135786 | 67135799 | 14 | 2 | 1.32E-08 | 3.13E-08 | 6.16E-06 | 3.17E-08 | -0.02098166 | SMAD3 |
| chr5 | 154754947 | 154756694 | 1748 | 9 | 7.44E-07 | 2.16E-07 | 0.000891 | 3.17E-08 | -0.00131031 | LARP1 |
| chr2 | 218672252 | 218673489 | 1238 | 6 | 3.69E-10 | 6.29E-08 | 0.000267 | 3.18E-08 | -0.00922238 | STK36 |
| chr13 | 49401247 | 49402974 | 1728 | 10 | 3.92E-06 | 4.28E-07 | 0.000281 | 3.18E-08 | -0.00577897 | CAB39L |
| chr17 | 80885945 | 80886805 | 861 | 6 | 9.04E-10 | 0.0002117 | 6.95E-05 | 3.20E-08 | -0.01175514 | RPTOR |
| chr6 | 165853094 | 165853112 | 19 | 2 | 1.21E-09 | 3.05E-07 | 3.93E-07 | 3.20E-08 | -0.01962419 | PDE10A |
| chr11 | 63986293 | 63987487 | 1195 | 8 | 5.59E-10 | 8.62E-07 | 0.000431 | 3.20E-08 | -0.00218187 | AP000721.1, OTUB1 |
| chr19 | 7963151 | 7964898 | 1748 | 6 | 2.46E-06 | 1.32E-06 | 0.000181 | 3.20E-08 | 0.002517738 | ELAVL1 |
| chr7 | 158704641 | 158705956 | 1316 | 10 | 1.70E-06 | 3.37E-07 | 0.00058 | 3.20E-08 | -0.00370972 | NCAPG2 |
| chr4 | 156970786 | 156971613 | 828 | 12 | 7.64E-09 | 7.75E-07 | 0.001586 | 3.20E-08 | 0.001704761 | PDGFC |
| chr19 | 14556197 | 14556740 | 544 | 4 | 7.31E-09 | 1.12E-06 | 0.000172 | 3.21E-08 | -0.01010254 | TECR, DNAJB1 |
| chr17 | 49361082 | 49362855 | 1774 | 14 | 4.16E-05 | 2.29E-07 | 0.005112 | 3.21E-08 | -0.00115803 | AC091180.2, ZNF652 |
| chr5 | 133934312 | 133935284 | 973 | 3 | 6.09E-09 | 1.06E-08 | 0.000248 | 3.21E-08 | -0.00971968 | NA |
| chr19 | 3626082 | 3627582 | 1501 | 13 | 1.76E-08 | 2.37E-07 | 0.000575 | 3.21E-08 | 0.000345673 | CACTIN |
| chr2 | 131034164 | 131034951 | 788 | 6 | 3.41E-11 | 7.43E-06 | 1.62E-06 | 3.21E-08 | -0.00342889 | ARHGEF4 |
| chr16 | 81635136 | 81635714 | 579 | 3 | 2.50E-10 | 1.33E-06 | 4.47E-06 | 3.22E-08 | -0.01338388 | CMIP |
| chr1 | 202928403 | 202929778 | 1376 | 6 | 7.77E-08 | 3.45E-07 | 5.53E-05 | 3.22E-08 | -0.0079896 | KLHL12 |
| chr6 | 4134251 | 4136330 | 2080 | 16 | 1.12E-06 | 5.73E-07 | 0.000952 | 3.22E-08 | -0.00058322 | AL136309.3, ECI2-DT, ECI2 |
| chr12 | 130953257 | 130954756 | 1500 | 15 | 2.31E-08 | 1.79E-07 | 0.000185 | 3.23E-08 | 0.001930511 | ADGRD1 |
| chr1 | 219173768 | 219174897 | 1130 | 12 | 7.16E-08 | 6.07E-07 | 0.000108 | 3.23E-08 | -0.00170582 | LYPLAL1, LYPLAL1-DT |
| chr18 | 32772334 | 32773736 | 1403 | 12 | 6.18E-09 | 2.42E-07 | 0.000647 | 3.23E-08 | 0.007788605 | AC012123.1, KLHL14 |
| chr1 | 205917558 | 205918002 | 445 | 2 | 3.72E-09 | 6.92E-08 | 1.75E-06 | 3.24E-08 | -0.00237583 | SLC26A9 |
| chr14 | 96383638 | 96384070 | 433 | 3 | 1.32E-09 | 2.79E-05 | 2.01E-06 | 3.25E-08 | -0.00216096 | GSKIP |
| chr20 | 18632052 | 18632407 | 356 | 2 | 1.02E-08 | 4.56E-08 | 3.21E-06 | 3.25E-08 | -0.01154957 | AL121900.1, DTD1 |
| chr19 | 2986837 | 2988005 | 1169 | 4 | 1.91E-09 | 1.94E-07 | 4.63E-05 | 3.25E-08 | -0.011045 | TLE6 |
| chr1 | 1173440 | 1174460 | 1021 | 9 | 2.09E-08 | 5.91E-08 | 0.001629 | 3.25E-08 | 0.020468069 | TTLL10, TTLL10-AS1 |
| chr6 | 75284412 | 75285427 | 1016 | 14 | 6.99E-08 | 1.20E-06 | 0.000456 | 3.25E-08 | 0.00083304 | TMEM30A-DT, TMEM30A |
| chr12 | 64403433 | 64404959 | 1527 | 11 | 4.98E-09 | 4.61E-06 | 7.00E-05 | 3.25E-08 | 5.53E-05 | XPOT |
| chr6 | 50713035 | 50714002 | 968 | 5 | 6.24E-11 | 1.23E-05 | 4.04E-06 | 3.26E-08 | 0.008616338 | TFAP2D |
| chr1 | 27492549 | 27492888 | 340 | 2 | 2.27E-10 | 0.000253 | 5.39E-08 | 3.26E-08 | 0.003501823 | NA |
| chr3 | 9396744 | 9398075 | 1332 | 14 | 3.02E-07 | 1.11E-06 | 0.000468 | 3.26E-08 | 0.000324434 | SETD5, THUMPD3-AS1 |
| chr14 | 52949814 | 52951394 | 1581 | 10 | 4.20E-07 | 2.84E-07 | 0.000656 | 3.26E-08 | 0.002530371 | FERMT2 |
| chr1 | 156927270 | 156928138 | 869 | 7 | 4.80E-09 | 5.13E-07 | 1.80E-06 | 3.26E-08 | -0.00096339 | LRRC71 |
| chr14 | 75699197 | 75699247 | 51 | 2 | 2.85E-09 | 1.40E-07 | 7.90E-07 | 3.26E-08 | 0.004866792 | TTLL5 |
| chr8 | 142315638 | 142317015 | 1378 | 7 | 1.01E-08 | 9.01E-08 | 0.000808 | 3.26E-08 | -0.00100843 | TSNARE1 |
| chr16 | 4015396 | 4016141 | 746 | 5 | 1.36E-07 | 6.15E-07 | 0.000207 | 3.26E-08 | -0.01084449 | ADCY9 |
| chr1 | 36858850 | 36860076 | 1227 | 8 | 5.32E-07 | 8.35E-09 | 0.000975 | 3.26E-08 | -0.00416489 | GRIK3 |
| chr9 | 130663033 | 130664721 | 1689 | 4 | 3.74E-09 | 1.06E-07 | 1.21E-05 | 3.27E-08 | 0.001063756 | PRDM12 |
| chr18 | 31649224 | 31649259 | 36 | 2 | 3.37E-08 | 2.09E-08 | 2.51E-05 | 3.27E-08 | 0.023366597 | B4GALT6 |
| chr22 | 41597353 | 41597718 | 366 | 3 | 5.53E-10 | 1.13E-06 | 5.98E-07 | 3.28E-08 | -0.01327851 | NA |
| chr19 | 46146583 | 46148296 | 1714 | 6 | 6.04E-06 | 9.18E-08 | 0.000113 | 3.28E-08 | -0.00249592 | AC007785.1, IGFL2 |
| chr12 | 70637422 | 70638813 | 1392 | 7 | 2.09E-09 | 2.35E-06 | 2.16E-05 | 3.30E-08 | -0.00480906 | PTPRB, PTPRR |
| chr6 | 131199557 | 131199963 | 407 | 6 | 6.19E-09 | 3.47E-06 | 0.000173 | 3.30E-08 | 0.002351082 | AKAP7 |
| chr20 | 52670799 | 52671797 | 999 | 4 | 1.72E-06 | 1.54E-08 | 0.000182 | 3.30E-08 | -0.00991283 | AL031674.1 |
| chr7 | 48454195 | 48455736 | 1542 | 9 | 1.44E-09 | 4.03E-05 | 8.08E-05 | 3.31E-08 | -0.00666466 | ABCA13 |
| chr11 | 130068488 | 130069015 | 528 | 3 | 3.30E-12 | 6.45E-06 | 1.30E-07 | 3.31E-08 | -0.00731862 | APLP2 |
| chr6 | 44245976 | 44248026 | 2051 | 19 | 2.86E-07 | 4.29E-06 | 0.000368 | 3.31E-08 | -6.50E-05 | HSP90AB1 |
| chr1 | 171652592 | 171654096 | 1505 | 6 | 1.69E-07 | 1.24E-06 | 0.000107 | 3.31E-08 | -0.00405869 | MYOC |
| chr2 | 235507029 | 235507420 | 392 | 4 | 1.39E-08 | 1.38E-07 | 2.88E-05 | 3.32E-08 | -0.0041565 | AGAP1, AGAP1-IT1 |
| chr18 | 50560210 | 50561992 | 1783 | 7 | 1.76E-09 | 1.16E-05 | 8.60E-06 | 3.33E-08 | 0.009257289 | MAPK4 |
| chr11 | 72040750 | 72041676 | 927 | 5 | 2.89E-12 | 9.04E-06 | 2.50E-07 | 3.34E-08 | 0.002258641 | NUMA1 |
| chr1 | 63329608 | 63330815 | 1208 | 8 | 6.08E-08 | 3.25E-07 | 8.55E-05 | 3.34E-08 | -0.00313465 | NA |
| chr7 | 111108812 | 111109518 | 707 | 3 | 5.54E-10 | 1.24E-06 | 1.61E-05 | 3.36E-08 | -0.0124043 | LRRN3, IMMP2L |
| chr6 | 35485728 | 35486845 | 1118 | 5 | 3.82E-08 | 8.66E-07 | 3.17E-05 | 3.36E-08 | -0.00510579 | TEAD3 |
| chr11 | 62805120 | 62806408 | 1289 | 15 | 1.58E-07 | 1.39E-06 | 0.001584 | 3.36E-08 | -0.00175967 | NXF1 |
| chr6 | 4610462 | 4611830 | 1369 | 6 | 2.36E-07 | 1.18E-06 | 0.000258 | 3.36E-08 | 0.000418312 | KU-MEL-3 |
| chr20 | 35625940 | 35627725 | 1786 | 4 | 1.51E-06 | 2.92E-07 | 2.37E-05 | 3.36E-08 | -0.00294799 | CPNE1 |
| chr5 | 10249411 | 10250469 | 1059 | 12 | 1.51E-09 | 7.47E-07 | 0.000107 | 3.36E-08 | 0.001332844 | AC012640.1, CCT5, ATPSCKMT |
| chr19 | 506588 | 508078 | 1491 | 13 | 1.65E-09 | 1.32E-06 | 2.01E-05 | 3.36E-08 | 0.001422388 | TPGS1, AC005775.1 |
| chr6 | 52087037 | 52088550 | 1514 | 6 | 4.21E-07 | 1.21E-06 | 9.23E-05 | 3.37E-08 | -0.00858327 | PKHD1 |
| chr13 | 40187448 | 40188332 | 885 | 7 | 7.32E-09 | 1.53E-06 | 5.03E-05 | 3.37E-08 | -0.00504471 | LINC00332, LINC00598 |
| chr4 | 48779894 | 48780823 | 930 | 12 | 1.75E-09 | 1.22E-07 | 0.001556 | 3.38E-08 | -7.34E-05 | FRYL |
| chr4 | 28686529 | 28686632 | 104 | 2 | 1.25E-08 | 1.68E-07 | 6.92E-07 | 3.38E-08 | -0.01837348 | NA |
| chr19 | 46716298 | 46717350 | 1053 | 6 | 9.34E-08 | 1.41E-08 | 0.000578 | 3.38E-08 | -0.00038952 | PRKD2 |
| chr13 | 97959715 | 97960168 | 454 | 4 | 1.68E-08 | 8.50E-08 | 5.34E-06 | 3.38E-08 | 0.001581537 | IPO5 |
| chr12 | 42483083 | 42484390 | 1308 | 16 | 2.08E-08 | 6.13E-06 | 9.08E-05 | 3.39E-08 | 0.001307919 | PRICKLE1 |
| chr19 | 39412329 | 39413762 | 1434 | 15 | 4.55E-10 | 1.42E-05 | 4.71E-06 | 3.40E-08 | 0.000936374 | PLEKHG2 |
| chr19 | 845662 | 846354 | 693 | 5 | 3.51E-08 | 1.14E-07 | 0.000176 | 3.40E-08 | -0.01026871 | PRTN3 |
| chr3 | 53110040 | 53110600 | 561 | 8 | 1.28E-08 | 1.63E-06 | 0.000158 | 3.40E-08 | -0.00787916 | RFT1 |
| chr3 | 137971014 | 137971485 | 472 | 4 | 3.03E-11 | 0.0044697 | 8.50E-09 | 3.40E-08 | -0.00752646 | NA |
| chr8 | 38468207 | 38469436 | 1230 | 13 | 5.83E-09 | 1.28E-06 | 6.35E-05 | 3.41E-08 | -0.00119099 | FGFR1 |
| chr2 | 127193057 | 127194468 | 1412 | 7 | 3.22E-06 | 1.24E-08 | 0.000957 | 3.42E-08 | -0.00253426 | CYP27C1 |
| chr20 | 34458612 | 34458902 | 291 | 2 | 1.35E-08 | 4.84E-08 | 3.24E-06 | 3.42E-08 | 0.000510056 | ITCH |
| chr8 | 41621150 | 41621381 | 232 | 3 | 6.03E-09 | 1.91E-08 | 4.94E-05 | 3.43E-08 | -0.00705155 | GPAT4, AC009630.3 |
| chr2 | 237124747 | 237125466 | 720 | 3 | 5.16E-08 | 1.56E-06 | 1.96E-05 | 3.43E-08 | 0.006646521 | NA |
| chr11 | 107928187 | 107929253 | 1067 | 9 | 4.63E-08 | 2.26E-08 | 0.001151 | 3.43E-08 | 0.002465209 | RAB39A, SLC35F2 |
| chr14 | 99944021 | 99944797 | 777 | 3 | 2.15E-07 | 6.96E-08 | 4.79E-05 | 3.43E-08 | -0.00140045 | NA |
| chr3 | 9843912 | 9844679 | 768 | 11 | 1.95E-11 | 4.81E-06 | 0.00011 | 3.44E-08 | -0.0028495 | TTLL3, RPUSD3 |
| chr1 | 24931440 | 24932615 | 1176 | 12 | 2.42E-11 | 1.34E-06 | 0.00013 | 3.44E-08 | -0.00453151 | RUNX3 |
| chr18 | 61746662 | 61747724 | 1063 | 3 | 3.60E-09 | 6.64E-08 | 9.20E-06 | 3.44E-08 | -0.00552133 | AC105094.2 |
| chr2 | 165573762 | 165574175 | 414 | 5 | 6.73E-10 | 7.47E-08 | 8.86E-05 | 3.44E-08 | -0.01251098 | CSRNP3 |
| chr6 | 43075953 | 43077449 | 1497 | 15 | 1.32E-07 | 5.02E-07 | 0.00026 | 3.45E-08 | 0.001010309 | PTK7 |
| chr5 | 132767909 | 132768237 | 329 | 2 | 3.49E-11 | 3.50E-05 | 6.63E-08 | 3.45E-08 | -0.00911446 | SEPTIN8 |
| chr1 | 43791532 | 43791541 | 10 | 2 | 1.93E-08 | 8.95E-08 | 1.39E-06 | 3.45E-08 | -0.01907649 | AL451062.2, ST3GAL3 |
| chr10 | 89414054 | 89416214 | 2161 | 15 | 1.11E-05 | 4.51E-07 | 0.000658 | 3.46E-08 | 0.001777275 | IFIT5, LIPA |
| chr11 | 111976170 | 111977675 | 1506 | 10 | 7.46E-10 | 3.92E-06 | 2.66E-05 | 3.46E-08 | -0.00262402 | DIXDC1 |
| chr8 | 11853402 | 11854871 | 1470 | 7 | 1.70E-12 | 4.37E-05 | 7.54E-06 | 3.46E-08 | -0.00518599 | CTSB |
| chr1 | 202009821 | 202011186 | 1366 | 10 | 4.96E-09 | 1.36E-06 | 4.11E-05 | 3.46E-08 | -0.00265259 | ELF3, ELF3-AS1 |
| chr8 | 85249823 | 85250245 | 423 | 5 | 1.96E-08 | 1.66E-06 | 7.34E-05 | 3.47E-08 | -0.00736987 | CA13, AC011773.2 |
| chr11 | 124864851 | 124866944 | 2094 | 16 | 9.60E-06 | 7.48E-08 | 0.000534 | 3.47E-08 | 0.000411427 | ROBO3 |
| chr16 | 67850265 | 67850570 | 306 | 2 | 4.04E-09 | 2.50E-07 | 5.11E-07 | 3.47E-08 | -0.01551156 | NUTF2 |
| chr2 | 240920477 | 240921578 | 1102 | 5 | 8.46E-11 | 4.26E-07 | 1.61E-06 | 3.47E-08 | -0.00678451 | CROCC2 |
| chr8 | 37625299 | 37626138 | 840 | 4 | 4.92E-09 | 6.58E-08 | 2.73E-05 | 3.47E-08 | -0.00878788 | AC137579.2, AC137579.1 |
| chr1 | 150233695 | 150233897 | 203 | 3 | 2.87E-10 | 4.53E-07 | 1.15E-06 | 3.47E-08 | 0.010383688 | ANP32E |
| chr9 | 88995547 | 88996321 | 775 | 4 | 1.89E-08 | 1.04E-07 | 0.000209 | 3.47E-08 | -0.0058994 | S1PR3 |
| chr6 | 57205824 | 57206797 | 974 | 3 | 1.36E-10 | 1.07E-06 | 3.31E-07 | 3.48E-08 | 0.001870625 | RAB23 |
| chr17 | 50833780 | 50834966 | 1187 | 12 | 2.55E-09 | 1.54E-07 | 0.000613 | 3.48E-08 | -0.00103199 | WFIKKN2 |
| chr3 | 70748995 | 70749734 | 740 | 2 | 4.94E-10 | 1.10E-07 | 1.11E-06 | 3.48E-08 | -0.00920156 | NA |
| chr6 | 107730299 | 107730889 | 591 | 2 | 6.13E-10 | 2.09E-07 | 5.99E-07 | 3.49E-08 | -0.01132323 | SCML4 |
| chr2 | 180978456 | 180980149 | 1694 | 8 | 8.92E-10 | 3.75E-06 | 3.77E-05 | 3.49E-08 | -0.00105029 | UBE2E3, AC104076.1 |
| chr13 | 25866907 | 25867740 | 834 | 4 | 2.74E-10 | 1.32E-05 | 4.90E-06 | 3.49E-08 | -0.00450611 | ATP8A2 |
| chr1 | 45521458 | 45522683 | 1226 | 13 | 3.23E-09 | 2.64E-06 | 0.00021 | 3.49E-08 | -0.00283031 | PRDX1 |
| chr12 | 90937806 | 90938702 | 897 | 5 | 2.45E-08 | 1.35E-07 | 0.000138 | 3.49E-08 | -0.00515107 | LINC00615, CCER1 |
| chr19 | 18557042 | 18558425 | 1384 | 12 | 7.90E-08 | 8.98E-07 | 0.001292 | 3.49E-08 | 0.000852339 | KXD1, AC005253.1 |
| chr17 | 28405818 | 28406565 | 748 | 12 | 2.37E-08 | 3.31E-08 | 0.002468 | 3.50E-08 | 0.003284241 | AC015917.2, SLC46A1 |
| chr18 | 69469070 | 69469946 | 877 | 6 | 6.34E-08 | 7.37E-09 | 0.001148 | 3.50E-08 | -0.008246 | DOK6, AC119868.1 |
| chr12 | 48183073 | 48184293 | 1221 | 8 | 1.46E-08 | 2.55E-08 | 8.80E-05 | 3.50E-08 | 0.00017885 | CCDC184 |
| chr3 | 111609111 | 111609744 | 634 | 2 | 2.96E-13 | 0.000998 | 5.43E-08 | 3.51E-08 | -0.00772436 | CD96 |
| chr9 | 98192005 | 98192937 | 933 | 7 | 1.29E-09 | 3.07E-06 | 2.62E-05 | 3.51E-08 | 0.001970759 | CORO2A |
| chr18 | 48933103 | 48933750 | 648 | 4 | 4.14E-09 | 1.24E-07 | 6.46E-05 | 3.51E-08 | 0.006452468 | SMAD7 |
| chr9 | 74951865 | 74953197 | 1333 | 14 | 1.57E-07 | 3.50E-07 | 7.87E-05 | 3.51E-08 | -0.00087661 | CARNMT1-AS1, C9orf40 |
| chr17 | 82626622 | 82627585 | 964 | 6 | 1.11E-07 | 2.30E-08 | 0.000615 | 3.51E-08 | 0.004025747 | FOXK2, WDR45B |
| chr15 | 88238769 | 88238912 | 144 | 2 | 2.86E-08 | 3.12E-08 | 8.37E-06 | 3.52E-08 | -0.01485456 | NTRK3 |
| chr10 | 119724843 | 119726694 | 1852 | 8 | 3.51E-06 | 1.31E-06 | 0.000741 | 3.52E-08 | 0.003692161 | INPP5F |
| chr8 | 60660435 | 60660815 | 381 | 4 | 3.64E-08 | 2.07E-08 | 0.000159 | 3.52E-08 | -0.01169999 | NA |
| chr6 | 160990628 | 160992023 | 1396 | 6 | 1.12E-10 | 5.98E-06 | 1.51E-07 | 3.53E-08 | -0.00094663 | MAP3K4, MAP3K4-AS1 |
| chr15 | 31338533 | 31339638 | 1106 | 6 | 2.16E-07 | 1.92E-07 | 0.000212 | 3.53E-08 | 0.00357076 | KLF13 |
| chr12 | 54294560 | 54296132 | 1573 | 4 | 5.20E-06 | 3.51E-08 | 0.000115 | 3.53E-08 | -0.01165979 | AC078778.1, NFE2 |
| chr22 | 29689896 | 29690963 | 1068 | 4 | 7.77E-09 | 6.68E-06 | 1.29E-05 | 3.53E-08 | -0.00797212 | NF2 |
| chr1 | 179080893 | 179082758 | 1866 | 19 | 1.39E-06 | 5.57E-07 | 0.001198 | 3.54E-08 | 0.001108722 | TOR3A |
| chr19 | 7636212 | 7637881 | 1670 | 14 | 3.74E-08 | 4.57E-07 | 0.001444 | 3.54E-08 | 0.000540992 | AC008763.2, STXBP2, AC008763.1 |
| chr2 | 11215527 | 11216192 | 666 | 5 | 3.19E-09 | 1.21E-05 | 2.07E-05 | 3.56E-08 | -0.00457138 | ROCK2 |
| chr3 | 48268715 | 48269407 | 693 | 4 | 4.73E-09 | 1.05E-07 | 9.60E-05 | 3.57E-08 | 0.000893607 | ZNF589 |
| chr16 | 11915982 | 11917010 | 1029 | 13 | 5.86E-08 | 4.17E-09 | 0.012995 | 3.57E-08 | -0.00966877 | GSPT1 |
| chr7 | 91940140 | 91941559 | 1420 | 10 | 1.54E-10 | 1.28E-05 | 2.00E-05 | 3.58E-08 | 0.001319873 | AKAP9 |
| chr8 | 38465111 | 38467095 | 1985 | 9 | 1.05E-06 | 7.50E-06 | 0.00025 | 3.58E-08 | 0.003510434 | FGFR1 |
| chr1 | 18290933 | 18291297 | 365 | 5 | 4.44E-08 | 1.96E-08 | 0.000125 | 3.58E-08 | 0.016933948 | IGSF21 |
| chr8 | 38457512 | 38458630 | 1119 | 4 | 1.03E-06 | 8.78E-08 | 3.43E-05 | 3.59E-08 | -0.00210831 | FGFR1 |
| chr11 | 60786282 | 60787493 | 1212 | 5 | 1.02E-06 | 9.59E-09 | 0.001244 | 3.59E-08 | -0.00708469 | MS4A10 |
| chr11 | 114058055 | 114059088 | 1034 | 7 | 3.50E-09 | 4.12E-07 | 0.000154 | 3.59E-08 | -0.01554974 | NA |
| chr2 | 158381368 | 158381958 | 591 | 2 | 5.40E-08 | 2.48E-08 | 1.86E-05 | 3.61E-08 | 0.007256956 | CCDC148 |
| chr4 | 902775 | 903799 | 1025 | 4 | 2.60E-09 | 1.91E-06 | 9.73E-06 | 3.61E-08 | -0.00760365 | GAK |
| chr8 | 42409413 | 42410143 | 731 | 3 | 6.58E-09 | 7.03E-08 | 3.40E-05 | 3.61E-08 | -0.00321328 | NA |
| chr3 | 10766870 | 10767196 | 327 | 2 | 1.30E-10 | 9.05E-06 | 9.28E-08 | 3.62E-08 | -0.01226747 | NA |

| | | | | | | | | | | |
|---|---|---|---|---|---|---|---|---|---|---|
| chr1 | 9835470 | 9836251 | 782 | 3 | 1.61E-08 | 3.81E-08 | 2.03E-05 | 3.63E-08 | 0.000610877 | NA |
| chr16 | 22947684 | 22948968 | 1285 | 8 | 1.26E-07 | 3.49E-08 | 0.000855 | 3.63E-08 | -0.00762073 | NA |
| chr7 | 95596027 | 95596996 | 970 | 9 | 8.92E-12 | 3.21E-05 | 4.62E-05 | 3.64E-08 | -0.00690204 | AC002451.1, PDK4 |
| chr20 | 13344780 | 13344782 | 3 | 2 | 6.16E-09 | 2.39E-06 | 1.46E-07 | 3.64E-08 | -0.0048921 | NA |
| chr17 | 36590430 | 36592164 | 1735 | 10 | 3.97E-08 | 4.91E-06 | 7.47E-05 | 3.66E-08 | 0.002245096 | DHRS11 |
| chr4 | 183096637 | 183097972 | 1336 | 4 | 5.39E-11 | 3.06E-06 | 3.28E-05 | 3.67E-08 | -0.01744708 | WWC2-AS2 |
| chr8 | 85220193 | 85221062 | 870 | 13 | 8.32E-10 | 6.19E-05 | 0.000271 | 3.67E-08 | 1.10E-06 | CA13, RBIS |
| chr8 | 141759722 | 141760669 | 948 | 5 | 3.61E-08 | 1.34E-08 | 0.000274 | 3.67E-08 | -0.00412224 | NA |
| chr3 | 129439340 | 129441311 | 1972 | 24 | 1.93E-06 | 2.76E-07 | 0.003503 | 3.67E-08 | -0.0007965 | IFT122, MBD4 |
| chr2 | 74648126 | 74648679 | 554 | 11 | 2.75E-09 | 1.64E-07 | 0.000235 | 3.68E-08 | -0.00845169 | M1AP |
| chr15 | 72057648 | 72058512 | 865 | 3 | 3.26E-09 | 5.47E-07 | 2.87E-05 | 3.68E-08 | -0.00568423 | MYO9A |
| chr2 | 241153351 | 241153873 | 523 | 4 | 1.15E-10 | 1.68E-05 | 1.01E-07 | 3.68E-08 | -0.00088181 | PPP1R7 |
| chr1 | 228060312 | 228061084 | 773 | 4 | 1.06E-09 | 4.77E-07 | 9.38E-07 | 3.69E-08 | -5.08E-06 | WNT3A |
| chr16 | 79505080 | 79505577 | 498 | 3 | 6.81E-09 | 1.66E-05 | 1.84E-06 | 3.69E-08 | -0.0090167 | NA |
| chr5 | 69043262 | 69045125 | 1864 | 5 | 2.65E-06 | 5.73E-08 | 0.000742 | 3.69E-08 | 0.002896584 | AC010273.3 |
| chr12 | 124989475 | 124990468 | 994 | 3 | 6.01E-09 | 4.45E-08 | 5.32E-05 | 3.69E-08 | -0.00241678 | NA |
| chr17 | 81251509 | 81252174 | 666 | 6 | 3.58E-08 | 6.17E-09 | 0.001426 | 3.70E-08 | 0.014629196 | AC027601.3, SLC38A10 |
| chr17 | 12664775 | 12666584 | 1810 | 17 | 3.01E-07 | 7.67E-08 | 0.013102 | 3.70E-08 | -0.00247472 | MYOCD |
| chr4 | 158231771 | 158232752 | 982 | 3 | 2.78E-08 | 3.62E-08 | 1.35E-05 | 3.71E-08 | 0.00531745 | TMEM144 |
| chr8 | 98064018 | 98064821 | 804 | 11 | 3.26E-08 | 1.10E-06 | 1.17E-05 | 3.71E-08 | -0.00192364 | ERICH5 |
| chr6 | 26234833 | 26235443 | 611 | 8 | 8.44E-11 | 1.39E-05 | 9.56E-08 | 3.72E-08 | -0.00666927 | H1-3 |
| chr15 | 98076192 | 98076299 | 108 | 2 | 3.69E-08 | 2.57E-08 | 1.86E-05 | 3.73E-08 | -0.00141666 | LINC02251, AC022523.1 |
| chr17 | 59105716 | 59107216 | 1501 | 17 | 2.29E-07 | 2.45E-06 | 0.002953 | 3.73E-08 | -0.00117602 | AC099850.1, TRIM37 |
| chr9 | 135301876 | 135302765 | 890 | 4 | 8.02E-08 | 2.16E-07 | 1.51E-05 | 3.74E-08 | 0.006450243 | NA |
| chr20 | 2799468 | 2801039 | 1572 | 13 | 3.50E-08 | 1.78E-07 | 0.005773 | 3.74E-08 | -0.00356539 | CPXM1 |
| chr11 | 34623008 | 34623677 | 670 | 3 | 4.35E-10 | 6.33E-08 | 1.37E-05 | 3.74E-08 | 0.000920693 | EHF |
| chr16 | 77971049 | 77971811 | 763 | 3 | 1.28E-09 | 5.91E-07 | 1.17E-06 | 3.75E-08 | -0.00087205 | VAT1L |
| chr11 | 62753213 | 62754893 | 1681 | 15 | 1.03E-06 | 1.98E-06 | 1.45E-05 | 3.76E-08 | -0.00128409 | ZBTB3 |
| chr2 | 10060580 | 10061396 | 817 | 3 | 4.76E-09 | 9.86E-08 | 4.70E-06 | 3.78E-08 | -0.00737622 | CYS1 |
| chr3 | 190322392 | 190323264 | 873 | 11 | 7.57E-10 | 2.81E-05 | 3.06E-05 | 3.78E-08 | 0.002927438 | CLDN16, CLDN1 |
| chr3 | 120093045 | 120093966 | 922 | 6 | 4.85E-09 | 7.16E-07 | 1.60E-05 | 3.78E-08 | -0.00376996 | GSK3B |
| chr20 | 59082116 | 59082423 | 308 | 2 | 1.84E-08 | 2.27E-08 | 4.01E-05 | 3.78E-08 | -0.02072699 | NA |
| chr5 | 132864645 | 132866459 | 1815 | 17 | 5.51E-08 | 6.94E-08 | 0.000104 | 3.79E-08 | -0.00114749 | GDF9 |
| chr21 | 46111375 | 46113283 | 1909 | 11 | 1.04E-06 | 9.57E-07 | 0.003027 | 3.79E-08 | -0.00184149 | COL6A2 |
| chr5 | 13783575 | 13783847 | 273 | 3 | 2.90E-08 | 4.67E-08 | 9.31E-06 | 3.79E-08 | -0.00434275 | DNAH5 |
| chr6 | 129710162 | 129710764 | 603 | 11 | 7.58E-09 | 2.88E-07 | 0.000262 | 3.80E-08 | -0.00734741 | ARHGAP18 |
| chr14 | 24298916 | 24300198 | 1283 | 19 | 1.22E-08 | 5.75E-08 | 0.001637 | 3.80E-08 | 0.001167409 | NOP9, DHRS1 |
| chr12 | 123465627 | 123466353 | 727 | 4 | 9.62E-10 | 8.50E-07 | 2.45E-05 | 3.80E-08 | -0.00587288 | SNRNP35 |
| chr15 | 98548181 | 98549185 | 1005 | 7 | 7.19E-09 | 4.30E-06 | 1.80E-05 | 3.80E-08 | 0.000627511 | NA |
| chr15 | 85618749 | 85619996 | 1248 | 6 | 1.59E-06 | 1.04E-08 | 0.000902 | 3.80E-08 | -0.01289235 | AKAP13, AC087286.2 |
| chr17 | 38677507 | 38677753 | 247 | 2 | 3.31E-08 | 2.28E-08 | 4.08E-05 | 3.80E-08 | -0.01616015 | NA |
| chr7 | 112205127 | 112206860 | 1734 | 17 | 1.05E-06 | 1.63E-06 | 0.00411 | 3.81E-08 | 0.000129649 | ZNF277, DOCK4 |
| chr10 | 130135828 | 130136892 | 1065 | 11 | 2.49E-08 | 1.69E-07 | 5.08E-05 | 3.81E-08 | -0.00344646 | GLRX3 |
| chr11 | 64855209 | 64856464 | 1256 | 5 | 9.99E-07 | 2.67E-06 | 3.33E-05 | 3.81E-08 | -0.00964889 | EHD1 |
| chr21 | 43826371 | 43827157 | 787 | 5 | 1.54E-08 | 1.59E-07 | 7.14E-05 | 3.82E-08 | -0.01269732 | NA |
| chr7 | 73521856 | 73522841 | 986 | 12 | 2.00E-08 | 7.60E-08 | 0.002551 | 3.83E-08 | -9.40E-05 | BAZ1B |
| chr2 | 24048916 | 24050584 | 1669 | 15 | 5.08E-08 | 1.55E-06 | 0.000288 | 3.83E-08 | -0.00322936 | MFSD2B, FKBP1B, WDCP |
| chr1 | 206555706 | 206557130 | 1425 | 14 | 1.07E-07 | 1.54E-06 | 0.00082 | 3.83E-08 | -0.00302101 | RASSF5 |
| chr7 | 151115195 | 151116109 | 915 | 2 | 1.38E-07 | 3.13E-08 | 1.08E-05 | 3.83E-08 | 0.006678396 | AGAP3 |
| chr2 | 148934355 | 148934966 | 612 | 4 | 1.16E-07 | 1.38E-08 | 0.000134 | 3.83E-08 | -0.00434238 | KIF5C |
| chr17 | 44014345 | 44015783 | 1439 | 17 | 6.57E-08 | 5.72E-07 | 0.001106 | 3.83E-08 | 0.000115314 | TMEM101 |
| chr14 | 101048305 | 101049542 | 1238 | 8 | 1.50E-07 | 1.37E-05 | 7.74E-05 | 3.83E-08 | -0.0039912 | MIR381HG, MIR544A |
| chr1 | 23691453 | 23692501 | 1049 | 15 | 8.59E-08 | 8.74E-08 | 0.000134 | 3.83E-08 | 0.001529574 | RPL11 |
| chr11 | 124301788 | 124302855 | 1068 | 3 | 4.38E-08 | 2.29E-06 | 6.55E-06 | 3.83E-08 | -0.00125509 | OR8D1 |
| chr11 | 77147602 | 77148913 | 1312 | 6 | 4.53E-08 | 9.77E-07 | 0.00025 | 3.84E-08 | -0.00889685 | MYO7A |
| chr14 | 100109480 | 100110434 | 955 | 4 | 1.43E-06 | 9.94E-09 | 0.000444 | 3.85E-08 | -0.00180759 | EVL, MIR342, MIR151B |
| chr12 | 55331695 | 55332207 | 513 | 3 | 5.59E-08 | 1.28E-08 | 0.000258 | 3.85E-08 | -0.01820974 | OR6C3 |
| chr2 | 40453147 | 40453471 | 325 | 2 | 1.03E-10 | 1.27E-05 | 8.97E-08 | 3.86E-08 | -0.00404897 | SLC8A1 |
| chr16 | 1351194 | 1352744 | 1551 | 13 | 1.58E-09 | 5.70E-07 | 0.000721 | 3.86E-08 | 0.000848217 | GNPTG, TSR3 |
| chr16 | 2154380 | 2155643 | 1264 | 13 | 6.79E-08 | 3.49E-06 | 0.00085 | 3.86E-08 | -0.00046535 | SNHG19, SNORD60, AC009065.9 |
| chr8 | 9259953 | 9260566 | 614 | 4 | 3.70E-09 | 8.20E-08 | 1.40E-05 | 3.86E-08 | 0.002848994 | AC022784.1, AC022784.3 |
| chr2 | 199955276 | 199957138 | 1863 | 14 | 1.07E-06 | 5.31E-08 | 0.006509 | 3.86E-08 | -0.00100532 | C2orf69, MAIP1, TYW5 |
| chr16 | 5034026 | 5034831 | 806 | 8 | 1.65E-08 | 3.45E-07 | 7.83E-05 | 3.86E-08 | -0.00034933 | NAGPA-AS1, ALG1, NAGPA |
| chr22 | 19659758 | 19660420 | 663 | 2 | 2.45E-08 | 3.94E-08 | 6.36E-06 | 3.86E-08 | 0.00364995 | NA |
| chr17 | 66229228 | 66229985 | 758 | 5 | 8.06E-09 | 2.39E-06 | 3.24E-06 | 3.87E-08 | 0.008869204 | APOH |
| chr19 | 18588015 | 18589743 | 1729 | 14 | 2.93E-10 | 0.0006251 | 1.86E-07 | 3.88E-08 | 0.002390141 | REX1BD, CRLF1 |
| chr1 | 173205511 | 173206189 | 679 | 8 | 2.39E-08 | 4.78E-08 | 0.000428 | 3.88E-08 | 0.002718966 | TNFSF4 |
| chr17 | 74776201 | 74777520 | 1320 | 15 | 1.94E-09 | 4.12E-06 | 2.90E-05 | 3.88E-08 | -0.00209323 | TMEM104, NAT9 |
| chr5 | 2186257 | 2186890 | 634 | 5 | 7.70E-08 | 4.64E-07 | 2.19E-05 | 3.89E-08 | 0.001269955 | NA |
| chr15 | 52652036 | 52653494 | 1459 | 9 | 2.19E-05 | 2.60E-09 | 0.00752 | 3.89E-08 | -0.01025093 | FAM214A |
| chr2 | 240234203 | 240235111 | 909 | 5 | 1.07E-10 | 1.38E-05 | 3.38E-07 | 3.89E-08 | -0.00578582 | AC124861.1 |
| chr1 | 100249292 | 100251343 | 2052 | 13 | 1.77E-07 | 1.91E-05 | 3.79E-05 | 3.89E-08 | -0.00085465 | AL445928.2, AL445928.1, DBT |
| chr15 | 40771731 | 40772302 | 572 | 4 | 9.79E-09 | 1.35E-07 | 9.74E-05 | 3.89E-08 | -0.00040915 | C15orf62, DNAJC17 |
| chr13 | 76952132 | 76952616 | 485 | 4 | 1.97E-10 | 6.11E-05 | 3.41E-06 | 3.89E-08 | 0.00385216 | ACOD1 |
| chr16 | 28091329 | 28092115 | 787 | 3 | 3.22E-09 | 4.77E-08 | 9.81E-06 | 3.89E-08 | -0.00490151 | NA |
| chr22 | 38699893 | 38701330 | 1438 | 14 | 3.23E-07 | 9.60E-07 | 0.000299 | 3.89E-08 | -0.00120407 | JOSD1 |
| chr13 | 22802476 | 22803076 | 601 | 4 | 1.24E-08 | 6.49E-08 | 1.05E-05 | 3.89E-08 | -0.01404673 | NA |
| chr14 | 90818890 | 90820062 | 1173 | 5 | 7.10E-07 | 2.60E-07 | 0.000151 | 3.89E-08 | -0.00531204 | NA |
| chr15 | 23645446 | 23646633 | 1188 | 8 | 2.03E-10 | 4.37E-07 | 3.35E-06 | 3.92E-08 | 0.002928505 | MAGEL2 |
| chr18 | 59454871 | 59455514 | 644 | 2 | 2.76E-07 | 2.36E-08 | 4.15E-05 | 3.93E-08 | -0.02221901 | CCBE1 |
| chr6 | 42928563 | 42929860 | 1298 | 16 | 2.07E-07 | 1.55E-07 | 0.003362 | 3.93E-08 | 0.001541209 | CNPY3, AL035587.2 |
| chr1 | 119482656 | 119483649 | 994 | 3 | 1.26E-09 | 1.37E-07 | 4.29E-06 | 3.93E-08 | -0.00730732 | NA |
| chr20 | 58466238 | 58467248 | 1011 | 5 | 3.00E-08 | 2.94E-07 | 0.000194 | 3.93E-08 | -0.00733868 | APCDD1L |
| chr10 | 98031759 | 98031906 | 148 | 2 | 1.52E-09 | 4.64E-08 | 4.85E-06 | 3.93E-08 | -0.03550602 | NA |
| chr17 | 8479623 | 8481491 | 1869 | 8 | 1.20E-06 | 2.75E-06 | 0.000252 | 3.94E-08 | 0.000468178 | NDEL1, MYH10 |
| chr3 | 42512814 | 42513759 | 946 | 6 | 1.31E-07 | 4.38E-08 | 0.001144 | 3.95E-08 | -0.00133945 | VIPR1, VIPR1-AS1 |
| chr2 | 238260896 | 238261212 | 317 | 3 | 2.41E-08 | 2.38E-07 | 3.11E-05 | 3.95E-08 | 0.000552283 | PER2 |
| chr1 | 159535225 | 159535643 | 419 | 2 | 8.92E-09 | 3.45E-08 | 9.26E-06 | 3.95E-08 | -0.01248546 | OR10J5 |
| chr1 | 7455021 | 7456591 | 1571 | 5 | 1.67E-06 | 2.57E-08 | 0.000243 | 3.95E-08 | -0.00556637 | CAMTA1 |
| chr7 | 158121049 | 158121787 | 739 | 2 | 2.54E-08 | 2.45E-08 | 3.42E-05 | 3.96E-08 | 0.000500659 | PTPRN2 |
| chr5 | 92660925 | 92661806 | 882 | 5 | 2.73E-07 | 1.29E-08 | 0.000299 | 3.96E-08 | -0.00171657 | AC026780.1 |
| chr16 | 8866976 | 8869194 | 2219 | 22 | 1.68E-05 | 5.30E-08 | 0.003041 | 3.96E-08 | -0.00404841 | CARHSP1 |
| chr19 | 6001977 | 6002903 | 927 | 5 | 2.39E-07 | 3.59E-08 | 0.000369 | 3.97E-08 | 0.002630436 | AC011444.1, RFX2 |
| chr21 | 42566673 | 42567895 | 1223 | 3 | 2.71E-09 | 7.26E-08 | 1.17E-05 | 3.99E-08 | -0.00669419 | SLC37A1 |
| chr22 | 49460989 | 49461606 | 618 | 2 | 1.27E-09 | 2.33E-07 | 6.65E-07 | 3.99E-08 | 0.014408931 | C22orf34 |
| chr6 | 53927190 | 53927891 | 702 | 2 | 1.59E-11 | 1.45E-05 | 8.97E-08 | 3.99E-08 | -0.01459558 | NA |
| chr12 | 132686815 | 132688139 | 1325 | 11 | 3.73E-08 | 4.14E-05 | 0.000197 | 3.99E-08 | -0.00082198 | PXMP2, AC135586.2, POLE |
| chr6 | 149577151 | 149577290 | 140 | 3 | 1.23E-08 | 2.94E-07 | 2.33E-05 | 4.01E-08 | -0.00371238 | GINM1, AL078581.4 |
| chr17 | 16426844 | 16427454 | 611 | 5 | 1.55E-09 | 7.40E-07 | 4.87E-06 | 4.02E-08 | -0.00022171 | TRPV2 |
| chr6 | 165987006 | 165988212 | 1207 | 15 | 6.15E-09 | 3.99E-08 | 0.000585 | 4.05E-08 | 0.002150862 | LINC00602, PDE10A |
| chr4 | 55852154 | 55854061 | 1908 | 14 | 3.45E-06 | 9.00E-06 | 0.000231 | 4.05E-08 | -0.00097791 | EXOC1 |
| chr4 | 38109973 | 38111284 | 1312 | 10 | 3.82E-11 | 2.76E-05 | 3.43E-05 | 4.05E-08 | -0.00228034 | TBC1D1 |
| chr16 | 67479565 | 67481441 | 1877 | 13 | 3.36E-07 | 8.49E-07 | 4.54E-05 | 4.05E-08 | -0.00136432 | AC009061.2, ATP6V0D1 |
| chr21 | 20997167 | 20998546 | 1380 | 10 | 6.05E-09 | 0.0002478 | 1.46E-05 | 4.06E-08 | -0.00049733 | NCAM2 |
| chr19 | 18504534 | 18504634 | 101 | 2 | 2.03E-08 | 5.40E-08 | 3.92E-06 | 4.06E-08 | -0.02450502 | ELL |
| chr5 | 1015686 | 1016653 | 968 | 5 | 1.80E-09 | 7.44E-07 | 2.87E-05 | 4.06E-08 | -0.00674794 | NKD2 |
| chr11 | 74493060 | 74494038 | 979 | 14 | 2.94E-09 | 3.35E-05 | 3.64E-05 | 4.07E-08 | -0.00089771 | AP001372.2, POLD3, LIPT2 |
| chr19 | 17117417 | 17117616 | 200 | 2 | 3.83E-08 | 2.60E-08 | 2.82E-05 | 4.07E-08 | 0.011653222 | MYO9B |
| chr2 | 14633264 | 14634778 | 1515 | 12 | 2.25E-10 | 7.11E-06 | 1.48E-05 | 4.07E-08 | -0.00076873 | LRATD1 |
| chr9 | 121808059 | 121809110 | 1052 | 4 | 3.90E-07 | 1.64E-06 | 2.19E-05 | 4.07E-08 | -0.00019393 | NA |
| chr2 | 86790830 | 86793254 | 2425 | 15 | 3.25E-07 | 2.49E-06 | 7.01E-05 | 4.08E-08 | -0.00272493 | CD8A |
| chr4 | 22752934 | 22754403 | 1470 | 5 | 2.61E-06 | 4.04E-07 | 5.41E-05 | 4.08E-08 | -0.00635538 | GBA3 |
| chr5 | 83714774 | 83715547 | 774 | 2 | 1.15E-08 | 7.60E-08 | 2.32E-06 | 4.09E-08 | -0.01446516 | HAPLN1 |
| chr2 | 109126508 | 109126858 | 351 | 3 | 2.40E-08 | 1.96E-08 | 0.000138 | 4.09E-08 | 0.000979413 | NA |
| chr11 | 64567492 | 64568261 | 770 | 4 | 3.93E-09 | 1.61E-07 | 6.30E-05 | 4.10E-08 | -0.00592871 | SLC22A11 |
| chr2 | 137963733 | 137964756 | 1024 | 10 | 8.54E-09 | 5.90E-07 | 0.000278 | 4.10E-08 | -0.00857122 | HNMT |
| chr14 | 105042727 | 105043039 | 313 | 3 | 1.47E-08 | 4.82E-07 | 1.18E-05 | 4.10E-08 | -0.00522128 | NA |
| chr4 | 76948111 | 76949620 | 1510 | 8 | 3.21E-09 | 1.63E-07 | 6.09E-05 | 4.10E-08 | -0.00410343 | NA |
| chr1 | 166452807 | 166453086 | 280 | 2 | 2.92E-08 | 3.35E-08 | 1.13E-05 | 4.10E-08 | -0.0091422 | NA |
| chr2 | 100241307 | 100242575 | 1269 | 5 | 1.33E-09 | 7.04E-07 | 8.49E-05 | 4.11E-08 | -0.00493887 | LINC01104 |
| chr3 | 160038082 | 160039340 | 1259 | 6 | 8.22E-07 | 8.34E-08 | 0.001747 | 4.13E-08 | -0.00530276 | IL12A-AS1 |
| chr7 | 26199619 | 26201506 | 1888 | 24 | 2.00E-07 | 6.93E-06 | 0.000899 | 4.14E-08 | -0.00029892 | CBX3, HNRNPA2B1 |
| chr1 | 8944996 | 8947189 | 2194 | 10 | 7.28E-10 | 3.65E-07 | 0.000123 | 4.14E-08 | -0.00457995 | CA6 |
| chr18 | 74407975 | 74408462 | 488 | 3 | 2.45E-09 | 2.37E-06 | 2.90E-07 | 4.14E-08 | -0.00681889 | NA |
| chr12 | 52258593 | 52259853 | 1261 | 7 | 6.79E-07 | 1.52E-08 | 0.00079 | 4.15E-08 | -0.0005493 | KRT86, KRT87P |
| chr10 | 17229007 | 17230617 | 1611 | 16 | 8.10E-06 | 1.43E-07 | 0.002596 | 4.15E-08 | 0.001261108 | VIM, VIM-AS1 |
| chr13 | 35941932 | 35942133 | 202 | 2 | 3.01E-09 | 4.77E-06 | 1.29E-07 | 4.15E-08 | -0.0199212 | DCLK1 |
| chr7 | 69432139 | 69432179 | 41 | 2 | 1.82E-08 | 8.79E-08 | 1.96E-06 | 4.15E-08 | -0.01168822 | NA |
| chr4 | 184473834 | 184475144 | 1311 | 15 | 1.69E-10 | 0.0001497 | 8.82E-05 | 4.17E-08 | 0.000381869 | IRF2-DT, IRF2 |
| chr9 | 89661626 | 89662600 | 975 | 5 | 1.32E-08 | 7.39E-08 | 0.00045 | 4.17E-08 | -0.01485472 | UNQ6494 |
| chr8 | 81841442 | 81842876 | 1435 | 13 | 3.89E-07 | 7.67E-07 | 0.000137 | 4.19E-08 | 0.00158498 | LINC02235, SNX16 |
| chr11 | 61055218 | 61055712 | 495 | 4 | 3.05E-10 | 2.19E-05 | 2.05E-07 | 4.20E-08 | -0.00460626 | AP003721.2 |
| chr8 | 17964570 | 17964653 | 84 | 2 | 2.40E-08 | 3.21E-08 | 1.39E-05 | 4.20E-08 | -0.01206253 | PCM1 |
| chr8 | 11017736 | 11018024 | 289 | 2 | 5.30E-09 | 8.59E-08 | 2.07E-06 | 4.20E-08 | 0.000965316 | XKR6 |
| chr1 | 75127265 | 75128907 | 1643 | 15 | 4.40E-07 | 3.03E-07 | 0.001683 | 4.20E-08 | -0.00953389 | LHX8, AC099786.1 |
| chr16 | 1675174 | 1675360 | 187 | 2 | 3.76E-09 | 1.21E-07 | 1.37E-06 | 4.21E-08 | -0.00819377 | CRAMP1, AL031708.1 |
| chr2 | 218259540 | 218261295 | 1756 | 9 | 6.42E-07 | 2.75E-07 | 9.76E-05 | 4.21E-08 | 0.001932313 | GPBAR1 |
| chr14 | 23407921 | 23408986 | 1066 | 4 | 2.20E-11 | 5.96E-06 | 1.43E-06 | 4.22E-08 | -0.00973584 | MYH6 |
| chr5 | 497282 | 498310 | 1029 | 6 | 2.12E-08 | 6.10E-08 | 6.67E-05 | 4.23E-08 | -0.01351725 | SLC9A3 |
| chr9 | 20622358 | 20624152 | 1795 | 13 | 1.11E-07 | 4.53E-08 | 0.002857 | 4.23E-08 | -0.00016508 | MLLT3 |
| chr10 | 79397070 | 79397918 | 849 | 3 | 3.49E-07 | 2.44E-08 | 0.000111 | 4.23E-08 | -0.00837699 | AL133481.1, ZCCHC24 |
| chr14 | 54566026 | 54568497 | 2472 | 17 | 3.31E-06 | 2.42E-06 | 5.42E-05 | 4.23E-08 | -0.00013736 | SAMD4A |
| chr2 | 239162445 | 239163172 | 728 | 4 | 1.34E-09 | 2.88E-07 | 6.72E-06 | 4.24E-08 | -0.00252458 | HDAC4 |
| chr16 | 10689962 | 10690805 | 844 | 4 | 5.51E-08 | 1.24E-07 | 5.03E-05 | 4.25E-08 | -0.00854825 | TEKT5 |
| chr2 | 171893453 | 171894735 | 1283 | 13 | 6.34E-08 | 9.10E-06 | 0.000374 | 4.25E-08 | 0.00237195 | SLC25A12 |
| chr3 | 185552310 | 185552770 | 461 | 6 | 1.28E-08 | 7.99E-07 | 0.001044 | 4.26E-08 | -0.01230655 | LIPH |
| chr3 | 110419314 | 110419341 | 28 | 2 | 2.99E-08 | 7.18E-08 | 2.74E-06 | 4.26E-08 | -0.01883651 | NA |
| chr10 | 125682973 | 125683898 | 926 | 8 | 2.51E-09 | 2.78E-07 | 8.02E-05 | 4.27E-08 | -0.00666217 | AL158835.1, TEX36 |
| chr20 | 3663216 | 3663653 | 438 | 8 | 2.76E-08 | 2.49E-06 | 7.54E-05 | 4.27E-08 | -0.01185368 | GFRA4 |
| chr1 | 22642499 | 22643846 | 1348 | 7 | 2.47E-07 | 6.86E-07 | 0.000392 | 4.27E-08 | -0.00363516 | C1QC |
| chr1 | 58248269 | 58248780 | 512 | 3 | 3.69E-10 | 8.15E-08 | 8.83E-06 | 4.29E-08 | 0.006828453 | DAB1 |
| chr17 | 1511466 | 1512256 | 791 | 3 | 4.65E-08 | 2.54E-08 | 0.000112 | 4.30E-08 | -0.01214476 | INPP5K |
| chr14 | 73460841 | 73461714 | 874 | 4 | 5.77E-09 | 2.46E-06 | 3.43E-06 | 4.31E-08 | 0.006151044 | AC005280.3, AC005280.2 |
| chr13 | 33284307 | 33286324 | 2018 | 14 | 3.34E-07 | 1.11E-06 | 4.17E-05 | 4.31E-08 | -0.00334157 | AL627232.1, STARD13 |
| chr19 | 55442005 | 55443606 | 1602 | 9 | 3.90E-09 | 4.56E-06 | 0.000233 | 4.31E-08 | 0.00046296 | SHISA7 |
| chr6 | 31648311 | 31649829 | 1519 | 15 | 4.36E-08 | 7.69E-06 | 0.000288 | 4.31E-08 | -0.00197326 | BAG6 |
| chr1 | 9733613 | 9735164 | 1552 | 5 | 3.77E-08 | 2.26E-08 | 0.000378 | 4.32E-08 | 0.00553245 | CLSTN1 |
| chr16 | 1401277 | 1401655 | 379 | 4 | 3.01E-08 | 1.16E-07 | 2.56E-05 | 4.32E-08 | -0.00471072 | UNKL |

| | | | | | | | | | | |
|---|---|---|---|---|---|---|---|---|---|---|
| chr8 | 60908707 | 60910260 | 1554 | 10 | 3.59E-07 | 2.65E-07 | 4.37E-05 | 4.32E-08 | -0.0027258 | AC022182.1 |
| chr1 | 83860502 | 83861743 | 1242 | 12 | 3.81E-08 | 5.50E-07 | 0.001873 | 4.33E-08 | 0.003015245 | LINC01725 |
| chr2 | 206423375 | 206423497 | 123 | 2 | 1.21E-08 | 4.27E-07 | 4.52E-07 | 4.34E-08 | -0.00038587 | NA |
| chr10 | 71822213 | 71822458 | 246 | 2 | 1.85E-10 | 3.33E-05 | 8.14E-08 | 4.34E-08 | 0.006343141 | PSAP |
| chr17 | 41499800 | 41501053 | 1254 | 6 | 1.61E-08 | 9.25E-07 | 1.74E-06 | 4.35E-08 | -0.00770236 | AC019349.1, KRT13 |
| chr5 | 143810819 | 143811350 | 532 | 2 | 2.05E-11 | 2.94E-07 | 6.10E-07 | 4.35E-08 | 0.00060072 | AC008696.2 |
| chr10 | 1596499 | 1596895 | 397 | 3 | 9.05E-09 | 2.77E-07 | 2.30E-06 | 4.35E-08 | -0.01536639 | ADARB2 |
| chr17 | 18038693 | 18039812 | 1120 | 15 | 5.54E-09 | 1.19E-06 | 0.001543 | 4.36E-08 | 0.003249614 | GID4, ATPAF2 |
| chr16 | 1045673 | 1046754 | 1082 | 5 | 9.17E-08 | 5.92E-05 | 1.03E-05 | 4.36E-08 | -0.00210222 | NA |
| chr20 | 40754499 | 40754583 | 85 | 2 | 4.39E-08 | 3.21E-08 | 1.62E-05 | 4.37E-08 | -0.01598947 | AL035665.2 |
| chr19 | 19515442 | 19517113 | 1672 | 19 | 1.11E-08 | 1.71E-05 | 3.66E-05 | 4.39E-08 | -1.41E-05 | NDUFA13, AC011448.1, TSSK6 |
| chr8 | 143926800 | 143928354 | 1555 | 7 | 2.13E-07 | 3.10E-06 | 7.09E-05 | 4.39E-08 | -0.01375739 | PLEC |
| chr11 | 66420248 | 66421826 | 1579 | 9 | 1.07E-06 | 1.59E-06 | 0.000242 | 4.39E-08 | 0.000682527 | NPAS4 |
| chr6 | 137223478 | 137224161 | 684 | 3 | 2.57E-09 | 3.76E-07 | 5.26E-06 | 4.39E-08 | 0.007989216 | NA |
| chr1 | 3623513 | 3624385 | 873 | 4 | 8.09E-08 | 1.48E-07 | 6.93E-05 | 4.42E-08 | -0.00876011 | AL513320.1 |
| chr18 | 58998848 | 58999007 | 160 | 2 | 3.67E-09 | 2.87E-07 | 6.44E-07 | 4.43E-08 | -0.00862632 | OACYLP |
| chr6 | 161435001 | 161435345 | 345 | 2 | 1.79E-09 | 2.98E-07 | 6.20E-07 | 4.43E-08 | -0.01816431 | AL138716.1, PRKN |
| chr6 | 134342284 | 134343277 | 994 | 2 | 6.32E-06 | 2.66E-08 | 4.28E-05 | 4.44E-08 | 0.013701836 | NA |
| chr11 | 1760275 | 1761660 | 1386 | 8 | 4.01E-06 | 1.21E-06 | 0.000384 | 4.45E-08 | -0.00742095 | AC068580.3, AC068580.4, CTSD |
| chr17 | 1569525 | 1569690 | 166 | 3 | 2.50E-08 | 8.00E-07 | 1.82E-05 | 4.45E-08 | -0.00470581 | SLC43A2 |
| chr7 | 118223524 | 118225058 | 1535 | 10 | 6.10E-10 | 3.28E-06 | 0.000132 | 4.45E-08 | -0.00185273 | ANKRD7 |
| chr3 | 46833736 | 46834864 | 1129 | 9 | 7.27E-11 | 0.0002563 | 1.84E-05 | 4.45E-08 | -0.00222687 | PRSS42P |
| chr10 | 48251757 | 48252506 | 750 | 4 | 2.52E-10 | 3.56E-06 | 2.48E-06 | 4.45E-08 | -0.00116651 | FRMPD2 |
| chr14 | 99575095 | 99575442 | 348 | 2 | 1.54E-09 | 2.07E-07 | 8.71E-07 | 4.45E-08 | -0.010413 | CCDC85C |
| chr18 | 59502103 | 59502775 | 673 | 2 | 6.07E-11 | 1.26E-05 | 1.02E-07 | 4.46E-08 | -0.01356616 | CCBE1 |
| chr7 | 87344649 | 87346218 | 1570 | 16 | 2.83E-10 | 1.28E-05 | 6.63E-05 | 4.46E-08 | 0.001263376 | AC005045.1, AC005045.2, CROT, TP53TG1 |
| chr17 | 50397380 | 50398399 | 1020 | 12 | 3.70E-09 | 7.52E-07 | 0.004166 | 4.46E-08 | 0.008647831 | AC004707.1, LRRC59 |
| chr19 | 49762115 | 49763079 | 965 | 2 | 7.67E-08 | 5.71E-08 | 4.40E-06 | 4.46E-08 | -0.01589104 | TSKS |
| chr12 | 53507245 | 53507774 | 530 | 5 | 1.09E-08 | 1.62E-07 | 0.000155 | 4.47E-08 | -0.00111924 | ATF7-NPFF, NPFF |
| chr8 | 73406819 | 73407741 | 923 | 2 | 1.49E-07 | 4.65E-08 | 6.52E-06 | 4.47E-08 | -0.0070764 | NA |
| chr1 | 77370715 | 77371292 | 578 | 2 | 1.20E-07 | 2.76E-08 | 3.65E-05 | 4.49E-08 | -0.00207555 | AK5 |
| chr20 | 4182515 | 4183225 | 711 | 3 | 2.61E-08 | 4.42E-08 | 1.52E-05 | 4.51E-08 | -0.00099284 | SMOX |
| chr16 | 86554538 | 86555677 | 1140 | 14 | 1.70E-08 | 5.12E-07 | 0.000245 | 4.51E-08 | 0.003309291 | FLJ30679, MTHFSD |
| chr17 | 34355751 | 34356462 | 712 | 7 | 5.77E-08 | 1.56E-07 | 0.000764 | 4.51E-08 | -0.00157683 | NA |
| chr16 | 87780227 | 87780572 | 346 | 3 | 1.19E-08 | 1.44E-08 | 0.000354 | 4.53E-08 | -0.01001999 | AC126696.1 |
| chr6 | 43283560 | 43285415 | 1856 | 10 | 2.85E-08 | 2.96E-06 | 0.000149 | 4.53E-08 | -0.00420055 | TTBK1 |
| chr4 | 3516052 | 3516394 | 343 | 4 | 1.90E-08 | 1.79E-08 | 0.000257 | 4.53E-08 | 0.014680052 | LRPAP1 |
| chr2 | 90068730 | 90069522 | 793 | 5 | 4.28E-08 | 3.62E-08 | 0.000384 | 4.54E-08 | -0.00324037 | NA |
| chr3 | 127337994 | 127338306 | 313 | 4 | 3.30E-08 | 1.86E-08 | 0.000154 | 4.55E-08 | -0.03564874 | LINC02016 |
| chr7 | 4714871 | 4715784 | 914 | 8 | 4.08E-09 | 2.72E-06 | 8.25E-06 | 4.55E-08 | -0.00178544 | FOXK1 |
| chr17 | 76671194 | 76672170 | 977 | 7 | 1.54E-08 | 4.18E-07 | 5.79E-05 | 4.55E-08 | 0.001439426 | AC005837.1 |
| chr12 | 31234511 | 31235279 | 769 | 3 | 1.93E-09 | 2.82E-06 | 2.19E-05 | 4.56E-08 | -0.01397632 | NA |
| chr5 | 143532091 | 143532313 | 223 | 3 | 1.20E-08 | 1.13E-07 | 7.27E-06 | 4.56E-08 | 0.002961221 | AC016598.2 |
| chr18 | 54358423 | 54359462 | 1040 | 11 | 7.48E-08 | 1.76E-07 | 0.001946 | 4.57E-08 | -0.00136691 | C18orf54 |
| chr14 | 92890389 | 92891420 | 1032 | 4 | 5.09E-09 | 2.56E-07 | 2.73E-06 | 4.57E-08 | -0.00821953 | LINC02833 |
| chr6 | 40524083 | 40525277 | 1195 | 4 | 1.40E-06 | 1.72E-08 | 0.000588 | 4.57E-08 | -0.01061751 | LRFN2 |
| chr12 | 59594963 | 59596639 | 1677 | 14 | 5.03E-07 | 5.76E-07 | 0.005016 | 4.57E-08 | 0.002717555 | SLC16A7 |
| chr14 | 100423705 | 100424026 | 322 | 2 | 2.56E-09 | 1.40E-07 | 1.33E-06 | 4.59E-08 | -0.00987333 | WDR25 |
| chr4 | 5887605 | 5889024 | 1420 | 17 | 1.29E-07 | 8.08E-07 | 0.000261 | 4.59E-08 | -0.00102335 | CRMP1 |
| chr5 | 158497988 | 158499104 | 1117 | 3 | 4.06E-09 | 1.87E-05 | 1.07E-05 | 4.60E-08 | -0.00862835 | AC091939.1 |
| chr12 | 560986 | 561861 | 876 | 5 | 1.86E-09 | 7.07E-06 | 1.69E-05 | 4.61E-08 | 0.003554523 | B4GALNT3 |
| chr3 | 172537343 | 172537739 | 397 | 2 | 3.62E-08 | 3.36E-08 | 1.70E-05 | 4.62E-08 | 0.021215204 | NA |
| chr10 | 126902428 | 126902560 | 133 | 3 | 7.02E-08 | 1.58E-08 | 0.000244 | 4.62E-08 | -0.00454694 | NA |
| chr11 | 83070440 | 83072310 | 1871 | 17 | 9.69E-07 | 3.45E-06 | 0.000743 | 4.62E-08 | -0.00120829 | RAB30-DT, RAB30 |
| chr18 | 8704168 | 8704724 | 557 | 4 | 2.45E-08 | 1.44E-07 | 4.19E-05 | 4.62E-08 | -0.01470483 | GACAT2 |
| chr1 | 54017680 | 54019121 | 1442 | 9 | 8.96E-07 | 7.28E-08 | 0.002098 | 4.62E-08 | -0.00666459 | LDLRAD1 |
| chr14 | 64796042 | 64797030 | 989 | 3 | 2.94E-10 | 6.49E-06 | 1.39E-07 | 4.62E-08 | -0.00772897 | SPTB |
| chr22 | 38655812 | 38657536 | 1725 | 20 | 3.77E-07 | 1.52E-07 | 0.00042 | 4.64E-08 | -0.00232266 | CBY1, FAM227A |
| chr2 | 260907 | 261362 | 456 | 3 | 3.49E-08 | 1.58E-07 | 1.80E-05 | 4.65E-08 | 0.00139987 | SH3YL1 |
| chr11 | 83285631 | 83287407 | 1777 | 17 | 4.92E-11 | 0.0032405 | 2.40E-06 | 4.65E-08 | -0.00326309 | AP000446.1, CCDC90B |
| chr7 | 116576783 | 116577845 | 1063 | 3 | 5.91E-08 | 1.10E-07 | 8.74E-06 | 4.65E-08 | -0.01092056 | COMETT |
| chr9 | 19379120 | 19380498 | 1379 | 8 | 6.90E-07 | 2.39E-06 | 0.000235 | 4.66E-08 | -0.00247063 | RPS6 |
| chr20 | 44717550 | 44718173 | 624 | 2 | 2.57E-09 | 1.79E-07 | 1.06E-06 | 4.67E-08 | -0.01022525 | CCN5, KCNK15-AS1 |
| chr19 | 3584595 | 3585611 | 1017 | 6 | 8.03E-10 | 1.93E-06 | 9.35E-06 | 4.67E-08 | 0.000903407 | GIPC3 |
| chr9 | 137225676 | 137227017 | 1342 | 5 | 6.71E-06 | 1.16E-08 | 0.000652 | 4.67E-08 | 2.24E-05 | CYSRT1 |
| chr11 | 2459976 | 2461505 | 1530 | 22 | 1.40E-07 | 1.53E-06 | 0.002863 | 4.68E-08 | -0.00184787 | KCNQ1 |
| chr4 | 83334759 | 83335881 | 1123 | 14 | 8.26E-08 | 2.84E-06 | 0.000135 | 4.68E-08 | -0.00209149 | HPSE |
| chr10 | 1048489 | 1050350 | 1862 | 15 | 6.59E-07 | 6.67E-06 | 0.002067 | 4.68E-08 | -0.001896 | WDR37, IDI1 |
| chr19 | 12627700 | 12628330 | 631 | 4 | 3.17E-09 | 8.67E-07 | 0.000101 | 4.69E-08 | 0.000547736 | ZNF791, ZNF490 |
| chr6 | 31859167 | 31861554 | 2388 | 32 | 1.68E-06 | 4.20E-08 | 0.019789 | 4.69E-08 | -0.00025595 | NEU1 |
| chr5 | 139680709 | 139682908 | 2200 | 10 | 2.48E-06 | 0.0001676 | 2.25E-05 | 4.69E-08 | -0.00468705 | CXXC5 |
| chr9 | 128321933 | 128324270 | 2338 | 21 | 6.21E-06 | 2.88E-07 | 0.002946 | 4.70E-08 | 0.000325674 | COQ4, TRUB2 |
| chr12 | 2672302 | 2673190 | 889 | 4 | 7.26E-08 | 2.86E-07 | 4.53E-05 | 4.71E-08 | -0.00066864 | CACNA1C |
| chr15 | 101347758 | 101348943 | 1186 | 6 | 2.26E-08 | 1.08E-06 | 6.82E-05 | 4.71E-08 | -0.00121603 | PCSK6 |
| chr9 | 109816344 | 109816563 | 220 | 2 | 1.06E-09 | 2.84E-05 | 8.97E-08 | 4.72E-08 | -0.00376276 | PALM2AKAP2 |
| chr15 | 72685411 | 72686113 | 703 | 10 | 2.54E-09 | 2.21E-05 | 0.000664 | 4.72E-08 | -0.00689135 | HIGD2B |
| chr19 | 3975390 | 3976715 | 1326 | 4 | 1.11E-06 | 2.57E-08 | 0.000384 | 4.73E-08 | 0.004423628 | EEF2 |
| chr2 | 208318235 | 208318736 | 502 | 3 | 3.25E-09 | 8.68E-06 | 1.43E-05 | 4.73E-08 | -0.01264265 | PIKFYVE |
| chr2 | 204664980 | 204665505 | 526 | 2 | 5.70E-08 | 3.23E-08 | 2.21E-05 | 4.73E-08 | -0.00028315 | PARD3B |
| chr8 | 1552201 | 1553801 | 1601 | 11 | 9.94E-07 | 7.80E-06 | 0.000118 | 4.73E-08 | -0.00029777 | DLGAP2 |
| chr5 | 1118555 | 1119533 | 979 | 4 | 2.09E-08 | 2.48E-06 | 2.60E-05 | 4.74E-08 | -0.00766551 | NA |
| chr20 | 3247802 | 3249345 | 1544 | 8 | 2.51E-08 | 1.18E-07 | 0.000715 | 4.74E-08 | -0.0026693 | C20orf194 |
| chr1 | 222465219 | 222465682 | 464 | 7 | 3.28E-08 | 6.52E-08 | 4.96E-05 | 4.74E-08 | -0.00088635 | NA |
| chr10 | 133072465 | 133073723 | 1259 | 5 | 1.03E-07 | 8.63E-06 | 3.72E-05 | 4.74E-08 | -0.00281658 | ADGRA1 |
| chr19 | 10290648 | 10291849 | 1202 | 5 | 2.10E-07 | 2.27E-08 | 0.000491 | 4.74E-08 | 0.003552365 | ICAM5 |
| chr3 | 149086291 | 149087922 | 1632 | 26 | 6.01E-07 | 1.63E-07 | 0.005194 | 4.74E-08 | -0.00129374 | HLTF-AS1, HLTF |
| chr5 | 141806289 | 141806345 | 57 | 2 | 5.19E-08 | 3.05E-08 | 2.89E-05 | 4.74E-08 | -0.02582637 | NA |
| chr11 | 6269721 | 6270721 | 1001 | 8 | 5.20E-09 | 2.40E-07 | 0.000433 | 4.76E-08 | -0.02162458 | CCKBR |
| chr6 | 40391391 | 40392332 | 942 | 5 | 8.76E-08 | 7.22E-08 | 0.000238 | 4.76E-08 | -0.01038532 | LRFN2 |
| chr2 | 239308757 | 239309360 | 604 | 6 | 2.22E-08 | 1.15E-06 | 3.19E-05 | 4.76E-08 | -0.00646491 | HDAC4 |
| chr22 | 17774086 | 17775898 | 1813 | 11 | 2.27E-07 | 3.80E-07 | 0.000466 | 4.76E-08 | -0.00233144 | BID |
| chr17 | 73191714 | 73192664 | 951 | 9 | 3.17E-11 | 1.24E-06 | 4.29E-05 | 4.76E-08 | -0.00851319 | AC097641.1, COG1 |
| chr7 | 96992011 | 96993397 | 1387 | 5 | 2.70E-06 | 4.02E-08 | 0.000296 | 4.77E-08 | -0.00911302 | DLX6-AS1 |
| chr16 | 3267157 | 3267428 | 272 | 2 | 3.65E-08 | 3.79E-08 | 1.31E-05 | 4.78E-08 | -0.058192 | LINC00921, ZNF263 |
| chr1 | 247078801 | 247079883 | 1083 | 8 | 1.14E-10 | 5.55E-07 | 4.49E-05 | 4.79E-08 | -0.01250291 | ZNF670-ZNF695, ZNF670 |
| chr1 | 165352504 | 165353547 | 1044 | 5 | 1.20E-09 | 8.19E-06 | 2.28E-06 | 4.79E-08 | 0.003988369 | LMX1A |
| chr19 | 14405733 | 14406635 | 903 | 6 | 8.34E-09 | 5.85E-08 | 2.67E-05 | 4.80E-08 | -0.00869854 | ADGRE5, AC008569.1 |
| chr4 | 154416570 | 154417014 | 445 | 6 | 8.99E-09 | 2.52E-06 | 6.59E-05 | 4.81E-08 | 0.002396223 | DCHS2 |
| chr10 | 133560143 | 133560298 | 156 | 2 | 3.59E-08 | 3.03E-08 | 3.30E-05 | 4.81E-08 | -0.02362369 | CYP2E1, SYCE1 |
| chr1 | 27871863 | 27872945 | 1083 | 12 | 1.54E-08 | 6.72E-07 | 0.000532 | 4.82E-08 | -0.00091888 | THEMIS2 |
| chr2 | 43371820 | 43372353 | 534 | 3 | 5.22E-09 | 1.03E-06 | 6.83E-07 | 4.82E-08 | 0.001217827 | THADA |
| chr1 | 35470943 | 35471972 | 1030 | 4 | 4.78E-08 | 1.89E-08 | 0.000392 | 4.82E-08 | -0.00644353 | KIAA0319L |
| chr20 | 36378136 | 36378865 | 730 | 2 | 2.46E-09 | 2.26E-07 | 8.98E-07 | 4.82E-08 | -0.02234602 | DLGAP4 |
| chr12 | 105106864 | 105108126 | 1263 | 18 | 3.91E-08 | 4.72E-06 | 0.001844 | 4.84E-08 | 0.001178836 | WASHC4, ALDH1L2 |
| chr17 | 16491304 | 16493183 | 1880 | 12 | 9.92E-07 | 1.86E-06 | 0.000253 | 4.84E-08 | 0.000623262 | LRRC75A |
| chr2 | 43158705 | 43159814 | 1110 | 4 | 1.52E-07 | 2.78E-07 | 3.71E-05 | 4.84E-08 | 0.002038403 | NA |
| chr1 | 3225966 | 3227244 | 1279 | 5 | 1.37E-08 | 7.11E-08 | 0.000384 | 4.84E-08 | 0.010662727 | PRDM16 |
| chr11 | 61555270 | 61556733 | 1464 | 9 | 5.14E-08 | 2.28E-08 | 0.000368 | 4.84E-08 | 0.002564541 | SYT7 |
| chr22 | 27438478 | 27438668 | 191 | 3 | 5.45E-08 | 1.58E-08 | 0.000325 | 4.84E-08 | 0.031701721 | AL050402.2 |
| chr20 | 31833806 | 31833971 | 166 | 2 | 1.35E-08 | 3.44E-07 | 6.22E-07 | 4.84E-08 | -0.0056069 | MYLK2 |
| chr4 | 37068659 | 37068779 | 121 | 2 | 3.33E-08 | 7.04E-08 | 3.65E-06 | 4.85E-08 | 0.028688358 | NA |
| chr11 | 65169394 | 65169589 | 196 | 4 | 5.80E-09 | 7.18E-08 | 0.000275 | 4.85E-08 | -0.03067494 | NA |
| chr17 | 351884 | 353601 | 1718 | 13 | 1.38E-07 | 3.02E-06 | 0.000465 | 4.85E-08 | 0.006371532 | AC129507.2, RPH3AL |
| chr8 | 6406635 | 6407469 | 835 | 6 | 2.53E-10 | 3.61E-05 | 4.82E-07 | 4.86E-08 | -0.00714502 | MCPH1, AC016065.1 |
| chr22 | 39316628 | 39317081 | 454 | 9 | 4.12E-08 | 1.20E-06 | 0.000284 | 4.87E-08 | -0.00524921 | RPL3 |
| chr19 | 15194627 | 15195127 | 501 | 2 | 3.07E-10 | 2.41E-07 | 8.62E-07 | 4.87E-08 | -0.01246901 | NOTCH3 |
| chr2 | 197310806 | 197311709 | 904 | 8 | 5.94E-11 | 3.35E-05 | 1.79E-07 | 4.88E-08 | 0.004147 | AC010746.2, ANKRD44 |
| chr1 | 3011581 | 3013751 | 2171 | 9 | 3.92E-06 | 2.87E-07 | 0.000738 | 4.88E-08 | -0.00285536 | NA |
| chr16 | 23754791 | 23755873 | 1083 | 5 | 4.48E-10 | 7.86E-05 | 2.94E-06 | 4.88E-08 | 0.000163669 | CHP2, AC012317.2 |
| chr5 | 421202 | 422518 | 1317 | 7 | 2.96E-07 | 1.15E-05 | 0.000148 | 4.89E-08 | 0.009642993 | PDCD6-AHRR, AHRR |
| chr6 | 169173655 | 169174019 | 365 | 3 | 6.06E-10 | 4.05E-06 | 3.23E-07 | 4.90E-08 | -0.00918416 | NA |
| chr17 | 57605041 | 57606328 | 1288 | 4 | 1.18E-08 | 2.55E-06 | 5.31E-05 | 4.90E-08 | 0.009245097 | MSI2, AC007431.1 |
| chr17 | 41177785 | 41179086 | 1302 | 5 | 1.07E-06 | 6.21E-08 | 0.000181 | 4.91E-08 | -0.00240206 | KRTAP4-2 |
| chr17 | 74370903 | 74372487 | 1585 | 8 | 1.14E-06 | 4.55E-07 | 0.000246 | 4.91E-08 | -0.00132499 | GPR142 |
| chr10 | 13271037 | 13271693 | 657 | 4 | 2.82E-08 | 2.96E-08 | 4.89E-05 | 4.91E-08 | -0.00498805 | NA |
| chr19 | 29714647 | 29716445 | 1799 | 11 | 2.40E-06 | 2.37E-07 | 0.001556 | 4.92E-08 | -0.00037306 | C19orf12 |
| chr4 | 110088832 | 110089235 | 404 | 2 | 2.48E-09 | 5.11E-07 | 4.69E-07 | 4.92E-08 | 0.001159082 | ELOVL6 |
| chr13 | 109297557 | 109297659 | 103 | 2 | 7.39E-09 | 3.42E-06 | 1.68E-07 | 4.92E-08 | -0.01232923 | LINC00370 |
| chr3 | 124032070 | 124034351 | 2282 | 8 | 3.44E-05 | 5.21E-08 | 0.002085 | 4.93E-08 | -0.00206292 | NA |
| chr14 | 91060119 | 91061337 | 1219 | 14 | 1.77E-08 | 4.64E-07 | 0.000185 | 4.94E-08 | 0.000499704 | DGLUCY, RPS6KA5 |
| chr12 | 15221523 | 15222847 | 1325 | 11 | 2.38E-13 | 7.87E-06 | 4.88E-07 | 4.95E-08 | -0.00287675 | RERG |
| chr6 | 154509864 | 154510916 | 1053 | 10 | 8.98E-08 | 3.83E-08 | 0.001359 | 4.95E-08 | 0.002561877 | AL357075.3, AL357075.4, CNKSR3 |
| chr1 | 24063624 | 24064434 | 811 | 2 | 9.03E-08 | 4.84E-08 | 7.77E-06 | 4.95E-08 | -0.01345169 | AL591178.1, MYOM3 |
| chr11 | 99020481 | 99021553 | 1073 | 14 | 6.42E-08 | 1.06E-07 | 0.001329 | 4.96E-08 | 0.000180102 | CNTN5 |
| chr15 | 79922700 | 79923999 | 1300 | 12 | 6.00E-07 | 8.40E-08 | 0.001025 | 4.96E-08 | -0.00138903 | ST20-AS1, ST20-MTHFS, ST20, AC015871.8 |
| chr4 | 1361533 | 1361636 | 104 | 3 | 2.45E-08 | 5.01E-08 | 6.31E-05 | 4.96E-08 | 0.002036675 | UVSSA |
| chr1 | 9270415 | 9270555 | 141 | 2 | 3.07E-08 | 5.12E-08 | 6.97E-06 | 4.97E-08 | -0.0084819 | H6PD |
| chr1 | 119711554 | 119713489 | 1936 | 18 | 3.15E-06 | 3.29E-06 | 0.00079 | 4.99E-08 | -0.00368346 | PHGDH |
| chr12 | 52032596 | 52033785 | 1190 | 7 | 1.08E-09 | 1.37E-06 | 8.80E-06 | 4.99E-08 | -0.00091139 | NR4A1 |
| chr7 | 79454131 | 79454880 | 750 | 13 | 5.38E-08 | 3.94E-07 | 0.002452 | 4.99E-08 | -0.00298548 | MAGI2-AS3 |
| chr11 | 72722014 | 72722821 | 808 | 9 | 1.15E-08 | 5.53E-06 | 0.000204 | 5.00E-08 | -0.00085148 | ARAP1 |
| chr10 | 133113306 | 133113373 | 68 | 2 | 1.09E-09 | 9.36E-06 | 1.24E-07 | 5.01E-08 | -0.01614064 | ADGRA1 |
| chr4 | 184283820 | 184284498 | 679 | 5 | 3.13E-09 | 1.22E-07 | 0.000113 | 5.01E-08 | -0.00241224 | NA |
| chr2 | 113888852 | 113890771 | 1920 | 17 | 6.48E-07 | 3.65E-06 | 0.000192 | 5.02E-08 | -0.00041436 | ACTR3, AC110769.2 |
| chr22 | 27275035 | 27275541 | 507 | 3 | 2.18E-09 | 8.07E-08 | 7.01E-06 | 5.03E-08 | -0.01772456 | NA |
| chr17 | 21043020 | 21044337 | 1318 | 10 | 4.54E-08 | 5.37E-09 | 0.002259 | 5.04E-08 | 0.001377454 | USP22 |
| chr6 | 149861463 | 149863002 | 1540 | 5 | 8.46E-08 | 6.58E-07 | 1.62E-05 | 5.04E-08 | -0.01007086 | AL355312.5, LRP11 |
| chr13 | 106151304 | 106152147 | 844 | 2 | 2.17E-07 | 4.15E-08 | 1.21E-05 | 5.05E-08 | -0.01919877 | NA |
| chr8 | 143794333 | 143795710 | 1378 | 7 | 1.19E-06 | 2.87E-07 | 0.000135 | 5.05E-08 | 0.000485211 | SCRIB |
| chr3 | 123285703 | 123287178 | 1476 | 4 | 9.67E-08 | 1.34E-05 | 4.05E-06 | 5.05E-08 | 0.000138154 | ADCY5 |
| chr10 | 50739157 | 50739802 | 646 | 3 | 8.95E-09 | 4.40E-08 | 7.51E-05 | 5.06E-08 | -0.01129536 | ASAH2B, AL589794.2 |
| chr8 | 35543541 | 35544390 | 850 | 5 | 5.52E-08 | 6.59E-07 | 3.96E-06 | 5.07E-08 | -0.00919716 | UNC5D |
| chr5 | 170232147 | 170233730 | 1584 | 17 | 1.79E-08 | 9.07E-06 | 4.64E-05 | 5.07E-08 | -0.00176915 | C5orf58 |
| chr18 | 48700785 | 48700879 | 95 | 2 | 2.05E-08 | 1.01E-07 | 2.33E-06 | 5.07E-08 | -0.01331787 | CTIF |
| chr16 | 78021178 | 78022729 | 1552 | 7 | 1.02E-08 | 9.15E-07 | 0.000367 | 5.08E-08 | -0.00745684 | CLEC3A, AC079414.1 |
| chr20 | 35146948 | 35148286 | 1339 | 13 | 2.85E-07 | 1.43E-05 | 0.000614 | 5.09E-08 | -0.00054769 | EDEM2 |
| chr1 | 236736316 | 236737857 | 1542 | 6 | 3.92E-06 | 1.16E-06 | 8.69E-05 | 5.09E-08 | -0.00148532 | ACTN2 |
| chr3 | 12898787 | 12899627 | 841 | 7 | 1.04E-08 | 1.85E-07 | 0.000192 | 5.09E-08 | -0.00160684 | IQSEC1 |

| | | | | | | | | | | |
|---|---|---|---|---|---|---|---|---|---|---|
| chr11 | 119594905 | 119595571 | 667 | 3 | 5.23E-08 | 2.69E-08 | 5.29E-05 | 5.10E-08 | -0.01486976 | NA |
| chr8 | 125951263 | 125952307 | 1045 | 5 | 1.98E-06 | 1.14E-07 | 0.000201 | 5.11E-08 | -0.00047378 | AC016074.2, LINC00861, SOD1P3 |
| chr6 | 30335349 | 30336125 | 777 | 13 | 1.35E-08 | 2.41E-06 | 0.000638 | 5.11E-08 | -0.00128216 | TRIM39, TRIM39-RPP21 |
| chr1 | 212546183 | 212546401 | 219 | 2 | 2.66E-08 | 3.10E-08 | 4.46E-05 | 5.11E-08 | -0.02505503 | LINC01740 |
| chr11 | 11502001 | 11502722 | 722 | 3 | 1.21E-12 | 1.41E-05 | 1.47E-07 | 5.12E-08 | 0.002999444 | GALNT18 |
| chr4 | 884493 | 885644 | 1152 | 4 | 1.38E-06 | 1.78E-07 | 9.58E-05 | 5.14E-08 | -0.00334666 | GAK |
| chr3 | 52278174 | 52279006 | 833 | 13 | 1.58E-09 | 4.00E-06 | 2.82E-06 | 5.15E-08 | 0.001896212 | WDR82 |
| chr5 | 134844666 | 134845837 | 1172 | 11 | 4.59E-09 | 2.33E-06 | 0.000332 | 5.17E-08 | -0.00149555 | DDX46, C5orf24 |
| chr15 | 71546259 | 71547308 | 1050 | 7 | 6.30E-08 | 1.34E-07 | 0.00017 | 5.18E-08 | -0.00579297 | THSD4, AC108861.1 |
| chr14 | 24310753 | 24311720 | 968 | 13 | 1.84E-07 | 5.14E-09 | 0.002491 | 5.18E-08 | -0.01855453 | LTB4R2, LTB4R, CIDEB |
| chr1 | 117121454 | 117123253 | 1800 | 16 | 4.20E-07 | 1.52E-06 | 0.00148 | 5.18E-08 | -0.0030532 | TRIM45 |
| chr9 | 138044922 | 138045567 | 646 | 3 | 3.01E-09 | 2.29E-06 | 2.12E-05 | 5.19E-08 | -0.00933917 | CACNA1B |
| chr6 | 83852607 | 83853725 | 1119 | 17 | 6.27E-09 | 2.41E-05 | 1.47E-05 | 5.19E-08 | -0.0018133 | RIPPLY2, AL139232.1 |
| chr4 | 7433027 | 7433379 | 353 | 2 | 9.22E-10 | 3.13E-07 | 7.46E-07 | 5.19E-08 | -0.01683729 | SORCS2, PSAPL1 |
| chr13 | 113364549 | 113365484 | 936 | 5 | 1.47E-08 | 3.21E-08 | 0.000461 | 5.19E-08 | -0.00895878 | NA |
| chr5 | 32489751 | 32490110 | 360 | 3 | 7.59E-08 | 2.84E-08 | 7.48E-05 | 5.20E-08 | 0.005675391 | NA |
| chr18 | 26549302 | 26551151 | 1850 | 16 | 1.15E-06 | 1.98E-07 | 0.001578 | 5.20E-08 | -0.00037892 | KCTD1 |
| chr7 | 157387079 | 157387432 | 354 | 6 | 1.67E-08 | 2.28E-06 | 0.000106 | 5.21E-08 | -0.00931886 | DNAJB6 |
| chr4 | 1002387 | 1003752 | 1366 | 9 | 1.94E-07 | 6.21E-09 | 0.005551 | 5.23E-08 | -0.00154009 | IDUA |
| chr6 | 166648397 | 166648738 | 342 | 4 | 1.68E-08 | 4.46E-07 | 2.59E-05 | 5.24E-08 | -0.00160943 | RPS6KA2 |
| chr20 | 46573839 | 46574476 | 638 | 3 | 2.04E-09 | 1.35E-06 | 8.39E-07 | 5.25E-08 | 0.014915608 | SLC13A3 |
| chr15 | 32640740 | 32642635 | 1896 | 10 | 6.14E-08 | 1.65E-05 | 0.00012 | 5.25E-08 | -0.00440719 | SCG5, AC123768.4 |
| chr21 | 33069764 | 33072278 | 2515 | 23 | 1.46E-05 | 2.12E-05 | 0.00016 | 5.25E-08 | -0.00095901 | OLIG1, AP000282.1 |
| chr2 | 241680316 | 241681704 | 1389 | 4 | 1.52E-06 | 1.77E-08 | 0.000472 | 5.26E-08 | -0.00282613 | DTYMK |
| chr18 | 24428938 | 24429503 | 566 | 5 | 3.88E-09 | 1.91E-06 | 3.08E-06 | 5.27E-08 | -0.00615548 | IMPACT |
| chr10 | 132115046 | 132116232 | 1187 | 6 | 4.17E-08 | 3.87E-08 | 0.000218 | 5.28E-08 | 0.000950518 | JAKMIP3 |
| chr11 | 48216744 | 48217215 | 472 | 2 | 6.37E-11 | 5.78E-06 | 1.50E-07 | 5.29E-08 | -0.03663394 | OR4B1 |
| chr8 | 57828390 | 57828705 | 316 | 2 | 9.17E-10 | 3.70E-05 | 9.44E-08 | 5.29E-08 | -0.02232967 | NA |
| chr14 | 100980758 | 100982248 | 1491 | 6 | 1.03E-07 | 5.19E-08 | 0.000345 | 5.31E-08 | -0.00703709 | MEG8, SNORD114-20, SNORD114-21 |
| chr4 | 880592 | 881711 | 1120 | 4 | 9.79E-08 | 5.59E-07 | 3.20E-05 | 5.31E-08 | -0.00505599 | GAK |
| chr1 | 145719365 | 145719950 | 586 | 5 | 1.49E-09 | 1.21E-07 | 6.63E-05 | 5.31E-08 | -0.00288443 | CD160 |
| chr12 | 120116668 | 120118235 | 1568 | 10 | 4.55E-07 | 3.21E-05 | 0.000289 | 5.32E-08 | -0.00033102 | AC004812.2, RAB35 |
| chr2 | 208395345 | 208395473 | 129 | 2 | 3.25E-08 | 9.70E-08 | 2.71E-06 | 5.32E-08 | -0.02232478 | PTH2R |
| chr1 | 111503249 | 111504695 | 1447 | 11 | 4.09E-08 | 3.81E-07 | 0.000794 | 5.34E-08 | 0.005174342 | TMIGD3, ADORA3 |
| chr7 | 16421154 | 16422173 | 1020 | 8 | 8.11E-09 | 0.000346 | 2.46E-06 | 5.34E-08 | 0.000362888 | CRPPA |
| chr1 | 180533633 | 180534375 | 743 | 3 | 1.69E-08 | 8.82E-08 | 1.62E-05 | 5.34E-08 | -0.01846884 | OVAAL |
| chr12 | 130336908 | 130338273 | 1366 | 13 | 4.81E-07 | 2.13E-08 | 0.005689 | 5.34E-08 | 0.022376865 | PIWIL1 |
| chr11 | 69496909 | 69497434 | 526 | 2 | 2.59E-09 | 2.45E-07 | 9.72E-07 | 5.35E-08 | -0.02044375 | NA |
| chr3 | 131715492 | 131716224 | 733 | 2 | 1.79E-08 | 8.34E-08 | 3.41E-06 | 5.35E-08 | 0.015472793 | CPNE4 |
| chr10 | 1215929 | 1217267 | 1339 | 5 | 1.91E-07 | 3.24E-07 | 5.07E-05 | 5.36E-08 | -0.00722541 | LINC00200, ADARB2 |
| chr20 | 59165014 | 59166003 | 990 | 3 | 4.63E-07 | 1.97E-08 | 0.000176 | 5.36E-08 | 0.01037647 | ZNF831 |
| chr12 | 7018419 | 7019675 | 1257 | 14 | 1.70E-07 | 6.74E-08 | 0.000726 | 5.37E-08 | -0.00146654 | C1S, LPCAT3 |
| chr20 | 58978755 | 58980068 | 1314 | 5 | 5.21E-08 | 4.54E-05 | 1.52E-05 | 5.37E-08 | -0.00048274 | AL109840.2 |
| chr7 | 2314047 | 2314849 | 803 | 11 | 9.28E-09 | 5.17E-07 | 0.000189 | 5.37E-08 | -0.00058346 | SNX8 |
| chr16 | 3401006 | 3401969 | 964 | 12 | 2.08E-08 | 1.16E-07 | 0.001153 | 5.38E-08 | -0.00102725 | AC025283.2, ZNF174, ZSCAN32 |
| chr16 | 89320390 | 89321988 | 1599 | 16 | 1.27E-06 | 2.92E-07 | 0.004648 | 5.38E-08 | -0.00087962 | AC137932.2, ANKRD11 |
| chr12 | 123269344 | 123270836 | 1493 | 14 | 7.52E-08 | 1.92E-07 | 0.000409 | 5.39E-08 | 0.00019663 | CDK2AP1 |
| chr9 | 130692400 | 130693025 | 626 | 5 | 3.18E-09 | 3.86E-08 | 0.000653 | 5.39E-08 | -0.01326466 | NA |
| chr8 | 104618668 | 104618829 | 162 | 2 | 1.52E-08 | 4.19E-07 | 6.15E-07 | 5.40E-08 | -0.01249011 | ZFPM2 |
| chr16 | 73231984 | 73232886 | 903 | 11 | 4.20E-09 | 9.84E-07 | 3.32E-05 | 5.41E-08 | -0.00379949 | AC140912.1, ZFHX3 |
| chr14 | 76153703 | 76154763 | 1061 | 5 | 1.25E-06 | 5.23E-08 | 0.000232 | 5.41E-08 | -0.0008671 | GPATCH2L |
| chr4 | 2467881 | 2470000 | 2120 | 17 | 1.84E-06 | 7.04E-07 | 0.002005 | 5.41E-08 | -0.00180818 | RNF4 |
| chr16 | 85986916 | 85987577 | 662 | 4 | 4.59E-12 | 1.20E-05 | 2.92E-07 | 5.41E-08 | -0.00597494 | AC092723.3 |
| chr7 | 139713779 | 139714774 | 996 | 3 | 2.55E-07 | 1.38E-07 | 8.68E-06 | 5.41E-08 | -0.00714718 | HIPK2 |
| chr3 | 57959550 | 57959968 | 419 | 3 | 6.80E-08 | 2.40E-08 | 0.000111 | 5.41E-08 | 0.039513733 | NA |
| chr12 | 133104630 | 133105296 | 667 | 3 | 6.02E-08 | 4.34E-07 | 6.38E-06 | 5.42E-08 | -0.01076606 | ZNF140 |
| chr17 | 60421616 | 60422825 | 1210 | 12 | 1.76E-09 | 2.40E-05 | 0.000382 | 5.42E-08 | -0.00597771 | C17orf64, USP32 |
| chr11 | 64561911 | 64563219 | 1309 | 7 | 1.44E-06 | 9.01E-07 | 0.000141 | 5.42E-08 | -0.00071913 | SLC22A11 |
| chr1 | 165540012 | 165540481 | 470 | 2 | 8.46E-10 | 1.75E-06 | 2.53E-07 | 5.43E-08 | -0.00685929 | LRRC52-AS1 |
| chr2 | 26785491 | 26786516 | 1026 | 14 | 1.38E-09 | 1.05E-05 | 8.29E-05 | 5.43E-08 | 0.00060627 | CENPA |
| chr16 | 70482465 | 70482810 | 346 | 4 | 5.80E-08 | 7.61E-08 | 5.75E-05 | 5.43E-08 | 0.001209275 | COG4 |
| chr13 | 113172240 | 113173214 | 975 | 2 | 3.86E-07 | 4.45E-08 | 1.28E-05 | 5.43E-08 | -0.00413056 | PROZ |
| chr8 | 30095139 | 30096085 | 947 | 7 | 1.66E-07 | 5.53E-08 | 0.001225 | 5.44E-08 | -0.00141317 | LEPROTL1 |
| chr3 | 164609068 | 164609585 | 518 | 2 | 2.94E-08 | 3.37E-08 | 4.00E-05 | 5.45E-08 | -0.01863641 | NA |
| chr6 | 29011433 | 29011721 | 289 | 11 | 3.02E-08 | 2.94E-07 | 3.01E-05 | 5.45E-08 | 0.002572372 | NA |
| chr7 | 158432391 | 158433204 | 814 | 5 | 3.35E-08 | 8.20E-08 | 2.61E-05 | 5.45E-08 | -0.00185327 | PTPRN2 |
| chr13 | 48302783 | 48304330 | 1548 | 21 | 1.58E-08 | 7.54E-06 | 0.004061 | 5.46E-08 | 0.000842023 | RB1, RB1-DT |
| chr1 | 154220079 | 154221573 | 1495 | 14 | 3.33E-07 | 1.02E-06 | 0.003726 | 5.46E-08 | 0.001155889 | UBAP2L, C1orf43 |
| chr13 | 78595737 | 78597168 | 1432 | 12 | 2.97E-07 | 1.06E-07 | 0.000923 | 5.47E-08 | -0.00145486 | OBI1-AS1, AL445209.1 |
| chr6 | 32077546 | 32078886 | 1341 | 6 | 2.22E-07 | 5.15E-08 | 9.20E-05 | 5.47E-08 | 0.001632821 | TNXB, RNA5SP206 |
| chr11 | 67425125 | 67425797 | 673 | 2 | 1.06E-08 | 4.13E-08 | 1.64E-05 | 5.47E-08 | -0.0078562 | CARNS1 |
| chr21 | 46647830 | 46647913 | 84 | 2 | 3.31E-10 | 7.50E-08 | 4.26E-06 | 5.48E-08 | -0.00574765 | PRMT2 |
| chr17 | 7023423 | 7024105 | 683 | 3 | 1.86E-09 | 3.83E-08 | 9.29E-05 | 5.49E-08 | -0.01781702 | BCL6B |
| chr12 | 45217306 | 45218040 | 735 | 2 | 3.83E-10 | 2.35E-07 | 1.05E-06 | 5.50E-08 | -0.01685714 | ANO6 |
| chr20 | 21106122 | 21106859 | 738 | 6 | 2.60E-08 | 7.73E-08 | 0.000251 | 5.50E-08 | -0.00244995 | LINC00237 |
| chr12 | 131710377 | 131710896 | 520 | 6 | 1.84E-09 | 1.21E-06 | 1.21E-05 | 5.50E-08 | -0.00866584 | NA |
| chr20 | 19210501 | 19211350 | 850 | 3 | 1.30E-09 | 2.11E-06 | 2.93E-05 | 5.51E-08 | -0.01018002 | AL136090.2 |
| chr1 | 45339850 | 45341542 | 1693 | 22 | 3.46E-08 | 7.19E-07 | 0.005071 | 5.52E-08 | -0.00079662 | TOE1, MUTYH, AL451136.1 |
| chr3 | 56076911 | 56077166 | 256 | 2 | 4.78E-08 | 4.10E-08 | 1.73E-05 | 5.52E-08 | -0.01551258 | ERC2 |
| chr1 | 160158281 | 160158798 | 518 | 3 | 2.81E-08 | 5.47E-08 | 4.30E-05 | 5.53E-08 | -0.0072759 | ATP1A4 |
| chr5 | 132664299 | 132665029 | 731 | 4 | 4.22E-09 | 2.49E-07 | 4.58E-05 | 5.53E-08 | -0.0078831 | NA |
| chr22 | 46541303 | 46542670 | 1368 | 6 | 5.76E-07 | 5.93E-08 | 8.73E-05 | 5.54E-08 | 0.001632981 | AL021392.1 |
| chr3 | 184154393 | 184155189 | 797 | 5 | 1.04E-08 | 3.69E-07 | 4.26E-05 | 5.54E-08 | -0.00443788 | NA |
| chr6 | 170250027 | 170251069 | 1043 | 13 | 1.76E-08 | 2.29E-08 | 0.002235 | 5.55E-08 | 0.002288769 | NA |
| chr11 | 69979173 | 69979629 | 457 | 3 | 3.46E-10 | 0.0001261 | 5.24E-08 | 5.55E-08 | -0.00891722 | NA |
| chr22 | 39556996 | 39558181 | 1186 | 5 | 6.30E-08 | 1.20E-06 | 1.66E-05 | 5.56E-08 | -0.0022024 | NA |
| chr21 | 41071535 | 41071776 | 242 | 2 | 3.17E-08 | 6.99E-08 | 5.09E-06 | 5.58E-08 | -0.00369995 | NA |
| chr16 | 68909363 | 68909597 | 235 | 2 | 4.15E-08 | 3.96E-08 | 2.04E-05 | 5.58E-08 | -0.01295123 | TANGO6 |
| chr17 | 81346673 | 81346852 | 180 | 2 | 8.20E-11 | 8.62E-05 | 8.71E-08 | 5.58E-08 | -0.01231811 | NA |
| chr2 | 98369851 | 98370508 | 658 | 4 | 8.37E-11 | 1.11E-06 | 1.71E-06 | 5.59E-08 | -0.00513116 | CNGA3 |
| chr21 | 41562267 | 41563834 | 1568 | 7 | 4.61E-06 | 1.09E-07 | 0.000179 | 5.59E-08 | -0.00305932 | AP001610.3 |
| chr3 | 64688063 | 64688790 | 728 | 6 | 1.74E-08 | 6.57E-07 | 0.000255 | 5.60E-08 | -0.00130415 | ADAMTS9-AS2 |
| chr9 | 30996290 | 30996789 | 500 | 2 | 1.17E-10 | 2.30E-06 | 2.31E-07 | 5.60E-08 | 0.015059156 | NA |
| chr2 | 96843616 | 96844760 | 1145 | 6 | 2.28E-10 | 3.18E-05 | 9.61E-06 | 5.60E-08 | 9.87E-05 | ANKRD23, ANKRD39 |
| chr3 | 49020610 | 49022054 | 1445 | 19 | 7.03E-09 | 4.79E-07 | 0.000793 | 5.60E-08 | 0.001170993 | NDUFAF3, DALRD3, MIR191 |
| chr16 | 843259 | 844919 | 1661 | 7 | 5.39E-08 | 5.25E-06 | 4.05E-05 | 5.61E-08 | -0.0050483 | AL023882.1 |
| chr11 | 115054782 | 115055083 | 302 | 2 | 1.89E-09 | 7.95E-06 | 1.44E-07 | 5.61E-08 | 0.007781636 | NA |
| chr15 | 81323528 | 81324994 | 1467 | 14 | 2.48E-09 | 1.22E-05 | 4.12E-06 | 5.61E-08 | 0.000221077 | TMC3-AS1, STARD5 |
| chr3 | 61250873 | 61252413 | 1541 | 16 | 4.62E-07 | 3.46E-08 | 0.01241 | 5.62E-08 | -0.00246415 | FHIT |
| chr19 | 50769803 | 50769849 | 47 | 2 | 1.23E-08 | 9.76E-07 | 3.60E-07 | 5.62E-08 | 0.010675152 | NA |
| chr3 | 100113928 | 100114949 | 1022 | 6 | 1.05E-08 | 2.42E-07 | 0.000117 | 5.62E-08 | -0.00032062 | CMSS1, FILIP1L |
| chr11 | 8128276 | 8129530 | 1255 | 3 | 1.21E-07 | 0.0001458 | 7.53E-06 | 5.62E-08 | -0.00264963 | RIC3 |
| chr17 | 67068937 | 67070610 | 1674 | 4 | 8.44E-06 | 5.77E-08 | 0.0003 | 5.63E-08 | -0.00783972 | HELZ |
| chr17 | 7211648 | 7212205 | 558 | 3 | 4.88E-09 | 3.64E-07 | 4.01E-05 | 5.63E-08 | -0.02844099 | DLG4 |
| chr14 | 39483942 | 39484818 | 877 | 4 | 2.29E-07 | 2.69E-06 | 6.52E-05 | 5.63E-08 | -0.00583392 | AL049828.1 |
| chr2 | 161414378 | 161415788 | 1411 | 11 | 9.43E-07 | 1.67E-07 | 0.000513 | 5.64E-08 | -0.00034858 | NA |
| chr14 | 94293099 | 94294074 | 976 | 10 | 9.44E-08 | 4.47E-08 | 0.001406 | 5.66E-08 | -0.00606817 | SERPINA10 |
| chr3 | 14588915 | 14589706 | 792 | 3 | 7.03E-08 | 4.72E-08 | 7.19E-05 | 5.67E-08 | -0.00511781 | NA |
| chr2 | 237387685 | 237388119 | 435 | 4 | 8.97E-09 | 3.78E-06 | 1.76E-06 | 5.67E-08 | -0.0027629 | COL6A3 |
| chr12 | 47754870 | 47756080 | 1211 | 7 | 1.77E-07 | 3.74E-08 | 0.001697 | 5.67E-08 | -0.00540517 | SLC48A1, RAPGEF3 |
| chr16 | 80569020 | 80569824 | 805 | 7 | 5.18E-09 | 3.21E-07 | 0.000473 | 5.68E-08 | -0.00418724 | LINC01227 |
| chr10 | 125027263 | 125028018 | 756 | 7 | 2.74E-09 | 4.71E-07 | 3.26E-05 | 5.68E-08 | -0.00116114 | CTBP2 |
| chr10 | 122909235 | 122911046 | 1812 | 7 | 1.74E-05 | 4.65E-07 | 0.000281 | 5.68E-08 | -0.00377676 | FAM24A |
| chr13 | 61319392 | 61319408 | 17 | 2 | 2.35E-08 | 4.13E-07 | 6.68E-07 | 5.69E-08 | 0.004225982 | NA |
| chr22 | 30692977 | 30694760 | 1784 | 10 | 3.18E-06 | 9.68E-08 | 0.001373 | 5.69E-08 | 0.002410163 | OSBP2 |
| chr11 | 114295718 | 114296773 | 1056 | 5 | 8.00E-11 | 2.39E-05 | 1.39E-05 | 5.71E-08 | -0.00250638 | NNMT, AP002518.2 |
| chr9 | 87702510 | 87703072 | 563 | 2 | 1.39E-09 | 5.31E-07 | 5.49E-07 | 5.71E-08 | -0.01659125 | DAPK1 |
| chr1 | 1551079 | 1552662 | 1584 | 7 | 1.50E-06 | 1.00E-06 | 0.000385 | 5.71E-08 | -0.0018619 | SSU72 |
| chr7 | 28723364 | 28724228 | 865 | 2 | 1.10E-06 | 3.70E-08 | 3.16E-05 | 5.72E-08 | -0.00802386 | CREB5 |
| chr22 | 41234978 | 41235797 | 820 | 5 | 3.32E-08 | 7.44E-08 | 2.11E-05 | 5.72E-08 | -0.00034353 | CHADL |
| chr4 | 20699952 | 20700754 | 803 | 11 | 2.73E-08 | 2.62E-07 | 0.000404 | 5.73E-08 | 0.002000462 | PACRGL |
| chr7 | 87416194 | 87416807 | 614 | 2 | 3.06E-10 | 1.61E-06 | 2.83E-07 | 5.73E-08 | 0.006744127 | ABCB4 |
| chr21 | 36319449 | 36320542 | 1094 | 15 | 7.27E-08 | 2.96E-07 | 0.001983 | 5.73E-08 | 0.000401932 | MORC3, AP000692.2 |
| chr12 | 115866791 | 115867152 | 362 | 4 | 6.43E-08 | 5.98E-08 | 0.000104 | 5.74E-08 | 0.01108301 | LINC02463 |
| chr13 | 60215021 | 60215444 | 424 | 5 | 4.47E-09 | 6.31E-05 | 1.46E-05 | 5.74E-08 | 0.00261946 | NA |
| chr2 | 96507976 | 96509522 | 1547 | 15 | 2.67E-08 | 7.31E-07 | 0.000431 | 5.74E-08 | -0.00100621 | NEURL3 |
| chr14 | 96663115 | 96663430 | 316 | 3 | 3.32E-08 | 5.12E-08 | 2.36E-05 | 5.74E-08 | -0.00649303 | NA |
| chr11 | 2149549 | 2150868 | 1320 | 12 | 1.57E-09 | 7.78E-06 | 4.35E-05 | 5.76E-08 | -0.00351187 | AC132217.2, INS-IGF2 |
| chr17 | 78473871 | 78474341 | 471 | 3 | 3.41E-10 | 5.63E-07 | 2.42E-06 | 5.76E-08 | -0.00241418 | DNAH17 |
| chr5 | 139439096 | 139440053 | 958 | 16 | 1.21E-08 | 4.71E-07 | 0.002761 | 5.77E-08 | -0.00024485 | DNAJC18 |
| chr21 | 17737860 | 17738813 | 954 | 5 | 9.48E-08 | 4.27E-07 | 8.69E-05 | 5.77E-08 | -0.00480104 | NA |
| chr9 | 76713606 | 76713728 | 123 | 2 | 6.15E-08 | 4.01E-08 | 2.28E-05 | 5.77E-08 | 0.010233002 | PCA3, PRUNE2 |
| chr3 | 53811260 | 53811903 | 644 | 2 | 7.98E-11 | 1.08E-06 | 3.51E-07 | 5.77E-08 | -0.00280213 | CACNA1D |
| chr17 | 2388340 | 2388745 | 406 | 3 | 1.77E-08 | 2.48E-07 | 5.70E-06 | 5.78E-08 | -0.0050578 | MNT |
| chr17 | 39684704 | 39685817 | 1114 | 3 | 9.89E-07 | 2.57E-08 | 0.000137 | 5.78E-08 | 0.000290844 | PGAP3 |
| chr9 | 34328754 | 34330592 | 1839 | 13 | 1.65E-06 | 4.57E-06 | 0.000276 | 5.78E-08 | -0.00282235 | NUDT2 |
| chr9 | 20806055 | 20807342 | 1288 | 4 | 1.49E-07 | 3.39E-08 | 0.000119 | 5.79E-08 | -0.00622867 | FOCAD |
| chr13 | 113429726 | 113431417 | 1692 | 9 | 2.05E-05 | 4.58E-08 | 0.001112 | 5.79E-08 | 0.000108241 | ADPRHL1 |
| chr16 | 30558098 | 30558657 | 560 | 9 | 6.02E-09 | 3.82E-06 | 1.34E-06 | 5.79E-08 | -0.00104381 | ZNF764 |
| chr12 | 113358092 | 113359041 | 950 | 9 | 2.38E-08 | 1.63E-06 | 2.41E-05 | 5.79E-08 | 0.001276169 | PLBD2, SLC8B1 |
| chr7 | 157902053 | 157903972 | 1920 | 9 | 2.78E-07 | 3.69E-07 | 0.000101 | 5.80E-08 | -0.00675397 | PTPRN2 |
| chr22 | 37328668 | 37329212 | 545 | 4 | 1.10E-08 | 7.06E-07 | 0.000172 | 5.80E-08 | -0.01508071 | NA |
| chr6 | 5260431 | 5261784 | 1354 | 13 | 4.04E-09 | 2.47E-05 | 3.32E-05 | 5.81E-08 | 0.000479412 | FARS2, LYRM4 |
| chr19 | 3707463 | 3708511 | 1049 | 13 | 1.88E-10 | 2.15E-05 | 4.83E-05 | 5.82E-08 | -0.00257257 | TJP3 |
| chr2 | 11669555 | 11671102 | 1548 | 14 | 6.20E-10 | 6.34E-05 | 7.77E-06 | 5.82E-08 | 0.003312016 | NTSR2 |
| chr17 | 47802069 | 47803521 | 1453 | 3 | 2.53E-06 | 1.01E-06 | 2.25E-05 | 5.82E-08 | -0.00795098 | NA |
| chr10 | 72272972 | 72273856 | 885 | 10 | 2.63E-11 | 9.42E-05 | 2.36E-06 | 5.82E-08 | 0.001813412 | NA |
| chr6 | 3279748 | 3280401 | 654 | 4 | 4.64E-09 | 2.71E-07 | 1.63E-05 | 5.82E-08 | 9.77E-07 | PSMG4, SLC22A23 |
| chr19 | 41875912 | 41877188 | 1277 | 5 | 5.39E-08 | 4.91E-07 | 8.27E-05 | 5.83E-08 | 0.007990681 | NA |
| chr15 | 45634430 | 45635356 | 927 | 16 | 1.03E-07 | 1.11E-06 | 0.000257 | 5.84E-08 | -0.00247824 | AC090527.2, SQOR |
| chr6 | 144064535 | 144065711 | 1177 | 19 | 5.10E-08 | 1.78E-07 | 0.002422 | 5.85E-08 | -0.0037663 | PLAGL1 |
| chr9 | 83544262 | 83544682 | 421 | 2 | 2.50E-08 | 6.31E-08 | 6.95E-06 | 5.86E-08 | -0.01587156 | NA |
| chr13 | 45712829 | 45714378 | 1550 | 7 | 3.61E-08 | 8.42E-07 | 0.000172 | 5.86E-08 | -0.00387772 | CBY2 |
| chr1 | 94863169 | 94864150 | 982 | 3 | 5.27E-07 | 1.20E-07 | 4.83E-05 | 5.86E-08 | -0.00927712 | SLC44A3 |
| chr5 | 912706 | 913896 | 1191 | 6 | 2.56E-06 | 1.08E-07 | 0.001046 | 5.87E-08 | -0.0003963 | TRIP13 |
| chr1 | 20175240 | 20177401 | 2162 | 6 | 2.60E-06 | 1.09E-06 | 0.000567 | 5.89E-08 | -0.00519036 | PLA2G2C |
| chr7 | 50434526 | 50434636 | 111 | 3 | 3.03E-08 | 1.14E-06 | 3.44E-05 | 5.89E-08 | -0.0086162 | NA |
| chr5 | 174751101 | 174752285 | 1185 | 7 | 6.72E-08 | 1.94E-06 | 7.26E-05 | 5.89E-08 | -4.47E-05 | AC113346.2, MIR4634 |

| | | | | | | | | | | |
|---|---|---|---|---|---|---|---|---|---|---|
| chr16 | 72947304 | 72948217 | 914 | 6 | 1.37E-09 | 2.81E-06 | 4.31E-05 | 5.90E-08 | -0.00335666 | ZFHX3 |
| chr9 | 68685252 | 68685919 | 668 | 3 | 8.56E-08 | 2.64E-08 | 0.000179 | 5.90E-08 | -0.00667644 | NA |
| chr7 | 50142019 | 50143473 | 1455 | 4 | 1.29E-05 | 1.50E-08 | 0.000443 | 5.90E-08 | -0.00429196 | SPATA48, AC020743.1 |
| chr22 | 36819492 | 36820023 | 532 | 7 | 7.00E-08 | 3.27E-07 | 0.000118 | 5.90E-08 | -0.01111442 | Z82185.1 |
| chr14 | 90060568 | 90062027 | 1460 | 13 | 3.07E-08 | 2.23E-06 | 4.56E-05 | 5.90E-08 | -0.00112483 | KCNK13 |
| chr13 | 113104424 | 113105729 | 1306 | 7 | 2.59E-07 | 1.73E-08 | 0.001499 | 5.91E-08 | -0.00400661 | NA |
| chr1 | 169112290 | 169112656 | 367 | 4 | 2.97E-08 | 7.57E-07 | 0.000139 | 5.92E-08 | 0.012859611 | ATP1B1 |
| chr17 | 5191586 | 5192950 | 1365 | 13 | 9.53E-07 | 1.73E-08 | 0.002398 | 5.93E-08 | -0.00221554 | AC087500.1, ZNF594 |
| chr7 | 127244222 | 127244947 | 726 | 4 | 1.57E-09 | 2.46E-06 | 1.83E-06 | 5.93E-08 | -0.00986348 | GRM8 |
| chr7 | 2734148 | 2734780 | 633 | 4 | 6.09E-10 | 1.08E-06 | 5.07E-05 | 5.93E-08 | 0.011714115 | AMZ1, GNA12 |
| chr1 | 213015380 | 213016481 | 1102 | 16 | 1.89E-07 | 3.76E-08 | 0.003383 | 5.94E-08 | 0.001577357 | ANGEL2 |
| chr19 | 16124211 | 16125041 | 831 | 3 | 2.96E-07 | 6.47E-07 | 6.07E-06 | 5.94E-08 | -0.00530885 | RAB8A, AC008894.2 |
| chr1 | 183249134 | 183249531 | 398 | 2 | 3.99E-08 | 4.81E-08 | 1.39E-05 | 5.95E-08 | -0.00567932 | NMNAT2 |
| chr12 | 14938767 | 14939500 | 734 | 5 | 1.66E-08 | 3.36E-05 | 2.01E-05 | 5.95E-08 | -0.00841694 | NA |
| chr16 | 2264360 | 2265778 | 1419 | 6 | 3.36E-06 | 1.35E-07 | 0.000459 | 5.97E-08 | 0.002878558 | RNPS1 |
| chr19 | 39445203 | 39446056 | 854 | 17 | 6.45E-09 | 5.55E-06 | 0.00106 | 5.97E-08 | 0.000466443 | SUPT5H |
| chr16 | 21277947 | 21279148 | 1202 | 9 | 3.58E-10 | 3.10E-05 | 1.71E-05 | 5.98E-08 | 0.001937194 | CRYM |
| chr2 | 212537709 | 212539528 | 1820 | 14 | 4.61E-09 | 9.06E-07 | 5.39E-05 | 5.98E-08 | -0.00185102 | ERBB4 |
| chr19 | 58511338 | 58512549 | 1212 | 4 | 9.22E-08 | 6.39E-08 | 0.000131 | 5.98E-08 | -0.00230785 | SLC27A5 |
| chr2 | 28990734 | 28990876 | 143 | 2 | 2.49E-09 | 2.98E-06 | 2.20E-07 | 5.99E-08 | -0.0034464 | TOGARAM2 |
| chr10 | 5445619 | 5447222 | 1604 | 12 | 2.30E-08 | 1.65E-06 | 0.000176 | 6.00E-08 | -0.0038177 | NET1 |
| chr3 | 52245387 | 52246686 | 1300 | 17 | 1.35E-07 | 1.20E-06 | 0.001355 | 6.00E-08 | -0.00241937 | PPM1M, TWF2 |
| chr9 | 93201220 | 93202325 | 1106 | 7 | 7.33E-11 | 4.10E-05 | 6.78E-06 | 6.00E-08 | -0.00595539 | WNK2 |
| chr15 | 88257279 | 88258243 | 965 | 8 | 3.36E-09 | 7.62E-05 | 4.67E-05 | 6.02E-08 | -0.00500115 | NTRK3-AS1 |
| chr14 | 74294411 | 74295287 | 877 | 6 | 2.04E-08 | 5.94E-07 | 0.00084 | 6.03E-08 | -0.00551014 | ABCD4, AC005519.1 |
| chr12 | 95470273 | 95470598 | 326 | 2 | 4.50E-08 | 4.81E-08 | 1.45E-05 | 6.03E-08 | 0.009625749 | NA |
| chr20 | 31604727 | 31606221 | 1495 | 15 | 1.30E-06 | 2.23E-07 | 0.000553 | 6.03E-08 | 0.001601172 | ID1 |
| chr7 | 5495320 | 5496303 | 984 | 12 | 3.19E-08 | 6.54E-07 | 0.000822 | 6.04E-08 | -0.00549389 | FBXL18, MIR589 |
| chr19 | 18856134 | 18856467 | 334 | 2 | 2.93E-10 | 3.67E-07 | 8.11E-07 | 6.04E-08 | -0.00210989 | UPF1 |
| chr11 | 6497033 | 6498223 | 1191 | 14 | 4.64E-08 | 3.76E-05 | 0.00016 | 6.04E-08 | -0.00028501 | AC084337.2, DNHD1 |
| chr12 | 47896219 | 47896944 | 726 | 4 | 1.48E-07 | 6.82E-08 | 0.000238 | 6.05E-08 | -0.00641632 | AC121338.1, VDR |
| chr9 | 124022131 | 124022818 | 688 | 3 | 2.33E-08 | 5.27E-06 | 2.68E-06 | 6.05E-08 | 0.001352955 | LHX2 |
| chr8 | 63012720 | 63013040 | 321 | 4 | 1.25E-08 | 2.53E-06 | 1.66E-05 | 6.05E-08 | 0.000140796 | NKAIN3 |
| chr17 | 7314650 | 7315640 | 991 | 10 | 9.13E-10 | 7.37E-06 | 6.54E-05 | 6.07E-08 | 0.001003269 | GPS2, AC026954.2, NEURL4 |
| chr11 | 67432440 | 67433759 | 1320 | 12 | 1.17E-08 | 4.80E-05 | 3.02E-05 | 6.08E-08 | 0.002636261 | RPS6KB2, RPS6KB2-AS1 |
| chr8 | 835815 | 837298 | 1484 | 10 | 9.53E-07 | 5.91E-06 | 0.000138 | 6.09E-08 | -0.00188439 | DLGAP2 |
| chr7 | 74532072 | 74532571 | 500 | 2 | 1.91E-08 | 7.55E-08 | 5.32E-06 | 6.09E-08 | -0.0028355 | GTF2IRD1 |
| chr11 | 12540381 | 12541793 | 1413 | 5 | 7.13E-08 | 1.08E-06 | 9.00E-06 | 6.09E-08 | -0.00684766 | AC009806.1 |
| chr12 | 45050025 | 45050840 | 816 | 8 | 5.22E-09 | 2.05E-07 | 0.000475 | 6.11E-08 | 0.002414839 | DBX2 |
| chr17 | 83151544 | 83151582 | 39 | 2 | 4.15E-08 | 3.76E-08 | 4.59E-05 | 6.12E-08 | -0.00737908 | AC139099.2 |
| chr8 | 10825534 | 10826237 | 704 | 6 | 1.25E-09 | 1.45E-05 | 4.42E-07 | 6.12E-08 | -0.006781 | AC105001.2, PINX1 |
| chr5 | 122844082 | 122845328 | 1247 | 12 | 3.08E-08 | 8.88E-07 | 0.000586 | 6.13E-08 | -0.00257507 | SNX24 |
| chr16 | 85812020 | 85812578 | 559 | 5 | 1.38E-07 | 5.89E-08 | 0.000519 | 6.13E-08 | -0.02015738 | NA |
| chr1 | 161177188 | 161179090 | 1903 | 18 | 2.51E-09 | 0.0001448 | 4.03E-05 | 6.16E-08 | -0.00238369 | PPOX, B4GALT3 |
| chr14 | 34035296 | 34035844 | 549 | 2 | 6.65E-10 | 1.26E-06 | 3.46E-07 | 6.17E-08 | -0.00716401 | EGLN3 |
| chr22 | 37198893 | 37199493 | 601 | 4 | 1.28E-09 | 2.17E-05 | 2.38E-06 | 6.17E-08 | 0.000829863 | C1QTNF6 |
| chr12 | 62438405 | 62438760 | 356 | 3 | 6.61E-08 | 3.28E-08 | 0.000142 | 6.18E-08 | -0.01526222 | NA |
| chr6 | 159788454 | 159790445 | 1992 | 22 | 4.86E-07 | 4.34E-05 | 4.55E-05 | 6.18E-08 | 7.82E-05 | MRPL18, TCP1 |
| chr8 | 29286682 | 29287121 | 440 | 2 | 2.85E-09 | 5.79E-07 | 5.75E-07 | 6.18E-08 | -0.00151557 | NA |
| chr3 | 46749987 | 46750972 | 986 | 6 | 1.61E-09 | 7.10E-07 | 2.01E-06 | 6.19E-08 | -0.00848867 | AC109583.1, PRSS45P, AC109583.4 |
| chr3 | 40505627 | 40506213 | 587 | 15 | 2.71E-07 | 2.49E-08 | 0.001036 | 6.19E-08 | -0.00178652 | ZNF620 |
| chr19 | 45691509 | 45692922 | 1414 | 16 | 3.92E-09 | 2.97E-05 | 3.23E-05 | 6.19E-08 | -0.00011276 | QPCTL, SNRPD2 |
| chr4 | 2046793 | 2047508 | 716 | 3 | 2.22E-08 | 3.99E-07 | 3.02E-06 | 6.19E-08 | -0.00835079 | NA |
| chr16 | 85659258 | 85660241 | 984 | 5 | 6.28E-07 | 6.08E-06 | 3.98E-05 | 6.20E-08 | -0.00142332 | GSE1, RN7SL381P |
| chr17 | 76584315 | 76584801 | 487 | 3 | 1.78E-09 | 3.04E-06 | 4.81E-07 | 6.21E-08 | 0.007050177 | ST6GALNAC2 |
| chr11 | 66509584 | 66510887 | 1304 | 12 | 9.93E-08 | 1.69E-07 | 0.000851 | 6.21E-08 | 3.59E-05 | DPP3, AP002748.5, BBS1 |
| chr16 | 88730711 | 88730987 | 277 | 3 | 1.36E-09 | 0.0002545 | 2.50E-07 | 6.21E-08 | -0.00282744 | PIEZO1 |
| chr8 | 126504087 | 126504633 | 547 | 4 | 4.95E-08 | 6.95E-08 | 2.75E-05 | 6.21E-08 | -0.00340024 | NA |
| chr12 | 101209551 | 101210642 | 1092 | 16 | 8.81E-08 | 4.48E-07 | 0.000453 | 6.22E-08 | 0.004699374 | SLC5A8 |
| chr1 | 109213792 | 109214461 | 670 | 4 | 6.92E-09 | 4.18E-08 | 0.000134 | 6.22E-08 | -0.00079132 | SARS1 |
| chr8 | 10334473 | 10335433 | 961 | 12 | 4.53E-08 | 1.91E-06 | 0.000305 | 6.22E-08 | -0.00100333 | MSRA |
| chr15 | 45510701 | 45511143 | 443 | 4 | 4.81E-09 | 8.50E-07 | 1.03E-06 | 6.23E-08 | -0.00718632 | AC025580.2, HMGN2P46, SLC30A4 |
| chr11 | 68839233 | 68840269 | 1037 | 7 | 2.75E-10 | 3.96E-06 | 1.90E-06 | 6.23E-08 | -0.00050431 | CPT1A |
| chr1 | 4804017 | 4804599 | 583 | 4 | 1.22E-07 | 1.71E-07 | 4.58E-05 | 6.23E-08 | -0.00556002 | NA |
| chr7 | 48009065 | 48009989 | 925 | 6 | 9.24E-08 | 2.33E-06 | 0.00022 | 6.23E-08 | -0.00526318 | SUN3 |
| chr20 | 49045288 | 49046699 | 1412 | 15 | 9.54E-08 | 7.74E-07 | 0.000819 | 6.24E-08 | -0.00025335 | CSE1L, CSE1L-AS1 |
| chr19 | 54189904 | 54191319 | 1416 | 16 | 2.13E-09 | 5.96E-06 | 1.83E-05 | 6.25E-08 | -0.0012119 | TSEN34 |
| chr6 | 41163475 | 41164317 | 843 | 5 | 8.96E-09 | 7.58E-06 | 4.70E-06 | 6.25E-08 | -0.00462242 | NA |
| chr20 | 9889877 | 9890774 | 898 | 2 | 6.84E-07 | 4.60E-08 | 1.91E-05 | 6.25E-08 | 0.005144051 | NA |
| chr20 | 47356103 | 47358302 | 2200 | 18 | 2.50E-06 | 4.06E-06 | 0.000736 | 6.25E-08 | 0.003257825 | ZMYND8 |
| chr8 | 139096031 | 139096446 | 416 | 3 | 4.37E-08 | 4.84E-08 | 8.83E-05 | 6.28E-08 | -0.02050285 | AC027541.1 |
| chr11 | 16612035 | 16613557 | 1523 | 8 | 1.98E-06 | 7.02E-08 | 0.000723 | 6.28E-08 | -0.00032524 | C11orf58, SOX6 |
| chr11 | 47247988 | 47249357 | 1370 | 20 | 1.70E-08 | 3.48E-06 | 0.000493 | 6.28E-08 | -0.0004143 | NR1H3, ACP2 |
| chr8 | 22140796 | 22142805 | 2010 | 18 | 4.67E-06 | 2.81E-06 | 0.001787 | 6.29E-08 | -0.0011332 | REEP4 |
| chr19 | 29677283 | 29677615 | 333 | 2 | 2.57E-08 | 3.88E-08 | 4.48E-05 | 6.30E-08 | -0.01039915 | NA |
| chr19 | 35084506 | 35084558 | 53 | 2 | 1.54E-08 | 1.90E-06 | 2.92E-07 | 6.33E-08 | -0.00684723 | HPN-AS1 |
| chr14 | 21030270 | 21030873 | 604 | 5 | 8.02E-09 | 2.45E-07 | 0.000423 | 6.34E-08 | -0.00146397 | TPPP2, NDRG2 |
| chr11 | 1778532 | 1779607 | 1076 | 4 | 1.09E-08 | 4.01E-07 | 1.04E-05 | 6.35E-08 | -0.00306457 | NA |
| chr11 | 72793572 | 72794562 | 991 | 13 | 1.67E-07 | 6.96E-08 | 0.006384 | 6.36E-08 | 0.000129446 | ARAP1, STARD10, AP002381.2 |
| chr12 | 6850307 | 6851930 | 1624 | 17 | 6.91E-12 | 0.0001613 | 4.88E-07 | 6.36E-08 | -0.00041457 | CDCA3 |
| chr15 | 75453037 | 75455409 | 2373 | 9 | 2.23E-05 | 2.58E-08 | 0.002443 | 6.36E-08 | -0.00057203 | SIN3A, AC105137.2 |
| chr5 | 40681342 | 40682231 | 890 | 6 | 4.87E-08 | 6.55E-09 | 0.004075 | 6.36E-08 | -0.00199925 | PTGER4, TTC33 |
| chr1 | 47765804 | 47767427 | 1624 | 4 | 3.90E-10 | 3.20E-06 | 3.47E-05 | 6.38E-08 | -0.00842412 | TRABD2B |
| chr16 | 67516608 | 67516670 | 63 | 2 | 4.50E-08 | 4.13E-08 | 3.30E-05 | 6.38E-08 | -0.01538303 | NA |
| chr16 | 72092522 | 72094847 | 2326 | 29 | 1.19E-05 | 1.88E-07 | 0.002587 | 6.39E-08 | 0.000575987 | DHX38, TXNL4B |
| chr17 | 4554540 | 4555888 | 1349 | 13 | 1.96E-08 | 8.41E-08 | 0.003941 | 6.39E-08 | 2.31E-05 | MYBBP1A |
| chr16 | 78098287 | 78100070 | 1784 | 15 | 2.01E-10 | 0.0002312 | 1.75E-06 | 6.39E-08 | -0.00125656 | WWOX |
| chr1 | 2069023 | 2070212 | 1190 | 6 | 2.82E-08 | 3.64E-07 | 6.73E-05 | 6.39E-08 | -0.00271915 | PRKCZ |
| chr10 | 100713265 | 100714053 | 789 | 5 | 1.37E-08 | 8.37E-07 | 1.48E-05 | 6.39E-08 | 0.008132625 | NA |
| chr1 | 160654619 | 160654863 | 245 | 2 | 1.61E-08 | 1.89E-07 | 1.68E-06 | 6.41E-08 | -0.00426439 | NA |
| chr4 | 7595845 | 7597009 | 1165 | 4 | 1.58E-06 | 9.51E-08 | 0.000172 | 6.41E-08 | -0.00745207 | SORCS2 |
| chr6 | 168381687 | 168382753 | 1067 | 7 | 6.28E-08 | 8.81E-08 | 0.000125 | 6.41E-08 | -0.0015034 | NA |
| chr8 | 40054773 | 40054934 | 162 | 2 | 1.22E-08 | 1.37E-06 | 3.46E-07 | 6.42E-08 | -0.03379614 | NA |
| chr15 | 88546378 | 88547279 | 902 | 11 | 7.37E-08 | 1.54E-07 | 0.00046 | 6.42E-08 | 0.000566123 | AC013489.1, DET1 |
| chr4 | 125324059 | 125324375 | 317 | 2 | 7.79E-08 | 3.96E-08 | 4.43E-05 | 6.42E-08 | -0.01615982 | FAT4 |
| chr12 | 100472399 | 100473980 | 1582 | 10 | 9.19E-08 | 1.14E-05 | 0.000689 | 6.43E-08 | 0.001688156 | NR1H4 |
| chr12 | 52068636 | 52069584 | 949 | 8 | 1.61E-09 | 7.73E-07 | 0.000234 | 6.44E-08 | -0.01907777 | ATG101 |
| chr1 | 197359534 | 197360394 | 861 | 4 | 9.89E-09 | 1.11E-06 | 1.14E-06 | 6.45E-08 | -0.00574879 | CRB1 |
| chr2 | 241229904 | 241231484 | 1581 | 11 | 1.05E-07 | 3.89E-06 | 4.05E-05 | 6.45E-08 | -0.00151528 | HDLBP |
| chr3 | 122564156 | 122566163 | 2008 | 19 | 1.01E-06 | 0.0001141 | 8.94E-05 | 6.45E-08 | 0.001228522 | DTX3L, PARP9 |
| chr13 | 32246376 | 32246738 | 363 | 2 | 3.02E-08 | 7.17E-08 | 6.78E-06 | 6.46E-08 | -0.0104974 | FRY |
| chr1 | 147756774 | 147757654 | 881 | 4 | 3.15E-08 | 1.02E-07 | 0.000108 | 6.46E-08 | -0.00984391 | AC241644.3, GJA5 |
| chr4 | 185395940 | 185397202 | 1263 | 13 | 1.83E-09 | 1.71E-06 | 0.000421 | 6.49E-08 | 0.000381047 | ANKRD37 |
| chr8 | 735830 | 736755 | 926 | 5 | 6.99E-07 | 2.08E-08 | 0.00065 | 6.49E-08 | -0.02046371 | ERICH1 |
| chr13 | 22926699 | 22927318 | 620 | 3 | 1.06E-09 | 2.31E-06 | 2.25E-05 | 6.49E-08 | -0.02292034 | NA |
| chr22 | 47292662 | 47292775 | 114 | 2 | 6.49E-08 | 5.25E-08 | 1.43E-05 | 6.49E-08 | -0.04270682 | NA |
| chr1 | 77978156 | 77979596 | 1441 | 16 | 6.34E-08 | 3.24E-06 | 0.001615 | 6.50E-08 | 0.000760751 | DNAJB4, GIPC2, FUBP1 |
| chr6 | 118787364 | 118787820 | 457 | 3 | 5.41E-09 | 2.33E-07 | 4.35E-06 | 6.51E-08 | 0.009813597 | NA |
| chr12 | 9115783 | 9117146 | 1364 | 10 | 5.25E-08 | 1.74E-05 | 2.92E-05 | 6.51E-08 | 0.001642211 | A2M |
| chr4 | 41880146 | 41881476 | 1331 | 10 | 3.44E-10 | 2.00E-05 | 0.000283 | 6.52E-08 | -0.00307574 | LINC00682 |
| chr2 | 47833108 | 47834100 | 993 | 2 | 6.68E-08 | 1.06E-07 | 3.56E-06 | 6.53E-08 | -0.00453715 | FBXO11 |
| chr5 | 2252764 | 2253015 | 252 | 4 | 4.89E-09 | 0.0003806 | 1.66E-06 | 6.53E-08 | -0.00736942 | NA |
| chr12 | 57549224 | 57550276 | 1053 | 10 | 9.21E-08 | 1.96E-06 | 0.001107 | 6.54E-08 | 0.001359366 | KIF5A |
| chr1 | 166988496 | 166989793 | 1298 | 15 | 3.13E-08 | 4.57E-06 | 0.000213 | 6.54E-08 | -0.00149846 | MAEL |
| chr12 | 24918335 | 24919314 | 980 | 3 | 3.71E-09 | 6.06E-06 | 1.69E-05 | 6.54E-08 | -0.00807216 | BCAT1 |
| chr1 | 153658175 | 153659854 | 1680 | 16 | 1.42E-06 | 2.82E-05 | 0.000921 | 6.54E-08 | -0.00155619 | SNAPIN |
| chr11 | 16604562 | 16605780 | 1219 | 6 | 8.45E-10 | 1.60E-05 | 2.53E-05 | 6.54E-08 | 0.001222812 | SOX6 |
| chr6 | 22568976 | 22569917 | 942 | 11 | 1.90E-08 | 9.13E-07 | 0.00032 | 6.54E-08 | -0.00177807 | CASC15, HDGFL1 |
| chr7 | 47557194 | 47557840 | 647 | 3 | 6.48E-08 | 3.41E-08 | 7.42E-05 | 6.54E-08 | -0.00315913 | TNS3 |
| chr5 | 94618127 | 94619375 | 1249 | 22 | 6.44E-09 | 1.22E-06 | 0.001021 | 6.55E-08 | 0.000729463 | SLF1, KIAA0825 |
| chr16 | 3735926 | 3737054 | 1129 | 4 | 3.84E-09 | 1.04E-06 | 7.79E-06 | 6.56E-08 | 0.000280335 | CREBBP |
| chr4 | 2041326 | 2042687 | 1362 | 15 | 1.58E-07 | 5.11E-07 | 0.002586 | 6.57E-08 | 0.000467035 | C4orf48, NELFA |
| chr20 | 38835536 | 38836682 | 1147 | 8 | 4.50E-09 | 2.26E-06 | 0.000108 | 6.58E-08 | -0.00348725 | PPP1R16B |
| chr21 | 44994124 | 44994671 | 548 | 5 | 8.43E-09 | 1.24E-07 | 4.91E-05 | 6.58E-08 | -0.00613082 | LINC00165 |
| chr20 | 21704763 | 21706090 | 1328 | 16 | 1.56E-09 | 4.72E-05 | 4.26E-05 | 6.60E-08 | -0.00063131 | PAX1 |
| chr3 | 193592237 | 193593955 | 1719 | 14 | 3.03E-06 | 9.10E-08 | 0.001002 | 6.63E-08 | -0.00058589 | OPA1, ATP13A4, AC048351.1 |
| chr6 | 4078399 | 4079820 | 1422 | 15 | 1.51E-07 | 6.83E-06 | 0.000135 | 6.63E-08 | 0.001152066 | C6orf201, FAM217A |
| chr2 | 232884703 | 232885879 | 1177 | 10 | 1.82E-09 | 6.76E-06 | 5.21E-06 | 6.63E-08 | 6.86E-05 | NGEF |
| chr14 | 36523028 | 36524312 | 1285 | 3 | 3.23E-11 | 3.36E-05 | 8.00E-07 | 6.64E-08 | 0.019140137 | AL132857.1 |
| chr3 | 113012283 | 113012572 | 290 | 2 | 2.39E-08 | 4.55E-08 | 2.61E-05 | 6.65E-08 | -0.01658658 | GTPBP8, NEPRO |
| chr4 | 7611106 | 7611767 | 662 | 2 | 1.03E-09 | 8.66E-07 | 4.79E-07 | 6.66E-08 | -0.00660407 | SORCS2 |
| chr1 | 42924724 | 42925034 | 311 | 3 | 2.25E-09 | 3.44E-07 | 4.65E-05 | 6.67E-08 | 0.00269047 | NA |
| chr3 | 9791430 | 9793071 | 1642 | 27 | 4.52E-08 | 4.38E-06 | 0.001688 | 6.68E-08 | 0.001534537 | ARPC4, TADA3 |
| chr5 | 34502683 | 34503075 | 393 | 2 | 2.28E-09 | 7.28E-08 | 7.12E-06 | 6.68E-08 | -0.01745432 | NA |
| chr5 | 135307261 | 135307401 | 141 | 3 | 4.26E-08 | 2.70E-07 | 1.33E-05 | 6.68E-08 | -0.00175574 | C5orf66 |
| chr17 | 81636487 | 81637952 | 1466 | 13 | 4.02E-09 | 2.24E-06 | 1.96E-06 | 6.68E-08 | 0.000725824 | TSPAN10, NPLOC4 |
| chr1 | 222011623 | 222011831 | 209 | 2 | 1.80E-09 | 0.0001942 | 9.44E-08 | 6.69E-08 | -0.01317821 | LINC01705 |
| chr16 | 68642375 | 68643461 | 1087 | 9 | 3.63E-08 | 1.02E-07 | 0.000209 | 6.70E-08 | -0.00469827 | CDH3 |
| chr4 | 6200364 | 6201833 | 1470 | 10 | 1.31E-08 | 1.49E-06 | 0.000137 | 6.70E-08 | 0.001812299 | LINC02495, C4orf50, JAKMIP1 |
| chr2 | 151923479 | 151923809 | 331 | 2 | 5.23E-08 | 5.25E-08 | 1.62E-05 | 6.72E-08 | 0.01650093 | AC068547.1, CACNB4 |
| chr10 | 50990181 | 50991170 | 990 | 10 | 6.45E-11 | 2.17E-06 | 0.000102 | 6.72E-08 | -0.00784973 | PRKG1 |
| chr8 | 125544463 | 125544814 | 352 | 3 | 2.95E-09 | 5.19E-06 | 6.05E-06 | 6.72E-08 | -0.00035703 | NA |
| chr3 | 33659198 | 33660215 | 1018 | 12 | 4.22E-07 | 8.50E-07 | 0.000846 | 6.73E-08 | -0.00255554 | CLASP2 |
| chr10 | 18400106 | 18401019 | 914 | 11 | 1.87E-08 | 1.45E-06 | 0.001124 | 6.77E-08 | 0.013921658 | CACNB2 |
| chr12 | 120099114 | 120100446 | 1333 | 6 | 2.00E-07 | 9.32E-08 | 0.00102 | 6.77E-08 | 0.001585449 | RAB35 |
| chr21 | 31334170 | 31334937 | 768 | 3 | 1.02E-08 | 1.95E-07 | 3.97E-05 | 6.78E-08 | 0.003453745 | TIAM1 |
| chr8 | 99893932 | 99895318 | 1387 | 10 | 8.31E-10 | 5.43E-07 | 3.13E-05 | 6.78E-08 | -0.00207323 | AC105328.1 |
| chr16 | 52950 | 54539 | 1590 | 11 | 1.13E-08 | 1.09E-05 | 3.11E-05 | 6.78E-08 | 0.001427349 | SNRNP25, POLR3K |
| chr2 | 23690224 | 23691042 | 819 | 5 | 1.38E-07 | 1.00E-07 | 0.000164 | 6.78E-08 | -0.00278988 | KLHL29 |
| chr17 | 79801888 | 79804668 | 2781 | 15 | 2.26E-07 | 0.0020424 | 2.96E-05 | 6.78E-08 | -0.00219961 | AC100791.2 |
| chr11 | 129363826 | 129364965 | 1140 | 7 | 5.05E-08 | 2.44E-07 | 0.000169 | 6.79E-08 | -0.00453529 | NA |
| chr16 | 4474051 | 4474774 | 724 | 12 | 2.44E-09 | 9.81E-07 | 2.90E-05 | 6.81E-08 | -0.003552 | HMOX2, NMRAL1 |
| chr7 | 102457557 | 102458024 | 468 | 4 | 3.00E-08 | 2.71E-07 | 1.27E-05 | 6.82E-08 | 7.91E-06 | ALKBH4 |
| chr16 | 11734813 | 11735633 | 821 | 4 | 8.81E-08 | 1.29E-07 | 3.44E-05 | 6.82E-08 | 0.001400497 | TXNDC11 |
| chr3 | 13072459 | 13073668 | 1210 | 10 | 2.51E-08 | 1.64E-06 | 0.001308 | 6.83E-08 | 0.005443569 | IQSEC1 |
| chr13 | 60612742 | 60613578 | 837 | 4 | 2.57E-07 | 2.57E-08 | 0.000167 | 6.84E-08 | -0.01719446 | NA |
| chr11 | 36510204 | 36510875 | 672 | 14 | 6.38E-08 | 7.00E-06 | 5.24E-05 | 6.84E-08 | -0.00189075 | AC061999.1, RAG1, TRAF6 |
| chr3 | 59064078 | 59064471 | 394 | 3 | 2.45E-08 | 5.92E-08 | 2.65E-05 | 6.84E-08 | 0.002114397 | AC126121.3 |

| | | | | | | | | | | |
|---|---|---|---|---|---|---|---|---|---|---|
| chr11 | 62391431 | 62391782 | 352 | 2 | 8.39E-09 | 1.50E-07 | 2.43E-06 | 6.86E-08 | 0.003090305 | ASRGL1, AP003064.1 |
| chr2 | 238403856 | 238404416 | 561 | 5 | 1.50E-07 | 1.61E-07 | 0.00067 | 6.86E-08 | -0.00059184 | NA |
| chr12 | 43937993 | 43938544 | 552 | 2 | 1.70E-08 | 9.95E-08 | 4.33E-06 | 6.86E-08 | -0.00921127 | TMEM117 |
| chr6 | 45663054 | 45664188 | 1135 | 7 | 1.72E-06 | 3.79E-08 | 0.000996 | 6.86E-08 | -0.00235354 | RUNX2 |
| chr10 | 815809 | 816697 | 889 | 4 | 1.94E-07 | 2.95E-08 | 0.000268 | 6.86E-08 | 0.002938384 | LARP4B |
| chr12 | 31958323 | 31959504 | 1182 | 11 | 6.88E-08 | 2.09E-08 | 0.001621 | 6.87E-08 | 0.001052348 | RESF1, LINC02422 |
| chr12 | 57631202 | 57633330 | 2129 | 16 | 5.45E-08 | 0.0078047 | 3.72E-05 | 6.87E-08 | -0.00242473 | B4GALNT1 |
| chr3 | 42852280 | 42853010 | 731 | 4 | 7.88E-11 | 6.90E-05 | 6.88E-07 | 6.89E-08 | -0.00800736 | ACKR2, AC099329.2, AC092042.3, KRBOX1, AC099329.1 |
| chr1 | 220751360 | 220752173 | 814 | 2 | 2.76E-07 | 4.81E-08 | 2.44E-05 | 6.90E-08 | 0.008810324 | MTARC2 |
| chr5 | 32016969 | 32017108 | 140 | 2 | 5.11E-08 | 8.33E-08 | 5.89E-06 | 6.90E-08 | -0.00798729 | PDZD2 |
| chr10 | 1210399 | 1210711 | 313 | 4 | 6.48E-09 | 1.11E-06 | 9.00E-06 | 6.91E-08 | -0.01120202 | LINC00200, ADARB2 |
| chr5 | 139872249 | 139873186 | 938 | 3 | 4.19E-07 | 4.48E-08 | 5.18E-05 | 6.91E-08 | -0.00231015 | NRG2 |
| chr2 | 76771608 | 76772231 | 624 | 3 | 4.13E-09 | 1.46E-07 | 8.59E-06 | 6.91E-08 | -0.00766928 | LRRTM4 |
| chr7 | 98228044 | 98228980 | 937 | 9 | 4.96E-08 | 1.43E-07 | 0.000292 | 6.91E-08 | 0.000876364 | TECPR1 |
| chr4 | 158201452 | 158201863 | 412 | 3 | 2.57E-09 | 2.30E-05 | 8.11E-07 | 6.92E-08 | 0.006563965 | FAM198B-AS1, TMEM144 |
| chr5 | 1335020 | 1335850 | 831 | 6 | 9.46E-09 | 2.94E-07 | 1.00E-05 | 6.92E-08 | -0.00231341 | CLPTM1L |
| chr22 | 30286807 | 30287954 | 1148 | 8 | 8.96E-08 | 8.91E-07 | 0.000452 | 6.93E-08 | -0.00455403 | CASTOR1, AC004997.1 |
| chr6 | 37819635 | 37820395 | 761 | 4 | 1.06E-08 | 7.17E-08 | 0.000201 | 6.94E-08 | -0.00360261 | ZFAND3 |
| chr20 | 59452852 | 59453664 | 813 | 3 | 2.81E-08 | 7.25E-08 | 2.32E-05 | 6.95E-08 | -0.00497946 | NA |
| chr2 | 241065117 | 241065685 | 569 | 4 | 4.58E-09 | 1.10E-05 | 5.46E-06 | 6.95E-08 | -0.00353708 | SNED1 |
| chr14 | 97789557 | 97790376 | 820 | 3 | 6.03E-08 | 3.11E-08 | 0.000189 | 6.95E-08 | -0.00253279 | NA |
| chr17 | 74592273 | 74592971 | 699 | 5 | 1.31E-07 | 1.34E-07 | 2.19E-05 | 6.96E-08 | -0.00583578 | C17orf77, CD300LD |
| chr6 | 108260861 | 108261867 | 1007 | 11 | 8.69E-08 | 8.98E-06 | 4.64E-05 | 6.96E-08 | -0.00247811 | Z98742.4, SNX3 |
| chr18 | 76979313 | 76980911 | 1599 | 9 | 1.06E-07 | 3.76E-06 | 0.000781 | 6.97E-08 | -0.00222021 | AC093330.1, MBP |
| chr14 | 20413281 | 20414800 | 1520 | 13 | 4.02E-06 | 5.13E-08 | 0.001826 | 6.97E-08 | 0.001393705 | TEP1 |
| chr1 | 151944258 | 151945067 | 810 | 3 | 6.89E-08 | 4.15E-08 | 0.000131 | 6.98E-08 | 0.005396054 | AL450992.3 |
| chr4 | 37889755 | 37891341 | 1587 | 6 | 1.74E-10 | 1.13E-05 | 3.20E-06 | 6.98E-08 | -0.00871279 | TBC1D1 |
| chr8 | 141392321 | 141393474 | 1154 | 5 | 8.24E-10 | 2.24E-06 | 3.32E-06 | 6.99E-08 | -0.00784626 | PTP4A3, AC100803.2 |
| chr13 | 43875347 | 43876708 | 1362 | 3 | 1.61E-12 | 0.0003003 | 7.31E-08 | 6.99E-08 | -0.00250382 | CCDC122 |
| chr17 | 8287352 | 8288706 | 1355 | 12 | 1.09E-09 | 2.13E-07 | 0.000145 | 6.99E-08 | -0.00338032 | RANGRF, SLC25A35 |
| chr18 | 79862587 | 79863598 | 1012 | 10 | 2.11E-07 | 1.38E-07 | 0.002547 | 6.99E-08 | 0.010360915 | NA |
| chr8 | 26040675 | 26041597 | 923 | 7 | 2.60E-08 | 4.68E-08 | 0.001051 | 6.99E-08 | 0.003992485 | EBF2 |
| chr13 | 111853330 | 111853904 | 575 | 9 | 6.67E-09 | 0.0001006 | 5.79E-05 | 7.00E-08 | -0.00054624 | NA |
| chr5 | 146337447 | 146338928 | 1482 | 10 | 1.32E-07 | 2.67E-06 | 4.63E-05 | 7.00E-08 | 0.001797337 | AC091959.1, POU4F3, AC011396.2 |
| chr7 | 158879672 | 158880067 | 396 | 3 | 1.33E-10 | 6.97E-05 | 7.18E-08 | 7.01E-08 | 0.00779048 | DYNC2I1 |
| chr9 | 35673721 | 35674003 | 283 | 4 | 2.85E-08 | 1.38E-07 | 2.01E-05 | 7.01E-08 | -0.00520805 | CA9, ARHGEF39 |
| chr19 | 43802463 | 43803297 | 835 | 6 | 3.05E-08 | 2.72E-06 | 0.000135 | 7.03E-08 | -0.01242627 | LYPD5 |
| chr12 | 48993788 | 48995204 | 1417 | 5 | 1.15E-07 | 2.75E-07 | 0.000298 | 7.03E-08 | -0.01040466 | AC011603.3, DDN |
| chr1 | 43384511 | 43385069 | 559 | 5 | 9.87E-08 | 5.11E-07 | 2.09E-05 | 7.04E-08 | -0.00183959 | MED8 |
| chr11 | 69671718 | 69672295 | 578 | 4 | 5.91E-09 | 4.31E-06 | 1.11E-05 | 7.06E-08 | -0.00341324 | LTO1 |
| chr17 | 78315787 | 78316374 | 588 | 3 | 9.83E-08 | 3.27E-08 | 0.000114 | 7.07E-08 | -0.01056709 | AC087645.2 |
| chr3 | 138959744 | 138961404 | 1661 | 6 | 1.89E-06 | 4.80E-08 | 0.000839 | 7.07E-08 | 0.000921914 | NA |
| chr16 | 89392162 | 89392849 | 688 | 2 | 2.98E-09 | 3.68E-07 | 1.01E-06 | 7.07E-08 | -0.00762015 | ANKRD11 |
| chr13 | 79480841 | 79481934 | 1094 | 13 | 5.77E-08 | 2.86E-07 | 0.000131 | 7.10E-08 | 0.000132525 | NDFIP2, NDFIP2-AS1 |
| chr7 | 25858287 | 25859574 | 1288 | 10 | 7.99E-08 | 1.81E-06 | 0.000178 | 7.13E-08 | -0.00129032 | NA |
| chr15 | 85768923 | 85769343 | 421 | 4 | 1.49E-08 | 2.37E-06 | 4.80E-05 | 7.15E-08 | -0.00107268 | KLHL25 |
| chr6 | 46651951 | 46654119 | 2169 | 17 | 5.15E-08 | 1.92E-05 | 0.000449 | 7.16E-08 | -0.00229692 | SLC25A27, CYP39A1 |
| chr22 | 45670896 | 45672116 | 1221 | 6 | 4.09E-07 | 7.19E-08 | 0.001101 | 7.16E-08 | -0.00465236 | Z95331.1, ATXN10 |
| chr5 | 114669455 | 114669544 | 90 | 2 | 6.34E-09 | 2.88E-06 | 2.70E-07 | 7.16E-08 | -0.01040731 | NA |
| chr1 | 237003789 | 237004975 | 1187 | 5 | 1.15E-08 | 6.92E-08 | 0.000393 | 7.18E-08 | -0.00732799 | MT1HL1 |
| chr3 | 194537641 | 194537855 | 215 | 2 | 1.07E-08 | 1.55E-06 | 3.69E-07 | 7.20E-08 | 0.009855428 | NA |
| chr1 | 19076295 | 19076812 | 518 | 4 | 8.57E-09 | 1.30E-05 | 4.17E-05 | 7.21E-08 | -0.00029085 | UBR4 |
| chr2 | 196613343 | 196613808 | 466 | 2 | 5.16E-09 | 3.58E-07 | 1.06E-06 | 7.22E-08 | -0.0081776 | NA |
| chr6 | 158312193 | 158313730 | 1538 | 10 | 1.99E-09 | 0.0001065 | 6.72E-07 | 7.24E-08 | -0.00183944 | TULP4, AL360169.2 |
| chr17 | 80214605 | 80215105 | 501 | 5 | 6.98E-08 | 6.96E-08 | 0.000558 | 7.24E-08 | -0.00392832 | SGSH |
| chr9 | 17134571 | 17134972 | 402 | 6 | 4.94E-08 | 1.33E-06 | 0.000413 | 7.25E-08 | -0.00208886 | NA |
| chr10 | 129031365 | 129033157 | 1793 | 13 | 6.96E-06 | 3.57E-06 | 0.00037 | 7.25E-08 | -0.00331615 | NA |
| chr17 | 4263552 | 4264798 | 1247 | 15 | 1.86E-08 | 3.23E-07 | 0.000297 | 7.26E-08 | -0.00025909 | ANKFY1 |
| chr11 | 67302693 | 67304175 | 1483 | 15 | 1.58E-08 | 3.94E-06 | 2.14E-05 | 7.26E-08 | -0.00316493 | SSH3 |
| chr17 | 47831108 | 47832162 | 1055 | 14 | 1.02E-09 | 3.65E-05 | 3.26E-05 | 7.26E-08 | -0.00048075 | LRRC46, MRPL10 |
| chr10 | 102151113 | 102152822 | 1710 | 16 | 3.07E-08 | 9.92E-05 | 0.000133 | 7.27E-08 | -0.00327603 | NOLC1 |
| chr12 | 121398754 | 121400206 | 1453 | 12 | 2.60E-09 | 1.87E-06 | 0.00088 | 7.27E-08 | 0.000571458 | AC048337.1, RNF34, ANAPC5 |
| chr16 | 2954038 | 2954362 | 325 | 2 | 1.54E-08 | 9.89E-08 | 4.94E-06 | 7.27E-08 | 0.006545101 | AC004034.1 |
| chr17 | 62679773 | 62681440 | 1668 | 10 | 1.25E-06 | 4.63E-05 | 9.70E-05 | 7.27E-08 | -0.00116393 | MRC2, AC080038.3 |
| chr21 | 29073032 | 29074129 | 1098 | 14 | 1.22E-08 | 3.40E-06 | 0.000106 | 7.30E-08 | 0.000324038 | CCT8 |
| chr19 | 36775846 | 36776124 | 279 | 2 | 2.20E-08 | 5.25E-08 | 2.20E-05 | 7.30E-08 | 0.033961713 | AC020928.1, AC020928.2 |
| chr11 | 65781213 | 65781970 | 758 | 7 | 1.44E-09 | 5.52E-06 | 5.25E-05 | 7.36E-08 | -0.00024034 | NA |
| chr8 | 141770904 | 141771849 | 946 | 5 | 8.85E-10 | 6.12E-05 | 4.95E-07 | 7.36E-08 | 0.002766666 | NA |
| chr1 | 248360369 | 248360934 | 566 | 2 | 4.25E-09 | 3.18E-07 | 1.22E-06 | 7.37E-08 | -0.03024654 | NA |
| chr9 | 124504938 | 124506040 | 1103 | 3 | 8.16E-08 | 3.68E-08 | 7.23E-05 | 7.37E-08 | -0.00440762 | NR5A1 |
| chr8 | 8664713 | 8665226 | 514 | 3 | 4.12E-09 | 2.04E-06 | 1.09E-06 | 7.38E-08 | -0.00776229 | NA |
| chr12 | 89019656 | 89020614 | 959 | 6 | 2.08E-08 | 7.38E-07 | 0.000233 | 7.38E-08 | -0.00470046 | LINC02458 |
| chr17 | 28985837 | 28986481 | 645 | 7 | 9.75E-08 | 1.37E-06 | 0.000879 | 7.38E-08 | -0.01227492 | PIPOX, SEZ6 |
| chr2 | 38751052 | 38752460 | 1409 | 13 | 9.53E-08 | 6.37E-08 | 0.003574 | 7.41E-08 | -0.00258544 | GEMIN6, SRSF7 |
| chr16 | 90019314 | 90020192 | 879 | 7 | 4.42E-10 | 2.13E-05 | 3.53E-05 | 7.42E-08 | -0.00117375 | GAS8, DBNDD1 |
| chr7 | 135182111 | 135182364 | 254 | 3 | 1.75E-08 | 1.27E-05 | 1.76E-05 | 7.43E-08 | -0.00762605 | NA |
| chr1 | 3292955 | 3293860 | 906 | 6 | 4.83E-08 | 9.92E-08 | 0.000121 | 7.43E-08 | 0.001152775 | PRDM16 |
| chr10 | 50624305 | 50625840 | 1536 | 9 | 2.30E-06 | 7.56E-07 | 0.000859 | 7.43E-08 | 0.001976821 | SGMS1-AS1, SGMS1 |
| chr5 | 178232228 | 178233508 | 1281 | 14 | 4.26E-07 | 2.47E-07 | 0.00164 | 7.43E-08 | 0.00042564 | PHYKPL |
| chr4 | 99868336 | 99869204 | 869 | 2 | 1.80E-06 | 4.71E-08 | 4.04E-05 | 7.45E-08 | -0.01103239 | DAPP1 |
| chr10 | 1475696 | 1476898 | 1203 | 4 | 2.49E-07 | 5.00E-08 | 5.58E-05 | 7.46E-08 | -0.00705976 | ADARB2 |
| chr14 | 100944545 | 100946228 | 1684 | 3 | 1.79E-07 | 1.73E-07 | 2.63E-05 | 7.48E-08 | -0.01942348 | AL117190.1, MEG8, SNORD113-9 |
| chr10 | 31693588 | 31693751 | 164 | 2 | 2.94E-08 | 2.92E-07 | 1.36E-06 | 7.48E-08 | -0.01751259 | MACORIS |
| chr7 | 156616364 | 156617512 | 1149 | 6 | 7.24E-08 | 4.07E-06 | 7.81E-05 | 7.48E-08 | -0.00987168 | LINC01006 |
| chr12 | 63150051 | 63151003 | 953 | 10 | 1.20E-08 | 6.68E-07 | 3.46E-05 | 7.50E-08 | -0.00387117 | AVPR1A |
| chr6 | 35737045 | 35738074 | 1030 | 6 | 2.18E-09 | 1.97E-06 | 5.82E-06 | 7.51E-08 | -0.00088055 | ARMC12 |
| chr11 | 104904817 | 104905714 | 898 | 2 | 3.64E-07 | 6.62E-08 | 1.22E-05 | 7.52E-08 | -0.01300646 | CASP4LP |
| chr6 | 10390014 | 10391179 | 1166 | 7 | 5.26E-10 | 4.27E-05 | 8.81E-06 | 7.54E-08 | -0.00042901 | NA |
| chr19 | 38387099 | 38389103 | 2005 | 12 | 5.11E-05 | 8.78E-07 | 0.003703 | 7.54E-08 | 0.0002898 | SPRED3, GGN |
| chr10 | 76887420 | 76888114 | 695 | 5 | 5.73E-08 | 9.70E-08 | 6.00E-05 | 7.54E-08 | 0.005633052 | KCNMA1-AS1, KCNMA1 |
| chr20 | 33731306 | 33732552 | 1247 | 10 | 5.54E-08 | 2.75E-06 | 0.000844 | 7.55E-08 | -0.00177525 | ZNF341, AL050349.1 |
| chr7 | 135557447 | 135558721 | 1275 | 15 | 5.53E-09 | 4.60E-06 | 0.001405 | 7.55E-08 | 0.001298473 | NUP205 |
| chr5 | 1216611 | 1217927 | 1317 | 8 | 1.07E-06 | 4.22E-07 | 0.00021 | 7.55E-08 | -0.00213328 | SLC6A19 |
| chr4 | 1050698 | 1051009 | 312 | 4 | 1.43E-08 | 3.13E-08 | 0.000175 | 7.56E-08 | -0.03501961 | NA |
| chr20 | 34714677 | 34715352 | 676 | 4 | 2.96E-09 | 1.35E-06 | 1.95E-06 | 7.58E-08 | -0.00167193 | NCOA6 |
| chr10 | 60389571 | 60391219 | 1649 | 8 | 1.39E-06 | 6.71E-05 | 1.22E-05 | 7.58E-08 | -0.00357601 | ANK3 |
| chr22 | 25068332 | 25069594 | 1263 | 3 | 9.84E-12 | 4.29E-06 | 8.43E-07 | 7.61E-08 | -0.00900901 | KIAA1671 |
| chr16 | 87777806 | 87778049 | 244 | 5 | 3.19E-08 | 3.91E-07 | 3.66E-05 | 7.62E-08 | -0.00116621 | AC126696.3 |
| chr1 | 209106036 | 209106588 | 553 | 4 | 1.70E-08 | 8.31E-07 | 5.67E-06 | 7.64E-08 | -0.00609252 | NA |
| chr4 | 159343103 | 159343509 | 407 | 2 | 3.10E-08 | 8.89E-08 | 6.56E-06 | 7.64E-08 | -0.00029211 | RAPGEF2 |
| chr7 | 5144361 | 5144905 | 545 | 3 | 1.06E-08 | 3.29E-07 | 4.98E-05 | 7.68E-08 | -0.04397406 | ZNF890P |
| chr3 | 149704313 | 149704479 | 167 | 2 | 3.10E-08 | 5.12E-08 | 3.05E-05 | 7.68E-08 | -0.01751157 | WWTR1 |
| chr6 | 44056949 | 44058016 | 1068 | 4 | 2.05E-08 | 4.47E-06 | 8.83E-05 | 7.68E-08 | -0.005334 | AL109615.3 |
| chr11 | 45846479 | 45847861 | 1383 | 14 | 1.38E-08 | 3.29E-05 | 0.000227 | 7.68E-08 | -0.00566771 | CRY2 |
| chr4 | 74109492 | 74109594 | 103 | 2 | 9.42E-08 | 4.74E-08 | 4.74E-05 | 7.68E-08 | -0.00206187 | NA |
| chr8 | 1837667 | 1838571 | 905 | 7 | 1.57E-07 | 5.94E-07 | 0.001991 | 7.71E-08 | -0.00145597 | AC019257.8, ARHGEF10 |
| chr2 | 235534173 | 235535763 | 1591 | 12 | 1.37E-08 | 0.0002384 | 1.78E-06 | 7.71E-08 | 0.000656028 | AGAP1 |
| chr11 | 78676145 | 78676524 | 380 | 6 | 6.92E-08 | 6.87E-06 | 0.000201 | 7.71E-08 | 0.003150839 | TENM4 |
| chr11 | 39221583 | 39221887 | 305 | 3 | 7.74E-08 | 4.83E-08 | 0.000122 | 7.71E-08 | -0.01201038 | NA |
| chr6 | 100296486 | 100297268 | 783 | 3 | 1.49E-09 | 3.50E-07 | 2.63E-06 | 7.72E-08 | 0.00330959 | NA |
| chr3 | 31981271 | 31982575 | 1305 | 13 | 3.45E-08 | 6.37E-07 | 0.00089 | 7.74E-08 | -0.00338731 | ZNF860, OSBPL10 |
| chr20 | 50509845 | 50510624 | 780 | 12 | 3.48E-08 | 6.13E-06 | 0.000981 | 7.78E-08 | 9.03E-05 | PTPN1 |
| chr10 | 49633819 | 49633902 | 84 | 2 | 4.55E-08 | 2.32E-07 | 1.83E-06 | 7.78E-08 | -0.01918558 | CHAT |
| chr6 | 39091882 | 39092293 | 412 | 2 | 2.54E-08 | 1.08E-07 | 4.94E-06 | 7.79E-08 | -0.01202757 | NA |
| chr1 | 159439058 | 159440212 | 1155 | 4 | 2.56E-08 | 2.27E-05 | 0.000158 | 7.80E-08 | -0.01113748 | OR10J1, AL513323.1 |
| chr3 | 25425746 | 25426043 | 298 | 2 | 1.17E-08 | 1.96E-07 | 2.22E-06 | 7.80E-08 | -0.01127282 | RARB |
| chr6 | 108557658 | 108559205 | 1548 | 7 | 1.66E-06 | 2.85E-07 | 0.000723 | 7.81E-08 | 0.00096874 | AL139106.1 |
| chr9 | 35689038 | 35690462 | 1425 | 10 | 2.90E-09 | 6.79E-07 | 0.000971 | 7.81E-08 | -0.00452974 | TPM2 |
| chr6 | 32761085 | 32762522 | 1438 | 25 | 3.13E-08 | 8.45E-07 | 0.000181 | 7.81E-08 | -0.00851926 | HLA-DQB2 |
| chr21 | 44903893 | 44904009 | 117 | 3 | 4.64E-08 | 6.87E-08 | 4.23E-05 | 7.82E-08 | -0.00136244 | ITGB2 |
| chr10 | 131773958 | 131774697 | 740 | 5 | 2.34E-09 | 6.60E-06 | 8.89E-05 | 7.83E-08 | 0.000226817 | LINC01164 |
| chr1 | 8025133 | 8027361 | 2229 | 11 | 8.07E-08 | 1.48E-06 | 0.000111 | 7.84E-08 | -0.0017574 | AL034417.2, ERRFI1 |
| chr17 | 19670548 | 19671053 | 506 | 2 | 2.13E-08 | 4.75E-08 | 5.56E-05 | 7.85E-08 | -0.00150833 | ALDH3A2 |
| chr12 | 57152094 | 57152314 | 221 | 2 | 1.20E-08 | 6.84E-08 | 1.30E-05 | 7.89E-08 | -0.01198669 | LRP1 |
| chr7 | 158442359 | 158443468 | 1110 | 6 | 1.74E-07 | 1.73E-06 | 2.23E-05 | 7.90E-08 | -0.00212523 | PTPRN2 |
| chr4 | 99821330 | 99822804 | 1475 | 3 | 1.88E-06 | 2.64E-08 | 0.000376 | 7.91E-08 | 0.007420192 | DAPP1 |
| chr10 | 73466600 | 73467190 | 591 | 2 | 1.85E-11 | 0.0084414 | 9.62E-08 | 7.92E-08 | -0.00358147 | PPP3CB |
| chr6 | 31797790 | 31798462 | 673 | 4 | 6.33E-09 | 6.28E-08 | 7.58E-05 | 7.92E-08 | -0.00419356 | LSM2 |
| chr5 | 42549314 | 42550103 | 790 | 5 | 1.70E-08 | 1.88E-07 | 9.04E-05 | 7.93E-08 | -0.00571558 | GHR |
| chr6 | 5003673 | 5005254 | 1582 | 14 | 1.59E-08 | 1.57E-05 | 5.84E-05 | 7.94E-08 | 0.002513421 | LYRM4-AS1, RPP40 |
| chr3 | 14432072 | 14432602 | 531 | 2 | 1.19E-08 | 4.82E-08 | 5.37E-05 | 7.94E-08 | 7.00E-05 | SLC6A6 |
| chr6 | 3075118 | 3076898 | 1781 | 8 | 9.59E-06 | 5.35E-07 | 0.000464 | 7.94E-08 | -0.00170868 | RIPK1 |
| chr8 | 143950469 | 143951010 | 542 | 9 | 1.81E-08 | 1.96E-06 | 7.30E-05 | 7.94E-08 | -0.00503643 | PLEC |
| chr21 | 37916045 | 37916963 | 919 | 10 | 7.04E-08 | 4.64E-07 | 0.000581 | 7.96E-08 | 0.001592158 | KCNJ6 |
| chr7 | 100160031 | 100160177 | 147 | 4 | 1.18E-07 | 1.02E-07 | 0.000235 | 7.97E-08 | -0.00102635 | GAL3ST4 |
| chr8 | 101540876 | 101541908 | 1033 | 4 | 1.99E-07 | 5.06E-08 | 0.000278 | 7.98E-08 | -0.00501583 | GRHL2 |
| chr1 | 244652621 | 244654110 | 1490 | 13 | 1.18E-06 | 6.35E-06 | 0.000387 | 8.01E-08 | 0.001057756 | DESI2 |
| chr11 | 10645452 | 10645458 | 7 | 2 | 9.70E-08 | 5.29E-08 | 3.24E-05 | 8.03E-08 | -0.01842803 | IRAG1 |
| chr13 | 24222285 | 24223499 | 1215 | 6 | 8.06E-08 | 1.87E-08 | 0.001404 | 8.03E-08 | -0.00209481 | AL359736.1, SPATA13 |
| chr7 | 627548 | 628655 | 1108 | 4 | 4.90E-08 | 2.52E-08 | 0.000599 | 8.04E-08 | -0.00283085 | PRKAR1B |
| chr22 | 47629619 | 47631135 | 1517 | 4 | 6.76E-06 | 1.79E-07 | 7.62E-05 | 8.05E-08 | -0.00423112 | AL117329.1, LINC00898 |
| chr2 | 227327376 | 227328065 | 690 | 6 | 1.19E-08 | 3.18E-06 | 2.93E-05 | 8.05E-08 | -0.00837478 | MFF |
| chr13 | 113074176 | 113075063 | 888 | 7 | 1.30E-09 | 1.79E-06 | 0.000249 | 8.05E-08 | 0.001316121 | MCF2L |
| chr11 | 75600301 | 75601668 | 1368 | 4 | 9.37E-06 | 9.87E-07 | 4.29E-05 | 8.06E-08 | -0.00733437 | MAP6 |
| chr15 | 92624838 | 92624911 | 74 | 2 | 4.22E-08 | 1.67E-07 | 2.84E-06 | 8.06E-08 | -0.01846353 | FAM174B |
| chr6 | 37696675 | 37697916 | 1242 | 8 | 6.49E-11 | 1.64E-05 | 1.35E-05 | 8.07E-08 | 0.001199025 | MDGA1 |
| chr21 | 33170457 | 33171663 | 1207 | 5 | 4.76E-09 | 3.25E-05 | 3.61E-05 | 8.10E-08 | -0.00737685 | LINC01548 |
| chr1 | 153678013 | 153679462 | 1450 | 14 | 1.87E-08 | 1.12E-05 | 3.10E-05 | 8.10E-08 | -0.00038767 | NPR1 |
| chr6 | 149723997 | 149724368 | 372 | 2 | 7.19E-08 | 5.83E-08 | 2.28E-05 | 8.10E-08 | -0.00397119 | NUP43 |
| chr1 | 34176110 | 34177497 | 1388 | 13 | 5.71E-08 | 9.77E-07 | 0.000274 | 8.10E-08 | -0.00196848 | C1orf94 |
| chr19 | 50513866 | 50515689 | 1824 | 12 | 1.03E-06 | 1.10E-06 | 0.00139 | 8.10E-08 | 0.007141898 | ASPDH |
| chr17 | 39659878 | 39661617 | 1740 | 6 | 1.98E-08 | 3.13E-05 | 3.79E-05 | 8.12E-08 | 0.001710249 | STARD3 |
| chr13 | 113121363 | 113121938 | 576 | 5 | 8.36E-10 | 5.45E-07 | 9.37E-05 | 8.13E-08 | -0.00334752 | NA |
| chr16 | 29811894 | 29813278 | 1385 | 10 | 1.43E-09 | 1.08E-05 | 0.000139 | 8.15E-08 | 0.000674706 | AC009133.5, PRRT2, AC009133.6, AC009133.1 |
| chr1 | 77778965 | 77779884 | 920 | 12 | 1.42E-07 | 1.10E-06 | 0.004208 | 8.15E-08 | 7.65E-05 | MIGA1 |
| chr15 | 63504214 | 63505823 | 1610 | 17 | 7.73E-09 | 1.86E-06 | 0.001002 | 8.16E-08 | 0.000306394 | USP3 |
| chr12 | 51172596 | 51173527 | 932 | 13 | 1.19E-08 | 1.78E-05 | 0.000432 | 8.16E-08 | 0.001772433 | TFCP2 |
| chr4 | 125314639 | 125316831 | 2193 | 16 | 1.51E-06 | 1.20E-06 | 0.001128 | 8.17E-08 | 3.66E-05 | FAT4 |

| | | | | | | | | | | |
|---|---|---|---|---|---|---|---|---|---|---|
| chr3 | 120095253 | 120095846 | 594 | 3 | 1.22E-11 | 1.50E-05 | 1.85E-07 | 8.17E-08 | 0.003618025 | AC092910.3 |
| chr10 | 132455538 | 132455736 | 199 | 2 | 3.07E-08 | 4.98E-08 | 5.20E-05 | 8.17E-08 | -0.00938579 | NA |
| chr2 | 80304358 | 80305812 | 1455 | 18 | 3.08E-08 | 2.96E-06 | 0.000463 | 8.17E-08 | 0.003484343 | CTNNA2, LRRTM1 |
| chr6 | 151764992 | 151765618 | 627 | 4 | 2.84E-10 | 1.60E-06 | 1.49E-06 | 8.19E-08 | -0.00381259 | ESR1 |
| chr2 | 29097003 | 29098039 | 1037 | 11 | 5.23E-09 | 5.26E-05 | 0.000189 | 8.19E-08 | -0.00047844 | CLIP4, AC105398.1 |
| chr12 | 89354523 | 89355600 | 1078 | 9 | 9.50E-09 | 2.69E-07 | 0.00056 | 8.19E-08 | -0.01581676 | AC010201.3 |
| chr12 | 3747678 | 3748855 | 1178 | 3 | 1.57E-06 | 6.97E-08 | 0.000104 | 8.19E-08 | -0.00898553 | CRACR2A |
| chr6 | 166626952 | 166628161 | 1210 | 10 | 1.09E-08 | 4.21E-05 | 4.47E-05 | 8.20E-08 | 0.000430827 | RPS6KA2 |
| chr5 | 140799905 | 140800713 | 809 | 5 | 1.80E-08 | 4.38E-07 | 0.000221 | 8.20E-08 | -0.01683228 | PCDHA1, PCDHA2, AC005609.5 |
| chr1 | 28087908 | 28088910 | 1003 | 14 | 1.39E-07 | 3.94E-08 | 0.00563 | 8.20E-08 | -0.00081105 | EYA3 |
| chr14 | 67532673 | 67533915 | 1243 | 13 | 2.65E-08 | 2.98E-06 | 5.08E-05 | 8.21E-08 | -0.00081521 | PLEKHH1, TMEM229B |
| chr6 | 37903544 | 37904243 | 700 | 2 | 7.86E-11 | 0.000287 | 1.08E-07 | 8.22E-08 | 0.012753971 | ZFAND3 |
| chr2 | 72354326 | 72354806 | 481 | 3 | 7.91E-09 | 6.01E-07 | 6.55E-06 | 8.22E-08 | -0.00966683 | EXOC6B |
| chr15 | 100113425 | 100114602 | 1178 | 5 | 1.13E-07 | 1.88E-07 | 0.000375 | 8.22E-08 | -0.0050165 | ADAMTS17 |
| chr3 | 129973993 | 129975822 | 1830 | 18 | 2.85E-06 | 2.27E-06 | 0.002242 | 8.22E-08 | 0.00030981 | TRH |
| chr2 | 85772375 | 85772750 | 376 | 3 | 4.38E-08 | 5.32E-08 | 5.48E-05 | 8.22E-08 | -0.01735149 | ATOH8 |
| chr15 | 89398925 | 89400200 | 1276 | 6 | 8.26E-08 | 1.32E-07 | 0.001171 | 8.23E-08 | -0.00206123 | NA |
| chr2 | 26970343 | 26971568 | 1226 | 11 | 1.17E-07 | 1.16E-06 | 0.003209 | 8.23E-08 | -0.00263393 | MAPRE3, AC013472.1 |
| chr19 | 10061443 | 10062223 | 781 | 4 | 1.00E-07 | 3.01E-08 | 0.0006 | 8.23E-08 | -0.02022556 | C3P1 |
| chr15 | 36818122 | 36818628 | 507 | 5 | 2.51E-09 | 8.93E-06 | 2.78E-05 | 8.24E-08 | -0.00352217 | CSNK1A1P1 |
| chr19 | 7354545 | 7354667 | 123 | 3 | 3.91E-08 | 7.59E-08 | 6.31E-05 | 8.25E-08 | -0.01051506 | AC119396.1 |
| chr2 | 42046942 | 42047479 | 538 | 5 | 1.64E-09 | 5.23E-05 | 5.10E-05 | 8.25E-08 | -0.0075328 | NA |
| chr1 | 150875972 | 150877329 | 1358 | 9 | 4.52E-09 | 1.20E-05 | 6.41E-07 | 8.27E-08 | 0.001230012 | ARNT |
| chr12 | 124535210 | 124535933 | 724 | 8 | 2.41E-09 | 2.12E-05 | 0.000175 | 8.27E-08 | 0.000971931 | NCOR2 |
| chr5 | 168600045 | 168600102 | 58 | 2 | 2.45E-09 | 0.0001344 | 1.15E-07 | 8.28E-08 | 0.007810413 | NA |
| chr9 | 126692644 | 126693218 | 575 | 4 | 3.98E-08 | 2.03E-07 | 7.32E-06 | 8.29E-08 | -0.00039881 | LMX1B |
| chr11 | 66045263 | 66046021 | 759 | 4 | 7.65E-09 | 1.55E-07 | 2.86E-05 | 8.31E-08 | -0.00318052 | GAL3ST3 |
| chr3 | 171692351 | 171692460 | 110 | 3 | 1.99E-08 | 4.66E-06 | 4.13E-07 | 8.31E-08 | -0.00471599 | PLD1 |
| chr13 | 97336486 | 97336705 | 220 | 3 | 8.02E-08 | 1.20E-07 | 5.44E-05 | 8.31E-08 | 0.000453738 | MBNL2 |
| chr4 | 119683822 | 119684364 | 543 | 2 | 1.07E-09 | 2.23E-06 | 3.56E-07 | 8.31E-08 | -0.01395 | NA |
| chr2 | 100976385 | 100977859 | 1475 | 10 | 9.55E-08 | 1.42E-07 | 0.00046 | 8.31E-08 | -0.00528541 | NPAS2, NPAS2-AS1 |
| chr16 | 65071029 | 65072382 | 1354 | 7 | 4.35E-06 | 7.41E-08 | 0.001296 | 8.31E-08 | -0.00305866 | CDH11 |
| chr11 | 1836212 | 1837921 | 1710 | 9 | 7.34E-07 | 2.96E-08 | 0.002258 | 8.32E-08 | -0.0011663 | SYT8 |
| chr6 | 83192779 | 83194372 | 1594 | 26 | 4.35E-09 | 1.55E-06 | 0.000467 | 8.33E-08 | -0.00092918 | RWDD2A, PGM3 |
| chr6 | 119349368 | 119351210 | 1843 | 8 | 2.86E-06 | 2.42E-06 | 0.002561 | 8.33E-08 | 0.002375849 | AL078600.1, MAN1A1 |
| chr8 | 35237844 | 35238445 | 602 | 2 | 3.86E-08 | 7.37E-08 | 1.24E-05 | 8.34E-08 | 0.023369883 | UNC5D |
| chr7 | 157552396 | 157553530 | 1135 | 8 | 3.70E-10 | 0.0002252 | 8.43E-06 | 8.34E-08 | -0.00545326 | PTPRN2 |
| chr9 | 15432381 | 15433172 | 792 | 3 | 2.69E-09 | 3.64E-06 | 3.71E-06 | 8.34E-08 | 0.000228098 | SNAPC3 |
| chr6 | 27091936 | 27092633 | 698 | 5 | 1.11E-08 | 1.01E-06 | 2.20E-05 | 8.34E-08 | -0.00822448 | NA |
| chr11 | 113232263 | 113233169 | 907 | 3 | 9.44E-09 | 2.12E-07 | 6.21E-05 | 8.37E-08 | -0.00686706 | NCAM1 |
| chr17 | 55783255 | 55784356 | 1102 | 5 | 3.83E-07 | 4.58E-08 | 0.000122 | 8.38E-08 | 0.009478469 | PCTP |
| chr7 | 55475506 | 55476005 | 500 | 2 | 3.91E-10 | 2.18E-06 | 3.64E-07 | 8.39E-08 | -0.00929459 | VOPP1 |
| chr3 | 193970069 | 193970849 | 781 | 3 | 3.05E-09 | 5.83E-05 | 4.52E-07 | 8.39E-08 | -0.01499418 | LINC02026 |
| chr1 | 66353275 | 66354940 | 1666 | 6 | 8.03E-07 | 4.75E-06 | 5.85E-05 | 8.40E-08 | 0.00629368 | PDE4B |
| chr7 | 1240404 | 1241564 | 1161 | 7 | 2.31E-10 | 0.0003458 | 3.03E-07 | 8.41E-08 | -0.00445137 | NA |
| chr13 | 39600548 | 39601144 | 597 | 3 | 1.06E-09 | 5.67E-07 | 1.75E-06 | 8.41E-08 | 0.008097782 | LHFPL6 |
| chr3 | 18745237 | 18745731 | 495 | 3 | 8.60E-08 | 1.08E-07 | 4.86E-05 | 8.43E-08 | 0.013049364 | SATB1-AS1 |
| chr6 | 10632469 | 10633224 | 756 | 3 | 4.54E-07 | 1.21E-06 | 1.09E-05 | 8.43E-08 | -0.00106534 | NA |
| chr11 | 108292245 | 108293424 | 1180 | 4 | 8.74E-07 | 2.15E-07 | 0.000138 | 8.45E-08 | -0.00106383 | ATM |
| chr1 | 150151300 | 150152893 | 1594 | 4 | 1.64E-06 | 6.19E-07 | 4.64E-05 | 8.46E-08 | 0.000227803 | PLEKHO1 |
| chr4 | 79210707 | 79211741 | 1035 | 5 | 1.51E-07 | 4.35E-08 | 0.000718 | 8.47E-08 | 0.002370403 | LINC01088 |
| chr19 | 56812472 | 56812927 | 456 | 3 | 1.01E-07 | 1.62E-07 | 1.49E-05 | 8.48E-08 | 0.003819822 | AC006115.2, ZIM2, PEG3 |
| chr9 | 123678304 | 123678660 | 357 | 4 | 3.36E-08 | 0.0008043 | 1.55E-05 | 8.48E-08 | -0.02057409 | DENND1A |
| chr9 | 98097682 | 98098208 | 527 | 3 | 8.21E-08 | 4.54E-08 | 0.000157 | 8.49E-08 | -0.00661455 | TRIM14 |
| chr7 | 100093183 | 100094319 | 1137 | 15 | 1.13E-07 | 4.26E-06 | 0.00054 | 8.49E-08 | -0.00329887 | MCM7, MIR25, MIR93, MIR106B |
| chr9 | 129482236 | 129483643 | 1408 | 12 | 3.37E-06 | 1.70E-06 | 0.000348 | 8.49E-08 | -0.00146558 | LINC00963, AL353803.4 |
| chr17 | 29568400 | 29569155 | 756 | 7 | 1.02E-08 | 3.51E-06 | 0.000132 | 8.50E-08 | 0.001700774 | ABHD15-AS1, TP53I13 |
| chr17 | 82733395 | 82735071 | 1677 | 6 | 1.58E-08 | 1.40E-05 | 1.72E-05 | 8.51E-08 | -0.00347782 | AC024361.3 |
| chr13 | 19673917 | 19674528 | 612 | 3 | 1.59E-08 | 7.55E-07 | 4.45E-06 | 8.53E-08 | 0.002702704 | NA |
| chr9 | 98086992 | 98088236 | 1245 | 6 | 4.82E-06 | 1.49E-08 | 0.001892 | 8.54E-08 | -0.01470382 | TRIM14 |
| chr5 | 14409181 | 14410543 | 1363 | 6 | 6.80E-07 | 1.94E-05 | 0.000111 | 8.55E-08 | -0.00351588 | TRIO |
| chr18 | 51390750 | 51391206 | 457 | 2 | 1.86E-08 | 1.53E-07 | 3.51E-06 | 8.55E-08 | -0.01987878 | LINC01630 |
| chr16 | 275826 | 276728 | 903 | 11 | 1.50E-09 | 5.94E-05 | 1.80E-05 | 8.56E-08 | -0.00081005 | Z69667.1, RGS11 |
| chr7 | 36189732 | 36190849 | 1118 | 3 | 1.29E-07 | 1.95E-07 | 3.31E-05 | 8.56E-08 | -0.00336022 | EEPD1 |
| chr16 | 49452244 | 49453726 | 1483 | 4 | 3.89E-12 | 3.55E-05 | 9.09E-07 | 8.57E-08 | -0.00502137 | AC007861.1 |
| chr10 | 69743036 | 69743564 | 529 | 2 | 4.05E-08 | 8.74E-08 | 8.99E-06 | 8.58E-08 | -0.01391972 | NA |
| chr21 | 37220311 | 37221453 | 1143 | 9 | 1.16E-07 | 2.04E-05 | 0.00021 | 8.58E-08 | -0.00105192 | DSCR9, AP001432.1 |
| chr1 | 17010357 | 17011888 | 1532 | 7 | 1.22E-07 | 2.42E-05 | 7.47E-05 | 8.58E-08 | 0.000388049 | ATP13A2, AL049569.3 |
| chr19 | 11108939 | 11108962 | 24 | 2 | 3.45E-08 | 8.20E-07 | 6.83E-07 | 8.59E-08 | 0.007466703 | LDLR |
| chr11 | 134387136 | 134388598 | 1463 | 12 | 2.93E-07 | 2.91E-07 | 0.002557 | 8.60E-08 | -0.00256033 | B3GAT1 |
| chr10 | 71850988 | 71852492 | 1505 | 13 | 1.81E-10 | 1.72E-06 | 0.000187 | 8.61E-08 | -0.00530979 | PSAP |
| chr2 | 200854206 | 200855012 | 807 | 3 | 1.36E-08 | 3.75E-05 | 6.00E-06 | 8.62E-08 | 0.000894068 | CLK1 |
| chr19 | 50791193 | 50791821 | 629 | 3 | 2.12E-08 | 3.14E-08 | 0.000248 | 8.62E-08 | -0.00992903 | ACP4, AC010325.1 |
| chr2 | 73212692 | 73214370 | 1679 | 13 | 1.51E-06 | 3.56E-06 | 0.000247 | 8.63E-08 | -0.00079982 | SMYD5 |
| chr10 | 93495338 | 93497002 | 1665 | 14 | 6.91E-07 | 3.76E-06 | 0.000979 | 8.63E-08 | -9.59E-05 | CEP55 |
| chr11 | 47714580 | 47716154 | 1575 | 14 | 1.04E-07 | 1.41E-06 | 0.000573 | 8.64E-08 | -0.00067875 | AGBL2 |
| chr4 | 158671466 | 158672889 | 1424 | 17 | 4.79E-09 | 1.78E-06 | 5.88E-05 | 8.64E-08 | -0.00092618 | ETFDH, C4orf46 |
| chr17 | 58417603 | 58418678 | 1076 | 6 | 9.95E-09 | 3.76E-05 | 1.26E-05 | 8.65E-08 | -0.00452754 | NA |
| chr2 | 169693913 | 169695787 | 1875 | 23 | 2.49E-07 | 8.97E-05 | 0.001339 | 8.66E-08 | 0.000275907 | PHOSPHO2, KLHL23, CCDC173 |
| chr9 | 19517516 | 19517633 | 118 | 2 | 6.42E-08 | 1.38E-07 | 4.18E-06 | 8.67E-08 | -0.01157255 | SLC24A2 |
| chr10 | 116669601 | 116670839 | 1239 | 9 | 9.39E-08 | 1.23E-06 | 0.000578 | 8.68E-08 | -0.00976711 | AC016825.1, C10orf82 |
| chr9 | 136855008 | 136856863 | 1856 | 6 | 1.20E-07 | 2.62E-07 | 3.73E-05 | 8.68E-08 | -0.00328375 | MAMDC4 |
| chr9 | 17018196 | 17018511 | 316 | 2 | 3.11E-09 | 1.14E-05 | 1.92E-07 | 8.68E-08 | -0.00412956 | AL162725.2 |
| chr20 | 46220568 | 46221542 | 975 | 4 | 1.48E-08 | 8.98E-07 | 6.69E-06 | 8.69E-08 | -0.00687013 | CDH22 |
| chr2 | 489332 | 489360 | 29 | 2 | 2.63E-08 | 9.20E-08 | 8.33E-06 | 8.69E-08 | -0.01014993 | NA |
| chr16 | 8572193 | 8573192 | 1000 | 8 | 6.38E-08 | 2.58E-07 | 0.002428 | 8.69E-08 | 0.003840621 | TMEM114 |
| chr3 | 50450228 | 50450855 | 628 | 5 | 1.61E-07 | 3.80E-08 | 0.000637 | 8.70E-08 | 0.023098966 | CACNA2D2 |
| chr6 | 154938033 | 154938331 | 299 | 2 | 2.07E-08 | 3.19E-07 | 1.57E-06 | 8.71E-08 | -0.00072601 | TIAM2 |
| chr17 | 36463898 | 36464870 | 973 | 8 | 3.81E-10 | 2.36E-05 | 5.40E-07 | 8.73E-08 | -0.00641965 | NA |
| chr7 | 20646123 | 20647089 | 967 | 5 | 7.68E-09 | 1.35E-05 | 3.20E-06 | 8.74E-08 | 0.00299623 | ABCB5 |
| chr9 | 132236301 | 132236391 | 91 | 3 | 6.65E-08 | 5.29E-07 | 8.40E-06 | 8.74E-08 | -0.01051741 | NTNG2 |
| chr22 | 46408629 | 46409781 | 1153 | 6 | 8.83E-09 | 6.46E-05 | 0.000158 | 8.75E-08 | -0.0035235 | CELSR1 |
| chr6 | 3444886 | 3445774 | 889 | 5 | 3.68E-07 | 1.76E-07 | 0.000125 | 8.75E-08 | -0.00193611 | SLC22A23 |
| chr7 | 2978973 | 2980286 | 1314 | 6 | 3.23E-09 | 2.62E-07 | 0.000458 | 8.76E-08 | 0.013466955 | CARD11 |
| chr1 | 29134305 | 29134558 | 254 | 4 | 1.24E-07 | 3.53E-08 | 0.000378 | 8.76E-08 | -0.02140513 | NA |
| chr10 | 97582736 | 97584407 | 1672 | 9 | 2.72E-06 | 1.79E-07 | 0.000892 | 8.76E-08 | -0.00316648 | ANKRD2, HOGA1, AL355315.1 |
| chr1 | 180970482 | 180971254 | 773 | 5 | 1.62E-07 | 6.97E-07 | 0.000549 | 8.76E-08 | 0.004500023 | AL162431.3, AL162431.1 |
| chr9 | 131189827 | 131190154 | 328 | 4 | 1.08E-08 | 3.77E-06 | 4.83E-06 | 8.76E-08 | -0.00430196 | NUP214, AL157938.3 |
| chr10 | 69953074 | 69953122 | 49 | 2 | 2.85E-08 | 1.07E-07 | 6.52E-06 | 8.76E-08 | -0.01198809 | COL13A1 |
| chr12 | 8066326 | 8067406 | 1081 | 5 | 2.63E-07 | 1.73E-08 | 0.001769 | 8.76E-08 | 0.008208377 | C3AR1 |
| chr17 | 64145547 | 64146464 | 918 | 13 | 1.11E-08 | 4.63E-07 | 0.000825 | 8.77E-08 | 0.001494701 | SNHG25, SNORD104, AC025362.1, SNORA50C |
| chr7 | 144437571 | 144438151 | 581 | 2 | 3.70E-08 | 9.89E-08 | 7.48E-06 | 8.78E-08 | -0.01131144 | NA |
| chr3 | 129045981 | 129047056 | 1076 | 5 | 2.50E-09 | 1.43E-06 | 7.19E-05 | 8.80E-08 | -0.01067288 | AC108673.1 |
| chr5 | 1594167 | 1594618 | 452 | 4 | 2.86E-07 | 2.96E-08 | 0.000633 | 8.82E-08 | -0.02029009 | SDHAP3 |
| chr2 | 219612025 | 219613188 | 1164 | 4 | 4.59E-07 | 4.29E-07 | 2.75E-05 | 8.83E-08 | 0.003239397 | STK11IP |
| chr5 | 14405523 | 14406623 | 1101 | 11 | 9.88E-08 | 7.71E-07 | 0.001675 | 8.83E-08 | -0.00114343 | TRIO |
| chr9 | 95552350 | 95553436 | 1087 | 5 | 9.33E-08 | 8.43E-08 | 0.001004 | 8.84E-08 | 0.014135203 | NA |
| chr10 | 42367428 | 42368146 | 719 | 6 | 1.39E-07 | 2.58E-08 | 0.002215 | 8.87E-08 | -0.03197256 | BX322639.1 |
| chr1 | 15756606 | 15758742 | 2137 | 12 | 1.71E-10 | 1.70E-06 | 0.00018 | 8.87E-08 | 1.22E-05 | FBLIM1 |
| chr12 | 93448866 | 93449352 | 487 | 3 | 6.46E-08 | 7.80E-08 | 2.94E-05 | 8.90E-08 | 0.001629099 | NA |
| chr2 | 78087302 | 78087736 | 435 | 3 | 1.40E-09 | 1.68E-05 | 1.76E-07 | 8.91E-08 | -0.00867877 | AC012494.1 |
| chr19 | 46040982 | 46042347 | 1366 | 6 | 2.83E-07 | 2.27E-07 | 6.33E-05 | 8.92E-08 | -0.00939039 | IGFL4 |
| chr17 | 78358540 | 78360180 | 1641 | 13 | 1.43E-06 | 1.42E-07 | 0.001828 | 8.93E-08 | -0.00628752 | SOCS3 |
| chr9 | 115112728 | 115113728 | 1001 | 6 | 2.92E-08 | 1.62E-07 | 1.35E-05 | 8.94E-08 | -0.00270403 | DELEC1, TNC |
| chr8 | 1095262 | 1096358 | 1097 | 4 | 1.48E-09 | 4.19E-06 | 1.81E-05 | 8.95E-08 | -0.00707442 | DLGAP2 |
| chr4 | 185710039 | 185710783 | 745 | 3 | 2.24E-07 | 2.97E-08 | 0.00043 | 8.95E-08 | 0.000485097 | SORBS2 |
| chr5 | 78627282 | 78628335 | 1054 | 4 | 1.90E-06 | 1.54E-06 | 3.22E-05 | 8.96E-08 | -0.01088769 | LHFPL2 |
| chr20 | 51904602 | 51904835 | 234 | 5 | 1.29E-08 | 2.00E-05 | 2.99E-05 | 8.97E-08 | -0.00134427 | NA |
| chr10 | 131259906 | 131260725 | 820 | 7 | 2.80E-09 | 9.50E-05 | 3.31E-07 | 8.97E-08 | 0.006023224 | TCERG1L |
| chr16 | 48276350 | 48276676 | 327 | 2 | 8.33E-09 | 9.06E-08 | 9.38E-06 | 8.97E-08 | 0.008502804 | LONP2 |
| chr1 | 222618172 | 222619245 | 1074 | 9 | 1.41E-08 | 8.98E-07 | 0.000947 | 8.97E-08 | -0.00231279 | MIA3 |
| chr13 | 106491266 | 106492622 | 1357 | 6 | 4.06E-07 | 1.26E-06 | 0.000248 | 9.00E-08 | 0.004316221 | AL138689.2, EFNB2 |
| chr4 | 116925932 | 116926862 | 931 | 5 | 7.33E-10 | 0.0003809 | 1.28E-05 | 9.01E-08 | -0.00766852 | AC093765.3 |
| chr2 | 231378244 | 231378479 | 236 | 2 | 4.48E-08 | 1.31E-07 | 4.96E-06 | 9.01E-08 | -0.01912743 | NA |
| chr16 | 72664467 | 72665319 | 853 | 12 | 1.15E-08 | 1.37E-06 | 1.79E-05 | 9.03E-08 | 0.0006176 | AC004943.2, LINC01572 |
| chr2 | 241149433 | 241150530 | 1098 | 19 | 9.73E-09 | 1.09E-06 | 1.05E-05 | 9.03E-08 | 0.000225038 | PPP1R7, PASK |
| chr12 | 52380046 | 52380363 | 318 | 3 | 3.99E-08 | 8.56E-08 | 5.03E-05 | 9.03E-08 | 0.056388331 | KRT84 |
| chr22 | 42367809 | 42368540 | 732 | 6 | 2.26E-08 | 6.70E-08 | 0.0005 | 9.04E-08 | -0.01579212 | LINC01315 |
| chr1 | 112395599 | 112396570 | 972 | 11 | 4.87E-09 | 1.38E-05 | 4.87E-05 | 9.06E-08 | 0.001878602 | CTTNBP2NL |
| chr1 | 202811300 | 202812823 | 1524 | 5 | 6.13E-10 | 1.53E-06 | 9.00E-06 | 9.06E-08 | -0.00410569 | PCAT6 |
| chr19 | 40575454 | 40576850 | 1397 | 12 | 2.06E-08 | 3.83E-06 | 6.00E-05 | 9.06E-08 | -0.00103756 | SPTBN4 |
| chr8 | 860491 | 861439 | 949 | 8 | 5.22E-07 | 1.89E-05 | 3.14E-05 | 9.06E-08 | -0.00533828 | DLGAP2 |
| chr11 | 487083 | 487916 | 834 | 5 | 6.81E-09 | 0.0001134 | 1.47E-06 | 9.06E-08 | -0.00438085 | PTDSS2 |
| chr22 | 20298730 | 20299322 | 593 | 3 | 6.75E-09 | 1.50E-07 | 2.04E-05 | 9.09E-08 | -0.01377244 | NA |
| chr2 | 127145739 | 127146391 | 653 | 2 | 5.58E-08 | 8.82E-08 | 1.03E-05 | 9.09E-08 | -0.00432241 | NA |
| chr10 | 112989035 | 112990365 | 1331 | 4 | 2.17E-09 | 6.43E-05 | 6.45E-07 | 9.10E-08 | -0.00102936 | TCF7L2 |
| chr10 | 97633006 | 97633876 | 871 | 13 | 1.46E-09 | 1.50E-06 | 2.41E-05 | 9.10E-08 | -0.00179492 | AL355315.1, MORN4 |
| chr8 | 80874627 | 80875952 | 1326 | 10 | 2.22E-07 | 1.88E-05 | 0.00035 | 9.11E-08 | -0.00333403 | ZNF704 |
| chr3 | 43879143 | 43879253 | 111 | 2 | 1.02E-07 | 6.84E-08 | 2.09E-05 | 9.11E-08 | -0.02265584 | NA |
| chr12 | 22544023 | 22545334 | 1312 | 15 | 7.73E-09 | 3.69E-06 | 0.000999 | 9.11E-08 | -0.00146785 | AC053513.1, C2CD5 |
| chr17 | 72556939 | 72557112 | 174 | 2 | 6.86E-09 | 4.04E-05 | 1.47E-07 | 9.11E-08 | -0.0212037 | LINC00511 |
| chr17 | 79080010 | 79080532 | 523 | 5 | 5.23E-08 | 1.46E-07 | 6.80E-05 | 9.12E-08 | -0.00076999 | ENGASE |
| chr15 | 40955683 | 40955851 | 169 | 4 | 9.93E-08 | 1.58E-07 | 2.48E-05 | 9.12E-08 | -0.00341117 | CHAC1 |
| chr1 | 32361242 | 32362590 | 1349 | 12 | 1.66E-08 | 0.0001893 | 0.000128 | 9.13E-08 | -0.00013832 | TSSK3, FAM229A |
| chr2 | 45004243 | 45005749 | 1507 | 8 | 1.49E-06 | 4.90E-07 | 0.001022 | 9.18E-08 | 0.003375313 | SIX2 |
| chr2 | 75315759 | 75316222 | 464 | 2 | 1.92E-08 | 1.71E-07 | 3.48E-06 | 9.18E-08 | -0.02137115 | NA |
| chr1 | 109240000 | 109240881 | 882 | 2 | 5.94E-08 | 1.59E-07 | 3.83E-06 | 9.19E-08 | -0.01611511 | NA |
| chr11 | 124897824 | 124898949 | 1126 | 11 | 2.51E-08 | 2.48E-06 | 0.001268 | 9.19E-08 | -0.00220831 | ROBO4 |
| chr4 | 176237326 | 176237745 | 420 | 3 | 2.32E-09 | 2.45E-06 | 2.08E-06 | 9.20E-08 | -0.01278324 | ASB5 |
| chr14 | 96210480 | 96211149 | 670 | 2 | 4.53E-09 | 2.46E-07 | 2.26E-06 | 9.20E-08 | -0.00014568 | BDKRB2, AL355102.2, AL355102.1 |
| chr14 | 93184170 | 93185546 | 1377 | 19 | 4.16E-09 | 0.0004231 | 0.000359 | 9.21E-08 | 0.000252711 | TMEM251, AL110118.1, MOAP1 |
| chr12 | 47706436 | 47707059 | 624 | 5 | 1.57E-08 | 2.34E-06 | 4.90E-05 | 9.21E-08 | -0.00610429 | AC004241.1 |
| chr11 | 94128613 | 94129583 | 971 | 15 | 6.51E-09 | 0.0009795 | 0.00048 | 9.21E-08 | -0.00065588 | PANX1 |
| chr11 | 68049842 | 68051297 | 1456 | 8 | 2.60E-09 | 4.19E-07 | 8.17E-05 | 9.21E-08 | -0.00023191 | TCIRG1, AP002807.1 |
| chr2 | 15993337 | 15993769 | 433 | 5 | 3.79E-08 | 8.40E-08 | 6.82E-05 | 9.21E-08 | 0.006010389 | NA |
| chr9 | 121374920 | 121375042 | 123 | 2 | 5.27E-08 | 9.48E-08 | 9.15E-06 | 9.22E-08 | -0.01946425 | AL359644.1 |
| chr4 | 37826424 | 37826878 | 455 | 8 | 3.64E-08 | 6.13E-06 | 0.001176 | 9.24E-08 | 2.25E-05 | PGM2 |
| chr9 | 38734800 | 38734847 | 48 | 2 | 3.50E-08 | 1.07E-06 | 6.26E-07 | 9.25E-08 | -0.00526658 | NA |

| | | | | | | | | | | |
|---|---|---|---|---|---|---|---|---|---|---|
| chr1 | 232151371 | 232151404 | 34 | 2 | 9.47E-08 | 6.39E-08 | 2.86E-05 | 9.25E-08 | -0.02720236 | NA |
| chr8 | 21788662 | 21790853 | 2192 | 10 | 2.30E-06 | 2.30E-06 | 0.001413 | 9.26E-08 | -0.00088908 | GFRA2 |
| chr22 | 43516029 | 43516199 | 171 | 4 | 3.88E-08 | 4.40E-07 | 7.13E-05 | 9.29E-08 | -0.02082942 | EFCAB6-AS1 |
| chr6 | 123637202 | 123638576 | 1375 | 6 | 1.66E-05 | 1.70E-06 | 7.98E-05 | 9.32E-08 | -0.00766846 | AL133257.1 |
| chr4 | 52721871 | 52722683 | 813 | 6 | 3.25E-08 | 1.76E-07 | 4.38E-05 | 9.33E-08 | 0.001467462 | DANCR, LINC01618, AC104066.4, ERVMER34-1 |
| chr6 | 169167360 | 169169645 | 2286 | 12 | 7.79E-06 | 7.52E-07 | 0.000299 | 9.33E-08 | -0.00050767 | NA |
| chr4 | 87391171 | 87392514 | 1344 | 12 | 2.38E-09 | 0.0001133 | 1.34E-05 | 9.34E-08 | -0.000322 | HSD17B11 |
| chr4 | 845740 | 845910 | 171 | 3 | 1.30E-07 | 7.21E-08 | 2.97E-05 | 9.34E-08 | 0.009589245 | AC139887.5 |
| chr7 | 39410594 | 39411977 | 1384 | 4 | 4.50E-06 | 7.92E-07 | 6.33E-05 | 9.34E-08 | -0.00400294 | POU6F2 |
| chr19 | 11353628 | 11355349 | 1722 | 10 | 2.70E-06 | 2.99E-05 | 0.000235 | 9.35E-08 | -0.00205267 | CCDC159 |
| chr4 | 87798625 | 87799488 | 864 | 6 | 9.77E-09 | 5.16E-06 | 5.40E-06 | 9.38E-08 | -0.00678934 | NA |
| chr6 | 117272790 | 117273537 | 748 | 2 | 8.35E-07 | 5.91E-08 | 4.57E-05 | 9.39E-08 | -0.02129554 | VGLL2 |
| chr19 | 10640583 | 10640641 | 59 | 2 | 9.87E-09 | 8.63E-05 | 1.36E-07 | 9.41E-08 | 0.00709084 | SLC44A2 |
| chr17 | 2815221 | 2816129 | 909 | 3 | 1.17E-09 | 6.20E-07 | 4.99E-06 | 9.42E-08 | -0.00861415 | RAP1GAP2 |
| chr8 | 29270409 | 29271208 | 800 | 3 | 9.61E-10 | 8.06E-06 | 6.66E-07 | 9.42E-08 | -0.00765982 | NA |
| chr1 | 39958812 | 39958991 | 180 | 2 | 2.04E-08 | 8.82E-08 | 1.14E-05 | 9.43E-08 | 0.009236096 | MFSD2A |
| chr2 | 241499265 | 241500297 | 1033 | 6 | 1.88E-07 | 3.07E-07 | 0.000174 | 9.44E-08 | -0.00102749 | STK25 |
| chr12 | 116263453 | 116264003 | 551 | 2 | 1.00E-08 | 2.65E-07 | 2.17E-06 | 9.45E-08 | 0.007453927 | MED13L |
| chr19 | 55639990 | 55641255 | 1266 | 10 | 4.01E-08 | 3.07E-08 | 0.001659 | 9.45E-08 | -0.0062648 | ZNF580, ZNF581, CCDC106 |
| chr1 | 3454156 | 3455022 | 867 | 9 | 2.59E-09 | 1.56E-05 | 4.82E-05 | 9.51E-08 | 1.04E-05 | ARHGEF16 |
| chr4 | 154626655 | 154628117 | 1463 | 7 | 2.76E-14 | 0.0005723 | 1.26E-08 | 9.51E-08 | -0.01023837 | LRAT |
| chr1 | 228102589 | 228104361 | 1773 | 13 | 1.18E-06 | 4.90E-08 | 0.002283 | 9.51E-08 | 0.006476533 | C1orf35 |
| chr18 | 5629250 | 5630952 | 1703 | 11 | 2.69E-06 | 2.23E-07 | 0.00083 | 9.51E-08 | -0.00037377 | EPB41L3 |
| chr17 | 64496618 | 64497714 | 1097 | 9 | 2.56E-07 | 3.01E-08 | 0.003721 | 9.52E-08 | 0.000714344 | POLG2 |
| chr9 | 113654362 | 113655411 | 1050 | 4 | 2.50E-09 | 2.00E-06 | 5.78E-06 | 9.53E-08 | 0.002228523 | AL157702.2 |
| chr7 | 106284205 | 106285093 | 889 | 8 | 1.59E-08 | 2.39E-06 | 0.000277 | 9.56E-08 | -0.00143035 | NAMPT |
| chr5 | 79235265 | 79235864 | 600 | 4 | 8.43E-16 | 0.0004195 | 5.20E-08 | 9.56E-08 | 0.014122865 | DMGDH |
| chr12 | 119676298 | 119677205 | 908 | 3 | 9.64E-08 | 1.47E-07 | 6.44E-05 | 9.61E-08 | -0.00877075 | PRKAB1 |
| chr12 | 57127488 | 57128814 | 1327 | 14 | 1.05E-07 | 1.25E-05 | 0.002667 | 9.61E-08 | -9.22E-05 | LRP1, STAT6 |
| chr19 | 48404845 | 48405300 | 456 | 4 | 7.29E-09 | 1.27E-06 | 7.16E-06 | 9.64E-08 | 0.000683385 | GRIN2D |
| chr9 | 93263131 | 93263749 | 619 | 2 | 6.38E-11 | 0.0001829 | 1.28E-07 | 9.64E-08 | 0.005019306 | WNK2 |
| chr7 | 1620554 | 1621273 | 720 | 9 | 2.01E-07 | 8.41E-07 | 0.000202 | 9.65E-08 | -0.00183737 | AC074389.2 |
| chr2 | 196810151 | 196811548 | 1398 | 6 | 1.67E-06 | 1.89E-07 | 0.000517 | 9.65E-08 | -0.00741038 | NA |
| chr14 | 103712195 | 103712823 | 629 | 7 | 1.04E-07 | 1.50E-07 | 0.000813 | 9.67E-08 | -0.00505179 | KLC1, XRCC3 |
| chr13 | 78277553 | 78278706 | 1154 | 5 | 3.35E-06 | 1.63E-08 | 0.001762 | 9.67E-08 | -0.00631769 | OBI1-AS1 |
| chr1 | 3152937 | 3153773 | 837 | 2 | 9.10E-07 | 5.92E-08 | 5.48E-05 | 9.67E-08 | -0.00949392 | PRDM16 |
| chr21 | 39866361 | 39867562 | 1202 | 11 | 9.07E-08 | 2.40E-06 | 0.000317 | 9.70E-08 | -3.73E-06 | PCP4 |
| chr20 | 8657655 | 8658602 | 948 | 5 | 1.31E-06 | 2.82E-06 | 0.000362 | 9.71E-08 | -0.00370353 | PLCB1 |
| chr19 | 33683439 | 33684648 | 1210 | 21 | 1.24E-07 | 2.69E-06 | 0.010745 | 9.72E-08 | 0.001806966 | CHST8 |
| chr12 | 54159205 | 54159931 | 727 | 3 | 5.31E-09 | 3.58E-07 | 3.61E-06 | 9.73E-08 | -0.01204881 | SMUG1 |
| chr6 | 29850186 | 29850523 | 338 | 7 | 3.65E-07 | 1.38E-08 | 0.002345 | 9.73E-08 | 0.028243018 | AL645929.2 |
| chr11 | 111454347 | 111455802 | 1456 | 6 | 1.71E-07 | 6.03E-08 | 5.70E-05 | 9.74E-08 | -0.00648298 | AP002008.2, POU2AF1 |
| chr1 | 100719361 | 100719755 | 395 | 6 | 5.55E-08 | 1.56E-07 | 0.000506 | 9.75E-08 | -0.00773516 | VCAM1 |
| chr7 | 45460517 | 45460785 | 269 | 3 | 7.45E-10 | 7.73E-05 | 1.61E-07 | 9.77E-08 | -0.00496086 | AC073325.1 |
| chr19 | 54434597 | 54435813 | 1217 | 6 | 5.67E-08 | 9.31E-08 | 9.22E-05 | 9.77E-08 | -0.03367522 | TTYH1, AC245884.9, AC245884.1 |
| chr21 | 45894154 | 45895175 | 1022 | 6 | 6.64E-10 | 4.10E-05 | 6.17E-06 | 9.77E-08 | -0.00437402 | PCBP3 |
| chr12 | 2884791 | 2885487 | 697 | 4 | 1.12E-07 | 6.53E-08 | 0.00029 | 9.77E-08 | -0.01271446 | RHNO1, TULP3 |
| chr5 | 178626469 | 178627694 | 1226 | 11 | 8.87E-09 | 6.16E-07 | 0.000258 | 9.78E-08 | 0.000843644 | CLK4 |
| chr7 | 2038401 | 2039623 | 1223 | 8 | 1.77E-08 | 2.07E-07 | 6.70E-05 | 9.78E-08 | 0.007156655 | MAD1L1, AC069288.1 |
| chr16 | 23492356 | 23493609 | 1254 | 5 | 2.00E-06 | 2.07E-07 | 0.000117 | 9.79E-08 | -0.0008587 | GGA2 |
| chr9 | 113649572 | 113650940 | 1369 | 6 | 5.36E-07 | 1.86E-07 | 3.78E-05 | 9.79E-08 | -0.00558755 | NA |
| chr20 | 45362706 | 45363773 | 1068 | 16 | 5.06E-08 | 4.13E-07 | 0.0018 | 9.81E-08 | -0.00133445 | SYS1, SYS1-DBNDD2 |
| chr3 | 11601793 | 11603030 | 1238 | 8 | 4.13E-06 | 7.10E-07 | 0.000775 | 9.81E-08 | 0.007186884 | VGLL4 |
| chr16 | 1443539 | 1445834 | 2296 | 16 | 8.95E-06 | 2.06E-06 | 0.002133 | 9.82E-08 | -0.00431674 | AL031600.3, CCDC154, CLCN7 |
| chr8 | 54174763 | 54175258 | 496 | 3 | 4.61E-09 | 5.04E-07 | 4.22E-05 | 9.82E-08 | -0.03348687 | NA |
| chr11 | 64826429 | 64826995 | 567 | 3 | 8.23E-09 | 9.49E-08 | 2.43E-05 | 9.84E-08 | -0.00047471 | CDC42BPG |
| chr1 | 246796580 | 246798188 | 1609 | 4 | 3.78E-07 | 1.80E-06 | 1.82E-05 | 9.84E-08 | -0.00064741 | KIF28P |
| chr17 | 48526873 | 48527412 | 540 | 4 | 1.09E-07 | 6.87E-08 | 0.000287 | 9.88E-08 | -0.01572187 | NA |
| chr1 | 35576832 | 35577401 | 570 | 4 | 1.71E-10 | 2.82E-05 | 1.34E-05 | 9.89E-08 | -0.01611388 | TFAP2E, TFAP2E-AS1 |
| chr12 | 56001403 | 56001655 | 253 | 4 | 5.73E-08 | 1.05E-06 | 4.22E-05 | 9.89E-08 | -0.01354568 | SUOX |
| chr8 | 48555271 | 48556553 | 1283 | 10 | 8.29E-08 | 1.61E-07 | 0.001089 | 9.89E-08 | -0.00153547 | AC022915.2, AC026904.2 |
| chr5 | 131944804 | 131945947 | 1144 | 10 | 3.09E-07 | 1.22E-08 | 0.010167 | 9.89E-08 | 0.003630364 | AC034228.2, MEIKIN, AC034228.3 |
| chr17 | 30116024 | 30117555 | 1532 | 18 | 9.63E-08 | 1.81E-06 | 0.000149 | 9.90E-08 | 4.76E-05 | NSRP1, MIR423, AC104984.2, hsa-mir-423, MIR3184 |
| chr16 | 88875168 | 88875950 | 783 | 5 | 6.26E-11 | 3.55E-05 | 3.94E-07 | 9.91E-08 | -0.00288237 | CBFA2T3 |
| chr13 | 20721389 | 20722838 | 1450 | 9 | 3.75E-07 | 2.51E-08 | 0.002382 | 9.91E-08 | -0.00847724 | IL17D |
| chr10 | 11012061 | 11013071 | 1011 | 3 | 2.70E-07 | 3.55E-08 | 0.000298 | 9.91E-08 | -0.02042017 | CELF2 |
| chr19 | 35039350 | 35040955 | 1606 | 13 | 9.27E-08 | 2.39E-06 | 0.002373 | 9.92E-08 | -0.0036759 | SCN1B, HPN |
| chr8 | 37780598 | 37781866 | 1269 | 3 | 9.07E-11 | 2.31E-06 | 8.15E-07 | 9.92E-08 | 0.008753818 | NA |
| chr17 | 17392272 | 17393174 | 903 | 2 | 1.53E-07 | 9.15E-08 | 1.19E-05 | 9.92E-08 | -0.00938564 | NA |
| chr9 | 129624085 | 129625260 | 1176 | 6 | 4.18E-11 | 8.70E-06 | 5.74E-06 | 9.94E-08 | -0.00653221 | NTMT1 |
| chr5 | 73336561 | 73337139 | 579 | 3 | 1.16E-08 | 1.73E-06 | 3.48E-05 | 9.95E-08 | -0.00016042 | NA |
| chr17 | 40218421 | 40219495 | 1075 | 13 | 1.44E-07 | 5.80E-07 | 0.000425 | 9.97E-08 | -0.00111868 | WIPF2 |
| chr11 | 968423 | 969000 | 578 | 6 | 5.21E-08 | 6.21E-06 | 3.71E-06 | 9.98E-08 | -0.00062972 | AP2A2 |
| chr12 | 9064483 | 9065263 | 781 | 8 | 1.06E-07 | 9.31E-08 | 0.006569 | 9.98E-08 | -0.03194111 | A2M-AS1, LINC00612 |
| chr6 | 89638290 | 89640141 | 1852 | 18 | 1.11E-07 | 3.27E-06 | 0.000179 | 9.98E-08 | -0.00277679 | LYRM2 |
| chr7 | 36692242 | 36692736 | 495 | 2 | 1.63E-07 | 6.35E-08 | 4.38E-05 | 9.99E-08 | -0.02186749 | AOAH |
| chr11 | 2996628 | 2997130 | 503 | 2 | 2.55E-10 | 0.0016654 | 1.18E-07 | 1.00E-07 | 0.003972532 | NA |
| chr12 | 132176410 | 132177094 | 685 | 5 | 1.50E-07 | 2.22E-07 | 0.000186 | 1.00E-07 | -0.00631195 | NA |
| chr10 | 132878902 | 132880163 | 1262 | 7 | 1.56E-09 | 8.21E-05 | 8.30E-05 | 1.00E-07 | 0.011281847 | CFAP46 |
| chr13 | 35174047 | 35174560 | 514 | 2 | 3.16E-08 | 1.32E-07 | 6.10E-06 | 1.00E-07 | -0.01682514 | NBEA |
| chr3 | 191330519 | 191331496 | 978 | 10 | 9.43E-08 | 5.35E-06 | 0.000122 | 1.01E-07 | 0.001296255 | CCDC50, UTS2B |
| chr20 | 24815553 | 24816033 | 481 | 2 | 4.77E-11 | 1.52E-05 | 2.01E-07 | 1.01E-07 | 0.003985227 | NA |
| chr19 | 28709867 | 28710989 | 1123 | 3 | 7.02E-12 | 0.0001798 | 1.09E-07 | 1.01E-07 | 0.017134524 | AC005394.2, AC005524.1 |
| chr17 | 19554595 | 19555754 | 1160 | 3 | 8.07E-12 | 9.10E-06 | 2.89E-07 | 1.01E-07 | 0.003004275 | SLC47A1 |
| chr19 | 4182524 | 4183453 | 930 | 11 | 5.88E-08 | 1.91E-06 | 3.39E-05 | 1.01E-07 | -0.00075273 | ANKRD24, SIRT6 |
| chr12 | 913311 | 914536 | 1226 | 7 | 2.79E-07 | 4.26E-08 | 0.001498 | 1.01E-07 | -0.00201129 | RAD52 |
| chr18 | 50281133 | 50283305 | 2173 | 18 | 1.61E-05 | 2.36E-07 | 0.002539 | 1.01E-07 | 0.000440256 | MBD1, CXXC1 |
| chr5 | 637673 | 638134 | 462 | 2 | 1.43E-09 | 2.82E-07 | 2.26E-06 | 1.01E-07 | -0.00188207 | CEP72 |
| chr10 | 99781605 | 99783226 | 1622 | 6 | 9.24E-08 | 7.29E-07 | 0.000123 | 1.01E-07 | 0.001149932 | ABCC2 |
| chr1 | 31880738 | 31881221 | 484 | 3 | 1.85E-09 | 5.66E-06 | 3.46E-07 | 1.01E-07 | 0.003907993 | AL136115.2 |
| chr1 | 43268982 | 43269503 | 522 | 5 | 5.43E-08 | 6.69E-07 | 0.000168 | 1.01E-07 | -0.00135732 | EBNA1BP2 |
| chr19 | 55339648 | 55341480 | 1833 | 14 | 3.89E-06 | 1.99E-06 | 7.95E-05 | 1.01E-07 | 0.001049875 | KMT5C |
| chr22 | 30265889 | 30267327 | 1439 | 12 | 4.54E-06 | 4.89E-07 | 0.003963 | 1.01E-07 | -0.00156379 | OSM |
| chr4 | 186588449 | 186589104 | 656 | 5 | 1.74E-08 | 2.92E-06 | 3.55E-05 | 1.01E-07 | 0.002602917 | FAT1 |
| chr18 | 69895806 | 69895812 | 7 | 2 | 1.22E-07 | 7.25E-08 | 2.48E-05 | 1.01E-07 | -0.00921824 | CD226 |
| chr11 | 74019924 | 74019979 | 56 | 2 | 3.27E-08 | 6.21E-07 | 1.05E-06 | 1.01E-07 | -0.01151527 | C2CD3 |
| chr22 | 37186013 | 37186280 | 268 | 2 | 1.17E-07 | 6.49E-08 | 4.18E-05 | 1.01E-07 | 0.012356713 | C1QTNF6 |
| chr3 | 71246184 | 71246533 | 350 | 4 | 3.55E-08 | 3.69E-07 | 2.71E-05 | 1.01E-07 | 0.001572273 | FOXP1, AC097634.4 |
| chr10 | 92847667 | 92849281 | 1615 | 14 | 8.29E-08 | 7.83E-06 | 0.000599 | 1.01E-07 | 0.000932905 | EXOC6 |
| chr14 | 22544066 | 22544882 | 817 | 4 | 2.57E-07 | 5.73E-08 | 0.000276 | 1.01E-07 | 0.00086341 | TRAJ2 |
| chr5 | 44807439 | 44808278 | 840 | 4 | 9.62E-09 | 1.77E-07 | 0.000173 | 1.01E-07 | -0.01068361 | MRPS30-DT |
| chr11 | 65110331 | 65111926 | 1596 | 15 | 1.15E-07 | 3.32E-06 | 0.000875 | 1.01E-07 | -0.00178155 | VPS51, TM7SF2, AP003068.1 |
| chr2 | 74482504 | 74483540 | 1037 | 18 | 8.56E-08 | 2.23E-06 | 0.000347 | 1.02E-07 | -0.00027783 | TTC31, CCDC142 |
| chr15 | 25076074 | 25076981 | 908 | 5 | 2.59E-08 | 5.18E-05 | 0.000122 | 1.02E-07 | -0.00695915 | SNHG14 |
| chr11 | 96155048 | 96156290 | 1243 | 4 | 1.15E-07 | 1.13E-07 | 0.00022 | 1.02E-07 | -0.00137658 | MAML2 |
| chr13 | 46553605 | 46554752 | 1148 | 4 | 3.27E-08 | 1.26E-05 | 3.17E-06 | 1.02E-07 | 0.002867751 | LRCH1 |
| chr3 | 169328358 | 169328799 | 442 | 2 | 1.26E-07 | 6.72E-08 | 3.70E-05 | 1.02E-07 | -0.0135374 | MECOM |
| chr3 | 18402893 | 18403220 | 328 | 3 | 4.92E-08 | 1.63E-06 | 1.16E-05 | 1.02E-07 | 0.002064652 | TBC1D5, SATB1 |
| chr17 | 7479289 | 7480445 | 1157 | 13 | 1.66E-07 | 1.40E-07 | 0.003559 | 1.02E-07 | 0.001639653 | ZBTB4 |
| chr17 | 38578228 | 38579331 | 1104 | 7 | 4.11E-08 | 2.46E-06 | 0.000751 | 1.02E-07 | 0.003345963 | SRCIN1 |
| chr14 | 51088440 | 51088802 | 363 | 4 | 1.32E-08 | 5.85E-06 | 2.48E-05 | 1.03E-07 | -0.01730679 | TRIM9 |
| chr19 | 39506515 | 39507650 | 1136 | 6 | 8.25E-07 | 1.75E-08 | 0.00252 | 1.03E-07 | 0.025611972 | DLL3 |
| chr11 | 2671911 | 2672775 | 865 | 3 | 2.41E-07 | 3.88E-08 | 0.00027 | 1.03E-07 | -0.00685251 | KCNQ1, KCNQ1OT1 |
| chr19 | 1255284 | 1257078 | 1795 | 8 | 3.77E-08 | 3.22E-07 | 0.000857 | 1.03E-07 | 3.66E-05 | MIDN |
| chr21 | 36783637 | 36783978 | 342 | 3 | 7.17E-08 | 2.13E-07 | 8.56E-06 | 1.03E-07 | -0.00647126 | HLCS |
| chr4 | 3514338 | 3515031 | 694 | 4 | 6.40E-08 | 3.93E-08 | 0.000165 | 1.03E-07 | 0.022966935 | LRPAP1 |
| chr6 | 2841234 | 2842707 | 1474 | 12 | 5.83E-07 | 3.84E-07 | 0.000501 | 1.03E-07 | 0.000854484 | SERPINB1 |
| chr6 | 31655492 | 31656872 | 1381 | 9 | 3.83E-08 | 2.31E-06 | 0.000345 | 1.03E-07 | 0.000135061 | APOM |
| chr7 | 27122675 | 27124201 | 1527 | 8 | 2.49E-08 | 5.72E-06 | 2.55E-05 | 1.03E-07 | -0.00493489 | HOXA-AS2, HOXA3 |
| chr6 | 46692449 | 46692457 | 9 | 2 | 8.40E-08 | 8.66E-08 | 1.53E-05 | 1.03E-07 | 0.001412437 | TDRD6 |
| chr19 | 41926758 | 41928112 | 1355 | 4 | 2.22E-07 | 6.36E-06 | 1.13E-05 | 1.03E-07 | -0.01480356 | ARHGEF1, ERFL |
| chr13 | 49109665 | 49110396 | 732 | 8 | 1.61E-08 | 9.62E-06 | 8.78E-05 | 1.03E-07 | 0.003077456 | FNDC3A |
| chr10 | 114751792 | 114752335 | 544 | 2 | 7.92E-08 | 8.28E-08 | 1.73E-05 | 1.03E-07 | -0.02862046 | ABLIM1 |
| chr16 | 1338165 | 1339573 | 1409 | 6 | 1.30E-07 | 1.70E-06 | 1.59E-05 | 1.03E-07 | -0.01082833 | BAIAP3 |
| chr22 | 36434865 | 36435626 | 762 | 4 | 8.87E-09 | 8.34E-07 | 6.38E-05 | 1.03E-07 | -0.0019884 | NA |
| chr12 | 2891204 | 2891737 | 534 | 2 | 5.64E-15 | 4.07E-05 | 1.61E-07 | 1.03E-07 | -0.02058233 | TULP3 |
| chr20 | 34874557 | 34875026 | 470 | 5 | 6.70E-09 | 6.41E-07 | 4.62E-05 | 1.03E-07 | 0.006093842 | ACSS2 |
| chr18 | 3924173 | 3924605 | 433 | 2 | 7.64E-09 | 6.97E-07 | 9.99E-07 | 1.03E-07 | -0.01284487 | DLGAP1 |
| chr6 | 168515465 | 168516544 | 1080 | 3 | 7.27E-08 | 2.62E-07 | 2.99E-05 | 1.04E-07 | -0.01309941 | SMOC2 |
| chr18 | 12777646 | 12778030 | 385 | 4 | 1.29E-07 | 8.89E-08 | 6.16E-05 | 1.04E-07 | 0.026530382 | NA |
| chr20 | 13638452 | 13639470 | 1019 | 15 | 1.95E-07 | 1.23E-07 | 0.002058 | 1.04E-07 | 0.002310052 | TASP1 |
| chr2 | 237256782 | 237257274 | 493 | 4 | 3.71E-09 | 4.10E-05 | 1.01E-06 | 1.04E-07 | 0.007173983 | AC112715.1 |
| chr18 | 50748404 | 50748738 | 335 | 3 | 7.31E-08 | 1.87E-06 | 9.78E-06 | 1.04E-07 | -0.01479891 | NA |
| chr13 | 21175833 | 21177094 | 1262 | 17 | 2.50E-09 | 3.31E-05 | 1.77E-05 | 1.04E-07 | 0.000677336 | MRPL57, SKA3 |
| chr14 | 96391707 | 96392654 | 948 | 9 | 1.61E-08 | 2.96E-07 | 0.000684 | 1.04E-07 | -0.00361304 | AK7 |
| chr10 | 118850658 | 118851113 | 456 | 2 | 2.42E-08 | 1.54E-07 | 5.11E-06 | 1.04E-07 | -0.00883181 | AL157388.1 |
| chr4 | 168848538 | 168849255 | 718 | 4 | 1.78E-07 | 3.49E-08 | 0.000642 | 1.04E-07 | 0.053363476 | PALLD |
| chr18 | 47149708 | 47151288 | 1581 | 16 | 1.45E-09 | 2.23E-05 | 2.95E-05 | 1.04E-07 | 0.00288017 | HDHD2, AC012254.2 |
| chr9 | 121334603 | 121335597 | 995 | 3 | 1.62E-06 | 2.62E-07 | 2.09E-05 | 1.04E-07 | -0.00401095 | NA |
| chr1 | 184539772 | 184539987 | 216 | 3 | 1.17E-07 | 9.95E-08 | 8.58E-05 | 1.04E-07 | -0.01204757 | C1orf21 |
| chr19 | 51593662 | 51595187 | 1526 | 7 | 4.98E-06 | 1.58E-07 | 0.001269 | 1.04E-07 | -0.00368797 | AC018755.3 |
| chr3 | 13896060 | 13897007 | 948 | 6 | 7.51E-09 | 1.73E-05 | 2.09E-05 | 1.04E-07 | -0.0066624 | NA |
| chr1 | 175146652 | 175147092 | 441 | 2 | 5.30E-08 | 7.11E-08 | 3.13E-05 | 1.04E-07 | -0.0197845 | TNN |
| chr10 | 131733611 | 131734238 | 628 | 2 | 7.96E-10 | 4.08E-06 | 3.39E-07 | 1.04E-07 | -0.01769377 | NA |
| chr17 | 31549200 | 31549725 | 526 | 5 | 3.27E-09 | 5.80E-06 | 3.97E-05 | 1.04E-07 | -0.00170696 | NA |
| chr9 | 112749998 | 112751476 | 1479 | 15 | 9.84E-08 | 3.80E-07 | 0.000224 | 1.04E-07 | 0.001533791 | SNX30, AL390067.1 |
| chr17 | 38906687 | 38907396 | 710 | 2 | 2.71E-09 | 6.41E-07 | 1.08E-06 | 1.04E-07 | -0.01519592 | LASP1 |
| chr16 | 81960037 | 81961272 | 1236 | 5 | 1.53E-10 | 3.35E-05 | 7.12E-07 | 1.04E-07 | -0.00427905 | PLCG2 |
| chr4 | 652103 | 653533 | 1431 | 8 | 1.01E-08 | 2.07E-07 | 9.55E-05 | 1.04E-07 | -0.00260891 | PDE6B, PDE6B-AS1 |
| chr14 | 88006093 | 88006694 | 602 | 3 | 6.97E-09 | 3.44E-06 | 3.30E-05 | 1.05E-07 | 0.012887054 | GPR65 |
| chr14 | 105900645 | 105901821 | 1177 | 6 | 1.95E-06 | 9.29E-08 | 0.001078 | 1.05E-07 | 0.011162394 | IGHD1-14, IGHD6-13 |
| chr3 | 193378808 | 193380190 | 1383 | 7 | 1.58E-12 | 1.03E-05 | 1.03E-06 | 1.05E-07 | -0.00710114 | ATP13A5 |
| chr6 | 28479017 | 28479338 | 322 | 5 | 1.61E-07 | 1.63E-08 | 0.002471 | 1.05E-07 | -0.03338542 | Z98745.2 |
| chr15 | 56463973 | 56464910 | 938 | 2 | 2.44E-09 | 1.32E-07 | 6.69E-06 | 1.05E-07 | -0.00698753 | MNS1 |
| chr2 | 73823843 | 73824059 | 217 | 2 | 1.60E-08 | 2.71E-06 | 4.17E-07 | 1.05E-07 | 0.003235137 | AC073046.4 |
| chr13 | 31923354 | 31923828 | 475 | 2 | 5.15E-08 | 1.07E-07 | 1.00E-05 | 1.05E-07 | 0.021875339 | FRY |
| chr6 | 38614300 | 38614998 | 699 | 2 | 2.31E-09 | 1.12E-06 | 7.15E-07 | 1.05E-07 | -0.00850968 | BTBD9 |
| chr16 | 3214442 | 3215971 | 1530 | 5 | 1.37E-06 | 2.07E-07 | 0.000103 | 1.05E-07 | -0.01182829 | AJ003147.2, OR1F2P |

| | | | | | | | | | | |
|---|---|---|---|---|---|---|---|---|---|---|
| chr3 | 187247233 | 187247362 | 130 | 2 | 7.05E-08 | 1.25E-07 | 7.42E-06 | 1.06E-07 | -0.00055918 | MASP1 |
| chr1 | 20696036 | 20697330 | 1295 | 5 | 4.74E-06 | 7.39E-07 | 6.88E-05 | 1.06E-07 | -0.00234697 | KIF17 |
| chr10 | 119655677 | 119656529 | 853 | 3 | 2.28E-08 | 7.08E-07 | 5.04E-06 | 1.06E-07 | -0.01043273 | BAG3 |
| chr1 | 26169383 | 26170900 | 1518 | 14 | 3.31E-07 | 1.91E-06 | 0.000533 | 1.06E-07 | 0.001583443 | ZNF593, AL391650.1 |
| chr2 | 224001694 | 224001766 | 73 | 5 | 8.25E-08 | 4.17E-06 | 5.39E-05 | 1.06E-07 | 0.000253761 | SERPINE2 |
| chr9 | 133120668 | 133121034 | 367 | 2 | 3.99E-09 | 1.29E-07 | 7.13E-06 | 1.06E-07 | 0.006615858 | RALGDS, AL162417.1 |
| chr10 | 132924794 | 132926430 | 1637 | 8 | 1.24E-06 | 1.45E-07 | 5.58E-05 | 1.06E-07 | -0.00135871 | CFAP46 |
| chr16 | 84193536 | 84195354 | 1819 | 6 | 1.01E-06 | 1.07E-07 | 0.00072 | 1.06E-07 | 0.003198074 | ADAD2, AC009123.1 |
| chr6 | 151239442 | 151240891 | 1450 | 15 | 4.02E-08 | 2.13E-06 | 0.005857 | 1.07E-07 | -0.00108178 | AKAP12 |
| chr22 | 20066093 | 20067104 | 1012 | 5 | 6.20E-08 | 1.84E-07 | 0.000145 | 1.07E-07 | -0.00018539 | TANGO2, AC006547.1 |
| chr4 | 88467147 | 88467773 | 627 | 4 | 5.65E-09 | 4.42E-07 | 3.56E-05 | 1.07E-07 | -0.00517657 | HERC5 |
| chr10 | 19488325 | 19489077 | 753 | 3 | 1.93E-07 | 6.68E-08 | 0.000152 | 1.07E-07 | -0.01467274 | MALRD1 |
| chr19 | 38710806 | 38711690 | 885 | 3 | 6.20E-11 | 0.0003113 | 2.99E-07 | 1.07E-07 | -0.00266602 | ACTN4 |
| chr20 | 844407 | 845414 | 1008 | 14 | 4.17E-08 | 2.35E-06 | 0.001192 | 1.07E-07 | 0.0074335 | FAM110A |
| chr5 | 38257905 | 38259296 | 1392 | 16 | 1.12E-07 | 4.78E-06 | 1.23E-05 | 1.07E-07 | -0.00040399 | EGFLAM |
| chr12 | 124385566 | 124386274 | 709 | 6 | 2.11E-07 | 2.70E-07 | 0.000376 | 1.07E-07 | -0.00179497 | NCOR2 |
| chr19 | 2278452 | 2278848 | 397 | 5 | 4.04E-08 | 1.89E-05 | 5.09E-06 | 1.07E-07 | -0.00335294 | AC005258.1, PEAK3 |
| chr1 | 119367582 | 119368829 | 1248 | 6 | 1.17E-07 | 6.60E-07 | 9.08E-05 | 1.07E-07 | -0.00594646 | HAO2 |
| chr19 | 1490195 | 1491290 | 1096 | 13 | 7.99E-09 | 2.17E-05 | 1.44E-05 | 1.08E-07 | 0.000111717 | REEP6, PCSK4 |
| chr3 | 16900779 | 16901036 | 258 | 3 | 1.45E-07 | 6.43E-08 | 5.68E-05 | 1.08E-07 | 0.017136599 | PLCL2 |
| chr6 | 56314836 | 56314993 | 158 | 2 | 8.61E-08 | 1.37E-07 | 6.66E-06 | 1.08E-07 | -0.02177354 | COL21A1 |
| chr15 | 74245173 | 74245689 | 517 | 5 | 3.00E-08 | 3.63E-06 | 5.65E-05 | 1.08E-07 | -0.00602447 | CCDC33 |
| chr3 | 187507800 | 187508142 | 343 | 3 | 5.69E-08 | 3.39E-07 | 6.45E-05 | 1.08E-07 | -0.01101412 | NA |
| chr5 | 172748318 | 172748852 | 535 | 5 | 2.41E-09 | 1.08E-06 | 5.78E-06 | 1.08E-07 | -0.00853718 | NA |
| chr11 | 67367136 | 67368131 | 996 | 5 | 4.37E-09 | 2.18E-06 | 7.56E-05 | 1.08E-07 | 0.009782742 | RAD9A, AP003419.1, CLCF1 |
| chr10 | 7475253 | 7475871 | 619 | 5 | 1.11E-07 | 6.07E-08 | 0.000299 | 1.09E-07 | -0.02319979 | NA |
| chr19 | 4338772 | 4339220 | 449 | 8 | 2.78E-08 | 7.31E-06 | 5.70E-06 | 1.09E-07 | -0.00631504 | STAP2 |
| chr2 | 174341124 | 174342516 | 1393 | 6 | 3.52E-08 | 5.71E-07 | 0.00024 | 1.09E-07 | 0.000450999 | NA |
| chr5 | 77629082 | 77630254 | 1173 | 7 | 2.62E-07 | 3.47E-06 | 0.000597 | 1.09E-07 | -0.00050375 | OTP |
| chr3 | 27729562 | 27730542 | 981 | 8 | 3.72E-10 | 1.54E-05 | 3.29E-07 | 1.09E-07 | 0.00071576 | NA |
| chr4 | 8096543 | 8097306 | 764 | 5 | 7.24E-09 | 9.03E-07 | 5.39E-05 | 1.09E-07 | 0.006435308 | ABLIM2 |
| chr8 | 143788779 | 143789538 | 760 | 10 | 7.71E-08 | 8.91E-07 | 0.000345 | 1.09E-07 | -0.00157414 | IQANK1 |
| chr16 | 1954051 | 1955061 | 1011 | 8 | 2.74E-07 | 1.84E-08 | 0.003727 | 1.09E-07 | -0.00569877 | RPL3L |
| chr9 | 133421545 | 133422289 | 745 | 10 | 5.30E-08 | 1.03E-06 | 0.000168 | 1.09E-07 | -0.00385463 | ADAMTS13 |
| chr11 | 1401089 | 1401717 | 629 | 5 | 3.00E-09 | 2.33E-06 | 4.37E-05 | 1.09E-07 | 0.006950251 | BRSK2 |
| chr1 | 3969068 | 3969740 | 673 | 2 | 2.69E-07 | 7.29E-08 | 3.42E-05 | 1.09E-07 | -0.0291091 | NA |
| chr2 | 136829999 | 136830123 | 125 | 4 | 1.17E-07 | 8.88E-07 | 0.000112 | 1.10E-07 | 0.000808998 | THSD7B |
| chr19 | 46660722 | 46661885 | 1164 | 12 | 1.69E-09 | 1.53E-05 | 4.95E-05 | 1.10E-07 | 5.90E-05 | DACT3-AS1, DACT3 |
| chr12 | 121858993 | 121859896 | 904 | 5 | 2.19E-09 | 4.68E-05 | 1.75E-05 | 1.10E-07 | -0.00786756 | HPD |
| chr10 | 33210920 | 33211303 | 384 | 6 | 4.05E-08 | 2.50E-05 | 4.73E-05 | 1.10E-07 | 0.001102309 | AL121748.1, NRP1 |
| chr9 | 86063906 | 86063918 | 13 | 2 | 1.42E-07 | 7.14E-08 | 4.05E-05 | 1.10E-07 | -0.00787776 | GOLM1 |
| chr4 | 1683012 | 1683828 | 817 | 7 | 2.36E-07 | 2.36E-05 | 3.65E-05 | 1.10E-07 | 0.002944056 | FAM53A, Y RNA |
| chr6 | 143449958 | 143451591 | 1634 | 19 | 5.89E-07 | 1.26E-06 | 0.000298 | 1.10E-07 | 0.000716792 | PEX3, ADAT2 |
| chr6 | 33609108 | 33609934 | 827 | 2 | 1.01E-08 | 3.54E-07 | 2.04E-06 | 1.10E-07 | -0.01523991 | NA |
| chr5 | 515236 | 515434 | 199 | 2 | 4.53E-08 | 3.47E-07 | 2.08E-06 | 1.10E-07 | -0.03507141 | SLC9A3 |
| chr10 | 48508285 | 48510212 | 1928 | 6 | 4.22E-05 | 2.11E-07 | 0.001919 | 1.11E-07 | 0.003546966 | ARHGAP22 |
| chr11 | 69704320 | 69705650 | 1331 | 14 | 1.03E-07 | 3.05E-05 | 0.000197 | 1.11E-07 | 0.002646003 | NA |
| chr5 | 1752553 | 1753235 | 683 | 4 | 5.28E-08 | 2.86E-06 | 2.17E-05 | 1.11E-07 | 0.001369135 | NA |
| chr2 | 214809940 | 214810913 | 974 | 6 | 8.57E-07 | 2.99E-08 | 0.001519 | 1.11E-07 | -0.00933332 | SNHG31 |
| chr15 | 70702065 | 70703537 | 1473 | 10 | 9.31E-10 | 1.83E-05 | 4.46E-05 | 1.11E-07 | -0.00296392 | UACA |
| chr4 | 82741448 | 82742327 | 880 | 2 | 2.19E-07 | 1.23E-07 | 8.73E-06 | 1.12E-07 | 0.011565544 | SCD5 |
| chr4 | 6659044 | 6659249 | 206 | 3 | 4.31E-08 | 1.14E-07 | 3.32E-05 | 1.12E-07 | 0.027675013 | LINC02482 |
| chr2 | 239608803 | 239608875 | 73 | 2 | 6.16E-09 | 3.43E-07 | 2.14E-06 | 1.12E-07 | -0.01687363 | NA |
| chr1 | 201189732 | 201190860 | 1129 | 8 | 1.79E-08 | 0.0001092 | 0.000322 | 1.12E-07 | -0.00825404 | IGFN1 |
| chr14 | 105033957 | 105034749 | 793 | 3 | 1.34E-08 | 7.74E-07 | 4.71E-05 | 1.12E-07 | 0.008389906 | NA |
| chr16 | 68509876 | 68510501 | 626 | 5 | 2.60E-09 | 9.09E-06 | 2.96E-05 | 1.12E-07 | -0.0010331 | NA |
| chr1 | 185734745 | 185735417 | 673 | 2 | 7.58E-13 | 4.48E-07 | 1.65E-06 | 1.12E-07 | -0.01332461 | HMCN1 |
| chr20 | 13220038 | 13221197 | 1160 | 25 | 5.67E-08 | 3.31E-06 | 0.012225 | 1.12E-07 | 0.000663406 | NA |
| chr3 | 143831676 | 143831885 | 210 | 2 | 1.23E-07 | 7.79E-08 | 2.89E-05 | 1.12E-07 | -0.00103439 | SLC9A9 |
| chr13 | 26956526 | 26957046 | 521 | 2 | 3.58E-10 | 5.43E-05 | 1.64E-07 | 1.12E-07 | -0.01107401 | NA |
| chr9 | 32550314 | 32551766 | 1453 | 8 | 1.95E-07 | 1.11E-06 | 0.001305 | 1.12E-07 | 0.003844789 | SMIM27, TOPORS |
| chr19 | 57320181 | 57320615 | 435 | 9 | 7.71E-10 | 0.0003561 | 2.50E-07 | 1.12E-07 | -0.00068245 | ZNF543 |
| chr21 | 31343593 | 31344505 | 913 | 6 | 1.96E-09 | 5.74E-06 | 2.93E-06 | 1.13E-07 | 0.00433744 | TIAM1 |
| chr8 | 103296991 | 103298152 | 1162 | 5 | 3.60E-10 | 2.14E-06 | 6.58E-05 | 1.13E-07 | -0.00601397 | BAALC-AS1 |
| chr6 | 1378442 | 1379560 | 1119 | 5 | 2.30E-08 | 4.88E-05 | 3.36E-06 | 1.13E-07 | 0.001056756 | NA |
| chr1 | 227763651 | 227764093 | 443 | 3 | 1.41E-07 | 7.14E-08 | 8.69E-05 | 1.13E-07 | -0.00466282 | SNAP47 |
| chr1 | 35192422 | 35194077 | 1656 | 19 | 4.53E-07 | 2.49E-06 | 0.000845 | 1.13E-07 | 0.001339503 | SFPQ |
| chr13 | 94465691 | 94466649 | 959 | 2 | 1.36E-06 | 8.60E-08 | 2.09E-05 | 1.13E-07 | 0.014011927 | DCT |
| chr2 | 26502848 | 26504368 | 1521 | 8 | 1.90E-08 | 5.17E-06 | 1.19E-05 | 1.13E-07 | 0.004734443 | OTOF |
| chr19 | 19203483 | 19204540 | 1058 | 11 | 1.13E-08 | 1.48E-05 | 9.23E-06 | 1.13E-07 | -0.0032373 | NA |
| chr11 | 68212789 | 68214277 | 1489 | 12 | 1.93E-07 | 5.36E-07 | 0.001211 | 1.13E-07 | 2.74E-05 | KMT5B |
| chr17 | 4607277 | 4607837 | 561 | 6 | 2.96E-08 | 3.38E-06 | 1.82E-05 | 1.13E-07 | -0.00611205 | SMTNL2 |
| chr5 | 140174278 | 140175231 | 954 | 13 | 9.77E-09 | 2.13E-05 | 0.000177 | 1.13E-07 | 0.00095092 | CYSTM1 |
| chr10 | 124619465 | 124619993 | 529 | 5 | 4.26E-08 | 8.00E-07 | 1.47E-05 | 1.13E-07 | 0.007974747 | AC068896.1, FAM53B |
| chr2 | 227717357 | 227718104 | 748 | 10 | 7.27E-10 | 1.94E-06 | 0.000118 | 1.13E-07 | -0.00301856 | SLC19A3 |
| chr12 | 15712952 | 15713681 | 730 | 6 | 1.31E-07 | 2.83E-06 | 0.001465 | 1.14E-07 | -0.00403295 | EPS8 |
| chr21 | 15728047 | 15729420 | 1374 | 4 | 1.11E-09 | 4.52E-07 | 1.47E-05 | 1.14E-07 | -0.00878414 | NA |
| chr3 | 12287157 | 12288246 | 1090 | 15 | 1.94E-10 | 3.27E-05 | 6.42E-05 | 1.14E-07 | -0.00141675 | PPARG |
| chr6 | 3387581 | 3387630 | 50 | 2 | 1.79E-08 | 1.27E-07 | 8.61E-06 | 1.14E-07 | 0.004523585 | SLC22A23 |
| chr6 | 41625601 | 41626107 | 507 | 2 | 2.85E-09 | 4.98E-07 | 1.53E-06 | 1.14E-07 | -0.00923367 | NA |
| chr7 | 4774523 | 4775321 | 799 | 5 | 1.57E-08 | 1.94E-07 | 0.001227 | 1.14E-07 | -0.00586693 | NA |
| chr10 | 14087105 | 14087713 | 609 | 2 | 1.35E-07 | 8.61E-08 | 2.19E-05 | 1.14E-07 | -0.00181807 | AL157896.1, FRMD4A |
| chr11 | 70485756 | 70486515 | 760 | 3 | 1.25E-07 | 4.33E-08 | 0.00024 | 1.15E-07 | 0.002729048 | SHANK2 |
| chr2 | 240256065 | 240257470 | 1406 | 7 | 2.58E-06 | 9.19E-08 | 0.000746 | 1.15E-07 | -0.00539434 | NA |
| chr10 | 73110482 | 73111523 | 1042 | 7 | 5.62E-08 | 1.55E-07 | 5.71E-05 | 1.15E-07 | -0.00630079 | NUDT13 |
| chr17 | 50273107 | 50274834 | 1728 | 10 | 1.42E-06 | 1.75E-05 | 6.63E-05 | 1.15E-07 | -8.09E-05 | TMEM92 |
| chr5 | 173616596 | 173617408 | 813 | 13 | 8.37E-08 | 1.66E-06 | 0.001021 | 1.15E-07 | 0.003568433 | BOD1 |
| chr5 | 168968168 | 168968283 | 116 | 2 | 1.51E-07 | 7.07E-08 | 5.66E-05 | 1.15E-07 | -0.02889317 | SLIT3 |
| chr10 | 114179204 | 114180769 | 1566 | 12 | 2.89E-07 | 5.11E-05 | 8.22E-05 | 1.15E-07 | 0.002776682 | TDRD1 |
| chr3 | 64067309 | 64067987 | 679 | 6 | 1.60E-08 | 1.80E-05 | 1.41E-05 | 1.15E-07 | -0.00810592 | PRICKLE2-AS1 |
| chr8 | 142173047 | 142174106 | 1060 | 7 | 5.33E-07 | 1.39E-07 | 0.00078 | 1.16E-07 | -0.0077183 | AC103758.1 |
| chr8 | 71854435 | 71854860 | 426 | 2 | 1.19E-09 | 2.78E-05 | 1.96E-07 | 1.16E-07 | 0.008149205 | MSC-AS1 |
| chr13 | 27162050 | 27162408 | 359 | 2 | 8.28E-08 | 9.95E-08 | 1.53E-05 | 1.16E-07 | 0.015001322 | USP12 |
| chr4 | 990981 | 991818 | 838 | 7 | 1.87E-09 | 2.98E-05 | 5.28E-06 | 1.16E-07 | -0.00064106 | IDUA, SLC26A1 |
| chr3 | 52485650 | 52486606 | 957 | 2 | 1.03E-06 | 8.37E-08 | 2.55E-05 | 1.16E-07 | -0.00430262 | NISCH |
| chr13 | 26557025 | 26558294 | 1270 | 13 | 7.04E-07 | 7.02E-07 | 0.00041 | 1.16E-07 | -0.00016878 | WASF3 |
| chr3 | 113837870 | 113839085 | 1216 | 12 | 4.41E-09 | 2.49E-05 | 0.000466 | 1.16E-07 | 0.000148878 | GRAMD1C |
| chr16 | 56344326 | 56345236 | 911 | 4 | 3.55E-08 | 1.19E-06 | 1.43E-05 | 1.16E-07 | -0.00636337 | GNAO1 |
| chr14 | 93332637 | 93333809 | 1173 | 18 | 1.70E-08 | 2.44E-05 | 6.77E-05 | 1.16E-07 | 0.000816669 | UNC79, BTBD7 |
| chr3 | 48847464 | 48848270 | 807 | 13 | 1.23E-07 | 4.47E-08 | 0.004873 | 1.16E-07 | -0.00263313 | PRKAR2A-AS1, PRKAR2A |
| chr5 | 140564251 | 140565522 | 1272 | 13 | 1.37E-08 | 9.49E-06 | 4.50E-05 | 1.16E-07 | -0.0013914 | SLC35A4, APBB3, AC116353.5 |
| chr8 | 3375142 | 3375904 | 763 | 3 | 2.76E-07 | 3.99E-08 | 0.00042 | 1.16E-07 | -0.00347154 | CSMD1, AC026991.1 |
| chr6 | 149701908 | 149702467 | 560 | 5 | 1.49E-07 | 1.21E-05 | 5.15E-05 | 1.17E-07 | -0.00097621 | LATS1 |
| chr15 | 52018591 | 52019434 | 844 | 10 | 7.51E-08 | 3.05E-08 | 0.001444 | 1.17E-07 | -0.00084992 | MAPK6, MAPK6-DT |
| chr8 | 40099277 | 40100231 | 955 | 2 | 6.28E-07 | 8.11E-08 | 2.95E-05 | 1.17E-07 | -0.01521583 | NA |
| chr5 | 137146283 | 137146633 | 351 | 2 | 1.42E-07 | 7.41E-08 | 4.49E-05 | 1.17E-07 | -0.00246198 | SPOCK1 |
| chr16 | 66578871 | 66580067 | 1197 | 13 | 2.40E-07 | 3.05E-07 | 0.004114 | 1.17E-07 | -0.00029255 | CKLF-CMTM1, CMTM1, CMTM2, AC010542.1 |
| chr2 | 86196821 | 86197095 | 275 | 3 | 4.15E-10 | 1.04E-07 | 2.89E-05 | 1.17E-07 | -0.01097815 | NA |
| chr12 | 124310300 | 124311863 | 1564 | 6 | 2.16E-06 | 8.43E-07 | 9.52E-05 | 1.17E-07 | -0.007358 | RFLNA |
| chr4 | 7915767 | 7915959 | 193 | 2 | 8.81E-08 | 8.52E-08 | 2.47E-05 | 1.17E-07 | 0.026823552 | AFAP1 |
| chr2 | 96108744 | 96109490 | 747 | 2 | 1.57E-07 | 1.10E-07 | 1.24E-05 | 1.17E-07 | -0.01615191 | NA |
| chr11 | 44763798 | 44764716 | 919 | 5 | 3.54E-10 | 4.06E-05 | 1.17E-07 | 1.17E-07 | -0.00514683 | TSPAN18 |
| chr3 | 132592683 | 132593399 | 717 | 2 | 5.49E-07 | 7.67E-08 | 3.79E-05 | 1.17E-07 | 0.00225669 | ACAD11, NPHP3-ACAD11 |
| chr12 | 109621395 | 109622243 | 849 | 3 | 8.54E-08 | 7.90E-08 | 0.000113 | 1.17E-07 | 0.024428102 | NA |
| chr1 | 957324 | 958034 | 711 | 4 | 4.52E-09 | 7.07E-07 | 0.000122 | 1.18E-07 | -0.01714116 | NOC2L |
| chr1 | 108721661 | 108722765 | 1105 | 3 | 1.07E-06 | 8.91E-08 | 5.11E-05 | 1.18E-07 | -0.00227151 | FNDC7 |
| chr6 | 37555458 | 37556847 | 1390 | 4 | 1.46E-07 | 4.72E-08 | 0.000178 | 1.18E-07 | -0.00093307 | NA |
| chr8 | 1296026 | 1296883 | 858 | 4 | 1.02E-06 | 6.78E-08 | 0.000249 | 1.18E-07 | -0.00261745 | DLGAP2, AF067845.2 |
| chr7 | 151023415 | 151023675 | 261 | 2 | 1.60E-08 | 1.09E-07 | 1.29E-05 | 1.18E-07 | -0.00759325 | ATG9B |
| chr1 | 981483 | 983035 | 1553 | 11 | 6.37E-08 | 3.18E-07 | 0.001813 | 1.18E-07 | -0.0073381 | PERM1 |
| chr8 | 127920529 | 127921185 | 657 | 2 | 8.06E-11 | 0.0003487 | 1.43E-07 | 1.18E-07 | -0.00696797 | PVT1 |
| chr5 | 1268661 | 1269194 | 534 | 7 | 1.67E-08 | 4.76E-06 | 0.000314 | 1.18E-07 | -0.00343441 | TERT |
| chr9 | 98872322 | 98872915 | 594 | 5 | 9.54E-08 | 7.71E-08 | 0.00044 | 1.18E-07 | 0.004010065 | AL136084.1 |
| chr2 | 186598871 | 186599344 | 474 | 4 | 1.20E-07 | 1.60E-06 | 3.39E-05 | 1.18E-07 | 4.55E-06 | ITGAV |
| chr4 | 84492593 | 84493720 | 1128 | 6 | 2.67E-07 | 3.67E-07 | 0.000166 | 1.18E-07 | -0.00401425 | NKX6-1 |
| chr1 | 3141635 | 3143262 | 1628 | 11 | 1.49E-09 | 5.46E-06 | 1.69E-05 | 1.18E-07 | 0.001401127 | PRDM16 |
| chr10 | 2985953 | 2986616 | 664 | 3 | 2.96E-07 | 6.21E-08 | 8.69E-05 | 1.18E-07 | 0.019305664 | NA |
| chr6 | 149922767 | 149923935 | 1169 | 9 | 1.52E-09 | 0.0002389 | 4.08E-05 | 1.18E-07 | 0.000156835 | RAET1G |
| chr3 | 170622843 | 170623223 | 381 | 4 | 5.36E-08 | 4.84E-06 | 0.000209 | 1.19E-07 | -0.0139142 | AC026316.4, SLC7A14-AS1 |
| chr20 | 38441942 | 38442005 | 64 | 3 | 6.57E-08 | 1.99E-07 | 1.10E-05 | 1.19E-07 | -0.01838606 | NA |
| chr7 | 19772856 | 19773961 | 1106 | 17 | 2.20E-08 | 6.88E-07 | 9.76E-05 | 1.19E-07 | -0.00331472 | TMEM196 |
| chr2 | 70767626 | 70768889 | 1264 | 18 | 1.61E-08 | 5.87E-07 | 0.002476 | 1.19E-07 | -0.0002281 | ADD2 |
| chr2 | 151973675 | 151975076 | 1402 | 10 | 1.78E-08 | 8.91E-07 | 0.003682 | 1.19E-07 | -0.00817099 | AC068547.1, CACNB4 |
| chr3 | 112671754 | 112672899 | 1146 | 3 | 2.65E-07 | 6.42E-08 | 0.000108 | 1.19E-07 | 0.004460404 | NA |
| chr1 | 56510017 | 56511507 | 1491 | 3 | 2.69E-06 | 1.90E-07 | 5.54E-05 | 1.19E-07 | -0.00042886 | AC119674.2, PLPP3 |
| chr1 | 162377675 | 162378474 | 800 | 5 | 1.67E-09 | 4.65E-07 | 2.60E-05 | 1.19E-07 | -0.00786671 | AL512785.2 |
| chr9 | 99182618 | 99183343 | 726 | 5 | 1.05E-08 | 2.55E-06 | 1.82E-06 | 1.19E-07 | 0.00349675 | NA |
| chr6 | 161415493 | 161416355 | 863 | 4 | 1.13E-07 | 2.64E-07 | 9.23E-05 | 1.19E-07 | -0.01495024 | PRKN |
| chr12 | 34335794 | 34337060 | 1267 | 5 | 6.27E-06 | 2.10E-06 | 0.000275 | 1.19E-07 | -0.01343838 | NA |
| chr16 | 23348500 | 23348717 | 218 | 3 | 1.81E-07 | 4.64E-08 | 0.000303 | 1.19E-07 | -0.01486516 | SCNN1B |
| chr3 | 120907806 | 120908977 | 1172 | 13 | 3.30E-08 | 1.04E-06 | 0.003447 | 1.20E-07 | -0.00348384 | STXBP5L, AC072026.2 |
| chr6 | 52876753 | 52877279 | 527 | 3 | 9.25E-08 | 1.18E-06 | 4.82E-05 | 1.20E-07 | -0.01061911 | GSTA10P |
| chr14 | 100295420 | 100296566 | 1147 | 8 | 2.73E-08 | 3.98E-06 | 4.38E-05 | 1.20E-07 | -0.00558392 | SLC25A29 |
| chr2 | 14401055 | 14402186 | 1132 | 4 | 7.03E-07 | 8.31E-07 | 9.92E-05 | 1.20E-07 | -0.00480659 | NA |
| chr17 | 49414170 | 49415521 | 1352 | 17 | 5.73E-07 | 3.44E-06 | 0.000124 | 1.20E-07 | -0.00027115 | AC091180.5, PHB |
| chr19 | 38606889 | 38607874 | 986 | 3 | 1.82E-06 | 5.29E-07 | 1.34E-05 | 1.20E-07 | 0.000256669 | MAP4K1 |
| chr20 | 38434883 | 38435861 | 979 | 11 | 2.03E-07 | 5.66E-06 | 0.00012 | 1.20E-07 | -0.00326041 | SNHG17 |
| chr11 | 61551594 | 61552195 | 602 | 4 | 2.96E-08 | 4.61E-07 | 7.29E-05 | 1.21E-07 | 0.000761284 | SYT7 |
| chr5 | 140826609 | 140828051 | 1443 | 12 | 8.16E-09 | 3.20E-05 | 0.000387 | 1.21E-07 | -0.00649767 | PCDHA1, PCDHA2, PCDHA3, PCDHA4, PCDHA5, PCDHA6, AC005609.5 |
| chr8 | 2098668 | 2099180 | 513 | 4 | 1.43E-07 | 1.19E-05 | 8.35E-05 | 1.21E-07 | -0.00956169 | MYOM2 |
| chr6 | 32814359 | 32815774 | 1416 | 8 | 9.21E-09 | 1.70E-06 | 1.45E-05 | 1.21E-07 | 0.001365321 | HLA-DOB, AL669918.1 |
| chr2 | 130839525 | 130840071 | 547 | 3 | 1.76E-07 | 4.82E-08 | 0.000194 | 1.21E-07 | -0.01288652 | ARHGEF4 |
| chr1 | 26787017 | 26788340 | 1324 | 20 | 3.83E-09 | 0.0002432 | 9.75E-05 | 1.21E-07 | 0.000184048 | PIGV |
| chr14 | 74616731 | 74617263 | 533 | 4 | 1.49E-08 | 4.54E-07 | 1.73E-05 | 1.21E-07 | -0.00825615 | NA |
| chr12 | 53727092 | 53727987 | 896 | 12 | 1.40E-08 | 0.0001068 | 2.04E-05 | 1.21E-07 | -0.00079145 | CALCOCO1 |
| chr15 | 98916213 | 98917100 | 888 | 5 | 2.38E-10 | 4.32E-05 | 1.72E-07 | 1.21E-07 | 0.001154255 | IGF1R |
| chr5 | 141441677 | 141442308 | 632 | 4 | 1.26E-09 | 9.80E-06 | 6.40E-07 | 1.21E-07 | -0.01361304 | PCDHGA1, PCDHGA2, PCDHGA3, PCDHGB1, PCDHGA4, PCDHGB2, PCDHGA5, PCDHGB3, PCDHGA6, PCDHGA7, PCDHGB4, PCDHGA8, PCDHGB5, PCDHGA9, PCDHGB6, PCDHGA10, PCDHGB7, PCDHGA11, PCDHGA12, PCDHGB9P |
| chr19 | 14529230 | 14530227 | 998 | 21 | 3.55E-08 | 6.86E-06 | 0.002142 | 1.21E-07 | 0.000171637 | TECR, MIR639, DNAJB1 |

| | | | | | | | | | | |
|---|---|---|---|---|---|---|---|---|---|---|
| chr20 | 35771310 | 35772274 | 965 | 13 | 1.60E-07 | 1.69E-05 | 0.006697 | 1.21E-07 | 0.000247243 | PHF20 |
| chr16 | 84144456 | 84145726 | 1271 | 14 | 7.28E-08 | 8.08E-06 | 0.004334 | 1.21E-07 | -0.00104877 | DNAAF1, HSDL1 |
| chr20 | 60308164 | 60308405 | 242 | 5 | 4.14E-08 | 1.43E-06 | 3.91E-06 | 1.22E-07 | -0.01396636 | MIR646HG |
| chr17 | 42608624 | 42609800 | 1177 | 21 | 4.72E-07 | 3.87E-08 | 0.007959 | 1.22E-07 | 0.00038865 | TUBG1, RETREG3 |
| chr5 | 177370511 | 177371048 | 538 | 5 | 2.17E-07 | 1.57E-08 | 0.003571 | 1.22E-07 | -0.05534991 | RGS14 |
| chr6 | 144330999 | 144331445 | 447 | 3 | 1.63E-07 | 5.42E-08 | 0.000245 | 1.22E-07 | -0.00204662 | UTRN, AL024474.2 |
| chr2 | 230960767 | 230961866 | 1100 | 4 | 1.30E-06 | 4.63E-08 | 0.000556 | 1.22E-07 | -0.00178224 | GPR55 |
| chr4 | 95620719 | 95621187 | 469 | 2 | 1.86E-08 | 3.19E-07 | 2.68E-06 | 1.22E-07 | -0.01051457 | NA |
| chr21 | 45455228 | 45455586 | 359 | 8 | 3.04E-07 | 1.77E-08 | 0.005061 | 1.22E-07 | -0.01351057 | COL18A1 |
| chr17 | 42107246 | 42108530 | 1285 | 8 | 1.80E-08 | 1.52E-06 | 0.001002 | 1.22E-07 | -0.01408537 | DHX58 |
| chr21 | 41598904 | 41599288 | 385 | 3 | 1.95E-07 | 5.08E-08 | 0.000203 | 1.22E-07 | 0.051655422 | NA |
| chr19 | 50639323 | 50640815 | 1493 | 13 | 1.30E-08 | 5.02E-06 | 8.16E-05 | 1.23E-07 | -0.00142447 | SYT3 |
| chr16 | 58249018 | 58250394 | 1377 | 15 | 2.19E-07 | 3.78E-06 | 0.000644 | 1.23E-07 | 0.000614149 | CCDC113 |
| chr17 | 32281036 | 32281348 | 313 | 3 | 7.75E-09 | 2.20E-05 | 9.43E-07 | 1.23E-07 | -0.00778828 | RHBDL3 |
| chr5 | 100817256 | 100817920 | 665 | 3 | 1.54E-08 | 1.01E-06 | 3.74E-06 | 1.24E-07 | 0.010106277 | ST8SIA4 |
| chr16 | 89308705 | 89309117 | 413 | 4 | 6.89E-08 | 1.07E-06 | 2.72E-05 | 1.24E-07 | -0.00108553 | ANKRD11 |
| chr3 | 10930610 | 10931206 | 597 | 2 | 8.02E-08 | 1.16E-07 | 1.27E-05 | 1.24E-07 | -0.00256089 | SLC6A11, AC027128.1 |
| chr7 | 2551636 | 2552705 | 1070 | 4 | 1.05E-06 | 4.62E-06 | 0.000139 | 1.24E-07 | 0.003928837 | BRAT1 |
| chr7 | 98211530 | 98212686 | 1157 | 13 | 1.90E-08 | 1.50E-05 | 0.000407 | 1.24E-07 | -0.00132412 | BHLHA15 |
| chr5 | 6822717 | 6823128 | 412 | 2 | 7.15E-08 | 7.41E-08 | 6.70E-05 | 1.24E-07 | -0.02361622 | LINC02236 |
| chr2 | 75558380 | 75560214 | 1835 | 6 | 2.82E-08 | 0.000659 | 1.98E-05 | 1.24E-07 | -0.00117347 | EVA1A |
| chr5 | 5139345 | 5140533 | 1189 | 15 | 3.06E-07 | 1.81E-06 | 0.003069 | 1.24E-07 | -0.00234883 | ADAMTS16, CTD-2297D10.2 |
| chr7 | 156104742 | 156105741 | 1000 | 5 | 1.03E-07 | 5.98E-08 | 0.000159 | 1.24E-07 | -0.00492096 | NA |
| chr4 | 7823818 | 7824530 | 713 | 2 | 5.88E-07 | 8.06E-08 | 4.03E-05 | 1.24E-07 | -0.01152454 | AFAP1 |
| chr1 | 230424085 | 230425047 | 963 | 3 | 7.55E-08 | 1.05E-07 | 2.80E-05 | 1.24E-07 | 0.00522569 | PGBD5 |
| chr14 | 73057580 | 73059491 | 1912 | 16 | 5.32E-06 | 2.23E-06 | 0.004751 | 1.24E-07 | -0.00048132 | RBM25, AL442663.3 |
| chr10 | 29119637 | 29120779 | 1143 | 4 | 6.43E-08 | 2.35E-07 | 1.31E-05 | 1.24E-07 | -0.00458288 | NA |
| chr3 | 45765798 | 45766220 | 423 | 6 | 1.41E-07 | 6.93E-08 | 0.000119 | 1.24E-07 | -0.00524919 | SLC6A20 |
| chr9 | 113590784 | 113591839 | 1056 | 4 | 6.75E-09 | 8.86E-06 | 8.18E-05 | 1.25E-07 | -0.01317544 | RGS3 |
| chr14 | 68401142 | 68401554 | 413 | 2 | 3.29E-09 | 4.95E-06 | 3.77E-07 | 1.25E-07 | 0.04829641 | RAD51B |
| chr16 | 89947561 | 89948633 | 1073 | 9 | 5.47E-07 | 4.51E-07 | 0.002553 | 1.25E-07 | 0.006352372 | NA |
| chr11 | 13313312 | 13314012 | 701 | 3 | 9.90E-10 | 1.35E-05 | 2.62E-07 | 1.25E-07 | 0.00432958 | ARNTL |
| chr20 | 21192073 | 21192303 | 231 | 2 | 1.74E-07 | 7.47E-08 | 6.76E-05 | 1.25E-07 | -0.02020811 | KIZ, KIZ-AS1 |
| chr18 | 2570889 | 2572966 | 2078 | 18 | 1.14E-05 | 1.40E-05 | 0.001037 | 1.25E-07 | 0.000522779 | NDC80, METTL4 |
| chr10 | 126225502 | 126226789 | 1288 | 4 | 2.28E-06 | 2.12E-07 | 0.00045 | 1.25E-07 | -0.00200374 | ADAM12 |
| chr3 | 49469024 | 49470411 | 1388 | 13 | 3.13E-07 | 2.04E-06 | 0.000283 | 1.25E-07 | -0.0006292 | DAG1 |
| chr1 | 160307517 | 160308339 | 823 | 2 | 1.91E-06 | 7.57E-08 | 6.24E-05 | 1.25E-07 | 0.02965462 | COPA |
| chr7 | 4610878 | 4611779 | 902 | 4 | 1.69E-09 | 2.31E-06 | 1.29E-06 | 1.25E-07 | 0.004958453 | NA |
| chr19 | 9584256 | 9584879 | 624 | 10 | 2.47E-07 | 1.15E-05 | 0.000775 | 1.25E-07 | -0.00292589 | ZNF121 |
| chr18 | 59219683 | 59220553 | 871 | 10 | 5.49E-08 | 6.16E-06 | 0.000143 | 1.26E-07 | 0.000690348 | GRP |
| chr7 | 569183 | 570700 | 1518 | 7 | 7.98E-07 | 2.18E-07 | 0.000863 | 1.26E-07 | -0.00054803 | PRKAR1B |
| chr6 | 73143336 | 73144488 | 1153 | 5 | 6.97E-07 | 2.08E-08 | 0.002027 | 1.26E-07 | -0.00191392 | KCNQ5, KCNQ5-AS1 |
| chr17 | 17226253 | 17226806 | 554 | 2 | 2.21E-08 | 1.03E-07 | 1.82E-05 | 1.26E-07 | 3.01E-05 | AC055811.2, FLCN |
| chr14 | 101556176 | 101557156 | 981 | 5 | 3.07E-07 | 5.29E-07 | 0.000278 | 1.26E-07 | -0.00706997 | DIO3OS |
| chr5 | 66843115 | 66843885 | 771 | 3 | 2.35E-09 | 3.57E-06 | 3.11E-06 | 1.26E-07 | -0.01190508 | MAST4 |
| chr14 | 72563441 | 72563958 | 518 | 3 | 3.71E-08 | 9.12E-08 | 0.000124 | 1.26E-07 | -0.00366416 | RGS6, AC004828.1 |
| chr9 | 92327079 | 92327588 | 510 | 2 | 2.84E-09 | 1.48E-07 | 8.21E-06 | 1.26E-07 | -0.00886536 | CENPP |
| chr3 | 114351122 | 114351893 | 772 | 12 | 1.67E-07 | 1.62E-07 | 0.00115 | 1.26E-07 | 9.50E-05 | ZBTB20-AS1, ZBTB20 |
| chr4 | 128287570 | 128289431 | 1862 | 14 | 7.75E-06 | 8.03E-07 | 0.001512 | 1.27E-07 | -0.00206953 | PGRMC2 |
| chr1 | 36309477 | 36309739 | 263 | 2 | 1.73E-07 | 7.64E-08 | 6.45E-05 | 1.27E-07 | 0.005946531 | SH3D21 |
| chr17 | 76278875 | 76279851 | 977 | 6 | 1.21E-06 | 8.11E-08 | 0.000283 | 1.27E-07 | -0.00190525 | QRICH2 |
| chr10 | 79346895 | 79348625 | 1731 | 16 | 1.94E-09 | 3.50E-05 | 0.000141 | 1.27E-07 | -0.00110259 | PPIF |
| chr2 | 6877440 | 6878022 | 583 | 6 | 6.72E-10 | 9.35E-06 | 2.78E-06 | 1.27E-07 | 0.007480142 | RSAD2 |
| chr8 | 22628155 | 22628208 | 54 | 2 | 4.90E-08 | 7.66E-08 | 6.40E-05 | 1.27E-07 | -0.02267401 | BIN3 |
| chr16 | 353314 | 354540 | 1227 | 6 | 8.21E-08 | 1.79E-06 | 0.000115 | 1.27E-07 | 0.00126316 | NA |
| chr1 | 109238149 | 109238984 | 836 | 2 | 2.33E-07 | 8.46E-08 | 3.65E-05 | 1.27E-07 | -0.00843449 | SARS1 |
| chr17 | 42194261 | 42194780 | 520 | 10 | 2.66E-07 | 1.56E-08 | 0.001291 | 1.28E-07 | -0.0037315 | GHDC |
| chr4 | 169619762 | 169621020 | 1259 | 15 | 4.71E-08 | 3.93E-07 | 0.001735 | 1.28E-07 | 0.000411476 | CLCN3 |
| chr1 | 24143074 | 24144037 | 964 | 8 | 3.86E-07 | 4.34E-06 | 0.000209 | 1.28E-07 | -0.00644825 | IL22RA1 |
| chr15 | 43105644 | 43106628 | 985 | 13 | 5.17E-07 | 2.66E-07 | 0.000411 | 1.28E-07 | -7.41E-05 | UBR1, AC068724.3 |
| chr1 | 21952350 | 21953209 | 860 | 2 | 1.06E-06 | 9.11E-08 | 2.76E-05 | 1.28E-07 | -0.01311559 | AL590556.2 |
| chr4 | 76251098 | 76252044 | 947 | 14 | 1.33E-07 | 9.76E-07 | 0.002766 | 1.28E-07 | -0.00266718 | FAM47E, FAM47E-STBD1, AC034139.1 |
| chr5 | 177447705 | 177448402 | 698 | 8 | 3.76E-08 | 9.11E-07 | 1.24E-05 | 1.28E-07 | 0.003118115 | PRR7 |
| chr8 | 144471964 | 144472733 | 770 | 4 | 8.22E-08 | 2.43E-07 | 0.000253 | 1.28E-07 | -0.01034038 | KIFC2 |
| chr2 | 167870097 | 167871199 | 1103 | 4 | 1.42E-07 | 5.91E-08 | 0.00063 | 1.28E-07 | -0.00632127 | B3GALT1, B3GALT1-AS1 |
| chr17 | 63479012 | 63479943 | 932 | 4 | 1.92E-06 | 1.28E-07 | 0.000399 | 1.28E-07 | 0.001928927 | ACE |
| chr4 | 79964125 | 79964448 | 324 | 3 | 5.34E-09 | 7.96E-07 | 3.25E-06 | 1.28E-07 | -0.00072565 | ANTXR2 |
| chr14 | 63203993 | 63205019 | 1027 | 13 | 6.34E-07 | 5.75E-05 | 0.001705 | 1.29E-07 | 0.025158768 | RHOJ |
| chr2 | 240591412 | 240591974 | 563 | 2 | 2.20E-09 | 8.03E-07 | 1.19E-06 | 1.29E-07 | -0.00467966 | CAPN10 |
| chr7 | 130379662 | 130380487 | 826 | 6 | 6.55E-08 | 1.41E-06 | 0.000267 | 1.29E-07 | -0.00333836 | CPA1 |
| chr2 | 74457753 | 74458827 | 1075 | 13 | 4.81E-08 | 3.47E-06 | 0.000241 | 1.29E-07 | 0.001101946 | INO80B, INO80B-WBP1, WBP1 |
| chr11 | 68750693 | 68752267 | 1575 | 10 | 2.86E-08 | 3.79E-07 | 0.00013 | 1.29E-07 | -0.00316446 | TESMIN |
| chr17 | 1533330 | 1534136 | 807 | 5 | 2.03E-08 | 1.92E-06 | 6.55E-05 | 1.30E-07 | -0.0025362 | PITPNA |
| chr12 | 57454777 | 57455487 | 711 | 4 | 2.32E-09 | 4.64E-05 | 1.24E-06 | 1.30E-07 | -0.0020152 | INHBE |
| chr8 | 40059189 | 40059696 | 508 | 2 | 1.01E-07 | 8.14E-08 | 4.89E-05 | 1.30E-07 | -0.01367483 | NA |
| chr15 | 98131985 | 98133081 | 1097 | 3 | 1.39E-05 | 5.13E-08 | 0.000222 | 1.30E-07 | -0.00471188 | AC022523.1 |
| chr10 | 132726314 | 132727469 | 1156 | 9 | 1.41E-08 | 8.75E-07 | 1.62E-05 | 1.30E-07 | -0.00024432 | INPP5A |
| chr11 | 70424342 | 70425429 | 1088 | 4 | 2.31E-11 | 0.0005575 | 1.64E-07 | 1.30E-07 | -0.00526519 | CTTN |
| chr19 | 4037611 | 4038958 | 1348 | 7 | 1.42E-06 | 0.0007657 | 1.58E-05 | 1.30E-07 | -0.00315251 | PIAS4 |
| chr17 | 17351789 | 17352457 | 669 | 6 | 1.85E-08 | 4.14E-06 | 1.94E-05 | 1.30E-07 | -0.0056249 | NA |
| chr12 | 51083289 | 51084479 | 1191 | 12 | 2.29E-07 | 2.19E-05 | 0.00019 | 1.30E-07 | -0.0017127 | CSRNP2 |
| chr11 | 123741520 | 123742504 | 985 | 6 | 2.36E-08 | 3.26E-07 | 3.03E-05 | 1.30E-07 | 0.000235663 | ZNF202 |
| chr20 | 12934999 | 12935443 | 445 | 3 | 5.03E-08 | 3.16E-06 | 3.20E-06 | 1.30E-07 | -0.00426231 | LINC01722, AL078623.1 |
| chr10 | 5194892 | 5195918 | 1027 | 6 | 4.11E-07 | 1.35E-05 | 7.07E-06 | 1.30E-07 | -0.00743687 | AKR1C4 |
| chr21 | 45528451 | 45528788 | 338 | 2 | 4.15E-09 | 2.95E-06 | 5.03E-07 | 1.30E-07 | 0.001486139 | SLC19A1 |
| chr5 | 123098045 | 123099249 | 1205 | 9 | 2.30E-08 | 4.33E-06 | 2.99E-05 | 1.30E-07 | -0.00499738 | PRDM6 |
| chr19 | 19597727 | 19598837 | 1111 | 4 | 1.34E-06 | 1.82E-07 | 5.42E-05 | 1.31E-07 | -0.01044559 | PBX4 |
| chr9 | 32782403 | 32782827 | 425 | 3 | 2.20E-10 | 2.75E-06 | 1.10E-06 | 1.31E-07 | -0.02094179 | NA |
| chr6 | 108157290 | 108158190 | 901 | 7 | 5.37E-08 | 7.42E-08 | 0.000533 | 1.31E-07 | -0.0002027 | OSTM1-AS1, OSTM1 |
| chr15 | 52803626 | 52805098 | 1473 | 10 | 1.28E-07 | 1.92E-07 | 0.001491 | 1.31E-07 | 0.000769897 | AC016044.1 |
| chr11 | 755588 | 755608 | 21 | 2 | 6.81E-08 | 8.45E-07 | 1.17E-06 | 1.31E-07 | -0.00065378 | TALDO1 |
| chr1 | 213988365 | 213989717 | 1353 | 10 | 2.55E-07 | 4.53E-07 | 0.000201 | 1.31E-07 | 0.002136109 | PROX1, PROX1-AS1 |
| chr6 | 108758717 | 108759385 | 669 | 4 | 2.41E-08 | 1.24E-07 | 0.000116 | 1.31E-07 | -0.00459673 | LINC00222 |
| chr3 | 13379163 | 13379461 | 299 | 4 | 1.85E-07 | 7.59E-08 | 8.94E-05 | 1.32E-07 | 0.009608663 | NUP210 |
| chr2 | 240030758 | 240031490 | 733 | 5 | 3.14E-07 | 3.50E-08 | 0.000401 | 1.32E-07 | -0.01037595 | NA |
| chr5 | 78614039 | 78614214 | 176 | 2 | 1.53E-07 | 8.86E-08 | 3.49E-05 | 1.32E-07 | -0.01254587 | LHFPL2 |
| chr1 | 248826606 | 248827552 | 947 | 6 | 8.62E-10 | 7.54E-06 | 2.25E-05 | 1.32E-07 | 0.006389291 | NA |
| chr7 | 42934293 | 42935095 | 803 | 3 | 8.59E-07 | 1.27E-07 | 0.000106 | 1.32E-07 | -0.0046668 | MRPL32 |
| chr8 | 42697131 | 42697576 | 446 | 8 | 2.99E-08 | 2.83E-07 | 8.22E-05 | 1.32E-07 | -0.00226562 | CHRNB3 |
| chr16 | 30006149 | 30006784 | 636 | 4 | 4.76E-09 | 1.20E-05 | 4.61E-06 | 1.32E-07 | 3.97E-06 | DOC2A |
| chr2 | 48372944 | 48373205 | 262 | 4 | 1.35E-08 | 0.0010336 | 1.26E-05 | 1.32E-07 | -0.01345259 | FOXN2 |
| chr11 | 7513446 | 7514752 | 1307 | 10 | 1.60E-06 | 9.03E-09 | 0.009826 | 1.32E-07 | -0.00426245 | PPFIBP2, AC107884.1 |
| chr3 | 98657546 | 98657730 | 185 | 2 | 1.70E-07 | 8.24E-08 | 5.18E-05 | 1.32E-07 | -0.02755866 | NA |
| chr12 | 12562877 | 12563864 | 988 | 5 | 5.46E-08 | 7.20E-05 | 1.05E-05 | 1.32E-07 | -0.00485761 | NA |
| chr17 | 8133485 | 8133861 | 377 | 3 | 4.49E-08 | 3.92E-07 | 7.07E-06 | 1.33E-07 | -0.00975893 | NA |
| chr20 | 25507905 | 25508514 | 610 | 4 | 1.17E-07 | 4.09E-07 | 2.28E-05 | 1.33E-07 | 0.000606375 | NINL |
| chr11 | 69667275 | 69668466 | 1192 | 6 | 6.21E-09 | 1.53E-05 | 0.000216 | 1.33E-07 | -0.00310175 | LTO1 |
| chr8 | 80029811 | 80031112 | 1302 | 13 | 9.97E-09 | 3.35E-07 | 0.00148 | 1.33E-07 | 0.00029443 | MRPS28, AC036214.3 |
| chr10 | 128470321 | 128470766 | 446 | 4 | 1.04E-07 | 3.50E-07 | 2.79E-05 | 1.33E-07 | -0.00747142 | NA |
| chr12 | 124423566 | 124424385 | 820 | 7 | 3.50E-08 | 1.48E-07 | 0.000249 | 1.33E-07 | -0.00838254 | NCOR2 |
| chr5 | 91379957 | 91381191 | 1235 | 10 | 1.55E-09 | 4.88E-06 | 0.000186 | 1.33E-07 | -4.84E-05 | ARRDC3-AS1, ARRDC3 |
| chr14 | 53613560 | 53614143 | 584 | 4 | 2.20E-08 | 1.22E-06 | 1.11E-05 | 1.33E-07 | -0.01690056 | AL365295.1, AL163953.1 |
| chr7 | 29145929 | 29147013 | 1085 | 14 | 4.38E-08 | 3.56E-06 | 0.001259 | 1.33E-07 | -0.00144755 | AC004593.2, CHN2, CPVL |
| chr7 | 561646 | 562191 | 546 | 7 | 9.08E-09 | 6.78E-06 | 6.31E-07 | 1.33E-07 | -0.00120583 | PRKAR1B-AS2, PRKAR1B |
| chr13 | 27968590 | 27969394 | 805 | 11 | 1.28E-07 | 1.29E-07 | 0.001985 | 1.33E-07 | 0.000599029 | CDX2 |
| chr16 | 4475917 | 4476444 | 528 | 11 | 1.11E-08 | 4.52E-06 | 0.000251 | 1.33E-07 | -0.00276164 | HMOX2, NMRAL1 |
| chr6 | 33670536 | 33671145 | 610 | 5 | 3.91E-07 | 2.35E-07 | 0.000189 | 1.33E-07 | -0.00416467 | ITPR3 |
| chr14 | 21318919 | 21319075 | 157 | 2 | 3.37E-08 | 1.98E-07 | 5.81E-06 | 1.34E-07 | -0.00667959 | RPGRIP1 |
| chr1 | 1421185 | 1422916 | 1732 | 14 | 3.67E-07 | 2.33E-06 | 0.00138 | 1.34E-07 | -0.00042945 | AL391244.1, ANKRD65 |
| chr10 | 97179683 | 97180748 | 1066 | 4 | 2.00E-09 | 7.79E-07 | 4.95E-06 | 1.34E-07 | -0.00995138 | SLIT1, ARHGAP19-SLIT1 |
| chr15 | 69390511 | 69391404 | 894 | 6 | 3.06E-08 | 2.22E-07 | 0.0003 | 1.34E-07 | -0.00216263 | PAQR5, AC027237.4 |
| chr12 | 71663878 | 71665513 | 1636 | 17 | 1.26E-11 | 0.0001275 | 1.51E-05 | 1.34E-07 | 0.000204255 | THAP2, ZFC3H1 |
| chr19 | 16807648 | 16808833 | 1186 | 5 | 4.75E-07 | 0.0001187 | 7.39E-05 | 1.34E-07 | -0.00371867 | NWD1 |
| chr3 | 42652947 | 42654088 | 1142 | 8 | 1.06E-08 | 2.84E-05 | 1.93E-06 | 1.34E-07 | -0.00379951 | ZBTB47, AC006059.1 |
| chr1 | 183634760 | 183636432 | 1673 | 18 | 1.11E-06 | 7.95E-07 | 0.00555 | 1.34E-07 | 0.001193573 | RGL1, ARPC5 |
| chr10 | 13382851 | 13383688 | 838 | 6 | 1.60E-07 | 4.44E-08 | 0.000409 | 1.34E-07 | 0.001196942 | NA |
| chr10 | 3172134 | 3173871 | 1738 | 15 | 2.26E-05 | 2.87E-07 | 0.009663 | 1.35E-07 | -0.00143672 | PITRM1 |
| chr1 | 221741234 | 221742454 | 1221 | 14 | 8.55E-07 | 1.76E-06 | 0.003615 | 1.35E-07 | -0.00042482 | DUSP10 |
| chr7 | 95275826 | 95276431 | 606 | 4 | 1.79E-07 | 1.26E-06 | 6.38E-05 | 1.35E-07 | -0.00473947 | PPP1R9A |
| chr17 | 77247335 | 77248164 | 830 | 4 | 2.70E-08 | 4.39E-07 | 9.28E-06 | 1.35E-07 | -0.00504007 | NA |
| chr8 | 48466615 | 48467074 | 460 | 3 | 3.36E-07 | 6.63E-08 | 0.000162 | 1.36E-07 | -0.01771621 | NA |
| chr13 | 74230403 | 74231453 | 1051 | 4 | 2.62E-08 | 4.89E-07 | 5.61E-05 | 1.36E-07 | -0.00731218 | NA |
| chr17 | 9115489 | 9116019 | 531 | 5 | 1.03E-07 | 5.00E-08 | 0.00154 | 1.36E-07 | 0.02311738 | NTN1 |
| chr4 | 3678994 | 3679243 | 250 | 3 | 1.67E-08 | 2.72E-06 | 2.31E-06 | 1.36E-07 | -0.00333707 | NA |
| chr4 | 150583573 | 150584315 | 743 | 5 | 4.26E-09 | 1.45E-05 | 2.16E-05 | 1.36E-07 | -0.00084884 | MAB21L2, LRBA |
| chr2 | 71276352 | 71276959 | 608 | 10 | 3.52E-10 | 6.25E-05 | 3.89E-06 | 1.36E-07 | 0.001864632 | ZNF638 |
| chr14 | 105150651 | 105152479 | 1829 | 9 | 6.27E-06 | 3.19E-06 | 0.000209 | 1.36E-07 | -0.00372693 | JAG2, MIR6765, AL512356.1 |
| chr16 | 89037801 | 89039086 | 1286 | 7 | 1.69E-08 | 9.09E-07 | 3.65E-05 | 1.36E-07 | 0.004063679 | NA |
| chr13 | 21583402 | 21583689 | 288 | 3 | 3.55E-08 | 9.81E-06 | 4.14E-05 | 1.36E-07 | 0.003106181 | MICU2 |
| chr21 | 42469517 | 42469721 | 205 | 2 | 8.11E-08 | 2.52E-07 | 4.35E-06 | 1.36E-07 | -0.00820528 | NA |
| chr1 | 236141499 | 236143011 | 1513 | 13 | 9.18E-07 | 0.0002717 | 0.0001 | 1.36E-07 | 0.000535109 | GPR137B |
| chr22 | 46239028 | 46239592 | 565 | 4 | 1.30E-08 | 4.69E-07 | 1.85E-05 | 1.36E-07 | 0.003906169 | PPARA |
| chr6 | 122421798 | 122422046 | 249 | 3 | 6.64E-08 | 5.23E-07 | 6.60E-05 | 1.37E-07 | -0.02532139 | HSF2 |
| chr2 | 217756535 | 217756857 | 323 | 9 | 4.39E-08 | 4.33E-06 | 1.18E-05 | 1.37E-07 | -0.00383892 | DIRC3 |
| chr9 | 119494374 | 119495303 | 930 | 4 | 5.42E-07 | 3.55E-08 | 0.000764 | 1.37E-07 | -0.00953864 | NA |
| chr4 | 22712656 | 22713710 | 1055 | 5 | 3.18E-09 | 8.00E-07 | 4.00E-05 | 1.37E-07 | -0.00531829 | GBA3 |
| chr21 | 45335990 | 45336733 | 744 | 6 | 1.63E-07 | 7.09E-07 | 0.000197 | 1.37E-07 | -0.00688635 | BX322557.1 |
| chr17 | 75779195 | 75780295 | 1101 | 14 | 5.26E-07 | 8.89E-07 | 0.00516 | 1.37E-07 | 0.003781159 | H3-3B |
| chr4 | 103099792 | 103100916 | 1125 | 6 | 7.28E-07 | 9.05E-08 | 0.00146 | 1.37E-07 | 8.95E-05 | BDH2 |
| chr14 | 102944626 | 102945562 | 937 | 8 | 1.60E-07 | 4.56E-07 | 0.001594 | 1.37E-07 | -0.00255088 | CDC42BPB |
| chr1 | 1116714 | 1117647 | 934 | 6 | 1.32E-08 | 1.71E-07 | 0.000176 | 1.37E-07 | -0.00551696 | NA |
| chr2 | 30807866 | 30808889 | 1024 | 5 | 1.20E-07 | 1.32E-07 | 0.000939 | 1.37E-07 | -0.00252185 | CAPN13 |
| chr10 | 93068848 | 93069333 | 486 | 3 | 4.94E-08 | 1.20E-07 | 3.14E-05 | 1.37E-07 | -0.00994428 | CYP26C1, AL358613.3 |
| chr15 | 25596684 | 25597389 | 706 | 5 | 2.42E-08 | 2.75E-06 | 5.71E-05 | 1.38E-07 | 0.000455984 | NA |
| chr5 | 115840959 | 115842434 | 1476 | 14 | 2.65E-07 | 9.20E-07 | 0.00462 | 1.38E-07 | -0.00035736 | AP3S1, ATG12 |
| chr3 | 184727158 | 184727773 | 616 | 3 | 1.21E-08 | 1.05E-05 | 3.29E-06 | 1.38E-07 | -0.01308734 | AC107294.2, LINC02069 |
| chr9 | 12922804 | 12923423 | 620 | 2 | 1.27E-08 | 4.39E-07 | 2.30E-06 | 1.38E-07 | 0.006844873 | NA |
| chr15 | 74051135 | 74051689 | 555 | 2 | 3.13E-10 | 6.45E-06 | 3.75E-07 | 1.38E-07 | -0.01109044 | NA |

| | | | | | | | | | | |
|---|---|---|---|---|---|---|---|---|---|---|
| chr3 | 139044828 | 139045530 | 703 | 10 | 1.11E-08 | 1.13E-06 | 0.000193 | 1.38E-07 | -0.01777529 | MRPS22, PRR23C |
| chr5 | 167934543 | 167934850 | 308 | 4 | 1.44E-07 | 1.07E-07 | 8.42E-05 | 1.38E-07 | -0.00391288 | TENM2 |
| chr4 | 2941793 | 2942965 | 1173 | 9 | 1.88E-07 | 3.26E-05 | 6.65E-05 | 1.39E-07 | -0.00381718 | NOP14-AS1, NOP14 |
| chr8 | 134713631 | 134714352 | 722 | 4 | 1.39E-08 | 3.56E-07 | 0.000154 | 1.39E-07 | 0.007517179 | NA |
| chr20 | 49385774 | 49385890 | 117 | 2 | 6.66E-09 | 2.07E-05 | 2.51E-07 | 1.39E-07 | -0.02844161 | AL035685.1, KCNB1 |
| chr8 | 144827229 | 144828281 | 1053 | 11 | 1.32E-06 | 3.11E-07 | 0.001156 | 1.39E-07 | 0.003398257 | ZNF7 |
| chr9 | 976802 | 977770 | 969 | 7 | 2.62E-08 | 9.80E-06 | 4.54E-05 | 1.39E-07 | 0.00032885 | DMRT3 |
| chr4 | 20850662 | 20850832 | 171 | 4 | 3.81E-07 | 3.67E-08 | 0.000793 | 1.40E-07 | -0.0003882 | KCNIP4 |
| chr22 | 37674994 | 37676094 | 1101 | 14 | 1.17E-09 | 7.51E-05 | 4.01E-05 | 1.40E-07 | -0.00711641 | LGALS1 |
| chr19 | 3556720 | 3558419 | 1700 | 11 | 9.18E-06 | 1.29E-05 | 0.000767 | 1.40E-07 | -0.00104498 | AC005786.3, AC005786.2, MFSD12 |
| chr8 | 53250409 | 53252319 | 1911 | 12 | 2.96E-06 | 1.32E-05 | 6.44E-05 | 1.40E-07 | -0.00128034 | OPRK1 |
| chr4 | 54332091 | 54332826 | 736 | 5 | 2.06E-08 | 6.38E-07 | 2.14E-05 | 1.40E-07 | -0.00075948 | NA |
| chr3 | 181723247 | 181724349 | 1103 | 9 | 3.84E-09 | 1.59E-05 | 0.000222 | 1.40E-07 | -0.00400409 | SOX2-OT |
| chr17 | 44777373 | 44777758 | 386 | 5 | 8.63E-08 | 3.70E-07 | 0.000591 | 1.41E-07 | -0.00506334 | ADAM11 |
| chr16 | 1524589 | 1526068 | 1480 | 9 | 1.63E-08 | 7.12E-08 | 0.002105 | 1.41E-07 | 0.000273901 | IFT140 |
| chr1 | 17574553 | 17575610 | 1058 | 3 | 7.22E-08 | 5.41E-08 | 0.000349 | 1.41E-07 | -0.00969005 | ARHGEF10L |
| chr12 | 2782812 | 2783671 | 860 | 2 | 2.43E-06 | 8.54E-08 | 6.36E-05 | 1.41E-07 | -0.01414646 | ITFG2-AS1 |
| chr14 | 57865398 | 57866666 | 1269 | 12 | 3.68E-07 | 4.93E-06 | 0.000103 | 1.41E-07 | 0.002934255 | SLC35F4 |
| chr3 | 194633140 | 194633959 | 820 | 13 | 3.70E-08 | 4.03E-06 | 8.94E-06 | 1.42E-07 | 0.001392694 | AC046143.1, TMEM44 |
| chr2 | 197499391 | 197500947 | 1557 | 21 | 5.12E-08 | 1.54E-06 | 0.003646 | 1.42E-07 | 0.000721524 | HSPE1, HSPE1-MOB4, HSPD1 |
| chr11 | 35616850 | 35618447 | 1598 | 10 | 6.80E-06 | 9.45E-07 | 0.002564 | 1.42E-07 | -0.00271992 | NA |
| chr8 | 95196013 | 95196834 | 822 | 2 | 7.93E-08 | 1.43E-07 | 1.17E-05 | 1.42E-07 | -0.00277497 | NA |
| chr13 | 113152085 | 113152448 | 364 | 2 | 4.23E-09 | 3.41E-07 | 3.17E-06 | 1.42E-07 | -0.01040885 | AL137002.2 |
| chr8 | 143246330 | 143247705 | 1376 | 18 | 5.40E-09 | 3.88E-06 | 0.000127 | 1.42E-07 | -0.00073811 | ZFP41, AC138696.1 |
| chr9 | 70157720 | 70158192 | 473 | 3 | 4.60E-08 | 1.95E-07 | 1.36E-05 | 1.42E-07 | -0.01458703 | MAMDC2, MAMDC2-AS1 |
| chr15 | 59456511 | 59457102 | 592 | 2 | 4.25E-09 | 3.70E-07 | 2.91E-06 | 1.42E-07 | -0.01607808 | FAM81A |
| chr10 | 125755966 | 125756708 | 743 | 6 | 1.27E-08 | 1.91E-06 | 2.07E-05 | 1.43E-07 | -0.00191845 | EDRF1, MMP21 |
| chr6 | 46901990 | 46903436 | 1447 | 5 | 1.43E-06 | 4.68E-07 | 7.01E-05 | 1.43E-07 | -0.00714055 | ADGRF5 |
| chr3 | 16536771 | 16537153 | 383 | 2 | 2.89E-09 | 2.20E-07 | 5.71E-06 | 1.43E-07 | 0.007633447 | LINC00690 |
| chr22 | 42552514 | 42553968 | 1455 | 10 | 1.54E-06 | 2.29E-07 | 0.00151 | 1.43E-07 | 0.004927145 | SERHL2 |
| chr11 | 33254977 | 33256436 | 1460 | 6 | 2.36E-09 | 4.03E-06 | 1.18E-05 | 1.43E-07 | -0.001005 | NA |
| chr20 | 45236130 | 45236427 | 298 | 3 | 1.11E-08 | 0.0001055 | 2.49E-07 | 1.44E-07 | -0.00818876 | NA |
| chr5 | 119353598 | 119355161 | 1564 | 9 | 1.88E-06 | 1.19E-06 | 0.001239 | 1.44E-07 | -0.00347916 | TNFAIP8 |
| chr16 | 88910870 | 88911479 | 610 | 3 | 2.61E-08 | 3.24E-06 | 4.45E-06 | 1.44E-07 | 0.012229945 | CBFA2T3 |
| chr12 | 107760429 | 107761502 | 1074 | 10 | 1.82E-07 | 4.06E-06 | 7.10E-05 | 1.44E-07 | 0.00156598 | PRDM4 |
| chr11 | 64796850 | 64797362 | 513 | 3 | 1.53E-08 | 2.75E-07 | 9.24E-06 | 1.44E-07 | -0.00664433 | MAP4K2 |
| chr9 | 123549501 | 123550097 | 597 | 3 | 2.51E-07 | 1.46E-07 | 5.75E-05 | 1.44E-07 | -0.01529108 | DENND1A |
| chr3 | 123758042 | 123758686 | 645 | 2 | 1.92E-09 | 2.97E-06 | 5.65E-07 | 1.44E-07 | 0.005316608 | MYLK |
| chr7 | 1432790 | 1433194 | 405 | 3 | 1.02E-07 | 5.22E-08 | 0.000356 | 1.44E-07 | -0.02014015 | MICALL2 |
| chr3 | 184386267 | 184386900 | 634 | 8 | 5.52E-09 | 2.84E-06 | 5.49E-06 | 1.44E-07 | -0.0090125 | CHRD |
| chr2 | 100818149 | 100820465 | 2317 | 14 | 1.96E-06 | 7.72E-05 | 0.000264 | 1.44E-07 | 0.000410294 | NPAS2 |
| chr10 | 124642276 | 124643503 | 1228 | 10 | 6.62E-08 | 4.86E-07 | 5.96E-05 | 1.44E-07 | -0.00074154 | AC068896.1, FAM53B |
| chr1 | 113978950 | 113980048 | 1099 | 11 | 2.81E-07 | 6.03E-07 | 4.15E-05 | 1.44E-07 | -0.00243524 | OLFML3 |
| chr17 | 83104819 | 83106069 | 1251 | 8 | 8.27E-08 | 5.37E-05 | 0.000174 | 1.45E-07 | -0.00495806 | AC144831.1 |
| chr8 | 98942653 | 98944646 | 1994 | 15 | 2.67E-05 | 1.13E-07 | 0.003478 | 1.45E-07 | -0.00189603 | AC016877.3, OSR2, STK3 |
| chr17 | 75289100 | 75290201 | 1102 | 11 | 1.92E-09 | 0.0026236 | 3.12E-05 | 1.46E-07 | -0.00032989 | SLC25A19 |
| chr2 | 212910954 | 212911398 | 445 | 2 | 3.18E-08 | 3.00E-07 | 3.86E-06 | 1.46E-07 | -0.01446293 | AC108066.1, AC093865.1 |
| chr17 | 17495648 | 17497318 | 1671 | 10 | 1.83E-08 | 1.57E-06 | 1.59E-05 | 1.46E-07 | 0.001643665 | RASD1 |
| chr21 | 38468845 | 38469319 | 475 | 2 | 5.14E-09 | 6.29E-07 | 1.75E-06 | 1.46E-07 | -0.00724265 | ERG |
| chr16 | 67545205 | 67545512 | 308 | 2 | 3.29E-08 | 1.77E-07 | 8.35E-06 | 1.46E-07 | 0.012371223 | RIPOR1 |
| chr1 | 161225373 | 161226103 | 731 | 12 | 6.55E-09 | 4.78E-06 | 0.000496 | 1.46E-07 | -0.0023331 | TOMM40L |
| chr10 | 34728890 | 34728954 | 65 | 3 | 5.59E-08 | 4.68E-07 | 5.82E-05 | 1.46E-07 | 0.006283373 | PARD3 |
| chr6 | 31576917 | 31578121 | 1205 | 9 | 1.43E-07 | 3.57E-06 | 0.001643 | 1.47E-07 | -0.00306465 | TNF |
| chr4 | 8586989 | 8587586 | 598 | 6 | 4.05E-08 | 3.74E-06 | 0.000275 | 1.47E-07 | -0.00334242 | GPR78 |
| chr8 | 98904656 | 98905699 | 1044 | 5 | 2.52E-07 | 4.93E-07 | 0.000223 | 1.47E-07 | -0.00135689 | STK3, AC016877.2 |
| chr16 | 88866547 | 88867588 | 1042 | 9 | 7.65E-09 | 9.23E-06 | 1.55E-06 | 1.47E-07 | -0.00140645 | PABPN1L |
| chr20 | 63499434 | 63500607 | 1174 | 8 | 8.21E-07 | 2.81E-07 | 0.000967 | 1.47E-07 | -0.00089585 | NA |
| chr17 | 28714873 | 28716128 | 1256 | 5 | 1.13E-07 | 7.06E-07 | 8.32E-05 | 1.47E-07 | -0.00071119 | RAB34 |
| chr15 | 48810983 | 48812076 | 1094 | 14 | 5.85E-07 | 1.30E-06 | 0.004324 | 1.47E-07 | 0.003334152 | AC012379.2, CEP152 |
| chr12 | 185707 | 186785 | 1079 | 5 | 3.75E-13 | 0.0034765 | 1.62E-08 | 1.47E-07 | 0.003336088 | NA |
| chr6 | 13014259 | 13014989 | 731 | 7 | 5.56E-08 | 8.18E-06 | 0.000703 | 1.47E-07 | -0.00746548 | PHACTR1 |
| chr22 | 45212635 | 45214599 | 1965 | 11 | 3.73E-06 | 6.28E-06 | 8.21E-05 | 1.48E-07 | 0.000860413 | KIAA0930 |
| chr2 | 215356079 | 215357337 | 1259 | 3 | 1.65E-06 | 8.74E-08 | 8.18E-05 | 1.48E-07 | -0.00864725 | NA |
| chr3 | 56467552 | 56468786 | 1235 | 11 | 5.32E-08 | 1.68E-06 | 0.00033 | 1.48E-07 | -0.00133326 | ERC2 |
| chr14 | 75935267 | 75935360 | 94 | 2 | 5.76E-08 | 1.63E-06 | 8.50E-07 | 1.48E-07 | 0.013395718 | TTLL5, IFT43 |
| chr14 | 44913670 | 44913740 | 71 | 2 | 4.06E-08 | 1.40E-07 | 1.36E-05 | 1.48E-07 | -0.01793 | NA |
| chr1 | 24322494 | 24323004 | 511 | 5 | 2.21E-08 | 5.26E-07 | 0.000474 | 1.48E-07 | -0.01191576 | GRHL3 |
| chr3 | 165837315 | 165838144 | 830 | 5 | 5.54E-08 | 1.68E-06 | 0.001052 | 1.48E-07 | -0.01842424 | LINC01322, BCHE |
| chr11 | 74948754 | 74950180 | 1427 | 18 | 2.78E-06 | 1.34E-06 | 0.002306 | 1.48E-07 | -0.00149685 | SPCS2, XRRA1 |
| chr11 | 33774049 | 33775535 | 1487 | 12 | 9.65E-07 | 1.69E-06 | 1.81E-05 | 1.48E-07 | -0.00055952 | FBXO3-DT, FBXO3 |
| chr2 | 178673314 | 178674214 | 901 | 3 | 7.61E-07 | 7.07E-08 | 0.000179 | 1.48E-07 | -0.0049728 | TTN-AS1, TTN |
| chr17 | 39461217 | 39462429 | 1213 | 12 | 1.13E-07 | 3.06E-06 | 0.000673 | 1.48E-07 | 0.000559765 | CDK12 |
| chr20 | 59364895 | 59365164 | 270 | 2 | 3.78E-08 | 9.09E-08 | 5.85E-05 | 1.48E-07 | 0.001557016 | NA |
| chr1 | 116763422 | 116764047 | 626 | 3 | 1.14E-09 | 5.12E-05 | 3.48E-06 | 1.48E-07 | -0.00932117 | CD2 |
| chr7 | 2145915 | 2146709 | 795 | 5 | 1.28E-07 | 4.74E-06 | 3.04E-05 | 1.48E-07 | -0.00448628 | MAD1L1, AC069288.1 |
| chr17 | 67245483 | 67247003 | 1521 | 10 | 3.18E-08 | 5.64E-05 | 1.39E-05 | 1.48E-07 | -0.00178325 | AC007448.5, AC007448.4, HELZ |
| chr12 | 31985078 | 31986003 | 926 | 4 | 3.29E-08 | 3.13E-07 | 1.22E-05 | 1.48E-07 | -0.0082237 | RESF1 |
| chr1 | 185156524 | 185157299 | 776 | 15 | 2.45E-09 | 3.50E-06 | 0.000773 | 1.49E-07 | 0.001820964 | SWT1, TRMT1L |
| chr9 | 136662349 | 136663468 | 1120 | 7 | 1.36E-07 | 1.12E-07 | 0.000137 | 1.49E-07 | -0.00850682 | EGFL7 |
| chr14 | 49619865 | 49620826 | 962 | 12 | 2.07E-08 | 4.29E-06 | 0.001247 | 1.49E-07 | -0.00056831 | MGAT2, RPL36AL, AL139099.1 |
| chr3 | 184389211 | 184389746 | 536 | 2 | 2.80E-07 | 9.39E-08 | 4.91E-05 | 1.49E-07 | -0.01321148 | CHRD |
| chr1 | 45550451 | 45551495 | 1045 | 16 | 2.49E-09 | 1.29E-05 | 0.000749 | 1.49E-07 | 0.000766217 | AKR1A1 |
| chr11 | 2839759 | 2840247 | 489 | 4 | 1.38E-07 | 4.83E-08 | 0.000357 | 1.49E-07 | -0.00573446 | KCNQ1, KCNQ1-AS1 |
| chr11 | 64267389 | 64268316 | 928 | 4 | 2.19E-08 | 2.11E-06 | 1.61E-06 | 1.49E-07 | -0.01117461 | PLCB3 |
| chr3 | 196785180 | 196786000 | 821 | 6 | 4.77E-07 | 4.57E-08 | 0.000269 | 1.49E-07 | -0.00133468 | PAK2 |
| chr2 | 201694175 | 201695358 | 1184 | 4 | 1.28E-09 | 9.49E-06 | 2.19E-06 | 1.50E-07 | -0.00281759 | MPP4 |
| chr5 | 126542956 | 126542990 | 35 | 2 | 1.98E-07 | 9.27E-08 | 5.51E-05 | 1.50E-07 | -0.00715661 | ALDH7A1 |
| chr11 | 46542279 | 46543175 | 897 | 6 | 1.79E-07 | 1.01E-06 | 0.000219 | 1.50E-07 | -0.00632677 | AMBRA1 |
| chr7 | 63925848 | 63926769 | 922 | 7 | 4.95E-07 | 5.76E-08 | 0.001487 | 1.50E-07 | -0.01575294 | AC092634.8, AC092634.3 |
| chr2 | 88186225 | 88187588 | 1364 | 5 | 1.30E-05 | 1.89E-07 | 0.000253 | 1.50E-07 | -0.00011878 | THNSL2 |
| chr20 | 43537704 | 43538356 | 653 | 2 | 5.84E-08 | 1.06E-07 | 3.07E-05 | 1.51E-07 | -0.01533112 | AL031681.2, L3MBTL1 |
| chr4 | 3301933 | 3302339 | 407 | 2 | 2.45E-07 | 9.09E-08 | 6.61E-05 | 1.51E-07 | 0.018277021 | RGS12 |
| chr17 | 1895975 | 1896433 | 459 | 4 | 1.20E-08 | 5.84E-06 | 9.00E-07 | 1.51E-07 | 0.003358938 | RPA1 |
| chr1 | 42743248 | 42743935 | 688 | 3 | 1.39E-07 | 1.07E-07 | 6.57E-05 | 1.51E-07 | 0.00211609 | NA |
| chr12 | 20508263 | 20509043 | 781 | 3 | 1.61E-07 | 2.59E-07 | 1.75E-05 | 1.51E-07 | -0.00515167 | PDE3A |
| chr3 | 177648006 | 177648020 | 15 | 2 | 9.22E-08 | 9.18E-08 | 6.23E-05 | 1.51E-07 | 0.022165397 | LINC00578 |
| chr1 | 170663844 | 170665189 | 1346 | 8 | 3.01E-07 | 9.79E-07 | 0.000735 | 1.51E-07 | 0.002502842 | PRRX1 |
| chr12 | 104138113 | 104139129 | 1017 | 7 | 3.02E-08 | 9.52E-06 | 2.32E-05 | 1.51E-07 | 0.001117296 | NFYB |
| chr11 | 14358784 | 14359688 | 905 | 8 | 5.40E-07 | 1.09E-07 | 0.001146 | 1.52E-07 | -0.00572027 | RRAS2 |
| chr16 | 58244155 | 58244319 | 165 | 2 | 6.92E-08 | 1.07E-07 | 3.11E-05 | 1.52E-07 | -0.00638097 | CCDC113 |
| chr4 | 151891386 | 151892325 | 940 | 4 | 1.29E-06 | 8.16E-08 | 0.000329 | 1.52E-07 | -0.01107098 | NA |
| chr4 | 83034071 | 83035240 | 1170 | 16 | 1.75E-08 | 4.35E-06 | 0.004804 | 1.52E-07 | -0.00377402 | COPS4 |
| chr7 | 47621364 | 47621691 | 328 | 3 | 4.02E-08 | 1.36E-06 | 3.06E-06 | 1.53E-07 | 0.004567805 | LINC01447 |
| chr12 | 99390573 | 99391258 | 686 | 2 | 9.83E-08 | 1.69E-07 | 9.98E-06 | 1.53E-07 | -0.0140923 | ANKS1B |
| chr4 | 664053 | 665192 | 1140 | 7 | 3.08E-07 | 5.45E-05 | 0.000122 | 1.53E-07 | 0.002724479 | PDE6B |
| chr10 | 133357142 | 133358527 | 1386 | 10 | 4.95E-07 | 5.32E-08 | 0.003537 | 1.53E-07 | -0.00395725 | FUOM |
| chr12 | 95173182 | 95174143 | 962 | 4 | 1.97E-08 | 2.08E-05 | 1.27E-05 | 1.53E-07 | 0.004303929 | FGD6 |
| chr10 | 73655946 | 73657086 | 1141 | 8 | 2.01E-06 | 1.37E-07 | 0.000211 | 1.53E-07 | 0.004682662 | AC073389.2, SYNPO2L |
| chr18 | 78792204 | 78792701 | 498 | 2 | 9.13E-09 | 5.61E-07 | 2.10E-06 | 1.53E-07 | -0.01139603 | NA |
| chr20 | 3749574 | 3750687 | 1114 | 6 | 5.48E-07 | 1.56E-07 | 0.000432 | 1.53E-07 | -0.0077602 | HSPA12B |
| chr7 | 156610329 | 156611782 | 1454 | 4 | 5.41E-06 | 4.44E-08 | 0.000904 | 1.53E-07 | -0.00546816 | LINC01006 |
| chr2 | 2662342 | 2663074 | 733 | 3 | 9.38E-09 | 1.05E-06 | 2.24E-06 | 1.54E-07 | -0.00802922 | AC018685.2 |
| chr8 | 141042677 | 141043858 | 1182 | 4 | 2.72E-06 | 2.14E-06 | 7.15E-05 | 1.54E-07 | 0.008710851 | NA |
| chr8 | 22010605 | 22011157 | 553 | 3 | 3.62E-09 | 1.32E-06 | 2.55E-06 | 1.54E-07 | -0.01572375 | NA |
| chr16 | 89854640 | 89856131 | 1492 | 8 | 1.14E-09 | 5.11E-06 | 0.000443 | 1.54E-07 | -0.00364177 | SPIRE2 |
| chr9 | 134816867 | 134818315 | 1449 | 5 | 1.83E-06 | 2.20E-07 | 0.000728 | 1.54E-07 | -0.00038103 | COL5A1 |
| chr11 | 118634379 | 118635634 | 1256 | 7 | 2.07E-07 | 7.25E-07 | 0.000627 | 1.55E-07 | -0.01521682 | PHLDB1 |
| chr19 | 3407063 | 3407725 | 663 | 2 | 2.31E-07 | 9.44E-08 | 6.09E-05 | 1.55E-07 | -0.00126607 | NFIC |
| chr18 | 79471298 | 79471654 | 357 | 2 | 1.66E-09 | 3.34E-07 | 3.75E-06 | 1.55E-07 | 0.003936395 | NFATC1 |
| chr17 | 60149620 | 60151121 | 1502 | 8 | 2.50E-06 | 1.57E-06 | 0.000171 | 1.55E-07 | 0.000565726 | CA4 |
| chr17 | 57762040 | 57762680 | 641 | 2 | 2.77E-10 | 0.0003475 | 1.77E-07 | 1.55E-07 | -0.01672821 | NA |
| chr1 | 182028829 | 182030233 | 1405 | 7 | 1.17E-07 | 1.69E-07 | 0.000445 | 1.55E-07 | -0.00207437 | NA |
| chr20 | 50138784 | 50138862 | 79 | 2 | 2.77E-08 | 1.32E-06 | 1.04E-06 | 1.56E-07 | -0.01058262 | PEDS1-UBE2V1, PEDS1 |
| chr2 | 94993209 | 94994348 | 1140 | 4 | 2.20E-08 | 1.26E-07 | 8.17E-05 | 1.56E-07 | -0.00393815 | AC103563.8 |
| chr17 | 76715124 | 76716347 | 1224 | 3 | 6.01E-10 | 2.44E-05 | 1.03E-06 | 1.56E-07 | 0.01184845 | JMJD6 |
| chr2 | 196799584 | 196800427 | 844 | 10 | 7.05E-08 | 2.28E-06 | 1.66E-05 | 1.56E-07 | 0.001253788 | GTF3C3 |
| chr1 | 1178244 | 1179199 | 956 | 3 | 4.05E-09 | 5.96E-06 | 3.39E-06 | 1.56E-07 | -0.00784895 | TTLL10, TTLL10-AS1 |
| chr22 | 38766380 | 38766741 | 362 | 4 | 6.83E-08 | 1.79E-06 | 0.000244 | 1.56E-07 | -0.01107795 | SUN2 |
| chr3 | 8962980 | 8963853 | 874 | 10 | 1.12E-07 | 6.98E-07 | 0.000128 | 1.57E-07 | -6.32E-05 | RAD18 |
| chr3 | 177521438 | 177522143 | 706 | 4 | 4.55E-10 | 1.79E-06 | 3.34E-06 | 1.57E-07 | -0.00262697 | LINC00578 |
| chr1 | 42830364 | 42830855 | 492 | 4 | 9.53E-08 | 4.21E-07 | 3.02E-05 | 1.57E-07 | -0.00341116 | ERMAP |
| chr7 | 93574383 | 93575427 | 1045 | 15 | 1.30E-08 | 7.38E-07 | 0.00098 | 1.57E-07 | -0.00056108 | CALCR |
| chr8 | 13275458 | 13277060 | 1603 | 9 | 1.32E-06 | 1.44E-06 | 0.001264 | 1.57E-07 | 0.002631046 | DLC1 |
| chr5 | 1389818 | 1390120 | 303 | 2 | 2.05E-07 | 9.71E-08 | 5.72E-05 | 1.57E-07 | 0.010400811 | NA |
| chr10 | 127738130 | 127739044 | 915 | 5 | 4.30E-07 | 1.15E-07 | 0.001229 | 1.57E-07 | -0.02040897 | FOXI2 |
| chr1 | 19643762 | 19644682 | 921 | 11 | 7.39E-08 | 5.74E-06 | 0.000408 | 1.57E-07 | 0.00133436 | NBL1, MICOS10-NBL1 |
| chr1 | 4666697 | 4666699 | 3 | 2 | 1.84E-07 | 1.45E-07 | 1.48E-05 | 1.58E-07 | -0.00410795 | AJAP1 |
| chr8 | 4994902 | 4995915 | 1014 | 10 | 7.12E-08 | 8.82E-07 | 0.000782 | 1.58E-07 | 0.010220882 | CSMD1 |
| chr7 | 108569153 | 108570112 | 960 | 13 | 1.58E-07 | 1.46E-07 | 0.002313 | 1.58E-07 | -0.00059174 | DNAJB9, PNPLA8, THAP5 |
| chr5 | 181120193 | 181120979 | 787 | 3 | 3.75E-08 | 1.93E-07 | 1.70E-05 | 1.58E-07 | 0.001615681 | NA |
| chr7 | 65252027 | 65252841 | 815 | 5 | 2.03E-08 | 1.28E-05 | 8.60E-05 | 1.58E-07 | 0.00211687 | NA |
| chr5 | 36066870 | 36067589 | 720 | 7 | 6.58E-09 | 4.76E-07 | 0.000103 | 1.58E-07 | -0.00822703 | UGT3A2 |
| chr12 | 6534638 | 6535736 | 1099 | 11 | 2.06E-08 | 4.64E-06 | 0.001446 | 1.58E-07 | 0.000567518 | GAPDH |
| chr17 | 50063048 | 50063375 | 328 | 2 | 4.93E-08 | 1.07E-07 | 3.72E-05 | 1.59E-07 | -0.01399625 | ITGA3 |
| chr6 | 31715274 | 31715575 | 302 | 6 | 6.03E-08 | 9.54E-08 | 0.000378 | 1.59E-07 | 0.009126084 | LY6G6F-LY6G6D, LY6G6D |
| chr10 | 1567188 | 1567558 | 371 | 3 | 1.26E-07 | 5.06E-07 | 3.28E-05 | 1.59E-07 | -0.01388526 | ADARB2 |
| chr7 | 157768318 | 157769233 | 916 | 8 | 1.72E-07 | 3.89E-06 | 0.001182 | 1.59E-07 | -0.00254016 | PTPRN2 |
| chr2 | 36966547 | 36967891 | 1345 | 6 | 2.57E-08 | 4.14E-06 | 2.02E-05 | 1.59E-07 | -0.00134811 | NA |
| chr1 | 64246586 | 64246900 | 315 | 2 | 5.68E-08 | 1.39E-07 | 1.67E-05 | 1.59E-07 | -0.00861792 | UBE2U |
| chr8 | 29991063 | 29991348 | 286 | 2 | 1.24E-08 | 8.78E-06 | 3.79E-07 | 1.59E-07 | 0.008854637 | NA |
| chr5 | 505870 | 506505 | 636 | 5 | 1.64E-09 | 2.84E-05 | 4.72E-07 | 1.59E-07 | -0.00214442 | SLC9A3 |
| chr6 | 145904692 | 145904849 | 158 | 2 | 1.80E-07 | 1.24E-07 | 2.27E-05 | 1.59E-07 | 0.007217202 | SHPRH |
| chr2 | 235570002 | 235570738 | 737 | 3 | 2.21E-08 | 1.35E-05 | 1.34E-05 | 1.59E-07 | -0.01660378 | AGAP1 |
| chr8 | 75426250 | 75426771 | 522 | 2 | 5.65E-09 | 6.03E-07 | 2.08E-06 | 1.60E-07 | 0.002054484 | HNF4G |
| chr21 | 44349458 | 44350415 | 958 | 5 | 3.28E-08 | 4.03E-06 | 0.000125 | 1.60E-07 | 0.004327268 | TRPM2 |
| chr18 | 32017515 | 32019161 | 1647 | 15 | 3.55E-07 | 1.86E-05 | 8.54E-05 | 1.60E-07 | -0.00261187 | RNF125, AC011825.3 |
| chr10 | 131114521 | 131114992 | 472 | 5 | 1.60E-08 | 1.87E-05 | 1.54E-05 | 1.60E-07 | -0.00054333 | TCERG1L |
| chr2 | 230828118 | 230828764 | 647 | 4 | 2.17E-07 | 2.69E-07 | 6.60E-05 | 1.60E-07 | -0.00267851 | NA |

| | | | | | | | | | | |
|---|---|---|---|---|---|---|---|---|---|---|
| chr20 | 59422214 | 59422907 | 694 | 3 | 5.90E-07 | 2.04E-07 | 9.94E-05 | 1.60E-07 | -0.00230947 | NA |
| chr14 | 67359292 | 67360888 | 1597 | 18 | 2.32E-08 | 7.97E-07 | 0.001113 | 1.60E-07 | 0.000365735 | EIF2S1, ATP6V1D |
| chr4 | 32775057 | 32775269 | 213 | 2 | 1.29E-07 | 1.82E-07 | 9.86E-06 | 1.61E-07 | -0.01500588 | NA |
| chr3 | 52454888 | 52455858 | 971 | 10 | 1.39E-09 | 0.0003144 | 2.69E-06 | 1.61E-07 | 8.85E-05 | NISCH |
| chr19 | 49688843 | 49690600 | 1758 | 9 | 4.43E-09 | 3.32E-07 | 0.000998 | 1.61E-07 | -0.01407169 | PRMT1, ADM5, AC011495.3 |
| chr12 | 11947522 | 11947726 | 205 | 2 | 2.33E-08 | 7.67E-06 | 4.06E-07 | 1.61E-07 | 0.008569442 | NA |
| chr8 | 54393124 | 54393764 | 641 | 2 | 1.15E-09 | 8.50E-06 | 3.89E-07 | 1.61E-07 | 0.0083109 | AC027250.2 |
| chr12 | 95548212 | 95549355 | 1144 | 13 | 5.87E-08 | 5.33E-06 | 0.00037 | 1.62E-07 | 0.000252287 | USP44 |
| chr5 | 140881376 | 140882719 | 1344 | 8 | 2.48E-08 | 4.71E-06 | 4.23E-05 | 1.62E-07 | -0.00651476 | PCDHA1, PCDHA2, PCDHA3, PCDHA4, PCDHA5, PCDHA6, PCDHA7, PCDHA8, PCDHA9, PCDHA10, PCDHA11, PCDHA12, PCDHA13, AC005609.1 |
| chr19 | 17334203 | 17335787 | 1585 | 12 | 2.55E-08 | 4.90E-07 | 0.00184 | 1.62E-07 | -0.00445849 | GTPBP3, ANO8 |
| chr1 | 945703 | 945889 | 187 | 2 | 7.34E-09 | 9.76E-05 | 2.06E-07 | 1.62E-07 | 0.002579313 | NOC2L |
| chr10 | 21387951 | 21388098 | 148 | 3 | 5.43E-08 | 6.66E-06 | 2.79E-06 | 1.62E-07 | -0.00861288 | NA |
| chr22 | 31973299 | 31974067 | 769 | 2 | 2.15E-08 | 4.81E-07 | 2.66E-06 | 1.62E-07 | -0.01242439 | LINC02558 |
| chr2 | 170927108 | 170927432 | 325 | 2 | 2.78E-10 | 9.05E-06 | 3.83E-07 | 1.63E-07 | 0.001900005 | NA |
| chr7 | 967110 | 967921 | 812 | 5 | 1.49E-09 | 2.09E-05 | 4.77E-07 | 1.63E-07 | -0.00163821 | COX19 |
| chr10 | 7620798 | 7621068 | 271 | 5 | 7.03E-08 | 7.42E-07 | 0.000128 | 1.63E-07 | -0.00480391 | ITIH5 |
| chr1 | 89575809 | 89576210 | 402 | 2 | 3.92E-08 | 3.67E-07 | 3.62E-06 | 1.63E-07 | 0.006469087 | LRRC8B |
| chr7 | 157653400 | 157654842 | 1443 | 8 | 4.08E-07 | 2.42E-06 | 0.001778 | 1.63E-07 | -0.00581036 | PTPRN2 |
| chr12 | 6942902 | 6944060 | 1159 | 13 | 5.11E-10 | 6.02E-07 | 0.000746 | 1.63E-07 | -0.00114392 | C12orf57, RNU7-1, U47924.2 |
| chr3 | 49723482 | 49724699 | 1218 | 10 | 2.04E-08 | 1.04E-05 | 3.45E-05 | 1.63E-07 | 0.000586857 | GMPPB, IP6K1 |
| chr1 | 32870230 | 32871010 | 781 | 3 | 3.71E-08 | 8.26E-08 | 0.000119 | 1.63E-07 | -0.00350624 | FNDC5 |
| chr16 | 1183262 | 1183640 | 379 | 2 | 3.22E-08 | 1.88E-07 | 9.62E-06 | 1.63E-07 | -0.01619863 | CACNA1H |
| chr7 | 80918373 | 80918991 | 619 | 4 | 1.46E-07 | 1.14E-07 | 0.000166 | 1.63E-07 | 0.006654103 | SEMA3C |
| chr11 | 2562475 | 2562771 | 297 | 2 | 1.69E-09 | 4.39E-05 | 2.41E-07 | 1.63E-07 | -0.00665 | KCNQ1 |
| chr5 | 50663869 | 50664740 | 872 | 5 | 1.81E-08 | 9.19E-06 | 1.62E-05 | 1.63E-07 | 0.006597268 | NA |
| chr19 | 17449956 | 17450386 | 431 | 3 | 3.16E-08 | 9.29E-08 | 9.25E-05 | 1.63E-07 | -0.01599745 | AC010319.2 |
| chr19 | 12658633 | 12659426 | 794 | 4 | 7.03E-07 | 4.17E-08 | 0.000868 | 1.64E-07 | -0.00205623 | MAN2B1 |
| chr19 | 39408736 | 39409870 | 1135 | 3 | 2.82E-06 | 4.76E-07 | 5.95E-05 | 1.64E-07 | -0.00116629 | ZFP36, MIR4530 |
| chr8 | 131039798 | 131041016 | 1219 | 11 | 7.04E-09 | 2.03E-06 | 0.000463 | 1.64E-07 | -0.00226448 | ADCY8 |
| chr16 | 68310172 | 68311479 | 1308 | 17 | 3.06E-08 | 1.70E-06 | 0.000381 | 1.64E-07 | -0.00030195 | PRMT7, SLC7A6OS |
| chr21 | 45409685 | 45410619 | 935 | 8 | 3.43E-08 | 1.37E-05 | 0.000421 | 1.64E-07 | -0.0007368 | COL18A1, COL18A1-AS2 |
| chr17 | 1022906 | 1023198 | 293 | 3 | 4.26E-08 | 6.76E-07 | 5.73E-06 | 1.64E-07 | -0.00816772 | ABR |
| chr21 | 25607467 | 25608302 | 836 | 11 | 4.80E-07 | 8.54E-08 | 0.00082 | 1.64E-07 | 0.001265843 | MRPL39 |
| chr1 | 228144562 | 228145967 | 1406 | 11 | 6.47E-08 | 3.62E-05 | 0.000407 | 1.64E-07 | -0.00133184 | GUK1 |
| chr10 | 80463781 | 80464648 | 868 | 3 | 7.15E-07 | 6.57E-08 | 0.000336 | 1.64E-07 | -0.00987471 | TSPAN14 |
| chr10 | 133446061 | 133447511 | 1451 | 8 | 1.70E-06 | 1.81E-06 | 0.000174 | 1.65E-07 | -0.01679788 | NA |
| chr13 | 113064139 | 113065695 | 1557 | 6 | 1.51E-09 | 2.14E-05 | 8.23E-06 | 1.65E-07 | -0.00098004 | MCF2L |
| chr8 | 6052358 | 6052539 | 182 | 3 | 2.81E-07 | 1.17E-07 | 8.04E-05 | 1.65E-07 | -0.00034551 | AC009435.1 |
| chr10 | 84241301 | 84241804 | 504 | 8 | 5.10E-08 | 7.99E-07 | 0.000201 | 1.65E-07 | -0.00303505 | RGR, LRIT1 |
| chr6 | 27864402 | 27865777 | 1376 | 11 | 7.91E-09 | 1.61E-05 | 8.86E-05 | 1.65E-07 | -0.00216684 | H2BC15, H2AC16 |
| chr10 | 31827126 | 31827529 | 404 | 3 | 1.52E-07 | 4.11E-07 | 7.98E-05 | 1.65E-07 | -0.00897311 | ARHGAP12 |
| chr9 | 137165093 | 137165816 | 724 | 4 | 2.70E-09 | 4.38E-06 | 1.41E-05 | 1.65E-07 | -0.00870372 | GRIN1 |
| chr11 | 19345107 | 19346219 | 1113 | 11 | 5.85E-08 | 3.15E-07 | 0.00219 | 1.65E-07 | 0.006342911 | NA |
| chr9 | 91423850 | 91425513 | 1664 | 14 | 8.69E-08 | 5.20E-05 | 0.000161 | 1.65E-07 | 0.002154065 | NA |
| chr10 | 15585057 | 15585317 | 261 | 2 | 1.43E-07 | 1.47E-07 | 1.62E-05 | 1.65E-07 | -0.02594755 | ITGA8 |
| chr9 | 4338969 | 4339595 | 627 | 10 | 9.10E-08 | 9.35E-06 | 0.000334 | 1.65E-07 | -0.00238483 | GLIS3 |
| chr1 | 38783590 | 38783992 | 403 | 7 | 2.04E-07 | 2.12E-07 | 0.000977 | 1.65E-07 | 0.021415909 | AL354702.1 |
| chr9 | 97711562 | 97711568 | 7 | 2 | 6.28E-08 | 1.84E-07 | 1.03E-05 | 1.65E-07 | -0.02094065 | PTCSC2 |
| chr12 | 128932144 | 128933273 | 1130 | 8 | 7.91E-09 | 6.16E-05 | 3.95E-05 | 1.65E-07 | -0.00904962 | GLT1D1 |
| chr11 | 65315321 | 65316503 | 1183 | 4 | 1.82E-07 | 3.70E-07 | 3.80E-05 | 1.65E-07 | -0.00051215 | AP000944.5, CDC42EP2 |
| chr22 | 30471686 | 30472851 | 1166 | 8 | 7.72E-07 | 7.81E-06 | 7.56E-05 | 1.65E-07 | -0.00311504 | SEC14L3 |
| chr21 | 41929220 | 41929790 | 571 | 3 | 1.25E-08 | 5.48E-07 | 4.76E-06 | 1.65E-07 | 0.011748225 | C2CD2 |
| chr8 | 140549132 | 140549642 | 511 | 6 | 3.66E-07 | 5.58E-07 | 0.000285 | 1.65E-07 | -0.00105649 | AGO2 |
| chr13 | 75374175 | 75374757 | 583 | 2 | 1.47E-10 | 1.15E-05 | 3.56E-07 | 1.66E-07 | -0.01400351 | TBC1D4 |
| chr18 | 8616151 | 8616553 | 403 | 4 | 3.98E-08 | 4.64E-07 | 2.74E-05 | 1.66E-07 | 0.003763314 | RAB12 |
| chr6 | 29976316 | 29977378 | 1063 | 11 | 3.64E-07 | 1.63E-08 | 0.006256 | 1.66E-07 | -0.0053302 | HCG9 |
| chr3 | 154324236 | 154324989 | 754 | 14 | 1.26E-06 | 8.82E-08 | 0.005942 | 1.66E-07 | 0.002381861 | DHX36 |
| chr1 | 230112975 | 230114609 | 1635 | 6 | 3.00E-06 | 4.97E-06 | 0.000122 | 1.66E-07 | 0.000777479 | GALNT2 |
| chr1 | 25167682 | 25169234 | 1553 | 4 | 4.00E-05 | 1.45E-07 | 0.000297 | 1.66E-07 | -0.00612 | NA |
| chr12 | 58919473 | 58920485 | 1013 | 14 | 8.62E-09 | 0.0007221 | 0.000359 | 1.66E-07 | 0.000836475 | LRIG3 |
| chr14 | 25041607 | 25042557 | 951 | 5 | 3.86E-08 | 3.35E-06 | 0.000365 | 1.66E-07 | -0.01077308 | STXBP6 |
| chr5 | 180324165 | 180324817 | 653 | 4 | 9.47E-09 | 8.03E-06 | 9.37E-06 | 1.66E-07 | -0.00710228 | GFPT2 |
| chr1 | 79006299 | 79007232 | 934 | 12 | 1.02E-07 | 1.94E-06 | 0.000251 | 1.67E-07 | -0.00535183 | ADGRL4 |
| chr6 | 85678192 | 85679315 | 1124 | 12 | 1.02E-08 | 4.77E-05 | 1.22E-05 | 1.67E-07 | -0.00122526 | SNHG5 |
| chr5 | 177311671 | 177312925 | 1255 | 13 | 7.11E-10 | 0.0001896 | 3.35E-05 | 1.67E-07 | 0.001351422 | MXD3 |
| chr17 | 63535700 | 63535906 | 207 | 4 | 3.66E-08 | 5.95E-06 | 1.67E-06 | 1.67E-07 | -0.00048085 | KCNH6 |
| chr4 | 186636438 | 186637691 | 1254 | 5 | 1.81E-06 | 6.61E-08 | 0.000848 | 1.67E-07 | 0.002503185 | FAT1 |
| chr17 | 22703275 | 22704162 | 888 | 5 | 3.96E-08 | 6.68E-06 | 4.97E-05 | 1.67E-07 | -0.0204523 | AC131274.3 |
| chr15 | 40806638 | 40807706 | 1069 | 15 | 1.56E-09 | 1.55E-05 | 8.40E-05 | 1.68E-07 | 0.001009557 | ZFYVE19, DNAJC17 |
| chr9 | 133133496 | 133134808 | 1313 | 5 | 2.56E-10 | 0.0008802 | 2.16E-06 | 1.68E-07 | -0.00087305 | RALGDS, AL162417.1 |
| chr1 | 42315592 | 42316083 | 492 | 3 | 4.05E-09 | 1.46E-05 | 5.75E-07 | 1.68E-07 | 0.002119062 | FOXJ3 |
| chr9 | 126421402 | 126422376 | 975 | 5 | 6.10E-08 | 5.74E-06 | 0.00016 | 1.68E-07 | -0.00792255 | MVB12B |
| chr2 | 147323402 | 147323889 | 488 | 3 | 3.26E-09 | 0.0001099 | 3.20E-07 | 1.68E-07 | -0.01179328 | NA |
| chr7 | 155811260 | 155812966 | 1707 | 14 | 7.18E-07 | 1.40E-06 | 0.000805 | 1.68E-07 | -0.0021522 | SHH |
| chr5 | 176810303 | 176811374 | 1072 | 6 | 1.03E-07 | 5.18E-07 | 2.24E-05 | 1.68E-07 | -0.00254982 | UNC5A |
| chr12 | 5433989 | 5434103 | 115 | 2 | 1.18E-08 | 1.10E-07 | 4.28E-05 | 1.68E-07 | -0.02702522 | NTF3 |
| chr6 | 31005074 | 31005785 | 712 | 4 | 1.39E-06 | 7.14E-08 | 0.000403 | 1.68E-07 | 0.01695544 | NA |
| chr2 | 26171649 | 26172990 | 1342 | 11 | 2.49E-07 | 2.56E-07 | 0.001463 | 1.68E-07 | -0.00173173 | NA |
| chr18 | 45212206 | 45213191 | 986 | 11 | 9.98E-09 | 0.0004171 | 9.34E-06 | 1.68E-07 | -0.00642544 | SLC14A2 |
| chr6 | 89217514 | 89218233 | 720 | 8 | 1.03E-08 | 5.11E-07 | 2.78E-05 | 1.68E-07 | -0.00448548 | GABRR1 |
| chr5 | 177094549 | 177094930 | 382 | 2 | 8.53E-11 | 2.94E-07 | 5.18E-06 | 1.69E-07 | -0.01586092 | FGFR4 |
| chr19 | 49487198 | 49488595 | 1398 | 15 | 3.43E-08 | 4.80E-06 | 0.000263 | 1.69E-07 | 7.98E-05 | AC010619.2, RPL13A |
| chr1 | 3242519 | 3243081 | 563 | 5 | 2.88E-09 | 8.07E-06 | 3.42E-05 | 1.69E-07 | -0.00502938 | PRDM16 |
| chr2 | 68973877 | 68974353 | 477 | 3 | 7.17E-09 | 5.02E-06 | 1.85E-05 | 1.69E-07 | -0.00390201 | NA |
| chr1 | 2928306 | 2928949 | 644 | 5 | 2.75E-08 | 1.25E-06 | 4.56E-05 | 1.69E-07 | -0.00242344 | NA |
| chr22 | 46762017 | 46763211 | 1195 | 16 | 6.28E-09 | 5.22E-06 | 0.000876 | 1.69E-07 | 0.000467224 | TBC1D22A, AL118516.1 |
| chr2 | 21636348 | 21636471 | 124 | 3 | 1.98E-08 | 9.70E-06 | 8.24E-07 | 1.69E-07 | -0.00544356 | AC018742.1, AC009411.2 |
| chr2 | 148876800 | 148877410 | 611 | 5 | 1.07E-07 | 9.86E-07 | 0.000175 | 1.69E-07 | -0.00492817 | KIF5C, KIF5C-AS1 |
| chr19 | 12830136 | 12831061 | 926 | 12 | 2.11E-09 | 3.52E-05 | 0.000298 | 1.70E-07 | 0.00507146 | AC020934.1, HOOK2, RTBDN |
| chr11 | 5343446 | 5344147 | 702 | 5 | 1.46E-07 | 2.10E-07 | 0.000299 | 1.70E-07 | -0.00573189 | HBG2, AC104389.5, HBE1, OR51B5 |
| chr10 | 44398259 | 44398654 | 396 | 3 | 2.39E-07 | 9.71E-08 | 6.96E-05 | 1.70E-07 | -0.02426498 | NA |
| chr18 | 13433934 | 13433939 | 6 | 2 | 1.22E-07 | 5.31E-07 | 2.59E-06 | 1.70E-07 | 0.004838372 | LDLRAD4 |
| chr1 | 63522654 | 63523665 | 1012 | 16 | 1.46E-07 | 5.64E-07 | 0.002478 | 1.70E-07 | 0.000913793 | EFCAB7, ITGB3BP |
| chr1 | 43590028 | 43591428 | 1401 | 7 | 1.26E-06 | 1.26E-05 | 0.000411 | 1.70E-07 | -0.00234307 | PTPRF |
| chr10 | 132589853 | 132590560 | 708 | 3 | 2.98E-10 | 3.97E-07 | 1.70E-05 | 1.70E-07 | 0.000536994 | INPP5A |
| chr21 | 41147978 | 41148732 | 755 | 6 | 3.91E-07 | 1.15E-07 | 0.000159 | 1.70E-07 | -0.00547028 | LINC00323 |
| chr6 | 7388327 | 7389316 | 990 | 4 | 6.27E-09 | 3.94E-06 | 3.06E-05 | 1.70E-07 | 0.004055614 | CAGE1 |
| chr8 | 46104018 | 46104246 | 229 | 4 | 1.05E-07 | 4.46E-07 | 0.000484 | 1.71E-07 | -0.00652654 | NA |
| chr7 | 105196504 | 105197571 | 1068 | 3 | 7.61E-06 | 6.88E-07 | 7.09E-05 | 1.71E-07 | -0.00062317 | SRPK2 |
| chr2 | 148874143 | 148875543 | 1401 | 9 | 2.00E-06 | 2.21E-06 | 0.001277 | 1.71E-07 | 0.000792897 | KIF5C, KIF5C-AS1 |
| chr20 | 35618408 | 35619029 | 622 | 3 | 4.93E-09 | 6.03E-07 | 6.36E-06 | 1.71E-07 | -0.0095027 | SPAG4 |
| chr5 | 95962083 | 95962969 | 887 | 10 | 7.56E-08 | 2.64E-05 | 7.38E-05 | 1.71E-07 | -0.00267411 | AC104123.1 |
| chr8 | 144416498 | 144417072 | 575 | 10 | 2.05E-09 | 4.02E-05 | 2.55E-05 | 1.71E-07 | -0.00085001 | SLC39A4 |
| chr15 | 51244648 | 51244885 | 238 | 2 | 2.54E-08 | 1.12E-07 | 4.43E-05 | 1.71E-07 | -0.04805547 | MIR4713HG, CYP19A1 |
| chr16 | 50553999 | 50554377 | 379 | 5 | 1.53E-07 | 9.78E-07 | 5.41E-05 | 1.72E-07 | -0.00529478 | NKD1 |
| chr1 | 210327044 | 210329160 | 2117 | 18 | 4.04E-07 | 9.56E-05 | 0.000357 | 1.72E-07 | 0.000806694 | HHAT |
| chr14 | 99188101 | 99189339 | 1239 | 5 | 8.62E-11 | 1.47E-05 | 5.12E-06 | 1.72E-07 | -0.0108555 | BCL11B |
| chr11 | 884165 | 884807 | 643 | 2 | 1.44E-07 | 1.04E-07 | 6.75E-05 | 1.72E-07 | -0.00637921 | CHID1 |
| chr21 | 43903921 | 43904575 | 655 | 4 | 4.40E-10 | 1.15E-06 | 1.91E-05 | 1.72E-07 | 0.000557094 | AGPAT3 |
| chr16 | 88714966 | 88716446 | 1481 | 7 | 2.39E-07 | 1.42E-05 | 6.93E-05 | 1.72E-07 | -0.003494 | CTU2, PIEZO1, MIR4722 |
| chr16 | 57479852 | 57480518 | 667 | 5 | 8.15E-09 | 4.60E-07 | 3.46E-05 | 1.72E-07 | -0.00311168 | DOK4 |
| chr15 | 25214276 | 25215388 | 1113 | 4 | 3.45E-08 | 1.02E-05 | 4.66E-05 | 1.72E-07 | -0.00262344 | SNHG14 |
| chr17 | 2016735 | 2017815 | 1081 | 7 | 3.86E-06 | 6.49E-07 | 0.000735 | 1.72E-07 | -0.00199413 | AC099684.1, RTN4RL1 |
| chr6 | 85593090 | 85594226 | 1137 | 15 | 2.46E-08 | 0.0005281 | 0.000251 | 1.72E-07 | -0.00040686 | SNX14, AL589666.1 |
| chr11 | 46762725 | 46763602 | 878 | 4 | 2.54E-08 | 7.03E-06 | 4.14E-05 | 1.73E-07 | -0.00416428 | CKAP5 |
| chr17 | 61443625 | 61444061 | 437 | 5 | 2.94E-07 | 1.08E-07 | 0.000117 | 1.73E-07 | -0.00779368 | NA |
| chr11 | 66616207 | 66617397 | 1191 | 13 | 1.67E-07 | 2.24E-07 | 0.008858 | 1.73E-07 | 0.000676219 | RBM14, RBM14-RBM4 |
| chr17 | 41786560 | 41787293 | 734 | 11 | 2.32E-10 | 0.0002413 | 8.31E-06 | 1.73E-07 | -0.00206828 | JUP |
| chr1 | 162317928 | 162319079 | 1152 | 5 | 4.35E-08 | 2.04E-05 | 1.72E-05 | 1.74E-07 | 0.004135096 | NOS1AP |
| chr9 | 137287297 | 137287307 | 11 | 2 | 2.41E-07 | 1.13E-07 | 4.56E-05 | 1.74E-07 | 0.022660406 | NA |
| chr21 | 26845357 | 26846327 | 971 | 4 | 2.27E-07 | 9.05E-07 | 0.000148 | 1.74E-07 | -0.00185466 | ADAMTS1 |
| chr3 | 42818130 | 42818262 | 133 | 3 | 9.70E-08 | 1.12E-06 | 2.06E-05 | 1.74E-07 | -0.00941915 | ACKR2, AC099329.2, AC092042.3, KRBOX1, AC099329.1 |
| chr15 | 43776048 | 43777665 | 1618 | 20 | 2.75E-07 | 3.86E-06 | 0.000849 | 1.74E-07 | -0.00250352 | SERF2, ELL3, AC018512.1 |
| chr8 | 85523216 | 85523870 | 655 | 4 | 8.54E-08 | 2.50E-07 | 9.30E-05 | 1.74E-07 | -0.00910371 | AC084734.1 |
| chr8 | 144461504 | 144463153 | 1650 | 9 | 1.00E-06 | 5.07E-07 | 0.000279 | 1.75E-07 | -4.41E-05 | CYHR1 |
| chr8 | 132000407 | 132000758 | 352 | 3 | 1.03E-07 | 1.95E-07 | 0.000102 | 1.75E-07 | -0.00255503 | EFR3A |
| chr9 | 125189388 | 125190702 | 1315 | 13 | 2.00E-06 | 5.81E-06 | 0.001164 | 1.75E-07 | -0.00055353 | PPP6C |
| chr5 | 135577869 | 135579398 | 1530 | 14 | 1.14E-05 | 3.50E-07 | 0.001778 | 1.75E-07 | -0.00023227 | AC034206.1, SLC25A48, CXCL14 |
| chr5 | 160399987 | 160400985 | 999 | 10 | 6.70E-10 | 6.56E-06 | 6.57E-06 | 1.75E-07 | 0.000366511 | ZBED8 |
| chr19 | 855536 | 856145 | 610 | 6 | 2.83E-07 | 9.18E-08 | 0.000538 | 1.75E-07 | -0.01308538 | ELANE |
| chr8 | 96059832 | 96060430 | 599 | 3 | 8.96E-07 | 5.96E-08 | 0.000532 | 1.75E-07 | -0.00622343 | NA |
| chr10 | 116324189 | 116325075 | 887 | 12 | 1.70E-07 | 5.41E-07 | 0.002014 | 1.75E-07 | -0.00842326 | CCDC172 |
| chr4 | 7893745 | 7894424 | 680 | 3 | 3.07E-09 | 2.18E-05 | 5.60E-06 | 1.75E-07 | -0.00743002 | AFAP1 |
| chr11 | 27700121 | 27701227 | 1107 | 16 | 1.19E-07 | 1.90E-07 | 0.004978 | 1.75E-07 | 0.000300638 | BDNF, AC103796.1 |
| chr2 | 9409148 | 9409207 | 60 | 2 | 1.99E-07 | 1.69E-07 | 1.41E-05 | 1.75E-07 | 0.019557685 | ITGB1BP1 |
| chr17 | 8108651 | 8110362 | 1712 | 8 | 4.36E-06 | 7.90E-08 | 0.003037 | 1.75E-07 | -0.00443616 | ALOXE3 |
| chr1 | 30878321 | 30879407 | 1087 | 5 | 3.55E-07 | 4.78E-08 | 0.001125 | 1.76E-07 | -0.00883735 | SDC3 |
| chr7 | 157825235 | 157825610 | 376 | 3 | 7.06E-08 | 1.36E-07 | 5.27E-05 | 1.76E-07 | -0.01174447 | PTPRN2 |
| chr4 | 73944195 | 73944593 | 399 | 3 | 1.85E-07 | 1.08E-07 | 0.000191 | 1.76E-07 | -0.00787304 | CXCL1P1 |
| chr19 | 48673020 | 48673711 | 692 | 6 | 5.90E-08 | 1.94E-06 | 0.000339 | 1.76E-07 | -0.00839664 | SEC1P, NTN5 |
| chr6 | 106085863 | 106086539 | 677 | 11 | 2.80E-08 | 2.36E-07 | 0.000529 | 1.76E-07 | -0.0003733 | PRDM1, ATG5 |
| chr22 | 31080684 | 31081768 | 1085 | 15 | 4.87E-07 | 2.87E-07 | 0.005529 | 1.76E-07 | -0.00135023 | SMTN |
| chr11 | 47176387 | 47177568 | 1182 | 14 | 2.04E-08 | 3.29E-05 | 0.000606 | 1.76E-07 | 0.001614907 | ARFGAP2, PACSIN3 |
| chr6 | 170583817 | 170585053 | 1237 | 12 | 1.95E-07 | 6.55E-06 | 0.00044 | 1.77E-07 | 0.002655348 | PDCD2 |
| chr8 | 54282750 | 54283219 | 470 | 2 | 8.94E-08 | 1.87E-07 | 1.19E-05 | 1.77E-07 | 0.005147846 | NA |
| chr3 | 188946843 | 188948141 | 1299 | 12 | 3.00E-07 | 2.33E-06 | 0.000263 | 1.77E-07 | -0.00525337 | TPRG1, TPRG1-AS1 |
| chr14 | 73825949 | 73825987 | 39 | 2 | 1.77E-07 | 2.11E-07 | 9.56E-06 | 1.77E-07 | -0.01790163 | LINC02274 |
| chr12 | 3252559 | 3252830 | 272 | 3 | 2.07E-08 | 0.0001002 | 2.07E-06 | 1.77E-07 | 0.002854531 | TSPAN9 |
| chr5 | 92908902 | 92909777 | 876 | 2 | 1.49E-06 | 1.20E-07 | 3.79E-05 | 1.77E-07 | -0.01390201 | LINC02058 |
| chr1 | 117210691 | 117211343 | 653 | 8 | 6.71E-08 | 4.01E-07 | 0.00148 | 1.77E-07 | -0.0046709 | VTCN1 |
| chr11 | 62856393 | 62857805 | 1413 | 5 | 5.86E-09 | 1.69E-06 | 2.71E-05 | 1.77E-07 | -0.00833446 | SLC3A2 |
| chr11 | 6480361 | 6482571 | 2211 | 16 | 9.10E-09 | 1.60E-05 | 0.000324 | 1.77E-07 | -0.00085094 | AC084337.2, TIMM10B, ARFIP2 |
| chr2 | 24360252 | 24361212 | 961 | 11 | 6.05E-09 | 1.80E-05 | 0.000135 | 1.78E-07 | -0.00043848 | ITSN2 |
| chr1 | 153549183 | 153550107 | 925 | 9 | 5.77E-08 | 1.29E-06 | 4.31E-05 | 1.78E-07 | 9.84E-05 | S100A4, S100A3 |
| chr22 | 17580673 | 17581458 | 786 | 6 | 3.38E-07 | 7.18E-08 | 0.000692 | 1.78E-07 | -0.01273539 | SLC25A18, AC007666.1 |
| chr20 | 3204292 | 3205446 | 1155 | 12 | 2.69E-08 | 3.84E-05 | 7.68E-05 | 1.78E-07 | -0.00066803 | DDRGK1 |
| chr19 | 33811477 | 33812093 | 617 | 3 | 5.67E-08 | 3.46E-07 | 2.43E-05 | 1.78E-07 | 0.007026857 | KCTD15 |
| chr20 | 16728712 | 16730196 | 1485 | 14 | 8.01E-11 | 5.08E-06 | 1.46E-06 | 1.78E-07 | -0.00234254 | SNRPB2, AL034428.1 |

| | | | | | | | | | | |
|---|---|---|---|---|---|---|---|---|---|---|
| chr8 | 123202713 | 123203134 | 422 | 6 | 4.54E-08 | 1.59E-06 | 0.000102 | 1.78E-07 | -0.00578169 | FAM83A, FAM83A-AS1 |
| chr6 | 31009752 | 31010469 | 718 | 4 | 3.30E-08 | 5.21E-07 | 1.67E-05 | 1.78E-07 | 0.006258505 | NA |
| chr9 | 135342282 | 135343456 | 1175 | 6 | 1.72E-07 | 9.58E-08 | 0.001005 | 1.79E-07 | -0.00625406 | C9orf62 |
| chr9 | 113274753 | 113276116 | 1364 | 15 | 1.75E-08 | 1.02E-05 | 1.38E-05 | 1.79E-07 | 3.01E-05 | PRPF4, CDC26 |
| chr19 | 3762180 | 3762975 | 796 | 9 | 6.92E-09 | 1.15E-05 | 6.29E-06 | 1.79E-07 | 0.000254233 | MRPL54 |
| chr11 | 90488600 | 90488982 | 383 | 2 | 1.33E-08 | 2.50E-06 | 8.17E-07 | 1.79E-07 | 0.005591805 | DISC1FP1 |
| chr19 | 18248113 | 18248762 | 650 | 8 | 1.23E-07 | 3.00E-07 | 7.20E-05 | 1.79E-07 | -0.00713815 | AC008397.2, PDE4C |
| chr21 | 43728451 | 43729492 | 1042 | 8 | 4.52E-07 | 5.01E-07 | 0.002619 | 1.79E-07 | 0.002022326 | PDXK |
| chr9 | 131848522 | 131849849 | 1328 | 4 | 4.46E-09 | 6.02E-06 | 5.38E-06 | 1.79E-07 | -0.00739789 | NA |
| chr4 | 1974493 | 1974634 | 142 | 3 | 2.06E-08 | 3.00E-06 | 4.12E-05 | 1.79E-07 | -0.00572975 | NSD2 |
| chr5 | 180829781 | 180831265 | 1485 | 7 | 9.59E-06 | 9.41E-07 | 0.000193 | 1.79E-07 | 0.001777894 | LINC00847, HEIH |
| chr14 | 77248804 | 77249700 | 897 | 2 | 9.59E-07 | 1.49E-07 | 2.02E-05 | 1.79E-07 | -0.00342604 | TMEM63C |
| chr6 | 41075114 | 41076037 | 924 | 2 | 1.01E-06 | 1.38E-07 | 2.51E-05 | 1.80E-07 | 0.013483715 | NFYA, OARD1 |
| chr16 | 64983457 | 64984101 | 645 | 3 | 1.88E-07 | 1.24E-07 | 5.36E-05 | 1.80E-07 | -0.01345354 | CDH11 |
| chr16 | 2967592 | 2969191 | 1600 | 13 | 2.17E-07 | 1.01E-06 | 0.004655 | 1.80E-07 | -0.00593352 | KREMEN2, PKMYT1 |
| chr1 | 19250542 | 19252311 | 1770 | 18 | 3.46E-06 | 3.15E-06 | 0.006172 | 1.80E-07 | 0.001546155 | MRTO4, EMC1 |
| chr1 | 2220868 | 2222015 | 1148 | 6 | 1.24E-06 | 7.82E-08 | 0.000428 | 1.80E-07 | -0.003304 | NA |
| chr6 | 33685560 | 33685729 | 170 | 4 | 5.72E-08 | 5.47E-06 | 2.92E-06 | 1.80E-07 | -0.0054606 | ITPR3 |
| chr18 | 74248934 | 74249462 | 529 | 2 | 4.67E-09 | 2.69E-06 | 7.92E-07 | 1.81E-07 | 0.022742991 | NA |
| chr12 | 22624475 | 22625551 | 1077 | 14 | 3.30E-09 | 1.30E-05 | 0.000187 | 1.81E-07 | -0.00088621 | ETNK1, AC087241.2 |
| chr10 | 87607627 | 87607949 | 323 | 2 | 1.29E-09 | 9.31E-07 | 1.70E-06 | 1.81E-07 | 0.003277149 | NA |
| chr11 | 134477141 | 134477446 | 306 | 2 | 4.45E-08 | 4.79E-07 | 3.20E-06 | 1.81E-07 | 0.002959089 | B3GAT1-DT |
| chr10 | 78366552 | 78367672 | 1121 | 7 | 1.41E-07 | 1.14E-06 | 0.000153 | 1.81E-07 | -0.0020668 | AC012560.1, LINC00595, SNORA71 |
| chr17 | 751868 | 752941 | 1074 | 15 | 5.96E-08 | 2.26E-06 | 0.00445 | 1.82E-07 | -0.00074896 | DBIL5P, GEMIN4 |
| chr8 | 27837579 | 27839014 | 1436 | 13 | 3.26E-07 | 4.26E-06 | 0.000765 | 1.82E-07 | 0.004250125 | AC104997.1, PBK |
| chr4 | 40585712 | 40585773 | 62 | 2 | 8.57E-08 | 1.61E-07 | 1.75E-05 | 1.82E-07 | 0.008083704 | RBM47 |
| chr3 | 185328179 | 185329193 | 1015 | 6 | 3.67E-08 | 1.86E-06 | 0.000161 | 1.83E-07 | -0.00040335 | MAP3K13 |
| chr1 | 86422902 | 86423045 | 144 | 4 | 7.97E-08 | 1.82E-06 | 1.16E-05 | 1.83E-07 | 0.016866121 | NA |
| chr5 | 69332413 | 69333029 | 617 | 8 | 8.18E-07 | 5.42E-07 | 0.000483 | 1.83E-07 | -0.00742933 | CCDC125 |
| chr9 | 72474258 | 72474558 | 301 | 2 | 1.32E-08 | 1.15E-06 | 1.46E-06 | 1.83E-07 | -0.01299824 | NA |
| chr10 | 62236456 | 62236822 | 367 | 4 | 1.66E-08 | 4.21E-06 | 1.04E-06 | 1.83E-07 | -0.0121483 | RTKN2 |
| chr3 | 187196212 | 187196932 | 721 | 3 | 3.33E-08 | 1.66E-07 | 2.88E-05 | 1.83E-07 | -0.00212564 | NA |
| chr1 | 55580670 | 55580816 | 147 | 3 | 1.44E-07 | 7.58E-07 | 5.70E-06 | 1.83E-07 | 0.007677397 | AL603840.1 |
| chr8 | 1032157 | 1032926 | 770 | 4 | 1.27E-06 | 6.17E-08 | 0.00051 | 1.83E-07 | 0.004466243 | DLGAP2 |
| chr20 | 57113821 | 57114340 | 520 | 2 | 4.77E-08 | 1.09E-07 | 8.03E-05 | 1.84E-07 | -0.00996212 | AL157414.1 |
| chr10 | 75409841 | 75410848 | 1008 | 4 | 5.46E-08 | 0.0001012 | 4.94E-06 | 1.84E-07 | -0.0007677 | AC010997.4 |
| chr7 | 151889781 | 151890464 | 684 | 3 | 4.25E-07 | 2.67E-07 | 3.43E-05 | 1.84E-07 | 0.003745779 | NA |
| chr7 | 25568663 | 25569774 | 1112 | 9 | 1.38E-07 | 3.85E-07 | 0.000918 | 1.84E-07 | -0.0104266 | NA |
| chr14 | 64749096 | 64749952 | 857 | 6 | 1.20E-07 | 3.56E-05 | 0.000133 | 1.84E-07 | 0.003796793 | PLEKHG3, SPTB |
| chr9 | 81412237 | 81412372 | 136 | 2 | 9.15E-08 | 9.04E-07 | 1.78E-06 | 1.85E-07 | -0.01374038 | NA |
| chr2 | 203582867 | 203583284 | 418 | 3 | 3.26E-07 | 7.18E-08 | 0.000259 | 1.85E-07 | -0.00524836 | NA |
| chr20 | 31547126 | 31548526 | 1401 | 9 | 2.29E-06 | 1.31E-06 | 0.002515 | 1.85E-07 | 0.005923369 | HM13, MCTS2P |
| chr17 | 48310865 | 48311103 | 239 | 4 | 3.32E-07 | 8.55E-08 | 0.000164 | 1.85E-07 | -0.01188514 | SKAP1 |
| chr11 | 11841061 | 11841797 | 737 | 8 | 3.14E-08 | 2.36E-07 | 0.000323 | 1.85E-07 | 0.008008682 | USP47 |
| chr18 | 6414331 | 6415923 | 1593 | 12 | 1.67E-07 | 1.55E-06 | 0.001629 | 1.86E-07 | -0.00298071 | L3MBTL4 |
| chr4 | 183758233 | 183759111 | 879 | 5 | 2.77E-08 | 3.57E-06 | 5.06E-05 | 1.86E-07 | -0.01246855 | NA |
| chr8 | 63167962 | 63168852 | 891 | 12 | 1.97E-07 | 6.62E-07 | 0.00285 | 1.87E-07 | 0.000737457 | YTHDF3, YTHDF3-AS1 |
| chr19 | 44986942 | 44987765 | 824 | 2 | 1.06E-06 | 1.28E-07 | 3.68E-05 | 1.87E-07 | -0.01273033 | CLPTM1 |
| chr12 | 123456501 | 123457188 | 688 | 3 | 3.34E-08 | 7.76E-07 | 8.02E-06 | 1.87E-07 | -0.00935254 | NA |
| chr7 | 7566970 | 7567853 | 884 | 6 | 1.51E-10 | 0.0011068 | 3.02E-06 | 1.87E-07 | -0.0053754 | MIOS, AC004982.2 |
| chr17 | 48002752 | 48003550 | 799 | 5 | 1.58E-07 | 1.34E-07 | 0.000214 | 1.87E-07 | -0.010261 | NA |
| chr17 | 80999291 | 81000414 | 1124 | 9 | 1.31E-06 | 2.92E-07 | 0.001306 | 1.87E-07 | -6.99E-05 | CHMP6, AC127496.6 |
| chr9 | 35111369 | 35111933 | 565 | 8 | 2.10E-09 | 0.0005482 | 3.56E-07 | 1.87E-07 | 0.00093975 | FAM214B |
| chr16 | 11205456 | 11206324 | 869 | 5 | 4.34E-08 | 1.68E-05 | 4.55E-05 | 1.88E-07 | -0.0085349 | AC007220.1 |
| chr15 | 40926786 | 40928067 | 1282 | 5 | 9.30E-09 | 4.85E-07 | 0.000356 | 1.88E-07 | 0.00497251 | NA |
| chr6 | 86936669 | 86937926 | 1258 | 15 | 8.04E-07 | 1.04E-05 | 0.000142 | 1.88E-07 | -0.00279955 | HTR1E |
| chr16 | 81644892 | 81645363 | 472 | 7 | 9.85E-08 | 4.55E-06 | 1.20E-05 | 1.89E-07 | 0.001166058 | CMIP |
| chr14 | 24953252 | 24954576 | 1325 | 3 | 1.59E-11 | 0.0001316 | 1.57E-07 | 1.89E-07 | -0.00269615 | STXBP6 |
| chr12 | 49055094 | 49056343 | 1250 | 8 | 3.14E-08 | 3.81E-05 | 7.54E-05 | 1.89E-07 | -0.00269779 | KMT2D |
| chr22 | 43155302 | 43155947 | 646 | 4 | 5.46E-09 | 2.32E-06 | 1.55E-05 | 1.89E-07 | -0.00992604 | TSPO |
| chr12 | 79407766 | 79408325 | 560 | 2 | 2.60E-07 | 1.31E-07 | 3.65E-05 | 1.89E-07 | -0.03905631 | SYT1, AC027288.3 |
| chr12 | 56724889 | 56726006 | 1118 | 17 | 1.02E-07 | 1.35E-05 | 0.000172 | 1.89E-07 | 0.000118366 | NACA, AC117378.1 |
| chr7 | 11613428 | 11613574 | 147 | 2 | 2.48E-07 | 1.26E-07 | 4.27E-05 | 1.89E-07 | 0.010873061 | THSD7A |
| chr1 | 156175179 | 156175427 | 249 | 2 | 5.57E-08 | 4.76E-07 | 3.46E-06 | 1.89E-07 | -0.00063455 | SEMA4A |
| chr13 | 98961311 | 98961871 | 561 | 4 | 3.99E-07 | 5.15E-08 | 0.00094 | 1.89E-07 | -0.00366896 | DOCK9 |
| chr1 | 197915004 | 197916477 | 1474 | 11 | 4.79E-09 | 1.85E-05 | 8.97E-06 | 1.90E-07 | -0.00095981 | LHX9 |
| chr5 | 111757148 | 111758718 | 1571 | 20 | 1.03E-07 | 1.13E-05 | 0.003776 | 1.90E-07 | -0.0012264 | NREP |
| chr15 | 26774550 | 26775193 | 644 | 5 | 2.86E-09 | 1.62E-06 | 8.40E-06 | 1.90E-07 | -0.00327535 | GABRB3 |
| chr1 | 238607028 | 238607128 | 101 | 2 | 7.18E-08 | 1.13E-07 | 8.10E-05 | 1.90E-07 | 0.013295109 | NA |
| chr19 | 3960365 | 3962152 | 1788 | 14 | 5.54E-07 | 6.35E-05 | 5.85E-05 | 1.90E-07 | -0.00128766 | DAPK3, MIR637 |
| chr18 | 58205635 | 58206116 | 482 | 3 | 3.20E-08 | 1.58E-07 | 3.28E-05 | 1.90E-07 | -0.01026399 | NEDD4L |
| chr2 | 183078073 | 183079320 | 1248 | 16 | 5.74E-07 | 3.21E-08 | 0.012088 | 1.90E-07 | -0.00014491 | DUSP19 |
| chr4 | 83524260 | 83524264 | 5 | 2 | 5.25E-08 | 1.02E-05 | 4.28E-07 | 1.90E-07 | -0.01557727 | NA |
| chr13 | 50843333 | 50844478 | 1146 | 13 | 1.94E-08 | 1.93E-06 | 0.002064 | 1.90E-07 | -0.00696592 | DLEU1, DLEU7 |
| chr9 | 5185586 | 5186522 | 937 | 7 | 1.04E-07 | 1.26E-06 | 3.44E-05 | 1.91E-07 | 0.002269543 | INSL6 |
| chr5 | 151567791 | 151567793 | 3 | 2 | 3.33E-08 | 5.72E-07 | 2.90E-06 | 1.91E-07 | 0.001286425 | FAT2 |
| chr8 | 126472805 | 126473108 | 304 | 2 | 5.04E-09 | 0.0009335 | 1.97E-07 | 1.91E-07 | -0.01651118 | NA |
| chr17 | 13000957 | 13001230 | 274 | 4 | 1.08E-07 | 4.87E-07 | 3.35E-05 | 1.91E-07 | -0.00677886 | ELAC2 |
| chr6 | 25041684 | 25042818 | 1135 | 13 | 1.77E-07 | 6.02E-07 | 0.003578 | 1.91E-07 | -0.00071026 | AL133268.3, RIPOR2 |
| chr3 | 9769723 | 9770496 | 774 | 11 | 2.50E-09 | 1.65E-05 | 1.51E-05 | 1.91E-07 | -0.0080851 | OGG1, CAMK1 |
| chr11 | 15629408 | 15629967 | 560 | 3 | 1.61E-08 | 9.70E-07 | 1.11E-05 | 1.91E-07 | -0.01447044 | AC087379.2 |
| chr16 | 2328801 | 2330241 | 1441 | 4 | 3.80E-05 | 7.10E-08 | 0.00063 | 1.91E-07 | -0.00708105 | ABCA3 |
| chr1 | 156381921 | 156382132 | 212 | 3 | 1.79E-07 | 8.60E-08 | 0.00027 | 1.92E-07 | -0.00648913 | RHBG |
| chr17 | 35981520 | 35982541 | 1022 | 8 | 6.61E-08 | 2.16E-07 | 0.002152 | 1.92E-07 | -0.00571429 | NA |
| chr20 | 47786616 | 47787115 | 500 | 10 | 1.46E-07 | 8.50E-08 | 0.001241 | 1.92E-07 | -0.00330635 | SULF2 |
| chr8 | 105321720 | 105322154 | 435 | 3 | 1.77E-07 | 1.56E-07 | 8.03E-05 | 1.92E-07 | -0.02049965 | ZFPM2 |
| chr11 | 46382067 | 46382671 | 605 | 3 | 1.63E-11 | 9.42E-05 | 7.66E-07 | 1.92E-07 | -0.00725253 | MDK |
| chr22 | 24774401 | 24775405 | 1005 | 7 | 6.77E-08 | 3.56E-07 | 5.03E-05 | 1.92E-07 | -0.006518 | PIWIL3 |
| chr1 | 45750523 | 45751615 | 1093 | 11 | 1.30E-06 | 4.76E-07 | 0.001943 | 1.92E-07 | -0.00040708 | IPP |
| chr21 | 30429514 | 30430112 | 599 | 5 | 6.74E-07 | 3.68E-08 | 0.002073 | 1.92E-07 | -0.00802093 | NA |
| chr5 | 95082188 | 95083855 | 1668 | 4 | 6.05E-06 | 8.67E-07 | 0.000112 | 1.92E-07 | -0.01254601 | MCTP1 |
| chr12 | 124738956 | 124739320 | 365 | 5 | 1.17E-07 | 1.27E-06 | 6.64E-05 | 1.93E-07 | -0.00823587 | NA |
| chr7 | 2018068 | 2019577 | 1510 | 8 | 1.43E-07 | 2.76E-06 | 0.003484 | 1.93E-07 | 0.001817336 | MAD1L1, AC069288.1 |
| chr4 | 56656596 | 56658038 | 1443 | 10 | 6.70E-10 | 0.0003445 | 0.000248 | 1.94E-07 | 0.016846851 | HOPX |
| chr5 | 119451630 | 119453085 | 1456 | 17 | 3.56E-08 | 6.30E-06 | 0.001018 | 1.94E-07 | 0.000173981 | HSD17B4 |
| chr14 | 94080955 | 94081642 | 688 | 16 | 1.61E-07 | 3.73E-06 | 0.003718 | 1.94E-07 | -0.00083002 | IFI27L1, DDX24 |
| chr10 | 1203786 | 1203903 | 118 | 2 | 1.10E-08 | 2.21E-07 | 1.09E-05 | 1.94E-07 | -0.01008994 | LINC00200, ADARB2, AL392083.2 |
| chr12 | 15156809 | 15157118 | 310 | 3 | 1.76E-07 | 1.75E-07 | 3.75E-05 | 1.94E-07 | -0.01447064 | RERG |
| chr1 | 228470670 | 228471698 | 1029 | 6 | 3.07E-08 | 1.34E-06 | 0.000411 | 1.94E-07 | -0.02096288 | NA |
| chr1 | 35031282 | 35033125 | 1844 | 12 | 1.34E-06 | 4.74E-05 | 0.000348 | 1.94E-07 | -0.0021464 | ZMYM1, AC114490.2, ZMYM6 |
| chr2 | 65556460 | 65556961 | 502 | 3 | 1.50E-07 | 2.14E-07 | 2.32E-05 | 1.95E-07 | -0.00462011 | AC007389.1 |
| chr8 | 139745121 | 139745592 | 472 | 2 | 1.27E-07 | 1.28E-07 | 4.56E-05 | 1.95E-07 | -0.00838395 | TRAPPC9 |
| chr14 | 96520090 | 96520851 | 762 | 2 | 1.93E-07 | 2.14E-07 | 1.16E-05 | 1.95E-07 | -0.00050934 | PAPOLA |
| chr15 | 84781339 | 84781644 | 306 | 3 | 1.02E-07 | 9.17E-06 | 4.99E-05 | 1.95E-07 | -0.00378585 | ZNF592 |
| chr1 | 169087465 | 169087820 | 356 | 3 | 1.82E-07 | 1.15E-07 | 0.000169 | 1.95E-07 | -0.00463935 | AL135926.1 |
| chr13 | 113321653 | 113322379 | 727 | 5 | 6.42E-11 | 0.0027633 | 4.59E-08 | 1.95E-07 | 0.000723328 | LAMP1 |
| chr20 | 31738564 | 31739693 | 1130 | 15 | 7.17E-08 | 9.69E-07 | 0.001419 | 1.96E-07 | 0.000842286 | TPX2 |
| chr11 | 93498676 | 93499665 | 990 | 5 | 9.21E-08 | 2.72E-06 | 4.05E-05 | 1.96E-07 | -0.00369963 | SMCO4 |
| chr12 | 684136 | 684670 | 535 | 6 | 2.64E-08 | 3.54E-07 | 0.000111 | 1.96E-07 | -0.0011462 | NA |
| chr3 | 88138876 | 88139135 | 260 | 8 | 1.13E-07 | 6.12E-06 | 0.001879 | 1.96E-07 | -0.00271496 | ZNF654, CGGBP1 |
| chr17 | 73369460 | 73370062 | 603 | 3 | 2.35E-10 | 1.80E-05 | 6.09E-07 | 1.96E-07 | 0.001644851 | SDK2 |
| chr20 | 3208240 | 3209556 | 1317 | 13 | 4.69E-08 | 1.58E-06 | 0.001192 | 1.96E-07 | 0.000588772 | ITPA |
| chr9 | 18776342 | 18776942 | 601 | 2 | 8.62E-09 | 7.27E-07 | 2.37E-06 | 1.96E-07 | 0.007331967 | ADAMTSL1 |
| chr1 | 226407633 | 226408967 | 1335 | 11 | 1.61E-07 | 4.75E-06 | 0.000164 | 1.96E-07 | 7.72E-05 | PARP1 |
| chr10 | 329308 | 329890 | 583 | 5 | 9.69E-07 | 4.19E-08 | 0.002083 | 1.96E-07 | 0.020685932 | DIP2C |
| chr17 | 17190297 | 17191429 | 1133 | 3 | 1.36E-05 | 2.50E-07 | 0.000106 | 1.96E-07 | 0.001485356 | MPRIP |
| chr12 | 54364580 | 54365512 | 933 | 8 | 2.78E-08 | 4.38E-06 | 4.66E-05 | 1.97E-07 | 0.000679561 | AC079313.2, AC079313.1 |
| chr1 | 155244634 | 155245367 | 734 | 8 | 6.68E-08 | 5.62E-07 | 7.87E-05 | 1.98E-07 | -0.00105745 | GBA |
| chr16 | 87053179 | 87053431 | 253 | 4 | 2.97E-07 | 1.43E-07 | 0.000149 | 1.98E-07 | -0.01121739 | NA |
| chr7 | 23205676 | 23205965 | 290 | 4 | 2.62E-09 | 3.74E-05 | 3.46E-07 | 1.98E-07 | -0.01873053 | AC005082.1, AC005082.2 |
| chr2 | 27274887 | 27275995 | 1109 | 14 | 2.25E-07 | 2.92E-07 | 0.0071 | 1.98E-07 | -0.00258713 | DNAJC5G, SLC30A3 |
| chr11 | 6682591 | 6683751 | 1161 | 14 | 1.06E-07 | 6.20E-06 | 9.94E-05 | 1.98E-07 | 0.000209739 | MRPL17 |
| chr16 | 88443168 | 88443449 | 282 | 4 | 1.85E-07 | 4.40E-07 | 5.65E-05 | 1.98E-07 | 0.004390177 | NA |
| chr1 | 18902122 | 18903309 | 1188 | 18 | 5.72E-08 | 6.65E-05 | 0.000341 | 1.98E-07 | 0.000437983 | ALDH4A1 |
| chr19 | 35328872 | 35329278 | 407 | 6 | 6.22E-08 | 1.03E-06 | 0.000157 | 1.98E-07 | -0.00473847 | CD22 |
| chr1 | 92298060 | 92299885 | 1826 | 20 | 2.57E-10 | 3.30E-06 | 0.000146 | 1.99E-07 | 0.000516326 | RPAP2, GLMN |
| chr5 | 501748 | 502805 | 1058 | 7 | 1.34E-10 | 3.01E-05 | 5.46E-06 | 1.99E-07 | 0.015627172 | SLC9A3 |
| chr6 | 33909514 | 33911051 | 1538 | 10 | 2.13E-07 | 9.79E-06 | 0.000879 | 1.99E-07 | 0.004242859 | AL138889.1 |
| chr5 | 35159125 | 35159847 | 723 | 3 | 2.15E-08 | 1.64E-06 | 3.73E-06 | 1.99E-07 | -0.00729412 | PRLR |
| chr10 | 75097772 | 75099166 | 1395 | 6 | 4.70E-07 | 2.55E-05 | 8.85E-06 | 1.99E-07 | -0.00697861 | DUSP13 |
| chr6 | 163404003 | 163405140 | 1138 | 4 | 4.43E-06 | 5.29E-08 | 0.001065 | 1.99E-07 | -0.00670237 | NA |
| chr22 | 47995708 | 47996593 | 886 | 3 | 4.10E-08 | 4.45E-07 | 9.05E-06 | 2.00E-07 | -0.00010574 | AL117329.1 |
| chr10 | 109884092 | 109884760 | 669 | 5 | 3.32E-09 | 2.88E-05 | 8.21E-06 | 2.01E-07 | -0.00437306 | XPNPEP1 |
| chr17 | 74961040 | 74962274 | 1235 | 6 | 2.92E-06 | 1.02E-06 | 0.00015 | 2.01E-07 | -0.00596964 | HID1 |
| chr3 | 49120336 | 49121703 | 1368 | 14 | 1.09E-07 | 3.98E-07 | 0.003017 | 2.01E-07 | 0.000604202 | USP19, LAMB2 |
| chr2 | 119543411 | 119544923 | 1513 | 12 | 4.23E-07 | 5.88E-06 | 5.80E-05 | 2.02E-07 | -0.00187758 | CFAP221 |
| chr21 | 37044163 | 37044198 | 36 | 2 | 1.29E-08 | 1.31E-05 | 4.13E-07 | 2.02E-07 | 0.008260979 | NA |
| chr4 | 103197395 | 103198482 | 1088 | 9 | 1.31E-09 | 0.0002429 | 3.07E-05 | 2.02E-07 | -0.00077762 | CENPE |
| chr5 | 1073355 | 1073770 | 416 | 4 | 3.86E-08 | 4.49E-07 | 1.66E-05 | 2.02E-07 | -0.00173323 | SLC12A7 |
| chr13 | 25300721 | 25302197 | 1477 | 13 | 1.14E-07 | 0.0003799 | 9.17E-05 | 2.03E-07 | -0.00545328 | NUP58, AL590787.1 |
| chr17 | 79987920 | 79988883 | 964 | 5 | 6.38E-08 | 1.00E-06 | 6.75E-06 | 2.03E-07 | 0.006388952 | TBC1D16 |
| chr2 | 25040938 | 25041924 | 987 | 8 | 7.99E-10 | 6.25E-06 | 1.97E-05 | 2.03E-07 | -0.00245253 | NA |
| chr16 | 23834958 | 23836495 | 1538 | 10 | 1.75E-07 | 5.67E-06 | 0.001484 | 2.03E-07 | 0.002461907 | PRKCB |
| chr8 | 138722200 | 138723281 | 1082 | 5 | 2.30E-06 | 1.99E-06 | 7.49E-05 | 2.03E-07 | 0.000695842 | COL22A1 |
| chr2 | 219851583 | 219852073 | 491 | 3 | 3.60E-07 | 6.82E-08 | 0.000597 | 2.03E-07 | -0.03902787 | NA |
| chr1 | 68818876 | 68819391 | 516 | 3 | 1.34E-07 | 2.78E-07 | 8.29E-05 | 2.03E-07 | 0.016026395 | AL033530.1 |
| chr3 | 78048020 | 78049000 | 981 | 2 | 6.89E-07 | 2.26E-07 | 1.16E-05 | 2.03E-07 | 0.002565556 | NA |
| chr21 | 44455240 | 44456544 | 1305 | 7 | 6.11E-07 | 5.29E-08 | 0.001316 | 2.04E-07 | -0.00126266 | LRRC3, LRRC3-DT |
| chr12 | 50351079 | 50351824 | 746 | 4 | 1.85E-07 | 2.27E-05 | 0.000369 | 2.04E-07 | -0.00578442 | FAM186A |
| chr17 | 18246279 | 18247640 | 1362 | 7 | 1.48E-06 | 5.76E-07 | 0.000914 | 2.04E-07 | -0.00723652 | FLII |
| chr8 | 1677229 | 1678531 | 1303 | 7 | 6.55E-07 | 0.0001168 | 6.28E-05 | 2.04E-07 | -0.00241445 | DLGAP2 |
| chr3 | 53235745 | 53236334 | 590 | 2 | 1.18E-08 | 3.72E-07 | 5.34E-06 | 2.05E-07 | -0.0092262 | TKT |
| chr18 | 48974460 | 48975826 | 1367 | 5 | 1.70E-10 | 7.44E-05 | 3.96E-06 | 2.05E-07 | -0.00912118 | NA |
| chr19 | 42408244 | 42408476 | 233 | 4 | 5.83E-09 | 0.0003512 | 4.08E-07 | 2.05E-07 | 0.003025012 | LIPE-AS1, LIPE |
| chr11 | 72553941 | 72554353 | 413 | 2 | 3.74E-07 | 1.22E-07 | 7.97E-05 | 2.05E-07 | -0.0227539 | NA |
| chr20 | 36790944 | 36791474 | 531 | 2 | 2.47E-08 | 6.23E-07 | 2.96E-06 | 2.05E-07 | -0.0016859 | SOGA1 |
| chr7 | 73849148 | 73849415 | 268 | 2 | 2.46E-08 | 1.21E-06 | 1.63E-06 | 2.06E-07 | -0.01012631 | NA |
| chr1 | 110082603 | 110082650 | 48 | 2 | 6.56E-08 | 1.24E-07 | 7.53E-05 | 2.06E-07 | -0.00106797 | NA |

| | | | | | | | | | | |
|---|---|---|---|---|---|---|---|---|---|---|
| chr19 | 6503873 | 6504019 | 147 | 3 | 2.45E-08 | 2.79E-07 | 3.73E-05 | 2.06E-07 | -0.00292934 | NA |
| chr3 | 120594456 | 120596017 | 1562 | 6 | 6.74E-11 | 0.0012745 | 7.41E-07 | 2.07E-07 | -0.00461995 | NA |
| chr19 | 40425790 | 40426369 | 580 | 11 | 3.86E-07 | 5.64E-08 | 0.001471 | 2.07E-07 | 0.000700071 | AC010271.2, SERTAD1 |
| chr1 | 17754505 | 17755508 | 1004 | 7 | 5.89E-08 | 2.53E-05 | 0.000285 | 2.07E-07 | 0.005114462 | ACTL8 |
| chr20 | 57262427 | 57263952 | 1526 | 6 | 4.13E-07 | 1.92E-05 | 6.66E-06 | 2.07E-07 | 0.006907018 | BMP7 |
| chr11 | 69673650 | 69675206 | 1557 | 6 | 4.17E-08 | 1.85E-06 | 0.000207 | 2.07E-07 | -0.00130361 | LTO1 |
| chr8 | 130401905 | 130402368 | 464 | 5 | 4.40E-08 | 8.22E-06 | 1.86E-05 | 2.07E-07 | -0.00180412 | ASAP1 |
| chr17 | 40704233 | 40704843 | 611 | 4 | 2.87E-08 | 2.31E-07 | 0.000596 | 2.07E-07 | -0.01240174 | NA |
| chr3 | 23399956 | 23400308 | 353 | 3 | 7.71E-08 | 9.02E-07 | 1.06E-05 | 2.08E-07 | -0.01119995 | UBE2E2 |
| chr1 | 245107124 | 245107184 | 61 | 2 | 2.40E-07 | 2.11E-07 | 1.39E-05 | 2.08E-07 | 0.027733588 | EFCAB2 |
| chr9 | 115058559 | 115059198 | 640 | 2 | 1.31E-06 | 1.23E-07 | 8.40E-05 | 2.08E-07 | 0.009508272 | DELEC1, TNC |
| chr2 | 210324545 | 210324717 | 173 | 4 | 7.04E-09 | 0.0005404 | 1.79E-07 | 2.08E-07 | 0.000597889 | NA |
| chr19 | 14230663 | 14231346 | 684 | 2 | 1.38E-07 | 2.31E-07 | 1.17E-05 | 2.08E-07 | -0.00557001 | NA |
| chr12 | 57078291 | 57079184 | 894 | 14 | 1.86E-07 | 2.33E-06 | 6.81E-05 | 2.08E-07 | 0.000440861 | NEMP1 |
| chr13 | 94327374 | 94327505 | 132 | 2 | 1.28E-07 | 6.78E-07 | 2.78E-06 | 2.08E-07 | 0.004124312 | GPC6 |
| chr1 | 155272530 | 155274016 | 1487 | 15 | 4.86E-09 | 3.01E-06 | 4.33E-05 | 2.08E-07 | -0.0006911 | CLK2 |
| chr7 | 44787187 | 44788268 | 1082 | 5 | 1.49E-07 | 1.21E-05 | 5.88E-05 | 2.08E-07 | -0.00453957 | AC013436.1 |
| chr8 | 95305240 | 95305806 | 567 | 3 | 7.07E-09 | 1.20E-05 | 4.82E-07 | 2.08E-07 | -0.01296534 | C8orf37-AS1 |
| chr11 | 69936966 | 69937495 | 530 | 2 | 1.56E-09 | 4.00E-06 | 7.41E-07 | 2.09E-07 | 0.014016019 | NA |
| chr17 | 43994451 | 43995254 | 804 | 6 | 1.98E-08 | 5.20E-05 | 6.26E-05 | 2.09E-07 | -0.00261723 | PYY |
| chr19 | 50625513 | 50626336 | 824 | 2 | 2.71E-08 | 1.80E-07 | 1.97E-05 | 2.09E-07 | -0.01144949 | SYT3 |
| chr3 | 107006757 | 107007395 | 639 | 3 | 1.77E-07 | 7.14E-06 | 5.25E-06 | 2.09E-07 | -0.00646563 | LINC00882 |
| chr4 | 158722708 | 158723867 | 1160 | 11 | 4.46E-07 | 1.53E-06 | 0.002723 | 2.09E-07 | 0.000691364 | PPID |
| chr3 | 126654791 | 126655995 | 1205 | 10 | 3.26E-07 | 0.0002381 | 2.51E-05 | 2.09E-07 | 0.002525479 | TXNRD3 |
| chr9 | 129933226 | 129933820 | 595 | 3 | 1.97E-08 | 8.04E-07 | 6.32E-05 | 2.09E-07 | 0.00077995 | AL158207.2, FNBP1 |
| chr4 | 2417724 | 2418934 | 1211 | 9 | 1.87E-09 | 1.55E-06 | 3.20E-05 | 2.10E-07 | 0.000247206 | ZFYVE28 |
| chr5 | 174617516 | 174618597 | 1082 | 8 | 1.89E-07 | 3.31E-05 | 9.58E-05 | 2.10E-07 | -0.00217222 | AC108112.1 |
| chr3 | 41961213 | 41962544 | 1332 | 11 | 3.78E-06 | 1.90E-07 | 0.006622 | 2.11E-07 | 5.46E-05 | ULK4 |
| chr1 | 150009705 | 150010905 | 1201 | 12 | 3.88E-08 | 3.28E-07 | 4.57E-05 | 2.11E-07 | -0.00040689 | OTUD7B |
| chr1 | 247948373 | 247948835 | 463 | 4 | 1.58E-07 | 5.64E-07 | 1.72E-05 | 2.11E-07 | -0.00039171 | NA |
| chr5 | 178994313 | 178996359 | 2047 | 13 | 1.14E-06 | 2.65E-07 | 0.003385 | 2.11E-07 | 0.003926782 | GRM6 |
| chr19 | 850485 | 851045 | 561 | 4 | 6.73E-10 | 3.35E-05 | 7.92E-07 | 2.11E-07 | -0.00395541 | ELANE |
| chr6 | 52910411 | 52911184 | 774 | 3 | 1.33E-07 | 1.37E-06 | 4.36E-05 | 2.11E-07 | -0.00638836 | NA |
| chr1 | 182788821 | 182789927 | 1107 | 12 | 5.39E-08 | 4.26E-05 | 0.000443 | 2.12E-07 | 0.000585016 | NPL |
| chr6 | 158078103 | 158078142 | 40 | 3 | 1.34E-07 | 1.33E-06 | 6.25E-06 | 2.12E-07 | 0.00107213 | SYNJ2 |
| chr17 | 53798889 | 53799240 | 352 | 2 | 1.91E-07 | 1.66E-07 | 2.53E-05 | 2.12E-07 | -0.01210135 | AC034268.2 |
| chr2 | 239792124 | 239792739 | 616 | 3 | 7.91E-07 | 1.89E-07 | 0.00015 | 2.12E-07 | -0.00706075 | AC093802.1 |
| chr3 | 12608856 | 12609324 | 469 | 5 | 5.41E-09 | 0.0001839 | 2.53E-06 | 2.13E-07 | 0.001606408 | RAF1 |
| chr18 | 6284001 | 6284897 | 897 | 6 | 3.69E-07 | 1.43E-07 | 0.000636 | 2.13E-07 | 0.000308348 | L3MBTL4 |
| chr6 | 170272414 | 170273455 | 1042 | 8 | 2.60E-08 | 4.89E-06 | 0.000164 | 2.13E-07 | -0.00426061 | LINC01624, AL109910.2 |
| chr22 | 29719359 | 29720339 | 981 | 5 | 6.18E-07 | 1.04E-07 | 0.00043 | 2.13E-07 | 0.000482925 | CABP7, AC004882.1, AC004882.2 |
| chr16 | 17972551 | 17973530 | 980 | 6 | 2.91E-06 | 4.09E-07 | 0.001733 | 2.13E-07 | -0.01200613 | AC010601.1 |
| chr13 | 109197177 | 109197485 | 309 | 2 | 9.58E-08 | 3.85E-07 | 5.51E-06 | 2.13E-07 | -0.01179738 | MYO16, MYO16-AS1 |
| chr15 | 66356770 | 66357523 | 754 | 9 | 2.35E-09 | 1.19E-05 | 0.000236 | 2.14E-07 | -5.62E-05 | TIPIN |
| chr11 | 117177813 | 117179517 | 1705 | 15 | 1.33E-05 | 1.65E-05 | 0.000738 | 2.14E-07 | -0.0013058 | SIDT2 |
| chr12 | 6962651 | 6964101 | 1451 | 13 | 6.01E-08 | 2.88E-05 | 0.000462 | 2.14E-07 | -0.0007323 | MIR200CHG, MIR200C, MIR141 |
| chr5 | 77850373 | 77851317 | 945 | 7 | 6.50E-07 | 1.15E-07 | 0.000584 | 2.14E-07 | 0.024073741 | TBCA |
| chr8 | 9556187 | 9556786 | 600 | 5 | 3.32E-08 | 9.68E-08 | 0.000748 | 2.14E-07 | -0.00309023 | TNKS, AC021242.3 |
| chr14 | 53615391 | 53615509 | 119 | 3 | 4.30E-08 | 4.44E-05 | 7.04E-06 | 2.14E-07 | 0.013923789 | AL365295.1 |
| chr17 | 5785998 | 5786424 | 427 | 4 | 9.74E-08 | 3.16E-07 | 2.96E-05 | 2.14E-07 | -0.00568556 | AC007846.1, AC007846.2 |
| chr11 | 115194092 | 115194125 | 34 | 2 | 3.09E-07 | 1.33E-07 | 6.29E-05 | 2.14E-07 | -0.02443704 | CADM1 |
| chr5 | 134544393 | 134545502 | 1110 | 4 | 1.96E-06 | 0.0001841 | 1.86E-05 | 2.14E-07 | 0.009143526 | JADE2 |
| chr16 | 85349587 | 85350229 | 643 | 2 | 6.41E-10 | 6.55E-07 | 3.01E-06 | 2.15E-07 | -0.01145378 | GSE1, AC092377.1 |
| chr19 | 17346442 | 17347317 | 876 | 2 | 2.94E-07 | 2.61E-07 | 1.01E-05 | 2.15E-07 | -0.01198319 | NA |
| chr13 | 22040910 | 22041195 | 286 | 5 | 2.77E-07 | 1.84E-07 | 0.000221 | 2.15E-07 | -0.02449936 | LINC00540 |
| chr1 | 10211133 | 10211666 | 534 | 6 | 2.63E-07 | 4.26E-08 | 0.0018 | 2.15E-07 | -0.00480383 | KIF1B |
| chr19 | 43798906 | 43799706 | 801 | 2 | 1.89E-11 | 3.65E-06 | 8.12E-07 | 2.15E-07 | -0.00890773 | LYPD5 |
| chr19 | 19366861 | 19366938 | 78 | 2 | 3.65E-08 | 5.40E-05 | 2.99E-07 | 2.15E-07 | -0.01622428 | NA |
| chr17 | 10656006 | 10656220 | 215 | 5 | 1.06E-07 | 6.09E-07 | 1.76E-05 | 2.16E-07 | 0.002041986 | MYH3 |
| chr17 | 71975642 | 71976107 | 466 | 2 | 1.47E-08 | 1.65E-06 | 1.36E-06 | 2.16E-07 | -0.01762287 | NA |
| chr11 | 121671503 | 121671929 | 427 | 3 | 3.54E-08 | 4.73E-06 | 2.22E-05 | 2.16E-07 | -0.00352945 | NA |
| chr14 | 53196121 | 53196776 | 656 | 2 | 8.36E-08 | 2.98E-07 | 8.17E-06 | 2.16E-07 | -0.00973662 | AL365295.1 |
| chr9 | 2857660 | 2858119 | 460 | 3 | 3.23E-09 | 5.71E-06 | 1.10E-06 | 2.16E-07 | -0.01370707 | NA |
| chr5 | 132747413 | 132748376 | 964 | 8 | 1.00E-08 | 1.78E-06 | 0.000191 | 2.16E-07 | -0.00537525 | CCNI2 |
| chr17 | 78705861 | 78707240 | 1380 | 4 | 1.16E-06 | 3.23E-07 | 4.95E-05 | 2.16E-07 | -2.58E-05 | CYTH1 |
| chr14 | 96638541 | 96638693 | 153 | 2 | 4.84E-09 | 1.23E-05 | 4.53E-07 | 2.16E-07 | -0.00640192 | NA |
| chr2 | 38601934 | 38603608 | 1675 | 13 | 3.38E-06 | 5.49E-06 | 0.005039 | 2.16E-07 | 0.000679205 | AC011247.1, HNRNPLL |
| chr7 | 25296698 | 25297105 | 408 | 2 | 2.53E-09 | 1.22E-05 | 4.58E-07 | 2.17E-07 | 0.004608827 | NA |
| chr6 | 56091517 | 56091811 | 295 | 2 | 1.63E-07 | 1.38E-07 | 5.70E-05 | 2.17E-07 | -0.02698845 | COL21A1 |
| chr7 | 131106250 | 131107846 | 1597 | 8 | 1.52E-07 | 5.19E-06 | 9.70E-05 | 2.17E-07 | 0.001655876 | AC016831.6, LINC-PINT |
| chr11 | 118526135 | 118527152 | 1018 | 4 | 2.69E-06 | 3.02E-07 | 0.000165 | 2.17E-07 | -0.00463413 | KMT2A, AP001267.3 |
| chr6 | 84022560 | 84022984 | 425 | 4 | 3.88E-08 | 1.52E-06 | 0.000138 | 2.17E-07 | -0.00695869 | LINC02857 |
| chr11 | 72658104 | 72658714 | 611 | 4 | 7.13E-08 | 3.91E-07 | 0.000137 | 2.18E-07 | -0.00646216 | PDE2A-AS1, PDE2A |
| chr17 | 16438287 | 16439149 | 863 | 9 | 7.71E-09 | 2.65E-07 | 0.000536 | 2.18E-07 | -0.00242464 | SNHG29 |
| chr2 | 43219848 | 43221421 | 1574 | 5 | 7.49E-06 | 3.58E-06 | 6.82E-05 | 2.18E-07 | -0.00036553 | AC010883.3 |
| chr19 | 43518401 | 43518796 | 396 | 2 | 2.00E-07 | 1.35E-07 | 6.61E-05 | 2.18E-07 | 0.0004925 | ETHE1 |
| chr3 | 46876401 | 46877376 | 976 | 5 | 3.56E-07 | 3.68E-07 | 0.000156 | 2.18E-07 | -0.00051883 | MYL3 |
| chr1 | 58700571 | 58701185 | 615 | 5 | 7.07E-10 | 3.74E-06 | 4.53E-06 | 2.19E-07 | 0.004827617 | NA |
| chr1 | 153536047 | 153537159 | 1113 | 13 | 1.08E-07 | 6.93E-06 | 0.000764 | 2.19E-07 | -0.00026965 | S100A6, S100A5 |
| chr2 | 232632939 | 232633972 | 1034 | 14 | 1.92E-08 | 2.84E-05 | 1.97E-05 | 2.19E-07 | 0.001894195 | EFHD1 |
| chr9 | 3180374 | 3181007 | 634 | 3 | 2.54E-08 | 3.43E-07 | 1.93E-05 | 2.19E-07 | 0.024642788 | NA |
| chr1 | 29349955 | 29350770 | 816 | 6 | 1.16E-07 | 3.58E-06 | 6.45E-05 | 2.19E-07 | -0.00296226 | LINC01756 |
| chr9 | 79570222 | 79571236 | 1015 | 5 | 1.34E-10 | 0.0001166 | 9.21E-07 | 2.19E-07 | -0.00726124 | NA |
| chr15 | 62110209 | 62110805 | 597 | 2 | 7.53E-08 | 2.62E-07 | 1.05E-05 | 2.19E-07 | -0.00375599 | NA |
| chr6 | 138217680 | 138218569 | 890 | 3 | 6.66E-07 | 1.86E-07 | 6.13E-05 | 2.19E-07 | -0.00529511 | ARFGEF3, PBOV1 |
| chr1 | 158547699 | 158548059 | 361 | 2 | 4.74E-08 | 1.88E-07 | 2.05E-05 | 2.20E-07 | -0.00024283 | OR6Y1 |
| chr6 | 36268803 | 36270453 | 1651 | 8 | 8.31E-07 | 5.80E-05 | 6.51E-05 | 2.20E-07 | -0.00587795 | PNPLA1 |
| chr17 | 81416926 | 81417271 | 346 | 3 | 1.10E-08 | 2.84E-06 | 4.21E-06 | 2.20E-07 | -0.00236415 | BAHCC1 |
| chr22 | 46085368 | 46086729 | 1362 | 13 | 7.62E-07 | 1.08E-06 | 0.000566 | 2.20E-07 | -0.00197282 | MIRLET7BHG |
| chr9 | 109167267 | 109167917 | 651 | 8 | 1.21E-07 | 1.40E-06 | 2.44E-05 | 2.20E-07 | -0.00233194 | NA |
| chr19 | 54231213 | 54231888 | 676 | 4 | 6.57E-08 | 8.18E-07 | 1.41E-05 | 2.20E-07 | 0.004296811 | RPS9 |
| chr11 | 106076662 | 106078379 | 1718 | 27 | 8.97E-07 | 8.84E-06 | 0.00419 | 2.20E-07 | -5.53E-05 | AASDHPPT, KBTBD3 |
| chr1 | 50749854 | 50750203 | 350 | 4 | 1.17E-07 | 3.28E-06 | 4.64E-05 | 2.20E-07 | 0.001574921 | FAF1 |
| chr7 | 30751067 | 30751678 | 612 | 3 | 1.06E-07 | 2.92E-07 | 1.95E-05 | 2.20E-07 | -0.00599658 | INMT |
| chr2 | 181656192 | 181657199 | 1008 | 10 | 2.26E-08 | 1.44E-06 | 0.001695 | 2.21E-07 | -0.00226026 | CERKL |
| chr7 | 157529905 | 157530750 | 846 | 7 | 1.47E-07 | 1.99E-06 | 4.56E-05 | 2.21E-07 | -0.00568641 | NA |
| chr2 | 120344856 | 120345784 | 929 | 5 | 1.02E-07 | 2.93E-06 | 0.00022 | 2.21E-07 | 0.000629114 | NA |
| chr15 | 48120348 | 48121147 | 800 | 8 | 1.25E-07 | 1.04E-06 | 0.001344 | 2.21E-07 | -0.00785106 | SLC24A5 |
| chr13 | 30617016 | 30617967 | 952 | 12 | 3.73E-07 | 1.52E-06 | 0.005084 | 2.21E-07 | 0.001416882 | USPL1, HMGB1 |
| chr19 | 44760680 | 44761131 | 452 | 3 | 6.63E-08 | 1.27E-06 | 3.27E-06 | 2.21E-07 | 0.006147345 | NA |
| chr16 | 25110887 | 25112542 | 1656 | 12 | 7.74E-08 | 7.02E-06 | 0.000508 | 2.21E-07 | -0.00064227 | LCMT1, LCMT1-AS1 |
| chr6 | 30647562 | 30648695 | 1134 | 22 | 2.06E-08 | 6.47E-06 | 0.001413 | 2.21E-07 | -0.00028289 | C6orf136 |
| chr21 | 25176911 | 25177145 | 235 | 2 | 1.70E-08 | 5.80E-05 | 3.03E-07 | 2.22E-07 | -0.01572328 | AP001341.1 |
| chr13 | 48037303 | 48038159 | 857 | 16 | 3.32E-08 | 2.98E-06 | 0.00316 | 2.22E-07 | -0.00030914 | NUDT15, SUCLA2 |
| chr12 | 52848949 | 52849705 | 757 | 7 | 2.48E-07 | 8.76E-07 | 0.000922 | 2.22E-07 | -0.00475612 | KRT78 |
| chr5 | 131270661 | 131271021 | 361 | 2 | 1.53E-09 | 8.91E-05 | 2.78E-07 | 2.22E-07 | 0.019204247 | CDC42SE2 |
| chr19 | 38989101 | 38989205 | 105 | 2 | 2.95E-07 | 1.59E-07 | 3.41E-05 | 2.22E-07 | -0.03933627 | AC011455.8 |
| chr5 | 31747747 | 31749370 | 1624 | 6 | 2.59E-06 | 0.0001937 | 6.00E-05 | 2.22E-07 | -0.00530195 | PDZD2, AC022447.7 |
| chr11 | 69708807 | 69709523 | 717 | 3 | 4.97E-08 | 5.10E-07 | 1.41E-05 | 2.22E-07 | -0.00786625 | NA |
| chr16 | 30075710 | 30076634 | 925 | 8 | 1.85E-07 | 9.24E-07 | 0.000109 | 2.22E-07 | 0.000494514 | PPP4C |
| chr12 | 109967749 | 109967843 | 95 | 2 | 3.11E-07 | 1.47E-07 | 4.70E-05 | 2.22E-07 | -0.01156436 | TCHP, GIT2 |
| chr10 | 121734810 | 121735358 | 549 | 3 | 2.10E-10 | 3.71E-05 | 3.33E-07 | 2.22E-07 | -0.00541079 | NA |
| chr16 | 1963572 | 1965702 | 2131 | 15 | 1.52E-06 | 2.18E-06 | 0.004263 | 2.23E-07 | -0.00294968 | SNHG9, SNORA78, RPS2, AC005363.1 |
| chr1 | 2483760 | 2484476 | 717 | 4 | 3.52E-08 | 5.81E-07 | 1.05E-05 | 2.23E-07 | -0.01244082 | PLCH2 |
| chr17 | 44988377 | 44988508 | 132 | 3 | 1.61E-07 | 7.35E-08 | 0.000647 | 2.23E-07 | -0.02462265 | NMT1 |
| chr5 | 83685384 | 83686245 | 862 | 6 | 7.71E-09 | 5.77E-05 | 3.84E-05 | 2.23E-07 | -0.00574278 | RN7SKP295, HAPLN1 |
| chr15 | 85730253 | 85730462 | 210 | 3 | 1.99E-07 | 8.52E-08 | 0.000292 | 2.23E-07 | -0.00757389 | AKAP13 |
| chr17 | 42900526 | 42901211 | 686 | 6 | 1.80E-08 | 3.61E-07 | 3.81E-05 | 2.23E-07 | -0.00353413 | G6PC |
| chr2 | 27204295 | 27205181 | 887 | 6 | 4.61E-08 | 5.74E-07 | 3.77E-05 | 2.24E-07 | 0.00061586 | SLC5A6 |
| chr1 | 7621335 | 7621977 | 643 | 3 | 1.73E-07 | 2.12E-06 | 1.52E-05 | 2.24E-07 | -0.01149649 | CAMTA1 |
| chr1 | 173970065 | 173970580 | 516 | 3 | 2.47E-08 | 4.92E-07 | 1.00E-05 | 2.24E-07 | -0.01000373 | RC3H1 |
| chr2 | 230124467 | 230125279 | 813 | 6 | 2.65E-08 | 1.32E-06 | 3.36E-05 | 2.24E-07 | -0.00231593 | AC009950.1 |
| chr20 | 57711335 | 57712238 | 904 | 11 | 2.54E-08 | 1.48E-05 | 0.001876 | 2.24E-07 | -0.00448765 | NKILA, PMEPA1 |
| chr6 | 91299617 | 91299966 | 350 | 2 | 1.13E-07 | 1.52E-07 | 4.25E-05 | 2.24E-07 | -0.01776869 | NA |
| chr22 | 42073752 | 42074490 | 739 | 7 | 2.83E-08 | 1.52E-06 | 0.000214 | 2.24E-07 | -0.01024679 | PHETA2 |
| chr1 | 3217598 | 3218519 | 922 | 5 | 6.93E-09 | 9.13E-06 | 3.36E-05 | 2.24E-07 | 0.007954886 | PRDM16 |
| chr2 | 213189182 | 213189825 | 644 | 3 | 9.21E-08 | 1.06E-07 | 0.000149 | 2.24E-07 | -0.01336948 | NA |
| chr19 | 15238125 | 15239366 | 1242 | 14 | 1.57E-06 | 1.29E-06 | 0.005392 | 2.24E-07 | 2.36E-05 | BRD4 |
| chr5 | 158255204 | 158255706 | 503 | 3 | 7.27E-08 | 5.12E-07 | 8.71E-06 | 2.24E-07 | -0.00770737 | NA |
| chr7 | 30937584 | 30938535 | 952 | 2 | 2.77E-06 | 1.54E-07 | 4.03E-05 | 2.24E-07 | -0.00256415 | NA |
| chr17 | 50188907 | 50189878 | 972 | 7 | 6.88E-09 | 4.87E-05 | 2.59E-06 | 2.24E-07 | -0.00363214 | COL1A1 |
| chr17 | 58528655 | 58529481 | 827 | 5 | 3.06E-08 | 7.78E-06 | 2.33E-05 | 2.24E-07 | -0.00438093 | SEPTIN4-AS1, SEPTIN4 |
| chr1 | 62069146 | 62069529 | 384 | 4 | 4.85E-07 | 5.10E-08 | 0.001668 | 2.24E-07 | -0.00659468 | PATJ |
| chr2 | 154589095 | 154590375 | 1281 | 6 | 2.51E-05 | 7.19E-07 | 0.000347 | 2.24E-07 | -0.00791894 | NA |
| chr18 | 46398662 | 46399456 | 795 | 3 | 9.32E-08 | 3.92E-07 | 1.19E-05 | 2.24E-07 | -0.00146886 | RNF165 |
| chr12 | 51814113 | 51815115 | 1003 | 5 | 2.49E-07 | 7.87E-08 | 0.001252 | 2.24E-07 | -0.010283 | AC068987.2, TMDD1 |
| chr15 | 77641346 | 77641650 | 305 | 5 | 2.00E-07 | 2.31E-06 | 5.47E-05 | 2.24E-07 | -0.0017034 | LINGO1 |
| chr2 | 105815084 | 105816458 | 1375 | 5 | 1.12E-06 | 4.80E-07 | 0.000506 | 2.25E-07 | 9.89E-05 | NCK2 |
| chr17 | 18152980 | 18153866 | 887 | 5 | 2.19E-06 | 1.47E-06 | 7.78E-05 | 2.25E-07 | 0.002892257 | MYO15A |
| chr14 | 101083424 | 101083940 | 517 | 2 | 1.48E-07 | 2.02E-07 | 1.90E-05 | 2.25E-07 | -0.00986416 | NA |
| chr20 | 25213365 | 25214636 | 1272 | 6 | 1.30E-06 | 1.20E-07 | 0.00068 | 2.25E-07 | -0.00022375 | ENTPD6 |
| chr15 | 74228830 | 74229636 | 807 | 3 | 3.11E-06 | 7.65E-08 | 0.000558 | 2.26E-07 | -0.01222859 | CCDC33 |
| chr1 | 243482609 | 243483485 | 877 | 8 | 1.95E-08 | 7.00E-06 | 9.32E-05 | 2.26E-07 | 0.0033343 | SDCCAG8 |
| chr1 | 223143914 | 223144508 | 595 | 7 | 1.53E-07 | 1.42E-07 | 0.003721 | 2.26E-07 | -0.00982185 | AL359979.2 |
| chr18 | 58412831 | 58413417 | 587 | 2 | 1.06E-06 | 1.45E-07 | 5.58E-05 | 2.26E-07 | -0.00713316 | NA |
| chr10 | 182255 | 182861 | 607 | 2 | 1.58E-07 | 2.22E-07 | 1.57E-05 | 2.26E-07 | -0.01576838 | ZMYND11 |
| chr8 | 122951791 | 122952698 | 908 | 6 | 1.15E-08 | 7.42E-06 | 0.000126 | 2.26E-07 | 2.58E-05 | ZHX2 |
| chr6 | 90218320 | 90219213 | 894 | 3 | 1.10E-07 | 1.64E-05 | 1.47E-05 | 2.26E-07 | -0.01165164 | BACH2 |
| chr1 | 115088894 | 115090106 | 1213 | 13 | 3.12E-07 | 1.06E-07 | 0.003897 | 2.27E-07 | -0.00061133 | TSPAN2 |
| chr11 | 72573518 | 72573740 | 223 | 2 | 2.43E-08 | 2.65E-05 | 3.71E-07 | 2.28E-07 | -0.01047705 | NA |
| chr9 | 36572414 | 36573075 | 662 | 11 | 1.63E-07 | 1.53E-05 | 0.002177 | 2.28E-07 | -0.00039738 | MELK |
| chr19 | 10478903 | 10479486 | 584 | 6 | 3.14E-07 | 3.29E-07 | 0.002508 | 2.28E-07 | 0.009147802 | NA |
| chr14 | 104222270 | 104222843 | 574 | 3 | 1.40E-08 | 5.95E-07 | 2.43E-05 | 2.28E-07 | -0.00230193 | NA |
| chr5 | 6337295 | 6338681 | 1387 | 7 | 9.78E-07 | 7.97E-06 | 8.45E-05 | 2.28E-07 | -0.0070903 | LINC02145 |
| chr1 | 1042129 | 1043018 | 890 | 4 | 1.30E-07 | 2.37E-07 | 0.000553 | 2.28E-07 | 0.00070956 | AGRN |
| chr8 | 139911987 | 139912493 | 507 | 2 | 1.17E-07 | 2.02E-07 | 1.97E-05 | 2.28E-07 | -0.01154736 | AC021744.1, TRAPPC9 |
| chr17 | 81034793 | 81035934 | 1142 | 14 | 1.16E-07 | 6.55E-07 | 0.004354 | 2.29E-07 | 0.000810955 | BAIAP2, BAIAP2-DT |
| chr6 | 1624151 | 1625521 | 1371 | 9 | 4.49E-07 | 1.83E-07 | 0.001552 | 2.29E-07 | -0.00432622 | GMDS |

| | | | | | | | | | | |
|---|---|---|---|---|---|---|---|---|---|---|
| chr20 | 3082472 | 3083369 | 898 | 4 | 1.80E-06 | 5.42E-07 | 0.000242 | 2.29E-07 | 0.000654234 | AVP |
| chr18 | 79529104 | 79529526 | 423 | 4 | 1.04E-07 | 1.53E-06 | 4.08E-05 | 2.29E-07 | -0.00353073 | NFATC1 |
| chr1 | 156241624 | 156242787 | 1164 | 11 | 1.35E-06 | 1.12E-05 | 0.001678 | 2.29E-07 | 0.006255067 | PMF1-BGLAP, BGLAP |
| chr16 | 4424937 | 4426537 | 1601 | 13 | 2.17E-08 | 0.0054275 | 2.42E-05 | 2.29E-07 | -0.00091526 | DNAJA3, CORO7 |
| chr18 | 74291669 | 74292598 | 930 | 14 | 6.14E-09 | 9.14E-05 | 2.98E-06 | 2.29E-07 | -0.00033719 | CYB5A |
| chr8 | 141147005 | 141147768 | 764 | 6 | 4.74E-08 | 5.72E-05 | 2.62E-05 | 2.30E-07 | -0.00174224 | DENND3 |
| chr2 | 42168533 | 42169742 | 1210 | 10 | 1.51E-06 | 2.03E-07 | 0.003501 | 2.30E-07 | -0.00211719 | EML4, EML4-AS1 |
| chr17 | 19673884 | 19674404 | 521 | 2 | 3.14E-09 | 1.32E-05 | 4.74E-07 | 2.30E-07 | -0.00813222 | ALDH3A2 |
| chr6 | 41638903 | 41639799 | 897 | 6 | 6.10E-08 | 1.11E-06 | 3.47E-05 | 2.31E-07 | -0.00363721 | MDFI |
| chr2 | 54866904 | 54867070 | 167 | 2 | 7.96E-08 | 2.40E-06 | 1.14E-06 | 2.31E-07 | -0.00521724 | EML6 |
| chr6 | 4705936 | 4706193 | 258 | 4 | 2.40E-08 | 0.0008417 | 1.64E-06 | 2.31E-07 | -0.0052128 | CDYL |
| chr11 | 17564498 | 17565031 | 534 | 2 | 3.04E-09 | 1.22E-05 | 4.88E-07 | 2.31E-07 | -0.01100236 | OTOG |
| chr22 | 20721452 | 20721729 | 278 | 4 | 2.98E-07 | 1.18E-07 | 0.000197 | 2.31E-07 | 0.002039634 | PI4KA |
| chr1 | 52725374 | 52726963 | 1590 | 12 | 1.07E-05 | 1.47E-07 | 0.002615 | 2.31E-07 | 0.001323486 | ZYG11B |
| chr5 | 131488317 | 131489290 | 974 | 2 | 1.81E-05 | 1.38E-07 | 8.56E-05 | 2.32E-07 | -0.00960471 | RAPGEF6, AC008695.1 |
| chr9 | 137235684 | 137235816 | 133 | 3 | 2.34E-07 | 1.19E-07 | 0.000221 | 2.32E-07 | 0.015712163 | SLC34A3 |
| chr19 | 18464596 | 18464963 | 368 | 2 | 7.26E-08 | 5.05E-07 | 4.58E-06 | 2.32E-07 | 0.024942398 | ELL |
| chr5 | 135953184 | 135953916 | 733 | 3 | 3.04E-09 | 1.78E-05 | 5.31E-07 | 2.33E-07 | 0.006840084 | LECT2 |
| chr12 | 33575441 | 33575570 | 130 | 3 | 4.74E-07 | 1.24E-07 | 0.00022 | 2.33E-07 | 0.027692462 | NA |
| chr12 | 124419968 | 124421125 | 1158 | 8 | 2.62E-07 | 3.68E-05 | 9.36E-06 | 2.33E-07 | 0.000903728 | NCOR2 |
| chr6 | 169162332 | 169163370 | 1039 | 9 | 3.75E-07 | 4.76E-07 | 0.002445 | 2.33E-07 | 0.001596156 | LINC01615 |
| chr7 | 66076760 | 66077788 | 1029 | 4 | 1.03E-06 | 2.54E-07 | 0.000178 | 2.33E-07 | 0.004064984 | ASL |
| chr17 | 57243128 | 57243983 | 856 | 5 | 1.61E-09 | 0.0010644 | 1.30E-06 | 2.34E-07 | -0.00540567 | NA |
| chr17 | 82022735 | 82024033 | 1299 | 17 | 5.58E-08 | 0.0001471 | 7.01E-05 | 2.34E-07 | 0.000894537 | LRRC45, CENPX |
| chr1 | 211206533 | 211207065 | 533 | 4 | 8.33E-08 | 0.0001392 | 1.17E-05 | 2.34E-07 | -0.00284055 | AC092017.3 |
| chr17 | 4967354 | 4968919 | 1566 | 13 | 7.39E-08 | 2.78E-05 | 3.64E-05 | 2.34E-07 | -0.00019248 | AC004771.3, SPAG7, CAMTA2 |
| chr14 | 25050278 | 25051515 | 1238 | 7 | 1.65E-06 | 1.56E-05 | 0.000764 | 2.34E-07 | -0.00015341 | STXBP6 |
| chr16 | 10939044 | 10939813 | 770 | 3 | 1.37E-07 | 7.07E-06 | 1.03E-05 | 2.34E-07 | 0.006548433 | CIITA, DEXI |
| chr6 | 33594732 | 33595534 | 803 | 2 | 7.67E-07 | 1.45E-07 | 6.72E-05 | 2.34E-07 | -0.00572609 | NA |
| chr3 | 126180558 | 126181981 | 1424 | 16 | 4.73E-09 | 9.17E-05 | 1.70E-05 | 2.34E-07 | -0.00265318 | ALDH1L1-AS2, ALDH1L1 |
| chr21 | 29332920 | 29333074 | 155 | 2 | 3.30E-07 | 1.49E-07 | 5.85E-05 | 2.35E-07 | -0.00162943 | BACH1 |
| chr1 | 163070416 | 163071249 | 834 | 6 | 2.51E-09 | 3.12E-06 | 0.000142 | 2.35E-07 | -0.01096578 | RGS4 |
| chr19 | 17859423 | 17860652 | 1230 | 16 | 8.53E-07 | 5.44E-06 | 0.002598 | 2.35E-07 | 0.000743836 | RPL18A |
| chr18 | 12254242 | 12255837 | 1596 | 14 | 2.84E-09 | 4.15E-05 | 0.000512 | 2.35E-07 | -0.00477883 | CIDEA |
| chr5 | 73153926 | 73154998 | 1073 | 3 | 4.09E-08 | 4.34E-07 | 1.11E-05 | 2.35E-07 | -0.0025043 | NA |
| chr3 | 194672110 | 194673089 | 980 | 13 | 1.37E-06 | 2.09E-06 | 0.003132 | 2.35E-07 | 0.000141721 | LSG1 |
| chr13 | 25881915 | 25882201 | 287 | 2 | 6.77E-08 | 2.90E-07 | 1.03E-05 | 2.35E-07 | -0.01117257 | ATP8A2 |
| chr19 | 1157684 | 1158360 | 677 | 4 | 6.52E-09 | 0.0001002 | 8.03E-06 | 2.35E-07 | -0.00967445 | SBNO2 |
| chr2 | 238420377 | 238421530 | 1154 | 4 | 2.55E-07 | 2.28E-07 | 0.000204 | 2.35E-07 | -0.00889609 | NA |
| chr4 | 70838784 | 70839967 | 1184 | 17 | 1.82E-08 | 8.87E-05 | 6.56E-05 | 2.36E-07 | 0.00098225 | GRSF1 |
| chr17 | 78241817 | 78242504 | 688 | 4 | 6.34E-08 | 1.03E-06 | 1.18E-05 | 2.36E-07 | -0.00048359 | NA |
| chr18 | 23951182 | 23952066 | 885 | 6 | 7.27E-08 | 0.0001072 | 0.000205 | 2.36E-07 | 0.00240478 | LAMA3 |
| chr9 | 69516739 | 69517257 | 519 | 2 | 5.35E-11 | 0.000214 | 2.63E-07 | 2.36E-07 | 0.003745791 | APBA1 |
| chr4 | 1856504 | 1857535 | 1032 | 5 | 2.05E-09 | 3.34E-06 | 4.06E-06 | 2.37E-07 | -0.006158 | NA |
| chr14 | 99125061 | 99125986 | 926 | 3 | 9.86E-07 | 6.66E-07 | 6.52E-05 | 2.37E-07 | -0.00343697 | NA |
| chr8 | 109974142 | 109975842 | 1701 | 14 | 8.67E-07 | 0.0001194 | 0.000102 | 2.37E-07 | 0.00032364 | AC027451.1, KCNV1 |
| chr14 | 70783358 | 70784464 | 1107 | 7 | 9.19E-09 | 7.50E-05 | 1.33E-06 | 2.37E-07 | -0.00257507 | MAP3K9 |
| chr3 | 196198933 | 196199203 | 271 | 2 | 1.20E-08 | 6.52E-06 | 6.66E-07 | 2.37E-07 | -0.00971317 | ZDHHC19 |
| chr16 | 3485674 | 3486023 | 350 | 2 | 5.77E-09 | 4.48E-07 | 5.51E-06 | 2.37E-07 | 0.006381926 | NAA60, AC004224.2 |
| chr7 | 99028897 | 99030217 | 1321 | 5 | 2.34E-07 | 8.56E-07 | 0.000124 | 2.37E-07 | -0.01495778 | TRRAP, SMURF1 |
| chr17 | 7213473 | 7214365 | 893 | 6 | 6.28E-08 | 1.55E-07 | 0.001382 | 2.38E-07 | -0.00838348 | DLG4 |
| chr12 | 3870554 | 3871608 | 1055 | 5 | 5.65E-07 | 3.47E-07 | 8.11E-05 | 2.38E-07 | -0.00701404 | PARP11-AS1, PARP11 |
| chr4 | 145907320 | 145908038 | 719 | 5 | 9.54E-08 | 4.42E-07 | 4.39E-05 | 2.38E-07 | 0.003204308 | ZNF827 |
| chr22 | 29132753 | 29132970 | 218 | 2 | 1.43E-07 | 4.92E-07 | 4.94E-06 | 2.38E-07 | -0.02267073 | KREMEN1 |
| chr18 | 45838183 | 45839536 | 1354 | 7 | 3.94E-07 | 4.32E-07 | 0.002036 | 2.38E-07 | -0.01688478 | SIGLEC15 |
| chr19 | 12806272 | 12806954 | 683 | 13 | 3.65E-08 | 2.45E-06 | 0.000359 | 2.38E-07 | -0.00030357 | THSD8, RNASEH2A, HOOK2 |
| chr11 | 67427438 | 67429035 | 1598 | 14 | 4.29E-07 | 2.50E-06 | 0.000756 | 2.38E-07 | -0.00010763 | RPS6KB2 |
| chr2 | 41876779 | 41877519 | 741 | 6 | 8.00E-08 | 2.84E-05 | 7.62E-05 | 2.39E-07 | -0.0028804 | LINC01913 |
| chr9 | 137578732 | 137579423 | 692 | 9 | 1.52E-08 | 6.82E-06 | 2.72E-05 | 2.40E-07 | -0.0060326 | DPH7 |
| chr12 | 10169132 | 10169821 | 690 | 5 | 2.31E-08 | 1.30E-05 | 9.42E-06 | 2.40E-07 | -0.00591227 | OLR1 |
| chr5 | 178465757 | 178466291 | 535 | 4 | 5.30E-07 | 3.68E-07 | 0.000273 | 2.40E-07 | 0.003344494 | COL23A1 |
| chr12 | 52993054 | 52993444 | 391 | 7 | 9.10E-07 | 3.54E-08 | 0.002246 | 2.40E-07 | -0.0014877 | NA |
| chr7 | 74172665 | 74173923 | 1259 | 5 | 2.92E-09 | 8.75E-07 | 0.00012 | 2.40E-07 | -0.01121697 | NA |
| chr18 | 13372158 | 13372299 | 142 | 2 | 1.86E-07 | 4.73E-07 | 5.25E-06 | 2.40E-07 | 0.017751666 | LDLRAD4 |
| chr20 | 226503 | 227217 | 715 | 4 | 6.87E-09 | 6.03E-06 | 1.00E-06 | 2.40E-07 | -0.00031895 | NA |
| chr12 | 39625730 | 39626276 | 547 | 11 | 3.83E-07 | 5.07E-07 | 0.000487 | 2.41E-07 | -0.00162326 | C12orf40 |
| chr8 | 68095205 | 68095978 | 774 | 3 | 1.67E-07 | 2.13E-07 | 3.36E-05 | 2.41E-07 | -0.00342031 | PREX2 |
| chr17 | 32341732 | 32342475 | 744 | 10 | 1.53E-07 | 5.56E-07 | 0.001845 | 2.41E-07 | 0.000211047 | AC005899.4, C17orf75 |
| chr14 | 102008972 | 102010012 | 1041 | 4 | 1.20E-05 | 5.09E-07 | 0.000111 | 2.41E-07 | -0.0001502 | DYNC1H1 |
| chr17 | 40501590 | 40502887 | 1298 | 11 | 4.93E-06 | 2.67E-07 | 0.00393 | 2.41E-07 | -0.00215608 | TNS4 |
| chr18 | 78426350 | 78427365 | 1016 | 4 | 4.71E-10 | 7.64E-06 | 1.17E-06 | 2.41E-07 | -0.00820573 | NA |
| chr8 | 140361445 | 140361981 | 537 | 2 | 6.34E-09 | 4.15E-06 | 8.67E-07 | 2.41E-07 | 0.003142507 | TRAPPC9 |
| chr2 | 85614950 | 85616257 | 1308 | 15 | 3.37E-07 | 2.74E-06 | 0.000464 | 2.41E-07 | -0.00089804 | USP39 |
| chr2 | 189783737 | 189784974 | 1238 | 18 | 3.74E-07 | 2.58E-06 | 0.003084 | 2.41E-07 | 0.000342528 | PMS1, ORMDL1 |
| chr1 | 46540056 | 46541001 | 946 | 6 | 1.84E-08 | 7.97E-06 | 2.35E-05 | 2.41E-07 | -0.00519424 | MKNK1-AS1, TMEM275 |
| chr17 | 75876503 | 75877843 | 1341 | 6 | 9.42E-06 | 4.98E-07 | 0.000592 | 2.41E-07 | 0.006281802 | AC087289.5, TRIM47 |
| chr4 | 119322097 | 119322584 | 488 | 4 | 6.96E-08 | 9.08E-06 | 6.62E-06 | 2.41E-07 | -0.00683724 | FABP2 |
| chr14 | 22318910 | 22319948 | 1039 | 3 | 1.80E-09 | 0.0001126 | 1.78E-06 | 2.42E-07 | -0.00223703 | AC244502.1 |
| chr2 | 219626708 | 219627462 | 755 | 9 | 8.48E-10 | 5.78E-05 | 2.01E-05 | 2.42E-07 | -0.0046073 | SLC4A3 |
| chr6 | 7051070 | 7051521 | 452 | 4 | 7.09E-08 | 1.37E-06 | 0.000323 | 2.42E-07 | -0.01782602 | NA |
| chr16 | 57794463 | 57795138 | 676 | 2 | 1.84E-07 | 2.25E-07 | 1.82E-05 | 2.43E-07 | -0.00929733 | KIFC3 |
| chr2 | 38868191 | 38868285 | 95 | 3 | 4.45E-07 | 1.56E-07 | 0.000145 | 2.43E-07 | -0.00214336 | DHX57 |
| chr1 | 218284339 | 218285527 | 1189 | 14 | 5.33E-09 | 0.0038472 | 4.96E-06 | 2.43E-07 | -0.00248951 | RRP15 |
| chr7 | 1596552 | 1596747 | 196 | 2 | 7.78E-08 | 1.78E-07 | 3.42E-05 | 2.43E-07 | -0.01079168 | NA |
| chr3 | 182793093 | 182793663 | 571 | 7 | 2.28E-08 | 3.79E-07 | 3.77E-05 | 2.44E-07 | -0.00261575 | ATP11B, AC069431.1 |
| chr20 | 45415688 | 45417145 | 1458 | 13 | 1.29E-06 | 3.33E-05 | 0.000879 | 2.44E-07 | -0.00115627 | PIGT |
| chr7 | 27114017 | 27115615 | 1599 | 18 | 5.40E-09 | 0.0004635 | 0.000138 | 2.44E-07 | -0.00139201 | HOXA-AS2, HOXA3 |
| chr2 | 5938439 | 5938606 | 168 | 2 | 1.72E-07 | 5.11E-07 | 4.94E-06 | 2.44E-07 | -0.02042854 | SILC1 |
| chr3 | 89106968 | 89107852 | 885 | 12 | 7.52E-07 | 7.28E-08 | 0.005614 | 2.44E-07 | 0.003607798 | EPHA3 |
| chr10 | 91909414 | 91910095 | 682 | 3 | 7.26E-10 | 2.75E-06 | 3.47E-06 | 2.45E-07 | 0.007869408 | FGFBP3 |
| chr19 | 55544953 | 55546020 | 1068 | 13 | 2.14E-07 | 2.81E-06 | 0.000238 | 2.45E-07 | -0.00215466 | SBK3 |
| chr7 | 140602880 | 140603828 | 949 | 5 | 8.46E-07 | 4.54E-06 | 2.96E-05 | 2.45E-07 | -0.00679974 | DENND2A |
| chr11 | 118343580 | 118344397 | 818 | 6 | 9.91E-08 | 1.43E-06 | 0.000646 | 2.45E-07 | -0.00432708 | CD3G |
| chr9 | 86255707 | 86256846 | 1140 | 7 | 9.37E-08 | 2.32E-06 | 0.000117 | 2.45E-07 | -0.00301267 | C9orf153 |
| chr1 | 2519600 | 2520728 | 1129 | 4 | 1.59E-06 | 9.58E-08 | 0.000357 | 2.45E-07 | -0.00165305 | PANK4 |
| chr11 | 77101550 | 77102318 | 769 | 4 | 3.78E-09 | 1.25E-05 | 2.73E-06 | 2.46E-07 | -0.00257216 | CAPN5 |
| chr17 | 282123 | 283610 | 1488 | 7 | 1.25E-06 | 7.01E-08 | 0.001292 | 2.46E-07 | -0.00766115 | RPH3AL |
| chr8 | 26655723 | 26656345 | 623 | 3 | 3.64E-08 | 2.15E-06 | 1.88E-05 | 2.46E-07 | -0.00183206 | DPYSL2 |
| chr2 | 112140823 | 112140916 | 94 | 2 | 1.73E-07 | 1.50E-07 | 7.62E-05 | 2.46E-07 | 0.036285555 | FBLN7 |
| chr17 | 61416632 | 61417683 | 1052 | 5 | 1.94E-07 | 1.18E-05 | 6.76E-05 | 2.46E-07 | -0.0061916 | NA |
| chr11 | 102011251 | 102011407 | 157 | 2 | 3.72E-07 | 1.46E-07 | 9.11E-05 | 2.46E-07 | -0.02494177 | NA |
| chr1 | 29237990 | 29239503 | 1514 | 4 | 4.01E-07 | 1.05E-06 | 0.000495 | 2.46E-07 | -0.01039366 | PTPRU |
| chr16 | 81686613 | 81686867 | 255 | 2 | 1.09E-07 | 5.05E-07 | 5.08E-06 | 2.46E-07 | -0.01832998 | CMIP |
| chr2 | 230719653 | 230720934 | 1282 | 3 | 1.75E-06 | 1.25E-07 | 0.0002 | 2.46E-07 | -0.00777407 | CAB39 |
| chr3 | 37573315 | 37573580 | 266 | 3 | 1.60E-07 | 1.04E-07 | 0.000334 | 2.47E-07 | -0.0051388 | ITGA9 |
| chr12 | 67269266 | 67270570 | 1305 | 9 | 1.13E-07 | 1.09E-05 | 0.000357 | 2.47E-07 | 0.001957616 | CAND1 |
| chr12 | 57756481 | 57757117 | 637 | 3 | 3.92E-09 | 0.0001146 | 5.77E-06 | 2.48E-07 | -0.01321333 | MARCHF9 |
| chr9 | 2621037 | 2621606 | 570 | 9 | 4.52E-09 | 5.50E-05 | 4.05E-05 | 2.48E-07 | -0.00519811 | VLDLR-AS1 |
| chr8 | 54476816 | 54477034 | 219 | 2 | 7.51E-08 | 2.05E-06 | 1.40E-06 | 2.48E-07 | -0.0156259 | NA |
| chr1 | 203272820 | 203273364 | 545 | 5 | 5.24E-10 | 0.0005591 | 1.91E-06 | 2.48E-07 | -0.00560771 | LINC01353, CHIT1 |
| chr13 | 102595634 | 102596979 | 1346 | 11 | 2.83E-09 | 2.30E-05 | 0.000214 | 2.48E-07 | -0.00021362 | NA |
| chr1 | 25099432 | 25100765 | 1334 | 4 | 3.28E-06 | 7.45E-07 | 0.000265 | 2.48E-07 | 0.007600733 | AL050344.1 |
| chr7 | 155509039 | 155509647 | 609 | 3 | 1.21E-07 | 1.16E-07 | 0.000167 | 2.49E-07 | -0.00887735 | CNPY1, AC009403.2 |
| chr12 | 126726875 | 126727982 | 1108 | 5 | 5.45E-07 | 1.56E-06 | 0.000344 | 2.49E-07 | -8.04E-05 | LINC00943 |
| chr7 | 157876131 | 157876865 | 735 | 4 | 2.88E-07 | 5.91E-07 | 0.000257 | 2.49E-07 | -0.01120691 | PTPRN2 |
| chr10 | 86312936 | 86312975 | 40 | 2 | 2.12E-07 | 5.95E-07 | 4.26E-06 | 2.50E-07 | -0.01769109 | GRID1 |
| chr11 | 3189402 | 3189742 | 341 | 3 | 8.36E-08 | 1.54E-06 | 5.77E-05 | 2.50E-07 | -0.01291257 | AC108448.1 |
| chr16 | 89738287 | 89740108 | 1822 | 11 | 3.29E-06 | 9.31E-06 | 0.000448 | 2.50E-07 | -0.00121577 | ZNF276, FANCA |
| chr16 | 87684355 | 87685055 | 701 | 2 | 5.97E-08 | 2.65E-07 | 1.41E-05 | 2.50E-07 | -0.00065536 | JPH3, AC010536.3 |
| chr14 | 73287178 | 73287262 | 85 | 2 | 7.98E-08 | 7.44E-06 | 6.65E-07 | 2.50E-07 | 0.004441043 | NUMB |
| chr15 | 89095342 | 89096806 | 1465 | 4 | 4.34E-06 | 1.03E-06 | 0.00014 | 2.50E-07 | -0.00793832 | ABHD2 |
| chr15 | 51095374 | 51096229 | 856 | 6 | 9.22E-09 | 8.03E-05 | 1.86E-05 | 2.51E-07 | -0.00753535 | MIR4713HG, TNFAIP8L3 |
| chr6 | 117262730 | 117263652 | 923 | 6 | 1.72E-07 | 8.28E-07 | 0.000345 | 2.51E-07 | -0.01262898 | Z98880.1 |
| chr1 | 17538839 | 17539560 | 722 | 6 | 3.08E-07 | 3.86E-06 | 0.000571 | 2.51E-07 | -0.0033045 | NA |
| chr6 | 27229324 | 27230457 | 1134 | 3 | 2.22E-05 | 4.16E-07 | 0.000108 | 2.51E-07 | 0.017802478 | NA |
| chr15 | 83446619 | 83447399 | 781 | 12 | 7.65E-08 | 9.28E-06 | 3.00E-05 | 2.51E-07 | 0.002832181 | SH3GL3 |
| chr6 | 34555634 | 34556560 | 927 | 7 | 4.79E-07 | 8.44E-08 | 0.002289 | 2.52E-07 | -0.00698269 | SPDEF |
| chr6 | 32204230 | 32205755 | 1526 | 12 | 2.02E-07 | 1.13E-06 | 5.85E-05 | 2.52E-07 | -0.00357524 | NOTCH4 |
| chr17 | 77560962 | 77561183 | 222 | 3 | 5.18E-07 | 9.51E-08 | 0.000328 | 2.52E-07 | -0.00496008 | AC021683.2 |
| chr17 | 28744551 | 28745233 | 683 | 5 | 7.68E-10 | 1.88E-05 | 1.56E-06 | 2.52E-07 | 0.000604052 | TRAF4 |
| chr3 | 33559027 | 33559408 | 382 | 2 | 2.95E-07 | 1.69E-07 | 4.67E-05 | 2.52E-07 | 0.019593035 | CLASP2 |
| chr19 | 1851167 | 1852166 | 1000 | 6 | 8.10E-06 | 1.15E-07 | 0.001765 | 2.52E-07 | -0.01394978 | NA |
| chr4 | 857597 | 858778 | 1182 | 6 | 3.63E-06 | 4.38E-06 | 0.001056 | 2.52E-07 | -0.00208292 | GAK |
| chr3 | 194299914 | 194300456 | 543 | 3 | 2.29E-09 | 7.08E-06 | 7.84E-06 | 2.52E-07 | 0.009631169 | AC117469.1, LINC00887 |
| chr20 | 32894306 | 32894745 | 440 | 3 | 2.32E-07 | 2.23E-07 | 4.83E-05 | 2.52E-07 | -0.01409801 | EFCAB8 |
| chr17 | 6712329 | 6713805 | 1477 | 14 | 8.73E-07 | 5.10E-06 | 0.001198 | 2.53E-07 | -0.00180513 | SLC13A5 |
| chr15 | 27902782 | 27903285 | 504 | 3 | 1.50E-07 | 2.05E-07 | 7.41E-05 | 2.53E-07 | -0.02244637 | OCA2 |
| chr3 | 157137304 | 157137626 | 323 | 2 | 1.56E-07 | 1.66E-07 | 5.19E-05 | 2.53E-07 | -0.0133777 | NA |
| chr4 | 172962756 | 172963233 | 478 | 2 | 5.74E-07 | 1.52E-07 | 8.62E-05 | 2.54E-07 | -0.01282658 | GALNTL6 |
| chr11 | 2909092 | 2909935 | 844 | 4 | 5.89E-08 | 0.0002904 | 6.54E-05 | 2.54E-07 | -0.02198624 | SLC22A18 |
| chr19 | 49616075 | 49616743 | 669 | 4 | 2.11E-12 | 0.0005631 | 7.61E-08 | 2.54E-07 | -0.01848578 | PRR12 |
| chr6 | 29603297 | 29604602 | 1306 | 16 | 5.05E-07 | 2.65E-05 | 0.001183 | 2.54E-07 | -0.00110429 | GABBR1 |
| chr4 | 47913684 | 47914623 | 940 | 10 | 1.37E-10 | 6.76E-05 | 1.55E-05 | 2.55E-07 | 0.001087509 | NIPAL1, NFXL1 |
| chr5 | 60487802 | 60488717 | 916 | 9 | 3.28E-07 | 2.54E-07 | 0.000326 | 2.55E-07 | -0.00072358 | AC109486.2, PART1, AC109486.3, PDE4D |
| chr13 | 97432837 | 97433695 | 859 | 5 | 8.44E-07 | 2.33E-07 | 0.000443 | 2.55E-07 | 0.000366114 | AL442067.3 |
| chr17 | 75667054 | 75667817 | 764 | 12 | 1.06E-06 | 1.99E-07 | 0.011238 | 2.55E-07 | 0.000703536 | SAP30BP, RECQL5 |
| chr2 | 127664539 | 127665407 | 869 | 12 | 4.24E-07 | 2.89E-05 | 0.000122 | 2.55E-07 | -0.00497484 | LIMS2 |
| chr18 | 54948736 | 54948779 | 44 | 2 | 1.36E-07 | 1.99E-07 | 2.89E-05 | 2.55E-07 | -0.02886061 | CCDC68 |
| chr1 | 51877728 | 51879306 | 1579 | 24 | 1.63E-06 | 2.79E-07 | 0.000541 | 2.55E-07 | 0.000239198 | NRDC |
| chr6 | 73393504 | 73394678 | 1175 | 10 | 2.17E-08 | 2.50E-06 | 0.000693 | 2.55E-07 | 0.001597725 | OOEP |
| chr10 | 132817093 | 132818198 | 1106 | 8 | 2.16E-07 | 5.93E-07 | 0.000361 | 2.56E-07 | 0.000428152 | CFAP46 |
| chr12 | 6540183 | 6540828 | 646 | 5 | 6.80E-08 | 4.39E-07 | 0.000117 | 2.56E-07 | -0.00783011 | IFFO1 |
| chr19 | 12835651 | 12836721 | 1071 | 7 | 3.34E-08 | 4.67E-07 | 0.002014 | 2.56E-07 | -0.00439198 | MAST1, HOOK2 |
| chr21 | 37005638 | 37006736 | 1099 | 8 | 7.99E-08 | 1.48E-05 | 0.001685 | 2.57E-07 | -0.00408415 | RIPPLY3 |
| chr18 | 34048518 | 34049281 | 764 | 4 | 2.16E-07 | 3.29E-06 | 0.000248 | 2.58E-07 | 0.03413814 | NOL4 |
| chr8 | 33805956 | 33806108 | 153 | 2 | 5.65E-08 | 1.09E-05 | 5.75E-07 | 2.58E-07 | -0.02187005 | AF279873.3 |
| chr17 | 1220518 | 1220897 | 380 | 4 | 2.82E-07 | 2.57E-07 | 6.96E-05 | 2.58E-07 | -0.00216274 | ABR |
| chr20 | 39225626 | 39226209 | 584 | 3 | 3.89E-09 | 1.52E-05 | 2.12E-06 | 2.58E-07 | -0.01287134 | NA |

| | | | | | | | | | | |
|---|---|---|---|---|---|---|---|---|---|---|
| chr3 | 12885236 | 12886452 | 1217 | 6 | 3.65E-12 | 8.74E-05 | 3.93E-07 | 2.58E-07 | 0.001534547 | NA |
| chr1 | 85276851 | 85278049 | 1199 | 10 | 1.34E-06 | 1.61E-07 | 0.005643 | 2.59E-07 | 0.002693919 | AL078459.1 |
| chr14 | 91292046 | 91292730 | 685 | 6 | 2.81E-09 | 5.47E-05 | 7.65E-06 | 2.59E-07 | -0.006531 | CCDC88C |
| chr8 | 22274386 | 22275563 | 1178 | 14 | 3.11E-08 | 6.14E-07 | 7.65E-05 | 2.59E-07 | -0.00697364 | PIWIL2, AC105206.2 |
| chr8 | 75484971 | 75485002 | 32 | 2 | 1.84E-07 | 4.15E-07 | 7.14E-06 | 2.59E-07 | -0.01386029 | HNF4G |
| chr7 | 158109513 | 158109716 | 204 | 2 | 4.75E-08 | 7.85E-06 | 6.71E-07 | 2.60E-07 | -0.01195805 | PTPRN2 |
| chr22 | 35551562 | 35552119 | 558 | 5 | 1.03E-07 | 1.14E-06 | 0.000176 | 2.60E-07 | -0.00320688 | RASD2 |
| chr10 | 86262987 | 86263486 | 500 | 6 | 9.47E-07 | 8.11E-08 | 0.002519 | 2.60E-07 | -0.0162954 | GRID1 |
| chr13 | 99971688 | 99972711 | 1024 | 10 | 2.71E-07 | 8.60E-07 | 0.000406 | 2.60E-07 | -0.00329348 | ZIC5 |
| chr12 | 52496428 | 52496908 | 481 | 2 | 7.08E-08 | 3.74E-07 | 8.40E-06 | 2.60E-07 | -0.02752436 | NA |
| chr13 | 20566083 | 20567625 | 1543 | 15 | 8.35E-09 | 0.0007557 | 4.00E-05 | 2.61E-07 | -0.00071685 | IFT88, AL590096.1 |
| chr2 | 159027749 | 159027970 | 222 | 3 | 8.95E-09 | 0.006997 | 9.62E-08 | 2.61E-07 | -0.00479293 | TANC1 |
| chr16 | 85213705 | 85214150 | 446 | 2 | 1.66E-07 | 2.15E-07 | 2.49E-05 | 2.61E-07 | 0.000132143 | GSE1 |
| chr1 | 151281483 | 151282774 | 1292 | 14 | 1.44E-06 | 1.13E-07 | 0.007097 | 2.61E-07 | 0.000921528 | ZNF687, ZNF687-AS1 |
| chr12 | 113576705 | 113577121 | 417 | 2 | 7.92E-09 | 1.34E-05 | 5.32E-07 | 2.61E-07 | 0.025445854 | NA |
| chr19 | 45440405 | 45441038 | 634 | 3 | 3.03E-08 | 4.18E-07 | 0.000103 | 2.61E-07 | -0.00768763 | ERCC1 |
| chr16 | 53703594 | 53704527 | 934 | 16 | 3.56E-08 | 2.91E-05 | 0.000109 | 2.61E-07 | -0.00014886 | FTO, RPGRIP1L |
| chr11 | 695322 | 696675 | 1354 | 18 | 3.61E-07 | 3.01E-06 | 0.002855 | 2.61E-07 | 0.000777248 | EPS8L2, TMEM80, DEAF1 |
| chr18 | 23871599 | 23872931 | 1333 | 10 | 6.21E-06 | 7.24E-07 | 0.002839 | 2.61E-07 | 0.00080011 | LAMA3 |
| chr17 | 13024261 | 13024657 | 397 | 3 | 4.33E-09 | 1.70E-05 | 5.00E-07 | 2.62E-07 | -0.00241842 | NA |
| chr1 | 44219710 | 44220350 | 641 | 3 | 1.40E-07 | 2.83E-07 | 2.48E-05 | 2.62E-07 | 0.000804767 | DMAP1 |
| chr1 | 53195150 | 53196374 | 1225 | 8 | 7.23E-07 | 4.67E-07 | 0.002656 | 2.62E-07 | -3.64E-05 | NA |
| chr21 | 31657376 | 31658467 | 1092 | 3 | 5.88E-06 | 3.12E-07 | 9.97E-05 | 2.62E-07 | 0.004764465 | AP000253.1 |
| chr12 | 8032080 | 8033335 | 1256 | 14 | 3.99E-07 | 8.30E-08 | 0.00378 | 2.63E-07 | 0.00068481 | FOXJ2 |
| chr11 | 20158790 | 20160517 | 1728 | 10 | 8.71E-07 | 5.01E-07 | 0.0002 | 2.63E-07 | 0.001348704 | DBX1 |
| chr10 | 123377004 | 123377231 | 228 | 2 | 1.43E-08 | 4.15E-05 | 3.77E-07 | 2.63E-07 | -0.01970253 | LINC02641 |
| chr19 | 30122649 | 30123492 | 844 | 2 | 1.24E-06 | 2.07E-07 | 2.86E-05 | 2.63E-07 | -0.0071373 | NA |
| chr1 | 64469486 | 64470174 | 689 | 5 | 2.11E-10 | 0.0003424 | 2.71E-06 | 2.63E-07 | -0.00732512 | CACHD1 |
| chr1 | 156193834 | 156194414 | 581 | 12 | 5.40E-08 | 2.63E-05 | 0.000711 | 2.63E-07 | -0.00015493 | SLC25A44 |
| chr21 | 34036097 | 34036444 | 348 | 2 | 3.64E-08 | 2.26E-06 | 1.41E-06 | 2.64E-07 | -0.00217761 | NA |
| chr22 | 39900403 | 39900763 | 361 | 2 | 2.35E-09 | 1.77E-06 | 1.69E-06 | 2.64E-07 | 0.016095691 | NA |
| chr10 | 132749265 | 132749646 | 382 | 3 | 7.22E-08 | 6.90E-07 | 5.18E-05 | 2.64E-07 | -0.00465687 | INPP5A |
| chr1 | 229753403 | 229753650 | 248 | 2 | 1.73E-07 | 4.10E-07 | 7.54E-06 | 2.64E-07 | -0.00438092 | NA |
| chr17 | 77477262 | 77477887 | 626 | 6 | 7.57E-09 | 0.0001415 | 3.27E-05 | 2.65E-07 | -0.00539559 | SEPTIN9 |
| chr17 | 76697803 | 76698156 | 354 | 4 | 2.18E-07 | 1.52E-06 | 0.000497 | 2.65E-07 | -0.00275312 | MXRA7 |
| chr2 | 27662565 | 27664021 | 1457 | 19 | 2.24E-07 | 1.12E-06 | 0.000511 | 2.65E-07 | 0.000183068 | SLC4A1AP, SUPT7L |
| chr3 | 120284445 | 120285420 | 976 | 6 | 2.94E-08 | 6.35E-06 | 0.00014 | 2.65E-07 | -0.00571753 | GPR156 |
| chr6 | 117547864 | 117548694 | 831 | 5 | 3.48E-07 | 2.65E-06 | 0.000108 | 2.65E-07 | 0.007963598 | DCBLD1, AL132671.2 |
| chr3 | 16173301 | 16175099 | 1799 | 11 | 2.40E-06 | 3.50E-06 | 0.000648 | 2.65E-07 | -0.00730238 | GALNT15 |
| chr10 | 111079225 | 111079688 | 464 | 3 | 1.79E-07 | 1.98E-07 | 6.95E-05 | 2.66E-07 | -0.00058214 | ADRA2A |
| chr6 | 157181109 | 157182105 | 997 | 2 | 1.22E-05 | 1.64E-07 | 7.25E-05 | 2.66E-07 | 0.001752461 | ARID1B |
| chr3 | 45510647 | 45511006 | 360 | 3 | 2.05E-07 | 9.58E-08 | 0.000498 | 2.67E-07 | -0.00161666 | LARS2 |
| chr18 | 34818060 | 34819023 | 964 | 7 | 1.58E-07 | 7.26E-06 | 0.000664 | 2.67E-07 | -0.00561368 | DTNA |
| chr9 | 95426500 | 95427099 | 600 | 4 | 2.75E-11 | 0.0017938 | 6.25E-08 | 2.67E-07 | 0.011827092 | FANCC |
| chr13 | 29595957 | 29597044 | 1088 | 7 | 1.24E-08 | 3.27E-06 | 6.01E-05 | 2.67E-07 | 0.002661351 | NA |
| chr4 | 83116109 | 83116183 | 75 | 2 | 5.91E-08 | 3.33E-06 | 1.10E-06 | 2.68E-07 | -0.0118833 | PLAC8 |
| chr19 | 58182083 | 58183403 | 1321 | 6 | 6.43E-07 | 1.15E-06 | 0.000106 | 2.68E-07 | -0.00853681 | ZNF274 |
| chr3 | 2156335 | 2156405 | 71 | 2 | 2.51E-07 | 4.86E-07 | 6.15E-06 | 2.68E-07 | -0.02562228 | CNTN4 |
| chr16 | 85395916 | 85396489 | 574 | 3 | 2.57E-09 | 7.55E-06 | 4.14E-05 | 2.69E-07 | -0.0099189 | GSE1 |
| chr2 | 238319895 | 238321087 | 1193 | 13 | 1.27E-07 | 2.75E-05 | 0.000151 | 2.69E-07 | 0.001980019 | TRAF3IP1 |
| chr20 | 2328061 | 2329277 | 1217 | 6 | 1.97E-07 | 1.25E-05 | 8.16E-05 | 2.69E-07 | -0.00286947 | AL121899.2, TGM3 |
| chr9 | 92659349 | 92660470 | 1122 | 4 | 3.41E-06 | 7.74E-08 | 0.001007 | 2.69E-07 | -0.00194153 | IPPK |
| chr16 | 72959338 | 72960699 | 1362 | 6 | 2.24E-08 | 1.06E-06 | 0.000329 | 2.69E-07 | -0.00333587 | ZFHX3 |
| chr4 | 587866 | 588845 | 980 | 6 | 1.90E-07 | 6.58E-07 | 0.000632 | 2.69E-07 | -0.01236366 | AC116565.1 |
| chr4 | 110628724 | 110629510 | 787 | 6 | 4.47E-08 | 4.17E-06 | 5.46E-05 | 2.69E-07 | -0.0043576 | PITX2 |
| chr1 | 53945344 | 53946562 | 1219 | 13 | 2.44E-07 | 5.84E-05 | 5.16E-05 | 2.69E-07 | 0.001135343 | LRRC42, HSPB11 |
| chr3 | 52445082 | 52446237 | 1156 | 8 | 7.98E-08 | 1.07E-05 | 0.000471 | 2.69E-07 | 0.001276751 | SEMA3G |
| chr22 | 18109789 | 18111469 | 1681 | 13 | 1.25E-08 | 1.81E-06 | 5.84E-05 | 2.69E-07 | 0.000668154 | PEX26, TUBA8, AC008079.1 |
| chr3 | 179251607 | 179252148 | 542 | 6 | 2.83E-07 | 5.23E-07 | 3.54E-05 | 2.69E-07 | -0.00052345 | KCNMB3 |
| chr1 | 17192847 | 17193721 | 875 | 3 | 4.59E-09 | 2.94E-06 | 1.99E-06 | 2.69E-07 | -0.00080017 | LINC02783, AL590644.2 |
| chr12 | 55743999 | 55744569 | 571 | 2 | 2.05E-07 | 1.64E-07 | 8.22E-05 | 2.70E-07 | -0.00519039 | GDF11 |
| chr9 | 89448622 | 89448694 | 73 | 3 | 5.85E-07 | 1.25E-07 | 0.00027 | 2.70E-07 | 0.006023135 | SEMA4D |
| chr1 | 43358985 | 43360233 | 1249 | 5 | 4.63E-11 | 3.64E-06 | 1.73E-05 | 2.71E-07 | -0.00735737 | CDC20 |
| chr4 | 42152823 | 42154165 | 1343 | 10 | 1.41E-09 | 0.0001366 | 4.50E-05 | 2.71E-07 | 0.001486689 | AC111006.1, BEND4 |
| chr14 | 105087877 | 105088745 | 869 | 5 | 3.34E-07 | 7.34E-08 | 0.001549 | 2.71E-07 | -0.01579615 | NA |
| chr20 | 2535856 | 2536967 | 1112 | 10 | 4.93E-08 | 2.13E-05 | 5.95E-05 | 2.71E-07 | -0.00590647 | TMC2 |
| chr15 | 80164851 | 80164901 | 51 | 3 | 1.33E-07 | 1.77E-05 | 4.82E-06 | 2.71E-07 | 0.00924575 | FAH |
| chr2 | 176131557 | 176132979 | 1423 | 4 | 1.26E-05 | 3.93E-07 | 0.000216 | 2.71E-07 | 0.005595053 | HOXD8, HOXD-AS2 |
| chr9 | 128454824 | 128456265 | 1442 | 16 | 3.00E-07 | 1.07E-05 | 0.004907 | 2.71E-07 | -8.60E-05 | ODF2 |
| chr11 | 65625827 | 65626753 | 927 | 5 | 1.29E-08 | 7.81E-05 | 1.60E-06 | 2.71E-07 | -0.00440963 | PCNX3 |
| chr12 | 131785018 | 131786284 | 1267 | 10 | 4.96E-07 | 3.99E-05 | 2.75E-05 | 2.71E-07 | -0.00100925 | SFSWAP |
| chr19 | 18155399 | 18156078 | 680 | 4 | 6.64E-09 | 1.51E-06 | 7.22E-05 | 2.72E-07 | -0.00306845 | AC007192.1, PIK3R2 |
| chr9 | 134049341 | 134049379 | 39 | 2 | 4.61E-08 | 1.92E-07 | 4.06E-05 | 2.72E-07 | -0.00988108 | BRD3 |
| chr22 | 50072953 | 50073357 | 405 | 4 | 2.54E-07 | 1.01E-07 | 0.00031 | 2.72E-07 | -0.00413506 | MLC1 |
| chr18 | 50267861 | 50268625 | 765 | 4 | 1.79E-08 | 9.76E-07 | 5.88E-05 | 2.72E-07 | -0.01199093 | MBD1 |
| chr1 | 6519524 | 6520465 | 942 | 9 | 2.59E-07 | 1.49E-06 | 0.002088 | 2.72E-07 | -0.00545813 | PLEKHG5 |
| chr6 | 168887170 | 168887192 | 23 | 2 | 5.60E-08 | 1.71E-07 | 6.79E-05 | 2.73E-07 | 0.001296539 | NA |
| chr19 | 37550254 | 37551883 | 1630 | 13 | 2.93E-06 | 9.46E-06 | 0.000473 | 2.73E-07 | 0.000289549 | ZNF571-AS1, ZNF540 |
| chr17 | 20983539 | 20983907 | 369 | 2 | 1.20E-07 | 4.17E-07 | 7.84E-06 | 2.73E-07 | -0.00634598 | CCDC144NL-AS1, AC087393.1 |
| chr6 | 158663338 | 158664066 | 729 | 3 | 1.99E-07 | 1.27E-07 | 0.000244 | 2.73E-07 | 0.016827473 | SYTL3 |
| chr15 | 77785314 | 77787029 | 1716 | 8 | 1.41E-06 | 5.74E-05 | 0.000299 | 2.73E-07 | -0.00716824 | LINGO1 |
| chr5 | 126679983 | 126680461 | 479 | 2 | 6.10E-09 | 1.21E-05 | 5.84E-07 | 2.73E-07 | -0.00650331 | NA |
| chr3 | 50550445 | 50550611 | 167 | 2 | 1.48E-08 | 1.18E-06 | 2.46E-06 | 2.74E-07 | -0.01922586 | NA |
| chr2 | 96869587 | 96870550 | 964 | 9 | 3.54E-08 | 3.19E-05 | 0.000965 | 2.74E-07 | 0.000707438 | SEMA4C |
| chr5 | 112204206 | 112204434 | 229 | 4 | 1.98E-07 | 6.49E-06 | 3.48E-05 | 2.74E-07 | 0.002436732 | AC010261.2, EPB41L4A |
| chr8 | 102388713 | 102389345 | 633 | 5 | 7.37E-07 | 1.28E-07 | 0.000345 | 2.74E-07 | -0.00262166 | UBR5 |
| chr6 | 34465362 | 34466548 | 1187 | 12 | 8.34E-08 | 1.27E-06 | 0.000149 | 2.74E-07 | -0.00129396 | PACSIN1 |
| chr11 | 124424882 | 124425804 | 923 | 6 | 1.23E-07 | 8.58E-06 | 3.95E-05 | 2.74E-07 | -0.00391734 | OR8B4 |
| chr5 | 1193388 | 1194011 | 624 | 6 | 6.86E-08 | 1.31E-05 | 2.84E-05 | 2.74E-07 | -0.00275182 | NA |
| chr2 | 64768411 | 64769007 | 597 | 3 | 6.89E-10 | 9.14E-06 | 8.11E-07 | 2.74E-07 | 0.003341118 | NA |
| chr1 | 155077624 | 155078680 | 1057 | 6 | 4.25E-09 | 5.99E-05 | 5.48E-06 | 2.74E-07 | -0.00266797 | AL691442.1 |
| chr3 | 44554686 | 44555999 | 1314 | 14 | 1.82E-07 | 4.99E-07 | 0.001695 | 2.74E-07 | -0.0024032 | ZKSCAN7 |
| chr9 | 135238836 | 135239682 | 847 | 4 | 4.16E-07 | 8.89E-07 | 3.77E-05 | 2.74E-07 | -0.00480743 | NA |
| chr15 | 78601660 | 78602297 | 638 | 8 | 2.78E-07 | 9.93E-07 | 0.000511 | 2.75E-07 | -0.00116039 | CHRNA3 |
| chr16 | 87951420 | 87952721 | 1302 | 5 | 1.56E-11 | 1.46E-05 | 3.13E-06 | 2.75E-07 | 0.002214274 | BANP |
| chr10 | 4383395 | 4384407 | 1013 | 8 | 1.89E-07 | 3.12E-06 | 0.000343 | 2.75E-07 | 0.000845843 | LINC00703 |
| chr2 | 12106525 | 12106578 | 54 | 2 | 1.85E-08 | 7.99E-07 | 3.61E-06 | 2.75E-07 | -0.01749346 | MIR3681HG |
| chr14 | 39431875 | 39432800 | 926 | 13 | 4.02E-08 | 3.33E-05 | 9.27E-05 | 2.75E-07 | 0.000170024 | AL049828.1, FBXO33 |
| chr16 | 75239130 | 75239692 | 563 | 5 | 1.93E-08 | 1.55E-05 | 8.39E-06 | 2.75E-07 | 0.00161202 | BCAR1 |
| chr8 | 47888804 | 47889411 | 608 | 3 | 1.02E-07 | 6.65E-07 | 4.45E-05 | 2.75E-07 | -0.00036446 | PRKDC |
| chr3 | 113273723 | 113274986 | 1264 | 4 | 3.51E-08 | 3.65E-05 | 2.83E-06 | 2.75E-07 | -0.00692747 | BOC |
| chr1 | 154324795 | 154326067 | 1273 | 13 | 3.49E-07 | 4.28E-06 | 0.001153 | 2.76E-07 | 0.001404212 | AQP10, ATP8B2 |
| chr8 | 133190224 | 133191598 | 1375 | 8 | 1.46E-09 | 8.23E-05 | 1.60E-05 | 2.76E-07 | -0.00168141 | CCN4 |
| chr3 | 194351041 | 194351687 | 647 | 11 | 3.70E-11 | 0.0013988 | 5.34E-07 | 2.76E-07 | -0.00196278 | CPN2 |
| chr19 | 932759 | 933173 | 415 | 3 | 5.14E-08 | 1.60E-06 | 3.36E-06 | 2.76E-07 | 0.005741074 | ARID3A |
| chr3 | 170908693 | 170909773 | 1081 | 9 | 7.71E-08 | 5.49E-06 | 0.000159 | 2.76E-07 | -0.00544773 | AC061708.1 |
| chr13 | 114234012 | 114234693 | 682 | 6 | 3.71E-09 | 1.19E-06 | 0.000128 | 2.76E-07 | 0.003049481 | NA |
| chr3 | 64511002 | 64511666 | 665 | 3 | 2.02E-09 | 3.88E-06 | 5.07E-06 | 2.76E-07 | -0.00884067 | NA |
| chr14 | 23071912 | 23072677 | 766 | 5 | 5.45E-08 | 3.36E-06 | 1.96E-05 | 2.77E-07 | -0.00639368 | ACIN1 |
| chr17 | 2265978 | 2266695 | 718 | 9 | 3.28E-07 | 1.40E-07 | 0.002608 | 2.77E-07 | -0.00948373 | SMG6 |
| chr5 | 55230009 | 55230708 | 700 | 3 | 2.58E-08 | 3.52E-07 | 9.09E-05 | 2.77E-07 | 0.004896357 | NA |
| chr2 | 188291889 | 188293805 | 1917 | 19 | 5.64E-09 | 8.99E-05 | 0.001244 | 2.77E-07 | -0.00539431 | GULP1, LINC01090 |
| chr15 | 35545839 | 35546878 | 1040 | 11 | 6.67E-08 | 2.23E-05 | 0.000432 | 2.78E-07 | -4.45E-05 | DPH6-DT, DPH6 |
| chr1 | 109621925 | 109622619 | 695 | 5 | 1.00E-07 | 7.74E-06 | 0.00052 | 2.78E-07 | 0.00829356 | AMPD2 |
| chr15 | 38695109 | 38695309 | 201 | 2 | 5.92E-09 | 7.89E-05 | 3.44E-07 | 2.78E-07 | -0.00799755 | LINC02694 |
| chr11 | 6728197 | 6728453 | 257 | 3 | 8.14E-08 | 3.78E-05 | 1.57E-05 | 2.78E-07 | -0.0203622 | GVINP1 |
| chr1 | 9718755 | 9719274 | 520 | 5 | 5.78E-09 | 8.86E-06 | 0.0002 | 2.78E-07 | -0.00841646 | PIK3CD |
| chr6 | 166908178 | 166908180 | 3 | 2 | 8.39E-08 | 1.34E-05 | 5.74E-07 | 2.78E-07 | -0.00666961 | AL159163.1 |
| chr8 | 60713626 | 60714273 | 648 | 5 | 1.78E-07 | 1.28E-07 | 0.00016 | 2.79E-07 | -0.01337334 | CHD7 |
| chr1 | 118989002 | 118990150 | 1149 | 16 | 7.36E-09 | 1.17E-05 | 0.000437 | 2.79E-07 | 0.000491639 | TBX15 |
| chr17 | 79085060 | 79085678 | 619 | 3 | 1.42E-08 | 0.0021298 | 3.25E-05 | 2.79E-07 | -0.00568697 | ENGASE |
| chr16 | 10580600 | 10581173 | 574 | 7 | 3.74E-08 | 7.09E-06 | 0.000118 | 2.79E-07 | -0.00528911 | EMP2 |
| chr15 | 48877568 | 48878503 | 936 | 15 | 1.11E-07 | 1.32E-06 | 0.001568 | 2.79E-07 | -0.0012497 | EID1, SHC4 |
| chr16 | 66477380 | 66478041 | 662 | 4 | 6.83E-08 | 1.92E-06 | 8.32E-05 | 2.79E-07 | -0.00780278 | BEAN1, BEAN1-AS1, AC010542.3 |
| chr8 | 134837849 | 134838678 | 830 | 5 | 3.67E-07 | 6.95E-07 | 0.000123 | 2.80E-07 | -0.00026253 | NCRNA00250 |
| chr22 | 19177873 | 19179045 | 1173 | 10 | 2.33E-07 | 1.72E-06 | 0.000387 | 2.80E-07 | -0.00087463 | SLC25A1 |
| chr6 | 10434110 | 10435219 | 1110 | 9 | 2.70E-06 | 3.30E-08 | 0.004671 | 2.80E-07 | -0.00171258 | MIR5689HG, LINC00518 |
| chr9 | 37484890 | 37486472 | 1583 | 11 | 4.04E-06 | 1.33E-05 | 0.000185 | 2.80E-07 | 0.00030896 | POLR1E |
| chr2 | 84870346 | 84870520 | 175 | 3 | 2.53E-07 | 9.01E-07 | 8.99E-06 | 2.81E-07 | 0.004444592 | TRABD2A |
| chr18 | 12396282 | 12396937 | 656 | 2 | 5.29E-07 | 2.09E-07 | 3.50E-05 | 2.81E-07 | 0.005746136 | NA |
| chr1 | 154807826 | 154809284 | 1459 | 5 | 4.65E-06 | 5.62E-07 | 0.000212 | 2.82E-07 | -0.00597941 | KCNN3 |
| chr7 | 148804740 | 148805359 | 620 | 3 | 4.78E-07 | 1.12E-07 | 0.000432 | 2.83E-07 | -0.00453897 | NA |
| chr12 | 10111593 | 10111611 | 19 | 2 | 4.36E-07 | 1.76E-07 | 7.29E-05 | 2.83E-07 | -0.00533387 | CLEC1A |
| chr17 | 78644828 | 78645630 | 803 | 9 | 6.16E-09 | 9.34E-06 | 1.81E-05 | 2.83E-07 | -0.00110268 | NA |
| chr1 | 109483379 | 109484163 | 785 | 8 | 4.77E-08 | 4.86E-06 | 6.01E-05 | 2.83E-07 | 8.93E-05 | ATXN7L2 |
| chr22 | 27639099 | 27639934 | 836 | 6 | 1.62E-06 | 7.25E-07 | 0.00032 | 2.83E-07 | -0.00191892 | NA |
| chr19 | 18469533 | 18470254 | 722 | 3 | 5.33E-08 | 7.47E-07 | 1.45E-05 | 2.84E-07 | -0.00872805 | ELL |
| chr10 | 132920630 | 132922389 | 1760 | 6 | 2.31E-05 | 4.95E-06 | 4.43E-05 | 2.84E-07 | -3.20E-05 | CFAP46 |
| chr16 | 2766367 | 2767979 | 1613 | 9 | 6.63E-08 | 9.78E-06 | 0.00016 | 2.84E-07 | -0.0006855 | SRRM2 |
| chr11 | 72243938 | 72245101 | 1164 | 11 | 1.55E-09 | 2.34E-05 | 5.90E-05 | 2.84E-07 | 0.001068239 | PHOX2A |
| chr3 | 64291387 | 64291513 | 127 | 2 | 4.17E-07 | 1.84E-07 | 6.03E-05 | 2.84E-07 | -0.02306361 | PRICKLE2 |
| chr1 | 228360068 | 228361141 | 1074 | 7 | 9.11E-08 | 5.83E-06 | 0.000282 | 2.84E-07 | -0.00275184 | OBSCN, AL670729.3 |
| chr11 | 627968 | 629523 | 1556 | 5 | 1.25E-06 | 1.97E-06 | 0.000506 | 2.84E-07 | 0.009576852 | NA |
| chr2 | 161623337 | 161624127 | 791 | 4 | 1.06E-12 | 5.91E-05 | 9.84E-07 | 2.84E-07 | -0.00711717 | SLC4A10, AC062022.2 |
| chr21 | 36927802 | 36927974 | 173 | 2 | 1.25E-07 | 1.69E-06 | 1.93E-06 | 2.84E-07 | -0.01395761 | HLCS |
| chr14 | 24235311 | 24236194 | 884 | 3 | 5.73E-07 | 2.30E-07 | 5.14E-05 | 2.84E-07 | 0.002607914 | GMPR2 |
| chr2 | 24008152 | 24010471 | 2320 | 16 | 9.47E-06 | 0.0001248 | 0.000368 | 2.84E-07 | 0.00051622 | MFSD2B |
| chr11 | 114439210 | 114440483 | 1274 | 11 | 5.90E-08 | 1.71E-05 | 0.00033 | 2.84E-07 | -0.00122377 | AP002373.1, REXO2 |
| chr2 | 229879998 | 229880513 | 516 | 6 | 2.46E-07 | 4.48E-06 | 0.000181 | 2.84E-07 | 0.000351025 | TRIP12 |
| chr13 | 37004148 | 37004709 | 562 | 2 | 4.00E-07 | 2.05E-07 | 3.88E-05 | 2.84E-07 | -0.02007464 | EXOSC8 |
| chr10 | 14449575 | 14450027 | 453 | 2 | 3.62E-07 | 1.95E-07 | 4.68E-05 | 2.85E-07 | -0.00896415 | FRMD4A |
| chr11 | 45668904 | 45669409 | 506 | 3 | 5.90E-07 | 1.16E-07 | 0.000407 | 2.85E-07 | -0.00623426 | NA |
| chr2 | 108987356 | 108987410 | 55 | 2 | 3.22E-07 | 3.40E-07 | 1.20E-05 | 2.85E-07 | -0.01197111 | EDAR |
| chr20 | 44449022 | 44449223 | 202 | 2 | 6.89E-09 | 1.64E-06 | 1.99E-06 | 2.85E-07 | -0.01015517 | LINC01620 |
| chr7 | 127587088 | 127588478 | 1391 | 10 | 2.99E-07 | 0.0002804 | 0.000172 | 2.85E-07 | -0.00170802 | ARF5, GCC1 |
| chr6 | 29886963 | 29887859 | 897 | 7 | 1.71E-07 | 4.32E-06 | 0.000408 | 2.85E-07 | -0.00741607 | HLA-H, AL645929.1 |
| chr1 | 114976244 | 114976603 | 360 | 3 | 6.37E-08 | 3.21E-06 | 1.90E-06 | 2.87E-07 | -0.01163137 | SYCP1 |
| chr11 | 125495912 | 125497216 | 1305 | 10 | 2.03E-08 | 9.45E-06 | 1.21E-05 | 2.88E-07 | -0.00479934 | AP000708.1, FEZ1 |

| | | | | | | | | | | |
|---|---|---|---|---|---|---|---|---|---|---|
| chr16 | 89167455 | 89168358 | 904 | 3 | 2.18E-07 | 2.09E-07 | 0.000151 | 2.88E-07 | 0.00573993 | LINC02138 |
| chr8 | 42377096 | 42378737 | 1642 | 12 | 9.35E-06 | 9.01E-07 | 0.002976 | 2.88E-07 | 0.003766162 | DKK4 |
| chr4 | 1718084 | 1718712 | 629 | 5 | 1.29E-07 | 3.71E-06 | 0.000228 | 2.88E-07 | -0.00766536 | TACC3, TMEM129 |
| chr13 | 113724499 | 113725197 | 699 | 19 | 3.58E-08 | 3.07E-06 | 0.003553 | 2.89E-07 | 0.001166625 | GRK1 |
| chr7 | 4730307 | 4731351 | 1045 | 6 | 5.60E-07 | 3.80E-07 | 8.93E-05 | 2.89E-07 | 0.008364737 | FOXK1 |
| chr3 | 121660267 | 121661686 | 1420 | 9 | 7.47E-10 | 6.49E-05 | 9.18E-07 | 2.90E-07 | 0.000377669 | HCLS1 |
| chr14 | 22515721 | 22516134 | 414 | 3 | 6.79E-07 | 1.02E-07 | 0.000536 | 2.90E-07 | 0.005473461 | NA |
| chr13 | 96052869 | 96054409 | 1541 | 12 | 4.87E-09 | 0.0002025 | 0.000307 | 2.91E-07 | -0.00117269 | UGGT2 |
| chr19 | 49676377 | 49677846 | 1470 | 14 | 6.33E-07 | 5.25E-05 | 0.000752 | 2.91E-07 | -4.66E-05 | PRMT1 |
| chr11 | 424617 | 425289 | 673 | 3 | 3.51E-08 | 2.42E-06 | 8.45E-06 | 2.91E-07 | -0.00052645 | ANO9 |
| chr10 | 44347817 | 44347927 | 111 | 2 | 4.58E-07 | 1.74E-07 | 9.74E-05 | 2.91E-07 | -0.00018488 | NA |
| chr1 | 47532965 | 47533844 | 880 | 6 | 1.61E-06 | 6.29E-08 | 0.001854 | 2.92E-07 | -0.00379362 | NA |
| chr7 | 36441231 | 36441512 | 282 | 2 | 1.10E-07 | 9.22E-07 | 3.42E-06 | 2.92E-07 | 0.011224037 | ANLN |
| chr2 | 10029104 | 10029548 | 445 | 2 | 2.47E-08 | 2.25E-06 | 1.63E-06 | 2.92E-07 | 0.008728382 | NA |
| chr1 | 38858950 | 38860507 | 1558 | 15 | 2.27E-07 | 1.70E-05 | 3.82E-05 | 2.92E-07 | 4.85E-05 | AL139260.1, RRAGC, AL139260.2 |
| chr22 | 40407124 | 40407325 | 202 | 2 | 2.03E-08 | 2.32E-07 | 2.93E-05 | 2.93E-07 | -0.01021375 | AL022238.3, SGSM3 |
| chr11 | 61752215 | 61753012 | 798 | 7 | 4.28E-08 | 3.58E-05 | 2.13E-05 | 2.93E-07 | -0.00092509 | MYRF, MYRF-AS1 |
| chr1 | 1303056 | 1303879 | 824 | 6 | 2.17E-08 | 1.78E-05 | 4.98E-05 | 2.93E-07 | 0.002990766 | ACAP3 |
| chr17 | 79379261 | 79379760 | 500 | 6 | 2.14E-07 | 1.29E-05 | 8.81E-05 | 2.93E-07 | -0.0032306 | RBFOX3 |
| chr15 | 52600619 | 52601178 | 560 | 2 | 8.62E-08 | 4.27E-07 | 8.74E-06 | 2.93E-07 | -0.02450579 | FAM214A |
| chr13 | 39654895 | 39656080 | 1186 | 9 | 1.02E-07 | 3.48E-05 | 2.70E-06 | 2.93E-07 | 0.000425821 | COG6 |
| chr1 | 163259455 | 163259732 | 278 | 2 | 5.18E-08 | 2.06E-06 | 1.74E-06 | 2.93E-07 | -0.02453767 | RGS5, RGS5 |
| chr15 | 79432446 | 79433300 | 855 | 9 | 1.50E-08 | 2.88E-06 | 0.000304 | 2.94E-07 | -0.0023327 | MINAR1 |
| chr14 | 103737826 | 103738962 | 1137 | 6 | 1.65E-08 | 2.62E-06 | 2.74E-05 | 2.94E-07 | 0.000709302 | PPP1R13B |
| chr2 | 55047842 | 55048244 | 403 | 2 | 1.91E-09 | 1.82E-07 | 7.72E-05 | 2.94E-07 | 0.015702253 | RTN4 |
| chr18 | 62324924 | 62325751 | 828 | 7 | 4.33E-07 | 6.78E-07 | 0.001831 | 2.94E-07 | -0.00205042 | TNFRSF11A |
| chr4 | 158818927 | 158819665 | 739 | 2 | 1.04E-06 | 1.83E-07 | 7.56E-05 | 2.94E-07 | -0.00672725 | FNIP2 |
| chr10 | 88064934 | 88065279 | 346 | 2 | 4.84E-09 | 2.59E-05 | 4.79E-07 | 2.94E-07 | -0.00240936 | NA |
| chr4 | 118941837 | 118942096 | 260 | 2 | 3.87E-08 | 1.12E-05 | 6.56E-07 | 2.94E-07 | -0.01921108 | SYNPO2 |
| chr13 | 112696165 | 112697356 | 1192 | 8 | 4.07E-10 | 8.95E-05 | 9.55E-06 | 2.94E-07 | -0.00130023 | ATP11A |
| chr19 | 48452683 | 48453632 | 950 | 6 | 8.80E-08 | 6.91E-06 | 0.000222 | 2.94E-07 | -0.00209616 | GRWD1 |
| chr1 | 158330943 | 158331248 | 306 | 3 | 2.77E-08 | 3.26E-06 | 1.96E-06 | 2.95E-07 | -0.01060935 | CD1B |
| chr3 | 119240332 | 119241138 | 807 | 11 | 2.07E-07 | 2.07E-06 | 0.000191 | 2.95E-07 | -0.00199468 | B4GALT4-AS1, B4GALT4 |
| chr11 | 75716517 | 75717865 | 1349 | 6 | 1.53E-09 | 1.98E-06 | 2.38E-05 | 2.96E-07 | -0.00516885 | MOGAT2 |
| chr1 | 66445510 | 66445986 | 477 | 2 | 2.84E-07 | 2.00E-07 | 5.05E-05 | 2.96E-07 | 0.011688488 | NA |
| chr8 | 80197655 | 80198065 | 411 | 2 | 2.67E-09 | 0.0001523 | 3.31E-07 | 2.96E-07 | -0.00445791 | TPD52 |
| chr11 | 45771059 | 45771972 | 914 | 8 | 6.03E-07 | 2.08E-07 | 0.003399 | 2.96E-07 | -0.00074059 | LINC02716 |
| chr15 | 74995040 | 74995948 | 909 | 9 | 1.09E-07 | 3.79E-06 | 0.000568 | 2.96E-07 | -0.00497569 | SCAMP5 |
| chr15 | 24954966 | 24956077 | 1112 | 9 | 5.15E-10 | 0.0003687 | 1.14E-06 | 2.96E-07 | -0.00604401 | SNRPN, SNURF, AC124312.1 |
| chr9 | 135573893 | 135574854 | 962 | 5 | 7.64E-07 | 1.91E-07 | 0.000651 | 2.97E-07 | -0.00699325 | NA |
| chr20 | 38633620 | 38634513 | 894 | 4 | 1.84E-09 | 0.0001286 | 1.89E-07 | 2.97E-07 | -0.00580442 | ARHGAP40 |
| chr1 | 223801971 | 223802828 | 858 | 6 | 3.11E-06 | 1.95E-07 | 0.000243 | 2.97E-07 | -0.00313089 | TP53BP2 |
| chr11 | 42239317 | 42240401 | 1085 | 4 | 4.84E-08 | 3.51E-07 | 5.71E-05 | 2.97E-07 | -0.00123683 | LINC02740 |
| chr9 | 72364587 | 72365824 | 1238 | 10 | 2.12E-06 | 6.73E-07 | 6.37E-05 | 2.98E-07 | -0.00176458 | ZFAND5 |
| chr10 | 112958385 | 112958657 | 273 | 2 | 6.94E-08 | 1.82E-07 | 8.63E-05 | 2.98E-07 | 0.005170202 | TCF7L2 |
| chr16 | 30122747 | 30123561 | 815 | 9 | 2.35E-07 | 1.73E-06 | 0.001445 | 2.98E-07 | -0.00072868 | MAPK3 |
| chr10 | 28559042 | 28559730 | 689 | 3 | 7.57E-08 | 5.24E-07 | 1.42E-05 | 2.99E-07 | -0.01318531 | WAC |
| chr8 | 38120954 | 38121325 | 372 | 3 | 2.45E-07 | 1.13E-07 | 0.00036 | 2.99E-07 | -0.00565305 | ASH2L |
| chr8 | 18209822 | 18211195 | 1374 | 9 | 3.33E-08 | 4.98E-06 | 0.001339 | 2.99E-07 | -0.00240243 | NAT1 |
| chr17 | 16653245 | 16654639 | 1395 | 14 | 9.20E-07 | 3.87E-07 | 0.008026 | 2.99E-07 | -0.00103212 | AC098850.2, ZNF624 |
| chr10 | 34792051 | 34793142 | 1092 | 4 | 1.08E-05 | 3.93E-07 | 0.000248 | 3.00E-07 | -0.00491522 | PARD3 |
| chr1 | 43974105 | 43975527 | 1423 | 16 | 1.43E-07 | 2.13E-06 | 0.000207 | 3.00E-07 | -8.72E-05 | ATP6V0B, AL357079.2 |
| chr5 | 180215786 | 180216824 | 1039 | 7 | 7.68E-09 | 0.0005901 | 3.65E-06 | 3.00E-07 | -0.00343374 | NA |
| chr14 | 78177944 | 78178627 | 684 | 4 | 3.04E-08 | 6.01E-05 | 2.28E-05 | 3.00E-07 | -0.00642636 | NRXN3 |
| chr2 | 219029836 | 219030695 | 860 | 5 | 5.05E-08 | 1.40E-05 | 4.49E-05 | 3.00E-07 | -0.0027549 | CFAP65 |
| chr1 | 2071759 | 2073529 | 1771 | 12 | 2.71E-08 | 9.38E-06 | 0.001062 | 3.00E-07 | -0.00314945 | PRKCZ |
| chr13 | 44268334 | 44268783 | 450 | 2 | 5.23E-07 | 1.87E-07 | 7.68E-05 | 3.00E-07 | -0.01069355 | NA |
| chr19 | 36972352 | 36973296 | 945 | 6 | 3.21E-09 | 1.33E-05 | 3.69E-05 | 3.01E-07 | -0.01359312 | ZNF568 |
| chr10 | 73784282 | 73786150 | 1869 | 14 | 8.49E-08 | 0.0019465 | 6.22E-05 | 3.01E-07 | -0.00118208 | ZSWIM8 |
| chr11 | 122116028 | 122116557 | 530 | 8 | 5.44E-08 | 0.0002688 | 0.000195 | 3.01E-07 | -0.00514433 | MIR100HG, BLID |
| chr5 | 624598 | 625589 | 992 | 6 | 1.44E-08 | 4.24E-06 | 2.22E-05 | 3.01E-07 | -0.00328747 | CEP72 |
| chr22 | 45003703 | 45004019 | 317 | 5 | 1.90E-07 | 1.19E-06 | 2.88E-05 | 3.01E-07 | -0.00086832 | PHF21B |
| chr4 | 113781671 | 113781941 | 271 | 2 | 1.55E-08 | 0.0003071 | 3.11E-07 | 3.02E-07 | -0.01912043 | NA |
| chr7 | 45887783 | 45888964 | 1182 | 10 | 1.37E-07 | 2.56E-06 | 1.12E-05 | 3.02E-07 | -0.00474756 | IGFBP1 |
| chr2 | 65227296 | 65228402 | 1107 | 12 | 4.17E-08 | 0.0001955 | 8.69E-05 | 3.02E-07 | 0.000700142 | ACTR2, AC007318.1 |
| chr8 | 84184445 | 84185194 | 750 | 9 | 1.93E-07 | 1.21E-06 | 0.001773 | 3.02E-07 | -0.00076174 | RALYL |
| chr17 | 81282273 | 81283417 | 1145 | 9 | 1.97E-07 | 7.37E-07 | 0.00015 | 3.02E-07 | -0.00033273 | SLC38A10 |
| chr15 | 85703526 | 85704264 | 739 | 4 | 1.98E-07 | 1.37E-07 | 0.000146 | 3.02E-07 | -0.01444433 | AKAP13 |
| chr1 | 168225588 | 168226595 | 1008 | 12 | 7.00E-07 | 7.18E-05 | 2.48E-05 | 3.02E-07 | 0.001467312 | SFT2D2 |
| chr11 | 17412721 | 17414155 | 1435 | 4 | 1.77E-07 | 1.45E-06 | 3.36E-05 | 3.02E-07 | -0.005305 | ABCC8 |
| chr2 | 112480799 | 112481796 | 998 | 7 | 4.17E-07 | 5.23E-07 | 0.000926 | 3.02E-07 | -0.00098043 | NA |
| chr14 | 75452422 | 75453495 | 1074 | 5 | 1.02E-05 | 6.39E-08 | 0.0022 | 3.03E-07 | -0.00842302 | JDP2 |
| chr2 | 46698614 | 46699871 | 1258 | 11 | 3.87E-07 | 3.57E-06 | 9.99E-05 | 3.03E-07 | 0.001398466 | LINC01118, SOCS5 |
| chr9 | 89096132 | 89096757 | 626 | 3 | 8.87E-07 | 1.41E-07 | 0.000331 | 3.03E-07 | -0.00608517 | SHC3, AL353150.1 |
| chr10 | 15096657 | 15098123 | 1467 | 19 | 4.11E-06 | 1.16E-06 | 0.008588 | 3.03E-07 | -0.00059433 | RPP38, RPP38-DT |
| chr7 | 34209101 | 34209379 | 279 | 4 | 3.75E-08 | 1.01E-05 | 4.28E-06 | 3.03E-07 | 0.008013493 | NA |
| chr1 | 206771689 | 206772821 | 1133 | 8 | 2.26E-07 | 7.37E-06 | 0.001729 | 3.04E-07 | 0.003034076 | IL19, IL10 |
| chr13 | 56910230 | 56910364 | 135 | 3 | 7.08E-07 | 1.15E-07 | 0.00039 | 3.04E-07 | -0.00148708 | NA |
| chr6 | 466893 | 467681 | 789 | 5 | 5.56E-09 | 5.57E-06 | 4.20E-06 | 3.05E-07 | -0.0019492 | AL512308.1 |
| chr15 | 67254253 | 67255737 | 1485 | 18 | 1.11E-07 | 6.68E-06 | 0.00011 | 3.05E-07 | 0.00185676 | IQCH, AAGAB |
| chr19 | 1854024 | 1854820 | 797 | 6 | 6.55E-07 | 2.15E-07 | 0.001705 | 3.05E-07 | -0.00427642 | KLF16 |
| chr19 | 58366489 | 58367692 | 1204 | 6 | 7.45E-07 | 1.36E-06 | 8.97E-05 | 3.05E-07 | -0.00625503 | AC012313.6, ZNF837 |
| chr15 | 84653931 | 84655346 | 1416 | 16 | 2.23E-06 | 4.47E-06 | 0.002115 | 3.05E-07 | -6.59E-05 | WDR73, NMB |
| chr17 | 42017229 | 42018604 | 1376 | 16 | 2.57E-07 | 9.69E-08 | 0.003149 | 3.06E-07 | 0.000572002 | NKIRAS2, DNAJC7 |
| chr9 | 128410133 | 128411280 | 1148 | 3 | 3.15E-07 | 3.01E-05 | 1.62E-05 | 3.06E-07 | -0.00039363 | NA |
| chr2 | 134448500 | 134449244 | 745 | 4 | 1.32E-07 | 4.16E-06 | 1.06E-05 | 3.06E-07 | -0.00485925 | MGAT5 |
| chr5 | 1515722 | 1517513 | 1792 | 4 | 1.22E-06 | 6.81E-07 | 8.26E-05 | 3.07E-07 | 0.008948442 | LPCAT1 |
| chr3 | 58121045 | 58121481 | 437 | 2 | 5.08E-09 | 1.53E-06 | 2.32E-06 | 3.07E-07 | -0.0046032 | FLNB |
| chr15 | 28099127 | 28100358 | 1232 | 12 | 4.19E-08 | 5.10E-06 | 0.000186 | 3.07E-07 | 0.000869988 | OCA2 |
| chr3 | 38164196 | 38165630 | 1435 | 13 | 1.98E-08 | 0.0083433 | 0.000185 | 3.07E-07 | 4.63E-05 | OXSR1 |
| chr2 | 143697870 | 143698470 | 601 | 2 | 1.55E-06 | 1.84E-07 | 9.99E-05 | 3.07E-07 | -0.00192954 | ARHGAP15, AC092652.1 |
| chr14 | 72643551 | 72644183 | 633 | 3 | 1.71E-08 | 1.15E-06 | 6.29E-06 | 3.07E-07 | -0.01142406 | DPF3 |
| chr7 | 43309453 | 43310318 | 866 | 3 | 1.50E-07 | 1.72E-07 | 0.000109 | 3.07E-07 | 0.003814172 | HECW1 |
| chr15 | 77821936 | 77822546 | 611 | 2 | 1.13E-07 | 3.44E-07 | 1.40E-05 | 3.07E-07 | 0.004581447 | NA |
| chr19 | 18257558 | 18258272 | 715 | 5 | 1.19E-07 | 1.29E-06 | 1.70E-05 | 3.07E-07 | -0.00425431 | IQCN |
| chr18 | 10524913 | 10525884 | 972 | 9 | 2.27E-06 | 2.69E-06 | 0.000408 | 3.07E-07 | -1.45E-05 | NA |
| chr11 | 118910441 | 118911376 | 936 | 10 | 2.87E-08 | 1.52E-06 | 0.000613 | 3.07E-07 | -0.00486774 | MIR4492, BCL9L |
| chr10 | 71294060 | 71294589 | 530 | 3 | 6.09E-08 | 3.11E-06 | 5.33E-05 | 3.07E-07 | -0.00992047 | UNC5B |
| chr14 | 69879084 | 69879760 | 677 | 12 | 2.21E-07 | 6.71E-07 | 0.003237 | 3.08E-07 | -0.00048373 | SMOC1 |
| chr19 | 1815426 | 1816628 | 1203 | 14 | 4.08E-06 | 1.25E-07 | 0.000582 | 3.08E-07 | -0.0016795 | AC012615.2, REXO1, MIR1909 |
| chr4 | 188333117 | 188333500 | 384 | 2 | 1.25E-07 | 2.26E-07 | 3.88E-05 | 3.09E-07 | -0.01505242 | NA |
| chr11 | 70116252 | 70116673 | 422 | 4 | 2.58E-07 | 6.96E-07 | 0.000361 | 3.10E-07 | -0.00872583 | ANO1 |
| chr7 | 141550028 | 141551833 | 1806 | 12 | 1.99E-06 | 2.01E-06 | 0.000353 | 3.11E-07 | 0.000713184 | AGK, AC073878.1 |
| chr8 | 691466 | 691497 | 32 | 2 | 1.48E-07 | 7.24E-07 | 4.94E-06 | 3.11E-07 | -0.02059956 | ERICH1 |
| chr17 | 1203259 | 1204166 | 908 | 6 | 1.20E-06 | 6.74E-07 | 7.78E-05 | 3.11E-07 | -0.01284605 | ABR |
| chr11 | 70181715 | 70182615 | 901 | 4 | 2.08E-06 | 7.89E-08 | 0.001366 | 3.12E-07 | -0.00412842 | ANO1 |
| chr1 | 23783746 | 23784807 | 1062 | 3 | 1.24E-09 | 1.53E-06 | 4.18E-06 | 3.12E-07 | -0.00378293 | PITHD1 |
| chr6 | 99567734 | 99568962 | 1229 | 13 | 1.77E-09 | 0.0005581 | 0.000106 | 3.12E-07 | 0.001019048 | TSTD3, CCNC |
| chr10 | 70232904 | 70234014 | 1111 | 12 | 4.20E-08 | 0.0005932 | 0.000142 | 3.12E-07 | -0.00105689 | PPA1 |
| chr1 | 182793415 | 182793970 | 556 | 3 | 1.48E-07 | 2.88E-07 | 7.44E-05 | 3.12E-07 | -6.39E-05 | NPL |
| chr2 | 95295265 | 95296538 | 1274 | 7 | 1.96E-07 | 2.16E-07 | 0.000683 | 3.12E-07 | -0.00157543 | NA |
| chr12 | 11161778 | 11162254 | 477 | 3 | 2.10E-09 | 0.0004801 | 1.54E-07 | 3.12E-07 | -0.00675958 | PRH1, AC018630.4, TAS2R14 |
| chr10 | 129402520 | 129402570 | 51 | 3 | 2.85E-07 | 1.13E-06 | 5.71E-06 | 3.12E-07 | -0.00291514 | NA |
| chr10 | 121310063 | 121310878 | 816 | 4 | 3.44E-07 | 3.66E-07 | 0.000222 | 3.12E-07 | 0.027155531 | NA |
| chr16 | 89089777 | 89090267 | 491 | 2 | 9.86E-09 | 2.15E-06 | 1.83E-06 | 3.13E-07 | -0.0216045 | ACSF3 |
| chr2 | 236517728 | 236517741 | 14 | 2 | 4.84E-07 | 2.05E-07 | 6.12E-05 | 3.14E-07 | -0.01564472 | NA |
| chr1 | 977689 | 978084 | 396 | 2 | 4.03E-09 | 2.77E-06 | 1.54E-06 | 3.14E-07 | 0.021919796 | PERM1 |
| chr12 | 2845282 | 2846128 | 847 | 3 | 8.45E-08 | 1.09E-06 | 8.96E-06 | 3.14E-07 | -0.00676657 | ITFG2 |
| chr3 | 105882523 | 105882990 | 468 | 2 | 6.23E-07 | 1.93E-07 | 8.44E-05 | 3.14E-07 | -0.05989434 | NA |
| chr8 | 43017321 | 43017785 | 465 | 3 | 4.04E-08 | 5.25E-06 | 5.05E-06 | 3.15E-07 | 0.004534245 | HOOK3 |
| chr21 | 42351011 | 42351814 | 804 | 7 | 9.86E-09 | 3.34E-05 | 1.07E-05 | 3.15E-07 | -0.00340757 | NA |
| chr9 | 2241249 | 2242173 | 925 | 9 | 8.92E-08 | 3.38E-06 | 0.002866 | 3.15E-07 | -0.01522575 | NA |
| chr2 | 240896253 | 240897087 | 835 | 7 | 6.77E-08 | 4.18E-07 | 0.002121 | 3.15E-07 | -0.01450603 | MAB21L4 |
| chr3 | 49898904 | 49899860 | 957 | 6 | 3.12E-07 | 1.46E-07 | 0.00037 | 3.15E-07 | -0.00735304 | AC105935.2, MST1R |
| chr2 | 134917355 | 134919325 | 1971 | 11 | 5.69E-07 | 5.65E-06 | 0.000797 | 3.15E-07 | -0.00246877 | CCNT2, CCNT2-AS1 |
| chr5 | 141359934 | 141361040 | 1107 | 12 | 1.10E-06 | 2.20E-06 | 0.000143 | 3.16E-07 | -0.00652331 | PCDHGA1, PCDHGA2, PCDHGA3, PCDHGB1, PCDHGA4, PCDHGB2 |
| chr2 | 19356305 | 19357440 | 1136 | 7 | 3.79E-09 | 0.0001645 | 2.35E-06 | 3.16E-07 | -0.00095905 | OSR1 |
| chr21 | 43946702 | 43947301 | 600 | 6 | 2.48E-07 | 3.42E-06 | 0.000306 | 3.16E-07 | 0.000331185 | AGPAT3, AP001054.1 |
| chr3 | 4515753 | 4516676 | 924 | 6 | 3.50E-07 | 4.41E-06 | 0.000331 | 3.16E-07 | -0.0004442 | ITPR1 |
| chr19 | 48630164 | 48630848 | 685 | 6 | 4.00E-08 | 7.98E-06 | 5.83E-05 | 3.18E-07 | -0.00624341 | SPHK2, DBP |
| chr3 | 32105120 | 32105631 | 512 | 3 | 5.81E-09 | 1.13E-06 | 1.03E-05 | 3.18E-07 | 0.005563598 | NA |
| chr5 | 139131374 | 139131904 | 531 | 3 | 1.88E-08 | 1.23E-06 | 5.86E-06 | 3.18E-07 | 0.012044175 | SIL1 |
| chr2 | 201140043 | 201140479 | 437 | 5 | 2.13E-07 | 7.73E-06 | 0.000513 | 3.19E-07 | -0.00157048 | CFLAR, CFLAR-AS1 |
| chr11 | 26993926 | 26994444 | 519 | 8 | 1.63E-07 | 1.69E-07 | 0.002026 | 3.19E-07 | -0.01438303 | FIBIN |
| chr9 | 16575762 | 16576173 | 412 | 3 | 1.49E-07 | 3.42E-07 | 8.03E-05 | 3.19E-07 | -0.02042663 | BNC2 |
| chr11 | 122367925 | 122369430 | 1506 | 6 | 7.28E-05 | 3.28E-07 | 0.000823 | 3.19E-07 | -0.00010186 | MIR100HG |
| chr6 | 116047292 | 116047558 | 267 | 2 | 5.20E-07 | 1.92E-07 | 9.92E-05 | 3.19E-07 | -0.0096026 | FRK |
| chr16 | 28597855 | 28598210 | 356 | 2 | 5.54E-08 | 1.93E-06 | 2.04E-06 | 3.19E-07 | 0.026035103 | AC009093.11, AC020765.6 |
| chr22 | 41533399 | 41533703 | 305 | 3 | 1.96E-07 | 5.94E-07 | 1.25E-05 | 3.20E-07 | -0.01097651 | POLR3H |
| chr18 | 3261973 | 3263084 | 1112 | 13 | 3.68E-08 | 5.65E-05 | 0.000264 | 3.20E-07 | 0.001311359 | MYL12B |
| chr10 | 131956101 | 131957128 | 1028 | 6 | 7.60E-09 | 1.83E-05 | 8.39E-06 | 3.20E-07 | -0.00070345 | PPP2R2D |
| chr20 | 44465255 | 44466179 | 925 | 3 | 2.35E-06 | 1.36E-07 | 0.000389 | 3.20E-07 | -0.00834526 | LINC01620 |
| chr12 | 131008349 | 131009699 | 1351 | 6 | 9.24E-09 | 1.77E-05 | 4.26E-06 | 3.21E-07 | -0.00130924 | ADGRD1 |
| chr7 | 29208813 | 29209793 | 981 | 3 | 5.34E-06 | 1.35E-07 | 0.000259 | 3.21E-07 | 0.001039626 | AC004593.2, CHN2, CHN2-AS1 |
| chr10 | 126476471 | 126476590 | 120 | 2 | 2.61E-07 | 7.25E-07 | 5.19E-06 | 3.21E-07 | -0.02621502 | C10orf90 |
| chr17 | 28576854 | 28578062 | 1209 | 10 | 2.93E-06 | 3.19E-06 | 0.000344 | 3.21E-07 | -8.00E-05 | AC005726.1, ALDOC, SPAG5 |
| chr17 | 7264205 | 7264685 | 481 | 4 | 9.45E-09 | 8.46E-07 | 1.60E-05 | 3.21E-07 | -0.00775226 | NA |
| chr14 | 79278648 | 79280016 | 1369 | 15 | 7.02E-08 | 3.09E-05 | 0.000106 | 3.21E-07 | 0.001387951 | NRXN3 |
| chr7 | 149773206 | 149773731 | 526 | 7 | 8.25E-09 | 0.0003676 | 5.79E-06 | 3.21E-07 | 0.001998247 | ZNF467 |
| chr12 | 11648816 | 11650265 | 1450 | 11 | 3.07E-06 | 2.82E-06 | 0.000987 | 3.21E-07 | 0.000919281 | ETV6 |
| chr6 | 18307418 | 18308071 | 654 | 2 | 1.72E-09 | 4.96E-05 | 4.34E-07 | 3.22E-07 | -0.00341118 | NA |
| chr17 | 3534732 | 3535748 | 1017 | 6 | 1.01E-07 | 4.01E-07 | 0.00072 | 3.22E-07 | -0.0026572 | TRPV3 |
| chr7 | 48336711 | 48337108 | 398 | 2 | 3.34E-07 | 2.42E-07 | 3.61E-05 | 3.23E-07 | -0.00667475 | ABCA13 |
| chr16 | 49277572 | 49278632 | 1061 | 9 | 2.98E-07 | 5.08E-06 | 0.000352 | 3.23E-07 | 0.000780535 | CBLN1 |
| chr3 | 132035831 | 132036485 | 655 | 4 | 4.81E-08 | 2.28E-06 | 2.34E-05 | 3.23E-07 | 0.01332273 | CPNE4 |
| chr3 | 39382653 | 39384610 | 1958 | 14 | 1.69E-05 | 2.13E-05 | 0.001788 | 3.24E-07 | -0.00155178 | SLC25A38, AC104850.2 |
| chr4 | 186144182 | 186145351 | 1170 | 12 | 8.12E-07 | 4.70E-07 | 0.001265 | 3.24E-07 | 0.001471516 | FAM149A |
| chr7 | 986674 | 987397 | 724 | 4 | 5.04E-09 | 4.60E-07 | 0.00023 | 3.24E-07 | -0.01022847 | CYP2W1 |

| | | | | | | | | | | |
|---|---|---|---|---|---|---|---|---|---|---|
| chr17 | 56737763 | 56738053 | 291 | 3 | 4.82E-07 | 2.87E-07 | 8.48E-05 | 3.24E-07 | 0.016627275 | NA |
| chr6 | 151451763 | 151453125 | 1363 | 20 | 2.00E-06 | 2.61E-05 | 0.000651 | 3.25E-07 | 0.001608212 | ARMT1, RMND1 |
| chr1 | 161090210 | 161090832 | 623 | 4 | 2.42E-08 | 1.82E-06 | 8.04E-05 | 3.25E-07 | 0.001327485 | NA |
| chr11 | 55809936 | 55810299 | 364 | 2 | 1.65E-07 | 4.42E-07 | 1.03E-05 | 3.25E-07 | -0.02370875 | NA |
| chr17 | 1586729 | 1587263 | 535 | 3 | 7.21E-07 | 1.54E-07 | 0.00034 | 3.26E-07 | -0.01244627 | SLC43A2 |
| chr11 | 134031783 | 134032615 | 833 | 6 | 1.73E-06 | 5.74E-06 | 0.001516 | 3.26E-07 | -0.00992556 | LINC02731 |
| chr19 | 39124512 | 39126178 | 1667 | 13 | 1.02E-05 | 2.61E-06 | 0.000179 | 3.26E-07 | -0.00121969 | PAK4 |
| chr3 | 32950666 | 32951673 | 1008 | 5 | 2.51E-07 | 1.06E-07 | 0.0005 | 3.26E-07 | 0.002408138 | CCR4 |
| chr2 | 208039844 | 208041215 | 1372 | 3 | 2.42E-07 | 1.15E-05 | 1.12E-05 | 3.26E-07 | -0.00383424 | NA |
| chr1 | 11898269 | 11899255 | 987 | 5 | 4.90E-06 | 2.64E-07 | 0.000607 | 3.26E-07 | -0.0058933 | NA |
| chr13 | 46796783 | 46797852 | 1070 | 14 | 1.30E-07 | 1.29E-05 | 0.000825 | 3.27E-07 | -0.00249688 | ESD |
| chr4 | 153309316 | 153309923 | 608 | 2 | 1.96E-07 | 3.50E-07 | 1.55E-05 | 3.27E-07 | -0.006035 | TRIM2, AC013477.1 |
| chr1 | 16207042 | 16208084 | 1043 | 6 | 4.10E-08 | 1.53E-05 | 0.000825 | 3.27E-07 | -0.013615 | ARHGEF19 |
| chr16 | 1984024 | 1984531 | 508 | 12 | 2.85E-07 | 1.93E-06 | 0.001195 | 3.27E-07 | 0.001225941 | GFER, NOXO1 |
| chr12 | 13000703 | 13002228 | 1526 | 7 | 2.51E-07 | 5.48E-06 | 0.000385 | 3.27E-07 | 0.001065929 | HTR7P1, AC023790.2 |
| chr2 | 85067146 | 85068111 | 966 | 9 | 5.67E-08 | 5.54E-07 | 5.24E-05 | 3.27E-07 | -0.00082719 | LINC01964 |
| chr1 | 84077473 | 84078601 | 1129 | 12 | 6.10E-08 | 0.0005501 | 9.44E-05 | 3.27E-07 | 9.72E-05 | PRKACB, AL359504.1 |
| chr2 | 53859717 | 53860415 | 699 | 12 | 2.39E-07 | 9.57E-07 | 0.008526 | 3.28E-07 | -0.01220872 | ASB3, GPR75 |
| chr12 | 188665 | 189318 | 654 | 5 | 8.22E-07 | 1.48E-07 | 0.001611 | 3.28E-07 | -0.01527085 | NA |
| chr10 | 1401491 | 1403122 | 1632 | 9 | 1.62E-05 | 5.44E-06 | 0.001068 | 3.28E-07 | -0.00298807 | ADARB2 |
| chr9 | 135175506 | 135175834 | 329 | 2 | 3.70E-09 | 2.44E-06 | 1.76E-06 | 3.28E-07 | -0.0176263 | NA |
| chr10 | 116620884 | 116621862 | 979 | 4 | 1.52E-08 | 1.68E-06 | 1.40E-05 | 3.28E-07 | -0.00089527 | PNLIPRP2 |
| chr9 | 121566048 | 121567235 | 1188 | 8 | 1.64E-06 | 1.02E-05 | 0.001002 | 3.28E-07 | -0.00478713 | DAB2IP |
| chr13 | 30956698 | 30956946 | 249 | 3 | 9.73E-09 | 0.0012206 | 1.33E-07 | 3.28E-07 | 0.002869204 | TEX26 |
| chr11 | 1298884 | 1300300 | 1417 | 8 | 8.26E-09 | 3.04E-07 | 0.000125 | 3.28E-07 | -0.00312489 | TOLLIP |
| chr11 | 125464633 | 125465039 | 407 | 2 | 4.23E-09 | 6.76E-05 | 4.13E-07 | 3.28E-07 | -0.00547747 | FEZ1 |
| chr15 | 26890585 | 26892315 | 1731 | 21 | 7.86E-06 | 1.08E-06 | 0.005947 | 3.28E-07 | 0.002875478 | GABRA5, GABRB3 |
| chr13 | 75536971 | 75538096 | 1126 | 11 | 5.25E-08 | 1.87E-06 | 0.000121 | 3.29E-07 | 0.002154878 | COMMD6 |
| chr14 | 71319604 | 71320498 | 895 | 5 | 4.50E-08 | 2.92E-06 | 7.97E-05 | 3.29E-07 | 0.002933528 | SIPA1L1, AC005476.2 |
| chr10 | 23438018 | 23438643 | 626 | 2 | 4.11E-08 | 3.28E-07 | 1.79E-05 | 3.29E-07 | -0.00058495 | NA |
| chr19 | 33175627 | 33177002 | 1376 | 5 | 5.17E-07 | 4.36E-07 | 0.000742 | 3.29E-07 | 0.004721862 | WDR88 |
| chr6 | 108114371 | 108116480 | 2110 | 12 | 6.13E-07 | 3.42E-06 | 3.06E-05 | 3.29E-07 | 0.003855386 | OSTM1 |
| chr1 | 38498800 | 38499254 | 455 | 2 | 2.95E-07 | 2.68E-07 | 2.91E-05 | 3.29E-07 | 0.024101502 | LINC01685 |
| chr7 | 17559244 | 17560605 | 1362 | 4 | 2.77E-10 | 4.80E-05 | 3.09E-06 | 3.29E-07 | 0.008713625 | NA |
| chr11 | 795307 | 796434 | 1128 | 7 | 8.11E-07 | 2.62E-07 | 0.00195 | 3.31E-07 | 0.002492775 | SLC25A22 |
| chr4 | 113340937 | 113341763 | 827 | 6 | 1.44E-06 | 9.06E-08 | 0.00112 | 3.32E-07 | -0.00532358 | ANK2 |
| chr3 | 196639566 | 196640684 | 1119 | 12 | 7.26E-09 | 4.48E-05 | 2.08E-05 | 3.32E-07 | 0.001235843 | NRROS, PIGX |
| chr4 | 188031938 | 188032815 | 878 | 8 | 2.73E-06 | 8.86E-07 | 0.000345 | 3.32E-07 | 0.001523224 | NA |
| chr3 | 152268362 | 152269600 | 1239 | 14 | 6.13E-08 | 1.06E-06 | 0.000342 | 3.33E-07 | 0.000361009 | MBNL1, MBNL1-AS1 |
| chr11 | 6570836 | 6571515 | 680 | 4 | 8.48E-07 | 8.36E-08 | 0.001643 | 3.33E-07 | 0.051664947 | DNHD1 |
| chr15 | 31391461 | 31392296 | 836 | 4 | 2.55E-09 | 2.56E-05 | 9.35E-07 | 3.33E-07 | -0.00013294 | KLF13 |
| chr16 | 21598583 | 21599801 | 1219 | 18 | 2.16E-06 | 2.91E-07 | 0.012091 | 3.33E-07 | 0.002015247 | METTL9 |
| chr15 | 26969459 | 26971158 | 1700 | 17 | 9.12E-06 | 1.74E-06 | 0.002004 | 3.34E-07 | 0.003233534 | TVP23BP1 |
| chr4 | 153700656 | 153701331 | 676 | 2 | 2.58E-06 | 2.00E-07 | 0.000102 | 3.34E-07 | -0.0236145 | TLR2 |
| chr20 | 3174253 | 3174843 | 591 | 5 | 3.39E-08 | 3.16E-05 | 0.000349 | 3.34E-07 | -0.01056716 | NA |
| chr2 | 27226558 | 27227331 | 774 | 4 | 2.81E-08 | 1.98E-06 | 0.000138 | 3.34E-07 | -0.002336 | CAD |
| chr12 | 71839014 | 71839792 | 779 | 12 | 4.47E-07 | 2.40E-07 | 0.005525 | 3.35E-07 | -0.00201218 | TBC1D15 |
| chr1 | 81349189 | 81349715 | 527 | 2 | 3.18E-09 | 5.71E-05 | 4.34E-07 | 3.35E-07 | -0.01114609 | ADGRL2 |
| chr4 | 56504608 | 56505761 | 1154 | 9 | 1.15E-07 | 5.71E-07 | 0.002479 | 3.35E-07 | -0.00518929 | ARL9 |
| chr14 | 23579319 | 23580171 | 853 | 4 | 3.28E-06 | 2.56E-07 | 0.000145 | 3.35E-07 | -0.00382605 | NA |
| chr3 | 146314032 | 146314799 | 768 | 3 | 6.74E-07 | 1.62E-07 | 0.000157 | 3.35E-07 | -0.01284148 | NA |
| chr1 | 48237242 | 48238283 | 1042 | 4 | 5.15E-06 | 1.85E-07 | 0.000424 | 3.35E-07 | -0.00864256 | SLC5A9 |
| chr21 | 34813663 | 34813760 | 98 | 3 | 4.31E-07 | 2.92E-07 | 4.20E-05 | 3.36E-07 | -0.01121722 | RUNX1 |
| chr18 | 2168308 | 2168394 | 87 | 2 | 3.41E-07 | 5.29E-07 | 8.31E-06 | 3.36E-07 | 0.004586874 | AP005230.1 |
| chr18 | 35267464 | 35268052 | 589 | 5 | 9.36E-07 | 1.46E-07 | 0.000747 | 3.36E-07 | -0.00661137 | ZSCAN30 |
| chr19 | 58097902 | 58098764 | 863 | 13 | 2.85E-08 | 5.55E-05 | 0.00093 | 3.37E-07 | -0.00233065 | ZSCAN18 |
| chr1 | 245563825 | 245564356 | 532 | 6 | 3.98E-07 | 4.64E-07 | 0.001508 | 3.37E-07 | -0.00127834 | KIF26B |
| chr10 | 119542756 | 119543538 | 783 | 4 | 1.96E-07 | 6.98E-07 | 4.60E-05 | 3.38E-07 | 0.003918468 | NA |
| chr5 | 80711340 | 80712018 | 679 | 3 | 1.27E-08 | 6.44E-06 | 1.37E-06 | 3.40E-07 | 0.001849096 | MSH3 |
| chr10 | 24465567 | 24466430 | 864 | 8 | 2.67E-09 | 0.0001609 | 2.58E-05 | 3.40E-07 | -0.00286011 | KIAA1217 |
| chr1 | 223543805 | 223544407 | 603 | 2 | 1.43E-08 | 2.65E-06 | 1.74E-06 | 3.40E-07 | -0.00884668 | CAPN8 |
| chr22 | 30308487 | 30309079 | 593 | 3 | 8.44E-10 | 0.0001625 | 1.25E-06 | 3.40E-07 | 0.007743402 | TBC1D10A |
| chr8 | 143125416 | 143125844 | 429 | 4 | 3.40E-08 | 2.59E-06 | 4.65E-05 | 3.40E-07 | -0.00832301 | NA |
| chr3 | 134249851 | 134251316 | 1466 | 11 | 2.70E-07 | 0.0001113 | 0.000427 | 3.40E-07 | 0.001282752 | RYK |
| chr2 | 172427851 | 172429365 | 1515 | 11 | 1.24E-09 | 0.0024573 | 3.59E-06 | 3.40E-07 | -0.00204538 | ITGA6, AC078883.2 |
| chr18 | 24425803 | 24426662 | 860 | 12 | 1.14E-09 | 1.66E-05 | 0.00054 | 3.40E-07 | -0.00108876 | IMPACT |
| chr3 | 197790651 | 197791840 | 1190 | 13 | 1.44E-08 | 3.13E-05 | 1.05E-05 | 3.40E-07 | -0.00223321 | LRCH3 |
| chr7 | 30914907 | 30915267 | 361 | 2 | 4.14E-07 | 2.19E-07 | 6.82E-05 | 3.41E-07 | -0.00189726 | AC004691.2, AQP1 |
| chr12 | 127596837 | 127597148 | 312 | 2 | 7.77E-09 | 2.45E-05 | 5.62E-07 | 3.41E-07 | 0.00398714 | NA |
| chr11 | 68002227 | 68003195 | 969 | 3 | 1.18E-06 | 1.94E-06 | 5.58E-05 | 3.41E-07 | -0.00904418 | UNC93B1 |
| chr8 | 138497429 | 138498285 | 857 | 4 | 1.29E-07 | 2.64E-06 | 5.62E-05 | 3.41E-07 | -0.00874028 | NA |
| chr20 | 32535593 | 32536187 | 595 | 4 | 5.31E-10 | 8.98E-05 | 4.02E-06 | 3.42E-07 | 0.005216224 | NOL4L |
| chr2 | 101251894 | 101253137 | 1244 | 10 | 6.01E-07 | 6.87E-07 | 0.001371 | 3.42E-07 | -0.00198696 | CNOT11, TBC1D8 |
| chr17 | 43813658 | 43814869 | 1212 | 3 | 3.57E-09 | 9.47E-06 | 4.11E-06 | 3.43E-07 | 0.000942651 | MPP3 |
| chr11 | 26721856 | 26722937 | 1082 | 5 | 1.39E-07 | 2.44E-07 | 0.000174 | 3.43E-07 | -2.22E-05 | SLC5A12 |
| chr8 | 120444294 | 120445594 | 1301 | 15 | 8.64E-11 | 0.0008501 | 1.40E-05 | 3.43E-07 | 0.00125065 | MTBP, MRPL13 |
| chr17 | 43758569 | 43759680 | 1112 | 8 | 1.59E-08 | 9.77E-06 | 2.01E-05 | 3.43E-07 | -0.001659 | SOST |
| chr5 | 1282242 | 1282631 | 390 | 5 | 4.84E-08 | 7.68E-05 | 3.02E-06 | 3.43E-07 | -0.00152888 | TERT |
| chr1 | 1293610 | 1294766 | 1157 | 7 | 1.62E-06 | 6.10E-07 | 0.002708 | 3.43E-07 | -0.01010573 | ACAP3 |
| chr22 | 21713218 | 21714382 | 1165 | 3 | 2.48E-05 | 1.56E-07 | 0.000369 | 3.44E-07 | -0.01118243 | AP000553.9, YPEL1 |
| chr11 | 78046241 | 78047297 | 1057 | 7 | 1.51E-07 | 8.43E-06 | 1.44E-05 | 3.44E-07 | -0.00588302 | NDUFC2-KCTD14 |
| chr5 | 168844014 | 168845285 | 1272 | 9 | 1.17E-06 | 5.68E-05 | 7.80E-05 | 3.44E-07 | -0.00011436 | SLIT3 |
| chr19 | 43880118 | 43881222 | 1105 | 6 | 4.49E-07 | 7.87E-06 | 0.001384 | 3.44E-07 | -0.00526258 | ZNF404 |
| chr6 | 158850752 | 158851219 | 468 | 2 | 1.36E-07 | 2.58E-07 | 3.73E-05 | 3.44E-07 | -0.00927769 | OSTCP1 |
| chr12 | 67671630 | 67672017 | 388 | 2 | 1.84E-07 | 4.29E-07 | 1.21E-05 | 3.45E-07 | 0.005330017 | NA |
| chr7 | 98866662 | 98867591 | 930 | 5 | 9.28E-08 | 1.07E-05 | 1.05E-05 | 3.45E-07 | -0.00547774 | TMEM130 |
| chr7 | 113118447 | 113118899 | 453 | 7 | 1.84E-07 | 2.95E-06 | 0.000344 | 3.45E-07 | 0.000495692 | AC073346.1, SMIM30 |
| chr7 | 27179248 | 27180130 | 883 | 7 | 1.81E-07 | 3.15E-07 | 0.00148 | 3.45E-07 | 0.00656874 | AC004080.3, HOXA10 |
| chr22 | 49738155 | 49738675 | 521 | 5 | 4.51E-08 | 2.50E-05 | 6.71E-06 | 3.45E-07 | -0.00489216 | NA |
| chr20 | 15986677 | 15986775 | 99 | 2 | 3.26E-07 | 6.24E-07 | 7.02E-06 | 3.45E-07 | 0.012404066 | MACROD2 |
| chr15 | 44710399 | 44711634 | 1236 | 13 | 1.26E-09 | 0.0001764 | 0.000327 | 3.45E-07 | -0.0019023 | B2M, PATL2 |
| chr6 | 15256005 | 15256676 | 672 | 2 | 2.37E-08 | 1.67E-06 | 2.53E-06 | 3.45E-07 | 0.022080247 | JARID2 |
| chr15 | 72560891 | 72561390 | 500 | 2 | 1.54E-07 | 4.23E-07 | 1.25E-05 | 3.45E-07 | -0.01357894 | ARIH1 |
| chr16 | 89273880 | 89274177 | 298 | 2 | 1.74E-07 | 2.47E-07 | 4.41E-05 | 3.45E-07 | 0.006599189 | ANKRD11 |
| chr19 | 646591 | 647766 | 1176 | 6 | 4.27E-08 | 7.41E-06 | 0.000128 | 3.45E-07 | 1.35E-07 | RNF126 |
| chr5 | 149357534 | 149358461 | 928 | 13 | 4.42E-07 | 2.90E-06 | 0.002761 | 3.46E-07 | 0.000533142 | PCYOX1L, GRPEL2-AS1 |
| chr9 | 93568363 | 93568635 | 273 | 2 | 1.81E-07 | 2.08E-07 | 0.000102 | 3.46E-07 | -0.00922963 | NA |
| chr22 | 39310382 | 39310974 | 593 | 2 | 6.21E-09 | 1.31E-06 | 3.15E-06 | 3.47E-07 | -0.02042446 | NA |
| chr5 | 77138445 | 77139663 | 1219 | 7 | 3.30E-07 | 8.97E-06 | 3.28E-05 | 3.48E-07 | -0.00304567 | AC008581.2, ZBED3-AS1, AC022414.1 |
| chr17 | 75951486 | 75952021 | 536 | 2 | 8.81E-09 | 7.83E-06 | 9.35E-07 | 3.48E-07 | 0.001738956 | ACOX1 |
| chr12 | 866658 | 866861 | 204 | 3 | 1.90E-07 | 8.18E-07 | 1.09E-05 | 3.48E-07 | -0.01479626 | WNK1, AC004765.1 |
| chr10 | 98463209 | 98463297 | 89 | 2 | 8.00E-08 | 1.56E-05 | 6.82E-07 | 3.49E-07 | 0.004027526 | HPSE2 |
| chr11 | 76782335 | 76783898 | 1564 | 12 | 9.51E-07 | 8.42E-05 | 0.000152 | 3.49E-07 | -0.00112161 | TSKU, AP003119.2 |
| chr8 | 22052125 | 22052879 | 755 | 5 | 1.70E-08 | 4.33E-05 | 9.29E-06 | 3.49E-07 | -0.00507083 | DMTN |
| chr6 | 35605737 | 35606383 | 647 | 2 | 4.29E-07 | 2.94E-07 | 2.70E-05 | 3.50E-07 | -0.01051183 | FKBP5 |
| chr12 | 56354428 | 56354580 | 153 | 2 | 6.48E-08 | 1.90E-06 | 2.32E-06 | 3.50E-07 | -0.00301899 | STAT2 |
| chr7 | 53087273 | 53087528 | 256 | 3 | 4.22E-07 | 3.99E-07 | 4.82E-05 | 3.50E-07 | -0.00116254 | NA |
| chr2 | 98345642 | 98347305 | 1664 | 13 | 3.13E-09 | 0.0010195 | 1.44E-05 | 3.51E-07 | -0.00361643 | CNGA3, AC092675.2, AC092675.1 |
| chr6 | 168440577 | 168441622 | 1046 | 7 | 1.44E-07 | 1.05E-06 | 0.000494 | 3.51E-07 | 0.007163881 | SMOC2 |
| chr20 | 40119550 | 40120123 | 574 | 3 | 1.04E-08 | 1.81E-06 | 6.87E-06 | 3.52E-07 | -0.00182916 | NA |
| chr2 | 3648275 | 3649158 | 884 | 4 | 6.03E-06 | 1.42E-07 | 0.000741 | 3.52E-07 | -0.00338991 | NA |
| chr6 | 162443429 | 162444226 | 798 | 7 | 5.71E-07 | 3.88E-05 | 3.32E-05 | 3.52E-07 | -0.00084469 | PRKN |
| chr8 | 20254664 | 20255677 | 1014 | 13 | 3.31E-07 | 2.42E-06 | 0.000662 | 3.52E-07 | 0.002645635 | LZTS1 |
| chr11 | 78062683 | 78063291 | 609 | 3 | 5.19E-08 | 4.84E-07 | 0.000135 | 3.52E-07 | 0.008487503 | NDUFC2-KCTD14 |
| chr1 | 20428754 | 20429041 | 288 | 4 | 6.81E-07 | 5.17E-07 | 0.000243 | 3.53E-07 | -0.00308035 | LINC01141 |
| chr22 | 36328024 | 36328978 | 955 | 3 | 1.38E-07 | 4.90E-07 | 2.09E-05 | 3.53E-07 | 0.00781253 | MYH9 |
| chr12 | 122966219 | 122967327 | 1109 | 13 | 2.94E-07 | 3.07E-06 | 0.003061 | 3.53E-07 | 0.000379323 | ABCB9 |
| chr16 | 55664196 | 55664210 | 15 | 2 | 5.13E-07 | 2.77E-07 | 3.31E-05 | 3.53E-07 | -0.01131547 | SLC6A2 |
| chr15 | 85329690 | 85330390 | 701 | 4 | 9.10E-09 | 3.26E-06 | 1.73E-05 | 3.53E-07 | 0.009599177 | ADAMTS7P4 |
| chr1 | 245687924 | 245688500 | 577 | 2 | 3.27E-07 | 2.57E-07 | 4.22E-05 | 3.53E-07 | 0.035352504 | KIF26B |
| chr17 | 2723697 | 2724954 | 1258 | 6 | 1.71E-07 | 2.51E-05 | 0.000628 | 3.54E-07 | -0.00090065 | AC005696.4, CCDC92B |
| chr11 | 10351621 | 10352463 | 843 | 3 | 1.02E-06 | 2.40E-07 | 6.81E-05 | 3.55E-07 | -0.00204642 | AMPD3 |
| chr3 | 4633367 | 4633693 | 327 | 2 | 1.78E-07 | 5.89E-07 | 7.92E-06 | 3.55E-07 | -0.01687249 | ITPR1 |
| chr17 | 27748384 | 27748571 | 188 | 2 | 4.71E-07 | 2.60E-07 | 4.10E-05 | 3.55E-07 | -0.01307372 | LGALS9DP |
| chr11 | 7905716 | 7906689 | 974 | 3 | 1.97E-06 | 2.80E-07 | 0.000192 | 3.56E-07 | -0.01939823 | AC044810.2 |
| chr9 | 133531985 | 133533043 | 1059 | 4 | 2.99E-08 | 5.49E-07 | 0.000158 | 3.56E-07 | -0.00181808 | ADAMTSL2 |
| chr14 | 87992573 | 87994007 | 1435 | 16 | 1.17E-06 | 2.51E-05 | 0.00313 | 3.56E-07 | -0.00014893 | GALC |
| chr21 | 41166461 | 41167626 | 1166 | 7 | 2.69E-06 | 1.55E-06 | 0.000556 | 3.56E-07 | -0.00701976 | MIR3197 |
| chr8 | 67721190 | 67722473 | 1284 | 4 | 2.40E-09 | 5.44E-06 | 7.83E-06 | 3.56E-07 | -6.86E-06 | CPA6 |
| chr14 | 103551547 | 103552398 | 852 | 4 | 7.04E-09 | 8.28E-06 | 8.55E-06 | 3.56E-07 | -0.00393283 | NA |
| chr14 | 101953318 | 101953659 | 342 | 2 | 4.22E-08 | 4.10E-06 | 1.38E-06 | 3.56E-07 | -0.00430581 | NA |
| chr4 | 76166500 | 76166885 | 386 | 2 | 2.26E-07 | 3.85E-07 | 1.57E-05 | 3.56E-07 | 0.003962056 | AC110795.1, SCARB2 |
| chr6 | 33182959 | 33184231 | 1273 | 8 | 3.15E-09 | 1.17E-05 | 0.000221 | 3.57E-07 | 0.003939958 | COL11A2 |
| chr9 | 138008855 | 138009504 | 650 | 3 | 8.54E-07 | 1.19E-07 | 0.000754 | 3.57E-07 | -0.00852413 | CACNA1B |
| chr12 | 120227510 | 120228454 | 945 | 4 | 2.40E-08 | 9.93E-07 | 2.83E-05 | 3.57E-07 | -0.00296823 | PXN |
| chr6 | 37202703 | 37203061 | 359 | 3 | 7.58E-07 | 1.34E-07 | 0.000468 | 3.57E-07 | 0.014804228 | NA |
| chr17 | 79986005 | 79986890 | 886 | 6 | 3.17E-08 | 0.000211 | 1.54E-06 | 3.58E-07 | -0.00540848 | TBC1D16 |
| chr11 | 76369363 | 76370010 | 648 | 3 | 2.51E-08 | 5.75E-06 | 3.70E-05 | 3.58E-07 | -0.00790332 | THAP12 |
| chr7 | 47406705 | 47407517 | 813 | 3 | 1.41E-07 | 2.20E-07 | 0.000233 | 3.58E-07 | -0.00419689 | TNS3 |
| chr11 | 36593801 | 36594815 | 1015 | 14 | 1.64E-07 | 5.47E-07 | 0.003742 | 3.58E-07 | -0.00022068 | IFTAP, RAG2 |
| chr20 | 53412943 | 53413405 | 463 | 5 | 2.59E-07 | 2.26E-06 | 2.43E-05 | 3.58E-07 | -0.00109953 | TSHZ2, AL109930.1 |
| chr5 | 42138 | 42385 | 248 | 2 | 3.06E-07 | 4.11E-07 | 1.42E-05 | 3.59E-07 | -0.02602177 | NA |
| chr10 | 129768463 | 129769757 | 1295 | 10 | 4.50E-07 | 1.34E-06 | 0.00338 | 3.59E-07 | 0.00658503 | MGMT, AL157832.1 |
| chr6 | 22250973 | 22251808 | 836 | 3 | 9.18E-08 | 6.14E-07 | 1.65E-05 | 3.59E-07 | -0.0080179 | CASC15 |
| chr11 | 65499011 | 65500092 | 1082 | 7 | 1.39E-06 | 2.23E-07 | 0.002066 | 3.59E-07 | 0.003006695 | MALAT1 |
| chr5 | 139463191 | 139463964 | 774 | 3 | 7.95E-08 | 3.19E-07 | 5.04E-05 | 3.60E-07 | -0.00409239 | NA |
| chr12 | 1592835 | 1594015 | 1181 | 8 | 2.02E-07 | 1.17E-05 | 0.000396 | 3.60E-07 | -0.00404346 | WNT5B, FBXL14 |
| chr19 | 10835987 | 10837219 | 1233 | 13 | 8.66E-07 | 3.33E-06 | 0.002957 | 3.60E-07 | 0.001002994 | C19orf38, TMED1 |
| chr4 | 8493896 | 8493966 | 71 | 2 | 5.34E-07 | 2.60E-07 | 4.33E-05 | 3.60E-07 | -0.01402326 | AC105345.1 |
| chr5 | 136058354 | 136059173 | 820 | 6 | 9.90E-08 | 1.59E-06 | 4.05E-05 | 3.61E-07 | -0.00813414 | TGFBI |
| chr6 | 7882710 | 7883186 | 477 | 3 | 2.28E-07 | 7.24E-07 | 1.19E-05 | 3.61E-07 | 0.000103274 | TXNDC5, BLOC1S5-TXNDC5 |
| chr2 | 157874559 | 157875674 | 1116 | 6 | 6.33E-08 | 2.54E-06 | 0.00013 | 3.61E-07 | 1.96E-05 | ACVR1 |
| chr6 | 16332249 | 16332763 | 515 | 3 | 1.59E-09 | 0.0005986 | 1.83E-07 | 3.61E-07 | 0.003722806 | ATXN1 |
| chr3 | 153483065 | 153484486 | 1422 | 6 | 2.61E-06 | 5.87E-06 | 8.01E-05 | 3.62E-07 | -0.0046529 | LINC02877, LINC02006 |
| chr4 | 25863753 | 25864774 | 1022 | 5 | 1.54E-07 | 6.82E-08 | 0.002435 | 3.62E-07 | -0.00576355 | SMIM20, SEL1L3 |
| chr16 | 951653 | 951998 | 346 | 3 | 8.32E-07 | 1.27E-07 | 0.000623 | 3.62E-07 | -0.01582695 | LMF1 |
| chr20 | 52863890 | 52864091 | 202 | 2 | 5.13E-08 | 7.92E-06 | 9.72E-07 | 3.63E-07 | -0.0121608 | NA |
| chr6 | 57013337 | 57013867 | 531 | 5 | 7.85E-07 | 3.88E-07 | 0.000362 | 3.63E-07 | 0.000407582 | BEND6 |

| | | | | | | | | | | |
|---|---|---|---|---|---|---|---|---|---|---|
| chr5 | 59768409 | 59769263 | 855 | 13 | 3.57E-07 | 1.13E-06 | 0.000105 | 3.63E-07 | -0.00054285 | PDE4D |
| chr22 | 50216938 | 50218172 | 1235 | 5 | 5.96E-10 | 0.0005707 | 1.71E-05 | 3.63E-07 | -0.00269211 | SELENOO, TUBGCP6 |
| chr17 | 55732960 | 55733485 | 526 | 4 | 2.62E-07 | 5.77E-06 | 0.000135 | 3.64E-07 | -0.00486367 | NA |
| chr6 | 149836266 | 149837012 | 747 | 5 | 6.42E-08 | 2.56E-05 | 0.000211 | 3.64E-07 | -0.00150961 | AL355312.5, LRP11 |
| chr11 | 57390035 | 57390888 | 854 | 6 | 1.89E-07 | 2.40E-06 | 0.000204 | 3.64E-07 | -0.00216301 | PRG2, AP000781.2 |
| chr12 | 105330035 | 105330756 | 722 | 11 | 2.95E-07 | 1.50E-06 | 0.003207 | 3.64E-07 | -0.00184665 | C12orf75 |
| chr20 | 49290455 | 49290757 | 303 | 2 | 1.43E-08 | 0.0007674 | 3.38E-07 | 3.65E-07 | -0.00514736 | ZFAS1 |
| chr3 | 186722480 | 186722831 | 352 | 2 | 2.20E-08 | 2.06E-05 | 6.45E-07 | 3.65E-07 | -0.01636116 | KNG1, AC068631.1 |
| chr1 | 245510554 | 245510985 | 432 | 2 | 7.43E-09 | 4.74E-06 | 1.30E-06 | 3.65E-07 | -0.00020528 | KIF26B |
| chr17 | 7021402 | 7022297 | 896 | 5 | 4.92E-07 | 5.36E-07 | 0.001357 | 3.65E-07 | -0.00335776 | NA |
| chr3 | 179071438 | 179072991 | 1554 | 17 | 1.60E-06 | 1.76E-06 | 0.003615 | 3.65E-07 | -0.00141506 | ZMAT3 |
| chr8 | 139405598 | 139406109 | 512 | 2 | 8.85E-07 | 2.23E-07 | 9.31E-05 | 3.65E-07 | -0.01777152 | NA |
| chr21 | 43023296 | 43024723 | 1428 | 6 | 1.10E-06 | 3.78E-05 | 5.95E-05 | 3.67E-07 | 0.001994686 | PKNOX1 |
| chr20 | 40892757 | 40893172 | 416 | 3 | 1.08E-07 | 8.25E-07 | 2.04E-05 | 3.67E-07 | -0.00188479 | NA |
| chr22 | 41927270 | 41928277 | 1008 | 6 | 5.75E-07 | 4.35E-06 | 0.001501 | 3.67E-07 | -0.00571621 | NA |
| chr6 | 144095428 | 144096400 | 973 | 11 | 3.58E-07 | 4.06E-07 | 0.003135 | 3.68E-07 | -0.0036944 | SF3B5 |
| chr9 | 106844371 | 106845501 | 1131 | 4 | 5.67E-06 | 9.25E-08 | 0.00138 | 3.68E-07 | 0.003849326 | NA |
| chr1 | 61005552 | 61006174 | 623 | 4 | 1.38E-07 | 1.73E-06 | 0.000486 | 3.68E-07 | -0.01602467 | NFIA, NFIA-AS2 |
| chr19 | 6074621 | 6075482 | 862 | 2 | 2.69E-06 | 2.68E-07 | 4.25E-05 | 3.68E-07 | -0.00773848 | AC011444.2, RFX2 |
| chr2 | 38917967 | 38918591 | 625 | 5 | 4.02E-08 | 9.65E-07 | 0.000194 | 3.69E-07 | 0.002736467 | MORN2, ARHGEF33 |
| chr21 | 46368461 | 46369033 | 573 | 3 | 4.96E-08 | 4.35E-06 | 3.51E-05 | 3.69E-07 | -0.00467699 | PCNT |
| chr5 | 141149204 | 141150886 | 1683 | 15 | 4.32E-06 | 1.52E-05 | 0.002022 | 3.69E-07 | -0.01422503 | PCDHB6, AC244517.2, AC244517.11 |
| chr17 | 38914512 | 38915128 | 617 | 3 | 3.09E-07 | 6.79E-07 | 0.00012 | 3.69E-07 | -0.00386366 | LASP1, MIR6779 |
| chr19 | 4277325 | 4278058 | 734 | 5 | 5.10E-07 | 1.09E-07 | 0.001569 | 3.70E-07 | -0.01282289 | NA |
| chr22 | 46649702 | 46650333 | 632 | 3 | 1.27E-07 | 9.15E-07 | 1.03E-05 | 3.70E-07 | 0.004438587 | GRAMD4 |
| chr8 | 29109015 | 29110544 | 1530 | 7 | 1.02E-05 | 1.31E-05 | 0.001103 | 3.70E-07 | -0.00394397 | KIF13B |
| chr5 | 79228540 | 79229369 | 830 | 2 | 1.01E-06 | 2.58E-07 | 4.97E-05 | 3.70E-07 | 0.003942413 | DMGDH |
| chr18 | 22932876 | 22934068 | 1193 | 16 | 1.40E-06 | 1.44E-07 | 0.003007 | 3.71E-07 | -0.0002013 | RBBP8, MIR4741, AC090912.1 |
| chr1 | 31788994 | 31790000 | 1007 | 5 | 2.16E-08 | 1.07E-06 | 3.27E-05 | 3.71E-07 | -0.00309862 | AL354919.2 |
| chr6 | 170403976 | 170404950 | 975 | 5 | 4.64E-07 | 8.96E-06 | 0.000334 | 3.71E-07 | 0.000338146 | FAM120B |
| chr5 | 179285050 | 179285635 | 586 | 3 | 1.03E-07 | 1.14E-06 | 1.04E-05 | 3.71E-07 | -0.00476481 | ADAMTS2 |
| chr14 | 103384743 | 103386080 | 1338 | 17 | 2.87E-06 | 2.32E-06 | 0.002972 | 3.72E-07 | 0.001285193 | MARK3 |
| chr9 | 133394416 | 133394648 | 233 | 2 | 4.22E-07 | 3.08E-07 | 2.87E-05 | 3.72E-07 | -0.01267826 | STKLD1 |
| chr15 | 59218182 | 59218432 | 251 | 3 | 4.58E-07 | 9.37E-07 | 0.000111 | 3.73E-07 | 0.000166453 | MYO1E |
| chr9 | 110206971 | 110208211 | 1241 | 8 | 9.72E-07 | 1.28E-05 | 0.000146 | 3.73E-07 | -0.00580615 | C9orf152 |
| chr2 | 38409382 | 38409599 | 218 | 2 | 3.73E-07 | 3.81E-07 | 1.79E-05 | 3.73E-07 | -0.01723227 | AC016995.1, LINC02613 |
| chr1 | 13828032 | 13828872 | 841 | 4 | 3.34E-08 | 7.50E-06 | 0.000224 | 3.73E-07 | -0.00201776 | NA |
| chr4 | 7266885 | 7267028 | 144 | 2 | 6.08E-07 | 2.24E-07 | 0.000108 | 3.73E-07 | 0.002401629 | SORCS2 |
| chr11 | 93330136 | 93331068 | 933 | 8 | 1.57E-06 | 4.10E-06 | 0.000509 | 3.73E-07 | -0.00369552 | DEUP1 |
| chr11 | 18287916 | 18288104 | 189 | 3 | 7.06E-08 | 9.55E-06 | 1.74E-06 | 3.74E-07 | 0.005400814 | HPS5 |
| chr17 | 62967783 | 62968700 | 918 | 3 | 6.02E-07 | 3.03E-05 | 6.27E-05 | 3.74E-07 | -0.00879005 | AC005972.3 |
| chr2 | 119158441 | 119160179 | 1739 | 12 | 8.48E-09 | 0.0001707 | 3.04E-05 | 3.75E-07 | 0.001006475 | C1QL2 |
| chr17 | 80918157 | 80919465 | 1309 | 4 | 3.86E-08 | 2.18E-05 | 1.32E-05 | 3.75E-07 | -0.00131407 | RPTOR |
| chr6 | 26239767 | 26240711 | 945 | 9 | 5.68E-08 | 1.69E-06 | 0.000122 | 3.75E-07 | -0.00322592 | H4C6 |
| chr3 | 81643480 | 81644126 | 647 | 2 | 1.52E-07 | 5.19E-07 | 1.07E-05 | 3.77E-07 | -0.00749251 | GBE1 |
| chr8 | 20311554 | 20312877 | 1324 | 5 | 5.05E-06 | 1.30E-06 | 0.000176 | 3.77E-07 | -0.00932145 | NA |
| chr5 | 43477326 | 43477993 | 668 | 3 | 3.53E-07 | 2.54E-07 | 0.000202 | 3.77E-07 | 0.00096654 | TMEM267 |
| chr20 | 62302011 | 62302749 | 739 | 10 | 1.09E-07 | 1.15E-06 | 0.000842 | 3.77E-07 | 0.001314728 | ADRM1 |
| chr11 | 68462739 | 68462779 | 41 | 2 | 3.27E-07 | 8.91E-07 | 5.31E-06 | 3.78E-07 | -0.00961356 | PPP6R3 |
| chr2 | 74497498 | 74498798 | 1301 | 7 | 2.80E-07 | 6.10E-06 | 0.000104 | 3.78E-07 | -0.00355371 | LBX2 |
| chr4 | 8506235 | 8506877 | 643 | 6 | 7.89E-09 | 9.13E-06 | 1.14E-05 | 3.78E-07 | -0.00822679 | AC105345.1 |
| chr6 | 100446637 | 100448031 | 1395 | 8 | 7.05E-07 | 1.22E-06 | 0.0014 | 3.78E-07 | -0.00283216 | SIM1 |
| chr19 | 43715523 | 43716185 | 663 | 5 | 4.36E-08 | 1.29E-05 | 0.000121 | 3.79E-07 | -0.00366301 | IRGC |
| chr4 | 168183606 | 168183908 | 303 | 2 | 3.36E-08 | 2.19E-05 | 6.55E-07 | 3.79E-07 | -0.00379588 | ANXA10, AC068989.1 |
| chr18 | 74196256 | 74196545 | 290 | 2 | 9.54E-08 | 1.06E-06 | 4.46E-06 | 3.79E-07 | -0.0027564 | NA |
| chr20 | 63735756 | 63737212 | 1457 | 14 | 4.05E-07 | 1.03E-06 | 0.005982 | 3.79E-07 | 0.008915964 | ZGPAT, AL121845.3, LIME1 |
| chr6 | 106098062 | 106099335 | 1274 | 13 | 8.54E-07 | 1.23E-05 | 3.14E-05 | 3.79E-07 | -0.0047198 | PRDM1, ATG5 |
| chr7 | 157466591 | 157468123 | 1533 | 4 | 4.28E-06 | 3.30E-07 | 8.81E-05 | 3.79E-07 | 0.00865668 | AC006372.3 |
| chr20 | 43470452 | 43471541 | 1090 | 3 | 8.35E-06 | 1.37E-07 | 0.000588 | 3.80E-07 | -0.00373126 | AL031681.2 |
| chr2 | 241752024 | 241752453 | 430 | 4 | 2.72E-07 | 4.55E-05 | 1.39E-05 | 3.80E-07 | -0.00183186 | D2HGDH |
| chr1 | 2352583 | 2353091 | 509 | 4 | 1.23E-08 | 2.92E-05 | 1.26E-05 | 3.80E-07 | -0.00431571 | MORN1, AL513477.1 |
| chr3 | 184314318 | 184315494 | 1177 | 13 | 9.66E-07 | 4.95E-07 | 0.003643 | 3.80E-07 | 0.001739108 | EIF4G1 |
| chr12 | 34376710 | 34377445 | 736 | 2 | 6.44E-07 | 2.83E-07 | 4.03E-05 | 3.81E-07 | -0.02798981 | NA |
| chr7 | 151992521 | 151992892 | 372 | 2 | 5.05E-09 | 0.0002673 | 3.82E-07 | 3.81E-07 | -0.01081563 | GALNTL5 |
| chr9 | 4305355 | 4305588 | 234 | 2 | 1.06E-07 | 2.28E-07 | 0.000112 | 3.82E-07 | 0.013917108 | AL162419.1, GLIS3 |
| chr2 | 229409020 | 229409845 | 826 | 2 | 3.73E-08 | 4.85E-07 | 1.23E-05 | 3.82E-07 | 0.00821009 | DNER |
| chr6 | 24490870 | 24491089 | 220 | 4 | 4.01E-08 | 2.35E-05 | 2.16E-06 | 3.82E-07 | -0.00300846 | GPLD1 |
| chr12 | 122991833 | 122992600 | 768 | 5 | 7.37E-09 | 2.48E-05 | 1.10E-05 | 3.82E-07 | -0.00264682 | PITPNM2 |
| chr5 | 160120392 | 160120877 | 486 | 2 | 4.73E-09 | 5.12E-07 | 1.13E-05 | 3.83E-07 | -0.01079114 | NA |
| chr2 | 219417521 | 219418720 | 1200 | 9 | 3.76E-06 | 2.48E-05 | 0.001324 | 3.83E-07 | -0.00800855 | DES |
| chr6 | 145963660 | 145964540 | 881 | 12 | 2.40E-08 | 5.00E-05 | 0.001632 | 3.84E-07 | 0.000321914 | SHPRH |
| chr19 | 35647683 | 35648772 | 1090 | 10 | 1.23E-07 | 1.59E-05 | 0.000307 | 3.84E-07 | 0.002543115 | COX6B1 |
| chr2 | 20052493 | 20053191 | 699 | 3 | 4.55E-09 | 1.49E-06 | 6.25E-06 | 3.84E-07 | -0.0019924 | LAPTM4A-DT |
| chr2 | 71668972 | 71669122 | 151 | 2 | 1.37E-07 | 5.49E-06 | 1.26E-06 | 3.84E-07 | -0.01042001 | DYSF |
| chr6 | 46735246 | 46736340 | 1095 | 13 | 3.49E-09 | 6.49E-05 | 4.51E-06 | 3.84E-07 | 0.0038261 | PLA2G7 |
| chr7 | 141909906 | 141911038 | 1133 | 3 | 2.29E-07 | 4.85E-07 | 3.63E-05 | 3.84E-07 | -0.01741838 | MGAM |
| chr10 | 17701584 | 17701690 | 107 | 2 | 3.76E-07 | 2.57E-07 | 5.98E-05 | 3.85E-07 | -0.00141968 | STAM |
| chr12 | 3290272 | 3290294 | 23 | 2 | 1.68E-07 | 2.84E-07 | 4.10E-05 | 3.85E-07 | -0.01227387 | NA |
| chr2 | 95120558 | 95121851 | 1294 | 6 | 6.76E-10 | 7.77E-06 | 1.47E-06 | 3.85E-07 | 0.001131582 | MRPS5 |
| chr8 | 69447980 | 69448328 | 349 | 3 | 8.82E-08 | 6.33E-07 | 3.52E-05 | 3.86E-07 | 0.0104051 | LINC01603 |
| chr1 | 46176580 | 46177713 | 1134 | 4 | 5.12E-07 | 5.43E-06 | 0.000106 | 3.86E-07 | -0.00880965 | TSPAN1 |
| chr10 | 97413136 | 97413283 | 148 | 2 | 3.16E-07 | 2.39E-07 | 8.79E-05 | 3.86E-07 | -0.01550865 | AL355490.2, RRP12 |
| chr11 | 2409944 | 2410373 | 430 | 3 | 2.10E-07 | 3.81E-07 | 5.96E-05 | 3.86E-07 | -0.01313587 | TRPM5 |
| chr10 | 34994792 | 34995296 | 505 | 4 | 1.87E-07 | 1.35E-06 | 0.000518 | 3.86E-07 | -0.00503985 | NA |
| chr17 | 41480424 | 41481429 | 1006 | 4 | 5.46E-08 | 1.04E-06 | 0.000125 | 3.87E-07 | -0.00012781 | KRT35 |
| chr20 | 45827283 | 45827845 | 563 | 6 | 9.44E-10 | 0.0044401 | 5.52E-08 | 3.87E-07 | -0.00107015 | TNNC2 |
| chr15 | 82808901 | 82809659 | 759 | 11 | 1.16E-06 | 2.19E-07 | 0.005804 | 3.87E-07 | -0.00306873 | WHAMM |
| chr12 | 64982980 | 64983341 | 362 | 2 | 6.73E-07 | 2.35E-07 | 0.000101 | 3.88E-07 | -0.00338711 | LINC02389 |
| chr7 | 159105780 | 159106201 | 422 | 4 | 2.77E-07 | 1.18E-05 | 5.74E-05 | 3.88E-07 | -0.00377221 | VIPR2 |
| chr14 | 74913777 | 74914154 | 378 | 2 | 3.92E-08 | 5.02E-06 | 1.36E-06 | 3.89E-07 | -0.00080783 | RPS6KL1 |
| chr12 | 123944342 | 123945397 | 1056 | 8 | 6.64E-09 | 0.0007382 | 9.63E-06 | 3.90E-07 | -0.00947616 | CCDC92 |
| chr20 | 24533636 | 24533827 | 192 | 2 | 4.17E-07 | 3.97E-07 | 1.84E-05 | 3.91E-07 | -0.00994559 | SYNDIG1 |
| chr12 | 26226323 | 26227059 | 737 | 5 | 3.55E-08 | 1.78E-06 | 6.93E-05 | 3.91E-07 | -0.00329263 | SSPN, AC022509.2 |
| chr22 | 36296963 | 36297816 | 854 | 3 | 2.88E-06 | 1.89E-07 | 0.00034 | 3.91E-07 | 0.000414097 | MYH9 |
| chr9 | 72952567 | 72953967 | 1401 | 7 | 1.03E-06 | 4.57E-07 | 0.001565 | 3.91E-07 | 0.009311379 | ALDH1A1 |
| chr10 | 12924805 | 12925729 | 925 | 5 | 2.54E-06 | 5.83E-08 | 0.004528 | 3.91E-07 | -0.00317339 | AL353586.1, CCDC3 |
| chr13 | 48749601 | 48749832 | 232 | 3 | 1.43E-07 | 5.20E-06 | 5.34E-06 | 3.91E-07 | -0.01155869 | NA |
| chr19 | 44111963 | 44113353 | 1391 | 13 | 1.84E-07 | 2.52E-05 | 0.000826 | 3.91E-07 | -0.00349132 | ZNF225, AC021092.1 |
| chr2 | 165469104 | 165469710 | 607 | 7 | 2.73E-07 | 1.34E-05 | 0.000357 | 3.91E-07 | -0.01254054 | CSRNP3 |
| chr6 | 165891119 | 165891400 | 282 | 3 | 2.82E-07 | 6.21E-07 | 0.000138 | 3.91E-07 | -0.00266193 | PDE10A |
| chr1 | 223756744 | 223757425 | 682 | 4 | 9.19E-09 | 1.05E-05 | 1.25E-06 | 3.92E-07 | -0.00198817 | CAPN2 |
| chr16 | 75464510 | 75465518 | 1009 | 9 | 3.42E-07 | 4.22E-07 | 0.000506 | 3.92E-07 | -0.00026157 | AC009163.5, TMEM170A |
| chr7 | 17939469 | 17940967 | 1499 | 9 | 4.88E-07 | 3.19E-07 | 0.002652 | 3.92E-07 | 0.002430473 | AC080080.1, SNX13 |
| chr1 | 162337182 | 162337438 | 257 | 3 | 4.82E-07 | 1.24E-06 | 3.66E-05 | 3.92E-07 | -0.00799194 | NOS1AP |
| chr12 | 122050948 | 122051639 | 692 | 3 | 5.30E-07 | 1.26E-06 | 3.39E-05 | 3.92E-07 | -0.00377397 | BCL7A |
| chr11 | 118086794 | 118087389 | 596 | 4 | 8.98E-07 | 3.84E-07 | 0.000226 | 3.93E-07 | -0.00236349 | TMPRSS4, SMIM35 |
| chr16 | 20899420 | 20900403 | 984 | 8 | 3.23E-08 | 1.58E-06 | 0.000701 | 3.93E-07 | -0.00040192 | LYRM1, ERI2, DCUN1D3 |
| chr6 | 27312138 | 27312943 | 806 | 12 | 7.63E-08 | 1.35E-05 | 0.000256 | 3.93E-07 | -1.49E-05 | POM121L2 |
| chr12 | 120079757 | 120080935 | 1179 | 4 | 7.33E-08 | 1.70E-06 | 3.48E-05 | 3.93E-07 | -0.00688304 | BICDL1 |
| chr5 | 158807719 | 158808115 | 397 | 4 | 9.35E-08 | 5.61E-06 | 1.18E-05 | 3.93E-07 | 0.006077495 | EBF1 |
| chr6 | 24721010 | 24721832 | 823 | 7 | 6.02E-07 | 1.93E-07 | 0.002561 | 3.93E-07 | -0.002893 | LINC02828 |
| chr4 | 47981436 | 47982377 | 942 | 7 | 1.13E-06 | 1.34E-06 | 0.001629 | 3.93E-07 | -0.00621323 | NIPAL1, CNGA1 |
| chr3 | 128650633 | 128651620 | 988 | 12 | 4.85E-08 | 1.39E-05 | 1.33E-05 | 3.94E-07 | -0.00337571 | RPN1 |
| chr5 | 179797562 | 179798558 | 997 | 6 | 1.52E-06 | 6.76E-08 | 0.004478 | 3.94E-07 | -0.00133667 | MGAT4B, MIR1229 |
| chr8 | 139789934 | 139790374 | 441 | 6 | 1.56E-07 | 2.47E-05 | 2.13E-05 | 3.94E-07 | -0.00301175 | TRAPPC9 |
| chr11 | 70661720 | 70662554 | 835 | 10 | 1.62E-08 | 2.70E-05 | 0.001063 | 3.95E-07 | -0.00800816 | SHANK2 |
| chr4 | 186031974 | 186032438 | 465 | 2 | 9.67E-07 | 2.37E-07 | 0.000111 | 3.95E-07 | -0.01194691 | NA |
| chr17 | 50550098 | 50550586 | 489 | 3 | 7.79E-08 | 1.45E-07 | 0.000585 | 3.95E-07 | -0.00287496 | SPATA20 |
| chr6 | 36487160 | 36487862 | 703 | 2 | 5.20E-08 | 1.20E-06 | 4.16E-06 | 3.95E-07 | -0.0053976 | KCTD20 |
| chr17 | 7173607 | 7174390 | 784 | 6 | 8.78E-07 | 1.06E-07 | 0.001072 | 3.95E-07 | -0.00154596 | ASGR1 |
| chr13 | 28100314 | 28101373 | 1060 | 8 | 4.38E-07 | 2.03E-06 | 0.001098 | 3.95E-07 | -0.00168853 | FLT3 |
| chr9 | 95475531 | 95476236 | 706 | 4 | 7.79E-07 | 9.29E-08 | 0.001622 | 3.95E-07 | -0.00336228 | PTCH1 |
| chr14 | 92952342 | 92953209 | 868 | 7 | 5.70E-08 | 3.72E-06 | 8.65E-05 | 3.95E-07 | -0.00339282 | ITPK1 |
| chr15 | 58864876 | 58865658 | 783 | 5 | 2.79E-08 | 2.83E-06 | 0.000428 | 3.96E-07 | -0.01036181 | ZNF444P1, RNF111, AC090515.4 |
| chr15 | 92571765 | 92572655 | 891 | 5 | 8.03E-08 | 1.58E-06 | 0.000106 | 3.96E-07 | -0.00578201 | LINC00930 |
| chr11 | 67468756 | 67470147 | 1392 | 14 | 2.22E-08 | 2.85E-05 | 0.00017 | 3.96E-07 | -0.0003528 | TMEM134 |
| chr11 | 119018068 | 119018931 | 864 | 15 | 1.42E-06 | 3.44E-07 | 0.012828 | 3.97E-07 | 0.001297354 | TRAPPC4, RPS25 |
| chr3 | 135635515 | 135636019 | 505 | 3 | 4.49E-07 | 2.03E-07 | 0.000152 | 3.97E-07 | -0.01109211 | NA |
| chr1 | 247936157 | 247937312 | 1156 | 13 | 2.07E-07 | 3.43E-06 | 0.00011 | 3.97E-07 | -0.01472833 | CLK3P2 |
| chr17 | 36948170 | 36949563 | 1394 | 12 | 6.35E-06 | 1.62E-06 | 0.001214 | 3.97E-07 | 0.000868384 | AATF |
| chr1 | 159587156 | 159588096 | 941 | 7 | 2.68E-07 | 2.78E-06 | 2.95E-05 | 3.98E-07 | -0.00368252 | APCS |
| chr18 | 9959089 | 9959403 | 315 | 2 | 1.11E-07 | 1.58E-06 | 3.24E-06 | 3.99E-07 | 0.005978062 | VAPA |
| chr20 | 62351931 | 62352950 | 1020 | 4 | 4.70E-07 | 1.19E-06 | 3.52E-05 | 3.99E-07 | -0.00592139 | LAMA5 |
| chr20 | 40582641 | 40583886 | 1246 | 3 | 2.57E-06 | 3.35E-07 | 9.55E-05 | 3.99E-07 | -0.0090673 | NA |
| chr13 | 98435932 | 98436469 | 538 | 5 | 5.63E-07 | 1.54E-07 | 0.00062 | 4.00E-07 | 0.004312118 | FARP1 |
| chr11 | 35138764 | 35139798 | 1035 | 14 | 7.63E-08 | 7.18E-06 | 0.000622 | 4.00E-07 | 0.000517243 | CD44 |
| chr6 | 4728149 | 4728233 | 85 | 2 | 5.92E-07 | 2.94E-07 | 4.18E-05 | 4.00E-07 | -0.01052778 | CDYL |
| chr8 | 90649386 | 90650142 | 757 | 2 | 9.28E-07 | 2.80E-07 | 5.06E-05 | 4.00E-07 | -0.00984691 | LINC00534, AC106038.1, TMEM64 |
| chr6 | 167121467 | 167123095 | 1629 | 10 | 1.13E-05 | 6.99E-07 | 0.014596 | 4.00E-07 | 0.006379573 | Z94721.2, CCR6, AL121935.1 |
| chr11 | 2418802 | 2419156 | 355 | 3 | 1.90E-07 | 1.54E-07 | 0.000404 | 4.01E-07 | 0.004282286 | TRPM5 |
| chr7 | 142720407 | 142720605 | 199 | 3 | 8.46E-07 | 1.68E-07 | 0.000455 | 4.01E-07 | 0.020184123 | NA |
| chr1 | 5862626 | 5863249 | 624 | 7 | 3.82E-08 | 2.42E-06 | 0.000214 | 4.02E-07 | -0.00286026 | MIR4689, NPHP4 |
| chr9 | 130042931 | 130044224 | 1294 | 11 | 6.60E-08 | 4.92E-05 | 5.50E-05 | 4.02E-07 | 2.96E-05 | FNBP1 |
| chr12 | 130704835 | 130705500 | 666 | 5 | 1.40E-07 | 5.70E-06 | 0.000179 | 4.02E-07 | -0.00458589 | RIMBP2 |
| chr12 | 124278549 | 124279254 | 706 | 4 | 8.26E-07 | 2.49E-07 | 0.000866 | 4.02E-07 | 0.001392367 | RFLNA |
| chr8 | 135456242 | 135457034 | 793 | 6 | 1.21E-07 | 7.57E-07 | 0.000539 | 4.03E-07 | -0.00089134 | AC040914.1 |
| chr8 | 52586993 | 52587364 | 372 | 4 | 1.30E-07 | 8.05E-06 | 2.15E-05 | 4.03E-07 | -0.00366005 | NA |
| chr13 | 49893835 | 49894154 | 320 | 2 | 2.34E-07 | 5.94E-07 | 1.01E-05 | 4.04E-07 | -0.00147387 | NA |
| chr2 | 160472447 | 160473096 | 650 | 2 | 1.12E-08 | 2.21E-06 | 2.49E-06 | 4.04E-07 | -0.00908832 | RBMS1 |
| chr2 | 55958614 | 55958654 | 41 | 2 | 3.73E-07 | 4.18E-07 | 1.81E-05 | 4.04E-07 | 0.012728231 | AC011306.1 |
| chr9 | 10612567 | 10614067 | 1501 | 11 | 9.46E-07 | 5.34E-05 | 0.000119 | 4.04E-07 | -0.00282375 | PTPRD-AS2, PTPRD |
| chr1 | 2207318 | 2208508 | 1191 | 11 | 1.43E-06 | 7.67E-07 | 0.002786 | 4.05E-07 | -0.00062407 | FAAP20 |
| chr10 | 70381619 | 70382907 | 1289 | 15 | 7.94E-09 | 0.0002662 | 0.000205 | 4.05E-07 | 0.000661498 | LRRC20 |
| chr1 | 244460651 | 244461483 | 833 | 10 | 5.80E-07 | 7.74E-06 | 0.00146 | 4.05E-07 | -0.00094024 | CATSPERE |
| chr12 | 48783810 | 48784999 | 1190 | 5 | 2.51E-09 | 0.0001004 | 2.51E-05 | 4.06E-07 | -0.00562105 | ADCY6 |
| chr19 | 405795 | 406560 | 766 | 4 | 1.42E-07 | 1.11E-05 | 6.58E-06 | 4.06E-07 | -0.0072037 | C2CD4C |
| chr2 | 102443125 | 102443527 | 403 | 3 | 2.17E-07 | 7.18E-07 | 2.22E-05 | 4.07E-07 | 0.002923088 | IL18RAP |
| chr19 | 17419029 | 17420079 | 1051 | 13 | 1.30E-06 | 4.01E-07 | 0.002956 | 4.07E-07 | -0.00219591 | BISPR, MVB12A, AC010319.4, AC010319.3 |

| | | | | | | | | | | |
|---|---|---|---|---|---|---|---|---|---|---|
| chr7 | 93890860 | 93891884 | 1025 | 16 | 3.42E-09 | 1.47E-05 | 0.000149 | 4.08E-07 | -4.04E-05 | GNGT1, TFPI2-DT |
| chr2 | 216774483 | 216774809 | 327 | 2 | 1.33E-07 | 1.27E-06 | 4.12E-06 | 4.08E-07 | -0.01174697 | AC007563.2 |
| chr6 | 131627870 | 131628603 | 734 | 9 | 1.50E-07 | 3.10E-06 | 0.000348 | 4.09E-07 | -0.00230819 | ENPP3, MED23 |
| chr7 | 67438966 | 67439176 | 211 | 2 | 2.92E-07 | 2.70E-07 | 6.44E-05 | 4.09E-07 | -0.02701041 | NA |
| chr20 | 45098448 | 45099005 | 558 | 3 | 3.80E-08 | 1.41E-06 | 6.82E-06 | 4.09E-07 | -0.00093606 | KCNS1 |
| chr21 | 42496049 | 42497303 | 1255 | 13 | 1.74E-07 | 6.42E-07 | 0.00994 | 4.10E-07 | 0.002033916 | SLC37A1, AP001625.3, RSPH1 |
| chr12 | 109477277 | 109478191 | 915 | 14 | 3.56E-07 | 3.48E-05 | 8.44E-05 | 4.10E-07 | 0.001087948 | UBE3B, KCTD10 |
| chr10 | 47310070 | 47310412 | 343 | 3 | 3.45E-08 | 3.11E-05 | 5.81E-07 | 4.10E-07 | 0.004899124 | GDF10 |
| chr14 | 76111570 | 76111641 | 72 | 2 | 2.26E-07 | 7.25E-07 | 7.84E-06 | 4.10E-07 | -0.00803175 | NA |
| chr20 | 44813318 | 44813482 | 165 | 2 | 2.60E-07 | 2.52E-07 | 9.69E-05 | 4.10E-07 | -0.00653018 | NA |
| chr10 | 126176084 | 126176182 | 99 | 2 | 6.26E-07 | 2.83E-07 | 5.44E-05 | 4.11E-07 | -0.01643015 | ADAM12 |
| chr8 | 22221469 | 22222369 | 901 | 8 | 7.11E-08 | 3.25E-05 | 2.18E-05 | 4.11E-07 | -0.00033189 | PHYHIP |
| chr9 | 13491932 | 13492140 | 209 | 2 | 5.34E-07 | 3.07E-07 | 4.01E-05 | 4.11E-07 | -0.01805089 | AL583785.1 |
| chr10 | 97780417 | 97781128 | 712 | 3 | 1.71E-07 | 0.0003848 | 4.96E-05 | 4.11E-07 | -0.01182549 | NA |
| chr12 | 55935512 | 55936464 | 953 | 6 | 1.63E-06 | 4.14E-07 | 0.00102 | 4.11E-07 | -0.00659568 | DGKA |
| chr1 | 75135256 | 75135651 | 396 | 4 | 2.67E-09 | 3.23E-05 | 9.26E-07 | 4.11E-07 | -0.00365866 | LHX8 |
| chr10 | 19986214 | 19987116 | 903 | 4 | 8.00E-06 | 7.68E-07 | 0.000158 | 4.12E-07 | -0.01246659 | PLXDC2 |
| chr10 | 34539964 | 34540194 | 231 | 2 | 2.45E-07 | 9.30E-07 | 5.84E-06 | 4.13E-07 | -0.00524736 | PARD3 |
| chr1 | 54413520 | 54413721 | 202 | 2 | 5.04E-08 | 9.58E-05 | 4.76E-07 | 4.13E-07 | -0.00477378 | NA |
| chr1 | 200279627 | 200279919 | 293 | 3 | 6.91E-08 | 4.40E-05 | 1.90E-06 | 4.13E-07 | -0.00824188 | LINC00862 |
| chr14 | 37598760 | 37600230 | 1471 | 13 | 5.18E-07 | 6.60E-06 | 0.004038 | 4.14E-07 | -0.00057861 | TTC6 |
| chr1 | 2412841 | 2414649 | 1809 | 19 | 2.71E-06 | 4.94E-05 | 0.001138 | 4.14E-07 | -0.00214035 | PEX10 |
| chr3 | 107240219 | 107241367 | 1149 | 15 | 2.98E-06 | 7.69E-06 | 0.003375 | 4.14E-07 | -0.00089761 | DUBR, LINC00882 |
| chr17 | 46969684 | 46969986 | 303 | 2 | 7.04E-08 | 2.76E-07 | 6.27E-05 | 4.14E-07 | 0.00030751 | GOSR2, AC005670.2 |
| chr1 | 22711828 | 22712432 | 605 | 3 | 7.63E-08 | 5.49E-07 | 2.48E-05 | 4.15E-07 | -0.00261181 | EPHB2 |
| chr5 | 1724777 | 1725708 | 932 | 7 | 3.01E-07 | 6.69E-08 | 0.003166 | 4.15E-07 | 0.017430337 | AC112176.1 |
| chr12 | 47905506 | 47906017 | 512 | 4 | 5.59E-07 | 3.01E-07 | 7.80E-05 | 4.16E-07 | 6.33E-05 | AC121338.2, VDR |
| chr5 | 136135769 | 136136194 | 426 | 2 | 2.65E-08 | 3.36E-07 | 3.23E-05 | 4.16E-07 | -0.00682027 | SMAD5 |
| chr15 | 90665139 | 90665696 | 558 | 6 | 3.03E-07 | 1.26E-06 | 0.00071 | 4.16E-07 | -0.01155771 | CRTC3-AS1 |
| chr13 | 108326406 | 108326790 | 385 | 3 | 2.91E-08 | 3.09E-05 | 6.44E-07 | 4.17E-07 | 0.005247893 | NA |
| chr5 | 171387283 | 171388991 | 1709 | 18 | 2.81E-05 | 5.10E-07 | 0.012532 | 4.17E-07 | 0.000445345 | NPM1 |
| chr5 | 134246968 | 134247130 | 163 | 2 | 2.47E-07 | 3.81E-07 | 2.42E-05 | 4.20E-07 | -0.00416805 | AC104109.4 |
| chr7 | 23250471 | 23251506 | 1036 | 3 | 1.06E-05 | 5.01E-07 | 9.22E-05 | 4.21E-07 | -0.00708522 | GPNMB |
| chr22 | 34834127 | 34834384 | 258 | 2 | 3.55E-07 | 2.72E-07 | 7.13E-05 | 4.21E-07 | -0.02588546 | LINC02885 |
| chr17 | 80965306 | 80966712 | 1407 | 11 | 6.13E-07 | 7.28E-05 | 7.96E-05 | 4.21E-07 | -0.00145285 | RPTOR, AC127496.1 |
| chr7 | 4216058 | 4216148 | 91 | 2 | 3.18E-08 | 1.36E-06 | 4.05E-06 | 4.21E-07 | -0.02939666 | SDK1 |
| chr19 | 42280063 | 42280441 | 379 | 5 | 8.62E-08 | 8.99E-07 | 6.71E-05 | 4.21E-07 | -0.00121464 | CIC |
| chr22 | 37905570 | 37906497 | 928 | 5 | 1.52E-07 | 2.00E-06 | 0.000214 | 4.22E-07 | -0.00435206 | MICALL1 |
| chr18 | 78544299 | 78544675 | 377 | 5 | 4.94E-07 | 3.55E-07 | 0.000732 | 4.22E-07 | 0.009042992 | NA |
| chr8 | 54464715 | 54465345 | 631 | 2 | 1.07E-06 | 2.53E-07 | 0.000117 | 4.22E-07 | 0.025891665 | NA |
| chr15 | 69669185 | 69670108 | 924 | 7 | 2.31E-06 | 3.84E-07 | 0.001162 | 4.23E-07 | 0.003021276 | DRAIC |
| chr9 | 137074174 | 137074798 | 625 | 4 | 9.10E-08 | 1.12E-06 | 7.76E-05 | 4.23E-07 | -0.00698232 | NA |
| chr10 | 1361725 | 1361743 | 19 | 2 | 1.92E-07 | 7.67E-07 | 7.73E-06 | 4.24E-07 | -0.01396759 | ADARB2 |
| chr1 | 26695741 | 26697960 | 2220 | 6 | 3.33E-05 | 2.85E-05 | 7.95E-05 | 4.24E-07 | -0.0027304 | ARID1A |
| chr8 | 105798630 | 105798915 | 286 | 3 | 6.01E-07 | 2.67E-07 | 0.000119 | 4.24E-07 | 0.002208094 | ZFPM2, ZFPM2-AS1 |
| chr4 | 156642324 | 156642855 | 532 | 6 | 1.91E-07 | 2.34E-06 | 0.000357 | 4.24E-07 | -0.00225244 | AC096736.2, LINC02272 |
| chr10 | 1736390 | 1738069 | 1680 | 17 | 1.03E-06 | 0.0003353 | 0.000234 | 4.24E-07 | -8.75E-05 | ADARB2 |
| chr1 | 225474648 | 225475064 | 417 | 5 | 3.48E-07 | 1.29E-06 | 0.000108 | 4.25E-07 | -0.00580515 | NA |
| chr5 | 160133265 | 160133685 | 421 | 3 | 4.56E-07 | 2.83E-07 | 9.89E-05 | 4.25E-07 | -0.01800252 | NA |
| chr3 | 188568013 | 188568985 | 973 | 7 | 3.91E-07 | 4.49E-06 | 0.001377 | 4.25E-07 | 0.000550272 | LPP, LPP-AS1 |
| chr4 | 185183813 | 185184237 | 425 | 3 | 2.39E-08 | 1.01E-05 | 1.13E-05 | 4.25E-07 | -0.01028998 | CFAP97 |
| chr2 | 227471153 | 227472212 | 1060 | 11 | 6.63E-07 | 1.08E-05 | 0.000268 | 4.25E-07 | -0.00285806 | MIR5703, AGFG1 |
| chr21 | 36990330 | 36991281 | 952 | 10 | 8.90E-07 | 2.16E-06 | 0.002407 | 4.25E-07 | -0.01097918 | NA |
| chr1 | 21293078 | 21294319 | 1242 | 6 | 1.62E-10 | 5.45E-05 | 3.46E-05 | 4.26E-07 | 0.005237973 | ECE1-AS1, ECE1 |
| chr1 | 83998401 | 83999326 | 926 | 6 | 3.04E-08 | 4.38E-06 | 0.000625 | 4.26E-07 | -0.00536829 | TTLL7 |
| chr7 | 73767906 | 73768576 | 671 | 2 | 3.19E-10 | 7.84E-06 | 1.17E-06 | 4.27E-07 | -0.01366596 | NA |
| chr10 | 84566922 | 84567480 | 559 | 2 | 2.16E-09 | 0.0009631 | 3.80E-07 | 4.27E-07 | -0.01238613 | NA |
| chr1 | 223628338 | 223628974 | 637 | 3 | 1.69E-06 | 4.71E-07 | 0.000138 | 4.27E-07 | -0.0064353 | CAPN8 |
| chr5 | 131796237 | 131796731 | 495 | 2 | 1.30E-10 | 2.21E-06 | 2.70E-06 | 4.27E-07 | 0.003538314 | AC008695.1, FNIP1 |
| chr3 | 150763500 | 150764127 | 628 | 6 | 2.90E-08 | 5.60E-06 | 0.000345 | 4.27E-07 | 0.002002208 | SIAH2-AS1 |
| chr17 | 44946846 | 44948287 | 1442 | 16 | 1.20E-06 | 4.28E-07 | 0.004536 | 4.27E-07 | 0.000902417 | AC015936.1, KIF18B |
| chr8 | 96233765 | 96234018 | 254 | 2 | 1.74E-08 | 0.0002278 | 4.29E-07 | 4.27E-07 | -0.00885388 | UQCRB |
| chr20 | 23034557 | 23035678 | 1122 | 8 | 6.78E-07 | 1.99E-06 | 0.000173 | 4.27E-07 | -0.00270538 | AL049651.2, SSTR4 |
| chr19 | 40989963 | 40990171 | 209 | 4 | 4.27E-07 | 2.43E-06 | 8.86E-05 | 4.27E-07 | 0.003633389 | NA |
| chr22 | 29267129 | 29269051 | 1923 | 18 | 4.13E-06 | 8.14E-07 | 0.012523 | 4.27E-07 | 0.00082806 | EWSR1, RHBDD3 |
| chr1 | 22915044 | 22915770 | 727 | 3 | 4.20E-08 | 8.25E-07 | 1.51E-05 | 4.28E-07 | -0.01163229 | EPHB2 |
| chr16 | 73062943 | 73063067 | 125 | 4 | 4.09E-07 | 6.53E-07 | 0.000203 | 4.28E-07 | -0.00555233 | ZFHX3 |
| chr2 | 232376471 | 232377620 | 1150 | 5 | 4.15E-06 | 6.52E-07 | 0.000733 | 4.28E-07 | 0.001161418 | NA |
| chr6 | 901014 | 901302 | 289 | 7 | 9.20E-08 | 7.40E-05 | 3.09E-05 | 4.28E-07 | 0.006656892 | NA |
| chr2 | 27118590 | 27119456 | 867 | 9 | 7.03E-08 | 2.62E-05 | 0.000128 | 4.28E-07 | -0.00488527 | CGREF1 |
| chr14 | 68795165 | 68796787 | 1623 | 17 | 2.34E-07 | 2.05E-06 | 0.000581 | 4.29E-07 | 0.001833118 | ZFP36L1 |
| chr19 | 41930233 | 41930420 | 188 | 2 | 2.92E-07 | 2.57E-07 | 0.000118 | 4.29E-07 | -0.02929932 | NA |
| chr15 | 42005586 | 42006118 | 533 | 5 | 2.46E-07 | 5.61E-06 | 0.000149 | 4.29E-07 | 0.000216357 | PLA2G4E |
| chr15 | 72796613 | 72797302 | 690 | 5 | 7.99E-09 | 2.81E-05 | 1.78E-06 | 4.29E-07 | 0.001630102 | ADPGK-AS1 |
| chr5 | 98455135 | 98455415 | 281 | 2 | 3.66E-07 | 4.59E-07 | 1.74E-05 | 4.29E-07 | -0.01804857 | NA |
| chr21 | 44928391 | 44929579 | 1189 | 9 | 1.18E-08 | 4.88E-05 | 0.000122 | 4.29E-07 | -0.00406929 | ITGB2-AS1, ITGB2 |
| chr2 | 6311182 | 6311415 | 234 | 2 | 1.32E-07 | 2.60E-06 | 2.38E-06 | 4.29E-07 | 0.007709077 | AC017053.1 |
| chr7 | 109597648 | 109597864 | 217 | 3 | 6.90E-07 | 1.73E-07 | 0.000337 | 4.30E-07 | -0.01265449 | NA |
| chr19 | 1400418 | 1401782 | 1365 | 13 | 8.10E-06 | 6.94E-06 | 0.002519 | 4.30E-07 | -0.00052768 | GAMT |
| chr20 | 59403179 | 59404271 | 1093 | 5 | 5.22E-07 | 1.34E-07 | 0.001667 | 4.30E-07 | -0.00286406 | NA |
| chr6 | 28616511 | 28616687 | 177 | 3 | 9.07E-06 | 3.76E-06 | 1.03E-05 | 4.30E-07 | -0.02247342 | NA |
| chr6 | 3175515 | 3176113 | 599 | 2 | 3.13E-08 | 9.04E-07 | 6.51E-06 | 4.31E-07 | -0.00994294 | NA |
| chr13 | 27450239 | 27451457 | 1219 | 20 | 1.11E-06 | 6.56E-07 | 0.00433 | 4.31E-07 | -0.00018643 | MTIF3 |
| chr1 | 111628585 | 111629122 | 538 | 2 | 2.24E-09 | 0.0109841 | 3.61E-07 | 4.33E-07 | -0.0227407 | RAP1A |
| chr4 | 180058167 | 180059531 | 1365 | 7 | 1.12E-07 | 1.90E-05 | 6.08E-05 | 4.33E-07 | 0.001801234 | AC108169.1 |
| chr18 | 79356151 | 79356310 | 160 | 3 | 1.13E-07 | 2.47E-05 | 7.47E-07 | 4.33E-07 | -0.00562999 | ATP9B |
| chr2 | 151048816 | 151048858 | 43 | 3 | 1.48E-07 | 1.43E-05 | 2.10E-05 | 4.34E-07 | -0.00479764 | NA |
| chr1 | 226810172 | 226810948 | 777 | 2 | 2.14E-06 | 3.02E-07 | 5.20E-05 | 4.34E-07 | 0.002472758 | NA |
| chr11 | 61949055 | 61950308 | 1254 | 10 | 5.36E-06 | 3.56E-06 | 0.000945 | 4.34E-07 | -0.00058914 | BEST1 |
| chr16 | 12089932 | 12090450 | 519 | 9 | 8.17E-08 | 7.03E-06 | 2.59E-05 | 4.34E-07 | -0.00637391 | SNX29, AC007601.1 |
| chr11 | 88097396 | 88098725 | 1330 | 6 | 1.27E-06 | 5.84E-06 | 9.46E-05 | 4.34E-07 | -0.00351664 | AP005436.2, AP005436.1 |
| chr11 | 65636934 | 65638289 | 1356 | 12 | 2.09E-06 | 3.65E-06 | 0.000941 | 4.34E-07 | 0.000947123 | PCNX3, SIPA1 |
| chr15 | 68578640 | 68579743 | 1104 | 8 | 4.33E-07 | 3.96E-07 | 0.003825 | 4.34E-07 | 0.001222906 | CORO2B |
| chr1 | 28232519 | 28233518 | 1000 | 11 | 2.25E-08 | 0.0001941 | 7.86E-06 | 4.34E-07 | 0.000429682 | DNAJC8 |
| chr20 | 2705099 | 2706717 | 1619 | 7 | 2.62E-05 | 3.61E-06 | 0.001322 | 4.35E-07 | -0.0028567 | EBF4 |
| chr1 | 3285294 | 3285389 | 96 | 2 | 3.68E-07 | 7.09E-07 | 9.03E-06 | 4.35E-07 | -0.01179283 | PRDM16 |
| chr17 | 4739247 | 4741400 | 2154 | 13 | 1.04E-08 | 4.16E-05 | 4.17E-06 | 4.35E-07 | -0.00331074 | ZMYND15, CXCL16 |
| chr19 | 15268237 | 15269019 | 783 | 4 | 6.75E-08 | 1.55E-05 | 5.15E-05 | 4.36E-07 | -0.01631216 | BRD4 |
| chr1 | 1073901 | 1074759 | 859 | 5 | 1.29E-08 | 5.72E-06 | 1.80E-05 | 4.36E-07 | -0.00488253 | RNF223 |
| chr4 | 73224169 | 73224230 | 62 | 2 | 7.92E-08 | 3.67E-07 | 2.96E-05 | 4.37E-07 | -0.00355044 | ANKRD17 |
| chr3 | 71753120 | 71754988 | 1869 | 16 | 1.54E-05 | 1.17E-05 | 0.000513 | 4.37E-07 | -0.00384557 | GPR27, AC097634.4, EIF4E3 |
| chr2 | 128090235 | 128091635 | 1401 | 11 | 9.16E-06 | 2.13E-07 | 0.007007 | 4.37E-07 | -0.00085215 | UGGT1 |
| chr4 | 177580369 | 177580748 | 380 | 4 | 1.39E-06 | 1.09E-07 | 0.001556 | 4.37E-07 | -0.01844577 | AC078881.1 |
| chr14 | 58827677 | 58828519 | 843 | 8 | 2.40E-07 | 1.48E-06 | 0.001746 | 4.38E-07 | -0.00087924 | LINC01500, AL049873.2 |
| chr17 | 81940856 | 81941753 | 898 | 5 | 2.13E-07 | 2.38E-07 | 0.000653 | 4.38E-07 | -0.00625475 | PYCR1, MYADML2 |
| chr21 | 45171121 | 45171624 | 504 | 4 | 3.54E-07 | 1.86E-06 | 3.10E-05 | 4.38E-07 | 0.001331051 | ADARB1 |
| chr7 | 85122519 | 85123119 | 601 | 4 | 2.47E-09 | 7.34E-05 | 3.56E-07 | 4.38E-07 | -0.00275693 | SEMA3D |
| chr15 | 100261573 | 100262245 | 673 | 6 | 5.19E-07 | 6.82E-06 | 0.00062 | 4.38E-07 | -0.00281041 | ADAMTS17 |
| chr19 | 1617028 | 1618612 | 1585 | 6 | 8.28E-07 | 1.27E-05 | 0.000372 | 4.38E-07 | -0.00203586 | TCF3 |
| chr18 | 78994611 | 78995898 | 1288 | 6 | 9.50E-07 | 5.16E-05 | 4.12E-05 | 4.38E-07 | -0.00350549 | SALL3 |
| chr15 | 90571110 | 90571174 | 65 | 2 | 4.21E-07 | 9.50E-07 | 6.31E-06 | 4.39E-07 | -0.02288572 | CRTC3 |
| chr2 | 241411366 | 241411986 | 621 | 2 | 1.39E-09 | 0.0001378 | 4.73E-07 | 4.39E-07 | -0.01073258 | FARP2 |
| chr1 | 67193962 | 67194918 | 957 | 2 | 1.41E-06 | 2.94E-07 | 6.21E-05 | 4.39E-07 | 0.00510257 | IL23R, C1orf141 |
| chr5 | 159209627 | 159210676 | 1050 | 11 | 2.27E-08 | 8.41E-05 | 3.52E-05 | 4.39E-07 | -0.00414578 | AC134043.2, RNF145 |
| chr7 | 154892341 | 154893253 | 913 | 9 | 8.79E-08 | 9.08E-07 | 0.003335 | 4.40E-07 | -0.01625163 | DPP6 |
| chr12 | 98592403 | 98593469 | 1067 | 9 | 5.68E-08 | 3.57E-06 | 3.18E-05 | 4.40E-07 | -0.00257932 | NA |
| chr8 | 54022357 | 54023045 | 689 | 10 | 5.41E-07 | 1.68E-06 | 0.005183 | 4.40E-07 | -0.00052118 | TCEA1 |
| chr2 | 237749233 | 237749911 | 679 | 3 | 1.52E-07 | 6.24E-05 | 5.90E-05 | 4.41E-07 | -0.00119307 | LRRFIP1 |
| chr20 | 17619830 | 17620598 | 769 | 6 | 9.34E-09 | 0.0001754 | 5.99E-07 | 4.41E-07 | -0.00251538 | RRBP1 |
| chr7 | 69373832 | 69374478 | 647 | 4 | 3.44E-08 | 8.25E-05 | 6.91E-07 | 4.42E-07 | 0.008399788 | AC092100.1 |
| chr14 | 88823868 | 88824893 | 1026 | 14 | 5.89E-07 | 8.88E-07 | 0.000246 | 4.42E-07 | 0.00177941 | TTC8 |
| chr12 | 69612396 | 69612729 | 334 | 2 | 1.40E-08 | 1.13E-06 | 5.30E-06 | 4.43E-07 | -0.01142279 | NA |
| chr17 | 8965163 | 8967038 | 1876 | 15 | 3.42E-06 | 1.84E-05 | 0.000233 | 4.43E-07 | -0.00116853 | AC002091.2, PIK3R5 |
| chr3 | 73403702 | 73404639 | 938 | 3 | 1.51E-08 | 1.34E-06 | 8.03E-06 | 4.44E-07 | -0.01060921 | PDZRN3 |
| chr8 | 41896493 | 41897410 | 918 | 7 | 6.43E-08 | 4.68E-05 | 1.94E-05 | 4.44E-07 | -0.0007041 | ANK1 |
| chr19 | 4669766 | 4671346 | 1581 | 14 | 8.96E-06 | 2.28E-06 | 0.008802 | 4.44E-07 | -0.0005634 | MYDGF |
| chr19 | 29212862 | 29213539 | 678 | 10 | 3.09E-07 | 2.48E-06 | 0.000177 | 4.45E-07 | -0.00029313 | AC007786.1, UQCRFS1 |
| chr16 | 27461018 | 27462023 | 1006 | 5 | 6.80E-06 | 4.53E-07 | 0.000455 | 4.46E-07 | -0.0015128 | GTF3C1 |
| chr17 | 82205298 | 82206371 | 1074 | 5 | 2.91E-08 | 1.73E-05 | 1.07E-05 | 4.46E-07 | -0.00169605 | CCDC57 |
| chr10 | 1488839 | 1489210 | 372 | 3 | 4.58E-07 | 2.44E-07 | 0.000134 | 4.46E-07 | 0.012243461 | ADARB2 |
| chr13 | 111923343 | 111923566 | 224 | 2 | 5.71E-07 | 2.94E-07 | 6.70E-05 | 4.46E-07 | -0.00400705 | NA |
| chr8 | 11444734 | 11446285 | 1552 | 7 | 3.39E-07 | 3.27E-06 | 0.000153 | 4.47E-07 | -0.00271222 | FAM167A |
| chr20 | 59249182 | 59250381 | 1200 | 6 | 7.21E-07 | 1.92E-05 | 3.02E-05 | 4.47E-07 | -0.00496648 | ZNF831 |
| chr5 | 32584455 | 32585789 | 1335 | 14 | 5.82E-07 | 9.26E-05 | 0.000336 | 4.47E-07 | -0.00423547 | SUB1 |
| chr15 | 24856480 | 24857230 | 751 | 4 | 2.58E-06 | 4.89E-07 | 0.000252 | 4.47E-07 | -0.00545265 | AC090983.2, SNRPN |
| chr3 | 50570356 | 50572120 | 1765 | 9 | 9.53E-07 | 0.0001446 | 0.000186 | 4.47E-07 | -0.0016486 | HEMK1, C3orf18 |
| chr10 | 35822534 | 35822550 | 17 | 2 | 7.03E-08 | 6.01E-05 | 5.65E-07 | 4.47E-07 | -0.00650829 | NA |
| chr22 | 36783868 | 36784241 | 374 | 2 | 1.89E-07 | 7.75E-07 | 8.40E-06 | 4.47E-07 | -0.00142232 | NA |
| chr2 | 66495970 | 66496216 | 247 | 2 | 6.25E-07 | 2.84E-07 | 8.21E-05 | 4.47E-07 | -0.01244182 | MEIS1 |
| chr2 | 218658659 | 218659853 | 1195 | 13 | 2.01E-07 | 9.29E-06 | 0.00374 | 4.47E-07 | 0.000132903 | BCS1L, ZNF142 |
| chr2 | 99255112 | 99255804 | 693 | 8 | 4.03E-09 | 0.0002989 | 2.46E-06 | 4.48E-07 | -0.00485533 | AC079447.1, LYG2 |
| chr6 | 111700163 | 111700473 | 311 | 5 | 6.15E-07 | 9.48E-07 | 0.000206 | 4.48E-07 | 0.000191996 | FYN |
| chr4 | 99144295 | 99144395 | 101 | 2 | 3.87E-07 | 2.93E-07 | 6.91E-05 | 4.48E-07 | 0.129660298 | AP002026.1, ADH4 |
| chr10 | 91589923 | 91590548 | 626 | 4 | 6.05E-07 | 1.26E-06 | 0.000542 | 4.48E-07 | -3.05E-05 | NA |
| chr11 | 115214667 | 115214731 | 65 | 2 | 4.26E-07 | 7.69E-07 | 8.58E-06 | 4.49E-07 | -0.00205177 | CADM1 |
| chr1 | 118185603 | 118186368 | 766 | 2 | 6.54E-09 | 3.14E-07 | 5.26E-05 | 4.50E-07 | -0.01026138 | AL391557.1 |
| chr3 | 148697656 | 148698521 | 866 | 8 | 1.53E-08 | 3.10E-05 | 0.00059 | 4.51E-07 | -0.00463751 | AGTR1 |
| chr10 | 91633629 | 91634501 | 873 | 3 | 6.32E-12 | 7.26E-06 | 1.79E-06 | 4.51E-07 | 0.009008603 | NA |
| chr16 | 89490960 | 89491616 | 657 | 10 | 3.30E-08 | 1.83E-05 | 0.000587 | 4.51E-07 | 0.000209639 | SPG7 |
| chr8 | 94951666 | 94952123 | 458 | 2 | 5.42E-08 | 2.26E-06 | 2.87E-06 | 4.51E-07 | 0.004534156 | NDUFAF6 |
| chr5 | 168529906 | 168531228 | 1323 | 3 | 6.53E-08 | 5.25E-06 | 3.90E-05 | 4.52E-07 | 0.005054985 | FBLL1 |
| chr3 | 194622263 | 194623085 | 823 | 5 | 2.56E-06 | 2.13E-07 | 0.001657 | 4.52E-07 | -0.01046122 | TMEM44 |
| chr6 | 29374115 | 29375038 | 924 | 10 | 1.06E-09 | 0.0046993 | 4.06E-05 | 4.52E-07 | 0.002711684 | OR5V1, OR12D3 |
| chr2 | 3133128 | 3133303 | 176 | 3 | 3.98E-07 | 1.09E-06 | 2.76E-05 | 4.52E-07 | 0.001171565 | AC019118.2 |
| chr7 | 143904975 | 143904978 | 4 | 2 | 4.61E-07 | 8.68E-07 | 7.42E-06 | 4.52E-07 | 0.002741893 | NA |
| chr7 | 101943481 | 101943996 | 516 | 4 | 7.57E-09 | 9.05E-05 | 1.20E-06 | 4.52E-07 | -0.00319064 | CUX1 |
| chr15 | 68729963 | 68730039 | 77 | 2 | 1.94E-07 | 2.83E-07 | 8.98E-05 | 4.53E-07 | -0.01687526 | NA |
| chr4 | 7635114 | 7635806 | 693 | 4 | 2.24E-07 | 3.26E-07 | 0.000116 | 4.53E-07 | -0.00077622 | SORCS2 |

| | | | | | | | | | | |
|---|---|---|---|---|---|---|---|---|---|---|
| chr11 | 125592256 | 125593544 | 1289 | 12 | 1.41E-07 | 1.93E-06 | 0.000271 | 4.53E-07 | -0.00040791 | STT3A, FEZ1 |
| chr2 | 190648125 | 190648966 | 842 | 8 | 1.88E-06 | 1.05E-07 | 0.004805 | 4.54E-07 | 0.001013171 | NAB1, AC006460.2 |
| chr12 | 1566803 | 1567090 | 288 | 3 | 1.77E-07 | 1.44E-05 | 6.63E-05 | 4.54E-07 | 0.002719945 | WNT5B, FBXL14 |
| chr3 | 122515287 | 122516271 | 985 | 4 | 2.53E-10 | 5.04E-05 | 7.24E-07 | 4.54E-07 | -0.00035181 | AC096861.2 |
| chr3 | 40456984 | 40458559 | 1576 | 15 | 3.86E-09 | 0.0001473 | 1.18E-05 | 4.54E-07 | -0.00094937 | RPL14 |
| chr17 | 1932441 | 1932543 | 103 | 2 | 5.43E-07 | 5.03E-07 | 1.68E-05 | 4.54E-07 | -0.00613516 | NA |
| chr4 | 1328294 | 1329645 | 1352 | 9 | 1.09E-06 | 0.0003434 | 0.000449 | 4.55E-07 | 0.001655695 | MAEA |
| chr1 | 115654849 | 115655530 | 682 | 3 | 9.97E-09 | 8.71E-05 | 5.55E-07 | 4.55E-07 | -0.00909715 | VANGL1 |
| chr1 | 247841584 | 247842421 | 838 | 8 | 9.90E-08 | 3.43E-06 | 5.17E-05 | 4.55E-07 | 0.00339639 | OR11L1 |
| chr17 | 47229833 | 47230380 | 548 | 5 | 4.74E-08 | 3.81E-05 | 2.03E-06 | 4.55E-07 | 0.003558694 | NA |
| chr16 | 67872512 | 67873470 | 959 | 12 | 5.44E-07 | 3.10E-06 | 0.00074 | 4.56E-07 | 0.001389085 | NUTF2, EDC4 |
| chr13 | 33829986 | 33830165 | 180 | 3 | 6.67E-07 | 2.63E-07 | 0.000311 | 4.56E-07 | 0.000819733 | RFC3 |
| chr1 | 36463673 | 36464781 | 1109 | 10 | 1.32E-07 | 2.87E-06 | 0.000322 | 4.56E-07 | 0.000682765 | MRPS15 |
| chr15 | 74797811 | 74799175 | 1365 | 5 | 7.59E-06 | 7.36E-07 | 0.000329 | 4.56E-07 | 0.000791099 | CSK |
| chr2 | 201261656 | 201261769 | 114 | 3 | 2.09E-07 | 1.54E-07 | 0.00082 | 4.57E-07 | 0.021056169 | CASP8 |
| chr14 | 94560551 | 94561768 | 1218 | 8 | 4.10E-08 | 6.80E-06 | 0.000674 | 4.57E-07 | -0.00467194 | SERPINA4, SERPINA5 |
| chr14 | 61228352 | 61228707 | 356 | 2 | 6.20E-08 | 1.01E-06 | 6.31E-06 | 4.58E-07 | -0.01355812 | PRKCH |
| chr12 | 53005309 | 53006419 | 1111 | 15 | 5.13E-06 | 4.45E-08 | 0.014112 | 4.58E-07 | -0.00132895 | EIF4B |
| chr10 | 101111386 | 101112008 | 623 | 2 | 8.21E-07 | 3.32E-07 | 4.66E-05 | 4.58E-07 | -0.0130572 | TLX1NB |
| chr2 | 54657328 | 54657890 | 563 | 3 | 1.43E-06 | 1.93E-07 | 0.000319 | 4.58E-07 | -0.0066405 | SPTBN1 |
| chr21 | 44338471 | 44339619 | 1149 | 13 | 1.87E-07 | 4.66E-06 | 0.001467 | 4.59E-07 | 0.001213713 | AP001062.2, CFAP410 |
| chr11 | 69669665 | 69670467 | 803 | 6 | 5.29E-07 | 2.58E-06 | 0.000125 | 4.59E-07 | -0.00488965 | LTO1 |
| chr1 | 159781562 | 159782389 | 828 | 2 | 1.00E-06 | 2.75E-07 | 0.000122 | 4.59E-07 | -0.00686827 | DUSP23 |
| chr12 | 49821587 | 49822030 | 444 | 4 | 4.70E-08 | 0.0002295 | 1.99E-06 | 4.59E-07 | 0.000144396 | NCKAP5L |
| chr1 | 2004981 | 2005272 | 292 | 3 | 1.38E-07 | 1.53E-07 | 0.000851 | 4.59E-07 | 0.005337435 | NA |
| chr1 | 213978733 | 213980302 | 1570 | 11 | 2.60E-06 | 0.0002049 | 0.000158 | 4.59E-07 | 0.005300319 | PROX1-AS1 |
| chr7 | 30282830 | 30284302 | 1473 | 8 | 5.49E-05 | 9.64E-07 | 0.000712 | 4.59E-07 | 0.000180838 | NA |
| chr2 | 219571502 | 219572596 | 1095 | 9 | 5.76E-08 | 2.54E-05 | 0.000419 | 4.59E-07 | -0.00621123 | INHA, OBSL1 |
| chr14 | 66507201 | 66508335 | 1135 | 15 | 1.09E-07 | 1.39E-06 | 0.000846 | 4.59E-07 | -0.00033632 | GPHN, AL359232.1 |
| chr2 | 127619590 | 127619805 | 216 | 2 | 1.19E-07 | 3.46E-07 | 4.08E-05 | 4.59E-07 | -0.00783521 | MYO7B |
| chr12 | 88534847 | 88535565 | 719 | 4 | 1.57E-08 | 1.62E-06 | 1.55E-05 | 4.59E-07 | -0.00610145 | KITLG |
| chr10 | 104141808 | 104142565 | 758 | 3 | 3.31E-07 | 3.89E-07 | 0.000102 | 4.59E-07 | -0.00517913 | CFAP43 |
| chr2 | 29919720 | 29920873 | 1154 | 7 | 2.08E-09 | 1.16E-05 | 1.04E-05 | 4.59E-07 | -0.00057942 | ALK, AC106870.3 |
| chr2 | 27059211 | 27060416 | 1206 | 4 | 7.23E-10 | 0.0001913 | 2.26E-07 | 4.60E-07 | -0.00490408 | AGBL5 |
| chr9 | 130053423 | 130054754 | 1332 | 14 | 6.36E-09 | 1.41E-05 | 1.19E-05 | 4.60E-07 | 0.001051826 | GPR107 |
| chr6 | 138545728 | 138545988 | 261 | 7 | 9.73E-08 | 5.26E-05 | 1.01E-05 | 4.60E-07 | 0.002995851 | NHSL1 |
| chr6 | 13327949 | 13328852 | 904 | 9 | 1.11E-06 | 6.76E-07 | 0.002224 | 4.60E-07 | 0.000374768 | TBC1D7 |
| chr2 | 237579391 | 237580062 | 672 | 3 | 2.80E-07 | 3.88E-07 | 6.96E-05 | 4.61E-07 | 0.00816547 | RAB17 |
| chr2 | 195796074 | 195796477 | 404 | 3 | 1.09E-06 | 1.62E-07 | 0.000632 | 4.61E-07 | -0.00382264 | DNAH7 |
| chr3 | 57692389 | 57693686 | 1298 | 13 | 5.78E-08 | 5.05E-05 | 0.000816 | 4.62E-07 | -0.00042418 | DENND6A-DT, DENND6A |
| chr3 | 113215597 | 113216243 | 647 | 3 | 7.28E-08 | 1.93E-05 | 3.89E-06 | 4.62E-07 | -0.00521892 | BOC |
| chr1 | 85259409 | 85260373 | 965 | 9 | 2.03E-07 | 2.93E-05 | 0.001444 | 4.62E-07 | -0.00300373 | C1orf52 |
| chr13 | 20189805 | 20190720 | 916 | 6 | 6.99E-08 | 7.21E-07 | 0.000177 | 4.62E-07 | 0.008227864 | GJB2 |
| chr19 | 45405182 | 45407190 | 2009 | 18 | 5.68E-09 | 0.0007601 | 1.10E-05 | 4.62E-07 | 0.001031021 | POLR1G, PPP1R13L |
| chr12 | 65602477 | 65603949 | 1473 | 7 | 1.09E-06 | 4.03E-05 | 3.56E-05 | 4.62E-07 | -0.00313609 | LINC02454, AC025419.1 |
| chr5 | 140351606 | 140351856 | 251 | 2 | 2.13E-07 | 1.57E-06 | 4.07E-06 | 4.62E-07 | 0.01065365 | AC008438.2 |
| chr9 | 121537398 | 121538257 | 860 | 3 | 2.09E-06 | 3.17E-07 | 8.29E-05 | 4.63E-07 | -0.00522905 | NA |
| chr4 | 159358387 | 159358877 | 491 | 2 | 1.00E-07 | 1.01E-06 | 6.50E-06 | 4.63E-07 | -0.00374136 | RAPGEF2 |
| chr10 | 27251884 | 27252688 | 805 | 6 | 1.68E-08 | 0.0002998 | 1.34E-05 | 4.64E-07 | -0.01012886 | LRRC37A6P |
| chr6 | 91816521 | 91816598 | 78 | 3 | 6.10E-07 | 8.81E-07 | 1.82E-05 | 4.64E-07 | -0.00548789 | NA |
| chr5 | 58422471 | 58422807 | 337 | 2 | 2.75E-08 | 1.06E-05 | 1.11E-06 | 4.66E-07 | 0.003259862 | NA |
| chr5 | 135930028 | 135930446 | 419 | 9 | 3.87E-07 | 5.67E-07 | 0.001107 | 4.66E-07 | 0.000390125 | FBXL21P, LECT2 |
| chr15 | 25072130 | 25072334 | 205 | 2 | 4.32E-08 | 3.98E-07 | 2.92E-05 | 4.66E-07 | 0.005810156 | SNHG14 |
| chr1 | 179575938 | 179577150 | 1213 | 10 | 1.05E-07 | 2.89E-07 | 0.001478 | 4.67E-07 | -0.00469785 | RNU5F-2P, NPHS2 |
| chr12 | 109707151 | 109707357 | 207 | 2 | 2.29E-07 | 7.65E-07 | 9.33E-06 | 4.67E-07 | -0.00523587 | NA |
| chr6 | 111086999 | 111088019 | 1021 | 13 | 8.22E-08 | 1.18E-05 | 0.000132 | 4.68E-07 | 0.001455876 | SLC16A10 |
| chr8 | 79327121 | 79327260 | 140 | 2 | 4.12E-07 | 3.07E-07 | 6.93E-05 | 4.68E-07 | -0.0067139 | NA |
| chr12 | 27523288 | 27524205 | 918 | 12 | 7.08E-07 | 7.76E-06 | 0.000725 | 4.68E-07 | 0.001795166 | PPFIBP1 |
| chr12 | 131456881 | 131457047 | 167 | 3 | 5.52E-07 | 7.36E-07 | 2.40E-05 | 4.68E-07 | -0.0091736 | AC073578.5 |
| chr2 | 1621244 | 1621920 | 677 | 5 | 3.25E-08 | 9.53E-05 | 1.71E-06 | 4.68E-07 | 0.000980393 | AC144450.1 |
| chr1 | 7847936 | 7848045 | 110 | 2 | 6.11E-07 | 2.95E-07 | 8.47E-05 | 4.68E-07 | -0.00776263 | UTS2 |
| chr4 | 18022010 | 18023306 | 1297 | 10 | 5.88E-06 | 1.45E-05 | 0.000186 | 4.69E-07 | -0.00135424 | NA |
| chr21 | 37267399 | 37268372 | 974 | 16 | 1.51E-06 | 1.20E-05 | 0.00032 | 4.69E-07 | 0.001573267 | AP001412.1, VPS26C |
| chr19 | 49867735 | 49868710 | 976 | 6 | 7.95E-10 | 5.55E-06 | 1.02E-05 | 4.69E-07 | -0.00693698 | PNKP |
| chr12 | 80762330 | 80762975 | 646 | 2 | 2.03E-09 | 6.12E-06 | 1.52E-06 | 4.70E-07 | -0.01200371 | NA |
| chr11 | 97943189 | 97943756 | 568 | 3 | 3.64E-08 | 3.86E-05 | 1.73E-06 | 4.71E-07 | 0.003096074 | LINC02713 |
| chr17 | 40100159 | 40101683 | 1525 | 13 | 1.96E-08 | 7.96E-05 | 0.000332 | 4.71E-07 | -0.00082842 | NR1D1 |
| chr20 | 38217883 | 38218524 | 642 | 3 | 1.19E-08 | 1.57E-05 | 1.64E-06 | 4.71E-07 | -0.00910486 | KIAA1755 |
| chr20 | 58832692 | 58833740 | 1049 | 4 | 1.76E-08 | 2.85E-06 | 8.80E-06 | 4.71E-07 | -0.00985596 | GNAS-AS1 |
| chr3 | 108223902 | 108225059 | 1158 | 4 | 4.11E-07 | 2.26E-05 | 7.14E-06 | 4.71E-07 | -0.00048388 | NA |
| chr17 | 82657045 | 82657816 | 772 | 3 | 8.05E-08 | 3.68E-07 | 5.74E-05 | 4.72E-07 | -0.0031564 | RAB40B |
| chr7 | 142963113 | 142964009 | 897 | 3 | 3.50E-09 | 0.0004447 | 2.31E-07 | 4.72E-07 | -0.00244026 | NA |
| chr6 | 127459918 | 127460728 | 811 | 4 | 9.08E-08 | 3.12E-06 | 8.39E-05 | 4.72E-07 | -0.0130007 | AL096711.2 |
| chr19 | 12773961 | 12774397 | 437 | 2 | 2.59E-07 | 2.99E-07 | 8.31E-05 | 4.72E-07 | -0.00505287 | HOOK2 |
| chr7 | 28955842 | 28957023 | 1182 | 9 | 1.52E-06 | 3.19E-07 | 0.00166 | 4.72E-07 | 0.002291349 | TRIL |
| chr7 | 102406087 | 102406647 | 561 | 3 | 8.96E-08 | 2.96E-05 | 3.16E-05 | 4.73E-07 | -0.00091721 | PRKRIP1 |
| chr1 | 203440784 | 203440952 | 169 | 2 | 2.04E-07 | 4.04E-06 | 1.97E-06 | 4.73E-07 | -0.01068319 | NA |
| chr19 | 5784173 | 5785241 | 1069 | 8 | 4.31E-08 | 2.72E-06 | 0.000271 | 4.73E-07 | 0.002165583 | PRR22, AC011499.1, DUS3L |
| chr6 | 125956009 | 125957007 | 999 | 11 | 2.08E-08 | 4.70E-05 | 0.000155 | 4.73E-07 | -0.00414993 | HINT3 |
| chr5 | 9631005 | 9631686 | 682 | 4 | 3.17E-08 | 5.55E-06 | 6.56E-06 | 4.73E-07 | -0.00275332 | AC026787.1, TAS2R1 |
| chr6 | 107847996 | 107849143 | 1148 | 8 | 1.63E-06 | 4.73E-07 | 0.000943 | 4.73E-07 | -0.00112459 | NA |
| chr16 | 20326877 | 20328098 | 1222 | 7 | 1.63E-07 | 7.16E-07 | 0.001061 | 4.74E-07 | -0.00628519 | GP2 |
| chr10 | 110609799 | 110610227 | 429 | 3 | 6.66E-08 | 7.76E-06 | 3.51E-05 | 4.74E-07 | -0.01987858 | NA |
| chr20 | 37402997 | 37404249 | 1253 | 6 | 7.43E-06 | 1.15E-06 | 0.000618 | 4.74E-07 | -0.00062538 | SRC |
| chr14 | 21100966 | 21102097 | 1132 | 5 | 2.60E-07 | 8.37E-06 | 3.59E-05 | 4.74E-07 | -0.00311383 | TMEM253, ZNF219 |
| chr21 | 15730445 | 15730680 | 236 | 3 | 2.07E-07 | 8.91E-07 | 1.73E-05 | 4.74E-07 | 0.001197525 | USP25 |
| chr17 | 75940645 | 75941335 | 691 | 11 | 5.06E-08 | 7.66E-06 | 0.000689 | 4.74E-07 | 0.001297502 | FBF1 |
| chr22 | 41672693 | 41673455 | 763 | 2 | 1.25E-06 | 3.76E-07 | 3.57E-05 | 4.74E-07 | -0.00528669 | NA |
| chr14 | 103057179 | 103058652 | 1474 | 9 | 1.69E-06 | 1.15E-06 | 0.000677 | 4.74E-07 | -0.00534942 | CDC42BPB |
| chr11 | 120118119 | 120118941 | 823 | 2 | 1.70E-06 | 3.75E-07 | 3.62E-05 | 4.75E-07 | 0.000738373 | TRIM29 |
| chr1 | 1342061 | 1342615 | 555 | 5 | 3.07E-08 | 0.0005395 | 1.71E-06 | 4.75E-07 | -0.00310437 | DVL1 |
| chr12 | 6974317 | 6974585 | 269 | 4 | 1.65E-07 | 1.94E-05 | 6.54E-06 | 4.75E-07 | 7.66E-05 | EMG1 |
| chr19 | 35533315 | 35533974 | 660 | 11 | 1.68E-07 | 2.65E-06 | 0.000443 | 4.75E-07 | -0.0102068 | GAPDHS |
| chr22 | 33163174 | 33164039 | 866 | 5 | 4.46E-08 | 0.0002086 | 1.66E-05 | 4.76E-07 | -0.00961282 | LARGE1 |
| chr16 | 2811031 | 2812019 | 989 | 2 | 3.18E-05 | 2.88E-07 | 0.000117 | 4.77E-07 | -0.01032426 | NA |
| chr13 | 25287512 | 25288677 | 1166 | 9 | 8.79E-08 | 3.13E-06 | 3.59E-05 | 4.77E-07 | -0.00832864 | MTMR6 |
| chr17 | 3471915 | 3472695 | 781 | 9 | 1.63E-06 | 1.13E-07 | 0.003947 | 4.78E-07 | 0.000525306 | ASPA, SPATA22 |
| chr12 | 117027793 | 117028044 | 252 | 2 | 1.24E-07 | 2.36E-06 | 3.00E-06 | 4.79E-07 | -0.00473105 | FBXW8 |
| chr4 | 8396863 | 8397682 | 820 | 6 | 5.40E-07 | 6.56E-07 | 0.000165 | 4.79E-07 | 0.001212096 | ACOX3 |
| chr16 | 86513022 | 86513924 | 903 | 7 | 4.60E-08 | 1.94E-05 | 0.001992 | 4.79E-07 | -0.00011943 | FOXF1 |
| chr1 | 168136489 | 168138507 | 2019 | 18 | 1.09E-05 | 2.62E-05 | 0.000733 | 4.79E-07 | -0.00042529 | GPR161 |
| chr16 | 85622045 | 85623072 | 1028 | 5 | 1.79E-07 | 3.32E-06 | 2.81E-05 | 4.79E-07 | -0.00143684 | GSE1 |
| chr13 | 42963695 | 42964199 | 505 | 2 | 7.15E-07 | 3.28E-07 | 5.89E-05 | 4.79E-07 | -0.0193157 | EPSTI1 |
| chr2 | 192645310 | 192646009 | 700 | 6 | 2.55E-08 | 8.68E-06 | 5.91E-06 | 4.80E-07 | -0.00211626 | AC062039.1, AC013401.1 |
| chr22 | 23179555 | 23180561 | 1007 | 9 | 1.91E-07 | 3.09E-06 | 1.34E-05 | 4.80E-07 | -0.00310102 | BCR, BCRP8 |
| chr1 | 200302327 | 200303087 | 761 | 6 | 1.02E-06 | 5.71E-07 | 0.001282 | 4.81E-07 | 0.017811618 | LINC00862 |
| chr1 | 25780646 | 25781026 | 381 | 2 | 6.66E-07 | 3.20E-07 | 6.65E-05 | 4.81E-07 | 0.006139279 | MAN1C1 |
| chr2 | 16726951 | 16727499 | 549 | 3 | 1.22E-06 | 2.78E-07 | 0.000174 | 4.81E-07 | 0.008001352 | NA |
| chr4 | 1414769 | 1415769 | 1001 | 6 | 1.44E-11 | 0.0009859 | 6.70E-07 | 4.81E-07 | 0.008704296 | NA |
| chr22 | 43142851 | 43144242 | 1392 | 13 | 3.33E-07 | 8.37E-06 | 0.001543 | 4.82E-07 | 0.000655725 | MCAT |
| chr14 | 101183596 | 101183937 | 342 | 2 | 4.12E-08 | 1.75E-05 | 9.21E-07 | 4.82E-07 | -0.01090177 | NA |
| chr20 | 25223409 | 25224019 | 611 | 2 | 1.39E-08 | 2.74E-06 | 2.69E-06 | 4.83E-07 | 0.006552467 | ENTPD6 |
| chr19 | 12401150 | 12401640 | 491 | 7 | 1.54E-07 | 8.41E-05 | 0.000171 | 4.83E-07 | 0.003456682 | AC008758.4, ZNF799, AC008758.5 |
| chr1 | 224457048 | 224457674 | 627 | 3 | 1.79E-07 | 4.68E-05 | 6.29E-05 | 4.83E-07 | -0.0014772 | CNIH3 |
| chr1 | 27098904 | 27100064 | 1161 | 5 | 1.00E-08 | 2.00E-05 | 8.25E-07 | 4.83E-07 | 0.004227769 | SLC9A1 |
| chr16 | 1528023 | 1528806 | 784 | 8 | 1.05E-08 | 5.94E-05 | 0.000548 | 4.83E-07 | -0.00591101 | TMEM204, IFT140 |
| chr1 | 6260157 | 6260700 | 544 | 5 | 2.53E-08 | 3.73E-06 | 4.81E-05 | 4.83E-07 | 0.001127484 | GPR153 |
| chr3 | 52063562 | 52063887 | 326 | 3 | 2.08E-07 | 6.68E-06 | 7.95E-05 | 4.83E-07 | -0.0042165 | NA |
| chr4 | 100189266 | 100190485 | 1220 | 10 | 4.17E-09 | 2.19E-05 | 0.000392 | 4.84E-07 | 0.000169336 | H2AZ1-DT, AP001961.1, DDIT4L |
| chr3 | 66453857 | 66453946 | 90 | 2 | 6.98E-08 | 0.004066 | 4.02E-07 | 4.84E-07 | -0.00898412 | LRIG1 |
| chr5 | 161684435 | 161685781 | 1347 | 7 | 1.22E-05 | 2.38E-05 | 0.000143 | 4.84E-07 | -0.00870686 | GABRA6 |
| chr16 | 22087136 | 22087172 | 37 | 3 | 3.89E-07 | 2.45E-06 | 4.91E-05 | 4.85E-07 | 0.010662564 | MOSMO, AC009019.1 |
| chr10 | 124092495 | 124094513 | 2019 | 15 | 3.29E-06 | 9.11E-06 | 0.001852 | 4.85E-07 | -0.0009331 | CHST15 |
| chr5 | 17302035 | 17302166 | 132 | 2 | 1.42E-07 | 1.45E-06 | 4.76E-06 | 4.85E-07 | -0.00736625 | NA |
| chr13 | 45340790 | 45341882 | 1093 | 12 | 2.59E-06 | 2.93E-06 | 0.001571 | 4.85E-07 | 8.35E-05 | AL138963.3, TPT1-AS1, TPT1 |
| chr12 | 132595879 | 132596193 | 315 | 2 | 9.06E-07 | 2.92E-07 | 0.000126 | 4.88E-07 | -0.02256561 | NA |
| chr5 | 139293474 | 139294813 | 1340 | 15 | 1.38E-07 | 5.17E-06 | 0.001473 | 4.88E-07 | 0.000704298 | MATR3, MATR3, SIL1 |
| chr3 | 172447727 | 172448727 | 1001 | 10 | 3.31E-07 | 2.05E-05 | 0.001572 | 4.88E-07 | 0.00419403 | GHSR |
| chr16 | 81785150 | 81785709 | 560 | 3 | 1.15E-08 | 5.91E-06 | 3.37E-06 | 4.88E-07 | -0.00500029 | PLCG2 |
| chr14 | 104017203 | 104017456 | 254 | 2 | 5.45E-07 | 4.18E-07 | 2.97E-05 | 4.88E-07 | 0.014174208 | TDRD9 |
| chr19 | 5208369 | 5209007 | 639 | 3 | 7.94E-07 | 2.23E-06 | 2.04E-05 | 4.89E-07 | -0.00372957 | PTPRS |
| chr19 | 40798167 | 40800195 | 2029 | 11 | 7.28E-08 | 2.43E-05 | 0.000656 | 4.90E-07 | 0.000791535 | RAB4B-EGLN2, EGLN2 |
| chr4 | 129093832 | 129094965 | 1134 | 6 | 2.42E-07 | 7.07E-06 | 0.00015 | 4.90E-07 | -0.00515729 | C4orf33 |
| chr3 | 138328520 | 138329560 | 1041 | 5 | 2.73E-06 | 4.85E-07 | 0.00117 | 4.90E-07 | -0.0031463 | NME9 |
| chr11 | 118971416 | 118971774 | 359 | 7 | 2.02E-07 | 2.50E-05 | 0.00012 | 4.90E-07 | -0.00685515 | FOXR1 |
| chr20 | 40962320 | 40963147 | 828 | 4 | 8.35E-07 | 5.36E-07 | 0.000155 | 4.91E-07 | -0.00704679 | NA |
| chr4 | 8844763 | 8844767 | 5 | 2 | 8.44E-07 | 2.93E-07 | 0.000127 | 4.91E-07 | -0.04391759 | NA |
| chr1 | 222711570 | 222713166 | 1597 | 15 | 1.02E-07 | 1.10E-05 | 0.00105 | 4.92E-07 | 0.000148091 | BROX, AIDA |
| chr7 | 37448559 | 37449571 | 1013 | 16 | 1.19E-07 | 5.33E-06 | 0.000905 | 4.92E-07 | 0.003295654 | ELMO1 |
| chr6 | 169808195 | 169808210 | 16 | 2 | 6.79E-07 | 4.14E-07 | 3.11E-05 | 4.93E-07 | -0.04070638 | NA |
| chr22 | 37608215 | 37609343 | 1129 | 21 | 9.92E-07 | 6.91E-07 | 0.008154 | 4.93E-07 | 0.001397586 | GGA1 |
| chr11 | 64240856 | 64242441 | 1586 | 16 | 1.29E-09 | 3.03E-05 | 0.000672 | 4.93E-07 | -0.00124322 | FKBP2, AP001453.3 |
| chr5 | 134226243 | 134227525 | 1283 | 13 | 4.30E-06 | 1.81E-06 | 0.001519 | 4.94E-07 | 0.001553552 | AC104109.2, AC104109.4 |
| chr15 | 99467105 | 99467356 | 252 | 2 | 1.35E-07 | 5.05E-06 | 1.81E-06 | 4.96E-07 | -0.00993898 | NA |
| chr20 | 46708317 | 46709757 | 1441 | 12 | 1.25E-05 | 8.47E-07 | 0.003636 | 4.96E-07 | -0.00044377 | SLC2A10 |
| chr6 | 119078155 | 119079315 | 1161 | 8 | 1.95E-06 | 1.88E-06 | 0.002701 | 4.96E-07 | 0.000365341 | AL137009.1, FAM184A |
| chr1 | 213491648 | 213492240 | 593 | 5 | 2.60E-08 | 2.63E-06 | 6.77E-05 | 4.97E-07 | -0.00130465 | NA |
| chr2 | 237590280 | 237591159 | 880 | 11 | 3.90E-07 | 2.72E-07 | 0.006188 | 4.97E-07 | -0.00342237 | AC104667.2, RAB17 |
| chr8 | 29348038 | 29349393 | 1356 | 9 | 9.98E-07 | 4.76E-07 | 0.001884 | 4.97E-07 | 0.002844405 | DUSP4 |
| chr4 | 7393063 | 7393885 | 823 | 7 | 7.82E-08 | 1.41E-05 | 4.81E-06 | 4.97E-07 | -0.00532443 | SORCS2 |
| chr8 | 144095484 | 144096382 | 899 | 6 | 5.38E-08 | 1.27E-06 | 0.000115 | 4.97E-07 | -0.00163693 | CYC1 |
| chr7 | 6409984 | 6410350 | 367 | 2 | 4.61E-09 | 1.13E-05 | 1.16E-06 | 4.98E-07 | 0.008916087 | DAGLB |
| chr16 | 84222430 | 84223259 | 830 | 4 | 1.40E-07 | 1.67E-05 | 8.08E-05 | 4.98E-07 | -0.0057532 | KCNG4 |
| chr1 | 176206521 | 176208030 | 1510 | 15 | 1.70E-07 | 5.05E-06 | 0.00049 | 4.98E-07 | 0.000914841 | AL591043.2, COP1 |
| chr10 | 124525189 | 124525317 | 129 | 2 | 4.56E-07 | 3.02E-07 | 0.000111 | 4.98E-07 | -0.01464766 | LHPP |
| chr22 | 17159102 | 17160082 | 981 | 8 | 2.91E-06 | 1.09E-07 | 0.009516 | 5.01E-07 | -0.00885257 | HDHD5-AS1, HDHD5 |
| chr12 | 117888187 | 117889554 | 1368 | 7 | 1.94E-05 | 9.27E-07 | 0.000979 | 5.01E-07 | -0.00395529 | AC084291.1, KSR2 |
| chr1 | 223652514 | 223653272 | 759 | 4 | 2.35E-07 | 3.43E-06 | 9.28E-05 | 5.02E-07 | -0.00757292 | CAPN8 |

| | | | | | | | | | | |
|---|---|---|---|---|---|---|---|---|---|---|
| chr6 | 28896528 | 28897055 | 528 | 17 | 7.87E-07 | 4.13E-07 | 0.003557 | 5.02E-07 | 5.32E-06 | HCG14 |
| chr5 | 140868805 | 140869102 | 298 | 6 | 2.97E-07 | 1.79E-06 | 3.09E-05 | 5.02E-07 | -0.00803558 | PCDHA1, PCDHA2, PCDHA3, PCDHA4, PCDHA5, PCDHA6, PCDHA7, PCDHA8, PCDHA9, PCDHA10, PCDHA11 |
| chr6 | 159978668 | 159979287 | 620 | 2 | 1.68E-06 | 3.06E-07 | 0.000109 | 5.04E-07 | 0.004051317 | IGF2R |
| chr6 | 151807695 | 151809053 | 1359 | 11 | 2.82E-06 | 2.94E-07 | 0.014862 | 5.04E-07 | -0.0036455 | ESR1 |
| chr6 | 30333650 | 30334165 | 516 | 3 | 5.99E-07 | 5.20E-07 | 6.31E-05 | 5.05E-07 | -0.00839084 | TRIM39, TRIM39-RPP21 |
| chr19 | 45037508 | 45038225 | 718 | 7 | 7.93E-08 | 1.72E-05 | 0.00012 | 5.05E-07 | 0.00061626 | RELB |
| chr8 | 129269488 | 129269841 | 354 | 2 | 3.24E-07 | 3.71E-07 | 4.58E-05 | 5.06E-07 | -0.0196417 | CCDC26 |
| chr3 | 136818327 | 136819527 | 1201 | 12 | 1.76E-07 | 4.48E-07 | 0.007191 | 5.06E-07 | 0.001175137 | SLC35G2 |
| chr15 | 79287080 | 79287856 | 777 | 2 | 5.07E-06 | 3.14E-07 | 9.50E-05 | 5.06E-07 | -0.0155605 | NA |
| chr4 | 24584256 | 24585535 | 1280 | 9 | 2.22E-06 | 0.0001038 | 0.000125 | 5.06E-07 | 0.00055342 | DHX15 |
| chr2 | 43831627 | 43832603 | 977 | 8 | 5.64E-08 | 4.86E-06 | 5.24E-05 | 5.06E-07 | -0.00340137 | ABCG8, ABCG5 |
| chr10 | 29710964 | 29712196 | 1233 | 3 | 3.79E-09 | 4.88E-06 | 3.73E-06 | 5.07E-07 | -0.0084292 | RNU6-908P, SVIL |
| chr16 | 65283552 | 65283921 | 370 | 2 | 2.32E-07 | 6.42E-07 | 1.42E-05 | 5.08E-07 | -0.01628079 | AC009055.1 |
| chr12 | 6354729 | 6355620 | 892 | 3 | 6.06E-07 | 8.91E-07 | 4.75E-05 | 5.09E-07 | -0.00595346 | SCNN1A |
| chr7 | 36986191 | 36987388 | 1198 | 11 | 2.08E-06 | 5.58E-07 | 0.000271 | 5.09E-07 | -0.00321388 | ELMO1 |
| chr4 | 90127406 | 90128577 | 1172 | 10 | 3.89E-07 | 1.22E-06 | 0.003352 | 5.09E-07 | 0.000660153 | CCSER1 |
| chr11 | 289441 | 290447 | 1007 | 7 | 7.46E-07 | 1.67E-07 | 0.003541 | 5.09E-07 | -0.04384298 | PGGHG |
| chr2 | 135231239 | 135231832 | 594 | 2 | 1.87E-06 | 3.09E-07 | 0.000109 | 5.10E-07 | 0.003384069 | ZRANB3 |
| chr7 | 17304924 | 17305597 | 674 | 2 | 1.09E-07 | 6.55E-07 | 1.39E-05 | 5.10E-07 | -0.00357326 | AHR, AC019117.3 |
| chr20 | 609599 | 610155 | 557 | 2 | 7.79E-13 | 0.0004187 | 4.72E-07 | 5.10E-07 | -0.01583488 | TCF15 |
| chr15 | 51622095 | 51623216 | 1122 | 10 | 1.25E-07 | 1.20E-05 | 3.57E-05 | 5.10E-07 | 0.000259644 | DMXL2 |
| chr17 | 7592424 | 7592851 | 428 | 4 | 2.12E-07 | 2.91E-06 | 7.30E-05 | 5.10E-07 | -0.00585915 | MPDU1, FXR2 |
| chr14 | 89711850 | 89712335 | 486 | 3 | 8.64E-07 | 2.24E-07 | 0.000322 | 5.10E-07 | -0.01106531 | AL137230.2 |
| chr7 | 55569430 | 55570537 | 1108 | 3 | 1.02E-06 | 2.44E-07 | 0.000412 | 5.10E-07 | 0.000473332 | VOPP1 |
| chr2 | 112085325 | 112085909 | 585 | 3 | 1.14E-06 | 1.91E-07 | 0.000543 | 5.10E-07 | 0.055236176 | TMEM87B |
| chr17 | 39867227 | 39868141 | 915 | 7 | 2.62E-09 | 7.43E-05 | 1.70E-06 | 5.11E-07 | -0.00962264 | NA |
| chr16 | 30697841 | 30698457 | 617 | 7 | 9.61E-08 | 7.41E-06 | 0.00029 | 5.11E-07 | -0.00216369 | SRCAP, AC093249.6 |
| chr5 | 138458018 | 138458620 | 603 | 2 | 2.21E-09 | 0.0007265 | 4.51E-07 | 5.12E-07 | -0.00897797 | NA |
| chr10 | 102460618 | 102461841 | 1224 | 10 | 1.18E-06 | 1.46E-07 | 0.005543 | 5.12E-07 | 0.001812615 | RPARP-AS1, MFSD13A |
| chr2 | 234422349 | 234423079 | 731 | 3 | 1.72E-06 | 3.40E-07 | 9.90E-05 | 5.12E-07 | -0.01183565 | NA |
| chr20 | 50636827 | 50637173 | 347 | 4 | 5.72E-08 | 9.97E-06 | 5.23E-06 | 5.12E-07 | -0.00853544 | RIPOR3 |
| chr11 | 58204698 | 58205141 | 444 | 2 | 3.93E-07 | 4.72E-07 | 2.56E-05 | 5.14E-07 | 0.01659891 | NA |
| chr6 | 166691113 | 166691847 | 735 | 6 | 1.06E-07 | 6.98E-06 | 0.000262 | 5.14E-07 | -0.00252824 | RPS6KA2 |
| chr2 | 206937729 | 206938615 | 887 | 4 | 1.54E-06 | 4.18E-07 | 0.000367 | 5.14E-07 | -0.00279893 | NA |
| chr15 | 84633410 | 84634561 | 1152 | 7 | 3.29E-08 | 9.12E-06 | 1.17E-05 | 5.16E-07 | -0.00055365 | SCAND2P, EGLN1P1, AC048382.5 |
| chr19 | 13763147 | 13764827 | 1681 | 12 | 1.50E-07 | 1.62E-07 | 0.00286 | 5.16E-07 | 0.011440927 | CCDC130, MRI1 |
| chr12 | 55726413 | 55727701 | 1289 | 8 | 6.41E-08 | 1.49E-05 | 0.000132 | 5.16E-07 | -0.00157078 | CD63 |
| chr3 | 50611721 | 50612979 | 1259 | 13 | 1.36E-06 | 1.46E-05 | 0.001458 | 5.17E-07 | -0.00119345 | MAPKAPK3, CISH |
| chr5 | 180689385 | 180689520 | 136 | 2 | 4.99E-08 | 2.33E-05 | 8.86E-07 | 5.17E-07 | -0.00848534 | NA |
| chr7 | 135388036 | 135388675 | 640 | 4 | 1.55E-07 | 2.23E-06 | 0.000188 | 5.18E-07 | 0.005150115 | CNOT4 |
| chr9 | 34710138 | 34710660 | 523 | 2 | 1.03E-07 | 1.23E-06 | 6.21E-06 | 5.19E-07 | -0.01482116 | AL162231.5 |
| chr5 | 119071005 | 119071830 | 826 | 12 | 6.44E-07 | 2.86E-06 | 0.007642 | 5.19E-07 | -0.00019823 | DMXL1 |
| chr11 | 75814480 | 75815864 | 1385 | 14 | 4.89E-07 | 7.75E-07 | 0.001547 | 5.19E-07 | 0.001414058 | UVRAG, UVRAG-DT |
| chr3 | 102673867 | 102674506 | 640 | 6 | 1.53E-06 | 3.03E-06 | 0.000217 | 5.20E-07 | -0.00528156 | AC063938.1 |
| chr15 | 32671954 | 32672862 | 909 | 7 | 7.86E-08 | 1.67E-06 | 0.000308 | 5.20E-07 | -0.00210174 | SCG5, SCG5-AS1 |
| chr9 | 8340450 | 8340565 | 116 | 2 | 6.30E-07 | 5.45E-07 | 1.97E-05 | 5.20E-07 | -0.01359662 | PTPRD |
| chr10 | 121927865 | 121928905 | 1041 | 16 | 1.18E-06 | 2.36E-06 | 0.007789 | 5.21E-07 | 0.009876073 | ATE1-AS1, ATE1 |
| chr6 | 46793107 | 46793466 | 360 | 6 | 8.80E-08 | 4.40E-05 | 3.17E-06 | 5.22E-07 | -0.00656199 | MEP1A |
| chr19 | 41363783 | 41365269 | 1487 | 19 | 8.39E-08 | 7.39E-06 | 0.001864 | 5.23E-07 | 0.001466206 | AC011462.1, TMEM91, B9D2 |
| chr12 | 116525048 | 116525666 | 619 | 3 | 2.45E-07 | 5.69E-07 | 7.18E-05 | 5.23E-07 | -0.00748838 | NA |
| chr15 | 65326415 | 65327225 | 811 | 2 | 4.53E-06 | 3.34E-07 | 8.22E-05 | 5.23E-07 | -0.01129163 | IGDCC3 |
| chr20 | 56596126 | 56596593 | 468 | 2 | 4.76E-07 | 3.82E-07 | 4.74E-05 | 5.24E-07 | -0.01801446 | LINC01716 |
| chr17 | 10469655 | 10470774 | 1120 | 6 | 1.32E-08 | 0.0001249 | 3.57E-05 | 5.24E-07 | -0.00162202 | AC005323.2, MYHAS |
| chr4 | 168741648 | 168741927 | 280 | 2 | 6.00E-08 | 3.10E-05 | 8.11E-07 | 5.24E-07 | -0.01301748 | PALLD |
| chr12 | 71032538 | 71033075 | 538 | 3 | 2.38E-08 | 5.11E-05 | 8.03E-06 | 5.26E-07 | -0.00358652 | NA |
| chr18 | 74484840 | 74485079 | 240 | 3 | 1.34E-06 | 1.81E-07 | 0.000778 | 5.26E-07 | -0.02737748 | NA |
| chr17 | 40363589 | 40364401 | 813 | 5 | 2.12E-09 | 0.0001627 | 0.000238 | 5.27E-07 | -0.01538337 | AC080112.1, GJD3 |
| chr22 | 48980374 | 48981074 | 701 | 5 | 2.38E-06 | 1.42E-05 | 0.000377 | 5.28E-07 | 0.001661986 | NA |
| chr7 | 1726418 | 1726937 | 520 | 2 | 1.48E-08 | 4.98E-07 | 2.46E-05 | 5.29E-07 | -0.01183269 | ELFN1 |
| chr21 | 45041545 | 45041821 | 277 | 2 | 8.85E-07 | 3.17E-07 | 0.000122 | 5.29E-07 | -0.01492517 | NA |
| chr22 | 26550214 | 26550776 | 563 | 2 | 1.93E-06 | 3.16E-07 | 0.000124 | 5.29E-07 | 0.003203634 | TPST2 |
| chr11 | 18394094 | 18396026 | 1933 | 17 | 4.01E-06 | 2.97E-05 | 0.001832 | 5.29E-07 | -0.00136278 | LDHA |
| chr11 | 128548656 | 128549617 | 962 | 6 | 6.17E-08 | 2.85E-05 | 1.77E-05 | 5.30E-07 | -0.00390485 | ETS1 |
| chr9 | 101613630 | 101614279 | 650 | 2 | 1.53E-06 | 3.51E-07 | 6.89E-05 | 5.30E-07 | -0.00623291 | GRIN3A |
| chr17 | 49514132 | 49515318 | 1187 | 5 | 2.80E-07 | 6.71E-06 | 7.51E-05 | 5.31E-07 | -0.00274718 | NGFR, AC006487.1 |
| chr1 | 77874894 | 77874948 | 55 | 2 | 2.37E-07 | 1.10E-05 | 1.26E-06 | 5.31E-07 | -0.00630988 | MIGA1 |
| chr6 | 40344376 | 40344605 | 230 | 2 | 3.11E-07 | 1.24E-06 | 6.40E-06 | 5.31E-07 | -0.02960383 | TDRG1, LINC00951 |
| chr7 | 100463988 | 100464440 | 453 | 3 | 2.65E-07 | 6.41E-06 | 1.09E-05 | 5.31E-07 | -0.00641643 | C7orf61, TSC22D4 |
| chr3 | 193852467 | 193852621 | 155 | 3 | 2.86E-07 | 9.72E-06 | 6.56E-06 | 5.31E-07 | -0.01254897 | NA |
| chr1 | 184036762 | 184037838 | 1077 | 8 | 7.51E-07 | 1.52E-06 | 0.000779 | 5.33E-07 | -0.00113264 | COLGALT2 |
| chr17 | 77088089 | 77089308 | 1220 | 17 | 4.19E-08 | 2.26E-05 | 0.000548 | 5.33E-07 | 0.000249962 | SNHG20, SEC14L1, SCARNA16 |
| chr19 | 13275312 | 13275849 | 538 | 2 | 1.32E-08 | 7.44E-06 | 1.56E-06 | 5.33E-07 | 0.003610302 | CACNA1A |
| chr4 | 127631843 | 127633172 | 1330 | 14 | 5.04E-08 | 9.71E-06 | 0.000232 | 5.34E-07 | -0.00011984 | INTU |
| chr17 | 43024452 | 43026007 | 1556 | 11 | 6.61E-07 | 9.02E-07 | 0.000189 | 5.34E-07 | 0.000434333 | RND2, VAT1 |
| chr16 | 1612298 | 1613676 | 1379 | 9 | 3.71E-07 | 1.06E-06 | 0.000145 | 5.34E-07 | 0.001796835 | CRAMP1 |
| chr6 | 132401631 | 132402908 | 1278 | 9 | 4.58E-07 | 3.10E-06 | 2.59E-05 | 5.34E-07 | -0.00207548 | NA |
| chr18 | 37566020 | 37567502 | 1483 | 25 | 1.14E-10 | 0.0001052 | 2.67E-05 | 5.34E-07 | 0.000837098 | AC009899.1 |
| chr11 | 120137317 | 120138290 | 974 | 9 | 1.60E-10 | 0.0001343 | 2.74E-07 | 5.34E-07 | 0.001118998 | TRIM29 |
| chr10 | 61661600 | 61663083 | 1484 | 12 | 6.81E-09 | 2.29E-06 | 0.000517 | 5.35E-07 | -0.00348466 | CABCOCO1 |
| chr1 | 151047251 | 151048257 | 1007 | 9 | 1.04E-06 | 1.15E-06 | 0.002435 | 5.35E-07 | -0.00022129 | BNIPL, C1orf56 |
| chr5 | 176861497 | 176862124 | 628 | 2 | 4.56E-09 | 3.43E-06 | 2.58E-06 | 5.36E-07 | -0.00305113 | UNC5A |
| chr6 | 31545421 | 31546964 | 1544 | 16 | 2.36E-07 | 1.69E-06 | 0.003857 | 5.37E-07 | -0.00148467 | NFKBIL1, ATP6V1G2-DDX39B, ATP6V1G2 |
| chr4 | 1010737 | 1011617 | 881 | 20 | 3.22E-07 | 3.35E-07 | 0.004781 | 5.37E-07 | 0.001161164 | FGFRL1 |
| chr16 | 75930495 | 75931130 | 636 | 2 | 2.90E-08 | 3.65E-06 | 2.47E-06 | 5.38E-07 | -0.02182488 | NA |
| chr11 | 94542585 | 94544024 | 1440 | 10 | 3.14E-12 | 0.0041236 | 7.86E-08 | 5.38E-07 | -0.003354 | PIWIL4, FUT4 |
| chr11 | 18046380 | 18046808 | 429 | 2 | 2.47E-07 | 3.21E-07 | 0.000129 | 5.39E-07 | -0.01699179 | NA |
| chr7 | 127414025 | 127414478 | 454 | 2 | 2.00E-07 | 7.75E-07 | 1.19E-05 | 5.39E-07 | 0.005711517 | ZNF800 |
| chr22 | 33657248 | 33658054 | 807 | 6 | 3.16E-09 | 4.16E-05 | 1.95E-06 | 5.40E-07 | -0.00655533 | LARGE1 |
| chr1 | 93163717 | 93164662 | 946 | 2 | 1.36E-05 | 3.42E-07 | 8.58E-05 | 5.40E-07 | -0.01182995 | TMED5 |
| chr1 | 55081417 | 55081420 | 4 | 2 | 6.99E-07 | 6.58E-07 | 1.55E-05 | 5.40E-07 | -0.00122022 | USP24 |
| chr6 | 26027866 | 26028553 | 688 | 5 | 2.71E-07 | 1.82E-06 | 4.65E-05 | 5.41E-07 | 0.001013087 | NA |
| chr12 | 1860566 | 1861004 | 439 | 2 | 1.85E-09 | 0.0023388 | 4.45E-07 | 5.41E-07 | 0.003849549 | CACNA2D4 |
| chr11 | 133305744 | 133305988 | 245 | 2 | 1.82E-07 | 3.89E-06 | 2.38E-06 | 5.41E-07 | -0.02724284 | OPCML |
| chr1 | 205749772 | 205750436 | 665 | 6 | 1.24E-07 | 1.63E-06 | 4.10E-05 | 5.41E-07 | 0.001421684 | NUCKS1 |
| chr11 | 34570363 | 34571308 | 946 | 6 | 2.02E-07 | 1.91E-06 | 0.000673 | 5.41E-07 | -0.01080525 | LINC02707 |
| chr3 | 9124977 | 9125621 | 645 | 4 | 2.38E-08 | 7.97E-06 | 2.41E-06 | 5.41E-07 | -0.00708281 | SRGAP3 |
| chr16 | 19167909 | 19168890 | 982 | 10 | 6.55E-08 | 2.14E-05 | 0.002161 | 5.41E-07 | -0.00251151 | AC099518.3, SYT17 |
| chr22 | 18028717 | 18029315 | 599 | 3 | 8.96E-08 | 1.40E-06 | 2.00E-05 | 5.42E-07 | -0.01544254 | NA |
| chr11 | 334251 | 334561 | 311 | 5 | 7.20E-07 | 5.33E-07 | 0.000308 | 5.42E-07 | -0.00811824 | NA |
| chr7 | 116614734 | 116615629 | 896 | 7 | 2.24E-06 | 5.75E-07 | 0.00114 | 5.43E-07 | -0.00152911 | COMETT |
| chr5 | 147509065 | 147510454 | 1390 | 15 | 1.82E-06 | 0.0001231 | 0.001396 | 5.43E-07 | -0.00144408 | DPYSL3 |
| chr3 | 44992777 | 44993418 | 642 | 2 | 3.30E-08 | 3.23E-06 | 2.75E-06 | 5.43E-07 | -0.00904896 | EXOSC7 |
| chr8 | 141176394 | 141176674 | 281 | 3 | 7.81E-09 | 2.78E-05 | 8.61E-07 | 5.43E-07 | -0.00307678 | DENND3 |
| chr6 | 28709164 | 28710072 | 909 | 6 | 5.68E-07 | 4.67E-06 | 2.43E-05 | 5.43E-07 | -0.00583316 | NA |
| chr19 | 53520478 | 53521395 | 918 | 15 | 4.42E-08 | 4.03E-05 | 0.000377 | 5.43E-07 | 0.00106043 | ZNF331 |
| chr21 | 41458752 | 41458879 | 128 | 4 | 2.38E-07 | 4.10E-05 | 2.32E-05 | 5.44E-07 | -0.00388532 | MX1 |
| chr2 | 741806 | 742427 | 622 | 7 | 4.04E-07 | 7.97E-06 | 0.000387 | 5.44E-07 | -0.00673879 | AC116609.2 |
| chr2 | 74178249 | 74179128 | 880 | 11 | 2.94E-07 | 2.01E-05 | 0.000217 | 5.44E-07 | 9.41E-05 | MOB1A |
| chr12 | 13095614 | 13097264 | 1651 | 7 | 4.71E-05 | 5.11E-07 | 0.001832 | 5.44E-07 | 0.002143235 | FAM234B, GSG1 |
| chr9 | 14847275 | 14847782 | 508 | 3 | 8.87E-08 | 2.30E-06 | 1.23E-05 | 5.44E-07 | -0.00331718 | FREM1 |
| chr2 | 129059237 | 129060076 | 840 | 4 | 1.15E-06 | 1.59E-07 | 0.001452 | 5.45E-07 | -0.003725 | NA |
| chr22 | 38289841 | 38290143 | 303 | 2 | 3.72E-08 | 2.95E-05 | 8.58E-07 | 5.45E-07 | -0.01183746 | NA |
| chr6 | 24910492 | 24911387 | 896 | 15 | 2.23E-08 | 0.0007018 | 4.60E-06 | 5.45E-07 | 0.000578545 | RIPOR2 |
| chr1 | 24400725 | 24401637 | 913 | 4 | 3.31E-08 | 3.88E-05 | 1.62E-05 | 5.45E-07 | -0.00129985 | STPG1 |
| chr5 | 177351471 | 177352376 | 906 | 16 | 5.81E-07 | 1.28E-05 | 0.000492 | 5.45E-07 | -0.00010805 | LMAN2 |
| chr2 | 42369080 | 42369408 | 329 | 3 | 2.45E-07 | 3.71E-06 | 9.68E-05 | 5.47E-07 | -0.01001433 | COX7A2L |
| chr9 | 125143655 | 125144659 | 1005 | 7 | 1.84E-07 | 4.66E-05 | 4.58E-05 | 5.48E-07 | -0.00336603 | NA |
| chr22 | 22043969 | 22044825 | 857 | 5 | 3.90E-06 | 1.11E-07 | 0.003707 | 5.48E-07 | -0.00784767 | IGLV10-54 |
| chr16 | 51150294 | 51151550 | 1257 | 23 | 5.93E-07 | 3.29E-06 | 0.00319 | 5.49E-07 | 0.000585503 | AC087564.1, SALL1 |
| chr11 | 66438523 | 66439423 | 901 | 7 | 1.72E-06 | 2.68E-07 | 0.001177 | 5.49E-07 | 0.003143928 | MRPL11 |
| chr2 | 27377021 | 27377456 | 436 | 2 | 3.04E-08 | 1.97E-06 | 4.23E-06 | 5.49E-07 | 0.004340769 | SNX17, ZNF513 |
| chr15 | 93982268 | 93982452 | 185 | 2 | 5.61E-08 | 2.60E-05 | 9.01E-07 | 5.49E-07 | -0.00913556 | LINC01580, LINC01579, LINC01581 |
| chr9 | 94156743 | 94157688 | 946 | 4 | 3.11E-07 | 5.34E-06 | 1.05E-05 | 5.49E-07 | 0.0087602 | AL158152.1 |
| chr16 | 83137046 | 83137694 | 649 | 4 | 5.88E-07 | 2.59E-07 | 0.000534 | 5.49E-07 | -0.02732201 | CDH13 |
| chr20 | 63244894 | 63244943 | 50 | 2 | 8.18E-07 | 4.37E-07 | 3.74E-05 | 5.50E-07 | -0.01062391 | NKAIN4 |
| chr4 | 1746379 | 1747514 | 1136 | 5 | 2.59E-06 | 1.49E-06 | 0.000206 | 5.50E-07 | -0.01000523 | NA |
| chr2 | 106841300 | 106841724 | 425 | 3 | 6.14E-08 | 3.69E-06 | 5.86E-05 | 5.50E-07 | -0.00524804 | ST6GAL2 |
| chr21 | 43811727 | 43812720 | 994 | 5 | 7.81E-07 | 1.26E-05 | 4.29E-05 | 5.51E-07 | 0.005659247 | AATBC |
| chr17 | 80743084 | 80743332 | 249 | 3 | 2.75E-07 | 8.12E-06 | 3.79E-05 | 5.51E-07 | -0.01665995 | RPTOR |
| chr5 | 472101 | 472892 | 792 | 7 | 2.23E-07 | 5.23E-06 | 0.000625 | 5.51E-07 | 0.009608807 | PP7080, SLC9A3 |
| chr3 | 135627426 | 135627812 | 387 | 2 | 2.54E-08 | 4.32E-05 | 7.66E-07 | 5.51E-07 | 0.004521324 | NA |
| chr11 | 65595420 | 65596646 | 1227 | 8 | 9.82E-09 | 2.97E-06 | 7.78E-05 | 5.51E-07 | -0.00105007 | KCNK7 |
| chr16 | 1186356 | 1187145 | 790 | 6 | 3.01E-07 | 3.19E-06 | 0.000379 | 5.51E-07 | -0.00072351 | CACNA1H |
| chr18 | 48538171 | 48539396 | 1226 | 11 | 5.67E-08 | 4.74E-05 | 0.000742 | 5.51E-07 | -0.0035777 | CTIF |
| chr7 | 148819657 | 148820679 | 1023 | 4 | 3.05E-07 | 2.41E-07 | 0.000222 | 5.51E-07 | -0.0098399 | EZH2 |
| chr12 | 3655662 | 3656079 | 418 | 2 | 5.23E-07 | 4.43E-07 | 3.65E-05 | 5.51E-07 | 0.012533826 | CRACR2A |
| chr12 | 53298861 | 53300234 | 1374 | 17 | 1.32E-09 | 0.0016893 | 1.76E-05 | 5.51E-07 | -0.00323224 | PFDN5, AC073611.1, MYG1, MYG1-AS1 |
| chr20 | 46215994 | 46216049 | 56 | 2 | 4.50E-07 | 1.80E-06 | 4.65E-06 | 5.52E-07 | -0.01040634 | CDH22 |
| chr19 | 50777693 | 50778228 | 536 | 3 | 1.88E-07 | 1.68E-06 | 8.71E-05 | 5.52E-07 | -0.00660551 | AC010325.1 |
| chr1 | 247490838 | 247491509 | 672 | 5 | 1.18E-07 | 9.50E-07 | 0.000304 | 5.52E-07 | 0.006040145 | OR2W5 |
| chr9 | 104535944 | 104537400 | 1457 | 7 | 2.00E-05 | 1.49E-06 | 0.000343 | 5.52E-07 | -0.00081218 | AL450426.1, OR13C3 |
| chr12 | 43572182 | 43572463 | 282 | 2 | 8.86E-07 | 3.43E-07 | 9.87E-05 | 5.53E-07 | -0.02162055 | NA |
| chr17 | 42963873 | 42964780 | 908 | 12 | 7.19E-07 | 6.79E-06 | 0.002407 | 5.53E-07 | -0.00067708 | AARSD1, PTGES3L-AARSD1 |
| chr7 | 74610329 | 74610630 | 302 | 2 | 9.58E-07 | 3.34E-07 | 0.000119 | 5.53E-07 | 0.002849338 | NA |
| chr13 | 81282369 | 81283144 | 776 | 3 | 1.82E-06 | 6.02E-07 | 0.000151 | 5.54E-07 | -0.00880627 | AL353633.1 |
| chr14 | 105382703 | 105383492 | 790 | 6 | 2.69E-08 | 0.0005703 | 3.55E-06 | 5.54E-07 | -0.00297634 | PACS2 |
| chr4 | 10461632 | 10462125 | 494 | 6 | 2.78E-07 | 2.11E-05 | 3.78E-05 | 5.55E-07 | 0.004654802 | AC005599.1 |
| chr16 | 54238812 | 54239543 | 732 | 3 | 3.10E-06 | 3.89E-07 | 0.000213 | 5.55E-07 | -0.00249415 | NA |
| chr10 | 407178 | 407378 | 201 | 3 | 2.08E-07 | 1.78E-05 | 6.33E-06 | 5.55E-07 | -0.00163574 | DIP2C |
| chr9 | 101738672 | 101739760 | 1089 | 8 | 2.17E-08 | 0.0004844 | 2.51E-05 | 5.56E-07 | -0.00461629 | NA |
| chr5 | 142621468 | 142622683 | 1216 | 4 | 2.94E-07 | 3.37E-05 | 0.000185 | 5.56E-07 | -0.00697749 | SPRY4-AS1, FGF1 |
| chr7 | 29193852 | 29194646 | 795 | 6 | 1.45E-09 | 4.42E-06 | 2.81E-05 | 5.56E-07 | -0.00621067 | AC004593.2, CHN2, CPVL, AC004593.1 |
| chr7 | 99990650 | 99990712 | 63 | 2 | 7.02E-07 | 3.61E-07 | 7.79E-05 | 5.56E-07 | -0.021828 | NA |
| chr10 | 13690625 | 13691814 | 1190 | 3 | 2.96E-05 | 2.58E-07 | 0.000449 | 5.57E-07 | -0.00768881 | FRMD4A |
| chr3 | 42803715 | 42804029 | 315 | 2 | 8.02E-10 | 7.68E-06 | 1.62E-06 | 5.57E-07 | -0.00526905 | AC006059.2, HIGD1A, AC099329.1 |
| chr13 | 25980052 | 25980733 | 682 | 3 | 9.69E-08 | 2.65E-06 | 6.57E-06 | 5.57E-07 | -0.00801061 | ATP8A2 |
| chr5 | 116446031 | 116447033 | 1003 | 4 | 1.22E-06 | 1.63E-07 | 0.000804 | 5.57E-07 | -0.00411535 | SEMA6A |
| chr2 | 99154001 | 99155521 | 1521 | 17 | 1.93E-08 | 3.08E-05 | 0.000997 | 5.58E-07 | -0.00031975 | AC079447.1, LIPT1, AC092587.1, TSGA10 |

| | | | | | | | | | | |
|---|---|---|---|---|---|---|---|---|---|---|
| chr2 | 28927029 | 28928009 | 981 | 5 | 1.25E-07 | 2.56E-06 | 1.35E-05 | 5.58E-07 | -0.00573998 | WDR43, SNORD53, SNORD53B, Y_RNA |
| chr1 | 20313693 | 20314646 | 954 | 3 | 3.60E-06 | 2.15E-07 | 0.000478 | 5.58E-07 | -0.00575118 | VWA5B1, AL020998.1 |
| chr5 | 170626228 | 170626763 | 536 | 2 | 1.35E-07 | 1.09E-06 | 8.03E-06 | 5.58E-07 | -0.00891591 | KCNIP1 |
| chr15 | 90638566 | 90638568 | 3 | 2 | 4.31E-07 | 4.03E-07 | 5.08E-05 | 5.59E-07 | -0.02999825 | CRTC3, CRTC3-AS1 |
| chr19 | 33226009 | 33227242 | 1234 | 8 | 7.26E-09 | 0.0001009 | 0.000202 | 5.59E-07 | -0.005857 | NA |
| chr15 | 69986620 | 69986872 | 253 | 3 | 8.48E-07 | 3.99E-07 | 0.000137 | 5.59E-07 | -0.02207516 | NA |
| chr1 | 47161840 | 47162470 | 631 | 2 | 2.67E-08 | 4.42E-06 | 2.30E-06 | 5.59E-07 | -0.01018674 | CYP4A22-AS1 |
| chr8 | 144620188 | 144621322 | 1135 | 4 | 6.22E-09 | 1.65E-05 | 2.44E-06 | 5.60E-07 | -0.00486667 | NA |
| chr12 | 58907083 | 58907574 | 492 | 2 | 4.35E-07 | 4.97E-07 | 2.89E-05 | 5.60E-07 | -0.00674311 | LRIG3 |
| chr4 | 186706619 | 186707367 | 749 | 8 | 1.82E-09 | 8.57E-05 | 3.37E-05 | 5.60E-07 | -0.0032171 | FAT1 |
| chr2 | 241871482 | 241872368 | 887 | 4 | 4.63E-08 | 2.56E-06 | 0.000137 | 5.60E-07 | -0.0088946 | RTP5 |
| chr1 | 243255178 | 243256228 | 1051 | 15 | 4.83E-08 | 0.0005236 | 4.56E-06 | 5.60E-07 | 0.000451515 | SDCCAG8, CEP170 |
| chr10 | 122489427 | 122490478 | 1052 | 5 | 4.41E-06 | 3.38E-06 | 0.000233 | 5.61E-07 | -0.01291185 | HTRA1 |
| chr15 | 85758756 | 85759740 | 985 | 6 | 1.32E-07 | 2.99E-06 | 0.000125 | 5.61E-07 | 0.001476802 | KLHL25 |
| chr3 | 49013392 | 49013829 | 438 | 2 | 1.07E-07 | 7.72E-07 | 1.31E-05 | 5.61E-07 | 0.005373861 | WDR6 |
| chr17 | 82923960 | 82925094 | 1135 | 5 | 2.50E-06 | 4.52E-07 | 0.000715 | 5.61E-07 | 0.002148418 | TBCD |
| chr1 | 184890177 | 184890944 | 768 | 3 | 6.91E-08 | 1.58E-05 | 2.05E-06 | 5.61E-07 | -0.00947757 | NIBAN1 |
| chr2 | 213150196 | 213151647 | 1452 | 10 | 7.20E-06 | 3.79E-06 | 0.000618 | 5.61E-07 | 0.000817355 | IKZF2 |
| chr20 | 45344682 | 45345593 | 912 | 3 | 1.12E-07 | 4.48E-07 | 0.000249 | 5.62E-07 | -0.00274449 | SDC4, AL021578.1 |
| chr10 | 1668045 | 1668831 | 787 | 3 | 3.22E-07 | 7.46E-06 | 4.38E-06 | 5.62E-07 | -0.00895795 | ADARB2 |
| chr12 | 69024050 | 69025062 | 1013 | 3 | 4.48E-06 | 2.64E-07 | 0.000228 | 5.62E-07 | -0.00775949 | NA |
| chr12 | 56244750 | 56245513 | 764 | 6 | 2.15E-07 | 5.33E-05 | 6.24E-05 | 5.62E-07 | -0.00312315 | ANKRD52 |
| chr21 | 33402420 | 33403153 | 734 | 9 | 3.32E-07 | 2.14E-05 | 0.000684 | 5.62E-07 | -0.00353025 | NA |
| chr9 | 77019623 | 77020529 | 907 | 4 | 1.97E-05 | 4.82E-07 | 0.000216 | 5.63E-07 | 0.005286575 | FOXB2 |
| chr1 | 8512648 | 8513093 | 446 | 4 | 1.12E-07 | 1.94E-05 | 5.87E-05 | 5.63E-07 | 0.004947676 | AL096855.2, RERE |
| chr11 | 68870100 | 68870632 | 533 | 2 | 4.41E-08 | 1.33E-06 | 6.54E-06 | 5.64E-07 | 0.029461101 | NA |
| chr2 | 239529812 | 239530200 | 389 | 2 | 7.02E-08 | 5.31E-06 | 2.06E-06 | 5.64E-07 | -0.00918608 | AC023787.2 |
| chr4 | 74308162 | 74308487 | 326 | 5 | 8.02E-08 | 1.25E-05 | 2.73E-06 | 5.64E-07 | 0.006716172 | EPGN |
| chr6 | 99152327 | 99152672 | 346 | 2 | 1.15E-07 | 3.60E-06 | 2.68E-06 | 5.64E-07 | 0.011228655 | NA |
| chr16 | 84828121 | 84828147 | 27 | 3 | 1.62E-07 | 9.83E-07 | 7.68E-05 | 5.64E-07 | 0.007003751 | CRISPLD2, AC025280.3 |
| chr19 | 43901281 | 43902089 | 809 | 6 | 7.03E-07 | 1.88E-07 | 0.002673 | 5.64E-07 | -0.00631956 | AC006213.3, ZNF404, AC006213.2 |
| chr6 | 167997689 | 167998516 | 828 | 3 | 3.77E-08 | 5.60E-07 | 0.000208 | 5.64E-07 | -0.00646225 | KIF25 |
| chr11 | 10742948 | 10743523 | 576 | 3 | 2.05E-07 | 1.16E-06 | 0.000118 | 5.64E-07 | 0.027129199 | NA |
| chr8 | 38231218 | 38232402 | 1185 | 19 | 6.34E-07 | 4.02E-06 | 0.00253 | 5.65E-07 | 0.000488889 | DDHD2, AC084024.4 |
| chr2 | 144365272 | 144365573 | 302 | 3 | 1.40E-07 | 8.95E-06 | 7.49E-06 | 5.65E-07 | -0.00309972 | ZEB2 |
| chr22 | 28441598 | 28442643 | 1046 | 5 | 5.19E-08 | 6.12E-06 | 0.000149 | 5.65E-07 | -0.00157685 | TTC28 |
| chr15 | 96361494 | 96363125 | 1632 | 7 | 7.74E-06 | 8.22E-06 | 0.001234 | 5.66E-07 | -0.00414088 | AC087477.2 |
| chr11 | 132331606 | 132332695 | 1090 | 4 | 1.63E-07 | 2.94E-06 | 8.70E-06 | 5.66E-07 | -0.00832272 | NTM |
| chr11 | 122851604 | 122851760 | 157 | 2 | 1.10E-07 | 5.05E-05 | 7.52E-07 | 5.66E-07 | -0.0098657 | CRTAM, AP002762.2 |
| chr6 | 44276013 | 44276853 | 841 | 4 | 2.06E-06 | 2.13E-07 | 0.000654 | 5.66E-07 | -0.01427084 | TMEM151B, AL353588.1 |
| chr17 | 49022278 | 49022645 | 368 | 2 | 5.05E-07 | 3.44E-07 | 0.000116 | 5.68E-07 | -0.00947074 | IGF2BP1 |
| chr18 | 3612508 | 3612576 | 69 | 2 | 9.14E-07 | 4.01E-07 | 5.54E-05 | 5.68E-07 | -0.01360737 | DLGAP1 |
| chr10 | 91410044 | 91411245 | 1202 | 8 | 4.68E-08 | 0.0002168 | 4.45E-05 | 5.68E-07 | -0.00278049 | HECTD2 |
| chr17 | 81966085 | 81966666 | 582 | 5 | 3.54E-07 | 2.98E-07 | 0.000531 | 5.68E-07 | 0.005705512 | AC145207.7 |
| chr8 | 76712507 | 76713518 | 1012 | 3 | 7.12E-06 | 2.59E-07 | 0.000433 | 5.68E-07 | -0.00244635 | ZFHX4 |
| chr17 | 40092096 | 40093476 | 1381 | 9 | 1.11E-06 | 2.48E-06 | 0.001327 | 5.69E-07 | -0.00337444 | THRA, NR1D1 |
| chr2 | 96641842 | 96641956 | 115 | 2 | 5.26E-09 | 2.39E-05 | 9.66E-07 | 5.69E-07 | -0.00709016 | KANSL3 |
| chr12 | 130761522 | 130761999 | 478 | 4 | 3.98E-07 | 3.19E-07 | 0.00017 | 5.70E-07 | 0.014112084 | NA |
| chr19 | 7845174 | 7846234 | 1061 | 5 | 6.57E-07 | 8.68E-06 | 2.84E-05 | 5.70E-07 | -0.0034271 | EVI5L |
| chr12 | 103161788 | 103162854 | 1067 | 4 | 7.03E-06 | 1.36E-07 | 0.002191 | 5.70E-07 | -0.00943542 | AC068643.2, AC068643.1 |
| chr1 | 47415584 | 47416065 | 482 | 4 | 1.84E-07 | 7.08E-06 | 3.78E-05 | 5.71E-07 | -0.00875063 | NA |
| chr1 | 3206490 | 3207290 | 801 | 6 | 5.03E-08 | 4.23E-05 | 2.00E-05 | 5.71E-07 | 0.00101472 | PRDM16, AL512383.1 |
| chr10 | 7817660 | 7818909 | 1250 | 14 | 3.69E-07 | 2.67E-06 | 0.000235 | 5.72E-07 | -0.00205988 | TAF3 |
| chr12 | 2876127 | 2877431 | 1305 | 12 | 4.87E-10 | 0.0002654 | 5.90E-06 | 5.72E-07 | -0.0016118 | RHNO1, TULP3, FOXM1 |
| chr7 | 25012898 | 25013386 | 489 | 2 | 1.50E-07 | 1.17E-06 | 7.74E-06 | 5.72E-07 | -0.01363421 | NA |
| chr9 | 37465026 | 37466351 | 1326 | 7 | 1.58E-07 | 1.52E-05 | 0.000549 | 5.72E-07 | -0.00045537 | ZBTB5 |
| chr18 | 46104066 | 46105394 | 1329 | 19 | 3.98E-07 | 4.11E-05 | 0.000189 | 5.72E-07 | -0.00115092 | HAUS1, ATP5F1A |
| chr8 | 128818464 | 128819034 | 571 | 3 | 3.24E-10 | 0.0002003 | 5.26E-07 | 5.73E-07 | -0.00474539 | CCDC26 |
| chr6 | 30159588 | 30160781 | 1194 | 14 | 1.61E-08 | 4.43E-06 | 4.66E-05 | 5.73E-07 | -0.00153321 | TRIM10 |
| chr21 | 15246579 | 15246598 | 20 | 2 | 1.27E-07 | 0.0002745 | 5.45E-07 | 5.75E-07 | -0.0020925 | NA |
| chr9 | 36738768 | 36739784 | 1017 | 3 | 6.80E-07 | 2.86E-07 | 0.000193 | 5.75E-07 | -0.0065981 | NA |
| chr17 | 40201939 | 40202172 | 234 | 2 | 6.03E-07 | 4.54E-07 | 3.92E-05 | 5.75E-07 | -0.00998685 | NA |
| chr5 | 141208273 | 141208852 | 580 | 8 | 1.05E-06 | 2.79E-07 | 0.001913 | 5.75E-07 | -0.01471469 | PCDHB12, AC244517.11 |
| chr3 | 146250455 | 146251271 | 817 | 8 | 1.64E-07 | 5.00E-06 | 0.00039 | 5.76E-07 | 0.001430265 | PLSCR4 |
| chr7 | 155506929 | 155507151 | 223 | 3 | 6.91E-07 | 7.23E-07 | 2.89E-05 | 5.77E-07 | -0.01020306 | CNPY1, AC009403.2 |
| chr2 | 27765709 | 27765991 | 283 | 4 | 1.26E-06 | 3.08E-07 | 0.000554 | 5.77E-07 | -0.01764021 | NA |
| chr11 | 130094830 | 130095613 | 784 | 3 | 3.00E-06 | 1.13E-06 | 5.89E-05 | 5.77E-07 | 0.003805379 | APLP2 |
| chr7 | 2638299 | 2639514 | 1216 | 7 | 1.38E-07 | 0.0001462 | 1.75E-05 | 5.77E-07 | -0.0043574 | TTYH3 |
| chr12 | 95231020 | 95231391 | 372 | 2 | 1.05E-06 | 3.51E-07 | 0.000117 | 5.78E-07 | -0.00066535 | VEZT |
| chr19 | 45141305 | 45141436 | 132 | 2 | 8.82E-07 | 4.19E-07 | 5.08E-05 | 5.79E-07 | -0.00206177 | MARK4, PPP1R37 |
| chr16 | 2339101 | 2339706 | 606 | 2 | 1.95E-08 | 9.75E-07 | 9.96E-06 | 5.79E-07 | 0.002092733 | ABCA17P, ABCA3 |
| chr1 | 177898157 | 177898348 | 192 | 3 | 3.17E-07 | 2.07E-05 | 8.72E-06 | 5.79E-07 | -0.01706885 | AL136114.1 |
| chr14 | 26871765 | 26872702 | 938 | 2 | 1.26E-05 | 3.90E-07 | 6.70E-05 | 5.80E-07 | -0.02748613 | AL110292.1, AL132718.1 |
| chr15 | 101065654 | 101066692 | 1039 | 6 | 5.33E-07 | 9.48E-07 | 9.97E-05 | 5.80E-07 | -0.00350825 | LRRK1, AC090907.2 |
| chr17 | 55265113 | 55266040 | 928 | 11 | 1.06E-06 | 2.24E-06 | 0.001112 | 5.80E-07 | 0.001669755 | HLF |
| chr14 | 77121040 | 77121964 | 925 | 3 | 5.90E-07 | 2.98E-07 | 0.000228 | 5.80E-07 | -0.02006906 | AC007375.2, TMEM63C |
| chr3 | 156509449 | 156509977 | 529 | 3 | 2.45E-07 | 5.23E-07 | 4.50E-05 | 5.81E-07 | 0.004923865 | KCNAB1, AC091607.2 |
| chr15 | 65102232 | 65102729 | 498 | 4 | 1.65E-07 | 1.84E-06 | 0.000107 | 5.81E-07 | 0.003106216 | UBAP1L |
| chr7 | 101804094 | 101804625 | 532 | 2 | 3.36E-07 | 5.42E-07 | 2.65E-05 | 5.82E-07 | -0.02155213 | NA |
| chr20 | 20550606 | 20550725 | 120 | 3 | 1.46E-06 | 2.13E-07 | 0.000719 | 5.82E-07 | 0.003670298 | RALGAPA2 |
| chr17 | 18266999 | 18267606 | 608 | 3 | 1.89E-09 | 3.05E-06 | 5.26E-06 | 5.82E-07 | -0.00280127 | NA |
| chr1 | 42152370 | 42153492 | 1123 | 11 | 1.17E-07 | 3.06E-05 | 2.40E-05 | 5.83E-07 | -0.0034824 | GUCA2B |
| chr7 | 50300622 | 50301614 | 993 | 3 | 4.45E-06 | 2.10E-07 | 0.000662 | 5.83E-07 | -0.00092816 | NA |
| chr1 | 179364852 | 179366463 | 1612 | 10 | 1.67E-05 | 2.44E-06 | 0.001214 | 5.84E-07 | -0.0031867 | AXDND1 |
| chr1 | 203054108 | 203054799 | 692 | 3 | 3.16E-09 | 9.78E-06 | 2.62E-06 | 5.84E-07 | -0.00375736 | PPFIA4 |
| chr2 | 219513937 | 219515246 | 1310 | 13 | 2.38E-06 | 5.23E-07 | 0.007511 | 5.85E-07 | 0.001559102 | ASIC4, ASIC4-AS1 |
| chr7 | 39013653 | 39014491 | 839 | 3 | 1.04E-06 | 3.84E-07 | 8.27E-05 | 5.85E-07 | -0.00359119 | POU6F2 |
| chr12 | 132335980 | 132336699 | 720 | 2 | 3.56E-06 | 3.48E-07 | 0.000138 | 5.85E-07 | -0.01102529 | NA |
| chr10 | 13630646 | 13631462 | 817 | 3 | 3.93E-06 | 4.00E-05 | 7.57E-05 | 5.86E-07 | -0.00158401 | PRPF18, AL157392.5, AL157392.3 |
| chr4 | 8066491 | 8066521 | 31 | 2 | 1.83E-07 | 4.94E-05 | 7.87E-07 | 5.86E-07 | 0.005509681 | ABLIM2 |
| chr7 | 104804955 | 104805488 | 534 | 3 | 1.67E-08 | 2.68E-06 | 3.94E-05 | 5.86E-07 | -0.00902052 | LHFPL3 |
| chr1 | 229221431 | 229222003 | 573 | 2 | 2.86E-09 | 0.0006418 | 5.10E-07 | 5.87E-07 | 0.004936535 | NA |
| chr22 | 25027096 | 25028227 | 1132 | 9 | 1.09E-07 | 2.08E-05 | 0.000499 | 5.87E-07 | -0.00085606 | KIAA1671 |
| chr7 | 50316519 | 50316587 | 69 | 2 | 3.25E-07 | 3.93E-07 | 6.91E-05 | 5.88E-07 | -0.01293239 | IKZF1 |
| chr13 | 30734224 | 30735360 | 1137 | 5 | 5.18E-06 | 1.04E-06 | 0.000734 | 5.88E-07 | 0.00451263 | ALOX5AP |
| chr16 | 67978187 | 67978370 | 184 | 4 | 6.64E-07 | 1.98E-06 | 0.000526 | 5.88E-07 | -0.00184807 | DPEP3 |
| chr10 | 4254214 | 4254889 | 676 | 2 | 1.55E-06 | 4.10E-07 | 5.86E-05 | 5.88E-07 | -0.01029952 | NA |
| chr13 | 24307961 | 24308876 | 916 | 4 | 8.05E-08 | 2.09E-07 | 0.000933 | 5.88E-07 | -0.03188119 | AL359736.1, C1QTNF9 |
| chr5 | 154858782 | 154859954 | 1173 | 9 | 9.11E-06 | 9.74E-06 | 0.000373 | 5.88E-07 | 0.002624361 | CNOT8, FAXDC2 |
| chr8 | 57278827 | 57279506 | 680 | 9 | 1.88E-07 | 0.0001017 | 0.000428 | 5.89E-07 | 0.000881267 | NA |
| chr12 | 89543860 | 89544950 | 1091 | 4 | 2.93E-08 | 2.93E-06 | 9.41E-06 | 5.89E-07 | -0.00556686 | POC1B-AS1, AC025034.2 |
| chr20 | 62775434 | 62776189 | 756 | 5 | 1.02E-07 | 9.57E-06 | 3.89E-05 | 5.89E-07 | -0.00488354 | LINC00659 |
| chr6 | 109428938 | 109429387 | 450 | 2 | 2.59E-08 | 1.79E-05 | 1.12E-06 | 5.89E-07 | -0.01190164 | PPIL6 |
| chr16 | 13454027 | 13454227 | 201 | 2 | 2.45E-07 | 4.51E-06 | 2.41E-06 | 5.89E-07 | -0.01520773 | U91319.1 |
| chr1 | 94298977 | 94299674 | 698 | 3 | 8.12E-07 | 1.04E-06 | 4.96E-05 | 5.90E-07 | -0.01371919 | ARHGAP29-AS1 |
| chr3 | 43031566 | 43031660 | 95 | 2 | 8.51E-07 | 5.13E-07 | 3.11E-05 | 5.90E-07 | 0.040017308 | AC092042.3, GASK1A |
| chr11 | 115627773 | 115627980 | 208 | 4 | 4.20E-07 | 2.60E-06 | 0.000104 | 5.90E-07 | -0.00675441 | NA |
| chr4 | 4688800 | 4689602 | 803 | 3 | 5.89E-07 | 6.94E-07 | 6.02E-05 | 5.90E-07 | -0.00450928 | STX18-AS1 |
| chr5 | 176662802 | 176663139 | 338 | 2 | 2.79E-08 | 2.28E-05 | 1.02E-06 | 5.90E-07 | -0.01108685 | NA |
| chr12 | 122974103 | 122975285 | 1183 | 21 | 1.88E-07 | 6.90E-05 | 0.000344 | 5.90E-07 | -0.00055324 | OGFOD2, ABCB9 |
| chr12 | 109773516 | 109774506 | 991 | 9 | 3.34E-06 | 1.58E-07 | 0.001979 | 5.91E-07 | 0.010064055 | NA |
| chr10 | 33772385 | 33773306 | 922 | 8 | 2.92E-07 | 2.74E-05 | 0.002056 | 5.91E-07 | 0.013609575 | LINC00838 |
| chr4 | 3121241 | 3121405 | 165 | 3 | 4.52E-07 | 1.34E-06 | 7.46E-05 | 5.91E-07 | -0.0145228 | HTT |
| chr12 | 90954465 | 90955948 | 1484 | 11 | 2.02E-07 | 1.44E-06 | 0.00012 | 5.91E-07 | -0.00250279 | CCER1 |
| chr3 | 193980147 | 193980629 | 483 | 5 | 1.78E-07 | 2.84E-06 | 0.000989 | 5.91E-07 | 0.002529563 | AC024559.1, LINC02026 |
| chr12 | 8662712 | 8664037 | 1326 | 7 | 1.42E-05 | 2.98E-06 | 0.000204 | 5.91E-07 | -0.00070707 | AC092490.2, MFAP5 |
| chr8 | 140458368 | 140458834 | 467 | 8 | 9.69E-08 | 4.13E-05 | 1.68E-05 | 5.92E-07 | -0.00342389 | TRAPPC9 |
| chr20 | 58753827 | 58754364 | 538 | 2 | 8.59E-09 | 4.00E-06 | 2.64E-06 | 5.92E-07 | -0.01076317 | AL132655.3, PIEZO1P2 |
| chr3 | 42875848 | 42876025 | 178 | 3 | 1.76E-07 | 2.13E-06 | 6.25E-05 | 5.92E-07 | -0.0070116 | ACKR2, AC099329.2, AC092042.3, KRBOX1, CYP8B1 |
| chr7 | 1964450 | 1965712 | 1263 | 6 | 4.30E-06 | 4.37E-06 | 0.000384 | 5.93E-07 | 0.000948578 | MAD1L1, AC069288.1 |
| chr17 | 78793085 | 78793722 | 638 | 3 | 1.27E-06 | 6.46E-05 | 3.49E-05 | 5.93E-07 | -0.00323753 | USP36 |
| chr19 | 48461553 | 48462718 | 1166 | 9 | 3.25E-07 | 2.56E-05 | 0.000974 | 5.94E-07 | -0.00433697 | KCNJ14 |
| chr22 | 39654925 | 39655591 | 667 | 2 | 4.13E-08 | 7.57E-07 | 1.52E-05 | 5.94E-07 | -0.01423278 | CACNA1I |
| chr14 | 105072144 | 105072797 | 654 | 5 | 7.57E-08 | 2.75E-05 | 5.03E-06 | 5.94E-07 | -0.00459526 | NA |
| chr9 | 124868410 | 124868807 | 398 | 4 | 4.12E-08 | 3.55E-06 | 0.000107 | 5.94E-07 | -0.00074382 | ARPC5L |
| chr17 | 42744706 | 42746160 | 1455 | 14 | 1.06E-06 | 1.87E-05 | 0.000536 | 5.95E-07 | 0.000111114 | EZH1 |
| chr20 | 12961293 | 12961574 | 282 | 4 | 1.33E-07 | 4.60E-05 | 7.92E-06 | 5.95E-07 | -0.00379181 | LINC01723 |
| chr16 | 58733222 | 58734106 | 885 | 9 | 1.52E-07 | 1.80E-06 | 0.000491 | 5.95E-07 | -0.00366594 | AC106793.1, GOT2 |
| chr19 | 49458832 | 49459757 | 926 | 4 | 3.14E-08 | 0.0001087 | 3.20E-05 | 5.95E-07 | 0.000396265 | ALDH16A1 |
| chr17 | 34081057 | 34081429 | 373 | 2 | 9.23E-08 | 4.76E-06 | 2.36E-06 | 5.95E-07 | -0.00105604 | ASIC2, AC004147.4, AC004147.1 |
| chr6 | 143511003 | 143512284 | 1282 | 16 | 2.36E-07 | 0.0006869 | 2.53E-05 | 5.95E-07 | 0.000893634 | FUCA2 |
| chr9 | 134050505 | 134051051 | 547 | 3 | 1.07E-07 | 1.57E-06 | 1.61E-05 | 5.95E-07 | -0.00300586 | BRD3 |
| chr3 | 49703486 | 49703928 | 443 | 3 | 3.06E-07 | 5.85E-07 | 0.000182 | 5.95E-07 | -0.00951529 | RNF123 |
| chr5 | 180628982 | 180630303 | 1322 | 6 | 9.09E-06 | 3.72E-07 | 0.000496 | 5.95E-07 | -0.00038442 | FLT4 |
| chr14 | 74426283 | 74426973 | 691 | 5 | 1.63E-09 | 8.95E-06 | 1.09E-05 | 5.96E-07 | -0.00871752 | NA |
| chr2 | 96897996 | 96898945 | 950 | 9 | 4.65E-08 | 2.75E-05 | 0.00034 | 5.96E-07 | -0.00327972 | FAM178B |
| chr4 | 78939353 | 78940244 | 892 | 11 | 2.74E-07 | 2.45E-05 | 0.000617 | 5.96E-07 | 0.001392655 | LINC01088, PAQR3 |
| chr17 | 80247444 | 80247610 | 167 | 3 | 6.16E-07 | 3.88E-07 | 0.000206 | 5.99E-07 | -0.00145682 | SLC26A11 |
| chr17 | 8899066 | 8899548 | 483 | 3 | 4.82E-08 | 0.0019733 | 1.54E-06 | 6.00E-07 | -0.00783279 | PIK3R5 |
| chr15 | 79005867 | 79006427 | 561 | 12 | 1.30E-07 | 3.51E-05 | 0.000107 | 6.01E-07 | -0.00148964 | RASGRF1 |
| chr2 | 121567852 | 121568200 | 349 | 2 | 6.69E-08 | 1.24E-05 | 1.37E-06 | 6.01E-07 | -0.00774777 | CLASP1 |
| chr6 | 128499476 | 128499572 | 97 | 3 | 3.71E-07 | 6.68E-06 | 3.29E-06 | 6.01E-07 | -0.01104067 | PTPRK |
| chr8 | 65643172 | 65644266 | 1095 | 9 | 2.82E-07 | 1.54E-06 | 0.000243 | 6.01E-07 | 0.004407388 | NA |
| chr15 | 40952417 | 40953718 | 1302 | 16 | 3.43E-07 | 3.76E-06 | 0.002561 | 6.01E-07 | 0.000787973 | CHAC1 |
| chr10 | 34739970 | 34740109 | 140 | 3 | 5.18E-07 | 5.32E-06 | 3.05E-05 | 6.02E-07 | -0.00781246 | PARD3 |
| chr3 | 126156683 | 126157326 | 644 | 5 | 1.14E-07 | 1.15E-06 | 5.90E-05 | 6.02E-07 | 0.001562684 | ALDH1L1 |
| chr16 | 1790004 | 1791495 | 1492 | 5 | 2.49E-08 | 2.89E-06 | 0.000378 | 6.02E-07 | -0.0010737 | SPSB3, IGFALS |
| chr3 | 185581986 | 185582788 | 803 | 5 | 3.74E-07 | 2.42E-05 | 0.000176 | 6.02E-07 | -0.00518387 | SENP2 |
| chr11 | 64018363 | 64019284 | 922 | 4 | 2.56E-06 | 1.81E-07 | 0.001379 | 6.03E-07 | 0.005790281 | MACROD1 |
| chr9 | 126616766 | 126617040 | 275 | 2 | 2.98E-08 | 6.96E-05 | 7.34E-07 | 6.04E-07 | 0.021694095 | LMX1B |
| chr3 | 136861798 | 136862347 | 550 | 9 | 1.28E-07 | 2.48E-06 | 0.000525 | 6.04E-07 | 0.00031338 | NCK1, NCK1-DT |
| chr3 | 161104480 | 161105993 | 1514 | 25 | 8.55E-07 | 0.0003297 | 0.000838 | 6.04E-07 | -0.00080649 | NMD3, B3GALNT1 |
| chr2 | 64341687 | 64341697 | 11 | 2 | 7.05E-07 | 3.60E-07 | 0.000141 | 6.04E-07 | -0.00392757 | NA |
| chr12 | 109995757 | 109996536 | 780 | 9 | 1.06E-07 | 2.01E-05 | 0.000134 | 6.05E-07 | 0.001688075 | GIT2 |
| chr16 | 3157966 | 3159179 | 1214 | 6 | 2.66E-07 | 4.27E-07 | 0.000412 | 6.05E-07 | 0.001166323 | NA |
| chr6 | 36340612 | 36341170 | 559 | 3 | 3.26E-07 | 7.67E-07 | 0.000169 | 6.06E-07 | -0.01249179 | NA |
| chr6 | 162705897 | 162706292 | 396 | 2 | 8.00E-07 | 4.18E-07 | 6.23E-05 | 6.06E-07 | -0.01314679 | PRKN |
| chr12 | 121654221 | 121654443 | 223 | 3 | 2.98E-07 | 2.22E-06 | 8.03E-06 | 6.06E-07 | -0.01007645 | MORN3 |

| | | | | | | | | | | |
|---|---|---|---|---|---|---|---|---|---|---|
| chr5 | 181059155 | 181060371 | 1217 | 6 | 2.52E-06 | 1.34E-05 | 0.000565 | 6.07E-07 | -0.00464301 | BTNL9 |
| chr1 | 205246106 | 205247339 | 1234 | 7 | 3.16E-07 | 1.87E-06 | 1.56E-05 | 6.07E-07 | -0.00235061 | TMCC2 |
| chr11 | 42066165 | 42066181 | 17 | 2 | 4.58E-07 | 3.30E-06 | 3.15E-06 | 6.07E-07 | -0.00658133 | LINC02745 |
| chr8 | 65705626 | 65707132 | 1507 | 4 | 2.95E-05 | 1.22E-06 | 8.01E-05 | 6.07E-07 | -0.00971883 | MTFR1 |
| chr10 | 121668551 | 121668861 | 311 | 2 | 9.29E-07 | 3.89E-07 | 8.62E-05 | 6.07E-07 | -0.0213975 | NA |
| chr1 | 54195688 | 54196411 | 724 | 4 | 5.70E-08 | 4.92E-05 | 1.53E-05 | 6.08E-07 | -0.00144033 | MRPL37, AL357673.1, CYB5RL |
| chr19 | 54415089 | 54416354 | 1266 | 12 | 2.47E-07 | 3.55E-07 | 0.00582 | 6.08E-07 | 0.000324073 | TTYH1 |
| chr4 | 140305106 | 140305753 | 648 | 3 | 3.31E-07 | 3.35E-07 | 0.000206 | 6.08E-07 | -0.00418254 | SCOC, SCOC-AS1 |
| chr9 | 35834282 | 35835018 | 737 | 3 | 1.15E-06 | 5.57E-07 | 0.000137 | 6.08E-07 | -0.00342431 | TMEM8B |
| chr22 | 38505549 | 38507020 | 1472 | 15 | 2.60E-06 | 1.03E-06 | 0.006172 | 6.09E-07 | 1.46E-06 | DDX17 |
| chr19 | 9214876 | 9215485 | 610 | 3 | 3.61E-07 | 7.97E-07 | 9.58E-05 | 6.10E-07 | -0.01407525 | OR7D4 |
| chr11 | 109421571 | 109422888 | 1318 | 11 | 1.50E-06 | 1.20E-06 | 0.003079 | 6.10E-07 | -0.0010647 | C11orf87, AP003049.2 |
| chr10 | 5523866 | 5524899 | 1034 | 11 | 7.66E-07 | 7.09E-08 | 0.015995 | 6.10E-07 | -0.00235851 | CALML3-AS1 |
| chr3 | 133895734 | 133897036 | 1303 | 13 | 2.51E-07 | 2.80E-06 | 0.0032 | 6.10E-07 | -0.00244712 | RAB6B |
| chr19 | 1237454 | 1238539 | 1086 | 11 | 3.36E-07 | 3.54E-05 | 0.000544 | 6.11E-07 | 0.001376371 | AC004221.1, CBARP |
| chr20 | 31355523 | 31355603 | 81 | 2 | 6.11E-07 | 1.33E-06 | 7.47E-06 | 6.11E-07 | -0.03231639 | NA |
| chr11 | 114762518 | 114762955 | 438 | 2 | 8.08E-07 | 4.25E-07 | 6.05E-05 | 6.11E-07 | -0.02446466 | NA |
| chr17 | 9532873 | 9533168 | 296 | 2 | 3.69E-07 | 4.23E-07 | 6.18E-05 | 6.11E-07 | -0.00115359 | STX8 |
| chr21 | 44558864 | 44559737 | 874 | 6 | 4.43E-07 | 4.74E-06 | 0.000876 | 6.12E-07 | 0.001805166 | TSPEAR |
| chr20 | 54840875 | 54841157 | 283 | 2 | 1.04E-06 | 3.74E-07 | 0.000116 | 6.12E-07 | -0.00010999 | NA |
| chr5 | 178443176 | 178443406 | 231 | 6 | 1.26E-06 | 6.07E-07 | 0.000371 | 6.12E-07 | 0.015376616 | COL23A1 |
| chr19 | 48754874 | 48756195 | 1322 | 9 | 6.70E-06 | 8.17E-06 | 0.00109 | 6.12E-07 | -0.00468011 | FGF21, FUT1 |
| chr19 | 58227968 | 58229495 | 1528 | 18 | 7.65E-06 | 1.18E-05 | 0.000293 | 6.12E-07 | -0.0020754 | ZNF544, AC020915.4 |
| chr19 | 48271757 | 48272132 | 376 | 3 | 7.87E-08 | 1.30E-06 | 3.53E-05 | 6.13E-07 | -0.00526247 | ZNF114 |
| chr2 | 127620859 | 127621085 | 227 | 2 | 2.42E-07 | 3.66E-07 | 0.00014 | 6.13E-07 | -0.09324477 | MYO7B |
| chr9 | 137187464 | 137188509 | 1046 | 8 | 1.50E-10 | 1.18E-05 | 1.52E-06 | 6.13E-07 | 0.00214697 | ANAPC2 |
| chr17 | 81483359 | 81484630 | 1272 | 7 | 7.96E-06 | 1.64E-05 | 0.001006 | 6.14E-07 | 0.00314679 | NA |
| chr16 | 69742136 | 69743145 | 1010 | 3 | 1.93E-06 | 2.46E-07 | 0.000419 | 6.15E-07 | -0.0130069 | AC092115.3, NOB1 |
| chr1 | 6235308 | 6236570 | 1263 | 11 | 1.84E-06 | 8.64E-07 | 0.006086 | 6.15E-07 | -0.00305988 | LINC00337, ICMT |
| chr9 | 98998564 | 98999205 | 642 | 3 | 7.37E-07 | 5.30E-07 | 5.06E-05 | 6.16E-07 | 0.017811608 | COL15A1 |
| chr15 | 99975632 | 99976425 | 794 | 4 | 6.93E-07 | 2.07E-05 | 0.000254 | 6.16E-07 | -0.00811865 | ADAMTS17 |
| chr12 | 105212583 | 105212778 | 196 | 2 | 1.04E-07 | 5.16E-06 | 2.34E-06 | 6.16E-07 | -0.01089446 | APPL2 |
| chr18 | 79933953 | 79934028 | 76 | 3 | 1.17E-06 | 4.14E-07 | 0.000296 | 6.17E-07 | -0.00210749 | SLC66A2 |
| chr14 | 69258468 | 69259680 | 1213 | 12 | 8.61E-10 | 0.0003021 | 3.24E-05 | 6.17E-07 | -0.0016137 | GALNT16, AL359317.1 |
| chr4 | 14360089 | 14360615 | 527 | 2 | 4.63E-09 | 0.0026279 | 4.95E-07 | 6.17E-07 | -0.00951145 | AC006296.3 |
| chr11 | 130197911 | 130198553 | 643 | 5 | 3.95E-07 | 1.65E-06 | 0.000449 | 6.17E-07 | -0.00194242 | ST14 |
| chr20 | 62728434 | 62728762 | 329 | 2 | 8.96E-07 | 4.07E-07 | 7.57E-05 | 6.17E-07 | -0.02717544 | NTSR1 |
| chr15 | 88638194 | 88639445 | 1252 | 9 | 1.90E-06 | 4.65E-06 | 0.003139 | 6.18E-07 | -0.00168921 | ISG20 |
| chr11 | 1279128 | 1280204 | 1077 | 6 | 8.88E-10 | 0.0001464 | 1.54E-06 | 6.18E-07 | -0.001159 | TOLLIP |
| chr6 | 70566371 | 70567423 | 1053 | 14 | 2.56E-07 | 6.14E-06 | 0.004585 | 6.18E-07 | -0.00129263 | SDHAF4 |
| chr8 | 102864345 | 102865147 | 803 | 8 | 5.36E-09 | 2.19E-05 | 4.23E-05 | 6.19E-07 | 0.002240622 | MAILR, AZIN1 |
| chr17 | 82772356 | 82773204 | 849 | 3 | 1.24E-06 | 0.0001107 | 1.43E-05 | 6.19E-07 | -0.01160409 | TBCD |
| chr11 | 120210072 | 120211530 | 1459 | 18 | 4.09E-07 | 1.47E-05 | 0.00125 | 6.19E-07 | 0.001127789 | OAF |
| chr20 | 44521770 | 44522617 | 848 | 12 | 7.72E-07 | 4.34E-06 | 0.000849 | 6.19E-07 | 0.001939855 | SERINC3 |
| chr2 | 236052421 | 236052545 | 125 | 3 | 2.90E-07 | 1.06E-05 | 6.11E-05 | 6.19E-07 | 0.00729395 | AGAP1 |
| chr17 | 53821908 | 53823173 | 1266 | 12 | 1.08E-05 | 5.91E-06 | 0.001217 | 6.19E-07 | 0.000993232 | KIF2B, AC034268.2 |
| chr13 | 102401059 | 102401806 | 748 | 8 | 3.00E-08 | 3.33E-05 | 2.77E-05 | 6.19E-07 | 0.001052388 | FGF14 |
| chr22 | 46444727 | 46444941 | 215 | 2 | 7.12E-09 | 4.66E-06 | 2.52E-06 | 6.21E-07 | -0.00843536 | CELSR1 |
| chr4 | 164929925 | 164930707 | 783 | 2 | 2.02E-06 | 4.78E-07 | 4.41E-05 | 6.21E-07 | -0.00390117 | AC106872.12, FAM218BP |
| chr11 | 123061961 | 123062982 | 1022 | 12 | 2.00E-07 | 4.20E-06 | 0.000652 | 6.21E-07 | 0.00214635 | AP000926.2, HSPA8 |
| chr8 | 22545246 | 22546120 | 875 | 7 | 8.03E-08 | 8.61E-06 | 0.000104 | 6.22E-07 | -0.00097195 | SORBS3, AC037459.4, AC087854.1 |
| chr20 | 58817517 | 58818222 | 706 | 4 | 9.77E-08 | 1.47E-06 | 0.000147 | 6.22E-07 | -0.00083011 | AL132655.2, MIR296 |
| chr10 | 129869686 | 129869730 | 45 | 2 | 1.74E-07 | 5.22E-06 | 2.35E-06 | 6.22E-07 | -0.00372289 | EBF3 |
| chr19 | 4694760 | 4695671 | 912 | 5 | 6.41E-07 | 4.30E-06 | 0.000236 | 6.22E-07 | -0.00477157 | DPP9 |
| chr11 | 64149204 | 64149219 | 16 | 2 | 5.98E-08 | 0.0001227 | 6.69E-07 | 6.23E-07 | -0.00204318 | MACROD1 |
| chr3 | 100260075 | 100261132 | 1058 | 19 | 1.85E-07 | 1.64E-05 | 0.000685 | 6.23E-07 | -0.00175627 | TBC1D23, AC129803.1 |
| chr19 | 34905571 | 34906202 | 632 | 3 | 4.66E-07 | 2.94E-07 | 0.000241 | 6.24E-07 | -0.01789354 | LINC01838 |
| chr17 | 57994384 | 57995002 | 619 | 4 | 1.85E-07 | 1.67E-06 | 8.29E-05 | 6.24E-07 | 0.007169687 | AC015813.2, AC015813.1 |
| chr6 | 117906569 | 117907708 | 1140 | 12 | 3.87E-07 | 3.18E-06 | 0.000238 | 6.24E-07 | -0.00192784 | SLC35F1 |
| chr14 | 22469683 | 22470365 | 683 | 5 | 1.21E-06 | 3.08E-07 | 0.000631 | 6.24E-07 | 0.015357521 | AC244502.1, TRDV3 |
| chr1 | 28668188 | 28668683 | 496 | 10 | 2.43E-06 | 9.41E-07 | 0.010382 | 6.25E-07 | -0.00132937 | NA |
| chr3 | 9748759 | 9750000 | 1242 | 11 | 1.21E-08 | 1.19E-05 | 0.001065 | 6.25E-07 | -0.00348084 | OGG1 |
| chr14 | 102966053 | 102966852 | 800 | 7 | 9.99E-08 | 1.88E-06 | 0.000735 | 6.25E-07 | 0.002556021 | CDC42BPB |
| chr12 | 14879258 | 14880118 | 861 | 3 | 4.48E-08 | 4.69E-06 | 5.26E-06 | 6.25E-07 | 0.002384407 | C12orf60 |
| chr1 | 202861698 | 202862968 | 1271 | 4 | 4.14E-10 | 6.69E-05 | 1.82E-05 | 6.26E-07 | -0.00391852 | ACTG1P25 |
| chr17 | 67030482 | 67030984 | 503 | 5 | 2.97E-08 | 1.37E-05 | 2.85E-05 | 6.26E-07 | -0.00208917 | CACNG4 |
| chr4 | 109702818 | 109703759 | 942 | 10 | 1.01E-07 | 3.39E-06 | 0.001376 | 6.26E-07 | 0.001958782 | CASP6 |
| chr14 | 93041065 | 93042234 | 1170 | 4 | 3.86E-08 | 8.60E-07 | 5.34E-05 | 6.26E-07 | -0.00828731 | ITPK1 |
| chr17 | 63537308 | 63538848 | 1541 | 4 | 2.78E-05 | 2.71E-05 | 0.00062 | 6.27E-07 | -0.00249872 | KCNH6 |
| chr2 | 173354198 | 173355472 | 1275 | 10 | 1.34E-07 | 6.17E-06 | 9.40E-05 | 6.27E-07 | -0.0015558 | CDCA7, AC092573.1 |
| chr12 | 57693174 | 57694224 | 1051 | 13 | 3.74E-08 | 7.59E-05 | 0.000164 | 6.27E-07 | 0.000403431 | OS9, AC025165.2 |
| chr1 | 147670062 | 147671124 | 1063 | 16 | 9.36E-08 | 3.55E-06 | 3.89E-05 | 6.27E-07 | 0.001121386 | ACP6 |
| chr2 | 25976917 | 25977132 | 216 | 2 | 3.57E-08 | 0.0002635 | 5.98E-07 | 6.28E-07 | 0.002907935 | KIF3C |
| chr5 | 69530967 | 69531814 | 848 | 3 | 7.22E-09 | 5.28E-05 | 7.52E-07 | 6.28E-07 | -0.01421298 | OCLN |
| chr15 | 74209859 | 74211142 | 1284 | 5 | 7.84E-09 | 1.83E-05 | 3.96E-05 | 6.28E-07 | -0.00020573 | CCDC33, STRA6 |
| chr22 | 33108193 | 33108454 | 262 | 4 | 1.16E-06 | 1.48E-06 | 0.000325 | 6.29E-07 | -0.01003913 | LINC01640 |
| chr4 | 108620237 | 108620988 | 752 | 16 | 2.54E-06 | 6.21E-07 | 0.016285 | 6.29E-07 | -0.00095401 | RPL34, RPL34-DT |
| chr2 | 67323490 | 67324650 | 1161 | 5 | 2.09E-07 | 1.74E-06 | 0.000216 | 6.29E-07 | -0.00394028 | AC023115.1, LINC01829 |
| chr13 | 99967915 | 99968746 | 832 | 5 | 7.31E-07 | 5.74E-07 | 0.000394 | 6.29E-07 | -0.00617879 | ZIC5 |
| chr12 | 64222045 | 64222605 | 561 | 12 | 2.47E-07 | 5.01E-06 | 0.001357 | 6.29E-07 | 0.000340105 | AC012158.1, C12orf66 |
| chr20 | 31879060 | 31880352 | 1293 | 8 | 4.29E-06 | 6.47E-05 | 0.000166 | 6.30E-07 | 3.54E-05 | TTLL9 |
| chr14 | 59262892 | 59263587 | 696 | 8 | 1.44E-09 | 0.0073858 | 4.77E-07 | 6.31E-07 | -0.00254997 | DAAM1 |
| chr9 | 98057260 | 98057646 | 387 | 2 | 8.47E-09 | 3.85E-07 | 0.000119 | 6.31E-07 | -0.00707959 | NANS |
| chr1 | 27635052 | 27635901 | 850 | 11 | 1.41E-07 | 3.55E-05 | 0.000373 | 6.31E-07 | -0.00367536 | FGR |
| chr15 | 55993063 | 55994138 | 1076 | 10 | 6.73E-08 | 1.66E-05 | 0.000225 | 6.31E-07 | 0.000186211 | NEDD4 |
| chr12 | 121297780 | 121298575 | 796 | 4 | 2.30E-05 | 3.47E-06 | 0.00027 | 6.31E-07 | 0.000708092 | CAMKK2 |
| chr15 | 89057751 | 89057847 | 97 | 2 | 5.07E-07 | 2.29E-06 | 4.57E-06 | 6.31E-07 | -0.01038978 | AC013565.1 |
| chr1 | 152825610 | 152826679 | 1070 | 6 | 1.62E-06 | 1.26E-05 | 3.55E-05 | 6.31E-07 | 0.004154446 | NA |
| chr8 | 61496445 | 61496834 | 390 | 2 | 1.57E-07 | 2.20E-06 | 4.77E-06 | 6.32E-07 | 0.013116667 | CLVS1 |
| chr8 | 86618089 | 86618774 | 686 | 2 | 7.46E-07 | 5.80E-07 | 2.88E-05 | 6.33E-07 | -0.02236122 | CNGB3 |
| chr12 | 124288040 | 124289122 | 1083 | 6 | 3.26E-08 | 1.15E-06 | 0.000573 | 6.33E-07 | -0.00237884 | RFLNA |
| chr11 | 32829541 | 32830404 | 864 | 11 | 2.37E-08 | 0.0020196 | 4.70E-05 | 6.34E-07 | 0.001891366 | PRRG4 |
| chr19 | 41912556 | 41913571 | 1016 | 4 | 9.74E-08 | 7.93E-05 | 2.52E-05 | 6.34E-07 | -0.00607536 | ARHGEF1, ERFL |
| chr2 | 136990412 | 136991263 | 852 | 6 | 7.95E-08 | 1.61E-05 | 1.12E-05 | 6.34E-07 | -0.0050488 | THSD7B |
| chr9 | 129219235 | 129220090 | 856 | 2 | 3.09E-06 | 4.64E-07 | 5.18E-05 | 6.34E-07 | 0.003289043 | AL158151.3, AL158151.1 |
| chr7 | 2701993 | 2702799 | 807 | 7 | 3.39E-08 | 0.0001388 | 6.87E-05 | 6.35E-07 | -0.00173014 | AMZ1 |
| chr3 | 97764135 | 97765075 | 941 | 12 | 3.93E-08 | 7.02E-05 | 0.000186 | 6.35E-07 | 0.000829626 | ARL6 |
| chr6 | 32223702 | 32224573 | 872 | 6 | 5.79E-08 | 3.67E-06 | 0.000172 | 6.35E-07 | -0.00800154 | NOTCH4 |
| chr8 | 113614281 | 113614735 | 455 | 3 | 1.14E-07 | 7.87E-06 | 9.47E-06 | 6.36E-07 | -0.00179002 | AC103993.1 |
| chr10 | 43904490 | 43905084 | 595 | 2 | 4.98E-07 | 5.82E-07 | 2.90E-05 | 6.37E-07 | -0.01175774 | LINC02659 |
| chr11 | 1409370 | 1410603 | 1234 | 6 | 3.79E-06 | 8.12E-06 | 9.38E-05 | 6.37E-07 | 0.006992925 | BRSK2 |
| chr13 | 107451212 | 107451495 | 284 | 2 | 9.16E-07 | 4.32E-07 | 6.90E-05 | 6.38E-07 | 0.024051135 | FAM155A |
| chr7 | 982462 | 983520 | 1059 | 13 | 9.61E-07 | 3.39E-05 | 0.002351 | 6.38E-07 | -7.42E-05 | CYP2W1 |
| chr10 | 33683647 | 33684263 | 617 | 3 | 5.71E-08 | 1.63E-05 | 1.50E-06 | 6.38E-07 | -0.00700373 | NA |
| chr3 | 158097883 | 158098885 | 1003 | 7 | 1.72E-07 | 1.05E-06 | 0.000584 | 6.38E-07 | 0.0018742 | SHOX2 |
| chr22 | 33106079 | 33106397 | 319 | 4 | 1.15E-06 | 8.10E-07 | 0.000903 | 6.39E-07 | 0.000898407 | LINC01640 |
| chr11 | 62517892 | 62518774 | 883 | 3 | 2.33E-07 | 7.52E-07 | 0.000154 | 6.39E-07 | -0.0022034 | AHNAK |
| chr4 | 1842703 | 1843451 | 749 | 7 | 1.19E-07 | 2.09E-05 | 0.000166 | 6.40E-07 | 0.000404 | LETM1 |
| chr5 | 136132057 | 136133289 | 1233 | 12 | 7.04E-06 | 5.81E-07 | 0.002377 | 6.40E-07 | 0.004658933 | SMAD5, SMAD5-AS1 |
| chr2 | 67826055 | 67826745 | 691 | 3 | 2.44E-06 | 3.51E-07 | 0.000198 | 6.42E-07 | -0.00207974 | NA |
| chr19 | 58470469 | 58471140 | 672 | 4 | 4.09E-07 | 2.84E-07 | 0.000646 | 6.42E-07 | -0.00182194 | ZNF324 |
| chr16 | 75251189 | 75253121 | 1933 | 16 | 2.19E-06 | 0.0028828 | 0.000248 | 6.42E-07 | 0.000382539 | BCAR1 |
| chr10 | 74883066 | 74883199 | 134 | 2 | 9.96E-07 | 4.70E-07 | 5.23E-05 | 6.42E-07 | 0.009509813 | KAT6B |
| chr12 | 104358441 | 104358445 | 5 | 2 | 1.30E-07 | 0.0001248 | 6.84E-07 | 6.42E-07 | -0.00429181 | NA |
| chr6 | 157981476 | 157982488 | 1013 | 8 | 6.36E-08 | 2.43E-05 | 0.000253 | 6.43E-07 | -0.00234053 | SYNJ2 |
| chr20 | 21397407 | 21398550 | 1144 | 16 | 9.08E-08 | 2.47E-05 | 0.00037 | 6.45E-07 | 0.000792203 | AL158013.1, NKX2-4 |
| chr3 | 150703034 | 150703978 | 945 | 12 | 7.04E-08 | 4.38E-06 | 0.000791 | 6.45E-07 | 0.005508809 | ERICH6-AS1, ERICH6 |
| chr7 | 139541838 | 139542419 | 582 | 4 | 7.33E-07 | 1.83E-06 | 0.000161 | 6.45E-07 | -0.00031477 | CLEC2L |
| chr10 | 48266746 | 48266883 | 138 | 2 | 1.12E-06 | 3.97E-07 | 0.000111 | 6.45E-07 | -0.00662081 | FRMPD2 |
| chr7 | 27141269 | 27141799 | 531 | 3 | 2.13E-08 | 4.34E-06 | 5.34E-06 | 6.45E-07 | -0.00302216 | HOXA-AS3, HOXA3, AC004080.6, HOXA5 |
| chr12 | 116697026 | 116698096 | 1071 | 5 | 4.12E-10 | 0.0006786 | 3.02E-06 | 6.45E-07 | -0.00353001 | AC125603.2 |
| chr16 | 23182337 | 23183412 | 1076 | 14 | 4.96E-07 | 1.30E-06 | 0.000593 | 6.45E-07 | 0.000379025 | SCNN1G, AC099482.2 |
| chr1 | 225677171 | 225677785 | 615 | 4 | 1.43E-06 | 3.46E-07 | 0.000252 | 6.45E-07 | -0.01235441 | NA |
| chr11 | 67284506 | 67285521 | 1016 | 5 | 3.40E-09 | 0.0001138 | 6.44E-07 | 6.46E-07 | -0.00699208 | GRK2 |
| chr12 | 64142456 | 64143287 | 832 | 3 | 8.03E-07 | 2.98E-06 | 3.53E-05 | 6.46E-07 | -0.00684098 | SRGAP1 |
| chr15 | 43067936 | 43068607 | 672 | 2 | 1.67E-07 | 1.26E-06 | 8.74E-06 | 6.46E-07 | -0.01385835 | UBR1 |
| chr1 | 39214565 | 39215066 | 502 | 3 | 1.31E-07 | 2.23E-06 | 1.84E-05 | 6.46E-07 | -0.00937357 | MACF1 |
| chr16 | 17368087 | 17368279 | 193 | 2 | 2.24E-07 | 6.02E-07 | 2.81E-05 | 6.46E-07 | 0.025796378 | XYLT1 |
| chr1 | 3238136 | 3239401 | 1266 | 4 | 4.76E-06 | 2.30E-06 | 0.000311 | 6.46E-07 | -0.00186525 | PRDM16 |
| chr2 | 55268473 | 55269943 | 1471 | 13 | 7.66E-07 | 0.0004155 | 0.000183 | 6.46E-07 | 0.001692738 | MTIF2 |
| chr17 | 38415021 | 38415768 | 748 | 4 | 1.04E-06 | 3.07E-07 | 0.00019 | 6.46E-07 | -0.00660869 | NA |
| chr10 | 46097783 | 46098458 | 676 | 2 | 1.89E-06 | 4.43E-07 | 6.67E-05 | 6.47E-07 | 0.006309503 | NA |
| chr3 | 52022289 | 52022815 | 527 | 4 | 2.13E-06 | 1.74E-07 | 0.001297 | 6.47E-07 | -0.00726859 | NA |
| chr22 | 45201846 | 45203268 | 1423 | 10 | 6.09E-09 | 0.0003289 | 0.000855 | 6.47E-07 | -0.00273383 | KIAA0930 |
| chr1 | 65419681 | 65421213 | 1533 | 18 | 1.48E-08 | 0.0008302 | 3.43E-05 | 6.47E-07 | 0.002007757 | LEPROT, LEPR |
| chr3 | 156290981 | 156291891 | 911 | 11 | 1.79E-06 | 1.63E-06 | 0.009245 | 6.47E-07 | -0.00040075 | KCNAB1, AC091607.2 |
| chr22 | 50285795 | 50286420 | 626 | 5 | 7.46E-09 | 4.76E-05 | 0.000138 | 6.48E-07 | -0.00337348 | PLXNB2 |
| chr1 | 113140899 | 113141393 | 495 | 3 | 5.25E-07 | 5.32E-07 | 8.13E-05 | 6.48E-07 | -0.01222304 | NA |
| chr16 | 89918754 | 89920137 | 1384 | 7 | 1.74E-06 | 1.12E-05 | 0.000353 | 6.48E-07 | 0.000826225 | MC1R, AC092143.1, AC092143.2 |
| chr2 | 31233024 | 31234421 | 1398 | 14 | 1.20E-07 | 8.23E-05 | 0.000473 | 6.48E-07 | 0.000926569 | EHD3, CAPN14 |
| chr6 | 28489847 | 28490551 | 705 | 5 | 2.01E-07 | 1.97E-06 | 0.00015 | 6.48E-07 | 0.001601312 | Z98745.1 |
| chr4 | 41359517 | 41360836 | 1320 | 12 | 1.57E-08 | 3.88E-06 | 0.001051 | 6.49E-07 | -0.00342532 | LIMCH1 |
| chr11 | 10652250 | 10653419 | 1170 | 7 | 1.66E-05 | 6.12E-06 | 0.000226 | 6.49E-07 | -0.00053174 | IRAG1 |
| chr11 | 208399 | 209287 | 889 | 11 | 7.32E-08 | 1.81E-06 | 0.001685 | 6.50E-07 | 0.000617584 | RIC8A |
| chr3 | 107429525 | 107430549 | 1025 | 6 | 1.68E-07 | 0.0001321 | 0.000256 | 6.50E-07 | -0.00222975 | NA |
| chr3 | 158799528 | 158799673 | 146 | 2 | 9.57E-08 | 4.62E-05 | 8.92E-07 | 6.51E-07 | -0.0051145 | MFSD1 |
| chr18 | 59268214 | 59269202 | 989 | 3 | 3.50E-09 | 2.89E-06 | 6.84E-06 | 6.51E-07 | -0.00499924 | RAX |
| chr1 | 201530492 | 201531265 | 774 | 4 | 1.09E-07 | 3.03E-05 | 5.48E-05 | 6.51E-07 | -0.01240498 | AC096677.1, RPS10P7 |
| chr17 | 57096063 | 57097089 | 1027 | 5 | 5.69E-08 | 6.49E-05 | 3.87E-06 | 6.52E-07 | -0.00365362 | AKAP1, AC007114.2 |
| chr11 | 94495474 | 94496188 | 715 | 3 | 1.54E-07 | 1.95E-06 | 9.31E-05 | 6.52E-07 | -0.00656193 | ANKRD49 |
| chr16 | 51134574 | 51135526 | 953 | 4 | 1.90E-07 | 7.15E-06 | 2.98E-05 | 6.52E-07 | 0.011365342 | NA |
| chr2 | 219332854 | 219333684 | 831 | 6 | 5.68E-07 | 2.83E-06 | 0.001411 | 6.52E-07 | -0.01650689 | RESP18 |
| chr4 | 142379812 | 142380229 | 418 | 3 | 1.18E-07 | 3.27E-05 | 2.33E-05 | 6.53E-07 | -0.0002734 | INPP4B |
| chr17 | 80023399 | 80024198 | 800 | 10 | 1.72E-07 | 3.11E-05 | 0.000385 | 6.53E-07 | -0.0052432 | AC116025.1, TBC1D16 |
| chr14 | 105269159 | 105269560 | 402 | 4 | 7.80E-07 | 9.63E-07 | 0.000255 | 6.53E-07 | 0.003060457 | BRF1 |
| chr10 | 43138309 | 43139043 | 735 | 12 | 1.31E-07 | 0.0002586 | 0.000186 | 6.54E-07 | 0.000158963 | CSGALNACT2, AC010864.1 |
| chr12 | 54324445 | 54325568 | 1124 | 17 | 1.80E-07 | 9.39E-06 | 0.001122 | 6.54E-07 | -0.00145069 | AC078778.1, COPZ1 |
| chr11 | 6608215 | 6609401 | 1187 | 7 | 1.76E-06 | 1.12E-05 | 0.000306 | 6.54E-07 | -0.00087428 | ILK, TAF10, AC091564.3 |

| | | | | | | | | | | |
|---|---|---|---|---|---|---|---|---|---|---|
| chr13 | 27917189 | 27918288 | 1100 | 8 | 1.02E-07 | 1.20E-05 | 4.42E-05 | 6.54E-07 | -0.00119124 | PLUT |
| chr12 | 62192486 | 62193111 | 626 | 11 | 6.38E-07 | 1.20E-06 | 0.000694 | 6.55E-07 | 0.000871429 | TAFA2 |
| chr11 | 64185628 | 64186755 | 1128 | 12 | 1.26E-06 | 1.43E-06 | 0.002142 | 6.55E-07 | 0.000414835 | STIP1 |
| chr10 | 697538 | 698807 | 1270 | 5 | 4.73E-08 | 6.79E-05 | 1.81E-06 | 6.56E-07 | -0.00462348 | NA |
| chr14 | 77330690 | 77331556 | 867 | 2 | 4.26E-06 | 4.85E-07 | 5.16E-05 | 6.56E-07 | -0.01152646 | GSTZ1 |
| chr5 | 1544507 | 1544867 | 361 | 2 | 1.03E-06 | 4.21E-07 | 9.02E-05 | 6.56E-07 | -0.00759148 | AC091849.2 |
| chr10 | 22210474 | 22211292 | 819 | 4 | 7.99E-08 | 2.74E-06 | 1.14E-05 | 6.56E-07 | -0.00171026 | NA |
| chr13 | 20232137 | 20233086 | 950 | 10 | 2.61E-06 | 1.00E-07 | 0.00491 | 6.57E-07 | -0.00099072 | GJB6 |
| chr13 | 113489633 | 113490288 | 656 | 5 | 6.45E-08 | 5.20E-06 | 5.09E-05 | 6.58E-07 | -0.00754255 | DCUN1D2 |
| chr1 | 243506361 | 243506950 | 590 | 3 | 6.59E-08 | 1.01E-05 | 3.24E-06 | 6.58E-07 | 0.002990338 | AKT3 |
| chr9 | 130915068 | 130915105 | 38 | 2 | 1.46E-07 | 0.000107 | 7.26E-07 | 6.58E-07 | -0.01566123 | FIBCD1 |
| chr17 | 81696009 | 81696934 | 926 | 5 | 4.32E-07 | 1.33E-05 | 0.000805 | 6.58E-07 | -0.00156737 | HGS |
| chr13 | 113929052 | 113930029 | 978 | 10 | 1.48E-06 | 1.08E-05 | 0.001692 | 6.59E-07 | -0.00172216 | NA |
| chr13 | 95432987 | 95434021 | 1035 | 14 | 1.00E-06 | 1.20E-06 | 0.00268 | 6.59E-07 | -0.00386966 | CLDN10 |
| chr6 | 44147390 | 44148531 | 1142 | 3 | 1.98E-05 | 1.21E-06 | 0.000143 | 6.60E-07 | -0.00535247 | TMEM63B |
| chr2 | 204089714 | 204089790 | 77 | 3 | 7.86E-07 | 7.44E-07 | 5.71E-05 | 6.60E-07 | -0.00652026 | NA |
| chr14 | 91339040 | 91339907 | 868 | 8 | 3.98E-07 | 5.17E-06 | 0.000181 | 6.60E-07 | -0.00248499 | CCDC88C |
| chr7 | 30933158 | 30933670 | 513 | 2 | 1.34E-06 | 3.93E-07 | 0.000147 | 6.60E-07 | -0.00931341 | NA |
| chr2 | 111535026 | 111535690 | 665 | 3 | 1.26E-06 | 1.78E-06 | 8.88E-05 | 6.60E-07 | -0.01058749 | AC017002.2, SOCAR, AC017002.3 |
| chr10 | 110871704 | 110872843 | 1140 | 10 | 1.52E-09 | 0.0007465 | 1.33E-06 | 6.60E-07 | 0.001796295 | PDCD4, PDCD4-AS1 |
| chr9 | 131482856 | 131483364 | 509 | 2 | 5.22E-08 | 4.33E-06 | 2.91E-06 | 6.61E-07 | 0.001393236 | PRRC2B |
| chr17 | 66958340 | 66958441 | 102 | 2 | 8.14E-07 | 8.38E-07 | 1.63E-05 | 6.61E-07 | -0.00370256 | NA |
| chr1 | 35180714 | 35181410 | 697 | 6 | 8.70E-07 | 1.46E-06 | 0.002222 | 6.63E-07 | -0.00205775 | SFPQ |
| chr1 | 103108144 | 103109063 | 920 | 12 | 2.97E-06 | 2.50E-07 | 0.009296 | 6.63E-07 | -0.00525638 | COL11A1 |
| chr17 | 82293059 | 82293821 | 763 | 6 | 1.54E-07 | 0.0088994 | 3.27E-05 | 6.64E-07 | -0.00643548 | AC132872.1 |
| chr5 | 93595467 | 93596775 | 1309 | 10 | 1.50E-06 | 1.48E-06 | 0.001563 | 6.65E-07 | -0.0011247 | NA |
| chr5 | 93620768 | 93621694 | 927 | 10 | 1.54E-07 | 0.000132 | 4.39E-05 | 6.66E-07 | -0.00121923 | AC106818.2, FAM172A, MIR2277 |
| chr9 | 130903995 | 130904300 | 306 | 3 | 1.88E-07 | 2.85E-06 | 6.73E-06 | 6.66E-07 | -0.00444507 | FIBCD1 |
| chr13 | 110409433 | 110409940 | 508 | 6 | 1.58E-07 | 7.55E-07 | 4.60E-05 | 6.66E-07 | 0.013359221 | COL4A2 |
| chr15 | 99454620 | 99455178 | 559 | 2 | 6.69E-08 | 2.10E-06 | 5.36E-06 | 6.66E-07 | -0.01099224 | NA |
| chr11 | 64629861 | 64630665 | 805 | 5 | 3.23E-07 | 3.26E-06 | 0.00034 | 6.67E-07 | -0.00811449 | NRXN2 |
| chr1 | 57171971 | 57172203 | 233 | 2 | 5.23E-07 | 1.14E-06 | 1.06E-05 | 6.67E-07 | -0.00770687 | DAB1 |
| chr12 | 56080189 | 56081280 | 1092 | 6 | 1.61E-08 | 8.68E-06 | 0.00014 | 6.67E-07 | -0.00550549 | ERBB3 |
| chr17 | 63742478 | 63743353 | 876 | 4 | 6.32E-09 | 8.31E-05 | 2.51E-05 | 6.67E-07 | 0.000469764 | AC046185.1 |
| chr1 | 221487932 | 221488341 | 410 | 3 | 2.38E-07 | 7.59E-06 | 2.83E-05 | 6.67E-07 | 0.010267965 | NA |
| chr19 | 13865796 | 13866559 | 764 | 6 | 4.48E-08 | 0.0002313 | 8.58E-07 | 6.67E-07 | 0.000687734 | NANOS3 |
| chr11 | 33014724 | 33016370 | 1647 | 12 | 4.12E-06 | 0.000121 | 0.000363 | 6.67E-07 | -0.00014015 | DEPDC7 |
| chr10 | 99534886 | 99536090 | 1205 | 6 | 9.94E-08 | 6.91E-05 | 2.03E-05 | 6.68E-07 | -0.00367509 | NKX2-3 |
| chr17 | 82577522 | 82578027 | 506 | 3 | 1.10E-08 | 0.0002213 | 5.32E-07 | 6.68E-07 | -0.00558991 | FOXK2, AC124283.3 |
| chr5 | 122775470 | 122776714 | 1245 | 5 | 3.92E-10 | 3.94E-05 | 3.43E-05 | 6.68E-07 | 0.005861973 | SNX2 |
| chr22 | 41285650 | 41287370 | 1721 | 18 | 1.58E-07 | 0.0034192 | 0.000147 | 6.69E-07 | -0.00033766 | RANGAP1 |
| chr17 | 42653488 | 42654175 | 688 | 6 | 4.93E-07 | 2.77E-06 | 0.000115 | 6.69E-07 | -0.01046077 | NA |
| chr6 | 144354224 | 144354860 | 637 | 3 | 1.72E-06 | 2.96E-07 | 0.000288 | 6.69E-07 | 0.011022669 | UTRN |
| chr17 | 17872707 | 17873308 | 602 | 3 | 4.26E-07 | 3.84E-07 | 0.000189 | 6.69E-07 | -0.01180582 | TOM1L2 |
| chr11 | 130122402 | 130122473 | 72 | 4 | 5.39E-07 | 1.97E-07 | 0.001087 | 6.70E-07 | 0.009060913 | APLP2 |
| chr17 | 43395380 | 43396058 | 679 | 2 | 9.89E-09 | 6.24E-07 | 2.88E-05 | 6.70E-07 | -0.01312628 | RNU6-1137P |
| chr1 | 46594973 | 46595584 | 612 | 4 | 5.76E-07 | 2.74E-06 | 0.000422 | 6.70E-07 | -0.00372505 | MKNK1 |
| chr5 | 39231337 | 39232442 | 1106 | 3 | 1.49E-06 | 2.20E-06 | 2.19E-05 | 6.70E-07 | -0.00083173 | FYB1 |
| chr20 | 33810617 | 33811716 | 1100 | 10 | 1.11E-07 | 8.44E-06 | 0.000104 | 6.70E-07 | -0.00012284 | CHMP4B, ZNF341-AS1 |
| chr1 | 156251436 | 156252307 | 872 | 5 | 2.36E-08 | 0.0002155 | 1.92E-06 | 6.71E-07 | 0.006192803 | SMG5 |
| chr13 | 51563946 | 51564162 | 217 | 2 | 2.05E-07 | 5.88E-07 | 3.29E-05 | 6.71E-07 | -0.00294398 | NA |
| chr22 | 24589884 | 24590735 | 852 | 2 | 2.20E-06 | 4.63E-07 | 6.62E-05 | 6.71E-07 | -0.01096103 | AP000356.5, LRRC75B |
| chr19 | 2935610 | 2936514 | 905 | 3 | 1.57E-05 | 5.17E-07 | 0.000177 | 6.71E-07 | 0.00304959 | ZNF77 |
| chr6 | 132963011 | 132963364 | 354 | 2 | 2.94E-08 | 8.40E-06 | 1.92E-06 | 6.72E-07 | -0.01934321 | LINC00326 |
| chr12 | 101876945 | 101877977 | 1033 | 11 | 2.20E-06 | 4.17E-06 | 0.003871 | 6.72E-07 | 0.000792142 | DRAM1 |
| chr2 | 120899242 | 120899771 | 530 | 2 | 5.60E-08 | 3.62E-06 | 3.39E-06 | 6.72E-07 | 0.01716895 | GLI2 |
| chr2 | 241902450 | 241903511 | 1062 | 7 | 1.14E-07 | 2.25E-06 | 6.23E-05 | 6.74E-07 | -0.01135907 | LINC01237, FAM240C |
| chr5 | 176188827 | 176188934 | 108 | 2 | 5.17E-07 | 5.56E-07 | 3.81E-05 | 6.74E-07 | -0.00898101 | AC139493.2 |
| chr6 | 36336679 | 36337378 | 700 | 5 | 6.53E-08 | 7.28E-06 | 2.68E-05 | 6.74E-07 | -0.00774902 | BNIP5 |
| chr9 | 26892071 | 26893205 | 1135 | 12 | 3.47E-08 | 1.46E-05 | 2.11E-05 | 6.74E-07 | 0.000391217 | CAAP1 |
| chr13 | 75877447 | 75877509 | 63 | 2 | 1.55E-07 | 1.35E-06 | 8.75E-06 | 6.74E-07 | -0.01018398 | LMO7DN, LMO7DN-IT1 |
| chr1 | 223712736 | 223713810 | 1075 | 6 | 3.05E-08 | 0.0001079 | 0.000134 | 6.74E-07 | -0.00255731 | CAPN2 |
| chr4 | 981033 | 981218 | 186 | 3 | 3.33E-07 | 6.33E-06 | 5.24E-06 | 6.75E-07 | 0.002236634 | DGKQ, SLC26A1 |
| chr20 | 63235148 | 63235997 | 850 | 9 | 1.05E-08 | 5.38E-05 | 0.001123 | 6.75E-07 | -0.00602528 | BIRC7 |
| chr6 | 21441513 | 21441549 | 37 | 2 | 8.81E-07 | 8.94E-07 | 1.53E-05 | 6.75E-07 | 0.014405592 | NA |
| chr17 | 83031189 | 83031955 | 767 | 3 | 1.83E-06 | 8.27E-06 | 5.82E-05 | 6.75E-07 | -0.00648133 | B3GNTL1 |
| chr5 | 111222929 | 111224266 | 1338 | 10 | 7.14E-07 | 6.52E-06 | 0.004936 | 6.75E-07 | -0.00221915 | CAMK4 |
| chr1 | 166040901 | 166041576 | 676 | 2 | 1.50E-06 | 4.53E-07 | 7.50E-05 | 6.75E-07 | 0.005199314 | NA |
| chr19 | 12441062 | 12441123 | 62 | 2 | 1.05E-06 | 5.71E-07 | 3.61E-05 | 6.76E-07 | 0.000897354 | AC008758.5, AC008758.6 |
| chr6 | 31794778 | 31796096 | 1319 | 29 | 9.21E-08 | 0.0003201 | 0.001665 | 6.77E-07 | -0.00040883 | VARS1 |
| chr6 | 144582103 | 144583425 | 1323 | 8 | 1.58E-05 | 0.0002765 | 0.00019 | 6.77E-07 | 0.00083631 | UTRN |
| chr11 | 118892150 | 118893782 | 1633 | 11 | 2.07E-05 | 3.54E-07 | 0.007269 | 6.77E-07 | 0.003921481 | CXCR5 |
| chr4 | 56861143 | 56861573 | 431 | 2 | 2.55E-08 | 1.84E-05 | 1.31E-06 | 6.78E-07 | 0.002215748 | NA |
| chr14 | 81533119 | 81534514 | 1396 | 14 | 1.34E-06 | 1.28E-05 | 0.000203 | 6.78E-07 | -0.00151588 | SEL1L |
| chr7 | 24763006 | 24763485 | 480 | 2 | 1.98E-06 | 4.05E-07 | 0.000149 | 6.79E-07 | 0.003555554 | NA |
| chr2 | 233379977 | 233380811 | 835 | 6 | 7.57E-07 | 2.66E-06 | 0.000119 | 6.80E-07 | -0.00212305 | DGKD |
| chr12 | 54047271 | 54048291 | 1021 | 5 | 3.02E-08 | 6.51E-06 | 4.46E-05 | 6.81E-07 | -0.00603462 | HOXC4 |
| chr17 | 75087331 | 75088883 | 1553 | 17 | 5.78E-06 | 1.15E-06 | 0.004045 | 6.81E-07 | 0.002098465 | SLC16A5 |
| chr4 | 174282976 | 174284338 | 1363 | 23 | 5.86E-07 | 3.58E-05 | 0.000954 | 6.81E-07 | 0.000282442 | CEP44, FBXO8 |
| chr1 | 2177699 | 2178484 | 786 | 5 | 4.10E-06 | 4.72E-07 | 0.000245 | 6.81E-07 | -0.00317329 | PRKCZ |
| chr8 | 53176653 | 53177489 | 837 | 4 | 5.36E-07 | 1.21E-06 | 5.85E-05 | 6.82E-07 | 0.015033092 | NA |
| chr16 | 28491226 | 28492521 | 1296 | 14 | 3.35E-06 | 3.73E-05 | 9.51E-05 | 6.83E-07 | -0.00013676 | AC009093.11, AC138894.1, CLN3 |
| chr8 | 41665203 | 41665962 | 760 | 8 | 7.95E-07 | 7.77E-06 | 4.32E-05 | 6.83E-07 | -0.00426033 | AC113133.1, ANK1 |
| chr17 | 37046099 | 37046683 | 585 | 3 | 1.25E-06 | 3.67E-07 | 0.000152 | 6.84E-07 | 0.005398789 | AATF, AC244093.5 |
| chr3 | 187701502 | 187702920 | 1419 | 8 | 5.99E-07 | 1.42E-06 | 0.00228 | 6.85E-07 | -0.00248208 | AC072022.1, RTP2 |
| chr15 | 98528476 | 98528491 | 16 | 2 | 4.67E-07 | 7.92E-07 | 1.93E-05 | 6.85E-07 | 0.000142425 | FAM169B |
| chr12 | 131066193 | 131066323 | 131 | 2 | 4.41E-07 | 4.48E-07 | 8.43E-05 | 6.85E-07 | -0.00696058 | ADGRD1 |
| chr8 | 132681867 | 132682288 | 422 | 2 | 7.00E-08 | 7.18E-06 | 2.17E-06 | 6.85E-07 | 0.00431257 | NA |
| chr9 | 124045232 | 124045827 | 596 | 3 | 3.22E-10 | 0.0006377 | 3.13E-07 | 6.86E-07 | -0.00766895 | NA |
| chr9 | 23825761 | 23826995 | 1235 | 12 | 1.75E-05 | 3.13E-05 | 0.001435 | 6.86E-07 | 0.001399241 | AL161628.1, ELAVL2 |
| chr1 | 37634202 | 37635594 | 1393 | 13 | 4.09E-06 | 1.75E-06 | 0.006397 | 6.86E-07 | -0.00379003 | RSPO1 |
| chr11 | 108121278 | 108122571 | 1294 | 18 | 6.42E-08 | 0.0002006 | 0.001631 | 6.86E-07 | 3.94E-05 | ACAT1, AP002433.1 |
| chr5 | 76951822 | 76953269 | 1448 | 11 | 5.35E-09 | 0.0003575 | 5.02E-06 | 6.87E-07 | -0.00114156 | CRHBP |
| chr6 | 117319965 | 117320449 | 485 | 2 | 7.11E-08 | 1.88E-06 | 6.27E-06 | 6.87E-07 | -0.00860585 | ROS1, AL132671.2 |
| chr4 | 1289300 | 1290542 | 1243 | 13 | 4.36E-06 | 2.83E-05 | 0.000343 | 6.87E-07 | -0.00011817 | MAEA |
| chr2 | 191791870 | 191792977 | 1108 | 4 | 4.10E-07 | 5.84E-07 | 0.000745 | 6.88E-07 | 0.005547016 | AC098872.1 |
| chr1 | 150268702 | 150269725 | 1024 | 15 | 4.98E-07 | 6.14E-06 | 0.000388 | 6.89E-07 | 0.001803902 | C1orf54, APH1A |
| chr12 | 52014736 | 52015794 | 1059 | 3 | 2.17E-06 | 3.28E-07 | 0.000445 | 6.89E-07 | -0.00765438 | TAMALIN |
| chr12 | 56152026 | 56152779 | 754 | 13 | 1.91E-08 | 0.0002983 | 7.03E-05 | 6.91E-07 | -0.00065774 | MYL6B, AC034102.5 |
| chr8 | 71547264 | 71547884 | 621 | 7 | 2.15E-07 | 3.05E-07 | 0.001296 | 6.91E-07 | -0.00646998 | EYA1 |
| chr2 | 127654846 | 127655435 | 590 | 2 | 1.12E-08 | 4.70E-06 | 2.92E-06 | 6.91E-07 | -0.01022538 | LIMS2 |
| chr13 | 46388657 | 46388828 | 172 | 2 | 3.25E-09 | 7.73E-07 | 2.05E-05 | 6.91E-07 | 0.001353951 | RUBCNL |
| chr6 | 27693787 | 27694446 | 660 | 7 | 3.43E-07 | 5.74E-07 | 0.000467 | 6.91E-07 | 0.000683455 | LINC01012 |
| chr2 | 241225894 | 241226103 | 210 | 3 | 4.82E-08 | 0.0002431 | 5.34E-07 | 6.92E-07 | -0.00537675 | NA |
| chr1 | 46340353 | 46341151 | 799 | 15 | 5.95E-07 | 5.96E-06 | 0.00125 | 6.92E-07 | -0.00065537 | NSUN4 |
| chr6 | 105609675 | 105609830 | 156 | 2 | 6.43E-07 | 4.13E-07 | 0.000152 | 6.93E-07 | -0.02562166 | NA |
| chr15 | 27270618 | 27271334 | 717 | 3 | 1.71E-06 | 7.99E-07 | 0.000113 | 6.93E-07 | -0.01545183 | GABRG3 |
| chr22 | 32799671 | 32801048 | 1378 | 11 | 1.29E-05 | 1.86E-06 | 0.002216 | 6.93E-07 | -0.00210769 | SYN3 |
| chr5 | 14290469 | 14290872 | 404 | 3 | 1.03E-07 | 5.98E-06 | 3.82E-06 | 6.94E-07 | 0.006294664 | TRIO |
| chr11 | 330872 | 331288 | 417 | 5 | 8.19E-08 | 4.48E-05 | 4.60E-05 | 6.94E-07 | -0.00755021 | NA |
| chr14 | 81324153 | 81324914 | 762 | 2 | 1.86E-05 | 4.94E-07 | 6.07E-05 | 6.95E-07 | 0.003972438 | STON2 |
| chr12 | 1690443 | 1691743 | 1301 | 13 | 2.02E-06 | 8.97E-07 | 0.006293 | 6.95E-07 | -0.00174162 | ADIPOR2 |
| chr17 | 67800587 | 67800699 | 113 | 2 | 5.94E-07 | 2.12E-06 | 5.70E-06 | 6.95E-07 | -0.01995894 | NA |
| chr6 | 22008590 | 22008862 | 273 | 2 | 5.41E-07 | 1.00E-06 | 1.37E-05 | 6.95E-07 | -0.01305229 | CASC15 |
| chr16 | 4261627 | 4262618 | 992 | 7 | 3.91E-08 | 0.0001069 | 2.03E-05 | 6.96E-07 | -0.00190417 | TFAP4 |
| chr15 | 92586577 | 92587805 | 1229 | 5 | 3.94E-06 | 3.67E-06 | 0.000174 | 6.96E-07 | -0.00233975 | AC091544.2 |
| chr20 | 59993871 | 59994510 | 640 | 5 | 1.64E-06 | 1.15E-06 | 0.000896 | 6.96E-07 | -0.00776661 | CDH26 |
| chr5 | 1910127 | 1910579 | 453 | 3 | 1.56E-07 | 8.25E-06 | 2.19E-05 | 6.96E-07 | -0.00508877 | NA |
| chr20 | 2840662 | 2841578 | 917 | 21 | 1.19E-06 | 2.39E-06 | 0.001778 | 6.97E-07 | 0.000420934 | VPS16, PCED1A |
| chr8 | 64806363 | 64806511 | 149 | 3 | 1.66E-06 | 2.96E-07 | 0.000466 | 6.98E-07 | -0.01056155 | AC104232.3, AC104232.1 |
| chr8 | 37747999 | 37749061 | 1063 | 6 | 1.59E-05 | 2.39E-06 | 0.001095 | 6.98E-07 | 0.006830104 | ERLIN2, AC138356.2 |
| chr16 | 1520064 | 1520880 | 817 | 3 | 2.76E-07 | 1.57E-06 | 0.000146 | 6.99E-07 | -0.00540967 | IFT140 |
| chr2 | 176763349 | 176764519 | 1171 | 4 | 1.64E-07 | 5.88E-07 | 0.000129 | 7.00E-07 | -0.00310009 | LINC01117, AC092162.2 |
| chr4 | 39700281 | 39700850 | 570 | 2 | 1.95E-08 | 1.19E-05 | 1.68E-06 | 7.00E-07 | 0.014533908 | UBE2K |
| chr14 | 102038514 | 102039474 | 961 | 7 | 6.65E-07 | 1.45E-05 | 0.000284 | 7.00E-07 | 0.000538331 | DYNC1H1, AL118558.1 |
| chr3 | 15522683 | 15523237 | 555 | 3 | 1.18E-06 | 7.45E-07 | 0.000135 | 7.00E-07 | 0.009167222 | NA |
| chr19 | 7675985 | 7677711 | 1727 | 10 | 1.79E-06 | 1.20E-05 | 0.002253 | 7.01E-07 | 0.001197238 | MCEMP1 |
| chr17 | 8470310 | 8471030 | 721 | 3 | 4.99E-07 | 3.65E-07 | 0.000172 | 7.01E-07 | -0.00262089 | NDEL1 |
| chr2 | 200533791 | 200535079 | 1289 | 4 | 2.98E-07 | 6.76E-07 | 0.000122 | 7.01E-07 | -0.00317951 | SGO2 |
| chr7 | 99843936 | 99843962 | 27 | 2 | 1.20E-06 | 4.95E-07 | 6.29E-05 | 7.01E-07 | -0.00760876 | CYP3A43 |
| chr21 | 43740348 | 43741478 | 1131 | 5 | 1.39E-08 | 2.92E-06 | 0.000188 | 7.02E-07 | 0.028694971 | PDXK |
| chr2 | 99554304 | 99554674 | 371 | 5 | 5.70E-07 | 1.69E-05 | 0.00017 | 7.02E-07 | -0.00164299 | AFF3 |
| chr16 | 2186772 | 2187220 | 449 | 4 | 2.52E-07 | 2.49E-06 | 0.000267 | 7.02E-07 | -0.00691257 | CASKIN1 |
| chr20 | 34363604 | 34364239 | 636 | 3 | 1.28E-09 | 1.86E-05 | 2.43E-06 | 7.02E-07 | -0.00566114 | ITCH |
| chr20 | 1184421 | 1185059 | 639 | 5 | 3.24E-07 | 7.67E-06 | 8.31E-05 | 7.03E-07 | -0.00719385 | PSMF1, TMEM74B |
| chr5 | 3121839 | 3122048 | 210 | 3 | 1.73E-06 | 2.83E-07 | 0.000458 | 7.03E-07 | -0.00937643 | NA |
| chr17 | 42572482 | 42572544 | 63 | 2 | 1.81E-07 | 1.07E-05 | 1.79E-06 | 7.03E-07 | -0.00828488 | MLX, PSMC3IP |
| chr6 | 111258691 | 111259736 | 1046 | 17 | 2.00E-07 | 2.93E-05 | 0.002153 | 7.04E-07 | -0.00103595 | MFSD4B, AL360227.1 |
| chr13 | 36818436 | 36819853 | 1418 | 14 | 6.22E-07 | 7.73E-06 | 0.005021 | 7.04E-07 | 0.000503844 | RFXAP |
| chr17 | 47695434 | 47696314 | 881 | 5 | 1.18E-08 | 6.05E-06 | 1.40E-05 | 7.05E-07 | -0.00802727 | TBKBP1 |
| chr19 | 14015892 | 14016673 | 782 | 2 | 9.63E-06 | 4.21E-07 | 0.000152 | 7.05E-07 | -0.00879126 | NA |
| chr10 | 111136931 | 111137114 | 184 | 3 | 4.60E-07 | 1.24E-05 | 2.37E-05 | 7.06E-07 | 0.000166199 | NA |
| chr16 | 50602638 | 50603181 | 544 | 3 | 7.15E-08 | 3.75E-05 | 8.55E-05 | 7.07E-07 | 0.012783833 | NKD1 |
| chr10 | 117375470 | 117376471 | 1002 | 8 | 8.60E-06 | 1.23E-06 | 0.000456 | 7.07E-07 | -0.00156206 | NA |
| chr16 | 71537029 | 71538147 | 1119 | 5 | 3.90E-06 | 7.32E-07 | 0.001162 | 7.07E-07 | -0.00600491 | CHST4, ZNF19 |
| chr14 | 38290521 | 38290977 | 457 | 5 | 7.32E-07 | 6.83E-06 | 0.000447 | 7.09E-07 | -0.02082072 | AL355835.1 |
| chr17 | 79146066 | 79146422 | 357 | 3 | 1.26E-07 | 3.21E-06 | 8.44E-05 | 7.09E-07 | -0.00899752 | RBFOX3 |
| chr10 | 31602626 | 31603262 | 637 | 5 | 4.07E-07 | 1.48E-06 | 8.94E-05 | 7.09E-07 | -0.00984842 | AL161935.1 |
| chr8 | 17313145 | 17313156 | 12 | 2 | 1.48E-07 | 3.74E-05 | 1.04E-06 | 7.10E-07 | 0.004399889 | MTMR7 |
| chr2 | 109185701 | 109185834 | 134 | 2 | 1.09E-06 | 4.56E-07 | 9.42E-05 | 7.10E-07 | -0.00938317 | SH3RF3 |
| chr21 | 34893233 | 34893447 | 215 | 2 | 1.10E-06 | 4.59E-07 | 9.13E-05 | 7.11E-07 | -0.01667132 | RUNX1 |
| chr8 | 70668582 | 70669494 | 913 | 12 | 1.39E-06 | 4.16E-06 | 0.002134 | 7.11E-07 | 0.001457693 | XKR9, AC015687.1, LACTB2 |
| chr4 | 183397773 | 183399263 | 1491 | 8 | 2.82E-08 | 5.64E-06 | 0.000789 | 7.11E-07 | -0.00079424 | NA |
| chr15 | 37110436 | 37111041 | 606 | 6 | 2.04E-06 | 4.96E-07 | 0.000829 | 7.11E-07 | -0.00332208 | NA |
| chr6 | 41636652 | 41637605 | 954 | 6 | 1.12E-07 | 2.54E-06 | 0.000335 | 7.12E-07 | -0.00737629 | MDFI |
| chr2 | 74653723 | 74654653 | 931 | 12 | 9.63E-09 | 0.0026792 | 3.61E-05 | 7.12E-07 | -0.00113976 | SEMA4F |
| chr14 | 88209422 | 88209509 | 88 | 2 | 1.27E-06 | 4.47E-07 | 0.000108 | 7.12E-07 | -0.02407507 | KCNK10 |
| chr17 | 49677614 | 49678472 | 859 | 10 | 1.87E-06 | 4.41E-06 | 8.30E-05 | 7.12E-07 | -0.00104835 | SPOP |

| | | | | | | | | | | |
|---|---|---|---|---|---|---|---|---|---|---|
| chr16 | 82453342 | 82454092 | 751 | 3 | 7.90E-07 | 2.99E-05 | 6.21E-05 | 7.13E-07 | 0.002547622 | NA |
| chr2 | 241188275 | 241188835 | 561 | 6 | 5.60E-08 | 2.59E-05 | 1.13E-05 | 7.13E-07 | -0.00529499 | ANO7 |
| chr2 | 161238185 | 161239151 | 967 | 9 | 5.28E-07 | 1.53E-06 | 0.00079 | 7.13E-07 | 0.002751056 | AC009299.2 |
| chr17 | 40308427 | 40309258 | 832 | 12 | 8.13E-07 | 3.01E-07 | 0.007642 | 7.13E-07 | -0.00213998 | RARA |
| chr11 | 65146632 | 65147353 | 722 | 2 | 5.62E-06 | 4.38E-07 | 0.000125 | 7.14E-07 | -0.01505509 | NA |
| chr8 | 38267386 | 38268582 | 1197 | 5 | 6.67E-06 | 5.62E-07 | 0.00084 | 7.14E-07 | -0.00107974 | DDHD2, PLPP5 |
| chr12 | 57766216 | 57767128 | 913 | 5 | 5.84E-08 | 7.98E-06 | 8.30E-05 | 7.14E-07 | -0.00296573 | CYP27B1 |
| chr2 | 20822671 | 20823282 | 612 | 10 | 6.13E-07 | 5.57E-07 | 0.000299 | 7.15E-07 | 0.00039895 | LDAH |
| chr19 | 50162981 | 50163295 | 315 | 6 | 3.06E-06 | 8.84E-08 | 0.006731 | 7.15E-07 | -0.02077286 | IZUMO2 |
| chr6 | 31901666 | 31903213 | 1548 | 27 | 8.00E-06 | 0.0001225 | 0.002167 | 7.16E-07 | -0.00032606 | C2, ZBTB12 |
| chr6 | 31834730 | 31835941 | 1212 | 23 | 2.81E-10 | 0.0018548 | 2.08E-05 | 7.16E-07 | -0.00023953 | AL671762.1, SNHG32, SNORD48 |
| chr8 | 94265914 | 94266501 | 588 | 3 | 1.83E-06 | 4.01E-07 | 0.000145 | 7.16E-07 | -0.01039758 | NA |
| chr1 | 18103860 | 18104747 | 888 | 3 | 3.26E-06 | 5.12E-07 | 0.000156 | 7.17E-07 | -0.01760921 | NA |
| chr7 | 99570016 | 99570301 | 286 | 3 | 1.27E-06 | 3.99E-07 | 0.000167 | 7.17E-07 | -0.011287 | ZNF655, AC005020.2 |
| chr7 | 74514125 | 74514449 | 325 | 2 | 7.09E-08 | 1.63E-06 | 7.80E-06 | 7.18E-07 | 0.002580842 | GTF2IRD1 |
| chr1 | 247300376 | 247301056 | 681 | 5 | 8.53E-08 | 2.06E-05 | 3.87E-05 | 7.19E-07 | -0.00058512 | ZNF496 |
| chr10 | 104580411 | 104580480 | 70 | 2 | 1.52E-07 | 3.03E-06 | 4.32E-06 | 7.20E-07 | -0.01273184 | AL161646.1 |
| chr2 | 101059304 | 101060304 | 1001 | 3 | 5.19E-06 | 2.82E-06 | 9.94E-05 | 7.21E-07 | -0.00078262 | TBC1D8 |
| chr12 | 49538206 | 49540246 | 2041 | 11 | 9.66E-06 | 1.99E-05 | 0.001141 | 7.21E-07 | -0.00302686 | KCNH3, AC020612.4 |
| chr21 | 42845596 | 42845798 | 203 | 3 | 4.11E-07 | 5.99E-06 | 1.29E-05 | 7.22E-07 | -0.00093031 | WDR4 |
| chr2 | 85663158 | 85663693 | 536 | 3 | 5.53E-07 | 5.78E-07 | 0.000273 | 7.22E-07 | -0.00306121 | SFTPB |
| chr18 | 62743519 | 62744241 | 723 | 2 | 7.74E-07 | 7.39E-07 | 2.48E-05 | 7.22E-07 | 0.008719022 | PHLPP1 |
| chr10 | 131475587 | 131476056 | 470 | 6 | 1.86E-06 | 1.40E-06 | 0.000807 | 7.22E-07 | -0.00387529 | NA |
| chr6 | 30818700 | 30818775 | 76 | 2 | 4.03E-07 | 5.64E-06 | 2.72E-06 | 7.22E-07 | -0.0158021 | LINC00243 |
| chr22 | 49323991 | 49324267 | 277 | 2 | 7.53E-08 | 9.31E-05 | 8.24E-07 | 7.23E-07 | -0.01320913 | NA |
| chr10 | 27088333 | 27088660 | 328 | 3 | 5.91E-07 | 6.57E-07 | 4.97E-05 | 7.23E-07 | 0.003731546 | ANKRD26, RNU6-490P |
| chr15 | 86062404 | 86062583 | 180 | 3 | 1.62E-06 | 3.35E-07 | 0.000354 | 7.25E-07 | -0.01385174 | NA |
| chr7 | 40833643 | 40834354 | 712 | 3 | 1.81E-06 | 3.19E-07 | 0.000363 | 7.25E-07 | -0.00716058 | SUGCT |
| chr10 | 118595115 | 118596522 | 1408 | 13 | 2.98E-06 | 2.52E-06 | 0.004475 | 7.25E-07 | -0.00171694 | PRLHR |
| chr13 | 112449757 | 112450783 | 1027 | 5 | 5.81E-07 | 1.02E-06 | 0.001017 | 7.26E-07 | -0.00015068 | NA |
| chr8 | 141560491 | 141560497 | 7 | 2 | 2.21E-07 | 4.07E-05 | 1.03E-06 | 7.26E-07 | 0.019684262 | NA |
| chr12 | 53379145 | 53380531 | 1387 | 11 | 1.01E-06 | 0.0004147 | 0.000126 | 7.26E-07 | -0.00153168 | SP1 |
| chr2 | 20595685 | 20596152 | 468 | 3 | 1.91E-06 | 2.90E-07 | 0.000652 | 7.26E-07 | -0.01341665 | HS1BP3 |
| chr8 | 29498642 | 29499512 | 871 | 3 | 2.95E-07 | 2.44E-06 | 7.03E-05 | 7.27E-07 | -0.00932811 | NA |
| chr16 | 48243648 | 48244743 | 1096 | 16 | 3.65E-06 | 2.98E-06 | 0.009892 | 7.27E-07 | 0.00032619 | LONP2, ABCC11 |
| chr22 | 23772410 | 23773381 | 972 | 10 | 1.02E-07 | 0.0004659 | 0.00011 | 7.27E-07 | -0.0026475 | MMP11 |
| chr1 | 31373057 | 31374566 | 1510 | 12 | 8.03E-06 | 6.54E-05 | 0.000463 | 7.28E-07 | 5.19E-05 | FABP3 |
| chr3 | 100400920 | 100402117 | 1198 | 15 | 1.79E-08 | 5.30E-05 | 0.000537 | 7.29E-07 | 0.000357476 | LNP1, TOMM70 |
| chr1 | 24411729 | 24412095 | 367 | 2 | 1.57E-07 | 9.99E-07 | 1.51E-05 | 7.29E-07 | -0.01122418 | STPG1 |
| chr21 | 45100320 | 45101366 | 1047 | 5 | 1.11E-07 | 5.50E-06 | 5.61E-05 | 7.29E-07 | 0.002992296 | ADARB1, AL133499.1 |
| chr20 | 591467 | 592040 | 574 | 2 | 5.23E-07 | 7.39E-07 | 2.55E-05 | 7.30E-07 | -0.00981554 | NA |
| chr10 | 110205646 | 110206336 | 691 | 5 | 1.65E-06 | 3.15E-06 | 0.001452 | 7.30E-07 | 0.015662903 | NA |
| chr8 | 29671297 | 29671695 | 399 | 2 | 1.90E-08 | 0.0004602 | 6.47E-07 | 7.30E-07 | 0.002951701 | NA |
| chr14 | 58350623 | 58351076 | 454 | 3 | 9.18E-08 | 2.63E-05 | 1.62E-06 | 7.30E-07 | 0.007797193 | ARID4A |
| chr1 | 230867330 | 230869509 | 2180 | 19 | 1.79E-08 | 0.002743 | 9.05E-05 | 7.30E-07 | -0.00012301 | AL118511.1, C1orf198 |
| chr11 | 119167999 | 119168731 | 733 | 11 | 1.54E-07 | 8.03E-05 | 7.82E-05 | 7.30E-07 | 0.001265138 | NLRX1 |
| chr12 | 62702654 | 62703484 | 831 | 3 | 1.44E-06 | 2.92E-07 | 0.000506 | 7.31E-07 | -0.01151809 | PPM1H |
| chr16 | 87350993 | 87351157 | 165 | 2 | 1.28E-06 | 4.51E-07 | 0.00012 | 7.31E-07 | -0.00144078 | FBXO31 |
| chr12 | 95473790 | 95474617 | 828 | 12 | 2.08E-06 | 1.88E-07 | 0.009028 | 7.31E-07 | 0.000128125 | METAP2 |
| chr9 | 127802230 | 127803099 | 870 | 14 | 8.38E-07 | 1.86E-05 | 0.0024 | 7.32E-07 | 0.001569285 | FPGS |
| chr7 | 101012308 | 101012902 | 595 | 3 | 1.58E-06 | 3.13E-07 | 0.000499 | 7.32E-07 | 0.007708471 | MUC12 |
| chr4 | 158769200 | 158770122 | 923 | 6 | 2.48E-06 | 1.58E-07 | 0.003592 | 7.33E-07 | -0.00310125 | FNIP2 |
| chr9 | 33290402 | 33290951 | 550 | 8 | 1.22E-06 | 1.55E-06 | 0.000774 | 7.33E-07 | 0.00182302 | NFX1 |
| chr2 | 8693486 | 8693634 | 149 | 2 | 1.33E-06 | 6.75E-07 | 3.12E-05 | 7.34E-07 | -0.01666726 | NA |
| chr19 | 38663186 | 38664192 | 1007 | 5 | 6.64E-08 | 8.20E-06 | 5.51E-06 | 7.34E-07 | -0.00548194 | ACTN4 |
| chr10 | 12832207 | 12832786 | 580 | 4 | 1.92E-08 | 7.67E-05 | 6.22E-07 | 7.34E-07 | 0.000912318 | CAMK1D |
| chr6 | 15663237 | 15664000 | 764 | 6 | 3.56E-10 | 1.18E-05 | 6.91E-06 | 7.35E-07 | -0.00117184 | NA |
| chr7 | 1437555 | 1438484 | 930 | 3 | 2.13E-09 | 4.04E-05 | 1.05E-06 | 7.36E-07 | -0.00558436 | MICALL2 |
| chr3 | 196632331 | 196633118 | 788 | 7 | 1.80E-07 | 7.54E-06 | 0.001 | 7.36E-07 | 0.001756273 | LINC01063 |
| chr16 | 55018706 | 55019141 | 436 | 4 | 4.35E-07 | 3.87E-06 | 7.70E-06 | 7.37E-07 | -0.00480921 | NA |
| chr2 | 40450460 | 40451946 | 1487 | 8 | 1.94E-05 | 2.59E-06 | 0.003228 | 7.37E-07 | -0.00271265 | SLC8A1 |
| chr11 | 64313686 | 64314402 | 717 | 5 | 1.73E-07 | 0.0002106 | 3.79E-06 | 7.38E-07 | -0.00482212 | ESRRA |
| chr6 | 40359438 | 40360073 | 636 | 2 | 1.70E-06 | 5.13E-07 | 6.75E-05 | 7.38E-07 | -0.01960466 | TDRG1, AL139275.1 |
| chr16 | 23131976 | 23132782 | 807 | 2 | 1.15E-05 | 4.49E-07 | 0.000135 | 7.39E-07 | -0.00842521 | USP31 |
| chr19 | 17376967 | 17377636 | 670 | 8 | 1.13E-07 | 1.20E-05 | 0.001438 | 7.39E-07 | -0.00171454 | PLVAP |
| chr6 | 44126039 | 44126879 | 841 | 7 | 2.97E-08 | 4.74E-06 | 5.17E-05 | 7.39E-07 | -0.00490126 | MRPL14 |
| chr11 | 62770029 | 62771063 | 1035 | 5 | 8.64E-08 | 1.04E-06 | 0.000149 | 7.39E-07 | -0.00309271 | NA |
| chr17 | 80414720 | 80415474 | 755 | 16 | 2.72E-07 | 7.08E-07 | 0.012797 | 7.39E-07 | 0.001335843 | ENDOV, RNF213-AS1 |
| chr22 | 19066059 | 19067049 | 991 | 4 | 1.43E-06 | 2.01E-06 | 0.000393 | 7.40E-07 | -0.00075097 | DGCR2 |
| chr16 | 50245826 | 50246812 | 987 | 5 | 4.24E-06 | 2.89E-07 | 0.0013 | 7.40E-07 | -8.15E-05 | ADCY7 |
| chr4 | 53682503 | 53683005 | 503 | 2 | 1.80E-06 | 4.55E-07 | 0.000125 | 7.40E-07 | 0.001116458 | AC058822.1, AC124017.1, LNX1 |
| chr14 | 68580528 | 68580887 | 360 | 2 | 4.25E-07 | 1.11E-06 | 1.35E-05 | 7.41E-07 | -0.00034413 | RAD51B |
| chr1 | 228108157 | 228109574 | 1418 | 18 | 3.51E-06 | 1.59E-05 | 0.004885 | 7.41E-07 | 0.000801576 | MRPL55 |
| chr8 | 101473664 | 101474505 | 842 | 2 | 1.62E-05 | 4.48E-07 | 0.000141 | 7.41E-07 | -0.00015864 | AP001207.3 |
| chr1 | 229249315 | 229249618 | 304 | 2 | 1.43E-07 | 1.52E-06 | 8.97E-06 | 7.41E-07 | 0.01157094 | NA |
| chr1 | 206671815 | 206672550 | 736 | 3 | 3.50E-07 | 2.83E-05 | 6.55E-06 | 7.42E-07 | -0.00625672 | DYRK3 |
| chr4 | 25988112 | 25988360 | 249 | 3 | 4.64E-07 | 3.72E-06 | 1.06E-05 | 7.42E-07 | -0.01015082 | NA |
| chr14 | 100938395 | 100939505 | 1111 | 9 | 3.62E-06 | 4.11E-06 | 0.000425 | 7.42E-07 | -0.00490849 | AL117190.1, MEG8 |
| chr12 | 94616082 | 94616535 | 454 | 5 | 4.39E-07 | 3.25E-06 | 4.02E-05 | 7.42E-07 | -0.00490984 | TMCC3 |
| chr12 | 47908408 | 47908471 | 64 | 2 | 9.00E-07 | 1.13E-06 | 1.32E-05 | 7.42E-07 | -0.00867319 | VDR |
| chr20 | 18612443 | 18612541 | 99 | 3 | 1.32E-06 | 4.13E-07 | 0.000381 | 7.42E-07 | 0.002669388 | AL121900.1, DTD1 |
| chr10 | 43194736 | 43194755 | 20 | 2 | 6.25E-07 | 4.56E-07 | 0.000127 | 7.42E-07 | 0.000719314 | RASGEF1A |
| chr16 | 24943831 | 24944566 | 736 | 3 | 2.89E-09 | 1.49E-05 | 3.53E-06 | 7.43E-07 | 0.003455902 | ARHGAP17 |
| chr2 | 124024540 | 124025009 | 470 | 6 | 6.23E-07 | 6.97E-07 | 7.79E-05 | 7.43E-07 | -0.00474469 | AC079154.1 |
| chr2 | 112720428 | 112721573 | 1146 | 7 | 3.94E-07 | 6.76E-05 | 0.000405 | 7.44E-07 | -0.00502876 | NT5DC4 |
| chr2 | 19049068 | 19049381 | 314 | 2 | 2.06E-07 | 4.42E-06 | 3.34E-06 | 7.44E-07 | -0.00707354 | LINC01376 |
| chr17 | 7018904 | 7020207 | 1304 | 6 | 3.10E-06 | 0.0001053 | 0.001133 | 7.44E-07 | -0.0066215 | AC040977.2, MIR497HG |
| chr1 | 26372143 | 26372860 | 718 | 2 | 5.49E-06 | 4.55E-07 | 0.00013 | 7.44E-07 | -0.00949146 | ZNF683 |
| chr3 | 146968609 | 146968655 | 47 | 2 | 9.65E-07 | 1.05E-06 | 1.48E-05 | 7.45E-07 | -0.01832959 | AC092957.1 |
| chr1 | 208039761 | 208040389 | 629 | 3 | 1.99E-07 | 3.62E-05 | 2.22E-05 | 7.45E-07 | -0.00154535 | PLXNA2 |
| chr4 | 55952127 | 55952194 | 68 | 2 | 8.91E-07 | 1.21E-06 | 1.21E-05 | 7.45E-07 | -0.00329141 | CEP135 |
| chr14 | 102047737 | 102048611 | 875 | 5 | 3.71E-06 | 1.15E-06 | 0.000161 | 7.45E-07 | 0.001452094 | DYNC1H1, AL118558.1 |
| chr1 | 161838537 | 161838790 | 254 | 2 | 8.96E-07 | 5.80E-07 | 4.73E-05 | 7.45E-07 | -0.01210418 | ATF6 |
| chr22 | 22554094 | 22554573 | 480 | 4 | 1.02E-06 | 3.55E-07 | 0.000241 | 7.45E-07 | 0.00281203 | PRAME |
| chr2 | 233386751 | 233388096 | 1346 | 9 | 7.77E-07 | 1.54E-06 | 0.003712 | 7.46E-07 | -0.00063441 | DGKD |
| chr6 | 30147546 | 30148601 | 1056 | 3 | 5.73E-09 | 7.48E-06 | 4.62E-06 | 7.47E-07 | -0.00631273 | TRIM40 |
| chr10 | 103561959 | 103562676 | 718 | 2 | 1.01E-07 | 9.19E-07 | 1.81E-05 | 7.47E-07 | -0.01533473 | NEURL1 |
| chr1 | 201887635 | 201889533 | 1899 | 18 | 6.25E-07 | 5.75E-05 | 0.000211 | 7.47E-07 | -0.00114338 | SHISA4 |
| chr11 | 1852527 | 1853090 | 564 | 6 | 2.41E-07 | 0.0026179 | 7.68E-05 | 7.48E-07 | 0.007259811 | LSP1 |
| chr9 | 130936429 | 130936571 | 143 | 3 | 6.90E-07 | 4.61E-07 | 0.000303 | 7.48E-07 | -0.01095594 | AL161733.1, FIBCD1 |
| chr16 | 58294305 | 58295552 | 1248 | 8 | 1.26E-05 | 3.97E-07 | 0.002638 | 7.49E-07 | -0.00583165 | GINS3, PRSS54 |
| chr1 | 10672006 | 10672973 | 968 | 11 | 3.27E-07 | 1.92E-05 | 0.000303 | 7.49E-07 | -0.00340176 | CASZ1 |
| chr19 | 40532005 | 40532673 | 669 | 2 | 4.03E-07 | 6.56E-07 | 3.52E-05 | 7.49E-07 | -0.01447296 | SPTBN4 |
| chr14 | 94517803 | 94518418 | 616 | 7 | 1.35E-07 | 3.30E-06 | 0.000133 | 7.49E-07 | -0.00532071 | SERPINA12 |
| chr16 | 67999606 | 68000099 | 494 | 5 | 5.57E-08 | 1.48E-05 | 5.33E-06 | 7.49E-07 | 0.002681886 | DUS2, DPEP2 |
| chr4 | 13537475 | 13537974 | 500 | 6 | 1.48E-06 | 2.50E-07 | 0.002673 | 7.49E-07 | -0.00435844 | NA |
| chr12 | 109820411 | 109820984 | 574 | 2 | 3.65E-08 | 1.10E-06 | 1.40E-05 | 7.49E-07 | -0.01421467 | TRPV4 |
| chr8 | 144099415 | 144100448 | 1034 | 4 | 6.16E-06 | 2.40E-06 | 0.000226 | 7.51E-07 | -0.00163544 | SHARPIN |
| chr4 | 187613294 | 187613758 | 465 | 5 | 7.49E-07 | 2.30E-06 | 4.94E-05 | 7.51E-07 | -0.0012127 | AC097521.1, LINC02492 |
| chr12 | 47988191 | 47989440 | 1250 | 6 | 1.47E-05 | 8.85E-07 | 0.001651 | 7.51E-07 | 0.002324873 | COL2A1 |
| chr19 | 52028185 | 52029167 | 983 | 11 | 6.57E-07 | 8.60E-07 | 0.000621 | 7.51E-07 | 0.000497563 | ZNF614 |
| chr3 | 30732820 | 30733389 | 570 | 2 | 2.69E-07 | 9.17E-07 | 1.85E-05 | 7.52E-07 | 0.016420955 | GADL1 |
| chr11 | 1235148 | 1235812 | 665 | 3 | 1.16E-06 | 6.40E-06 | 9.21E-05 | 7.52E-07 | -0.00965157 | MUC5B |
| chr1 | 233295079 | 233296207 | 1129 | 8 | 3.35E-07 | 1.51E-05 | 0.000265 | 7.53E-07 | 0.001726784 | PCNX2, AL133380.1 |
| chr19 | 45178729 | 45180005 | 1277 | 5 | 1.80E-08 | 4.61E-07 | 0.000413 | 7.53E-07 | -0.00494986 | MARK4, BLOC1S3 |
| chr2 | 108785598 | 108787236 | 1639 | 16 | 4.42E-07 | 2.29E-05 | 0.001683 | 7.54E-07 | 0.000541296 | RANBP2, CCDC138 |
| chr13 | 33918323 | 33918864 | 542 | 2 | 1.17E-07 | 2.40E-06 | 5.72E-06 | 7.54E-07 | -0.00772208 | RFC3 |
| chr3 | 38038712 | 38039434 | 723 | 9 | 1.21E-07 | 1.94E-06 | 0.001341 | 7.54E-07 | -0.00076638 | DLEC1 |
| chr16 | 84294785 | 84296132 | 1348 | 5 | 7.05E-07 | 3.72E-06 | 0.000439 | 7.54E-07 | 0.005361501 | WFDC1, AC010551.2 |
| chr20 | 37344133 | 37345904 | 1772 | 10 | 2.04E-05 | 5.84E-06 | 0.003705 | 7.54E-07 | 0.001141496 | SRC |
| chr16 | 89004239 | 89004516 | 278 | 5 | 7.24E-08 | 1.93E-05 | 7.96E-05 | 7.55E-07 | 0.002629869 | NA |
| chr22 | 26452322 | 26453040 | 719 | 4 | 2.37E-07 | 9.77E-06 | 3.26E-05 | 7.55E-07 | -9.47E-05 | HPS4 |
| chr19 | 47886178 | 47886857 | 680 | 7 | 3.10E-07 | 2.92E-06 | 0.000399 | 7.55E-07 | -0.0047719 | SULT2A1 |
| chr1 | 4781880 | 4782658 | 779 | 6 | 5.49E-07 | 7.67E-07 | 0.00061 | 7.55E-07 | -0.00419349 | AJAP1 |
| chr11 | 47414739 | 47415124 | 386 | 3 | 1.68E-07 | 1.13E-06 | 3.79E-05 | 7.56E-07 | -0.00458648 | SLC39A13 |
| chr2 | 235864256 | 235864616 | 361 | 4 | 5.65E-07 | 1.20E-06 | 0.000618 | 7.57E-07 | -0.01240321 | AGAP1 |
| chr11 | 134583537 | 134583924 | 388 | 2 | 6.51E-08 | 1.79E-05 | 1.51E-06 | 7.57E-07 | 2.38E-05 | NA |
| chr13 | 111901509 | 111901673 | 165 | 3 | 3.17E-07 | 1.06E-05 | 4.30E-05 | 7.57E-07 | -0.00894366 | NA |
| chr13 | 60326214 | 60326726 | 513 | 2 | 2.16E-06 | 4.58E-07 | 0.000144 | 7.58E-07 | 0.001152916 | NA |
| chr12 | 106188770 | 106189476 | 707 | 3 | 7.85E-07 | 3.60E-07 | 0.000246 | 7.59E-07 | 0.003668004 | NA |
| chr5 | 126857778 | 126858601 | 824 | 2 | 3.00E-06 | 5.97E-07 | 4.64E-05 | 7.59E-07 | 0.005131724 | NA |
| chr3 | 197960462 | 197961511 | 1050 | 6 | 1.37E-09 | 0.0005999 | 7.96E-05 | 7.59E-07 | -0.00797042 | LMLN |
| chr8 | 118994783 | 118995391 | 609 | 3 | 7.01E-07 | 1.55E-06 | 2.61E-05 | 7.60E-07 | -0.00325404 | NA |
| chr1 | 40039925 | 40041203 | 1279 | 18 | 5.00E-08 | 0.0002042 | 2.27E-05 | 7.60E-07 | -0.00025172 | CAP1 |
| chr7 | 25847555 | 25847965 | 411 | 2 | 1.56E-06 | 4.63E-07 | 0.000139 | 7.61E-07 | 0.001073766 | NA |
| chr2 | 130354946 | 130356294 | 1349 | 7 | 1.73E-08 | 1.76E-05 | 3.27E-06 | 7.62E-07 | -0.00375841 | PTPN18 |
| chr10 | 26436971 | 26438996 | 2026 | 16 | 2.05E-06 | 4.45E-05 | 0.000421 | 7.62E-07 | 0.002473298 | APBB1IP |
| chr11 | 66728677 | 66729144 | 468 | 3 | 3.46E-08 | 8.67E-07 | 4.97E-05 | 7.62E-07 | -0.0098174 | SPTBN2 |
| chr15 | 70779116 | 70779769 | 654 | 3 | 2.24E-08 | 0.0002605 | 1.77E-05 | 7.62E-07 | 0.001460825 | NA |
| chr19 | 1507933 | 1508556 | 624 | 4 | 2.37E-06 | 3.21E-06 | 0.000152 | 7.62E-07 | -0.00779952 | PLK5, AC027307.1, ADAMTSL5 |
| chr2 | 218357592 | 218358801 | 1210 | 5 | 9.48E-06 | 4.98E-07 | 0.000838 | 7.62E-07 | -0.0034464 | CATIP, CATIP-AS2 |
| chr9 | 128771518 | 128772494 | 977 | 14 | 3.75E-07 | 9.72E-06 | 0.000394 | 7.62E-07 | -0.00036406 | ZER1 |
| chr7 | 5501710 | 5501935 | 226 | 3 | 1.07E-08 | 6.55E-05 | 1.20E-06 | 7.63E-07 | -0.00400205 | FBXL18 |
| chr3 | 187027459 | 187027854 | 396 | 2 | 1.23E-06 | 4.86E-07 | 0.000102 | 7.63E-07 | 0.021578986 | ST6GAL1 |
| chr12 | 48936252 | 48936375 | 124 | 2 | 4.94E-07 | 5.39E-07 | 6.57E-05 | 7.64E-07 | -0.00750543 | AC073610.2, ARF3 |
| chr3 | 47474753 | 47475511 | 759 | 4 | 4.00E-08 | 1.12E-06 | 6.12E-05 | 7.65E-07 | 0.002295457 | SCAP |
| chr6 | 32830402 | 32832326 | 1925 | 12 | 7.47E-08 | 3.13E-05 | 8.04E-05 | 7.65E-07 | -0.0001394 | AL669918.1, TAP2 |
| chr1 | 197774465 | 197775657 | 1193 | 16 | 3.46E-09 | 0.0004604 | 9.39E-05 | 7.66E-07 | 6.95E-05 | DENND1B |
| chr1 | 15465300 | 15465886 | 587 | 3 | 4.61E-07 | 8.67E-07 | 0.000187 | 7.67E-07 | 0.003632332 | CELA2A |
| chr3 | 129605658 | 129606874 | 1217 | 9 | 2.75E-06 | 3.55E-07 | 0.003064 | 7.67E-07 | 0.000583929 | PLXND1 |
| chr16 | 84616596 | 84617564 | 969 | 6 | 2.84E-06 | 1.96E-07 | 0.00514 | 7.67E-07 | -0.0097016 | COTL1 |
| chr13 | 25171268 | 25172506 | 1239 | 16 | 1.71E-07 | 2.90E-05 | 0.000217 | 7.68E-07 | 0.005096448 | LINC01053, AMER2 |
| chr16 | 17557496 | 17558193 | 698 | 2 | 1.14E-06 | 6.69E-07 | 3.61E-05 | 7.68E-07 | -0.00526732 | NA |
| chr2 | 219229446 | 219230612 | 1167 | 18 | 5.14E-08 | 8.00E-05 | 0.000428 | 7.68E-07 | -0.00105793 | ANKZF1, ATG9A |
| chr9 | 113885164 | 113885416 | 253 | 2 | 9.51E-08 | 9.67E-06 | 2.10E-06 | 7.68E-07 | 0.007531799 | ZNF618 |
| chr5 | 6868242 | 6868717 | 476 | 5 | 1.98E-06 | 1.36E-07 | 0.003847 | 7.68E-07 | -0.00062225 | LINC02196, AC122710.1 |
| chr20 | 45833663 | 45834589 | 927 | 9 | 6.07E-07 | 1.84E-06 | 0.004411 | 7.68E-07 | -0.00130381 | SNX21, TNNC2 |
| chr8 | 126368658 | 126369321 | 664 | 3 | 2.10E-06 | 5.39E-06 | 8.22E-05 | 7.68E-07 | -0.00976392 | NA |

| | | | | | | | | | | |
|---|---|---|---|---|---|---|---|---|---|---|
| chr5 | 122155793 | 122156157 | 365 | 3 | 1.33E-07 | 2.40E-05 | 1.11E-05 | 7.68E-07 | -0.00578246 | AC010255.2, ZNF474-AS1 |
| chr10 | 33337064 | 33337870 | 807 | 4 | 3.76E-07 | 1.12E-06 | 0.000563 | 7.70E-07 | -0.01033532 | NA |
| chr17 | 11769021 | 11769152 | 132 | 2 | 1.24E-06 | 5.70E-07 | 5.63E-05 | 7.70E-07 | -0.00609269 | DNAH9 |
| chr9 | 137014301 | 137014709 | 409 | 4 | 2.52E-07 | 1.84E-05 | 1.71E-05 | 7.70E-07 | -0.00473395 | ABCA2 |
| chr17 | 42098499 | 42098985 | 487 | 3 | 5.75E-09 | 2.14E-05 | 1.59E-06 | 7.71E-07 | 0.004393192 | NA |
| chr21 | 29757944 | 29759023 | 1080 | 6 | 9.91E-09 | 4.33E-05 | 5.36E-06 | 7.74E-07 | -0.00652085 | GRIK1-AS1, GRIK1 |
| chr20 | 1265449 | 1266414 | 966 | 9 | 1.44E-07 | 4.75E-06 | 2.74E-05 | 7.74E-07 | -0.00138509 | RAD21L1, SNPH |
| chr15 | 71146728 | 71147655 | 928 | 4 | 1.70E-05 | 2.44E-07 | 0.000886 | 7.74E-07 | -0.00350429 | THSD4, THSD4-AS1 |
| chr3 | 69541095 | 69541861 | 767 | 5 | 2.33E-06 | 6.52E-07 | 0.000342 | 7.74E-07 | -0.00879981 | FRMD4B |
| chr4 | 156791995 | 156792665 | 671 | 3 | 2.96E-09 | 0.0261149 | 2.56E-07 | 7.75E-07 | -0.01121524 | PDGFC |
| chr7 | 21076564 | 21076789 | 226 | 2 | 1.25E-06 | 5.12E-07 | 8.66E-05 | 7.75E-07 | -0.00934166 | NA |
| chr2 | 11023183 | 11024461 | 1279 | 7 | 5.79E-08 | 1.40E-05 | 0.001459 | 7.76E-07 | -0.00902153 | NA |
| chr4 | 2059207 | 2060714 | 1508 | 10 | 2.21E-05 | 2.65E-07 | 0.005102 | 7.76E-07 | -0.00467861 | NAT8L |
| chr14 | 51243562 | 51243945 | 384 | 3 | 3.44E-09 | 0.000171 | 6.83E-07 | 7.76E-07 | 0.006022206 | TMX1 |
| chr12 | 132306828 | 132307484 | 657 | 3 | 7.61E-07 | 3.06E-07 | 0.000509 | 7.76E-07 | 0.01311609 | GALNT9 |
| chr2 | 239378215 | 239378465 | 251 | 4 | 1.16E-06 | 5.93E-07 | 0.000606 | 7.76E-07 | -0.01043035 | HDAC4 |
| chr17 | 76307358 | 76308538 | 1181 | 10 | 3.26E-06 | 3.97E-07 | 0.002373 | 7.76E-07 | -4.68E-05 | QRICH2 |
| chr11 | 134149188 | 134149545 | 358 | 5 | 9.94E-07 | 5.30E-07 | 0.001373 | 7.77E-07 | -0.00070821 | JAM3 |
| chr12 | 132006164 | 132006253 | 90 | 2 | 1.31E-06 | 5.61E-07 | 6.16E-05 | 7.79E-07 | -0.00039119 | EP400 |
| chr1 | 19889616 | 19890431 | 816 | 4 | 7.99E-09 | 0.0067046 | 2.37E-07 | 7.79E-07 | 0.002334609 | OTUD3 |
| chr13 | 45311756 | 45312168 | 413 | 3 | 2.35E-08 | 9.79E-06 | 2.59E-05 | 7.79E-07 | -0.01035472 | NA |
| chr7 | 47949526 | 47949815 | 290 | 4 | 4.53E-07 | 1.96E-05 | 4.32E-05 | 7.79E-07 | 0.005160341 | NA |
| chr18 | 54269342 | 54270083 | 742 | 13 | 2.65E-06 | 1.20E-06 | 0.00306 | 7.79E-07 | 0.000263998 | POLI, AC093462.1 |
| chr19 | 45370036 | 45370895 | 860 | 16 | 3.27E-07 | 2.53E-05 | 0.000834 | 7.80E-07 | 0.000779716 | ERCC2 |
| chr20 | 57329058 | 57330020 | 963 | 12 | 2.66E-07 | 3.74E-05 | 0.000482 | 7.80E-07 | -0.00200712 | SPO11 |
| chr5 | 136365734 | 136366645 | 912 | 4 | 5.07E-11 | 0.0003205 | 3.63E-06 | 7.81E-07 | -0.00969345 | NA |
| chr8 | 1197747 | 1198043 | 297 | 2 | 4.74E-07 | 5.07E-07 | 9.44E-05 | 7.81E-07 | -0.00076842 | DLGAP2 |
| chr4 | 83484358 | 83485809 | 1452 | 13 | 4.60E-06 | 1.82E-05 | 0.000738 | 7.82E-07 | 0.001148839 | ABRAXAS1 |
| chr2 | 176324948 | 176325507 | 560 | 2 | 2.94E-07 | 1.18E-06 | 1.38E-05 | 7.82E-07 | -0.00843838 | MTX2 |
| chr10 | 129652268 | 129652498 | 231 | 2 | 7.87E-07 | 9.52E-07 | 1.89E-05 | 7.82E-07 | -0.00022594 | MGMT |
| chr4 | 112520452 | 112521648 | 1197 | 10 | 1.96E-08 | 0.0002313 | 6.01E-05 | 7.83E-07 | -0.0026013 | NEUROG2-AS1 |
| chr6 | 27815448 | 27815558 | 111 | 3 | 1.09E-06 | 4.34E-07 | 0.000248 | 7.84E-07 | -0.02185822 | H2BC14 |
| chr7 | 77696024 | 77697525 | 1502 | 12 | 2.57E-06 | 6.55E-05 | 0.000411 | 7.84E-07 | -0.00072107 | RSBN1L, APTR |
| chr11 | 10494824 | 10495327 | 504 | 2 | 5.33E-08 | 7.59E-06 | 2.47E-06 | 7.85E-07 | -0.0044537 | AMPD3 |
| chr10 | 132240298 | 132241293 | 996 | 6 | 3.40E-07 | 1.71E-05 | 5.76E-05 | 7.85E-07 | 0.000243683 | STK32C |
| chr19 | 3807789 | 3808864 | 1076 | 5 | 1.73E-06 | 4.25E-07 | 0.001859 | 7.85E-07 | -0.00343817 | ZFR2 |
| chr3 | 36714558 | 36714594 | 37 | 3 | 1.14E-06 | 1.74E-06 | 4.84E-05 | 7.85E-07 | 0.002954002 | DCLK3 |
| chr7 | 155349016 | 155350053 | 1038 | 6 | 1.38E-07 | 9.00E-06 | 2.91E-05 | 7.87E-07 | -0.01804547 | NA |
| chr3 | 37398215 | 37398824 | 610 | 5 | 7.24E-08 | 6.31E-06 | 2.90E-05 | 7.88E-07 | -0.00239173 | C3orf35 |
| chr7 | 90594844 | 90595572 | 729 | 5 | 7.06E-07 | 2.97E-07 | 0.001767 | 7.89E-07 | -0.00202839 | CDK14, AC002456.1 |
| chr2 | 186693628 | 186694403 | 776 | 9 | 5.19E-08 | 0.0031938 | 0.00029 | 7.89E-07 | 8.31E-05 | FAM171B, AC017101.1 |
| chr14 | 104571014 | 104571348 | 335 | 3 | 3.42E-07 | 3.51E-07 | 0.000379 | 7.89E-07 | -0.00473293 | TMEM179 |
| chr22 | 35617260 | 35618309 | 1050 | 13 | 2.69E-06 | 2.95E-07 | 0.0062 | 7.90E-07 | -0.00222394 | MB |
| chr17 | 47200032 | 47200500 | 469 | 3 | 1.08E-06 | 4.90E-07 | 0.000135 | 7.90E-07 | -0.01188051 | MYL4 |
| chr5 | 176152470 | 176153024 | 555 | 2 | 1.29E-07 | 9.34E-07 | 1.99E-05 | 7.90E-07 | -0.01655326 | AC139491.5, AC139491.6, AC139491.2 |
| chr2 | 240560145 | 240561925 | 1781 | 13 | 3.62E-05 | 4.50E-07 | 0.00642 | 7.90E-07 | 0.00055299 | DUSP28, ANKMY1 |
| chr6 | 42722253 | 42723304 | 1052 | 6 | 5.39E-07 | 1.51E-05 | 2.21E-05 | 7.91E-07 | -0.00023584 | PRPH2 |
| chr5 | 42547010 | 42548035 | 1026 | 3 | 2.34E-08 | 7.84E-06 | 3.48E-06 | 7.91E-07 | -0.0101165 | GHR |
| chr11 | 118436749 | 118437067 | 319 | 3 | 1.22E-07 | 3.73E-06 | 6.66E-06 | 7.92E-07 | 0.000499492 | AP001267.5, KMT2A |
| chr12 | 8954311 | 8954319 | 9 | 2 | 1.21E-06 | 7.87E-07 | 2.77E-05 | 7.93E-07 | -0.01395137 | KLRG1 |
| chr11 | 73245642 | 73245717 | 76 | 2 | 4.69E-07 | 8.67E-06 | 2.32E-06 | 7.93E-07 | -0.00631131 | NA |
| chr10 | 1755507 | 1755918 | 412 | 2 | 6.89E-08 | 2.37E-06 | 6.25E-06 | 7.93E-07 | 0.014559204 | NA |
| chr8 | 92094607 | 92096328 | 1722 | 14 | 6.17E-07 | 0.0024513 | 8.90E-05 | 7.93E-07 | -0.00339051 | RUNX1T1 |
| chr16 | 82137440 | 82138834 | 1395 | 4 | 1.26E-06 | 9.98E-07 | 0.000624 | 7.93E-07 | 0.011394234 | AC092142.1 |
| chr9 | 126375867 | 126376321 | 455 | 3 | 1.28E-07 | 5.13E-05 | 2.77E-05 | 7.93E-07 | -0.00320301 | MVB12B |
| chr2 | 11406450 | 11406872 | 423 | 2 | 7.12E-08 | 1.12E-05 | 2.01E-06 | 7.94E-07 | 0.008046498 | AC099344.3 |
| chr6 | 31954584 | 31955435 | 852 | 8 | 3.24E-06 | 2.03E-06 | 0.001856 | 7.95E-07 | -0.00293701 | NELFE |
| chr12 | 110123538 | 110124944 | 1407 | 12 | 8.97E-06 | 3.70E-06 | 0.001457 | 7.95E-07 | -0.00049299 | IFT81 |
| chr11 | 67647712 | 67648317 | 606 | 4 | 9.77E-08 | 2.04E-05 | 0.000451 | 7.95E-07 | 0.016307799 | ACY3 |
| chr4 | 184453612 | 184453762 | 151 | 2 | 1.30E-06 | 5.61E-07 | 6.73E-05 | 7.95E-07 | 0.006291223 | IRF2 |
| chr15 | 101048363 | 101049659 | 1297 | 6 | 6.64E-07 | 0.000145 | 4.31E-05 | 7.96E-07 | -0.00448483 | LRRK1, AC090907.1 |
| chr6 | 24877354 | 24878603 | 1250 | 5 | 1.13E-05 | 3.32E-06 | 0.001477 | 7.96E-07 | -0.00472874 | RIPOR2 |
| chr10 | 35195605 | 35196224 | 620 | 9 | 1.52E-08 | 0.0001282 | 4.58E-05 | 7.96E-07 | 0.001046067 | CREM |
| chr5 | 138178693 | 138179872 | 1180 | 14 | 5.20E-09 | 5.78E-06 | 0.005136 | 7.98E-07 | -0.00019776 | KIF20A, BRD8 |
| chr2 | 46850053 | 46850249 | 197 | 2 | 4.00E-07 | 5.08E-07 | 0.000107 | 8.00E-07 | 0.026399892 | LINC01119 |
| chr16 | 2458717 | 2459197 | 481 | 2 | 1.56E-09 | 4.25E-06 | 3.80E-06 | 8.00E-07 | -0.00940576 | CCNF, AC106820.5 |
| chr5 | 81069179 | 81070010 | 832 | 3 | 6.01E-06 | 3.84E-07 | 0.000345 | 8.00E-07 | -0.00510357 | RASGRF2 |
| chr10 | 124801518 | 124802192 | 675 | 10 | 3.55E-07 | 3.53E-05 | 0.000256 | 8.00E-07 | -0.00078462 | ABRAXAS2 |
| chr9 | 124503069 | 124503710 | 642 | 2 | 3.85E-07 | 5.21E-07 | 9.35E-05 | 8.01E-07 | -0.01299008 | NR5A1 |
| chr18 | 48620713 | 48621477 | 765 | 3 | 5.82E-09 | 1.81E-05 | 1.34E-05 | 8.02E-07 | -0.00880649 | CTIF |
| chr21 | 43776358 | 43776779 | 422 | 6 | 4.11E-07 | 2.99E-06 | 0.000308 | 8.02E-07 | 0.001364025 | NA |
| chr12 | 114680972 | 114681661 | 690 | 4 | 2.44E-07 | 1.70E-06 | 3.32E-05 | 8.03E-07 | -0.00058989 | TBX3 |
| chr12 | 130212718 | 130213573 | 856 | 3 | 1.13E-08 | 0.0001963 | 9.20E-07 | 8.03E-07 | -0.02623494 | NA |
| chr6 | 36848558 | 36849272 | 715 | 2 | 4.18E-06 | 5.17E-07 | 0.000101 | 8.06E-07 | -0.02044812 | NA |
| chr5 | 151080539 | 151081650 | 1112 | 14 | 1.58E-05 | 1.44E-06 | 0.003325 | 8.06E-07 | -0.00058971 | TNIP1 |
| chr7 | 73667563 | 73668139 | 577 | 12 | 1.78E-07 | 4.89E-05 | 3.32E-05 | 8.06E-07 | -0.00024977 | VPS37D |
| chr4 | 67892541 | 67892928 | 388 | 3 | 1.45E-06 | 7.33E-07 | 0.000269 | 8.06E-07 | -0.00080663 | UBA6-AS1 |
| chr17 | 5032819 | 5033560 | 742 | 3 | 9.52E-08 | 6.46E-05 | 1.80E-06 | 8.07E-07 | -0.00410422 | SLC52A1 |
| chr7 | 64182899 | 64183580 | 682 | 2 | 5.73E-07 | 9.25E-07 | 2.12E-05 | 8.07E-07 | -0.02743114 | NA |
| chr7 | 99140991 | 99142159 | 1169 | 7 | 3.48E-08 | 2.96E-05 | 1.29E-05 | 8.08E-07 | -0.00136589 | SMURF1 |
| chr10 | 377967 | 378920 | 954 | 6 | 2.54E-07 | 3.10E-07 | 0.001326 | 8.08E-07 | -0.00861885 | DIP2C |
| chr14 | 81617105 | 81617137 | 33 | 2 | 7.27E-07 | 7.37E-07 | 3.34E-05 | 8.09E-07 | -0.00461456 | LINC01467 |
| chr6 | 146769888 | 146771106 | 1219 | 7 | 1.35E-07 | 7.73E-06 | 0.00102 | 8.09E-07 | -0.00646824 | ADGB |
| chr3 | 184275292 | 184276454 | 1163 | 13 | 2.78E-08 | 8.53E-05 | 0.000572 | 8.10E-07 | -0.00506582 | EEF1AKMT4-ECE2, ECE2 |
| chr19 | 57102786 | 57103888 | 1103 | 3 | 4.93E-05 | 2.63E-06 | 0.000106 | 8.10E-07 | -0.00069277 | NA |
| chr7 | 549459 | 550722 | 1264 | 7 | 1.12E-05 | 1.93E-06 | 0.000677 | 8.11E-07 | -0.00152976 | PRKAR1B |
| chr7 | 131327347 | 131328448 | 1102 | 16 | 1.56E-06 | 1.86E-06 | 0.003954 | 8.11E-07 | 0.000832466 | MKLN1, MKLN1-AS |
| chr12 | 117452191 | 117453369 | 1179 | 6 | 1.24E-05 | 4.50E-07 | 0.001372 | 8.11E-07 | -0.00158688 | AC073864.1, KSR2 |
| chr12 | 6384601 | 6386139 | 1539 | 8 | 6.33E-07 | 1.54E-05 | 0.000191 | 8.13E-07 | -0.00290394 | LTBR |
| chr7 | 66088828 | 66089677 | 850 | 6 | 1.22E-07 | 0.0003136 | 3.46E-05 | 8.13E-07 | -0.00322979 | ASL, AC068533.4 |
| chr22 | 50085257 | 50086339 | 1083 | 16 | 8.60E-06 | 7.72E-06 | 0.002846 | 8.13E-07 | 0.002405486 | MLC1 |
| chr9 | 137986711 | 137987334 | 624 | 2 | 9.95E-08 | 2.88E-06 | 5.37E-06 | 8.13E-07 | -0.01784456 | CACNA1B |
| chr3 | 38346585 | 38347317 | 733 | 8 | 1.18E-08 | 7.20E-06 | 9.38E-06 | 8.13E-07 | -0.0033519 | XYLB |
| chr1 | 26899811 | 26900892 | 1082 | 11 | 4.89E-07 | 1.53E-05 | 0.001989 | 8.13E-07 | -0.00123413 | NUDC, GPATCH3 |
| chr17 | 39606202 | 39608040 | 1839 | 15 | 4.09E-06 | 1.23E-05 | 0.005364 | 8.13E-07 | 0.000274772 | NEUROD2 |
| chr5 | 180815568 | 180816026 | 459 | 4 | 8.58E-09 | 0.0003598 | 7.26E-07 | 8.14E-07 | 0.00802668 | MGAT1 |
| chr9 | 122674869 | 122675375 | 507 | 4 | 5.80E-07 | 1.64E-05 | 0.000331 | 8.14E-07 | -0.00223546 | OR1L3 |
| chr15 | 94743677 | 94743789 | 113 | 2 | 1.08E-06 | 9.48E-07 | 2.08E-05 | 8.15E-07 | -0.01893769 | NA |
| chr2 | 17793430 | 17794012 | 583 | 3 | 8.00E-08 | 4.91E-05 | 1.95E-05 | 8.16E-07 | -0.00465139 | SMC6 |
| chr7 | 158719689 | 158719980 | 292 | 4 | 1.40E-06 | 5.07E-07 | 0.000565 | 8.16E-07 | -0.01222388 | AC019084.1 |
| chr2 | 218430969 | 218431027 | 59 | 2 | 1.02E-07 | 0.0001142 | 8.87E-07 | 8.17E-07 | -0.00977627 | VIL1 |
| chr7 | 1320993 | 1321595 | 603 | 2 | 2.61E-07 | 1.37E-06 | 1.21E-05 | 8.18E-07 | -0.00522938 | NA |
| chr19 | 5829161 | 5829844 | 684 | 4 | 2.46E-08 | 4.14E-06 | 0.000128 | 8.18E-07 | -0.00745089 | NA |
| chr5 | 172754316 | 172755031 | 716 | 5 | 4.87E-06 | 2.62E-07 | 0.000899 | 8.20E-07 | -0.00771616 | NA |
| chr2 | 233566247 | 233566820 | 574 | 12 | 5.28E-07 | 9.89E-06 | 0.000117 | 8.20E-07 | -0.00140117 | USP40 |
| chr8 | 25685294 | 25685766 | 473 | 5 | 4.67E-07 | 4.07E-06 | 1.52E-05 | 8.21E-07 | -0.004724 | AC009623.1 |
| chr5 | 154093416 | 154094573 | 1158 | 6 | 6.75E-05 | 2.26E-06 | 0.000233 | 8.22E-07 | 0.002154192 | MFAP3 |
| chr6 | 31877333 | 31878472 | 1140 | 11 | 2.59E-07 | 6.83E-05 | 0.000131 | 8.22E-07 | -0.00297469 | EHMT2-AS1, SLC44A4 |
| chr7 | 93149512 | 93149650 | 139 | 2 | 1.27E-06 | 5.03E-07 | 0.000137 | 8.23E-07 | -0.00269884 | NA |
| chr10 | 130203283 | 130204323 | 1041 | 4 | 3.05E-06 | 4.16E-07 | 0.000823 | 8.23E-07 | -0.00315487 | NA |
| chr8 | 143527535 | 143528384 | 850 | 4 | 3.97E-08 | 5.83E-06 | 1.39E-05 | 8.24E-07 | 0.002721653 | ZC3H3 |
| chr17 | 8117789 | 8119257 | 1469 | 16 | 2.79E-07 | 0.0004152 | 4.93E-05 | 8.24E-07 | -0.00057714 | ALOXE3 |
| chr1 | 2428020 | 2428635 | 616 | 5 | 1.37E-07 | 0.0005622 | 5.92E-05 | 8.24E-07 | 0.004130265 | PLCH2 |
| chr10 | 11757637 | 11758294 | 658 | 4 | 7.75E-07 | 6.59E-07 | 0.000108 | 8.24E-07 | 0.002365056 | ECHDC3 |
| chr21 | 19301017 | 19301099 | 83 | 2 | 6.47E-07 | 4.01E-06 | 4.15E-06 | 8.24E-07 | -0.03278717 | NA |
| chr1 | 117643153 | 117643343 | 191 | 3 | 3.55E-07 | 9.40E-06 | 3.20E-06 | 8.24E-07 | -0.00510956 | NA |
| chr1 | 206634473 | 206635824 | 1352 | 13 | 7.49E-07 | 1.19E-05 | 0.000448 | 8.24E-07 | 0.001766875 | DYRK3, DYRK3-AS1 |
| chr1 | 6138044 | 6138607 | 564 | 2 | 5.91E-08 | 1.68E-06 | 9.60E-06 | 8.25E-07 | -0.00735437 | CHD5 |
| chr4 | 38014425 | 38014992 | 568 | 6 | 8.19E-07 | 1.82E-06 | 3.59E-05 | 8.25E-07 | -2.82E-05 | TBC1D1 |
| chr16 | 85290741 | 85291644 | 904 | 3 | 9.90E-06 | 3.79E-07 | 0.000279 | 8.25E-07 | 0.004203337 | GSE1, AC092275.1 |
| chr11 | 66288212 | 66289473 | 1262 | 8 | 2.62E-08 | 6.47E-06 | 0.000252 | 8.25E-07 | -0.00244957 | YIF1A |
| chr10 | 2266757 | 2266778 | 22 | 2 | 2.55E-07 | 0.0001182 | 8.90E-07 | 8.25E-07 | -0.0057313 | NA |
| chr6 | 4350125 | 4350551 | 427 | 2 | 7.28E-07 | 7.07E-07 | 3.89E-05 | 8.26E-07 | -0.01060858 | AL162718.1 |
| chr5 | 112191206 | 112191258 | 53 | 2 | 3.14E-08 | 8.70E-07 | 2.52E-05 | 8.27E-07 | -0.01263687 | EPB41L4A |
| chr9 | 136544771 | 136545728 | 958 | 4 | 9.80E-07 | 1.90E-05 | 0.000715 | 8.27E-07 | 0.000804921 | NOTCH1 |
| chr3 | 71305797 | 71306152 | 356 | 4 | 1.29E-06 | 9.69E-07 | 0.000412 | 8.28E-07 | 0.015403594 | FOXP1-AS1, FOXP1, AC097634.4 |
| chr2 | 162243120 | 162244002 | 883 | 6 | 3.02E-06 | 8.15E-06 | 0.000669 | 8.28E-07 | -0.00303235 | FAP |
| chr16 | 79229445 | 79229842 | 398 | 6 | 3.42E-07 | 0.0001348 | 2.92E-05 | 8.29E-07 | -0.00349794 | AC092376.1 |
| chr11 | 81388893 | 81388953 | 61 | 2 | 1.34E-06 | 5.27E-07 | 0.000107 | 8.29E-07 | 0.006553293 | AP003464.1 |
| chr2 | 208025249 | 208026078 | 830 | 13 | 6.66E-08 | 0.000143 | 5.50E-05 | 8.29E-07 | -0.00109145 | PLEKHM3 |
| chr14 | 103890978 | 103891497 | 520 | 2 | 9.22E-07 | 6.51E-07 | 4.93E-05 | 8.30E-07 | -0.01407946 | NA |
| chr17 | 2071924 | 2072583 | 660 | 6 | 1.05E-06 | 4.65E-07 | 0.003504 | 8.30E-07 | -0.00572676 | SMG6 |
| chr19 | 41225778 | 41226763 | 986 | 8 | 1.12E-06 | 3.27E-06 | 0.002677 | 8.30E-07 | -0.00327392 | AXL |
| chr18 | 72750497 | 72750561 | 65 | 3 | 3.87E-07 | 1.18E-05 | 2.80E-06 | 8.30E-07 | -0.00784661 | NETO1 |
| chr13 | 48479850 | 48480012 | 163 | 2 | 5.05E-07 | 4.56E-06 | 3.78E-06 | 8.31E-07 | -0.00185621 | RB1 |
| chr20 | 63272143 | 63273871 | 1729 | 13 | 9.61E-06 | 2.73E-05 | 0.006961 | 8.32E-07 | 0.001525484 | ARFGAP1, NKAIN4 |
| chr6 | 29606448 | 29607368 | 921 | 7 | 4.38E-07 | 5.49E-06 | 0.001093 | 8.32E-07 | 0.000694064 | GABBR1 |
| chr11 | 62393322 | 62393621 | 300 | 4 | 1.51E-06 | 2.21E-07 | 0.001863 | 8.32E-07 | 0.04040102 | ASRGL1, AP003064.1 |
| chr12 | 12915572 | 12916475 | 904 | 3 | 6.82E-06 | 3.01E-07 | 0.00089 | 8.33E-07 | 0.003338209 | GPRC5A, MIR614 |
| chr6 | 108713737 | 108714671 | 935 | 3 | 4.19E-07 | 4.51E-07 | 0.000424 | 8.33E-07 | 0.030732413 | NA |
| chr6 | 45576857 | 45577841 | 985 | 5 | 4.13E-08 | 4.21E-05 | 5.40E-06 | 8.35E-07 | 0.002691915 | RUNX2 |
| chr8 | 143982622 | 143983764 | 1143 | 5 | 1.23E-05 | 7.41E-07 | 0.000946 | 8.35E-07 | -0.00369309 | PARP10 |
| chr14 | 91241857 | 91242133 | 277 | 3 | 7.82E-07 | 1.28E-06 | 2.50E-05 | 8.37E-07 | 0.001987519 | GPR68 |
| chr17 | 17025789 | 17026073 | 285 | 3 | 6.90E-07 | 2.18E-06 | 5.66E-05 | 8.37E-07 | 0.00042791 | NA |
| chr11 | 57335491 | 57336070 | 580 | 8 | 4.66E-08 | 4.50E-06 | 3.71E-05 | 8.39E-07 | 0.001474349 | SSRP1 |
| chr9 | 97468219 | 97468690 | 472 | 4 | 1.59E-06 | 6.06E-07 | 0.000508 | 8.39E-07 | 0.002440964 | TDRD7 |
| chr11 | 134046713 | 134047825 | 1113 | 5 | 3.90E-07 | 7.63E-07 | 0.000105 | 8.41E-07 | 0.000637512 | LINC02731 |
| chr6 | 34053739 | 34053787 | 49 | 3 | 1.34E-06 | 1.96E-06 | 0.000154 | 8.41E-07 | -0.00531692 | GRM4 |
| chr22 | 43129324 | 43129673 | 350 | 3 | 3.04E-08 | 0.0002875 | 5.83E-07 | 8.41E-07 | -0.00915013 | BIK |
| chr1 | 95370966 | 95371033 | 68 | 2 | 3.28E-07 | 2.78E-06 | 5.84E-06 | 8.42E-07 | -0.01080989 | AL445218.1 |
| chr20 | 63042785 | 63043727 | 943 | 6 | 5.95E-09 | 0.0017426 | 2.42E-07 | 8.42E-07 | -0.00877507 | LINC01749, LINC01056 |
| chr12 | 122997027 | 122997506 | 480 | 4 | 1.10E-06 | 8.34E-07 | 0.000578 | 8.42E-07 | -0.00263457 | PITPNM2 |
| chr13 | 111584292 | 111585132 | 841 | 6 | 6.71E-06 | 2.41E-06 | 0.000523 | 8.43E-07 | -0.00615976 | NA |
| chr1 | 10429905 | 10431247 | 1343 | 18 | 4.34E-07 | 2.11E-05 | 0.007409 | 8.43E-07 | -0.00059642 | CENPS-CORT, CENPS, AL139424.1 |
| chr8 | 100490520 | 100491155 | 636 | 3 | 2.88E-08 | 4.48E-05 | 1.31E-05 | 8.43E-07 | -0.00286122 | AP000424.1 |
| chr9 | 92536464 | 92536687 | 224 | 2 | 5.99E-08 | 2.41E-06 | 6.74E-06 | 8.43E-07 | 0.017681034 | CENPP, ECM2 |
| chr13 | 33236328 | 33236768 | 441 | 2 | 8.48E-08 | 8.33E-06 | 2.55E-06 | 8.43E-07 | -0.01075711 | STARD13-AS, STARD13 |
| chr4 | 41865398 | 41866186 | 789 | 6 | 9.20E-07 | 1.04E-05 | 7.62E-05 | 8.43E-07 | 0.002972524 | NA |

| | | | | | | | | | | |
|---|---|---|---|---|---|---|---|---|---|---|
| chr9 | 127170178 | 127170273 | 96 | 2 | 1.13E-06 | 1.02E-06 | 1.99E-05 | 8.43E-07 | -0.00769733 | RALGPS1 |
| chr8 | 105189589 | 105189665 | 77 | 3 | 1.26E-06 | 3.59E-07 | 0.000464 | 8.43E-07 | -0.01680436 | ZFPM2 |
| chr2 | 105912308 | 105912687 | 380 | 2 | 1.51E-07 | 2.69E-06 | 6.07E-06 | 8.43E-07 | -0.00470889 | AC009505.1 |
| chr14 | 99971614 | 99972620 | 1007 | 7 | 1.67E-06 | 7.03E-07 | 0.001713 | 8.44E-07 | -0.00075493 | EVL |
| chr5 | 90558476 | 90559162 | 687 | 12 | 3.97E-08 | 7.38E-05 | 0.001052 | 8.44E-07 | 0.000495773 | ADGRV1 |
| chr16 | 30922616 | 30923406 | 791 | 12 | 4.65E-06 | 2.58E-07 | 0.002762 | 8.44E-07 | 0.000556204 | FBXL19, FBXL19-AS1 |
| chr15 | 55372955 | 55373038 | 84 | 2 | 3.20E-07 | 7.71E-06 | 2.69E-06 | 8.45E-07 | 0.007429906 | CCPG1, DNAAF4-CCPG1, MIR628 |
| chr1 | 28914451 | 28915350 | 900 | 8 | 1.49E-06 | 7.73E-06 | 0.000374 | 8.46E-07 | 0.000808458 | EPB41 |
| chr7 | 143881938 | 143883054 | 1117 | 6 | 3.16E-06 | 5.25E-07 | 0.000876 | 8.47E-07 | 0.003821387 | TCAF1 |
| chr9 | 122799476 | 122800350 | 875 | 6 | 2.32E-07 | 9.60E-06 | 4.88E-05 | 8.47E-07 | -0.00401105 | OR1K1, PDCL |
| chr22 | 28020252 | 28021051 | 800 | 6 | 1.37E-07 | 1.91E-05 | 0.000112 | 8.48E-07 | -0.00986797 | TTC28 |
| chr22 | 19978011 | 19978850 | 840 | 2 | 1.08E-07 | 7.51E-07 | 3.64E-05 | 8.48E-07 | -0.00690483 | ARVCF |
| chr19 | 11639503 | 11640289 | 787 | 7 | 1.53E-07 | 1.26E-05 | 1.79E-05 | 8.49E-07 | 0.000572375 | AC008543.1 |
| chr9 | 116372876 | 116373103 | 228 | 3 | 2.07E-06 | 3.56E-07 | 0.000621 | 8.50E-07 | -0.00642053 | PAPPA |
| chr12 | 51197942 | 51198649 | 708 | 9 | 2.03E-07 | 1.05E-05 | 7.27E-05 | 8.50E-07 | -0.00114681 | POU6F1 |
| chr8 | 113377127 | 113378150 | 1024 | 8 | 3.00E-06 | 2.70E-05 | 0.001201 | 8.50E-07 | -0.00408162 | AC107890.1, CSMD3 |
| chr7 | 97003148 | 97004454 | 1307 | 9 | 3.52E-06 | 1.26E-05 | 0.000429 | 8.51E-07 | -0.00581172 | DLX6-AS1 |
| chr10 | 78301544 | 78303291 | 1748 | 5 | 3.61E-07 | 3.83E-06 | 5.05E-05 | 8.51E-07 | 0.001952617 | AC012560.1, LINC00595 |
| chr3 | 87796475 | 87797030 | 556 | 2 | 1.54E-06 | 5.84E-07 | 7.62E-05 | 8.51E-07 | -0.00855398 | HTR1F |
| chr5 | 19988091 | 19989410 | 1320 | 14 | 4.11E-06 | 4.85E-07 | 0.00924 | 8.51E-07 | -0.00229464 | CDH18 |
| chr4 | 74364674 | 74365880 | 1207 | 11 | 1.10E-07 | 0.0009242 | 0.000113 | 8.52E-07 | 0.001653344 | EREG |
| chr8 | 23457917 | 23458945 | 1029 | 7 | 2.46E-07 | 3.49E-05 | 0.000154 | 8.52E-07 | 0.003932732 | AC104561.1 |
| chr3 | 138433995 | 138434597 | 603 | 8 | 7.71E-08 | 2.12E-05 | 0.001571 | 8.53E-07 | -0.01913526 | ESYT3 |
| chr5 | 172983438 | 172984157 | 720 | 13 | 6.53E-07 | 3.92E-05 | 0.000174 | 8.53E-07 | 0.001209211 | ATP6V0E1 |
| chr12 | 53177181 | 53177580 | 400 | 2 | 1.40E-07 | 5.22E-06 | 3.54E-06 | 8.53E-07 | 0.006751717 | CSAD |
| chr15 | 57640840 | 57641038 | 199 | 2 | 3.93E-07 | 6.64E-06 | 3.01E-06 | 8.54E-07 | -0.00344473 | MYZAP, GCOM1 |
| chr4 | 17975675 | 17975820 | 146 | 2 | 1.16E-06 | 8.73E-07 | 2.70E-05 | 8.54E-07 | -0.00749365 | LCORL |
| chr10 | 117164831 | 117167104 | 2274 | 9 | 4.36E-05 | 4.24E-05 | 0.000817 | 8.55E-07 | 0.001374611 | MIR3663HG |
| chr7 | 1505749 | 1507124 | 1376 | 7 | 2.10E-06 | 5.01E-07 | 0.004683 | 8.55E-07 | 0.003801233 | NA |
| chr1 | 244302524 | 244303126 | 603 | 2 | 1.12E-07 | 2.94E-06 | 5.70E-06 | 8.55E-07 | -0.00173818 | NA |
| chr14 | 60514455 | 60515917 | 1463 | 10 | 1.13E-07 | 6.51E-05 | 0.001303 | 8.55E-07 | 0.000119239 | AL049874.3, C14orf39 |
| chr8 | 97868632 | 97869395 | 764 | 11 | 3.17E-07 | 3.24E-05 | 0.000466 | 8.56E-07 | -0.00068285 | MATN2 |
| chr19 | 52492830 | 52493364 | 535 | 4 | 1.85E-06 | 2.79E-07 | 0.00112 | 8.57E-07 | -0.02096782 | ZNF578 |
| chr3 | 130929798 | 130930422 | 625 | 5 | 2.24E-07 | 1.28E-06 | 7.63E-05 | 8.57E-07 | -0.01379282 | ATP2C1, AC055733.2 |
| chr9 | 133107587 | 133108195 | 609 | 2 | 1.15E-06 | 5.64E-07 | 9.14E-05 | 8.57E-07 | 0.000294633 | RALGDS, AL162417.1 |
| chr20 | 32801062 | 32801328 | 267 | 2 | 4.55E-08 | 3.09E-06 | 5.46E-06 | 8.57E-07 | -0.00552673 | DNMT3B |
| chr10 | 1180705 | 1180946 | 242 | 4 | 2.26E-06 | 2.80E-07 | 0.001638 | 8.57E-07 | -0.01456586 | LINC00200, ADARB2 |
| chr18 | 79968011 | 79968073 | 63 | 2 | 7.71E-07 | 3.05E-06 | 5.52E-06 | 8.57E-07 | -0.00116148 | HSBP1L1 |
| chr14 | 70045866 | 70046319 | 454 | 3 | 1.10E-06 | 4.09E-07 | 0.000393 | 8.57E-07 | -0.02392285 | SLC8A3 |
| chr9 | 95316847 | 95317930 | 1084 | 15 | 1.97E-09 | 0.0008102 | 5.05E-07 | 8.57E-07 | 0.000345579 | FANCC |
| chr11 | 12725249 | 12725999 | 751 | 3 | 8.33E-07 | 3.76E-06 | 9.55E-05 | 8.57E-07 | -0.0139451 | TEAD1 |
| chr10 | 95039008 | 95039108 | 101 | 2 | 5.79E-07 | 5.88E-06 | 3.29E-06 | 8.59E-07 | -0.00476744 | CYP2C8 |
| chr1 | 42681733 | 42683446 | 1714 | 15 | 1.64E-06 | 8.36E-06 | 0.005291 | 8.59E-07 | 0.000208979 | YBX1, AC098484.4 |
| chr3 | 130850190 | 130850988 | 799 | 8 | 9.19E-07 | 3.04E-05 | 4.18E-05 | 8.59E-07 | 0.003159824 | ATP2C1 |
| chr2 | 240319966 | 240320693 | 728 | 3 | 1.16E-06 | 8.24E-07 | 0.000262 | 8.59E-07 | -0.00172721 | NA |
| chr13 | 71977093 | 71977370 | 278 | 3 | 1.37E-06 | 3.84E-07 | 0.000317 | 8.59E-07 | -0.00066984 | NA |
| chr11 | 20120753 | 20120878 | 126 | 3 | 1.83E-06 | 4.91E-07 | 0.000151 | 8.59E-07 | -0.00109776 | NAV2 |
| chr2 | 96116152 | 96117001 | 850 | 7 | 1.91E-06 | 3.51E-06 | 0.001918 | 8.60E-07 | -0.00172568 | ADRA2B |
| chr14 | 64213054 | 64214278 | 1225 | 6 | 9.11E-08 | 1.22E-06 | 0.001441 | 8.60E-07 | -0.00052831 | SYNE2, ESR2 |
| chr15 | 50816201 | 50816603 | 403 | 2 | 1.13E-06 | 6.21E-07 | 6.46E-05 | 8.60E-07 | -0.01222422 | NA |
| chr8 | 1098844 | 1099477 | 634 | 6 | 2.88E-09 | 0.0062939 | 1.00E-07 | 8.61E-07 | -0.00407576 | DLGAP2 |
| chr6 | 31722219 | 31723626 | 1408 | 14 | 2.59E-06 | 6.01E-06 | 0.004198 | 8.61E-07 | 0.003160093 | MPIG6B |
| chr10 | 126070680 | 126070762 | 83 | 2 | 4.56E-08 | 0.0008276 | 7.12E-07 | 8.61E-07 | -0.01458046 | ADAM12 |
| chr19 | 12899880 | 12900498 | 619 | 2 | 2.58E-07 | 8.99E-07 | 2.60E-05 | 8.62E-07 | -0.01064875 | GCDH, SYCE2 |
| chr17 | 40647003 | 40648059 | 1057 | 10 | 4.29E-06 | 5.67E-07 | 0.002226 | 8.62E-07 | -0.00045629 | SMARCE1, AC073508.2 |
| chr13 | 112101549 | 112102335 | 787 | 3 | 2.38E-08 | 2.10E-05 | 2.42E-05 | 8.64E-07 | 0.019952574 | SOX1-OT |
| chr9 | 113620660 | 113621685 | 1026 | 4 | 2.08E-05 | 5.42E-07 | 0.000602 | 8.64E-07 | -0.00930966 | AL162727.1 |
| chr3 | 53495628 | 53496820 | 1193 | 5 | 1.70E-07 | 5.72E-06 | 0.001331 | 8.64E-07 | 0.006802203 | CACNA1D |
| chr4 | 84581512 | 84582900 | 1389 | 12 | 1.23E-07 | 4.92E-05 | 0.0002 | 8.64E-07 | 0.001601228 | NA |
| chr13 | 44219135 | 44219148 | 14 | 2 | 8.40E-07 | 5.17E-07 | 0.000167 | 8.65E-07 | 0.001565238 | SMIM2-AS1 |
| chr1 | 84141752 | 84141870 | 119 | 2 | 6.97E-07 | 2.44E-06 | 6.92E-06 | 8.65E-07 | 0.017961653 | PRKACB |
| chr1 | 183417396 | 183418616 | 1221 | 9 | 1.23E-06 | 4.42E-06 | 0.002345 | 8.65E-07 | 0.000339379 | NMNAT2 |
| chr20 | 3226294 | 3226526 | 233 | 3 | 2.10E-07 | 2.28E-05 | 3.05E-06 | 8.67E-07 | -0.00052069 | NA |
| chr1 | 233326829 | 233327757 | 929 | 7 | 1.07E-11 | 0.0292627 | 5.24E-08 | 8.67E-07 | -0.0057694 | MAP3K21 |
| chr19 | 11102705 | 11102888 | 184 | 2 | 2.54E-07 | 2.80E-05 | 1.44E-06 | 8.68E-07 | 0.002993749 | LDLR |
| chr2 | 240870620 | 240871069 | 450 | 2 | 2.37E-07 | 9.44E-07 | 2.42E-05 | 8.68E-07 | -0.00752153 | AGXT |
| chr16 | 87636226 | 87636962 | 737 | 6 | 9.81E-07 | 5.16E-07 | 0.001563 | 8.69E-07 | -0.00557556 | JPH3 |
| chr15 | 40252519 | 40253593 | 1075 | 14 | 9.22E-08 | 8.53E-05 | 3.36E-05 | 8.69E-07 | 0.000434033 | PAK6, BUB1B-PAK6, C15orf56 |
| chr6 | 3787303 | 3787616 | 314 | 2 | 5.57E-08 | 6.93E-05 | 1.05E-06 | 8.69E-07 | -0.00279354 | NA |
| chr1 | 235842357 | 235843389 | 1033 | 4 | 6.77E-08 | 0.0001459 | 1.33E-06 | 8.70E-07 | 0.005937506 | LYST |
| chr12 | 53197265 | 53197982 | 718 | 6 | 2.95E-07 | 1.36E-06 | 0.000248 | 8.70E-07 | -0.01209204 | ITGB7 |
| chr8 | 882917 | 883724 | 808 | 5 | 1.21E-06 | 2.67E-07 | 0.001742 | 8.70E-07 | -0.021627 | DLGAP2 |
| chr1 | 236686173 | 236687061 | 889 | 9 | 4.16E-07 | 4.99E-05 | 0.001558 | 8.70E-07 | -0.00418881 | ACTN2 |
| chr2 | 106825053 | 106826011 | 959 | 4 | 2.34E-06 | 7.45E-05 | 5.74E-05 | 8.70E-07 | 0.003057782 | ST6GAL2 |
| chr1 | 214377160 | 214378515 | 1356 | 4 | 1.53E-05 | 5.76E-07 | 0.000813 | 8.71E-07 | 0.000189464 | PTPN14 |
| chr1 | 78004428 | 78005681 | 1254 | 10 | 3.17E-06 | 2.52E-06 | 0.005611 | 8.72E-07 | -4.45E-05 | DNAJB4, GIPC2, AC103591.3 |
| chr19 | 6590095 | 6591489 | 1395 | 7 | 1.66E-05 | 6.39E-07 | 0.001977 | 8.72E-07 | 0.000242554 | CD70 |
| chr17 | 4127290 | 4127875 | 586 | 2 | 4.29E-08 | 9.08E-06 | 2.52E-06 | 8.73E-07 | -0.00435129 | ZZEF1 |
| chr12 | 95740040 | 95740537 | 498 | 5 | 4.01E-07 | 4.00E-05 | 0.000318 | 8.73E-07 | -0.00488314 | NTN4 |
| chr2 | 241733555 | 241734564 | 1010 | 11 | 4.03E-07 | 2.83E-06 | 0.004295 | 8.73E-07 | -0.00331175 | NA |
| chr7 | 24722576 | 24722943 | 368 | 2 | 5.60E-07 | 5.55E-07 | 0.00011 | 8.73E-07 | 0.009024576 | GSDME |
| chr9 | 126672767 | 126673858 | 1092 | 3 | 2.83E-05 | 3.78E-07 | 0.000609 | 8.74E-07 | 0.014559188 | LMX1B |
| chr2 | 29114731 | 29115566 | 836 | 12 | 6.32E-07 | 8.52E-06 | 0.000475 | 8.74E-07 | -0.00134104 | CLIP4 |
| chr1 | 183478325 | 183478841 | 517 | 2 | 1.43E-06 | 7.35E-07 | 4.19E-05 | 8.74E-07 | 0.000934315 | SMG7 |
| chr5 | 139788738 | 139789765 | 1028 | 3 | 5.00E-07 | 5.87E-07 | 9.68E-05 | 8.74E-07 | -0.00430125 | NA |
| chr12 | 18246760 | 18247607 | 848 | 5 | 1.47E-06 | 1.02E-06 | 0.000129 | 8.74E-07 | -0.007407 | PIK3C2G, RERGL |
| chr19 | 52068879 | 52068958 | 80 | 2 | 2.19E-07 | 2.65E-05 | 1.49E-06 | 8.76E-07 | 0.007519373 | ZNF432, ZNF841 |
| chr20 | 33644365 | 33645019 | 655 | 7 | 4.74E-07 | 7.06E-06 | 0.000119 | 8.76E-07 | 0.00212106 | CBFA2T2 |
| chr17 | 76271749 | 76272501 | 753 | 4 | 1.43E-08 | 0.0001619 | 6.24E-05 | 8.77E-07 | 0.011160877 | NA |
| chr17 | 182066 | 182712 | 647 | 6 | 1.40E-06 | 1.77E-07 | 0.00278 | 8.78E-07 | -0.02177621 | NA |
| chr15 | 75367868 | 75368899 | 1032 | 14 | 4.57E-06 | 9.02E-07 | 0.004812 | 8.78E-07 | -0.00053154 | AC068338.2, MAN2C1 |
| chr17 | 40461378 | 40461482 | 105 | 3 | 4.86E-07 | 5.15E-06 | 6.31E-06 | 8.79E-07 | -0.00213669 | NA |
| chr3 | 177351674 | 177351890 | 217 | 2 | 7.04E-07 | 1.68E-06 | 1.08E-05 | 8.81E-07 | 0.011746446 | NA |
| chr12 | 122167053 | 122167747 | 695 | 7 | 3.20E-06 | 1.38E-06 | 0.002777 | 8.81E-07 | -0.00332296 | LRRC43 |
| chr1 | 27300567 | 27300929 | 363 | 3 | 1.58E-06 | 1.52E-06 | 8.56E-05 | 8.81E-07 | 0.004077481 | WDTC1 |
| chr17 | 8072461 | 8072967 | 507 | 3 | 1.23E-07 | 1.84E-06 | 0.000144 | 8.82E-07 | -0.0091339 | ALOXE3P1, ALOX12B |
| chr3 | 49103338 | 49104857 | 1520 | 10 | 1.19E-05 | 1.98E-05 | 0.000826 | 8.83E-07 | -0.00097513 | QARS1 |
| chr16 | 74984563 | 74985869 | 1307 | 14 | 1.05E-07 | 2.69E-05 | 1.79E-05 | 8.83E-07 | -0.0006399 | WDR59 |
| chr2 | 201118309 | 201118985 | 677 | 11 | 5.73E-07 | 1.39E-05 | 0.000763 | 8.83E-07 | 0.000130807 | CFLAR |
| chr3 | 160026004 | 160026755 | 752 | 6 | 2.82E-07 | 3.20E-06 | 2.20E-05 | 8.84E-07 | -0.00168856 | LINC01100, IL12A-AS1 |
| chr1 | 6268377 | 6268895 | 519 | 4 | 3.94E-07 | 3.37E-06 | 2.05E-05 | 8.84E-07 | -0.00648123 | ACOT7 |
| chr9 | 136590903 | 136591350 | 448 | 2 | 1.76E-08 | 1.32E-05 | 2.08E-06 | 8.84E-07 | -0.00905943 | NA |
| chr9 | 128838076 | 128838506 | 431 | 2 | 7.81E-07 | 5.73E-07 | 0.000101 | 8.85E-07 | -0.00142982 | AL441992.2, KYAT1 |
| chr1 | 113949462 | 113950013 | 552 | 3 | 9.17E-07 | 2.08E-06 | 2.85E-05 | 8.87E-07 | 0.001193835 | HIPK1 |
| chr1 | 205430675 | 205431069 | 395 | 4 | 1.78E-07 | 9.35E-06 | 9.36E-06 | 8.89E-07 | -0.00410339 | LEMD1 |
| chr2 | 71380217 | 71381179 | 963 | 2 | 5.58E-05 | 5.68E-07 | 0.000108 | 8.89E-07 | 0.008663853 | ZNF638 |
| chr10 | 77715267 | 77715874 | 608 | 2 | 4.68E-07 | 1.12E-06 | 1.93E-05 | 8.89E-07 | -0.01388698 | NA |
| chr12 | 49619228 | 49619611 | 384 | 2 | 2.11E-07 | 3.96E-06 | 4.67E-06 | 8.90E-07 | -0.00522523 | PRPF40B, HIGD1AP9 |
| chr20 | 63665872 | 63666395 | 524 | 3 | 1.40E-07 | 2.87E-05 | 2.09E-06 | 8.90E-07 | -0.00995883 | RTEL1, RTEL1-TNFRSF6B |
| chr7 | 31793105 | 31794117 | 1013 | 4 | 2.05E-06 | 5.91E-07 | 0.000262 | 8.91E-07 | -0.00299195 | PDE1C |
| chr8 | 72536155 | 72537289 | 1135 | 9 | 1.68E-05 | 5.22E-05 | 0.000179 | 8.91E-07 | 0.002962963 | KCNB2 |
| chr1 | 48498510 | 48498645 | 136 | 3 | 1.45E-07 | 3.23E-05 | 1.43E-06 | 8.92E-07 | -0.0110789 | NA |
| chr3 | 143736515 | 143736715 | 201 | 2 | 1.54E-06 | 5.73E-07 | 0.000105 | 8.92E-07 | -0.01092887 | SLC9A9 |
| chr10 | 129556609 | 129557038 | 430 | 7 | 1.56E-07 | 0.000701 | 0.00026 | 8.92E-07 | -0.00211839 | MGMT |
| chr2 | 11544767 | 11545109 | 343 | 2 | 4.67E-08 | 0.0004102 | 7.92E-07 | 8.92E-07 | -0.00941743 | GREB1 |
| chr21 | 45509586 | 45510045 | 460 | 5 | 1.31E-06 | 7.72E-07 | 0.000214 | 8.92E-07 | -0.00110676 | COL18A1, SLC19A1 |
| chr16 | 10694533 | 10695125 | 593 | 5 | 2.24E-06 | 4.76E-07 | 0.001 | 8.92E-07 | -0.00481601 | TEKT5 |
| chr7 | 777502 | 778725 | 1224 | 6 | 7.97E-09 | 0.0006665 | 9.21E-07 | 8.92E-07 | -0.00019186 | DNAAF5 |
| chr4 | 7460365 | 7461411 | 1047 | 7 | 1.70E-07 | 2.90E-06 | 0.000843 | 8.94E-07 | 0.00813478 | SORCS2 |
| chr11 | 1435889 | 1436116 | 228 | 3 | 5.74E-07 | 1.13E-06 | 3.74E-05 | 8.94E-07 | 0.053475523 | BRSK2 |
| chr7 | 5569523 | 5570633 | 1111 | 6 | 2.25E-08 | 2.97E-05 | 2.96E-06 | 8.95E-07 | 0.000744334 | NA |
| chr4 | 151918185 | 151918530 | 346 | 3 | 6.32E-07 | 2.15E-06 | 0.000164 | 8.95E-07 | -0.00870381 | AC097375.2 |
| chr4 | 127964850 | 127966091 | 1242 | 22 | 5.88E-07 | 0.0001853 | 0.001202 | 8.96E-07 | 0.000783693 | ABHD18, MFSD8 |
| chr15 | 40871992 | 40872206 | 215 | 3 | 7.23E-08 | 0.0029469 | 9.54E-07 | 8.96E-07 | 0.008947459 | NA |
| chr2 | 26178729 | 26179449 | 721 | 9 | 2.15E-07 | 3.84E-05 | 3.24E-05 | 8.98E-07 | -0.01151409 | GAREM2 |
| chr21 | 17890787 | 17891340 | 554 | 3 | 6.45E-07 | 8.83E-07 | 0.000265 | 8.98E-07 | -0.00714599 | NA |
| chr11 | 61447993 | 61448708 | 716 | 2 | 1.70E-06 | 7.13E-07 | 4.93E-05 | 8.98E-07 | -0.05257499 | AP003108.2 |
| chr14 | 104866147 | 104866997 | 851 | 3 | 2.41E-07 | 7.51E-06 | 7.84E-06 | 8.99E-07 | -0.00686782 | CEP170B |
| chr11 | 44957093 | 44957775 | 683 | 5 | 1.06E-06 | 1.40E-06 | 0.000606 | 9.00E-07 | -0.00956595 | NA |
| chr7 | 14747772 | 14747778 | 7 | 3 | 1.22E-06 | 2.40E-06 | 2.12E-05 | 9.00E-07 | 0.02728853 | DGKB |
| chr12 | 120469498 | 120470421 | 924 | 10 | 4.16E-07 | 1.10E-05 | 0.000133 | 9.01E-07 | -0.00095839 | DYNLL1, SRSF9 |
| chr20 | 15496957 | 15497005 | 49 | 2 | 2.79E-07 | 0.0001272 | 9.53E-07 | 9.02E-07 | -0.00554163 | MACROD2 |
| chr9 | 5459738 | 5460696 | 959 | 3 | 1.01E-05 | 3.53E-07 | 0.000591 | 9.02E-07 | -0.00704276 | CD274, AL162253.2 |
| chr19 | 3025267 | 3025831 | 565 | 8 | 6.03E-08 | 5.34E-06 | 2.47E-05 | 9.03E-07 | -0.01137789 | TLE2 |
| chr2 | 86440305 | 86441415 | 1111 | 16 | 1.24E-07 | 2.74E-06 | 0.000357 | 9.03E-07 | -0.00033419 | KDM3A |
| chr1 | 231338824 | 231339599 | 776 | 3 | 7.08E-08 | 9.70E-07 | 9.27E-05 | 9.03E-07 | -0.02096059 | SPRTN, AL117352.1 |
| chr21 | 30562047 | 30562159 | 113 | 2 | 2.44E-07 | 0.0001198 | 9.65E-07 | 9.03E-07 | -0.01310378 | NA |
| chr3 | 46368680 | 46368931 | 252 | 4 | 4.52E-07 | 3.58E-06 | 0.000446 | 9.05E-07 | -0.01017117 | CCR5AS |
| chr3 | 152647035 | 152647321 | 287 | 2 | 3.01E-07 | 2.04E-06 | 8.98E-06 | 9.05E-07 | -0.01029172 | NA |
| chr7 | 35982854 | 35983231 | 378 | 4 | 2.51E-06 | 3.44E-07 | 0.000925 | 9.05E-07 | -0.01155689 | AC083864.5 |
| chr13 | 27352597 | 27353353 | 757 | 3 | 1.66E-07 | 4.46E-06 | 8.11E-06 | 9.07E-07 | -0.01328855 | NA |
| chr13 | 114027268 | 114028155 | 888 | 6 | 2.06E-07 | 8.92E-07 | 0.000625 | 9.08E-07 | 0.000761097 | RASA3 |
| chr7 | 101602491 | 101602612 | 122 | 2 | 4.85E-07 | 1.05E-05 | 2.43E-06 | 9.08E-07 | 0.000285609 | NA |
| chr5 | 1503144 | 1503864 | 721 | 6 | 3.37E-07 | 1.99E-05 | 0.000309 | 9.08E-07 | 0.00635407 | LPCAT1 |
| chr1 | 7106736 | 7107305 | 570 | 3 | 1.67E-07 | 1.68E-06 | 5.07E-05 | 9.09E-07 | -0.00957007 | CAMTA1 |
| chr16 | 31115628 | 31115972 | 345 | 2 | 7.50E-08 | 1.58E-06 | 1.23E-05 | 9.09E-07 | -0.00491473 | KAT8 |
| chr11 | 709627 | 710392 | 766 | 4 | 1.16E-08 | 7.85E-06 | 3.08E-05 | 9.10E-07 | -0.01083889 | EPS8L2, AP006621.4 |
| chr21 | 36460928 | 36461233 | 306 | 3 | 6.04E-07 | 1.64E-06 | 2.03E-05 | 9.10E-07 | 0.000915011 | AP000695.1, AP000695.2, CLDN14 |
| chr11 | 13352307 | 13353108 | 802 | 4 | 6.29E-06 | 4.40E-07 | 0.00066 | 9.10E-07 | -0.00160065 | ARNTL |
| chr8 | 2044604 | 2045778 | 1175 | 10 | 1.20E-06 | 1.09E-06 | 0.002949 | 9.11E-07 | 0.004678179 | MYOM2 |
| chr4 | 13546712 | 13547731 | 1020 | 6 | 1.42E-07 | 4.79E-07 | 0.001454 | 9.11E-07 | 0.001405975 | LINC01096 |
| chr4 | 2462287 | 2462852 | 566 | 4 | 3.31E-07 | 4.42E-07 | 0.000457 | 9.11E-07 | 0.000795345 | CFAP99, RNF4 |
| chr7 | 121312954 | 121313012 | 59 | 2 | 4.35E-08 | 1.26E-06 | 1.68E-05 | 9.11E-07 | -0.00903862 | NA |
| chr5 | 61265824 | 61266057 | 234 | 3 | 2.15E-08 | 0.0206303 | 2.25E-07 | 9.12E-07 | -0.00437584 | LINC02057 |
| chr11 | 117428405 | 117428755 | 351 | 3 | 5.40E-07 | 2.13E-05 | 2.84E-05 | 9.12E-07 | 0.000544162 | DSCAML1 |
| chr12 | 123382836 | 123383763 | 928 | 6 | 5.58E-06 | 8.94E-06 | 0.00015 | 9.14E-07 | -0.00424544 | NA |
| chr7 | 129210759 | 129212089 | 1331 | 14 | 3.51E-09 | 2.80E-05 | 0.000473 | 9.15E-07 | -0.00337933 | SMO, AC011005.4 |
| chr6 | 41779629 | 41780805 | 1177 | 9 | 4.54E-06 | 1.45E-06 | 0.002195 | 9.15E-07 | 0.000784085 | AL365205.1, PRICKLE4, FRS3 |

| | | | | | | | | | | |
|---|---|---|---|---|---|---|---|---|---|---|
| chr22 | 23893382 | 23894537 | 1156 | 14 | 3.18E-06 | 1.34E-06 | 0.005382 | 9.16E-07 | -0.00074438 | AP000350.4, MIF, MIF-AS1 |
| chr17 | 44689408 | 44690345 | 938 | 11 | 6.02E-06 | 6.05E-05 | 0.001791 | 9.16E-07 | 0.002449606 | MEIOC, AC091152.4, CCDC43 |
| chr16 | 85092999 | 85094059 | 1061 | 3 | 3.84E-05 | 7.80E-07 | 0.000282 | 9.16E-07 | -0.00180238 | KIAA0513 |
| chr2 | 113052667 | 113053748 | 1082 | 6 | 7.21E-07 | 1.80E-06 | 0.001942 | 9.16E-07 | -0.0043033 | IL36B |
| chr10 | 71713478 | 71714036 | 559 | 3 | 1.56E-08 | 1.04E-05 | 3.28E-06 | 9.16E-07 | -0.00659086 | CDH23, C10orf105 |
| chr4 | 8023243 | 8023952 | 710 | 4 | 5.67E-07 | 1.19E-06 | 0.00032 | 9.17E-07 | -0.00662429 | ABLIM2 |
| chr2 | 28682995 | 28683326 | 332 | 2 | 4.22E-08 | 1.37E-06 | 1.50E-05 | 9.17E-07 | 0.004333916 | NA |
| chr3 | 169107631 | 169107814 | 184 | 3 | 1.43E-06 | 1.71E-06 | 0.000154 | 9.18E-07 | -0.00778419 | MECOM |
| chr8 | 71557653 | 71558747 | 1095 | 6 | 6.21E-07 | 9.51E-07 | 0.001409 | 9.18E-07 | 0.00016807 | EYA1 |
| chr12 | 70609796 | 70610592 | 797 | 8 | 1.53E-06 | 6.75E-06 | 0.001423 | 9.19E-07 | -0.00044242 | PTPRB |
| chr5 | 113679121 | 113679429 | 309 | 3 | 2.05E-07 | 7.89E-06 | 2.17E-05 | 9.19E-07 | 0.001459386 | NA |
| chr14 | 89160588 | 89160651 | 64 | 2 | 2.44E-07 | 1.04E-05 | 2.48E-06 | 9.21E-07 | 0.003225799 | FOXN3 |
| chr4 | 8640580 | 8641050 | 471 | 4 | 5.28E-07 | 7.52E-05 | 0.000253 | 9.21E-07 | -0.00328545 | NA |
| chr10 | 68559544 | 68560775 | 1232 | 16 | 9.31E-07 | 5.97E-07 | 0.007852 | 9.21E-07 | 0.003005775 | TET1 |
| chr3 | 48989501 | 48990395 | 895 | 16 | 2.34E-06 | 1.20E-06 | 0.004532 | 9.21E-07 | 0.001122903 | P4HTM, AC137630.1 |
| chr19 | 17455817 | 17456132 | 316 | 2 | 4.45E-07 | 8.07E-07 | 3.88E-05 | 9.21E-07 | -0.02099263 | AC010319.2, NXNL1 |
| chr13 | 99727326 | 99727681 | 356 | 2 | 6.80E-07 | 7.05E-07 | 5.61E-05 | 9.22E-07 | -0.00796123 | CLYBL, CLYBL-AS1 |
| chr1 | 85047274 | 85048209 | 936 | 5 | 7.42E-07 | 4.02E-06 | 4.33E-05 | 9.24E-07 | -0.00822211 | DNAI3, MCOLN3 |
| chr12 | 96377972 | 96378027 | 56 | 2 | 6.22E-07 | 8.37E-06 | 2.86E-06 | 9.24E-07 | -0.01149386 | CDK17 |
| chr9 | 22682273 | 22683215 | 943 | 4 | 2.02E-08 | 0.0001127 | 1.38E-06 | 9.24E-07 | -0.01865318 | LINC01239 |
| chr10 | 3137629 | 3138768 | 1140 | 9 | 3.20E-06 | 1.36E-06 | 0.001796 | 9.24E-07 | -0.00042723 | PFKP, AL451164.2, PITRM1 |
| chr9 | 121074135 | 121075567 | 1433 | 11 | 1.85E-05 | 3.83E-06 | 0.002386 | 9.25E-07 | -0.00037976 | CNTRL |
| chr11 | 130600763 | 130601403 | 641 | 3 | 2.14E-07 | 1.82E-06 | 5.07E-05 | 9.25E-07 | -0.0204423 | NA |
| chr22 | 30418794 | 30419277 | 484 | 3 | 1.00E-06 | 5.15E-07 | 0.000205 | 9.25E-07 | -0.00157964 | SEC14L2, AC004832.3, RNF215 |
| chr12 | 57205934 | 57206721 | 788 | 3 | 1.00E-05 | 4.26E-07 | 0.000574 | 9.26E-07 | 0.006312288 | LRP1 |
| chr11 | 114559563 | 114560578 | 1016 | 5 | 6.69E-06 | 7.63E-07 | 0.000929 | 9.26E-07 | -0.00833956 | NXPE1 |
| chr1 | 26943858 | 26945113 | 1256 | 4 | 3.03E-06 | 2.74E-07 | 0.001654 | 9.26E-07 | 0.003763471 | NUDC |
| chr3 | 171771162 | 171771835 | 674 | 3 | 1.09E-07 | 0.0005133 | 4.93E-05 | 9.28E-07 | -0.01501923 | PLD1 |
| chr10 | 8642894 | 8643165 | 272 | 2 | 4.96E-08 | 6.89E-05 | 1.12E-06 | 9.28E-07 | -0.01278392 | NA |
| chr19 | 7380082 | 7380964 | 883 | 6 | 1.36E-05 | 7.22E-07 | 0.00064 | 9.28E-07 | -0.00759203 | AC119396.1 |
| chr6 | 41501164 | 41501398 | 235 | 2 | 8.45E-08 | 1.40E-06 | 1.49E-05 | 9.28E-07 | -0.01327515 | LINC01276, FOXP4-AS1 |
| chr7 | 6058867 | 6060134 | 1268 | 14 | 8.97E-07 | 3.31E-05 | 0.006548 | 9.28E-07 | 0.001067208 | EIF2AK1 |
| chr5 | 55101764 | 55102727 | 964 | 5 | 1.49E-07 | 9.10E-06 | 1.10E-05 | 9.29E-07 | 0.011693711 | GZMA |
| chr5 | 154851607 | 154852264 | 658 | 3 | 1.60E-07 | 8.33E-06 | 4.31E-05 | 9.29E-07 | 0.000912134 | FAXDC2 |
| chr8 | 144853048 | 144853980 | 933 | 12 | 1.96E-07 | 5.28E-05 | 0.000302 | 9.29E-07 | -0.00026296 | COMMD5 |
| chr5 | 56278815 | 56279376 | 562 | 2 | 2.29E-06 | 5.90E-07 | 0.000115 | 9.29E-07 | -0.0036556 | NA |
| chr3 | 169134002 | 169134586 | 585 | 6 | 7.72E-07 | 6.78E-07 | 0.000807 | 9.30E-07 | -0.00938408 | MECOM |
| chr14 | 76227190 | 76227594 | 405 | 3 | 1.37E-06 | 5.62E-07 | 0.000218 | 9.30E-07 | -0.00871091 | GPATCH2L |
| chr20 | 46954683 | 46955276 | 594 | 4 | 4.50E-07 | 2.91E-06 | 0.000162 | 9.31E-07 | -0.00231449 | EYA2 |
| chr9 | 136268684 | 136269151 | 468 | 3 | 2.22E-09 | 0.0001236 | 7.24E-07 | 9.31E-07 | 0.002366776 | CCDC187 |
| chr11 | 107719701 | 107720191 | 491 | 5 | 5.69E-08 | 0.0062347 | 7.20E-05 | 9.31E-07 | 0.00277703 | NA |
| chr13 | 114064780 | 114065667 | 888 | 6 | 6.01E-08 | 1.26E-05 | 7.69E-05 | 9.32E-07 | -0.00671898 | RASA3 |
| chr5 | 6581661 | 6582429 | 769 | 4 | 1.70E-07 | 1.12E-06 | 0.000771 | 9.32E-07 | -0.00173402 | LINC01018 |
| chr3 | 64501351 | 64501653 | 303 | 3 | 8.65E-07 | 1.63E-06 | 2.79E-05 | 9.33E-07 | 0.013694288 | NA |
| chr17 | 9540276 | 9540614 | 339 | 5 | 7.42E-07 | 2.84E-06 | 5.72E-05 | 9.33E-07 | -0.00166067 | STX8 |
| chr12 | 80598306 | 80598505 | 200 | 2 | 1.67E-06 | 5.88E-07 | 0.000123 | 9.35E-07 | -0.00913179 | PTPRQ |
| chr19 | 14031235 | 14032151 | 917 | 13 | 1.37E-06 | 1.58E-06 | 0.006951 | 9.36E-07 | -0.00107847 | RLN3, IL27RA, AC022098.2 |
| chr14 | 97298545 | 97298850 | 306 | 2 | 3.50E-07 | 1.99E-06 | 9.76E-06 | 9.36E-07 | -0.01106736 | NA |
| chr14 | 99644508 | 99645604 | 1097 | 11 | 9.77E-08 | 0.0001514 | 0.000121 | 9.36E-07 | 0.001739781 | HHIPL1 |
| chr1 | 230701426 | 230701770 | 345 | 3 | 1.72E-07 | 7.18E-05 | 1.38E-06 | 9.36E-07 | -0.00601364 | NA |
| chr12 | 6035332 | 6035631 | 300 | 2 | 7.44E-07 | 1.14E-06 | 2.08E-05 | 9.36E-07 | 0.007601539 | VWF |
| chr2 | 239692628 | 239693575 | 948 | 4 | 2.22E-06 | 3.83E-07 | 0.000603 | 9.36E-07 | -0.00226639 | NA |
| chr8 | 10510346 | 10511167 | 822 | 6 | 1.74E-06 | 4.88E-05 | 0.000253 | 9.36E-07 | -0.00742452 | PRSS51 |
| chr9 | 111601786 | 111601881 | 96 | 2 | 1.85E-07 | 1.43E-06 | 1.48E-05 | 9.38E-07 | -0.02984901 | NA |
| chr15 | 83207802 | 83208562 | 761 | 9 | 2.96E-08 | 7.68E-05 | 1.25E-06 | 9.39E-07 | 0.000755575 | AC103876.1, HDGFL3 |
| chr2 | 218072449 | 218072912 | 464 | 5 | 2.82E-07 | 2.18E-05 | 0.000354 | 9.39E-07 | -0.00941787 | RUFY4 |
| chr2 | 1422296 | 1423073 | 778 | 5 | 3.95E-06 | 3.06E-07 | 0.002728 | 9.39E-07 | -0.00311904 | TPO, AC105450.1 |
| chr9 | 38026079 | 38026425 | 347 | 3 | 2.09E-06 | 3.72E-07 | 0.000722 | 9.40E-07 | 0.015022809 | AL138752.2, SHB |
| chr12 | 109110959 | 109111292 | 334 | 7 | 1.56E-06 | 9.36E-07 | 0.003772 | 9.41E-07 | -0.01237617 | UNG, AC007637.1 |
| chr4 | 8005166 | 8005804 | 639 | 5 | 4.72E-07 | 6.55E-07 | 0.000343 | 9.42E-07 | -0.00449158 | ABLIM2, MIR95 |
| chr12 | 57840369 | 57840902 | 534 | 2 | 5.10E-08 | 1.08E-05 | 2.49E-06 | 9.43E-07 | 0.008414927 | AC083805.2, CTDSP2 |
| chr20 | 20860240 | 20860326 | 87 | 2 | 7.61E-07 | 4.52E-06 | 4.55E-06 | 9.43E-07 | -0.00361753 | NA |
| chr5 | 33277629 | 33278390 | 762 | 5 | 1.39E-07 | 0.00013 | 3.07E-05 | 9.43E-07 | -0.00737216 | LINC02160, AC010343.3 |
| chr11 | 105610044 | 105611261 | 1218 | 20 | 2.02E-07 | 8.89E-05 | 6.25E-05 | 9.43E-07 | -0.00015443 | GRIA4 |
| chr16 | 83717996 | 83718296 | 301 | 3 | 5.55E-07 | 5.67E-07 | 0.000133 | 9.43E-07 | -0.01788989 | CDH13 |
| chr13 | 41194353 | 41195349 | 997 | 9 | 5.89E-07 | 3.88E-06 | 0.001269 | 9.44E-07 | -0.00060126 | AL354696.2, KBTBD7 |
| chr1 | 156646762 | 156647282 | 521 | 6 | 9.35E-08 | 1.37E-05 | 0.000355 | 9.44E-07 | -0.00932383 | BCAN, AL590666.2 |
| chr11 | 57325356 | 57326144 | 789 | 7 | 4.66E-07 | 6.99E-07 | 0.000746 | 9.44E-07 | 0.009267439 | AP000781.1, SSRP1 |
| chr19 | 52614602 | 52615466 | 865 | 5 | 8.67E-06 | 4.76E-06 | 0.000638 | 9.44E-07 | -0.00330036 | ZNF83 |
| chr1 | 24902091 | 24902701 | 611 | 9 | 2.68E-07 | 3.92E-06 | 0.000638 | 9.44E-07 | -0.00068938 | RUNX3 |
| chr9 | 100352850 | 100353836 | 987 | 7 | 1.53E-06 | 2.85E-06 | 0.000367 | 9.44E-07 | 0.00049503 | TEX10 |
| chr3 | 9135763 | 9136565 | 803 | 4 | 1.57E-08 | 0.0001123 | 1.28E-05 | 9.45E-07 | -0.00915095 | SRGAP3 |
| chr5 | 151303478 | 151304259 | 782 | 6 | 2.35E-07 | 1.28E-05 | 4.92E-05 | 9.45E-07 | 0.000663975 | SLC36A3 |
| chr10 | 812877 | 812983 | 107 | 3 | 1.20E-06 | 4.46E-06 | 6.99E-05 | 9.45E-07 | -0.00570517 | LARP4B |
| chr19 | 12929614 | 12930819 | 1206 | 5 | 2.14E-09 | 4.08E-05 | 5.42E-06 | 9.46E-07 | 0.004574192 | FARSA-AS1, FARSA |
| chr1 | 19377868 | 19379058 | 1191 | 4 | 4.05E-06 | 1.45E-06 | 9.09E-05 | 9.47E-07 | 0.003224337 | CAPZB |
| chr7 | 41028794 | 41029127 | 334 | 2 | 5.50E-08 | 2.38E-05 | 1.70E-06 | 9.47E-07 | -0.00709463 | NA |
| chr9 | 121621345 | 121621575 | 231 | 2 | 1.67E-06 | 5.95E-07 | 0.000124 | 9.48E-07 | -0.01171856 | DAB2IP |
| chr2 | 144332589 | 144333289 | 701 | 10 | 5.36E-07 | 0.0003464 | 0.000331 | 9.48E-07 | -0.00189249 | NA |
| chr14 | 21711553 | 21711563 | 11 | 2 | 9.69E-07 | 5.83E-07 | 0.000144 | 9.48E-07 | 0.020769632 | NA |
| chr9 | 128243389 | 128243397 | 9 | 2 | 1.33E-06 | 1.10E-06 | 2.28E-05 | 9.49E-07 | 0.011426856 | DNM1 |
| chr2 | 170819796 | 170820415 | 620 | 4 | 1.78E-08 | 0.0007529 | 7.45E-05 | 9.49E-07 | 0.013243905 | GAD1, AC007405.4 |
| chr1 | 37734579 | 37735451 | 873 | 4 | 1.63E-06 | 1.36E-06 | 0.000344 | 9.49E-07 | -0.01492041 | EPHA10 |
| chr9 | 128946339 | 128947192 | 854 | 4 | 5.97E-08 | 7.72E-06 | 0.000237 | 9.49E-07 | 0.000109168 | AL672142.1, DOLK |
| chr6 | 52143915 | 52144356 | 442 | 6 | 5.60E-07 | 5.26E-06 | 6.74E-05 | 9.49E-07 | -0.00457363 | MIR206 |
| chr19 | 2327681 | 2329404 | 1724 | 20 | 3.04E-05 | 1.74E-05 | 0.001554 | 9.49E-07 | -0.00311612 | AC005258.1, SPPL2B, LSM7 |
| chr16 | 2787635 | 2788121 | 487 | 6 | 1.38E-06 | 7.08E-07 | 0.001904 | 9.50E-07 | -0.01277495 | PRSS33 |
| chr6 | 127120559 | 127121503 | 945 | 3 | 2.39E-06 | 1.63E-06 | 0.000107 | 9.50E-07 | -0.00434098 | RSPO3, AL356534.1 |
| chr10 | 43255037 | 43255064 | 28 | 2 | 8.02E-07 | 1.07E-06 | 2.41E-05 | 9.51E-07 | -0.01082799 | RASGEF1A |
| chr11 | 12182699 | 12183325 | 627 | 2 | 9.62E-07 | 6.77E-07 | 7.22E-05 | 9.52E-07 | -0.00817781 | MICAL2 |
| chr10 | 127424304 | 127424673 | 370 | 2 | 1.33E-07 | 1.26E-05 | 2.32E-06 | 9.52E-07 | -0.02231111 | DOCK1 |
| chr2 | 41733739 | 41734367 | 629 | 5 | 4.98E-07 | 2.20E-06 | 0.000974 | 9.52E-07 | -0.00438599 | NA |
| chr5 | 115150619 | 115150907 | 289 | 2 | 7.66E-07 | 8.75E-07 | 3.57E-05 | 9.53E-07 | -0.01780404 | TRIM36 |
| chr10 | 6918472 | 6918893 | 422 | 2 | 2.41E-08 | 2.05E-05 | 1.82E-06 | 9.54E-07 | 9.99E-05 | AL392086.3 |
| chr10 | 129328094 | 129329005 | 912 | 3 | 4.49E-06 | 3.32E-07 | 0.001121 | 9.54E-07 | -0.01157117 | NA |
| chr16 | 76194396 | 76194436 | 41 | 3 | 2.21E-06 | 6.26E-07 | 0.00031 | 9.54E-07 | -0.0245838 | NA |
| chr22 | 19990803 | 19990991 | 189 | 3 | 9.36E-07 | 1.01E-06 | 0.000132 | 9.56E-07 | -0.00028482 | ARVCF |
| chr6 | 30678930 | 30680243 | 1314 | 17 | 6.07E-08 | 0.0001403 | 7.81E-05 | 9.57E-07 | -0.00399904 | PPP1R18 |
| chr10 | 131518752 | 131519938 | 1187 | 6 | 2.14E-07 | 1.94E-06 | 8.79E-05 | 9.57E-07 | -0.00391876 | NA |
| chr6 | 116279475 | 116280643 | 1169 | 12 | 8.03E-07 | 2.14E-06 | 0.003081 | 9.57E-07 | 0.001061141 | DSE, TSPYL1 |
| chr5 | 69234504 | 69235402 | 899 | 17 | 1.81E-06 | 1.73E-07 | 0.005498 | 9.57E-07 | 5.11E-05 | CDK7 |
| chr17 | 1197799 | 1198301 | 503 | 4 | 2.80E-06 | 2.92E-07 | 0.001216 | 9.57E-07 | 0.003524764 | ABR |
| chr19 | 35909070 | 35909535 | 466 | 5 | 3.88E-07 | 4.64E-06 | 0.000169 | 9.58E-07 | 0.005191074 | NA |
| chr8 | 68022387 | 68022680 | 294 | 2 | 2.44E-07 | 9.97E-06 | 2.69E-06 | 9.58E-07 | -0.00804945 | PREX2 |
| chr3 | 119557839 | 119558488 | 650 | 4 | 8.80E-07 | 5.00E-07 | 0.000877 | 9.59E-07 | -0.00857495 | CD80 |
| chr11 | 1006564 | 1007447 | 884 | 8 | 2.43E-06 | 9.09E-08 | 0.009315 | 9.59E-07 | -0.00273057 | AP2A2 |
| chr5 | 57499371 | 57499561 | 191 | 2 | 1.24E-06 | 9.72E-07 | 2.91E-05 | 9.59E-07 | -0.00426159 | AC025470.2 |
| chr14 | 22536919 | 22537519 | 601 | 3 | 6.57E-07 | 1.75E-06 | 2.23E-05 | 9.60E-07 | -0.00844187 | NA |
| chr16 | 11586544 | 11587951 | 1408 | 13 | 8.43E-07 | 2.69E-05 | 0.001469 | 9.61E-07 | -0.00175242 | LITAF |
| chr12 | 95995916 | 95997474 | 1559 | 11 | 2.43E-08 | 9.06E-05 | 2.56E-05 | 9.61E-07 | 0.000725243 | AC007298.2, HAL |
| chr9 | 91824616 | 91824789 | 174 | 2 | 8.50E-07 | 7.98E-07 | 4.56E-05 | 9.61E-07 | 0.004848395 | ROR2 |
| chr17 | 67994313 | 67995259 | 947 | 6 | 4.12E-06 | 3.38E-07 | 0.003522 | 9.61E-07 | 0.005430962 | C17orf58 |
| chr15 | 76139507 | 76140644 | 1138 | 3 | 2.22E-05 | 0.0001103 | 9.57E-05 | 9.62E-07 | -0.00889991 | TMEM266 |
| chr1 | 22051919 | 22052557 | 639 | 7 | 5.48E-08 | 1.28E-05 | 0.000147 | 9.62E-07 | -0.00190731 | CDC42, CDC42-AS1 |
| chr5 | 180388975 | 180390084 | 1110 | 7 | 6.31E-06 | 3.30E-06 | 0.000966 | 9.63E-07 | -0.00343591 | NA |
| chr7 | 72404479 | 72405629 | 1151 | 4 | 1.21E-05 | 1.05E-05 | 7.31E-05 | 9.63E-07 | 0.004064744 | CALN1 |
| chr16 | 15034069 | 15034818 | 750 | 3 | 2.02E-07 | 2.52E-06 | 6.95E-05 | 9.63E-07 | -0.01558239 | PDXDC1 |
| chr6 | 26103168 | 26104332 | 1165 | 12 | 3.42E-07 | 1.05E-05 | 0.00017 | 9.64E-07 | -0.00160061 | H4C3 |
| chr11 | 1917704 | 1918495 | 792 | 5 | 2.41E-06 | 2.93E-07 | 0.001968 | 9.64E-07 | 0.017270442 | NA |
| chr14 | 37369523 | 37369811 | 289 | 4 | 2.15E-06 | 3.95E-07 | 0.000531 | 9.65E-07 | -0.00936184 | MIPOL1 |
| chr14 | 24213710 | 24214485 | 776 | 5 | 4.71E-08 | 1.86E-06 | 4.05E-05 | 9.66E-07 | -4.42E-05 | CHMP4A, AL096870.2, MDP1, NEDD8-MDP1 |
| chr9 | 97369252 | 97369803 | 552 | 2 | 1.56E-06 | 6.76E-07 | 7.79E-05 | 9.66E-07 | 0.001996369 | STRA6LP, CCDC180 |
| chr2 | 33726580 | 33727356 | 777 | 2 | 8.03E-08 | 4.76E-06 | 4.53E-06 | 9.67E-07 | -0.12488454 | LINC01320, MYADML |
| chr1 | 1019703 | 1020919 | 1217 | 10 | 2.75E-05 | 6.34E-05 | 0.000651 | 9.67E-07 | 0.000550876 | AGRN |
| chr2 | 9095843 | 9096339 | 497 | 2 | 5.66E-07 | 7.49E-07 | 5.56E-05 | 9.67E-07 | -0.01504218 | NA |
| chr18 | 77111698 | 77112457 | 760 | 3 | 1.10E-07 | 3.94E-05 | 4.09E-05 | 9.68E-07 | -0.01872082 | MBP |
| chr9 | 6757146 | 6758140 | 995 | 16 | 1.15E-06 | 3.13E-06 | 0.006678 | 9.68E-07 | 0.000716826 | KDM4C |
| chr15 | 58987357 | 58988520 | 1164 | 16 | 4.74E-06 | 6.50E-06 | 0.000986 | 9.68E-07 | -0.00062707 | RNF111 |
| chr14 | 104932838 | 104933258 | 421 | 3 | 5.78E-08 | 4.42E-06 | 6.50E-05 | 9.68E-07 | -0.01347431 | PLD4 |
| chr20 | 2403594 | 2403818 | 225 | 4 | 3.18E-07 | 0.0001659 | 4.33E-06 | 9.68E-07 | -0.00382888 | TGM6 |
| chr5 | 132673938 | 132675242 | 1305 | 7 | 1.51E-06 | 9.30E-06 | 6.98E-05 | 9.68E-07 | -0.00531266 | IL4 |
| chr6 | 167746649 | 167747338 | 690 | 7 | 1.77E-06 | 3.29E-06 | 0.001634 | 9.68E-07 | -0.00523873 | NA |
| chr10 | 7301211 | 7301330 | 120 | 2 | 3.73E-08 | 0.0015271 | 7.66E-07 | 9.69E-07 | -0.00725644 | SFMBT2 |
| chr17 | 74641307 | 74642166 | 860 | 3 | 5.51E-08 | 1.07E-05 | 3.57E-06 | 9.69E-07 | -0.00428973 | NA |
| chr6 | 32940790 | 32941746 | 957 | 13 | 6.21E-08 | 0.0014727 | 0.000881 | 9.69E-07 | 0.001024139 | HLA-DMB, AL645941.2 |
| chr2 | 135023266 | 135024225 | 960 | 3 | 9.09E-06 | 7.16E-06 | 0.000122 | 9.69E-07 | -0.00761672 | MAP3K19 |
| chr19 | 54257227 | 54257964 | 738 | 6 | 1.76E-07 | 5.82E-05 | 3.41E-05 | 9.69E-07 | -0.0023415 | LILRB5 |
| chr5 | 1147258 | 1147414 | 157 | 3 | 1.97E-07 | 0.000194 | 6.42E-07 | 9.70E-07 | -0.00678188 | NA |
| chr19 | 49490608 | 49491228 | 621 | 7 | 8.58E-07 | 4.94E-06 | 0.000451 | 9.70E-07 | -0.00326203 | RPL13A, SNORD33, SNORD34, SNORD35A |
| chr1 | 15243179 | 15243981 | 803 | 3 | 4.98E-06 | 3.67E-07 | 0.000844 | 9.71E-07 | -0.00333247 | NA |
| chr14 | 78402794 | 78403889 | 1096 | 8 | 1.63E-07 | 1.85E-05 | 0.000105 | 9.71E-07 | 0.006154741 | NRXN3 |
| chr21 | 31509347 | 31509895 | 549 | 3 | 7.30E-08 | 0.0001072 | 4.20E-05 | 9.71E-07 | -0.00795933 | TIAM1 |
| chr1 | 62319408 | 62320411 | 1004 | 6 | 5.20E-07 | 2.35E-06 | 0.000926 | 9.71E-07 | -0.00171252 | KANK4 |
| chr13 | 51283900 | 51284359 | 460 | 2 | 1.93E-06 | 6.11E-07 | 0.000127 | 9.71E-07 | -0.00708579 | FAM124A |
| chr5 | 4441114 | 4441637 | 524 | 3 | 6.19E-08 | 6.93E-05 | 3.63E-06 | 9.72E-07 | -0.00691056 | NA |
| chr6 | 143060206 | 143061361 | 1156 | 11 | 1.68E-07 | 6.30E-05 | 3.27E-06 | 9.72E-07 | 0.001544939 | AIG1 |
| chr6 | 170207000 | 170207423 | 424 | 2 | 6.81E-09 | 5.38E-05 | 1.28E-06 | 9.72E-07 | -0.00920227 | NA |
| chr12 | 13441072 | 13441677 | 606 | 3 | 7.03E-07 | 7.74E-07 | 0.00015 | 9.72E-07 | -0.01534367 | GRIN2B |
| chr7 | 17402416 | 17402692 | 277 | 2 | 1.12E-07 | 0.0001381 | 1.01E-06 | 9.74E-07 | 0.013078911 | AC019117.3, AC019117.2 |
| chr3 | 170221903 | 170222408 | 506 | 11 | 9.53E-08 | 0.0001105 | 0.00038 | 9.75E-07 | -0.00326379 | NA |
| chr11 | 70397737 | 70398585 | 849 | 10 | 3.38E-06 | 1.97E-06 | 0.003666 | 9.76E-07 | 0.000268728 | CTTN, AP000487.1 |
| chr1 | 209724269 | 209725160 | 892 | 4 | 1.81E-05 | 2.86E-07 | 0.001448 | 9.77E-07 | -0.00777691 | HSD11B1 |
| chr12 | 109845004 | 109845791 | 788 | 6 | 7.10E-07 | 1.84E-05 | 5.07E-05 | 9.77E-07 | -0.00046001 | NA |
| chr10 | 132548660 | 132549281 | 622 | 4 | 1.78E-07 | 3.89E-06 | 2.16E-05 | 9.77E-07 | -0.01272424 | INPP5A |
| chr1 | 3358067 | 3358719 | 653 | 5 | 1.46E-07 | 4.28E-05 | 6.37E-05 | 9.77E-07 | -0.0044212 | PRDM16 |
| chr16 | 29784801 | 29785804 | 1004 | 7 | 3.54E-07 | 0.0001055 | 0.002661 | 9.78E-07 | -0.0074728 | NA |
| chr1 | 64526450 | 64526750 | 301 | 2 | 1.74E-06 | 6.14E-07 | 0.00013 | 9.78E-07 | 0.017435423 | CACHD1 |
| chr3 | 153881342 | 153881384 | 43 | 2 | 7.21E-07 | 7.16E-07 | 6.71E-05 | 9.79E-07 | -0.00291967 | LINC02006 |

| | | | | | | | | | | |
|---|---|---|---|---|---|---|---|---|---|---|
| chr9 | 126587356 | 126587681 | 326 | 3 | 1.27E-06 | 1.29E-06 | 8.18E-05 | 9.80E-07 | -0.00948105 | NA |
| chr6 | 31702418 | 31704414 | 1997 | 38 | 3.27E-06 | 8.68E-06 | 0.004399 | 9.81E-07 | 0.000207443 | ABHD16A, AL662899.1 |
| chr7 | 27663817 | 27664446 | 630 | 4 | 2.80E-07 | 5.52E-06 | 4.32E-05 | 9.81E-07 | -0.00482749 | NA |
| chr14 | 46882208 | 46882900 | 693 | 5 | 5.52E-08 | 1.61E-05 | 4.12E-05 | 9.82E-07 | -0.01217876 | MDGA2 |
| chr17 | 76539669 | 76540187 | 519 | 8 | 6.89E-07 | 3.24E-06 | 8.11E-05 | 9.82E-07 | -0.00203266 | PRCD, CYGB |
| chr21 | 45465816 | 45466198 | 383 | 2 | 1.47E-06 | 6.34E-07 | 0.000111 | 9.82E-07 | -0.01183857 | COL18A1 |
| chr2 | 1891871 | 1893448 | 1578 | 8 | 1.23E-05 | 7.51E-05 | 0.000116 | 9.82E-07 | -0.00367036 | MYT1L |
| chr14 | 105818742 | 105819419 | 678 | 2 | 1.70E-06 | 6.37E-07 | 0.000109 | 9.82E-07 | 0.010882682 | NA |
| chr19 | 8363260 | 8363610 | 351 | 4 | 1.35E-07 | 3.39E-06 | 0.000153 | 9.82E-07 | -0.01540588 | ANGPTL4 |
| chr4 | 113436043 | 113436445 | 403 | 2 | 4.85E-07 | 1.55E-06 | 1.47E-05 | 9.84E-07 | -0.02904394 | NA |
| chr12 | 55990024 | 55990724 | 701 | 4 | 2.84E-07 | 1.37E-05 | 0.000633 | 9.85E-07 | -0.02070405 | RAB5B |
| chr19 | 7170573 | 7170849 | 277 | 2 | 6.59E-07 | 1.84E-06 | 1.18E-05 | 9.85E-07 | -0.00290111 | INSR |
| chr20 | 33318027 | 33318330 | 304 | 3 | 4.84E-07 | 1.09E-05 | 3.57E-06 | 9.86E-07 | -0.0072896 | BPIFB5P |
| chr4 | 122924435 | 122925101 | 667 | 3 | 8.46E-08 | 2.56E-05 | 1.85E-06 | 9.86E-07 | -0.00074743 | SPATA5 |
| chr1 | 50421770 | 50423770 | 2001 | 17 | 1.28E-05 | 0.000139 | 0.000305 | 9.86E-07 | -0.00218976 | AL049637.1, DMRTA2 |
| chr11 | 84720314 | 84721269 | 956 | 6 | 3.80E-08 | 0.0002224 | 1.17E-05 | 9.87E-07 | 0.00147773 | AP001825.1, DLG2 |
| chr13 | 28750877 | 28751700 | 824 | 3 | 7.58E-07 | 4.78E-07 | 0.000336 | 9.87E-07 | 0.013727007 | NA |
| chr1 | 153633239 | 153634140 | 902 | 10 | 2.85E-08 | 1.40E-05 | 7.75E-06 | 9.88E-07 | -0.00556329 | CHTOP, AL162258.1, AL162258.2 |
| chr10 | 49679166 | 49679888 | 723 | 10 | 1.65E-08 | 0.0001686 | 3.26E-06 | 9.88E-07 | -0.00434313 | C10orf53 |
| chr6 | 19837980 | 19839144 | 1165 | 5 | 8.10E-08 | 4.86E-06 | 0.000273 | 9.90E-07 | 0.000849518 | ID4, LNC-LBCS |
| chr4 | 20252699 | 20253723 | 1025 | 15 | 2.24E-06 | 5.58E-05 | 9.31E-05 | 9.90E-07 | -0.00033313 | SLIT2 |
| chr4 | 94451045 | 94452252 | 1208 | 12 | 2.03E-07 | 2.26E-06 | 0.002396 | 9.91E-07 | 0.00070895 | PDLIM5 |
| chr17 | 441226 | 441712 | 487 | 5 | 1.47E-07 | 3.38E-06 | 0.000246 | 9.92E-07 | -0.0035648 | RFLNB |
| chr7 | 2519783 | 2520470 | 688 | 9 | 1.06E-07 | 1.13E-05 | 0.000381 | 9.92E-07 | 0.000719937 | LFNG |
| chr4 | 44725809 | 44726942 | 1134 | 14 | 9.98E-07 | 3.28E-05 | 0.00012 | 9.96E-07 | 0.000637192 | GNPDA2 |
| chr2 | 180692585 | 180693332 | 748 | 4 | 1.53E-07 | 2.76E-05 | 4.52E-05 | 9.96E-07 | -0.01361553 | SCHLAP1 |
| chr9 | 127423175 | 127424267 | 1093 | 8 | 2.11E-09 | 0.0022961 | 6.55E-06 | 9.96E-07 | -0.0042384 | NA |
| chr2 | 60495178 | 60495667 | 490 | 4 | 1.26E-07 | 6.45E-06 | 9.09E-06 | 9.98E-07 | 0.005732595 | BCL11A |
| chr4 | 101159085 | 101159187 | 103 | 2 | 6.35E-07 | 2.51E-06 | 8.44E-06 | 9.99E-07 | -0.00725635 | PPP3CA |
| chr13 | 98860214 | 98860633 | 420 | 2 | 2.42E-07 | 3.54E-06 | 6.09E-06 | 1.00E-06 | -0.00074817 | DOCK9 |
| chr3 | 128553880 | 128554666 | 787 | 6 | 3.07E-07 | 1.63E-05 | 0.000165 | 1.00E-06 | -0.00189066 | NA |
| chr16 | 12662939 | 12663568 | 630 | 2 | 4.31E-07 | 1.36E-06 | 1.81E-05 | 1.00E-06 | -0.0019694 | CPPED1 |
| chr9 | 114287390 | 114288466 | 1077 | 4 | 1.59E-06 | 2.45E-06 | 0.000173 | 1.00E-06 | -0.0100626 | COL27A1 |
| chr16 | 83963056 | 83963298 | 243 | 2 | 4.55E-07 | 1.03E-06 | 2.92E-05 | 1.00E-06 | -0.00542417 | OSGIN1 |
| chr12 | 92540576 | 92540785 | 210 | 2 | 1.70E-06 | 6.47E-07 | 0.000113 | 1.00E-06 | 0.001461282 | NA |
| chr7 | 37211413 | 37211768 | 356 | 3 | 1.84E-06 | 6.54E-07 | 0.000112 | 1.00E-06 | -0.00170618 | ELMO1 |
| chr11 | 134223603 | 134225173 | 1571 | 17 | 4.22E-07 | 0.0007884 | 0.001659 | 1.00E-06 | 0.000697362 | VPS26B, NCAPD3 |
| chr1 | 9764965 | 9765116 | 152 | 2 | 6.07E-07 | 2.66E-06 | 8.01E-06 | 1.00E-06 | 0.000314977 | CLSTN1 |
| chr5 | 137889215 | 137889820 | 606 | 8 | 1.58E-06 | 5.57E-07 | 0.003556 | 1.00E-06 | 0.006584345 | PKD2L2, AC106791.1 |
| chr15 | 44667150 | 44667474 | 325 | 2 | 3.56E-07 | 1.80E-06 | 1.25E-05 | 1.00E-06 | -0.00121291 | PATL2 |
| chr2 | 151734222 | 151735211 | 990 | 5 | 1.31E-07 | 4.63E-05 | 5.00E-05 | 1.00E-06 | -0.00704521 | NEB |
| chr4 | 167102742 | 167103763 | 1022 | 4 | 1.26E-05 | 0.0002731 | 2.60E-05 | 1.00E-06 | -4.13E-05 | SPOCK3 |
| chr11 | 64224445 | 64225071 | 627 | 5 | 5.72E-07 | 4.49E-07 | 0.00041 | 1.01E-06 | -0.00033819 | TRPT1 |
| chr18 | 12910905 | 12911529 | 625 | 4 | 8.22E-08 | 2.52E-06 | 9.68E-05 | 1.01E-06 | 0.011062847 | STK25P1, PTPN2 |
| chr14 | 69619943 | 69620568 | 626 | 3 | 8.50E-07 | 8.73E-07 | 0.000309 | 1.01E-06 | -0.00317229 | SUSD6 |
| chr6 | 81767081 | 81767781 | 701 | 2 | 9.26E-07 | 1.09E-06 | 2.69E-05 | 1.01E-06 | 0.000980643 | NA |
| chr20 | 57664002 | 57665399 | 1398 | 6 | 3.61E-07 | 5.87E-06 | 9.03E-05 | 1.01E-06 | -0.00358377 | PMEPA1 |
| chr4 | 151760827 | 151761929 | 1103 | 15 | 1.22E-06 | 0.000429 | 0.001026 | 1.01E-06 | 0.00140616 | GATB |
| chr17 | 78138636 | 78139180 | 545 | 3 | 2.45E-06 | 3.75E-07 | 0.000813 | 1.01E-06 | -0.00748459 | TMC8 |
| chr7 | 100646328 | 100647323 | 996 | 2 | 4.48E-05 | 6.18E-07 | 0.000158 | 1.01E-06 | -0.01277489 | ACTL6B |
| chr10 | 34723686 | 34724090 | 405 | 3 | 8.43E-07 | 8.14E-07 | 0.000101 | 1.01E-06 | -0.01286428 | PARD3 |
| chr9 | 127875030 | 127875980 | 951 | 3 | 5.98E-06 | 6.28E-07 | 0.000366 | 1.01E-06 | -0.00482356 | AK1, AL157935.2 |
| chr5 | 157116642 | 157116726 | 85 | 2 | 3.09E-07 | 0.0001313 | 1.05E-06 | 1.01E-06 | -0.0088983 | HAVCR2 |
| chr12 | 120291767 | 120292179 | 413 | 9 | 1.76E-06 | 1.30E-06 | 0.001641 | 1.01E-06 | -0.00203389 | RNU4-2 |
| chr8 | 48550833 | 48551507 | 675 | 4 | 1.38E-07 | 2.65E-05 | 0.000143 | 1.01E-06 | -0.0049834 | NA |
| chr5 | 34929061 | 34929914 | 854 | 9 | 1.40E-07 | 3.66E-06 | 0.000109 | 1.01E-06 | 0.000140083 | DNAJC21 |
| chr10 | 78408889 | 78409115 | 227 | 3 | 1.49E-06 | 1.12E-06 | 4.29E-05 | 1.01E-06 | -0.01636121 | AC012560.1, LINC00595 |
| chr6 | 45988396 | 45988770 | 375 | 3 | 1.69E-06 | 5.17E-07 | 0.000396 | 1.01E-06 | -0.00717202 | CLIC5 |
| chr7 | 92131567 | 92132538 | 972 | 4 | 1.40E-07 | 1.29E-05 | 1.21E-05 | 1.01E-06 | -0.00852302 | CYP51A1, AC000120.2, AC000120.3 |
| chr17 | 19362866 | 19363779 | 914 | 8 | 8.45E-10 | 6.10E-05 | 0.000112 | 1.01E-06 | -0.00477274 | AC124066.1, B9D1 |
| chr2 | 241633193 | 241634557 | 1365 | 7 | 1.37E-07 | 2.52E-05 | 0.000531 | 1.01E-06 | -0.00093004 | THAP4 |
| chr2 | 143519200 | 143519705 | 506 | 3 | 2.14E-07 | 8.36E-06 | 7.48E-06 | 1.01E-06 | -0.00535971 | ARHGAP15, AC079793.1 |
| chr8 | 58844731 | 58844875 | 145 | 2 | 6.10E-07 | 7.61E-06 | 3.43E-06 | 1.02E-06 | 0.003774034 | TOX |
| chr13 | 23465952 | 23467112 | 1161 | 9 | 1.45E-06 | 1.12E-05 | 0.00026 | 1.02E-06 | 0.002620861 | LINC00327 |
| chr6 | 4663561 | 4664195 | 635 | 2 | 5.89E-07 | 1.20E-06 | 2.31E-05 | 1.02E-06 | 0.003767186 | NA |
| chr6 | 134317604 | 134318344 | 741 | 11 | 8.51E-07 | 8.27E-06 | 0.0051 | 1.02E-06 | -0.00101662 | SGK1 |
| chr9 | 133738421 | 133739480 | 1060 | 8 | 2.59E-07 | 1.23E-06 | 0.000148 | 1.02E-06 | -0.00130209 | SARDH |
| chr16 | 89699916 | 89700789 | 874 | 5 | 5.75E-08 | 2.32E-06 | 0.000332 | 1.02E-06 | -0.00130548 | SPATA2L |
| chr22 | 38487828 | 38489703 | 1876 | 6 | 1.91E-05 | 0.0001101 | 4.72E-05 | 1.02E-06 | -0.00191949 | DDX17 |
| chr12 | 68483666 | 68484357 | 692 | 2 | 1.42E-06 | 8.95E-07 | 4.08E-05 | 1.02E-06 | 0.008966575 | NA |
| chr2 | 64993014 | 64993968 | 955 | 4 | 5.02E-07 | 1.29E-06 | 9.07E-05 | 1.02E-06 | -0.00549101 | SLC1A4, LINC02245 |
| chr10 | 84327810 | 84328925 | 1116 | 12 | 1.53E-07 | 3.40E-05 | 6.68E-06 | 1.02E-06 | 0.00100392 | CCSER2 |
| chr6 | 41920811 | 41921799 | 989 | 12 | 2.14E-07 | 1.27E-05 | 6.02E-05 | 1.02E-06 | 0.000920057 | BYSL, MED20 |
| chr4 | 1410340 | 1410819 | 480 | 4 | 7.76E-08 | 0.0005253 | 3.69E-06 | 1.02E-06 | -0.00591303 | NA |
| chr16 | 1347790 | 1348225 | 436 | 4 | 9.78E-07 | 1.14E-06 | 8.56E-05 | 1.02E-06 | -0.0028501 | BAIAP3 |
| chr7 | 24829498 | 24830101 | 604 | 2 | 2.27E-07 | 2.35E-06 | 9.45E-06 | 1.02E-06 | -0.02134353 | OSBPL3 |
| chr17 | 6705682 | 6706785 | 1104 | 3 | 3.49E-05 | 1.90E-06 | 0.000132 | 1.02E-06 | -0.00341906 | SLC13A5 |
| chr6 | 32876892 | 32876898 | 7 | 2 | 4.08E-07 | 3.78E-05 | 1.53E-06 | 1.02E-06 | -0.00586411 | NA |
| chr18 | 36296164 | 36296640 | 477 | 2 | 3.39E-09 | 7.27E-05 | 1.23E-06 | 1.02E-06 | -0.01952358 | NA |
| chr1 | 234742751 | 234742786 | 36 | 2 | 2.51E-07 | 0.0004716 | 8.86E-07 | 1.02E-06 | 0.002733139 | NA |
| chr22 | 39456276 | 39456857 | 582 | 5 | 1.09E-06 | 4.29E-07 | 0.001393 | 1.03E-06 | -0.00795289 | NA |
| chr19 | 19667607 | 19668667 | 1061 | 5 | 2.85E-09 | 2.94E-05 | 6.93E-06 | 1.03E-06 | 0.000704671 | NA |
| chr1 | 81920166 | 81920679 | 514 | 2 | 3.72E-08 | 3.15E-05 | 1.65E-06 | 1.03E-06 | -0.00593966 | ADGRL2 |
| chr9 | 117377164 | 117377171 | 8 | 2 | 5.58E-07 | 6.56E-07 | 0.000121 | 1.03E-06 | 0.025461974 | ASTN2 |
| chr7 | 38177907 | 38178939 | 1033 | 13 | 2.15E-06 | 4.79E-06 | 0.000127 | 1.03E-06 | 0.001147129 | STARD3NL |
| chr1 | 109185692 | 109186544 | 853 | 6 | 5.20E-07 | 0.0002925 | 9.54E-05 | 1.03E-06 | -0.00810787 | ELAPOR1 |
| chr8 | 17800433 | 17801559 | 1127 | 13 | 2.00E-06 | 6.93E-06 | 0.002316 | 1.03E-06 | 0.000424262 | AC027117.2, MTUS1 |
| chr18 | 48562532 | 48563076 | 545 | 2 | 4.88E-08 | 1.52E-05 | 2.31E-06 | 1.03E-06 | -0.00485754 | CTIF |
| chr5 | 181170246 | 181170642 | 397 | 4 | 1.51E-06 | 2.26E-06 | 0.000331 | 1.03E-06 | -0.01912859 | NA |
| chr1 | 202822079 | 202822389 | 311 | 3 | 1.98E-07 | 2.47E-06 | 2.16E-05 | 1.03E-06 | -0.00935879 | MGAT4EP |
| chr8 | 61292385 | 61292681 | 297 | 2 | 3.26E-07 | 6.90E-06 | 3.72E-06 | 1.03E-06 | -0.01863165 | CLVS1 |
| chr15 | 90233229 | 90234467 | 1239 | 11 | 2.76E-06 | 9.69E-06 | 0.000577 | 1.03E-06 | 0.000842728 | GDPGP1, AC091167.7, CIB1 |
| chr11 | 122746359 | 122746383 | 25 | 2 | 4.63E-07 | 1.44E-06 | 1.77E-05 | 1.03E-06 | 0.013024126 | UBASH3B |
| chr10 | 96128746 | 96130133 | 1388 | 18 | 1.21E-08 | 2.09E-05 | 0.000573 | 1.03E-06 | -0.00245186 | ZNF518A, AC021037.1 |
| chr5 | 140806381 | 140807152 | 772 | 7 | 1.70E-07 | 6.03E-06 | 6.26E-05 | 1.03E-06 | -0.00492325 | PCDHA1, PCDHA2, PCDHA3, PCDHA4, AC005609.5 |
| chr19 | 34346071 | 34346533 | 463 | 3 | 2.99E-06 | 3.87E-07 | 0.000857 | 1.03E-06 | -0.01678122 | GARRE1 |
| chr14 | 24085330 | 24085954 | 625 | 3 | 1.02E-06 | 1.13E-06 | 0.000204 | 1.03E-06 | -0.00576144 | NRL |
| chr22 | 30667505 | 30668590 | 1086 | 13 | 1.45E-06 | 1.57E-05 | 0.002509 | 1.03E-06 | 0.000168315 | SLC35E4, AC003072.2, DUSP18 |
| chr2 | 112232045 | 112232074 | 30 | 3 | 1.64E-06 | 9.41E-07 | 5.83E-05 | 1.03E-06 | -0.0165261 | ZC3H8 |
| chr16 | 50592258 | 50592667 | 410 | 2 | 1.76E-06 | 6.78E-07 | 0.000106 | 1.03E-06 | 0.012841241 | NKD1 |
| chr7 | 494258 | 494317 | 60 | 3 | 1.95E-06 | 3.81E-07 | 0.000971 | 1.04E-06 | 0.038882524 | AC147651.1 |
| chr14 | 37986085 | 37986593 | 509 | 2 | 3.23E-06 | 6.26E-07 | 0.000178 | 1.04E-06 | -0.02606757 | TTC6 |
| chr16 | 58032574 | 58033080 | 507 | 3 | 3.20E-07 | 1.24E-05 | 2.53E-05 | 1.04E-06 | -0.00569754 | MMP15 |
| chr17 | 74432228 | 74433497 | 1270 | 6 | 8.27E-08 | 5.98E-06 | 0.000236 | 1.04E-06 | -0.00740551 | GPRC5C |
| chr7 | 73683362 | 73684288 | 927 | 15 | 2.18E-07 | 8.64E-05 | 0.003457 | 1.04E-06 | 0.00063993 | BUD23, DNAJC30 |
| chr1 | 231240457 | 231241864 | 1408 | 16 | 1.51E-06 | 1.00E-04 | 0.000163 | 1.04E-06 | -0.00146026 | GNPAT, C1orf131 |
| chr1 | 3611063 | 3612544 | 1482 | 11 | 7.16E-06 | 1.64E-05 | 0.001948 | 1.04E-06 | -0.0015886 | MEGF6 |
| chr5 | 139954976 | 139955657 | 682 | 2 | 5.10E-06 | 6.60E-07 | 0.000127 | 1.04E-06 | -0.01314431 | NRG2 |
| chr5 | 150639934 | 150641438 | 1505 | 6 | 1.64E-07 | 4.68E-06 | 4.85E-05 | 1.04E-06 | -0.00648072 | SYNPO |
| chr3 | 84289852 | 84290326 | 475 | 2 | 1.86E-08 | 6.09E-06 | 4.15E-06 | 1.04E-06 | -0.01927901 | NA |
| chr8 | 33389043 | 33389382 | 340 | 2 | 1.23E-06 | 8.60E-07 | 4.83E-05 | 1.04E-06 | -0.00080489 | FUT10 |
| chr7 | 907514 | 908930 | 1417 | 5 | 5.23E-06 | 8.41E-07 | 0.000585 | 1.04E-06 | 0.006051444 | ADAP1, COX19 |
| chr3 | 72847912 | 72848918 | 1007 | 16 | 7.61E-06 | 3.00E-06 | 0.00607 | 1.04E-06 | 0.00079682 | SHQ1 |
| chr3 | 42223131 | 42223293 | 163 | 2 | 1.53E-07 | 1.55E-06 | 1.63E-05 | 1.04E-06 | -0.00101825 | TRAK1 |
| chr3 | 45958545 | 45958608 | 64 | 2 | 3.65E-07 | 8.10E-07 | 5.77E-05 | 1.05E-06 | 0.005919532 | FYCO1 |
| chr1 | 220813199 | 220813426 | 228 | 3 | 1.11E-06 | 3.94E-06 | 4.74E-05 | 1.05E-06 | -0.00301716 | MTARC1, AL445423.3 |
| chr11 | 12258231 | 12258530 | 300 | 2 | 7.90E-07 | 1.16E-06 | 2.60E-05 | 1.05E-06 | 0.005309069 | MICAL2 |
| chr17 | 40818635 | 40819197 | 563 | 8 | 1.75E-06 | 4.32E-07 | 0.006051 | 1.05E-06 | 0.000813538 | TMEM99, KRT10 |
| chr20 | 450114 | 450437 | 324 | 2 | 1.35E-06 | 8.22E-07 | 5.57E-05 | 1.05E-06 | -0.00745261 | TBC1D20 |
| chr16 | 49868114 | 49868525 | 412 | 3 | 1.90E-06 | 9.78E-07 | 0.000199 | 1.05E-06 | -0.00653203 | NA |
| chr16 | 18849982 | 18850394 | 413 | 2 | 4.06E-08 | 2.37E-05 | 1.89E-06 | 1.05E-06 | -0.00520913 | SMG1 |
| chr1 | 217800297 | 217800385 | 89 | 2 | 1.51E-06 | 1.23E-06 | 2.37E-05 | 1.05E-06 | -0.00763226 | SPATA17 |
| chr8 | 72542733 | 72542996 | 264 | 2 | 1.05E-06 | 1.26E-06 | 2.29E-05 | 1.05E-06 | 0.026594785 | KCNB2 |
| chr6 | 19734123 | 19735248 | 1126 | 4 | 3.76E-06 | 4.96E-07 | 0.00062 | 1.05E-06 | -0.00390837 | LNC-LBCS |
| chr11 | 94545232 | 94545902 | 671 | 9 | 2.71E-07 | 8.74E-07 | 0.002808 | 1.05E-06 | -0.00151818 | PIWIL4, FUT4, AP000943.2 |
| chr5 | 172095625 | 172096221 | 597 | 3 | 4.69E-07 | 1.24E-06 | 0.000115 | 1.05E-06 | 0.004394622 | STK10 |
| chr4 | 76585529 | 76586498 | 970 | 8 | 1.19E-07 | 5.45E-06 | 1.59E-05 | 1.05E-06 | -0.00534815 | SHROOM3 |
| chr5 | 55994356 | 55995176 | 821 | 7 | 3.03E-07 | 2.75E-06 | 0.00357 | 1.06E-06 | 0.001675264 | AC008892.1, FLJ31104, IL6ST |
| chr11 | 64126928 | 64127330 | 403 | 4 | 8.12E-07 | 3.39E-06 | 0.00028 | 1.06E-06 | -0.00248987 | MACROD1 |
| chr3 | 5298770 | 5299347 | 578 | 2 | 2.03E-07 | 2.79E-06 | 8.24E-06 | 1.06E-06 | -0.01012598 | NA |
| chr2 | 149002713 | 149003218 | 506 | 3 | 2.27E-07 | 6.97E-06 | 3.10E-05 | 1.06E-06 | -0.00405972 | KIF5C |
| chr7 | 107742699 | 107743800 | 1102 | 14 | 3.31E-07 | 0.0023237 | 2.67E-05 | 1.06E-06 | -0.00104391 | AC002467.1 |
| chr12 | 124914551 | 124915418 | 868 | 12 | 3.66E-06 | 4.28E-07 | 0.006609 | 1.06E-06 | -0.00110978 | UBC |
| chr10 | 62091295 | 62091816 | 522 | 3 | 9.90E-09 | 6.49E-05 | 1.33E-06 | 1.06E-06 | 0.001467189 | ARID5B |
| chr17 | 82309008 | 82309223 | 216 | 4 | 9.74E-07 | 1.63E-05 | 0.000316 | 1.06E-06 | -0.00707026 | NA |
| chr8 | 141429389 | 141430150 | 762 | 2 | 2.85E-10 | 5.67E-05 | 1.37E-06 | 1.06E-06 | -0.00740453 | PTP4A3 |
| chr8 | 133571488 | 133572903 | 1416 | 10 | 3.49E-08 | 7.25E-05 | 6.12E-05 | 1.06E-06 | -0.00101161 | ST3GAL1 |
| chr18 | 59068383 | 59068896 | 514 | 3 | 4.30E-07 | 2.35E-06 | 3.91E-05 | 1.06E-06 | -0.01509271 | OACYLP |
| chr7 | 6169595 | 6170605 | 1011 | 5 | 8.44E-07 | 2.15E-06 | 0.000372 | 1.06E-06 | -0.00764318 | CYTH3 |
| chr1 | 166920552 | 166921407 | 856 | 6 | 7.53E-06 | 1.77E-06 | 0.001764 | 1.06E-06 | -0.0009858 | ILDR2 |
| chr8 | 143937238 | 143937720 | 483 | 2 | 5.82E-07 | 8.44E-07 | 5.33E-05 | 1.06E-06 | -0.01079892 | PLEC |
| chr1 | 2031810 | 2032355 | 546 | 3 | 1.37E-07 | 8.21E-06 | 2.51E-05 | 1.06E-06 | -0.01013782 | NA |
| chr11 | 65966156 | 65967487 | 1332 | 6 | 7.41E-06 | 1.58E-06 | 0.00261 | 1.06E-06 | -0.00285285 | SART1 |
| chr2 | 191409797 | 191411236 | 1440 | 4 | 4.66E-06 | 5.00E-05 | 7.88E-05 | 1.06E-06 | -0.00903187 | MYO1B |
| chr5 | 141363598 | 141364882 | 1285 | 12 | 1.65E-05 | 8.04E-07 | 0.000806 | 1.06E-06 | -0.00401855 | PCDHGA1, PCDHGA2, PCDHGA3, PCDHGB1, PCDHGA4, PCDHGB2, PCDHGA5 |
| chr1 | 234964026 | 234965043 | 1018 | 6 | 3.08E-07 | 9.37E-06 | 0.00023 | 1.06E-06 | -0.0139921 | AL391832.2 |
| chr9 | 122125539 | 122125982 | 444 | 3 | 1.36E-06 | 5.71E-07 | 0.0005 | 1.06E-06 | -0.00956523 | NA |
| chr19 | 13502277 | 13503544 | 1268 | 4 | 1.23E-09 | 4.69E-05 | 2.32E-06 | 1.06E-06 | 0.006429031 | CACNA1A |
| chr10 | 27081029 | 27081881 | 853 | 3 | 1.49E-05 | 5.88E-07 | 0.000466 | 1.06E-06 | -0.00717888 | ANKRD26 |
| chr6 | 166998832 | 166999933 | 1102 | 15 | 6.14E-06 | 1.76E-06 | 0.00198 | 1.06E-06 | 0.000748845 | CEP43, Z94721.2, Z94721.1, Z94721.3 |
| chr10 | 131146526 | 131146641 | 116 | 3 | 9.36E-07 | 1.27E-06 | 3.84E-05 | 1.06E-06 | 0.001866266 | TCERG1L |
| chr7 | 6157883 | 6159215 | 1333 | 9 | 4.64E-07 | 4.16E-05 | 0.000319 | 1.06E-06 | 0.00174859 | USP42 |
| chr12 | 57617981 | 57618248 | 268 | 4 | 1.01E-06 | 4.23E-07 | 0.001785 | 1.07E-06 | -0.02007552 | AC025165.6, AC025165.1 |
| chr4 | 105930722 | 105931017 | 296 | 2 | 8.85E-07 | 8.00E-07 | 6.38E-05 | 1.07E-06 | -0.00233601 | NPNT, AC109361.2 |
| chr5 | 111976801 | 111978251 | 1451 | 4 | 3.68E-05 | 5.30E-07 | 0.000513 | 1.07E-06 | 0.004747993 | NREP-AS1, NREP |
| chr1 | 29251285 | 29251663 | 379 | 3 | 2.96E-07 | 4.89E-05 | 5.04E-05 | 1.07E-06 | -0.01115905 | PTPRU |
| chr2 | 172674135 | 172674534 | 400 | 3 | 5.11E-07 | 4.79E-07 | 0.000399 | 1.07E-06 | -0.03683161 | AC018712.1 |
| chr6 | 21666323 | 21667584 | 1262 | 10 | 3.26E-06 | 2.06E-05 | 0.001162 | 1.07E-06 | 0.001092494 | CASC15 |

| | | | | | | | | | | |
|---|---|---|---|---|---|---|---|---|---|---|
| chr11 | 67239934 | 67241288 | 1355 | 15 | 6.77E-06 | 0.0001852 | 0.001123 | 1.07E-06 | -8.75E-05 | KDM2A |
| chr13 | 113749847 | 113750112 | 266 | 4 | 1.73E-06 | 2.11E-06 | 0.000674 | 1.07E-06 | 0.003515856 | LINC00552 |
| chr3 | 78729084 | 78729533 | 450 | 2 | 8.19E-07 | 1.11E-06 | 3.00E-05 | 1.07E-06 | 0.005952646 | ROBO1 |
| chr1 | 201946318 | 201946871 | 554 | 9 | 2.61E-08 | 0.0002276 | 3.57E-06 | 1.07E-06 | 0.004754446 | LMOD1 |
| chr9 | 137956913 | 137957262 | 350 | 2 | 8.46E-07 | 7.84E-07 | 6.95E-05 | 1.07E-06 | -0.0104309 | CACNA1B |
| chr22 | 39360905 | 39361105 | 201 | 2 | 4.08E-07 | 1.47E-05 | 2.45E-06 | 1.07E-06 | -0.0063016 | SYNGR1 |
| chr9 | 87107398 | 87107517 | 120 | 2 | 2.60E-07 | 0.0002972 | 9.75E-07 | 1.07E-06 | -0.0101748 | NA |
| chr7 | 895920 | 896954 | 1035 | 4 | 6.38E-07 | 3.27E-06 | 8.27E-05 | 1.07E-06 | -0.00410747 | SUN1, GET4 |
| chr8 | 55879675 | 55880516 | 842 | 5 | 3.15E-07 | 1.66E-06 | 0.000502 | 1.07E-06 | 0.000488392 | LYN |
| chr4 | 186555172 | 186555911 | 740 | 11 | 1.46E-06 | 3.33E-07 | 0.00429 | 1.07E-06 | 4.19E-05 | AC018709.1, MTNR1A |
| chr22 | 50582531 | 50583404 | 874 | 12 | 4.62E-07 | 9.50E-06 | 0.000132 | 1.07E-06 | -0.00031172 | CHKB-DT, CHKB-CPT1B, CHKB |
| chr14 | 104187732 | 104188508 | 777 | 3 | 2.65E-06 | 1.00E-06 | 7.29E-05 | 1.07E-06 | -0.01186131 | NA |
| chr22 | 39222775 | 39223394 | 620 | 2 | 7.08E-07 | 7.82E-07 | 7.05E-05 | 1.07E-06 | -0.01127658 | PDGFB |
| chr17 | 2965593 | 2966009 | 417 | 4 | 9.98E-07 | 5.77E-07 | 0.000198 | 1.08E-06 | -0.00032333 | RAP1GAP2, AC015921.1 |
| chr1 | 40508048 | 40509262 | 1215 | 18 | 1.44E-06 | 2.47E-05 | 0.000404 | 1.08E-06 | -0.00053249 | EXO5, AL603839.3 |
| chr17 | 49574091 | 49575153 | 1063 | 7 | 2.03E-07 | 2.26E-06 | 0.000234 | 1.08E-06 | -0.00140565 | AC006487.2 |
| chr8 | 127854279 | 127855266 | 988 | 3 | 8.25E-07 | 1.30E-06 | 4.11E-05 | 1.08E-06 | -0.00633563 | PVT1 |
| chr7 | 150992451 | 150993881 | 1431 | 8 | 4.02E-05 | 7.67E-05 | 0.000251 | 1.08E-06 | -0.00094579 | NOS3 |
| chr1 | 31585026 | 31585716 | 691 | 2 | 1.59E-06 | 7.44E-07 | 8.65E-05 | 1.08E-06 | -0.00665195 | TINAGL1 |
| chr19 | 36121469 | 36122009 | 541 | 4 | 1.93E-06 | 8.62E-07 | 0.000457 | 1.08E-06 | 0.008670354 | TBCB |
| chr1 | 239297767 | 239298222 | 456 | 3 | 4.14E-07 | 2.04E-06 | 2.75E-05 | 1.08E-06 | -0.02025461 | NA |
| chr14 | 33061949 | 33062671 | 723 | 3 | 1.66E-07 | 1.11E-05 | 1.61E-05 | 1.08E-06 | -0.00966547 | NPAS3 |
| chr18 | 45062600 | 45063971 | 1372 | 7 | 8.49E-07 | 6.17E-05 | 0.000118 | 1.08E-06 | 0.001123377 | SETBP1 |
| chr5 | 10489915 | 10490223 | 309 | 2 | 1.48E-06 | 8.20E-07 | 6.20E-05 | 1.08E-06 | -0.00521771 | NA |
| chr16 | 66746610 | 66747419 | 810 | 3 | 1.47E-05 | 3.71E-07 | 0.001263 | 1.08E-06 | -0.00798938 | DYNC1LI2 |
| chr4 | 76743949 | 76744480 | 532 | 2 | 1.11E-06 | 9.25E-07 | 4.46E-05 | 1.08E-06 | -0.00679913 | SHROOM3, SHROOM3-AS1 |
| chr19 | 2137181 | 2137789 | 609 | 3 | 1.26E-08 | 1.11E-05 | 3.93E-06 | 1.08E-06 | -0.00917377 | AP3D1 |
| chr4 | 6695857 | 6696441 | 585 | 2 | 1.05E-07 | 5.44E-06 | 4.78E-06 | 1.08E-06 | 0.04761517 | S100P |
| chr3 | 196247106 | 196247946 | 841 | 6 | 9.36E-07 | 3.70E-06 | 3.89E-05 | 1.08E-06 | -0.00561164 | PCYT1A |
| chr11 | 121892222 | 121892844 | 623 | 2 | 1.04E-06 | 8.93E-07 | 4.90E-05 | 1.08E-06 | 0.00189108 | AP001977.1 |
| chr2 | 222655692 | 222656837 | 1146 | 14 | 1.00E-08 | 0.0003209 | 0.000114 | 1.08E-06 | 0.000329455 | FARSB |
| chr8 | 124610410 | 124610533 | 124 | 2 | 1.40E-06 | 6.61E-07 | 0.000171 | 1.08E-06 | -0.01254707 | MTSS1 |
| chr14 | 63388875 | 63389888 | 1014 | 4 | 1.09E-05 | 6.49E-05 | 0.000204 | 1.08E-06 | -0.00858631 | PPP2R5E |
| chr14 | 35045705 | 35046652 | 948 | 12 | 3.33E-07 | 2.75E-05 | 0.000196 | 1.09E-06 | 0.000374729 | FAM177A1 |
| chr12 | 25958764 | 25959985 | 1222 | 15 | 1.69E-06 | 3.99E-06 | 0.000758 | 1.09E-06 | 0.003631501 | RASSF8, RASSF8-AS1 |
| chr21 | 38751958 | 38752924 | 967 | 5 | 2.41E-07 | 2.26E-06 | 0.000344 | 1.09E-06 | -0.00940337 | NA |
| chr6 | 157530583 | 157531284 | 702 | 2 | 1.34E-08 | 6.77E-06 | 4.07E-06 | 1.09E-06 | -0.01089627 | ZDHHC14 |
| chr16 | 34133954 | 34134160 | 207 | 2 | 5.04E-07 | 3.14E-06 | 7.70E-06 | 1.09E-06 | -0.0305582 | NA |
| chr10 | 99527302 | 99528089 | 788 | 8 | 5.10E-07 | 1.02E-06 | 0.001483 | 1.09E-06 | -0.00599041 | AL513542.1, LINC01475 |
| chr15 | 88910137 | 88910576 | 440 | 3 | 7.32E-07 | 1.76E-06 | 2.74E-05 | 1.09E-06 | -0.01027382 | MFGE8 |
| chr20 | 11810432 | 11811640 | 1209 | 4 | 8.58E-07 | 1.08E-05 | 4.54E-05 | 1.09E-06 | -0.0063314 | LINC00687 |
| chr9 | 128829503 | 128830534 | 1032 | 10 | 1.50E-06 | 5.75E-06 | 0.001509 | 1.09E-06 | 4.13E-05 | SPOUT1, AL441992.2 |
| chr12 | 93380991 | 93380996 | 6 | 2 | 1.34E-06 | 6.59E-07 | 0.000184 | 1.09E-06 | -0.01865858 | NUDT4 |
| chr2 | 16319177 | 16320075 | 899 | 4 | 1.95E-06 | 2.64E-06 | 0.000171 | 1.09E-06 | 0.002815807 | AC010745.2, AC010745.4 |
| chr11 | 33808369 | 33809283 | 915 | 4 | 1.41E-07 | 1.77E-05 | 6.27E-06 | 1.09E-06 | -0.00903491 | NA |
| chr17 | 10518048 | 10518901 | 854 | 6 | 8.12E-07 | 8.91E-07 | 0.001945 | 1.09E-06 | -0.00866634 | AC005323.2, MYHAS, MYH1 |
| chr2 | 115341177 | 115341411 | 235 | 2 | 1.73E-06 | 7.87E-07 | 7.42E-05 | 1.09E-06 | -0.00039295 | DPP10 |
| chr11 | 61757481 | 61758450 | 970 | 8 | 2.80E-07 | 2.22E-05 | 0.00058 | 1.09E-06 | -0.00029838 | MYRF, MYRF-AS1 |
| chr12 | 111842357 | 111843167 | 811 | 13 | 2.39E-06 | 3.03E-07 | 0.006967 | 1.10E-06 | 0.000194505 | MAPKAPK5, MAPKAPK5-AS1 |
| chr6 | 31870997 | 31871552 | 556 | 4 | 5.48E-08 | 1.17E-05 | 0.000128 | 1.10E-06 | -0.00574965 | SLC44A4 |
| chr3 | 155766131 | 155766508 | 378 | 2 | 2.51E-07 | 5.96E-06 | 4.53E-06 | 1.10E-06 | -0.00196029 | C3orf33, AC104472.3 |
| chr1 | 163321697 | 163322450 | 754 | 15 | 6.90E-08 | 4.56E-06 | 0.003175 | 1.10E-06 | 0.00049235 | NUF2, RGS5, RGS5 |
| chr2 | 150911834 | 150912009 | 176 | 2 | 1.83E-06 | 7.96E-07 | 7.24E-05 | 1.10E-06 | -0.03379078 | NA |
| chr2 | 3423447 | 3423940 | 494 | 3 | 1.63E-08 | 1.62E-05 | 3.04E-06 | 1.10E-06 | -0.00835803 | TRAPPC12 |
| chr16 | 89646295 | 89646931 | 637 | 3 | 3.83E-07 | 8.84E-07 | 7.48E-05 | 1.10E-06 | -4.02E-05 | CHMP1A |
| chr2 | 231041364 | 231041693 | 330 | 2 | 1.90E-06 | 6.98E-07 | 0.000131 | 1.10E-06 | -0.01013467 | C2orf72 |
| chr1 | 52637099 | 52637622 | 524 | 2 | 7.41E-07 | 1.20E-06 | 2.76E-05 | 1.10E-06 | -0.0032679 | SHISAL2A |
| chr3 | 38453842 | 38454825 | 984 | 7 | 9.81E-06 | 5.24E-07 | 0.001841 | 1.10E-06 | -0.00074698 | ACVR2B, ACVR2B-AS1 |
| chr10 | 124681713 | 124682133 | 421 | 5 | 5.77E-07 | 2.59E-05 | 0.000164 | 1.10E-06 | -0.00023921 | AC068896.1, FAM53B |
| chr17 | 11506841 | 11506844 | 4 | 2 | 1.90E-06 | 9.44E-07 | 4.49E-05 | 1.10E-06 | -0.01356885 | SHISA6 |
| chr1 | 31390355 | 31390551 | 197 | 2 | 1.03E-06 | 2.10E-06 | 1.22E-05 | 1.10E-06 | -0.00642108 | NA |
| chr8 | 126588538 | 126589211 | 674 | 6 | 1.13E-06 | 3.19E-06 | 0.001125 | 1.10E-06 | -0.00150336 | PCAT1 |
| chr6 | 2992591 | 2992848 | 258 | 2 | 6.09E-07 | 1.94E-06 | 1.35E-05 | 1.10E-06 | 2.77E-05 | NQO2, NQO2-AS1 |
| chr1 | 208239240 | 208239774 | 535 | 2 | 3.31E-07 | 1.10E-06 | 3.25E-05 | 1.10E-06 | -0.01087951 | PLXNA2 |
| chr16 | 87367563 | 87368517 | 955 | 2 | 5.04E-05 | 6.69E-07 | 0.000181 | 1.10E-06 | 0.041367401 | FBXO31 |
| chr9 | 127981277 | 127982028 | 752 | 7 | 2.75E-08 | 8.43E-05 | 7.70E-06 | 1.10E-06 | 0.000340757 | NA |
| chr15 | 75676063 | 75677294 | 1232 | 8 | 5.97E-07 | 6.72E-06 | 0.000111 | 1.11E-06 | -0.00328852 | AC105020.4, CSPG4 |
| chr8 | 37359737 | 37360564 | 828 | 3 | 5.05E-06 | 4.42E-07 | 0.000586 | 1.11E-06 | -0.00847101 | NA |
| chr3 | 196812034 | 196812249 | 216 | 2 | 1.52E-06 | 9.75E-07 | 4.20E-05 | 1.11E-06 | -0.02987894 | PAK2 |
| chr10 | 131201384 | 131202208 | 825 | 7 | 2.41E-07 | 0.0016657 | 0.000318 | 1.11E-06 | -0.00376973 | TCERG1L |
| chr7 | 157409554 | 157410430 | 877 | 3 | 7.31E-06 | 3.67E-06 | 7.86E-05 | 1.11E-06 | -0.00255465 | DNAJB6 |
| chr15 | 63331882 | 63332743 | 862 | 5 | 7.34E-07 | 2.97E-06 | 0.000103 | 1.11E-06 | -0.0043434 | CA12 |
| chr12 | 115508068 | 115508217 | 150 | 3 | 4.45E-07 | 5.14E-05 | 2.12E-06 | 1.11E-06 | -0.00159822 | NA |
| chr4 | 174522092 | 174522716 | 625 | 7 | 5.00E-07 | 0.0001324 | 8.88E-05 | 1.11E-06 | 0.002263707 | AC096751.2, HPGD |
| chr17 | 35764186 | 35764882 | 697 | 5 | 5.17E-07 | 1.12E-05 | 0.000271 | 1.11E-06 | 7.40E-05 | TAF15, C17orf50, MMP28 |
| chr14 | 76761202 | 76762347 | 1146 | 12 | 6.71E-07 | 3.57E-06 | 0.000489 | 1.11E-06 | 0.002068794 | VASH1, AC007376.2 |
| chr3 | 197498331 | 197499643 | 1313 | 6 | 9.38E-08 | 1.75E-05 | 4.67E-05 | 1.11E-06 | 0.004057958 | NA |
| chr13 | 110635597 | 110636694 | 1098 | 7 | 1.83E-06 | 1.49E-06 | 0.003426 | 1.11E-06 | -0.00119126 | NAXD |
| chr14 | 35331970 | 35332430 | 461 | 2 | 1.41E-08 | 6.94E-05 | 1.36E-06 | 1.11E-06 | -0.00632714 | NA |
| chr1 | 154995861 | 154996305 | 445 | 2 | 1.92E-08 | 0.00017 | 1.10E-06 | 1.11E-06 | -0.01730853 | NA |
| chr15 | 63041277 | 63043481 | 2205 | 16 | 1.01E-05 | 1.24E-05 | 0.007771 | 1.11E-06 | -0.00306048 | TPM1 |
| chr10 | 103810748 | 103811317 | 570 | 2 | 1.11E-08 | 0.0003014 | 1.01E-06 | 1.11E-06 | -0.00890897 | SH3PXD2A |
| chr15 | 101179318 | 101180139 | 822 | 4 | 2.56E-08 | 0.0001277 | 7.44E-05 | 1.11E-06 | 0.008257963 | CHSY1 |
| chr12 | 13103732 | 13104958 | 1227 | 7 | 9.68E-08 | 2.85E-06 | 0.00013 | 1.11E-06 | -0.00103477 | FAM234B |
| chr19 | 4854678 | 4855362 | 685 | 2 | 7.51E-07 | 1.36E-06 | 2.27E-05 | 1.11E-06 | -0.002128 | PLIN3 |
| chr12 | 109093067 | 109094430 | 1364 | 10 | 4.20E-07 | 2.37E-05 | 0.000836 | 1.11E-06 | -0.0038562 | ALKBH2 |
| chr17 | 47987553 | 47987961 | 409 | 2 | 2.22E-07 | 1.23E-06 | 2.71E-05 | 1.11E-06 | 0.007925437 | AC018521.4 |
| chr5 | 148382737 | 148383978 | 1242 | 13 | 3.20E-06 | 7.63E-06 | 0.002769 | 1.11E-06 | -0.00066158 | FBXO38, AC011346.1 |
| chr1 | 115545525 | 115546290 | 766 | 3 | 3.05E-08 | 7.23E-05 | 1.18E-06 | 1.12E-06 | -0.00751402 | NA |
| chr21 | 46664818 | 46665605 | 788 | 2 | 1.74E-05 | 6.76E-07 | 0.000182 | 1.12E-06 | -0.0081812 | PRMT2 |
| chr13 | 52652009 | 52652675 | 667 | 11 | 3.76E-07 | 4.18E-05 | 9.58E-05 | 1.12E-06 | -0.00068391 | SUGT1-DT |
| chr9 | 706082 | 706836 | 755 | 7 | 2.51E-08 | 8.32E-06 | 0.000771 | 1.12E-06 | 0.01353255 | KANK1 |
| chr8 | 132865713 | 132866803 | 1091 | 5 | 1.43E-05 | 4.05E-05 | 0.000202 | 1.12E-06 | -0.0015982 | NA |
| chr19 | 48605138 | 48606031 | 894 | 4 | 3.32E-07 | 8.42E-07 | 0.00068 | 1.12E-06 | -0.00119085 | FAM83E |
| chr10 | 23363664 | 23363920 | 257 | 2 | 5.46E-07 | 3.89E-06 | 6.56E-06 | 1.12E-06 | 0.007984948 | NA |
| chr14 | 101065066 | 101065833 | 768 | 14 | 4.35E-08 | 4.33E-05 | 0.001248 | 1.12E-06 | -0.00140427 | MIR409, MIR412, MIR369 |
| chr14 | 93433021 | 93433468 | 448 | 2 | 3.47E-07 | 2.62E-06 | 9.73E-06 | 1.12E-06 | 0.008757711 | UNC79 |
| chr13 | 100544382 | 100545416 | 1035 | 6 | 8.08E-08 | 0.0004628 | 4.38E-05 | 1.12E-06 | -0.00079438 | GGACT, AL356966.1 |
| chr15 | 48373656 | 48374221 | 566 | 2 | 1.02E-08 | 3.65E-06 | 6.99E-06 | 1.12E-06 | -0.00315472 | NA |
| chr3 | 195913102 | 195913372 | 271 | 2 | 3.61E-07 | 6.97E-07 | 0.000153 | 1.12E-06 | -0.0170046 | TNK2-AS1, AC124944.2 |
| chr12 | 116533051 | 116534128 | 1078 | 11 | 2.44E-06 | 1.02E-06 | 0.008097 | 1.12E-06 | -0.00042841 | LINC00173 |
| chr1 | 115112823 | 115113812 | 990 | 3 | 7.71E-07 | 5.97E-06 | 2.15E-05 | 1.12E-06 | -0.00722464 | NA |
| chr8 | 134685470 | 134686212 | 743 | 4 | 2.42E-06 | 3.21E-07 | 0.002064 | 1.12E-06 | 0.012808194 | ZFAT |
| chr20 | 36113828 | 36114307 | 480 | 3 | 5.57E-07 | 6.87E-06 | 3.97E-05 | 1.12E-06 | -0.00960387 | EPB41L1 |
| chr12 | 16359961 | 16360075 | 115 | 3 | 1.68E-06 | 3.34E-06 | 0.000117 | 1.12E-06 | -0.01072483 | MGST1 |
| chr4 | 140967959 | 140969067 | 1109 | 3 | 2.06E-06 | 7.98E-07 | 0.00013 | 1.13E-06 | 0.001755017 | RNF150 |
| chr15 | 37984077 | 37984450 | 374 | 2 | 3.64E-08 | 0.0003033 | 1.02E-06 | 1.13E-06 | 0.005148244 | LINC02345 |
| chr10 | 3502866 | 3503683 | 818 | 5 | 1.03E-07 | 4.83E-06 | 5.81E-05 | 1.13E-06 | -0.00594826 | LINC02669 |
| chr12 | 110689334 | 110690442 | 1109 | 12 | 1.29E-07 | 3.32E-05 | 7.65E-05 | 1.13E-06 | 0.000325964 | HVCN1 |
| chr17 | 61486954 | 61487593 | 640 | 6 | 5.14E-07 | 6.98E-05 | 5.65E-05 | 1.13E-06 | -0.00137724 | NA |
| chr7 | 100836361 | 100836888 | 528 | 2 | 4.29E-08 | 5.68E-06 | 4.90E-06 | 1.13E-06 | -0.00019455 | SLC12A9 |
| chr19 | 9785421 | 9786114 | 694 | 3 | 5.32E-08 | 4.74E-05 | 2.02E-05 | 1.13E-06 | 0.002862082 | ZNF846 |
| chr19 | 53868383 | 53869789 | 1407 | 14 | 9.99E-08 | 5.82E-05 | 5.59E-05 | 1.13E-06 | -4.97E-05 | MYADM, MYADM-AS1 |
| chr1 | 224355811 | 224357062 | 1252 | 13 | 3.41E-06 | 2.08E-05 | 0.000514 | 1.13E-06 | -0.00071669 | CNIH4 |
| chr1 | 154056903 | 154057268 | 366 | 3 | 1.33E-07 | 2.11E-05 | 9.23E-06 | 1.13E-06 | -0.00807672 | NUP210L |
| chr1 | 58929527 | 58929981 | 455 | 2 | 1.11E-06 | 6.80E-07 | 0.000195 | 1.13E-06 | -0.00930591 | LINC02777 |
| chr2 | 3127316 | 3127600 | 285 | 5 | 1.47E-06 | 2.94E-06 | 0.002405 | 1.13E-06 | 0.012475111 | NA |
| chr14 | 104179350 | 104180718 | 1369 | 6 | 5.33E-07 | 1.22E-06 | 0.00089 | 1.13E-06 | -0.00096898 | KIF26A |
| chr1 | 3063253 | 3064373 | 1121 | 7 | 3.01E-06 | 1.39E-05 | 0.000819 | 1.13E-06 | -0.00023598 | PRDM16-DT |
| chr13 | 99355539 | 99356703 | 1165 | 6 | 1.10E-06 | 5.58E-07 | 0.004104 | 1.13E-06 | -0.00325183 | UBAC2, MIR623 |
| chr1 | 151909498 | 151910303 | 806 | 11 | 9.68E-07 | 6.23E-07 | 0.003089 | 1.13E-06 | -0.00449243 | THEM4 |
| chr9 | 134544404 | 134545532 | 1129 | 4 | 7.34E-06 | 2.47E-05 | 0.000258 | 1.13E-06 | -0.00259196 | AL669970.3 |
| chr6 | 13574039 | 13575217 | 1179 | 10 | 4.65E-07 | 1.58E-05 | 0.000533 | 1.13E-06 | 0.000652842 | SIRT5 |
| chr10 | 99876500 | 99877043 | 544 | 2 | 2.66E-07 | 2.69E-06 | 9.63E-06 | 1.13E-06 | -0.00168824 | DNMBP |
| chr17 | 3810195 | 3810964 | 770 | 4 | 5.41E-07 | 2.33E-06 | 0.000268 | 1.13E-06 | -0.0028611 | NCBP3 |
| chr17 | 80084905 | 80084979 | 75 | 3 | 2.10E-06 | 8.80E-07 | 0.000325 | 1.13E-06 | -0.0089479 | CCDC40 |
| chr4 | 87821433 | 87821708 | 276 | 2 | 1.01E-06 | 7.39E-07 | 0.000116 | 1.14E-06 | 0.010025296 | MEPE |
| chr14 | 70907582 | 70908632 | 1051 | 6 | 1.21E-05 | 6.44E-06 | 0.001487 | 1.14E-06 | 0.001710405 | PCNX1 |
| chr18 | 66424263 | 66424714 | 452 | 2 | 5.49E-07 | 1.23E-06 | 2.82E-05 | 1.14E-06 | -0.02350703 | NA |
| chr6 | 5994002 | 5994917 | 916 | 8 | 1.39E-06 | 1.18E-05 | 0.000301 | 1.14E-06 | 0.001115041 | NA |
| chr10 | 31769876 | 31770528 | 653 | 2 | 2.59E-06 | 8.26E-07 | 7.41E-05 | 1.14E-06 | -0.01103364 | NA |
| chr12 | 53944898 | 53945869 | 972 | 9 | 8.41E-07 | 1.21E-05 | 0.000462 | 1.14E-06 | -0.00293075 | HOXC13 |
| chr2 | 219729216 | 219729488 | 273 | 3 | 9.63E-07 | 5.15E-07 | 0.000554 | 1.14E-06 | -0.0150433 | LINC02832 |
| chr21 | 46138300 | 46138679 | 380 | 2 | 5.12E-07 | 9.91E-07 | 4.44E-05 | 1.14E-06 | 0.014721535 | FTCD |
| chr1 | 22911943 | 22912122 | 180 | 2 | 1.58E-06 | 9.75E-07 | 4.62E-05 | 1.14E-06 | -0.00370924 | EPHB2 |
| chr1 | 66542917 | 66543822 | 906 | 2 | 2.28E-05 | 7.24E-07 | 0.000136 | 1.14E-06 | -0.00878796 | SGIP1 |
| chr9 | 135961546 | 135962511 | 966 | 7 | 1.11E-07 | 2.47E-05 | 1.58E-05 | 1.14E-06 | -0.00627436 | NA |
| chr13 | 111305040 | 111305501 | 462 | 3 | 8.43E-07 | 7.61E-07 | 0.00012 | 1.15E-06 | -0.00010455 | ARHGEF7 |
| chr3 | 64267858 | 64268436 | 579 | 7 | 1.19E-06 | 7.52E-05 | 0.000191 | 1.15E-06 | -0.00669898 | PRICKLE2 |
| chr11 | 132024670 | 132025705 | 1036 | 3 | 1.73E-05 | 5.76E-07 | 0.000487 | 1.15E-06 | -0.00420182 | NTM |
| chr5 | 96590629 | 96590840 | 212 | 2 | 1.04E-07 | 1.59E-05 | 2.55E-06 | 1.15E-06 | -0.00862498 | AC104123.1, CAST |
| chr1 | 1600674 | 1601016 | 343 | 3 | 9.28E-08 | 3.42E-06 | 0.000123 | 1.15E-06 | -0.00958683 | AL691432.4 |
| chr17 | 30476540 | 30477661 | 1122 | 10 | 3.19E-06 | 7.30E-06 | 0.004263 | 1.15E-06 | -0.00387008 | GOSR1 |
| chr19 | 56403395 | 56404836 | 1442 | 14 | 4.18E-06 | 0.0001846 | 6.24E-05 | 1.15E-06 | -0.00287924 | ZNF583 |
| chr2 | 148887019 | 148887825 | 807 | 4 | 6.31E-06 | 7.27E-07 | 0.000469 | 1.15E-06 | -0.0081944 | KIF5C, KIF5C-AS1 |
| chr12 | 166913 | 168993 | 2081 | 8 | 1.22E-05 | 2.47E-05 | 0.000225 | 1.15E-06 | 0.002748658 | IQSEC3, AC007406.3 |
| chr16 | 2205649 | 2206715 | 1067 | 8 | 5.08E-08 | 7.59E-06 | 0.002934 | 1.15E-06 | -0.00893468 | MLST8 |
| chr17 | 77674598 | 77674867 | 270 | 2 | 1.33E-06 | 9.11E-07 | 5.66E-05 | 1.15E-06 | -0.00836221 | AC021683.5 |
| chr14 | 51239444 | 51241257 | 1814 | 11 | 2.24E-05 | 7.40E-05 | 0.000759 | 1.15E-06 | -0.00183666 | TMX1 |
| chr17 | 75261202 | 75262190 | 989 | 13 | 6.64E-08 | 0.0001111 | 6.41E-05 | 1.15E-06 | 0.000173577 | MRPS7, GGA3 |
| chr11 | 72434311 | 72434800 | 490 | 9 | 6.64E-07 | 2.05E-05 | 0.000186 | 1.15E-06 | 0.000153888 | CLPB |
| chr21 | 36698683 | 36699886 | 1204 | 11 | 6.02E-08 | 2.51E-05 | 0.000514 | 1.16E-06 | 0.001663806 | SIM2, AP000697.1 |
| chr3 | 46496930 | 46497528 | 599 | 7 | 1.92E-06 | 2.69E-07 | 0.003124 | 1.16E-06 | -0.00526586 | NA |
| chr14 | 102593054 | 102593649 | 596 | 5 | 1.19E-06 | 6.56E-07 | 0.001964 | 1.16E-06 | -0.00123804 | RCOR1 |
| chr1 | 169893368 | 169894391 | 1024 | 14 | 2.58E-06 | 1.82E-06 | 0.007472 | 1.16E-06 | -0.00080758 | SCYL3 |
| chr10 | 123274254 | 123275302 | 1049 | 9 | 1.99E-05 | 6.20E-06 | 0.000375 | 1.16E-06 | 0.008518214 | NA |
| chr22 | 50360870 | 50361343 | 474 | 4 | 1.57E-07 | 5.14E-05 | 1.33E-06 | 1.16E-06 | 0.003041293 | PPP6R2 |
| chr1 | 87334231 | 87335328 | 1098 | 5 | 3.13E-06 | 4.69E-05 | 0.000223 | 1.16E-06 | -0.0083355 | LMO4 |

| | | | | | | | | | | |
|---|---|---|---|---|---|---|---|---|---|---|
| chr17 | 17557591 | 17558040 | 450 | 2 | 5.28E-07 | 1.74E-06 | 1.70E-05 | 1.16E-06 | 0.000321647 | PEMT |
| chr1 | 150158185 | 150158259 | 75 | 2 | 4.73E-07 | 1.55E-06 | 2.01E-05 | 1.16E-06 | 0.015994881 | PLEKHO1 |
| chr14 | 102139881 | 102141009 | 1129 | 9 | 3.13E-07 | 0.0003128 | 5.88E-05 | 1.16E-06 | -0.00298001 | WDR20 |
| chr6 | 28940790 | 28941559 | 770 | 6 | 4.87E-08 | 0.0001517 | 9.11E-06 | 1.16E-06 | 0.000155251 | NA |
| chr3 | 124268976 | 124269819 | 844 | 3 | 7.91E-08 | 4.79E-06 | 5.44E-05 | 1.16E-06 | -0.00695013 | KALRN |
| chr22 | 41444288 | 41444445 | 158 | 2 | 4.51E-08 | 0.0005226 | 9.79E-07 | 1.16E-06 | -0.00430579 | TOB2 |
| chr10 | 86693921 | 86694066 | 146 | 2 | 2.16E-06 | 7.59E-07 | 0.000115 | 1.16E-06 | -0.01539078 | LDB3 |
| chr15 | 43648669 | 43648874 | 206 | 4 | 1.98E-06 | 1.45E-06 | 0.000117 | 1.16E-06 | -0.00687919 | CATSPER2 |
| chr1 | 34791874 | 34793332 | 1459 | 9 | 1.32E-06 | 5.93E-05 | 0.000282 | 1.16E-06 | -0.00263699 | GJA4, SMIM12 |
| chr19 | 18931911 | 18932530 | 620 | 5 | 2.99E-07 | 1.98E-06 | 6.14E-05 | 1.16E-06 | -0.0045863 | AC002985.1, HOMER3 |
| chr2 | 88350388 | 88351624 | 1237 | 9 | 5.13E-07 | 3.07E-05 | 0.000555 | 1.16E-06 | -0.00050583 | NA |
| chr6 | 30163584 | 30164694 | 1111 | 17 | 5.73E-07 | 3.96E-05 | 0.001004 | 1.16E-06 | -0.00058162 | TRIM15 |
| chr19 | 17540045 | 17540239 | 195 | 3 | 3.35E-07 | 7.57E-06 | 7.04E-05 | 1.16E-06 | -0.00905134 | NIBAN3 |
| chr5 | 150671490 | 150672004 | 515 | 5 | 2.42E-08 | 1.12E-05 | 0.000336 | 1.16E-06 | -0.00212389 | MYOZ3, AC008453.1 |
| chr21 | 42237820 | 42238503 | 684 | 4 | 3.05E-06 | 1.63E-06 | 0.000295 | 1.16E-06 | -0.00460798 | ABCG1 |
| chr11 | 86069804 | 86070677 | 874 | 5 | 1.22E-07 | 4.16E-06 | 6.71E-05 | 1.16E-06 | -0.00452095 | PICALM |
| chr16 | 57831006 | 57831752 | 747 | 4 | 9.43E-07 | 6.34E-07 | 0.000707 | 1.17E-06 | -0.00451783 | KIFC3 |
| chr4 | 11414334 | 11414648 | 315 | 5 | 1.56E-06 | 1.95E-06 | 0.000383 | 1.17E-06 | -4.89E-05 | HS3ST1 |
| chr3 | 11725863 | 11726230 | 368 | 2 | 7.61E-07 | 7.02E-07 | 0.000199 | 1.17E-06 | -0.00048936 | VGLL4 |
| chr19 | 12969310 | 12969595 | 286 | 4 | 8.73E-07 | 3.89E-06 | 2.77E-05 | 1.17E-06 | -0.00237494 | DAND5 |
| chr22 | 41575050 | 41575959 | 910 | 4 | 7.28E-06 | 3.54E-05 | 2.56E-05 | 1.17E-06 | -0.00228151 | CSDC2 |
| chr10 | 96618798 | 96619342 | 545 | 5 | 5.72E-07 | 6.97E-07 | 0.000216 | 1.17E-06 | -0.00190161 | PIK3AP1 |
| chr8 | 98891167 | 98891757 | 591 | 2 | 1.30E-06 | 1.01E-06 | 4.48E-05 | 1.17E-06 | 0.00016485 | STK3 |
| chr20 | 1924938 | 1925300 | 363 | 2 | 7.46E-07 | 9.72E-07 | 4.97E-05 | 1.17E-06 | -0.00011048 | SIRPA |
| chr19 | 44945156 | 44946044 | 889 | 11 | 7.53E-06 | 1.80E-05 | 0.005153 | 1.17E-06 | -0.01534947 | APOC4, APOC4-APOC2, APOC2 |
| chr16 | 75861508 | 75862051 | 544 | 2 | 1.26E-06 | 7.08E-07 | 0.000188 | 1.17E-06 | -0.01939342 | NA |
| chr22 | 49918314 | 49919638 | 1325 | 11 | 1.27E-05 | 3.70E-05 | 0.000514 | 1.17E-06 | 0.003308364 | CRELD2, ALG12 |
| chr1 | 38153305 | 38153896 | 592 | 3 | 2.29E-08 | 7.05E-05 | 3.44E-06 | 1.17E-06 | -0.02682 | AL390839.1 |
| chr3 | 127591448 | 127592326 | 879 | 4 | 1.09E-08 | 8.38E-06 | 1.86E-05 | 1.17E-06 | -0.01363098 | TPRA1 |
| chr1 | 99715665 | 99716013 | 349 | 2 | 1.29E-07 | 1.76E-06 | 1.71E-05 | 1.17E-06 | -0.01465466 | FRRS1 |
| chr15 | 77709403 | 77710575 | 1173 | 3 | 2.69E-05 | 0.0002533 | 9.89E-05 | 1.17E-06 | -0.01034891 | LINGO1 |
| chr9 | 126054517 | 126054621 | 105 | 2 | 1.62E-06 | 8.34E-07 | 8.03E-05 | 1.17E-06 | 0.000830805 | NA |
| chr9 | 123303431 | 123304043 | 613 | 2 | 4.08E-08 | 1.24E-05 | 3.01E-06 | 1.17E-06 | -0.00263463 | NA |
| chr5 | 86410006 | 86410335 | 330 | 5 | 1.21E-06 | 1.88E-05 | 0.00032 | 1.17E-06 | -0.00291823 | NA |
| chr1 | 158253723 | 158254144 | 422 | 4 | 1.34E-07 | 7.61E-06 | 1.46E-05 | 1.17E-06 | -0.00555183 | NA |
| chr5 | 172235231 | 172236550 | 1320 | 3 | 2.93E-09 | 3.50E-05 | 1.89E-06 | 1.17E-06 | -0.00609652 | UBTD2 |
| chr18 | 24137722 | 24139322 | 1601 | 17 | 9.54E-10 | 0.0057039 | 1.69E-05 | 1.17E-06 | -0.00048284 | CABYR |
| chr7 | 1232289 | 1233074 | 786 | 7 | 4.12E-07 | 1.15E-06 | 0.000301 | 1.17E-06 | -6.32E-05 | UNCX |
| chr7 | 45239576 | 45240309 | 734 | 2 | 3.36E-06 | 8.80E-07 | 6.74E-05 | 1.17E-06 | 0.007229981 | NA |
| chr8 | 33472776 | 33473449 | 674 | 13 | 5.12E-07 | 3.30E-05 | 0.001859 | 1.18E-06 | -0.0001773 | FUT10, TTI2 |
| chr5 | 10701327 | 10702234 | 908 | 4 | 4.51E-08 | 2.18E-05 | 1.71E-05 | 1.18E-06 | -0.00499081 | DAP |
| chr1 | 6056951 | 6057099 | 149 | 2 | 1.72E-06 | 1.12E-06 | 3.70E-05 | 1.18E-06 | -0.00917431 | KCNAB2 |
| chr14 | 101917671 | 101917740 | 70 | 2 | 6.71E-07 | 7.25E-07 | 0.000168 | 1.18E-06 | -0.00477212 | PPP2R5C |
| chr1 | 47776740 | 47776985 | 246 | 2 | 2.28E-06 | 7.23E-07 | 0.000172 | 1.18E-06 | -0.00191568 | TRABD2B |
| chr8 | 40152924 | 40153827 | 904 | 6 | 4.63E-06 | 7.44E-06 | 0.001072 | 1.18E-06 | -0.00526575 | TCIM |
| chr15 | 34238540 | 34239517 | 978 | 4 | 7.47E-07 | 5.52E-06 | 9.50E-05 | 1.18E-06 | -0.00246138 | SLC12A6 |
| chr4 | 153131688 | 153131756 | 69 | 2 | 2.37E-06 | 7.21E-07 | 0.00018 | 1.18E-06 | -0.00665507 | NA |
| chr21 | 39727535 | 39727664 | 130 | 3 | 6.97E-07 | 7.99E-06 | 7.76E-06 | 1.18E-06 | -0.00692466 | NA |
| chr16 | 66924747 | 66926448 | 1702 | 12 | 9.27E-06 | 5.57E-05 | 0.002144 | 1.18E-06 | -0.00233504 | RRAD |
| chr11 | 76704640 | 76705465 | 826 | 4 | 3.10E-08 | 2.03E-05 | 3.96E-06 | 1.18E-06 | -0.00710983 | GUCY2EP |
| chr7 | 4265147 | 4266096 | 950 | 6 | 2.64E-06 | 1.92E-05 | 0.000936 | 1.19E-06 | 0.015798481 | SDK1 |
| chr12 | 132329025 | 132330477 | 1453 | 9 | 4.31E-06 | 0.0001054 | 0.000415 | 1.19E-06 | 0.000875331 | AC148477.1, GALNT9 |
| chr5 | 35118095 | 35118757 | 663 | 5 | 2.09E-07 | 3.93E-06 | 5.34E-05 | 1.19E-06 | -0.00814475 | PRLR |
| chr13 | 21558184 | 21558511 | 328 | 2 | 1.52E-06 | 9.48E-07 | 5.62E-05 | 1.19E-06 | -0.02060642 | MICU2 |
| chr11 | 78257196 | 78257637 | 442 | 3 | 5.01E-07 | 4.26E-06 | 3.01E-05 | 1.19E-06 | -0.01052782 | GAB2 |
| chr3 | 194313565 | 194313729 | 165 | 3 | 3.43E-07 | 1.73E-06 | 0.000113 | 1.19E-06 | -0.00665096 | LINC00887 |
| chr19 | 37718479 | 37719858 | 1380 | 12 | 1.75E-08 | 0.0002195 | 0.000554 | 1.19E-06 | -0.0037637 | AC093227.2, ZNF607 |
| chr17 | 82184733 | 82185307 | 575 | 3 | 1.51E-06 | 7.47E-07 | 0.000443 | 1.19E-06 | -0.00515689 | CCDC57 |
| chr2 | 176490344 | 176490876 | 533 | 4 | 6.42E-08 | 1.18E-05 | 4.71E-06 | 1.19E-06 | -0.00460884 | NA |
| chr6 | 33711360 | 33712784 | 1425 | 9 | 4.24E-06 | 1.87E-06 | 0.006217 | 1.19E-06 | -0.00398426 | UQCC2 |
| chr6 | 7728443 | 7729561 | 1119 | 6 | 1.13E-05 | 2.44E-07 | 0.003529 | 1.19E-06 | -0.00231046 | BMP6 |
| chr19 | 58450726 | 58451638 | 913 | 11 | 3.51E-08 | 2.59E-05 | 0.000715 | 1.19E-06 | -0.00244883 | ZNF324B |
| chr8 | 139854858 | 139855471 | 614 | 2 | 2.58E-06 | 7.59E-07 | 0.000135 | 1.19E-06 | -0.00463164 | TRAPPC9 |
| chr13 | 95552239 | 95553706 | 1468 | 15 | 3.70E-06 | 9.55E-07 | 0.006202 | 1.19E-06 | -0.0042722 | CLDN10 |
| chr8 | 119247919 | 119248345 | 427 | 3 | 1.28E-06 | 7.36E-07 | 0.000155 | 1.19E-06 | -0.01437022 | NA |
| chr10 | 18658535 | 18659470 | 936 | 10 | 1.25E-06 | 7.32E-07 | 0.000932 | 1.20E-06 | 0.005779061 | ARL5B, NSUN6 |
| chr21 | 15905562 | 15905883 | 322 | 2 | 1.05E-07 | 7.77E-05 | 1.42E-06 | 1.20E-06 | 0.018093769 | NA |
| chr11 | 47477312 | 47477827 | 516 | 2 | 8.52E-07 | 9.86E-07 | 5.17E-05 | 1.20E-06 | 0.001449565 | CELF1 |
| chr19 | 12736588 | 12737911 | 1324 | 15 | 1.70E-05 | 1.40E-06 | 0.004154 | 1.20E-06 | -0.00029281 | GET3 |
| chr7 | 108037509 | 108037986 | 478 | 4 | 1.26E-07 | 1.65E-05 | 6.97E-06 | 1.20E-06 | -0.00111666 | LAMB4 |
| chr20 | 25065418 | 25065817 | 400 | 2 | 1.38E-07 | 1.81E-05 | 2.50E-06 | 1.20E-06 | -0.00849786 | NA |
| chr11 | 196824 | 197507 | 684 | 3 | 3.48E-06 | 1.50E-06 | 8.48E-05 | 1.20E-06 | -0.01051312 | ODF3, BET1L |
| chr4 | 55125776 | 55126522 | 747 | 6 | 9.43E-07 | 3.30E-05 | 6.54E-05 | 1.20E-06 | 0.002367776 | NA |
| chr11 | 29444906 | 29445206 | 301 | 2 | 9.64E-07 | 1.74E-06 | 1.82E-05 | 1.20E-06 | -0.00564947 | LINC02755 |
| chr12 | 129872667 | 129872778 | 112 | 2 | 4.07E-07 | 8.77E-05 | 1.38E-06 | 1.20E-06 | -0.02633167 | TMEM132D |
| chr2 | 151408832 | 151410068 | 1237 | 13 | 9.23E-06 | 3.35E-06 | 0.001945 | 1.20E-06 | -0.00252115 | RIF1 |
| chr11 | 75513614 | 75514020 | 407 | 2 | 1.14E-06 | 1.12E-06 | 3.89E-05 | 1.20E-06 | -0.00423577 | GDPD5 |
| chr16 | 78925216 | 78925498 | 283 | 2 | 4.46E-07 | 7.41E-06 | 4.36E-06 | 1.20E-06 | -0.01375232 | WWOX |
| chr19 | 7899689 | 7900149 | 461 | 2 | 2.42E-07 | 1.93E-06 | 1.59E-05 | 1.20E-06 | 0.004931003 | LRRC8E, AC010336.1 |
| chr17 | 82636319 | 82637234 | 916 | 5 | 1.03E-07 | 1.30E-05 | 2.62E-05 | 1.20E-06 | -0.00659304 | FOXK2, WDR45B |
| chr17 | 43952359 | 43952919 | 561 | 2 | 2.47E-07 | 3.01E-06 | 9.43E-06 | 1.20E-06 | -0.01235409 | PYY |
| chr16 | 84301632 | 84302908 | 1277 | 7 | 1.03E-05 | 1.86E-05 | 0.000838 | 1.20E-06 | -0.0010973 | WFDC1 |
| chr8 | 143512793 | 143512829 | 37 | 2 | 4.12E-07 | 0.0001721 | 1.19E-06 | 1.21E-06 | 0.002986187 | ZC3H3 |
| chr17 | 18332429 | 18332773 | 345 | 3 | 1.20E-06 | 1.53E-05 | 5.51E-05 | 1.21E-06 | 0.000484769 | SHMT1 |
| chr10 | 10173008 | 10173218 | 211 | 2 | 1.00E-06 | 2.77E-06 | 1.04E-05 | 1.21E-06 | -0.00542988 | NA |
| chr17 | 67528680 | 67529119 | 440 | 2 | 7.03E-07 | 1.61E-06 | 2.08E-05 | 1.21E-06 | -0.01137377 | PITPNC1 |
| chr3 | 110893522 | 110894461 | 940 | 4 | 1.11E-07 | 1.30E-05 | 4.36E-06 | 1.21E-06 | -0.00648765 | NECTIN3-AS1 |
| chr16 | 15501997 | 15502566 | 570 | 10 | 8.12E-07 | 5.29E-05 | 0.000103 | 1.21E-06 | 0.001871977 | AC140504.1, BMERB1 |
| chr1 | 212284082 | 212284820 | 739 | 7 | 7.21E-09 | 0.0003851 | 5.31E-07 | 1.21E-06 | -0.0028587 | NA |
| chr7 | 838403 | 838877 | 475 | 5 | 1.12E-07 | 2.64E-05 | 4.15E-06 | 1.21E-06 | 0.000978357 | SUN1 |
| chr1 | 17638851 | 17639671 | 821 | 2 | 1.10E-05 | 7.75E-07 | 0.00013 | 1.21E-06 | -0.01263289 | ARHGEF10L |
| chr11 | 124611094 | 124612159 | 1066 | 5 | 1.04E-05 | 5.94E-07 | 0.001629 | 1.21E-06 | -0.00069508 | PANX3 |
| chr1 | 46543774 | 46544551 | 778 | 7 | 1.35E-07 | 8.50E-05 | 4.18E-05 | 1.21E-06 | -0.00433096 | MKNK1-AS1, TMEM275 |
| chr15 | 63246872 | 63247219 | 348 | 2 | 8.44E-07 | 1.67E-06 | 1.98E-05 | 1.21E-06 | -0.00757801 | RAB8B |
| chr6 | 116366866 | 116367061 | 196 | 2 | 5.10E-07 | 8.03E-06 | 4.15E-06 | 1.21E-06 | -0.0072254 | DSE |
| chr3 | 187519208 | 187519514 | 307 | 3 | 4.92E-07 | 3.58E-05 | 3.20E-05 | 1.21E-06 | 0.010927575 | NA |
| chr19 | 43774122 | 43774875 | 754 | 5 | 4.23E-07 | 0.0010091 | 0.000107 | 1.21E-06 | -0.01010373 | KCNN4 |
| chr14 | 104117761 | 104118695 | 935 | 4 | 5.97E-07 | 3.44E-07 | 0.001826 | 1.22E-06 | -0.00653806 | NA |
| chr7 | 44285730 | 44286437 | 708 | 3 | 3.24E-06 | 2.52E-06 | 0.000158 | 1.22E-06 | -3.19E-06 | CAMK2B |
| chr22 | 26164311 | 26164489 | 179 | 3 | 4.18E-07 | 1.20E-05 | 4.87E-06 | 1.22E-06 | -0.01024653 | AL022337.1 |
| chr5 | 173455652 | 173456340 | 689 | 2 | 1.04E-06 | 1.01E-06 | 5.16E-05 | 1.22E-06 | -0.00096413 | NA |
| chr11 | 44557175 | 44557251 | 77 | 2 | 1.73E-06 | 7.33E-07 | 0.000203 | 1.22E-06 | -0.00966118 | NA |
| chr3 | 195906942 | 195908094 | 1153 | 7 | 3.52E-05 | 4.38E-07 | 0.003347 | 1.22E-06 | 0.000147642 | TNK2-AS1, TNK2 |
| chr9 | 70869023 | 70869771 | 749 | 8 | 1.54E-07 | 8.59E-05 | 4.94E-05 | 1.22E-06 | -0.00497869 | TRPM3 |
| chr13 | 110711505 | 110713392 | 1888 | 16 | 1.19E-05 | 2.29E-05 | 0.001587 | 1.22E-06 | -0.00136566 | ING1, CARS2 |
| chr12 | 55829618 | 55831332 | 1715 | 14 | 1.16E-06 | 1.71E-05 | 0.001655 | 1.22E-06 | 0.000115324 | TMEM198B, DNAJC14 |
| chr16 | 662147 | 663101 | 955 | 10 | 2.35E-06 | 0.0002773 | 0.001412 | 1.22E-06 | -0.00110153 | WDR90 |
| chr17 | 3635916 | 3637046 | 1131 | 15 | 2.31E-07 | 1.24E-05 | 0.000614 | 1.22E-06 | 0.000801919 | CTNS, AC027796.3, SHPK |
| chr16 | 405931 | 406384 | 454 | 3 | 3.54E-08 | 0.0001811 | 2.98E-06 | 1.22E-06 | -7.97E-05 | NME4, DECR2 |
| chr1 | 156496937 | 156497396 | 460 | 3 | 1.24E-07 | 1.23E-05 | 6.12E-06 | 1.22E-06 | 0.008748798 | MEF2D |
| chr1 | 154182819 | 154183944 | 1126 | 11 | 5.31E-07 | 0.0007239 | 7.11E-05 | 1.22E-06 | 0.001990799 | TPM3 |
| chr7 | 45027051 | 45027720 | 670 | 8 | 3.36E-06 | 4.45E-07 | 0.005514 | 1.22E-06 | -0.00015555 | CCM2 |
| chr8 | 94262459 | 94263347 | 889 | 5 | 2.29E-05 | 3.30E-07 | 0.001606 | 1.22E-06 | -0.00300832 | NA |
| chr15 | 44955744 | 44956734 | 991 | 7 | 4.71E-07 | 5.18E-06 | 0.000108 | 1.22E-06 | -0.00346664 | TERB2 |
| chr15 | 60374376 | 60374901 | 526 | 3 | 5.05E-09 | 0.0009896 | 8.02E-07 | 1.23E-06 | -0.00354217 | ANXA2 |
| chr11 | 19447331 | 19447694 | 364 | 2 | 6.45E-08 | 0.0005135 | 1.03E-06 | 1.23E-06 | -0.02214934 | NAV2 |
| chr16 | 31487777 | 31489637 | 1861 | 10 | 1.09E-06 | 8.92E-06 | 0.003791 | 1.23E-06 | -0.0041175 | SLC5A2, AC026471.3, RUSF1 |
| chr18 | 304733 | 305256 | 524 | 2 | 8.59E-07 | 1.32E-06 | 2.97E-05 | 1.23E-06 | 0.005364662 | NA |
| chr4 | 37000395 | 37000932 | 538 | 3 | 1.17E-06 | 4.96E-07 | 0.00077 | 1.23E-06 | 0.010531784 | NA |
| chr11 | 86726307 | 86727223 | 917 | 4 | 8.47E-06 | 2.77E-05 | 0.000215 | 1.23E-06 | -0.00847773 | NA |
| chr8 | 138700563 | 138700641 | 79 | 2 | 3.22E-07 | 0.0005155 | 1.03E-06 | 1.23E-06 | -0.01306196 | COL22A1 |
| chr7 | 151805236 | 151806183 | 948 | 2 | 4.26E-05 | 7.45E-07 | 0.000193 | 1.23E-06 | 0.005682345 | PRKAG2 |
| chr14 | 51093416 | 51095094 | 1679 | 8 | 3.00E-07 | 1.12E-05 | 0.000165 | 1.23E-06 | 0.000525976 | TRIM9 |
| chr9 | 92974381 | 92975153 | 773 | 8 | 9.64E-08 | 8.20E-05 | 1.21E-05 | 1.23E-06 | 4.84E-05 | FGD3 |
| chr20 | 32856175 | 32856977 | 803 | 4 | 1.29E-06 | 2.36E-06 | 0.000135 | 1.23E-06 | 0.002254444 | AL035071.1 |
| chr6 | 41691622 | 41691920 | 299 | 4 | 1.86E-06 | 1.86E-06 | 0.000186 | 1.23E-06 | -0.00593168 | TFEB |
| chr8 | 132906193 | 132906449 | 257 | 3 | 1.41E-06 | 2.79E-06 | 0.000125 | 1.23E-06 | -0.00562138 | TG |
| chr9 | 133239331 | 133239911 | 581 | 4 | 2.13E-06 | 6.10E-07 | 0.001341 | 1.23E-06 | -0.00582522 | NA |
| chr3 | 50224813 | 50224976 | 164 | 2 | 6.63E-07 | 3.35E-06 | 8.77E-06 | 1.23E-06 | -0.00773731 | NA |
| chr12 | 81598315 | 81599158 | 844 | 4 | 3.18E-06 | 3.58E-07 | 0.001415 | 1.23E-06 | -0.01896389 | PPFIA2 |
| chr11 | 90319124 | 90319219 | 96 | 2 | 2.46E-06 | 7.61E-07 | 0.000171 | 1.23E-06 | -0.01025001 | DISC1FP1 |
| chr10 | 18260449 | 18261694 | 1246 | 14 | 2.38E-07 | 1.61E-05 | 0.00753 | 1.23E-06 | 0.009052346 | CACNB2, AL390783.1 |
| chr2 | 239734508 | 239734917 | 410 | 3 | 8.99E-07 | 1.80E-06 | 8.11E-05 | 1.24E-06 | -0.00740808 | NA |
| chr6 | 25652019 | 25652587 | 569 | 12 | 9.06E-07 | 3.85E-05 | 0.001263 | 1.24E-06 | -0.00157084 | SCGN |
| chr10 | 133456085 | 133457327 | 1243 | 7 | 1.40E-06 | 1.12E-05 | 0.00025 | 1.24E-06 | -0.0029087 | SCART1 |
| chr2 | 75701390 | 75702387 | 998 | 3 | 1.01E-05 | 3.85E-05 | 3.23E-05 | 1.24E-06 | 0.000203181 | GCFC2 |
| chr9 | 134594976 | 134594984 | 9 | 2 | 3.31E-07 | 0.0006134 | 1.02E-06 | 1.24E-06 | -0.00515456 | NA |
| chr13 | 30667958 | 30668565 | 608 | 2 | 5.76E-07 | 1.74E-06 | 1.93E-05 | 1.24E-06 | -0.00248495 | NA |
| chr19 | 7862084 | 7863180 | 1097 | 5 | 9.67E-07 | 1.29E-06 | 0.001491 | 1.24E-06 | -0.00204781 | EVI5L |
| chr22 | 47033188 | 47033761 | 574 | 3 | 1.90E-07 | 1.55E-06 | 7.73E-05 | 1.24E-06 | 0.001874082 | TBC1D22A |
| chr7 | 157849045 | 157849133 | 89 | 2 | 1.08E-06 | 2.39E-06 | 1.29E-05 | 1.24E-06 | -0.00576542 | PTPRN2 |
| chr3 | 46743340 | 46744467 | 1128 | 3 | 1.05E-06 | 4.39E-06 | 2.95E-05 | 1.24E-06 | -4.05E-05 | AC109583.1, PRSS45P, AC109583.4 |
| chr8 | 144408903 | 144409980 | 1078 | 12 | 1.09E-06 | 2.43E-05 | 0.002804 | 1.24E-06 | 0.00181384 | AC233992.3, CPSF1, SLC39A4 |
| chr13 | 107531377 | 107531930 | 554 | 3 | 1.53E-06 | 6.70E-07 | 0.000263 | 1.25E-06 | -0.00611598 | FAM155A |
| chr12 | 94009594 | 94009632 | 39 | 2 | 2.13E-07 | 1.21E-05 | 3.28E-06 | 1.25E-06 | 0.002572896 | NA |
| chr14 | 100663082 | 100663808 | 727 | 3 | 9.55E-08 | 1.49E-05 | 3.79E-06 | 1.25E-06 | 0.005094362 | LINC00523 |
| chr10 | 102484559 | 102485141 | 583 | 3 | 1.22E-06 | 1.22E-06 | 0.000198 | 1.25E-06 | -0.00032672 | ACTR1A |
| chr12 | 4224328 | 4224705 | 378 | 3 | 1.98E-06 | 1.13E-06 | 6.64E-05 | 1.25E-06 | 0.009707509 | NA |
| chr18 | 77016882 | 77017595 | 714 | 14 | 3.14E-06 | 1.12E-06 | 0.012693 | 1.25E-06 | 0.00069892 | MBP |
| chr20 | 46088824 | 46090089 | 1266 | 8 | 2.11E-05 | 0.0011671 | 0.000367 | 1.25E-06 | 0.000816738 | NCOA5 |
| chr8 | 23353704 | 23354409 | 706 | 6 | 2.75E-07 | 6.05E-06 | 9.21E-05 | 1.25E-06 | -0.00118535 | AC090197.1, LOXL2 |
| chr19 | 2151585 | 2152007 | 423 | 9 | 1.26E-06 | 3.80E-06 | 0.000683 | 1.25E-06 | -0.00076966 | AP3D1 |
| chr4 | 973644 | 974815 | 1172 | 10 | 1.19E-07 | 9.41E-05 | 9.12E-05 | 1.25E-06 | -0.00271391 | DGKQ |
| chr14 | 91332778 | 91333013 | 236 | 2 | 1.27E-07 | 8.73E-06 | 4.08E-06 | 1.25E-06 | -0.0099707 | CCDC88C |
| chr7 | 100886250 | 100887157 | 908 | 7 | 1.43E-06 | 1.14E-05 | 0.000551 | 1.25E-06 | -0.00322462 | SRRT |
| chr1 | 204928916 | 204929295 | 380 | 2 | 4.35E-07 | 7.84E-07 | 0.000153 | 1.25E-06 | -0.01246671 | NFASC |
| chr7 | 74258605 | 74258874 | 270 | 4 | 5.49E-07 | 0.0001627 | 8.78E-05 | 1.25E-06 | 0.00420721 | NA |
| chr14 | 68363583 | 68364096 | 514 | 4 | 1.39E-07 | 4.67E-06 | 0.000201 | 1.26E-06 | 0.017413183 | RAD51B |

| | | | | | | | | | | |
|---|---|---|---|---|---|---|---|---|---|---|
| chr9 | 112869899 | 112870457 | 559 | 3 | 1.17E-06 | 9.93E-07 | 0.000273 | 1.26E-06 | -0.00147343 | SNX30 |
| chr20 | 45355361 | 45355607 | 247 | 2 | 1.61E-06 | 1.18E-06 | 3.88E-05 | 1.26E-06 | -0.00768471 | NA |
| chr10 | 125343064 | 125343709 | 646 | 2 | 1.90E-06 | 1.03E-06 | 5.41E-05 | 1.26E-06 | -0.00145953 | NA |
| chr6 | 43307936 | 43309413 | 1478 | 13 | 8.78E-06 | 5.11E-07 | 0.003645 | 1.26E-06 | -0.00334987 | CRIP3, ZNF318 |
| chr9 | 133255796 | 133256320 | 525 | 2 | 6.75E-08 | 6.31E-06 | 5.21E-06 | 1.26E-06 | 0.006836902 | ABO |
| chr4 | 186012302 | 186012739 | 438 | 4 | 2.84E-07 | 1.34E-05 | 8.77E-06 | 1.26E-06 | -0.00388683 | NA |
| chr12 | 58874794 | 58875476 | 683 | 2 | 5.74E-06 | 7.84E-07 | 0.000158 | 1.26E-06 | 0.00288202 | LRIG3 |
| chr2 | 20538511 | 20539149 | 639 | 2 | 5.04E-06 | 7.63E-07 | 0.000197 | 1.26E-06 | -0.02607668 | NA |
| chr7 | 122315840 | 122317167 | 1328 | 6 | 4.46E-09 | 0.000331 | 0.00011 | 1.26E-06 | -0.00218519 | NA |
| chr11 | 78418618 | 78419411 | 794 | 4 | 6.28E-09 | 0.0003877 | 6.84E-07 | 1.26E-06 | -0.0048811 | AP003086.1 |
| chr15 | 72376009 | 72376808 | 800 | 12 | 3.22E-08 | 6.72E-05 | 0.000242 | 1.26E-06 | 0.001383782 | HEXA-AS1, HEXA |
| chr1 | 45510808 | 45511326 | 519 | 2 | 1.28E-07 | 1.74E-06 | 2.02E-05 | 1.26E-06 | 0.002436532 | MMACHC, PRDX1 |
| chr17 | 61411274 | 61412491 | 1218 | 10 | 2.57E-07 | 8.94E-05 | 0.000327 | 1.26E-06 | 0.001054925 | LINC02875, TBX2-AS1 |
| chr3 | 20185472 | 20186448 | 977 | 15 | 5.33E-07 | 5.21E-05 | 0.002547 | 1.26E-06 | 0.000572243 | SGO1-AS1, SGO1 |
| chr10 | 3119925 | 3119999 | 75 | 3 | 1.68E-06 | 8.10E-07 | 0.000193 | 1.26E-06 | -0.00360109 | PFKP |
| chr6 | 52670463 | 52671261 | 799 | 12 | 1.01E-06 | 3.31E-05 | 0.000168 | 1.27E-06 | 0.001826845 | TMEM14A |
| chr17 | 3673306 | 3673774 | 469 | 3 | 1.00E-06 | 1.36E-06 | 0.000291 | 1.27E-06 | -0.01071029 | P2RX5-TAX1BP3, P2RX5 |
| chr6 | 32212085 | 32213248 | 1164 | 8 | 1.35E-09 | 3.52E-05 | 5.09E-06 | 1.27E-06 | -0.00115433 | NOTCH4 |
| chr16 | 14435857 | 14435980 | 124 | 2 | 3.10E-07 | 0.0004433 | 1.08E-06 | 1.27E-06 | 0.005586253 | PARN |
| chr6 | 152953770 | 152954318 | 549 | 2 | 2.57E-06 | 7.71E-07 | 0.000193 | 1.27E-06 | -0.00255346 | NA |
| chr6 | 88145266 | 88146401 | 1136 | 7 | 6.29E-07 | 5.83E-06 | 0.001071 | 1.27E-06 | -0.00255398 | CNR1 |
| chr12 | 44972703 | 44973166 | 464 | 2 | 8.78E-07 | 1.43E-06 | 2.80E-05 | 1.27E-06 | -0.00051297 | NA |
| chr7 | 30910598 | 30912392 | 1795 | 16 | 5.32E-09 | 0.0018046 | 8.33E-05 | 1.27E-06 | -0.00138317 | AC004691.2, AQP1 |
| chr6 | 3350667 | 3351511 | 845 | 3 | 1.35E-07 | 2.43E-05 | 7.88E-06 | 1.27E-06 | 0.001540729 | SLC22A23 |
| chr22 | 50456922 | 50457933 | 1012 | 5 | 2.73E-07 | 1.09E-05 | 9.64E-05 | 1.27E-06 | 0.001837011 | SBF1 |
| chr20 | 37178974 | 37180225 | 1252 | 18 | 3.03E-06 | 7.36E-05 | 0.001111 | 1.27E-06 | 9.17E-05 | RPN2, MROH8 |
| chr1 | 10917869 | 10918539 | 671 | 2 | 1.52E-06 | 8.38E-07 | 0.000115 | 1.27E-06 | 0.017459417 | NA |
| chr11 | 94763501 | 94764308 | 808 | 2 | 4.78E-06 | 8.20E-07 | 0.000129 | 1.27E-06 | -0.00340736 | AMOTL1 |
| chr16 | 22813168 | 22814448 | 1281 | 12 | 4.73E-07 | 6.19E-06 | 0.00133 | 1.27E-06 | -0.00108845 | HS3ST2, AC130466.1 |
| chr1 | 207052767 | 207053731 | 965 | 15 | 3.18E-07 | 0.0002825 | 0.001349 | 1.27E-06 | 0.00097283 | PFKFB2, YOD1 |
| chr10 | 73966003 | 73966329 | 327 | 2 | 4.48E-07 | 5.76E-06 | 5.70E-06 | 1.27E-06 | -0.00921984 | NA |
| chr2 | 118309976 | 118310921 | 946 | 8 | 2.42E-06 | 8.96E-06 | 0.000199 | 1.28E-06 | 0.000820159 | NA |
| chr4 | 67545679 | 67546247 | 569 | 5 | 3.36E-07 | 1.41E-06 | 0.001029 | 1.28E-06 | -0.0080919 | NA |
| chr2 | 241573531 | 241573943 | 413 | 3 | 6.29E-07 | 8.25E-06 | 1.28E-05 | 1.28E-06 | -0.00376545 | BOK |
| chr3 | 123800269 | 123801180 | 912 | 4 | 1.09E-06 | 6.42E-06 | 0.000287 | 1.28E-06 | -0.00715557 | MYLK |
| chr1 | 9449362 | 9449422 | 61 | 2 | 6.79E-07 | 3.39E-06 | 9.16E-06 | 1.28E-06 | 0.00943689 | NA |
| chr3 | 149464323 | 149464595 | 273 | 2 | 2.98E-07 | 2.64E-05 | 2.25E-06 | 1.28E-06 | -0.00461333 | AC108751.3 |
| chr19 | 1244376 | 1244965 | 590 | 4 | 4.74E-08 | 4.04E-05 | 5.77E-05 | 1.28E-06 | 0.01097834 | ATP5F1D |
| chr5 | 32713358 | 32714419 | 1062 | 6 | 2.53E-06 | 1.10E-06 | 0.000328 | 1.28E-06 | 0.000468326 | NPR3 |
| chr15 | 85588396 | 85588960 | 565 | 3 | 2.43E-07 | 8.13E-06 | 7.34E-06 | 1.28E-06 | -0.01722999 | AKAP13 |
| chr2 | 27071230 | 27072252 | 1023 | 10 | 1.88E-06 | 1.57E-06 | 0.002951 | 1.28E-06 | 0.000665426 | OST4 |
| chr6 | 166803371 | 166803733 | 363 | 5 | 3.17E-07 | 0.0002747 | 8.27E-05 | 1.28E-06 | -0.00522875 | RPS6KA2 |
| chr15 | 99434127 | 99435129 | 1003 | 6 | 1.75E-05 | 1.77E-06 | 0.001775 | 1.28E-06 | 0.010851416 | AC015660.1 |
| chr9 | 128120411 | 128120421 | 11 | 2 | 1.18E-07 | 1.92E-05 | 2.63E-06 | 1.28E-06 | -0.00466252 | NA |
| chr2 | 44929503 | 44930474 | 972 | 4 | 4.50E-07 | 1.79E-05 | 0.00015 | 1.28E-06 | -0.00529151 | LINC01833 |
| chr7 | 143959060 | 143960084 | 1025 | 4 | 2.17E-07 | 2.68E-06 | 5.89E-05 | 1.29E-06 | -0.00067281 | OR2F1 |
| chr3 | 158732283 | 158733569 | 1287 | 10 | 2.59E-05 | 2.94E-05 | 0.000392 | 1.29E-06 | -0.00199734 | MFSD1, AC080013.1, RARRES1 |
| chr1 | 3313015 | 3313860 | 846 | 6 | 1.48E-06 | 2.70E-07 | 0.003705 | 1.29E-06 | 0.012230849 | PRDM16, AL354743.2 |
| chr17 | 61615871 | 61616140 | 270 | 2 | 2.98E-07 | 2.87E-05 | 2.18E-06 | 1.29E-06 | -0.0334467 | NA |
| chr1 | 26031465 | 26031761 | 297 | 2 | 7.05E-07 | 1.77E-06 | 2.03E-05 | 1.29E-06 | -0.00342526 | EXTL1 |
| chr3 | 196142188 | 196143138 | 951 | 6 | 2.01E-06 | 8.32E-06 | 0.000133 | 1.29E-06 | -0.00030288 | LINC00885 |
| chr3 | 184230270 | 184231155 | 886 | 8 | 2.10E-07 | 9.48E-05 | 0.000111 | 1.29E-06 | 0.002873337 | VWA5B2 |
| chr9 | 136540612 | 136541719 | 1108 | 3 | 1.66E-05 | 9.67E-07 | 0.000389 | 1.29E-06 | -0.00698038 | NOTCH1 |
| chr5 | 179678621 | 179679332 | 712 | 5 | 1.34E-06 | 1.07E-06 | 0.000807 | 1.29E-06 | -0.00348529 | CANX, HMGB3P22, CBY3 |
| chr5 | 132470901 | 132471182 | 282 | 2 | 9.51E-07 | 1.23E-06 | 3.87E-05 | 1.29E-06 | -0.0048187 | IRF1-AS1, AC116366.2, AC116366.1 |
| chr8 | 141161138 | 141161757 | 620 | 2 | 4.48E-07 | 1.40E-06 | 3.03E-05 | 1.30E-06 | -0.01611974 | DENND3 |
| chr1 | 226827717 | 226828323 | 607 | 9 | 1.65E-07 | 0.0006842 | 5.87E-05 | 1.30E-06 | -0.00133722 | AL359732.1 |
| chr12 | 122182113 | 122183490 | 1378 | 9 | 2.13E-05 | 1.05E-05 | 0.001723 | 1.30E-06 | 0.000299692 | LRRC43 |
| chr1 | 31408709 | 31409788 | 1080 | 6 | 1.47E-05 | 2.08E-05 | 0.00058 | 1.30E-06 | -1.68E-05 | SERINC2 |
| chr5 | 41112100 | 41112451 | 352 | 3 | 1.62E-07 | 3.99E-05 | 1.66E-05 | 1.30E-06 | -0.0055756 | NA |
| chr7 | 155931533 | 155931566 | 34 | 2 | 2.66E-06 | 7.82E-07 | 0.000205 | 1.30E-06 | -0.02031368 | NA |
| chr3 | 9403245 | 9403704 | 460 | 2 | 4.61E-07 | 1.94E-06 | 1.81E-05 | 1.30E-06 | -0.00890059 | SETD5 |
| chr1 | 225444944 | 225445187 | 244 | 3 | 1.00E-06 | 3.94E-06 | 1.64E-05 | 1.30E-06 | 0.00017282 | NA |
| chr8 | 26189876 | 26190898 | 1023 | 7 | 1.18E-06 | 1.14E-06 | 0.003213 | 1.30E-06 | -0.00827928 | NA |
| chr6 | 32971415 | 32972881 | 1467 | 38 | 4.64E-07 | 1.42E-05 | 0.000992 | 1.30E-06 | 0.000466805 | BRD2, AL645941.3 |
| chr11 | 17542878 | 17543149 | 272 | 2 | 5.01E-09 | 3.99E-05 | 1.92E-06 | 1.30E-06 | -0.0064969 | USH1C |
| chr2 | 200864110 | 200864960 | 851 | 14 | 2.34E-07 | 6.37E-05 | 5.07E-05 | 1.30E-06 | -0.00083947 | CLK1 |
| chr20 | 21516542 | 21517462 | 921 | 4 | 7.22E-08 | 4.44E-05 | 3.18E-05 | 1.30E-06 | 0.001106162 | NA |
| chr5 | 54049860 | 54049934 | 75 | 2 | 1.37E-06 | 3.86E-06 | 8.27E-06 | 1.30E-06 | -0.01614373 | ARL15 |
| chr7 | 83016592 | 83016785 | 194 | 3 | 2.72E-06 | 1.00E-06 | 0.000383 | 1.30E-06 | -0.00354915 | PCLO |
| chr1 | 39491050 | 39491728 | 679 | 6 | 1.92E-06 | 2.29E-06 | 0.000711 | 1.30E-06 | 1.27E-05 | BMP8A |
| chr19 | 4607045 | 4607358 | 314 | 7 | 4.57E-06 | 4.25E-07 | 0.001757 | 1.30E-06 | -0.00620176 | NA |
| chr5 | 66209353 | 66210079 | 727 | 3 | 2.61E-06 | 5.27E-07 | 0.000631 | 1.30E-06 | -0.00631464 | LINC02065 |
| chr12 | 13100808 | 13101739 | 932 | 7 | 1.08E-06 | 5.25E-07 | 0.001548 | 1.31E-06 | 0.000710597 | FAM234B, GSG1 |
| chr10 | 97640036 | 97640711 | 676 | 12 | 2.37E-08 | 3.51E-05 | 0.000109 | 1.31E-06 | 6.02E-05 | AL355315.1, PI4K2A |
| chr13 | 23411528 | 23411985 | 458 | 3 | 4.88E-07 | 1.03E-05 | 6.16E-06 | 1.31E-06 | -0.0009386 | SACS |
| chr8 | 38359563 | 38359725 | 163 | 2 | 2.42E-07 | 2.10E-05 | 2.56E-06 | 1.31E-06 | 0.016086521 | NSD3 |
| chr4 | 2425223 | 2425647 | 425 | 3 | 3.51E-07 | 1.57E-06 | 4.31E-05 | 1.31E-06 | -0.00581172 | CFAP99 |
| chr6 | 47203822 | 47204079 | 258 | 3 | 5.80E-07 | 2.02E-06 | 3.16E-05 | 1.31E-06 | -0.00121695 | NA |
| chr12 | 132493191 | 132494711 | 1521 | 5 | 1.13E-05 | 9.58E-05 | 6.16E-05 | 1.31E-06 | -0.00391013 | FBRSL1 |
| chr1 | 10530424 | 10530615 | 192 | 2 | 1.05E-06 | 2.70E-06 | 1.22E-05 | 1.31E-06 | -0.02069866 | PEX14 |
| chr22 | 31947464 | 31947738 | 275 | 2 | 2.87E-07 | 5.33E-06 | 6.29E-06 | 1.31E-06 | -0.00961522 | Z82190.2, YWHAH |
| chr12 | 124779275 | 124779572 | 298 | 3 | 3.74E-07 | 1.38E-05 | 6.71E-06 | 1.31E-06 | -5.46E-05 | SCARB1 |
| chr4 | 135888098 | 135888732 | 635 | 2 | 2.29E-08 | 4.69E-05 | 1.83E-06 | 1.31E-06 | -0.01409268 | LINC00613 |
| chr3 | 14297421 | 14298200 | 780 | 4 | 3.77E-06 | 3.73E-07 | 0.002138 | 1.31E-06 | -0.02034203 | AC093496.1 |
| chr13 | 41136716 | 41136784 | 69 | 3 | 1.66E-06 | 6.23E-06 | 3.66E-05 | 1.31E-06 | -0.00344122 | AL354696.2 |
| chr17 | 81265772 | 81266103 | 332 | 3 | 4.10E-07 | 8.51E-05 | 2.73E-05 | 1.31E-06 | -0.00230382 | SLC38A10 |
| chr5 | 7746256 | 7746777 | 522 | 2 | 1.12E-07 | 2.75E-06 | 1.20E-05 | 1.31E-06 | 0.007444833 | ADCY2, AC093305.1 |
| chr12 | 119592168 | 119593929 | 1762 | 9 | 3.73E-06 | 9.21E-05 | 9.50E-05 | 1.31E-06 | -0.00033618 | TMEM233, AC002070.1 |
| chr7 | 151350897 | 151351240 | 344 | 4 | 7.10E-07 | 7.26E-06 | 8.56E-05 | 1.31E-06 | -0.00942228 | NUB1 |
| chr5 | 139647606 | 139648569 | 964 | 7 | 3.46E-08 | 3.24E-05 | 6.24E-05 | 1.32E-06 | 0.002170553 | CXXC5 |
| chr7 | 157057798 | 157059080 | 1283 | 6 | 3.14E-05 | 1.33E-06 | 0.003488 | 1.32E-06 | -0.00527838 | AC006967.2 |
| chr19 | 35139747 | 35140269 | 523 | 3 | 1.22E-07 | 6.58E-05 | 5.88E-05 | 1.32E-06 | -0.01024477 | FXYD1, AC020907.2, LGI4 |
| chr2 | 42926277 | 42927045 | 769 | 5 | 1.60E-06 | 4.85E-07 | 0.002048 | 1.32E-06 | 0.006723137 | NA |
| chr7 | 16645648 | 16646686 | 1039 | 14 | 3.25E-06 | 6.88E-07 | 0.005982 | 1.32E-06 | -0.00051342 | BZW2, ANKMY2 |
| chr14 | 104474032 | 104474602 | 571 | 4 | 1.54E-06 | 6.95E-07 | 0.00144 | 1.32E-06 | -0.02278734 | NA |
| chr21 | 36245124 | 36246158 | 1035 | 4 | 3.29E-05 | 3.24E-06 | 0.000132 | 1.32E-06 | -0.00216894 | DOP1B |
| chr11 | 122885369 | 122886257 | 889 | 5 | 2.00E-06 | 5.83E-07 | 0.000866 | 1.32E-06 | -0.00614849 | JHY |
| chr19 | 58198032 | 58198518 | 487 | 2 | 6.72E-07 | 1.84E-06 | 2.02E-05 | 1.32E-06 | -0.00938288 | ZNF274 |
| chr16 | 48384007 | 48384522 | 516 | 3 | 2.79E-08 | 2.33E-05 | 3.85E-05 | 1.32E-06 | -0.00769549 | SIAH1 |
| chr1 | 44949451 | 44949737 | 287 | 2 | 2.48E-06 | 7.97E-07 | 0.000208 | 1.32E-06 | -0.00112885 | EIF2B3 |
| chr11 | 134416078 | 134417121 | 1044 | 3 | 1.82E-10 | 0.0016491 | 5.24E-07 | 1.33E-06 | -0.00617303 | B3GAT1-DT |
| chr13 | 34101841 | 34102301 | 461 | 3 | 1.04E-07 | 0.0003307 | 6.98E-06 | 1.33E-06 | -0.00230175 | NA |
| chr2 | 47525375 | 47525845 | 471 | 2 | 8.34E-07 | 9.15E-07 | 9.40E-05 | 1.33E-06 | 0.000933621 | MSH2, KCNK12 |
| chr7 | 100157284 | 100158124 | 841 | 9 | 5.59E-07 | 5.17E-06 | 0.000971 | 1.33E-06 | -0.00107475 | MAP11 |
| chr5 | 57482175 | 57483293 | 1119 | 7 | 1.09E-06 | 4.51E-05 | 2.75E-05 | 1.33E-06 | -0.00390503 | AC025470.2, ACTBL2 |
| chr17 | 43766599 | 43767048 | 450 | 4 | 6.39E-08 | 0.0114235 | 6.70E-07 | 1.33E-06 | -0.0068272 | DUSP3 |
| chr6 | 43297601 | 43298108 | 508 | 4 | 5.13E-07 | 1.00E-06 | 0.000585 | 1.33E-06 | -0.00819838 | SLC22A7 |
| chr1 | 164320930 | 164321596 | 667 | 8 | 2.40E-07 | 8.57E-05 | 7.64E-05 | 1.33E-06 | 0.004626422 | NA |
| chr6 | 34358697 | 34359101 | 405 | 2 | 6.00E-08 | 0.0002807 | 1.20E-06 | 1.33E-06 | 0.006787655 | NUDT3, RPS10-NUDT3 |
| chr19 | 18791351 | 18792729 | 1379 | 9 | 1.06E-08 | 0.0001266 | 0.000926 | 1.33E-06 | 0.000170156 | NA |
| chr19 | 40466127 | 40467464 | 1338 | 8 | 1.79E-07 | 4.29E-06 | 0.000176 | 1.33E-06 | 0.003937686 | SPTBN4 |
| chr17 | 20156098 | 20157144 | 1047 | 6 | 3.80E-09 | 0.0011659 | 0.000119 | 1.34E-06 | -0.00024274 | SPECC1 |
| chr1 | 178544765 | 178544903 | 139 | 2 | 1.75E-07 | 9.64E-05 | 1.50E-06 | 1.34E-06 | -0.00579098 | TEX35, C1orf220 |
| chr9 | 120928308 | 120928885 | 578 | 6 | 3.58E-08 | 1.19E-05 | 0.001047 | 1.34E-06 | -0.00845415 | TRAF1 |
| chr1 | 14959060 | 14959285 | 226 | 2 | 9.09E-07 | 8.68E-07 | 0.000126 | 1.34E-06 | 0.018095492 | KAZN |
| chr8 | 22134135 | 22134847 | 713 | 3 | 5.94E-07 | 6.91E-07 | 0.000393 | 1.34E-06 | -0.00806851 | NA |
| chr9 | 21402507 | 21403133 | 627 | 6 | 2.09E-06 | 1.09E-05 | 0.000225 | 1.34E-06 | 0.00151535 | IFNA12P |
| chr19 | 58581245 | 58581905 | 661 | 5 | 3.09E-06 | 4.90E-07 | 0.002401 | 1.34E-06 | -0.00975002 | CENPBD1P1, AC016629.3 |
| chr12 | 45214908 | 45216006 | 1099 | 10 | 2.18E-08 | 3.30E-05 | 4.29E-05 | 1.34E-06 | 0.000259025 | ANO6, PLEKHA8P1 |
| chr2 | 73284549 | 73285060 | 512 | 5 | 2.11E-07 | 3.06E-05 | 1.77E-05 | 1.34E-06 | 0.001988194 | NA |
| chr3 | 42025304 | 42025824 | 521 | 2 | 4.77E-08 | 4.40E-06 | 7.64E-06 | 1.34E-06 | 0.019282941 | TRAK1 |
| chr6 | 40951318 | 40951320 | 3 | 2 | 9.33E-07 | 1.58E-05 | 3.07E-06 | 1.34E-06 | -0.01518563 | NA |
| chr6 | 30323572 | 30323991 | 420 | 3 | 1.02E-07 | 8.13E-06 | 7.21E-06 | 1.34E-06 | -0.00642325 | HCG17, HCG18 |
| chr2 | 67846467 | 67846506 | 40 | 2 | 2.24E-06 | 1.32E-06 | 3.62E-05 | 1.34E-06 | -0.00831342 | NA |
| chr12 | 112125291 | 112126261 | 971 | 12 | 1.56E-07 | 0.0005451 | 3.11E-05 | 1.34E-06 | 0.000647811 | TRAFD1 |
| chr3 | 196708950 | 196709588 | 639 | 4 | 3.07E-06 | 3.78E-07 | 0.002116 | 1.34E-06 | 0.002121043 | PIGX, CEP19, RNU6-646P |
| chr1 | 20358686 | 20359125 | 440 | 3 | 2.50E-07 | 1.33E-05 | 4.08E-05 | 1.34E-06 | -0.01548232 | NA |
| chr16 | 14957732 | 14957889 | 158 | 3 | 1.56E-06 | 1.98E-06 | 0.00017 | 1.35E-06 | -0.00119508 | NA |
| chr8 | 115563407 | 115563675 | 269 | 2 | 2.45E-06 | 8.62E-07 | 0.000137 | 1.35E-06 | 0.025303288 | TRPS1 |
| chr6 | 33413903 | 33414543 | 641 | 6 | 2.57E-07 | 0.0002441 | 2.51E-05 | 1.35E-06 | 0.001142073 | PHF1 |
| chr14 | 105498170 | 105499042 | 873 | 7 | 7.32E-07 | 6.12E-06 | 0.001931 | 1.35E-06 | -0.00141386 | TEDC1 |
| chr22 | 40358302 | 40358961 | 660 | 5 | 2.50E-06 | 2.79E-06 | 0.000657 | 1.35E-06 | -0.0028434 | ADSL, AL022238.3 |
| chr2 | 113062137 | 113062516 | 380 | 6 | 2.20E-06 | 1.15E-05 | 0.00025 | 1.35E-06 | -0.00183752 | IL36RN |
| chr17 | 81082737 | 81083692 | 956 | 6 | 2.21E-07 | 1.13E-05 | 0.000304 | 1.35E-06 | -0.00438525 | BAIAP2 |
| chr2 | 165890895 | 165891328 | 434 | 4 | 1.88E-06 | 4.45E-07 | 0.002174 | 1.35E-06 | -0.01095584 | TTC21B |
| chr13 | 90119911 | 90120744 | 834 | 3 | 9.28E-08 | 2.01E-05 | 3.80E-06 | 1.35E-06 | 0.003721653 | NA |
| chr1 | 16977939 | 16978767 | 829 | 5 | 9.49E-08 | 0.0001577 | 4.92E-05 | 1.35E-06 | -0.00525367 | MFAP2 |
| chr3 | 169966181 | 169966895 | 715 | 13 | 1.66E-07 | 3.68E-05 | 0.000718 | 1.36E-06 | -0.00543625 | SEC62, AC008040.1 |
| chr9 | 89359241 | 89359253 | 13 | 2 | 1.16E-06 | 1.03E-06 | 6.91E-05 | 1.36E-06 | -0.0141598 | SECISBP2 |
| chr1 | 1434413 | 1435395 | 983 | 9 | 1.15E-06 | 8.38E-07 | 0.001934 | 1.36E-06 | -0.00196709 | VWA1, LINC01770 |
| chr17 | 63838214 | 63838776 | 563 | 3 | 2.17E-07 | 1.03E-05 | 4.87E-05 | 1.36E-06 | -0.00685215 | SMARCD2 |
| chr7 | 142320364 | 142320557 | 194 | 2 | 2.12E-06 | 1.11E-06 | 5.61E-05 | 1.36E-06 | 0.005079033 | NA |
| chr3 | 179376215 | 179377015 | 801 | 3 | 4.85E-06 | 1.58E-06 | 6.93E-05 | 1.36E-06 | 0.000779953 | MFN1 |
| chr7 | 157539682 | 157541468 | 1787 | 13 | 2.88E-07 | 0.0032785 | 1.44E-05 | 1.36E-06 | -0.00029467 | PTPRN2 |
| chr3 | 159992492 | 159992919 | 428 | 2 | 1.19E-06 | 9.65E-07 | 8.62E-05 | 1.36E-06 | 0.000901532 | IL12A, IL12A-AS1 |
| chr14 | 103725841 | 103726742 | 902 | 6 | 4.92E-07 | 1.93E-05 | 0.000259 | 1.36E-06 | -0.0003711 | ZFYVE21 |
| chr7 | 108007305 | 108007740 | 436 | 2 | 2.32E-06 | 9.09E-07 | 0.000109 | 1.36E-06 | -3.13E-06 | NA |
| chr1 | 210025218 | 210025724 | 507 | 2 | 2.10E-08 | 0.0021898 | 1.01E-06 | 1.36E-06 | -0.01150664 | SYT14 |
| chr6 | 10410683 | 10412253 | 1571 | 13 | 1.35E-07 | 9.49E-06 | 3.69E-05 | 1.36E-06 | -0.00096805 | TFAP2A-AS1, TFAP2A |
| chr18 | 61333265 | 61333925 | 661 | 7 | 1.58E-07 | 5.32E-06 | 0.000374 | 1.36E-06 | -0.00244771 | CDH20 |
| chr14 | 102544357 | 102544995 | 639 | 4 | 2.51E-07 | 2.87E-05 | 0.000166 | 1.36E-06 | -0.00798215 | NA |
| chr19 | 37345587 | 37346288 | 702 | 5 | 5.98E-06 | 0.0001094 | 0.000457 | 1.36E-06 | 0.003544783 | ZNF875 |
| chr18 | 52499204 | 52499403 | 200 | 2 | 2.63E-07 | 0.0002917 | 1.22E-06 | 1.36E-06 | -0.0064504 | DCC |
| chr17 | 79027025 | 79027184 | 160 | 2 | 5.82E-07 | 2.21E-06 | 1.68E-05 | 1.37E-06 | -0.0120543 | C1QTNF1, C1QTNF1-AS1 |
| chr17 | 74545793 | 74546779 | 987 | 6 | 1.18E-05 | 1.49E-07 | 0.010736 | 1.37E-06 | -0.00864096 | CD300C |
| chr6 | 167974764 | 167975645 | 882 | 5 | 1.97E-06 | 2.48E-06 | 7.49E-05 | 1.37E-06 | -0.00155863 | NA |
| chr1 | 14928702 | 14929985 | 1284 | 9 | 4.14E-07 | 0.0015773 | 3.46E-05 | 1.37E-06 | -0.00267866 | KAZN |

| | | | | | | | | | | |
|---|---|---|---|---|---|---|---|---|---|---|
| chr6 | 6057823 | 6058430 | 608 | 6 | 8.45E-07 | 2.87E-06 | 0.00041 | 1.37E-06 | 0.007748428 | NA |
| chr16 | 30112344 | 30113641 | 1298 | 6 | 3.80E-06 | 1.49E-05 | 9.25E-05 | 1.38E-06 | -0.00308712 | GDPD3 |
| chr16 | 2934406 | 2934767 | 362 | 5 | 5.89E-07 | 1.48E-05 | 8.76E-05 | 1.38E-06 | -0.00556756 | FLYWCH1 |
| chr2 | 174927717 | 174928097 | 381 | 3 | 3.83E-07 | 9.37E-06 | 7.01E-06 | 1.38E-06 | -0.00701306 | CHN1 |
| chr1 | 89150864 | 89150874 | 11 | 2 | 1.58E-06 | 1.17E-06 | 5.08E-05 | 1.38E-06 | -0.0006666 | GBP7 |
| chr6 | 132512660 | 132513775 | 1116 | 14 | 5.05E-08 | 0.0003685 | 7.81E-06 | 1.38E-06 | -0.00064709 | STX7 |
| chr3 | 66997227 | 66998830 | 1604 | 12 | 1.38E-05 | 2.88E-05 | 0.001711 | 1.38E-06 | -0.00317179 | KBTBD8, AC020655.1 |
| chr8 | 41490003 | 41490686 | 684 | 8 | 1.25E-06 | 1.87E-05 | 0.000427 | 1.38E-06 | 0.000346267 | GOLGA7 |
| chr22 | 30356318 | 30357153 | 836 | 10 | 4.67E-07 | 2.36E-05 | 0.002283 | 1.38E-06 | 0.001007241 | CCDC157, SF3A1 |
| chr19 | 6463938 | 6464874 | 937 | 11 | 2.80E-07 | 5.34E-06 | 0.000713 | 1.38E-06 | -0.00286633 | CRB3, SLC25A23 |
| chr14 | 94174785 | 94175444 | 660 | 2 | 6.30E-06 | 8.49E-07 | 0.000183 | 1.38E-06 | -0.00352042 | PPP4R4 |
| chr12 | 131217241 | 131217744 | 504 | 5 | 5.82E-07 | 1.29E-05 | 9.47E-06 | 1.38E-06 | -0.00494749 | LINC01257 |
| chr13 | 60012863 | 60013435 | 573 | 6 | 2.26E-06 | 6.02E-07 | 0.001763 | 1.38E-06 | -0.00125934 | DIAPH3-AS1, DIAPH3 |
| chr7 | 27175215 | 27175991 | 777 | 5 | 1.86E-05 | 6.99E-07 | 0.001181 | 1.39E-06 | -0.00298866 | AC004080.3, HOXA10 |
| chr18 | 35040565 | 35041784 | 1220 | 12 | 3.54E-06 | 4.60E-06 | 0.002252 | 1.39E-06 | 0.000574235 | MAPRE2 |
| chr8 | 132661697 | 132662328 | 632 | 4 | 1.78E-06 | 7.39E-05 | 5.14E-05 | 1.39E-06 | 0.002817492 | LRRC6 |
| chr1 | 3434415 | 3435980 | 1566 | 8 | 2.20E-08 | 4.18E-05 | 2.97E-05 | 1.39E-06 | -0.00073436 | PRDM16 |
| chr15 | 92821923 | 92822723 | 801 | 2 | 1.25E-05 | 8.60E-07 | 0.000174 | 1.39E-06 | -0.00317083 | CHASERR |
| chr14 | 24081636 | 24082080 | 445 | 7 | 1.01E-06 | 1.18E-06 | 0.000699 | 1.39E-06 | -0.006474 | NRL |
| chr1 | 5749028 | 5749401 | 374 | 4 | 2.83E-07 | 0.0009651 | 5.23E-06 | 1.39E-06 | -0.00583537 | NA |
| chr8 | 38757289 | 38758038 | 750 | 5 | 2.63E-07 | 5.76E-06 | 0.000262 | 1.39E-06 | -0.00450709 | TACC1 |
| chr19 | 30479106 | 30479385 | 280 | 2 | 3.05E-06 | 8.35E-07 | 0.000222 | 1.39E-06 | -0.01684736 | ZNF536 |
| chr3 | 36856903 | 36856959 | 57 | 3 | 4.67E-07 | 0.0001218 | 1.19E-06 | 1.39E-06 | -0.00113473 | TRANK1 |
| chr17 | 63621393 | 63622485 | 1093 | 10 | 1.97E-05 | 7.94E-06 | 0.003991 | 1.39E-06 | -0.00046716 | MAP3K3 |
| chr12 | 96303136 | 96304060 | 925 | 2 | 6.39E-05 | 8.35E-07 | 0.000222 | 1.39E-06 | -0.00449129 | CDK17, RNU4-24P |
| chr4 | 26161802 | 26162256 | 455 | 2 | 1.65E-07 | 1.20E-05 | 3.78E-06 | 1.40E-06 | -0.00902767 | NA |
| chr14 | 23539542 | 23539920 | 379 | 3 | 3.63E-07 | 4.19E-06 | 7.56E-05 | 1.40E-06 | -0.00669413 | ZFHX2-AS1, ZFHX2 |
| chr14 | 62116670 | 62117948 | 1279 | 15 | 1.38E-05 | 0.0001347 | 0.000407 | 1.40E-06 | -0.00357741 | LINC00643, AL390816.2 |
| chr10 | 35306999 | 35307367 | 369 | 2 | 6.27E-08 | 0.002504 | 1.02E-06 | 1.40E-06 | 0.01068047 | CCNY |
| chr10 | 43229708 | 43230694 | 987 | 9 | 3.20E-06 | 7.64E-06 | 0.000197 | 1.40E-06 | -0.00382438 | RASGEF1A |
| chr15 | 89017647 | 89018353 | 707 | 5 | 2.49E-07 | 6.20E-05 | 6.73E-05 | 1.40E-06 | -0.00663006 | NA |
| chr6 | 13634024 | 13634193 | 170 | 3 | 2.05E-06 | 2.81E-06 | 0.0001 | 1.40E-06 | -0.00629799 | RANBP9 |
| chr11 | 125454125 | 125454630 | 506 | 3 | 2.28E-07 | 2.03E-05 | 0.000103 | 1.40E-06 | -0.0014794 | FEZ1 |
| chr4 | 872064 | 873023 | 960 | 9 | 1.30E-07 | 2.07E-05 | 1.78E-05 | 1.40E-06 | -0.00023189 | GAK |
| chr4 | 183322561 | 183323678 | 1118 | 7 | 7.42E-08 | 0.000101 | 4.36E-06 | 1.40E-06 | -0.00145479 | NA |
| chr9 | 84340828 | 84341658 | 831 | 5 | 1.92E-06 | 1.36E-06 | 0.000508 | 1.40E-06 | 0.000633332 | AL157886.1 |
| chr10 | 13474135 | 13474746 | 612 | 2 | 6.83E-06 | 8.49E-07 | 0.000204 | 1.40E-06 | -0.00208499 | BEND7 |
| chr1 | 150561214 | 150561789 | 576 | 7 | 1.24E-07 | 6.52E-06 | 0.000602 | 1.40E-06 | 0.001647859 | ADAMTSL4-AS1, MCL1 |
| chr2 | 112884943 | 112885362 | 420 | 2 | 1.73E-07 | 4.05E-06 | 8.84E-06 | 1.40E-06 | -0.0160065 | NA |
| chr17 | 50133069 | 50133375 | 307 | 2 | 3.34E-07 | 1.54E-06 | 3.05E-05 | 1.40E-06 | 0.009609548 | NA |
| chr4 | 185015324 | 185016251 | 928 | 3 | 7.43E-07 | 1.16E-06 | 0.000101 | 1.41E-06 | 0.015138725 | NA |
| chr3 | 49276722 | 49277975 | 1254 | 13 | 1.23E-06 | 3.55E-05 | 0.001648 | 1.41E-06 | 0.001215051 | C3orf62, USP4 |
| chr2 | 111883820 | 111884586 | 767 | 10 | 7.43E-07 | 0.0001257 | 0.00057 | 1.41E-06 | 0.000285916 | ANAPC1 |
| chr5 | 14508206 | 14508836 | 631 | 5 | 9.99E-09 | 2.86E-05 | 5.54E-05 | 1.41E-06 | -0.00126712 | TRIO |
| chr12 | 133036847 | 133037728 | 882 | 9 | 3.48E-07 | 1.11E-05 | 0.00025 | 1.41E-06 | -0.00354905 | ZNF84, ZNF84-DT |
| chr22 | 50280996 | 50281057 | 62 | 3 | 4.77E-07 | 1.47E-06 | 7.31E-05 | 1.41E-06 | -0.00134031 | PLXNB2 |
| chr16 | 2798408 | 2799857 | 1450 | 8 | 1.29E-05 | 2.41E-05 | 0.00032 | 1.41E-06 | -0.01095046 | PRSS41 |
| chr16 | 20348122 | 20348649 | 528 | 3 | 2.30E-08 | 6.27E-05 | 0.000121 | 1.41E-06 | -0.00433657 | UMOD |
| chr3 | 9317177 | 9317230 | 54 | 2 | 1.62E-06 | 3.67E-06 | 9.81E-06 | 1.41E-06 | -0.01624249 | SRGAP3, AC026191.1 |
| chr15 | 80586403 | 80587153 | 751 | 2 | 1.16E-05 | 8.77E-07 | 0.000166 | 1.41E-06 | -0.00063866 | ARNT2 |
| chr9 | 4904699 | 4905203 | 505 | 2 | 5.06E-06 | 8.44E-07 | 0.000223 | 1.41E-06 | 0.016699234 | NA |
| chr11 | 125574558 | 125574580 | 23 | 2 | 2.75E-06 | 9.29E-07 | 0.000118 | 1.41E-06 | 0.000225688 | EI24, FEZ1 |
| chr1 | 101235808 | 101236963 | 1156 | 14 | 2.93E-06 | 3.78E-05 | 0.001237 | 1.41E-06 | 0.000336005 | S1PR1, AL109741.1 |
| chr15 | 59994940 | 59996205 | 1266 | 8 | 5.59E-07 | 5.94E-05 | 0.000172 | 1.41E-06 | 0.003093834 | NA |
| chr3 | 151340950 | 151341049 | 100 | 3 | 3.48E-06 | 7.46E-07 | 0.000238 | 1.41E-06 | 0.018468048 | MED12L, P2RY12 |
| chr1 | 16366984 | 16368380 | 1397 | 14 | 1.87E-05 | 0.0004236 | 6.35E-05 | 1.41E-06 | 0.001590843 | SZRD1 |
| chr6 | 12573274 | 12573795 | 522 | 2 | 9.90E-08 | 1.60E-05 | 3.24E-06 | 1.42E-06 | 0.007397476 | NA |
| chr8 | 139711341 | 139711795 | 455 | 3 | 1.70E-07 | 4.31E-05 | 4.45E-06 | 1.42E-06 | -0.00760578 | NA |
| chr19 | 51691976 | 51692306 | 331 | 4 | 4.17E-07 | 9.95E-05 | 0.000191 | 1.42E-06 | -0.00417995 | SPACA6, SPACA6P-AS |
| chr15 | 88881416 | 88881638 | 223 | 3 | 1.54E-06 | 1.38E-06 | 0.000123 | 1.42E-06 | -0.00248658 | HAPLN3 |
| chr20 | 56347771 | 56348152 | 382 | 2 | 2.87E-06 | 8.78E-07 | 0.000171 | 1.42E-06 | -0.06769333 | NA |
| chr5 | 177599189 | 177600588 | 1400 | 12 | 5.68E-07 | 4.33E-05 | 0.000688 | 1.42E-06 | 0.00137017 | B4GALT7 |
| chr11 | 116829333 | 116830328 | 996 | 6 | 5.72E-07 | 1.83E-05 | 0.000155 | 1.42E-06 | -0.00417703 | APOC3 |
| chr14 | 102316188 | 102316883 | 696 | 3 | 2.33E-07 | 7.74E-07 | 0.000246 | 1.42E-06 | 0.002199305 | NA |
| chr17 | 5184064 | 5185179 | 1116 | 5 | 2.90E-06 | 3.83E-06 | 9.87E-05 | 1.42E-06 | 0.002274681 | ZNF594 |
| chr15 | 58013967 | 58014339 | 373 | 3 | 6.95E-07 | 1.36E-06 | 0.000326 | 1.42E-06 | -0.01239213 | ALDH1A2 |
| chr11 | 32307898 | 32307982 | 85 | 2 | 8.51E-07 | 2.07E-05 | 2.86E-06 | 1.42E-06 | -0.00535204 | THEM7P |
| chr15 | 82950285 | 82950771 | 487 | 2 | 1.67E-08 | 1.11E-05 | 4.07E-06 | 1.42E-06 | -0.01289976 | AC022558.2, HOMER2 |
| chr17 | 38886870 | 38887510 | 641 | 3 | 8.24E-07 | 8.78E-07 | 0.00026 | 1.42E-06 | -0.00290826 | LASP1 |
| chr13 | 24042489 | 24042519 | 31 | 2 | 1.27E-06 | 7.32E-06 | 5.46E-06 | 1.42E-06 | 0.009793793 | AL359736.1, SPATA13 |
| chr1 | 168178361 | 168179816 | 1456 | 17 | 7.18E-06 | 1.59E-05 | 0.00202 | 1.43E-06 | 0.000988031 | TIPRL |
| chr3 | 143331508 | 143331649 | 142 | 2 | 5.15E-07 | 6.55E-05 | 1.78E-06 | 1.43E-06 | -0.00716279 | SLC9A9 |
| chr11 | 72583860 | 72585094 | 1235 | 8 | 4.82E-07 | 6.37E-05 | 0.000127 | 1.43E-06 | -0.00332849 | PDE2A-AS2, PDE2A |
| chr11 | 28757739 | 28758455 | 717 | 4 | 1.33E-07 | 3.44E-06 | 0.000183 | 1.43E-06 | -0.00496899 | LINC02742 |
| chr3 | 49015775 | 49016883 | 1109 | 4 | 1.88E-07 | 2.06E-06 | 6.46E-05 | 1.43E-06 | 0.004314133 | WDR6, DALRD3 |
| chr10 | 71318821 | 71319592 | 772 | 8 | 4.78E-06 | 9.41E-07 | 0.000909 | 1.43E-06 | -0.00249995 | SLC29A3 |
| chr16 | 75555674 | 75556447 | 774 | 8 | 2.32E-07 | 1.23E-05 | 0.00098 | 1.43E-06 | -0.00174484 | AC025287.2, TMEM231, AC025287.1 |
| chr19 | 47778751 | 47779978 | 1228 | 5 | 1.71E-07 | 0.0025402 | 2.47E-05 | 1.43E-06 | 0.000806334 | SELENOW |
| chr6 | 131183143 | 131183641 | 499 | 2 | 2.39E-06 | 9.94E-07 | 9.50E-05 | 1.43E-06 | -0.00708472 | AKAP7 |
| chr16 | 87346587 | 87347156 | 570 | 5 | 3.55E-08 | 2.56E-06 | 0.000328 | 1.43E-06 | -0.00235852 | FBXO31 |
| chr1 | 232180289 | 232180714 | 426 | 2 | 1.70E-07 | 6.05E-06 | 6.42E-06 | 1.44E-06 | -0.01148548 | AL353052.1 |
| chr10 | 61900311 | 61901737 | 1427 | 17 | 2.36E-06 | 1.15E-05 | 0.002659 | 1.44E-06 | -0.00020469 | ARID5B |
| chr22 | 29818228 | 29818230 | 3 | 2 | 1.34E-06 | 8.71E-07 | 0.000203 | 1.44E-06 | -0.00044571 | ASCC2 |
| chr12 | 45990501 | 45990883 | 383 | 8 | 3.15E-07 | 0.0002793 | 9.71E-05 | 1.44E-06 | -0.00052933 | SCAF11 |
| chr12 | 122986369 | 122987603 | 1235 | 5 | 8.53E-09 | 3.09E-05 | 4.96E-06 | 1.44E-06 | 0.001680867 | PITPNM2 |
| chr11 | 4693392 | 4693449 | 58 | 3 | 1.61E-06 | 4.30E-06 | 1.77E-05 | 1.44E-06 | -0.00798466 | OR51E2, OR51C1P |
| chr17 | 35734578 | 35735554 | 977 | 5 | 1.79E-06 | 1.69E-05 | 4.36E-05 | 1.44E-06 | 0.000222383 | TAF15, RASL10B |
| chr16 | 30052828 | 30053576 | 749 | 8 | 2.37E-08 | 0.0003719 | 1.83E-05 | 1.44E-06 | 0.001712958 | AC093512.2, TLCD3B |
| chr22 | 50251038 | 50252081 | 1044 | 13 | 1.89E-07 | 0.0001318 | 0.000158 | 1.44E-06 | -0.00132158 | HDAC10, MAPK12 |
| chr15 | 43302045 | 43303076 | 1032 | 9 | 6.01E-07 | 1.18E-05 | 5.79E-05 | 1.44E-06 | -7.27E-05 | AC009852.1, TGM7 |
| chr1 | 23845184 | 23845630 | 447 | 2 | 5.02E-08 | 0.000271 | 1.31E-06 | 1.45E-06 | 0.004893143 | FUCA1 |
| chr19 | 46784521 | 46785658 | 1138 | 8 | 1.59E-06 | 6.11E-06 | 0.000121 | 1.45E-06 | -0.00336945 | SLC1A5 |
| chr3 | 4878758 | 4878976 | 219 | 2 | 1.27E-06 | 9.52E-07 | 0.00012 | 1.45E-06 | -0.00135071 | AC018816.1 |
| chr12 | 65947939 | 65948588 | 650 | 3 | 3.87E-06 | 1.20E-06 | 0.000364 | 1.45E-06 | -0.01096333 | HMGA2, AC090673.1 |
| chr4 | 8371711 | 8372970 | 1260 | 7 | 2.87E-06 | 4.46E-06 | 0.004378 | 1.45E-06 | -0.00064082 | ACOX3 |
| chr5 | 95542084 | 95542403 | 320 | 2 | 9.27E-07 | 9.84E-07 | 0.000103 | 1.45E-06 | -0.0136385 | TTC37 |
| chr15 | 68559346 | 68560086 | 741 | 6 | 1.28E-06 | 3.47E-05 | 0.000188 | 1.45E-06 | 0.001939389 | NA |
| chr1 | 169366939 | 169368622 | 1684 | 14 | 4.42E-06 | 7.62E-06 | 0.001376 | 1.45E-06 | 0.001579215 | BLZF1, NME7 |
| chr1 | 89994875 | 89995581 | 707 | 11 | 7.97E-08 | 2.97E-05 | 0.000538 | 1.45E-06 | 0.002359878 | ZNF326 |
| chr16 | 56558556 | 56559319 | 764 | 2 | 2.03E-05 | 8.73E-07 | 0.000217 | 1.45E-06 | -0.00930126 | NA |
| chr7 | 116327989 | 116328406 | 418 | 5 | 1.90E-06 | 1.66E-06 | 0.000307 | 1.45E-06 | -0.00447022 | CAV2, AC002066.1 |
| chr4 | 89129747 | 89130184 | 438 | 2 | 4.25E-07 | 8.79E-07 | 0.000208 | 1.46E-06 | 0.01292294 | NA |
| chr12 | 52675408 | 52676149 | 742 | 3 | 2.65E-06 | 9.01E-07 | 0.000175 | 1.46E-06 | -0.00772603 | KRT1 |
| chr16 | 85709891 | 85710622 | 732 | 7 | 3.29E-06 | 3.41E-07 | 0.007201 | 1.46E-06 | -0.00338953 | AC018695.9, C16orf74 |
| chr19 | 3210431 | 3210673 | 243 | 2 | 5.88E-07 | 9.09E-07 | 0.000163 | 1.46E-06 | -0.01243751 | NA |
| chr1 | 110893221 | 110893613 | 393 | 3 | 2.00E-07 | 2.08E-05 | 4.69E-05 | 1.46E-06 | -0.00128228 | CD53 |
| chr11 | 71086827 | 71086883 | 57 | 2 | 2.94E-06 | 9.31E-07 | 0.000142 | 1.46E-06 | -0.01697764 | SHANK2 |
| chr1 | 163068455 | 163069394 | 940 | 14 | 2.27E-06 | 2.75E-05 | 0.003136 | 1.46E-06 | -0.00604155 | RGS4 |
| chr14 | 102359514 | 102360194 | 681 | 3 | 4.54E-06 | 5.49E-07 | 0.001055 | 1.46E-06 | -0.00238329 | CINP |
| chr19 | 42204233 | 42204412 | 180 | 2 | 1.88E-06 | 1.76E-06 | 2.66E-05 | 1.47E-06 | -0.00777953 | DEDD2 |
| chr22 | 30607140 | 30608380 | 1241 | 9 | 4.37E-10 | 6.51E-05 | 4.88E-05 | 1.47E-06 | -0.00179264 | TCN2 |
| chr15 | 67542799 | 67543492 | 694 | 7 | 3.95E-07 | 7.39E-06 | 4.47E-05 | 1.47E-06 | 0.001568074 | MAP2K5 |
| chr3 | 194396289 | 194396956 | 668 | 6 | 4.23E-07 | 7.02E-06 | 9.69E-05 | 1.47E-06 | 0.003358037 | GP5 |
| chr5 | 180684381 | 180684966 | 586 | 5 | 4.71E-07 | 1.36E-05 | 0.000356 | 1.47E-06 | -0.00362943 | LINC02222 |
| chr5 | 121961339 | 121962683 | 1345 | 13 | 8.64E-07 | 7.16E-05 | 0.005058 | 1.47E-06 | 0.000500037 | SRFBP1 |
| chr16 | 71808390 | 71809744 | 1355 | 15 | 2.17E-07 | 4.52E-06 | 0.002191 | 1.47E-06 | 0.000353231 | AP1G1 |
| chr7 | 102321329 | 102321800 | 472 | 4 | 1.60E-06 | 1.29E-06 | 0.001227 | 1.47E-06 | -0.018025 | SH2B2 |
| chr8 | 23243350 | 23243403 | 54 | 2 | 1.93E-06 | 9.09E-07 | 0.000173 | 1.47E-06 | 0.024734584 | NA |
| chr13 | 79446383 | 79446511 | 129 | 2 | 5.10E-07 | 7.42E-05 | 1.76E-06 | 1.47E-06 | -0.00738628 | NA |
| chr19 | 3116217 | 3116718 | 502 | 2 | 2.84E-07 | 2.66E-06 | 1.51E-05 | 1.47E-06 | -0.00857168 | GNA11 |
| chr6 | 52573892 | 52575129 | 1238 | 4 | 3.44E-05 | 7.53E-07 | 0.000427 | 1.47E-06 | 0.000959796 | TRAM2 |
| chr10 | 72892643 | 72893691 | 1049 | 5 | 4.16E-05 | 3.42E-06 | 0.001886 | 1.47E-06 | 0.003780436 | OIT3 |
| chr4 | 26859324 | 26859639 | 316 | 2 | 3.14E-08 | 9.04E-07 | 0.000183 | 1.47E-06 | -0.01426212 | STIM2 |
| chr22 | 43094240 | 43094394 | 155 | 2 | 9.32E-07 | 1.08E-05 | 4.33E-06 | 1.47E-06 | -0.0109714 | NA |
| chr20 | 36862098 | 36862952 | 855 | 3 | 1.74E-06 | 1.45E-05 | 3.78E-05 | 1.48E-06 | -0.00698636 | SOGA1 |
| chr12 | 89709466 | 89710287 | 822 | 4 | 4.67E-08 | 7.41E-05 | 3.92E-06 | 1.48E-06 | 0.001463801 | ATP2B1-AS1 |
| chr15 | 89647406 | 89648085 | 680 | 4 | 8.20E-07 | 9.49E-05 | 3.32E-05 | 1.48E-06 | -0.01176342 | KIF7 |
| chr5 | 160370157 | 160370944 | 788 | 14 | 1.49E-06 | 4.13E-06 | 0.002592 | 1.48E-06 | -0.00109643 | C1QTNF2 |
| chr2 | 169583436 | 169584830 | 1395 | 11 | 4.65E-07 | 0.0016576 | 6.48E-06 | 1.48E-06 | 5.04E-05 | PPIG |
| chr21 | 34821489 | 34822617 | 1129 | 4 | 2.03E-06 | 3.16E-06 | 0.000748 | 1.48E-06 | -0.00417374 | RUNX1 |
| chr19 | 18385449 | 18386391 | 943 | 8 | 2.00E-06 | 1.00E-06 | 0.003028 | 1.48E-06 | 0.010027662 | GDF15 |
| chr12 | 6778377 | 6778950 | 574 | 4 | 3.17E-07 | 1.36E-05 | 2.01E-05 | 1.48E-06 | -0.00698156 | LAG3 |
| chr22 | 38793554 | 38794444 | 891 | 11 | 9.57E-08 | 8.40E-05 | 0.000188 | 1.48E-06 | -0.00082229 | SUN2, DNAL4 |
| chr11 | 13338549 | 13339248 | 700 | 3 | 2.11E-07 | 7.98E-07 | 0.000227 | 1.48E-06 | -0.01734036 | ARNTL |
| chr5 | 140400699 | 140401800 | 1102 | 14 | 1.11E-07 | 0.0004187 | 7.19E-05 | 1.48E-06 | 0.000181447 | AC008438.1 |
| chr12 | 132413859 | 132414691 | 833 | 6 | 5.95E-07 | 7.86E-06 | 0.000732 | 1.48E-06 | -0.00645447 | NA |
| chr9 | 91809728 | 91810245 | 518 | 2 | 1.68E-06 | 1.23E-06 | 5.58E-05 | 1.48E-06 | -0.00012329 | ROR2 |
| chr11 | 64809866 | 64811062 | 1197 | 15 | 2.16E-07 | 0.0001483 | 1.94E-05 | 1.49E-06 | 0.00189667 | MEN1 |
| chr5 | 88888010 | 88888034 | 25 | 2 | 1.64E-07 | 3.64E-06 | 1.08E-05 | 1.49E-06 | 0.01343049 | MEF2C-AS1, MEF2C |
| chr3 | 52770928 | 52771622 | 695 | 10 | 4.08E-06 | 4.18E-07 | 0.005292 | 1.49E-06 | 0.000496567 | NEK4 |
| chr14 | 28765345 | 28767015 | 1671 | 21 | 2.42E-05 | 2.06E-05 | 0.0113 | 1.49E-06 | -0.00139145 | AL049777.1, FOXG1 |
| chr19 | 36378475 | 36379363 | 889 | 9 | 3.96E-07 | 6.03E-06 | 0.000278 | 1.49E-06 | 0.001238054 | AC092296.1, ZFP14, AC092296.4 |
| chr14 | 59819091 | 59819592 | 502 | 2 | 4.04E-08 | 0.0001698 | 1.47E-06 | 1.49E-06 | -0.00364758 | RTN1 |
| chr12 | 89840264 | 89840614 | 351 | 2 | 7.93E-07 | 3.11E-06 | 1.28E-05 | 1.49E-06 | 0.013604318 | AC009522.1 |
| chr15 | 68337767 | 68338403 | 637 | 3 | 5.34E-06 | 1.12E-06 | 0.000356 | 1.49E-06 | -0.00286242 | ITGA11 |
| chr7 | 133427705 | 133428306 | 602 | 2 | 5.65E-06 | 9.17E-07 | 0.000178 | 1.49E-06 | -0.00814336 | EXOC4 |
| chr1 | 2307493 | 2308329 | 837 | 4 | 1.51E-05 | 3.23E-06 | 0.00038 | 1.49E-06 | 0.003770436 | SKI |
| chr20 | 642543 | 643215 | 673 | 5 | 1.16E-06 | 1.62E-06 | 0.000222 | 1.49E-06 | -0.01164512 | NA |
| chr1 | 206798260 | 206798917 | 658 | 7 | 4.32E-06 | 5.30E-07 | 0.003568 | 1.49E-06 | -0.00050271 | IL19 |
| chr2 | 33475782 | 33476638 | 857 | 10 | 7.29E-07 | 0.0001251 | 6.73E-05 | 1.49E-06 | 0.000220992 | RASGRP3 |
| chr1 | 25998156 | 25999379 | 1224 | 7 | 1.90E-09 | 9.20E-05 | 7.77E-07 | 1.49E-06 | -0.00556619 | NA |
| chr5 | 138559813 | 138560235 | 423 | 3 | 1.66E-09 | 0.0025459 | 5.08E-07 | 1.49E-06 | 0.006296431 | HSPA9 |
| chr3 | 49814006 | 49814509 | 504 | 4 | 5.86E-07 | 3.69E-06 | 0.000522 | 1.49E-06 | -0.00686311 | NA |
| chr5 | 176361231 | 176362611 | 1381 | 17 | 9.14E-07 | 3.55E-05 | 0.002596 | 1.49E-06 | 0.000448785 | KIAA1191 |
| chr8 | 11801611 | 11802461 | 851 | 7 | 1.95E-10 | 5.06E-05 | 5.08E-06 | 1.49E-06 | -0.00225962 | FDFT1, AC069185.1 |
| chr2 | 176637291 | 176637656 | 366 | 6 | 8.40E-08 | 2.05E-05 | 1.03E-05 | 1.49E-06 | -0.0051473 | LINC01117, LINC01116 |
| chr19 | 49793376 | 49793878 | 503 | 3 | 1.02E-07 | 2.01E-05 | 3.71E-06 | 1.49E-06 | -0.00686126 | AP2A1 |
| chr10 | 123357851 | 123358198 | 348 | 4 | 3.50E-07 | 0.0003482 | 2.79E-05 | 1.49E-06 | 0.00630373 | LINC02641 |

| | | | | | | | | | | |
|---|---|---|---|---|---|---|---|---|---|---|
| chr7 | 73433532 | 73434373 | 842 | 5 | 7.78E-08 | 1.73E-05 | 7.64E-06 | 1.50E-06 | 0.001073378 | FZD9 |
| chr20 | 63604918 | 63605643 | 726 | 4 | 6.62E-07 | 8.86E-06 | 7.94E-06 | 1.50E-06 | 0.000753257 | GMEB2 |
| chr16 | 49455118 | 49455448 | 331 | 2 | 2.03E-07 | 1.19E-06 | 6.29E-05 | 1.50E-06 | -0.00837964 | AC007861.2 |
| chr8 | 23528392 | 23529219 | 828 | 13 | 4.68E-07 | 0.0001456 | 0.001426 | 1.50E-06 | -0.00086527 | SLC25A37 |
| chr12 | 103632281 | 103632773 | 493 | 2 | 3.72E-06 | 9.25E-07 | 0.000175 | 1.50E-06 | -0.01282713 | STAB2 |
| chr16 | 85553813 | 85554745 | 933 | 5 | 4.02E-06 | 8.05E-07 | 0.001419 | 1.50E-06 | 0.000309464 | GSE1 |
| chr10 | 97445586 | 97447184 | 1599 | 16 | 5.79E-07 | 2.97E-06 | 0.000183 | 1.50E-06 | 0.000151668 | ZDHHC16, EXOSC1 |
| chr14 | 66510408 | 66510520 | 113 | 2 | 6.44E-08 | 4.76E-05 | 2.09E-06 | 1.50E-06 | -0.00993954 | GPHN |
| chr9 | 136500632 | 136500752 | 121 | 4 | 1.50E-07 | 0.0011819 | 3.14E-06 | 1.50E-06 | 0.000709038 | NOTCH1 |
| chr9 | 34501973 | 34502004 | 32 | 2 | 5.84E-07 | 9.55E-07 | 0.000147 | 1.50E-06 | -0.01481211 | DNAI1 |
| chr19 | 48495768 | 48495785 | 18 | 2 | 1.91E-06 | 1.48E-06 | 3.84E-05 | 1.50E-06 | -0.01328949 | LMTK3 |
| chr2 | 239249161 | 239250052 | 892 | 5 | 5.56E-06 | 2.64E-05 | 0.000685 | 1.50E-06 | -0.00538369 | HDAC4 |
| chr10 | 5551423 | 5551986 | 564 | 5 | 7.93E-08 | 3.83E-05 | 0.000294 | 1.50E-06 | -0.00820836 | AL732437.3 |
| chr6 | 48830201 | 48830781 | 581 | 2 | 1.56E-07 | 2.02E-06 | 2.28E-05 | 1.50E-06 | -0.01724798 | NA |
| chr7 | 128864982 | 128865714 | 733 | 4 | 1.60E-07 | 1.04E-06 | 0.000568 | 1.50E-06 | -0.00659045 | ATP6V1F, KCP |
| chr9 | 21965074 | 21966006 | 933 | 10 | 6.55E-07 | 3.42E-05 | 1.80E-05 | 1.50E-06 | -0.0018342 | AL359922.1 |
| chr9 | 133332336 | 133333457 | 1122 | 7 | 1.01E-07 | 1.84E-05 | 6.10E-05 | 1.51E-06 | -0.00547417 | SURF6 |
| chr1 | 3484851 | 3485133 | 283 | 3 | 6.10E-08 | 0.0013891 | 8.72E-07 | 1.51E-06 | -0.00360641 | NA |
| chr11 | 57711825 | 57712922 | 1098 | 15 | 1.58E-06 | 2.17E-06 | 0.005892 | 1.51E-06 | 3.92E-05 | AP001931.2, TMX2, MED19 |
| chr17 | 17722207 | 17722705 | 499 | 2 | 6.57E-08 | 4.03E-06 | 9.91E-06 | 1.51E-06 | -0.03990611 | RAI1 |
| chr17 | 36535412 | 36536495 | 1084 | 13 | 6.07E-08 | 1.31E-05 | 0.000111 | 1.51E-06 | 0.002460393 | PIGW, MYO19 |
| chr13 | 94188875 | 94188990 | 116 | 2 | 5.15E-07 | 0.0001318 | 1.56E-06 | 1.51E-06 | 0.008486872 | GPC6 |
| chr16 | 88726704 | 88727635 | 932 | 5 | 1.78E-08 | 0.0008297 | 3.72E-05 | 1.51E-06 | -0.00122368 | PIEZO1 |
| chr11 | 6557984 | 6558609 | 626 | 4 | 1.61E-07 | 3.36E-05 | 4.30E-06 | 1.51E-06 | 0.000594479 | DNHD1 |
| chr10 | 126391451 | 126392110 | 660 | 3 | 1.84E-06 | 7.67E-07 | 0.000628 | 1.51E-06 | -0.01673715 | NA |
| chr3 | 108123031 | 108124024 | 994 | 2 | 1.49E-05 | 9.02E-07 | 0.000231 | 1.51E-06 | 0.01134661 | NA |
| chr11 | 88326359 | 88326705 | 347 | 2 | 1.03E-06 | 2.38E-06 | 1.83E-05 | 1.51E-06 | -0.0075235 | CTSC |
| chr2 | 120239477 | 120240049 | 573 | 5 | 9.05E-07 | 2.65E-06 | 0.000112 | 1.52E-06 | -0.00869755 | NA |
| chr12 | 47770670 | 47771536 | 867 | 5 | 2.85E-06 | 5.94E-06 | 0.000879 | 1.52E-06 | 0.003389829 | SLC48A1, RAPGEF3 |
| chr8 | 60865206 | 60865729 | 524 | 5 | 3.18E-07 | 1.01E-05 | 0.000416 | 1.52E-06 | -0.00542134 | CHD7 |
| chr4 | 85147731 | 85147992 | 262 | 2 | 1.75E-06 | 1.65E-06 | 3.23E-05 | 1.52E-06 | -0.00466714 | NA |
| chr2 | 39436990 | 39437964 | 975 | 10 | 3.07E-06 | 5.93E-05 | 0.00298 | 1.52E-06 | -0.00063562 | MAP4K3-DT, MAP4K3 |
| chr9 | 134703285 | 134703314 | 30 | 2 | 1.50E-06 | 1.66E-06 | 3.19E-05 | 1.52E-06 | -0.01722478 | COL5A1 |
| chr10 | 84424825 | 84425220 | 396 | 4 | 2.72E-07 | 9.35E-05 | 7.40E-05 | 1.52E-06 | 0.006907265 | CCSER2 |
| chr10 | 128098598 | 128098981 | 384 | 2 | 1.06E-07 | 0.0001344 | 1.57E-06 | 1.52E-06 | -0.0062808 | MKI67 |
| chr7 | 150957284 | 150958045 | 762 | 5 | 5.68E-07 | 1.75E-05 | 0.000367 | 1.52E-06 | 0.001872992 | KCNH2 |
| chr1 | 3732271 | 3733326 | 1056 | 6 | 3.44E-06 | 2.14E-05 | 0.001941 | 1.52E-06 | -0.01064318 | TP73 |
| chr5 | 90474043 | 90474882 | 840 | 14 | 1.04E-06 | 1.38E-05 | 0.012415 | 1.52E-06 | 0.000411571 | POLR3G, MBLAC2 |
| chr15 | 55907597 | 55907889 | 293 | 2 | 1.10E-07 | 0.0013183 | 1.14E-06 | 1.52E-06 | 0.004423462 | NEDD4 |
| chr17 | 58679141 | 58679701 | 561 | 3 | 9.51E-07 | 8.41E-07 | 0.000274 | 1.52E-06 | -0.04162607 | TEX14, IGBP1P2, RNVU1-34 |
| chr1 | 94542485 | 94542923 | 439 | 4 | 1.22E-08 | 3.33E-06 | 2.54E-05 | 1.52E-06 | -0.00105692 | NA |
| chr5 | 119332082 | 119333015 | 934 | 5 | 1.85E-07 | 1.63E-05 | 5.45E-05 | 1.52E-06 | 0.003108921 | TNFAIP8 |
| chr17 | 63496974 | 63497383 | 410 | 4 | 3.49E-06 | 2.85E-06 | 0.000262 | 1.53E-06 | -0.00324184 | ACE, AC113554.1 |
| chr4 | 118684980 | 118685768 | 789 | 9 | 2.56E-08 | 0.0323457 | 0.000214 | 1.53E-06 | 1.91E-05 | METTL14, METTL14-DT |
| chr11 | 64960131 | 64961087 | 957 | 6 | 1.99E-06 | 1.17E-05 | 0.0006 | 1.53E-06 | -0.00474517 | MAJIN |
| chr1 | 152110449 | 152110816 | 368 | 4 | 3.43E-06 | 1.70E-06 | 0.00017 | 1.53E-06 | 0.011075603 | TCHH |
| chr5 | 77644531 | 77644624 | 94 | 2 | 1.29E-07 | 9.22E-07 | 0.000212 | 1.53E-06 | -0.0110206 | NA |
| chr13 | 109867908 | 109868114 | 207 | 2 | 2.52E-06 | 1.12E-06 | 8.22E-05 | 1.53E-06 | -0.0021497 | NA |
| chr21 | 32297678 | 32298948 | 1271 | 7 | 2.74E-06 | 0.0002296 | 7.57E-05 | 1.53E-06 | -0.00124088 | MRAP |
| chr3 | 36860125 | 36861547 | 1423 | 6 | 6.06E-07 | 1.22E-05 | 0.000196 | 1.53E-06 | -0.00217508 | TRANK1 |
| chr16 | 47143737 | 47145046 | 1310 | 10 | 5.25E-06 | 9.07E-07 | 0.00831 | 1.53E-06 | -0.0024307 | ITFG1-AS1, NETO2 |
| chr3 | 107522552 | 107523378 | 827 | 11 | 2.56E-07 | 3.91E-05 | 0.00012 | 1.53E-06 | 0.00068888 | BBX |
| chr10 | 129707498 | 129708415 | 918 | 8 | 2.81E-06 | 6.68E-06 | 0.00131 | 1.53E-06 | -0.00074438 | MGMT |
| chr13 | 112831346 | 112831734 | 389 | 3 | 5.80E-07 | 2.06E-06 | 0.00027 | 1.54E-06 | 0.000175064 | ATP11A |
| chr7 | 82506371 | 82506961 | 591 | 3 | 1.69E-06 | 1.25E-06 | 0.000112 | 1.54E-06 | -0.00077474 | NA |
| chr17 | 73305144 | 73305460 | 317 | 2 | 6.11E-07 | 1.10E-06 | 8.79E-05 | 1.54E-06 | -0.02129447 | CDC42EP4 |
| chr16 | 88484042 | 88485014 | 973 | 7 | 8.80E-06 | 6.07E-06 | 0.003216 | 1.54E-06 | 0.00091268 | ZFPM1 |
| chr2 | 38168734 | 38168971 | 238 | 2 | 9.92E-07 | 3.73E-06 | 1.12E-05 | 1.54E-06 | -0.04238043 | CYP1B1-AS1 |
| chr20 | 23403845 | 23404380 | 536 | 2 | 7.54E-07 | 1.82E-06 | 2.81E-05 | 1.54E-06 | -0.01563164 | NAPB |
| chr1 | 226965130 | 226965658 | 529 | 5 | 8.22E-07 | 2.02E-05 | 5.07E-05 | 1.54E-06 | -0.00086925 | AL391628.1, COQ8A |
| chr4 | 103019554 | 103020524 | 971 | 13 | 4.89E-06 | 6.03E-07 | 0.005311 | 1.54E-06 | -0.00526115 | SLC9B1, SLC9B2 |
| chr5 | 2110383 | 2111344 | 962 | 3 | 3.47E-06 | 7.89E-07 | 0.000554 | 1.54E-06 | 0.009673104 | AC124852.1 |
| chr5 | 72248378 | 72248979 | 602 | 2 | 1.90E-08 | 4.96E-05 | 2.12E-06 | 1.54E-06 | -0.01814077 | MRPS27 |
| chr10 | 129811194 | 129811607 | 414 | 2 | 1.92E-06 | 1.22E-06 | 6.51E-05 | 1.54E-06 | -0.0121224 | NA |
| chr22 | 30942533 | 30943567 | 1035 | 5 | 1.79E-05 | 0.0036838 | 0.000249 | 1.55E-06 | 0.001595878 | MORC2 |
| chr2 | 238518346 | 238518378 | 33 | 2 | 3.25E-07 | 0.000301 | 1.37E-06 | 1.55E-06 | -0.00723103 | LINC01107 |
| chr10 | 70511754 | 70512204 | 451 | 2 | 5.54E-07 | 2.65E-06 | 1.66E-05 | 1.55E-06 | -0.00692528 | PALD1 |
| chr2 | 241509128 | 241510350 | 1223 | 12 | 1.79E-07 | 3.68E-05 | 0.003822 | 1.55E-06 | -0.00411393 | STK25 |
| chr5 | 73298908 | 73300131 | 1224 | 10 | 2.13E-07 | 2.06E-06 | 0.000689 | 1.55E-06 | -0.00256332 | NA |
| chr2 | 233311242 | 233311472 | 231 | 2 | 1.41E-06 | 2.19E-06 | 2.14E-05 | 1.55E-06 | -0.01383497 | SAG |
| chr20 | 25623826 | 25624542 | 717 | 16 | 1.51E-06 | 4.16E-06 | 0.000909 | 1.55E-06 | -0.0002308 | ZNF337-AS1, NANP |
| chr14 | 65102277 | 65103535 | 1259 | 16 | 1.60E-06 | 9.05E-06 | 0.002052 | 1.55E-06 | 0.002081568 | MAX |
| chr16 | 71581886 | 71582068 | 183 | 3 | 3.28E-06 | 9.81E-07 | 0.000183 | 1.55E-06 | -0.00028308 | NA |
| chr11 | 2422958 | 2424212 | 1255 | 17 | 4.16E-08 | 0.0008625 | 0.00036 | 1.55E-06 | -0.00088988 | TRPM5 |
| chr2 | 224221606 | 224222515 | 910 | 4 | 1.27E-06 | 0.0002151 | 4.58E-05 | 1.55E-06 | -0.00257213 | NA |
| chr1 | 147171892 | 147173040 | 1149 | 14 | 6.44E-08 | 0.0003741 | 0.000375 | 1.55E-06 | 0.001430236 | PDIA3P1, AC242426.2, PRKAB2 |
| chr18 | 59056761 | 59057160 | 400 | 2 | 4.00E-06 | 9.28E-07 | 0.000234 | 1.56E-06 | 0.011317576 | OACYLP |
| chr2 | 218269781 | 218271079 | 1299 | 16 | 1.98E-06 | 6.64E-06 | 8.53E-05 | 1.56E-06 | -0.00011903 | PNKD, AAMP |
| chr12 | 51983226 | 51983609 | 384 | 2 | 2.22E-07 | 2.18E-05 | 3.06E-06 | 1.56E-06 | 0.003194925 | ACVR1B |
| chr13 | 67574853 | 67574869 | 17 | 2 | 3.12E-06 | 1.05E-06 | 0.00011 | 1.56E-06 | -0.01162966 | NA |
| chr5 | 118988068 | 118988932 | 865 | 9 | 8.04E-06 | 1.88E-06 | 0.003366 | 1.56E-06 | -0.00589857 | DTWD2 |
| chr13 | 48531929 | 48533113 | 1185 | 7 | 3.88E-09 | 7.42E-05 | 2.91E-05 | 1.56E-06 | -0.0066291 | RB1, AL157813.2, RCBTB2, AL157813.1 |
| chr5 | 14869974 | 14871627 | 1654 | 7 | 7.80E-08 | 3.35E-06 | 0.000392 | 1.56E-06 | -0.01098235 | ANKH |
| chr22 | 29772791 | 29773366 | 576 | 2 | 4.11E-06 | 1.04E-06 | 0.000117 | 1.56E-06 | -0.00815836 | NA |
| chr20 | 62437330 | 62437547 | 218 | 2 | 7.73E-07 | 9.19E-06 | 5.22E-06 | 1.56E-06 | -0.0074207 | AL121832.1 |
| chr2 | 106832966 | 106834104 | 1139 | 3 | 3.98E-05 | 9.66E-07 | 0.000502 | 1.56E-06 | 0.005332962 | ST6GAL2 |
| chr1 | 15975737 | 15976521 | 785 | 11 | 3.69E-07 | 1.37E-05 | 0.000585 | 1.56E-06 | 0.000462901 | ZBTB17 |
| chr14 | 21020793 | 21021429 | 637 | 3 | 2.81E-06 | 9.19E-07 | 0.000507 | 1.56E-06 | -0.00370859 | NDRG2 |
| chr1 | 16003909 | 16004358 | 450 | 7 | 1.45E-06 | 3.13E-05 | 0.000158 | 1.56E-06 | -0.0048453 | SRARP |
| chr4 | 3422965 | 3423654 | 690 | 4 | 3.05E-07 | 1.74E-06 | 0.000173 | 1.56E-06 | -0.00219491 | RGS12 |
| chr5 | 154581898 | 154582433 | 536 | 2 | 7.69E-07 | 2.17E-06 | 2.22E-05 | 1.56E-06 | -0.00965928 | NA |
| chr5 | 5057143 | 5057606 | 464 | 6 | 2.56E-06 | 4.07E-06 | 0.000405 | 1.56E-06 | 0.004215674 | LINC01020 |
| chr9 | 121201854 | 121202384 | 531 | 11 | 4.87E-06 | 7.11E-07 | 0.009586 | 1.56E-06 | -0.00261404 | RAB14 |
| chr13 | 98577384 | 98578136 | 753 | 5 | 5.48E-07 | 3.09E-06 | 0.000536 | 1.57E-06 | -0.00336807 | STK24-AS1, STK24 |
| chr5 | 173562203 | 173562306 | 104 | 3 | 6.75E-07 | 2.96E-05 | 3.09E-06 | 1.57E-06 | -0.0084331 | NA |
| chr5 | 140591889 | 140592349 | 461 | 3 | 5.43E-08 | 1.25E-05 | 2.28E-05 | 1.57E-06 | -0.00187846 | AC116353.5, AC116353.6 |
| chr5 | 78359953 | 78361097 | 1145 | 12 | 4.19E-07 | 6.15E-05 | 1.43E-05 | 1.57E-06 | 9.73E-05 | SCAMP1, SCAMP1-AS1 |
| chr7 | 148589228 | 148590374 | 1147 | 5 | 6.61E-07 | 1.59E-05 | 4.12E-05 | 1.57E-06 | 6.95E-05 | AC005229.5 |
| chr12 | 118980817 | 118981981 | 1165 | 13 | 3.01E-06 | 3.26E-05 | 0.000121 | 1.57E-06 | -0.00210955 | SRRM4 |
| chr14 | 22475655 | 22476592 | 938 | 6 | 5.56E-07 | 5.00E-06 | 0.000386 | 1.57E-06 | -0.00471619 | TRAJ60, TRAJ59, AC244502.1 |
| chr16 | 1224873 | 1225475 | 603 | 4 | 1.29E-06 | 5.89E-06 | 0.000199 | 1.57E-06 | -0.00531724 | TPSG1 |
| chr18 | 62526264 | 62527119 | 856 | 4 | 2.50E-06 | 1.30E-06 | 0.000101 | 1.57E-06 | 0.001406909 | ZCCHC2, AC064801.1 |
| chr7 | 116738861 | 116738948 | 88 | 3 | 3.82E-06 | 7.19E-07 | 0.000679 | 1.57E-06 | 0.001523971 | MET |
| chr5 | 16936260 | 16937270 | 1011 | 11 | 8.21E-08 | 0.000132 | 0.000134 | 1.57E-06 | -0.00120044 | MYO10 |
| chr21 | 34909691 | 34910128 | 438 | 2 | 2.47E-06 | 1.10E-06 | 9.76E-05 | 1.57E-06 | -0.01903887 | RUNX1 |
| chr6 | 137874891 | 137876111 | 1221 | 5 | 8.37E-06 | 2.38E-05 | 0.000173 | 1.57E-06 | 0.002768358 | TNFAIP3 |
| chr10 | 49298248 | 49298851 | 604 | 6 | 9.12E-06 | 3.42E-07 | 0.005556 | 1.57E-06 | -0.0012052 | C10orf71-AS1 |
| chr2 | 152433099 | 152433965 | 867 | 2 | 4.19E-05 | 9.42E-07 | 0.000229 | 1.57E-06 | -0.01339632 | FMNL2 |
| chr2 | 63060153 | 63060551 | 399 | 2 | 7.15E-08 | 1.34E-05 | 4.08E-06 | 1.57E-06 | -0.01322719 | NA |
| chr20 | 62236909 | 62237288 | 380 | 2 | 1.30E-07 | 2.52E-06 | 1.82E-05 | 1.57E-06 | -0.01109394 | OSBPL2 |
| chr2 | 593345 | 593391 | 47 | 2 | 9.86E-08 | 0.0006874 | 1.25E-06 | 1.57E-06 | -0.02007425 | NA |
| chr8 | 90984773 | 90985757 | 985 | 8 | 6.10E-08 | 0.0004656 | 1.19E-06 | 1.58E-06 | -0.00128946 | C8orf88 |
| chr10 | 77926674 | 77927921 | 1248 | 11 | 3.12E-06 | 6.42E-06 | 0.000491 | 1.58E-06 | -0.00194816 | DLG5-AS1, DLG5 |
| chr7 | 2721058 | 2721445 | 388 | 3 | 1.69E-07 | 1.61E-06 | 0.000303 | 1.58E-06 | 0.056777189 | AMZ1 |
| chr4 | 112145135 | 112145915 | 781 | 6 | 9.69E-08 | 0.0001045 | 9.95E-06 | 1.58E-06 | -0.00315276 | FAM241A |
| chr22 | 49999078 | 49999400 | 323 | 2 | 6.12E-08 | 8.86E-05 | 1.80E-06 | 1.58E-06 | -0.01365294 | IL17REL |
| chr3 | 148946914 | 148947052 | 139 | 3 | 2.67E-06 | 6.13E-07 | 0.000806 | 1.58E-06 | -0.02562737 | AC092979.2, AC092979.1 |
| chr19 | 1066968 | 1068488 | 1521 | 10 | 1.83E-05 | 0.0001643 | 0.001772 | 1.58E-06 | -0.00089005 | ARHGAP45 |
| chr6 | 46325672 | 46326401 | 730 | 9 | 3.99E-07 | 9.34E-05 | 0.000342 | 1.58E-06 | 0.001193658 | RCAN2 |
| chr1 | 20552047 | 20553151 | 1105 | 10 | 1.77E-07 | 9.81E-06 | 2.01E-05 | 1.58E-06 | -0.00134882 | FAM43B |
| chr2 | 11746127 | 11747196 | 1070 | 9 | 5.04E-08 | 0.000432 | 9.05E-06 | 1.58E-06 | 0.001072947 | LPIN1 |
| chr11 | 64593252 | 64593548 | 297 | 6 | 1.60E-06 | 7.39E-06 | 0.000449 | 1.58E-06 | -0.00272099 | SLC22A12 |
| chr4 | 3645185 | 3646058 | 874 | 3 | 2.03E-06 | 1.02E-06 | 0.000162 | 1.58E-06 | 0.005919454 | NA |
| chr9 | 107491519 | 107491685 | 167 | 2 | 3.68E-07 | 2.85E-06 | 1.58E-05 | 1.59E-06 | 0.013161449 | NA |
| chr2 | 45013234 | 45014439 | 1206 | 9 | 3.50E-07 | 3.05E-05 | 3.86E-05 | 1.59E-06 | 0.000598908 | AC093702.1 |
| chr9 | 5449663 | 5449890 | 228 | 3 | 1.61E-08 | 0.0002562 | 4.07E-06 | 1.59E-06 | -0.00333767 | NA |
| chr10 | 67905439 | 67905900 | 462 | 4 | 2.81E-08 | 9.49E-05 | 3.02E-06 | 1.59E-06 | -0.00184881 | SIRT1 |
| chr9 | 134170267 | 134170934 | 668 | 4 | 3.07E-08 | 2.76E-05 | 5.12E-05 | 1.59E-06 | -0.00416321 | NA |
| chr5 | 179060122 | 179060825 | 704 | 11 | 8.36E-07 | 1.44E-05 | 0.002958 | 1.59E-06 | 0.000532717 | ZNF354C |
| chr8 | 55102506 | 55103225 | 720 | 5 | 2.39E-06 | 4.14E-05 | 0.000577 | 1.59E-06 | -0.00117154 | XKR4 |
| chr4 | 177357120 | 177357828 | 709 | 5 | 2.70E-06 | 9.05E-06 | 0.00039 | 1.59E-06 | -0.00471169 | NEIL3 |
| chr2 | 27808230 | 27809444 | 1215 | 3 | 5.62E-05 | 9.68E-07 | 0.000516 | 1.59E-06 | 0.002985951 | MRPL33, RBKS |
| chr12 | 53954834 | 53955787 | 954 | 8 | 1.16E-07 | 9.75E-06 | 3.72E-05 | 1.59E-06 | -0.00410539 | HOXC12 |
| chr2 | 150120508 | 150120927 | 420 | 2 | 2.08E-06 | 1.22E-06 | 7.22E-05 | 1.59E-06 | 0.001842063 | NA |
| chr13 | 19601769 | 19601787 | 19 | 2 | 1.46E-06 | 8.80E-06 | 5.51E-06 | 1.59E-06 | -0.01224707 | NA |
| chr5 | 179624567 | 179625006 | 440 | 7 | 1.82E-07 | 5.54E-05 | 3.95E-05 | 1.59E-06 | -0.00231135 | HNRNPH1 |
| chr14 | 36508314 | 36509428 | 1115 | 6 | 2.61E-07 | 1.86E-05 | 1.97E-05 | 1.59E-06 | 0.00064984 | AL132857.1, SFTA3, SFTA3 |
| chr19 | 37403079 | 37404070 | 992 | 6 | 3.17E-07 | 1.27E-05 | 7.17E-05 | 1.59E-06 | -0.01884491 | AC008806.1 |
| chr16 | 1826590 | 1827337 | 748 | 11 | 1.10E-06 | 2.68E-06 | 0.002835 | 1.60E-06 | 0.001171425 | FAHD1, HAGH |
| chr1 | 27489789 | 27490893 | 1105 | 16 | 1.16E-05 | 5.01E-05 | 0.00031 | 1.60E-06 | -0.00038905 | WASF2 |
| chr10 | 18551738 | 18551938 | 201 | 3 | 5.26E-07 | 2.86E-05 | 7.92E-06 | 1.60E-06 | -0.00222167 | NSUN6 |
| chr2 | 229893856 | 229894278 | 423 | 4 | 1.24E-06 | 1.87E-05 | 0.00046 | 1.60E-06 | -0.00912043 | TRIP12 |
| chr17 | 46769854 | 46770061 | 208 | 3 | 3.08E-06 | 1.03E-06 | 0.000177 | 1.60E-06 | -0.02421635 | WNT3 |
| chr10 | 132104186 | 132104883 | 698 | 7 | 7.50E-06 | 3.97E-06 | 0.001837 | 1.60E-06 | -0.00278469 | JAKMIP3 |
| chr10 | 113165407 | 113166585 | 1179 | 5 | 3.48E-05 | 6.98E-06 | 0.000692 | 1.60E-06 | 0.000141142 | TCF7L2 |
| chr6 | 88165517 | 88166525 | 1009 | 10 | 1.52E-06 | 9.09E-06 | 0.000763 | 1.60E-06 | -0.00195046 | CNR1 |
| chr8 | 28778027 | 28778512 | 486 | 2 | 1.51E-07 | 3.50E-06 | 1.27E-05 | 1.60E-06 | 0.016383279 | INTS9 |
| chr12 | 53045716 | 53046708 | 993 | 8 | 3.33E-07 | 1.06E-05 | 0.001576 | 1.60E-06 | -0.00785859 | AC068888.1 |
| chr8 | 132187249 | 132188171 | 923 | 4 | 3.90E-07 | 5.80E-06 | 0.000639 | 1.61E-06 | -0.00254572 | KCNQ3 |
| chr3 | 193554772 | 193555686 | 915 | 9 | 4.80E-06 | 2.41E-07 | 0.012152 | 1.61E-06 | -0.01540333 | ATP13A4-AS1, ATP13A4 |
| chr12 | 2229779 | 2230448 | 670 | 5 | 1.72E-06 | 8.57E-07 | 0.002338 | 1.61E-06 | -0.01132722 | CACNA1C |
| chr1 | 39691414 | 39692404 | 991 | 15 | 1.01E-07 | 6.20E-05 | 0.0004 | 1.61E-06 | 0.001412492 | PPIE, HPCAL4 |
| chr17 | 82840005 | 82840830 | 826 | 12 | 1.68E-05 | 2.03E-07 | 0.023364 | 1.61E-06 | -4.02E-05 | TBCD, ZNF750 |
| chr13 | 27944545 | 27945182 | 638 | 3 | 3.85E-08 | 3.23E-05 | 1.33E-05 | 1.62E-06 | -0.00874638 | NA |
| chr19 | 57614018 | 57614685 | 668 | 14 | 9.24E-07 | 2.37E-05 | 0.007283 | 1.62E-06 | 0.000532336 | ZNF134 |
| chr13 | 43356226 | 43356782 | 557 | 4 | 1.75E-06 | 6.71E-06 | 0.000122 | 1.62E-06 | -0.00749627 | ENOX1 |
| chr19 | 36849308 | 36849792 | 485 | 3 | 1.13E-07 | 1.52E-06 | 0.000355 | 1.62E-06 | 0.000812655 | ZNF790 |
| chr11 | 108222291 | 108223384 | 1094 | 25 | 1.20E-07 | 0.0001083 | 0.002141 | 1.62E-06 | -0.00067656 | ATM, NPAT |

| | | | | | | | | | | |
|---|---|---|---|---|---|---|---|---|---|---|
| chr3 | 100635401 | 100635777 | 377 | 3 | 4.05E-07 | 5.94E-06 | 0.000159 | 1.62E-06 | -0.00891146 | ADGRG7 |
| chr8 | 23573311 | 23573501 | 191 | 4 | 8.68E-07 | 5.75E-06 | 1.86E-05 | 1.62E-06 | -0.00072091 | SLC25A37 |
| chr10 | 133094233 | 133095118 | 886 | 3 | 1.29E-06 | 1.02E-06 | 0.000154 | 1.62E-06 | -0.0046425 | ADGRA1 |
| chr11 | 118126213 | 118126741 | 529 | 2 | 2.94E-06 | 1.12E-06 | 0.000102 | 1.62E-06 | 0.002096166 | NA |
| chr5 | 142235804 | 142236500 | 697 | 3 | 2.21E-06 | 1.30E-06 | 0.000124 | 1.62E-06 | -0.0025692 | NA |
| chr4 | 79408130 | 79409198 | 1069 | 9 | 4.16E-05 | 5.44E-06 | 0.002385 | 1.62E-06 | -0.00188074 | GK2 |
| chr8 | 115648279 | 115648836 | 558 | 3 | 9.12E-07 | 9.29E-06 | 1.42E-05 | 1.62E-06 | 0.002577972 | TRPS1 |
| chr1 | 200254308 | 200254664 | 357 | 2 | 1.10E-07 | 0.0004354 | 1.36E-06 | 1.62E-06 | -0.01014671 | LINC00862 |
| chr2 | 38484004 | 38484559 | 556 | 3 | 5.94E-07 | 1.93E-05 | 1.64E-05 | 1.62E-06 | -0.00583304 | LINC02613 |
| chr1 | 46485619 | 46486382 | 764 | 5 | 5.15E-06 | 1.09E-06 | 0.002463 | 1.63E-06 | 0.003543394 | NA |
| chr11 | 129061448 | 129061659 | 212 | 2 | 2.16E-06 | 1.72E-06 | 3.48E-05 | 1.63E-06 | -0.01793039 | ARHGAP32 |
| chr20 | 31608166 | 31608911 | 746 | 5 | 8.70E-07 | 4.75E-06 | 0.000227 | 1.63E-06 | -0.00872485 | NA |
| chr2 | 95025076 | 95025510 | 435 | 5 | 6.42E-08 | 3.38E-05 | 2.01E-05 | 1.63E-06 | -0.00457623 | AC103563.7 |
| chr8 | 103819143 | 103819418 | 276 | 4 | 2.43E-06 | 2.65E-06 | 0.000103 | 1.63E-06 | -0.00241205 | RIMS2 |
| chr2 | 203014055 | 203015302 | 1248 | 14 | 8.72E-07 | 6.17E-06 | 0.000668 | 1.63E-06 | -0.00045694 | NBEAL1, WDR12 |
| chr18 | 35826065 | 35826758 | 694 | 3 | 1.25E-06 | 1.08E-06 | 0.000141 | 1.63E-06 | 0.006844897 | NA |
| chr11 | 1881879 | 1882103 | 225 | 3 | 2.33E-06 | 6.58E-07 | 0.000923 | 1.63E-06 | 0.018664547 | LSP1 |
| chr12 | 98417623 | 98418103 | 481 | 2 | 4.94E-08 | 1.59E-05 | 3.85E-06 | 1.63E-06 | 0.004817814 | RNU4-41P |
| chr12 | 104985385 | 104986132 | 748 | 6 | 5.77E-07 | 3.12E-05 | 5.30E-05 | 1.63E-06 | -0.00239881 | NA |
| chr7 | 102933658 | 102934523 | 866 | 5 | 1.18E-05 | 1.16E-06 | 0.002182 | 1.64E-06 | 0.000933576 | LRRC17, FBXL13 |
| chr22 | 45268736 | 45269655 | 920 | 7 | 7.18E-07 | 9.53E-05 | 0.000652 | 1.64E-06 | 0.000502767 | NA |
| chr6 | 33175159 | 33176294 | 1136 | 7 | 1.86E-06 | 3.42E-05 | 0.001205 | 1.64E-06 | -0.00281318 | COL11A2 |
| chr1 | 226675206 | 226676103 | 898 | 5 | 5.01E-06 | 6.87E-06 | 0.000606 | 1.64E-06 | 0.001651066 | ITPKB-AS1, ITPKB |
| chr5 | 124644390 | 124645685 | 1296 | 6 | 2.88E-05 | 9.24E-06 | 0.000158 | 1.64E-06 | -0.00351662 | ZNF608 |
| chr10 | 29409319 | 29410341 | 1023 | 9 | 1.73E-06 | 2.43E-06 | 0.002738 | 1.64E-06 | -0.00528169 | SVIL-AS1, AL158167.1 |
| chr11 | 469636 | 470429 | 794 | 5 | 3.29E-07 | 0.000101 | 6.54E-06 | 1.64E-06 | -0.00281385 | PTDSS2 |
| chr1 | 44760327 | 44760828 | 502 | 3 | 3.31E-07 | 0.0002831 | 6.48E-06 | 1.64E-06 | 0.00166755 | KIF2C, AL592166.1 |
| chr22 | 49043858 | 49044329 | 472 | 2 | 9.15E-07 | 2.25E-06 | 2.33E-05 | 1.64E-06 | -0.01252072 | NA |
| chr20 | 12964251 | 12964431 | 181 | 3 | 9.29E-07 | 1.25E-05 | 6.87E-06 | 1.64E-06 | -0.01034146 | LINC01723 |
| chr5 | 135834311 | 135835490 | 1180 | 11 | 4.63E-06 | 4.30E-06 | 0.006462 | 1.64E-06 | 0.00244503 | SLC25A48 |
| chr19 | 728176 | 728385 | 210 | 2 | 2.21E-06 | 1.31E-06 | 6.52E-05 | 1.65E-06 | -0.0266547 | PALM |
| chr1 | 53092254 | 53093329 | 1076 | 7 | 5.63E-06 | 6.55E-07 | 0.004688 | 1.65E-06 | -0.00795718 | SLC1A7 |
| chr5 | 945525 | 946441 | 917 | 6 | 2.49E-06 | 1.84E-05 | 0.000569 | 1.65E-06 | 0.005669371 | AC122719.1 |
| chr9 | 122510273 | 122510369 | 97 | 2 | 8.12E-07 | 8.02E-06 | 6.17E-06 | 1.65E-06 | -0.00062251 | AL359636.2 |
| chr5 | 123063416 | 123063567 | 152 | 2 | 1.79E-06 | 3.48E-06 | 1.34E-05 | 1.65E-06 | -0.00722 | NA |
| chr11 | 36818091 | 36818451 | 361 | 2 | 1.58E-06 | 1.07E-06 | 0.000138 | 1.65E-06 | -0.00231219 | NA |
| chr19 | 54011915 | 54012557 | 643 | 5 | 2.11E-06 | 3.50E-06 | 0.000968 | 1.65E-06 | 0.018201139 | CACNG6 |
| chr2 | 100470044 | 100471113 | 1070 | 6 | 6.35E-06 | 7.25E-07 | 0.003597 | 1.65E-06 | -0.00676661 | NMS |
| chr6 | 41424063 | 41424665 | 603 | 2 | 1.50E-08 | 0.0114687 | 1.12E-06 | 1.65E-06 | 0.001861721 | NA |
| chr12 | 55940101 | 55940779 | 679 | 4 | 1.71E-06 | 5.96E-06 | 9.24E-05 | 1.66E-06 | 2.25E-05 | DGKA |
| chr3 | 104717850 | 104717852 | 3 | 2 | 3.47E-06 | 1.03E-06 | 0.00018 | 1.66E-06 | 0.015890411 | NA |
| chr17 | 81888609 | 81889810 | 1202 | 7 | 1.72E-07 | 6.76E-06 | 0.000161 | 1.66E-06 | -0.00122862 | ALYREF |
| chr3 | 123420111 | 123420313 | 203 | 2 | 8.65E-08 | 3.64E-05 | 2.58E-06 | 1.66E-06 | -0.00386848 | ADCY5 |
| chr5 | 523315 | 523679 | 365 | 4 | 7.99E-08 | 2.14E-05 | 9.51E-06 | 1.66E-06 | 0.008745794 | SLC9A3 |
| chr10 | 22927253 | 22928244 | 992 | 10 | 3.52E-06 | 1.05E-05 | 0.002376 | 1.66E-06 | 0.001097116 | ARMC3 |
| chr16 | 50369485 | 50370375 | 891 | 3 | 5.19E-06 | 6.29E-07 | 0.00093 | 1.66E-06 | -0.00417395 | AC007493.2 |
| chr2 | 38359611 | 38359710 | 100 | 2 | 1.56E-06 | 1.64E-06 | 4.03E-05 | 1.67E-06 | -0.01477443 | ATL2 |
| chr8 | 100105674 | 100106320 | 647 | 11 | 8.77E-08 | 7.77E-05 | 0.001108 | 1.67E-06 | -0.00395109 | RGS22 |
| chr8 | 1495202 | 1495302 | 101 | 2 | 2.17E-07 | 1.16E-05 | 4.85E-06 | 1.67E-06 | -0.00391942 | DLGAP2 |
| chr8 | 94474526 | 94475253 | 728 | 9 | 4.08E-07 | 9.04E-06 | 3.45E-05 | 1.67E-06 | 0.002525752 | RAD54B |
| chr16 | 1561341 | 1562219 | 879 | 5 | 3.31E-06 | 3.03E-07 | 0.005475 | 1.67E-06 | 0.000223613 | IFT140 |
| chr10 | 52313287 | 52314118 | 832 | 7 | 7.83E-08 | 9.71E-05 | 0.000196 | 1.67E-06 | 0.000573946 | PRKG1-AS1 |
| chr10 | 68755016 | 68755487 | 472 | 3 | 6.46E-08 | 0.0004852 | 7.18E-06 | 1.68E-06 | 0.002669667 | CCAR1, SNORD98 |
| chr11 | 76860414 | 76860566 | 153 | 5 | 6.26E-07 | 1.49E-05 | 0.000159 | 1.68E-06 | -0.0066567 | NA |
| chr10 | 70814812 | 70815849 | 1038 | 8 | 6.21E-06 | 2.73E-05 | 0.000173 | 1.68E-06 | -0.00190644 | NA |
| chr4 | 114601572 | 114602003 | 432 | 2 | 6.49E-07 | 2.64E-06 | 1.92E-05 | 1.68E-06 | -0.01668458 | UGT8 |
| chr10 | 96720234 | 96721279 | 1046 | 11 | 4.61E-06 | 2.75E-05 | 0.000489 | 1.68E-06 | 0.000513046 | PIK3AP1 |
| chr1 | 53508000 | 53508054 | 55 | 2 | 2.89E-06 | 1.62E-06 | 4.21E-05 | 1.68E-06 | 0.000583168 | GLIS1 |
| chr22 | 50479932 | 50480518 | 587 | 3 | 5.68E-08 | 1.93E-05 | 0.000122 | 1.68E-06 | -0.01809918 | NA |
| chr16 | 52654657 | 52655258 | 602 | 4 | 6.03E-06 | 4.12E-07 | 0.003155 | 1.68E-06 | -0.00713649 | AC026462.4 |
| chr17 | 48593248 | 48594499 | 1252 | 17 | 9.23E-07 | 5.09E-06 | 0.004125 | 1.69E-06 | -0.00051124 | HOXB-AS3, HOXB3, HOXB5 |
| chr17 | 77422261 | 77422591 | 331 | 3 | 1.01E-06 | 1.18E-06 | 0.000233 | 1.69E-06 | -0.00204171 | SEPTIN9 |
| chr1 | 2642592 | 2643256 | 665 | 5 | 2.70E-06 | 7.99E-06 | 0.000611 | 1.69E-06 | 0.003209069 | TTC34 |
| chr11 | 2362549 | 2363297 | 749 | 4 | 1.35E-06 | 4.41E-06 | 0.000593 | 1.69E-06 | -0.00242468 | CD81-AS1 |
| chr5 | 138351977 | 138353005 | 1029 | 12 | 1.72E-06 | 4.41E-06 | 0.000172 | 1.69E-06 | -0.00141654 | KDM3B |
| chr6 | 27669523 | 27670612 | 1090 | 5 | 1.33E-05 | 3.45E-06 | 0.002761 | 1.69E-06 | -0.01035209 | NA |
| chr1 | 160400050 | 160401665 | 1616 | 16 | 4.09E-07 | 5.14E-05 | 1.55E-05 | 1.69E-06 | -0.00223911 | VANGL2 |
| chr13 | 106567777 | 106568704 | 928 | 11 | 8.02E-07 | 8.05E-05 | 0.001089 | 1.69E-06 | -0.00021583 | LINC00551, ARGLU1 |
| chr19 | 19305942 | 19306671 | 730 | 5 | 1.23E-05 | 1.83E-06 | 0.000617 | 1.69E-06 | -0.00100748 | SUGP1 |
| chr2 | 176159774 | 176161131 | 1358 | 7 | 3.97E-08 | 0.0001042 | 9.22E-05 | 1.69E-06 | -0.00267224 | HOXD3 |
| chr2 | 43838120 | 43839155 | 1036 | 20 | 5.56E-07 | 0.0001141 | 0.000759 | 1.69E-06 | -0.00202811 | ABCG8, ABCG5 |
| chr11 | 64715646 | 64716219 | 574 | 2 | 8.36E-07 | 1.42E-06 | 5.79E-05 | 1.69E-06 | -0.01212964 | NRXN2 |
| chr3 | 132285220 | 132285756 | 537 | 4 | 9.41E-07 | 2.55E-06 | 5.86E-05 | 1.69E-06 | 0.002794165 | CPNE4 |
| chr4 | 8043774 | 8045066 | 1293 | 5 | 4.81E-08 | 5.26E-05 | 5.15E-06 | 1.69E-06 | -0.00426401 | ABLIM2 |
| chr1 | 46603460 | 46604606 | 1147 | 12 | 1.34E-07 | 6.32E-05 | 0.002498 | 1.69E-06 | 0.000429551 | MKNK1 |
| chr1 | 84300841 | 84302195 | 1355 | 8 | 1.86E-05 | 0.0003718 | 0.000142 | 1.69E-06 | 0.006878002 | SAMD13 |
| chr15 | 88466978 | 88467914 | 937 | 18 | 1.04E-06 | 4.55E-05 | 0.002422 | 1.69E-06 | 0.001999148 | MRPS11, MRPL46, AC013489.1 |
| chr19 | 49876542 | 49878200 | 1659 | 20 | 2.50E-06 | 7.80E-07 | 0.001766 | 1.70E-06 | 0.001017391 | TBC1D17, PNKP, AKT1S1 |
| chr2 | 33706483 | 33707256 | 774 | 4 | 2.13E-06 | 3.25E-05 | 0.00035 | 1.70E-06 | 0.000733564 | LINC01320 |
| chr16 | 49528323 | 49528832 | 510 | 3 | 3.54E-06 | 6.53E-07 | 0.000894 | 1.70E-06 | -0.01467557 | ZNF423 |
| chr22 | 41151796 | 41152324 | 529 | 4 | 1.93E-06 | 1.04E-06 | 0.000687 | 1.70E-06 | -0.0055729 | EP300 |
| chr4 | 104555483 | 104555527 | 45 | 2 | 1.01E-06 | 3.52E-06 | 1.39E-05 | 1.70E-06 | -0.00641288 | CXXC4-AS1 |
| chr12 | 132247936 | 132248561 | 626 | 4 | 2.39E-06 | 1.19E-06 | 0.001225 | 1.70E-06 | -0.00677772 | GALNT9 |
| chr12 | 110246958 | 110248021 | 1064 | 4 | 4.51E-05 | 2.20E-05 | 0.000151 | 1.70E-06 | -0.0047278 | NA |
| chr10 | 60352226 | 60352427 | 202 | 2 | 4.08E-07 | 1.72E-05 | 3.86E-06 | 1.70E-06 | -0.01616151 | ANK3 |
| chr5 | 14439762 | 14440965 | 1204 | 7 | 1.34E-05 | 8.72E-06 | 0.001421 | 1.70E-06 | -0.00231743 | TRIO |
| chr12 | 15162911 | 15163178 | 268 | 2 | 1.05E-06 | 5.14E-06 | 9.45E-06 | 1.70E-06 | -0.01215288 | RERG |
| chr15 | 56733036 | 56734397 | 1362 | 12 | 2.80E-07 | 2.97E-05 | 0.000746 | 1.71E-06 | -0.0046233 | ZNF280D, AC090517.4 |
| chr10 | 58868882 | 58869090 | 209 | 4 | 8.97E-07 | 1.26E-05 | 6.09E-05 | 1.71E-06 | -0.00809943 | NA |
| chr2 | 65231995 | 65232327 | 333 | 3 | 2.06E-08 | 0.0167765 | 3.93E-07 | 1.71E-06 | 0.00491306 | ACTR2 |
| chr1 | 115479304 | 115479470 | 167 | 4 | 2.58E-06 | 7.62E-06 | 0.00044 | 1.71E-06 | -0.0155423 | AL512638.2 |
| chr4 | 8908700 | 8908863 | 164 | 5 | 1.04E-06 | 2.62E-05 | 5.31E-06 | 1.71E-06 | -0.01751746 | NA |
| chr18 | 76348641 | 76349417 | 777 | 3 | 5.08E-07 | 2.69E-06 | 3.18E-05 | 1.71E-06 | -0.01136672 | NA |
| chr7 | 148482367 | 148482408 | 42 | 2 | 3.62E-06 | 1.02E-06 | 0.000238 | 1.71E-06 | -0.00182796 | AC006974.1 |
| chr7 | 2589666 | 2590350 | 685 | 5 | 6.29E-06 | 1.32E-06 | 0.000455 | 1.71E-06 | 0.000983764 | IQCE |
| chr13 | 27402638 | 27403642 | 1005 | 4 | 6.39E-09 | 0.0056482 | 7.70E-07 | 1.71E-06 | -0.00352195 | NA |
| chr5 | 35032678 | 35033507 | 830 | 3 | 2.42E-07 | 3.76E-05 | 7.29E-06 | 1.71E-06 | -0.01196086 | AGXT2 |
| chr4 | 184455684 | 184456763 | 1080 | 7 | 1.60E-06 | 6.03E-07 | 0.006524 | 1.71E-06 | 0.001576488 | IRF2 |
| chr12 | 45926207 | 45927107 | 901 | 6 | 4.08E-06 | 5.19E-06 | 6.46E-05 | 1.71E-06 | -0.00111722 | SCAF11 |
| chr20 | 256361 | 256659 | 299 | 3 | 2.56E-07 | 1.76E-05 | 1.89E-05 | 1.71E-06 | -0.01187023 | NA |
| chr19 | 2650729 | 2651148 | 420 | 5 | 4.71E-07 | 2.36E-05 | 8.99E-06 | 1.71E-06 | -0.00125733 | GNG7 |
| chr14 | 101564276 | 101565166 | 891 | 6 | 9.39E-06 | 3.92E-06 | 0.000624 | 1.71E-06 | -0.00019004 | NA |
| chr18 | 21682858 | 21683249 | 392 | 2 | 7.54E-08 | 6.52E-05 | 2.14E-06 | 1.71E-06 | -0.01430904 | ABHD3 |
| chr1 | 39133040 | 39133515 | 476 | 2 | 1.15E-07 | 1.35E-05 | 4.53E-06 | 1.72E-06 | -0.00948891 | MACF1 |
| chr5 | 141320332 | 141321109 | 778 | 15 | 5.12E-10 | 0.0018883 | 1.57E-05 | 1.72E-06 | -0.00011717 | AC005618.4, TAF7 |
| chr11 | 57540669 | 57541513 | 845 | 6 | 6.61E-08 | 0.0002671 | 3.92E-05 | 1.72E-06 | 0.000995424 | NA |
| chr4 | 75674316 | 75675264 | 949 | 4 | 5.40E-06 | 0.000192 | 5.93E-05 | 1.72E-06 | -0.00161539 | G3BP2 |
| chr8 | 85106666 | 85107628 | 963 | 15 | 1.98E-07 | 8.01E-06 | 0.002703 | 1.72E-06 | 0.000976564 | LRRCC1 |
| chr22 | 43424882 | 43425109 | 228 | 3 | 8.87E-07 | 6.24E-05 | 4.78E-06 | 1.72E-06 | -0.000656 | MPPED1 |
| chr21 | 41973300 | 41974436 | 1137 | 5 | 5.03E-08 | 0.0015008 | 2.84E-06 | 1.72E-06 | -0.00723283 | NA |
| chr22 | 20085186 | 20086473 | 1288 | 13 | 4.03E-07 | 0.0002582 | 4.17E-05 | 1.72E-06 | -0.00103435 | DGCR8, MIR3618, MIR1306 |
| chr12 | 117897540 | 117898063 | 524 | 2 | 1.36E-07 | 1.75E-05 | 3.89E-06 | 1.73E-06 | 0.005153015 | KSR2 |
| chr3 | 52552076 | 52552327 | 252 | 3 | 2.34E-06 | 8.57E-07 | 0.000555 | 1.73E-06 | -0.01032808 | SMIM4, PBRM1 |
| chr2 | 24046996 | 24047856 | 861 | 12 | 6.14E-07 | 7.03E-06 | 0.000506 | 1.73E-06 | 0.000392832 | MFSD2B, WDCP |
| chr8 | 60792773 | 60793148 | 376 | 2 | 2.18E-06 | 1.39E-06 | 6.51E-05 | 1.73E-06 | 0.006785209 | CHD7 |
| chr17 | 39688112 | 39689413 | 1302 | 12 | 2.57E-05 | 6.88E-05 | 0.000108 | 1.73E-06 | -0.00162318 | ERBB2, PGAP3 |
| chr18 | 26752116 | 26752427 | 312 | 2 | 3.97E-06 | 1.03E-06 | 0.000249 | 1.73E-06 | -0.01813951 | AQP4-AS1 |
| chr2 | 219495205 | 219495659 | 455 | 2 | 8.61E-07 | 1.16E-06 | 0.000118 | 1.73E-06 | -0.00217009 | AC053503.6, ASIC4-AS1 |
| chr14 | 68824907 | 68825139 | 233 | 3 | 9.03E-07 | 0.0001346 | 3.65E-05 | 1.73E-06 | -0.01088162 | NA |
| chr12 | 65169549 | 65170615 | 1067 | 6 | 4.87E-09 | 0.0001747 | 1.10E-05 | 1.73E-06 | 0.00162137 | LEMD3 |
| chr3 | 49783907 | 49784360 | 454 | 2 | 4.56E-06 | 1.04E-06 | 0.000227 | 1.73E-06 | -0.00538713 | IP6K1 |
| chr1 | 156848800 | 156849601 | 802 | 6 | 1.97E-06 | 8.14E-06 | 0.000157 | 1.73E-06 | 0.001348587 | NTRK1, INSRR |
| chr8 | 51014233 | 51014461 | 229 | 2 | 6.60E-07 | 2.34E-05 | 3.34E-06 | 1.74E-06 | 0.001605382 | NA |
| chr12 | 27013863 | 27014879 | 1017 | 12 | 6.86E-07 | 2.58E-05 | 0.00046 | 1.74E-06 | 0.00105204 | TM7SF3 |
| chr1 | 245692020 | 245692239 | 220 | 2 | 1.69E-06 | 2.11E-06 | 2.86E-05 | 1.74E-06 | 0.007587511 | KIF26B |
| chr3 | 51710974 | 51711305 | 332 | 3 | 2.80E-07 | 2.71E-05 | 9.52E-06 | 1.74E-06 | -0.00273638 | GRM2 |
| chr5 | 69189259 | 69189927 | 669 | 11 | 2.09E-06 | 8.74E-06 | 0.002089 | 1.74E-06 | 0.001791296 | CENPH, AC022107.1 |
| chr5 | 109846 | 110749 | 904 | 2 | 5.14E-05 | 1.04E-06 | 0.000251 | 1.74E-06 | 0.004375939 | PLEKHG4B |
| chr6 | 73695156 | 73696517 | 1362 | 18 | 2.64E-06 | 3.79E-06 | 0.013087 | 1.74E-06 | 0.000564513 | CD109, AL590428.1 |
| chr7 | 38317128 | 38317446 | 319 | 5 | 3.36E-06 | 9.91E-07 | 0.000546 | 1.74E-06 | 0.013456705 | TRGV9 |
| chr1 | 25875262 | 25876449 | 1188 | 9 | 1.94E-07 | 0.0001537 | 0.000195 | 1.74E-06 | 0.001629028 | PAQR7 |
| chr7 | 158740513 | 158741199 | 687 | 3 | 2.25E-06 | 7.06E-07 | 0.000745 | 1.74E-06 | -0.01754342 | ESYT2 |
| chr19 | 56839954 | 56841216 | 1263 | 18 | 5.61E-08 | 9.03E-06 | 0.006238 | 1.75E-06 | -0.00475852 | MIMT1, ZIM2, PEG3 |
| chr10 | 119003905 | 119004001 | 97 | 2 | 2.40E-06 | 1.66E-06 | 4.42E-05 | 1.75E-06 | -0.01089814 | AL157388.1 |
| chr4 | 1577560 | 1578650 | 1091 | 8 | 1.29E-07 | 2.87E-05 | 0.000842 | 1.75E-06 | -0.01395301 | AC147067.1 |
| chr6 | 42226817 | 42226953 | 137 | 2 | 2.28E-06 | 1.23E-06 | 9.98E-05 | 1.75E-06 | 0.003593591 | TRERF1 |
| chr15 | 77696164 | 77696942 | 779 | 5 | 4.82E-06 | 5.17E-07 | 0.001253 | 1.75E-06 | -0.00915427 | LINGO1 |
| chr19 | 12722249 | 12723846 | 1598 | 10 | 1.37E-07 | 0.0001667 | 4.84E-06 | 1.75E-06 | 0.001189133 | TNPO2 |
| chr19 | 40780235 | 40780625 | 391 | 2 | 7.54E-07 | 4.07E-06 | 1.24E-05 | 1.75E-06 | -0.00526586 | MIA-RAB4B, RAB4B, RAB4B-EGLN2, AC008537.4 |
| chr16 | 71640604 | 71641035 | 432 | 3 | 9.58E-07 | 5.72E-06 | 0.000182 | 1.75E-06 | 0.002398972 | MARVELD3, PHLPP2 |
| chr21 | 42833005 | 42833501 | 497 | 4 | 4.41E-06 | 4.49E-07 | 0.002283 | 1.75E-06 | -0.00424874 | AP001628.1 |
| chr10 | 48305986 | 48306985 | 1000 | 8 | 1.59E-07 | 0.0016068 | 0.000212 | 1.75E-06 | -0.00475915 | MAPK8 |
| chr1 | 185629457 | 185630006 | 550 | 3 | 7.18E-07 | 3.10E-06 | 7.38E-05 | 1.75E-06 | -9.18E-05 | NA |
| chr10 | 93059802 | 93060619 | 818 | 8 | 7.30E-07 | 1.87E-05 | 0.000401 | 1.75E-06 | -0.01106781 | AL358613.1 |
| chr6 | 26246042 | 26247042 | 1001 | 9 | 4.58E-07 | 3.44E-05 | 0.000721 | 1.75E-06 | -0.00188856 | H4C7 |
| chr9 | 130861082 | 130861239 | 158 | 3 | 5.01E-06 | 6.37E-07 | 0.001192 | 1.75E-06 | -0.01540406 | ABL1 |
| chr1 | 193112556 | 193113634 | 1079 | 3 | 1.25E-05 | 7.12E-07 | 0.000971 | 1.75E-06 | -0.00613339 | NA |
| chr3 | 47581313 | 47582291 | 979 | 4 | 8.30E-07 | 0.0010479 | 0.000367 | 1.75E-06 | -0.00539123 | NA |
| chr8 | 144994542 | 144994772 | 231 | 3 | 4.20E-06 | 7.62E-07 | 0.00053 | 1.75E-06 | -0.00449391 | AF235103.1, ZNF252P |
| chr17 | 11882788 | 11883632 | 845 | 3 | 5.81E-10 | 0.0080631 | 5.74E-07 | 1.75E-06 | -0.01367437 | DNAH9 |
| chr12 | 30927118 | 30927489 | 372 | 2 | 3.82E-08 | 1.31E-06 | 8.11E-05 | 1.76E-06 | -0.00920778 | TSPAN11 |
| chr16 | 2989445 | 2990131 | 687 | 4 | 1.32E-06 | 8.31E-06 | 9.16E-05 | 1.76E-06 | -0.00642532 | LINC00514 |
| chr12 | 8698123 | 8698644 | 522 | 4 | 8.72E-07 | 1.39E-06 | 0.000362 | 1.76E-06 | -0.00296112 | RIMKLB |
| chr17 | 2232456 | 2233093 | 638 | 4 | 1.41E-07 | 5.45E-05 | 9.86E-05 | 1.76E-06 | -0.00124439 | AL450226.1, SMG6 |
| chr11 | 133076560 | 133077039 | 480 | 3 | 8.37E-07 | 5.34E-06 | 1.91E-05 | 1.76E-06 | -0.00171526 | OPCML |
| chr2 | 201806435 | 201807560 | 1126 | 6 | 3.44E-06 | 0.0002351 | 6.67E-06 | 1.76E-06 | -0.00438158 | CDK15 |
| chr19 | 42005649 | 42005941 | 293 | 4 | 1.21E-07 | 0.0001596 | 1.17E-06 | 1.76E-06 | -0.00150032 | GRIK5 |
| chr1 | 114715906 | 114716946 | 1041 | 10 | 9.07E-07 | 0.0001025 | 0.000343 | 1.76E-06 | -0.00175819 | NRAS, CSDE1 |

| | | | | | | | | | | |
|---|---|---|---|---|---|---|---|---|---|---|
| chr17 | 76500871 | 76501769 | 899 | 9 | 1.96E-07 | 1.89E-05 | 0.000135 | 1.76E-06 | -0.00059172 | RHBDF2 |
| chr5 | 2100547 | 2101063 | 517 | 3 | 3.82E-07 | 1.50E-05 | 2.72E-05 | 1.76E-06 | 0.019769614 | AC124852.1 |
| chr8 | 144326426 | 144327718 | 1293 | 11 | 2.59E-07 | 0.00054 | 0.000319 | 1.76E-06 | 0.002242431 | DGAT1 |
| chr2 | 96759943 | 96761296 | 1354 | 12 | 1.68E-06 | 7.23E-06 | 0.002216 | 1.76E-06 | 0.000632672 | CNNM4 |
| chr9 | 128341728 | 128342146 | 419 | 3 | 3.19E-07 | 7.11E-06 | 4.90E-05 | 1.77E-06 | -0.00222091 | SLC27A4 |
| chr20 | 34190119 | 34190806 | 688 | 5 | 1.48E-06 | 5.97E-06 | 8.32E-05 | 1.77E-06 | -0.0070126 | NA |
| chr22 | 32761374 | 32762250 | 877 | 4 | 2.27E-08 | 0.0071365 | 3.99E-06 | 1.77E-06 | -0.01007579 | SYN3 |
| chr6 | 27110648 | 27111053 | 406 | 2 | 2.39E-07 | 8.27E-06 | 6.62E-06 | 1.77E-06 | -0.01515308 | NA |
| chr2 | 120771611 | 120772276 | 666 | 3 | 1.10E-06 | 1.77E-06 | 0.000239 | 1.77E-06 | -0.00169876 | GLI2 |
| chr10 | 126421777 | 126422209 | 433 | 5 | 1.29E-06 | 3.03E-06 | 5.08E-05 | 1.77E-06 | -0.00757674 | LINC00601 |
| chr10 | 47325205 | 47326198 | 994 | 8 | 1.49E-07 | 3.76E-05 | 0.000502 | 1.78E-06 | -0.00045836 | GDF2 |
| chr1 | 108963064 | 108964183 | 1120 | 12 | 1.55E-07 | 0.0004337 | 0.000534 | 1.78E-06 | 0.000694296 | AKNAD1, CLCC1 |
| chr20 | 35547019 | 35547522 | 504 | 6 | 2.55E-06 | 1.79E-05 | 0.001954 | 1.78E-06 | -0.00240062 | ERGIC3 |
| chr6 | 28986047 | 28986985 | 939 | 6 | 4.08E-06 | 2.14E-06 | 0.001756 | 1.78E-06 | -0.00762024 | HCG15, AL662791.1 |
| chr7 | 2195934 | 2196770 | 837 | 3 | 2.96E-07 | 7.30E-06 | 2.04E-05 | 1.78E-06 | -0.00376888 | MAD1L1, AC069288.1 |
| chr15 | 38251008 | 38251802 | 795 | 4 | 1.09E-06 | 6.95E-07 | 0.001231 | 1.78E-06 | -0.00447714 | NA |
| chr14 | 95157347 | 95157956 | 610 | 10 | 2.46E-06 | 4.49E-07 | 0.005439 | 1.78E-06 | 0.001429923 | DICER1-AS1, DICER1 |
| chr11 | 65060727 | 65061330 | 604 | 2 | 2.21E-06 | 1.52E-06 | 5.71E-05 | 1.78E-06 | -0.02025216 | NA |
| chr2 | 237207796 | 237208113 | 318 | 2 | 7.28E-07 | 7.67E-06 | 7.10E-06 | 1.78E-06 | -0.00314608 | NA |
| chr19 | 57167437 | 57168222 | 786 | 10 | 4.59E-06 | 1.16E-05 | 0.000135 | 1.78E-06 | 0.006682708 | DUXA |
| chr9 | 133054360 | 133055742 | 1383 | 5 | 9.21E-08 | 0.0001435 | 2.06E-06 | 1.79E-06 | 0.002832495 | GTF3C5 |
| chr14 | 23195870 | 23195976 | 107 | 2 | 2.03E-06 | 4.38E-06 | 1.19E-05 | 1.79E-06 | 0.002266898 | RNF212B |
| chr3 | 113530715 | 113531385 | 671 | 4 | 1.61E-07 | 9.82E-06 | 0.000105 | 1.79E-06 | -0.0057298 | NA |
| chr8 | 88297836 | 88298665 | 830 | 3 | 7.52E-06 | 7.07E-06 | 6.41E-05 | 1.79E-06 | -0.00185086 | MMP16 |
| chr2 | 25323680 | 25324470 | 791 | 3 | 2.71E-05 | 9.41E-07 | 0.000463 | 1.79E-06 | 0.000175743 | DNMT3A |
| chr15 | 93900067 | 93900870 | 804 | 9 | 6.62E-07 | 1.03E-05 | 0.004347 | 1.79E-06 | -0.00093073 | LINC01580, LINC01579 |
| chr11 | 9385078 | 9385842 | 765 | 5 | 2.43E-06 | 9.29E-06 | 4.67E-05 | 1.79E-06 | -0.00360295 | IPO7 |
| chr1 | 110963399 | 110964538 | 1140 | 18 | 4.11E-06 | 3.63E-05 | 0.001472 | 1.79E-06 | 0.002078775 | AL360270.3, LRIF1 |
| chr6 | 46491401 | 46492734 | 1334 | 16 | 3.72E-06 | 2.49E-06 | 0.008615 | 1.79E-06 | -0.00471642 | AL035670.1, RCAN2 |
| chr8 | 69833393 | 69834431 | 1039 | 15 | 4.52E-07 | 4.84E-06 | 0.001437 | 1.79E-06 | 0.000555181 | AC079089.1, SLCO5A1 |
| chr17 | 34318729 | 34319137 | 409 | 5 | 1.09E-06 | 2.67E-05 | 0.00019 | 1.79E-06 | -0.00885548 | NA |
| chr16 | 54038085 | 54038313 | 229 | 2 | 1.35E-06 | 5.12E-06 | 1.03E-05 | 1.79E-06 | 0.005843915 | FTO, AC007347.1 |
| chr17 | 80194439 | 80195040 | 602 | 2 | 3.74E-07 | 4.12E-06 | 1.28E-05 | 1.79E-06 | -0.00827124 | CARD14 |
| chr17 | 62134596 | 62134669 | 74 | 3 | 1.27E-06 | 1.84E-05 | 5.23E-06 | 1.79E-06 | -0.00308155 | NA |
| chr5 | 96702522 | 96702910 | 389 | 11 | 3.35E-06 | 5.99E-05 | 0.002153 | 1.79E-06 | 0.00203197 | CAST |
| chr20 | 46588156 | 46588420 | 265 | 4 | 2.06E-06 | 2.42E-06 | 6.19E-05 | 1.79E-06 | -0.0018319 | SLC13A3 |
| chr1 | 156133847 | 156134868 | 1022 | 7 | 1.54E-06 | 2.09E-05 | 0.000754 | 1.79E-06 | -0.00460455 | LMNA |
| chr21 | 41601636 | 41601942 | 307 | 2 | 2.05E-06 | 1.78E-06 | 4.12E-05 | 1.79E-06 | -0.02352571 | NA |
| chr8 | 96492731 | 96493899 | 1169 | 9 | 1.18E-06 | 1.06E-05 | 0.002511 | 1.80E-06 | -0.00018338 | SDC2 |
| chr17 | 8581997 | 8582647 | 651 | 2 | 3.79E-07 | 3.41E-06 | 1.57E-05 | 1.80E-06 | -0.00486849 | MYH10 |
| chr3 | 12005118 | 12005502 | 385 | 2 | 3.41E-08 | 2.09E-06 | 3.12E-05 | 1.80E-06 | -0.01352948 | SYN2 |
| chr21 | 44939203 | 44940529 | 1327 | 11 | 1.91E-05 | 0.0001389 | 0.000646 | 1.80E-06 | 0.00020052 | FAM207A, LINC01547 |
| chr9 | 558572 | 558625 | 54 | 2 | 3.45E-06 | 1.40E-06 | 7.30E-05 | 1.80E-06 | -0.00835952 | KANK1 |
| chr22 | 30238044 | 30238164 | 121 | 2 | 3.64E-08 | 0.0001501 | 1.80E-06 | 1.80E-06 | 0.00331918 | NA |
| chr1 | 206946439 | 206947283 | 845 | 7 | 4.93E-06 | 3.32E-06 | 0.001171 | 1.80E-06 | -0.00023353 | PIGR |
| chr1 | 32702381 | 32703626 | 1246 | 8 | 6.67E-08 | 0.0007849 | 8.75E-06 | 1.80E-06 | 0.001453144 | SYNC |
| chr4 | 6366986 | 6367573 | 588 | 2 | 2.59E-08 | 0.0005501 | 1.46E-06 | 1.80E-06 | -0.01796123 | PPP2R2C |
| chr2 | 101418840 | 101419490 | 651 | 4 | 5.50E-07 | 3.10E-05 | 5.95E-05 | 1.80E-06 | -0.00695996 | RFX8 |
| chr20 | 49507516 | 49507577 | 62 | 2 | 2.26E-06 | 2.16E-06 | 2.98E-05 | 1.80E-06 | -0.00632829 | PTGIS |
| chr12 | 131030574 | 131030889 | 316 | 3 | 4.43E-06 | 8.68E-07 | 0.000429 | 1.80E-06 | -0.00612792 | ADGRD1, AC078925.1 |
| chr2 | 241040000 | 241040464 | 465 | 2 | 8.34E-07 | 1.76E-06 | 4.29E-05 | 1.80E-06 | -0.0083853 | SNED1, SNED1-AS1 |
| chr6 | 36024025 | 36024914 | 890 | 11 | 5.86E-06 | 1.17E-06 | 0.001348 | 1.81E-06 | -1.15E-05 | SLC26A8 |
| chr6 | 117776209 | 117776582 | 374 | 2 | 3.53E-06 | 1.14E-06 | 0.00017 | 1.81E-06 | -0.0214385 | NA |
| chr3 | 197298709 | 197300032 | 1324 | 14 | 2.90E-07 | 0.0003484 | 0.000114 | 1.81E-06 | 0.000757066 | DLG1-AS1, DLG1 |
| chr13 | 30832983 | 30833077 | 95 | 2 | 2.04E-06 | 1.10E-06 | 0.000224 | 1.81E-06 | 0.017864531 | NA |
| chr4 | 57164315 | 57165032 | 718 | 4 | 7.94E-11 | 0.0008901 | 8.17E-07 | 1.81E-06 | 0.001454831 | IGFBP7-AS1 |
| chr16 | 89158946 | 89159514 | 569 | 7 | 1.06E-06 | 9.10E-06 | 0.000152 | 1.81E-06 | -0.00096936 | ACSF3, LINC00304 |
| chr8 | 28203034 | 28203343 | 310 | 2 | 2.05E-06 | 1.81E-06 | 4.10E-05 | 1.81E-06 | -0.0127147 | NA |
| chr4 | 38662025 | 38663033 | 1009 | 4 | 3.46E-08 | 0.0006276 | 3.68E-06 | 1.81E-06 | -0.01337872 | KLF3-AS1 |
| chr16 | 767098 | 767463 | 366 | 2 | 1.13E-08 | 0.0002456 | 1.65E-06 | 1.81E-06 | 0.03645281 | MSLN |
| chr1 | 42849299 | 42850167 | 869 | 4 | 2.49E-07 | 3.38E-05 | 8.94E-06 | 1.82E-06 | -0.00055876 | ZNF691 |
| chr6 | 31880322 | 31881495 | 1174 | 5 | 2.15E-07 | 0.0003736 | 8.55E-06 | 1.82E-06 | -0.00151414 | EHMT2-AS1, EHMT2 |
| chr19 | 30450951 | 30451189 | 239 | 3 | 4.73E-07 | 4.35E-06 | 2.67E-05 | 1.82E-06 | 0.014261011 | ZNF536 |
| chr1 | 39459994 | 39460761 | 768 | 5 | 1.74E-06 | 4.67E-06 | 5.58E-05 | 1.82E-06 | -0.00127939 | MACF1 |
| chr17 | 49032337 | 49032583 | 247 | 2 | 1.40E-06 | 4.34E-06 | 1.23E-05 | 1.82E-06 | 0.005372185 | IGF2BP1 |
| chr2 | 231057238 | 231058445 | 1208 | 3 | 2.71E-08 | 6.09E-06 | 1.39E-05 | 1.82E-06 | 0.009143778 | PSMD1 |
| chr3 | 8769318 | 8770072 | 755 | 9 | 1.02E-06 | 6.08E-05 | 0.002929 | 1.82E-06 | 0.00906833 | CAV3, OXTR |
| chr1 | 44250270 | 44250624 | 355 | 5 | 1.05E-06 | 6.69E-06 | 1.89E-05 | 1.82E-06 | -0.00198702 | ERI3 |
| chr17 | 75130404 | 75131373 | 970 | 4 | 3.92E-08 | 6.16E-05 | 1.19E-05 | 1.82E-06 | -0.00308214 | AC022211.4, NT5C |
| chr19 | 16355487 | 16355825 | 339 | 5 | 1.59E-06 | 2.76E-05 | 0.000296 | 1.82E-06 | -0.00078325 | EPS15L1 |
| chr15 | 78951084 | 78951184 | 101 | 2 | 9.25E-07 | 2.20E-06 | 2.97E-05 | 1.82E-06 | -0.01613948 | NA |
| chr22 | 36470464 | 36470857 | 394 | 2 | 8.30E-08 | 0.0013921 | 1.34E-06 | 1.82E-06 | 0.002696441 | TXN2 |
| chr2 | 218962938 | 218963911 | 974 | 6 | 4.28E-08 | 3.40E-05 | 4.08E-05 | 1.82E-06 | 0.002302211 | NA |
| chr2 | 219447299 | 219448132 | 834 | 3 | 9.63E-08 | 3.76E-06 | 2.37E-05 | 1.82E-06 | -0.0151245 | SPEG |
| chr17 | 9771500 | 9771895 | 396 | 5 | 7.26E-07 | 9.44E-06 | 2.87E-05 | 1.83E-06 | -0.00179738 | AC027045.2, DHRS7C |
| chr7 | 135562898 | 135562946 | 49 | 2 | 2.28E-06 | 2.91E-06 | 1.97E-05 | 1.83E-06 | -0.00085931 | NUP205 |
| chr13 | 96140840 | 96141201 | 362 | 2 | 1.31E-07 | 0.0003906 | 1.54E-06 | 1.83E-06 | -0.01616949 | HS6ST3 |
| chr16 | 67458772 | 67460316 | 1545 | 5 | 1.24E-06 | 1.36E-05 | 6.82E-05 | 1.83E-06 | 0.000763335 | ATP6V0D1 |
| chr16 | 78145540 | 78145743 | 204 | 4 | 1.26E-06 | 1.72E-05 | 1.84E-05 | 1.83E-06 | -0.00326125 | WWOX |
| chr3 | 161226191 | 161226471 | 281 | 2 | 5.81E-07 | 1.57E-05 | 4.46E-06 | 1.83E-06 | -0.00614795 | NMD3 |
| chr13 | 52851500 | 52851897 | 398 | 6 | 2.09E-06 | 4.71E-06 | 0.000629 | 1.83E-06 | 0.000686958 | NA |
| chr8 | 143219724 | 143220374 | 651 | 3 | 1.96E-06 | 1.80E-05 | 5.16E-05 | 1.83E-06 | -0.0129217 | NA |
| chr4 | 70687837 | 70688846 | 1010 | 14 | 1.29E-05 | 4.89E-07 | 0.007838 | 1.83E-06 | 0.000580907 | UTP3 |
| chr9 | 135021228 | 135021677 | 450 | 2 | 2.41E-07 | 1.57E-05 | 4.47E-06 | 1.83E-06 | -0.01064827 | NA |
| chr20 | 17651082 | 17651551 | 470 | 2 | 1.66E-06 | 1.61E-06 | 5.37E-05 | 1.83E-06 | -0.03024145 | RRBP1 |
| chr1 | 226939748 | 226940836 | 1089 | 10 | 6.89E-08 | 8.03E-05 | 1.98E-05 | 1.83E-06 | 0.000264508 | AL391628.1, COQ8A |
| chr17 | 17577015 | 17578078 | 1064 | 12 | 1.56E-06 | 1.18E-05 | 0.002347 | 1.84E-06 | -0.00370166 | PEMT |
| chr1 | 6681130 | 6681714 | 585 | 8 | 3.17E-06 | 5.23E-06 | 0.002685 | 1.84E-06 | -0.00390316 | DNAJC11 |
| chr11 | 27506378 | 27507156 | 779 | 11 | 4.83E-06 | 1.86E-05 | 0.002376 | 1.84E-06 | -0.00084025 | BDNF-AS, LIN7C |
| chr21 | 44507078 | 44508509 | 1432 | 3 | 1.91E-05 | 7.93E-07 | 0.000569 | 1.84E-06 | 0.001365377 | TSPEAR-AS1, TSPEAR |
| chr2 | 27771151 | 27772212 | 1062 | 12 | 3.60E-06 | 2.60E-06 | 0.017717 | 1.84E-06 | 0.000741012 | MRPL33 |
| chr21 | 36980350 | 36981018 | 669 | 7 | 1.14E-06 | 1.06E-05 | 0.001228 | 1.84E-06 | -0.00379809 | HLCS |
| chr8 | 27771869 | 27772988 | 1120 | 8 | 1.74E-06 | 0.0002968 | 0.000303 | 1.84E-06 | -0.0098192 | ESCO2, CCDC25 |
| chr7 | 149396781 | 149397394 | 614 | 2 | 4.93E-09 | 0.00017 | 1.79E-06 | 1.85E-06 | -0.00755994 | NA |
| chr15 | 40038940 | 40039665 | 726 | 12 | 2.16E-07 | 0.0008131 | 6.36E-05 | 1.85E-06 | 0.000383122 | SRP14-AS1, SRP14 |
| chr19 | 17026394 | 17027271 | 878 | 10 | 4.76E-08 | 0.0002771 | 4.72E-05 | 1.85E-06 | -0.00077732 | CPAMD8 |
| chr9 | 111691682 | 111692012 | 331 | 2 | 1.58E-07 | 0.0005497 | 1.49E-06 | 1.85E-06 | -0.00897329 | SHOC1 |
| chr6 | 89561620 | 89562502 | 883 | 8 | 3.73E-06 | 4.74E-05 | 0.000354 | 1.85E-06 | -0.00204771 | ANKRD6, AL159174.1 |
| chr2 | 240721903 | 240722706 | 804 | 3 | 2.70E-07 | 3.33E-05 | 2.97E-05 | 1.85E-06 | -0.00222169 | KIF1A |
| chr15 | 96778621 | 96779902 | 1282 | 7 | 1.30E-06 | 5.08E-06 | 0.000369 | 1.85E-06 | -0.00582711 | SPATA8-AS1 |
| chr8 | 926130 | 927074 | 945 | 4 | 1.89E-05 | 9.12E-06 | 0.000296 | 1.85E-06 | 0.001247582 | DLGAP2 |
| chr11 | 111878476 | 111879916 | 1441 | 20 | 1.12E-05 | 3.09E-05 | 0.006087 | 1.85E-06 | 0.000276905 | C11orf1, AP001781.2, FDXACB1 |
| chr11 | 68405772 | 68405930 | 159 | 2 | 5.78E-07 | 8.50E-06 | 6.88E-06 | 1.85E-06 | -0.00471569 | LRP5 |
| chr3 | 23944559 | 23945715 | 1157 | 7 | 1.57E-06 | 1.22E-06 | 0.00455 | 1.85E-06 | 0.001980671 | NR1D2, NKIRAS1 |
| chr6 | 27545070 | 27545127 | 58 | 2 | 1.01E-06 | 7.63E-06 | 7.53E-06 | 1.85E-06 | -0.00647957 | NA |
| chr3 | 195178470 | 195178993 | 524 | 2 | 7.13E-08 | 1.70E-05 | 4.33E-06 | 1.86E-06 | -0.01316985 | XXYLT1 |
| chr18 | 23585017 | 23585463 | 447 | 2 | 2.62E-07 | 3.60E-06 | 1.57E-05 | 1.86E-06 | -0.00943881 | NPC1 |
| chr17 | 35795165 | 35796322 | 1158 | 13 | 1.71E-07 | 5.11E-05 | 0.001064 | 1.86E-06 | -0.00016339 | TAF15, MMP28 |
| chr22 | 43530083 | 43530592 | 510 | 2 | 9.01E-07 | 2.05E-06 | 3.48E-05 | 1.86E-06 | 0.00642702 | EFCAB6-AS1, EFCAB6 |
| chr10 | 90834633 | 90835052 | 420 | 2 | 6.59E-07 | 4.88E-06 | 1.14E-05 | 1.86E-06 | -0.00883078 | HTR7 |
| chr6 | 31717653 | 31717798 | 146 | 6 | 2.86E-06 | 6.72E-06 | 0.000445 | 1.86E-06 | -0.00084091 | LY6G6F-LY6G6D, LY6G6D |
| chr22 | 42719931 | 42721229 | 1299 | 13 | 7.68E-06 | 4.23E-06 | 0.007209 | 1.86E-06 | -0.00071019 | A4GALT |
| chr12 | 113097557 | 113097927 | 371 | 3 | 2.29E-06 | 8.67E-07 | 0.000781 | 1.86E-06 | -0.01144853 | DTX1 |
| chr19 | 4902564 | 4902887 | 324 | 6 | 1.67E-06 | 1.02E-05 | 3.90E-05 | 1.86E-06 | 0.004529342 | ARRDC5 |
| chr2 | 40745486 | 40746307 | 822 | 3 | 1.52E-06 | 5.24E-05 | 0.000144 | 1.87E-06 | 0.01047273 | NA |
| chr7 | 154899394 | 154900147 | 754 | 4 | 6.38E-06 | 4.98E-07 | 0.002576 | 1.87E-06 | -0.00574453 | NA |
| chr4 | 108350348 | 108350683 | 336 | 2 | 7.64E-07 | 6.99E-06 | 8.17E-06 | 1.87E-06 | -0.00509713 | NA |
| chr10 | 116827013 | 116827045 | 33 | 2 | 4.05E-06 | 1.12E-06 | 0.000261 | 1.87E-06 | -0.00474716 | HSPA12A |
| chr16 | 89885294 | 89885541 | 248 | 3 | 7.11E-07 | 2.66E-05 | 6.63E-05 | 1.87E-06 | -0.00597784 | TCF25 |
| chr2 | 85186290 | 85186374 | 85 | 3 | 8.78E-07 | 4.28E-06 | 6.76E-05 | 1.87E-06 | -0.00914335 | TCF7L1 |
| chr7 | 32489928 | 32491133 | 1206 | 19 | 3.84E-06 | 1.45E-05 | 0.001023 | 1.87E-06 | 0.00094488 | LSM5 |
| chr2 | 45812313 | 45812639 | 327 | 2 | 8.27E-07 | 6.60E-06 | 8.61E-06 | 1.87E-06 | 0.001363129 | PRKCE |
| chr12 | 31354021 | 31354179 | 159 | 2 | 3.13E-06 | 1.61E-06 | 5.71E-05 | 1.87E-06 | -0.01371723 | NA |
| chr7 | 74453127 | 74454540 | 1414 | 13 | 1.78E-07 | 0.0015317 | 0.000685 | 1.87E-06 | 0.000741828 | GTF2IRD1 |
| chr4 | 140237657 | 140238269 | 613 | 2 | 4.57E-07 | 1.31E-06 | 0.000104 | 1.87E-06 | 0.003913784 | NA |
| chr1 | 40096892 | 40097733 | 842 | 10 | 1.21E-06 | 3.17E-06 | 0.000644 | 1.88E-06 | 0.000749351 | PPT1 |
| chr18 | 59567598 | 59567739 | 142 | 2 | 3.22E-06 | 1.61E-06 | 5.81E-05 | 1.88E-06 | -0.00185116 | AC090213.1, CCBE1 |
| chr19 | 55132714 | 55132861 | 148 | 2 | 2.00E-06 | 2.51E-06 | 2.56E-05 | 1.88E-06 | -0.0327752 | TNNT1 |
| chr14 | 90555951 | 90556064 | 114 | 2 | 2.52E-06 | 2.74E-06 | 2.26E-05 | 1.88E-06 | -0.00763496 | TTC7B |
| chr11 | 63635829 | 63636347 | 519 | 4 | 3.78E-06 | 7.52E-06 | 0.000239 | 1.88E-06 | -0.00546425 | ATL3 |
| chr11 | 39861726 | 39862887 | 1162 | 3 | 1.84E-09 | 0.0008412 | 8.00E-07 | 1.88E-06 | -0.01315198 | NA |
| chr14 | 73236242 | 73236455 | 214 | 3 | 1.97E-07 | 0.0001137 | 0.00013 | 1.88E-06 | -0.0111813 | NA |
| chr20 | 48231596 | 48232016 | 421 | 2 | 4.68E-07 | 5.50E-06 | 1.03E-05 | 1.88E-06 | -0.00049551 | NA |
| chr4 | 140012643 | 140012847 | 205 | 2 | 3.16E-06 | 1.46E-06 | 7.54E-05 | 1.88E-06 | -0.01273882 | MAML3 |
| chr4 | 87610411 | 87610761 | 351 | 3 | 4.63E-07 | 0.0006358 | 4.12E-06 | 1.88E-06 | 0.00297632 | DSPP, AC093895.1 |
| chr16 | 971074 | 972008 | 935 | 6 | 1.01E-06 | 3.76E-06 | 0.000597 | 1.88E-06 | -0.00601333 | LMF1 |
| chr1 | 1084985 | 1085919 | 935 | 5 | 1.94E-08 | 7.58E-05 | 2.61E-05 | 1.88E-06 | -0.00255836 | C1orf159 |
| chr12 | 120063747 | 120064613 | 867 | 4 | 1.52E-05 | 4.61E-06 | 0.00014 | 1.88E-06 | -0.00518177 | BICDL1 |
| chr16 | 87708305 | 87709680 | 1376 | 8 | 1.06E-07 | 0.0001509 | 1.98E-05 | 1.88E-06 | -0.00100988 | KLHDC4 |
| chr15 | 45377870 | 45379510 | 1641 | 17 | 1.30E-07 | 6.57E-05 | 0.000841 | 1.89E-06 | 0.005248633 | AC025580.3, GATM |
| chr17 | 696589 | 697346 | 758 | 4 | 3.00E-06 | 2.78E-06 | 9.78E-05 | 1.89E-06 | 0.003447643 | VPS53 |
| chr10 | 13889052 | 13890304 | 1253 | 3 | 1.08E-05 | 6.70E-07 | 0.001431 | 1.89E-06 | -0.00836534 | FRMD4A |
| chr4 | 178196841 | 178197202 | 362 | 3 | 1.49E-06 | 4.15E-06 | 0.00011 | 1.89E-06 | -0.00986258 | NA |
| chr10 | 7702759 | 7703491 | 733 | 4 | 2.29E-06 | 1.77E-06 | 0.000308 | 1.89E-06 | -0.00640054 | ITIH2 |
| chr2 | 39782361 | 39782734 | 374 | 2 | 2.41E-07 | 4.49E-05 | 2.73E-06 | 1.89E-06 | 0.003275523 | NA |
| chr5 | 78294487 | 78295596 | 1110 | 12 | 8.75E-07 | 9.89E-06 | 0.001729 | 1.89E-06 | 0.000543044 | AP3B1 |
| chr19 | 31363166 | 31363269 | 104 | 2 | 2.93E-06 | 2.26E-06 | 3.06E-05 | 1.89E-06 | -0.01473286 | AC025809.1 |
| chr15 | 98906865 | 98907560 | 696 | 3 | 5.81E-07 | 2.42E-06 | 0.00013 | 1.89E-06 | 0.006123662 | IGF1R |
| chr6 | 106325820 | 106326690 | 871 | 9 | 7.80E-08 | 0.0017368 | 0.000165 | 1.89E-06 | -0.00869171 | NA |
| chr18 | 5073871 | 5074288 | 418 | 2 | 8.69E-07 | 1.17E-06 | 0.000202 | 1.90E-06 | 0.020780922 | NA |
| chr16 | 2911484 | 2912050 | 567 | 7 | 5.26E-07 | 2.09E-05 | 7.70E-06 | 1.90E-06 | 0.000173712 | FLYWCH1 |
| chr16 | 88880497 | 88881535 | 1039 | 12 | 6.36E-06 | 8.00E-07 | 0.010626 | 1.90E-06 | -0.00191366 | AC092384.2, CBFA2T3 |
| chr17 | 18889166 | 18889538 | 373 | 3 | 1.56E-06 | 4.39E-06 | 5.34E-05 | 1.90E-06 | -0.00917302 | PRPSAP2 |
| chr13 | 42485381 | 42485572 | 192 | 2 | 5.85E-07 | 9.09E-05 | 2.15E-06 | 1.90E-06 | -0.00705871 | LINC02341 |
| chr6 | 39715483 | 39716002 | 520 | 2 | 1.31E-06 | 2.16E-06 | 3.34E-05 | 1.90E-06 | -0.01154957 | KIF6 |
| chr2 | 24707406 | 24707526 | 121 | 2 | 1.38E-06 | 1.09E-05 | 5.95E-06 | 1.90E-06 | 0.004220698 | NCOA1 |
| chr6 | 159786453 | 159786866 | 414 | 2 | 5.92E-08 | 2.56E-05 | 3.54E-06 | 1.90E-06 | -0.0134486 | TCP1 |
| chr22 | 46059219 | 46059790 | 572 | 4 | 5.79E-07 | 0.0001513 | 0.000111 | 1.90E-06 | 0.002227407 | MIRLET7BHG |

| | | | | | | | | | | |
|---|---|---|---|---|---|---|---|---|---|---|
| chr16 | 6018830 | 6019730 | 901 | 13 | 1.12E-05 | 6.09E-07 | 0.015524 | 1.90E-06 | 0.000447867 | RBFOX1 |
| chr13 | 114161029 | 114161560 | 532 | 6 | 5.63E-07 | 7.86E-05 | 0.000572 | 1.91E-06 | 0.003607194 | CFAP97D2 |
| chr15 | 90388220 | 90388758 | 539 | 4 | 4.06E-06 | 1.45E-06 | 0.000142 | 1.91E-06 | 0.000800061 | IQGAP1 |
| chr22 | 33873322 | 33873433 | 112 | 2 | 5.77E-07 | 1.48E-05 | 4.88E-06 | 1.91E-06 | -0.00886274 | LARGE1 |
| chr2 | 241482608 | 241482686 | 79 | 2 | 8.89E-07 | 5.94E-05 | 2.47E-06 | 1.91E-06 | -0.00789249 | FARP2 |
| chr6 | 150831690 | 150831780 | 91 | 2 | 2.85E-06 | 2.59E-06 | 2.52E-05 | 1.91E-06 | -0.00798567 | PLEKHG1 |
| chr22 | 28771707 | 28772400 | 694 | 4 | 9.85E-07 | 1.36E-06 | 0.000734 | 1.91E-06 | -0.00221732 | NA |
| chr11 | 504301 | 504933 | 633 | 6 | 2.97E-07 | 1.25E-06 | 0.002191 | 1.91E-06 | -0.00338039 | RNH1 |
| chr17 | 3803296 | 3804062 | 767 | 5 | 1.45E-06 | 2.79E-06 | 0.000571 | 1.91E-06 | -0.00014519 | NCBP3 |
| chr8 | 43281019 | 43281685 | 667 | 5 | 7.57E-07 | 7.17E-06 | 8.17E-05 | 1.91E-06 | 0.000974976 | NA |
| chr10 | 84194197 | 84195202 | 1006 | 9 | 1.32E-07 | 3.35E-06 | 0.000789 | 1.91E-06 | -0.00129613 | CDHR1 |
| chr17 | 8758684 | 8759977 | 1294 | 4 | 8.86E-06 | 1.41E-05 | 0.0003 | 1.91E-06 | -0.00556988 | NA |
| chr20 | 3785615 | 3786314 | 700 | 5 | 1.59E-06 | 1.15E-06 | 0.001535 | 1.92E-06 | -0.00683733 | CENPB |
| chr4 | 6269774 | 6270560 | 787 | 9 | 3.56E-07 | 4.30E-06 | 0.000793 | 1.92E-06 | 0.00182911 | WFS1 |
| chr1 | 70720386 | 70721082 | 697 | 4 | 6.51E-07 | 7.42E-05 | 0.000217 | 1.92E-06 | -0.00686988 | LINC01788 |
| chr6 | 93785341 | 93785507 | 167 | 2 | 2.74E-06 | 1.14E-06 | 0.000265 | 1.92E-06 | -0.00951699 | NA |
| chr20 | 12244440 | 12244677 | 238 | 4 | 3.07E-06 | 3.42E-06 | 7.62E-05 | 1.92E-06 | -0.00574515 | AL109838.1 |
| chr5 | 112627723 | 112628262 | 540 | 2 | 2.55E-07 | 3.09E-06 | 2.00E-05 | 1.92E-06 | -0.01367984 | NA |
| chr2 | 65070113 | 65070718 | 606 | 5 | 4.42E-06 | 4.61E-07 | 0.002488 | 1.92E-06 | 0.006418667 | CEP68, RAB1A |
| chr19 | 55345791 | 55346669 | 879 | 6 | 1.19E-06 | 1.08E-05 | 0.000241 | 1.92E-06 | 0.00075986 | KMT5C |
| chr3 | 4752336 | 4752859 | 524 | 4 | 3.40E-06 | 2.57E-06 | 0.000243 | 1.92E-06 | 0.008540575 | ITPR1 |
| chr18 | 10374593 | 10375089 | 497 | 5 | 5.09E-06 | 6.08E-06 | 0.001031 | 1.92E-06 | 0.001952949 | NA |
| chr4 | 118836140 | 118837038 | 899 | 10 | 1.19E-05 | 1.33E-06 | 0.002303 | 1.93E-06 | 0.000657879 | SEC24D |
| chr22 | 50076894 | 50077818 | 925 | 5 | 3.20E-06 | 4.17E-06 | 0.001351 | 1.93E-06 | -0.00077079 | MLC1 |
| chr6 | 155084871 | 155085164 | 294 | 3 | 8.25E-07 | 5.68E-06 | 1.63E-05 | 1.93E-06 | -0.00600043 | TIAM2 |
| chr7 | 16804982 | 16805596 | 615 | 6 | 4.40E-08 | 0.0002058 | 8.03E-06 | 1.93E-06 | 0.003359967 | AGR2 |
| chr6 | 145815235 | 145815846 | 612 | 5 | 4.53E-08 | 1.21E-05 | 0.000488 | 1.93E-06 | -0.00468069 | FBXO30-DT |
| chr7 | 226444 | 227051 | 608 | 5 | 5.25E-08 | 0.0007576 | 3.00E-06 | 1.93E-06 | -8.94E-07 | FAM20C |
| chr14 | 39347016 | 39347575 | 560 | 3 | 1.52E-07 | 0.0002598 | 1.33E-06 | 1.93E-06 | 0.005640529 | MIA2 |
| chr17 | 16281380 | 16281986 | 607 | 3 | 1.28E-06 | 2.61E-06 | 5.74E-05 | 1.93E-06 | -0.00392626 | PIGL |
| chr7 | 99785601 | 99785627 | 27 | 2 | 4.08E-06 | 1.25E-06 | 0.000156 | 1.93E-06 | -0.00661967 | NA |
| chr8 | 47697593 | 47697752 | 160 | 2 | 1.66E-07 | 0.0001682 | 1.88E-06 | 1.93E-06 | -0.01626424 | SPIDR |
| chr1 | 245673533 | 245674420 | 888 | 2 | 2.54E-06 | 1.58E-06 | 6.68E-05 | 1.94E-06 | -0.00448144 | KIF26B, AC104462.1 |
| chr5 | 180860685 | 180861855 | 1171 | 11 | 7.79E-07 | 4.98E-05 | 0.001523 | 1.94E-06 | 0.000470751 | ZFP62 |
| chr9 | 131836575 | 131837362 | 788 | 3 | 1.14E-05 | 9.57E-07 | 0.000692 | 1.94E-06 | -0.01299073 | NA |
| chr8 | 144787041 | 144787844 | 804 | 9 | 1.60E-06 | 7.80E-06 | 0.000232 | 1.94E-06 | -0.00116864 | ZNF34 |
| chr4 | 25655743 | 25656505 | 763 | 9 | 5.53E-06 | 1.52E-05 | 0.004334 | 1.94E-06 | 0.003575119 | SLC34A2 |
| chr11 | 128939017 | 128939244 | 228 | 4 | 2.26E-06 | 4.63E-06 | 0.00018 | 1.94E-06 | -0.00113978 | TP53AIP1 |
| chr17 | 50055350 | 50056322 | 973 | 11 | 8.09E-08 | 4.87E-05 | 0.00057 | 1.94E-06 | 0.000823518 | ITGA3, PICART1 |
| chr2 | 201115265 | 201116144 | 880 | 8 | 1.46E-07 | 0.0001003 | 2.24E-05 | 1.94E-06 | -0.00165737 | NA |
| chr8 | 140953232 | 140953545 | 314 | 2 | 4.71E-07 | 2.59E-05 | 3.60E-06 | 1.94E-06 | 0.005327352 | PTK2 |
| chr14 | 96100236 | 96100334 | 99 | 3 | 1.08E-06 | 2.74E-05 | 1.10E-05 | 1.94E-06 | -0.00362459 | NA |
| chr12 | 132322917 | 132324352 | 1436 | 10 | 8.40E-07 | 3.40E-05 | 0.001046 | 1.95E-06 | 0.006965911 | GALNT9 |
| chr15 | 81374171 | 81375096 | 926 | 6 | 9.20E-06 | 1.12E-05 | 0.000195 | 1.95E-06 | -0.00073908 | TMC3-AS1, TMC3 |
| chr5 | 67095166 | 67095610 | 445 | 2 | 6.19E-08 | 0.0001137 | 2.08E-06 | 1.95E-06 | -0.01197686 | MAST4 |
| chr11 | 63894754 | 63895154 | 401 | 4 | 2.26E-06 | 8.06E-06 | 5.20E-05 | 1.95E-06 | 0.000581604 | MARK2 |
| chr16 | 71577616 | 71578229 | 614 | 2 | 3.13E-08 | 4.57E-05 | 2.81E-06 | 1.95E-06 | -0.01218822 | TAT-AS1 |
| chr7 | 1693794 | 1694753 | 960 | 8 | 2.80E-06 | 5.48E-06 | 0.002255 | 1.95E-06 | -0.0057044 | ELFN1, AC074389.1 |
| chr2 | 55050485 | 55051291 | 807 | 9 | 1.41E-06 | 1.27E-06 | 0.001886 | 1.95E-06 | -0.00018984 | RTN4 |
| chr2 | 159347601 | 159347813 | 213 | 2 | 1.53E-06 | 5.97E-06 | 1.01E-05 | 1.95E-06 | -0.00143853 | BAZ2B |
| chr11 | 66637979 | 66639273 | 1295 | 10 | 1.17E-07 | 0.0001151 | 0.000149 | 1.95E-06 | 9.00E-05 | RBM14-RBM4, RBM4 |
| chr1 | 156060868 | 156061704 | 837 | 8 | 3.03E-06 | 1.46E-05 | 0.000434 | 1.95E-06 | -0.00179418 | RAB25 |
| chr5 | 57780365 | 57780447 | 83 | 2 | 2.68E-07 | 3.53E-05 | 3.12E-06 | 1.95E-06 | 0.005708705 | NA |
| chr17 | 41929865 | 41930801 | 937 | 13 | 3.92E-06 | 2.43E-05 | 0.001105 | 1.96E-06 | 0.001068068 | TTC25, ACLY |
| chr15 | 68277098 | 68278011 | 914 | 11 | 6.27E-07 | 3.82E-05 | 0.000867 | 1.96E-06 | -0.00096633 | FEM1B, AC107871.2 |
| chr10 | 6735912 | 6736200 | 289 | 3 | 9.92E-07 | 1.43E-05 | 1.01E-05 | 1.96E-06 | -0.00067958 | AL158210.1, LINP1 |
| chr10 | 102802538 | 102803272 | 735 | 4 | 4.26E-06 | 9.88E-05 | 0.000617 | 1.96E-06 | -0.00259961 | WBP1L |
| chr16 | 76234887 | 76235286 | 400 | 2 | 1.79E-06 | 2.04E-06 | 3.95E-05 | 1.96E-06 | -0.00048598 | AC010528.1, RPL18P13 |
| chr12 | 16608485 | 16610546 | 2062 | 12 | 4.22E-06 | 0.0001297 | 0.00041 | 1.96E-06 | 0.00710321 | MGST1, LMO3 |
| chr10 | 43025969 | 43026895 | 927 | 7 | 1.27E-06 | 5.73E-06 | 0.000262 | 1.96E-06 | -0.00915704 | NA |
| chr18 | 58135279 | 58135726 | 448 | 2 | 1.10E-06 | 1.86E-06 | 4.72E-05 | 1.96E-06 | 0.006256275 | NEDD4L |
| chr20 | 18059100 | 18059445 | 346 | 6 | 1.90E-06 | 6.40E-06 | 0.000579 | 1.97E-06 | -0.00194046 | OVOL2 |
| chr5 | 31832183 | 31832334 | 152 | 4 | 2.05E-06 | 9.11E-06 | 2.70E-05 | 1.97E-06 | -0.00095115 | PDZD2 |
| chr1 | 9708484 | 9709619 | 1136 | 4 | 7.72E-08 | 1.05E-05 | 1.33E-05 | 1.97E-06 | 0.00655864 | PIK3CD |
| chr19 | 14518428 | 14519105 | 678 | 6 | 1.89E-07 | 1.56E-06 | 0.00417 | 1.97E-06 | 0.001332696 | TECR, DNAJB1 |
| chr10 | 100996679 | 100997633 | 955 | 15 | 1.78E-06 | 7.48E-06 | 4.73E-05 | 1.97E-06 | -0.0011136 | LZTS2 |
| chr11 | 76680209 | 76680958 | 750 | 4 | 3.54E-06 | 1.77E-06 | 0.000531 | 1.97E-06 | -0.00197908 | AP001189.4 |
| chr8 | 98949317 | 98950119 | 803 | 6 | 5.13E-07 | 7.90E-06 | 0.000758 | 1.97E-06 | -0.00697649 | OSR2 |
| chr11 | 61332780 | 61333821 | 1042 | 11 | 1.44E-06 | 3.63E-06 | 0.003477 | 1.97E-06 | -0.00024609 | TKFC, DDB1 |
| chr12 | 101283508 | 101283583 | 76 | 2 | 1.15E-06 | 4.28E-05 | 2.93E-06 | 1.97E-06 | -0.00611848 | UTP20 |
| chr11 | 33379028 | 33379943 | 916 | 4 | 3.68E-07 | 5.12E-05 | 4.06E-06 | 1.98E-06 | -0.00096615 | KIAA1549L |
| chr20 | 63156809 | 63157631 | 823 | 3 | 3.39E-08 | 5.75E-06 | 1.66E-05 | 1.98E-06 | 0.002626003 | NA |
| chr19 | 43926403 | 43926886 | 484 | 4 | 8.12E-08 | 3.42E-05 | 7.98E-06 | 1.98E-06 | 0.00385181 | AC006213.3, ZNF45 |
| chr6 | 158167731 | 158168825 | 1095 | 16 | 1.19E-05 | 1.22E-06 | 0.005157 | 1.98E-06 | 0.000530069 | GTF2H5, SERAC1 |
| chr14 | 23366003 | 23366838 | 836 | 5 | 5.27E-08 | 0.00014 | 0.000261 | 1.98E-06 | -0.00981932 | NA |
| chr4 | 121696152 | 121696852 | 701 | 5 | 2.84E-07 | 3.39E-05 | 2.05E-05 | 1.98E-06 | 0.002439278 | ANXA5 |
| chr10 | 71816516 | 71816653 | 138 | 2 | 4.77E-07 | 2.60E-05 | 3.66E-06 | 1.98E-06 | 0.004640602 | PSAP |
| chr8 | 753273 | 754148 | 876 | 6 | 6.42E-07 | 5.03E-06 | 0.001138 | 1.98E-06 | 0.011011766 | DLGAP2 |
| chr19 | 12156649 | 12157618 | 970 | 10 | 2.81E-06 | 1.27E-05 | 0.006086 | 1.98E-06 | -0.00139323 | ZNF625-ZNF20, ZNF625 |
| chr12 | 72273927 | 72275111 | 1185 | 4 | 8.76E-06 | 9.21E-06 | 0.000122 | 1.98E-06 | -0.00934512 | TRHDE, TRHDE-AS1 |
| chr16 | 637604 | 639107 | 1504 | 6 | 4.67E-05 | 8.61E-06 | 0.001123 | 1.98E-06 | -0.00561218 | MCRIP2 |
| chr3 | 100185374 | 100185806 | 433 | 6 | 9.67E-07 | 3.58E-05 | 8.41E-05 | 1.98E-06 | -0.00023239 | TMEM30CP, AC129803.1 |
| chr11 | 73875731 | 73876415 | 685 | 5 | 2.79E-08 | 1.75E-05 | 2.78E-05 | 1.98E-06 | -0.00780221 | COA4 |
| chr2 | 241686801 | 241688006 | 1206 | 12 | 1.28E-05 | 2.45E-06 | 0.007871 | 1.98E-06 | -0.00220122 | ING5, DTYMK |
| chr21 | 44626125 | 44626476 | 352 | 4 | 2.59E-06 | 1.73E-05 | 8.51E-05 | 1.98E-06 | -0.00152835 | KRTAP10-4, TSPEAR |
| chr1 | 6306247 | 6306598 | 352 | 4 | 6.20E-06 | 9.81E-07 | 0.001117 | 1.98E-06 | -0.00039583 | ACOT7 |
| chr10 | 78267018 | 78267375 | 358 | 5 | 5.63E-07 | 2.56E-05 | 2.28E-05 | 1.98E-06 | -0.00637477 | AC012560.1, LINC00595 |
| chr3 | 9554798 | 9555165 | 368 | 2 | 5.80E-08 | 6.61E-06 | 9.37E-06 | 1.98E-06 | -0.00989097 | NA |
| chr18 | 69949294 | 69950053 | 760 | 2 | 1.53E-06 | 1.61E-06 | 6.75E-05 | 1.98E-06 | -0.00772118 | CD226 |
| chr10 | 129207432 | 129207514 | 83 | 2 | 9.69E-07 | 7.55E-05 | 2.37E-06 | 1.98E-06 | -0.0031195 | NA |
| chr10 | 109411988 | 109412070 | 83 | 2 | 2.42E-06 | 4.71E-06 | 1.31E-05 | 1.98E-06 | 0.004228395 | NA |
| chr3 | 3485002 | 3485609 | 608 | 4 | 5.74E-06 | 1.32E-06 | 0.000295 | 1.98E-06 | 0.007376506 | AC034195.1 |
| chr18 | 78362391 | 78362611 | 221 | 2 | 1.14E-07 | 4.81E-06 | 1.28E-05 | 1.98E-06 | -0.00440564 | NA |
| chr2 | 241198877 | 241199126 | 250 | 3 | 3.01E-07 | 8.31E-05 | 2.12E-06 | 1.98E-06 | -0.00228989 | ANO7 |
| chr4 | 99563471 | 99564292 | 822 | 14 | 2.93E-06 | 2.21E-06 | 0.005148 | 1.98E-06 | -0.00102884 | MTTP, TRMT10A |
| chr1 | 151190124 | 151191299 | 1176 | 9 | 3.08E-05 | 1.51E-05 | 0.000768 | 1.98E-06 | -0.00041202 | VPS72 |
| chr18 | 79711434 | 79711861 | 428 | 3 | 1.79E-07 | 3.09E-05 | 1.30E-05 | 1.98E-06 | -0.03017721 | CTDP1 |
| chr11 | 1306732 | 1307423 | 692 | 7 | 3.00E-07 | 0.0002185 | 0.00017 | 1.98E-06 | 0.004579661 | TOLLIP |
| chr10 | 84235438 | 84236006 | 569 | 2 | 6.51E-06 | 1.21E-06 | 0.000226 | 1.98E-06 | -0.00926281 | RGR, LRIT1 |
| chr12 | 51855998 | 51856068 | 71 | 2 | 3.23E-06 | 2.36E-06 | 3.17E-05 | 1.98E-06 | -0.01328181 | NA |
| chr14 | 28876790 | 28876803 | 14 | 2 | 3.78E-07 | 2.31E-06 | 3.28E-05 | 1.99E-06 | -0.0184388 | LINC02327, LINC02326 |
| chr1 | 7664473 | 7664978 | 506 | 3 | 3.40E-08 | 0.0002885 | 5.48E-06 | 1.99E-06 | 0.000706925 | CAMTA1 |
| chr17 | 50668294 | 50668955 | 662 | 2 | 8.13E-06 | 1.20E-06 | 0.000244 | 1.99E-06 | -0.0143493 | ABCC3 |
| chr8 | 38417102 | 38418088 | 987 | 6 | 2.46E-06 | 3.43E-05 | 0.000156 | 1.99E-06 | -0.00251139 | FGFR1 |
| chr10 | 5943937 | 5945254 | 1318 | 3 | 1.65E-08 | 1.14E-05 | 8.53E-06 | 1.99E-06 | 0.008248437 | AL137186.2, IL15RA |
| chr12 | 132080588 | 132081015 | 428 | 2 | 5.91E-08 | 1.87E-05 | 4.45E-06 | 1.99E-06 | 0.002913957 | NA |
| chr19 | 1163486 | 1164146 | 661 | 3 | 4.49E-07 | 7.92E-06 | 1.85E-05 | 1.99E-06 | -0.00776751 | SBNO2 |
| chr15 | 63601213 | 63602097 | 885 | 6 | 8.74E-06 | 3.47E-07 | 0.007128 | 1.99E-06 | -0.00525142 | FBXL22, USP3-AS1 |
| chr10 | 132557129 | 132557138 | 10 | 2 | 1.04E-06 | 1.20E-06 | 0.000243 | 1.99E-06 | -0.00036707 | INPP5A |
| chr3 | 56556914 | 56557746 | 833 | 12 | 1.11E-06 | 7.31E-05 | 0.001474 | 1.99E-06 | 0.000592624 | CCDC66 |
| chr5 | 149999570 | 150000793 | 1224 | 14 | 1.57E-06 | 2.90E-05 | 0.000978 | 1.99E-06 | -0.00212682 | HMGXB3, TIGD6 |
| chr22 | 36379707 | 36380064 | 358 | 3 | 1.48E-06 | 6.25E-06 | 0.000101 | 1.99E-06 | -0.00024958 | MYH9 |
| chr9 | 131545505 | 131546130 | 626 | 3 | 3.68E-06 | 6.96E-07 | 0.001602 | 1.99E-06 | -0.00731997 | PRRT1B |
| chr14 | 94479831 | 94480366 | 536 | 3 | 4.84E-07 | 1.56E-05 | 0.000152 | 1.99E-06 | -0.00061164 | NA |
| chr14 | 93115533 | 93116434 | 902 | 8 | 8.52E-07 | 6.04E-06 | 0.000383 | 2.00E-06 | -4.67E-05 | ITPK1 |
| chr10 | 34954895 | 34955463 | 569 | 2 | 4.50E-08 | 1.74E-05 | 4.70E-06 | 2.00E-06 | -0.00895815 | NA |
| chr9 | 127785546 | 127786837 | 1292 | 10 | 7.62E-07 | 7.16E-06 | 0.000371 | 2.00E-06 | -0.0013804 | CDK9, MIR3960, MIR2861 |
| chr5 | 141094273 | 141094994 | 722 | 7 | 8.19E-08 | 1.65E-05 | 0.000114 | 2.00E-06 | -0.0125815 | PCDHB2, AC244517.5 |
| chr5 | 10324363 | 10324455 | 93 | 2 | 1.43E-06 | 6.76E-06 | 9.33E-06 | 2.00E-06 | -0.00934057 | NA |
| chr3 | 177440420 | 177440963 | 544 | 3 | 2.51E-07 | 8.16E-06 | 3.19E-05 | 2.00E-06 | 0.009849251 | NA |
| chr4 | 158209958 | 158210633 | 676 | 10 | 2.15E-06 | 4.46E-06 | 0.004557 | 2.00E-06 | -0.00084981 | TMEM144 |
| chr11 | 70816452 | 70816942 | 491 | 2 | 4.46E-06 | 1.21E-06 | 0.00025 | 2.00E-06 | -0.0085418 | SHANK2 |
| chr5 | 14011307 | 14011925 | 619 | 4 | 2.95E-06 | 1.67E-06 | 0.00016 | 2.00E-06 | -0.00281131 | NA |
| chr12 | 45379225 | 45379601 | 377 | 2 | 4.15E-07 | 1.54E-05 | 5.05E-06 | 2.00E-06 | 0.00311498 | ANO6 |
| chr11 | 67389490 | 67389835 | 346 | 3 | 2.01E-06 | 8.67E-07 | 0.00081 | 2.00E-06 | -0.00369071 | RAD9A |
| chr17 | 58007453 | 58008048 | 596 | 11 | 3.14E-07 | 6.34E-05 | 0.000369 | 2.00E-06 | -0.00022428 | AC015813.5 |
| chr3 | 135514028 | 135514303 | 276 | 4 | 1.16E-07 | 0.0039176 | 3.85E-07 | 2.00E-06 | -0.00927854 | NA |
| chr3 | 111541909 | 111542456 | 548 | 8 | 7.55E-07 | 2.51E-05 | 0.000367 | 2.00E-06 | 0.002441885 | CD96 |
| chr19 | 1262968 | 1263081 | 114 | 2 | 9.88E-07 | 7.00E-06 | 9.08E-06 | 2.01E-06 | 0.014708868 | CIRBP |
| chr2 | 314573 | 315136 | 564 | 4 | 5.38E-07 | 1.11E-05 | 4.53E-05 | 2.01E-06 | 0.000497255 | NA |
| chr1 | 36154621 | 36155605 | 985 | 5 | 1.88E-05 | 2.01E-05 | 0.00096 | 2.01E-06 | -0.00295518 | MAP7D1, TRAPPC3 |
| chr12 | 46825843 | 46826414 | 572 | 12 | 6.13E-06 | 1.94E-07 | 0.009389 | 2.01E-06 | -0.01437563 | AC008014.1, SLC38A4 |
| chr3 | 53283638 | 53284053 | 416 | 3 | 2.87E-07 | 4.26E-05 | 3.48E-06 | 2.01E-06 | -0.00448693 | DCP1A |
| chr16 | 27209203 | 27210122 | 920 | 5 | 1.24E-05 | 4.96E-06 | 0.001763 | 2.01E-06 | -0.00034638 | KDM8 |
| chr6 | 148972523 | 148972956 | 434 | 3 | 6.69E-07 | 2.09E-05 | 2.57E-05 | 2.01E-06 | -0.01627271 | UST |
| chr19 | 17405425 | 17406559 | 1135 | 11 | 3.74E-07 | 4.59E-06 | 0.001229 | 2.01E-06 | -0.00206408 | BISPR, MVB12A, BST2 |
| chr12 | 113465080 | 113466571 | 1492 | 8 | 1.54E-07 | 0.0004308 | 0.001215 | 2.02E-06 | -0.00112736 | LHX5 |
| chr17 | 6444213 | 6444905 | 693 | 13 | 2.46E-07 | 0.0002224 | 0.000196 | 2.02E-06 | 0.00227468 | PIMREG |
| chr9 | 135905018 | 135905183 | 166 | 2 | 1.50E-06 | 2.77E-06 | 2.56E-05 | 2.02E-06 | -0.00698448 | CAMSAP1 |
| chr2 | 237096545 | 237097169 | 625 | 2 | 5.01E-08 | 5.13E-05 | 2.79E-06 | 2.02E-06 | -0.00564713 | COPS8 |
| chr8 | 95269276 | 95269787 | 512 | 6 | 2.18E-07 | 0.0005227 | 0.000408 | 2.02E-06 | -0.00144307 | C8orf37-AS1 |
| chr16 | 2183797 | 2184570 | 774 | 6 | 2.41E-06 | 8.60E-07 | 0.000583 | 2.02E-06 | 0.000641463 | CASKIN1 |
| chr2 | 235026301 | 235027493 | 1193 | 5 | 1.86E-07 | 2.34E-05 | 0.000108 | 2.03E-06 | -0.00039586 | SH3BP4 |
| chr16 | 635686 | 636584 | 899 | 8 | 1.22E-06 | 5.09E-06 | 0.001648 | 2.03E-06 | 0.002662416 | METTL26 |
| chr2 | 11539458 | 11540018 | 561 | 8 | 1.26E-07 | 0.0003839 | 2.61E-05 | 2.03E-06 | -0.00443443 | GREB1 |
| chr5 | 180440108 | 180440607 | 500 | 3 | 1.43E-08 | 3.72E-05 | 4.88E-06 | 2.03E-06 | -0.01109672 | AC034213.1 |
| chr11 | 7484704 | 7485709 | 1006 | 6 | 1.55E-06 | 2.39E-06 | 0.001283 | 2.03E-06 | -0.00683484 | OLFML1, AC107884.1 |
| chr14 | 104700963 | 104701390 | 428 | 5 | 1.87E-05 | 4.01E-07 | 0.004948 | 2.03E-06 | -0.00870544 | INF2 |
| chr14 | 89629807 | 89630468 | 662 | 2 | 3.21E-06 | 1.67E-06 | 6.75E-05 | 2.03E-06 | -0.0099147 | AL137230.2, AL137230.1 |
| chr15 | 67863399 | 67864243 | 845 | 5 | 3.37E-06 | 5.28E-05 | 0.000177 | 2.04E-06 | -0.0053464 | AC009292.2 |
| chr7 | 41732564 | 41733219 | 656 | 3 | 1.90E-06 | 8.50E-07 | 0.000852 | 2.04E-06 | -0.0147644 | INHBA-AS1 |
| chr10 | 75109702 | 75110602 | 901 | 5 | 4.82E-08 | 0.0001548 | 3.62E-06 | 2.04E-06 | -0.00212593 | SAMD8 |
| chr7 | 156940413 | 156940880 | 468 | 3 | 2.88E-07 | 0.0002086 | 3.88E-05 | 2.04E-06 | -0.00928395 | NA |
| chr1 | 61424507 | 61424515 | 9 | 2 | 1.29E-06 | 4.38E-05 | 3.00E-06 | 2.04E-06 | 0.020018028 | NFIA |
| chr4 | 102614365 | 102614959 | 595 | 2 | 1.80E-07 | 1.23E-05 | 5.99E-06 | 2.04E-06 | -0.00986698 | NFKB1 |
| chr2 | 169827111 | 169827945 | 835 | 9 | 7.49E-07 | 1.40E-05 | 0.000749 | 2.04E-06 | -0.00129678 | UBR3 |
| chr13 | 24746250 | 24747356 | 1107 | 7 | 1.24E-05 | 1.39E-06 | 0.001415 | 2.04E-06 | -0.00296369 | NA |
| chr14 | 95071908 | 95072143 | 236 | 3 | 1.77E-06 | 6.55E-06 | 2.08E-05 | 2.05E-06 | 0.013964159 | NA |

| | | | | | | | | | | |
|---|---|---|---|---|---|---|---|---|---|---|
| chr15 | 66953207 | 66953347 | 141 | 2 | 3.62E-06 | 1.71E-06 | 6.46E-05 | 2.05E-06 | 0.001114066 | LINC02206 |
| chr10 | 131410223 | 131410721 | 499 | 5 | 2.40E-07 | 9.19E-06 | 4.25E-05 | 2.05E-06 | 0.002540759 | NA |
| chr3 | 141864681 | 141865299 | 619 | 2 | 2.67E-07 | 2.79E-06 | 2.60E-05 | 2.05E-06 | -0.00031347 | HMGN2P25 |
| chr19 | 16015215 | 16015508 | 294 | 3 | 6.76E-07 | 4.15E-05 | 8.74E-06 | 2.05E-06 | -0.005572 | LINC00661 |
| chr11 | 1425254 | 1426078 | 825 | 4 | 7.32E-08 | 1.58E-05 | 9.35E-05 | 2.05E-06 | -0.00767323 | BRSK2 |
| chr12 | 121240671 | 121240917 | 247 | 3 | 2.02E-06 | 1.52E-06 | 0.000485 | 2.05E-06 | 0.006583185 | CAMKK2 |
| chr19 | 2782605 | 2783158 | 554 | 4 | 4.27E-07 | 1.32E-06 | 0.000267 | 2.05E-06 | -0.00104342 | SGTA |
| chr10 | 96089724 | 96090642 | 919 | 8 | 3.73E-06 | 6.00E-06 | 0.002206 | 2.05E-06 | -0.00351985 | ENTPD1-AS1 |
| chr3 | 148279656 | 148280045 | 390 | 2 | 9.06E-08 | 0.0016383 | 1.48E-06 | 2.06E-06 | -0.00663969 | LINC02046, LINC02045 |
| chr3 | 14532361 | 14532906 | 546 | 3 | 2.03E-06 | 1.48E-06 | 0.000173 | 2.06E-06 | -0.01048855 | GRIP2 |
| chr3 | 148036481 | 148036676 | 196 | 2 | 4.52E-06 | 1.23E-06 | 0.00027 | 2.06E-06 | -0.01983034 | AC092958.4 |
| chr12 | 24709583 | 24709593 | 11 | 2 | 4.24E-06 | 1.46E-06 | 0.000106 | 2.06E-06 | -0.00914323 | AC023796.1 |
| chr1 | 88992352 | 88993166 | 815 | 11 | 6.90E-07 | 1.75E-05 | 0.00016 | 2.06E-06 | 0.002030141 | KYAT3, RBMXL1 |
| chr2 | 205975893 | 205976537 | 645 | 2 | 2.52E-06 | 1.34E-06 | 0.000157 | 2.06E-06 | -0.00766297 | NA |
| chr6 | 7230014 | 7231226 | 1213 | 3 | 8.92E-08 | 2.32E-06 | 5.61E-05 | 2.06E-06 | -0.00303652 | RREB1 |
| chr7 | 91556871 | 91557475 | 605 | 4 | 1.18E-06 | 3.82E-06 | 0.000693 | 2.06E-06 | -0.01790907 | NA |
| chr1 | 3630959 | 3630970 | 12 | 2 | 9.76E-07 | 3.39E-05 | 3.39E-06 | 2.06E-06 | 0.012383358 | WRAP73 |
| chr19 | 1272854 | 1274061 | 1208 | 4 | 2.80E-07 | 0.0006065 | 1.02E-05 | 2.06E-06 | -0.00613847 | CIRBP |
| chr16 | 17541129 | 17541711 | 583 | 2 | 8.81E-09 | 0.000498 | 1.67E-06 | 2.07E-06 | -0.01002599 | NA |
| chr2 | 171521471 | 171522176 | 706 | 8 | 8.67E-07 | 5.68E-06 | 0.002495 | 2.07E-06 | 0.007738879 | NA |
| chr2 | 10141032 | 10141866 | 835 | 7 | 2.46E-06 | 3.23E-06 | 0.000409 | 2.07E-06 | -0.0039306 | RRM2 |
| chr10 | 10795405 | 10796125 | 721 | 3 | 2.51E-07 | 9.25E-05 | 1.05E-05 | 2.07E-06 | -0.01365127 | NA |
| chr10 | 23190481 | 23191308 | 828 | 7 | 1.23E-06 | 3.19E-05 | 0.001413 | 2.07E-06 | 0.001801214 | NA |
| chr17 | 78838696 | 78839258 | 563 | 4 | 2.83E-08 | 0.0099406 | 4.03E-07 | 2.07E-06 | 0.001522344 | USP36 |
| chr7 | 142312711 | 142312788 | 78 | 2 | 3.32E-06 | 1.24E-06 | 0.000257 | 2.07E-06 | 0.020276249 | NA |
| chr16 | 1125678 | 1125947 | 270 | 2 | 3.49E-07 | 0.0002208 | 1.90E-06 | 2.07E-06 | 0.010386991 | NA |
| chr11 | 22432606 | 22433115 | 510 | 5 | 1.07E-06 | 1.46E-05 | 0.000305 | 2.07E-06 | -0.01076861 | NA |
| chr11 | 118481910 | 118482457 | 548 | 3 | 2.08E-07 | 0.0001051 | 5.51E-06 | 2.07E-06 | 0.00292045 | AP001267.5, KMT2A |
| chr3 | 133926579 | 133927660 | 1082 | 4 | 3.06E-06 | 1.96E-05 | 6.16E-05 | 2.08E-06 | -0.00630422 | NA |
| chr21 | 45477267 | 45477319 | 53 | 2 | 2.90E-07 | 7.91E-06 | 8.56E-06 | 2.08E-06 | -0.01071419 | COL18A1 |
| chr10 | 73801407 | 73802490 | 1084 | 5 | 1.84E-07 | 3.50E-06 | 0.000227 | 2.08E-06 | -0.00395407 | ZSWIM8, AC022400.5, NDST2 |
| chr14 | 50532201 | 50533340 | 1140 | 16 | 1.92E-06 | 3.89E-05 | 0.000103 | 2.08E-06 | 0.000426124 | ATL1, MAP4K5 |
| chr4 | 70225495 | 70225665 | 171 | 3 | 8.74E-07 | 3.40E-05 | 5.23E-06 | 2.08E-06 | -0.01326386 | NA |
| chr19 | 38335009 | 38335888 | 880 | 10 | 6.10E-07 | 7.66E-05 | 2.18E-05 | 2.08E-06 | 0.002878962 | CATSPERG |
| chr5 | 112706150 | 112706172 | 23 | 2 | 2.02E-08 | 0.0001834 | 1.99E-06 | 2.08E-06 | 0.003681802 | NA |
| chr10 | 24385979 | 24385986 | 8 | 2 | 4.51E-06 | 1.30E-06 | 0.000198 | 2.08E-06 | -0.03064495 | KIAA1217 |
| chr9 | 114657784 | 114658181 | 398 | 5 | 2.91E-06 | 5.50E-06 | 0.000255 | 2.08E-06 | -0.00145854 | TEX53 |
| chr6 | 29016058 | 29016939 | 882 | 11 | 1.88E-07 | 5.94E-06 | 0.004768 | 2.09E-06 | -0.00852312 | NA |
| chr19 | 44304477 | 44305210 | 734 | 10 | 1.44E-06 | 3.28E-05 | 7.92E-05 | 2.09E-06 | 0.000523532 | ZNF235 |
| chr15 | 86631879 | 86632481 | 603 | 3 | 2.38E-06 | 0.0002554 | 6.82E-05 | 2.09E-06 | 0.009634112 | AGBL1 |
| chr19 | 47256143 | 47256554 | 412 | 11 | 2.52E-06 | 5.19E-06 | 0.007175 | 2.09E-06 | -0.00667133 | CCDC9, AC008532.1 |
| chr15 | 98969537 | 98969661 | 125 | 4 | 2.97E-06 | 2.61E-06 | 0.000436 | 2.09E-06 | 0.001492189 | AC036108.1, PGPEP1L |
| chr7 | 6659891 | 6659895 | 5 | 2 | 1.01E-06 | 1.79E-06 | 6.12E-05 | 2.09E-06 | 0.013280432 | NA |
| chr17 | 81064353 | 81064912 | 560 | 2 | 1.31E-07 | 2.61E-05 | 3.92E-06 | 2.09E-06 | -0.00425043 | BAIAP2 |
| chr9 | 109317946 | 109319166 | 1221 | 4 | 9.61E-07 | 2.78E-06 | 0.00054 | 2.09E-06 | 0.003301009 | EPB41L4B |
| chr17 | 28861419 | 28861858 | 440 | 6 | 4.44E-06 | 9.65E-07 | 0.000552 | 2.09E-06 | 0.002973117 | MIR451B, AC024267.1, MIR451A, MIR144, MIR4732 |
| chr3 | 73434707 | 73435352 | 646 | 3 | 4.88E-08 | 0.000306 | 2.69E-05 | 2.09E-06 | 0.001814449 | PDZRN3 |
| chr16 | 85463979 | 85465311 | 1333 | 6 | 3.04E-06 | 0.0006363 | 3.21E-05 | 2.09E-06 | -0.00287696 | GSE1 |
| chr16 | 80892573 | 80893139 | 567 | 5 | 8.05E-06 | 7.11E-07 | 0.001947 | 2.09E-06 | -0.00610235 | ARLNC1, AC009070.1 |
| chr3 | 196528761 | 196529352 | 592 | 5 | 2.01E-06 | 2.03E-05 | 0.000351 | 2.09E-06 | -0.01344493 | AC092933.1 |
| chr2 | 21123347 | 21124020 | 674 | 7 | 1.58E-06 | 7.84E-06 | 0.000138 | 2.09E-06 | -0.00213114 | TDRD15 |
| chr14 | 101965085 | 101965767 | 683 | 4 | 3.08E-07 | 5.82E-05 | 1.17E-05 | 2.09E-06 | 0.00038793 | DYNC1H1 |
| chr9 | 127263134 | 127263572 | 439 | 7 | 4.17E-06 | 6.80E-07 | 0.00354 | 2.09E-06 | -0.00405421 | GARNL3 |
| chr3 | 188686549 | 188687172 | 624 | 2 | 1.23E-06 | 2.54E-06 | 3.15E-05 | 2.09E-06 | -0.00326248 | LPP |
| chr2 | 106281773 | 106281930 | 158 | 2 | 3.59E-06 | 1.80E-06 | 6.10E-05 | 2.10E-06 | -0.0021011 | NA |
| chr2 | 165506131 | 165506751 | 621 | 2 | 4.62E-06 | 1.33E-06 | 0.000182 | 2.10E-06 | -0.00035524 | CSRNP3 |
| chr1 | 24364686 | 24365430 | 745 | 3 | 3.55E-06 | 1.26E-06 | 0.000277 | 2.10E-06 | -0.00652086 | STPG1 |
| chr12 | 2057396 | 2057665 | 270 | 2 | 2.98E-07 | 0.0005454 | 1.68E-06 | 2.10E-06 | -0.0086464 | CACNA1C |
| chr1 | 28328837 | 28329636 | 800 | 9 | 4.72E-07 | 0.0002195 | 0.000119 | 2.10E-06 | 0.000912662 | MED18 |
| chr17 | 28357511 | 28358410 | 900 | 15 | 1.36E-07 | 4.05E-05 | 0.00164 | 2.10E-06 | -0.0034114 | TMEM199, AC002094.1, POLDIP2 |
| chr13 | 44244101 | 44244441 | 341 | 3 | 1.01E-06 | 4.00E-06 | 3.19E-05 | 2.10E-06 | 0.007110143 | AL589745.1 |
| chr22 | 25628501 | 25628652 | 152 | 2 | 1.20E-06 | 4.74E-06 | 1.42E-05 | 2.10E-06 | 0.00413486 | GRK3 |
| chr6 | 168570455 | 168571758 | 1304 | 7 | 1.80E-08 | 0.0008276 | 1.03E-05 | 2.10E-06 | 0.002625077 | SMOC2 |
| chr11 | 850296 | 851048 | 753 | 3 | 3.17E-06 | 5.29E-06 | 0.000142 | 2.11E-06 | -0.01362035 | TSPAN4 |
| chr11 | 1068018 | 1068600 | 583 | 2 | 1.12E-06 | 2.83E-06 | 2.69E-05 | 2.11E-06 | 0.007718718 | NA |
| chr19 | 7122657 | 7123103 | 447 | 4 | 1.63E-07 | 0.0001657 | 1.96E-06 | 2.11E-06 | -0.00182318 | INSR |
| chr12 | 6863331 | 6864102 | 772 | 3 | 1.41E-06 | 7.16E-05 | 3.86E-05 | 2.11E-06 | -0.00343297 | USP5 |
| chr6 | 138693734 | 138694366 | 633 | 5 | 2.91E-07 | 2.35E-05 | 3.31E-05 | 2.11E-06 | -0.00346392 | AL590617.2 |
| chr1 | 111961244 | 111961374 | 131 | 2 | 4.67E-06 | 1.26E-06 | 0.000279 | 2.11E-06 | -0.01336799 | KCND3 |
| chr9 | 21675187 | 21675753 | 567 | 2 | 1.89E-06 | 2.08E-06 | 4.56E-05 | 2.11E-06 | -0.00114227 | NA |
| chr1 | 33709520 | 33710136 | 617 | 5 | 3.69E-06 | 1.65E-06 | 0.00324 | 2.11E-06 | -0.00314516 | CSMD2 |
| chr3 | 25885573 | 25886116 | 544 | 2 | 5.09E-08 | 0.0002809 | 1.85E-06 | 2.12E-06 | -0.00499751 | NA |
| chr7 | 39720823 | 39720844 | 22 | 2 | 4.68E-06 | 1.26E-06 | 0.000272 | 2.12E-06 | -0.00725109 | NA |
| chr1 | 7388997 | 7389897 | 901 | 5 | 1.05E-06 | 3.81E-06 | 6.16E-05 | 2.12E-06 | -0.00911769 | CAMTA1, CAMTA1-AS2 |
| chr7 | 139716068 | 139716755 | 688 | 6 | 6.49E-07 | 0.0104551 | 5.34E-05 | 2.12E-06 | -0.00089789 | HIPK2 |
| chr18 | 59358909 | 59359536 | 628 | 11 | 2.99E-07 | 0.0002292 | 5.60E-05 | 2.12E-06 | -0.0012398 | LMAN1 |
| chr17 | 31392005 | 31392764 | 760 | 5 | 1.85E-07 | 2.99E-05 | 1.30E-05 | 2.12E-06 | 0.007129747 | RAB11FIP4 |
| chr5 | 135325412 | 135325813 | 402 | 2 | 5.06E-08 | 1.85E-05 | 4.86E-06 | 2.12E-06 | -0.00333635 | C5orf66 |
| chr12 | 117034415 | 117034999 | 585 | 3 | 1.99E-07 | 3.24E-05 | 0.000102 | 2.12E-06 | -0.00456285 | NA |
| chr10 | 33226175 | 33226308 | 134 | 4 | 4.97E-06 | 2.94E-06 | 0.000496 | 2.12E-06 | -0.0007889 | NRP1 |
| chr3 | 100951242 | 100951565 | 324 | 2 | 1.49E-06 | 3.91E-06 | 1.79E-05 | 2.12E-06 | -0.00700892 | ABI3BP |
| chr11 | 74726362 | 74726694 | 333 | 3 | 8.55E-07 | 5.76E-06 | 2.39E-05 | 2.12E-06 | -0.0040154 | CHRDL2 |
| chr2 | 239178024 | 239179302 | 1279 | 5 | 2.14E-07 | 0.0001408 | 1.35E-05 | 2.13E-06 | 0.001607709 | HDAC4 |
| chr4 | 6562913 | 6563319 | 407 | 3 | 2.71E-07 | 2.47E-06 | 5.37E-05 | 2.13E-06 | 0.010321217 | PPP2R2C |
| chr4 | 95839492 | 95840447 | 956 | 11 | 8.32E-06 | 2.01E-06 | 0.0052 | 2.13E-06 | -0.00246033 | PDHA2 |
| chr1 | 2293239 | 2293860 | 622 | 4 | 1.34E-06 | 3.81E-06 | 0.001056 | 2.13E-06 | -0.00037364 | SKI |
| chr12 | 114818178 | 114818247 | 70 | 3 | 3.44E-06 | 8.23E-06 | 0.000177 | 2.13E-06 | -0.0138267 | NA |
| chr19 | 18677983 | 18678533 | 551 | 2 | 4.11E-06 | 1.40E-06 | 0.00015 | 2.13E-06 | -0.01676452 | NA |
| chr15 | 36878180 | 36878974 | 795 | 5 | 3.16E-06 | 7.33E-07 | 0.00481 | 2.13E-06 | 0.004395113 | AC018563.1 |
| chr19 | 24001398 | 24002131 | 734 | 8 | 4.54E-06 | 1.66E-06 | 0.003038 | 2.13E-06 | -0.0048439 | NA |
| chr3 | 45594201 | 45594894 | 694 | 9 | 5.42E-06 | 4.58E-06 | 0.00084 | 2.13E-06 | -0.00471215 | LIMD1 |
| chr11 | 118572122 | 118573151 | 1030 | 13 | 3.07E-06 | 6.18E-05 | 0.000411 | 2.13E-06 | 3.52E-05 | ARCN1, IFT46 |
| chr17 | 52159405 | 52160484 | 1080 | 16 | 3.92E-07 | 2.26E-05 | 0.000176 | 2.13E-06 | -0.00193806 | CA10 |
| chr7 | 101952851 | 101953124 | 274 | 5 | 2.09E-06 | 1.26E-05 | 0.00013 | 2.13E-06 | -0.00609449 | CUX1 |
| chr16 | 4233374 | 4233650 | 277 | 2 | 4.76E-06 | 1.28E-06 | 0.000268 | 2.13E-06 | -0.00538879 | SRL |
| chr6 | 43773317 | 43774182 | 866 | 3 | 7.02E-07 | 0.0001393 | 6.70E-06 | 2.13E-06 | -0.01060231 | VEGFA |
| chr6 | 52724327 | 52724586 | 260 | 2 | 2.69E-07 | 0.0025238 | 1.49E-06 | 2.13E-06 | -0.00359314 | NA |
| chr9 | 126482967 | 126483865 | 899 | 2 | 9.84E-06 | 1.27E-06 | 0.00028 | 2.13E-06 | 0.008393051 | MVB12B |
| chr19 | 15299183 | 15299909 | 727 | 3 | 4.94E-06 | 2.00E-05 | 9.39E-05 | 2.14E-06 | 0.005497389 | BRD4 |
| chr17 | 43645757 | 43646651 | 895 | 5 | 1.29E-06 | 2.07E-05 | 0.000268 | 2.14E-06 | -0.00044832 | MEOX1 |
| chr3 | 126062425 | 126063104 | 680 | 2 | 5.39E-07 | 2.59E-06 | 3.19E-05 | 2.14E-06 | -0.00738488 | SLC41A3 |
| chr19 | 14441201 | 14441538 | 338 | 6 | 1.47E-06 | 1.00E-05 | 0.000205 | 2.14E-06 | -0.00032435 | PKN1 |
| chr20 | 10173338 | 10173396 | 59 | 3 | 2.54E-07 | 0.0004663 | 2.72E-06 | 2.14E-06 | -0.0051109 | AL354824.1, AL354824.2, SNAP25-AS1 |
| chr7 | 154609762 | 154610058 | 297 | 3 | 1.28E-06 | 2.82E-05 | 7.67E-05 | 2.14E-06 | -0.00292545 | DPP6 |
| chr16 | 69482210 | 69482802 | 593 | 4 | 3.94E-06 | 1.07E-06 | 0.000857 | 2.14E-06 | 0.016891008 | NA |
| chr20 | 36807449 | 36807654 | 206 | 2 | 9.70E-07 | 1.19E-05 | 6.54E-06 | 2.14E-06 | -0.00108821 | SOGA1 |
| chr10 | 43209948 | 43210556 | 609 | 3 | 2.25E-07 | 3.41E-06 | 0.000307 | 2.15E-06 | -0.00643668 | RASGEF1A |
| chr5 | 122703094 | 122703218 | 125 | 2 | 1.57E-06 | 2.74E-06 | 2.98E-05 | 2.16E-06 | -0.01195611 | LINC02201 |
| chr15 | 82571158 | 82572041 | 884 | 8 | 1.08E-07 | 0.0003529 | 3.37E-05 | 2.16E-06 | -0.00603677 | AC245033.1, CPEB1 |
| chr4 | 75513930 | 75514883 | 954 | 16 | 5.38E-06 | 3.99E-06 | 0.017168 | 2.16E-06 | 0.000375893 | THAP6, RCHY1 |
| chr16 | 75594285 | 75594328 | 44 | 2 | 4.24E-06 | 1.75E-06 | 7.23E-05 | 2.17E-06 | -0.01607923 | AC025287.1 |
| chr2 | 23670193 | 23670355 | 163 | 2 | 1.30E-06 | 2.14E-05 | 4.57E-06 | 2.17E-06 | -0.00504781 | KLHL29, AC009242.1 |
| chr4 | 3648641 | 3649744 | 1104 | 5 | 8.09E-08 | 0.00019 | 5.64E-06 | 2.17E-06 | -0.007797 | NA |
| chr1 | 56645197 | 56645838 | 642 | 7 | 1.93E-07 | 1.59E-05 | 6.79E-05 | 2.17E-06 | -0.00215799 | PRKAA2, PLPP3 |
| chr7 | 6015462 | 6015951 | 490 | 3 | 8.80E-08 | 1.14E-05 | 1.31E-05 | 2.17E-06 | -0.00971125 | AIMP2 |
| chr16 | 69410195 | 69410545 | 351 | 2 | 3.42E-06 | 1.37E-06 | 0.00019 | 2.18E-06 | 0.004672247 | NA |
| chr18 | 50819536 | 50820473 | 938 | 11 | 3.12E-06 | 0.0003775 | 5.52E-05 | 2.18E-06 | -0.00192316 | MRO |
| chr12 | 2690889 | 2691753 | 865 | 10 | 2.75E-06 | 1.39E-06 | 0.001093 | 2.18E-06 | 0.001065505 | CACNA1C, CACNA1C-AS1 |
| chr6 | 149648364 | 149648964 | 601 | 8 | 1.81E-07 | 2.86E-05 | 4.22E-05 | 2.18E-06 | 0.001633638 | KATNA1 |
| chr19 | 50375530 | 50376706 | 1177 | 10 | 1.00E-06 | 7.47E-05 | 0.001524 | 2.18E-06 | -0.00213706 | NR1H2 |
| chr1 | 42784740 | 42785582 | 843 | 7 | 1.60E-06 | 1.41E-05 | 0.004173 | 2.18E-06 | -0.00730608 | AC098484.3, TMEM269 |
| chr17 | 28661589 | 28662906 | 1318 | 16 | 1.26E-06 | 5.90E-06 | 0.004236 | 2.18E-06 | 0.001412846 | SUPT6H, SDF2 |
| chr19 | 1871381 | 1871484 | 104 | 2 | 1.48E-06 | 1.01E-05 | 7.53E-06 | 2.18E-06 | 0.005423777 | NA |
| chr19 | 47499745 | 47500443 | 699 | 3 | 6.30E-07 | 1.47E-05 | 1.11E-05 | 2.18E-06 | -0.00361025 | NAPA-AS1, NAPA |
| chr7 | 2906288 | 2906705 | 418 | 4 | 2.41E-06 | 2.99E-06 | 0.000159 | 2.18E-06 | 0.000486886 | CARD11 |
| chr1 | 31655395 | 31656003 | 609 | 5 | 3.60E-06 | 8.15E-07 | 0.001779 | 2.18E-06 | -0.00331792 | PEF1-AS1, COL16A1 |
| chr5 | 172639961 | 172640300 | 340 | 3 | 1.65E-08 | 7.06E-05 | 2.48E-06 | 2.18E-06 | -0.00069351 | NA |
| chr2 | 149320277 | 149320763 | 487 | 5 | 4.04E-06 | 1.80E-06 | 0.000678 | 2.18E-06 | -0.0183159 | FAM8A3P |
| chr2 | 9378195 | 9378622 | 428 | 2 | 8.52E-08 | 0.0005628 | 1.72E-06 | 2.18E-06 | 0.011930266 | ASAP2 |
| chr15 | 47824387 | 47825313 | 927 | 3 | 3.29E-07 | 5.26E-06 | 2.77E-05 | 2.18E-06 | -0.01340341 | AC092078.2, LINC01491 |
| chr3 | 22989864 | 22990768 | 905 | 6 | 1.18E-06 | 4.50E-06 | 9.35E-05 | 2.19E-06 | -0.00258691 | SALL4P5 |
| chr19 | 11986769 | 11988002 | 1234 | 9 | 3.82E-07 | 0.0001223 | 0.000456 | 2.19E-06 | -0.00371865 | ZNF433-AS1, AC008770.4 |
| chr1 | 230873708 | 230874619 | 912 | 2 | 0.000119562 | 1.30E-06 | 0.000281 | 2.19E-06 | 0.012095272 | AL118511.1 |
| chr7 | 155380302 | 155380337 | 36 | 2 | 2.36E-06 | 2.27E-06 | 4.20E-05 | 2.19E-06 | -0.00577371 | NA |
| chr17 | 45961520 | 45962183 | 664 | 4 | 2.52E-07 | 1.80E-05 | 1.86E-05 | 2.19E-06 | -0.0047216 | MAPT |
| chr19 | 45497484 | 45498292 | 809 | 4 | 2.84E-06 | 2.90E-06 | 0.000201 | 2.19E-06 | -0.0055014 | PPM1N |
| chr1 | 25775264 | 25775535 | 272 | 2 | 4.48E-06 | 1.36E-06 | 0.00021 | 2.19E-06 | 0.013811983 | MAN1C1 |
| chr21 | 35720472 | 35721042 | 571 | 6 | 7.53E-07 | 7.59E-06 | 0.00015 | 2.19E-06 | 0.001204352 | MIR802, RUNX1, AF015720.1 |
| chr2 | 169824154 | 169825153 | 1000 | 11 | 4.05E-06 | 0.0005657 | 0.001818 | 2.19E-06 | -0.00238699 | METTL5 |
| chr10 | 22481788 | 22482318 | 531 | 2 | 4.45E-06 | 1.48E-06 | 0.000134 | 2.19E-06 | -0.00283667 | NA |
| chr22 | 48207624 | 48207899 | 276 | 2 | 3.72E-07 | 0.00012 | 2.31E-06 | 2.19E-06 | -0.00555471 | NA |
| chr22 | 38209891 | 38210089 | 199 | 2 | 8.26E-07 | 3.64E-05 | 3.51E-06 | 2.19E-06 | -0.00548328 | MAFF, PLA2G6 |
| chr11 | 47184972 | 47186447 | 1476 | 12 | 8.47E-07 | 0.0050088 | 0.000318 | 2.20E-06 | -0.00223547 | PACSIN3 |
| chr19 | 57745924 | 57746957 | 1034 | 9 | 1.94E-07 | 7.56E-05 | 0.000124 | 2.20E-06 | -0.00518257 | AC003006.1, ZNF776 |
| chr16 | 883619 | 883739 | 121 | 3 | 1.09E-06 | 0.0001319 | 6.27E-06 | 2.20E-06 | -0.01320446 | LMF1 |
| chr2 | 43596128 | 43596988 | 861 | 12 | 2.46E-09 | 0.0021898 | 4.51E-07 | 2.20E-06 | 0.003945611 | NA |
| chr16 | 1714710 | 1715122 | 413 | 2 | 1.04E-07 | 1.96E-06 | 5.78E-05 | 2.20E-06 | -0.00330766 | MAPK8IP3 |
| chr17 | 555746 | 556702 | 957 | 3 | 4.82E-06 | 1.03E-06 | 0.000625 | 2.20E-06 | -0.01012912 | VPS53 |
| chr1 | 3320739 | 3321588 | 850 | 2 | 2.06E-05 | 1.33E-06 | 0.000263 | 2.20E-06 | -0.00823476 | PRDM16, AL354743.2 |
| chr7 | 43907030 | 43907948 | 919 | 8 | 1.40E-06 | 4.30E-06 | 0.000375 | 2.20E-06 | -0.00377547 | URGCP |
| chr2 | 183107886 | 183108537 | 652 | 4 | 2.95E-06 | 3.47E-05 | 4.74E-05 | 2.20E-06 | -0.00462511 | AC064871.2 |
| chr11 | 2088434 | 2089381 | 948 | 5 | 5.56E-06 | 3.13E-06 | 0.000734 | 2.21E-06 | -0.00180434 | NA |
| chr6 | 77463233 | 77464003 | 771 | 11 | 3.35E-06 | 6.99E-06 | 0.000404 | 2.21E-06 | -0.00029339 | HTR1B |
| chr9 | 87888589 | 87889177 | 589 | 3 | 1.32E-06 | 3.79E-06 | 0.00029 | 2.21E-06 | -0.00790712 | SPATA31E1 |
| chr16 | 21188318 | 21188809 | 492 | 4 | 4.47E-06 | 8.05E-07 | 0.001552 | 2.21E-06 | -0.00686323 | NA |
| chr17 | 51153221 | 51153868 | 648 | 14 | 7.91E-07 | 7.05E-05 | 0.00012 | 2.21E-06 | 0.00154121 | NME1, NME1-NME2 |
| chr22 | 17950763 | 17951286 | 524 | 4 | 1.60E-06 | 3.25E-06 | 6.10E-05 | 2.21E-06 | -0.00762049 | MICAL3 |
| chr16 | 89906185 | 89907308 | 1124 | 7 | 3.29E-08 | 4.17E-05 | 4.66E-05 | 2.21E-06 | 0.001615713 | TCF25, AC092143.3 |

| | | | | | | | | | | |
|---|---|---|---|---|---|---|---|---|---|---|
| chr14 | 21269686 | 21270314 | 629 | 7 | 3.52E-08 | 3.16E-05 | 7.27E-05 | 2.21E-06 | -0.00200592 | NA |
| chr12 | 2052964 | 2053527 | 564 | 11 | 6.34E-07 | 4.82E-05 | 0.00171 | 2.21E-06 | 0.001552294 | CACNA1C |
| chr6 | 90025262 | 90025740 | 479 | 2 | 5.17E-06 | 1.39E-06 | 0.000196 | 2.21E-06 | -0.01043378 | BACH2 |
| chr10 | 100744892 | 100746110 | 1219 | 9 | 1.53E-06 | 8.37E-06 | 8.28E-05 | 2.21E-06 | 0.001129575 | PAX2 |
| chr4 | 70591459 | 70592189 | 731 | 7 | 1.40E-07 | 0.0100742 | 8.45E-06 | 2.21E-06 | -0.00728077 | NA |
| chr14 | 54627344 | 54627728 | 385 | 2 | 2.85E-07 | 1.20E-05 | 6.78E-06 | 2.21E-06 | 0.016511005 | SAMD4A |
| chr1 | 206479681 | 206480472 | 792 | 5 | 6.49E-07 | 8.00E-06 | 3.71E-05 | 2.21E-06 | -0.0006765 | IKBKE |
| chr4 | 9893893 | 9893914 | 22 | 2 | 1.02E-06 | 0.0001376 | 2.25E-06 | 2.21E-06 | 0.003346651 | SLC2A9 |
| chr11 | 102421428 | 102421540 | 113 | 2 | 1.10E-06 | 2.30E-05 | 4.49E-06 | 2.21E-06 | -0.01590598 | TMEM123 |
| chr1 | 10032571 | 10033766 | 1196 | 17 | 1.79E-05 | 7.60E-06 | 0.002683 | 2.21E-06 | -0.001152 | UBE4B |
| chr15 | 89893872 | 89895004 | 1133 | 13 | 1.03E-07 | 0.0002929 | 0.000405 | 2.21E-06 | -0.00357324 | AP3S2, ARPIN-AP3S2 |
| chr8 | 144991904 | 144992127 | 224 | 2 | 2.07E-07 | 0.0007654 | 1.69E-06 | 2.22E-06 | -0.01094887 | ZNF252P |
| chr6 | 3156890 | 3157736 | 847 | 6 | 2.59E-06 | 0.0001158 | 0.000663 | 2.22E-06 | 0.000983087 | TUBB2A |
| chr7 | 1043436 | 1044526 | 1091 | 8 | 1.97E-07 | 3.90E-06 | 0.001668 | 2.22E-06 | -0.00479768 | AC073957.1, C7orf50 |
| chr9 | 34255096 | 34255151 | 56 | 2 | 2.69E-07 | 0.0005565 | 1.75E-06 | 2.22E-06 | -0.00965377 | KIF24 |
| chr6 | 159762069 | 159763009 | 941 | 13 | 1.71E-06 | 4.26E-05 | 0.000899 | 2.23E-06 | -2.32E-05 | ACAT2, SOD2, SOD2-OT1 |
| chr2 | 26605351 | 26606264 | 914 | 4 | 1.16E-06 | 6.63E-06 | 0.000451 | 2.23E-06 | -0.00465585 | CIB4 |
| chr2 | 157256716 | 157256807 | 92 | 2 | 1.85E-07 | 4.80E-05 | 3.17E-06 | 2.23E-06 | -0.01233423 | NA |
| chr1 | 7218398 | 7219329 | 932 | 6 | 3.53E-06 | 0.0001579 | 3.30E-05 | 2.23E-06 | 0.001364553 | CAMTA1 |
| chr10 | 49077303 | 49077563 | 261 | 4 | 6.22E-06 | 1.74E-06 | 0.000964 | 2.23E-06 | -0.01562067 | VSTM4 |
| chr10 | 73414738 | 73415378 | 641 | 3 | 7.17E-09 | 0.0001602 | 1.70E-06 | 2.23E-06 | 0.008008248 | NA |
| chr9 | 101184894 | 101185466 | 573 | 3 | 3.77E-07 | 1.02E-05 | 1.16E-05 | 2.23E-06 | -0.01865613 | PLPPR1 |
| chr19 | 57934944 | 57935620 | 677 | 10 | 1.10E-05 | 1.90E-06 | 0.015325 | 2.23E-06 | -0.0046396 | ZNF418 |
| chr17 | 17813692 | 17814653 | 962 | 11 | 3.28E-06 | 2.36E-05 | 0.000137 | 2.23E-06 | -0.00270534 | SREBF1, MIR33B |
| chr6 | 157382344 | 157383007 | 664 | 2 | 5.40E-09 | 0.0009212 | 1.67E-06 | 2.23E-06 | -0.01102267 | ZDHHC14 |
| chr19 | 10194629 | 10195367 | 739 | 9 | 7.00E-07 | 3.25E-05 | 0.000161 | 2.23E-06 | -0.00313474 | DNMT1 |
| chr1 | 1126141 | 1126857 | 717 | 4 | 5.91E-06 | 2.04E-06 | 0.000435 | 2.24E-06 | 0.000545693 | NA |
| chr11 | 116224749 | 116225057 | 309 | 2 | 2.50E-06 | 2.38E-06 | 4.05E-05 | 2.24E-06 | 0.024459348 | NA |
| chr20 | 59603970 | 59605302 | 1333 | 13 | 2.35E-05 | 2.11E-05 | 0.00397 | 2.24E-06 | 0.001439464 | PHACTR3 |
| chr19 | 36838078 | 36838957 | 880 | 18 | 1.22E-06 | 2.67E-06 | 0.002253 | 2.24E-06 | -0.00066534 | ZNF790 |
| chr3 | 133398576 | 133398860 | 285 | 3 | 3.62E-06 | 1.04E-06 | 0.000543 | 2.24E-06 | -0.00733734 | NA |
| chr18 | 9987700 | 9987971 | 272 | 3 | 7.71E-07 | 8.53E-06 | 1.36E-05 | 2.24E-06 | -0.00763255 | NA |
| chr17 | 65594002 | 65594457 | 456 | 2 | 3.03E-08 | 0.0001803 | 2.14E-06 | 2.24E-06 | -0.01094678 | AC004805.1 |
| chr8 | 29904940 | 29905618 | 679 | 4 | 1.51E-06 | 6.40E-05 | 3.78E-05 | 2.24E-06 | -0.00492013 | NA |
| chr1 | 183921996 | 183922431 | 436 | 4 | 7.99E-07 | 5.31E-05 | 9.11E-05 | 2.24E-06 | 0.000123315 | RGL1 |
| chr22 | 43881364 | 43881382 | 19 | 2 | 2.79E-06 | 1.63E-06 | 0.000101 | 2.24E-06 | -0.00109 | PNPLA5 |
| chr16 | 89442204 | 89442601 | 398 | 3 | 4.94E-07 | 8.08E-06 | 1.42E-05 | 2.25E-06 | 0.01028315 | AC092120.3, ANKRD11 |
| chr13 | 110044473 | 110044634 | 162 | 2 | 4.27E-06 | 1.64E-06 | 0.0001 | 2.25E-06 | -0.02493009 | AL390755.1 |
| chr14 | 100775903 | 100776111 | 209 | 3 | 6.07E-06 | 1.01E-06 | 0.000536 | 2.25E-06 | -0.00184741 | NA |
| chr2 | 46973589 | 46973731 | 143 | 2 | 7.15E-07 | 4.30E-06 | 1.76E-05 | 2.25E-06 | -0.00187897 | TTC7A |
| chr15 | 22926080 | 22926153 | 74 | 2 | 6.99E-07 | 0.0013302 | 1.62E-06 | 2.25E-06 | 0.002566228 | CYFIP1 |
| chr2 | 216109083 | 216110131 | 1049 | 6 | 1.71E-07 | 1.85E-05 | 3.53E-05 | 2.25E-06 | 0.000495903 | XRCC5 |
| chr7 | 51028860 | 51029274 | 415 | 6 | 3.83E-06 | 5.47E-05 | 0.000886 | 2.25E-06 | 0.002120326 | COBL |
| chr2 | 55133708 | 55134174 | 467 | 3 | 3.88E-06 | 9.10E-07 | 0.001108 | 2.25E-06 | 0.003271107 | NA |
| chr5 | 127088658 | 127089105 | 448 | 2 | 4.17E-06 | 1.51E-06 | 0.000144 | 2.25E-06 | 0.000712508 | NA |
| chr17 | 9965092 | 9965950 | 859 | 2 | 5.03E-05 | 1.36E-06 | 0.000266 | 2.25E-06 | -0.01634863 | GAS7 |
| chr6 | 104859686 | 104860964 | 1279 | 12 | 2.43E-06 | 0.0004363 | 0.000879 | 2.25E-06 | -0.000868 | AL357315.2, HACE1 |
| chr15 | 63050089 | 63050600 | 512 | 4 | 8.90E-08 | 2.12E-05 | 0.000124 | 2.25E-06 | -0.01353648 | TPM1 |
| chr20 | 43513365 | 43514375 | 1011 | 21 | 5.24E-07 | 4.86E-05 | 0.004748 | 2.25E-06 | 0.002045521 | AL031681.2, L3MBTL1 |
| chr19 | 54200284 | 54200974 | 691 | 9 | 1.35E-06 | 1.04E-05 | 0.000914 | 2.25E-06 | -0.0002227 | RPS9 |
| chr1 | 47232276 | 47233304 | 1029 | 10 | 7.24E-07 | 1.03E-05 | 0.002434 | 2.25E-06 | -0.00203197 | NA |
| chr17 | 36662812 | 36663675 | 864 | 2 | 2.97E-05 | 1.40E-06 | 0.000218 | 2.26E-06 | -0.00174747 | NA |
| chr5 | 42993338 | 42994021 | 684 | 5 | 2.74E-07 | 0.0002515 | 0.000335 | 2.26E-06 | -0.00219941 | NA |
| chr16 | 23072721 | 23073169 | 449 | 4 | 4.73E-07 | 7.91E-06 | 1.64E-05 | 2.26E-06 | -0.0042255 | USP31 |
| chr9 | 34380892 | 34382301 | 1410 | 10 | 2.01E-06 | 1.59E-05 | 0.002157 | 2.26E-06 | -0.00351571 | C9orf24 |
| chr6 | 136037008 | 136037908 | 901 | 4 | 9.54E-07 | 8.44E-06 | 0.000265 | 2.26E-06 | -0.0059294 | PDE7B, AL138828.1 |
| chr10 | 32542027 | 32542120 | 94 | 2 | 3.90E-06 | 1.35E-06 | 0.000283 | 2.27E-06 | -0.01225771 | CCDC7 |
| chr11 | 132860850 | 132861102 | 253 | 2 | 1.54E-06 | 1.37E-06 | 0.000262 | 2.27E-06 | -0.01594845 | OPCML |
| chr1 | 16404106 | 16404803 | 698 | 2 | 1.33E-05 | 1.35E-06 | 0.000286 | 2.27E-06 | -0.00315623 | SPATA21 |
| chr18 | 811510 | 812263 | 754 | 4 | 4.69E-07 | 6.61E-06 | 9.12E-05 | 2.27E-06 | 0.005500407 | YES1 |
| chr16 | 30354519 | 30355526 | 1008 | 14 | 1.03E-07 | 0.0001131 | 4.26E-05 | 2.27E-06 | -0.00017022 | CD2BP2-DT, CD2BP2 |
| chr17 | 7327500 | 7327869 | 370 | 2 | 3.74E-08 | 8.42E-05 | 2.63E-06 | 2.27E-06 | -0.00584898 | NEURL4 |
| chr6 | 73369596 | 73370537 | 942 | 12 | 1.18E-05 | 0.000113 | 0.002638 | 2.27E-06 | 7.14E-05 | OOEP-AS1, OOEP |
| chr17 | 4395934 | 4396925 | 992 | 3 | 2.28E-09 | 0.000516 | 1.12E-06 | 2.27E-06 | -0.00577512 | NA |
| chr9 | 88829008 | 88829580 | 573 | 2 | 5.76E-07 | 5.54E-06 | 1.37E-05 | 2.27E-06 | -0.01071136 | NA |
| chr20 | 38032182 | 38033046 | 865 | 4 | 1.62E-07 | 8.71E-05 | 2.96E-05 | 2.27E-06 | 0.002161966 | TTI1 |
| chr17 | 82090935 | 82091471 | 537 | 5 | 3.22E-06 | 2.18E-06 | 0.001391 | 2.27E-06 | -0.00132105 | FASN |
| chr19 | 10491344 | 10492037 | 694 | 4 | 1.10E-06 | 1.03E-06 | 0.000712 | 2.27E-06 | -0.00289159 | KEAP1 |
| chr8 | 32264921 | 32265075 | 155 | 2 | 3.29E-06 | 2.85E-06 | 3.12E-05 | 2.28E-06 | -0.00577287 | NRG1 |
| chr17 | 74117233 | 74117836 | 604 | 2 | 4.39E-06 | 1.54E-06 | 0.000136 | 2.28E-06 | 0.001391581 | LINC02074 |
| chr9 | 137175293 | 137175774 | 482 | 4 | 1.80E-06 | 2.80E-06 | 0.000377 | 2.28E-06 | -0.01448918 | ANAPC2 |
| chr1 | 70410809 | 70411698 | 890 | 11 | 3.85E-06 | 5.97E-05 | 0.00033 | 2.28E-06 | -0.00071973 | CTH |
| chr18 | 62186683 | 62187889 | 1207 | 10 | 8.36E-07 | 2.20E-05 | 0.000413 | 2.28E-06 | 0.000416356 | RELCH, PIGN |
| chr11 | 62671382 | 62672330 | 949 | 17 | 1.07E-06 | 1.48E-05 | 0.00029 | 2.28E-06 | -0.00036775 | UQCC3, LBHD1 |
| chr13 | 42883221 | 42883858 | 638 | 2 | 1.99E-06 | 2.41E-06 | 4.15E-05 | 2.28E-06 | 0.024538601 | NA |
| chr5 | 174044723 | 174045729 | 1007 | 9 | 4.34E-07 | 3.19E-05 | 0.001745 | 2.28E-06 | 0.001311839 | NSG2 |
| chr4 | 139775641 | 139776504 | 864 | 2 | 3.32E-05 | 1.42E-06 | 0.000211 | 2.28E-06 | -0.0089544 | MAML3 |
| chr12 | 24949128 | 24949844 | 717 | 12 | 1.01E-06 | 0.0005647 | 0.00095 | 2.28E-06 | -0.00341414 | AC026310.1 |
| chr6 | 29462319 | 29463633 | 1315 | 8 | 1.51E-05 | 2.08E-05 | 0.000615 | 2.28E-06 | -0.0014749 | OR2H1 |
| chr1 | 40665781 | 40666350 | 570 | 6 | 1.28E-06 | 0.0002225 | 0.000285 | 2.28E-06 | -0.00114889 | AL031289.2 |
| chr8 | 142989363 | 142989543 | 181 | 2 | 1.19E-06 | 2.42E-06 | 4.14E-05 | 2.29E-06 | 0.001608007 | LY6E-DT |
| chr1 | 155884260 | 155884640 | 381 | 2 | 6.52E-08 | 4.14E-05 | 3.47E-06 | 2.29E-06 | -0.00347824 | SYT11 |
| chr6 | 100228869 | 100229269 | 401 | 6 | 2.39E-07 | 0.000774 | 3.53E-06 | 2.29E-06 | 0.000630783 | NA |
| chr7 | 2165614 | 2166110 | 497 | 5 | 3.85E-08 | 0.0011663 | 1.84E-06 | 2.29E-06 | -0.00420916 | MAD1L1, AC069288.1 |
| chr6 | 49858683 | 49859070 | 388 | 2 | 1.97E-06 | 1.68E-06 | 9.88E-05 | 2.29E-06 | -0.00488134 | CRISP1 |
| chr6 | 153002228 | 153003121 | 894 | 14 | 4.25E-07 | 0.0001667 | 0.000348 | 2.29E-06 | 0.000407972 | MTRF1L |
| chr6 | 166703710 | 166704410 | 701 | 2 | 2.31E-06 | 1.86E-06 | 7.43E-05 | 2.30E-06 | 0.011578896 | RPS6KA2 |
| chr6 | 79077844 | 79079047 | 1204 | 16 | 5.75E-06 | 0.0017349 | 0.000388 | 2.30E-06 | -0.00025607 | AL356776.2, PHIP |
| chr20 | 56968472 | 56968589 | 118 | 2 | 4.70E-06 | 1.57E-06 | 0.00013 | 2.30E-06 | 0.030299413 | NA |
| chr15 | 42358026 | 42358888 | 863 | 5 | 2.48E-07 | 3.14E-05 | 9.25E-06 | 2.30E-06 | -0.00337552 | AC012651.1 |
| chr1 | 162560401 | 162561633 | 1233 | 12 | 3.81E-06 | 1.95E-06 | 0.008684 | 2.30E-06 | 0.003727003 | UAP1, AL596325.1 |
| chr19 | 2607728 | 2607905 | 178 | 4 | 4.20E-07 | 0.0001941 | 1.61E-06 | 2.30E-06 | -0.00462601 | GNG7 |
| chr17 | 17860389 | 17860948 | 560 | 2 | 2.99E-07 | 1.13E-05 | 7.43E-06 | 2.30E-06 | -0.00846625 | TOM1L2 |
| chr7 | 4762292 | 4763245 | 954 | 7 | 1.08E-07 | 5.15E-05 | 0.001054 | 2.30E-06 | -0.00132144 | FOXK1 |
| chr15 | 39641760 | 39642266 | 507 | 3 | 2.54E-06 | 3.07E-06 | 4.57E-05 | 2.30E-06 | 0.009939758 | FSIP1 |
| chr12 | 66631331 | 66631597 | 267 | 2 | 1.96E-06 | 4.43E-06 | 1.77E-05 | 2.30E-06 | -0.00473762 | GRIP1 |
| chr3 | 32371614 | 32372370 | 757 | 6 | 3.25E-06 | 2.66E-05 | 0.002192 | 2.30E-06 | 0.010364581 | NA |
| chr4 | 79596098 | 79596432 | 335 | 2 | 7.68E-07 | 1.37E-05 | 6.50E-06 | 2.30E-06 | 0.002038546 | LINC00989 |
| chr18 | 31683778 | 31684542 | 765 | 8 | 6.40E-06 | 5.75E-05 | 0.002581 | 2.31E-06 | 0.002694661 | B4GALT6 |
| chr17 | 74359856 | 74360115 | 260 | 4 | 6.61E-06 | 5.64E-07 | 0.003212 | 2.31E-06 | -0.00196432 | BTBD17 |
| chr2 | 55224507 | 55225227 | 721 | 2 | 1.75E-06 | 2.09E-06 | 5.74E-05 | 2.31E-06 | 0.00729433 | CDPF1P1, CLHC1 |
| chr4 | 58565219 | 58565570 | 352 | 3 | 1.75E-07 | 0.0009379 | 1.05E-06 | 2.31E-06 | -0.00578003 | NA |
| chr1 | 210239423 | 210239934 | 512 | 2 | 1.81E-07 | 3.01E-05 | 4.08E-06 | 2.31E-06 | 0.011443086 | SERTAD4 |
| chr20 | 50115923 | 50116537 | 615 | 10 | 3.50E-07 | 0.0001992 | 9.17E-05 | 2.31E-06 | 0.00177731 | UBE2V1, PEDS1-UBE2V1 |
| chr2 | 23030201 | 23030683 | 483 | 3 | 3.83E-07 | 2.79E-06 | 5.35E-05 | 2.31E-06 | 0.008187544 | AC018467.1 |
| chr6 | 30340876 | 30341990 | 1115 | 17 | 1.11E-06 | 6.68E-05 | 0.004028 | 2.31E-06 | -0.00063818 | TRIM39, TRIM39-RPP21 |
| chr10 | 133340385 | 133340468 | 84 | 4 | 1.68E-06 | 3.09E-05 | 5.23E-06 | 2.31E-06 | -0.00620946 | ZNF511-PRAP1 |
| chr1 | 20661208 | 20661744 | 537 | 9 | 1.69E-06 | 4.78E-06 | 0.000567 | 2.31E-06 | -0.00011098 | DDOST |
| chr10 | 124035620 | 124036400 | 781 | 4 | 8.29E-06 | 2.59E-06 | 0.000193 | 2.31E-06 | -0.00742944 | CHST15 |
| chr2 | 26002844 | 26003148 | 305 | 3 | 6.73E-07 | 2.57E-05 | 3.20E-05 | 2.31E-06 | -0.00721857 | NA |
| chr10 | 128125695 | 128126507 | 813 | 10 | 3.20E-06 | 1.99E-06 | 0.001561 | 2.31E-06 | -0.00089865 | MKI67 |
| chr6 | 37657253 | 37657734 | 482 | 2 | 1.44E-06 | 1.76E-06 | 8.79E-05 | 2.31E-06 | -0.0246895 | MDGA1 |
| chr11 | 67983621 | 67983692 | 72 | 2 | 2.73E-06 | 1.68E-06 | 0.000103 | 2.31E-06 | 0.00474828 | NA |
| chr15 | 90889066 | 90889750 | 685 | 7 | 4.01E-06 | 1.13E-06 | 0.00395 | 2.31E-06 | 0.001114171 | FES |
| chr1 | 1540889 | 1541721 | 833 | 6 | 8.45E-07 | 7.27E-06 | 0.000135 | 2.31E-06 | -0.00396532 | SSU72 |
| chr8 | 136046132 | 136046174 | 43 | 2 | 8.20E-07 | 1.54E-05 | 6.06E-06 | 2.32E-06 | 0.002863432 | LINC02055 |
| chr5 | 486368 | 486736 | 369 | 3 | 8.34E-07 | 1.82E-06 | 0.000183 | 2.32E-06 | 0.004082614 | SLC9A3 |
| chr19 | 55503934 | 55504853 | 920 | 4 | 2.00E-06 | 1.95E-06 | 0.000231 | 2.32E-06 | -0.00678697 | SSC5D |
| chr5 | 36606000 | 36606963 | 964 | 15 | 6.82E-07 | 2.15E-05 | 0.001766 | 2.32E-06 | 0.001519697 | SLC1A3 |
| chr17 | 83020541 | 83021164 | 624 | 3 | 3.77E-07 | 8.16E-06 | 2.91E-05 | 2.32E-06 | -0.00360543 | B3GNTL1 |
| chr19 | 10876363 | 10876977 | 615 | 2 | 5.84E-07 | 5.22E-06 | 1.51E-05 | 2.32E-06 | -0.00556133 | CARM1 |
| chr22 | 32562563 | 32563117 | 555 | 3 | 6.73E-07 | 1.18E-05 | 1.13E-05 | 2.33E-06 | -0.00606498 | SYN3 |
| chr13 | 37869646 | 37870542 | 897 | 12 | 5.53E-06 | 9.04E-06 | 0.003867 | 2.33E-06 | 0.003361085 | AL356495.1, TRPC4 |
| chr1 | 57877329 | 57877342 | 14 | 2 | 3.32E-06 | 1.39E-06 | 0.000296 | 2.33E-06 | -0.01262469 | DAB1-AS1, DAB1 |
| chr3 | 14178350 | 14179335 | 986 | 19 | 1.66E-06 | 3.06E-05 | 0.001118 | 2.33E-06 | 0.001570333 | LSM3, XPC |
| chr2 | 70899809 | 70901061 | 1253 | 17 | 8.25E-07 | 2.97E-05 | 0.001855 | 2.33E-06 | 0.000938896 | AC007040.3, VAX2 |
| chr15 | 25405464 | 25406135 | 672 | 3 | 1.11E-08 | 0.0002591 | 1.64E-06 | 2.33E-06 | -0.00764786 | SNHG14, UBE3A |
| chr11 | 64015887 | 64016654 | 768 | 5 | 7.00E-07 | 0.0004182 | 0.000173 | 2.33E-06 | -0.000783 | MACROD1 |
| chr1 | 222927682 | 222928506 | 825 | 3 | 3.45E-05 | 1.12E-06 | 0.000807 | 2.33E-06 | -0.00166995 | DISP1 |
| chr10 | 129542248 | 129542851 | 604 | 4 | 3.98E-06 | 1.33E-06 | 0.000493 | 2.33E-06 | -0.00160169 | MGMT |
| chr21 | 26801459 | 26801660 | 202 | 2 | 8.35E-07 | 7.08E-05 | 2.90E-06 | 2.34E-06 | -0.01054939 | NA |
| chr5 | 149728891 | 149730561 | 1671 | 11 | 1.91E-06 | 3.92E-05 | 0.001248 | 2.34E-06 | -0.00373749 | PPARGC1B |
| chr17 | 4566088 | 4567322 | 1235 | 4 | 2.11E-08 | 0.0001933 | 0.000103 | 2.34E-06 | -0.00984264 | AC118754.2 |
| chr8 | 65789671 | 65790465 | 795 | 5 | 7.36E-07 | 5.73E-06 | 0.000593 | 2.34E-06 | 0.001123869 | PDE7A |
| chr12 | 10612901 | 10613129 | 229 | 3 | 8.81E-07 | 1.33E-06 | 0.000573 | 2.34E-06 | -0.00572629 | MAGOHB |
| chr15 | 80856596 | 80856882 | 287 | 2 | 5.14E-07 | 7.31E-05 | 2.86E-06 | 2.34E-06 | -0.01063792 | CEMIP |
| chr1 | 36575937 | 36576853 | 917 | 3 | 3.52E-06 | 2.98E-05 | 3.09E-05 | 2.34E-06 | -0.00025915 | NA |
| chr14 | 105218619 | 105219323 | 705 | 5 | 7.54E-07 | 5.60E-06 | 0.001818 | 2.34E-06 | -0.00225905 | BRF1 |
| chr5 | 37839987 | 37840737 | 751 | 7 | 1.31E-06 | 2.05E-06 | 0.000693 | 2.34E-06 | 0.001725791 | GDNF-AS1, GDNF |
| chr2 | 67694643 | 67695043 | 401 | 2 | 6.42E-07 | 1.08E-05 | 7.88E-06 | 2.34E-06 | 0.007560652 | NA |
| chr4 | 96352265 | 96352439 | 175 | 2 | 4.65E-07 | 0.0088341 | 1.54E-06 | 2.34E-06 | -0.01544058 | LINC02267 |
| chr4 | 142474027 | 142474703 | 677 | 3 | 9.30E-07 | 2.21E-06 | 0.000329 | 2.34E-06 | -0.00626296 | INPP4B |
| chr9 | 95606100 | 95606648 | 549 | 4 | 5.68E-07 | 3.67E-05 | 2.36E-05 | 2.34E-06 | 0.007456248 | AL392185.1 |
| chr12 | 106301818 | 106302895 | 1078 | 11 | 1.87E-06 | 6.17E-05 | 0.000364 | 2.34E-06 | -0.00064768 | TCP11L2, CKAP4 |
| chr9 | 133370810 | 133371516 | 707 | 2 | 2.59E-06 | 2.37E-06 | 4.58E-05 | 2.34E-06 | 0.022186291 | SURF4 |
| chr2 | 70293183 | 70294460 | 1278 | 16 | 1.92E-06 | 7.16E-06 | 0.000594 | 2.34E-06 | -0.0002023 | SNRPG |
| chr8 | 143707653 | 143708147 | 495 | 5 | 3.69E-06 | 4.34E-07 | 0.006341 | 2.35E-06 | 0.018566834 | ZNF707, CCDC166 |
| chr19 | 10461490 | 10461686 | 197 | 2 | 1.37E-06 | 1.82E-05 | 5.53E-06 | 2.35E-06 | 0.005607886 | PDE4A |
| chr11 | 533457 | 534269 | 813 | 5 | 3.92E-07 | 2.63E-06 | 0.001325 | 2.35E-06 | -0.00345331 | HRAS |
| chr17 | 49959032 | 49959319 | 288 | 2 | 2.77E-06 | 1.61E-06 | 0.000131 | 2.35E-06 | -0.01350271 | NA |
| chr13 | 113817820 | 113819110 | 1291 | 12 | 1.67E-06 | 7.06E-06 | 0.001616 | 2.35E-06 | -0.00092408 | GAS6-AS1 |
| chr1 | 240823212 | 240823598 | 387 | 5 | 1.42E-06 | 8.85E-05 | 0.000164 | 2.35E-06 | -0.00065721 | RGS7, RFKP1 |
| chr3 | 126051856 | 126052000 | 145 | 2 | 2.31E-06 | 7.73E-06 | 1.05E-05 | 2.36E-06 | -0.0041184 | SLC41A3 |
| chr14 | 54396658 | 54397526 | 869 | 12 | 6.98E-06 | 2.39E-06 | 0.003463 | 2.36E-06 | -0.00014814 | CDKN3 |
| chr5 | 1959736 | 1959802 | 67 | 2 | 4.27E-06 | 2.25E-06 | 5.18E-05 | 2.36E-06 | 0.006346914 | AC124852.1 |
| chr5 | 76325793 | 76326571 | 779 | 2 | 1.61E-05 | 1.54E-06 | 0.000165 | 2.36E-06 | -0.00967176 | SV2C |
| chr6 | 4458832 | 4459435 | 604 | 2 | 4.36E-06 | 1.71E-06 | 0.000105 | 2.36E-06 | -0.01001746 | AL162718.2, AL162718.1 |
| chr1 | 164576316 | 164576906 | 591 | 7 | 3.93E-07 | 0.0003764 | 0.000576 | 2.36E-06 | -0.00634771 | PBX1 |

| | | | | | | | | | | |
|---|---|---|---|---|---|---|---|---|---|---|
| chr13 | 95641223 | 95641877 | 655 | 4 | 6.68E-07 | 2.67E-05 | 0.000117 | 2.37E-06 | -0.0093518 | DZIP1 |
| chr9 | 136991852 | 136992765 | 914 | 9 | 2.29E-07 | 3.38E-05 | 1.17E-05 | 2.37E-06 | -0.00093653 | PAXX |
| chr7 | 4171317 | 4172123 | 807 | 8 | 3.79E-06 | 2.65E-06 | 0.000881 | 2.37E-06 | 0.000426657 | SDK1 |
| chr9 | 36035991 | 36037343 | 1353 | 9 | 5.41E-08 | 3.00E-05 | 5.08E-06 | 2.37E-06 | -0.00200286 | RECK |
| chr3 | 49799915 | 49800605 | 691 | 4 | 1.67E-06 | 3.38E-06 | 8.66E-05 | 2.37E-06 | -0.00019014 | NA |
| chr7 | 712193 | 713823 | 1631 | 18 | 8.12E-07 | 0.0001886 | 5.02E-05 | 2.37E-06 | -0.00056214 | PRKAR1B |
| chr3 | 13420359 | 13421403 | 1045 | 6 | 6.52E-07 | 2.07E-05 | 0.000981 | 2.37E-06 | -0.00825083 | NA |
| chr19 | 55464248 | 55464460 | 213 | 2 | 2.36E-06 | 2.52E-06 | 4.18E-05 | 2.37E-06 | -0.01708851 | C19orf85 |
| chr19 | 35563617 | 35564111 | 495 | 5 | 1.29E-06 | 6.97E-05 | 2.22E-05 | 2.37E-06 | -0.00694407 | AD000090.1, ATP4A |
| chr13 | 73849482 | 73849587 | 106 | 2 | 8.16E-07 | 0.0003032 | 2.05E-06 | 2.37E-06 | 0.002554986 | KLF12 |
| chr11 | 70628311 | 70628910 | 600 | 2 | 2.58E-07 | 2.91E-06 | 3.29E-05 | 2.37E-06 | -0.01450609 | SHANK2-AS1, SHANK2 |
| chr14 | 91285379 | 91285429 | 51 | 3 | 4.18E-06 | 1.12E-06 | 0.00049 | 2.37E-06 | 0.014378753 | CCDC88C |
| chr13 | 19184606 | 19185280 | 675 | 3 | 4.69E-06 | 2.88E-05 | 4.62E-05 | 2.37E-06 | 0.011348712 | AL139327.1 |
| chr11 | 95230230 | 95231653 | 1424 | 14 | 9.63E-07 | 0.0005942 | 0.002718 | 2.37E-06 | 0.001589917 | AP000787.1, SESN3 |
| chr10 | 120975520 | 120976076 | 557 | 2 | 8.45E-07 | 4.44E-06 | 1.86E-05 | 2.38E-06 | -0.00431972 | AC010998.1 |
| chr7 | 121327820 | 121328234 | 415 | 3 | 4.39E-08 | 5.26E-05 | 5.85E-06 | 2.38E-06 | -0.00634847 | WNT16 |
| chr6 | 36782873 | 36783015 | 143 | 2 | 1.69E-06 | 1.54E-06 | 0.000176 | 2.38E-06 | -0.01457822 | CPNE5 |
| chr3 | 167734935 | 167736551 | 1617 | 11 | 1.02E-05 | 6.76E-06 | 0.001581 | 2.38E-06 | -0.00198032 | SERPINI1, PDCD10 |
| chr17 | 81253440 | 81253514 | 75 | 2 | 3.83E-07 | 0.0017855 | 1.67E-06 | 2.38E-06 | 0.018773523 | SLC38A10 |
| chr7 | 4746733 | 4747355 | 623 | 4 | 1.08E-06 | 5.57E-06 | 0.000385 | 2.38E-06 | 0.00330701 | FOXK1 |
| chr5 | 116084466 | 116085497 | 1032 | 9 | 3.43E-06 | 6.60E-06 | 0.002187 | 2.38E-06 | 0.001161984 | COMMD10, AC034236.2 |
| chr16 | 4344135 | 4345424 | 1290 | 6 | 5.05E-05 | 3.96E-06 | 0.002785 | 2.38E-06 | -0.00293345 | PAM16, CORO7-PAM16 |
| chr11 | 69772800 | 69773422 | 623 | 2 | 3.41E-07 | 1.70E-06 | 0.000111 | 2.38E-06 | 0.024751239 | FGF4 |
| chr20 | 3043461 | 3044197 | 737 | 6 | 1.78E-08 | 0.0012284 | 1.83E-06 | 2.38E-06 | -0.00313397 | GNRH2 |
| chr12 | 13172706 | 13173094 | 389 | 4 | 1.11E-06 | 2.83E-05 | 6.40E-05 | 2.38E-06 | -0.00531262 | NA |
| chr11 | 108467435 | 108468482 | 1048 | 11 | 4.04E-06 | 4.50E-06 | 0.006 | 2.38E-06 | -0.00115259 | C11orf65 |
| chr22 | 17165199 | 17165373 | 175 | 4 | 4.25E-07 | 4.48E-05 | 0.000281 | 2.39E-06 | -0.00328757 | HDHD5-AS1, HDHD5 |
| chr12 | 91678895 | 91679044 | 150 | 2 | 1.03E-06 | 8.27E-05 | 2.80E-06 | 2.39E-06 | -0.00685247 | NA |
| chr5 | 42912418 | 42912638 | 221 | 2 | 2.22E-06 | 5.32E-06 | 1.54E-05 | 2.39E-06 | 0.008904676 | AC008945.2 |
| chr1 | 6459863 | 6461430 | 1568 | 10 | 1.66E-05 | 0.000674 | 0.000281 | 2.39E-06 | -0.00138859 | ESPN, TNFRSF25 |
| chr6 | 26108615 | 26109035 | 421 | 4 | 6.47E-07 | 1.76E-06 | 0.000145 | 2.39E-06 | -0.00071825 | NA |
| chr6 | 149425259 | 149425637 | 379 | 2 | 2.49E-06 | 1.74E-06 | 0.000103 | 2.39E-06 | -0.0019336 | NA |
| chr8 | 141018526 | 141019042 | 517 | 2 | 2.25E-06 | 1.97E-06 | 7.27E-05 | 2.39E-06 | -0.01555209 | NA |
| chr10 | 129899447 | 129899716 | 270 | 2 | 6.41E-07 | 1.74E-06 | 0.000103 | 2.39E-06 | -0.0229951 | EBF3 |
| chr16 | 89324201 | 89324560 | 360 | 4 | 5.95E-06 | 7.51E-07 | 0.002023 | 2.39E-06 | -0.00936364 | AC137932.2, ANKRD11 |
| chr11 | 103798857 | 103799441 | 585 | 2 | 2.52E-07 | 1.05E-05 | 8.28E-06 | 2.39E-06 | -0.01254395 | AP002989.1 |
| chr1 | 174975933 | 174977041 | 1109 | 3 | 9.48E-06 | 9.41E-07 | 0.000995 | 2.39E-06 | 2.62E-05 | RABGAP1L |
| chr2 | 231529991 | 231530615 | 625 | 6 | 2.16E-07 | 6.21E-05 | 0.000108 | 2.39E-06 | -0.00205191 | NMUR1 |
| chr5 | 31530334 | 31530833 | 500 | 3 | 2.13E-07 | 1.35E-06 | 0.000602 | 2.39E-06 | -0.00067094 | DROSHA |
| chr10 | 97885585 | 97886007 | 423 | 2 | 6.17E-07 | 6.20E-06 | 1.33E-05 | 2.39E-06 | -0.00357382 | CRTAC1 |
| chr12 | 103178435 | 103178922 | 488 | 3 | 1.89E-07 | 0.0032472 | 4.24E-06 | 2.39E-06 | -0.01281656 | AC068643.2, AC068643.1 |
| chr3 | 58214294 | 58215867 | 1574 | 8 | 9.41E-06 | 6.27E-06 | 0.000335 | 2.40E-06 | 0.004105087 | DNASE1L3 |
| chr22 | 30204700 | 30204718 | 19 | 2 | 3.59E-06 | 2.09E-06 | 6.36E-05 | 2.40E-06 | -0.00927308 | AC002378.1 |
| chr19 | 45882772 | 45883704 | 933 | 2 | 2.16E-05 | 1.43E-06 | 0.0003 | 2.40E-06 | -0.01510449 | IRF2BP1 |
| chr17 | 78997190 | 78997616 | 427 | 6 | 1.85E-07 | 0.0001113 | 9.59E-06 | 2.40E-06 | 0.000613357 | CANT1 |
| chr2 | 95761137 | 95761425 | 289 | 2 | 9.50E-07 | 1.72E-05 | 5.90E-06 | 2.40E-06 | -0.00669564 | NA |
| chr14 | 80959568 | 80960233 | 666 | 5 | 2.74E-06 | 5.44E-06 | 0.000869 | 2.41E-06 | 0.006389411 | TSHR |
| chr6 | 117448416 | 117449037 | 622 | 5 | 3.50E-06 | 1.57E-06 | 0.000312 | 2.41E-06 | -0.0074609 | AL132671.2 |
| chr6 | 24774485 | 24775352 | 868 | 10 | 7.30E-07 | 1.96E-05 | 0.000135 | 2.41E-06 | -0.00107371 | GMNN |
| chr16 | 89136163 | 89136745 | 583 | 7 | 8.14E-06 | 2.95E-06 | 0.000813 | 2.41E-06 | 0.001988285 | ACSF3 |
| chr4 | 1394035 | 1394525 | 491 | 3 | 4.46E-06 | 1.44E-06 | 0.000337 | 2.41E-06 | -0.00049363 | UVSSA |
| chr2 | 231051517 | 231052647 | 1131 | 5 | 1.12E-06 | 9.83E-07 | 0.003112 | 2.42E-06 | -0.00898337 | AC009407.1 |
| chr3 | 72123874 | 72124156 | 283 | 4 | 7.22E-07 | 4.79E-05 | 7.52E-06 | 2.42E-06 | -0.00300131 | LINC00877 |
| chr15 | 72398299 | 72398658 | 360 | 2 | 1.53E-07 | 2.06E-05 | 5.32E-06 | 2.42E-06 | -0.00746049 | TMEM202 |
| chr16 | 906062 | 906700 | 639 | 3 | 1.84E-07 | 4.52E-05 | 4.03E-06 | 2.42E-06 | -0.00278971 | LMF1 |
| chr9 | 35616247 | 35616729 | 483 | 2 | 4.97E-07 | 2.87E-06 | 3.50E-05 | 2.42E-06 | -0.01300941 | CD72 |
| chr6 | 96649620 | 96650170 | 551 | 4 | 1.00E-06 | 1.10E-05 | 2.47E-05 | 2.42E-06 | -0.00844095 | NA |
| chr7 | 158428678 | 158429084 | 407 | 3 | 1.34E-06 | 1.06E-06 | 0.000613 | 2.42E-06 | 0.008005105 | PTPRN2 |
| chr17 | 44223425 | 44223942 | 518 | 4 | 9.98E-07 | 1.41E-05 | 4.22E-05 | 2.42E-06 | 0.000676544 | AC003102.1 |
| chr19 | 53553954 | 53554831 | 878 | 8 | 2.70E-06 | 0.000116 | 0.006833 | 2.42E-06 | -0.00308716 | ZNF331 |
| chr2 | 43398110 | 43398566 | 457 | 2 | 1.17E-07 | 0.0002436 | 2.17E-06 | 2.42E-06 | -0.00645747 | THADA |
| chr20 | 49916586 | 49917463 | 878 | 4 | 1.28E-07 | 5.03E-06 | 3.56E-05 | 2.42E-06 | -0.00039152 | NA |
| chr2 | 171921831 | 171922601 | 771 | 12 | 4.06E-06 | 3.98E-06 | 0.002028 | 2.42E-06 | -0.00033552 | HAT1, SLC25A12 |
| chr15 | 62326650 | 62327267 | 618 | 2 | 5.22E-06 | 1.74E-06 | 0.000111 | 2.42E-06 | -0.01032272 | NA |
| chr9 | 131950215 | 131950489 | 275 | 2 | 3.09E-07 | 0.0015979 | 1.71E-06 | 2.42E-06 | -0.01051348 | MED27 |
| chr10 | 19178855 | 19179084 | 230 | 2 | 5.80E-07 | 0.0002088 | 2.25E-06 | 2.43E-06 | 0.009602867 | MALRD1 |
| chr8 | 1369850 | 1370040 | 191 | 4 | 7.33E-07 | 4.73E-05 | 0.000133 | 2.43E-06 | -0.00519181 | DLGAP2 |
| chr10 | 23345992 | 23346271 | 280 | 3 | 3.55E-07 | 3.34E-05 | 5.14E-06 | 2.43E-06 | 0.003762186 | NA |
| chr1 | 10860430 | 10861301 | 872 | 3 | 2.59E-05 | 3.65E-06 | 0.000335 | 2.43E-06 | -0.0138574 | NA |
| chr18 | 23133795 | 23134563 | 769 | 8 | 2.11E-06 | 3.45E-05 | 0.002195 | 2.43E-06 | 0.003825598 | NA |
| chr21 | 44261340 | 44261951 | 612 | 2 | 4.70E-06 | 1.64E-06 | 0.000144 | 2.43E-06 | 0.003121091 | DNMT3L |
| chr20 | 46020885 | 46021514 | 630 | 4 | 3.51E-07 | 1.09E-05 | 0.000434 | 2.43E-06 | -0.00294746 | SLC12A5-AS1 |
| chr9 | 114305856 | 114306252 | 397 | 4 | 3.51E-07 | 8.28E-05 | 3.47E-06 | 2.43E-06 | -0.00905121 | COL27A1 |
| chr17 | 40797033 | 40797260 | 228 | 2 | 2.00E-06 | 1.46E-06 | 0.000293 | 2.43E-06 | -0.02372639 | KRT28 |
| chr2 | 37436414 | 37436587 | 174 | 2 | 5.10E-06 | 1.55E-06 | 0.000191 | 2.43E-06 | 0.007038804 | NA |
| chr22 | 43419944 | 43420277 | 334 | 2 | 1.42E-06 | 1.46E-06 | 0.0003 | 2.44E-06 | -0.01231494 | MPPED1 |
| chr19 | 8444451 | 8444949 | 499 | 11 | 5.68E-06 | 3.94E-06 | 0.001943 | 2.44E-06 | 0.001037995 | HNRNPM, AC136469.2 |
| chr2 | 61184286 | 61184357 | 72 | 2 | 5.97E-07 | 1.74E-05 | 5.97E-06 | 2.44E-06 | 0.007911197 | AHSA2P |
| chr11 | 134441020 | 134441335 | 316 | 2 | 3.48E-06 | 1.92E-06 | 8.27E-05 | 2.44E-06 | 0.003662857 | B3GAT1-DT |
| chr15 | 100849067 | 100849528 | 462 | 6 | 2.95E-06 | 1.40E-06 | 0.003398 | 2.44E-06 | 0.025028931 | NA |
| chr8 | 51663417 | 51663614 | 198 | 2 | 6.33E-07 | 0.0003247 | 2.08E-06 | 2.44E-06 | 0.01283738 | PXDNL |
| chr17 | 66876376 | 66877476 | 1101 | 9 | 8.61E-06 | 1.14E-06 | 0.002653 | 2.44E-06 | 0.000396779 | CACNG5 |
| chr5 | 173056277 | 173057409 | 1133 | 13 | 5.17E-07 | 3.19E-05 | 0.000245 | 2.44E-06 | 0.000585418 | CREBRF |
| chr10 | 59362196 | 59363338 | 1143 | 15 | 7.46E-06 | 8.71E-06 | 0.007578 | 2.44E-06 | -0.00077592 | AL355474.1, FAM13C |
| chr17 | 9901757 | 9902261 | 505 | 2 | 2.37E-06 | 1.52E-06 | 0.000226 | 2.44E-06 | -0.02940393 | RCVRN |
| chr8 | 26230335 | 26230396 | 62 | 2 | 5.48E-06 | 1.50E-06 | 0.000253 | 2.45E-06 | -0.01241346 | NA |
| chr6 | 167712488 | 167713061 | 574 | 3 | 2.94E-07 | 8.11E-05 | 2.95E-06 | 2.45E-06 | -0.01564379 | NA |
| chr17 | 42154682 | 42156233 | 1552 | 12 | 7.60E-06 | 0.000787 | 4.42E-05 | 2.45E-06 | -0.00148804 | AC099811.3, AC099811.2, RAB5C |
| chr16 | 24496382 | 24496897 | 516 | 2 | 8.62E-07 | 1.57E-06 | 0.000187 | 2.45E-06 | 0.000298993 | NA |
| chr5 | 139759295 | 139759741 | 447 | 4 | 7.61E-06 | 6.68E-07 | 0.002417 | 2.45E-06 | -0.02162028 | NA |
| chr17 | 3571325 | 3571656 | 332 | 5 | 1.25E-07 | 0.0003051 | 9.75E-07 | 2.45E-06 | -0.00198912 | TRPV1 |
| chr7 | 98209347 | 98209423 | 77 | 3 | 4.40E-06 | 1.62E-06 | 0.000261 | 2.45E-06 | 0.001627247 | LMTK2 |
| chr1 | 90729947 | 90730931 | 985 | 6 | 1.78E-07 | 6.40E-06 | 0.000147 | 2.45E-06 | -0.00169736 | NA |
| chr7 | 2211361 | 2211913 | 553 | 2 | 8.19E-09 | 3.27E-05 | 4.20E-06 | 2.45E-06 | -0.00330045 | MAD1L1, AC069288.1 |
| chr10 | 124713779 | 124714056 | 278 | 2 | 4.37E-07 | 0.0002189 | 2.25E-06 | 2.46E-06 | -0.01031038 | FAM53B-AS1, AC068896.1, FAM53B |
| chr11 | 94135032 | 94135233 | 202 | 2 | 5.25E-06 | 1.52E-06 | 0.00023 | 2.46E-06 | -0.01616942 | PANX1 |
| chr5 | 132181504 | 132181863 | 360 | 2 | 2.54E-08 | 0.0005129 | 1.94E-06 | 2.46E-06 | -0.00927273 | NA |
| chr2 | 204473852 | 204474116 | 265 | 4 | 3.32E-06 | 3.00E-06 | 8.93E-05 | 2.46E-06 | -0.00371742 | AC016903.2 |
| chr10 | 117240572 | 117240761 | 190 | 4 | 1.01E-06 | 1.51E-06 | 0.001046 | 2.46E-06 | -0.01149242 | AL731557.1 |
| chr1 | 183471569 | 183472738 | 1170 | 14 | 5.14E-06 | 0.000122 | 0.000954 | 2.46E-06 | -3.19E-05 | SMG7, SMG7-AS1 |
| chr7 | 26636572 | 26637755 | 1184 | 7 | 1.62E-08 | 0.0003601 | 1.57E-06 | 2.46E-06 | -0.00404522 | NA |
| chr12 | 120219017 | 120219511 | 495 | 4 | 3.26E-07 | 2.73E-05 | 0.000358 | 2.46E-06 | -0.00279709 | AC004263.2, PXN |
| chr12 | 121352013 | 121352808 | 796 | 12 | 3.26E-06 | 1.04E-05 | 0.004467 | 2.46E-06 | -0.00064417 | ANAPC5 |
| chr3 | 142447361 | 142448298 | 938 | 6 | 9.98E-08 | 0.0001181 | 1.76E-05 | 2.47E-06 | -0.00033822 | XRN1 |
| chr4 | 4290148 | 4291352 | 1205 | 12 | 6.08E-10 | 0.0006086 | 3.28E-06 | 2.47E-06 | 0.000246926 | ZBTB49, LYAR |
| chr10 | 33507427 | 33508194 | 768 | 2 | 5.34E-06 | 1.70E-06 | 0.00013 | 2.47E-06 | 0.005734759 | NA |
| chr17 | 9791155 | 9792281 | 1127 | 9 | 5.00E-06 | 6.56E-07 | 0.003985 | 2.47E-06 | -0.00258549 | AC027045.2, DHRS7C, AC027045.1 |
| chr16 | 545486 | 546009 | 524 | 7 | 2.24E-06 | 5.35E-06 | 0.000653 | 2.47E-06 | -0.00221755 | CAPN15 |
| chr6 | 8435245 | 8436235 | 991 | 12 | 1.58E-06 | 8.69E-06 | 0.007468 | 2.47E-06 | -0.00459107 | AL355499.2, HULC, SLC35B3 |
| chr14 | 60091175 | 60092482 | 1308 | 14 | 4.83E-05 | 1.37E-05 | 0.001235 | 2.47E-06 | 0.001010258 | PCNX4, AL157911.1 |
| chr4 | 188003563 | 188003598 | 36 | 2 | 5.17E-06 | 1.48E-06 | 0.000295 | 2.47E-06 | -0.0079138 | ZFP42 |
| chr1 | 226723968 | 226725147 | 1180 | 3 | 7.12E-07 | 4.26E-06 | 6.80E-05 | 2.47E-06 | 0.009240865 | ITPKB |
| chr10 | 13001313 | 13001986 | 674 | 10 | 1.02E-06 | 4.64E-06 | 0.000293 | 2.47E-06 | -0.00127923 | CCDC3 |
| chr17 | 39924125 | 39924576 | 452 | 3 | 6.67E-07 | 2.00E-05 | 3.50E-05 | 2.47E-06 | 0.001618348 | ORMDL3 |
| chr2 | 36732348 | 36733187 | 840 | 3 | 1.14E-05 | 2.03E-06 | 0.000296 | 2.47E-06 | -0.00654285 | VIT |
| chr12 | 132701380 | 132702303 | 924 | 3 | 3.98E-07 | 3.96E-05 | 1.34E-05 | 2.47E-06 | -0.00401748 | PXMP2, AC135586.2 |
| chr1 | 84754336 | 84754447 | 112 | 3 | 9.53E-07 | 3.47E-05 | 2.21E-05 | 2.48E-06 | -0.00198672 | NA |
| chr7 | 111581285 | 111581438 | 154 | 2 | 1.97E-06 | 1.35E-05 | 7.19E-06 | 2.48E-06 | -0.01596178 | NA |
| chr18 | 51569288 | 51569709 | 422 | 2 | 7.03E-06 | 1.48E-06 | 0.000307 | 2.48E-06 | 0.014498962 | LINC01630 |
| chr14 | 74239030 | 74239633 | 604 | 7 | 4.71E-07 | 0.0001115 | 0.000413 | 2.48E-06 | -0.00172381 | VSX2 |
| chr8 | 123231306 | 123232279 | 974 | 3 | 2.81E-06 | 1.30E-06 | 0.000579 | 2.48E-06 | -0.00976778 | C8orf76, ZHX1-C8orf76 |
| chr11 | 45701780 | 45702235 | 456 | 5 | 3.61E-07 | 9.13E-05 | 3.16E-05 | 2.48E-06 | -0.0099986 | NA |
| chr6 | 42021428 | 42021432 | 5 | 2 | 1.55E-06 | 3.96E-06 | 2.33E-05 | 2.48E-06 | 0.000528684 | CCND3 |
| chr14 | 23369592 | 23370258 | 667 | 2 | 9.49E-06 | 1.57E-06 | 0.000204 | 2.48E-06 | -0.01316137 | NA |
| chr1 | 234676257 | 234676428 | 172 | 3 | 1.68E-06 | 6.60E-06 | 2.21E-05 | 2.49E-06 | -0.00506094 | AL160408.1, AL160408.3 |
| chr3 | 15477610 | 15478071 | 462 | 3 | 4.26E-06 | 1.30E-06 | 0.000771 | 2.49E-06 | 0.016122686 | COLQ |
| chr19 | 45145538 | 45146494 | 957 | 3 | 2.39E-05 | 3.87E-06 | 0.000226 | 2.49E-06 | -0.00540789 | MARK4, PPP1R37 |
| chr11 | 131760570 | 131760754 | 185 | 3 | 5.46E-06 | 1.63E-06 | 0.000446 | 2.49E-06 | -0.03481621 | NTM |
| chr7 | 26486062 | 26486474 | 413 | 2 | 1.09E-06 | 5.71E-06 | 1.53E-05 | 2.49E-06 | -0.00743455 | AC004540.1 |
| chr7 | 92786056 | 92786209 | 154 | 2 | 9.48E-07 | 0.0001421 | 2.50E-06 | 2.49E-06 | -0.00117803 | CDK6 |
| chr15 | 58979728 | 58980187 | 460 | 4 | 2.67E-09 | 0.0027999 | 1.46E-06 | 2.49E-06 | -0.00480362 | RNF111 |
| chr11 | 135074176 | 135074331 | 156 | 2 | 3.91E-06 | 2.42E-06 | 5.13E-05 | 2.49E-06 | -0.0004132 | LINC02684 |
| chr2 | 38073394 | 38074295 | 902 | 6 | 8.33E-07 | 1.61E-05 | 0.001205 | 2.49E-06 | -0.0069551 | CYP1B1-AS1, CYP1B1 |
| chr17 | 48530466 | 48531512 | 1047 | 15 | 2.18E-06 | 7.36E-06 | 0.000902 | 2.49E-06 | -0.00115397 | HOXB1 |
| chr18 | 59664601 | 59665196 | 596 | 2 | 1.38E-05 | 1.50E-06 | 0.000305 | 2.50E-06 | -0.01203778 | CCBE1 |
| chr10 | 63268983 | 63269408 | 426 | 6 | 4.18E-07 | 9.92E-05 | 0.000227 | 2.50E-06 | 0.003032353 | JMJD1C |
| chr9 | 113503243 | 113503943 | 701 | 2 | 1.74E-05 | 1.53E-06 | 0.000256 | 2.50E-06 | 0.003056565 | RGS3 |
| chr2 | 238925023 | 238925954 | 932 | 4 | 1.65E-07 | 0.0003138 | 7.86E-05 | 2.50E-06 | -0.00353489 | LINC01940 |
| chr3 | 134406484 | 134407539 | 1056 | 6 | 4.60E-07 | 1.38E-05 | 1.72E-05 | 2.50E-06 | -0.00387617 | NA |
| chr10 | 1129815 | 1130209 | 395 | 4 | 1.07E-07 | 1.70E-05 | 1.62E-05 | 2.50E-06 | 0.004507754 | WDR37 |
| chr12 | 120327915 | 120328420 | 506 | 3 | 3.79E-07 | 3.26E-06 | 5.03E-05 | 2.51E-06 | -0.01003521 | NA |
| chr6 | 57313815 | 57314252 | 438 | 5 | 2.33E-06 | 1.22E-05 | 0.000281 | 2.51E-06 | -0.00848638 | NA |
| chr22 | 32477539 | 32477974 | 436 | 4 | 5.78E-07 | 3.23E-05 | 0.000163 | 2.51E-06 | 0.001180944 | FBXO7 |
| chr11 | 94658604 | 94658612 | 9 | 2 | 5.56E-06 | 1.62E-06 | 0.000184 | 2.51E-06 | -0.00989138 | AP000943.2 |
| chr2 | 177166305 | 177166593 | 289 | 2 | 2.76E-07 | 2.38E-05 | 5.14E-06 | 2.51E-06 | -0.01194352 | AC079305.2 |
| chr1 | 156641402 | 156642348 | 947 | 8 | 8.27E-06 | 0.0001062 | 0.000488 | 2.51E-06 | 0.006835454 | BCAN, AL365181.3 |
| chr5 | 143058475 | 143059098 | 624 | 2 | 7.25E-06 | 1.66E-06 | 0.000159 | 2.51E-06 | 0.001126893 | ARHGAP26 |
| chr9 | 114524630 | 114525174 | 545 | 2 | 6.35E-08 | 2.55E-05 | 4.95E-06 | 2.51E-06 | -0.01471465 | NA |
| chr11 | 64317494 | 64318548 | 1055 | 20 | 6.66E-07 | 0.0024721 | 0.000271 | 2.51E-06 | -0.00014883 | PRDX5, TRMT112 |
| chr7 | 121149050 | 121149455 | 406 | 2 | 5.30E-06 | 1.59E-06 | 0.000205 | 2.51E-06 | 0.00015408 | CPED1 |
| chr3 | 197509497 | 197510283 | 787 | 6 | 2.22E-06 | 7.17E-06 | 0.001232 | 2.51E-06 | 0.007417943 | BDH1 |
| chr15 | 62884404 | 62884804 | 401 | 4 | 3.73E-06 | 5.95E-06 | 0.00032 | 2.51E-06 | 0.002611738 | NA |
| chr5 | 154606347 | 154606624 | 278 | 3 | 2.40E-06 | 2.41E-05 | 0.000125 | 2.52E-06 | -0.00876884 | NA |
| chr2 | 1875218 | 1875608 | 391 | 3 | 2.14E-07 | 1.49E-05 | 0.000175 | 2.52E-06 | -0.0048734 | MYT1L |
| chr12 | 10030573 | 10031251 | 679 | 7 | 4.29E-06 | 7.43E-07 | 0.001695 | 2.52E-06 | -0.01613748 | CLEC9A, AC024224.2 |
| chr3 | 192408234 | 192409060 | 827 | 5 | 2.20E-05 | 1.10E-05 | 6.89E-05 | 2.52E-06 | 0.002722336 | FGF12 |
| chr1 | 2189539 | 2190082 | 544 | 5 | 9.71E-06 | 5.17E-07 | 0.004416 | 2.52E-06 | -0.02862577 | FAAP20 |
| chr19 | 3979391 | 3979990 | 600 | 5 | 1.01E-06 | 0.0001093 | 0.000131 | 2.52E-06 | 0.000350199 | EEF2 |

| | | | | | | | | | | |
|---|---|---|---|---|---|---|---|---|---|---|
| chr13 | 32588809 | 32589038 | 230 | 2 | 5.34E-06 | 1.54E-06 | 0.000267 | 2.52E-06 | -0.01450153 | PDS5B |
| chr1 | 184974429 | 184975156 | 728 | 9 | 1.79E-07 | 0.0008164 | 6.97E-06 | 2.52E-06 | -0.0025854 | NIBAN1 |
| chr1 | 99766126 | 99767265 | 1140 | 7 | 5.89E-07 | 5.66E-05 | 0.000185 | 2.52E-06 | 0.000237791 | FRRS1 |
| chr11 | 34513538 | 34514634 | 1097 | 13 | 3.60E-07 | 0.0002301 | 0.000143 | 2.52E-06 | -9.55E-05 | ELF5 |
| chr6 | 36660310 | 36660326 | 17 | 2 | 1.61E-06 | 1.66E-06 | 0.000165 | 2.53E-06 | -0.01527293 | NA |
| chr5 | 124127760 | 124127890 | 131 | 2 | 5.75E-06 | 1.52E-06 | 0.000306 | 2.53E-06 | -0.0017416 | LINC01170 |
| chr5 | 317725 | 318354 | 630 | 2 | 1.49E-05 | 1.78E-06 | 0.000122 | 2.53E-06 | -0.01704372 | PDCD6-AHRR |
| chr12 | 95209029 | 95209517 | 489 | 3 | 4.40E-06 | 1.25E-06 | 0.000564 | 2.53E-06 | 0.000644963 | FGD6 |
| chr7 | 149146528 | 149147512 | 985 | 8 | 7.36E-07 | 5.10E-05 | 0.000342 | 2.53E-06 | -0.00209103 | ZNF398 |
| chr22 | 39490512 | 39490543 | 32 | 2 | 1.41E-06 | 1.66E-06 | 0.000167 | 2.53E-06 | -0.01646361 | MGAT3 |
| chr12 | 120504839 | 120505178 | 340 | 4 | 1.55E-06 | 0.0002227 | 0.000172 | 2.53E-06 | -0.00409219 | COQ5 |
| chr7 | 157017791 | 157018823 | 1033 | 9 | 9.40E-06 | 1.75E-05 | 0.00511 | 2.53E-06 | 0.000732812 | NA |
| chr12 | 132828534 | 132829723 | 1190 | 11 | 1.85E-06 | 2.64E-05 | 0.002076 | 2.53E-06 | 0.001631246 | GOLGA3, CHFR |
| chr1 | 212791391 | 212792406 | 1016 | 17 | 6.30E-07 | 2.87E-05 | 0.001905 | 2.54E-06 | 0.000200355 | TATDN3, NSL1 |
| chr3 | 70108258 | 70108333 | 76 | 2 | 3.45E-06 | 5.33E-06 | 1.70E-05 | 2.54E-06 | 0.025761262 | SAMMSON |
| chr22 | 21774451 | 21775295 | 845 | 2 | 2.78E-05 | 1.58E-06 | 0.000231 | 2.54E-06 | -0.00438088 | MAPK1 |
| chr2 | 97663899 | 97664937 | 1039 | 15 | 3.57E-06 | 2.18E-06 | 0.010659 | 2.54E-06 | 0.00182778 | C2orf92, ACTR1B, RNU4-8P |
| chr8 | 37479491 | 37479942 | 452 | 2 | 1.39E-06 | 2.42E-06 | 5.39E-05 | 2.54E-06 | -0.01118426 | LINC01605 |
| chr12 | 54829886 | 54830119 | 234 | 3 | 2.06E-06 | 9.68E-06 | 1.41E-05 | 2.54E-06 | -0.01037735 | NA |
| chr12 | 91361682 | 91362352 | 671 | 6 | 4.10E-06 | 2.27E-06 | 0.001466 | 2.55E-06 | 0.004345317 | LINC02823 |
| chr11 | 111913150 | 111913846 | 697 | 5 | 1.43E-06 | 7.98E-05 | 3.40E-05 | 2.55E-06 | -0.00348578 | HSPB2, HSPB2-C11orf52, CRYAB |
| chr7 | 108527686 | 108528959 | 1274 | 9 | 7.33E-07 | 3.66E-05 | 0.001786 | 2.55E-06 | -0.00122261 | PNPLA8 |
| chr2 | 134253186 | 134253318 | 133 | 3 | 1.72E-06 | 1.00E-05 | 7.40E-05 | 2.55E-06 | 0.003321059 | MGAT5 |
| chr16 | 47121061 | 47121301 | 241 | 6 | 3.85E-06 | 6.62E-06 | 0.000103 | 2.55E-06 | -0.00411597 | NETO2 |
| chr2 | 201451614 | 201452879 | 1266 | 11 | 3.50E-08 | 6.03E-06 | 0.003819 | 2.56E-06 | -0.0011086 | STRADB |
| chr2 | 61065650 | 61066590 | 941 | 14 | 2.51E-07 | 0.0004511 | 4.97E-06 | 2.56E-06 | 0.000992372 | KIAA1841 |
| chr14 | 22614254 | 22614313 | 60 | 2 | 1.90E-06 | 8.30E-06 | 1.12E-05 | 2.56E-06 | -0.00474263 | NA |
| chr1 | 26828008 | 26828578 | 571 | 2 | 2.25E-08 | 6.37E-05 | 3.30E-06 | 2.56E-06 | 0.014648102 | ZDHHC18 |
| chr1 | 19758267 | 19758677 | 411 | 2 | 7.52E-07 | 1.03E-05 | 9.31E-06 | 2.56E-06 | 0.005804133 | TMCO4 |
| chr6 | 28531602 | 28532048 | 447 | 4 | 4.19E-06 | 8.08E-07 | 0.001728 | 2.57E-06 | -0.00655334 | GPX5 |
| chr19 | 54474203 | 54474216 | 14 | 2 | 1.12E-06 | 2.35E-06 | 5.92E-05 | 2.57E-06 | -0.01793414 | NA |
| chr17 | 50531971 | 50532894 | 924 | 6 | 3.98E-07 | 9.46E-05 | 0.000343 | 2.57E-06 | -0.00267464 | EPN3, AC021491.2 |
| chr3 | 50370273 | 50371179 | 907 | 4 | 4.76E-07 | 0.0207626 | 3.85E-05 | 2.57E-06 | -0.00489452 | CACNA2D2 |
| chr5 | 77632803 | 77633239 | 437 | 3 | 2.59E-07 | 0.000258 | 7.08E-06 | 2.57E-06 | -0.00802455 | OTP |
| chr22 | 46077841 | 46078856 | 1016 | 7 | 1.92E-07 | 0.0008385 | 0.000138 | 2.57E-06 | -0.00081636 | MIRLET7BHG |
| chr8 | 48583485 | 48583831 | 347 | 3 | 2.49E-06 | 8.00E-06 | 2.08E-05 | 2.58E-06 | 0.002720432 | AC022915.2 |
| chr10 | 121531764 | 121532436 | 673 | 3 | 4.36E-06 | 1.04E-05 | 0.000166 | 2.58E-06 | -0.01196827 | FGFR2 |
| chr10 | 99906800 | 99906836 | 37 | 2 | 4.44E-06 | 1.56E-06 | 0.000285 | 2.58E-06 | 0.001215761 | DNMBP |
| chr12 | 49268622 | 49268822 | 201 | 3 | 1.92E-06 | 2.08E-06 | 0.000514 | 2.59E-06 | -0.01496726 | TUBA1C, AC125611.3 |
| chr2 | 235106525 | 235106905 | 381 | 2 | 4.63E-07 | 8.87E-06 | 1.07E-05 | 2.59E-06 | -0.01273207 | NA |
| chr20 | 18811726 | 18812415 | 690 | 4 | 1.21E-06 | 1.88E-06 | 0.000284 | 2.59E-06 | -0.00234428 | SCP2D1-AS1 |
| chr4 | 950348 | 950532 | 185 | 3 | 3.65E-06 | 1.59E-06 | 0.000213 | 2.59E-06 | -0.00329436 | TMEM175 |
| chr5 | 149549609 | 149550811 | 1203 | 10 | 1.94E-07 | 1.74E-05 | 0.000367 | 2.59E-06 | 0.001711759 | CSNK1A1 |
| chr2 | 170504089 | 170504354 | 266 | 2 | 1.73E-06 | 3.16E-06 | 3.47E-05 | 2.59E-06 | -0.01736942 | MYO3B |
| chr1 | 50330628 | 50330970 | 343 | 2 | 1.33E-07 | 0.0053808 | 1.70E-06 | 2.60E-06 | 0.010308744 | NA |
| chr14 | 56810071 | 56812011 | 1941 | 13 | 2.67E-05 | 0.0005196 | 0.001601 | 2.60E-06 | -0.0037202 | OTX2 |
| chr7 | 106867891 | 106868975 | 1085 | 6 | 1.34E-05 | 7.40E-05 | 0.000302 | 2.60E-06 | 0.001588217 | PIK3CG |
| chr1 | 149887554 | 149889254 | 1701 | 12 | 7.77E-08 | 0.0002661 | 3.92E-05 | 2.60E-06 | -0.00154983 | BOLA1, H2AC21 |
| chr21 | 36745343 | 36746037 | 695 | 5 | 1.77E-06 | 3.20E-06 | 0.000421 | 2.60E-06 | -0.00139898 | SIM2 |
| chr4 | 99651836 | 99652693 | 858 | 6 | 3.01E-07 | 0.0001288 | 4.15E-05 | 2.60E-06 | -0.00549203 | C4orf54 |
| chr19 | 13253050 | 13253420 | 371 | 4 | 2.44E-06 | 4.11E-06 | 0.000881 | 2.61E-06 | -0.00090104 | CACNA1A |
| chr6 | 50818044 | 50818957 | 914 | 3 | 1.10E-08 | 0.0010096 | 1.38E-06 | 2.61E-06 | -0.00268313 | TFAP2B |
| chr1 | 205522290 | 205522413 | 124 | 3 | 2.17E-06 | 4.68E-06 | 3.25E-05 | 2.61E-06 | -0.00027576 | CDK18 |
| chr19 | 5851111 | 5852487 | 1377 | 10 | 3.34E-07 | 0.0001732 | 4.12E-05 | 2.61E-06 | -0.00326605 | AC024592.2, FUT3 |
| chr17 | 57337459 | 57337532 | 74 | 2 | 2.73E-06 | 4.01E-06 | 2.50E-05 | 2.61E-06 | 0.003830796 | MSI2 |
| chr16 | 15996747 | 15996906 | 160 | 2 | 5.03E-06 | 1.81E-06 | 0.000132 | 2.61E-06 | -0.00879664 | ABCC1 |
| chr11 | 65888424 | 65889018 | 595 | 8 | 2.20E-08 | 0.0009439 | 3.03E-06 | 2.61E-06 | -0.00066538 | FIBP |
| chr12 | 57087826 | 57088991 | 1166 | 10 | 5.87E-07 | 9.15E-06 | 0.000597 | 2.61E-06 | 0.000614787 | NEMP1 |
| chr16 | 88882675 | 88883789 | 1115 | 8 | 2.95E-06 | 1.58E-06 | 0.001079 | 2.61E-06 | -0.00244531 | AC092384.2, CBFA2T3 |
| chr6 | 114748007 | 114748374 | 368 | 2 | 7.13E-07 | 1.71E-05 | 6.56E-06 | 2.62E-06 | -0.01973489 | NA |
| chr17 | 75012042 | 75013117 | 1076 | 12 | 2.04E-06 | 5.35E-06 | 0.002271 | 2.62E-06 | 3.57E-06 | MRPL58 |
| chr5 | 639861 | 640267 | 407 | 2 | 3.27E-08 | 4.61E-05 | 3.86E-06 | 2.62E-06 | -0.00347582 | CEP72 |
| chr19 | 3490715 | 3491215 | 501 | 3 | 5.16E-07 | 0.0001829 | 4.33E-05 | 2.62E-06 | 0.001809967 | DOHH |
| chr10 | 3537174 | 3538301 | 1128 | 4 | 1.53E-06 | 1.64E-05 | 0.000128 | 2.62E-06 | -0.00277739 | AL357833.1 |
| chr5 | 500087 | 500450 | 364 | 5 | 7.60E-07 | 0.0003946 | 9.19E-05 | 2.62E-06 | 0.001085758 | SLC9A3 |
| chr10 | 43670972 | 43671780 | 809 | 4 | 2.58E-06 | 2.87E-06 | 0.000771 | 2.62E-06 | 0.004984182 | ZNF32-AS3 |
| chr8 | 23198784 | 23198988 | 205 | 2 | 4.86E-07 | 4.20E-05 | 4.03E-06 | 2.62E-06 | 0.00381273 | TNFRSF10A |
| chr15 | 79209149 | 79209795 | 647 | 5 | 2.69E-06 | 4.88E-06 | 5.03E-05 | 2.62E-06 | -0.00621071 | MIR184, ANKRD34C-AS1 |
| chr4 | 171812609 | 171813692 | 1084 | 9 | 1.79E-05 | 2.83E-05 | 0.000447 | 2.62E-06 | 0.002171847 | GALNTL6 |
| chr15 | 59999681 | 60000620 | 940 | 5 | 2.34E-06 | 6.99E-06 | 4.08E-05 | 2.62E-06 | 0.000323709 | NA |
| chr13 | 110397063 | 110397698 | 636 | 3 | 1.43E-05 | 9.51E-07 | 0.001489 | 2.62E-06 | -0.01551307 | COL4A2 |
| chr9 | 135352018 | 135352706 | 689 | 4 | 1.07E-06 | 3.36E-05 | 2.59E-05 | 2.62E-06 | 0.001546327 | AL353615.1 |
| chr8 | 28337910 | 28338729 | 820 | 6 | 3.26E-07 | 5.17E-05 | 8.06E-05 | 2.62E-06 | -0.00035681 | PNOC, AC021678.2 |
| chr16 | 22027599 | 22027988 | 390 | 2 | 3.56E-07 | 5.38E-05 | 3.60E-06 | 2.62E-06 | -0.01148049 | MOSMO |
| chr17 | 2745845 | 2747194 | 1350 | 4 | 7.36E-07 | 2.98E-06 | 8.77E-05 | 2.62E-06 | -0.00276499 | CCDC92B |
| chr5 | 159331131 | 159331345 | 215 | 2 | 2.24E-05 | 1.63E-06 | 0.000236 | 2.62E-06 | 0.001785211 | AC008691.1 |
| chr3 | 165206603 | 165206912 | 310 | 4 | 1.33E-05 | 2.02E-06 | 0.000196 | 2.62E-06 | 0.004389327 | NA |
| chr4 | 1581506 | 1582171 | 666 | 2 | 6.01E-07 | 6.24E-06 | 1.52E-05 | 2.63E-06 | -0.00099383 | NA |
| chr10 | 101310352 | 101311098 | 747 | 6 | 8.58E-07 | 0.0003319 | 0.000461 | 2.63E-06 | 0.014507277 | AL133387.1 |
| chr3 | 14401928 | 14403064 | 1137 | 13 | 3.83E-06 | 1.06E-06 | 0.001777 | 2.63E-06 | -0.00111513 | SLC6A6 |
| chr1 | 6645702 | 6646317 | 616 | 3 | 2.67E-06 | 2.14E-05 | 0.000177 | 2.63E-06 | -0.00067804 | DNAJC11 |
| chr10 | 133206863 | 133207318 | 456 | 3 | 1.26E-06 | 1.57E-06 | 0.000504 | 2.63E-06 | -0.01186003 | KNDC1 |
| chr10 | 123843775 | 123844223 | 449 | 3 | 6.34E-06 | 1.58E-06 | 0.000673 | 2.63E-06 | -0.00348339 | CPXM2 |
| chr6 | 155314067 | 155315119 | 1053 | 13 | 4.43E-06 | 5.14E-06 | 0.000512 | 2.63E-06 | -8.78E-06 | TFB1M |
| chr7 | 143476458 | 143477981 | 1524 | 4 | 5.72E-07 | 0.0001694 | 4.68E-05 | 2.64E-06 | -0.00497719 | EPHA1-AS1, TAS2R41 |
| chr13 | 30905615 | 30906509 | 895 | 10 | 5.96E-06 | 1.70E-06 | 0.002268 | 2.64E-06 | -0.01371035 | MEDAG, TEX26-AS1 |
| chr11 | 88169663 | 88169768 | 106 | 2 | 5.42E-06 | 1.91E-06 | 0.000112 | 2.64E-06 | 0.026175578 | RAB38 |
| chr5 | 876612 | 876961 | 350 | 3 | 8.32E-08 | 0.0004752 | 2.16E-06 | 2.64E-06 | 0.000777082 | BRD9 |
| chr1 | 90716571 | 90717494 | 924 | 12 | 6.54E-06 | 9.51E-05 | 0.000479 | 2.64E-06 | -0.00238283 | BARHL2 |
| chr6 | 125300649 | 125301842 | 1194 | 6 | 1.54E-05 | 9.41E-06 | 0.00039 | 2.64E-06 | -0.00164654 | HDDC2 |
| chr1 | 84689865 | 84690041 | 177 | 3 | 1.64E-07 | 1.06E-06 | 0.000992 | 2.64E-06 | 0.000669791 | SSX2IP |
| chr7 | 157044079 | 157044886 | 808 | 5 | 1.67E-07 | 0.0001283 | 2.21E-05 | 2.64E-06 | 0.004918625 | NA |
| chr2 | 218216662 | 218217573 | 912 | 16 | 7.65E-07 | 0.0001785 | 0.00025 | 2.64E-06 | 0.001158073 | ARPC2 |
| chr1 | 234613029 | 234613722 | 694 | 5 | 2.20E-06 | 8.08E-06 | 0.000217 | 2.65E-06 | 0.007061713 | NA |
| chr16 | 85360900 | 85362055 | 1156 | 3 | 7.67E-08 | 6.88E-05 | 4.17E-06 | 2.65E-06 | -0.00950323 | GSE1 |
| chr13 | 86059367 | 86059452 | 86 | 2 | 1.18E-06 | 1.71E-06 | 0.000184 | 2.65E-06 | 0.00358735 | NA |
| chr1 | 28258884 | 28260222 | 1339 | 10 | 6.01E-05 | 0.0009255 | 0.001722 | 2.65E-06 | -0.00287188 | SESN2 |
| chr3 | 124503970 | 124504641 | 672 | 4 | 6.32E-06 | 1.18E-06 | 0.001167 | 2.65E-06 | 0.015132979 | KALRN |
| chr10 | 122331196 | 122332450 | 1255 | 7 | 6.05E-05 | 3.99E-06 | 0.003791 | 2.65E-06 | -0.00094994 | BTBD16 |
| chr16 | 88419021 | 88419248 | 228 | 2 | 4.26E-06 | 2.29E-06 | 6.87E-05 | 2.65E-06 | -0.00166202 | ZNF469 |
| chr1 | 174998526 | 174999320 | 795 | 10 | 1.92E-07 | 6.36E-06 | 0.001241 | 2.65E-06 | -0.00650598 | CACYBP, Z99127.3 |
| chr5 | 132490173 | 132491242 | 1070 | 9 | 8.19E-07 | 4.16E-06 | 0.00558 | 2.65E-06 | 0.001937687 | AC116366.2, IRF1 |
| chr13 | 94959457 | 94959811 | 355 | 3 | 1.71E-06 | 3.25E-05 | 0.000152 | 2.65E-06 | -0.00862433 | NA |
| chr2 | 102355218 | 102355871 | 654 | 6 | 4.79E-06 | 9.30E-06 | 0.000226 | 2.65E-06 | 0.005381312 | IL18R1 |
| chr14 | 105855010 | 105855446 | 437 | 3 | 5.48E-06 | 1.14E-06 | 0.000733 | 2.66E-06 | 0.001523683 | IGHM |
| chr11 | 64807138 | 64807640 | 503 | 3 | 9.91E-06 | 9.92E-07 | 0.001453 | 2.66E-06 | -0.00136525 | MEN1 |
| chr2 | 197399042 | 197399525 | 484 | 2 | 2.35E-07 | 1.45E-05 | 7.52E-06 | 2.66E-06 | -0.00029196 | SF3B1 |
| chr11 | 45693222 | 45694454 | 1233 | 8 | 4.26E-07 | 0.0002302 | 0.000104 | 2.66E-06 | -0.00026714 | NA |
| chr19 | 51947952 | 51948841 | 890 | 2 | 1.69E-08 | 4.37E-05 | 4.03E-06 | 2.66E-06 | 0.000183204 | ZNF613 |
| chr7 | 2815367 | 2816125 | 759 | 8 | 2.04E-06 | 1.23E-05 | 0.001831 | 2.66E-06 | 0.000130486 | GNA12 |
| chr19 | 11738716 | 11739198 | 483 | 9 | 3.51E-06 | 7.29E-06 | 0.004454 | 2.66E-06 | 0.001043833 | ZNF823 |
| chr8 | 1362285 | 1363410 | 1126 | 6 | 3.92E-07 | 0.0001571 | 0.000459 | 2.66E-06 | -0.00313967 | DLGAP2 |
| chr9 | 130913286 | 130913439 | 154 | 2 | 4.49E-06 | 2.12E-06 | 8.43E-05 | 2.67E-06 | 0.025707601 | FIBCD1 |
| chr6 | 116087533 | 116088648 | 1116 | 3 | 3.23E-08 | 0.0003194 | 4.30E-06 | 2.67E-06 | -0.00832965 | NA |
| chr5 | 135035707 | 135035744 | 38 | 2 | 1.28E-07 | 1.59E-06 | 0.000316 | 2.67E-06 | 0.017855439 | C5orf66, AC008406.3 |
| chr1 | 228463652 | 228464147 | 496 | 3 | 3.60E-07 | 9.90E-06 | 0.00019 | 2.67E-06 | 0.005399248 | H2BU2P |
| chr4 | 2811629 | 2812337 | 709 | 6 | 3.00E-06 | 2.62E-06 | 0.002011 | 2.67E-06 | -0.00184986 | SH3BP2 |
| chr3 | 58572053 | 58572648 | 596 | 4 | 1.41E-05 | 2.37E-06 | 0.000516 | 2.67E-06 | -0.00503056 | FAM107A |
| chr22 | 44032472 | 44032969 | 498 | 3 | 2.00E-06 | 3.11E-06 | 6.16E-05 | 2.67E-06 | -0.00537029 | PARVB |
| chr4 | 97496218 | 97496630 | 413 | 2 | 4.96E-06 | 1.78E-06 | 0.000158 | 2.67E-06 | -0.00641851 | STPG2, AC034154.1 |
| chr8 | 8324408 | 8325231 | 824 | 6 | 5.32E-06 | 1.61E-06 | 0.002805 | 2.68E-06 | -0.00294058 | PRAG1 |
| chr10 | 13528012 | 13528550 | 539 | 8 | 6.15E-07 | 3.02E-05 | 0.000102 | 2.68E-06 | 0.000700628 | AL590677.1, BEND7 |
| chr11 | 62796101 | 62797150 | 1050 | 3 | 2.17E-05 | 4.33E-05 | 0.000112 | 2.68E-06 | -0.00822557 | NXF1 |
| chr16 | 1896175 | 1897054 | 880 | 4 | 2.50E-06 | 3.94E-06 | 5.52E-05 | 2.68E-06 | -0.0096554 | NA |
| chr17 | 46013496 | 46013696 | 201 | 2 | 5.27E-06 | 1.83E-06 | 0.000143 | 2.68E-06 | -0.00702026 | MAPT |
| chr17 | 37727035 | 37727643 | 609 | 3 | 2.09E-07 | 0.0001899 | 4.12E-06 | 2.69E-06 | -0.00727281 | HNF1B |
| chr3 | 190701148 | 190701334 | 187 | 3 | 7.39E-06 | 1.08E-06 | 0.001209 | 2.69E-06 | -0.0051685 | NA |
| chr7 | 73860741 | 73861480 | 740 | 9 | 2.93E-06 | 5.20E-05 | 0.000849 | 2.69E-06 | -0.00047043 | TMEM270 |
| chr12 | 121005506 | 121005697 | 192 | 3 | 7.01E-06 | 1.30E-06 | 0.000534 | 2.69E-06 | -0.00979754 | C12orf43 |
| chr6 | 30561250 | 30561275 | 26 | 2 | 1.87E-06 | 6.97E-06 | 1.42E-05 | 2.70E-06 | 0.000563104 | PRR3 |
| chr14 | 75046519 | 75047006 | 488 | 3 | 1.50E-07 | 4.72E-05 | 1.10E-05 | 2.70E-06 | -0.0040326 | MLH3 |
| chr1 | 65484186 | 65485284 | 1099 | 3 | 1.06E-05 | 4.72E-05 | 6.91E-05 | 2.70E-06 | 0.003752346 | LEPR |
| chr10 | 22713585 | 22714850 | 1266 | 8 | 1.39E-05 | 1.20E-05 | 0.000133 | 2.70E-06 | -0.00135344 | PIP4K2A |
| chr10 | 70928317 | 70928366 | 50 | 2 | 1.54E-06 | 4.99E-06 | 2.03E-05 | 2.70E-06 | -0.01099807 | AC073176.2 |
| chr7 | 20533054 | 20533185 | 132 | 2 | 6.15E-06 | 1.62E-06 | 0.000301 | 2.70E-06 | -0.0106325 | NA |
| chr11 | 22491558 | 22492070 | 513 | 6 | 2.21E-07 | 1.67E-05 | 0.000385 | 2.70E-06 | 0.003945592 | LINC01495 |
| chr12 | 6627672 | 6627730 | 59 | 2 | 1.31E-06 | 9.27E-06 | 1.10E-05 | 2.70E-06 | -0.01733402 | LPAR5 |
| chr1 | 180946221 | 180946496 | 276 | 3 | 4.72E-06 | 2.13E-06 | 0.000301 | 2.70E-06 | -0.01080506 | KIAA1614, AL162431.1 |
| chr9 | 133485104 | 133485296 | 193 | 2 | 1.45E-06 | 1.57E-05 | 7.25E-06 | 2.70E-06 | -0.00929293 | NA |
| chr15 | 74869832 | 74870214 | 383 | 2 | 2.74E-07 | 6.17E-06 | 1.61E-05 | 2.70E-06 | 0.01030528 | SCAMP2 |
| chr11 | 4607804 | 4608682 | 879 | 12 | 5.01E-06 | 5.26E-06 | 0.006932 | 2.70E-06 | -0.00470879 | TRIM68 |
| chr12 | 95956485 | 95957017 | 533 | 6 | 8.95E-06 | 7.59E-07 | 0.001777 | 2.71E-06 | -0.01506602 | AMDHD1 |
| chr10 | 19793413 | 19793828 | 416 | 3 | 3.39E-06 | 2.09E-06 | 0.00025 | 2.71E-06 | -0.00854121 | NA |
| chr17 | 50346493 | 50347653 | 1161 | 5 | 3.11E-06 | 2.32E-06 | 0.001445 | 2.71E-06 | -0.00693881 | XYLT2 |
| chr1 | 112507917 | 112508709 | 793 | 8 | 3.31E-06 | 1.77E-06 | 0.002238 | 2.71E-06 | 5.81E-05 | WNT2B |
| chr13 | 70924400 | 70924507 | 108 | 2 | 4.74E-06 | 2.42E-06 | 6.40E-05 | 2.71E-06 | -0.00617938 | MTCL1P1 |
| chr3 | 11037442 | 11038193 | 752 | 7 | 5.23E-08 | 0.0003033 | 0.000135 | 2.71E-06 | -0.00356828 | SLC6A1 |
| chr19 | 46225703 | 46226244 | 542 | 2 | 3.06E-08 | 0.0198925 | 1.72E-06 | 2.72E-06 | -0.00680183 | NA |
| chr15 | 74906299 | 74907690 | 1392 | 19 | 7.47E-08 | 0.0097262 | 2.49E-06 | 2.72E-06 | 0.001223838 | FAM219B |
| chr15 | 66328845 | 66329101 | 257 | 5 | 2.73E-06 | 3.58E-05 | 0.000583 | 2.72E-06 | 0.000165692 | DIS3L, AC055855.1 |
| chr5 | 26904750 | 26905522 | 773 | 5 | 4.28E-06 | 1.70E-06 | 0.001353 | 2.72E-06 | -0.006078 | CDH9 |
| chr17 | 44328451 | 44328816 | 366 | 2 | 3.10E-07 | 0.0001132 | 2.92E-06 | 2.72E-06 | -0.00914287 | AC003043.1 |
| chr1 | 44631827 | 44633371 | 1545 | 6 | 4.04E-07 | 8.39E-06 | 2.96E-05 | 2.72E-06 | 0.00545354 | RNF220 |
| chr11 | 65121460 | 65123068 | 1609 | 22 | 6.38E-06 | 4.16E-06 | 0.004502 | 2.72E-06 | 0.000116571 | MRPL49, FAU, SYVN1 |
| chr6 | 7129481 | 7130373 | 893 | 3 | 6.50E-07 | 9.97E-05 | 2.36E-05 | 2.72E-06 | 0.004587064 | RREB1 |
| chr21 | 45392630 | 45393081 | 452 | 2 | 2.92E-08 | 0.0079676 | 1.75E-06 | 2.72E-06 | -0.00454839 | NA |
| chr19 | 57769433 | 57770480 | 1048 | 10 | 3.22E-05 | 5.60E-06 | 0.001521 | 2.72E-06 | -0.00684026 | ZNF586 |
| chr19 | 42145386 | 42145507 | 122 | 3 | 5.83E-06 | 2.62E-06 | 0.000161 | 2.72E-06 | -0.00450117 | POU2F2 |

| | | | | | | | | | | |
|---|---|---|---|---|---|---|---|---|---|---|
| chr9 | 132894015 | 132894555 | 541 | 2 | 7.90E-06 | 1.71E-06 | 0.000229 | 2.73E-06 | -0.00639041 | TSC1 |
| chr1 | 153140786 | 153141639 | 854 | 3 | 8.27E-06 | 1.76E-06 | 0.000545 | 2.73E-06 | 0.005569989 | NA |
| chr4 | 7287755 | 7288167 | 413 | 7 | 3.63E-07 | 7.25E-05 | 3.37E-06 | 2.74E-06 | 0.002724092 | SORCS2 |
| chr4 | 82284639 | 82285451 | 813 | 6 | 3.98E-06 | 2.76E-06 | 0.001956 | 2.74E-06 | 0.002417304 | VAMP9P |
| chr18 | 9707558 | 9708923 | 1366 | 9 | 7.17E-05 | 8.67E-07 | 0.004341 | 2.74E-06 | -0.00299592 | RAB31 |
| chr17 | 28226541 | 28227584 | 1044 | 5 | 4.29E-08 | 0.0002321 | 2.28E-06 | 2.74E-06 | -0.0078604 | PYY2 |
| chr8 | 131859902 | 131860311 | 410 | 2 | 4.57E-08 | 0.0008706 | 2.02E-06 | 2.74E-06 | 0.014388462 | NA |
| chr9 | 131233635 | 131234102 | 468 | 4 | 3.40E-06 | 3.85E-06 | 7.14E-05 | 2.74E-06 | -0.0046048 | NUP214 |
| chr1 | 18637464 | 18637906 | 443 | 3 | 1.42E-07 | 0.0001849 | 0.000127 | 2.74E-06 | -0.00049165 | PAX7 |
| chr14 | 89351638 | 89352281 | 644 | 4 | 1.99E-07 | 0.0002443 | 5.37E-06 | 2.74E-06 | 0.005562101 | AL357093.2, FOXN3 |
| chr22 | 44676575 | 44677285 | 711 | 11 | 1.76E-06 | 4.09E-06 | 0.002987 | 2.74E-06 | 0.00113332 | PRR5 |
| chr17 | 63826414 | 63828036 | 1623 | 19 | 1.99E-05 | 9.47E-05 | 0.002208 | 2.74E-06 | -6.14E-06 | PSMC5, FTSJ3 |
| chr15 | 42716492 | 42717027 | 536 | 2 | 5.57E-07 | 9.46E-06 | 1.10E-05 | 2.75E-06 | -0.00446739 | STARD9 |
| chr12 | 57613839 | 57614519 | 681 | 4 | 3.66E-06 | 9.65E-07 | 0.00156 | 2.75E-06 | -0.00721525 | ARHGEF25, AC025165.1 |
| chr8 | 125008418 | 125009262 | 845 | 2 | 3.06E-05 | 1.75E-06 | 0.000202 | 2.75E-06 | 0.001805104 | SQLE |
| chr22 | 37274767 | 37275395 | 629 | 2 | 5.91E-06 | 1.64E-06 | 0.000322 | 2.75E-06 | -0.01404038 | NA |
| chr11 | 72726686 | 72727357 | 672 | 6 | 2.07E-05 | 3.42E-07 | 0.012066 | 2.75E-06 | -0.00307858 | ARAP1 |
| chr15 | 51230079 | 51230387 | 309 | 2 | 1.38E-06 | 2.70E-06 | 5.26E-05 | 2.75E-06 | -0.01143995 | MIR4713HG, CYP19A1 |
| chr7 | 81483396 | 81483924 | 529 | 2 | 3.78E-07 | 1.07E-05 | 9.94E-06 | 2.75E-06 | -0.000101 | NA |
| chr20 | 57959212 | 57959261 | 50 | 2 | 2.63E-06 | 1.67E-06 | 0.000289 | 2.76E-06 | -0.00637028 | AL354984.1 |
| chr15 | 38564327 | 38565494 | 1168 | 12 | 6.39E-07 | 2.85E-05 | 0.003711 | 2.76E-06 | 0.000810743 | RASGRP1 |
| chr19 | 16283555 | 16283963 | 409 | 5 | 2.70E-06 | 1.02E-05 | 0.000162 | 2.76E-06 | 0.003487889 | AC020911.2 |
| chr7 | 149861247 | 149861455 | 209 | 2 | 1.11E-07 | 0.0002039 | 2.55E-06 | 2.76E-06 | -0.00278326 | ZNF862, AC004877.1 |
| chr3 | 133818978 | 133819248 | 271 | 2 | 1.44E-06 | 3.54E-06 | 3.32E-05 | 2.76E-06 | 0.008092293 | SRPRB |
| chr20 | 46514786 | 46514872 | 87 | 2 | 4.38E-07 | 1.59E-05 | 7.40E-06 | 2.77E-06 | 0.004154499 | NA |
| chr3 | 46959419 | 46960105 | 687 | 3 | 7.25E-06 | 1.02E-06 | 0.001446 | 2.77E-06 | 0.012430692 | CCDC12 |
| chr16 | 88570036 | 88571066 | 1031 | 7 | 1.05E-06 | 1.60E-06 | 0.000435 | 2.77E-06 | 0.000193756 | ZC3H18 |
| chr8 | 127838653 | 127838805 | 153 | 2 | 3.48E-06 | 4.02E-06 | 2.79E-05 | 2.77E-06 | -0.00209116 | PVT1 |
| chr11 | 77994442 | 77995437 | 996 | 9 | 4.47E-07 | 0.0001861 | 6.46E-05 | 2.78E-06 | 0.000925828 | INTS4 |
| chr20 | 50909704 | 50910395 | 692 | 8 | 4.23E-08 | 0.0056918 | 4.42E-05 | 2.78E-06 | -0.00340662 | ADNP |
| chr11 | 45650979 | 45651860 | 882 | 5 | 3.23E-07 | 2.48E-05 | 4.22E-05 | 2.78E-06 | -0.00255317 | AC087442.1, CHST1 |
| chr20 | 36734001 | 36734590 | 590 | 2 | 1.62E-06 | 3.13E-06 | 4.10E-05 | 2.78E-06 | -0.00882441 | NDRG3 |
| chr1 | 225824799 | 225825983 | 1185 | 8 | 4.00E-06 | 0.0006456 | 0.000197 | 2.78E-06 | -0.00342076 | EPHX1 |
| chr20 | 34946460 | 34947051 | 592 | 4 | 3.88E-06 | 1.23E-06 | 0.001023 | 2.78E-06 | 0.000513027 | GSS |
| chr12 | 68627502 | 68627556 | 55 | 2 | 7.91E-07 | 0.0001466 | 2.79E-06 | 2.78E-06 | -0.00266218 | RAP1B |
| chr2 | 127043040 | 127043695 | 656 | 3 | 9.98E-06 | 1.34E-06 | 0.00049 | 2.78E-06 | -4.09E-05 | NA |
| chr2 | 219567344 | 219568150 | 807 | 7 | 5.50E-06 | 0.0001069 | 0.000185 | 2.78E-06 | -0.00046625 | OBSL1 |
| chr5 | 1537496 | 1538409 | 914 | 5 | 5.94E-06 | 7.62E-07 | 0.002875 | 2.78E-06 | -0.00366627 | NA |
| chr22 | 42687933 | 42688979 | 1047 | 4 | 1.55E-07 | 1.71E-05 | 1.25E-05 | 2.78E-06 | -0.0122624 | NA |
| chr17 | 7137259 | 7137541 | 283 | 2 | 2.03E-07 | 0.0029519 | 1.84E-06 | 2.78E-06 | -0.01236646 | NA |
| chr10 | 101319518 | 101319728 | 211 | 2 | 1.61E-06 | 4.30E-06 | 2.58E-05 | 2.79E-06 | 0.006720892 | NA |
| chr1 | 18956188 | 18957267 | 1080 | 7 | 1.28E-06 | 3.53E-06 | 0.003933 | 2.79E-06 | 0.001294485 | IFFO2 |
| chr12 | 65882208 | 65883194 | 987 | 9 | 8.11E-06 | 1.03E-05 | 0.001892 | 2.79E-06 | -0.00435514 | HMGA2 |
| chr5 | 141191597 | 141192210 | 614 | 3 | 3.80E-07 | 2.72E-05 | 1.03E-05 | 2.79E-06 | -0.02435628 | AC244517.11, AC244517.4, AC244517.7 |
| chr2 | 17815149 | 17816512 | 1364 | 7 | 1.87E-06 | 6.46E-06 | 0.001076 | 2.79E-06 | 0.001244713 | MSGN1 |
| chr10 | 132756553 | 132756854 | 302 | 3 | 5.31E-06 | 2.35E-06 | 0.00012 | 2.79E-06 | -0.00623329 | INPP5A |
| chr6 | 109382310 | 109383312 | 1003 | 13 | 3.54E-06 | 3.03E-06 | 0.009052 | 2.79E-06 | 1.07E-05 | AL359711.2, CD164 |
| chr16 | 56515845 | 56516222 | 378 | 2 | 1.44E-06 | 3.13E-06 | 4.13E-05 | 2.79E-06 | -0.01450318 | BBS2 |
| chr5 | 174583939 | 174584487 | 549 | 2 | 1.06E-06 | 5.16E-06 | 2.07E-05 | 2.79E-06 | -0.00210349 | NA |
| chr13 | 49939019 | 49939099 | 81 | 2 | 1.59E-06 | 4.59E-05 | 4.15E-06 | 2.79E-06 | -0.00956182 | NA |
| chr8 | 133449382 | 133450304 | 923 | 6 | 1.71E-06 | 1.06E-05 | 0.000216 | 2.79E-06 | 0.004423131 | NA |
| chr1 | 19664131 | 19664744 | 614 | 6 | 5.40E-07 | 0.0001107 | 2.45E-05 | 2.79E-06 | -0.00380041 | NA |
| chr8 | 32605427 | 32605575 | 149 | 2 | 4.06E-06 | 3.84E-06 | 3.02E-05 | 2.79E-06 | -0.00934579 | NRG1 |
| chr14 | 68564459 | 68565054 | 596 | 3 | 5.37E-06 | 1.26E-06 | 0.000606 | 2.79E-06 | 0.004765178 | RAD51B |
| chr7 | 10973340 | 10974538 | 1199 | 13 | 1.51E-05 | 6.65E-07 | 0.022931 | 2.80E-06 | 2.25E-05 | PHF14 |
| chr8 | 61714433 | 61715333 | 901 | 10 | 9.73E-08 | 0.0009929 | 1.98E-05 | 2.80E-06 | -0.00017312 | MIR4470, ASPH |
| chr11 | 908683 | 908781 | 99 | 2 | 1.10E-06 | 7.36E-06 | 1.42E-05 | 2.80E-06 | -0.00897225 | CHID1 |
| chr18 | 13218444 | 13219226 | 783 | 8 | 1.15E-06 | 5.47E-05 | 0.000487 | 2.80E-06 | 0.000305986 | LDLRAD4 |
| chr16 | 50681224 | 50681789 | 566 | 10 | 1.47E-06 | 6.89E-05 | 0.002129 | 2.80E-06 | -0.00164355 | SNX20 |
| chr12 | 95551344 | 95551991 | 648 | 8 | 4.98E-06 | 1.28E-05 | 0.000817 | 2.80E-06 | -0.00207346 | PGAM1P5, USP44 |
| chr15 | 31223547 | 31224278 | 732 | 9 | 1.75E-06 | 4.14E-06 | 0.001624 | 2.80E-06 | 0.013528837 | LINC02352, AC012236.1 |
| chr19 | 21493249 | 21493817 | 569 | 4 | 9.38E-06 | 1.02E-06 | 0.001538 | 2.80E-06 | -0.01556231 | LINC00664 |
| chr1 | 237890528 | 237891202 | 675 | 5 | 9.14E-08 | 3.54E-05 | 1.67E-05 | 2.80E-06 | -0.00604782 | AL359924.1, ZP4 |
| chr7 | 128915842 | 128916716 | 875 | 3 | 5.86E-06 | 0.0003226 | 6.64E-05 | 2.81E-06 | -0.01533365 | NA |
| chr10 | 79334055 | 79334525 | 471 | 2 | 6.43E-06 | 1.78E-06 | 0.000212 | 2.81E-06 | 0.016090891 | NA |
| chr20 | 31909433 | 31909504 | 72 | 2 | 4.29E-06 | 4.55E-06 | 2.43E-05 | 2.81E-06 | -0.0020137 | TTLL9 |
| chr22 | 37027423 | 37027661 | 239 | 2 | 1.45E-06 | 3.94E-06 | 2.96E-05 | 2.81E-06 | -0.00507652 | MPST |
| chr3 | 192406879 | 192407055 | 177 | 2 | 3.49E-06 | 2.07E-06 | 0.00011 | 2.82E-06 | 0.001483797 | FGF12 |
| chr1 | 205448410 | 205449282 | 873 | 5 | 2.58E-05 | 2.26E-06 | 0.002146 | 2.82E-06 | -0.00154167 | LEMD1, BLACAT1 |
| chr2 | 216014180 | 216014376 | 197 | 4 | 5.59E-07 | 1.43E-05 | 6.20E-05 | 2.82E-06 | -0.00293601 | MREG, PECR |
| chr6 | 87701810 | 87702554 | 745 | 9 | 2.10E-06 | 3.42E-06 | 0.004334 | 2.82E-06 | 0.00070153 | AKIRIN2 |
| chr13 | 89554398 | 89554504 | 107 | 3 | 2.56E-06 | 7.11E-06 | 0.000119 | 2.82E-06 | 0.0058068 | LINC02336 |
| chr8 | 66869987 | 66870871 | 885 | 9 | 1.35E-06 | 2.04E-05 | 0.000139 | 2.83E-06 | -0.00239053 | MCMDC2 |
| chr2 | 96138508 | 96139383 | 876 | 4 | 1.20E-05 | 1.32E-06 | 0.001508 | 2.83E-06 | -0.01826386 | NA |
| chr16 | 88899938 | 88900495 | 558 | 5 | 6.12E-08 | 1.62E-05 | 2.81E-05 | 2.83E-06 | -0.00303952 | CBFA2T3 |
| chr2 | 219492792 | 219492886 | 95 | 2 | 4.34E-06 | 1.87E-06 | 0.00017 | 2.83E-06 | -0.02785629 | SPEG, AC053503.6, ASIC4-AS1 |
| chr18 | 7753488 | 7754311 | 824 | 3 | 4.68E-07 | 6.37E-06 | 6.08E-05 | 2.83E-06 | -0.00975806 | PTPRM, AP000897.2 |
| chr10 | 132770026 | 132771156 | 1131 | 4 | 3.69E-07 | 2.00E-05 | 1.57E-05 | 2.83E-06 | -0.00187054 | INPP5A |
| chr16 | 2216564 | 2216806 | 243 | 2 | 6.74E-07 | 2.10E-06 | 0.000106 | 2.83E-06 | 0.001652543 | NA |
| chr16 | 31195354 | 31195813 | 460 | 2 | 1.43E-07 | 0.0002921 | 2.43E-06 | 2.83E-06 | -0.0041074 | NA |
| chr12 | 124927820 | 124928618 | 799 | 6 | 1.74E-06 | 4.04E-06 | 0.000196 | 2.84E-06 | -0.00737664 | NA |
| chr3 | 53346776 | 53347838 | 1063 | 10 | 1.57E-05 | 4.76E-05 | 0.001353 | 2.84E-06 | -5.91E-05 | CACNA1D, DCP1A |
| chr6 | 70303062 | 70304172 | 1111 | 7 | 1.26E-07 | 4.57E-05 | 9.29E-05 | 2.84E-06 | -0.00311885 | COL9A1 |
| chr17 | 825465 | 826021 | 557 | 5 | 3.13E-08 | 0.0002483 | 5.87E-06 | 2.84E-06 | -0.00603279 | NXN |
| chr6 | 146026969 | 146028995 | 2027 | 16 | 4.65E-06 | 0.0001075 | 0.001534 | 2.84E-06 | 0.003035811 | GRM1 |
| chr19 | 42428356 | 42428872 | 517 | 4 | 2.71E-06 | 3.18E-06 | 0.000913 | 2.84E-06 | -0.00254975 | LIPE-AS1, CXCL17 |
| chr17 | 62749841 | 62750164 | 324 | 2 | 5.30E-08 | 0.0002653 | 2.48E-06 | 2.85E-06 | -0.00588375 | AC005821.1, MARCHF10 |
| chr2 | 9004117 | 9004635 | 519 | 5 | 1.20E-07 | 0.0020278 | 2.50E-05 | 2.85E-06 | -0.0043463 | NA |
| chr7 | 2511879 | 2512362 | 484 | 6 | 1.24E-07 | 2.90E-05 | 0.000102 | 2.85E-06 | 0.006781179 | NA |
| chr1 | 24501606 | 24502380 | 775 | 8 | 2.92E-06 | 3.54E-06 | 0.001937 | 2.85E-06 | 0.000119756 | RCAN3, RCAN3AS |
| chr6 | 73520051 | 73521331 | 1281 | 13 | 7.77E-06 | 3.10E-05 | 0.000707 | 2.85E-06 | 0.001219639 | EEF1A1 |
| chr8 | 141020103 | 141020470 | 368 | 2 | 2.78E-07 | 1.72E-05 | 7.30E-06 | 2.85E-06 | 0.008261516 | NA |
| chr5 | 146400310 | 146400515 | 206 | 2 | 8.68E-07 | 2.57E-05 | 5.70E-06 | 2.86E-06 | -0.0149584 | NA |
| chr12 | 69584871 | 69585842 | 972 | 12 | 7.01E-06 | 1.84E-05 | 0.011335 | 2.86E-06 | 0.000196166 | CCT2 |
| chr17 | 49435144 | 49435532 | 389 | 2 | 4.58E-06 | 2.07E-06 | 0.000117 | 2.86E-06 | -0.01315168 | AC091180.5 |
| chr1 | 243931466 | 243932113 | 648 | 6 | 8.79E-06 | 4.08E-07 | 0.009386 | 2.86E-06 | -0.01148085 | LINC02774 |
| chr3 | 72571849 | 72571871 | 23 | 2 | 1.24E-06 | 5.94E-05 | 3.81E-06 | 2.86E-06 | -0.00262617 | NA |
| chr6 | 24359686 | 24360375 | 690 | 8 | 5.79E-06 | 5.54E-06 | 0.002545 | 2.86E-06 | -0.00745683 | NA |
| chr9 | 129080459 | 129081354 | 896 | 12 | 8.81E-08 | 0.0003343 | 7.51E-05 | 2.86E-06 | 0.000996355 | DOLPP1, AL592211.2 |
| chr20 | 36575568 | 36575818 | 251 | 2 | 2.23E-06 | 1.73E-06 | 0.000293 | 2.86E-06 | 0.004466386 | TGIF2, TGIF2-RAB5IF |
| chr1 | 47332797 | 47334162 | 1366 | 10 | 3.65E-08 | 0.0004557 | 0.000208 | 2.87E-06 | -0.003875 | CMPK1 |
| chr5 | 95162102 | 95162992 | 891 | 3 | 1.74E-05 | 1.42E-06 | 0.000877 | 2.87E-06 | -0.02193002 | MCTP1 |
| chr3 | 38031120 | 38031459 | 340 | 2 | 3.88E-07 | 1.95E-06 | 0.000149 | 2.87E-06 | -0.00400142 | NA |
| chr8 | 11458012 | 11458053 | 42 | 2 | 2.15E-06 | 1.85E-06 | 0.000197 | 2.87E-06 | -0.01051453 | FAM167A |
| chr12 | 129698244 | 129698296 | 53 | 3 | 4.53E-07 | 2.19E-05 | 1.14E-05 | 2.87E-06 | 0.001438822 | AC117373.2, TMEM132D |
| chr9 | 134661834 | 134662245 | 412 | 2 | 2.83E-08 | 8.91E-06 | 1.24E-05 | 2.87E-06 | -0.00349787 | COL5A1 |
| chr14 | 21536859 | 21537523 | 665 | 6 | 1.02E-07 | 0.0004443 | 0.000103 | 2.87E-06 | 0.003686034 | SALL2 |
| chr4 | 4480431 | 4481099 | 669 | 2 | 4.92E-06 | 1.92E-06 | 0.000164 | 2.87E-06 | -0.00209761 | STX18, STX18-IT1 |
| chr12 | 124626517 | 124627169 | 653 | 2 | 7.61E-07 | 2.69E-06 | 5.92E-05 | 2.87E-06 | -0.00044892 | NA |
| chr16 | 1679794 | 1680578 | 785 | 6 | 4.82E-06 | 8.85E-07 | 0.002268 | 2.88E-06 | 0.001543214 | AL031708.1, JPT2 |
| chr5 | 136023730 | 136024064 | 335 | 2 | 5.02E-06 | 1.73E-06 | 0.00031 | 2.88E-06 | -0.01392385 | NA |
| chr15 | 49851427 | 49852103 | 677 | 2 | 1.50E-05 | 3.33E-06 | 3.99E-05 | 2.88E-06 | -0.0239381 | NA |
| chr12 | 110917614 | 110918188 | 575 | 2 | 2.32E-06 | 3.05E-06 | 4.63E-05 | 2.88E-06 | 0.000158049 | MYL2 |
| chr18 | 55303103 | 55303982 | 880 | 5 | 9.54E-06 | 2.10E-06 | 0.002256 | 2.88E-06 | -0.00577316 | TCF4 |
| chr17 | 81929899 | 81930113 | 215 | 2 | 1.42E-06 | 2.77E-06 | 5.63E-05 | 2.88E-06 | -0.00615153 | MAFG-DT |
| chr6 | 149717400 | 149718631 | 1232 | 14 | 5.63E-07 | 0.0005255 | 0.000504 | 2.89E-06 | -0.0015455 | AL358852.1, LATS1 |
| chr17 | 48892861 | 48893736 | 876 | 8 | 1.32E-06 | 2.98E-06 | 0.000412 | 2.89E-06 | 0.000917343 | ATP5MC1, SUMO2P17 |
| chr10 | 47299752 | 47300145 | 394 | 9 | 4.09E-06 | 4.03E-06 | 0.001963 | 2.89E-06 | -0.00395909 | NA |
| chr1 | 161258665 | 161259619 | 955 | 8 | 2.05E-06 | 6.92E-06 | 0.00012 | 2.89E-06 | -0.00068258 | PCP4L1 |
| chr7 | 151122833 | 151124270 | 1438 | 6 | 2.87E-05 | 5.16E-06 | 0.000737 | 2.89E-06 | -0.00539282 | AGAP3 |
| chr20 | 63537110 | 63537602 | 493 | 10 | 6.55E-06 | 7.01E-06 | 0.00352 | 2.89E-06 | -0.00458649 | PTK6 |
| chr22 | 20967146 | 20967775 | 630 | 4 | 5.49E-07 | 3.18E-06 | 0.000187 | 2.89E-06 | -0.00601217 | AIFM3 |
| chr8 | 81695764 | 81696353 | 590 | 4 | 3.22E-07 | 9.07E-06 | 0.000348 | 2.89E-06 | -0.00068807 | NA |
| chr15 | 65285623 | 65286492 | 870 | 4 | 3.51E-06 | 9.01E-06 | 0.000758 | 2.89E-06 | -0.00020702 | PARP16 |
| chr12 | 111182395 | 111182509 | 115 | 3 | 1.07E-07 | 2.76E-05 | 1.76E-05 | 2.89E-06 | -0.00385271 | CUX2 |
| chr9 | 133939315 | 133940076 | 762 | 6 | 2.91E-07 | 4.26E-05 | 0.000823 | 2.89E-06 | -0.00698076 | VAV2 |
| chr7 | 98976763 | 98977042 | 280 | 2 | 1.40E-06 | 5.85E-06 | 1.90E-05 | 2.90E-06 | -0.00035644 | TRRAP |
| chr20 | 64038010 | 64038454 | 445 | 6 | 9.41E-08 | 8.11E-05 | 2.79E-06 | 2.90E-06 | 0.003827101 | C20orf204 |
| chr1 | 213994089 | 213995089 | 1001 | 5 | 1.64E-07 | 0.0009215 | 3.76E-05 | 2.90E-06 | -0.00687595 | PROX1 |
| chr9 | 125613186 | 125613416 | 231 | 2 | 1.57E-06 | 2.15E-05 | 6.46E-06 | 2.90E-06 | -0.00499286 | MAPKAP1 |
| chr21 | 45748221 | 45749309 | 1089 | 5 | 2.20E-08 | 0.0003187 | 5.08E-06 | 2.90E-06 | -0.01446599 | PCBP3 |
| chr16 | 11430212 | 11430214 | 3 | 2 | 5.48E-06 | 2.98E-06 | 4.92E-05 | 2.90E-06 | 0.01996569 | AC099489.1 |
| chr10 | 23165966 | 23166768 | 803 | 3 | 8.14E-08 | 4.21E-05 | 1.11E-05 | 2.90E-06 | -0.01448521 | NA |
| chr10 | 133442846 | 133443440 | 595 | 5 | 6.71E-07 | 0.0003021 | 6.66E-05 | 2.90E-06 | 0.003878987 | NA |
| chr21 | 45984195 | 45984354 | 160 | 3 | 7.24E-06 | 1.09E-06 | 0.001493 | 2.90E-06 | 0.002799902 | COL6A1 |
| chr4 | 39032664 | 39033397 | 734 | 3 | 1.00E-05 | 3.52E-05 | 0.000155 | 2.91E-06 | 0.002459898 | TMEM156 |
| chr13 | 23615647 | 23615984 | 338 | 2 | 8.35E-07 | 2.53E-05 | 5.86E-06 | 2.91E-06 | -0.00374911 | TNFRSF19 |
| chr1 | 116769090 | 116769463 | 374 | 2 | 1.56E-07 | 0.0001004 | 3.22E-06 | 2.91E-06 | -0.00686697 | CD2 |
| chr3 | 190513878 | 190515071 | 1194 | 11 | 1.83E-05 | 1.84E-06 | 0.009343 | 2.91E-06 | 0.001602654 | IL1RAP |
| chr4 | 27635113 | 27635631 | 519 | 3 | 1.90E-07 | 0.0001553 | 4.41E-06 | 2.91E-06 | -0.00265144 | NA |
| chr19 | 9435576 | 9436456 | 881 | 11 | 2.21E-07 | 1.12E-05 | 0.005982 | 2.91E-06 | -0.0063579 | NA |
| chr12 | 56714163 | 56714366 | 204 | 2 | 2.65E-06 | 1.81E-06 | 0.000247 | 2.91E-06 | -0.00714578 | NACA |
| chr13 | 77990667 | 77991211 | 545 | 5 | 6.24E-06 | 2.38E-06 | 0.001212 | 2.91E-06 | -0.01242352 | OBI1-AS1, LINC01069 |
| chr1 | 156054068 | 156055317 | 1250 | 16 | 2.57E-05 | 1.52E-05 | 0.004072 | 2.91E-06 | -4.59E-05 | LAMTOR2 |
| chr2 | 162058986 | 162059392 | 407 | 4 | 1.35E-06 | 3.49E-05 | 2.75E-05 | 2.91E-06 | -0.00759259 | DPP4 |
| chr9 | 128346382 | 128347098 | 717 | 2 | 2.75E-05 | 1.75E-06 | 0.000323 | 2.92E-06 | -0.00763785 | SLC27A4 |
| chr3 | 178429349 | 178429415 | 67 | 3 | 3.75E-06 | 2.40E-06 | 0.000177 | 2.92E-06 | -0.00405976 | KCNMB2, AC117457.1, LINC01014 |
| chr4 | 127843927 | 127844447 | 521 | 4 | 5.72E-06 | 5.73E-06 | 0.000286 | 2.92E-06 | -0.00397438 | AC093591.2 |
| chr16 | 29926678 | 29927415 | 738 | 6 | 1.13E-05 | 1.65E-05 | 0.001124 | 2.92E-06 | 0.003766242 | AC120114.1 |
| chr11 | 2796261 | 2797286 | 1026 | 6 | 5.49E-07 | 7.38E-05 | 9.55E-05 | 2.92E-06 | 0.000703862 | KCNQ1 |
| chr7 | 1874437 | 1874887 | 451 | 3 | 5.16E-06 | 1.28E-06 | 0.001053 | 2.92E-06 | 0.003227187 | MAD1L1, AC069288.1 |
| chr8 | 10425039 | 10426285 | 1247 | 7 | 4.05E-07 | 2.26E-05 | 0.000327 | 2.92E-06 | -0.00578233 | MSRA |
| chr17 | 4987205 | 4988247 | 1043 | 14 | 5.52E-08 | 6.08E-05 | 1.13E-05 | 2.93E-06 | 0.001097969 | AC004771.4, AC004771.2, CAMTA2, INCA1 |
| chr16 | 1065517 | 1066397 | 881 | 4 | 2.02E-07 | 6.60E-06 | 0.000945 | 2.93E-06 | -0.01521126 | AC009041.2, SSTR5-AS1 |
| chr10 | 102503715 | 102504266 | 552 | 10 | 1.73E-06 | 7.29E-06 | 0.001553 | 2.93E-06 | -0.0004918 | SUFU |
| chr18 | 79875987 | 79876561 | 575 | 3 | 4.39E-06 | 1.81E-06 | 0.000296 | 2.93E-06 | -0.00662852 | KCNG2 |
| chr12 | 129110029 | 129110849 | 821 | 3 | 1.83E-05 | 4.11E-06 | 0.000249 | 2.93E-06 | -0.01203571 | TMEM132D-AS1, TMEM132D |
| chr11 | 64367745 | 64368791 | 1047 | 11 | 1.28E-06 | 3.56E-05 | 0.000141 | 2.93E-06 | -0.00144115 | RPS6KA4, MIR1237 |

| | | | | | | | | | | |
|---|---|---|---|---|---|---|---|---|---|---|
| chr22 | 35379966 | 35380726 | 761 | 6 | 5.51E-07 | 3.61E-06 | 0.000767 | 2.93E-06 | -0.00301178 | HMOX1 |
| chr6 | 143502533 | 143503541 | 1009 | 4 | 2.29E-05 | 7.09E-05 | 0.00064 | 2.93E-06 | -0.0002117 | AL031320.2, VDAC1P8, FUCA2 |
| chr13 | 112835056 | 112835698 | 643 | 7 | 1.23E-07 | 0.0001013 | 1.20E-05 | 2.94E-06 | -0.00195017 | ATP11A |
| chr7 | 2482691 | 2483202 | 512 | 2 | 2.23E-06 | 3.30E-06 | 4.23E-05 | 2.94E-06 | -0.00457436 | NA |
| chr17 | 49509592 | 49510684 | 1093 | 4 | 2.72E-05 | 2.02E-05 | 0.001281 | 2.94E-06 | -0.00984726 | NGFR, AC006487.1 |
| chr17 | 82418996 | 82419996 | 1001 | 6 | 1.06E-06 | 2.62E-05 | 0.000147 | 2.95E-06 | -0.00713133 | HEXD |
| chr9 | 83201270 | 83201568 | 299 | 3 | 2.02E-06 | 2.12E-05 | 8.50E-06 | 2.95E-06 | 0.003056093 | NA |
| chr15 | 74461028 | 74461671 | 644 | 14 | 5.56E-07 | 7.75E-06 | 0.002997 | 2.95E-06 | -0.00019497 | UBL7-AS1, UBL7 |
| chr15 | 101086105 | 101086142 | 38 | 3 | 1.09E-06 | 6.09E-05 | 7.34E-06 | 2.95E-06 | -0.00306173 | NA |
| chr17 | 72192679 | 72192782 | 104 | 2 | 2.38E-06 | 2.47E-05 | 6.06E-06 | 2.95E-06 | 0.010907748 | AC007461.1, SOX9-AS1 |
| chr20 | 56629286 | 56630169 | 884 | 6 | 2.37E-08 | 0.0002747 | 3.22E-05 | 2.95E-06 | 0.000411386 | TFAP2C |
| chr2 | 216995058 | 216995288 | 231 | 3 | 3.96E-06 | 2.08E-06 | 0.000198 | 2.95E-06 | -0.0052213 | NA |
| chr8 | 94553021 | 94554248 | 1228 | 13 | 3.63E-07 | 4.25E-05 | 2.51E-05 | 2.95E-06 | 0.000101504 | AC023632.2, VIRMA |
| chr8 | 868662 | 868875 | 214 | 3 | 4.11E-06 | 4.42E-06 | 7.22E-05 | 2.95E-06 | 0.006238805 | DLGAP2 |
| chr12 | 48044126 | 48044473 | 348 | 2 | 1.57E-06 | 7.87E-06 | 1.44E-05 | 2.95E-06 | 0.001259301 | SENP1 |
| chr22 | 39689896 | 39690435 | 540 | 2 | 7.23E-07 | 2.88E-06 | 5.50E-05 | 2.95E-06 | -0.00872727 | NA |
| chr1 | 43057887 | 43058304 | 418 | 2 | 1.19E-06 | 2.08E-06 | 0.000133 | 2.95E-06 | -0.0076407 | NA |
| chr17 | 40354391 | 40356510 | 2120 | 10 | 1.09E-05 | 2.09E-05 | 0.001462 | 2.95E-06 | -0.0020127 | RARA |
| chr3 | 188431344 | 188431429 | 86 | 2 | 2.82E-06 | 2.57E-06 | 7.07E-05 | 2.95E-06 | -0.00706503 | LPP |
| chr20 | 45578649 | 45578904 | 256 | 2 | 4.67E-06 | 2.22E-06 | 0.000106 | 2.96E-06 | -0.00385858 | WFDC8 |
| chr8 | 126302744 | 126303279 | 536 | 2 | 8.47E-07 | 6.35E-06 | 1.80E-05 | 2.96E-06 | -0.01480378 | NA |
| chr4 | 177308497 | 177309024 | 528 | 2 | 5.37E-07 | 8.91E-06 | 1.30E-05 | 2.96E-06 | -0.01172681 | NA |
| chr22 | 42565773 | 42566246 | 474 | 2 | 1.23E-07 | 0.0004969 | 2.32E-06 | 2.96E-06 | 0.004383272 | SERHL2, RRP7BP |
| chr17 | 40026917 | 40028393 | 1477 | 8 | 2.57E-05 | 1.37E-06 | 0.012951 | 2.96E-06 | 0.004132118 | MED24, SNORD124 |
| chr1 | 84619630 | 84619654 | 25 | 2 | 9.78E-07 | 1.67E-05 | 7.80E-06 | 2.96E-06 | -0.02178866 | NA |
| chr12 | 66916058 | 66916477 | 420 | 3 | 1.24E-07 | 9.59E-05 | 1.96E-05 | 2.97E-06 | -0.00050642 | GRIP1 |
| chr1 | 77888096 | 77889540 | 1445 | 15 | 6.51E-07 | 0.0005776 | 8.70E-05 | 2.97E-06 | -0.00273295 | NEXN, NEXN-AS1 |
| chr14 | 21556887 | 21557381 | 495 | 2 | 4.21E-07 | 8.48E-06 | 1.36E-05 | 2.97E-06 | -0.01659943 | NA |
| chr1 | 60925306 | 60925514 | 209 | 2 | 8.65E-07 | 0.0002355 | 2.67E-06 | 2.98E-06 | -0.00322565 | NFIA, NFIA-AS2 |
| chr5 | 151115499 | 151115658 | 160 | 2 | 6.44E-06 | 1.92E-06 | 0.000201 | 2.98E-06 | -0.0091607 | ANXA6 |
| chr11 | 94512058 | 94512629 | 572 | 10 | 1.09E-07 | 0.0004772 | 0.00012 | 2.98E-06 | 0.000851482 | C11orf97 |
| chr20 | 36604550 | 36605539 | 990 | 8 | 5.69E-07 | 4.23E-06 | 0.002333 | 2.98E-06 | -0.00524616 | TGIF2-RAB5IF |
| chr1 | 246834753 | 246834834 | 82 | 2 | 3.35E-06 | 2.79E-06 | 6.02E-05 | 2.98E-06 | -0.015678 | KIF28P |
| chr21 | 45427200 | 45427983 | 784 | 5 | 3.94E-07 | 0.0001344 | 1.02E-05 | 2.98E-06 | -0.00416055 | COL18A1 |
| chr1 | 192555135 | 192555183 | 49 | 3 | 4.87E-06 | 1.22E-05 | 0.000221 | 2.98E-06 | -0.00539574 | AL390957.1 |
| chr12 | 132405289 | 132406191 | 903 | 7 | 5.79E-06 | 1.22E-06 | 0.005195 | 2.99E-06 | 0.005742234 | NA |
| chr1 | 206498075 | 206498725 | 651 | 4 | 2.47E-06 | 1.04E-05 | 0.000283 | 2.99E-06 | -0.00282216 | NA |
| chr15 | 51586194 | 51586431 | 238 | 2 | 3.59E-06 | 4.06E-06 | 3.18E-05 | 2.99E-06 | -0.00180472 | DMXL2 |
| chr14 | 105508098 | 105508243 | 146 | 3 | 1.67E-06 | 1.22E-06 | 0.000931 | 2.99E-06 | -0.00524651 | NA |
| chr10 | 117685208 | 117685638 | 431 | 2 | 6.02E-08 | 1.78E-05 | 7.56E-06 | 2.99E-06 | -0.01706977 | NA |
| chr2 | 95279765 | 95279774 | 10 | 3 | 3.69E-06 | 1.14E-05 | 7.31E-05 | 2.99E-06 | -0.00382297 | PROM2 |
| chr7 | 84701463 | 84701969 | 507 | 3 | 3.11E-07 | 0.0014846 | 1.32E-05 | 2.99E-06 | -0.00132797 | NA |
| chr2 | 85887553 | 85888468 | 916 | 4 | 5.67E-07 | 1.67E-06 | 0.000439 | 2.99E-06 | 0.003316091 | ST3GAL5 |
| chr1 | 9094191 | 9094195 | 5 | 2 | 6.99E-06 | 1.79E-06 | 0.000325 | 2.99E-06 | 0.016463707 | NA |
| chr10 | 110407359 | 110408303 | 945 | 4 | 1.84E-05 | 3.57E-05 | 0.00015 | 2.99E-06 | -0.00452799 | NA |
| chr6 | 40754260 | 40754777 | 518 | 4 | 4.67E-08 | 0.0062539 | 2.83E-05 | 3.00E-06 | -0.00811506 | NA |
| chr15 | 40893602 | 40894586 | 985 | 11 | 4.33E-07 | 0.0002726 | 1.60E-05 | 3.00E-06 | -0.0006935 | VPS18 |
| chr2 | 33378862 | 33379505 | 644 | 2 | 2.63E-06 | 2.95E-06 | 5.45E-05 | 3.00E-06 | -0.00602301 | LTBP1 |
| chr6 | 83229635 | 83230105 | 471 | 3 | 1.59E-06 | 8.48E-06 | 6.49E-05 | 3.00E-06 | -0.01093506 | ME1 |
| chr19 | 37217982 | 37218805 | 824 | 6 | 2.92E-06 | 1.00E-05 | 0.000301 | 3.00E-06 | 0.002647194 | ZNF383, ZNF585B |
| chr7 | 23105180 | 23106109 | 930 | 14 | 8.68E-06 | 3.02E-06 | 0.006368 | 3.00E-06 | -0.00083578 | KLHL7, KLHL7-DT |
| chr17 | 19457917 | 19458341 | 425 | 2 | 4.73E-07 | 1.18E-05 | 1.04E-05 | 3.00E-06 | -0.03720443 | AC004448.1, AC004448.2 |
| chr10 | 121736877 | 121737407 | 531 | 2 | 1.92E-06 | 2.03E-06 | 0.000162 | 3.00E-06 | -0.02644713 | AC025947.1 |
| chr17 | 73261841 | 73262897 | 1057 | 9 | 2.36E-05 | 0.0001437 | 0.000574 | 3.00E-06 | -0.00029951 | CPSF4L |
| chr11 | 68658923 | 68659552 | 630 | 2 | 8.25E-07 | 3.40E-06 | 4.24E-05 | 3.01E-06 | 0.002857307 | NA |
| chr19 | 49503225 | 49503542 | 318 | 3 | 6.20E-07 | 6.68E-05 | 5.76E-06 | 3.01E-06 | -0.00645321 | NA |
| chr11 | 64249619 | 64250837 | 1219 | 5 | 9.32E-09 | 5.20E-05 | 1.08E-05 | 3.01E-06 | -0.00508425 | NA |
| chr2 | 77629818 | 77629957 | 140 | 2 | 1.74E-06 | 5.26E-05 | 4.26E-06 | 3.01E-06 | -0.01220937 | NA |
| chr12 | 9932337 | 9932575 | 239 | 3 | 4.13E-06 | 4.63E-06 | 0.00015 | 3.01E-06 | -0.00320644 | CLEC2A |
| chr2 | 240986090 | 240986520 | 431 | 4 | 1.52E-07 | 0.0426849 | 1.49E-06 | 3.01E-06 | 0.001008366 | CROCC2 |
| chr12 | 108598001 | 108598372 | 372 | 6 | 5.15E-07 | 0.0001584 | 6.89E-05 | 3.01E-06 | -0.00099852 | TMEM119 |
| chr10 | 133039306 | 133039840 | 535 | 5 | 9.55E-07 | 5.91E-06 | 0.000292 | 3.01E-06 | -0.01424129 | NA |
| chr3 | 137035113 | 137035615 | 503 | 3 | 5.28E-06 | 3.76E-06 | 0.000103 | 3.01E-06 | -0.00642713 | NA |
| chr2 | 36857912 | 36857945 | 34 | 2 | 6.28E-06 | 2.30E-06 | 0.000102 | 3.02E-06 | 0.002407218 | STRN |
| chr10 | 73252385 | 73253319 | 935 | 17 | 2.99E-06 | 3.58E-06 | 0.007506 | 3.02E-06 | -0.00010553 | DNAJC9-AS1, DNAJC9-AS1, MRPS16 |
| chr17 | 83085669 | 83085906 | 238 | 3 | 2.45E-06 | 2.85E-06 | 0.00042 | 3.02E-06 | -0.00423072 | METRNL |
| chr10 | 119667971 | 119668270 | 300 | 3 | 1.50E-06 | 9.02E-05 | 3.85E-05 | 3.02E-06 | -0.00149657 | BAG3 |
| chr15 | 89912403 | 89913387 | 985 | 12 | 2.97E-05 | 5.59E-06 | 0.000429 | 3.03E-06 | -0.00200013 | ARPIN-AP3S2, ARPIN |
| chr10 | 101582800 | 101583365 | 566 | 4 | 9.82E-06 | 8.14E-07 | 0.003578 | 3.03E-06 | -0.0025656 | DPCD, POLL |
| chr7 | 129865526 | 129865563 | 38 | 3 | 8.36E-06 | 1.60E-06 | 0.000544 | 3.03E-06 | -0.00124631 | UBE2H |
| chr19 | 3700334 | 3700886 | 553 | 8 | 3.69E-06 | 3.07E-06 | 0.00378 | 3.03E-06 | -0.00048344 | PIP5K1C |
| chr20 | 47705155 | 47705339 | 185 | 2 | 4.71E-06 | 2.68E-06 | 6.95E-05 | 3.03E-06 | -0.00688763 | SULF2 |
| chr11 | 75149578 | 75149940 | 363 | 3 | 2.00E-07 | 0.0001073 | 5.10E-06 | 3.03E-06 | -0.00606563 | SLCO2B1 |
| chr1 | 206744560 | 206744897 | 338 | 2 | 1.58E-06 | 8.37E-06 | 1.42E-05 | 3.03E-06 | -0.01312465 | NA |
| chr15 | 52564785 | 52565505 | 721 | 2 | 1.39E-05 | 1.88E-06 | 0.000258 | 3.04E-06 | -0.01247125 | ARPP19 |
| chr16 | 86072800 | 86073530 | 731 | 3 | 1.40E-05 | 3.23E-06 | 0.000148 | 3.04E-06 | -0.0101763 | NA |
| chr1 | 61724976 | 61726223 | 1248 | 10 | 1.16E-06 | 0.0001519 | 0.000107 | 3.04E-06 | 0.001558562 | TM2D1 |
| chr16 | 6482763 | 6483788 | 1026 | 12 | 1.60E-06 | 3.65E-05 | 0.000917 | 3.04E-06 | 0.004522487 | RBFOX1 |
| chr15 | 48730158 | 48730915 | 758 | 3 | 1.71E-05 | 4.72E-05 | 0.000199 | 3.04E-06 | 0.000192509 | CEP152 |
| chr12 | 110280506 | 110281236 | 731 | 9 | 7.48E-06 | 8.61E-07 | 0.005483 | 3.04E-06 | 0.001466904 | ATP2A2 |
| chr1 | 9249830 | 9250198 | 369 | 3 | 1.39E-06 | 2.02E-05 | 3.02E-05 | 3.04E-06 | -0.00858134 | H6PD |
| chr6 | 27399003 | 27399123 | 121 | 2 | 5.53E-06 | 2.91E-06 | 5.82E-05 | 3.04E-06 | -0.01579557 | ZNF391 |
| chr20 | 39358051 | 39358443 | 393 | 2 | 7.84E-06 | 1.83E-06 | 0.000319 | 3.05E-06 | -0.01752816 | NA |
| chr6 | 168950652 | 168951384 | 733 | 6 | 1.37E-05 | 7.82E-07 | 0.004107 | 3.05E-06 | -0.00032811 | AL513210.1 |
| chr4 | 107862130 | 107862902 | 773 | 5 | 6.66E-07 | 7.39E-05 | 0.000338 | 3.05E-06 | -0.00339877 | SGMS2 |
| chr17 | 45974127 | 45974238 | 112 | 2 | 1.87E-06 | 1.84E-06 | 0.000313 | 3.05E-06 | -0.01736118 | MAPT |
| chr17 | 50558492 | 50559285 | 794 | 4 | 1.91E-06 | 1.93E-06 | 0.001668 | 3.05E-06 | -0.00213225 | CACNA1G-AS1 |
| chr15 | 77482843 | 77483281 | 439 | 2 | 4.70E-06 | 1.89E-06 | 0.000261 | 3.05E-06 | -0.00583499 | HMG20A |
| chr1 | 211975514 | 211976081 | 568 | 2 | 7.42E-06 | 1.90E-06 | 0.000261 | 3.06E-06 | 0.000622069 | INTS7 |
| chr18 | 12102287 | 12102700 | 414 | 4 | 3.61E-06 | 7.18E-06 | 0.000193 | 3.07E-06 | 0.00388164 | ANKRD62, SDHDP1 |
| chr17 | 40287397 | 40288371 | 975 | 14 | 4.86E-07 | 0.0002601 | 0.000433 | 3.07E-06 | -0.00043598 | CDC6 |
| chr20 | 41293027 | 41293518 | 492 | 3 | 1.91E-06 | 1.00E-05 | 1.80E-05 | 3.07E-06 | -0.00646055 | ZHX3 |
| chr11 | 2929938 | 2930751 | 814 | 19 | 3.53E-05 | 3.21E-06 | 0.026756 | 3.07E-06 | 0.000515078 | NA |
| chr17 | 64378032 | 64378034 | 3 | 2 | 6.73E-07 | 1.22E-05 | 1.04E-05 | 3.08E-06 | -0.01153985 | PECAM1 |
| chr12 | 113077128 | 113077634 | 507 | 3 | 3.23E-06 | 1.65E-06 | 0.000583 | 3.08E-06 | -0.01146414 | DTX1 |
| chr6 | 1615001 | 1615874 | 874 | 5 | 6.13E-07 | 0.000157 | 1.21E-05 | 3.08E-06 | 0.008576473 | NA |
| chr9 | 35079654 | 35080662 | 1009 | 9 | 2.57E-07 | 3.82E-05 | 0.000354 | 3.08E-06 | 0.001000798 | VCP, FANCG |
| chr13 | 112182847 | 112183838 | 992 | 6 | 4.20E-06 | 0.0006539 | 0.000313 | 3.08E-06 | -0.00270325 | NA |
| chr10 | 132928862 | 132930047 | 1186 | 5 | 7.40E-08 | 2.74E-05 | 7.01E-05 | 3.08E-06 | 0.000290242 | CFAP46 |
| chr13 | 113654304 | 113654434 | 131 | 3 | 8.42E-06 | 1.57E-06 | 0.000581 | 3.08E-06 | -0.0115932 | ATP4B |
| chr6 | 36206891 | 36207241 | 351 | 2 | 4.34E-06 | 3.41E-06 | 4.46E-05 | 3.09E-06 | -0.01085532 | BRPF3 |
| chr17 | 16406983 | 16407548 | 566 | 4 | 2.05E-06 | 5.61E-06 | 0.000766 | 3.09E-06 | 0.003194877 | NA |
| chr10 | 101119047 | 101119214 | 168 | 2 | 7.08E-06 | 1.87E-06 | 0.000308 | 3.09E-06 | -0.01019592 | TLX1NB |
| chr14 | 60204306 | 60204849 | 544 | 3 | 1.19E-07 | 5.60E-05 | 9.42E-06 | 3.09E-06 | 0.006835173 | NA |
| chr7 | 157736643 | 157737324 | 682 | 3 | 4.49E-08 | 1.91E-05 | 0.000213 | 3.09E-06 | -0.00717385 | PTPRN2 |
| chr3 | 142964041 | 142964487 | 447 | 8 | 4.22E-07 | 0.0006028 | 4.14E-05 | 3.09E-06 | -0.00502822 | PAQR9-AS1 |
| chr8 | 101453681 | 101453707 | 27 | 2 | 4.26E-07 | 0.0001112 | 3.33E-06 | 3.09E-06 | 0.003970299 | NA |
| chr18 | 79969909 | 79970650 | 742 | 2 | 2.50E-07 | 1.12E-05 | 1.13E-05 | 3.09E-06 | 0.00383984 | HSBP1L1 |
| chr8 | 11844210 | 11844905 | 696 | 2 | 4.76E-07 | 8.48E-06 | 1.44E-05 | 3.09E-06 | -0.00023554 | CTSB |
| chr7 | 143287063 | 143287926 | 864 | 7 | 6.13E-08 | 0.0001781 | 0.000116 | 3.09E-06 | 0.001519686 | TMEM139, TMEM139-AS1 |
| chr19 | 58353989 | 58354996 | 1008 | 5 | 1.45E-05 | 4.33E-06 | 0.000559 | 3.09E-06 | -0.00290132 | A1BG-AS1, AC012313.3, ZNF497 |
| chr1 | 35078869 | 35079751 | 883 | 13 | 1.27E-05 | 2.17E-05 | 0.017939 | 3.09E-06 | -0.00046369 | ZMYM1 |
| chr17 | 10037335 | 10037778 | 444 | 3 | 5.20E-07 | 3.63E-06 | 6.67E-05 | 3.10E-06 | -0.00420549 | GAS7 |
| chr12 | 17372604 | 17373234 | 631 | 3 | 1.35E-06 | 1.64E-06 | 0.000662 | 3.10E-06 | -0.01243699 | NA |
| chr1 | 200974808 | 200975179 | 372 | 2 | 4.31E-06 | 2.53E-06 | 8.50E-05 | 3.10E-06 | -0.00722841 | KIF21B |
| chr13 | 101876350 | 101877163 | 814 | 3 | 4.30E-05 | 3.01E-06 | 0.000427 | 3.11E-06 | 0.001614868 | FGF14 |
| chr10 | 96765353 | 96765820 | 468 | 3 | 3.55E-06 | 2.31E-06 | 0.000413 | 3.11E-06 | -0.00841862 | NA |
| chr9 | 136944911 | 136945980 | 1070 | 13 | 1.03E-07 | 0.0059519 | 0.0003 | 3.11E-06 | -0.00187654 | C8G |
| chr3 | 13645029 | 13645898 | 870 | 3 | 1.79E-06 | 4.87E-06 | 6.75E-05 | 3.11E-06 | -0.00697207 | NA |
| chr19 | 39439396 | 39439434 | 39 | 2 | 1.88E-06 | 1.48E-05 | 9.09E-06 | 3.11E-06 | -0.00541568 | SUPT5H |
| chr2 | 45650927 | 45651838 | 912 | 7 | 6.93E-08 | 0.0002796 | 2.07E-05 | 3.11E-06 | 0.000625479 | PRKCE |
| chr10 | 114043081 | 114044060 | 980 | 10 | 1.06E-05 | 1.62E-05 | 0.000279 | 3.11E-06 | 0.000949353 | ADRB1 |
| chr8 | 101903059 | 101903086 | 28 | 2 | 2.42E-06 | 7.99E-06 | 1.54E-05 | 3.11E-06 | -0.02187541 | NCALD |
| chr10 | 96359947 | 96360485 | 539 | 4 | 2.05E-07 | 4.85E-06 | 0.001509 | 3.12E-06 | -0.00647687 | NA |
| chr1 | 243843537 | 243844797 | 1261 | 6 | 9.08E-06 | 8.00E-06 | 0.003115 | 3.12E-06 | -0.00765152 | AKT3 |
| chr22 | 30408760 | 30409228 | 469 | 6 | 2.05E-06 | 5.91E-06 | 8.29E-05 | 3.12E-06 | -0.00719347 | SEC14L2, RNF215 |
| chr6 | 27019796 | 27020510 | 715 | 10 | 4.40E-06 | 4.12E-05 | 0.000487 | 3.12E-06 | -0.00373602 | LINC00240, AL590062.1 |
| chr12 | 81369629 | 81369713 | 85 | 2 | 2.52E-07 | 0.0012005 | 2.19E-06 | 3.12E-06 | 0.013656823 | PPFIA2 |
| chr14 | 37197284 | 37198027 | 744 | 15 | 1.63E-06 | 4.43E-06 | 0.000568 | 3.12E-06 | 0.000515523 | MIPOL1 |
| chr5 | 107670298 | 107671310 | 1013 | 6 | 4.43E-07 | 0.0002272 | 1.64E-05 | 3.12E-06 | -0.0014948 | EFNA5 |
| chr8 | 67950281 | 67952089 | 1809 | 8 | 1.24E-06 | 1.13E-05 | 5.19E-05 | 3.12E-06 | 0.001301835 | PREX2 |
| chr13 | 110706169 | 110706538 | 370 | 9 | 1.37E-05 | 6.21E-07 | 0.016548 | 3.12E-06 | 0.001915611 | CARS2 |
| chr6 | 159232996 | 159233383 | 388 | 2 | 1.87E-06 | 2.84E-06 | 6.59E-05 | 3.12E-06 | -0.02291262 | FNDC1 |
| chr11 | 116869745 | 116870359 | 615 | 3 | 3.87E-08 | 2.48E-05 | 7.98E-06 | 3.12E-06 | 0.004484157 | SIK3 |
| chr7 | 76317944 | 76318428 | 485 | 4 | 3.57E-07 | 3.92E-05 | 4.25E-05 | 3.12E-06 | 6.21E-05 | NA |
| chr22 | 39095898 | 39096298 | 401 | 5 | 5.61E-06 | 2.02E-05 | 0.001687 | 3.12E-06 | -0.00084912 | NA |
| chr22 | 27142604 | 27143080 | 477 | 2 | 4.80E-06 | 2.28E-06 | 0.000119 | 3.12E-06 | -0.01435551 | NA |
| chr2 | 55960982 | 55961452 | 471 | 2 | 9.61E-06 | 1.88E-06 | 0.000328 | 3.12E-06 | -0.00059751 | AC011306.1 |
| chr5 | 95861679 | 95861772 | 94 | 3 | 5.60E-06 | 1.62E-06 | 0.000797 | 3.13E-06 | -0.00367858 | NA |
| chr11 | 73759575 | 73760529 | 955 | 6 | 3.01E-06 | 0.0001567 | 4.54E-05 | 3.13E-06 | -0.00505929 | RAB6A |
| chr2 | 65075423 | 65075902 | 480 | 2 | 4.62E-07 | 1.41E-05 | 9.54E-06 | 3.13E-06 | -0.01131024 | CEP68, RAB1A |
| chr19 | 35500417 | 35500679 | 263 | 3 | 1.73E-06 | 7.36E-06 | 0.000313 | 3.13E-06 | -0.00878919 | DMKN |
| chr18 | 79346342 | 79346426 | 85 | 2 | 1.02E-06 | 3.73E-06 | 3.98E-05 | 3.13E-06 | 0.002698471 | ATP9B |
| chr5 | 55222839 | 55223858 | 1020 | 8 | 1.36E-05 | 1.03E-06 | 0.006012 | 3.14E-06 | -0.00023629 | MCIDAS |
| chr7 | 30470847 | 30471104 | 258 | 2 | 7.03E-07 | 0.0001196 | 3.31E-06 | 3.14E-06 | 0.009420285 | NOD1 |
| chr17 | 9701035 | 9701645 | 611 | 4 | 5.84E-07 | 7.27E-05 | 8.43E-06 | 3.14E-06 | -0.00418705 | USP43 |
| chr7 | 152136444 | 152137017 | 574 | 3 | 1.28E-06 | 3.17E-06 | 8.86E-05 | 3.14E-06 | -0.00295195 | KMT2C |
| chr7 | 47293336 | 47293487 | 152 | 2 | 1.15E-06 | 3.62E-06 | 4.19E-05 | 3.14E-06 | -0.00441166 | TNS3 |
| chr3 | 56930418 | 56930912 | 495 | 2 | 6.22E-07 | 1.37E-05 | 9.80E-06 | 3.14E-06 | -0.00878477 | ARHGEF3 |
| chr3 | 139280287 | 139280897 | 611 | 3 | 8.61E-07 | 4.00E-06 | 5.63E-05 | 3.14E-06 | 0.006774249 | MRPS22 |
| chr1 | 50369959 | 50370406 | 448 | 3 | 8.93E-06 | 1.32E-06 | 0.000825 | 3.14E-06 | -0.00642733 | NA |
| chr10 | 132685326 | 132686248 | 923 | 11 | 5.16E-05 | 1.18E-05 | 0.001896 | 3.14E-06 | -0.00238601 | INPP5A |
| chr8 | 66112416 | 66113298 | 883 | 9 | 2.67E-07 | 5.23E-05 | 1.22E-05 | 3.15E-06 | -0.00169361 | AC084082.1 |
| chr5 | 58969989 | 58970631 | 643 | 2 | 3.45E-06 | 3.04E-06 | 5.81E-05 | 3.15E-06 | -0.02044959 | PDE4D |
| chr12 | 132955341 | 132956412 | 1072 | 5 | 4.15E-07 | 5.68E-06 | 0.000866 | 3.15E-06 | -0.00125037 | CHFR, ZNF605 |
| chr7 | 926933 | 927641 | 709 | 8 | 5.03E-07 | 0.0002955 | 2.74E-05 | 3.15E-06 | -0.0017324 | ADAP1, COX19 |
| chr18 | 59042224 | 59042304 | 81 | 2 | 7.39E-06 | 1.90E-06 | 0.000326 | 3.15E-06 | -0.01050753 | OACYLP |
| chr3 | 157130214 | 157131065 | 852 | 6 | 1.44E-05 | 5.05E-06 | 0.002779 | 3.16E-06 | -0.00015631 | LINC00881 |

| | | | | | | | | | | |
|---|---|---|---|---|---|---|---|---|---|---|
| chr11 | 44939303 | 44939808 | 506 | 3 | 2.33E-06 | 2.21E-06 | 0.000258 | 3.16E-06 | -0.00980295 | TP53I11 |
| chr6 | 87472389 | 87472884 | 496 | 10 | 8.57E-08 | 0.0001792 | 0.000207 | 3.16E-06 | 0.008148204 | AL049697.1, SLC35A1 |
| chr6 | 150640678 | 150641285 | 608 | 2 | 1.18E-05 | 1.98E-06 | 0.000248 | 3.16E-06 | -0.01603157 | PLEKHG1 |
| chr2 | 239052217 | 239052958 | 742 | 8 | 9.18E-07 | 0.0001448 | 0.000254 | 3.16E-06 | -0.00095133 | HDAC4 |
| chr9 | 70810963 | 70811342 | 380 | 2 | 2.67E-07 | 2.33E-05 | 6.84E-06 | 3.16E-06 | -0.01398366 | TRPM3 |
| chr2 | 37083848 | 37084963 | 1116 | 13 | 1.65E-07 | 0.000745 | 0.000869 | 3.16E-06 | 0.000961318 | GPATCH11, HEATR5B |
| chr15 | 80969899 | 80970694 | 796 | 5 | 2.80E-06 | 1.19E-05 | 0.00116 | 3.16E-06 | -0.00226824 | MESD |
| chr15 | 58750043 | 58750959 | 917 | 10 | 2.05E-06 | 5.71E-06 | 0.000546 | 3.16E-06 | -0.00350819 | AC090515.6 |
| chr6 | 15517112 | 15517900 | 789 | 4 | 5.54E-06 | 4.71E-05 | 2.81E-05 | 3.17E-06 | -0.0042298 | JARID2 |
| chr9 | 419673 | 420082 | 410 | 2 | 8.95E-08 | 0.0006568 | 2.38E-06 | 3.17E-06 | 0.002769389 | DOCK8 |
| chr7 | 6212870 | 6213153 | 284 | 3 | 3.15E-06 | 4.44E-05 | 0.000201 | 3.17E-06 | -0.00383951 | CYTH3 |
| chr16 | 2778301 | 2779414 | 1114 | 4 | 1.35E-05 | 1.63E-05 | 0.000194 | 3.17E-06 | -0.00212041 | AC092117.1 |
| chr10 | 94156200 | 94156276 | 77 | 2 | 5.87E-06 | 1.89E-06 | 0.00035 | 3.17E-06 | -0.0081105 | PLCE1 |
| chr16 | 553568 | 554188 | 621 | 7 | 1.40E-06 | 0.0001069 | 0.000127 | 3.17E-06 | -0.00271484 | CAPN15, Z97986.1 |
| chr4 | 55793756 | 55794294 | 539 | 7 | 5.43E-06 | 3.78E-06 | 0.002032 | 3.17E-06 | -0.01059574 | NA |
| chr6 | 5086797 | 5087515 | 719 | 5 | 3.73E-06 | 4.34E-06 | 0.000903 | 3.17E-06 | 0.005177807 | PPP1R3G |
| chr12 | 123350270 | 123350322 | 53 | 2 | 1.43E-06 | 2.14E-06 | 0.000168 | 3.18E-06 | 0.001002541 | SBNO1 |
| chr2 | 233691036 | 233691777 | 742 | 7 | 2.83E-06 | 2.52E-06 | 0.000556 | 3.18E-06 | 0.000601775 | UGT1A8, UGT1A10, UGT1A9, UGT1A7, UGT1A6 |
| chr12 | 122028628 | 122029322 | 695 | 3 | 4.93E-06 | 4.03E-06 | 6.85E-05 | 3.18E-06 | 0.010444738 | BCL7A |
| chr2 | 233337863 | 233338490 | 628 | 4 | 2.85E-06 | 8.09E-06 | 6.13E-05 | 3.18E-06 | -0.00528178 | SAG |
| chr6 | 35435354 | 35435961 | 608 | 3 | 4.97E-06 | 3.18E-06 | 9.09E-05 | 3.18E-06 | 0.000526892 | NA |
| chr15 | 93802837 | 93803700 | 864 | 6 | 4.69E-06 | 4.14E-06 | 0.00138 | 3.18E-06 | -0.00405123 | LINC01579 |
| chr3 | 184378625 | 184379652 | 1028 | 6 | 1.22E-05 | 2.62E-05 | 0.000171 | 3.18E-06 | -0.00307371 | THPO |
| chr20 | 48763240 | 48763377 | 138 | 3 | 4.74E-06 | 1.87E-06 | 0.000261 | 3.19E-06 | 0.008387512 | PREX1 |
| chr17 | 82848259 | 82848559 | 301 | 4 | 4.76E-06 | 9.71E-06 | 0.000242 | 3.19E-06 | -0.00909367 | TBCD |
| chr6 | 26536722 | 26538443 | 1722 | 15 | 9.11E-06 | 0.0002175 | 0.005219 | 3.19E-06 | 0.000136247 | HMGN4 |
| chr21 | 15514275 | 15514841 | 567 | 3 | 2.31E-07 | 0.0005458 | 6.02E-05 | 3.19E-06 | 0.009135622 | AJ009632.2 |
| chr22 | 24011866 | 24012366 | 501 | 4 | 3.67E-07 | 2.62E-06 | 0.001205 | 3.19E-06 | -0.00826317 | CABIN1 |
| chr16 | 2759730 | 2760374 | 645 | 5 | 3.64E-06 | 2.99E-06 | 0.002479 | 3.19E-06 | -0.00254459 | SRRM2 |
| chr19 | 42320048 | 42320428 | 381 | 3 | 1.86E-06 | 8.71E-05 | 0.00017 | 3.19E-06 | -0.00026931 | TMEM145 |
| chr13 | 112850341 | 112851240 | 900 | 3 | 6.95E-06 | 2.27E-06 | 0.000181 | 3.19E-06 | -0.00529036 | ATP11A |
| chr4 | 1985056 | 1985928 | 873 | 5 | 2.08E-07 | 9.84E-05 | 2.63E-05 | 3.20E-06 | -0.00088828 | NELFA |
| chr12 | 112695310 | 112695779 | 470 | 3 | 2.87E-06 | 4.34E-06 | 0.000374 | 3.20E-06 | -0.01368428 | RPH3A |
| chr8 | 139830430 | 139830956 | 527 | 2 | 7.77E-08 | 0.0004992 | 2.49E-06 | 3.20E-06 | 0.002854958 | TRAPPC9 |
| chr15 | 51782406 | 51782798 | 393 | 2 | 5.62E-06 | 2.23E-06 | 0.000147 | 3.20E-06 | -0.00567628 | TMOD2 |
| chr11 | 114228364 | 114229295 | 932 | 4 | 9.51E-08 | 3.52E-05 | 7.78E-06 | 3.20E-06 | 0.002206352 | ZBTB16, AP002518.2 |
| chr6 | 39114826 | 39115429 | 604 | 8 | 3.90E-06 | 5.31E-05 | 0.000157 | 3.20E-06 | -0.0003452 | SAYSD1 |
| chr16 | 87132795 | 87132825 | 31 | 2 | 3.53E-06 | 2.77E-06 | 7.46E-05 | 3.20E-06 | -0.00199135 | AC136285.3, C16orf95 |
| chr21 | 15693148 | 15693194 | 47 | 2 | 7.69E-07 | 5.78E-05 | 4.38E-06 | 3.20E-06 | -0.01841314 | NA |
| chr1 | 155207770 | 155208764 | 995 | 13 | 6.53E-06 | 6.06E-05 | 0.001247 | 3.20E-06 | -0.00220579 | MTX1, THBS3 |
| chr12 | 122979215 | 122980362 | 1148 | 14 | 5.87E-06 | 2.48E-06 | 0.01279 | 3.20E-06 | -0.00548602 | OGFOD2, AC026362.1, ARL6IP4, ABCB9 |
| chr1 | 114581586 | 114581894 | 309 | 10 | 2.49E-07 | 0.0079582 | 3.17E-05 | 3.20E-06 | 0.000122739 | BCAS2 |
| chr14 | 103334135 | 103334846 | 712 | 11 | 2.30E-05 | 1.44E-05 | 0.005542 | 3.21E-06 | 0.000816399 | EIF5 |
| chr8 | 93258901 | 93259550 | 650 | 3 | 1.66E-06 | 4.98E-06 | 0.000317 | 3.21E-06 | -0.01296736 | LINC00535 |
| chr4 | 4866996 | 4867445 | 450 | 5 | 7.77E-07 | 7.40E-06 | 0.000158 | 3.21E-06 | -0.00039726 | NA |
| chr1 | 110067830 | 110069090 | 1261 | 6 | 7.48E-07 | 9.93E-06 | 0.000228 | 3.21E-06 | -0.00389405 | STRIP1, ALX3 |
| chr15 | 99100543 | 99100855 | 313 | 5 | 2.60E-06 | 1.73E-05 | 0.000197 | 3.21E-06 | 0.001436964 | SYNM, AC036108.1 |
| chr3 | 49117202 | 49117391 | 190 | 2 | 2.39E-07 | 8.18E-06 | 1.58E-05 | 3.21E-06 | 0.000876485 | USP19 |
| chr1 | 178055368 | 178055965 | 598 | 2 | 3.33E-08 | 0.0002224 | 2.93E-06 | 3.21E-06 | -0.00696281 | NA |
| chr1 | 160638097 | 160638394 | 298 | 2 | 9.71E-07 | 4.56E-05 | 4.91E-06 | 3.21E-06 | 0.01711463 | SLAMF1 |
| chr2 | 70820517 | 70821228 | 712 | 6 | 4.52E-06 | 1.03E-05 | 0.004528 | 3.22E-06 | -0.0074971 | CLEC4F |
| chr5 | 79514350 | 79515074 | 725 | 6 | 8.90E-09 | 0.0001755 | 2.18E-06 | 3.22E-06 | 0.000615576 | NA |
| chr8 | 640487 | 640650 | 164 | 2 | 3.09E-06 | 2.00E-06 | 0.000261 | 3.22E-06 | -0.01387555 | ERICH1 |
| chr11 | 5269516 | 5270068 | 553 | 3 | 2.18E-07 | 2.28E-05 | 1.86E-05 | 3.22E-06 | 0.004389309 | HBG2, AC104389.5, HBE1 |
| chr8 | 103140920 | 103141601 | 682 | 10 | 4.38E-06 | 2.17E-05 | 0.00493 | 3.22E-06 | -8.48E-05 | BAALC, BAALC-AS2 |
| chr12 | 125255385 | 125255862 | 478 | 3 | 5.54E-07 | 9.89E-06 | 2.50E-05 | 3.22E-06 | 0.007900663 | TMEM132B |
| chr17 | 55314365 | 55315247 | 883 | 4 | 2.30E-06 | 1.55E-05 | 4.29E-05 | 3.23E-06 | -0.00461619 | HLF |
| chr4 | 75034905 | 75035421 | 517 | 3 | 2.29E-06 | 2.43E-06 | 0.000147 | 3.23E-06 | -0.00534396 | PARM1 |
| chr3 | 183697336 | 183698280 | 945 | 7 | 1.36E-05 | 4.48E-06 | 0.001752 | 3.23E-06 | 0.00231984 | YEATS2 |
| chr9 | 91775227 | 91776185 | 959 | 3 | 2.79E-07 | 7.06E-05 | 9.47E-06 | 3.23E-06 | -0.00559135 | ROR2 |
| chr6 | 73461627 | 73462019 | 393 | 10 | 9.86E-08 | 0.0002213 | 1.55E-06 | 3.23E-06 | -0.00057181 | MTO1 |
| chr19 | 49451737 | 49452609 | 873 | 6 | 8.31E-07 | 5.96E-06 | 0.000706 | 3.23E-06 | -0.00360943 | PIH1D1 |
| chr18 | 3879595 | 3880558 | 964 | 7 | 1.25E-06 | 9.56E-06 | 0.000373 | 3.24E-06 | -0.00318297 | DLGAP1-AS3, DLGAP1 |
| chr9 | 128294458 | 128294484 | 27 | 2 | 9.41E-07 | 3.28E-05 | 5.78E-06 | 3.24E-06 | -0.00687779 | SWI5 |
| chr10 | 62555037 | 62555105 | 69 | 2 | 4.55E-06 | 7.10E-06 | 1.85E-05 | 3.24E-06 | -0.00887101 | AC024598.1, AC067752.1 |
| chr6 | 37691836 | 37692627 | 792 | 4 | 9.65E-06 | 1.55E-06 | 0.00135 | 3.24E-06 | -0.01289983 | MDGA1 |
| chr3 | 197519966 | 197520349 | 384 | 4 | 7.23E-06 | 5.11E-06 | 0.001569 | 3.24E-06 | -0.00117645 | BDH1 |
| chr14 | 105136816 | 105137123 | 308 | 3 | 1.01E-06 | 2.35E-05 | 1.63E-05 | 3.24E-06 | -0.01160758 | AL512356.5 |
| chr15 | 95326689 | 95327761 | 1073 | 11 | 2.02E-05 | 6.78E-05 | 0.001155 | 3.24E-06 | 0.001726765 | LINC00924, LINC01197 |
| chr17 | 82882945 | 82883258 | 314 | 2 | 1.68E-07 | 1.63E-05 | 8.95E-06 | 3.24E-06 | 0.007477392 | TBCD |
| chr1 | 111747034 | 111747369 | 336 | 2 | 4.22E-06 | 2.96E-06 | 6.65E-05 | 3.24E-06 | -0.00540393 | INKA2-AS1, INKA2, AL049557.1 |
| chr1 | 15874023 | 15874046 | 24 | 2 | 7.69E-07 | 5.78E-06 | 2.35E-05 | 3.24E-06 | -0.01349368 | SPEN |
| chr4 | 7985346 | 7986107 | 762 | 4 | 6.41E-07 | 8.11E-06 | 0.000369 | 3.24E-06 | 0.006221964 | ABLIM2 |
| chr11 | 72676989 | 72677238 | 250 | 3 | 1.42E-06 | 6.75E-06 | 0.000334 | 3.24E-06 | -0.00249349 | NA |
| chr1 | 38047573 | 38047969 | 397 | 4 | 8.96E-08 | 6.74E-05 | 4.61E-05 | 3.24E-06 | -0.00680398 | MIR3659HG |
| chr10 | 46000847 | 46001541 | 695 | 3 | 3.85E-07 | 1.21E-05 | 7.54E-05 | 3.24E-06 | -0.00162308 | TIMM23 |
| chr12 | 114933609 | 114934088 | 480 | 3 | 1.49E-06 | 1.47E-05 | 0.000233 | 3.24E-06 | -0.01266454 | NA |
| chr2 | 129071839 | 129071885 | 47 | 3 | 2.49E-06 | 3.58E-05 | 0.00011 | 3.25E-06 | -0.00160659 | NA |
| chr19 | 10459453 | 10459746 | 294 | 2 | 2.96E-07 | 0.000154 | 3.20E-06 | 3.25E-06 | -0.00328665 | PDE4A |
| chr1 | 54390504 | 54390742 | 239 | 2 | 1.71E-06 | 2.50E-05 | 6.78E-06 | 3.25E-06 | -0.00320848 | SSBP3 |
| chr5 | 2415731 | 2415911 | 181 | 3 | 6.62E-06 | 2.93E-06 | 0.00014 | 3.25E-06 | 0.023301871 | NA |
| chr2 | 98454331 | 98454566 | 236 | 2 | 5.50E-07 | 2.65E-06 | 8.74E-05 | 3.25E-06 | -0.01184225 | INPP4A |
| chr17 | 1878961 | 1879390 | 430 | 4 | 1.07E-05 | 1.72E-06 | 0.001934 | 3.26E-06 | 5.81E-05 | RPA1 |
| chr14 | 76777471 | 76777972 | 502 | 4 | 8.46E-07 | 4.75E-06 | 0.000561 | 3.26E-06 | -0.01065268 | VASH1, AF111169.3 |
| chr10 | 128860886 | 128860981 | 96 | 2 | 4.77E-06 | 2.48E-06 | 0.000106 | 3.26E-06 | -0.0109965 | NA |
| chr16 | 67931853 | 67932718 | 866 | 11 | 5.56E-06 | 4.73E-05 | 0.002815 | 3.26E-06 | 0.000280299 | CTRL, AC040162.1 |
| chr22 | 45972145 | 45973573 | 1429 | 7 | 7.58E-07 | 3.66E-05 | 0.000165 | 3.27E-06 | 0.007615739 | WNT7B |
| chr8 | 141374780 | 141374801 | 22 | 2 | 2.63E-06 | 3.89E-05 | 5.37E-06 | 3.27E-06 | -0.00647841 | NA |
| chr1 | 10662881 | 10663573 | 693 | 3 | 4.39E-07 | 1.85E-05 | 1.18E-05 | 3.27E-06 | -0.00572294 | CASZ1 |
| chr8 | 26389685 | 26390286 | 602 | 3 | 4.55E-06 | 2.41E-06 | 0.000155 | 3.27E-06 | -0.01071409 | BNIP3L |
| chr8 | 30383404 | 30384145 | 742 | 8 | 3.64E-07 | 6.51E-05 | 0.000104 | 3.27E-06 | 0.004698902 | RBPMS-AS1 |
| chr11 | 111698261 | 111698961 | 701 | 2 | 2.34E-05 | 2.00E-06 | 0.000301 | 3.27E-06 | -0.01425765 | SIK2 |
| chr17 | 40497557 | 40498155 | 599 | 2 | 6.53E-06 | 2.06E-06 | 0.000249 | 3.28E-06 | -0.01031316 | TNS4 |
| chr3 | 51387050 | 51387510 | 461 | 2 | 7.30E-06 | 1.96E-06 | 0.000357 | 3.28E-06 | -0.00337388 | MANF |
| chr8 | 63030957 | 63031605 | 649 | 3 | 1.34E-06 | 1.34E-05 | 0.000102 | 3.28E-06 | -0.00389982 | GGH |
| chr9 | 121282649 | 121282985 | 337 | 3 | 3.29E-06 | 3.32E-06 | 0.0001 | 3.28E-06 | -0.00950892 | GSN, AL513122.2, GSN-AS1 |
| chr12 | 4909063 | 4910333 | 1271 | 14 | 6.59E-06 | 4.21E-06 | 0.000905 | 3.28E-06 | 0.000227561 | AC005906.2, KCNA1, AC006063.4 |
| chr9 | 136171241 | 136172330 | 1090 | 3 | 1.62E-05 | 1.19E-06 | 0.001717 | 3.28E-06 | -0.01365369 | NA |
| chr9 | 127912581 | 127913159 | 579 | 4 | 9.41E-08 | 0.0001639 | 2.98E-06 | 3.28E-06 | 0.00262555 | ST6GALNAC4 |
| chr1 | 30411681 | 30412472 | 792 | 3 | 3.08E-05 | 2.34E-06 | 0.000637 | 3.28E-06 | -0.00797307 | NA |
| chr22 | 32882127 | 32882563 | 437 | 2 | 2.94E-06 | 3.57E-06 | 4.75E-05 | 3.28E-06 | -0.00775918 | SYN3 |
| chr1 | 14924773 | 14925385 | 613 | 4 | 8.31E-08 | 0.0001478 | 8.65E-06 | 3.28E-06 | -0.00707005 | KAZN |
| chr2 | 152097641 | 152098456 | 816 | 4 | 1.90E-06 | 1.21E-06 | 0.001788 | 3.28E-06 | -0.00947203 | AC079790.1, CACNB4 |
| chr14 | 100292527 | 100293478 | 952 | 5 | 9.30E-07 | 2.82E-06 | 0.000302 | 3.28E-06 | 0.000114125 | AL157871.1, SLC25A29 |
| chr4 | 86107346 | 86108188 | 843 | 4 | 2.12E-06 | 4.51E-05 | 3.03E-05 | 3.29E-06 | -0.00278224 | MAPK10 |
| chr6 | 158016624 | 158017387 | 764 | 11 | 7.19E-06 | 1.76E-05 | 0.001415 | 3.29E-06 | -0.00271821 | SYNJ2 |
| chr4 | 2464976 | 2465148 | 173 | 2 | 1.15E-06 | 4.80E-05 | 4.92E-06 | 3.29E-06 | -0.01266496 | RNF4 |
| chr22 | 46861672 | 46861762 | 91 | 2 | 4.67E-06 | 6.56E-06 | 2.07E-05 | 3.29E-06 | 0.002842285 | TBC1D22A |
| chr11 | 618993 | 619080 | 88 | 4 | 2.35E-06 | 3.59E-05 | 0.000211 | 3.29E-06 | -0.00030109 | CDHR5 |
| chr11 | 61429075 | 61430012 | 938 | 9 | 4.89E-08 | 0.0001067 | 0.000239 | 3.29E-06 | 0.000317983 | AP003108.2, CPSF7 |
| chr10 | 1172823 | 1174039 | 1217 | 7 | 7.18E-07 | 0.0001005 | 0.000695 | 3.29E-06 | -0.00488778 | LINC00200 |
| chr7 | 27160832 | 27161340 | 509 | 4 | 1.47E-06 | 8.38E-06 | 0.001114 | 3.29E-06 | -0.00063191 | NA |
| chr12 | 111599582 | 111600064 | 483 | 11 | 1.32E-05 | 1.04E-06 | 0.004928 | 3.29E-06 | 0.000762024 | ATXN2-AS, ATXN2 |
| chr11 | 71447879 | 71448859 | 981 | 9 | 1.41E-06 | 8.45E-05 | 0.000315 | 3.30E-06 | -0.00063809 | AP002387.1, DHCR7 |
| chr20 | 37902518 | 37903013 | 496 | 6 | 3.18E-06 | 5.58E-06 | 0.000418 | 3.30E-06 | 0.006123125 | NA |
| chr6 | 4950074 | 4950439 | 366 | 3 | 6.05E-07 | 1.70E-05 | 1.20E-05 | 3.30E-06 | -0.00319002 | CDYL |
| chr1 | 29245371 | 29245966 | 596 | 3 | 9.27E-07 | 9.73E-05 | 1.39E-05 | 3.30E-06 | 0.001309375 | PTPRU |
| chr7 | 97871915 | 97872596 | 682 | 10 | 3.89E-06 | 2.12E-05 | 0.001317 | 3.30E-06 | 0.000916042 | ASNS, AC079781.5 |
| chr11 | 56029096 | 56030152 | 1057 | 5 | 1.03E-05 | 0.0001228 | 0.000181 | 3.30E-06 | 0.000128689 | OR5AS1 |
| chr9 | 134536391 | 134536399 | 9 | 2 | 1.37E-06 | 4.31E-06 | 3.55E-05 | 3.30E-06 | -0.00768377 | AL669970.3 |
| chr1 | 9025699 | 9026589 | 891 | 6 | 1.19E-05 | 0.0001242 | 2.98E-05 | 3.30E-06 | 0.001362154 | SLC2A7 |
| chr10 | 45453458 | 45454472 | 1015 | 5 | 3.09E-05 | 1.09E-06 | 0.00528 | 3.31E-06 | -0.01105391 | NA |
| chr15 | 27694990 | 27695501 | 512 | 4 | 5.08E-06 | 1.58E-06 | 0.000688 | 3.31E-06 | 0.005083733 | NA |
| chr19 | 46228684 | 46229607 | 924 | 5 | 2.17E-05 | 3.24E-06 | 0.001118 | 3.31E-06 | -0.0035481 | AC006262.3 |
| chr19 | 48501266 | 48501577 | 312 | 4 | 4.62E-06 | 7.24E-06 | 0.000484 | 3.31E-06 | -0.01462261 | LMTK3 |
| chr15 | 83107519 | 83108330 | 812 | 8 | 6.79E-06 | 1.77E-06 | 0.002104 | 3.31E-06 | -0.00131244 | TM6SF1, AC024270.4 |
| chr7 | 99469493 | 99470484 | 992 | 5 | 2.28E-05 | 1.43E-05 | 0.001541 | 3.32E-06 | -0.01575018 | NA |
| chr12 | 48472336 | 48472607 | 272 | 5 | 2.40E-06 | 0.0005797 | 3.61E-05 | 3.32E-06 | 0.004462016 | ANP32D |
| chr19 | 17932601 | 17933323 | 723 | 11 | 7.73E-06 | 8.55E-06 | 0.008989 | 3.32E-06 | 0.004203779 | CCDC124 |
| chr1 | 64895206 | 64895954 | 749 | 4 | 2.41E-05 | 1.32E-06 | 0.002321 | 3.32E-06 | 0.003722829 | JAK1 |
| chr13 | 48000993 | 48001791 | 799 | 13 | 5.19E-07 | 0.0003549 | 0.00025 | 3.32E-06 | -0.00340522 | SUCLA2-AS1, SUCLA2 |
| chr6 | 75085279 | 75085963 | 685 | 3 | 3.53E-06 | 1.26E-06 | 0.001521 | 3.32E-06 | 0.01085384 | COL12A1 |
| chr10 | 123135427 | 123136613 | 1187 | 13 | 1.65E-06 | 0.0010097 | 0.000815 | 3.32E-06 | 0.001434855 | HMX3 |
| chr17 | 75128543 | 75129058 | 516 | 3 | 1.21E-06 | 0.0007776 | 0.000135 | 3.33E-06 | -0.00360192 | ARMC7 |
| chr17 | 68247443 | 68248055 | 613 | 10 | 3.02E-06 | 2.27E-05 | 0.005424 | 3.33E-06 | 0.000145374 | AMZ2, AC005332.1 |
| chr11 | 2575070 | 2575238 | 169 | 2 | 4.25E-07 | 0.0001333 | 3.42E-06 | 3.33E-06 | -0.00338434 | KCNQ1 |
| chr2 | 173075027 | 173076317 | 1291 | 12 | 5.94E-07 | 7.04E-05 | 0.002443 | 3.33E-06 | -0.00165793 | MAP3K20 |
| chr4 | 78542412 | 78542910 | 499 | 2 | 3.39E-07 | 3.89E-05 | 5.49E-06 | 3.33E-06 | -0.00280275 | FRAS1 |
| chr6 | 165665325 | 165665355 | 31 | 3 | 4.30E-06 | 1.41E-05 | 0.000125 | 3.33E-06 | -0.00735688 | PDE10A |
| chr3 | 114309012 | 114309039 | 28 | 2 | 7.06E-06 | 2.39E-06 | 0.000132 | 3.33E-06 | -0.01536427 | TIGIT |
| chr11 | 83292944 | 83292967 | 24 | 2 | 3.63E-06 | 1.71E-05 | 8.99E-06 | 3.33E-06 | -0.01901028 | AP000446.1 |
| chr7 | 48088328 | 48089165 | 838 | 16 | 7.46E-07 | 0.0009165 | 0.000924 | 3.33E-06 | -0.00111285 | UPP1 |
| chr8 | 52009080 | 52009691 | 612 | 3 | 2.81E-07 | 0.0003722 | 4.40E-05 | 3.34E-06 | -0.01260234 | AC064807.2 |
| chr3 | 15686091 | 15686464 | 374 | 4 | 7.16E-06 | 2.23E-06 | 0.000193 | 3.34E-06 | -0.00384212 | BTD, ANKRD28 |
| chr17 | 39152575 | 39153319 | 745 | 7 | 1.38E-07 | 3.49E-05 | 9.01E-05 | 3.34E-06 | -0.00022032 | PLXDC1 |
| chr1 | 220045821 | 220046833 | 1013 | 13 | 2.50E-06 | 0.0003677 | 3.44E-05 | 3.34E-06 | 0.000129121 | EPRS1 |
| chr20 | 62475504 | 62476739 | 1236 | 22 | 4.48E-06 | 8.20E-05 | 0.005436 | 3.34E-06 | -0.00115122 | GATA5 |
| chr22 | 26576189 | 26576251 | 63 | 2 | 2.90E-06 | 3.07E-05 | 6.25E-06 | 3.35E-06 | -0.01566646 | TPST2 |
| chr12 | 10214659 | 10215268 | 610 | 4 | 2.05E-06 | 0.0001775 | 8.53E-05 | 3.35E-06 | -0.00831385 | GABARAPL1, GABARAPL1-AS1 |
| chr7 | 102185990 | 102186759 | 770 | 5 | 1.02E-06 | 1.49E-05 | 1.79E-05 | 3.35E-06 | -0.00166404 | CUX1 |
| chr2 | 233820740 | 233821020 | 281 | 2 | 1.07E-06 | 3.28E-06 | 5.87E-05 | 3.35E-06 | -0.00891853 | MROH2A |
| chr4 | 68404054 | 68404371 | 318 | 2 | 6.94E-07 | 8.81E-05 | 3.91E-06 | 3.35E-06 | -0.01440283 | NA |
| chr8 | 6621030 | 6621642 | 613 | 6 | 1.24E-06 | 1.47E-05 | 5.88E-05 | 3.35E-06 | 0.000493778 | MCPH1, MCPH1-AS1 |
| chr4 | 176195425 | 176196097 | 673 | 11 | 2.66E-06 | 1.47E-05 | 8.15E-05 | 3.36E-06 | -0.0010329 | SPATA4 |
| chr12 | 94173008 | 94173027 | 20 | 2 | 1.78E-06 | 4.45E-06 | 3.50E-05 | 3.36E-06 | 0.006543992 | PLXNC1, AC123567.2 |
| chr5 | 14325828 | 14326577 | 750 | 7 | 9.10E-06 | 2.22E-06 | 0.009434 | 3.36E-06 | 0.013104085 | TRIO |
| chr15 | 101174930 | 101175614 | 685 | 4 | 4.15E-06 | 4.09E-06 | 0.000519 | 3.36E-06 | 0.002510722 | NA |
| chr10 | 125718700 | 125719996 | 1297 | 14 | 1.62E-05 | 4.51E-05 | 0.005458 | 3.36E-06 | -0.00163387 | EDRF1, EDRF1-DT, AL158835.3, AL158835.2 |

| | | | | | | | | | | |
|---|---|---|---|---|---|---|---|---|---|---|
| chr11 | 18680733 | 18680902 | 170 | 2 | 3.64E-06 | 8.80E-06 | 1.58E-05 | 3.36E-06 | -0.00690654 | NA |
| chr9 | 131429415 | 131430245 | 831 | 7 | 6.00E-06 | 4.61E-06 | 0.000691 | 3.36E-06 | 0.002100114 | PRRC2B |
| chr9 | 126514720 | 126515384 | 665 | 4 | 2.45E-06 | 6.16E-06 | 0.00037 | 3.37E-06 | 0.001443519 | NA |
| chr12 | 123037956 | 123037972 | 17 | 2 | 6.14E-06 | 3.99E-06 | 4.15E-05 | 3.37E-06 | 0.000567493 | PITPNM2 |
| chr15 | 70763365 | 70764150 | 786 | 12 | 4.41E-06 | 8.82E-06 | 0.001931 | 3.37E-06 | 0.000596222 | UACA |
| chr12 | 52519747 | 52520817 | 1071 | 9 | 4.54E-07 | 0.0002626 | 0.000111 | 3.37E-06 | 0.002559163 | KRT5 |
| chr1 | 21783064 | 21784550 | 1487 | 13 | 1.31E-05 | 0.0008008 | 0.004842 | 3.37E-06 | -0.00214627 | USP48 |
| chr16 | 22614285 | 22614878 | 594 | 6 | 7.29E-07 | 6.71E-05 | 4.25E-05 | 3.37E-06 | -0.00281435 | NA |
| chr2 | 120984554 | 120985265 | 712 | 4 | 1.03E-05 | 9.34E-05 | 0.000851 | 3.37E-06 | -0.00148426 | GLI2 |
| chr17 | 1347211 | 1347896 | 686 | 2 | 1.95E-05 | 2.09E-06 | 0.000276 | 3.37E-06 | -0.00201743 | YWHAE |
| chr16 | 84117069 | 84117695 | 627 | 7 | 1.09E-06 | 1.84E-05 | 5.88E-05 | 3.37E-06 | -0.00306264 | AC040169.1 |
| chr5 | 77869323 | 77869852 | 530 | 3 | 4.54E-06 | 2.21E-06 | 0.000619 | 3.37E-06 | 0.001394418 | NA |
| chr22 | 26796396 | 26797395 | 1000 | 4 | 1.52E-05 | 1.00E-05 | 0.000777 | 3.38E-06 | -0.00424057 | NA |
| chr16 | 19698081 | 19699292 | 1212 | 5 | 1.97E-08 | 0.0013942 | 5.89E-07 | 3.38E-06 | -0.00307808 | VPS35L |
| chr15 | 96039010 | 96039165 | 156 | 2 | 2.18E-06 | 3.87E-05 | 5.60E-06 | 3.38E-06 | 0.009274297 | AC012409.2 |
| chr1 | 37989664 | 37990315 | 652 | 14 | 2.91E-06 | 2.63E-05 | 0.004139 | 3.38E-06 | 0.001035996 | SF3A3 |
| chr12 | 130752892 | 130753163 | 272 | 4 | 7.98E-06 | 2.15E-06 | 0.000554 | 3.38E-06 | -0.01155214 | NA |
| chr16 | 66366026 | 66366696 | 671 | 10 | 1.81E-06 | 0.0004245 | 0.000377 | 3.38E-06 | -0.00131886 | CDH5 |
| chr17 | 42459575 | 42459977 | 403 | 2 | 2.24E-07 | 8.25E-06 | 1.69E-05 | 3.38E-06 | -0.01178601 | ATP6V0A1 |
| chr6 | 155147870 | 155148396 | 527 | 3 | 1.92E-06 | 2.59E-06 | 0.000595 | 3.38E-06 | 0.001695389 | TIAM2 |
| chr16 | 86252317 | 86252871 | 555 | 2 | 7.55E-06 | 2.30E-06 | 0.000168 | 3.39E-06 | -0.01093583 | LINC01081 |
| chr10 | 5449197 | 5450060 | 864 | 3 | 2.96E-07 | 5.46E-05 | 4.84E-05 | 3.39E-06 | -0.02072189 | NET1 |
| chr11 | 1499401 | 1500383 | 983 | 7 | 2.35E-06 | 4.14E-05 | 0.000123 | 3.39E-06 | -0.00575888 | MOB2 |
| chr4 | 52712144 | 52713186 | 1043 | 13 | 1.13E-06 | 5.02E-05 | 0.000749 | 3.39E-06 | 0.00103139 | DANCR, LINC01618, MIR4449, AC104066.4 |
| chr4 | 79325559 | 79326474 | 916 | 10 | 4.54E-06 | 6.38E-05 | 4.93E-05 | 3.39E-06 | -4.48E-05 | NAA11 |
| chr10 | 43648358 | 43649288 | 931 | 12 | 9.46E-08 | 0.0009201 | 2.30E-06 | 3.39E-06 | 0.001517648 | ZNF32-AS3, ZNF32 |
| chr2 | 105811489 | 105812293 | 805 | 2 | 2.15E-05 | 2.03E-06 | 0.000364 | 3.39E-06 | 0.00049982 | NCK2 |
| chr10 | 101569583 | 101570452 | 870 | 7 | 7.39E-06 | 1.64E-06 | 0.000887 | 3.40E-06 | 0.002857397 | NA |
| chr13 | 26010141 | 26010824 | 684 | 2 | 2.93E-05 | 2.05E-06 | 0.000343 | 3.40E-06 | -0.00691868 | ATP8A2 |
| chr21 | 46393110 | 46393576 | 467 | 5 | 1.42E-05 | 1.19E-06 | 0.001964 | 3.40E-06 | -0.00440181 | PCNT |
| chr14 | 56578874 | 56579757 | 884 | 9 | 3.12E-06 | 2.45E-05 | 0.000672 | 3.40E-06 | 0.000754814 | TMEM260 |
| chr22 | 35311018 | 35311295 | 278 | 2 | 2.37E-07 | 0.0022286 | 2.25E-06 | 3.40E-06 | -0.00773803 | TOM1 |
| chr20 | 45338627 | 45339296 | 670 | 4 | 1.67E-07 | 0.0001424 | 0.000173 | 3.40E-06 | -0.00595722 | SDC4 |
| chr17 | 10026211 | 10026877 | 667 | 9 | 4.02E-06 | 1.54E-05 | 0.001163 | 3.40E-06 | -0.00502591 | GAS7 |
| chr2 | 51581819 | 51581885 | 67 | 2 | 8.05E-06 | 2.07E-06 | 0.00032 | 3.40E-06 | -0.01107679 | AC007402.1 |
| chr10 | 99234624 | 99236340 | 1717 | 10 | 1.31E-05 | 0.0003623 | 0.000145 | 3.40E-06 | 0.000227224 | HPSE2 |
| chr16 | 46836475 | 46836518 | 44 | 2 | 7.94E-06 | 2.16E-06 | 0.000245 | 3.40E-06 | 0.010714797 | NA |
| chr5 | 146342569 | 146343346 | 778 | 8 | 4.44E-06 | 2.88E-06 | 0.001346 | 3.41E-06 | -0.00041442 | AC011396.2 |
| chr12 | 52870756 | 52871306 | 551 | 4 | 6.73E-06 | 1.55E-06 | 0.001363 | 3.41E-06 | -0.01397426 | NA |
| chr7 | 127615265 | 127615916 | 652 | 5 | 2.18E-06 | 1.34E-05 | 0.000143 | 3.41E-06 | -0.01232399 | PAX4 |
| chr2 | 45778914 | 45779232 | 319 | 4 | 3.48E-06 | 5.73E-06 | 6.05E-05 | 3.41E-06 | -0.00783801 | PRKCE |
| chr1 | 207910910 | 207911698 | 789 | 14 | 2.42E-05 | 1.13E-06 | 0.015374 | 3.41E-06 | 0.001621793 | AL356275.1, CD34 |
| chr6 | 49532098 | 49532856 | 759 | 3 | 5.21E-06 | 2.67E-06 | 0.0005 | 3.41E-06 | -0.00389799 | NA |
| chr5 | 2135749 | 2136063 | 315 | 2 | 1.14E-06 | 2.22E-06 | 0.000216 | 3.42E-06 | -0.01473544 | NA |
| chr19 | 9092627 | 9093580 | 954 | 6 | 6.98E-08 | 0.0003487 | 5.93E-05 | 3.42E-06 | -0.00641332 | OR1M1 |
| chr9 | 136059802 | 136060137 | 336 | 2 | 2.49E-07 | 3.74E-05 | 5.75E-06 | 3.42E-06 | -0.01103893 | NACC2 |
| chr17 | 12019945 | 12019972 | 28 | 2 | 2.22E-07 | 2.24E-06 | 0.000202 | 3.42E-06 | -0.01086633 | AC005410.1 |
| chr8 | 142742316 | 142743168 | 853 | 11 | 4.46E-06 | 4.03E-06 | 0.004014 | 3.42E-06 | -0.00296738 | SLURP1 |
| chr8 | 29070755 | 29071431 | 677 | 2 | 0.000115983 | 2.05E-06 | 0.000368 | 3.42E-06 | 0.004212428 | KIF13B |
| chr7 | 2517328 | 2518462 | 1135 | 11 | 2.32E-06 | 0.0020203 | 0.000498 | 3.43E-06 | -0.0017423 | LFNG |
| chr4 | 183676441 | 183677054 | 614 | 2 | 2.12E-06 | 4.46E-06 | 3.63E-05 | 3.43E-06 | -0.0202959 | TRAPPC11 |
| chr20 | 18095647 | 18095683 | 37 | 2 | 1.38E-06 | 7.98E-05 | 4.15E-06 | 3.43E-06 | -0.00853332 | NA |
| chr9 | 83281720 | 83282360 | 641 | 4 | 8.41E-06 | 2.21E-06 | 0.00117 | 3.43E-06 | -0.00814218 | FRMD3 |
| chr22 | 21665228 | 21666268 | 1041 | 10 | 6.47E-08 | 0.002604 | 1.73E-05 | 3.43E-06 | 0.003587039 | AP000553.1, PPIL2 |
| chr7 | 47536982 | 47537492 | 511 | 4 | 3.93E-06 | 1.52E-06 | 0.001056 | 3.43E-06 | -0.00338659 | TNS3 |
| chr5 | 44723895 | 44723977 | 83 | 2 | 4.01E-06 | 2.05E-06 | 0.000366 | 3.43E-06 | -0.01544204 | NA |
| chr6 | 166470529 | 166470942 | 414 | 2 | 2.25E-06 | 3.86E-06 | 4.61E-05 | 3.43E-06 | 0.001291705 | RPS6KA2 |
| chr21 | 32021801 | 32022572 | 772 | 5 | 4.66E-05 | 9.93E-07 | 0.004475 | 3.43E-06 | -0.00668735 | HUNK |
| chr10 | 113119211 | 113119904 | 694 | 5 | 4.08E-07 | 0.0001519 | 2.55E-05 | 3.44E-06 | -0.00210226 | TCF7L2 |
| chr2 | 135531880 | 135532182 | 303 | 5 | 7.46E-07 | 1.08E-05 | 0.001341 | 3.44E-06 | 0.001604575 | R3HDM1 |
| chr16 | 1958722 | 1959359 | 638 | 9 | 5.69E-07 | 5.19E-05 | 0.000196 | 3.44E-06 | -0.00561695 | NA |
| chr15 | 22942454 | 22943181 | 728 | 5 | 3.20E-06 | 6.48E-06 | 0.000429 | 3.44E-06 | -0.00095744 | CYFIP1 |
| chr2 | 118858666 | 118859552 | 887 | 5 | 9.54E-06 | 1.78E-05 | 0.000348 | 3.44E-06 | -0.00468512 | NA |
| chr1 | 145926778 | 145927977 | 1200 | 14 | 7.88E-07 | 4.16E-06 | 0.000916 | 3.44E-06 | 0.001340686 | LIX1L-AS1, AC243547.3, RBM8A |
| chr11 | 73948819 | 73950184 | 1366 | 6 | 8.89E-07 | 3.03E-05 | 0.000186 | 3.44E-06 | -0.00725295 | NA |
| chr6 | 12335620 | 12336026 | 407 | 2 | 4.10E-06 | 2.11E-06 | 0.000298 | 3.44E-06 | 0.001062233 | NA |
| chr11 | 5107147 | 5107652 | 506 | 2 | 8.15E-06 | 2.06E-06 | 0.000359 | 3.44E-06 | 0.000638487 | AC113331.1 |
| chr5 | 1886714 | 1887895 | 1182 | 16 | 1.31E-05 | 1.59E-05 | 0.002101 | 3.44E-06 | 0.000114222 | CTD-2194D22.4, IRX4 |
| chr9 | 16120876 | 16121844 | 969 | 6 | 4.16E-05 | 5.46E-06 | 0.005047 | 3.44E-06 | -0.00116233 | NA |
| chr7 | 130921469 | 130921786 | 318 | 4 | 6.73E-06 | 2.39E-06 | 0.000417 | 3.44E-06 | -0.00082782 | LINC00513, AC016831.6 |
| chr21 | 44304587 | 44305258 | 672 | 6 | 2.19E-07 | 0.0007452 | 1.89E-06 | 3.44E-06 | -0.00648558 | PFKL |
| chr12 | 68844471 | 68844528 | 58 | 2 | 3.92E-06 | 4.60E-06 | 3.51E-05 | 3.44E-06 | -0.00855508 | MDM2, CPM |
| chr12 | 123966748 | 123967238 | 491 | 2 | 4.06E-06 | 3.00E-06 | 7.65E-05 | 3.45E-06 | -0.01118868 | CCDC92 |
| chr4 | 88284448 | 88285458 | 1011 | 12 | 5.02E-06 | 5.81E-05 | 0.003777 | 3.45E-06 | 0.00184167 | PPM1K-DT, PPM1K |
| chr13 | 112837955 | 112838462 | 508 | 4 | 2.53E-06 | 1.41E-06 | 0.001998 | 3.45E-06 | 0.002790012 | ATP11A |
| chr2 | 118371537 | 118371920 | 384 | 2 | 2.28E-06 | 5.50E-06 | 2.79E-05 | 3.45E-06 | -0.01142413 | NA |
| chr4 | 174828958 | 174829850 | 893 | 13 | 2.14E-06 | 1.55E-05 | 0.000522 | 3.46E-06 | -0.00213696 | ADAM29, GLRA3 |
| chr22 | 32533619 | 32533676 | 58 | 2 | 9.72E-07 | 0.0003328 | 2.90E-06 | 3.46E-06 | -0.00688594 | SYN3 |
| chr10 | 132753071 | 132754255 | 1185 | 9 | 4.92E-07 | 3.83E-05 | 5.07E-05 | 3.46E-06 | -0.00026693 | INPP5A |
| chr12 | 13877446 | 13877503 | 58 | 2 | 4.44E-06 | 1.03E-05 | 1.42E-05 | 3.46E-06 | 0.001538743 | GRIN2B |
| chr16 | 1399307 | 1399938 | 632 | 6 | 2.09E-06 | 3.63E-06 | 0.001749 | 3.46E-06 | -0.00071864 | UNKL |
| chr16 | 87125321 | 87125668 | 348 | 3 | 2.91E-06 | 1.52E-06 | 0.00114 | 3.46E-06 | -0.00415245 | C16orf95 |
| chr3 | 112033998 | 112034837 | 840 | 4 | 7.53E-06 | 5.53E-06 | 0.001678 | 3.46E-06 | -0.00418532 | NA |
| chr15 | 81279024 | 81279712 | 689 | 2 | 2.78E-05 | 2.07E-06 | 0.000366 | 3.46E-06 | -0.00676666 | IL16 |
| chr19 | 57727067 | 57728304 | 1238 | 13 | 3.16E-06 | 1.15E-05 | 0.002915 | 3.46E-06 | -0.00035005 | AC003006.1, ZNF671 |
| chr5 | 190707 | 191988 | 1282 | 14 | 2.03E-05 | 6.20E-06 | 0.009716 | 3.46E-06 | -0.00122047 | LRRC14B |
| chr1 | 3425380 | 3425743 | 364 | 3 | 1.78E-06 | 1.34E-06 | 0.001461 | 3.46E-06 | 0.000290405 | PRDM16 |
| chr17 | 1684594 | 1685151 | 558 | 10 | 1.89E-06 | 3.36E-05 | 0.000937 | 3.46E-06 | 0.000490903 | AC130343.3, PRPF8 |
| chr12 | 15151103 | 15151395 | 293 | 2 | 5.95E-06 | 2.57E-06 | 0.00012 | 3.47E-06 | 0.007223167 | RERG |
| chr19 | 33235749 | 33236719 | 971 | 5 | 1.99E-06 | 1.38E-05 | 4.71E-05 | 3.47E-06 | -0.00751287 | NA |
| chr9 | 131478012 | 131479052 | 1041 | 3 | 1.09E-08 | 0.0014057 | 1.23E-06 | 3.47E-06 | -0.00837878 | PRRC2B |
| chr10 | 129415945 | 129417205 | 1261 | 5 | 5.10E-06 | 4.14E-05 | 0.000379 | 3.47E-06 | -0.00201317 | NA |
| chr17 | 75153909 | 75154857 | 949 | 12 | 1.16E-05 | 6.31E-06 | 0.012419 | 3.47E-06 | 0.000429989 | JPT1 |
| chr17 | 55284447 | 55285261 | 815 | 2 | 1.28E-05 | 2.38E-06 | 0.000166 | 3.47E-06 | -0.00126643 | HLF |
| chr6 | 71411648 | 71412144 | 497 | 2 | 9.51E-07 | 8.34E-06 | 1.75E-05 | 3.47E-06 | -0.00393756 | AL136164.2, LINC00472 |
| chr8 | 2022465 | 2023318 | 854 | 5 | 4.39E-06 | 4.04E-06 | 0.001097 | 3.48E-06 | -0.01092726 | NA |
| chr8 | 51256762 | 51257014 | 253 | 2 | 3.18E-07 | 1.70E-05 | 9.56E-06 | 3.48E-06 | -0.00722909 | NA |
| chr16 | 84176735 | 84176936 | 202 | 2 | 4.63E-07 | 7.45E-05 | 4.33E-06 | 3.48E-06 | 0.00240749 | DNAAF1 |
| chr13 | 112768954 | 112769964 | 1011 | 5 | 2.62E-07 | 1.87E-05 | 0.000187 | 3.48E-06 | -0.00235981 | ATP11A |
| chr20 | 17227052 | 17228786 | 1735 | 6 | 4.74E-07 | 2.77E-05 | 0.001177 | 3.48E-06 | 0.002969532 | PCSK2 |
| chr7 | 27156936 | 27158810 | 1875 | 16 | 4.87E-05 | 1.23E-06 | 0.020435 | 3.48E-06 | -0.00525278 | AC004080.4, HOXA7 |
| chr3 | 71893348 | 71893701 | 354 | 2 | 1.97E-06 | 8.67E-06 | 1.69E-05 | 3.48E-06 | -0.01138538 | NA |
| chr1 | 23531040 | 23531809 | 770 | 10 | 4.98E-07 | 0.0001349 | 0.000218 | 3.48E-06 | 0.000948137 | E2F2 |
| chr19 | 47108057 | 47108183 | 127 | 2 | 2.47E-06 | 2.14E-06 | 0.000307 | 3.49E-06 | 0.008860571 | ZC3H4 |
| chr7 | 75966703 | 75966889 | 187 | 4 | 9.00E-07 | 0.0002723 | 4.08E-05 | 3.49E-06 | 0.002062254 | POR |
| chr20 | 57109373 | 57109708 | 336 | 2 | 1.50E-07 | 0.0003746 | 2.86E-06 | 3.49E-06 | -0.01681204 | NA |
| chr16 | 2291441 | 2291627 | 187 | 2 | 7.28E-07 | 0.0010396 | 2.46E-06 | 3.49E-06 | -0.0038548 | ABCA3 |
| chr6 | 24799353 | 24799529 | 177 | 3 | 1.97E-05 | 2.14E-06 | 0.000401 | 3.49E-06 | 0.004671514 | AL512428.1 |
| chr22 | 29663522 | 29663737 | 216 | 2 | 7.81E-06 | 2.09E-06 | 0.000369 | 3.49E-06 | -0.0190122 | NF2 |
| chr10 | 61001742 | 61002095 | 354 | 5 | 2.18E-06 | 8.58E-06 | 0.000102 | 3.50E-06 | -0.00598658 | NA |
| chr1 | 223123648 | 223123832 | 185 | 2 | 5.47E-06 | 3.91E-06 | 4.68E-05 | 3.50E-06 | 0.009037815 | TLR5 |
| chr12 | 13739768 | 13739862 | 95 | 2 | 4.59E-06 | 8.69E-06 | 1.70E-05 | 3.50E-06 | 0.009147741 | GRIN2B |
| chr16 | 87662175 | 87662981 | 807 | 3 | 4.45E-05 | 2.37E-06 | 0.000409 | 3.50E-06 | -0.00298393 | JPH3 |
| chr7 | 92591268 | 92591571 | 304 | 3 | 1.26E-07 | 5.57E-05 | 8.00E-06 | 3.50E-06 | -0.00279819 | NA |
| chr8 | 64579349 | 64580379 | 1031 | 13 | 2.31E-05 | 1.70E-05 | 0.004696 | 3.50E-06 | -0.00204773 | BHLHE22, AC090136.3 |
| chr3 | 50292419 | 50293670 | 1252 | 15 | 4.95E-07 | 9.14E-06 | 0.001078 | 3.50E-06 | 0.002181468 | IFRD2, HYAL3 |
| chr5 | 113434506 | 113435024 | 519 | 6 | 6.30E-06 | 1.06E-05 | 0.0024 | 3.50E-06 | -0.00194397 | AC079465.1, MCC, TSSK1B |
| chr19 | 30444270 | 30445198 | 929 | 4 | 9.91E-08 | 1.92E-05 | 2.64E-05 | 3.50E-06 | -0.00287667 | ZNF536 |
| chr3 | 8997075 | 8997152 | 78 | 2 | 7.61E-06 | 2.59E-06 | 0.000123 | 3.50E-06 | -0.01161003 | SRGAP3 |
| chr20 | 34311620 | 34312476 | 857 | 8 | 4.23E-06 | 1.64E-05 | 0.001056 | 3.51E-06 | 0.00213482 | AHCY |
| chr12 | 112575559 | 112575903 | 345 | 3 | 1.94E-06 | 3.66E-06 | 8.02E-05 | 3.51E-06 | -0.0083705 | RPH3A |
| chr1 | 22216317 | 22216651 | 335 | 2 | 8.54E-06 | 2.13E-06 | 0.000339 | 3.51E-06 | -0.01549258 | NA |
| chr11 | 66277579 | 66278654 | 1076 | 16 | 1.13E-05 | 2.26E-05 | 0.006551 | 3.51E-06 | 0.000551816 | CNIH2, AP001107.1 |
| chr15 | 92883756 | 92884508 | 753 | 7 | 3.88E-06 | 1.53E-05 | 0.002308 | 3.51E-06 | 0.003791755 | CHASERR, AC013394.1 |
| chr1 | 168279704 | 168280529 | 826 | 3 | 1.03E-07 | 2.55E-05 | 9.94E-06 | 3.51E-06 | 0.000108722 | NA |
| chr14 | 52548609 | 52548927 | 319 | 2 | 2.94E-06 | 6.01E-06 | 2.57E-05 | 3.51E-06 | -0.01534464 | TXNDC16 |
| chr4 | 70721723 | 70722597 | 875 | 8 | 6.29E-07 | 0.0001895 | 0.00016 | 3.52E-06 | 0.00110826 | RUFY3 |
| chr13 | 110070931 | 110071266 | 336 | 2 | 5.71E-06 | 2.68E-06 | 0.000111 | 3.52E-06 | 0.009555131 | AL390755.1 |
| chr15 | 64162628 | 64163863 | 1236 | 10 | 1.34E-06 | 8.41E-05 | 0.000334 | 3.52E-06 | 0.001087238 | PPIB |
| chr19 | 18903364 | 18903946 | 583 | 3 | 6.11E-06 | 1.43E-06 | 0.001384 | 3.53E-06 | 0.003010119 | COPE |
| chr1 | 161231331 | 161231410 | 80 | 3 | 2.24E-07 | 0.0014231 | 2.10E-06 | 3.53E-06 | 0.005738774 | NR1I3 |
| chr6 | 169641324 | 169641932 | 609 | 5 | 4.16E-06 | 2.43E-05 | 0.000292 | 3.54E-06 | -0.0019421 | AL031315.1, WDR27 |
| chr9 | 137180771 | 137180839 | 69 | 2 | 5.22E-06 | 2.16E-06 | 0.000322 | 3.54E-06 | 0.001331985 | ANAPC2 |
| chr5 | 75511126 | 75512378 | 1253 | 14 | 1.32E-06 | 3.02E-05 | 0.001516 | 3.54E-06 | 0.000905147 | POLK, CERT1 |
| chr4 | 56757616 | 56757747 | 132 | 2 | 4.30E-06 | 3.74E-06 | 5.21E-05 | 3.54E-06 | 0.02296807 | NA |
| chr16 | 912818 | 913219 | 402 | 5 | 2.22E-07 | 0.0019235 | 9.18E-05 | 3.54E-06 | -0.00382792 | LMF1 |
| chr8 | 48950734 | 48951240 | 507 | 2 | 5.67E-07 | 9.36E-06 | 1.62E-05 | 3.54E-06 | 0.005935837 | NA |
| chr11 | 70269553 | 70270008 | 456 | 3 | 1.19E-07 | 1.13E-05 | 2.00E-05 | 3.54E-06 | -0.00421872 | NA |
| chr4 | 97366509 | 97366911 | 403 | 3 | 3.60E-06 | 8.70E-05 | 0.000168 | 3.54E-06 | -0.0007453 | STPG2-AS1, STPG2, AC034154.1 |
| chr5 | 152433374 | 152433520 | 147 | 2 | 4.19E-06 | 3.31E-06 | 6.65E-05 | 3.55E-06 | -0.01084342 | AC008571.2 |
| chr12 | 12449038 | 12449940 | 903 | 4 | 7.27E-06 | 5.62E-06 | 0.000182 | 3.55E-06 | -0.00668202 | BORCS5 |
| chr5 | 67350444 | 67350887 | 444 | 3 | 5.73E-06 | 2.09E-06 | 0.000282 | 3.55E-06 | -0.02196672 | NA |
| chr17 | 36679976 | 36680654 | 679 | 3 | 2.42E-05 | 1.58E-06 | 0.001147 | 3.55E-06 | -0.00816458 | NA |
| chr4 | 3034407 | 3034512 | 106 | 2 | 1.98E-06 | 0.0001186 | 3.77E-06 | 3.55E-06 | -0.01281033 | GRK4 |
| chr17 | 18263861 | 18264522 | 662 | 5 | 8.98E-07 | 5.03E-06 | 0.000343 | 3.55E-06 | -0.00232615 | MIEF2 |
| chr17 | 48614458 | 48615497 | 1040 | 15 | 1.30E-06 | 2.45E-05 | 0.001718 | 3.56E-06 | -0.0023996 | HOXB7, HOXB8 |
| chr22 | 37259574 | 37259734 | 161 | 2 | 6.34E-06 | 2.89E-06 | 9.22E-05 | 3.56E-06 | -0.00842917 | NA |
| chr19 | 39314946 | 39316527 | 1582 | 10 | 1.65E-05 | 5.16E-06 | 0.001486 | 3.56E-06 | -0.00471726 | LRFN1 |
| chr11 | 8677196 | 8677992 | 797 | 4 | 9.01E-07 | 0.0001431 | 0.000142 | 3.56E-06 | -0.00240468 | TRIM66 |
| chr12 | 101358514 | 101359039 | 526 | 3 | 4.52E-07 | 2.70E-05 | 0.000184 | 3.56E-06 | -0.00957874 | UTP20 |
| chr20 | 59933021 | 59934187 | 1167 | 11 | 7.50E-07 | 0.0001208 | 0.000519 | 3.57E-06 | 0.006297342 | FAM217B, SYCP2 |
| chr6 | 111568510 | 111568736 | 227 | 2 | 2.26E-08 | 0.0057581 | 2.24E-06 | 3.57E-06 | 0.004921177 | TRAF3IP2-AS1, TRAF3IP2 |
| chr3 | 11271762 | 11272649 | 888 | 10 | 1.43E-06 | 0.0001319 | 1.27E-05 | 3.57E-06 | 0.001808456 | ATG7 |
| chr3 | 122936416 | 122936590 | 175 | 2 | 9.45E-07 | 0.0020672 | 2.36E-06 | 3.57E-06 | -0.01026685 | SEMA5B |
| chr9 | 74834165 | 74834382 | 218 | 2 | 1.53E-06 | 6.91E-05 | 4.60E-06 | 3.57E-06 | 0.002437365 | TRPM6 |
| chr5 | 140668483 | 140668928 | 446 | 6 | 1.31E-06 | 5.94E-05 | 0.001348 | 3.58E-06 | -0.00037068 | WDR55 |
| chr1 | 153734260 | 153734602 | 343 | 2 | 7.82E-07 | 1.59E-05 | 1.05E-05 | 3.58E-06 | 0.004053364 | INTS3 |
| chr17 | 80013503 | 80013683 | 181 | 3 | 3.06E-06 | 3.06E-05 | 8.34E-06 | 3.59E-06 | -0.00374091 | TBC1D16 |

| | | | | | | | | | | |
|---|---|---|---|---|---|---|---|---|---|---|
| chr3 | 58053854 | 58053943 | 90 | 2 | 5.46E-06 | 4.33E-06 | 4.18E-05 | 3.59E-06 | -0.00225365 | FLNB |
| chr19 | 11172669 | 11173012 | 344 | 2 | 7.56E-06 | 2.20E-06 | 0.000314 | 3.59E-06 | -0.0011463 | KANK2 |
| chr1 | 1919596 | 1920799 | 1204 | 17 | 7.77E-07 | 5.92E-06 | 0.017936 | 3.59E-06 | -0.00097117 | NA |
| chr11 | 134942818 | 134942868 | 51 | 2 | 5.98E-06 | 2.26E-06 | 0.000266 | 3.59E-06 | -0.00343652 | NA |
| chr11 | 60993282 | 60993903 | 622 | 2 | 1.32E-05 | 2.15E-06 | 0.000378 | 3.59E-06 | -0.01871343 | CD6 |
| chr3 | 196215301 | 196216403 | 1103 | 7 | 4.46E-10 | 0.0248683 | 3.84E-05 | 3.59E-06 | -0.00026679 | SLC51A, PCYT1A |
| chr5 | 6737665 | 6738294 | 630 | 3 | 5.85E-06 | 1.65E-06 | 0.000896 | 3.59E-06 | -0.00765011 | TENT4A |
| chr10 | 69788916 | 69789207 | 292 | 2 | 3.58E-06 | 5.46E-06 | 3.03E-05 | 3.59E-06 | -0.01580759 | NA |
| chr10 | 123146495 | 123147944 | 1450 | 10 | 3.32E-07 | 2.30E-05 | 0.000266 | 3.60E-06 | 0.000199963 | NA |
| chr7 | 129550516 | 129551000 | 485 | 3 | 1.56E-05 | 1.45E-06 | 0.001369 | 3.60E-06 | 0.000443448 | NA |
| chr9 | 95082280 | 95082619 | 340 | 2 | 4.26E-07 | 2.34E-05 | 8.00E-06 | 3.60E-06 | -0.00300602 | AOPEP |
| chr8 | 121068138 | 121068470 | 333 | 2 | 2.66E-06 | 6.92E-06 | 2.28E-05 | 3.60E-06 | -0.00194798 | AC068413.1 |
| chr16 | 84593901 | 84595249 | 1349 | 7 | 3.24E-07 | 4.18E-05 | 5.63E-05 | 3.60E-06 | 0.001660578 | AC092145.1, COTL1 |
| chr10 | 99834172 | 99834808 | 637 | 4 | 1.98E-06 | 8.66E-05 | 3.12E-05 | 3.60E-06 | -0.00092019 | ABCC2 |
| chr7 | 28570430 | 28570854 | 425 | 2 | 1.51E-06 | 9.14E-06 | 1.69E-05 | 3.60E-06 | 0.001051576 | CREB5 |
| chr10 | 68720974 | 68722358 | 1385 | 12 | 9.07E-06 | 1.10E-06 | 0.006492 | 3.60E-06 | 0.00136687 | CCAR1 |
| chr6 | 151615991 | 151616518 | 528 | 4 | 7.02E-06 | 2.57E-05 | 0.000691 | 3.60E-06 | 0.005851038 | CCDC170 |
| chr19 | 10535826 | 10535971 | 146 | 2 | 6.64E-06 | 3.33E-06 | 6.84E-05 | 3.60E-06 | -0.00990148 | NA |
| chr3 | 39154408 | 39155478 | 1071 | 6 | 1.97E-06 | 1.56E-05 | 0.000608 | 3.61E-06 | -0.00568379 | AC092053.3, AC092053.2, CSRNP1 |
| chr7 | 142655835 | 142656526 | 692 | 2 | 1.32E-05 | 2.44E-06 | 0.000176 | 3.61E-06 | 0.007737102 | NA |
| chr5 | 61636754 | 61636961 | 208 | 2 | 4.17E-06 | 6.62E-06 | 2.40E-05 | 3.61E-06 | -0.00569227 | C5orf64 |
| chr3 | 49413472 | 49413784 | 313 | 2 | 1.92E-07 | 2.59E-05 | 7.54E-06 | 3.61E-06 | 0.005888699 | TCTA |
| chr17 | 9974245 | 9974777 | 533 | 2 | 1.97E-07 | 0.0001089 | 3.96E-06 | 3.61E-06 | 0.004050882 | GAS7 |
| chr13 | 111115676 | 111116734 | 1059 | 6 | 1.99E-05 | 5.96E-06 | 0.000554 | 3.61E-06 | 0.003110371 | ARHGEF7, ARHGEF7-AS2 |
| chr10 | 102432209 | 102432762 | 554 | 8 | 4.25E-06 | 1.18E-05 | 0.002436 | 3.61E-06 | 0.000103833 | CUEDC2 |
| chr9 | 26243627 | 26244011 | 385 | 3 | 3.06E-06 | 8.09E-06 | 0.000257 | 3.62E-06 | -0.00910386 | NA |
| chr13 | 31200385 | 31200896 | 512 | 4 | 1.53E-07 | 2.86E-05 | 9.73E-06 | 3.62E-06 | 0.001095138 | B3GLCT |
| chr22 | 17769176 | 17769675 | 500 | 2 | 1.12E-06 | 7.61E-06 | 2.06E-05 | 3.62E-06 | -0.01543941 | BID |
| chr6 | 36289402 | 36289780 | 379 | 2 | 2.85E-06 | 4.17E-06 | 4.55E-05 | 3.62E-06 | -0.0008989 | PNPLA1 |
| chr3 | 33412748 | 33412796 | 49 | 2 | 1.52E-06 | 4.76E-06 | 3.70E-05 | 3.62E-06 | -0.00297829 | UBP1 |
| chr2 | 72916405 | 72917380 | 976 | 7 | 2.20E-07 | 0.0006744 | 9.11E-05 | 3.62E-06 | 0.00062223 | EMX1 |
| chr3 | 185498523 | 185499342 | 820 | 12 | 1.83E-07 | 0.0001845 | 0.002248 | 3.63E-06 | 0.000163701 | AC099661.1, TMEM41A |
| chr14 | 50233398 | 50234109 | 712 | 3 | 2.33E-06 | 1.60E-06 | 0.000738 | 3.63E-06 | -0.0010584 | NA |
| chr22 | 33746451 | 33746522 | 72 | 2 | 6.45E-06 | 3.48E-06 | 6.37E-05 | 3.63E-06 | 0.004977449 | LARGE-AS1, LARGE1 |
| chr10 | 43077173 | 43078446 | 1274 | 6 | 5.08E-08 | 0.0015909 | 3.61E-05 | 3.63E-06 | -0.00378307 | RET |
| chr8 | 141239603 | 141240158 | 556 | 3 | 2.90E-07 | 9.80E-06 | 6.96E-05 | 3.63E-06 | -0.01142172 | SLC45A4 |
| chr2 | 74552672 | 74553490 | 819 | 9 | 5.44E-07 | 5.55E-05 | 1.21E-05 | 3.63E-06 | 0.005255781 | DOK1, LOXL3 |
| chr15 | 74422122 | 74422764 | 643 | 2 | 2.86E-05 | 2.18E-06 | 0.000381 | 3.64E-06 | -0.00957947 | SEMA7A |
| chr5 | 170434991 | 170435735 | 745 | 3 | 1.06E-05 | 2.59E-06 | 0.00035 | 3.64E-06 | -0.00163198 | KCNIP1 |
| chr20 | 21391824 | 21392225 | 402 | 4 | 1.25E-07 | 9.50E-05 | 8.09E-06 | 3.64E-06 | -0.00471347 | NA |
| chr10 | 131574570 | 131575099 | 530 | 5 | 1.30E-06 | 7.42E-05 | 1.84E-05 | 3.64E-06 | -0.00069893 | NA |
| chr3 | 71007045 | 71007400 | 356 | 2 | 1.97E-06 | 9.74E-06 | 1.62E-05 | 3.64E-06 | -0.01587917 | FOXP1, AC097634.4 |
| chr16 | 27066667 | 27067323 | 657 | 6 | 8.35E-07 | 8.47E-05 | 0.000296 | 3.65E-06 | -0.00440258 | C16orf82, AC092725.1 |
| chr12 | 21657039 | 21658119 | 1081 | 19 | 1.81E-06 | 3.74E-05 | 0.00244 | 3.65E-06 | 0.00061291 | AC010197.2, LDHB |
| chr13 | 28322678 | 28323284 | 607 | 5 | 2.87E-06 | 6.10E-06 | 0.000432 | 3.65E-06 | -0.00817231 | FLT1 |
| chr3 | 193572729 | 193572749 | 21 | 2 | 8.52E-06 | 2.39E-06 | 0.00021 | 3.65E-06 | -0.03222154 | ATP13A4 |
| chr15 | 52111832 | 52112690 | 859 | 4 | 9.12E-07 | 1.35E-05 | 0.001034 | 3.65E-06 | -0.00906179 | BCL2L10 |
| chr6 | 1417074 | 1417131 | 58 | 2 | 4.67E-06 | 9.07E-06 | 1.74E-05 | 3.65E-06 | -0.0147079 | NA |
| chr8 | 142004389 | 142005098 | 710 | 6 | 3.98E-06 | 1.90E-05 | 0.001339 | 3.66E-06 | -4.25E-05 | NA |
| chr11 | 5132272 | 5132604 | 333 | 2 | 8.37E-07 | 8.44E-06 | 1.87E-05 | 3.66E-06 | -0.00704022 | OR52A5 |
| chr17 | 40022405 | 40022588 | 184 | 2 | 4.18E-06 | 2.24E-06 | 0.000325 | 3.67E-06 | -0.02262827 | MED24 |
| chr7 | 2903689 | 2904493 | 805 | 4 | 7.62E-06 | 7.17E-06 | 9.64E-05 | 3.67E-06 | -0.00579174 | NA |
| chr2 | 217869294 | 217869671 | 378 | 2 | 4.34E-07 | 0.000207 | 3.37E-06 | 3.67E-06 | -0.0064654 | TNS1 |
| chr17 | 67577868 | 67578083 | 216 | 2 | 8.05E-06 | 2.32E-06 | 0.00026 | 3.67E-06 | 0.001248151 | PITPNC1 |
| chr6 | 32220410 | 32221833 | 1424 | 11 | 1.08E-07 | 9.85E-05 | 4.67E-05 | 3.67E-06 | 0.00109212 | NOTCH4 |
| chr17 | 74893685 | 74894452 | 768 | 6 | 7.10E-07 | 3.77E-05 | 1.91E-05 | 3.67E-06 | -0.00126999 | FADS6 |
| chr20 | 9050987 | 9051555 | 569 | 3 | 7.89E-07 | 4.15E-05 | 6.39E-05 | 3.67E-06 | 0.001420884 | NA |
| chr2 | 86788690 | 86789597 | 908 | 5 | 1.07E-06 | 4.43E-06 | 0.000133 | 3.68E-06 | -0.00679281 | CD8A |
| chr6 | 423875 | 424126 | 252 | 3 | 2.01E-06 | 1.31E-05 | 1.79E-05 | 3.68E-06 | -0.00158915 | NA |
| chr6 | 108075747 | 108075810 | 64 | 2 | 2.79E-07 | 7.74E-05 | 4.53E-06 | 3.68E-06 | 0.00338816 | OSTM1 |
| chr21 | 31476598 | 31477086 | 489 | 2 | 1.39E-06 | 8.19E-06 | 1.95E-05 | 3.68E-06 | -0.00051311 | TIAM1 |
| chr2 | 25161424 | 25161940 | 517 | 4 | 4.75E-06 | 1.70E-06 | 0.000708 | 3.68E-06 | -0.01007151 | POMC |
| chr17 | 75317823 | 75318370 | 548 | 2 | 8.05E-06 | 2.49E-06 | 0.000179 | 3.68E-06 | -0.01136028 | GRB2 |
| chr20 | 52191576 | 52192461 | 886 | 16 | 7.19E-06 | 4.18E-05 | 0.006713 | 3.68E-06 | 0.000501849 | LINC01524, ZFP64 |
| chr4 | 3462926 | 3463337 | 412 | 9 | 4.75E-06 | 2.08E-06 | 0.000328 | 3.69E-06 | -0.00252021 | DOK7 |
| chr11 | 115240575 | 115241254 | 680 | 5 | 5.03E-06 | 1.32E-05 | 9.67E-05 | 3.69E-06 | 0.002506096 | CADM1 |
| chr2 | 100417795 | 100418567 | 773 | 7 | 1.66E-06 | 1.71E-05 | 0.0012 | 3.69E-06 | 0.004544719 | NA |
| chr17 | 82632749 | 82632894 | 146 | 2 | 1.15E-06 | 0.001623 | 2.47E-06 | 3.69E-06 | -0.00503645 | FOXK2, WDR45B |
| chr4 | 165982951 | 165983385 | 435 | 2 | 5.47E-06 | 2.85E-06 | 0.000109 | 3.70E-06 | 0.001183281 | TLL1 |
| chr17 | 50707828 | 50708241 | 414 | 7 | 7.97E-06 | 4.78E-06 | 0.001395 | 3.70E-06 | 0.002185685 | ANKRD40 |
| chr11 | 70738147 | 70738846 | 700 | 3 | 7.79E-07 | 1.10E-05 | 2.44E-05 | 3.70E-06 | -0.00689944 | SHANK2 |
| chr2 | 241962587 | 241962642 | 56 | 3 | 1.15E-05 | 1.69E-06 | 0.001065 | 3.70E-06 | 0.010705743 | LINC01237 |
| chr3 | 13936330 | 13936386 | 57 | 2 | 1.46E-06 | 1.59E-05 | 1.09E-05 | 3.70E-06 | 0.000798034 | FGD5P1 |
| chr11 | 4181731 | 4182312 | 582 | 2 | 2.09E-06 | 5.14E-06 | 3.46E-05 | 3.70E-06 | 0.001972257 | LINC02749 |
| chr15 | 41847952 | 41848517 | 566 | 2 | 2.09E-07 | 4.31E-06 | 4.50E-05 | 3.70E-06 | 0.000628575 | JMJD7-PLA2G4B, PLA2G4B, SPTBN5 |
| chr20 | 19074784 | 19075695 | 912 | 4 | 1.46E-06 | 1.56E-05 | 2.34E-05 | 3.70E-06 | -0.00144238 | NA |
| chr8 | 57805200 | 57805293 | 94 | 2 | 9.26E-07 | 4.36E-05 | 5.84E-06 | 3.70E-06 | -0.01050191 | NA |
| chr14 | 50883213 | 50883579 | 367 | 3 | 2.68E-07 | 0.0001455 | 3.34E-06 | 3.70E-06 | 0.007595792 | ABHD12B, PYGL |
| chr8 | 69671975 | 69672597 | 623 | 2 | 1.13E-05 | 2.22E-06 | 0.000383 | 3.70E-06 | -0.01691544 | SLCO5A1 |
| chr2 | 24489788 | 24490712 | 925 | 5 | 5.51E-07 | 2.46E-05 | 0.000295 | 3.71E-06 | -0.01016871 | NA |
| chr9 | 95084452 | 95085151 | 700 | 3 | 1.95E-05 | 4.27E-06 | 0.000134 | 3.71E-06 | -0.00050551 | AOPEP |
| chr6 | 129546775 | 129547214 | 440 | 4 | 3.81E-06 | 1.20E-05 | 3.68E-05 | 3.71E-06 | 0.001340913 | AL356124.1 |
| chr11 | 7596382 | 7597842 | 1461 | 7 | 1.68E-05 | 9.32E-06 | 0.001521 | 3.71E-06 | -0.00375492 | PPFIBP2 |
| chr7 | 137354671 | 137354704 | 34 | 2 | 3.97E-06 | 2.10E-05 | 8.96E-06 | 3.71E-06 | -0.00342113 | NA |
| chr8 | 20187381 | 20187449 | 69 | 2 | 6.55E-06 | 4.74E-06 | 3.92E-05 | 3.72E-06 | -0.01671754 | NA |
| chr18 | 78502286 | 78503059 | 774 | 3 | 3.08E-06 | 6.27E-05 | 0.000199 | 3.72E-06 | -0.00011815 | NA |
| chr12 | 49548974 | 49549823 | 850 | 6 | 3.29E-08 | 0.0049666 | 2.41E-06 | 3.72E-06 | -0.00445495 | KCNH3 |
| chr20 | 11540522 | 11540933 | 412 | 3 | 2.97E-06 | 1.73E-05 | 1.66E-05 | 3.72E-06 | -0.00813383 | AL109837.2 |
| chr10 | 89331924 | 89332265 | 342 | 3 | 1.64E-07 | 0.0332718 | 1.32E-06 | 3.73E-06 | 0.006783355 | IFIT3, LIPA |
| chr2 | 218002662 | 218002813 | 152 | 2 | 5.14E-06 | 2.24E-06 | 0.000371 | 3.73E-06 | 0.014274006 | TNS1 |
| chr9 | 122940500 | 122941561 | 1062 | 10 | 2.53E-06 | 9.79E-06 | 4.73E-05 | 3.73E-06 | -0.00113446 | RABGAP1, AC007066.3 |
| chr16 | 4242199 | 4242676 | 478 | 5 | 3.37E-06 | 2.27E-05 | 0.000157 | 3.73E-06 | -0.00526709 | NA |
| chr17 | 20206118 | 20206728 | 611 | 3 | 3.99E-06 | 3.15E-06 | 0.000382 | 3.74E-06 | -0.00970123 | SPECC1, AC004702.1 |
| chr2 | 129252866 | 129253547 | 682 | 2 | 3.27E-06 | 4.06E-06 | 5.07E-05 | 3.74E-06 | -0.00282378 | LINC01854 |
| chr7 | 55120093 | 55120939 | 847 | 3 | 5.30E-05 | 2.50E-06 | 0.00064 | 3.74E-06 | 0.003311926 | EGFR |
| chr6 | 125819839 | 125820019 | 181 | 2 | 1.66E-06 | 9.85E-06 | 1.66E-05 | 3.74E-06 | 0.012390466 | NCOA7 |
| chr14 | 69013985 | 69014800 | 816 | 4 | 1.86E-05 | 1.69E-06 | 0.001667 | 3.74E-06 | -0.0034464 | NA |
| chr17 | 68198993 | 68199166 | 174 | 2 | 1.12E-06 | 2.42E-06 | 0.000223 | 3.74E-06 | -0.00964358 | ARHGAP27P2 |
| chr8 | 142441932 | 142442026 | 95 | 2 | 9.04E-06 | 2.25E-06 | 0.000372 | 3.74E-06 | -0.01159582 | NA |
| chr8 | 30441166 | 30441687 | 522 | 2 | 4.06E-07 | 1.81E-05 | 1.01E-05 | 3.75E-06 | 0.008782999 | RBPMS |
| chr5 | 135190434 | 135191062 | 629 | 4 | 5.64E-06 | 1.13E-06 | 0.002288 | 3.75E-06 | -0.01230989 | C5orf66 |
| chr17 | 63972678 | 63973359 | 682 | 8 | 6.97E-06 | 9.71E-06 | 0.000989 | 3.75E-06 | -0.00248565 | SCN4A, AC127029.2 |
| chr5 | 2562188 | 2562716 | 529 | 3 | 1.12E-05 | 1.59E-06 | 0.001048 | 3.75E-06 | -0.00578615 | NA |
| chr13 | 109067121 | 109067542 | 422 | 2 | 5.31E-07 | 5.91E-05 | 5.18E-06 | 3.76E-06 | -0.00611426 | MYO16 |
| chr17 | 76318572 | 76318825 | 254 | 3 | 3.75E-06 | 1.06E-05 | 0.000308 | 3.76E-06 | 0.004062032 | PRPSAP1 |
| chr7 | 5989514 | 5990473 | 960 | 3 | 1.79E-07 | 0.0001116 | 7.13E-06 | 3.76E-06 | -0.00618084 | PMS2 |
| chr6 | 7909855 | 7911007 | 1153 | 8 | 2.89E-08 | 6.38E-05 | 0.00081 | 3.76E-06 | 3.71E-05 | TXNDC5, BLOC1S5-TXNDC5 |
| chr2 | 210476402 | 210477537 | 1136 | 12 | 2.43E-07 | 0.0007323 | 0.00039 | 3.76E-06 | -0.00062533 | LANCL1 |
| chr8 | 60653654 | 60654122 | 469 | 6 | 7.29E-07 | 2.80E-05 | 0.0002 | 3.76E-06 | -0.00054392 | NA |
| chr12 | 57475248 | 57476158 | 911 | 5 | 2.52E-06 | 4.56E-06 | 0.001312 | 3.76E-06 | -0.00261802 | MARS1, ARHGAP9 |
| chr8 | 15238023 | 15238552 | 530 | 6 | 9.04E-06 | 1.80E-06 | 0.004615 | 3.76E-06 | 0.001341259 | SGCZ |
| chr2 | 9105904 | 9106570 | 667 | 4 | 2.09E-06 | 5.03E-06 | 0.000229 | 3.76E-06 | -0.00560164 | NA |
| chr9 | 128383076 | 128384006 | 931 | 3 | 5.63E-09 | 0.0004491 | 2.83E-06 | 3.77E-06 | 0.006499354 | URM1 |
| chr14 | 92923127 | 92923546 | 420 | 8 | 1.15E-07 | 0.000174 | 5.14E-05 | 3.77E-06 | -0.00061154 | CHGA |
| chr12 | 62603323 | 62603851 | 529 | 11 | 7.43E-06 | 2.32E-05 | 0.003992 | 3.77E-06 | -0.0003879 | AC048341.1, MIRLET7I, LINC01465 |
| chr9 | 98796048 | 98797495 | 1448 | 9 | 4.87E-05 | 6.68E-05 | 0.000232 | 3.77E-06 | 0.002074532 | ANKS6 |
| chr21 | 17526902 | 17527592 | 691 | 3 | 2.67E-06 | 8.30E-06 | 0.000284 | 3.77E-06 | -0.01259041 | CXADR, Y RNA |
| chr10 | 45377287 | 45378425 | 1139 | 3 | 2.17E-08 | 0.0008439 | 2.36E-06 | 3.77E-06 | -0.00558704 | ALOX5 |
| chr10 | 116756525 | 116756673 | 149 | 2 | 2.53E-06 | 2.27E-06 | 0.00036 | 3.77E-06 | 0.000167356 | HSPA12A |
| chr12 | 4643891 | 4644047 | 157 | 2 | 4.81E-06 | 3.40E-06 | 7.39E-05 | 3.77E-06 | 0.004584017 | AC005832.4, AKAP3 |
| chr3 | 70803745 | 70804002 | 258 | 2 | 6.26E-07 | 0.0031439 | 2.41E-06 | 3.77E-06 | -0.00551502 | NA |
| chr10 | 123997767 | 123998088 | 322 | 3 | 2.34E-06 | 2.12E-05 | 2.99E-05 | 3.77E-06 | 0.004572696 | NA |
| chr11 | 113951454 | 113951664 | 211 | 2 | 3.77E-06 | 9.37E-06 | 1.77E-05 | 3.78E-06 | 0.010682091 | NA |
| chr16 | 3129327 | 3130173 | 847 | 7 | 3.90E-06 | 6.89E-05 | 8.45E-05 | 3.78E-06 | -0.00056525 | ZNF213, ZNF213-AS1 |
| chr6 | 150069758 | 150069827 | 70 | 3 | 1.89E-06 | 3.10E-06 | 0.000173 | 3.78E-06 | -0.02016942 | NA |
| chr14 | 24146427 | 24147554 | 1128 | 11 | 1.07E-05 | 5.07E-05 | 0.005106 | 3.78E-06 | -0.00036109 | RNF31, AL136295.5, PSME2 |
| chr2 | 172462219 | 172462642 | 424 | 3 | 3.75E-06 | 1.65E-06 | 0.001236 | 3.78E-06 | 0.007072775 | ITGA6 |
| chr12 | 95003411 | 95004033 | 623 | 12 | 2.47E-05 | 3.31E-06 | 0.005502 | 3.78E-06 | -0.00471194 | NDUFA12 |
| chr8 | 127937937 | 127938406 | 470 | 3 | 3.38E-06 | 4.44E-05 | 4.91E-05 | 3.78E-06 | -0.0012564 | PVT1 |
| chr18 | 63936445 | 63937484 | 1040 | 7 | 1.24E-05 | 0.0002594 | 0.000122 | 3.78E-06 | -0.00456584 | NA |
| chr11 | 70571328 | 70572067 | 740 | 4 | 8.02E-06 | 2.49E-06 | 0.001087 | 3.79E-06 | -0.00276179 | SHANK2 |
| chr1 | 15943100 | 15944066 | 967 | 5 | 3.94E-06 | 9.85E-07 | 0.004487 | 3.79E-06 | -0.0037757 | ZBTB17 |
| chr6 | 109690483 | 109691285 | 803 | 12 | 2.68E-05 | 1.70E-06 | 0.003228 | 3.79E-06 | 0.00010779 | FIG4, AK9 |
| chr3 | 133573447 | 133574655 | 1209 | 18 | 1.85E-05 | 9.24E-05 | 0.002052 | 3.79E-06 | 0.00081508 | CDV3 |
| chr1 | 112693784 | 112694508 | 725 | 2 | 3.81E-05 | 2.28E-06 | 0.000368 | 3.79E-06 | -7.95E-05 | MOV10, AL603832.1 |
| chr1 | 7536625 | 7537350 | 726 | 3 | 1.46E-06 | 6.83E-06 | 4.62E-05 | 3.79E-06 | -0.00768776 | CAMTA1 |
| chr10 | 93893621 | 93894401 | 781 | 11 | 1.59E-06 | 1.61E-05 | 0.001642 | 3.79E-06 | -0.00018402 | SLC35G1 |
| chr1 | 151198134 | 151199186 | 1053 | 12 | 3.39E-06 | 1.15E-05 | 0.004123 | 3.79E-06 | -0.00028654 | PIP5K1A |
| chr9 | 135886516 | 135886838 | 323 | 3 | 7.04E-06 | 6.75E-06 | 0.000157 | 3.80E-06 | -0.00759207 | CAMSAP1 |
| chr6 | 75749794 | 75749926 | 133 | 2 | 1.92E-07 | 0.0005037 | 2.94E-06 | 3.80E-06 | -0.01673565 | MYO6 |
| chr4 | 113761452 | 113762666 | 1215 | 12 | 2.61E-06 | 0.0001778 | 0.000971 | 3.80E-06 | 0.00075058 | CAMK2D |
| chr11 | 108500383 | 108500973 | 591 | 2 | 2.13E-06 | 2.79E-06 | 0.000131 | 3.80E-06 | -0.01128832 | NA |
| chr17 | 31558768 | 31560030 | 1263 | 8 | 4.96E-06 | 4.61E-06 | 0.000505 | 3.80E-06 | -0.00579688 | MIR193A |
| chr11 | 122926587 | 122927024 | 438 | 3 | 4.41E-06 | 2.91E-06 | 0.000175 | 3.81E-06 | -0.00455708 | JHY |
| chr4 | 7659223 | 7660002 | 780 | 4 | 2.92E-06 | 6.56E-05 | 0.000172 | 3.81E-06 | -0.0041555 | SORCS2 |
| chr22 | 39925662 | 39926747 | 1086 | 8 | 1.88E-06 | 6.97E-05 | 0.000142 | 3.81E-06 | -0.00168856 | GRAP2 |
| chr12 | 132615073 | 132615351 | 279 | 3 | 2.79E-06 | 2.39E-05 | 2.41E-05 | 3.81E-06 | -0.00856298 | NA |
| chr4 | 152485356 | 152485890 | 535 | 2 | 9.55E-06 | 2.48E-06 | 0.000223 | 3.81E-06 | -0.01572142 | AC023424.2, FBXW7 |
| chr13 | 33610526 | 33611093 | 568 | 8 | 1.66E-06 | 0.0001618 | 0.00014 | 3.81E-06 | -0.00602222 | AL161719.1, AL139383.1 |
| chr1 | 4170700 | 4171160 | 461 | 3 | 6.25E-06 | 2.20E-06 | 0.000556 | 3.81E-06 | -0.00142179 | NA |
| chr13 | 98804182 | 98804729 | 548 | 2 | 2.29E-06 | 5.14E-06 | 3.67E-05 | 3.82E-06 | -0.00378569 | DOCK9 |
| chr12 | 32679035 | 32679527 | 493 | 7 | 2.11E-06 | 1.19E-05 | 0.001494 | 3.82E-06 | -0.0008519 | DNM1L |
| chr9 | 129637420 | 129637937 | 518 | 3 | 4.56E-06 | 3.90E-06 | 8.92E-05 | 3.82E-06 | 0.007008536 | ASB6 |
| chr10 | 1205799 | 1206564 | 766 | 6 | 2.09E-06 | 3.65E-05 | 0.000411 | 3.82E-06 | -0.00445395 | LINC00200, ADARB2, AL392083.2 |
| chr12 | 47753251 | 47753692 | 442 | 3 | 1.84E-07 | 0.0003289 | 6.42E-06 | 3.83E-06 | -0.01126317 | RAPGEF3 |
| chr16 | 81519742 | 81519881 | 140 | 2 | 6.97E-06 | 2.33E-06 | 0.000336 | 3.83E-06 | -0.00936394 | CMIP, AC092139.2 |
| chr17 | 50079840 | 50080277 | 438 | 4 | 2.58E-06 | 1.13E-05 | 0.00026 | 3.83E-06 | -0.00541896 | ITGA3 |
| chr18 | 5543802 | 5544662 | 861 | 14 | 1.38E-06 | 4.75E-05 | 0.000238 | 3.83E-06 | 0.000708156 | EPB41L3 |

| | | | | | | | | | | |
|---|---|---|---|---|---|---|---|---|---|---|
| chr1 | 203286715 | 203287187 | 473 | 5 | 8.74E-07 | 3.91E-05 | 1.47E-05 | 3.83E-06 | 0.00026396 | LINC01353 |
| chr13 | 114049167 | 114049663 | 497 | 6 | 2.66E-06 | 0.0040663 | 0.000143 | 3.83E-06 | -0.0013945 | RASA3 |
| chr6 | 161375753 | 161375889 | 137 | 5 | 5.29E-06 | 1.59E-05 | 4.14E-05 | 3.84E-06 | -0.00314215 | PRKN |
| chr3 | 172578149 | 172578242 | 94 | 2 | 7.91E-06 | 3.20E-06 | 8.93E-05 | 3.84E-06 | -0.02275198 | LINC02068 |
| chr7 | 44999578 | 45000722 | 1145 | 11 | 4.71E-07 | 0.0001513 | 0.000159 | 3.84E-06 | -0.00117175 | CCM2 |
| chr5 | 57834387 | 57835137 | 751 | 3 | 2.39E-06 | 3.39E-06 | 0.000114 | 3.84E-06 | -0.00576027 | NA |
| chr15 | 74337896 | 74338689 | 794 | 2 | 1.21E-05 | 2.93E-06 | 0.000115 | 3.84E-06 | -0.00374943 | CYP11A1 |
| chr12 | 27589391 | 27589955 | 565 | 3 | 8.81E-06 | 4.38E-06 | 0.000487 | 3.84E-06 | -0.01120115 | PPFIBP1 |
| chr7 | 151794340 | 151794827 | 488 | 3 | 2.41E-06 | 2.26E-06 | 0.00071 | 3.84E-06 | -0.00357314 | PRKAG2 |
| chr9 | 37467448 | 37468241 | 794 | 2 | 1.08E-05 | 2.75E-06 | 0.000146 | 3.84E-06 | -0.00885968 | NA |
| chr3 | 9809488 | 9810668 | 1181 | 15 | 1.40E-06 | 0.0008162 | 0.000128 | 3.84E-06 | 0.000163574 | ARPC4-TTLL3, TTLL3 |
| chr4 | 68246016 | 68246705 | 690 | 5 | 2.92E-06 | 2.72E-05 | 0.000344 | 3.85E-06 | -0.01048126 | NA |
| chr11 | 70544993 | 70545601 | 609 | 4 | 5.81E-06 | 5.99E-05 | 0.000199 | 3.85E-06 | -0.0067181 | SHANK2 |
| chr21 | 36125554 | 36125813 | 260 | 2 | 6.32E-06 | 2.85E-06 | 0.000128 | 3.85E-06 | -0.01728209 | AP000688.1 |
| chr10 | 113131533 | 113132003 | 471 | 3 | 4.11E-06 | 4.67E-06 | 0.000385 | 3.85E-06 | 0.002875068 | TCF7L2 |
| chr11 | 130672160 | 130673002 | 843 | 5 | 1.83E-05 | 6.78E-05 | 0.000156 | 3.85E-06 | -0.00682463 | LINC02873 |
| chr22 | 32457160 | 32457984 | 825 | 2 | 7.20E-09 | 6.90E-05 | 5.00E-06 | 3.86E-06 | -0.01337959 | BPIFC |
| chr1 | 226980638 | 226981044 | 407 | 2 | 3.41E-06 | 2.33E-06 | 0.000359 | 3.86E-06 | 0.00171477 | AL391628.1, COQ8A |
| chr1 | 94856125 | 94856262 | 138 | 2 | 3.66E-06 | 5.23E-06 | 3.65E-05 | 3.86E-06 | -0.00910542 | SLC44A3 |
| chr6 | 83215390 | 83215585 | 196 | 2 | 1.28E-06 | 1.46E-05 | 1.24E-05 | 3.86E-06 | -0.00473998 | ME1 |
| chr17 | 16483386 | 16483490 | 105 | 2 | 8.99E-06 | 2.35E-06 | 0.000337 | 3.86E-06 | -0.00902035 | LRRC75A |
| chr5 | 178777355 | 178777765 | 411 | 3 | 4.82E-06 | 1.62E-05 | 0.000123 | 3.86E-06 | -0.00994746 | AACSP1 |
| chr19 | 42075998 | 42076389 | 392 | 5 | 8.56E-08 | 0.0010037 | 1.66E-06 | 3.86E-06 | -0.00063223 | ZNF574 |
| chr8 | 66774532 | 66775239 | 708 | 12 | 7.10E-07 | 0.0002004 | 0.000146 | 3.86E-06 | 0.000962162 | C8orf44-SGK3, SGK3 |
| chr6 | 138107430 | 138108174 | 745 | 10 | 6.83E-06 | 2.87E-06 | 0.000436 | 3.87E-06 | 0.001014115 | NA |
| chr17 | 82900953 | 82901263 | 311 | 5 | 2.22E-06 | 6.65E-05 | 0.000666 | 3.87E-06 | -0.00565754 | TBCD |
| chr15 | 64149825 | 64150379 | 555 | 3 | 8.10E-07 | 2.61E-05 | 6.40E-05 | 3.87E-06 | 0.000863718 | NA |
| chr13 | 92059765 | 92059878 | 114 | 2 | 1.37E-06 | 2.31E-06 | 0.000394 | 3.87E-06 | 0.00022325 | GPC5, RNU4ATAC3P |
| chr15 | 23025402 | 23025475 | 74 | 2 | 3.54E-06 | 3.47E-06 | 7.55E-05 | 3.87E-06 | -0.00553202 | TUBGCP5 |
| chr9 | 132233781 | 132233940 | 160 | 2 | 2.05E-06 | 9.02E-05 | 4.53E-06 | 3.88E-06 | -0.00508945 | NTNG2 |
| chr5 | 177553152 | 177553908 | 757 | 3 | 1.88E-07 | 4.35E-05 | 7.05E-06 | 3.88E-06 | -0.00543065 | FAM193B |
| chr14 | 105364269 | 105364522 | 254 | 3 | 3.59E-06 | 2.38E-05 | 2.28E-05 | 3.88E-06 | 0.013274974 | PACS2 |
| chr15 | 89368745 | 89369064 | 320 | 4 | 4.58E-07 | 0.0002393 | 2.69E-05 | 3.88E-06 | 0.004348149 | MIR9-3HG |
| chr19 | 1472937 | 1473180 | 244 | 3 | 7.80E-06 | 1.68E-06 | 0.000918 | 3.88E-06 | -0.01651691 | APC2, C19orf25 |
| chr17 | 57253477 | 57253481 | 5 | 2 | 7.69E-06 | 3.94E-06 | 5.91E-05 | 3.88E-06 | -0.00489699 | AC091181.2 |
| chr11 | 130478203 | 130478909 | 707 | 3 | 5.83E-07 | 5.44E-05 | 7.98E-05 | 3.89E-06 | -0.00927515 | NA |
| chr1 | 17272867 | 17273314 | 448 | 2 | 2.28E-06 | 6.03E-06 | 3.06E-05 | 3.89E-06 | -0.00617423 | PADI3 |
| chr13 | 33818026 | 33818732 | 707 | 5 | 1.19E-06 | 5.82E-06 | 0.000284 | 3.89E-06 | 0.000309595 | RFC3 |
| chr11 | 86607832 | 86608433 | 602 | 3 | 1.26E-05 | 2.05E-05 | 0.000225 | 3.89E-06 | -0.00458721 | AP001831.1, ME3 |
| chr8 | 31252271 | 31252889 | 619 | 2 | 1.46E-05 | 2.45E-06 | 0.000273 | 3.89E-06 | 0.003663242 | NA |
| chr3 | 193774677 | 193775244 | 568 | 2 | 4.42E-07 | 2.16E-05 | 9.35E-06 | 3.90E-06 | -0.00703063 | AC069421.3 |
| chr3 | 127673560 | 127674098 | 539 | 4 | 6.03E-07 | 1.98E-06 | 0.002491 | 3.90E-06 | -0.00947227 | ABTB1 |
| chr8 | 23187876 | 23188209 | 334 | 3 | 8.93E-06 | 8.98E-06 | 0.000276 | 3.90E-06 | -0.01151548 | NA |
| chr6 | 10464247 | 10464669 | 423 | 2 | 6.90E-07 | 1.47E-05 | 1.25E-05 | 3.90E-06 | 0.01390144 | NA |
| chr1 | 247330948 | 247331948 | 1001 | 8 | 5.01E-07 | 0.0002352 | 5.84E-05 | 3.90E-06 | 0.000918312 | ZNF496 |
| chr8 | 98357683 | 98358217 | 535 | 2 | 6.88E-07 | 8.33E-06 | 2.10E-05 | 3.90E-06 | 0.002476985 | NA |
| chr1 | 172443902 | 172444472 | 571 | 8 | 2.37E-05 | 1.02E-06 | 0.004107 | 3.90E-06 | -0.00060483 | C1orf105, PIGC |
| chr22 | 45448761 | 45448895 | 135 | 3 | 9.64E-06 | 2.51E-06 | 0.000614 | 3.90E-06 | -0.00227603 | NA |
| chr15 | 32730369 | 32731036 | 668 | 4 | 1.80E-07 | 6.29E-05 | 0.000161 | 3.91E-06 | -0.00814899 | GREM1 |
| chr5 | 169583263 | 169584170 | 908 | 13 | 3.34E-06 | 2.66E-05 | 0.002172 | 3.91E-06 | -0.00074412 | SPDL1 |
| chr12 | 57459613 | 57460453 | 841 | 8 | 8.53E-06 | 2.13E-06 | 0.00462 | 3.91E-06 | -0.00174599 | GLI1, AC022506.2 |
| chr15 | 68352612 | 68353631 | 1020 | 3 | 9.52E-07 | 8.69E-06 | 4.26E-05 | 3.91E-06 | -0.00240946 | ITGA11 |
| chr7 | 91880455 | 91881283 | 829 | 12 | 1.44E-05 | 8.42E-06 | 0.003685 | 3.91E-06 | -0.00385497 | AC003086.1, MTERF1 |
| chr11 | 6419226 | 6419738 | 513 | 12 | 3.88E-07 | 9.10E-05 | 0.000606 | 3.91E-06 | -0.00160298 | APBB1 |
| chr17 | 41506471 | 41507519 | 1049 | 5 | 5.99E-06 | 8.98E-06 | 9.53E-05 | 3.92E-06 | -0.00375599 | NA |
| chr7 | 123655155 | 123656044 | 890 | 5 | 3.57E-05 | 0.0004036 | 0.00017 | 3.92E-06 | -0.00633714 | LMOD2 |
| chr17 | 43544981 | 43546342 | 1362 | 9 | 2.05E-06 | 7.67E-05 | 0.002534 | 3.92E-06 | -0.00332741 | DHX8, ETV4 |
| chr12 | 123534446 | 123534762 | 317 | 4 | 1.51E-06 | 4.36E-05 | 0.000162 | 3.92E-06 | -0.00681333 | NA |
| chr15 | 90716453 | 90717416 | 964 | 14 | 5.71E-07 | 0.0003939 | 0.000278 | 3.93E-06 | 0.00013047 | BLM, CRTC3-AS1 |
| chr5 | 169581304 | 169581711 | 408 | 3 | 1.59E-06 | 0.0001346 | 7.43E-06 | 3.94E-06 | -0.00947322 | NA |
| chr22 | 37433947 | 37433992 | 46 | 2 | 8.16E-06 | 3.52E-06 | 7.68E-05 | 3.94E-06 | -0.00887054 | NA |
| chr10 | 125786752 | 125787137 | 386 | 2 | 1.68E-06 | 1.25E-05 | 1.44E-05 | 3.94E-06 | -0.007672 | UROS |
| chr4 | 82797012 | 82797799 | 788 | 3 | 1.19E-05 | 1.34E-05 | 0.000275 | 3.94E-06 | 0.000132999 | SCD5 |
| chr16 | 66740459 | 66740723 | 265 | 2 | 1.56E-06 | 2.26E-05 | 9.22E-06 | 3.94E-06 | -0.01159513 | AC018557.3, DYNC1LI2 |
| chr13 | 74575696 | 74576002 | 307 | 4 | 4.56E-06 | 4.32E-05 | 0.000123 | 3.94E-06 | -0.00074788 | NA |
| chr12 | 94649451 | 94650246 | 796 | 9 | 5.48E-06 | 8.86E-06 | 0.002188 | 3.95E-06 | 0.000980278 | TMCC3 |
| chr12 | 108851255 | 108851806 | 552 | 2 | 1.24E-06 | 2.53E-06 | 0.000244 | 3.95E-06 | -0.0076271 | SSH1 |
| chr17 | 81653748 | 81654242 | 495 | 3 | 7.04E-06 | 2.23E-05 | 0.000266 | 3.95E-06 | 0.001898492 | PDE6G |
| chr5 | 55306955 | 55308063 | 1109 | 19 | 1.28E-05 | 2.77E-06 | 0.015258 | 3.95E-06 | -0.00016219 | MTREX, DHX29 |
| chr14 | 76825959 | 76826390 | 432 | 5 | 1.91E-06 | 7.48E-06 | 0.000115 | 3.95E-06 | -0.00013413 | LRRC74A, ANGEL1 |
| chr3 | 41194781 | 41195129 | 349 | 4 | 1.46E-06 | 5.83E-05 | 6.13E-06 | 3.95E-06 | 0.001298621 | CTNNB1 |
| chr10 | 22345649 | 22346214 | 566 | 4 | 6.50E-09 | 0.0001275 | 2.46E-05 | 3.95E-06 | -0.01484954 | SPAG6 |
| chr4 | 147733219 | 147734077 | 859 | 3 | 2.43E-08 | 9.16E-05 | 7.77E-06 | 3.95E-06 | 0.007813112 | ARHGAP10 |
| chr19 | 40145032 | 40145100 | 69 | 2 | 7.87E-07 | 0.0015761 | 2.64E-06 | 3.95E-06 | -0.00810691 | NA |
| chr4 | 6008348 | 6008437 | 90 | 2 | 7.29E-06 | 2.42E-06 | 0.000327 | 3.95E-06 | 0.042699169 | C4orf50 |
| chr1 | 161748917 | 161749711 | 795 | 9 | 6.76E-07 | 2.74E-05 | 0.000787 | 3.95E-06 | -0.00030737 | NA |
| chr3 | 100603649 | 100603785 | 137 | 2 | 7.58E-06 | 2.79E-06 | 0.000155 | 3.96E-06 | 0.016466147 | NA |
| chr3 | 136026020 | 136026944 | 925 | 5 | 4.00E-06 | 0.0001715 | 0.00056 | 3.96E-06 | -0.0006802 | PPP2R3A |
| chr15 | 82699322 | 82699913 | 592 | 4 | 8.05E-06 | 2.22E-05 | 0.000484 | 3.96E-06 | -0.01006348 | AP3B2 |
| chr16 | 67305837 | 67305901 | 65 | 2 | 9.69E-06 | 2.37E-06 | 0.000395 | 3.96E-06 | -0.00671013 | KCTD19 |
| chr12 | 53972976 | 53974038 | 1063 | 10 | 2.59E-08 | 0.0003696 | 0.000242 | 3.96E-06 | 0.003544135 | HOXC11, HOTAIR |
| chr18 | 34976140 | 34976868 | 729 | 9 | 5.17E-06 | 1.84E-05 | 0.001529 | 3.96E-06 | -0.00251778 | NA |
| chr4 | 103076730 | 103077560 | 831 | 11 | 6.86E-07 | 5.38E-05 | 0.001679 | 3.96E-06 | 0.01086431 | SLC9B2 |
| chr2 | 179428916 | 179429283 | 368 | 2 | 5.55E-06 | 3.34E-06 | 8.88E-05 | 3.96E-06 | -0.02672641 | NA |
| chr10 | 17562830 | 17562837 | 8 | 2 | 9.63E-06 | 2.42E-06 | 0.000335 | 3.96E-06 | 0.051262665 | NA |
| chr6 | 33069157 | 33069885 | 729 | 3 | 2.31E-06 | 8.61E-06 | 0.000131 | 3.97E-06 | 0.003416421 | HLA-DPA1 |
| chr5 | 141172524 | 141173510 | 987 | 12 | 1.70E-06 | 4.82E-05 | 0.004298 | 3.97E-06 | -0.01216659 | PCDHB7, AC244517.2, AC244517.11 |
| chr5 | 139561832 | 139563078 | 1247 | 6 | 3.51E-07 | 0.0003854 | 0.000311 | 3.97E-06 | -0.00155898 | UBE2D2 |
| chr22 | 21982637 | 21983071 | 435 | 11 | 2.47E-06 | 0.0004006 | 0.00043 | 3.97E-06 | 0.000344895 | TOP3B |
| chr10 | 132124389 | 132125389 | 1001 | 8 | 1.05E-05 | 3.29E-05 | 0.000399 | 3.97E-06 | -0.00057089 | JAKMIP3 |
| chr9 | 6761072 | 6761547 | 476 | 2 | 3.21E-06 | 4.60E-06 | 4.73E-05 | 3.98E-06 | -0.00796584 | KDM4C |
| chr10 | 4469711 | 4470231 | 521 | 2 | 6.24E-06 | 3.03E-06 | 0.000117 | 3.98E-06 | -0.00943471 | NA |
| chr19 | 11374076 | 11374760 | 685 | 10 | 7.12E-06 | 1.17E-05 | 0.007129 | 3.98E-06 | -0.00029056 | SWSAP1, AC024575.1 |
| chr16 | 84747092 | 84747134 | 43 | 4 | 7.78E-06 | 1.30E-05 | 0.000227 | 3.98E-06 | -0.00405872 | USP10 |
| chr18 | 13382616 | 13383399 | 784 | 4 | 1.01E-06 | 1.56E-05 | 2.04E-05 | 3.98E-06 | 0.008426164 | LDLRAD4 |
| chr1 | 247855288 | 247855932 | 645 | 3 | 3.03E-07 | 1.50E-05 | 6.75E-05 | 3.98E-06 | -0.00783337 | NA |
| chr5 | 132394901 | 132395023 | 123 | 2 | 9.66E-06 | 2.39E-06 | 0.000394 | 3.99E-06 | 0.002603228 | SLC22A5 |
| chr7 | 87475408 | 87476100 | 693 | 10 | 2.04E-06 | 1.90E-05 | 0.003009 | 3.99E-06 | -0.00327369 | ABCB4 |
| chr6 | 35321264 | 35321698 | 435 | 2 | 2.48E-06 | 6.03E-06 | 3.21E-05 | 3.99E-06 | -0.00167145 | DEF6 |
| chr15 | 93092464 | 93092511 | 48 | 2 | 2.11E-06 | 3.26E-06 | 9.65E-05 | 3.99E-06 | -0.00951172 | NA |
| chr2 | 233117078 | 233117175 | 98 | 2 | 6.67E-06 | 4.31E-06 | 5.34E-05 | 3.99E-06 | -0.01119481 | INPP5D |
| chr10 | 6239172 | 6239365 | 194 | 2 | 8.97E-06 | 2.55E-06 | 0.000253 | 3.99E-06 | -0.0261408 | PFKFB3 |
| chr9 | 136646011 | 136646370 | 360 | 2 | 2.01E-07 | 1.17E-05 | 1.57E-05 | 4.00E-06 | 0.012349039 | NA |
| chr12 | 118361120 | 118361242 | 123 | 3 | 4.19E-06 | 9.73E-06 | 0.000137 | 4.00E-06 | -0.01418278 | TAOK3 |
| chr17 | 19004369 | 19005610 | 1242 | 13 | 2.41E-07 | 0.0304659 | 0.000239 | 4.00E-06 | -0.00074106 | SLC5A10, FAM83G |
| chr7 | 157286161 | 157286190 | 30 | 2 | 3.87E-06 | 3.20E-05 | 7.53E-06 | 4.00E-06 | -0.00685518 | NA |
| chr22 | 49586454 | 49586557 | 104 | 2 | 5.76E-06 | 7.94E-06 | 2.31E-05 | 4.00E-06 | 0.001821706 | C22orf34 |
| chr6 | 47780960 | 47782306 | 1347 | 8 | 2.69E-07 | 0.0030509 | 6.31E-06 | 4.01E-06 | -0.00249572 | OPN5 |
| chr16 | 57260444 | 57260727 | 284 | 3 | 4.19E-06 | 1.52E-05 | 0.000274 | 4.01E-06 | -0.01404297 | PLLP |
| chr20 | 38913794 | 38914579 | 786 | 4 | 9.55E-06 | 0.000208 | 0.000104 | 4.01E-06 | -0.00565608 | PPP1R16B |
| chr13 | 46587032 | 46587292 | 261 | 2 | 6.69E-07 | 0.0006327 | 2.99E-06 | 4.01E-06 | -0.00451004 | LRCH1 |
| chr7 | 100586287 | 100587133 | 847 | 12 | 8.10E-06 | 1.09E-06 | 0.012883 | 4.01E-06 | 0.000381977 | FBXO24 |
| chr7 | 77416570 | 77417246 | 677 | 6 | 1.59E-06 | 1.80E-06 | 0.001173 | 4.01E-06 | -0.00665134 | AC004921.1 |
| chr3 | 11878983 | 11879140 | 158 | 2 | 8.85E-06 | 2.71E-06 | 0.00019 | 4.02E-06 | -0.0153926 | FANCD2P2, CYCSP12 |
| chr3 | 146921496 | 146921938 | 443 | 5 | 1.62E-05 | 2.01E-06 | 0.002186 | 4.02E-06 | 0.000852976 | LINC02010, AC092957.1 |
| chr11 | 65266150 | 65266289 | 140 | 2 | 3.15E-06 | 2.83E-05 | 8.12E-06 | 4.02E-06 | -0.01521475 | POLA2, AP000944.5 |
| chr20 | 41204154 | 41204829 | 676 | 2 | 4.00E-06 | 4.19E-06 | 5.71E-05 | 4.02E-06 | -0.00555585 | ZHX3 |
| chr8 | 93837679 | 93837994 | 316 | 2 | 7.90E-06 | 2.71E-06 | 0.000192 | 4.02E-06 | -0.01064694 | AC084346.1 |
| chr4 | 84279791 | 84280282 | 492 | 2 | 4.92E-07 | 4.06E-05 | 6.68E-06 | 4.02E-06 | -0.01946004 | AC079160.1 |
| chr9 | 108415939 | 108416208 | 270 | 2 | 7.68E-06 | 2.86E-06 | 0.000152 | 4.02E-06 | -0.00546313 | NA |
| chr12 | 16236887 | 16237337 | 451 | 3 | 8.25E-06 | 7.38E-06 | 0.00018 | 4.02E-06 | 0.001894786 | SLC15A5 |
| chr10 | 80276355 | 80276564 | 210 | 3 | 1.50E-06 | 0.0006339 | 1.03E-05 | 4.02E-06 | -0.00335194 | MAT1A |
| chr11 | 66069723 | 66070766 | 1044 | 5 | 2.09E-06 | 2.27E-06 | 0.001515 | 4.03E-06 | 0.001437997 | PACS1 |
| chr2 | 237669659 | 237670084 | 426 | 5 | 2.33E-06 | 0.000164 | 0.00017 | 4.03E-06 | -0.00699401 | LRRFIP1 |
| chr8 | 132702219 | 132702754 | 536 | 2 | 3.37E-07 | 5.41E-05 | 5.80E-06 | 4.03E-06 | -0.0073573 | TMEM71 |
| chr10 | 80458621 | 80459654 | 1034 | 5 | 9.26E-07 | 4.08E-05 | 5.41E-05 | 4.03E-06 | 4.08E-06 | TSPAN14 |
| chr2 | 67626545 | 67626927 | 383 | 2 | 6.91E-06 | 2.90E-06 | 0.000145 | 4.03E-06 | -0.00791122 | AC007422.1 |
| chr13 | 111893145 | 111893992 | 848 | 6 | 3.38E-06 | 2.87E-06 | 0.000622 | 4.04E-06 | 0.016384986 | LINC00354 |
| chr3 | 46395400 | 46395523 | 124 | 2 | 4.25E-06 | 9.10E-06 | 2.03E-05 | 4.04E-06 | -0.00912298 | CCR5AS |
| chr10 | 132537154 | 132538261 | 1108 | 8 | 6.15E-07 | 0.0004491 | 6.29E-05 | 4.04E-06 | 8.08E-05 | INPP5A |
| chr16 | 87364190 | 87364434 | 245 | 2 | 9.37E-06 | 2.48E-06 | 0.000323 | 4.04E-06 | -0.05478696 | AC010531.5, FBXO31 |
| chr8 | 109086008 | 109086694 | 687 | 7 | 5.36E-06 | 6.38E-06 | 0.000772 | 4.05E-06 | 0.002656894 | TRHR |
| chr11 | 70645477 | 70646142 | 666 | 6 | 4.59E-06 | 9.66E-05 | 0.000288 | 4.05E-06 | -0.00483744 | SHANK2 |
| chr12 | 5431536 | 5432648 | 1113 | 8 | 1.59E-07 | 0.0004359 | 3.04E-05 | 4.05E-06 | -0.00058154 | NTF3 |
| chr1 | 1988672 | 1989228 | 557 | 3 | 7.01E-06 | 2.35E-06 | 0.000468 | 4.05E-06 | 0.007487227 | CFAP74 |
| chr13 | 112977429 | 112978330 | 902 | 4 | 4.80E-07 | 7.87E-06 | 0.000152 | 4.05E-06 | -0.00472286 | MCF2L |
| chr3 | 183252719 | 183253520 | 802 | 12 | 1.09E-05 | 0.0001923 | 0.002452 | 4.05E-06 | -0.00122514 | B3GNT5, MCF2L2 |
| chr1 | 1622992 | 1623775 | 784 | 5 | 2.55E-06 | 7.75E-05 | 0.000349 | 4.06E-06 | -0.00241883 | MIB2 |
| chr8 | 102528236 | 102528786 | 551 | 5 | 1.90E-06 | 0.0004859 | 0.000154 | 4.06E-06 | 0.016895767 | AP002852.1 |
| chr2 | 1138179 | 1138261 | 83 | 2 | 1.68E-06 | 0.000949 | 2.87E-06 | 4.06E-06 | 0.002532971 | SNTG2 |
| chr8 | 52714376 | 52715068 | 693 | 12 | 4.19E-06 | 6.41E-05 | 0.000563 | 4.06E-06 | 0.000848597 | RB1CC1 |
| chr1 | 231274239 | 231274736 | 498 | 4 | 7.61E-07 | 0.0150035 | 3.76E-05 | 4.06E-06 | 0.000616189 | GNPAT |
| chr15 | 82943336 | 82943688 | 353 | 2 | 2.52E-06 | 5.28E-06 | 4.01E-05 | 4.07E-06 | -0.00757979 | AC022558.2, HOMER2 |
| chr1 | 98044772 | 98046073 | 1302 | 8 | 1.54E-06 | 5.42E-05 | 0.0019 | 4.07E-06 | -0.00433302 | MIR137HG, MIR2682, MIR137 |
| chr2 | 239296787 | 239297110 | 324 | 2 | 6.13E-07 | 0.0002755 | 3.50E-06 | 4.07E-06 | -0.00704519 | HDAC4 |
| chr6 | 41052711 | 41053422 | 712 | 6 | 3.87E-06 | 0.0002846 | 0.00078 | 4.08E-06 | -0.00539867 | APOBEC2, OARD1 |
| chr10 | 110148871 | 110149335 | 465 | 2 | 2.39E-07 | 0.0004039 | 3.25E-06 | 4.08E-06 | -0.00608412 | NA |
| chr20 | 6123206 | 6124239 | 1034 | 10 | 2.86E-05 | 4.64E-06 | 0.008369 | 4.08E-06 | -0.00581252 | NA |
| chr2 | 131011528 | 131011609 | 82 | 2 | 3.36E-07 | 5.05E-05 | 6.10E-06 | 4.09E-06 | -0.00642771 | ARHGEF4 |
| chr1 | 153684366 | 153684924 | 559 | 3 | 1.02E-06 | 2.49E-05 | 1.31E-05 | 4.09E-06 | -0.00580281 | NPR1 |
| chr13 | 112981584 | 112982365 | 782 | 3 | 1.76E-06 | 1.01E-05 | 5.45E-05 | 4.10E-06 | -0.00480525 | MCF2L |
| chr11 | 70048777 | 70049396 | 620 | 4 | 5.38E-06 | 3.90E-05 | 0.000161 | 4.10E-06 | 0.002945661 | NA |
| chr4 | 138182800 | 138183224 | 425 | 2 | 1.36E-07 | 0.0004952 | 3.15E-06 | 4.10E-06 | -0.01071684 | SLC7A11 |
| chr2 | 27363402 | 27364197 | 796 | 2 | 2.20E-05 | 2.46E-06 | 0.000384 | 4.10E-06 | -0.00755351 | AC074117.1 |
| chr12 | 53989450 | 53989908 | 459 | 5 | 4.49E-06 | 7.47E-06 | 0.002234 | 4.10E-06 | 0.005333136 | HOXC10, AC012531.3 |
| chr6 | 143606733 | 143607565 | 833 | 5 | 5.40E-06 | 2.42E-05 | 6.02E-05 | 4.10E-06 | 0.008598289 | PHACTR2 |
| chr15 | 74812284 | 74812945 | 662 | 8 | 5.20E-07 | 2.04E-05 | 6.63E-05 | 4.10E-06 | -0.00286342 | LMAN1L |
| chr2 | 50961168 | 50961543 | 376 | 2 | 5.79E-07 | 1.40E-05 | 1.40E-05 | 4.11E-06 | -0.01486994 | NRXN1 |

| | | | | | | | | | | |
|---|---|---|---|---|---|---|---|---|---|---|
| chr11 | 69813030 | 69813403 | 374 | 2 | 1.03E-06 | 1.47E-05 | 1.34E-05 | 4.11E-06 | 0.014008413 | FGF3 |
| chr16 | 75575162 | 75575488 | 327 | 3 | 5.82E-06 | 1.56E-06 | 0.00164 | 4.11E-06 | -0.00038638 | GABARAPL2, AC025287.1 |
| chr17 | 17780696 | 17780926 | 231 | 2 | 3.51E-06 | 2.52E-06 | 0.000326 | 4.11E-06 | -0.00289917 | RAI1 |
| chr11 | 3641737 | 3642858 | 1122 | 14 | 2.86E-05 | 1.50E-05 | 0.004631 | 4.11E-06 | -0.0016482 | ART5 |
| chr8 | 43191107 | 43191446 | 340 | 3 | 6.63E-06 | 6.55E-05 | 0.000149 | 4.11E-06 | -0.00196694 | HGSNAT |
| chr14 | 94114214 | 94114787 | 574 | 3 | 4.86E-06 | 5.19E-06 | 7.11E-05 | 4.11E-06 | -0.00306582 | IFI27 |
| chr2 | 100285667 | 100285783 | 117 | 2 | 6.53E-06 | 5.11E-06 | 4.30E-05 | 4.11E-06 | 0.032505458 | LONRF2 |
| chr15 | 89720550 | 89720978 | 429 | 4 | 3.97E-06 | 1.20E-05 | 0.000461 | 4.11E-06 | -0.00760911 | WDR93 |
| chr10 | 69179515 | 69180903 | 1389 | 14 | 1.76E-05 | 7.81E-06 | 0.000163 | 4.11E-06 | -0.00133706 | SUPV3L1 |
| chr19 | 42325100 | 42326444 | 1345 | 14 | 7.07E-06 | 9.75E-06 | 0.012546 | 4.11E-06 | -0.00093169 | MEGF8 |
| chr9 | 16112068 | 16112637 | 570 | 2 | 9.57E-06 | 2.80E-06 | 0.000184 | 4.11E-06 | -0.00500631 | NA |
| chr15 | 25708137 | 25708367 | 231 | 3 | 2.83E-06 | 9.80E-06 | 6.83E-05 | 4.12E-06 | -0.00062672 | ATP10A |
| chr17 | 1405322 | 1405433 | 112 | 5 | 5.25E-06 | 2.70E-05 | 0.000285 | 4.12E-06 | -0.00227778 | NA |
| chr15 | 50354976 | 50355950 | 975 | 21 | 3.17E-06 | 2.11E-05 | 0.006713 | 4.12E-06 | 0.000946646 | GABPB1-AS1, GABPB1 |
| chr1 | 235853700 | 235854025 | 326 | 4 | 2.37E-06 | 2.88E-05 | 0.000221 | 4.12E-06 | -0.01993027 | LYST |
| chr11 | 95696753 | 95697506 | 754 | 3 | 3.49E-07 | 1.86E-05 | 1.68E-05 | 4.12E-06 | -0.00143725 | AP000820.2 |
| chr22 | 39318188 | 39319151 | 964 | 6 | 3.16E-07 | 5.92E-05 | 0.000987 | 4.12E-06 | -0.00138858 | RPL3, SNORD43 |
| chr19 | 12513652 | 12514235 | 584 | 9 | 2.17E-06 | 1.29E-05 | 0.001939 | 4.12E-06 | -0.00747469 | ZNF709, AC008758.1 |
| chr3 | 158672285 | 158673251 | 967 | 13 | 2.54E-07 | 0.0004319 | 3.31E-05 | 4.13E-06 | -0.00066413 | GFM1, LXN |
| chr14 | 24373240 | 24373955 | 716 | 4 | 1.26E-06 | 0.0019761 | 5.78E-05 | 4.13E-06 | -0.00367453 | NFATC4 |
| chr16 | 4752468 | 4752989 | 522 | 5 | 9.10E-07 | 0.0003234 | 0.000174 | 4.14E-06 | -0.00023799 | AC020663.1, ZNF500 |
| chr9 | 134839179 | 134840034 | 856 | 4 | 1.97E-05 | 1.73E-06 | 0.001422 | 4.14E-06 | -0.00300246 | COL5A1 |
| chr11 | 62605965 | 62606287 | 323 | 4 | 6.26E-07 | 5.98E-06 | 0.000137 | 4.14E-06 | -0.00407465 | AP001458.1, EML3 |
| chr14 | 19874894 | 19875225 | 332 | 2 | 1.04E-07 | 0.00085 | 2.96E-06 | 4.14E-06 | 0.022507627 | OR4K2 |
| chr6 | 33073441 | 33073920 | 480 | 8 | 1.26E-07 | 3.09E-05 | 2.40E-05 | 4.14E-06 | -0.00177965 | HLA-DPA1 |
| chr18 | 48054031 | 48054651 | 621 | 3 | 2.87E-06 | 3.57E-05 | 2.92E-05 | 4.15E-06 | -0.00059117 | ZBTB7C |
| chr12 | 132052452 | 132053230 | 779 | 7 | 1.11E-05 | 1.49E-05 | 0.001948 | 4.15E-06 | 0.000325309 | EP400 |
| chr9 | 89364234 | 89364821 | 588 | 3 | 2.57E-06 | 2.73E-05 | 0.00018 | 4.15E-06 | -0.00531363 | SEMA4D |
| chr19 | 18939755 | 18940938 | 1184 | 11 | 5.88E-06 | 1.00E-06 | 0.010223 | 4.15E-06 | -0.00131583 | HOMER3-AS1, HOMER3 |
| chr7 | 122886617 | 122887468 | 852 | 13 | 3.81E-07 | 7.72E-06 | 0.001279 | 4.15E-06 | 0.000655143 | CADPS2 |
| chr12 | 131222446 | 131222950 | 505 | 3 | 6.07E-08 | 6.55E-05 | 6.05E-06 | 4.15E-06 | 0.005902388 | LINC01257 |
| chr12 | 6039740 | 6040564 | 825 | 4 | 9.07E-06 | 1.43E-06 | 0.001998 | 4.16E-06 | -0.00338692 | VWF |
| chr6 | 38599216 | 38599565 | 350 | 3 | 2.65E-06 | 5.02E-05 | 1.33E-05 | 4.16E-06 | -0.00353663 | BTBD9 |
| chr1 | 206790706 | 206791079 | 374 | 2 | 4.07E-06 | 4.88E-06 | 4.73E-05 | 4.16E-06 | 0.004038503 | IL19 |
| chr13 | 52616134 | 52616911 | 778 | 3 | 1.59E-08 | 0.006114 | 1.10E-06 | 4.16E-06 | 0.004267749 | AL137058.2, MRPS31P4 |
| chr18 | 77342316 | 77342871 | 556 | 2 | 1.59E-06 | 4.22E-06 | 6.12E-05 | 4.16E-06 | -0.00228368 | NA |
| chr13 | 28033950 | 28034145 | 196 | 3 | 1.03E-05 | 2.55E-06 | 0.000291 | 4.16E-06 | -0.00380646 | FLT3 |
| chr6 | 170221577 | 170222580 | 1004 | 6 | 9.49E-08 | 1.86E-05 | 0.000271 | 4.16E-06 | -0.00455813 | NA |
| chr20 | 59645740 | 59646076 | 337 | 2 | 8.10E-06 | 2.81E-06 | 0.000192 | 4.16E-06 | 0.007571124 | PHACTR3 |
| chr11 | 9138049 | 9138774 | 726 | 3 | 4.86E-06 | 1.54E-06 | 0.001862 | 4.16E-06 | -0.00810684 | SCUBE2 |
| chr7 | 35261458 | 35262250 | 793 | 4 | 1.26E-07 | 0.0011822 | 4.50E-06 | 4.16E-06 | -0.00153031 | NA |
| chr11 | 59866401 | 59866803 | 403 | 4 | 1.02E-06 | 3.52E-05 | 3.06E-05 | 4.17E-06 | -0.01077286 | TCN1 |
| chr2 | 73385174 | 73385765 | 592 | 12 | 1.98E-06 | 6.88E-05 | 0.00061 | 4.17E-06 | -2.85E-05 | ALMS1, AC074008.2 |
| chr13 | 23569196 | 23569551 | 356 | 3 | 1.91E-08 | 0.0001159 | 4.22E-06 | 4.18E-06 | -0.01692849 | NA |
| chr11 | 15864859 | 15865162 | 304 | 2 | 2.62E-06 | 1.33E-05 | 1.49E-05 | 4.18E-06 | -0.01004812 | AC009869.1 |
| chr12 | 58172654 | 58172834 | 181 | 3 | 3.26E-06 | 3.99E-05 | 1.24E-05 | 4.18E-06 | 5.32E-05 | NA |
| chr12 | 110500060 | 110500464 | 405 | 3 | 6.60E-09 | 0.0050957 | 1.10E-06 | 4.18E-06 | -0.01045188 | VPS29 |
| chr2 | 10838019 | 10838844 | 826 | 4 | 3.81E-06 | 6.78E-06 | 0.0003 | 4.18E-06 | -0.00445302 | NA |
| chr19 | 19179730 | 19180157 | 428 | 3 | 1.48E-05 | 1.48E-06 | 0.002199 | 4.19E-06 | -0.00884879 | MEF2B, BORCS8-MEF2B, BORCS8 |
| chr17 | 57365482 | 57365653 | 172 | 2 | 1.93E-06 | 7.62E-06 | 2.62E-05 | 4.19E-06 | 0.007789302 | MSI2 |
| chr17 | 74147307 | 74147882 | 576 | 2 | 1.94E-06 | 6.88E-06 | 2.97E-05 | 4.19E-06 | -0.00389177 | LINC02074 |
| chr11 | 124697009 | 124697116 | 108 | 2 | 4.50E-06 | 1.84E-05 | 1.16E-05 | 4.19E-06 | -0.00208868 | SPA17 |
| chr17 | 35568209 | 35568590 | 382 | 3 | 1.93E-06 | 8.13E-06 | 8.75E-05 | 4.19E-06 | 0.003217188 | SNHG30 |
| chr22 | 23856821 | 23857832 | 1012 | 12 | 2.54E-06 | 0.0001124 | 0.003245 | 4.19E-06 | 0.002439247 | SLC2A11, AP000350.5 |
| chr22 | 36609994 | 36610669 | 676 | 2 | 5.52E-08 | 0.0002637 | 3.62E-06 | 4.19E-06 | -0.00936434 | CACNG2 |
| chr6 | 3357388 | 3357828 | 441 | 2 | 2.43E-06 | 3.56E-06 | 8.97E-05 | 4.19E-06 | -0.00501154 | SLC22A23 |
| chr16 | 30014043 | 30015026 | 984 | 4 | 2.26E-06 | 0.0006901 | 1.32E-05 | 4.19E-06 | 0.000517258 | DOC2A |
| chr11 | 133532289 | 133533316 | 1028 | 9 | 1.81E-07 | 0.0008203 | 2.95E-05 | 4.20E-06 | 0.002022916 | AP004606.1, OPCML |
| chr21 | 45017562 | 45017761 | 200 | 2 | 4.48E-06 | 7.91E-06 | 2.51E-05 | 4.20E-06 | 0.00780231 | NA |
| chr17 | 40984114 | 40984558 | 445 | 4 | 4.56E-06 | 2.38E-06 | 0.000429 | 4.20E-06 | -0.01833936 | KRT40 |
| chr11 | 68029876 | 68031065 | 1190 | 15 | 1.80E-05 | 1.52E-06 | 0.023445 | 4.20E-06 | 0.001075847 | NDUFS8, AC004923.4 |
| chr11 | 45253760 | 45253892 | 133 | 4 | 5.33E-06 | 8.12E-06 | 4.53E-05 | 4.20E-06 | -0.00248013 | AC103681.1, SYT13 |
| chr17 | 49848002 | 49848877 | 876 | 5 | 4.70E-06 | 1.09E-05 | 0.001344 | 4.21E-06 | -0.0052916 | AC027801.1, TAC4 |
| chr5 | 177467689 | 177468167 | 479 | 3 | 2.39E-07 | 0.000299 | 4.64E-06 | 4.21E-06 | 0.003276565 | DBN1 |
| chr6 | 36840638 | 36841118 | 481 | 4 | 1.06E-06 | 3.93E-06 | 0.000705 | 4.21E-06 | -0.00745163 | NA |
| chr11 | 115153391 | 115154223 | 833 | 5 | 1.42E-06 | 6.93E-06 | 0.000498 | 4.21E-06 | 0.004459621 | NA |
| chr9 | 89352857 | 89353433 | 577 | 3 | 1.02E-06 | 9.91E-06 | 8.86E-05 | 4.21E-06 | -0.00507663 | SECISBP2 |
| chr16 | 49529848 | 49530459 | 612 | 4 | 1.49E-05 | 2.84E-06 | 0.001505 | 4.21E-06 | -0.01229984 | ZNF423 |
| chr2 | 45849775 | 45850104 | 330 | 2 | 1.97E-06 | 3.12E-06 | 0.000134 | 4.21E-06 | -0.025291 | PRKCE |
| chr1 | 15764223 | 15764723 | 501 | 6 | 2.44E-05 | 1.21E-06 | 0.005609 | 4.22E-06 | -0.0057749 | FBLIM1 |
| chr3 | 146085872 | 146086169 | 298 | 2 | 2.77E-06 | 2.99E-06 | 0.000156 | 4.22E-06 | -0.01384684 | AC107021.1, PLOD2 |
| chr17 | 49016127 | 49016890 | 764 | 3 | 2.38E-05 | 1.79E-06 | 0.00093 | 4.22E-06 | -0.01818174 | IGF2BP1 |
| chr6 | 162726390 | 162727258 | 869 | 8 | 4.96E-06 | 6.75E-06 | 0.000686 | 4.22E-06 | 0.007505307 | PACRG, PRKN |
| chr12 | 6361988 | 6362185 | 198 | 3 | 9.56E-06 | 1.91E-06 | 0.000763 | 4.22E-06 | -0.01151833 | SCNN1A |
| chr2 | 74384373 | 74385095 | 723 | 4 | 5.84E-07 | 2.09E-05 | 0.000121 | 4.22E-06 | -0.00119145 | DCTN1 |
| chr7 | 139105168 | 139105695 | 528 | 2 | 1.70E-07 | 0.0002333 | 3.76E-06 | 4.22E-06 | 0.00461474 | ZC3HAV1 |
| chr22 | 35607287 | 35607343 | 57 | 2 | 2.24E-06 | 4.18E-05 | 6.96E-06 | 4.22E-06 | -0.00652487 | MB |
| chr3 | 120742032 | 120742947 | 916 | 15 | 7.82E-06 | 2.18E-05 | 0.001214 | 4.22E-06 | 0.001024861 | GTF2E1, RABL3 |
| chr3 | 179027787 | 179028138 | 352 | 2 | 5.88E-07 | 0.0003154 | 3.53E-06 | 4.23E-06 | -0.03250325 | ZMAT3 |
| chr10 | 99358483 | 99358516 | 34 | 2 | 1.17E-06 | 3.23E-06 | 0.000121 | 4.23E-06 | -0.00347113 | CNNM1 |
| chr1 | 94274919 | 94275127 | 209 | 2 | 6.80E-06 | 2.58E-06 | 0.000349 | 4.23E-06 | -0.00342485 | ARHGAP29-AS1, ARHGAP29 |
| chr1 | 44043810 | 44044301 | 492 | 4 | 2.54E-06 | 2.26E-06 | 0.001423 | 4.23E-06 | 0.00700844 | AL139220.2 |
| chr1 | 222814379 | 222815097 | 719 | 7 | 2.43E-06 | 1.93E-05 | 0.002108 | 4.24E-06 | 0.007216407 | DISP1 |
| chr1 | 3139135 | 3139978 | 844 | 4 | 9.97E-08 | 2.69E-05 | 3.53E-05 | 4.24E-06 | 0.004031801 | PRDM16 |
| chr12 | 27433267 | 27434242 | 976 | 2 | 9.89E-05 | 2.55E-06 | 0.000389 | 4.24E-06 | -0.01399477 | ARNTL2-AS1 |
| chr6 | 159906313 | 159906328 | 16 | 2 | 4.18E-06 | 4.16E-06 | 6.56E-05 | 4.24E-06 | -0.0046386 | MAS1 |
| chr13 | 33312524 | 33312875 | 352 | 2 | 7.90E-06 | 2.95E-06 | 0.000173 | 4.25E-06 | -0.01066391 | STARD13 |
| chr6 | 36697463 | 36697493 | 31 | 2 | 2.09E-05 | 4.25E-06 | 6.36E-05 | 4.25E-06 | -0.01010732 | NA |
| chr20 | 34515779 | 34516658 | 880 | 11 | 1.22E-05 | 5.14E-06 | 0.00333 | 4.26E-06 | 3.27E-05 | ITCH, DYNLRB1 |
| chr19 | 43781416 | 43781924 | 509 | 6 | 1.16E-06 | 6.41E-06 | 0.000382 | 4.26E-06 | -0.00430933 | NA |
| chr11 | 134393094 | 134393797 | 704 | 7 | 7.45E-06 | 2.62E-05 | 0.00427 | 4.26E-06 | -0.00746964 | B3GAT1 |
| chr1 | 3533261 | 3533663 | 403 | 2 | 1.32E-06 | 5.60E-06 | 4.04E-05 | 4.26E-06 | -0.00901576 | MEGF6 |
| chr1 | 171780977 | 171781786 | 810 | 14 | 7.66E-06 | 1.92E-05 | 0.005378 | 4.26E-06 | 0.000108818 | EEF1AKNMT |
| chr9 | 21077880 | 21078540 | 661 | 3 | 3.71E-06 | 2.71E-06 | 0.000265 | 4.26E-06 | -0.00862245 | IFNB1 |
| chr14 | 61322092 | 61323378 | 1287 | 15 | 6.55E-07 | 1.27E-05 | 0.005857 | 4.27E-06 | -0.00170977 | PRKCH, AL359220.1 |
| chr16 | 88778091 | 88778796 | 706 | 5 | 1.21E-06 | 5.42E-05 | 9.60E-05 | 4.27E-06 | 0.005708238 | PIEZO1 |
| chr5 | 73381403 | 73382059 | 657 | 5 | 4.91E-06 | 2.87E-06 | 0.00107 | 4.27E-06 | 0.002958297 | NA |
| chr4 | 21205928 | 21206273 | 346 | 3 | 7.74E-06 | 2.84E-06 | 0.000221 | 4.27E-06 | -0.00944611 | KCNIP4 |
| chr22 | 43919385 | 43919863 | 479 | 3 | 1.25E-05 | 1.75E-06 | 0.001082 | 4.27E-06 | -0.00774051 | NA |
| chr9 | 137199225 | 137200618 | 1394 | 5 | 9.74E-08 | 0.0002717 | 3.40E-05 | 4.27E-06 | -0.00470965 | TPRN |
| chr11 | 2198792 | 2199155 | 364 | 2 | 1.60E-07 | 4.74E-06 | 5.27E-05 | 4.27E-06 | -0.00434256 | NA |
| chr5 | 6758855 | 6759016 | 162 | 3 | 6.22E-06 | 3.09E-05 | 0.000152 | 4.27E-06 | 0.000112455 | NA |
| chr4 | 123547685 | 123548209 | 525 | 2 | 4.63E-08 | 0.0003898 | 3.43E-06 | 4.28E-06 | -0.00830441 | LINC01091 |
| chr17 | 50686804 | 50687605 | 802 | 5 | 1.36E-06 | 0.000182 | 0.000357 | 4.28E-06 | -0.00550856 | ABCC3 |
| chr11 | 579119 | 579621 | 503 | 3 | 3.50E-06 | 3.43E-06 | 0.000145 | 4.28E-06 | -0.00772422 | PHRF1 |
| chr1 | 92970717 | 92971151 | 435 | 3 | 4.29E-06 | 1.81E-06 | 0.00133 | 4.28E-06 | 0.00424003 | NA |
| chr21 | 28537969 | 28538440 | 472 | 3 | 6.22E-08 | 5.48E-05 | 7.93E-06 | 4.28E-06 | -0.00085344 | AF165147.1 |
| chr19 | 44954416 | 44955555 | 1140 | 18 | 2.60E-06 | 0.0011486 | 0.004634 | 4.28E-06 | -0.0008899 | CLPTM1 |
| chr1 | 111289487 | 111289971 | 485 | 4 | 7.20E-06 | 5.10E-06 | 0.000209 | 4.29E-06 | -0.00687164 | NA |
| chr6 | 33450879 | 33452450 | 1572 | 20 | 1.43E-05 | 0.0020839 | 0.000951 | 4.29E-06 | -0.0014051 | SYNGAP1, SYNGAP1-AS1 |
| chr13 | 75548950 | 75550288 | 1339 | 14 | 2.27E-05 | 5.72E-05 | 0.007131 | 4.29E-06 | 0.000156687 | UCHL3, AL137782.1, COMMD6 |
| chr17 | 1170907 | 1171571 | 665 | 3 | 9.73E-06 | 2.08E-06 | 0.000978 | 4.29E-06 | -0.00539033 | ABR |
| chr12 | 56193302 | 56193742 | 441 | 2 | 3.03E-06 | 5.99E-06 | 3.70E-05 | 4.29E-06 | -0.00984329 | NA |
| chr2 | 42810487 | 42811259 | 773 | 5 | 4.08E-06 | 3.93E-05 | 0.000171 | 4.29E-06 | 0.001951595 | NA |
| chr20 | 57555851 | 57556013 | 163 | 4 | 5.09E-06 | 2.08E-05 | 2.43E-05 | 4.29E-06 | -0.01770889 | NA |
| chr13 | 114226214 | 114226965 | 752 | 5 | 1.06E-06 | 0.0001492 | 3.02E-05 | 4.29E-06 | -0.00790072 | NA |
| chr22 | 48835751 | 48835801 | 51 | 2 | 1.24E-06 | 0.0003991 | 3.43E-06 | 4.29E-06 | 0.008626231 | TAFA5 |
| chr9 | 111759173 | 111759793 | 621 | 5 | 4.23E-07 | 0.0027534 | 7.45E-06 | 4.29E-06 | 0.002130595 | SHOC1 |
| chr22 | 30425517 | 30426522 | 1006 | 8 | 7.98E-06 | 1.13E-05 | 0.000615 | 4.30E-06 | -0.00189696 | AC004832.3, MTFP1 |
| chr1 | 2274331 | 2274463 | 133 | 3 | 1.42E-05 | 2.59E-06 | 0.000851 | 4.30E-06 | -0.00466563 | SKI |
| chr14 | 22819784 | 22819905 | 122 | 3 | 2.81E-06 | 4.06E-06 | 0.00011 | 4.30E-06 | -0.00822534 | SLC7A7 |
| chr18 | 35497533 | 35498755 | 1223 | 13 | 1.36E-06 | 4.01E-05 | 0.000255 | 4.30E-06 | -0.00210537 | AC007998.2, INO80C |
| chr16 | 85282317 | 85283005 | 689 | 7 | 1.08E-05 | 2.58E-06 | 0.009578 | 4.30E-06 | -0.00377562 | GSE1, AC092275.1, LINC00311 |
| chr15 | 47523951 | 47524195 | 245 | 2 | 3.19E-06 | 3.46E-06 | 0.000104 | 4.30E-06 | -0.00968985 | SEMA6D |
| chr12 | 57147672 | 57148438 | 767 | 3 | 4.43E-06 | 5.30E-06 | 0.000311 | 4.31E-06 | 0.003662136 | LRP1 |
| chr11 | 82901365 | 82902108 | 744 | 12 | 1.43E-06 | 1.67E-05 | 0.008989 | 4.31E-06 | -0.00030684 | DDIAS, PRCP |
| chr9 | 101027731 | 101028450 | 720 | 5 | 4.06E-08 | 6.58E-05 | 0.000244 | 4.31E-06 | -0.00846572 | NA |
| chr11 | 113858541 | 113859244 | 704 | 3 | 7.53E-07 | 0.0005146 | 7.16E-06 | 4.31E-06 | -0.0072538 | USP28 |
| chr17 | 38455447 | 38456665 | 1219 | 5 | 1.29E-06 | 0.000224 | 1.93E-05 | 4.31E-06 | -0.00563373 | ARHGAP23 |
| chr3 | 69013611 | 69015024 | 1414 | 13 | 4.80E-06 | 7.73E-06 | 0.000295 | 4.31E-06 | 0.000890621 | AC109587.1, EOGT |
| chr1 | 6084494 | 6085071 | 578 | 3 | 1.45E-06 | 2.72E-05 | 1.50E-05 | 4.31E-06 | -8.63E-05 | KCNAB2 |
| chr19 | 10502504 | 10503335 | 832 | 9 | 4.26E-06 | 5.41E-06 | 0.004555 | 4.31E-06 | 0.000390463 | KEAP1 |
| chr20 | 59987491 | 59988385 | 895 | 3 | 1.96E-05 | 1.23E-05 | 8.06E-05 | 4.32E-06 | -0.00590207 | CDH26 |
| chr1 | 169629761 | 169630516 | 756 | 4 | 1.98E-06 | 0.0001932 | 2.04E-05 | 4.32E-06 | -0.00702999 | SELP |
| chr8 | 57978000 | 57978098 | 99 | 2 | 7.54E-06 | 5.62E-06 | 4.14E-05 | 4.33E-06 | -0.0095273 | LINC01602 |
| chr3 | 141095077 | 141095291 | 215 | 3 | 1.04E-05 | 2.45E-06 | 0.000876 | 4.33E-06 | -0.01749822 | SPSB4 |
| chr12 | 55704858 | 55705630 | 773 | 3 | 9.57E-06 | 1.63E-06 | 0.001642 | 4.33E-06 | -0.00247896 | ITGA7 |
| chr15 | 88126966 | 88127221 | 256 | 2 | 1.23E-06 | 4.73E-06 | 5.48E-05 | 4.33E-06 | -0.01792665 | NTRK3 |
| chr1 | 117366400 | 117366987 | 588 | 5 | 1.75E-07 | 0.0049688 | 3.08E-06 | 4.33E-06 | -0.00059951 | NA |
| chr8 | 93166763 | 93167155 | 393 | 2 | 2.07E-06 | 5.00E-06 | 4.98E-05 | 4.33E-06 | -0.00496289 | C8orf87 |
| chr14 | 101526081 | 101527161 | 1081 | 5 | 1.67E-06 | 1.31E-05 | 0.000823 | 4.33E-06 | -0.00371448 | NA |
| chr19 | 19655135 | 19655852 | 718 | 5 | 4.28E-06 | 3.89E-06 | 0.000318 | 4.34E-06 | -7.68E-05 | ATP13A1 |
| chr10 | 80356129 | 80356852 | 724 | 12 | 2.72E-06 | 4.11E-05 | 0.001168 | 4.34E-06 | -0.00157811 | DYDC2, DYDC1 |
| chr4 | 10021360 | 10021577 | 218 | 4 | 4.72E-06 | 2.87E-05 | 0.000102 | 4.34E-06 | -0.0066236 | SLC2A9 |
| chr1 | 24083979 | 24084525 | 547 | 6 | 1.45E-06 | 0.0003405 | 2.11E-05 | 4.34E-06 | -0.0036377 | AL591178.1, MYOM3 |
| chr15 | 74819805 | 74820316 | 512 | 3 | 6.02E-06 | 1.80E-06 | 0.001219 | 4.35E-06 | -0.00482389 | LMAN1L, AC091230.1 |
| chr16 | 47122745 | 47122989 | 245 | 2 | 2.00E-06 | 4.27E-05 | 7.11E-06 | 4.35E-06 | -0.00477759 | NETO2 |
| chr8 | 1462744 | 1463533 | 790 | 2 | 1.47E-05 | 2.61E-06 | 0.000395 | 4.35E-06 | -0.00481318 | DLGAP2 |
| chr7 | 129208845 | 129209418 | 574 | 4 | 3.07E-06 | 2.64E-06 | 0.001131 | 4.35E-06 | 0.001422802 | SMO |
| chr5 | 181221409 | 181222174 | 766 | 4 | 9.70E-08 | 4.95E-06 | 0.000884 | 4.35E-06 | -0.01382055 | NA |
| chr11 | 30918407 | 30918913 | 507 | 2 | 1.25E-07 | 0.0459153 | 2.55E-06 | 4.35E-06 | -0.0087919 | DCDC1 |
| chr7 | 103148949 | 103149721 | 773 | 8 | 8.95E-06 | 3.33E-05 | 0.000895 | 4.35E-06 | -0.00147885 | NAPEPLD |
| chr1 | 2627881 | 2628911 | 1031 | 6 | 3.63E-07 | 0.0002534 | 1.02E-05 | 4.36E-06 | -0.00800564 | MMEL1 |
| chr11 | 119701853 | 119702294 | 442 | 3 | 2.77E-06 | 7.77E-06 | 0.000164 | 4.36E-06 | -0.00344736 | NECTIN1 |
| chr9 | 81605177 | 81605448 | 272 | 2 | 5.35E-06 | 5.55E-06 | 4.29E-05 | 4.36E-06 | -0.01978393 | TLE1 |
| chr19 | 35213923 | 35214089 | 167 | 2 | 1.02E-05 | 2.70E-06 | 0.00032 | 4.36E-06 | 0.001998975 | NA |
| chr4 | 173334343 | 173335252 | 910 | 15 | 4.94E-06 | 4.93E-05 | 0.005805 | 4.36E-06 | 0.000306946 | HMGB2 |
| chr5 | 68290342 | 68291100 | 759 | 6 | 7.91E-06 | 8.12E-06 | 0.000104 | 4.36E-06 | -0.00321182 | PIK3R1 |

| | | | | | | | | | | |
|---|---|---|---|---|---|---|---|---|---|---|
| chr5 | 85112839 | 85113386 | 548 | 4 | 1.03E-05 | 3.32E-06 | 0.001108 | 4.36E-06 | -0.01202593 | NA |
| chr14 | 26413184 | 26413458 | 275 | 5 | 8.37E-06 | 3.82E-06 | 0.000323 | 4.36E-06 | 0.001636071 | NA |
| chr14 | 28771485 | 28771786 | 302 | 3 | 6.10E-07 | 0.0025025 | 1.35E-06 | 4.38E-06 | -0.01153277 | AL049777.1, LINC01551 |
| chr5 | 133294987 | 133295728 | 742 | 2 | 6.22E-05 | 2.61E-06 | 0.000423 | 4.39E-06 | -0.00861646 | FSTL4 |
| chr10 | 10959478 | 10960099 | 622 | 5 | 4.72E-07 | 0.0002077 | 1.64E-05 | 4.39E-06 | -0.00687378 | CELF2 |
| chr12 | 123149181 | 123149612 | 432 | 2 | 9.75E-07 | 5.88E-06 | 3.98E-05 | 4.39E-06 | -0.00705731 | PITPNM2 |
| chr14 | 29896318 | 29896548 | 231 | 2 | 6.03E-06 | 5.56E-06 | 4.33E-05 | 4.39E-06 | 0.002165185 | PRKD1 |
| chr7 | 140478167 | 140479823 | 1657 | 15 | 2.81E-06 | 4.97E-05 | 0.00657 | 4.39E-06 | 0.001142525 | MKRN1 |
| chr17 | 45639149 | 45639251 | 103 | 3 | 7.67E-06 | 5.23E-06 | 7.38E-05 | 4.39E-06 | -0.00142384 | LINC02210, LINC02210-CRHR1 |
| chr2 | 38956289 | 38956962 | 674 | 4 | 1.24E-05 | 8.33E-06 | 0.001244 | 4.39E-06 | -0.00073909 | ARHGEF33 |
| chr8 | 145051814 | 145052617 | 804 | 8 | 9.91E-06 | 2.48E-06 | 0.003464 | 4.39E-06 | 0.000483594 | C8orf33 |
| chr2 | 95277377 | 95277734 | 358 | 2 | 6.78E-08 | 7.79E-05 | 5.46E-06 | 4.39E-06 | -0.00521391 | PROM2 |
| chr16 | 68698201 | 68698671 | 471 | 4 | 3.35E-06 | 2.76E-05 | 0.000221 | 4.39E-06 | -0.00060979 | CDH3 |
| chr7 | 106057151 | 106057903 | 753 | 3 | 5.64E-06 | 4.82E-06 | 0.000224 | 4.39E-06 | -0.00688774 | NA |
| chr6 | 35643777 | 35643834 | 58 | 3 | 2.03E-06 | 2.02E-05 | 1.84E-05 | 4.40E-06 | 0.005271579 | FKBP5 |
| chr1 | 26343883 | 26344456 | 574 | 4 | 2.75E-06 | 0.0001648 | 6.23E-05 | 4.40E-06 | -0.00093376 | CRYBG2 |
| chr5 | 170502952 | 170504385 | 1434 | 11 | 1.08E-05 | 8.87E-05 | 0.001573 | 4.40E-06 | -0.00232962 | KCNIP1 |
| chr20 | 25248740 | 25249588 | 849 | 3 | 1.26E-06 | 3.62E-06 | 0.000144 | 4.41E-06 | -0.01030162 | PYGB |
| chr12 | 114399303 | 114400554 | 1252 | 6 | 1.37E-07 | 0.0001172 | 0.000198 | 4.41E-06 | -0.00125456 | TBX5 |
| chr10 | 124667946 | 124668495 | 550 | 2 | 4.56E-07 | 5.78E-06 | 4.14E-05 | 4.41E-06 | 0.001629615 | AC068896.1, FAM53B |
| chr12 | 513451 | 514011 | 561 | 3 | 1.13E-05 | 3.30E-06 | 0.000285 | 4.41E-06 | 0.000246924 | B4GALNT3 |
| chr7 | 3371724 | 3371871 | 148 | 2 | 6.07E-06 | 8.38E-06 | 2.56E-05 | 4.42E-06 | 0.010980467 | SDK1 |
| chr20 | 44684842 | 44685301 | 460 | 4 | 7.05E-06 | 6.48E-06 | 0.000805 | 4.42E-06 | -0.00481919 | AL139352.1 |
| chr7 | 153835474 | 153836138 | 665 | 2 | 2.18E-05 | 2.76E-06 | 0.000305 | 4.42E-06 | 0.015704495 | NA |
| chr6 | 49877462 | 49878247 | 786 | 4 | 1.30E-06 | 6.55E-05 | 0.000262 | 4.42E-06 | -0.01359148 | NA |
| chr17 | 47840855 | 47841638 | 784 | 10 | 6.52E-08 | 0.002001 | 0.000141 | 4.42E-06 | -3.79E-05 | SCRN2 |
| chr12 | 98745523 | 98745793 | 271 | 3 | 4.42E-06 | 6.19E-06 | 7.66E-05 | 4.42E-06 | -0.00963164 | ANKS1B |
| chr7 | 76265317 | 76265358 | 42 | 2 | 2.53E-06 | 3.01E-06 | 0.00019 | 4.42E-06 | -0.00541713 | SRRM3 |
| chr19 | 16076030 | 16076821 | 792 | 6 | 4.75E-06 | 2.19E-06 | 0.001341 | 4.43E-06 | 0.000675518 | TPM4, AC008894.3 |
| chr5 | 29947416 | 29947423 | 8 | 2 | 8.44E-06 | 5.40E-06 | 4.63E-05 | 4.43E-06 | 0.007251792 | NA |
| chr8 | 60516616 | 60517268 | 653 | 10 | 8.61E-06 | 1.77E-05 | 0.000765 | 4.43E-06 | 0.000673633 | RAB2A, LINC01301 |
| chr15 | 90184131 | 90184925 | 795 | 11 | 5.06E-06 | 1.13E-05 | 0.00267 | 4.44E-06 | -0.00646115 | SEMA4B |
| chr20 | 50133679 | 50133697 | 19 | 2 | 4.03E-06 | 9.97E-06 | 2.14E-05 | 4.44E-06 | 0.003279128 | PEDS1-UBE2V1, PEDS1 |
| chr14 | 69483326 | 69484178 | 853 | 3 | 7.89E-07 | 2.14E-06 | 0.000616 | 4.44E-06 | -0.01035311 | AL157996.2 |
| chr17 | 31805229 | 31805837 | 609 | 2 | 2.07E-06 | 6.68E-06 | 3.42E-05 | 4.44E-06 | -0.00836658 | NA |
| chr7 | 82444023 | 82444864 | 842 | 7 | 1.82E-07 | 0.0006 | 2.58E-05 | 4.44E-06 | -0.0050621 | NA |
| chr2 | 191150788 | 191151748 | 961 | 13 | 6.70E-08 | 0.0001048 | 4.99E-05 | 4.45E-06 | 0.001023594 | STAT4 |
| chr19 | 4457570 | 4458455 | 886 | 10 | 2.83E-05 | 1.47E-05 | 0.01327 | 4.45E-06 | -0.00035295 | UBXN6, AC011498.1 |
| chr11 | 76721756 | 76722256 | 501 | 11 | 2.90E-06 | 0.0003391 | 0.00074 | 4.45E-06 | -0.00081433 | NA |
| chr15 | 69461957 | 69462375 | 419 | 6 | 3.99E-08 | 0.0007046 | 2.17E-06 | 4.45E-06 | -0.00314539 | NA |
| chr8 | 142798311 | 142798695 | 385 | 3 | 2.01E-06 | 2.17E-05 | 7.62E-05 | 4.45E-06 | -0.01438476 | AC083841.2, AC083841.4 |
| chr15 | 80398085 | 80398719 | 635 | 3 | 1.83E-06 | 1.86E-05 | 0.000189 | 4.45E-06 | 0.003469297 | AC016705.2 |
| chr19 | 45766065 | 45767064 | 1000 | 4 | 6.58E-05 | 0.00031 | 0.000167 | 4.45E-06 | -0.0051739 | AC074212.1, SIX5 |
| chr10 | 7728801 | 7728896 | 96 | 3 | 1.44E-05 | 1.81E-06 | 0.001345 | 4.46E-06 | -0.00468872 | ITIH2 |
| chr12 | 121185749 | 121186974 | 1226 | 4 | 5.20E-05 | 2.38E-06 | 0.00252 | 4.46E-06 | 0.001115919 | P2RX7, AC069209.2 |
| chr19 | 7167810 | 7168061 | 252 | 2 | 4.39E-06 | 6.81E-06 | 3.36E-05 | 4.46E-06 | 0.007657867 | INSR |
| chr1 | 36083424 | 36083596 | 173 | 4 | 2.54E-06 | 0.0001364 | 1.24E-05 | 4.46E-06 | 0.008252849 | AL138787.1 |
| chr17 | 6077584 | 6077749 | 166 | 2 | 8.94E-06 | 2.69E-06 | 0.000387 | 4.47E-06 | -0.00842038 | AC007846.2, WSCD1 |
| chr1 | 212607630 | 212609094 | 1465 | 13 | 1.95E-05 | 0.0002054 | 0.000616 | 4.47E-06 | 0.001121123 | ATF3 |
| chr7 | 1326109 | 1326232 | 124 | 3 | 4.75E-06 | 7.27E-06 | 0.000433 | 4.47E-06 | -0.00717468 | NA |
| chr19 | 10416523 | 10416912 | 390 | 8 | 5.60E-06 | 4.67E-05 | 0.001179 | 4.47E-06 | -0.00226022 | PDE4A, CDC37 |
| chr6 | 36727591 | 36727634 | 44 | 2 | 7.13E-06 | 2.67E-06 | 0.000419 | 4.47E-06 | 0.00494914 | RAB44 |
| chr14 | 49596677 | 49596808 | 132 | 2 | 6.26E-06 | 2.76E-06 | 0.000333 | 4.47E-06 | 0.010854421 | RPS29 |
| chr12 | 114658425 | 114658622 | 198 | 2 | 6.48E-06 | 4.08E-06 | 7.86E-05 | 4.47E-06 | -0.01218614 | NA |
| chr1 | 5504490 | 5504791 | 302 | 2 | 7.98E-06 | 3.33E-06 | 0.000136 | 4.47E-06 | -0.01777773 | NA |
| chr9 | 126833194 | 126833301 | 108 | 4 | 7.26E-06 | 1.48E-05 | 0.000177 | 4.47E-06 | -0.00224932 | ZBTB43 |
| chr20 | 5190322 | 5190732 | 411 | 2 | 1.07E-06 | 1.69E-05 | 1.35E-05 | 4.48E-06 | -0.00233415 | CDS2 |
| chr8 | 118280850 | 118280988 | 139 | 3 | 7.37E-06 | 4.78E-06 | 0.000262 | 4.48E-06 | -0.00498192 | SAMD12 |
| chr19 | 1075068 | 1075596 | 529 | 3 | 7.67E-06 | 2.64E-06 | 0.000317 | 4.48E-06 | 0.003579603 | ARHGAP45 |
| chr7 | 92645544 | 92645861 | 318 | 2 | 7.61E-06 | 3.19E-06 | 0.00016 | 4.48E-06 | -0.00278946 | CDK6 |
| chr3 | 183427092 | 183428112 | 1021 | 6 | 9.46E-06 | 2.03E-05 | 0.001472 | 4.48E-06 | 0.001872401 | MCF2L2 |
| chr9 | 94800030 | 94800058 | 29 | 2 | 9.32E-07 | 5.33E-06 | 4.84E-05 | 4.48E-06 | -0.01996772 | AOPEP |
| chr4 | 94338989 | 94339419 | 431 | 2 | 2.25E-06 | 9.42E-06 | 2.32E-05 | 4.49E-06 | 0.011031315 | AC109925.1, HPGDS |
| chr16 | 53502901 | 53503691 | 791 | 10 | 1.67E-06 | 2.44E-05 | 3.59E-05 | 4.49E-06 | 0.000206652 | AKTIP |
| chr9 | 95874616 | 95875604 | 989 | 11 | 1.71E-06 | 1.53E-06 | 0.005723 | 4.49E-06 | 0.000922006 | ERCC6L2, LINC00476 |
| chr21 | 33588277 | 33588999 | 723 | 9 | 1.60E-06 | 4.14E-05 | 5.61E-05 | 4.50E-06 | -7.19E-05 | DONSON, AP000311.1 |
| chr7 | 19145327 | 19145784 | 458 | 9 | 9.27E-07 | 6.40E-05 | 7.47E-05 | 4.50E-06 | 0.001350564 | AC003986.1, FERD3L |
| chr1 | 209859963 | 209860540 | 578 | 4 | 2.77E-06 | 3.52E-06 | 0.000184 | 4.50E-06 | -0.0078383 | NA |
| chr14 | 22111747 | 22112468 | 722 | 5 | 7.41E-06 | 1.70E-05 | 0.000368 | 4.50E-06 | -0.01031737 | TRAV25 |
| chr20 | 38748221 | 38748973 | 753 | 8 | 9.58E-06 | 1.14E-05 | 0.004528 | 4.50E-06 | -0.00131143 | ACTR5 |
| chr9 | 23829191 | 23829437 | 247 | 2 | 1.89E-06 | 9.59E-06 | 2.29E-05 | 4.51E-06 | -0.01883679 | NA |
| chr12 | 62335054 | 62335340 | 287 | 3 | 1.31E-05 | 1.81E-06 | 0.00137 | 4.51E-06 | -0.00744184 | USP15 |
| chr3 | 16085134 | 16085603 | 470 | 2 | 2.43E-06 | 7.72E-06 | 2.93E-05 | 4.51E-06 | 0.037607996 | NA |
| chr17 | 82083098 | 82083948 | 851 | 5 | 4.16E-07 | 3.09E-05 | 4.03E-05 | 4.51E-06 | -0.0022217 | FASN |
| chr21 | 26111948 | 26112269 | 322 | 2 | 3.46E-06 | 9.31E-06 | 2.37E-05 | 4.51E-06 | -0.0092388 | APP |
| chr11 | 65872108 | 65873106 | 999 | 11 | 1.52E-06 | 8.19E-05 | 0.001023 | 4.51E-06 | -0.00223449 | EFEMP2 |
| chr13 | 46571317 | 46571962 | 646 | 2 | 2.55E-05 | 2.70E-06 | 0.000418 | 4.51E-06 | -0.00419753 | LRCH1 |
| chr1 | 225427967 | 225428966 | 1000 | 13 | 2.87E-06 | 0.0004591 | 0.000404 | 4.51E-06 | 0.000982093 | LBR |
| chr11 | 31805536 | 31807167 | 1632 | 13 | 5.97E-06 | 1.63E-05 | 0.000831 | 4.52E-06 | 0.000699105 | PAX6 |
| chr11 | 19240704 | 19241544 | 841 | 9 | 3.46E-06 | 5.87E-06 | 0.004447 | 4.52E-06 | 0.00079112 | CSRP3-AS1, E2F8 |
| chr19 | 3936584 | 3936878 | 295 | 2 | 8.17E-07 | 8.22E-05 | 5.51E-06 | 4.52E-06 | -0.00961826 | NMRK2 |
| chr7 | 44884709 | 44886205 | 1497 | 9 | 2.05E-05 | 9.53E-05 | 0.000209 | 4.53E-06 | 0.003251353 | AC004854.2, PURB |
| chr16 | 85750697 | 85751524 | 828 | 4 | 1.17E-06 | 3.74E-06 | 0.000166 | 4.53E-06 | -0.00803788 | C16orf74 |
| chr6 | 3160268 | 3160940 | 673 | 6 | 1.94E-06 | 0.0013963 | 0.000283 | 4.53E-06 | 0.007488996 | NA |
| chr7 | 33107196 | 33107525 | 330 | 2 | 2.17E-06 | 1.06E-05 | 2.07E-05 | 4.53E-06 | -0.01565731 | RP9 |
| chr15 | 34655875 | 34656259 | 385 | 3 | 1.22E-06 | 3.93E-05 | 0.000109 | 4.53E-06 | -0.0045776 | LINC02252 |
| chr4 | 70334606 | 70335034 | 429 | 4 | 1.14E-05 | 6.62E-06 | 0.000366 | 4.53E-06 | -0.00786745 | CABS1 |
| chr22 | 29683485 | 29684284 | 800 | 4 | 2.37E-06 | 3.54E-06 | 0.000384 | 4.53E-06 | -0.00149323 | NF2 |
| chr13 | 27827619 | 27828118 | 500 | 4 | 2.26E-06 | 2.34E-05 | 0.00039 | 4.53E-06 | 0.001239936 | PLUT |
| chr14 | 90256438 | 90257118 | 681 | 8 | 2.30E-05 | 5.05E-06 | 0.004131 | 4.53E-06 | -0.00078684 | PSMC1 |
| chr17 | 50386669 | 50386944 | 276 | 3 | 9.53E-06 | 4.30E-06 | 0.000581 | 4.53E-06 | -0.0010282 | LRRC59 |
| chr22 | 21761184 | 21761385 | 202 | 2 | 1.75E-06 | 1.05E-05 | 2.10E-05 | 4.54E-06 | -0.0106517 | MAPK1 |
| chr5 | 147877656 | 147877914 | 259 | 5 | 7.30E-06 | 8.30E-06 | 0.002515 | 4.54E-06 | -0.00301565 | SCGB3A2 |
| chr10 | 87817775 | 87818908 | 1134 | 13 | 3.46E-06 | 0.0004982 | 0.001471 | 4.55E-06 | -0.00035526 | CFL1P1, ATAD1 |
| chr13 | 105505871 | 105506709 | 839 | 5 | 3.18E-06 | 7.22E-06 | 0.000316 | 4.55E-06 | 0.0021603 | NA |
| chr12 | 100573352 | 100574365 | 1014 | 9 | 1.06E-06 | 0.0001351 | 0.000589 | 4.55E-06 | 0.000453729 | GAS2L3 |
| chr8 | 23403448 | 23404646 | 1199 | 11 | 2.91E-06 | 0.0001032 | 0.000483 | 4.55E-06 | -0.0015433 | LOXL2, ENTPD4 |
| chr5 | 134039208 | 134039727 | 520 | 3 | 2.13E-07 | 0.0033348 | 7.36E-06 | 4.55E-06 | 0.002162397 | NA |
| chr20 | 2754263 | 2754477 | 215 | 2 | 9.14E-06 | 3.02E-06 | 0.000219 | 4.55E-06 | -0.00615556 | EBF4 |
| chr1 | 206792495 | 206793117 | 623 | 3 | 5.29E-07 | 0.0002411 | 6.33E-06 | 4.56E-06 | 0.00018521 | IL19 |
| chr11 | 9625417 | 9625628 | 212 | 2 | 2.77E-06 | 2.46E-05 | 1.05E-05 | 4.56E-06 | 0.0019616 | NA |
| chr6 | 11503878 | 11503914 | 37 | 2 | 7.73E-06 | 7.53E-06 | 3.09E-05 | 4.56E-06 | -0.02428268 | AL445430.2 |
| chr10 | 119140866 | 119141181 | 316 | 2 | 2.70E-06 | 8.84E-06 | 2.55E-05 | 4.56E-06 | -0.01029163 | SFXN4 |
| chr1 | 112720667 | 112721142 | 476 | 2 | 2.63E-07 | 3.99E-05 | 7.79E-06 | 4.56E-06 | -0.00516558 | TAFA3 |
| chr22 | 50170898 | 50171831 | 934 | 5 | 5.31E-07 | 9.52E-06 | 0.000387 | 4.57E-06 | -0.00580849 | PANX2 |
| chr2 | 168705783 | 168706051 | 269 | 2 | 3.17E-06 | 1.67E-05 | 1.40E-05 | 4.57E-06 | -0.00531652 | CERS6 |
| chr2 | 102418767 | 102419665 | 899 | 6 | 1.65E-06 | 1.84E-05 | 7.12E-05 | 4.57E-06 | -0.00357289 | IL18RAP |
| chr4 | 5895496 | 5896164 | 669 | 3 | 8.39E-07 | 0.0001393 | 1.94E-05 | 4.57E-06 | -0.01022803 | NA |
| chr7 | 99543168 | 99543589 | 422 | 4 | 1.11E-06 | 7.31E-05 | 1.90E-05 | 4.57E-06 | -0.00216626 | NA |
| chr19 | 41911151 | 41911513 | 363 | 2 | 4.54E-07 | 9.84E-06 | 2.28E-05 | 4.57E-06 | -0.00986696 | ARHGEF1, ERFL |
| chr19 | 8697667 | 8698313 | 647 | 4 | 4.95E-07 | 2.13E-05 | 0.000496 | 4.57E-06 | -0.00340319 | ACTL9 |
| chr1 | 18597616 | 18598439 | 824 | 3 | 5.07E-06 | 2.91E-05 | 5.44E-05 | 4.57E-06 | -0.0165545 | NA |
| chr20 | 51607580 | 51608208 | 629 | 2 | 9.60E-06 | 2.98E-06 | 0.000241 | 4.58E-06 | -0.00916191 | ATP9A |
| chr6 | 32074322 | 32075784 | 1463 | 10 | 7.08E-07 | 1.20E-05 | 0.00029 | 4.58E-06 | -0.00156337 | TNXB |
| chr19 | 44724872 | 44725591 | 720 | 3 | 1.76E-06 | 1.93E-06 | 0.001021 | 4.59E-06 | -0.01129456 | AC243964.3 |
| chr1 | 77531319 | 77532036 | 718 | 2 | 1.56E-05 | 3.18E-06 | 0.000181 | 4.59E-06 | 0.007293283 | AK5 |
| chr12 | 120157079 | 120157240 | 162 | 2 | 7.00E-06 | 5.17E-06 | 5.36E-05 | 4.59E-06 | -0.01007491 | GCN1 |
| chr9 | 98993340 | 98993911 | 572 | 3 | 5.03E-06 | 3.96E-06 | 0.000321 | 4.59E-06 | -0.01914821 | COL15A1 |
| chr1 | 43198892 | 43199460 | 569 | 2 | 2.14E-05 | 2.73E-06 | 0.000434 | 4.59E-06 | -0.00290672 | CFAP57, EBNA1BP2 |
| chr17 | 7419396 | 7420774 | 1379 | 12 | 3.17E-06 | 0.0001044 | 6.62E-05 | 4.60E-06 | -0.00304157 | NLGN2, AC113189.2, SPEM1 |
| chr12 | 132887359 | 132888427 | 1069 | 27 | 6.61E-06 | 1.17E-05 | 0.026322 | 4.60E-06 | 0.001116269 | AC127070.1, CHFR |
| chr13 | 21998185 | 21998319 | 135 | 2 | 5.37E-06 | 1.43E-05 | 1.61E-05 | 4.60E-06 | 0.000880995 | NA |
| chr12 | 26523846 | 26523976 | 131 | 3 | 1.37E-05 | 1.65E-06 | 0.002177 | 4.60E-06 | 0.006903726 | ITPR2 |
| chr8 | 123002238 | 123002953 | 716 | 3 | 1.56E-06 | 5.80E-06 | 8.70E-05 | 4.61E-06 | -0.0112672 | AC104316.1 |
| chr14 | 22984095 | 22984111 | 17 | 2 | 2.13E-06 | 2.75E-06 | 0.000429 | 4.61E-06 | -0.0117135 | AL132780.2 |
| chr14 | 99758723 | 99759017 | 295 | 2 | 5.64E-06 | 2.78E-06 | 0.000396 | 4.61E-06 | -0.00382663 | EML1 |
| chr7 | 116222449 | 116222972 | 524 | 7 | 6.97E-06 | 1.76E-05 | 0.001341 | 4.62E-06 | -0.00121986 | TES |
| chr11 | 8030825 | 8030989 | 165 | 2 | 4.68E-06 | 2.75E-06 | 0.000436 | 4.62E-06 | -0.00969637 | TUB |
| chr13 | 25371117 | 25372259 | 1143 | 8 | 7.61E-08 | 0.0274337 | 5.09E-06 | 4.62E-06 | 0.003362521 | ATP8A2 |
| chr6 | 32207361 | 32207756 | 396 | 2 | 2.32E-06 | 1.15E-05 | 1.98E-05 | 4.62E-06 | -0.01652667 | NOTCH4 |
| chr12 | 105235342 | 105236390 | 1049 | 10 | 1.39E-06 | 0.0002722 | 0.000815 | 4.63E-06 | 0.000952914 | C12orf75, AC078874.1, APPL2 |
| chr13 | 109141350 | 109141868 | 519 | 2 | 6.48E-07 | 4.39E-06 | 7.32E-05 | 4.63E-06 | -0.01881808 | MYO16 |
| chr2 | 216003171 | 216003386 | 216 | 3 | 1.46E-05 | 1.90E-06 | 0.001525 | 4.63E-06 | 0.003357965 | MREG, PECR |
| chr9 | 28956472 | 28957209 | 738 | 3 | 2.55E-05 | 3.33E-06 | 0.000283 | 4.63E-06 | -0.00933316 | NA |
| chr13 | 66377842 | 66378495 | 654 | 2 | 2.62E-06 | 2.78E-06 | 0.000406 | 4.63E-06 | -0.00168251 | PCDH9 |
| chr13 | 40648943 | 40649626 | 684 | 3 | 4.56E-06 | 0.0003886 | 3.84E-05 | 4.63E-06 | 0.005812987 | FOXO1 |
| chr3 | 52189935 | 52190232 | 298 | 2 | 1.07E-06 | 1.39E-05 | 1.66E-05 | 4.63E-06 | -0.00206517 | NA |
| chr2 | 85577353 | 85577609 | 257 | 7 | 3.06E-06 | 1.26E-05 | 3.35E-05 | 4.63E-06 | -0.00241747 | VAMP8 |
| chr1 | 1535456 | 1535637 | 182 | 3 | 6.62E-06 | 2.43E-06 | 0.000482 | 4.63E-06 | -0.00078198 | TMEM240 |
| chr8 | 58346551 | 58346706 | 156 | 2 | 1.10E-05 | 2.89E-06 | 0.000309 | 4.63E-06 | -0.00877794 | NA |
| chr7 | 45148303 | 45148845 | 543 | 5 | 2.18E-06 | 1.43E-05 | 0.000268 | 4.63E-06 | 0.000354892 | NA |
| chr7 | 14902699 | 14903427 | 729 | 8 | 7.49E-06 | 4.04E-05 | 0.002108 | 4.63E-06 | 0.006133496 | DGKB |
| chr9 | 134299815 | 134300222 | 408 | 2 | 3.72E-06 | 3.41E-06 | 0.000145 | 4.64E-06 | -0.0027872 | AL354796.1 |
| chr1 | 1330798 | 1331401 | 604 | 7 | 1.47E-06 | 0.0006544 | 0.001831 | 4.64E-06 | -0.00628979 | TAS1R3 |
| chr8 | 24076966 | 24077324 | 359 | 3 | 7.24E-06 | 2.18E-05 | 0.00027 | 4.64E-06 | -0.00855209 | RNU1-148P |
| chr11 | 3220636 | 3221839 | 1204 | 5 | 5.91E-05 | 1.68E-06 | 0.002798 | 4.64E-06 | -0.00489173 | MRGPRG-AS1 |
| chr10 | 130787774 | 130787789 | 16 | 2 | 2.60E-06 | 8.11E-05 | 5.70E-06 | 4.64E-06 | 0.015133886 | NA |
| chr22 | 40177200 | 40177873 | 674 | 4 | 2.85E-06 | 3.09E-06 | 0.000456 | 4.64E-06 | -0.00358498 | TNRC6B |
| chr19 | 36686532 | 36687538 | 1007 | 7 | 1.72E-05 | 1.65E-06 | 0.008746 | 4.64E-06 | -0.00546807 | LINC01534 |
| chr2 | 46616724 | 46617594 | 871 | 14 | 5.01E-06 | 6.65E-05 | 0.000126 | 4.64E-06 | 0.000166856 | CRIPT, PIGF |
| chr9 | 136851670 | 136852776 | 1107 | 8 | 3.24E-06 | 3.58E-06 | 0.003063 | 4.64E-06 | -0.00199558 | MAMDC4 |
| chr2 | 27135284 | 27135978 | 695 | 4 | 6.89E-07 | 0.0002247 | 0.000365 | 4.64E-06 | -0.00340338 | NA |
| chr16 | 2009558 | 2010326 | 769 | 14 | 5.13E-06 | 6.68E-06 | 0.004379 | 4.64E-06 | -0.00078071 | NPW, ZNF598 |
| chr14 | 64294113 | 64295104 | 992 | 11 | 1.12E-05 | 1.66E-05 | 0.003 | 4.64E-06 | 0.000451932 | ESR2 |
| chr7 | 157005887 | 157006699 | 813 | 8 | 2.39E-06 | 0.0006615 | 4.63E-05 | 4.64E-06 | -0.00372097 | MNX1-AS2, MNX1 |
| chr16 | 716396 | 716888 | 493 | 4 | 1.59E-06 | 2.30E-06 | 0.001967 | 4.65E-06 | 0.014590023 | METRN |
| chr16 | 68824326 | 68824394 | 69 | 2 | 4.12E-06 | 7.86E-06 | 3.02E-05 | 4.65E-06 | 0.007315173 | CDH1 |
| chr20 | 37524411 | 37525240 | 830 | 6 | 9.01E-07 | 2.48E-05 | 0.000387 | 4.65E-06 | -0.00546979 | BLCAP |

| | | | | | | | | | | |
|---|---|---|---|---|---|---|---|---|---|---|
| chr1 | 24719916 | 24720119 | 204 | 2 | 4.66E-06 | 4.01E-06 | 9.09E-05 | 4.65E-06 | -0.01393581 | NA |
| chr6 | 98980844 | 98981009 | 166 | 2 | 1.02E-05 | 3.25E-06 | 0.000176 | 4.65E-06 | 0.001575333 | NA |
| chr10 | 1463277 | 1463980 | 704 | 5 | 3.12E-05 | 8.72E-07 | 0.007745 | 4.65E-06 | 0.025964894 | ADARB2 |
| chr1 | 4161328 | 4162022 | 695 | 6 | 3.90E-06 | 3.15E-05 | 0.000667 | 4.65E-06 | -0.0043936 | NA |
| chr9 | 137298868 | 137299448 | 581 | 3 | 7.81E-08 | 2.68E-05 | 1.29E-05 | 4.66E-06 | -0.00250197 | BX255925.4 |
| chr7 | 157645081 | 157645924 | 844 | 5 | 5.33E-06 | 3.89E-06 | 0.000767 | 4.66E-06 | -0.00318728 | PTPRN2 |
| chr2 | 23636070 | 23636478 | 409 | 2 | 3.94E-06 | 2.99E-06 | 0.000265 | 4.66E-06 | -0.00824613 | KLHL29 |
| chr2 | 183038370 | 183039338 | 969 | 12 | 1.67E-06 | 5.86E-05 | 0.002346 | 4.66E-06 | -0.00113362 | NCKAP1 |
| chr16 | 2222589 | 2223559 | 971 | 11 | 1.97E-05 | 5.03E-06 | 0.013873 | 4.66E-06 | -0.00056533 | NA |
| chr10 | 88095314 | 88095510 | 197 | 2 | 3.63E-06 | 7.21E-06 | 3.38E-05 | 4.66E-06 | 0.006326802 | NA |
| chr15 | 27541203 | 27542083 | 881 | 9 | 7.00E-06 | 8.68E-06 | 0.014229 | 4.67E-06 | 0.002186741 | GABRG3, AC104002.3 |
| chr14 | 99480756 | 99481952 | 1197 | 17 | 4.30E-07 | 0.0001441 | 3.97E-05 | 4.68E-06 | -0.00052943 | CCNK, SETD3 |
| chr2 | 127061662 | 127061879 | 218 | 2 | 1.30E-06 | 1.27E-05 | 1.83E-05 | 4.68E-06 | -0.01725485 | BIN1 |
| chr22 | 49203301 | 49203777 | 477 | 2 | 1.51E-05 | 2.81E-06 | 0.000408 | 4.68E-06 | 0.004507318 | NA |
| chr10 | 113679108 | 113679873 | 766 | 14 | 1.45E-05 | 1.95E-06 | 0.011598 | 4.68E-06 | 0.000422909 | CASP7 |
| chr4 | 173522215 | 173523156 | 942 | 6 | 3.17E-06 | 8.11E-05 | 0.000175 | 4.68E-06 | -0.00372365 | NA |
| chr1 | 215523659 | 215523828 | 170 | 2 | 1.15E-05 | 2.82E-06 | 0.0004 | 4.68E-06 | -0.01804339 | NA |
| chr10 | 129896411 | 129896850 | 440 | 5 | 7.06E-06 | 0.0001623 | 0.000868 | 4.69E-06 | -0.00690454 | EBF3 |
| chr4 | 139730798 | 139731175 | 378 | 5 | 1.68E-06 | 2.80E-05 | 0.000629 | 4.69E-06 | -0.00482363 | MGST2, MAML3 |
| chr18 | 49561639 | 49562410 | 772 | 10 | 2.17E-05 | 2.15E-06 | 0.011857 | 4.69E-06 | 0.000911478 | LIPG |
| chr5 | 4100977 | 4101423 | 447 | 6 | 6.45E-06 | 2.92E-06 | 0.000467 | 4.69E-06 | 0.006330501 | NA |
| chr2 | 23532982 | 23533116 | 135 | 3 | 9.22E-06 | 1.09E-05 | 0.000274 | 4.70E-06 | -0.00914697 | KLHL29 |
| chr11 | 22677299 | 22677412 | 114 | 2 | 8.32E-06 | 5.78E-06 | 4.69E-05 | 4.70E-06 | -0.01819646 | GAS2 |
| chr8 | 81685905 | 81686266 | 362 | 3 | 9.62E-07 | 6.09E-05 | 7.22E-06 | 4.70E-06 | -0.00261694 | IMPA1 |
| chr20 | 61923659 | 61924113 | 455 | 2 | 7.63E-06 | 3.09E-06 | 0.000236 | 4.70E-06 | -0.0099899 | CDH4 |
| chr4 | 140373368 | 140374207 | 840 | 12 | 5.08E-06 | 2.42E-05 | 0.002894 | 4.71E-06 | 0.001144171 | SCOC, SCOC-AS1 |
| chr19 | 45280400 | 45280716 | 317 | 3 | 6.09E-06 | 1.73E-05 | 0.000332 | 4.71E-06 | -0.00951305 | MARK4 |
| chr7 | 1459005 | 1459421 | 417 | 2 | 1.96E-08 | 7.15E-05 | 6.11E-06 | 4.71E-06 | 0.003175092 | MICALL2 |
| chr3 | 57948945 | 57949296 | 352 | 2 | 2.02E-07 | 0.0001399 | 4.85E-06 | 4.71E-06 | 0.003882898 | NA |
| chr6 | 6940433 | 6940990 | 558 | 2 | 5.65E-07 | 3.37E-05 | 8.94E-06 | 4.71E-06 | -0.00862704 | NA |
| chr11 | 65539260 | 65539535 | 276 | 3 | 2.12E-06 | 1.76E-06 | 0.001752 | 4.71E-06 | -0.00621683 | LTBP3 |
| chr6 | 87784460 | 87784831 | 372 | 3 | 6.63E-06 | 6.08E-06 | 0.00044 | 4.71E-06 | -0.0031703 | NA |
| chr16 | 11414111 | 11414684 | 574 | 3 | 3.69E-06 | 1.01E-05 | 3.94E-05 | 4.71E-06 | -0.00567921 | AC099489.1 |
| chr11 | 61920110 | 61920661 | 552 | 6 | 3.14E-06 | 0.00023 | 5.05E-05 | 4.72E-06 | -0.00488622 | RAB3IL1 |
| chr1 | 226222496 | 226222697 | 202 | 2 | 1.92E-07 | 1.15E-05 | 2.04E-05 | 4.72E-06 | -0.00063092 | NA |
| chr4 | 63127705 | 63128174 | 470 | 3 | 9.85E-06 | 2.15E-06 | 0.001278 | 4.74E-06 | -0.01291876 | NA |
| chr14 | 103099530 | 103100251 | 722 | 8 | 1.79E-06 | 0.0001525 | 0.000132 | 4.74E-06 | 0.000999057 | EXOC3L4 |
| chr17 | 1013032 | 1013919 | 888 | 5 | 1.24E-05 | 5.83E-06 | 0.000763 | 4.74E-06 | -0.00274188 | ABR |
| chr19 | 50024839 | 50025761 | 923 | 13 | 5.19E-06 | 0.000146 | 0.002844 | 4.74E-06 | -0.00051774 | ZNF473, VRK3 |
| chr18 | 55585950 | 55586678 | 729 | 5 | 5.58E-07 | 4.50E-05 | 0.001122 | 4.74E-06 | -0.01145395 | TCF4 |
| chr2 | 73176283 | 73177081 | 799 | 4 | 5.04E-08 | 3.42E-05 | 0.000286 | 4.74E-06 | -0.00852576 | NA |
| chr4 | 121165543 | 121165729 | 187 | 2 | 6.65E-07 | 4.19E-05 | 7.94E-06 | 4.74E-06 | -0.00517247 | TNIP3 |
| chr15 | 45198677 | 45199012 | 336 | 4 | 2.86E-06 | 3.38E-05 | 0.00077 | 4.74E-06 | 0.000890375 | AC051619.3, SHF, AC051619.6 |
| chr9 | 131621958 | 131622179 | 222 | 2 | 6.69E-06 | 6.10E-06 | 4.41E-05 | 4.75E-06 | -0.00353594 | RAPGEF1 |
| chr13 | 79571763 | 79572270 | 508 | 3 | 4.70E-06 | 1.62E-05 | 3.65E-05 | 4.75E-06 | -0.00840055 | NA |
| chr10 | 25083708 | 25083968 | 261 | 2 | 4.84E-06 | 8.69E-06 | 2.78E-05 | 4.75E-06 | -0.00163818 | AL512598.2 |
| chr2 | 101001342 | 101001932 | 591 | 7 | 8.89E-06 | 4.19E-06 | 0.001319 | 4.76E-06 | -0.00753714 | AC016738.1 |
| chr21 | 39317595 | 39317688 | 94 | 2 | 5.15E-06 | 1.08E-05 | 2.21E-05 | 4.76E-06 | -0.01375658 | BRWD1-AS1, BRWD1 |
| chr18 | 58202703 | 58203003 | 301 | 2 | 7.17E-06 | 2.91E-06 | 0.00036 | 4.76E-06 | -0.00783803 | NEDD4L |
| chr12 | 6326350 | 6327387 | 1038 | 4 | 3.60E-07 | 0.0012059 | 5.24E-06 | 4.76E-06 | -0.00482532 | PLEKHG6 |
| chr16 | 21704539 | 21704726 | 188 | 3 | 1.45E-06 | 0.000254 | 1.28E-05 | 4.76E-06 | -0.00393829 | OTOA |
| chr4 | 48493662 | 48494387 | 726 | 5 | 1.13E-05 | 1.48E-05 | 0.003533 | 4.76E-06 | 0.002580722 | ZAR1 |
| chr12 | 1649898 | 1650555 | 658 | 2 | 1.83E-05 | 2.84E-06 | 0.000442 | 4.76E-06 | -0.01186407 | NA |
| chr14 | 103732565 | 103733095 | 531 | 2 | 7.74E-06 | 2.98E-06 | 0.000311 | 4.77E-06 | -0.00572489 | ZFYVE21 |
| chr11 | 1053885 | 1054538 | 654 | 3 | 8.85E-07 | 2.04E-05 | 8.31E-05 | 4.77E-06 | -0.00729607 | LINC02688 |
| chr7 | 103344097 | 103344943 | 847 | 10 | 1.10E-06 | 0.0002801 | 6.19E-05 | 4.77E-06 | 0.000388862 | PSMC2, DNAJC2 |
| chr12 | 56361042 | 56361715 | 674 | 4 | 2.91E-06 | 3.56E-05 | 0.000531 | 4.77E-06 | -0.0023421 | APOF |
| chr7 | 65430505 | 65430919 | 415 | 2 | 1.80E-06 | 5.98E-06 | 4.60E-05 | 4.78E-06 | -0.0134871 | NA |
| chr4 | 133163069 | 133163087 | 19 | 2 | 4.79E-06 | 3.90E-05 | 8.34E-06 | 4.78E-06 | -0.00530309 | PCDH10 |
| chr4 | 1770335 | 1770424 | 90 | 2 | 4.85E-06 | 3.02E-06 | 0.000293 | 4.78E-06 | 0.012301605 | NA |
| chr14 | 24139706 | 24140043 | 338 | 2 | 1.97E-06 | 3.41E-06 | 0.000165 | 4.79E-06 | 0.010289086 | AL136295.2, EMC9 |
| chr4 | 64726517 | 64726889 | 373 | 4 | 1.27E-06 | 0.000259 | 4.90E-05 | 4.79E-06 | -0.00180913 | NA |
| chr6 | 33279118 | 33279732 | 615 | 11 | 3.78E-06 | 1.48E-05 | 0.004391 | 4.79E-06 | -0.001132 | B3GALT4, WDR46 |
| chr20 | 5112155 | 5113393 | 1239 | 13 | 1.54E-06 | 0.0003493 | 0.00011 | 4.79E-06 | 0.000149467 | TMEM230 |
| chr5 | 10353459 | 10354451 | 993 | 7 | 1.30E-05 | 0.0001306 | 0.000565 | 4.79E-06 | 0.000522351 | MARCHF6, AC012640.2 |
| chr2 | 170928969 | 170929806 | 838 | 9 | 5.87E-06 | 3.80E-05 | 8.07E-05 | 4.80E-06 | -0.00133207 | GORASP2 |
| chr2 | 3517487 | 3519134 | 1648 | 5 | 8.68E-05 | 0.000473 | 0.000125 | 4.80E-06 | -0.00049231 | ADI1 |
| chr10 | 16951959 | 16952443 | 485 | 2 | 1.01E-05 | 2.89E-06 | 0.000402 | 4.80E-06 | 0.007218943 | CUBN |
| chr12 | 13079744 | 13079799 | 56 | 2 | 1.21E-05 | 2.86E-06 | 0.000437 | 4.80E-06 | 0.004581858 | FAM234B |
| chr21 | 42010274 | 42011162 | 889 | 8 | 1.44E-05 | 1.30E-05 | 0.003396 | 4.80E-06 | 0.001385999 | ZNF295-AS1, ZBTB21 |
| chr13 | 112974935 | 112975003 | 69 | 2 | 3.53E-06 | 3.68E-06 | 0.000129 | 4.80E-06 | -0.00867232 | MCF2L |
| chr7 | 151900154 | 151900686 | 533 | 2 | 1.85E-06 | 4.01E-06 | 0.0001 | 4.80E-06 | -0.01546544 | RNU6-604P |
| chr5 | 10479108 | 10479618 | 511 | 3 | 5.18E-06 | 4.26E-06 | 0.000124 | 4.81E-06 | 0.002734724 | LINC01513 |
| chr14 | 72892202 | 72893104 | 903 | 11 | 4.28E-07 | 0.0005606 | 0.000134 | 4.81E-06 | 0.000851314 | DPF3 |
| chr1 | 67760030 | 67760523 | 494 | 3 | 1.05E-06 | 0.0016677 | 4.89E-06 | 4.81E-06 | -0.00523268 | GNG12 |
| chr16 | 81958889 | 81959025 | 137 | 2 | 3.26E-06 | 3.05E-06 | 0.000287 | 4.81E-06 | -0.01366213 | PLCG2 |
| chr6 | 89819024 | 89819575 | 552 | 6 | 3.08E-06 | 2.16E-06 | 0.005094 | 4.81E-06 | -0.0011569 | MDN1 |
| chr3 | 126353492 | 126354565 | 1074 | 4 | 0.000117095 | 3.29E-06 | 0.001086 | 4.81E-06 | -0.0072215 | KLF15 |
| chr7 | 150405325 | 150406157 | 833 | 8 | 5.62E-07 | 0.0002993 | 0.000875 | 4.82E-06 | 0.004057306 | AC073111.2, AC073111.3 |
| chr2 | 23383901 | 23385130 | 1230 | 5 | 3.72E-08 | 0.0009406 | 9.66E-07 | 4.82E-06 | 0.002167986 | NA |
| chr6 | 131894661 | 131894893 | 233 | 2 | 5.23E-06 | 9.63E-06 | 2.54E-05 | 4.82E-06 | -0.00841627 | ENPP1 |
| chr4 | 54588660 | 54589007 | 348 | 2 | 6.98E-06 | 4.17E-06 | 9.21E-05 | 4.82E-06 | -0.01105299 | NA |
| chr2 | 231205382 | 231206067 | 686 | 4 | 2.97E-05 | 2.89E-06 | 0.000721 | 4.83E-06 | -0.00096016 | ARMC9 |
| chr1 | 202750771 | 202751478 | 708 | 3 | 3.24E-06 | 0.0008502 | 2.19E-05 | 4.83E-06 | -0.00045337 | KDM5B |
| chr19 | 18312209 | 18312749 | 541 | 2 | 1.46E-05 | 3.01E-06 | 0.00032 | 4.83E-06 | -0.00379441 | LSM4 |
| chr3 | 178434342 | 178434588 | 247 | 2 | 7.73E-06 | 3.58E-06 | 0.000145 | 4.83E-06 | -0.02082904 | KCNMB2, AC117457.1, LINC01014 |
| chr6 | 44074714 | 44075455 | 742 | 5 | 1.06E-05 | 2.63E-06 | 0.003853 | 4.83E-06 | -0.00571351 | AL109615.2 |
| chr17 | 59331242 | 59332334 | 1093 | 8 | 6.43E-07 | 3.58E-05 | 0.002499 | 4.84E-06 | -0.00038047 | YPEL2 |
| chr1 | 241355913 | 241356456 | 544 | 4 | 7.18E-07 | 7.42E-06 | 0.00056 | 4.84E-06 | -0.00243839 | RGS7 |
| chr11 | 3702462 | 3702691 | 230 | 2 | 5.05E-06 | 9.28E-06 | 2.67E-05 | 4.84E-06 | -0.00521137 | NUP98 |
| chr11 | 71997439 | 71997858 | 420 | 3 | 1.46E-06 | 2.37E-06 | 0.000624 | 4.85E-06 | -0.00200419 | RNF121 |
| chr1 | 4774900 | 4775238 | 339 | 2 | 2.48E-06 | 1.57E-05 | 1.60E-05 | 4.86E-06 | 0.003358472 | AJAP1 |
| chr16 | 85062445 | 85062827 | 383 | 5 | 9.73E-06 | 1.10E-05 | 0.000929 | 4.86E-06 | -0.00186037 | KIAA0513 |
| chr16 | 77787890 | 77788874 | 985 | 10 | 5.79E-07 | 0.0006906 | 7.87E-05 | 4.86E-06 | 0.000279671 | VAT1L |
| chr19 | 40709001 | 40709060 | 60 | 2 | 9.17E-06 | 5.96E-06 | 4.79E-05 | 4.86E-06 | -0.00173078 | COQ8B |
| chr16 | 84788894 | 84789618 | 725 | 8 | 1.62E-05 | 5.63E-06 | 0.002713 | 4.86E-06 | -0.00406679 | NA |
| chr5 | 175680769 | 175681426 | 658 | 7 | 2.86E-06 | 4.91E-06 | 0.000223 | 4.86E-06 | 0.002087115 | HRH2 |
| chr2 | 29330271 | 29330488 | 218 | 3 | 1.10E-05 | 4.76E-06 | 0.000172 | 4.86E-06 | -0.00415622 | AC074096.1, ALK |
| chr2 | 192193902 | 192195173 | 1272 | 8 | 1.45E-05 | 5.07E-05 | 0.002175 | 4.86E-06 | -0.00209105 | TMEFF2 |
| chr1 | 11839580 | 11840595 | 1016 | 4 | 2.93E-05 | 3.43E-06 | 0.001103 | 4.86E-06 | -0.00220906 | CLCN6 |
| chr17 | 58332157 | 58332583 | 427 | 5 | 1.49E-07 | 0.0003998 | 2.44E-06 | 4.86E-06 | 0.00045317 | TSPOAP1-AS1, AC004687.1 |
| chr1 | 192575122 | 192575586 | 465 | 2 | 1.97E-07 | 8.44E-05 | 5.91E-06 | 4.86E-06 | -0.009551 | AL390957.1 |
| chr10 | 29859834 | 29860327 | 494 | 3 | 1.12E-06 | 6.81E-05 | 0.000194 | 4.86E-06 | -0.00936624 | NA |
| chr4 | 138851592 | 138851981 | 390 | 2 | 1.09E-05 | 2.90E-06 | 0.000444 | 4.86E-06 | -0.02001056 | AC109927.1 |
| chr20 | 12949833 | 12950305 | 473 | 3 | 2.10E-06 | 9.54E-05 | 1.03E-05 | 4.87E-06 | -0.00598644 | LINC01723, LINC01722 |
| chr19 | 7918721 | 7919286 | 566 | 7 | 4.64E-06 | 1.31E-06 | 0.006254 | 4.87E-06 | -0.01775863 | TGFBR3L, AC010336.2 |
| chr7 | 142799565 | 142799710 | 146 | 2 | 9.67E-06 | 3.09E-06 | 0.000291 | 4.87E-06 | 0.014760113 | NA |
| chr15 | 30073385 | 30073772 | 388 | 2 | 3.28E-05 | 3.34E-06 | 0.000198 | 4.88E-06 | -0.01306936 | NA |
| chr5 | 1032846 | 1033961 | 1116 | 5 | 1.26E-07 | 0.000251 | 6.64E-06 | 4.88E-06 | -0.00144624 | NKD2 |
| chr8 | 143579747 | 143580755 | 1009 | 4 | 3.90E-06 | 1.55E-05 | 3.53E-05 | 4.88E-06 | -0.00109891 | AC067930.5, AC067930.1, EEF1D |
| chr13 | 98411419 | 98411884 | 466 | 3 | 3.77E-06 | 6.39E-06 | 7.55E-05 | 4.88E-06 | 0.000110818 | FARP1 |
| chr21 | 14501036 | 14501441 | 406 | 3 | 9.33E-06 | 3.13E-06 | 0.000714 | 4.88E-06 | 0.004255693 | SAMSN1 |
| chr22 | 46003098 | 46003551 | 454 | 3 | 2.75E-06 | 2.47E-05 | 1.47E-05 | 4.88E-06 | 3.99E-05 | NA |
| chr4 | 56170132 | 56170237 | 106 | 2 | 8.77E-06 | 3.43E-06 | 0.000176 | 4.88E-06 | -0.00121768 | CRACD |
| chr8 | 56557770 | 56558732 | 963 | 5 | 2.78E-05 | 1.18E-06 | 0.004347 | 4.88E-06 | -0.00486884 | LINC00968 |
| chr14 | 35369034 | 35370019 | 986 | 7 | 4.53E-06 | 1.09E-05 | 0.002681 | 4.89E-06 | 0.01603211 | NA |
| chr1 | 161399172 | 161400180 | 1009 | 10 | 2.38E-06 | 0.0010602 | 0.000237 | 4.89E-06 | -0.00054771 | AL592295.4, AL592295.5 |
| chr21 | 15379810 | 15379927 | 118 | 2 | 1.22E-05 | 2.96E-06 | 0.000398 | 4.89E-06 | -0.02411588 | AJ009632.2 |
| chr13 | 33187989 | 33188655 | 667 | 2 | 1.00E-05 | 3.11E-06 | 0.00029 | 4.89E-06 | -0.00925661 | STARD13-AS, STARD13 |
| chr17 | 79752438 | 79752458 | 21 | 2 | 7.07E-06 | 1.29E-05 | 1.93E-05 | 4.90E-06 | 0.016561199 | NA |
| chr16 | 945827 | 946172 | 346 | 2 | 6.60E-07 | 0.0001518 | 4.94E-06 | 4.90E-06 | -0.01569581 | LMF1 |
| chr6 | 168072374 | 168072612 | 239 | 3 | 7.11E-07 | 0.0008652 | 3.27E-06 | 4.90E-06 | -0.00714693 | FRMD1 |
| chr8 | 20700033 | 20700627 | 595 | 4 | 1.00E-05 | 2.48E-05 | 0.000603 | 4.90E-06 | -0.00232265 | AC018541.1 |
| chr6 | 154436830 | 154437045 | 216 | 2 | 1.01E-05 | 3.01E-06 | 0.000357 | 4.91E-06 | -0.01108231 | AL357075.4, CNKSR3 |
| chr7 | 102393589 | 102393831 | 243 | 2 | 4.53E-06 | 1.27E-05 | 1.97E-05 | 4.91E-06 | -0.01406348 | PRKRIP1 |
| chr13 | 114124055 | 114125091 | 1037 | 9 | 2.13E-07 | 0.0001215 | 0.0002 | 4.92E-06 | -0.00250289 | RASA3 |
| chr11 | 2381161 | 2381788 | 628 | 4 | 4.56E-07 | 2.82E-05 | 1.62E-05 | 4.92E-06 | -0.00399777 | CD81 |
| chr2 | 38875927 | 38876231 | 305 | 6 | 8.67E-05 | 2.11E-06 | 0.00185 | 4.93E-06 | 0.001073474 | MORN2, DHX57 |
| chr9 | 5608645 | 5609143 | 499 | 2 | 4.96E-06 | 4.89E-06 | 6.96E-05 | 4.93E-06 | 0.013266676 | AL162253.2 |
| chr20 | 58837890 | 58839107 | 1218 | 6 | 2.21E-06 | 5.56E-06 | 0.000742 | 4.93E-06 | -0.00473523 | GNAS-AS1 |
| chr17 | 44623384 | 44623497 | 114 | 2 | 9.16E-06 | 5.52E-06 | 5.63E-05 | 4.93E-06 | -0.01520849 | NA |
| chr12 | 47948364 | 47948594 | 231 | 2 | 4.65E-06 | 4.01E-06 | 0.000109 | 4.93E-06 | -0.00054773 | NA |
| chr1 | 111222116 | 111222235 | 120 | 2 | 2.28E-06 | 1.68E-05 | 1.55E-05 | 4.93E-06 | -0.01578232 | CHI3L2 |
| chr1 | 31581972 | 31582528 | 557 | 2 | 2.09E-07 | 5.89E-06 | 5.04E-05 | 4.93E-06 | -0.00693429 | LINC01226, TINAGL1 |
| chr3 | 113514706 | 113515246 | 541 | 8 | 9.45E-06 | 2.09E-05 | 0.000279 | 4.93E-06 | 0.001264912 | AC112128.1, SPICE1 |
| chr5 | 11026908 | 11027157 | 250 | 5 | 8.90E-06 | 5.24E-06 | 0.000752 | 4.93E-06 | -0.00071768 | CTNND2 |
| chr2 | 106062093 | 106062602 | 510 | 2 | 4.02E-05 | 2.94E-06 | 0.000442 | 4.93E-06 | 0.010874471 | NA |
| chr16 | 84341723 | 84342028 | 306 | 2 | 8.04E-07 | 0.0004115 | 3.91E-06 | 4.94E-06 | 0.004066276 | NA |
| chr19 | 51926937 | 51927518 | 582 | 10 | 1.20E-06 | 4.35E-05 | 2.42E-05 | 4.94E-06 | -0.00053681 | ZNF613 |
| chr2 | 72886838 | 72887127 | 290 | 4 | 7.58E-05 | 1.70E-06 | 0.001543 | 4.94E-06 | -0.00743906 | NA |
| chr22 | 44689221 | 44689543 | 323 | 3 | 6.29E-06 | 7.41E-06 | 8.89E-05 | 4.94E-06 | -0.00481738 | PRR5 |
| chr2 | 17518421 | 17519254 | 834 | 9 | 5.38E-06 | 2.44E-05 | 0.001302 | 4.94E-06 | -0.00087782 | RAD51AP2 |
| chr1 | 32291881 | 32292699 | 819 | 10 | 1.20E-06 | 2.82E-05 | 0.000327 | 4.94E-06 | 0.000473842 | HDAC1 |
| chr7 | 137849611 | 137849824 | 214 | 2 | 4.01E-06 | 1.87E-05 | 1.43E-05 | 4.94E-06 | 0.004757534 | NA |
| chr7 | 138089683 | 138090358 | 676 | 2 | 2.41E-06 | 5.57E-06 | 5.59E-05 | 4.94E-06 | 0.012770211 | AKR1D1 |
| chr10 | 70598603 | 70599832 | 1230 | 9 | 1.34E-07 | 0.0001886 | 0.000127 | 4.95E-06 | -0.00254054 | PRF1 |
| chr19 | 1632633 | 1633074 | 442 | 2 | 1.38E-07 | 0.0006541 | 3.60E-06 | 4.95E-06 | -0.00288751 | TCF3 |
| chr9 | 122343107 | 122343385 | 279 | 2 | 2.91E-06 | 3.17E-06 | 0.000276 | 4.95E-06 | -0.00635067 | NA |
| chr2 | 133262469 | 133262880 | 412 | 3 | 8.96E-06 | 2.99E-06 | 0.000765 | 4.95E-06 | 0.001706993 | NCKAP5 |
| chr16 | 15600550 | 15601134 | 585 | 2 | 1.02E-06 | 4.26E-06 | 9.48E-05 | 4.95E-06 | 0.004542388 | BMERB1, MARF1 |
| chr18 | 66604104 | 66605014 | 911 | 8 | 1.94E-06 | 0.0001683 | 0.000448 | 4.95E-06 | -0.00034108 | CDH19 |
| chr2 | 100207874 | 100208468 | 595 | 6 | 1.49E-05 | 2.67E-06 | 0.006599 | 4.95E-06 | -0.00065355 | LINC01104 |
| chr2 | 109201997 | 109202056 | 60 | 2 | 1.22E-05 | 3.16E-06 | 0.000288 | 4.96E-06 | -0.00731554 | SH3RF3 |
| chr1 | 43073778 | 43073810 | 33 | 2 | 2.17E-06 | 6.25E-06 | 4.64E-05 | 4.96E-06 | -0.00382304 | NA |
| chr2 | 33563429 | 33563454 | 26 | 2 | 1.58E-06 | 2.98E-06 | 0.000421 | 4.97E-06 | -0.00192778 | RASGRP3, RASGRP3-AS1 |
| chr3 | 38137807 | 38139085 | 1279 | 15 | 3.10E-06 | 0.0002168 | 0.001076 | 4.97E-06 | -0.0001114 | MYD88 |
| chr1 | 95072626 | 95073281 | 656 | 13 | 2.60E-06 | 0.0013339 | 0.000606 | 4.97E-06 | 0.00145106 | ALG14 |
| chr2 | 98604970 | 98605232 | 263 | 2 | 5.31E-06 | 3.47E-06 | 0.000183 | 4.97E-06 | -0.01217003 | COA5 |

| | | | | | | | | | | |
|---|---|---|---|---|---|---|---|---|---|---|
| chr6 | 151154749 | 151155049 | 301 | 2 | 1.78E-06 | 5.99E-05 | 7.02E-06 | 4.97E-06 | 0.005179427 | AL138733.1 |
| chr22 | 39152126 | 39153094 | 969 | 11 | 1.31E-05 | 0.0001548 | 0.001274 | 4.98E-06 | -0.00104795 | CBX7 |
| chr2 | 230995231 | 230996503 | 1273 | 7 | 1.16E-05 | 0.0001207 | 0.000865 | 4.98E-06 | -0.00214901 | SPATA3, SPATA3-AS1 |
| chr4 | 98659379 | 98659698 | 320 | 2 | 2.00E-06 | 3.01E-06 | 0.000399 | 4.98E-06 | -0.01545255 | AC114811.2 |
| chr1 | 3360123 | 3361059 | 937 | 8 | 3.36E-06 | 1.69E-05 | 0.000259 | 4.98E-06 | -0.00022554 | PRDM16 |
| chr2 | 234913404 | 234913967 | 564 | 2 | 1.27E-06 | 4.75E-06 | 7.59E-05 | 4.98E-06 | -0.00496132 | NA |
| chr8 | 347359 | 347436 | 78 | 2 | 1.10E-06 | 0.0019745 | 3.19E-06 | 4.99E-06 | -0.01483918 | NA |
| chr11 | 117143633 | 117144675 | 1043 | 10 | 3.65E-06 | 0.0006582 | 0.000149 | 4.99E-06 | -0.00218397 | PAFAH1B2, AP005018.1 |
| chr14 | 74302498 | 74303578 | 1081 | 10 | 6.15E-06 | 0.0006225 | 0.000126 | 4.99E-06 | 0.001149801 | VRTN, ABCD4 |
| chr5 | 152972908 | 152972967 | 60 | 2 | 3.99E-06 | 4.27E-06 | 9.62E-05 | 4.99E-06 | 0.007372657 | LINC01470 |
| chr16 | 56657310 | 56658165 | 856 | 8 | 5.03E-06 | 2.82E-05 | 6.63E-05 | 5.00E-06 | -0.00190508 | MT1F |
| chr3 | 55177434 | 55178096 | 663 | 5 | 1.78E-05 | 1.74E-05 | 0.000986 | 5.00E-06 | 0.003548916 | NA |
| chr7 | 158730591 | 158730700 | 110 | 2 | 3.21E-06 | 8.39E-05 | 6.10E-06 | 5.00E-06 | -0.00702681 | NA |
| chr2 | 96245671 | 96246287 | 617 | 3 | 1.60E-05 | 0.0001186 | 0.000235 | 5.00E-06 | -0.00047814 | NA |
| chr3 | 180036451 | 180037497 | 1047 | 13 | 2.20E-06 | 0.0001234 | 0.002093 | 5.00E-06 | 0.001269979 | PEX5L |
| chr4 | 38565347 | 38565863 | 517 | 3 | 1.10E-05 | 8.14E-06 | 0.000144 | 5.00E-06 | -0.00253972 | LINC02278 |
| chr17 | 10696703 | 10697659 | 957 | 10 | 1.22E-05 | 2.66E-05 | 0.001612 | 5.01E-06 | -0.00077992 | ADPRM, SCO1 |
| chr3 | 170344197 | 170344723 | 527 | 2 | 1.61E-05 | 3.05E-06 | 0.000377 | 5.01E-06 | 0.003062019 | NA |
| chr15 | 40322899 | 40323088 | 190 | 2 | 1.36E-06 | 3.40E-06 | 0.00021 | 5.01E-06 | -0.0128555 | NA |
| chr11 | 75723426 | 75723459 | 34 | 2 | 7.33E-06 | 3.24E-06 | 0.000265 | 5.01E-06 | -0.01033623 | MOGAT2 |
| chr9 | 138057648 | 138057795 | 148 | 2 | 9.56E-06 | 3.74E-06 | 0.000144 | 5.01E-06 | -0.01658001 | CACNA1B |
| chr17 | 77958334 | 77958612 | 279 | 2 | 1.76E-07 | 4.98E-05 | 7.75E-06 | 5.01E-06 | 0.00249255 | NA |
| chr7 | 158743085 | 158743518 | 434 | 5 | 8.66E-07 | 0.000108 | 1.15E-05 | 5.02E-06 | -2.12E-05 | ESYT2 |
| chr1 | 226882929 | 226883266 | 338 | 2 | 2.14E-06 | 2.99E-06 | 0.000456 | 5.02E-06 | 0.005422577 | PSEN2, AL391628.1 |
| chr5 | 179314003 | 179314373 | 371 | 4 | 9.06E-07 | 0.000245 | 6.62E-06 | 5.02E-06 | -0.00262881 | ADAMTS2 |
| chr13 | 96994385 | 96994830 | 446 | 9 | 1.51E-05 | 5.72E-06 | 0.003756 | 5.03E-06 | -0.00624427 | OXGR1 |
| chr19 | 35959456 | 35960014 | 559 | 3 | 1.12E-06 | 5.22E-06 | 0.000108 | 5.03E-06 | -0.01163716 | NA |
| chr1 | 205849828 | 205850481 | 654 | 11 | 3.90E-05 | 2.30E-07 | 0.039063 | 5.03E-06 | -0.07123778 | AC119673.2, PM20D1 |
| chr13 | 75789526 | 75789715 | 190 | 3 | 6.36E-06 | 6.45E-06 | 7.09E-05 | 5.03E-06 | -0.01088493 | AL137782.1, LMO7 |
| chr17 | 13786491 | 13786530 | 40 | 2 | 1.92E-06 | 0.0002135 | 4.62E-06 | 5.03E-06 | -0.00529962 | COX10-AS1 |
| chr3 | 132694870 | 132695292 | 423 | 2 | 1.83E-06 | 6.40E-06 | 4.63E-05 | 5.04E-06 | -0.01655514 | NPHP3-ACAD11, NPHP3 |
| chr4 | 25108000 | 25108038 | 39 | 2 | 4.71E-06 | 5.17E-05 | 7.65E-06 | 5.04E-06 | -0.00542635 | NA |
| chr1 | 154936764 | 154937515 | 752 | 15 | 8.28E-06 | 1.63E-05 | 0.009993 | 5.04E-06 | 0.000342959 | AL451085.1 |
| chr7 | 30504793 | 30505458 | 666 | 12 | 1.13E-05 | 2.65E-05 | 0.000905 | 5.04E-06 | 0.000274958 | GGCT, AC005154.5 |
| chr8 | 30155793 | 30156751 | 959 | 10 | 7.42E-06 | 4.80E-06 | 0.000359 | 5.04E-06 | 0.001893841 | LEPROTL1, DCTN6, AC026979.2 |
| chr11 | 33039003 | 33040170 | 1168 | 12 | 1.46E-05 | 0.0004142 | 0.000275 | 5.04E-06 | -0.0002784 | TCP11L1 |
| chr5 | 2904590 | 2904996 | 407 | 2 | 1.51E-05 | 3.01E-06 | 0.00045 | 5.04E-06 | 0.00230447 | NA |
| chr5 | 14463955 | 14464381 | 427 | 2 | 1.14E-07 | 6.31E-06 | 4.74E-05 | 5.04E-06 | -0.01395682 | TRIO |
| chr6 | 129894710 | 129895058 | 349 | 2 | 2.75E-06 | 1.57E-05 | 1.70E-05 | 5.05E-06 | -0.01232468 | NA |
| chr19 | 55437182 | 55437675 | 494 | 4 | 9.92E-06 | 6.27E-06 | 0.001749 | 5.05E-06 | -0.00250397 | SHISA7 |
| chr19 | 782861 | 783593 | 733 | 2 | 0.000141263 | 3.65E-06 | 0.000162 | 5.05E-06 | -0.00071669 | LINC01836 |
| chr2 | 23092054 | 23092244 | 191 | 3 | 4.57E-06 | 0.0001624 | 0.000107 | 5.05E-06 | 0.023440638 | AC018467.1 |
| chr9 | 18460171 | 18460504 | 334 | 2 | 3.92E-06 | 9.92E-06 | 2.66E-05 | 5.05E-06 | 0.003598145 | ADAMTSL1 |
| chr12 | 11187465 | 11187750 | 286 | 3 | 2.97E-06 | 2.04E-05 | 8.01E-05 | 5.05E-06 | -0.01815924 | NA |
| chr16 | 75647839 | 75648429 | 591 | 8 | 2.05E-06 | 0.0002611 | 0.000631 | 5.05E-06 | 0.000947109 | TERF2IP, KARS1 |
| chr2 | 97733839 | 97735085 | 1247 | 11 | 3.04E-06 | 1.08E-05 | 0.000471 | 5.05E-06 | -0.00115902 | ZAP70 |
| chr14 | 37809859 | 37810421 | 563 | 3 | 2.12E-05 | 2.39E-06 | 0.001252 | 5.06E-06 | -8.14E-05 | TTC6 |
| chr5 | 181195072 | 181195643 | 572 | 7 | 1.12E-05 | 1.52E-06 | 0.006839 | 5.06E-06 | -0.00868508 | TRIM7-AS1, TRIM7 |
| chr16 | 77861837 | 77862183 | 347 | 3 | 1.68E-06 | 7.64E-05 | 0.000129 | 5.06E-06 | -0.00350342 | VAT1L |
| chr9 | 94110597 | 94110726 | 130 | 2 | 1.65E-06 | 0.0096204 | 2.98E-06 | 5.06E-06 | -0.00338919 | NA |
| chr2 | 20651270 | 20652027 | 758 | 9 | 5.44E-06 | 5.22E-05 | 0.002035 | 5.06E-06 | 0.001479008 | NA |
| chr16 | 86991975 | 86992630 | 656 | 2 | 1.16E-05 | 3.01E-06 | 0.00046 | 5.07E-06 | -0.00883538 | NA |
| chr17 | 21052183 | 21052810 | 628 | 2 | 6.26E-06 | 4.69E-06 | 8.16E-05 | 5.07E-06 | -0.01411101 | NA |
| chr12 | 116441580 | 116441591 | 12 | 2 | 8.50E-06 | 9.31E-06 | 2.87E-05 | 5.07E-06 | -0.0077127 | NA |
| chr15 | 36895348 | 36895900 | 553 | 2 | 7.76E-07 | 1.30E-05 | 2.01E-05 | 5.07E-06 | -0.00505748 | MEIS2 |
| chr15 | 65070684 | 65071176 | 493 | 2 | 5.05E-07 | 7.37E-05 | 6.55E-06 | 5.07E-06 | 0.008678683 | RASL12 |
| chr20 | 58673260 | 58673574 | 315 | 5 | 1.13E-05 | 1.26E-05 | 0.002089 | 5.07E-06 | -0.00374808 | STX16, STX16-NPEPL1 |
| chr6 | 128930118 | 128930300 | 183 | 2 | 2.73E-06 | 4.65E-06 | 8.33E-05 | 5.07E-06 | -0.0035815 | LAMA2 |
| chr2 | 2682026 | 2682325 | 300 | 2 | 7.92E-07 | 1.42E-05 | 1.86E-05 | 5.07E-06 | 0.006651101 | AC018685.2 |
| chr18 | 24530773 | 24530887 | 115 | 2 | 1.87E-06 | 0.0031857 | 3.12E-06 | 5.08E-06 | 0.00337084 | AC007922.3 |
| chr6 | 136993476 | 136994173 | 698 | 3 | 4.20E-06 | 1.11E-05 | 7.47E-05 | 5.08E-06 | -0.00667001 | NA |
| chr12 | 53254239 | 53254790 | 552 | 2 | 5.46E-07 | 3.84E-05 | 9.08E-06 | 5.08E-06 | -0.00412521 | MFSD5 |
| chr10 | 132858100 | 132859413 | 1314 | 10 | 1.42E-06 | 0.0010462 | 0.000146 | 5.08E-06 | -0.00036856 | CFAP46 |
| chr18 | 13726492 | 13727231 | 740 | 14 | 6.42E-06 | 4.28E-05 | 0.0042 | 5.08E-06 | 0.000690969 | RNMT, FAM210A |
| chr1 | 112923340 | 112923867 | 528 | 5 | 3.28E-07 | 8.64E-05 | 0.000157 | 5.08E-06 | -0.00275015 | AKR7A2P1, SLC16A1 |
| chr2 | 190781432 | 190782006 | 575 | 2 | 7.45E-06 | 4.17E-06 | 0.000108 | 5.08E-06 | -0.02093647 | AC006460.1 |
| chr1 | 198858876 | 198859373 | 498 | 7 | 6.57E-06 | 0.0001101 | 0.000231 | 5.09E-06 | -0.00162481 | MIR181A1HG, MIR181B1, MIR181A1 |
| chr1 | 21820217 | 21820870 | 654 | 2 | 8.38E-07 | 8.17E-06 | 3.37E-05 | 5.09E-06 | -0.00571743 | LDLRAD2 |
| chr12 | 47837342 | 47837354 | 13 | 2 | 2.03E-06 | 0.0004676 | 3.93E-06 | 5.09E-06 | 0.006600239 | LINC02354 |
| chr4 | 86593718 | 86594858 | 1141 | 18 | 2.31E-06 | 5.46E-05 | 0.000297 | 5.09E-06 | 0.000966415 | PTPN13, MAPK10 |
| chr2 | 120528672 | 120528811 | 140 | 2 | 9.46E-06 | 3.04E-06 | 0.00045 | 5.10E-06 | -0.01257175 | NA |
| chr7 | 6057030 | 6057339 | 310 | 2 | 1.53E-06 | 2.88E-05 | 1.09E-05 | 5.10E-06 | -0.01921381 | EIF2AK1 |
| chr1 | 21111342 | 21111554 | 213 | 3 | 3.03E-06 | 0.0008553 | 9.22E-05 | 5.10E-06 | -0.00354978 | EIF4G3 |
| chr14 | 95319443 | 95320464 | 1022 | 8 | 2.01E-05 | 5.44E-06 | 0.004577 | 5.10E-06 | -0.00259716 | LINC02292, CLMN |
| chr14 | 94066022 | 94066161 | 140 | 2 | 4.30E-06 | 4.03E-05 | 8.89E-06 | 5.10E-06 | -0.0075394 | DDX24 |
| chr17 | 41030323 | 41030417 | 95 | 3 | 1.20E-05 | 2.54E-06 | 0.00067 | 5.11E-06 | -0.0097701 | NA |
| chr20 | 19340684 | 19341280 | 597 | 4 | 3.62E-06 | 4.31E-05 | 0.000325 | 5.11E-06 | -0.00196023 | SLC24A3 |
| chr17 | 44758320 | 44758807 | 488 | 4 | 1.48E-06 | 8.16E-06 | 0.00032 | 5.11E-06 | -0.00269308 | NA |
| chr6 | 38177304 | 38177688 | 385 | 4 | 2.56E-06 | 1.30E-05 | 0.000551 | 5.12E-06 | -0.00431041 | BTBD9 |
| chr22 | 28051933 | 28052373 | 441 | 2 | 1.98E-06 | 1.48E-05 | 1.82E-05 | 5.12E-06 | -0.01099658 | TTC28 |
| chr1 | 17409212 | 17410056 | 845 | 4 | 2.26E-05 | 2.74E-06 | 0.001327 | 5.12E-06 | 0.000189329 | RCC2 |
| chr12 | 50532020 | 50532874 | 855 | 2 | 7.45E-05 | 3.16E-06 | 0.000359 | 5.12E-06 | -0.00666679 | DIP2B |
| chr7 | 2222465 | 2222844 | 380 | 5 | 1.40E-07 | 0.0066574 | 6.48E-07 | 5.12E-06 | -0.00178511 | MAD1L1, AC069288.1 |
| chr6 | 144150061 | 144150848 | 788 | 5 | 2.97E-06 | 2.27E-05 | 0.000107 | 5.12E-06 | -0.00143779 | STX11 |
| chr19 | 6693258 | 6693427 | 170 | 2 | 1.24E-05 | 3.18E-06 | 0.000338 | 5.12E-06 | -0.00530224 | C3 |
| chr20 | 63919062 | 63919387 | 326 | 6 | 9.90E-06 | 1.39E-06 | 0.004634 | 5.13E-06 | 0.001803484 | DNAJC5 |
| chr11 | 63368942 | 63369926 | 985 | 6 | 6.66E-06 | 0.0003561 | 0.000318 | 5.13E-06 | -0.00814063 | SLC22A10, SLC22A9 |
| chr6 | 44674314 | 44674790 | 477 | 2 | 5.89E-07 | 2.13E-05 | 1.36E-05 | 5.13E-06 | -0.00927979 | NA |
| chr3 | 58588111 | 58588315 | 205 | 2 | 1.19E-07 | 7.64E-06 | 3.75E-05 | 5.14E-06 | -0.01042765 | FAM107A |
| chr14 | 94617544 | 94617560 | 17 | 3 | 3.76E-06 | 6.31E-06 | 0.000494 | 5.14E-06 | -0.0001378 | AL049839.2, SERPINA3 |
| chr11 | 62726455 | 62727107 | 653 | 6 | 4.59E-06 | 7.31E-05 | 0.001815 | 5.15E-06 | 0.000885337 | HNRNPUL2-BSCL2, HNRNPUL2 |
| chr12 | 88545786 | 88545814 | 29 | 2 | 1.24E-05 | 3.58E-06 | 0.000191 | 5.15E-06 | -0.00296675 | KITLG |
| chr1 | 31066635 | 31066767 | 133 | 2 | 2.43E-07 | 2.83E-05 | 1.12E-05 | 5.15E-06 | -0.00503778 | NA |
| chr4 | 3444678 | 3445812 | 1135 | 5 | 3.84E-08 | 0.0001341 | 7.79E-06 | 5.15E-06 | -0.0049607 | HGFAC |
| chr17 | 79368214 | 79368456 | 243 | 2 | 3.53E-06 | 1.67E-05 | 1.66E-05 | 5.15E-06 | -0.01318068 | RBFOX3 |
| chr4 | 166053322 | 166053732 | 411 | 4 | 3.13E-06 | 0.0001271 | 0.000229 | 5.15E-06 | 0.003554917 | TLL1 |
| chr9 | 137302333 | 137303101 | 769 | 7 | 2.23E-06 | 1.84E-05 | 0.00118 | 5.15E-06 | -0.00061717 | NA |
| chr1 | 228424902 | 228425559 | 658 | 7 | 4.18E-06 | 1.49E-05 | 0.000269 | 5.15E-06 | -0.00133149 | H3-4 |
| chr7 | 143362944 | 143363443 | 500 | 5 | 3.74E-06 | 3.23E-06 | 0.001112 | 5.15E-06 | -0.01180234 | NA |
| chr18 | 74796932 | 74797599 | 668 | 3 | 2.90E-05 | 6.12E-06 | 0.00048 | 5.16E-06 | -0.00578834 | ZNF407 |
| chr13 | 19782451 | 19783561 | 1111 | 9 | 8.36E-07 | 5.26E-05 | 7.81E-05 | 5.16E-06 | -0.00231016 | PSPC1 |
| chr5 | 50441515 | 50441939 | 425 | 2 | 1.90E-07 | 0.0001941 | 4.86E-06 | 5.16E-06 | 0.021707478 | EMB |
| chr1 | 192492050 | 192492593 | 544 | 3 | 2.38E-06 | 1.55E-05 | 6.69E-05 | 5.16E-06 | -0.0024387 | AL390957.1 |
| chr19 | 19628364 | 19629011 | 648 | 10 | 9.12E-06 | 2.22E-06 | 0.009672 | 5.16E-06 | 0.003526263 | LPAR2 |
| chr11 | 64765367 | 64766151 | 785 | 7 | 1.38E-06 | 0.0017012 | 9.71E-06 | 5.17E-06 | 0.003487292 | SF1 |
| chr16 | 51392995 | 51393000 | 6 | 2 | 1.32E-05 | 3.11E-06 | 0.000428 | 5.17E-06 | -0.0139711 | AC087564.1, AC007344.1 |
| chr16 | 24419782 | 24419947 | 166 | 2 | 1.32E-06 | 7.92E-05 | 6.50E-06 | 5.17E-06 | 0.01522888 | NA |
| chr5 | 138962397 | 138962904 | 508 | 2 | 7.29E-07 | 1.15E-05 | 2.36E-05 | 5.18E-06 | -0.00400375 | SIL1 |
| chr11 | 5789795 | 5789954 | 160 | 2 | 5.88E-06 | 4.10E-06 | 0.000121 | 5.18E-06 | -0.002852 | TRIM5, OR52N1 |
| chr13 | 112700746 | 112701042 | 297 | 2 | 6.29E-06 | 6.95E-06 | 4.34E-05 | 5.18E-06 | -0.00222489 | ATP11A |
| chr3 | 195104281 | 195104586 | 306 | 2 | 3.89E-06 | 4.95E-06 | 7.76E-05 | 5.18E-06 | -0.00828961 | XXYLT1 |
| chr2 | 10846596 | 10847555 | 960 | 4 | 3.18E-06 | 5.56E-05 | 9.13E-05 | 5.18E-06 | -0.0064594 | LINC01954 |
| chr11 | 93936215 | 93936852 | 638 | 3 | 3.13E-05 | 5.47E-06 | 0.000424 | 5.18E-06 | -0.00514782 | NA |
| chr7 | 30465325 | 30466063 | 739 | 3 | 5.98E-08 | 0.0191151 | 1.20E-06 | 5.18E-06 | 0.004944507 | NOD1 |
| chr14 | 20469413 | 20470406 | 994 | 6 | 7.01E-08 | 4.88E-05 | 0.000113 | 5.18E-06 | 0.000256685 | PNP |
| chr5 | 72106814 | 72108163 | 1350 | 9 | 1.13E-05 | 0.0003553 | 0.000167 | 5.19E-06 | 0.000528032 | MAP1B, AC025774.1 |
| chr8 | 140764394 | 140765273 | 880 | 6 | 3.53E-05 | 4.05E-06 | 0.002849 | 5.19E-06 | -0.011556 | PTK2 |
| chr3 | 42515519 | 42516302 | 784 | 4 | 6.28E-06 | 0.0002148 | 0.00085 | 5.20E-06 | -0.002764 | VIPR1, VIPR1-AS1 |
| chr1 | 173660294 | 173660658 | 365 | 2 | 1.83E-06 | 2.06E-05 | 1.42E-05 | 5.20E-06 | -0.01282271 | AL139142.2, ANKRD45 |
| chr5 | 1813658 | 1814087 | 430 | 5 | 8.35E-07 | 0.0004843 | 4.00E-06 | 5.20E-06 | 0.000832487 | NDUFS6 |
| chr12 | 96044183 | 96044806 | 624 | 4 | 3.94E-06 | 4.28E-06 | 0.000312 | 5.20E-06 | -0.00879317 | NA |
| chr5 | 150449373 | 150450621 | 1249 | 14 | 1.43E-05 | 7.56E-05 | 0.008214 | 5.20E-06 | 0.001501576 | RPS14 |
| chr8 | 141002418 | 141002894 | 477 | 3 | 6.21E-06 | 3.30E-05 | 0.000219 | 5.21E-06 | -0.00032601 | NA |
| chr5 | 15934791 | 15935178 | 388 | 4 | 1.05E-06 | 0.0002255 | 4.73E-05 | 5.21E-06 | -0.00561355 | FBXL7 |
| chr4 | 105594293 | 105594731 | 439 | 2 | 2.29E-06 | 1.29E-05 | 2.12E-05 | 5.21E-06 | 0.015303624 | ARHGEF38 |
| chr9 | 137836354 | 137837100 | 747 | 4 | 1.97E-05 | 5.28E-06 | 0.000578 | 5.21E-06 | -0.00383193 | EHMT1 |
| chr2 | 68799665 | 68799907 | 243 | 2 | 1.03E-06 | 0.0087393 | 3.06E-06 | 5.21E-06 | -0.00306352 | ARHGAP25 |
| chr10 | 22700462 | 22701199 | 738 | 2 | 5.86E-06 | 4.33E-06 | 0.000106 | 5.21E-06 | 0.008900924 | PIP4K2A |
| chr10 | 68301945 | 68302431 | 487 | 2 | 1.55E-07 | 0.001239 | 3.48E-06 | 5.21E-06 | -0.01440998 | PBLD |
| chr12 | 111024513 | 111024560 | 48 | 2 | 1.96E-06 | 0.0007009 | 3.75E-06 | 5.21E-06 | -0.00741643 | NA |
| chr16 | 85353950 | 85354733 | 784 | 2 | 4.44E-05 | 3.20E-06 | 0.000377 | 5.21E-06 | -0.01240167 | GSE1 |
| chr7 | 63114547 | 63114720 | 174 | 2 | 1.30E-05 | 3.10E-06 | 0.000468 | 5.22E-06 | -0.02256885 | SAPCD2P4 |
| chr3 | 127287444 | 127287960 | 517 | 4 | 1.51E-05 | 1.73E-06 | 0.002245 | 5.22E-06 | -0.00316463 | PRR20G |
| chr1 | 162885737 | 162886113 | 377 | 2 | 3.11E-06 | 1.20E-05 | 2.31E-05 | 5.22E-06 | -0.00915843 | NA |
| chr2 | 223837550 | 223838713 | 1164 | 8 | 7.17E-07 | 0.0001716 | 0.000125 | 5.22E-06 | -0.00092477 | AP1S3, AC093884.1 |
| chr15 | 101008471 | 101009102 | 632 | 3 | 2.01E-05 | 3.07E-06 | 0.000838 | 5.23E-06 | 0.0059114 | LRRK1 |
| chr20 | 41233629 | 41234632 | 1004 | 3 | 6.78E-07 | 3.22E-05 | 1.57E-05 | 5.23E-06 | -0.00513641 | ZHX3 |
| chr13 | 57631526 | 57632338 | 813 | 6 | 7.07E-06 | 3.63E-06 | 0.001566 | 5.23E-06 | -4.10E-05 | PCDH17 |
| chr5 | 2206091 | 2207062 | 972 | 6 | 1.75E-06 | 2.28E-06 | 0.002281 | 5.23E-06 | -0.01038346 | NA |
| chr4 | 47844401 | 47844694 | 294 | 4 | 3.82E-06 | 8.65E-06 | 6.54E-05 | 5.24E-06 | 0.000643346 | AC107068.2 |
| chr8 | 143824206 | 143824640 | 435 | 5 | 3.84E-06 | 1.02E-05 | 0.002001 | 5.24E-06 | -0.0058962 | PUF60 |
| chr10 | 2889884 | 2890428 | 545 | 4 | 1.84E-05 | 2.57E-06 | 0.002708 | 5.24E-06 | -0.00354586 | NA |
| chr15 | 55217791 | 55217805 | 15 | 2 | 1.02E-05 | 6.50E-06 | 4.91E-05 | 5.24E-06 | -0.00449914 | RAB27A |
| chr21 | 34791822 | 34792451 | 630 | 5 | 4.00E-06 | 2.43E-06 | 0.000573 | 5.24E-06 | -0.00334871 | RUNX1 |
| chr3 | 44113154 | 44113645 | 492 | 5 | 6.65E-06 | 4.84E-06 | 0.003088 | 5.24E-06 | -0.00137351 | NA |
| chr10 | 113550029 | 113550852 | 824 | 5 | 4.20E-05 | 1.37E-06 | 0.004928 | 5.24E-06 | -0.00170449 | HABP2 |
| chr14 | 72670265 | 72671185 | 921 | 5 | 9.79E-08 | 0.0003751 | 0.000103 | 5.24E-06 | 0.006447945 | DPF3 |
| chr13 | 111588223 | 111588730 | 508 | 3 | 3.45E-07 | 0.0007444 | 3.25E-06 | 5.24E-06 | -0.00666813 | AL359649.1 |
| chr3 | 123143871 | 123144365 | 495 | 6 | 1.61E-05 | 8.32E-06 | 0.002173 | 5.25E-06 | 0.000322467 | PDIA5 |
| chr7 | 13931353 | 13931744 | 392 | 2 | 1.21E-05 | 3.33E-06 | 0.000308 | 5.25E-06 | -0.0070113 | ETV1 |
| chr20 | 7346165 | 7346252 | 88 | 3 | 1.02E-05 | 1.25E-05 | 0.000385 | 5.25E-06 | 0.000730666 | NA |
| chr2 | 165846098 | 165846559 | 462 | 5 | 6.53E-06 | 6.69E-06 | 0.000292 | 5.25E-06 | 0.002971553 | NA |
| chr5 | 180650359 | 180650670 | 312 | 2 | 2.26E-06 | 3.48E-06 | 0.000245 | 5.26E-06 | -0.01428594 | NA |
| chr3 | 48564152 | 48565361 | 1210 | 8 | 4.60E-07 | 0.0011647 | 0.000122 | 5.26E-06 | -0.00321227 | COL7A1 |
| chr4 | 140642642 | 140643117 | 476 | 3 | 2.63E-06 | 1.51E-05 | 0.000259 | 5.26E-06 | 0.003003608 | TBC1D9, TNRC18P1 |
| chr11 | 79315628 | 79315943 | 316 | 2 | 4.46E-06 | 1.02E-05 | 2.78E-05 | 5.27E-06 | -0.02012817 | TENM4 |
| chr2 | 130819852 | 130820389 | 538 | 3 | 1.15E-05 | 2.61E-06 | 0.00074 | 5.27E-06 | -0.00984093 | NA |
| chr1 | 247112042 | 247112794 | 753 | 10 | 3.31E-06 | 1.35E-05 | 0.000157 | 5.27E-06 | -0.0048595 | C1orf229 |

| | | | | | | | | | | |
|---|---|---|---|---|---|---|---|---|---|---|
| chr2 | 205323780 | 205324163 | 384 | 2 | 7.34E-06 | 4.56E-06 | 9.64E-05 | 5.27E-06 | -0.00064921 | PARD3B |
| chr3 | 25292788 | 25293419 | 632 | 7 | 1.42E-05 | 1.18E-05 | 0.000576 | 5.27E-06 | 0.003433805 | RARB |
| chr21 | 46643108 | 46643513 | 406 | 2 | 1.84E-06 | 5.53E-06 | 6.57E-05 | 5.27E-06 | 0.007131309 | PRMT2 |
| chr3 | 182500544 | 182500777 | 234 | 2 | 3.09E-06 | 2.02E-05 | 1.47E-05 | 5.27E-06 | -0.02107645 | LINC01995 |
| chr14 | 68733099 | 68733254 | 156 | 2 | 3.12E-06 | 5.25E-05 | 8.02E-06 | 5.28E-06 | 0.013566934 | AL132986.1 |
| chr1 | 77292847 | 77293533 | 687 | 4 | 2.39E-06 | 2.52E-05 | 8.50E-05 | 5.28E-06 | -0.00879541 | AK5 |
| chr2 | 216426777 | 216427205 | 429 | 6 | 1.15E-05 | 2.14E-05 | 0.000375 | 5.28E-06 | -0.00015679 | SMARCAL1 |
| chr8 | 130086094 | 130086365 | 272 | 3 | 1.09E-05 | 6.83E-06 | 0.000109 | 5.28E-06 | -0.0043998 | ASAP1 |
| chr13 | 31119578 | 31119934 | 357 | 2 | 1.42E-06 | 1.12E-05 | 2.52E-05 | 5.28E-06 | -0.01505671 | NA |
| chr15 | 94354900 | 94355889 | 990 | 6 | 7.62E-06 | 0.0014534 | 0.00026 | 5.28E-06 | -0.00276015 | MCTP2 |
| chr12 | 22963457 | 22964403 | 947 | 4 | 1.29E-06 | 5.82E-05 | 5.82E-05 | 5.28E-06 | -0.0068785 | AC084816.1 |
| chr11 | 18742230 | 18742533 | 304 | 2 | 4.13E-07 | 7.50E-05 | 6.81E-06 | 5.29E-06 | -0.00638602 | PTPN5 |
| chr5 | 93579580 | 93579875 | 296 | 2 | 4.13E-07 | 5.68E-06 | 6.32E-05 | 5.29E-06 | -0.00487287 | NR2F1-AS1 |
| chr10 | 126071867 | 126072078 | 212 | 2 | 1.37E-06 | 5.17E-06 | 7.53E-05 | 5.30E-06 | -0.00347026 | ADAM12 |
| chr20 | 61881240 | 61882093 | 854 | 5 | 6.06E-06 | 2.53E-05 | 0.00022 | 5.30E-06 | 0.001647971 | CDH4 |
| chr8 | 53743773 | 53744104 | 332 | 2 | 2.73E-06 | 2.02E-05 | 1.48E-05 | 5.30E-06 | 0.000544632 | ATP6V1H |
| chr5 | 140875616 | 140876194 | 579 | 4 | 8.18E-07 | 5.99E-05 | 1.16E-05 | 5.30E-06 | -0.0088649 | PCDHA1, PCDHA2, PCDHA3, PCDHA4, PCDHA5, PCDHA6, PCDHA7, PCDHA8, PCDHA9, PCDHA10, PCDHA11, PCDHA12, AC005609.3 |
| chr12 | 102480788 | 102481523 | 736 | 6 | 1.52E-06 | 0.0001282 | 1.51E-05 | 5.30E-06 | -0.00620511 | IGF1 |
| chr8 | 66504952 | 66505640 | 689 | 6 | 5.18E-06 | 1.78E-05 | 0.00339 | 5.30E-06 | -0.00177702 | AC009879.3, VXN |
| chr16 | 31032789 | 31033438 | 650 | 15 | 6.85E-06 | 7.11E-05 | 0.00373 | 5.30E-06 | 0.00060661 | STX4 |
| chr10 | 121483644 | 121483981 | 338 | 4 | 1.73E-06 | 9.97E-05 | 2.14E-05 | 5.30E-06 | 0.005249259 | FGFR2 |
| chr5 | 8839497 | 8839610 | 114 | 2 | 3.87E-06 | 7.98E-06 | 3.75E-05 | 5.31E-06 | -0.01086919 | NA |
| chr16 | 86664101 | 86664553 | 453 | 3 | 1.92E-06 | 8.92E-05 | 0.00019 | 5.31E-06 | -0.00040783 | AC009154.1 |
| chr10 | 98251812 | 98251967 | 156 | 4 | 1.42E-06 | 6.66E-05 | 2.75E-05 | 5.31E-06 | -0.00126779 | LOXL4 |
| chr6 | 41409078 | 41409550 | 473 | 4 | 2.80E-06 | 4.84E-06 | 0.00229 | 5.31E-06 | -0.01865874 | NA |
| chr22 | 46112114 | 46112724 | 611 | 6 | 2.33E-07 | 0.0003128 | 5.31E-05 | 5.31E-06 | -0.00117405 | MIRLET7BHG |
| chr1 | 204196981 | 204197860 | 880 | 5 | 3.15E-07 | 0.0010404 | 4.53E-06 | 5.32E-06 | 0.004194296 | NA |
| chr17 | 74203227 | 74204421 | 1195 | 14 | 2.97E-05 | 2.72E-05 | 0.001461 | 5.32E-06 | -0.00102869 | RPL38 |
| chr17 | 74584319 | 74584942 | 624 | 6 | 3.62E-06 | 6.68E-06 | 0.002144 | 5.32E-06 | -0.0050815 | C17orf77, CD300LD |
| chr1 | 9724112 | 9724887 | 776 | 3 | 3.66E-06 | 1.28E-05 | 0.000337 | 5.32E-06 | 0.000241256 | PIK3CD |
| chr10 | 133419313 | 133419905 | 593 | 5 | 1.74E-05 | 9.35E-06 | 0.00181 | 5.32E-06 | -0.00078008 | AL360181.3, MTG1 |
| chr16 | 3203043 | 3204207 | 1165 | 6 | 6.95E-06 | 1.73E-05 | 0.003193 | 5.32E-06 | 0.002327639 | AJ003147.2 |
| chr1 | 111138986 | 111139659 | 674 | 11 | 2.09E-06 | 0.0005734 | 0.000656 | 5.32E-06 | -0.00077164 | CEPT1, DRAM2 |
| chr3 | 48223077 | 48223263 | 187 | 2 | 1.90E-07 | 2.12E-05 | 1.44E-05 | 5.33E-06 | -0.01021439 | NA |
| chr14 | 72975305 | 72975986 | 682 | 3 | 4.00E-07 | 0.0002979 | 4.13E-06 | 5.33E-06 | -0.00242018 | ZFYVE1 |
| chr12 | 30930457 | 30931163 | 707 | 3 | 7.70E-06 | 3.66E-06 | 0.000497 | 5.33E-06 | -0.0079184 | TSPAN11 |
| chr1 | 965755 | 966645 | 891 | 10 | 1.41E-05 | 1.17E-05 | 0.006299 | 5.33E-06 | -0.0069756 | PLEKHN1 |
| chr17 | 56153058 | 56153635 | 578 | 10 | 9.69E-06 | 2.53E-05 | 0.00075 | 5.34E-06 | -0.00151001 | ANKFN1 |
| chr1 | 162334717 | 162335203 | 487 | 2 | 9.70E-07 | 3.42E-05 | 1.04E-05 | 5.34E-06 | -0.01051033 | NOS1AP |
| chr2 | 16228350 | 16228875 | 526 | 5 | 6.22E-06 | 1.88E-05 | 0.000772 | 5.34E-06 | 0.000401764 | AC010745.2 |
| chr19 | 48445623 | 48446206 | 584 | 8 | 6.85E-08 | 0.0006662 | 0.00031 | 5.34E-06 | -0.00073472 | GRWD1 |
| chr1 | 39676784 | 39677417 | 634 | 4 | 6.81E-06 | 5.80E-06 | 0.000467 | 5.34E-06 | -0.00978197 | NA |
| chr15 | 84881769 | 84881808 | 40 | 2 | 9.90E-06 | 7.26E-06 | 4.35E-05 | 5.35E-06 | -0.00752239 | NA |
| chr6 | 157460776 | 157461175 | 400 | 2 | 1.10E-05 | 3.51E-06 | 0.00026 | 5.36E-06 | 0.003064477 | ZDHHC14 |
| chr8 | 94940123 | 94941025 | 903 | 2 | 6.31E-05 | 3.34E-06 | 0.00034 | 5.36E-06 | -0.00143712 | NDUFAF6, TP53INP1 |
| chr6 | 32202101 | 32202717 | 617 | 5 | 1.23E-07 | 0.0002137 | 2.09E-05 | 5.36E-06 | -0.01899502 | NOTCH4 |
| chr16 | 73385400 | 73386075 | 676 | 4 | 4.44E-06 | 5.99E-06 | 0.000907 | 5.36E-06 | -0.00483717 | ZFHX3 |
| chr7 | 1243619 | 1244345 | 727 | 5 | 2.31E-06 | 8.37E-06 | 0.001741 | 5.36E-06 | -0.00131301 | NA |
| chr17 | 924718 | 925288 | 571 | 3 | 1.26E-06 | 4.84E-05 | 7.38E-05 | 5.36E-06 | -0.00750824 | NXN |
| chr5 | 60700010 | 60701114 | 1105 | 10 | 1.88E-05 | 0.0004104 | 0.001926 | 5.37E-06 | -0.00218101 | AC109133.1, DEPDC1B |
| chr16 | 14487742 | 14488110 | 369 | 3 | 3.31E-06 | 5.61E-05 | 0.000287 | 5.37E-06 | 0.008465938 | PARN |
| chr15 | 31347455 | 31347608 | 154 | 2 | 8.16E-06 | 3.47E-06 | 0.000281 | 5.37E-06 | -0.0084847 | KLF13 |
| chr7 | 77498728 | 77498979 | 252 | 3 | 1.15E-05 | 2.77E-06 | 0.001129 | 5.38E-06 | 0.003269356 | AC004921.2 |
| chr5 | 67166056 | 67166834 | 779 | 4 | 8.44E-06 | 3.71E-06 | 0.0009 | 5.38E-06 | -0.01781328 | MAST4 |
| chr6 | 5905133 | 5905539 | 407 | 2 | 9.68E-06 | 7.28E-06 | 4.38E-05 | 5.38E-06 | -0.01279222 | NA |
| chr10 | 1096154 | 1096723 | 570 | 5 | 1.36E-05 | 1.07E-05 | 0.001932 | 5.38E-06 | 0.001176927 | WDR37 |
| chr17 | 82157060 | 82157175 | 116 | 2 | 8.35E-06 | 9.33E-06 | 3.18E-05 | 5.38E-06 | -0.00678975 | CCDC57 |
| chr13 | 106859073 | 106859199 | 127 | 2 | 1.90E-06 | 6.68E-06 | 4.98E-05 | 5.38E-06 | -0.00393447 | NA |
| chr2 | 172061035 | 172061164 | 130 | 3 | 5.30E-06 | 3.64E-05 | 1.30E-05 | 5.38E-06 | -0.00211176 | METAP1D |
| chr9 | 133578462 | 133579006 | 545 | 2 | 6.65E-09 | 0.0092141 | 3.14E-06 | 5.38E-06 | 0.013431967 | FAM163B |
| chr7 | 6654742 | 6655461 | 720 | 5 | 4.92E-06 | 9.97E-06 | 0.001562 | 5.38E-06 | -0.00084058 | ZNF316 |
| chr1 | 19003562 | 19003661 | 100 | 2 | 8.71E-06 | 9.28E-06 | 3.21E-05 | 5.39E-06 | -0.01203081 | NA |
| chr11 | 18371515 | 18371644 | 130 | 2 | 1.28E-06 | 0.0016857 | 3.47E-06 | 5.39E-06 | -0.01188542 | NA |
| chr15 | 48719866 | 48720509 | 644 | 2 | 9.20E-06 | 3.23E-06 | 0.000461 | 5.39E-06 | -0.01376272 | CEP152 |
| chr3 | 44761057 | 44761752 | 696 | 12 | 4.51E-05 | 5.72E-06 | 0.004879 | 5.39E-06 | -0.0031978 | KIF15, KIAA1143 |
| chr17 | 78993369 | 78993883 | 515 | 6 | 5.01E-06 | 1.09E-05 | 0.001257 | 5.39E-06 | -0.00337616 | CANT1 |
| chr21 | 45472918 | 45473241 | 324 | 2 | 2.35E-06 | 1.39E-05 | 2.09E-05 | 5.40E-06 | 0.003546466 | COL18A1 |
| chr21 | 42108550 | 42108758 | 209 | 4 | 7.84E-06 | 1.28E-06 | 0.005778 | 5.40E-06 | 0.035474971 | UMODL1 |
| chr15 | 77568391 | 77569112 | 722 | 3 | 1.01E-07 | 0.0003798 | 3.17E-06 | 5.40E-06 | 0.001405518 | AC046168.2 |
| chr4 | 109815107 | 109815511 | 405 | 12 | 1.11E-05 | 2.73E-06 | 0.002265 | 5.40E-06 | 0.000741808 | GAR1, AC126283.1 |
| chr7 | 77200528 | 77201279 | 752 | 5 | 8.99E-06 | 2.22E-06 | 0.001301 | 5.40E-06 | 0.008956452 | CCDC146 |
| chr11 | 12376451 | 12376588 | 138 | 3 | 3.38E-06 | 7.70E-06 | 6.21E-05 | 5.40E-06 | 0.00171285 | NA |
| chr3 | 186806262 | 186807070 | 809 | 18 | 4.89E-06 | 4.56E-05 | 0.002174 | 5.41E-06 | 0.000111378 | RFC4 |
| chr5 | 132294429 | 132295467 | 1039 | 11 | 7.38E-06 | 1.12E-05 | 0.000502 | 5.41E-06 | -0.00033366 | SLC22A4, P4HA2 |
| chr7 | 2082468 | 2082942 | 475 | 3 | 1.66E-06 | 2.22E-05 | 4.51E-05 | 5.42E-06 | -0.00016872 | MAD1L1, AC069288.1 |
| chr19 | 18192518 | 18193232 | 715 | 11 | 9.52E-06 | 1.19E-05 | 0.004247 | 5.42E-06 | 0.00036873 | MPV17L2 |
| chr2 | 69913654 | 69914157 | 504 | 2 | 6.63E-07 | 4.23E-06 | 0.00013 | 5.42E-06 | -0.00997708 | MXD1 |
| chr2 | 46555114 | 46555908 | 795 | 4 | 7.66E-06 | 7.28E-05 | 0.000224 | 5.43E-06 | -0.00243354 | RHOQ |
| chr6 | 158972523 | 158973080 | 558 | 2 | 2.17E-05 | 3.29E-06 | 0.000423 | 5.44E-06 | 0.016347891 | RSPH3 |
| chr2 | 203238588 | 203239536 | 949 | 10 | 1.10E-05 | 4.37E-06 | 0.005643 | 5.44E-06 | 0.001578917 | CYP20A1 |
| chr2 | 71068031 | 71068972 | 942 | 15 | 4.65E-06 | 9.98E-06 | 0.002384 | 5.45E-06 | -0.00035592 | NAGK, AC007881.3 |
| chr11 | 116703127 | 116703563 | 437 | 2 | 2.19E-06 | 1.55E-05 | 1.91E-05 | 5.45E-06 | 0.00177501 | NA |
| chr16 | 56563597 | 56564036 | 440 | 4 | 4.90E-06 | 1.15E-05 | 0.000721 | 5.45E-06 | -0.00297221 | NA |
| chr17 | 70168773 | 70170100 | 1328 | 11 | 8.21E-07 | 9.89E-05 | 0.001204 | 5.45E-06 | 0.001340698 | KCNJ2, KCNJ2-AS1 |
| chr11 | 19524586 | 19524609 | 24 | 2 | 4.63E-06 | 3.26E-05 | 1.10E-05 | 5.46E-06 | -0.006312 | NAV2 |
| chr19 | 10489300 | 10489741 | 442 | 4 | 6.81E-06 | 8.80E-06 | 0.000327 | 5.46E-06 | -0.00067314 | KEAP1 |
| chr18 | 62605733 | 62606163 | 431 | 2 | 9.16E-06 | 3.69E-06 | 0.000225 | 5.46E-06 | -0.0049063 | NA |
| chr10 | 68736649 | 68737320 | 672 | 3 | 2.86E-05 | 2.60E-06 | 0.001215 | 5.46E-06 | -0.00757201 | CCAR1 |
| chr9 | 124032327 | 124033206 | 880 | 7 | 1.43E-06 | 2.62E-05 | 0.000128 | 5.46E-06 | -0.00494378 | LHX2, AC006450.2 |
| chr12 | 21774491 | 21774889 | 399 | 7 | 3.80E-06 | 3.30E-05 | 0.000131 | 5.46E-06 | 0.000732125 | KCNJ8 |
| chr7 | 146654164 | 146654315 | 152 | 2 | 1.77E-06 | 0.0050577 | 3.25E-06 | 5.46E-06 | -0.00925553 | CNTNAP2 |
| chr2 | 201643506 | 201644068 | 563 | 8 | 1.50E-07 | 3.43E-05 | 6.78E-05 | 5.46E-06 | 0.003609586 | AC007279.1, TMEM237 |
| chr8 | 41276906 | 41277116 | 211 | 2 | 7.82E-06 | 3.27E-06 | 0.000463 | 5.47E-06 | -0.00941942 | SFRP1, AC104393.1 |
| chr10 | 78905257 | 78906203 | 947 | 3 | 2.22E-06 | 3.21E-06 | 0.000549 | 5.47E-06 | 0.001074429 | NA |
| chr19 | 4791666 | 4791687 | 22 | 3 | 0.00015276 | 2.52E-05 | 1.76E-05 | 5.47E-06 | -5.16E-07 | NA |
| chr17 | 43577049 | 43577523 | 475 | 2 | 2.59E-06 | 3.79E-06 | 0.000202 | 5.47E-06 | -0.00850425 | DHX8, ETV4 |
| chr9 | 136333242 | 136333295 | 54 | 2 | 2.15E-06 | 4.31E-06 | 0.000127 | 5.47E-06 | -0.00811312 | GPSM1 |
| chr12 | 98644268 | 98645524 | 1257 | 21 | 7.80E-06 | 0.0003721 | 0.01058 | 5.47E-06 | 0.001105623 | APAF1, IKBIP |
| chr7 | 128892159 | 128892724 | 566 | 2 | 3.75E-07 | 1.31E-05 | 2.26E-05 | 5.47E-06 | -0.00390948 | KCP |
| chr12 | 26127324 | 26128108 | 785 | 2 | 2.43E-05 | 3.42E-06 | 0.000341 | 5.47E-06 | -0.01550652 | SSPN |
| chr12 | 50530498 | 50530761 | 264 | 2 | 3.73E-06 | 1.15E-05 | 2.60E-05 | 5.48E-06 | -0.00354225 | DIP2B |
| chr1 | 84997681 | 84998607 | 927 | 5 | 7.81E-06 | 2.09E-05 | 0.000235 | 5.48E-06 | 0.004695256 | NA |
| chr3 | 152374548 | 152374806 | 259 | 2 | 1.52E-06 | 0.0003993 | 4.37E-06 | 5.49E-06 | -0.00527541 | MBNL1 |
| chr16 | 70646201 | 70646914 | 714 | 8 | 5.96E-06 | 2.90E-05 | 0.000622 | 5.49E-06 | -0.00125491 | IL34 |
| chr12 | 112997633 | 112998175 | 543 | 3 | 1.69E-06 | 7.01E-06 | 0.000134 | 5.49E-06 | -0.00297516 | OAS2, AC004551.1 |
| chr10 | 97422564 | 97422576 | 13 | 2 | 5.16E-06 | 5.34E-06 | 7.74E-05 | 5.49E-06 | -0.00174482 | RRP12 |
| chr9 | 121597985 | 121598268 | 284 | 4 | 8.46E-06 | 4.50E-06 | 0.000232 | 5.49E-06 | -0.0081031 | DAB2IP |
| chr15 | 34987737 | 34988513 | 777 | 8 | 8.70E-06 | 3.63E-05 | 0.00042 | 5.49E-06 | 0.001356636 | ZNF770 |
| chr1 | 228174532 | 228174808 | 277 | 6 | 1.25E-05 | 1.10E-06 | 0.008298 | 5.49E-06 | 0.000888478 | IBA57 |
| chr11 | 43895235 | 43896067 | 833 | 3 | 4.94E-05 | 3.63E-06 | 0.000813 | 5.49E-06 | 0.00048391 | ALKBH3 |
| chr14 | 73644372 | 73645191 | 820 | 11 | 4.07E-06 | 0.0008847 | 0.001178 | 5.50E-06 | 0.001475408 | DNAL1 |
| chr7 | 87713548 | 87714192 | 645 | 2 | 1.71E-05 | 3.64E-06 | 0.000252 | 5.50E-06 | 0.022136323 | RUNDC3B |
| chr10 | 86338137 | 86338364 | 228 | 2 | 7.86E-06 | 3.32E-06 | 0.000436 | 5.50E-06 | 0.001764504 | GRID1 |
| chr12 | 5511930 | 5512290 | 361 | 3 | 1.36E-06 | 1.42E-05 | 7.13E-05 | 5.50E-06 | -0.02355674 | NTF3 |
| chr20 | 35740586 | 35742312 | 1727 | 14 | 7.53E-07 | 2.38E-05 | 0.000115 | 5.50E-06 | -0.00090549 | RBM39 |
| chr14 | 100321954 | 100323172 | 1219 | 5 | 1.44E-05 | 2.05E-06 | 0.003296 | 5.51E-06 | -0.00352778 | NA |
| chr14 | 102500544 | 102500556 | 13 | 2 | 1.10E-05 | 3.39E-06 | 0.000376 | 5.51E-06 | -0.01009679 | TECPR2 |
| chr19 | 50607928 | 50608131 | 204 | 2 | 5.75E-06 | 1.54E-05 | 1.96E-05 | 5.51E-06 | -0.01618747 | NA |
| chr6 | 37646459 | 37647592 | 1134 | 4 | 9.27E-06 | 2.88E-06 | 0.001836 | 5.51E-06 | -0.00575826 | MDGA1 |
| chr22 | 32204959 | 32205667 | 709 | 6 | 8.45E-07 | 3.18E-06 | 0.001249 | 5.51E-06 | -0.02056531 | SLC5A4-AS1, RFPL2 |
| chr9 | 74615232 | 74615593 | 362 | 4 | 3.38E-06 | 8.15E-05 | 6.04E-05 | 5.51E-06 | -0.0030385 | RORB |
| chr11 | 69537529 | 69537727 | 199 | 2 | 5.20E-06 | 4.14E-06 | 0.00015 | 5.51E-06 | -0.00621492 | NA |
| chr12 | 3367744 | 3367987 | 244 | 4 | 8.54E-06 | 3.97E-06 | 0.002258 | 5.51E-06 | -0.00014721 | LINC02417 |
| chr16 | 59755000 | 59755581 | 582 | 6 | 1.09E-07 | 0.0006057 | 1.53E-05 | 5.52E-06 | -0.00044339 | APOOP5 |
| chr16 | 90021948 | 90022628 | 681 | 10 | 1.00E-06 | 5.07E-05 | 0.000356 | 5.52E-06 | -0.00697073 | GAS8 |
| chr12 | 55973426 | 55974034 | 609 | 14 | 8.24E-06 | 2.85E-05 | 0.008159 | 5.53E-06 | -0.0002903 | RAB5B |
| chr10 | 129908480 | 129908615 | 136 | 3 | 1.66E-05 | 2.38E-06 | 0.0015 | 5.53E-06 | -0.00115545 | EBF3 |
| chr16 | 3881414 | 3881896 | 483 | 4 | 1.85E-06 | 5.22E-06 | 0.000128 | 5.53E-06 | -0.01178901 | NA |
| chr1 | 35883336 | 35883942 | 607 | 5 | 8.01E-06 | 1.10E-05 | 0.002059 | 5.53E-06 | 0.000870637 | AGO1 |
| chr9 | 97855717 | 97855833 | 117 | 3 | 3.55E-06 | 0.0001412 | 6.19E-06 | 5.54E-06 | -0.00330953 | FOXE1 |
| chr2 | 9799460 | 9800159 | 700 | 4 | 1.01E-05 | 1.02E-05 | 0.000268 | 5.55E-06 | -7.27E-05 | NA |
| chr19 | 33293053 | 33293513 | 461 | 2 | 9.81E-06 | 3.41E-06 | 0.000385 | 5.55E-06 | -0.0023908 | NA |
| chr19 | 55362967 | 55363483 | 517 | 6 | 8.12E-07 | 8.84E-06 | 0.002104 | 5.55E-06 | -0.00617622 | FAM71E2 |
| chr11 | 128709582 | 128710151 | 570 | 2 | 3.03E-06 | 7.16E-06 | 4.78E-05 | 5.55E-06 | 0.000131921 | FLI1 |
| chr17 | 81445111 | 81445557 | 447 | 2 | 1.33E-07 | 7.79E-05 | 7.07E-06 | 5.56E-06 | -0.00560013 | BAHCC1 |
| chr1 | 29283553 | 29283913 | 361 | 3 | 2.09E-06 | 0.0001097 | 0.000256 | 5.56E-06 | -0.01028902 | PTPRU |
| chr1 | 156081650 | 156082405 | 756 | 7 | 6.03E-07 | 0.0008661 | 1.61E-05 | 5.56E-06 | 0.000724386 | MEX3A |
| chr1 | 95889073 | 95889987 | 915 | 4 | 1.12E-05 | 1.57E-06 | 0.003433 | 5.57E-06 | -0.01089883 | NA |
| chr4 | 7766313 | 7766933 | 621 | 5 | 3.39E-05 | 1.83E-06 | 0.003667 | 5.57E-06 | -0.00569642 | AFAP1-AS1, AFAP1 |
| chr11 | 44726224 | 44727058 | 835 | 4 | 1.99E-06 | 3.90E-06 | 0.000513 | 5.57E-06 | 0.003314749 | TSPAN18, TSPAN18-AS1 |
| chr9 | 5085741 | 5086105 | 365 | 3 | 9.71E-06 | 2.94E-06 | 0.000493 | 5.57E-06 | -0.00400116 | JAK2, PDSS1P1 |
| chr19 | 51647294 | 51647668 | 375 | 5 | 3.97E-06 | 4.11E-06 | 0.000307 | 5.57E-06 | 0.002309085 | NA |
| chr5 | 56233581 | 56234427 | 847 | 5 | 2.68E-07 | 5.39E-05 | 0.000245 | 5.58E-06 | -0.00560771 | NA |
| chr16 | 24853663 | 24854042 | 380 | 2 | 1.06E-05 | 3.74E-06 | 0.000236 | 5.58E-06 | -0.0254513 | SLC5A11 |
| chr20 | 16573127 | 16573886 | 760 | 11 | 5.06E-06 | 2.11E-05 | 0.003008 | 5.58E-06 | -0.00331591 | KIF16B |
| chr2 | 63883435 | 63883573 | 139 | 2 | 4.07E-05 | 5.44E-06 | 7.78E-05 | 5.58E-06 | 0.004785253 | UGP2 |
| chr3 | 195817684 | 195818585 | 902 | 5 | 3.68E-05 | 0.0008142 | 0.000208 | 5.58E-06 | -0.0065652 | NA |
| chr3 | 125279193 | 125279493 | 301 | 2 | 2.05E-06 | 6.86E-05 | 7.53E-06 | 5.58E-06 | -0.01503405 | ZNF148 |
| chr6 | 138044545 | 138044781 | 237 | 2 | 3.26E-06 | 4.73E-05 | 9.07E-06 | 5.58E-06 | -0.0059893 | NA |
| chr14 | 55339782 | 55340615 | 834 | 4 | 5.81E-07 | 0.0001232 | 6.73E-06 | 5.58E-06 | -0.00433826 | FBXO34 |
| chr22 | 25488187 | 25488570 | 384 | 2 | 6.96E-06 | 5.39E-06 | 7.92E-05 | 5.58E-06 | 0.012124715 | CRYBB2P1 |
| chr2 | 74454428 | 74455073 | 646 | 13 | 1.84E-07 | 0.0003883 | 0.002684 | 5.59E-06 | -0.00364667 | NA |
| chr14 | 29354499 | 29354781 | 283 | 3 | 1.39E-05 | 2.87E-06 | 0.000943 | 5.59E-06 | -0.00286935 | AL135878.1, AL133166.1 |
| chr9 | 33418344 | 33418881 | 538 | 3 | 3.80E-06 | 6.47E-06 | 0.000255 | 5.59E-06 | -0.00354291 | NA |
| chr2 | 206201986 | 206202615 | 630 | 2 | 3.92E-06 | 6.72E-06 | 5.36E-05 | 5.60E-06 | -0.00057114 | GPR1 |
| chr11 | 46741018 | 46741342 | 325 | 2 | 4.78E-06 | 1.02E-05 | 3.05E-05 | 5.60E-06 | -0.00288615 | NA |
| chr6 | 165710100 | 165710174 | 75 | 3 | 3.77E-07 | 0.0006631 | 2.74E-06 | 5.60E-06 | -0.00556561 | PDE10A |
| chr15 | 57607677 | 57608301 | 625 | 2 | 7.85E-06 | 4.91E-06 | 9.72E-05 | 5.60E-06 | -0.00168917 | MYZAP, GCOM1 |

| | | | | | | | | | | |
|---|---|---|---|---|---|---|---|---|---|---|
| chr19 | 12940612 | 12940830 | 219 | 4 | 1.71E-06 | 9.75E-06 | 0.000398 | 5.61E-06 | -0.00873219 | CALR |
| chr11 | 93939331 | 93939846 | 516 | 2 | 2.32E-07 | 0.0013198 | 3.69E-06 | 5.61E-06 | 0.00414526 | NA |
| chr11 | 94647365 | 94647432 | 68 | 2 | 6.90E-06 | 2.34E-05 | 1.43E-05 | 5.61E-06 | 0.013645396 | LINC02700, AP000943.2 |
| chr5 | 39215088 | 39215405 | 318 | 2 | 1.15E-06 | 0.0003391 | 4.63E-06 | 5.62E-06 | 0.011681403 | FYB1 |
| chr12 | 80068549 | 80069367 | 819 | 4 | 6.20E-06 | 1.75E-05 | 0.000321 | 5.63E-06 | -0.01020547 | RNU7-106P |
| chr3 | 150761973 | 150762493 | 521 | 5 | 3.52E-07 | 3.76E-05 | 0.000116 | 5.64E-06 | 0.001781382 | SIAH2-AS1, SIAH2 |
| chr19 | 17724010 | 17724014 | 5 | 2 | 3.09E-06 | 5.87E-05 | 8.19E-06 | 5.64E-06 | -0.00476036 | MAP1S |
| chr7 | 3175654 | 3175910 | 257 | 3 | 3.55E-06 | 1.64E-05 | 0.000117 | 5.64E-06 | -0.0086229 | NA |
| chr6 | 26087379 | 26088218 | 840 | 7 | 1.68E-07 | 0.0004603 | 4.89E-05 | 5.64E-06 | 0.002046405 | HFE |
| chr6 | 121435414 | 121435805 | 392 | 3 | 6.00E-07 | 0.0012957 | 1.38E-05 | 5.64E-06 | -0.00535952 | GJA1 |
| chr1 | 54356430 | 54357249 | 820 | 4 | 1.62E-06 | 2.68E-05 | 0.000112 | 5.65E-06 | 0.007563246 | SSBP3 |
| chr12 | 80706983 | 80707910 | 928 | 5 | 3.84E-06 | 2.18E-06 | 0.001953 | 5.65E-06 | -0.00756143 | MYF6 |
| chr13 | 114067732 | 114068417 | 686 | 3 | 5.23E-06 | 1.00E-05 | 0.000112 | 5.65E-06 | -0.00450169 | RASA3 |
| chr3 | 93959718 | 93959864 | 147 | 2 | 1.94E-06 | 0.0036029 | 3.42E-06 | 5.65E-06 | -0.01768467 | PROS1 |
| chr7 | 16526476 | 16527259 | 784 | 10 | 1.40E-06 | 0.000472 | 0.000779 | 5.65E-06 | 0.000450775 | LRRC72, SOSTDC1 |
| chr15 | 83715968 | 83716795 | 828 | 3 | 5.32E-05 | 1.33E-05 | 0.000364 | 5.65E-06 | -0.00144467 | ADAMTSL3 |
| chr13 | 27040092 | 27040238 | 147 | 2 | 2.48E-06 | 0.0002539 | 4.98E-06 | 5.66E-06 | -0.00890093 | NA |
| chr19 | 48982332 | 48982953 | 622 | 3 | 1.11E-05 | 2.14E-06 | 0.002073 | 5.66E-06 | -0.00173155 | GYS1 |
| chr6 | 62775300 | 62775871 | 572 | 2 | 4.14E-06 | 6.95E-06 | 5.20E-05 | 5.66E-06 | -0.00581471 | NA |
| chr19 | 5621961 | 5622859 | 899 | 7 | 6.71E-06 | 2.17E-05 | 0.001096 | 5.66E-06 | -0.00030308 | SAFB2 |
| chr11 | 57791505 | 57791784 | 280 | 4 | 3.06E-06 | 0.0001014 | 9.98E-06 | 5.66E-06 | 0.001649676 | AP001931.2, AP001931.1, CTNND1 |
| chr22 | 24582154 | 24582368 | 215 | 3 | 5.11E-06 | 2.69E-06 | 0.000714 | 5.66E-06 | -0.00392486 | AP000356.5 |
| chr4 | 37434636 | 37435044 | 409 | 2 | 9.12E-07 | 6.25E-06 | 6.22E-05 | 5.67E-06 | -0.00675519 | NWD2 |
| chr1 | 2304744 | 2305464 | 721 | 4 | 2.67E-07 | 0.0010065 | 9.69E-05 | 5.67E-06 | 0.003178445 | SKI |
| chr1 | 162023574 | 162024573 | 1000 | 15 | 1.06E-06 | 0.0006123 | 0.000176 | 5.67E-06 | 0.001794131 | OLFML2B |
| chr11 | 63969156 | 63969216 | 61 | 2 | 4.07E-06 | 0.0001004 | 6.55E-06 | 5.67E-06 | -0.00642601 | NA |
| chr6 | 26532686 | 26533521 | 836 | 6 | 2.24E-06 | 4.10E-05 | 2.11E-05 | 5.67E-06 | 0.002222692 | NA |
| chr22 | 43879584 | 43880133 | 550 | 3 | 1.47E-05 | 2.63E-06 | 0.001373 | 5.68E-06 | -0.01558194 | PNPLA5 |
| chr16 | 88473453 | 88473988 | 536 | 5 | 2.21E-06 | 3.24E-05 | 1.92E-05 | 5.68E-06 | -0.00431254 | ZFPM1 |
| chr6 | 36871548 | 36871798 | 251 | 4 | 1.29E-05 | 9.30E-06 | 0.000432 | 5.68E-06 | -0.00033554 | PPIL1 |
| chr7 | 72446556 | 72447426 | 871 | 6 | 2.43E-05 | 9.66E-05 | 0.000139 | 5.69E-06 | 0.006283785 | CALN1 |
| chr13 | 113823285 | 113823680 | 396 | 4 | 2.87E-06 | 6.11E-06 | 0.001575 | 5.69E-06 | 0.000586821 | GAS6-AS1, GAS6 |
| chr9 | 36855134 | 36855652 | 519 | 4 | 6.12E-06 | 6.07E-06 | 0.000636 | 5.70E-06 | -0.01136796 | PAX5 |
| chr16 | 87839783 | 87840786 | 1004 | 5 | 5.04E-08 | 0.0028526 | 2.01E-06 | 5.70E-06 | -0.00298871 | SLC7A5 |
| chr17 | 57744405 | 57745284 | 880 | 5 | 1.98E-06 | 8.48E-05 | 3.68E-05 | 5.70E-06 | -0.01094766 | CCDC182 |
| chr1 | 31270336 | 31270621 | 286 | 3 | 1.85E-05 | 3.44E-06 | 0.000838 | 5.70E-06 | 0.003471012 | SNRNP40 |
| chr3 | 46880996 | 46881943 | 948 | 4 | 3.26E-07 | 0.0009689 | 4.84E-05 | 5.70E-06 | -0.0073459 | PTH1R, MYL3 |
| chr15 | 99478135 | 99478374 | 240 | 2 | 4.22E-06 | 3.40E-06 | 0.000486 | 5.71E-06 | 0.004009426 | NA |
| chr5 | 6404115 | 6404312 | 198 | 2 | 2.67E-06 | 4.05E-05 | 1.02E-05 | 5.71E-06 | -0.01057471 | AC093307.1 |
| chr22 | 29048986 | 29049915 | 930 | 5 | 2.27E-05 | 0.0056175 | 9.12E-05 | 5.71E-06 | -0.00224388 | ZNRF3 |
| chr2 | 69829015 | 69830347 | 1333 | 11 | 6.04E-05 | 0.0003321 | 0.000194 | 5.71E-06 | 0.001145384 | GMCL1 |
| chr11 | 62611591 | 62612641 | 1051 | 7 | 2.50E-07 | 0.0005277 | 0.000191 | 5.71E-06 | -0.00056559 | ROM1, EML3 |
| chr1 | 1625069 | 1625514 | 446 | 4 | 2.33E-06 | 3.27E-05 | 0.000461 | 5.71E-06 | -0.00111983 | MIB2 |
| chr22 | 28916853 | 28917304 | 452 | 6 | 9.82E-07 | 2.89E-05 | 1.85E-05 | 5.71E-06 | -0.00098403 | ZNRF3 |
| chr1 | 197904300 | 197904947 | 648 | 3 | 1.05E-05 | 5.64E-06 | 0.000138 | 5.72E-06 | -0.01947041 | C1orf53 |
| chr12 | 94459810 | 94460693 | 884 | 14 | 3.09E-07 | 0.0052529 | 4.95E-05 | 5.72E-06 | -0.00024031 | CEP83-DT, CEP83 |
| chr4 | 2276534 | 2277083 | 550 | 5 | 2.94E-06 | 4.05E-06 | 0.000433 | 5.72E-06 | 0.00048979 | ZFYVE28 |
| chr17 | 79851590 | 79851999 | 410 | 3 | 1.44E-06 | 1.53E-05 | 0.000279 | 5.72E-06 | -0.00344778 | NA |
| chr4 | 42656935 | 42657758 | 824 | 10 | 1.04E-06 | 0.0011628 | 0.000134 | 5.73E-06 | -0.00153654 | AC096734.2, ATP8A1 |
| chr18 | 75213416 | 75213724 | 309 | 4 | 7.99E-06 | 9.37E-06 | 9.82E-05 | 5.73E-06 | -0.00727581 | TSHZ1 |
| chr19 | 57240223 | 57241387 | 1165 | 14 | 2.01E-05 | 5.27E-05 | 0.005113 | 5.73E-06 | 0.000209572 | ZNF805 |
| chr8 | 144469457 | 144469632 | 176 | 3 | 6.70E-06 | 4.04E-06 | 0.000795 | 5.73E-06 | -0.00224202 | KIFC2 |
| chr17 | 50199681 | 50200725 | 1045 | 5 | 1.26E-07 | 4.74E-05 | 3.19E-05 | 5.73E-06 | -0.00378099 | AC015909.1, COL1A1 |
| chr10 | 100286072 | 100287011 | 940 | 23 | 2.62E-07 | 0.0025479 | 8.77E-05 | 5.73E-06 | 0.001446475 | BLOC1S2 |
| chr16 | 48235194 | 48235908 | 715 | 6 | 7.01E-06 | 4.08E-06 | 0.001967 | 5.74E-06 | -0.00227659 | ABCC11 |
| chr10 | 133087695 | 133088471 | 777 | 10 | 1.01E-05 | 5.11E-06 | 0.004551 | 5.74E-06 | 0.002373332 | ADGRA1, ADGRA1-AS1 |
| chr18 | 9771523 | 9772092 | 570 | 3 | 1.20E-05 | 9.67E-06 | 0.000198 | 5.74E-06 | -0.00544065 | RAB31 |
| chr1 | 207866068 | 207866908 | 841 | 3 | 2.16E-05 | 5.88E-06 | 0.000602 | 5.74E-06 | 0.001253264 | MIR29B2CHG |
| chr10 | 118259068 | 118259470 | 403 | 2 | 2.47E-06 | 1.97E-05 | 1.69E-05 | 5.75E-06 | 0.016644713 | AL354863.1 |
| chr10 | 27503679 | 27504512 | 834 | 10 | 8.25E-06 | 6.62E-05 | 0.003493 | 5.75E-06 | 0.001843052 | RAB18 |
| chr11 | 134330250 | 134331568 | 1319 | 5 | 1.43E-09 | 0.0018449 | 6.77E-06 | 5.75E-06 | -0.00995616 | NA |
| chr12 | 121831044 | 121831769 | 726 | 5 | 2.59E-06 | 0.0005818 | 0.000315 | 5.75E-06 | -0.0034836 | SETD1B |
| chr1 | 116277819 | 116278766 | 948 | 4 | 2.45E-06 | 1.11E-05 | 7.45E-05 | 5.76E-06 | -0.00052484 | U3 |
| chr16 | 2087579 | 2088614 | 1036 | 9 | 1.04E-06 | 0.0001116 | 0.00012 | 5.76E-06 | 8.29E-05 | TSC2 |
| chr7 | 102390623 | 102390807 | 185 | 2 | 1.18E-05 | 4.64E-06 | 0.000123 | 5.76E-06 | -0.01148713 | PRKRIP1 |
| chr5 | 179698824 | 179699857 | 1034 | 9 | 9.80E-06 | 0.0007263 | 0.006847 | 5.76E-06 | 0.001542936 | CANX |
| chr12 | 121625100 | 121626062 | 963 | 5 | 1.86E-07 | 3.51E-06 | 0.000729 | 5.76E-06 | -0.00223212 | NA |
| chr16 | 15726049 | 15727046 | 998 | 6 | 2.50E-07 | 0.000168 | 7.69E-06 | 5.76E-06 | -0.00214766 | NDE1, AF001548.2, MYH11 |
| chr2 | 79874189 | 79874275 | 87 | 2 | 1.47E-05 | 3.55E-06 | 0.000387 | 5.76E-06 | -0.00571824 | CTNNA2 |
| chr15 | 57540823 | 57541417 | 595 | 3 | 1.13E-05 | 5.04E-05 | 0.000307 | 5.76E-06 | -0.00244323 | CGNL1 |
| chr15 | 89356498 | 89357224 | 727 | 3 | 6.33E-06 | 4.31E-06 | 0.000215 | 5.77E-06 | 0.00316987 | NA |
| chr4 | 173185274 | 173185780 | 507 | 2 | 4.24E-07 | 0.0001997 | 5.38E-06 | 5.77E-06 | 0.006785001 | GALNT7 |
| chr9 | 135693884 | 135694465 | 582 | 2 | 3.50E-06 | 7.77E-06 | 4.57E-05 | 5.77E-06 | 0.002900253 | SOHLH1 |
| chr14 | 77477335 | 77477592 | 258 | 2 | 5.79E-06 | 7.83E-06 | 4.52E-05 | 5.77E-06 | -0.00612485 | ISM2 |
| chr6 | 168574471 | 168574928 | 458 | 2 | 4.34E-07 | 0.0002251 | 5.23E-06 | 5.77E-06 | -0.00650978 | SMOC2 |
| chr13 | 105410789 | 105410823 | 35 | 3 | 1.62E-05 | 3.92E-06 | 0.000252 | 5.78E-06 | -0.02115457 | AL138954.1 |
| chr10 | 67956898 | 67957306 | 409 | 2 | 9.03E-06 | 3.60E-06 | 0.000364 | 5.78E-06 | -0.01962634 | HERC4 |
| chr11 | 1252431 | 1252910 | 480 | 5 | 3.19E-06 | 4.05E-05 | 0.00022 | 5.79E-06 | -0.00308697 | MUC5B |
| chr5 | 180873777 | 180874165 | 389 | 2 | 8.64E-06 | 4.77E-06 | 0.000115 | 5.79E-06 | 0.0174828 | NA |
| chr16 | 537568 | 538482 | 915 | 5 | 9.18E-07 | 0.0001633 | 5.66E-05 | 5.79E-06 | 0.000629608 | CAPN15 |
| chr12 | 68584455 | 68585003 | 549 | 2 | 1.78E-06 | 1.50E-05 | 2.16E-05 | 5.79E-06 | 0.001497222 | NA |
| chr5 | 143434671 | 143435369 | 699 | 6 | 6.60E-06 | 0.0002042 | 0.000212 | 5.79E-06 | 0.006113498 | NR3C1 |
| chr15 | 57387703 | 57388281 | 579 | 2 | 1.62E-05 | 3.80E-06 | 0.000269 | 5.79E-06 | -0.01449855 | CGNL1 |
| chr17 | 44911416 | 44911779 | 364 | 5 | 1.18E-05 | 4.45E-06 | 0.0007 | 5.79E-06 | 0.001166406 | GFAP |
| chr5 | 120780662 | 120780776 | 115 | 2 | 6.05E-06 | 6.38E-06 | 6.32E-05 | 5.80E-06 | -0.02524694 | NA |
| chr7 | 44679024 | 44679329 | 306 | 3 | 1.31E-06 | 0.0002563 | 4.17E-06 | 5.80E-06 | 0.004120085 | OGDH |
| chr3 | 112000444 | 112000635 | 192 | 3 | 5.35E-06 | 2.49E-06 | 0.001199 | 5.80E-06 | -0.00276953 | TAGLN3 |
| chr1 | 100132603 | 100133613 | 1011 | 12 | 6.58E-06 | 1.30E-05 | 0.000495 | 5.80E-06 | 0.001353077 | TRMT13, SASS6 |
| chr12 | 130916386 | 130917334 | 949 | 8 | 3.28E-06 | 1.19E-05 | 0.00016 | 5.81E-06 | 0.008904848 | NA |
| chr20 | 2470562 | 2471486 | 925 | 10 | 9.23E-07 | 0.0001013 | 2.64E-05 | 5.81E-06 | -0.00165084 | SNRPB, AL049650.1 |
| chr14 | 99262602 | 99262944 | 343 | 6 | 1.41E-05 | 1.08E-05 | 0.000487 | 5.81E-06 | 0.009743549 | BCL11B |
| chr10 | 1747818 | 1747991 | 174 | 2 | 3.37E-06 | 7.72E-06 | 4.69E-05 | 5.82E-06 | -0.01518406 | NA |
| chr19 | 38417783 | 38418446 | 664 | 4 | 1.27E-05 | 1.97E-06 | 0.002689 | 5.82E-06 | -0.00186705 | RASGRP4 |
| chr17 | 55238217 | 55238836 | 620 | 6 | 1.53E-05 | 4.27E-06 | 0.003408 | 5.83E-06 | 0.002961296 | NA |
| chr15 | 65415130 | 65415370 | 241 | 2 | 1.15E-05 | 4.34E-06 | 0.000158 | 5.83E-06 | -0.00893747 | IGDCC4 |
| chr11 | 66431865 | 66432662 | 798 | 4 | 2.70E-05 | 6.85E-06 | 0.001119 | 5.83E-06 | -0.0049532 | NA |
| chr2 | 14274829 | 14275401 | 573 | 3 | 7.79E-06 | 4.68E-06 | 0.00017 | 5.84E-06 | -0.01419368 | LINC00276 |
| chr2 | 239578106 | 239578260 | 155 | 3 | 2.36E-06 | 1.94E-05 | 3.68E-05 | 5.84E-06 | 0.008412122 | NA |
| chr14 | 105466145 | 105466946 | 802 | 5 | 2.65E-06 | 4.57E-06 | 0.001201 | 5.84E-06 | -0.00012299 | MTA1 |
| chr4 | 158787366 | 158788033 | 668 | 2 | 1.47E-05 | 3.58E-06 | 0.000398 | 5.84E-06 | 0.006419039 | FNIP2 |
| chr4 | 9898894 | 9898901 | 8 | 2 | 1.34E-05 | 4.92E-06 | 0.00011 | 5.84E-06 | 0.031030696 | SLC2A9 |
| chr6 | 125790976 | 125791765 | 790 | 5 | 5.00E-06 | 5.00E-06 | 0.001551 | 5.85E-06 | 0.000483716 | NCOA7 |
| chr19 | 30003382 | 30004170 | 789 | 6 | 2.55E-06 | 0.0002361 | 3.64E-05 | 5.85E-06 | -0.00525709 | URI1 |
| chr5 | 77847434 | 77848192 | 759 | 2 | 6.29E-06 | 5.63E-06 | 8.19E-05 | 5.85E-06 | 0.023634161 | TBCA |
| chr17 | 31487364 | 31488018 | 655 | 3 | 2.67E-06 | 1.31E-05 | 0.000302 | 5.85E-06 | 0.000911923 | RAB11FIP4 |
| chr13 | 51974777 | 51975451 | 675 | 6 | 1.52E-05 | 4.68E-05 | 0.001756 | 5.85E-06 | -0.00376121 | ATP7B |
| chr6 | 31441431 | 31441798 | 368 | 7 | 1.23E-05 | 8.81E-06 | 0.000568 | 5.85E-06 | -0.01083619 | AL645933.3, LINC01149 |
| chr1 | 181207509 | 181208319 | 811 | 3 | 1.50E-05 | 2.22E-06 | 0.002103 | 5.86E-06 | -0.00899141 | NA |
| chr5 | 93603 | 93613 | 11 | 2 | 7.85E-06 | 5.60E-06 | 8.30E-05 | 5.86E-06 | 0.001063139 | PLEKHG4B |
| chr17 | 50202896 | 50203071 | 176 | 4 | 4.06E-06 | 2.19E-05 | 0.00025 | 5.86E-06 | -0.0056728 | AC015909.1 |
| chr17 | 3885242 | 3885912 | 671 | 3 | 1.62E-06 | 4.75E-05 | 1.32E-05 | 5.87E-06 | -0.00575072 | CAMKK1 |
| chr18 | 13269698 | 13269812 | 115 | 2 | 1.28E-05 | 3.57E-06 | 0.000434 | 5.87E-06 | 0.0176029 | LDLRAD4 |
| chr14 | 92088465 | 92088720 | 256 | 2 | 3.39E-06 | 4.26E-05 | 1.03E-05 | 5.87E-06 | -0.00153018 | ATXN3 |
| chr1 | 204996872 | 204997384 | 513 | 7 | 1.48E-05 | 4.90E-06 | 0.002396 | 5.88E-06 | -0.00102717 | NFASC |
| chr1 | 192009452 | 192010379 | 928 | 5 | 4.68E-05 | 1.33E-05 | 0.000456 | 5.88E-06 | -0.01006267 | LINC02770 |
| chr17 | 38718917 | 38719715 | 799 | 3 | 3.06E-07 | 5.39E-06 | 0.000133 | 5.88E-06 | 0.006935572 | MLLT6, MIR4726, AC006449.3 |
| chr19 | 55947560 | 55948188 | 629 | 6 | 7.70E-06 | 1.18E-05 | 0.000425 | 5.88E-06 | -0.0020582 | NLRP8 |
| chr6 | 29612563 | 29613451 | 889 | 10 | 1.71E-05 | 0.0006963 | 0.002469 | 5.88E-06 | 0.000502506 | GABBR1 |
| chr3 | 183447248 | 183448016 | 769 | 8 | 7.59E-06 | 6.19E-06 | 0.002397 | 5.88E-06 | 0.001039316 | LINC00888 |
| chr12 | 58007911 | 58007970 | 60 | 2 | 1.47E-05 | 3.83E-06 | 0.000282 | 5.88E-06 | -0.00481843 | NA |
| chr16 | 71230379 | 71231117 | 739 | 12 | 9.59E-06 | 6.13E-06 | 0.001411 | 5.88E-06 | 0.00185399 | HYDIN |
| chr15 | 27545574 | 27545710 | 137 | 2 | 2.79E-06 | 0.0001866 | 5.61E-06 | 5.89E-06 | -0.01351961 | NA |
| chr6 | 83536018 | 83536449 | 432 | 3 | 8.60E-06 | 3.72E-06 | 0.000305 | 5.89E-06 | 0.006253571 | NA |
| chr2 | 66435366 | 66436002 | 637 | 4 | 5.37E-06 | 2.24E-06 | 0.001978 | 5.89E-06 | 0.004532841 | MEIS1 |
| chr17 | 847001 | 847608 | 608 | 6 | 1.27E-05 | 2.21E-05 | 0.00301 | 5.89E-06 | 0.01047225 | NXN |
| chr10 | 129840862 | 129841362 | 501 | 3 | 4.26E-07 | 1.09E-05 | 5.74E-05 | 5.90E-06 | -0.00744941 | EBF3 |
| chr1 | 167219348 | 167220386 | 1039 | 4 | 6.48E-07 | 2.84E-06 | 0.001106 | 5.90E-06 | -0.00038608 | AL451050.2 |
| chr11 | 61007480 | 61008359 | 880 | 5 | 3.13E-06 | 2.34E-06 | 0.003154 | 5.90E-06 | -0.01708026 | CD6 |
| chr10 | 86536607 | 86537052 | 446 | 5 | 6.02E-06 | 5.14E-06 | 0.000258 | 5.90E-06 | 0.006452403 | NA |
| chr5 | 42745266 | 42745527 | 262 | 2 | 1.30E-05 | 3.91E-06 | 0.000259 | 5.90E-06 | -0.01857636 | NA |
| chr19 | 48825295 | 48825570 | 276 | 2 | 7.33E-06 | 6.68E-06 | 6.08E-05 | 5.90E-06 | -0.00932252 | HSD17B14 |
| chr4 | 99946942 | 99947702 | 761 | 5 | 1.05E-05 | 5.23E-06 | 0.000506 | 5.90E-06 | -0.00269311 | NA |
| chr11 | 122747581 | 122747712 | 132 | 2 | 4.87E-06 | 3.55E-06 | 0.000469 | 5.91E-06 | -0.00065251 | UBASH3B |
| chr7 | 156471133 | 156471467 | 335 | 4 | 1.12E-05 | 1.36E-05 | 0.0003 | 5.91E-06 | -0.00041963 | LINC01006 |
| chr3 | 47417211 | 47417333 | 123 | 2 | 3.68E-06 | 4.92E-06 | 0.000115 | 5.91E-06 | 0.000234826 | SCAP |
| chr9 | 127835331 | 127835423 | 93 | 2 | 3.45E-06 | 0.0001408 | 6.12E-06 | 5.92E-06 | 0.002292439 | ENG |
| chr2 | 55232065 | 55232725 | 661 | 13 | 1.42E-05 | 1.16E-05 | 0.000487 | 5.92E-06 | 0.000578372 | RPS27A, CLHC1 |
| chr10 | 1366099 | 1366650 | 552 | 3 | 8.85E-06 | 6.45E-06 | 0.000151 | 5.92E-06 | -0.00950386 | ADARB2 |
| chr19 | 1577596 | 1578730 | 1135 | 5 | 8.52E-06 | 2.88E-06 | 0.000671 | 5.92E-06 | 0.002550015 | MBD3, AC005943.1 |
| chr1 | 58903432 | 58904282 | 851 | 9 | 2.68E-07 | 0.0024289 | 7.14E-06 | 5.93E-06 | -0.00011191 | LINC02777 |
| chr12 | 52056429 | 52056520 | 92 | 2 | 1.35E-06 | 0.0001886 | 5.64E-06 | 5.93E-06 | 0.004849642 | NR4A1 |
| chr19 | 444385 | 444523 | 139 | 3 | 1.63E-05 | 3.69E-06 | 0.000363 | 5.93E-06 | -0.00723338 | SHC2 |
| chr12 | 113965862 | 113966891 | 1030 | 11 | 4.32E-06 | 6.77E-05 | 0.004175 | 5.93E-06 | 0.001770019 | RBM19 |
| chr11 | 68932939 | 68933552 | 614 | 5 | 2.60E-06 | 7.64E-06 | 0.000897 | 5.94E-06 | 0.001633477 | IGHMBP2 |
| chr21 | 42199200 | 42199541 | 342 | 4 | 3.00E-06 | 2.30E-06 | 0.0023 | 5.94E-06 | 0.002275713 | NA |
| chr22 | 50331109 | 50331509 | 401 | 3 | 1.63E-06 | 5.84E-05 | 9.67E-06 | 5.95E-06 | -0.00671547 | NA |
| chr4 | 147480121 | 147480751 | 631 | 6 | 1.98E-06 | 4.33E-06 | 0.001478 | 5.95E-06 | -0.00100862 | NA |
| chr8 | 53880730 | 53881623 | 894 | 9 | 4.30E-06 | 7.74E-06 | 0.002977 | 5.95E-06 | -0.00073346 | RGS20 |
| chr3 | 68004341 | 68004806 | 466 | 2 | 5.59E-07 | 5.05E-06 | 0.00011 | 5.96E-06 | 0.002714947 | TAFA1 |
| chr13 | 100580148 | 100580430 | 283 | 5 | 4.88E-07 | 0.0026845 | 2.59E-06 | 5.96E-06 | 0.002182855 | GGACT, AL356966.1 |
| chr9 | 122214587 | 122214624 | 38 | 2 | 9.23E-07 | 5.59E-06 | 8.71E-05 | 5.96E-06 | -0.01128819 | LHX6 |
| chr11 | 7963426 | 7963729 | 304 | 5 | 5.29E-06 | 7.88E-05 | 0.000273 | 5.96E-06 | -0.00499504 | NLRP10 |
| chr5 | 15614981 | 15615324 | 344 | 3 | 3.53E-06 | 1.18E-05 | 5.05E-05 | 5.97E-06 | -0.00328528 | FBXL7, CTD-2350J17.1 |
| chr17 | 2416156 | 2416612 | 457 | 5 | 7.73E-06 | 5.45E-06 | 0.002103 | 5.98E-06 | -0.00485657 | METTL16 |
| chr12 | 75699873 | 75700181 | 309 | 3 | 8.85E-06 | 5.78E-06 | 0.000178 | 5.98E-06 | -0.0041543 | AC078923.1 |
| chr21 | 36126891 | 36127361 | 471 | 2 | 1.19E-07 | 0.0032199 | 3.60E-06 | 5.98E-06 | 0.002639194 | NA |
| chr2 | 103161158 | 103161538 | 381 | 2 | 1.38E-06 | 7.71E-05 | 7.71E-06 | 5.99E-06 | 0.013162357 | AC073987.1 |
| chr2 | 114419423 | 114419598 | 176 | 3 | 1.35E-05 | 2.98E-06 | 0.000669 | 5.99E-06 | 0.002367435 | NA |

| | | | | | | | | | | |
|---|---|---|---|---|---|---|---|---|---|---|
| chr1 | 2808019 | 2808261 | 243 | 3 | 7.38E-06 | 1.19E-05 | 0.000143 | 5.99E-06 | -0.00717533 | NA |
| chr20 | 48016481 | 48016574 | 94 | 2 | 1.39E-05 | 4.49E-06 | 0.000157 | 5.99E-06 | -0.006577 | AL139351.2 |
| chr10 | 68526896 | 68527740 | 845 | 15 | 1.44E-05 | 2.34E-05 | 0.000306 | 5.99E-06 | -0.00027073 | SLC25A16 |
| chr16 | 87581911 | 87582642 | 732 | 4 | 4.22E-06 | 1.00E-05 | 0.00022 | 6.00E-06 | -0.01030752 | NA |
| chr22 | 19805959 | 19806633 | 675 | 3 | 4.06E-06 | 4.51E-05 | 4.05E-05 | 6.00E-06 | -0.00576454 | GNB1L |
| chr19 | 55157244 | 55158920 | 1677 | 6 | 1.12E-05 | 2.14E-05 | 0.000571 | 6.00E-06 | -0.01531375 | TNNI3, AC010327.2, DNAAF3 |
| chr6 | 28881123 | 28881443 | 321 | 3 | 1.98E-05 | 2.90E-06 | 0.000883 | 6.00E-06 | 0.01456538 | NA |
| chr10 | 3924390 | 3924435 | 46 | 2 | 6.01E-06 | 5.43E-05 | 9.20E-06 | 6.00E-06 | -0.00453249 | LINC02660 |
| chr9 | 27715150 | 27715218 | 69 | 2 | 1.47E-05 | 4.13E-06 | 0.000221 | 6.01E-06 | -0.01107802 | NA |
| chr11 | 2273860 | 2274499 | 640 | 4 | 5.82E-06 | 3.63E-05 | 0.000368 | 6.01E-06 | 0.000477444 | NA |
| chr13 | 27113576 | 27113661 | 86 | 2 | 1.57E-05 | 3.59E-06 | 0.000496 | 6.02E-06 | 0.00841753 | USP12 |
| chr14 | 51508130 | 51508523 | 394 | 2 | 1.70E-05 | 3.62E-06 | 0.000469 | 6.02E-06 | 0.055233668 | FRMD6, FRMD6-AS2 |
| chr4 | 2939269 | 2940033 | 765 | 4 | 2.39E-06 | 5.27E-05 | 0.0003 | 6.02E-06 | -0.00163 | NOP14-AS1, NOP14 |
| chr18 | 79448807 | 79449372 | 566 | 4 | 3.11E-06 | 1.93E-05 | 0.000111 | 6.02E-06 | 0.000575415 | NFATC1 |
| chr12 | 102062117 | 102062709 | 593 | 11 | 4.78E-06 | 5.02E-05 | 0.00148 | 6.03E-06 | 0.004886942 | WASHC3 |
| chr18 | 31497164 | 31497900 | 737 | 5 | 1.79E-05 | 4.32E-06 | 0.000725 | 6.03E-06 | 0.002406705 | AC021549.1 |
| chr2 | 37573407 | 37574423 | 1017 | 3 | 5.10E-07 | 3.62E-05 | 2.46E-05 | 6.03E-06 | 0.001200366 | AC006369.1 |
| chr6 | 169232256 | 169233229 | 974 | 5 | 1.70E-05 | 0.0111411 | 0.000115 | 6.03E-06 | -0.00057079 | BX322234.1, THBS2 |
| chr7 | 158160809 | 158161335 | 527 | 3 | 3.47E-07 | 0.0001107 | 6.46E-06 | 6.03E-06 | -0.01045006 | PTPRN2 |
| chr16 | 57025650 | 57025837 | 188 | 3 | 3.33E-06 | 3.09E-06 | 0.001221 | 6.03E-06 | 0.00975745 | NLRC5 |
| chr2 | 171160906 | 171161426 | 521 | 7 | 3.67E-06 | 0.0003038 | 0.000305 | 6.04E-06 | 0.001280444 | TLK1 |
| chr2 | 58462005 | 58462383 | 379 | 3 | 7.12E-06 | 2.42E-05 | 0.000112 | 6.05E-06 | -0.00349186 | LINC01122, AC007238.1 |
| chr3 | 51670335 | 51671134 | 800 | 8 | 4.37E-06 | 4.64E-05 | 0.000701 | 6.05E-06 | -0.00293817 | TEX264, AC099050.1 |
| chr3 | 129897856 | 129898293 | 438 | 2 | 5.67E-07 | 0.0003954 | 4.83E-06 | 6.05E-06 | 0.003525582 | TMCC1-AS1 |
| chr12 | 117360822 | 117361642 | 821 | 6 | 3.06E-06 | 1.32E-05 | 0.006678 | 6.05E-06 | 0.000405246 | NOS1 |
| chr10 | 132975538 | 132976310 | 773 | 4 | 1.74E-05 | 2.56E-06 | 0.000999 | 6.05E-06 | 0.001295034 | LINC01168 |
| chr6 | 142146920 | 142147519 | 600 | 12 | 2.65E-06 | 0.0004706 | 0.001009 | 6.06E-06 | 0.001068784 | VTA1 |
| chr11 | 126940199 | 126940837 | 639 | 3 | 2.65E-06 | 1.26E-05 | 0.000457 | 6.06E-06 | -0.01111257 | KIRREL3-AS2, KIRREL3, AP002833.1 |
| chr21 | 41245232 | 41245917 | 686 | 3 | 9.51E-08 | 0.0043414 | 5.39E-06 | 6.07E-06 | -0.00131715 | BACE2 |
| chr20 | 35431901 | 35432251 | 351 | 3 | 1.22E-06 | 0.0002757 | 1.75E-05 | 6.07E-06 | -0.00500337 | NA |
| chr3 | 100561492 | 100562358 | 867 | 3 | 2.99E-05 | 0.0001739 | 6.66E-05 | 6.07E-06 | -0.00716424 | TMEM45A |
| chr1 | 178512672 | 178513193 | 522 | 9 | 3.33E-06 | 9.39E-05 | 0.00024 | 6.07E-06 | -0.00132529 | TEX35, AL513013.1 |
| chr1 | 35443365 | 35444234 | 870 | 4 | 1.21E-07 | 0.0017667 | 3.47E-06 | 6.07E-06 | -0.00438909 | KIAA0319L |
| chr14 | 104954761 | 104955507 | 747 | 3 | 3.85E-06 | 3.78E-06 | 0.000326 | 6.08E-06 | -0.00667138 | AHNAK2 |
| chr17 | 41028626 | 41028635 | 10 | 2 | 5.98E-06 | 4.79E-06 | 0.000136 | 6.08E-06 | 4.61E-05 | NA |
| chr12 | 46536731 | 46537356 | 626 | 3 | 7.73E-07 | 0.0156591 | 5.12E-06 | 6.08E-06 | -0.00093247 | AC008014.1 |
| chr11 | 116732228 | 116732382 | 155 | 2 | 3.95E-06 | 1.44E-05 | 2.43E-05 | 6.08E-06 | -0.01395094 | NA |
| chr13 | 40573804 | 40573858 | 55 | 2 | 1.54E-05 | 3.65E-06 | 0.000477 | 6.08E-06 | -0.00686481 | AL133318.1, FOXO1 |
| chr1 | 30037655 | 30037884 | 230 | 2 | 3.26E-06 | 7.56E-05 | 7.91E-06 | 6.08E-06 | -0.00337034 | NA |
| chr13 | 28657942 | 28658662 | 721 | 5 | 3.40E-06 | 2.26E-06 | 0.001764 | 6.08E-06 | 0.004410096 | NA |
| chr12 | 26156187 | 26156256 | 70 | 2 | 5.96E-06 | 5.43E-05 | 9.34E-06 | 6.08E-06 | 0.00734231 | SSPN, AC022509.5 |
| chr9 | 18437919 | 18438151 | 233 | 2 | 3.33E-07 | 5.22E-06 | 0.000107 | 6.08E-06 | -0.00012422 | ADAMTSL1 |
| chr2 | 217249708 | 217250283 | 576 | 3 | 4.82E-06 | 5.58E-06 | 0.000406 | 6.08E-06 | -0.00464347 | NA |
| chr17 | 32384726 | 32385370 | 645 | 3 | 3.26E-05 | 2.26E-06 | 0.002152 | 6.08E-06 | -0.00058813 | NA |
| chr5 | 74866777 | 74867898 | 1122 | 10 | 8.43E-06 | 8.03E-06 | 0.005608 | 6.08E-06 | -0.00322729 | AC010501.2, FAM169A |
| chr13 | 28015231 | 28016104 | 874 | 4 | 1.75E-06 | 0.0001309 | 3.19E-05 | 6.09E-06 | 0.000703697 | FLT3 |
| chr4 | 146578880 | 146579138 | 259 | 2 | 7.19E-06 | 5.71E-06 | 8.80E-05 | 6.09E-06 | 0.010720801 | NA |
| chr22 | 49046154 | 49046603 | 450 | 3 | 1.98E-05 | 2.34E-06 | 0.001836 | 6.09E-06 | -0.0093594 | NA |
| chr19 | 15640064 | 15640802 | 739 | 5 | 6.12E-06 | 7.04E-06 | 0.000163 | 6.09E-06 | -0.00478061 | NA |
| chr11 | 111564796 | 111565307 | 512 | 2 | 1.04E-05 | 4.57E-06 | 0.000159 | 6.09E-06 | -0.00082262 | NA |
| chr4 | 168840624 | 168841586 | 963 | 3 | 1.84E-05 | 2.23E-06 | 0.002395 | 6.09E-06 | 0.017847053 | PALLD |
| chr5 | 84384813 | 84385745 | 933 | 8 | 9.05E-06 | 0.0001004 | 0.000621 | 6.09E-06 | -0.00300095 | EDIL3-DT, EDIL3 |
| chr15 | 70799877 | 70800464 | 588 | 3 | 9.19E-06 | 2.54E-05 | 4.50E-05 | 6.10E-06 | 0.004685306 | RPL29P30 |
| chr3 | 99999038 | 99999341 | 304 | 3 | 1.26E-05 | 4.82E-06 | 0.000184 | 6.10E-06 | -0.00116869 | CMSS1, FILIP1L |
| chr7 | 99143734 | 99144575 | 842 | 12 | 5.01E-06 | 0.0001941 | 0.001656 | 6.10E-06 | 0.004057003 | SMURF1 |
| chr15 | 79064695 | 79064969 | 275 | 5 | 1.79E-05 | 2.56E-06 | 0.004524 | 6.10E-06 | 0.000862611 | RASGRF1 |
| chr4 | 139084493 | 139085235 | 743 | 6 | 4.48E-06 | 7.69E-05 | 0.000644 | 6.11E-06 | -0.00394595 | ELF2 |
| chr2 | 113990177 | 113990249 | 73 | 2 | 3.91E-06 | 3.76E-06 | 0.000403 | 6.12E-06 | 0.007907331 | LINC01191 |
| chr5 | 58454012 | 58454472 | 461 | 2 | 3.29E-07 | 1.94E-05 | 1.87E-05 | 6.12E-06 | -0.0090277 | PLK2 |
| chr2 | 189581768 | 189582066 | 299 | 3 | 1.02E-06 | 2.84E-06 | 0.001025 | 6.12E-06 | 0.010835912 | SLC40A1 |
| chr11 | 126456516 | 126456801 | 286 | 2 | 7.18E-06 | 7.62E-06 | 5.33E-05 | 6.12E-06 | -0.00685533 | KIRREL3 |
| chr1 | 44617032 | 44617606 | 575 | 7 | 1.05E-05 | 2.02E-06 | 0.00751 | 6.12E-06 | -0.0015124 | RNF220 |
| chr10 | 124059716 | 124060377 | 662 | 3 | 3.55E-05 | 2.52E-06 | 0.001756 | 6.12E-06 | -0.00591772 | CHST15 |
| chr7 | 19119331 | 19119946 | 616 | 4 | 2.26E-08 | 0.0001399 | 9.37E-05 | 6.13E-06 | -0.00840795 | AC003986.3 |
| chr1 | 39632923 | 39633698 | 776 | 7 | 1.09E-05 | 3.62E-06 | 0.002614 | 6.13E-06 | -0.00732369 | AL035404.2, HEYL |
| chr9 | 134652926 | 134653359 | 434 | 5 | 2.85E-06 | 7.96E-05 | 0.000563 | 6.13E-06 | -0.00566932 | COL5A1, COL5A1-AS1 |
| chr9 | 100472659 | 100473254 | 596 | 6 | 1.65E-05 | 2.68E-06 | 0.0035 | 6.13E-06 | -0.0030478 | MSANTD3-TMEFF1, TMEFF1 |
| chr5 | 69175928 | 69176024 | 97 | 2 | 5.78E-07 | 0.0001005 | 7.14E-06 | 6.14E-06 | -0.00756224 | CCNB1 |
| chr11 | 65430062 | 65430666 | 605 | 7 | 6.49E-07 | 0.0004361 | 5.93E-05 | 6.15E-06 | 0.000422273 | NEAT1 |
| chr6 | 792305 | 792778 | 474 | 6 | 2.62E-06 | 8.52E-06 | 0.000161 | 6.15E-06 | -0.01295963 | AL357054.2 |
| chr4 | 967717 | 968336 | 620 | 3 | 7.59E-06 | 3.37E-06 | 0.001122 | 6.15E-06 | 0.032983744 | DGKQ |
| chr17 | 82012259 | 82012437 | 179 | 3 | 9.51E-06 | 8.88E-06 | 0.000163 | 6.15E-06 | 0.016280139 | ASPSCR1 |
| chr9 | 86259523 | 86260473 | 951 | 7 | 1.44E-05 | 2.42E-05 | 0.001509 | 6.15E-06 | 0.000972389 | C9orf153 |
| chr16 | 3092206 | 3093134 | 929 | 8 | 8.82E-05 | 3.15E-06 | 0.006315 | 6.15E-06 | 8.98E-05 | ZSCAN10 |
| chr7 | 135231004 | 135231645 | 642 | 4 | 6.17E-06 | 5.29E-05 | 0.000898 | 6.15E-06 | -0.00293659 | NA |
| chr21 | 38259024 | 38259908 | 885 | 4 | 8.67E-06 | 7.58E-05 | 0.000213 | 6.15E-06 | -0.0029819 | KCNJ15 |
| chr2 | 176147389 | 176147978 | 590 | 7 | 4.94E-06 | 1.08E-05 | 0.001329 | 6.15E-06 | 0.005320443 | HOXD3 |
| chr8 | 8286063 | 8286692 | 630 | 2 | 9.19E-06 | 3.82E-06 | 0.000383 | 6.16E-06 | -0.01253525 | NA |
| chr14 | 104084633 | 104085547 | 915 | 6 | 2.85E-06 | 1.72E-05 | 0.000696 | 6.16E-06 | -0.00631852 | NA |
| chr4 | 7425468 | 7425586 | 119 | 2 | 5.91E-06 | 4.39E-05 | 1.07E-05 | 6.16E-06 | 0.01002645 | SORCS2 |
| chr8 | 22232859 | 22234284 | 1426 | 7 | 6.33E-08 | 0.0015091 | 3.09E-05 | 6.17E-06 | 0.003694898 | NA |
| chr4 | 121070190 | 121071243 | 1054 | 6 | 6.88E-07 | 0.0012224 | 0.000104 | 6.17E-06 | -0.01082553 | NDNF |
| chr7 | 6374565 | 6375903 | 1339 | 5 | 7.17E-07 | 1.65E-05 | 8.06E-05 | 6.17E-06 | -0.00650412 | RAC1 |
| chr5 | 2141659 | 2141938 | 280 | 2 | 5.44E-06 | 9.43E-06 | 4.00E-05 | 6.17E-06 | -0.00836093 | NA |
| chr1 | 3389883 | 3390651 | 769 | 6 | 6.18E-06 | 1.75E-05 | 0.003198 | 6.17E-06 | -0.00244526 | PRDM16 |
| chr1 | 206932068 | 206932748 | 681 | 3 | 2.78E-06 | 2.01E-05 | 7.94E-05 | 6.17E-06 | -0.00367459 | PIGR |
| chr12 | 34303696 | 34304137 | 442 | 2 | 4.96E-06 | 4.16E-06 | 0.000246 | 6.18E-06 | -0.01189103 | NA |
| chr6 | 42911193 | 42911671 | 479 | 3 | 2.86E-05 | 2.56E-06 | 0.001619 | 6.18E-06 | -0.01252066 | NA |
| chr4 | 153152219 | 153153003 | 785 | 12 | 8.56E-06 | 0.0011443 | 0.002265 | 6.18E-06 | -0.00117298 | TRIM2, AC013477.1 |
| chr11 | 10497807 | 10498331 | 525 | 5 | 2.38E-05 | 4.06E-06 | 0.004308 | 6.19E-06 | -0.00059556 | AMPD3 |
| chr2 | 135168439 | 135168798 | 360 | 2 | 3.14E-06 | 6.82E-06 | 6.53E-05 | 6.19E-06 | 0.010918324 | RAB3GAP1, ZRANB3 |
| chr6 | 72097024 | 72097125 | 102 | 3 | 1.32E-05 | 9.39E-06 | 0.000109 | 6.19E-06 | -0.00531745 | RIMS1 |
| chr10 | 12656882 | 12656959 | 78 | 2 | 2.58E-06 | 0.006034 | 3.59E-06 | 6.19E-06 | -0.01756768 | CAMK1D |
| chr11 | 3095635 | 3095951 | 317 | 3 | 4.76E-06 | 3.08E-05 | 4.00E-05 | 6.19E-06 | -0.00569389 | OSBPL5 |
| chr10 | 511169 | 511996 | 828 | 8 | 1.61E-05 | 2.11E-05 | 0.000757 | 6.19E-06 | -0.002787 | DIP2C |
| chr19 | 53511930 | 53512352 | 423 | 3 | 2.77E-06 | 1.36E-05 | 4.10E-05 | 6.19E-06 | -0.00743897 | AC011487.1, AC011487.3 |
| chr13 | 20435357 | 20435722 | 366 | 2 | 1.70E-05 | 4.29E-06 | 0.00022 | 6.19E-06 | -0.02057363 | CRYL1 |
| chr2 | 177923528 | 177924124 | 597 | 3 | 2.79E-05 | 4.01E-06 | 0.000822 | 6.20E-06 | -0.00761377 | PDE11A, PDE11A |
| chr7 | 4127779 | 4128414 | 636 | 6 | 1.04E-05 | 1.55E-05 | 0.00033 | 6.20E-06 | -0.00395027 | SDK1 |
| chr20 | 63010102 | 63010641 | 540 | 2 | 4.56E-06 | 4.38E-06 | 0.0002 | 6.20E-06 | -0.01851176 | LINC01749 |
| chr15 | 94287818 | 94288370 | 553 | 2 | 4.15E-06 | 8.22E-06 | 4.86E-05 | 6.20E-06 | 0.004930033 | MCTP2 |
| chr17 | 1154737 | 1155271 | 535 | 2 | 2.17E-07 | 7.02E-05 | 8.39E-06 | 6.20E-06 | -0.00478506 | ABR |
| chr19 | 10732630 | 10732672 | 43 | 2 | 4.33E-06 | 3.10E-05 | 1.34E-05 | 6.21E-06 | 0.006757207 | DNM2 |
| chr20 | 38201456 | 38202014 | 559 | 6 | 9.16E-06 | 3.02E-05 | 0.001035 | 6.21E-06 | -0.00176287 | NA |
| chr5 | 11384469 | 11384955 | 487 | 5 | 2.09E-06 | 0.0001088 | 0.000272 | 6.21E-06 | -0.00548678 | CTNND2 |
| chr14 | 100836170 | 100836943 | 774 | 2 | 4.81E-06 | 4.10E-06 | 0.000271 | 6.21E-06 | 0.002679122 | MEG3, AL117190.1, AL117190.2 |
| chr10 | 70462187 | 70462872 | 686 | 3 | 4.76E-06 | 8.55E-06 | 0.000559 | 6.21E-06 | -0.00693584 | NA |
| chr7 | 128409630 | 128410528 | 899 | 14 | 3.21E-06 | 0.0010293 | 0.000686 | 6.22E-06 | -0.00055315 | IMPDH1 |
| chr22 | 39149888 | 39150271 | 384 | 5 | 3.57E-06 | 0.0002133 | 6.49E-05 | 6.22E-06 | -0.00178705 | CBX7 |
| chr17 | 3473940 | 3474503 | 564 | 6 | 1.17E-06 | 4.11E-05 | 0.000287 | 6.22E-06 | -0.00488771 | ASPA, SPATA22 |
| chr1 | 160189880 | 160190976 | 1097 | 12 | 3.48E-06 | 2.94E-05 | 0.005556 | 6.22E-06 | -0.00241697 | CASQ1 |
| chr22 | 41240937 | 41241463 | 527 | 5 | 9.57E-06 | 7.85E-06 | 0.001346 | 6.22E-06 | -0.00255077 | NA |
| chr9 | 74774072 | 74774363 | 292 | 2 | 1.05E-05 | 5.31E-06 | 0.000111 | 6.22E-06 | 0.011568233 | TRPM6 |
| chr17 | 20127587 | 20127599 | 13 | 2 | 7.90E-06 | 8.68E-06 | 4.53E-05 | 6.22E-06 | -0.00435948 | SPECC1 |
| chr6 | 36442491 | 36443439 | 949 | 14 | 1.59E-06 | 0.0003224 | 0.000334 | 6.23E-06 | -0.00016617 | KCTD20, PXT1 |
| chr11 | 61385007 | 61385424 | 418 | 2 | 1.33E-05 | 4.82E-06 | 0.000146 | 6.23E-06 | -0.00597898 | NA |
| chr17 | 49454443 | 49454524 | 82 | 2 | 1.58E-05 | 3.76E-06 | 0.00047 | 6.23E-06 | -0.00280767 | AC091180.5 |
| chr15 | 45704142 | 45704589 | 448 | 5 | 1.57E-06 | 1.15E-05 | 7.24E-05 | 6.24E-06 | -0.01026027 | NA |
| chr7 | 26191138 | 26191781 | 644 | 2 | 5.36E-06 | 3.99E-06 | 0.000323 | 6.24E-06 | -0.01253329 | HNRNPA2B1 |
| chr6 | 10838051 | 10838848 | 798 | 9 | 2.16E-05 | 5.63E-06 | 0.006988 | 6.24E-06 | -0.00054062 | TMEM14B, AL024498.2, MAK |
| chr10 | 4654699 | 4655213 | 515 | 3 | 8.79E-06 | 5.91E-06 | 0.000701 | 6.24E-06 | -0.00269461 | MANCR |
| chr3 | 147411991 | 147412749 | 759 | 5 | 4.07E-06 | 1.30E-05 | 0.000477 | 6.25E-06 | -0.00054929 | ZIC1 |
| chr16 | 55670496 | 55671317 | 822 | 4 | 5.08E-06 | 2.98E-06 | 0.002024 | 6.25E-06 | 0.000156136 | SLC6A2 |
| chr2 | 10475052 | 10476111 | 1060 | 5 | 3.78E-05 | 1.53E-05 | 0.000677 | 6.25E-06 | 0.001743622 | NA |
| chr11 | 34186297 | 34186790 | 494 | 3 | 5.84E-06 | 5.29E-06 | 0.000278 | 6.25E-06 | -0.0033515 | ABTB2 |
| chr17 | 62653893 | 62654524 | 632 | 2 | 1.85E-05 | 4.12E-06 | 0.000276 | 6.25E-06 | -0.00052925 | MRC2 |
| chr17 | 40986987 | 40987635 | 649 | 6 | 2.47E-05 | 4.04E-06 | 0.0045 | 6.26E-06 | -0.00942816 | KRT40 |
| chr12 | 6960422 | 6961320 | 899 | 5 | 1.93E-07 | 0.0001581 | 2.41E-05 | 6.26E-06 | -0.00402969 | PTPN6 |
| chr7 | 129054962 | 129056126 | 1165 | 14 | 2.55E-06 | 8.64E-05 | 0.002343 | 6.26E-06 | -0.0023529 | TPI1P2, TNPO3 |
| chr4 | 57192262 | 57192526 | 265 | 2 | 1.06E-05 | 3.77E-06 | 0.000481 | 6.26E-06 | -0.01953126 | IGFBP7-AS1 |
| chr8 | 106268842 | 106269593 | 752 | 4 | 1.35E-06 | 0.0001205 | 5.39E-05 | 6.26E-06 | -0.01197523 | NA |
| chr20 | 1841728 | 1841740 | 13 | 2 | 8.03E-06 | 4.15E-06 | 0.00027 | 6.27E-06 | -0.00963411 | NA |
| chr7 | 99390943 | 99391106 | 164 | 3 | 1.57E-05 | 2.85E-06 | 0.000884 | 6.27E-06 | -0.00238287 | AC004922.1, ARPC1B |
| chr12 | 6867467 | 6868301 | 835 | 11 | 1.59E-06 | 0.0001783 | 0.000548 | 6.27E-06 | -0.00045136 | TPI1 |
| chr11 | 35077508 | 35077943 | 436 | 2 | 6.97E-07 | 0.0002535 | 5.52E-06 | 6.27E-06 | 0.008083921 | NA |
| chr3 | 132653421 | 132654554 | 1134 | 6 | 2.21E-06 | 0.0003514 | 0.000771 | 6.27E-06 | -0.0006382 | UBA5, ACAD11, NPHP3-ACAD11 |
| chr1 | 245244843 | 245245016 | 174 | 2 | 7.73E-06 | 3.98E-06 | 0.00034 | 6.27E-06 | -0.00014028 | KIF26B |
| chr6 | 42263422 | 42263464 | 43 | 2 | 5.77E-06 | 7.66E-05 | 8.16E-06 | 6.28E-06 | -0.00505961 | TRERF1 |
| chr16 | 73891260 | 73891349 | 90 | 2 | 2.52E-06 | 0.0085279 | 3.59E-06 | 6.28E-06 | 0.005231982 | ZFHX3 |
| chr6 | 44301426 | 44302379 | 954 | 4 | 1.22E-05 | 6.95E-06 | 0.001103 | 6.28E-06 | -0.00294825 | TMEM151B, AL353588.1, AARS2 |
| chr1 | 242449383 | 242449943 | 561 | 7 | 1.02E-06 | 0.0005271 | 2.69E-05 | 6.28E-06 | -1.94E-05 | PLD5 |
| chr18 | 35289876 | 35290804 | 929 | 14 | 2.39E-05 | 3.14E-05 | 0.003894 | 6.28E-06 | 0.00067494 | ZNF271P, ZSCAN30, AC011815.1 |
| chr9 | 136907592 | 136907981 | 390 | 2 | 1.47E-07 | 4.59E-05 | 1.07E-05 | 6.29E-06 | -0.00138586 | TRAF2 |
| chr7 | 51327749 | 51328237 | 489 | 2 | 1.45E-05 | 7.17E-06 | 6.25E-05 | 6.30E-06 | -0.0016551 | NA |
| chr11 | 1460946 | 1461333 | 388 | 2 | 2.61E-06 | 1.02E-05 | 3.73E-05 | 6.30E-06 | -0.00609053 | BRSK2 |
| chr22 | 31159684 | 31160581 | 898 | 12 | 4.01E-06 | 2.27E-05 | 0.000713 | 6.30E-06 | 0.00038228 | RNF185, MIR3928 |
| chr11 | 119686855 | 119687487 | 633 | 3 | 2.60E-06 | 1.66E-05 | 0.000427 | 6.30E-06 | -0.00923916 | NECTIN1 |
| chr6 | 34082879 | 34083352 | 474 | 3 | 6.01E-06 | 1.45E-05 | 0.000448 | 6.30E-06 | -0.00890545 | GRM4 |
| chr3 | 23916473 | 23917217 | 745 | 13 | 1.13E-05 | 4.70E-05 | 0.002678 | 6.30E-06 | -3.85E-05 | RPL15, NKIRAS1 |
| chr5 | 126342741 | 126342994 | 254 | 2 | 1.69E-05 | 3.76E-06 | 0.000516 | 6.31E-06 | -0.01623112 | AC116362.1 |
| chr21 | 42449966 | 42450127 | 162 | 2 | 9.59E-06 | 4.09E-06 | 0.000303 | 6.31E-06 | -0.0047487 | NA |
| chr11 | 119030329 | 119031641 | 1313 | 15 | 1.05E-05 | 8.54E-06 | 0.007601 | 6.32E-06 | 5.89E-05 | SLC37A4 |
| chr2 | 23926990 | 23927752 | 763 | 9 | 2.76E-06 | 1.29E-05 | 0.002008 | 6.32E-06 | 0.000815104 | UBXN2A, ATAD2B |
| chr8 | 17832295 | 17832578 | 284 | 2 | 4.70E-06 | 2.05E-05 | 1.87E-05 | 6.32E-06 | -0.01061212 | AC027117.2 |
| chr1 | 147242843 | 147243864 | 1022 | 4 | 2.43E-07 | 5.26E-05 | 0.000579 | 6.32E-06 | 0.000172192 | AC242426.2, CHD1L, FMO5 |
| chr2 | 203866345 | 203866926 | 582 | 2 | 3.46E-06 | 3.93E-06 | 0.000384 | 6.32E-06 | 0.007758112 | NA |
| chr20 | 47993270 | 47993752 | 483 | 2 | 1.17E-05 | 4.18E-06 | 0.000274 | 6.32E-06 | -0.00657393 | NA |
| chr17 | 4845128 | 4845449 | 322 | 2 | 1.23E-06 | 0.0005142 | 4.80E-06 | 6.32E-06 | 0.003604638 | MINK1 |

| | | | | | | | | | | |
|---|---|---|---|---|---|---|---|---|---|---|
| chr1 | 20405830 | 20406061 | 232 | 3 | 9.72E-06 | 1.37E-05 | 0.000432 | 6.33E-06 | 0.013419557 | LINC01141 |
| chr1 | 225998876 | 225999777 | 902 | 10 | 3.11E-07 | 0.0001917 | 7.92E-06 | 6.33E-06 | 0.00216005 | SDE2 |
| chr6 | 12957190 | 12957923 | 734 | 7 | 1.58E-05 | 4.46E-06 | 0.003066 | 6.33E-06 | 0.00167553 | PHACTR1 |
| chr1 | 168302496 | 168302934 | 439 | 2 | 9.25E-06 | 5.22E-06 | 0.000123 | 6.33E-06 | 0.003246649 | TBX19 |
| chr20 | 36746178 | 36746483 | 306 | 9 | 2.14E-05 | 1.61E-06 | 0.007608 | 6.33E-06 | 0.002161848 | NA |
| chr12 | 112456089 | 112456294 | 206 | 2 | 1.29E-05 | 5.08E-06 | 0.000133 | 6.33E-06 | -0.0042905 | PTPN11 |
| chr12 | 119000286 | 119000990 | 705 | 4 | 6.83E-06 | 0.0039511 | 0.000154 | 6.33E-06 | -0.00832183 | SRRM4 |
| chr3 | 196502712 | 196503437 | 726 | 4 | 5.54E-07 | 0.0004905 | 2.38E-05 | 6.34E-06 | 0.000571473 | RNF168 |
| chr11 | 46700551 | 46701472 | 922 | 13 | 6.77E-06 | 7.34E-06 | 0.003324 | 6.34E-06 | 0.000710368 | ZNF408, ARHGAP1 |
| chr19 | 47021226 | 47021932 | 707 | 5 | 2.51E-06 | 9.80E-06 | 0.000212 | 6.34E-06 | -0.00107898 | NPAS1 |
| chr5 | 170267346 | 170267743 | 398 | 3 | 1.35E-06 | 2.44E-05 | 0.000241 | 6.35E-06 | -0.01119201 | LCP2 |
| chr6 | 21556855 | 21557217 | 363 | 2 | 6.80E-07 | 0.0001979 | 5.97E-06 | 6.35E-06 | 0.004230452 | AL512380.2 |
| chr7 | 100431683 | 100432193 | 511 | 2 | 1.15E-06 | 3.79E-06 | 0.000521 | 6.35E-06 | -0.00207903 | MEPCE |
| chr10 | 15627404 | 15627497 | 94 | 2 | 2.99E-06 | 0.0014678 | 4.12E-06 | 6.35E-06 | -0.00799178 | ITGA8 |
| chr5 | 122159492 | 122160243 | 752 | 6 | 2.50E-05 | 1.29E-06 | 0.009082 | 6.36E-06 | -0.00470159 | AC010255.2 |
| chr4 | 155953450 | 155954509 | 1060 | 9 | 1.53E-06 | 0.0007324 | 0.000231 | 6.36E-06 | 0.001270658 | CTSO |
| chr1 | 34759125 | 34759725 | 601 | 6 | 1.69E-05 | 1.69E-06 | 0.003322 | 6.36E-06 | -0.01017018 | SMIM12 |
| chr3 | 49557507 | 49557844 | 338 | 2 | 7.67E-07 | 0.0021463 | 3.96E-06 | 6.36E-06 | -0.00787561 | BSN |
| chr1 | 160789899 | 160789991 | 93 | 2 | 1.33E-05 | 6.40E-06 | 7.84E-05 | 6.36E-06 | 0.012891756 | NA |
| chr4 | 188555459 | 188555591 | 133 | 2 | 1.64E-05 | 3.90E-06 | 0.000429 | 6.37E-06 | -0.00135693 | LINC01060 |
| chr3 | 161107819 | 161108341 | 523 | 2 | 9.39E-06 | 5.17E-06 | 0.00013 | 6.37E-06 | 0.005404852 | NMD3 |
| chr8 | 82959111 | 82959842 | 732 | 3 | 1.03E-05 | 6.10E-06 | 0.000538 | 6.37E-06 | -0.01607627 | AC060765.2 |
| chr18 | 23137255 | 23138337 | 1083 | 7 | 4.56E-07 | 4.15E-05 | 4.05E-05 | 6.38E-06 | 0.003488684 | CABLES1 |
| chr20 | 46558711 | 46558784 | 74 | 2 | 1.05E-05 | 4.75E-06 | 0.000169 | 6.38E-06 | -0.00917351 | SLC13A3 |
| chr9 | 129110801 | 129111907 | 1107 | 12 | 3.19E-06 | 0.0003383 | 0.002584 | 6.39E-06 | -0.00105554 | PTPA, CRAT |
| chr11 | 85919701 | 85919947 | 247 | 2 | 1.84E-06 | 0.000704 | 4.58E-06 | 6.39E-06 | -0.02087274 | CCDC83, AP003128.1 |
| chr3 | 51982812 | 51983718 | 907 | 16 | 1.03E-06 | 0.000254 | 0.000407 | 6.40E-06 | -0.00037406 | ABHD14A-ACY1, ACY1, ABHD14B |
| chr6 | 56540744 | 56541286 | 543 | 3 | 1.46E-05 | 7.07E-06 | 0.000113 | 6.41E-06 | -0.01069377 | DST |
| chr2 | 240445877 | 240446181 | 305 | 3 | 6.92E-06 | 7.68E-06 | 8.57E-05 | 6.41E-06 | -0.00252096 | GPC1 |
| chr20 | 44921147 | 44921847 | 701 | 2 | 1.12E-05 | 4.75E-06 | 0.000173 | 6.41E-06 | -0.0090583 | PABPC1L |
| chr16 | 69852938 | 69853145 | 208 | 2 | 1.32E-05 | 5.04E-06 | 0.000143 | 6.41E-06 | 0.00055677 | WWP2 |
| chr11 | 8080903 | 8081554 | 652 | 6 | 4.94E-06 | 1.48E-05 | 0.000296 | 6.41E-06 | 0.000121612 | TUB |
| chr1 | 225422395 | 225422520 | 126 | 2 | 7.20E-06 | 7.43E-06 | 6.14E-05 | 6.41E-06 | -0.01061217 | LBR |
| chr19 | 50310315 | 50310659 | 345 | 2 | 1.30E-05 | 4.06E-06 | 0.000346 | 6.41E-06 | 0.024090957 | MYH14, AC008655.1 |
| chr11 | 69083407 | 69084464 | 1058 | 9 | 1.48E-05 | 0.0001498 | 0.000329 | 6.42E-06 | 1.69E-05 | TPCN2, AP003071.5 |
| chr19 | 55173514 | 55174006 | 493 | 3 | 1.91E-07 | 0.0162493 | 1.84E-06 | 6.42E-06 | -0.00136391 | AC010327.4, SYT5 |
| chr6 | 105726869 | 105727052 | 184 | 2 | 2.47E-06 | 0.0007982 | 4.52E-06 | 6.43E-06 | -0.01258454 | AL591518.1 |
| chr2 | 24174352 | 24175301 | 950 | 10 | 9.74E-07 | 5.02E-05 | 0.000191 | 6.43E-06 | -0.00728777 | AC008073.3, FAM228A, AC008073.2 |
| chr12 | 109573138 | 109574263 | 1126 | 23 | 1.61E-06 | 0.0001344 | 0.000232 | 6.43E-06 | 0.0004665 | MVK, MMAB |
| chr1 | 160839704 | 160839849 | 146 | 3 | 9.14E-06 | 5.62E-06 | 0.000177 | 6.43E-06 | -0.00764119 | CD244 |
| chr1 | 33337462 | 33338204 | 743 | 3 | 2.27E-05 | 2.35E-06 | 0.002444 | 6.43E-06 | 0.000266258 | AL513327.1, PHC2 |
| chr8 | 141257768 | 141258187 | 420 | 2 | 2.44E-06 | 2.33E-05 | 1.73E-05 | 6.44E-06 | -0.01380757 | SLC45A4 |
| chr18 | 79006007 | 79006747 | 741 | 4 | 1.06E-06 | 2.00E-05 | 0.000156 | 6.44E-06 | -0.01145455 | NA |
| chr10 | 130028191 | 130028625 | 435 | 2 | 6.14E-06 | 3.84E-06 | 0.00053 | 6.44E-06 | -0.00571433 | C10orf143 |
| chr11 | 57183753 | 57184092 | 340 | 2 | 1.68E-06 | 6.76E-05 | 8.95E-06 | 6.44E-06 | -0.008185 | LRRC55 |
| chr6 | 151445244 | 151445787 | 544 | 3 | 4.86E-06 | 1.21E-05 | 6.78E-05 | 6.45E-06 | -0.00561508 | RMND1 |
| chr10 | 7541119 | 7541252 | 134 | 3 | 5.01E-06 | 7.89E-05 | 1.18E-05 | 6.45E-06 | -0.00567204 | NA |
| chr7 | 27249933 | 27250210 | 278 | 3 | 9.83E-06 | 2.83E-05 | 0.00012 | 6.45E-06 | -0.00757278 | EVX1 |
| chr10 | 81873923 | 81874520 | 598 | 4 | 1.00E-06 | 0.0013737 | 6.23E-05 | 6.45E-06 | 0.001904173 | AL096706.1 |
| chr2 | 176189412 | 176190440 | 1029 | 4 | 1.21E-05 | 0.0005293 | 0.000297 | 6.45E-06 | -0.00501439 | HOXD1, AC009336.2 |
| chr1 | 157100883 | 157101284 | 402 | 4 | 4.71E-06 | 2.13E-05 | 0.00028 | 6.45E-06 | -0.00361527 | ETV3L |
| chr11 | 66857382 | 66857807 | 426 | 5 | 2.41E-05 | 2.63E-06 | 0.004295 | 6.45E-06 | -0.00415333 | LRFN4, PC |
| chr5 | 127042510 | 127042879 | 370 | 2 | 6.91E-06 | 6.55E-06 | 7.78E-05 | 6.46E-06 | 0.003199408 | C5orf63 |
| chr17 | 5501188 | 5502081 | 894 | 6 | 1.18E-06 | 7.67E-06 | 0.003878 | 6.46E-06 | 0.000458995 | NLRP1 |
| chr10 | 102135674 | 102136311 | 638 | 2 | 5.44E-07 | 3.33E-05 | 1.35E-05 | 6.47E-06 | -0.01001587 | PPRC1 |
| chr8 | 118621605 | 118622395 | 791 | 10 | 6.12E-06 | 6.59E-06 | 0.001286 | 6.47E-06 | 0.001550446 | SAMD12-AS1, SAMD12 |
| chr5 | 161548786 | 161549411 | 626 | 9 | 4.21E-06 | 0.0002254 | 0.000953 | 6.47E-06 | -0.00025635 | GABRA6, GABRB2 |
| chr20 | 57246161 | 57246259 | 99 | 2 | 1.29E-05 | 3.86E-06 | 0.000526 | 6.47E-06 | -0.00742792 | BMP7 |
| chr14 | 54737704 | 54738282 | 579 | 2 | 8.49E-06 | 5.74E-06 | 0.000103 | 6.47E-06 | 0.02261149 | SAMD4A |
| chr8 | 3997893 | 3998055 | 163 | 3 | 1.80E-05 | 4.01E-06 | 0.000842 | 6.47E-06 | -0.00575174 | CSMD1 |
| chr8 | 25060754 | 25061348 | 595 | 3 | 4.28E-06 | 1.89E-05 | 0.000289 | 6.47E-06 | -0.01237141 | NA |
| chr2 | 1813491 | 1813637 | 147 | 3 | 6.04E-06 | 2.47E-06 | 0.002183 | 6.48E-06 | 0.057233727 | MYT1L |
| chr6 | 97925137 | 97925534 | 398 | 2 | 9.65E-06 | 5.43E-06 | 0.00012 | 6.48E-06 | 0.002976899 | AL589740.1 |
| chr10 | 112949214 | 112950001 | 788 | 5 | 9.88E-10 | 0.0014254 | 1.11E-05 | 6.48E-06 | -0.00143679 | NA |
| chr19 | 54035974 | 54036039 | 66 | 2 | 6.69E-06 | 1.81E-05 | 2.15E-05 | 6.48E-06 | -0.01924715 | NA |
| chr1 | 37514442 | 37515177 | 736 | 12 | 7.80E-06 | 1.89E-05 | 0.002476 | 6.48E-06 | 0.001108022 | MEAF6 |
| chr10 | 35089998 | 35090799 | 802 | 11 | 1.28E-07 | 0.0041102 | 3.53E-06 | 6.48E-06 | 0.001114753 | CUL2 |
| chr4 | 105894932 | 105895226 | 295 | 4 | 1.38E-05 | 3.17E-06 | 0.000873 | 6.49E-06 | 0.009193012 | NPNT, INTS12 |
| chr17 | 4691344 | 4691902 | 559 | 3 | 1.52E-06 | 9.93E-05 | 7.53E-06 | 6.49E-06 | -0.00393663 | AC091153.2, PELP1 |
| chr3 | 124821458 | 124821655 | 198 | 2 | 9.88E-06 | 9.01E-06 | 4.68E-05 | 6.49E-06 | -0.00507673 | ITGB5 |
| chr16 | 77709850 | 77710214 | 365 | 3 | 1.19E-05 | 1.52E-05 | 0.000448 | 6.49E-06 | 0.000629267 | NA |
| chr15 | 74815069 | 74815748 | 680 | 3 | 3.67E-07 | 4.91E-05 | 3.23E-05 | 6.49E-06 | -0.0079479 | LMAN1L |
| chr1 | 228372738 | 228373407 | 670 | 3 | 8.73E-07 | 5.98E-05 | 1.10E-05 | 6.49E-06 | 0.007571933 | OBSCN |
| chr10 | 13709379 | 13710399 | 1021 | 5 | 6.39E-06 | 6.55E-06 | 0.000263 | 6.49E-06 | -0.00129754 | FRMD4A |
| chr6 | 27205795 | 27206212 | 418 | 5 | 8.87E-06 | 1.51E-05 | 0.001018 | 6.49E-06 | -0.00158062 | NA |
| chr2 | 210225393 | 210225960 | 568 | 9 | 1.22E-05 | 8.73E-05 | 0.001195 | 6.49E-06 | -0.00395445 | AC006994.1, ACADL |
| chr14 | 75968085 | 75968436 | 352 | 2 | 3.44E-06 | 1.84E-05 | 2.14E-05 | 6.49E-06 | 0.00737029 | IFT43, TGFB3 |
| chr12 | 54388998 | 54389270 | 273 | 2 | 5.26E-06 | 9.00E-06 | 4.69E-05 | 6.49E-06 | 0.005735149 | AC079313.2, AC079313.1, ZNF385A |
| chr9 | 136652072 | 136652103 | 32 | 2 | 7.09E-06 | 4.85E-05 | 1.08E-05 | 6.49E-06 | 0.00128167 | AL590226.1 |
| chr11 | 103334002 | 103334341 | 340 | 2 | 8.62E-07 | 0.0026321 | 3.97E-06 | 6.49E-06 | -0.01630756 | DYNC2H1 |
| chr9 | 88586665 | 88587320 | 656 | 3 | 7.82E-06 | 3.52E-06 | 0.000625 | 6.49E-06 | -0.00121327 | NA |
| chr9 | 128684549 | 128685716 | 1168 | 7 | 1.64E-07 | 0.0011962 | 2.31E-05 | 6.50E-06 | -0.00353571 | SET |
| chr19 | 15281021 | 15281646 | 626 | 5 | 1.95E-07 | 1.77E-05 | 6.48E-05 | 6.50E-06 | 0.005304652 | BRD4 |
| chr6 | 41656058 | 41656312 | 255 | 3 | 6.31E-06 | 3.12E-05 | 0.000196 | 6.50E-06 | -0.00693719 | NA |
| chr17 | 78159291 | 78159985 | 695 | 6 | 1.80E-05 | 2.08E-05 | 0.002017 | 6.50E-06 | -0.00779719 | C17orf99 |
| chr12 | 238025 | 238521 | 497 | 3 | 4.92E-06 | 3.56E-06 | 0.000683 | 6.50E-06 | -0.00782419 | SLC6A13 |
| chr12 | 108810040 | 108810980 | 941 | 3 | 7.54E-06 | 2.68E-05 | 9.74E-05 | 6.51E-06 | -0.00169665 | SSH1 |
| chr10 | 21184550 | 21185291 | 742 | 2 | 0.000139482 | 4.17E-06 | 0.000331 | 6.51E-06 | -0.00594097 | NEBL |
| chr15 | 47718395 | 47719269 | 875 | 7 | 2.01E-07 | 0.0005291 | 7.85E-06 | 6.51E-06 | -0.00057066 | SEMA6D |
| chr14 | 64764601 | 64764775 | 175 | 4 | 3.65E-06 | 1.64E-05 | 0.00041 | 6.51E-06 | -0.008718 | SPTB |
| chr8 | 117883808 | 117884009 | 202 | 2 | 8.44E-07 | 0.000412 | 5.15E-06 | 6.52E-06 | 0.007668856 | EXT1 |
| chr1 | 183817311 | 183817882 | 572 | 2 | 9.66E-06 | 5.44E-06 | 0.000122 | 6.52E-06 | -0.01237861 | RGL1 |
| chr8 | 36266052 | 36266177 | 126 | 2 | 1.52E-05 | 4.79E-06 | 0.000181 | 6.52E-06 | -0.02116706 | NA |
| chr3 | 161220737 | 161221658 | 922 | 9 | 3.78E-07 | 0.0024045 | 1.70E-05 | 6.52E-06 | -0.00037646 | NMD3 |
| chr11 | 133924838 | 133924874 | 37 | 2 | 1.42E-05 | 4.04E-06 | 0.000407 | 6.52E-06 | -0.00463251 | IGSF9B |
| chr2 | 3171829 | 3172377 | 549 | 4 | 5.98E-07 | 0.0007994 | 2.72E-05 | 6.52E-06 | 0.002327773 | NA |
| chr11 | 22703137 | 22703927 | 791 | 2 | 8.36E-05 | 3.94E-06 | 0.00048 | 6.52E-06 | -0.01183969 | GAS2 |
| chr18 | 24459532 | 24460041 | 510 | 4 | 2.85E-05 | 1.85E-06 | 0.004409 | 6.52E-06 | 0.007675544 | NA |
| chr2 | 231255244 | 231255588 | 345 | 2 | 3.40E-06 | 1.03E-05 | 3.96E-05 | 6.53E-06 | -0.00618645 | ARMC9 |
| chr18 | 74315456 | 74316387 | 932 | 10 | 4.52E-06 | 7.81E-06 | 0.004654 | 6.53E-06 | -0.00037859 | C18orf63 |
| chr3 | 157174240 | 157175246 | 1007 | 8 | 1.60E-06 | 9.34E-06 | 0.001408 | 6.53E-06 | 0.000926072 | AC092944.1 |
| chr19 | 28998311 | 28999022 | 712 | 3 | 2.78E-05 | 6.30E-06 | 0.000344 | 6.53E-06 | -0.01038935 | NA |
| chr16 | 70576705 | 70577653 | 949 | 3 | 1.18E-05 | 6.16E-05 | 5.89E-05 | 6.53E-06 | 0.002304103 | SF3B3 |
| chr6 | 33632743 | 33633492 | 750 | 6 | 5.07E-06 | 5.94E-06 | 0.000412 | 6.54E-06 | -0.01182265 | ITPR3 |
| chr12 | 114656631 | 114657220 | 590 | 3 | 2.07E-05 | 1.83E-05 | 0.000423 | 6.54E-06 | -0.0132748 | NA |
| chr10 | 47047886 | 47047945 | 60 | 2 | 1.08E-05 | 1.35E-05 | 2.92E-05 | 6.55E-06 | -0.01772605 | NA |
| chr10 | 132746049 | 132746856 | 808 | 7 | 3.48E-05 | 8.16E-05 | 0.00145 | 6.55E-06 | -0.00282177 | INPP5A |
| chr7 | 73869248 | 73869748 | 501 | 3 | 7.79E-07 | 0.0006841 | 1.19E-05 | 6.55E-06 | -0.00155038 | NA |
| chr15 | 62843548 | 62844092 | 545 | 3 | 6.23E-07 | 0.0001098 | 1.39E-05 | 6.55E-06 | 0.000760817 | TLN2, AC103740.1 |
| chr12 | 6635891 | 6636541 | 651 | 11 | 1.10E-06 | 0.0001092 | 0.002497 | 6.55E-06 | 0.006302065 | LPAR5 |
| chr1 | 84724704 | 84724717 | 14 | 2 | 1.20E-05 | 1.08E-05 | 3.76E-05 | 6.55E-06 | -0.02463309 | NA |
| chr9 | 125240976 | 125241863 | 888 | 11 | 9.34E-06 | 2.49E-05 | 0.004544 | 6.56E-06 | 0.000597488 | AL354710.2, HSPA5 |
| chr9 | 79609869 | 79610835 | 967 | 6 | 3.33E-06 | 2.61E-05 | 0.000239 | 6.56E-06 | 0.002272076 | TLE4 |
| chr2 | 69386975 | 69387813 | 839 | 8 | 1.11E-06 | 4.31E-05 | 0.000329 | 6.56E-06 | 0.000349227 | GFPT1 |
| chr21 | 33023913 | 33024827 | 915 | 7 | 2.06E-06 | 0.0002328 | 0.00011 | 6.56E-06 | -0.00502433 | AP000282.1 |
| chr6 | 33746854 | 33747333 | 480 | 4 | 6.94E-06 | 1.14E-05 | 0.000628 | 6.56E-06 | -0.00665947 | IP6K3 |
| chr1 | 183609676 | 183610335 | 660 | 2 | 4.09E-05 | 3.92E-06 | 0.000528 | 6.56E-06 | -0.00385025 | NA |
| chr7 | 57179912 | 57180311 | 400 | 6 | 1.59E-06 | 0.001882 | 4.01E-05 | 6.56E-06 | -0.00745848 | GUSBP10 |
| chr8 | 40119086 | 40119397 | 312 | 3 | 1.54E-05 | 4.96E-06 | 0.000272 | 6.56E-06 | -0.00390579 | LINC02866 |
| chr1 | 17231849 | 17232853 | 1005 | 3 | 1.52E-06 | 4.51E-06 | 0.000315 | 6.56E-06 | -0.00597554 | PADI1 |
| chr10 | 120857614 | 120857631 | 18 | 2 | 3.68E-06 | 0.0001346 | 6.96E-06 | 6.56E-06 | -0.00723171 | WDR11 |
| chr20 | 57672246 | 57673071 | 826 | 4 | 7.69E-07 | 0.0006713 | 8.22E-05 | 6.57E-06 | -0.00546807 | PMEPA1 |
| chr19 | 18611471 | 18612681 | 1211 | 8 | 3.58E-06 | 1.80E-05 | 0.000302 | 6.57E-06 | -0.00016661 | TMEM59L |
| chr9 | 86722030 | 86722095 | 66 | 2 | 7.75E-06 | 9.85E-06 | 4.25E-05 | 6.57E-06 | -0.01399039 | NA |
| chr10 | 131078435 | 131079025 | 591 | 3 | 8.73E-06 | 6.09E-06 | 0.000275 | 6.57E-06 | -0.00152857 | NA |
| chr1 | 205356660 | 205357309 | 650 | 9 | 1.56E-05 | 0.0002883 | 0.003737 | 6.57E-06 | -0.00547169 | KLHDC8A |
| chr2 | 101136809 | 101136941 | 133 | 3 | 9.18E-06 | 1.56E-05 | 6.75E-05 | 6.58E-06 | -0.00192948 | TBC1D8 |
| chr12 | 112380797 | 112381604 | 808 | 5 | 8.10E-06 | 7.08E-06 | 0.001694 | 6.58E-06 | 0.001083816 | HECTD4 |
| chr12 | 19546570 | 19546898 | 329 | 3 | 4.65E-06 | 0.0008419 | 5.61E-05 | 6.59E-06 | 0.009413153 | AEBP2 |
| chr10 | 17151540 | 17151592 | 53 | 2 | 1.01E-05 | 5.43E-06 | 0.000128 | 6.59E-06 | -0.00918051 | TRDMT1 |
| chr13 | 112806991 | 112807731 | 741 | 8 | 5.69E-06 | 7.92E-05 | 0.00173 | 6.61E-06 | -2.65E-05 | ATP11A |
| chr2 | 37231214 | 37232183 | 970 | 17 | 5.28E-06 | 0.0001059 | 0.007169 | 6.61E-06 | 0.000908567 | NDUFAF7, CEBPZ |
| chr11 | 67021268 | 67021830 | 563 | 4 | 5.46E-07 | 7.48E-06 | 0.000432 | 6.61E-06 | -0.00878547 | SYT12 |
| chr12 | 120668564 | 120669239 | 676 | 4 | 2.39E-05 | 8.20E-05 | 0.000238 | 6.61E-06 | -0.00789388 | NA |
| chr6 | 169663791 | 169664237 | 447 | 2 | 6.52E-07 | 4.63E-05 | 1.14E-05 | 6.61E-06 | -0.00455356 | AL031315.1, WDR27 |
| chr8 | 86098050 | 86098727 | 678 | 3 | 5.20E-06 | 1.56E-05 | 7.87E-05 | 6.61E-06 | -0.00852255 | ATP6V0D2 |
| chr7 | 4838240 | 4839070 | 831 | 4 | 1.62E-07 | 0.0005352 | 1.11E-05 | 6.61E-06 | -0.00448264 | RADIL |
| chr11 | 66744152 | 66745122 | 971 | 6 | 3.60E-06 | 1.78E-05 | 0.000281 | 6.62E-06 | 0.00194586 | C11orf80 |
| chr2 | 136025533 | 136025786 | 254 | 2 | 1.30E-05 | 4.15E-06 | 0.000381 | 6.62E-06 | -0.01346101 | NA |
| chr8 | 120518679 | 120518785 | 107 | 2 | 3.42E-06 | 0.0007559 | 4.70E-06 | 6.62E-06 | -0.01125985 | MTBP |
| chr2 | 16223803 | 16224524 | 722 | 3 | 3.01E-06 | 6.41E-06 | 0.000523 | 6.62E-06 | 0.002109832 | AC010745.2 |
| chr20 | 44355087 | 44356238 | 1152 | 12 | 6.64E-06 | 0.0013044 | 0.001046 | 6.62E-06 | 0.000484606 | HNF4A, AL117382.1 |
| chr16 | 69040081 | 69040429 | 349 | 2 | 1.93E-05 | 3.95E-06 | 0.000534 | 6.62E-06 | 0.004009749 | TANGO6 |
| chr1 | 171783800 | 171784488 | 689 | 6 | 2.31E-07 | 0.0023057 | 0.000107 | 6.62E-06 | -0.0015249 | EEF1AKNMT |
| chr20 | 41340291 | 41341409 | 1119 | 11 | 1.75E-06 | 0.0003133 | 0.000114 | 6.62E-06 | 0.004824697 | LPIN3 |
| chr17 | 35495602 | 35495985 | 384 | 3 | 2.48E-06 | 8.36E-06 | 0.000284 | 6.63E-06 | -0.00667635 | SLFN12L |
| chr1 | 44425569 | 44425626 | 58 | 2 | 3.01E-06 | 0.0029535 | 4.02E-06 | 6.63E-06 | -0.00947338 | RNF220 |
| chr10 | 101588222 | 101589110 | 889 | 11 | 2.23E-06 | 7.73E-05 | 0.00248 | 6.63E-06 | -0.00264481 | DPCD, POLL |
| chr11 | 17496559 | 17496935 | 377 | 2 | 2.62E-06 | 2.29E-05 | 1.83E-05 | 6.63E-06 | -0.01239856 | USH1C |
| chr4 | 83535518 | 83536669 | 1152 | 25 | 3.88E-06 | 0.0001519 | 0.008739 | 6.63E-06 | 0.000154761 | GPAT3 |
| chr5 | 178859734 | 178860543 | 810 | 7 | 2.98E-06 | 0.0005181 | 0.000905 | 6.64E-06 | 0.001166775 | ZNF354B |
| chr6 | 6656824 | 6657063 | 240 | 3 | 6.65E-06 | 0.000121 | 0.000277 | 6.64E-06 | -0.00092986 | NA |
| chr4 | 95549099 | 95550027 | 929 | 15 | 2.20E-06 | 0.0003343 | 0.005866 | 6.64E-06 | -3.81E-05 | AC106881.1, UNC5C |
| chr4 | 133148438 | 133149286 | 849 | 10 | 2.65E-05 | 1.38E-05 | 0.000595 | 6.64E-06 | -1.90E-05 | AC105383.1 |
| chr20 | 36366875 | 36367287 | 413 | 6 | 5.07E-06 | 1.04E-05 | 0.002215 | 6.64E-06 | -0.0068104 | DLGAP4 |
| chr2 | 228181690 | 228182985 | 1296 | 10 | 1.23E-09 | 0.0018532 | 0.00013 | 6.65E-06 | -0.00210126 | NA |
| chr5 | 176642769 | 176643734 | 966 | 4 | 1.41E-06 | 6.13E-05 | 4.24E-05 | 6.65E-06 | -0.00193393 | EIF4E1B |
| chr11 | 71501356 | 71501902 | 547 | 2 | 1.52E-07 | 1.79E-05 | 2.28E-05 | 6.65E-06 | 0.001852448 | NADSYN1 |

| | | | | | | | | | | |
|---|---|---|---|---|---|---|---|---|---|---|
| chr10 | 69238962 | 69239336 | 375 | 4 | 1.97E-05 | 3.90E-06 | 0.002249 | 6.66E-06 | -0.0057608 | HKDC1 |
| chr16 | 84031619 | 84031996 | 378 | 4 | 2.29E-06 | 2.67E-05 | 3.83E-05 | 6.66E-06 | 0.000278978 | SLC38A8 |
| chr15 | 71802441 | 71802957 | 517 | 3 | 1.67E-05 | 3.57E-06 | 0.000644 | 6.66E-06 | -0.01011724 | NR2E3 |
| chr1 | 204142989 | 204143765 | 777 | 3 | 1.71E-05 | 1.56E-05 | 0.000359 | 6.66E-06 | -0.0066751 | ERLNC1, ETNK2 |
| chr3 | 53882264 | 53882973 | 710 | 6 | 3.60E-06 | 3.81E-05 | 0.000457 | 6.67E-06 | -0.00225837 | NA |
| chr2 | 108612433 | 108612881 | 449 | 2 | 1.45E-05 | 4.05E-06 | 0.000471 | 6.67E-06 | -0.0122382 | LIMS1 |
| chr18 | 79400212 | 79401214 | 1003 | 7 | 2.56E-06 | 3.08E-05 | 0.000623 | 6.67E-06 | -0.00327546 | NFATC1 |
| chr19 | 3223917 | 3224398 | 482 | 3 | 5.63E-07 | 3.55E-05 | 0.000351 | 6.67E-06 | 0.004399902 | NA |
| chr7 | 47481472 | 47482340 | 869 | 2 | 5.95E-05 | 4.01E-06 | 0.000514 | 6.68E-06 | 0.004325293 | TNS3 |
| chr11 | 134527456 | 134527832 | 377 | 3 | 1.24E-05 | 2.10E-05 | 0.000197 | 6.68E-06 | 0.00385587 | NA |
| chr1 | 46171611 | 46172052 | 442 | 2 | 1.26E-05 | 4.08E-06 | 0.000455 | 6.68E-06 | -0.0092437 | P3R3URF-PIK3R3 |
| chr7 | 55364319 | 55364999 | 681 | 5 | 2.61E-06 | 2.20E-05 | 6.49E-05 | 6.68E-06 | 0.005529154 | NA |
| chr19 | 50910559 | 50910775 | 217 | 5 | 5.79E-06 | 6.72E-05 | 0.000239 | 6.68E-06 | 0.00090688 | KLK4 |
| chr17 | 34580113 | 34580978 | 866 | 11 | 1.21E-05 | 4.37E-05 | 0.000169 | 6.69E-06 | 0.001174174 | TMEM132E |
| chr11 | 134519127 | 134519749 | 623 | 3 | 3.42E-05 | 3.23E-06 | 0.001338 | 6.69E-06 | -0.00318262 | NA |
| chr7 | 2251900 | 2252325 | 426 | 2 | 2.17E-06 | 1.01E-05 | 4.29E-05 | 6.70E-06 | -0.0212157 | SNX8 |
| chr10 | 102773417 | 102774545 | 1129 | 4 | 6.92E-05 | 0.0001581 | 0.000817 | 6.70E-06 | 0.00443978 | WBP1L |
| chr2 | 178450011 | 178450350 | 340 | 2 | 2.19E-06 | 4.01E-06 | 0.000538 | 6.70E-06 | -0.01860537 | PRKRA |
| chr7 | 69597096 | 69597635 | 540 | 5 | 2.24E-05 | 1.77E-06 | 0.002859 | 6.71E-06 | 0.00197929 | CT66 |
| chr3 | 5169876 | 5169914 | 39 | 2 | 3.33E-06 | 0.0024529 | 4.12E-06 | 6.71E-06 | 0.015150221 | ARL8B, AC026202.2 |
| chr8 | 52388606 | 52389249 | 644 | 5 | 1.20E-05 | 3.11E-05 | 0.000353 | 6.71E-06 | -0.00593554 | ST18 |
| chr1 | 23817577 | 23817932 | 356 | 2 | 9.01E-07 | 3.90E-05 | 1.29E-05 | 6.72E-06 | -0.00245673 | HMGCL |
| chr16 | 30417883 | 30418827 | 945 | 6 | 6.18E-05 | 1.16E-05 | 0.001103 | 6.72E-06 | -0.00311583 | ZNF771 |
| chr16 | 57752901 | 57753236 | 336 | 3 | 9.97E-07 | 4.87E-05 | 1.31E-05 | 6.72E-06 | -0.00476556 | KATNB1 |
| chr5 | 176365120 | 176365877 | 758 | 13 | 4.89E-06 | 7.63E-05 | 0.00045 | 6.72E-06 | 0.001059383 | ARL10 |
| chr18 | 13542551 | 13543049 | 499 | 2 | 4.15E-07 | 6.12E-06 | 9.94E-05 | 6.72E-06 | 0.000771672 | LDLRAD4 |
| chr16 | 46844252 | 46844641 | 390 | 2 | 1.47E-06 | 2.06E-05 | 2.04E-05 | 6.72E-06 | -0.02731701 | CKBP1 |
| chr2 | 206085694 | 206086434 | 741 | 8 | 2.35E-05 | 8.45E-06 | 0.001937 | 6.73E-06 | 0.000996988 | AC007383.1, INO80D |
| chr10 | 59622433 | 59622978 | 546 | 2 | 8.87E-06 | 5.90E-06 | 0.000109 | 6.73E-06 | 0.011832993 | AC026391.1 |
| chr20 | 50818537 | 50818746 | 210 | 3 | 2.20E-06 | 0.0001432 | 1.12E-05 | 6.73E-06 | -0.00534343 | BCAS4 |
| chr12 | 11885241 | 11885968 | 728 | 4 | 1.39E-05 | 1.01E-05 | 0.000376 | 6.73E-06 | 0.000285217 | ETV6 |
| chr2 | 110115454 | 110116164 | 711 | 7 | 4.36E-06 | 2.29E-05 | 0.002545 | 6.73E-06 | 0.006488455 | MALL |
| chr22 | 19479256 | 19479919 | 664 | 14 | 3.10E-05 | 2.84E-05 | 0.003056 | 6.74E-06 | -0.0006177 | CDC45 |
| chr9 | 10232563 | 10232876 | 314 | 2 | 1.16E-05 | 5.54E-06 | 0.00013 | 6.74E-06 | -0.01353227 | PTPRD |
| chr19 | 33101105 | 33101391 | 287 | 3 | 4.62E-06 | 0.0001636 | 3.05E-05 | 6.74E-06 | -0.00331977 | GPATCH1 |
| chr2 | 241279560 | 241279955 | 396 | 2 | 1.66E-05 | 4.24E-06 | 0.000379 | 6.74E-06 | 0.004443868 | HDLBP |
| chr11 | 119106669 | 119107798 | 1130 | 15 | 1.06E-06 | 0.0009018 | 5.86E-05 | 6.74E-06 | -0.00067304 | C2CD2L, DPAGT1 |
| chr3 | 171026751 | 171027175 | 425 | 4 | 7.81E-07 | 2.65E-05 | 0.000273 | 6.74E-06 | -0.00461262 | AC061708.1 |
| chr11 | 45145295 | 45145942 | 648 | 4 | 7.75E-06 | 2.39E-06 | 0.0021 | 6.76E-06 | -0.00687481 | PRDM11 |
| chr9 | 16039200 | 16039923 | 724 | 3 | 4.40E-07 | 0.0001309 | 7.88E-06 | 6.76E-06 | -0.01251226 | CCDC171 |
| chr21 | 41853689 | 41854725 | 1037 | 5 | 1.06E-05 | 1.95E-06 | 0.005836 | 6.76E-06 | 0.001288513 | PRDM15 |
| chr14 | 102461625 | 102462100 | 476 | 2 | 1.53E-05 | 4.50E-06 | 0.000281 | 6.77E-06 | -0.01965254 | TECPR2 |
| chr5 | 92689575 | 92689694 | 120 | 2 | 3.03E-06 | 1.33E-05 | 3.14E-05 | 6.78E-06 | 0.003636267 | NA |
| chr1 | 36398789 | 36399291 | 503 | 2 | 7.07E-08 | 7.70E-05 | 8.95E-06 | 6.78E-06 | -0.00365895 | NA |
| chr1 | 32768886 | 32769723 | 838 | 2 | 1.72E-07 | 1.34E-05 | 3.12E-05 | 6.78E-06 | -0.00122887 | KIAA1522 |
| chr22 | 19447622 | 19448335 | 714 | 11 | 2.02E-05 | 5.13E-05 | 0.000408 | 6.78E-06 | -0.00014248 | AC000068.1, C22orf39 |
| chr15 | 58887235 | 58887385 | 151 | 3 | 3.49E-05 | 6.89E-06 | 0.00014 | 6.78E-06 | -0.00351573 | RNF111, SLTM |
| chr6 | 32948829 | 32949878 | 1050 | 10 | 8.72E-07 | 0.0003683 | 2.97E-05 | 6.78E-06 | 0.002776271 | AL645941.2, HLA-DMA |
| chr1 | 240596810 | 240597377 | 568 | 3 | 1.64E-06 | 0.0001071 | 0.00018 | 6.79E-06 | -0.01700372 | GREM2 |
| chr15 | 74302640 | 74302933 | 294 | 3 | 4.66E-06 | 5.37E-05 | 0.000282 | 6.79E-06 | -0.00856506 | CCDC33, AC023300.3 |
| chr6 | 139375634 | 139376174 | 541 | 8 | 6.28E-07 | 0.0002217 | 2.87E-05 | 6.80E-06 | 0.001347916 | AL592429.2 |
| chr19 | 34359040 | 34359883 | 844 | 5 | 3.87E-05 | 0.0001113 | 0.000241 | 6.80E-06 | -0.00364286 | GPI, AC010504.1 |
| chr7 | 121302330 | 121302951 | 622 | 3 | 1.27E-05 | 3.87E-06 | 0.000737 | 6.81E-06 | -0.00299142 | NA |
| chr17 | 61481141 | 61481672 | 532 | 2 | 1.03E-05 | 5.01E-06 | 0.000187 | 6.81E-06 | -0.00886345 | TBX4 |
| chr5 | 179150497 | 179150541 | 45 | 2 | 1.01E-05 | 5.01E-06 | 0.000187 | 6.81E-06 | 0.004170263 | ADAMTS2 |
| chr2 | 235088749 | 235089147 | 399 | 4 | 3.20E-06 | 0.0003308 | 0.000229 | 6.81E-06 | -0.00994625 | NA |
| chr16 | 529607 | 529627 | 21 | 2 | 4.99E-06 | 1.76E-05 | 2.40E-05 | 6.81E-06 | 0.004998197 | CAPN15 |
| chr3 | 109355749 | 109356262 | 514 | 2 | 1.90E-05 | 4.33E-06 | 0.000358 | 6.82E-06 | -0.01886466 | NA |
| chr1 | 28812407 | 28813246 | 840 | 7 | 1.09E-06 | 0.0148141 | 9.32E-06 | 6.82E-06 | 0.003922311 | OPRD1 |
| chr14 | 24339216 | 24340553 | 1338 | 13 | 6.98E-06 | 0.0002493 | 0.000236 | 6.82E-06 | -0.00011143 | RIPK3 |
| chr4 | 54289044 | 54289467 | 424 | 2 | 4.24E-08 | 0.000152 | 6.97E-06 | 6.82E-06 | -0.00750164 | AC058822.1, PDGFRA |
| chr14 | 36654473 | 36655229 | 757 | 7 | 3.44E-06 | 2.81E-05 | 0.000239 | 6.82E-06 | -0.00044375 | AL132857.1, AL079303.1 |
| chr19 | 55069484 | 55070192 | 709 | 8 | 1.74E-06 | 0.0004556 | 0.000165 | 6.83E-06 | 0.00649793 | RDH13 |
| chr19 | 58575271 | 58575834 | 564 | 7 | 2.62E-06 | 0.0003802 | 9.50E-05 | 6.84E-06 | 0.001062457 | CENPBD1P1, AC016629.3 |
| chr17 | 64505920 | 64506447 | 528 | 9 | 2.02E-06 | 1.50E-06 | 0.016008 | 6.84E-06 | 0.0006517 | DDX5 |
| chr1 | 154870241 | 154871071 | 831 | 10 | 3.02E-05 | 5.53E-05 | 0.002962 | 6.84E-06 | -0.00280892 | KCNN3 |
| chr1 | 46530874 | 46531571 | 698 | 4 | 1.65E-06 | 1.11E-05 | 0.001404 | 6.84E-06 | -0.00582416 | NA |
| chr19 | 37920781 | 37921217 | 437 | 2 | 7.81E-06 | 6.85E-06 | 8.27E-05 | 6.84E-06 | -0.01149868 | SIPA1L3 |
| chr17 | 47220016 | 47220640 | 625 | 4 | 2.64E-05 | 6.24E-05 | 0.00032 | 6.85E-06 | -0.00587388 | MYL4 |
| chr7 | 128343601 | 128344403 | 803 | 14 | 1.14E-05 | 2.11E-05 | 0.002283 | 6.86E-06 | -0.00084371 | RNU7-27P, RBM28 |
| chr4 | 152779519 | 152780808 | 1290 | 8 | 1.78E-05 | 0.0001248 | 0.000909 | 6.86E-06 | 0.001050631 | ARFIP1, TIGD4 |
| chr9 | 5509131 | 5509695 | 565 | 3 | 1.83E-05 | 4.12E-06 | 0.000899 | 6.86E-06 | 0.005254306 | AL162253.2 |
| chr1 | 44117930 | 44118439 | 510 | 5 | 6.48E-06 | 2.68E-06 | 0.001731 | 6.86E-06 | -0.00242822 | NA |
| chr2 | 3574167 | 3574288 | 122 | 2 | 3.77E-08 | 5.98E-05 | 1.03E-05 | 6.86E-06 | -0.00504979 | AC108488.3 |
| chr9 | 89720095 | 89720375 | 281 | 2 | 7.66E-06 | 4.23E-06 | 0.000436 | 6.87E-06 | -0.0094932 | NA |
| chr2 | 218751139 | 218751699 | 561 | 2 | 3.92E-06 | 6.78E-06 | 8.52E-05 | 6.88E-06 | -0.00580443 | TTLL4 |
| chr1 | 9910199 | 9911057 | 859 | 15 | 1.89E-05 | 2.06E-05 | 0.007291 | 6.88E-06 | 0.001070355 | CTNNBIP1 |
| chr9 | 35658013 | 35658455 | 443 | 7 | 1.92E-05 | 5.20E-06 | 0.000984 | 6.88E-06 | -0.00090165 | CCDC107, RMRP, RMRP |
| chr11 | 45699612 | 45700303 | 692 | 3 | 1.77E-07 | 0.0006754 | 4.04E-06 | 6.88E-06 | -0.00405435 | NA |
| chr14 | 73633651 | 73634284 | 634 | 8 | 2.04E-06 | 0.0002292 | 1.88E-05 | 6.88E-06 | 0.002304754 | AC005225.2 |
| chr15 | 80408419 | 80408478 | 60 | 2 | 3.86E-06 | 0.0008949 | 4.75E-06 | 6.88E-06 | 0.00316141 | ARNT2 |
| chr2 | 241755063 | 241756188 | 1126 | 6 | 1.25E-05 | 1.75E-06 | 0.006515 | 6.88E-06 | 0.001305839 | D2HGDH, AC114730.2 |
| chr1 | 151841303 | 151841595 | 293 | 4 | 1.90E-06 | 7.72E-05 | 0.000112 | 6.89E-06 | 0.003369699 | NA |
| chr9 | 127965733 | 127966215 | 483 | 3 | 3.18E-06 | 6.68E-05 | 0.000246 | 6.89E-06 | 0.004637485 | FAM102A |
| chr4 | 185838902 | 185839224 | 323 | 3 | 3.47E-06 | 4.27E-05 | 1.77E-05 | 6.89E-06 | -0.00238488 | SORBS2 |
| chr2 | 206609861 | 206610237 | 377 | 5 | 6.62E-06 | 2.11E-05 | 8.46E-05 | 6.89E-06 | 0.008361999 | ADAM23 |
| chr11 | 128686487 | 128687370 | 884 | 6 | 2.95E-06 | 3.28E-06 | 0.001642 | 6.89E-06 | -0.00218366 | FLI1, AP001122.1 |
| chr16 | 67282009 | 67282256 | 248 | 2 | 3.13E-06 | 1.67E-05 | 2.56E-05 | 6.90E-06 | -0.00471283 | PLEKHG4 |
| chr2 | 9740821 | 9741880 | 1060 | 5 | 1.19E-06 | 0.0001624 | 3.58E-05 | 6.90E-06 | -0.00069104 | NA |
| chr7 | 36299395 | 36299796 | 402 | 4 | 6.04E-06 | 5.91E-05 | 0.000144 | 6.90E-06 | 0.000393144 | EEPD1 |
| chr17 | 76140677 | 76141992 | 1316 | 10 | 1.26E-05 | 9.39E-05 | 0.000387 | 6.90E-06 | 0.00044577 | RNF157-AS1, FOXJ1 |
| chr1 | 203339912 | 203340304 | 393 | 2 | 4.24E-07 | 0.0019353 | 4.33E-06 | 6.91E-06 | -0.00828043 | NA |
| chr5 | 140338403 | 140338901 | 499 | 2 | 7.38E-07 | 0.000128 | 7.48E-06 | 6.91E-06 | -0.00902545 | HBEGF |
| chr8 | 100008551 | 100008720 | 170 | 3 | 4.54E-06 | 0.0001317 | 7.29E-06 | 6.91E-06 | -0.00833924 | RGS22 |
| chr17 | 3093507 | 3094111 | 605 | 4 | 1.18E-05 | 0.0006084 | 0.000774 | 6.91E-06 | -0.00155241 | OR1D2 |
| chr12 | 111037324 | 111037434 | 111 | 3 | 2.25E-06 | 0.000121 | 2.46E-05 | 6.91E-06 | 0.016637845 | CUX2 |
| chr1 | 1232656 | 1233604 | 949 | 6 | 2.28E-06 | 0.0001593 | 5.15E-05 | 6.91E-06 | -0.00426458 | B3GALT6 |
| chr1 | 46174583 | 46175439 | 857 | 5 | 3.46E-07 | 0.0008641 | 7.81E-06 | 6.91E-06 | -0.0045893 | TSPAN1, P3R3URF-PIK3R3 |
| chr3 | 67990345 | 67990735 | 391 | 3 | 2.19E-05 | 3.28E-06 | 0.001012 | 6.91E-06 | -0.01363144 | NA |
| chr12 | 27710701 | 27711328 | 628 | 8 | 4.99E-05 | 1.97E-06 | 0.010496 | 6.92E-06 | -0.00016227 | MRPS35, AC009509.1 |
| chr17 | 79132363 | 79132541 | 179 | 3 | 2.60E-06 | 4.11E-05 | 3.69E-05 | 6.92E-06 | -0.00866358 | RBFOX3 |
| chr16 | 89095788 | 89096477 | 690 | 5 | 5.86E-06 | 5.49E-05 | 0.001745 | 6.92E-06 | -0.00089815 | ACSF3 |
| chr2 | 32277157 | 32277900 | 744 | 9 | 1.55E-07 | 5.53E-05 | 0.000817 | 6.92E-06 | -0.00223326 | NA |
| chr1 | 3560604 | 3561126 | 523 | 6 | 9.22E-06 | 1.49E-05 | 0.001241 | 6.92E-06 | -0.00380984 | MEGF6, MIR551A |
| chr11 | 19591347 | 19591887 | 541 | 3 | 9.78E-07 | 2.80E-05 | 8.80E-05 | 6.92E-06 | -0.00687697 | NAV2 |
| chr22 | 36814913 | 36815476 | 564 | 4 | 1.47E-06 | 0.0001455 | 1.01E-05 | 6.92E-06 | -0.0156788 | PVALB |
| chr11 | 134036798 | 134037602 | 805 | 7 | 4.49E-06 | 1.88E-05 | 0.000754 | 6.92E-06 | 0.002752805 | LINC02731, AP000911.1 |
| chr2 | 11656660 | 11656891 | 232 | 4 | 3.94E-06 | 0.0023776 | 0.000283 | 6.93E-06 | 0.01343315 | NA |
| chr1 | 212665423 | 212666247 | 825 | 8 | 4.40E-06 | 0.0001166 | 0.000313 | 6.93E-06 | -0.00244603 | LINC02773 |
| chr12 | 80456267 | 80456659 | 393 | 2 | 4.94E-06 | 1.27E-05 | 3.45E-05 | 6.93E-06 | -0.00931186 | PTPRQ |
| chr21 | 14455287 | 14455290 | 4 | 2 | 4.77E-06 | 0.0003666 | 5.63E-06 | 6.94E-06 | -0.00546348 | NA |
| chr10 | 86374199 | 86374696 | 498 | 4 | 5.05E-06 | 1.51E-05 | 0.000214 | 6.94E-06 | -0.00788381 | NA |
| chr11 | 125456074 | 125456551 | 478 | 4 | 4.38E-06 | 1.73E-05 | 0.000313 | 6.95E-06 | -0.00740664 | FEZ1 |
| chr14 | 105525761 | 105526282 | 522 | 3 | 3.44E-06 | 9.45E-06 | 0.000197 | 6.95E-06 | -0.01211866 | NA |
| chr9 | 22147984 | 22148136 | 153 | 2 | 7.08E-06 | 3.51E-05 | 1.43E-05 | 6.96E-06 | 0.005587152 | NA |
| chr16 | 88976763 | 88977299 | 537 | 9 | 1.68E-06 | 0.0002105 | 0.000392 | 6.96E-06 | 0.000499048 | CBFA2T3 |
| chr19 | 7699663 | 7700417 | 755 | 5 | 9.69E-06 | 1.62E-05 | 0.001151 | 6.96E-06 | 0.0017265 | FCER2 |
| chr19 | 55590260 | 55590550 | 291 | 2 | 9.26E-06 | 4.19E-06 | 0.000525 | 6.96E-06 | -0.00996173 | NA |
| chr11 | 119481516 | 119482418 | 903 | 6 | 1.20E-05 | 4.06E-05 | 0.000157 | 6.96E-06 | -0.00592843 | USP2-AS1 |
| chr17 | 44957387 | 44957927 | 541 | 2 | 6.73E-06 | 7.18E-06 | 7.92E-05 | 6.97E-06 | 0.003840781 | NA |
| chr8 | 125091326 | 125092298 | 973 | 10 | 2.17E-06 | 0.0011617 | 0.000561 | 6.97E-06 | 0.001529883 | NSMCE2, WASHC5 |
| chr4 | 113372582 | 113373161 | 580 | 3 | 2.34E-06 | 6.03E-05 | 1.17E-05 | 6.97E-06 | 0.004142645 | ANK2 |
| chr8 | 1865797 | 1865987 | 191 | 4 | 5.34E-06 | 2.63E-06 | 0.001777 | 6.98E-06 | -0.00010082 | AC019257.8, ARHGEF10 |
| chr5 | 14580454 | 14581112 | 659 | 2 | 4.71E-07 | 5.26E-06 | 0.000174 | 6.98E-06 | -0.00176589 | NA |
| chr10 | 127012220 | 127013019 | 800 | 6 | 4.69E-05 | 9.67E-07 | 0.014174 | 6.99E-06 | 0.021806931 | DOCK1, AL359094.1 |
| chr11 | 17772411 | 17772931 | 521 | 3 | 3.15E-06 | 5.02E-06 | 0.000831 | 6.99E-06 | -0.00526608 | KCNC1 |
| chr7 | 102152871 | 102153687 | 817 | 3 | 2.32E-06 | 9.19E-05 | 1.98E-05 | 7.00E-06 | -0.00357 | CUX1, AC005086.2 |
| chr8 | 109691851 | 109692060 | 210 | 3 | 6.25E-05 | 4.38E-06 | 0.000683 | 7.00E-06 | 0.001940696 | NA |
| chr7 | 24406451 | 24406938 | 488 | 2 | 1.71E-06 | 3.26E-05 | 1.52E-05 | 7.01E-06 | -0.01604511 | AC003044.1 |
| chr8 | 48137174 | 48137324 | 151 | 2 | 5.07E-06 | 0.0001042 | 8.18E-06 | 7.01E-06 | -0.00750734 | NA |
| chr10 | 594798 | 595701 | 904 | 7 | 1.07E-05 | 2.44E-05 | 0.002916 | 7.01E-06 | 0.001030703 | DIP2C |
| chr17 | 17222627 | 17223384 | 758 | 5 | 2.12E-06 | 5.44E-05 | 0.001907 | 7.01E-06 | 9.93E-05 | AC055811.2, FLCN |
| chr3 | 11568664 | 11569237 | 574 | 9 | 7.15E-06 | 0.0001375 | 0.000592 | 7.01E-06 | -0.00068163 | VGLL4 |
| chr5 | 74819910 | 74820472 | 563 | 2 | 1.65E-05 | 4.71E-06 | 0.000276 | 7.01E-06 | 0.003852716 | FAM169A |
| chr3 | 77015131 | 77015281 | 151 | 2 | 1.39E-05 | 7.08E-06 | 8.27E-05 | 7.02E-06 | 0.008694662 | ROBO2 |
| chr7 | 70694229 | 70694848 | 620 | 4 | 2.00E-06 | 1.60E-05 | 4.66E-05 | 7.02E-06 | 0.000958877 | AUTS2 |
| chr14 | 101045824 | 101046736 | 913 | 13 | 6.47E-06 | 6.54E-06 | 0.003591 | 7.02E-06 | -0.01019307 | MIR381HG, MIR381, MIR487B |
| chr3 | 120091750 | 120091996 | 247 | 2 | 2.78E-06 | 1.76E-05 | 2.52E-05 | 7.03E-06 | 0.007793851 | GSK3B |
| chr21 | 45643039 | 45644215 | 1177 | 9 | 2.97E-07 | 0.0053154 | 0.000576 | 7.03E-06 | 0.001357504 | PCBP3 |
| chr2 | 206442820 | 206443651 | 832 | 7 | 1.20E-05 | 7.61E-06 | 0.004629 | 7.03E-06 | -0.00023133 | ADAM23 |
| chr11 | 15114513 | 15114665 | 153 | 3 | 6.13E-05 | 1.19E-05 | 0.00049 | 7.03E-06 | -0.00175713 | INSC |
| chr10 | 59176470 | 59177113 | 644 | 11 | 1.67E-05 | 0.0003728 | 0.00082 | 7.03E-06 | 0.00254381 | PHYHIPL |
| chr7 | 157661083 | 157661515 | 433 | 6 | 1.34E-06 | 0.0041775 | 5.83E-05 | 7.04E-06 | -0.00548407 | PTPRN2 |
| chr5 | 1406062 | 1406890 | 829 | 3 | 3.19E-06 | 2.66E-06 | 0.002232 | 7.04E-06 | -0.00011786 | SLC6A3 |
| chr10 | 29881632 | 29881672 | 41 | 2 | 1.35E-05 | 1.04E-05 | 4.53E-05 | 7.04E-06 | 0.003465305 | NA |
| chr9 | 36976475 | 36976737 | 263 | 2 | 8.71E-06 | 1.06E-05 | 4.38E-05 | 7.04E-06 | -0.00371721 | PAX5 |
| chr4 | 146241611 | 146242015 | 405 | 3 | 7.02E-06 | 3.91E-06 | 0.000618 | 7.05E-06 | -0.01018967 | AC097372.1 |
| chr3 | 127002288 | 127002743 | 456 | 3 | 6.75E-06 | 4.83E-05 | 0.000295 | 7.05E-06 | -0.00662565 | PLXNA1 |
| chr8 | 23951334 | 23951737 | 404 | 2 | 1.29E-05 | 5.17E-06 | 0.000194 | 7.06E-06 | -0.00337164 | NA |
| chr5 | 131532210 | 131532521 | 312 | 3 | 1.10E-05 | 1.39E-05 | 0.000247 | 7.06E-06 | -0.01453484 | RAPGEF6, AC008695.1 |
| chr8 | 143786576 | 143787352 | 777 | 2 | 8.77E-05 | 4.26E-06 | 0.000518 | 7.06E-06 | -0.00199187 | IQANK1 |
| chr18 | 79443371 | 79444561 | 1191 | 5 | 3.16E-05 | 0.0002548 | 0.00078 | 7.06E-06 | -0.00085652 | NFATC1 |
| chr16 | 19969489 | 19969771 | 283 | 2 | 1.26E-05 | 5.99E-06 | 0.000122 | 7.07E-06 | -0.00823937 | NA |
| chr15 | 27123616 | 27124068 | 453 | 2 | 3.69E-06 | 1.71E-05 | 2.60E-05 | 7.07E-06 | -0.00361365 | GABRG3, GABRG3-AS1 |
| chr6 | 24666345 | 24667628 | 1284 | 13 | 2.24E-05 | 9.37E-05 | 0.003902 | 7.07E-06 | 0.000668027 | ACOT13, TDP2 |
| chr10 | 132356758 | 132357198 | 441 | 2 | 1.74E-05 | 4.28E-06 | 0.000507 | 7.07E-06 | -0.01552185 | LRRC27 |
| chr9 | 88970055 | 88970195 | 141 | 2 | 1.26E-06 | 1.97E-05 | 2.29E-05 | 7.08E-06 | -0.0026725 | NA |
| chr12 | 68911367 | 68911881 | 515 | 4 | 3.86E-06 | 0.0001215 | 0.000207 | 7.08E-06 | -0.00033142 | CPM |

| | | | | | | | | | | |
|---|---|---|---|---|---|---|---|---|---|---|
| chr11 | 13440235 | 13440716 | 482 | 8 | 8.38E-06 | 6.83E-05 | 0.001743 | 7.08E-06 | -0.00066699 | BTBD10 |
| chr3 | 44058571 | 44058888 | 318 | 2 | 1.04E-05 | 4.52E-06 | 0.000355 | 7.08E-06 | -0.02374744 | NA |
| chr3 | 46558754 | 46559224 | 471 | 2 | 8.27E-07 | 1.23E-05 | 3.69E-05 | 7.08E-06 | -0.02451978 | LRRC2-AS1, LRRC2 |
| chr11 | 110249373 | 110249957 | 585 | 3 | 2.15E-05 | 3.05E-06 | 0.00133 | 7.08E-06 | -0.00670963 | RDX |
| chr13 | 21060806 | 21061984 | 1179 | 12 | 1.81E-07 | 0.000195 | 0.00014 | 7.09E-06 | 0.000287117 | LATS2 |
| chr9 | 129406573 | 129407035 | 463 | 2 | 1.62E-05 | 4.70E-06 | 0.000296 | 7.09E-06 | -0.00337475 | NA |
| chr6 | 20534119 | 20534888 | 770 | 15 | 7.21E-07 | 0.0001979 | 0.000122 | 7.09E-06 | 0.000232071 | CDKAL1 |
| chr1 | 166875814 | 166876789 | 976 | 14 | 2.45E-06 | 0.0001412 | 0.002047 | 7.09E-06 | 5.75E-05 | TADA1 |
| chr6 | 116002779 | 116003384 | 606 | 5 | 1.06E-05 | 0.0020356 | 0.000118 | 7.09E-06 | 0.004173508 | FRK |
| chr2 | 174585346 | 174585458 | 113 | 2 | 2.44E-06 | 6.64E-05 | 1.02E-05 | 7.10E-06 | 0.004023549 | AC010894.2, AC010894.3, WIPF1 |
| chr4 | 103719403 | 103720739 | 1337 | 13 | 2.03E-07 | 0.0002189 | 0.001483 | 7.10E-06 | -0.0013051 | TACR3 |
| chr16 | 88784887 | 88785561 | 675 | 6 | 4.36E-06 | 2.36E-05 | 0.002299 | 7.11E-06 | 0.000955086 | PIEZO1 |
| chr14 | 75660821 | 75661660 | 840 | 11 | 9.16E-06 | 0.0002586 | 0.002598 | 7.11E-06 | 0.000722748 | FLVCR2, TTLL5, ERG28 |
| chr4 | 154332791 | 154333174 | 384 | 5 | 1.24E-05 | 1.50E-05 | 0.001179 | 7.12E-06 | 0.007725889 | DCHS2 |
| chr7 | 53035001 | 53035883 | 883 | 8 | 6.52E-06 | 3.47E-05 | 0.001744 | 7.13E-06 | -0.00291899 | POM121L12 |
| chr20 | 60482119 | 60482407 | 289 | 2 | 2.61E-06 | 0.000136 | 7.57E-06 | 7.13E-06 | -0.01500099 | MIR646HG |
| chr8 | 37851091 | 37851497 | 407 | 5 | 6.48E-06 | 2.80E-05 | 0.000591 | 7.13E-06 | -0.00335977 | NA |
| chr1 | 36156847 | 36157711 | 865 | 5 | 7.16E-06 | 3.90E-05 | 0.000735 | 7.13E-06 | -0.00257702 | MAP7D1 |
| chr16 | 22001083 | 22001635 | 553 | 10 | 9.41E-06 | 0.00014 | 0.001346 | 7.13E-06 | -0.00122157 | PDZD9 |
| chr7 | 137317514 | 137318004 | 491 | 2 | 6.22E-07 | 4.06E-05 | 1.35E-05 | 7.13E-06 | -0.00952554 | PTN |
| chr4 | 153759757 | 153760467 | 711 | 8 | 6.42E-06 | 5.08E-06 | 0.001563 | 7.13E-06 | -0.00100816 | RNF175 |
| chr1 | 94417663 | 94418504 | 842 | 10 | 3.80E-06 | 8.60E-05 | 0.006421 | 7.14E-06 | 0.000687364 | ABCD3, AC118469.2 |
| chr12 | 15214825 | 15215308 | 484 | 3 | 9.00E-06 | 3.03E-06 | 0.001321 | 7.14E-06 | -0.00433855 | RERG |
| chr15 | 94475568 | 94476362 | 795 | 5 | 1.86E-05 | 8.45E-05 | 0.000188 | 7.14E-06 | 0.000545776 | MCTP2, AC009432.2 |
| chr20 | 5545424 | 5546173 | 750 | 2 | 6.72E-05 | 4.34E-06 | 0.000495 | 7.14E-06 | -0.00529248 | GPCPD1 |
| chr8 | 125602592 | 125602776 | 185 | 2 | 1.33E-05 | 4.29E-06 | 0.000547 | 7.15E-06 | -0.00281522 | NA |
| chr9 | 136202006 | 136203031 | 1026 | 5 | 5.94E-05 | 1.45E-05 | 0.000675 | 7.15E-06 | 0.001935136 | LHX3 |
| chr2 | 109377485 | 109377973 | 489 | 2 | 4.32E-06 | 1.16E-05 | 4.05E-05 | 7.15E-06 | 0.000939045 | SH3RF3 |
| chr17 | 1160601 | 1160916 | 316 | 4 | 2.04E-05 | 2.84E-06 | 0.001819 | 7.15E-06 | 0.005063945 | ABR |
| chr16 | 57938735 | 57939985 | 1251 | 3 | 2.74E-07 | 0.0010361 | 1.08E-05 | 7.16E-06 | -0.00635861 | CNGB1 |
| chr12 | 9667502 | 9668243 | 742 | 3 | 2.64E-06 | 1.38E-05 | 0.000441 | 7.16E-06 | -0.01120004 | CLEC2D |
| chr3 | 48415842 | 48416377 | 536 | 2 | 5.41E-06 | 6.49E-06 | 0.000104 | 7.16E-06 | -0.00587389 | PLXNB1 |
| chr15 | 43285937 | 43286012 | 76 | 2 | 4.91E-06 | 0.0002845 | 6.15E-06 | 7.16E-06 | -0.00902838 | TGM7 |
| chr5 | 177445316 | 177446207 | 892 | 4 | 5.04E-07 | 0.000249 | 0.000776 | 7.16E-06 | -0.00758403 | PRR7-AS1 |
| chr16 | 85480141 | 85480372 | 232 | 3 | 6.20E-06 | 6.01E-05 | 1.52E-05 | 7.17E-06 | -0.00929043 | GSE1 |
| chr3 | 14278484 | 14279023 | 540 | 2 | 1.50E-05 | 4.76E-06 | 0.000296 | 7.17E-06 | -0.01010367 | AC093496.1 |
| chr8 | 32711106 | 32711591 | 486 | 3 | 1.50E-06 | 0.0002173 | 1.28E-05 | 7.17E-06 | 0.000768133 | NRG1 |
| chr3 | 155869872 | 155870923 | 1052 | 11 | 5.08E-06 | 0.0002175 | 4.79E-05 | 7.17E-06 | 0.000222068 | GMPS |
| chr1 | 209784256 | 209784405 | 150 | 5 | 2.27E-05 | 4.14E-05 | 2.91E-05 | 7.17E-06 | -0.00046991 | C1orf74 |
| chr2 | 9944776 | 9945089 | 314 | 4 | 1.03E-05 | 3.16E-05 | 0.001658 | 7.17E-06 | -0.00625119 | AC010969.3 |
| chr3 | 112640641 | 112641599 | 959 | 9 | 7.22E-06 | 4.27E-05 | 0.00196 | 7.17E-06 | -0.00920126 | CCDC80 |
| chr5 | 3203062 | 3203961 | 900 | 3 | 7.89E-05 | 6.76E-05 | 0.000242 | 7.18E-06 | -0.00117106 | NA |
| chr9 | 133554403 | 133555213 | 811 | 6 | 1.04E-06 | 0.0003152 | 9.16E-06 | 7.18E-06 | 0.001416308 | ADAMTSL2 |
| chr17 | 75621682 | 75622204 | 523 | 2 | 2.24E-05 | 4.47E-06 | 0.000417 | 7.18E-06 | -0.00104685 | MYO15B |
| chr3 | 50100540 | 50101039 | 500 | 2 | 2.03E-05 | 4.48E-06 | 0.000414 | 7.18E-06 | -0.01199947 | RBM5, RBM5-AS1 |
| chr3 | 139837002 | 139837214 | 213 | 3 | 6.47E-06 | 1.24E-05 | 0.000263 | 7.18E-06 | -0.00576294 | NA |
| chr8 | 142454076 | 142454530 | 455 | 7 | 2.90E-06 | 0.0005214 | 0.000233 | 7.19E-06 | 0.004259418 | ADGRB1 |
| chr11 | 2094133 | 2094343 | 211 | 2 | 6.73E-06 | 1.23E-05 | 3.79E-05 | 7.19E-06 | -0.01019295 | NA |
| chr7 | 23478516 | 23478617 | 102 | 2 | 1.29E-05 | 1.08E-05 | 4.46E-05 | 7.19E-06 | -0.00858989 | NA |
| chr6 | 12289630 | 12290546 | 917 | 12 | 2.21E-06 | 0.0217538 | 0.003435 | 7.19E-06 | 0.000712609 | EDN1 |
| chr20 | 44965798 | 44966594 | 797 | 13 | 8.77E-06 | 5.57E-05 | 0.00689 | 7.19E-06 | 9.71E-05 | STK4, STK4-AS1 |
| chr14 | 105199331 | 105199501 | 171 | 2 | 1.19E-05 | 4.48E-06 | 0.000422 | 7.20E-06 | -0.01460077 | NA |
| chr1 | 153609018 | 153609470 | 453 | 7 | 4.37E-06 | 3.59E-06 | 0.004302 | 7.21E-06 | 0.000127768 | S100A16 |
| chr17 | 29091266 | 29091873 | 608 | 5 | 9.97E-06 | 4.05E-06 | 0.004265 | 7.21E-06 | -0.00074547 | MYO18A |
| chr8 | 32439544 | 32440162 | 619 | 3 | 4.57E-06 | 1.40E-05 | 0.000506 | 7.21E-06 | -0.00392559 | NRG1 |
| chr7 | 55143163 | 55143692 | 530 | 4 | 4.80E-06 | 1.95E-06 | 0.004561 | 7.21E-06 | -0.00601367 | EGFR |
| chr21 | 42692085 | 42692368 | 284 | 2 | 5.40E-06 | 1.26E-05 | 3.69E-05 | 7.21E-06 | -0.00305728 | PDE9A |
| chr10 | 1324193 | 1324220 | 28 | 2 | 1.05E-05 | 1.23E-05 | 3.81E-05 | 7.21E-06 | 0.021102693 | ADARB2 |
| chr2 | 61502273 | 61502296 | 24 | 2 | 1.23E-05 | 1.51E-05 | 3.04E-05 | 7.21E-06 | 0.017994171 | XPO1 |
| chr1 | 2278518 | 2278896 | 379 | 2 | 1.49E-06 | 9.25E-06 | 5.62E-05 | 7.22E-06 | -0.00624136 | SKI |
| chr7 | 128352262 | 128352580 | 319 | 3 | 3.26E-06 | 6.44E-05 | 1.18E-05 | 7.22E-06 | -0.01213817 | PRRT4 |
| chr10 | 128518020 | 128518134 | 115 | 2 | 4.93E-06 | 0.000213 | 6.71E-06 | 7.23E-06 | -0.00704608 | NA |
| chr7 | 101085062 | 101086131 | 1070 | 10 | 6.57E-06 | 4.52E-05 | 0.000225 | 7.23E-06 | 0.000136466 | TRIM56 |
| chr15 | 76311316 | 76312476 | 1161 | 14 | 2.80E-05 | 9.52E-05 | 0.002767 | 7.23E-06 | 0.002789072 | ETFA |
| chr19 | 7903028 | 7903978 | 951 | 10 | 1.76E-05 | 5.26E-06 | 0.007566 | 7.23E-06 | 0.003586141 | MAP2K7, AC010336.1 |
| chr6 | 41940043 | 41941446 | 1404 | 6 | 4.17E-05 | 6.95E-06 | 0.000851 | 7.24E-06 | -0.00156876 | CCND3 |
| chr16 | 51671921 | 51671929 | 9 | 2 | 1.02E-05 | 6.42E-06 | 0.00011 | 7.24E-06 | 0.010078133 | NA |
| chr19 | 39390568 | 39391554 | 987 | 15 | 2.39E-05 | 2.69E-06 | 0.029261 | 7.24E-06 | -0.00018392 | MED29, PAF1 |
| chr4 | 8251508 | 8251980 | 473 | 4 | 8.43E-06 | 7.43E-06 | 0.001361 | 7.25E-06 | -0.00112874 | NA |
| chr17 | 80477055 | 80478161 | 1107 | 6 | 3.48E-07 | 0.0007037 | 2.70E-05 | 7.25E-06 | 0.003557834 | NPTX1 |
| chr11 | 116792762 | 116793088 | 327 | 3 | 4.27E-07 | 0.0001885 | 6.10E-06 | 7.25E-06 | 0.00178049 | NA |
| chr8 | 96645031 | 96646097 | 1067 | 12 | 4.04E-07 | 0.0002386 | 0.000367 | 7.25E-06 | 0.000819437 | CPQ |
| chr17 | 41764959 | 41764979 | 21 | 2 | 7.47E-06 | 2.00E-05 | 2.35E-05 | 7.25E-06 | -0.00644635 | JUP |
| chr2 | 241720009 | 241721309 | 1301 | 7 | 6.16E-06 | 4.44E-05 | 0.000781 | 7.26E-06 | -0.0009541 | ING5 |
| chr5 | 75953474 | 75953732 | 259 | 3 | 5.64E-06 | 0.0002513 | 3.24E-05 | 7.26E-06 | 0.004521421 | NA |
| chr17 | 62379755 | 62380040 | 286 | 3 | 2.03E-05 | 2.76E-06 | 0.002333 | 7.27E-06 | -0.01005345 | EFCAB3 |
| chr1 | 202341696 | 202342364 | 669 | 12 | 1.02E-06 | 0.0030363 | 1.20E-05 | 7.28E-06 | -0.00114208 | UBE2T |
| chr14 | 75051094 | 75051851 | 758 | 8 | 4.22E-05 | 8.32E-06 | 0.003657 | 7.28E-06 | -0.00116888 | MLH3 |
| chr15 | 74369010 | 74369491 | 482 | 4 | 6.30E-06 | 8.20E-06 | 0.001461 | 7.28E-06 | -0.00099948 | AC090826.1 |
| chr2 | 232503405 | 232503833 | 429 | 4 | 1.82E-05 | 9.80E-06 | 0.000682 | 7.29E-06 | 0.00114783 | NA |
| chr5 | 175895459 | 175895628 | 170 | 2 | 4.56E-06 | 3.00E-05 | 1.70E-05 | 7.29E-06 | 0.009573724 | NA |
| chr2 | 1494043 | 1494293 | 251 | 3 | 1.49E-05 | 1.20E-05 | 0.000116 | 7.29E-06 | 0.000914949 | TPO, AC105450.1 |
| chr10 | 1665375 | 1665922 | 548 | 3 | 2.89E-06 | 2.00E-05 | 3.28E-05 | 7.30E-06 | -0.00327439 | ADARB2 |
| chr6 | 159836865 | 159837239 | 375 | 4 | 9.86E-06 | 2.47E-05 | 0.000188 | 7.30E-06 | 0.006546785 | NA |
| chr17 | 72634675 | 72635487 | 813 | 4 | 2.37E-05 | 1.41E-05 | 0.000514 | 7.30E-06 | -0.00313499 | LINC00511 |
| chr19 | 12847524 | 12848099 | 576 | 7 | 1.89E-06 | 0.0003588 | 4.69E-05 | 7.30E-06 | -0.00291113 | MAST1, HOOK2 |
| chr19 | 5341584 | 5342090 | 507 | 2 | 2.38E-06 | 4.43E-06 | 0.000499 | 7.30E-06 | 0.001915841 | NA |
| chr16 | 84864467 | 84864544 | 78 | 2 | 1.39E-05 | 1.33E-05 | 3.55E-05 | 7.30E-06 | -0.00037781 | CRISPLD2 |
| chr1 | 110165739 | 110166431 | 693 | 3 | 7.55E-06 | 4.72E-06 | 0.000906 | 7.30E-06 | -0.00599973 | SLC6A17, SLC6A17-AS1 |
| chr9 | 68584480 | 68585048 | 569 | 3 | 2.13E-05 | 9.44E-06 | 0.000309 | 7.30E-06 | -0.0022812 | TMEM252-DT |
| chr19 | 20039022 | 20039892 | 871 | 11 | 1.96E-05 | 9.52E-06 | 0.001642 | 7.31E-06 | 0.00064927 | ZNF682 |
| chr11 | 1354074 | 1354427 | 354 | 3 | 1.79E-05 | 1.11E-05 | 0.000362 | 7.31E-06 | -0.00874354 | LINC02689 |
| chr2 | 219160008 | 219160878 | 871 | 6 | 2.15E-06 | 9.83E-05 | 0.000154 | 7.31E-06 | 0.00128381 | NHEJ1, AC068946.1 |
| chr8 | 8273078 | 8273555 | 478 | 3 | 1.35E-06 | 0.0001677 | 6.52E-06 | 7.32E-06 | 0.001825045 | NA |
| chr11 | 126112660 | 126112662 | 3 | 2 | 7.70E-06 | 5.94E-06 | 0.000142 | 7.32E-06 | -0.0075842 | NA |
| chr14 | 94026330 | 94026950 | 621 | 7 | 3.52E-06 | 3.78E-06 | 0.006944 | 7.32E-06 | 0.003357334 | OTUB2 |
| chr18 | 54959245 | 54960102 | 858 | 11 | 1.16E-05 | 1.20E-05 | 0.009415 | 7.33E-06 | -0.00114925 | CCDC68 |
| chr10 | 48400514 | 48400909 | 396 | 2 | 4.17E-06 | 7.11E-06 | 9.11E-05 | 7.33E-06 | -0.0007706 | MAPK8 |
| chr3 | 171457924 | 171458227 | 304 | 4 | 2.49E-06 | 2.95E-05 | 0.000398 | 7.33E-06 | 0.005563788 | TNIK |
| chr6 | 131629991 | 131630061 | 71 | 2 | 3.72E-06 | 0.0003183 | 6.12E-06 | 7.33E-06 | -0.00926778 | ENPP3 |
| chr1 | 3598357 | 3598630 | 274 | 4 | 1.74E-05 | 5.71E-06 | 0.000465 | 7.33E-06 | 0.018020058 | MEGF6 |
| chr5 | 1053659 | 1054312 | 654 | 4 | 1.16E-06 | 0.0001153 | 1.63E-05 | 7.33E-06 | -0.0023762 | SLC12A7 |
| chr17 | 79948710 | 79948976 | 267 | 3 | 3.09E-06 | 1.67E-05 | 0.000359 | 7.33E-06 | -0.00517898 | TBC1D16 |
| chr1 | 6094021 | 6094888 | 868 | 6 | 1.47E-06 | 0.001571 | 0.000534 | 7.33E-06 | -0.00257863 | KCNAB2 |
| chr2 | 121331039 | 121331085 | 47 | 2 | 3.64E-06 | 0.0031853 | 4.38E-06 | 7.34E-06 | 0.003574252 | NA |
| chr6 | 167094080 | 167094682 | 603 | 6 | 3.71E-05 | 1.87E-06 | 0.0083 | 7.34E-06 | 0.005347339 | CEP43, Z94721.2 |
| chr8 | 139730793 | 139731220 | 428 | 2 | 9.94E-07 | 3.46E-05 | 1.55E-05 | 7.34E-06 | -0.00103633 | TRAPPC9 |
| chr8 | 140527035 | 140527354 | 320 | 3 | 1.94E-06 | 1.60E-05 | 4.17E-05 | 7.34E-06 | -0.00184646 | AGO2 |
| chr1 | 188217774 | 188217783 | 10 | 2 | 8.01E-06 | 7.59E-06 | 8.12E-05 | 7.35E-06 | 0.002857047 | NA |
| chr17 | 82596094 | 82596896 | 803 | 6 | 1.69E-06 | 1.40E-05 | 0.000473 | 7.35E-06 | -0.00786272 | FOXK2 |
| chr11 | 86951926 | 86952376 | 451 | 5 | 1.62E-06 | 0.0005914 | 0.000191 | 7.35E-06 | -0.00388921 | PRSS23, FZD4 |
| chr4 | 212329 | 213088 | 760 | 6 | 1.47E-05 | 5.87E-06 | 0.000585 | 7.36E-06 | 0.002958949 | ZNF876P |
| chr13 | 81226986 | 81227640 | 655 | 3 | 1.22E-06 | 0.0001097 | 2.92E-05 | 7.36E-06 | -0.00113681 | NA |
| chr4 | 1608666 | 1609045 | 380 | 2 | 3.11E-06 | 3.16E-05 | 1.66E-05 | 7.36E-06 | -0.00442641 | NA |
| chr2 | 144319900 | 144320008 | 109 | 2 | 3.05E-06 | 5.50E-06 | 0.000187 | 7.36E-06 | -0.00663061 | GTDC1 |
| chr11 | 61710361 | 61710930 | 570 | 2 | 4.51E-06 | 5.04E-06 | 0.000263 | 7.37E-06 | 0.000629642 | DAGLA |
| chr17 | 7351718 | 7352417 | 700 | 6 | 1.13E-05 | 5.73E-06 | 0.0019 | 7.37E-06 | -0.00461431 | KCTD11 |
| chr10 | 71323466 | 71324458 | 993 | 4 | 6.02E-07 | 2.45E-05 | 3.35E-05 | 7.37E-06 | 0.003242693 | SLC29A3 |
| chr17 | 74479992 | 74480489 | 498 | 3 | 1.47E-05 | 3.18E-06 | 0.001748 | 7.37E-06 | -0.0059628 | CD300A |
| chr2 | 205719101 | 205719329 | 229 | 2 | 5.63E-06 | 3.76E-05 | 1.47E-05 | 7.37E-06 | 0.004130988 | NRP2 |
| chr1 | 244788967 | 244789092 | 126 | 3 | 1.55E-05 | 1.01E-05 | 0.000107 | 7.37E-06 | -0.00763737 | NA |
| chr20 | 44626506 | 44627655 | 1150 | 5 | 8.08E-05 | 0.0002304 | 5.07E-05 | 7.38E-06 | -0.00333928 | ADA |
| chr5 | 75381652 | 75381744 | 93 | 2 | 1.94E-05 | 4.65E-06 | 0.000398 | 7.38E-06 | -0.01619623 | CERT1 |
| chr10 | 84244525 | 84245365 | 841 | 8 | 5.25E-06 | 0.0001043 | 0.002838 | 7.38E-06 | -0.00589642 | RGR |
| chr3 | 9447116 | 9448321 | 1206 | 7 | 8.63E-06 | 1.01E-05 | 0.001715 | 7.38E-06 | -0.00034019 | SETD5 |
| chr6 | 131135629 | 131136366 | 738 | 6 | 1.28E-05 | 5.54E-05 | 0.001245 | 7.38E-06 | -0.00393704 | AKAP7 |
| chr1 | 51576360 | 51577000 | 641 | 2 | 2.28E-05 | 4.41E-06 | 0.000574 | 7.38E-06 | 0.001233926 | CALR4P |
| chr17 | 73584332 | 73584924 | 593 | 3 | 1.36E-05 | 3.59E-06 | 0.001394 | 7.39E-06 | -0.0048621 | SDK2 |
| chr8 | 123206837 | 123207485 | 649 | 5 | 3.70E-06 | 1.99E-05 | 0.001575 | 7.39E-06 | 0.016001819 | FAM83A |
| chr20 | 31368195 | 31368527 | 333 | 4 | 5.66E-06 | 2.19E-05 | 0.00015 | 7.39E-06 | 0.010349631 | NA |
| chr12 | 113389074 | 113389361 | 288 | 4 | 2.03E-05 | 3.00E-06 | 0.001562 | 7.39E-06 | -0.00166816 | PLBD2 |
| chr12 | 128117712 | 128118857 | 1146 | 10 | 8.77E-07 | 0.0002708 | 2.67E-05 | 7.39E-06 | -0.00828853 | LINC02368, LINC02369 |
| chr11 | 132712593 | 132712913 | 321 | 4 | 7.13E-06 | 4.87E-05 | 4.56E-05 | 7.40E-06 | -0.00396667 | OPCML |
| chr5 | 93092045 | 93092306 | 262 | 2 | 9.98E-06 | 1.00E-05 | 5.22E-05 | 7.40E-06 | -0.00384723 | NA |
| chr1 | 212558327 | 212559114 | 788 | 4 | 4.64E-06 | 7.81E-06 | 0.000954 | 7.40E-06 | -0.00335599 | AC092803.1 |
| chr5 | 134568542 | 134568984 | 443 | 3 | 1.58E-06 | 0.0002448 | 4.74E-05 | 7.40E-06 | -0.00119447 | JADE2 |
| chr10 | 114763752 | 114764466 | 715 | 3 | 1.45E-06 | 8.13E-05 | 1.57E-05 | 7.40E-06 | -0.00452833 | ABLIM1 |
| chr16 | 3500662 | 3501320 | 659 | 11 | 4.34E-07 | 0.0005075 | 0.00026 | 7.40E-06 | -0.00384276 | AC004224.2, CLUAP1 |
| chr21 | 24694611 | 24694990 | 380 | 2 | 9.24E-06 | 7.61E-06 | 8.25E-05 | 7.40E-06 | -0.00182611 | NA |
| chr11 | 67400355 | 67401146 | 792 | 3 | 2.24E-06 | 5.11E-06 | 0.000301 | 7.40E-06 | -0.0008144 | PPP1CA |
| chr12 | 14614971 | 14615045 | 75 | 2 | 1.30E-05 | 4.47E-06 | 0.000526 | 7.40E-06 | 0.002638515 | PLBD1-AS1, GUCY2C |
| chr4 | 24980073 | 24980551 | 479 | 11 | 5.83E-06 | 0.0001092 | 0.001109 | 7.41E-06 | -0.0014213 | CCDC149 |
| chr7 | 44606342 | 44606618 | 277 | 7 | 0.000131367 | 5.10E-06 | 0.003519 | 7.41E-06 | -0.00048754 | OGDH |
| chr20 | 50009478 | 50009900 | 423 | 2 | 1.10E-06 | 1.35E-05 | 3.60E-05 | 7.41E-06 | 0.001225858 | NA |
| chr16 | 16190158 | 16190229 | 72 | 2 | 1.88E-06 | 0.0002376 | 6.68E-06 | 7.41E-06 | -0.00414954 | ABCC6 |
| chr14 | 99270050 | 99270641 | 592 | 5 | 2.97E-05 | 2.91E-05 | 0.000922 | 7.41E-06 | 0.003301343 | BCL11B |
| chr10 | 59904869 | 59905799 | 931 | 4 | 3.57E-05 | 8.86E-05 | 0.000337 | 7.41E-06 | 0.008085423 | CCDC6 |
| chr15 | 98946832 | 98947169 | 338 | 2 | 4.20E-06 | 2.12E-05 | 2.31E-05 | 7.42E-06 | -0.00342757 | IGF1R |
| chr9 | 100104471 | 100104505 | 35 | 2 | 5.27E-06 | 8.60E-05 | 9.47E-06 | 7.42E-06 | 0.003236173 | INVS |
| chr2 | 238912746 | 238913435 | 690 | 2 | 4.37E-05 | 4.52E-06 | 0.000495 | 7.42E-06 | -0.00108188 | NA |
| chr5 | 74766856 | 74767549 | 694 | 15 | 6.35E-07 | 0.000896 | 0.001333 | 7.43E-06 | 0.000433476 | NSA2, GFM2 |
| chr7 | 87600236 | 87601028 | 793 | 10 | 3.36E-06 | 1.47E-05 | 0.001354 | 7.43E-06 | 0.000308697 | ABCB1 |
| chr1 | 22563228 | 22563555 | 328 | 2 | 4.99E-06 | 8.03E-06 | 7.48E-05 | 7.43E-06 | -0.00049642 | EPHA8 |
| chr20 | 17626884 | 17626912 | 29 | 2 | 1.48E-05 | 1.02E-05 | 5.18E-05 | 7.44E-06 | -0.01901168 | RRBP1 |
| chr5 | 141210018 | 141210041 | 24 | 2 | 2.80E-06 | 2.17E-05 | 2.26E-05 | 7.44E-06 | -0.02846891 | PCDHB12, AC244517.11 |
| chr20 | 31900181 | 31900685 | 505 | 3 | 2.31E-06 | 3.17E-06 | 0.001484 | 7.44E-06 | 0.005328103 | TTLL9 |
| chr9 | 135260697 | 135260919 | 223 | 3 | 1.39E-05 | 5.52E-06 | 0.000799 | 7.45E-06 | -0.02101494 | NA |
| chr20 | 63238061 | 63238185 | 125 | 3 | 1.59E-05 | 3.48E-06 | 0.001565 | 7.45E-06 | 0.016496462 | BIRC7 |

| | | | | | | | | | | |
|---|---|---|---|---|---|---|---|---|---|---|
| chr1 | 1955426 | 1955832 | 407 | 3 | 1.17E-06 | 3.98E-05 | 6.30E-05 | 7.45E-06 | -0.00416916 | CFAP74 |
| chr10 | 128476172 | 128476644 | 473 | 4 | 2.18E-06 | 0.003181 | 8.48E-06 | 7.45E-06 | 0.004933679 | NA |
| chr12 | 104456441 | 104457394 | 954 | 13 | 7.46E-06 | 2.05E-05 | 0.003937 | 7.46E-06 | 0.000177648 | CHST11 |
| chr4 | 8724905 | 8725297 | 393 | 2 | 6.02E-06 | 6.52E-06 | 0.000117 | 7.47E-06 | -0.03055825 | NA |
| chr22 | 20135700 | 20135822 | 123 | 2 | 1.54E-05 | 7.61E-06 | 8.42E-05 | 7.47E-06 | -0.00084937 | ZDHHC8 |
| chr13 | 99606270 | 99606700 | 431 | 8 | 2.70E-05 | 9.33E-05 | 0.001701 | 7.47E-06 | -0.00014221 | CLYBL |
| chr19 | 2511111 | 2511477 | 367 | 2 | 1.94E-05 | 4.62E-06 | 0.000454 | 7.47E-06 | -0.00664826 | GNG7 |
| chr16 | 68363887 | 68364630 | 744 | 4 | 3.92E-06 | 1.61E-05 | 0.000275 | 7.47E-06 | -0.00452043 | SMPD3 |
| chr3 | 69081419 | 69081767 | 349 | 3 | 8.78E-07 | 0.0002605 | 7.53E-06 | 7.48E-06 | -0.00878164 | NA |
| chr9 | 122913397 | 122914568 | 1172 | 5 | 1.83E-07 | 2.80E-05 | 3.91E-05 | 7.49E-06 | -0.00814699 | NA |
| chr9 | 33249250 | 33249548 | 299 | 2 | 4.92E-06 | 6.97E-06 | 0.000101 | 7.49E-06 | -0.01700093 | BAG1 |
| chr7 | 156963011 | 156964164 | 1154 | 8 | 3.40E-06 | 0.0002485 | 0.00056 | 7.50E-06 | 7.03E-05 | NOM1 |
| chr20 | 2477036 | 2477460 | 425 | 2 | 8.09E-06 | 8.11E-06 | 7.50E-05 | 7.50E-06 | 0.000831644 | AL049650.1 |
| chr16 | 89371794 | 89372484 | 691 | 7 | 7.25E-08 | 0.0006059 | 2.52E-05 | 7.50E-06 | -0.00279556 | ANKRD11 |
| chr17 | 76710469 | 76711403 | 935 | 6 | 6.09E-05 | 5.95E-05 | 0.002288 | 7.50E-06 | -0.00277725 | MXRA7 |
| chr13 | 35715094 | 35715256 | 163 | 2 | 3.08E-06 | 4.81E-05 | 1.30E-05 | 7.50E-06 | -0.00198185 | NA |
| chr13 | 52129192 | 52129704 | 513 | 8 | 8.35E-06 | 2.46E-05 | 0.001811 | 7.51E-06 | 0.000573168 | AL139082.1 |
| chr1 | 9181556 | 9182281 | 726 | 6 | 1.99E-06 | 5.67E-05 | 0.000187 | 7.51E-06 | 0.005627748 | LNCTAM34A, MIR34AHG |
| chr8 | 143829501 | 143830652 | 1152 | 8 | 1.23E-06 | 0.0001449 | 3.12E-05 | 7.51E-06 | 0.000480723 | AC234917.3 |
| chr6 | 96562195 | 96563027 | 833 | 3 | 5.93E-08 | 0.001027 | 5.08E-06 | 7.51E-06 | -0.0102065 | FHL5 |
| chr1 | 155071085 | 155071869 | 785 | 6 | 5.53E-06 | 2.15E-05 | 0.004832 | 7.52E-06 | -0.00608912 | AL691442.1 |
| chr12 | 26051642 | 26052125 | 484 | 3 | 6.44E-06 | 1.05E-05 | 0.000148 | 7.52E-06 | -0.00879965 | RASSF8 |
| chr17 | 40071387 | 40071545 | 159 | 2 | 4.59E-06 | 8.13E-06 | 7.53E-05 | 7.52E-06 | 0.011795947 | THRA |
| chr9 | 37603184 | 37603218 | 35 | 2 | 1.99E-06 | 0.0028258 | 4.53E-06 | 7.52E-06 | -0.00690515 | AL138752.2 |
| chr4 | 36158838 | 36159818 | 981 | 5 | 3.83E-06 | 6.74E-05 | 0.000432 | 7.52E-06 | -0.00772188 | ARAP2 |
| chr5 | 174383595 | 174383890 | 296 | 2 | 6.25E-06 | 1.08E-05 | 4.88E-05 | 7.53E-06 | -0.00973094 | LINC01411 |
| chr1 | 165444952 | 165445933 | 982 | 15 | 3.22E-05 | 0.0001477 | 0.01608 | 7.53E-06 | -0.00203695 | RXRG |
| chr19 | 37362727 | 37362928 | 202 | 2 | 1.95E-05 | 4.65E-06 | 0.000461 | 7.53E-06 | 0.005754292 | ZNF875 |
| chr19 | 46377759 | 46378176 | 418 | 2 | 1.37E-05 | 5.57E-06 | 0.000196 | 7.54E-06 | -0.01968801 | PPP5C |
| chr4 | 16508100 | 16508612 | 513 | 2 | 9.71E-06 | 5.52E-06 | 0.000204 | 7.54E-06 | -0.0020521 | AC097515.1, LDB2 |
| chr21 | 45379756 | 45379871 | 116 | 3 | 6.92E-06 | 0.0001393 | 3.59E-05 | 7.54E-06 | -0.01143306 | NA |
| chr6 | 32953744 | 32954028 | 285 | 3 | 1.08E-06 | 1.25E-05 | 0.000565 | 7.55E-06 | 0.019215603 | HLA-DMA |
| chr11 | 10450126 | 10450918 | 793 | 9 | 5.72E-06 | 3.35E-05 | 0.00249 | 7.55E-06 | 0.00089421 | AMPD3 |
| chr10 | 35167135 | 35167544 | 410 | 3 | 1.55E-05 | 2.85E-06 | 0.002451 | 7.55E-06 | 0.005835364 | CREM |
| chr14 | 93206377 | 93207075 | 699 | 8 | 5.67E-05 | 4.78E-05 | 0.00123 | 7.55E-06 | -0.00161473 | AL110118.1, GON7 |
| chr7 | 134365650 | 134365826 | 177 | 3 | 5.32E-06 | 0.0051405 | 1.64E-05 | 7.55E-06 | -0.0093385 | NA |
| chr8 | 10725816 | 10726317 | 502 | 2 | 2.62E-05 | 4.81E-06 | 0.000382 | 7.56E-06 | -0.01014299 | SOX7, AC105001.2 |
| chr22 | 45974764 | 45975272 | 509 | 2 | 4.60E-08 | 0.0069719 | 4.29E-06 | 7.56E-06 | 0.008719689 | WNT7B |
| chr16 | 81891367 | 81891527 | 161 | 4 | 5.84E-06 | 0.0004828 | 0.000145 | 7.56E-06 | -0.00083092 | PLCG2 |
| chr1 | 85404501 | 85404970 | 470 | 5 | 1.09E-05 | 2.13E-05 | 0.000711 | 7.56E-06 | -0.00146769 | AL078459.1, DDAH1 |
| chr4 | 153448655 | 153448682 | 28 | 2 | 4.91E-06 | 0.0002017 | 7.13E-06 | 7.56E-06 | 0.006237227 | NA |
| chr10 | 14073689 | 14073715 | 27 | 2 | 2.32E-06 | 5.43E-05 | 1.22E-05 | 7.57E-06 | -0.00782357 | FRMD4A |
| chr3 | 129002401 | 129002885 | 485 | 2 | 1.28E-05 | 4.53E-06 | 0.000571 | 7.57E-06 | -0.01335349 | EFCC1, CFAP92 |
| chr16 | 69132563 | 69133448 | 886 | 10 | 1.04E-06 | 3.46E-05 | 0.000131 | 7.57E-06 | -0.00037793 | UTP4, CHTF8 |
| chr2 | 64019700 | 64019896 | 197 | 4 | 5.24E-06 | 0.0011428 | 0.000165 | 7.58E-06 | -0.00033302 | NA |
| chr1 | 9355157 | 9356120 | 964 | 6 | 1.45E-07 | 0.0017145 | 3.96E-05 | 7.58E-06 | 0.001513576 | SPSB1 |
| chr20 | 45469076 | 45469718 | 643 | 7 | 5.67E-05 | 9.55E-06 | 0.003752 | 7.58E-06 | -0.00264744 | NA |
| chr18 | 58065249 | 58065394 | 146 | 2 | 7.96E-06 | 4.56E-06 | 0.000549 | 7.58E-06 | 0.005979226 | NEDD4L |
| chr19 | 53197176 | 53197735 | 560 | 3 | 7.03E-07 | 6.94E-05 | 0.000313 | 7.58E-06 | -0.07250618 | AC092070.2 |
| chr6 | 31403326 | 31404043 | 718 | 20 | 4.51E-06 | 1.79E-05 | 0.001829 | 7.58E-06 | 0.000711702 | MICA, AL645933.3 |
| chr6 | 33784199 | 33784750 | 552 | 2 | 8.19E-06 | 5.02E-06 | 0.000316 | 7.58E-06 | -0.00558113 | LEMD2 |
| chr4 | 7708450 | 7708636 | 187 | 3 | 1.15E-06 | 0.0009431 | 7.13E-06 | 7.58E-06 | 0.001403888 | SORCS2 |
| chr6 | 36986036 | 36987150 | 1115 | 15 | 1.77E-05 | 2.27E-06 | 0.026821 | 7.59E-06 | -0.00115813 | MTCH1 |
| chr10 | 606950 | 607627 | 678 | 4 | 5.20E-06 | 0.0006542 | 9.04E-05 | 7.59E-06 | -0.00488911 | DIP2C |
| chr15 | 50765220 | 50765944 | 725 | 10 | 1.24E-05 | 9.82E-06 | 0.007085 | 7.60E-06 | -0.00028616 | SPPL2A |
| chr7 | 44210403 | 44210888 | 486 | 3 | 2.47E-07 | 0.0406763 | 1.37E-06 | 7.60E-06 | 0.004164432 | YKT6 |
| chr19 | 24004392 | 24004756 | 365 | 4 | 2.05E-05 | 3.39E-05 | 0.000834 | 7.60E-06 | -0.0040028 | RNA5-8SP4 |
| chr7 | 149375438 | 149375890 | 453 | 2 | 1.67E-05 | 5.14E-06 | 0.000285 | 7.60E-06 | -0.0127648 | NA |
| chr17 | 39612530 | 39612551 | 22 | 2 | 4.78E-06 | 1.48E-05 | 3.40E-05 | 7.61E-06 | -0.00897905 | NA |
| chr5 | 50711149 | 50711545 | 397 | 2 | 7.44E-06 | 9.57E-06 | 5.93E-05 | 7.61E-06 | 0.001713638 | PARP8 |
| chr7 | 12570758 | 12571611 | 854 | 9 | 5.63E-07 | 1.06E-05 | 0.001348 | 7.61E-06 | -0.00640981 | SCIN, AC005281.1 |
| chr21 | 39984635 | 39985037 | 403 | 2 | 1.43E-05 | 5.53E-06 | 0.00021 | 7.61E-06 | 0.006268266 | NA |
| chr1 | 162671215 | 162671829 | 615 | 2 | 2.77E-05 | 4.57E-06 | 0.000568 | 7.62E-06 | -0.00754317 | DDR2 |
| chr6 | 163415213 | 163415686 | 474 | 2 | 1.06E-06 | 5.55E-06 | 0.000209 | 7.62E-06 | 0.000389188 | QKI |
| chr3 | 12158458 | 12159167 | 710 | 5 | 1.04E-05 | 5.90E-06 | 0.000567 | 7.62E-06 | -0.01252444 | SYN2, TIMP4 |
| chr12 | 95790385 | 95791286 | 902 | 10 | 6.36E-06 | 5.73E-05 | 0.001875 | 7.63E-06 | 0.000942103 | NTN4 |
| chr22 | 29602682 | 29603969 | 1288 | 16 | 2.56E-06 | 1.79E-05 | 7.17E-05 | 7.63E-06 | 0.000530596 | NF2 |
| chr1 | 91884873 | 91886084 | 1212 | 7 | 2.05E-06 | 3.38E-05 | 0.001404 | 7.63E-06 | -0.0016244 | TGFBR3 |
| chr2 | 108533266 | 108533875 | 610 | 3 | 1.75E-06 | 1.47E-05 | 0.000267 | 7.63E-06 | -0.0042481 | GCC2-AS1 |
| chr2 | 47403008 | 47404006 | 999 | 13 | 1.50E-06 | 6.51E-06 | 0.002911 | 7.63E-06 | 0.000824043 | MSH2 |
| chr5 | 177050714 | 177051046 | 333 | 3 | 6.59E-06 | 1.19E-05 | 6.49E-05 | 7.63E-06 | 0.000296348 | ZNF346 |
| chr15 | 45714651 | 45714728 | 78 | 2 | 9.50E-06 | 1.07E-05 | 5.08E-05 | 7.63E-06 | -0.01740065 | AC068722.1 |
| chr7 | 139415571 | 139415897 | 327 | 2 | 1.28E-05 | 4.59E-06 | 0.000558 | 7.63E-06 | -0.00739527 | LUC7L2, FMC1-LUC7L2 |
| chr11 | 62728858 | 62729328 | 471 | 4 | 3.68E-06 | 1.42E-05 | 7.68E-05 | 7.64E-06 | 0.002664659 | TTC9C |
| chr8 | 1512050 | 1512552 | 503 | 6 | 3.12E-05 | 1.92E-06 | 0.007345 | 7.64E-06 | -0.00267438 | DLGAP2 |
| chr6 | 28542504 | 28542963 | 460 | 13 | 1.89E-06 | 4.39E-05 | 1.36E-05 | 7.65E-06 | 0.000234092 | NA |
| chr12 | 111098860 | 111100038 | 1179 | 8 | 1.95E-07 | 0.0016832 | 7.77E-06 | 7.65E-06 | 0.001475785 | CUX2 |
| chr3 | 113156684 | 113157088 | 405 | 2 | 2.05E-06 | 1.36E-05 | 3.76E-05 | 7.65E-06 | -0.0127413 | AC078785.1, LINC02044 |
| chr11 | 78436486 | 78436752 | 267 | 2 | 5.14E-06 | 4.03E-05 | 1.48E-05 | 7.66E-06 | 0.004690095 | AP003086.1, NARS2 |
| chr1 | 153827814 | 153828223 | 410 | 2 | 1.24E-06 | 0.0002088 | 7.15E-06 | 7.66E-06 | 0.01208298 | GATAD2B |
| chr22 | 44312789 | 44313734 | 946 | 8 | 1.78E-06 | 7.25E-05 | 0.000155 | 7.66E-06 | -0.00394096 | SHISAL1 |
| chr19 | 7476874 | 7477305 | 432 | 4 | 3.72E-07 | 0.0001375 | 1.23E-05 | 7.66E-06 | -0.00560433 | PEX11G |
| chr14 | 66110404 | 66110796 | 393 | 2 | 4.16E-06 | 5.87E-06 | 0.000175 | 7.66E-06 | -0.00331244 | NA |
| chr7 | 135211475 | 135212228 | 754 | 12 | 2.85E-05 | 8.07E-05 | 0.001386 | 7.67E-06 | 0.003187183 | WDR91 |
| chr1 | 25922063 | 25922406 | 344 | 2 | 3.65E-07 | 9.85E-05 | 9.31E-06 | 7.68E-06 | 0.0071653 | NA |
| chr6 | 118285194 | 118285242 | 49 | 2 | 1.88E-05 | 5.62E-06 | 0.000206 | 7.68E-06 | 0.001133799 | SLC35F1 |
| chr5 | 179802932 | 179803096 | 165 | 4 | 7.60E-07 | 0.0001748 | 4.04E-05 | 7.70E-06 | -0.00768725 | MGAT4B |
| chr5 | 138337212 | 138338213 | 1002 | 12 | 4.25E-06 | 4.70E-05 | 0.00081 | 7.70E-06 | -0.00033147 | FAM53C, CDC25C |
| chr1 | 15876221 | 15876607 | 387 | 6 | 9.50E-07 | 0.0039766 | 1.50E-06 | 7.70E-06 | -0.00195612 | SPEN |
| chr9 | 87727320 | 87727723 | 404 | 2 | 2.08E-06 | 5.47E-06 | 0.000233 | 7.70E-06 | -0.00455811 | CTSL |
| chr2 | 232662228 | 232662717 | 490 | 5 | 2.86E-05 | 5.48E-06 | 0.001851 | 7.70E-06 | -0.00313639 | EFHD1 |
| chr11 | 120710572 | 120710646 | 75 | 2 | 3.69E-06 | 0.0036594 | 4.54E-06 | 7.71E-06 | -0.01376012 | GRIK4 |
| chr1 | 20488407 | 20488420 | 14 | 2 | 1.08E-05 | 7.68E-06 | 8.93E-05 | 7.71E-06 | -0.02415899 | NA |
| chr3 | 183318448 | 183319053 | 606 | 3 | 2.81E-06 | 0.0002649 | 1.11E-05 | 7.72E-06 | -0.00707918 | MCF2L2 |
| chr2 | 127024659 | 127025237 | 579 | 6 | 1.93E-05 | 2.59E-06 | 0.008264 | 7.73E-06 | -0.01272957 | AC012508.3, AC012508.1, AC012508.2 |
| chr10 | 13666083 | 13666265 | 183 | 8 | 2.45E-05 | 8.41E-06 | 0.007362 | 7.73E-06 | 0.001902367 | AL157392.3, FRMD4A |
| chr11 | 104163330 | 104164254 | 925 | 6 | 2.22E-06 | 0.001271 | 0.000399 | 7.73E-06 | -0.00096229 | PDGFD |
| chr1 | 51329565 | 51330709 | 1145 | 5 | 3.42E-09 | 0.0001997 | 2.35E-05 | 7.73E-06 | 0.003144098 | TTC39A-AS1, TTC39A |
| chr16 | 87303784 | 87304526 | 743 | 2 | 8.56E-07 | 1.50E-05 | 3.44E-05 | 7.73E-06 | -0.00941081 | C16orf95, AC010531.1 |
| chr2 | 232292892 | 232293122 | 231 | 2 | 8.70E-06 | 1.78E-05 | 2.88E-05 | 7.73E-06 | -0.01160205 | DIS3L2 |
| chr1 | 220689921 | 220690424 | 504 | 10 | 3.08E-05 | 1.03E-05 | 0.002401 | 7.73E-06 | -0.00026308 | C1orf115 |
| chr11 | 55819268 | 55819628 | 361 | 4 | 1.60E-06 | 0.0011895 | 2.12E-05 | 7.73E-06 | 0.002648545 | OR5D18 |
| chr15 | 71817389 | 71817998 | 610 | 4 | 7.45E-06 | 0.0001437 | 0.000158 | 7.74E-06 | 0.000874495 | NR2E3 |
| chr12 | 62145711 | 62146513 | 803 | 5 | 3.41E-07 | 0.0024723 | 1.74E-06 | 7.74E-06 | -0.00128436 | TAFA2 |
| chr15 | 74265186 | 74265677 | 492 | 3 | 9.23E-08 | 0.0004818 | 4.85E-06 | 7.74E-06 | 0.00471253 | CCDC33 |
| chr3 | 41955406 | 41955615 | 210 | 3 | 1.39E-05 | 1.55E-05 | 0.000111 | 7.75E-06 | -0.00369595 | RPL36P20, ULK4 |
| chr9 | 5773894 | 5774228 | 335 | 2 | 3.45E-06 | 4.94E-05 | 1.33E-05 | 7.75E-06 | 0.003584103 | RIC1, ERMP1 |
| chr6 | 43181303 | 43182465 | 1163 | 9 | 1.14E-06 | 0.0003439 | 2.52E-05 | 7.75E-06 | -0.0036131 | SRF, CUL9 |
| chr4 | 10054882 | 10055083 | 202 | 3 | 3.08E-06 | 8.08E-05 | 2.77E-05 | 7.75E-06 | -0.00435032 | SLC2A9 |
| chr21 | 45302668 | 45303252 | 585 | 3 | 1.46E-06 | 0.0003145 | 1.53E-05 | 7.75E-06 | -0.00656295 | BX322557.2, LINC00315 |
| chr19 | 19642937 | 19644100 | 1164 | 11 | 1.10E-06 | 0.0001489 | 0.005926 | 7.76E-06 | -0.00065324 | GMIP |
| chr8 | 38995801 | 38996910 | 1110 | 15 | 9.44E-06 | 5.21E-06 | 0.011597 | 7.76E-06 | 0.000118395 | ADAM9, TM2D2 |
| chr17 | 7042191 | 7043208 | 1018 | 5 | 3.31E-06 | 1.05E-05 | 0.002093 | 7.76E-06 | -0.00234111 | SLC16A11 |
| chr12 | 25655831 | 25656070 | 240 | 2 | 1.66E-05 | 5.77E-06 | 0.000196 | 7.76E-06 | -0.00749691 | NA |
| chr12 | 108302532 | 108303165 | 634 | 4 | 1.53E-05 | 5.83E-06 | 0.001734 | 7.76E-06 | -0.00717506 | CMKLR1 |
| chr11 | 120236290 | 120237121 | 832 | 3 | 2.41E-07 | 2.73E-05 | 3.23E-05 | 7.77E-06 | -0.00809302 | POU2F3 |
| chr11 | 126228943 | 126229391 | 449 | 2 | 4.57E-07 | 0.0016684 | 4.92E-06 | 7.77E-06 | -0.0052087 | FAM118B |
| chr2 | 238140909 | 238141447 | 539 | 3 | 2.33E-06 | 2.79E-05 | 4.66E-05 | 7.77E-06 | 0.000587872 | KLHL30 |
| chr10 | 48461670 | 48462509 | 840 | 2 | 7.99E-08 | 0.0003702 | 6.29E-06 | 7.77E-06 | -0.00515843 | ARHGAP22 |
| chr2 | 121754353 | 121754405 | 53 | 3 | 8.78E-06 | 1.45E-05 | 0.000154 | 7.77E-06 | -0.00297802 | TSN |
| chr17 | 289501 | 290038 | 538 | 3 | 8.04E-07 | 8.31E-05 | 1.97E-05 | 7.77E-06 | -0.00761302 | RPH3AL |
| chr17 | 83063543 | 83063894 | 352 | 5 | 3.43E-06 | 5.44E-05 | 0.000433 | 7.77E-06 | -0.00137874 | NA |
| chr8 | 743891 | 744857 | 967 | 4 | 4.23E-07 | 7.71E-05 | 1.79E-05 | 7.78E-06 | -0.0016231 | DLGAP2 |
| chr8 | 47959150 | 47960066 | 917 | 6 | 3.91E-06 | 3.51E-06 | 0.004725 | 7.78E-06 | -0.00528717 | PRKDC |
| chr4 | 102868764 | 102869537 | 774 | 8 | 5.11E-06 | 3.09E-05 | 0.003885 | 7.78E-06 | 0.000449914 | CISD2, UBE2D3 |
| chr15 | 45175419 | 45175430 | 12 | 2 | 1.76E-05 | 6.52E-06 | 0.000134 | 7.78E-06 | -0.0062115 | AC051619.3, SHF |
| chr9 | 133928418 | 133928425 | 8 | 2 | 1.17E-05 | 5.83E-06 | 0.000191 | 7.79E-06 | -0.00240869 | VAV2 |
| chr11 | 68457896 | 68458275 | 380 | 2 | 1.03E-06 | 0.0001406 | 8.25E-06 | 7.79E-06 | 0.005518907 | NA |
| chr9 | 94594169 | 94594693 | 525 | 3 | 1.07E-05 | 6.37E-06 | 0.000863 | 7.79E-06 | -0.00057225 | PCAT7 |
| chr16 | 1655666 | 1656200 | 535 | 7 | 2.80E-06 | 1.30E-05 | 0.001092 | 7.80E-06 | -0.0021737 | CRAMP1, AL031708.1 |
| chr2 | 88127508 | 88128308 | 801 | 6 | 6.39E-06 | 8.32E-06 | 0.001902 | 7.80E-06 | -0.00686329 | FABP1 |
| chr22 | 38734741 | 38734850 | 110 | 2 | 8.34E-06 | 4.68E-06 | 0.000581 | 7.80E-06 | -0.01166312 | GTPBP1, AL021707.5, AL021707.3, SUN2 |
| chr7 | 30169302 | 30169897 | 596 | 3 | 6.16E-06 | 7.19E-06 | 0.000797 | 7.80E-06 | -0.00371644 | NA |
| chr16 | 30709899 | 30710394 | 496 | 3 | 3.12E-05 | 3.03E-06 | 0.002269 | 7.80E-06 | 0.009067248 | SRCAP, AC106886.5 |
| chr6 | 134182730 | 134182753 | 24 | 2 | 7.85E-06 | 1.10E-05 | 5.12E-05 | 7.81E-06 | 0.001097251 | SGK1 |
| chr14 | 50944549 | 50945374 | 826 | 11 | 8.49E-06 | 7.50E-05 | 0.008096 | 7.81E-06 | -0.0048286 | NA |
| chr6 | 39322383 | 39323101 | 719 | 7 | 1.20E-06 | 0.0002155 | 0.000117 | 7.81E-06 | -0.0032166 | KCNK16 |
| chr1 | 205042778 | 205043321 | 544 | 7 | 1.30E-06 | 0.0006728 | 3.25E-05 | 7.81E-06 | -0.00174589 | CNTN2 |
| chr2 | 219253551 | 219254771 | 1221 | 16 | 3.40E-06 | 0.0001683 | 0.003092 | 7.81E-06 | -0.00042919 | TUBA4B, TUBA4A |
| chr3 | 183386055 | 183386486 | 432 | 2 | 2.53E-06 | 3.66E-05 | 1.62E-05 | 7.81E-06 | -0.01192003 | MCF2L2 |
| chr4 | 138282262 | 138282551 | 290 | 3 | 5.69E-06 | 8.35E-05 | 1.35E-05 | 7.81E-06 | -0.008194 | NA |
| chr9 | 33816426 | 33816965 | 540 | 6 | 9.29E-06 | 4.46E-05 | 0.001391 | 7.81E-06 | 0.001676912 | UBE2R2-AS1 |
| chr20 | 61928251 | 61928393 | 143 | 3 | 5.51E-06 | 2.59E-05 | 4.21E-05 | 7.82E-06 | -0.00350707 | CDH4 |
| chr17 | 27883955 | 27884364 | 410 | 2 | 4.06E-06 | 2.02E-05 | 2.60E-05 | 7.82E-06 | -0.00726194 | AC005697.1, LYRM9 |
| chr5 | 175517371 | 175517861 | 491 | 4 | 2.18E-05 | 3.67E-06 | 0.00172 | 7.82E-06 | -0.004303 | SFXN1 |
| chr8 | 57193535 | 57193961 | 427 | 4 | 1.95E-05 | 2.44E-05 | 0.000744 | 7.82E-06 | -0.00143017 | LINC01606 |
| chr6 | 136234644 | 136235325 | 682 | 2 | 0.000109068 | 5.10E-06 | 0.000351 | 7.83E-06 | 0.00900337 | MTFR2 |
| chr5 | 139543528 | 139543682 | 155 | 2 | 1.68E-05 | 6.94E-06 | 0.000117 | 7.83E-06 | -0.00913137 | UBE2D2 |
| chr15 | 89680481 | 89680867 | 387 | 2 | 5.13E-07 | 0.0001885 | 7.54E-06 | 7.83E-06 | 0.003666248 | PEX11A |
| chr21 | 33753100 | 33753244 | 145 | 3 | 3.52E-06 | 0.00018 | 2.26E-05 | 7.83E-06 | 0.00325055 | ITSN1, AP000311.1 |
| chr17 | 8466699 | 8467230 | 532 | 4 | 4.46E-06 | 2.57E-05 | 5.11E-05 | 7.83E-06 | -0.00175515 | NDEL1 |
| chr12 | 117189645 | 117190784 | 1140 | 13 | 7.57E-06 | 6.22E-06 | 0.000427 | 7.83E-06 | -0.00016622 | FBXO21 |
| chr1 | 245577361 | 245577901 | 541 | 3 | 2.20E-05 | 3.37E-06 | 0.001829 | 7.84E-06 | -0.00062044 | KIF26B |
| chr6 | 25961540 | 25961749 | 210 | 3 | 3.65E-06 | 6.77E-05 | 0.00024 | 7.84E-06 | -0.00227395 | NA |
| chr3 | 9940999 | 9941024 | 26 | 3 | 1.43E-05 | 3.40E-05 | 0.000228 | 7.84E-06 | -6.56E-05 | AC018809.3, CRELD1 |
| chr3 | 147355257 | 147355331 | 75 | 2 | 1.26E-05 | 6.17E-06 | 0.000163 | 7.85E-06 | -0.01091532 | AC092957.1 |
| chr7 | 156178260 | 156178704 | 445 | 6 | 2.65E-06 | 1.49E-05 | 0.000242 | 7.85E-06 | 0.009214744 | NA |
| chr16 | 74449796 | 74450377 | 582 | 6 | 1.54E-06 | 2.08E-05 | 0.005895 | 7.85E-06 | -0.00087276 | GLG1 |

| | | | | | | | | | | |
|---|---|---|---|---|---|---|---|---|---|---|
| chr5 | 176628854 | 176629681 | 828 | 4 | 4.58E-06 | 1.97E-05 | 9.07E-05 | 7.85E-06 | -0.00921332 | SNCB, MIR4281 |
| chr19 | 11105185 | 11105688 | 504 | 5 | 4.44E-06 | 0.0002123 | 2.86E-05 | 7.85E-06 | -0.00211225 | LDLR |
| chr22 | 20880524 | 20880753 | 230 | 2 | 6.63E-06 | 9.46E-06 | 6.47E-05 | 7.86E-06 | -0.00440752 | SNAP29 |
| chr1 | 184805672 | 184806311 | 640 | 3 | 7.52E-06 | 9.82E-06 | 0.000108 | 7.86E-06 | -0.0073798 | NIBAN1 |
| chr3 | 124635271 | 124635719 | 449 | 2 | 1.85E-06 | 4.65E-05 | 1.41E-05 | 7.87E-06 | -0.01162989 | KALRN |
| chr4 | 14995505 | 14995669 | 165 | 3 | 4.44E-06 | 0.000115 | 1.05E-05 | 7.87E-06 | -0.00417673 | CPEB2-DT |
| chr21 | 30492621 | 30492754 | 134 | 2 | 2.12E-06 | 1.16E-05 | 4.84E-05 | 7.87E-06 | -0.00156148 | NA |
| chr19 | 46750269 | 46750347 | 79 | 2 | 9.28E-06 | 2.16E-05 | 2.47E-05 | 7.87E-06 | -0.01123927 | FKRP |
| chr7 | 109960342 | 109960676 | 335 | 4 | 1.11E-05 | 2.65E-05 | 0.000336 | 7.87E-06 | 0.002020594 | NA |
| chr17 | 82050665 | 82051589 | 925 | 5 | 4.63E-08 | 0.0009475 | 5.81E-06 | 7.87E-06 | -0.00655529 | GPS1, RFNG |
| chr22 | 50612489 | 50612784 | 296 | 2 | 1.61E-06 | 0.0015975 | 5.00E-06 | 7.87E-06 | -0.01426539 | MAPK8IP2 |
| chr8 | 96333264 | 96333845 | 582 | 8 | 2.63E-05 | 3.31E-06 | 0.003087 | 7.87E-06 | -0.00130282 | PTDSS1 |
| chr15 | 89496382 | 89497046 | 665 | 11 | 5.91E-05 | 3.42E-06 | 0.010123 | 7.87E-06 | -0.00272503 | RHCG |
| chr1 | 77219280 | 77219812 | 533 | 11 | 1.98E-05 | 1.79E-05 | 0.00115 | 7.88E-06 | 0.001272595 | PIGK |
| chr3 | 122993176 | 122994178 | 1003 | 6 | 9.16E-06 | 6.98E-06 | 0.000449 | 7.88E-06 | -0.0123394 | SEMA5B |
| chr14 | 89575857 | 89576236 | 380 | 7 | 1.98E-05 | 1.94E-06 | 0.007567 | 7.89E-06 | 0.000111469 | FOXN3-AS2, FOXN3, AL137230.2 |
| chr12 | 7936222 | 7936818 | 597 | 6 | 2.29E-05 | 0.0001005 | 0.000414 | 7.89E-06 | -0.00377561 | NA |
| chr16 | 24036883 | 24036953 | 71 | 2 | 9.57E-06 | 3.80E-05 | 1.61E-05 | 7.89E-06 | -0.01475655 | PRKCB |
| chr11 | 46663471 | 46663846 | 376 | 2 | 3.02E-06 | 1.47E-05 | 3.63E-05 | 7.89E-06 | 0.011512448 | ATG13 |
| chr17 | 50432194 | 50432612 | 419 | 2 | 1.79E-05 | 5.24E-06 | 0.00032 | 7.89E-06 | -0.00604942 | ACSF2 |
| chr17 | 28599439 | 28599850 | 412 | 6 | 9.96E-06 | 6.98E-05 | 0.001247 | 7.89E-06 | 0.001339934 | SPAG5-AS1, AC005726.1 |
| chr4 | 38782957 | 38783317 | 361 | 4 | 1.79E-07 | 0.0033386 | 9.20E-06 | 7.89E-06 | -0.00193909 | TLR10 |
| chr11 | 69493276 | 69494000 | 725 | 4 | 2.29E-07 | 0.0031244 | 1.71E-06 | 7.89E-06 | -0.00213675 | LINC01488 |
| chr2 | 61604701 | 61605596 | 896 | 5 | 3.71E-06 | 7.41E-05 | 0.000502 | 7.89E-06 | 0.001997926 | NA |
| chr18 | 9966263 | 9966654 | 392 | 2 | 1.48E-05 | 5.81E-06 | 0.000207 | 7.89E-06 | -0.00578143 | NA |
| chr19 | 7394236 | 7395175 | 940 | 11 | 8.31E-07 | 4.64E-05 | 0.002745 | 7.90E-06 | -0.00363179 | AC008878.3, ARHGEF18, AC119396.2 |
| chr19 | 35843879 | 35844418 | 540 | 5 | 1.27E-05 | 2.11E-05 | 0.000397 | 7.90E-06 | -0.00182952 | NPHS1 |
| chr12 | 132145514 | 132145830 | 317 | 2 | 7.07E-07 | 5.23E-06 | 0.000325 | 7.90E-06 | -0.00153325 | NOC4L |
| chr11 | 59846446 | 59846739 | 294 | 4 | 4.54E-06 | 0.001669 | 0.000191 | 7.90E-06 | -0.00115461 | NA |
| chr10 | 13727634 | 13728043 | 410 | 4 | 2.24E-06 | 0.0007299 | 0.000219 | 7.91E-06 | 0.000409284 | RNA5SP301, FRMD4A |
| chr2 | 24363449 | 24363751 | 303 | 3 | 4.69E-06 | 7.59E-05 | 1.16E-05 | 7.91E-06 | -0.0001048 | NA |
| chr2 | 236014187 | 236015115 | 929 | 5 | 1.80E-05 | 2.03E-06 | 0.005106 | 7.91E-06 | -0.00811501 | AGAP1 |
| chr6 | 136792638 | 136793375 | 738 | 6 | 2.69E-06 | 0.0002885 | 6.43E-05 | 7.92E-06 | -0.00268059 | NA |
| chr14 | 67573813 | 67573846 | 34 | 2 | 4.31E-06 | 0.0010073 | 5.33E-06 | 7.92E-06 | 0.00221547 | PLEKHH1 |
| chr8 | 119873120 | 119874210 | 1091 | 12 | 4.79E-05 | 3.78E-05 | 0.005405 | 7.93E-06 | -0.00198778 | DEPTOR, AP005717.1 |
| chr4 | 48804753 | 48804945 | 193 | 2 | 2.95E-06 | 0.0015388 | 5.06E-06 | 7.94E-06 | -0.00352917 | NA |
| chr12 | 132028074 | 132028397 | 324 | 4 | 4.25E-06 | 5.43E-05 | 5.01E-05 | 7.94E-06 | -0.00430342 | EP400 |
| chr19 | 15150604 | 15150710 | 107 | 3 | 1.39E-05 | 2.61E-05 | 0.000119 | 7.94E-06 | -0.0030814 | NA |
| chr14 | 26926212 | 26926526 | 315 | 3 | 1.65E-05 | 2.24E-05 | 0.000473 | 7.95E-06 | -0.00188532 | AL110292.1 |
| chr20 | 62799547 | 62799959 | 413 | 2 | 4.97E-06 | 6.15E-06 | 0.000173 | 7.95E-06 | 0.002533359 | MRGBP |
| chr19 | 36035120 | 36035349 | 230 | 2 | 8.93E-06 | 1.60E-05 | 3.35E-05 | 7.95E-06 | 0.005096195 | AC002116.2, THAP8 |
| chr2 | 79522262 | 79522509 | 248 | 4 | 2.61E-05 | 2.26E-06 | 0.004857 | 7.95E-06 | -0.00186397 | CTNNA2 |
| chr10 | 94683408 | 94683626 | 219 | 3 | 2.21E-05 | 4.25E-06 | 0.000664 | 7.95E-06 | -0.00221768 | NA |
| chr3 | 9301324 | 9302283 | 960 | 3 | 9.53E-06 | 2.01E-05 | 0.000134 | 7.96E-06 | -0.00769515 | SRGAP3, AC026191.1 |
| chr16 | 57263532 | 57265225 | 1694 | 6 | 4.83E-05 | 4.04E-05 | 0.000619 | 7.97E-06 | -0.00358086 | PLLP |
| chr2 | 232550191 | 232550943 | 753 | 16 | 2.12E-05 | 6.36E-05 | 0.009264 | 7.97E-06 | 0.001179586 | EIF4E2, TIGD1, MIR5001 |
| chr15 | 65107283 | 65107841 | 559 | 3 | 2.23E-05 | 4.06E-06 | 0.000708 | 7.97E-06 | 0.006744204 | UBAP1L |
| chr7 | 92793991 | 92794595 | 605 | 3 | 1.12E-05 | 7.97E-06 | 0.000135 | 7.99E-06 | -0.00587696 | CDK6 |
| chr5 | 140558396 | 140558757 | 362 | 6 | 1.43E-06 | 5.21E-06 | 0.001684 | 7.99E-06 | 0.000730783 | APBB3 |
| chr5 | 90529677 | 90530091 | 415 | 6 | 6.13E-07 | 0.0036464 | 6.85E-06 | 7.99E-06 | -0.00098835 | ADGRV1 |
| chr1 | 121519571 | 121519606 | 36 | 2 | 3.79E-06 | 7.22E-05 | 1.12E-05 | 8.00E-06 | 0.004263264 | EMBP1 |
| chr22 | 49575208 | 49575436 | 229 | 2 | 1.26E-06 | 5.51E-05 | 1.30E-05 | 8.00E-06 | 0.003765411 | C22orf34, Z97192.2 |
| chr10 | 93702404 | 93702942 | 539 | 13 | 2.66E-05 | 1.01E-05 | 0.006902 | 8.00E-06 | 0.001669829 | FRA10AC1 |
| chr17 | 79041856 | 79041883 | 28 | 2 | 4.33E-06 | 2.04E-05 | 2.67E-05 | 8.00E-06 | -0.00266891 | C1QTNF1 |
| chr6 | 37432124 | 37432530 | 407 | 2 | 4.89E-08 | 0.000383 | 6.43E-06 | 8.01E-06 | -0.00250199 | NA |
| chr8 | 41577527 | 41578185 | 659 | 7 | 1.22E-05 | 1.53E-05 | 0.003102 | 8.01E-06 | 0.001449153 | GPAT4, AC009630.1 |
| chr21 | 40146448 | 40146696 | 249 | 2 | 2.18E-05 | 4.82E-06 | 0.000577 | 8.01E-06 | -0.01460941 | DSCAM |
| chr19 | 1417237 | 1417655 | 419 | 2 | 4.36E-07 | 0.0006645 | 5.75E-06 | 8.01E-06 | 0.005366259 | DAZAP1 |
| chr17 | 72412164 | 72412823 | 660 | 2 | 1.92E-05 | 5.97E-06 | 0.0002 | 8.02E-06 | -0.0075764 | LINC00511 |
| chr13 | 33016135 | 33016355 | 221 | 5 | 4.32E-05 | 2.56E-06 | 0.004631 | 8.02E-06 | -0.00011813 | NA |
| chr9 | 133113072 | 133113141 | 70 | 2 | 3.87E-06 | 0.0001189 | 9.09E-06 | 8.03E-06 | 0.00897563 | RALGDS, AL162417.1 |
| chr2 | 230873026 | 230873243 | 218 | 4 | 2.39E-05 | 7.32E-06 | 0.000723 | 8.03E-06 | -0.01364564 | ITM2C |
| chr6 | 135189818 | 135190160 | 343 | 2 | 1.69E-06 | 0.0003617 | 6.54E-06 | 8.04E-06 | -0.00899879 | MYB |
| chr2 | 2825951 | 2825987 | 37 | 3 | 1.19E-05 | 7.45E-05 | 8.86E-05 | 8.04E-06 | -0.00270727 | AC011995.2 |
| chr16 | 67184681 | 67185534 | 854 | 8 | 2.21E-07 | 3.69E-05 | 0.000685 | 8.04E-06 | -0.00566126 | EXOC3L1 |
| chr16 | 743677 | 744222 | 546 | 3 | 3.45E-06 | 0.0005292 | 8.18E-05 | 8.05E-06 | -0.00618541 | NA |
| chr6 | 19143903 | 19143935 | 33 | 2 | 1.86E-05 | 7.73E-06 | 9.84E-05 | 8.05E-06 | 0.00215387 | AL589647.1 |
| chr12 | 108129632 | 108130568 | 937 | 6 | 1.31E-05 | 2.53E-05 | 0.0008 | 8.06E-06 | -0.00213522 | WSCD2 |
| chr19 | 39131286 | 39131320 | 35 | 2 | 3.97E-06 | 5.11E-06 | 0.000413 | 8.06E-06 | -0.0123122 | PAK4 |
| chr14 | 102438132 | 102438269 | 138 | 3 | 8.78E-06 | 0.0001125 | 2.41E-05 | 8.06E-06 | -0.00859718 | TECPR2 |
| chr5 | 147782361 | 147783018 | 658 | 8 | 8.11E-06 | 4.92E-05 | 0.002207 | 8.06E-06 | 0.003121805 | JAKMIP2 |
| chr10 | 100065163 | 100065428 | 266 | 5 | 8.69E-06 | 0.0002273 | 0.000155 | 8.06E-06 | -0.0122391 | CPN1 |
| chr10 | 22325347 | 22326004 | 658 | 5 | 7.07E-06 | 7.83E-05 | 0.000229 | 8.06E-06 | -0.00018312 | COMMD3-BMI1, BMI1 |
| chr2 | 170813352 | 170814468 | 1117 | 7 | 4.28E-06 | 0.0003951 | 0.000505 | 8.06E-06 | -0.00194785 | GAD1, AC007405.1 |
| chr17 | 39995406 | 39995652 | 247 | 2 | 4.85E-06 | 5.21E-06 | 0.000373 | 8.07E-06 | 0.001705902 | PSMD3 |
| chr1 | 60979156 | 60979407 | 252 | 2 | 2.00E-06 | 0.0037877 | 4.73E-06 | 8.07E-06 | -0.0143177 | NFIA, NFIA-AS2 |
| chr16 | 2744900 | 2745389 | 490 | 4 | 2.26E-06 | 0.0002601 | 0.000953 | 8.07E-06 | -0.00439862 | SRRM2-AS1 |
| chr2 | 64453564 | 64454003 | 440 | 4 | 2.84E-06 | 1.85E-05 | 6.36E-05 | 8.08E-06 | 0.001030643 | LGALSL, LGALSL-DT |
| chr2 | 242047705 | 242048294 | 590 | 6 | 1.08E-05 | 1.60E-05 | 0.001216 | 8.08E-06 | 0.006436491 | LINC01237, LINC01880 |
| chr5 | 134579716 | 134580415 | 700 | 2 | 6.51E-07 | 5.23E-06 | 0.000367 | 8.08E-06 | -0.00438465 | JADE2 |
| chr19 | 58572656 | 58573575 | 920 | 8 | 1.01E-05 | 0.0010149 | 0.002288 | 8.08E-06 | 0.001199367 | MZF1-AS1, CENPBD1P1, MZF1 |
| chr11 | 133923046 | 133923349 | 304 | 2 | 8.01E-07 | 0.0027916 | 4.85E-06 | 8.08E-06 | -0.0058032 | IGSF9B |
| chr20 | 59247447 | 59247856 | 410 | 2 | 2.13E-05 | 4.84E-06 | 0.000606 | 8.08E-06 | -0.00679411 | ZNF831 |
| chr3 | 71130552 | 71131302 | 751 | 13 | 2.43E-05 | 1.09E-05 | 0.008454 | 8.09E-06 | 0.003622347 | FOXP1, AC097634.4 |
| chr22 | 49156773 | 49156827 | 55 | 2 | 3.53E-06 | 0.0067083 | 4.58E-06 | 8.09E-06 | -0.00938202 | NA |
| chr1 | 29230309 | 29231308 | 1000 | 14 | 2.49E-05 | 1.25E-05 | 0.002501 | 8.10E-06 | -0.00192054 | MECR |
| chr10 | 376041 | 376390 | 350 | 4 | 5.06E-06 | 7.33E-05 | 2.74E-05 | 8.10E-06 | -0.00344042 | DIP2C |
| chr2 | 219562126 | 219562620 | 495 | 5 | 6.62E-07 | 0.0004556 | 2.27E-05 | 8.10E-06 | -0.00515737 | OBSL1 |
| chr12 | 24562316 | 24563270 | 955 | 12 | 4.84E-06 | 1.78E-05 | 0.003479 | 8.10E-06 | 0.000334112 | SOX5 |
| chr14 | 88630663 | 88630890 | 228 | 2 | 2.98E-06 | 0.000663 | 5.81E-06 | 8.10E-06 | 0.002957286 | EML5 |
| chr17 | 79747832 | 79748719 | 888 | 6 | 1.64E-06 | 7.62E-05 | 0.000188 | 8.11E-06 | 9.25E-06 | NA |
| chr6 | 30894477 | 30894694 | 218 | 2 | 1.04E-05 | 1.15E-05 | 5.15E-05 | 8.11E-06 | -0.00112125 | DDR1 |
| chr3 | 197778351 | 197778360 | 10 | 2 | 6.23E-06 | 5.14E-06 | 0.00042 | 8.11E-06 | -0.02334505 | FYTTD1 |
| chr6 | 14921397 | 14922109 | 713 | 2 | 8.70E-05 | 4.87E-06 | 0.000603 | 8.12E-06 | -0.0114162 | AL138720.1 |
| chr9 | 134745199 | 134745466 | 268 | 2 | 2.20E-06 | 6.54E-05 | 1.21E-05 | 8.12E-06 | -0.00655232 | COL5A1 |
| chr7 | 149813642 | 149814005 | 364 | 2 | 1.38E-05 | 6.54E-06 | 0.000155 | 8.12E-06 | 0.006080824 | SSPOP |
| chr18 | 35345050 | 35345338 | 289 | 3 | 5.07E-06 | 9.49E-06 | 0.000146 | 8.12E-06 | 0.002179772 | ZNF24 |
| chr9 | 105386319 | 105386514 | 196 | 2 | 1.58E-05 | 7.81E-06 | 9.83E-05 | 8.12E-06 | -0.00018467 | SLC44A1 |
| chr6 | 1526393 | 1526837 | 445 | 2 | 3.59E-06 | 7.87E-06 | 9.69E-05 | 8.13E-06 | 0.022218322 | NA |
| chr10 | 33134013 | 33134214 | 202 | 2 | 5.72E-06 | 5.22E-06 | 0.000386 | 8.13E-06 | -0.01280326 | NA |
| chr7 | 157020786 | 157021418 | 633 | 4 | 7.48E-06 | 4.28E-06 | 0.001052 | 8.13E-06 | -0.00077713 | NA |
| chr13 | 95676303 | 95676697 | 395 | 5 | 2.92E-06 | 5.13E-05 | 9.61E-05 | 8.13E-06 | 0.003346278 | DNAJC3-DT |
| chr17 | 27219308 | 27219327 | 20 | 2 | 7.78E-06 | 0.0001138 | 9.38E-06 | 8.14E-06 | -0.0127412 | NA |
| chr11 | 131234316 | 131234589 | 274 | 5 | 8.92E-06 | 0.0004866 | 0.000266 | 8.14E-06 | -0.00468979 | AP002856.4 |
| chr13 | 91250410 | 91250719 | 310 | 2 | 2.23E-05 | 4.94E-06 | 0.000552 | 8.14E-06 | 0.00541839 | NA |
| chr12 | 56115819 | 56116871 | 1053 | 14 | 3.05E-05 | 5.24E-06 | 0.023913 | 8.14E-06 | 0.001030714 | RPL41 |
| chr2 | 110995929 | 110996139 | 211 | 2 | 1.53E-05 | 8.46E-06 | 8.46E-05 | 8.14E-06 | -0.00190128 | ACOXL |
| chr16 | 86466461 | 86466932 | 472 | 3 | 2.51E-05 | 1.42E-05 | 0.000289 | 8.15E-06 | 0.000550348 | NA |
| chr13 | 102799869 | 102800865 | 997 | 8 | 2.09E-06 | 0.0005021 | 0.001678 | 8.16E-06 | 0.001789717 | BIVM-ERCC5, BIVM |
| chr20 | 31871355 | 31871663 | 309 | 2 | 4.00E-07 | 6.60E-06 | 0.000154 | 8.17E-06 | 0.001287999 | TTLL9 |
| chr4 | 78181756 | 78182136 | 381 | 2 | 2.90E-06 | 5.28E-05 | 1.37E-05 | 8.17E-06 | -0.00408873 | FRAS1, SERBP1P5 |
| chr20 | 35453882 | 35454191 | 310 | 5 | 1.53E-05 | 5.41E-06 | 0.00086 | 8.17E-06 | -0.00298799 | GDF5, MIR1289-1 |
| chr5 | 39274660 | 39275490 | 831 | 3 | 2.58E-05 | 3.29E-06 | 0.001793 | 8.18E-06 | -0.00405141 | NA |
| chr7 | 100538545 | 100539145 | 601 | 9 | 3.86E-05 | 3.35E-05 | 0.009298 | 8.18E-06 | -9.77E-05 | NA |
| chr6 | 25191488 | 25191601 | 114 | 2 | 7.28E-06 | 1.58E-05 | 3.55E-05 | 8.18E-06 | -0.00974403 | CMAHP |
| chr1 | 154954417 | 154955215 | 799 | 3 | 1.77E-06 | 5.46E-05 | 1.79E-05 | 8.19E-06 | 0.004493996 | PBXIP1 |
| chr6 | 11877297 | 11877381 | 85 | 2 | 6.68E-06 | 2.12E-05 | 2.66E-05 | 8.19E-06 | -0.00708422 | NA |
| chr1 | 3134273 | 3134965 | 693 | 4 | 2.30E-05 | 2.13E-05 | 0.00058 | 8.19E-06 | 0.000454263 | PRDM16 |
| chr8 | 22156486 | 22157233 | 748 | 10 | 1.08E-06 | 0.0097924 | 0.001228 | 8.19E-06 | 0.001668923 | SFTPC, LGI3 |
| chr21 | 25054716 | 25054864 | 149 | 2 | 1.04E-05 | 8.05E-06 | 9.44E-05 | 8.19E-06 | -0.01048927 | LINC01692 |
| chr22 | 29305539 | 29307190 | 1652 | 16 | 3.50E-05 | 0.0028699 | 0.000943 | 8.19E-06 | -0.00248458 | GAS2L1 |
| chr9 | 133918318 | 133918643 | 326 | 2 | 3.68E-06 | 5.91E-05 | 1.29E-05 | 8.20E-06 | 0.01727468 | VAV2 |
| chr14 | 104151691 | 104152451 | 761 | 3 | 5.86E-05 | 1.66E-05 | 0.000463 | 8.20E-06 | 0.000605608 | KIF26A |
| chr7 | 150400349 | 150400674 | 326 | 3 | 8.26E-06 | 1.40E-05 | 0.000555 | 8.20E-06 | 0.00463638 | AC073111.2 |
| chr13 | 22428525 | 22428661 | 137 | 3 | 1.58E-05 | 1.98E-05 | 0.000486 | 8.20E-06 | 0.00859196 | NA |
| chr1 | 9849787 | 9850079 | 293 | 3 | 2.93E-05 | 3.94E-06 | 0.001438 | 8.21E-06 | 0.008334425 | AL357140.1, CTNNBIP1 |
| chr10 | 132490395 | 132490524 | 130 | 2 | 3.12E-06 | 6.14E-06 | 0.000198 | 8.22E-06 | 0.005132134 | NA |
| chr20 | 48164941 | 48164957 | 17 | 2 | 6.14E-06 | 5.42E-05 | 1.36E-05 | 8.22E-06 | -0.00988139 | NA |
| chr17 | 45241393 | 45242015 | 623 | 5 | 1.88E-05 | 1.08E-05 | 0.002083 | 8.22E-06 | -0.00025389 | FMNL1, AC008105.1 |
| chr8 | 73092787 | 73093578 | 792 | 11 | 3.58E-06 | 0.0001482 | 0.001753 | 8.22E-06 | 0.001868657 | SBSPON |
| chr4 | 88018196 | 88018694 | 499 | 2 | 4.94E-07 | 9.43E-05 | 1.03E-05 | 8.23E-06 | 0.006921914 | PKD2 |
| chr22 | 44365231 | 44365524 | 294 | 4 | 2.81E-06 | 0.0005192 | 1.72E-05 | 8.23E-06 | -0.00662738 | NA |
| chr11 | 3099719 | 3100170 | 452 | 4 | 6.13E-07 | 0.00035 | 0.000103 | 8.23E-06 | 0.001406738 | OSBPL5 |
| chr1 | 90706780 | 90707675 | 896 | 8 | 6.15E-06 | 6.40E-05 | 0.00311 | 8.23E-06 | 0.000831014 | NA |
| chr2 | 27427611 | 27428268 | 658 | 6 | 9.35E-08 | 4.75E-05 | 0.000344 | 8.23E-06 | 0.005595706 | NRBP1 |
| chr17 | 82439571 | 82440086 | 516 | 5 | 3.90E-06 | 1.99E-05 | 0.001785 | 8.24E-06 | 0.001418659 | HEXD |
| chr17 | 2365146 | 2365644 | 499 | 5 | 1.27E-06 | 0.0001004 | 0.000576 | 8.24E-06 | -0.00141016 | SGSM2 |
| chr19 | 6745310 | 6745706 | 397 | 2 | 1.44E-05 | 7.40E-06 | 0.000116 | 8.24E-06 | -0.00161682 | TRIP10 |
| chr18 | 36732620 | 36732676 | 57 | 2 | 6.09E-06 | 0.0003374 | 6.81E-06 | 8.25E-06 | 0.005640273 | FHOD3 |
| chr11 | 66393338 | 66393352 | 15 | 2 | 2.86E-06 | 1.10E-05 | 5.70E-05 | 8.25E-06 | -0.00745652 | NA |
| chr18 | 34708978 | 34709652 | 675 | 5 | 7.63E-06 | 3.01E-05 | 0.000876 | 8.25E-06 | 0.005687294 | DTNA |
| chr12 | 114480495 | 114481337 | 843 | 5 | 2.59E-06 | 3.42E-06 | 0.002864 | 8.26E-06 | 0.001511117 | NA |
| chr16 | 75012143 | 75012694 | 552 | 2 | 5.93E-06 | 7.15E-06 | 0.000128 | 8.26E-06 | -0.01770867 | ZNRF1 |
| chr22 | 28989803 | 28990281 | 479 | 3 | 2.05E-05 | 3.11E-06 | 0.002459 | 8.26E-06 | -0.00401497 | ZNRF3 |
| chr19 | 5998701 | 5998917 | 217 | 2 | 1.03E-05 | 5.20E-06 | 0.000439 | 8.26E-06 | 0.003521365 | AC011444.1, RFX2 |
| chr2 | 7725068 | 7725522 | 455 | 2 | 3.29E-06 | 2.42E-05 | 2.41E-05 | 8.26E-06 | 0.020890696 | NA |
| chr14 | 45268668 | 45269046 | 379 | 2 | 2.35E-05 | 5.04E-06 | 0.000544 | 8.27E-06 | -0.01459766 | NA |
| chr15 | 63218675 | 63218743 | 69 | 2 | 9.98E-06 | 4.19E-05 | 1.60E-05 | 8.27E-06 | -0.01107189 | RAB8B |
| chr17 | 19724422 | 19725108 | 687 | 5 | 6.44E-06 | 1.39E-05 | 0.000551 | 8.27E-06 | 0.016754473 | NA |
| chr4 | 80184669 | 80186031 | 1363 | 9 | 2.02E-05 | 3.07E-05 | 0.001104 | 8.28E-06 | -0.00375721 | PRDM8, AC021127.1 |
| chr10 | 99731841 | 99732885 | 1045 | 16 | 9.95E-07 | 0.0010709 | 0.000508 | 8.28E-06 | 0.000536507 | CUTC, AL133353.2, COX15 |
| chr7 | 1348635 | 1349081 | 447 | 5 | 1.02E-05 | 7.83E-05 | 0.001514 | 8.28E-06 | -0.00842444 | NA |
| chr7 | 76379333 | 76379963 | 631 | 2 | 1.07E-05 | 4.99E-06 | 0.000595 | 8.28E-06 | 0.005245721 | NA |
| chr2 | 28810195 | 28810486 | 292 | 5 | 2.49E-05 | 7.26E-06 | 0.000819 | 8.28E-06 | 0.000685981 | SPDYA, AC097724.1 |
| chr7 | 22488414 | 22488919 | 506 | 2 | 2.57E-05 | 5.17E-06 | 0.000464 | 8.28E-06 | -0.00727108 | STEAP1B |
| chr2 | 60461908 | 60461978 | 71 | 2 | 6.31E-06 | 6.02E-06 | 0.000221 | 8.28E-06 | -0.00616798 | BCL11A |
| chr7 | 12685929 | 12686806 | 878 | 9 | 7.26E-06 | 1.62E-06 | 0.017546 | 8.28E-06 | -0.00571758 | AC011891.3 |
| chr3 | 35744243 | 35744988 | 746 | 5 | 1.68E-06 | 6.37E-05 | 0.000396 | 8.28E-06 | -0.00743602 | ARPP21, MIR128-2 |

| | | | | | | | | | | |
|---|---|---|---|---|---|---|---|---|---|---|
| chr19 | 17235280 | 17235698 | 419 | 3 | 1.21E-06 | 1.71E-05 | 8.38E-05 | 8.29E-06 | -0.00322414 | NR2F6 |
| chr18 | 60144683 | 60145270 | 588 | 2 | 1.37E-05 | 6.76E-06 | 0.000151 | 8.29E-06 | 0.004988209 | AC090771.2 |
| chr14 | 49987983 | 49988403 | 421 | 2 | 3.76E-06 | 2.57E-05 | 2.30E-05 | 8.29E-06 | 0.004640248 | LINC01588 |
| chr16 | 89305126 | 89305574 | 449 | 5 | 1.03E-05 | 2.10E-05 | 0.00086 | 8.29E-06 | -0.00175467 | ANKRD11 |
| chr12 | 10806320 | 10807122 | 803 | 4 | 2.13E-05 | 0.0053165 | 2.23E-05 | 8.30E-06 | -0.00589181 | TAS2R8 |
| chr17 | 18183523 | 18184266 | 744 | 10 | 1.13E-08 | 0.0013515 | 3.53E-06 | 8.30E-06 | 0.00120981 | ALKBH5, AC087164.1 |
| chr6 | 27872290 | 27873344 | 1055 | 10 | 1.39E-05 | 1.18E-05 | 0.002145 | 8.30E-06 | 0.001168958 | H3C11, H4C13 |
| chr8 | 67481500 | 67481672 | 173 | 2 | 1.11E-05 | 1.99E-05 | 2.90E-05 | 8.30E-06 | 0.001954668 | CPA6 |
| chr10 | 131014596 | 131014773 | 178 | 3 | 1.51E-05 | 1.47E-05 | 0.000108 | 8.30E-06 | -0.00675632 | NA |
| chr1 | 9073294 | 9073355 | 62 | 2 | 1.58E-05 | 5.96E-06 | 0.000233 | 8.31E-06 | 0.012738745 | SLC2A5 |
| chr10 | 71677577 | 71677606 | 30 | 2 | 4.53E-06 | 0.0001114 | 9.69E-06 | 8.31E-06 | -0.00386815 | CDH23 |
| chr1 | 147560999 | 147561301 | 303 | 2 | 1.05E-05 | 1.09E-05 | 5.88E-05 | 8.31E-06 | 0.004349163 | BCL9 |
| chr8 | 100867318 | 100867484 | 167 | 3 | 7.51E-06 | 9.38E-06 | 0.000116 | 8.31E-06 | -0.00234985 | NA |
| chr3 | 10812581 | 10812661 | 81 | 2 | 2.11E-05 | 6.01E-06 | 0.000226 | 8.31E-06 | -0.00403023 | NA |
| chr7 | 25385644 | 25386373 | 730 | 4 | 2.32E-05 | 8.69E-06 | 0.001069 | 8.32E-06 | 0.00038617 | AC005100.1 |
| chr20 | 32277045 | 32277699 | 655 | 10 | 1.77E-05 | 2.05E-05 | 0.006043 | 8.32E-06 | 0.001186993 | KIF3B, AL121897.1 |
| chr19 | 48468692 | 48469744 | 1053 | 14 | 1.50E-06 | 0.0001343 | 0.001574 | 8.32E-06 | -0.00024095 | AC008403.1, CYTH2, AC008403.2 |
| chr21 | 44289793 | 44290757 | 965 | 7 | 6.02E-07 | 0.0017626 | 1.54E-05 | 8.32E-06 | -0.00173625 | AIRE |
| chr8 | 750220 | 750734 | 515 | 5 | 3.41E-06 | 2.00E-05 | 5.34E-05 | 8.32E-06 | -0.00451933 | DLGAP2 |
| chr4 | 48830814 | 48831570 | 757 | 17 | 4.87E-06 | 4.79E-05 | 0.002227 | 8.32E-06 | 0.000894798 | OCIAD1 |
| chr14 | 58635001 | 58635555 | 555 | 2 | 9.50E-06 | 6.09E-06 | 0.000217 | 8.32E-06 | -0.0108193 | DACT1 |
| chr11 | 31305436 | 31305669 | 234 | 4 | 2.99E-06 | 0.0003612 | 6.52E-06 | 8.32E-06 | -0.0051679 | DCDC1 |
| chr20 | 17531181 | 17532150 | 970 | 9 | 7.63E-06 | 3.44E-05 | 0.000924 | 8.33E-06 | -0.0036026 | BFSP1 |
| chr13 | 29848587 | 29849745 | 1159 | 3 | 4.94E-07 | 3.63E-05 | 0.000397 | 8.33E-06 | 0.008410964 | UBL3 |
| chr12 | 52789724 | 52790493 | 770 | 7 | 4.00E-06 | 6.21E-05 | 3.45E-05 | 8.33E-06 | 0.010273475 | KRT3 |
| chr19 | 10352984 | 10353644 | 661 | 5 | 9.57E-06 | 1.70E-05 | 0.000181 | 8.34E-06 | -0.00185735 | TYK2 |
| chr7 | 46969297 | 46969668 | 372 | 4 | 9.57E-06 | 4.67E-05 | 4.96E-05 | 8.34E-06 | -0.00317928 | AC004870.2, AC004870.4 |
| chr16 | 54193878 | 54194446 | 569 | 6 | 3.12E-05 | 2.72E-05 | 0.000455 | 8.34E-06 | -0.00353165 | NA |
| chr14 | 87989069 | 87989818 | 750 | 5 | 2.28E-05 | 2.67E-06 | 0.005348 | 8.35E-06 | -0.00683286 | GALC |
| chr9 | 36091581 | 36091891 | 311 | 2 | 3.46E-07 | 0.0111166 | 4.63E-06 | 8.35E-06 | 0.013610253 | RECK |
| chr9 | 77431619 | 77432524 | 906 | 5 | 2.56E-06 | 0.0001191 | 0.003402 | 8.36E-06 | -0.00049103 | GNA14 |
| chr8 | 102810581 | 102811034 | 454 | 3 | 1.25E-06 | 0.0001406 | 3.47E-05 | 8.36E-06 | 0.003110187 | NA |
| chr22 | 39533929 | 39534330 | 402 | 3 | 3.84E-06 | 4.47E-06 | 0.000614 | 8.36E-06 | -0.00074454 | NA |
| chr22 | 31345298 | 31345897 | 600 | 5 | 7.31E-06 | 5.44E-06 | 0.001034 | 8.36E-06 | 0.003115418 | PATZ1 |
| chr1 | 6157850 | 6158056 | 207 | 3 | 2.03E-05 | 4.56E-06 | 0.000592 | 8.36E-06 | -0.00969284 | CHD5 |
| chr1 | 185433212 | 185433805 | 594 | 3 | 7.30E-06 | 2.51E-05 | 8.95E-05 | 8.37E-06 | -0.00168575 | NA |
| chr20 | 35285265 | 35285860 | 596 | 5 | 1.89E-07 | 0.0001809 | 0.000468 | 8.37E-06 | -0.00351461 | FAM83C-AS1, FAM83C |
| chr2 | 25362750 | 25363224 | 475 | 5 | 3.59E-06 | 0.0001022 | 3.45E-05 | 8.38E-06 | 0.002065869 | NA |
| chr7 | 37354956 | 37355102 | 147 | 4 | 2.17E-05 | 7.35E-06 | 0.00121 | 8.38E-06 | -0.00837442 | ELMO1 |
| chr17 | 58992045 | 58992659 | 615 | 2 | 3.12E-05 | 5.38E-06 | 0.000397 | 8.38E-06 | 0.006174243 | TRIM37 |
| chr15 | 24772985 | 24773302 | 318 | 4 | 1.28E-05 | 3.84E-06 | 0.002111 | 8.38E-06 | -0.00250347 | PWRN1, AC090983.2 |
| chr8 | 10760756 | 10761228 | 473 | 3 | 2.21E-06 | 3.58E-05 | 2.33E-05 | 8.38E-06 | 0.004498647 | AC105001.1, AC105001.2 |
| chr12 | 110026974 | 110027305 | 332 | 2 | 2.97E-07 | 0.0001482 | 8.81E-06 | 8.39E-06 | -0.01322215 | ANKRD13A, C12orf76 |
| chr2 | 28566082 | 28567310 | 1229 | 8 | 9.23E-07 | 0.000212 | 5.75E-06 | 8.39E-06 | -0.00039605 | PLB1 |
| chr10 | 132994398 | 132994655 | 258 | 2 | 6.27E-07 | 1.48E-05 | 4.02E-05 | 8.39E-06 | -0.02217745 | NA |
| chr5 | 56816395 | 56817081 | 687 | 4 | 2.75E-05 | 1.51E-05 | 0.002086 | 8.39E-06 | 0.002952358 | MAP3K1 |
| chr8 | 133552269 | 133552434 | 166 | 2 | 2.49E-06 | 0.0010839 | 5.58E-06 | 8.40E-06 | -0.00838301 | ST3GAL1 |
| chr15 | 89317876 | 89318479 | 604 | 2 | 1.24E-05 | 6.58E-06 | 0.000172 | 8.40E-06 | -0.00867545 | POLG |
| chr4 | 86371444 | 86371566 | 123 | 2 | 1.53E-05 | 6.46E-06 | 0.000183 | 8.40E-06 | -0.00175714 | MAPK10 |
| chr15 | 41852068 | 41852382 | 315 | 4 | 1.45E-05 | 1.02E-05 | 0.000756 | 8.40E-06 | -0.0017331 | SPTBN5 |
| chr5 | 163502607 | 163503319 | 713 | 10 | 3.54E-05 | 3.36E-06 | 0.01154 | 8.40E-06 | -0.00311247 | MAT2B |
| chr13 | 41962504 | 41963036 | 533 | 2 | 1.06E-07 | 2.67E-05 | 2.27E-05 | 8.41E-06 | 0.004998864 | VWA8-AS1 |
| chr11 | 116822819 | 116823656 | 838 | 6 | 6.43E-07 | 0.0003504 | 0.000114 | 8.41E-06 | -0.00305097 | APOA4 |
| chr3 | 154224559 | 154224643 | 85 | 3 | 1.22E-05 | 4.20E-05 | 3.61E-05 | 8.41E-06 | 0.01412498 | ARHGEF26 |
| chr10 | 66925185 | 66925729 | 545 | 3 | 3.73E-06 | 1.62E-05 | 0.000346 | 8.41E-06 | -0.01570004 | CTNNA3 |
| chr18 | 13813607 | 13813640 | 34 | 2 | 1.53E-05 | 5.55E-06 | 0.000347 | 8.41E-06 | 0.018353094 | NA |
| chr9 | 34635591 | 34636246 | 656 | 3 | 1.35E-05 | 4.81E-06 | 0.000514 | 8.42E-06 | 0.001187776 | SIGMAR1 |
| chr14 | 51283906 | 51284040 | 135 | 2 | 1.17E-05 | 9.00E-06 | 8.25E-05 | 8.42E-06 | 0.004565953 | NA |
| chr1 | 234709736 | 234710023 | 288 | 2 | 2.64E-06 | 6.95E-06 | 0.000148 | 8.42E-06 | 0.000158149 | AL160408.6 |
| chr11 | 9314067 | 9315298 | 1232 | 16 | 1.68E-06 | 2.21E-05 | 0.005429 | 8.42E-06 | 0.000934913 | TMEM41B |
| chr5 | 1879907 | 1880799 | 893 | 4 | 1.74E-05 | 8.62E-05 | 0.000543 | 8.43E-06 | -0.0078679 | IRX4 |
| chr17 | 34386777 | 34387314 | 538 | 2 | 5.37E-06 | 1.25E-05 | 5.01E-05 | 8.43E-06 | -0.00467467 | NA |
| chr22 | 37330635 | 37331164 | 530 | 2 | 2.45E-05 | 5.41E-06 | 0.000403 | 8.43E-06 | -0.01050351 | NA |
| chr11 | 18526755 | 18527742 | 988 | 14 | 1.35E-06 | 0.0001751 | 0.000384 | 8.43E-06 | -0.00225325 | TSG101 |
| chr1 | 47223521 | 47224112 | 592 | 3 | 1.02E-06 | 4.78E-05 | 0.000104 | 8.44E-06 | -0.00386083 | TAL1 |
| chr8 | 56929397 | 56929767 | 371 | 2 | 9.43E-06 | 7.13E-06 | 0.000139 | 8.44E-06 | -0.0082898 | NA |
| chr14 | 86712921 | 86713024 | 104 | 2 | 1.68E-05 | 1.06E-05 | 6.38E-05 | 8.44E-06 | -0.01185709 | NA |
| chr15 | 58770535 | 58771179 | 645 | 10 | 1.22E-05 | 3.71E-06 | 0.001431 | 8.44E-06 | 0.000914636 | AC090515.2 |
| chr3 | 52494939 | 52495508 | 570 | 8 | 1.29E-05 | 3.97E-05 | 0.000871 | 8.44E-06 | 0.001872066 | STAB1 |
| chr8 | 143177869 | 143178353 | 485 | 2 | 8.78E-07 | 0.0001382 | 9.11E-06 | 8.44E-06 | -0.00351283 | NA |
| chr7 | 143434854 | 143434936 | 83 | 2 | 2.20E-05 | 5.44E-06 | 0.000394 | 8.45E-06 | -0.00340448 | EPHA1-AS1 |
| chr5 | 74499698 | 74500399 | 702 | 3 | 6.32E-07 | 2.07E-05 | 9.91E-05 | 8.46E-06 | -0.01368751 | LINC01331 |
| chr1 | 228902466 | 228903128 | 663 | 3 | 1.89E-05 | 5.63E-06 | 0.001058 | 8.46E-06 | 0.00623659 | NA |
| chr12 | 45232652 | 45233155 | 504 | 2 | 2.72E-06 | 1.57E-05 | 3.80E-05 | 8.46E-06 | -0.00889906 | ANO6 |
| chr11 | 118322847 | 118322904 | 58 | 2 | 1.29E-05 | 2.54E-05 | 2.39E-05 | 8.47E-06 | -0.00291233 | AP001582.1 |
| chr8 | 101263063 | 101263869 | 807 | 4 | 6.89E-06 | 9.32E-05 | 6.67E-05 | 8.47E-06 | 0.000742578 | NA |
| chr19 | 3825143 | 3825443 | 301 | 4 | 1.96E-05 | 9.77E-06 | 0.000598 | 8.47E-06 | -0.00772286 | ZFR2 |
| chr20 | 4592553 | 4592925 | 373 | 4 | 1.01E-05 | 1.48E-05 | 9.09E-05 | 8.47E-06 | 0.000347934 | NA |
| chr2 | 100641991 | 100642530 | 540 | 2 | 2.69E-05 | 5.30E-06 | 0.00047 | 8.47E-06 | -0.01055622 | AC092845.1 |
| chr8 | 5509132 | 5509388 | 257 | 4 | 4.17E-06 | 0.000789 | 5.95E-05 | 8.48E-06 | -0.00195483 | NA |
| chr5 | 81750589 | 81751427 | 839 | 6 | 3.26E-07 | 0.0002545 | 2.74E-05 | 8.48E-06 | -0.001418 | SSBP2 |
| chr17 | 20300679 | 20301116 | 438 | 2 | 1.46E-05 | 6.45E-06 | 0.000192 | 8.48E-06 | 0.009636897 | SPECC1, AC004702.1 |
| chr8 | 28621760 | 28622122 | 363 | 4 | 1.70E-05 | 1.78E-05 | 0.001474 | 8.49E-06 | -0.00215718 | EXTL3 |
| chr21 | 41740201 | 41741152 | 952 | 5 | 6.24E-06 | 6.27E-05 | 0.001019 | 8.50E-06 | 4.73E-05 | AP001615.1, RIPK4 |
| chr3 | 87006448 | 87006868 | 421 | 2 | 1.76E-05 | 5.92E-06 | 0.00027 | 8.50E-06 | -0.01153552 | NA |
| chr5 | 58455872 | 58456295 | 424 | 3 | 1.88E-06 | 3.62E-06 | 0.001609 | 8.50E-06 | -0.00319675 | PLK2 |
| chr13 | 31846342 | 31846683 | 342 | 5 | 1.40E-05 | 9.00E-06 | 0.003679 | 8.50E-06 | 0.001064529 | AL138708.1 |
| chr6 | 153044098 | 153044285 | 188 | 2 | 4.81E-06 | 2.85E-05 | 2.19E-05 | 8.51E-06 | -0.01045131 | RGS17 |
| chr2 | 18041498 | 18042019 | 522 | 3 | 1.63E-06 | 2.32E-05 | 0.000242 | 8.51E-06 | -0.01317061 | KCNS3 |
| chr4 | 169200371 | 169200732 | 362 | 2 | 2.28E-06 | 5.19E-05 | 1.45E-05 | 8.51E-06 | -0.00416379 | SH3RF1 |
| chr19 | 46124862 | 46125833 | 972 | 5 | 1.16E-06 | 0.0001326 | 1.19E-05 | 8.51E-06 | -0.00701555 | AC007785.1 |
| chr2 | 19574727 | 19575006 | 280 | 3 | 8.59E-06 | 5.24E-05 | 0.000329 | 8.51E-06 | -0.01314662 | NA |
| chr9 | 131358803 | 131358949 | 147 | 3 | 1.38E-05 | 3.28E-06 | 0.002286 | 8.51E-06 | -0.00882856 | PLPP7 |
| chr12 | 108732147 | 108732965 | 819 | 5 | 1.64E-06 | 7.69E-05 | 0.000385 | 8.51E-06 | -0.00600637 | NA |
| chr15 | 78625878 | 78626218 | 341 | 3 | 1.88E-05 | 4.34E-06 | 0.001362 | 8.51E-06 | 0.001765977 | AC067863.1, CHRNB4 |
| chr19 | 1407377 | 1408141 | 765 | 7 | 1.13E-05 | 9.56E-06 | 0.003576 | 8.52E-06 | 0.000805662 | DAZAP1 |
| chr7 | 27238951 | 27239710 | 760 | 3 | 7.55E-06 | 1.12E-05 | 8.77E-05 | 8.52E-06 | -0.0007197 | AC004080.1 |
| chr7 | 75881942 | 75882226 | 285 | 2 | 1.99E-06 | 6.41E-05 | 1.30E-05 | 8.53E-06 | 0.003724237 | RHBDD2 |
| chr2 | 169068405 | 169068930 | 526 | 3 | 1.12E-06 | 4.16E-05 | 3.78E-05 | 8.53E-06 | 0.001109167 | DHRS9 |
| chr2 | 54113797 | 54113879 | 83 | 2 | 9.56E-06 | 5.47E-06 | 0.000409 | 8.54E-06 | 0.003819106 | ACYP2 |
| chr1 | 109718226 | 109718505 | 280 | 2 | 1.78E-06 | 0.0001001 | 1.05E-05 | 8.54E-06 | 0.00728732 | GSTM5 |
| chr21 | 45377617 | 45377984 | 368 | 2 | 4.43E-06 | 9.47E-06 | 7.79E-05 | 8.54E-06 | -0.00059458 | NA |
| chr2 | 134301745 | 134301768 | 24 | 2 | 1.86E-05 | 9.98E-06 | 7.15E-05 | 8.55E-06 | 0.002376492 | MGAT5 |
| chr2 | 226802011 | 226802853 | 843 | 4 | 6.50E-05 | 2.27E-05 | 0.002091 | 8.55E-06 | -0.00343354 | AC010735.2 |
| chr7 | 1570157 | 1570584 | 428 | 6 | 5.00E-06 | 0.0002668 | 5.85E-05 | 8.55E-06 | -0.00031649 | PSMG3-AS1, PSMG3 |
| chr7 | 103421542 | 103421677 | 136 | 2 | 1.55E-05 | 1.21E-05 | 5.36E-05 | 8.55E-06 | -0.00966735 | SLC26A5 |
| chr7 | 55526994 | 55527175 | 182 | 2 | 2.97E-05 | 9.97E-06 | 7.16E-05 | 8.55E-06 | 0.019992762 | VOPP1 |
| chr21 | 44370030 | 44370638 | 609 | 3 | 1.15E-06 | 1.90E-05 | 0.000448 | 8.55E-06 | 0.009935821 | TRPM2 |
| chr2 | 74501496 | 74502583 | 1088 | 7 | 7.90E-07 | 4.74E-05 | 5.38E-05 | 8.55E-06 | -0.00758691 | AC005041.3, LBX2 |
| chr1 | 153261561 | 153261911 | 351 | 4 | 4.96E-06 | 1.25E-05 | 0.000437 | 8.56E-06 | 0.000915023 | LORICRIN |
| chr21 | 45340201 | 45340247 | 47 | 2 | 5.94E-06 | 0.0003894 | 6.85E-06 | 8.56E-06 | -0.00633369 | LINC00316 |
| chr9 | 72256598 | 72257268 | 671 | 4 | 5.49E-05 | 3.81E-06 | 0.003252 | 8.56E-06 | -0.00710362 | GDA |
| chr6 | 43741290 | 43741688 | 399 | 2 | 7.19E-06 | 1.12E-05 | 5.97E-05 | 8.56E-06 | -0.00907888 | NA |
| chr11 | 108593375 | 108593771 | 397 | 6 | 3.46E-05 | 0.0004654 | 8.93E-05 | 8.56E-06 | 0.001721824 | EXPH5 |
| chr18 | 48777038 | 48777218 | 181 | 2 | 6.99E-06 | 6.90E-05 | 1.25E-05 | 8.57E-06 | -0.0081441 | CTIF |
| chr1 | 35361659 | 35362040 | 382 | 2 | 7.03E-07 | 5.67E-05 | 1.40E-05 | 8.57E-06 | 0.009780114 | ZMYM4, ZMYM4-AS1 |
| chr1 | 1081639 | 1082003 | 365 | 4 | 2.15E-06 | 0.0002972 | 2.11E-05 | 8.57E-06 | -0.00321348 | C1orf159 |
| chr4 | 3385554 | 3386267 | 714 | 7 | 1.08E-05 | 7.19E-06 | 0.002474 | 8.57E-06 | 0.001690401 | RGS12 |
| chr11 | 102109624 | 102110457 | 834 | 12 | 2.59E-06 | 0.0006566 | 5.20E-05 | 8.57E-06 | 0.000153889 | AP001527.1, YAP1, AP001527.2 |
| chr13 | 42271364 | 42271558 | 195 | 4 | 2.98E-06 | 4.05E-06 | 0.002159 | 8.57E-06 | -0.01539647 | NA |
| chr12 | 95244540 | 95244558 | 19 | 2 | 8.67E-06 | 2.37E-05 | 2.59E-05 | 8.57E-06 | 0.001477535 | VEZT |
| chr12 | 54018904 | 54020152 | 1249 | 9 | 1.30E-05 | 1.68E-06 | 0.01583 | 8.58E-06 | 0.008339168 | AC012531.3, HOXC6, HOXC4, AC012531.2, AC012531.1 |
| chr11 | 133228529 | 133228562 | 34 | 2 | 9.02E-06 | 1.78E-05 | 3.41E-05 | 8.59E-06 | 0.011741304 | OPCML |
| chr7 | 45459391 | 45459479 | 89 | 2 | 6.70E-07 | 2.90E-05 | 2.19E-05 | 8.59E-06 | -0.00114115 | NA |
| chr6 | 161154197 | 161154635 | 439 | 7 | 1.09E-06 | 0.0007349 | 1.01E-05 | 8.60E-06 | 0.000455815 | AGPAT4 |
| chr12 | 64101281 | 64101400 | 120 | 2 | 2.13E-05 | 6.25E-06 | 0.000229 | 8.61E-06 | -0.01926325 | SRGAP1, AC025576.3 |
| chr5 | 173989271 | 173989964 | 694 | 5 | 8.27E-06 | 1.83E-05 | 0.000226 | 8.61E-06 | -0.00610155 | C5orf47 |
| chr4 | 105399792 | 105400103 | 312 | 2 | 4.70E-06 | 4.86E-05 | 1.53E-05 | 8.61E-06 | -0.00942007 | PPA2 |
| chr9 | 128169610 | 128170142 | 533 | 3 | 2.37E-05 | 6.98E-06 | 0.000269 | 8.61E-06 | -0.00213399 | CIZ1 |
| chr16 | 27549474 | 27550238 | 765 | 13 | 3.48E-06 | 0.0003681 | 0.006004 | 8.61E-06 | 0.000333367 | KATNIP, GTF3C1 |
| chr7 | 64563146 | 64564361 | 1216 | 9 | 2.47E-05 | 6.34E-05 | 0.003217 | 8.62E-06 | -0.00158921 | NA |
| chr10 | 86880056 | 86880545 | 490 | 2 | 6.45E-07 | 0.0007661 | 6.05E-06 | 8.62E-06 | -0.00984303 | BMPR1A |
| chr9 | 21385441 | 21385837 | 397 | 2 | 7.48E-07 | 0.0012843 | 5.59E-06 | 8.62E-06 | -0.01790112 | NA |
| chr11 | 82718386 | 82719307 | 922 | 4 | 4.19E-07 | 0.0001384 | 3.44E-05 | 8.62E-06 | 0.003381453 | NA |
| chr4 | 38688289 | 38688936 | 648 | 6 | 2.41E-05 | 4.62E-06 | 0.001493 | 8.63E-06 | -0.00113341 | KLF3 |
| chr21 | 42403495 | 42404153 | 659 | 10 | 1.32E-05 | 0.0010221 | 0.000167 | 8.63E-06 | -0.00152086 | UBASH3A |
| chr11 | 33376133 | 33376820 | 688 | 6 | 1.51E-05 | 0.0002143 | 0.000597 | 8.63E-06 | 0.002238684 | KIAA1549L |
| chr17 | 42185005 | 42185874 | 870 | 11 | 4.21E-06 | 0.0011785 | 0.000426 | 8.65E-06 | -0.00220431 | HCRT |
| chr1 | 76764934 | 76765274 | 341 | 4 | 1.24E-05 | 1.20E-05 | 0.000365 | 8.66E-06 | -0.0239563 | LINC02567 |
| chr1 | 76074425 | 76075314 | 890 | 12 | 7.55E-07 | 0.0001151 | 0.001939 | 8.67E-06 | 0.001198291 | ST6GALNAC3 |
| chr4 | 3206573 | 3206951 | 379 | 6 | 1.87E-07 | 0.1344019 | 5.98E-07 | 8.67E-06 | 0.001555636 | HTT |
| chr8 | 61550690 | 61550808 | 119 | 2 | 7.57E-06 | 8.63E-06 | 9.53E-05 | 8.67E-06 | 0.003372192 | ASPH |
| chr1 | 15585718 | 15586339 | 622 | 4 | 4.85E-06 | 8.14E-05 | 7.45E-05 | 8.68E-06 | -0.04339008 | DNAJC16, AL121992.1 |
| chr15 | 92082366 | 92082929 | 564 | 3 | 6.98E-06 | 6.21E-05 | 9.00E-05 | 8.68E-06 | -0.00030279 | SLCO3A1 |
| chr1 | 13541812 | 13542296 | 485 | 2 | 3.02E-05 | 5.25E-06 | 0.000602 | 8.68E-06 | 0.002070116 | NA |
| chr13 | 110371819 | 110372348 | 530 | 3 | 3.40E-06 | 1.61E-05 | 5.86E-05 | 8.69E-06 | -0.00513839 | COL4A2 |
| chr7 | 86786317 | 86786979 | 663 | 8 | 5.05E-06 | 0.001836 | 0.000286 | 8.69E-06 | -0.0018528 | GRM3, GRM3-AS1 |
| chr12 | 54301141 | 54301425 | 285 | 6 | 8.68E-06 | 0.0015868 | 0.000217 | 8.69E-06 | -0.00383555 | AC078778.1, COPZ1 |
| chr12 | 52953150 | 52953547 | 398 | 2 | 1.99E-06 | 1.75E-05 | 3.53E-05 | 8.69E-06 | 0.001580991 | NA |
| chr3 | 66153945 | 66154120 | 176 | 2 | 2.10E-05 | 6.19E-06 | 0.000249 | 8.69E-06 | -0.02130325 | SLC25A26 |
| chr16 | 30814208 | 30814630 | 423 | 4 | 4.50E-06 | 3.38E-05 | 7.84E-05 | 8.70E-06 | 0.001539676 | NA |
| chr10 | 102232716 | 102233271 | 556 | 3 | 1.05E-07 | 0.0003729 | 5.75E-06 | 8.71E-06 | -0.00723659 | PITX3 |
| chr16 | 323931 | 324570 | 640 | 3 | 1.69E-06 | 9.08E-06 | 0.000757 | 8.71E-06 | 0.001132326 | AXIN1 |
| chr14 | 45253329 | 45254167 | 839 | 9 | 6.49E-06 | 3.50E-05 | 0.000167 | 8.71E-06 | -0.00577166 | MIS18BP1 |
| chr12 | 5964610 | 5965146 | 537 | 3 | 1.01E-05 | 4.12E-06 | 0.001378 | 8.71E-06 | -0.01429501 | VWF |
| chr22 | 39312541 | 39312578 | 38 | 2 | 6.68E-06 | 0.000109 | 1.03E-05 | 8.71E-06 | -0.00805462 | NA |
| chr11 | 123577133 | 123577993 | 861 | 5 | 4.27E-06 | 0.0001644 | 0.000261 | 8.71E-06 | -0.00444194 | GRAMD1B |
| chr4 | 118981433 | 118982203 | 771 | 3 | 9.79E-05 | 1.47E-05 | 0.000525 | 8.72E-06 | -0.00694846 | SYNPO2 |

| | | | | | | | | | | |
|---|---|---|---|---|---|---|---|---|---|---|
| chr12 | 113312368 | 113312703 | 336 | 2 | 6.60E-06 | 1.85E-05 | 3.36E-05 | 8.72E-06 | -0.00891305 | SLC8B1 |
| chr2 | 6731213 | 6731786 | 574 | 3 | 9.52E-07 | 0.0005393 | 4.88E-06 | 8.72E-06 | -0.00389414 | LINC00487 |
| chr19 | 3969524 | 3969952 | 429 | 4 | 1.56E-06 | 0.0004259 | 1.14E-05 | 8.72E-06 | -0.00507588 | DAPK3 |
| chr8 | 890173 | 890770 | 598 | 4 | 2.87E-05 | 2.10E-05 | 0.001858 | 8.72E-06 | -0.00368216 | DLGAP2 |
| chr2 | 98775339 | 98775641 | 303 | 2 | 1.34E-05 | 8.92E-06 | 9.13E-05 | 8.73E-06 | -0.02053945 | LINC02611 |
| chr14 | 60658204 | 60658732 | 529 | 6 | 2.83E-05 | 3.03E-05 | 0.000705 | 8.73E-06 | -0.00512171 | SIX1 |
| chr13 | 99308431 | 99308703 | 273 | 3 | 3.67E-06 | 0.0001639 | 1.13E-05 | 8.73E-06 | -0.00797202 | UBAC2 |
| chr6 | 42782714 | 42783506 | 793 | 6 | 2.24E-05 | 9.37E-06 | 0.006669 | 8.73E-06 | 0.004530831 | BICRAL |
| chr2 | 177392614 | 177393697 | 1084 | 8 | 2.67E-05 | 2.86E-06 | 0.011129 | 8.73E-06 | 0.002875353 | AGPS, NFE2L2, AC019080.1 |
| chr10 | 14509050 | 14509379 | 330 | 4 | 8.10E-07 | 0.0107451 | 1.37E-06 | 8.73E-06 | -0.00419381 | NA |
| chr1 | 156368822 | 156369495 | 674 | 10 | 5.64E-06 | 4.71E-05 | 0.001114 | 8.74E-06 | -0.00214207 | RHBG |
| chr14 | 96093135 | 96093571 | 437 | 2 | 1.15E-06 | 0.0003261 | 7.31E-06 | 8.74E-06 | -0.0100293 | C14orf132 |
| chr21 | 44423617 | 44423980 | 364 | 3 | 1.03E-05 | 5.32E-06 | 0.000551 | 8.74E-06 | -0.00810276 | TRPM2, TRPM2-AS |
| chr12 | 95633416 | 95633810 | 395 | 2 | 1.34E-05 | 7.28E-06 | 0.000148 | 8.75E-06 | -0.01368712 | PGAM1P5 |
| chr10 | 68404052 | 68404579 | 528 | 2 | 1.29E-05 | 9.35E-06 | 8.42E-05 | 8.75E-06 | 0.001476454 | RUFY2 |
| chr15 | 88913350 | 88914010 | 661 | 11 | 1.12E-05 | 1.06E-05 | 0.006312 | 8.76E-06 | -0.00365917 | MFGE8 |
| chr17 | 77304067 | 77304202 | 136 | 2 | 1.17E-05 | 9.08E-06 | 8.91E-05 | 8.76E-06 | -0.00187257 | SEPTIN9 |
| chr7 | 23298224 | 23299278 | 1055 | 13 | 3.45E-06 | 0.0001159 | 0.002055 | 8.76E-06 | -0.00068827 | MALSU1 |
| chr15 | 75023383 | 75023982 | 600 | 10 | 4.36E-06 | 0.0004319 | 8.80E-05 | 8.76E-06 | 0.000526681 | PPCDC |
| chr21 | 44956579 | 44957155 | 577 | 3 | 9.98E-07 | 1.12E-05 | 0.00052 | 8.77E-06 | -0.00253076 | FAM207A |
| chr7 | 93220145 | 93220773 | 629 | 4 | 7.02E-06 | 3.93E-06 | 0.001851 | 8.77E-06 | -0.00485398 | HEPACAM2 |
| chr1 | 37473822 | 37474887 | 1066 | 11 | 6.65E-07 | 0.0021457 | 0.000134 | 8.77E-06 | 0.001787081 | ZC3H12A, LINC01137 |
| chr22 | 28953160 | 28953642 | 483 | 3 | 5.56E-06 | 2.09E-05 | 0.000133 | 8.77E-06 | 0.000227562 | ZNRF3 |
| chr10 | 28534104 | 28534162 | 59 | 2 | 2.71E-06 | 1.06E-05 | 6.92E-05 | 8.78E-06 | 0.000322215 | WAC |
| chr10 | 103446130 | 103446879 | 750 | 3 | 2.82E-05 | 1.52E-05 | 0.000413 | 8.79E-06 | -0.00536603 | PDCD11, CALHM2 |
| chr13 | 113084937 | 113085263 | 327 | 4 | 6.51E-06 | 5.90E-05 | 8.48E-05 | 8.80E-06 | -0.00161908 | MCF2L |
| chr17 | 7670363 | 7670711 | 349 | 2 | 1.97E-05 | 5.32E-06 | 0.000607 | 8.80E-06 | -0.00044405 | TP53 |
| chr19 | 10817496 | 10817963 | 468 | 8 | 6.40E-06 | 6.13E-05 | 0.000335 | 8.80E-06 | -0.00080179 | DNM2, MIR199A1 |
| chr1 | 203487563 | 203487690 | 128 | 2 | 6.45E-05 | 6.63E-06 | 0.000204 | 8.81E-06 | -0.00377205 | PRELP |
| chr17 | 81719403 | 81720045 | 643 | 4 | 2.73E-07 | 0.0002787 | 4.98E-05 | 8.81E-06 | 0.000181971 | AC139530.2, SLC25A10 |
| chr14 | 68934300 | 68934352 | 53 | 2 | 1.47E-05 | 1.77E-05 | 3.57E-05 | 8.81E-06 | 0.020008774 | ACTN1 |
| chr14 | 68248745 | 68249132 | 388 | 2 | 4.42E-06 | 8.35E-06 | 0.000107 | 8.82E-06 | 0.002714062 | RAD51B |
| chr12 | 9221687 | 9222002 | 316 | 2 | 5.10E-06 | 4.27E-05 | 1.72E-05 | 8.82E-06 | -0.01807827 | NA |
| chr2 | 201131006 | 201131550 | 545 | 2 | 4.64E-06 | 6.45E-06 | 0.000226 | 8.82E-06 | -0.02514628 | CFLAR |
| chr9 | 135074738 | 135075174 | 437 | 4 | 3.09E-07 | 9.28E-05 | 0.00018 | 8.82E-06 | -0.01305288 | NA |
| chr7 | 1103129 | 1103936 | 808 | 3 | 4.98E-05 | 1.16E-05 | 0.000257 | 8.82E-06 | -0.00756192 | C7orf50 |
| chr2 | 45610672 | 45611442 | 771 | 9 | 9.11E-06 | 0.000104 | 0.001035 | 8.82E-06 | 0.000234399 | SRBD1 |
| chr18 | 76491528 | 76492179 | 652 | 5 | 8.05E-06 | 9.78E-05 | 0.000367 | 8.83E-06 | 0.001611783 | AC018413.1, ZNF516 |
| chr12 | 94872934 | 94873093 | 160 | 2 | 1.14E-05 | 5.34E-06 | 0.000616 | 8.85E-06 | -0.00348897 | NA |
| chr19 | 45548468 | 45548719 | 252 | 2 | 5.00E-06 | 9.58E-05 | 1.11E-05 | 8.85E-06 | -0.00841792 | OPA3 |
| chr1 | 7582219 | 7582295 | 77 | 2 | 1.42E-05 | 7.24E-06 | 0.000157 | 8.85E-06 | -0.00747405 | CAMTA1 |
| chr11 | 119056966 | 119057630 | 665 | 8 | 2.50E-05 | 4.19E-06 | 0.006361 | 8.85E-06 | 0.000714473 | HYOU1 |
| chr5 | 148268392 | 148268929 | 538 | 3 | 5.65E-06 | 6.20E-06 | 0.000926 | 8.85E-06 | 0.000988273 | SPINK13, AC011346.1 |
| chr1 | 171248050 | 171248618 | 569 | 5 | 7.51E-06 | 1.98E-05 | 0.000462 | 8.85E-06 | -0.00398148 | FMO1, BX284613.2 |
| chr8 | 98427457 | 98428051 | 595 | 5 | 7.14E-06 | 1.49E-05 | 0.000879 | 8.86E-06 | 0.002746503 | KCNS2, STK3 |
| chr5 | 43125438 | 43125448 | 11 | 2 | 1.41E-05 | 2.62E-05 | 2.50E-05 | 8.88E-06 | -0.00322671 | ZNF131 |
| chr1 | 51263570 | 51263931 | 362 | 3 | 1.84E-05 | 5.06E-06 | 0.000643 | 8.88E-06 | 0.002830305 | RNF11 |
| chr12 | 75478357 | 75478597 | 241 | 2 | 4.96E-06 | 3.10E-05 | 2.18E-05 | 8.89E-06 | 0.005536896 | NA |
| chr1 | 37765474 | 37766096 | 623 | 4 | 1.17E-05 | 7.51E-06 | 0.000692 | 8.89E-06 | -0.00057096 | NA |
| chr16 | 29661847 | 29662710 | 864 | 5 | 2.50E-07 | 7.00E-05 | 0.000302 | 8.89E-06 | -0.004048 | NA |
| chr3 | 129248550 | 129249569 | 1020 | 8 | 1.09E-05 | 4.29E-05 | 0.001446 | 8.90E-06 | 0.001561548 | NA |
| chr7 | 102454917 | 102455300 | 384 | 2 | 3.47E-06 | 1.78E-05 | 3.61E-05 | 8.90E-06 | -0.00669759 | ORAI2 |
| chr20 | 8829603 | 8829854 | 252 | 2 | 5.57E-06 | 4.71E-05 | 1.63E-05 | 8.91E-06 | -0.00876429 | PLCB1 |
| chr3 | 66004073 | 66004793 | 721 | 5 | 1.41E-05 | 0.0002271 | 0.000348 | 8.91E-06 | -0.00412127 | MAGI1 |
| chr17 | 79925179 | 79925409 | 231 | 2 | 1.83E-05 | 5.78E-06 | 0.000386 | 8.91E-06 | -0.02447716 | LINC01978, LINC01979 |
| chr3 | 98195553 | 98195717 | 165 | 2 | 1.83E-05 | 8.69E-06 | 0.000101 | 8.91E-06 | -0.0284561 | NA |
| chr16 | 51650913 | 51651131 | 219 | 3 | 2.62E-05 | 5.57E-06 | 0.000417 | 8.91E-06 | -0.01362435 | NA |
| chr22 | 35351638 | 35352287 | 650 | 3 | 6.53E-06 | 3.65E-06 | 0.001866 | 8.91E-06 | -0.01406618 | NA |
| chr19 | 1025543 | 1026208 | 666 | 3 | 5.35E-07 | 0.0002714 | 6.62E-06 | 8.91E-06 | -0.00664039 | NA |
| chr12 | 93081273 | 93081325 | 53 | 2 | 3.61E-06 | 0.0009407 | 6.03E-06 | 8.91E-06 | 0.007972327 | AC138123.1 |
| chr12 | 48882512 | 48883153 | 642 | 3 | 1.16E-05 | 0.0005636 | 6.52E-05 | 8.91E-06 | -0.00304841 | NA |
| chr17 | 49512232 | 49512964 | 733 | 5 | 2.78E-05 | 8.37E-06 | 0.00417 | 8.92E-06 | -0.00764971 | NGFR, AC006487.1 |
| chr11 | 68460170 | 68460760 | 591 | 10 | 1.60E-06 | 0.0007952 | 6.97E-06 | 8.92E-06 | 0.000259079 | PPP6R3 |
| chr13 | 77623386 | 77623867 | 482 | 3 | 2.33E-05 | 2.34E-05 | 0.000374 | 8.92E-06 | -0.00289162 | SCEL, RNY3P7 |
| chr9 | 135216829 | 135216854 | 26 | 2 | 2.45E-06 | 2.44E-05 | 2.67E-05 | 8.92E-06 | -0.00898841 | AL390778.1 |
| chr15 | 62245761 | 62245960 | 200 | 2 | 4.17E-05 | 3.72E-05 | 1.91E-05 | 8.93E-06 | -0.00788397 | AC126323.1 |
| chr10 | 132305557 | 132306494 | 938 | 8 | 3.03E-06 | 3.31E-05 | 0.004261 | 8.93E-06 | -0.0020151 | STK32C |
| chr22 | 28741539 | 28742283 | 745 | 13 | 6.57E-06 | 5.04E-05 | 0.006418 | 8.93E-06 | 0.000621106 | HSCB, CHEK2 |
| chr19 | 42287074 | 42287566 | 493 | 5 | 6.50E-07 | 6.87E-05 | 0.00021 | 8.93E-06 | -0.00289494 | CIC |
| chr12 | 133130730 | 133130993 | 264 | 5 | 4.86E-05 | 4.47E-05 | 0.000687 | 8.94E-06 | 0.000807616 | ZNF10, AC026786.1 |
| chr13 | 30373172 | 30373985 | 814 | 5 | 2.17E-05 | 4.63E-06 | 0.004017 | 8.94E-06 | 0.002910815 | LINC00426 |
| chr21 | 43190741 | 43191270 | 530 | 2 | 2.34E-06 | 2.27E-05 | 2.87E-05 | 8.94E-06 | -0.00581001 | NA |
| chr10 | 127986995 | 127987273 | 279 | 4 | 4.52E-06 | 0.0002622 | 5.48E-06 | 8.95E-06 | -0.00562584 | PTPRE |
| chr15 | 78077905 | 78077984 | 80 | 3 | 0.000148362 | 4.43E-06 | 0.0013 | 8.95E-06 | 0.013411594 | SH2D7 |
| chr12 | 111792533 | 111792778 | 246 | 2 | 1.08E-05 | 1.73E-05 | 3.76E-05 | 8.96E-06 | -0.00589207 | ALDH2 |
| chr21 | 46455406 | 46455763 | 358 | 3 | 7.91E-07 | 0.0007377 | 6.10E-06 | 8.97E-06 | 0.004996198 | NA |
| chr8 | 66513595 | 66513971 | 377 | 3 | 6.54E-06 | 3.68E-05 | 2.82E-05 | 8.97E-06 | -0.00455199 | AC009879.3, VXN |
| chr2 | 50348186 | 50348489 | 304 | 3 | 1.20E-06 | 6.38E-06 | 0.000634 | 8.97E-06 | -0.02559465 | NRXN1 |
| chr2 | 15180422 | 15180790 | 369 | 3 | 3.27E-06 | 0.0004575 | 1.74E-05 | 8.97E-06 | -0.00271575 | NBAS |
| chr1 | 15748882 | 15750017 | 1136 | 4 | 5.99E-05 | 2.82E-05 | 0.000634 | 8.97E-06 | 1.47E-05 | SLC25A34-AS1 |
| chr19 | 32719260 | 32719945 | 686 | 11 | 2.19E-06 | 0.0002314 | 0.001251 | 8.97E-06 | 0.003749451 | TDRD12, AC008736.2 |
| chr5 | 138128510 | 138128813 | 304 | 2 | 5.95E-06 | 3.23E-05 | 2.14E-05 | 8.97E-06 | -0.01515347 | NME5 |
| chr3 | 183984047 | 183985098 | 1052 | 4 | 8.91E-06 | 4.73E-06 | 0.001863 | 8.98E-06 | -0.00563091 | ABCC5 |
| chr15 | 72800845 | 72801352 | 508 | 3 | 1.54E-05 | 7.64E-05 | 7.17E-05 | 8.98E-06 | -0.01454197 | NA |
| chr4 | 1278009 | 1278394 | 386 | 3 | 6.61E-07 | 0.0024803 | 3.35E-06 | 8.99E-06 | -0.00500618 | CTBP1-DT |
| chr19 | 41447951 | 41447970 | 20 | 2 | 2.37E-05 | 6.23E-06 | 0.000283 | 8.99E-06 | -0.01370981 | NA |
| chr9 | 108934473 | 108935365 | 893 | 9 | 8.61E-07 | 7.59E-05 | 0.001092 | 8.99E-06 | -0.00252704 | ABITRAM |
| chr6 | 168161334 | 168161728 | 395 | 3 | 1.20E-05 | 1.24E-05 | 0.000179 | 8.99E-06 | -0.00427209 | NA |
| chr16 | 67876969 | 67877602 | 634 | 6 | 1.23E-05 | 4.98E-05 | 0.000548 | 8.99E-06 | 0.000922308 | EDC4 |
| chr3 | 98901972 | 98902612 | 641 | 8 | 4.28E-07 | 0.0005571 | 0.000332 | 9.00E-06 | -0.00318173 | AC091212.1 |
| chr14 | 36477791 | 36477859 | 69 | 2 | 2.96E-05 | 8.14E-06 | 0.00012 | 9.00E-06 | -0.01515886 | AL132857.1, SFTA3, SFTA3 |
| chr16 | 87177308 | 87177674 | 367 | 3 | 9.22E-06 | 2.00E-05 | 8.67E-05 | 9.00E-06 | -0.00540575 | C16orf95 |
| chr2 | 1448488 | 1448893 | 406 | 3 | 1.10E-05 | 3.21E-05 | 0.000175 | 9.02E-06 | -9.60E-05 | TPO, AC105450.1 |
| chr19 | 1479219 | 1479574 | 356 | 10 | 1.35E-05 | 0.0002035 | 0.00183 | 9.02E-06 | 0.00086002 | C19orf25 |
| chr7 | 143051361 | 143052445 | 1085 | 7 | 6.63E-06 | 9.99E-06 | 0.001892 | 9.03E-06 | -0.00118889 | OR6V1 |
| chr19 | 1021258 | 1021717 | 460 | 4 | 2.11E-06 | 8.78E-06 | 0.000206 | 9.03E-06 | -0.0010267 | RNU6-2 |
| chr11 | 121023283 | 121023637 | 355 | 3 | 5.06E-06 | 4.02E-05 | 0.000109 | 9.03E-06 | -0.00514258 | NA |
| chr11 | 125345922 | 125346140 | 219 | 3 | 3.10E-06 | 0.0005396 | 9.45E-06 | 9.03E-06 | -0.0049925 | PKNOX2 |
| chr20 | 57157211 | 57157217 | 7 | 2 | 3.05E-06 | 5.46E-06 | 0.000617 | 9.03E-06 | 0.000463871 | NA |
| chr18 | 54220388 | 54220853 | 466 | 2 | 1.69E-05 | 6.68E-06 | 0.000221 | 9.03E-06 | -0.00480166 | MBD2 |
| chr6 | 125985403 | 125986332 | 930 | 7 | 5.54E-07 | 0.0001572 | 0.001284 | 9.04E-06 | 0.005209587 | NA |
| chr10 | 49524450 | 49525150 | 701 | 4 | 6.60E-06 | 0.0013055 | 0.000228 | 9.04E-06 | -0.00128802 | ERCC6 |
| chr1 | 17125939 | 17126316 | 378 | 2 | 9.46E-06 | 1.18E-05 | 6.26E-05 | 9.04E-06 | -0.00779539 | NA |
| chr12 | 69037273 | 69037290 | 18 | 2 | 1.26E-05 | 4.05E-05 | 1.84E-05 | 9.04E-06 | -0.00402355 | NA |
| chr1 | 180275327 | 180275929 | 603 | 3 | 4.94E-05 | 3.32E-06 | 0.002956 | 9.05E-06 | -0.00016285 | LHX4, ACBD6 |
| chr16 | 50391418 | 50391503 | 86 | 3 | 2.86E-06 | 0.0017778 | 9.53E-06 | 9.05E-06 | 0.003482293 | LINC02178 |
| chr6 | 152749633 | 152750425 | 793 | 3 | 1.06E-05 | 3.48E-05 | 4.58E-05 | 9.05E-06 | -0.01420793 | NA |
| chr1 | 3779617 | 3780005 | 389 | 3 | 3.89E-06 | 1.21E-05 | 9.05E-05 | 9.06E-06 | 0.000367794 | LRRC47 |
| chr17 | 1716714 | 1717551 | 838 | 7 | 8.92E-07 | 2.07E-05 | 0.004446 | 9.06E-06 | 0.000398626 | WDR81, MIR22HG |
| chr19 | 9796573 | 9796893 | 321 | 3 | 1.64E-05 | 1.66E-05 | 0.000283 | 9.06E-06 | -0.01040495 | AC008752.1 |
| chr1 | 226982735 | 226982962 | 228 | 4 | 4.17E-06 | 1.40E-05 | 8.91E-05 | 9.06E-06 | 0.000371544 | AL391628.1, COQ8A |
| chr10 | 3141386 | 3141945 | 560 | 6 | 7.64E-07 | 0.0004123 | 0.000266 | 9.07E-06 | 0.001195391 | AL451164.2, PITRM1-AS1, PITRM1 |
| chr22 | 43811733 | 43812669 | 937 | 13 | 2.83E-06 | 0.0002296 | 0.002797 | 9.07E-06 | -4.80E-05 | Z97055.2, EFCAB6 |
| chr5 | 174455825 | 174456358 | 534 | 2 | 9.99E-08 | 0.0005109 | 6.84E-06 | 9.07E-06 | -0.00415356 | LINC01411 |
| chr5 | 118594928 | 118595148 | 221 | 2 | 7.34E-06 | 5.14E-05 | 1.58E-05 | 9.07E-06 | -0.00404259 | AC114311.1 |
| chr19 | 47540874 | 47541143 | 270 | 2 | 1.36E-05 | 5.47E-06 | 0.000634 | 9.07E-06 | -0.00234544 | ZNF541 |
| chr4 | 40334238 | 40335320 | 1083 | 6 | 2.52E-06 | 1.75E-05 | 0.000943 | 9.08E-06 | -0.00512928 | NA |
| chr3 | 72262516 | 72262895 | 380 | 4 | 1.88E-05 | 8.29E-06 | 0.000449 | 9.08E-06 | 0.000513777 | LINC00877 |
| chr17 | 4007745 | 4008121 | 377 | 4 | 2.98E-05 | 2.52E-06 | 0.004423 | 9.08E-06 | -0.00318484 | ZZEF1 |
| chr17 | 27471389 | 27472421 | 1033 | 8 | 2.29E-05 | 1.69E-06 | 0.015614 | 9.09E-06 | -0.00459516 | KSR1 |
| chr20 | 35438686 | 35439518 | 833 | 5 | 2.42E-05 | 3.16E-06 | 0.001776 | 9.09E-06 | -0.01069057 | GDF5 |
| chr1 | 49048047 | 49048185 | 139 | 2 | 5.67E-06 | 0.0003987 | 7.25E-06 | 9.10E-06 | -0.00658579 | AC099788.1, AGBL4 |
| chr3 | 33492072 | 33492776 | 705 | 5 | 1.46E-06 | 0.0018167 | 1.18E-05 | 9.10E-06 | -0.00015358 | RNA5SP128 |
| chr7 | 43758240 | 43758608 | 369 | 4 | 1.12E-05 | 7.41E-06 | 0.001553 | 9.10E-06 | -0.00225286 | NA |
| chr14 | 76491070 | 76491601 | 532 | 5 | 1.11E-05 | 0.0012305 | 0.000628 | 9.11E-06 | -0.00077739 | ESRRB |
| chr11 | 100340718 | 100341163 | 446 | 2 | 2.00E-06 | 8.30E-05 | 1.23E-05 | 9.12E-06 | 0.005363436 | CNTN5 |
| chr16 | 2074102 | 2074498 | 397 | 5 | 9.90E-06 | 4.80E-05 | 0.002166 | 9.12E-06 | -0.00327093 | TSC2 |
| chr1 | 233520481 | 233520751 | 271 | 2 | 2.40E-05 | 5.66E-06 | 0.000515 | 9.12E-06 | -0.01589634 | NA |
| chr13 | 111152967 | 111153583 | 617 | 8 | 2.84E-05 | 1.18E-05 | 0.003612 | 9.12E-06 | 0.003895916 | ARHGEF7, AL353704.1 |
| chr3 | 10164035 | 10164729 | 695 | 7 | 5.15E-06 | 2.74E-05 | 0.003954 | 9.12E-06 | -0.00319039 | NA |
| chr9 | 93144237 | 93144795 | 559 | 3 | 1.03E-05 | 7.40E-06 | 0.000543 | 9.12E-06 | -0.0055651 | NA |
| chr1 | 165938615 | 165938622 | 8 | 2 | 2.56E-05 | 5.59E-06 | 0.000569 | 9.13E-06 | 0.015296715 | NA |
| chr6 | 157673417 | 157673887 | 471 | 4 | 7.86E-06 | 5.30E-06 | 0.001092 | 9.13E-06 | -0.00681765 | ZDHHC14 |
| chr16 | 85639003 | 85639993 | 991 | 3 | 1.51E-05 | 5.23E-06 | 0.000777 | 9.13E-06 | -0.00210358 | GSE1 |
| chr2 | 128485790 | 128486615 | 826 | 5 | 3.44E-05 | 0.0019202 | 0.00011 | 9.13E-06 | -0.001492 | NA |
| chr22 | 38754869 | 38755634 | 766 | 8 | 3.40E-09 | 0.0029831 | 3.66E-06 | 9.13E-06 | 0.002066017 | SUN2 |
| chr11 | 57752979 | 57753516 | 538 | 7 | 3.19E-07 | 0.0001399 | 1.33E-05 | 9.14E-06 | 0.000817369 | AP001931.2, AP001931.1, CTNND1 |
| chr5 | 138138669 | 138139011 | 343 | 2 | 2.18E-06 | 1.18E-05 | 6.38E-05 | 9.15E-06 | -0.01834836 | NME5 |
| chr11 | 35682852 | 35683095 | 244 | 3 | 9.50E-06 | 0.0001253 | 2.92E-05 | 9.15E-06 | -0.00161789 | TRIM44 |
| chr19 | 51502339 | 51502851 | 513 | 4 | 4.76E-06 | 3.75E-05 | 0.000339 | 9.15E-06 | 0.000335252 | NA |
| chr1 | 62316066 | 62316535 | 470 | 2 | 3.04E-06 | 2.55E-05 | 2.67E-05 | 9.15E-06 | -0.0055551 | KANK4 |
| chr3 | 32934516 | 32934648 | 133 | 2 | 5.05E-06 | 0.0008302 | 6.31E-06 | 9.15E-06 | -0.02954387 | NA |
| chr22 | 20032854 | 20033437 | 584 | 4 | 1.34E-06 | 0.0004127 | 4.32E-05 | 9.16E-06 | 0.001968027 | TANGO2, MIR185 |
| chr8 | 52409761 | 52410217 | 457 | 7 | 6.75E-06 | 0.0002669 | 0.000159 | 9.16E-06 | -0.00255542 | ST18 |
| chr14 | 77307564 | 77307681 | 118 | 2 | 1.78E-06 | 0.0001491 | 9.71E-06 | 9.16E-06 | 0.003605978 | POMT2 |
| chr1 | 247452131 | 247452994 | 864 | 6 | 3.36E-06 | 0.0001252 | 1.85E-05 | 9.16E-06 | -0.0046318 | OR2B11 |
| chr16 | 3762612 | 3762687 | 76 | 3 | 1.28E-05 | 8.62E-06 | 0.000571 | 9.16E-06 | -0.00122632 | CREBBP |
| chr1 | 91546851 | 91547179 | 329 | 4 | 2.68E-05 | 3.57E-06 | 0.003447 | 9.16E-06 | 0.019421073 | NA |
| chr5 | 132387547 | 132388354 | 808 | 2 | 6.68E-05 | 5.53E-06 | 0.000634 | 9.16E-06 | -0.01100349 | SLC22A5 |
| chr11 | 72642005 | 72642560 | 556 | 9 | 1.26E-05 | 3.65E-05 | 0.001504 | 9.16E-06 | -0.00062246 | PDE2A |
| chr3 | 183288933 | 183289348 | 416 | 2 | 9.21E-07 | 0.0016459 | 5.73E-06 | 9.17E-06 | -0.00687736 | B3GNT5, MCF2L2 |
| chr1 | 24470415 | 24470843 | 429 | 5 | 2.00E-05 | 7.14E-06 | 0.000954 | 9.17E-06 | -0.00772103 | NIPAL3 |
| chr8 | 143540987 | 143542055 | 1069 | 8 | 3.33E-06 | 3.91E-05 | 0.000437 | 9.17E-06 | 0.001237553 | AC067930.9, ZC3H3, AC067930.3 |
| chr6 | 106195162 | 106195346 | 185 | 2 | 2.29E-05 | 6.23E-06 | 0.000317 | 9.18E-06 | 0.001723461 | ATG5 |
| chr6 | 166950540 | 166950577 | 38 | 2 | 1.00E-05 | 6.69E-06 | 0.000236 | 9.18E-06 | -0.01281045 | AL159163.1, RNASET2 |
| chr6 | 52284709 | 52285462 | 754 | 11 | 1.05E-05 | 2.74E-05 | 0.007001 | 9.18E-06 | 0.001252711 | MCM3 |
| chr20 | 63707277 | 63708160 | 884 | 14 | 8.38E-06 | 0.0002666 | 0.00684 | 9.18E-06 | 0.001187676 | ZGPAT, ARFRP1 |
| chr5 | 72109705 | 72110361 | 657 | 5 | 6.55E-06 | 4.48E-05 | 9.22E-05 | 9.20E-06 | -0.00453668 | MAP1B |

| | | | | | | | | | | |
|---|---|---|---|---|---|---|---|---|---|---|
| chr2 | 37005179 | 37005453 | 275 | 2 | 2.21E-05 | 6.09E-06 | 0.000358 | 9.20E-06 | -0.05758553 | HEATR5B |
| chr4 | 169244508 | 169244747 | 240 | 3 | 2.50E-05 | 4.91E-06 | 0.000735 | 9.21E-06 | 0.008262608 | SH3RF1 |
| chr12 | 118103780 | 118104601 | 822 | 9 | 2.61E-05 | 4.46E-06 | 0.003525 | 9.21E-06 | 0.000124295 | VSIG10 |
| chr3 | 30696074 | 30696312 | 239 | 2 | 1.82E-06 | 0.0006011 | 6.73E-06 | 9.22E-06 | 0.002887456 | NA |
| chr5 | 141879144 | 141879691 | 548 | 5 | 3.59E-05 | 4.47E-06 | 0.005521 | 9.22E-06 | 0.002168382 | PCDH1 |
| chr12 | 8604257 | 8604312 | 56 | 2 | 1.96E-05 | 6.94E-06 | 0.00021 | 9.23E-06 | -0.03270384 | AICDA |
| chr10 | 70132837 | 70133403 | 567 | 10 | 1.78E-06 | 0.0001285 | 0.000148 | 9.23E-06 | -0.00473126 | AIFM2 |
| chr8 | 131935413 | 131936017 | 605 | 3 | 1.63E-06 | 6.63E-05 | 5.20E-05 | 9.23E-06 | -0.00464509 | EFR3A |
| chr5 | 57488368 | 57488727 | 360 | 2 | 5.85E-06 | 2.66E-05 | 2.60E-05 | 9.23E-06 | -0.00773317 | AC025470.2 |
| chr4 | 1490157 | 1490297 | 141 | 3 | 5.33E-06 | 0.0004091 | 5.73E-06 | 9.23E-06 | -0.00632498 | NA |
| chr10 | 97465934 | 97466218 | 285 | 2 | 6.04E-06 | 2.87E-05 | 2.45E-05 | 9.23E-06 | -0.0028877 | MMS19 |
| chr12 | 49202171 | 49202219 | 49 | 2 | 1.39E-05 | 9.42E-06 | 9.43E-05 | 9.23E-06 | -0.00264013 | TUBA1C |
| chr1 | 169794321 | 169795067 | 747 | 9 | 1.43E-05 | 1.78E-05 | 0.003262 | 9.23E-06 | 0.00050225 | C1orf112, METTL18 |
| chr7 | 70631278 | 70631392 | 115 | 2 | 8.08E-06 | 7.93E-05 | 1.28E-05 | 9.24E-06 | 0.011064932 | AUTS2 |
| chr2 | 10326846 | 10327083 | 238 | 2 | 1.55E-06 | 0.0017389 | 5.73E-06 | 9.24E-06 | 0.006325094 | HPCAL1 |
| chr6 | 27247015 | 27247597 | 583 | 4 | 1.14E-06 | 2.00E-05 | 0.000309 | 9.24E-06 | -0.00095023 | NA |
| chr2 | 126655116 | 126655954 | 839 | 11 | 3.45E-05 | 1.20E-05 | 0.011764 | 9.24E-06 | 0.001955378 | NA |
| chr9 | 95516973 | 95518071 | 1099 | 8 | 1.53E-06 | 3.93E-05 | 0.001522 | 9.25E-06 | 0.00865817 | PTCH1 |
| chr1 | 69569397 | 69569796 | 400 | 4 | 2.35E-06 | 8.69E-06 | 0.000763 | 9.25E-06 | -0.00199559 | LRRC7 |
| chr10 | 33145653 | 33146185 | 533 | 4 | 1.03E-05 | 2.06E-05 | 0.001839 | 9.25E-06 | 0.000694713 | NA |
| chr6 | 143286227 | 143286598 | 372 | 3 | 2.02E-06 | 0.0048821 | 1.12E-05 | 9.25E-06 | 0.000787173 | AIG1 |
| chr17 | 82707815 | 82708121 | 307 | 3 | 1.60E-05 | 1.04E-05 | 0.000483 | 9.25E-06 | 0.006071993 | NA |
| chr1 | 156625372 | 156626055 | 684 | 3 | 1.09E-05 | 4.33E-06 | 0.001106 | 9.26E-06 | -0.00354655 | HAPLN2 |
| chr20 | 19824026 | 19824249 | 224 | 2 | 2.51E-05 | 5.69E-06 | 0.00056 | 9.26E-06 | -0.00157991 | RIN2 |
| chr17 | 9768707 | 9768957 | 251 | 2 | 2.67E-05 | 5.57E-06 | 0.000653 | 9.26E-06 | 0.000785163 | AC027045.2 |
| chr8 | 142324302 | 142324831 | 530 | 4 | 1.05E-05 | 1.25E-05 | 0.001896 | 9.26E-06 | -0.00316405 | TSNARE1 |
| chr12 | 11310910 | 11311673 | 764 | 3 | 2.91E-05 | 1.80E-05 | 0.000119 | 9.26E-06 | -0.00195532 | NA |
| chr16 | 87334007 | 87334599 | 593 | 6 | 3.91E-06 | 0.0022931 | 2.65E-05 | 9.26E-06 | 0.00171047 | AC010531.1, FBXO31 |
| chr2 | 133844195 | 133844283 | 89 | 2 | 1.77E-05 | 5.68E-06 | 0.000572 | 9.27E-06 | -0.01442328 | NA |
| chr1 | 162881506 | 162881724 | 219 | 3 | 1.83E-05 | 1.16E-05 | 0.000306 | 9.27E-06 | -0.002464 | NA |
| chr7 | 44146920 | 44147711 | 792 | 4 | 4.52E-05 | 7.05E-06 | 0.001301 | 9.27E-06 | -0.00485649 | GCK |
| chr21 | 42672833 | 42673411 | 579 | 2 | 3.85E-05 | 5.58E-06 | 0.000658 | 9.27E-06 | -0.00273987 | PDE9A |
| chr21 | 44461222 | 44461624 | 403 | 2 | 8.01E-06 | 1.07E-05 | 7.67E-05 | 9.27E-06 | -0.0021733 | LRRC3 |
| chr6 | 68652171 | 68652326 | 156 | 3 | 7.62E-06 | 0.0001649 | 5.64E-05 | 9.28E-06 | 0.000815757 | ADGRB3 |
| chr5 | 112861000 | 112861652 | 653 | 11 | 1.85E-06 | 0.0003696 | 0.000687 | 9.28E-06 | -0.00039796 | AC008575.1, SRP19 |
| chr3 | 188212419 | 188212916 | 498 | 5 | 2.32E-06 | 1.87E-05 | 0.002781 | 9.29E-06 | -0.0017613 | LPP |
| chr7 | 39084457 | 39084860 | 404 | 2 | 1.60E-07 | 2.95E-05 | 2.42E-05 | 9.29E-06 | 0.008187515 | POU6F2 |
| chr1 | 171787771 | 171787846 | 76 | 3 | 1.28E-05 | 4.64E-06 | 0.001212 | 9.29E-06 | -0.00014192 | EEF1AKNMT |
| chr15 | 75647960 | 75648885 | 926 | 9 | 1.11E-06 | 0.0004852 | 0.002162 | 9.30E-06 | -0.00359901 | SNX33, IMP3 |
| chr16 | 30655532 | 30656066 | 535 | 2 | 6.80E-07 | 7.24E-05 | 1.35E-05 | 9.30E-06 | 0.002049542 | PRR14 |
| chr2 | 32926490 | 32926682 | 193 | 2 | 1.03E-05 | 3.25E-05 | 2.25E-05 | 9.30E-06 | -0.00233151 | LINC00486 |
| chr9 | 69034649 | 69035456 | 808 | 4 | 2.94E-06 | 8.87E-06 | 0.000237 | 9.31E-06 | 0.002173275 | NA |
| chr5 | 170297586 | 170298096 | 511 | 7 | 3.48E-06 | 0.001906 | 0.000112 | 9.31E-06 | -2.89E-05 | LCP2 |
| chr8 | 143148803 | 143149213 | 411 | 3 | 3.92E-06 | 0.0006736 | 1.21E-05 | 9.31E-06 | -0.00401136 | NA |
| chr1 | 39560089 | 39560848 | 760 | 5 | 2.65E-07 | 0.0016432 | 3.47E-06 | 9.31E-06 | 0.002842625 | PABPC4 |
| chr6 | 1608161 | 1608988 | 828 | 6 | 8.28E-06 | 7.39E-05 | 0.000593 | 9.31E-06 | 0.001505251 | NA |
| chr21 | 45019321 | 45019555 | 235 | 2 | 2.51E-06 | 0.0001215 | 1.07E-05 | 9.31E-06 | -0.00827175 | NA |
| chr9 | 8887341 | 8888118 | 778 | 4 | 2.43E-05 | 6.75E-06 | 0.001684 | 9.31E-06 | -0.00935856 | PTPRD |
| chr2 | 85391562 | 85392044 | 483 | 2 | 3.59E-05 | 6.08E-06 | 0.000389 | 9.32E-06 | -0.00846821 | ELMOD3 |
| chr10 | 129968610 | 129969123 | 514 | 3 | 5.00E-06 | 2.51E-05 | 0.000485 | 9.33E-06 | 0.004511685 | EBF3 |
| chr3 | 50358219 | 50358821 | 603 | 3 | 7.20E-06 | 6.15E-05 | 0.000294 | 9.33E-06 | -0.00709723 | CYB561D2, Z84492.2, TMEM115 |
| chr22 | 36702414 | 36703019 | 606 | 5 | 5.72E-07 | 0.0002769 | 0.000366 | 9.33E-06 | 0.011889192 | CACNG2 |
| chr4 | 181796623 | 181796839 | 217 | 3 | 2.48E-05 | 6.61E-06 | 0.000327 | 9.34E-06 | -0.01808245 | NA |
| chr19 | 48170695 | 48172048 | 1354 | 9 | 2.28E-08 | 0.00045 | 0.000372 | 9.34E-06 | -0.00192845 | ZSWIM9 |
| chr16 | 74843909 | 74843973 | 65 | 2 | 1.84E-05 | 1.41E-05 | 5.17E-05 | 9.34E-06 | -0.00705163 | NA |
| chr20 | 31717824 | 31718311 | 488 | 2 | 3.18E-05 | 5.62E-06 | 0.00066 | 9.34E-06 | 0.007331192 | BCL2L1 |
| chr19 | 2507306 | 2507426 | 121 | 3 | 3.19E-05 | 4.25E-06 | 0.001446 | 9.35E-06 | -0.0060974 | NA |
| chr3 | 46241807 | 46242280 | 474 | 2 | 3.38E-05 | 5.81E-06 | 0.000523 | 9.35E-06 | -0.00387396 | CCR3 |
| chr8 | 22023334 | 22024040 | 707 | 8 | 1.50E-07 | 7.27E-05 | 0.001722 | 9.37E-06 | -0.00296198 | NA |
| chr8 | 101205321 | 101206216 | 896 | 14 | 7.17E-06 | 6.81E-05 | 0.000857 | 9.37E-06 | 0.000902543 | ZNF706 |
| chr3 | 129530161 | 129530580 | 420 | 2 | 6.11E-06 | 1.66E-05 | 4.26E-05 | 9.38E-06 | -0.00261116 | RHO |
| chr1 | 20168657 | 20168976 | 320 | 2 | 9.14E-06 | 1.71E-05 | 4.14E-05 | 9.38E-06 | -0.00965646 | PLA2G2C |
| chr10 | 46028997 | 46029619 | 623 | 3 | 3.71E-06 | 4.51E-06 | 0.001638 | 9.38E-06 | -0.0131023 | NCOA4 |
| chr14 | 105463083 | 105463847 | 765 | 6 | 6.91E-07 | 0.0003691 | 0.00029 | 9.39E-06 | -0.0048078 | MTA1 |
| chr14 | 20452032 | 20452415 | 384 | 4 | 1.34E-05 | 9.37E-06 | 0.001086 | 9.39E-06 | -0.00070197 | OSGEP |
| chr7 | 101912700 | 101913005 | 306 | 5 | 2.21E-05 | 3.53E-06 | 0.003531 | 9.39E-06 | 0.000887216 | CUX1 |
| chr2 | 38943750 | 38944689 | 940 | 2 | 2.31E-05 | 6.72E-06 | 0.000256 | 9.39E-06 | -0.00531323 | ARHGEF33 |
| chr19 | 2983597 | 2983747 | 151 | 2 | 8.01E-06 | 6.44E-06 | 0.000307 | 9.39E-06 | -0.02443389 | TLE6 |
| chr9 | 120877646 | 120878196 | 551 | 4 | 1.88E-06 | 2.44E-05 | 0.000579 | 9.39E-06 | -0.00497139 | PHF19 |
| chr14 | 68792923 | 68794007 | 1085 | 11 | 3.44E-07 | 0.012279 | 1.03E-05 | 9.39E-06 | 0.000688441 | ZFP36L1 |
| chr6 | 166308271 | 166308574 | 304 | 3 | 5.34E-06 | 0.0001559 | 7.89E-05 | 9.39E-06 | 0.004198258 | PRR18 |
| chr6 | 41469938 | 41470569 | 632 | 3 | 4.84E-06 | 6.91E-06 | 0.000467 | 9.40E-06 | -0.00440721 | NA |
| chr16 | 85234451 | 85235254 | 804 | 3 | 1.37E-05 | 6.00E-06 | 0.00041 | 9.40E-06 | -0.00813781 | GSE1, AC092275.1 |
| chr1 | 112456006 | 112456267 | 262 | 2 | 2.57E-07 | 0.0001998 | 9.05E-06 | 9.40E-06 | -0.00597804 | CTTNBP2NL |
| chr4 | 4861555 | 4862252 | 698 | 5 | 1.28E-05 | 3.68E-05 | 0.000806 | 9.40E-06 | -0.00338305 | MSX1 |
| chr8 | 9918241 | 9918523 | 283 | 2 | 9.91E-07 | 0.0023626 | 5.65E-06 | 9.41E-06 | 0.010213828 | NA |
| chr1 | 41382764 | 41383531 | 768 | 5 | 1.27E-05 | 7.89E-05 | 0.001615 | 9.41E-06 | 0.000724314 | FOXO6 |
| chr22 | 38035716 | 38035719 | 4 | 2 | 1.38E-05 | 5.66E-06 | 0.000663 | 9.41E-06 | -0.00900762 | POLR2F |
| chr20 | 1329078 | 1329807 | 730 | 4 | 1.83E-06 | 7.87E-05 | 0.000128 | 9.41E-06 | -0.00591964 | SDCBP2-AS1, SDCBP2, AL136531.2 |
| chr5 | 65623850 | 65624813 | 964 | 18 | 6.08E-06 | 9.35E-06 | 0.009743 | 9.41E-06 | 0.000431786 | TRIM23 |
| chr3 | 187186821 | 187186868 | 48 | 2 | 1.02E-05 | 5.09E-05 | 1.67E-05 | 9.41E-06 | 0.004264525 | NA |
| chr20 | 63540855 | 63541236 | 382 | 3 | 2.04E-06 | 0.0001361 | 1.97E-05 | 9.41E-06 | -0.00118232 | SRMS |
| chr18 | 9886067 | 9886408 | 342 | 2 | 5.84E-07 | 2.13E-05 | 3.30E-05 | 9.42E-06 | -0.00635041 | TXNDC2 |
| chr20 | 13956794 | 13956801 | 8 | 2 | 2.44E-06 | 0.0022188 | 5.70E-06 | 9.42E-06 | -0.01981637 | SEL1L2 |
| chr6 | 121437834 | 121438118 | 285 | 2 | 3.77E-06 | 1.30E-05 | 5.87E-05 | 9.43E-06 | -0.01125084 | GJA1 |
| chr9 | 137757324 | 137757339 | 16 | 2 | 9.10E-07 | 0.0054518 | 5.31E-06 | 9.44E-06 | -0.00308446 | EHMT1 |
| chr12 | 110631966 | 110632723 | 758 | 6 | 1.00E-05 | 3.05E-05 | 0.00024 | 9.44E-06 | -0.00169775 | TCTN1, HVCN1 |
| chr13 | 114062018 | 114062789 | 772 | 4 | 9.88E-07 | 0.0006115 | 0.000138 | 9.45E-06 | -0.0142287 | RASA3 |
| chr13 | 51356222 | 51356553 | 332 | 2 | 2.18E-05 | 6.33E-06 | 0.000344 | 9.45E-06 | -0.00414302 | SERPINE3, INTS6 |
| chr8 | 96329917 | 96330128 | 212 | 2 | 1.97E-05 | 7.41E-06 | 0.000187 | 9.46E-06 | 0.00347069 | PTDSS1 |
| chr6 | 7691890 | 7692473 | 584 | 4 | 3.46E-06 | 5.77E-05 | 0.000244 | 9.46E-06 | -0.01478395 | NA |
| chr11 | 122795922 | 122796481 | 560 | 3 | 2.09E-05 | 4.59E-06 | 0.001348 | 9.46E-06 | -0.00731753 | UBASH3B, AP002762.2 |
| chr17 | 82907996 | 82908491 | 496 | 5 | 2.67E-06 | 0.0001325 | 6.19E-05 | 9.46E-06 | 0.00451948 | TBCD |
| chr1 | 234591962 | 234592517 | 556 | 3 | 2.12E-06 | 0.0002966 | 2.08E-05 | 9.46E-06 | -0.00036274 | NA |
| chr22 | 45878236 | 45878600 | 365 | 2 | 6.92E-07 | 1.55E-05 | 4.71E-05 | 9.46E-06 | 0.007592193 | BX324167.2, BX324167.1 |
| chr3 | 190699050 | 190699344 | 295 | 2 | 9.57E-07 | 0.0009456 | 6.38E-06 | 9.47E-06 | -0.00478441 | LINC02013 |
| chr13 | 110812207 | 110812681 | 475 | 3 | 2.63E-05 | 3.44E-06 | 0.003182 | 9.47E-06 | -0.01865143 | LINC00567 |
| chr5 | 132815355 | 132816110 | 756 | 2 | 7.67E-05 | 5.70E-06 | 0.000663 | 9.47E-06 | -0.00535414 | SOWAHA |
| chr14 | 60603531 | 60603972 | 442 | 3 | 3.19E-05 | 4.78E-06 | 0.00147 | 9.48E-06 | -0.00233134 | NA |
| chr3 | 98086696 | 98087250 | 555 | 3 | 7.22E-06 | 1.77E-05 | 0.000264 | 9.48E-06 | 0.003452225 | OR5AC2 |
| chr16 | 72950492 | 72950865 | 374 | 4 | 4.94E-06 | 0.0170137 | 0.000185 | 9.48E-06 | 0.002409828 | ZFHX3 |
| chr1 | 91165386 | 91165765 | 380 | 3 | 1.18E-05 | 1.40E-05 | 8.78E-05 | 9.48E-06 | -0.00391706 | NA |
| chr11 | 68616258 | 68616917 | 660 | 8 | 7.52E-06 | 7.00E-05 | 0.002502 | 9.48E-06 | -0.00083246 | AP003096.1 |
| chr22 | 31944948 | 31945794 | 847 | 13 | 1.43E-06 | 0.000362 | 0.000523 | 9.49E-06 | 0.001236309 | Z82190.2, YWHAH, C22orf24 |
| chr17 | 73324298 | 73324623 | 326 | 3 | 7.60E-06 | 2.39E-05 | 0.000457 | 9.50E-06 | -0.00430016 | NA |
| chr3 | 49028778 | 49029718 | 941 | 12 | 6.26E-07 | 0.0006971 | 3.26E-05 | 9.51E-06 | 0.001764717 | AC137630.3, IMPDH2, QRICH1 |
| chr9 | 133497152 | 133497251 | 100 | 2 | 2.54E-05 | 6.34E-06 | 0.000353 | 9.51E-06 | -0.00139981 | NA |
| chr10 | 100659452 | 100659860 | 409 | 7 | 2.74E-06 | 0.0045822 | 0.000691 | 9.51E-06 | -0.00175126 | NA |
| chr7 | 50540198 | 50541430 | 1233 | 3 | 4.06E-05 | 3.85E-06 | 0.002084 | 9.51E-06 | -0.00404617 | DDC-AS1, FIGNL1, DDC |
| chr14 | 99277378 | 99278066 | 689 | 3 | 1.38E-06 | 0.0003954 | 6.36E-06 | 9.51E-06 | -0.0165074 | NA |
| chr1 | 11526202 | 11526806 | 605 | 2 | 3.52E-07 | 0.0001806 | 9.49E-06 | 9.51E-06 | -0.00758353 | DISP3 |
| chr4 | 158914091 | 158914549 | 459 | 3 | 2.97E-05 | 3.87E-06 | 0.002195 | 9.51E-06 | 0.026343983 | C4orf45 |
| chr12 | 131264938 | 131265420 | 483 | 2 | 2.47E-06 | 7.64E-06 | 0.000174 | 9.51E-06 | -0.00703018 | NA |
| chr4 | 153576533 | 153576961 | 429 | 2 | 1.12E-06 | 0.0007478 | 6.68E-06 | 9.52E-06 | -0.00869326 | TMEM131L |
| chr6 | 74535146 | 74535265 | 120 | 2 | 4.29E-06 | 5.82E-05 | 1.57E-05 | 9.53E-06 | -0.01032819 | AL357507.1, AL356277.3 |
| chr13 | 42405891 | 42406626 | 736 | 2 | 0.000103131 | 5.76E-06 | 0.000645 | 9.53E-06 | -0.01104544 | LINC02341 |
| chr5 | 35939508 | 35939935 | 428 | 3 | 1.23E-05 | 7.92E-06 | 0.000893 | 9.53E-06 | -0.01585387 | AC112204.2 |
| chr10 | 127374234 | 127374422 | 189 | 4 | 1.64E-05 | 0.0001157 | 0.000464 | 9.53E-06 | -0.00272781 | DOCK1 |
| chr9 | 137069233 | 137069848 | 616 | 4 | 1.47E-06 | 8.96E-06 | 0.001062 | 9.53E-06 | -0.00169995 | SAPCD2 |
| chr1 | 6253702 | 6254053 | 352 | 4 | 1.09E-05 | 0.0001843 | 0.000114 | 9.53E-06 | -0.0027613 | GPR153 |
| chr11 | 127739101 | 127739408 | 308 | 2 | 1.53E-05 | 9.27E-06 | 0.000106 | 9.54E-06 | -0.01443572 | NA |
| chr5 | 150133062 | 150133638 | 577 | 2 | 5.55E-05 | 6.37E-06 | 0.000352 | 9.54E-06 | -0.00591877 | PDGFRB |
| chr13 | 109671998 | 109672451 | 454 | 2 | 5.40E-06 | 1.99E-05 | 3.62E-05 | 9.54E-06 | -0.00850595 | AL163541.1 |
| chr17 | 40885172 | 40886311 | 1140 | 5 | 1.11E-08 | 0.0015178 | 1.78E-05 | 9.55E-06 | -0.00882977 | KRT20 |
| chr12 | 3999842 | 4000358 | 517 | 3 | 1.14E-05 | 2.81E-05 | 9.06E-05 | 9.55E-06 | 4.87E-05 | NA |
| chr2 | 237015358 | 237015605 | 248 | 3 | 1.43E-05 | 7.89E-06 | 0.000268 | 9.55E-06 | -0.00377469 | AC012063.1 |
| chr1 | 179953887 | 179955181 | 1295 | 13 | 1.65E-07 | 0.0027484 | 2.31E-06 | 9.55E-06 | 0.000576854 | CEP350, AL353708.1 |
| chr7 | 100420537 | 100420665 | 129 | 3 | 1.28E-05 | 0.0001406 | 0.000203 | 9.55E-06 | -0.00491553 | ZCWPW1 |
| chr12 | 6492701 | 6493021 | 321 | 3 | 5.37E-06 | 1.85E-05 | 0.000508 | 9.55E-06 | -0.00036236 | MRPL51 |
| chr3 | 69293134 | 69293374 | 241 | 2 | 2.24E-06 | 0.0004717 | 7.35E-06 | 9.56E-06 | 0.002752339 | FRMD4B |
| chr3 | 189806384 | 189806681 | 298 | 2 | 1.82E-05 | 7.39E-06 | 0.000198 | 9.57E-06 | -0.00505832 | TP63 |
| chr16 | 3262601 | 3263282 | 682 | 4 | 3.49E-06 | 1.06E-05 | 0.001269 | 9.57E-06 | -0.00480842 | NA |
| chr4 | 47838033 | 47839297 | 1265 | 10 | 6.80E-08 | 0.0001201 | 0.000479 | 9.58E-06 | 0.001899615 | AC107068.2, CORIN |
| chr2 | 183123830 | 183124490 | 661 | 11 | 3.03E-06 | 0.0010875 | 0.000127 | 9.58E-06 | 0.000440864 | NUP35 |
| chr15 | 89335959 | 89336284 | 326 | 2 | 2.55E-07 | 0.0017094 | 5.95E-06 | 9.58E-06 | 0.016350257 | AC124068.2 |
| chr16 | 65577466 | 65577639 | 174 | 2 | 2.43E-06 | 1.46E-05 | 5.19E-05 | 9.58E-06 | -0.01054527 | NA |
| chr19 | 33906534 | 33907359 | 826 | 6 | 9.52E-06 | 0.0001294 | 0.000275 | 9.59E-06 | -0.00710119 | AC008556.1 |
| chr16 | 68842895 | 68843811 | 917 | 7 | 3.83E-06 | 8.78E-05 | 0.000184 | 9.60E-06 | 0.002788173 | TANGO6 |
| chr1 | 228280203 | 228280923 | 721 | 7 | 1.26E-05 | 4.14E-05 | 0.000496 | 9.60E-06 | -7.23E-06 | OBSCN |
| chr6 | 37783843 | 37784297 | 455 | 4 | 1.61E-05 | 6.85E-05 | 0.000626 | 9.60E-06 | 0.00344911 | NA |
| chr3 | 146161176 | 146161923 | 748 | 15 | 3.07E-05 | 1.21E-05 | 0.010917 | 9.60E-06 | 0.005848008 | PLOD2 |
| chr16 | 89720378 | 89721152 | 775 | 9 | 3.83E-06 | 0.0001144 | 0.000788 | 9.60E-06 | 0.000213973 | ZNF276, VPS9D1 |
| chr1 | 202007789 | 202008262 | 474 | 4 | 4.94E-06 | 0.001071 | 0.000148 | 9.60E-06 | -0.0065368 | ELF3, ELF3-AS1 |
| chr7 | 158818935 | 158819606 | 672 | 2 | 5.20E-05 | 5.91E-06 | 0.000569 | 9.60E-06 | 0.001632329 | ESYT2 |
| chr7 | 19754896 | 19755348 | 453 | 2 | 3.07E-06 | 4.39E-05 | 1.89E-05 | 9.60E-06 | 0.004227863 | TMEM196 |
| chr9 | 87119335 | 87119889 | 555 | 3 | 2.23E-05 | 4.68E-06 | 0.000936 | 9.61E-06 | -0.01346567 | NA |
| chr8 | 37267250 | 37267290 | 41 | 2 | 1.19E-05 | 6.14E-05 | 1.54E-05 | 9.61E-06 | -0.00439787 | NA |
| chr16 | 86377616 | 86377850 | 235 | 5 | 4.75E-06 | 0.019417 | 0.000111 | 9.62E-06 | -0.00535442 | NA |
| chr2 | 114281349 | 114281410 | 62 | 2 | 1.03E-05 | 9.46E-06 | 0.000104 | 9.62E-06 | 0.005465048 | NA |
| chr1 | 155282568 | 155282745 | 178 | 3 | 2.62E-05 | 2.04E-05 | 0.000422 | 9.62E-06 | -0.00318271 | HCN3 |
| chr7 | 106042663 | 106042954 | 292 | 2 | 3.54E-06 | 6.11E-05 | 1.54E-05 | 9.62E-06 | -0.0081773 | NA |
| chr19 | 5237235 | 5237608 | 374 | 4 | 2.43E-05 | 3.46E-06 | 0.003675 | 9.63E-06 | -4.16E-05 | PTPRS |
| chr7 | 100127165 | 100127500 | 336 | 3 | 1.26E-06 | 7.98E-06 | 0.000371 | 9.63E-06 | 0.00481614 | MBLAC1 |
| chr14 | 104882549 | 104883439 | 891 | 6 | 6.10E-06 | 6.24E-05 | 0.000457 | 9.63E-06 | -0.00349386 | CEP170B |
| chr3 | 10530165 | 10530836 | 672 | 3 | 1.26E-05 | 8.62E-06 | 0.000204 | 9.63E-06 | -0.01028527 | ATP2B2 |
| chr9 | 69820617 | 69820915 | 299 | 6 | 4.42E-05 | 1.55E-05 | 0.008133 | 9.64E-06 | -0.00685874 | C9orf135, C9orf135-DT |
| chr11 | 5151999 | 5152375 | 377 | 3 | 9.37E-06 | 9.89E-06 | 0.00014 | 9.65E-06 | -0.01286923 | OR52A1 |
| chr11 | 125139686 | 125139789 | 104 | 3 | 4.33E-06 | 0.0002744 | 7.53E-06 | 9.65E-06 | -0.0099482 | NA |

| | | | | | | | | | | |
|---|---|---|---|---|---|---|---|---|---|---|
| chr7 | 1160205 | 1160898 | 694 | 12 | 4.67E-06 | 0.0001625 | 0.005427 | 9.65E-06 | -0.00196575 | ZFAND2A-DT, ZFAND2A |
| chr7 | 45974224 | 45974525 | 302 | 2 | 2.71E-05 | 5.87E-06 | 0.000623 | 9.66E-06 | -0.02675254 | AC073115.2 |
| chr1 | 153778320 | 153779189 | 870 | 4 | 2.42E-07 | 8.65E-05 | 9.84E-05 | 9.66E-06 | -0.00598591 | SLC27A3 |
| chr12 | 48865632 | 48866214 | 583 | 8 | 2.18E-06 | 6.22E-05 | 0.004611 | 9.66E-06 | 0.000356631 | RND1 |
| chr17 | 82037454 | 82038037 | 584 | 9 | 6.94E-06 | 5.07E-05 | 0.000144 | 9.67E-06 | 0.000237378 | DCXR-DT, DCXR |
| chr12 | 108633079 | 108633310 | 232 | 4 | 1.85E-05 | 5.39E-06 | 0.001931 | 9.67E-06 | -0.00716275 | AC007569.1, SELPLG |
| chr17 | 57379051 | 57379174 | 124 | 2 | 2.45E-05 | 6.95E-06 | 0.00026 | 9.68E-06 | 0.009794695 | MSI2 |
| chr7 | 20798904 | 20799012 | 109 | 4 | 2.64E-05 | 7.19E-06 | 0.000692 | 9.68E-06 | -0.00371512 | NA |
| chr2 | 233461141 | 233461424 | 284 | 4 | 2.36E-05 | 6.79E-06 | 0.002289 | 9.68E-06 | -0.00275811 | DGKD |
| chr6 | 160679542 | 160679640 | 99 | 3 | 3.32E-05 | 1.06E-05 | 0.000296 | 9.68E-06 | -0.00833532 | NA |
| chr14 | 102221063 | 102221508 | 446 | 4 | 1.68E-05 | 1.12E-05 | 0.001005 | 9.68E-06 | 0.001463881 | WDR20 |
| chr1 | 152970687 | 152970830 | 144 | 3 | 1.67E-05 | 4.28E-06 | 0.00182 | 9.68E-06 | -0.00851165 | SPRR4 |
| chr9 | 69421716 | 69422372 | 657 | 3 | 2.04E-06 | 0.0007643 | 1.24E-05 | 9.69E-06 | -0.00750359 | NA |
| chr11 | 130212331 | 130212582 | 252 | 2 | 1.40E-05 | 5.83E-06 | 0.000677 | 9.69E-06 | -0.00862128 | NA |
| chr10 | 114547727 | 114547955 | 229 | 3 | 2.05E-05 | 6.15E-06 | 0.001031 | 9.70E-06 | -0.01608592 | ABLIM1 |
| chr7 | 178613 | 179032 | 420 | 4 | 2.31E-05 | 3.01E-06 | 0.004662 | 9.70E-06 | -0.00136335 | NA |
| chr4 | 10730122 | 10730371 | 250 | 2 | 1.06E-05 | 6.67E-06 | 0.000311 | 9.70E-06 | -0.01117354 | NA |
| chr15 | 36880073 | 36880816 | 744 | 5 | 5.63E-05 | 1.50E-05 | 0.000447 | 9.71E-06 | -0.00450322 | AC018563.1 |
| chr13 | 110758437 | 110758819 | 383 | 2 | 5.59E-07 | 0.0008138 | 6.71E-06 | 9.71E-06 | -0.01085933 | NA |
| chr4 | 134327031 | 134327467 | 437 | 4 | 2.79E-06 | 0.0002402 | 3.67E-05 | 9.71E-06 | 0.007351737 | PES1P1, AC015631.1 |
| chr11 | 63613672 | 63614753 | 1082 | 12 | 3.96E-06 | 0.000147 | 0.003095 | 9.72E-06 | -0.00015656 | PLAAT3 |
| chr9 | 93826268 | 93826773 | 506 | 4 | 3.57E-06 | 0.0008139 | 0.000196 | 9.72E-06 | -0.00968819 | AL442224.1 |
| chr16 | 27775367 | 27776070 | 704 | 3 | 1.31E-06 | 2.89E-05 | 4.54E-05 | 9.72E-06 | -0.00570514 | KATNIP |
| chr8 | 142879725 | 142880098 | 374 | 6 | 2.30E-05 | 1.51E-05 | 0.002626 | 9.73E-06 | -0.00694358 | GML, CYP11B1 |
| chr19 | 6669838 | 6670379 | 542 | 3 | 1.07E-05 | 1.50E-05 | 0.000432 | 9.74E-06 | -0.01397596 | TNFSF14 |
| chr11 | 69599825 | 69600155 | 331 | 2 | 6.25E-06 | 2.34E-05 | 3.18E-05 | 9.74E-06 | 0.004385894 | NA |
| chr14 | 34649766 | 34649815 | 50 | 2 | 2.05E-05 | 1.29E-05 | 6.32E-05 | 9.75E-06 | -0.01596905 | NA |
| chr17 | 42823961 | 42824758 | 798 | 8 | 1.01E-05 | 9.53E-06 | 0.001583 | 9.75E-06 | 0.001071836 | PSME3, BECN1 |
| chr15 | 78449307 | 78449778 | 472 | 2 | 3.47E-06 | 1.79E-05 | 4.18E-05 | 9.75E-06 | -0.00832052 | IREB2 |
| chr1 | 160210265 | 160211234 | 970 | 4 | 7.14E-07 | 0.000265 | 0.000271 | 9.76E-06 | -0.00196503 | PEA15 |
| chr7 | 2863743 | 2864403 | 661 | 3 | 4.48E-05 | 4.23E-06 | 0.001843 | 9.76E-06 | 0.005341692 | NA |
| chr10 | 132414067 | 132414226 | 160 | 2 | 2.44E-06 | 0.0001876 | 9.63E-06 | 9.76E-06 | 0.003461741 | PWWP2B |
| chr3 | 17740396 | 17740719 | 324 | 2 | 6.15E-06 | 1.07E-05 | 8.57E-05 | 9.76E-06 | -0.01394419 | TBC1D5 |
| chr12 | 129926659 | 129926789 | 131 | 3 | 3.40E-05 | 4.44E-06 | 0.001234 | 9.77E-06 | -0.00851553 | NA |
| chr9 | 129874123 | 129875043 | 921 | 4 | 1.27E-05 | 7.19E-06 | 0.001123 | 9.77E-06 | -0.00146347 | USP20 |
| chr11 | 123939972 | 123940403 | 432 | 2 | 2.64E-06 | 3.01E-05 | 2.56E-05 | 9.78E-06 | 0.002792681 | OR4D5 |
| chr2 | 148885440 | 148885762 | 323 | 4 | 1.39E-05 | 0.0009101 | 0.000202 | 9.79E-06 | 0.000508562 | KIF5C, KIF5C-AS1 |
| chr5 | 61162349 | 61163067 | 719 | 9 | 1.10E-05 | 2.91E-05 | 0.002443 | 9.79E-06 | -0.00335652 | SMIM15-AS1, SMIM15 |
| chr16 | 2284255 | 2284595 | 341 | 4 | 2.09E-05 | 3.89E-06 | 0.00198 | 9.79E-06 | 0.001249678 | ABCA3 |
| chr12 | 31217777 | 31217794 | 18 | 2 | 2.52E-05 | 7.80E-06 | 0.000182 | 9.80E-06 | -0.03954578 | NA |
| chr15 | 82921395 | 82921441 | 47 | 2 | 2.72E-05 | 5.92E-06 | 0.00066 | 9.80E-06 | -0.00153142 | HOMER2 |
| chr20 | 32972246 | 32972493 | 248 | 2 | 5.08E-06 | 1.67E-05 | 4.57E-05 | 9.80E-06 | 0.003652475 | NA |
| chr11 | 55664108 | 55664150 | 43 | 2 | 5.14E-06 | 0.0073094 | 5.41E-06 | 9.80E-06 | 0.014081265 | OR4C6 |
| chr15 | 54012095 | 54012581 | 487 | 2 | 8.62E-05 | 6.01E-06 | 0.000583 | 9.81E-06 | -0.01749974 | UNC13C |
| chr15 | 48204893 | 48205456 | 564 | 5 | 2.53E-05 | 2.46E-06 | 0.0067 | 9.81E-06 | -0.00121676 | SLC12A1 |
| chr19 | 29724194 | 29724861 | 668 | 5 | 4.87E-05 | 5.87E-06 | 0.002488 | 9.81E-06 | 0.004355261 | NA |
| chr8 | 27048980 | 27049289 | 310 | 3 | 1.65E-06 | 0.0001532 | 1.75E-05 | 9.82E-06 | -0.00260441 | NA |
| chr10 | 73871830 | 73872806 | 977 | 4 | 8.46E-08 | 0.003901 | 3.79E-06 | 9.82E-06 | 0.003613925 | CAMK2G |
| chr14 | 99234996 | 99235079 | 84 | 3 | 1.32E-05 | 4.99E-06 | 0.001498 | 9.82E-06 | -0.00791261 | BCL11B |
| chr9 | 136436117 | 136436958 | 842 | 3 | 1.49E-06 | 4.73E-05 | 2.56E-05 | 9.82E-06 | -0.00342431 | INPP5E |
| chr1 | 52295295 | 52295692 | 398 | 2 | 1.65E-06 | 3.93E-05 | 2.10E-05 | 9.83E-06 | -0.00312453 | ZFYVE9 |
| chr3 | 45007278 | 45007402 | 125 | 3 | 1.38E-05 | 1.86E-05 | 7.43E-05 | 9.83E-06 | 0.001371519 | EXOSC7, CLEC3B |
| chr2 | 11555546 | 11556271 | 726 | 4 | 6.93E-06 | 1.73E-05 | 0.000443 | 9.83E-06 | -0.00459615 | GREB1 |
| chr18 | 79713568 | 79714091 | 524 | 3 | 8.18E-06 | 7.74E-06 | 0.000272 | 9.83E-06 | -0.00735989 | CTDP1 |
| chr11 | 134941060 | 134941496 | 437 | 2 | 6.72E-06 | 1.01E-05 | 9.65E-05 | 9.84E-06 | -0.01357625 | NA |
| chr2 | 177039279 | 177039281 | 3 | 2 | 2.81E-05 | 5.94E-06 | 0.000668 | 9.84E-06 | -0.00981887 | AC074286.1 |
| chr12 | 1590654 | 1590865 | 212 | 3 | 5.61E-06 | 0.0005292 | 0.000342 | 9.84E-06 | -0.00521792 | WNT5B, FBXL14 |
| chr22 | 31891577 | 31891903 | 327 | 2 | 2.04E-06 | 0.0012709 | 6.34E-06 | 9.84E-06 | 0.010192398 | DEPDC5, Z82190.2 |
| chr5 | 126750331 | 126750811 | 481 | 2 | 2.93E-05 | 6.17E-06 | 0.000509 | 9.84E-06 | -0.00797102 | NA |
| chr18 | 76374812 | 76374847 | 36 | 3 | 1.13E-05 | 2.10E-05 | 0.000134 | 9.84E-06 | -0.01563749 | AC009716.1, ZNF516 |
| chr9 | 110257429 | 110258302 | 874 | 3 | 1.63E-06 | 6.96E-06 | 0.001079 | 9.85E-06 | -0.00437023 | NA |
| chr4 | 99219317 | 99219748 | 432 | 2 | 2.63E-06 | 1.45E-05 | 5.54E-05 | 9.85E-06 | 0.012413498 | AP002026.1, ADH6 |
| chr1 | 17520212 | 17520938 | 727 | 6 | 1.21E-05 | 4.38E-05 | 0.000884 | 9.86E-06 | 0.000677054 | NA |
| chr12 | 108321532 | 108321594 | 63 | 2 | 1.10E-05 | 7.64E-06 | 0.000201 | 9.88E-06 | -0.00409117 | CMKLR1 |
| chr16 | 346164 | 346560 | 397 | 4 | 1.20E-05 | 1.65E-05 | 0.000144 | 9.88E-06 | -0.00253437 | AXIN1 |
| chr12 | 23585406 | 23585921 | 516 | 4 | 7.84E-06 | 3.24E-05 | 0.000362 | 9.88E-06 | -0.01243779 | SOX5 |
| chr19 | 3659349 | 3660028 | 680 | 7 | 9.89E-06 | 2.81E-05 | 0.000708 | 9.89E-06 | 0.00075386 | PIP5K1C |
| chr16 | 3731319 | 3731829 | 511 | 5 | 1.42E-06 | 2.90E-05 | 0.000529 | 9.90E-06 | 0.003385719 | CREBBP |
| chr20 | 58700139 | 58701004 | 866 | 2 | 4.68E-05 | 6.71E-06 | 0.000334 | 9.90E-06 | 0.000358551 | STX16-NPEPL1, NPEPL1 |
| chr16 | 1180137 | 1180935 | 799 | 4 | 7.34E-06 | 5.10E-06 | 0.000784 | 9.90E-06 | 0.000473424 | CACNA1H |
| chr20 | 35614706 | 35614799 | 94 | 2 | 9.08E-06 | 7.19E-06 | 0.000252 | 9.90E-06 | -0.01167512 | NA |
| chr16 | 54924247 | 54924353 | 107 | 2 | 1.01E-05 | 7.74E-06 | 0.000194 | 9.91E-06 | -0.01739784 | CRNDE |
| chr4 | 87209630 | 87210263 | 634 | 3 | 8.73E-06 | 5.17E-05 | 0.000401 | 9.93E-06 | -0.00907463 | KLHL8 |
| chr8 | 124608781 | 124609151 | 371 | 2 | 1.48E-05 | 6.08E-06 | 0.000595 | 9.93E-06 | 0.00390354 | MTSS1 |
| chr13 | 102591991 | 102592788 | 798 | 8 | 7.21E-05 | 6.92E-06 | 0.012955 | 9.95E-06 | -0.00064721 | NA |
| chr7 | 9961039 | 9961404 | 366 | 2 | 6.17E-06 | 2.95E-05 | 2.67E-05 | 9.95E-06 | -0.01369555 | AC010991.1 |
| chr5 | 141199473 | 141200071 | 599 | 9 | 5.37E-06 | 7.78E-05 | 0.000173 | 9.95E-06 | -0.01477683 | PCDHB11, AC244517.11, AC244517.4 |
| chr4 | 15938239 | 15938715 | 477 | 6 | 1.60E-05 | 2.04E-05 | 0.000243 | 9.96E-06 | -0.00768994 | FGFBP1 |
| chr19 | 7474824 | 7475752 | 929 | 2 | 0.000128719 | 6.05E-06 | 0.000625 | 9.96E-06 | 0.008861763 | NA |
| chr6 | 30488030 | 30488299 | 270 | 5 | 8.85E-06 | 5.53E-06 | 0.003194 | 9.96E-06 | 0.003522454 | NA |
| chr21 | 37749277 | 37749347 | 71 | 2 | 2.50E-05 | 8.06E-06 | 0.000173 | 9.96E-06 | 0.004980716 | KCNJ6 |
| chr3 | 127616928 | 127617379 | 452 | 3 | 3.16E-06 | 2.86E-05 | 5.27E-05 | 9.96E-06 | -0.00595836 | MCM2 |
| chr11 | 94566904 | 94567635 | 732 | 8 | 1.95E-05 | 4.39E-05 | 0.00044 | 9.96E-06 | -0.00679615 | PIWIL4, AP000943.2 |
| chr1 | 998305 | 998656 | 352 | 3 | 1.80E-05 | 1.05E-05 | 0.000241 | 9.96E-06 | 0.004113421 | NA |
| chr3 | 72348196 | 72348340 | 145 | 2 | 2.72E-05 | 6.21E-06 | 0.000531 | 9.96E-06 | 0.015924074 | NA |
| chr8 | 143617460 | 143618197 | 738 | 6 | 3.50E-05 | 1.93E-05 | 0.000934 | 9.97E-06 | -0.00111205 | GFUS |
| chr7 | 40792212 | 40792503 | 292 | 2 | 2.62E-05 | 6.25E-06 | 0.000511 | 9.97E-06 | -0.00121268 | SUGCT |
| chr6 | 65838049 | 65838702 | 654 | 4 | 8.00E-06 | 0.0001883 | 4.84E-05 | 9.97E-06 | -0.00758629 | NA |
| chr13 | 106861440 | 106861458 | 19 | 2 | 2.77E-05 | 6.00E-06 | 0.000677 | 9.97E-06 | -0.01146664 | NA |
| chr1 | 230153971 | 230154342 | 372 | 3 | 3.70E-05 | 3.73E-06 | 0.002882 | 9.97E-06 | -0.00801685 | GALNT2 |
| chr8 | 28386753 | 28387252 | 500 | 5 | 7.34E-07 | 2.53E-05 | 0.000989 | 9.97E-06 | -0.00508056 | ZNF395, FBXO16 |
| chr7 | 44080055 | 44080259 | 205 | 2 | 1.81E-05 | 7.26E-06 | 0.000251 | 9.97E-06 | -0.00574537 | POLM |
| chr18 | 4444736 | 4445094 | 359 | 2 | 1.98E-05 | 7.41E-06 | 0.000234 | 9.98E-06 | -0.01547716 | DLGAP1 |
| chr10 | 97292218 | 97293393 | 1176 | 11 | 5.30E-06 | 3.11E-05 | 0.00142 | 9.99E-06 | 0.000658057 | ARHGAP19-SLIT1, ARHGAP19 |
| chr7 | 35800389 | 35801117 | 729 | 12 | 8.68E-06 | 0.0003762 | 0.002726 | 9.99E-06 | 0.000750085 | SEPTIN7, SEPTIN7-DT |
| chr11 | 79691813 | 79691928 | 116 | 3 | 1.14E-05 | 0.000119 | 2.56E-05 | 9.99E-06 | 0.004457433 | NA |
| chr18 | 79738474 | 79738956 | 483 | 2 | 1.15E-05 | 6.39E-06 | 0.000457 | 9.99E-06 | -0.01625824 | CTDP1 |
| chr19 | 10259279 | 10260244 | 966 | 6 | 1.56E-05 | 0.0002753 | 0.000265 | 9.99E-06 | -0.00403789 | MRPL4, AC011511.2, AC011511.3 |
| chr9 | 124453161 | 124453768 | 608 | 6 | 1.22E-07 | 0.0129882 | 1.14E-06 | 1.00E-05 | 0.00314169 | ADGRD2 |
| chr15 | 29003922 | 29003988 | 67 | 2 | 1.18E-05 | 1.58E-05 | 5.08E-05 | 1.00E-05 | 0.01024535 | APBA2 |
| chr17 | 28061124 | 28061446 | 323 | 2 | 2.81E-05 | 6.01E-06 | 0.000684 | 1.00E-05 | -0.00269002 | NLK |
| chr17 | 49845416 | 49846113 | 698 | 4 | 1.06E-06 | 0.0003973 | 2.28E-05 | 1.00E-05 | -0.00339744 | AC027801.1, TAC4 |
| chr5 | 146291293 | 146291635 | 343 | 2 | 3.35E-06 | 0.0001449 | 1.09E-05 | 1.00E-05 | 0.006951519 | AC091959.1 |
| chr1 | 45332445 | 45333108 | 664 | 3 | 3.10E-05 | 4.05E-06 | 0.002397 | 1.00E-05 | -0.00481484 | MUTYH, AL451136.1 |
| chr10 | 94088892 | 94089306 | 415 | 4 | 2.05E-06 | 0.0001886 | 1.03E-05 | 1.00E-05 | -0.00440785 | PLCE1, PLCE1-AS2 |
| chr9 | 127402309 | 127402939 | 631 | 3 | 1.50E-05 | 7.50E-06 | 0.000299 | 1.00E-05 | -0.00419676 | SLC2A8 |
| chr22 | 37607113 | 37607200 | 88 | 4 | 4.39E-06 | 4.32E-05 | 4.52E-05 | 1.00E-05 | 0.000989442 | NA |
| chr1 | 226122021 | 226122774 | 754 | 3 | 2.84E-07 | 0.0013638 | 4.90E-06 | 1.00E-05 | -0.003672 | NA |
| chr2 | 241776692 | 241776973 | 282 | 6 | 3.89E-05 | 1.41E-05 | 0.002476 | 1.00E-05 | -0.00538801 | GAL3ST2 |
| chr20 | 43584590 | 43585089 | 500 | 3 | 1.46E-05 | 8.16E-06 | 0.000231 | 1.00E-05 | -0.00761071 | SGK2 |
| chr1 | 228752340 | 228752827 | 488 | 3 | 3.42E-05 | 7.64E-06 | 0.000265 | 1.00E-05 | -0.01058842 | NA |
| chr14 | 53946500 | 53947490 | 991 | 8 | 2.38E-06 | 0.0002201 | 0.000117 | 1.00E-05 | 0.00264199 | NA |
| chr15 | 51280779 | 51281389 | 611 | 3 | 2.18E-05 | 5.26E-06 | 0.000771 | 1.00E-05 | -0.00495457 | MIR4713HG, CYP19A1 |
| chr5 | 92775625 | 92776026 | 402 | 2 | 2.52E-06 | 8.04E-05 | 1.40E-05 | 1.00E-05 | -0.01174557 | NA |
| chr18 | 14747662 | 14748299 | 638 | 8 | 1.10E-06 | 0.0014962 | 8.20E-05 | 1.01E-05 | -0.01069997 | ANKRD30B |
| chr22 | 38056859 | 38057482 | 624 | 12 | 1.25E-05 | 1.85E-05 | 0.003874 | 1.01E-05 | 0.000718022 | PICK1, AL031587.2 |
| chr2 | 173271687 | 173272161 | 475 | 4 | 1.26E-06 | 0.0006447 | 5.08E-05 | 1.01E-05 | 0.006518614 | MAP3K20-AS1 |
| chr18 | 9334490 | 9335104 | 615 | 8 | 4.86E-06 | 0.0002398 | 0.0002 | 1.01E-05 | 0.000613115 | TWSG1 |
| chr5 | 123511072 | 123512272 | 1201 | 13 | 7.76E-05 | 0.0002431 | 0.007864 | 1.01E-05 | -0.00095961 | CSNK1G3 |
| chr3 | 11016752 | 11017364 | 613 | 9 | 1.85E-06 | 0.0001013 | 0.00116 | 1.01E-05 | -0.00408174 | SLC6A1, SLC6A1-AS1 |
| chr10 | 28280716 | 28280773 | 58 | 2 | 6.35E-06 | 1.90E-05 | 4.14E-05 | 1.01E-05 | 0.000574714 | MPP7 |
| chr3 | 157158356 | 157159091 | 736 | 3 | 2.55E-06 | 0.0001747 | 2.27E-05 | 1.01E-05 | -0.01540007 | CCNL1 |
| chr12 | 120980938 | 120981473 | 536 | 3 | 1.77E-05 | 6.29E-06 | 0.001 | 1.01E-05 | -0.00368286 | HNF1A, HNF1A-AS1 |
| chr6 | 22303240 | 22303568 | 329 | 2 | 2.34E-06 | 0.0001105 | 1.22E-05 | 1.01E-05 | -0.00682175 | CASC15 |
| chr1 | 94599353 | 94599418 | 66 | 2 | 2.10E-05 | 8.54E-06 | 0.000154 | 1.01E-05 | 0.015705981 | NA |
| chr16 | 69837787 | 69838749 | 963 | 6 | 6.57E-06 | 9.96E-06 | 0.002326 | 1.01E-05 | -0.00396902 | WWP2 |
| chr15 | 25705921 | 25706016 | 96 | 2 | 6.76E-06 | 0.0006413 | 7.29E-06 | 1.01E-05 | 0.003218505 | ATP10A |
| chr10 | 101036015 | 101037210 | 1196 | 6 | 4.73E-07 | 0.0030462 | 0.000245 | 1.01E-05 | -0.00467203 | SFXN3 |
| chr4 | 5885998 | 5886323 | 326 | 2 | 6.15E-07 | 0.0017763 | 6.23E-06 | 1.01E-05 | -0.01412338 | CRMP1 |
| chr11 | 70100908 | 70101040 | 133 | 3 | 6.53E-06 | 0.0008145 | 1.14E-05 | 1.01E-05 | -0.00892472 | ANO1 |
| chr5 | 40679353 | 40679937 | 585 | 12 | 2.55E-05 | 1.32E-05 | 0.010705 | 1.01E-05 | 0.001253606 | PTGER4, TTC33 |
| chr7 | 157336736 | 157337597 | 862 | 11 | 1.42E-06 | 0.003412 | 0.000464 | 1.01E-05 | 0.000974275 | DNAJB6 |
| chr2 | 70790079 | 70790912 | 834 | 8 | 2.80E-06 | 5.97E-06 | 0.008384 | 1.01E-05 | -0.00349645 | FIGLA |
| chr15 | 81304306 | 81304407 | 102 | 2 | 1.25E-05 | 0.000104 | 1.26E-05 | 1.01E-05 | 0.001512064 | IL16, AC103858.1 |
| chr14 | 34232979 | 34232995 | 17 | 2 | 1.49E-05 | 1.12E-05 | 8.61E-05 | 1.01E-05 | 0.008068775 | EGLN3 |
| chr3 | 196211177 | 196211565 | 389 | 6 | 1.32E-05 | 2.93E-05 | 0.001943 | 1.01E-05 | -0.00659578 | SLC51A, ZDHHC19 |
| chr4 | 168790074 | 168790607 | 534 | 2 | 2.90E-05 | 6.39E-06 | 0.000503 | 1.02E-05 | 0.007057436 | PALLD |
| chr5 | 129903074 | 129903580 | 507 | 3 | 7.38E-07 | 3.60E-05 | 3.09E-05 | 1.02E-05 | 0.01099616 | AC008591.1 |
| chr3 | 11770563 | 11770598 | 36 | 2 | 1.62E-05 | 1.32E-05 | 6.66E-05 | 1.02E-05 | -0.00308139 | VGLL4 |
| chr21 | 33265704 | 33266443 | 740 | 12 | 1.26E-06 | 7.66E-05 | 0.00014 | 1.02E-05 | 0.002607161 | AP000295.1, IL10RB, IL10RB-DT |
| chr12 | 8564303 | 8564891 | 589 | 5 | 6.90E-06 | 0.0006168 | 0.001698 | 1.02E-05 | -0.00685189 | AC092746.1 |
| chr1 | 119675073 | 119675735 | 663 | 3 | 9.61E-06 | 1.57E-05 | 9.55E-05 | 1.02E-05 | 0.030443847 | PHGDH |
| chr10 | 96396275 | 96396363 | 89 | 3 | 1.38E-05 | 4.73E-06 | 0.001745 | 1.02E-05 | -0.01106964 | TLL2 |
| chr22 | 49049916 | 49050023 | 108 | 3 | 1.02E-05 | 0.0001092 | 1.56E-05 | 1.02E-05 | -0.00411114 | AL078622.1 |
| chr16 | 31458811 | 31459632 | 822 | 7 | 2.98E-07 | 7.07E-05 | 0.00022 | 1.02E-05 | 0.001101217 | ARMC5, AC026471.1 |
| chr10 | 45235496 | 45236006 | 511 | 2 | 1.12E-05 | 9.24E-06 | 0.00013 | 1.02E-05 | 0.025888851 | CUBNP2 |
| chr7 | 6416873 | 6417497 | 625 | 4 | 1.88E-05 | 9.11E-06 | 0.000487 | 1.02E-05 | -0.00034376 | DAGLB |
| chr1 | 1672408 | 1672525 | 118 | 3 | 4.83E-06 | 0.0003099 | 0.000216 | 1.02E-05 | -0.00353445 | AL691432.1, SLC35E2B |
| chr1 | 68050105 | 68051030 | 926 | 14 | 3.58E-05 | 5.16E-05 | 0.009227 | 1.02E-05 | -0.00314581 | GNG12-AS1, DIRAS3 |
| chr20 | 46705506 | 46705559 | 54 | 2 | 2.87E-05 | 6.45E-06 | 0.00049 | 1.02E-05 | -0.0029445 | NA |
| chr12 | 2624942 | 2625425 | 484 | 5 | 3.98E-05 | 5.26E-06 | 0.001628 | 1.02E-05 | -0.00864895 | CACNA1C |
| chr20 | 61560122 | 61560711 | 590 | 3 | 2.05E-05 | 4.90E-06 | 0.001149 | 1.02E-05 | -0.00021385 | CDH4 |
| chr12 | 57186314 | 57187333 | 1020 | 3 | 1.59E-07 | 0.003016 | 3.08E-05 | 1.02E-05 | -0.00338237 | LRP1 |
| chr18 | 57629468 | 57630145 | 678 | 4 | 8.53E-06 | 2.72E-06 | 0.005795 | 1.02E-05 | -0.00026035 | NA |
| chr3 | 3126458 | 3126641 | 184 | 3 | 4.84E-05 | 0.000106 | 0.000315 | 1.02E-05 | -0.00730895 | IL5RA |
| chr8 | 123852033 | 123852094 | 62 | 2 | 1.12E-05 | 6.17E-06 | 0.00067 | 1.02E-05 | -0.00555514 | FER1L6 |
| chr11 | 35040217 | 35040699 | 483 | 3 | 2.45E-05 | 3.78E-06 | 0.003125 | 1.02E-05 | -0.00997925 | NA |
| chr19 | 53647931 | 53648345 | 415 | 5 | 6.46E-06 | 0.002028 | 2.89E-05 | 1.02E-05 | 0.001976184 | NA |

| | | | | | | | | | | |
|---|---|---|---|---|---|---|---|---|---|---|
| chr2 | 20392447 | 20392684 | 238 | 2 | 2.53E-05 | 6.87E-06 | 0.000355 | 1.02E-05 | -0.00575575 | SLC7A15P |
| chr1 | 46203150 | 46203673 | 524 | 7 | 3.38E-05 | 9.38E-06 | 0.000294 | 1.02E-05 | 0.000455673 | LURAP1, POMGNT1 |
| chr8 | 107497528 | 107498086 | 559 | 5 | 5.10E-06 | 3.59E-05 | 0.001106 | 1.02E-05 | -0.0007357 | ANGPT1 |
| chr1 | 26773456 | 26774728 | 1273 | 5 | 2.57E-05 | 0.0004326 | 0.001704 | 1.02E-05 | 0.001158524 | ARID1A |
| chr5 | 164353339 | 164353616 | 278 | 2 | 1.81E-05 | 6.67E-06 | 0.000413 | 1.02E-05 | -0.02417442 | AC109466.1 |
| chr5 | 37833867 | 37834069 | 203 | 3 | 3.02E-06 | 1.57E-05 | 0.000192 | 1.02E-05 | -0.00203017 | GDNF-AS1, GDNF |
| chr7 | 774754 | 775211 | 458 | 6 | 6.15E-06 | 4.92E-05 | 0.000118 | 1.02E-05 | -0.00324984 | DNAAF5 |
| chr11 | 2846016 | 2846518 | 503 | 2 | 2.97E-05 | 6.55E-06 | 0.000463 | 1.02E-05 | -0.00053123 | KCNQ1, KCNQ1-AS1 |
| chr5 | 134737740 | 134737889 | 150 | 5 | 7.81E-07 | 0.0065521 | 8.90E-06 | 1.02E-05 | -0.00340686 | NA |
| chr6 | 135497602 | 135498214 | 613 | 6 | 1.66E-05 | 0.0001166 | 0.001711 | 1.02E-05 | 0.001041655 | LINC00271, AHI1 |
| chr9 | 121550907 | 121551040 | 134 | 3 | 1.55E-05 | 3.90E-06 | 0.002746 | 1.02E-05 | -0.00258847 | NA |
| chr2 | 35000363 | 35000463 | 101 | 2 | 9.48E-06 | 3.07E-05 | 2.70E-05 | 1.03E-05 | -0.01776369 | AC012593.1 |
| chr17 | 54172601 | 54172814 | 214 | 2 | 2.31E-05 | 8.00E-06 | 0.000199 | 1.03E-05 | 0.028895314 | AC034268.2 |
| chr15 | 70535227 | 70535298 | 72 | 2 | 1.31E-05 | 1.67E-05 | 4.96E-05 | 1.03E-05 | 0.01630602 | NA |
| chr1 | 35448181 | 35448480 | 300 | 4 | 4.19E-05 | 2.84E-06 | 0.006201 | 1.03E-05 | -0.01129947 | KIAA0319L |
| chr4 | 3430110 | 3430615 | 506 | 3 | 3.27E-06 | 1.66E-05 | 0.000481 | 1.03E-05 | -0.00535615 | RGS12 |
| chr5 | 135233003 | 135233148 | 146 | 2 | 2.30E-05 | 6.57E-06 | 0.00046 | 1.03E-05 | -0.00761558 | C5orf66 |
| chr6 | 42322518 | 42322783 | 266 | 3 | 3.20E-05 | 5.76E-06 | 0.000915 | 1.03E-05 | 0.001374389 | TRERF1 |
| chr8 | 1743941 | 1744560 | 620 | 3 | 1.53E-05 | 1.28E-05 | 0.000115 | 1.03E-05 | -0.00456898 | NA |
| chr6 | 108174181 | 108174781 | 601 | 5 | 8.39E-06 | 0.0001697 | 0.000694 | 1.03E-05 | 0.007687085 | NR2E1 |
| chr22 | 43570971 | 43571328 | 358 | 3 | 1.49E-05 | 4.43E-06 | 0.001514 | 1.03E-05 | -0.01740515 | EFCAB6 |
| chr3 | 156301275 | 156301425 | 151 | 3 | 2.24E-05 | 1.99E-05 | 0.000403 | 1.03E-05 | 0.017402082 | KCNAB1, AC091607.2 |
| chr7 | 30852168 | 30852221 | 54 | 2 | 1.83E-06 | 7.17E-06 | 0.000305 | 1.03E-05 | -0.00842056 | INMT-MINDY4, MINDY4 |
| chr8 | 87376201 | 87376579 | 379 | 2 | 1.11E-05 | 1.26E-05 | 7.39E-05 | 1.03E-05 | -0.012972 | CNBD1 |
| chr16 | 74107767 | 74107905 | 139 | 2 | 2.18E-05 | 1.09E-05 | 9.39E-05 | 1.03E-05 | -0.00720879 | AC138627.1 |
| chr5 | 125492166 | 125492646 | 481 | 2 | 2.09E-05 | 6.23E-06 | 0.000664 | 1.03E-05 | -0.01603781 | LINC02240, AC116362.1 |
| chr4 | 73987616 | 73987768 | 153 | 2 | 6.78E-06 | 3.19E-05 | 2.63E-05 | 1.03E-05 | -0.00735043 | PPBP |
| chr22 | 21032509 | 21033208 | 700 | 10 | 9.28E-06 | 0.0001108 | 0.000741 | 1.03E-05 | -0.00239578 | AC002472.1, SLC7A4 |
| chr5 | 75236844 | 75237454 | 611 | 10 | 3.16E-05 | 3.84E-05 | 0.007015 | 1.03E-05 | 0.00078358 | ANKRD31 |
| chr8 | 125445743 | 125446326 | 584 | 2 | 3.90E-05 | 6.45E-06 | 0.000529 | 1.03E-05 | -0.00614961 | NA |
| chr7 | 51446218 | 51446783 | 566 | 2 | 4.32E-05 | 6.33E-06 | 0.000601 | 1.03E-05 | -0.02506875 | NA |
| chr15 | 43791571 | 43792248 | 678 | 12 | 2.48E-05 | 1.00E-06 | 0.030753 | 1.03E-05 | 0.001388563 | SERF2, AC018512.1 |
| chr6 | 27238724 | 27239132 | 409 | 4 | 1.13E-05 | 3.75E-05 | 0.000676 | 1.03E-05 | -0.01887119 | NA |
| chr6 | 87154710 | 87155596 | 887 | 7 | 1.26E-06 | 0.0001677 | 8.38E-06 | 1.03E-05 | -0.00028406 | ZNF292, AL139274.2 |
| chr2 | 168890150 | 168890717 | 568 | 13 | 3.31E-06 | 9.56E-06 | 0.009344 | 1.03E-05 | 0.000529878 | SPC25 |
| chr6 | 168555638 | 168555738 | 101 | 2 | 3.85E-06 | 0.0001404 | 1.14E-05 | 1.03E-05 | -0.00424293 | SMOC2 |
| chr10 | 120607753 | 120608259 | 507 | 2 | 5.48E-07 | 0.006059 | 5.73E-06 | 1.03E-05 | -0.00846937 | NA |
| chr17 | 77779777 | 77780299 | 523 | 3 | 1.97E-06 | 0.0044794 | 0.000133 | 1.03E-05 | 0.008686159 | NA |
| chr7 | 64037755 | 64038420 | 666 | 3 | 4.45E-05 | 4.50E-06 | 0.001539 | 1.03E-05 | -0.02273663 | AC115220.2 |
| chr5 | 66584663 | 66585047 | 385 | 3 | 2.68E-05 | 8.13E-06 | 0.0003 | 1.03E-05 | -0.00510835 | NA |
| chr9 | 8188954 | 8189263 | 310 | 2 | 4.53E-06 | 0.0001194 | 1.22E-05 | 1.04E-05 | -0.00201241 | NA |
| chr22 | 46292807 | 46292926 | 120 | 2 | 2.75E-05 | 7.04E-06 | 0.000342 | 1.04E-05 | 0.000836576 | TTC38 |
| chr7 | 158314111 | 158314690 | 580 | 6 | 3.27E-05 | 1.10E-05 | 0.008989 | 1.04E-05 | -0.00224065 | PTPRN2 |
| chr20 | 38246029 | 38246066 | 38 | 2 | 2.83E-06 | 0.0003963 | 8.29E-06 | 1.04E-05 | -0.00651278 | KIAA1755 |
| chr10 | 129137045 | 129137170 | 126 | 3 | 1.94E-05 | 3.14E-05 | 0.000244 | 1.04E-05 | -0.00242534 | NA |
| chr6 | 13423400 | 13423489 | 90 | 2 | 2.09E-05 | 7.73E-06 | 0.000235 | 1.04E-05 | -0.0005492 | GFOD1 |
| chr19 | 57855519 | 57855850 | 332 | 3 | 4.65E-06 | 0.0001013 | 3.31E-05 | 1.04E-05 | 0.007430259 | AC010522.1, ZNF587, ZNF814 |
| chr4 | 153033707 | 153034170 | 464 | 3 | 1.70E-06 | 0.0004064 | 1.85E-05 | 1.04E-05 | -0.00859491 | NA |
| chr1 | 159955170 | 159955626 | 457 | 3 | 3.12E-05 | 4.22E-06 | 0.002366 | 1.04E-05 | -0.00465147 | NA |
| chr11 | 77515546 | 77515951 | 406 | 2 | 1.03E-05 | 1.38E-05 | 6.57E-05 | 1.04E-05 | -0.00672459 | CLNS1A |
| chr8 | 57994161 | 57994828 | 668 | 7 | 1.79E-05 | 1.70E-05 | 0.003041 | 1.04E-05 | 0.004373461 | FAM110B |
| chr2 | 72149873 | 72150629 | 757 | 4 | 1.20E-06 | 0.0002404 | 7.30E-05 | 1.04E-05 | 8.38E-05 | NA |
| chr7 | 36291263 | 36291724 | 462 | 4 | 3.18E-05 | 7.17E-06 | 0.000817 | 1.04E-05 | -0.00599959 | EEPD1 |
| chr12 | 106955166 | 106955863 | 698 | 10 | 3.28E-05 | 3.90E-05 | 0.0031 | 1.04E-05 | -0.00162363 | TMEM263, AC007541.1 |
| chr15 | 74433541 | 74435075 | 1535 | 18 | 3.61E-06 | 4.61E-05 | 0.002949 | 1.04E-05 | -0.00131725 | SEMA7A |
| chr7 | 143315867 | 143316521 | 655 | 4 | 1.42E-05 | 2.12E-05 | 0.000273 | 1.04E-05 | 0.00461959 | CLCN1 |
| chr22 | 45199243 | 45199484 | 242 | 2 | 1.04E-05 | 6.33E-06 | 0.000652 | 1.04E-05 | -0.00346102 | KIAA0930 |
| chr7 | 122301956 | 122303233 | 1278 | 4 | 2.84E-05 | 3.33E-06 | 0.004375 | 1.04E-05 | -0.00135961 | FEZF1 |
| chr12 | 25885808 | 25886095 | 288 | 5 | 1.02E-06 | 0.0424022 | 1.26E-06 | 1.04E-05 | -0.00187177 | AC019209.1 |
| chr21 | 45283582 | 45283835 | 254 | 2 | 2.64E-06 | 0.0001236 | 1.21E-05 | 1.04E-05 | -0.00346712 | POFUT2 |
| chr19 | 51009574 | 51009639 | 66 | 2 | 1.91E-05 | 6.35E-05 | 1.67E-05 | 1.04E-05 | -0.01732146 | AC011483.1, AC011473.4, KLK9 |
| chr5 | 56058400 | 56058520 | 121 | 3 | 1.15E-05 | 0.0002068 | 4.04E-05 | 1.04E-05 | -0.01259058 | AC008892.1 |
| chr12 | 183045 | 183058 | 14 | 2 | 2.01E-05 | 6.82E-06 | 0.000414 | 1.04E-05 | -0.01166118 | NA |
| chr3 | 42638732 | 42639207 | 476 | 2 | 6.83E-06 | 1.77E-05 | 4.80E-05 | 1.04E-05 | -0.00167149 | NKTR, AC006059.1 |
| chr17 | 59619154 | 59620128 | 975 | 14 | 8.14E-06 | 0.0005424 | 0.001297 | 1.05E-05 | 0.001598049 | CLTC, AC091271.1 |
| chr5 | 172114823 | 172115323 | 501 | 3 | 2.16E-07 | 0.0433581 | 1.83E-06 | 1.05E-05 | -0.00469691 | STK10 |
| chr19 | 7639046 | 7639137 | 92 | 3 | 1.26E-05 | 4.06E-05 | 6.90E-05 | 1.05E-05 | -0.0023089 | AC008763.2, STXBP2 |
| chr13 | 111076401 | 111076563 | 163 | 2 | 2.20E-05 | 1.05E-05 | 0.000104 | 1.05E-05 | 0.002383135 | NA |
| chr2 | 189763201 | 189764219 | 1019 | 7 | 1.22E-05 | 3.15E-05 | 0.000859 | 1.05E-05 | -0.00011165 | OSGEPL1-AS1, OSGEPL1, AC013468.1 |
| chr15 | 86261948 | 86262455 | 508 | 3 | 3.56E-06 | 0.0002798 | 0.000148 | 1.05E-05 | 0.002213702 | AGBL1 |
| chr10 | 3008701 | 3009289 | 589 | 3 | 3.29E-06 | 0.0002929 | 0.000296 | 1.05E-05 | 0.012436588 | NA |
| chr2 | 46429120 | 46430245 | 1126 | 3 | 1.20E-05 | 4.96E-06 | 0.001815 | 1.05E-05 | -0.00788596 | LINC02583, AC016912.2, AC016912.1 |
| chr13 | 45418310 | 45419277 | 968 | 13 | 2.94E-05 | 6.51E-05 | 0.002878 | 1.05E-05 | 0.001999177 | SLC25A30-AS1, SLC25A30 |
| chr14 | 94367878 | 94368426 | 549 | 5 | 1.79E-05 | 2.40E-05 | 0.002632 | 1.05E-05 | -0.00103007 | NA |
| chr19 | 51704067 | 51704539 | 473 | 6 | 3.36E-06 | 6.27E-05 | 0.001045 | 1.05E-05 | 0.001120879 | SPACA6 |
| chr17 | 81955427 | 81955555 | 129 | 4 | 7.16E-06 | 8.48E-05 | 1.65E-05 | 1.05E-05 | -0.00226161 | NOTUM |
| chr10 | 23686779 | 23686810 | 32 | 2 | 1.30E-05 | 7.53E-05 | 1.53E-05 | 1.05E-05 | 0.009382951 | NA |
| chr4 | 6662736 | 6663600 | 865 | 3 | 5.49E-06 | 2.85E-05 | 0.00056 | 1.05E-05 | 0.00369189 | AC093323.1, LINC02482 |
| chr11 | 90134217 | 90134936 | 720 | 14 | 4.49E-05 | 2.07E-05 | 0.006288 | 1.05E-05 | -0.00392328 | NAALAD2 |
| chr8 | 143572717 | 143573383 | 667 | 9 | 1.33E-05 | 2.48E-05 | 0.003202 | 1.05E-05 | -0.00750997 | MROH6 |
| chr16 | 66534075 | 66534331 | 257 | 2 | 2.87E-05 | 6.51E-06 | 0.000572 | 1.05E-05 | -0.00286817 | TK2 |
| chr14 | 20435251 | 20436010 | 760 | 12 | 8.82E-07 | 0.0004095 | 0.004788 | 1.05E-05 | -0.00145369 | KLHL33 |
| chr11 | 119123701 | 119123931 | 231 | 2 | 1.28E-06 | 0.0007724 | 7.32E-06 | 1.05E-05 | -0.00818414 | HINFP |
| chr8 | 6901094 | 6901266 | 173 | 2 | 2.09E-05 | 1.14E-05 | 9.11E-05 | 1.05E-05 | 0.028360502 | NA |
| chr1 | 207323917 | 207324635 | 719 | 5 | 2.28E-07 | 0.0023019 | 3.43E-06 | 1.05E-05 | 0.000382232 | CD55 |
| chr17 | 80189469 | 80189991 | 523 | 4 | 6.37E-06 | 2.42E-05 | 0.000664 | 1.05E-05 | -0.0066512 | CARD14 |
| chr12 | 51889440 | 51890018 | 579 | 2 | 1.23E-05 | 9.75E-06 | 0.000127 | 1.05E-05 | -0.00764025 | ANKRD33 |
| chr20 | 61161937 | 61162573 | 637 | 3 | 2.25E-05 | 3.90E-06 | 0.003165 | 1.05E-05 | 0.003289179 | NA |
| chr10 | 132185117 | 132185409 | 293 | 4 | 3.37E-06 | 4.69E-05 | 0.000515 | 1.05E-05 | 0.013298434 | AL512622.1 |
| chr9 | 132409808 | 132409820 | 13 | 3 | 1.26E-05 | 3.51E-05 | 3.26E-05 | 1.05E-05 | -0.01103467 | NA |
| chr3 | 16828640 | 16829078 | 439 | 3 | 9.34E-06 | 2.21E-05 | 0.00014 | 1.06E-05 | 0.001675932 | PLCL2 |
| chr2 | 109305050 | 109305661 | 612 | 4 | 7.25E-06 | 1.06E-05 | 0.001094 | 1.06E-05 | 0.001775578 | SH3RF3 |
| chr19 | 7561290 | 7561464 | 175 | 3 | 3.18E-05 | 6.00E-06 | 0.000963 | 1.06E-05 | -0.00474629 | PNPLA6 |
| chr19 | 57099168 | 57099603 | 436 | 5 | 1.50E-05 | 4.19E-05 | 0.000514 | 1.06E-05 | -0.00691182 | NA |
| chr4 | 3107541 | 3108194 | 654 | 2 | 4.02E-05 | 6.39E-06 | 0.000692 | 1.06E-05 | 0.003723538 | HTT |
| chr5 | 157651565 | 157651814 | 250 | 4 | 3.11E-06 | 7.19E-06 | 0.000887 | 1.06E-05 | -0.0115245 | SOX30 |
| chr10 | 42888837 | 42888843 | 7 | 2 | 8.11E-06 | 0.0006078 | 7.70E-06 | 1.06E-05 | -0.00910825 | NA |
| chr11 | 78188519 | 78189594 | 1076 | 14 | 1.16E-06 | 0.0003869 | 1.56E-05 | 1.06E-05 | 1.72E-05 | USP35, KCTD21 |
| chr12 | 32501560 | 32502258 | 699 | 5 | 1.36E-05 | 8.27E-05 | 0.000225 | 1.06E-05 | -0.00197179 | FGD4 |
| chr3 | 48983895 | 48984482 | 588 | 3 | 2.10E-05 | 0.0002358 | 0.000145 | 1.06E-05 | 0.003046824 | ARIH2, AC137630.2 |
| chr2 | 85298720 | 85298970 | 251 | 5 | 1.70E-05 | 0.0003289 | 0.00044 | 1.06E-05 | -0.00185611 | TCF7L1 |
| chr17 | 4004055 | 4004700 | 646 | 5 | 1.10E-06 | 0.0035743 | 4.14E-05 | 1.06E-05 | 0.007851633 | ZZEF1 |
| chr16 | 4331660 | 4332222 | 563 | 6 | 7.31E-06 | 7.99E-05 | 0.00109 | 1.06E-05 | -0.00104218 | GLIS2, PAM16 |
| chr20 | 297082 | 297456 | 375 | 7 | 1.24E-05 | 2.49E-05 | 8.22E-05 | 1.06E-05 | 0.000681358 | NA |
| chr7 | 41692450 | 41693349 | 900 | 5 | 2.21E-05 | 4.39E-05 | 0.002632 | 1.06E-05 | 0.001603343 | INHBA |
| chr12 | 27973597 | 27974040 | 444 | 3 | 2.47E-07 | 0.017298 | 2.12E-06 | 1.06E-05 | 0.011072971 | NA |
| chr8 | 8989499 | 8989843 | 345 | 2 | 3.22E-06 | 0.0002126 | 1.02E-05 | 1.06E-05 | -0.01374838 | NA |
| chr22 | 40370465 | 40371214 | 750 | 10 | 1.33E-05 | 0.0001434 | 0.000577 | 1.06E-05 | 0.001020305 | ADSL, AL022238.3, SGSM3 |
| chr22 | 37098979 | 37099041 | 63 | 2 | 9.00E-06 | 5.68E-05 | 1.82E-05 | 1.06E-05 | -0.01849164 | TMPRSS6 |
| chr15 | 43464067 | 43464544 | 478 | 2 | 7.35E-07 | 0.0004971 | 8.04E-06 | 1.06E-05 | -0.00107379 | TP53BP1 |
| chr17 | 76568739 | 76568927 | 189 | 3 | 1.67E-05 | 5.50E-06 | 0.001265 | 1.06E-05 | 0.012283572 | AC015802.6, ST6GALNAC2 |
| chr12 | 49544214 | 49544235 | 22 | 2 | 2.47E-05 | 6.93E-06 | 0.000426 | 1.06E-05 | -0.01260094 | KCNH3 |
| chr2 | 215840133 | 215840428 | 296 | 2 | 2.87E-05 | 6.61E-06 | 0.00056 | 1.06E-05 | 0.007765432 | LINC00607 |
| chr16 | 788502 | 789166 | 665 | 7 | 4.94E-06 | 0.0006766 | 0.000317 | 1.06E-05 | 0.005802937 | CHTF18 |
| chr5 | 139794622 | 139794980 | 359 | 4 | 1.52E-05 | 1.20E-05 | 0.002495 | 1.06E-05 | -0.00866366 | NA |
| chr12 | 53748164 | 53748219 | 56 | 2 | 2.05E-05 | 6.95E-06 | 0.000422 | 1.06E-05 | 0.012340401 | AC076968.2, CISTR |
| chr7 | 893662 | 893670 | 9 | 3 | 2.00E-06 | 0.0002267 | 2.61E-05 | 1.06E-05 | -0.00546429 | SUN1, GET4 |
| chr6 | 42821093 | 42821315 | 223 | 2 | 8.07E-07 | 0.0010159 | 7.06E-06 | 1.06E-05 | -0.00555209 | BICRAL |
| chr17 | 63440805 | 63441190 | 386 | 7 | 6.23E-07 | 0.0043056 | 0.000208 | 1.06E-05 | -0.00077714 | CYB561 |
| chr7 | 56092716 | 56093327 | 612 | 10 | 6.63E-05 | 2.60E-06 | 0.008019 | 1.06E-05 | -0.00899076 | PHKG1 |
| chr17 | 48048936 | 48049414 | 479 | 2 | 1.94E-06 | 6.56E-06 | 0.000598 | 1.07E-05 | -0.00825067 | NFE2L1 |
| chr1 | 2496831 | 2497639 | 809 | 9 | 5.21E-06 | 0.0001346 | 0.000899 | 1.07E-05 | -0.00371247 | PLCH2 |
| chr10 | 103244877 | 103245192 | 316 | 3 | 1.02E-05 | 1.35E-05 | 0.000446 | 1.07E-05 | 0.004404711 | NT5C2 |
| chr15 | 69090400 | 69090935 | 536 | 2 | 4.56E-05 | 6.42E-06 | 0.000715 | 1.07E-05 | -0.01278536 | EWSAT1, AC027088.2 |
| chr1 | 116359926 | 116360291 | 366 | 2 | 1.92E-06 | 0.0009679 | 7.14E-06 | 1.07E-05 | -0.00514591 | NA |
| chr5 | 153497128 | 153497487 | 360 | 2 | 5.72E-06 | 4.46E-05 | 2.15E-05 | 1.07E-05 | -0.00174995 | GRIA1 |
| chr8 | 41505698 | 41506159 | 462 | 2 | 3.78E-05 | 6.48E-06 | 0.000679 | 1.07E-05 | -0.00300829 | GOLGA7 |
| chr2 | 9318796 | 9319467 | 672 | 8 | 5.10E-06 | 0.0004483 | 0.000113 | 1.07E-05 | 0.001144164 | ASAP2 |
| chr9 | 34989141 | 34989699 | 559 | 8 | 1.00E-05 | 0.0011043 | 5.28E-05 | 1.07E-05 | -8.09E-05 | DNAJB5, DNAJB5-DT |
| chr1 | 1980626 | 1980696 | 71 | 2 | 1.90E-05 | 1.75E-05 | 5.08E-05 | 1.07E-05 | -0.001937 | CFAP74 |
| chr10 | 124389225 | 124389678 | 454 | 3 | 3.88E-07 | 0.0004846 | 9.23E-06 | 1.07E-05 | -0.00764428 | NA |
| chr21 | 21686660 | 21687098 | 439 | 3 | 4.41E-06 | 0.0001196 | 1.40E-05 | 1.07E-05 | 0.006207222 | NA |
| chr10 | 71553198 | 71553675 | 478 | 2 | 2.17E-05 | 7.76E-06 | 0.000267 | 1.07E-05 | -0.00891902 | CDH23 |
| chr12 | 113276784 | 113277248 | 465 | 3 | 1.00E-05 | 1.46E-05 | 0.000143 | 1.07E-05 | 0.003884273 | TPCN1 |
| chr17 | 18107877 | 18108199 | 323 | 3 | 5.04E-06 | 5.76E-06 | 0.000878 | 1.07E-05 | -0.00215789 | DRG2, AC087164.2 |
| chr17 | 1032772 | 1033310 | 539 | 2 | 2.76E-05 | 6.74E-06 | 0.000528 | 1.07E-05 | 0.003426328 | ABR |
| chr14 | 74922577 | 74923698 | 1122 | 13 | 2.38E-06 | 1.35E-05 | 0.009701 | 1.07E-05 | 0.003334878 | RPS6KL1 |
| chr8 | 9182534 | 9182939 | 406 | 3 | 5.87E-06 | 3.19E-05 | 6.83E-05 | 1.07E-05 | -0.00264599 | AC022784.1 |
| chr3 | 71658302 | 71658648 | 347 | 2 | 7.22E-06 | 4.85E-05 | 2.04E-05 | 1.07E-05 | -0.00160753 | AC097634.4 |
| chr3 | 158644392 | 158644741 | 350 | 7 | 1.68E-06 | 0.0001938 | 0.000146 | 1.07E-05 | -0.00045514 | GFM1 |
| chr6 | 29670721 | 29671141 | 421 | 3 | 7.94E-06 | 1.06E-05 | 0.000785 | 1.07E-05 | -0.01187591 | MOG |
| chr12 | 62519293 | 62519698 | 406 | 2 | 1.87E-06 | 3.88E-05 | 2.41E-05 | 1.08E-05 | -0.00173249 | MON2 |
| chr8 | 23984637 | 23985090 | 454 | 3 | 2.24E-05 | 1.17E-05 | 0.000163 | 1.08E-05 | -0.01689048 | NA |
| chr8 | 43092654 | 43093409 | 756 | 10 | 1.67E-05 | 0.0001628 | 0.003256 | 1.08E-05 | -0.00108167 | NA |
| chr15 | 89574943 | 89575906 | 964 | 11 | 7.03E-07 | 0.0001664 | 0.006036 | 1.08E-05 | 0.000970775 | AC013391.2, TICRR |
| chr5 | 139862777 | 139863368 | 592 | 2 | 1.46E-05 | 7.51E-06 | 0.000314 | 1.08E-05 | -0.00626158 | NRG2 |
| chr4 | 100193985 | 100194027 | 43 | 2 | 1.75E-05 | 3.17E-05 | 2.83E-05 | 1.08E-05 | -0.01031134 | H2AZ1-DT, AP001961.1 |
| chr11 | 130156803 | 130156903 | 101 | 2 | 1.55E-05 | 1.05E-05 | 0.000114 | 1.08E-05 | -0.00746892 | NA |
| chr21 | 17196122 | 17196617 | 496 | 2 | 1.46E-06 | 2.19E-05 | 3.99E-05 | 1.08E-05 | 0.005191833 | NA |
| chr20 | 63084388 | 63084526 | 139 | 2 | 2.56E-05 | 6.76E-06 | 0.000539 | 1.08E-05 | -0.00920787 | LINC01749 |
| chr20 | 4823218 | 4824263 | 1046 | 7 | 9.73E-06 | 5.87E-06 | 0.007564 | 1.08E-05 | -0.00633202 | RASSF2 |
| chr10 | 54801185 | 54801783 | 599 | 6 | 5.82E-05 | 5.88E-06 | 0.005481 | 1.08E-05 | 0.004711933 | PCDH15 |
| chr14 | 103536036 | 103536317 | 282 | 2 | 2.25E-06 | 0.0001554 | 1.16E-05 | 1.08E-05 | -0.00401055 | TRMT61A |
| chr5 | 142644209 | 142645050 | 842 | 5 | 2.24E-05 | 2.69E-06 | 0.008464 | 1.08E-05 | -0.00766863 | SPRY4-AS1, FGF1 |
| chr6 | 158819045 | 158819782 | 738 | 9 | 5.85E-05 | 1.84E-05 | 0.006771 | 1.08E-05 | -0.00296596 | EZR-AS1, EZR |
| chr1 | 7333626 | 7335092 | 1467 | 4 | 1.84E-06 | 0.0007966 | 3.79E-05 | 1.08E-05 | -0.00130539 | CAMTA1 |
| chr12 | 113831647 | 113831666 | 20 | 2 | 6.76E-06 | 0.0001718 | 1.11E-05 | 1.08E-05 | 0.001770079 | RBM19 |
| chr7 | 97599290 | 97599500 | 211 | 3 | 3.38E-05 | 5.77E-06 | 0.001539 | 1.08E-05 | -0.00615086 | NA |

| | | | | | | | | | | |
|---|---|---|---|---|---|---|---|---|---|---|
| chr5 | 431039 | 431248 | 210 | 2 | 2.51E-06 | 1.29E-05 | 7.87E-05 | 1.08E-05 | -0.02860897 | PDCD6-AHRR, AHRR |
| chr1 | 232293043 | 232293122 | 80 | 2 | 3.09E-05 | 6.51E-06 | 0.000714 | 1.08E-05 | 0.016051972 | NA |
| chr7 | 127892308 | 127892727 | 420 | 2 | 5.09E-06 | 3.56E-05 | 2.58E-05 | 1.08E-05 | 0.006620971 | SND1 |
| chr2 | 237564827 | 237565230 | 404 | 5 | 8.17E-06 | 1.78E-05 | 0.000847 | 1.08E-05 | -0.00173636 | NA |
| chr1 | 66924666 | 66925408 | 743 | 12 | 4.79E-06 | 2.36E-05 | 0.008294 | 1.08E-05 | -0.00031105 | MIER1, DNAI4 |
| chr17 | 6474378 | 6474726 | 349 | 2 | 8.61E-06 | 1.42E-05 | 6.85E-05 | 1.08E-05 | 0.00176795 | PITPNM3 |
| chr6 | 31177525 | 31178445 | 921 | 5 | 1.10E-05 | 0.0001137 | 0.000255 | 1.08E-05 | -0.00250459 | POU5F1, PSORS1C3 |
| chr6 | 166465194 | 166465873 | 680 | 6 | 1.24E-05 | 4.58E-05 | 0.000757 | 1.08E-05 | 0.002632939 | RPS6KA2, RPS6KA2-IT1 |
| chr5 | 122948033 | 122948557 | 525 | 2 | 1.61E-05 | 9.27E-06 | 0.000157 | 1.08E-05 | -0.01310726 | SNX24 |
| chr2 | 237056707 | 237056884 | 178 | 2 | 3.19E-06 | 6.92E-05 | 1.66E-05 | 1.08E-05 | -0.00349629 | AC012063.1 |
| chr2 | 120828846 | 120829208 | 363 | 3 | 1.99E-05 | 8.80E-06 | 0.000292 | 1.08E-05 | -0.01159945 | GLI2 |
| chr9 | 130367003 | 130367417 | 415 | 2 | 1.39E-06 | 0.0006649 | 7.74E-06 | 1.08E-05 | -0.00382868 | HMCN2 |
| chr19 | 55945925 | 55946009 | 85 | 2 | 2.95E-05 | 6.53E-06 | 0.000701 | 1.08E-05 | 0.008245304 | NA |
| chr20 | 141286 | 141988 | 703 | 3 | 7.12E-06 | 1.30E-05 | 0.000722 | 1.08E-05 | -0.02258576 | NA |
| chr3 | 126056570 | 126057311 | 742 | 6 | 1.75E-05 | 3.00E-05 | 0.000356 | 1.08E-05 | -0.00298342 | AC117422.1, SLC41A3 |
| chr7 | 130373260 | 130373895 | 636 | 2 | 7.99E-06 | 1.34E-05 | 7.47E-05 | 1.08E-05 | 0.015163476 | NA |
| chr8 | 90000948 | 90001469 | 522 | 10 | 8.43E-06 | 0.000214 | 0.00033 | 1.08E-05 | -9.10E-05 | DECR1, NBN |
| chr7 | 24980132 | 24980379 | 248 | 6 | 4.16E-05 | 3.23E-06 | 0.007958 | 1.08E-05 | 0.003323999 | OSBPL3 |
| chr3 | 72345796 | 72346298 | 503 | 2 | 3.80E-05 | 6.62E-06 | 0.000643 | 1.08E-05 | 0.015726516 | NA |
| chr22 | 39598150 | 39598830 | 681 | 4 | 2.78E-05 | 5.03E-05 | 0.000348 | 1.09E-05 | -0.004962 | CACNA1I |
| chr6 | 133439782 | 133440476 | 695 | 3 | 3.30E-05 | 9.84E-06 | 0.000437 | 1.09E-05 | -0.00420779 | EYA4 |
| chr8 | 6891552 | 6892011 | 460 | 2 | 8.52E-06 | 9.53E-06 | 0.000147 | 1.09E-05 | -0.00383326 | NA |
| chr11 | 110185616 | 110185986 | 371 | 3 | 8.07E-06 | 5.78E-05 | 8.52E-05 | 1.09E-05 | -0.00039423 | RDX |
| chr19 | 6110302 | 6110977 | 676 | 9 | 1.76E-05 | 1.70E-05 | 0.001017 | 1.09E-05 | 0.000372413 | AC011444.3, RFX2 |
| chr9 | 89816091 | 89816474 | 384 | 2 | 3.27E-05 | 6.54E-06 | 0.000718 | 1.09E-05 | 0.020261088 | BX470209.1 |
| chr19 | 45354725 | 45354991 | 267 | 3 | 1.74E-05 | 1.88E-05 | 9.02E-05 | 1.09E-05 | 0.000819517 | ERCC2 |
| chr5 | 314438 | 314527 | 90 | 4 | 1.16E-05 | 0.0001499 | 2.58E-05 | 1.09E-05 | 0.002133259 | AC021087.5, PDCD6, PDCD6-AHRR |
| chr20 | 21709121 | 21709398 | 278 | 3 | 1.19E-05 | 7.24E-06 | 0.000558 | 1.09E-05 | -0.00222009 | PAX1 |
| chr22 | 18970165 | 18970367 | 203 | 7 | 1.79E-05 | 3.94E-05 | 0.000372 | 1.09E-05 | -0.0003946 | NA |
| chr10 | 47406809 | 47407071 | 263 | 2 | 9.18E-06 | 4.12E-05 | 2.34E-05 | 1.09E-05 | -0.01004129 | NA |
| chr2 | 1373670 | 1374263 | 594 | 6 | 1.14E-05 | 4.46E-05 | 0.000207 | 1.09E-05 | -0.00210475 | TPO, AC105450.1 |
| chr6 | 135114026 | 135114544 | 519 | 2 | 2.21E-05 | 7.97E-06 | 0.000258 | 1.09E-05 | -0.00741355 | NA |
| chr6 | 57221495 | 57222643 | 1149 | 16 | 3.87E-06 | 3.15E-05 | 0.007601 | 1.09E-05 | 0.000334176 | RAB23 |
| chr22 | 37166378 | 37166714 | 337 | 4 | 1.03E-05 | 3.53E-05 | 0.000781 | 1.09E-05 | 0.005403462 | IL2RB |
| chr1 | 240179990 | 240180345 | 356 | 3 | 1.48E-05 | 1.61E-05 | 8.90E-05 | 1.09E-05 | -0.01240281 | FMN2 |
| chr5 | 1155738 | 1156338 | 601 | 2 | 1.67E-05 | 8.85E-06 | 0.000183 | 1.09E-05 | -0.00552812 | NA |
| chr16 | 88682114 | 88682154 | 41 | 2 | 3.44E-06 | 7.11E-05 | 1.65E-05 | 1.09E-05 | 0.014663604 | SNAI3-AS1, SNAI3 |
| chr10 | 2328776 | 2329244 | 469 | 5 | 2.29E-06 | 0.0006824 | 0.0005 | 1.09E-05 | -0.00044489 | NA |
| chr1 | 42787904 | 42788011 | 108 | 2 | 4.52E-06 | 0.0002664 | 9.76E-06 | 1.09E-05 | -0.00438204 | AC098484.3, TMEM269 |
| chr1 | 234744301 | 234744990 | 690 | 3 | 1.90E-06 | 9.69E-05 | 2.01E-05 | 1.09E-05 | -0.00592555 | NA |
| chr10 | 113854075 | 113854953 | 879 | 18 | 3.07E-05 | 8.85E-06 | 0.021233 | 1.09E-05 | 0.000564261 | NHLRC2, DCLRE1A |
| chr8 | 143988909 | 143989194 | 286 | 4 | 7.48E-07 | 0.000131 | 1.45E-05 | 1.09E-05 | -0.01214376 | PARP10 |
| chr11 | 1763750 | 1764574 | 825 | 14 | 2.61E-06 | 0.0001559 | 0.001461 | 1.09E-05 | -0.00107917 | AC068580.4, CTSD |
| chr18 | 49048214 | 49048419 | 206 | 2 | 6.28E-06 | 0.0002977 | 9.46E-06 | 1.09E-05 | -0.0159945 | AC016866.1, DYM |
| chr2 | 241125996 | 241126483 | 488 | 3 | 2.71E-06 | 2.89E-05 | 0.000144 | 1.09E-05 | -0.00163958 | PASK |
| chr5 | 137736110 | 137737246 | 1137 | 5 | 1.01E-05 | 3.70E-05 | 0.001051 | 1.09E-05 | 0.000160046 | NA |
| chr12 | 54561065 | 54561359 | 295 | 4 | 1.62E-05 | 5.04E-06 | 0.002291 | 1.09E-05 | -0.01021707 | PDE1B |
| chr9 | 134367461 | 134368301 | 841 | 4 | 2.98E-06 | 0.0015782 | 1.44E-05 | 1.09E-05 | -0.00281641 | RXRA |
| chr12 | 132910845 | 132911536 | 692 | 3 | 6.23E-05 | 2.28E-05 | 0.000514 | 1.09E-05 | 0.001399781 | AC127070.2, CHFR |
| chr7 | 134552448 | 134552551 | 104 | 2 | 3.16E-05 | 6.63E-06 | 0.000683 | 1.09E-05 | -0.00612608 | AKR1B15 |
| chr18 | 48825353 | 48825476 | 124 | 2 | 1.22E-05 | 7.14E-05 | 1.66E-05 | 1.09E-05 | -0.00472493 | CTIF |
| chr12 | 129697086 | 129697153 | 68 | 2 | 9.74E-07 | 0.0011868 | 7.08E-06 | 1.10E-05 | 0.00932718 | AC117373.2, TMEM132D, AC117373.1 |
| chr7 | 151130211 | 151130793 | 583 | 2 | 5.12E-05 | 9.96E-06 | 0.000136 | 1.10E-05 | -0.00456333 | AGAP3 |
| chr20 | 710271 | 710567 | 297 | 3 | 1.12E-05 | 3.99E-05 | 0.000114 | 1.10E-05 | -0.00437285 | NA |
| chr1 | 207064497 | 207064991 | 495 | 5 | 1.86E-05 | 2.32E-06 | 0.007774 | 1.10E-05 | -0.00048617 | PFKFB2 |
| chr2 | 10526459 | 10526736 | 278 | 4 | 1.96E-05 | 7.63E-06 | 0.001576 | 1.10E-05 | -0.00652472 | NA |
| chr7 | 156641709 | 156641960 | 252 | 4 | 3.25E-06 | 8.03E-05 | 0.000161 | 1.10E-05 | -0.00037127 | RNF32 |
| chr22 | 21919598 | 21920262 | 665 | 3 | 4.88E-05 | 7.64E-06 | 0.001067 | 1.10E-05 | -0.0037924 | PPM1F |
| chr22 | 49823171 | 49823786 | 616 | 6 | 7.93E-06 | 8.77E-06 | 0.00181 | 1.10E-05 | -0.00186744 | BRD1 |
| chr1 | 73191952 | 73192036 | 85 | 2 | 3.16E-05 | 6.71E-06 | 0.000648 | 1.10E-05 | -0.00465066 | NA |
| chr1 | 157238241 | 157238357 | 117 | 2 | 3.15E-05 | 6.70E-06 | 0.000656 | 1.10E-05 | -0.01722849 | NA |
| chr19 | 58059422 | 58059712 | 291 | 4 | 1.26E-05 | 3.04E-06 | 0.005208 | 1.10E-05 | -0.00278446 | ZNF135 |
| chr6 | 43006557 | 43006981 | 425 | 2 | 2.47E-05 | 1.02E-05 | 0.000129 | 1.10E-05 | 0.00300427 | PPP2R5D |
| chr20 | 46178943 | 46179004 | 62 | 2 | 4.44E-06 | 2.34E-05 | 3.84E-05 | 1.10E-05 | -0.00755972 | CDH22 |
| chr4 | 3871691 | 3872416 | 726 | 6 | 6.73E-05 | 6.41E-05 | 0.004887 | 1.10E-05 | -0.00255773 | NA |
| chr17 | 33779117 | 33779513 | 397 | 2 | 2.38E-05 | 7.71E-06 | 0.000311 | 1.10E-05 | -0.01318929 | ASIC2 |
| chr11 | 126063260 | 126063803 | 544 | 8 | 5.31E-05 | 2.09E-06 | 0.010734 | 1.10E-05 | -0.00014361 | CDON |
| chr8 | 144180048 | 144180507 | 460 | 5 | 1.54E-05 | 9.33E-06 | 0.003091 | 1.10E-05 | -0.00076791 | MROH1 |
| chr1 | 109548055 | 109549003 | 949 | 15 | 3.52E-06 | 0.0003526 | 0.001418 | 1.10E-05 | 0.000326447 | GPR61, GNAI3 |
| chr15 | 89415319 | 89415713 | 395 | 2 | 7.55E-07 | 0.0006324 | 7.95E-06 | 1.10E-05 | -0.01861468 | NA |
| chr2 | 96195842 | 96196147 | 306 | 2 | 2.29E-06 | 0.0001076 | 1.37E-05 | 1.10E-05 | 0.007410006 | STARD7 |
| chr1 | 24187728 | 24188627 | 900 | 8 | 1.29E-05 | 5.15E-05 | 0.004545 | 1.10E-05 | -0.00029054 | IFNLR1 |
| chr1 | 161201421 | 161202501 | 1081 | 16 | 7.27E-06 | 0.0001101 | 0.000989 | 1.10E-05 | -0.00391814 | NDUFS2 |
| chr11 | 130473182 | 130473638 | 457 | 3 | 3.24E-05 | 5.00E-06 | 0.00135 | 1.11E-05 | -0.00485628 | ADAMTS15 |
| chr5 | 179895739 | 179896035 | 297 | 2 | 9.35E-06 | 2.24E-05 | 4.05E-05 | 1.11E-05 | -0.00113447 | TBC1D9B |
| chr19 | 5875438 | 5875736 | 299 | 3 | 7.85E-06 | 5.50E-05 | 0.000129 | 1.11E-05 | 0.000389738 | AC024592.3 |
| chr14 | 70359717 | 70360725 | 1009 | 10 | 2.67E-06 | 0.0001804 | 0.001505 | 1.11E-05 | -0.00735203 | COX16, SYNJ2BP-COX16 |
| chr3 | 121644209 | 121644460 | 252 | 4 | 4.85E-06 | 0.0009743 | 0.000183 | 1.11E-05 | -0.00482357 | HCLS1 |
| chr9 | 96080361 | 96080800 | 440 | 2 | 2.39E-05 | 6.65E-06 | 0.000728 | 1.11E-05 | -0.00156536 | ERCC6L2, AL449403.1 |
| chr15 | 25230308 | 25230909 | 602 | 5 | 1.15E-05 | 0.00019 | 0.000412 | 1.11E-05 | -0.00652203 | SNHG14, SNORD115-33 |
| chr13 | 26046474 | 26046814 | 341 | 3 | 1.27E-06 | 0.0078382 | 2.98E-06 | 1.11E-05 | -0.00242983 | SHISA2 |
| chr15 | 47400180 | 47400934 | 755 | 3 | 3.84E-05 | 7.85E-06 | 0.00087 | 1.11E-05 | 0.008266463 | SEMA6D |
| chr9 | 109817607 | 109817996 | 390 | 3 | 1.24E-05 | 5.50E-06 | 0.001423 | 1.11E-05 | -0.00958736 | PALM2AKAP2 |
| chr8 | 133051094 | 133051706 | 613 | 2 | 2.46E-07 | 3.20E-05 | 2.92E-05 | 1.11E-05 | -0.00408369 | TG, SLA |
| chr2 | 202911800 | 202912274 | 475 | 9 | 2.82E-05 | 3.24E-05 | 0.001819 | 1.11E-05 | -0.00063029 | CARF, WDR12 |
| chr10 | 29659111 | 29659395 | 285 | 2 | 9.73E-06 | 2.31E-05 | 3.96E-05 | 1.11E-05 | -0.0092896 | SVIL |
| chr3 | 55615419 | 55615770 | 352 | 2 | 4.85E-06 | 1.09E-05 | 0.000115 | 1.11E-05 | -0.00656488 | ERC2 |
| chr7 | 55964133 | 55964671 | 539 | 9 | 1.98E-05 | 0.0001896 | 0.002598 | 1.11E-05 | -0.00032598 | NIPSNAP2 |
| chr5 | 43019558 | 43019771 | 214 | 3 | 2.38E-05 | 4.21E-06 | 0.002858 | 1.11E-05 | 0.00548393 | AC025171.3, AC025171.1 |
| chr4 | 138918741 | 138919360 | 620 | 4 | 5.09E-05 | 3.58E-06 | 0.00362 | 1.11E-05 | 0.020398743 | AC109927.1 |
| chr12 | 29388452 | 29388496 | 45 | 2 | 6.65E-06 | 0.0001478 | 1.21E-05 | 1.11E-05 | -0.01559141 | NA |
| chr21 | 32413104 | 32413810 | 707 | 4 | 3.92E-07 | 7.36E-05 | 0.000478 | 1.11E-05 | -0.00247132 | EVA1C, CFAP298-TCP10L |
| chr13 | 96229278 | 96229418 | 141 | 2 | 2.37E-05 | 1.19E-05 | 9.64E-05 | 1.11E-05 | -0.00337797 | HS6ST3 |
| chr1 | 9936458 | 9936479 | 22 | 2 | 4.09E-05 | 7.98E-06 | 0.000283 | 1.11E-05 | 0.013126506 | LZIC |
| chr5 | 132059990 | 132060664 | 675 | 10 | 2.34E-05 | 0.000212 | 0.002394 | 1.11E-05 | 0.002261163 | IL3 |
| chr9 | 133355996 | 133356415 | 420 | 5 | 3.68E-05 | 1.21E-05 | 0.001248 | 1.11E-05 | 0.001312679 | SURF1 |
| chr2 | 224564315 | 224564575 | 261 | 2 | 1.35E-05 | 1.09E-05 | 0.000116 | 1.11E-05 | -0.00541337 | CUL3 |
| chr6 | 54266219 | 54266568 | 350 | 2 | 2.71E-05 | 7.07E-06 | 0.000508 | 1.11E-05 | -0.0025534 | MLIP |
| chr5 | 139383123 | 139383781 | 659 | 6 | 1.38E-05 | 8.54E-06 | 0.001092 | 1.11E-05 | 0.003076228 | SLC23A1 |
| chr11 | 66371086 | 66372398 | 1313 | 12 | 1.58E-06 | 0.0001332 | 0.003592 | 1.12E-05 | 0.001553588 | SLC29A2 |
| chr10 | 71795991 | 71796270 | 280 | 2 | 6.35E-06 | 8.92E-06 | 0.000194 | 1.12E-05 | -0.0015916 | CDH23 |
| chr2 | 105081131 | 105081319 | 189 | 3 | 1.16E-05 | 6.66E-05 | 2.01E-05 | 1.12E-05 | -0.00046784 | MRPS9 |
| chr1 | 60073463 | 60074003 | 541 | 9 | 2.80E-05 | 9.66E-05 | 0.002981 | 1.12E-05 | -0.00428454 | C1orf87 |
| chr12 | 45065302 | 45065484 | 183 | 4 | 3.22E-06 | 0.0026762 | 2.67E-06 | 1.12E-05 | -0.00161467 | AC008127.1, RACGAP1P1 |
| chr7 | 39949463 | 39950413 | 951 | 10 | 3.22E-07 | 0.0002155 | 1.61E-05 | 1.12E-05 | 0.000925522 | CDK13, AC072061.1 |
| chr19 | 50718346 | 50718348 | 3 | 2 | 2.16E-06 | 7.25E-06 | 0.000449 | 1.12E-05 | -0.00074255 | SHANK1 |
| chr4 | 40190165 | 40190579 | 415 | 3 | 1.24E-05 | 4.57E-06 | 0.002347 | 1.12E-05 | -0.00047958 | AC095057.4 |
| chr1 | 24555662 | 24556500 | 839 | 10 | 9.29E-06 | 1.86E-05 | 0.00175 | 1.12E-05 | -0.00042333 | NCMAP, NCMAP-DT |
| chr12 | 6982851 | 6983403 | 553 | 2 | 1.14E-05 | 6.75E-06 | 0.000704 | 1.12E-05 | -0.01084661 | EMG1, LPCAT3 |
| chr17 | 56833824 | 56834496 | 673 | 5 | 2.00E-06 | 0.0001558 | 0.000155 | 1.12E-05 | -0.00039082 | DGKE, C17orf67 |
| chr11 | 32583638 | 32583893 | 256 | 8 | 5.40E-05 | 2.29E-05 | 0.005031 | 1.12E-05 | -0.00058019 | EIF3M |
| chr2 | 231706428 | 231707193 | 766 | 7 | 3.77E-06 | 9.99E-05 | 0.000478 | 1.12E-05 | -0.00138001 | PTMA |
| chr19 | 47660541 | 47660562 | 22 | 2 | 3.10E-05 | 7.58E-06 | 0.000367 | 1.12E-05 | -0.00056897 | AC010519.1, BICRA |
| chr6 | 13923681 | 13924703 | 1023 | 8 | 2.88E-06 | 3.18E-05 | 0.004133 | 1.12E-05 | -0.00396593 | RNF182 |
| chr11 | 2741941 | 2742403 | 463 | 2 | 5.24E-06 | 2.97E-05 | 3.16E-05 | 1.12E-05 | -0.00455085 | KCNQ1 |
| chr16 | 1580025 | 1580466 | 442 | 4 | 1.10E-05 | 1.76E-05 | 0.000632 | 1.12E-05 | -0.00736329 | IFT140, AL133297.1 |
| chr1 | 58432880 | 58433121 | 242 | 3 | 3.08E-05 | 6.63E-06 | 0.00121 | 1.12E-05 | 0.036951855 | DAB1, OMA1 |
| chr16 | 74632588 | 74632771 | 184 | 3 | 3.94E-05 | 4.65E-06 | 0.002426 | 1.12E-05 | -0.00410234 | RFWD3 |
| chr16 | 28974728 | 28975508 | 781 | 8 | 2.40E-05 | 0.0009885 | 0.000194 | 1.12E-05 | -4.61E-05 | AC009093.11, SPNS1, AC109460.3, AC109460.1 |
| chr10 | 73791920 | 73792566 | 647 | 4 | 2.00E-05 | 2.51E-05 | 7.89E-05 | 1.12E-05 | -0.00243189 | ZSWIM8 |
| chr9 | 134746575 | 134746871 | 297 | 2 | 1.20E-05 | 1.26E-05 | 8.92E-05 | 1.12E-05 | -0.00115504 | COL5A1 |
| chr4 | 2823958 | 2824693 | 736 | 7 | 2.58E-05 | 6.03E-05 | 0.000768 | 1.12E-05 | 5.56E-05 | SH3BP2 |
| chr5 | 89271098 | 89271434 | 337 | 2 | 2.60E-05 | 7.65E-06 | 0.000356 | 1.12E-05 | 0.012747737 | MEF2C-AS1 |
| chr11 | 1195940 | 1196302 | 363 | 3 | 2.22E-05 | 5.16E-06 | 0.001812 | 1.12E-05 | -0.00191452 | MUC5AC |
| chr3 | 12187501 | 12187520 | 20 | 2 | 9.21E-06 | 8.55E-06 | 0.000228 | 1.12E-05 | 0.006005041 | SYN2 |
| chr2 | 236048219 | 236049102 | 884 | 4 | 5.21E-06 | 5.15E-05 | 0.000173 | 1.12E-05 | 0.00017986 | AGAP1 |
| chr12 | 122500233 | 122501096 | 864 | 10 | 7.98E-07 | 0.0002248 | 0.000584 | 1.12E-05 | 0.000700241 | ZCCHC8 |
| chr3 | 45884624 | 45885177 | 554 | 3 | 6.54E-06 | 3.36E-05 | 0.000437 | 1.13E-05 | -0.00780434 | LZTFL1 |
| chr1 | 32961139 | 32961540 | 402 | 3 | 1.30E-05 | 3.74E-05 | 4.13E-05 | 1.13E-05 | -0.01041164 | RNF19B |
| chr9 | 89700027 | 89700335 | 309 | 2 | 2.88E-06 | 0.0002103 | 1.09E-05 | 1.13E-05 | 0.005752751 | UNQ6494 |
| chr6 | 35719533 | 35719680 | 148 | 3 | 2.31E-05 | 1.15E-05 | 0.000166 | 1.13E-05 | -0.00063718 | FKBP5 |
| chr15 | 47942758 | 47943120 | 363 | 2 | 7.97E-07 | 0.0271831 | 5.87E-06 | 1.13E-05 | -0.01169326 | AC092078.2 |
| chr17 | 49578002 | 49578479 | 478 | 3 | 1.49E-05 | 1.49E-05 | 0.000219 | 1.13E-05 | -0.00377382 | NXPH3 |
| chr3 | 16315669 | 16316084 | 416 | 3 | 1.18E-05 | 2.41E-05 | 6.55E-05 | 1.13E-05 | 0.005676239 | OXNAD1, RFTN1 |
| chr8 | 495647 | 496004 | 358 | 2 | 4.10E-07 | 0.0017486 | 6.90E-06 | 1.13E-05 | -0.00970576 | TDRP |
| chr16 | 1986529 | 1986893 | 365 | 3 | 3.76E-06 | 0.0001896 | 4.14E-05 | 1.13E-05 | -0.00740377 | GFER, AC005606.2 |
| chr22 | 42344238 | 42344848 | 611 | 4 | 1.46E-05 | 5.40E-06 | 0.001514 | 1.13E-05 | -0.00460923 | NA |
| chr5 | 141634495 | 141634956 | 462 | 2 | 3.68E-08 | 0.0005272 | 8.45E-06 | 1.13E-05 | -0.00647672 | HDAC3 |
| chr10 | 14173306 | 14174101 | 796 | 6 | 2.20E-06 | 0.0004713 | 5.58E-05 | 1.13E-05 | -0.00432492 | FRMD4A |
| chr15 | 63155789 | 63155876 | 88 | 2 | 1.21E-05 | 7.96E-06 | 0.000304 | 1.13E-05 | 0.004822545 | RPS27L |
| chr3 | 52153775 | 52154752 | 978 | 10 | 8.77E-06 | 0.0009442 | 2.56E-05 | 1.13E-05 | -0.0013652 | POC1A |
| chr6 | 30480707 | 30481737 | 1031 | 10 | 2.11E-05 | 0.0003259 | 0.00226 | 1.13E-05 | 0.000936629 | NA |
| chr6 | 144351751 | 144351863 | 113 | 2 | 8.49E-06 | 0.0001804 | 1.15E-05 | 1.13E-05 | 0.013449134 | UTRN |
| chr2 | 60550771 | 60551838 | 1068 | 6 | 9.51E-06 | 6.03E-06 | 0.001338 | 1.13E-05 | 0.002196797 | BCL11A |
| chr17 | 50538526 | 50539045 | 520 | 3 | 1.09E-05 | 7.55E-06 | 0.001068 | 1.13E-05 | -0.00133527 | EPN3, AC021491.2 |
| chr17 | 45120757 | 45121243 | 487 | 4 | 2.25E-05 | 5.54E-06 | 0.001719 | 1.13E-05 | -0.01470201 | PLCD3 |
| chr8 | 13365091 | 13365519 | 429 | 2 | 2.90E-06 | 0.0001064 | 1.41E-05 | 1.13E-05 | -0.01005955 | DLC1 |
| chr12 | 115752941 | 115753255 | 315 | 3 | 6.47E-06 | 2.35E-05 | 0.000141 | 1.13E-05 | -0.01405446 | NA |
| chr1 | 204149712 | 204150067 | 356 | 3 | 9.99E-06 | 7.64E-05 | 6.74E-05 | 1.13E-05 | -0.005588 | ETNK2 |
| chr1 | 69929844 | 69930011 | 168 | 2 | 2.46E-05 | 9.74E-06 | 0.000159 | 1.13E-05 | -0.01857887 | LRRC7 |
| chr14 | 103156866 | 103158196 | 1331 | 3 | 5.19E-05 | 6.40E-06 | 0.000735 | 1.13E-05 | -0.00621774 | NA |
| chr16 | 50121960 | 50122745 | 786 | 5 | 3.16E-06 | 0.0009497 | 0.000206 | 1.13E-05 | -0.00711894 | AC007610.5 |
| chr20 | 8334706 | 8335326 | 621 | 3 | 2.14E-05 | 9.46E-06 | 0.000271 | 1.13E-05 | -0.00039859 | PLCB1 |
| chr12 | 113307719 | 113307970 | 252 | 2 | 1.82E-05 | 1.25E-05 | 9.21E-05 | 1.13E-05 | -0.00104306 | SLC8B1 |
| chr22 | 46043867 | 46044514 | 648 | 3 | 1.00E-06 | 6.11E-06 | 0.001541 | 1.13E-05 | 0.01062707 | LINC00899 |
| chr2 | 39546336 | 39546671 | 336 | 2 | 2.47E-05 | 7.52E-06 | 0.00041 | 1.14E-05 | -0.00318708 | MAP4K3-DT |
| chr12 | 121672561 | 121673274 | 714 | 6 | 3.68E-05 | 1.16E-05 | 0.001458 | 1.14E-05 | -0.00164756 | MORN3 |
| chr19 | 6686015 | 6686566 | 552 | 2 | 2.35E-05 | 7.89E-06 | 0.000327 | 1.14E-05 | 0.010402648 | C3 |

| | | | | | | | | | | |
|---|---|---|---|---|---|---|---|---|---|---|
| chr13 | 98406764 | 98406771 | 8 | 2 | 4.88E-06 | 5.19E-05 | 2.10E-05 | 1.14E-05 | -0.00634439 | FARP1 |
| chr13 | 30963929 | 30964074 | 146 | 2 | 1.14E-05 | 8.95E-05 | 1.54E-05 | 1.14E-05 | -0.01816047 | TEX26 |
| chr11 | 76719789 | 76719843 | 55 | 3 | 0.000129224 | 8.58E-06 | 0.000903 | 1.14E-05 | -0.00141163 | GUCY2EP |
| chr12 | 1811764 | 1811949 | 186 | 3 | 1.45E-05 | 5.78E-06 | 0.001532 | 1.14E-05 | -0.01319657 | CACNA2D4 |
| chr12 | 123900787 | 123901453 | 667 | 2 | 6.49E-05 | 7.03E-06 | 0.000605 | 1.14E-05 | 0.007201134 | DNAH10 |
| chr12 | 25011072 | 25011378 | 307 | 4 | 9.47E-06 | 6.77E-05 | 7.23E-05 | 1.14E-05 | -0.00311023 | IRAG2 |
| chr20 | 45100769 | 45101600 | 832 | 12 | 4.68E-05 | 3.74E-06 | 0.004829 | 1.14E-05 | -0.00284901 | KCNS1 |
| chr15 | 89378441 | 89379371 | 931 | 10 | 1.20E-06 | 5.71E-05 | 0.001404 | 1.14E-05 | 0.003257909 | MIR9-3HG |
| chr20 | 46562636 | 46562845 | 210 | 2 | 6.66E-06 | 5.54E-05 | 2.03E-05 | 1.14E-05 | -0.00569721 | SLC13A3 |
| chr12 | 6637806 | 6638847 | 1042 | 5 | 1.99E-05 | 2.53E-06 | 0.007977 | 1.14E-05 | -0.00597442 | ACRBP |
| chr6 | 73450483 | 73451194 | 712 | 3 | 3.87E-07 | 0.0002736 | 1.02E-05 | 1.14E-05 | -0.01114979 | CGAS |
| chr10 | 116273778 | 116274845 | 1068 | 8 | 1.39E-07 | 0.0007432 | 2.08E-05 | 1.14E-05 | -0.00170371 | AC012470.1 |
| chr11 | 119323449 | 119324027 | 579 | 5 | 4.49E-06 | 2.51E-05 | 5.53E-05 | 1.14E-05 | -0.00347838 | NA |
| chr2 | 98821779 | 98822440 | 662 | 4 | 1.96E-05 | 2.40E-05 | 0.00012 | 1.14E-05 | -0.00554472 | CRACDL |
| chr17 | 7445334 | 7445878 | 545 | 5 | 9.29E-09 | 0.0004021 | 1.18E-05 | 1.14E-05 | 0.001900411 | AC113189.4, CHRNB1 |
| chr18 | 6943342 | 6943623 | 282 | 2 | 7.23E-06 | 7.62E-05 | 1.68E-05 | 1.14E-05 | -0.00843841 | LAMA1 |
| chr13 | 32031117 | 32031965 | 849 | 11 | 3.22E-05 | 5.86E-05 | 0.001066 | 1.14E-05 | 0.001715231 | FRY, FRY-AS1 |
| chr5 | 140111645 | 140112781 | 1137 | 5 | 8.63E-07 | 1.89E-05 | 0.00051 | 1.14E-05 | 0.009244079 | PURA |
| chr3 | 69387640 | 69387721 | 82 | 2 | 1.83E-06 | 5.26E-05 | 2.10E-05 | 1.14E-05 | -0.00588984 | FRMD4B |
| chr1 | 158831083 | 158831481 | 399 | 5 | 1.89E-05 | 1.29E-05 | 0.00075 | 1.14E-05 | 0.010663337 | MNDA |
| chr2 | 205925565 | 205926174 | 610 | 6 | 2.55E-06 | 0.0005266 | 6.81E-06 | 1.14E-05 | 0.001711109 | NA |
| chr2 | 417421 | 417731 | 311 | 2 | 6.49E-06 | 4.50E-05 | 2.35E-05 | 1.14E-05 | 0.003937743 | NA |
| chr14 | 41348229 | 41348563 | 335 | 3 | 7.95E-06 | 0.0001622 | 7.80E-05 | 1.14E-05 | -0.01050979 | NA |
| chr22 | 29813715 | 29814252 | 538 | 2 | 4.87E-05 | 6.95E-06 | 0.000696 | 1.15E-05 | -0.00228462 | ASCC2 |
| chr17 | 58517633 | 58518852 | 1220 | 10 | 7.55E-06 | 0.000277 | 0.006069 | 1.15E-05 | 0.001404102 | MTMR4 |
| chr1 | 107530921 | 107531502 | 582 | 2 | 2.13E-05 | 8.39E-06 | 0.000267 | 1.15E-05 | -0.00725942 | NA |
| chr5 | 128537920 | 128538894 | 975 | 12 | 1.26E-05 | 3.46E-05 | 0.001025 | 1.15E-05 | -0.00016357 | SLC27A6, FBN2 |
| chr17 | 66192030 | 66192899 | 870 | 11 | 3.45E-06 | 2.43E-05 | 8.68E-05 | 1.15E-05 | 0.000687948 | CEP112 |
| chr22 | 42934563 | 42934812 | 250 | 3 | 7.61E-06 | 9.95E-05 | 1.68E-05 | 1.15E-05 | -0.0033619 | PACSIN2 |
| chr11 | 111436706 | 111437041 | 336 | 2 | 2.20E-05 | 8.58E-06 | 0.000248 | 1.15E-05 | -0.02059153 | POU2AF1 |
| chr1 | 10615518 | 10616233 | 716 | 3 | 1.69E-05 | 7.62E-06 | 0.000414 | 1.15E-05 | -0.00892419 | PEX14, AL139423.2 |
| chr18 | 74082112 | 74082547 | 436 | 3 | 8.85E-06 | 3.53E-05 | 5.39E-05 | 1.15E-05 | 0.003636659 | FBXO15 |
| chr16 | 1457264 | 1457651 | 388 | 2 | 1.52E-05 | 1.11E-05 | 0.000122 | 1.15E-05 | -0.01397515 | CLCN7 |
| chr2 | 197293950 | 197294395 | 446 | 2 | 3.58E-05 | 7.09E-06 | 0.000625 | 1.15E-05 | 0.01268681 | ANKRD44, ANKRD44-IT1 |
| chr22 | 50464126 | 50464885 | 760 | 4 | 1.53E-06 | 0.0007892 | 8.10E-06 | 1.15E-05 | -0.00206207 | SBF1 |
| chr7 | 18086836 | 18087880 | 1045 | 8 | 2.34E-07 | 0.0011192 | 0.000535 | 1.15E-05 | -0.00653074 | HDAC9 |
| chr15 | 26546063 | 26546521 | 459 | 3 | 1.18E-05 | 1.85E-05 | 0.000713 | 1.15E-05 | -0.0012175 | GABRB3 |
| chr16 | 12289833 | 12290276 | 444 | 2 | 4.07E-06 | 1.68E-05 | 6.13E-05 | 1.15E-05 | -0.00429352 | SNX29 |
| chr4 | 87006503 | 87007438 | 936 | 14 | 3.06E-06 | 0.0013265 | 0.001688 | 1.15E-05 | 0.001409673 | AFF1 |
| chr6 | 34110745 | 34111045 | 301 | 3 | 3.26E-05 | 6.69E-06 | 0.000578 | 1.15E-05 | -0.00558232 | GRM4 |
| chr2 | 2932651 | 2933038 | 388 | 3 | 3.83E-05 | 7.02E-06 | 0.001366 | 1.16E-05 | -0.00475189 | LINC01250 |
| chr4 | 6986579 | 6987857 | 1279 | 18 | 2.45E-06 | 0.0004315 | 0.000414 | 1.16E-05 | -0.00100206 | TBC1D14, AC097382.3 |
| chr2 | 229920334 | 229920981 | 648 | 4 | 1.02E-06 | 0.0001496 | 2.39E-05 | 1.16E-05 | 0.001636965 | TRIP12 |
| chr6 | 26033166 | 26033917 | 752 | 8 | 1.49E-06 | 0.0002017 | 0.000203 | 1.16E-05 | -0.00044164 | H2AC4 |
| chr11 | 64352505 | 64352841 | 337 | 4 | 3.17E-05 | 2.37E-05 | 0.000684 | 1.16E-05 | -0.00206774 | CCDC88B |
| chr9 | 133092635 | 133093030 | 396 | 3 | 7.04E-06 | 0.0001051 | 2.02E-05 | 1.16E-05 | -0.00308203 | NA |
| chr11 | 2213745 | 2214220 | 476 | 4 | 6.32E-05 | 5.62E-06 | 0.003656 | 1.16E-05 | 0.006243313 | NA |
| chr5 | 119340967 | 119341696 | 730 | 4 | 4.32E-05 | 3.59E-06 | 0.005361 | 1.16E-05 | 0.008363068 | TNFAIP8 |
| chr1 | 15742519 | 15743360 | 842 | 6 | 4.86E-07 | 0.0009434 | 0.000118 | 1.16E-05 | -0.00372473 | TMEM82, SLC25A34-AS1 |
| chr20 | 63123517 | 63123693 | 177 | 3 | 2.60E-05 | 8.60E-06 | 0.001148 | 1.16E-05 | -0.00703149 | NA |
| chr2 | 44941368 | 44942542 | 1175 | 10 | 6.50E-06 | 0.001146 | 0.000408 | 1.16E-05 | -0.00162858 | SIX3, SIX3-AS1 |
| chr9 | 100411356 | 100411861 | 506 | 6 | 1.25E-05 | 3.41E-05 | 5.86E-05 | 1.16E-05 | -0.00466861 | NA |
| chr7 | 128030517 | 128031208 | 692 | 9 | 9.39E-08 | 0.0007593 | 2.28E-05 | 1.16E-05 | -0.00176826 | SND1, LRRC4 |
| chr12 | 12983648 | 12983921 | 274 | 2 | 2.76E-05 | 8.01E-06 | 0.000337 | 1.16E-05 | -0.01534077 | GPRC5D-AS1, HEBP1 |
| chr20 | 58388666 | 58389397 | 732 | 12 | 1.02E-05 | 3.15E-05 | 0.000468 | 1.16E-05 | 0.001354432 | VAPB |
| chr19 | 34327904 | 34328150 | 247 | 2 | 3.14E-05 | 7.04E-06 | 0.000697 | 1.16E-05 | -0.00301224 | GARRE1 |
| chr4 | 112705210 | 112705339 | 130 | 2 | 8.18E-06 | 1.38E-05 | 8.22E-05 | 1.16E-05 | 0.006949674 | AC106864.1 |
| chr9 | 87974615 | 87975512 | 898 | 10 | 1.45E-05 | 0.0001378 | 0.002209 | 1.16E-05 | 0.000572035 | CDK20 |
| chr5 | 111997496 | 111997556 | 61 | 3 | 1.21E-05 | 0.0002885 | 1.78E-05 | 1.16E-05 | 0.01174204 | NREP-AS1 |
| chr5 | 150086503 | 150087152 | 650 | 7 | 4.23E-06 | 0.0013244 | 0.000911 | 1.16E-05 | -0.00549061 | CSF1R |
| chr8 | 37778219 | 37778581 | 363 | 2 | 2.96E-06 | 0.0001394 | 1.31E-05 | 1.16E-05 | 0.003188219 | PLPBP |
| chr2 | 45675236 | 45676229 | 994 | 5 | 1.02E-06 | 0.001242 | 4.93E-06 | 1.17E-05 | 0.005438834 | PRKCE, PRKCE-AS1 |
| chr12 | 12787676 | 12788067 | 392 | 2 | 8.29E-07 | 3.76E-05 | 2.75E-05 | 1.17E-05 | -0.00022678 | APOLD1 |
| chr1 | 211212102 | 211212114 | 13 | 2 | 1.09E-05 | 8.44E-06 | 0.000279 | 1.17E-05 | 0.001608029 | NA |
| chr2 | 8138125 | 8138176 | 52 | 2 | 3.00E-05 | 7.96E-06 | 0.000359 | 1.17E-05 | -0.00413976 | LINC00298, LINC00299 |
| chr2 | 201071251 | 201072303 | 1053 | 18 | 1.37E-05 | 0.0003348 | 0.001672 | 1.17E-05 | 0.000163965 | NDUFB3, FAM126B |
| chr8 | 1829311 | 1829600 | 290 | 3 | 1.77E-06 | 0.0007128 | 8.54E-06 | 1.17E-05 | 0.005593938 | AC019257.8, ARHGEF10 |
| chr1 | 25338366 | 25338983 | 618 | 5 | 4.47E-05 | 6.43E-06 | 0.002707 | 1.17E-05 | -0.00016193 | TMEM50A |
| chr17 | 3732272 | 3732286 | 15 | 2 | 2.06E-05 | 2.91E-05 | 3.43E-05 | 1.17E-05 | 0.000311972 | ITGAE |
| chr12 | 131872217 | 131872443 | 227 | 3 | 1.49E-05 | 4.10E-05 | 6.93E-05 | 1.17E-05 | -0.00813857 | NA |
| chr22 | 38019499 | 38019551 | 53 | 2 | 1.16E-05 | 3.48E-05 | 2.94E-05 | 1.17E-05 | -0.00677734 | POLR2F |
| chr3 | 192566663 | 192566949 | 287 | 2 | 4.86E-06 | 0.0002692 | 1.05E-05 | 1.17E-05 | 0.002218066 | FGF12 |
| chr15 | 90881416 | 90881488 | 73 | 3 | 6.26E-06 | 0.0001321 | 8.61E-05 | 1.17E-05 | 0.00385888 | FURIN |
| chr19 | 8595874 | 8596197 | 324 | 2 | 2.79E-05 | 7.73E-06 | 0.000421 | 1.17E-05 | 0.007298213 | ADAMTS10 |
| chr12 | 68804963 | 68805257 | 295 | 3 | 6.10E-07 | 0.0064781 | 2.90E-06 | 1.17E-05 | -0.00201861 | AC025423.3 |
| chr11 | 15110870 | 15111541 | 672 | 3 | 1.13E-07 | 0.0011069 | 5.52E-06 | 1.17E-05 | -0.00247514 | NA |
| chr19 | 40923818 | 40924284 | 467 | 4 | 8.76E-06 | 6.53E-05 | 6.30E-05 | 1.17E-05 | 0.001822815 | CYP2B7P |
| chr7 | 144403537 | 144403926 | 390 | 5 | 1.10E-05 | 0.0003989 | 0.000694 | 1.17E-05 | -0.00340664 | NOBOX |
| chr8 | 50397884 | 50398346 | 463 | 3 | 2.07E-06 | 0.0006171 | 3.65E-05 | 1.17E-05 | -0.00238163 | SNTG1 |
| chr4 | 14163095 | 14163399 | 305 | 2 | 2.48E-05 | 7.18E-06 | 0.00065 | 1.17E-05 | 0.004317546 | NA |
| chr14 | 96276583 | 96277156 | 574 | 5 | 5.26E-06 | 0.00186 | 0.000135 | 1.17E-05 | 0.003210428 | AL355102.3 |
| chr3 | 197749311 | 197750367 | 1057 | 18 | 5.64E-06 | 1.47E-05 | 0.005469 | 1.17E-05 | 0.000827008 | FYTTD1, RUBCN |
| chr4 | 42704229 | 42704671 | 443 | 2 | 8.19E-06 | 2.10E-05 | 4.80E-05 | 1.17E-05 | 0.005197357 | NA |
| chr19 | 3805862 | 3806100 | 239 | 3 | 2.92E-05 | 6.91E-06 | 0.001463 | 1.17E-05 | -0.00248478 | ZFR2 |
| chr1 | 25233054 | 25233252 | 199 | 3 | 8.59E-07 | 0.0001406 | 1.36E-05 | 1.17E-05 | 0.007115982 | AL031432.4 |
| chr8 | 132747590 | 132748028 | 439 | 3 | 6.76E-06 | 6.19E-05 | 4.73E-05 | 1.17E-05 | 0.002558974 | TMEM71 |
| chr4 | 72567954 | 72568827 | 874 | 9 | 7.75E-06 | 6.29E-05 | 0.000332 | 1.17E-05 | -0.0002562 | ADAMTS3 |
| chr1 | 3414516 | 3414714 | 199 | 3 | 5.40E-06 | 0.0004401 | 0.000169 | 1.18E-05 | -0.00439189 | PRDM16 |
| chr10 | 94622183 | 94622394 | 212 | 2 | 2.49E-05 | 9.90E-06 | 0.000172 | 1.18E-05 | -0.0120592 | NA |
| chr2 | 85328117 | 85328896 | 780 | 13 | 5.73E-06 | 0.0006625 | 0.000406 | 1.18E-05 | 0.000947359 | TGOLN2 |
| chr7 | 859838 | 860400 | 563 | 5 | 8.18E-06 | 0.0014144 | 6.58E-05 | 1.18E-05 | -0.00325181 | SUN1 |
| chr19 | 49890396 | 49891041 | 646 | 3 | 1.01E-05 | 6.00E-06 | 0.001227 | 1.18E-05 | -0.01197778 | IL4I1 |
| chr5 | 32312911 | 32313388 | 478 | 12 | 1.60E-05 | 0.001672 | 0.001333 | 1.18E-05 | 0.000289147 | MTMR12 |
| chr2 | 37318164 | 37318525 | 362 | 5 | 2.54E-05 | 1.02E-05 | 0.001887 | 1.18E-05 | -0.0035782 | PRKD3 |
| chr7 | 1946610 | 1947005 | 396 | 5 | 1.15E-05 | 4.57E-05 | 0.001529 | 1.18E-05 | -0.02002704 | MAD1L1, AC069288.1 |
| chr22 | 49543996 | 49544122 | 127 | 2 | 1.25E-05 | 9.24E-05 | 1.58E-05 | 1.18E-05 | 0.002931794 | C22orf34 |
| chr2 | 242011603 | 242012134 | 532 | 5 | 3.20E-05 | 7.27E-06 | 0.00275 | 1.18E-05 | 0.00566481 | LINC01237 |
| chr19 | 46600461 | 46601226 | 766 | 9 | 8.65E-06 | 1.80E-05 | 0.00147 | 1.18E-05 | -0.00017813 | CALM3, PPP5D1 |
| chr20 | 49982755 | 49983666 | 912 | 9 | 1.82E-05 | 5.51E-05 | 0.005972 | 1.18E-05 | -0.00071447 | SNAI1 |
| chr1 | 100345007 | 100345015 | 9 | 2 | 1.20E-05 | 1.21E-05 | 0.000108 | 1.18E-05 | -0.00845727 | AC104457.2, CDC14A |
| chr16 | 70251505 | 70252527 | 1023 | 11 | 7.26E-06 | 1.14E-05 | 0.000766 | 1.18E-05 | 0.000284145 | EXOSC6, AARS1 |
| chr8 | 43246260 | 43246479 | 220 | 2 | 2.86E-05 | 8.45E-06 | 0.000293 | 1.18E-05 | -0.03262058 | NA |
| chr5 | 179211012 | 179211025 | 14 | 2 | 1.56E-05 | 9.17E-06 | 0.000221 | 1.18E-05 | -0.01169339 | ADAMTS2 |
| chr3 | 56702478 | 56702937 | 460 | 2 | 1.85E-05 | 9.95E-06 | 0.000173 | 1.18E-05 | -0.01532754 | NA |
| chr6 | 40622062 | 40622103 | 42 | 2 | 1.22E-05 | 7.29E-05 | 1.79E-05 | 1.18E-05 | 0.007746137 | NA |
| chr8 | 125576019 | 125576310 | 292 | 2 | 3.47E-06 | 0.0002339 | 1.11E-05 | 1.18E-05 | -0.01029197 | NA |
| chr2 | 5132511 | 5132953 | 443 | 4 | 7.06E-06 | 3.48E-05 | 0.000396 | 1.18E-05 | 0.000351235 | NA |
| chr2 | 113278987 | 113279483 | 497 | 6 | 6.26E-06 | 7.16E-05 | 0.001814 | 1.18E-05 | -0.00509751 | NA |
| chr17 | 31155058 | 31155843 | 786 | 3 | 5.04E-06 | 6.90E-06 | 0.001271 | 1.18E-05 | -0.00254718 | NF1 |
| chr15 | 74889789 | 74890545 | 757 | 11 | 3.91E-05 | 1.78E-05 | 0.022767 | 1.18E-05 | -0.00072804 | MPI |
| chr12 | 12696043 | 12697148 | 1106 | 12 | 1.42E-06 | 0.0002852 | 5.83E-05 | 1.18E-05 | 0.003994635 | GPR19 |
| chr2 | 73381048 | 73381710 | 663 | 3 | 5.34E-05 | 2.36E-05 | 0.000572 | 1.18E-05 | -0.00123387 | AC074008.2 |
| chr6 | 41936606 | 41936745 | 140 | 3 | 2.14E-05 | 2.54E-05 | 0.000285 | 1.18E-05 | -0.01231588 | CCND3 |
| chr13 | 31952746 | 31953034 | 289 | 2 | 7.58E-06 | 7.23E-05 | 1.81E-05 | 1.18E-05 | 0.003325505 | FRY, EEF1DP3 |
| chr11 | 932687 | 932760 | 74 | 2 | 3.87E-06 | 0.0003937 | 9.54E-06 | 1.18E-05 | -0.01438013 | AP2A2 |
| chr20 | 51488168 | 51488407 | 240 | 2 | 1.34E-05 | 3.11E-05 | 3.30E-05 | 1.18E-05 | -0.00121119 | NFATC2 |
| chr16 | 89791768 | 89792219 | 452 | 4 | 1.71E-05 | 2.60E-05 | 0.000433 | 1.18E-05 | 0.001917238 | FANCA |
| chr8 | 60776394 | 60776728 | 335 | 2 | 2.45E-06 | 0.0017555 | 7.23E-06 | 1.18E-05 | -0.00180636 | CHD7 |
| chr21 | 39534992 | 39535842 | 851 | 6 | 4.44E-06 | 0.002004 | 0.0002 | 1.19E-05 | -0.00243747 | NA |
| chr16 | 75069955 | 75070252 | 298 | 2 | 8.00E-06 | 5.86E-05 | 2.06E-05 | 1.19E-05 | 0.003226647 | ZNRF1 |
| chr5 | 41071303 | 41072042 | 740 | 4 | 1.16E-05 | 0.0003387 | 0.001455 | 1.19E-05 | -0.006574 | MROH2B |
| chr13 | 97257662 | 97258104 | 443 | 3 | 2.80E-05 | 1.33E-05 | 0.000578 | 1.19E-05 | 0.002910632 | MBNL2 |
| chr5 | 61769309 | 61769589 | 281 | 2 | 3.31E-06 | 0.0020308 | 7.10E-06 | 1.19E-05 | -0.00615975 | C5orf64 |
| chr17 | 7717311 | 7717800 | 490 | 6 | 3.89E-05 | 2.68E-06 | 0.008987 | 1.19E-05 | 0.002019336 | DNAH2 |
| chr6 | 19714704 | 19714720 | 17 | 2 | 2.64E-05 | 1.54E-05 | 7.32E-05 | 1.19E-05 | -0.02101324 | LNC-LBCS |
| chr15 | 63878862 | 63879296 | 435 | 2 | 1.61E-06 | 0.0001191 | 1.42E-05 | 1.19E-05 | -0.00335915 | NA |
| chr10 | 22563679 | 22564283 | 605 | 4 | 6.57E-05 | 3.00E-06 | 0.007298 | 1.19E-05 | -0.03310095 | PIP4K2A |
| chr2 | 239641524 | 239642044 | 521 | 2 | 2.47E-05 | 8.67E-06 | 0.000276 | 1.19E-05 | 0.003611761 | NA |
| chr19 | 1219836 | 1221227 | 1392 | 9 | 7.34E-06 | 1.98E-05 | 0.00853 | 1.19E-05 | -0.0004342 | STK11 |
| chr22 | 24460909 | 24461224 | 316 | 2 | 1.98E-05 | 9.59E-06 | 0.000197 | 1.19E-05 | -0.00796069 | ADORA2A-AS1 |
| chr8 | 143126895 | 143127268 | 374 | 4 | 1.91E-06 | 3.50E-05 | 0.000196 | 1.19E-05 | -0.00418081 | NA |
| chr3 | 52360655 | 52361182 | 528 | 4 | 2.36E-06 | 0.0014517 | 4.13E-05 | 1.19E-05 | -0.00195527 | DNAH1 |
| chr5 | 179326431 | 179326802 | 372 | 2 | 6.28E-06 | 9.29E-06 | 0.00022 | 1.19E-05 | -0.02483048 | ADAMTS2 |
| chr17 | 81664423 | 81664520 | 98 | 2 | 2.79E-06 | 3.04E-05 | 3.40E-05 | 1.19E-05 | -0.01051104 | NA |
| chr5 | 173163308 | 173163740 | 433 | 2 | 1.44E-05 | 1.26E-05 | 0.000103 | 1.19E-05 | -0.005715 | BNIP1 |
| chr5 | 31981344 | 31982238 | 895 | 4 | 6.10E-06 | 6.77E-05 | 3.87E-05 | 1.19E-05 | -0.00327141 | PDZD2 |
| chr13 | 44815913 | 44816534 | 622 | 4 | 1.36E-05 | 6.01E-05 | 9.53E-05 | 1.20E-05 | 0.002506614 | NA |
| chr16 | 4783705 | 4784031 | 327 | 4 | 1.70E-05 | 1.61E-05 | 0.000813 | 1.20E-05 | -0.00411673 | SEPTIN12, AC020663.4 |
| chr19 | 43728735 | 43728748 | 14 | 2 | 1.18E-05 | 1.38E-05 | 8.81E-05 | 1.20E-05 | -0.00123233 | SMG9 |
| chr12 | 31645931 | 31646184 | 254 | 4 | 2.63E-05 | 4.77E-06 | 0.002683 | 1.20E-05 | -0.01234422 | AC068774.1 |
| chr5 | 37872745 | 37872761 | 17 | 2 | 1.36E-05 | 7.25E-06 | 0.000714 | 1.20E-05 | 0.009406123 | GDNF-AS1 |
| chr12 | 120325684 | 120326093 | 410 | 6 | 1.78E-05 | 9.94E-06 | 0.00056 | 1.20E-05 | -0.00038134 | PLA2G1B |
| chr1 | 67653060 | 67653216 | 157 | 2 | 2.94E-05 | 9.35E-06 | 0.000219 | 1.20E-05 | 0.010547036 | NA |
| chr19 | 45792332 | 45793027 | 696 | 7 | 4.27E-05 | 1.25E-05 | 0.003119 | 1.20E-05 | 0.001129092 | DMWD |
| chr12 | 124847448 | 124847637 | 190 | 3 | 9.50E-06 | 0.0001816 | 0.000253 | 1.20E-05 | -0.00530748 | SCARB1 |
| chr4 | 3689101 | 3689670 | 570 | 4 | 1.11E-05 | 2.35E-05 | 0.001376 | 1.20E-05 | 0.00720507 | NA |
| chr1 | 204226021 | 204226336 | 316 | 3 | 1.64E-05 | 9.92E-06 | 0.000372 | 1.20E-05 | -0.00403712 | PLEKHA6 |
| chr1 | 213996474 | 213996898 | 425 | 8 | 5.98E-06 | 0.0002779 | 0.000438 | 1.20E-05 | 0.00301327 | PROX1 |
| chr19 | 42225376 | 42226141 | 766 | 5 | 1.64E-07 | 0.0004426 | 8.06E-05 | 1.20E-05 | 0.00157841 | ZNF526, AC006486.3 |
| chr10 | 73301969 | 73302184 | 216 | 2 | 2.28E-06 | 0.0012662 | 7.64E-06 | 1.20E-05 | -0.00450815 | CFAP70 |
| chr11 | 88337612 | 88338592 | 981 | 11 | 2.16E-05 | 2.50E-06 | 0.015422 | 1.20E-05 | 0.000318251 | CTSC |
| chr4 | 8124578 | 8124931 | 354 | 3 | 4.63E-06 | 4.80E-05 | 2.98E-05 | 1.20E-05 | -0.00407801 | ABLIM2 |
| chr17 | 34156541 | 34157240 | 700 | 13 | 1.84E-05 | 5.55E-05 | 0.002018 | 1.20E-05 | -0.00014108 | AC004147.2, ASIC2 |
| chr5 | 72269344 | 72269703 | 360 | 2 | 1.90E-05 | 1.11E-05 | 0.000136 | 1.20E-05 | -0.01512823 | MRPS27 |
| chr5 | 68434768 | 68435816 | 1049 | 3 | 6.40E-06 | 0.0001138 | 7.47E-05 | 1.20E-05 | -7.88E-05 | NA |
| chr4 | 182246759 | 182247005 | 247 | 3 | 1.18E-05 | 5.23E-06 | 0.002288 | 1.20E-05 | -0.01223814 | TENM3 |
| chr15 | 74854173 | 74854322 | 150 | 3 | 3.77E-05 | 7.16E-06 | 0.000717 | 1.20E-05 | -0.00216793 | SCAMP2 |
| chr1 | 36087358 | 36088130 | 773 | 5 | 1.01E-05 | 7.79E-05 | 0.000318 | 1.21E-05 | -0.00555126 | TEKT2 |

| | | | | | | | | | | |
|---|---|---|---|---|---|---|---|---|---|---|
| chr17 | 17678365 | 17678693 | 329 | 2 | 1.17E-05 | 2.28E-05 | 4.57E-05 | 1.21E-05 | -0.00563564 | NA |
| chr22 | 20405916 | 20406632 | 717 | 4 | 8.87E-06 | 1.42E-05 | 0.001002 | 1.21E-05 | 0.000714054 | ZNF74 |
| chr12 | 102961159 | 102961548 | 390 | 6 | 2.82E-05 | 4.33E-06 | 0.004277 | 1.21E-05 | -0.0008994 | NA |
| chr16 | 14070163 | 14071003 | 841 | 4 | 8.46E-07 | 8.95E-05 | 0.000178 | 1.21E-05 | -0.00067472 | NA |
| chr3 | 12769395 | 12769855 | 461 | 2 | 1.07E-05 | 1.60E-05 | 7.14E-05 | 1.21E-05 | -0.00989746 | TMEM40 |
| chr1 | 1143521 | 1144499 | 979 | 5 | 1.21E-06 | 6.48E-05 | 0.000179 | 1.21E-05 | 0.00015076 | LINC01342 |
| chr11 | 124761770 | 124763285 | 1516 | 9 | 1.40E-05 | 0.000202 | 0.000533 | 1.21E-05 | 0.001452374 | AP000866.2, ESAM |
| chr6 | 30907809 | 30908115 | 307 | 12 | 4.86E-05 | 3.80E-05 | 0.011642 | 1.21E-05 | 0.001707413 | NA |
| chr3 | 45647268 | 45647618 | 351 | 2 | 5.64E-06 | 7.68E-05 | 1.79E-05 | 1.21E-05 | -0.0099977 | LIMD1 |
| chr4 | 186251799 | 186252297 | 499 | 6 | 3.36E-05 | 7.98E-06 | 0.00889 | 1.21E-05 | -0.00028399 | KLKB1 |
| chr5 | 177950831 | 177951230 | 400 | 3 | 1.68E-05 | 1.98E-05 | 8.42E-05 | 1.21E-05 | -0.00126947 | AC106795.3, AC106795.2 |
| chr16 | 74296298 | 74297181 | 884 | 14 | 6.07E-07 | 0.0003063 | 7.87E-05 | 1.21E-05 | 0.000288762 | PSMD7, AC138627.1 |
| chr12 | 52013450 | 52013512 | 63 | 2 | 9.41E-06 | 3.32E-05 | 3.21E-05 | 1.21E-05 | -0.00323449 | TAMALIN |
| chr2 | 130930455 | 130930933 | 479 | 3 | 9.41E-06 | 2.60E-05 | 0.000651 | 1.21E-05 | -0.01088829 | ARHGEF4 |
| chr8 | 21912184 | 21912722 | 539 | 4 | 6.61E-08 | 0.0093311 | 1.76E-05 | 1.21E-05 | -0.00613951 | DOK2 |
| chr12 | 131084603 | 131084862 | 260 | 3 | 6.45E-06 | 6.54E-06 | 0.001364 | 1.21E-05 | -0.00449615 | ADGRD1 |
| chr14 | 104989463 | 104989686 | 224 | 5 | 1.31E-05 | 0.0003052 | 0.000123 | 1.21E-05 | -0.00515972 | CLBA1 |
| chr11 | 70327250 | 70327406 | 157 | 3 | 9.47E-06 | 7.77E-06 | 0.00045 | 1.21E-05 | -0.00314454 | PPFIA1, AP002336.2 |
| chr10 | 43418026 | 43418174 | 149 | 2 | 1.91E-05 | 2.63E-05 | 3.98E-05 | 1.21E-05 | 0.001092743 | NA |
| chr2 | 3482450 | 3482622 | 173 | 4 | 2.20E-05 | 1.26E-05 | 0.000169 | 1.21E-05 | -0.00182836 | TRAPPC12 |
| chr11 | 45751631 | 45752277 | 647 | 8 | 6.99E-06 | 0.0003325 | 0.004761 | 1.21E-05 | 0.009192699 | LINC02690 |
| chr3 | 50355125 | 50355613 | 489 | 2 | 2.24E-06 | 3.80E-05 | 2.88E-05 | 1.21E-05 | 0.004230012 | CYB561D2, Z84492.2, TMEM115 |
| chr1 | 26179609 | 26179864 | 256 | 3 | 1.42E-05 | 4.59E-05 | 0.000222 | 1.21E-05 | -0.00371481 | CNKSR1 |
| chr12 | 71219182 | 71219744 | 563 | 5 | 4.99E-06 | 6.86E-05 | 8.95E-05 | 1.21E-05 | -0.00897088 | TSPAN8 |
| chr17 | 17382991 | 17383161 | 171 | 2 | 9.57E-06 | 2.75E-05 | 3.81E-05 | 1.21E-05 | -0.0133638 | NA |
| chr11 | 78532378 | 78533156 | 779 | 4 | 2.94E-05 | 8.04E-05 | 0.000707 | 1.21E-05 | 0.002550706 | NARS2 |
| chr8 | 59138797 | 59138940 | 144 | 2 | 1.13E-05 | 1.54E-05 | 7.64E-05 | 1.21E-05 | -0.0079982 | NA |
| chr17 | 522822 | 523296 | 475 | 6 | 6.63E-06 | 0.0001348 | 0.001469 | 1.21E-05 | 0.000990589 | AC015853.3, VPS53 |
| chr18 | 49967204 | 49967423 | 220 | 2 | 6.78E-06 | 4.69E-05 | 2.47E-05 | 1.21E-05 | -0.0142287 | MYO5B |
| chr13 | 110298678 | 110298906 | 229 | 3 | 6.43E-05 | 5.35E-06 | 0.002128 | 1.22E-05 | 0.009583357 | COL4A1 |
| chr10 | 132545855 | 132546536 | 682 | 4 | 3.70E-05 | 1.08E-05 | 0.001072 | 1.22E-05 | -0.00081789 | INPP5A |
| chr2 | 173964414 | 173965322 | 909 | 5 | 1.23E-05 | 0.0009491 | 0.000188 | 1.22E-05 | 0.002806874 | SP3 |
| chr19 | 4952834 | 4953221 | 388 | 2 | 1.26E-05 | 1.66E-05 | 6.91E-05 | 1.22E-05 | -0.00248816 | UHRF1 |
| chr1 | 26233589 | 26234834 | 1246 | 15 | 2.99E-06 | 0.0539878 | 5.90E-06 | 1.22E-05 | 0.000917131 | CEP85 |
| chr11 | 113447884 | 113448379 | 496 | 2 | 1.38E-05 | 9.20E-06 | 0.000249 | 1.22E-05 | -0.00222297 | DRD2 |
| chr1 | 43221488 | 43221739 | 252 | 2 | 3.19E-05 | 7.90E-06 | 0.000475 | 1.22E-05 | -0.00866281 | CFAP57, EBNA1BP2 |
| chr8 | 141643555 | 141644291 | 737 | 6 | 1.77E-05 | 0.0001857 | 0.000561 | 1.22E-05 | -0.00343036 | AC025839.1 |
| chr20 | 21139170 | 21139233 | 64 | 2 | 2.40E-05 | 7.47E-06 | 0.000681 | 1.22E-05 | -0.01591355 | KIZ |
| chr2 | 71746742 | 71747165 | 424 | 3 | 3.89E-05 | 8.23E-06 | 0.000897 | 1.22E-05 | -0.00132724 | NA |
| chr19 | 49950093 | 49950285 | 193 | 2 | 3.13E-06 | 1.72E-05 | 6.64E-05 | 1.22E-05 | 0.000440173 | AC011452.1, SIGLEC11 |
| chr19 | 44776508 | 44776927 | 420 | 5 | 1.92E-05 | 3.85E-05 | 0.000496 | 1.22E-05 | -0.00167354 | NA |
| chr2 | 151504256 | 151504906 | 651 | 3 | 6.92E-06 | 8.34E-05 | 3.02E-05 | 1.22E-05 | 0.002559094 | RIF1, NEB |
| chr20 | 62707925 | 62708187 | 263 | 2 | 2.26E-06 | 8.00E-06 | 0.000448 | 1.22E-05 | -0.01086531 | NA |
| chr1 | 179744214 | 179744815 | 602 | 3 | 3.52E-05 | 4.52E-06 | 0.00345 | 1.22E-05 | -0.02360131 | FAM163A |
| chr9 | 129485440 | 129485710 | 271 | 2 | 1.37E-05 | 1.69E-05 | 6.81E-05 | 1.22E-05 | 0.012687285 | LINC00963 |
| chr19 | 56508462 | 56508989 | 528 | 3 | 7.41E-07 | 0.0003693 | 0.00012 | 1.22E-05 | -0.00596527 | ZNF471 |
| chr2 | 60391712 | 60392085 | 374 | 3 | 6.37E-05 | 1.54E-05 | 0.000738 | 1.22E-05 | -0.00647969 | MIR4432HG |
| chr19 | 57279773 | 57280442 | 670 | 12 | 1.59E-06 | 0.009764 | 0.000167 | 1.22E-05 | 0.000739882 | ZNF460, ZNF460-AS1 |
| chr19 | 45994791 | 45995131 | 341 | 7 | 3.64E-06 | 0.0010147 | 0.001131 | 1.22E-05 | 0.002994532 | NA |
| chr17 | 77886206 | 77886527 | 322 | 3 | 1.22E-05 | 1.42E-05 | 0.000168 | 1.23E-05 | -0.00010876 | NA |
| chr16 | 58487819 | 58488033 | 215 | 3 | 1.18E-05 | 0.0001501 | 1.48E-05 | 1.23E-05 | -0.0016652 | NDRG4 |
| chr5 | 35090664 | 35090724 | 61 | 2 | 7.82E-06 | 8.09E-05 | 1.77E-05 | 1.23E-05 | -0.00311201 | PRLR |
| chr16 | 56332097 | 56332993 | 897 | 4 | 3.92E-07 | 0.0039449 | 4.18E-06 | 1.23E-05 | -0.00504622 | GNAO1 |
| chr11 | 36512173 | 36512375 | 203 | 2 | 2.59E-06 | 0.0004766 | 9.45E-06 | 1.23E-05 | -0.00886942 | RAG1 |
| chr13 | 50000655 | 50000816 | 162 | 2 | 0.000106904 | 7.39E-06 | 0.000772 | 1.23E-05 | -0.01063721 | TRIM13, DLEU2 |
| chr16 | 49483848 | 49484503 | 656 | 5 | 1.91E-05 | 4.19E-06 | 0.002246 | 1.23E-05 | 0.015331426 | NA |
| chr17 | 35106488 | 35107074 | 587 | 4 | 6.13E-05 | 5.50E-06 | 0.001747 | 1.23E-05 | 0.000821295 | AC004223.3, RAD51D |
| chr16 | 86542735 | 86543224 | 490 | 2 | 2.30E-06 | 7.39E-06 | 0.000776 | 1.23E-05 | -0.00609881 | MTHFSD |
| chr12 | 57193931 | 57194567 | 637 | 7 | 5.61E-06 | 1.25E-05 | 0.000502 | 1.23E-05 | -0.00398295 | LRP1, MIR1228 |
| chr4 | 1763670 | 1763823 | 154 | 2 | 1.70E-05 | 3.25E-05 | 3.36E-05 | 1.23E-05 | 0.010926975 | NA |
| chr6 | 30665549 | 30665681 | 133 | 6 | 7.45E-06 | 0.0001888 | 3.44E-05 | 1.23E-05 | -0.0010859 | DHX16 |
| chr20 | 63700781 | 63700923 | 143 | 3 | 0.000119407 | 2.27E-05 | 8.04E-05 | 1.23E-05 | 0.002640839 | ARFRP1 |
| chr19 | 12035132 | 12036199 | 1068 | 14 | 5.46E-06 | 0.0002577 | 0.001776 | 1.23E-05 | 0.000626934 | ZNF433-AS1, AC008770.4, ZNF433, AC008770.1 |
| chr10 | 3105189 | 3106134 | 946 | 5 | 4.91E-06 | 0.0021817 | 0.000179 | 1.23E-05 | -0.00206307 | PFKP |
| chr12 | 106086768 | 106087046 | 279 | 5 | 3.67E-05 | 6.76E-06 | 0.00095 | 1.23E-05 | -0.00084588 | NUAK1 |
| chr8 | 141241192 | 141241276 | 85 | 2 | 4.31E-06 | 1.20E-05 | 0.000124 | 1.24E-05 | -0.01074611 | SLC45A4 |
| chr12 | 10636363 | 10636615 | 253 | 2 | 5.50E-06 | 0.0004581 | 9.60E-06 | 1.24E-05 | 0.009574039 | STYK1 |
| chr15 | 42725288 | 42725639 | 352 | 3 | 1.97E-05 | 6.93E-06 | 0.000707 | 1.24E-05 | 0.00185498 | CDAN1 |
| chr12 | 113899871 | 113900341 | 471 | 4 | 4.53E-05 | 3.79E-06 | 0.004294 | 1.24E-05 | -0.00152204 | RBM19 |
| chr11 | 132114090 | 132114291 | 202 | 2 | 1.21E-05 | 6.28E-05 | 2.08E-05 | 1.24E-05 | -0.01673978 | NTM |
| chr13 | 91350697 | 91351164 | 468 | 4 | 3.85E-06 | 4.26E-05 | 7.32E-05 | 1.24E-05 | -0.00850363 | MIR17HG, MIR18A, MIR19A, MIR20A |
| chr8 | 13515581 | 13516138 | 558 | 3 | 2.89E-05 | 6.14E-06 | 0.001767 | 1.24E-05 | -0.02414064 | DLC1 |
| chr22 | 29899864 | 29899969 | 106 | 3 | 1.35E-05 | 0.0001359 | 1.71E-05 | 1.24E-05 | -0.01515109 | MTMR3 |
| chr1 | 223393105 | 223393831 | 727 | 9 | 4.56E-05 | 8.10E-07 | 0.035404 | 1.24E-05 | 0.022823752 | CCDC185 |
| chr6 | 79671699 | 79671920 | 222 | 4 | 2.38E-05 | 3.48E-05 | 0.00105 | 1.24E-05 | -0.01070391 | SH3BGRL2 |
| chr10 | 120502231 | 120502664 | 434 | 2 | 1.46E-06 | 0.0001577 | 1.34E-05 | 1.24E-05 | -0.00421492 | PLPP4 |
| chr14 | 102900650 | 102901521 | 872 | 5 | 3.15E-06 | 0.004294 | 0.000587 | 1.24E-05 | 0.01882632 | TRAF3 |
| chr8 | 144831261 | 144831459 | 199 | 2 | 1.84E-05 | 1.42E-05 | 9.09E-05 | 1.24E-05 | -0.00399906 | ZNF7 |
| chr3 | 187138652 | 187139530 | 879 | 4 | 6.09E-07 | 0.0002348 | 7.83E-05 | 1.24E-05 | -0.00764207 | RPL39L |
| chr6 | 168242760 | 168242983 | 224 | 2 | 2.58E-06 | 1.02E-05 | 0.000191 | 1.24E-05 | -0.01101095 | AL606970.3 |
| chr4 | 88378617 | 88379004 | 388 | 8 | 3.09E-05 | 1.31E-05 | 0.00037 | 1.24E-05 | -0.00072566 | HERC6 |
| chr14 | 77549161 | 77549652 | 492 | 3 | 2.47E-05 | 1.04E-05 | 0.00027 | 1.24E-05 | 0.000403973 | SPTLC2 |
| chr6 | 159726194 | 159727365 | 1172 | 16 | 8.23E-06 | 2.86E-05 | 0.001066 | 1.24E-05 | -0.00129609 | WTAP, SOD2 |
| chr11 | 112073696 | 112074276 | 581 | 11 | 1.60E-05 | 2.34E-05 | 0.00582 | 1.24E-05 | 0.00075033 | NKAPD1, PIH1D2 |
| chr16 | 70294370 | 70294733 | 364 | 6 | 0.000124861 | 7.86E-05 | 0.000178 | 1.24E-05 | -0.00173325 | DDX19B |
| chr19 | 50896941 | 50897096 | 156 | 3 | 1.12E-05 | 6.04E-05 | 3.19E-05 | 1.24E-05 | -0.01213297 | NA |
| chr8 | 66178823 | 66180327 | 1505 | 7 | 2.15E-06 | 4.74E-05 | 0.000228 | 1.25E-05 | -0.00489112 | NA |
| chr7 | 2253642 | 2254133 | 492 | 4 | 2.86E-06 | 0.0005295 | 1.87E-05 | 1.25E-05 | 9.31E-05 | SNX8 |
| chr16 | 50712033 | 50712152 | 120 | 3 | 3.70E-06 | 0.0001152 | 6.91E-05 | 1.25E-05 | -0.00522655 | NOD2 |
| chr18 | 63366385 | 63367091 | 707 | 3 | 4.20E-06 | 1.56E-05 | 0.000245 | 1.25E-05 | 0.002305963 | KDSR |
| chr7 | 73597876 | 73598749 | 874 | 2 | 2.11E-05 | 7.70E-06 | 0.000645 | 1.25E-05 | -0.01090398 | MLXIPL |
| chr12 | 6331234 | 6331445 | 212 | 2 | 7.50E-06 | 7.52E-06 | 0.000766 | 1.25E-05 | -0.006411 | TNFRSF1A |
| chr6 | 26284638 | 26285130 | 493 | 5 | 9.78E-06 | 3.45E-05 | 8.75E-05 | 1.25E-05 | -0.00295696 | H4C8 |
| chr10 | 100806486 | 100806719 | 234 | 2 | 2.79E-05 | 9.96E-06 | 0.000212 | 1.25E-05 | -0.00947756 | PAX2 |
| chr22 | 38872746 | 38873211 | 466 | 5 | 8.67E-07 | 0.0002687 | 0.000193 | 1.25E-05 | -0.00256784 | NA |
| chr14 | 22417085 | 22417806 | 722 | 3 | 1.70E-05 | 7.54E-06 | 0.000713 | 1.25E-05 | -0.01644027 | AC244502.3, AC244502.1 |
| chr2 | 47904656 | 47905272 | 617 | 4 | 3.37E-06 | 8.23E-05 | 0.000145 | 1.25E-05 | 0.008024304 | FBXO11 |
| chr2 | 181620373 | 181620385 | 13 | 2 | 5.31E-06 | 0.0038213 | 7.01E-06 | 1.25E-05 | -0.0027051 | CERKL |
| chr12 | 18261081 | 18261517 | 437 | 7 | 1.45E-05 | 0.000297 | 0.00087 | 1.25E-05 | -0.00471095 | PIK3C2G, RERGL |
| chr2 | 128359222 | 128359231 | 10 | 2 | 2.98E-05 | 1.38E-05 | 9.80E-05 | 1.25E-05 | -0.00868016 | NA |
| chr13 | 28529945 | 28530292 | 348 | 2 | 6.06E-06 | 9.86E-06 | 0.00022 | 1.25E-05 | 0.011793947 | NA |
| chr14 | 87805306 | 87805832 | 527 | 2 | 5.16E-05 | 7.61E-06 | 0.000729 | 1.25E-05 | -0.01496093 | AL359237.1 |
| chr8 | 43377569 | 43378084 | 516 | 2 | 3.29E-06 | 1.08E-05 | 0.000171 | 1.25E-05 | 0.035921636 | NA |
| chr7 | 151226629 | 151227981 | 1353 | 14 | 4.11E-06 | 0.0002705 | 0.001409 | 1.25E-05 | 0.00042721 | ABCF2-H2BE1, ABCF2 |
| chr16 | 55956021 | 55956417 | 397 | 4 | 8.43E-07 | 0.0010164 | 7.53E-06 | 1.26E-05 | -0.00810007 | CES5A |
| chr16 | 76263505 | 76263551 | 47 | 2 | 2.68E-05 | 7.62E-06 | 0.000735 | 1.26E-05 | -0.01767074 | NA |
| chr22 | 37751703 | 37751774 | 72 | 2 | 2.14E-05 | 1.52E-05 | 8.42E-05 | 1.26E-05 | -0.00548262 | TRIOBP |
| chr17 | 19336851 | 19337619 | 769 | 3 | 1.53E-07 | 0.000939 | 5.67E-06 | 1.26E-05 | -0.02356968 | B9D1 |
| chr14 | 23551537 | 23552268 | 732 | 12 | 7.21E-06 | 0.000119 | 0.000918 | 1.26E-05 | 0.000745351 | ZFHX2-AS1, ZFHX2 |
| chr19 | 33535464 | 33535627 | 164 | 2 | 1.58E-05 | 2.43E-05 | 4.61E-05 | 1.26E-05 | -0.02178288 | NA |
| chr20 | 62402489 | 62402966 | 478 | 3 | 1.22E-05 | 0.000297 | 0.000308 | 1.26E-05 | -0.00264597 | CABLES2, AL121832.3 |
| chr7 | 157713297 | 157713774 | 478 | 3 | 6.52E-07 | 0.0065235 | 7.84E-06 | 1.26E-05 | 0.006128941 | PTPRN2 |
| chr2 | 148999436 | 148999954 | 519 | 3 | 5.73E-06 | 0.0002664 | 2.38E-05 | 1.26E-05 | -0.00605861 | KIF5C |
| chr9 | 104927366 | 104927794 | 429 | 6 | 1.10E-05 | 6.05E-05 | 0.001251 | 1.26E-05 | 0.001072007 | AL359182.1, ABCA1 |
| chr1 | 242604425 | 242604640 | 216 | 2 | 2.44E-05 | 9.03E-06 | 0.000307 | 1.26E-05 | -0.00125763 | NA |
| chr3 | 11898366 | 11898547 | 182 | 3 | 1.57E-05 | 0.000105 | 2.98E-05 | 1.26E-05 | -0.00563641 | NA |
| chr19 | 1299756 | 1300086 | 331 | 2 | 4.17E-06 | 2.45E-05 | 4.59E-05 | 1.26E-05 | 0.017392068 | EFNA2 |
| chr19 | 17719836 | 17720279 | 444 | 4 | 1.01E-06 | 0.0006882 | 4.85E-06 | 1.26E-05 | 0.002528854 | MAP1S |
| chr5 | 404563 | 405373 | 811 | 5 | 6.99E-06 | 0.001312 | 9.92E-05 | 1.26E-05 | -0.00296034 | PDCD6-AHRR, AHRR |
| chr12 | 120359044 | 120359514 | 471 | 2 | 1.65E-06 | 0.000426 | 1.00E-05 | 1.26E-05 | -0.00551718 | MSI1 |
| chr19 | 4586766 | 4586789 | 24 | 2 | 1.91E-05 | 3.62E-05 | 3.17E-05 | 1.26E-05 | -0.01069935 | NA |
| chr17 | 63484109 | 63484413 | 305 | 5 | 2.16E-05 | 0.0001389 | 0.000204 | 1.26E-05 | -0.00409323 | ACE |
| chr11 | 45810987 | 45811578 | 592 | 6 | 2.65E-05 | 3.88E-05 | 0.003918 | 1.26E-05 | 0.000277828 | SLC35C1 |
| chr13 | 99657908 | 99658587 | 680 | 3 | 9.45E-06 | 1.05E-05 | 0.000283 | 1.26E-05 | 0.004593656 | CLYBL |
| chr8 | 40901428 | 40901942 | 515 | 3 | 6.72E-06 | 3.69E-05 | 4.56E-05 | 1.26E-05 | 0.000698142 | AC048387.1 |
| chr1 | 19081376 | 19081877 | 502 | 4 | 2.43E-05 | 5.53E-05 | 0.000308 | 1.26E-05 | -0.00353582 | UBR4 |
| chr4 | 931864 | 932742 | 879 | 14 | 5.87E-05 | 5.11E-05 | 0.004339 | 1.26E-05 | 0.000276079 | TMEM175, GAK |
| chr6 | 138238693 | 138238844 | 152 | 2 | 9.90E-05 | 1.25E-05 | 0.000122 | 1.27E-05 | -0.00779278 | ARFGEF3 |
| chr6 | 52422925 | 52423279 | 355 | 3 | 1.59E-05 | 3.10E-05 | 0.000629 | 1.27E-05 | -0.00627723 | EFHC1 |
| chr16 | 86761792 | 86762247 | 456 | 4 | 3.10E-06 | 0.0001621 | 0.000189 | 1.27E-05 | 0.022662257 | NA |
| chr9 | 134437391 | 134437495 | 105 | 2 | 4.88E-05 | 8.00E-06 | 0.000559 | 1.27E-05 | -0.00210535 | RXRA |
| chr2 | 109335809 | 109336518 | 710 | 3 | 2.85E-05 | 5.84E-06 | 0.001912 | 1.27E-05 | 0.006804744 | SH3RF3 |
| chr14 | 99205511 | 99205811 | 301 | 2 | 1.56E-05 | 1.99E-05 | 5.91E-05 | 1.27E-05 | -0.00876793 | BCL11B |
| chr14 | 104887875 | 104887930 | 56 | 3 | 1.43E-05 | 5.92E-05 | 5.50E-05 | 1.27E-05 | -0.00106847 | CEP170B |
| chr19 | 40222348 | 40222710 | 363 | 2 | 1.50E-06 | 3.58E-05 | 3.23E-05 | 1.27E-05 | -0.00193452 | CCNP |
| chr2 | 153479079 | 153479888 | 810 | 8 | 2.55E-06 | 7.49E-05 | 0.001433 | 1.27E-05 | -0.00853816 | AC012501.2 |
| chr6 | 148869308 | 148869747 | 440 | 2 | 2.40E-05 | 8.01E-06 | 0.00056 | 1.27E-05 | -0.00995211 | UST |
| chr11 | 113706664 | 113706938 | 275 | 2 | 2.92E-06 | 2.81E-05 | 4.03E-05 | 1.27E-05 | -0.00239369 | NA |
| chr10 | 68471640 | 68472315 | 676 | 12 | 1.11E-05 | 0.0001268 | 0.000228 | 1.27E-05 | -0.00042317 | DNA2 |
| chr14 | 34761936 | 34761999 | 64 | 2 | 2.42E-05 | 2.45E-05 | 4.63E-05 | 1.27E-05 | 0.005660955 | BAZ1A |
| chr8 | 36764082 | 36764160 | 79 | 2 | 5.05E-05 | 1.04E-05 | 0.000194 | 1.27E-05 | -0.01545552 | AC090809.1 |
| chr2 | 19474309 | 19474344 | 36 | 2 | 5.41E-05 | 1.35E-05 | 0.000105 | 1.27E-05 | 0.001299158 | LINC01808 |
| chr9 | 135817898 | 135818481 | 584 | 4 | 3.35E-07 | 8.94E-05 | 0.000392 | 1.27E-05 | -0.00241527 | CAMSAP1 |
| chr19 | 50728300 | 50728750 | 451 | 5 | 4.50E-05 | 5.92E-06 | 0.00255 | 1.27E-05 | -0.01244258 | NA |
| chr8 | 133150430 | 133150812 | 383 | 3 | 1.69E-05 | 1.47E-05 | 0.000254 | 1.27E-05 | -0.0037717 | NA |
| chr20 | 5065911 | 5066132 | 222 | 2 | 1.12E-05 | 7.55E-05 | 1.93E-05 | 1.27E-05 | -0.02232711 | AL121890.4 |
| chr11 | 63888870 | 63889393 | 524 | 2 | 1.38E-08 | 0.000621 | 9.25E-06 | 1.27E-05 | -0.00421634 | MARK2 |
| chr10 | 13161349 | 13161598 | 250 | 7 | 2.87E-05 | 2.37E-05 | 0.002809 | 1.28E-05 | 0.00048415 | MCM10 |
| chr9 | 128339743 | 128340360 | 618 | 5 | 3.10E-06 | 0.0001007 | 5.83E-05 | 1.28E-05 | -0.00088064 | NA |
| chr5 | 178112360 | 178113242 | 883 | 7 | 1.42E-05 | 4.48E-05 | 0.000299 | 1.28E-05 | -0.00239055 | NA |
| chr17 | 7046080 | 7046275 | 196 | 2 | 4.42E-06 | 1.97E-05 | 6.03E-05 | 1.28E-05 | 0.000866551 | NA |
| chr18 | 70288301 | 70288732 | 432 | 7 | 2.19E-05 | 0.0001381 | 0.000421 | 1.28E-05 | -0.00185432 | NA |
| chr19 | 32607350 | 32607562 | 213 | 2 | 2.39E-05 | 1.06E-05 | 0.000192 | 1.28E-05 | -0.00373459 | ANKRD27 |
| chr2 | 109475700 | 109476062 | 363 | 2 | 1.99E-05 | 8.99E-06 | 0.000332 | 1.28E-05 | 0.010835064 | SH3RF3 |
| chr6 | 36692073 | 36692372 | 300 | 2 | 2.73E-06 | 0.0135496 | 6.67E-06 | 1.28E-05 | 0.00514514 | NA |
| chr10 | 100724086 | 100724742 | 657 | 5 | 1.54E-05 | 0.0006179 | 0.000352 | 1.28E-05 | -0.0005271 | NA |
| chr15 | 96325706 | 96326801 | 1096 | 8 | 5.15E-05 | 4.60E-06 | 0.013338 | 1.28E-05 | 0.00135059 | NR2F2, NR2F2-AS1 |
| chr20 | 64055718 | 64056201 | 484 | 3 | 3.52E-07 | 7.06E-05 | 0.000271 | 1.28E-05 | -0.00645753 | TCEA2 |

| | | | | | | | | | | |
|---|---|---|---|---|---|---|---|---|---|---|
| chr15 | 99361399 | 99361797 | 399 | 2 | 7.26E-06 | 3.03E-05 | 3.78E-05 | 1.28E-05 | -0.01248899 | LRRC28 |
| chr3 | 112653697 | 112654152 | 456 | 4 | 4.67E-06 | 5.77E-05 | 0.000159 | 1.28E-05 | -0.00465265 | NA |
| chr5 | 172878239 | 172879133 | 895 | 7 | 7.73E-06 | 3.10E-06 | 0.008819 | 1.28E-05 | -0.00053533 | ERGIC1 |
| chr1 | 245609492 | 245609882 | 391 | 2 | 6.80E-06 | 4.59E-05 | 2.70E-05 | 1.28E-05 | -0.00201185 | KIF26B |
| chr5 | 160438857 | 160439465 | 609 | 5 | 2.97E-05 | 3.21E-06 | 0.007584 | 1.28E-05 | -0.00086316 | MIR3142HG |
| chr15 | 92809829 | 92810192 | 364 | 5 | 3.45E-05 | 6.13E-06 | 0.005592 | 1.28E-05 | 0.002176505 | FAM174B |
| chr18 | 22559020 | 22559479 | 460 | 2 | 7.74E-08 | 0.0003657 | 1.06E-05 | 1.28E-05 | -0.00523136 | NA |
| chr20 | 22579164 | 22579843 | 680 | 3 | 9.87E-06 | 3.83E-05 | 0.000442 | 1.28E-05 | -0.00832645 | NA |
| chr9 | 137555008 | 137555010 | 3 | 2 | 2.56E-05 | 1.67E-05 | 7.61E-05 | 1.28E-05 | -0.00734148 | DPH7 |
| chr18 | 48073377 | 48073723 | 347 | 2 | 3.53E-06 | 0.0002214 | 1.23E-05 | 1.28E-05 | -0.00451144 | ZBTB7C |
| chr12 | 124512843 | 124513391 | 549 | 4 | 1.30E-05 | 2.13E-05 | 0.002677 | 1.28E-05 | 0.00359035 | AC073592.1, NCOR2 |
| chr14 | 92576915 | 92577192 | 278 | 2 | 4.39E-05 | 8.03E-06 | 0.000597 | 1.28E-05 | -0.01124384 | RIN3 |
| chr1 | 245134183 | 245134381 | 199 | 3 | 3.42E-05 | 1.39E-05 | 0.00072 | 1.29E-05 | 0.001341589 | NA |
| chr1 | 63326251 | 63326568 | 318 | 6 | 1.54E-05 | 0.0003853 | 9.14E-05 | 1.29E-05 | -0.00592128 | NA |
| chr13 | 49447199 | 49447917 | 719 | 3 | 5.70E-06 | 5.29E-05 | 4.80E-05 | 1.29E-05 | 0.012959663 | SETDB2 |
| chr13 | 113098646 | 113098930 | 285 | 3 | 1.43E-05 | 4.48E-05 | 0.000213 | 1.29E-05 | -1.30E-06 | MCF2L |
| chr8 | 22683963 | 22684207 | 245 | 4 | 1.97E-05 | 9.95E-06 | 0.001401 | 1.29E-05 | -0.00396431 | AC105046.1 |
| chr4 | 2261334 | 2262470 | 1137 | 11 | 4.50E-06 | 7.94E-05 | 0.000122 | 1.29E-05 | 0.000524091 | MXD4 |
| chr12 | 117874753 | 117875088 | 336 | 2 | 7.19E-06 | 7.75E-06 | 0.000795 | 1.29E-05 | -0.00248876 | KSR2 |
| chr2 | 232669336 | 232669388 | 53 | 2 | 2.67E-05 | 2.02E-05 | 5.95E-05 | 1.29E-05 | -0.01880921 | EFHD1 |
| chr3 | 139709897 | 139710168 | 272 | 2 | 1.42E-05 | 9.98E-06 | 0.000236 | 1.29E-05 | -0.01105422 | AC046134.2 |
| chr10 | 16436835 | 16437193 | 359 | 7 | 4.98E-05 | 3.47E-05 | 0.000226 | 1.29E-05 | -0.00142661 | PTER |
| chr7 | 131013443 | 131013911 | 469 | 3 | 1.04E-05 | 1.96E-05 | 0.000494 | 1.29E-05 | -0.01404933 | AC016831.6, LINC-PINT |
| chr2 | 39504381 | 39504532 | 152 | 2 | 4.56E-06 | 0.0001342 | 1.49E-05 | 1.29E-05 | -0.01554615 | MAP4K3-DT |
| chr4 | 6573802 | 6573972 | 171 | 2 | 2.32E-06 | 7.94E-06 | 0.00067 | 1.29E-05 | 0.012345053 | NA |
| chr2 | 197452844 | 197453622 | 779 | 18 | 3.91E-06 | 0.0005637 | 0.013122 | 1.29E-05 | -0.00053492 | COQ10B |
| chr7 | 72337589 | 72338448 | 860 | 5 | 8.33E-07 | 0.0041212 | 0.00172 | 1.29E-05 | -0.01127042 | CALN1 |
| chr11 | 130426956 | 130427633 | 678 | 4 | 1.98E-05 | 5.01E-06 | 0.005031 | 1.29E-05 | 0.003186556 | ADAMTS8 |
| chr10 | 112823436 | 112823836 | 401 | 4 | 6.43E-06 | 0.0005835 | 0.000331 | 1.29E-05 | -0.0021085 | AL158212.3 |
| chr7 | 1987330 | 1987896 | 567 | 4 | 3.12E-05 | 1.21E-05 | 0.001076 | 1.30E-05 | -0.00250302 | MAD1L1, AC069288.1 |
| chr15 | 78064657 | 78064919 | 263 | 2 | 1.84E-06 | 8.83E-05 | 1.81E-05 | 1.30E-05 | 0.006392761 | TBC1D2B |
| chr6 | 90677302 | 90677573 | 272 | 2 | 1.71E-05 | 1.79E-05 | 7.07E-05 | 1.30E-05 | 0.003661437 | NA |
| chr4 | 152975829 | 152976553 | 725 | 7 | 3.41E-05 | 7.51E-06 | 0.002437 | 1.30E-05 | -0.00291321 | FHDC1 |
| chr7 | 141076702 | 141076853 | 152 | 2 | 3.26E-05 | 9.96E-06 | 0.000243 | 1.30E-05 | -0.00680232 | TMEM178B |
| chr10 | 7964683 | 7964796 | 114 | 3 | 9.61E-06 | 0.0004945 | 9.87E-06 | 1.30E-05 | -0.00249831 | TAF3 |
| chr1 | 115338418 | 115338923 | 506 | 7 | 8.94E-07 | 2.46E-05 | 0.000117 | 1.30E-05 | -0.00121639 | NGF-AS1 |
| chr3 | 170062343 | 170062950 | 608 | 4 | 8.27E-06 | 4.22E-05 | 0.000111 | 1.30E-05 | 0.000827906 | GPR160, KMT5AP3 |
| chr1 | 217275472 | 217276046 | 575 | 3 | 8.74E-06 | 0.0009161 | 2.66E-05 | 1.30E-05 | 0.001514965 | NA |
| chr11 | 46593783 | 46594489 | 707 | 15 | 9.32E-06 | 0.0003671 | 0.000649 | 1.30E-05 | -0.0003659 | AMBRA1 |
| chr4 | 94344114 | 94344499 | 386 | 3 | 1.17E-05 | 1.92E-05 | 0.00059 | 1.30E-05 | -8.05E-05 | NA |
| chr1 | 54781194 | 54781735 | 542 | 5 | 3.50E-05 | 2.82E-06 | 0.010019 | 1.30E-05 | -0.01454132 | TTC22 |
| chr1 | 27383085 | 27383780 | 696 | 15 | 5.25E-06 | 0.0004201 | 0.00096 | 1.30E-05 | -0.00374363 | CD164L2 |
| chr22 | 39434527 | 39434587 | 61 | 2 | 3.88E-05 | 7.80E-06 | 0.000804 | 1.30E-05 | -0.00664987 | TAB1 |
| chr5 | 119358030 | 119358069 | 40 | 2 | 2.19E-05 | 3.71E-05 | 3.25E-05 | 1.30E-05 | -0.01394814 | TNFAIP8 |
| chr5 | 137753216 | 137753673 | 458 | 2 | 1.10E-05 | 7.81E-06 | 0.000797 | 1.30E-05 | 0.004864005 | HNRNPA0 |
| chr12 | 122995486 | 122995761 | 276 | 2 | 2.21E-05 | 7.84E-06 | 0.000776 | 1.30E-05 | 0.013197424 | PITPNM2 |
| chr4 | 110475720 | 110476425 | 706 | 4 | 4.72E-06 | 9.27E-05 | 0.000568 | 1.30E-05 | -0.0108641 | ENPEP |
| chr15 | 64844741 | 64845384 | 644 | 4 | 6.89E-05 | 5.75E-06 | 0.002242 | 1.30E-05 | -0.00825328 | PLEKHO2, AC069368.1 |
| chr12 | 54393743 | 54394507 | 765 | 3 | 7.79E-05 | 4.80E-05 | 0.000298 | 1.30E-05 | -0.00873212 | AC079313.2, AC079313.1 |
| chr10 | 73495742 | 73496178 | 437 | 9 | 5.78E-05 | 2.94E-06 | 0.017502 | 1.30E-05 | 0.000821298 | PPP3CB-AS1, PPP3CB |
| chr2 | 219073582 | 219073784 | 203 | 2 | 1.47E-05 | 4.84E-05 | 2.66E-05 | 1.30E-05 | -0.00646146 | NHEJ1 |
| chr3 | 138594169 | 138594859 | 691 | 13 | 2.98E-05 | 0.0001431 | 0.007037 | 1.31E-05 | -0.00023506 | CEP70 |
| chr18 | 6850001 | 6850603 | 603 | 3 | 4.20E-06 | 9.30E-05 | 0.000336 | 1.31E-05 | -0.00806279 | ARHGAP28 |
| chr12 | 25385010 | 25385446 | 437 | 5 | 1.65E-05 | 2.00E-05 | 0.000512 | 1.31E-05 | -0.01178641 | NA |
| chr10 | 92687759 | 92688097 | 339 | 4 | 7.85E-06 | 4.41E-05 | 0.000867 | 1.31E-05 | 0.000864524 | NA |
| chr20 | 60116321 | 60116732 | 412 | 2 | 9.86E-06 | 9.56E-06 | 0.000288 | 1.31E-05 | -0.01219986 | MIR646HG |
| chr17 | 49588879 | 49588996 | 118 | 2 | 2.17E-05 | 3.04E-05 | 3.89E-05 | 1.31E-05 | 0.006400997 | NA |
| chr4 | 4758766 | 4759153 | 388 | 2 | 3.29E-06 | 0.0002177 | 1.27E-05 | 1.31E-05 | -0.00948579 | STX18-AS1 |
| chr17 | 46875331 | 46875339 | 9 | 2 | 2.22E-05 | 7.89E-06 | 0.000767 | 1.31E-05 | -0.00951565 | WNT9B |
| chr1 | 10686424 | 10687050 | 627 | 2 | 2.83E-05 | 8.52E-06 | 0.000474 | 1.31E-05 | -0.01019218 | CASZ1 |
| chr13 | 48316936 | 48317408 | 473 | 2 | 9.54E-08 | 0.0009639 | 8.70E-06 | 1.31E-05 | -0.0168559 | RB1, PPP1R26P1 |
| chr20 | 63047354 | 63047770 | 417 | 2 | 6.48E-06 | 1.68E-05 | 7.89E-05 | 1.31E-05 | -0.00257747 | LINC01749, LINC01056 |
| chr8 | 37966461 | 37967247 | 787 | 10 | 9.17E-06 | 1.47E-05 | 0.002293 | 1.31E-05 | -0.00020287 | ADRB3 |
| chr12 | 120699531 | 120699768 | 238 | 6 | 5.50E-06 | 0.0107876 | 9.00E-05 | 1.31E-05 | 0.00272108 | MLEC, AC069234.2 |
| chr4 | 6715560 | 6716254 | 695 | 8 | 3.67E-05 | 5.01E-05 | 0.004819 | 1.31E-05 | -0.00298023 | BLOC1S4 |
| chr16 | 510441 | 510855 | 415 | 3 | 3.62E-06 | 0.0001905 | 1.43E-05 | 1.31E-05 | -0.00522993 | RAB11FIP3 |
| chr16 | 10681849 | 10682040 | 192 | 3 | 1.65E-06 | 0.0013062 | 5.25E-06 | 1.31E-05 | -0.00778727 | TEKT5 |
| chr8 | 22865223 | 22865695 | 473 | 3 | 7.53E-06 | 1.69E-05 | 0.000119 | 1.31E-05 | -0.00122843 | PEBP4 |
| chr5 | 25448456 | 25448975 | 520 | 2 | 2.90E-05 | 9.34E-06 | 0.000321 | 1.31E-05 | -0.00375407 | NA |
| chr7 | 84085986 | 84086085 | 100 | 2 | 3.02E-05 | 1.40E-05 | 0.000106 | 1.31E-05 | -0.00282683 | SEMA3A |
| chr14 | 22871924 | 22872378 | 455 | 6 | 3.83E-05 | 5.50E-05 | 0.001853 | 1.31E-05 | 0.002271644 | LRP10 |
| chr9 | 135087132 | 135087561 | 430 | 7 | 1.01E-05 | 8.37E-06 | 0.003703 | 1.31E-05 | -0.00172596 | OLFM1 |
| chr13 | 41463165 | 41463297 | 133 | 2 | 4.07E-06 | 0.0001102 | 1.66E-05 | 1.31E-05 | -0.00874082 | RGCC |
| chr17 | 48566365 | 48566813 | 449 | 2 | 2.67E-05 | 8.04E-06 | 0.000684 | 1.31E-05 | -0.00834364 | HOXB-AS3, HOXB3 |
| chr10 | 86355575 | 86355975 | 401 | 3 | 9.95E-06 | 2.94E-05 | 0.000513 | 1.31E-05 | 0.003538813 | GRID1 |
| chr16 | 58024725 | 58025245 | 521 | 7 | 2.57E-05 | 0.0007661 | 0.001976 | 1.31E-05 | -0.00174254 | NA |
| chr3 | 134312171 | 134312843 | 673 | 4 | 2.70E-05 | 6.55E-06 | 0.002103 | 1.31E-05 | -0.01179255 | NA |
| chr16 | 11275635 | 11276538 | 904 | 7 | 2.76E-06 | 0.0003775 | 3.61E-05 | 1.31E-05 | 5.01E-05 | RMI2, PRM2 |
| chr5 | 42811850 | 42812742 | 893 | 7 | 2.92E-06 | 0.0002011 | 8.16E-05 | 1.31E-05 | -0.00626737 | SELENOP |
| chr19 | 57947549 | 57947990 | 442 | 11 | 1.32E-05 | 3.98E-05 | 0.001621 | 1.31E-05 | 0.000504671 | ZNF256 |
| chr22 | 23890598 | 23890612 | 15 | 2 | 3.39E-05 | 8.63E-06 | 0.000459 | 1.32E-05 | -0.00619764 | AP000350.4 |
| chr22 | 49961776 | 49962431 | 656 | 3 | 4.02E-08 | 0.0001757 | 3.78E-05 | 1.32E-05 | 0.002245178 | PIM3 |
| chr2 | 33599103 | 33600005 | 903 | 16 | 6.83E-05 | 2.00E-05 | 0.008284 | 1.32E-05 | 0.000597086 | AC017050.1, FAM98A |
| chr12 | 79805595 | 79805880 | 286 | 2 | 7.89E-06 | 0.000107 | 1.68E-05 | 1.32E-05 | -0.00208848 | PPP1R12A |
| chr18 | 49559379 | 49559866 | 488 | 2 | 4.57E-06 | 1.06E-05 | 0.00021 | 1.32E-05 | -0.01373099 | NA |
| chr15 | 92655788 | 92656306 | 519 | 10 | 2.41E-05 | 5.74E-05 | 0.00261 | 1.32E-05 | -0.00070548 | FAM174B |
| chr13 | 94443683 | 94443846 | 164 | 3 | 2.41E-05 | 3.02E-05 | 5.79E-05 | 1.32E-05 | -0.0054375 | DCT |
| chr10 | 92830216 | 92831039 | 824 | 4 | 4.90E-06 | 2.57E-05 | 0.00081 | 1.32E-05 | -0.00539374 | EXOC6 |
| chr7 | 159058573 | 159058878 | 306 | 2 | 7.97E-06 | 1.79E-05 | 7.24E-05 | 1.32E-05 | -0.00349079 | VIPR2 |
| chr1 | 16115884 | 16116135 | 252 | 2 | 1.19E-05 | 3.90E-05 | 3.18E-05 | 1.32E-05 | -0.00892243 | NA |
| chr6 | 157184275 | 157184862 | 588 | 6 | 7.37E-05 | 1.01E-05 | 0.003331 | 1.32E-05 | -0.00089711 | ARID1B |
| chr5 | 178730771 | 178731085 | 315 | 9 | 6.33E-06 | 0.001208 | 0.000448 | 1.32E-05 | -0.00615805 | NA |
| chr8 | 75030642 | 75030671 | 30 | 2 | 1.83E-05 | 8.58E-06 | 0.000486 | 1.32E-05 | -0.0208032 | CRISPLD1 |
| chr9 | 134234268 | 134234270 | 3 | 2 | 3.95E-05 | 7.93E-06 | 0.000798 | 1.32E-05 | -0.00546548 | NA |
| chr1 | 1001517 | 1002029 | 513 | 4 | 7.65E-07 | 0.000834 | 8.81E-05 | 1.32E-05 | 0.009114759 | ISG15 |
| chr6 | 46586829 | 46587161 | 333 | 3 | 3.26E-05 | 1.14E-05 | 0.00035 | 1.32E-05 | -0.01037619 | AL035670.1, CYP39A1 |
| chr12 | 132030313 | 132031523 | 1211 | 4 | 1.04E-06 | 0.0011486 | 0.000172 | 1.32E-05 | -0.00384853 | EP400, SNORA49 |
| chr17 | 419551 | 419560 | 10 | 2 | 6.00E-06 | 0.0035861 | 7.40E-06 | 1.32E-05 | 0.002852436 | C17orf97 |
| chr20 | 47013628 | 47013805 | 178 | 2 | 2.25E-05 | 8.11E-06 | 0.000685 | 1.32E-05 | 0.004946828 | EYA2 |
| chr19 | 17747292 | 17748020 | 729 | 11 | 1.65E-05 | 7.09E-05 | 0.000583 | 1.32E-05 | 0.000115492 | FCHO1 |
| chr16 | 66517957 | 66518006 | 50 | 2 | 2.30E-06 | 9.01E-05 | 1.84E-05 | 1.32E-05 | 0.007225507 | TK2 |
| chr4 | 127880490 | 127881149 | 660 | 13 | 1.80E-05 | 0.0004038 | 0.004187 | 1.32E-05 | 0.000749663 | PLK4 |
| chr3 | 128607782 | 128608095 | 314 | 4 | 3.64E-05 | 6.55E-06 | 0.001876 | 1.32E-05 | -0.00863589 | NA |
| chr13 | 114314017 | 114314492 | 476 | 4 | 5.66E-06 | 4.49E-05 | 0.000236 | 1.32E-05 | 0.003669663 | CHAMP1 |
| chr15 | 94856838 | 94856930 | 93 | 2 | 1.65E-06 | 0.0025715 | 7.66E-06 | 1.32E-05 | -0.00794346 | AC087633.2, AC107976.1 |
| chr6 | 107115292 | 107116026 | 735 | 6 | 2.39E-06 | 0.0004275 | 0.000201 | 1.33E-05 | 0.001810897 | BEND3 |
| chr1 | 8883674 | 8883822 | 149 | 2 | 2.38E-05 | 1.33E-05 | 0.000121 | 1.33E-05 | -0.01198074 | NA |
| chr11 | 116276863 | 116277212 | 350 | 4 | 2.67E-05 | 0.0002025 | 0.000709 | 1.33E-05 | -0.0074599 | NA |
| chr6 | 30453314 | 30453878 | 565 | 9 | 4.70E-05 | 1.12E-05 | 0.005557 | 1.33E-05 | -0.00766949 | AL662873.1 |
| chr10 | 123142833 | 123143798 | 966 | 6 | 3.74E-06 | 0.0051753 | 0.000114 | 1.33E-05 | -0.00126812 | NA |
| chr3 | 17140413 | 17140442 | 30 | 2 | 3.09E-05 | 9.05E-06 | 0.000389 | 1.33E-05 | 0.001260938 | NA |
| chr19 | 2321627 | 2321990 | 364 | 5 | 1.32E-05 | 3.49E-05 | 0.000118 | 1.33E-05 | -0.00084866 | AC005258.1, LSM7 |
| chr9 | 127570494 | 127570790 | 297 | 2 | 3.12E-06 | 9.47E-05 | 1.80E-05 | 1.33E-05 | 0.002021141 | NIBAN2 |
| chr3 | 99817063 | 99818209 | 1147 | 18 | 6.65E-06 | 0.0004011 | 0.00087 | 1.33E-05 | -0.00034166 | CMSS1 |
| chr3 | 56987247 | 56987546 | 300 | 2 | 1.79E-05 | 1.82E-05 | 7.23E-05 | 1.33E-05 | 0.006882162 | ARHGEF3 |
| chr1 | 47192975 | 47193273 | 299 | 2 | 1.52E-06 | 0.0001406 | 1.51E-05 | 1.33E-05 | -0.00887021 | NA |
| chr1 | 229343169 | 229343748 | 580 | 7 | 9.06E-06 | 7.80E-05 | 0.000515 | 1.33E-05 | 0.002079346 | CCSAP |
| chr4 | 83219568 | 83219861 | 294 | 3 | 2.19E-05 | 1.55E-05 | 0.000611 | 1.33E-05 | 0.008231253 | NA |
| chr22 | 24711823 | 24712236 | 414 | 3 | 2.83E-05 | 9.35E-06 | 0.001308 | 1.33E-05 | 0.001062854 | NA |
| chr1 | 11339411 | 11339855 | 445 | 2 | 3.29E-05 | 8.50E-06 | 0.000533 | 1.33E-05 | -0.01043469 | NA |
| chr17 | 82927482 | 82927806 | 325 | 2 | 3.74E-06 | 1.55E-05 | 9.22E-05 | 1.33E-05 | -0.00192567 | TBCD |
| chr1 | 196650501 | 196651298 | 798 | 4 | 1.97E-05 | 2.23E-05 | 0.000121 | 1.33E-05 | 0.001507434 | NA |
| chr20 | 3782680 | 3783166 | 487 | 3 | 1.26E-06 | 0.000183 | 1.64E-05 | 1.33E-05 | -0.00562091 | NA |
| chr2 | 67179047 | 67179208 | 162 | 2 | 1.89E-05 | 1.12E-05 | 0.00019 | 1.33E-05 | -0.00278637 | LINC01828, LINC01829 |
| chr4 | 123871495 | 123871799 | 305 | 3 | 1.94E-05 | 3.30E-05 | 0.000292 | 1.33E-05 | -0.00691731 | LINC01091 |
| chr3 | 141656979 | 141657496 | 518 | 2 | 4.15E-05 | 8.00E-06 | 0.000801 | 1.33E-05 | 2.06E-05 | NA |
| chr6 | 7543224 | 7544144 | 921 | 3 | 2.12E-05 | 0.0001188 | 9.05E-05 | 1.33E-05 | 0.010255017 | DSP |
| chr18 | 79022346 | 79022895 | 550 | 7 | 1.82E-06 | 0.0006967 | 2.91E-05 | 1.33E-05 | -0.00251206 | NA |
| chr16 | 49575145 | 49575541 | 397 | 4 | 1.65E-05 | 5.40E-05 | 5.62E-05 | 1.33E-05 | -0.0003256 | ZNF423 |
| chr19 | 48240962 | 48241227 | 266 | 3 | 1.02E-05 | 0.0001231 | 1.97E-05 | 1.33E-05 | -0.01296199 | CARD8 |
| chr18 | 48410051 | 48410701 | 651 | 4 | 7.24E-06 | 2.60E-05 | 0.000376 | 1.33E-05 | -0.00305048 | ZBTB7C |
| chr6 | 43687230 | 43688062 | 833 | 12 | 4.49E-06 | 0.0030848 | 1.75E-05 | 1.33E-05 | 0.002534711 | MRPS18A |
| chr17 | 75320199 | 75320880 | 682 | 12 | 4.21E-05 | 2.59E-05 | 0.007492 | 1.33E-05 | -0.00049123 | GRB2 |
| chr6 | 170043721 | 170044171 | 451 | 4 | 7.75E-06 | 0.0001902 | 0.000134 | 1.33E-05 | -0.00434839 | NA |
| chr2 | 85853168 | 85853372 | 205 | 2 | 1.33E-05 | 1.77E-05 | 7.61E-05 | 1.33E-05 | 0.009898928 | ST3GAL5 |
| chr8 | 73292998 | 73294296 | 1299 | 14 | 7.32E-06 | 4.56E-05 | 0.006679 | 1.34E-05 | 0.000355153 | RPL7 |
| chr10 | 11725024 | 11725312 | 289 | 2 | 8.34E-06 | 9.51E-05 | 1.81E-05 | 1.34E-05 | -0.0120314 | NA |
| chr11 | 48305849 | 48306580 | 732 | 3 | 6.51E-05 | 0.0003331 | 0.000377 | 1.34E-05 | -0.01787294 | OR4S1 |
| chr10 | 45003456 | 45003685 | 230 | 2 | 3.36E-06 | 0.0002004 | 1.34E-05 | 1.34E-05 | -0.00369956 | ZNF22, CEP164P1 |
| chr12 | 6774629 | 6775095 | 467 | 5 | 6.06E-06 | 0.000158 | 5.09E-05 | 1.34E-05 | 0.002523328 | LAG3 |
| chr12 | 15534326 | 15534774 | 449 | 3 | 1.28E-05 | 5.74E-05 | 0.00014 | 1.34E-05 | 0.010443296 | PTPRO |
| chr5 | 44952037 | 44952530 | 494 | 2 | 3.24E-05 | 9.11E-06 | 0.000393 | 1.34E-05 | 0.002009134 | NA |
| chr4 | 154399461 | 154399474 | 14 | 2 | 2.96E-05 | 1.86E-05 | 7.12E-05 | 1.34E-05 | 0.007799059 | DCHS2 |
| chr7 | 99616522 | 99617104 | 583 | 13 | 2.21E-05 | 3.48E-05 | 0.010644 | 1.34E-05 | 0.00032887 | ZSCAN25 |
| chr22 | 41468501 | 41469096 | 596 | 12 | 8.89E-06 | 0.0010496 | 0.000584 | 1.34E-05 | 0.000533752 | ACO2, PHF5A |
| chr4 | 165209020 | 165209854 | 835 | 3 | 1.06E-06 | 0.0307425 | 3.02E-06 | 1.34E-05 | 0.000635436 | KLHL2 |
| chr1 | 42816528 | 42817760 | 1233 | 17 | 7.62E-06 | 0.0043697 | 9.42E-05 | 1.34E-05 | 0.001453541 | TMEM269, ERMAP, SVBP |
| chr10 | 61993791 | 61994453 | 663 | 2 | 7.67E-05 | 8.03E-06 | 0.000821 | 1.34E-05 | -0.00757711 | ARID5B |
| chr20 | 36864318 | 36864975 | 658 | 4 | 1.49E-06 | 0.0004373 | 0.000732 | 1.34E-05 | -0.0061078 | NA |
| chr3 | 187739340 | 187740842 | 1503 | 9 | 1.16E-06 | 1.63E-05 | 0.005932 | 1.34E-05 | -0.00170948 | BCL6 |
| chr12 | 62621169 | 62621634 | 466 | 2 | 1.90E-05 | 1.12E-05 | 0.00019 | 1.34E-05 | 0.003448046 | AC048341.1 |
| chr3 | 157504043 | 157504612 | 570 | 4 | 4.27E-05 | 4.66E-06 | 0.004249 | 1.34E-05 | -0.00014718 | VEPH1 |
| chr20 | 35674125 | 35674795 | 671 | 5 | 3.85E-05 | 1.02E-05 | 0.001165 | 1.34E-05 | -0.00158192 | AL109827.1, NFS1 |
| chr12 | 81125812 | 81125927 | 116 | 2 | 2.09E-06 | 0.0022887 | 7.85E-06 | 1.35E-05 | -0.00953933 | ACSS3, AC078955.1 |
| chr11 | 17014087 | 17014943 | 857 | 9 | 2.00E-05 | 0.0005184 | 0.000447 | 1.35E-05 | 0.000132648 | OR7E14P, PLEKHA7 |
| chr11 | 36385512 | 36385873 | 362 | 3 | 2.13E-05 | 9.57E-06 | 0.000546 | 1.35E-05 | -0.00705916 | PRR5L |
| chr2 | 161426923 | 161428133 | 1211 | 9 | 1.17E-05 | 4.59E-05 | 0.001205 | 1.35E-05 | -0.00134513 | AC009487.2, SLC4A10 |
| chr3 | 100338644 | 100339092 | 449 | 3 | 1.23E-05 | 8.88E-05 | 0.000182 | 1.35E-05 | 0.001689895 | NIT2 |

| | | | | | | | | | | |
|---|---|---|---|---|---|---|---|---|---|---|
| chr22 | 48745981 | 48746077 | 97 | 2 | 2.38E-05 | 1.05E-05 | 0.000234 | 1.35E-05 | -0.01005159 | TAFA5 |
| chr1 | 160415846 | 160416066 | 221 | 3 | 6.61E-06 | 0.0001133 | 2.01E-05 | 1.35E-05 | -0.00363971 | VANGL2 |
| chr9 | 84265856 | 84266017 | 162 | 2 | 7.65E-06 | 0.0011501 | 8.68E-06 | 1.35E-05 | -0.00523146 | NA |
| chr4 | 12311247 | 12311385 | 139 | 2 | 1.20E-05 | 3.03E-05 | 4.10E-05 | 1.35E-05 | 0.000736788 | NA |
| chr9 | 4678949 | 4679784 | 836 | 13 | 6.66E-06 | 0.0001558 | 0.005744 | 1.35E-05 | 0.001067795 | CDC37L1, CDC37L1-DT |
| chr12 | 99092405 | 99092653 | 249 | 3 | 4.61E-06 | 0.0073036 | 2.00E-05 | 1.35E-05 | 0.006208838 | ANKS1B |
| chr8 | 143198215 | 143198241 | 27 | 2 | 9.44E-06 | 8.58E-06 | 0.000551 | 1.35E-05 | -0.00734679 | NA |
| chr10 | 124523135 | 124523445 | 311 | 4 | 2.84E-05 | 2.16E-05 | 0.000385 | 1.35E-05 | -0.01164598 | LHPP |
| chr1 | 67267767 | 67267845 | 79 | 2 | 3.57E-05 | 8.69E-06 | 0.000514 | 1.35E-05 | -0.0011203 | NA |
| chr8 | 6877117 | 6877440 | 324 | 2 | 1.13E-06 | 0.0001176 | 1.66E-05 | 1.35E-05 | -0.00860154 | GS1-24F4.2, DEFB1 |
| chr12 | 123634435 | 123634703 | 269 | 3 | 6.49E-07 | 0.0003779 | 4.46E-05 | 1.35E-05 | -0.01045993 | GTF2H3 |
| chr12 | 57463733 | 57464244 | 512 | 3 | 8.24E-06 | 8.59E-06 | 0.00083 | 1.35E-05 | -0.00295857 | GLI1 |
| chr13 | 95493898 | 95494172 | 275 | 2 | 3.67E-06 | 9.43E-06 | 0.000355 | 1.35E-05 | -0.00368242 | CLDN10, CLDN10-AS1 |
| chr4 | 3724528 | 3724974 | 447 | 3 | 1.34E-05 | 4.25E-05 | 8.22E-05 | 1.35E-05 | 0.013947526 | NA |
| chr10 | 132948585 | 132948875 | 291 | 2 | 2.85E-05 | 1.06E-05 | 0.000231 | 1.35E-05 | -0.00961758 | LINC01166 |
| chr22 | 23898937 | 23899108 | 172 | 2 | 3.00E-06 | 0.0012303 | 8.61E-06 | 1.35E-05 | -0.00978137 | NA |
| chr8 | 23287919 | 23288451 | 533 | 9 | 4.28E-06 | 3.25E-05 | 0.000801 | 1.35E-05 | 0.001413528 | R3HCC1 |
| chr1 | 162663656 | 162664099 | 444 | 2 | 1.49E-06 | 0.0003266 | 1.16E-05 | 1.35E-05 | -0.02089753 | DDR2 |
| chr11 | 9663562 | 9664221 | 660 | 9 | 7.40E-06 | 0.0005847 | 0.000629 | 1.35E-05 | -0.00043971 | SWAP70 |
| chr16 | 49465702 | 49466098 | 397 | 4 | 3.60E-05 | 0.0011939 | 0.00022 | 1.35E-05 | -0.00082619 | NA |
| chr1 | 202348599 | 202349067 | 469 | 6 | 1.68E-06 | 3.51E-05 | 0.000132 | 1.35E-05 | -0.00080454 | PPP1R12B |
| chr1 | 116409974 | 116410143 | 170 | 2 | 4.28E-06 | 0.0016934 | 8.22E-06 | 1.35E-05 | 0.00143957 | ATP1A1, ATP1A1-AS1 |
| chr9 | 124834944 | 124835307 | 364 | 3 | 1.56E-05 | 1.90E-05 | 9.89E-05 | 1.35E-05 | -0.00476205 | NA |
| chr15 | 65529975 | 65530748 | 774 | 11 | 1.36E-05 | 9.70E-06 | 0.00463 | 1.35E-05 | -0.00066267 | HACD3 |
| chr16 | 3011834 | 3011986 | 153 | 3 | 5.00E-05 | 1.72E-05 | 0.000126 | 1.35E-05 | 0.007613959 | NA |
| chr11 | 20085615 | 20085726 | 112 | 2 | 9.40E-06 | 0.0009062 | 9.12E-06 | 1.36E-05 | -0.00423192 | NAV2 |
| chr5 | 150309716 | 150309998 | 283 | 2 | 2.86E-06 | 2.62E-05 | 4.82E-05 | 1.36E-05 | -0.00858361 | ARSI |
| chr1 | 220653936 | 220654035 | 100 | 2 | 2.69E-05 | 1.66E-05 | 8.65E-05 | 1.36E-05 | -0.0093024 | MARK1 |
| chr16 | 28931698 | 28932356 | 659 | 7 | 2.44E-05 | 3.89E-06 | 0.009529 | 1.36E-05 | 0.001026746 | AC009093.11, CD19, RABEP2 |
| chr17 | 67366625 | 67367197 | 573 | 10 | 6.44E-05 | 5.08E-05 | 0.004705 | 1.36E-05 | -0.00225195 | NA |
| chr2 | 3643645 | 3644061 | 417 | 5 | 1.30E-05 | 0.0006094 | 0.000409 | 1.36E-05 | 0.000489491 | COLEC11 |
| chr16 | 1263492 | 1264164 | 673 | 6 | 3.81E-06 | 0.0001211 | 0.000539 | 1.36E-05 | -0.00036807 | PRSS29P |
| chr10 | 101617083 | 101617148 | 66 | 2 | 9.18E-06 | 0.0001828 | 1.40E-05 | 1.36E-05 | 0.001430573 | FBXW4 |
| chr11 | 72674654 | 72675496 | 843 | 5 | 1.29E-06 | 0.0005406 | 3.19E-05 | 1.36E-05 | -0.00621541 | NA |
| chr10 | 58792531 | 58792964 | 434 | 2 | 8.43E-06 | 1.02E-05 | 0.000272 | 1.36E-05 | -0.00829725 | BICC1 |
| chr1 | 197735644 | 197735855 | 212 | 4 | 2.88E-05 | 3.24E-05 | 0.000148 | 1.36E-05 | -0.00518785 | AL365258.2, DENND1B |
| chr14 | 51397711 | 51398337 | 627 | 3 | 9.62E-06 | 0.000163 | 0.000466 | 1.36E-05 | -0.00875401 | NA |
| chr3 | 4302977 | 4303754 | 778 | 17 | 2.71E-05 | 2.57E-05 | 0.000737 | 1.36E-05 | -0.00242311 | SETMAR, SUMF1, AC023483.1 |
| chr3 | 47892770 | 47893303 | 534 | 6 | 3.47E-05 | 3.09E-05 | 0.001459 | 1.36E-05 | 0.000945786 | MAP4 |
| chr22 | 27311933 | 27312062 | 130 | 2 | 2.42E-05 | 9.19E-06 | 0.000418 | 1.36E-05 | -0.0176291 | LINC02554, AL020994.1 |
| chr8 | 140667881 | 140668347 | 467 | 3 | 6.36E-07 | 0.0012582 | 1.11E-05 | 1.36E-05 | -0.00720532 | PTK2 |
| chr3 | 47007111 | 47007295 | 185 | 2 | 1.42E-05 | 2.39E-05 | 5.39E-05 | 1.36E-05 | 0.011811746 | NBEAL2 |
| chr9 | 135888263 | 135888770 | 508 | 2 | 2.74E-05 | 9.93E-06 | 0.000304 | 1.36E-05 | -0.00145738 | CAMSAP1 |
| chr9 | 126671353 | 126671500 | 148 | 3 | 2.78E-05 | 1.21E-05 | 0.000721 | 1.36E-05 | 0.017835786 | LMX1B |
| chr1 | 34783496 | 34783726 | 231 | 3 | 1.09E-05 | 1.63E-05 | 0.000144 | 1.37E-05 | -0.00084773 | GJB3, SMIM12, AL121988.1 |
| chr12 | 122197167 | 122197199 | 33 | 2 | 4.03E-05 | 8.60E-06 | 0.000592 | 1.37E-05 | 0.012834733 | LRRC43 |
| chr5 | 125201818 | 125202142 | 325 | 2 | 1.54E-05 | 1.07E-05 | 0.000234 | 1.37E-05 | -0.00668161 | LINC02240 |
| chr4 | 7792919 | 7793397 | 479 | 2 | 4.07E-07 | 0.0019667 | 8.12E-06 | 1.37E-05 | -0.01438651 | AFAP1 |
| chr4 | 38791559 | 38791815 | 257 | 2 | 1.77E-05 | 1.23E-05 | 0.00016 | 1.37E-05 | -0.01035264 | TLR1 |
| chr4 | 183852795 | 183853390 | 596 | 5 | 2.58E-05 | 8.93E-05 | 0.00189 | 1.37E-05 | -0.00409061 | STOX2 |
| chr11 | 70667336 | 70668194 | 859 | 4 | 1.46E-07 | 0.0011764 | 4.75E-06 | 1.37E-05 | -0.00740061 | SHANK2 |
| chr16 | 72025039 | 72025510 | 472 | 2 | 1.07E-05 | 8.62E-06 | 0.000585 | 1.37E-05 | 0.005982439 | DHODH |
| chr3 | 134408611 | 134408744 | 134 | 2 | 7.73E-06 | 9.62E-05 | 1.85E-05 | 1.37E-05 | -0.0055042 | NA |
| chr19 | 35496172 | 35496327 | 156 | 3 | 3.74E-05 | 6.14E-06 | 0.001573 | 1.37E-05 | -0.00894999 | NA |
| chr13 | 114115719 | 114116081 | 363 | 4 | 5.70E-05 | 1.04E-05 | 0.00234 | 1.37E-05 | 0.002733187 | RASA3 |
| chr2 | 28747716 | 28748221 | 506 | 2 | 3.55E-05 | 9.15E-06 | 0.000435 | 1.37E-05 | -0.00819627 | AC092164.1 |
| chr19 | 5456250 | 5456853 | 604 | 2 | 6.71E-07 | 4.87E-05 | 2.83E-05 | 1.37E-05 | 0.000472181 | ZNRF4 |
| chr1 | 64471662 | 64471845 | 184 | 3 | 1.03E-05 | 1.22E-05 | 0.000236 | 1.37E-05 | -0.00864988 | CACHD1 |
| chr6 | 21592898 | 21594089 | 1192 | 12 | 4.88E-06 | 0.0009273 | 0.002464 | 1.37E-05 | 0.000361947 | SOX4, AL512380.2 |
| chr10 | 123995116 | 123995635 | 520 | 3 | 9.07E-06 | 3.83E-05 | 0.000299 | 1.37E-05 | -0.01595619 | NA |
| chr11 | 65711318 | 65712207 | 890 | 12 | 2.60E-05 | 4.97E-05 | 0.013099 | 1.37E-05 | -0.00116319 | KAT5 |
| chr19 | 54419187 | 54419766 | 580 | 2 | 1.08E-05 | 8.25E-06 | 0.000801 | 1.37E-05 | 0.00100427 | TTYH1 |
| chr19 | 17211718 | 17212481 | 764 | 5 | 3.60E-06 | 0.0005377 | 2.25E-05 | 1.37E-05 | -0.00089812 | MYO9B |
| chr1 | 4407991 | 4408029 | 39 | 2 | 4.09E-05 | 8.39E-06 | 0.000708 | 1.37E-05 | 0.005156815 | NA |
| chr10 | 120850791 | 120851629 | 839 | 9 | 2.58E-05 | 4.66E-06 | 0.003945 | 1.37E-05 | 0.003555292 | WDR11, WDR11-AS1 |
| chr1 | 204126900 | 204127287 | 388 | 2 | 3.56E-06 | 1.85E-05 | 7.53E-05 | 1.37E-05 | -0.00877949 | SOX13 |
| chr1 | 6961981 | 6962224 | 244 | 4 | 1.51E-05 | 8.97E-05 | 0.000173 | 1.37E-05 | -0.00396336 | CAMTA1 |
| chr14 | 35826051 | 35826823 | 773 | 10 | 7.90E-06 | 7.32E-05 | 0.006241 | 1.37E-05 | -9.50E-05 | BRMS1L, AL162311.3 |
| chr9 | 116154214 | 116154830 | 617 | 3 | 7.17E-07 | 8.78E-05 | 2.19E-05 | 1.37E-05 | -0.00251377 | PAPPA |
| chr3 | 58577370 | 58578180 | 811 | 10 | 1.58E-05 | 2.78E-05 | 0.005503 | 1.37E-05 | -0.0042793 | FAM107A |
| chr12 | 12979543 | 12980197 | 655 | 8 | 4.79E-05 | 7.89E-06 | 0.001874 | 1.37E-05 | -0.00198755 | GPRC5D-AS1, HEBP1 |
| chr11 | 15650045 | 15650712 | 668 | 8 | 2.56E-05 | 0.000103 | 0.003436 | 1.38E-05 | -0.00139993 | AC087379.2 |
| chr7 | 158529015 | 158529460 | 446 | 2 | 2.76E-06 | 0.0001127 | 1.73E-05 | 1.38E-05 | -0.01582867 | PTPRN2 |
| chr15 | 31482570 | 31483203 | 634 | 3 | 3.39E-07 | 0.0071898 | 3.56E-06 | 1.38E-05 | -0.00403144 | OTUD7A |
| chr1 | 225810530 | 225810582 | 53 | 2 | 1.11E-05 | 1.42E-05 | 0.000117 | 1.38E-05 | 0.003674076 | EPHX1 |
| chr21 | 37700926 | 37700966 | 41 | 3 | 2.77E-05 | 5.09E-06 | 0.003741 | 1.38E-05 | -0.00778969 | KCNJ6 |
| chr1 | 33875451 | 33876009 | 559 | 2 | 4.09E-06 | 1.52E-05 | 0.000103 | 1.38E-05 | -0.01339364 | CSMD2-AS1, CSMD2 |
| chr20 | 50252731 | 50253046 | 316 | 2 | 4.03E-05 | 8.25E-06 | 0.000834 | 1.38E-05 | -0.00784135 | PELATON |
| chr3 | 44584500 | 44585368 | 869 | 9 | 0.000103617 | 5.90E-06 | 0.008665 | 1.38E-05 | -0.00125778 | ZKSCAN7, ZNF660, ZNF197, ZKSCAN7-AS1 |
| chr7 | 102203713 | 102203772 | 60 | 2 | 2.50E-06 | 5.40E-05 | 2.65E-05 | 1.38E-05 | 0.001416002 | CUX1 |
| chr2 | 68465092 | 68465316 | 225 | 2 | 2.31E-05 | 9.46E-06 | 0.000386 | 1.38E-05 | -0.01077129 | FBXO48 |
| chr7 | 99624258 | 99624484 | 227 | 2 | 1.53E-05 | 6.76E-05 | 2.29E-05 | 1.38E-05 | -0.02054819 | ZSCAN25 |
| chr12 | 57225507 | 57225618 | 112 | 3 | 8.86E-05 | 5.72E-06 | 0.002639 | 1.38E-05 | 0.006490748 | NXPH4 |
| chr8 | 144104831 | 144105313 | 483 | 4 | 1.72E-06 | 1.86E-05 | 0.000207 | 1.38E-05 | -0.00508407 | MAF1, SHARPIN |
| chr15 | 66708433 | 66708617 | 185 | 5 | 1.85E-05 | 4.27E-05 | 0.000145 | 1.38E-05 | -0.00028711 | SMAD6 |
| chr4 | 129779534 | 129779809 | 276 | 2 | 2.92E-05 | 8.26E-06 | 0.000834 | 1.38E-05 | -0.01271973 | LINC02465 |
| chr9 | 136505716 | 136506147 | 432 | 2 | 4.89E-06 | 8.68E-06 | 0.000604 | 1.38E-05 | -0.01569447 | NOTCH1 |
| chr17 | 82179199 | 82179787 | 589 | 4 | 9.44E-06 | 2.13E-05 | 0.000558 | 1.38E-05 | -0.00517892 | CCDC57 |
| chr15 | 88805517 | 88805617 | 101 | 2 | 1.82E-05 | 6.35E-05 | 2.39E-05 | 1.38E-05 | 0.023972742 | ACAN |
| chr11 | 19725913 | 19726042 | 130 | 2 | 6.51E-06 | 0.0279278 | 7.01E-06 | 1.38E-05 | -0.00259953 | NAV2 |
| chr10 | 132522495 | 132522880 | 386 | 4 | 1.46E-05 | 6.27E-05 | 0.001496 | 1.38E-05 | -0.00615425 | NA |
| chr10 | 30354060 | 30354346 | 287 | 2 | 1.38E-05 | 2.71E-05 | 4.78E-05 | 1.38E-05 | 0.004052304 | DNM1P17, MTPAP |
| chr20 | 1207008 | 1207345 | 338 | 2 | 5.74E-06 | 0.0001588 | 1.51E-05 | 1.38E-05 | 0.00372537 | AL031665.1, C20orf202 |
| chr1 | 109394520 | 109394569 | 50 | 4 | 4.09E-05 | 1.01E-05 | 0.001048 | 1.38E-05 | -0.00431068 | SORT1 |
| chr2 | 219565189 | 219565507 | 319 | 2 | 3.18E-05 | 9.15E-06 | 0.000462 | 1.38E-05 | -0.00360045 | OBSL1 |
| chr1 | 149884970 | 149885941 | 972 | 7 | 2.28E-06 | 0.0004139 | 0.000129 | 1.38E-05 | 0.001583226 | H2BC21 |
| chr5 | 180207383 | 180208152 | 770 | 4 | 2.17E-06 | 0.0003208 | 0.000366 | 1.39E-05 | -0.0132061 | RASGEF1C |
| chr10 | 69738739 | 69738942 | 204 | 2 | 1.89E-05 | 1.55E-05 | 0.000101 | 1.39E-05 | -0.00843508 | NA |
| chr7 | 154304806 | 154305533 | 728 | 10 | 2.12E-05 | 2.58E-05 | 0.003741 | 1.39E-05 | 0.003053747 | DPP6 |
| chr11 | 111335036 | 111335776 | 741 | 2 | 1.71E-05 | 1.21E-05 | 0.000175 | 1.39E-05 | -0.00100528 | NA |
| chr11 | 19905506 | 19905585 | 80 | 2 | 1.03E-05 | 0.0001271 | 1.66E-05 | 1.39E-05 | -0.013995 | NAV2 |
| chr7 | 116672543 | 116672850 | 308 | 3 | 1.66E-05 | 1.25E-05 | 0.000339 | 1.39E-05 | -0.01176002 | MET |
| chr19 | 1082613 | 1083194 | 582 | 4 | 2.11E-05 | 6.99E-06 | 0.00325 | 1.39E-05 | -0.00372579 | ARHGAP45 |
| chr18 | 74266414 | 74267042 | 629 | 2 | 6.39E-05 | 8.36E-06 | 0.000795 | 1.39E-05 | -0.00472366 | CYB5A |
| chr4 | 54224645 | 54225011 | 367 | 2 | 2.77E-05 | 9.30E-06 | 0.000436 | 1.39E-05 | -0.00433383 | AC058822.1 |
| chr17 | 27559403 | 27560198 | 796 | 5 | 1.40E-06 | 0.0001681 | 0.000302 | 1.39E-05 | -0.00278171 | KSR1 |
| chr10 | 103794561 | 103795131 | 571 | 2 | 1.48E-06 | 6.09E-05 | 2.47E-05 | 1.39E-05 | -0.00686614 | SH3PXD2A |
| chr16 | 11345676 | 11345928 | 253 | 4 | 4.02E-06 | 0.0001198 | 0.00035 | 1.39E-05 | -0.0001917 | RMI2 |
| chr20 | 49189768 | 49189864 | 97 | 2 | 9.02E-06 | 2.51E-05 | 5.23E-05 | 1.39E-05 | -0.00038456 | NA |
| chr12 | 26970524 | 26970768 | 245 | 3 | 4.77E-05 | 7.76E-06 | 0.001477 | 1.39E-05 | -0.00112853 | NA |
| chr5 | 175843181 | 175843404 | 224 | 2 | 1.29E-05 | 6.51E-05 | 2.37E-05 | 1.39E-05 | -0.00994015 | CPLX2 |
| chr10 | 1101690 | 1102181 | 492 | 4 | 2.57E-06 | 0.0008874 | 2.84E-05 | 1.39E-05 | 0.003915069 | WDR37 |
| chr16 | 31042168 | 31043090 | 923 | 8 | 2.90E-06 | 0.0001546 | 0.000259 | 1.39E-05 | 0.000463833 | STX4 |
| chr16 | 57461965 | 57463098 | 1134 | 11 | 2.40E-06 | 0.0001408 | 0.000338 | 1.39E-05 | -0.00177496 | AC009052.1, POLR2C |
| chr12 | 132672343 | 132672692 | 350 | 5 | 1.90E-05 | 1.05E-05 | 0.001073 | 1.39E-05 | -0.0022596 | POLE |
| chr20 | 34111672 | 34112474 | 803 | 7 | 9.07E-06 | 4.79E-05 | 0.000286 | 1.39E-05 | 0.000669658 | EIF2S2 |
| chr14 | 105146416 | 105146765 | 350 | 3 | 9.75E-06 | 1.81E-05 | 0.000659 | 1.39E-05 | -0.00299197 | JAG2 |
| chr3 | 11350285 | 11350472 | 188 | 2 | 2.41E-05 | 1.26E-05 | 0.00016 | 1.39E-05 | -0.00156505 | ATG7 |
| chr7 | 158266704 | 158267098 | 395 | 3 | 3.16E-05 | 5.75E-06 | 0.002721 | 1.39E-05 | 0.011052827 | PTPRN2 |
| chr3 | 69999226 | 69999521 | 296 | 3 | 1.24E-05 | 6.27E-05 | 3.54E-05 | 1.39E-05 | -0.00763169 | NA |
| chr4 | 112756649 | 112756671 | 23 | 2 | 3.55E-05 | 1.31E-05 | 0.000144 | 1.39E-05 | -0.00291746 | NA |
| chr11 | 44780621 | 44781014 | 394 | 2 | 1.01E-05 | 1.30E-05 | 0.000147 | 1.39E-05 | 0.012149387 | TSPAN18 |
| chr12 | 132062114 | 132062734 | 621 | 4 | 3.56E-05 | 4.05E-05 | 0.000636 | 1.39E-05 | -0.0004951 | EP400 |
| chr12 | 94075451 | 94075736 | 286 | 2 | 2.83E-05 | 1.08E-05 | 0.000245 | 1.39E-05 | -0.01169875 | NA |
| chr13 | 32311293 | 32312003 | 711 | 6 | 2.23E-05 | 1.07E-05 | 0.003244 | 1.39E-05 | -0.00060789 | ZAR1L |
| chr20 | 63034806 | 63035202 | 397 | 2 | 1.03E-06 | 1.08E-05 | 0.000245 | 1.39E-05 | 0.000500848 | LINC01749, LINC00029 |
| chr17 | 75065273 | 75065753 | 481 | 3 | 5.23E-06 | 0.0001732 | 0.000247 | 1.39E-05 | -0.00216085 | KCTD2 |
| chr8 | 141920906 | 141921180 | 275 | 2 | 3.82E-05 | 1.61E-05 | 9.56E-05 | 1.39E-05 | 0.01070734 | NA |
| chr5 | 112818932 | 112819274 | 343 | 2 | 1.06E-05 | 8.63E-06 | 0.000668 | 1.39E-05 | -0.00069551 | APC |
| chr20 | 18137367 | 18138068 | 702 | 13 | 1.64E-05 | 6.87E-05 | 0.002515 | 1.39E-05 | -0.00033572 | KAT14, PET117 |
| chr11 | 60658346 | 60658540 | 195 | 2 | 3.42E-05 | 1.07E-05 | 0.00026 | 1.39E-05 | -0.00507951 | LINC00301 |
| chr2 | 134504650 | 134504707 | 58 | 2 | 4.10E-05 | 8.92E-06 | 0.000557 | 1.40E-05 | -0.00046731 | TMEM163 |
| chr5 | 62412341 | 62413307 | 967 | 12 | 3.96E-05 | 0.0001273 | 0.003168 | 1.40E-05 | 0.00137961 | AC114982.2, IPO11 |
| chr5 | 54319852 | 54320108 | 257 | 3 | 3.77E-05 | 8.86E-06 | 0.001474 | 1.40E-05 | -0.00761335 | LINC01033 |
| chr6 | 33292486 | 33293074 | 589 | 3 | 6.42E-05 | 3.70E-05 | 5.08E-05 | 1.40E-05 | 0.005366762 | PFDN6, RGL2 |
| chr2 | 20224662 | 20225919 | 1258 | 15 | 9.62E-06 | 0.0008671 | 0.001441 | 1.40E-05 | -0.00043241 | SDC1 |
| chr15 | 90248991 | 90249081 | 91 | 3 | 7.00E-06 | 4.61E-05 | 0.000387 | 1.40E-05 | 0.013328544 | AC091167.7 |
| chr17 | 42679016 | 42679347 | 332 | 3 | 2.11E-06 | 0.0002432 | 9.19E-05 | 1.40E-05 | 0.005217537 | CCR10 |
| chr14 | 93100381 | 93100784 | 404 | 2 | 1.72E-06 | 0.0001208 | 1.71E-05 | 1.40E-05 | -0.00018962 | ITPK1 |
| chr11 | 7555193 | 7555570 | 378 | 3 | 3.69E-06 | 0.0007216 | 2.08E-05 | 1.40E-05 | 0.001472728 | PPFIBP2 |
| chr11 | 121100533 | 121100752 | 220 | 2 | 1.62E-05 | 1.31E-05 | 0.000146 | 1.40E-05 | -0.01109366 | TBCEL-TECTA |
| chr3 | 55490906 | 55491549 | 644 | 4 | 1.13E-05 | 1.34E-05 | 0.00174 | 1.40E-05 | -0.01239184 | NA |
| chr22 | 41445487 | 41445815 | 329 | 3 | 4.20E-06 | 0.0001587 | 0.000434 | 1.40E-05 | 0.002530734 | TOB2 |
| chr1 | 114407132 | 114407559 | 428 | 3 | 1.21E-05 | 0.0001224 | 0.000433 | 1.40E-05 | -0.01029184 | TRIM33 |
| chr9 | 132586548 | 132587135 | 588 | 4 | 3.38E-05 | 6.45E-06 | 0.002202 | 1.40E-05 | -0.0021494 | BARHL1 |
| chr6 | 33902522 | 33902924 | 403 | 3 | 1.26E-05 | 1.23E-05 | 0.000383 | 1.40E-05 | 0.011531241 | AL138889.1 |
| chr1 | 164558568 | 164558864 | 297 | 4 | 3.70E-07 | 4.23E-05 | 0.000214 | 1.40E-05 | 0.000679182 | PBX1 |
| chr15 | 45430238 | 45430721 | 484 | 6 | 3.10E-05 | 3.59E-05 | 0.001836 | 1.40E-05 | 0.000691861 | C15orf48, AC025580.1 |
| chr16 | 52237701 | 52238041 | 341 | 3 | 3.82E-05 | 6.30E-06 | 0.00216 | 1.40E-05 | -0.00181671 | NA |
| chr12 | 190483 | 190541 | 59 | 2 | 7.64E-06 | 8.40E-06 | 0.000838 | 1.40E-05 | -0.00173606 | SLC6A12 |
| chr17 | 5444330 | 5444653 | 324 | 3 | 2.36E-05 | 2.13E-05 | 9.31E-05 | 1.40E-05 | 0.00018042 | C1QBP, DHX33 |
| chr7 | 364619 | 364954 | 336 | 2 | 0.000109868 | 1.11E-05 | 0.000231 | 1.40E-05 | 0.002239053 | NA |
| chr12 | 110345884 | 110346374 | 491 | 5 | 1.79E-05 | 5.58E-05 | 0.000721 | 1.40E-05 | 0.001888854 | ATP2A2 |
| chr19 | 40205959 | 40206180 | 222 | 5 | 2.17E-05 | 6.19E-05 | 3.56E-05 | 1.40E-05 | -0.00102142 | MAP3K10 |
| chr7 | 2376761 | 2377398 | 638 | 4 | 1.38E-05 | 7.76E-05 | 0.000381 | 1.40E-05 | -0.00323648 | EIF3B |
| chr12 | 97686850 | 97686984 | 135 | 2 | 1.41E-05 | 1.81E-05 | 8.17E-05 | 1.40E-05 | -0.01604836 | NA |
| chr3 | 194090794 | 194091360 | 567 | 4 | 5.06E-05 | 2.31E-05 | 0.001572 | 1.40E-05 | -0.00422692 | NA |
| chr1 | 162268199 | 162268593 | 395 | 2 | 3.72E-05 | 8.99E-06 | 0.000551 | 1.40E-05 | 0.005115337 | NOS1AP |

| | | | | | | | | | | |
|---|---|---|---|---|---|---|---|---|---|---|
| chr19 | 7910489 | 7911033 | 545 | 4 | 2.30E-07 | 0.0061373 | 2.33E-06 | 1.40E-05 | -0.00263937 | MAP2K7 |
| chr20 | 57688079 | 57688107 | 29 | 2 | 3.21E-06 | 1.40E-05 | 0.000128 | 1.40E-05 | -0.00104378 | PMEPA1 |
| chr22 | 20858879 | 20859837 | 959 | 7 | 9.78E-06 | 7.03E-06 | 0.003837 | 1.40E-05 | -0.00604141 | SNAP29, PI4KA |
| chr6 | 37368514 | 37368596 | 83 | 2 | 3.86E-05 | 1.03E-05 | 0.000301 | 1.40E-05 | 0.003017625 | RNF8 |
| chr13 | 101751717 | 101751725 | 9 | 2 | 1.12E-05 | 0.0009696 | 9.34E-06 | 1.40E-05 | 0.001972007 | FGF14 |
| chr21 | 31251585 | 31251990 | 406 | 5 | 3.13E-05 | 8.48E-06 | 0.004894 | 1.40E-05 | -0.00208903 | TIAM1 |
| chr11 | 93850343 | 93850807 | 465 | 6 | 9.78E-05 | 2.43E-05 | 0.000782 | 1.40E-05 | 0.009252844 | VSTM5 |
| chr10 | 113720501 | 113720945 | 445 | 6 | 4.91E-06 | 9.21E-05 | 0.001029 | 1.41E-05 | 0.002615878 | CASP7 |
| chr1 | 240697796 | 240698271 | 476 | 3 | 1.88E-05 | 5.23E-06 | 0.003648 | 1.41E-05 | -0.01150617 | NA |
| chr8 | 6139254 | 6139790 | 537 | 2 | 8.83E-05 | 9.90E-06 | 0.000353 | 1.41E-05 | 0.004094399 | AC009435.1 |
| chr4 | 54447292 | 54447351 | 60 | 2 | 2.37E-05 | 9.53E-06 | 0.000415 | 1.41E-05 | -0.00620284 | NA |
| chr5 | 139701367 | 139701588 | 222 | 5 | 2.05E-05 | 7.35E-05 | 0.000998 | 1.41E-05 | 0.001622405 | AC008667.1 |
| chr11 | 47243431 | 47243552 | 122 | 2 | 2.24E-05 | 1.04E-05 | 0.000294 | 1.41E-05 | -0.00681108 | ACP2 |
| chr13 | 75226171 | 75226337 | 167 | 2 | 6.55E-06 | 5.43E-05 | 2.72E-05 | 1.41E-05 | -0.00552565 | NA |
| chr17 | 2485777 | 2486033 | 257 | 3 | 1.60E-05 | 2.55E-05 | 0.000108 | 1.41E-05 | -0.00595101 | METTL16 |
| chr7 | 128830163 | 128831546 | 1384 | 8 | 1.68E-07 | 0.0196278 | 0.000165 | 1.41E-05 | -0.00282214 | FLNC |
| chr9 | 93300072 | 93300331 | 260 | 3 | 1.31E-05 | 7.34E-05 | 5.93E-05 | 1.41E-05 | -0.00746251 | WNK2 |
| chr10 | 122190434 | 122190688 | 255 | 2 | 1.04E-05 | 4.80E-05 | 2.99E-05 | 1.41E-05 | 0.012894191 | TACC2 |
| chr17 | 980536 | 981248 | 713 | 3 | 2.70E-06 | 0.0002471 | 0.000468 | 1.41E-05 | 0.008069314 | NA |
| chr2 | 177137906 | 177138063 | 158 | 2 | 2.76E-05 | 9.74E-06 | 0.000388 | 1.41E-05 | 0.001990505 | AC074286.1, AC079305.2 |
| chr20 | 16864227 | 16864263 | 37 | 2 | 2.11E-05 | 2.14E-05 | 6.57E-05 | 1.41E-05 | -0.00351863 | NA |
| chr5 | 90967658 | 90967907 | 250 | 2 | 2.59E-05 | 1.50E-05 | 0.000113 | 1.41E-05 | 0.022339933 | ADGRV1 |
| chr2 | 128319340 | 128319830 | 491 | 3 | 4.17E-05 | 5.41E-06 | 0.00339 | 1.41E-05 | -0.01562232 | NA |
| chr12 | 48326841 | 48327221 | 381 | 2 | 1.87E-05 | 9.52E-06 | 0.000429 | 1.41E-05 | -0.00351195 | NA |
| chr8 | 21129290 | 21129642 | 353 | 2 | 1.61E-05 | 2.05E-05 | 6.95E-05 | 1.41E-05 | -0.01560294 | AC021613.1 |
| chr3 | 48232716 | 48232845 | 130 | 2 | 1.66E-05 | 3.27E-05 | 4.12E-05 | 1.41E-05 | -0.01534572 | NA |
| chr18 | 12423139 | 12423173 | 35 | 2 | 9.64E-06 | 4.20E-05 | 3.32E-05 | 1.41E-05 | 0.010970048 | PRELID3A |
| chr9 | 2024342 | 2024728 | 387 | 3 | 2.50E-05 | 8.21E-06 | 0.001388 | 1.41E-05 | 0.010896317 | SMARCA2 |
| chr17 | 79073987 | 79074711 | 725 | 4 | 2.03E-05 | 5.15E-05 | 0.000638 | 1.41E-05 | 0.002620029 | NA |
| chr6 | 15432923 | 15433342 | 420 | 3 | 1.35E-05 | 2.75E-05 | 0.000734 | 1.41E-05 | 0.004921584 | JARID2 |
| chr10 | 86668056 | 86668672 | 617 | 9 | 7.23E-06 | 3.13E-05 | 0.001253 | 1.41E-05 | -0.0068426 | LDB3 |
| chr1 | 19367290 | 19367794 | 505 | 3 | 6.75E-06 | 0.0002398 | 0.000405 | 1.42E-05 | -0.00271367 | CAPZB |
| chr3 | 94062809 | 94063336 | 528 | 12 | 1.87E-05 | 0.0001193 | 0.008841 | 1.42E-05 | -0.00074931 | NSUN3, DHFR2 |
| chr19 | 11580338 | 11580465 | 128 | 3 | 3.99E-06 | 0.000432 | 0.000335 | 1.42E-05 | 0.001050184 | ZNF627 |
| chr10 | 61780266 | 61780565 | 300 | 2 | 1.69E-05 | 2.45E-05 | 5.59E-05 | 1.42E-05 | 0.015449942 | NA |
| chr17 | 31574787 | 31575383 | 597 | 8 | 1.46E-06 | 0.0032247 | 5.36E-05 | 1.42E-05 | -0.00065343 | AC003101.1, MIR4725 |
| chr12 | 97757601 | 97757908 | 308 | 2 | 5.70E-06 | 4.63E-05 | 3.09E-05 | 1.42E-05 | -0.02267036 | PAFAH1B2P2 |
| chr19 | 2339164 | 2339510 | 347 | 2 | 2.00E-05 | 9.05E-06 | 0.00057 | 1.42E-05 | 0.000545572 | AC005258.1, SPPL2B |
| chr11 | 126268082 | 126269568 | 1487 | 20 | 8.20E-06 | 0.0003077 | 0.000639 | 1.42E-05 | 0.000656752 | FOXRED1, SRPRA |
| chr16 | 86078220 | 86078413 | 194 | 3 | 4.32E-05 | 7.63E-06 | 0.000825 | 1.42E-05 | -0.00732419 | NA |
| chr9 | 82496460 | 82496491 | 32 | 2 | 3.79E-05 | 1.01E-05 | 0.00034 | 1.42E-05 | -0.02582443 | AL162726.3 |
| chr9 | 36868135 | 36868364 | 230 | 2 | 1.94E-05 | 2.55E-05 | 5.36E-05 | 1.42E-05 | -0.01422597 | PAX5 |
| chr12 | 43154542 | 43155139 | 598 | 3 | 2.51E-05 | 9.66E-06 | 0.000601 | 1.42E-05 | -0.01553447 | NA |
| chr16 | 75148137 | 75148293 | 157 | 3 | 3.76E-05 | 0.0002353 | 1.28E-05 | 1.42E-05 | -0.00154585 | NA |
| chr16 | 88906058 | 88906457 | 400 | 4 | 3.43E-06 | 2.48E-05 | 0.000151 | 1.42E-05 | -0.00402169 | CBFA2T3 |
| chr19 | 40512537 | 40513378 | 842 | 6 | 1.06E-05 | 7.16E-05 | 5.24E-05 | 1.42E-05 | -0.00116294 | SPTBN4 |
| chr11 | 58404235 | 58404621 | 387 | 2 | 9.34E-06 | 1.53E-05 | 0.000109 | 1.42E-05 | -0.01198551 | OR5B3 |
| chr12 | 56599171 | 56599760 | 590 | 2 | 4.82E-05 | 8.86E-06 | 0.000649 | 1.42E-05 | -0.00382792 | BAZ2A |
| chr8 | 122828433 | 122828505 | 73 | 2 | 2.18E-05 | 5.07E-05 | 2.89E-05 | 1.42E-05 | -0.00364156 | ZHX2 |
| chr13 | 109880980 | 109881477 | 498 | 3 | 7.45E-05 | 6.81E-06 | 0.001402 | 1.42E-05 | 0.007767399 | NA |
| chr10 | 69630120 | 69630629 | 510 | 8 | 2.64E-05 | 1.21E-05 | 0.001928 | 1.42E-05 | -0.00030508 | FAM241B |
| chr6 | 43021358 | 43021862 | 505 | 10 | 1.84E-05 | 0.0006485 | 0.006019 | 1.42E-05 | 0.000825615 | RRP36 |
| chr10 | 1380238 | 1380670 | 433 | 3 | 7.76E-06 | 1.48E-05 | 0.000168 | 1.42E-05 | -0.00373759 | ADARB2 |
| chr11 | 624603 | 625274 | 672 | 8 | 2.29E-05 | 0.0001345 | 0.003275 | 1.42E-05 | -0.00244376 | CDHR5 |
| chr7 | 46754773 | 46755280 | 508 | 2 | 6.21E-06 | 3.05E-05 | 4.46E-05 | 1.42E-05 | 0.021999322 | AC011294.1 |
| chr9 | 113401322 | 113401465 | 144 | 4 | 0.000113559 | 0.0001372 | 0.00046 | 1.42E-05 | -0.00256941 | NA |
| chr22 | 30830391 | 30830548 | 158 | 2 | 0.000106692 | 1.15E-05 | 0.00022 | 1.42E-05 | -0.00656252 | OSBP2 |
| chr17 | 42166285 | 42166472 | 188 | 5 | 1.76E-05 | 0.0005875 | 0.000147 | 1.42E-05 | -0.00052834 | KCNH4 |
| chr17 | 39632825 | 39633789 | 965 | 4 | 1.19E-07 | 0.001284 | 8.53E-06 | 1.42E-05 | -0.00315848 | PPP1R1B |
| chr12 | 57831221 | 57831752 | 532 | 3 | 5.72E-06 | 2.09E-05 | 0.000719 | 1.43E-05 | -0.00553942 | CTDSP2 |
| chr14 | 24052634 | 24053145 | 512 | 3 | 6.65E-06 | 2.14E-05 | 0.000511 | 1.43E-05 | -0.00558033 | CARMIL3 |
| chr5 | 181187561 | 181188650 | 1090 | 7 | 1.11E-05 | 4.18E-05 | 0.003837 | 1.43E-05 | -0.00501994 | LINC01962 |
| chr1 | 239952972 | 239952976 | 5 | 2 | 3.11E-05 | 8.86E-06 | 0.00067 | 1.43E-05 | -0.00811762 | NA |
| chr11 | 82267561 | 82267571 | 11 | 2 | 1.17E-05 | 0.0008651 | 9.69E-06 | 1.43E-05 | 0.005885245 | MIR4300HG |
| chr18 | 74242650 | 74242990 | 341 | 2 | 2.10E-05 | 1.05E-05 | 0.000301 | 1.43E-05 | 0.007603178 | NA |
| chr18 | 10305101 | 10305215 | 115 | 2 | 3.19E-05 | 1.64E-05 | 9.86E-05 | 1.43E-05 | 0.008218236 | AP005209.2 |
| chr16 | 23394345 | 23395088 | 744 | 3 | 2.86E-05 | 6.92E-06 | 0.001257 | 1.43E-05 | 0.000619405 | COG7 |
| chr3 | 181699296 | 181699825 | 530 | 3 | 1.73E-05 | 6.39E-06 | 0.002098 | 1.43E-05 | -0.01586594 | SOX2-OT, AC117415.1, AC125613.1 |
| chr14 | 29382424 | 29382464 | 41 | 3 | 3.51E-06 | 0.0082284 | 3.90E-06 | 1.43E-05 | 0.01294472 | AL133166.1 |
| chr7 | 86945128 | 86945235 | 108 | 2 | 1.13E-05 | 0.0001455 | 1.62E-05 | 1.43E-05 | -0.0050373 | ELAPOR2 |
| chr3 | 141777161 | 141778479 | 1319 | 8 | 1.28E-05 | 1.50E-05 | 0.001456 | 1.43E-05 | -0.00197982 | AC112504.2, GRK7 |
| chr17 | 48872011 | 48872342 | 332 | 3 | 2.70E-05 | 7.65E-06 | 0.001066 | 1.43E-05 | 0.003236202 | NA |
| chr2 | 10329115 | 10329262 | 148 | 3 | 5.11E-06 | 0.0002638 | 0.000261 | 1.43E-05 | 0.002718316 | HPCAL1 |
| chr12 | 122174609 | 122174706 | 98 | 5 | 3.71E-05 | 0.0001225 | 0.000607 | 1.43E-05 | -0.00125404 | LRRC43 |
| chr18 | 35950034 | 35950597 | 564 | 5 | 5.77E-05 | 7.48E-06 | 0.002748 | 1.43E-05 | -0.00253168 | NA |
| chr2 | 235767323 | 235767428 | 106 | 3 | 2.49E-05 | 9.10E-06 | 0.000511 | 1.43E-05 | -0.00274602 | AGAP1 |
| chr19 | 2493742 | 2494597 | 856 | 5 | 7.67E-07 | 0.0003113 | 1.06E-05 | 1.43E-05 | -0.00716368 | NA |
| chr1 | 25872699 | 25873263 | 565 | 2 | 9.08E-07 | 3.07E-05 | 4.48E-05 | 1.43E-05 | -0.00648988 | PAQR7 |
| chr2 | 96694920 | 96695527 | 608 | 5 | 1.72E-06 | 0.0006855 | 1.64E-05 | 1.43E-05 | 0.002179055 | FER1L5 |
| chr5 | 123094271 | 123095126 | 856 | 9 | 3.69E-06 | 3.96E-05 | 0.000552 | 1.43E-05 | -0.00455643 | PRDM6 |
| chr17 | 44325514 | 44325871 | 358 | 4 | 3.67E-06 | 8.75E-05 | 3.74E-05 | 1.43E-05 | 0.001982924 | NA |
| chr14 | 24058260 | 24058899 | 640 | 3 | 3.24E-05 | 2.93E-05 | 0.000429 | 1.43E-05 | 0.02452253 | CARMIL3 |
| chr11 | 93523345 | 93523376 | 32 | 2 | 8.04E-06 | 2.67E-05 | 5.18E-05 | 1.43E-05 | 0.004685788 | SMCO4 |
| chr15 | 90474149 | 90474435 | 287 | 2 | 1.23E-05 | 1.10E-05 | 0.000263 | 1.43E-05 | 0.007326005 | IQGAP1 |
| chr1 | 23140428 | 23140875 | 448 | 2 | 3.51E-05 | 9.65E-06 | 0.000437 | 1.43E-05 | -0.00964481 | AL161672.1, LUZP1 |
| chr1 | 8427434 | 8427810 | 377 | 3 | 2.63E-06 | 0.0008699 | 7.00E-06 | 1.43E-05 | 0.012198026 | RERE-AS1, RERE |
| chr7 | 74093870 | 74094664 | 795 | 8 | 1.58E-05 | 0.0001931 | 0.001389 | 1.43E-05 | 0.003681825 | LIMK1 |
| chr19 | 11456094 | 11456654 | 561 | 2 | 1.76E-05 | 1.35E-05 | 0.000148 | 1.43E-05 | 0.00133414 | ELAVL3 |
| chr8 | 123348957 | 123349369 | 413 | 3 | 8.54E-06 | 1.77E-05 | 0.000127 | 1.44E-05 | -0.00207587 | ATAD2 |
| chr2 | 238786911 | 238787373 | 463 | 4 | 1.08E-05 | 0.0001268 | 2.25E-05 | 1.44E-05 | -0.00495976 | NA |
| chr19 | 13154409 | 13154524 | 116 | 2 | 7.16E-06 | 8.63E-06 | 0.000848 | 1.44E-05 | 0.005169864 | IER2 |
| chr1 | 229436381 | 229436959 | 579 | 2 | 3.80E-06 | 2.14E-05 | 6.75E-05 | 1.44E-05 | -0.00298034 | NA |
| chr11 | 63015651 | 63016372 | 722 | 9 | 1.03E-05 | 6.17E-05 | 0.000112 | 1.44E-05 | -0.00437923 | SLC22A8 |
| chr14 | 56797275 | 56797569 | 295 | 3 | 2.96E-05 | 1.02E-05 | 0.000492 | 1.44E-05 | -0.00017874 | NA |
| chr15 | 58623004 | 58623506 | 503 | 2 | 2.76E-05 | 9.59E-06 | 0.000457 | 1.44E-05 | 0.013097361 | ADAM10 |
| chr22 | 50532015 | 50532680 | 666 | 7 | 6.90E-06 | 2.22E-05 | 0.002359 | 1.44E-05 | 0.00234456 | ODF3B |
| chr21 | 46195841 | 46196406 | 566 | 3 | 3.14E-05 | 5.90E-06 | 0.002383 | 1.44E-05 | -0.00318318 | LSS |
| chr6 | 118388519 | 118388599 | 81 | 2 | 2.49E-05 | 3.75E-05 | 3.75E-05 | 1.44E-05 | -0.00835081 | NA |
| chr19 | 5031240 | 5031749 | 510 | 2 | 4.16E-06 | 1.52E-05 | 0.000116 | 1.44E-05 | 0.00277209 | KDM4B |
| chr3 | 12965363 | 12965687 | 325 | 2 | 7.76E-06 | 1.20E-05 | 0.000206 | 1.45E-05 | -0.00391364 | IQSEC1 |
| chr3 | 177815638 | 177815697 | 60 | 2 | 2.21E-05 | 8.97E-06 | 0.000683 | 1.45E-05 | -0.0061484 | NA |
| chr22 | 37386741 | 37387066 | 326 | 4 | 2.85E-05 | 3.02E-05 | 0.002088 | 1.45E-05 | 0.004200214 | ELFN2, ELFN2 |
| chr13 | 31044558 | 31044606 | 49 | 2 | 4.09E-05 | 3.60E-05 | 3.90E-05 | 1.45E-05 | -0.00323961 | NA |
| chr12 | 132268750 | 132269439 | 690 | 5 | 1.13E-05 | 6.69E-05 | 0.000183 | 1.45E-05 | -0.00706309 | GALNT9 |
| chr22 | 22022408 | 22022857 | 450 | 3 | 3.70E-06 | 0.001784 | 6.89E-05 | 1.45E-05 | 0.014246268 | PRAMENP |
| chr14 | 64879772 | 64880157 | 386 | 6 | 2.22E-05 | 3.27E-05 | 0.002367 | 1.45E-05 | 0.002143382 | SPTB |
| chr6 | 2999677 | 3000196 | 520 | 9 | 1.72E-05 | 6.23E-05 | 0.001717 | 1.45E-05 | -9.05E-05 | NQO2 |
| chr4 | 1027155 | 1027503 | 349 | 4 | 1.63E-05 | 2.62E-05 | 0.000134 | 1.45E-05 | -0.00313676 | NA |
| chr16 | 1475168 | 1475697 | 530 | 9 | 3.68E-07 | 0.0079863 | 4.64E-06 | 1.45E-05 | 0.003771168 | NA |
| chr6 | 148964588 | 148965239 | 652 | 6 | 3.35E-05 | 0.0001237 | 0.000695 | 1.45E-05 | -0.00202829 | UST, UST-AS1 |
| chr7 | 98107541 | 98108239 | 699 | 3 | 1.10E-07 | 0.0011095 | 1.20E-05 | 1.45E-05 | -0.01046928 | LMTK2 |
| chr2 | 100106004 | 100106967 | 964 | 8 | 8.98E-06 | 1.75E-05 | 0.002206 | 1.45E-05 | -0.00237368 | AC092667.1, AFF3 |
| chr6 | 137218243 | 137218700 | 458 | 4 | 6.92E-06 | 7.68E-06 | 0.001324 | 1.46E-05 | 0.000482435 | IFNGR1 |
| chr8 | 52941865 | 52941935 | 71 | 2 | 1.37E-05 | 1.41E-05 | 0.000139 | 1.46E-05 | -0.00078949 | NPBWR1 |
| chr19 | 8148699 | 8149548 | 850 | 4 | 1.07E-06 | 0.0002203 | 3.34E-05 | 1.46E-05 | 0.002346561 | FBN3 |
| chr12 | 53251418 | 53252409 | 992 | 12 | 1.86E-05 | 5.90E-05 | 0.000198 | 1.46E-05 | 0.001264011 | MFSD5 |
| chr19 | 13008509 | 13008516 | 8 | 2 | 3.61E-05 | 9.79E-06 | 0.000445 | 1.46E-05 | -0.00437101 | NFIX |
| chr22 | 50015008 | 50015707 | 700 | 3 | 3.98E-05 | 3.33E-05 | 0.000483 | 1.46E-05 | -0.0080057 | TTLL8 |
| chr8 | 81193767 | 81194220 | 454 | 3 | 3.83E-05 | 0.000165 | 0.000494 | 1.46E-05 | 0.003823632 | NA |
| chr14 | 88189641 | 88189784 | 144 | 2 | 3.08E-05 | 1.21E-05 | 0.000209 | 1.46E-05 | -0.01339885 | KCNK10 |
| chr1 | 222622819 | 222622886 | 68 | 2 | 3.50E-05 | 1.50E-05 | 0.000123 | 1.46E-05 | -0.00535967 | MIA3 |
| chr22 | 38820104 | 38820244 | 141 | 2 | 2.15E-05 | 1.04E-05 | 0.000345 | 1.46E-05 | -0.01291111 | NPTXR |
| chr20 | 4721612 | 4721935 | 324 | 5 | 1.33E-05 | 0.0003558 | 0.000553 | 1.46E-05 | -0.00198958 | PRND |
| chr9 | 5465479 | 5465685 | 207 | 3 | 1.67E-05 | 3.07E-05 | 0.000279 | 1.46E-05 | -0.00182657 | CD274, AL162253.2 |
| chr2 | 85583621 | 85584348 | 728 | 10 | 7.18E-06 | 3.48E-05 | 0.001135 | 1.46E-05 | -0.00709916 | NA |
| chr3 | 125445610 | 125445952 | 343 | 4 | 2.47E-05 | 6.85E-05 | 0.000341 | 1.46E-05 | -0.00298684 | NA |
| chr22 | 48731075 | 48731431 | 357 | 2 | 2.10E-05 | 1.58E-05 | 0.000111 | 1.46E-05 | 0.001409472 | TAFA5 |
| chr19 | 41327198 | 41327318 | 121 | 2 | 3.85E-06 | 9.53E-05 | 2.03E-05 | 1.47E-05 | 0.000681723 | TGFB1 |
| chr3 | 2865972 | 2866805 | 834 | 4 | 7.65E-06 | 7.56E-05 | 0.000254 | 1.47E-05 | 0.005272746 | CNTN4 |
| chr16 | 87085282 | 87085646 | 365 | 2 | 3.67E-05 | 9.39E-06 | 0.000567 | 1.47E-05 | -0.00239977 | C16orf95 |
| chr2 | 29580561 | 29580757 | 197 | 2 | 2.41E-05 | 1.07E-05 | 0.00032 | 1.47E-05 | -0.0018119 | ALK |
| chr7 | 150978428 | 150979441 | 1014 | 8 | 7.92E-05 | 0.0003545 | 0.005543 | 1.47E-05 | 0.001207994 | NA |
| chr16 | 4972642 | 4972747 | 106 | 2 | 2.37E-05 | 4.14E-05 | 3.54E-05 | 1.47E-05 | -0.01491811 | SEC14L5 |
| chr13 | 26987730 | 26987933 | 204 | 2 | 6.89E-05 | 1.47E-05 | 0.000131 | 1.47E-05 | -0.01584692 | AL160035.1 |
| chr19 | 19193933 | 19194218 | 286 | 3 | 3.93E-06 | 9.03E-05 | 6.89E-05 | 1.47E-05 | 0.006949029 | RFXANK |
| chr1 | 44218275 | 44218651 | 377 | 2 | 6.47E-07 | 0.0015792 | 8.99E-06 | 1.47E-05 | -0.00168303 | DMAP1 |
| chr5 | 150290192 | 150291262 | 1071 | 4 | 4.92E-06 | 0.0002813 | 0.000121 | 1.47E-05 | 0.004530099 | CAMK2A |
| chr10 | 84543208 | 84543396 | 189 | 2 | 2.16E-05 | 1.54E-05 | 0.000118 | 1.47E-05 | -0.01445331 | NA |
| chr16 | 67252556 | 67252771 | 216 | 2 | 3.62E-05 | 9.37E-06 | 0.000582 | 1.47E-05 | 0.003530689 | SLC9A5 |
| chr7 | 155816498 | 155816741 | 244 | 2 | 2.34E-05 | 1.02E-05 | 0.000392 | 1.47E-05 | -0.01454332 | NA |
| chr4 | 77193714 | 77194197 | 484 | 4 | 4.86E-05 | 5.62E-06 | 0.004631 | 1.47E-05 | 0.006329325 | CCNG2 |
| chr8 | 106858177 | 106858597 | 421 | 2 | 2.52E-05 | 1.18E-05 | 0.000232 | 1.47E-05 | 0.014969265 | NA |
| chr9 | 27297091 | 27297408 | 318 | 3 | 4.10E-05 | 1.14E-05 | 0.000409 | 1.47E-05 | -0.00574143 | EQTN |
| chr12 | 132560774 | 132561293 | 520 | 2 | 9.14E-07 | 4.67E-05 | 3.24E-05 | 1.47E-05 | 0.003251094 | FBRSL1 |
| chr8 | 54165993 | 54166216 | 224 | 2 | 1.95E-05 | 3.38E-05 | 4.25E-05 | 1.47E-05 | -0.00347287 | NA |
| chr19 | 908894 | 909533 | 640 | 4 | 5.92E-05 | 1.19E-05 | 0.002233 | 1.47E-05 | 0.005238361 | R3HDM4 |
| chr3 | 190586171 | 190586507 | 337 | 3 | 1.63E-06 | 0.0006492 | 5.86E-05 | 1.48E-05 | 0.008780428 | IL1RAP |
| chr3 | 8570928 | 8571702 | 775 | 4 | 2.09E-06 | 0.0028495 | 1.88E-05 | 1.48E-05 | -0.0032046 | LMCD1, LMCD1-AS1 |
| chr2 | 233712794 | 233712926 | 133 | 2 | 7.38E-06 | 2.00E-05 | 7.80E-05 | 1.48E-05 | -0.0079295 | UGT1A8, UGT1A10, UGT1A9, UGT1A7, UGT1A6 |
| chr12 | 48975837 | 48976446 | 610 | 2 | 3.26E-05 | 9.65E-06 | 0.000508 | 1.48E-05 | -0.01226849 | NA |
| chr5 | 178153210 | 178154071 | 862 | 10 | 7.36E-05 | 2.47E-05 | 0.006392 | 1.48E-05 | -0.00192105 | NHP2 |
| chr3 | 186611753 | 186612278 | 526 | 5 | 2.42E-06 | 0.00121 | 0.000228 | 1.48E-05 | -0.00315306 | AC068631.1 |
| chr3 | 39013503 | 39013973 | 471 | 2 | 1.37E-05 | 1.92E-05 | 8.27E-05 | 1.48E-05 | -0.01329973 | SCN11A |
| chr4 | 76589850 | 76590344 | 495 | 2 | 4.86E-05 | 8.85E-06 | 0.000868 | 1.48E-05 | -0.01158341 | SHROOM3 |
| chr12 | 2874260 | 2874489 | 230 | 3 | 1.19E-05 | 7.83E-05 | 3.11E-05 | 1.48E-05 | -0.00567709 | FOXM1 |
| chr7 | 2200168 | 2200741 | 574 | 4 | 1.51E-05 | 0.0017673 | 0.000159 | 1.48E-05 | 0.001907145 | MAD1L1, AC069288.1 |
| chr22 | 42609761 | 42609909 | 149 | 2 | 8.17E-06 | 9.35E-06 | 0.00061 | 1.48E-05 | 0.007707347 | POLDIP3 |
| chr6 | 8707600 | 8708118 | 519 | 3 | 3.78E-05 | 9.25E-06 | 0.000569 | 1.48E-05 | 0.008424694 | AL355499.2, HULC |

| | | | | | | | | | | |
|---|---|---|---|---|---|---|---|---|---|---|
| chr16 | 84082363 | 84082557 | 195 | 2 | 1.93E-05 | 9.63E-06 | 0.000517 | 1.48E-05 | -0.01065697 | MBTPS1 |
| chr3 | 182364462 | 182365121 | 660 | 6 | 5.28E-05 | 3.18E-05 | 0.001703 | 1.48E-05 | -0.00800764 | NA |
| chr17 | 42009397 | 42009543 | 147 | 2 | 2.96E-05 | 9.11E-06 | 0.000721 | 1.48E-05 | 0.010489794 | DNAJC7 |
| chr13 | 112879037 | 112880201 | 1165 | 6 | 1.06E-07 | 0.000303 | 2.16E-05 | 1.48E-05 | -0.0029433 | ATP11A |
| chr11 | 44859393 | 44859893 | 501 | 3 | 7.80E-05 | 1.13E-05 | 0.000916 | 1.48E-05 | -0.00409425 | TSPAN18 |
| chr11 | 126405733 | 126406697 | 965 | 6 | 3.43E-06 | 0.0004806 | 0.000103 | 1.48E-05 | -0.00675347 | ST3GAL4 |
| chr22 | 37376053 | 37376441 | 389 | 3 | 1.60E-06 | 9.22E-05 | 0.000126 | 1.48E-05 | 0.000152484 | ELFN2, ELFN2 |
| chr1 | 9238382 | 9238896 | 515 | 4 | 5.18E-05 | 7.39E-06 | 0.002998 | 1.48E-05 | -0.00754167 | H6PD |
| chr5 | 150161733 | 150161999 | 267 | 2 | 9.59E-06 | 1.96E-05 | 8.11E-05 | 1.48E-05 | -0.00932374 | NA |
| chr10 | 825286 | 825597 | 312 | 2 | 1.03E-05 | 6.83E-05 | 2.50E-05 | 1.48E-05 | -0.00326755 | LARP4B |
| chr9 | 38141588 | 38142078 | 491 | 2 | 4.39E-05 | 9.48E-06 | 0.000579 | 1.48E-05 | 0.001688453 | NA |
| chr6 | 113611447 | 113611700 | 254 | 2 | 4.77E-06 | 0.0050486 | 8.00E-06 | 1.49E-05 | 0.008858082 | NA |
| chr8 | 125696933 | 125697395 | 463 | 2 | 7.39E-06 | 3.34E-05 | 4.37E-05 | 1.49E-05 | -0.0131663 | AC016074.2 |
| chr8 | 126301143 | 126301165 | 23 | 2 | 1.48E-05 | 3.35E-05 | 4.35E-05 | 1.49E-05 | -0.01308272 | NA |
| chr17 | 7990026 | 7990549 | 524 | 6 | 7.31E-06 | 0.0002848 | 2.69E-05 | 1.49E-05 | -0.00397324 | NA |
| chr12 | 110718704 | 110719180 | 477 | 3 | 5.01E-06 | 0.0013991 | 0.000305 | 1.49E-05 | -0.00043737 | NA |
| chr2 | 115163036 | 115163682 | 647 | 4 | 5.80E-06 | 1.47E-05 | 0.00086 | 1.49E-05 | 0.006538958 | DPP10 |
| chr10 | 116255818 | 116256049 | 232 | 3 | 4.01E-05 | 6.23E-06 | 0.002475 | 1.49E-05 | -0.01553306 | GFRA1 |
| chr8 | 78798249 | 78798608 | 360 | 4 | 2.61E-05 | 2.02E-05 | 0.000281 | 1.49E-05 | -0.00617085 | IL7 |
| chr15 | 70802349 | 70802946 | 598 | 5 | 8.32E-06 | 0.0003426 | 0.001464 | 1.49E-05 | -0.0020032 | RPL29P30 |
| chr17 | 8249126 | 8249965 | 840 | 9 | 2.95E-05 | 0.0043407 | 0.000522 | 1.49E-05 | -0.00203675 | PFAS |
| chr3 | 30518456 | 30518815 | 360 | 2 | 2.22E-05 | 1.31E-05 | 0.000177 | 1.49E-05 | -0.00121743 | NA |
| chr16 | 54075973 | 54075979 | 7 | 2 | 3.45E-05 | 1.18E-05 | 0.000241 | 1.49E-05 | -0.01534049 | FTO |
| chr15 | 78148895 | 78149818 | 924 | 10 | 4.34E-06 | 0.0002949 | 0.001356 | 1.49E-05 | -0.00105831 | IDH3A |
| chr17 | 75703291 | 75703317 | 27 | 2 | 3.11E-05 | 9.92E-06 | 0.000468 | 1.49E-05 | 0.000535943 | SAP30BP |
| chr17 | 63989212 | 63989461 | 250 | 2 | 2.30E-06 | 0.0001479 | 1.69E-05 | 1.49E-05 | -0.00395719 | AC127029.2 |
| chr6 | 44298526 | 44298735 | 210 | 3 | 1.91E-05 | 7.42E-06 | 0.001222 | 1.49E-05 | -0.0027867 | TMEM151B, AL353588.1, AARS2 |
| chr7 | 24572799 | 24573016 | 218 | 3 | 1.04E-05 | 6.28E-06 | 0.002467 | 1.49E-05 | -0.00347894 | NA |
| chr3 | 96776530 | 96777291 | 762 | 5 | 2.56E-05 | 1.78E-05 | 0.000992 | 1.49E-05 | -0.00978409 | CDV3P1 |
| chr9 | 137582638 | 137583205 | 568 | 4 | 2.24E-06 | 0.0016745 | 3.89E-06 | 1.49E-05 | -0.00189349 | ZMYND19 |
| chr3 | 4806596 | 4806669 | 74 | 2 | 4.51E-05 | 9.13E-06 | 0.000767 | 1.49E-05 | 0.004737529 | ITPR1 |
| chr12 | 62346042 | 62346048 | 7 | 2 | 3.10E-05 | 2.65E-05 | 5.62E-05 | 1.49E-05 | 0.001406798 | USP15 |
| chr12 | 31659547 | 31659860 | 314 | 7 | 1.60E-05 | 0.000676 | 0.001467 | 1.50E-05 | -0.00502796 | ETFBKMT |
| chr11 | 67642682 | 67643183 | 502 | 2 | 1.74E-05 | 1.19E-05 | 0.000239 | 1.50E-05 | -0.00367989 | ACY3 |
| chr20 | 290259 | 291031 | 773 | 11 | 4.93E-05 | 3.82E-05 | 0.00927 | 1.50E-05 | -0.00036942 | C20orf96 |
| chr2 | 176085545 | 176086115 | 571 | 4 | 1.33E-06 | 0.0001857 | 7.24E-05 | 1.50E-05 | 0.000950907 | NA |
| chr20 | 61519737 | 61519799 | 63 | 2 | 1.35E-05 | 0.0004336 | 1.19E-05 | 1.50E-05 | -0.00979559 | CDH4 |
| chr12 | 12725277 | 12725953 | 677 | 10 | 7.61E-06 | 0.0001093 | 4.25E-05 | 1.50E-05 | 0.000263974 | APOLD1 |
| chr16 | 48622670 | 48623353 | 684 | 5 | 4.04E-06 | 2.64E-05 | 0.001104 | 1.50E-05 | -0.00847236 | NA |
| chr17 | 35778395 | 35778902 | 508 | 3 | 3.37E-05 | 0.0004736 | 0.000167 | 1.50E-05 | -0.00523086 | TAF15, MMP28 |
| chr2 | 235614703 | 235614718 | 16 | 2 | 3.75E-05 | 1.04E-05 | 0.000383 | 1.50E-05 | -0.01044221 | AGAP1 |
| chr17 | 70129221 | 70129553 | 333 | 2 | 3.14E-06 | 1.67E-05 | 0.000106 | 1.50E-05 | -0.00699361 | KCNJ16 |
| chr14 | 105450788 | 105450876 | 89 | 3 | 3.83E-05 | 8.30E-06 | 0.001754 | 1.50E-05 | -0.00305143 | MTA1 |
| chr19 | 34428506 | 34429186 | 681 | 6 | 2.13E-05 | 6.17E-05 | 0.000301 | 1.50E-05 | 0.003405785 | UBA2 |
| chr9 | 130449801 | 130449811 | 11 | 2 | 1.05E-05 | 3.32E-05 | 4.46E-05 | 1.50E-05 | -0.00194508 | ASS1 |
| chr19 | 40715194 | 40715613 | 420 | 5 | 6.27E-08 | 0.0005276 | 8.08E-06 | 1.50E-05 | -0.00189077 | COQ8B |
| chr16 | 88406694 | 88407305 | 612 | 3 | 7.49E-06 | 7.14E-06 | 0.001328 | 1.50E-05 | -0.00233441 | ZNF469 |
| chr13 | 100510694 | 100510945 | 252 | 2 | 4.09E-05 | 9.35E-06 | 0.000671 | 1.50E-05 | -0.00623161 | PCCA |
| chr3 | 187291787 | 187291914 | 128 | 2 | 5.54E-06 | 6.41E-05 | 2.64E-05 | 1.50E-05 | -0.01193048 | AC007920.2, MASP1 |
| chr2 | 88172833 | 88173029 | 197 | 2 | 2.25E-05 | 0.0001045 | 1.99E-05 | 1.50E-05 | -0.00482569 | THNSL2 |
| chr15 | 70323460 | 70323470 | 11 | 2 | 2.14E-06 | 0.000463 | 1.18E-05 | 1.50E-05 | 0.001933212 | AC048383.1 |
| chr4 | 173399227 | 173399621 | 395 | 2 | 1.19E-06 | 0.0038645 | 8.27E-06 | 1.50E-05 | 0.002826927 | AC093849.4, SCRG1 |
| chr16 | 31442085 | 31442270 | 186 | 3 | 1.04E-05 | 8.65E-06 | 0.001664 | 1.50E-05 | -0.01789398 | ZNF843 |
| chr2 | 180521124 | 180521454 | 331 | 3 | 1.62E-05 | 6.41E-05 | 3.36E-05 | 1.50E-05 | 0.007045041 | NA |
| chr16 | 80628469 | 80629107 | 639 | 3 | 2.64E-06 | 1.56E-05 | 0.00035 | 1.50E-05 | -0.00810963 | CDYL2 |
| chr15 | 91947376 | 91948348 | 973 | 4 | 8.48E-07 | 0.0001246 | 6.14E-05 | 1.50E-05 | -0.00597806 | SLCO3A1 |
| chr11 | 2805098 | 2805758 | 661 | 4 | 3.39E-05 | 4.13E-06 | 0.006036 | 1.50E-05 | -0.00476923 | KCNQ1 |
| chr2 | 222859979 | 222860331 | 353 | 4 | 4.96E-06 | 7.95E-05 | 0.000182 | 1.50E-05 | -0.00899326 | NA |
| chr1 | 150250171 | 150250286 | 116 | 2 | 1.73E-05 | 1.71E-05 | 0.000103 | 1.50E-05 | -0.01051421 | NA |
| chr14 | 31106760 | 31107000 | 241 | 3 | 3.69E-05 | 9.81E-06 | 0.000974 | 1.50E-05 | -0.0010516 | AP4S1, HECTD1 |
| chr19 | 35946870 | 35947056 | 187 | 2 | 2.05E-05 | 1.32E-05 | 0.000182 | 1.50E-05 | -0.01203339 | NA |
| chr6 | 41286019 | 41286481 | 463 | 2 | 1.07E-06 | 1.97E-05 | 8.30E-05 | 1.50E-05 | -0.0062601 | TREM1 |
| chr13 | 20999320 | 20999531 | 212 | 3 | 6.01E-05 | 1.26E-05 | 0.000625 | 1.51E-05 | -0.00966525 | LATS2 |
| chr13 | 34943017 | 34943497 | 481 | 4 | 3.13E-05 | 2.89E-05 | 0.000436 | 1.51E-05 | -0.00481328 | NBEA |
| chr8 | 69073793 | 69074186 | 394 | 2 | 2.12E-05 | 1.53E-05 | 0.000128 | 1.51E-05 | 0.00331455 | LINC01592 |
| chr9 | 68779886 | 68780251 | 366 | 6 | 3.14E-05 | 2.30E-05 | 0.002585 | 1.51E-05 | -9.60E-05 | PIP5K1B, FAM122A |
| chr17 | 68955470 | 68955762 | 293 | 3 | 1.71E-05 | 0.0001602 | 0.000448 | 1.51E-05 | 0.001538156 | ABCA9-AS1 |
| chr5 | 92146865 | 92147423 | 559 | 3 | 1.77E-05 | 2.12E-05 | 0.000125 | 1.51E-05 | -0.00030552 | AC114316.1 |
| chr1 | 29253743 | 29254171 | 429 | 4 | 2.78E-05 | 1.67E-05 | 0.000546 | 1.51E-05 | -0.00368307 | PTPRU |
| chr1 | 211680931 | 211681801 | 871 | 6 | 4.00E-06 | 0.0004781 | 4.49E-05 | 1.51E-05 | 0.008664231 | AC096637.2 |
| chr4 | 28752099 | 28752689 | 591 | 3 | 8.81E-06 | 4.84E-05 | 4.10E-05 | 1.51E-05 | -0.00233347 | NA |
| chr1 | 25126832 | 25126934 | 103 | 2 | 2.81E-05 | 2.59E-05 | 5.88E-05 | 1.51E-05 | -0.00491811 | NA |
| chr12 | 90623142 | 90623146 | 5 | 2 | 1.32E-05 | 2.17E-05 | 7.29E-05 | 1.51E-05 | -0.01125943 | LINC02822, AC112481.2 |
| chr12 | 2284505 | 2285017 | 513 | 4 | 9.72E-06 | 0.0002123 | 0.000109 | 1.51E-05 | -0.00595062 | CACNA1C, CACNA1C-IT3 |
| chr11 | 102613016 | 102613079 | 64 | 2 | 1.60E-05 | 3.93E-05 | 3.87E-05 | 1.51E-05 | 0.000360424 | AP000851.1, MMP20 |
| chr14 | 94963118 | 94963597 | 480 | 4 | 2.17E-05 | 4.17E-05 | 0.000375 | 1.51E-05 | 0.002542839 | NA |
| chr2 | 6770262 | 6771010 | 749 | 6 | 5.31E-07 | 0.0030084 | 5.01E-06 | 1.51E-05 | -0.00607286 | LINC00487 |
| chr5 | 81232611 | 81234124 | 1514 | 13 | 6.11E-05 | 8.18E-06 | 0.010981 | 1.51E-05 | 0.002971769 | CKMT2, CKMT2-AS1 |
| chr12 | 21936626 | 21936867 | 242 | 2 | 2.01E-05 | 1.02E-05 | 0.000453 | 1.51E-05 | -0.01434223 | ABCC9 |
| chr1 | 115668957 | 115669018 | 62 | 2 | 1.52E-05 | 0.0002597 | 1.40E-05 | 1.51E-05 | 0.010255055 | VANGL1 |
| chr19 | 2905985 | 2905987 | 3 | 2 | 2.22E-05 | 5.48E-05 | 2.99E-05 | 1.52E-05 | 0.023868479 | ZNF57 |
| chr11 | 124441235 | 124441654 | 420 | 7 | 3.98E-06 | 0.0035413 | 2.80E-05 | 1.52E-05 | -0.00124607 | OR8B8 |
| chr11 | 118339037 | 118339302 | 266 | 2 | 8.93E-06 | 7.00E-05 | 2.53E-05 | 1.52E-05 | -0.01112894 | CD3D |
| chr17 | 82601183 | 82602138 | 956 | 4 | 4.45E-06 | 2.12E-05 | 0.000408 | 1.52E-05 | 0.000999785 | FOXK2 |
| chr17 | 57068460 | 57068598 | 139 | 2 | 1.52E-05 | 0.0005235 | 1.16E-05 | 1.52E-05 | -0.00836424 | NA |
| chr1 | 52033805 | 52034279 | 475 | 6 | 2.80E-06 | 0.0006354 | 9.14E-05 | 1.52E-05 | -0.00337305 | AL445685.1, TXNDC12, KTI12 |
| chr2 | 11775585 | 11776134 | 550 | 5 | 9.55E-06 | 0.0001675 | 0.000101 | 1.52E-05 | -0.00124517 | LPIN1 |
| chr1 | 10682995 | 10683737 | 743 | 3 | 2.14E-05 | 0.017298 | 9.86E-05 | 1.52E-05 | -0.0004325 | CASZ1 |
| chr7 | 151009040 | 151009267 | 228 | 3 | 1.73E-05 | 3.02E-05 | 0.000253 | 1.52E-05 | -0.01509963 | NOS3 |
| chr14 | 77273536 | 77274018 | 483 | 3 | 1.96E-05 | 6.06E-05 | 0.000227 | 1.52E-05 | -0.00307231 | NA |
| chr5 | 883445 | 883918 | 474 | 7 | 6.29E-06 | 0.0011942 | 0.000139 | 1.52E-05 | -0.00042419 | BRD9 |
| chr7 | 114921438 | 114922792 | 1355 | 11 | 2.27E-05 | 1.22E-05 | 0.003593 | 1.52E-05 | -0.00467898 | MDFIC |
| chr7 | 92134536 | 92135025 | 490 | 13 | 2.81E-05 | 2.60E-05 | 0.000505 | 1.52E-05 | -0.00012419 | CYP51A1-AS1, CYP51A1, AC000120.2, AC000120.3 |
| chr12 | 130161477 | 130162668 | 1192 | 9 | 1.11E-05 | 0.0006769 | 0.001205 | 1.52E-05 | 0.001430428 | FZD10, FZD10-AS1 |
| chr3 | 128354804 | 128355023 | 220 | 2 | 1.27E-05 | 1.54E-05 | 0.00013 | 1.52E-05 | -0.00720936 | EEFSEC |
| chr3 | 8575379 | 8575566 | 188 | 2 | 3.08E-05 | 1.73E-05 | 0.000104 | 1.52E-05 | -0.00324114 | LMCD1-AS1, AC034187.1 |
| chr7 | 31052974 | 31053561 | 588 | 4 | 7.86E-06 | 3.52E-05 | 8.06E-05 | 1.52E-05 | 8.51E-05 | ADCYAP1R1 |
| chr7 | 107953818 | 107954036 | 219 | 2 | 3.59E-05 | 1.04E-05 | 0.000418 | 1.53E-05 | 0.008676718 | LAMB1 |
| chr12 | 109273336 | 109273488 | 153 | 2 | 3.16E-05 | 1.88E-05 | 9.15E-05 | 1.53E-05 | -0.01867194 | NA |
| chr14 | 22982118 | 22982902 | 785 | 8 | 4.92E-05 | 3.08E-06 | 0.014613 | 1.53E-05 | 0.000340492 | AL132780.2, AL132780.3, AJUBA |
| chr16 | 56589039 | 56589637 | 599 | 11 | 1.18E-06 | 0.0001448 | 0.001015 | 1.53E-05 | -0.00187827 | MT3 |
| chr1 | 227779259 | 227779484 | 226 | 2 | 3.70E-06 | 2.13E-05 | 7.65E-05 | 1.53E-05 | -0.00646056 | SNAP47 |
| chr16 | 87183958 | 87184034 | 77 | 2 | 9.33E-06 | 0.0022051 | 8.94E-06 | 1.53E-05 | -0.00278053 | C16orf95 |
| chr15 | 89689552 | 89689856 | 305 | 2 | 5.63E-07 | 0.0001187 | 1.91E-05 | 1.53E-05 | -0.00411937 | PEX11A |
| chr12 | 111402988 | 111403425 | 438 | 5 | 4.83E-05 | 8.00E-06 | 0.004555 | 1.53E-05 | -0.00440031 | LINC02356 |
| chr9 | 97982823 | 97983693 | 871 | 7 | 6.98E-06 | 5.94E-05 | 0.000768 | 1.53E-05 | 0.000104528 | ANP32B |
| chr8 | 143018883 | 143019155 | 273 | 4 | 2.60E-06 | 0.0006882 | 8.33E-06 | 1.53E-05 | 6.68E-05 | LY6E |
| chr1 | 10449625 | 10450339 | 715 | 10 | 5.63E-06 | 0.0001232 | 0.004202 | 1.53E-05 | -0.00242811 | CENPS-CORT, CORT |
| chr5 | 176798591 | 176798713 | 123 | 2 | 7.48E-05 | 1.07E-05 | 0.000387 | 1.53E-05 | -0.00375616 | NA |
| chr10 | 49818019 | 49818043 | 25 | 2 | 8.81E-06 | 0.005993 | 8.12E-06 | 1.53E-05 | -0.00777995 | NA |
| chr5 | 4116058 | 4116447 | 390 | 4 | 1.72E-05 | 0.0023736 | 0.000349 | 1.53E-05 | 0.000914955 | NA |
| chr6 | 166553640 | 166554150 | 511 | 4 | 2.02E-05 | 2.72E-05 | 0.000116 | 1.53E-05 | -0.00112037 | RPS6KA2 |
| chr5 | 93599420 | 93599603 | 184 | 3 | 1.07E-05 | 3.46E-05 | 0.000115 | 1.53E-05 | 0.00106946 | NA |
| chr3 | 138047419 | 138047885 | 467 | 2 | 1.25E-06 | 0.000909 | 1.03E-05 | 1.53E-05 | 0.006317871 | NA |
| chr10 | 119512180 | 119512372 | 193 | 4 | 2.24E-05 | 7.35E-05 | 6.97E-05 | 1.53E-05 | 0.004522567 | RGS10 |
| chr3 | 68931860 | 68932947 | 1088 | 13 | 3.55E-05 | 2.16E-05 | 0.005071 | 1.53E-05 | -0.00147919 | TAFA4 |
| chr2 | 152421410 | 152422289 | 880 | 4 | 7.74E-07 | 0.0003971 | 3.63E-05 | 1.53E-05 | 0.000904056 | FMNL2 |
| chr10 | 77866392 | 77867077 | 686 | 5 | 5.23E-06 | 0.0003333 | 2.56E-05 | 1.53E-05 | -0.00282841 | AL391421.1, DLG5 |
| chr1 | 160261584 | 160262226 | 643 | 6 | 9.09E-06 | 3.74E-06 | 0.010841 | 1.53E-05 | 0.002695877 | AL139011.1, DCAF8, AL139011.2 |
| chr2 | 218333573 | 218334214 | 642 | 3 | 4.27E-05 | 1.01E-05 | 0.001288 | 1.53E-05 | 0.014046954 | PNKD, CATIP-AS2 |
| chr4 | 147731283 | 147731698 | 416 | 4 | 1.79E-06 | 0.0002385 | 1.35E-05 | 1.54E-05 | -0.00394174 | NA |
| chr15 | 100671587 | 100671955 | 369 | 2 | 4.13E-05 | 9.84E-06 | 0.000584 | 1.54E-05 | -0.01244877 | NA |
| chr10 | 103570201 | 103570320 | 120 | 2 | 8.98E-06 | 8.19E-05 | 2.34E-05 | 1.54E-05 | -0.01178189 | NEURL1 |
| chr19 | 4493645 | 4494087 | 443 | 3 | 1.18E-05 | 4.82E-05 | 4.44E-05 | 1.54E-05 | 0.001190075 | HDGFL2 |
| chr20 | 34268941 | 34269421 | 481 | 5 | 6.05E-05 | 6.96E-06 | 0.007583 | 1.54E-05 | -0.01317082 | ASIP, AL035458.2 |
| chr19 | 1109155 | 1109253 | 99 | 2 | 5.08E-06 | 1.65E-05 | 0.000116 | 1.54E-05 | -0.00470348 | SBNO2 |
| chr17 | 17972427 | 17973521 | 1095 | 14 | 1.61E-06 | 0.0004445 | 0.002195 | 1.54E-05 | 9.55E-05 | DRC3 |
| chr4 | 111323570 | 111323974 | 405 | 2 | 3.65E-06 | 0.0003483 | 1.31E-05 | 1.54E-05 | 0.003095979 | NA |
| chr4 | 30795069 | 30795086 | 18 | 2 | 2.63E-05 | 9.37E-06 | 0.000802 | 1.54E-05 | -0.0125016 | PCDH7 |
| chr1 | 10824342 | 10824807 | 466 | 2 | 3.10E-05 | 1.01E-05 | 0.000527 | 1.54E-05 | -0.00176348 | NA |
| chr12 | 31303344 | 31303946 | 603 | 5 | 5.73E-05 | 7.00E-05 | 0.000572 | 1.54E-05 | -0.00396983 | SINHCAF |
| chr17 | 81475011 | 81475180 | 170 | 3 | 2.56E-06 | 0.0044414 | 5.32E-06 | 1.54E-05 | -0.00434477 | NA |
| chr7 | 74062007 | 74062345 | 339 | 3 | 1.12E-05 | 3.22E-05 | 6.66E-05 | 1.54E-05 | 0.009708023 | ELN, ELN-AS1 |
| chr17 | 7631260 | 7631619 | 360 | 3 | 1.87E-05 | 0.0001545 | 0.000237 | 1.54E-05 | -0.00582279 | SHBG |
| chr5 | 172462508 | 172462627 | 120 | 2 | 3.83E-05 | 1.39E-05 | 0.00017 | 1.54E-05 | -0.0051856 | NA |
| chr6 | 32113486 | 32113836 | 351 | 4 | 3.08E-05 | 0.0002205 | 0.000303 | 1.54E-05 | -0.00315247 | TNXB, AL662884.2 |
| chr15 | 75727238 | 75727652 | 415 | 3 | 3.08E-06 | 9.16E-05 | 0.000387 | 1.54E-05 | -0.00750816 | ODF3L1 |
| chr13 | 46146690 | 46146895 | 206 | 2 | 2.82E-05 | 1.22E-05 | 0.000246 | 1.54E-05 | -0.01261894 | CPB2-AS1, LCP1 |
| chr15 | 41011313 | 41011818 | 506 | 3 | 3.48E-05 | 9.67E-06 | 0.000552 | 1.54E-05 | 0.000137866 | AC020661.2, INO80 |
| chr6 | 30661387 | 30661839 | 453 | 3 | 6.99E-06 | 0.0001148 | 6.57E-05 | 1.54E-05 | -0.00317737 | DHX16 |
| chr22 | 47783898 | 47784162 | 265 | 2 | 7.12E-06 | 0.000524 | 1.17E-05 | 1.54E-05 | 0.011405763 | AL117329.1 |
| chr1 | 111190676 | 111191046 | 371 | 2 | 2.40E-05 | 1.19E-05 | 0.000267 | 1.54E-05 | -0.03899654 | DENND2D |
| chr4 | 4677901 | 4678382 | 482 | 2 | 6.41E-05 | 1.05E-05 | 0.000445 | 1.54E-05 | -0.01571013 | STX18-AS1 |
| chr22 | 38179196 | 38180080 | 885 | 3 | 1.21E-05 | 8.89E-06 | 0.000703 | 1.54E-05 | -0.00670624 | PLA2G6 |
| chr1 | 20652289 | 20652914 | 626 | 4 | 1.63E-05 | 0.000103 | 0.000118 | 1.54E-05 | -0.00417615 | DDOST |
| chr17 | 28882396 | 28882607 | 212 | 3 | 3.67E-05 | 1.37E-05 | 0.000368 | 1.54E-05 | -1.60E-05 | FLOT2 |
| chr11 | 13276486 | 13277136 | 651 | 6 | 1.11E-06 | 0.0020226 | 4.53E-06 | 1.54E-05 | 0.002462977 | ARNTL |
| chr17 | 1454940 | 1455430 | 491 | 4 | 4.99E-06 | 0.0013268 | 0.000102 | 1.55E-05 | -0.0082227 | CRK |
| chr13 | 72849951 | 72850587 | 637 | 3 | 2.00E-05 | 0.0001121 | 0.000378 | 1.55E-05 | -0.00580647 | PIBF1 |
| chr15 | 41511230 | 41511652 | 423 | 3 | 5.81E-05 | 8.11E-06 | 0.001922 | 1.55E-05 | -0.00989841 | LTK |
| chr11 | 61281758 | 61282185 | 428 | 5 | 1.94E-06 | 0.0054408 | 6.27E-05 | 1.55E-05 | -0.01145024 | VWCE |
| chr1 | 157155193 | 157155416 | 224 | 2 | 4.28E-06 | 0.0008661 | 1.05E-05 | 1.55E-05 | -0.0059667 | NA |
| chr12 | 111404969 | 111405781 | 813 | 8 | 1.67E-05 | 3.66E-05 | 0.003026 | 1.55E-05 | 0.001096347 | NA |
| chr7 | 92809943 | 92810448 | 506 | 2 | 5.68E-05 | 9.35E-06 | 0.000849 | 1.55E-05 | -0.00770148 | CDK6 |
| chr17 | 45740205 | 45740232 | 28 | 2 | 3.10E-05 | 3.08E-05 | 5.07E-05 | 1.55E-05 | 0.000791862 | LINC02210-CRHR1 |
| chr15 | 79896786 | 79897479 | 694 | 9 | 2.75E-05 | 2.26E-05 | 0.001777 | 1.55E-05 | 0.001406983 | MTHFS, ST20-MTHFS |
| chr7 | 157601645 | 157601969 | 325 | 4 | 1.65E-05 | 0.0001225 | 0.000308 | 1.55E-05 | -0.00210051 | PTPRN2 |
| chr1 | 190098730 | 190098962 | 233 | 4 | 1.41E-05 | 0.0004363 | 4.46E-05 | 1.55E-05 | -0.00182619 | BRINP3 |

| | | | | | | | | | | |
|---|---|---|---|---|---|---|---|---|---|---|
| chr11 | 2862299 | 2862908 | 610 | 6 | 4.94E-06 | 3.13E-05 | 0.000928 | 1.55E-05 | 0.000239346 | NA |
| chr2 | 70102499 | 70102516 | 18 | 2 | 5.87E-06 | 0.0003234 | 1.35E-05 | 1.55E-05 | -0.01060371 | PCBP1-AS1 |
| chr17 | 82936075 | 82936367 | 293 | 4 | 2.73E-05 | 9.75E-06 | 0.001747 | 1.55E-05 | 0.002528615 | TBCD |
| chr11 | 128765000 | 128765050 | 51 | 2 | 2.41E-05 | 8.70E-05 | 2.29E-05 | 1.55E-05 | 0.00561818 | FLI1 |
| chr17 | 6084520 | 6084929 | 410 | 3 | 9.00E-06 | 0.0001439 | 2.09E-05 | 1.55E-05 | 0.002081886 | WSCD1 |
| chr15 | 72113666 | 72113901 | 236 | 2 | 8.64E-05 | 1.19E-05 | 0.000279 | 1.55E-05 | -0.011517 | MYO9A |
| chr9 | 137778114 | 137778607 | 494 | 3 | 1.29E-06 | 0.000736 | 8.23E-06 | 1.55E-05 | 0.000527299 | EHMT1, AL590627.1 |
| chr7 | 133252248 | 133253228 | 981 | 12 | 1.31E-05 | 5.71E-05 | 0.002958 | 1.55E-05 | -0.00065865 | EXOC4 |
| chr1 | 235650709 | 235651119 | 411 | 10 | 9.87E-06 | 2.10E-05 | 0.007006 | 1.55E-05 | -0.00016224 | GNG4 |
| chr1 | 24582152 | 24582599 | 448 | 2 | 1.55E-05 | 2.01E-05 | 8.66E-05 | 1.55E-05 | -0.00631277 | NCMAP |
| chr2 | 2708897 | 2709137 | 241 | 2 | 2.10E-05 | 2.73E-05 | 5.84E-05 | 1.55E-05 | -0.00851089 | AC011995.2 |
| chr11 | 2655217 | 2655222 | 6 | 2 | 2.58E-05 | 1.72E-05 | 0.000111 | 1.55E-05 | -0.00323129 | KCNQ1, KCNQ1OT1 |
| chr19 | 30371143 | 30371269 | 127 | 2 | 5.99E-06 | 9.91E-06 | 0.000609 | 1.56E-05 | 0.008207682 | ZNF536 |
| chr21 | 46219724 | 46220237 | 514 | 4 | 2.59E-05 | 4.54E-05 | 0.000328 | 1.56E-05 | 0.005927809 | LSS |
| chr3 | 196338235 | 196338486 | 252 | 7 | 1.94E-05 | 0.0006197 | 9.30E-05 | 1.56E-05 | -0.00438864 | TM4SF19-TCTEX1D2, TM4SF19 |
| chr1 | 227541762 | 227541979 | 218 | 2 | 3.32E-05 | 9.37E-06 | 0.000882 | 1.56E-05 | -0.01209333 | NA |
| chr12 | 130972029 | 130972207 | 179 | 2 | 1.67E-06 | 0.0002259 | 1.52E-05 | 1.56E-05 | -0.009861 | ADGRD1 |
| chr8 | 30563877 | 30564310 | 434 | 2 | 5.36E-05 | 9.68E-06 | 0.000706 | 1.56E-05 | -6.38E-05 | RBPMS |
| chr9 | 110148491 | 110148518 | 28 | 2 | 8.35E-05 | 9.81E-06 | 0.000654 | 1.56E-05 | 0.022266824 | PALM2AKAP2 |
| chr20 | 56466001 | 56466489 | 489 | 3 | 3.29E-05 | 5.98E-06 | 0.003471 | 1.56E-05 | 0.015879969 | NA |
| chr19 | 36114790 | 36115493 | 704 | 9 | 3.18E-05 | 4.35E-06 | 0.010116 | 1.56E-05 | 0.000770058 | TBCB, AD001527.2, POLR2I |
| chr16 | 67438460 | 67438959 | 500 | 5 | 1.86E-05 | 7.77E-05 | 0.000875 | 1.56E-05 | 0.001411786 | ATP6V0D1 |
| chr16 | 1420502 | 1421085 | 584 | 9 | 6.64E-05 | 0.0001366 | 0.00751 | 1.56E-05 | 2.77E-05 | C16orf91 |
| chr11 | 103091405 | 103092486 | 1082 | 12 | 3.18E-06 | 0.0002168 | 0.000229 | 1.56E-05 | 0.000442596 | DCUN1D5 |
| chr6 | 32834821 | 32835723 | 903 | 12 | 2.29E-05 | 0.0001812 | 0.0041 | 1.56E-05 | 0.000192113 | AL669918.1, TAP2 |
| chr11 | 33742974 | 33743513 | 540 | 2 | 5.81E-05 | 9.70E-06 | 0.000712 | 1.56E-05 | -0.00166912 | FBXO3 |
| chr2 | 128472917 | 128473627 | 711 | 6 | 1.50E-05 | 3.68E-05 | 0.001209 | 1.56E-05 | -0.00077145 | NA |
| chr1 | 31666273 | 31666905 | 633 | 3 | 3.75E-05 | 0.0007014 | 0.000156 | 1.56E-05 | -0.00791982 | COL16A1 |
| chr3 | 127822329 | 127822686 | 358 | 6 | 4.26E-06 | 0.0029069 | 8.72E-06 | 1.57E-05 | -0.0027809 | MGLL |
| chr22 | 21769311 | 21769466 | 156 | 4 | 5.05E-05 | 1.23E-05 | 0.003741 | 1.57E-05 | -0.0097026 | MAPK1 |
| chr17 | 811493 | 811695 | 203 | 3 | 5.33E-05 | 6.96E-06 | 0.002341 | 1.57E-05 | 0.005616331 | NXN |
| chr4 | 51844439 | 51845105 | 667 | 5 | 6.46E-06 | 3.28E-05 | 0.000101 | 1.57E-05 | -0.00040148 | DCUN1D4 |
| chr2 | 235042079 | 235042128 | 50 | 2 | 1.84E-05 | 1.35E-05 | 0.000195 | 1.57E-05 | -0.00723021 | SH3BP4 |
| chr14 | 100149264 | 100149608 | 345 | 3 | 1.54E-05 | 8.45E-06 | 0.001452 | 1.57E-05 | -0.00279393 | DEGS2 |
| chr16 | 88766077 | 88766164 | 88 | 3 | 2.84E-05 | 3.00E-05 | 0.000129 | 1.57E-05 | -0.00645848 | PIEZO1 |
| chr6 | 73339802 | 73340241 | 440 | 2 | 1.83E-05 | 1.77E-05 | 0.000107 | 1.57E-05 | -0.00786642 | NA |
| chr1 | 243891380 | 243891906 | 527 | 2 | 1.43E-05 | 1.23E-05 | 0.000259 | 1.57E-05 | -0.01048829 | NA |
| chr3 | 101821649 | 101821729 | 81 | 3 | 4.23E-05 | 1.75E-05 | 0.00029 | 1.57E-05 | -0.00094651 | NXPE3 |
| chr2 | 159907953 | 159908407 | 455 | 2 | 2.25E-05 | 1.45E-05 | 0.000165 | 1.57E-05 | -0.00254847 | NA |
| chr9 | 91588505 | 91588658 | 154 | 2 | 1.20E-05 | 9.94E-06 | 0.000642 | 1.57E-05 | -0.00094599 | ROR2 |
| chr18 | 46343964 | 46344096 | 133 | 2 | 3.40E-05 | 1.89E-05 | 9.71E-05 | 1.57E-05 | -0.00331789 | RNF165 |
| chr5 | 155419348 | 155419461 | 114 | 2 | 2.03E-05 | 9.02E-05 | 2.29E-05 | 1.58E-05 | 0.010681154 | NA |
| chr16 | 88387171 | 88387531 | 361 | 5 | 6.84E-05 | 5.75E-06 | 0.004278 | 1.58E-05 | 0.008492936 | ZNF469 |
| chr5 | 109690012 | 109690340 | 329 | 3 | 3.34E-06 | 1.74E-05 | 0.000221 | 1.58E-05 | -0.00308082 | MAN2A1 |
| chr15 | 68229937 | 68231341 | 1405 | 7 | 1.17E-05 | 3.54E-05 | 0.006845 | 1.58E-05 | 0.003037272 | CLN6 |
| chr6 | 25987690 | 25988041 | 352 | 2 | 1.61E-05 | 2.76E-05 | 5.89E-05 | 1.58E-05 | 0.005614205 | TRIM38, U91328.2 |
| chr19 | 16066839 | 16067289 | 451 | 5 | 5.27E-07 | 0.0001012 | 0.000101 | 1.58E-05 | 0.000943648 | TPM4 |
| chr12 | 109931609 | 109932241 | 633 | 3 | 1.66E-06 | 0.0003368 | 2.14E-05 | 1.58E-05 | 0.002694093 | TCHP, GIT2 |
| chr21 | 33479800 | 33480550 | 751 | 9 | 9.30E-06 | 0.0001178 | 0.000214 | 1.58E-05 | 5.96E-05 | TMEM50B |
| chr9 | 91888442 | 91888623 | 182 | 2 | 2.45E-05 | 9.46E-06 | 0.000905 | 1.58E-05 | -0.0015793 | ROR2 |
| chr16 | 85175551 | 85175706 | 156 | 3 | 2.63E-05 | 2.30E-05 | 0.000209 | 1.58E-05 | -0.00215408 | GSE1 |
| chr19 | 12669509 | 12670143 | 635 | 10 | 8.64E-06 | 0.0001061 | 0.001346 | 1.58E-05 | 0.001058322 | WDR83, AC010422.8, AC010422.7 |
| chr8 | 143280873 | 143282128 | 1256 | 8 | 7.66E-06 | 0.000229 | 0.000567 | 1.58E-05 | -0.00191975 | MINCR |
| chr6 | 43383677 | 43383905 | 229 | 2 | 2.55E-05 | 2.26E-05 | 7.55E-05 | 1.58E-05 | -0.00182624 | AL359813.1 |
| chr20 | 60137004 | 60137044 | 41 | 2 | 1.87E-05 | 1.76E-05 | 0.000111 | 1.58E-05 | -0.00148909 | MIR646HG |
| chr13 | 101408229 | 101408439 | 211 | 2 | 1.53E-05 | 9.37E-05 | 2.26E-05 | 1.58E-05 | -0.0119707 | NALCN |
| chr5 | 151252965 | 151253101 | 137 | 5 | 5.16E-05 | 1.33E-05 | 0.002879 | 1.58E-05 | -0.00225011 | GM2A |
| chr11 | 13961726 | 13962530 | 805 | 10 | 3.70E-05 | 3.50E-06 | 0.024482 | 1.58E-05 | -0.01769876 | NA |
| chr12 | 56087513 | 56087606 | 94 | 2 | 8.93E-05 | 2.57E-05 | 6.47E-05 | 1.58E-05 | 0.002504369 | ERBB3 |
| chr3 | 196197251 | 196197767 | 517 | 2 | 1.02E-06 | 1.56E-05 | 0.000141 | 1.58E-05 | -0.00884585 | ZDHHC19 |
| chr8 | 66036434 | 66036493 | 60 | 2 | 4.19E-05 | 1.24E-05 | 0.000262 | 1.58E-05 | -0.01453198 | DNAJC5B |
| chr17 | 35087918 | 35088423 | 506 | 3 | 6.12E-06 | 1.10E-05 | 0.000463 | 1.58E-05 | 0.000313637 | RFFL, AC004223.3 |
| chr10 | 42782402 | 42783487 | 1086 | 5 | 1.56E-06 | 0.0032057 | 3.19E-05 | 1.59E-05 | 0.000755522 | BMS1 |
| chr1 | 147271648 | 147271746 | 99 | 2 | 5.15E-06 | 1.79E-05 | 0.000109 | 1.59E-05 | 0.003466384 | AC242426.2, CHD1L, LINC00624 |
| chr6 | 26365559 | 26366155 | 597 | 2 | 2.78E-05 | 9.54E-06 | 0.000888 | 1.59E-05 | -0.0191608 | BTN3A2 |
| chr15 | 95494258 | 95494771 | 514 | 2 | 2.63E-05 | 9.60E-06 | 0.00085 | 1.59E-05 | -0.01579418 | LINC00924 |
| chr5 | 82107218 | 82107706 | 489 | 2 | 1.35E-05 | 9.57E-06 | 0.00087 | 1.59E-05 | -0.01427493 | ATG10 |
| chr7 | 76285609 | 76285987 | 379 | 5 | 1.47E-05 | 0.0001228 | 9.78E-05 | 1.59E-05 | -0.00213355 | SRRM3 |
| chr19 | 30402155 | 30402605 | 451 | 3 | 3.43E-05 | 1.51E-05 | 0.000226 | 1.59E-05 | 0.00111427 | ZNF536 |
| chr1 | 217925080 | 217925251 | 172 | 3 | 4.78E-05 | 1.19E-05 | 0.000726 | 1.59E-05 | 0.013516963 | NA |
| chr4 | 183571503 | 183571823 | 321 | 3 | 9.66E-06 | 0.0002787 | 5.41E-05 | 1.59E-05 | -0.0064512 | NA |
| chr7 | 2555893 | 2556259 | 367 | 4 | 2.18E-05 | 1.76E-05 | 0.001951 | 1.59E-05 | -0.00075356 | NA |
| chr9 | 125706991 | 125707504 | 514 | 7 | 1.14E-05 | 0.0002413 | 0.001182 | 1.59E-05 | -0.00131259 | MAPKAP1 |
| chr1 | 2082805 | 2083189 | 385 | 4 | 3.35E-05 | 1.07E-05 | 0.001296 | 1.59E-05 | -0.00470321 | PRKCZ |
| chr5 | 107300823 | 107301185 | 363 | 2 | 3.09E-05 | 1.02E-05 | 0.000592 | 1.59E-05 | 0.002682438 | NA |
| chr6 | 168104257 | 168104889 | 633 | 2 | 1.99E-05 | 1.50E-05 | 0.000158 | 1.59E-05 | 0.009523859 | NA |
| chr6 | 35509260 | 35509889 | 630 | 3 | 9.14E-06 | 0.0004577 | 6.70E-05 | 1.59E-05 | -0.00646037 | TULP1 |
| chr2 | 227796224 | 227796339 | 116 | 2 | 4.41E-05 | 1.15E-05 | 0.000344 | 1.59E-05 | -0.01069439 | NA |
| chr9 | 36141419 | 36141431 | 13 | 2 | 1.90E-05 | 1.09E-05 | 0.000439 | 1.59E-05 | -0.01645413 | GLIPR2 |
| chr8 | 37079218 | 37079406 | 189 | 2 | 2.87E-05 | 1.11E-05 | 0.000405 | 1.59E-05 | -0.01931185 | NA |
| chr2 | 49155020 | 49155284 | 265 | 5 | 1.39E-05 | 0.0013525 | 0.000135 | 1.59E-05 | 0.004218222 | AC009975.1 |
| chr11 | 75090942 | 75091253 | 312 | 3 | 1.01E-05 | 0.0004148 | 1.79E-05 | 1.59E-05 | -0.01194357 | AP003175.1, OR2AT4 |
| chr11 | 112806878 | 112807184 | 307 | 2 | 2.97E-05 | 1.03E-05 | 0.000568 | 1.59E-05 | -0.01963219 | NA |
| chr1 | 63621779 | 63621956 | 178 | 4 | 1.19E-05 | 0.0002008 | 0.000119 | 1.59E-05 | 0.000905659 | PGM1 |
| chr16 | 46620779 | 46621763 | 985 | 11 | 5.23E-05 | 2.80E-05 | 0.002488 | 1.59E-05 | -0.00027399 | SHCBP1 |
| chr16 | 4556072 | 4556417 | 346 | 4 | 6.10E-05 | 1.12E-05 | 0.00232 | 1.59E-05 | -0.00349699 | NA |
| chr6 | 83431274 | 83431828 | 555 | 7 | 7.93E-05 | 7.02E-06 | 0.00325 | 1.60E-05 | -0.00571601 | NA |
| chr1 | 163174803 | 163175167 | 365 | 11 | 4.57E-05 | 1.45E-05 | 0.003103 | 1.60E-05 | -0.00259197 | RGS5-AS1, RGS5 |
| chr2 | 74958025 | 74958980 | 956 | 10 | 6.73E-06 | 8.60E-05 | 0.001272 | 1.60E-05 | -0.00170772 | POLE4 |
| chr19 | 18671564 | 18671747 | 184 | 2 | 2.56E-05 | 2.13E-05 | 8.33E-05 | 1.60E-05 | -0.00713527 | KLHL26 |
| chr1 | 226703227 | 226703491 | 265 | 4 | 4.28E-05 | 8.02E-05 | 0.000441 | 1.60E-05 | 0.001421976 | ITPKB |
| chr2 | 112182658 | 112182832 | 175 | 3 | 2.27E-05 | 8.13E-06 | 0.001792 | 1.60E-05 | -0.00740559 | FBLN7 |
| chr9 | 21335127 | 21335741 | 615 | 9 | 3.28E-05 | 1.88E-05 | 0.003175 | 1.60E-05 | 0.0016746 | KLHL9 |
| chr8 | 99872813 | 99872828 | 16 | 2 | 1.13E-05 | 0.0002465 | 1.51E-05 | 1.60E-05 | -0.00190729 | VPS13B |
| chr11 | 15800314 | 15800787 | 474 | 2 | 3.02E-05 | 1.24E-05 | 0.000272 | 1.60E-05 | -0.00777864 | NA |
| chr20 | 19658037 | 19658232 | 196 | 2 | 4.29E-05 | 1.10E-05 | 0.000418 | 1.60E-05 | 0.016159548 | SLC24A3 |
| chr2 | 2787504 | 2787824 | 321 | 3 | 6.95E-06 | 0.0002864 | 3.38E-05 | 1.60E-05 | -0.00658031 | AC011995.2 |
| chr7 | 4758695 | 4759196 | 502 | 4 | 6.37E-06 | 4.20E-05 | 0.001255 | 1.60E-05 | 0.001308032 | FOXK1 |
| chr1 | 58896308 | 58896880 | 573 | 6 | 9.03E-06 | 0.0001632 | 0.000113 | 1.60E-05 | -0.00409929 | LINC01135, LINC02777 |
| chr19 | 12702059 | 12702121 | 63 | 2 | 3.31E-05 | 1.50E-05 | 0.00016 | 1.60E-05 | -0.00929412 | TNPO2 |
| chr18 | 74117371 | 74117572 | 202 | 2 | 2.68E-05 | 2.60E-05 | 6.49E-05 | 1.60E-05 | -0.01787977 | FBXO15 |
| chr12 | 42398174 | 42398496 | 323 | 3 | 8.96E-06 | 0.0002723 | 0.000259 | 1.60E-05 | -0.00740924 | PPHLN1 |
| chr14 | 81741049 | 81741126 | 78 | 2 | 1.43E-05 | 9.89E-06 | 0.000749 | 1.60E-05 | -0.01184248 | AL355838.1, LINC02311 |
| chr6 | 14276812 | 14277236 | 425 | 2 | 4.85E-05 | 9.97E-06 | 0.000713 | 1.60E-05 | 0.009013919 | NA |
| chr1 | 1183554 | 1184049 | 496 | 2 | 2.07E-06 | 2.16E-05 | 8.27E-05 | 1.60E-05 | 0.007012989 | TTLL10 |
| chr10 | 44974751 | 44975114 | 364 | 5 | 4.75E-05 | 5.58E-06 | 0.005283 | 1.60E-05 | -0.00232678 | RASSF4, DEPP1 |
| chr10 | 3891231 | 3891578 | 348 | 2 | 3.97E-06 | 9.54E-05 | 2.27E-05 | 1.60E-05 | -0.00425311 | NA |
| chr16 | 667621 | 668405 | 785 | 13 | 1.10E-05 | 0.001357 | 0.000385 | 1.60E-05 | 0.000567354 | WDR90, RHOT2 |
| chr18 | 268031 | 268601 | 571 | 10 | 2.06E-05 | 0.001464 | 0.002708 | 1.61E-05 | -0.00042632 | AP000845.1, THOC1 |
| chr12 | 41436264 | 41436518 | 255 | 3 | 6.63E-06 | 0.0014413 | 0.000108 | 1.61E-05 | 0.000417009 | PDZRN4, AC090531.1 |
| chr1 | 3557359 | 3557812 | 454 | 3 | 1.08E-05 | 4.64E-05 | 0.000369 | 1.61E-05 | 0.001623834 | MEGF6 |
| chr16 | 89393956 | 89394343 | 388 | 2 | 1.59E-06 | 8.51E-05 | 2.42E-05 | 1.61E-05 | -0.00843287 | ANKRD11 |
| chr8 | 85438049 | 85438789 | 741 | 9 | 1.44E-07 | 0.0027907 | 0.000371 | 1.61E-05 | -0.00426351 | CA3 |
| chr3 | 98594554 | 98595149 | 596 | 3 | 3.24E-06 | 7.33E-05 | 0.00049 | 1.61E-05 | 0.008260231 | NA |
| chr22 | 21568349 | 21568863 | 515 | 3 | 9.18E-07 | 0.0001014 | 2.46E-05 | 1.61E-05 | -0.00360983 | UBE2L3 |
| chr2 | 36485427 | 36485666 | 240 | 2 | 5.45E-06 | 6.18E-05 | 2.98E-05 | 1.61E-05 | -0.00323361 | CRIM1 |
| chr20 | 57259964 | 57260294 | 331 | 2 | 1.26E-05 | 1.59E-05 | 0.000143 | 1.61E-05 | 0.021697762 | BMP7 |
| chr22 | 43837105 | 43837371 | 267 | 2 | 1.50E-05 | 5.67E-05 | 3.17E-05 | 1.61E-05 | -0.01057312 | SULT4A1 |
| chr14 | 100841081 | 100841093 | 13 | 2 | 3.36E-05 | 2.91E-05 | 5.76E-05 | 1.61E-05 | -0.00437609 | MEG3, AL117190.1, AL117190.2 |
| chr17 | 56895695 | 56895881 | 187 | 2 | 4.83E-05 | 9.82E-06 | 0.000812 | 1.61E-05 | -0.00356714 | TRIM25 |
| chr19 | 7682334 | 7682859 | 526 | 3 | 3.82E-05 | 0.0004599 | 0.000486 | 1.61E-05 | -0.00731831 | AC008763.3, TRAPPC5 |
| chr3 | 100278663 | 100279230 | 568 | 3 | 3.19E-05 | 0.0011221 | 0.000176 | 1.61E-05 | -0.00845509 | TBC1D23 |
| chr18 | 70180947 | 70181289 | 343 | 2 | 3.20E-05 | 1.29E-05 | 0.000242 | 1.61E-05 | -0.00374764 | RTTN |
| chr6 | 168188939 | 168189254 | 316 | 3 | 2.87E-05 | 1.33E-05 | 0.000437 | 1.61E-05 | -0.00061168 | NA |
| chr4 | 174213394 | 174214278 | 885 | 6 | 8.64E-07 | 0.000677 | 2.38E-05 | 1.61E-05 | -0.00425871 | LINC02268 |
| chr5 | 64574356 | 64574867 | 512 | 4 | 2.39E-06 | 0.0006506 | 3.87E-05 | 1.61E-05 | -0.01052396 | RGS7BP |
| chr1 | 18294705 | 18295231 | 527 | 2 | 8.31E-06 | 9.68E-06 | 0.000918 | 1.61E-05 | -0.01970262 | IGSF21 |
| chr13 | 94713790 | 94714202 | 413 | 4 | 1.09E-05 | 0.0001262 | 0.000453 | 1.61E-05 | 0.005940309 | SOX21-AS1 |
| chr16 | 2991348 | 2991994 | 647 | 5 | 1.41E-05 | 5.70E-06 | 0.005442 | 1.61E-05 | -7.79E-05 | LINC00514 |
| chr18 | 70016051 | 70016356 | 306 | 2 | 8.31E-06 | 0.0001879 | 1.69E-05 | 1.61E-05 | -0.0052502 | RTTN |
| chr19 | 38183892 | 38184582 | 691 | 3 | 2.52E-06 | 0.0005578 | 9.29E-06 | 1.61E-05 | -0.0003703 | SIPA1L3, AC011479.3 |
| chr16 | 759476 | 760292 | 817 | 6 | 1.82E-06 | 0.0005391 | 0.000143 | 1.61E-05 | -0.00275226 | NA |
| chr10 | 117763957 | 117764046 | 90 | 2 | 1.01E-05 | 0.0015976 | 9.86E-06 | 1.61E-05 | 0.007558437 | NA |
| chr9 | 99239642 | 99239997 | 356 | 2 | 1.54E-05 | 3.03E-05 | 5.54E-05 | 1.62E-05 | 0.013166964 | NA |
| chr12 | 111886561 | 111886823 | 263 | 3 | 4.76E-05 | 6.18E-06 | 0.003612 | 1.62E-05 | 0.000914054 | MAPKAPK5 |
| chr16 | 81907346 | 81907705 | 360 | 2 | 4.77E-06 | 5.11E-05 | 3.45E-05 | 1.62E-05 | -0.00593462 | PLCG2 |
| chr17 | 21002167 | 21002474 | 308 | 4 | 5.45E-05 | 1.38E-05 | 0.001504 | 1.62E-05 | 0.004017815 | CCDC144NL-AS1, USP22 |
| chr10 | 14478612 | 14478742 | 131 | 2 | 4.65E-05 | 1.04E-05 | 0.000583 | 1.62E-05 | -0.01567811 | NA |
| chr16 | 67969746 | 67970004 | 259 | 3 | 2.17E-07 | 0.004266 | 4.48E-06 | 1.62E-05 | 0.00350433 | NA |
| chr3 | 47380782 | 47380968 | 187 | 7 | 7.76E-05 | 0.0002493 | 0.001024 | 1.62E-05 | 0.000405065 | AC099778.1 |
| chr15 | 41796164 | 41796720 | 557 | 2 | 8.82E-05 | 1.01E-05 | 0.00069 | 1.62E-05 | -0.01298135 | MAPKBP1 |
| chr14 | 75008920 | 75009198 | 279 | 2 | 2.37E-05 | 2.32E-05 | 7.67E-05 | 1.62E-05 | -0.0017476 | EIF2B2 |
| chr1 | 23387480 | 23387880 | 401 | 6 | 2.50E-05 | 1.96E-05 | 0.000315 | 1.62E-05 | -0.00206459 | TCEA3 |
| chr1 | 10836286 | 10836393 | 108 | 2 | 3.01E-05 | 1.06E-05 | 0.000541 | 1.62E-05 | -0.01021229 | NA |
| chr11 | 44693785 | 44694475 | 691 | 3 | 7.71E-06 | 6.49E-05 | 4.35E-05 | 1.62E-05 | -0.00634642 | NA |
| chr11 | 117929518 | 117930617 | 1100 | 5 | 9.01E-06 | 0.0002412 | 5.81E-05 | 1.62E-05 | -0.00453587 | NA |
| chr8 | 116874298 | 116875273 | 976 | 16 | 2.08E-05 | 0.0016772 | 0.001985 | 1.62E-05 | -0.00061978 | RAD21-AS1, RAD21, MIR3610 |
| chr20 | 44302961 | 44303518 | 558 | 3 | 1.07E-05 | 0.0001025 | 4.30E-05 | 1.62E-05 | -0.00239357 | FITM2 |
| chr20 | 33660820 | 33661189 | 370 | 3 | 7.30E-07 | 0.0015014 | 7.53E-06 | 1.62E-05 | -0.0047682 | NECAB3 |
| chr17 | 50071702 | 50072007 | 306 | 3 | 2.93E-05 | 1.56E-05 | 0.000214 | 1.62E-05 | 0.002744225 | ITGA3 |
| chr8 | 1605095 | 1605698 | 604 | 4 | 3.41E-05 | 4.63E-05 | 0.000314 | 1.63E-05 | -0.00133012 | DLGAP2, DLGAP2-AS1 |
| chr3 | 52697721 | 52697883 | 163 | 4 | 4.65E-06 | 0.0046022 | 1.28E-05 | 1.63E-05 | -0.00145094 | GLT8D1 |
| chr11 | 45802061 | 45802590 | 530 | 2 | 4.94E-05 | 9.76E-06 | 0.000922 | 1.63E-05 | -0.00488227 | NA |
| chr11 | 46242993 | 46243725 | 733 | 6 | 1.80E-05 | 0.0001569 | 0.000484 | 1.63E-05 | -0.00022006 | NA |
| chr19 | 5799329 | 5799456 | 128 | 3 | 2.80E-05 | 3.11E-05 | 8.44E-05 | 1.63E-05 | -0.0124662 | NA |
| chr1 | 3296867 | 3297104 | 238 | 2 | 3.31E-05 | 1.38E-05 | 0.00021 | 1.63E-05 | -0.00709952 | PRDM16 |
| chr19 | 49473905 | 49474416 | 512 | 10 | 4.88E-05 | 5.35E-05 | 0.002949 | 1.63E-05 | -9.09E-05 | AC010619.1, FLT3LG |
| chr5 | 35130283 | 35130445 | 163 | 2 | 3.44E-05 | 1.04E-05 | 0.000606 | 1.63E-05 | -0.00911087 | PRLR |

| | | | | | | | | | | |
|---|---|---|---|---|---|---|---|---|---|---|
| chr17 | 2690197 | 2690316 | 120 | 2 | 1.41E-05 | 0.0001174 | 2.08E-05 | 1.63E-05 | 0.00562379 | CLUH |
| chr16 | 17194069 | 17194437 | 369 | 3 | 4.12E-05 | 1.19E-05 | 0.00141 | 1.63E-05 | -0.01024047 | XYLT1, AC099494.1 |
| chr3 | 10290486 | 10290922 | 437 | 5 | 8.05E-05 | 3.67E-06 | 0.009201 | 1.63E-05 | 0.00208164 | GHRLOS, GHRL |
| chr20 | 42191342 | 42191727 | 386 | 3 | 1.08E-05 | 9.59E-05 | 0.000388 | 1.63E-05 | 0.001485191 | PTPRT |
| chr9 | 34628028 | 34628632 | 605 | 5 | 3.38E-06 | 4.56E-05 | 0.001831 | 1.63E-05 | -0.01553236 | ARID3C |
| chr17 | 28982095 | 28983009 | 915 | 7 | 1.01E-06 | 0.0004527 | 2.18E-05 | 1.63E-05 | -0.00065337 | PIPOX, SEZ6 |
| chr18 | 12434097 | 12434981 | 885 | 5 | 4.06E-06 | 9.27E-06 | 0.000762 | 1.63E-05 | -0.0004019 | AP001029.1 |
| chr12 | 20629915 | 20630206 | 292 | 3 | 1.08E-05 | 0.0003088 | 2.37E-05 | 1.63E-05 | -0.00383898 | PDE3A |
| chr17 | 38869674 | 38870388 | 715 | 8 | 1.14E-05 | 3.24E-05 | 0.000308 | 1.64E-05 | 0.001069134 | LASP1 |
| chr19 | 55665214 | 55665270 | 57 | 3 | 1.45E-05 | 3.00E-05 | 0.000128 | 1.64E-05 | 8.80E-05 | U2AF2, AC008735.2 |
| chr8 | 142988130 | 142988280 | 151 | 2 | 8.78E-06 | 4.82E-05 | 3.66E-05 | 1.64E-05 | -0.00337789 | LY6E-DT |
| chr14 | 56759287 | 56759303 | 17 | 3 | 9.52E-05 | 6.18E-06 | 0.003752 | 1.64E-05 | -0.01340778 | NA |
| chr2 | 129242905 | 129243435 | 531 | 2 | 3.63E-05 | 1.19E-05 | 0.000348 | 1.64E-05 | -0.00911401 | LINC01854 |
| chr2 | 50118722 | 50119993 | 1272 | 5 | 5.95E-05 | 0.0005137 | 0.00061 | 1.64E-05 | -0.01099936 | NRXN1 |
| chr5 | 87300351 | 87300807 | 457 | 3 | 4.83E-06 | 0.0007102 | 0.000163 | 1.64E-05 | 0.000324714 | RASA1 |
| chr10 | 49131311 | 49132128 | 818 | 4 | 1.19E-05 | 8.51E-05 | 4.48E-05 | 1.64E-05 | -0.00178954 | FAM170B-AS1, FAM170B |
| chr2 | 69520343 | 69520944 | 602 | 3 | 1.79E-05 | 7.69E-06 | 0.001452 | 1.64E-05 | -0.00159382 | AAK1 |
| chr6 | 53064685 | 53065949 | 1265 | 16 | 1.02E-05 | 5.22E-05 | 0.000271 | 1.64E-05 | 4.71E-05 | FBXO9, AL031178.2 |
| chr22 | 39683102 | 39683621 | 520 | 3 | 3.15E-05 | 2.88E-05 | 0.000149 | 1.64E-05 | -0.00054228 | CACNA1I |
| chr13 | 109568410 | 109568592 | 183 | 2 | 1.08E-05 | 0.000146 | 1.91E-05 | 1.64E-05 | -0.00899337 | AL163541.1 |
| chr13 | 46675698 | 46675788 | 91 | 3 | 3.89E-05 | 3.02E-05 | 0.000287 | 1.64E-05 | -0.01446917 | LRCH1 |
| chr9 | 128233549 | 128233937 | 389 | 3 | 2.21E-06 | 0.0021286 | 5.64E-06 | 1.65E-05 | -0.00425317 | DNM1 |
| chr3 | 126343107 | 126343805 | 699 | 4 | 4.58E-05 | 1.67E-05 | 0.000364 | 1.65E-05 | -0.00083602 | KLF15 |
| chr19 | 3572412 | 3573545 | 1134 | 11 | 5.66E-06 | 3.98E-05 | 0.001429 | 1.65E-05 | 0.000502858 | HMG20B, MFSD12 |
| chr15 | 93088349 | 93089390 | 1042 | 11 | 5.78E-06 | 0.0002009 | 0.000159 | 1.65E-05 | -0.00041985 | RGMA |
| chr12 | 125983013 | 125983917 | 905 | 8 | 9.63E-06 | 0.0001917 | 0.000403 | 1.65E-05 | 0.000316492 | LINC02826, LINC00939 |
| chr7 | 51316392 | 51316674 | 283 | 2 | 1.70E-05 | 1.23E-05 | 0.000313 | 1.65E-05 | -0.01036819 | COBL |
| chr1 | 99645319 | 99645459 | 141 | 3 | 1.85E-05 | 1.88E-05 | 0.000621 | 1.65E-05 | -0.0001045 | NA |
| chr8 | 66613044 | 66613614 | 571 | 7 | 5.36E-05 | 1.21E-05 | 0.005539 | 1.65E-05 | -8.19E-06 | MYBL1 |
| chr6 | 129861268 | 129861822 | 555 | 8 | 3.89E-06 | 0.0002121 | 9.84E-05 | 1.65E-05 | -0.00430315 | TMEM244 |
| chr19 | 9834343 | 9835236 | 894 | 12 | 1.33E-05 | 5.94E-06 | 0.008176 | 1.65E-05 | -0.00082979 | AC008752.2 |
| chr8 | 140098899 | 140099632 | 734 | 5 | 1.76E-05 | 1.25E-05 | 0.000854 | 1.65E-05 | -0.00326197 | TRAPPC9, PEG13 |
| chr16 | 3745350 | 3745431 | 82 | 3 | 0.00010629 | 2.75E-05 | 0.000409 | 1.65E-05 | 0.002747508 | CREBBP |
| chr19 | 35030868 | 35031516 | 649 | 4 | 2.18E-08 | 0.0004742 | 1.56E-05 | 1.65E-05 | -0.00232271 | SCN1B |
| chr2 | 88442642 | 88443073 | 432 | 2 | 4.63E-05 | 1.05E-05 | 0.000617 | 1.65E-05 | -0.01359971 | NA |
| chr3 | 186464579 | 186464888 | 310 | 2 | 1.17E-05 | 4.53E-05 | 3.90E-05 | 1.65E-05 | -0.00614381 | LINC02052 |
| chr18 | 50339978 | 50340461 | 484 | 6 | 9.57E-05 | 3.01E-06 | 0.010718 | 1.65E-05 | -0.0025721 | NA |
| chr2 | 150611277 | 150612210 | 934 | 3 | 1.48E-06 | 0.000103 | 0.000172 | 1.65E-05 | 0.008295994 | AC104777.1 |
| chr8 | 1202941 | 1203004 | 64 | 2 | 5.67E-05 | 1.06E-05 | 0.000597 | 1.65E-05 | -0.0036293 | DLGAP2 |
| chr7 | 18027814 | 18028330 | 517 | 9 | 9.37E-06 | 0.0002672 | 0.000859 | 1.65E-05 | 0.000275847 | PRPS1L1 |
| chr10 | 114925333 | 114925508 | 176 | 2 | 4.83E-05 | 1.05E-05 | 0.000659 | 1.65E-05 | -0.0126188 | NA |
| chr15 | 80071366 | 80071650 | 285 | 3 | 4.18E-05 | 1.07E-05 | 0.001211 | 1.65E-05 | 0.001995273 | ZFAND6 |
| chr3 | 127172937 | 127173252 | 316 | 3 | 1.18E-05 | 0.0001445 | 1.98E-05 | 1.66E-05 | -0.00688337 | AC112482.2 |
| chr19 | 18431834 | 18431853 | 20 | 2 | 3.24E-05 | 1.04E-05 | 0.000683 | 1.66E-05 | -0.01223429 | SSBP4 |
| chr18 | 31203965 | 31203990 | 26 | 2 | 3.20E-05 | 2.05E-05 | 9.52E-05 | 1.66E-05 | -0.0045409 | NA |
| chr5 | 88381966 | 88382327 | 362 | 3 | 7.96E-06 | 0.0007369 | 3.65E-05 | 1.66E-05 | -0.00790451 | TMEM161B-AS1, RPS3AP22 |
| chr7 | 151802186 | 151802412 | 227 | 2 | 3.92E-05 | 1.31E-05 | 0.00026 | 1.66E-05 | -0.00357838 | PRKAG2 |
| chr11 | 57598475 | 57598952 | 478 | 2 | 8.04E-08 | 0.0007518 | 1.16E-05 | 1.66E-05 | 0.013849465 | SERPING1 |
| chr1 | 153751237 | 153751877 | 641 | 2 | 5.44E-05 | 1.67E-05 | 0.000138 | 1.66E-05 | -0.0087507 | INTS3, AL513523.3 |
| chr1 | 222972752 | 222972776 | 25 | 2 | 1.27E-05 | 3.25E-05 | 5.37E-05 | 1.66E-05 | -0.00171881 | DISP1 |
| chr3 | 81173534 | 81173777 | 244 | 2 | 1.38E-05 | 0.0001087 | 2.21E-05 | 1.66E-05 | -0.01699664 | NA |
| chr15 | 23036126 | 23036248 | 123 | 2 | 2.55E-05 | 2.08E-05 | 9.39E-05 | 1.66E-05 | -0.01426129 | TUBGCP5 |
| chr4 | 66357484 | 66357611 | 128 | 2 | 5.16E-05 | 9.96E-06 | 0.000933 | 1.66E-05 | -0.01644824 | NA |
| chr4 | 1711829 | 1712672 | 844 | 8 | 2.38E-06 | 0.0001089 | 7.23E-05 | 1.66E-05 | 0.000425815 | SLBP |
| chr20 | 34634622 | 34635056 | 435 | 5 | 4.29E-05 | 3.11E-05 | 0.00177 | 1.66E-05 | 0.008679668 | PIGU |
| chr2 | 113627317 | 113627592 | 276 | 2 | 1.63E-05 | 5.25E-05 | 3.50E-05 | 1.66E-05 | -0.00114075 | RABL2A |
| chr9 | 93108578 | 93108960 | 383 | 2 | 2.12E-06 | 0.0001725 | 1.80E-05 | 1.66E-05 | -0.00634273 | CARD19 |
| chr8 | 142614466 | 142615170 | 705 | 9 | 3.01E-05 | 8.05E-05 | 0.001485 | 1.66E-05 | -0.00193852 | ARC |
| chr12 | 120904394 | 120904806 | 413 | 7 | 4.53E-05 | 1.39E-05 | 0.010063 | 1.66E-05 | -0.00058879 | AC069214.1 |
| chr15 | 80151566 | 80152887 | 1322 | 8 | 4.55E-06 | 5.19E-05 | 0.001511 | 1.66E-05 | -0.00207509 | FAH |
| chr14 | 39110069 | 39110301 | 233 | 2 | 1.20E-05 | 0.000204 | 1.69E-05 | 1.66E-05 | -0.00680914 | NA |
| chr11 | 1890317 | 1890956 | 640 | 3 | 1.92E-07 | 7.95E-05 | 4.63E-05 | 1.66E-05 | 0.005523143 | LSP1, PRR33 |
| chr16 | 88888754 | 88888816 | 63 | 2 | 7.29E-06 | 0.0002561 | 1.56E-05 | 1.67E-05 | -0.01113827 | CBFA2T3 |
| chr19 | 39204191 | 39204597 | 407 | 6 | 1.03E-05 | 0.0001039 | 0.000213 | 1.67E-05 | -0.0063567 | SYCN |
| chr3 | 32369374 | 32369913 | 540 | 6 | 1.31E-05 | 7.65E-05 | 0.000355 | 1.67E-05 | 0.001094979 | CMTM8 |
| chr7 | 318075 | 318550 | 476 | 4 | 2.51E-05 | 2.79E-05 | 0.002251 | 1.67E-05 | -0.00520668 | NA |
| chr9 | 90801746 | 90802321 | 576 | 6 | 2.06E-05 | 1.26E-05 | 0.004587 | 1.67E-05 | 0.000483763 | SYK |
| chr20 | 50850716 | 50850879 | 164 | 2 | 2.41E-05 | 3.40E-05 | 5.18E-05 | 1.67E-05 | 0.001879812 | BCAS4 |
| chr12 | 8949472 | 8950588 | 1117 | 13 | 1.61E-05 | 5.21E-05 | 0.000323 | 1.67E-05 | -0.00013859 | KLRG1, M6PR |
| chr13 | 99896161 | 99896763 | 603 | 5 | 6.16E-06 | 3.50E-05 | 0.001512 | 1.67E-05 | 0.00895521 | CLYBL, AL137139.2 |
| chr1 | 167439272 | 167439961 | 690 | 8 | 3.50E-05 | 5.95E-05 | 0.004924 | 1.67E-05 | -0.00853518 | CD247 |
| chr5 | 5886161 | 5886955 | 795 | 4 | 7.01E-07 | 0.000295 | 2.64E-05 | 1.67E-05 | -0.00421902 | NA |
| chr22 | 38681734 | 38682306 | 573 | 5 | 5.51E-05 | 2.07E-05 | 0.000978 | 1.67E-05 | -0.00026762 | TOMM22, AL021707.2 |
| chr12 | 101697441 | 101697609 | 169 | 6 | 0.000111642 | 4.84E-05 | 0.00175 | 1.67E-05 | 0.000742926 | CHPT1 |
| chr19 | 39336028 | 39336893 | 866 | 9 | 3.19E-05 | 5.21E-05 | 0.00276 | 1.67E-05 | -0.00027886 | GMFG |
| chr6 | 26602672 | 26602729 | 58 | 3 | 1.44E-05 | 6.63E-05 | 6.05E-05 | 1.67E-05 | -0.00216477 | NA |
| chr11 | 34251235 | 34251564 | 330 | 3 | 6.60E-05 | 6.56E-06 | 0.003174 | 1.67E-05 | 0.018391459 | ABTB2 |
| chr16 | 4218003 | 4218372 | 370 | 3 | 4.66E-05 | 3.80E-05 | 0.000454 | 1.67E-05 | -0.00441241 | SRL |
| chr8 | 71045271 | 71045643 | 373 | 3 | 3.67E-05 | 1.22E-05 | 0.000394 | 1.68E-05 | -0.00574637 | AC015687.1 |
| chr4 | 38752887 | 38753092 | 206 | 2 | 5.81E-05 | 1.15E-05 | 0.000434 | 1.68E-05 | 0.000312255 | NA |
| chr21 | 32537461 | 32537633 | 173 | 2 | 3.16E-05 | 1.29E-05 | 0.000286 | 1.68E-05 | 0.003752165 | CFAP298-TCP10L, TCP10L |
| chr4 | 153340571 | 153340579 | 9 | 2 | 5.19E-05 | 1.02E-05 | 0.000872 | 1.68E-05 | -0.00332469 | AC013477.1 |
| chr17 | 2928004 | 2928609 | 606 | 4 | 3.13E-05 | 2.39E-05 | 0.000427 | 1.68E-05 | -0.0034098 | RAP1GAP2 |
| chr2 | 317826 | 317938 | 113 | 2 | 0.00011412 | 0.0003527 | 1.42E-05 | 1.68E-05 | -0.0123011 | LINC01865 |
| chr18 | 78914571 | 78915070 | 500 | 4 | 1.69E-05 | 6.07E-05 | 0.000951 | 1.68E-05 | 0.018074096 | AC091027.2 |
| chr16 | 88688161 | 88688401 | 241 | 2 | 4.57E-06 | 4.80E-05 | 3.81E-05 | 1.68E-05 | -0.01172152 | NA |
| chr17 | 77097025 | 77097097 | 73 | 2 | 1.08E-05 | 0.0001108 | 2.23E-05 | 1.68E-05 | 0.010116281 | SNHG20, SEC14L1 |
| chr20 | 53701688 | 53701889 | 202 | 2 | 2.85E-05 | 2.70E-05 | 6.76E-05 | 1.68E-05 | 0.001011434 | NA |
| chr20 | 56248223 | 56248822 | 600 | 3 | 8.40E-06 | 0.0002919 | 2.92E-05 | 1.68E-05 | -0.00669648 | MC3R |
| chr8 | 144012132 | 144012546 | 415 | 3 | 8.42E-05 | 1.34E-05 | 0.001089 | 1.69E-05 | 0.000238992 | SPATC1, PARP10 |
| chr14 | 61810955 | 61811023 | 69 | 2 | 6.83E-06 | 1.11E-05 | 0.000525 | 1.69E-05 | 0.005912069 | NA |
| chr14 | 105066685 | 105066773 | 89 | 2 | 2.14E-06 | 1.02E-05 | 0.000896 | 1.69E-05 | -0.00875081 | NA |
| chr2 | 64523920 | 64524369 | 450 | 7 | 8.83E-06 | 0.0002529 | 0.001037 | 1.69E-05 | -7.71E-05 | AFTPH, AC008074.2 |
| chr19 | 3828613 | 3828910 | 298 | 2 | 4.84E-05 | 1.05E-05 | 0.000723 | 1.69E-05 | 0.000592752 | ZFR2 |
| chr19 | 8333330 | 8333793 | 464 | 4 | 3.04E-05 | 2.47E-05 | 0.001852 | 1.69E-05 | 0.01295275 | KANK3 |
| chr18 | 76690014 | 76690053 | 40 | 2 | 3.81E-05 | 1.88E-05 | 0.000116 | 1.69E-05 | -0.00289396 | LINC01879 |
| chr1 | 205928031 | 205928762 | 732 | 6 | 2.02E-05 | 2.92E-05 | 0.000221 | 1.69E-05 | -0.00327474 | SLC26A9 |
| chr5 | 86352737 | 86352742 | 6 | 2 | 9.80E-06 | 8.27E-05 | 2.62E-05 | 1.69E-05 | -0.01084717 | NA |
| chr7 | 73886270 | 73886578 | 309 | 2 | 3.03E-05 | 1.55E-05 | 0.000174 | 1.69E-05 | 0.005842271 | NA |
| chr2 | 143776205 | 143776713 | 509 | 6 | 4.78E-06 | 0.0006424 | 2.82E-05 | 1.69E-05 | 0.000319252 | NA |
| chr4 | 8607674 | 8607867 | 194 | 2 | 2.62E-05 | 1.41E-05 | 0.000226 | 1.69E-05 | -0.00064763 | GPR78, CPZ |
| chr14 | 89773400 | 89773780 | 381 | 3 | 3.78E-06 | 0.0022063 | 5.77E-06 | 1.69E-05 | -0.0027119 | AL137230.2 |
| chr14 | 24241876 | 24242867 | 992 | 15 | 1.99E-05 | 0.0001421 | 0.00079 | 1.69E-05 | -0.00075828 | TINF2 |
| chr10 | 116138541 | 116139017 | 477 | 2 | 4.31E-06 | 9.98E-05 | 2.37E-05 | 1.69E-05 | -0.00586514 | GFRA1 |
| chr14 | 70600629 | 70601471 | 843 | 9 | 4.39E-06 | 0.0001835 | 0.000348 | 1.70E-05 | -0.00045811 | MED6, AL357153.3 |
| chr11 | 64555418 | 64555836 | 419 | 5 | 1.62E-05 | 1.25E-05 | 0.005051 | 1.70E-05 | 0.003166137 | SLC22A11 |
| chr6 | 126643389 | 126643696 | 308 | 3 | 1.72E-05 | 3.52E-05 | 6.94E-05 | 1.70E-05 | -0.00639255 | PRELID1P1, AL356534.1 |
| chr2 | 8238708 | 8239046 | 339 | 2 | 1.77E-06 | 0.0002061 | 1.72E-05 | 1.70E-05 | 0.002184616 | LINC00298, LINC00299 |
| chr21 | 44299521 | 44299949 | 429 | 7 | 8.21E-06 | 6.07E-05 | 8.49E-05 | 1.70E-05 | 0.001192304 | NA |
| chr12 | 69476194 | 69476376 | 183 | 2 | 1.22E-05 | 0.0004518 | 1.35E-05 | 1.70E-05 | 0.012579947 | FRS2 |
| chr7 | 81367172 | 81367587 | 416 | 2 | 6.44E-05 | 1.02E-05 | 0.000946 | 1.70E-05 | -0.00679243 | NA |
| chr9 | 72525032 | 72525459 | 428 | 3 | 1.04E-06 | 0.0363699 | 3.08E-06 | 1.70E-05 | -0.01373228 | TMC1 |
| chr3 | 38479688 | 38480607 | 920 | 3 | 2.02E-06 | 6.23E-05 | 4.49E-05 | 1.70E-05 | 0.002262616 | ACVR2B |
| chr14 | 77029329 | 77029372 | 44 | 3 | 2.85E-05 | 7.96E-06 | 0.001491 | 1.70E-05 | -0.00961942 | AC007686.3 |
| chr1 | 182586755 | 182586978 | 224 | 2 | 1.46E-05 | 3.83E-05 | 4.74E-05 | 1.70E-05 | 0.006888455 | RNASEL |
| chr20 | 62553282 | 62554184 | 903 | 5 | 5.13E-06 | 1.00E-05 | 0.006837 | 1.70E-05 | -0.00405758 | MIR1-1HG |
| chr2 | 12728260 | 12728896 | 637 | 3 | 8.58E-06 | 0.0001004 | 0.000116 | 1.70E-05 | -0.00083599 | TRIB2 |
| chr14 | 75258945 | 75259458 | 514 | 7 | 4.13E-05 | 7.46E-06 | 0.002786 | 1.70E-05 | 0.002726414 | AF111167.1, AL691403.1 |
| chr2 | 97737760 | 97737829 | 70 | 3 | 2.88E-05 | 2.33E-05 | 0.000864 | 1.70E-05 | -0.00182638 | ZAP70 |
| chr17 | 68258743 | 68258907 | 165 | 2 | 5.08E-05 | 1.06E-05 | 0.00074 | 1.70E-05 | -0.01676559 | NA |
| chr17 | 50219874 | 50220166 | 293 | 3 | 1.03E-05 | 0.0004831 | 0.000114 | 1.70E-05 | -0.00422758 | NA |
| chr10 | 121409801 | 121410493 | 693 | 3 | 1.47E-05 | 6.01E-05 | 5.64E-05 | 1.70E-05 | 0.005204848 | NA |
| chr6 | 170421325 | 170421605 | 281 | 3 | 2.97E-05 | 1.58E-05 | 0.000234 | 1.70E-05 | 0.005257818 | NA |
| chr20 | 53939480 | 53939938 | 459 | 3 | 7.51E-05 | 5.18E-05 | 0.000451 | 1.70E-05 | -0.01502106 | BCAS1 |
| chr8 | 133113078 | 133113512 | 435 | 3 | 1.40E-05 | 3.30E-05 | 0.000255 | 1.70E-05 | -0.00328848 | TG |
| chr20 | 10524563 | 10524944 | 382 | 2 | 9.25E-07 | 0.0006192 | 1.25E-05 | 1.70E-05 | -0.00403851 | SLX4IP |
| chr20 | 54070468 | 54070855 | 388 | 5 | 2.47E-05 | 2.16E-05 | 0.001953 | 1.70E-05 | 0.016732986 | BCAS1 |
| chr2 | 126888828 | 126889016 | 189 | 3 | 6.25E-06 | 0.0022863 | 5.97E-06 | 1.70E-05 | -0.00480206 | NA |
| chr16 | 21653681 | 21653917 | 237 | 4 | 1.60E-05 | 0.0001534 | 1.89E-05 | 1.71E-05 | 0.003923455 | METTL9 |
| chr14 | 75430639 | 75431138 | 500 | 5 | 2.77E-05 | 4.45E-05 | 0.001679 | 1.71E-05 | 0.001177898 | JDP2 |
| chr10 | 46550360 | 46550713 | 354 | 2 | 5.92E-06 | 5.01E-05 | 3.77E-05 | 1.71E-05 | -0.00738944 | GPRIN2 |
| chr9 | 135184928 | 135185330 | 403 | 2 | 6.53E-06 | 1.98E-05 | 0.000108 | 1.71E-05 | 0.005040865 | NA |
| chr11 | 8168901 | 8169290 | 390 | 9 | 6.90E-05 | 2.74E-05 | 0.000885 | 1.71E-05 | 0.00300348 | RIC3-DT, RIC3 |
| chr8 | 143438683 | 143439437 | 755 | 8 | 4.33E-05 | 0.0001343 | 0.00185 | 1.71E-05 | -1.32E-05 | ZC3H3 |
| chr12 | 58087725 | 58087827 | 103 | 3 | 1.58E-05 | 2.51E-05 | 0.00039 | 1.71E-05 | -0.0105904 | NA |
| chr7 | 47088152 | 47088912 | 761 | 4 | 3.42E-05 | 0.0014271 | 0.000153 | 1.71E-05 | 0.00319575 | NA |
| chr2 | 227887016 | 227887090 | 75 | 2 | 4.34E-05 | 1.07E-05 | 0.000693 | 1.71E-05 | 0.00362862 | DAW1 |
| chr19 | 40646178 | 40646525 | 348 | 2 | 6.62E-06 | 0.0002551 | 1.61E-05 | 1.71E-05 | -0.01295613 | NA |
| chr1 | 1221496 | 1222161 | 666 | 3 | 6.66E-06 | 2.43E-05 | 0.000236 | 1.71E-05 | -0.00165787 | SDF4 |
| chr6 | 52116213 | 52116423 | 211 | 2 | 3.74E-05 | 1.61E-05 | 0.000164 | 1.71E-05 | -0.01161714 | NA |
| chr12 | 10803853 | 10804039 | 187 | 3 | 3.02E-06 | 0.000415 | 2.84E-05 | 1.71E-05 | -0.00540793 | NA |
| chr13 | 24743129 | 24743858 | 730 | 4 | 2.77E-06 | 0.0013974 | 2.54E-05 | 1.71E-05 | -0.0095982 | IRX1P1 |
| chr7 | 141917479 | 141917818 | 340 | 2 | 2.17E-05 | 1.49E-05 | 0.000198 | 1.71E-05 | 0.004856397 | MGAM, OR9A4 |
| chr15 | 73767841 | 73767974 | 134 | 3 | 4.38E-05 | 1.85E-05 | 0.000194 | 1.71E-05 | -0.01270337 | INSYN1-AS1 |
| chr14 | 75280032 | 75280629 | 598 | 4 | 8.40E-06 | 1.16E-05 | 0.003568 | 1.71E-05 | -0.00533357 | FOS |
| chr8 | 141169301 | 141170010 | 710 | 6 | 6.01E-07 | 0.0003826 | 4.03E-05 | 1.71E-05 | -0.00362229 | DENND3 |
| chr1 | 44405016 | 44405427 | 412 | 8 | 3.97E-05 | 8.59E-06 | 0.013158 | 1.71E-05 | 0.001558651 | RNF220 |
| chr8 | 26044884 | 26045590 | 707 | 6 | 5.19E-05 | 5.06E-05 | 0.0035 | 1.71E-05 | -0.00435717 | EBF2 |
| chr7 | 128444439 | 128444851 | 413 | 2 | 5.68E-06 | 0.0001184 | 2.21E-05 | 1.72E-05 | -0.00497148 | NA |
| chr3 | 130820831 | 130820968 | 138 | 2 | 5.64E-05 | 1.37E-05 | 0.000257 | 1.72E-05 | 0.011104317 | NA |
| chr14 | 21634875 | 21634926 | 52 | 2 | 9.54E-06 | 0.001117 | 1.11E-05 | 1.72E-05 | -0.00865685 | OR10G2 |
| chr13 | 33123819 | 33124231 | 413 | 2 | 3.97E-06 | 0.0004102 | 1.40E-05 | 1.72E-05 | -0.00862041 | STARD13 |
| chr11 | 8086951 | 8087284 | 334 | 2 | 5.18E-05 | 1.05E-05 | 0.00082 | 1.72E-05 | -0.01155192 | TUB |
| chr16 | 57175786 | 57176201 | 416 | 4 | 6.38E-05 | 5.43E-05 | 0.001517 | 1.72E-05 | -0.00601822 | PSME3IP1 |
| chr4 | 123747051 | 123747434 | 384 | 3 | 1.72E-05 | 0.0002765 | 5.12E-05 | 1.72E-05 | 0.006349411 | LINC01091 |
| chr17 | 17742444 | 17742615 | 172 | 2 | 2.17E-05 | 1.50E-05 | 0.000199 | 1.72E-05 | -0.01044236 | RAI1 |
| chr2 | 3702389 | 3702975 | 587 | 7 | 5.00E-06 | 0.0001017 | 0.001071 | 1.72E-05 | -0.00369336 | ALLC, AC010907.1 |
| chr1 | 7842712 | 7842793 | 82 | 2 | 1.23E-05 | 0.002098 | 1.01E-05 | 1.72E-05 | 0.005255143 | PER3 |

| | | | | | | | | | | |
|---|---|---|---|---|---|---|---|---|---|---|
| chr4 | 188208839 | 188208910 | 72 | 2 | 5.10E-05 | 1.15E-05 | 0.000509 | 1.72E-05 | 0.000165422 | NA |
| chr17 | 57369618 | 57370051 | 434 | 2 | 7.70E-05 | 1.10E-05 | 0.00062 | 1.72E-05 | 0.013583698 | MSI2 |
| chr2 | 9556476 | 9557422 | 947 | 5 | 1.41E-06 | 4.70E-05 | 9.84E-05 | 1.72E-05 | 0.000503674 | AC073195.1, ADAM17 |
| chr2 | 108448671 | 108449572 | 902 | 14 | 3.71E-05 | 0.0003333 | 0.000536 | 1.72E-05 | 0.00103211 | GCC2 |
| chr7 | 101733844 | 101733983 | 140 | 2 | 3.12E-05 | 3.27E-05 | 5.69E-05 | 1.72E-05 | -0.0134443 | NA |
| chr22 | 45891584 | 45891726 | 143 | 2 | 1.49E-05 | 1.10E-05 | 0.000642 | 1.72E-05 | 0.009070261 | NA |
| chr2 | 96145319 | 96146128 | 810 | 7 | 4.83E-05 | 6.60E-05 | 0.001075 | 1.72E-05 | 0.002503714 | DUSP2, AC012307.1 |
| chr13 | 99542935 | 99543004 | 70 | 2 | 4.19E-05 | 1.90E-05 | 0.000119 | 1.72E-05 | -0.01118351 | TM9SF2 |
| chr16 | 1911526 | 1912144 | 619 | 5 | 2.87E-05 | 2.91E-05 | 0.005107 | 1.72E-05 | -0.00665361 | HS3ST6 |
| chr5 | 11082818 | 11082830 | 13 | 2 | 1.29E-05 | 0.0025749 | 9.83E-06 | 1.72E-05 | -0.0026688 | CTNND2 |
| chr6 | 100455815 | 100456500 | 686 | 8 | 2.59E-05 | 2.39E-05 | 0.00178 | 1.72E-05 | -0.0029461 | SIM1 |
| chr19 | 4769519 | 4769678 | 160 | 7 | 3.42E-05 | 3.85E-06 | 0.012788 | 1.72E-05 | 0.001570372 | MIR7-3HG |
| chr18 | 35501173 | 35501425 | 253 | 2 | 1.08E-05 | 0.0002579 | 1.62E-05 | 1.72E-05 | -0.00250971 | NA |
| chr16 | 52552766 | 52552854 | 89 | 2 | 1.96E-05 | 2.12E-05 | 9.87E-05 | 1.72E-05 | -0.00147562 | CASC16 |
| chr14 | 23185300 | 23185490 | 191 | 2 | 2.81E-05 | 1.53E-05 | 0.000192 | 1.72E-05 | -0.02197089 | RNF212B |
| chr4 | 145097430 | 145098143 | 714 | 13 | 3.15E-05 | 2.64E-05 | 0.00385 | 1.73E-05 | 0.000476365 | AC098588.2, ANAPC10 |
| chr10 | 11443630 | 11444050 | 421 | 3 | 6.23E-06 | 0.0001774 | 0.000568 | 1.73E-05 | 0.000114921 | NA |
| chr19 | 1885143 | 1885718 | 576 | 5 | 5.30E-06 | 1.18E-05 | 0.001991 | 1.73E-05 | -0.001455 | ABHD17A |
| chr11 | 71369345 | 71369837 | 493 | 2 | 5.65E-05 | 1.07E-05 | 0.000761 | 1.73E-05 | -0.00089451 | NA |
| chr4 | 188270352 | 188270550 | 199 | 3 | 3.57E-05 | 2.61E-05 | 0.000123 | 1.73E-05 | -0.00023122 | NA |
| chr21 | 16539113 | 16539922 | 810 | 5 | 3.81E-05 | 4.36E-05 | 0.00095 | 1.73E-05 | 0.000417511 | MIR99AHG, MIR99A, MIRLET7C |
| chr1 | 3821661 | 3822046 | 386 | 6 | 3.29E-05 | 6.92E-05 | 0.005943 | 1.73E-05 | -0.0012529 | CEP104 |
| chr17 | 8189170 | 8189860 | 691 | 4 | 3.70E-06 | 8.82E-06 | 0.004608 | 1.73E-05 | -0.00412567 | AC129492.6, BORCS6 |
| chr17 | 79211048 | 79211342 | 295 | 3 | 1.43E-05 | 2.18E-05 | 0.000135 | 1.73E-05 | -0.0127779 | RBFOX3 |
| chr4 | 25858188 | 25858499 | 312 | 2 | 4.91E-05 | 1.06E-05 | 0.000839 | 1.73E-05 | -0.00494661 | SEL1L3 |
| chr13 | 99439774 | 99439827 | 54 | 3 | 2.80E-05 | 0.0001272 | 6.29E-05 | 1.73E-05 | -0.00147859 | NA |
| chr11 | 77287176 | 77287456 | 281 | 2 | 3.62E-05 | 9.16E-05 | 2.55E-05 | 1.73E-05 | -0.00748453 | GDPD4 |
| chr3 | 107674378 | 107674609 | 232 | 3 | 5.90E-06 | 0.0023011 | 5.99E-06 | 1.73E-05 | 0.003382708 | BBX |
| chr22 | 25199442 | 25200343 | 902 | 8 | 1.78E-05 | 0.0001005 | 0.000512 | 1.73E-05 | -0.00421842 | CRYBB3 |
| chr2 | 26034585 | 26035542 | 958 | 6 | 6.46E-05 | 1.69E-05 | 0.006806 | 1.73E-05 | -0.00052005 | RAB10 |
| chr1 | 209576617 | 209577094 | 478 | 3 | 6.36E-05 | 1.53E-05 | 0.001195 | 1.73E-05 | -0.0082649 | NA |
| chr9 | 96449227 | 96450078 | 852 | 7 | 7.29E-06 | 0.0002174 | 6.45E-05 | 1.73E-05 | 0.001001023 | NA |
| chr2 | 128503985 | 128504226 | 242 | 2 | 1.65E-05 | 1.15E-05 | 0.000514 | 1.73E-05 | 0.001570678 | NA |
| chr5 | 172188934 | 172189570 | 637 | 5 | 5.58E-06 | 1.30E-05 | 0.003155 | 1.73E-05 | 0.004757629 | NA |
| chr16 | 67004806 | 67005469 | 664 | 6 | 1.04E-05 | 0.0002278 | 2.54E-05 | 1.73E-05 | 6.28E-05 | CES4A |
| chr5 | 88660571 | 88661065 | 495 | 3 | 4.71E-07 | 2.00E-05 | 0.0004 | 1.73E-05 | -0.00385318 | LINC00461 |
| chr1 | 165144807 | 165145216 | 410 | 2 | 5.23E-06 | 0.0001554 | 1.98E-05 | 1.73E-05 | 0.010137388 | NA |
| chr2 | 9113807 | 9113941 | 135 | 2 | 4.80E-05 | 1.31E-05 | 0.000311 | 1.73E-05 | 0.023828773 | AC093904.3 |
| chr14 | 95876382 | 95876782 | 401 | 6 | 2.86E-05 | 0.0001112 | 0.002736 | 1.73E-05 | 0.000197704 | TUNAR |
| chr3 | 15809458 | 15810031 | 574 | 2 | 6.44E-05 | 1.10E-05 | 0.000673 | 1.73E-05 | -0.00435359 | ANKRD28 |
| chr17 | 81442471 | 81442739 | 269 | 2 | 1.27E-06 | 1.12E-05 | 0.000593 | 1.73E-05 | 0.001702258 | BAHCC1 |
| chr7 | 102464360 | 102465334 | 975 | 11 | 8.04E-05 | 7.18E-05 | 0.001205 | 1.73E-05 | 0.0009696 | LRWD1, ALKBH4 |
| chr1 | 10704455 | 10704839 | 385 | 3 | 3.05E-05 | 6.47E-06 | 0.004098 | 1.73E-05 | -0.01426962 | CASZ1 |
| chr19 | 779163 | 779556 | 394 | 4 | 6.07E-05 | 9.84E-06 | 0.002998 | 1.73E-05 | -0.0013379 | NA |
| chr2 | 200585783 | 200586580 | 798 | 12 | 2.22E-05 | 0.0004246 | 0.005073 | 1.74E-05 | 0.000161793 | AOX1 |
| chr2 | 118843122 | 118843680 | 559 | 3 | 2.80E-05 | 1.21E-05 | 0.000664 | 1.74E-05 | 0.000595647 | EN1 |
| chr14 | 105247816 | 105249127 | 1312 | 17 | 3.86E-05 | 0.0001894 | 0.008923 | 1.74E-05 | -0.00178023 | BTBD6, BRF1 |
| chr8 | 142356164 | 142356627 | 464 | 5 | 1.65E-05 | 6.54E-05 | 0.001248 | 1.74E-05 | -0.00169818 | TSNARE1 |
| chr2 | 37344035 | 37344589 | 555 | 7 | 5.72E-07 | 0.0005936 | 0.000272 | 1.74E-05 | -0.00326583 | QPCT |
| chr17 | 77307781 | 77307911 | 131 | 2 | 2.53E-05 | 1.07E-05 | 0.00081 | 1.74E-05 | -0.00720555 | SEPTIN9 |
| chr2 | 85753137 | 85753898 | 762 | 14 | 8.06E-06 | 0.0005853 | 0.004316 | 1.74E-05 | -0.00114034 | ATOH8 |
| chr15 | 74129989 | 74130736 | 748 | 13 | 2.67E-06 | 0.0017518 | 0.000377 | 1.74E-05 | 0.000694265 | ISLR2 |
| chr6 | 10305917 | 10306212 | 296 | 2 | 2.20E-05 | 3.02E-05 | 6.33E-05 | 1.74E-05 | -0.01546799 | NA |
| chr20 | 41717530 | 41717543 | 14 | 2 | 3.14E-05 | 2.48E-05 | 8.06E-05 | 1.74E-05 | 0.000873193 | NA |
| chr18 | 24662516 | 24662617 | 102 | 2 | 3.93E-06 | 0.0024882 | 1.00E-05 | 1.74E-05 | 0.004887112 | LINC01915 |
| chr11 | 68296244 | 68296489 | 246 | 2 | 9.36E-06 | 0.0004411 | 1.39E-05 | 1.74E-05 | -0.0049518 | NA |
| chr6 | 4491372 | 4491795 | 424 | 2 | 4.67E-06 | 0.0001761 | 1.89E-05 | 1.74E-05 | -0.00590983 | LINC02533, AL162718.1 |
| chr11 | 130897483 | 130897865 | 383 | 2 | 2.93E-05 | 1.58E-05 | 0.000185 | 1.75E-05 | -0.01365459 | SNX19 |
| chr18 | 77058446 | 77058859 | 414 | 4 | 1.98E-06 | 0.0011803 | 9.32E-06 | 1.75E-05 | -0.00261595 | MBP |
| chr1 | 157691663 | 157691747 | 85 | 2 | 2.64E-06 | 0.000387 | 1.45E-05 | 1.75E-05 | -0.00531183 | FCRL3 |
| chr7 | 133262821 | 133263078 | 258 | 2 | 9.59E-06 | 0.0001701 | 1.92E-05 | 1.75E-05 | -0.00999916 | EXOC4 |
| chr7 | 150799977 | 150800258 | 282 | 5 | 2.76E-05 | 0.0001095 | 0.001816 | 1.75E-05 | -0.01037386 | TMEM176B |
| chr7 | 64649812 | 64649874 | 63 | 2 | 5.41E-05 | 1.09E-05 | 0.000783 | 1.75E-05 | 0.000442589 | NA |
| chr22 | 39988352 | 39988895 | 544 | 2 | 1.11E-05 | 1.18E-05 | 0.000499 | 1.75E-05 | 0.012879905 | NA |
| chr5 | 13944980 | 13945881 | 902 | 4 | 1.12E-07 | 0.0004255 | 1.17E-05 | 1.76E-05 | -0.00240597 | NA |
| chr5 | 141421719 | 141421915 | 197 | 3 | 2.71E-06 | 2.18E-05 | 0.000516 | 1.76E-05 | -0.01339812 | PCDHGA1, PCDHGA2, PCDHGA3, PCDHGB1, PCDHGA4, PCDHGB2, PCDHGA5, PCDHGB3, PCDHGA6, PCDHGA7, PCDHGB4, PCDHGA8, PCDHGB5, PCDHGA9, PCDHGB6, PCDHGA10, PCDHGB7, PCDHGA11 |
| chr2 | 46689936 | 46689948 | 13 | 2 | 8.39E-06 | 0.0209987 | 8.73E-06 | 1.76E-05 | 0.004920576 | NA |
| chr2 | 207680802 | 207681359 | 558 | 3 | 2.46E-05 | 0.0004245 | 0.000381 | 1.76E-05 | -0.01502862 | NA |
| chr1 | 95509676 | 95510299 | 624 | 6 | 2.96E-05 | 8.41E-05 | 0.000702 | 1.76E-05 | -0.00178453 | LINC02607 |
| chr1 | 2443571 | 2444251 | 681 | 3 | 2.41E-05 | 6.87E-06 | 0.003481 | 1.76E-05 | 0.004722383 | PLCH2 |
| chr13 | 25067169 | 25067187 | 19 | 2 | 3.22E-05 | 4.53E-05 | 4.30E-05 | 1.76E-05 | -0.01426077 | NA |
| chr17 | 74936493 | 74936897 | 405 | 3 | 3.07E-06 | 0.0096887 | 0.000331 | 1.76E-05 | -0.00722659 | OTOP3 |
| chr11 | 62678083 | 62678649 | 567 | 4 | 9.94E-07 | 2.71E-05 | 0.000155 | 1.76E-05 | -0.00473087 | UBXN1 |
| chr4 | 141711006 | 141711327 | 322 | 2 | 1.15E-05 | 9.11E-05 | 2.61E-05 | 1.76E-05 | -0.00732636 | IL15 |
| chr12 | 12266818 | 12267201 | 384 | 6 | 1.89E-05 | 9.34E-06 | 0.007992 | 1.76E-05 | 0.0006244 | LRP6 |
| chr1 | 25858895 | 25860139 | 1245 | 10 | 4.17E-06 | 0.0005232 | 4.66E-05 | 1.76E-05 | -0.00082467 | AL033528.2, AUNIP |
| chr19 | 45456395 | 45456463 | 69 | 2 | 1.40E-05 | 0.0002056 | 1.80E-05 | 1.76E-05 | -0.00324156 | ERCC1 |
| chr2 | 230991344 | 230991540 | 197 | 2 | 9.42E-06 | 0.0001368 | 2.13E-05 | 1.76E-05 | -0.00768043 | SPATA3, SPATA3-AS1 |
| chr9 | 95048335 | 95048487 | 153 | 2 | 6.72E-05 | 1.16E-05 | 0.000557 | 1.76E-05 | -3.24E-05 | AOPEP |
| chr11 | 116826853 | 116827163 | 311 | 2 | 5.19E-05 | 1.09E-05 | 0.000794 | 1.76E-05 | -0.01100056 | NA |
| chr2 | 42492363 | 42493186 | 824 | 6 | 6.17E-06 | 0.0001632 | 0.000228 | 1.77E-05 | -0.005423 | KCNG3 |
| chr2 | 204325678 | 204325845 | 168 | 2 | 1.09E-05 | 0.0001552 | 2.02E-05 | 1.77E-05 | -0.00337373 | NA |
| chr2 | 47889742 | 47889985 | 244 | 4 | 1.84E-05 | 0.0006771 | 3.45E-05 | 1.77E-05 | -0.00458131 | FBXO11 |
| chr15 | 99014935 | 99015490 | 556 | 3 | 9.77E-06 | 0.0001112 | 3.61E-05 | 1.77E-05 | 0.00157753 | LUNAR1, AC036108.1 |
| chr10 | 95194196 | 95194495 | 300 | 2 | 1.46E-05 | 1.28E-05 | 0.000366 | 1.77E-05 | 0.005680359 | ACSM6 |
| chr19 | 38596703 | 38597127 | 425 | 2 | 5.04E-06 | 1.48E-05 | 0.000228 | 1.77E-05 | 0.000328518 | AC008649.1, MAP4K1 |
| chr3 | 45865006 | 45865364 | 359 | 2 | 2.01E-05 | 1.63E-05 | 0.000178 | 1.77E-05 | -0.01336864 | AC099782.1, LZTFL1 |
| chr6 | 158501753 | 158502748 | 996 | 5 | 2.05E-05 | 0.0001095 | 0.000357 | 1.77E-05 | 0.000937614 | TULP4 |
| chr9 | 36258117 | 36258974 | 858 | 14 | 1.96E-06 | 0.0014026 | 7.57E-05 | 1.77E-05 | -2.08E-05 | CLTA, GNE |
| chr8 | 133477746 | 133478023 | 278 | 2 | 1.13E-05 | 1.40E-05 | 0.000269 | 1.77E-05 | 0.000794654 | ST3GAL1 |
| chr1 | 240591317 | 240591402 | 86 | 3 | 1.19E-05 | 0.0027441 | 1.05E-05 | 1.77E-05 | -0.00649718 | GREM2 |
| chr10 | 71209051 | 71209583 | 533 | 4 | 1.21E-05 | 0.0003991 | 0.000238 | 1.77E-05 | 0.002137117 | NA |
| chr10 | 5233921 | 5234355 | 435 | 3 | 2.58E-05 | 2.07E-05 | 0.000881 | 1.77E-05 | -0.01074901 | NA |
| chr14 | 24240550 | 24240635 | 86 | 2 | 4.12E-05 | 1.25E-05 | 0.000408 | 1.77E-05 | 0.004655582 | TINF2 |
| chr19 | 2809057 | 2809434 | 378 | 5 | 2.05E-05 | 1.37E-05 | 0.000476 | 1.77E-05 | -0.00085813 | THOP1 |
| chr8 | 96161998 | 96162154 | 157 | 2 | 7.05E-06 | 7.24E-05 | 3.05E-05 | 1.77E-05 | -0.01609526 | NA |
| chr6 | 157875536 | 157875611 | 76 | 2 | 3.04E-05 | 4.96E-05 | 4.03E-05 | 1.77E-05 | -0.00028684 | SNX9 |
| chr22 | 39995348 | 39996067 | 720 | 7 | 8.64E-06 | 0.0003948 | 0.000971 | 1.78E-05 | -0.00494387 | FAM83F |
| chr5 | 7755186 | 7755687 | 502 | 3 | 1.96E-06 | 0.0012005 | 0.000223 | 1.78E-05 | -0.03333777 | ADCY2 |
| chr21 | 33976403 | 33976827 | 425 | 6 | 1.65E-05 | 7.06E-05 | 0.002949 | 1.78E-05 | -0.00597762 | LINC00649 |
| chr6 | 624306 | 624671 | 366 | 2 | 2.87E-05 | 1.09E-05 | 0.000811 | 1.78E-05 | 0.000428242 | EXOC2 |
| chr20 | 51862936 | 51863121 | 186 | 4 | 1.52E-05 | 0.0002677 | 1.67E-05 | 1.78E-05 | 0.000635871 | NA |
| chr9 | 95087652 | 95088068 | 417 | 2 | 1.45E-05 | 1.75E-05 | 0.000154 | 1.78E-05 | -0.00326143 | NA |
| chr6 | 29738038 | 29738101 | 64 | 2 | 5.14E-05 | 1.96E-05 | 0.000122 | 1.78E-05 | -0.0062405 | HLA-F, HLA-F-AS1 |
| chr1 | 31772322 | 31773059 | 738 | 4 | 6.53E-06 | 4.77E-05 | 0.000137 | 1.78E-05 | 0.003769236 | NA |
| chr11 | 3208708 | 3209065 | 358 | 2 | 1.96E-05 | 2.75E-05 | 7.33E-05 | 1.78E-05 | -0.00603786 | NA |
| chr11 | 49208484 | 49208616 | 133 | 4 | 3.77E-05 | 7.36E-05 | 4.99E-05 | 1.78E-05 | -0.00787203 | FOLH1 |
| chr3 | 132661346 | 132661733 | 388 | 2 | 1.53E-07 | 0.0008703 | 1.21E-05 | 1.78E-05 | -0.0163464 | UBA5, NPHP3-ACAD11 |
| chr6 | 160277669 | 160278088 | 420 | 3 | 1.01E-05 | 2.88E-05 | 0.000838 | 1.78E-05 | -0.00072118 | NA |
| chr6 | 29632059 | 29632865 | 807 | 14 | 3.05E-05 | 3.26E-05 | 0.013778 | 1.78E-05 | -0.00148636 | GABBR1 |
| chr14 | 30656545 | 30657082 | 538 | 2 | 6.11E-05 | 1.13E-05 | 0.00069 | 1.78E-05 | 0.003707401 | SCFD1 |
| chr19 | 49659515 | 49659641 | 127 | 2 | 1.50E-05 | 0.0005061 | 1.38E-05 | 1.78E-05 | -0.00312926 | IRF3 |
| chr2 | 37157380 | 37157702 | 323 | 3 | 4.41E-06 | 1.62E-05 | 0.000774 | 1.78E-05 | 0.01246503 | NA |
| chr1 | 95936589 | 95936602 | 14 | 2 | 2.60E-05 | 1.69E-05 | 0.000168 | 1.78E-05 | -0.00110217 | NA |
| chr1 | 154520471 | 154520476 | 6 | 2 | 9.69E-05 | 1.10E-05 | 0.0008 | 1.78E-05 | -0.00991714 | TDRD10 |
| chr11 | 12137976 | 12138579 | 604 | 5 | 1.22E-05 | 0.0001743 | 6.32E-05 | 1.78E-05 | -0.0042024 | MICAL2 |
| chr10 | 93062415 | 93062445 | 31 | 2 | 1.84E-05 | 1.10E-05 | 0.00079 | 1.78E-05 | 0.009397206 | CYP26C1 |
| chr17 | 81937130 | 81937708 | 579 | 17 | 1.07E-05 | 0.0001717 | 0.000156 | 1.78E-05 | -0.00031089 | PYCR1 |
| chr7 | 9736878 | 9737311 | 434 | 3 | 3.51E-05 | 7.14E-06 | 0.002981 | 1.79E-05 | 0.001216167 | AC060834.2 |
| chr22 | 40030597 | 40031119 | 523 | 2 | 8.80E-07 | 0.0005941 | 1.33E-05 | 1.79E-05 | -0.00385327 | FAM83F |
| chr11 | 60701195 | 60701357 | 163 | 3 | 0.000107279 | 1.01E-05 | 0.001175 | 1.79E-05 | -0.00705736 | MS4A8 |
| chr16 | 4733988 | 4734530 | 543 | 12 | 2.40E-05 | 0.0002195 | 0.00343 | 1.79E-05 | 0.000587771 | C16orf71, ANKS3, AC020663.1 |
| chr8 | 80697163 | 80697524 | 362 | 3 | 3.65E-05 | 1.02E-05 | 0.001113 | 1.79E-05 | -0.00116652 | ZNF704 |
| chr2 | 239049555 | 239049858 | 304 | 2 | 5.50E-06 | 3.36E-05 | 5.89E-05 | 1.79E-05 | 0.006916117 | HDAC4 |
| chr15 | 50120179 | 50120658 | 480 | 3 | 1.22E-05 | 1.11E-05 | 0.00066 | 1.79E-05 | -0.02007936 | ATP8B4 |
| chr4 | 54274549 | 54274917 | 369 | 3 | 1.72E-06 | 0.0021805 | 6.27E-06 | 1.79E-05 | -0.00328976 | AC058822.1, PDGFRA |
| chr8 | 116136461 | 116136759 | 299 | 2 | 7.94E-06 | 0.0004332 | 1.43E-05 | 1.79E-05 | -0.00594585 | LINC00536 |
| chr13 | 110305328 | 110306071 | 744 | 3 | 3.02E-05 | 7.34E-06 | 0.0032 | 1.79E-05 | 0.009334026 | COL4A2, COL4A1 |
| chr5 | 173233793 | 173234484 | 692 | 5 | 2.57E-06 | 0.0001118 | 4.33E-05 | 1.79E-05 | -0.00176134 | NKX2-5 |
| chr1 | 1519923 | 1519926 | 4 | 2 | 4.23E-05 | 1.48E-05 | 0.000242 | 1.79E-05 | 0.005803544 | ATAD3A |
| chr6 | 129292876 | 129292940 | 65 | 2 | 3.80E-05 | 3.00E-05 | 6.70E-05 | 1.79E-05 | -0.00034722 | LAMA2 |
| chr17 | 74604753 | 74605423 | 671 | 2 | 7.84E-05 | 1.08E-05 | 0.000948 | 1.79E-05 | -0.00852842 | AC064805.1 |
| chr3 | 139477649 | 139477971 | 323 | 3 | 5.80E-06 | 0.0011384 | 9.38E-06 | 1.79E-05 | -0.0091248 | AC046134.2, RBP2 |
| chr7 | 39330433 | 39330463 | 31 | 2 | 2.76E-05 | 3.04E-05 | 6.61E-05 | 1.79E-05 | -0.01199127 | POU6F2 |
| chr6 | 30649788 | 30650119 | 332 | 4 | 3.59E-07 | 0.0074017 | 3.54E-06 | 1.80E-05 | -0.00232979 | C6orf136 |
| chr7 | 102093635 | 102093972 | 338 | 3 | 3.90E-05 | 1.17E-05 | 0.000789 | 1.80E-05 | -0.00176536 | CUX1 |
| chr9 | 134843828 | 134843882 | 55 | 2 | 1.15E-05 | 0.0001245 | 2.28E-05 | 1.80E-05 | -0.00184574 | COL5A1 |
| chr10 | 132376555 | 132376688 | 134 | 2 | 1.82E-05 | 6.72E-05 | 3.26E-05 | 1.80E-05 | -0.00451865 | LRRC27 |
| chr11 | 69205439 | 69206174 | 736 | 6 | 6.00E-05 | 1.21E-05 | 0.004035 | 1.80E-05 | -0.0030997 | NA |
| chr17 | 76498131 | 76498708 | 578 | 3 | 1.08E-05 | 1.20E-05 | 0.001608 | 1.80E-05 | -0.00164907 | RHBDF2 |
| chr8 | 86069541 | 86070135 | 595 | 9 | 9.43E-06 | 0.0001428 | 2.32E-05 | 1.80E-05 | -0.00813416 | ATP6V0D2, PSKH2 |
| chr7 | 137344226 | 137344646 | 421 | 4 | 9.10E-07 | 4.82E-05 | 0.000102 | 1.80E-05 | 0.009547655 | NA |
| chr15 | 41557755 | 41558305 | 551 | 4 | 1.12E-05 | 0.0080165 | 9.15E-05 | 1.80E-05 | 0.000803037 | TYRO3 |
| chr12 | 110431232 | 110431552 | 321 | 4 | 2.41E-06 | 0.0109399 | 3.40E-05 | 1.80E-05 | -0.00920577 | AC144548.1 |
| chr7 | 1931287 | 1931831 | 545 | 4 | 7.78E-06 | 2.79E-05 | 0.001999 | 1.80E-05 | -0.00288075 | MAD1L1, AC069288.1 |
| chr19 | 50715543 | 50715910 | 368 | 2 | 4.39E-06 | 1.15E-05 | 0.000648 | 1.80E-05 | -0.00859941 | SHANK1 |
| chr15 | 45139456 | 45139492 | 37 | 2 | 3.39E-05 | 1.08E-05 | 0.00096 | 1.80E-05 | -0.00643085 | DUOX1 |
| chr15 | 78809639 | 78810879 | 1241 | 9 | 6.01E-06 | 0.0001023 | 0.001675 | 1.80E-05 | -0.0004361 | MORF4L1, ADAMTS7 |
| chr9 | 95477738 | 95478475 | 738 | 4 | 5.81E-06 | 0.0002807 | 6.66E-05 | 1.80E-05 | -0.00429283 | PTCH1 |
| chr17 | 21198258 | 21198647 | 390 | 4 | 7.80E-06 | 0.0001801 | 4.08E-05 | 1.80E-05 | -0.00045399 | TMEM11 |
| chr7 | 22822889 | 22823128 | 240 | 5 | 1.43E-05 | 4.84E-05 | 0.00015 | 1.80E-05 | 0.003841995 | AC005682.2 |
| chr1 | 960846 | 961383 | 538 | 5 | 8.28E-06 | 1.59E-05 | 0.003506 | 1.80E-05 | 0.000495372 | KLHL17 |
| chr19 | 19138600 | 19139258 | 659 | 5 | 8.96E-06 | 0.0001286 | 0.001326 | 1.80E-05 | 0.000895625 | NA |
| chr8 | 129415563 | 129416117 | 555 | 4 | 1.18E-05 | 9.94E-06 | 0.00344 | 1.80E-05 | 0.018999197 | AC011257.1, CCDC26 |
| chr12 | 109698210 | 109698239 | 30 | 2 | 4.82E-05 | 1.68E-05 | 0.000175 | 1.80E-05 | -0.00625135 | NA |
| chr20 | 4250509 | 4250897 | 389 | 2 | 2.71E-05 | 1.23E-05 | 0.000471 | 1.80E-05 | -0.01063777 | NA |
| chr10 | 23499745 | 23499803 | 59 | 2 | 5.26E-05 | 1.30E-05 | 0.000376 | 1.80E-05 | -0.00190038 | AL512603.2 |
| chr16 | 89711839 | 89712252 | 414 | 3 | 0.000123113 | 8.28E-06 | 0.001915 | 1.80E-05 | 0.003666243 | VPS9D1-AS1, VPS9D1 |

| | | | | | | | | | | |
|---|---|---|---|---|---|---|---|---|---|---|
| chr12 | 122859287 | 122860223 | 937 | 4 | 5.78E-06 | 3.91E-05 | 8.01E-05 | 1.81E-05 | -0.00225806 | HIP1R |
| chr1 | 228486893 | 228487969 | 1077 | 11 | 3.37E-05 | 2.61E-05 | 0.003515 | 1.81E-05 | 0.001080656 | RNF187, AL139288.2 |
| chr22 | 20182772 | 20183247 | 476 | 2 | 4.66E-05 | 1.90E-05 | 0.000134 | 1.81E-05 | -0.00680682 | NA |
| chr6 | 166463002 | 166463514 | 513 | 6 | 6.82E-05 | 2.47E-05 | 0.003221 | 1.81E-05 | 0.016423728 | RPS6KA2, RPS6KA2-IT1 |
| chr3 | 126266245 | 126266720 | 476 | 3 | 5.45E-06 | 5.31E-05 | 4.91E-05 | 1.81E-05 | -0.00896852 | NA |
| chr10 | 13439846 | 13440679 | 834 | 6 | 1.31E-05 | 0.003231 | 0.000414 | 1.81E-05 | -0.00329377 | BEND7 |
| chr2 | 199910282 | 199911150 | 869 | 9 | 2.56E-06 | 0.0001602 | 0.001834 | 1.81E-05 | -0.01887945 | AC097717.1 |
| chr20 | 61251688 | 61252156 | 469 | 3 | 2.96E-05 | 1.02E-05 | 0.000847 | 1.81E-05 | -0.00446449 | NA |
| chr6 | 31123152 | 31123606 | 455 | 4 | 7.63E-06 | 0.0003378 | 6.72E-05 | 1.81E-05 | -0.00486713 | PSORS1C1 |
| chr22 | 30423262 | 30424269 | 1008 | 10 | 8.22E-06 | 0.0010255 | 0.000141 | 1.81E-05 | -0.00078043 | SEC14L2, AC004832.3 |
| chr12 | 58543097 | 58543205 | 109 | 2 | 4.01E-05 | 2.38E-05 | 9.20E-05 | 1.81E-05 | -0.01050299 | NA |
| chr5 | 141176675 | 141176900 | 226 | 2 | 4.53E-06 | 4.66E-05 | 4.38E-05 | 1.81E-05 | -0.01087579 | AC244517.11 |
| chr12 | 26194663 | 26195244 | 582 | 6 | 6.06E-05 | 0.0002701 | 0.001031 | 1.81E-05 | -0.00473273 | SSPN |
| chr12 | 8841524 | 8842076 | 553 | 3 | 1.15E-05 | 2.13E-05 | 0.000289 | 1.81E-05 | -0.01319161 | A2ML1 |
| chr13 | 20394797 | 20395354 | 558 | 3 | 4.51E-06 | 0.0026449 | 0.000338 | 1.81E-05 | -0.01668392 | NA |
| chr10 | 32022179 | 32022502 | 324 | 2 | 2.10E-06 | 0.002075 | 1.06E-05 | 1.81E-05 | 0.01318836 | KIF5B |
| chr6 | 89353612 | 89354266 | 655 | 3 | 7.53E-06 | 7.46E-06 | 0.003187 | 1.81E-05 | 0.015509747 | NA |
| chr12 | 26622869 | 26623356 | 488 | 4 | 2.47E-05 | 8.32E-06 | 0.002967 | 1.81E-05 | -0.00676384 | ITPR2 |
| chr13 | 40770777 | 40771433 | 657 | 13 | 2.00E-05 | 0.0001617 | 0.003682 | 1.81E-05 | 0.000572548 | MRPS31 |
| chr10 | 48289763 | 48289809 | 47 | 2 | 1.21E-05 | 1.63E-05 | 0.000193 | 1.81E-05 | -0.01744705 | NA |
| chr17 | 74447896 | 74448036 | 141 | 2 | 5.16E-06 | 0.0015464 | 1.11E-05 | 1.81E-05 | -0.011034 | GPRC5C |
| chr20 | 46307174 | 46307489 | 316 | 3 | 3.99E-06 | 0.0007206 | 1.66E-05 | 1.81E-05 | -0.00351891 | CDH22 |
| chr8 | 119416284 | 119417278 | 995 | 4 | 2.11E-06 | 0.0005491 | 0.000364 | 1.81E-05 | -0.00284028 | CCN3, AC021733.4 |
| chr11 | 365961 | 366487 | 527 | 5 | 3.76E-05 | 0.0001043 | 0.001085 | 1.81E-05 | -0.0060327 | NA |
| chr9 | 136720241 | 136720804 | 564 | 5 | 1.32E-05 | 0.0002782 | 0.00011 | 1.82E-05 | -0.00275983 | DIPK1B |
| chr15 | 91926164 | 91926185 | 22 | 2 | 3.11E-05 | 2.64E-05 | 8.04E-05 | 1.82E-05 | -0.00801226 | SLCO3A1 |
| chr11 | 84712284 | 84712889 | 606 | 4 | 1.05E-05 | 0.0001428 | 0.000747 | 1.82E-05 | -0.00638874 | DLG2 |
| chr12 | 12685493 | 12685653 | 161 | 2 | 1.85E-05 | 1.20E-05 | 0.000555 | 1.82E-05 | -0.00208697 | GPR19 |
| chr17 | 41350825 | 41351412 | 588 | 7 | 1.53E-05 | 2.99E-05 | 0.005007 | 1.82E-05 | -0.00124651 | KRT33A |
| chr20 | 44954073 | 44954517 | 445 | 3 | 5.28E-06 | 0.0001882 | 1.89E-05 | 1.82E-05 | 0.001339264 | PABPC1L, TOMM34 |
| chr22 | 38818563 | 38818976 | 414 | 2 | 1.06E-05 | 4.65E-05 | 4.43E-05 | 1.82E-05 | 0.000313317 | NPTXR |
| chr11 | 69115888 | 69115966 | 79 | 2 | 5.84E-05 | 1.90E-05 | 0.000137 | 1.82E-05 | -0.00230524 | TPCN2, AP003071.5 |
| chr15 | 49879100 | 49879751 | 652 | 2 | 4.21E-05 | 1.79E-05 | 0.000156 | 1.82E-05 | -0.0119564 | AC025040.1, ATP8B4 |
| chr17 | 79845480 | 79845703 | 224 | 2 | 1.13E-05 | 6.10E-05 | 3.56E-05 | 1.82E-05 | 0.007562636 | NA |
| chr8 | 141367982 | 141368366 | 385 | 3 | 4.91E-05 | 7.77E-06 | 0.002359 | 1.82E-05 | 0.000355093 | NA |
| chr4 | 51995536 | 51995665 | 130 | 2 | 5.55E-05 | 1.13E-05 | 0.000776 | 1.82E-05 | -0.00430643 | LRRC66 |
| chr8 | 22696202 | 22696719 | 518 | 2 | 2.52E-05 | 1.22E-05 | 0.000511 | 1.82E-05 | -0.00173537 | AC055854.1 |
| chr7 | 155296612 | 155297320 | 709 | 5 | 1.91E-05 | 0.0001498 | 0.00032 | 1.82E-05 | -0.0013847 | AC144652.1 |
| chr3 | 12809870 | 12810077 | 208 | 2 | 5.04E-05 | 1.13E-05 | 0.000793 | 1.82E-05 | -0.00616108 | CAND2 |
| chr2 | 208146329 | 208147571 | 1243 | 5 | 7.10E-06 | 7.02E-05 | 0.000178 | 1.82E-05 | -0.00366419 | NA |
| chr14 | 90398188 | 90398558 | 371 | 4 | 2.13E-05 | 4.66E-05 | 0.000633 | 1.82E-05 | -0.00176943 | CALM1 |
| chr2 | 68467036 | 68467577 | 542 | 12 | 2.37E-05 | 0.0004119 | 0.000857 | 1.82E-05 | 0.000625973 | APLF, FBXO48 |
| chr3 | 184037719 | 184038056 | 338 | 2 | 2.88E-05 | 1.40E-05 | 0.000301 | 1.82E-05 | -0.01063558 | HTR3D |
| chr1 | 53685160 | 53685380 | 221 | 2 | 5.73E-05 | 1.10E-05 | 0.000965 | 1.82E-05 | -0.0241013 | GLIS1 |
| chr4 | 4693848 | 4693988 | 141 | 2 | 1.03E-05 | 2.39E-05 | 9.31E-05 | 1.82E-05 | -0.00634258 | STX18-AS1 |
| chr22 | 48334316 | 48334672 | 357 | 3 | 2.77E-05 | 4.77E-05 | 0.000142 | 1.82E-05 | -0.00839297 | NA |
| chr3 | 27826971 | 27827160 | 190 | 2 | 3.17E-05 | 3.08E-05 | 6.73E-05 | 1.82E-05 | 0.00257049 | LINC01980 |
| chr13 | 110361530 | 110362038 | 509 | 2 | 4.88E-06 | 3.75E-05 | 5.45E-05 | 1.82E-05 | 0.019113129 | COL4A2 |
| chr12 | 130424548 | 130425111 | 564 | 3 | 1.02E-05 | 0.0002399 | 3.30E-05 | 1.82E-05 | -0.004431 | AC063926.3, RIMBP2 |
| chr8 | 48921274 | 48921795 | 522 | 5 | 4.42E-05 | 1.28E-05 | 0.000752 | 1.83E-05 | 9.97E-05 | SNAI2 |
| chr1 | 30953765 | 30953918 | 154 | 2 | 5.69E-05 | 1.10E-05 | 0.000946 | 1.83E-05 | 0.000211766 | PUM1 |
| chr2 | 46296180 | 46296651 | 472 | 4 | 2.10E-05 | 6.95E-06 | 0.00353 | 1.83E-05 | -0.01262772 | EPAS1 |
| chr5 | 149958078 | 149958143 | 66 | 2 | 3.93E-05 | 2.03E-05 | 0.000122 | 1.83E-05 | 0.005275128 | NA |
| chr19 | 32676719 | 32677285 | 567 | 5 | 2.55E-05 | 0.0013418 | 0.000157 | 1.83E-05 | -0.00649165 | RGS9BP |
| chr19 | 19320450 | 19321038 | 589 | 12 | 1.02E-05 | 0.0005514 | 0.000758 | 1.83E-05 | -0.00018725 | MAU2, SUGP1 |
| chr12 | 2273702 | 2274065 | 364 | 2 | 1.04E-05 | 8.70E-05 | 2.83E-05 | 1.83E-05 | -0.00405364 | CACNA1C, CACNA1C-IT3 |
| chr17 | 44980905 | 44981199 | 295 | 2 | 1.44E-05 | 8.50E-05 | 2.87E-05 | 1.83E-05 | -0.02150816 | NMT1 |
| chr14 | 103867644 | 103867775 | 132 | 2 | 6.99E-06 | 0.006955 | 9.51E-06 | 1.83E-05 | -0.00498843 | AL132712.1 |
| chr17 | 44025086 | 44025323 | 238 | 3 | 8.10E-06 | 4.02E-05 | 6.84E-05 | 1.83E-05 | 0.001531714 | U3 |
| chr1 | 204101903 | 204102334 | 432 | 4 | 1.35E-05 | 3.34E-05 | 0.000165 | 1.83E-05 | 0.002922123 | SOX13 |
| chr2 | 932987 | 933272 | 286 | 5 | 3.91E-05 | 2.29E-05 | 0.001847 | 1.83E-05 | -0.01188148 | NA |
| chr14 | 81427927 | 81428357 | 431 | 4 | 3.76E-05 | 7.42E-05 | 0.000868 | 1.83E-05 | -0.00777708 | STON2 |
| chr10 | 124711341 | 124711407 | 67 | 2 | 3.48E-05 | 3.15E-05 | 6.59E-05 | 1.83E-05 | 5.57E-05 | FAM53B-AS1, AC068896.1, FAM53B |
| chr13 | 43785279 | 43785860 | 582 | 4 | 4.18E-05 | 5.71E-06 | 0.005899 | 1.83E-05 | -0.00351449 | ENOX1 |
| chr13 | 50528822 | 50529237 | 416 | 3 | 2.03E-06 | 0.0005704 | 1.53E-05 | 1.83E-05 | -0.00507341 | DLEU1, DLEU7 |
| chr10 | 326649 | 327218 | 570 | 5 | 8.03E-05 | 1.31E-05 | 0.003328 | 1.83E-05 | 0.000920612 | DIP2C |
| chr20 | 19688528 | 19688538 | 11 | 2 | 1.96E-05 | 1.90E-05 | 0.000141 | 1.83E-05 | -0.01210427 | SLC24A3 |
| chr4 | 122826378 | 122826894 | 517 | 9 | 9.07E-05 | 1.95E-05 | 0.008801 | 1.83E-05 | -0.00517391 | FGF2 |
| chr2 | 138712831 | 138712850 | 20 | 2 | 1.86E-05 | 0.000531 | 1.40E-05 | 1.83E-05 | -0.00478804 | NXPH2 |
| chr9 | 128058195 | 128058290 | 96 | 2 | 1.77E-05 | 1.49E-05 | 0.000259 | 1.83E-05 | -0.00293815 | NA |
| chr12 | 70365185 | 70366140 | 956 | 8 | 1.59E-06 | 9.13E-05 | 7.10E-05 | 1.83E-05 | -0.00240988 | NA |
| chr19 | 51986867 | 51988015 | 1149 | 10 | 3.24E-07 | 0.0005571 | 0.000195 | 1.84E-05 | -0.00155428 | NA |
| chr9 | 83970798 | 83971252 | 455 | 2 | 2.36E-06 | 0.0004789 | 1.44E-05 | 1.84E-05 | -0.00461449 | HNRNPK |
| chr17 | 49494601 | 49495248 | 648 | 5 | 2.25E-06 | 0.0002031 | 0.000126 | 1.84E-05 | -0.00081751 | AC015656.1 |
| chr14 | 91506388 | 91506777 | 390 | 2 | 1.46E-05 | 4.05E-05 | 5.10E-05 | 1.84E-05 | -0.0067139 | PPP4R3A |
| chr22 | 36775801 | 36776669 | 869 | 11 | 1.69E-05 | 0.0022471 | 3.83E-05 | 1.84E-05 | -1.06E-05 | IFT27 |
| chr17 | 81397138 | 81397881 | 744 | 5 | 7.56E-06 | 6.62E-05 | 0.001019 | 1.84E-05 | 0.003657008 | BAHCC1, AC110285.6 |
| chr4 | 17810332 | 17811002 | 671 | 13 | 1.70E-05 | 0.0004953 | 0.005737 | 1.84E-05 | 0.000462593 | NCAPG, DCAF16 |
| chr9 | 28720255 | 28720524 | 270 | 2 | 2.29E-05 | 1.13E-05 | 0.000854 | 1.84E-05 | -0.01503998 | NA |
| chr10 | 73625636 | 73626330 | 695 | 6 | 1.69E-05 | 1.62E-05 | 0.005788 | 1.84E-05 | 0.001336834 | AC073389.1, USP54 |
| chr17 | 38427728 | 38428497 | 770 | 7 | 1.39E-05 | 0.0002434 | 6.28E-05 | 1.84E-05 | 0.001200956 | ARHGAP23 |
| chr21 | 45534969 | 45535288 | 320 | 2 | 1.20E-05 | 1.12E-05 | 0.000906 | 1.84E-05 | -0.00467262 | SLC19A1 |
| chr19 | 17830035 | 17830179 | 145 | 3 | 6.85E-06 | 0.0009864 | 8.53E-06 | 1.84E-05 | -0.00296307 | JAK3 |
| chr16 | 49858093 | 49858785 | 693 | 7 | 1.86E-06 | 1.87E-05 | 0.000361 | 1.84E-05 | 0.001058355 | NA |
| chr20 | 24468500 | 24469280 | 781 | 7 | 1.87E-05 | 1.86E-05 | 0.003477 | 1.84E-05 | -0.00560977 | NA |
| chr11 | 59184950 | 59185234 | 285 | 2 | 1.12E-06 | 0.0464318 | 8.99E-06 | 1.84E-05 | -0.00964363 | DTX4 |
| chr1 | 243338636 | 243338837 | 202 | 2 | 7.78E-06 | 0.0114472 | 9.31E-06 | 1.84E-05 | -0.00744537 | SDCCAG8 |
| chr9 | 126618449 | 126618726 | 278 | 2 | 1.31E-05 | 9.66E-05 | 2.68E-05 | 1.84E-05 | -0.00582558 | LMX1B |
| chr5 | 172061174 | 172061353 | 180 | 2 | 2.86E-05 | 1.18E-05 | 0.000666 | 1.84E-05 | 0.004292179 | STK10 |
| chr1 | 3061297 | 3061766 | 470 | 2 | 3.07E-05 | 1.13E-05 | 0.000871 | 1.84E-05 | -0.0092369 | PRDM16-DT |
| chr9 | 133691736 | 133692019 | 284 | 2 | 3.60E-05 | 1.68E-05 | 0.000186 | 1.84E-05 | -0.00904146 | SARDH |
| chr6 | 17169231 | 17169479 | 249 | 2 | 2.64E-05 | 3.15E-05 | 6.68E-05 | 1.84E-05 | -0.0040265 | NA |
| chr6 | 44622391 | 44622883 | 493 | 2 | 9.76E-06 | 1.77E-05 | 0.000166 | 1.84E-05 | 0.000261303 | NA |
| chr4 | 38867497 | 38868158 | 662 | 12 | 8.29E-05 | 3.93E-05 | 0.004511 | 1.84E-05 | -0.00204205 | FAM114A1, MIR574 |
| chr4 | 121800737 | 121801458 | 722 | 9 | 5.33E-05 | 0.0003429 | 0.007698 | 1.84E-05 | 0.009237893 | EXOSC9 |
| chr6 | 32371431 | 32371734 | 304 | 2 | 7.67E-06 | 0.0004484 | 1.47E-05 | 1.84E-05 | -0.00705404 | TSBP1-AS1, TSBP1 |
| chr1 | 150264993 | 150265506 | 514 | 2 | 4.92E-05 | 1.21E-05 | 0.000583 | 1.84E-05 | 0.001142462 | CA14, APH1A |
| chr12 | 113861129 | 113861271 | 143 | 2 | 2.77E-05 | 4.19E-05 | 4.97E-05 | 1.84E-05 | 0.00257055 | RBM19 |
| chr5 | 1182922 | 1183309 | 388 | 5 | 1.02E-05 | 0.0003907 | 9.02E-05 | 1.84E-05 | -0.00355271 | NA |
| chr19 | 5718618 | 5718901 | 284 | 2 | 2.50E-06 | 1.35E-05 | 0.000361 | 1.84E-05 | -0.00830916 | LONP1 |
| chr1 | 247008128 | 247008584 | 457 | 9 | 3.01E-06 | 0.0001127 | 0.000626 | 1.84E-05 | 0.008458729 | ZNF670-ZNF695 |
| chr12 | 130120446 | 130120567 | 122 | 2 | 8.56E-06 | 6.04E-05 | 3.66E-05 | 1.84E-05 | 0.022983443 | NA |
| chr3 | 101196488 | 101196848 | 361 | 2 | 5.42E-05 | 1.15E-05 | 0.000763 | 1.84E-05 | -0.00382486 | NA |
| chr17 | 62807454 | 62807889 | 436 | 3 | 6.96E-06 | 0.0003277 | 1.78E-05 | 1.85E-05 | -0.00523455 | MARCHF10 |
| chr15 | 76932016 | 76932646 | 631 | 4 | 6.07E-06 | 1.51E-05 | 0.000844 | 1.85E-05 | 0.005006771 | RCN2 |
| chr17 | 57685083 | 57686029 | 947 | 5 | 4.69E-06 | 0.0080584 | 1.43E-05 | 1.85E-05 | -0.00033329 | RNU7-134P |
| chr18 | 58709391 | 58710135 | 745 | 3 | 9.22E-05 | 3.76E-05 | 0.000333 | 1.85E-05 | 0.002294831 | MALT1, AC104365.1 |
| chr4 | 103478345 | 103478858 | 514 | 2 | 5.58E-05 | 1.20E-05 | 0.000622 | 1.85E-05 | 0.014068153 | NA |
| chr9 | 126060973 | 126061101 | 129 | 2 | 2.30E-05 | 5.27E-05 | 4.09E-05 | 1.85E-05 | 0.01002905 | AL589923.1 |
| chr20 | 16028963 | 16029085 | 123 | 2 | 1.19E-05 | 0.0029553 | 1.04E-05 | 1.85E-05 | 0.008372227 | MACROD2 |
| chr3 | 129061321 | 129062050 | 730 | 4 | 1.46E-05 | 1.17E-05 | 0.00229 | 1.85E-05 | -0.00207047 | GP9 |
| chr11 | 122838763 | 122838843 | 81 | 2 | 4.69E-05 | 1.48E-05 | 0.000268 | 1.85E-05 | -0.00438812 | CRTAM, AP002762.2 |
| chr8 | 142196934 | 142198131 | 1198 | 5 | 3.67E-07 | 0.0007279 | 0.000154 | 1.85E-05 | 0.004616609 | NA |
| chr19 | 1088783 | 1088988 | 206 | 2 | 3.27E-06 | 8.92E-05 | 2.83E-05 | 1.85E-05 | 0.007181654 | POLR2E |
| chr1 | 2165143 | 2165719 | 577 | 4 | 8.01E-06 | 0.0001657 | 3.53E-05 | 1.85E-05 | 0.001749281 | PRKCZ |
| chr1 | 7671022 | 7671315 | 294 | 3 | 3.80E-06 | 8.59E-05 | 0.000108 | 1.85E-05 | -0.00062663 | CAMTA1 |
| chr13 | 110094907 | 110095105 | 199 | 2 | 5.06E-05 | 1.29E-05 | 0.000445 | 1.85E-05 | 0.027910202 | AL390755.1 |
| chr3 | 75752444 | 75752533 | 90 | 2 | 5.51E-05 | 1.25E-05 | 0.000513 | 1.85E-05 | -0.01310606 | ZNF717 |
| chr11 | 64651170 | 64651435 | 266 | 4 | 3.96E-05 | 4.76E-06 | 0.009212 | 1.85E-05 | 0.00322331 | NRXN2-AS1, NRXN2 |
| chr2 | 23293452 | 23294010 | 559 | 2 | 5.03E-05 | 1.20E-05 | 0.000622 | 1.85E-05 | 0.004602479 | NA |
| chr8 | 101847912 | 101848258 | 347 | 3 | 6.38E-05 | 8.85E-06 | 0.001724 | 1.85E-05 | 0.003525689 | NCALD |
| chr9 | 134405766 | 134406544 | 779 | 9 | 1.58E-05 | 5.57E-05 | 0.001647 | 1.85E-05 | -0.00223524 | RXRA |
| chr8 | 40849056 | 40849358 | 303 | 2 | 3.35E-05 | 1.26E-05 | 0.000489 | 1.85E-05 | -0.01438183 | ZMAT4 |
| chr21 | 46155220 | 46155986 | 767 | 8 | 2.71E-05 | 3.27E-05 | 0.001164 | 1.85E-05 | -0.00023783 | FTCD |
| chr6 | 3742584 | 3743295 | 712 | 3 | 1.87E-06 | 3.23E-05 | 0.000754 | 1.85E-05 | 0.025358087 | PXDC1 |
| chr1 | 3482322 | 3482754 | 433 | 3 | 7.89E-06 | 7.06E-05 | 3.92E-05 | 1.86E-05 | 0.010423673 | NA |
| chr3 | 195810562 | 195810637 | 76 | 2 | 1.57E-05 | 1.22E-05 | 0.000584 | 1.86E-05 | -0.00533148 | MUC4 |
| chr1 | 212034841 | 212035814 | 974 | 16 | 4.11E-05 | 3.24E-06 | 0.038073 | 1.86E-05 | -8.14E-05 | DTL, INTS7 |
| chr3 | 119034813 | 119035250 | 438 | 10 | 5.70E-05 | 3.21E-05 | 0.007213 | 1.86E-05 | 0.000442299 | IGSF11 |
| chr2 | 104582335 | 104582705 | 371 | 3 | 4.64E-06 | 3.03E-05 | 0.000874 | 1.86E-05 | -0.00524614 | NA |
| chr16 | 83400810 | 83401001 | 192 | 2 | 1.09E-05 | 6.28E-05 | 3.59E-05 | 1.86E-05 | -0.01682923 | CDH13 |
| chr6 | 75360715 | 75360837 | 123 | 2 | 2.01E-05 | 1.45E-05 | 0.000293 | 1.86E-05 | -0.01530911 | AL445465.1, FILIP1 |
| chr6 | 30058858 | 30059432 | 575 | 2 | 6.08E-06 | 1.12E-05 | 0.000982 | 1.86E-05 | 0.006422895 | POLR1H, ZNRD1ASP |
| chr9 | 135506542 | 135506636 | 95 | 3 | 1.89E-05 | 1.75E-05 | 0.00037 | 1.86E-05 | -0.0047545 | NA |
| chr20 | 10641763 | 10641799 | 37 | 3 | 1.15E-05 | 0.0005088 | 0.000425 | 1.86E-05 | 0.000406204 | JAG1 |
| chr10 | 132572194 | 132572586 | 393 | 2 | 2.57E-05 | 1.27E-05 | 0.000483 | 1.86E-05 | -0.01326645 | INPP5A |
| chr15 | 56393922 | 56393990 | 69 | 2 | 3.58E-05 | 1.56E-05 | 0.000236 | 1.86E-05 | -0.00685345 | TEX9 |
| chr3 | 41235041 | 41235630 | 590 | 3 | 1.28E-05 | 6.60E-05 | 5.06E-05 | 1.86E-05 | -0.00942017 | CTNNB1 |
| chr4 | 3711482 | 3711560 | 79 | 3 | 2.97E-05 | 8.53E-05 | 3.61E-05 | 1.86E-05 | -0.00512651 | NA |
| chr10 | 97570204 | 97570690 | 487 | 5 | 6.93E-06 | 0.0002677 | 0.000754 | 1.86E-05 | -0.0016729 | UBTD1 |
| chr22 | 46509049 | 46509441 | 393 | 3 | 1.53E-05 | 4.91E-05 | 5.92E-05 | 1.86E-05 | -0.00249932 | CELSR1 |
| chr7 | 133600746 | 133601139 | 394 | 3 | 7.27E-05 | 9.95E-06 | 0.001404 | 1.86E-05 | 0.001409601 | EXOC4 |
| chr8 | 141152832 | 141153708 | 877 | 3 | 1.35E-05 | 1.74E-05 | 0.000254 | 1.86E-05 | -0.00514243 | DENND3 |
| chr17 | 75476097 | 75476360 | 264 | 2 | 4.39E-06 | 0.0003741 | 1.57E-05 | 1.86E-05 | -0.006487 | TMEM94 |
| chr2 | 11482778 | 11483160 | 383 | 2 | 2.12E-05 | 1.15E-05 | 0.000822 | 1.86E-05 | -0.00083557 | GREB1 |
| chr16 | 35639854 | 35639901 | 48 | 2 | 1.40E-05 | 0.0001141 | 2.50E-05 | 1.87E-05 | -0.02078648 | KIF18BP1 |
| chr8 | 940966 | 941513 | 548 | 4 | 5.32E-05 | 0.0010303 | 0.000767 | 1.87E-05 | -0.00592068 | DLGAP2 |
| chr8 | 47659496 | 47659995 | 500 | 5 | 1.41E-05 | 0.0001206 | 0.00059 | 1.87E-05 | -0.01568072 | SPIDR |
| chr17 | 28571583 | 28572194 | 612 | 9 | 3.30E-05 | 0.0005436 | 0.005066 | 1.87E-05 | -0.00235015 | AC005726.1, PIGS |
| chr10 | 87743376 | 87743396 | 21 | 2 | 1.38E-05 | 0.0002373 | 1.82E-05 | 1.87E-05 | -0.00310789 | PAPSS2 |
| chr17 | 3812449 | 3812953 | 505 | 3 | 0.000101724 | 0.0005231 | 2.18E-05 | 1.87E-05 | -0.00602341 | NCBP3 |
| chr5 | 128536234 | 128536507 | 274 | 4 | 6.14E-05 | 9.78E-05 | 0.000268 | 1.87E-05 | -0.00293022 | FBN2 |
| chr12 | 113186125 | 113186453 | 329 | 2 | 2.53E-05 | 1.13E-05 | 0.000929 | 1.87E-05 | -0.00800493 | RITA1, AC089999.1 |
| chr2 | 133517570 | 133517977 | 408 | 4 | 2.56E-06 | 0.0018065 | 6.51E-06 | 1.87E-05 | -0.00370728 | NCKAP5 |
| chr13 | 112871168 | 112871760 | 593 | 6 | 1.26E-05 | 7.20E-05 | 0.002407 | 1.87E-05 | 0.000480219 | ATP11A |
| chr6 | 85126718 | 85127113 | 396 | 2 | 2.20E-05 | 2.41E-05 | 9.69E-05 | 1.87E-05 | -0.01065798 | NA |
| chr13 | 114121242 | 114121894 | 653 | 5 | 6.15E-06 | 0.0004922 | 5.20E-05 | 1.87E-05 | -0.0030925 | RASA3 |
| chr7 | 139821246 | 139821573 | 328 | 2 | 1.46E-06 | 0.00625 | 9.78E-06 | 1.87E-05 | -0.00377287 | TBXAS1 |

| | | | | | | | | | | |
|---|---|---|---|---|---|---|---|---|---|---|
| chr3 | 73474578 | 73474744 | 167 | 2 | 5.43E-06 | 1.49E-05 | 0.000277 | 1.87E-05 | -0.00782633 | PDZRN3 |
| chr3 | 48429736 | 48430113 | 378 | 4 | 5.00E-06 | 0.0001149 | 0.000393 | 1.87E-05 | 0.001003166 | PLXNB1 |
| chr19 | 40541045 | 40541537 | 493 | 3 | 1.04E-05 | 7.78E-05 | 0.000198 | 1.87E-05 | -0.00556152 | SPTBN4 |
| chr8 | 140590092 | 140590780 | 689 | 3 | 3.08E-05 | 3.91E-05 | 0.000687 | 1.87E-05 | 0.008810823 | AGO2 |
| chr5 | 167753928 | 167754514 | 587 | 3 | 8.31E-05 | 2.02E-05 | 0.000187 | 1.87E-05 | -0.00706678 | TENM2 |
| chr10 | 3066459 | 3066478 | 20 | 2 | 5.90E-06 | 1.36E-05 | 0.000384 | 1.87E-05 | -0.01321564 | PFKP |
| chr2 | 37196671 | 37197368 | 698 | 7 | 4.26E-06 | 0.0006488 | 0.001603 | 1.88E-05 | -0.00217215 | CEBPZOS |
| chr21 | 36481842 | 36481872 | 31 | 2 | 4.67E-06 | 0.0051337 | 9.96E-06 | 1.88E-05 | -0.00304388 | AP000695.2, CLDN14 |
| chr1 | 217630996 | 217631535 | 540 | 12 | 2.51E-05 | 0.0007702 | 0.006956 | 1.88E-05 | 0.002060725 | SPATA17, GPATCH2 |
| chr1 | 229558841 | 229559344 | 504 | 7 | 4.67E-05 | 7.74E-05 | 0.001587 | 1.88E-05 | 0.001145275 | NA |
| chr3 | 133041264 | 133041700 | 437 | 3 | 1.68E-05 | 2.95E-05 | 0.000102 | 1.88E-05 | 0.004126245 | TMEM108 |
| chr16 | 85087264 | 85087450 | 187 | 2 | 6.36E-06 | 1.32E-05 | 0.000426 | 1.88E-05 | -0.01093695 | KIAA0513 |
| chr9 | 99259514 | 99259935 | 422 | 3 | 5.17E-05 | 8.55E-06 | 0.001763 | 1.88E-05 | -0.01686137 | NA |
| chr19 | 38653867 | 38654299 | 433 | 2 | 5.94E-05 | 1.13E-05 | 0.001009 | 1.88E-05 | -0.00084812 | ACTN4 |
| chr2 | 196176766 | 196177275 | 510 | 2 | 0.000119778 | 1.43E-05 | 0.000326 | 1.88E-05 | -0.0012369 | NA |
| chr17 | 6635178 | 6635654 | 477 | 4 | 8.21E-06 | 0.0013713 | 6.66E-05 | 1.88E-05 | -0.00221812 | KIAA0753, AC004706.3 |
| chr19 | 32692371 | 32692979 | 609 | 2 | 2.71E-07 | 0.0001202 | 2.47E-05 | 1.88E-05 | 0.00954835 | NUDT19 |
| chr22 | 33320428 | 33320978 | 551 | 6 | 5.15E-05 | 3.56E-05 | 0.002486 | 1.88E-05 | -0.00163301 | Z82173.1, LARGE1 |
| chr1 | 64133023 | 64133655 | 633 | 3 | 4.01E-05 | 1.66E-05 | 0.000344 | 1.89E-05 | -0.0091901 | ROR1, ROR1-AS1 |
| chr21 | 46269711 | 46269772 | 62 | 3 | 7.06E-05 | 7.94E-06 | 0.002548 | 1.89E-05 | -0.00106572 | MCM3AP |
| chr15 | 25208877 | 25209192 | 316 | 3 | 8.59E-06 | 1.56E-05 | 0.000592 | 1.89E-05 | -0.00666672 | SNHG14 |
| chr20 | 50464144 | 50464496 | 353 | 2 | 1.55E-05 | 5.04E-05 | 4.37E-05 | 1.89E-05 | -0.00761083 | NA |
| chr8 | 1881356 | 1881899 | 544 | 3 | 5.52E-06 | 2.52E-05 | 0.000596 | 1.89E-05 | -0.00398764 | AC019257.8, ARHGEF10 |
| chr1 | 202889020 | 202889698 | 679 | 13 | 3.06E-05 | 2.71E-05 | 0.007526 | 1.89E-05 | 0.000281469 | RABIF |
| chr1 | 183647315 | 183647833 | 519 | 2 | 5.36E-05 | 1.25E-05 | 0.000563 | 1.89E-05 | -0.00573962 | RGL1, APOBEC4 |
| chr8 | 92017871 | 92018923 | 1053 | 6 | 1.15E-06 | 0.0015916 | 7.64E-05 | 1.89E-05 | -0.00744978 | RUNX1T1 |
| chr12 | 33407187 | 33407342 | 156 | 3 | 3.06E-05 | 7.16E-05 | 7.36E-05 | 1.89E-05 | 0.003869337 | SYT10 |
| chr1 | 35462525 | 35462620 | 96 | 2 | 5.56E-05 | 1.31E-05 | 0.000456 | 1.89E-05 | 0.009778137 | KIAA0319L |
| chr1 | 9320697 | 9321372 | 676 | 4 | 3.93E-05 | 5.51E-06 | 0.005953 | 1.89E-05 | -0.00703903 | SPSB1 |
| chr1 | 41639466 | 41639851 | 386 | 2 | 5.39E-06 | 0.0004247 | 1.53E-05 | 1.89E-05 | -0.00852159 | HIVEP3 |
| chr19 | 48394606 | 48394903 | 298 | 6 | 2.20E-06 | 0.0040368 | 5.43E-06 | 1.89E-05 | 0.002082024 | GRIN2D |
| chr9 | 135806103 | 135806113 | 11 | 2 | 2.30E-05 | 2.53E-05 | 9.23E-05 | 1.89E-05 | -0.01449351 | NA |
| chr10 | 76176344 | 76176357 | 14 | 2 | 4.66E-05 | 2.29E-05 | 0.000108 | 1.89E-05 | -0.00360707 | LRMDA |
| chr5 | 114418488 | 114418514 | 27 | 2 | 2.35E-05 | 0.0002307 | 1.87E-05 | 1.89E-05 | -0.00588351 | KCNN2 |
| chr3 | 149084336 | 149084698 | 363 | 2 | 2.84E-05 | 1.18E-05 | 0.000811 | 1.89E-05 | 0.003742622 | HLTF |
| chr16 | 24361105 | 24361398 | 294 | 2 | 8.04E-06 | 2.86E-05 | 7.89E-05 | 1.89E-05 | -0.00892283 | CACNG3 |
| chr5 | 17204415 | 17204500 | 86 | 2 | 3.47E-05 | 1.18E-05 | 0.000815 | 1.89E-05 | -0.01274789 | BASP1, BASP1-AS1 |
| chr7 | 50558575 | 50558601 | 27 | 2 | 0.000105447 | 1.14E-05 | 0.001013 | 1.89E-05 | -0.01061774 | DDC |
| chr5 | 104911128 | 104911333 | 206 | 2 | 1.07E-05 | 2.00E-05 | 0.00014 | 1.90E-05 | 0.007542938 | AC099520.1 |
| chr21 | 24685732 | 24685845 | 114 | 2 | 2.99E-05 | 2.94E-05 | 7.61E-05 | 1.90E-05 | -0.01292357 | NA |
| chr20 | 14337651 | 14338479 | 829 | 6 | 5.34E-06 | 2.15E-05 | 0.001399 | 1.90E-05 | -0.00417514 | MACROD2 |
| chr5 | 73473856 | 73474024 | 169 | 2 | 1.41E-05 | 0.0006303 | 1.39E-05 | 1.90E-05 | -0.00632765 | NA |
| chr2 | 227408078 | 227408310 | 233 | 2 | 3.74E-05 | 2.08E-05 | 0.000129 | 1.90E-05 | -0.01526112 | NA |
| chr16 | 21233145 | 21234141 | 997 | 7 | 5.71E-06 | 0.0002507 | 7.46E-05 | 1.90E-05 | -0.00110381 | ANKS4B |
| chr17 | 17515377 | 17515929 | 553 | 3 | 2.26E-05 | 2.75E-05 | 0.000781 | 1.90E-05 | 0.002232539 | PEMT |
| chr3 | 72447302 | 72447724 | 423 | 3 | 1.30E-06 | 1.84E-05 | 0.001209 | 1.90E-05 | -0.01036796 | NA |
| chr17 | 45429463 | 45429652 | 190 | 2 | 1.02E-05 | 0.0003439 | 1.64E-05 | 1.90E-05 | -0.00262261 | ARHGAP27 |
| chr1 | 243803637 | 243804079 | 443 | 2 | 2.25E-05 | 2.19E-05 | 0.000118 | 1.90E-05 | 0.011516285 | AKT3 |
| chr19 | 5790795 | 5791337 | 543 | 6 | 1.80E-05 | 4.52E-05 | 0.000167 | 1.90E-05 | -0.00079462 | DUS3L |
| chr2 | 148457862 | 148458315 | 454 | 4 | 2.20E-05 | 2.44E-05 | 0.000436 | 1.90E-05 | -0.00294296 | MBD5 |
| chr6 | 17015995 | 17016253 | 259 | 3 | 5.27E-05 | 1.21E-05 | 0.001116 | 1.90E-05 | -0.02398239 | AL133405.1 |
| chr17 | 12900567 | 12900811 | 245 | 2 | 4.87E-05 | 1.37E-05 | 0.000394 | 1.90E-05 | -0.01056692 | ARHGAP44 |
| chr6 | 46018111 | 46018264 | 154 | 2 | 6.29E-06 | 0.0046011 | 1.02E-05 | 1.90E-05 | -0.00884503 | CLIC5 |
| chr8 | 139829231 | 139829423 | 193 | 3 | 1.56E-05 | 0.000213 | 3.21E-05 | 1.90E-05 | 0.005198712 | TRAPPC9 |
| chr22 | 32203524 | 32203661 | 138 | 3 | 1.60E-05 | 1.02E-05 | 0.0013 | 1.90E-05 | -0.04125712 | RFPL2 |
| chr10 | 128157393 | 128157682 | 290 | 2 | 4.90E-06 | 1.86E-05 | 0.000164 | 1.90E-05 | -0.00865345 | NA |
| chr13 | 112723987 | 112724517 | 531 | 2 | 1.06E-05 | 2.16E-05 | 0.000121 | 1.91E-05 | 0.000923588 | ATP11A |
| chr17 | 61863086 | 61863705 | 620 | 9 | 6.89E-05 | 1.72E-05 | 0.001029 | 1.91E-05 | 0.000655113 | BRIP1 |
| chr14 | 105313800 | 105314393 | 594 | 8 | 6.78E-06 | 0.0002269 | 0.007121 | 1.91E-05 | 0.000751459 | PACS2, BRF1 |
| chr15 | 90954909 | 90955793 | 885 | 9 | 5.05E-06 | 2.41E-05 | 0.005432 | 1.91E-05 | -0.0040325 | RCCD1, RCCD1-AS1 |
| chr9 | 36819793 | 36820218 | 426 | 2 | 2.13E-06 | 0.0408851 | 9.31E-06 | 1.91E-05 | -0.00983505 | AL450267.2 |
| chr1 | 246005337 | 246005376 | 40 | 2 | 1.55E-05 | 0.0005016 | 1.48E-05 | 1.91E-05 | -0.00928764 | SMYD3 |
| chr1 | 53328539 | 53328641 | 103 | 2 | 6.50E-05 | 2.34E-05 | 0.000106 | 1.91E-05 | -0.01962542 | LINC01771 |
| chr1 | 11658118 | 11658402 | 285 | 2 | 2.71E-05 | 2.97E-05 | 7.64E-05 | 1.91E-05 | -0.00848619 | FBXO44 |
| chr16 | 83912019 | 83912682 | 664 | 6 | 7.18E-06 | 0.0001601 | 2.58E-05 | 1.91E-05 | -0.00036565 | MLYCD |
| chr12 | 100199645 | 100200808 | 1164 | 13 | 2.77E-06 | 1.47E-05 | 0.005227 | 1.91E-05 | 0.000169762 | ACTR6, AC010203.1 |
| chr7 | 151308985 | 151309259 | 275 | 2 | 5.48E-06 | 0.0010171 | 1.27E-05 | 1.91E-05 | -0.01909493 | NA |
| chr19 | 52369474 | 52370024 | 551 | 12 | 1.27E-05 | 0.0001126 | 0.006341 | 1.91E-05 | -0.00590152 | ZNF880 |
| chr5 | 3736504 | 3736702 | 199 | 3 | 3.56E-05 | 5.53E-05 | 5.17E-05 | 1.91E-05 | -0.00924777 | NA |
| chr10 | 17391563 | 17392051 | 489 | 2 | 3.64E-05 | 1.15E-05 | 0.001019 | 1.92E-05 | -0.00151569 | ST8SIA6-AS1, ST8SIA6 |
| chr3 | 195893104 | 195893276 | 173 | 2 | 4.03E-05 | 7.72E-05 | 3.24E-05 | 1.92E-05 | 0.011546926 | TNK2 |
| chr12 | 32327963 | 32328388 | 426 | 6 | 1.58E-05 | 0.000106 | 8.19E-05 | 1.92E-05 | 0.001051119 | BICD1 |
| chr4 | 108650246 | 108650570 | 325 | 9 | 3.11E-05 | 4.93E-05 | 0.001728 | 1.92E-05 | 2.82E-05 | NA |
| chr11 | 45964989 | 45965604 | 616 | 4 | 1.33E-05 | 0.000489 | 2.17E-05 | 1.92E-05 | -0.00196734 | PHF21A |
| chr6 | 98832319 | 98834101 | 1783 | 9 | 5.64E-05 | 8.47E-05 | 0.006071 | 1.92E-05 | 0.000926053 | AL589826.2 |
| chr2 | 165794259 | 165795061 | 803 | 13 | 0.000108958 | 1.24E-05 | 0.020143 | 1.92E-05 | -0.00178762 | AC009495.3, GALNT3, AC009495.1 |
| chr3 | 48898500 | 48899546 | 1047 | 10 | 2.89E-06 | 0.0016802 | 0.000111 | 1.92E-05 | -0.00027679 | SLC25A20 |
| chr6 | 30141009 | 30141431 | 423 | 3 | 2.44E-05 | 3.17E-05 | 0.00012 | 1.92E-05 | -0.00464087 | TRIM40 |
| chr6 | 167824759 | 167825545 | 787 | 6 | 1.38E-05 | 0.0001238 | 2.77E-05 | 1.92E-05 | -0.00820907 | AL009178.3, AFDN-DT |
| chr2 | 235776643 | 235777013 | 371 | 3 | 5.02E-06 | 2.01E-05 | 0.000226 | 1.92E-05 | -0.0017602 | AGAP1, AC064874.1 |
| chr3 | 195062545 | 195063237 | 693 | 6 | 6.62E-06 | 0.0003123 | 0.000779 | 1.92E-05 | -0.00208561 | NA |
| chr10 | 13100097 | 13100679 | 583 | 6 | 1.38E-05 | 0.0001469 | 0.000871 | 1.92E-05 | 0.000662587 | OPTN |
| chr2 | 200509724 | 200510477 | 754 | 11 | 3.25E-05 | 0.0002682 | 0.001655 | 1.92E-05 | 0.001058101 | SGO2, KCTD18 |
| chr22 | 44180989 | 44181385 | 397 | 9 | 1.98E-05 | 6.43E-06 | 0.017069 | 1.92E-05 | 0.002640919 | PARVG |
| chr6 | 10000564 | 10000652 | 89 | 2 | 1.99E-05 | 0.0003843 | 1.61E-05 | 1.92E-05 | -0.00488793 | OFCC1 |
| chr19 | 46731356 | 46731616 | 261 | 3 | 4.17E-05 | 1.36E-05 | 0.000499 | 1.93E-05 | -0.00696431 | AC008635.1, STRN4 |
| chr5 | 176406411 | 176406692 | 282 | 2 | 1.85E-05 | 1.53E-05 | 0.000285 | 1.93E-05 | 0.003725258 | CLTB |
| chr2 | 222306754 | 222307610 | 857 | 4 | 3.39E-06 | 6.18E-05 | 0.0003 | 1.93E-05 | -0.00382756 | NA |
| chr1 | 145918566 | 145919286 | 721 | 17 | 1.43E-05 | 0.0001565 | 0.000135 | 1.93E-05 | -0.00014644 | GNRHR2, PEX11B, RBM8A |
| chr15 | 96356914 | 96357414 | 501 | 4 | 5.55E-05 | 4.15E-05 | 0.000817 | 1.93E-05 | -0.00246279 | AC087477.2 |
| chr17 | 80825921 | 80826974 | 1054 | 6 | 3.56E-05 | 6.14E-05 | 0.007144 | 1.93E-05 | 0.001313805 | RPTOR, AC016245.2 |
| chr12 | 1616622 | 1617126 | 505 | 8 | 2.10E-06 | 0.0005474 | 2.34E-05 | 1.93E-05 | -0.00301741 | WNT5B |
| chr7 | 138173859 | 138173947 | 89 | 2 | 2.39E-05 | 1.25E-05 | 0.000645 | 1.93E-05 | -0.00338067 | NA |
| chr2 | 26244445 | 26245456 | 1012 | 15 | 8.05E-06 | 0.0009175 | 0.000367 | 1.93E-05 | 0.000956604 | HADHB, HADHA |
| chr7 | 794819 | 794928 | 110 | 2 | 2.07E-05 | 0.0002728 | 1.80E-05 | 1.93E-05 | -0.00306029 | NA |
| chr2 | 219055494 | 219055623 | 130 | 2 | 4.67E-05 | 5.66E-05 | 4.11E-05 | 1.93E-05 | -0.00960329 | IHH |
| chr3 | 50298901 | 50300063 | 1163 | 22 | 2.51E-05 | 0.0013887 | 0.0005 | 1.93E-05 | 0.000285862 | HYAL3, NAA80, HYAL1 |
| chr14 | 76697155 | 76697208 | 54 | 2 | 3.35E-05 | 3.15E-05 | 7.26E-05 | 1.93E-05 | -0.01195691 | AC008050.1 |
| chr14 | 76727559 | 76727948 | 390 | 3 | 3.08E-05 | 2.17E-05 | 0.000816 | 1.93E-05 | -0.00125975 | AC007376.2 |
| chr22 | 31623656 | 31623772 | 117 | 2 | 2.47E-05 | 1.34E-05 | 0.000463 | 1.94E-05 | -0.00533622 | PISD |
| chr18 | 76463064 | 76463252 | 189 | 5 | 2.84E-05 | 5.31E-05 | 0.00022 | 1.94E-05 | -0.00205432 | ZNF516 |
| chr2 | 207558579 | 207558938 | 360 | 3 | 3.37E-05 | 5.69E-05 | 7.46E-05 | 1.94E-05 | -0.00020883 | CREB1 |
| chr12 | 43278397 | 43278462 | 66 | 2 | 7.48E-06 | 0.0008596 | 1.33E-05 | 1.94E-05 | -0.01085178 | NA |
| chr7 | 129765866 | 129766284 | 419 | 2 | 1.66E-05 | 2.04E-05 | 0.000142 | 1.94E-05 | -0.00112419 | AC084864.2 |
| chr3 | 59049681 | 59050432 | 752 | 13 | 5.78E-05 | 3.73E-05 | 0.001621 | 1.94E-05 | 8.04E-05 | AC126121.3, CFAP20DC |
| chr12 | 122773894 | 122774783 | 890 | 16 | 8.24E-07 | 0.0067231 | 8.36E-06 | 1.94E-05 | 0.001894095 | CCDC62 |
| chr14 | 23050983 | 23051242 | 260 | 3 | 1.10E-05 | 0.0003504 | 3.85E-05 | 1.94E-05 | -0.00839769 | CDH24 |
| chr16 | 66750898 | 66751074 | 177 | 2 | 1.02E-06 | 0.0043329 | 1.04E-05 | 1.94E-05 | 0.004311013 | DYNC1LI2 |
| chr15 | 51715236 | 51715320 | 85 | 2 | 4.01E-05 | 3.49E-05 | 6.53E-05 | 1.94E-05 | -0.00323039 | SCG3 |
| chr12 | 56477999 | 56478454 | 456 | 2 | 2.30E-05 | 1.71E-05 | 0.000212 | 1.94E-05 | 0.001505981 | SPRYD4, AC097104.1, GLS2 |
| chr1 | 31647292 | 31647712 | 421 | 2 | 1.85E-05 | 2.11E-05 | 0.000133 | 1.94E-05 | -0.00089917 | PEF1-AS1, AC114488.2 |
| chr8 | 122846514 | 122847218 | 705 | 6 | 7.12E-06 | 0.0003907 | 4.25E-05 | 1.94E-05 | -0.00674868 | ZHX2 |
| chr16 | 10963967 | 10964301 | 335 | 2 | 8.42E-06 | 1.77E-05 | 0.000196 | 1.94E-05 | 2.89E-06 | CLEC16A |
| chr4 | 13524973 | 13525145 | 173 | 3 | 3.36E-06 | 0.0040175 | 5.61E-06 | 1.94E-05 | -0.00563871 | NA |
| chr2 | 151947593 | 151947941 | 349 | 3 | 2.71E-05 | 2.14E-05 | 0.000267 | 1.95E-05 | 0.004970808 | AC068547.1, CACNB4 |
| chr3 | 45043671 | 45043909 | 239 | 2 | 7.87E-06 | 0.0028641 | 1.10E-05 | 1.95E-05 | -0.0033144 | NA |
| chr16 | 58339651 | 58339903 | 253 | 2 | 2.86E-05 | 3.39E-05 | 6.76E-05 | 1.95E-05 | -0.01108691 | GINS3 |
| chr7 | 140063774 | 140063973 | 200 | 7 | 4.82E-05 | 2.69E-05 | 0.001958 | 1.95E-05 | -0.00393796 | NA |
| chr19 | 41195989 | 41196248 | 260 | 2 | 2.34E-05 | 4.92E-05 | 4.68E-05 | 1.95E-05 | 0.001504989 | CYP2S1 |
| chr8 | 48589592 | 48590607 | 1016 | 6 | 7.10E-06 | 3.37E-05 | 0.003312 | 1.95E-05 | -0.00655542 | AC022915.2, LINC02599 |
| chr2 | 218820861 | 218821285 | 425 | 2 | 1.04E-05 | 5.42E-05 | 4.32E-05 | 1.95E-05 | 0.001483889 | NA |
| chr10 | 230860 | 230897 | 38 | 3 | 3.54E-05 | 0.0002166 | 0.000535 | 1.95E-05 | -0.00922093 | ZMYND11 |
| chr1 | 3940065 | 3940684 | 620 | 8 | 2.33E-05 | 0.0007116 | 0.002489 | 1.95E-05 | -0.00390852 | LINC01346 |
| chr1 | 110488509 | 110488881 | 373 | 3 | 4.33E-05 | 1.15E-05 | 0.001693 | 1.95E-05 | 0.002541099 | AL358215.3, CYMP, CYMP-AS1 |
| chr19 | 56229594 | 56229650 | 57 | 2 | 3.86E-05 | 3.20E-05 | 7.27E-05 | 1.95E-05 | 0.000291033 | ZSCAN5A |
| chr6 | 34142840 | 34143306 | 467 | 3 | 3.29E-05 | 1.28E-05 | 0.001377 | 1.95E-05 | -0.01263251 | GRM4 |
| chr2 | 205685383 | 205686095 | 713 | 3 | 8.32E-06 | 3.92E-05 | 0.000179 | 1.95E-05 | 0.001672041 | NRP2 |
| chr7 | 157721934 | 157722130 | 197 | 3 | 3.36E-05 | 1.33E-05 | 0.000525 | 1.95E-05 | -0.01192643 | PTPRN2 |
| chr1 | 231425406 | 231425887 | 482 | 3 | 3.80E-05 | 1.06E-05 | 0.001047 | 1.95E-05 | -0.0078094 | AL445524.2 |
| chr1 | 202236220 | 202236866 | 647 | 4 | 3.85E-05 | 1.67E-05 | 0.000362 | 1.95E-05 | -0.00594352 | LGR6 |
| chr15 | 82088628 | 82088670 | 43 | 2 | 6.81E-06 | 2.95E-05 | 8.04E-05 | 1.96E-05 | -0.01654933 | LINC01583 |
| chr7 | 45684859 | 45685178 | 320 | 2 | 3.97E-05 | 1.22E-05 | 0.00079 | 1.96E-05 | -0.01122107 | ADCY1 |
| chr7 | 53811510 | 53812427 | 918 | 9 | 6.98E-06 | 0.0007205 | 0.001534 | 1.96E-05 | -0.00566171 | LINC01446 |
| chr21 | 45325975 | 45326208 | 234 | 2 | 3.72E-05 | 1.26E-05 | 0.000672 | 1.96E-05 | -0.01226326 | NA |
| chr9 | 98556192 | 98556423 | 232 | 3 | 6.16E-05 | 1.07E-05 | 0.001537 | 1.96E-05 | -0.00103234 | GABBR2 |
| chr6 | 44238176 | 44238559 | 384 | 4 | 3.14E-05 | 7.08E-06 | 0.002731 | 1.96E-05 | 0.001803154 | NA |
| chr7 | 77327245 | 77327669 | 425 | 2 | 3.71E-05 | 1.19E-05 | 0.000961 | 1.96E-05 | 0.033139931 | CCDC146, GSAP |
| chr12 | 117330755 | 117331232 | 478 | 6 | 3.47E-05 | 3.08E-05 | 0.000938 | 1.96E-05 | -0.00296231 | NOS1 |
| chr1 | 2028003 | 2028191 | 189 | 2 | 9.70E-05 | 1.21E-05 | 0.000856 | 1.96E-05 | -0.00111117 | GABRD |
| chr6 | 161574270 | 161574337 | 68 | 3 | 3.30E-05 | 9.52E-05 | 4.97E-05 | 1.96E-05 | -0.00318884 | PRKN |
| chr8 | 10960342 | 10960678 | 337 | 2 | 2.48E-05 | 2.90E-05 | 8.29E-05 | 1.97E-05 | -0.00139571 | XKR6 |
| chr16 | 30095785 | 30096938 | 1154 | 8 | 8.00E-06 | 0.0002825 | 0.001766 | 1.97E-05 | 0.002815507 | AC012645.1, YPEL3 |
| chr21 | 38768859 | 38769342 | 484 | 3 | 4.18E-05 | 1.85E-05 | 0.000345 | 1.97E-05 | -0.00970003 | NA |
| chr16 | 46748264 | 46748630 | 367 | 6 | 5.48E-05 | 4.87E-06 | 0.009517 | 1.97E-05 | -0.00181799 | MYLK3 |
| chr2 | 234994457 | 234994898 | 442 | 3 | 2.70E-06 | 0.0002852 | 1.68E-05 | 1.97E-05 | 0.01536442 | SH3BP4 |
| chr8 | 144547694 | 144548543 | 850 | 3 | 1.22E-05 | 7.70E-06 | 0.003615 | 1.97E-05 | -0.00606628 | ARHGAP39 |
| chr2 | 21317142 | 21317452 | 311 | 2 | 1.09E-05 | 0.0003856 | 1.65E-05 | 1.97E-05 | -0.00511976 | AC018742.1 |
| chr21 | 39183935 | 39184745 | 811 | 4 | 1.17E-05 | 5.73E-06 | 0.005795 | 1.97E-05 | 0.000616144 | AF129408.1, BRWD1 |
| chr8 | 141340911 | 141341866 | 956 | 3 | 5.69E-07 | 0.0007062 | 1.99E-05 | 1.97E-05 | -0.01136474 | LINC01300 |
| chr17 | 76029627 | 76029780 | 154 | 3 | 1.72E-05 | 0.0002941 | 4.05E-05 | 1.97E-05 | 0.000666629 | NA |
| chr2 | 26704269 | 26704825 | 557 | 3 | 8.74E-06 | 2.61E-05 | 0.000568 | 1.97E-05 | -0.00342973 | KCNK3 |
| chr8 | 38413815 | 38414599 | 785 | 5 | 2.95E-05 | 1.24E-05 | 0.001338 | 1.97E-05 | -0.00087742 | FGFR1 |
| chr17 | 17805693 | 17805970 | 278 | 2 | 1.64E-05 | 0.0001007 | 2.87E-05 | 1.97E-05 | -0.00381429 | RAI1 |
| chr1 | 229381454 | 229382115 | 662 | 2 | 6.29E-05 | 1.29E-05 | 0.000615 | 1.97E-05 | -0.00958689 | NA |
| chr8 | 70246381 | 70246686 | 306 | 2 | 1.57E-05 | 2.50E-05 | 0.000102 | 1.97E-05 | -0.00247948 | NCOA2 |
| chr6 | 34791241 | 34791387 | 147 | 3 | 3.44E-06 | 4.36E-05 | 0.000143 | 1.97E-05 | -0.00415749 | NA |
| chr10 | 5596118 | 5596619 | 502 | 5 | 6.75E-05 | 7.72E-06 | 0.006275 | 1.97E-05 | -0.00398676 | LASTR |
| chr8 | 117134526 | 117134924 | 399 | 3 | 3.30E-05 | 2.12E-05 | 0.000319 | 1.97E-05 | -0.00548936 | SLC30A8 |

| | | | | | | | | | | |
|---|---|---|---|---|---|---|---|---|---|---|
| chr1 | 161621698 | 161621801 | 104 | 2 | 5.94E-05 | 1.31E-05 | 0.00057 | 1.97E-05 | -0.00440628 | NA |
| chr6 | 132624376 | 132624738 | 363 | 6 | 2.72E-05 | 0.0004371 | 0.001259 | 1.97E-05 | -0.0052585 | NA |
| chr3 | 36993239 | 36994134 | 896 | 25 | 2.71E-06 | 5.13E-05 | 0.000126 | 1.97E-05 | 0.00030441 | MLH1 |
| chr12 | 120388588 | 120388884 | 297 | 2 | 9.61E-06 | 6.22E-05 | 3.93E-05 | 1.97E-05 | -0.0078897 | NA |
| chr5 | 172167749 | 172167994 | 246 | 2 | 2.74E-05 | 1.30E-05 | 0.000592 | 1.98E-05 | -0.01325883 | STK10 |
| chr4 | 184833479 | 184833878 | 400 | 3 | 8.13E-05 | 7.51E-06 | 0.004091 | 1.98E-05 | -0.00362598 | AC084871.3 |
| chr3 | 47009851 | 47010056 | 206 | 3 | 3.05E-05 | 1.06E-05 | 0.001027 | 1.98E-05 | -0.01102028 | NA |
| chr4 | 113035854 | 113036015 | 162 | 2 | 1.79E-05 | 5.28E-05 | 4.51E-05 | 1.98E-05 | 0.000918006 | ANK2, ANK2-AS1 |
| chr10 | 11091866 | 11091895 | 30 | 3 | 4.16E-05 | 8.82E-05 | 0.000369 | 1.98E-05 | -0.00910496 | CELF2, CELF2-AS2 |
| chr12 | 51162629 | 51162670 | 42 | 2 | 4.39E-05 | 2.80E-05 | 8.82E-05 | 1.98E-05 | -0.00720348 | TFCP2 |
| chr3 | 111048402 | 111048732 | 331 | 3 | 4.51E-05 | 5.14E-05 | 0.00037 | 1.98E-05 | -0.0047553 | NECTIN3-AS1 |
| chr2 | 221372613 | 221372642 | 30 | 2 | 5.97E-05 | 1.38E-05 | 0.000462 | 1.98E-05 | -0.01688653 | NA |
| chr12 | 132189807 | 132190472 | 666 | 6 | 3.07E-05 | 8.67E-05 | 0.00152 | 1.98E-05 | 0.005020825 | AC138466.1 |
| chr16 | 27470235 | 27470800 | 566 | 3 | 1.73E-05 | 8.60E-05 | 5.13E-05 | 1.98E-05 | -0.00263928 | GTF3C1 |
| chr6 | 3737596 | 3737722 | 127 | 3 | 2.13E-05 | 0.0001785 | 2.43E-05 | 1.98E-05 | -0.0024162 | PXDC1 |
| chr17 | 18018166 | 18018625 | 460 | 3 | 4.13E-05 | 1.21E-05 | 0.000721 | 1.98E-05 | -0.00032355 | ATPAF2 |
| chr14 | 64942941 | 64943761 | 821 | 7 | 1.15E-05 | 0.0013068 | 0.000113 | 1.98E-05 | 0.00385787 | CHURC1, CHURC1-FNTB |
| chr1 | 236794884 | 236795682 | 799 | 16 | 2.10E-05 | 0.0010192 | 0.00038 | 1.98E-05 | 0.000541571 | MTR |
| chr6 | 30016022 | 30016512 | 491 | 3 | 8.23E-05 | 7.70E-06 | 0.003888 | 1.98E-05 | 0.006131724 | ZNRD1ASP |
| chr22 | 46064010 | 46064458 | 449 | 2 | 1.70E-06 | 4.87E-05 | 4.85E-05 | 1.98E-05 | 1.19E-05 | MIRLET7BHG |
| chr2 | 239387828 | 239387986 | 159 | 3 | 2.90E-05 | 1.44E-05 | 0.000583 | 1.98E-05 | 0.004681249 | HDAC4 |
| chr1 | 239997672 | 239998147 | 476 | 4 | 4.54E-05 | 9.46E-06 | 0.001684 | 1.98E-05 | -0.00804167 | NA |
| chr13 | 40666366 | 40666624 | 259 | 3 | 9.40E-06 | 1.72E-05 | 0.000314 | 1.98E-05 | -0.00096299 | FOXO1 |
| chr17 | 77830529 | 77830631 | 103 | 2 | 4.75E-05 | 2.28E-05 | 0.000122 | 1.98E-05 | -0.01141784 | NA |
| chr13 | 29428197 | 29428794 | 598 | 8 | 5.14E-06 | 0.0002367 | 0.000336 | 1.98E-05 | 0.002441396 | MTUS2 |
| chr4 | 154492240 | 154492765 | 526 | 6 | 5.66E-06 | 0.0002482 | 0.001401 | 1.98E-05 | -0.01089903 | NA |
| chr2 | 62673914 | 62674040 | 127 | 2 | 1.22E-05 | 3.48E-05 | 6.80E-05 | 1.98E-05 | -0.00550635 | EHBP1 |
| chr15 | 34016741 | 34017189 | 449 | 3 | 3.56E-05 | 9.40E-05 | 0.000529 | 1.98E-05 | -0.00471689 | CHRM5, AVEN |
| chr2 | 10083720 | 10084105 | 386 | 5 | 4.51E-05 | 1.31E-05 | 0.000759 | 1.98E-05 | 0.002307821 | AC104794.2 |
| chr4 | 80263771 | 80264074 | 304 | 3 | 5.90E-06 | 3.36E-05 | 0.000101 | 1.98E-05 | -0.01064484 | NA |
| chr11 | 119274654 | 119274909 | 256 | 2 | 1.52E-05 | 1.40E-05 | 0.000435 | 1.98E-05 | -0.00677881 | CBL |
| chr1 | 7940892 | 7941249 | 358 | 5 | 1.96E-05 | 4.08E-05 | 0.000465 | 1.98E-05 | -0.00617135 | TNFRSF9 |
| chr11 | 924527 | 924646 | 120 | 2 | 2.34E-05 | 1.33E-05 | 0.000555 | 1.98E-05 | -0.00914602 | NA |
| chr15 | 71214629 | 71215356 | 728 | 5 | 4.10E-05 | 8.11E-06 | 0.004291 | 1.98E-05 | -0.01005843 | THSD4 |
| chr13 | 98890365 | 98890462 | 98 | 3 | 5.51E-05 | 1.88E-05 | 0.000734 | 1.99E-05 | -0.00811016 | DOCK9 |
| chr1 | 89610856 | 89611543 | 688 | 3 | 1.89E-06 | 0.0295818 | 7.54E-06 | 1.99E-05 | -0.00716984 | LRRC8C-DT |
| chr2 | 86105039 | 86106214 | 1176 | 13 | 1.59E-06 | 0.0001375 | 0.000943 | 1.99E-05 | -0.00105707 | POLR1A |
| chr9 | 95383309 | 95383516 | 208 | 2 | 3.08E-05 | 1.93E-05 | 0.000171 | 1.99E-05 | -0.00049088 | FANCC |
| chr2 | 114123079 | 114123338 | 260 | 2 | 7.56E-06 | 1.20E-05 | 0.001024 | 1.99E-05 | -0.00242164 | AC010982.1 |
| chr17 | 29622717 | 29623395 | 679 | 8 | 4.46E-05 | 2.96E-05 | 0.001536 | 1.99E-05 | -0.00248867 | ABHD15-AS1, CORO6 |
| chr17 | 699356 | 699832 | 477 | 4 | 9.27E-05 | 1.39E-05 | 0.004082 | 1.99E-05 | -0.00597821 | VPS53 |
| chr8 | 53807871 | 53808164 | 294 | 2 | 2.49E-05 | 1.21E-05 | 0.00096 | 1.99E-05 | 0.006232441 | ATP6V1H |
| chr14 | 63849719 | 63850443 | 725 | 3 | 5.13E-05 | 5.36E-05 | 0.000174 | 1.99E-05 | -0.0024464 | SYNE2 |
| chr5 | 177436128 | 177436530 | 403 | 5 | 6.10E-05 | 6.45E-06 | 0.007064 | 1.99E-05 | -0.00097843 | GRK6 |
| chr11 | 115173330 | 115173472 | 143 | 2 | 5.54E-05 | 3.71E-05 | 6.38E-05 | 1.99E-05 | 0.003362518 | CADM1 |
| chr7 | 80368858 | 80368870 | 13 | 2 | 1.59E-06 | 0.0021334 | 1.16E-05 | 1.99E-05 | -0.003875 | AC004862.1 |
| chr1 | 24905846 | 24906426 | 581 | 2 | 6.81E-05 | 1.24E-05 | 0.000824 | 1.99E-05 | 0.000444736 | RUNX3 |
| chr3 | 56801976 | 56802777 | 802 | 6 | 1.02E-07 | 0.0002372 | 0.00025 | 1.99E-05 | -0.00430064 | ARHGEF3 |
| chr17 | 75909682 | 75910594 | 913 | 5 | 1.42E-05 | 5.69E-05 | 0.006649 | 1.99E-05 | 0.021931626 | AC087289.3, FBF1 |
| chr16 | 68445650 | 68446019 | 370 | 2 | 3.81E-05 | 1.21E-05 | 0.000989 | 1.99E-05 | 0.005368288 | SMPD3 |
| chr20 | 25609785 | 25610063 | 279 | 3 | 2.24E-05 | 0.0001548 | 0.000194 | 1.99E-05 | 0.000228275 | NA |
| chr1 | 10644896 | 10644978 | 83 | 2 | 1.86E-05 | 8.86E-05 | 3.14E-05 | 1.99E-05 | -0.00424294 | AL139423.1, CASZ1 |
| chr12 | 55817114 | 55817599 | 486 | 4 | 1.61E-07 | 0.0046216 | 1.42E-05 | 1.99E-05 | 0.003351326 | SARNP, AC023055.1 |
| chr17 | 17591859 | 17592301 | 443 | 6 | 9.23E-05 | 0.0001133 | 4.59E-05 | 1.99E-05 | 0.001978576 | AC020558.1 |
| chr7 | 28279724 | 28280057 | 334 | 4 | 2.67E-05 | 0.0001158 | 0.000641 | 1.99E-05 | 1.89E-05 | NA |
| chr8 | 138477514 | 138477982 | 469 | 2 | 5.71E-05 | 1.20E-05 | 0.001037 | 1.99E-05 | -0.00354438 | FAM135B |
| chr11 | 65833731 | 65833956 | 226 | 8 | 2.13E-05 | 0.0001597 | 0.002012 | 2.00E-05 | 0.001094706 | SNX32, CFL1 |
| chr6 | 164085549 | 164086273 | 725 | 10 | 5.30E-05 | 1.01E-05 | 0.0082 | 2.00E-05 | -0.01268323 | Z97205.2 |
| chr4 | 37583332 | 37583672 | 341 | 2 | 5.29E-05 | 1.20E-05 | 0.001042 | 2.00E-05 | -0.00222573 | C4orf19 |
| chr11 | 2186225 | 2186753 | 529 | 3 | 3.47E-05 | 2.43E-05 | 0.001086 | 2.00E-05 | -0.00493965 | NA |
| chr18 | 51136810 | 51137053 | 244 | 2 | 2.28E-05 | 1.20E-05 | 0.001047 | 2.00E-05 | -0.00729209 | NA |
| chr12 | 9333608 | 9333946 | 339 | 3 | 3.37E-06 | 0.0292449 | 4.40E-06 | 2.00E-05 | 0.005806204 | NA |
| chr8 | 143590072 | 143590546 | 475 | 2 | 3.77E-05 | 1.25E-05 | 0.000789 | 2.00E-05 | -0.01337968 | EEF1D |
| chr4 | 25866823 | 25867045 | 223 | 2 | 1.00E-05 | 0.0001343 | 2.52E-05 | 2.00E-05 | 0.004315715 | SMIM20, AC133961.1 |
| chr22 | 38016317 | 38016458 | 142 | 2 | 4.16E-05 | 3.02E-05 | 8.17E-05 | 2.00E-05 | -0.00793525 | POLR2F |
| chr11 | 61965574 | 61966733 | 1160 | 4 | 8.97E-08 | 0.0024824 | 1.50E-05 | 2.00E-05 | -0.01144508 | FTH1 |
| chr10 | 8235286 | 8235560 | 275 | 5 | 6.38E-05 | 1.48E-05 | 0.000937 | 2.00E-05 | -0.00319228 | NA |
| chr10 | 63465784 | 63466619 | 836 | 11 | 1.04E-05 | 3.25E-05 | 0.001485 | 2.00E-05 | 0.001519037 | JMJD1C-AS1, JMJD1C |
| chr12 | 122063329 | 122063979 | 651 | 4 | 5.77E-06 | 8.02E-05 | 0.000142 | 2.00E-05 | -0.00896539 | AC156455.1 |
| chr20 | 62246319 | 62246357 | 39 | 2 | 4.15E-05 | 1.50E-05 | 0.000356 | 2.00E-05 | -0.00146738 | OSBPL2 |
| chr3 | 42532620 | 42532811 | 192 | 3 | 3.08E-05 | 7.81E-05 | 0.000257 | 2.00E-05 | -0.00439199 | VIPR1, VIPR1-AS1 |
| chr13 | 111970374 | 111970477 | 104 | 3 | 1.28E-05 | 0.0017969 | 9.04E-06 | 2.00E-05 | 0.003069813 | NA |
| chr7 | 23709988 | 23710466 | 479 | 10 | 8.49E-06 | 8.16E-05 | 0.00205 | 2.00E-05 | -0.00108205 | STK31 |
| chr11 | 46380501 | 46380814 | 314 | 6 | 1.17E-05 | 0.0007428 | 0.001315 | 2.00E-05 | 0.000972469 | DGKZ, MDK |
| chr14 | 91780686 | 91780776 | 91 | 2 | 6.22E-05 | 1.27E-05 | 0.000749 | 2.00E-05 | 0.019195568 | CATSPERB, TC2N |
| chr14 | 105442356 | 105442614 | 259 | 3 | 1.16E-05 | 0.0001981 | 4.52E-05 | 2.00E-05 | -0.00029525 | TEX22, MTA1 |
| chr8 | 131042309 | 131042489 | 181 | 4 | 5.61E-06 | 0.0003785 | 2.63E-05 | 2.01E-05 | -0.00076587 | NA |
| chr9 | 127789300 | 127789454 | 155 | 3 | 2.39E-05 | 2.63E-05 | 0.000145 | 2.01E-05 | -0.00330103 | CDK9 |
| chr2 | 8723096 | 8723436 | 341 | 3 | 3.95E-06 | 0.0005534 | 4.27E-05 | 2.01E-05 | -0.00105004 | KIDINS220 |
| chr8 | 142111588 | 142111770 | 183 | 3 | 1.27E-05 | 0.0019733 | 9.95E-06 | 2.01E-05 | 0.006404406 | AC103758.1 |
| chr17 | 50095154 | 50095870 | 717 | 13 | 3.03E-05 | 7.11E-05 | 0.011288 | 2.01E-05 | -0.00086086 | PDK2 |
| chr5 | 83471132 | 83472149 | 1018 | 16 | 5.52E-05 | 0.0010811 | 0.007131 | 2.01E-05 | 0.000200783 | VCAN |
| chr19 | 6375759 | 6376565 | 807 | 7 | 1.09E-05 | 7.18E-05 | 0.002496 | 2.01E-05 | -0.0066082 | PSPN |
| chr8 | 56303545 | 56303955 | 411 | 2 | 1.01E-05 | 8.60E-05 | 3.23E-05 | 2.01E-05 | -0.00855521 | SDR16C5 |
| chr11 | 6240181 | 6240624 | 444 | 6 | 3.23E-05 | 2.20E-05 | 0.000238 | 2.01E-05 | -0.00427199 | CNGA4 |
| chr1 | 3171073 | 3171960 | 888 | 6 | 9.81E-06 | 9.02E-05 | 0.001551 | 2.01E-05 | 4.40E-05 | PRDM16 |
| chr19 | 40219429 | 40219861 | 433 | 3 | 2.05E-07 | 0.0046324 | 7.23E-06 | 2.01E-05 | -0.01198677 | NA |
| chr21 | 39387608 | 39387768 | 161 | 6 | 4.16E-05 | 1.77E-05 | 0.001933 | 2.01E-05 | 0.023112151 | GET1, GET1-SH3BGR |
| chr1 | 84477776 | 84477820 | 45 | 2 | 2.72E-05 | 1.72E-05 | 0.000238 | 2.01E-05 | -0.003096 | AL844170.1 |
| chr2 | 38671610 | 38672705 | 1096 | 4 | 6.66E-07 | 0.0120337 | 4.55E-06 | 2.01E-05 | 0.004848155 | GALM |
| chr3 | 47083908 | 47084303 | 396 | 4 | 7.06E-05 | 1.83E-05 | 0.000951 | 2.01E-05 | -0.00172958 | SETD2 |
| chr14 | 55267975 | 55268266 | 292 | 2 | 4.81E-05 | 1.45E-05 | 0.000414 | 2.01E-05 | -0.00077919 | AL158801.2 |
| chr15 | 58261582 | 58261784 | 203 | 2 | 1.84E-05 | 6.54E-05 | 3.90E-05 | 2.02E-05 | -0.00065405 | ALDH1A2 |
| chr17 | 9836384 | 9836402 | 19 | 2 | 1.40E-05 | 0.009178 | 1.03E-05 | 2.02E-05 | -0.00418718 | GLP2R |
| chr17 | 46832830 | 46833729 | 900 | 6 | 1.01E-05 | 0.0288273 | 0.000124 | 2.02E-05 | -0.00300784 | WNT9B, WNT3 |
| chr5 | 164600063 | 164600490 | 428 | 3 | 2.19E-05 | 1.73E-05 | 0.000323 | 2.02E-05 | -0.00928497 | AC109466.1 |
| chr20 | 35954368 | 35955159 | 792 | 10 | 1.03E-05 | 0.0009784 | 0.000299 | 2.02E-05 | 0.001566144 | CNBD2, SCAND1 |
| chr15 | 67836295 | 67836731 | 437 | 2 | 1.58E-06 | 2.36E-05 | 0.000119 | 2.02E-05 | -0.00188922 | AC009292.2, AC009292.1 |
| chr5 | 132555982 | 132557031 | 1050 | 11 | 1.68E-06 | 0.0010851 | 0.003321 | 2.02E-05 | -0.00482691 | AC116366.2, RAD50, IL5 |
| chr19 | 1313408 | 1313528 | 121 | 3 | 1.54E-05 | 0.0002671 | 0.000301 | 2.02E-05 | -0.00887782 | NA |
| chr5 | 126812427 | 126812454 | 28 | 2 | 5.03E-05 | 1.25E-05 | 0.000869 | 2.02E-05 | -0.00828698 | LMNB1 |
| chr17 | 7304787 | 7304839 | 53 | 2 | 3.56E-05 | 2.32E-05 | 0.000123 | 2.02E-05 | 0.000717219 | NA |
| chr5 | 135111187 | 135111621 | 435 | 2 | 3.24E-06 | 4.53E-05 | 5.37E-05 | 2.02E-05 | -0.00533463 | C5orf66 |
| chr7 | 128167447 | 128168039 | 593 | 4 | 1.02E-05 | 4.14E-05 | 7.94E-05 | 2.02E-05 | -0.00015938 | NA |
| chr21 | 28517227 | 28517388 | 162 | 2 | 3.77E-05 | 3.50E-05 | 6.98E-05 | 2.02E-05 | -0.01083923 | AF165147.1 |
| chr3 | 149474348 | 149474590 | 243 | 3 | 1.97E-05 | 9.56E-06 | 0.001769 | 2.02E-05 | -0.00611445 | NA |
| chr16 | 55462362 | 55463174 | 813 | 3 | 3.15E-05 | 2.76E-05 | 0.000214 | 2.02E-05 | -0.00722435 | MMP2 |
| chr15 | 75178436 | 75179510 | 1075 | 6 | 1.57E-05 | 0.0006505 | 0.000556 | 2.02E-05 | -0.00114649 | NA |
| chr12 | 88932125 | 88932546 | 422 | 2 | 5.35E-06 | 0.0006805 | 1.45E-05 | 2.02E-05 | -0.0128271 | NA |
| chr22 | 48608612 | 48608945 | 334 | 3 | 3.71E-05 | 2.00E-05 | 0.000574 | 2.02E-05 | -0.00928175 | TAFA5 |
| chr12 | 27752804 | 27752961 | 158 | 3 | 2.71E-05 | 0.0001473 | 5.78E-05 | 2.02E-05 | 0.005489691 | MRPS35, Y RNA |
| chr17 | 64230427 | 64231103 | 677 | 3 | 2.93E-06 | 4.60E-05 | 6.88E-05 | 2.02E-05 | 0.000772973 | TEX2 |
| chr2 | 171434531 | 171435173 | 643 | 7 | 2.37E-05 | 7.59E-06 | 0.005989 | 2.02E-05 | 0.000758826 | DCAF17, METTL8 |
| chr9 | 136474297 | 136474702 | 406 | 2 | 2.31E-05 | 2.53E-05 | 0.000107 | 2.02E-05 | 0.011507761 | SEC16A |
| chr18 | 49613149 | 49613737 | 589 | 4 | 2.03E-05 | 0.0002933 | 0.000597 | 2.02E-05 | 0.002592523 | NA |
| chr11 | 1219856 | 1220111 | 256 | 3 | 5.68E-06 | 0.0036515 | 0.000238 | 2.03E-05 | -0.01120619 | AC061979.1 |
| chr8 | 133491820 | 133492218 | 399 | 2 | 4.12E-05 | 1.29E-05 | 0.000734 | 2.03E-05 | 0.011691804 | ST3GAL1 |
| chr4 | 159349945 | 159350199 | 255 | 2 | 2.58E-05 | 7.99E-05 | 3.42E-05 | 2.03E-05 | -0.00224184 | RAPGEF2 |
| chr14 | 64974333 | 64974689 | 357 | 2 | 8.94E-06 | 9.99E-05 | 3.00E-05 | 2.03E-05 | -0.01005656 | CHURC1-FNTB |
| chr7 | 4178537 | 4178579 | 43 | 2 | 1.37E-05 | 1.57E-05 | 0.000318 | 2.03E-05 | -0.00320838 | SDK1 |
| chr15 | 31605603 | 31605830 | 228 | 3 | 4.99E-05 | 2.09E-05 | 0.00035 | 2.03E-05 | -0.00221219 | OTUD7A |
| chr9 | 131788104 | 131788335 | 232 | 2 | 5.71E-05 | 1.34E-05 | 0.000604 | 2.03E-05 | -6.82E-06 | NA |
| chr2 | 662602 | 663024 | 423 | 3 | 5.47E-05 | 9.73E-06 | 0.001725 | 2.03E-05 | -0.0028615 | NA |
| chr15 | 99384010 | 99384075 | 66 | 3 | 4.06E-05 | 1.63E-05 | 0.000402 | 2.03E-05 | 0.001766527 | LRRC28 |
| chr20 | 5586865 | 5587057 | 193 | 2 | 1.62E-05 | 1.83E-05 | 0.00021 | 2.03E-05 | -0.01523019 | GPCPD1 |
| chr11 | 129895511 | 129896259 | 749 | 6 | 5.50E-06 | 6.04E-05 | 0.000563 | 2.03E-05 | -0.00296694 | NFRKB |
| chr3 | 63443072 | 63443231 | 160 | 3 | 3.45E-06 | 0.0013118 | 0.000347 | 2.03E-05 | 0.015706237 | SYNPR, SYNPR-AS1 |
| chr1 | 8877581 | 8878756 | 1176 | 10 | 7.26E-08 | 0.0006027 | 1.14E-05 | 2.03E-05 | 0.001386126 | ENO1 |
| chr2 | 120521907 | 120522530 | 624 | 4 | 1.33E-05 | 4.97E-05 | 0.000221 | 2.03E-05 | 0.01306962 | NA |
| chr5 | 135535772 | 135536625 | 854 | 18 | 1.59E-05 | 3.05E-05 | 0.006892 | 2.03E-05 | -0.00074611 | NEUROG1 |
| chr22 | 37925825 | 37926385 | 561 | 3 | 2.09E-06 | 0.0013992 | 4.08E-05 | 2.03E-05 | 0.002120846 | MICALL1 |
| chr1 | 247437696 | 247438176 | 481 | 2 | 5.41E-05 | 1.37E-05 | 0.000558 | 2.03E-05 | -0.03327039 | NLRP3 |
| chr5 | 1002954 | 1004035 | 1082 | 5 | 2.71E-06 | 0.0124013 | 5.48E-05 | 2.03E-05 | -0.00158206 | NA |
| chr15 | 93476998 | 93477109 | 112 | 2 | 6.98E-06 | 0.0755575 | 9.81E-06 | 2.04E-05 | -0.01789058 | AC091078.1 |
| chr4 | 39048593 | 39048931 | 339 | 2 | 8.14E-06 | 7.01E-05 | 3.77E-05 | 2.04E-05 | 0.014204383 | KLHL5 |
| chr10 | 119037164 | 119037239 | 76 | 2 | 4.82E-05 | 2.65E-05 | 0.000101 | 2.04E-05 | -0.0051804 | EIF3A |
| chr6 | 30998880 | 30999161 | 282 | 2 | 1.00E-05 | 0.0005592 | 1.54E-05 | 2.04E-05 | -0.01587204 | NA |
| chr2 | 40103293 | 40103958 | 666 | 2 | 8.56E-05 | 1.33E-05 | 0.000645 | 2.04E-05 | 0.013406947 | SLC8A1-AS1, SLC8A1 |
| chr9 | 136830287 | 136830312 | 26 | 2 | 4.25E-05 | 2.13E-05 | 0.000149 | 2.04E-05 | -0.00892732 | RABL6 |
| chr19 | 859065 | 860565 | 1501 | 10 | 7.56E-06 | 0.011514 | 6.83E-06 | 2.04E-05 | 0.002458653 | CFD |
| chr6 | 63441834 | 63441840 | 7 | 2 | 4.27E-05 | 4.21E-05 | 5.86E-05 | 2.04E-05 | -0.01666033 | AL121949.1 |
| chr10 | 133453826 | 133453963 | 138 | 3 | 5.54E-05 | 1.04E-05 | 0.001743 | 2.04E-05 | -0.00325473 | SCART1 |
| chr3 | 133399995 | 133400306 | 312 | 3 | 2.30E-06 | 0.1659317 | 5.17E-06 | 2.04E-05 | -0.00628801 | BFSP2 |
| chr3 | 140941493 | 140942458 | 966 | 11 | 2.30E-05 | 9.18E-05 | 0.001459 | 2.04E-05 | 0.000321918 | SLC25A36 |
| chr5 | 123400401 | 123400509 | 109 | 2 | 5.82E-05 | 1.51E-05 | 0.000376 | 2.04E-05 | -0.00872727 | CEP120 |
| chr3 | 15065280 | 15065802 | 523 | 10 | 3.53E-05 | 4.14E-06 | 0.018579 | 2.04E-05 | -0.00500834 | MRPS25 |
| chr11 | 41394240 | 41394403 | 164 | 2 | 2.84E-05 | 1.31E-05 | 0.000701 | 2.04E-05 | -0.01741009 | LRRC4C |
| chr20 | 44753813 | 44754090 | 278 | 2 | 2.05E-05 | 6.88E-05 | 3.84E-05 | 2.05E-05 | -0.00791979 | RIMS4 |
| chr14 | 32710700 | 32710710 | 11 | 2 | 3.74E-05 | 1.72E-05 | 0.000254 | 2.05E-05 | 0.008172418 | AKAP6 |
| chr16 | 20352389 | 20352962 | 574 | 3 | 8.90E-05 | 9.19E-06 | 0.002765 | 2.05E-05 | -6.60E-05 | UMOD |
| chr12 | 53676347 | 53676826 | 480 | 11 | 3.29E-05 | 0.0002447 | 0.008851 | 2.05E-05 | 0.003356996 | ATP5MC2 |
| chr16 | 27906804 | 27907296 | 493 | 2 | 5.32E-05 | 1.39E-05 | 0.000527 | 2.05E-05 | -0.00677116 | GSG1L |
| chr17 | 65055754 | 65055957 | 204 | 5 | 1.40E-05 | 8.69E-05 | 0.000394 | 2.05E-05 | 0.000692705 | GNA13 |
| chr9 | 132240195 | 132241001 | 807 | 4 | 5.16E-06 | 0.0003927 | 0.000388 | 2.05E-05 | -0.0051733 | NTNG2 |
| chr1 | 11529429 | 11529986 | 558 | 4 | 2.54E-05 | 2.80E-05 | 0.002698 | 2.05E-05 | -0.00360802 | DISP3 |
| chr10 | 17003335 | 17003801 | 467 | 3 | 3.69E-05 | 1.13E-05 | 0.000984 | 2.05E-05 | -0.00155009 | CUBN |
| chr4 | 1863138 | 1863540 | 403 | 2 | 2.51E-05 | 1.30E-05 | 0.000751 | 2.05E-05 | -5.54E-05 | NA |
| chr2 | 10404939 | 10405602 | 664 | 2 | 7.82E-07 | 5.55E-05 | 4.57E-05 | 2.05E-05 | -0.00397596 | HPCAL1 |

| | | | | | | | | | | |
|---|---|---|---|---|---|---|---|---|---|---|
| chr16 | 81061152 | 81061397 | 246 | 2 | 3.24E-05 | 3.32E-05 | 7.63E-05 | 2.05E-05 | 0.002029812 | AC092718.8, C16orf46, AC092718.3 |
| chr2 | 3490094 | 3490686 | 593 | 2 | 2.62E-05 | 1.42E-05 | 0.000492 | 2.05E-05 | 0.001268003 | NA |
| chr22 | 32531256 | 32531316 | 61 | 2 | 9.50E-06 | 1.31E-05 | 0.000728 | 2.05E-05 | 0.001358184 | SYN3 |
| chr2 | 238452795 | 238452847 | 53 | 2 | 7.56E-06 | 2.91E-05 | 9.04E-05 | 2.05E-05 | 0.0028332 | NA |
| chr7 | 74234127 | 74234574 | 448 | 3 | 1.52E-05 | 6.11E-05 | 5.55E-05 | 2.06E-05 | 5.45E-05 | RFC2 |
| chr18 | 58559785 | 58559962 | 178 | 2 | 5.52E-05 | 1.45E-05 | 0.000451 | 2.06E-05 | -0.00656547 | AC104971.2, ALPK2 |
| chr9 | 111838354 | 111838476 | 123 | 2 | 4.61E-05 | 1.76E-05 | 0.000241 | 2.06E-05 | 0.001512681 | NA |
| chr12 | 120630331 | 120630578 | 248 | 3 | 4.44E-05 | 2.36E-05 | 0.000501 | 2.06E-05 | -0.00075612 | AC125616.1 |
| chr13 | 111630134 | 111630356 | 223 | 3 | 7.23E-05 | 2.59E-05 | 0.001101 | 2.06E-05 | -0.00729285 | AL359649.1, LINC02337 |
| chr6 | 693728 | 694246 | 519 | 3 | 5.44E-06 | 1.70E-05 | 0.00077 | 2.06E-05 | 0.001247187 | AL357054.2 |
| chr7 | 157373762 | 157373765 | 4 | 2 | 4.92E-05 | 1.39E-05 | 0.000554 | 2.06E-05 | -0.00502143 | DNAJB6 |
| chr3 | 181711445 | 181712123 | 679 | 11 | 1.60E-05 | 0.0002412 | 0.003317 | 2.06E-05 | 0.000857669 | SOX2-OT, SOX2 |
| chr7 | 76400331 | 76400564 | 234 | 4 | 7.72E-05 | 8.75E-06 | 0.00574 | 2.06E-05 | -0.00355266 | ZP3, SSC4D |
| chr10 | 119198194 | 119198614 | 421 | 2 | 6.34E-05 | 1.26E-05 | 0.000927 | 2.06E-05 | -0.01743309 | NA |
| chr1 | 181318165 | 181318831 | 667 | 7 | 2.89E-05 | 4.39E-05 | 0.00328 | 2.06E-05 | 0.001870228 | CACNA1E |
| chr7 | 97731709 | 97732447 | 739 | 10 | 1.78E-05 | 0.0002564 | 0.000119 | 2.06E-05 | -0.00161561 | TAC1 |
| chr16 | 11401213 | 11401490 | 278 | 2 | 9.33E-06 | 3.04E-05 | 8.60E-05 | 2.06E-05 | -0.00963896 | AC099489.1 |
| chr10 | 122464196 | 122464954 | 759 | 3 | 5.58E-06 | 0.0002579 | 2.50E-05 | 2.06E-05 | -0.00128544 | HTRA1 |
| chr10 | 87932743 | 87932750 | 8 | 2 | 5.83E-05 | 1.72E-05 | 0.000259 | 2.06E-05 | -0.0047301 | PTEN |
| chr21 | 45265239 | 45265597 | 359 | 4 | 1.74E-06 | 0.0143807 | 2.95E-06 | 2.07E-05 | -0.00188027 | POFUT2 |
| chr16 | 67256680 | 67256946 | 267 | 2 | 7.85E-06 | 0.0027625 | 1.17E-05 | 2.07E-05 | -0.0053922 | SLC9A5 |
| chr20 | 18378594 | 18379006 | 413 | 5 | 1.09E-05 | 0.0001793 | 0.000475 | 2.07E-05 | 0.002081724 | NA |
| chr11 | 70717159 | 70718011 | 853 | 7 | 1.87E-05 | 0.0001363 | 0.000461 | 2.07E-05 | 0.00378484 | SHANK2 |
| chr2 | 88191799 | 88192106 | 308 | 2 | 6.93E-06 | 0.0017542 | 1.24E-05 | 2.07E-05 | -0.00951414 | NA |
| chr15 | 100310894 | 100311298 | 405 | 2 | 5.84E-05 | 1.33E-05 | 0.000691 | 2.07E-05 | -0.01198003 | ADAMTS17 |
| chr5 | 53478982 | 53479323 | 342 | 2 | 2.18E-05 | 1.25E-05 | 0.001043 | 2.07E-05 | 0.001660092 | NA |
| chr17 | 80087846 | 80087933 | 88 | 3 | 0.000105888 | 2.46E-05 | 0.001122 | 2.07E-05 | 0.006308487 | CCDC40 |
| chr14 | 69005592 | 69005935 | 344 | 2 | 1.05E-05 | 0.0001513 | 2.49E-05 | 2.07E-05 | -0.00642895 | NA |
| chr14 | 21099487 | 21099637 | 151 | 2 | 0.000110359 | 2.99E-05 | 8.89E-05 | 2.07E-05 | -0.010545 | TMEM253, ZNF219 |
| chr1 | 54403334 | 54403698 | 365 | 2 | 3.33E-05 | 1.92E-05 | 0.000198 | 2.07E-05 | 0.01124417 | SSBP3 |
| chr6 | 53153943 | 53154188 | 246 | 3 | 1.62E-05 | 8.73E-05 | 0.000214 | 2.08E-05 | -0.0064124 | NA |
| chr1 | 206539937 | 206540200 | 264 | 2 | 8.20E-06 | 8.62E-05 | 3.36E-05 | 2.08E-05 | 0.006027805 | RASSF5 |
| chr7 | 229225 | 229695 | 471 | 4 | 2.56E-06 | 9.13E-05 | 7.45E-05 | 2.08E-05 | -0.00492662 | FAM20C, AC145676.1 |
| chr1 | 228445574 | 228446564 | 991 | 7 | 5.80E-05 | 7.10E-06 | 0.012297 | 2.08E-05 | 0.00102478 | NA |
| chr2 | 108287496 | 108288047 | 552 | 3 | 3.18E-06 | 0.0004313 | 3.90E-05 | 2.08E-05 | -0.00824371 | NA |
| chr3 | 58496564 | 58496649 | 86 | 2 | 0.000151296 | 1.40E-05 | 0.000552 | 2.08E-05 | -0.00657275 | KCTD6 |
| chr6 | 3063813 | 3064522 | 710 | 7 | 4.81E-06 | 0.0009577 | 2.95E-05 | 2.08E-05 | -0.0005507 | RIPK1 |
| chr16 | 85149097 | 85149135 | 39 | 2 | 5.29E-06 | 0.0038539 | 1.13E-05 | 2.08E-05 | -0.00844626 | LINC02139 |
| chr6 | 167468371 | 167468398 | 28 | 2 | 2.76E-05 | 0.0002099 | 2.16E-05 | 2.08E-05 | -0.00726686 | NA |
| chr14 | 99437011 | 99437169 | 159 | 2 | 5.37E-05 | 1.77E-05 | 0.000245 | 2.08E-05 | -0.00703746 | SETD3 |
| chr1 | 11806873 | 11807251 | 379 | 3 | 1.11E-06 | 1.32E-05 | 0.00161 | 2.08E-05 | -0.00060182 | CLCN6 |
| chr22 | 40519742 | 40519813 | 72 | 2 | 4.94E-05 | 2.62E-05 | 0.000107 | 2.08E-05 | -0.00518397 | MRTFA |
| chr19 | 44025071 | 44025531 | 461 | 6 | 3.70E-05 | 0.0003538 | 0.000251 | 2.08E-05 | -0.00041432 | ZNF222, AC067968.1, AC067968.2 |
| chr8 | 57555325 | 57555776 | 452 | 3 | 3.86E-05 | 5.78E-05 | 0.000718 | 2.08E-05 | -0.00893344 | AC090796.1 |
| chr9 | 112379481 | 112379597 | 117 | 4 | 2.17E-05 | 1.51E-05 | 0.003733 | 2.08E-05 | 0.000476067 | NA |
| chr16 | 46832590 | 46832785 | 196 | 2 | 3.94E-06 | 0.0001807 | 2.31E-05 | 2.08E-05 | -0.00567671 | NA |
| chr11 | 68633008 | 68633023 | 16 | 2 | 6.64E-05 | 1.30E-05 | 0.00085 | 2.08E-05 | -0.01435485 | NA |
| chr17 | 80178370 | 80178667 | 298 | 6 | 9.49E-05 | 0.0005754 | 0.000623 | 2.08E-05 | -0.00490089 | CARD14 |
| chr2 | 239367021 | 239367131 | 111 | 2 | 2.67E-05 | 0.0001126 | 2.90E-05 | 2.08E-05 | 0.004750377 | HDAC4 |
| chr16 | 71707642 | 71708072 | 431 | 2 | 1.69E-05 | 3.80E-05 | 6.73E-05 | 2.08E-05 | -0.00261727 | PHLPP2 |
| chr13 | 114280583 | 114280862 | 280 | 3 | 2.49E-06 | 0.0004937 | 4.98E-05 | 2.08E-05 | -0.00963661 | NA |
| chr3 | 19346245 | 19346449 | 205 | 3 | 2.98E-05 | 0.0001859 | 0.000514 | 2.08E-05 | -0.00926855 | KCNH8 |
| chr8 | 102806408 | 102806691 | 284 | 4 | 5.84E-06 | 1.64E-05 | 0.002212 | 2.09E-05 | 0.001516483 | GASAL1 |
| chr17 | 81294814 | 81295238 | 425 | 4 | 2.00E-05 | 6.46E-05 | 0.00091 | 2.09E-05 | 0.002432227 | SLC38A10 |
| chr5 | 59790606 | 59791024 | 419 | 2 | 7.48E-05 | 1.26E-05 | 0.00104 | 2.09E-05 | -0.00136937 | PDE4D |
| chr1 | 3771240 | 3771693 | 454 | 4 | 5.32E-06 | 0.0017286 | 0.000348 | 2.09E-05 | -0.00455634 | CCDC27 |
| chr22 | 27797922 | 27798008 | 87 | 2 | 1.63E-05 | 8.99E-05 | 3.30E-05 | 2.09E-05 | 0.001925623 | MN1 |
| chr10 | 1154085 | 1154259 | 175 | 3 | 3.11E-05 | 3.54E-05 | 0.000142 | 2.09E-05 | -0.02021551 | NA |
| chr2 | 3448792 | 3449425 | 634 | 6 | 1.44E-05 | 0.0001783 | 0.000161 | 2.09E-05 | -0.00392186 | TRAPPC12 |
| chr3 | 192449394 | 192449573 | 180 | 2 | 3.37E-05 | 4.80E-05 | 5.36E-05 | 2.09E-05 | 0.021021691 | FGF12 |
| chr4 | 87974539 | 87974804 | 266 | 2 | 1.73E-05 | 1.76E-05 | 0.000254 | 2.09E-05 | 0.007871023 | AC131944.1 |
| chr11 | 60184084 | 60184347 | 264 | 3 | 1.15E-05 | 0.0008432 | 0.00017 | 2.09E-05 | 0.009403605 | MS4A6A |
| chr4 | 104490599 | 104491142 | 544 | 6 | 4.87E-06 | 0.0039431 | 2.00E-05 | 2.09E-05 | -0.00017762 | CXXC4-AS1, CXXC4 |
| chr1 | 19439070 | 19439818 | 749 | 3 | 1.26E-05 | 2.13E-05 | 0.000284 | 2.09E-05 | -0.00265396 | CAPZB |
| chr9 | 136052058 | 136052328 | 271 | 9 | 5.06E-05 | 4.92E-05 | 0.003647 | 2.09E-05 | 0.001041933 | NACC2 |
| chr7 | 75872489 | 75872678 | 190 | 2 | 9.62E-06 | 0.017756 | 1.03E-05 | 2.09E-05 | -0.00959197 | RHBDD2 |
| chr5 | 1385517 | 1386035 | 519 | 5 | 2.74E-05 | 1.96E-05 | 0.001706 | 2.09E-05 | -0.00010762 | AC026748.3 |
| chr5 | 36241411 | 36242797 | 1387 | 13 | 1.97E-05 | 0.0009888 | 0.003386 | 2.09E-05 | 0.002426212 | NADK2 |
| chr6 | 47098878 | 47098947 | 70 | 2 | 1.69E-05 | 0.0003702 | 1.79E-05 | 2.09E-05 | -0.01255532 | NA |
| chr8 | 140561945 | 140562043 | 99 | 2 | 3.26E-05 | 0.0001054 | 3.03E-05 | 2.10E-05 | -0.0183719 | AGO2 |
| chr17 | 77280346 | 77280706 | 361 | 3 | 4.46E-06 | 1.29E-05 | 0.001349 | 2.10E-05 | -0.00871395 | SEPTIN9, SEPTIN9-DT |
| chr19 | 38222410 | 38222733 | 324 | 3 | 1.67E-05 | 1.04E-05 | 0.001441 | 2.10E-05 | 0.022891605 | DPF1 |
| chr2 | 27715204 | 27715245 | 42 | 2 | 7.03E-05 | 0.0013175 | 1.32E-05 | 2.10E-05 | -0.00969902 | LINC01460 |
| chr19 | 58562002 | 58562063 | 62 | 3 | 6.46E-05 | 8.49E-06 | 0.003293 | 2.10E-05 | -0.01162744 | MZF1-AS1, MZF1 |
| chr1 | 37265250 | 37265255 | 6 | 2 | 6.39E-05 | 1.48E-05 | 0.000456 | 2.10E-05 | 0.002715021 | AL353604.1 |
| chr11 | 59715008 | 59715225 | 218 | 3 | 1.60E-05 | 0.0008445 | 2.65E-05 | 2.10E-05 | 0.001195289 | STX3 |
| chr9 | 23851772 | 23851856 | 85 | 2 | 1.92E-05 | 8.73E-05 | 3.39E-05 | 2.10E-05 | -0.01294419 | NA |
| chr5 | 166083756 | 166083799 | 44 | 2 | 6.16E-05 | 1.63E-05 | 0.00033 | 2.10E-05 | -0.01793742 | NA |
| chr6 | 163309764 | 163310946 | 1183 | 9 | 9.58E-07 | 0.0003805 | 0.000183 | 2.10E-05 | 0.000148131 | PACRG, PACRG-AS1 |
| chr1 | 26992192 | 26993087 | 896 | 4 | 4.53E-05 | 1.36E-05 | 0.000985 | 2.10E-05 | 3.56E-05 | NA |
| chr4 | 5863927 | 5863971 | 45 | 2 | 6.79E-05 | 1.27E-05 | 0.00107 | 2.10E-05 | -0.00189439 | CRMP1 |
| chr1 | 23016833 | 23017049 | 217 | 2 | 7.65E-05 | 2.78E-05 | 0.000101 | 2.10E-05 | -0.01333005 | NA |
| chr10 | 69219181 | 69220055 | 875 | 4 | 2.98E-06 | 6.26E-05 | 9.60E-05 | 2.10E-05 | -0.004736 | AL596223.1 |
| chr1 | 110150278 | 110150374 | 97 | 3 | 2.22E-05 | 6.12E-05 | 0.000153 | 2.10E-05 | -0.00190624 | NA |
| chr19 | 1174155 | 1174668 | 514 | 7 | 9.71E-06 | 0.0017139 | 0.000618 | 2.11E-05 | -0.00296446 | SBNO2 |
| chr4 | 107283576 | 107283944 | 369 | 4 | 6.90E-05 | 8.23E-06 | 0.003733 | 2.11E-05 | 0.000941374 | AC104663.1, DKK2 |
| chr15 | 63193964 | 63194065 | 102 | 2 | 2.47E-05 | 0.0002416 | 2.08E-05 | 2.11E-05 | 0.002369074 | RAB8B |
| chr2 | 71720799 | 71720969 | 171 | 2 | 4.16E-05 | 3.12E-05 | 8.69E-05 | 2.11E-05 | -0.01304882 | NA |
| chr17 | 81641864 | 81642320 | 457 | 5 | 4.95E-06 | 8.90E-05 | 8.93E-05 | 2.11E-05 | -0.00195031 | TSPAN10, NPLOC4 |
| chr22 | 42696602 | 42697116 | 515 | 3 | 8.50E-07 | 0.0012936 | 1.31E-05 | 2.11E-05 | -0.00512001 | A4GALT |
| chr5 | 138875746 | 138876220 | 475 | 4 | 7.17E-06 | 1.90E-05 | 0.002265 | 2.11E-05 | -0.00401071 | CTNNA1 |
| chr9 | 8734803 | 8735059 | 257 | 3 | 4.43E-06 | 0.000295 | 3.97E-05 | 2.11E-05 | -0.00218457 | PTPRD |
| chr8 | 1359523 | 1360730 | 1208 | 6 | 6.86E-06 | 2.65E-05 | 0.004051 | 2.11E-05 | -0.00152373 | DLGAP2 |
| chr12 | 124532110 | 124532588 | 479 | 3 | 4.84E-05 | 7.57E-05 | 0.000166 | 2.11E-05 | -0.00792264 | NCOR2 |
| chr1 | 29264789 | 29265214 | 426 | 2 | 5.05E-05 | 1.48E-05 | 0.000475 | 2.11E-05 | -0.00622336 | PTPRU |
| chr5 | 37868454 | 37868511 | 58 | 3 | 6.65E-05 | 9.33E-06 | 0.002912 | 2.11E-05 | 0.000215173 | GDNF-AS1 |
| chr9 | 136760446 | 136760701 | 256 | 4 | 3.53E-05 | 2.46E-05 | 0.001294 | 2.11E-05 | -0.00561835 | LCN15 |
| chr19 | 46856283 | 46856286 | 4 | 2 | 0.000107471 | 1.29E-05 | 0.000952 | 2.11E-05 | -0.00384252 | NA |
| chr3 | 39087811 | 39088220 | 410 | 2 | 2.16E-06 | 0.0002789 | 1.99E-05 | 2.12E-05 | 0.008313912 | WDR48 |
| chr16 | 1342587 | 1343102 | 516 | 3 | 1.54E-05 | 8.15E-05 | 0.000342 | 2.12E-05 | 0.000497084 | BAIAP3 |
| chr10 | 109963867 | 109964208 | 342 | 2 | 4.15E-05 | 1.81E-05 | 0.000247 | 2.12E-05 | -0.006752 | ADD3-AS1 |
| chr1 | 151611899 | 151612447 | 549 | 12 | 5.13E-05 | 1.04E-05 | 0.012266 | 2.12E-05 | -0.00031203 | SNX27, AL391335.1 |
| chr2 | 178795878 | 178796465 | 588 | 2 | 2.39E-05 | 2.16E-05 | 0.000161 | 2.12E-05 | -0.01563673 | TTN |
| chr12 | 113018252 | 113018374 | 123 | 2 | 6.77E-06 | 0.0005908 | 1.59E-05 | 2.12E-05 | -0.00387033 | NA |
| chr17 | 32486971 | 32487496 | 526 | 3 | 0.000101615 | 1.55E-05 | 0.00108 | 2.12E-05 | 0.001309883 | CDK5R1 |
| chr10 | 133008230 | 133008497 | 268 | 3 | 1.36E-05 | 0.0003336 | 1.71E-05 | 2.12E-05 | 0.001296765 | NA |
| chr2 | 55661535 | 55661636 | 102 | 2 | 5.06E-05 | 1.39E-05 | 0.000642 | 2.12E-05 | -0.0099711 | PNPT1 |
| chr14 | 22906257 | 22906366 | 110 | 3 | 3.96E-06 | 0.0140099 | 4.57E-06 | 2.12E-05 | 0.003656345 | RBM23 |
| chr13 | 44373457 | 44374016 | 560 | 6 | 2.64E-05 | 9.98E-05 | 0.000743 | 2.13E-05 | -0.00385134 | SERP2 |
| chr6 | 118757013 | 118757360 | 348 | 2 | 2.13E-05 | 1.65E-05 | 0.000328 | 2.13E-05 | 0.001093303 | NA |
| chr12 | 108768865 | 108768972 | 108 | 2 | 4.35E-05 | 0.0028258 | 1.19E-05 | 2.13E-05 | -0.00316863 | NA |
| chr7 | 886794 | 886986 | 193 | 2 | 2.40E-05 | 1.86E-05 | 0.000233 | 2.13E-05 | -0.00014422 | SUN1, GET4 |
| chr6 | 28408741 | 28409281 | 541 | 3 | 9.14E-05 | 1.37E-05 | 0.000942 | 2.13E-05 | -0.0001494 | NA |
| chr19 | 13872569 | 13872999 | 431 | 3 | 3.18E-06 | 0.0156038 | 0.000115 | 2.13E-05 | -0.0058875 | NANOS3 |
| chr3 | 136101763 | 136102098 | 336 | 4 | 4.77E-05 | 2.09E-05 | 0.000776 | 2.13E-05 | -0.00197616 | PPP2R3A |
| chr19 | 45002040 | 45002285 | 246 | 2 | 1.41E-05 | 1.29E-05 | 0.001051 | 2.13E-05 | 0.000434315 | RELB |
| chr18 | 28079731 | 28079834 | 104 | 3 | 3.83E-05 | 9.44E-05 | 9.14E-05 | 2.13E-05 | -0.0073796 | CDH2 |
| chr12 | 121202919 | 121203275 | 357 | 3 | 1.64E-05 | 0.000139 | 4.79E-05 | 2.13E-05 | 0.002621863 | NA |
| chr6 | 157822801 | 157823654 | 854 | 6 | 3.36E-06 | 6.03E-05 | 0.000212 | 2.13E-05 | 0.00118958 | SNX9 |
| chr17 | 8289980 | 8290134 | 155 | 2 | 3.89E-06 | 0.0003338 | 1.88E-05 | 2.13E-05 | -0.00603028 | RANGRF, SLC25A35 |
| chr1 | 13665772 | 13666046 | 275 | 2 | 6.69E-05 | 1.30E-05 | 0.000992 | 2.13E-05 | -0.00302909 | NA |
| chr1 | 2251926 | 2251999 | 74 | 2 | 6.77E-06 | 0.0003841 | 1.80E-05 | 2.13E-05 | -0.00707888 | SKI |
| chr15 | 78139761 | 78140058 | 298 | 2 | 1.26E-05 | 0.0001081 | 3.06E-05 | 2.13E-05 | -0.00948232 | IDH3A |
| chr8 | 73747541 | 73747737 | 197 | 3 | 8.21E-05 | 1.66E-05 | 0.00085 | 2.13E-05 | 0.007015247 | STAU2, AC022826.2 |
| chr19 | 17556345 | 17556827 | 483 | 2 | 2.10E-07 | 0.0032939 | 1.18E-05 | 2.13E-05 | -0.00899373 | COLGALT1 |
| chr10 | 125012067 | 125012364 | 298 | 3 | 1.19E-05 | 0.0003862 | 1.97E-05 | 2.13E-05 | -0.00206743 | CTBP2 |
| chr3 | 44873179 | 44873402 | 224 | 2 | 1.96E-05 | 1.28E-05 | 0.001089 | 2.13E-05 | 0.001240497 | KIF15 |
| chr16 | 3454484 | 3455017 | 534 | 2 | 1.23E-06 | 0.0013678 | 1.34E-05 | 2.14E-05 | -0.00424737 | AC025283.2, NAA60 |
| chr1 | 8525944 | 8526246 | 303 | 6 | 1.43E-05 | 0.0039161 | 0.001022 | 2.14E-05 | 0.007043036 | RERE |
| chr19 | 54502440 | 54502738 | 299 | 5 | 0.000139147 | 4.30E-06 | 0.012642 | 2.14E-05 | 0.016045214 | LAIR2 |
| chr12 | 69737912 | 69738936 | 1025 | 8 | 7.44E-06 | 3.82E-05 | 0.000318 | 2.14E-05 | 0.002722734 | RAB3IP, AC025263.1 |
| chr7 | 100694642 | 100694706 | 65 | 2 | 1.51E-05 | 7.43E-05 | 3.86E-05 | 2.14E-05 | 0.00389732 | NA |
| chr7 | 129503196 | 129503420 | 225 | 2 | 1.08E-06 | 0.001801 | 1.28E-05 | 2.14E-05 | -0.00929813 | SMKR1 |
| chr13 | 46127632 | 46127934 | 303 | 2 | 1.20E-05 | 5.67E-05 | 4.79E-05 | 2.14E-05 | 0.007509671 | CPB2-AS1, LCP1 |
| chr1 | 109493795 | 109494504 | 710 | 10 | 2.93E-05 | 3.08E-05 | 0.016807 | 2.14E-05 | 0.00071317 | CYB561D1 |
| chr1 | 248817027 | 248817590 | 564 | 5 | 1.78E-05 | 0.0001094 | 0.000294 | 2.14E-05 | -0.00133374 | SH3BP5L |
| chr11 | 44789671 | 44790142 | 472 | 2 | 7.16E-05 | 1.30E-05 | 0.001038 | 2.14E-05 | -0.0017207 | TSPAN18 |
| chr7 | 157598338 | 157598605 | 268 | 3 | 8.67E-06 | 0.0019278 | 7.77E-06 | 2.14E-05 | -0.00363109 | PTPRN2 |
| chr7 | 44796128 | 44796597 | 470 | 8 | 1.33E-05 | 0.0001108 | 0.002667 | 2.14E-05 | 0.001098656 | NA |
| chr12 | 110065936 | 110065945 | 10 | 2 | 4.07E-05 | 1.36E-05 | 0.000781 | 2.14E-05 | -0.00717354 | C12orf76, AC007546.3 |
| chr13 | 99205453 | 99205672 | 220 | 3 | 5.33E-05 | 2.35E-05 | 0.0008 | 2.14E-05 | 0.002582263 | UBAC2 |
| chr8 | 70835178 | 70835335 | 158 | 2 | 6.83E-05 | 1.30E-05 | 0.001009 | 2.14E-05 | -0.00066801 | AC015687.1 |
| chr7 | 156643329 | 156643790 | 462 | 2 | 6.81E-05 | 1.78E-05 | 0.00027 | 2.14E-05 | -0.00968239 | RNF32 |
| chr3 | 191568316 | 191568531 | 216 | 3 | 9.16E-05 | 8.11E-06 | 0.00442 | 2.15E-05 | -0.01084593 | AC073365.1 |
| chr8 | 120123775 | 120123901 | 127 | 2 | 2.52E-06 | 0.0145091 | 1.06E-05 | 2.15E-05 | -0.00674038 | COL14A1 |
| chr3 | 138947088 | 138947631 | 544 | 9 | 4.13E-06 | 0.0014325 | 0.000335 | 2.15E-05 | 0.000772264 | FOXL2NB, FOXL2 |
| chr18 | 59139392 | 59140316 | 925 | 9 | 8.38E-06 | 0.0005361 | 0.001545 | 2.15E-05 | 5.24E-05 | SEC11C |
| chr1 | 15171414 | 15172055 | 642 | 8 | 4.77E-05 | 8.42E-06 | 0.011321 | 2.15E-05 | -0.00711131 | TMEM51 |
| chr11 | 93721437 | 93721460 | 24 | 2 | 5.31E-05 | 1.84E-05 | 0.000246 | 2.15E-05 | -0.00586495 | CEP295 |
| chr21 | 46322890 | 46323940 | 1051 | 9 | 6.44E-06 | 1.84E-05 | 0.009135 | 2.15E-05 | -0.00028429 | C21orf58 |
| chr15 | 28123917 | 28124281 | 365 | 3 | 2.72E-05 | 4.81E-05 | 7.23E-05 | 2.15E-05 | -0.0034658 | HERC2 |
| chr13 | 113359659 | 113359750 | 92 | 2 | 4.13E-05 | 4.88E-05 | 5.53E-05 | 2.15E-05 | -0.00409711 | GRTP1-AS1, GRTP1 |
| chr13 | 98383081 | 98383302 | 222 | 3 | 2.53E-05 | 0.000249 | 8.48E-05 | 2.15E-05 | 0.004417281 | FARP1 |
| chr15 | 59168859 | 59169400 | 542 | 2 | 8.93E-05 | 1.32E-05 | 0.000938 | 2.15E-05 | 0.006974714 | MYO1E |
| chr8 | 48408258 | 48408585 | 328 | 2 | 4.41E-05 | 1.31E-05 | 0.00099 | 2.15E-05 | -0.00101367 | NA |
| chr1 | 215004085 | 215004120 | 36 | 2 | 2.45E-05 | 3.27E-05 | 8.50E-05 | 2.15E-05 | -0.00200164 | NA |
| chr6 | 168449172 | 168449679 | 508 | 2 | 5.90E-05 | 1.44E-05 | 0.000592 | 2.15E-05 | -0.00474983 | SMOC2 |
| chr19 | 847943 | 848071 | 129 | 4 | 4.29E-05 | 8.61E-05 | 0.000414 | 2.15E-05 | 0.00628914 | PRTN3 |

| | | | | | | | | | | |
|---|---|---|---|---|---|---|---|---|---|---|
| chr2 | 11345426 | 11345658 | 233 | 3 | 5.00E-05 | 1.46E-05 | 0.000608 | 2.15E-05 | -0.00440423 | ROCK2 |
| chr3 | 156816287 | 156816591 | 305 | 2 | 2.68E-05 | 1.64E-05 | 0.000352 | 2.15E-05 | -0.01212259 | LINC00886 |
| chr2 | 44676084 | 44676401 | 318 | 2 | 7.00E-06 | 0.0017349 | 1.30E-05 | 2.15E-05 | -0.00183835 | CAMKMT |
| chr3 | 38606333 | 38606709 | 377 | 2 | 1.11E-05 | 1.31E-05 | 0.000995 | 2.16E-05 | -0.00075976 | SCN5A |
| chr10 | 35151908 | 35152207 | 300 | 2 | 3.39E-05 | 2.78E-05 | 0.000107 | 2.16E-05 | 0.011014051 | CREM |
| chr10 | 129887839 | 129887944 | 106 | 3 | 6.57E-05 | 7.66E-05 | 0.000728 | 2.16E-05 | -0.00617409 | EBF3 |
| chr2 | 8500932 | 8501341 | 410 | 4 | 4.74E-05 | 3.37E-05 | 0.001469 | 2.16E-05 | -0.00533765 | AC092617.1, LINC01814 |
| chr6 | 163051270 | 163051539 | 270 | 3 | 3.46E-05 | 0.0007975 | 0.00032 | 2.16E-05 | -0.01774856 | PACRG, PACRG-AS2 |
| chr1 | 9650763 | 9651605 | 843 | 8 | 9.26E-06 | 0.000552 | 0.000179 | 2.16E-05 | 0.002687924 | NA |
| chr14 | 22818288 | 22818344 | 57 | 2 | 5.22E-05 | 1.37E-05 | 0.000788 | 2.16E-05 | -0.01157112 | SLC7A7 |
| chr21 | 46189805 | 46190182 | 378 | 3 | 2.37E-05 | 2.63E-05 | 0.000204 | 2.16E-05 | 0.005295968 | LSS |
| chr13 | 110528604 | 110529035 | 432 | 2 | 6.74E-06 | 3.69E-05 | 7.41E-05 | 2.16E-05 | -0.0072746 | RAB20 |
| chr1 | 244406402 | 244406442 | 41 | 2 | 6.44E-05 | 1.62E-05 | 0.00037 | 2.16E-05 | -0.01348557 | AL645465.1 |
| chr12 | 50504468 | 50505235 | 768 | 12 | 7.14E-05 | 1.60E-05 | 0.024485 | 2.16E-05 | -0.00219548 | DIP2B |
| chr1 | 30783181 | 30783403 | 223 | 2 | 3.56E-05 | 1.34E-05 | 0.000907 | 2.16E-05 | -0.05261707 | NA |
| chr16 | 56864874 | 56865271 | 398 | 5 | 4.51E-05 | 4.94E-05 | 0.000386 | 2.16E-05 | 0.002869018 | SLC12A3 |
| chr1 | 151540665 | 151541026 | 362 | 5 | 4.91E-05 | 1.75E-05 | 0.000873 | 2.16E-05 | 0.001685137 | TUFT1, AL365436.2 |
| chr5 | 59390252 | 59390658 | 407 | 2 | 5.62E-05 | 1.46E-05 | 0.000578 | 2.16E-05 | 0.010390081 | PDE4D |
| chr6 | 54307385 | 54307809 | 425 | 3 | 3.82E-05 | 4.39E-05 | 0.000615 | 2.16E-05 | -0.01325013 | NA |
| chr9 | 133875832 | 133876017 | 186 | 2 | 3.32E-05 | 1.83E-05 | 0.00026 | 2.16E-05 | -0.000174 | VAV2 |
| chr8 | 125352125 | 125352839 | 715 | 2 | 1.30E-05 | 1.47E-05 | 0.000556 | 2.17E-05 | 0.00688478 | NSMCE2 |
| chr16 | 85113162 | 85113540 | 379 | 4 | 1.03E-05 | 1.37E-05 | 0.00151 | 2.17E-05 | -0.00440837 | NA |
| chr13 | 112988143 | 112988470 | 328 | 5 | 3.72E-05 | 5.97E-05 | 0.000862 | 2.17E-05 | -0.00235214 | MCF2L |
| chr11 | 10934454 | 10934528 | 75 | 2 | 2.87E-05 | 9.15E-05 | 3.44E-05 | 2.17E-05 | -0.00594447 | NA |
| chr18 | 79679856 | 79680287 | 432 | 4 | 1.76E-06 | 8.99E-05 | 0.000113 | 2.17E-05 | 0.00225319 | CTDP1 |
| chr12 | 67222508 | 67222766 | 259 | 2 | 1.82E-05 | 0.0001511 | 2.64E-05 | 2.17E-05 | 0.001030826 | NA |
| chr11 | 130449796 | 130450324 | 529 | 3 | 1.32E-06 | 0.0001297 | 7.22E-05 | 2.17E-05 | -0.01004306 | ADAMTS15 |
| chr1 | 203117834 | 203117964 | 131 | 2 | 1.67E-05 | 9.62E-05 | 3.35E-05 | 2.17E-05 | -0.00833194 | ADORA1, AC105940.2 |
| chr6 | 143163523 | 143164046 | 524 | 2 | 1.93E-05 | 1.37E-05 | 0.000813 | 2.17E-05 | -0.00796338 | AIG1 |
| chr17 | 48595562 | 48595640 | 79 | 2 | 5.12E-06 | 0.0007653 | 1.53E-05 | 2.17E-05 | -0.00294305 | HOXB-AS3, HOXB3 |
| chr1 | 163363700 | 163363957 | 258 | 2 | 1.05E-05 | 0.0012018 | 1.39E-05 | 2.17E-05 | -0.00915777 | NA |
| chr10 | 119652314 | 119652974 | 661 | 3 | 1.94E-06 | 7.18E-05 | 6.83E-05 | 2.17E-05 | 0.00375027 | BAG3 |
| chr19 | 45937306 | 45937771 | 466 | 3 | 8.69E-06 | 0.0004442 | 0.000443 | 2.18E-05 | -0.01166126 | NOVA2 |
| chr17 | 80356647 | 80356677 | 31 | 2 | 2.06E-05 | 7.42E-05 | 3.97E-05 | 2.18E-05 | -0.00411229 | RNF213, RNF213-AS1 |
| chr7 | 2646518 | 2647210 | 693 | 4 | 2.20E-05 | 0.000106 | 0.001423 | 2.18E-05 | -0.00191962 | TTYH3 |
| chr8 | 41652360 | 41652680 | 321 | 2 | 6.38E-06 | 1.82E-05 | 0.000268 | 2.18E-05 | -0.00229756 | NA |
| chr8 | 37899424 | 37900353 | 930 | 10 | 3.96E-06 | 0.0007858 | 0.000146 | 2.18E-05 | 6.15E-05 | RAB11FIP1 |
| chr12 | 51291926 | 51292333 | 408 | 2 | 4.03E-05 | 1.84E-05 | 0.000263 | 2.18E-05 | -0.01174076 | BIN2 |
| chr5 | 132221798 | 132222225 | 428 | 3 | 5.85E-05 | 1.07E-05 | 0.002543 | 2.18E-05 | 0.004985395 | P4HA2 |
| chr14 | 54858784 | 54859022 | 239 | 4 | 6.83E-05 | 1.03E-05 | 0.001495 | 2.18E-05 | 6.36E-05 | GCH1 |
| chr6 | 25726230 | 25726728 | 499 | 7 | 7.44E-05 | 1.45E-05 | 0.005836 | 2.18E-05 | -3.58E-06 | H2AC1 |
| chr7 | 115910891 | 115911252 | 362 | 2 | 5.15E-05 | 3.08E-05 | 9.49E-05 | 2.19E-05 | -0.00772564 | NA |
| chr6 | 71449957 | 71449996 | 40 | 2 | 3.61E-05 | 1.71E-05 | 0.00033 | 2.19E-05 | 0.00272703 | NA |
| chr8 | 144510970 | 144511759 | 790 | 6 | 2.99E-06 | 0.0003535 | 0.000268 | 2.19E-05 | -0.00250924 | MFSD3, RECQL4 |
| chr1 | 92080341 | 92081009 | 669 | 8 | 4.38E-06 | 0.0004897 | 0.001204 | 2.19E-05 | 0.001828779 | BTBD8 |
| chr9 | 76181961 | 76181964 | 4 | 2 | 2.77E-05 | 6.19E-05 | 4.62E-05 | 2.19E-05 | 0.003283916 | PCSK5 |
| chr20 | 57560956 | 57561111 | 156 | 4 | 0.000101356 | 0.0007736 | 0.000241 | 2.19E-05 | 0.000442112 | PCK1 |
| chr11 | 120685224 | 120685429 | 206 | 2 | 6.52E-05 | 1.41E-05 | 0.000713 | 2.19E-05 | -0.00556389 | GRIK4 |
| chr13 | 95533756 | 95534007 | 252 | 3 | 2.66E-05 | 0.0007691 | 0.000385 | 2.19E-05 | -0.00745914 | CLDN10, CLDN10-AS1 |
| chr22 | 30555967 | 30556246 | 280 | 2 | 3.92E-06 | 0.0002939 | 2.03E-05 | 2.19E-05 | -0.00469665 | GAL3ST1 |
| chr3 | 64561634 | 64561670 | 37 | 3 | 0.000102475 | 1.26E-05 | 0.001704 | 2.19E-05 | -0.00133936 | ADAMTS9-AS1, ADAMTS9 |
| chr19 | 41887895 | 41888266 | 372 | 3 | 7.12E-06 | 0.0002275 | 2.48E-05 | 2.20E-05 | 0.005091538 | ARHGEF1 |
| chr12 | 30822392 | 30822815 | 424 | 5 | 3.37E-05 | 4.70E-05 | 0.00248 | 2.20E-05 | 0.000848583 | AC010198.2 |
| chr19 | 2866621 | 2866625 | 5 | 2 | 6.32E-06 | 8.13E-05 | 3.76E-05 | 2.20E-05 | -0.00365046 | NA |
| chr12 | 47844908 | 47844980 | 73 | 2 | 1.86E-05 | 2.38E-05 | 0.000146 | 2.20E-05 | 0.000924389 | VDR |
| chr14 | 21104880 | 21105249 | 370 | 4 | 2.21E-05 | 1.99E-05 | 0.001306 | 2.20E-05 | -0.0043494 | NA |
| chr10 | 48589214 | 48589325 | 112 | 2 | 5.90E-05 | 1.95E-05 | 0.000229 | 2.20E-05 | -0.01412825 | ARHGAP22 |
| chr4 | 48134435 | 48134829 | 395 | 3 | 1.97E-05 | 2.01E-05 | 0.000764 | 2.20E-05 | 0.00018975 | NA |
| chr9 | 135521166 | 135521462 | 297 | 4 | 2.49E-05 | 0.0025169 | 0.000378 | 2.20E-05 | -0.00725182 | LCN1 |
| chr12 | 80714211 | 80714255 | 45 | 3 | 3.58E-05 | 0.0003077 | 0.0002 | 2.20E-05 | -6.20E-05 | NA |
| chr6 | 112059477 | 112059911 | 435 | 2 | 2.66E-06 | 0.000254 | 2.15E-05 | 2.20E-05 | 0.001355759 | CCN6 |
| chr22 | 41678517 | 41678915 | 399 | 3 | 2.93E-05 | 8.83E-05 | 0.000466 | 2.20E-05 | -0.00145276 | SNU13 |
| chr8 | 39948780 | 39949182 | 403 | 4 | 1.18E-05 | 0.0007643 | 0.000268 | 2.20E-05 | 0.001920071 | IDO2, AC007991.3 |
| chr17 | 5331724 | 5332008 | 285 | 2 | 1.96E-05 | 1.55E-05 | 0.000474 | 2.20E-05 | 0.001537903 | RABEP1 |
| chr21 | 41187725 | 41187781 | 57 | 2 | 3.51E-05 | 1.35E-05 | 0.000962 | 2.20E-05 | -0.01443496 | BACE2 |
| chr19 | 49422813 | 49423819 | 1007 | 9 | 9.52E-06 | 0.0015155 | 3.63E-05 | 2.21E-05 | 0.001759159 | GFY, PTH2 |
| chr1 | 100085379 | 100085568 | 190 | 3 | 4.60E-05 | 4.14E-05 | 0.000268 | 2.21E-05 | -0.00309461 | SASS6 |
| chr8 | 48693661 | 48693845 | 185 | 2 | 3.57E-05 | 0.0007267 | 1.57E-05 | 2.21E-05 | -0.00749471 | AC022915.2 |
| chr1 | 235971849 | 235972582 | 734 | 6 | 1.36E-05 | 2.04E-05 | 0.001011 | 2.21E-05 | -0.00901356 | NA |
| chr10 | 3233305 | 3233347 | 43 | 3 | 2.30E-05 | 3.17E-05 | 0.00017 | 2.21E-05 | -0.00369852 | LINC02668 |
| chr13 | 19873771 | 19873967 | 197 | 3 | 3.63E-05 | 2.02E-05 | 0.000299 | 2.21E-05 | 0.005119867 | NA |
| chr1 | 44826241 | 44826532 | 292 | 3 | 8.22E-05 | 1.25E-05 | 0.001387 | 2.21E-05 | -0.00471979 | PTCH2 |
| chr17 | 58191809 | 58192049 | 241 | 3 | 2.20E-05 | 1.40E-05 | 0.00073 | 2.21E-05 | -0.00658356 | NA |
| chr9 | 92115062 | 92115620 | 559 | 6 | 5.57E-05 | 3.22E-05 | 0.007442 | 2.21E-05 | 0.001032541 | SPTLC1 |
| chr6 | 43016553 | 43016957 | 405 | 4 | 5.10E-05 | 8.88E-06 | 0.0045 | 2.21E-05 | -0.00384883 | KLHDC3, MEA1 |
| chr1 | 207068046 | 207068451 | 406 | 3 | 3.84E-06 | 6.63E-05 | 5.72E-05 | 2.21E-05 | -0.00231692 | PFKFB2 |
| chr7 | 140060575 | 140060871 | 297 | 2 | 5.58E-05 | 2.16E-05 | 0.000183 | 2.21E-05 | 0.013275628 | PARP12 |
| chr17 | 81200864 | 81201332 | 469 | 3 | 2.88E-05 | 8.59E-05 | 0.000721 | 2.21E-05 | -0.00541832 | CEP131 |
| chr11 | 67310548 | 67310942 | 395 | 2 | 2.98E-06 | 0.0003197 | 1.99E-05 | 2.21E-05 | -0.00591607 | SSH3 |
| chr12 | 109231827 | 109232512 | 686 | 4 | 8.19E-07 | 0.0009612 | 0.000149 | 2.21E-05 | -0.00328132 | ACACB |
| chr9 | 121268770 | 121269173 | 404 | 2 | 2.35E-06 | 1.69E-05 | 0.000354 | 2.21E-05 | -0.00991694 | GSN |
| chr9 | 37888613 | 37888623 | 11 | 2 | 4.61E-05 | 4.86E-05 | 5.83E-05 | 2.21E-05 | -0.01798922 | AL138752.2, SLC25A51 |
| chr15 | 89665684 | 89666095 | 412 | 4 | 6.60E-05 | 3.67E-05 | 0.000701 | 2.22E-05 | -0.00209062 | PLIN1 |
| chr3 | 196568347 | 196568655 | 309 | 7 | 0.000104052 | 0.0001736 | 0.00247 | 2.22E-05 | 0.000611238 | FBXO45, WDR53 |
| chr4 | 54231253 | 54231822 | 570 | 5 | 4.66E-05 | 1.00E-05 | 0.00327 | 2.22E-05 | 0.000233427 | AC058822.1, PDGFRA |
| chr6 | 32109967 | 32110847 | 881 | 11 | 3.17E-05 | 3.51E-05 | 0.006642 | 2.22E-05 | -0.00117706 | AL662884.5, TNXB, AL662884.2 |
| chr6 | 149594065 | 149594644 | 580 | 3 | 2.30E-05 | 2.46E-05 | 0.00034 | 2.22E-05 | -0.0028516 | RPS18P9, AL078581.1 |
| chr11 | 111924624 | 111924860 | 237 | 3 | 2.20E-05 | 2.78E-05 | 0.000599 | 2.22E-05 | 0.003182543 | HSPB2-C11orf52, C11orf52 |
| chr17 | 1485656 | 1486281 | 626 | 6 | 3.89E-06 | 6.10E-05 | 0.000138 | 2.23E-05 | -0.00085376 | MYO1C |
| chr11 | 108008451 | 108009211 | 761 | 6 | 1.88E-05 | 0.0002107 | 0.001681 | 2.23E-05 | -0.00213351 | CUL5, AP003307.1 |
| chr1 | 7150766 | 7150888 | 123 | 2 | 1.23E-05 | 3.71E-05 | 7.77E-05 | 2.23E-05 | -0.01864064 | CAMTA1 |
| chr14 | 54441917 | 54442491 | 575 | 4 | 6.69E-06 | 0.000232 | 2.64E-05 | 2.23E-05 | -0.00368278 | NA |
| chr3 | 50303578 | 50303938 | 361 | 5 | 3.64E-05 | 0.0001696 | 0.002438 | 2.23E-05 | 0.000436608 | HYAL1 |
| chr19 | 3683646 | 3683813 | 168 | 3 | 6.65E-05 | 8.91E-06 | 0.003771 | 2.23E-05 | 0.02494265 | PIP5K1C |
| chr20 | 10673299 | 10674640 | 1342 | 9 | 4.38E-05 | 0.0001207 | 0.002887 | 2.23E-05 | 0.000751634 | AL050403.2, JAG1 |
| chr7 | 8689353 | 8689486 | 134 | 2 | 7.41E-06 | 4.93E-05 | 5.82E-05 | 2.23E-05 | -0.00527072 | NXPH1 |
| chr7 | 3056647 | 3056927 | 281 | 4 | 9.21E-05 | 1.46E-05 | 0.003152 | 2.23E-05 | -0.00099969 | NA |
| chr18 | 48742515 | 48742636 | 122 | 2 | 2.31E-05 | 0.0001757 | 2.54E-05 | 2.23E-05 | -0.00463853 | CTIF |
| chr22 | 36933791 | 36933858 | 68 | 2 | 5.68E-05 | 2.47E-05 | 0.000142 | 2.23E-05 | -0.00675992 | CSF2RB |
| chr9 | 6327804 | 6328386 | 583 | 7 | 3.14E-05 | 0.000209 | 0.002628 | 2.23E-05 | -0.00393197 | TPD52L3 |
| chr5 | 96901508 | 96901730 | 223 | 2 | 1.94E-05 | 0.0002177 | 2.33E-05 | 2.23E-05 | 0.003256027 | ERAP2, AC009126.1 |
| chr8 | 66137213 | 66137805 | 593 | 2 | 0.000141282 | 1.34E-05 | 0.001117 | 2.23E-05 | 0.007459128 | TRIM55 |
| chr2 | 136645279 | 136645858 | 580 | 4 | 4.37E-06 | 0.0057098 | 0.000324 | 2.23E-05 | -0.00707847 | NA |
| chr6 | 36139573 | 36139875 | 303 | 2 | 1.26E-05 | 2.74E-05 | 0.000118 | 2.23E-05 | -0.01352577 | MAPK13 |
| chr15 | 47301736 | 47301741 | 6 | 2 | 7.13E-05 | 1.36E-05 | 0.001044 | 2.23E-05 | -0.01250111 | SEMA6D |
| chr6 | 14844643 | 14844812 | 170 | 3 | 6.30E-05 | 2.10E-05 | 0.000992 | 2.23E-05 | -0.0047701 | AL138720.1 |
| chr1 | 209416146 | 209416299 | 154 | 2 | 3.33E-05 | 1.35E-05 | 0.001079 | 2.23E-05 | -0.00937279 | NA |
| chr15 | 68058127 | 68058210 | 84 | 2 | 7.27E-05 | 1.35E-05 | 0.001086 | 2.24E-05 | -0.00325228 | PIAS1 |
| chr13 | 111018253 | 111018373 | 121 | 3 | 1.34E-05 | 0.0090588 | 1.16E-05 | 2.24E-05 | -0.00055731 | NA |
| chr22 | 20551269 | 20551438 | 170 | 3 | 9.09E-05 | 1.14E-05 | 0.001337 | 2.24E-05 | 0.023651869 | MED15 |
| chr3 | 9191380 | 9191730 | 351 | 3 | 9.60E-06 | 4.48E-05 | 0.000854 | 2.24E-05 | -0.00101304 | SRGAP3 |
| chr10 | 1609007 | 1609196 | 190 | 3 | 3.47E-05 | 0.0002107 | 0.000266 | 2.24E-05 | -0.00989267 | ADARB2 |
| chr20 | 63428039 | 63428206 | 168 | 5 | 3.49E-05 | 0.0002076 | 0.000148 | 2.24E-05 | -0.00283103 | KCNQ2 |
| chr16 | 48116851 | 48117408 | 558 | 3 | 2.19E-07 | 0.0113342 | 8.96E-06 | 2.24E-05 | -0.00190638 | ABCC12 |
| chr20 | 41137308 | 41138154 | 847 | 12 | 9.83E-06 | 0.0001002 | 0.000746 | 2.24E-05 | -2.25E-05 | PLCG1, PLCG1-AS1 |
| chr19 | 45584517 | 45585192 | 676 | 12 | 3.44E-05 | 0.0003067 | 0.001338 | 2.24E-05 | 0.000333555 | OPA3 |
| chr1 | 159926806 | 159927266 | 461 | 2 | 5.14E-06 | 9.77E-05 | 3.46E-05 | 2.24E-05 | -0.00643267 | IGSF9 |
| chr10 | 43361853 | 43362193 | 341 | 2 | 5.44E-05 | 3.00E-05 | 0.000103 | 2.24E-05 | 0.007710981 | NA |
| chr17 | 15943919 | 15944098 | 180 | 2 | 1.72E-05 | 2.02E-05 | 0.000226 | 2.24E-05 | -0.01525874 | NA |
| chr16 | 21300647 | 21300955 | 309 | 5 | 5.59E-05 | 4.07E-05 | 0.001541 | 2.24E-05 | 0.003654069 | CRYM-AS1, CRYM |
| chr9 | 134324364 | 134324687 | 324 | 2 | 2.14E-06 | 0.0001407 | 2.85E-05 | 2.25E-05 | -0.00994062 | RXRA |
| chr16 | 10528917 | 10528927 | 11 | 2 | 1.42E-05 | 0.0021444 | 1.31E-05 | 2.25E-05 | -0.00902217 | EMP2 |
| chr1 | 171565940 | 171566652 | 713 | 4 | 7.33E-05 | 3.83E-05 | 0.001391 | 2.25E-05 | -0.0027677 | PRRC2C |
| chr17 | 30926780 | 30926851 | 72 | 2 | 2.28E-05 | 2.91E-05 | 0.000108 | 2.25E-05 | -0.01035225 | ADAP2 |
| chr2 | 120437422 | 120437785 | 364 | 2 | 4.40E-05 | 1.90E-05 | 0.000267 | 2.25E-05 | -0.00309282 | NA |
| chr5 | 44001776 | 44002173 | 398 | 2 | 7.56E-05 | 1.38E-05 | 0.001006 | 2.25E-05 | -0.01338692 | NA |
| chr19 | 35225190 | 35225198 | 9 | 2 | 2.71E-05 | 1.41E-05 | 0.000843 | 2.25E-05 | -0.04859647 | FAM187B |
| chr2 | 8709832 | 8709936 | 105 | 3 | 4.02E-05 | 1.37E-05 | 0.001652 | 2.25E-05 | -0.00369426 | NA |
| chr5 | 10286820 | 10287195 | 376 | 2 | 4.81E-05 | 1.74E-05 | 0.000346 | 2.25E-05 | 0.000192646 | CMBL |
| chr3 | 124785173 | 124785253 | 81 | 2 | 5.77E-05 | 2.43E-05 | 0.00015 | 2.25E-05 | 0.00431916 | ITGB5-AS1, ITGB5 |
| chr3 | 25454384 | 25454652 | 269 | 2 | 3.26E-05 | 2.30E-05 | 0.000169 | 2.25E-05 | -0.00903105 | RARB |
| chr1 | 21883736 | 21884581 | 846 | 4 | 6.40E-06 | 0.0001673 | 0.000245 | 2.25E-05 | -0.00542504 | HSPG2 |
| chr3 | 18426119 | 18426324 | 206 | 2 | 3.90E-06 | 0.0001657 | 2.64E-05 | 2.25E-05 | 0.007377694 | TBC1D5, SATB1 |
| chr1 | 40754339 | 40754434 | 96 | 3 | 2.12E-05 | 0.0001719 | 4.73E-05 | 2.26E-05 | -0.00341878 | NFYC, MIR30E |
| chr20 | 35541393 | 35541640 | 248 | 2 | 2.89E-06 | 0.0092983 | 1.14E-05 | 2.26E-05 | 0.002483262 | NA |
| chr17 | 66953828 | 66953851 | 24 | 2 | 7.06E-05 | 1.38E-05 | 0.001001 | 2.26E-05 | -0.00541765 | NA |
| chr7 | 30931692 | 30931879 | 188 | 2 | 4.03E-05 | 1.84E-05 | 0.000295 | 2.26E-05 | -0.00315684 | NA |
| chr4 | 105632315 | 105632692 | 378 | 2 | 2.57E-05 | 2.32E-05 | 0.000167 | 2.26E-05 | -0.05555415 | ARHGEF38 |
| chr12 | 49323009 | 49323886 | 878 | 11 | 1.94E-05 | 0.0001139 | 0.000754 | 2.26E-05 | 0.000917047 | TROAP, AC125611.4 |
| chr6 | 125374356 | 125374744 | 389 | 2 | 2.12E-05 | 4.27E-05 | 6.83E-05 | 2.26E-05 | -0.01071862 | AL450332.1 |
| chr17 | 44123620 | 44124029 | 410 | 10 | 4.40E-05 | 0.0009659 | 0.00071 | 2.26E-05 | 0.001717966 | HDAC5 |
| chr15 | 77682435 | 77682845 | 411 | 2 | 2.56E-05 | 3.07E-05 | 0.000102 | 2.26E-05 | -0.01302778 | LINGO1 |
| chr19 | 43652338 | 43652349 | 12 | 2 | 5.81E-05 | 1.87E-05 | 0.000282 | 2.26E-05 | -0.019255 | PLAUR |
| chr8 | 79892565 | 79892680 | 116 | 2 | 4.14E-05 | 1.54E-05 | 0.00055 | 2.26E-05 | 0.004391055 | AC036214.4 |
| chr22 | 31835847 | 31836017 | 171 | 2 | 8.37E-06 | 1.49E-05 | 0.000654 | 2.26E-05 | -0.01652988 | DEPDC5, Z82190.2 |
| chr1 | 5871753 | 5872291 | 539 | 3 | 3.92E-06 | 8.86E-05 | 5.77E-05 | 2.26E-05 | -0.00340041 | NPHP4 |
| chr4 | 169008607 | 169009278 | 672 | 3 | 7.67E-06 | 1.87E-05 | 0.001065 | 2.26E-05 | 0.001995586 | CBR4 |
| chr1 | 23198642 | 23198751 | 110 | 2 | 4.99E-05 | 1.39E-05 | 0.000979 | 2.26E-05 | 0.008399426 | NA |
| chr1 | 207097677 | 207098001 | 325 | 4 | 1.81E-05 | 0.002076 | 0.000265 | 2.26E-05 | -0.00471995 | C4BPB |
| chr3 | 4693644 | 4694056 | 413 | 2 | 2.68E-05 | 2.78E-05 | 0.000119 | 2.26E-05 | 0.014858609 | ITPR1 |
| chr3 | 87552899 | 87553530 | 632 | 4 | 6.12E-05 | 1.57E-05 | 0.001461 | 2.26E-05 | -0.00886242 | NA |
| chr2 | 127057630 | 127057686 | 57 | 2 | 0.000162953 | 1.37E-05 | 0.001114 | 2.26E-05 | 0.00035972 | BIN1 |
| chr11 | 134406779 | 134407333 | 555 | 3 | 1.63E-05 | 0.0001018 | 5.23E-05 | 2.26E-05 | -0.0097532 | B3GAT1 |
| chr12 | 25405212 | 25405227 | 16 | 2 | 5.12E-05 | 3.91E-05 | 7.56E-05 | 2.27E-05 | -0.00450853 | NA |
| chr12 | 53173523 | 53174042 | 520 | 6 | 5.37E-05 | 0.0001209 | 0.001187 | 2.27E-05 | -0.00438539 | CSAD |
| chr14 | 89954105 | 89954648 | 544 | 6 | 5.90E-07 | 0.0006688 | 0.000135 | 2.27E-05 | -0.00864016 | AL137230.2, EFCAB11 |
| chr6 | 142474549 | 142474888 | 340 | 2 | 1.76E-05 | 2.42E-05 | 0.000154 | 2.27E-05 | -0.00029765 | NA |
| chr22 | 35423279 | 35423377 | 99 | 2 | 7.45E-06 | 1.66E-05 | 0.000417 | 2.27E-05 | -0.00123561 | MCM5 |

| | | | | | | | | | | |
|---|---|---|---|---|---|---|---|---|---|---|
| chr17 | 41185736 | 41186254 | 519 | 4 | 2.31E-05 | 1.30E-05 | 0.001155 | 2.27E-05 | -0.01161772 | NA |
| chr17 | 75078466 | 75078709 | 244 | 4 | 8.49E-06 | 0.0012492 | 0.000397 | 2.27E-05 | -0.00664709 | TRIM80P |
| chr8 | 123767773 | 123768611 | 839 | 13 | 1.12E-05 | 0.0006667 | 0.008847 | 2.27E-05 | 0.000630087 | FAM91A1 |
| chr2 | 237798366 | 237798946 | 581 | 10 | 7.53E-06 | 8.19E-05 | 0.002883 | 2.27E-05 | -0.00088102 | LRRFIP1, RBM44 |
| chr1 | 91737407 | 91737430 | 24 | 2 | 5.65E-05 | 1.69E-05 | 0.000394 | 2.27E-05 | -1.50E-05 | TGFBR3 |
| chr12 | 131000812 | 131000975 | 164 | 2 | 4.30E-06 | 6.63E-05 | 4.61E-05 | 2.27E-05 | -0.00586927 | ADGRD1 |
| chr4 | 385977 | 386607 | 631 | 4 | 1.10E-05 | 0.001609 | 0.00015 | 2.27E-05 | 0.005726297 | AC092574.2 |
| chr17 | 40612977 | 40613043 | 67 | 2 | 5.11E-05 | 1.56E-05 | 0.000541 | 2.28E-05 | 0.002829566 | NA |
| chr11 | 13008175 | 13009382 | 1208 | 14 | 4.20E-07 | 0.0476064 | 3.43E-05 | 2.28E-05 | -6.79E-05 | RASSF10, RASSF10-DT |
| chr11 | 14364581 | 14364936 | 356 | 5 | 0.000142095 | 6.81E-06 | 0.005134 | 2.28E-05 | 0.000762378 | NA |
| chr3 | 47501667 | 47502104 | 438 | 2 | 8.81E-07 | 1.95E-05 | 0.000257 | 2.28E-05 | -0.00010306 | ELP6 |
| chr5 | 132761480 | 132761536 | 57 | 2 | 7.13E-06 | 1.68E-05 | 0.000411 | 2.28E-05 | -0.00461438 | SEPTIN8 |
| chr3 | 49093049 | 49094229 | 1181 | 13 | 4.41E-05 | 5.26E-05 | 0.001986 | 2.28E-05 | 0.001219685 | QRICH1 |
| chr5 | 124806636 | 124806703 | 68 | 2 | 4.64E-05 | 4.91E-05 | 6.02E-05 | 2.28E-05 | 0.021737775 | NA |
| chr17 | 3890917 | 3891534 | 618 | 4 | 3.90E-05 | 5.48E-05 | 0.000773 | 2.28E-05 | -0.00633131 | CAMKK1 |
| chr7 | 148290358 | 148290865 | 508 | 2 | 7.76E-05 | 1.44E-05 | 0.000828 | 2.28E-05 | -0.01272958 | CNTNAP2 |
| chr7 | 1474725 | 1474987 | 263 | 3 | 4.17E-05 | 2.13E-05 | 0.000377 | 2.28E-05 | -0.00428476 | INTS1 |
| chr7 | 39700365 | 39701181 | 817 | 3 | 7.93E-06 | 9.84E-05 | 4.06E-05 | 2.28E-05 | -0.0055561 | RALA, AC004837.2 |
| chr14 | 60153992 | 60154200 | 209 | 3 | 4.18E-06 | 0.0012392 | 1.37E-05 | 2.29E-05 | 0.002193633 | PCNX4, DHRS7 |
| chr13 | 114043380 | 114043616 | 237 | 3 | 6.24E-06 | 0.0097762 | 3.04E-05 | 2.29E-05 | 0.005742521 | RASA3 |
| chr6 | 24645704 | 24646554 | 851 | 15 | 1.61E-05 | 0.0001071 | 0.000391 | 2.29E-05 | 0.001917781 | KIAA0319 |
| chr12 | 13061386 | 13062136 | 751 | 6 | 1.25E-05 | 0.0001906 | 0.000508 | 2.29E-05 | -0.00342074 | FAM234B |
| chr15 | 65119750 | 65119879 | 130 | 2 | 5.11E-05 | 2.29E-05 | 0.000177 | 2.29E-05 | 0.000377815 | PDCD7 |
| chr2 | 236776915 | 236777205 | 291 | 2 | 6.38E-05 | 1.51E-05 | 0.000649 | 2.29E-05 | 0.003391089 | NA |
| chr11 | 69966008 | 69966190 | 183 | 2 | 4.74E-06 | 0.0001841 | 2.56E-05 | 2.29E-05 | -0.01140702 | NA |
| chr8 | 143460842 | 143461781 | 940 | 6 | 2.00E-06 | 0.0002718 | 0.0001 | 2.29E-05 | -0.004262 | ZC3H3 |
| chr3 | 67384972 | 67385165 | 194 | 3 | 4.82E-05 | 4.98E-05 | 0.000749 | 2.30E-05 | -0.00880069 | SUCLG2 |
| chr20 | 53608101 | 53608310 | 210 | 3 | 4.97E-05 | 1.30E-05 | 0.001071 | 2.30E-05 | -0.01506198 | ZNF217 |
| chr7 | 23411000 | 23411185 | 186 | 2 | 2.82E-05 | 8.06E-05 | 4.03E-05 | 2.30E-05 | 0.002919572 | IGF2BP3 |
| chr6 | 168263308 | 168263823 | 516 | 4 | 3.08E-06 | 0.0010941 | 1.54E-05 | 2.30E-05 | 0.001892088 | NA |
| chr2 | 96265757 | 96266766 | 1010 | 12 | 2.78E-05 | 0.0002849 | 0.000712 | 2.30E-05 | 8.98E-05 | CIAO1, TMEM127 |
| chr2 | 20149714 | 20149774 | 61 | 2 | 2.97E-05 | 3.99E-05 | 7.58E-05 | 2.30E-05 | 0.001012239 | NA |
| chr1 | 230495913 | 230496090 | 178 | 2 | 2.35E-05 | 0.0002337 | 2.34E-05 | 2.30E-05 | -0.00704363 | NA |
| chr6 | 110981859 | 110982589 | 731 | 12 | 1.51E-05 | 0.0004629 | 0.001012 | 2.30E-05 | -3.29E-05 | RPF2 |
| chr16 | 8862350 | 8862589 | 240 | 5 | 0.000129722 | 7.54E-06 | 0.009755 | 2.30E-05 | 0.000753436 | CARHSP1 |
| chr13 | 113832177 | 113832465 | 289 | 5 | 8.13E-06 | 0.0002568 | 2.36E-05 | 2.30E-05 | -0.00308333 | GAS6-AS1, GAS6 |
| chr8 | 137186332 | 137186379 | 48 | 2 | 5.22E-05 | 3.80E-05 | 8.04E-05 | 2.30E-05 | 0.012705064 | NA |
| chr8 | 139737134 | 139737552 | 419 | 3 | 1.67E-06 | 0.0012692 | 2.90E-05 | 2.30E-05 | -0.00337701 | TRAPPC9 |
| chr8 | 143713601 | 143713935 | 335 | 6 | 3.41E-06 | 0.0027351 | 1.97E-05 | 2.30E-05 | -0.00283898 | ZNF707 |
| chr2 | 117814440 | 117814469 | 30 | 2 | 0.000118275 | 3.14E-05 | 0.000102 | 2.30E-05 | 0.003081507 | NA |
| chr16 | 88927784 | 88928332 | 549 | 4 | 1.19E-05 | 2.31E-05 | 0.001089 | 2.30E-05 | 0.000107745 | CBFA2T3 |
| chr17 | 42019722 | 42020630 | 909 | 13 | 1.77E-05 | 0.0001203 | 0.000544 | 2.30E-05 | -0.00094517 | NKIRAS2, DNAJC7 |
| chr5 | 134905414 | 134905848 | 435 | 7 | 1.18E-05 | 5.47E-05 | 0.00417 | 2.31E-05 | 0.001249793 | PCBD2, AC006077.2 |
| chr8 | 78805026 | 78806247 | 1222 | 11 | 5.35E-05 | 0.0042613 | 0.001618 | 2.31E-05 | -0.00125025 | AC083837.1, IL7 |
| chr1 | 190478102 | 190478875 | 774 | 5 | 2.86E-06 | 7.20E-05 | 0.00081 | 2.31E-05 | -0.00017321 | LINC01351, BRINP3 |
| chr4 | 3662513 | 3662744 | 232 | 3 | 1.41E-05 | 0.0008437 | 1.32E-05 | 2.31E-05 | -0.00158022 | NA |
| chr4 | 188236868 | 188237215 | 348 | 4 | 4.88E-05 | 2.12E-05 | 0.001593 | 2.31E-05 | -0.00296436 | NA |
| chr3 | 14261333 | 14261648 | 316 | 4 | 1.37E-05 | 0.0002061 | 6.86E-05 | 2.31E-05 | -0.00808969 | NA |
| chr15 | 69032931 | 69033429 | 499 | 5 | 5.36E-05 | 1.85E-05 | 0.002377 | 2.31E-05 | 0.010682937 | NOX5 |
| chr17 | 8913874 | 8913901 | 28 | 2 | 9.24E-06 | 0.0001252 | 3.13E-05 | 2.31E-05 | 0.003411094 | PIK3R5 |
| chr5 | 65076388 | 65076793 | 406 | 3 | 7.72E-06 | 0.0014196 | 2.92E-05 | 2.31E-05 | -0.00650542 | NA |
| chr1 | 115837695 | 115838325 | 631 | 5 | 1.11E-05 | 0.0065194 | 5.17E-05 | 2.31E-05 | -0.00341835 | NHLH2 |
| chr5 | 81301464 | 81302009 | 546 | 11 | 3.52E-06 | 0.0064866 | 0.000173 | 2.31E-05 | 0.000448232 | ZCCHC9, CKMT2-AS1 |
| chr4 | 40915693 | 40916174 | 482 | 2 | 3.25E-05 | 2.12E-05 | 0.000218 | 2.32E-05 | -0.00739067 | APBB2 |
| chr15 | 55109129 | 55109426 | 298 | 2 | 5.95E-06 | 2.29E-05 | 0.000182 | 2.32E-05 | -0.00810354 | NA |
| chr9 | 110143778 | 110144068 | 291 | 2 | 2.67E-05 | 3.09E-05 | 0.000106 | 2.32E-05 | 0.002972391 | PALM2AKAP2 |
| chr12 | 122704699 | 122704709 | 11 | 3 | 5.76E-05 | 4.65E-05 | 0.000217 | 2.32E-05 | -0.00338571 | AC026333.3 |
| chr2 | 147876757 | 147877453 | 697 | 4 | 2.14E-06 | 0.0194765 | 4.05E-06 | 2.32E-05 | -0.00297984 | ACVR2A, RNU6-1275P |
| chr4 | 84733262 | 84733608 | 347 | 2 | 9.25E-06 | 0.0004701 | 1.86E-05 | 2.32E-05 | 0.004276906 | WDFY3 |
| chr12 | 118164836 | 118165207 | 372 | 2 | 1.94E-06 | 0.0004555 | 1.87E-05 | 2.32E-05 | -0.0063738 | TAOK3 |
| chr16 | 4347872 | 4347894 | 23 | 2 | 2.49E-05 | 0.0005618 | 1.77E-05 | 2.33E-05 | 0.003991679 | PAM16, CORO7-PAM16, AC012676.5 |
| chr1 | 44784509 | 44785168 | 660 | 4 | 1.12E-05 | 0.0001193 | 5.55E-05 | 2.33E-05 | -0.0036316 | BEST4 |
| chr1 | 145720960 | 145721367 | 408 | 4 | 1.67E-06 | 0.0227568 | 3.07E-06 | 2.33E-05 | -0.00036173 | CD160 |
| chr4 | 188127413 | 188127581 | 169 | 2 | 3.56E-05 | 6.99E-05 | 4.57E-05 | 2.33E-05 | -0.01006395 | NA |
| chr11 | 115219017 | 115219565 | 549 | 3 | 2.44E-05 | 3.00E-05 | 0.001141 | 2.33E-05 | 0.002013976 | CADM1 |
| chr3 | 133038243 | 133038897 | 655 | 9 | 3.03E-05 | 2.49E-05 | 0.004335 | 2.33E-05 | 0.001237426 | TMEM108 |
| chr10 | 132631098 | 132631485 | 388 | 2 | 4.86E-06 | 0.0018044 | 1.39E-05 | 2.33E-05 | -0.00729271 | INPP5A |
| chr3 | 129542635 | 129543212 | 578 | 6 | 5.51E-05 | 1.75E-05 | 0.001746 | 2.33E-05 | -0.00365001 | NA |
| chr1 | 22952785 | 22953192 | 408 | 4 | 2.16E-05 | 3.77E-05 | 0.000127 | 2.33E-05 | -0.01088074 | LACTBL1 |
| chr1 | 38990746 | 38991598 | 853 | 11 | 1.05E-05 | 0.000208 | 0.003956 | 2.33E-05 | 0.000527087 | AKIRIN1 |
| chr17 | 28643898 | 28645036 | 1139 | 5 | 3.54E-06 | 0.0002025 | 0.00026 | 2.33E-05 | -0.00326015 | KIAA0100 |
| chr16 | 89557191 | 89557679 | 489 | 4 | 3.08E-05 | 7.53E-05 | 0.000367 | 2.33E-05 | -0.00660401 | SPG7 |
| chr6 | 36673777 | 36673807 | 31 | 2 | 2.88E-05 | 0.0001095 | 3.40E-05 | 2.33E-05 | 0.007875952 | PANDAR |
| chr10 | 324037 | 324069 | 33 | 2 | 8.97E-06 | 0.0001424 | 2.96E-05 | 2.33E-05 | 0.013538447 | DIP2C |
| chr6 | 128520476 | 128521173 | 698 | 11 | 1.91E-05 | 0.0001855 | 0.005109 | 2.34E-05 | 0.000390766 | PTPRK |
| chr3 | 126938003 | 126938317 | 315 | 2 | 4.90E-05 | 1.99E-05 | 0.000266 | 2.34E-05 | 0.003950445 | CHCHD6 |
| chr7 | 107550366 | 107550622 | 257 | 3 | 2.96E-05 | 4.57E-05 | 0.000202 | 2.34E-05 | -0.00270129 | COG5 |
| chr12 | 108515077 | 108515782 | 706 | 9 | 8.98E-07 | 0.0023411 | 6.61E-05 | 2.34E-05 | 3.20E-05 | FICD |
| chr2 | 229025697 | 229025886 | 190 | 3 | 9.93E-05 | 9.11E-06 | 0.00415 | 2.34E-05 | -0.00142638 | PID1 |
| chr16 | 84618823 | 84619448 | 626 | 4 | 5.38E-06 | 0.0001351 | 0.000525 | 2.34E-05 | -0.00760314 | NA |
| chr3 | 38275953 | 38276198 | 246 | 2 | 2.22E-05 | 2.47E-05 | 0.00016 | 2.34E-05 | -0.00446082 | SLC22A13 |
| chr8 | 100003953 | 100004053 | 101 | 3 | 6.96E-05 | 2.17E-05 | 0.000301 | 2.34E-05 | -0.0057006 | RGS22 |
| chr6 | 39215402 | 39215585 | 184 | 2 | 3.38E-05 | 7.31E-05 | 4.44E-05 | 2.34E-05 | -0.00643326 | KCNK5 |
| chr5 | 103451040 | 103451070 | 31 | 2 | 2.30E-05 | 0.0001106 | 3.40E-05 | 2.34E-05 | -0.0057865 | PDZPH1P |
| chr6 | 170280395 | 170280809 | 415 | 5 | 1.76E-05 | 4.46E-06 | 0.015377 | 2.34E-05 | 0.027081264 | NA |
| chr5 | 180432344 | 180432512 | 169 | 2 | 7.06E-05 | 1.54E-05 | 0.000673 | 2.34E-05 | -0.00385795 | AC034213.1 |
| chr17 | 79810870 | 79811392 | 523 | 3 | 2.56E-06 | 0.000168 | 3.97E-05 | 2.34E-05 | 0.003789104 | NA |
| chr11 | 1074682 | 1074869 | 188 | 5 | 2.27E-05 | 7.40E-05 | 0.000525 | 2.34E-05 | -0.00102702 | NA |
| chr9 | 16625558 | 16625587 | 30 | 3 | 1.53E-05 | 0.000201 | 2.56E-05 | 2.34E-05 | -0.00033025 | BNC2 |
| chr16 | 81496342 | 81496358 | 17 | 2 | 1.53E-05 | 4.14E-05 | 7.50E-05 | 2.34E-05 | -0.00308085 | CMIP |
| chr12 | 116066802 | 116067030 | 229 | 2 | 6.11E-05 | 1.45E-05 | 0.000929 | 2.34E-05 | -0.01418763 | MED13L |
| chr17 | 49788518 | 49789100 | 583 | 7 | 4.99E-05 | 0.000247 | 0.002569 | 2.34E-05 | -7.56E-05 | KAT7, FAM117A |
| chr15 | 32721771 | 32721820 | 50 | 2 | 4.60E-05 | 5.18E-05 | 5.98E-05 | 2.34E-05 | -0.00826098 | GREM1 |
| chr2 | 96335996 | 96336568 | 573 | 4 | 1.32E-06 | 0.0015649 | 1.19E-05 | 2.34E-05 | -0.00142279 | NCAPH |
| chr2 | 27217035 | 27217896 | 862 | 8 | 7.87E-06 | 0.0003605 | 1.44E-05 | 2.34E-05 | 0.001872219 | ATRAID, CAD |
| chr18 | 65725205 | 65725491 | 287 | 2 | 3.38E-05 | 3.80E-05 | 8.30E-05 | 2.34E-05 | -0.01930239 | NA |
| chr15 | 40051375 | 40051592 | 218 | 2 | 3.52E-05 | 5.31E-05 | 5.85E-05 | 2.34E-05 | 0.000388175 | SRP14-AS1 |
| chr5 | 150245118 | 150245681 | 564 | 5 | 4.94E-05 | 4.38E-05 | 0.001161 | 2.34E-05 | -0.0022119 | CAMK2A |
| chr16 | 30499503 | 30499817 | 315 | 3 | 2.39E-05 | 0.0001398 | 0.00063 | 2.35E-05 | -0.00843731 | ITGAL, AC116348.2 |
| chr6 | 154247475 | 154247970 | 496 | 5 | 7.44E-05 | 2.41E-05 | 0.000989 | 2.35E-05 | 0.006398002 | IPCEF1, AL357075.4 |
| chr3 | 130243243 | 130243521 | 279 | 2 | 2.77E-06 | 0.0009259 | 1.58E-05 | 2.35E-05 | 0.001643801 | COL6A4P2 |
| chr14 | 69924329 | 69924486 | 158 | 2 | 1.48E-05 | 0.0034377 | 1.28E-05 | 2.35E-05 | -0.0024451 | SMOC1 |
| chr21 | 44345528 | 44345866 | 339 | 2 | 5.11E-05 | 1.86E-05 | 0.000327 | 2.35E-05 | -0.00586296 | NA |
| chr1 | 116451665 | 116452416 | 752 | 3 | 4.82E-06 | 2.07E-05 | 0.001013 | 2.35E-05 | 0.000855727 | LINC01762 |
| chr18 | 79069138 | 79069834 | 697 | 6 | 1.11E-05 | 0.0003575 | 0.000347 | 2.35E-05 | -0.00047082 | ATP9B |
| chr9 | 130396424 | 130397073 | 650 | 2 | 7.99E-07 | 5.47E-05 | 5.73E-05 | 2.35E-05 | -0.00977199 | HMCN2 |
| chr8 | 9151171 | 9151989 | 819 | 9 | 2.69E-05 | 1.77E-05 | 0.0166 | 2.35E-05 | -0.01140782 | AC022784.1, PPP1R3B |
| chr16 | 89476401 | 89476533 | 133 | 2 | 2.41E-05 | 2.47E-05 | 0.000163 | 2.36E-05 | 0.010351218 | ANKRD11 |
| chr8 | 73185271 | 73185567 | 297 | 4 | 5.83E-05 | 3.86E-05 | 0.000407 | 2.36E-05 | -0.0044466 | NA |
| chr4 | 157220784 | 157221739 | 956 | 6 | 7.15E-06 | 5.18E-05 | 0.00098 | 2.36E-05 | -0.00540779 | GRIA2 |
| chr5 | 400086 | 400717 | 632 | 5 | 3.38E-05 | 5.57E-05 | 0.00015 | 2.36E-05 | -0.00683954 | PDCD6-AHRR, AHRR |
| chr19 | 18275120 | 18275611 | 492 | 2 | 1.74E-05 | 1.49E-05 | 0.000826 | 2.36E-05 | -0.00201858 | NA |
| chr3 | 71494477 | 71494533 | 57 | 2 | 4.79E-05 | 1.48E-05 | 0.000853 | 2.36E-05 | 0.003053713 | FOXP1, AC097634.4 |
| chr14 | 53153489 | 53153683 | 195 | 7 | 1.18E-05 | 0.0013971 | 0.000875 | 2.36E-05 | 0.004626302 | AL356020.1 |
| chr17 | 76917071 | 76917678 | 608 | 5 | 1.29E-05 | 0.0081953 | 5.63E-05 | 2.36E-05 | -0.00282774 | MGAT5B |
| chr20 | 19254573 | 19254575 | 3 | 2 | 5.47E-05 | 1.86E-05 | 0.000336 | 2.36E-05 | -0.03348614 | SLC24A3, SLC24A3-AS1 |
| chr22 | 42070749 | 42071248 | 500 | 9 | 2.99E-06 | 0.0010654 | 4.18E-05 | 2.36E-05 | -0.00030678 | Z99716.2, NAGA |
| chr11 | 61744102 | 61744637 | 536 | 4 | 4.48E-05 | 0.0026727 | 0.000333 | 2.36E-05 | -0.00346719 | DAGLA |
| chr17 | 877598 | 877835 | 238 | 6 | 4.89E-05 | 0.0003054 | 0.002819 | 2.36E-05 | -0.00794882 | NXN |
| chr5 | 180292171 | 180292502 | 332 | 7 | 1.84E-05 | 0.0001477 | 0.000445 | 2.36E-05 | -4.81E-05 | NA |
| chr6 | 26341397 | 26341795 | 399 | 2 | 2.92E-05 | 3.03E-05 | 0.000114 | 2.36E-05 | 0.003383472 | NA |
| chr18 | 10453627 | 10454088 | 462 | 5 | 3.19E-05 | 3.09E-05 | 0.000488 | 2.36E-05 | -0.00656402 | NA |
| chr17 | 78891229 | 78891794 | 566 | 2 | 4.65E-05 | 1.75E-05 | 0.000412 | 2.36E-05 | 0.004259723 | TIMP2, CEP295NL |
| chr12 | 85278033 | 85278845 | 813 | 5 | 6.26E-07 | 0.007125 | 0.000112 | 2.36E-05 | 0.003598787 | NA |
| chr4 | 107034384 | 107034678 | 295 | 3 | 1.19E-05 | 3.95E-05 | 0.001046 | 2.36E-05 | 0.002697787 | DKK2 |
| chr22 | 40347855 | 40348332 | 478 | 4 | 5.88E-06 | 0.0002177 | 0.000268 | 2.36E-05 | 0.004364294 | ADSL, AL022238.3 |
| chr11 | 73661233 | 73661631 | 399 | 3 | 1.20E-05 | 4.29E-05 | 0.000742 | 2.36E-05 | 0.00628711 | PLEKHB1 |
| chr7 | 36367100 | 36367634 | 535 | 13 | 4.66E-05 | 1.66E-05 | 0.0049 | 2.36E-05 | 0.001358315 | KIAA0895 |
| chr11 | 22341162 | 22341824 | 663 | 3 | 9.14E-05 | 2.31E-05 | 0.001033 | 2.37E-05 | -0.01004932 | SLC17A6 |
| chr1 | 21439912 | 21439914 | 3 | 2 | 6.22E-05 | 0.0002773 | 2.26E-05 | 2.37E-05 | 0.00352107 | NA |
| chr17 | 82315366 | 82315809 | 444 | 4 | 3.35E-06 | 0.0009296 | 0.000326 | 2.37E-05 | 0.009498119 | CD7 |
| chr3 | 181174965 | 181175120 | 156 | 2 | 3.97E-05 | 6.02E-05 | 5.31E-05 | 2.37E-05 | -0.01685398 | SOX2-OT |
| chr11 | 2959957 | 2960163 | 207 | 2 | 1.23E-05 | 0.0003758 | 2.03E-05 | 2.37E-05 | -0.00454644 | NAP1L4 |
| chr9 | 112747355 | 112747490 | 136 | 2 | 2.84E-05 | 0.0002138 | 2.51E-05 | 2.37E-05 | -0.0027072 | NA |
| chr8 | 127340400 | 127340527 | 128 | 2 | 2.65E-05 | 0.0001522 | 2.92E-05 | 2.37E-05 | -0.00416733 | PCAT1, POU5F1B, CASC8 |
| chr16 | 1303626 | 1303945 | 320 | 2 | 8.13E-05 | 1.78E-05 | 0.000393 | 2.37E-05 | -0.01539801 | NA |
| chr3 | 23712310 | 23712494 | 185 | 2 | 1.10E-05 | 0.0391944 | 1.13E-05 | 2.37E-05 | -0.00701621 | NA |
| chr11 | 34149679 | 34149904 | 226 | 2 | 2.16E-05 | 0.0001407 | 3.05E-05 | 2.37E-05 | -0.00786698 | NA |
| chr6 | 138377951 | 138378147 | 197 | 2 | 3.56E-05 | 2.29E-05 | 0.000196 | 2.37E-05 | -0.00542788 | SMIM28 |
| chr12 | 30518856 | 30518993 | 138 | 2 | 1.56E-05 | 0.0040266 | 1.27E-05 | 2.38E-05 | -0.01915674 | NA |
| chr10 | 24315100 | 24315235 | 136 | 2 | 1.34E-05 | 0.0003489 | 2.09E-05 | 2.38E-05 | -0.00659564 | KIAA1217 |
| chr17 | 62779245 | 62779496 | 252 | 2 | 3.74E-05 | 2.71E-05 | 0.00014 | 2.38E-05 | -0.00381959 | MARCHF10 |
| chr17 | 81865027 | 81865469 | 443 | 3 | 0.00010954 | 9.03E-06 | 0.004611 | 2.38E-05 | 0.002801243 | NA |
| chr20 | 58910136 | 58911021 | 886 | 10 | 1.42E-05 | 0.000213 | 0.007029 | 2.38E-05 | 0.000621114 | GNAS |
| chr7 | 1966833 | 1966883 | 51 | 2 | 3.70E-06 | 0.0006285 | 1.76E-05 | 2.38E-05 | -0.00257377 | MAD1L1, AC069288.1 |
| chr19 | 6713216 | 6713448 | 233 | 3 | 1.22E-06 | 0.0031485 | 4.77E-05 | 2.38E-05 | -0.00434668 | C3 |
| chr1 | 200053760 | 200053968 | 209 | 4 | 5.37E-05 | 0.0004075 | 0.002375 | 2.38E-05 | -0.00194669 | NR5A2 |
| chr6 | 42960318 | 42961182 | 865 | 22 | 2.57E-07 | 0.0005059 | 6.10E-05 | 2.38E-05 | -0.00350689 | GNMT |
| chr16 | 84263859 | 84263942 | 84 | 2 | 1.73E-05 | 0.0003647 | 2.06E-05 | 2.38E-05 | 0.001299504 | NA |
| chr9 | 2161155 | 2161868 | 714 | 4 | 1.30E-05 | 0.0002758 | 0.000209 | 2.38E-05 | -0.01021583 | SMARCA2 |
| chr13 | 42991263 | 42991978 | 716 | 3 | 6.20E-08 | 0.0001253 | 0.000136 | 2.38E-05 | 0.024146741 | EPSTI1 |
| chr12 | 67590583 | 67590653 | 71 | 2 | 4.87E-05 | 1.45E-05 | 0.001089 | 2.38E-05 | 0.013135711 | LINC02408 |
| chr8 | 28118860 | 28119121 | 262 | 2 | 6.88E-05 | 1.55E-05 | 0.000707 | 2.38E-05 | -0.00872884 | ELP3 |
| chr6 | 13815015 | 13815336 | 322 | 2 | 8.49E-07 | 0.0008319 | 1.65E-05 | 2.38E-05 | -0.01105881 | NA |
| chr4 | 124712476 | 124712485 | 10 | 2 | 3.65E-06 | 2.74E-05 | 0.000138 | 2.38E-05 | 0.002345049 | ANKRD50 |
| chr10 | 117458781 | 117458800 | 20 | 2 | 5.26E-05 | 4.52E-05 | 7.05E-05 | 2.38E-05 | -0.00484565 | AC005871.2 |
| chr1 | 208030357 | 208031205 | 849 | 5 | 1.19E-06 | 0.0008114 | 7.68E-06 | 2.39E-05 | -0.00603399 | PLXNA2 |
| chr8 | 141027403 | 141027749 | 347 | 2 | 5.41E-06 | 0.00016 | 2.88E-05 | 2.39E-05 | -0.01517514 | NA |

| | | | | | | | | | | |
|---|---|---|---|---|---|---|---|---|---|---|
| chr12 | 7444926 | 7445012 | 87 | 4 | 3.53E-05 | 2.90E-05 | 0.002648 | 2.39E-05 | -0.00173639 | CD163L1 |
| chr5 | 14203379 | 14203690 | 312 | 2 | 8.97E-06 | 0.0003788 | 2.05E-05 | 2.39E-05 | 0.007183343 | TRIO |
| chr2 | 158126314 | 158126677 | 364 | 3 | 3.88E-05 | 4.17E-05 | 0.000382 | 2.39E-05 | -0.00395396 | UPP2 |
| chr1 | 17439677 | 17440499 | 823 | 11 | 4.72E-05 | 3.37E-05 | 0.015273 | 2.39E-05 | -0.00046859 | RCC2 |
| chr12 | 98896126 | 98896600 | 475 | 2 | 9.61E-06 | 1.84E-05 | 0.000364 | 2.39E-05 | -0.01593114 | ANKS1B |
| chr19 | 14472467 | 14472756 | 290 | 3 | 6.92E-06 | 0.0001444 | 3.32E-05 | 2.39E-05 | -2.42E-05 | PTGER1 |
| chr11 | 68446512 | 68447412 | 901 | 8 | 4.18E-06 | 0.0084808 | 0.000195 | 2.39E-05 | -0.00107621 | LRP5 |
| chr15 | 40941503 | 40942192 | 690 | 4 | 6.37E-06 | 3.28E-05 | 0.000565 | 2.39E-05 | 0.00023892 | NA |
| chr13 | 24668770 | 24669142 | 373 | 2 | 5.14E-05 | 1.86E-05 | 0.000354 | 2.39E-05 | -0.00052912 | NA |
| chr1 | 15523961 | 15524006 | 46 | 2 | 4.03E-05 | 0.0001472 | 3.01E-05 | 2.39E-05 | 0.003388835 | CASP9 |
| chr7 | 99272985 | 99273212 | 228 | 4 | 7.26E-06 | 0.0024513 | 6.02E-06 | 2.39E-05 | -0.00228074 | MYH16, AC004834.1 |
| chr6 | 29223791 | 29223961 | 171 | 3 | 2.04E-05 | 0.0015908 | 2.06E-05 | 2.39E-05 | 5.12E-05 | NA |
| chr7 | 18296754 | 18297117 | 364 | 2 | 5.44E-06 | 3.38E-05 | 9.99E-05 | 2.40E-05 | -0.00540366 | HDAC9 |
| chr22 | 37385349 | 37385661 | 313 | 3 | 3.42E-05 | 1.26E-05 | 0.001409 | 2.40E-05 | -0.00713551 | ELFN2, ELFN2 |
| chr21 | 36691572 | 36691936 | 365 | 2 | 1.52E-05 | 2.90E-05 | 0.000127 | 2.40E-05 | -0.00852085 | NA |
| chr2 | 205707710 | 205708359 | 650 | 3 | 4.10E-05 | 1.11E-05 | 0.001942 | 2.40E-05 | -0.00099434 | NRP2 |
| chr13 | 106462965 | 106463121 | 157 | 2 | 7.51E-05 | 1.51E-05 | 0.000842 | 2.40E-05 | 0.009013938 | NA |
| chr10 | 103602275 | 103603042 | 768 | 6 | 1.09E-06 | 0.0018948 | 0.000256 | 2.40E-05 | -0.00293641 | SH3PXD2A |
| chr2 | 48398398 | 48399363 | 966 | 4 | 1.43E-07 | 0.0013052 | 8.80E-06 | 2.40E-05 | 0.00156292 | NA |
| chr11 | 19935445 | 19935827 | 383 | 2 | 7.21E-06 | 0.0002331 | 2.46E-05 | 2.40E-05 | -0.01506779 | NAV2 |
| chr10 | 42638244 | 42638854 | 611 | 5 | 7.49E-05 | 8.01E-06 | 0.002792 | 2.40E-05 | 0.003220376 | AL022345.4, ZNF33B |
| chr2 | 11155096 | 11155707 | 612 | 7 | 8.42E-05 | 0.0004253 | 0.001059 | 2.40E-05 | -0.0003043 | SLC66A3 |
| chr12 | 121827585 | 121828047 | 463 | 4 | 4.15E-05 | 2.86E-05 | 0.001698 | 2.40E-05 | -0.00104839 | SETD1B |
| chr11 | 118928529 | 118929145 | 617 | 8 | 7.70E-05 | 4.31E-05 | 0.009571 | 2.40E-05 | 0.000143706 | UPK2 |
| chr9 | 96875589 | 96875900 | 312 | 3 | 3.79E-05 | 3.58E-05 | 0.00016 | 2.40E-05 | 0.002580227 | ZNF782 |
| chr16 | 625116 | 625614 | 499 | 3 | 2.45E-05 | 7.56E-05 | 7.36E-05 | 2.40E-05 | -0.00213513 | RAB40C |
| chr18 | 49858917 | 49859211 | 295 | 3 | 7.01E-05 | 1.20E-05 | 0.00147 | 2.40E-05 | 0.001552255 | MYO5B |
| chr8 | 104497087 | 104497266 | 180 | 3 | 1.56E-05 | 4.55E-05 | 0.000172 | 2.40E-05 | -0.00412365 | LRP12 |
| chr1 | 224175747 | 224175873 | 127 | 2 | 6.97E-05 | 1.79E-05 | 0.000406 | 2.40E-05 | -0.01670336 | DEGS1 |
| chr20 | 38304787 | 38305354 | 568 | 2 | 2.11E-05 | 2.14E-05 | 0.00024 | 2.41E-05 | -0.0124247 | BPI, AL359555.1 |
| chr19 | 3505779 | 3506560 | 782 | 9 | 3.09E-05 | 6.49E-05 | 0.001999 | 2.41E-05 | 6.18E-05 | FZR1 |
| chr5 | 149062661 | 149063436 | 776 | 8 | 1.25E-05 | 0.0012277 | 0.00164 | 2.41E-05 | -0.0040749 | SH3TC2-DT, SH3TC2 |
| chr9 | 121500112 | 121500420 | 309 | 6 | 1.97E-05 | 7.80E-05 | 0.00313 | 2.41E-05 | -0.01907802 | NA |
| chr9 | 126460943 | 126461298 | 356 | 2 | 5.37E-06 | 0.0004787 | 1.92E-05 | 2.41E-05 | 0.00352484 | MVB12B |
| chr19 | 1972140 | 1972235 | 96 | 2 | 2.03E-05 | 0.0001574 | 2.93E-05 | 2.41E-05 | 0.000568107 | CSNK1G2 |
| chr14 | 22829675 | 22830347 | 673 | 12 | 7.43E-06 | 2.93E-05 | 0.000635 | 2.41E-05 | -1.16E-05 | MRPL52, SLC7A7 |
| chr16 | 81580842 | 81580877 | 36 | 2 | 1.09E-05 | 0.0006675 | 1.75E-05 | 2.41E-05 | -0.00654288 | CMIP |
| chr12 | 51677760 | 51678572 | 813 | 4 | 6.63E-06 | 0.0003578 | 3.58E-05 | 2.41E-05 | 0.00122695 | SCN8A |
| chr14 | 56730573 | 56731046 | 474 | 6 | 2.53E-06 | 0.0035333 | 1.13E-05 | 2.41E-05 | -0.00485388 | NA |
| chr7 | 130508813 | 130508861 | 49 | 2 | 5.47E-05 | 1.66E-05 | 0.000563 | 2.42E-05 | -0.00533024 | COPG2 |
| chr11 | 74170159 | 74171231 | 1073 | 17 | 9.37E-06 | 0.0001542 | 0.001477 | 2.42E-05 | 0.000538028 | C2CD3 |
| chr11 | 120302838 | 120303251 | 414 | 2 | 6.01E-06 | 7.70E-05 | 4.48E-05 | 2.42E-05 | 0.000551125 | POU2F3 |
| chr12 | 124807751 | 124808068 | 318 | 3 | 5.90E-06 | 0.0023284 | 9.36E-06 | 2.42E-05 | -0.00221478 | AC073593.2, SCARB1 |
| chr1 | 156881585 | 156881648 | 64 | 2 | 1.66E-05 | 0.0001558 | 2.97E-05 | 2.42E-05 | -0.00256999 | NTRK1 |
| chr1 | 230678569 | 230678952 | 384 | 2 | 0.000105252 | 1.83E-05 | 0.000392 | 2.42E-05 | -0.01240242 | COG2 |
| chr15 | 38759452 | 38759674 | 223 | 4 | 1.53E-05 | 0.0451498 | 6.32E-05 | 2.42E-05 | -0.00738364 | LINC02694 |
| chr7 | 112069907 | 112070320 | 414 | 2 | 2.23E-05 | 2.68E-05 | 0.00015 | 2.42E-05 | -0.00518593 | DOCK4 |
| chr2 | 121790028 | 121790430 | 403 | 5 | 0.000120449 | 5.62E-06 | 0.010326 | 2.42E-05 | -0.00347625 | LINC01823 |
| chr16 | 35748368 | 35749013 | 646 | 2 | 1.07E-06 | 2.78E-05 | 0.000141 | 2.42E-05 | 0.004158034 | LINC02167, RNA5SP414 |
| chr10 | 12566496 | 12566876 | 381 | 2 | 1.79E-05 | 7.12E-05 | 4.77E-05 | 2.42E-05 | 0.004320845 | CAMK1D, AL731559.1 |
| chr1 | 39298907 | 39299054 | 148 | 2 | 4.81E-05 | 4.95E-05 | 6.61E-05 | 2.42E-05 | -0.0032798 | MACF1 |
| chr3 | 50567332 | 50567967 | 636 | 9 | 7.03E-06 | 5.58E-05 | 0.002438 | 2.43E-05 | 0.000377676 | C3orf18 |
| chr7 | 140231772 | 140232332 | 561 | 2 | 5.12E-05 | 1.50E-05 | 0.00099 | 2.43E-05 | -0.01763545 | NA |
| chr4 | 56049181 | 56049734 | 554 | 5 | 7.03E-05 | 0.0054225 | 0.001205 | 2.43E-05 | -0.00056656 | CRACD |
| chr1 | 22666136 | 22666227 | 92 | 2 | 4.92E-05 | 2.30E-05 | 0.000207 | 2.43E-05 | -0.01254585 | NA |
| chr5 | 17340705 | 17340956 | 252 | 2 | 3.41E-05 | 5.17E-05 | 6.35E-05 | 2.43E-05 | -0.01627253 | NA |
| chr19 | 2078332 | 2078511 | 180 | 3 | 1.92E-05 | 0.000536 | 2.44E-05 | 2.43E-05 | -0.00228168 | MOB3A |
| chr16 | 57694261 | 57694720 | 460 | 2 | 1.58E-05 | 1.50E-05 | 0.000981 | 2.43E-05 | -0.01617456 | AC018552.2 |
| chr2 | 203484302 | 203484713 | 412 | 2 | 6.02E-05 | 1.69E-05 | 0.000538 | 2.43E-05 | 0.009240187 | RAPH1 |
| chr6 | 28001330 | 28001670 | 341 | 2 | 1.55E-05 | 0.0001424 | 3.12E-05 | 2.43E-05 | -0.00873531 | NA |
| chr14 | 53151560 | 53152014 | 455 | 3 | 2.72E-05 | 3.14E-05 | 0.001122 | 2.43E-05 | 0.009492004 | DDHD1 |
| chr4 | 74473327 | 74473791 | 465 | 2 | 8.60E-05 | 1.50E-05 | 0.000995 | 2.43E-05 | -0.00530711 | NA |
| chr11 | 4998537 | 4999352 | 816 | 7 | 1.53E-05 | 0.000273 | 0.000383 | 2.43E-05 | -0.00929615 | OR51L1 |
| chr9 | 128550479 | 128550910 | 432 | 2 | 9.16E-06 | 1.48E-05 | 0.001061 | 2.43E-05 | -0.01080666 | AL356481.1 |
| chr12 | 53214937 | 53215618 | 682 | 2 | 1.79E-05 | 1.90E-05 | 0.000348 | 2.43E-05 | -0.00780565 | RARG |
| chr19 | 3942160 | 3942332 | 173 | 4 | 8.34E-05 | 1.29E-05 | 0.004957 | 2.43E-05 | 0.002403147 | NMRK2 |
| chr5 | 14414838 | 14415547 | 710 | 5 | 9.71E-05 | 8.13E-05 | 0.001022 | 2.43E-05 | -0.00644469 | TRIO |
| chr10 | 97164479 | 97165134 | 656 | 2 | 3.93E-05 | 1.54E-05 | 0.000856 | 2.43E-05 | -0.01598949 | SLIT1, ARHGAP19-SLIT1 |
| chr11 | 1473478 | 1473898 | 421 | 4 | 5.90E-05 | 0.0003573 | 0.000881 | 2.44E-05 | -0.00284897 | MOB2 |
| chr6 | 25830561 | 25830624 | 64 | 2 | 2.00E-05 | 0.0009757 | 1.63E-05 | 2.44E-05 | -0.00655714 | SLC17A1 |
| chr2 | 238554605 | 238555075 | 471 | 3 | 6.20E-06 | 0.0008062 | 0.000513 | 2.44E-05 | 0.022294441 | LINC01937, LINC01107 |
| chr19 | 1799936 | 1800338 | 403 | 6 | 5.69E-05 | 0.0001946 | 0.001492 | 2.44E-05 | -0.00120903 | ATP8B3 |
| chr10 | 102409369 | 102410083 | 715 | 5 | 2.65E-05 | 0.0002007 | 0.000327 | 2.44E-05 | 0.002327668 | PSD |
| chr4 | 151909148 | 151909537 | 390 | 2 | 5.63E-06 | 0.0017649 | 1.45E-05 | 2.44E-05 | -0.00332194 | AC097375.2 |
| chr11 | 128890198 | 128890472 | 275 | 2 | 6.86E-06 | 2.79E-05 | 0.000142 | 2.44E-05 | -0.0111726 | NA |
| chr7 | 5235344 | 5235386 | 43 | 2 | 4.79E-05 | 1.55E-05 | 0.000828 | 2.44E-05 | 0.020970265 | NA |
| chr8 | 71155083 | 71155361 | 279 | 2 | 9.23E-05 | 1.55E-05 | 0.000833 | 2.44E-05 | -0.00247561 | AC015687.1 |
| chr4 | 148065199 | 148065594 | 396 | 3 | 6.37E-06 | 8.53E-05 | 5.35E-05 | 2.44E-05 | -0.01061686 | ARHGAP10 |
| chr10 | 129525496 | 129525925 | 430 | 5 | 2.10E-05 | 0.0001038 | 6.50E-05 | 2.45E-05 | -0.0001811 | MGMT |
| chr3 | 13637418 | 13638074 | 657 | 6 | 5.67E-06 | 0.0008771 | 1.73E-05 | 2.45E-05 | 0.000682502 | FBLN2 |
| chr7 | 120273522 | 120273717 | 196 | 3 | 1.14E-06 | 4.59E-05 | 0.000357 | 2.45E-05 | 0.003053228 | KCND2 |
| chr1 | 1548825 | 1549129 | 305 | 2 | 9.82E-06 | 4.54E-05 | 7.35E-05 | 2.45E-05 | 0.002609268 | SSU72 |
| chr1 | 28863143 | 28863573 | 431 | 5 | 7.08E-05 | 6.21E-05 | 0.005023 | 2.45E-05 | -0.012749 | OPRD1 |
| chr16 | 2486152 | 2486400 | 249 | 2 | 6.20E-06 | 0.0001445 | 3.13E-05 | 2.45E-05 | -0.00853509 | TBC1D24 |
| chr1 | 163160938 | 163161527 | 590 | 4 | 1.99E-05 | 6.11E-05 | 0.002609 | 2.45E-05 | -0.00695966 | RGS5 |
| chr12 | 114671647 | 114671699 | 53 | 2 | 8.48E-05 | 1.94E-05 | 0.000341 | 2.45E-05 | -0.00832305 | TBX3 |
| chr3 | 185253864 | 185254383 | 520 | 8 | 1.97E-05 | 0.0004389 | 0.001895 | 2.45E-05 | -0.00447177 | EHHADH |
| chr1 | 156070169 | 156070528 | 360 | 2 | 4.96E-06 | 0.0211498 | 1.18E-05 | 2.45E-05 | -0.01899579 | RAB25 |
| chr18 | 76502689 | 76502772 | 84 | 2 | 3.05E-05 | 0.0003006 | 2.29E-05 | 2.45E-05 | 0.019679411 | NA |
| chr17 | 80532857 | 80533334 | 478 | 2 | 8.60E-05 | 1.48E-05 | 0.001185 | 2.45E-05 | 0.001269706 | NA |
| chr5 | 34487564 | 34487838 | 275 | 3 | 2.34E-05 | 0.0001198 | 3.99E-05 | 2.45E-05 | 0.001692993 | NA |
| chr11 | 119521131 | 119521304 | 174 | 2 | 6.00E-05 | 2.19E-05 | 0.000241 | 2.45E-05 | -0.00628776 | USP2-AS1 |
| chr3 | 136752285 | 136752735 | 451 | 7 | 5.70E-05 | 0.0001838 | 0.004427 | 2.45E-05 | 0.001041746 | AC117382.2, STAG1 |
| chr9 | 33263102 | 33263465 | 364 | 2 | 4.82E-05 | 3.27E-05 | 0.00011 | 2.45E-05 | -0.01067207 | BAG1 |
| chr1 | 27959523 | 27960373 | 851 | 11 | 1.51E-05 | 0.0001685 | 8.42E-05 | 2.46E-05 | 9.29E-05 | XKR8, AL512288.1 |
| chr10 | 126387062 | 126387739 | 678 | 4 | 2.56E-07 | 0.0028619 | 8.36E-06 | 2.46E-05 | 0.000901559 | ADAM12 |
| chr7 | 149775845 | 149776089 | 245 | 3 | 2.52E-05 | 3.32E-05 | 0.00053 | 2.46E-05 | 0.01019593 | SSPOP |
| chr17 | 64708725 | 64708997 | 273 | 2 | 5.56E-05 | 2.08E-05 | 0.000279 | 2.46E-05 | 0.003334697 | NA |
| chr22 | 35399959 | 35400710 | 752 | 7 | 1.42E-05 | 0.000118 | 0.001156 | 2.46E-05 | 0.000264152 | MCM5 |
| chr19 | 8399744 | 8400214 | 471 | 4 | 9.64E-05 | 1.51E-05 | 0.004491 | 2.46E-05 | -0.00066261 | RAB11B |
| chr4 | 181150031 | 181150395 | 365 | 2 | 6.08E-05 | 1.73E-05 | 0.000512 | 2.46E-05 | -0.01519835 | LINC00290 |
| chr16 | 19858643 | 19858770 | 128 | 2 | 6.57E-05 | 1.48E-05 | 0.001157 | 2.46E-05 | -0.00758529 | GPRC5B |
| chr2 | 43136324 | 43136487 | 164 | 2 | 1.67E-05 | 5.80E-05 | 5.83E-05 | 2.46E-05 | 0.004708644 | NA |
| chr9 | 136523953 | 136524546 | 594 | 6 | 2.13E-05 | 0.000232 | 0.000232 | 2.46E-05 | 0.000528566 | NOTCH1 |
| chr6 | 26018306 | 26019157 | 852 | 6 | 7.52E-08 | 0.0128851 | 7.95E-06 | 2.46E-05 | -0.00351214 | NA |
| chr20 | 52456581 | 52456881 | 301 | 3 | 8.80E-05 | 1.06E-05 | 0.003421 | 2.47E-05 | -0.01076858 | LINC01524 |
| chr7 | 2585593 | 2585878 | 286 | 4 | 1.02E-05 | 0.0006749 | 0.000176 | 2.47E-05 | -0.00260132 | IQCE |
| chr14 | 24113847 | 24114230 | 384 | 4 | 1.91E-06 | 0.0002641 | 0.001613 | 2.47E-05 | -0.00075696 | DCAF11, NRL |
| chr15 | 39331764 | 39331832 | 69 | 2 | 6.32E-05 | 1.50E-05 | 0.001107 | 2.47E-05 | -0.00117097 | AC013652.1 |
| chr1 | 109619138 | 109620079 | 942 | 15 | 2.95E-05 | 2.12E-05 | 0.010439 | 2.47E-05 | 0.000721333 | AMPD2, GNAT2 |
| chr10 | 37784321 | 37784459 | 139 | 4 | 4.59E-05 | 0.0001307 | 0.000143 | 2.47E-05 | 0.004274027 | ZNF248 |
| chr4 | 1228050 | 1228504 | 455 | 5 | 2.82E-06 | 8.43E-05 | 0.000285 | 2.47E-05 | 0.003905591 | CTBP1 |
| chr18 | 58232446 | 58232558 | 113 | 2 | 6.72E-05 | 0.0002726 | 2.40E-05 | 2.47E-05 | -0.00258356 | NEDD4L |
| chr2 | 140349708 | 140349754 | 47 | 2 | 6.13E-05 | 3.25E-05 | 0.000113 | 2.47E-05 | -0.00924228 | LRP1B |
| chr21 | 46147362 | 46147974 | 613 | 3 | 5.24E-08 | 0.0055673 | 9.98E-06 | 2.48E-05 | -0.00251851 | FTCD |
| chr9 | 77648671 | 77649334 | 664 | 4 | 8.98E-07 | 0.0003157 | 0.000157 | 2.48E-05 | -0.00918426 | NA |
| chr7 | 100578711 | 100578741 | 31 | 2 | 2.79E-05 | 1.64E-05 | 0.00067 | 2.48E-05 | -0.00014669 | LRCH4 |
| chr12 | 30200099 | 30200361 | 263 | 3 | 1.89E-05 | 0.00096 | 3.82E-05 | 2.48E-05 | -0.00808308 | NA |
| chr2 | 217846509 | 217846911 | 403 | 2 | 2.11E-05 | 1.50E-05 | 0.00113 | 2.48E-05 | -0.01306676 | TNS1 |
| chr20 | 4835441 | 4835552 | 112 | 2 | 6.26E-05 | 2.68E-05 | 0.000159 | 2.48E-05 | -0.00829524 | NA |
| chr12 | 124466123 | 124466444 | 322 | 5 | 0.000104438 | 6.64E-06 | 0.008792 | 2.48E-05 | 0.005279639 | NCOR2 |
| chr7 | 116671334 | 116671442 | 109 | 2 | 4.72E-05 | 1.66E-05 | 0.000632 | 2.48E-05 | -0.00566054 | NA |
| chr1 | 175066366 | 175067551 | 1186 | 5 | 0.000127454 | 0.0002074 | 0.001201 | 2.48E-05 | -0.00534187 | NA |
| chr9 | 35072640 | 35072980 | 341 | 6 | 1.32E-05 | 0.000131 | 0.001253 | 2.48E-05 | 0.000189126 | VCP |
| chr19 | 10230908 | 10231680 | 773 | 9 | 2.25E-05 | 0.0006043 | 0.000158 | 2.48E-05 | 0.001029606 | DNMT1, S1PR2 |
| chr9 | 35791985 | 35792960 | 976 | 7 | 2.31E-06 | 0.0120962 | 2.87E-05 | 2.48E-05 | -0.00154253 | NPR2 |
| chr20 | 50033757 | 50034074 | 318 | 3 | 2.08E-05 | 0.0001976 | 0.000168 | 2.48E-05 | -0.00699912 | NA |
| chr14 | 33956964 | 33957412 | 449 | 2 | 7.92E-05 | 1.56E-05 | 0.000881 | 2.48E-05 | -0.00971683 | EGLN3 |
| chr6 | 79537320 | 79537569 | 250 | 5 | 0.000154839 | 6.03E-05 | 0.001013 | 2.48E-05 | 0.000678453 | AL391840.3, LCA5 |
| chr12 | 102478825 | 102479159 | 335 | 3 | 4.82E-05 | 1.83E-05 | 0.000708 | 2.48E-05 | 0.000535769 | IGF1 |
| chr5 | 135303994 | 135304248 | 255 | 2 | 5.90E-05 | 2.07E-05 | 0.000296 | 2.49E-05 | 0.003260977 | C5orf66 |
| chr5 | 157535406 | 157535814 | 409 | 2 | 7.89E-05 | 3.07E-05 | 0.000126 | 2.49E-05 | 0.003079085 | ADAM19 |
| chr1 | 64691579 | 64691721 | 143 | 2 | 4.18E-05 | 8.34E-05 | 4.40E-05 | 2.49E-05 | -0.00476587 | CACHD1 |
| chr14 | 102043944 | 102044400 | 457 | 6 | 8.39E-05 | 0.005845 | 0.000289 | 2.49E-05 | -0.00267732 | DYNC1H1, AL118558.1 |
| chr11 | 5690539 | 5690879 | 341 | 2 | 3.06E-05 | 1.85E-05 | 0.000418 | 2.49E-05 | -0.00118137 | TRIM22, TRIM5 |
| chr4 | 139454315 | 139454693 | 379 | 3 | 1.04E-05 | 5.60E-05 | 0.000102 | 2.49E-05 | 0.002939564 | RAB33B |
| chr13 | 38687597 | 38687659 | 63 | 2 | 2.40E-05 | 1.56E-05 | 0.000899 | 2.49E-05 | -0.00459864 | FREM2 |
| chr9 | 110540872 | 110540967 | 96 | 2 | 3.03E-05 | 0.0003122 | 2.30E-05 | 2.49E-05 | -0.01287058 | SVEP1 |
| chr3 | 111978541 | 111979418 | 878 | 14 | 1.88E-05 | 0.0013253 | 0.000312 | 2.49E-05 | -0.00492343 | ABHD10 |
| chr13 | 104007179 | 104007477 | 299 | 4 | 6.45E-05 | 1.37E-05 | 0.002118 | 2.49E-05 | -0.00601267 | NA |
| chr6 | 21664421 | 21664924 | 504 | 3 | 1.42E-05 | 3.12E-05 | 0.001219 | 2.49E-05 | -0.0119864 | CASC15 |
| chr20 | 17649270 | 17649541 | 272 | 3 | 5.99E-05 | 2.01E-05 | 0.000778 | 2.49E-05 | -0.02695857 | RRBP1 |
| chr11 | 117327708 | 117328231 | 524 | 13 | 2.82E-05 | 0.0001745 | 0.00275 | 2.49E-05 | 0.001217058 | CEP164, AP000892.1 |
| chr17 | 77935163 | 77935487 | 325 | 2 | 2.62E-05 | 3.57E-05 | 0.000101 | 2.50E-05 | -0.01150623 | NA |
| chr1 | 16018041 | 16018482 | 442 | 6 | 2.36E-05 | 2.92E-05 | 0.000707 | 2.50E-05 | -0.00372413 | HSPB7 |
| chr5 | 36422590 | 36422840 | 251 | 2 | 6.27E-05 | 1.94E-05 | 0.000365 | 2.50E-05 | -0.01444828 | NA |
| chr7 | 157717565 | 157717980 | 416 | 4 | 4.72E-05 | 4.05E-05 | 0.000279 | 2.50E-05 | -0.00114695 | PTPRN2 |
| chr19 | 5117247 | 5117548 | 302 | 2 | 6.08E-05 | 1.84E-05 | 0.000432 | 2.50E-05 | -0.00601673 | KDM4B |
| chr14 | 76131125 | 76131481 | 357 | 3 | 3.71E-05 | 6.11E-05 | 0.000208 | 2.50E-05 | -0.00332633 | NA |
| chr8 | 142726844 | 142727396 | 553 | 7 | 2.71E-05 | 2.68E-05 | 0.000951 | 2.50E-05 | -0.00089703 | THEM6, LNCOC1, AC108002.1 |
| chr1 | 203220132 | 203220539 | 408 | 3 | 1.30E-05 | 0.0013196 | 1.51E-05 | 2.50E-05 | -0.00160258 | CHIT1 |
| chr6 | 49498587 | 49498669 | 83 | 2 | 1.26E-05 | 0.0056643 | 1.30E-05 | 2.50E-05 | -0.0135843 | NA |
| chr11 | 78017080 | 78017134 | 55 | 3 | 4.21E-05 | 9.23E-05 | 8.96E-05 | 2.50E-05 | -0.00255354 | KCTD14, NDUFC2-KCTD14 |
| chr19 | 18915436 | 18915553 | 118 | 3 | 0.000128489 | 1.06E-05 | 0.003515 | 2.50E-05 | -0.00620291 | COPE, AC002985.1 |
| chr6 | 37174442 | 37174444 | 3 | 2 | 5.64E-05 | 1.55E-05 | 0.000993 | 2.50E-05 | -0.01934651 | PIM1 |
| chr6 | 30732757 | 30732839 | 83 | 2 | 4.13E-05 | 1.93E-05 | 0.000377 | 2.50E-05 | -0.00345825 | FLOT1 |
| chr2 | 235192044 | 235192107 | 64 | 3 | 5.15E-05 | 6.75E-05 | 6.52E-05 | 2.51E-05 | 0.004239212 | NA |
| chr3 | 11570389 | 11570404 | 16 | 2 | 1.63E-05 | 2.29E-05 | 0.000231 | 2.51E-05 | -0.00336211 | VGLL4 |

| | | | | | | | | | | |
|---|---|---|---|---|---|---|---|---|---|---|
| chr2 | 7357043 | 7357435 | 393 | 2 | 7.45E-05 | 1.59E-05 | 0.000832 | 2.51E-05 | -0.00248199 | NA |
| chr12 | 67413842 | 67413845 | 4 | 2 | 4.44E-05 | 9.16E-05 | 4.17E-05 | 2.51E-05 | 0.002110652 | NA |
| chr11 | 116074112 | 116074156 | 45 | 2 | 5.89E-05 | 3.33E-05 | 0.000112 | 2.51E-05 | -0.01434251 | NA |
| chr1 | 160681662 | 160681764 | 103 | 3 | 8.26E-05 | 1.80E-05 | 0.000545 | 2.51E-05 | -0.01630752 | AL121985.1, CD48 |
| chr2 | 24015997 | 24016267 | 271 | 2 | 2.51E-05 | 2.34E-05 | 0.000221 | 2.51E-05 | -0.01033855 | MFSD2B |
| chr15 | 82087314 | 82087606 | 293 | 3 | 7.30E-06 | 0.0008668 | 2.00E-05 | 2.51E-05 | -0.00420025 | NA |
| chr13 | 63844543 | 63844545 | 3 | 2 | 2.92E-05 | 0.0005603 | 1.92E-05 | 2.51E-05 | -0.01017608 | NA |
| chr16 | 83886569 | 83886680 | 112 | 2 | 6.49E-05 | 1.52E-05 | 0.001141 | 2.51E-05 | 0.004965971 | NA |
| chr2 | 152175543 | 152176229 | 687 | 10 | 1.65E-05 | 0.000163 | 4.86E-05 | 2.51E-05 | 0.000811111 | AC079790.2, STAM2 |
| chr11 | 85525885 | 85525943 | 59 | 2 | 6.60E-05 | 1.51E-05 | 0.001196 | 2.51E-05 | -0.01395691 | DLG2 |
| chr7 | 131558119 | 131558604 | 486 | 5 | 2.93E-05 | 0.003087 | 2.85E-05 | 2.51E-05 | -0.00483832 | PODXL |
| chr5 | 178840956 | 178840992 | 37 | 2 | 5.08E-05 | 1.54E-05 | 0.001062 | 2.51E-05 | -0.0073156 | AC126915.1 |
| chr3 | 158528382 | 158528465 | 84 | 3 | 2.24E-05 | 0.0009115 | 1.52E-05 | 2.51E-05 | 0.001280619 | RSRC1 |
| chr17 | 47717773 | 47718130 | 358 | 3 | 2.09E-06 | 0.0055695 | 1.18E-05 | 2.51E-05 | -0.0023323 | NA |
| chr8 | 64376582 | 64376670 | 89 | 3 | 3.56E-05 | 6.64E-05 | 0.00091 | 2.52E-05 | 0.014385281 | MIR124-2HG |
| chr16 | 50719277 | 50719362 | 86 | 3 | 0.000125402 | 2.76E-05 | 0.000326 | 2.52E-05 | 0.001854129 | NOD2 |
| chr13 | 23578421 | 23580000 | 1580 | 15 | 1.36E-06 | 0.001932 | 4.66E-05 | 2.52E-05 | 0.000726435 | TNFRSF19 |
| chr5 | 64165389 | 64166103 | 715 | 9 | 4.40E-05 | 0.0001594 | 0.001504 | 2.52E-05 | -0.00457309 | RNF180 |
| chr14 | 58297665 | 58298607 | 943 | 15 | 1.02E-05 | 0.0029307 | 2.89E-05 | 2.52E-05 | -0.00011996 | ARID4A, ARMH4, PSMA3-AS1 |
| chr12 | 27333552 | 27333847 | 296 | 2 | 1.12E-05 | 0.0001446 | 3.23E-05 | 2.52E-05 | 0.001230421 | ARNTL2 |
| chr6 | 108560252 | 108560981 | 730 | 6 | 7.25E-06 | 7.95E-05 | 0.001561 | 2.52E-05 | 0.002366015 | FOXO3 |
| chr2 | 70939479 | 70939496 | 18 | 2 | 3.05E-05 | 1.59E-05 | 0.000888 | 2.52E-05 | -0.01710172 | VAX2, ATP6V1B1, AC007040.2 |
| chr14 | 78754134 | 78754648 | 515 | 3 | 9.51E-06 | 0.0016553 | 2.24E-05 | 2.52E-05 | 0.006255349 | NRXN3 |
| chr1 | 202957994 | 202958893 | 900 | 19 | 3.74E-05 | 0.000226 | 0.000693 | 2.52E-05 | 0.000265644 | ADIPOR1 |
| chr2 | 68318828 | 68319556 | 729 | 7 | 2.86E-06 | 0.0002539 | 0.003001 | 2.52E-05 | -0.00732903 | CNRIP1 |
| chr2 | 25248014 | 25248039 | 26 | 2 | 5.65E-05 | 1.71E-05 | 0.000611 | 2.52E-05 | -0.00118195 | DNMT3A |
| chr10 | 35271748 | 35272024 | 277 | 2 | 1.62E-05 | 0.0003671 | 2.20E-05 | 2.52E-05 | -0.00326373 | CCNY |
| chr7 | 35831714 | 35832012 | 299 | 2 | 2.24E-05 | 2.36E-05 | 0.000221 | 2.52E-05 | -0.01382487 | SEPTIN7 |
| chr15 | 98295126 | 98295338 | 213 | 2 | 5.15E-05 | 2.86E-05 | 0.000148 | 2.52E-05 | -0.0096982 | NA |
| chr9 | 111411833 | 111412508 | 676 | 3 | 2.81E-05 | 1.49E-05 | 0.00175 | 2.52E-05 | 0.000706972 | ECPAS |
| chr1 | 59930062 | 59930086 | 25 | 2 | 2.77E-05 | 0.0007373 | 1.80E-05 | 2.53E-05 | -0.00508971 | NA |
| chr10 | 132637689 | 132637754 | 66 | 2 | 1.21E-05 | 1.58E-05 | 0.000908 | 2.53E-05 | -0.00624891 | INPP5A |
| chr1 | 68116191 | 68116461 | 271 | 2 | 2.22E-05 | 0.0001481 | 3.21E-05 | 2.53E-05 | 0.006452684 | GNG12-AS1, WLS |
| chr20 | 42685066 | 42685306 | 241 | 3 | 1.07E-05 | 0.0134619 | 1.73E-05 | 2.53E-05 | -0.01209207 | PTPRT |
| chr15 | 98540465 | 98540630 | 166 | 2 | 1.07E-05 | 0.0120072 | 1.24E-05 | 2.53E-05 | -0.0075356 | FAM169B |
| chr1 | 16176089 | 16176398 | 310 | 2 | 1.56E-05 | 1.00E-04 | 4.01E-05 | 2.53E-05 | -0.00810762 | NA |
| chr8 | 81462488 | 81462903 | 416 | 3 | 1.30E-05 | 2.95E-05 | 0.000213 | 2.53E-05 | -0.00976688 | AC018616.1 |
| chr14 | 66822878 | 66822989 | 112 | 3 | 5.84E-05 | 4.57E-05 | 0.000173 | 2.53E-05 | -0.00235986 | GPHN |
| chr5 | 42235662 | 42236109 | 448 | 2 | 6.85E-06 | 4.36E-05 | 8.17E-05 | 2.53E-05 | -0.00949702 | NA |
| chr19 | 22286415 | 22286566 | 152 | 5 | 3.38E-05 | 0.0001889 | 7.13E-05 | 2.53E-05 | -0.00247013 | ZNF729 |
| chr16 | 4639853 | 4640130 | 278 | 3 | 9.10E-06 | 0.0002529 | 0.000551 | 2.54E-05 | -0.00318048 | MGRN1, AC023830.1 |
| chr19 | 2090499 | 2091162 | 664 | 3 | 7.24E-06 | 0.0092141 | 6.23E-05 | 2.54E-05 | 0.006774179 | MOB3A |
| chr1 | 32851293 | 32851331 | 39 | 2 | 1.54E-05 | 0.0001209 | 3.58E-05 | 2.54E-05 | 0.003281609 | S100PBP |
| chr3 | 182642722 | 182642912 | 191 | 2 | 1.90E-05 | 6.04E-05 | 5.89E-05 | 2.54E-05 | -0.00708547 | NA |
| chr19 | 45065809 | 45066023 | 215 | 2 | 5.49E-05 | 2.43E-05 | 0.000209 | 2.54E-05 | -0.00306007 | CLASRP |
| chr1 | 6178556 | 6179097 | 542 | 2 | 0.000114665 | 1.62E-05 | 0.000823 | 2.54E-05 | 0.003238112 | CHD5, AL031847.1 |
| chr12 | 117359251 | 117359404 | 154 | 2 | 1.65E-05 | 1.67E-05 | 0.000711 | 2.54E-05 | 0.018603546 | NOS1 |
| chr7 | 30026578 | 30027388 | 811 | 14 | 5.45E-05 | 3.08E-05 | 0.018921 | 2.54E-05 | -2.45E-05 | FKBP14-AS1, FKBP14 |
| chr15 | 72271188 | 72271922 | 735 | 5 | 4.66E-05 | 2.72E-05 | 0.003227 | 2.54E-05 | -0.00037918 | PARP6, AC009690.3 |
| chr3 | 161101173 | 161101575 | 403 | 2 | 5.15E-05 | 1.66E-05 | 0.000723 | 2.54E-05 | -0.00091456 | B3GALNT1 |
| chr5 | 139555541 | 139555644 | 104 | 2 | 5.74E-05 | 3.09E-05 | 0.000131 | 2.54E-05 | -0.01849111 | UBE2D2 |
| chr6 | 110270023 | 110270434 | 412 | 2 | 4.42E-05 | 2.28E-05 | 0.000245 | 2.54E-05 | -0.00577102 | METTL24 |
| chr1 | 155014394 | 155014622 | 229 | 2 | 7.36E-06 | 0.0005448 | 1.96E-05 | 2.54E-05 | -0.00723637 | ZBTB7B |
| chr6 | 159315582 | 159315926 | 345 | 3 | 3.63E-05 | 0.0001591 | 0.000167 | 2.54E-05 | 0.006051164 | NA |
| chr9 | 99266347 | 99266590 | 244 | 2 | 1.50E-05 | 0.0009367 | 1.71E-05 | 2.55E-05 | -0.00290024 | NA |
| chr17 | 81994491 | 81995183 | 693 | 6 | 2.20E-05 | 3.38E-05 | 0.001324 | 2.55E-05 | 0.00146668 | ASPSCR1 |
| chr19 | 54452127 | 54452164 | 38 | 5 | 3.68E-05 | 9.88E-05 | 0.000977 | 2.55E-05 | -0.00276331 | LENG8 |
| chr14 | 96234186 | 96234895 | 710 | 6 | 1.94E-05 | 0.0007111 | 7.08E-05 | 2.55E-05 | -0.00262097 | BDKRB2, AL355102.2 |
| chr16 | 81403519 | 81404104 | 586 | 4 | 1.79E-05 | 0.0015243 | 0.000212 | 2.55E-05 | 0.002837295 | NA |
| chr9 | 998261 | 998547 | 287 | 3 | 1.70E-05 | 0.0006973 | 0.000667 | 2.56E-05 | 0.021085792 | NA |
| chr22 | 37158130 | 37158181 | 52 | 2 | 7.66E-05 | 1.68E-05 | 0.000726 | 2.56E-05 | -0.00334511 | IL2RB |
| chr18 | 596703 | 597111 | 409 | 5 | 0.00012916 | 1.79E-05 | 0.006565 | 2.56E-05 | -0.00383029 | CLUL1 |
| chr13 | 114020865 | 114021349 | 485 | 3 | 5.96E-05 | 1.09E-05 | 0.003007 | 2.56E-05 | -0.00011181 | RASA3 |
| chr4 | 1521347 | 1521599 | 253 | 3 | 7.61E-05 | 1.38E-05 | 0.001479 | 2.56E-05 | -0.02432055 | NA |
| chr17 | 28532913 | 28533101 | 189 | 2 | 4.66E-05 | 3.25E-05 | 0.000123 | 2.56E-05 | -0.00934913 | FOXN1, AC005726.1 |
| chr12 | 120264672 | 120264859 | 188 | 2 | 9.33E-06 | 2.64E-05 | 0.000178 | 2.56E-05 | -0.00935074 | PXN |
| chr11 | 60831052 | 60831782 | 731 | 4 | 5.42E-06 | 0.002251 | 2.64E-05 | 2.56E-05 | 0.003066523 | NA |
| chr12 | 78687667 | 78688085 | 419 | 2 | 1.32E-05 | 0.0002798 | 2.47E-05 | 2.56E-05 | -0.00764119 | NA |
| chr12 | 54280763 | 54281684 | 922 | 6 | 5.19E-06 | 1.99E-05 | 0.002073 | 2.56E-05 | 0.000917739 | AC078778.1, HNRNPA1 |
| chr19 | 3599931 | 3600597 | 667 | 4 | 5.66E-06 | 0.0001237 | 6.93E-05 | 2.57E-05 | 0.001424535 | TBXA2R |
| chr1 | 89566832 | 89567096 | 265 | 3 | 1.23E-05 | 0.0001777 | 3.28E-05 | 2.57E-05 | 0.006773397 | LRRC8B |
| chr11 | 70377937 | 70378277 | 341 | 2 | 6.59E-05 | 1.58E-05 | 0.001018 | 2.57E-05 | -0.00920923 | PPFIA1, AP000487.1 |
| chr5 | 98779625 | 98779699 | 75 | 2 | 8.55E-05 | 1.54E-05 | 0.001207 | 2.57E-05 | -0.00625487 | RGMB |
| chr8 | 143920667 | 143921078 | 412 | 3 | 6.75E-06 | 0.0003985 | 0.000416 | 2.57E-05 | 0.000332477 | PLEC |
| chr1 | 47996886 | 47997632 | 747 | 9 | 1.13E-05 | 0.0002219 | 0.005604 | 2.57E-05 | 0.001230792 | TRABD2B |
| chr1 | 39789883 | 39790231 | 349 | 2 | 8.10E-06 | 1.65E-05 | 0.000798 | 2.57E-05 | -0.00350745 | AL033527.2 |
| chr13 | 36370157 | 36370503 | 347 | 6 | 4.67E-05 | 7.54E-05 | 0.001224 | 2.57E-05 | 0.000815905 | SPART |
| chr20 | 45856847 | 45857123 | 277 | 4 | 9.38E-06 | 5.74E-05 | 0.000144 | 2.57E-05 | -0.00117877 | ACOT8 |
| chr11 | 69914157 | 69914654 | 498 | 3 | 3.23E-06 | 0.0005682 | 5.41E-05 | 2.57E-05 | -0.00655364 | NA |
| chr16 | 23729904 | 23730179 | 276 | 2 | 7.85E-05 | 1.61E-05 | 0.000943 | 2.57E-05 | -0.0057055 | NA |
| chr5 | 107466133 | 107466577 | 445 | 3 | 1.60E-05 | 0.0002312 | 2.66E-05 | 2.58E-05 | 0.00440766 | EFNA5 |
| chr21 | 34110689 | 34110919 | 231 | 2 | 4.78E-05 | 2.80E-05 | 0.000162 | 2.58E-05 | -0.00282157 | MRPS6, AP000317.2 |
| chr19 | 11528802 | 11529286 | 485 | 13 | 3.12E-05 | 0.0030134 | 0.010181 | 2.58E-05 | 0.000566462 | ECSIT |
| chr5 | 178388409 | 178388692 | 284 | 2 | 7.30E-05 | 1.65E-05 | 0.000813 | 2.58E-05 | -0.01224169 | COL23A1 |
| chr20 | 32509892 | 32510379 | 488 | 4 | 5.70E-06 | 0.0004086 | 0.000173 | 2.58E-05 | 0.006115341 | AL034550.2, NOL4L |
| chr1 | 27287541 | 27287797 | 257 | 4 | 5.11E-05 | 1.42E-05 | 0.001544 | 2.58E-05 | -0.00117467 | WDTC1 |
| chr21 | 28885439 | 28885929 | 491 | 7 | 7.31E-06 | 0.0003514 | 0.000166 | 2.58E-05 | 0.000285829 | NA |
| chr22 | 40346022 | 40346698 | 677 | 10 | 1.88E-05 | 0.0007459 | 0.000766 | 2.58E-05 | -0.00023928 | ADSL, AL022238.3 |
| chr6 | 31535013 | 31535479 | 467 | 8 | 5.48E-06 | 0.0006236 | 0.000728 | 2.58E-05 | -0.00136118 | DDX39B, ATP6V1G2-DDX39B |
| chr1 | 35202050 | 35202103 | 54 | 2 | 7.06E-05 | 2.59E-05 | 0.00019 | 2.58E-05 | -0.00519769 | NA |
| chr17 | 45159872 | 45160819 | 948 | 9 | 9.60E-06 | 5.74E-05 | 0.00413 | 2.58E-05 | 0.003590851 | HEXIM2, AC138150.1 |
| chr20 | 62158555 | 62158758 | 204 | 5 | 7.28E-05 | 2.36E-05 | 0.000883 | 2.58E-05 | -0.00203064 | SS18L1 |
| chr22 | 37568723 | 37569035 | 313 | 2 | 4.20E-05 | 2.41E-05 | 0.000226 | 2.58E-05 | -0.00688014 | CDC42EP1 |
| chr18 | 48925024 | 48925211 | 188 | 2 | 6.19E-05 | 1.89E-05 | 0.000459 | 2.59E-05 | -0.01453217 | SMAD7 |
| chr3 | 53845224 | 53845923 | 700 | 4 | 1.27E-06 | 2.39E-05 | 0.000897 | 2.59E-05 | 0.001064028 | CHDH |
| chr6 | 43204981 | 43205115 | 135 | 3 | 9.41E-06 | 7.75E-05 | 6.12E-05 | 2.59E-05 | -0.00013634 | CUL9 |
| chr4 | 6709865 | 6710268 | 404 | 9 | 1.55E-05 | 0.0001151 | 0.001576 | 2.59E-05 | 0.000369854 | MRFAP1L1 |
| chr9 | 4666266 | 4667099 | 834 | 12 | 1.60E-05 | 0.0003845 | 0.000203 | 2.59E-05 | -0.00330277 | SPATA6L |
| chr18 | 74534355 | 74534550 | 196 | 8 | 0.000100587 | 5.39E-05 | 0.005512 | 2.59E-05 | 0.000455228 | CNDP1 |
| chr2 | 169072348 | 169072623 | 276 | 2 | 2.20E-05 | 2.21E-05 | 0.000282 | 2.59E-05 | -0.01267115 | DHRS9 |
| chr11 | 119148217 | 119149135 | 919 | 6 | 9.00E-07 | 0.0005468 | 2.66E-05 | 2.59E-05 | -0.00570036 | ABCG4 |
| chr3 | 197736875 | 197736898 | 24 | 3 | 0.00013729 | 4.44E-05 | 0.000511 | 2.59E-05 | 0.000692556 | RUBCN |
| chr6 | 160092464 | 160092822 | 359 | 2 | 1.87E-05 | 4.81E-05 | 7.61E-05 | 2.59E-05 | 0.005470449 | IGF2R |
| chr16 | 1449394 | 1449811 | 418 | 3 | 1.07E-05 | 1.41E-05 | 0.001397 | 2.59E-05 | -0.00179783 | CLCN7 |
| chr1 | 152372532 | 152372956 | 425 | 2 | 1.39E-05 | 4.19E-05 | 8.90E-05 | 2.59E-05 | -0.01872153 | FLG-AS1 |
| chr7 | 100014906 | 100015582 | 677 | 9 | 6.64E-06 | 8.88E-05 | 0.001491 | 2.60E-05 | 0.002602323 | ZKSCAN1 |
| chr4 | 151701384 | 151701537 | 154 | 2 | 4.88E-05 | 5.20E-05 | 7.05E-05 | 2.60E-05 | -0.00376822 | GATB |
| chr12 | 9680549 | 9680824 | 276 | 2 | 4.15E-05 | 3.83E-05 | 9.94E-05 | 2.60E-05 | -0.00955889 | CLEC2D |
| chr6 | 44295018 | 44295238 | 221 | 2 | 5.29E-05 | 1.80E-05 | 0.000559 | 2.60E-05 | -0.01228328 | TMEM151B, AL353588.1, TCTE1 |
| chr15 | 65517225 | 65517866 | 642 | 11 | 7.07E-07 | 0.0002645 | 0.000156 | 2.60E-05 | 0.001369472 | DPP8 |
| chr17 | 35760134 | 35760263 | 130 | 2 | 0.000140415 | 1.61E-05 | 0.000998 | 2.60E-05 | -0.00705559 | TAF15, MMP28 |
| chr2 | 102182191 | 102182294 | 104 | 2 | 6.77E-05 | 1.64E-05 | 0.000892 | 2.60E-05 | 0.000617687 | NA |
| chr19 | 17718682 | 17718687 | 6 | 2 | 3.60E-05 | 2.35E-05 | 0.000245 | 2.60E-05 | -0.01340768 | NA |
| chr7 | 20791038 | 20791425 | 388 | 4 | 3.85E-05 | 0.0222078 | 0.0003 | 2.60E-05 | -0.00375372 | NA |
| chr16 | 19095831 | 19095968 | 138 | 2 | 1.33E-05 | 0.0009588 | 1.74E-05 | 2.60E-05 | -0.01121249 | AC099518.1 |
| chr1 | 152757551 | 152757755 | 205 | 2 | 1.55E-05 | 0.0025619 | 1.46E-05 | 2.60E-05 | -0.0168255 | NA |
| chr7 | 121314414 | 121314495 | 82 | 3 | 1.88E-05 | 2.09E-05 | 0.001365 | 2.60E-05 | -0.00938498 | NA |
| chr18 | 9079865 | 9080007 | 143 | 2 | 3.55E-05 | 0.0001496 | 3.31E-05 | 2.60E-05 | -0.01250111 | NA |
| chr11 | 89922632 | 89922703 | 72 | 2 | 3.41E-05 | 4.03E-05 | 9.39E-05 | 2.60E-05 | 0.004999486 | NA |
| chr4 | 55547687 | 55548542 | 856 | 5 | 2.03E-05 | 3.62E-05 | 0.000299 | 2.60E-05 | -0.00232015 | AC024243.1 |
| chr9 | 34659834 | 34660009 | 176 | 2 | 4.78E-05 | 2.00E-05 | 0.000389 | 2.60E-05 | 0.003439028 | IL11RA |
| chr5 | 141402710 | 141402943 | 234 | 7 | 4.44E-05 | 4.89E-05 | 0.003142 | 2.60E-05 | -0.01222779 | PCDHGA1, PCDHGA2, PCDHGA3, PCDHGB1, PCDHGA4, PCDHGB2, PCDHGA5, PCDHGB3, PCDHGA6, PCDHGA7, PCDHGB4, PCDHGA8, PCDHGB5, PCDHGA9 |
| chr9 | 35161629 | 35162154 | 526 | 9 | 4.64E-05 | 2.91E-05 | 0.00776 | 2.60E-05 | -0.00344287 | UNC13B |
| chr3 | 112409905 | 112410122 | 218 | 3 | 5.13E-05 | 1.97E-05 | 0.001455 | 2.60E-05 | 0.002771861 | NA |
| chr20 | 653139 | 654087 | 949 | 8 | 1.06E-05 | 9.59E-05 | 0.008242 | 2.61E-05 | -0.00618349 | SRXN1, AL121758.1 |
| chr5 | 73860244 | 73860480 | 237 | 4 | 5.44E-05 | 3.40E-05 | 0.001653 | 2.61E-05 | 0.00174664 | ARHGEF28 |
| chr1 | 96538496 | 96538746 | 251 | 2 | 1.44E-05 | 0.0011788 | 1.67E-05 | 2.61E-05 | -0.01146969 | NA |
| chr2 | 150538943 | 150539118 | 176 | 2 | 2.29E-05 | 2.45E-05 | 0.000223 | 2.61E-05 | -0.01209546 | RND3 |
| chr8 | 112888002 | 112888409 | 408 | 4 | 7.48E-05 | 1.48E-05 | 0.004648 | 2.61E-05 | -0.0088129 | CSMD3 |
| chr12 | 54040106 | 54040615 | 510 | 3 | 2.38E-05 | 1.64E-05 | 0.000759 | 2.61E-05 | 0.011182827 | HOXC4 |
| chr3 | 124887643 | 124887736 | 94 | 3 | 4.85E-05 | 0.0001501 | 7.80E-05 | 2.61E-05 | 0.002779695 | ITGB5 |
| chr6 | 15143002 | 15143554 | 553 | 2 | 0.000148754 | 1.57E-05 | 0.001235 | 2.61E-05 | -0.00550838 | NA |
| chr1 | 154192223 | 154192518 | 296 | 3 | 1.29E-06 | 0.008096 | 6.40E-06 | 2.61E-05 | -0.00350357 | TPM3 |
| chr9 | 109118604 | 109119433 | 830 | 4 | 2.39E-05 | 3.48E-05 | 0.003207 | 2.61E-05 | 0.007151234 | TMEM245 |
| chr3 | 141739031 | 141739548 | 518 | 3 | 2.33E-05 | 1.75E-05 | 0.000755 | 2.61E-05 | -0.00357588 | RNF7, AC112504.2 |
| chr6 | 10060547 | 10060705 | 159 | 2 | 5.73E-05 | 2.76E-05 | 0.000173 | 2.62E-05 | -0.00744181 | OFCC1 |
| chr11 | 9613202 | 9613764 | 563 | 5 | 6.09E-05 | 2.59E-05 | 0.00223 | 2.62E-05 | 0.002287573 | NA |
| chr12 | 127063442 | 127063449 | 8 | 2 | 7.37E-05 | 2.35E-05 | 0.00025 | 2.62E-05 | 0.004234391 | NA |
| chr1 | 6243668 | 6244439 | 772 | 11 | 6.70E-06 | 0.0006714 | 0.001329 | 2.62E-05 | -0.00189553 | HES3 |
| chr19 | 18426118 | 18426564 | 447 | 2 | 2.09E-05 | 1.86E-05 | 0.000516 | 2.62E-05 | 0.008551966 | SSBP4 |
| chr12 | 4556334 | 4556381 | 48 | 2 | 2.87E-05 | 5.70E-05 | 6.53E-05 | 2.62E-05 | -0.01177436 | RAD51AP1 |
| chr15 | 75636335 | 75636353 | 19 | 2 | 4.02E-05 | 0.0001018 | 4.14E-05 | 2.62E-05 | 0.006996015 | AC105020.2 |
| chr1 | 179942737 | 179942852 | 116 | 3 | 4.16E-05 | 0.0001096 | 4.50E-05 | 2.62E-05 | -0.01241811 | NA |
| chr10 | 89564399 | 89564614 | 216 | 2 | 2.09E-05 | 0.0004696 | 2.11E-05 | 2.62E-05 | -0.0146648 | NA |
| chr17 | 1628567 | 1629562 | 996 | 7 | 3.05E-06 | 0.0001581 | 0.004676 | 2.62E-05 | 0.000550128 | SLC43A2 |
| chr16 | 670309 | 670883 | 575 | 4 | 1.17E-05 | 0.0003544 | 0.000143 | 2.62E-05 | -2.20E-05 | RHOT2 |
| chr19 | 39480497 | 39481176 | 680 | 7 | 2.32E-06 | 0.0001604 | 0.000121 | 2.62E-05 | -0.00424945 | TIMM50 |
| chr11 | 30424053 | 30424497 | 445 | 3 | 2.44E-06 | 0.0004392 | 5.26E-05 | 2.62E-05 | -0.0040806 | MPPED2 |
| chr20 | 64267902 | 64268010 | 109 | 2 | 5.74E-05 | 1.88E-05 | 0.000493 | 2.62E-05 | -0.00083423 | PCMTD2 |
| chr15 | 62694567 | 62694716 | 150 | 2 | 5.78E-05 | 3.53E-05 | 0.000113 | 2.62E-05 | -0.0080212 | TLN2 |
| chr10 | 3494539 | 3494581 | 43 | 2 | 4.16E-05 | 0.0004481 | 2.15E-05 | 2.62E-05 | -0.00496746 | LINC02669 |
| chr6 | 38712988 | 38713317 | 330 | 3 | 5.47E-05 | 2.61E-05 | 0.000903 | 2.62E-05 | 0.005407525 | NA |
| chr6 | 33304594 | 33305700 | 1107 | 10 | 9.94E-07 | 0.0004542 | 0.001888 | 2.62E-05 | -0.00098048 | TAPBP |
| chr20 | 33370410 | 33370547 | 138 | 2 | 1.15E-05 | 0.0006584 | 1.93E-05 | 2.63E-05 | -0.00895967 | CDK5RAP1 |
| chr8 | 126824954 | 126825137 | 184 | 2 | 5.64E-05 | 1.70E-05 | 0.000782 | 2.63E-05 | -0.03215572 | PCAT1, AC024382.1 |
| chr3 | 129486227 | 129486495 | 269 | 2 | 7.21E-05 | 1.82E-05 | 0.000577 | 2.63E-05 | -0.01126831 | IFT122 |
| chr13 | 48706271 | 48706344 | 74 | 3 | 0.000175501 | 9.98E-05 | 0.000622 | 2.63E-05 | -0.00470573 | CYSLTR2 |
| chr10 | 68561911 | 68562685 | 775 | 5 | 1.55E-05 | 1.39E-05 | 0.003874 | 2.63E-05 | 0.018267588 | TET1 |

| | | | | | | | | | | |
|---|---|---|---|---|---|---|---|---|---|---|
| chr1 | 107805331 | 107805732 | 402 | 2 | 5.74E-06 | 0.002033 | 1.53E-05 | 2.63E-05 | -0.01208082 | VAV3 |
| chr3 | 48918385 | 48919504 | 1120 | 13 | 1.82E-05 | 0.0003005 | 0.003149 | 2.63E-05 | 0.001281283 | ARIH2, ARIH2OS |
| chr11 | 104036752 | 104037261 | 510 | 4 | 7.21E-05 | 1.01E-05 | 0.006093 | 2.63E-05 | -0.00361017 | DDI1, PDGFD |
| chr11 | 417540 | 418251 | 712 | 8 | 1.75E-05 | 0.0014149 | 0.0009 | 2.63E-05 | -0.00151446 | ANO9 |
| chr17 | 9465373 | 9465478 | 106 | 2 | 2.16E-05 | 0.0003237 | 2.42E-05 | 2.64E-05 | -0.01117371 | AC087501.1, AC087501.2, STX8 |
| chr1 | 110003377 | 110003640 | 264 | 2 | 2.40E-05 | 2.37E-05 | 0.000249 | 2.64E-05 | -0.00512022 | AHCYL1 |
| chr6 | 74879286 | 74879490 | 205 | 3 | 2.62E-05 | 0.0001733 | 3.26E-05 | 2.64E-05 | -0.00298806 | NA |
| chr16 | 84990444 | 84990649 | 206 | 4 | 9.69E-06 | 0.0024359 | 2.73E-05 | 2.64E-05 | -0.00263119 | ZDHHC7 |
| chr1 | 114751041 | 114751347 | 307 | 3 | 1.06E-05 | 0.0003062 | 2.34E-05 | 2.64E-05 | 0.001049738 | CSDE1 |
| chr15 | 70032627 | 70033008 | 382 | 2 | 4.11E-06 | 0.0009101 | 1.79E-05 | 2.64E-05 | -0.01040121 | NA |
| chr19 | 38003428 | 38003730 | 303 | 3 | 5.40E-05 | 1.71E-05 | 0.000847 | 2.64E-05 | 0.003847817 | SIPA1L3 |
| chr1 | 27626154 | 27626641 | 488 | 3 | 1.27E-05 | 5.59E-05 | 0.000409 | 2.64E-05 | -0.00426224 | FGR |
| chr17 | 10127989 | 10128165 | 177 | 2 | 1.41E-05 | 1.88E-05 | 0.000509 | 2.64E-05 | -0.04438696 | GAS7 |
| chr22 | 37150285 | 37150325 | 41 | 2 | 0.000112934 | 1.86E-05 | 0.000539 | 2.64E-05 | -0.01306269 | IL2RB |
| chr7 | 122145277 | 122145406 | 130 | 2 | 5.75E-07 | 0.0005981 | 1.99E-05 | 2.64E-05 | -0.01063673 | AC015983.2 |
| chr11 | 58968527 | 58968631 | 105 | 3 | 0.000112663 | 1.03E-05 | 0.0044 | 2.65E-05 | -0.0135744 | GLYATL1, AP001636.3 |
| chr8 | 66923213 | 66923618 | 406 | 2 | 1.08E-05 | 0.0001002 | 4.24E-05 | 2.65E-05 | -0.01981032 | SNHG6 |
| chr15 | 89999992 | 90000160 | 169 | 2 | 6.97E-05 | 2.90E-05 | 0.000162 | 2.65E-05 | -0.00740231 | NA |
| chr20 | 17611416 | 17611761 | 346 | 3 | 8.61E-06 | 2.08E-05 | 0.000443 | 2.65E-05 | -0.00639893 | NA |
| chr17 | 1720357 | 1720427 | 71 | 2 | 8.20E-05 | 1.69E-05 | 0.000841 | 2.65E-05 | 0.000497365 | WDR81 |
| chr17 | 48265726 | 48266105 | 380 | 2 | 9.50E-06 | 4.88E-05 | 7.76E-05 | 2.65E-05 | -0.01369982 | SKAP1 |
| chr3 | 50523025 | 50523458 | 434 | 2 | 1.54E-05 | 2.59E-05 | 0.000205 | 2.65E-05 | -0.01003465 | NA |
| chr2 | 44155291 | 44155792 | 502 | 2 | 0.00011437 | 1.67E-05 | 0.000909 | 2.65E-05 | 0.016003216 | NA |
| chr13 | 42573510 | 42574548 | 1039 | 9 | 4.49E-06 | 0.0008778 | 0.001061 | 2.65E-05 | 0.000937697 | TNFSF11 |
| chr5 | 2387924 | 2388035 | 112 | 2 | 4.31E-05 | 0.0002388 | 2.74E-05 | 2.65E-05 | 0.011623513 | NA |
| chr6 | 46658173 | 46658520 | 348 | 3 | 3.73E-05 | 4.99E-05 | 0.000184 | 2.65E-05 | -0.00395268 | SLC25A27 |
| chr20 | 57975429 | 57976278 | 850 | 5 | 3.09E-05 | 2.92E-05 | 0.001892 | 2.66E-05 | -0.00887182 | NA |
| chr11 | 72425351 | 72425667 | 317 | 2 | 6.49E-05 | 1.97E-05 | 0.000446 | 2.66E-05 | -0.00910939 | CLPB |
| chr10 | 101121084 | 101121788 | 705 | 7 | 2.09E-05 | 0.0005329 | 0.001141 | 2.66E-05 | -0.00184282 | TLX1NB |
| chr10 | 101066696 | 101067386 | 691 | 3 | 1.38E-05 | 1.10E-05 | 0.003719 | 2.66E-05 | 0.001020976 | KAZALD1 |
| chr19 | 372718 | 372978 | 261 | 2 | 2.53E-05 | 2.21E-05 | 0.000308 | 2.66E-05 | 0.020955452 | THEG |
| chr22 | 39943443 | 39943604 | 162 | 2 | 1.99E-05 | 4.35E-05 | 8.91E-05 | 2.66E-05 | -0.01298209 | GRAP2 |
| chr22 | 50576008 | 50576078 | 71 | 2 | 6.04E-05 | 1.61E-05 | 0.001151 | 2.66E-05 | -0.0059038 | CPT1B, CHKB-CPT1B |
| chr14 | 56808021 | 56808695 | 675 | 6 | 2.75E-05 | 0.0001224 | 0.001481 | 2.66E-05 | 0.002002793 | OTX2 |
| chr6 | 132751198 | 132751412 | 215 | 2 | 2.76E-05 | 0.0001325 | 3.62E-05 | 2.66E-05 | -0.01595217 | VNN2 |
| chr5 | 163565835 | 163566368 | 534 | 6 | 4.66E-05 | 1.57E-05 | 0.008001 | 2.66E-05 | -0.00109732 | NA |
| chr12 | 74624236 | 74624338 | 103 | 2 | 7.76E-05 | 2.12E-05 | 0.000349 | 2.66E-05 | -0.01601364 | NA |
| chr3 | 31708941 | 31709221 | 281 | 4 | 4.33E-05 | 4.54E-05 | 0.000644 | 2.66E-05 | 0.003069944 | OSBPL10-AS1, OSBPL10 |
| chr9 | 78288391 | 78288482 | 92 | 2 | 6.02E-05 | 4.24E-05 | 9.19E-05 | 2.66E-05 | 0.015114774 | NA |
| chr10 | 48745834 | 48746141 | 308 | 3 | 1.45E-05 | 0.0023728 | 1.57E-05 | 2.66E-05 | -0.00218131 | WDFY4, RPL13AP19 |
| chr1 | 99727922 | 99728281 | 360 | 2 | 3.58E-06 | 0.0001208 | 3.81E-05 | 2.66E-05 | 0.019898868 | FRRS1 |
| chr19 | 50943462 | 50943697 | 236 | 2 | 4.61E-05 | 4.19E-05 | 9.35E-05 | 2.67E-05 | -0.0048949 | KLK5 |
| chr13 | 99366729 | 99367393 | 665 | 4 | 0.000112635 | 0.0037644 | 0.000429 | 2.67E-05 | -0.00105014 | UBAC2 |
| chr1 | 31768810 | 31769226 | 417 | 2 | 6.33E-06 | 6.08E-05 | 6.33E-05 | 2.67E-05 | -0.00253871 | NA |
| chr13 | 77718338 | 77718557 | 220 | 2 | 5.45E-05 | 2.07E-05 | 0.000383 | 2.67E-05 | 0.016373749 | SLAIN1 |
| chr2 | 173184313 | 173184504 | 192 | 2 | 7.10E-05 | 2.15E-05 | 0.000338 | 2.67E-05 | -0.00861991 | MAP3K20, MAP3K20-AS1 |
| chr1 | 47592718 | 47593313 | 596 | 4 | 7.12E-06 | 0.0003151 | 0.000742 | 2.67E-05 | 0.001702452 | NA |
| chr7 | 33815775 | 33815846 | 72 | 2 | 4.39E-05 | 1.69E-05 | 0.000888 | 2.67E-05 | -0.01034146 | BBS9 |
| chr8 | 20233790 | 20234172 | 383 | 2 | 1.46E-05 | 7.20E-05 | 5.49E-05 | 2.68E-05 | -0.0124291 | NA |
| chr9 | 137251112 | 137251259 | 148 | 5 | 5.01E-05 | 5.83E-05 | 0.000768 | 2.68E-05 | -0.00546732 | STPG3-AS1 |
| chr9 | 74886619 | 74887529 | 911 | 6 | 1.68E-05 | 0.000806 | 0.000545 | 2.68E-05 | -0.00219771 | TRPM6 |
| chr10 | 3786142 | 3786740 | 599 | 2 | 8.11E-06 | 1.67E-05 | 0.000954 | 2.68E-05 | -0.00153215 | NA |
| chr2 | 108605996 | 108606422 | 427 | 3 | 1.36E-05 | 0.0002752 | 2.51E-05 | 2.68E-05 | -0.00457461 | LIMS1 |
| chr11 | 6451041 | 6451407 | 367 | 2 | 8.86E-06 | 5.61E-05 | 6.88E-05 | 2.68E-05 | -0.00506177 | TRIM3 |
| chr5 | 147227118 | 147227184 | 67 | 2 | 8.67E-05 | 4.99E-05 | 7.76E-05 | 2.68E-05 | -0.0053164 | STK32A-AS1 |
| chr6 | 166956003 | 166956970 | 968 | 10 | 1.44E-05 | 1.56E-05 | 0.005499 | 2.68E-05 | -6.22E-05 | AL159163.1, RNASET2 |
| chr21 | 15224034 | 15224437 | 404 | 3 | 1.62E-05 | 0.0001908 | 0.000727 | 2.68E-05 | -0.00368735 | AF222685.1 |
| chr1 | 70706340 | 70706803 | 464 | 3 | 8.17E-05 | 1.02E-05 | 0.004875 | 2.69E-05 | -0.01179298 | LINC01788 |
| chr19 | 47485099 | 47485582 | 484 | 5 | 1.37E-05 | 6.47E-05 | 0.000816 | 2.69E-05 | 0.007308093 | NAPA-AS1 |
| chr14 | 101447527 | 101447637 | 111 | 2 | 2.36E-05 | 6.02E-05 | 6.47E-05 | 2.69E-05 | -0.00781117 | NA |
| chr18 | 63752567 | 63752779 | 213 | 2 | 4.97E-05 | 0.0004241 | 2.25E-05 | 2.69E-05 | -0.00147451 | NA |
| chr11 | 71080337 | 71080556 | 220 | 2 | 6.83E-05 | 2.21E-05 | 0.000322 | 2.69E-05 | 0.016781224 | SHANK2 |
| chr7 | 98215079 | 98215635 | 557 | 4 | 4.33E-06 | 0.0012876 | 1.66E-05 | 2.69E-05 | -0.00269262 | BHLHA15, TECPR1 |
| chr1 | 48680689 | 48681222 | 534 | 3 | 5.92E-05 | 2.05E-05 | 0.000486 | 2.69E-05 | 0.000237598 | AGBL4 |
| chr2 | 230690430 | 230690556 | 127 | 3 | 8.69E-05 | 1.56E-05 | 0.000992 | 2.69E-05 | -0.00289986 | LINC01907 |
| chr5 | 151108771 | 151109073 | 303 | 2 | 4.12E-05 | 1.63E-05 | 0.001184 | 2.69E-05 | -0.00085216 | ANXA6 |
| chr6 | 37547293 | 37547683 | 391 | 2 | 1.63E-06 | 0.0002987 | 2.54E-05 | 2.69E-05 | -0.00368804 | AL353597.1 |
| chr2 | 233013147 | 233014004 | 858 | 8 | 3.86E-06 | 0.0010264 | 4.00E-05 | 2.69E-05 | 0.000112963 | AC106876.1, NGEF |
| chr3 | 4985490 | 4985931 | 442 | 3 | 1.09E-06 | 0.0070509 | 6.68E-06 | 2.70E-05 | 0.00241408 | NA |
| chr4 | 113335771 | 113336037 | 267 | 2 | 0.000112374 | 1.83E-05 | 0.000632 | 2.70E-05 | -0.00746704 | ANK2 |
| chr10 | 70938878 | 70939254 | 377 | 4 | 3.90E-05 | 1.07E-05 | 0.002549 | 2.70E-05 | 0.003145482 | AC073176.2, LINC02622 |
| chr15 | 43457441 | 43457487 | 47 | 2 | 1.88E-05 | 0.0001909 | 3.08E-05 | 2.70E-05 | -0.00362562 | TP53BP1 |
| chr17 | 3846886 | 3846992 | 107 | 2 | 3.60E-05 | 2.10E-05 | 0.000383 | 2.70E-05 | -0.00630966 | NA |
| chr5 | 133243837 | 133243930 | 94 | 2 | 0.000103561 | 2.49E-05 | 0.000239 | 2.70E-05 | -0.00669906 | AC010307.3, FSTL4 |
| chr12 | 11171017 | 11171412 | 396 | 3 | 2.29E-06 | 0.0011819 | 5.97E-05 | 2.70E-05 | -0.00181182 | SMIM10L1, PRH1, AC018630.4, TAS2R14, AC134349.1 |
| chr12 | 121133072 | 121133265 | 194 | 3 | 1.24E-05 | 0.0039114 | 9.50E-06 | 2.70E-05 | 0.001579647 | P2RX7 |
| chr17 | 49315774 | 49316184 | 411 | 2 | 2.63E-05 | 2.09E-05 | 0.000389 | 2.71E-05 | -0.00201015 | FLJ40194, ZNF652 |
| chr12 | 56153834 | 56153890 | 57 | 2 | 1.26E-05 | 1.63E-05 | 0.001264 | 2.71E-05 | 0.000653532 | MYL6B, AC034102.5 |
| chr7 | 44224243 | 44224791 | 549 | 2 | 5.74E-05 | 1.69E-05 | 0.000974 | 2.71E-05 | -0.01059323 | CAMK2B |
| chr22 | 42604095 | 42604733 | 639 | 4 | 1.48E-05 | 0.0002535 | 8.08E-05 | 2.71E-05 | 0.000560507 | POLDIP3 |
| chr14 | 50823944 | 50824033 | 90 | 2 | 1.28E-05 | 1.72E-05 | 0.00089 | 2.71E-05 | 0.016391137 | NIN |
| chr4 | 1305354 | 1305367 | 14 | 2 | 9.40E-05 | 4.52E-05 | 8.82E-05 | 2.71E-05 | -0.00396115 | MAEA |
| chr9 | 91681410 | 91681841 | 432 | 4 | 4.74E-05 | 1.50E-05 | 0.003416 | 2.71E-05 | 0.008066105 | ROR2 |
| chr1 | 8096258 | 8096606 | 349 | 3 | 1.28E-05 | 2.65E-05 | 0.000292 | 2.72E-05 | -0.00172835 | AL034417.2 |
| chr16 | 87705017 | 87705227 | 211 | 3 | 6.29E-06 | 0.0040012 | 9.31E-05 | 2.72E-05 | 0.003795486 | KLHDC4 |
| chr2 | 105851744 | 105852090 | 347 | 3 | 1.77E-06 | 0.0001938 | 3.19E-05 | 2.72E-05 | -0.0060324 | NCK2, AC010978.1 |
| chr17 | 82392529 | 82393511 | 983 | 5 | 2.94E-05 | 2.34E-05 | 0.003454 | 2.72E-05 | -0.00501258 | OGFOD3 |
| chr14 | 32808734 | 32809154 | 421 | 2 | 5.87E-05 | 2.07E-05 | 0.000411 | 2.72E-05 | -0.03096624 | AKAP6 |
| chr2 | 132671263 | 132672453 | 1191 | 6 | 6.98E-07 | 0.0002656 | 0.000272 | 2.72E-05 | -0.0088174 | LYPD1, NCKAP5 |
| chr8 | 30913101 | 30913517 | 417 | 2 | 3.41E-05 | 1.66E-05 | 0.001129 | 2.72E-05 | -0.00704681 | NA |
| chr13 | 22897625 | 22897627 | 3 | 2 | 0.000134882 | 1.89E-05 | 0.000566 | 2.72E-05 | 0.000632211 | LINC00621, BASP1P1 |
| chr2 | 161417417 | 161418288 | 872 | 8 | 7.09E-06 | 3.50E-05 | 0.000819 | 2.72E-05 | -0.00346636 | TBR1 |
| chr1 | 11131688 | 11131725 | 38 | 3 | 2.47E-05 | 0.0002325 | 0.000132 | 2.72E-05 | 0.000385546 | MTOR |
| chr2 | 207143328 | 207143726 | 399 | 2 | 7.17E-06 | 0.0003684 | 2.39E-05 | 2.72E-05 | 0.007201801 | KLF7 |
| chr8 | 141276939 | 141277165 | 227 | 4 | 1.99E-05 | 0.0005869 | 2.38E-05 | 2.72E-05 | 0.000209478 | SLC45A4 |
| chr12 | 132198063 | 132198335 | 273 | 2 | 1.07E-05 | 5.28E-05 | 7.48E-05 | 2.72E-05 | 0.00530676 | GALNT9 |
| chr2 | 207172837 | 207173244 | 408 | 3 | 1.33E-05 | 0.0014578 | 1.33E-05 | 2.72E-05 | -0.00032185 | MYOSLID |
| chr16 | 22296624 | 22296644 | 21 | 2 | 3.71E-05 | 2.70E-05 | 0.000202 | 2.72E-05 | -0.01933583 | NA |
| chr9 | 138061558 | 138061599 | 42 | 2 | 1.71E-05 | 5.26E-05 | 7.53E-05 | 2.73E-05 | -0.00105386 | CACNA1B |
| chr11 | 134155837 | 134156647 | 811 | 6 | 2.93E-05 | 4.53E-05 | 0.004662 | 2.73E-05 | -3.88E-05 | NCAPD3 |
| chr22 | 17628511 | 17629145 | 635 | 9 | 4.05E-05 | 6.48E-05 | 0.003313 | 2.73E-05 | 0.000122004 | BCL2L13, ATP6V1E1 |
| chr9 | 122402576 | 122403487 | 912 | 3 | 6.87E-07 | 0.0004919 | 3.44E-05 | 2.73E-05 | 0.002390133 | AL359636.2, AL359636.1 |
| chr16 | 84158329 | 84158623 | 295 | 2 | 9.35E-06 | 0.0059062 | 1.39E-05 | 2.73E-05 | -0.00973949 | DNAAF1 |
| chr18 | 80246354 | 80247297 | 944 | 4 | 6.40E-06 | 1.12E-05 | 0.003005 | 2.73E-05 | -0.00096864 | PARD6G |
| chr6 | 168396555 | 168396828 | 274 | 3 | 2.96E-05 | 8.04E-05 | 6.32E-05 | 2.73E-05 | -0.0021941 | NA |
| chr13 | 98021792 | 98022172 | 381 | 3 | 5.52E-05 | 0.0004972 | 0.000235 | 2.73E-05 | -0.00036218 | IPO5 |
| chr18 | 3247296 | 3247508 | 213 | 5 | 0.000105473 | 1.53E-05 | 0.005507 | 2.73E-05 | -0.00082511 | MYL12A |
| chr15 | 42084173 | 42084567 | 395 | 4 | 5.27E-05 | 3.49E-05 | 0.000201 | 2.73E-05 | 0.004653731 | PLA2G4D |
| chr13 | 36131860 | 36132296 | 437 | 4 | 5.77E-05 | 1.52E-05 | 0.003288 | 2.73E-05 | 0.003014282 | NA |
| chr11 | 46333080 | 46333733 | 654 | 6 | 2.52E-06 | 5.53E-05 | 0.000843 | 2.73E-05 | -0.00258231 | DGKZ |
| chr1 | 24266344 | 24266678 | 335 | 2 | 9.03E-06 | 0.0019551 | 1.60E-05 | 2.74E-05 | -0.00540256 | NA |
| chr9 | 115739278 | 115739283 | 6 | 2 | 0.00014783 | 3.67E-05 | 0.000117 | 2.74E-05 | 0.008729303 | AL691420.1 |
| chr2 | 39717193 | 39717330 | 138 | 2 | 3.60E-05 | 0.0001932 | 3.11E-05 | 2.74E-05 | -0.0042279 | TMEM178A |
| chr1 | 4689105 | 4689189 | 85 | 2 | 9.26E-06 | 6.52E-05 | 6.19E-05 | 2.74E-05 | 0.003901722 | AJAP1 |
| chr7 | 152187608 | 152187764 | 157 | 2 | 9.01E-05 | 1.66E-05 | 0.001173 | 2.74E-05 | -0.00689748 | KMT2C |
| chr22 | 25942639 | 25942884 | 246 | 2 | 2.29E-05 | 4.38E-05 | 9.30E-05 | 2.74E-05 | -0.00062644 | MYO18B |
| chr9 | 93134462 | 93134934 | 473 | 6 | 1.25E-05 | 0.0005798 | 3.13E-05 | 2.74E-05 | -0.00043078 | NA |
| chr11 | 57530608 | 57531121 | 514 | 8 | 4.72E-05 | 4.28E-05 | 0.005775 | 2.74E-05 | 0.000468288 | TIMM10 |
| chr19 | 49162203 | 49162683 | 481 | 2 | 7.83E-05 | 1.65E-05 | 0.00127 | 2.74E-05 | -0.00639192 | TRPM4 |
| chr19 | 53329058 | 53329417 | 360 | 4 | 7.00E-05 | 2.07E-05 | 0.000797 | 2.74E-05 | -0.0153674 | NA |
| chr11 | 122979432 | 122979837 | 406 | 5 | 8.03E-05 | 3.78E-05 | 0.001407 | 2.74E-05 | -0.00264541 | BSX |
| chr11 | 118171738 | 118171915 | 178 | 4 | 6.90E-05 | 7.23E-05 | 0.001129 | 2.74E-05 | 0.000775927 | SCN2B |
| chr15 | 27805001 | 27806116 | 1116 | 5 | 5.18E-06 | 0.0001722 | 0.000122 | 2.75E-05 | -0.00244219 | OCA2 |
| chr5 | 112160812 | 112161379 | 568 | 8 | 4.67E-05 | 0.0001243 | 0.000257 | 2.75E-05 | 0.002282614 | EPB41L4A-AS1, EPB41L4A |
| chr22 | 29337784 | 29337985 | 202 | 2 | 1.22E-05 | 0.0001953 | 3.11E-05 | 2.75E-05 | -0.00638977 | AP1B1 |
| chr21 | 36704742 | 36705373 | 632 | 7 | 3.34E-05 | 0.0003164 | 0.001665 | 2.75E-05 | -0.00293012 | SIM2 |
| chr9 | 131823190 | 131823539 | 350 | 2 | 1.12E-06 | 0.0023825 | 1.56E-05 | 2.75E-05 | -0.00552229 | NA |
| chr18 | 8717325 | 8717928 | 604 | 6 | 7.38E-06 | 0.040473 | 7.16E-05 | 2.75E-05 | -0.007113 | MTCL1 |
| chr10 | 31031267 | 31031409 | 143 | 2 | 2.30E-06 | 0.0083702 | 1.37E-05 | 2.75E-05 | -0.00888168 | ZNF438 |
| chr14 | 95363572 | 95363838 | 267 | 5 | 5.58E-05 | 4.97E-05 | 0.001134 | 2.75E-05 | -0.01005717 | NA |
| chr7 | 5189148 | 5189642 | 495 | 4 | 3.32E-06 | 0.0005332 | 1.60E-05 | 2.75E-05 | 0.000771271 | NA |
| chr14 | 95467939 | 95468405 | 467 | 2 | 1.18E-05 | 3.06E-05 | 0.000162 | 2.75E-05 | -0.00097784 | SYNE3 |
| chr13 | 31368183 | 31368407 | 225 | 3 | 6.37E-05 | 1.89E-05 | 0.001931 | 2.75E-05 | -0.00535925 | NA |
| chr8 | 90510310 | 90510338 | 29 | 2 | 6.87E-05 | 1.86E-05 | 0.000656 | 2.75E-05 | 0.001373327 | LINC00534, AC004083.1 |
| chr10 | 6114975 | 6115488 | 514 | 4 | 7.52E-06 | 0.0127905 | 6.48E-05 | 2.75E-05 | -0.0054743 | RBM17 |
| chr12 | 51005343 | 51005962 | 620 | 3 | 2.95E-05 | 3.55E-05 | 0.000202 | 2.75E-05 | -0.00638728 | SLC11A2 |
| chr10 | 18651362 | 18651990 | 629 | 14 | 4.24E-05 | 0.0001669 | 0.005322 | 2.75E-05 | 0.000120208 | NSUN6 |
| chr7 | 5214110 | 5214467 | 358 | 9 | 5.02E-05 | 0.0002333 | 0.003369 | 2.76E-05 | 0.000141995 | WIPI2 |
| chr12 | 42588579 | 42589127 | 549 | 5 | 4.62E-05 | 4.23E-05 | 0.002142 | 2.76E-05 | -0.00713148 | PRICKLE1 |
| chr3 | 134382033 | 134382039 | 7 | 2 | 6.17E-05 | 5.28E-05 | 7.66E-05 | 2.76E-05 | -0.00465226 | NA |
| chr8 | 139463046 | 139463369 | 324 | 2 | 3.53E-05 | 4.80E-05 | 8.51E-05 | 2.76E-05 | -0.01198221 | NA |
| chr9 | 114273722 | 114273884 | 163 | 3 | 3.62E-05 | 2.67E-05 | 0.001351 | 2.76E-05 | -0.00625847 | COL27A1 |
| chr2 | 169479945 | 169480327 | 383 | 2 | 1.22E-06 | 8.28E-05 | 5.12E-05 | 2.76E-05 | 0.016558007 | AC093899.2, BBS5 |
| chr20 | 13990418 | 13991165 | 748 | 4 | 8.94E-06 | 0.0177097 | 0.000243 | 2.76E-05 | -0.01522167 | SEL1L2 |
| chr14 | 95129604 | 95130071 | 468 | 3 | 4.96E-05 | 1.22E-05 | 0.002925 | 2.76E-05 | -0.00563676 | DICER1 |
| chr1 | 26189987 | 26190830 | 844 | 8 | 5.38E-05 | 1.30E-05 | 0.009997 | 2.77E-05 | -0.00287614 | CATSPER4 |
| chr6 | 7698141 | 7698370 | 230 | 2 | 8.52E-05 | 1.76E-05 | 0.000873 | 2.77E-05 | -0.003947 | NA |
| chr15 | 38040165 | 38040342 | 178 | 3 | 8.81E-05 | 4.49E-05 | 0.000629 | 2.77E-05 | -0.00538059 | LINC02345 |
| chr22 | 17942557 | 17942732 | 176 | 3 | 5.41E-05 | 1.20E-05 | 0.002743 | 2.77E-05 | -0.00221647 | MICAL3 |
| chr16 | 628127 | 629347 | 1221 | 5 | 3.31E-07 | 0.0009173 | 9.05E-05 | 2.77E-05 | -0.00268914 | RAB40C, WFIKKN1 |
| chr20 | 53061184 | 53061312 | 129 | 2 | 3.33E-05 | 6.35E-05 | 6.45E-05 | 2.77E-05 | 0.008019464 | TSHZ2 |
| chr2 | 219905531 | 219905697 | 167 | 3 | 8.01E-05 | 1.67E-05 | 0.001773 | 2.77E-05 | 0.000427899 | AC008281.1 |
| chr10 | 28303011 | 28303170 | 160 | 4 | 8.83E-05 | 0.0001079 | 0.002093 | 2.77E-05 | 0.002216607 | MPP7 |
| chr8 | 58993944 | 58994131 | 188 | 2 | 5.09E-05 | 3.16E-05 | 0.000155 | 2.77E-05 | 0.006549408 | TOX |
| chr2 | 230777996 | 230778291 | 296 | 2 | 6.71E-06 | 0.0003741 | 2.43E-05 | 2.77E-05 | -0.01087719 | CAB39 |
| chr6 | 4620069 | 4620446 | 378 | 2 | 1.33E-05 | 0.0002385 | 2.88E-05 | 2.77E-05 | -0.00083378 | NA |
| chr14 | 73885430 | 73886217 | 788 | 2 | 3.02E-08 | 0.0007116 | 2.00E-05 | 2.77E-05 | 0.00295541 | PTGR2, AC005520.1, AC005520.5 |
| chr20 | 58855608 | 58856147 | 540 | 2 | 3.54E-05 | 1.74E-05 | 0.000952 | 2.77E-05 | -0.01992154 | GNAS |

| | | | | | | | | | | |
|---|---|---|---|---|---|---|---|---|---|---|
| chr14 | 93778501 | 93778535 | 35 | 2 | 8.08E-05 | 1.73E-05 | 0.001005 | 2.77E-05 | -0.01154909 | PRIMA1 |
| chr10 | 59138226 | 59138327 | 102 | 3 | 6.14E-05 | 6.48E-05 | 8.91E-05 | 2.77E-05 | -0.01300125 | NA |
| chr16 | 70289464 | 70290012 | 549 | 13 | 3.58E-05 | 0.0001361 | 0.000363 | 2.77E-05 | -0.0009012 | DDX19B, AARS1 |
| chr11 | 71330277 | 71330645 | 369 | 2 | 5.85E-05 | 1.85E-05 | 0.000691 | 2.77E-05 | -0.00694956 | NA |
| chr11 | 8259194 | 8259488 | 295 | 2 | 1.16E-05 | 0.0016738 | 1.66E-05 | 2.77E-05 | 0.016200324 | LMO1 |
| chr19 | 36151818 | 36152630 | 813 | 4 | 1.30E-06 | 4.37E-05 | 0.000913 | 2.77E-05 | 0.0116205 | COX7A1 |
| chr6 | 32172981 | 32173814 | 834 | 6 | 6.49E-06 | 1.02E-05 | 0.007889 | 2.78E-05 | -0.00144054 | AGPAT1 |
| chr7 | 6673046 | 6673789 | 744 | 4 | 1.40E-05 | 0.0004328 | 0.000121 | 2.78E-05 | -0.00573948 | AC073343.2, AC073343.1 |
| chr2 | 8570912 | 8571126 | 215 | 5 | 6.08E-05 | 3.16E-05 | 0.003388 | 2.78E-05 | 0.001164626 | LINC01814 |
| chr16 | 87964004 | 87964475 | 472 | 3 | 2.57E-06 | 6.20E-05 | 8.51E-05 | 2.78E-05 | 0.006339811 | BANP |
| chr8 | 128855243 | 128855609 | 367 | 2 | 1.02E-05 | 0.0006406 | 2.06E-05 | 2.78E-05 | 0.003509233 | CCDC26 |
| chr1 | 232986027 | 232986389 | 363 | 5 | 7.03E-05 | 7.37E-05 | 0.000264 | 2.78E-05 | -0.00093337 | PCNX2 |
| chr19 | 54158998 | 54159548 | 551 | 4 | 1.17E-07 | 0.0001984 | 0.000174 | 2.78E-05 | -0.00154855 | LENG1 |
| chr19 | 58381981 | 58382319 | 339 | 3 | 6.44E-06 | 4.22E-05 | 0.001193 | 2.78E-05 | -0.00689165 | NA |
| chr8 | 94895161 | 94896391 | 1231 | 10 | 5.65E-07 | 0.000379 | 0.004354 | 2.79E-05 | 0.00075689 | NDUFAF6, CCNE2 |
| chr8 | 144311735 | 144312342 | 608 | 3 | 1.72E-06 | 0.0001945 | 3.48E-05 | 2.79E-05 | 0.002775531 | HSF1 |
| chr2 | 11465413 | 11465618 | 206 | 3 | 3.31E-06 | 0.0256903 | 9.89E-06 | 2.79E-05 | -0.00171539 | E2F6 |
| chr20 | 833395 | 834093 | 699 | 6 | 9.21E-06 | 0.0037263 | 0.000124 | 2.79E-05 | 0.001098139 | FAM110A |
| chr6 | 136628833 | 136629257 | 425 | 3 | 1.87E-05 | 0.0007687 | 0.000171 | 2.79E-05 | -0.00859403 | MAP3K5-AS1, MAP3K5 |
| chr10 | 121682493 | 121682812 | 320 | 3 | 3.32E-05 | 0.0002044 | 6.75E-05 | 2.79E-05 | 0.003257208 | NA |
| chr4 | 40908206 | 40908964 | 759 | 3 | 6.36E-07 | 0.030666 | 5.28E-06 | 2.79E-05 | -0.0033339 | APBB2 |
| chr1 | 117985607 | 117985939 | 333 | 2 | 8.23E-05 | 1.79E-05 | 0.00085 | 2.79E-05 | -0.00098082 | SPAG17 |
| chr6 | 131643414 | 131643574 | 161 | 3 | 8.85E-05 | 2.25E-05 | 0.000478 | 2.79E-05 | -0.00476195 | ENPP3 |
| chr7 | 42153207 | 42153887 | 681 | 2 | 3.19E-05 | 1.72E-05 | 0.001071 | 2.79E-05 | -0.00675179 | GLI3 |
| chr4 | 82718452 | 82718536 | 85 | 3 | 0.000109626 | 1.51E-05 | 0.002463 | 2.79E-05 | 0.002496238 | SCD5 |
| chr22 | 37549862 | 37550210 | 349 | 3 | 1.72E-05 | 6.23E-05 | 0.000528 | 2.79E-05 | -0.00356309 | AL022315.1 |
| chr12 | 80187147 | 80187282 | 136 | 2 | 2.10E-05 | 0.0007599 | 1.98E-05 | 2.79E-05 | 0.004375577 | OTOGL |
| chr9 | 110048120 | 110048674 | 555 | 6 | 4.91E-05 | 0.0001971 | 0.00096 | 2.79E-05 | 0.002197921 | PALM2AKAP2 |
| chr17 | 41536015 | 41536109 | 95 | 2 | 5.78E-05 | 2.81E-05 | 0.0002 | 2.80E-05 | -0.00845308 | NA |
| chr8 | 144538028 | 144538378 | 351 | 2 | 2.96E-05 | 5.76E-05 | 7.18E-05 | 2.80E-05 | 0.008227477 | ARHGAP39 |
| chr22 | 46673661 | 46673902 | 242 | 3 | 7.64E-06 | 8.73E-05 | 8.55E-05 | 2.80E-05 | 0.002963264 | GRAMD4 |
| chr5 | 132765011 | 132765068 | 58 | 2 | 9.07E-05 | 1.80E-05 | 0.000844 | 2.80E-05 | 0.001322146 | SEPTIN8 |
| chr9 | 26975799 | 26975927 | 129 | 2 | 3.70E-05 | 0.0001308 | 3.88E-05 | 2.80E-05 | -0.01113036 | IFT74 |
| chr8 | 11680736 | 11681277 | 542 | 3 | 3.31E-05 | 2.52E-05 | 0.000348 | 2.80E-05 | -0.00711858 | GATA4 |
| chr17 | 1829666 | 1830347 | 682 | 6 | 1.28E-05 | 4.85E-05 | 0.00324 | 2.80E-05 | 0.000811268 | RPA1, SMYD4 |
| chr3 | 178386254 | 178386545 | 292 | 3 | 8.97E-05 | 1.43E-05 | 0.001932 | 2.80E-05 | -0.00305393 | KCNMB2 |
| chr16 | 89770338 | 89770659 | 322 | 3 | 4.49E-06 | 3.25E-05 | 0.001353 | 2.80E-05 | -0.00850092 | FANCA |
| chr12 | 7060009 | 7060596 | 588 | 7 | 2.16E-05 | 9.71E-05 | 0.002781 | 2.80E-05 | -0.00564629 | C1S |
| chr13 | 50082023 | 50082541 | 519 | 6 | 2.46E-05 | 0.0008455 | 0.005018 | 2.80E-05 | -0.00239555 | DLEU1, AL137060.6, DLEU2 |
| chr17 | 2045285 | 2045341 | 57 | 2 | 8.14E-05 | 1.75E-05 | 0.000989 | 2.80E-05 | -0.00823493 | NA |
| chr10 | 78292641 | 78293063 | 423 | 3 | 1.02E-05 | 1.50E-05 | 0.00154 | 2.80E-05 | -0.00707076 | AC012560.1, LINC00595 |
| chr5 | 139273530 | 139273877 | 348 | 8 | 1.30E-05 | 0.0003078 | 0.00013 | 2.80E-05 | -0.00190814 | MATR3, SIL1 |
| chr13 | 24254406 | 24254554 | 149 | 2 | 2.97E-05 | 8.29E-05 | 5.23E-05 | 2.80E-05 | -0.00625551 | AL359736.1, SPATA13, SPATA13-AS1 |
| chr16 | 57542356 | 57542950 | 595 | 4 | 1.04E-05 | 0.000305 | 0.000158 | 2.80E-05 | 0.015137609 | ADGRG5 |
| chr4 | 183796893 | 183797248 | 356 | 3 | 2.56E-05 | 0.0001589 | 0.000199 | 2.80E-05 | -0.00863876 | NA |
| chr21 | 37564807 | 37565327 | 521 | 3 | 3.38E-05 | 1.10E-05 | 0.004396 | 2.80E-05 | -0.01439769 | NA |
| chr11 | 16925948 | 16926004 | 57 | 2 | 3.15E-05 | 4.52E-05 | 9.37E-05 | 2.80E-05 | 0.006212821 | PLEKHA7 |
| chr2 | 224646169 | 224646659 | 491 | 2 | 0.00011383 | 1.70E-05 | 0.001185 | 2.81E-05 | 0.000272992 | NA |
| chr11 | 69940220 | 69940408 | 189 | 2 | 4.28E-05 | 4.07E-05 | 0.000107 | 2.81E-05 | -0.01222787 | NA |
| chr19 | 1138907 | 1139148 | 242 | 2 | 2.01E-05 | 0.00054 | 2.19E-05 | 2.81E-05 | -0.0076873 | SBNO2 |
| chr10 | 28218034 | 28218051 | 18 | 2 | 8.16E-05 | 2.56E-05 | 0.000252 | 2.81E-05 | -0.00987396 | MPP7 |
| chr2 | 38960392 | 38960865 | 474 | 6 | 1.51E-05 | 0.0008697 | 0.000171 | 2.81E-05 | -0.00491154 | ARHGEF33 |
| chr6 | 44313295 | 44313611 | 317 | 6 | 4.01E-05 | 0.0005481 | 0.000309 | 2.81E-05 | -0.00030429 | AL353588.1, AARS2 |
| chr2 | 66236360 | 66236529 | 170 | 2 | 3.19E-05 | 1.69E-05 | 0.00126 | 2.81E-05 | -0.00058629 | NA |
| chr6 | 45423046 | 45423628 | 583 | 2 | 1.26E-05 | 1.96E-05 | 0.000585 | 2.81E-05 | 0.002478683 | RUNX2 |
| chr19 | 2926884 | 2927447 | 564 | 4 | 0.000139933 | 1.24E-05 | 0.003803 | 2.81E-05 | -0.00495586 | NA |
| chr5 | 40755568 | 40756410 | 843 | 7 | 9.16E-05 | 3.88E-05 | 0.003156 | 2.82E-05 | -0.00474829 | TTC33 |
| chr7 | 151790649 | 151790942 | 294 | 4 | 1.53E-05 | 1.49E-05 | 0.00354 | 2.82E-05 | 0.003244955 | PRKAG2 |
| chr2 | 120977801 | 120978551 | 751 | 7 | 4.73E-06 | 0.0069795 | 0.000192 | 2.82E-05 | -0.00073171 | GLI2 |
| chr3 | 147393873 | 147394754 | 882 | 7 | 1.26E-06 | 0.0025917 | 1.28E-05 | 2.82E-05 | -0.00361104 | ZIC1, ZIC4 |
| chr17 | 82605343 | 82605858 | 516 | 3 | 1.82E-05 | 2.95E-05 | 0.000327 | 2.82E-05 | -0.00428829 | FOXK2 |
| chr5 | 555569 | 555780 | 212 | 3 | 6.42E-05 | 1.21E-05 | 0.003291 | 2.82E-05 | -0.00159337 | NA |
| chr1 | 1937062 | 1937199 | 138 | 2 | 2.36E-05 | 1.93E-05 | 0.00063 | 2.82E-05 | 0.010232263 | CFAP74 |
| chr17 | 42281786 | 42282112 | 327 | 2 | 2.35E-05 | 3.50E-05 | 0.000136 | 2.82E-05 | -0.00493349 | NA |
| chr2 | 75710870 | 75711633 | 764 | 11 | 3.00E-05 | 4.62E-05 | 0.000441 | 2.82E-05 | 0.00476781 | AC005034.6, GCFC2 |
| chr10 | 4279368 | 4279813 | 446 | 3 | 0.000112432 | 1.26E-05 | 0.003408 | 2.82E-05 | -0.00486924 | NA |
| chr8 | 22433958 | 22433994 | 37 | 3 | 8.12E-05 | 0.000209 | 7.09E-05 | 2.82E-05 | -0.00106048 | SLC39A14 |
| chr20 | 45539335 | 45539545 | 211 | 3 | 7.89E-05 | 2.75E-05 | 0.000539 | 2.82E-05 | -0.00588153 | WFDC6, EPPIN-WFDC6 |
| chr8 | 143732819 | 143733479 | 661 | 5 | 9.57E-06 | 0.0002291 | 0.000634 | 2.82E-05 | 0.003875848 | FAM83H, MIR4664 |
| chr8 | 98046444 | 98046798 | 355 | 3 | 1.13E-06 | 0.00033 | 9.27E-05 | 2.82E-05 | -0.00443274 | RPL30 |
| chr19 | 18650322 | 18650754 | 433 | 7 | 1.38E-05 | 0.0013674 | 7.20E-05 | 2.83E-05 | 0.002157518 | KLHL26 |
| chr14 | 74610682 | 74611580 | 899 | 2 | 2.31E-05 | 2.03E-05 | 0.000515 | 2.83E-05 | -0.00030659 | LTBP2 |
| chr20 | 3916989 | 3917489 | 501 | 5 | 1.40E-06 | 0.0012492 | 7.47E-06 | 2.83E-05 | 0.00290189 | PANK2 |
| chr19 | 55483685 | 55484484 | 800 | 4 | 1.63E-07 | 4.44E-05 | 0.000144 | 2.83E-05 | -0.00619531 | ZNF628 |
| chr10 | 127177633 | 127178074 | 442 | 2 | 5.98E-05 | 1.89E-05 | 0.000694 | 2.83E-05 | 0.003106983 | DOCK1, INSYN2A |
| chr12 | 62466249 | 62466690 | 442 | 10 | 2.69E-06 | 0.0001867 | 0.001584 | 2.83E-05 | 0.001258566 | NA |
| chr2 | 8313016 | 8313399 | 384 | 3 | 1.27E-05 | 0.0001581 | 7.61E-05 | 2.83E-05 | -0.00709907 | LINC00299 |
| chr7 | 123607866 | 123608192 | 327 | 2 | 2.28E-06 | 0.0040467 | 1.49E-05 | 2.83E-05 | -0.01250519 | ASB15, ASB15-AS1 |
| chr19 | 9185151 | 9185491 | 341 | 3 | 3.13E-05 | 4.27E-05 | 0.000145 | 2.83E-05 | -0.00784069 | OR7D2 |
| chr14 | 89900061 | 89900593 | 533 | 2 | 9.20E-05 | 1.77E-05 | 0.001015 | 2.83E-05 | -0.00834069 | AL137230.2, EFCAB11 |
| chr7 | 74231393 | 74231636 | 244 | 5 | 0.000119509 | 1.42E-05 | 0.004049 | 2.84E-05 | -0.00390351 | RFC2 |
| chr11 | 77411461 | 77411884 | 424 | 4 | 3.53E-05 | 8.44E-06 | 0.007993 | 2.84E-05 | 0.002304028 | PAK1 |
| chr10 | 37975653 | 37976012 | 360 | 2 | 1.97E-07 | 0.0049737 | 1.46E-05 | 2.84E-05 | 0.003769466 | ZNF25 |
| chr12 | 131443331 | 131443510 | 180 | 2 | 5.64E-05 | 4.48E-05 | 9.66E-05 | 2.84E-05 | -0.00545507 | NA |
| chr5 | 156923999 | 156924255 | 257 | 4 | 6.41E-05 | 0.0001031 | 0.001047 | 2.84E-05 | -0.0094591 | APOOP1, TIMD4 |
| chr8 | 36780148 | 36780151 | 4 | 2 | 3.56E-05 | 3.65E-05 | 0.000129 | 2.84E-05 | 0.015308288 | NA |
| chr16 | 55876582 | 55876821 | 240 | 3 | 1.68E-05 | 3.80E-05 | 0.000987 | 2.84E-05 | 0.008437181 | CES5A |
| chr8 | 143367746 | 143368439 | 694 | 4 | 5.05E-08 | 0.0047444 | 7.26E-06 | 2.84E-05 | -0.00418799 | RHPN1-AS1 |
| chr9 | 19127699 | 19128184 | 486 | 6 | 4.81E-05 | 6.52E-05 | 0.006363 | 2.84E-05 | -0.00079422 | PLIN2 |
| chr1 | 1339040 | 1339757 | 718 | 2 | 6.66E-05 | 1.75E-05 | 0.001085 | 2.84E-05 | -0.00360735 | DVL1, MIR6808 |
| chr5 | 79774254 | 79774416 | 163 | 2 | 6.65E-05 | 3.30E-05 | 0.000153 | 2.84E-05 | -0.01570171 | CMYA5, AC008496.2 |
| chr7 | 33732123 | 33732248 | 126 | 2 | 0.000132033 | 0.0003645 | 2.52E-05 | 2.84E-05 | -0.00754528 | BBS9 |
| chr20 | 4148474 | 4148836 | 363 | 5 | 8.30E-05 | 4.59E-05 | 0.004906 | 2.84E-05 | 0.003134061 | SMOX |
| chr11 | 67435337 | 67435574 | 238 | 3 | 1.05E-05 | 2.49E-05 | 0.000362 | 2.84E-05 | 0.00591699 | RPS6KB2, RPS6KB2-AS1, PTPRCAP, CORO1B |
| chr2 | 229942244 | 229942709 | 466 | 5 | 5.55E-05 | 0.0003764 | 0.002027 | 2.84E-05 | 0.001273018 | FBXO36 |
| chr11 | 4393686 | 4394157 | 472 | 5 | 3.70E-05 | 2.44E-05 | 0.000519 | 2.84E-05 | -0.01309904 | OR52K3P, TRIM21 |
| chr3 | 143848189 | 143849240 | 1052 | 5 | 4.17E-06 | 0.0001145 | 0.000143 | 2.85E-05 | -0.00125231 | SLC9A9 |
| chr10 | 11162995 | 11163238 | 244 | 2 | 4.36E-05 | 2.21E-05 | 0.000397 | 2.85E-05 | -0.00557355 | CELF2 |
| chr1 | 246586570 | 246587005 | 436 | 2 | 4.54E-05 | 2.01E-05 | 0.000556 | 2.85E-05 | -0.01575511 | CNST |
| chr15 | 22952899 | 22953236 | 338 | 3 | 2.69E-05 | 0.001077 | 0.000257 | 2.85E-05 | -0.00503294 | CYFIP1 |
| chr14 | 30005617 | 30005790 | 174 | 2 | 1.73E-05 | 0.0003169 | 2.66E-05 | 2.85E-05 | -0.00420339 | PRKD1, AL133372.2 |
| chr19 | 54952076 | 54952250 | 175 | 2 | 5.12E-05 | 1.71E-05 | 0.001292 | 2.85E-05 | 0.000116095 | NLRP7 |
| chr6 | 35797438 | 35798284 | 847 | 6 | 8.49E-06 | 2.82E-05 | 0.002657 | 2.85E-05 | -0.00420171 | LHFPL5 |
| chr18 | 11148511 | 11149069 | 559 | 9 | 5.76E-07 | 0.0032731 | 6.51E-06 | 2.85E-05 | -0.00253776 | PIEZO2 |
| chr1 | 197267989 | 197268209 | 221 | 3 | 1.59E-05 | 2.69E-05 | 0.000398 | 2.85E-05 | -0.00236432 | CRB1 |
| chr19 | 7620353 | 7620715 | 363 | 2 | 1.10E-05 | 6.17E-05 | 6.95E-05 | 2.85E-05 | -0.00113574 | XAB2 |
| chr7 | 157594414 | 157594669 | 256 | 4 | 2.96E-05 | 5.08E-05 | 0.001149 | 2.85E-05 | -0.0090005 | PTPRN2 |
| chr20 | 51334422 | 51334817 | 396 | 2 | 1.27E-05 | 7.18E-05 | 6.06E-05 | 2.86E-05 | -0.00775451 | NA |
| chr18 | 52339695 | 52340989 | 1295 | 17 | 3.46E-06 | 0.0015312 | 0.003649 | 2.86E-05 | 0.001585876 | DCC |
| chr10 | 79090692 | 79090848 | 157 | 2 | 1.66E-05 | 0.0186028 | 1.36E-05 | 2.86E-05 | 0.004469993 | ZMIZ1 |
| chr10 | 104253776 | 104254107 | 332 | 3 | 6.53E-07 | 0.0001244 | 0.0008 | 2.86E-05 | 0.006504925 | GSTO1 |
| chr1 | 45622664 | 45623373 | 710 | 6 | 2.27E-05 | 3.69E-05 | 0.005784 | 2.86E-05 | -0.01015271 | CCDC17 |
| chr5 | 140346092 | 140346881 | 790 | 4 | 4.15E-06 | 6.19E-05 | 0.000131 | 2.86E-05 | 3.00E-05 | HBEGF |
| chr11 | 58229227 | 58229328 | 102 | 3 | 6.46E-05 | 4.79E-05 | 0.000883 | 2.86E-05 | 0.002489817 | NA |
| chr10 | 27981925 | 27982267 | 343 | 3 | 6.72E-05 | 2.06E-05 | 0.000558 | 2.86E-05 | -0.00235271 | ARMC4 |
| chr8 | 54328314 | 54328422 | 109 | 2 | 5.09E-05 | 2.64E-05 | 0.000246 | 2.86E-05 | -0.01071858 | NA |
| chr12 | 7066686 | 7067619 | 934 | 5 | 2.04E-06 | 9.17E-05 | 6.49E-05 | 2.86E-05 | -0.00450129 | C1S |
| chr15 | 27768894 | 27769374 | 481 | 4 | 1.05E-05 | 0.0021341 | 7.98E-05 | 2.86E-05 | -0.01362475 | OCA2 |
| chr15 | 52794660 | 52795290 | 631 | 6 | 3.93E-05 | 0.0001884 | 0.000347 | 2.86E-05 | 0.003034046 | NA |
| chr16 | 16014945 | 16015429 | 485 | 2 | 3.68E-06 | 0.0001795 | 3.41E-05 | 2.86E-05 | -0.00311565 | ABCC1 |
| chr21 | 22018891 | 22019121 | 231 | 2 | 7.17E-06 | 0.0441833 | 1.33E-05 | 2.86E-05 | -0.01056144 | LINC01687 |
| chr22 | 47062183 | 47062580 | 398 | 2 | 3.32E-05 | 1.97E-05 | 0.000626 | 2.86E-05 | -0.00088201 | TBC1D22A |
| chr14 | 49688543 | 49689053 | 511 | 7 | 1.89E-06 | 0.0021133 | 4.31E-05 | 2.86E-05 | 0.001681601 | NA |
| chr17 | 42273710 | 42274131 | 422 | 3 | 5.54E-06 | 0.0004541 | 0.000106 | 2.86E-05 | 0.004097099 | STAT5B, AC099811.5 |
| chr6 | 89985504 | 89986299 | 796 | 3 | 9.16E-06 | 5.00E-05 | 0.000158 | 2.86E-05 | -0.00092886 | BACH2 |
| chr1 | 153259086 | 153259984 | 899 | 7 | 6.43E-06 | 0.0040983 | 0.001149 | 2.87E-05 | -0.00182437 | LORICRIN |
| chr11 | 75573598 | 75573744 | 147 | 2 | 8.49E-06 | 1.93E-05 | 0.00068 | 2.87E-05 | -0.00665077 | NA |
| chr8 | 142368157 | 142368439 | 283 | 2 | 2.67E-05 | 0.0001239 | 4.15E-05 | 2.87E-05 | -0.0154086 | TSNARE1 |
| chr22 | 41485244 | 41485317 | 74 | 2 | 5.19E-05 | 0.0001007 | 4.72E-05 | 2.87E-05 | -0.00734307 | ACO2 |
| chr5 | 178679625 | 178679750 | 126 | 2 | 8.64E-05 | 2.31E-05 | 0.000358 | 2.87E-05 | 0.004135606 | NA |
| chr2 | 9720676 | 9720897 | 222 | 2 | 1.71E-05 | 6.76E-05 | 6.45E-05 | 2.87E-05 | 0.002527755 | NA |
| chr21 | 33125103 | 33125110 | 8 | 2 | 0.000172 | 2.10E-05 | 0.000488 | 2.87E-05 | -0.00202947 | NA |
| chr2 | 70469628 | 70469815 | 188 | 3 | 8.57E-05 | 2.56E-05 | 0.000394 | 2.87E-05 | 0.00327636 | TGFA |
| chr6 | 125302946 | 125303427 | 482 | 3 | 7.80E-06 | 0.0001623 | 4.10E-05 | 2.87E-05 | -0.00028203 | NA |
| chr11 | 69636347 | 69636877 | 531 | 3 | 6.39E-05 | 1.61E-05 | 0.002109 | 2.87E-05 | -0.00992598 | NA |
| chr17 | 74652138 | 74652434 | 297 | 3 | 2.51E-05 | 4.39E-05 | 0.000146 | 2.87E-05 | -0.00675149 | NA |
| chr16 | 90047485 | 90048155 | 671 | 7 | 7.26E-06 | 4.54E-05 | 0.00632 | 2.87E-05 | 0.001874544 | URAHP |
| chr21 | 33245580 | 33245632 | 53 | 2 | 6.39E-05 | 1.96E-05 | 0.000644 | 2.87E-05 | 0.004621646 | IFNAR2, AP000295.1 |
| chr10 | 62805882 | 62806012 | 131 | 4 | 5.89E-05 | 8.07E-06 | 0.009428 | 2.87E-05 | -0.0124076 | ADO, AC067751.1 |
| chr5 | 150623475 | 150624018 | 544 | 3 | 3.20E-06 | 0.0010954 | 2.74E-05 | 2.87E-05 | -0.00722608 | SYNPO, AC008453.2 |
| chr15 | 71801009 | 71801150 | 142 | 2 | 2.76E-05 | 7.46E-05 | 5.93E-05 | 2.88E-05 | -0.01227595 | NR2E3 |
| chr11 | 111255261 | 111256065 | 805 | 6 | 6.09E-06 | 0.0001364 | 0.002124 | 2.88E-05 | -0.00365787 | C11orf53 |
| chr1 | 167328353 | 167328379 | 27 | 2 | 2.56E-06 | 0.0001107 | 4.47E-05 | 2.88E-05 | -0.01734985 | POU2F1 |
| chr19 | 16570186 | 16570526 | 341 | 3 | 1.21E-05 | 0.0008509 | 1.61E-05 | 2.88E-05 | -0.00270016 | AC008764.1, SLC35E1 |
| chr10 | 30403684 | 30403736 | 53 | 3 | 6.10E-05 | 7.73E-05 | 7.28E-05 | 2.88E-05 | -0.00040019 | CCND3P1 |
| chr2 | 10490428 | 10490818 | 391 | 2 | 5.71E-06 | 0.0077238 | 1.43E-05 | 2.88E-05 | -0.00674708 | NA |
| chr16 | 17160694 | 17161093 | 400 | 3 | 5.20E-05 | 4.01E-05 | 0.000325 | 2.88E-05 | -0.00564357 | XYLT1 |
| chr1 | 167546239 | 167546800 | 562 | 3 | 9.47E-06 | 4.69E-05 | 0.000159 | 2.88E-05 | -0.00755716 | CREG1 |
| chr4 | 7843191 | 7843255 | 65 | 2 | 7.52E-05 | 1.82E-05 | 0.000964 | 2.88E-05 | -0.00474416 | AFAP1 |
| chr15 | 89223068 | 89223096 | 29 | 2 | 4.11E-05 | 7.46E-05 | 5.96E-05 | 2.88E-05 | -0.00298836 | NA |
| chr6 | 31713785 | 31714249 | 465 | 7 | 1.51E-06 | 0.0031076 | 0.000519 | 2.88E-05 | 0.000653008 | LY6G6F-LY6G6D, AL662899.1, LY6G6E |
| chr12 | 99647263 | 99647580 | 318 | 3 | 3.65E-05 | 2.33E-05 | 0.001065 | 2.88E-05 | 0.002110559 | ANKS1B |
| chr19 | 38681265 | 38681278 | 14 | 2 | 4.36E-05 | 4.01E-05 | 0.000116 | 2.88E-05 | -0.0028875 | ACTN4 |
| chr7 | 65239609 | 65240328 | 720 | 4 | 2.82E-05 | 9.56E-05 | 0.000898 | 2.89E-05 | -0.00076036 | NA |
| chr17 | 39199708 | 39200301 | 594 | 8 | 5.96E-05 | 0.0002659 | 0.003113 | 2.89E-05 | 0.001568984 | RPL19 |
| chr22 | 29855804 | 29856034 | 231 | 3 | 3.77E-05 | 0.0001743 | 4.68E-05 | 2.89E-05 | -0.00556298 | NA |
| chr4 | 84966229 | 84966616 | 388 | 5 | 7.24E-05 | 2.87E-05 | 0.002406 | 2.89E-05 | 0.002040109 | WDFY3-AS2, WDFY3 |
| chr1 | 161119167 | 161119549 | 383 | 4 | 1.86E-05 | 7.52E-05 | 0.0005 | 2.89E-05 | 0.00261016 | NIT1 |

| | | | | | | | | | | |
|---|---|---|---|---|---|---|---|---|---|---|
| chr21 | 46036401 | 46037002 | 602 | 6 | 5.23E-06 | 0.0062797 | 0.000688 | 2.89E-05 | -0.00307999 | NA |
| chr10 | 129915565 | 129915587 | 23 | 2 | 6.76E-05 | 5.04E-05 | 8.75E-05 | 2.89E-05 | -0.0049435 | EBF3 |
| chr3 | 126503798 | 126504300 | 503 | 2 | 3.11E-06 | 0.0001909 | 3.35E-05 | 2.89E-05 | -0.00678743 | UROC1 |
| chr11 | 119691711 | 119692823 | 1113 | 4 | 8.91E-07 | 0.0001068 | 0.000199 | 2.89E-05 | -0.0079937 | NECTIN1 |
| chr11 | 102588951 | 102589129 | 179 | 2 | 2.94E-05 | 0.0003737 | 2.55E-05 | 2.89E-05 | -0.01211427 | MMP20 |
| chr12 | 56424825 | 56425276 | 452 | 3 | 0.000115239 | 1.61E-05 | 0.002426 | 2.90E-05 | 0.007615275 | TIMELESS |
| chr19 | 41439447 | 41439919 | 473 | 10 | 0.000107661 | 0.0002052 | 0.001268 | 2.90E-05 | 0.001561267 | DMAC2 |
| chr16 | 27698199 | 27698380 | 182 | 2 | 2.55E-05 | 0.0005491 | 2.26E-05 | 2.90E-05 | -0.00252985 | KATNIP, AC016597.1 |
| chr2 | 26473170 | 26473657 | 488 | 4 | 3.96E-05 | 0.0016743 | 7.49E-05 | 2.90E-05 | -0.00252464 | OTOF |
| chr13 | 99414114 | 99414181 | 68 | 2 | 8.72E-05 | 2.34E-05 | 0.000359 | 2.90E-05 | -0.00886538 | CCR12P |
| chr17 | 40361389 | 40361661 | 273 | 2 | 2.66E-05 | 1.86E-05 | 0.000863 | 2.90E-05 | -0.01926628 | AC080112.1, GJD3 |
| chr11 | 2753526 | 2753792 | 267 | 2 | 2.38E-05 | 0.0002499 | 2.99E-05 | 2.90E-05 | -0.00783004 | KCNQ1 |
| chr16 | 1555066 | 1555338 | 273 | 3 | 1.70E-05 | 0.0010591 | 0.000106 | 2.90E-05 | -0.00492336 | TMEM204, IFT140 |
| chr14 | 92041169 | 92041434 | 266 | 2 | 7.31E-06 | 0.0002521 | 2.98E-05 | 2.90E-05 | 0.006292173 | NA |
| chr17 | 8376776 | 8377233 | 458 | 6 | 2.97E-05 | 4.51E-05 | 0.003796 | 2.90E-05 | -0.00395245 | AC135178.4, AC135178.3 |
| chr7 | 149474510 | 149475097 | 588 | 4 | 2.49E-05 | 1.24E-05 | 0.004946 | 2.91E-05 | -0.00112953 | ZNF746 |
| chr10 | 132635017 | 132635091 | 75 | 2 | 5.48E-05 | 5.92E-05 | 7.46E-05 | 2.91E-05 | -0.00851482 | INPP5A |
| chr5 | 65722029 | 65722682 | 654 | 11 | 5.11E-05 | 6.21E-05 | 0.005448 | 2.91E-05 | -0.00061878 | NLN, SGTB |
| chr7 | 54339281 | 54339311 | 31 | 2 | 0.00012041 | 0.0001753 | 3.51E-05 | 2.91E-05 | -0.00639319 | LINC01445 |
| chr16 | 88646044 | 88646342 | 299 | 2 | 4.23E-05 | 2.55E-05 | 0.000284 | 2.91E-05 | -0.01031549 | CYBA |
| chr10 | 79275320 | 79275831 | 512 | 2 | 4.49E-05 | 1.78E-05 | 0.001144 | 2.91E-05 | 0.0046889 | ZMIZ1 |
| chr8 | 86342516 | 86343396 | 881 | 10 | 2.33E-05 | 0.0016338 | 0.000745 | 2.91E-05 | 0.000380819 | WWP1, AC103760.1 |
| chr16 | 89265589 | 89265892 | 304 | 3 | 4.05E-05 | 4.90E-05 | 0.000138 | 2.91E-05 | 0.002452546 | NA |
| chr3 | 194097942 | 194097998 | 57 | 2 | 6.93E-05 | 2.68E-05 | 0.000252 | 2.91E-05 | 0.004332278 | AC080129.1 |
| chr10 | 43911421 | 43911765 | 345 | 3 | 3.79E-05 | 0.0002939 | 0.000529 | 2.91E-05 | -0.00533186 | LINC00841, LINC02659 |
| chr18 | 4124505 | 4124514 | 10 | 2 | 4.30E-05 | 3.26E-05 | 0.000168 | 2.91E-05 | -0.00553943 | DLGAP1 |
| chr19 | 33815558 | 33815986 | 429 | 3 | 9.77E-05 | 2.16E-05 | 0.001388 | 2.91E-05 | 0.004134045 | KCTD15 |
| chr1 | 109751277 | 109752029 | 753 | 3 | 9.56E-06 | 8.39E-05 | 7.89E-05 | 2.91E-05 | -0.00368347 | GSTM5, AL158847.1, EPS8L3 |
| chr17 | 43901122 | 43901467 | 346 | 4 | 1.45E-05 | 0.0003001 | 0.000691 | 2.92E-05 | -0.00205693 | MPP2 |
| chr13 | 29359400 | 29359428 | 29 | 2 | 0.000154988 | 2.37E-05 | 0.00035 | 2.92E-05 | -0.00644095 | MTUS2 |
| chr16 | 1158129 | 1158531 | 403 | 2 | 1.36E-05 | 1.96E-05 | 0.000695 | 2.92E-05 | 0.003887557 | CACNA1H |
| chr10 | 44958664 | 44959348 | 685 | 4 | 9.33E-06 | 0.001996 | 1.89E-05 | 2.92E-05 | 0.002025966 | TMEM72-AS1, AL353801.3 |
| chr2 | 17790972 | 17791325 | 354 | 3 | 0.000108734 | 1.68E-05 | 0.001357 | 2.92E-05 | -0.00891496 | SMC6 |
| chr15 | 77070511 | 77071295 | 785 | 9 | 1.50E-05 | 2.01E-05 | 0.007526 | 2.92E-05 | 0.001372801 | TSPAN3 |
| chr10 | 131675662 | 131675703 | 42 | 3 | 0.000134759 | 0.0003321 | 0.00013 | 2.92E-05 | -0.01219415 | NA |
| chr16 | 228853 | 229747 | 895 | 12 | 2.23E-05 | 2.95E-05 | 0.011665 | 2.92E-05 | -0.00092962 | LUC7L |
| chr17 | 409922 | 410267 | 346 | 10 | 0.000104901 | 3.35E-05 | 0.008064 | 2.92E-05 | 0.006827986 | AC141424.1 |
| chr20 | 44883332 | 44883675 | 344 | 2 | 2.49E-06 | 0.008475 | 1.44E-05 | 2.92E-05 | 0.005447032 | AL008725.1 |
| chr14 | 76400207 | 76400584 | 378 | 3 | 5.05E-05 | 0.001181 | 0.000183 | 2.92E-05 | 0.002763145 | ESRRB |
| chr12 | 124660900 | 124661393 | 494 | 3 | 5.06E-05 | 4.58E-05 | 0.001087 | 2.92E-05 | -0.00230153 | NA |
| chr2 | 191020231 | 191020294 | 64 | 2 | 5.24E-05 | 1.77E-05 | 0.001234 | 2.92E-05 | -0.00239161 | STAT1 |
| chr17 | 50966462 | 50966672 | 211 | 3 | 1.54E-05 | 0.006219 | 2.05E-05 | 2.93E-05 | -0.00597973 | SPAG9 |
| chr10 | 94590140 | 94590545 | 406 | 2 | 3.96E-05 | 1.81E-05 | 0.001106 | 2.93E-05 | 0.002497171 | HELLS, AL138759.1 |
| chr8 | 38785824 | 38786074 | 251 | 3 | 0.000124358 | 1.93E-05 | 0.000863 | 2.93E-05 | -0.00237977 | TACC1 |
| chr1 | 71080630 | 71081295 | 666 | 10 | 4.03E-05 | 4.75E-05 | 0.01239 | 2.93E-05 | 0.00050371 | ZRANB2 |
| chr5 | 75337097 | 75337702 | 606 | 8 | 4.21E-05 | 0.0003087 | 0.000144 | 2.93E-05 | 0.000565038 | HMGCR |
| chr2 | 190627928 | 190628309 | 382 | 2 | 7.80E-05 | 1.99E-05 | 0.000672 | 2.93E-05 | 0.013832834 | AC006460.2 |
| chr16 | 489845 | 490031 | 187 | 3 | 6.45E-05 | 1.78E-05 | 0.001419 | 2.94E-05 | 0.001703814 | RAB11FIP3 |
| chr10 | 75712389 | 75712793 | 405 | 3 | 0.000106429 | 1.40E-05 | 0.00192 | 2.94E-05 | -0.01470175 | LRMDA |
| chr1 | 205565860 | 205566104 | 245 | 3 | 3.98E-05 | 0.0014537 | 0.000708 | 2.94E-05 | -0.00585965 | NA |
| chr6 | 41649906 | 41650065 | 160 | 2 | 4.04E-05 | 1.81E-05 | 0.001101 | 2.94E-05 | -0.00231059 | MDFI |
| chr2 | 33279961 | 33280112 | 152 | 2 | 7.59E-05 | 2.87E-05 | 0.000221 | 2.94E-05 | -0.00689561 | LTBP1, AC019127.1 |
| chr16 | 79081658 | 79081979 | 322 | 3 | 1.68E-05 | 0.0004789 | 4.85E-05 | 2.94E-05 | -0.0074396 | WWOX |
| chr12 | 111217573 | 111217843 | 271 | 4 | 1.16E-05 | 0.0005972 | 2.79E-05 | 2.94E-05 | 0.002567921 | CUX2 |
| chr5 | 69217255 | 69217729 | 475 | 8 | 0.000113888 | 0.0006164 | 0.000884 | 2.94E-05 | -0.00247824 | NA |
| chr16 | 46688730 | 46689633 | 904 | 13 | 4.26E-05 | 0.000617 | 0.010965 | 2.94E-05 | 0.001006566 | VPS35 |
| chr3 | 127628500 | 127629312 | 813 | 6 | 2.75E-06 | 0.0421583 | 2.70E-05 | 2.94E-05 | -0.00401173 | PODXL2, AC023593.1 |
| chr2 | 112153219 | 112153328 | 110 | 2 | 5.20E-05 | 4.94E-05 | 9.22E-05 | 2.94E-05 | -0.0039903 | FBLN7 |
| chr8 | 385281 | 385563 | 283 | 3 | 0.000107975 | 1.35E-05 | 0.002526 | 2.95E-05 | -0.00069463 | NA |
| chr8 | 57702696 | 57702911 | 216 | 2 | 7.70E-06 | 0.0003234 | 2.75E-05 | 2.95E-05 | -0.01178198 | AC104051.1 |
| chr14 | 102683675 | 102683963 | 289 | 3 | 1.16E-05 | 0.0103032 | 7.61E-06 | 2.95E-05 | 0.00158179 | RCOR1 |
| chr6 | 158923778 | 158923983 | 206 | 2 | 7.23E-05 | 2.73E-05 | 0.000252 | 2.95E-05 | 0.010950162 | NA |
| chr16 | 89084935 | 89085621 | 687 | 5 | 7.66E-06 | 0.000514 | 0.002081 | 2.95E-05 | 0.003895596 | NA |
| chr11 | 28902018 | 28902249 | 232 | 4 | 5.85E-05 | 9.02E-05 | 0.000157 | 2.95E-05 | -0.00064796 | LINC02742 |
| chr12 | 132374290 | 132374509 | 220 | 2 | 5.68E-05 | 1.95E-05 | 0.000753 | 2.95E-05 | 0.017308214 | NA |
| chr2 | 96702316 | 96702609 | 294 | 2 | 1.89E-05 | 0.0003908 | 2.56E-05 | 2.95E-05 | -0.00434192 | FER1L5 |
| chr4 | 2949154 | 2949564 | 411 | 2 | 6.60E-05 | 3.34E-05 | 0.000166 | 2.95E-05 | 0.004824954 | NOP14-AS1, NOP14 |
| chr9 | 122905259 | 122905665 | 407 | 8 | 3.44E-05 | 0.000197 | 0.000442 | 2.95E-05 | 5.69E-05 | RC3H2 |
| chr1 | 156961895 | 156962146 | 252 | 2 | 1.41E-05 | 9.83E-05 | 5.01E-05 | 2.96E-05 | -0.00633299 | ARHGEF11 |
| chr10 | 132641929 | 132642508 | 580 | 5 | 4.52E-05 | 0.000162 | 4.92E-05 | 2.96E-05 | -0.00084872 | INPP5A |
| chr20 | 3745214 | 3745681 | 468 | 3 | 0.000138731 | 1.46E-05 | 0.002577 | 2.96E-05 | -0.00981949 | HSPA12B |
| chr15 | 92892382 | 92892571 | 190 | 2 | 9.51E-05 | 1.85E-05 | 0.001016 | 2.96E-05 | 0.003925287 | CHASERR, AC013394.1 |
| chr1 | 18795306 | 18795612 | 307 | 4 | 0.000154292 | 9.23E-06 | 0.007583 | 2.96E-05 | 0.014614354 | NA |
| chr12 | 71157078 | 71157250 | 173 | 2 | 4.43E-05 | 1.96E-05 | 0.000749 | 2.96E-05 | -0.01061313 | TSPAN8 |
| chr1 | 246681088 | 246681729 | 642 | 3 | 2.12E-05 | 6.00E-05 | 9.67E-05 | 2.96E-05 | 0.001060189 | NA |
| chr1 | 48760619 | 48761011 | 393 | 3 | 5.17E-05 | 7.38E-05 | 0.000476 | 2.96E-05 | -0.00555109 | AGBL4, BEND5 |
| chr1 | 170285228 | 170285256 | 29 | 2 | 5.76E-05 | 1.83E-05 | 0.001097 | 2.96E-05 | -0.01901015 | LINC01681 |
| chr2 | 8118462 | 8118622 | 161 | 4 | 5.58E-05 | 2.38E-05 | 0.000835 | 2.96E-05 | 0.003781461 | LINC00298, LINC00299 |
| chr13 | 113076953 | 113077561 | 609 | 4 | 1.60E-06 | 0.0060304 | 8.46E-06 | 2.96E-05 | -0.0022706 | MCF2L |
| chr6 | 33892257 | 33893136 | 880 | 5 | 1.99E-05 | 7.15E-05 | 0.00186 | 2.96E-05 | -0.00203002 | AL138889.1, LINC01016 |
| chr5 | 6598477 | 6598797 | 321 | 2 | 1.05E-05 | 0.0026007 | 1.65E-05 | 2.96E-05 | 0.004355543 | LINC01018 |
| chr15 | 77027607 | 77028043 | 437 | 5 | 4.40E-05 | 6.77E-05 | 0.001419 | 2.96E-05 | -0.00062143 | PSTPIP1 |
| chr15 | 83653525 | 83654402 | 878 | 7 | 3.51E-05 | 0.0001187 | 0.001574 | 2.96E-05 | 0.003995759 | ADAMTSL3 |
| chr11 | 62603951 | 62604124 | 174 | 3 | 3.60E-05 | 3.65E-05 | 0.000225 | 2.96E-05 | 0.003241929 | EML3 |
| chr7 | 99434905 | 99435220 | 316 | 5 | 4.64E-05 | 2.35E-05 | 0.004937 | 2.96E-05 | -0.00284479 | PTCD1, ATP5MF-PTCD1 |
| chr17 | 62693854 | 62694011 | 158 | 3 | 4.93E-05 | 1.51E-05 | 0.002476 | 2.97E-05 | -0.00287879 | NA |
| chr5 | 179606494 | 179607188 | 695 | 5 | 4.06E-06 | 0.0009318 | 1.31E-05 | 2.97E-05 | 0.002619346 | RUFY1 |
| chr11 | 67502631 | 67503329 | 699 | 2 | 3.54E-06 | 4.21E-05 | 0.000115 | 2.97E-05 | -0.00079428 | PITPNM1 |
| chr5 | 917776 | 918263 | 488 | 3 | 4.95E-05 | 8.49E-05 | 0.00015 | 2.97E-05 | 0.000170275 | TRIP13 |
| chr15 | 71216442 | 71216781 | 340 | 2 | 2.96E-06 | 0.0006997 | 2.16E-05 | 2.97E-05 | 0.005384708 | THSD4 |
| chr14 | 37434695 | 37434961 | 267 | 4 | 0.00010964 | 1.64E-05 | 0.005594 | 2.97E-05 | 0.000900707 | MIPOL1 |
| chr8 | 42768575 | 42768643 | 69 | 4 | 7.20E-05 | 7.19E-05 | 0.000601 | 2.97E-05 | 0.006684143 | CHRNA6 |
| chr10 | 1662775 | 1662891 | 117 | 3 | 0.000147083 | 6.41E-05 | 0.000117 | 2.97E-05 | 0.01128029 | ADARB2 |
| chr13 | 112452909 | 112453043 | 135 | 2 | 8.85E-06 | 5.28E-05 | 8.75E-05 | 2.98E-05 | -0.00725468 | NA |
| chr8 | 141149982 | 141150136 | 155 | 2 | 0.00012509 | 1.79E-05 | 0.001307 | 2.98E-05 | -0.0052987 | DENND3 |
| chr13 | 32639737 | 32639939 | 203 | 2 | 9.86E-05 | 1.82E-05 | 0.001199 | 2.98E-05 | 0.005225644 | PDS5B |
| chr5 | 39424828 | 39425837 | 1010 | 13 | 2.97E-05 | 8.29E-05 | 0.001012 | 2.98E-05 | -0.00134107 | C9, DAB2 |
| chr1 | 32638603 | 32638738 | 136 | 2 | 3.28E-05 | 0.0003654 | 2.66E-05 | 2.98E-05 | -0.00807471 | ZBTB8OS |
| chr17 | 58360214 | 58360569 | 356 | 3 | 6.31E-05 | 0.0001337 | 0.000195 | 2.98E-05 | -0.00069486 | TSPOAP1-AS1, AC004687.2, RNF43 |
| chr8 | 37878090 | 37878208 | 119 | 3 | 8.20E-06 | 0.0012276 | 2.30E-05 | 2.98E-05 | -0.00530506 | RAB11FIP1 |
| chr9 | 137670496 | 137670500 | 5 | 2 | 4.97E-05 | 2.16E-05 | 0.000518 | 2.98E-05 | -0.00819643 | AL590627.2, EHMT1 |
| chr14 | 70188665 | 70189399 | 735 | 8 | 3.17E-05 | 0.0001004 | 0.00234 | 2.98E-05 | -0.00286706 | AL160191.1, SLC8A3 |
| chr12 | 49903994 | 49904963 | 970 | 11 | 6.05E-06 | 0.0024686 | 0.000112 | 2.98E-05 | 0.002078108 | LINC02395, FAIM2 |
| chr8 | 133694580 | 133694733 | 154 | 2 | 1.75E-05 | 0.0233052 | 1.40E-05 | 2.98E-05 | -0.00284861 | NA |
| chr8 | 80080464 | 80080747 | 284 | 3 | 4.21E-06 | 0.0013277 | 2.85E-05 | 2.98E-05 | 0.005993621 | AC036214.3, TPD52 |
| chr19 | 38893416 | 38894127 | 712 | 2 | 8.94E-06 | 4.65E-05 | 0.000102 | 2.99E-05 | -0.00252409 | SIRT2 |
| chr4 | 3641677 | 3641748 | 72 | 2 | 6.88E-05 | 3.36E-05 | 0.000168 | 2.99E-05 | -0.00928611 | NA |
| chr16 | 84056751 | 84056917 | 167 | 2 | 2.82E-05 | 2.54E-05 | 0.000314 | 2.99E-05 | -0.00316385 | MBTPS1 |
| chr2 | 200963148 | 200963557 | 410 | 6 | 0.000104784 | 5.44E-05 | 0.003185 | 2.99E-05 | 0.001333636 | AC005037.1, ORC2 |
| chr2 | 218710064 | 218711273 | 1210 | 11 | 1.04E-05 | 0.0002975 | 0.009432 | 2.99E-05 | -0.00121007 | TTLL4 |
| chr17 | 41689292 | 41690023 | 732 | 4 | 1.24E-07 | 0.0191313 | 3.07E-06 | 2.99E-05 | -0.00166604 | EIF1 |
| chr20 | 46460366 | 46460479 | 114 | 2 | 4.66E-06 | 0.0003055 | 2.86E-05 | 2.99E-05 | -0.01196461 | AL031686.1 |
| chr19 | 7928888 | 7929560 | 673 | 3 | 9.91E-05 | 7.13E-05 | 0.00025 | 2.99E-05 | -0.00701069 | TIMM44 |
| chr14 | 67541952 | 67542247 | 296 | 2 | 7.92E-05 | 2.13E-05 | 0.00055 | 2.99E-05 | -0.0300364 | PLEKHH1 |
| chr21 | 27360457 | 27360618 | 162 | 2 | 0.000111112 | 1.80E-05 | 0.00132 | 2.99E-05 | -0.01069572 | AP001605.1 |
| chr2 | 73807977 | 73807983 | 7 | 2 | 2.41E-05 | 0.0093819 | 1.46E-05 | 2.99E-05 | -0.00504713 | C2orf78 |
| chr2 | 16202014 | 16202408 | 395 | 3 | 2.42E-05 | 0.0001075 | 0.000294 | 2.99E-05 | 0.000137827 | NA |
| chr15 | 88836177 | 88836458 | 282 | 2 | 5.88E-05 | 2.67E-05 | 0.000278 | 2.99E-05 | -0.00641567 | ACAN |
| chr9 | 69173843 | 69174157 | 315 | 2 | 4.13E-06 | 0.0001675 | 3.72E-05 | 2.99E-05 | 0.002089206 | AL358113.1, TJP2 |
| chr11 | 134640950 | 134641225 | 276 | 2 | 1.89E-05 | 2.29E-05 | 0.000435 | 2.99E-05 | -0.01030025 | NA |
| chr21 | 33229381 | 33229852 | 472 | 12 | 2.08E-05 | 0.0017825 | 0.000929 | 3.00E-05 | -0.00197836 | NA |
| chr9 | 136075181 | 136075501 | 321 | 3 | 2.72E-05 | 1.31E-05 | 0.00352 | 3.00E-05 | -0.01644836 | NACC2 |
| chr4 | 57424234 | 57424585 | 352 | 3 | 5.52E-05 | 1.41E-05 | 0.002098 | 3.00E-05 | -0.01338915 | LINC02380 |
| chr11 | 69702171 | 69702703 | 533 | 4 | 1.11E-05 | 0.0001537 | 0.001345 | 3.00E-05 | 0.00256486 | FGF19 |
| chr6 | 149941627 | 149942058 | 432 | 4 | 2.10E-05 | 0.0002452 | 9.05E-05 | 3.00E-05 | 0.002501616 | ULBP2 |
| chr8 | 134848668 | 134849075 | 408 | 3 | 1.13E-05 | 0.0024092 | 1.56E-05 | 3.00E-05 | -0.00466538 | NA |
| chr4 | 1035622 | 1036266 | 645 | 5 | 7.15E-05 | 8.85E-05 | 0.001219 | 3.00E-05 | -0.00337114 | NA |
| chr19 | 16525825 | 16526498 | 674 | 4 | 3.31E-05 | 2.36E-05 | 0.001066 | 3.00E-05 | 9.15E-05 | AC008764.1, CHERP |
| chr20 | 49058174 | 49058413 | 240 | 2 | 3.52E-05 | 8.22E-05 | 5.86E-05 | 3.01E-05 | 0.004111808 | CSE1L |
| chr1 | 8007292 | 8007298 | 7 | 2 | 9.90E-05 | 0.0001331 | 4.24E-05 | 3.01E-05 | -0.00707744 | ERRFI1 |
| chr9 | 132978484 | 132978859 | 376 | 7 | 0.000133741 | 7.60E-06 | 0.014896 | 3.01E-05 | 0.002059189 | GFI1B |
| chr11 | 65587937 | 65588330 | 394 | 3 | 0.000127668 | 1.43E-05 | 0.002521 | 3.01E-05 | 0.003976625 | EHBP1L1 |
| chr12 | 107320329 | 107320896 | 568 | 2 | 5.29E-06 | 2.75E-05 | 0.000261 | 3.01E-05 | 9.49E-05 | BTBD11 |
| chr9 | 87010520 | 87010832 | 313 | 3 | 8.15E-05 | 1.28E-05 | 0.003508 | 3.01E-05 | 0.000548968 | AL513318.1 |
| chr1 | 41010801 | 41011246 | 446 | 2 | 9.40E-05 | 1.81E-05 | 0.001321 | 3.01E-05 | -0.00218637 | CTPS1 |
| chr2 | 135050799 | 135051047 | 249 | 5 | 9.84E-06 | 0.0026283 | 2.12E-05 | 3.01E-05 | 0.001151896 | NA |
| chr12 | 65556909 | 65557205 | 297 | 4 | 3.55E-05 | 0.0001185 | 0.000532 | 3.01E-05 | -0.00778461 | AC090023.1, AC025419.1 |
| chr19 | 49461703 | 49462006 | 304 | 4 | 3.79E-05 | 0.0002296 | 0.000128 | 3.01E-05 | 0.000889298 | ALDH16A1 |
| chr2 | 8681528 | 8681922 | 395 | 5 | 2.76E-05 | 7.20E-05 | 0.002786 | 3.01E-05 | 0.003281512 | ID2, ID2-AS1 |
| chr22 | 44931492 | 44931515 | 24 | 2 | 7.15E-05 | 2.59E-05 | 0.000306 | 3.01E-05 | -0.00181399 | PHF21B |
| chr7 | 64888755 | 64889328 | 574 | 4 | 3.83E-05 | 0.0004522 | 0.00062 | 3.01E-05 | 0.000913204 | ZNF273, AC073349.1 |
| chr4 | 101790472 | 101791059 | 588 | 7 | 4.86E-05 | 2.85E-05 | 0.00161 | 3.01E-05 | 0.002895325 | BANK1 |
| chr9 | 114162451 | 114162660 | 210 | 5 | 1.43E-05 | 0.0023781 | 5.72E-05 | 3.01E-05 | -0.00222079 | COL27A1 |
| chr17 | 81128632 | 81129169 | 538 | 7 | 3.99E-05 | 0.0016405 | 0.002753 | 3.01E-05 | -0.00083865 | AATK |
| chr12 | 48978204 | 48978691 | 488 | 8 | 8.18E-05 | 1.84E-05 | 0.01159 | 3.01E-05 | -0.00034623 | WNT1 |
| chr3 | 27632301 | 27632310 | 10 | 2 | 8.48E-05 | 2.09E-05 | 0.000622 | 3.01E-05 | -0.00039721 | NA |
| chr2 | 11741099 | 11741336 | 238 | 4 | 6.62E-05 | 4.14E-05 | 0.001682 | 3.02E-05 | -0.00390084 | LPIN1, AC012456.2 |
| chr14 | 70010013 | 70010249 | 237 | 3 | 7.30E-05 | 3.26E-05 | 0.000371 | 3.02E-05 | 0.00182192 | SMOC1 |
| chr17 | 43745222 | 43745532 | 311 | 2 | 8.75E-05 | 1.99E-05 | 0.00079 | 3.02E-05 | -0.01329681 | NA |
| chr3 | 47356719 | 47356784 | 66 | 2 | 8.02E-05 | 2.79E-05 | 0.000256 | 3.02E-05 | -0.00765737 | NA |
| chr4 | 118352672 | 118353193 | 522 | 6 | 1.35E-05 | 0.0002085 | 0.001051 | 3.02E-05 | 0.002169159 | PRSS12 |
| chr4 | 93661613 | 93661967 | 355 | 2 | 4.16E-05 | 4.20E-05 | 0.00012 | 3.02E-05 | -0.00599419 | GRID2 |
| chr3 | 185489948 | 185490764 | 817 | 4 | 3.12E-06 | 0.0008953 | 4.44E-05 | 3.02E-05 | 0.000332068 | TMEM41A |
| chr20 | 51001471 | 51001972 | 502 | 2 | 3.93E-05 | 2.07E-05 | 0.000661 | 3.02E-05 | 0.000359735 | AL050404.1 |
| chr17 | 73092723 | 73093484 | 762 | 10 | 4.82E-05 | 0.0001238 | 0.002442 | 3.02E-05 | -0.00204672 | NA |
| chr17 | 18978594 | 18978826 | 233 | 3 | 8.01E-06 | 0.0143025 | 6.85E-06 | 3.02E-05 | -0.00514932 | SLC5A10, FAM83G |
| chr7 | 108157608 | 108158102 | 495 | 3 | 1.37E-05 | 2.37E-05 | 0.001705 | 3.02E-05 | -0.00592048 | NRCAM |
| chr19 | 17244976 | 17245501 | 526 | 5 | 4.37E-06 | 0.0005035 | 4.40E-05 | 3.02E-05 | -0.00419435 | NR2F6, AC010646.1 |

| | | | | | | | | | | |
|---|---|---|---|---|---|---|---|---|---|---|
| chr2 | 85185139 | 85185233 | 95 | 3 | 2.16E-05 | 4.96E-05 | 0.000286 | 3.03E-05 | 0.001088332 | TCF7L1 |
| chr6 | 32352931 | 32352938 | 8 | 2 | 9.65E-05 | 1.89E-05 | 0.001041 | 3.03E-05 | 0.003646854 | TSBP1-AS1, RNU6-603P, TSBP1 |
| chr5 | 156820137 | 156820404 | 268 | 2 | 9.25E-05 | 1.95E-05 | 0.000896 | 3.03E-05 | -0.01486505 | NA |
| chr22 | 38181571 | 38182172 | 602 | 13 | 3.66E-06 | 0.0032524 | 9.33E-05 | 3.03E-05 | -0.00072511 | PLA2G6 |
| chr12 | 108511934 | 108512241 | 308 | 3 | 6.57E-05 | 1.74E-05 | 0.002443 | 3.03E-05 | -0.00627073 | LINC01498 |
| chr12 | 57864705 | 57865299 | 595 | 2 | 7.80E-08 | 0.0006294 | 2.28E-05 | 3.03E-05 | -0.00861184 | NA |
| chr12 | 26335870 | 26336086 | 217 | 4 | 6.51E-06 | 0.0159051 | 9.42E-06 | 3.03E-05 | 0.005714939 | AC024145.1, ITPR2, AC055720.2 |
| chr19 | 10294407 | 10294550 | 144 | 2 | 1.36E-05 | 2.40E-05 | 0.000394 | 3.03E-05 | -0.02899439 | ICAM5 |
| chr1 | 16167507 | 16167648 | 142 | 2 | 8.08E-05 | 7.99E-05 | 6.08E-05 | 3.03E-05 | -0.00563179 | NA |
| chr3 | 10329869 | 10329907 | 39 | 2 | 1.58E-05 | 0.0001475 | 4.06E-05 | 3.04E-05 | -0.00912657 | ATP2B2 |
| chr16 | 88066096 | 88066134 | 39 | 3 | 9.07E-05 | 1.60E-05 | 0.001867 | 3.04E-05 | 0.000464003 | BANP |
| chr14 | 50321211 | 50321526 | 316 | 2 | 6.21E-05 | 2.01E-05 | 0.000771 | 3.04E-05 | -0.00338645 | DMAC2L |
| chr20 | 37941127 | 37941541 | 415 | 3 | 7.13E-05 | 3.59E-05 | 0.000461 | 3.04E-05 | -0.0087937 | VSTM2L |
| chr1 | 61081822 | 61082637 | 816 | 12 | 9.27E-07 | 0.0578716 | 2.01E-05 | 3.04E-05 | 0.000894187 | NFIA |
| chr19 | 38427489 | 38427613 | 125 | 5 | 4.18E-05 | 0.0002725 | 0.000234 | 3.04E-05 | -0.00678826 | NA |
| chr1 | 13730143 | 13730477 | 335 | 2 | 2.93E-05 | 1.96E-05 | 0.000873 | 3.04E-05 | -0.00570548 | PRDM2 |
| chr19 | 17870883 | 17871146 | 264 | 4 | 2.31E-06 | 0.0040626 | 6.67E-06 | 3.04E-05 | -0.00225228 | NA |
| chr1 | 10643174 | 10643536 | 363 | 2 | 5.83E-05 | 1.99E-05 | 0.000819 | 3.04E-05 | 0.0003166 | AL139423.1, CASZ1 |
| chr15 | 96782495 | 96782955 | 461 | 3 | 1.20E-06 | 0.0005308 | 3.55E-05 | 3.04E-05 | -0.00686395 | SPATA8-AS1 |
| chr7 | 157827993 | 157828890 | 898 | 4 | 3.36E-05 | 1.37E-05 | 0.004188 | 3.04E-05 | -0.00502812 | PTPRN2 |
| chr12 | 91184211 | 91184601 | 391 | 3 | 2.30E-05 | 6.76E-05 | 9.00E-05 | 3.04E-05 | -0.00603859 | NA |
| chr17 | 7627553 | 7628138 | 586 | 10 | 5.30E-05 | 3.46E-05 | 0.005746 | 3.04E-05 | 0.001104478 | SHBG, SAT2 |
| chr3 | 196947886 | 196948339 | 454 | 3 | 0.000115678 | 2.84E-05 | 0.000341 | 3.04E-05 | -0.00106788 | PIGZ |
| chr16 | 402067 | 402547 | 481 | 3 | 3.48E-08 | 0.0003876 | 2.94E-05 | 3.04E-05 | -0.00943438 | NME4, DECR2 |
| chr8 | 85031928 | 85032196 | 269 | 2 | 1.21E-05 | 0.0084688 | 1.50E-05 | 3.05E-05 | -0.00866402 | NA |
| chr1 | 40072880 | 40072933 | 54 | 2 | 4.15E-05 | 0.0001533 | 4.00E-05 | 3.05E-05 | 0.000286901 | PPT1 |
| chr19 | 49590284 | 49590945 | 662 | 4 | 6.76E-05 | 3.43E-05 | 0.00158 | 3.05E-05 | 0.002906718 | PRRG2 |
| chr2 | 170695794 | 170696111 | 318 | 2 | 9.40E-05 | 1.86E-05 | 0.001234 | 3.05E-05 | -0.01390113 | NA |
| chr1 | 47037959 | 47038356 | 398 | 2 | 8.97E-05 | 1.96E-05 | 0.000887 | 3.05E-05 | 0.000732142 | CYP4X1 |
| chr9 | 138075494 | 138075705 | 212 | 3 | 2.86E-05 | 0.0163739 | 6.36E-06 | 3.05E-05 | -0.01030341 | CACNA1B |
| chr4 | 6345787 | 6346629 | 843 | 4 | 1.80E-05 | 1.57E-05 | 0.004209 | 3.05E-05 | -0.00412968 | PPP2R2C |
| chr6 | 137870944 | 137871521 | 578 | 5 | 4.06E-05 | 1.33E-05 | 0.002058 | 3.05E-05 | 0.005567378 | TNFAIP3 |
| chr1 | 220267914 | 220268254 | 341 | 2 | 8.05E-06 | 0.0110116 | 1.47E-05 | 3.05E-05 | -0.00502166 | RAB3GAP2, AURKAP1 |
| chr1 | 3655766 | 3655903 | 138 | 2 | 4.68E-05 | 2.06E-05 | 0.000709 | 3.05E-05 | -0.0031379 | TP73 |
| chr17 | 10638084 | 10638447 | 364 | 4 | 1.45E-05 | 0.0021237 | 1.57E-05 | 3.05E-05 | -0.00015724 | MYH3 |
| chr1 | 204467398 | 204468592 | 1195 | 4 | 5.61E-06 | 7.26E-05 | 0.001914 | 3.05E-05 | -0.00112413 | PIK3C2B |
| chr11 | 118997536 | 118998675 | 1140 | 13 | 3.98E-06 | 0.0042017 | 0.000429 | 3.06E-05 | 0.000158794 | CENATAC, CENATAC-DT |
| chr11 | 19234445 | 19234903 | 459 | 3 | 2.78E-05 | 0.0001312 | 4.91E-05 | 3.06E-05 | -0.00212043 | CSRP3-AS1, E2F8 |
| chr4 | 2732007 | 2732243 | 237 | 4 | 4.13E-05 | 6.66E-05 | 0.000338 | 3.06E-05 | -0.00353862 | FAM193A |
| chr9 | 124941033 | 124941763 | 731 | 9 | 2.71E-05 | 0.0006999 | 0.001777 | 3.06E-05 | 0.000269573 | GOLGA1 |
| chr8 | 144527920 | 144528809 | 890 | 5 | 1.82E-05 | 1.38E-05 | 0.00133 | 3.06E-05 | -0.00712626 | C8orf82 |
| chr2 | 217146924 | 217147168 | 245 | 2 | 1.74E-05 | 0.0017353 | 1.82E-05 | 3.06E-05 | -0.0052292 | NA |
| chr10 | 29735399 | 29736364 | 966 | 7 | 1.42E-05 | 0.0001163 | 8.62E-05 | 3.06E-05 | 0.000986599 | SVIL |
| chr17 | 18984496 | 18984502 | 7 | 2 | 4.85E-05 | 4.46E-05 | 0.000114 | 3.06E-05 | -0.00246346 | SLC5A10, FAM83G |
| chr6 | 138403583 | 138404094 | 512 | 8 | 8.27E-05 | 2.85E-05 | 0.001654 | 3.06E-05 | 0.001731191 | HEBP2 |
| chr8 | 47739090 | 47739261 | 172 | 3 | 1.30E-05 | 1.44E-05 | 0.002662 | 3.06E-05 | 0.016532457 | NA |
| chr6 | 148502115 | 148502649 | 535 | 2 | 3.47E-05 | 2.82E-05 | 0.000263 | 3.07E-05 | 0.00439613 | SASH1 |
| chr11 | 17276185 | 17276800 | 616 | 10 | 8.08E-05 | 0.0001352 | 0.000732 | 3.07E-05 | 0.000613883 | NUCB2 |
| chr2 | 29074565 | 29074851 | 287 | 3 | 6.72E-05 | 6.22E-05 | 0.000229 | 3.07E-05 | -0.00078054 | NA |
| chr11 | 125079031 | 125079284 | 254 | 4 | 4.24E-05 | 4.06E-05 | 0.001115 | 3.07E-05 | -0.01226479 | SLC37A2 |
| chr7 | 8440956 | 8441907 | 952 | 4 | 1.89E-05 | 2.46E-05 | 0.003612 | 3.07E-05 | -0.00639568 | NXPH1 |
| chr8 | 125334287 | 125334598 | 312 | 3 | 6.87E-06 | 0.000663 | 2.12E-05 | 3.07E-05 | 0.004672024 | NSMCE2 |
| chr6 | 49333975 | 49334078 | 104 | 2 | 5.78E-05 | 2.53E-05 | 0.000349 | 3.07E-05 | -0.01421684 | NA |
| chr16 | 83995852 | 83995906 | 55 | 2 | 9.23E-05 | 2.47E-05 | 0.000377 | 3.07E-05 | 0.013268394 | NECAB2 |
| chr5 | 134501339 | 134501582 | 244 | 2 | 1.52E-05 | 0.0010544 | 2.02E-05 | 3.07E-05 | 0.003300619 | NA |
| chr16 | 24876423 | 24876778 | 356 | 2 | 2.71E-05 | 8.89E-05 | 5.71E-05 | 3.07E-05 | -0.00874851 | SLC5A11 |
| chr15 | 26052406 | 26052659 | 254 | 4 | 7.80E-05 | 4.06E-05 | 0.003578 | 3.07E-05 | -0.00575516 | LINC02346 |
| chr12 | 107645913 | 107646752 | 840 | 2 | 2.92E-07 | 0.0032063 | 1.66E-05 | 3.07E-05 | -0.00553264 | BTBD11 |
| chr11 | 93542553 | 93542660 | 108 | 2 | 2.87E-06 | 1.90E-05 | 0.001136 | 3.07E-05 | 0.00325822 | SMCO4 |
| chr8 | 66666972 | 66667381 | 410 | 7 | 6.42E-05 | 0.0002484 | 0.001268 | 3.07E-05 | 0.000949113 | VCPIP1 |
| chr18 | 70359126 | 70359448 | 323 | 2 | 9.54E-06 | 0.0001248 | 4.55E-05 | 3.07E-05 | -0.00280219 | NA |
| chr6 | 125192362 | 125192749 | 388 | 3 | 7.63E-06 | 0.0354428 | 7.85E-06 | 3.07E-05 | -0.00419192 | TPD52L1 |
| chr16 | 83933317 | 83933758 | 442 | 2 | 1.57E-05 | 2.09E-05 | 0.000693 | 3.08E-05 | -0.01187806 | MLYCD, OSGIN1 |
| chr4 | 36286281 | 36286501 | 221 | 3 | 9.45E-05 | 2.19E-05 | 0.00138 | 3.08E-05 | 0.01413247 | DTHD1 |
| chr3 | 194075184 | 194075298 | 115 | 2 | 4.69E-05 | 6.51E-05 | 7.47E-05 | 3.08E-05 | 0.002810141 | AC024559.2 |
| chr13 | 45970409 | 45970460 | 52 | 2 | 3.55E-05 | 3.45E-05 | 0.000174 | 3.08E-05 | -0.00546952 | ZC3H13 |
| chr16 | 67165772 | 67166589 | 818 | 8 | 2.41E-05 | 0.0002461 | 0.003539 | 3.08E-05 | -0.00453952 | AC074143.1, HSF4 |
| chr8 | 54381976 | 54382322 | 347 | 6 | 7.83E-05 | 6.21E-05 | 0.001477 | 3.08E-05 | -0.00495332 | AC027250.2 |
| chr10 | 93961875 | 93962756 | 882 | 3 | 2.65E-05 | 1.72E-05 | 0.002304 | 3.08E-05 | -0.00302369 | NA |
| chr1 | 44153555 | 44153747 | 193 | 3 | 5.09E-05 | 0.0001232 | 0.00039 | 3.08E-05 | -0.00591034 | NA |
| chr8 | 21780843 | 21781272 | 430 | 2 | 4.34E-05 | 1.85E-05 | 0.001365 | 3.08E-05 | -0.0085198 | GFRA2 |
| chr18 | 75370615 | 75370617 | 3 | 2 | 4.89E-05 | 0.0001028 | 5.17E-05 | 3.08E-05 | -0.00508906 | NA |
| chr2 | 44299404 | 44299980 | 577 | 3 | 1.35E-05 | 0.0010736 | 7.40E-05 | 3.08E-05 | -0.00798913 | AC013717.1, SLC3A1 |
| chr8 | 21919112 | 21920061 | 950 | 8 | 1.01E-05 | 0.0012827 | 0.000892 | 3.08E-05 | -0.0008734 | XPO7 |
| chr8 | 52564985 | 52566064 | 1080 | 8 | 6.04E-05 | 3.47E-05 | 0.005633 | 3.08E-05 | 0.00306439 | ALKAL1 |
| chr13 | 20987846 | 20988099 | 254 | 2 | 2.02E-05 | 2.21E-05 | 0.000558 | 3.08E-05 | 0.001110496 | LATS2 |
| chr1 | 155261975 | 155262588 | 614 | 13 | 3.74E-05 | 0.0004215 | 0.001495 | 3.08E-05 | 0.001063582 | SCAMP3 |
| chr1 | 205514569 | 205514718 | 150 | 2 | 7.17E-06 | 0.0241481 | 1.44E-05 | 3.09E-05 | 0.003576886 | CDK18 |
| chr5 | 120160081 | 120160262 | 182 | 2 | 1.68E-05 | 0.0234941 | 1.44E-05 | 3.09E-05 | -0.00526977 | NA |
| chr7 | 985037 | 985482 | 446 | 4 | 7.36E-06 | 0.0003608 | 3.86E-05 | 3.09E-05 | -0.00499352 | CYP2W1 |
| chr5 | 172849440 | 172849655 | 216 | 2 | 9.45E-05 | 2.48E-05 | 0.000376 | 3.09E-05 | 0.005390572 | ERGIC1 |
| chr12 | 6771084 | 6771474 | 391 | 3 | 2.26E-05 | 8.25E-05 | 0.000238 | 3.09E-05 | -0.0025203 | NA |
| chr7 | 71911122 | 71911396 | 275 | 2 | 2.72E-05 | 9.30E-05 | 5.58E-05 | 3.09E-05 | -0.00170151 | CALN1 |
| chr7 | 23679834 | 23680042 | 209 | 2 | 3.40E-05 | 0.0001807 | 3.73E-05 | 3.09E-05 | 0.000189756 | NA |
| chr2 | 240125984 | 240126248 | 265 | 2 | 2.45E-05 | 0.0001042 | 5.14E-05 | 3.09E-05 | -0.00080151 | NA |
| chr11 | 59143594 | 59144928 | 1335 | 13 | 7.22E-06 | 0.0109511 | 3.38E-05 | 3.09E-05 | 0.001090884 | FAM111A |
| chr14 | 60169892 | 60170526 | 635 | 3 | 0.000101717 | 0.0002234 | 0.000304 | 3.09E-05 | -0.00219269 | NA |
| chr22 | 48279308 | 48279649 | 342 | 2 | 2.64E-05 | 1.98E-05 | 0.000916 | 3.09E-05 | -0.00545983 | NA |
| chr14 | 68685707 | 68686119 | 413 | 3 | 2.06E-06 | 0.0486894 | 5.81E-06 | 3.09E-05 | 0.005527739 | RAD51B |
| chr16 | 88891903 | 88892194 | 292 | 4 | 2.48E-05 | 0.0004899 | 3.50E-05 | 3.09E-05 | -0.00010477 | CBFA2T3 |
| chr14 | 21779974 | 21780296 | 323 | 2 | 6.78E-05 | 1.98E-05 | 0.000942 | 3.09E-05 | -0.00861843 | NA |
| chr9 | 37576423 | 37576961 | 539 | 5 | 6.07E-05 | 1.27E-05 | 0.007289 | 3.09E-05 | 0.004063726 | AL513165.2 |
| chr12 | 122888459 | 122888626 | 168 | 2 | 5.05E-05 | 7.23E-05 | 6.85E-05 | 3.09E-05 | 0.008904738 | VPS37B, AC027290.2 |
| chr16 | 68247283 | 68248076 | 794 | 4 | 8.65E-06 | 0.001195 | 0.000124 | 3.10E-05 | -0.00259069 | PLA2G15 |
| chr7 | 100125539 | 100125561 | 23 | 2 | 0.000141605 | 0.0002464 | 3.25E-05 | 3.10E-05 | -0.00503684 | AC073842.2 |
| chr19 | 2050860 | 2051278 | 419 | 7 | 8.88E-06 | 0.0001926 | 0.00735 | 3.10E-05 | 0.001646965 | MKNK2 |
| chr2 | 26770762 | 26771110 | 349 | 3 | 3.04E-05 | 0.0001418 | 0.000142 | 3.10E-05 | -0.00364211 | SLC35F6, CENPA |
| chr3 | 71541655 | 71542278 | 624 | 2 | 0.000146338 | 1.87E-05 | 0.001338 | 3.10E-05 | -0.01197111 | FOXP1, AC097634.4, MIR1284 |
| chr8 | 73448683 | 73448815 | 133 | 4 | 7.94E-05 | 0.0002282 | 6.98E-05 | 3.10E-05 | -0.00201637 | STAU2 |
| chr11 | 120284635 | 120284929 | 295 | 2 | 0.00010432 | 1.89E-05 | 0.001257 | 3.10E-05 | -0.01115315 | POU2F3 |
| chr16 | 89858467 | 89858589 | 123 | 2 | 3.63E-05 | 2.68E-05 | 0.000311 | 3.10E-05 | -0.00613377 | SPIRE2 |
| chr21 | 38784485 | 38784532 | 48 | 2 | 7.89E-05 | 4.28E-05 | 0.000125 | 3.11E-05 | -0.00235809 | NA |
| chr5 | 66143692 | 66144283 | 592 | 10 | 1.62E-05 | 7.27E-05 | 0.00898 | 3.11E-05 | 0.000193959 | SREK1, AC025442.1 |
| chr20 | 62347406 | 62347647 | 242 | 2 | 2.46E-05 | 0.000462 | 2.57E-05 | 3.11E-05 | -0.0051679 | LAMA5 |
| chr5 | 126756595 | 126756891 | 297 | 2 | 2.61E-05 | 5.75E-05 | 8.62E-05 | 3.11E-05 | 0.001600726 | LMNB1-DT |
| chr16 | 66426816 | 66427172 | 357 | 5 | 6.81E-05 | 0.0005341 | 0.001825 | 3.11E-05 | 0.00403485 | NA |
| chr17 | 17224520 | 17225127 | 608 | 2 | 3.27E-08 | 0.0411112 | 1.43E-05 | 3.11E-05 | -0.0021597 | AC055811.2, FLCN |
| chr8 | 141874488 | 141874615 | 128 | 2 | 8.31E-05 | 3.07E-05 | 0.000226 | 3.11E-05 | 0.00478038 | NA |
| chr15 | 68621743 | 68622193 | 451 | 2 | 1.75E-05 | 2.91E-05 | 0.000255 | 3.11E-05 | 0.006724611 | CORO2B |
| chr15 | 39583454 | 39584047 | 594 | 3 | 1.07E-05 | 0.0008661 | 2.51E-05 | 3.11E-05 | -0.00444913 | THBS1 |
| chr12 | 124795205 | 124795272 | 68 | 2 | 7.85E-05 | 3.72E-05 | 0.000156 | 3.11E-05 | -0.00084507 | AC073593.2, SCARB1 |
| chr10 | 28668500 | 28668805 | 306 | 2 | 7.61E-05 | 1.96E-05 | 0.00104 | 3.12E-05 | 0.004263556 | NA |
| chr6 | 106694532 | 106694612 | 81 | 2 | 2.98E-05 | 3.33E-05 | 0.000192 | 3.12E-05 | -0.00125331 | NA |
| chr21 | 33858974 | 33859109 | 136 | 2 | 2.65E-05 | 0.0008202 | 2.18E-05 | 3.12E-05 | -0.00430817 | ITSN1, AP000311.1 |
| chr5 | 154037606 | 154038365 | 760 | 2 | 5.61E-05 | 1.90E-05 | 0.00127 | 3.12E-05 | -0.0006257 | FAM114A2 |
| chr5 | 140664397 | 140664477 | 81 | 3 | 0.000115049 | 2.84E-05 | 0.001683 | 3.12E-05 | 0.000618658 | NA |
| chr11 | 64000560 | 64000568 | 9 | 2 | 6.79E-05 | 6.71E-05 | 7.44E-05 | 3.12E-05 | 0.003309316 | OTUB1, MACROD1 |
| chr16 | 89115399 | 89115533 | 135 | 4 | 5.65E-05 | 4.73E-05 | 0.001794 | 3.13E-05 | -0.00014648 | ACSF3 |
| chr3 | 23805529 | 23805934 | 406 | 5 | 6.71E-05 | 1.26E-05 | 0.0065 | 3.13E-05 | -0.0001176 | UBE2E1-AS1 |
| chr9 | 136694196 | 136694961 | 766 | 15 | 3.86E-05 | 0.0002157 | 0.002573 | 3.13E-05 | -0.0044546 | NA |
| chr1 | 90003411 | 90003768 | 358 | 3 | 6.92E-06 | 0.0107317 | 6.95E-06 | 3.13E-05 | -0.00917431 | ZNF326 |
| chr4 | 117576845 | 117576868 | 24 | 2 | 9.75E-06 | 2.79E-05 | 0.000289 | 3.13E-05 | 0.014641909 | LINC01378 |
| chr16 | 85249983 | 85250391 | 409 | 5 | 1.97E-05 | 0.0004263 | 0.000681 | 3.13E-05 | -0.00338715 | GSE1, AC092275.1 |
| chr4 | 2006979 | 2008052 | 1074 | 4 | 4.70E-06 | 0.0004878 | 0.000732 | 3.13E-05 | -0.00215452 | NELFA |
| chr1 | 201222807 | 201223158 | 352 | 3 | 3.86E-05 | 1.73E-05 | 0.0015 | 3.13E-05 | -0.00348303 | IGFN1, AC103925.1 |
| chr1 | 37690379 | 37690980 | 602 | 11 | 1.28E-05 | 0.0001848 | 0.000103 | 3.14E-05 | -0.0086985 | C1orf109 |
| chr10 | 68333238 | 68334009 | 772 | 5 | 1.24E-06 | 0.0001284 | 0.000542 | 3.14E-05 | 0.004767453 | HNRNPH3 |
| chr11 | 33353866 | 33354219 | 354 | 3 | 6.98E-06 | 0.0076307 | 7.92E-06 | 3.14E-05 | -0.01022948 | HIPK3 |
| chr14 | 100961703 | 100962127 | 425 | 3 | 8.45E-05 | 2.36E-05 | 0.000642 | 3.14E-05 | -0.00216898 | MEG8 |
| chr7 | 3094923 | 3095308 | 386 | 3 | 8.12E-05 | 2.06E-05 | 0.00086 | 3.14E-05 | 0.008075161 | AC024028.1 |
| chr2 | 196316349 | 196316351 | 3 | 2 | 2.37E-05 | 0.0497985 | 1.44E-05 | 3.14E-05 | -0.00940066 | HECW2 |
| chr6 | 2764970 | 2765711 | 742 | 5 | 1.60E-05 | 0.0003718 | 7.35E-05 | 3.14E-05 | 0.000917851 | WRNIP1 |
| chr17 | 39926872 | 39927535 | 664 | 6 | 3.17E-05 | 3.90E-05 | 0.002024 | 3.14E-05 | -0.00222825 | ORMDL3 |
| chr6 | 49950608 | 49950622 | 15 | 2 | 1.75E-05 | 3.25E-05 | 0.000207 | 3.14E-05 | -0.01066866 | NA |
| chr10 | 26810257 | 26810742 | 486 | 2 | 0.000119711 | 1.91E-05 | 0.001303 | 3.15E-05 | 0.000794291 | ABI1 |
| chr11 | 96045986 | 96046290 | 305 | 2 | 0.000102299 | 1.95E-05 | 0.001145 | 3.15E-05 | -0.02703307 | MAML2 |
| chr19 | 42697970 | 42698160 | 191 | 2 | 6.52E-05 | 2.09E-05 | 0.000775 | 3.15E-05 | 0.031507483 | NA |
| chr11 | 112356008 | 112356057 | 50 | 2 | 0.000108449 | 1.89E-05 | 0.001382 | 3.15E-05 | 0.000121397 | LINC02762 |
| chr8 | 58513316 | 58513743 | 428 | 3 | 9.90E-05 | 1.67E-05 | 0.002282 | 3.16E-05 | -0.00908051 | NA |
| chr18 | 76822612 | 76823678 | 1067 | 5 | 6.02E-07 | 0.0022749 | 8.90E-05 | 3.16E-05 | -0.00266763 | ZNF236 |
| chr8 | 141142242 | 141142440 | 199 | 2 | 2.91E-05 | 5.00E-05 | 0.000103 | 3.16E-05 | -0.00663498 | DENND3 |
| chr6 | 85643969 | 85644300 | 332 | 4 | 2.84E-05 | 0.0070731 | 5.93E-05 | 3.16E-05 | -0.0016545 | SYNCRIP |
| chr20 | 19977248 | 19977548 | 301 | 2 | 3.26E-05 | 1.91E-05 | 0.001327 | 3.16E-05 | -0.00728367 | RIN2 |
| chr9 | 33222000 | 33222316 | 317 | 3 | 1.84E-05 | 3.63E-05 | 0.000287 | 3.16E-05 | 0.004248772 | SPINK4 |
| chr20 | 31967180 | 31968105 | 926 | 7 | 4.89E-06 | 0.0078293 | 0.000127 | 3.17E-05 | -0.00144208 | NA |
| chr21 | 27449035 | 27449242 | 208 | 2 | 5.24E-05 | 6.98E-05 | 7.31E-05 | 3.17E-05 | -0.00790123 | NA |
| chr2 | 196988089 | 196988180 | 92 | 2 | 0.000107685 | 1.93E-05 | 0.00125 | 3.17E-05 | 0.003452518 | ANKRD44 |
| chr13 | 19588033 | 19588231 | 199 | 2 | 9.01E-06 | 0.0001267 | 4.68E-05 | 3.17E-05 | -0.00581741 | LINC00350 |
| chr17 | 192808 | 193068 | 261 | 2 | 7.57E-05 | 1.97E-05 | 0.001102 | 3.17E-05 | 0.01416664 | NA |
| chr14 | 56676016 | 56676069 | 54 | 2 | 4.09E-05 | 0.0005077 | 2.55E-05 | 3.17E-05 | -0.00877406 | AL161757.5 |
| chr12 | 119177548 | 119177703 | 156 | 2 | 1.29E-05 | 2.21E-05 | 0.000624 | 3.17E-05 | -0.01296228 | HSPB8 |
| chr11 | 2792068 | 2792298 | 231 | 2 | 7.50E-05 | 2.30E-05 | 0.000539 | 3.17E-05 | -0.02473269 | KCNQ1 |
| chr1 | 196466403 | 196466448 | 46 | 2 | 0.000122729 | 3.41E-05 | 0.000192 | 3.17E-05 | -0.01722332 | KCNT2 |
| chr9 | 114348724 | 114349083 | 360 | 3 | 9.83E-06 | 0.0003512 | 0.000237 | 3.18E-05 | -0.00937325 | AKNA |
| chr17 | 7404078 | 7405284 | 1207 | 14 | 2.14E-07 | 0.0005381 | 0.000372 | 3.18E-05 | -0.00033026 | NLGN2, TMEM256-PLSCR3, TMEM256 |
| chr6 | 31826525 | 31826982 | 458 | 4 | 8.37E-06 | 6.24E-05 | 0.001446 | 3.18E-05 | -0.00769312 | NA |
| chr4 | 140755695 | 140756081 | 387 | 4 | 2.35E-06 | 0.011376 | 5.44E-06 | 3.18E-05 | -0.00680059 | TBC1D9 |
| chr9 | 133814536 | 133814863 | 328 | 2 | 2.42E-05 | 9.17E-05 | 5.86E-05 | 3.18E-05 | -0.00881739 | VAV2 |

| | | | | | | | | | | |
|---|---|---|---|---|---|---|---|---|---|---|
| chr2 | 16049999 | 16050694 | 696 | 5 | 5.19E-05 | 9.40E-06 | 0.004921 | 3.18E-05 | -0.00098005 | GACAT3, AC010145.1 |
| chr11 | 59181818 | 59181990 | 173 | 6 | 0.000121895 | 1.67E-05 | 0.005591 | 3.18E-05 | 0.001109759 | DTX4 |
| chr6 | 41027649 | 41028353 | 705 | 6 | 5.93E-06 | 0.0007978 | 0.000398 | 3.18E-05 | -0.00460625 | UNC5CL |
| chr16 | 2161632 | 2161983 | 352 | 2 | 2.45E-05 | 3.28E-05 | 0.000209 | 3.18E-05 | -0.00586775 | TRAF7 |
| chr17 | 82248340 | 82249044 | 705 | 3 | 2.69E-05 | 6.55E-05 | 0.000397 | 3.18E-05 | -0.00292843 | SLC16A3, CSNK1D |
| chr1 | 34111571 | 34111881 | 311 | 2 | 1.72E-05 | 0.00062 | 2.41E-05 | 3.18E-05 | -0.01285001 | CSMD2 |
| chr19 | 37468068 | 37469205 | 1138 | 10 | 1.79E-06 | 0.0012611 | 0.002081 | 3.18E-05 | -0.00150016 | ZNF570, ZNF569 |
| chr17 | 3896972 | 3897100 | 129 | 2 | 3.04E-05 | 0.0008491 | 2.21E-05 | 3.18E-05 | -0.00485068 | P2RX1 |
| chr1 | 15730668 | 15731534 | 867 | 2 | 4.53E-05 | 1.93E-05 | 0.001328 | 3.18E-05 | -0.005185 | PLEKHM2, AL450998.2 |
| chr3 | 119579236 | 119579585 | 350 | 8 | 1.39E-05 | 0.0005397 | 0.000292 | 3.19E-05 | -0.00430844 | ADPRH, AC073352.2 |
| chr14 | 91303708 | 91304074 | 367 | 4 | 3.53E-05 | 0.0021894 | 0.000119 | 3.19E-05 | -0.00448831 | CCDC88C |
| chr6 | 52163260 | 52163598 | 339 | 2 | 8.27E-05 | 2.26E-05 | 0.000587 | 3.19E-05 | 0.009201924 | NA |
| chr19 | 47331745 | 47332218 | 474 | 7 | 1.47E-05 | 0.0020003 | 8.33E-05 | 3.19E-05 | -0.00411046 | C5AR2 |
| chr20 | 46901846 | 46901874 | 29 | 2 | 2.78E-05 | 2.02E-05 | 0.000991 | 3.19E-05 | -0.00803347 | EYA2 |
| chr10 | 24618287 | 24618421 | 135 | 2 | 7.52E-05 | 4.12E-05 | 0.00014 | 3.19E-05 | -0.00677911 | ARHGAP21 |
| chr1 | 179818627 | 179818833 | 207 | 2 | 9.96E-06 | 0.0010605 | 2.09E-05 | 3.19E-05 | 0.002890571 | NA |
| chr20 | 64063821 | 64064781 | 961 | 5 | 1.35E-06 | 0.0007815 | 0.000866 | 3.20E-05 | 0.008910702 | TCEA2 |
| chr2 | 109568118 | 109568250 | 133 | 3 | 4.74E-05 | 0.0001902 | 4.23E-05 | 3.20E-05 | 0.001489387 | SEPTIN10 |
| chr11 | 123493872 | 123494112 | 241 | 2 | 9.24E-05 | 2.21E-05 | 0.000646 | 3.20E-05 | 0.008655989 | GRAMD1B |
| chr8 | 27430522 | 27430888 | 367 | 7 | 5.46E-05 | 8.21E-05 | 0.000657 | 3.20E-05 | 0.0013244 | PTK2B |
| chr3 | 157500014 | 157500470 | 457 | 4 | 1.07E-06 | 7.73E-05 | 0.000144 | 3.20E-05 | 0.003705711 | VEPH1 |
| chr6 | 157276295 | 157276741 | 447 | 3 | 3.93E-05 | 1.25E-05 | 0.004952 | 3.20E-05 | -0.00408562 | NA |
| chr11 | 1000888 | 1001652 | 765 | 4 | 4.98E-06 | 7.03E-05 | 0.002553 | 3.20E-05 | -8.51E-05 | AP2A2 |
| chr10 | 128431671 | 128431795 | 125 | 3 | 5.46E-05 | 0.0002098 | 0.00053 | 3.20E-05 | 0.010229361 | NA |
| chr6 | 116111765 | 116112116 | 352 | 2 | 8.95E-05 | 1.94E-05 | 0.001302 | 3.20E-05 | -0.00018486 | NT5DC1 |
| chr17 | 68734775 | 68734785 | 11 | 2 | 6.75E-05 | 8.29E-05 | 6.40E-05 | 3.20E-05 | 0.002520641 | LINC01482 |
| chr18 | 13801144 | 13801272 | 129 | 3 | 0.000161541 | 3.40E-05 | 0.000641 | 3.20E-05 | 0.005416055 | NA |
| chr14 | 21663733 | 21663807 | 75 | 3 | 4.93E-05 | 0.0002344 | 9.20E-05 | 3.20E-05 | -0.01011269 | OR4E2 |
| chr17 | 3768951 | 3768989 | 39 | 2 | 6.00E-05 | 8.56E-05 | 6.25E-05 | 3.20E-05 | 0.007319827 | ITGAE |
| chr5 | 14492266 | 14492711 | 446 | 4 | 0.000132939 | 0.000287 | 0.000585 | 3.21E-05 | 0.00115542 | TRIO |
| chr1 | 165530260 | 165530364 | 105 | 2 | 4.32E-05 | 0.0002884 | 3.17E-05 | 3.21E-05 | -0.01395159 | LRRC52-AS1 |
| chr3 | 50530414 | 50530825 | 412 | 2 | 5.54E-06 | 0.0039414 | 1.68E-05 | 3.21E-05 | -0.00646716 | NA |
| chr9 | 95289482 | 95290068 | 587 | 2 | 7.40E-05 | 1.97E-05 | 0.001208 | 3.21E-05 | 0.002976103 | FANCC |
| chr6 | 42452771 | 42453033 | 263 | 2 | 7.51E-05 | 2.31E-05 | 0.000556 | 3.21E-05 | 0.000477441 | NA |
| chr14 | 92908627 | 92908931 | 305 | 2 | 8.28E-05 | 1.95E-05 | 0.001302 | 3.21E-05 | -0.00119388 | NA |
| chr7 | 107961211 | 107961238 | 28 | 2 | 3.91E-05 | 0.0007061 | 2.34E-05 | 3.21E-05 | -0.00380427 | LAMB1 |
| chr2 | 37188487 | 37188604 | 118 | 2 | 6.92E-05 | 1.97E-05 | 0.001218 | 3.21E-05 | -4.51E-05 | SULT6B1 |
| chr11 | 3143401 | 3143621 | 221 | 3 | 6.17E-06 | 0.0110476 | 7.13E-06 | 3.21E-05 | -0.00388309 | OSBPL5 |
| chr11 | 22337518 | 22338401 | 884 | 10 | 9.51E-06 | 0.0003302 | 0.000748 | 3.21E-05 | -0.00241539 | SLC17A6, AC104009.1 |
| chr13 | 41645083 | 41645340 | 258 | 2 | 1.49E-05 | 6.48E-05 | 8.06E-05 | 3.21E-05 | 0.004822102 | VWA8 |
| chr1 | 153969213 | 153969428 | 216 | 3 | 4.87E-05 | 2.16E-05 | 0.001689 | 3.21E-05 | -0.00528503 | CREB3L4, AL358472.7 |
| chr22 | 43777565 | 43777842 | 278 | 2 | 2.06E-05 | 0.0006087 | 2.45E-05 | 3.21E-05 | 0.016772378 | EFCAB6 |
| chr2 | 203781086 | 203781268 | 183 | 3 | 4.74E-06 | 0.0116083 | 7.74E-06 | 3.21E-05 | 0.002223691 | NA |
| chr12 | 2927732 | 2927869 | 138 | 2 | 6.30E-05 | 0.0003685 | 2.89E-05 | 3.21E-05 | 0.005415456 | TULP3 |
| chr16 | 69120931 | 69121189 | 259 | 3 | 8.75E-05 | 1.37E-05 | 0.003983 | 3.21E-05 | -0.00082523 | DERPC, CHTF8 |
| chr1 | 109655234 | 109656109 | 876 | 9 | 2.03E-05 | 8.82E-05 | 0.009916 | 3.21E-05 | 0.001704398 | GSTM4 |
| chr15 | 59292496 | 59292749 | 254 | 2 | 1.85E-05 | 4.74E-05 | 0.000115 | 3.21E-05 | 0.000626818 | MYO1E |
| chr9 | 106588797 | 106588867 | 71 | 3 | 2.17E-05 | 0.0034489 | 1.02E-05 | 3.21E-05 | -0.00549305 | LINC01505, AL451140.1 |
| chr10 | 132977819 | 132978198 | 380 | 3 | 3.50E-06 | 0.002429 | 1.15E-05 | 3.21E-05 | -0.00936599 | NA |
| chr12 | 48787273 | 48787771 | 499 | 2 | 3.52E-07 | 0.0001709 | 4.04E-05 | 3.22E-05 | 0.005421502 | ADCY6 |
| chr18 | 37253376 | 37253975 | 600 | 3 | 1.12E-05 | 0.0001148 | 0.00012 | 3.22E-05 | -0.00184175 | CELF4 |
| chr1 | 237168053 | 237168249 | 197 | 2 | 4.60E-05 | 0.0001166 | 5.04E-05 | 3.22E-05 | -0.00796106 | RYR2 |
| chr15 | 62697736 | 62698271 | 536 | 6 | 1.94E-05 | 0.00195 | 0.000105 | 3.22E-05 | -0.00065874 | TLN2 |
| chr6 | 28137325 | 28137639 | 315 | 3 | 7.96E-06 | 0.0011464 | 0.000532 | 3.22E-05 | 0.000732466 | AL358933.1 |
| chr12 | 123787886 | 123788500 | 615 | 3 | 3.06E-05 | 2.10E-05 | 0.0008 | 3.22E-05 | 0.000673516 | DNAH10 |
| chr12 | 34365632 | 34366022 | 391 | 3 | 8.78E-06 | 4.90E-05 | 0.000166 | 3.22E-05 | -0.0102679 | NA |
| chr22 | 43197979 | 43198333 | 355 | 2 | 2.04E-05 | 1.94E-05 | 0.001382 | 3.22E-05 | -0.00643949 | SCUBE1 |
| chr2 | 201129072 | 201129373 | 302 | 4 | 2.91E-05 | 0.0002236 | 0.000664 | 3.22E-05 | 0.004006703 | CFLAR |
| chr2 | 106165276 | 106165368 | 93 | 2 | 3.11E-05 | 0.0010209 | 2.13E-05 | 3.22E-05 | 0.00774522 | UXS1 |
| chr2 | 11634016 | 11634726 | 711 | 6 | 2.20E-06 | 0.089228 | 4.15E-06 | 3.22E-05 | -0.00246501 | GREB1 |
| chr1 | 67463664 | 67463724 | 61 | 2 | 0.00010215 | 2.31E-05 | 0.000572 | 3.22E-05 | -0.00575262 | NA |
| chr7 | 140386942 | 140387236 | 295 | 2 | 0.000107895 | 1.97E-05 | 0.001258 | 3.22E-05 | 0.013080092 | SLC37A3 |
| chr16 | 58697813 | 58697880 | 68 | 2 | 3.50E-05 | 0.0010755 | 2.11E-05 | 3.22E-05 | -0.00714321 | NA |
| chr10 | 6138304 | 6138601 | 298 | 2 | 1.99E-05 | 6.01E-05 | 8.76E-05 | 3.22E-05 | -0.00688568 | NA |
| chr9 | 130199685 | 130200011 | 327 | 3 | 8.03E-06 | 0.0017894 | 0.000327 | 3.22E-05 | -0.00130252 | NCS1 |
| chr6 | 75473897 | 75474031 | 135 | 2 | 7.50E-05 | 2.04E-05 | 0.001014 | 3.22E-05 | 0.002304337 | FILIP1 |
| chr8 | 88121190 | 88121221 | 32 | 2 | 9.95E-06 | 0.0026406 | 1.79E-05 | 3.22E-05 | -0.01300815 | MMP16 |
| chr1 | 32464567 | 32465097 | 531 | 10 | 2.78E-05 | 0.0001827 | 0.002346 | 3.22E-05 | 0.001081459 | AL033529.1, ZBTB8B |
| chr16 | 89247707 | 89248274 | 568 | 3 | 2.70E-05 | 1.30E-05 | 0.004535 | 3.22E-05 | 0.006947428 | NA |
| chr20 | 45115951 | 45116237 | 287 | 3 | 1.56E-06 | 0.0013922 | 2.13E-05 | 3.23E-05 | -0.00638804 | NA |
| chr4 | 155376464 | 155377275 | 812 | 15 | 1.35E-05 | 0.005352 | 0.000836 | 3.23E-05 | -0.00160455 | AC097467.3, MAP9 |
| chr2 | 23693230 | 23693254 | 25 | 2 | 3.42E-05 | 0.0010601 | 2.12E-05 | 3.23E-05 | -0.00213009 | KLHL29 |
| chr1 | 25805402 | 25805692 | 291 | 2 | 3.09E-05 | 3.42E-05 | 0.0002 | 3.23E-05 | -0.00920924 | SELENON |
| chr4 | 5427089 | 5427256 | 168 | 3 | 6.65E-05 | 5.66E-05 | 0.000916 | 3.23E-05 | -0.0110882 | STK32B |
| chr19 | 3282160 | 3282511 | 352 | 2 | 8.77E-06 | 0.0062115 | 1.62E-05 | 3.23E-05 | -0.00866191 | CELF5 |
| chr16 | 67327805 | 67328065 | 261 | 2 | 5.73E-06 | 2.77E-05 | 0.000325 | 3.23E-05 | -0.01054082 | LRRC36 |
| chr1 | 223720789 | 223720947 | 159 | 2 | 0.000102732 | 2.09E-05 | 0.000891 | 3.23E-05 | 0.005772949 | CAPN2 |
| chr3 | 170869905 | 170870631 | 727 | 12 | 4.35E-05 | 0.0001526 | 0.001137 | 3.23E-05 | 4.33E-06 | RPL22L1 |
| chr3 | 197512824 | 197512827 | 4 | 2 | 1.81E-06 | 0.0118364 | 1.55E-05 | 3.23E-05 | -0.00644125 | BDH1 |
| chr15 | 73974631 | 73974827 | 197 | 2 | 6.27E-05 | 2.11E-05 | 0.000858 | 3.23E-05 | 0.032411873 | NA |
| chr3 | 25664300 | 25665333 | 1034 | 11 | 3.20E-05 | 0.0002701 | 0.000366 | 3.23E-05 | 0.001135305 | TOP2B, MIR4442 |
| chr17 | 50123765 | 50124300 | 536 | 4 | 4.24E-05 | 0.0003907 | 0.000867 | 3.23E-05 | -0.00547956 | SAMD14 |
| chr17 | 57114281 | 57114393 | 113 | 2 | 6.16E-05 | 2.91E-05 | 0.000287 | 3.24E-05 | -0.0105095 | AKAP1 |
| chr13 | 51335445 | 51335685 | 241 | 5 | 7.51E-05 | 6.48E-05 | 0.002837 | 3.24E-05 | -0.00230849 | NA |
| chr19 | 53537520 | 53537582 | 63 | 4 | 0.000119522 | 3.51E-05 | 0.004084 | 3.24E-05 | -0.01262792 | ZNF331 |
| chr6 | 87435610 | 87435648 | 39 | 2 | 1.79E-05 | 0.0006117 | 2.47E-05 | 3.24E-05 | 0.002456145 | CFAP206, AL049697.1 |
| chr22 | 46573134 | 46573370 | 237 | 2 | 3.19E-05 | 0.0001473 | 4.43E-05 | 3.24E-05 | -0.00309286 | NA |
| chr4 | 23766522 | 23766532 | 11 | 2 | 0.000107038 | 2.21E-05 | 0.000703 | 3.24E-05 | -0.01190743 | AC093607.1, PPARGC1A |
| chr3 | 38122356 | 38122663 | 308 | 2 | 2.25E-06 | 2.40E-05 | 0.000507 | 3.24E-05 | 0.00424137 | DLEC1, ACAA1 |
| chr6 | 17282053 | 17282882 | 830 | 12 | 8.63E-06 | 0.0001524 | 0.003233 | 3.24E-05 | -0.00451846 | RBM24 |
| chr2 | 86624909 | 86625355 | 447 | 3 | 3.52E-07 | 0.0069983 | 8.35E-06 | 3.24E-05 | 0.001542102 | RNF103-CHMP3 |
| chr6 | 157830573 | 157831162 | 590 | 4 | 4.11E-05 | 0.0001526 | 0.000228 | 3.24E-05 | 0.005091635 | SNX9, AL391863.2 |
| chr1 | 228141373 | 228141753 | 381 | 2 | 1.97E-05 | 1.95E-05 | 0.001406 | 3.24E-05 | 0.001814431 | GUK1 |
| chr3 | 128051480 | 128052248 | 769 | 9 | 2.39E-05 | 3.16E-05 | 0.005963 | 3.24E-05 | 0.001131225 | SEC61A1, MGLL |
| chr9 | 68705536 | 68705777 | 242 | 6 | 2.93E-05 | 0.000269 | 0.000131 | 3.24E-05 | -0.00166873 | PIP5K1B |
| chr17 | 43823219 | 43823307 | 89 | 2 | 7.81E-05 | 4.84E-05 | 0.000115 | 3.24E-05 | -0.01004482 | MPP3 |
| chr2 | 239155688 | 239155861 | 174 | 2 | 5.15E-05 | 1.96E-05 | 0.001381 | 3.24E-05 | -0.00855349 | HDAC4 |
| chr2 | 24972128 | 24972732 | 605 | 6 | 1.06E-05 | 5.97E-05 | 0.000281 | 3.25E-05 | 0.004688213 | DNAJC27-AS1 |
| chr16 | 67198557 | 67199099 | 543 | 4 | 3.29E-05 | 2.93E-05 | 0.002416 | 3.25E-05 | -0.01012678 | E2F4 |
| chr19 | 33807243 | 33807305 | 63 | 2 | 7.25E-05 | 3.12E-05 | 0.000248 | 3.25E-05 | -0.00031428 | KCTD15 |
| chr17 | 79893023 | 79893259 | 237 | 2 | 6.40E-05 | 3.21E-05 | 0.000234 | 3.25E-05 | -0.00872555 | NA |
| chr5 | 146245405 | 146245744 | 340 | 5 | 0.000114521 | 3.35E-05 | 0.005303 | 3.25E-05 | 0.001024077 | AC091959.1, RBM27 |
| chr7 | 47439654 | 47439913 | 260 | 3 | 9.23E-05 | 2.69E-05 | 0.001844 | 3.25E-05 | 0.000652794 | TNS3 |
| chr14 | 67414585 | 67414804 | 220 | 2 | 2.44E-05 | 0.0009039 | 2.23E-05 | 3.25E-05 | -0.00892549 | NA |
| chr10 | 100994071 | 100994519 | 449 | 2 | 4.04E-05 | 2.63E-05 | 0.000385 | 3.25E-05 | -0.01565314 | TWNK |
| chr2 | 30985853 | 30986149 | 297 | 2 | 6.65E-05 | 3.22E-05 | 0.000233 | 3.25E-05 | -0.0048832 | GALNT14 |
| chr5 | 90409188 | 90409762 | 575 | 5 | 1.93E-05 | 0.0011069 | 0.00056 | 3.25E-05 | -0.00198844 | CETN3 |
| chr17 | 79089733 | 79090306 | 574 | 4 | 0.000116685 | 9.64E-05 | 0.001515 | 3.25E-05 | -0.00358854 | RBFOX3 |
| chr5 | 122323504 | 122323631 | 128 | 3 | 7.05E-05 | 5.75E-05 | 0.000218 | 3.26E-05 | -0.00391714 | SNCAIP |
| chr17 | 40342564 | 40342844 | 281 | 4 | 1.03E-05 | 0.0004414 | 0.000624 | 3.26E-05 | 0.006936081 | RARA, RARA-AS1 |
| chr16 | 47941497 | 47942334 | 838 | 5 | 4.84E-06 | 0.0015338 | 7.10E-05 | 3.26E-05 | 0.000134154 | NA |
| chr16 | 75116558 | 75116982 | 425 | 8 | 8.20E-05 | 5.71E-06 | 0.020663 | 3.26E-05 | 0.01573806 | LDHD |
| chr15 | 99005715 | 99005849 | 135 | 3 | 6.56E-05 | 3.24E-05 | 0.001624 | 3.26E-05 | -0.01735169 | AC036108.1, PGPEP1L |
| chr3 | 141400420 | 141400699 | 280 | 2 | 1.71E-05 | 2.11E-05 | 0.0009 | 3.26E-05 | -0.00159071 | ZBTB38 |
| chr16 | 854510 | 854543 | 34 | 2 | 3.20E-05 | 0.000733 | 2.36E-05 | 3.26E-05 | -0.00458502 | AL023882.1, LMF1 |
| chr8 | 1809792 | 1809857 | 66 | 2 | 3.35E-05 | 2.15E-05 | 0.000823 | 3.26E-05 | 0.001223032 | AC019257.8 |
| chr5 | 54590764 | 54590881 | 118 | 2 | 0.000107528 | 2.11E-05 | 0.000908 | 3.27E-05 | -0.01257106 | NA |
| chr18 | 77132811 | 77132850 | 40 | 4 | 4.52E-05 | 4.27E-05 | 0.000907 | 3.27E-05 | 0.00097209 | MBP |
| chr22 | 42160876 | 42161176 | 301 | 2 | 3.70E-05 | 2.48E-05 | 0.000466 | 3.27E-05 | -0.00599569 | TCF20 |
| chr18 | 51284873 | 51284893 | 21 | 2 | 7.45E-05 | 4.24E-05 | 0.000142 | 3.27E-05 | -0.01350231 | NA |
| chr6 | 42142450 | 42142758 | 309 | 3 | 9.16E-05 | 7.40E-05 | 0.000385 | 3.27E-05 | 0.00035859 | C6orf132 |
| chr12 | 49911172 | 49911525 | 354 | 2 | 8.69E-06 | 8.90E-05 | 6.26E-05 | 3.27E-05 | -0.00056074 | LINC02395, LINC02396 |
| chr9 | 86631212 | 86631215 | 4 | 2 | 6.68E-05 | 9.61E-05 | 5.91E-05 | 3.27E-05 | -0.01002087 | NA |
| chr7 | 43764204 | 43764403 | 200 | 3 | 4.49E-05 | 0.0003879 | 0.000236 | 3.27E-05 | 0.006642332 | BLVRA |
| chr15 | 68651328 | 68651608 | 281 | 3 | 3.07E-05 | 0.0005238 | 0.000566 | 3.27E-05 | -0.01800316 | CORO2B |
| chr11 | 460587 | 460914 | 328 | 3 | 7.73E-05 | 2.65E-05 | 0.001041 | 3.27E-05 | 0.001823808 | PTDSS2 |
| chr9 | 93945942 | 93946006 | 65 | 2 | 7.01E-05 | 2.13E-05 | 0.000876 | 3.28E-05 | 0.000673636 | NA |
| chr12 | 127108565 | 127108802 | 238 | 2 | 2.22E-05 | 0.0010409 | 2.16E-05 | 3.28E-05 | 0.011266918 | NA |
| chr11 | 45928854 | 45929145 | 292 | 2 | 2.45E-05 | 2.74E-05 | 0.000354 | 3.28E-05 | 0.000523685 | LARGE2 |
| chr16 | 4538666 | 4539421 | 756 | 13 | 4.46E-05 | 0.0001185 | 0.001357 | 3.28E-05 | -0.00157722 | CDIP1 |
| chr11 | 95151711 | 95152233 | 523 | 4 | 1.65E-05 | 5.33E-05 | 0.000269 | 3.28E-05 | 0.004183472 | AP000787.1 |
| chr6 | 33266424 | 33267199 | 776 | 9 | 6.12E-05 | 6.17E-05 | 0.002966 | 3.28E-05 | -0.00220387 | VPS52 |
| chr12 | 118372631 | 118373269 | 639 | 9 | 1.82E-05 | 0.0001594 | 0.000574 | 3.28E-05 | -0.00115595 | TAOK3 |
| chr3 | 52012723 | 52012825 | 103 | 2 | 6.22E-05 | 9.14E-05 | 6.17E-05 | 3.28E-05 | 0.001734031 | NA |
| chr4 | 7043993 | 7044680 | 688 | 4 | 1.65E-05 | 3.33E-05 | 0.000379 | 3.28E-05 | -0.00495409 | TADA2B, AC097382.2 |
| chr8 | 119387870 | 119388323 | 454 | 2 | 0.000120405 | 1.97E-05 | 0.001421 | 3.28E-05 | 0.002257556 | AC021733.4 |
| chr19 | 45092829 | 45093570 | 742 | 9 | 2.80E-05 | 0.0001245 | 0.000501 | 3.28E-05 | -6.29E-05 | MARK4, PPP1R37 |
| chr16 | 84037282 | 84037451 | 170 | 2 | 8.53E-05 | 3.15E-05 | 0.000251 | 3.28E-05 | -0.00959216 | SLC38A8 |
| chr6 | 41445520 | 41445760 | 241 | 2 | 8.62E-05 | 2.03E-05 | 0.001173 | 3.29E-05 | 0.001354163 | NA |
| chr17 | 22521433 | 22521659 | 227 | 4 | 2.33E-05 | 0.0037089 | 2.41E-05 | 3.29E-05 | 0.003329003 | NA |
| chr9 | 129061140 | 129061476 | 337 | 2 | 2.20E-05 | 2.13E-05 | 0.00089 | 3.29E-05 | 0.001705993 | MIGA2 |
| chr6 | 13873693 | 13874020 | 328 | 4 | 0.000123474 | 1.22E-05 | 0.007665 | 3.29E-05 | -0.00789322 | NA |
| chr16 | 8776512 | 8776601 | 90 | 2 | 3.80E-05 | 0.000729 | 2.39E-05 | 3.29E-05 | -0.00691214 | ABAT |
| chr3 | 127013675 | 127014126 | 452 | 2 | 0.000122458 | 2.17E-05 | 0.000813 | 3.29E-05 | -0.0096829 | PLXNA1 |
| chr16 | 66886934 | 66886960 | 27 | 2 | 9.88E-05 | 2.70E-05 | 0.000372 | 3.29E-05 | -0.00849641 | PDP2 |
| chr13 | 112735697 | 112735899 | 203 | 3 | 5.33E-05 | 0.0002686 | 0.000119 | 3.30E-05 | 0.000532548 | ATP11A |
| chr12 | 92736271 | 92736475 | 205 | 3 | 0.000109608 | 0.0001041 | 6.66E-05 | 3.30E-05 | -0.00953694 | PLEKHG7 |
| chr14 | 23372284 | 23372916 | 633 | 4 | 2.48E-05 | 0.0026751 | 0.000189 | 3.30E-05 | -0.00534923 | IL25 |
| chr16 | 58418802 | 58419351 | 550 | 2 | 1.87E-05 | 2.03E-05 | 0.001237 | 3.30E-05 | -0.00759342 | NA |
| chr6 | 161408371 | 161408452 | 82 | 2 | 4.54E-05 | 0.0003599 | 3.01E-05 | 3.30E-05 | 0.003370078 | PRKN |
| chr11 | 115760382 | 115760734 | 353 | 7 | 7.36E-05 | 0.0004994 | 0.00037 | 3.31E-05 | 0.002581564 | LINC02698, LINC00900 |
| chr19 | 39915546 | 39915836 | 291 | 3 | 5.21E-05 | 4.97E-05 | 0.00023 | 3.31E-05 | 0.006315175 | FCGBP |
| chr6 | 30781799 | 30782038 | 240 | 4 | 7.39E-05 | 0.0001759 | 0.001708 | 3.31E-05 | -0.00621566 | HCG20 |
| chr12 | 54352136 | 54352392 | 257 | 4 | 5.84E-05 | 0.0003584 | 0.003198 | 3.31E-05 | -0.01131131 | NA |
| chr15 | 82047579 | 82047986 | 408 | 4 | 1.69E-05 | 3.17E-05 | 0.000531 | 3.31E-05 | 0.001058328 | NA |
| chr16 | 16122018 | 16122251 | 234 | 3 | 3.42E-05 | 1.91E-05 | 0.001226 | 3.31E-05 | -0.01658562 | ABCC1 |
| chr9 | 19472150 | 19472383 | 234 | 3 | 4.29E-05 | 0.0043274 | 0.000747 | 3.31E-05 | -0.01031443 | NA |
| chr4 | 183176947 | 183177026 | 80 | 2 | 6.95E-05 | 2.09E-05 | 0.001034 | 3.31E-05 | -0.00121153 | WWC2 |

| | | | | | | | | | | |
|---|---|---|---|---|---|---|---|---|---|---|
| chr17 | 67244214 | 67244441 | 228 | 2 | 1.58E-06 | 0.0107299 | 1.59E-05 | 3.31E-05 | -0.01666478 | AC007448.5, HELZ |
| chr15 | 65129149 | 65129200 | 52 | 2 | 2.50E-05 | 9.79E-05 | 5.95E-05 | 3.31E-05 | -0.00277323 | PDCD7 |
| chr12 | 52601269 | 52601918 | 650 | 9 | 1.48E-05 | 0.0025041 | 0.000182 | 3.31E-05 | -0.00624538 | KRT73-AS1, KRT72 |
| chr22 | 19515344 | 19515819 | 476 | 2 | 2.22E-06 | 0.0018123 | 1.95E-05 | 3.32E-05 | 0.002046846 | CDC45 |
| chr3 | 50607675 | 50607995 | 321 | 4 | 0.000116383 | 1.46E-05 | 0.004575 | 3.32E-05 | -0.00686802 | CISH |
| chr6 | 12355410 | 12355604 | 195 | 2 | 3.55E-05 | 2.01E-05 | 0.001352 | 3.32E-05 | 0.003290773 | NA |
| chr10 | 112444422 | 112444487 | 66 | 2 | 6.13E-05 | 2.36E-05 | 0.000596 | 3.32E-05 | -0.00010079 | ZDHHC6 |
| chr16 | 56905584 | 56905997 | 414 | 2 | 0.000113681 | 2.05E-05 | 0.001189 | 3.32E-05 | -0.00311835 | SLC12A3 |
| chr5 | 10884869 | 10884926 | 58 | 2 | 5.97E-05 | 0.0001421 | 4.66E-05 | 3.32E-05 | -0.00401672 | NA |
| chr12 | 10179408 | 10179600 | 193 | 2 | 3.17E-05 | 0.00829 | 1.62E-05 | 3.32E-05 | 0.003131004 | TMEM52B |
| chr15 | 31445049 | 31445256 | 208 | 2 | 0.000106219 | 2.12E-05 | 0.000976 | 3.32E-05 | -0.00882449 | NA |
| chr2 | 106542812 | 106542999 | 188 | 4 | 2.06E-05 | 0.0027816 | 1.40E-05 | 3.32E-05 | 0.005701182 | CD8B2, AC108868.2 |
| chr17 | 81372537 | 81372562 | 26 | 2 | 6.15E-05 | 6.05E-05 | 9.18E-05 | 3.33E-05 | -0.00022091 | AC110285.3 |
| chr13 | 113658448 | 113659099 | 652 | 3 | 1.26E-05 | 0.0008234 | 7.55E-05 | 3.33E-05 | -0.00338366 | NA |
| chr3 | 99808378 | 99808632 | 255 | 2 | 2.87E-05 | 2.53E-05 | 0.000473 | 3.33E-05 | -0.00812583 | NA |
| chr16 | 13935400 | 13935964 | 565 | 8 | 6.51E-06 | 0.0004329 | 9.56E-05 | 3.33E-05 | -0.00226509 | ERCC4, AC010401.1 |
| chr12 | 115755188 | 115755221 | 34 | 3 | 4.76E-05 | 6.09E-05 | 0.000189 | 3.33E-05 | -0.00418253 | NA |
| chr8 | 144950537 | 144951299 | 763 | 11 | 2.27E-05 | 0.0005181 | 0.006724 | 3.33E-05 | -0.0012609 | ZNF16 |
| chr1 | 205422753 | 205423113 | 361 | 3 | 1.49E-05 | 3.50E-05 | 0.00106 | 3.33E-05 | -0.00920656 | LEMD1 |
| chr11 | 368351 | 368712 | 362 | 12 | 2.82E-05 | 9.59E-05 | 0.003343 | 3.33E-05 | 0.006693991 | NA |
| chr11 | 1746545 | 1746563 | 19 | 2 | 9.87E-05 | 2.02E-05 | 0.001367 | 3.33E-05 | -0.01486484 | IFITM10, AC068580.4 |
| chr7 | 461573 | 461921 | 349 | 3 | 6.15E-05 | 3.93E-05 | 0.000267 | 3.33E-05 | -0.00843488 | NA |
| chr12 | 111753273 | 111753718 | 446 | 5 | 4.78E-06 | 0.0041906 | 1.09E-05 | 3.33E-05 | -0.00544347 | ACAD10 |
| chr5 | 126999440 | 126999609 | 170 | 2 | 8.00E-05 | 3.82E-05 | 0.000177 | 3.34E-05 | -0.00577112 | MARCHF3 |
| chr19 | 4555826 | 4555999 | 174 | 3 | 2.39E-05 | 0.0012846 | 5.13E-05 | 3.34E-05 | -0.00206172 | SEMA6B |
| chr19 | 15201179 | 15201431 | 253 | 4 | 9.74E-05 | 4.30E-05 | 0.000504 | 3.34E-05 | -0.0035513 | NA |
| chr11 | 69195990 | 69196270 | 281 | 2 | 1.41E-05 | 0.0046086 | 1.72E-05 | 3.34E-05 | -0.00373627 | NA |
| chr22 | 49854494 | 49855259 | 766 | 3 | 7.26E-06 | 6.43E-05 | 0.000923 | 3.34E-05 | 0.00218098 | ZBED4 |
| chr19 | 14097868 | 14098240 | 373 | 2 | 2.31E-05 | 2.65E-05 | 0.000413 | 3.34E-05 | 0.004249865 | PRKACA |
| chr21 | 32261585 | 32261757 | 173 | 2 | 0.000104833 | 2.31E-05 | 0.000669 | 3.34E-05 | -0.00382325 | AP000265.1 |
| chr1 | 100423918 | 100424116 | 199 | 2 | 4.24E-05 | 3.44E-05 | 0.000218 | 3.34E-05 | -0.0015683 | CDC14A |
| chr21 | 40576891 | 40576960 | 70 | 3 | 0.000144044 | 1.41E-05 | 0.003685 | 3.34E-05 | -0.0061846 | DSCAM |
| chr14 | 92644212 | 92644505 | 294 | 2 | 0.000116992 | 2.01E-05 | 0.001415 | 3.35E-05 | -0.00105252 | RIN3 |
| chr1 | 202121721 | 202122047 | 327 | 5 | 6.35E-06 | 0.0030533 | 0.000756 | 3.35E-05 | 0.000813256 | NA |
| chr9 | 100216041 | 100216253 | 213 | 2 | 2.09E-05 | 0.0028164 | 1.83E-05 | 3.35E-05 | -0.00605194 | INVS |
| chr12 | 7190025 | 7190424 | 400 | 9 | 0.000109768 | 1.28E-05 | 0.019446 | 3.35E-05 | -0.0036546 | PEX5 |
| chr1 | 43558479 | 43558561 | 83 | 3 | 6.36E-05 | 1.30E-05 | 0.004973 | 3.35E-05 | -0.01441953 | PTPRF |
| chr16 | 49586552 | 49586625 | 74 | 2 | 4.49E-05 | 0.0001408 | 4.74E-05 | 3.35E-05 | -0.00402925 | ZNF423 |
| chr21 | 46458368 | 46459080 | 713 | 10 | 3.30E-06 | 0.0024826 | 7.20E-05 | 3.35E-05 | 0.000272429 | DIP2A |
| chr3 | 32685597 | 32685697 | 101 | 2 | 1.55E-05 | 2.49E-05 | 0.000515 | 3.35E-05 | 0.000541132 | CNOT10 |
| chr5 | 38734569 | 38734659 | 91 | 2 | 0.000108178 | 2.33E-05 | 0.000656 | 3.35E-05 | -0.01354048 | OSMR-AS1 |
| chr21 | 39934652 | 39935083 | 432 | 2 | 4.15E-05 | 2.28E-05 | 0.000723 | 3.35E-05 | -0.00922929 | NA |
| chr20 | 57548573 | 57548844 | 272 | 2 | 0.000115934 | 2.03E-05 | 0.001386 | 3.36E-05 | -0.00683115 | NA |
| chr1 | 159829920 | 159829940 | 21 | 3 | 0.000131962 | 4.12E-05 | 0.000221 | 3.36E-05 | -0.00535146 | SLAMF8 |
| chr2 | 9244953 | 9245004 | 52 | 2 | 0.000106114 | 2.56E-05 | 0.000471 | 3.36E-05 | 0.003630648 | ASAP2 |
| chr1 | 165355321 | 165356596 | 1276 | 14 | 1.10E-05 | 0.0050839 | 0.000508 | 3.36E-05 | 0.000739623 | LMX1A |
| chr4 | 61142526 | 61142905 | 380 | 3 | 5.53E-05 | 2.21E-05 | 0.00088 | 3.36E-05 | -0.00119237 | NA |
| chr22 | 30201915 | 30202287 | 373 | 2 | 3.75E-05 | 2.11E-05 | 0.001083 | 3.36E-05 | 0.00421142 | AC002378.1 |
| chr19 | 48491381 | 48491620 | 240 | 3 | 2.28E-05 | 0.0002422 | 3.65E-05 | 3.36E-05 | 0.001629109 | LMTK3 |
| chr15 | 23686793 | 23687265 | 473 | 4 | 5.04E-05 | 0.0004612 | 0.000458 | 3.36E-05 | -0.00897668 | NDN |
| chr7 | 1909996 | 1911222 | 1227 | 8 | 4.88E-06 | 0.0001252 | 0.002408 | 3.37E-05 | 0.001341993 | MAD1L1, AC069288.1 |
| chr1 | 32537843 | 32538823 | 981 | 4 | 1.86E-05 | 0.0040259 | 0.00053 | 3.37E-05 | 0.001542911 | AL033529.1 |
| chr4 | 169635667 | 169635803 | 137 | 2 | 6.14E-05 | 2.61E-05 | 0.000447 | 3.37E-05 | -0.01489564 | CLCN3 |
| chr21 | 43332887 | 43333421 | 535 | 2 | 3.19E-05 | 2.14E-05 | 0.000999 | 3.37E-05 | -0.01603579 | NA |
| chr9 | 95450669 | 95451473 | 805 | 4 | 4.70E-05 | 5.76E-05 | 0.001745 | 3.37E-05 | -0.00314271 | PTCH1 |
| chr8 | 126383110 | 126383941 | 832 | 3 | 3.12E-06 | 5.48E-05 | 0.000585 | 3.37E-05 | -0.0143292 | NA |
| chr3 | 87792869 | 87794186 | 1318 | 5 | 8.12E-06 | 4.54E-05 | 0.001785 | 3.37E-05 | -0.00086178 | HTR1F, AC108749.1 |
| chr20 | 3230597 | 3230919 | 323 | 4 | 5.15E-06 | 0.0016464 | 2.34E-05 | 3.37E-05 | 0.000336166 | SLC4A11 |
| chr16 | 87461438 | 87461742 | 305 | 2 | 4.26E-05 | 2.07E-05 | 0.001248 | 3.37E-05 | -0.01048492 | ZCCHC14 |
| chr17 | 82345696 | 82345838 | 143 | 3 | 5.47E-05 | 0.0001754 | 9.42E-05 | 3.37E-05 | -0.00608392 | NA |
| chr2 | 236217592 | 236217730 | 139 | 2 | 3.19E-05 | 0.0004942 | 2.75E-05 | 3.37E-05 | -0.00163035 | AC019068.1, ASB18 |
| chr19 | 19543959 | 19544543 | 585 | 3 | 7.29E-06 | 7.24E-05 | 0.000196 | 3.37E-05 | -0.00722733 | CILP2 |
| chr16 | 88800049 | 88800219 | 171 | 2 | 0.000105113 | 2.30E-05 | 0.000718 | 3.37E-05 | 0.009061903 | NA |
| chr1 | 53913878 | 53914213 | 336 | 2 | 5.62E-05 | 2.82E-05 | 0.000358 | 3.38E-05 | 0.006430686 | NA |
| chr5 | 97096430 | 97096814 | 385 | 2 | 0.000103402 | 2.15E-05 | 0.000978 | 3.38E-05 | -0.00720592 | LIX1-AS1, LIX1 |
| chr19 | 35154000 | 35155247 | 1248 | 15 | 1.74E-05 | 0.0018536 | 0.00158 | 3.38E-05 | -0.00032438 | FXYD7, FXYD5 |
| chr13 | 31005150 | 31005263 | 114 | 3 | 2.77E-05 | 0.0026991 | 1.67E-05 | 3.38E-05 | 0.000415842 | NA |
| chr9 | 109183034 | 109183330 | 297 | 2 | 3.72E-06 | 0.0177563 | 1.58E-05 | 3.38E-05 | -0.0095212 | EPB41L4B |
| chr11 | 1197578 | 1198290 | 713 | 5 | 0.000138154 | 1.33E-05 | 0.006149 | 3.38E-05 | 0.003621353 | MUC5AC |
| chr1 | 27341567 | 27342065 | 499 | 9 | 4.15E-06 | 0.0105659 | 0.000263 | 3.38E-05 | 0.001051178 | SYTL1 |
| chr12 | 49094771 | 49095115 | 345 | 5 | 2.09E-05 | 2.29E-05 | 0.002626 | 3.39E-05 | -0.00482836 | DHH |
| chr4 | 27048261 | 27048411 | 151 | 2 | 3.99E-05 | 0.0002583 | 3.53E-05 | 3.39E-05 | 0.001951365 | NA |
| chr7 | 5356500 | 5356897 | 398 | 4 | 7.14E-05 | 0.0037767 | 0.00025 | 3.39E-05 | -0.00357133 | TNRC18 |
| chr19 | 14112708 | 14113381 | 674 | 5 | 2.98E-05 | 0.0001158 | 0.001126 | 3.39E-05 | -0.00424652 | PRKACA |
| chr2 | 9738950 | 9739609 | 660 | 3 | 1.32E-05 | 1.44E-05 | 0.003952 | 3.39E-05 | -0.04164192 | NA |
| chr22 | 21641827 | 21642420 | 594 | 8 | 3.22E-05 | 0.0001936 | 0.00015 | 3.39E-05 | 0.001018966 | SDF2L1 |
| chr16 | 67528521 | 67529394 | 874 | 8 | 1.85E-05 | 3.93E-05 | 0.010417 | 3.39E-05 | 0.002853166 | RIPOR1, AC027682.6 |
| chr3 | 184364771 | 184364971 | 201 | 2 | 1.10E-05 | 0.0002972 | 3.34E-05 | 3.39E-05 | 0.006203657 | POLR2H |
| chr17 | 81855534 | 81855631 | 98 | 3 | 3.48E-05 | 9.72E-05 | 0.000816 | 3.39E-05 | -0.00703863 | P4HB |
| chr9 | 105151598 | 105151873 | 276 | 2 | 4.17E-05 | 9.62E-05 | 6.24E-05 | 3.39E-05 | -0.00164177 | AL591506.1 |
| chr9 | 131111972 | 131112083 | 112 | 2 | 7.33E-06 | 2.04E-05 | 0.001429 | 3.40E-05 | 0.004261757 | AIF1L |
| chr7 | 45157652 | 45158010 | 359 | 7 | 4.84E-05 | 9.08E-05 | 0.002088 | 3.40E-05 | -0.00234607 | RAMP3 |
| chr9 | 97944782 | 97945115 | 334 | 4 | 2.32E-05 | 0.0003442 | 4.33E-05 | 3.40E-05 | -0.00451778 | HEMGN |
| chr1 | 34904896 | 34905037 | 142 | 2 | 2.19E-05 | 0.0004445 | 2.88E-05 | 3.40E-05 | -0.01455129 | DLGAP3 |
| chr20 | 32744547 | 32745262 | 716 | 4 | 4.78E-07 | 0.0003965 | 0.000216 | 3.40E-05 | 0.006804073 | NA |
| chr11 | 62513709 | 62513719 | 11 | 2 | 1.38E-05 | 0.001818 | 2.00E-05 | 3.40E-05 | -0.00345127 | AHNAK |
| chr4 | 184370027 | 184370380 | 354 | 3 | 4.93E-05 | 5.67E-05 | 0.000385 | 3.40E-05 | 0.001503291 | LINC02362 |
| chr12 | 45849731 | 45850016 | 286 | 2 | 5.00E-05 | 4.87E-05 | 0.000126 | 3.41E-05 | -0.00885552 | ARID2 |
| chr1 | 3373720 | 3373849 | 130 | 2 | 4.29E-05 | 0.0001074 | 5.81E-05 | 3.41E-05 | -0.00649426 | PRDM16, AL354743.1 |
| chr5 | 1549341 | 1549360 | 20 | 2 | 6.87E-06 | 0.0001001 | 6.09E-05 | 3.41E-05 | -0.00789345 | AC091849.2 |
| chr17 | 79930186 | 79930597 | 412 | 5 | 1.13E-05 | 0.0003101 | 4.01E-05 | 3.41E-05 | 0.001993095 | NA |
| chr6 | 686506 | 686818 | 313 | 2 | 2.42E-05 | 0.0002471 | 3.63E-05 | 3.41E-05 | -0.00788144 | EXOC2 |
| chr2 | 208164834 | 208165058 | 225 | 5 | 3.06E-05 | 0.0006196 | 0.000184 | 3.41E-05 | -0.00176729 | NA |
| chr11 | 112086442 | 112087148 | 707 | 18 | 1.27E-05 | 0.0046562 | 5.41E-05 | 3.41E-05 | 0.000204034 | SDHD, AP002884.2, TIMM8B |
| chr19 | 32608888 | 32609294 | 407 | 2 | 5.32E-05 | 3.48E-05 | 0.000224 | 3.41E-05 | 0.000618271 | ANKRD27 |
| chr16 | 71758384 | 71758402 | 19 | 2 | 4.18E-05 | 3.32E-05 | 0.000249 | 3.41E-05 | -0.00496237 | AP1G1, SNORD71 |
| chr17 | 7798228 | 7798263 | 36 | 2 | 0.000169406 | 5.93E-05 | 9.79E-05 | 3.41E-05 | -0.00555262 | DNAH2 |
| chr11 | 111876776 | 111876974 | 199 | 4 | 5.20E-05 | 6.82E-05 | 0.000941 | 3.41E-05 | -0.00721794 | AP001781.2, FDXACB1 |
| chr8 | 1817051 | 1817311 | 261 | 7 | 0.000124237 | 0.0002397 | 0.001813 | 3.41E-05 | 0.006575981 | AC019257.8, MIR596, AC100810.7 |
| chr2 | 88671959 | 88672170 | 212 | 2 | 0.000114956 | 2.07E-05 | 0.001343 | 3.41E-05 | -0.01868749 | NA |
| chr5 | 32030355 | 32030742 | 388 | 2 | 0.000119735 | 2.08E-05 | 0.001299 | 3.41E-05 | -0.00514037 | PDZD2 |
| chr17 | 4269594 | 4269938 | 345 | 2 | 6.62E-06 | 0.0106709 | 1.63E-05 | 3.41E-05 | 0.00275281 | UBE2G1 |
| chr17 | 29160550 | 29160710 | 161 | 2 | 4.20E-05 | 8.08E-05 | 7.22E-05 | 3.42E-05 | -0.00795881 | MYO18A |
| chr4 | 3313313 | 3313703 | 391 | 5 | 7.21E-05 | 3.31E-05 | 0.000678 | 3.42E-05 | -0.00252953 | RGS12 |
| chr6 | 169452480 | 169452747 | 268 | 3 | 7.44E-05 | 1.48E-05 | 0.003392 | 3.42E-05 | -0.00489552 | AL031315.1 |
| chr11 | 120337615 | 120337913 | 299 | 2 | 6.56E-06 | 6.63E-05 | 8.77E-05 | 3.43E-05 | -0.012369 | ARHGEF12 |
| chr1 | 22909494 | 22909708 | 215 | 2 | 2.50E-05 | 0.0001412 | 4.89E-05 | 3.43E-05 | -0.00704901 | EPHB2 |
| chr22 | 41253455 | 41254333 | 879 | 3 | 6.76E-05 | 5.31E-05 | 0.000226 | 3.43E-05 | -0.00852472 | RANGAP1 |
| chr2 | 20439525 | 20439678 | 154 | 2 | 8.50E-05 | 2.26E-05 | 0.000847 | 3.43E-05 | -0.00933019 | NA |
| chr14 | 76139509 | 76139861 | 353 | 3 | 2.08E-05 | 1.34E-05 | 0.005092 | 3.43E-05 | -0.02133366 | AC016526.4 |
| chr17 | 57860467 | 57860481 | 15 | 2 | 1.49E-05 | 2.29E-05 | 0.000799 | 3.43E-05 | -0.00967621 | NA |
| chr2 | 141557260 | 141557558 | 299 | 2 | 0.000103484 | 2.23E-05 | 0.000908 | 3.43E-05 | -0.01033164 | LRP1B |
| chr8 | 1334917 | 1335276 | 360 | 2 | 7.62E-05 | 2.35E-05 | 0.000704 | 3.43E-05 | -0.00040655 | DLGAP2 |
| chr6 | 33255249 | 33255410 | 162 | 3 | 0.000132914 | 3.99E-05 | 0.000878 | 3.43E-05 | 0.00501664 | VPS52 |
| chr15 | 43185095 | 43185978 | 884 | 15 | 6.78E-06 | 0.0147813 | 0.001477 | 3.44E-05 | 0.000300477 | TMEM62, CCNDBP1, AC068724.3, AC068724.4, AC068724.1 |
| chr5 | 31409257 | 31409602 | 346 | 2 | 1.30E-05 | 0.0003315 | 3.25E-05 | 3.44E-05 | 0.00894996 | DROSHA |
| chr4 | 38832699 | 38832934 | 236 | 2 | 8.02E-05 | 3.35E-05 | 0.00025 | 3.44E-05 | -0.00533235 | TLR1, TLR6 |
| chr3 | 58645958 | 58646273 | 316 | 2 | 6.46E-05 | 3.59E-05 | 0.000215 | 3.44E-05 | -0.00403602 | FAM3D |
| chr4 | 145651450 | 145651535 | 86 | 2 | 5.91E-05 | 2.77E-05 | 0.000406 | 3.44E-05 | 0.000586711 | MMAA |
| chr7 | 43209092 | 43209160 | 69 | 2 | 6.53E-05 | 0.0001193 | 5.47E-05 | 3.44E-05 | -0.00205885 | HECW1 |
| chr18 | 47749081 | 47749805 | 725 | 4 | 4.82E-05 | 3.14E-05 | 0.000462 | 3.44E-05 | -0.00256214 | NA |
| chr8 | 52499669 | 52499786 | 118 | 2 | 0.000117852 | 2.10E-05 | 0.0013 | 3.44E-05 | -0.00888071 | NA |
| chr7 | 100082655 | 100082967 | 313 | 5 | 9.30E-06 | 0.0005792 | 4.10E-05 | 3.45E-05 | -0.00078279 | NA |
| chr9 | 19154871 | 19155073 | 203 | 2 | 2.59E-05 | 0.0001925 | 4.15E-05 | 3.45E-05 | 0.006302929 | NA |
| chr2 | 240135785 | 240136298 | 514 | 3 | 5.06E-07 | 3.73E-05 | 0.000346 | 3.45E-05 | -0.00263145 | COPS9 |
| chr17 | 41869441 | 41869512 | 72 | 2 | 7.80E-05 | 6.69E-05 | 8.79E-05 | 3.45E-05 | -0.01223975 | ACLY |
| chr8 | 133644910 | 133645569 | 660 | 2 | 3.81E-07 | 4.85E-05 | 0.00013 | 3.45E-05 | 0.001290365 | NA |
| chr22 | 33743293 | 33743361 | 69 | 2 | 1.73E-05 | 0.0091135 | 1.67E-05 | 3.45E-05 | -0.00543057 | LARGE-AS1, LARGE1 |
| chr7 | 98619540 | 98619791 | 252 | 5 | 0.000108825 | 1.89E-05 | 0.00923 | 3.45E-05 | -0.00620862 | NPTX2 |
| chr4 | 183653772 | 183653955 | 184 | 3 | 3.23E-05 | 2.53E-05 | 0.000622 | 3.45E-05 | -0.00592631 | RWDD4 |
| chr21 | 44005553 | 44005616 | 64 | 2 | 2.52E-05 | 0.004304 | 1.79E-05 | 3.45E-05 | -0.00556606 | NA |
| chr6 | 36491563 | 36491821 | 259 | 2 | 2.21E-05 | 0.0010036 | 2.31E-05 | 3.45E-05 | -0.00563855 | NA |
| chr1 | 50048255 | 50048788 | 534 | 3 | 6.17E-07 | 0.0023666 | 7.52E-05 | 3.45E-05 | -0.00789941 | ELAVL4 |
| chr16 | 11316236 | 11316770 | 535 | 6 | 6.99E-05 | 0.0011298 | 0.000533 | 3.45E-05 | -0.0016914 | RMI2 |
| chr18 | 12237245 | 12237889 | 645 | 3 | 2.40E-05 | 0.0002058 | 0.00059 | 3.46E-05 | 0.004551348 | NA |
| chr13 | 111325617 | 111326003 | 387 | 3 | 2.21E-05 | 0.0009539 | 0.000647 | 3.46E-05 | -0.0052938 | TEX29 |
| chr12 | 102430800 | 102430816 | 17 | 2 | 0.000107407 | 2.86E-05 | 0.000374 | 3.46E-05 | 0.006182423 | LINC02456, IGF1 |
| chr1 | 16119443 | 16119617 | 175 | 2 | 2.58E-05 | 2.22E-05 | 0.000967 | 3.46E-05 | -0.0048188 | NA |
| chr5 | 181243058 | 181243379 | 322 | 4 | 5.33E-05 | 0.0001741 | 0.000233 | 3.46E-05 | 0.00159734 | RACK1, SNORD95 |
| chr10 | 34018455 | 34019101 | 647 | 2 | 5.15E-06 | 0.0002632 | 3.60E-05 | 3.47E-05 | 0.007475753 | NA |
| chr1 | 173917963 | 173918222 | 260 | 3 | 1.14E-05 | 0.0191776 | 7.13E-06 | 3.47E-05 | 0.002148048 | NA |
| chr8 | 132050604 | 132050727 | 124 | 2 | 2.40E-05 | 0.0001669 | 4.51E-05 | 3.47E-05 | 0.001477417 | OC90, AC100868.1 |
| chr16 | 740595 | 741252 | 658 | 8 | 5.90E-06 | 0.0001067 | 0.000256 | 3.47E-05 | -0.00175834 | CIAO3 |
| chr8 | 48260278 | 48260556 | 279 | 2 | 4.56E-05 | 2.12E-05 | 0.001271 | 3.47E-05 | -0.0061478 | NA |
| chr13 | 27621411 | 27622233 | 823 | 9 | 2.51E-06 | 0.0013192 | 0.000252 | 3.47E-05 | 3.52E-05 | POLR1D |
| chr1 | 26620253 | 26620360 | 108 | 2 | 3.34E-05 | 2.11E-05 | 0.001335 | 3.47E-05 | 0.004639298 | NA |
| chr2 | 145653131 | 145653289 | 159 | 2 | 2.60E-05 | 0.0033313 | 1.85E-05 | 3.47E-05 | 0.005866343 | AC079163.2 |
| chr19 | 48363449 | 48364555 | 1107 | 13 | 5.60E-06 | 0.0019749 | 0.000957 | 3.47E-05 | 0.000230265 | SYNGR4, TMEM143 |
| chr11 | 126440830 | 126441200 | 371 | 3 | 7.17E-06 | 4.57E-05 | 0.00027 | 3.47E-05 | -0.00571624 | KIRREL3 |
| chr1 | 205269030 | 205269405 | 376 | 5 | 8.07E-06 | 0.0030041 | 1.40E-05 | 3.47E-05 | -0.00126553 | TMCC2 |
| chr20 | 718015 | 718028 | 14 | 2 | 8.66E-05 | 5.34E-05 | 0.000115 | 3.48E-05 | -0.0124423 | NA |
| chr6 | 29727813 | 29728487 | 675 | 2 | 0.000133909 | 2.11E-05 | 0.001355 | 3.48E-05 | 0.005485543 | HLA-F, HLA-F-AS1 |
| chr5 | 140647340 | 140648279 | 940 | 14 | 4.72E-06 | 0.0026588 | 4.73E-05 | 3.48E-05 | -0.00106998 | IK, MIR3655, NDUFA2 |
| chr1 | 170530813 | 170531009 | 197 | 2 | 9.44E-06 | 0.0001773 | 4.40E-05 | 3.49E-05 | -0.01644246 | GORAB-AS1 |
| chr12 | 110937031 | 110937227 | 197 | 2 | 3.09E-05 | 2.49E-05 | 0.000606 | 3.49E-05 | -0.00827582 | LINC01405 |
| chr18 | 31718523 | 31718713 | 191 | 3 | 4.88E-05 | 0.0001714 | 0.000314 | 3.49E-05 | -0.00503419 | NA |
| chr3 | 119146105 | 119147080 | 976 | 9 | 5.20E-08 | 0.0128535 | 0.00011 | 3.49E-05 | -0.00143208 | TEX55 |
| chr8 | 25468190 | 25468236 | 47 | 2 | 5.88E-05 | 2.25E-05 | 0.000952 | 3.49E-05 | 0.00398769 | CDCA2 |

| | | | | | | | | | | |
|---|---|---|---|---|---|---|---|---|---|---|
| chr19 | 38615408 | 38615545 | 138 | 2 | 1.55E-05 | 5.26E-05 | 0.000119 | 3.49E-05 | -0.00710941 | MAP4K1 |
| chr4 | 33089986 | 33090084 | 99 | 3 | 9.90E-05 | 4.46E-05 | 0.000237 | 3.49E-05 | -0.01313946 | NA |
| chr4 | 5622678 | 5622844 | 167 | 3 | 7.27E-05 | 0.0001163 | 0.000541 | 3.49E-05 | -0.00345854 | EVC2 |
| chr16 | 8852702 | 8853186 | 485 | 4 | 2.68E-05 | 0.0001776 | 0.004093 | 3.50E-05 | -0.00512023 | AC022167.2, CARHSP1 |
| chr17 | 6978995 | 6979013 | 19 | 2 | 6.83E-05 | 2.10E-05 | 0.001471 | 3.50E-05 | -0.00944556 | ALOX12-AS1 |
| chr10 | 133226746 | 133226921 | 176 | 2 | 7.82E-05 | 8.92E-05 | 6.91E-05 | 3.50E-05 | -0.01417253 | NA |
| chr20 | 5610969 | 5611785 | 817 | 9 | 4.16E-06 | 0.003928 | 0.000142 | 3.50E-05 | -0.00096613 | GPCPD1 |
| chr7 | 94059807 | 94060298 | 492 | 2 | 0.000113479 | 2.10E-05 | 0.00147 | 3.50E-05 | -0.00779855 | AC003092.1 |
| chr17 | 15272296 | 15272361 | 66 | 3 | 4.59E-05 | 2.43E-05 | 0.000921 | 3.50E-05 | 0.002791784 | AC005703.6 |
| chr1 | 84398384 | 84398926 | 543 | 6 | 9.04E-05 | 3.94E-05 | 0.00065 | 3.50E-05 | -0.00161802 | DNASE2B |
| chr6 | 9503580 | 9503671 | 92 | 2 | 0.000109977 | 2.63E-05 | 0.000506 | 3.50E-05 | 0.02906133 | NA |
| chr18 | 79526052 | 79526260 | 209 | 3 | 5.65E-05 | 5.14E-05 | 0.001313 | 3.50E-05 | -0.00342944 | NFATC1 |
| chr22 | 47161808 | 47162241 | 434 | 3 | 1.65E-05 | 0.0005582 | 5.25E-05 | 3.51E-05 | 0.002670098 | TBC1D22A |
| chr1 | 15799719 | 15800235 | 517 | 3 | 9.42E-05 | 0.0005078 | 0.000386 | 3.51E-05 | -0.00395659 | NA |
| chr12 | 32397216 | 32397220 | 5 | 2 | 3.91E-05 | 0.0001526 | 4.81E-05 | 3.51E-05 | -0.01033365 | NA |
| chr19 | 46636134 | 46636239 | 106 | 2 | 4.13E-05 | 2.21E-05 | 0.001084 | 3.51E-05 | -0.02077098 | NA |
| chr4 | 72037991 | 72038149 | 159 | 3 | 0.00015475 | 0.0002278 | 7.92E-05 | 3.51E-05 | -0.00572243 | NPFFR2 |
| chr3 | 13573574 | 13573629 | 56 | 2 | 0.000119503 | 2.16E-05 | 0.001247 | 3.51E-05 | -0.00029389 | FBLN2 |
| chr14 | 100830410 | 100830912 | 503 | 5 | 3.35E-05 | 7.17E-05 | 0.002222 | 3.51E-05 | -0.00216174 | MEG3 |
| chr19 | 5103458 | 5103470 | 13 | 2 | 6.24E-05 | 4.34E-05 | 0.000161 | 3.51E-05 | 0.003389585 | KDM4B |
| chr15 | 31032825 | 31033062 | 238 | 2 | 3.35E-05 | 6.89E-05 | 8.78E-05 | 3.51E-05 | -0.00109273 | AC009562.1, TRPM1 |
| chr5 | 177433862 | 177434251 | 390 | 4 | 8.16E-05 | 2.37E-05 | 0.001861 | 3.51E-05 | -0.00212593 | GRK6 |
| chr15 | 65033004 | 65033251 | 248 | 2 | 3.77E-05 | 8.25E-05 | 7.40E-05 | 3.51E-05 | -0.01561908 | NA |
| chr19 | 15792928 | 15793300 | 373 | 4 | 4.29E-05 | 0.000168 | 0.000163 | 3.51E-05 | 0.004058311 | OR10H5 |
| chr19 | 3519685 | 3519807 | 123 | 2 | 6.28E-05 | 4.74E-05 | 0.00014 | 3.52E-05 | -0.00139061 | FZR1 |
| chr20 | 34859383 | 34859548 | 166 | 3 | 0.000126286 | 1.54E-05 | 0.003422 | 3.52E-05 | 0.000685581 | GGT7 |
| chr2 | 224401546 | 224402402 | 857 | 13 | 2.80E-05 | 0.0001848 | 0.003179 | 3.52E-05 | -0.00093552 | FAM124B |
| chr14 | 60165270 | 60165814 | 545 | 8 | 2.09E-05 | 0.000836 | 3.95E-05 | 3.52E-05 | 0.001298512 | PCNX4, DHRS7 |
| chr12 | 121452857 | 121453328 | 472 | 3 | 5.14E-05 | 1.48E-05 | 0.004293 | 3.52E-05 | -0.00614747 | KDM2B |
| chr15 | 61229711 | 61230156 | 446 | 6 | 2.67E-05 | 1.64E-05 | 0.010286 | 3.52E-05 | -0.00466038 | NA |
| chr8 | 41650592 | 41651099 | 508 | 4 | 3.70E-05 | 4.14E-05 | 0.002808 | 3.52E-05 | 0.003606375 | NKX6-3 |
| chr11 | 472948 | 473158 | 211 | 3 | 5.49E-05 | 0.0001573 | 6.90E-05 | 3.52E-05 | -0.00381322 | PTDSS2 |
| chr6 | 166383573 | 166383941 | 369 | 19 | 0.000102733 | 2.25E-05 | 0.021652 | 3.52E-05 | -0.00019832 | AL022069.1 |
| chr20 | 37608950 | 37608962 | 13 | 2 | 2.97E-05 | 5.63E-05 | 0.000111 | 3.53E-05 | -0.00775576 | NA |
| chr11 | 64409147 | 64409370 | 224 | 2 | 7.14E-05 | 2.30E-05 | 0.000901 | 3.53E-05 | -0.00569211 | NA |
| chr1 | 155911114 | 155911608 | 495 | 8 | 3.43E-05 | 0.0001022 | 0.003243 | 3.53E-05 | 0.000537639 | RIT1 |
| chr18 | 12145420 | 12145774 | 355 | 3 | 1.31E-06 | 0.003995 | 1.06E-05 | 3.53E-05 | -0.00513382 | NA |
| chr18 | 50784711 | 50785046 | 336 | 2 | 9.73E-05 | 2.42E-05 | 0.00072 | 3.53E-05 | -0.01995996 | NA |
| chr12 | 111429015 | 111429317 | 303 | 2 | 2.92E-05 | 0.0001791 | 4.44E-05 | 3.53E-05 | 0.002956874 | SH2B3 |
| chr9 | 37323874 | 37324743 | 870 | 5 | 6.03E-06 | 0.0040959 | 6.78E-05 | 3.53E-05 | 0.000860197 | ZCCHC7 |
| chr14 | 76486367 | 76486554 | 188 | 2 | 9.65E-05 | 3.24E-05 | 0.000292 | 3.53E-05 | -0.00598109 | ESRRB |
| chr22 | 18091324 | 18091726 | 403 | 2 | 7.40E-06 | 4.56E-05 | 0.000151 | 3.53E-05 | -0.00896296 | PEX26 |
| chr10 | 70674780 | 70674812 | 33 | 2 | 3.59E-05 | 0.0001706 | 4.57E-05 | 3.53E-05 | 0.00188331 | ADAMTS14 |
| chr16 | 74667510 | 74667523 | 14 | 2 | 5.40E-05 | 2.12E-05 | 0.001484 | 3.53E-05 | -0.0108602 | TMPOP2 |
| chr2 | 84905268 | 84906138 | 871 | 6 | 8.78E-06 | 0.0001265 | 0.000217 | 3.54E-05 | -0.00081942 | TMSB10, TRABD2A |
| chr19 | 7518041 | 7518180 | 140 | 2 | 2.88E-05 | 0.0002084 | 4.12E-05 | 3.54E-05 | -0.0106392 | AC008878.1, ZNF358 |
| chr14 | 91947362 | 91948081 | 720 | 10 | 2.96E-05 | 4.74E-05 | 0.004697 | 3.54E-05 | -0.00065047 | FBLN5 |
| chr1 | 90834658 | 90835002 | 345 | 6 | 2.21E-05 | 6.19E-05 | 0.003321 | 3.54E-05 | -0.0037901 | LINC02609 |
| chr6 | 30828422 | 30828630 | 209 | 6 | 5.49E-05 | 0.0002217 | 0.004262 | 3.54E-05 | -0.01461867 | LINC00243 |
| chr3 | 64351433 | 64351815 | 383 | 2 | 1.44E-05 | 0.0004242 | 3.06E-05 | 3.54E-05 | -0.00834252 | PRICKLE2 |
| chr11 | 1926513 | 1926719 | 207 | 2 | 5.94E-05 | 3.05E-05 | 0.000343 | 3.54E-05 | 0.021683977 | TNNT3 |
| chr11 | 18697941 | 18698811 | 871 | 8 | 6.23E-06 | 0.0016444 | 0.000519 | 3.54E-05 | -0.00308612 | TMEM86A |
| chr3 | 161224329 | 161224994 | 666 | 3 | 9.23E-05 | 0.0004798 | 0.000182 | 3.54E-05 | -0.00928767 | NMD3 |
| chr3 | 115721270 | 115721454 | 185 | 2 | 6.36E-05 | 7.43E-05 | 8.28E-05 | 3.54E-05 | -0.00418763 | GAP43 |
| chr10 | 12648978 | 12649148 | 171 | 2 | 0.000111649 | 2.40E-05 | 0.000764 | 3.54E-05 | -0.00938876 | CAMK1D |
| chr11 | 72669247 | 72669773 | 527 | 3 | 4.26E-05 | 2.84E-05 | 0.001863 | 3.55E-05 | -0.00686852 | PDE2A |
| chr13 | 111858051 | 111858371 | 321 | 3 | 2.82E-05 | 0.0003525 | 3.69E-05 | 3.55E-05 | -0.00672946 | NA |
| chr16 | 2092652 | 2093018 | 367 | 3 | 2.33E-05 | 5.55E-05 | 0.000325 | 3.55E-05 | -0.00312882 | AC009065.2, PKD1 |
| chr9 | 128279166 | 128279237 | 72 | 2 | 5.95E-05 | 0.0001957 | 4.26E-05 | 3.55E-05 | -0.00255955 | SWI5 |
| chr21 | 14753730 | 14753866 | 137 | 2 | 5.67E-05 | 2.19E-05 | 0.001233 | 3.55E-05 | -0.00203037 | AF127936.1 |
| chr20 | 17576612 | 17577012 | 401 | 3 | 7.21E-05 | 2.44E-05 | 0.001076 | 3.55E-05 | -0.00135945 | DSTN |
| chr2 | 101022390 | 101022634 | 245 | 3 | 7.54E-05 | 5.76E-05 | 0.000237 | 3.55E-05 | -0.00059271 | RPL31, TBC1D8 |
| chr10 | 101844465 | 101844771 | 307 | 2 | 5.97E-06 | 2.13E-05 | 0.001494 | 3.55E-05 | -0.01144365 | NA |
| chr13 | 111320507 | 111320796 | 290 | 9 | 3.66E-05 | 0.0003958 | 0.003546 | 3.55E-05 | 3.68E-05 | TEX29 |
| chr19 | 808116 | 808528 | 413 | 4 | 9.21E-06 | 5.99E-05 | 0.001471 | 3.55E-05 | 9.91E-05 | PTBP1 |
| chr8 | 141226025 | 141226818 | 794 | 4 | 2.50E-05 | 2.76E-05 | 0.003088 | 3.55E-05 | -0.00287272 | SLC45A4, AC011676.5 |
| chr16 | 85343135 | 85343158 | 24 | 4 | 0.000139656 | 1.47E-05 | 0.005023 | 3.55E-05 | -0.00192071 | GSE1 |
| chr17 | 5047622 | 5047665 | 44 | 2 | 0.000122719 | 2.27E-05 | 0.001008 | 3.55E-05 | -0.02526541 | SLC52A1 |
| chr20 | 58512123 | 58512542 | 420 | 2 | 1.79E-06 | 0.0119505 | 1.68E-05 | 3.55E-05 | -0.0038432 | APCDD1L |
| chr8 | 63584604 | 63584622 | 19 | 2 | 8.76E-05 | 2.39E-05 | 0.000782 | 3.56E-05 | -0.00399621 | NA |
| chr1 | 16048726 | 16048751 | 26 | 2 | 6.03E-05 | 8.37E-05 | 7.46E-05 | 3.56E-05 | -0.00576494 | CLCNKB |
| chr11 | 2915499 | 2916173 | 675 | 5 | 1.71E-06 | 0.0109936 | 2.31E-05 | 3.56E-05 | -0.00120926 | SLC22A18 |
| chr10 | 49133944 | 49134100 | 157 | 4 | 8.24E-05 | 0.0001201 | 0.00179 | 3.56E-05 | -0.00174493 | FAM170B-AS1, FAM170B |
| chr8 | 13116835 | 13116982 | 148 | 4 | 4.49E-05 | 2.07E-05 | 0.002259 | 3.56E-05 | -0.00117037 | DLC1 |
| chr3 | 50674232 | 50674458 | 227 | 2 | 1.75E-05 | 0.0007548 | 2.56E-05 | 3.56E-05 | -0.00928151 | NA |
| chr2 | 209652717 | 209653051 | 335 | 2 | 2.05E-05 | 2.67E-05 | 0.00052 | 3.56E-05 | -0.01511777 | MAP2 |
| chr7 | 158424463 | 158425169 | 707 | 6 | 4.22E-05 | 0.0004273 | 0.000527 | 3.56E-05 | -0.00037628 | PTPRN2 |
| chr1 | 6385572 | 6385856 | 285 | 7 | 2.30E-05 | 0.0002589 | 0.000718 | 3.57E-05 | -0.00330948 | ACOT7 |
| chr4 | 80266752 | 80267317 | 566 | 5 | 1.54E-05 | 9.31E-06 | 0.009697 | 3.57E-05 | 0.00016802 | FGF5 |
| chr15 | 89654693 | 89655295 | 603 | 4 | 4.12E-05 | 6.23E-05 | 0.00134 | 3.57E-05 | 1.13E-05 | KIF7 |
| chr7 | 50492826 | 50493101 | 276 | 4 | 6.91E-05 | 8.91E-05 | 0.002537 | 3.57E-05 | -0.00147912 | FIGNL1, DDC |
| chr3 | 130746728 | 130747217 | 490 | 8 | 4.11E-05 | 0.0001254 | 0.000487 | 3.58E-05 | -0.0006459 | PIK3R4 |
| chr19 | 4208037 | 4208786 | 750 | 2 | 3.14E-06 | 0.0001541 | 4.91E-05 | 3.58E-05 | -0.01308045 | ANKRD24 |
| chr14 | 54691705 | 54691936 | 232 | 3 | 5.75E-05 | 2.14E-05 | 0.001695 | 3.58E-05 | 0.01846272 | SAMD4A |
| chr1 | 3510843 | 3511034 | 192 | 3 | 4.50E-05 | 7.70E-05 | 0.000205 | 3.58E-05 | 0.007693145 | MEGF6 |
| chr2 | 120278775 | 120279215 | 441 | 5 | 0.000135025 | 9.99E-06 | 0.007527 | 3.58E-05 | -0.00026399 | RALB |
| chr9 | 81588041 | 81588277 | 237 | 3 | 8.51E-05 | 1.64E-05 | 0.003689 | 3.58E-05 | -0.01362559 | TLE1 |
| chr11 | 69744693 | 69744791 | 99 | 2 | 0.000111631 | 2.66E-05 | 0.000535 | 3.58E-05 | -0.00948251 | NA |
| chr12 | 48974128 | 48974624 | 497 | 3 | 0.000105538 | 1.69E-05 | 0.002661 | 3.58E-05 | -0.00679152 | NA |
| chr22 | 43823594 | 43823802 | 209 | 2 | 7.25E-05 | 2.98E-05 | 0.000378 | 3.58E-05 | -0.01007432 | NA |
| chr13 | 112983381 | 112983705 | 325 | 5 | 8.67E-05 | 1.70E-05 | 0.00704 | 3.58E-05 | -0.00223312 | MCF2L |
| chr17 | 73236399 | 73236455 | 57 | 2 | 6.92E-05 | 0.0001259 | 5.60E-05 | 3.58E-05 | 0.004385682 | C17orf80, FAM104A |
| chr5 | 55737124 | 55737786 | 663 | 3 | 7.05E-05 | 0.0001449 | 6.02E-05 | 3.58E-05 | -0.00044332 | SLC38A9 |
| chr4 | 144708022 | 144708190 | 169 | 2 | 5.07E-05 | 3.49E-05 | 0.000258 | 3.58E-05 | -0.00996808 | HHIP, AC098588.3, AC098588.2 |
| chr1 | 117001584 | 117001794 | 211 | 2 | 8.77E-05 | 2.15E-05 | 0.001482 | 3.58E-05 | -0.00911542 | CD101 |
| chr16 | 4192566 | 4192905 | 340 | 2 | 9.58E-06 | 0.002968 | 1.94E-05 | 3.59E-05 | 0.002441342 | SRL |
| chr3 | 184812179 | 184812807 | 629 | 7 | 2.44E-05 | 3.71E-05 | 0.000719 | 3.59E-05 | 0.000844461 | VPS8 |
| chr2 | 177177740 | 177178277 | 538 | 4 | 5.62E-05 | 6.22E-05 | 0.00021 | 3.59E-05 | 0.002727173 | STUB1P1, AC079305.2 |
| chr19 | 10395860 | 10396207 | 348 | 2 | 5.28E-05 | 2.68E-05 | 0.000525 | 3.59E-05 | -0.00472413 | CDC37 |
| chr20 | 22490892 | 22491121 | 230 | 2 | 3.41E-05 | 2.86E-05 | 0.00043 | 3.59E-05 | -0.01858945 | NA |
| chr19 | 970973 | 971130 | 158 | 2 | 8.01E-05 | 4.78E-05 | 0.000145 | 3.59E-05 | -0.00270336 | ARID3A |
| chr20 | 20052337 | 20052889 | 553 | 17 | 1.98E-05 | 0.0028468 | 0.0022 | 3.59E-05 | 0.000627305 | CFAP61, CRNKL1 |
| chr6 | 14542875 | 14543197 | 323 | 2 | 1.24E-05 | 0.0012827 | 2.27E-05 | 3.59E-05 | -0.00545812 | NA |
| chr11 | 67482575 | 67482838 | 264 | 5 | 8.60E-05 | 4.66E-05 | 0.000599 | 3.59E-05 | -0.00198042 | NA |
| chr15 | 41416516 | 41416719 | 204 | 4 | 5.93E-05 | 0.0001458 | 0.000249 | 3.60E-05 | 0.002166237 | RTF1 |
| chr11 | 65333604 | 65334371 | 768 | 7 | 6.82E-06 | 0.0029959 | 4.65E-05 | 3.60E-05 | -0.00056148 | DPF2 |
| chr1 | 13583027 | 13583339 | 313 | 5 | 9.21E-05 | 0.0002192 | 0.000798 | 3.60E-05 | 0.008642222 | NA |
| chr1 | 44654475 | 44654960 | 486 | 6 | 2.93E-06 | 0.0009033 | 1.55E-05 | 3.60E-05 | -0.0021227 | TMEM53 |
| chr8 | 96643765 | 96643892 | 128 | 2 | 2.00E-05 | 2.73E-05 | 0.000505 | 3.60E-05 | -0.00176457 | NA |
| chr8 | 85245135 | 85245506 | 372 | 10 | 3.06E-05 | 1.66E-05 | 0.012175 | 3.60E-05 | -0.00405491 | CA13, AC011773.3 |
| chr15 | 99443887 | 99443937 | 51 | 2 | 6.96E-05 | 0.0001284 | 5.57E-05 | 3.60E-05 | -0.0012301 | NA |
| chr10 | 2176562 | 2176677 | 116 | 2 | 0.000104265 | 2.17E-05 | 0.001464 | 3.60E-05 | -6.35E-05 | LINC02662 |
| chr7 | 25895527 | 25895603 | 77 | 2 | 2.95E-05 | 0.0098482 | 1.72E-05 | 3.60E-05 | 0.004661648 | NA |
| chr2 | 167288971 | 167289035 | 65 | 2 | 6.56E-05 | 0.0001541 | 4.97E-05 | 3.61E-05 | -0.00524099 | NA |
| chr14 | 85531648 | 85532325 | 678 | 6 | 1.48E-06 | 0.0019364 | 4.54E-05 | 3.61E-05 | 0.000656504 | FLRT2 |
| chr21 | 41804856 | 41805185 | 330 | 2 | 1.46E-05 | 0.0011223 | 2.35E-05 | 3.61E-05 | -0.0025213 | PRDM15 |
| chr3 | 128605153 | 128605489 | 337 | 2 | 1.13E-05 | 0.0009809 | 2.43E-05 | 3.61E-05 | -0.00950973 | NA |
| chr16 | 85746779 | 85747099 | 321 | 2 | 1.25E-05 | 0.0056673 | 1.81E-05 | 3.61E-05 | -0.01101582 | C16orf74 |
| chr8 | 119066934 | 119067386 | 453 | 2 | 5.65E-05 | 2.39E-05 | 0.000855 | 3.61E-05 | -0.02052815 | COLEC10, AC107953.2 |
| chr6 | 132060696 | 132060792 | 97 | 2 | 8.50E-05 | 5.89E-05 | 0.00011 | 3.61E-05 | -0.01164043 | AL133346.1 |
| chr19 | 5288203 | 5288620 | 418 | 2 | 0.000138587 | 2.18E-05 | 0.00144 | 3.61E-05 | -0.00160431 | PTPRS |
| chr13 | 113292705 | 113293371 | 667 | 2 | 9.11E-05 | 2.35E-05 | 0.000929 | 3.61E-05 | -0.00127525 | NA |
| chr4 | 41646415 | 41646603 | 189 | 2 | 9.67E-05 | 2.51E-05 | 0.000683 | 3.61E-05 | -0.00830146 | LIMCH1 |
| chr1 | 78685685 | 78685766 | 82 | 2 | 0.000101782 | 2.22E-05 | 0.00128 | 3.61E-05 | -0.00427694 | NA |
| chr22 | 45930146 | 45930795 | 650 | 3 | 7.60E-06 | 8.06E-05 | 0.000101 | 3.61E-05 | -0.01559901 | WNT7B |
| chr8 | 144515713 | 144516541 | 829 | 5 | 2.19E-06 | 0.0003561 | 0.000637 | 3.61E-05 | -0.0012647 | RECQL4 |
| chr15 | 70480124 | 70480421 | 298 | 2 | 5.40E-05 | 3.52E-05 | 0.000259 | 3.61E-05 | -0.00410664 | NA |
| chr16 | 64006 | 64116 | 111 | 2 | 6.85E-05 | 2.25E-05 | 0.001193 | 3.61E-05 | -0.00807863 | RHBDF1 |
| chr14 | 34982587 | 34983235 | 649 | 12 | 3.07E-05 | 0.0001583 | 0.002339 | 3.61E-05 | 0.000693388 | SRP54 |
| chr19 | 43413690 | 43414256 | 567 | 3 | 3.95E-06 | 0.000497 | 0.000767 | 3.61E-05 | -0.00435903 | TEX101 |
| chr2 | 106582316 | 106582680 | 365 | 5 | 6.81E-05 | 4.54E-05 | 0.002181 | 3.61E-05 | 0.014952209 | NA |
| chr14 | 55128509 | 55129638 | 1130 | 9 | 9.12E-05 | 2.77E-05 | 0.021569 | 3.61E-05 | -0.00120489 | LGALS3 |
| chr3 | 138694867 | 138694899 | 33 | 2 | 3.30E-05 | 0.0051535 | 1.83E-05 | 3.62E-05 | -0.00427113 | PIK3CB |
| chr7 | 99501411 | 99501622 | 212 | 2 | 9.03E-06 | 2.18E-05 | 0.001463 | 3.62E-05 | 0.003423263 | ZNF789, ZNF394 |
| chr3 | 36945064 | 36945549 | 486 | 7 | 1.76E-05 | 0.0001098 | 0.002432 | 3.62E-05 | 0.00011793 | NA |
| chr4 | 55068628 | 55068775 | 148 | 2 | 5.41E-05 | 0.0001896 | 4.46E-05 | 3.62E-05 | -0.00693884 | AC111194.2 |
| chr17 | 47117608 | 47117642 | 35 | 2 | 6.26E-05 | 3.61E-05 | 0.000247 | 3.62E-05 | 0.002645422 | NA |
| chr2 | 136116728 | 136118067 | 1340 | 8 | 4.07E-07 | 0.0002285 | 0.000558 | 3.62E-05 | 6.95E-05 | CXCR4 |
| chr5 | 134758392 | 134759182 | 791 | 11 | 2.18E-05 | 0.0077295 | 0.00034 | 3.62E-05 | 0.001687803 | DDX46 |
| chr5 | 3225742 | 3225808 | 67 | 2 | 4.36E-05 | 0.0009244 | 2.48E-05 | 3.62E-05 | -0.00440015 | NA |
| chr3 | 126988570 | 126989232 | 663 | 3 | 8.56E-06 | 0.0001906 | 0.000104 | 3.62E-05 | -0.00390093 | PLXNA1 |
| chr5 | 1911987 | 1912053 | 67 | 2 | 0.00010155 | 2.25E-05 | 0.0012 | 3.62E-05 | -0.00663791 | NA |
| chr6 | 168078942 | 168079333 | 392 | 5 | 2.53E-05 | 0.0004574 | 0.000125 | 3.62E-05 | -0.00354538 | FRMD1 |
| chr12 | 24704290 | 24704518 | 229 | 4 | 7.65E-05 | 7.10E-05 | 0.000129 | 3.62E-05 | -0.00535599 | AC023796.1 |
| chr17 | 47896413 | 47897053 | 641 | 2 | 7.82E-06 | 5.72E-05 | 0.000115 | 3.63E-05 | 0.005601764 | SP2 |
| chr7 | 151761709 | 151761712 | 4 | 2 | 3.27E-05 | 0.0062261 | 1.80E-05 | 3.63E-05 | 0.001860851 | PRKAG2 |
| chr2 | 183064596 | 183065196 | 601 | 3 | 4.63E-05 | 0.0007116 | 0.000114 | 3.63E-05 | -0.00183278 | NA |
| chr20 | 25354095 | 25354527 | 433 | 4 | 4.43E-05 | 0.0006055 | 6.46E-05 | 3.63E-05 | -0.00894968 | ABHD12 |
| chr16 | 75409632 | 75409670 | 39 | 2 | 0.00012682 | 2.20E-05 | 0.001408 | 3.63E-05 | -0.00306768 | CFDP1 |
| chr8 | 42411524 | 42411806 | 283 | 2 | 1.74E-05 | 0.0040759 | 1.88E-05 | 3.63E-05 | -0.00723554 | NA |
| chr7 | 143408502 | 143409037 | 536 | 9 | 7.76E-05 | 0.0002397 | 0.004379 | 3.63E-05 | 0.00164053 | EPHA1-AS1, EPHA1 |
| chr1 | 21629809 | 21630012 | 204 | 2 | 0.000111726 | 2.29E-05 | 0.001108 | 3.63E-05 | -0.01314303 | RAP1GAP |
| chr12 | 106251661 | 106252186 | 526 | 3 | 0.000145857 | 7.63E-05 | 0.000725 | 3.63E-05 | -0.00499033 | AC079174.2, CKAP4 |
| chr2 | 60569871 | 60570078 | 208 | 3 | 4.23E-05 | 0.0002489 | 4.22E-05 | 3.63E-05 | 0.000464747 | NA |
| chr8 | 51809952 | 51810885 | 934 | 5 | 1.09E-06 | 0.0004787 | 0.00018 | 3.63E-05 | -0.00209316 | AC090186.1 |
| chr9 | 107733217 | 107733723 | 507 | 3 | 4.30E-05 | 0.0001036 | 0.00013 | 3.63E-05 | -0.01131607 | NA |
| chr2 | 65388180 | 65388395 | 216 | 2 | 8.70E-06 | 0.0213815 | 1.67E-05 | 3.63E-05 | -0.00556088 | SPRED2 |
| chr14 | 101193763 | 101193944 | 182 | 2 | 7.10E-05 | 4.23E-05 | 0.000183 | 3.63E-05 | -0.00613679 | AL355836.3 |

| | | | | | | | | | | |
|---|---|---|---|---|---|---|---|---|---|---|
| chr8 | 140238330 | 140238995 | 666 | 6 | 3.83E-06 | 0.001721 | 2.45E-05 | 3.63E-05 | -1.29E-05 | TRAPPC9 |
| chr22 | 33579617 | 33579969 | 353 | 2 | 1.70E-05 | 0.0006518 | 2.74E-05 | 3.64E-05 | 0.012511773 | LARGE1 |
| chr5 | 176843729 | 176843935 | 207 | 3 | 0.000105424 | 2.01E-05 | 0.001404 | 3.64E-05 | -0.00227203 | UNC5A |
| chr19 | 12890750 | 12891176 | 427 | 11 | 4.03E-05 | 0.0004786 | 0.009575 | 3.64E-05 | 0.001182732 | GCDH |
| chr4 | 119070981 | 119071377 | 397 | 2 | 6.42E-06 | 0.0107877 | 1.73E-05 | 3.64E-05 | -0.01057689 | NA |
| chr4 | 153265807 | 153266086 | 280 | 4 | 4.81E-05 | 0.0007719 | 0.000216 | 3.64E-05 | 0.002953906 | TRIM2, AC013477.1 |
| chr2 | 214830862 | 214830955 | 94 | 2 | 0.000123592 | 2.32E-05 | 0.001044 | 3.64E-05 | -0.02106902 | SNHG31 |
| chr10 | 99540749 | 99540850 | 102 | 2 | 0.000144789 | 2.20E-05 | 0.001456 | 3.64E-05 | -1.83E-05 | NA |
| chr11 | 33327501 | 33327664 | 164 | 2 | 3.07E-05 | 0.001756 | 2.16E-05 | 3.64E-05 | 0.003838098 | HIPK3 |
| chr15 | 56365239 | 56365918 | 680 | 10 | 1.14E-05 | 0.0022753 | 0.002786 | 3.64E-05 | -0.00145524 | TEX9 |
| chr4 | 26069401 | 26069716 | 316 | 2 | 0.000116695 | 2.29E-05 | 0.001132 | 3.65E-05 | -0.00804188 | NA |
| chr19 | 49115095 | 49115471 | 377 | 5 | 7.81E-05 | 1.42E-05 | 0.00503 | 3.65E-05 | -5.20E-05 | LIN7B |
| chr5 | 34944848 | 34944879 | 32 | 2 | 0.000110584 | 3.30E-05 | 0.00031 | 3.65E-05 | -0.00518853 | DNAJC21 |
| chr3 | 66384106 | 66384212 | 107 | 2 | 1.73E-05 | 4.28E-05 | 0.000181 | 3.65E-05 | -0.00207347 | SLC25A26, LRIG1 |
| chr1 | 17247683 | 17247835 | 153 | 2 | 2.43E-05 | 0.0001473 | 5.20E-05 | 3.65E-05 | -0.00543245 | NA |
| chr19 | 53681017 | 53681354 | 338 | 3 | 1.31E-05 | 0.002164 | 1.63E-05 | 3.66E-05 | -0.00538022 | NA |
| chr11 | 4709224 | 4709561 | 338 | 2 | 0.000106996 | 3.71E-05 | 0.00024 | 3.66E-05 | 0.006820694 | MMP26 |
| chr16 | 31330158 | 31330554 | 397 | 4 | 6.34E-05 | 3.10E-05 | 0.001655 | 3.66E-05 | -0.00875165 | ITGAM |
| chr19 | 54662483 | 54662719 | 237 | 5 | 5.23E-05 | 0.000119 | 0.001722 | 3.66E-05 | -0.00461957 | LILRB4 |
| chr1 | 155046131 | 155046453 | 323 | 5 | 4.46E-06 | 0.0072535 | 6.17E-06 | 3.67E-05 | -0.00152266 | DCST1 |
| chr2 | 133567148 | 133567325 | 178 | 2 | 8.85E-06 | 0.0001938 | 4.49E-05 | 3.67E-05 | -0.00368663 | NCKAP5 |
| chr10 | 80699541 | 80699663 | 123 | 3 | 1.14E-05 | 0.0045243 | 1.58E-05 | 3.67E-05 | 0.002562872 | NA |
| chr2 | 16352996 | 16353142 | 147 | 2 | 0.000102107 | 3.36E-05 | 0.000303 | 3.67E-05 | 0.011011666 | NA |
| chr2 | 44244041 | 44244244 | 204 | 2 | 9.75E-05 | 2.38E-05 | 0.000964 | 3.67E-05 | -0.01278792 | PPM1B, AC013717.1 |
| chr3 | 129059717 | 129059895 | 179 | 6 | 1.34E-05 | 0.0006193 | 0.000397 | 3.68E-05 | 0.006260169 | NA |
| chr2 | 96819882 | 96820335 | 454 | 2 | 4.86E-06 | 9.30E-05 | 7.21E-05 | 3.68E-05 | -0.00597804 | CNNM3 |
| chr10 | 7717527 | 7717658 | 132 | 2 | 3.24E-05 | 0.002321 | 2.07E-05 | 3.68E-05 | 0.003586559 | ITIH2 |
| chr6 | 106359807 | 106359989 | 183 | 3 | 1.66E-05 | 1.50E-05 | 0.004722 | 3.68E-05 | 0.010347277 | NA |
| chr12 | 14814436 | 14814799 | 364 | 2 | 2.81E-05 | 0.0001186 | 6.04E-05 | 3.68E-05 | 0.005992054 | C12orf60 |
| chr20 | 64115644 | 64115855 | 212 | 2 | 3.56E-05 | 0.0069475 | 1.80E-05 | 3.68E-05 | -0.0054645 | MYT1 |
| chr2 | 231594203 | 231594959 | 757 | 3 | 2.13E-06 | 0.0320402 | 7.00E-06 | 3.68E-05 | -0.00140226 | TEX44 |
| chr16 | 31873378 | 31874160 | 783 | 11 | 6.90E-06 | 0.0008576 | 0.000207 | 3.68E-05 | 0.001132702 | ZNF267 |
| chr7 | 131692105 | 131692326 | 222 | 3 | 0.00010446 | 5.31E-05 | 0.000801 | 3.68E-05 | -0.00205957 | NA |
| chr18 | 24402591 | 24402656 | 66 | 2 | 0.000103338 | 3.96E-05 | 0.000214 | 3.68E-05 | -0.00701367 | AC023983.2 |
| chr19 | 10643944 | 10644460 | 517 | 2 | 4.50E-05 | 2.36E-05 | 0.001012 | 3.68E-05 | 0.036342144 | SLC44A2 |
| chr14 | 73273995 | 73274378 | 384 | 2 | 6.11E-06 | 0.0005681 | 2.90E-05 | 3.69E-05 | 0.002011853 | PAPLN, AC004846.2 |
| chr14 | 60213165 | 60213263 | 99 | 2 | 0.000111382 | 3.09E-05 | 0.00038 | 3.69E-05 | 0.002269829 | NA |
| chr3 | 87551520 | 87551708 | 189 | 2 | 5.94E-05 | 9.07E-05 | 7.40E-05 | 3.69E-05 | -0.00834018 | NA |
| chr6 | 35804793 | 35805446 | 654 | 10 | 0.000114705 | 8.34E-05 | 0.011272 | 3.70E-05 | -0.00253329 | LHFPL5 |
| chr20 | 47236391 | 47236479 | 89 | 2 | 1.42E-05 | 0.0001267 | 5.83E-05 | 3.70E-05 | 0.00291616 | ZMYND8 |
| chr11 | 67605612 | 67606742 | 1131 | 11 | 8.92E-06 | 0.0008552 | 0.004872 | 3.70E-05 | -0.00282654 | AP003385.3, NDUFV1, C11orf72 |
| chr10 | 13775804 | 13776133 | 330 | 2 | 2.76E-05 | 0.0001879 | 4.61E-05 | 3.70E-05 | -0.00804658 | FRMD4A |
| chr1 | 186178522 | 186178534 | 13 | 2 | 6.69E-05 | 3.00E-05 | 0.000417 | 3.70E-05 | 0.000261692 | HMCN1 |
| chr10 | 6255785 | 6255948 | 164 | 3 | 9.03E-05 | 0.00012 | 0.00049 | 3.70E-05 | -0.00299697 | NA |
| chr1 | 44228370 | 44228549 | 180 | 2 | 0.000104328 | 2.36E-05 | 0.001052 | 3.70E-05 | -0.00444515 | ERI3 |
| chr9 | 131817802 | 131817950 | 149 | 2 | 3.82E-05 | 2.54E-05 | 0.000744 | 3.70E-05 | -0.00404061 | AL160271.2 |
| chr10 | 17201827 | 17202193 | 367 | 4 | 0.000111214 | 2.47E-05 | 0.003926 | 3.70E-05 | -0.00580784 | TRDMT1 |
| chr18 | 23689808 | 23690384 | 577 | 4 | 1.75E-05 | 0.000426 | 0.001252 | 3.70E-05 | -0.0023983 | LAMA3 |
| chr19 | 55113761 | 55113816 | 56 | 3 | 1.46E-05 | 0.0045962 | 1.93E-05 | 3.71E-05 | -0.00573664 | PPP1R12C |
| chr15 | 94120146 | 94120432 | 287 | 2 | 9.58E-05 | 2.84E-05 | 0.000493 | 3.71E-05 | -0.00181423 | NA |
| chr8 | 1101510 | 1101895 | 386 | 3 | 1.05E-05 | 0.0003595 | 6.39E-05 | 3.71E-05 | 0.003528627 | DLGAP2 |
| chr1 | 183195277 | 183195561 | 285 | 2 | 0.000130563 | 2.23E-05 | 0.001478 | 3.71E-05 | -0.01330186 | LAMC2 |
| chr12 | 69690056 | 69690721 | 666 | 5 | 6.05E-06 | 6.01E-05 | 0.001968 | 3.71E-05 | 0.001831774 | BEST3 |
| chr5 | 120464539 | 120465447 | 909 | 6 | 2.51E-06 | 0.000118 | 0.000581 | 3.71E-05 | 0.004548385 | PRR16 |
| chr3 | 23671784 | 23671950 | 167 | 2 | 3.62E-05 | 0.0011416 | 2.41E-05 | 3.71E-05 | -0.01255634 | NA |
| chr12 | 130576730 | 130577214 | 485 | 3 | 5.49E-05 | 5.56E-05 | 0.00017 | 3.71E-05 | -0.00123089 | RIMBP2 |
| chr1 | 244036410 | 244036479 | 70 | 2 | 0.000129334 | 2.26E-05 | 0.001371 | 3.71E-05 | -0.00649784 | LINC02774 |
| chr17 | 62065130 | 62065814 | 685 | 5 | 1.14E-05 | 0.0003253 | 0.000746 | 3.71E-05 | -0.00367901 | MED13 |
| chr7 | 6272423 | 6272822 | 400 | 4 | 0.000141624 | 2.57E-05 | 0.00189 | 3.71E-05 | -0.00108829 | CYTH3 |
| chr10 | 128710393 | 128710531 | 139 | 2 | 6.41E-06 | 2.24E-05 | 0.001479 | 3.71E-05 | 0.020646493 | NA |
| chr15 | 74344920 | 74345217 | 298 | 3 | 5.98E-05 | 2.13E-05 | 0.001242 | 3.71E-05 | -0.00472374 | CYP11A1 |
| chr3 | 9216349 | 9216889 | 541 | 5 | 1.30E-05 | 0.0008152 | 2.54E-05 | 3.72E-05 | -0.00232176 | SRGAP3 |
| chr17 | 74261824 | 74262092 | 269 | 4 | 5.66E-05 | 1.17E-05 | 0.009254 | 3.72E-05 | -0.00232432 | TTYH2, AC100786.1 |
| chr17 | 48430328 | 48430743 | 416 | 10 | 2.76E-05 | 0.0066588 | 0.000306 | 3.72E-05 | -0.00037779 | NA |
| chr19 | 463460 | 463990 | 531 | 3 | 1.61E-06 | 0.0002312 | 5.36E-05 | 3.72E-05 | -0.00487542 | ODF3L2 |
| chr16 | 571764 | 571841 | 78 | 3 | 1.38E-05 | 0.0040731 | 1.98E-05 | 3.72E-05 | 0.006257256 | PIGQ, Z98883.1 |
| chr16 | 57627605 | 57628435 | 831 | 11 | 3.57E-06 | 0.0004334 | 0.00085 | 3.72E-05 | -0.00181342 | ADGRG1, AC018552.3 |
| chr8 | 64576083 | 64576358 | 276 | 3 | 0.00010188 | 6.38E-05 | 0.000451 | 3.72E-05 | -0.00309016 | AC090136.3 |
| chr2 | 240913684 | 240913977 | 294 | 2 | 1.72E-05 | 3.18E-05 | 0.000363 | 3.72E-05 | -0.00692926 | CROCC2 |
| chr15 | 67841140 | 67841634 | 495 | 3 | 4.55E-05 | 2.26E-05 | 0.001093 | 3.72E-05 | -0.01026787 | AC009292.2 |
| chr6 | 166765430 | 166765467 | 38 | 2 | 1.20E-05 | 0.0056669 | 1.86E-05 | 3.73E-05 | -0.00236136 | RPS6KA2 |
| chr14 | 67427447 | 67427546 | 100 | 2 | 6.96E-05 | 2.77E-05 | 0.000554 | 3.73E-05 | -0.0059223 | NA |
| chr1 | 53889599 | 53890233 | 635 | 9 | 4.85E-05 | 0.0005749 | 0.009259 | 3.73E-05 | 0.000457866 | YIPF1 |
| chr11 | 102335983 | 102336271 | 289 | 3 | 1.80E-05 | 0.0018635 | 9.08E-05 | 3.73E-05 | -0.00968335 | BIRC3 |
| chr15 | 98707211 | 98707778 | 568 | 7 | 5.65E-07 | 0.0107299 | 6.29E-05 | 3.73E-05 | 0.001710949 | IGF1R |
| chr1 | 155962663 | 155963003 | 341 | 2 | 2.74E-05 | 3.62E-05 | 0.000268 | 3.73E-05 | -0.0122504 | ARHGEF2 |
| chr18 | 25466642 | 25466884 | 243 | 2 | 1.68E-05 | 2.94E-05 | 0.000455 | 3.73E-05 | -0.00436395 | NA |
| chr12 | 118135726 | 118136463 | 738 | 11 | 4.91E-05 | 1.12E-05 | 0.011748 | 3.73E-05 | 0.001564935 | PEBP1, VSIG10 |
| chr1 | 70221099 | 70221876 | 778 | 8 | 3.69E-05 | 0.0017563 | 0.000172 | 3.74E-05 | -0.00208253 | SRSF11 |
| chr17 | 72110648 | 72111044 | 397 | 3 | 6.30E-05 | 2.48E-05 | 0.001607 | 3.74E-05 | 0.018367251 | AC007461.1, SOX9-AS1 |
| chr8 | 1500832 | 1501865 | 1034 | 8 | 1.03E-05 | 0.004303 | 0.000103 | 3.74E-05 | 0.000286783 | DLGAP2 |
| chr3 | 44948768 | 44948902 | 135 | 2 | 6.59E-05 | 0.0001257 | 5.94E-05 | 3.74E-05 | 0.002946441 | ZDHHC3 |
| chr16 | 84104205 | 84104352 | 148 | 2 | 4.76E-05 | 0.000194 | 4.60E-05 | 3.74E-05 | -0.01030407 | MBTPS1 |
| chr2 | 111919176 | 111919257 | 82 | 2 | 0.000110125 | 2.25E-05 | 0.001541 | 3.74E-05 | -0.00449082 | MERTK |
| chr20 | 63328263 | 63328359 | 97 | 2 | 4.17E-05 | 8.21E-05 | 8.24E-05 | 3.74E-05 | -0.00733965 | COL20A1 |
| chr6 | 118776791 | 118776972 | 182 | 2 | 0.000105538 | 3.08E-05 | 0.000404 | 3.74E-05 | -0.00127212 | SELENOKP3 |
| chr15 | 26131120 | 26131498 | 379 | 2 | 6.30E-05 | 2.40E-05 | 0.001025 | 3.75E-05 | -0.00119498 | LINC00929 |
| chr12 | 14843209 | 14843842 | 634 | 11 | 7.52E-05 | 0.0001196 | 0.001682 | 3.75E-05 | 0.001654257 | C12orf60, AC007655.1, ART4 |
| chr3 | 151066942 | 151067223 | 282 | 2 | 9.37E-05 | 3.02E-05 | 0.000427 | 3.75E-05 | 0.004900764 | CLRN1-AS1 |
| chr17 | 77319162 | 77319786 | 625 | 9 | 3.80E-05 | 7.51E-05 | 0.004835 | 3.75E-05 | 0.005282525 | SEPTIN9 |
| chr12 | 109739884 | 109739938 | 55 | 2 | 0.000122521 | 2.72E-05 | 0.000602 | 3.75E-05 | -0.01498102 | FAM222A, FAM222A-AS1 |
| chr8 | 59593701 | 59593909 | 209 | 4 | 3.50E-05 | 0.0003489 | 4.20E-05 | 3.75E-05 | -0.00449314 | AC087664.1 |
| chr19 | 12917994 | 12918561 | 568 | 2 | 1.35E-05 | 4.82E-05 | 0.000159 | 3.75E-05 | 0.00407385 | SYCE2 |
| chr6 | 125268211 | 125268512 | 302 | 2 | 0.000130954 | 2.27E-05 | 0.001444 | 3.75E-05 | -0.00017152 | AL121938.1, HDDC2 |
| chr12 | 106736564 | 106736590 | 27 | 2 | 0.000107571 | 4.07E-05 | 0.000214 | 3.76E-05 | 0.017878453 | RFX4, AC079385.1 |
| chr11 | 3032830 | 3032912 | 83 | 2 | 6.88E-05 | 0.0001495 | 5.37E-05 | 3.76E-05 | 0.00306713 | CARS1-AS1, CARS1 |
| chr19 | 1546207 | 1546386 | 180 | 4 | 0.000127519 | 4.65E-05 | 0.001417 | 3.76E-05 | 0.00271344 | NA |
| chr4 | 153538320 | 153538586 | 267 | 2 | 5.84E-05 | 2.99E-05 | 0.000448 | 3.76E-05 | -0.00785113 | TMEM131L |
| chr3 | 39466568 | 39466857 | 290 | 4 | 0.000117064 | 2.43E-05 | 0.002052 | 3.76E-05 | -0.00299977 | NA |
| chr17 | 835582 | 835629 | 48 | 2 | 1.83E-05 | 7.38E-05 | 9.22E-05 | 3.77E-05 | -0.00977773 | NXN |
| chr16 | 85422242 | 85422305 | 64 | 2 | 9.85E-05 | 2.33E-05 | 0.001273 | 3.77E-05 | 0.033212415 | GSE1 |
| chr12 | 13347380 | 13347396 | 17 | 2 | 0.000113544 | 3.53E-05 | 0.000291 | 3.77E-05 | -0.01395722 | NA |
| chr19 | 18097125 | 18097695 | 571 | 9 | 4.68E-06 | 0.003752 | 0.000103 | 3.77E-05 | -0.00316732 | IL12RB1 |
| chr12 | 12946746 | 12946950 | 205 | 2 | 8.24E-06 | 2.27E-05 | 0.001531 | 3.77E-05 | 0.001945754 | GPRC5D-AS1, GPRC5D |
| chr11 | 131636444 | 131636645 | 202 | 3 | 0.000138425 | 2.14E-05 | 0.001506 | 3.77E-05 | -0.00045414 | NTM, NTM-AS1 |
| chr19 | 53671352 | 53671933 | 582 | 4 | 6.59E-05 | 9.81E-06 | 0.012568 | 3.77E-05 | 0.001498363 | NA |
| chr14 | 104582716 | 104582783 | 68 | 2 | 7.70E-05 | 7.77E-05 | 8.78E-05 | 3.77E-05 | -0.00406724 | C14orf180, TMEM179 |
| chr17 | 78344834 | 78345123 | 290 | 2 | 1.63E-05 | 0.0036762 | 1.98E-05 | 3.77E-05 | -0.00889337 | AC087645.2 |
| chr17 | 74213059 | 74213663 | 605 | 10 | 2.70E-05 | 0.0005975 | 0.002029 | 3.77E-05 | 0.001136921 | TTYH2, MGC16275 |
| chr2 | 202588985 | 202589087 | 103 | 3 | 0.000127672 | 3.12E-05 | 0.000593 | 3.78E-05 | -0.00309414 | NA |
| chr16 | 85436879 | 85437068 | 190 | 3 | 5.37E-05 | 2.12E-05 | 0.002683 | 3.78E-05 | -0.01146984 | GSE1 |
| chr9 | 136094303 | 136094533 | 231 | 3 | 4.55E-05 | 8.76E-05 | 0.001173 | 3.78E-05 | 0.004082767 | NACC2 |
| chr15 | 54082700 | 54083005 | 306 | 2 | 2.75E-05 | 0.0003547 | 3.53E-05 | 3.78E-05 | -0.01370568 | UNC13C |
| chr18 | 62524171 | 62524438 | 268 | 3 | 1.32E-05 | 2.06E-05 | 0.002656 | 3.78E-05 | 0.002906612 | ZCCHC2 |
| chr1 | 53262506 | 53262611 | 106 | 2 | 8.27E-05 | 2.27E-05 | 0.001549 | 3.78E-05 | -0.00545554 | LRP8 |
| chr10 | 117133919 | 117134669 | 751 | 5 | 3.10E-05 | 0.000535 | 0.000586 | 3.78E-05 | 0.000217326 | VAX1 |
| chr1 | 2923198 | 2923743 | 546 | 5 | 1.82E-05 | 0.0002676 | 3.53E-05 | 3.78E-05 | 0.00596867 | NA |
| chr1 | 3111666 | 3112346 | 681 | 8 | 3.49E-06 | 0.0008638 | 3.78E-05 | 3.78E-05 | 0.002934794 | PRDM16 |
| chr16 | 50547980 | 50548542 | 563 | 7 | 4.34E-05 | 3.81E-05 | 0.011501 | 3.78E-05 | 0.000265028 | NKD1 |
| chr2 | 96489867 | 96490503 | 637 | 2 | 8.99E-07 | 0.0019198 | 2.21E-05 | 3.79E-05 | -0.01541816 | NA |
| chr2 | 241801858 | 241801992 | 135 | 3 | 3.38E-05 | 2.92E-05 | 0.001585 | 3.79E-05 | -0.00536758 | GAL3ST2, AC131097.1 |
| chr12 | 113471913 | 113472796 | 884 | 11 | 5.80E-05 | 8.54E-05 | 0.003622 | 3.79E-05 | 0.000786429 | LHX5-AS1, LHX5 |
| chr7 | 76357644 | 76358293 | 650 | 4 | 1.57E-05 | 1.59E-05 | 0.005555 | 3.79E-05 | -0.00086472 | YWHAG |
| chr1 | 110746720 | 110746818 | 99 | 2 | 5.05E-05 | 0.0004802 | 3.17E-05 | 3.79E-05 | -0.01200389 | NA |
| chr12 | 1601508 | 1601683 | 176 | 2 | 3.63E-05 | 0.0004138 | 3.34E-05 | 3.79E-05 | -0.00295513 | WNT5B |
| chr8 | 80404644 | 80404673 | 30 | 2 | 7.52E-05 | 0.0001348 | 5.81E-05 | 3.79E-05 | -0.00045363 | AC034114.2 |
| chr1 | 3718247 | 3718646 | 400 | 6 | 2.98E-05 | 0.0003582 | 0.001645 | 3.80E-05 | -0.00983187 | TP73 |
| chr6 | 165704021 | 165704188 | 168 | 2 | 0.00011395 | 2.84E-05 | 0.000544 | 3.80E-05 | -0.00624486 | PDE10A |
| chr2 | 71130115 | 71130670 | 556 | 16 | 8.85E-05 | 0.0017362 | 0.001166 | 3.80E-05 | 0.000439955 | MPHOSPH10, MCEE |
| chr4 | 168593694 | 168593718 | 25 | 2 | 3.69E-05 | 3.46E-05 | 0.000314 | 3.80E-05 | -0.01307725 | PALLD |
| chr3 | 48579089 | 48579650 | 562 | 3 | 3.27E-05 | 1.46E-05 | 0.005654 | 3.80E-05 | -0.00127965 | COL7A1 |
| chr14 | 99936132 | 99936438 | 307 | 3 | 7.50E-05 | 3.80E-05 | 0.001627 | 3.80E-05 | -0.00073737 | EML1 |
| chr7 | 128934340 | 128934705 | 366 | 3 | 9.59E-06 | 0.0124264 | 9.47E-06 | 3.80E-05 | -0.0106116 | NA |
| chr4 | 136282113 | 136282339 | 227 | 2 | 9.65E-05 | 2.44E-05 | 0.001042 | 3.80E-05 | -0.0205778 | AC018680.1 |
| chr1 | 15737744 | 15738843 | 1100 | 4 | 1.88E-05 | 1.66E-05 | 0.003581 | 3.81E-05 | -0.00348709 | SLC25A34 |
| chr4 | 87831508 | 87832027 | 520 | 3 | 2.47E-05 | 0.000157 | 6.37E-05 | 3.81E-05 | -0.00085359 | MEPE |
| chr11 | 130847631 | 130847849 | 219 | 4 | 6.46E-05 | 6.53E-05 | 0.000347 | 3.81E-05 | -0.0046385 | LINC02551 |
| chr10 | 1244427 | 1244616 | 190 | 2 | 3.08E-05 | 0.0001137 | 6.54E-05 | 3.81E-05 | -0.00513196 | LINC00200, ADARB2 |
| chr20 | 49914844 | 49915067 | 224 | 3 | 1.07E-06 | 0.0078349 | 2.68E-05 | 3.81E-05 | 0.002767114 | SPATA2 |
| chr9 | 136453988 | 136454698 | 711 | 4 | 3.56E-05 | 2.02E-05 | 0.003864 | 3.81E-05 | 0.002081457 | SEC16A |
| chr1 | 2183817 | 2184246 | 430 | 5 | 6.99E-05 | 0.0001041 | 0.001167 | 3.81E-05 | -0.00045678 | PRKCZ, PRKCZ-AS1 |
| chr12 | 117036198 | 117036276 | 79 | 2 | 3.48E-05 | 8.72E-05 | 8.05E-05 | 3.81E-05 | 0.005635301 | NA |
| chr1 | 149918737 | 149919103 | 367 | 4 | 4.09E-06 | 0.0024172 | 4.64E-05 | 3.82E-05 | -0.00364621 | NA |
| chr6 | 41097868 | 41098150 | 283 | 2 | 2.93E-05 | 2.29E-05 | 0.001557 | 3.82E-05 | 0.005000401 | NFYA, AL031778.1 |
| chr15 | 99029358 | 99029556 | 199 | 3 | 5.44E-05 | 0.0008134 | 0.000391 | 3.82E-05 | -0.00503752 | LUNAR1, AC036108.1 |
| chr8 | 137851589 | 137851663 | 75 | 2 | 4.58E-05 | 2.29E-05 | 0.001555 | 3.82E-05 | -0.01478767 | AC046195.1 |
| chr3 | 114056589 | 114056931 | 343 | 9 | 2.84E-05 | 0.0006999 | 0.000658 | 3.82E-05 | 0.000653234 | QTRT2, CCDC191 |
| chr12 | 3040931 | 3041397 | 467 | 4 | 1.90E-05 | 0.0003715 | 3.04E-05 | 3.82E-05 | 0.001443519 | NA |
| chr3 | 69122584 | 69123052 | 469 | 3 | 4.50E-05 | 1.92E-05 | 0.002091 | 3.82E-05 | -0.0036775 | LMOD3 |
| chr14 | 104636640 | 104637157 | 518 | 4 | 7.08E-05 | 0.0003224 | 0.000716 | 3.83E-05 | 0.000543003 | NA |
| chr19 | 4851563 | 4852493 | 931 | 7 | 9.98E-06 | 0.0019076 | 3.79E-05 | 3.83E-05 | -0.00089619 | PLIN3 |
| chr11 | 70575784 | 70576078 | 295 | 2 | 0.000106235 | 2.30E-05 | 0.001544 | 3.83E-05 | -0.00970568 | SHANK2 |
| chr1 | 23169361 | 23169504 | 144 | 2 | 7.18E-05 | 0.0010999 | 2.51E-05 | 3.83E-05 | -0.01316851 | LUZP1 |
| chr16 | 29825735 | 29825946 | 212 | 2 | 7.66E-05 | 2.60E-05 | 0.000796 | 3.83E-05 | -0.01453843 | AC120114.4, MVP |
| chr22 | 44442875 | 44443242 | 368 | 4 | 2.67E-05 | 0.0006853 | 0.000447 | 3.83E-05 | -0.01232067 | NA |
| chr16 | 88803393 | 88803896 | 504 | 4 | 2.87E-05 | 0.0001792 | 0.000581 | 3.84E-05 | -0.00402748 | CDT1 |
| chr12 | 70920807 | 70921484 | 678 | 8 | 3.06E-07 | 0.0087626 | 0.000343 | 3.84E-05 | -0.00496476 | NA |
| chr19 | 10113767 | 10114084 | 318 | 3 | 2.03E-05 | 0.0021515 | 0.000103 | 3.84E-05 | 0.005230944 | PPAN-P2RY11, P2RY11 |

| | | | | | | | | | | |
|---|---|---|---|---|---|---|---|---|---|---|
| chr5 | 150256131 | 150256869 | 739 | 4 | 1.05E-06 | 0.0074364 | 6.73E-06 | 3.84E-05 | -0.00757529 | CAMK2A |
| chr10 | 119165581 | 119166120 | 540 | 10 | 5.60E-05 | 0.0003159 | 0.000209 | 3.84E-05 | -0.00114904 | SFXN4 |
| chr5 | 393232 | 393251 | 20 | 2 | 0.000145833 | 2.64E-05 | 0.000747 | 3.84E-05 | -0.00892624 | PDCD6-AHRR, AHRR |
| chr6 | 30516060 | 30516557 | 498 | 11 | 1.58E-05 | 0.0013466 | 0.000119 | 3.84E-05 | 0.000413302 | LINC02569 |
| chr19 | 42219757 | 42220357 | 601 | 11 | 3.90E-05 | 0.0005874 | 0.001729 | 3.85E-05 | 0.000613045 | ZNF526, DEDD2, AC006486.3 |
| chr5 | 68214905 | 68215045 | 141 | 4 | 3.46E-05 | 6.26E-05 | 0.002318 | 3.85E-05 | 0.003793572 | NA |
| chr21 | 46431556 | 46432027 | 472 | 6 | 3.12E-05 | 0.0002467 | 0.002365 | 3.85E-05 | 0.002537147 | PCNT |
| chr1 | 39258462 | 39258523 | 62 | 2 | 0.00011826 | 4.84E-05 | 0.000168 | 3.85E-05 | 0.001406168 | MACF1 |
| chr1 | 3183240 | 3183933 | 694 | 6 | 2.55E-05 | 6.01E-05 | 0.007752 | 3.85E-05 | 0.001961528 | PRDM16 |
| chr19 | 53101968 | 53102178 | 211 | 2 | 4.10E-05 | 4.32E-05 | 0.000204 | 3.85E-05 | -0.00386398 | ZNF160 |
| chr19 | 47598180 | 47598793 | 614 | 2 | 1.26E-06 | 0.0004799 | 3.23E-05 | 3.85E-05 | -0.01423797 | NA |
| chr3 | 188385496 | 188385689 | 194 | 2 | 2.28E-05 | 0.0008351 | 2.72E-05 | 3.85E-05 | 0.010775721 | LPP |
| chr1 | 27109442 | 27109897 | 456 | 3 | 2.33E-05 | 0.0006943 | 0.00039 | 3.85E-05 | -0.00019493 | SLC9A1 |
| chr2 | 3470081 | 3470398 | 318 | 3 | 9.46E-06 | 0.0017345 | 1.97E-05 | 3.85E-05 | 0.002917623 | TRAPPC12 |
| chr19 | 894450 | 894654 | 205 | 4 | 2.81E-05 | 0.0029056 | 0.000336 | 3.85E-05 | -0.00044981 | NA |
| chr3 | 44511393 | 44511924 | 532 | 5 | 3.02E-05 | 0.0004359 | 0.002987 | 3.85E-05 | -0.00096876 | NA |
| chr11 | 31814336 | 31814706 | 371 | 3 | 5.82E-05 | 9.89E-05 | 0.000334 | 3.86E-05 | -0.00416529 | PAUPAR, AL035078.4, PAX6 |
| chr15 | 42094528 | 42094995 | 468 | 4 | 1.24E-05 | 5.03E-05 | 0.00132 | 3.86E-05 | -0.00499049 | PLA2G4D |
| chr2 | 97713367 | 97714030 | 664 | 5 | 1.12E-05 | 0.0007859 | 0.001249 | 3.86E-05 | 0.002740069 | ZAP70 |
| chr7 | 102164632 | 102164641 | 10 | 2 | 4.25E-05 | 0.0012596 | 2.45E-05 | 3.86E-05 | 0.003220422 | CUX1 |
| chr14 | 89986617 | 89986621 | 5 | 2 | 0.000126516 | 2.87E-05 | 0.000562 | 3.86E-05 | 0.008501717 | TDP1 |
| chr19 | 47788635 | 47789015 | 381 | 3 | 1.78E-05 | 0.0030267 | 3.29E-05 | 3.86E-05 | -0.01117185 | NA |
| chr1 | 160348927 | 160349030 | 104 | 2 | 3.39E-05 | 0.000118 | 6.50E-05 | 3.86E-05 | 0.000270479 | NCSTN |
| chr5 | 1356267 | 1356669 | 403 | 3 | 1.71E-05 | 0.0019466 | 2.10E-05 | 3.86E-05 | 0.002614944 | LINC01511 |
| chr2 | 75518075 | 75518470 | 396 | 4 | 1.59E-05 | 0.0005863 | 0.000341 | 3.86E-05 | -0.00621652 | EVA1A |
| chr22 | 32414297 | 32414404 | 108 | 2 | 3.87E-05 | 0.0004897 | 3.22E-05 | 3.87E-05 | -0.01297141 | BPIFC |
| chr6 | 71288414 | 71288695 | 282 | 5 | 1.88E-05 | 0.0001696 | 0.001728 | 3.87E-05 | 0.000565223 | AL589935.1 |
| chr1 | 24415059 | 24415651 | 593 | 8 | 2.37E-05 | 0.0001495 | 0.003557 | 3.87E-05 | 0.001963553 | STPG1 |
| chr6 | 151341516 | 151341658 | 143 | 3 | 8.72E-05 | 3.08E-05 | 0.000729 | 3.87E-05 | -0.00162631 | AKAP12 |
| chr22 | 42090739 | 42091536 | 798 | 12 | 2.60E-05 | 0.0066493 | 0.001585 | 3.87E-05 | 6.69E-05 | NDUFA6-DT, NDUFA6 |
| chr11 | 121767155 | 121767195 | 41 | 2 | 5.91E-05 | 0.0003776 | 3.54E-05 | 3.87E-05 | 0.010009333 | AP001977.1 |
| chr2 | 167868765 | 167869025 | 261 | 5 | 6.97E-05 | 5.83E-05 | 0.005731 | 3.87E-05 | -0.00310045 | B3GALT1, B3GALT1-AS1 |
| chr17 | 44762473 | 44762665 | 193 | 3 | 6.10E-05 | 0.0001641 | 0.000375 | 3.87E-05 | -0.00832133 | ADAM11 |
| chr14 | 36807549 | 36807942 | 394 | 2 | 5.90E-05 | 3.07E-05 | 0.000459 | 3.87E-05 | -0.0004652 | SLC25A21 |
| chr20 | 63553721 | 63554077 | 357 | 4 | 5.26E-05 | 4.82E-05 | 0.000828 | 3.87E-05 | 0.003204411 | FNDC11 |
| chr19 | 55400609 | 55401020 | 412 | 2 | 2.29E-05 | 0.0001017 | 7.28E-05 | 3.87E-05 | -0.00331392 | RPL28, UBE2S |
| chr4 | 105560248 | 105560329 | 82 | 2 | 0.000101497 | 2.49E-05 | 0.001037 | 3.87E-05 | -0.00088222 | ARHGEF38 |
| chr9 | 109428728 | 109429421 | 694 | 4 | 1.25E-05 | 0.0012174 | 0.000485 | 3.88E-05 | -0.00878918 | PTPN3 |
| chr4 | 87649989 | 87650501 | 513 | 4 | 2.45E-05 | 0.0014855 | 0.001336 | 3.88E-05 | 0.001651817 | DMP1, AC093895.1 |
| chr11 | 76444443 | 76445090 | 648 | 12 | 4.51E-05 | 8.52E-05 | 0.0023 | 3.88E-05 | 0.000975054 | EMSY, AP002360.1 |
| chr5 | 40798185 | 40798710 | 526 | 13 | 8.34E-05 | 0.0002112 | 0.003049 | 3.88E-05 | 2.80E-05 | PRKAA1 |
| chr8 | 116948583 | 116948891 | 309 | 2 | 7.93E-06 | 0.0017191 | 2.31E-05 | 3.89E-05 | 0.008640162 | NA |
| chr19 | 49434947 | 49435611 | 665 | 2 | 1.12E-05 | 2.85E-05 | 0.000591 | 3.89E-05 | -0.00048248 | SLC17A7 |
| chr9 | 109639990 | 109641021 | 1032 | 11 | 5.68E-06 | 0.0271137 | 2.09E-05 | 3.89E-05 | 0.001806301 | PALM2AKAP2 |
| chr6 | 49462883 | 49462900 | 18 | 2 | 9.09E-05 | 2.45E-05 | 0.00115 | 3.89E-05 | 0.000296601 | MMUT |
| chr18 | 22172331 | 22172703 | 373 | 5 | 2.12E-05 | 0.000258 | 0.0021 | 3.89E-05 | -0.00109728 | GATA6 |
| chr6 | 98825342 | 98825954 | 613 | 8 | 1.21E-05 | 0.0070848 | 0.001233 | 3.89E-05 | -0.00269671 | NA |
| chr9 | 33452951 | 33453245 | 295 | 2 | 2.46E-05 | 0.0007256 | 2.86E-05 | 3.89E-05 | -0.01061149 | NA |
| chr20 | 33166755 | 33167142 | 388 | 3 | 1.08E-05 | 0.0016752 | 0.000233 | 3.89E-05 | -0.00531199 | BPIFA2 |
| chr16 | 89625952 | 89626307 | 356 | 4 | 4.14E-05 | 0.0021798 | 6.13E-05 | 3.89E-05 | -0.00220443 | DPEP1 |
| chr22 | 43272766 | 43273138 | 373 | 2 | 1.44E-06 | 0.0353865 | 1.75E-05 | 3.89E-05 | -0.00392308 | SCUBE1 |
| chr19 | 48719635 | 48720021 | 387 | 6 | 7.04E-05 | 0.0002641 | 0.002883 | 3.90E-05 | -0.00687793 | MAMSTR |
| chr14 | 100656662 | 100657345 | 684 | 7 | 4.00E-05 | 0.0005814 | 0.000239 | 3.90E-05 | -0.00184671 | LINC00523 |
| chr22 | 27286346 | 27286400 | 55 | 2 | 0.000125477 | 2.99E-05 | 0.000513 | 3.90E-05 | -0.0064856 | AL020994.3 |
| chr14 | 88384272 | 88385040 | 769 | 5 | 6.90E-06 | 0.0001135 | 0.000932 | 3.90E-05 | -0.00831714 | SPATA7 |
| chr19 | 53432023 | 53432077 | 55 | 2 | 5.03E-05 | 2.57E-05 | 0.000908 | 3.90E-05 | -0.00219202 | ZNF761, TPM3P9 |
| chr11 | 118180545 | 118180682 | 138 | 2 | 0.000137041 | 2.35E-05 | 0.001522 | 3.90E-05 | -0.0101238 | NA |
| chr15 | 43493776 | 43494033 | 258 | 3 | 1.46E-05 | 7.50E-05 | 0.000175 | 3.91E-05 | -0.0065163 | TP53BP1 |
| chr1 | 33863468 | 33864187 | 720 | 5 | 1.19E-05 | 0.0021442 | 0.000436 | 3.91E-05 | -0.00606908 | HMGB4, CSMD2 |
| chr17 | 17440077 | 17440261 | 185 | 2 | 9.67E-05 | 6.68E-05 | 0.00011 | 3.91E-05 | -0.0229745 | NA |
| chr12 | 47577646 | 47577677 | 32 | 2 | 1.60E-05 | 2.64E-05 | 0.000815 | 3.91E-05 | -0.00773962 | NA |
| chr17 | 76005058 | 76005575 | 518 | 2 | 4.18E-05 | 2.36E-05 | 0.001551 | 3.91E-05 | 0.006679594 | TEN1-CDK3, CDK3, EVPL |
| chr11 | 47754623 | 47754662 | 40 | 2 | 5.67E-05 | 0.0004882 | 3.26E-05 | 3.91E-05 | -0.00245993 | FNBP4 |
| chr14 | 100973232 | 100973641 | 410 | 2 | 4.43E-05 | 6.95E-05 | 0.000105 | 3.92E-05 | -0.01511698 | MEG8, SNORD114-16 |
| chr3 | 142830011 | 142830369 | 359 | 2 | 9.51E-05 | 2.43E-05 | 0.001279 | 3.92E-05 | 0.001337957 | PCOLCE2 |
| chr21 | 41819839 | 41820203 | 365 | 4 | 2.67E-05 | 0.0006293 | 0.000559 | 3.92E-05 | -0.00184081 | PRDM15 |
| chr12 | 14934860 | 14935050 | 191 | 3 | 1.13E-05 | 0.0557763 | 6.99E-06 | 3.92E-05 | -0.00523808 | ERP27 |
| chr7 | 2541027 | 2541271 | 245 | 3 | 5.26E-05 | 9.62E-05 | 0.000108 | 3.92E-05 | -0.00064232 | BRAT1 |
| chr9 | 136403680 | 136404210 | 531 | 2 | 0.000100931 | 2.35E-05 | 0.001581 | 3.92E-05 | -0.0130211 | ENTR1 |
| chr14 | 24377173 | 24377196 | 24 | 2 | 4.10E-05 | 6.99E-05 | 0.000105 | 3.92E-05 | 0.000766585 | NFATC4 |
| chr16 | 17134309 | 17134582 | 274 | 3 | 2.51E-05 | 0.0008028 | 2.28E-05 | 3.92E-05 | -0.00742992 | AC109446.2, XYLT1 |
| chr22 | 17084104 | 17084259 | 156 | 2 | 6.32E-05 | 2.37E-05 | 0.001509 | 3.92E-05 | -0.01529802 | NA |
| chr7 | 193580 | 194719 | 1140 | 7 | 1.09E-06 | 0.0007827 | 2.27E-05 | 3.92E-05 | -0.0014462 | FAM20C, AC093627.3 |
| chr5 | 178591185 | 178592013 | 829 | 3 | 7.32E-06 | 5.58E-05 | 0.000271 | 3.93E-05 | 0.003016155 | NA |
| chr1 | 205031635 | 205031782 | 148 | 2 | 2.09E-05 | 0.0082757 | 1.89E-05 | 3.93E-05 | -0.00587822 | NA |
| chr11 | 95789670 | 95789782 | 113 | 2 | 0.000141191 | 0.0002204 | 4.58E-05 | 3.93E-05 | 0.001870042 | FAM76B |
| chr8 | 10474150 | 10474381 | 232 | 4 | 6.25E-05 | 0.0001968 | 0.000398 | 3.93E-05 | -0.00020186 | NA |
| chr1 | 59360256 | 59360382 | 127 | 2 | 7.03E-05 | 3.49E-05 | 0.000341 | 3.93E-05 | 0.002260805 | FGGY |
| chr10 | 110226271 | 110226364 | 94 | 3 | 6.01E-06 | 2.95E-05 | 0.001103 | 3.93E-05 | 5.53E-05 | MXI1 |
| chr11 | 118901665 | 118901822 | 158 | 3 | 5.24E-05 | 0.000213 | 0.000214 | 3.93E-05 | -0.00156492 | BCL9L |
| chr3 | 179321633 | 179322077 | 445 | 2 | 5.80E-05 | 2.41E-05 | 0.001358 | 3.93E-05 | -0.01885489 | NA |
| chr19 | 17971163 | 17971216 | 54 | 2 | 3.37E-05 | 3.47E-05 | 0.000348 | 3.94E-05 | -0.01134146 | KCNN1 |
| chr3 | 93980020 | 93980508 | 489 | 8 | 8.80E-05 | 1.81E-05 | 0.012024 | 3.94E-05 | 0.000123971 | ARL13B |
| chr11 | 75240314 | 75241745 | 1432 | 6 | 1.78E-05 | 0.0003432 | 0.000411 | 3.94E-05 | -0.00189541 | TPBGL, AP001972.3 |
| chr10 | 5285941 | 5285982 | 42 | 3 | 0.000117025 | 6.13E-05 | 0.000302 | 3.94E-05 | 0.001553521 | AKR1C7P |
| chr10 | 122021697 | 122022087 | 391 | 7 | 6.70E-07 | 0.0493322 | 8.87E-07 | 3.94E-05 | -0.00253456 | TACC2 |
| chr15 | 50076717 | 50076980 | 264 | 2 | 3.63E-05 | 0.0003216 | 3.85E-05 | 3.94E-05 | -0.01198944 | ATP8B4 |
| chr12 | 95406588 | 95406602 | 15 | 3 | 0.000105927 | 5.10E-05 | 0.000255 | 3.94E-05 | -0.00188137 | AC084879.1 |
| chr18 | 54828173 | 54828839 | 667 | 12 | 1.88E-06 | 0.0050249 | 7.09E-05 | 3.94E-05 | -0.00115282 | RAB27B |
| chr4 | 186533657 | 186533737 | 81 | 3 | 1.87E-05 | 0.0001761 | 6.28E-05 | 3.94E-05 | -0.00400163 | AC018709.1, MTNR1A |
| chr11 | 111309869 | 111310171 | 303 | 2 | 2.59E-05 | 0.0004115 | 3.50E-05 | 3.94E-05 | -0.00710886 | NA |
| chr9 | 137245800 | 137246418 | 619 | 6 | 1.59E-05 | 0.0001463 | 0.000986 | 3.95E-05 | -0.00333003 | FAM166A |
| chr12 | 54162675 | 54162962 | 288 | 2 | 1.76E-05 | 4.62E-05 | 0.000193 | 3.95E-05 | -0.00391134 | SMUG1, AC023794.4 |
| chr17 | 81672560 | 81672660 | 101 | 2 | 4.44E-05 | 0.000523 | 3.22E-05 | 3.95E-05 | 0.002875418 | CCDC137 |
| chr8 | 96235549 | 96236170 | 622 | 9 | 7.44E-05 | 3.53E-05 | 0.014446 | 3.95E-05 | -0.00158349 | AP003465.1 |
| chr3 | 151440606 | 151440942 | 337 | 5 | 5.41E-05 | 0.0004306 | 0.001987 | 3.95E-05 | -0.00700046 | IGSF10 |
| chr6 | 108251722 | 108252257 | 536 | 4 | 7.16E-06 | 0.0013959 | 0.00014 | 3.95E-05 | -0.00655658 | Z98742.2, SNX3 |
| chr10 | 68329840 | 68330226 | 387 | 2 | 1.13E-05 | 0.0002446 | 4.39E-05 | 3.95E-05 | 0.009386727 | PBLD |
| chr3 | 184317349 | 184317739 | 391 | 6 | 4.08E-05 | 0.0002183 | 0.000108 | 3.96E-05 | -0.00278319 | EIF4G1 |
| chr1 | 45491101 | 45492167 | 1067 | 11 | 4.40E-05 | 1.67E-05 | 0.01396 | 3.96E-05 | -8.52E-05 | AL451136.1, TESK2 |
| chr10 | 133336635 | 133337013 | 379 | 8 | 6.22E-05 | 0.0002897 | 0.0011 | 3.96E-05 | 0.002325889 | ZNF511-PRAP1, CALY |
| chr10 | 119413997 | 119414247 | 251 | 2 | 6.26E-05 | 2.37E-05 | 0.001599 | 3.96E-05 | -0.00188725 | GRK5 |
| chr19 | 5897405 | 5897417 | 13 | 2 | 7.62E-05 | 3.74E-05 | 0.000297 | 3.96E-05 | -0.0001429 | AC024592.3, NDUFA11 |
| chr12 | 64080415 | 64080535 | 121 | 2 | 0.000122231 | 2.99E-05 | 0.000543 | 3.96E-05 | -0.00860426 | SRGAP1, AC020611.2 |
| chr11 | 22673577 | 22673644 | 68 | 2 | 5.46E-05 | 4.07E-05 | 0.000249 | 3.96E-05 | -0.01296338 | GAS2 |
| chr5 | 76546557 | 76546728 | 172 | 3 | 0.00011992 | 3.96E-05 | 0.000541 | 3.96E-05 | 0.005566928 | IQGAP2 |
| chr2 | 121648896 | 121650064 | 1169 | 15 | 1.47E-05 | 0.0001418 | 0.000905 | 3.97E-05 | 0.000707798 | NIFK-AS1, CLASP1 |
| chr19 | 16369217 | 16369832 | 616 | 2 | 2.41E-06 | 3.64E-05 | 0.000318 | 3.97E-05 | -0.00050939 | EPS15L1 |
| chr21 | 30395993 | 30396001 | 9 | 2 | 1.92E-05 | 0.0002939 | 4.05E-05 | 3.97E-05 | 0.005430941 | NA |
| chr2 | 178038861 | 178038893 | 33 | 2 | 6.71E-05 | 0.0002678 | 4.22E-05 | 3.97E-05 | -0.00501174 | PDE11A, PDE11A |
| chr1 | 184753772 | 184754119 | 348 | 3 | 8.96E-06 | 0.0006808 | 0.000898 | 3.97E-05 | -0.00270342 | EDEM3 |
| chr1 | 3067362 | 3068266 | 905 | 9 | 0.000100857 | 2.79E-05 | 0.014251 | 3.97E-05 | 0.003629712 | PRDM16-DT |
| chr18 | 79409782 | 79410046 | 265 | 4 | 0.000137299 | 0.0001997 | 0.002645 | 3.97E-05 | 0.000232508 | NFATC1 |
| chr10 | 125135420 | 125135829 | 410 | 2 | 1.74E-06 | 0.0014478 | 2.45E-05 | 3.98E-05 | -0.00441691 | CTBP2 |
| chr16 | 25050486 | 25050894 | 409 | 2 | 5.20E-07 | 0.0349014 | 1.78E-05 | 3.98E-05 | 0.003449166 | AC133552.2 |
| chr7 | 16397326 | 16397751 | 426 | 3 | 7.82E-06 | 0.0022208 | 2.23E-05 | 3.98E-05 | -0.00308935 | CRPPA |
| chr8 | 1451803 | 1452099 | 297 | 3 | 5.64E-05 | 0.0001502 | 0.000632 | 3.98E-05 | -0.00158211 | DLGAP2 |
| chr7 | 152987224 | 152987390 | 167 | 2 | 0.000106795 | 3.84E-05 | 0.000284 | 3.98E-05 | 0.007919277 | NA |
| chr11 | 100190274 | 100190478 | 205 | 2 | 8.62E-06 | 0.0005896 | 3.13E-05 | 3.98E-05 | -0.01805821 | CNTN5 |
| chr8 | 96579971 | 96580251 | 281 | 4 | 4.92E-05 | 0.0001192 | 0.000688 | 3.98E-05 | -0.00043201 | SDC2 |
| chr4 | 1354514 | 1355156 | 643 | 4 | 4.52E-05 | 0.0030251 | 0.001569 | 3.99E-05 | -0.00226247 | UVSSA |
| chr18 | 58600555 | 58600800 | 246 | 2 | 5.32E-05 | 5.07E-05 | 0.000169 | 3.99E-05 | -0.00774804 | SNORA108, ALPK2 |
| chr4 | 128353986 | 128354370 | 385 | 2 | 5.18E-06 | 0.0025466 | 2.21E-05 | 3.99E-05 | 0.011602104 | LINC02615 |
| chr17 | 2336875 | 2337733 | 859 | 8 | 2.32E-05 | 0.0001438 | 0.008265 | 3.99E-05 | 1.91E-05 | SGSM2 |
| chr16 | 81707888 | 81707986 | 99 | 2 | 4.12E-05 | 0.0008102 | 2.85E-05 | 3.99E-05 | -0.00387488 | CMIP |
| chr15 | 36045640 | 36046066 | 427 | 4 | 7.10E-05 | 2.84E-05 | 0.00457 | 3.99E-05 | -0.00647202 | LINC02853, AC021351.1 |
| chr17 | 6206970 | 6206994 | 25 | 2 | 7.23E-05 | 7.44E-05 | 0.000101 | 3.99E-05 | 0.00257767 | NA |
| chr17 | 7589832 | 7590833 | 1002 | 11 | 1.67E-05 | 0.026364 | 7.08E-05 | 4.00E-05 | -0.00167088 | MPDU1, SOX15 |
| chr1 | 19311836 | 19312804 | 969 | 7 | 2.24E-05 | 8.73E-05 | 0.000926 | 4.00E-05 | 0.002708585 | SLC66A1, AKR7A2 |
| chr17 | 35051412 | 35051592 | 181 | 2 | 0.000137223 | 2.45E-05 | 0.001367 | 4.00E-05 | 0.001926792 | RFFL, AC004223.3 |
| chr8 | 11325470 | 11325505 | 36 | 3 | 0.000115118 | 1.73E-05 | 0.003634 | 4.00E-05 | -0.0015376 | MTMR9 |
| chr14 | 23953159 | 23953210 | 52 | 2 | 1.94E-05 | 0.0001239 | 6.61E-05 | 4.00E-05 | -0.01249771 | DHRS4-AS1 |
| chr19 | 15479258 | 15480232 | 975 | 8 | 2.72E-05 | 0.0006162 | 0.001355 | 4.00E-05 | -0.00209758 | PGLYRP2 |
| chr22 | 39946329 | 39946533 | 205 | 3 | 4.83E-05 | 5.16E-05 | 0.000918 | 4.00E-05 | -0.00030434 | GRAP2 |
| chr2 | 109606943 | 109607093 | 151 | 3 | 6.89E-05 | 8.78E-05 | 0.000107 | 4.00E-05 | -0.0076972 | SEPTIN10 |
| chr9 | 130883102 | 130883160 | 59 | 2 | 9.44E-05 | 6.94E-05 | 0.00011 | 4.00E-05 | -0.00237467 | ABL1 |
| chr16 | 49662066 | 49662087 | 22 | 2 | 0.00012193 | 7.73E-05 | 9.72E-05 | 4.00E-05 | -0.00504878 | ZNF423 |
| chr1 | 87154588 | 87154800 | 213 | 2 | 6.47E-05 | 2.41E-05 | 0.001598 | 4.01E-05 | 0.00837531 | AC093155.3, LINC01140 |
| chr1 | 13699559 | 13699987 | 429 | 5 | 6.36E-06 | 0.0006691 | 0.002584 | 4.01E-05 | -0.01253392 | NA |
| chr5 | 144170486 | 144171346 | 861 | 9 | 5.69E-05 | 0.000162 | 0.002599 | 4.01E-05 | -0.0004477 | KCTD16, YIPF5 |
| chr5 | 16428356 | 16428412 | 57 | 3 | 3.79E-05 | 8.68E-05 | 0.000163 | 4.01E-05 | -0.00745626 | LINC02150 |
| chr9 | 126564957 | 126565103 | 147 | 2 | 8.68E-05 | 7.09E-05 | 0.000107 | 4.01E-05 | -0.00717325 | NA |
| chr5 | 160418567 | 160419216 | 650 | 5 | 6.76E-06 | 3.71E-05 | 0.004794 | 4.01E-05 | -0.00032396 | SLU7 |
| chr1 | 183186231 | 183186928 | 698 | 4 | 2.73E-06 | 0.0004642 | 5.11E-05 | 4.02E-05 | -0.00578419 | LAMC2 |
| chr4 | 109847565 | 109848291 | 727 | 4 | 2.43E-05 | 5.82E-05 | 0.000744 | 4.02E-05 | -0.00051516 | RRH, LRIT3 |
| chr7 | 156843426 | 156843583 | 158 | 2 | 5.21E-05 | 0.0003534 | 3.79E-05 | 4.02E-05 | 0.00332584 | LMBR1 |
| chr4 | 1593567 | 1593788 | 222 | 3 | 8.08E-05 | 2.19E-05 | 0.002514 | 4.02E-05 | 0.00404589 | NA |
| chr7 | 1603662 | 1604242 | 581 | 3 | 5.72E-05 | 3.12E-05 | 0.001101 | 4.02E-05 | -0.0016947 | NA |
| chr13 | 98650229 | 98650562 | 334 | 2 | 1.37E-05 | 0.0055827 | 2.00E-05 | 4.02E-05 | 0.002027429 | NA |
| chr4 | 134758001 | 134758086 | 86 | 3 | 5.65E-05 | 0.000415 | 3.77E-05 | 4.02E-05 | 0.007428559 | NA |
| chr11 | 66472995 | 66473287 | 293 | 2 | 2.72E-05 | 5.26E-05 | 0.000162 | 4.02E-05 | -0.00350341 | PELI3 |
| chr6 | 137452915 | 137453060 | 146 | 2 | 4.60E-05 | 0.0008353 | 2.85E-05 | 4.02E-05 | -0.00332848 | NA |
| chr15 | 59752253 | 59752368 | 116 | 2 | 0.000126686 | 3.05E-05 | 0.000544 | 4.03E-05 | -0.01225912 | NA |
| chr1 | 7744759 | 7744883 | 125 | 2 | 4.83E-05 | 6.96E-05 | 0.000111 | 4.03E-05 | -0.00326081 | CAMTA1 |
| chr3 | 42263474 | 42264323 | 850 | 10 | 8.82E-05 | 3.22E-05 | 0.009323 | 4.03E-05 | 0.001170053 | CCK |
| chr3 | 50348958 | 50348971 | 14 | 2 | 4.20E-05 | 4.38E-05 | 0.000225 | 4.03E-05 | 0.000492005 | NPRL2 |
| chr20 | 14522043 | 14522595 | 553 | 2 | 1.03E-05 | 2.43E-05 | 0.001545 | 4.03E-05 | -0.00636387 | MACROD2, AL121582.1 |
| chr14 | 89026651 | 89027045 | 395 | 4 | 3.73E-05 | 0.0005632 | 0.000822 | 4.03E-05 | -0.00859778 | NA |

| | | | | | | | | | | |
|---|---|---|---|---|---|---|---|---|---|---|
| chr9 | 126380380 | 126380544 | 165 | 2 | 9.42E-05 | 5.39E-05 | 0.000157 | 4.03E-05 | -0.00215455 | MVB12B |
| chr19 | 2269147 | 2270299 | 1153 | 9 | 3.69E-06 | 0.0082906 | 3.53E-05 | 4.03E-05 | 2.64E-05 | OAZ1, AC005258.1, JSRP1 |
| chr19 | 3691726 | 3692093 | 368 | 3 | 1.39E-05 | 0.0008566 | 0.000135 | 4.03E-05 | 0.000389494 | PIP5K1C |
| chr6 | 31507406 | 31507615 | 210 | 3 | 6.23E-05 | 4.44E-05 | 0.000347 | 4.03E-05 | 0.003364188 | MICB |
| chr16 | 31073633 | 31073930 | 298 | 4 | 6.80E-05 | 1.53E-05 | 0.005258 | 4.03E-05 | -0.00017219 | ZNF668 |
| chr8 | 102738906 | 102739395 | 490 | 2 | 6.59E-07 | 8.18E-05 | 9.31E-05 | 4.03E-05 | 0.00424674 | NA |
| chr6 | 157013788 | 157014036 | 249 | 2 | 5.66E-05 | 9.42E-05 | 8.27E-05 | 4.04E-05 | 0.002200017 | ARID1B |
| chr10 | 22477932 | 22478147 | 216 | 2 | 2.83E-05 | 3.27E-05 | 0.000447 | 4.04E-05 | -0.02031127 | AL513128.3, AL513128.2 |
| chr3 | 133499141 | 133499429 | 289 | 2 | 3.70E-05 | 0.0002509 | 4.45E-05 | 4.04E-05 | -0.00488568 | NA |
| chr15 | 40002387 | 40002770 | 384 | 2 | 5.10E-05 | 4.47E-05 | 0.000218 | 4.04E-05 | -0.00429308 | EIF2AK4 |
| chr16 | 4925272 | 4925453 | 182 | 3 | 6.31E-05 | 1.81E-05 | 0.003106 | 4.04E-05 | -0.00922164 | PPL |
| chr8 | 142985426 | 142985433 | 8 | 2 | 8.64E-05 | 4.84E-05 | 0.000189 | 4.04E-05 | -8.07E-06 | LY6E-DT |
| chr16 | 72610309 | 72610472 | 164 | 2 | 8.01E-05 | 8.29E-05 | 9.23E-05 | 4.04E-05 | -0.00517681 | LINC01572 |
| chr2 | 47156383 | 47156581 | 199 | 3 | 3.06E-06 | 0.0007687 | 3.74E-05 | 4.04E-05 | 0.006023295 | AC073283.3 |
| chr20 | 51400258 | 51400566 | 309 | 2 | 5.71E-05 | 5.09E-05 | 0.000174 | 4.04E-05 | 0.002481711 | NFATC2 |
| chr14 | 60245605 | 60245845 | 241 | 2 | 6.56E-05 | 2.43E-05 | 0.001608 | 4.05E-05 | 0.002517327 | PPM1A, AL157756.1 |
| chr12 | 102957654 | 102958222 | 569 | 6 | 7.83E-05 | 6.15E-05 | 0.001277 | 4.05E-05 | 1.26E-05 | ASCL1, PAH |
| chr7 | 36777308 | 36777399 | 92 | 2 | 0.000116312 | 3.51E-05 | 0.00037 | 4.05E-05 | 0.005429477 | NA |
| chr4 | 30723219 | 30723293 | 75 | 2 | 7.64E-05 | 7.23E-05 | 0.000107 | 4.05E-05 | -0.01062113 | PCDH7 |
| chr19 | 14891337 | 14891390 | 54 | 2 | 0.000133689 | 2.95E-05 | 0.000624 | 4.05E-05 | -0.01322425 | NA |
| chr1 | 1626730 | 1627391 | 662 | 5 | 3.36E-07 | 0.0116043 | 5.11E-05 | 4.05E-05 | -0.00051946 | MIB2 |
| chr17 | 81468871 | 81468941 | 71 | 2 | 9.84E-05 | 6.94E-05 | 0.000113 | 4.05E-05 | -0.00154262 | NA |
| chr1 | 213992747 | 213992866 | 120 | 3 | 6.13E-05 | 0.0005896 | 0.000122 | 4.05E-05 | -0.00473029 | PROX1 |
| chr8 | 143524401 | 143524859 | 459 | 2 | 2.28E-05 | 2.65E-05 | 0.000966 | 4.05E-05 | -0.00383734 | ZC3H3 |
| chr15 | 67548038 | 67548416 | 379 | 5 | 3.08E-05 | 0.0003307 | 0.000867 | 4.05E-05 | 0.000946393 | MAP2K5 |
| chr17 | 29074126 | 29074594 | 469 | 4 | 3.46E-05 | 7.37E-05 | 0.00017 | 4.06E-05 | 0.004928465 | MYO18A, TIAF1 |
| chr3 | 43352316 | 43352454 | 139 | 2 | 9.91E-05 | 2.74E-05 | 0.000844 | 4.06E-05 | 0.000966207 | SNRK |
| chr9 | 122159054 | 122159309 | 256 | 4 | 1.28E-05 | 3.91E-05 | 0.00493 | 4.07E-05 | -0.00253079 | NDUFA8 |
| chr2 | 43124158 | 43124163 | 6 | 2 | 4.40E-05 | 3.91E-05 | 0.000292 | 4.07E-05 | 0.011638518 | LINC02580 |
| chr1 | 30847343 | 30847715 | 373 | 2 | 2.11E-05 | 0.0001634 | 5.68E-05 | 4.07E-05 | -0.00157969 | NA |
| chr16 | 4200317 | 4200547 | 231 | 2 | 0.00012143 | 2.87E-05 | 0.00071 | 4.07E-05 | -0.00864752 | SRL |
| chr9 | 136508090 | 136508435 | 346 | 3 | 1.70E-05 | 0.0002892 | 0.000922 | 4.07E-05 | -0.003897 | NOTCH1 |
| chr7 | 56051519 | 56051853 | 335 | 6 | 4.06E-05 | 0.0003775 | 0.00097 | 4.07E-05 | 0.000981583 | CCT6A, PSPH |
| chr7 | 2476560 | 2477310 | 751 | 7 | 1.56E-05 | 0.0005381 | 9.59E-05 | 4.07E-05 | 0.00156949 | GRIFIN |
| chr12 | 57520455 | 57520932 | 478 | 9 | 7.84E-06 | 0.0002137 | 0.003604 | 4.08E-05 | 6.90E-05 | MBD6, DDIT3, AC022506.1 |
| chr6 | 27810458 | 27811058 | 601 | 3 | 6.47E-05 | 0.0001912 | 0.000308 | 4.08E-05 | -0.00080256 | H3C10 |
| chr11 | 85663503 | 85663850 | 348 | 5 | 6.91E-06 | 0.0032079 | 1.25E-05 | 4.08E-05 | 0.00117267 | CREBZF |
| chr7 | 2125855 | 2126177 | 323 | 2 | 9.63E-05 | 2.57E-05 | 0.001179 | 4.08E-05 | 0.003842688 | MAD1L1, AC069288.1 |
| chr7 | 123566821 | 123566843 | 23 | 2 | 0.000106586 | 2.45E-05 | 0.001585 | 4.08E-05 | -0.0102518 | NA |
| chr14 | 95108342 | 95108675 | 334 | 2 | 1.97E-05 | 5.97E-05 | 0.00014 | 4.08E-05 | -0.00108751 | DICER1 |
| chr11 | 6128060 | 6128238 | 179 | 2 | 0.000105908 | 4.12E-05 | 0.000264 | 4.08E-05 | -0.01855662 | AC022762.1 |
| chr1 | 22089995 | 22090242 | 248 | 2 | 5.12E-05 | 0.0001568 | 5.86E-05 | 4.09E-05 | 0.004725744 | CDC42 |
| chr1 | 151008321 | 151009471 | 1151 | 13 | 2.95E-05 | 0.0200507 | 0.00061 | 4.09E-05 | 0.000450804 | PRUNE1, MINDY1 |
| chr15 | 59574841 | 59575279 | 439 | 3 | 3.51E-05 | 4.93E-05 | 0.001656 | 4.09E-05 | -0.00428611 | NA |
| chr4 | 88708596 | 88708993 | 398 | 3 | 4.41E-05 | 0.0001713 | 0.000121 | 4.09E-05 | -0.00240353 | NA |
| chr10 | 70376607 | 70376744 | 138 | 2 | 2.32E-05 | 0.0002488 | 4.55E-05 | 4.09E-05 | -0.00433956 | LRRC20 |
| chr17 | 69896891 | 69896938 | 48 | 2 | 0.000167159 | 0.0001383 | 6.35E-05 | 4.09E-05 | -0.00918768 | LINC01483 |
| chr14 | 53850972 | 53851497 | 526 | 3 | 2.11E-05 | 0.0001744 | 7.02E-05 | 4.09E-05 | -0.01081039 | AL365295.1 |
| chr11 | 14041354 | 14041516 | 163 | 2 | 0.000146596 | 2.45E-05 | 0.001632 | 4.10E-05 | 0.007437272 | SPON1 |
| chr10 | 119571533 | 119571782 | 250 | 2 | 1.04E-05 | 0.00929 | 1.95E-05 | 4.10E-05 | -0.01208454 | NA |
| chr19 | 2295766 | 2295840 | 75 | 2 | 6.93E-05 | 4.82E-05 | 0.000197 | 4.10E-05 | 0.005261282 | AC005258.1, AC104530.1 |
| chr16 | 19885771 | 19886231 | 461 | 4 | 4.99E-05 | 0.0011704 | 0.000216 | 4.10E-05 | -0.00642013 | GPRC5B |
| chr3 | 141343669 | 141343880 | 212 | 5 | 4.23E-05 | 0.0001783 | 0.002604 | 4.11E-05 | -0.00285611 | PXYLP1, ZBTB38, AC117383.1 |
| chr4 | 151014796 | 151015874 | 1079 | 11 | 2.44E-06 | 0.0043927 | 0.000148 | 4.11E-05 | 0.000610505 | LRBA |
| chr20 | 38723969 | 38724149 | 181 | 2 | 2.34E-05 | 2.59E-05 | 0.001186 | 4.11E-05 | 0.004772641 | NA |
| chr17 | 82767185 | 82767338 | 154 | 3 | 6.81E-05 | 0.000349 | 0.000915 | 4.11E-05 | -0.01250051 | TBCD |
| chr7 | 97014142 | 97014509 | 368 | 5 | 8.04E-05 | 1.24E-05 | 0.008315 | 4.11E-05 | 0.002548512 | NA |
| chr1 | 21610191 | 21610242 | 52 | 3 | 6.02E-05 | 2.02E-05 | 0.002121 | 4.11E-05 | -0.00311161 | RAP1GAP |
| chr17 | 58352139 | 58352520 | 382 | 10 | 2.46E-05 | 0.0032399 | 6.08E-05 | 4.11E-05 | 0.000533696 | TSPOAP1-AS1, SUPT4H1, AC004687.2 |
| chr1 | 26361489 | 26362009 | 521 | 2 | 5.22E-06 | 0.0001952 | 5.18E-05 | 4.11E-05 | -0.0042791 | ZNF683 |
| chr4 | 148443391 | 148443729 | 339 | 4 | 4.02E-06 | 0.0001882 | 0.001054 | 4.11E-05 | 0.001493213 | NR3C2 |
| chr6 | 73522951 | 73523556 | 606 | 4 | 2.92E-05 | 3.99E-05 | 0.003076 | 4.11E-05 | -0.00446547 | EEF1A1 |
| chr9 | 98833291 | 98833353 | 63 | 2 | 2.92E-05 | 0.0139193 | 1.90E-05 | 4.11E-05 | 0.002193283 | GALNT12 |
| chr13 | 99124821 | 99124847 | 27 | 2 | 0.0001131 | 2.70E-05 | 0.00097 | 4.11E-05 | 0.004118117 | NA |
| chr10 | 15210053 | 15210152 | 100 | 2 | 0.000115018 | 3.40E-05 | 0.000423 | 4.11E-05 | -0.00449281 | NA |
| chr10 | 97850079 | 97850673 | 595 | 6 | 3.80E-06 | 0.0102666 | 0.000468 | 4.11E-05 | -0.00175803 | GOLGA7B |
| chr4 | 2247249 | 2247418 | 170 | 3 | 5.84E-05 | 0.0001738 | 8.07E-05 | 4.11E-05 | -0.00229483 | NA |
| chr16 | 318524 | 318761 | 238 | 3 | 3.86E-05 | 4.10E-05 | 0.000579 | 4.11E-05 | -0.00072058 | AXIN1 |
| chr20 | 34391264 | 34391317 | 54 | 2 | 0.000157304 | 2.49E-05 | 0.001521 | 4.12E-05 | -0.01209836 | ITCH |
| chr5 | 74720201 | 74720316 | 116 | 2 | 3.66E-05 | 0.0014326 | 2.55E-05 | 4.12E-05 | -0.00804703 | HEXB |
| chr8 | 122926473 | 122926630 | 158 | 2 | 4.77E-05 | 2.51E-05 | 0.001464 | 4.12E-05 | 0.00408185 | ZHX2 |
| chr7 | 152872864 | 152872995 | 132 | 2 | 5.50E-05 | 0.0002012 | 5.11E-05 | 4.12E-05 | -0.00603978 | NA |
| chr11 | 24496510 | 24497025 | 516 | 13 | 5.12E-05 | 0.0006523 | 0.005473 | 4.12E-05 | 0.002575602 | LUZP2 |
| chr19 | 589070 | 589411 | 342 | 2 | 3.83E-06 | 0.0001428 | 6.28E-05 | 4.12E-05 | -0.00860108 | NA |
| chr22 | 38110582 | 38110774 | 193 | 6 | 6.27E-05 | 0.0001029 | 0.000382 | 4.12E-05 | -0.00249228 | BAIAP2L2 |
| chr1 | 97968963 | 97969181 | 219 | 3 | 9.60E-05 | 6.73E-05 | 0.000162 | 4.12E-05 | -0.00237988 | DPYD, MIR137HG |
| chr2 | 85311636 | 85311669 | 34 | 3 | 4.89E-05 | 0.0003631 | 3.68E-05 | 4.12E-05 | -0.0025709 | NA |
| chr1 | 17587575 | 17587844 | 270 | 2 | 4.30E-05 | 0.002077 | 2.37E-05 | 4.12E-05 | 0.00316636 | ARHGEF10L |
| chr1 | 1658982 | 1659256 | 275 | 6 | 4.81E-05 | 0.0027832 | 0.001569 | 4.12E-05 | -0.00131771 | CDK11B |
| chr20 | 44905579 | 44905720 | 142 | 2 | 8.47E-05 | 8.78E-05 | 9.08E-05 | 4.12E-05 | -0.00557733 | YWHAB |
| chr15 | 85718760 | 85718925 | 166 | 2 | 3.92E-05 | 0.0099697 | 1.95E-05 | 4.12E-05 | -0.00720877 | AKAP13 |
| chr3 | 46936926 | 46937120 | 195 | 2 | 3.06E-05 | 4.16E-05 | 0.000266 | 4.12E-05 | 0.001030774 | CCDC12 |
| chr12 | 49599484 | 49599772 | 289 | 6 | 1.49E-05 | 0.004731 | 4.12E-05 | 4.12E-05 | -0.00162693 | PRPF40B, FAM186B |
| chr16 | 28069945 | 28070235 | 291 | 4 | 2.87E-05 | 0.0170036 | 0.000205 | 4.13E-05 | -0.00110557 | NA |
| chr7 | 143397043 | 143397574 | 532 | 2 | 2.13E-05 | 2.98E-05 | 0.000652 | 4.13E-05 | -0.0075292 | EPHA1 |
| chr20 | 35174671 | 35174870 | 200 | 3 | 0.000123681 | 2.92E-05 | 0.001432 | 4.13E-05 | -0.00467788 | PROCR, AL356652.1 |
| chr17 | 82710735 | 82711030 | 296 | 3 | 3.87E-05 | 0.0026399 | 0.000458 | 4.13E-05 | -0.009846 | NA |
| chr1 | 235867796 | 235868127 | 332 | 2 | 4.70E-05 | 2.60E-05 | 0.001208 | 4.13E-05 | 0.029086355 | LYST |
| chr1 | 161357206 | 161357218 | 13 | 2 | 0.000109515 | 5.99E-05 | 0.000143 | 4.14E-05 | -0.00569363 | SDHC |
| chr9 | 135478072 | 135478451 | 380 | 3 | 2.09E-05 | 3.13E-05 | 0.000734 | 4.14E-05 | -0.00061951 | PPP1R26-AS1 |
| chr12 | 54390963 | 54391525 | 563 | 7 | 8.21E-05 | 6.80E-05 | 0.003971 | 4.14E-05 | 0.000954361 | AC079313.2, AC079313.1, ZNF385A |
| chr2 | 200608320 | 200608389 | 70 | 2 | 6.27E-05 | 0.0002353 | 4.73E-05 | 4.14E-05 | -0.01409259 | AOX1 |
| chr1 | 151035935 | 151036495 | 561 | 4 | 1.14E-05 | 0.0001441 | 0.000927 | 4.14E-05 | -0.00155772 | BNIPL |
| chr19 | 1319207 | 1319243 | 37 | 2 | 0.000110819 | 4.47E-05 | 0.000233 | 4.14E-05 | -0.01601817 | NA |
| chr6 | 22147853 | 22148509 | 657 | 3 | 6.46E-06 | 0.0004969 | 0.000329 | 4.14E-05 | -0.00675512 | CASC15 |
| chr19 | 1242099 | 1242324 | 226 | 3 | 6.25E-07 | 0.0048121 | 1.37E-05 | 4.15E-05 | -0.00927846 | ATP5F1D |
| chr17 | 36646218 | 36646280 | 63 | 2 | 7.44E-05 | 0.0001669 | 5.76E-05 | 4.15E-05 | 0.001924469 | NA |
| chr17 | 81049401 | 81049831 | 431 | 2 | 2.17E-05 | 2.53E-05 | 0.001463 | 4.15E-05 | 0.045478037 | BAIAP2 |
| chr22 | 39570648 | 39570916 | 269 | 3 | 1.27E-05 | 0.000719 | 2.55E-05 | 4.15E-05 | -0.0045831 | CACNA1I |
| chr20 | 58463407 | 58463416 | 10 | 2 | 4.75E-05 | 2.66E-05 | 0.001086 | 4.15E-05 | -0.02302746 | APCDD1L |
| chr15 | 26842168 | 26842407 | 240 | 2 | 6.58E-05 | 9.44E-05 | 8.63E-05 | 4.15E-05 | -0.0297621 | GABRB3 |
| chr12 | 75305637 | 75305845 | 209 | 3 | 5.09E-05 | 6.53E-05 | 0.000176 | 4.15E-05 | -0.0036604 | AC092552.1, CAPS2 |
| chr11 | 70290335 | 70290365 | 31 | 2 | 0.000147344 | 2.54E-05 | 0.001434 | 4.15E-05 | -0.00026195 | PPFIA1, AP002336.2 |
| chr1 | 193186078 | 193186260 | 183 | 3 | 4.17E-05 | 0.0001401 | 9.86E-05 | 4.15E-05 | 0.004871988 | CDC73, B3GALT2 |
| chr1 | 181235292 | 181235837 | 546 | 2 | 6.68E-05 | 2.61E-05 | 0.001217 | 4.15E-05 | -0.00668163 | NA |
| chr20 | 25246852 | 25247669 | 818 | 4 | 3.54E-06 | 3.16E-05 | 0.00062 | 4.16E-05 | 0.008223892 | NA |
| chr1 | 43144831 | 43145078 | 248 | 3 | 5.70E-05 | 6.39E-05 | 0.000203 | 4.16E-05 | -0.00434323 | NA |
| chr17 | 781250 | 782137 | 888 | 8 | 6.08E-05 | 4.48E-05 | 0.002941 | 4.16E-05 | -0.00090493 | GLOD4 |
| chr19 | 47232484 | 47232896 | 413 | 5 | 5.16E-06 | 0.0124136 | 1.36E-05 | 4.16E-05 | -0.00569485 | BBC3 |
| chr17 | 40177680 | 40178187 | 508 | 8 | 3.05E-05 | 0.0004161 | 0.001438 | 4.16E-05 | 0.00363398 | RAPGEFL1 |
| chr7 | 28178905 | 28179314 | 410 | 3 | 1.55E-05 | 0.0005332 | 0.000457 | 4.16E-05 | -0.0074845 | JAZF1 |
| chr15 | 55909298 | 55909490 | 193 | 2 | 8.84E-05 | 4.71E-05 | 0.000214 | 4.16E-05 | 0.006016365 | NEDD4 |
| chr3 | 170358240 | 170358491 | 252 | 5 | 2.55E-05 | 3.90E-05 | 0.00799 | 4.16E-05 | -0.00371541 | SKIL |
| chr19 | 55432864 | 55433237 | 374 | 3 | 3.44E-05 | 0.0002903 | 4.30E-05 | 4.17E-05 | 0.006191004 | SHISA7 |
| chr10 | 97332580 | 97332951 | 372 | 2 | 2.70E-05 | 9.25E-05 | 8.82E-05 | 4.17E-05 | -0.0093947 | FRAT2 |
| chr1 | 14733131 | 14733496 | 366 | 3 | 7.94E-05 | 2.17E-05 | 0.001796 | 4.17E-05 | -0.01827939 | KAZN |
| chr16 | 73228802 | 73228916 | 115 | 2 | 0.000127169 | 3.45E-05 | 0.000428 | 4.17E-05 | -0.01034859 | ZFHX3 |
| chr14 | 101914044 | 101914304 | 261 | 3 | 2.16E-05 | 0.0009013 | 2.81E-05 | 4.17E-05 | -0.00392733 | PPP2R5C |
| chr6 | 10585450 | 10585656 | 207 | 3 | 8.64E-05 | 0.0056425 | 0.000446 | 4.17E-05 | 0.003223502 | GCNT2 |
| chr1 | 205720375 | 205720598 | 224 | 2 | 9.74E-05 | 4.51E-05 | 0.000234 | 4.17E-05 | -0.01302526 | NUCKS1 |
| chr9 | 122930466 | 122930652 | 187 | 2 | 3.09E-05 | 4.47E-05 | 0.000238 | 4.17E-05 | -0.00861666 | ZBTB26 |
| chr7 | 67378533 | 67378886 | 354 | 3 | 8.97E-05 | 5.97E-05 | 0.000282 | 4.17E-05 | -0.00693575 | NA |
| chr19 | 54096020 | 54096190 | 171 | 2 | 4.80E-05 | 3.28E-05 | 0.000499 | 4.17E-05 | -0.00314153 | OSCAR |
| chr19 | 41218748 | 41219247 | 500 | 3 | 6.42E-05 | 2.97E-05 | 0.000798 | 4.17E-05 | -0.01195299 | AXL |
| chr15 | 69380420 | 69380844 | 425 | 2 | 9.04E-05 | 2.83E-05 | 0.000841 | 4.18E-05 | -0.00489578 | PAQR5 |
| chr6 | 32980427 | 32980749 | 323 | 5 | 4.94E-06 | 0.0009073 | 0.00013 | 4.18E-05 | -0.00015146 | BRD2 |
| chr9 | 137407041 | 137407665 | 625 | 3 | 3.40E-05 | 2.62E-05 | 0.001042 | 4.18E-05 | -0.00823547 | EXD3 |
| chr12 | 112905604 | 112905798 | 195 | 4 | 8.28E-05 | 0.0001224 | 0.001397 | 4.18E-05 | 0.000222973 | NA |
| chr10 | 101001929 | 101002644 | 716 | 4 | 4.01E-06 | 0.0006921 | 0.000123 | 4.18E-05 | -0.00380608 | LZTS2 |
| chr16 | 1978005 | 1978739 | 735 | 5 | 7.36E-06 | 0.0003553 | 0.000165 | 4.18E-05 | -0.00065431 | TBL3 |
| chr8 | 56210592 | 56211559 | 968 | 7 | 1.18E-05 | 2.92E-05 | 0.01232 | 4.19E-05 | -0.00023942 | PLAG1 |
| chr13 | 113053469 | 113053615 | 147 | 3 | 5.37E-05 | 0.0016332 | 1.78E-05 | 4.19E-05 | -0.00627853 | MCF2L |
| chr3 | 122615197 | 122615483 | 287 | 5 | 4.62E-05 | 6.23E-05 | 0.004852 | 4.19E-05 | -0.00641379 | PARP15 |
| chr4 | 67701243 | 67701456 | 214 | 9 | 0.000133136 | 3.59E-05 | 0.016944 | 4.19E-05 | -0.000363 | UBA6-AS1 |
| chr4 | 7811748 | 7812184 | 437 | 3 | 6.15E-05 | 2.94E-05 | 0.000809 | 4.19E-05 | 0.008903191 | AFAP1 |
| chr8 | 53843017 | 53843664 | 648 | 15 | 5.14E-06 | 0.0003348 | 0.001754 | 4.19E-05 | 0.000512961 | ATP6V1H |
| chr12 | 95224818 | 95225015 | 198 | 2 | 0.000109444 | 4.00E-05 | 0.000306 | 4.20E-05 | 0.0071538 | VEZT |
| chr22 | 29315464 | 29316150 | 687 | 5 | 3.67E-05 | 0.0002733 | 0.003507 | 4.20E-05 | 0.000141577 | RASL10A |
| chr1 | 2337034 | 2337223 | 190 | 2 | 3.17E-05 | 5.80E-05 | 0.000155 | 4.20E-05 | -0.00235616 | MORN1 |
| chr10 | 88911787 | 88912289 | 503 | 3 | 3.72E-05 | 3.68E-05 | 0.000466 | 4.21E-05 | -0.00242596 | STAMBPL1 |
| chr17 | 83195458 | 83196011 | 554 | 4 | 2.93E-05 | 2.10E-05 | 0.002894 | 4.21E-05 | 0.002993095 | AC139099.2 |
| chr13 | 113623119 | 113623199 | 81 | 2 | 5.64E-05 | 0.0002591 | 4.62E-05 | 4.21E-05 | 0.002014889 | TFDP1 |
| chr4 | 76087028 | 76087367 | 340 | 2 | 1.58E-05 | 0.003091 | 2.27E-05 | 4.21E-05 | -0.01813732 | ART3 |
| chr7 | 128274692 | 128274928 | 237 | 2 | 7.05E-05 | 7.04E-05 | 0.000119 | 4.21E-05 | 0.008792335 | NA |
| chr17 | 66448404 | 66448466 | 63 | 2 | 7.95E-05 | 2.52E-05 | 0.001661 | 4.21E-05 | -0.01018979 | PRKCA |
| chr17 | 79097324 | 79097784 | 461 | 5 | 4.05E-05 | 0.0021098 | 0.000481 | 4.21E-05 | -0.00662122 | RBFOX3 |
| chr19 | 19225110 | 19225612 | 503 | 6 | 4.65E-05 | 0.0006244 | 0.001361 | 4.21E-05 | -0.00709348 | NCAN |
| chr15 | 71294554 | 71294687 | 134 | 2 | 3.88E-05 | 0.0056364 | 2.09E-05 | 4.21E-05 | 0.00566693 | THSD4 |
| chr13 | 45463869 | 45463967 | 99 | 2 | 1.44E-05 | 2.54E-05 | 0.001609 | 4.22E-05 | 0.006901385 | NA |
| chr7 | 70766195 | 70766318 | 124 | 2 | 7.93E-05 | 0.0001085 | 7.92E-05 | 4.22E-05 | -0.00065201 | AUTS2 |
| chr22 | 44955213 | 44955454 | 242 | 2 | 4.75E-05 | 2.80E-05 | 0.000936 | 4.23E-05 | -0.01188965 | PHF21B |
| chr17 | 82872756 | 82873046 | 291 | 3 | 1.07E-05 | 0.0013457 | 2.23E-05 | 4.23E-05 | -0.00752542 | TBCD |
| chr12 | 123845882 | 123846100 | 219 | 2 | 3.62E-05 | 0.0009324 | 2.91E-05 | 4.23E-05 | -0.01122276 | DNAH10 |
| chr17 | 60154433 | 60154980 | 548 | 3 | 1.81E-05 | 0.0013312 | 0.000224 | 4.23E-05 | -0.00373247 | CA4 |
| chr7 | 136115163 | 136115748 | 586 | 3 | 1.16E-05 | 0.001617 | 1.87E-05 | 4.23E-05 | -0.00505504 | AC078845.1 |
| chr1 | 75788823 | 75789380 | 558 | 3 | 6.10E-06 | 0.0527645 | 1.31E-05 | 4.23E-05 | -0.01017302 | RABGGTB |
| chr1 | 3119604 | 3119794 | 191 | 2 | 2.22E-05 | 2.85E-05 | 0.000871 | 4.23E-05 | -0.00987504 | PRDM16 |
| chr5 | 116361749 | 116361999 | 251 | 3 | 7.02E-05 | 3.35E-05 | 0.000576 | 4.23E-05 | -0.01122695 | COMMD10 |

| | | | | | | | | | | |
|---|---|---|---|---|---|---|---|---|---|---|
| chr15 | 40900688 | 40900845 | 158 | 4 | 0.000111536 | 0.0003825 | 0.000898 | 4.23E-05 | 0.003366975 | VPS18 |
| chr6 | 29560093 | 29560108 | 16 | 2 | 0.000122242 | 2.84E-05 | 0.000897 | 4.23E-05 | -0.00178656 | GABBR1 |
| chr22 | 46198354 | 46198538 | 185 | 5 | 3.71E-05 | 0.0012744 | 0.000246 | 4.24E-05 | 0.001262172 | PPARA |
| chr1 | 24418648 | 24418905 | 258 | 3 | 1.25E-05 | 0.0004556 | 3.38E-05 | 4.24E-05 | -0.00161696 | NIPAL3 |
| chr16 | 27998513 | 27998515 | 3 | 2 | 7.27E-05 | 2.66E-05 | 0.001243 | 4.24E-05 | 0.001658545 | GSG1L |
| chr13 | 21818857 | 21818988 | 132 | 2 | 7.88E-05 | 0.0122602 | 1.97E-05 | 4.24E-05 | 0.001861909 | NA |
| chr21 | 39439187 | 39439190 | 4 | 2 | 7.63E-05 | 9.92E-05 | 8.58E-05 | 4.24E-05 | -0.0049375 | GET1-SH3BGR, LCA5L |
| chr2 | 179049900 | 179050306 | 407 | 5 | 5.82E-05 | 0.0002825 | 0.000388 | 4.24E-05 | 0.001553208 | CCDC141 |
| chr1 | 27542506 | 27542541 | 36 | 2 | 0.000120731 | 0.000449 | 3.68E-05 | 4.24E-05 | 0.005825189 | AHDC1 |
| chr14 | 103132560 | 103133215 | 656 | 4 | 3.46E-05 | 0.0014314 | 0.000419 | 4.24E-05 | 0.000874526 | TNFAIP2 |
| chr3 | 197428402 | 197428584 | 183 | 2 | 3.08E-05 | 0.0043073 | 2.17E-05 | 4.25E-05 | -0.00952692 | NA |
| chr17 | 7035607 | 7036030 | 424 | 10 | 5.35E-05 | 0.0005919 | 0.000458 | 4.25E-05 | 0.000253129 | SLC16A13 |
| chr1 | 10766450 | 10766597 | 148 | 2 | 1.88E-05 | 2.63E-05 | 0.001368 | 4.25E-05 | -0.00785102 | CASZ1 |
| chr4 | 10129956 | 10130080 | 125 | 2 | 3.93E-05 | 0.0019359 | 2.47E-05 | 4.25E-05 | 0.002736293 | NA |
| chr7 | 128321539 | 128321708 | 170 | 3 | 8.06E-05 | 6.85E-05 | 0.00017 | 4.25E-05 | 0.013263139 | RBM28 |
| chr1 | 54251958 | 54252063 | 106 | 2 | 3.57E-05 | 0.0057102 | 2.10E-05 | 4.25E-05 | -0.00273315 | SSBP3 |
| chr5 | 145872531 | 145873425 | 895 | 4 | 8.01E-06 | 5.35E-05 | 0.001936 | 4.25E-05 | 0.001562675 | GRXCR2 |
| chr3 | 10994851 | 10995212 | 362 | 3 | 4.84E-05 | 8.99E-05 | 0.000394 | 4.26E-05 | -0.00318007 | SLC6A1 |
| chr19 | 49127785 | 49127926 | 142 | 3 | 2.86E-05 | 0.0054691 | 2.13E-05 | 4.26E-05 | -0.0118756 | PPFIA3 |
| chr16 | 50024886 | 50025511 | 626 | 10 | 2.69E-05 | 0.0008529 | 0.000137 | 4.26E-05 | 0.000994224 | CNEP1R1 |
| chr9 | 91900243 | 91900396 | 154 | 4 | 0.000126987 | 4.86E-05 | 0.000386 | 4.26E-05 | 0.00721307 | ROR2 |
| chr19 | 15231213 | 15231938 | 726 | 3 | 7.25E-06 | 0.0002075 | 0.000116 | 4.26E-05 | -0.00393813 | EPHX3 |
| chr2 | 25115582 | 25116086 | 505 | 5 | 1.52E-06 | 0.0309091 | 1.79E-05 | 4.27E-05 | 0.000821092 | EFR3B |
| chr5 | 135337791 | 135337813 | 23 | 3 | 0.000126987 | 2.46E-05 | 0.001556 | 4.27E-05 | -0.00392054 | C5orf66, MACROH2A1 |
| chr6 | 138194398 | 138194419 | 22 | 2 | 0.000140044 | 0.0003005 | 4.38E-05 | 4.27E-05 | 0.003502614 | ARFGEF3 |
| chr10 | 129777087 | 129777832 | 746 | 3 | 4.28E-05 | 0.0002009 | 0.000849 | 4.27E-05 | -0.0034981 | NA |
| chr2 | 240298554 | 240298581 | 28 | 2 | 8.61E-05 | 4.81E-05 | 0.00022 | 4.27E-05 | -0.00108056 | NA |
| chr21 | 42892837 | 42893317 | 481 | 10 | 0.000107021 | 1.29E-05 | 0.00946 | 4.27E-05 | -0.00016554 | NDUFV3 |
| chr5 | 141050984 | 141051564 | 581 | 10 | 0.000123617 | 2.70E-05 | 0.017473 | 4.27E-05 | 0.003913796 | PCDHB1, AC244517.5 |
| chr8 | 30727772 | 30728184 | 413 | 10 | 6.37E-05 | 0.0001097 | 0.000506 | 4.27E-05 | 0.000779284 | GSR |
| chr2 | 25233203 | 25233629 | 427 | 2 | 1.33E-05 | 3.02E-05 | 0.000729 | 4.27E-05 | -0.00545072 | DNMT3A |
| chr14 | 21432661 | 21432912 | 252 | 3 | 5.02E-05 | 4.68E-05 | 0.000307 | 4.27E-05 | -0.00042901 | CHD8 |
| chr5 | 150114964 | 150115188 | 225 | 2 | 6.43E-05 | 3.55E-05 | 0.000432 | 4.27E-05 | 0.003450074 | PDGFRB |
| chr19 | 55062204 | 55063057 | 854 | 3 | 3.96E-07 | 0.0001075 | 0.000112 | 4.28E-05 | -0.02426143 | RDH13 |
| chr16 | 16076327 | 16076578 | 252 | 2 | 5.00E-05 | 2.81E-05 | 0.000986 | 4.28E-05 | -0.01491179 | ABCC1 |
| chr14 | 51988834 | 51989044 | 211 | 2 | 2.48E-05 | 7.28E-05 | 0.000118 | 4.28E-05 | -0.00284257 | NA |
| chr17 | 80983746 | 80983798 | 53 | 2 | 7.83E-05 | 5.47E-05 | 0.000177 | 4.28E-05 | -0.00525979 | NA |
| chr12 | 107774785 | 107775210 | 426 | 3 | 4.79E-05 | 0.0001741 | 0.00011 | 4.28E-05 | -0.00719065 | ASCL4 |
| chr10 | 126586439 | 126586465 | 27 | 3 | 4.88E-05 | 0.0003653 | 3.85E-05 | 4.28E-05 | -0.00378014 | C10orf90 |
| chr2 | 39265290 | 39266229 | 940 | 3 | 3.40E-05 | 6.00E-05 | 0.000965 | 4.28E-05 | -0.01762357 | MAP4K3 |
| chr17 | 68466079 | 68466136 | 58 | 2 | 4.93E-05 | 0.0016093 | 2.58E-05 | 4.28E-05 | -0.01139536 | AC007780.1 |
| chr18 | 37274337 | 37274973 | 637 | 3 | 2.20E-05 | 0.0003233 | 0.000922 | 4.29E-05 | -0.01104668 | AC090386.1, CELF4 |
| chr10 | 89207295 | 89207432 | 138 | 6 | 0.000120005 | 5.99E-05 | 0.000499 | 4.29E-05 | -0.00031218 | CH25H |
| chr12 | 63844827 | 63844939 | 113 | 2 | 8.52E-05 | 4.74E-05 | 0.000228 | 4.29E-05 | -0.00246172 | SRGAP1 |
| chr8 | 739326 | 739908 | 583 | 3 | 8.18E-05 | 1.64E-05 | 0.006051 | 4.29E-05 | -0.02216091 | DLGAP2, AC100797.4 |
| chr14 | 49551197 | 49551363 | 167 | 2 | 9.63E-05 | 2.76E-05 | 0.001095 | 4.29E-05 | -0.01915464 | NA |
| chr19 | 6990665 | 6991150 | 486 | 3 | 5.14E-05 | 7.48E-05 | 0.000175 | 4.29E-05 | -0.0070977 | ADGRE4P |
| chr3 | 184572601 | 184573006 | 406 | 5 | 1.78E-05 | 0.0022426 | 1.75E-05 | 4.29E-05 | -0.00250196 | EPHB3 |
| chr1 | 1273046 | 1273329 | 284 | 3 | 4.02E-05 | 5.04E-05 | 0.000621 | 4.29E-05 | 0.000339104 | UBE2J2 |
| chr8 | 144700563 | 144700704 | 142 | 4 | 0.000108877 | 4.84E-05 | 0.002028 | 4.29E-05 | -0.00456531 | AF186192.2 |
| chr5 | 157743405 | 157744179 | 775 | 11 | 2.49E-05 | 4.66E-05 | 0.001759 | 4.29E-05 | -0.00082931 | LSM11 |
| chr9 | 130433186 | 130433833 | 648 | 5 | 1.30E-05 | 0.0110225 | 0.000461 | 4.29E-05 | -0.00185834 | HMCN2 |
| chr4 | 1231719 | 1231941 | 223 | 3 | 6.26E-05 | 2.48E-05 | 0.001788 | 4.29E-05 | -0.00033748 | CTBP1 |
| chr16 | 2924780 | 2925208 | 429 | 2 | 3.78E-06 | 0.0009027 | 2.98E-05 | 4.29E-05 | 0.001648694 | FLYWCH1 |
| chr10 | 79286665 | 79286794 | 130 | 2 | 8.82E-05 | 2.58E-05 | 0.001669 | 4.29E-05 | -0.00279 | ZMIZ1 |
| chr11 | 119653202 | 119653533 | 332 | 3 | 1.59E-05 | 0.0020317 | 1.68E-05 | 4.29E-05 | -0.00140911 | AP003393.1, NECTIN1 |
| chr1 | 151058810 | 151059203 | 394 | 5 | 3.66E-05 | 1.23E-05 | 0.007855 | 4.30E-05 | 0.002165315 | CDC42SE1 |
| chr20 | 49906179 | 49906596 | 418 | 3 | 3.95E-05 | 0.0001123 | 0.00072 | 4.30E-05 | -0.00054567 | SPATA2 |
| chr14 | 105326516 | 105327217 | 702 | 3 | 1.07E-05 | 0.0002395 | 0.000267 | 4.30E-05 | -0.00324634 | PACS2 |
| chr12 | 57824276 | 57824460 | 185 | 2 | 3.26E-05 | 4.34E-05 | 0.000274 | 4.30E-05 | -0.00211917 | CTDSP2 |
| chr5 | 157730985 | 157731630 | 646 | 8 | 1.14E-05 | 0.0001954 | 0.000791 | 4.30E-05 | 0.001580249 | THG1L |
| chr19 | 55258669 | 55259364 | 696 | 10 | 3.45E-06 | 0.0003447 | 0.000777 | 4.31E-05 | 0.001532756 | PPP6R1 |
| chr5 | 80256314 | 80256651 | 338 | 4 | 6.06E-05 | 0.0004973 | 0.000261 | 4.31E-05 | -0.0002955 | NA |
| chr19 | 46426441 | 46426988 | 548 | 5 | 6.01E-05 | 0.0004633 | 0.000686 | 4.31E-05 | -0.01111673 | PNMA8C |
| chr8 | 70356663 | 70357091 | 429 | 3 | 1.00E-05 | 0.0088967 | 1.21E-05 | 4.31E-05 | -0.00746383 | NCOA2 |
| chr12 | 96327035 | 96327090 | 56 | 2 | 0.000120789 | 3.37E-05 | 0.000518 | 4.32E-05 | 0.012509698 | CDK17 |
| chr22 | 46777789 | 46777819 | 31 | 2 | 0.000105748 | 6.73E-05 | 0.000133 | 4.32E-05 | 0.009466834 | TBC1D22A |
| chr1 | 156338171 | 156338937 | 767 | 14 | 2.48E-05 | 0.0010901 | 0.000618 | 4.32E-05 | 0.000317026 | TSACC, CCT3 |
| chr18 | 23495788 | 23496011 | 224 | 2 | 0.000155579 | 2.59E-05 | 0.001671 | 4.32E-05 | 0.011729038 | NA |
| chr2 | 222423494 | 222423612 | 119 | 2 | 7.98E-05 | 2.67E-05 | 0.001362 | 4.32E-05 | 0.024259445 | NA |
| chr21 | 39681550 | 39681727 | 178 | 2 | 0.000114632 | 4.24E-05 | 0.000291 | 4.32E-05 | -0.01294186 | AF064860.1 |
| chr5 | 179241835 | 179241900 | 66 | 2 | 3.71E-05 | 2.63E-05 | 0.001538 | 4.33E-05 | -0.02795979 | ADAMTS2 |
| chr6 | 134821918 | 134821989 | 72 | 2 | 4.50E-05 | 9.34E-05 | 9.29E-05 | 4.33E-05 | -0.00590161 | NA |
| chr6 | 168861081 | 168861331 | 251 | 3 | 5.03E-05 | 0.0004471 | 0.000281 | 4.33E-05 | -0.00318195 | NA |
| chr8 | 38528717 | 38528918 | 202 | 3 | 0.000135286 | 0.0001612 | 0.000118 | 4.33E-05 | -0.00680977 | C8orf86 |
| chr18 | 74835194 | 74835324 | 131 | 3 | 6.88E-05 | 0.0003996 | 7.94E-05 | 4.33E-05 | -0.00249148 | ZNF407 |
| chr19 | 13024504 | 13024817 | 314 | 9 | 5.23E-05 | 1.38E-05 | 0.010885 | 4.33E-05 | 0.003727096 | NFIX |
| chr9 | 108156376 | 108156651 | 276 | 2 | 6.73E-05 | 8.23E-05 | 0.000105 | 4.34E-05 | 0.002236364 | NA |
| chr16 | 67517922 | 67518376 | 455 | 2 | 1.32E-05 | 3.64E-05 | 0.000422 | 4.34E-05 | 0.001895565 | AC027682.6 |
| chr5 | 10235532 | 10235548 | 17 | 2 | 0.000142186 | 3.37E-05 | 0.00053 | 4.34E-05 | -0.00634591 | ATPSCKMT |
| chr15 | 96764644 | 96764706 | 63 | 2 | 0.000142641 | 3.24E-05 | 0.000602 | 4.34E-05 | 0.036221422 | NA |
| chr5 | 141651040 | 141651421 | 382 | 6 | 1.31E-05 | 0.0091947 | 0.000809 | 4.34E-05 | 0.000899401 | FCHSD1 |
| chr16 | 27154947 | 27155153 | 207 | 2 | 7.33E-05 | 0.0001094 | 8.21E-05 | 4.34E-05 | -0.01449274 | NA |
| chr1 | 170200828 | 170200835 | 8 | 2 | 0.000154065 | 2.69E-05 | 0.001351 | 4.34E-05 | -0.02225369 | LINC01681 |
| chr12 | 120977703 | 120977956 | 254 | 2 | 2.13E-06 | 0.0001456 | 6.67E-05 | 4.34E-05 | -0.00267583 | HNF1A-AS1 |
| chr11 | 113973684 | 113974106 | 423 | 3 | 1.25E-05 | 0.0002871 | 0.000866 | 4.34E-05 | 0.009842681 | NA |
| chr8 | 129701452 | 129701610 | 159 | 2 | 3.38E-05 | 0.0048404 | 2.19E-05 | 4.35E-05 | -0.00577718 | AC103718.1 |
| chr1 | 75124798 | 75125344 | 547 | 4 | 0.000117218 | 1.40E-05 | 0.009024 | 4.35E-05 | -0.03195909 | NA |
| chr7 | 47783739 | 47784049 | 311 | 2 | 7.37E-05 | 3.10E-05 | 0.000705 | 4.35E-05 | -0.01000533 | PKD1L1 |
| chr1 | 53459313 | 53460279 | 967 | 13 | 2.55E-05 | 0.0004253 | 0.013608 | 4.35E-05 | -0.00010684 | DMRTB1 |
| chr1 | 228006201 | 228006335 | 135 | 2 | 7.72E-06 | 0.002175 | 2.47E-05 | 4.35E-05 | -0.00814466 | NA |
| chr7 | 80513048 | 80513318 | 271 | 2 | 4.48E-05 | 2.78E-05 | 0.001138 | 4.35E-05 | -0.00324488 | CD36 |
| chr11 | 129999759 | 130000370 | 612 | 2 | 1.05E-05 | 7.71E-05 | 0.000114 | 4.35E-05 | 0.003885829 | PRDM10 |
| chr17 | 32349983 | 32350200 | 218 | 7 | 0.000134807 | 0.0023749 | 0.000434 | 4.35E-05 | 0.000259632 | AC005899.4, MIR632, ZNF207, C17orf75 |
| chr6 | 136072878 | 136073364 | 487 | 5 | 6.25E-05 | 4.84E-05 | 0.001782 | 4.35E-05 | -0.00107887 | PDE7B, AL138828.1 |
| chr7 | 76327614 | 76328059 | 446 | 7 | 9.15E-05 | 9.34E-05 | 0.005688 | 4.36E-05 | -0.0017196 | YWHAG |
| chr16 | 1981216 | 1981236 | 21 | 2 | 0.000139769 | 4.52E-05 | 0.000261 | 4.36E-05 | -0.02087836 | TBL3, NOXO1 |
| chr16 | 85335432 | 85335536 | 105 | 2 | 5.62E-05 | 2.61E-05 | 0.001696 | 4.36E-05 | 0.040212403 | GSE1 |
| chr1 | 19446120 | 19446336 | 217 | 3 | 0.000105014 | 6.36E-05 | 0.000728 | 4.36E-05 | 0.00307276 | CAPZB |
| chr1 | 3890748 | 3891145 | 398 | 2 | 7.51E-05 | 2.93E-05 | 0.000891 | 4.36E-05 | -0.00969564 | C1orf174 |
| chr4 | 171025091 | 171025095 | 5 | 2 | 0.000106378 | 4.01E-05 | 0.00034 | 4.36E-05 | 0.011861239 | NA |
| chr2 | 102586714 | 102586765 | 52 | 2 | 0.000121108 | 5.36E-05 | 0.000191 | 4.36E-05 | 0.002707058 | NA |
| chr13 | 60396076 | 60396874 | 799 | 15 | 8.75E-05 | 0.001242 | 0.007652 | 4.36E-05 | 0.000360132 | TDRD3 |
| chr2 | 23524056 | 23524504 | 449 | 5 | 1.58E-05 | 0.0212748 | 5.97E-05 | 4.36E-05 | -0.00237435 | KLHL29, AC011239.1 |
| chr12 | 56643722 | 56643873 | 152 | 2 | 1.87E-05 | 5.03E-05 | 0.000214 | 4.36E-05 | 0.010936206 | ATP5F1B |
| chr13 | 112051322 | 112052063 | 742 | 4 | 6.62E-05 | 0.0006797 | 0.000249 | 4.36E-05 | -0.0145746 | NA |
| chr7 | 106112402 | 106112701 | 300 | 11 | 9.93E-05 | 0.003185 | 0.006414 | 4.37E-05 | 0.000438852 | SYPL1 |
| chr16 | 24790304 | 24791306 | 1003 | 3 | 5.82E-06 | 0.0001666 | 7.32E-05 | 4.37E-05 | -0.00350123 | TNRC6A |
| chr17 | 34613282 | 34613376 | 95 | 2 | 0.000123645 | 4.64E-05 | 0.00025 | 4.37E-05 | -0.03910356 | TMEM132E |
| chr1 | 15343956 | 15344126 | 171 | 4 | 4.46E-05 | 0.0003402 | 0.00122 | 4.37E-05 | -0.01348868 | FHAD1 |
| chr12 | 101039753 | 101040041 | 289 | 2 | 5.85E-06 | 0.0019486 | 2.54E-05 | 4.37E-05 | -0.00689918 | ANO4 |
| chr15 | 69591119 | 69591121 | 3 | 2 | 0.000157163 | 2.62E-05 | 0.001699 | 4.37E-05 | 0.00087164 | DRAIC |
| chr1 | 12202673 | 12202858 | 186 | 2 | 7.77E-05 | 3.69E-05 | 0.00042 | 4.37E-05 | -0.00022029 | TNFRSF1B |
| chr10 | 8332330 | 8332563 | 234 | 3 | 2.65E-05 | 3.64E-05 | 0.00063 | 4.37E-05 | 0.001646781 | NA |
| chr16 | 17162449 | 17162548 | 100 | 2 | 7.32E-05 | 2.74E-05 | 0.001291 | 4.37E-05 | -0.02594075 | XYLT1 |
| chr8 | 29525889 | 29525942 | 54 | 2 | 7.22E-05 | 3.33E-05 | 0.00057 | 4.37E-05 | -0.01021201 | NA |
| chr10 | 94403052 | 94403400 | 349 | 3 | 2.81E-06 | 0.0001454 | 0.001106 | 4.38E-05 | 0.0026602 | TBC1D12 |
| chr7 | 127997725 | 127997818 | 94 | 2 | 7.85E-05 | 9.90E-05 | 9.04E-05 | 4.38E-05 | 0.002585873 | SND1, SND1-IT1 |
| chr15 | 68590917 | 68591151 | 235 | 2 | 0.000105795 | 4.06E-05 | 0.000335 | 4.38E-05 | -0.00562055 | CORO2B |
| chr2 | 97710017 | 97710098 | 82 | 2 | 8.12E-05 | 4.78E-05 | 0.000239 | 4.38E-05 | -0.0089488 | NA |
| chr14 | 67567284 | 67567617 | 334 | 2 | 4.28E-05 | 3.68E-05 | 0.000426 | 4.38E-05 | -0.00483426 | PLEKHH1 |
| chr1 | 173868401 | 173868952 | 552 | 7 | 6.13E-05 | 0.0102252 | 0.000173 | 4.38E-05 | -0.00098676 | ZBTB37, GAS5 |
| chr12 | 113808278 | 113808408 | 131 | 2 | 8.33E-05 | 0.0001175 | 7.88E-05 | 4.38E-05 | -0.0035365 | NA |
| chr2 | 127575453 | 127575478 | 26 | 2 | 6.81E-05 | 3.24E-05 | 0.000629 | 4.39E-05 | -0.00619491 | MYO7B |
| chr12 | 131041949 | 131041990 | 42 | 3 | 0.000126538 | 8.63E-05 | 0.001217 | 4.39E-05 | 0.002614573 | ADGRD1 |
| chr8 | 123645695 | 123645903 | 209 | 4 | 8.33E-05 | 0.0001391 | 8.99E-05 | 4.39E-05 | 0.000958386 | AC135166.1, KLHL38 |
| chr17 | 80895478 | 80895727 | 250 | 2 | 2.96E-06 | 0.0007038 | 3.28E-05 | 4.39E-05 | -0.0040835 | RPTOR |
| chr10 | 102291644 | 102291728 | 85 | 3 | 6.50E-05 | 0.0005581 | 4.79E-05 | 4.39E-05 | 0.000421095 | GBF1 |
| chr10 | 7570285 | 7570452 | 168 | 2 | 0.000154823 | 2.66E-05 | 0.001579 | 4.39E-05 | -5.02E-05 | ITIH5 |
| chr16 | 4614853 | 4615306 | 454 | 7 | 5.22E-05 | 9.47E-05 | 0.005025 | 4.39E-05 | -0.00073121 | UBALD1 |
| chr1 | 95119716 | 95120054 | 339 | 2 | 8.03E-06 | 0.0002219 | 5.27E-05 | 4.39E-05 | -0.00827777 | TLCD4, TLCD4-RWDD3 |
| chr1 | 1631308 | 1631481 | 174 | 3 | 5.15E-05 | 1.73E-05 | 0.005498 | 4.39E-05 | 0.009093097 | NA |
| chr2 | 199471423 | 199471957 | 535 | 5 | 4.76E-05 | 1.37E-05 | 0.009339 | 4.40E-05 | -0.00220365 | SATB2-AS1 |
| chr3 | 97961740 | 97961914 | 175 | 2 | 3.11E-05 | 0.0070304 | 2.12E-05 | 4.40E-05 | -0.00456802 | RIOX2 |
| chr4 | 77075687 | 77076412 | 726 | 9 | 2.47E-05 | 0.0004118 | 0.00041 | 4.40E-05 | 0.002428973 | CCNI |
| chr22 | 36455418 | 36456027 | 610 | 3 | 2.03E-05 | 5.31E-05 | 0.000624 | 4.40E-05 | -0.00681161 | NA |
| chr7 | 31051440 | 31051630 | 191 | 2 | 3.14E-05 | 2.68E-05 | 0.001549 | 4.41E-05 | 0.007697159 | NA |
| chr11 | 57667214 | 57668331 | 1118 | 12 | 2.77E-07 | 0.0093906 | 2.94E-06 | 4.41E-05 | 0.000726771 | ZDHHC5 |
| chr10 | 86377451 | 86377594 | 144 | 3 | 0.000138445 | 0.0001345 | 0.000319 | 4.41E-05 | -0.0031209 | NA |
| chr1 | 111990599 | 111991127 | 529 | 3 | 5.63E-06 | 3.12E-05 | 0.000916 | 4.41E-05 | 0.00365743 | LINC01750 |
| chr3 | 49903386 | 49904180 | 795 | 13 | 1.90E-05 | 0.0011009 | 0.000407 | 4.41E-05 | -0.00040089 | AC105935.2, AC105935.1, MST1R |
| chr2 | 11374750 | 11375122 | 373 | 4 | 5.04E-05 | 0.0025011 | 0.000172 | 4.41E-05 | 0.003154445 | LINC00570 |
| chr12 | 110959085 | 110959158 | 74 | 2 | 5.14E-05 | 0.0001265 | 7.52E-05 | 4.41E-05 | -0.00976146 | LINC01405 |
| chr2 | 8324633 | 8324773 | 141 | 3 | 0.000116851 | 1.85E-05 | 0.00418 | 4.42E-05 | -0.03160065 | LINC00299 |
| chr6 | 165687117 | 165687205 | 89 | 3 | 4.25E-05 | 0.0001256 | 0.000153 | 4.42E-05 | 0.003285259 | PDE10A |
| chr1 | 156813999 | 156815496 | 1498 | 9 | 9.11E-05 | 3.80E-05 | 0.017662 | 4.42E-05 | -0.00136447 | SH2D2A |
| chr17 | 83018814 | 83019134 | 321 | 3 | 6.06E-05 | 4.70E-05 | 0.000495 | 4.42E-05 | -0.00289355 | B3GNTL1 |
| chr8 | 119969645 | 119969689 | 45 | 3 | 8.84E-05 | 9.05E-05 | 0.000607 | 4.42E-05 | -0.00443853 | DEPTOR |
| chr20 | 58179885 | 58179925 | 41 | 2 | 3.23E-05 | 0.0025313 | 2.45E-05 | 4.42E-05 | 0.001912652 | NA |
| chr19 | 58475894 | 58476815 | 922 | 8 | 1.55E-05 | 0.0006224 | 0.000169 | 4.42E-05 | 0.000977931 | ZNF446 |
| chr3 | 58236347 | 58236842 | 496 | 3 | 2.99E-05 | 2.91E-05 | 0.002607 | 4.42E-05 | 0.004239607 | NA |
| chr20 | 45506623 | 45506837 | 215 | 2 | 0.000111105 | 4.36E-05 | 0.000294 | 4.42E-05 | 0.002056719 | NA |
| chr12 | 52902078 | 52902416 | 339 | 2 | 3.80E-05 | 0.0001461 | 6.84E-05 | 4.42E-05 | 0.001345227 | KRT8 |
| chr20 | 33793632 | 33793797 | 166 | 2 | 1.01E-05 | 0.0054143 | 2.20E-05 | 4.43E-05 | -0.00678603 | ZNF341-AS1 |
| chr20 | 64161063 | 64161115 | 53 | 2 | 6.18E-05 | 3.11E-05 | 0.000761 | 4.43E-05 | -0.02513935 | MYT1 |
| chr8 | 73935663 | 73935812 | 150 | 2 | 4.85E-05 | 0.0009327 | 3.06E-05 | 4.43E-05 | 0.001921437 | NA |
| chr6 | 165896215 | 165896254 | 40 | 2 | 5.33E-05 | 0.0001396 | 7.07E-05 | 4.43E-05 | -0.00398204 | PDE10A |
| chr12 | 66024389 | 66024489 | 101 | 2 | 3.47E-05 | 0.0236193 | 1.98E-05 | 4.43E-05 | -0.01059159 | NA |

| | | | | | | | | | | |
|---|---|---|---|---|---|---|---|---|---|---|
| chr8 | 1776732 | 1776974 | 243 | 3 | 5.56E-05 | 0.000326 | 4.40E-05 | 4.43E-05 | 0.005333022 | CLN8, AC019257.8 |
| chr17 | 4143560 | 4144280 | 721 | 6 | 7.35E-06 | 0.0007161 | 0.000486 | 4.44E-05 | 0.000130873 | CYB5D2 |
| chr5 | 172711471 | 172711478 | 8 | 2 | 0.000128329 | 4.41E-05 | 0.000289 | 4.44E-05 | -0.00932737 | NA |
| chr10 | 69578835 | 69578918 | 84 | 2 | 2.98E-05 | 0.0001332 | 7.31E-05 | 4.44E-05 | 0.007443906 | NA |
| chr22 | 29767281 | 29768173 | 893 | 7 | 7.05E-08 | 0.0045149 | 1.32E-05 | 4.44E-05 | 0.000985949 | UQCR10, AC004882.3 |
| chr9 | 6689890 | 6690431 | 542 | 2 | 2.30E-06 | 0.0004001 | 4.06E-05 | 4.44E-05 | -0.00256013 | NA |
| chr22 | 19770644 | 19771052 | 409 | 3 | 0.00011366 | 7.04E-05 | 0.000511 | 4.44E-05 | -0.00146832 | TBX1 |
| chr14 | 35346824 | 35347123 | 300 | 2 | 1.91E-05 | 0.0012112 | 2.87E-05 | 4.44E-05 | -0.00625708 | NA |
| chr17 | 33519584 | 33519873 | 290 | 2 | 3.59E-05 | 5.82E-05 | 0.000175 | 4.44E-05 | -0.001213 | ASIC2 |
| chr6 | 161417965 | 161418435 | 471 | 4 | 5.28E-05 | 0.0150309 | 0.000483 | 4.44E-05 | -0.00379035 | PRKN |
| chr9 | 90898188 | 90898375 | 188 | 2 | 0.000132318 | 3.41E-05 | 0.000561 | 4.44E-05 | -0.00810602 | SYK |
| chr6 | 104955702 | 104956024 | 323 | 2 | 6.81E-06 | 0.0003617 | 4.23E-05 | 4.44E-05 | -0.00523264 | LIN28B |
| chr19 | 55279521 | 55280127 | 607 | 7 | 7.04E-06 | 0.0001263 | 0.000294 | 4.44E-05 | 0.003196185 | HSPBP1 |
| chr17 | 78228506 | 78228760 | 255 | 2 | 0.000139445 | 2.73E-05 | 0.001458 | 4.44E-05 | -0.00182775 | NA |
| chr14 | 77320638 | 77321428 | 791 | 14 | 2.29E-05 | 0.0008628 | 0.000984 | 4.44E-05 | 0.001161013 | GSTZ1, POMT2 |
| chr7 | 151047187 | 151047512 | 326 | 3 | 3.58E-05 | 2.68E-05 | 0.001644 | 4.45E-05 | 0.010123063 | ABCB8 |
| chr15 | 58065676 | 58066681 | 1006 | 15 | 0.00015283 | 0.0010286 | 0.001044 | 4.45E-05 | 0.001409367 | AC025431.1, ALDH1A2 |
| chr19 | 6268054 | 6268265 | 212 | 2 | 6.23E-05 | 0.0001542 | 6.66E-05 | 4.45E-05 | 0.004539518 | MLLT1 |
| chr13 | 111559728 | 111559919 | 192 | 4 | 5.17E-05 | 8.14E-05 | 0.001354 | 4.45E-05 | 0.007291539 | NA |
| chr4 | 132345038 | 132345167 | 130 | 2 | 3.60E-05 | 0.0153503 | 2.03E-05 | 4.46E-05 | -0.00494797 | AC093916.1 |
| chr8 | 22250539 | 22250710 | 172 | 2 | 4.91E-05 | 0.0005845 | 3.55E-05 | 4.46E-05 | -0.0079688 | POLR3D |
| chr4 | 150079083 | 150079532 | 450 | 2 | 5.37E-06 | 2.81E-05 | 0.00125 | 4.46E-05 | -0.00303962 | DCLK2 |
| chr10 | 62267990 | 62268786 | 797 | 7 | 1.68E-06 | 0.0001079 | 0.003993 | 4.46E-05 | -0.00359617 | RTKN2 |
| chr8 | 1161512 | 1162153 | 642 | 4 | 7.95E-06 | 0.0006959 | 0.000112 | 4.46E-05 | -0.00410373 | DLGAP2 |
| chr20 | 50673693 | 50674032 | 340 | 2 | 2.37E-05 | 0.0001615 | 6.49E-05 | 4.46E-05 | -0.00550083 | RIPOR3 |
| chr18 | 31041524 | 31042234 | 711 | 5 | 2.53E-05 | 7.88E-05 | 0.002125 | 4.46E-05 | -0.00407513 | DSC3 |
| chr21 | 38956211 | 38956752 | 542 | 5 | 2.84E-05 | 0.0001497 | 0.00242 | 4.46E-05 | -0.00371013 | AP001042.1 |
| chr20 | 9032112 | 9032136 | 25 | 2 | 0.000168933 | 0.0001022 | 9.05E-05 | 4.46E-05 | -0.00809606 | NA |
| chr11 | 95923820 | 95924260 | 441 | 7 | 0.000116449 | 0.0001279 | 0.010474 | 4.46E-05 | 0.00208411 | MTMR2 |
| chr6 | 164611035 | 164611091 | 57 | 3 | 0.000147801 | 4.51E-05 | 0.000733 | 4.46E-05 | -0.001274 | NA |
| chr19 | 55736926 | 55737152 | 227 | 3 | 0.000137875 | 1.83E-05 | 0.004835 | 4.46E-05 | 0.01036355 | NLRP9 |
| chr14 | 67617402 | 67617473 | 72 | 2 | 2.92E-05 | 2.87E-05 | 0.001136 | 4.46E-05 | -0.00471173 | Y RNA |
| chr2 | 20232750 | 20233021 | 272 | 2 | 4.79E-05 | 0.0001291 | 7.55E-05 | 4.47E-05 | 0.004084171 | NA |
| chr20 | 20056292 | 20056702 | 411 | 4 | 4.42E-05 | 7.40E-05 | 0.002858 | 4.47E-05 | -0.01593626 | CFAP61 |
| chr5 | 53922646 | 53922686 | 41 | 2 | 8.95E-05 | 0.0001432 | 7.03E-05 | 4.47E-05 | 0.008800798 | ARL15 |
| chr15 | 51163016 | 51163652 | 637 | 3 | 1.30E-05 | 0.0019976 | 2.09E-05 | 4.47E-05 | -0.00617891 | MIR4713HG |
| chr22 | 43015077 | 43016035 | 959 | 12 | 1.86E-05 | 0.002423 | 8.23E-05 | 4.47E-05 | -0.00029354 | PACSIN2 |
| chr20 | 45290698 | 45291028 | 331 | 3 | 9.90E-06 | 0.0044919 | 2.61E-05 | 4.48E-05 | 0.003380352 | NA |
| chr17 | 74860058 | 74861231 | 1174 | 9 | 1.84E-05 | 3.86E-05 | 0.016668 | 4.48E-05 | 0.001733969 | GRIN2C |
| chr11 | 43269124 | 43269660 | 537 | 5 | 0.000105771 | 9.38E-05 | 0.003157 | 4.48E-05 | -0.01833819 | HNRNPKP3 |
| chr11 | 73362380 | 73362669 | 290 | 3 | 5.48E-05 | 0.0003306 | 0.000226 | 4.48E-05 | -0.00437685 | ARHGEF17 |
| chr11 | 64891293 | 64891593 | 301 | 9 | 5.37E-05 | 0.0005306 | 0.002448 | 4.48E-05 | 0.000200359 | MIR194-2HG, MIR194-2 |
| chr7 | 32070219 | 32070627 | 409 | 4 | 4.08E-06 | 0.0024735 | 0.001111 | 4.49E-05 | 0.016843617 | PDE1C |
| chr19 | 40359340 | 40359576 | 237 | 6 | 6.17E-05 | 0.0032668 | 0.001127 | 4.49E-05 | -0.00397667 | PLD3, AC118344.2 |
| chr3 | 52777281 | 52777621 | 341 | 3 | 1.92E-05 | 0.0003411 | 4.38E-05 | 4.49E-05 | -0.00524657 | ITIH1 |
| chr20 | 32976369 | 32976635 | 267 | 2 | 5.97E-05 | 7.82E-05 | 0.000119 | 4.49E-05 | 0.006291825 | NA |
| chr12 | 132151530 | 132151757 | 228 | 6 | 8.03E-05 | 0.0046566 | 0.006205 | 4.49E-05 | 0.002130846 | NOC4L |
| chr11 | 2056077 | 2056292 | 216 | 2 | 0.00013426 | 3.29E-05 | 0.000662 | 4.49E-05 | 0.019130267 | NA |
| chr6 | 10425320 | 10425415 | 96 | 3 | 2.72E-05 | 4.62E-05 | 0.001833 | 4.49E-05 | -0.00664465 | AL138885.2 |
| chr1 | 176799397 | 176799405 | 9 | 2 | 0.000137218 | 4.39E-05 | 0.000304 | 4.49E-05 | 0.005419017 | PAPPA2 |
| chr6 | 149910806 | 149911067 | 262 | 4 | 6.72E-05 | 0.0002867 | 0.000242 | 4.49E-05 | -0.00157107 | RAET1E-AS1, RAET1E-AS1 |
| chr4 | 143336841 | 143337295 | 455 | 5 | 2.97E-05 | 0.0003446 | 0.004438 | 4.49E-05 | 0.001383545 | GAB1 |
| chr16 | 84461779 | 84461821 | 43 | 2 | 2.35E-05 | 3.34E-05 | 0.000631 | 4.50E-05 | -0.00471026 | ATP2C2, ATP2C2-AS1 |
| chr12 | 103769441 | 103769689 | 249 | 3 | 2.91E-05 | 0.0053797 | 3.17E-05 | 4.50E-05 | -0.00428379 | NA |
| chr20 | 62937504 | 62937983 | 480 | 7 | 9.24E-05 | 9.99E-05 | 0.00154 | 4.50E-05 | 0.001059843 | DIDO1 |
| chr17 | 38769725 | 38769754 | 30 | 2 | 1.61E-05 | 0.0040759 | 2.32E-05 | 4.50E-05 | -0.00639745 | PIP4K2B |
| chr22 | 30887406 | 30887688 | 283 | 3 | 7.96E-05 | 0.0001289 | 0.000718 | 4.50E-05 | -0.00212778 | OSBP2 |
| chr16 | 15889156 | 15889484 | 329 | 5 | 8.60E-06 | 0.0004282 | 4.63E-05 | 4.50E-05 | -0.00278896 | NA |
| chr10 | 60943667 | 60944449 | 783 | 9 | 3.51E-05 | 0.0151118 | 0.001774 | 4.50E-05 | 0.00068381 | RHOBTB1 |
| chr11 | 62645760 | 62646027 | 268 | 5 | 6.85E-06 | 0.0156058 | 6.69E-05 | 4.50E-05 | -0.0017848 | GANAB |
| chr12 | 42238799 | 42239529 | 731 | 4 | 2.02E-06 | 3.88E-05 | 0.003963 | 4.50E-05 | -0.00299646 | PPHLN1 |
| chr2 | 11586842 | 11587047 | 206 | 2 | 8.89E-05 | 7.79E-05 | 0.00012 | 4.51E-05 | -0.00735355 | GREB1 |
| chr3 | 49666870 | 49667096 | 227 | 2 | 2.79E-05 | 0.0003187 | 4.57E-05 | 4.51E-05 | -0.00912593 | BSN |
| chr11 | 33295469 | 33295547 | 79 | 3 | 0.00010581 | 0.000123 | 0.000158 | 4.51E-05 | -0.00252281 | HIPK3 |
| chr15 | 33465525 | 33465626 | 102 | 2 | 4.91E-05 | 2.71E-05 | 0.001728 | 4.51E-05 | -0.01937555 | RYR3 |
| chr12 | 25552927 | 25553019 | 93 | 2 | 0.000110578 | 3.52E-05 | 0.000537 | 4.51E-05 | -0.00802526 | LMNTD1 |
| chr10 | 129771789 | 129772483 | 695 | 4 | 8.47E-05 | 0.0001583 | 0.000482 | 4.52E-05 | -0.00051133 | NA |
| chr10 | 11171361 | 11171433 | 73 | 2 | 3.48E-05 | 0.0006086 | 3.55E-05 | 4.52E-05 | 0.006139564 | CELF2, AL136320.1 |
| chr1 | 160344271 | 160344956 | 686 | 7 | 4.35E-06 | 0.0016612 | 1.17E-05 | 4.52E-05 | -0.00079371 | NCSTN |
| chr1 | 232988672 | 232988933 | 262 | 3 | 1.02E-05 | 0.0141383 | 9.96E-06 | 4.52E-05 | 0.00067488 | PCNX2 |
| chr11 | 9245590 | 9246119 | 530 | 3 | 7.34E-05 | 7.75E-05 | 0.00126 | 4.52E-05 | -0.00584344 | DENND5A |
| chr16 | 8788748 | 8788830 | 83 | 3 | 8.14E-05 | 0.0004563 | 9.37E-05 | 4.53E-05 | -0.00295115 | PMM2, TMEM186 |
| chr8 | 1873989 | 1874402 | 414 | 3 | 0.000113686 | 2.16E-05 | 0.003324 | 4.53E-05 | -0.01660406 | AC019257.8, ARHGEF10 |
| chr1 | 3815079 | 3815271 | 193 | 4 | 6.39E-05 | 0.0001606 | 0.000223 | 4.53E-05 | -0.00328907 | CEP104 |
| chr17 | 80608589 | 80608847 | 259 | 3 | 0.000113279 | 5.43E-05 | 0.000864 | 4.53E-05 | 7.67E-06 | RPTOR |
| chr15 | 72306687 | 72307390 | 704 | 5 | 4.18E-06 | 0.0003468 | 0.00027 | 4.53E-05 | -0.00118512 | AC009690.2, AC009690.3, CELF6, AC009690.1 |
| chr9 | 35732715 | 35732885 | 171 | 3 | 0.000140265 | 7.09E-05 | 0.000967 | 4.54E-05 | -0.00175658 | CREB3 |
| chr16 | 2468285 | 2468496 | 212 | 3 | 7.68E-05 | 0.0001727 | 0.001181 | 4.54E-05 | 0.002315158 | NA |
| chr6 | 15509106 | 15509592 | 487 | 2 | 2.91E-06 | 0.0001893 | 6.04E-05 | 4.54E-05 | -0.00243601 | JARID2 |
| chr2 | 86160254 | 86160758 | 505 | 3 | 6.99E-05 | 8.73E-05 | 0.000205 | 4.54E-05 | 0.003919926 | IMMT |
| chr6 | 70221851 | 70221967 | 117 | 3 | 0.000119232 | 3.31E-05 | 0.000763 | 4.55E-05 | -0.01089571 | COL9A1 |
| chr4 | 123396676 | 123397368 | 693 | 8 | 2.15E-05 | 5.92E-05 | 0.010642 | 4.55E-05 | 0.001403414 | SPRY1 |
| chr5 | 528847 | 529143 | 297 | 2 | 3.02E-05 | 2.73E-05 | 0.001723 | 4.55E-05 | 0.020930143 | NA |
| chr18 | 8240750 | 8240753 | 4 | 2 | 0.000162776 | 2.82E-05 | 0.001408 | 4.56E-05 | -0.00878314 | PTPRM |
| chr16 | 8847711 | 8848483 | 773 | 6 | 2.67E-06 | 0.0113921 | 0.000455 | 4.56E-05 | 0.002206564 | PMM2, AC022167.2, AC012173.1 |
| chr8 | 82441518 | 82441663 | 146 | 2 | 0.000113199 | 4.51E-05 | 0.0003 | 4.56E-05 | 0.016159721 | AC060765.2 |
| chr17 | 41705615 | 41705717 | 103 | 2 | 0.000158788 | 2.91E-05 | 0.001185 | 4.57E-05 | -0.00306443 | NA |
| chr9 | 137790807 | 137790853 | 47 | 2 | 0.00010487 | 0.0001772 | 6.32E-05 | 4.57E-05 | -0.003967 | EHMT1 |
| chr14 | 101069655 | 101069833 | 179 | 3 | 2.43E-06 | 0.000477 | 4.53E-05 | 4.57E-05 | 0.001871989 | MEG9 |
| chr17 | 8383268 | 8383928 | 661 | 5 | 4.10E-06 | 0.0001062 | 0.004045 | 4.57E-05 | -0.00101514 | NA |
| chr5 | 36741199 | 36741376 | 178 | 2 | 5.88E-05 | 0.0012419 | 2.93E-05 | 4.57E-05 | 0.003620835 | NA |
| chr4 | 55345152 | 55345266 | 115 | 2 | 0.000105384 | 8.63E-05 | 0.00011 | 4.57E-05 | 0.001724232 | NA |
| chr14 | 67240589 | 67241594 | 1006 | 11 | 2.56E-05 | 0.0015082 | 0.000319 | 4.57E-05 | 0.000715178 | MPP5 |
| chr12 | 43096009 | 43096144 | 136 | 2 | 0.000164101 | 2.76E-05 | 0.001674 | 4.58E-05 | 0.007060296 | NA |
| chr14 | 75334320 | 75334388 | 69 | 2 | 0.000146225 | 2.75E-05 | 0.001741 | 4.58E-05 | 0.003828498 | NA |
| chr9 | 135085911 | 135086083 | 173 | 2 | 7.98E-06 | 0.000395 | 4.24E-05 | 4.58E-05 | -0.00930713 | OLFM1 |
| chr18 | 35377365 | 35378242 | 878 | 9 | 4.25E-05 | 0.0039529 | 0.001629 | 4.59E-05 | -0.00387117 | NA |
| chr11 | 64229049 | 64230114 | 1066 | 6 | 1.44E-06 | 0.0001018 | 0.000945 | 4.59E-05 | -0.00212238 | NUDT22, AP001453.1 |
| chr2 | 239304506 | 239304548 | 43 | 2 | 3.76E-05 | 3.85E-05 | 0.000444 | 4.59E-05 | -0.01307788 | HDAC4 |
| chr6 | 119069213 | 119069253 | 41 | 2 | 0.000149202 | 0.0001187 | 8.39E-05 | 4.59E-05 | -0.00862326 | AL137009.1, FAM184A |
| chr11 | 8707341 | 8707670 | 330 | 3 | 1.51E-05 | 0.0018882 | 1.88E-05 | 4.59E-05 | -0.00068361 | RPL27A, DENND2B |
| chr17 | 42675745 | 42676154 | 410 | 2 | 5.27E-05 | 3.07E-05 | 0.000955 | 4.59E-05 | 0.00338584 | PLEKHH3 |
| chr6 | 31712335 | 31712451 | 117 | 3 | 5.67E-05 | 0.0005637 | 8.80E-05 | 4.59E-05 | 0.010340352 | LY6G6F-LY6G6D, AL662899.1, LY6G6E |
| chr13 | 43390031 | 43390089 | 59 | 2 | 0.000121097 | 2.82E-05 | 0.001506 | 4.60E-05 | -0.02350641 | ENOX1 |
| chr1 | 32204178 | 32205094 | 917 | 4 | 1.37E-06 | 0.0001134 | 0.000455 | 4.61E-05 | -0.00184281 | CCDC28B, AL049795.1 |
| chr14 | 93019958 | 93020191 | 234 | 2 | 2.24E-05 | 2.79E-05 | 0.001643 | 4.61E-05 | -0.01280765 | ITPK1 |
| chr17 | 57772988 | 57773023 | 36 | 2 | 0.0001065 | 0.0001073 | 9.13E-05 | 4.61E-05 | -0.01459273 | AC015845.2 |
| chr9 | 137550441 | 137550554 | 114 | 4 | 7.32E-05 | 0.00019 | 0.000223 | 4.61E-05 | 0.002021588 | NA |
| chr8 | 143358887 | 143359903 | 1017 | 4 | 5.07E-06 | 2.93E-05 | 0.001038 | 4.61E-05 | 0.000475497 | TOP1MT |
| chr6 | 65787693 | 65787853 | 161 | 2 | 8.13E-05 | 3.99E-05 | 0.00041 | 4.61E-05 | 0.000676727 | NA |
| chr5 | 180686357 | 180686361 | 5 | 2 | 0.000142801 | 2.91E-05 | 0.001281 | 4.61E-05 | -0.00518153 | LINC02222 |
| chr16 | 2448951 | 2449222 | 272 | 3 | 2.49E-05 | 0.0010941 | 6.02E-05 | 4.61E-05 | -0.00237008 | CCNF |
| chr10 | 3935146 | 3935416 | 271 | 3 | 3.00E-05 | 0.0003474 | 0.000266 | 4.62E-05 | -0.00691284 | LINC02660 |
| chr2 | 1859843 | 1860379 | 537 | 2 | 4.13E-05 | 2.77E-05 | 0.001753 | 4.62E-05 | -0.0036994 | MYT1L |
| chr10 | 124461002 | 124461922 | 921 | 11 | 5.15E-05 | 0.0002267 | 0.00758 | 4.62E-05 | 0.00093467 | LHPP |
| chr3 | 49115027 | 49115307 | 281 | 2 | 6.58E-05 | 2.77E-05 | 0.001741 | 4.62E-05 | -0.00061964 | USP19 |
| chr6 | 163919493 | 163919582 | 90 | 2 | 1.64E-05 | 0.0008127 | 3.32E-05 | 4.62E-05 | -0.00302264 | AL078602.1 |
| chr9 | 106923299 | 106923466 | 168 | 2 | 4.05E-05 | 5.74E-05 | 0.000197 | 4.63E-05 | -0.00858949 | ZNF462 |
| chr1 | 157963798 | 157964104 | 307 | 2 | 9.71E-06 | 0.0885069 | 2.01E-05 | 4.63E-05 | -0.01129397 | NA |
| chr19 | 11694605 | 11694906 | 302 | 3 | 2.46E-05 | 0.0011855 | 0.000125 | 4.63E-05 | 0.003766904 | NA |
| chr9 | 136342211 | 136342325 | 115 | 3 | 5.84E-05 | 2.79E-05 | 0.002839 | 4.63E-05 | -0.00576405 | GPSM1 |
| chr5 | 39284712 | 39284815 | 104 | 2 | 0.000141738 | 2.78E-05 | 0.001763 | 4.64E-05 | -0.00075932 | C9 |
| chr8 | 47971280 | 47971366 | 87 | 2 | 3.14E-05 | 0.0001243 | 8.23E-05 | 4.64E-05 | 0.002369984 | MCM4 |
| chr6 | 168412654 | 168413482 | 829 | 5 | 3.46E-05 | 4.90E-05 | 0.002755 | 4.65E-05 | -0.00466506 | NA |
| chr14 | 73851459 | 73852071 | 613 | 13 | 5.72E-05 | 0.0003825 | 0.004288 | 4.65E-05 | -1.08E-05 | PTGR2, AC005520.1 |
| chr5 | 151753141 | 151753930 | 790 | 5 | 3.41E-05 | 0.0068872 | 7.81E-05 | 4.65E-05 | -0.00296196 | ATOX1 |
| chr5 | 103526712 | 103526717 | 6 | 2 | 0.00010396 | 0.0001261 | 8.16E-05 | 4.65E-05 | -0.00820714 | NA |
| chr12 | 96063494 | 96063695 | 202 | 2 | 8.65E-05 | 3.11E-05 | 0.000966 | 4.65E-05 | -0.00146246 | NA |
| chr22 | 39245867 | 39246276 | 410 | 3 | 3.86E-06 | 0.0006749 | 4.12E-05 | 4.65E-05 | -0.00778829 | AL031590.1 |
| chr12 | 132438797 | 132439048 | 252 | 3 | 4.47E-05 | 0.000232 | 0.001017 | 4.65E-05 | 0.00032945 | NA |
| chr2 | 157791527 | 157791612 | 86 | 2 | 5.67E-05 | 0.0003127 | 4.78E-05 | 4.65E-05 | 0.001419884 | ACVR1 |
| chr1 | 109169940 | 109170001 | 62 | 2 | 0.000153764 | 3.52E-05 | 0.000604 | 4.66E-05 | -0.01159769 | ELAPOR1 |
| chr13 | 110262227 | 110262378 | 152 | 2 | 7.68E-05 | 9.52E-05 | 0.000103 | 4.66E-05 | -0.01024698 | COL4A1 |
| chr5 | 115601917 | 115602451 | 535 | 9 | 5.69E-06 | 0.0057209 | 0.002495 | 4.66E-05 | -0.00014091 | AC010226.1, TICAM2, TMED7-TICAM2 |
| chr3 | 194687827 | 194688236 | 410 | 4 | 3.98E-05 | 2.27E-05 | 0.003505 | 4.66E-05 | -0.0027742 | FAM43A |
| chr17 | 81715921 | 81716727 | 807 | 4 | 3.87E-07 | 0.0007891 | 0.000123 | 4.66E-05 | -0.00326715 | AC139530.2, SLC25A10 |
| chr11 | 65976380 | 65976688 | 309 | 4 | 4.58E-05 | 0.0005092 | 7.98E-05 | 4.66E-05 | 0.001547221 | SART1 |
| chr18 | 23024166 | 23024176 | 11 | 2 | 0.000139293 | 4.92E-05 | 0.000265 | 4.66E-05 | -0.01218261 | RBBP8 |
| chr2 | 98296262 | 98296659 | 398 | 3 | 0.000103439 | 2.56E-05 | 0.002558 | 4.66E-05 | 0.002578532 | VWA3B |
| chr11 | 62462441 | 62462539 | 99 | 2 | 9.59E-06 | 0.0003774 | 4.42E-05 | 4.66E-05 | -0.00650156 | AHNAK |
| chr17 | 40665925 | 40665947 | 23 | 2 | 0.000141244 | 7.24E-05 | 0.000142 | 4.66E-05 | -0.00782065 | NA |
| chr16 | 75038345 | 75038738 | 394 | 3 | 4.54E-05 | 0.0004197 | 4.59E-05 | 4.66E-05 | 0.001689452 | ZNRF1 |
| chr10 | 5852209 | 5852436 | 228 | 2 | 9.96E-05 | 3.93E-05 | 0.000443 | 4.67E-05 | 0.006278615 | NA |
| chr2 | 232385660 | 232385861 | 202 | 2 | 8.89E-05 | 9.17E-05 | 0.000107 | 4.67E-05 | -0.0050774 | ECEL1P2 |
| chr3 | 27369479 | 27369574 | 96 | 4 | 2.71E-05 | 4.75E-05 | 0.000525 | 4.67E-05 | 0.00050534 | AC099535.2 |
| chr4 | 139030947 | 139031006 | 60 | 2 | 8.19E-05 | 0.0002873 | 5.00E-05 | 4.67E-05 | 0.007223992 | NOCT, ELF2 |
| chr10 | 34727669 | 34727745 | 77 | 2 | 1.52E-05 | 0.0005726 | 3.75E-05 | 4.67E-05 | -0.01392145 | PARD3 |
| chr22 | 47928749 | 47929023 | 275 | 2 | 4.71E-05 | 0.0002891 | 4.99E-05 | 4.67E-05 | -0.01298912 | AL117329.1 |
| chr18 | 55322290 | 55322784 | 495 | 4 | 2.39E-06 | 0.0067158 | 0.000195 | 4.67E-05 | 0.004547297 | TCF4 |
| chr6 | 168365622 | 168365986 | 365 | 3 | 1.35E-05 | 0.0214671 | 8.90E-06 | 4.67E-05 | -0.00271975 | NA |
| chr5 | 14185924 | 14186389 | 466 | 2 | 3.13E-05 | 3.27E-05 | 0.000807 | 4.68E-05 | 0.007102922 | TRIO |
| chr8 | 144730598 | 144730639 | 42 | 3 | 2.37E-05 | 2.66E-05 | 0.001596 | 4.68E-05 | 0.037197372 | ZNF251 |
| chr6 | 131951842 | 131952015 | 174 | 3 | 1.31E-05 | 4.35E-05 | 0.000449 | 4.68E-05 | -0.00068026 | AL133346.1 |
| chr12 | 95793843 | 95793880 | 38 | 2 | 0.00011536 | 2.89E-05 | 0.001469 | 4.69E-05 | -0.00918001 | AC090001.1 |
| chr6 | 158644748 | 158645150 | 403 | 11 | 0.000109136 | 2.92E-05 | 0.019908 | 4.69E-05 | 3.68E-05 | NA |
| chr7 | 99388089 | 99388176 | 88 | 2 | 3.83E-05 | 0.0016916 | 2.81E-05 | 4.69E-05 | -0.00158622 | AC004922.1, ARPC1B |
| chr4 | 185624512 | 185624684 | 173 | 2 | 3.67E-05 | 2.81E-05 | 0.001769 | 4.69E-05 | -0.01151192 | SORBS2 |
| chr6 | 35024978 | 35025500 | 523 | 2 | 9.76E-06 | 0.0003964 | 4.36E-05 | 4.69E-05 | -0.01852818 | ANKS1A |

| | | | | | | | | | | |
|---|---|---|---|---|---|---|---|---|---|---|
| chr19 | 39264077 | 39264334 | 258 | 2 | 0.00011667 | 4.11E-05 | 0.000404 | 4.70E-05 | -0.00555602 | NA |
| chr18 | 4117462 | 4117506 | 45 | 2 | 5.18E-05 | 3.60E-05 | 0.000585 | 4.70E-05 | 0.010929566 | DLGAP1 |
| chr1 | 939965 | 940500 | 536 | 3 | 2.67E-06 | 9.63E-05 | 0.000173 | 4.70E-05 | 0.001930725 | SAMD11 |
| chr4 | 7434966 | 7435429 | 464 | 5 | 4.29E-05 | 5.39E-05 | 0.001187 | 4.70E-05 | -0.0037742 | SORCS2 |
| chr4 | 80335339 | 80336115 | 777 | 12 | 1.39E-05 | 0.0040152 | 9.67E-05 | 4.70E-05 | -0.00029994 | FGF5, CFAP299 |
| chr4 | 73263457 | 73263493 | 37 | 2 | 5.24E-05 | 0.0005517 | 3.84E-05 | 4.70E-05 | -0.00344505 | AC053527.2 |
| chr1 | 30245734 | 30246275 | 542 | 3 | 8.46E-05 | 5.30E-05 | 0.000311 | 4.71E-05 | 0.001433997 | NA |
| chr19 | 11346344 | 11347023 | 680 | 10 | 0.000107749 | 0.0002714 | 0.003475 | 4.71E-05 | -0.00125773 | CCDC159, TMEM205 |
| chr19 | 10681939 | 10682167 | 229 | 3 | 3.81E-06 | 4.50E-05 | 0.000527 | 4.71E-05 | -0.00061189 | ILF3 |
| chr19 | 48513268 | 48513307 | 40 | 3 | 0.000123602 | 2.35E-05 | 0.00287 | 4.71E-05 | -0.01548722 | LMTK3 |
| chr1 | 32316731 | 32316816 | 86 | 2 | 0.000168902 | 2.88E-05 | 0.001548 | 4.72E-05 | -0.00476801 | HDAC1 |
| chr19 | 39514299 | 39515118 | 820 | 11 | 1.70E-05 | 0.0022876 | 0.00069 | 4.72E-05 | -0.00318475 | SELENOV |
| chr17 | 8163351 | 8163698 | 348 | 3 | 5.70E-05 | 2.55E-05 | 0.001658 | 4.72E-05 | -0.02828153 | VAMP2 |
| chr7 | 155448795 | 155449275 | 481 | 3 | 6.32E-05 | 2.04E-05 | 0.004758 | 4.72E-05 | 0.001709469 | AC008060.3 |
| chr19 | 39532641 | 39532928 | 288 | 8 | 0.000141891 | 9.95E-05 | 0.000136 | 4.72E-05 | 0.00087585 | AC011500.3, EID2B |
| chr12 | 14073447 | 14073927 | 481 | 4 | 1.05E-05 | 0.0082567 | 3.67E-05 | 4.72E-05 | 0.000210232 | NA |
| chr5 | 157521573 | 157521859 | 287 | 2 | 3.44E-05 | 0.0002178 | 5.87E-05 | 4.72E-05 | 0.006560069 | ADAM19 |
| chr6 | 116779975 | 116780228 | 254 | 2 | 4.32E-05 | 0.0005322 | 3.90E-05 | 4.73E-05 | -0.01286118 | NA |
| chr16 | 48609902 | 48610109 | 208 | 4 | 3.04E-05 | 2.14E-05 | 0.002469 | 4.73E-05 | 0.001069135 | N4BP1 |
| chr6 | 24949344 | 24949473 | 130 | 3 | 3.76E-05 | 0.0026267 | 4.42E-05 | 4.73E-05 | -0.0060047 | RIPOR2 |
| chr10 | 124046267 | 124046874 | 608 | 4 | 1.93E-05 | 0.0001436 | 0.000566 | 4.73E-05 | -1.12E-05 | CHST15 |
| chr12 | 113242158 | 113242690 | 533 | 2 | 1.36E-06 | 0.0002071 | 6.06E-05 | 4.74E-05 | -0.00751381 | TPCN1 |
| chr9 | 34617103 | 34617389 | 287 | 2 | 0.000123583 | 0.0001934 | 6.31E-05 | 4.74E-05 | -0.00156079 | DCTN3 |
| chr15 | 63946050 | 63946298 | 249 | 3 | 0.000124266 | 0.000139 | 0.000107 | 4.74E-05 | -0.00965264 | DAPK2 |
| chr16 | 3254261 | 3254449 | 189 | 3 | 0.000149493 | 4.37E-05 | 0.00098 | 4.74E-05 | -0.01425111 | MEFV |
| chr17 | 41397708 | 41397908 | 201 | 3 | 9.75E-05 | 8.10E-05 | 0.000168 | 4.74E-05 | 0.000114729 | NA |
| chr20 | 49270472 | 49270682 | 211 | 2 | 5.75E-05 | 4.94E-05 | 0.000274 | 4.74E-05 | -0.00771147 | ZNFX1 |
| chr5 | 39372205 | 39372309 | 105 | 2 | 3.48E-05 | 7.95E-05 | 0.00013 | 4.74E-05 | -0.00719242 | C9, DAB2 |
| chr2 | 236503644 | 236503796 | 153 | 4 | 6.49E-05 | 0.0004131 | 8.04E-05 | 4.74E-05 | 0.003408518 | IQCA1 |
| chr1 | 1758171 | 1758448 | 278 | 2 | 5.15E-05 | 0.0001581 | 7.16E-05 | 4.74E-05 | -0.00536688 | NADK |
| chr3 | 33113641 | 33114150 | 510 | 11 | 9.03E-05 | 0.003275 | 0.002456 | 4.74E-05 | 0.00064438 | CRTAP |
| chr3 | 179452829 | 179453071 | 243 | 2 | 2.96E-05 | 2.90E-05 | 0.001534 | 4.74E-05 | -0.00836635 | NA |
| chr2 | 60553280 | 60554082 | 803 | 12 | 1.60E-05 | 0.0002063 | 0.004459 | 4.74E-05 | 0.002276617 | BCL11A |
| chr1 | 42734955 | 42735230 | 276 | 2 | 3.26E-05 | 0.000204 | 6.13E-05 | 4.75E-05 | -0.00305038 | CLDN19 |
| chr13 | 110559767 | 110559827 | 61 | 2 | 6.72E-05 | 0.0006598 | 3.65E-05 | 4.75E-05 | -0.00474473 | RAB20 |
| chr16 | 75207166 | 75207407 | 242 | 7 | 7.89E-05 | 7.19E-05 | 0.000349 | 4.75E-05 | -0.00229459 | NA |
| chr12 | 5368726 | 5369116 | 391 | 3 | 0.000107413 | 2.19E-05 | 0.002914 | 4.75E-05 | -0.00560098 | AC006206.2 |
| chr11 | 122157295 | 122157338 | 44 | 2 | 0.000170915 | 2.89E-05 | 0.001603 | 4.75E-05 | -0.01870321 | MIR100HG |
| chr7 | 44250118 | 44250254 | 137 | 2 | 0.000141828 | 4.12E-05 | 0.000417 | 4.75E-05 | -0.00970477 | CAMK2B |
| chr4 | 75048098 | 75048250 | 153 | 2 | 0.000163795 | 3.01E-05 | 0.001267 | 4.75E-05 | -0.0072393 | PARM1 |
| chr1 | 206051904 | 206052343 | 440 | 2 | 2.44E-06 | 0.0003901 | 4.45E-05 | 4.75E-05 | 0.010137354 | BX571818.1 |
| chr10 | 5889690 | 5890737 | 1048 | 8 | 1.21E-06 | 0.0048996 | 0.000165 | 4.75E-05 | 0.000529347 | FBH1, ANKRD16 |
| chr1 | 3684171 | 3684315 | 145 | 3 | 0.000106799 | 2.47E-05 | 0.002337 | 4.75E-05 | -0.01824306 | TP73 |
| chr20 | 36578456 | 36578903 | 448 | 5 | 7.10E-05 | 0.0001796 | 0.000159 | 4.75E-05 | 4.72E-05 | TGIF2, TGIF2-RAB5IF |
| chr1 | 38044402 | 38044850 | 449 | 4 | 6.91E-05 | 6.66E-05 | 0.000454 | 4.76E-05 | 2.87E-05 | POU3F1 |
| chr11 | 495198 | 495908 | 711 | 2 | 1.61E-07 | 0.0010718 | 3.17E-05 | 4.76E-05 | 0.009336457 | RNH1 |
| chr8 | 130852560 | 130852628 | 69 | 2 | 9.55E-05 | 0.0001718 | 6.82E-05 | 4.76E-05 | 0.017233071 | ADCY8 |
| chr19 | 42067187 | 42067346 | 160 | 3 | 9.00E-05 | 2.26E-05 | 0.002781 | 4.76E-05 | 0.000965643 | GRIK5 |
| chr6 | 107745342 | 107745444 | 103 | 2 | 5.43E-05 | 0.0002213 | 5.89E-05 | 4.76E-05 | -0.00767001 | SCML4 |
| chr4 | 152956755 | 152956903 | 149 | 2 | 7.85E-05 | 0.0002175 | 5.94E-05 | 4.76E-05 | -0.00690499 | FHDC1 |
| chr1 | 186680387 | 186680853 | 467 | 10 | 5.44E-05 | 9.38E-05 | 0.007639 | 4.77E-05 | 0.001243312 | PACERR, PTGS2 |
| chr12 | 11556614 | 11556923 | 310 | 3 | 1.53E-05 | 6.42E-05 | 0.000248 | 4.77E-05 | 0.01051553 | LINC01252 |
| chr10 | 14953958 | 14954928 | 971 | 17 | 2.02E-05 | 0.0006014 | 0.010968 | 4.78E-05 | 0.001202875 | DCLRE1C |
| chr11 | 995292 | 995478 | 187 | 3 | 1.98E-05 | 0.0004558 | 0.000105 | 4.78E-05 | -0.00541712 | AP2A2 |
| chr11 | 1008880 | 1009438 | 559 | 7 | 3.60E-05 | 0.0001548 | 0.000216 | 4.78E-05 | 0.00084695 | AP2A2 |
| chr2 | 101272957 | 101273292 | 336 | 8 | 9.66E-05 | 0.0018979 | 0.001044 | 4.78E-05 | 0.000875881 | RNF149, SNORD89 |
| chr15 | 42155881 | 42156588 | 708 | 4 | 4.94E-05 | 0.0001236 | 0.001506 | 4.78E-05 | -0.00152228 | PLA2G4F |
| chr12 | 21526465 | 21527286 | 822 | 4 | 2.02E-05 | 4.74E-05 | 0.000834 | 4.79E-05 | -0.01146972 | SPX |
| chr11 | 117300357 | 117300578 | 222 | 4 | 4.97E-05 | 8.46E-05 | 0.002558 | 4.79E-05 | -0.00322621 | BACE1 |
| chr12 | 15672625 | 15672820 | 196 | 3 | 7.77E-05 | 0.0002649 | 5.64E-05 | 4.79E-05 | -0.00712414 | EPS8 |
| chr17 | 19384364 | 19384665 | 302 | 3 | 8.30E-05 | 3.94E-05 | 0.001002 | 4.79E-05 | 0.005216774 | MFAP4 |
| chr2 | 104053105 | 104053435 | 331 | 4 | 7.25E-05 | 0.0003118 | 8.83E-05 | 4.79E-05 | -0.00272865 | LINC01965 |
| chr10 | 71761849 | 71761998 | 150 | 4 | 9.09E-05 | 0.0002855 | 0.000693 | 4.79E-05 | 8.10E-05 | CDH23, VSIR |
| chr1 | 9875134 | 9875576 | 443 | 4 | 0.00013373 | 8.05E-05 | 0.000674 | 4.79E-05 | -0.00541998 | CTNNBIP1 |
| chr4 | 24469328 | 24469671 | 344 | 2 | 8.32E-05 | 3.10E-05 | 0.001143 | 4.79E-05 | -0.01833966 | NA |
| chr14 | 63641874 | 63642607 | 734 | 11 | 3.97E-06 | 0.0005877 | 0.000778 | 4.79E-05 | 0.000964868 | AL136038.4 |
| chr6 | 81561831 | 81561931 | 101 | 2 | 2.62E-05 | 0.0008118 | 3.46E-05 | 4.79E-05 | -0.00658129 | TENT5A |
| chr5 | 530848 | 531190 | 343 | 4 | 8.58E-05 | 0.0004984 | 0.000611 | 4.80E-05 | 0.010812093 | NA |
| chr3 | 13843100 | 13843150 | 51 | 2 | 5.60E-05 | 0.001655 | 2.88E-05 | 4.80E-05 | 0.003219498 | WNT7A |
| chr11 | 3061534 | 3061538 | 5 | 2 | 0.000174325 | 2.88E-05 | 0.001755 | 4.80E-05 | -0.00514361 | AC108448.3 |
| chr9 | 33043426 | 33044119 | 694 | 3 | 6.84E-05 | 3.38E-05 | 0.002281 | 4.80E-05 | -0.00471571 | SMU1 |
| chr4 | 41608927 | 41609092 | 166 | 2 | 8.48E-05 | 9.94E-05 | 0.000104 | 4.80E-05 | -0.00480116 | LIMCH1, AC108050.1 |
| chr14 | 69853791 | 69854317 | 527 | 4 | 0.000107878 | 0.0002536 | 0.0034 | 4.80E-05 | -0.00036259 | SMOC1 |
| chr10 | 104640199 | 104641122 | 924 | 8 | 5.67E-06 | 0.0004432 | 6.57E-05 | 4.80E-05 | 0.001877288 | NA |
| chr7 | 90707954 | 90708205 | 252 | 3 | 3.13E-05 | 0.0001715 | 9.52E-05 | 4.80E-05 | -0.00469413 | CDK14 |
| chr12 | 111457802 | 111457851 | 50 | 2 | 3.03E-05 | 0.000719 | 3.60E-05 | 4.80E-05 | -0.00645015 | ATXN2 |
| chr9 | 77455437 | 77455616 | 180 | 3 | 0.000133673 | 2.57E-05 | 0.003363 | 4.80E-05 | -0.00813 | GNA14 |
| chr22 | 30909441 | 30909474 | 34 | 2 | 0.000112651 | 0.0001098 | 9.58E-05 | 4.80E-05 | -0.00559374 | NA |
| chr1 | 181104598 | 181104873 | 276 | 2 | 3.22E-06 | 0.0010178 | 3.25E-05 | 4.81E-05 | 0.001988843 | NA |
| chr15 | 85592267 | 85592433 | 167 | 2 | 0.0001093 | 3.61E-05 | 0.000633 | 4.81E-05 | -0.01440086 | AKAP13 |
| chr16 | 85107626 | 85107756 | 131 | 3 | 0.000132951 | 3.55E-05 | 0.002209 | 4.81E-05 | -0.00741123 | CIBAR2 |
| chr2 | 8657740 | 8658030 | 291 | 3 | 5.86E-05 | 0.0002553 | 0.000724 | 4.82E-05 | 0.004345675 | NA |
| chr12 | 53930707 | 53931111 | 405 | 4 | 2.51E-05 | 0.0018421 | 2.27E-05 | 4.82E-05 | 0.006790645 | NA |
| chr3 | 183640879 | 183640891 | 13 | 2 | 9.37E-05 | 0.0002272 | 5.90E-05 | 4.82E-05 | 0.002074462 | KLHL24 |
| chr12 | 52243050 | 52243645 | 596 | 3 | 1.37E-05 | 0.0005783 | 5.52E-05 | 4.82E-05 | -0.00511813 | KRT7 |
| chr3 | 195543773 | 195544217 | 445 | 5 | 8.80E-05 | 1.74E-05 | 0.010269 | 4.83E-05 | -0.00594263 | AC069213.1 |
| chr16 | 62036842 | 62037591 | 750 | 10 | 2.80E-05 | 2.79E-05 | 0.01199 | 4.83E-05 | 0.003068882 | CDH8 |
| chr19 | 1248288 | 1248883 | 596 | 9 | 3.17E-05 | 0.0002246 | 0.001814 | 4.83E-05 | 0.00182667 | MIDN |
| chr15 | 34367472 | 34367875 | 404 | 6 | 0.000114047 | 0.0003235 | 0.000387 | 4.83E-05 | -0.00077655 | NA |
| chr5 | 140107430 | 140107511 | 82 | 3 | 7.78E-05 | 3.66E-05 | 0.00076 | 4.83E-05 | 0.003244093 | MALINC1 |
| chr1 | 3130778 | 3131113 | 336 | 2 | 1.93E-05 | 0.0036484 | 2.52E-05 | 4.83E-05 | -0.0043825 | PRDM16 |
| chr3 | 186635094 | 186635561 | 468 | 2 | 3.07E-06 | 3.65E-05 | 0.000625 | 4.83E-05 | 0.000883177 | AC068631.1 |
| chr1 | 154961697 | 154961920 | 224 | 3 | 0.000146862 | 0.0001573 | 0.001152 | 4.84E-05 | -0.00201948 | AL451085.2, PYGO2 |
| chr19 | 39502673 | 39503057 | 385 | 4 | 2.40E-05 | 0.0080626 | 1.93E-05 | 4.84E-05 | 0.007659369 | DLL3 |
| chr16 | 86531186 | 86531322 | 137 | 2 | 9.05E-06 | 3.94E-05 | 0.000501 | 4.84E-05 | 0.004091223 | MTHFSD |
| chr21 | 44324761 | 44325222 | 462 | 4 | 8.40E-05 | 3.86E-05 | 0.002281 | 4.84E-05 | -0.00026861 | PFKL |
| chr16 | 4797966 | 4798175 | 210 | 3 | 1.74E-05 | 0.0653844 | 9.36E-06 | 4.84E-05 | -0.00359441 | AC020663.4, ROGDI |
| chr20 | 25196271 | 25196704 | 434 | 4 | 4.34E-07 | 0.1008481 | 1.29E-05 | 4.84E-05 | 0.001813841 | ENTPD6 |
| chr1 | 234229794 | 234230010 | 217 | 2 | 3.99E-05 | 9.99E-05 | 0.000106 | 4.84E-05 | -0.00641797 | SLC35F3 |
| chr7 | 151726476 | 151726653 | 178 | 3 | 5.99E-05 | 0.0002475 | 0.000468 | 4.85E-05 | -0.003099 | PRKAG2 |
| chr17 | 41441561 | 41441728 | 168 | 3 | 4.94E-05 | 0.010402 | 0.000787 | 4.85E-05 | -0.00606777 | NA |
| chr19 | 11596372 | 11596657 | 286 | 4 | 3.45E-06 | 0.0003768 | 0.002683 | 4.85E-05 | 0.000596283 | ZNF627 |
| chr8 | 10352757 | 10352772 | 16 | 2 | 0.000141618 | 5.26E-05 | 0.000259 | 4.85E-05 | 0.009141029 | MSRA |
| chr20 | 48682979 | 48683056 | 78 | 2 | 3.82E-05 | 0.0005311 | 4.04E-05 | 4.85E-05 | -0.01541695 | PREX1 |
| chr16 | 30400415 | 30400728 | 314 | 2 | 1.32E-05 | 7.98E-05 | 0.000135 | 4.85E-05 | 0.000951076 | NA |
| chr8 | 143899470 | 143899663 | 194 | 2 | 3.67E-05 | 0.0037364 | 2.52E-05 | 4.86E-05 | -0.00598281 | AC109322.2 |
| chr5 | 138869192 | 138869745 | 554 | 2 | 9.67E-05 | 2.91E-05 | 0.001773 | 4.86E-05 | 0.00087916 | CTNNA1, LRRTM2 |
| chr4 | 139295616 | 139296158 | 543 | 7 | 4.21E-05 | 0.0001161 | 0.002844 | 4.86E-05 | 0.001508316 | NDUFC1 |
| chr1 | 18868202 | 18868388 | 187 | 2 | 0.000159762 | 0.0001644 | 7.22E-05 | 4.86E-05 | -0.01452855 | NA |
| chr1 | 12132531 | 12132563 | 33 | 2 | 0.000113626 | 0.0001538 | 7.57E-05 | 4.87E-05 | 0.000714012 | TNFRSF8 |
| chr5 | 132776757 | 132777509 | 753 | 12 | 3.67E-05 | 7.53E-05 | 0.004792 | 4.87E-05 | -0.00070542 | SEPTIN8 |
| chr19 | 6087846 | 6088013 | 168 | 2 | 3.43E-05 | 0.003019 | 2.61E-05 | 4.87E-05 | -0.02374801 | RFX2 |
| chr5 | 1093465 | 1093920 | 456 | 5 | 3.95E-05 | 0.0025117 | 0.000614 | 4.87E-05 | 0.00019102 | SLC12A7 |
| chr14 | 95573417 | 95573596 | 180 | 2 | 5.69E-05 | 8.92E-05 | 0.00012 | 4.87E-05 | 0.003714802 | LINC02318 |
| chr13 | 110106970 | 110107077 | 108 | 2 | 0.000100603 | 0.0001358 | 8.29E-05 | 4.87E-05 | -0.00282653 | AL390755.1 |
| chr14 | 95649924 | 95649970 | 47 | 2 | 7.95E-05 | 3.66E-05 | 0.00064 | 4.88E-05 | 0.006250311 | NA |
| chr3 | 49939673 | 49939995 | 323 | 10 | 0.000141735 | 0.0002608 | 0.001367 | 4.88E-05 | -1.07E-05 | NA |
| chr8 | 48634548 | 48634566 | 19 | 2 | 0.000174359 | 3.09E-05 | 0.0013 | 4.88E-05 | 0.004903317 | AC022915.2 |
| chr14 | 91352220 | 91352977 | 758 | 3 | 1.40E-06 | 9.02E-05 | 0.000165 | 4.88E-05 | -0.00700675 | CCDC88C |
| chr20 | 36173176 | 36173187 | 12 | 2 | 0.000112039 | 6.65E-05 | 0.000177 | 4.88E-05 | -0.00753223 | EPB41L1 |
| chr17 | 41973590 | 41973763 | 174 | 3 | 2.65E-05 | 0.0070387 | 1.68E-05 | 4.88E-05 | 0.004567999 | CNP |
| chr19 | 38229986 | 38230212 | 227 | 4 | 2.66E-05 | 0.0004833 | 9.02E-05 | 4.88E-05 | -0.00894128 | NA |
| chr19 | 45233950 | 45234534 | 585 | 7 | 4.48E-05 | 6.19E-05 | 0.001703 | 4.88E-05 | -0.00728648 | MARK4, EXOC3L2 |
| chr15 | 74873283 | 74873770 | 488 | 8 | 7.99E-05 | 0.0001769 | 0.007779 | 4.89E-05 | -0.00198738 | SCAMP2 |
| chr2 | 54256340 | 54256680 | 341 | 4 | 0.000100591 | 2.73E-05 | 0.004373 | 4.89E-05 | -0.00765113 | ACYP2 |
| chr4 | 183505606 | 183505664 | 59 | 2 | 0.000128162 | 0.000171 | 7.11E-05 | 4.89E-05 | 0.001963951 | ING2 |
| chr5 | 1807074 | 1807224 | 151 | 3 | 0.000117148 | 6.53E-05 | 0.00064 | 4.89E-05 | 0.006176426 | NDUFS6 |
| chr12 | 123233159 | 123234159 | 1001 | 18 | 1.53E-05 | 0.0005622 | 0.008111 | 4.89E-05 | -0.00037735 | C12orf65, MPHOSPH9 |
| chr9 | 127915270 | 127915740 | 471 | 2 | 3.51E-05 | 5.80E-05 | 0.000222 | 4.89E-05 | 0.008852604 | ST6GALNAC4 |
| chr4 | 8453364 | 8453373 | 10 | 2 | 9.46E-05 | 5.36E-05 | 0.000256 | 4.90E-05 | -0.00625228 | TRMT44 |
| chr10 | 72089348 | 72089868 | 521 | 4 | 2.32E-07 | 0.0058499 | 8.25E-05 | 4.90E-05 | 0.004627414 | NA |
| chr15 | 63288444 | 63288861 | 418 | 3 | 0.000134042 | 2.44E-05 | 0.002434 | 4.90E-05 | 0.006246968 | APH1B |
| chr14 | 38255050 | 38255740 | 691 | 5 | 1.17E-05 | 0.00088 | 0.000307 | 4.90E-05 | -0.00520838 | CLEC14A |
| chr5 | 142869808 | 142870166 | 359 | 2 | 1.22E-05 | 0.0001083 | 0.0001 | 4.91E-05 | 0.00653027 | ARHGAP26 |
| chr1 | 157690521 | 157690623 | 103 | 3 | 1.13E-05 | 0.0084166 | 1.74E-05 | 4.91E-05 | 0.001088248 | FCRL3 |
| chr5 | 35194960 | 35195335 | 376 | 2 | 2.22E-05 | 4.42E-05 | 0.000389 | 4.91E-05 | 0.001889428 | PRLR |
| chr1 | 89198862 | 89199099 | 238 | 5 | 7.39E-05 | 0.0003091 | 0.000401 | 4.91E-05 | 0.002064049 | AC099063.4, GBP4 |
| chr5 | 41160271 | 41160315 | 45 | 2 | 0.000139178 | 6.02E-05 | 0.000209 | 4.91E-05 | 0.002874197 | C6 |
| chr6 | 168308913 | 168309523 | 611 | 4 | 2.24E-05 | 0.0001262 | 0.00028 | 4.91E-05 | 0.000280867 | DACT2 |
| chr7 | 55132524 | 55132964 | 441 | 2 | 4.84E-07 | 0.0208155 | 2.18E-05 | 4.91E-05 | -0.00722021 | EGFR |
| chr10 | 60200427 | 60200823 | 397 | 2 | 2.61E-05 | 2.95E-05 | 0.001763 | 4.91E-05 | -0.00177613 | ANK3 |
| chr8 | 119832420 | 119832936 | 517 | 7 | 1.50E-05 | 3.39E-05 | 0.002725 | 4.91E-05 | -0.00129061 | AP005717.2, TAF2 |
| chr20 | 57696250 | 57696308 | 59 | 2 | 7.49E-05 | 4.57E-05 | 0.000361 | 4.91E-05 | -0.0129045 | PMEPA1 |
| chr7 | 150412941 | 150413138 | 198 | 3 | 8.48E-05 | 7.80E-05 | 0.000971 | 4.91E-05 | -0.00308738 | NA |
| chr3 | 51872289 | 51872601 | 313 | 2 | 0.000126361 | 3.39E-05 | 0.000879 | 4.92E-05 | -0.00873858 | RN7SL504P |
| chr2 | 71575874 | 71576281 | 408 | 5 | 4.70E-05 | 0.0009577 | 0.000962 | 4.92E-05 | 0.000863163 | DYSF |
| chr19 | 45008535 | 45008864 | 330 | 2 | 2.14E-05 | 0.0028648 | 2.66E-05 | 4.92E-05 | -0.01420382 | RELB |
| chr12 | 49929114 | 49929218 | 105 | 3 | 0.000109359 | 1.89E-05 | 0.006547 | 4.93E-05 | -0.00335622 | LINC02395 |
| chr8 | 37996628 | 37996790 | 163 | 2 | 9.22E-05 | 0.0011615 | 3.22E-05 | 4.93E-05 | -0.00588046 | NA |
| chr11 | 2582781 | 2583269 | 489 | 3 | 2.94E-05 | 0.0048457 | 0.000166 | 4.93E-05 | -0.00606019 | KCNQ1 |
| chr3 | 15049790 | 15050898 | 1109 | 3 | 5.70E-07 | 0.001141 | 3.23E-05 | 4.93E-05 | -0.0059588 | NR2C2, MRPS25 |
| chr3 | 156227982 | 156228064 | 83 | 2 | 0.000116402 | 4.46E-05 | 0.000387 | 4.93E-05 | -0.01293732 | KCNAB1, AC091607.2 |
| chr21 | 41479498 | 41480591 | 1094 | 3 | 1.02E-07 | 0.0126808 | 1.09E-05 | 4.93E-05 | -0.00859495 | TMPRSS2 |
| chr5 | 172174678 | 172174784 | 107 | 2 | 3.89E-05 | 8.54E-05 | 0.000129 | 4.93E-05 | 0.001987215 | STK10 |
| chr11 | 17390165 | 17390228 | 64 | 3 | 0.000147243 | 4.26E-05 | 0.001989 | 4.93E-05 | -0.00677747 | NA |
| chr20 | 45778156 | 45778242 | 87 | 2 | 2.41E-05 | 0.0064789 | 2.39E-05 | 4.94E-05 | 0.002185462 | WFDC3 |
| chr12 | 6946289 | 6946845 | 557 | 8 | 5.63E-05 | 1.09E-05 | 0.026215 | 4.94E-05 | 0.002079854 | PTPN6 |
| chr22 | 19942034 | 19943279 | 1246 | 7 | 3.24E-05 | 0.0002003 | 0.01087 | 4.94E-05 | 0.001897426 | COMT |

| | | | | | | | | | | |
|---|---|---|---|---|---|---|---|---|---|---|
| chr12 | 32364879 | 32364957 | 79 | 2 | 0.000121241 | 4.09E-05 | 0.000485 | 4.94E-05 | -0.00818006 | BICD1 |
| chr5 | 132370639 | 132371253 | 615 | 2 | 2.39E-05 | 6.11E-05 | 0.000208 | 4.94E-05 | -0.00072743 | SLC22A5 |
| chr11 | 132993175 | 132993324 | 150 | 2 | 3.14E-05 | 0.0005761 | 4.01E-05 | 4.95E-05 | -0.00971549 | OPCML |
| chr12 | 54587497 | 54587779 | 283 | 2 | 1.45E-05 | 8.01E-05 | 0.00014 | 4.95E-05 | -0.0043632 | PPP1R1A |
| chr6 | 20421038 | 20421055 | 18 | 2 | 0.000157116 | 2.97E-05 | 0.001784 | 4.95E-05 | 0.006045164 | E2F3 |
| chr22 | 47138149 | 47138170 | 22 | 2 | 5.88E-05 | 0.0003105 | 5.19E-05 | 4.96E-05 | -0.00208441 | TBC1D22A |
| chr19 | 41997246 | 41997311 | 66 | 2 | 7.77E-05 | 0.0002056 | 6.47E-05 | 4.96E-05 | 0.002782753 | ATP1A3 |
| chr9 | 97984998 | 97985485 | 488 | 3 | 1.67E-05 | 0.0005504 | 7.18E-05 | 4.96E-05 | 7.61E-05 | ANP32B |
| chr7 | 149719732 | 149720549 | 818 | 5 | 5.33E-05 | 0.0011431 | 0.000466 | 4.97E-05 | 0.001576626 | KRBA1 |
| chr1 | 11319085 | 11319319 | 235 | 3 | 2.66E-05 | 0.0070471 | 5.07E-05 | 4.97E-05 | -0.00798024 | AL031291.1 |
| chr6 | 101000202 | 101000490 | 289 | 2 | 2.62E-05 | 0.0031449 | 2.64E-05 | 4.97E-05 | 0.009377644 | NA |
| chr7 | 150797297 | 150797910 | 614 | 3 | 2.52E-06 | 0.0001555 | 0.000144 | 4.98E-05 | -0.00350245 | TMEM176B |
| chr20 | 57579743 | 57579841 | 99 | 3 | 4.60E-05 | 0.0014534 | 6.92E-05 | 4.98E-05 | -0.01467905 | NA |
| chr7 | 118461049 | 118461143 | 95 | 2 | 8.95E-05 | 0.0002482 | 5.86E-05 | 4.98E-05 | -0.0198397 | ANKRD7 |
| chr18 | 48841470 | 48841731 | 262 | 3 | 0.000136704 | 4.01E-05 | 0.00148 | 4.99E-05 | 0.004242148 | CTIF |
| chr14 | 20693138 | 20693460 | 323 | 2 | 1.46E-05 | 0.0107637 | 2.31E-05 | 4.99E-05 | -0.00502783 | ANG, RNASE4, AL163636.1 |
| chr11 | 46591578 | 46591584 | 7 | 3 | 0.000151285 | 0.0006261 | 5.87E-05 | 4.99E-05 | -0.00205539 | AMBRA1 |
| chr2 | 134714427 | 134714572 | 146 | 2 | 1.98E-05 | 0.0142704 | 2.27E-05 | 4.99E-05 | -0.00506728 | TMEM163 |
| chr13 | 73618332 | 73618726 | 395 | 3 | 0.000150517 | 8.10E-05 | 0.000516 | 5.00E-05 | -0.01300472 | LINC00393 |
| chr10 | 101292404 | 101292859 | 456 | 5 | 1.62E-05 | 0.0007112 | 0.000142 | 5.00E-05 | -0.00142438 | NA |
| chr2 | 205682420 | 205682755 | 336 | 6 | 1.30E-05 | 0.0005735 | 0.000511 | 5.01E-05 | 0.001329342 | NRP2 |
| chr8 | 99156529 | 99156609 | 81 | 2 | 4.71E-05 | 0.0004423 | 4.49E-05 | 5.01E-05 | -0.00265144 | VPS13B |
| chr13 | 76456509 | 76456639 | 131 | 2 | 8.20E-05 | 9.50E-05 | 0.000118 | 5.01E-05 | -0.01273081 | NA |
| chr8 | 457247 | 457482 | 236 | 2 | 9.83E-06 | 0.0013343 | 3.16E-05 | 5.01E-05 | -0.00633293 | FBXO25 |
| chr13 | 113840037 | 113841102 | 1066 | 12 | 1.24E-05 | 0.0001039 | 0.003858 | 5.01E-05 | 0.003339615 | GAS6-AS1, GAS6 |
| chr11 | 120324757 | 120325196 | 440 | 8 | 4.98E-05 | 0.0001736 | 0.005704 | 5.02E-05 | -0.00315643 | NA |
| chr1 | 203085594 | 203086534 | 941 | 11 | 1.32E-05 | 0.0027526 | 0.001205 | 5.02E-05 | -0.00199941 | MYOG |
| chr10 | 79238234 | 79238373 | 140 | 2 | 4.41E-05 | 3.12E-05 | 0.001469 | 5.03E-05 | -0.00709872 | ZMIZ1 |
| chr12 | 67647538 | 67647958 | 421 | 4 | 5.40E-05 | 0.0001211 | 0.000913 | 5.03E-05 | -0.00425434 | NA |
| chr20 | 61980134 | 61980157 | 24 | 2 | 3.38E-05 | 0.0008279 | 3.61E-05 | 5.03E-05 | 0.003076877 | TAF4 |
| chr2 | 60352632 | 60353424 | 793 | 5 | 1.07E-05 | 0.0017198 | 3.26E-05 | 5.03E-05 | -0.0016163 | MIR4432HG |
| chr12 | 2652198 | 2652383 | 186 | 4 | 4.17E-05 | 6.40E-05 | 0.000639 | 5.03E-05 | -0.00120321 | CACNA1C |
| chr1 | 40757839 | 40758173 | 335 | 2 | 8.65E-05 | 3.03E-05 | 0.001773 | 5.03E-05 | -0.00020317 | NFYC |
| chr10 | 117734983 | 117735504 | 522 | 6 | 5.96E-05 | 0.0011948 | 5.14E-05 | 5.04E-05 | 0.001126884 | LINC02674 |
| chr16 | 2882998 | 2883307 | 310 | 8 | 2.10E-05 | 0.0014983 | 0.001506 | 5.04E-05 | 0.000965392 | FLYWCH2 |
| chr21 | 33855317 | 33855343 | 27 | 2 | 0.000151637 | 5.31E-05 | 0.000281 | 5.04E-05 | -0.00119104 | ITSN1, AP000311.1 |
| chr3 | 180601340 | 180601930 | 591 | 5 | 2.60E-05 | 0.0005052 | 0.002248 | 5.04E-05 | -0.00859939 | AC125618.1 |
| chr3 | 49001445 | 49001509 | 65 | 2 | 0.000174001 | 3.44E-05 | 0.000923 | 5.04E-05 | -0.00541216 | P4HTM |
| chr9 | 37753359 | 37753951 | 593 | 5 | 2.11E-05 | 0.0020512 | 2.45E-05 | 5.05E-05 | -0.00064768 | TRMT10B, AL138752.2 |
| chr7 | 157670608 | 157670663 | 56 | 2 | 3.80E-05 | 4.84E-05 | 0.000341 | 5.05E-05 | -0.00296281 | PTPRN2 |
| chr8 | 143170703 | 143171042 | 340 | 2 | 3.69E-06 | 0.005996 | 2.46E-05 | 5.05E-05 | -0.01214535 | NA |
| chr17 | 18253891 | 18254164 | 274 | 4 | 2.84E-05 | 0.0037644 | 9.23E-05 | 5.05E-05 | -0.0015709 | FLII |
| chr5 | 42564949 | 42565764 | 816 | 6 | 2.76E-05 | 0.0057173 | 0.000833 | 5.05E-05 | -4.03E-05 | GHR |
| chr2 | 1974341 | 1974435 | 95 | 2 | 9.79E-05 | 0.0001861 | 7.05E-05 | 5.06E-05 | -0.00512516 | MYT1L |
| chr20 | 43915679 | 43916459 | 781 | 11 | 5.32E-05 | 0.0002083 | 0.003314 | 5.06E-05 | -0.00018806 | TOX2 |
| chr10 | 69879616 | 69879946 | 331 | 2 | 5.12E-05 | 4.86E-05 | 0.000341 | 5.06E-05 | -0.0008577 | COL13A1 |
| chr5 | 54982229 | 54982356 | 128 | 2 | 8.25E-05 | 0.0003012 | 5.41E-05 | 5.07E-05 | -0.00603044 | ESM1 |
| chr4 | 62709839 | 62710079 | 241 | 2 | 7.07E-05 | 0.0001813 | 7.19E-05 | 5.07E-05 | -0.00563801 | NA |
| chr5 | 138467413 | 138467680 | 268 | 4 | 3.29E-05 | 0.0031838 | 5.80E-05 | 5.07E-05 | -0.00117627 | EGR1 |
| chr5 | 176597649 | 176597969 | 321 | 2 | 3.73E-05 | 0.0001697 | 7.50E-05 | 5.07E-05 | -0.00837024 | GPRIN1 |
| chr1 | 6281841 | 6282375 | 535 | 3 | 2.57E-05 | 0.0003109 | 0.000566 | 5.07E-05 | -0.00589479 | ACOT7 |
| chr3 | 195094234 | 195094843 | 610 | 5 | 2.36E-05 | 0.0007031 | 5.30E-05 | 5.08E-05 | 0.003970547 | XXYLT1-AS1, XXYLT1 |
| chr9 | 133944963 | 133944993 | 31 | 2 | 6.35E-05 | 0.0003087 | 5.36E-05 | 5.08E-05 | 0.001160414 | VAV2 |
| chr2 | 109398793 | 109398892 | 100 | 2 | 5.32E-05 | 6.34E-05 | 0.000208 | 5.08E-05 | -0.00356177 | SH3RF3 |
| chr7 | 69401440 | 69401482 | 43 | 2 | 1.78E-05 | 0.000166 | 7.63E-05 | 5.08E-05 | -0.00099452 | AC092100.1 |
| chr19 | 3200936 | 3201682 | 747 | 6 | 1.47E-05 | 0.0007754 | 0.001589 | 5.08E-05 | -0.00095638 | NCLN |
| chr5 | 134177709 | 134177820 | 112 | 2 | 1.23E-05 | 6.06E-05 | 0.000225 | 5.08E-05 | -0.00537519 | AC104109.3 |
| chr1 | 114780738 | 114781829 | 1092 | 6 | 8.46E-07 | 0.0003265 | 4.48E-05 | 5.08E-05 | -0.00368116 | NA |
| chr9 | 135657885 | 135658231 | 347 | 2 | 4.40E-06 | 0.0032785 | 2.68E-05 | 5.08E-05 | 0.002617491 | NA |
| chr4 | 958110 | 958578 | 469 | 4 | 2.86E-05 | 0.0093782 | 2.26E-05 | 5.09E-05 | 0.001443873 | TMEM175 |
| chr17 | 74780099 | 74780147 | 49 | 2 | 7.73E-05 | 3.08E-05 | 0.001734 | 5.09E-05 | 0.011201295 | TMEM104 |
| chr15 | 101333584 | 101333715 | 132 | 2 | 9.18E-05 | 0.0001161 | 9.99E-05 | 5.09E-05 | 0.002391795 | PCSK6 |
| chr14 | 88550370 | 88550658 | 289 | 5 | 2.29E-05 | 0.0003739 | 0.00039 | 5.09E-05 | 0.000845653 | PTPN21 |
| chr11 | 17169688 | 17170070 | 383 | 2 | 3.47E-06 | 0.0021878 | 2.89E-05 | 5.09E-05 | 0.003242157 | PIK3C2A |
| chr10 | 42981830 | 42982147 | 318 | 2 | 9.32E-05 | 5.15E-05 | 0.000307 | 5.09E-05 | -0.00878712 | NA |
| chr10 | 67763725 | 67764514 | 790 | 8 | 1.48E-05 | 0.0010273 | 0.003116 | 5.10E-05 | 0.000789621 | NA |
| chr20 | 45971302 | 45972339 | 1038 | 12 | 1.26E-06 | 0.00118 | 0.000418 | 5.11E-05 | 0.002309364 | ZNF335 |
| chr1 | 61851418 | 61851574 | 157 | 3 | 7.26E-05 | 3.00E-05 | 0.001601 | 5.11E-05 | -0.0092907 | PATJ |
| chr1 | 13554836 | 13555047 | 212 | 3 | 8.75E-05 | 0.0006874 | 3.64E-05 | 5.11E-05 | 0.001877679 | BRWD1P1 |
| chr19 | 54460347 | 54460699 | 353 | 2 | 5.12E-05 | 5.52E-05 | 0.00027 | 5.11E-05 | 0.011998076 | LENG8 |
| chr17 | 72053663 | 72053806 | 144 | 2 | 2.35E-05 | 7.37E-05 | 0.000167 | 5.11E-05 | 0.016232018 | ROCR, AC007461.1 |
| chr11 | 70708250 | 70708435 | 186 | 2 | 4.50E-05 | 0.0020376 | 2.94E-05 | 5.11E-05 | -0.00718674 | SHANK2 |
| chr20 | 38316514 | 38317144 | 631 | 3 | 4.11E-06 | 0.0012458 | 4.22E-05 | 5.11E-05 | -0.00316397 | BPI, AL359555.1 |
| chr10 | 120431194 | 120431232 | 39 | 2 | 0.000162235 | 4.56E-05 | 0.000407 | 5.11E-05 | 0.000923514 | NA |
| chr22 | 20974103 | 20974452 | 350 | 3 | 5.32E-05 | 0.0001085 | 0.000145 | 5.11E-05 | -0.00115378 | AIFM3 |
| chr13 | 112246899 | 112247037 | 139 | 2 | 9.38E-05 | 3.45E-05 | 0.000983 | 5.11E-05 | -0.00479506 | NA |
| chr13 | 20193639 | 20194397 | 759 | 8 | 1.67E-05 | 0.0001944 | 0.000439 | 5.12E-05 | -0.00080165 | NA |
| chr2 | 1234541 | 1234654 | 114 | 4 | 8.53E-05 | 0.0062235 | 0.000198 | 5.12E-05 | -0.00176743 | SNTG2 |
| chr4 | 15779226 | 15779579 | 354 | 3 | 4.50E-05 | 9.69E-05 | 0.000408 | 5.12E-05 | 0.001264425 | CD38 |
| chr4 | 16397386 | 16397412 | 27 | 2 | 9.33E-05 | 4.06E-05 | 0.00056 | 5.12E-05 | -0.00605328 | NA |
| chr8 | 64798717 | 64799467 | 751 | 10 | 1.81E-05 | 0.000319 | 0.002753 | 5.12E-05 | -0.00452853 | AC104232.3, CYP7B1 |
| chr4 | 127780859 | 127780980 | 122 | 2 | 2.76E-05 | 0.0001603 | 7.92E-05 | 5.12E-05 | 0.006278647 | NA |
| chr8 | 37939940 | 37940114 | 175 | 2 | 6.49E-05 | 0.0001685 | 7.65E-05 | 5.13E-05 | -0.00344826 | GOT1L1, AC144573.1 |
| chr19 | 38912183 | 38912407 | 225 | 6 | 0.000119502 | 4.34E-05 | 0.008626 | 5.13E-05 | -0.01186532 | NA |
| chr11 | 212669 | 212842 | 174 | 2 | 1.98E-05 | 0.0004804 | 4.47E-05 | 5.13E-05 | -0.00347495 | RIC8A |
| chr14 | 50651097 | 50651152 | 56 | 2 | 7.03E-05 | 0.0009896 | 3.50E-05 | 5.13E-05 | -0.00338413 | SAV1 |
| chr1 | 145957948 | 145958216 | 269 | 7 | 0.000149181 | 0.0001173 | 0.000871 | 5.14E-05 | -0.00082353 | LIX1L-AS1, AC243547.3, LIX1L |
| chr4 | 6688975 | 6689366 | 392 | 3 | 0.000102217 | 0.001082 | 0.000936 | 5.14E-05 | 0.006247842 | LINC02481 |
| chr1 | 76866734 | 76867389 | 656 | 5 | 1.04E-05 | 0.0327104 | 1.05E-05 | 5.14E-05 | -0.00683769 | NA |
| chr16 | 50153665 | 50154161 | 497 | 6 | 3.14E-05 | 0.0049859 | 0.001125 | 5.14E-05 | -0.00147066 | TENT4B |
| chr1 | 43299946 | 43300535 | 590 | 5 | 3.57E-05 | 0.01159 | 0.000478 | 5.14E-05 | -0.0039497 | NA |
| chr16 | 68538989 | 68539337 | 349 | 6 | 2.56E-05 | 0.0089309 | 6.42E-05 | 5.15E-05 | -0.00289022 | ZFP90 |
| chr19 | 6135310 | 6135653 | 344 | 5 | 3.29E-05 | 0.0008111 | 0.000814 | 5.15E-05 | 0.004737859 | ACSBG2, RFX2 |
| chr5 | 850986 | 851343 | 358 | 7 | 0.000113304 | 0.000132 | 0.000462 | 5.15E-05 | 0.000209695 | ZDHHC11, BRD9 |
| chr16 | 67547534 | 67547772 | 239 | 2 | 2.12E-05 | 0.0003913 | 4.89E-05 | 5.15E-05 | 0.01140623 | NA |
| chr1 | 237862114 | 237862287 | 174 | 4 | 5.07E-05 | 0.0022581 | 0.000191 | 5.15E-05 | -0.00307284 | AL359924.1 |
| chr22 | 35447915 | 35448072 | 158 | 2 | 0.000134953 | 3.18E-05 | 0.001538 | 5.16E-05 | -0.01224542 | NA |
| chr1 | 63316968 | 63317905 | 938 | 9 | 1.03E-05 | 0.0040879 | 0.000454 | 5.16E-05 | -0.00212097 | LINC00466 |
| chr4 | 70471941 | 70472156 | 216 | 4 | 2.73E-05 | 0.0058525 | 2.31E-05 | 5.16E-05 | -0.0076557 | MUC7 |
| chr17 | 61415088 | 61415571 | 484 | 2 | 4.06E-05 | 3.10E-05 | 0.001833 | 5.16E-05 | -0.00096782 | NA |
| chr10 | 12043431 | 12044037 | 607 | 6 | 1.40E-05 | 2.43E-05 | 0.002833 | 5.17E-05 | -0.00473537 | NA |
| chr3 | 2144328 | 2144977 | 650 | 5 | 3.50E-05 | 5.65E-05 | 0.000511 | 5.17E-05 | 0.001342999 | CNTN4 |
| chr9 | 71919437 | 71919718 | 282 | 2 | 1.21E-05 | 0.0004178 | 4.77E-05 | 5.17E-05 | -0.01012597 | C9orf85 |
| chr16 | 28831971 | 28832028 | 58 | 2 | 4.06E-05 | 0.0032249 | 2.74E-05 | 5.17E-05 | -0.00483139 | AC009093.11, ATXN2L, AC133550.2 |
| chr4 | 139735253 | 139735772 | 520 | 4 | 9.47E-05 | 7.03E-05 | 0.000696 | 5.17E-05 | 0.00210268 | MGST2, MAML3 |
| chr17 | 76881011 | 76881331 | 321 | 2 | 1.72E-05 | 0.0001685 | 7.76E-05 | 5.17E-05 | -0.00908081 | MGAT5B |
| chr5 | 149271190 | 149271581 | 392 | 5 | 4.07E-05 | 0.0001094 | 0.000445 | 5.17E-05 | -0.0066891 | AC012613.1, AC012613.2 |
| chr4 | 6147477 | 6147570 | 94 | 2 | 0.000113727 | 5.73E-05 | 0.000261 | 5.17E-05 | -0.01482224 | C4orf50, JAKMIP1 |
| chr17 | 3941009 | 3941129 | 121 | 3 | 2.83E-05 | 0.0028706 | 6.69E-05 | 5.17E-05 | -0.00392384 | ATP2A3 |
| chr13 | 50125208 | 50126195 | 988 | 7 | 6.11E-06 | 0.0009129 | 0.000243 | 5.18E-05 | 0.001736992 | DLEU1, AL137060.3, DLEU2 |
| chr14 | 25121633 | 25121870 | 238 | 2 | 5.31E-05 | 0.0005224 | 4.37E-05 | 5.18E-05 | -0.00783662 | NA |
| chr12 | 50742482 | 50742633 | 152 | 2 | 8.16E-05 | 4.74E-05 | 0.000388 | 5.18E-05 | -0.01010727 | DIP2B |
| chr3 | 186785662 | 186785854 | 193 | 3 | 1.83E-05 | 0.000751 | 9.72E-05 | 5.18E-05 | -0.00832216 | EIF4A2 |
| chr15 | 66775999 | 66776040 | 42 | 2 | 0.000131515 | 3.62E-05 | 0.000853 | 5.18E-05 | -0.0019386 | SMAD6 |
| chr2 | 129877848 | 129878572 | 725 | 5 | 5.34E-06 | 0.0005025 | 0.000448 | 5.18E-05 | -0.00704297 | AC079776.5 |
| chr9 | 74798409 | 74798436 | 28 | 2 | 0.000133897 | 3.37E-05 | 0.001172 | 5.18E-05 | 0.011643639 | TRPM6 |
| chr3 | 195147698 | 195148114 | 417 | 6 | 2.29E-05 | 0.000226 | 0.000378 | 5.18E-05 | -0.00626071 | XXYLT1-AS2, XXYLT1 |
| chr10 | 132451982 | 132452559 | 578 | 4 | 3.17E-05 | 0.0001157 | 0.00149 | 5.18E-05 | -0.00406125 | NA |
| chr11 | 3152638 | 3152898 | 261 | 3 | 2.06E-05 | 0.0138826 | 1.59E-05 | 5.18E-05 | 0.000336397 | OSBPL5 |
| chr9 | 109463388 | 109463539 | 152 | 2 | 2.56E-05 | 0.0005844 | 4.19E-05 | 5.18E-05 | 0.002962014 | PTPN3 |
| chr18 | 62597014 | 62597321 | 308 | 2 | 8.46E-07 | 7.25E-05 | 0.000177 | 5.18E-05 | -0.01516506 | NA |
| chr1 | 231038407 | 231038604 | 198 | 3 | 7.51E-05 | 0.0001023 | 0.000763 | 5.18E-05 | -0.00149087 | FAM89A |
| chr3 | 185152349 | 185152686 | 338 | 5 | 5.45E-05 | 0.0002827 | 0.000752 | 5.18E-05 | -4.54E-05 | C3orf70 |
| chr10 | 114359448 | 114359530 | 83 | 2 | 9.12E-05 | 0.0003075 | 5.53E-05 | 5.19E-05 | -0.00632214 | AFAP1L2 |
| chr17 | 8744626 | 8745686 | 1061 | 7 | 4.17E-06 | 0.0006063 | 0.000401 | 5.19E-05 | 0.003166585 | CCDC42 |
| chr9 | 133425828 | 133426023 | 196 | 2 | 0.000158907 | 3.16E-05 | 0.001668 | 5.19E-05 | -0.00736492 | ADAMTS13 |
| chr15 | 90211560 | 90212226 | 667 | 3 | 3.34E-06 | 8.96E-05 | 0.000637 | 5.19E-05 | -0.00333302 | SEMA4B |
| chr9 | 122222942 | 122223477 | 536 | 3 | 3.78E-05 | 0.0002774 | 0.001184 | 5.20E-05 | -0.00303764 | LHX6 |
| chr2 | 27703421 | 27703463 | 43 | 2 | 6.59E-05 | 6.51E-05 | 0.000211 | 5.20E-05 | -0.01749307 | NA |
| chr19 | 41396753 | 41397105 | 353 | 3 | 1.67E-05 | 9.12E-05 | 0.000316 | 5.20E-05 | -0.00252678 | AC011462.1, EXOSC5 |
| chr1 | 156176817 | 156176961 | 145 | 3 | 9.78E-05 | 0.0001109 | 0.000745 | 5.20E-05 | -0.00248581 | SEMA4A |
| chr1 | 119611682 | 119611768 | 87 | 2 | 9.20E-05 | 3.22E-05 | 0.001522 | 5.20E-05 | -0.01215058 | NA |
| chr1 | 150235272 | 150236127 | 856 | 4 | 1.13E-06 | 1.64E-05 | 0.009713 | 5.21E-05 | 0.002631023 | ANP32E |
| chr15 | 88602348 | 88602369 | 22 | 2 | 0.000119823 | 0.0001306 | 9.43E-05 | 5.21E-05 | -0.00333416 | LINC01586 |
| chr16 | 81895753 | 81896112 | 360 | 10 | 6.67E-05 | 0.0008966 | 0.000588 | 5.21E-05 | -5.23E-05 | PLCG2 |
| chr5 | 1097550 | 1097679 | 130 | 2 | 8.95E-05 | 3.55E-05 | 0.000949 | 5.21E-05 | 0.010191968 | SLC12A7 |
| chr3 | 122929603 | 122930002 | 400 | 2 | 2.74E-06 | 0.0009547 | 3.60E-05 | 5.21E-05 | -0.00768447 | SEMA5B |
| chr18 | 10405567 | 10406052 | 486 | 2 | 2.48E-05 | 6.11E-05 | 0.000237 | 5.21E-05 | 0.001095484 | LINC01254 |
| chr15 | 91649860 | 91650118 | 259 | 2 | 2.58E-06 | 0.0031016 | 2.78E-05 | 5.21E-05 | -0.00409293 | NA |
| chr17 | 48999542 | 48999803 | 262 | 2 | 3.72E-05 | 3.19E-05 | 0.001641 | 5.22E-05 | -0.0223376 | IGF2BP1 |
| chr11 | 69558031 | 69558280 | 250 | 2 | 0.00011357 | 3.51E-05 | 0.001004 | 5.22E-05 | -0.00455529 | NA |
| chr11 | 71476462 | 71476844 | 383 | 3 | 6.46E-05 | 2.88E-05 | 0.001988 | 5.22E-05 | -0.00477126 | NADSYN1 |
| chr5 | 57468430 | 57468594 | 165 | 2 | 6.05E-05 | 0.0008434 | 3.74E-05 | 5.22E-05 | 0.00768698 | AC025470.2 |
| chr11 | 1305278 | 1305649 | 372 | 4 | 3.45E-05 | 0.0016925 | 4.65E-05 | 5.23E-05 | 0.000429824 | TOLLIP |
| chr16 | 4371097 | 4371602 | 506 | 6 | 1.04E-05 | 8.89E-05 | 0.000212 | 5.23E-05 | -0.00444157 | CORO7-PAM16, CORO7 |
| chr12 | 52216298 | 52216517 | 220 | 3 | 6.31E-05 | 3.16E-05 | 0.001281 | 5.23E-05 | -0.00354808 | LINC00592 |
| chr1 | 6447968 | 6447975 | 8 | 2 | 3.48E-05 | 0.0131722 | 2.37E-05 | 5.23E-05 | 0.002954108 | ESPN |
| chr11 | 125956314 | 125956451 | 138 | 2 | 0.000135788 | 6.00E-05 | 0.000246 | 5.23E-05 | 0.003931993 | VSIG10L2, CDON |
| chr1 | 209583631 | 209583762 | 132 | 2 | 0.000104643 | 9.32E-05 | 0.000131 | 5.23E-05 | 0.01575229 | CAMK1G |
| chr11 | 125931561 | 125931624 | 64 | 2 | 2.54E-05 | 0.0024972 | 2.89E-05 | 5.23E-05 | -0.0054864 | DDX25, AP000842.2 |
| chr14 | 60641509 | 60642089 | 581 | 3 | 1.32E-05 | 2.31E-05 | 0.003679 | 5.24E-05 | 0.000478894 | SALRNA1 |
| chr16 | 14370358 | 14370518 | 161 | 4 | 0.000150626 | 9.64E-05 | 0.004597 | 5.24E-05 | -0.00552905 | NA |
| chr1 | 247406468 | 247407082 | 615 | 3 | 2.86E-05 | 0.0003997 | 6.95E-05 | 5.24E-05 | -0.00495958 | NA |
| chr19 | 55599505 | 55600136 | 632 | 8 | 6.55E-05 | 0.0010069 | 0.002299 | 5.24E-05 | 0.00119958 | ZNF524, FIZ1 |
| chr2 | 113274214 | 113274277 | 64 | 2 | 4.64E-05 | 3.42E-05 | 0.001156 | 5.24E-05 | -0.04770227 | PAX8-AS1, PAX8 |
| chr20 | 48372801 | 48372966 | 166 | 2 | 0.000133415 | 5.52E-05 | 0.00029 | 5.24E-05 | -0.00764756 | NA |
| chr10 | 12388860 | 12388938 | 79 | 3 | 0.000106049 | 0.0001497 | 0.00011 | 5.25E-05 | 0.008604198 | CAMK1D |
| chr8 | 33601075 | 33601273 | 199 | 2 | 1.41E-05 | 3.19E-05 | 0.00173 | 5.25E-05 | -0.00533012 | NA |

| | | | | | | | | | | |
|---|---|---|---|---|---|---|---|---|---|---|
| chr5 | 180648862 | 180648875 | 14 | 2 | 0.000157138 | 6.00E-05 | 0.00025 | 5.25E-05 | -0.00884613 | FLT4 |
| chr9 | 109331026 | 109331466 | 441 | 2 | 1.09E-05 | 6.98E-05 | 0.000194 | 5.26E-05 | 0.001826066 | NA |
| chr11 | 57514899 | 57515681 | 783 | 6 | 1.13E-05 | 8.18E-05 | 0.000291 | 5.26E-05 | -0.00138034 | SLC43A1 |
| chr3 | 80789844 | 80789856 | 13 | 2 | 0.000159867 | 3.15E-05 | 0.001883 | 5.26E-05 | -0.01633388 | NA |
| chr2 | 6634962 | 6635472 | 511 | 3 | 7.70E-06 | 9.55E-05 | 0.000197 | 5.26E-05 | -0.00584194 | MIR7515HG |
| chr4 | 128553242 | 128553439 | 198 | 2 | 3.42E-05 | 0.0001501 | 8.62E-05 | 5.26E-05 | -0.00494425 | AC110609.1 |
| chr6 | 136525967 | 136526329 | 363 | 4 | 0.000126516 | 5.91E-05 | 0.003997 | 5.26E-05 | -0.00119512 | MAP7 |
| chr14 | 92728952 | 92729035 | 84 | 2 | 4.52E-05 | 3.47E-05 | 0.001108 | 5.26E-05 | -0.02072116 | LGMN |
| chr6 | 26383069 | 26383445 | 377 | 5 | 4.60E-05 | 7.73E-05 | 0.000546 | 5.26E-05 | 0.001619709 | BTN2A2 |
| chr4 | 7873665 | 7873712 | 48 | 2 | 7.26E-05 | 0.0001134 | 0.000108 | 5.26E-05 | -0.00299428 | AFAP1 |
| chr9 | 130405323 | 130405385 | 63 | 2 | 2.59E-05 | 8.31E-05 | 0.000151 | 5.26E-05 | -0.00467064 | HMCN2 |
| chr11 | 46360935 | 46361175 | 241 | 5 | 0.000140627 | 3.90E-05 | 0.004447 | 5.26E-05 | 0.000337743 | DGKZ |
| chr15 | 58889523 | 58889964 | 442 | 2 | 7.62E-05 | 4.01E-05 | 0.000643 | 5.26E-05 | -0.01424059 | RNF111, SLTM |
| chr11 | 36289223 | 36289889 | 667 | 10 | 1.86E-05 | 0.0528161 | 0.000223 | 5.26E-05 | -0.00180992 | COMMD9 |
| chr12 | 6987792 | 6988246 | 455 | 5 | 0.000119232 | 0.0002401 | 0.002067 | 5.26E-05 | -0.00289285 | EMG1, LPCAT3 |
| chr15 | 33194450 | 33194919 | 470 | 8 | 8.53E-05 | 0.0010503 | 0.00312 | 5.26E-05 | 0.002064759 | FMN1 |
| chr10 | 17646202 | 17646211 | 10 | 2 | 0.000103259 | 0.0001492 | 8.68E-05 | 5.26E-05 | 0.004108094 | STAM |
| chr9 | 93911962 | 93912051 | 90 | 2 | 0.000105394 | 3.77E-05 | 0.00079 | 5.26E-05 | -0.00655855 | NA |
| chr6 | 4087357 | 4087807 | 451 | 5 | 2.82E-05 | 0.0034466 | 0.000327 | 5.26E-05 | -0.00091272 | C6orf201 |
| chr13 | 110678087 | 110678757 | 671 | 5 | 9.03E-06 | 0.0206831 | 1.35E-05 | 5.27E-05 | 0.001344476 | CARS2 |
| chr22 | 23249978 | 23250008 | 31 | 2 | 6.22E-05 | 3.19E-05 | 0.001741 | 5.27E-05 | -0.00973743 | BCR |
| chr9 | 136571743 | 136571967 | 225 | 2 | 8.19E-05 | 3.48E-05 | 0.001101 | 5.27E-05 | -0.00632868 | NA |
| chr1 | 145997266 | 145997772 | 507 | 3 | 6.48E-07 | 0.0009112 | 3.13E-05 | 5.27E-05 | 0.003365441 | NA |
| chr1 | 3752355 | 3752378 | 24 | 3 | 4.48E-05 | 0.0045584 | 1.68E-05 | 5.27E-05 | -0.00700267 | CCDC27 |
| chr18 | 9449617 | 9449631 | 15 | 2 | 9.18E-05 | 0.0003787 | 5.10E-05 | 5.27E-05 | -0.00910859 | NA |
| chr14 | 65669114 | 65669390 | 277 | 3 | 0.000102457 | 9.45E-05 | 0.000344 | 5.27E-05 | -0.00162506 | FUT8 |
| chr16 | 88811682 | 88812188 | 507 | 9 | 8.53E-05 | 3.73E-05 | 0.005623 | 5.27E-05 | -0.00121455 | APRT |
| chr7 | 3687534 | 3687557 | 24 | 2 | 0.000115766 | 0.0001638 | 8.15E-05 | 5.27E-05 | -0.00333878 | SDK1 |
| chr18 | 59273433 | 59274014 | 582 | 6 | 4.15E-05 | 0.0003638 | 0.000207 | 5.27E-05 | 0.000695906 | RAX |
| chr13 | 24251589 | 24251835 | 247 | 3 | 1.51E-05 | 5.46E-05 | 0.000586 | 5.28E-05 | -0.00552357 | AL359736.1, SPATA13 |
| chr20 | 57935059 | 57935188 | 130 | 2 | 8.90E-05 | 0.0001767 | 7.76E-05 | 5.28E-05 | -0.00676909 | NA |
| chr15 | 25783640 | 25783901 | 262 | 4 | 6.68E-05 | 0.0020461 | 4.94E-05 | 5.28E-05 | -0.00862302 | ATP10A |
| chr5 | 42238149 | 42238344 | 196 | 2 | 6.31E-05 | 0.0001903 | 7.40E-05 | 5.28E-05 | -0.00809239 | NA |
| chr2 | 64641381 | 64641576 | 196 | 2 | 6.52E-05 | 8.00E-05 | 0.00016 | 5.28E-05 | 0.009483221 | SERTAD2 |
| chr17 | 78974775 | 78974851 | 77 | 3 | 8.67E-05 | 0.00024 | 8.19E-05 | 5.28E-05 | -0.00517712 | LGALS3BP |
| chr7 | 140641180 | 140641535 | 356 | 3 | 3.65E-05 | 0.0003315 | 0.0002 | 5.28E-05 | -0.00533729 | AC006452.1, DENND2A |
| chr5 | 141354696 | 141355154 | 459 | 4 | 8.55E-06 | 0.0002163 | 0.001279 | 5.29E-05 | -0.01310276 | PCDHGA1, PCDHGA2, PCDHGA3, PCDHGB1, PCDHGA4 |
| chr2 | 188798361 | 188798487 | 127 | 2 | 5.10E-05 | 5.34E-05 | 0.000317 | 5.29E-05 | 0.004009958 | AC092598.1 |
| chr11 | 4368887 | 4369033 | 147 | 4 | 0.000123258 | 0.0001077 | 0.000556 | 5.29E-05 | -0.00197139 | NA |
| chr7 | 150720255 | 150720929 | 675 | 5 | 2.96E-05 | 0.0024843 | 0.000324 | 5.29E-05 | -5.35E-05 | GIMAP1, GIMAP1-GIMAP5 |
| chr3 | 193241043 | 193241973 | 931 | 8 | 6.50E-06 | 0.005355 | 0.000666 | 5.29E-05 | -0.00045398 | PLAAT1 |
| chr6 | 89894580 | 89895058 | 479 | 2 | 3.28E-05 | 4.13E-05 | 0.000601 | 5.29E-05 | -0.00975791 | GJA10 |
| chr10 | 97498631 | 97499228 | 598 | 11 | 2.13E-05 | 0.0008771 | 0.000742 | 5.29E-05 | -0.00060128 | UBTD1, MMS19 |
| chr10 | 49188380 | 49188788 | 409 | 6 | 7.64E-05 | 0.0002043 | 0.000127 | 5.29E-05 | 0.003948074 | TMEM273 |
| chr21 | 26171365 | 26171734 | 370 | 3 | 5.67E-05 | 0.0002646 | 7.49E-05 | 5.29E-05 | -0.01021366 | AP001439.1, AP000229.1 |
| chr20 | 56987935 | 56988090 | 156 | 3 | 0.000148651 | 2.77E-05 | 0.003176 | 5.30E-05 | -0.01204825 | NA |
| chr17 | 7610722 | 7611173 | 452 | 3 | 2.22E-05 | 0.0006822 | 0.000154 | 5.30E-05 | -0.00344525 | FXR2 |
| chr9 | 111630821 | 111631698 | 878 | 11 | 7.19E-05 | 7.38E-05 | 0.012585 | 5.30E-05 | 0.000543785 | DNAJC25, DNAJC25-GNG10, LRRC37A5P |
| chr11 | 43489845 | 43489901 | 57 | 2 | 9.43E-05 | 0.0003444 | 5.38E-05 | 5.30E-05 | -0.00655287 | TTC17 |
| chr15 | 76191761 | 76192284 | 524 | 4 | 3.51E-05 | 0.0019945 | 0.000112 | 5.30E-05 | -0.01137795 | TMEM266 |
| chr7 | 41808607 | 41808628 | 22 | 2 | 3.96E-05 | 0.0041903 | 2.69E-05 | 5.30E-05 | -0.00714168 | NA |
| chr14 | 77822084 | 77822436 | 353 | 2 | 1.81E-05 | 0.0001123 | 0.000111 | 5.30E-05 | 0.004793091 | ADCK1 |
| chr2 | 232771263 | 232771701 | 439 | 5 | 7.37E-05 | 0.0006153 | 0.000652 | 5.30E-05 | -0.00255303 | GIGYF2, KCNJ13 |
| chr15 | 60519948 | 60519979 | 32 | 2 | 0.000119631 | 7.36E-05 | 0.000183 | 5.31E-05 | 0.000554258 | RORA-AS1, RORA |
| chr9 | 37967135 | 37967454 | 320 | 2 | 6.22E-06 | 0.0009046 | 3.73E-05 | 5.31E-05 | -0.01263459 | AL138752.2, SHB |
| chr19 | 9324375 | 9324731 | 357 | 3 | 1.05E-05 | 7.27E-05 | 0.000256 | 5.31E-05 | -0.01591516 | ZNF559, ZNF559-ZNF177 |
| chr13 | 51221544 | 51222192 | 649 | 9 | 3.88E-05 | 0.000221 | 0.001134 | 5.32E-05 | -0.00153166 | NA |
| chr6 | 166545956 | 166546166 | 211 | 4 | 0.000132434 | 3.55E-05 | 0.002413 | 5.32E-05 | -0.00330789 | RPS6KA2 |
| chr10 | 34024946 | 34025037 | 92 | 2 | 6.92E-05 | 0.0010845 | 3.55E-05 | 5.32E-05 | -0.01604812 | NA |
| chr12 | 120725891 | 120726843 | 953 | 8 | 5.67E-06 | 0.0089534 | 0.000294 | 5.32E-05 | -0.00055998 | ACADS |
| chr1 | 36319874 | 36319928 | 55 | 2 | 3.45E-05 | 3.24E-05 | 0.001708 | 5.32E-05 | -4.35E-06 | SH3D21 |
| chr1 | 114709625 | 114709736 | 112 | 2 | 0.000105113 | 3.57E-05 | 0.001034 | 5.32E-05 | 1.15E-05 | NRAS |
| chr22 | 41560311 | 41560436 | 126 | 2 | 8.52E-05 | 0.0023646 | 2.97E-05 | 5.32E-05 | 0.006744198 | AL023553.1 |
| chr4 | 128811110 | 128811820 | 711 | 11 | 7.38E-05 | 0.0005061 | 0.011036 | 5.33E-05 | -0.00039936 | JADE1 |
| chr11 | 62402401 | 62403109 | 709 | 2 | 4.97E-05 | 3.43E-05 | 0.001252 | 5.33E-05 | 0.002233842 | NA |
| chr19 | 35404168 | 35404193 | 26 | 2 | 1.38E-05 | 0.0110082 | 2.44E-05 | 5.33E-05 | -0.00359175 | LINC01531 |
| chr11 | 69646566 | 69646683 | 118 | 2 | 4.32E-05 | 5.11E-05 | 0.000358 | 5.33E-05 | -0.00548471 | CCND1 |
| chr15 | 41513937 | 41514390 | 454 | 8 | 6.70E-05 | 4.16E-05 | 0.01049 | 5.33E-05 | -0.00233273 | NA |
| chr1 | 54517500 | 54517830 | 331 | 3 | 4.65E-05 | 0.0065361 | 0.000792 | 5.33E-05 | -0.01119036 | NA |
| chr16 | 2707656 | 2707805 | 150 | 2 | 8.40E-06 | 0.000105 | 0.00012 | 5.34E-05 | 0.003878562 | KCTD5 |
| chr5 | 157215882 | 157216208 | 327 | 2 | 7.27E-05 | 3.25E-05 | 0.001729 | 5.34E-05 | -0.00748139 | ITK, AC010609.1 |
| chr20 | 1297003 | 1297170 | 168 | 2 | 2.61E-05 | 6.46E-05 | 0.000229 | 5.34E-05 | -0.00554541 | SNPH |
| chr15 | 65045087 | 65045579 | 493 | 6 | 3.04E-05 | 0.0003464 | 0.00017 | 5.34E-05 | -0.00083677 | SLC51B |
| chr16 | 68995544 | 68995555 | 12 | 2 | 0.0001599 | 5.91E-05 | 0.000269 | 5.34E-05 | -0.00455311 | TANGO6 |
| chr13 | 39786033 | 39786280 | 248 | 2 | 1.40E-05 | 0.0207101 | 2.36E-05 | 5.34E-05 | -0.00796369 | COG6 |
| chr2 | 170822241 | 170822444 | 204 | 2 | 0.000114635 | 3.20E-05 | 0.001922 | 5.35E-05 | 0.010788624 | GAD1 |
| chr15 | 67146460 | 67147094 | 635 | 4 | 1.83E-06 | 0.0014646 | 4.66E-05 | 5.36E-05 | 0.004712639 | SMAD3, AC012568.1 |
| chr2 | 26401360 | 26401997 | 638 | 10 | 3.01E-05 | 0.001272 | 3.31E-05 | 5.36E-05 | -0.00337334 | DRC1 |
| chr2 | 99140268 | 99140893 | 626 | 3 | 1.95E-05 | 3.23E-05 | 0.001459 | 5.36E-05 | 0.000736129 | TSGA10 |
| chr4 | 183503948 | 183504133 | 186 | 3 | 2.26E-05 | 0.0007778 | 3.52E-05 | 5.36E-05 | -0.00025655 | AC107214.1 |
| chr11 | 47594346 | 47594544 | 199 | 2 | 2.05E-06 | 0.0004678 | 4.76E-05 | 5.36E-05 | 0.008751303 | C1QTNF4 |
| chr4 | 39228383 | 39228464 | 82 | 2 | 0.000175537 | 3.66E-05 | 0.00097 | 5.37E-05 | -0.0013911 | WDR19 |
| chr8 | 133067009 | 133067198 | 190 | 2 | 0.000105874 | 0.0001067 | 0.000119 | 5.38E-05 | -0.003526 | TG, SLA |
| chr9 | 128203186 | 128203482 | 297 | 6 | 3.86E-05 | 0.0061846 | 0.003323 | 5.38E-05 | 0.002394033 | DNM1, CIZ1 |
| chr19 | 15049349 | 15049507 | 159 | 5 | 0.000160161 | 0.0002581 | 0.001642 | 5.38E-05 | -0.00278868 | CASP14 |
| chr17 | 74994207 | 74994352 | 146 | 2 | 0.00013069 | 7.93E-05 | 0.000169 | 5.38E-05 | -0.00069624 | CDR2L |
| chr1 | 2855281 | 2855647 | 367 | 4 | 7.96E-06 | 0.0049164 | 3.29E-05 | 5.38E-05 | -0.00157059 | NA |
| chr1 | 203765039 | 203765431 | 393 | 8 | 4.72E-05 | 5.01E-05 | 0.02158 | 5.38E-05 | -0.00721544 | LAX1 |
| chr17 | 63997964 | 63998395 | 432 | 5 | 3.29E-05 | 0.0051202 | 0.00018 | 5.39E-05 | 0.000441449 | PRR29, PRR29-AS1 |
| chr14 | 53954292 | 53954860 | 569 | 9 | 5.64E-05 | 4.33E-05 | 0.01526 | 5.39E-05 | -0.00084709 | AL138479.2, BMP4 |
| chr2 | 241249128 | 241249481 | 354 | 2 | 0.000121461 | 3.29E-05 | 0.00171 | 5.40E-05 | -0.00401569 | HDLBP |
| chr7 | 138001520 | 138002180 | 661 | 9 | 1.81E-06 | 0.0005658 | 0.000117 | 5.40E-05 | -0.00038192 | CREB3L2 |
| chr2 | 30732504 | 30732768 | 265 | 2 | 0.000100002 | 4.61E-05 | 0.000477 | 5.41E-05 | 0.001105061 | CAPN13 |
| chr3 | 180870988 | 180871253 | 266 | 2 | 7.69E-05 | 5.96E-05 | 0.000272 | 5.41E-05 | 0.007716783 | FXR1, AC108734.4 |
| chr16 | 50697100 | 50697204 | 105 | 2 | 0.000164204 | 6.36E-05 | 0.000242 | 5.41E-05 | -0.00176217 | NOD2 |
| chr11 | 58927447 | 58927704 | 258 | 4 | 1.95E-05 | 0.0002727 | 0.000408 | 5.41E-05 | 0.011510324 | GLYATL1, AP001652.1 |
| chr8 | 133455231 | 133455735 | 505 | 2 | 1.55E-06 | 0.0002387 | 6.68E-05 | 5.42E-05 | 0.005671611 | ST3GAL1 |
| chr1 | 231282021 | 231282310 | 290 | 3 | 1.00E-05 | 0.0163024 | 2.50E-05 | 5.42E-05 | 0.003636374 | NA |
| chr3 | 72150308 | 72150477 | 170 | 2 | 3.98E-05 | 0.0008777 | 3.85E-05 | 5.42E-05 | 0.004071025 | LINC00877 |
| chr18 | 23358130 | 23358245 | 116 | 2 | 4.23E-05 | 0.000155 | 8.82E-05 | 5.42E-05 | -0.01047795 | TMEM241 |
| chr5 | 598609 | 598791 | 183 | 3 | 8.29E-05 | 0.001254 | 0.000352 | 5.42E-05 | 0.0122294 | NA |
| chr11 | 811162 | 811954 | 793 | 7 | 5.12E-05 | 0.0003779 | 0.000744 | 5.43E-05 | -0.00110632 | RPLP2, SNORA52 |
| chr9 | 3898591 | 3898614 | 24 | 2 | 5.75E-05 | 0.0001045 | 0.000124 | 5.43E-05 | -0.02077728 | GLIS3 |
| chr3 | 134366934 | 134367214 | 281 | 2 | 8.73E-05 | 4.41E-05 | 0.000545 | 5.43E-05 | -0.0058555 | AMOTL2 |
| chr22 | 27377695 | 27377753 | 59 | 2 | 7.96E-05 | 6.47E-05 | 0.000238 | 5.43E-05 | -0.00242071 | AL020994.2 |
| chr10 | 134564 | 135348 | 785 | 11 | 5.21E-08 | 0.0018699 | 0.00012 | 5.44E-05 | -0.00089001 | ZMYND11 |
| chr9 | 34555044 | 34555097 | 54 | 2 | 4.64E-05 | 0.0008693 | 3.87E-05 | 5.44E-05 | -0.0134148 | CNTFR |
| chr19 | 40366235 | 40366820 | 586 | 3 | 5.24E-06 | 0.0009985 | 3.45E-05 | 5.44E-05 | -0.007842 | PLD3 |
| chr13 | 111538775 | 111538925 | 151 | 3 | 5.31E-05 | 0.0001266 | 0.000455 | 5.44E-05 | -0.00485849 | NA |
| chr10 | 71648428 | 71648663 | 236 | 2 | 8.49E-05 | 0.0001307 | 0.000101 | 5.44E-05 | -0.00588953 | CDH23 |
| chr17 | 58363111 | 58363581 | 471 | 4 | 0.000106269 | 0.0005069 | 0.003957 | 5.44E-05 | -0.00339681 | TSPOAP1-AS1, AC004687.2, RNF43 |
| chr12 | 1500079 | 1500575 | 497 | 9 | 1.11E-05 | 0.0100864 | 0.000145 | 5.45E-05 | 0.00278754 | LINC00942 |
| chr10 | 15248742 | 15248985 | 244 | 4 | 8.72E-06 | 0.027398 | 7.47E-06 | 5.45E-05 | -0.00188015 | FAM171A1 |
| chr17 | 17475795 | 17476438 | 644 | 5 | 6.70E-05 | 1.87E-05 | 0.010624 | 5.45E-05 | 0.002681047 | NA |
| chr13 | 113857413 | 113857886 | 474 | 7 | 1.59E-05 | 0.0013635 | 0.00186 | 5.45E-05 | -0.00643356 | GAS6 |
| chr1 | 210440347 | 210440497 | 151 | 2 | 5.21E-05 | 6.37E-05 | 0.000246 | 5.45E-05 | -4.58E-05 | HHAT |
| chr14 | 100986660 | 100987094 | 435 | 6 | 0.000124367 | 9.98E-05 | 0.002652 | 5.45E-05 | -0.00269343 | MEG8, SNORD114-26 |
| chr15 | 75450795 | 75451658 | 864 | 4 | 8.34E-06 | 2.39E-05 | 0.002671 | 5.45E-05 | 0.001897933 | SIN3A |
| chr15 | 34336530 | 34336999 | 470 | 5 | 4.36E-05 | 1.48E-05 | 0.015651 | 5.46E-05 | -0.00281783 | SLC12A6 |
| chr2 | 135809584 | 135809700 | 117 | 2 | 0.000101649 | 5.57E-05 | 0.000318 | 5.46E-05 | -0.00621324 | LCT |
| chr17 | 8832819 | 8832976 | 158 | 3 | 9.03E-05 | 0.0001863 | 8.78E-05 | 5.46E-05 | -0.00660044 | PIK3R6 |
| chr3 | 126433180 | 126433195 | 16 | 3 | 0.000113719 | 0.0004201 | 0.000241 | 5.46E-05 | -0.00217415 | CFAP100, AC073439.1 |
| chr4 | 10518557 | 10518775 | 219 | 2 | 2.80E-05 | 7.18E-05 | 0.000203 | 5.46E-05 | -0.00998382 | AC005599.1, CLNK |
| chr16 | 89493660 | 89493868 | 209 | 2 | 7.24E-05 | 0.0003187 | 5.82E-05 | 5.47E-05 | -0.02009157 | SPG7, AC092123.1 |
| chr2 | 239401334 | 239402019 | 686 | 9 | 1.78E-05 | 0.0003032 | 0.001547 | 5.47E-05 | -0.00030353 | HDAC4-AS1, HDAC4 |
| chr9 | 136117429 | 136118447 | 1019 | 4 | 4.89E-07 | 0.0039919 | 1.16E-05 | 5.47E-05 | 0.00055397 | TMEM250 |
| chr14 | 51208931 | 51209103 | 173 | 2 | 0.000101079 | 7.78E-05 | 0.00018 | 5.47E-05 | -0.01532988 | NA |
| chr17 | 8944907 | 8944956 | 50 | 2 | 0.000123746 | 0.0001407 | 9.60E-05 | 5.47E-05 | -0.00448189 | PIK3R5 |
| chr2 | 10344130 | 10344454 | 325 | 3 | 1.52E-05 | 0.0069334 | 5.35E-05 | 5.47E-05 | 0.007986742 | HPCAL1 |
| chr8 | 9436265 | 9436454 | 190 | 2 | 8.94E-05 | 5.73E-05 | 0.000303 | 5.47E-05 | -0.00068489 | NA |
| chr22 | 50017224 | 50017428 | 205 | 2 | 7.68E-05 | 0.0002168 | 7.18E-05 | 5.48E-05 | 0.000267308 | TTLL8 |
| chr13 | 41019249 | 41019446 | 198 | 4 | 0.00014335 | 6.79E-05 | 0.001139 | 5.48E-05 | 0.006986023 | ELF1 |
| chr4 | 95051315 | 95051639 | 325 | 8 | 9.75E-05 | 0.00061 | 0.001999 | 5.48E-05 | -0.00378513 | BMPR1B |
| chr18 | 2577213 | 2578085 | 873 | 5 | 2.85E-05 | 0.000919 | 0.000126 | 5.48E-05 | 0.000762945 | NDC80 |
| chr4 | 85929568 | 85930272 | 705 | 7 | 1.87E-05 | 0.0115225 | 3.57E-05 | 5.49E-05 | -0.00189472 | ARHGAP24 |
| chr16 | 89565936 | 89566341 | 406 | 4 | 3.93E-05 | 9.31E-05 | 0.005102 | 5.49E-05 | 0.009763545 | NA |
| chr15 | 50547129 | 50548020 | 892 | 5 | 5.76E-05 | 0.0011015 | 0.003567 | 5.49E-05 | 0.006685387 | NA |
| chr2 | 182715967 | 182716419 | 453 | 7 | 8.54E-05 | 7.98E-05 | 0.001777 | 5.49E-05 | 0.001653072 | DNAJC10 |
| chr20 | 1893036 | 1893231 | 196 | 2 | 5.26E-05 | 0.000155 | 9.00E-05 | 5.49E-05 | -0.00154399 | AL117335.1 |
| chr16 | 2977967 | 2978108 | 142 | 2 | 6.70E-05 | 4.82E-05 | 0.000448 | 5.49E-05 | -0.00152159 | PKMYT1 |
| chr22 | 38115517 | 38115760 | 244 | 3 | 7.68E-05 | 0.00241 | 0.001053 | 5.49E-05 | -0.0026363 | PLA2G6 |
| chr7 | 3244394 | 3244817 | 424 | 3 | 1.57E-05 | 0.0008198 | 3.44E-05 | 5.50E-05 | 0.007768687 | NA |
| chr2 | 216607062 | 216607378 | 317 | 4 | 0.000171238 | 5.46E-05 | 0.002139 | 5.50E-05 | 0.002905954 | AC073321.1 |
| chr7 | 100707046 | 100707440 | 395 | 3 | 5.37E-07 | 0.0020802 | 2.49E-05 | 5.51E-05 | -0.00351391 | POP7 |
| chr12 | 125178084 | 125178375 | 292 | 2 | 2.43E-05 | 0.0142221 | 2.47E-05 | 5.51E-05 | -0.0084731 | NA |
| chr2 | 74917818 | 74918260 | 443 | 4 | 3.65E-05 | 0.005283 | 8.55E-05 | 5.51E-05 | -0.00497241 | LINC01291 |
| chr3 | 32472829 | 32472998 | 170 | 2 | 5.00E-05 | 0.0027916 | 2.98E-05 | 5.51E-05 | -0.00605725 | CMTM7 |
| chr11 | 69100497 | 69100942 | 446 | 2 | 1.02E-05 | 0.0001254 | 0.000107 | 5.51E-05 | -0.01320716 | TPCN2, AP003071.5 |
| chr12 | 50953164 | 50953316 | 153 | 3 | 1.72E-05 | 0.0072862 | 1.75E-05 | 5.51E-05 | -0.00037688 | NA |
| chr8 | 10528712 | 10528807 | 96 | 2 | 4.08E-05 | 3.41E-05 | 0.001585 | 5.51E-05 | 0.025112853 | PRSS55, PRSS51 |
| chr1 | 34868285 | 34868928 | 644 | 4 | 1.98E-05 | 0.0027488 | 3.07E-05 | 5.52E-05 | -0.00093363 | DLGAP3 |
| chr21 | 43967883 | 43968098 | 216 | 2 | 6.61E-05 | 0.0004454 | 5.03E-05 | 5.52E-05 | -0.00791129 | AGPAT3 |
| chr1 | 37794536 | 37795385 | 850 | 6 | 2.54E-05 | 2.14E-05 | 0.004206 | 5.52E-05 | -0.00903068 | MANEAL |
| chr17 | 38749556 | 38749745 | 190 | 3 | 0.000119436 | 2.50E-05 | 0.004383 | 5.52E-05 | 0.001459757 | AC006449.5, PCGF2 |
| chr11 | 111936824 | 111937380 | 557 | 9 | 3.32E-05 | 0.0005355 | 0.000444 | 5.52E-05 | -0.00066959 | DIXDC1 |
| chr19 | 48111455 | 48111516 | 62 | 3 | 4.11E-05 | 6.57E-05 | 0.000652 | 5.52E-05 | -0.00940284 | NA |
| chr15 | 101308984 | 101309195 | 212 | 2 | 8.00E-05 | 3.55E-05 | 0.001297 | 5.52E-05 | -0.00702101 | PCSK6 |
| chr1 | 19596288 | 19596848 | 561 | 5 | 3.19E-06 | 0.000911 | 0.000188 | 5.53E-05 | -0.00471371 | MICOS10, AL031727.2 |

| | | | | | | | | | | |
|---|---|---|---|---|---|---|---|---|---|---|
| chr15 | 65356152 | 65356433 | 282 | 2 | 5.10E-06 | 0.0015361 | 3.38E-05 | 5.53E-05 | -0.01348399 | IGDCC3 |
| chr12 | 51589230 | 51589264 | 35 | 2 | 2.46E-06 | 0.000394 | 5.32E-05 | 5.53E-05 | -0.0030746 | NA |
| chr17 | 35931214 | 35931399 | 186 | 2 | 9.34E-05 | 3.69E-05 | 0.001076 | 5.53E-05 | -0.00371113 | NA |
| chr1 | 228735930 | 228736542 | 613 | 5 | 1.08E-05 | 0.0019637 | 3.54E-05 | 5.53E-05 | -0.00294506 | RHOU |
| chr11 | 6389847 | 6390239 | 393 | 6 | 7.53E-06 | 0.0036975 | 0.00135 | 5.53E-05 | -0.00571404 | NA |
| chr1 | 35715572 | 35715846 | 275 | 3 | 0.000124967 | 8.69E-05 | 0.000919 | 5.54E-05 | -0.00774901 | C1orf216 |
| chr11 | 8594147 | 8595419 | 1273 | 12 | 6.67E-07 | 0.0077403 | 9.65E-05 | 5.54E-05 | -0.00073727 | STK33 |
| chr7 | 64352905 | 64353067 | 163 | 2 | 0.000133879 | 7.74E-05 | 0.000186 | 5.54E-05 | -0.00451402 | ZNF736 |
| chr16 | 87305947 | 87306271 | 325 | 4 | 5.46E-05 | 0.0004362 | 0.000108 | 5.54E-05 | -0.00358649 | C16orf95, AC010531.1 |
| chr17 | 42567564 | 42567636 | 73 | 2 | 2.98E-05 | 0.0003009 | 6.08E-05 | 5.55E-05 | -5.99E-05 | MLX |
| chr22 | 41781214 | 41781679 | 466 | 3 | 3.92E-07 | 0.0043918 | 1.98E-05 | 5.55E-05 | 0.007084936 | MEI1 |
| chr11 | 809095 | 809449 | 355 | 4 | 6.18E-05 | 1.85E-05 | 0.007233 | 5.55E-05 | 0.001712443 | PIDD1 |
| chr12 | 57806323 | 57806616 | 294 | 4 | 0.000114564 | 7.96E-05 | 0.00194 | 5.56E-05 | -0.00014471 | TSFM, AVIL |
| chr9 | 93575353 | 93575700 | 348 | 4 | 2.24E-05 | 0.0010569 | 4.18E-05 | 5.56E-05 | -0.01116859 | NA |
| chr6 | 158767127 | 158767356 | 230 | 3 | 4.81E-05 | 0.0035109 | 5.69E-05 | 5.56E-05 | -0.00629252 | EZR |
| chr4 | 3740454 | 3740630 | 177 | 3 | 2.51E-05 | 9.84E-05 | 0.000176 | 5.56E-05 | 0.008991329 | NA |
| chr10 | 133345443 | 133345498 | 56 | 2 | 0.000175527 | 3.84E-05 | 0.000949 | 5.57E-05 | -0.00818655 | ZNF511-PRAP1 |
| chr12 | 6788250 | 6788565 | 316 | 2 | 1.44E-05 | 0.0001246 | 0.000109 | 5.57E-05 | -0.00542023 | CD4 |
| chr4 | 3829241 | 3829615 | 375 | 4 | 5.07E-05 | 0.0002943 | 0.000842 | 5.57E-05 | -0.00299983 | NA |
| chr3 | 184032273 | 184032902 | 630 | 5 | 7.55E-05 | 0.0002703 | 0.001938 | 5.57E-05 | -0.00276627 | HTR3D |
| chr4 | 142405332 | 142405588 | 257 | 4 | 0.000101612 | 3.72E-05 | 0.004577 | 5.57E-05 | -0.00040887 | INPP4B |
| chr9 | 12694273 | 12694940 | 668 | 3 | 9.99E-05 | 2.96E-05 | 0.002082 | 5.57E-05 | 0.001631485 | TYRP1 |
| chr18 | 3469010 | 3469153 | 144 | 2 | 0.000119622 | 0.0001184 | 0.000115 | 5.57E-05 | -0.01092889 | GAPLINC |
| chr1 | 40530277 | 40531392 | 1116 | 6 | 6.94E-08 | 0.0009039 | 0.000116 | 5.58E-05 | -0.00314988 | NA |
| chr14 | 20455863 | 20456579 | 717 | 3 | 1.43E-05 | 3.33E-05 | 0.003138 | 5.58E-05 | 0.003113081 | APEX1 |
| chr15 | 90245966 | 90246001 | 36 | 2 | 0.000132797 | 4.47E-05 | 0.000578 | 5.58E-05 | -0.01550942 | AC091167.7 |
| chr16 | 31180154 | 31180538 | 385 | 2 | 2.23E-05 | 3.79E-05 | 0.001008 | 5.58E-05 | -0.00238512 | FUS |
| chr19 | 18785583 | 18786043 | 461 | 4 | 7.62E-05 | 0.000584 | 0.000122 | 5.58E-05 | -0.00874097 | COMP |
| chr5 | 38555769 | 38556333 | 565 | 4 | 1.02E-05 | 0.0002361 | 0.000729 | 5.58E-05 | 0.002989467 | LIFR |
| chr17 | 59892660 | 59892800 | 141 | 4 | 0.000142523 | 5.39E-05 | 0.000509 | 5.58E-05 | 0.002786332 | TUBD1 |
| chr1 | 21811499 | 21811616 | 118 | 2 | 4.85E-05 | 0.0004134 | 5.27E-05 | 5.58E-05 | -0.00772791 | NA |
| chr12 | 93378435 | 93378991 | 557 | 3 | 4.01E-06 | 0.0174067 | 0.000111 | 5.59E-05 | 0.002218584 | NUDT4 |
| chr20 | 60123497 | 60123528 | 32 | 2 | 8.57E-05 | 0.0007639 | 4.16E-05 | 5.59E-05 | 0.004204155 | MIR646HG |
| chr1 | 210069861 | 210070155 | 295 | 2 | 3.33E-05 | 0.0020116 | 3.22E-05 | 5.59E-05 | -0.00567246 | SYT14 |
| chr14 | 69191173 | 69192218 | 1046 | 16 | 4.70E-05 | 0.0002545 | 0.009062 | 5.59E-05 | 0.000450943 | EXD2, AL359317.2 |
| chr16 | 88628623 | 88629019 | 397 | 2 | 2.56E-06 | 0.0017465 | 3.32E-05 | 5.59E-05 | -0.00779996 | ZC3H18 |
| chr13 | 24512504 | 24512952 | 449 | 3 | 6.48E-05 | 2.38E-05 | 0.004447 | 5.61E-05 | 0.002292791 | TPTE2P6, PARP4 |
| chr3 | 126464821 | 126464888 | 68 | 2 | 0.000109121 | 0.0002575 | 6.71E-05 | 5.62E-05 | 0.004307399 | ZXDC |
| chr9 | 120966346 | 120966562 | 217 | 2 | 9.20E-05 | 0.0001672 | 8.81E-05 | 5.62E-05 | -0.0014183 | C5 |
| chr6 | 43473608 | 43473750 | 143 | 2 | 4.50E-05 | 0.0009001 | 3.98E-05 | 5.62E-05 | 0.004193285 | NA |
| chr9 | 136949869 | 136950010 | 142 | 2 | 1.68E-05 | 5.10E-05 | 0.000419 | 5.62E-05 | -0.00024526 | LCN12 |
| chr11 | 115659432 | 115659713 | 282 | 5 | 4.45E-05 | 0.0010627 | 2.93E-05 | 5.62E-05 | 0.001862862 | AP000997.4, LINC02698 |
| chr9 | 119088143 | 119088200 | 58 | 2 | 0.00010568 | 0.0003057 | 6.15E-05 | 5.62E-05 | -0.00567944 | NA |
| chr9 | 17906340 | 17906563 | 224 | 2 | 5.28E-06 | 0.004387 | 2.83E-05 | 5.62E-05 | 0.003254072 | ADAMTSL1 |
| chr19 | 17100813 | 17100962 | 150 | 2 | 3.53E-05 | 7.39E-05 | 0.000207 | 5.62E-05 | 0.012360429 | MYO9B |
| chr8 | 95024595 | 95025432 | 838 | 8 | 6.59E-05 | 0.0039973 | 0.005262 | 5.63E-05 | -0.00082492 | NDUFAF6 |
| chr17 | 15257370 | 15257442 | 73 | 3 | 7.62E-05 | 2.75E-05 | 0.003464 | 5.63E-05 | 0.003215154 | PMP22 |
| chr10 | 125585223 | 125585245 | 23 | 2 | 9.66E-05 | 0.0002399 | 7.01E-05 | 5.63E-05 | 0.008156663 | TEX36 |
| chr22 | 42210257 | 42210370 | 114 | 3 | 0.000145508 | 2.49E-05 | 0.004953 | 5.63E-05 | 0.000662475 | TCF20 |
| chr17 | 81510827 | 81511147 | 321 | 3 | 1.23E-05 | 0.0001916 | 0.000127 | 5.64E-05 | -0.00862224 | ACTG1 |
| chr2 | 236633981 | 236634063 | 83 | 2 | 0.000150983 | 7.99E-05 | 0.000185 | 5.64E-05 | -0.00389173 | NA |
| chr1 | 28643042 | 28643475 | 434 | 12 | 2.19E-05 | 0.0079082 | 5.39E-05 | 5.64E-05 | 0.001279722 | LINC01715, TAF12 |
| chr15 | 78872372 | 78872883 | 512 | 4 | 3.24E-05 | 0.0002299 | 0.00011 | 5.64E-05 | -0.00373314 | MORF4L1 |
| chr19 | 14480221 | 14480533 | 313 | 5 | 5.46E-05 | 0.0021959 | 0.000652 | 5.65E-05 | -0.00928851 | GIPC1 |
| chr2 | 232487729 | 232488494 | 766 | 5 | 1.87E-05 | 0.0003936 | 8.78E-05 | 5.65E-05 | -0.00662434 | ECEL1 |
| chr2 | 229714719 | 229715378 | 660 | 5 | 2.37E-06 | 0.0140968 | 0.000446 | 5.65E-05 | 0.001555455 | NA |
| chr3 | 52314935 | 52315323 | 389 | 3 | 6.56E-06 | 0.0004683 | 0.000457 | 5.65E-05 | 0.001190915 | NA |
| chr11 | 67316670 | 67317816 | 1147 | 9 | 2.61E-06 | 0.0120408 | 0.000596 | 5.66E-05 | -0.00208717 | AP001885.2 |
| chr7 | 11252059 | 11252176 | 118 | 2 | 2.66E-05 | 3.51E-05 | 0.001573 | 5.66E-05 | 0.006746445 | AC004160.1 |
| chr14 | 103140911 | 103140928 | 18 | 2 | 8.56E-05 | 0.0004824 | 5.01E-05 | 5.66E-05 | -0.01491542 | NDUFB4P11 |
| chr2 | 7024396 | 7024455 | 60 | 2 | 0.000141617 | 9.39E-05 | 0.000152 | 5.66E-05 | -0.00071327 | RNF144A |
| chr8 | 22439595 | 22439907 | 313 | 2 | 3.19E-05 | 0.0001027 | 0.000137 | 5.66E-05 | 0.000452063 | NA |
| chr9 | 78439255 | 78439833 | 579 | 10 | 3.31E-05 | 0.005184 | 0.000156 | 5.66E-05 | -0.00339887 | NA |
| chr7 | 1989119 | 1989605 | 487 | 5 | 3.29E-05 | 0.0011804 | 0.002175 | 5.67E-05 | -0.00067242 | MAD1L1, AC069288.1 |
| chr6 | 7204547 | 7204560 | 14 | 2 | 0.000157395 | 5.97E-05 | 0.000307 | 5.67E-05 | 0.000690032 | RREB1 |
| chr12 | 132777261 | 132777824 | 564 | 3 | 2.20E-05 | 0.0099145 | 2.94E-05 | 5.67E-05 | 0.002721709 | GOLGA3 |
| chr20 | 56202630 | 56202892 | 263 | 2 | 2.92E-05 | 0.0133484 | 2.55E-05 | 5.67E-05 | -0.01504486 | NA |
| chr9 | 134421780 | 134421814 | 35 | 2 | 2.89E-05 | 0.0014133 | 3.54E-05 | 5.67E-05 | -0.00637949 | RXRA |
| chr6 | 43912925 | 43913161 | 237 | 2 | 4.46E-05 | 6.47E-05 | 0.000265 | 5.67E-05 | -0.0058371 | NA |
| chr20 | 36498850 | 36499070 | 221 | 2 | 1.61E-05 | 0.0068067 | 2.70E-05 | 5.68E-05 | 0.003857499 | DLGAP4 |
| chr10 | 112339641 | 112339697 | 57 | 3 | 0.000129527 | 0.0002285 | 0.000186 | 5.68E-05 | -0.00446011 | GUCY2GP |
| chr5 | 88680175 | 88680477 | 303 | 3 | 7.84E-05 | 3.99E-05 | 0.002841 | 5.68E-05 | -0.00198252 | MEF2C-AS2, LINC00461 |
| chr10 | 29535454 | 29535637 | 184 | 2 | 1.64E-05 | 0.0047264 | 2.83E-05 | 5.68E-05 | -0.01514299 | SVIL |
| chr1 | 242211708 | 242211978 | 271 | 2 | 2.59E-05 | 3.98E-05 | 0.000909 | 5.68E-05 | 0.016778546 | PLD5 |
| chr22 | 26784008 | 26784138 | 131 | 2 | 6.88E-05 | 0.0012042 | 3.70E-05 | 5.69E-05 | -0.01101924 | NA |
| chr3 | 186570117 | 186570654 | 538 | 9 | 3.57E-05 | 0.000387 | 0.002172 | 5.69E-05 | 0.000623394 | DNAJB11, TBCCD1 |
| chr17 | 82696795 | 82697046 | 252 | 4 | 4.90E-05 | 2.15E-05 | 0.007063 | 5.69E-05 | 0.000644428 | RAB40B |
| chr1 | 217785132 | 217785273 | 142 | 2 | 5.16E-05 | 0.0015227 | 3.49E-05 | 5.69E-05 | 0.010050174 | SPATA17 |
| chr14 | 36514489 | 36514927 | 439 | 6 | 9.28E-05 | 0.000157 | 0.000174 | 5.69E-05 | -0.0017819 | AL132857.1, SFTA3 |
| chr19 | 45615765 | 45616378 | 614 | 3 | 1.20E-05 | 0.0040191 | 2.06E-05 | 5.69E-05 | -0.01000735 | EML2 |
| chr2 | 43226731 | 43227489 | 759 | 14 | 2.74E-05 | 0.0004674 | 0.006944 | 5.69E-05 | 0.000193145 | LINC01126 |
| chr20 | 61897009 | 61897275 | 267 | 2 | 2.37E-05 | 0.0004355 | 5.27E-05 | 5.69E-05 | -0.01392438 | CDH4 |
| chr12 | 110339605 | 110339625 | 21 | 2 | 8.50E-05 | 0.0002114 | 7.69E-05 | 5.70E-05 | 0.00347531 | ATP2A2 |
| chr10 | 103715617 | 103715768 | 152 | 2 | 2.87E-05 | 0.0035125 | 2.97E-05 | 5.70E-05 | -0.00747926 | SH3PXD2A |
| chr2 | 70931493 | 70931831 | 339 | 3 | 0.000106414 | 0.0003666 | 0.001197 | 5.71E-05 | -0.00578137 | VAX2 |
| chr16 | 1990194 | 1990626 | 433 | 4 | 5.29E-05 | 2.90E-05 | 0.004644 | 5.71E-05 | -0.00074268 | SYNGR3, AC005606.2 |
| chr15 | 82853385 | 82853445 | 61 | 2 | 9.28E-05 | 4.05E-05 | 0.000873 | 5.72E-05 | -0.0182226 | HOMER2 |
| chr8 | 40898345 | 40898715 | 371 | 4 | 3.64E-06 | 0.0016874 | 5.28E-05 | 5.72E-05 | 0.004531043 | NA |
| chr3 | 133880350 | 133880418 | 69 | 2 | 0.000165568 | 6.66E-05 | 0.000256 | 5.72E-05 | 0.001737996 | RAB6B |
| chr16 | 84124480 | 84124658 | 179 | 2 | 9.24E-05 | 4.46E-05 | 0.000641 | 5.72E-05 | -0.00223537 | HSDL1 |
| chr16 | 88454340 | 88454879 | 540 | 4 | 2.20E-06 | 0.0021898 | 2.09E-05 | 5.73E-05 | -0.00099003 | ZFPM1 |
| chr1 | 1286804 | 1286990 | 187 | 2 | 4.59E-05 | 0.0001694 | 8.99E-05 | 5.73E-05 | 0.004923631 | SCNN1D |
| chr7 | 102170656 | 102170829 | 174 | 4 | 0.000102037 | 0.0033151 | 4.80E-05 | 5.74E-05 | -0.00300668 | CUX1 |
| chr8 | 100472147 | 100472158 | 12 | 2 | 7.78E-05 | 0.000143 | 0.000102 | 5.74E-05 | 0.006977494 | NA |
| chr8 | 29353063 | 29353561 | 499 | 6 | 6.72E-05 | 0.0001498 | 0.000289 | 5.74E-05 | 0.005238556 | AC084262.2, AC084262.1 |
| chr20 | 37262601 | 37262683 | 83 | 2 | 2.30E-05 | 0.0001065 | 0.000135 | 5.74E-05 | -0.00795652 | NA |
| chr20 | 36497454 | 36497687 | 234 | 2 | 5.11E-05 | 3.81E-05 | 0.00114 | 5.74E-05 | -0.00038642 | DLGAP4 |
| chr7 | 127374506 | 127374694 | 189 | 3 | 5.52E-05 | 0.0001498 | 0.000188 | 5.74E-05 | -0.00265942 | ZNF800 |
| chr2 | 199468702 | 199469414 | 713 | 6 | 6.72E-06 | 0.001999 | 0.002959 | 5.75E-05 | -0.0030858 | SATB2-AS1, SATB2 |
| chr18 | 47176174 | 47176525 | 352 | 6 | 9.93E-05 | 2.22E-05 | 0.008683 | 5.75E-05 | 0.001606036 | AC012254.2, IER3IP1 |
| chr21 | 22425108 | 22425239 | 132 | 2 | 0.000105519 | 3.46E-05 | 0.00192 | 5.75E-05 | -0.0022151 | NA |
| chr4 | 2242259 | 2242611 | 353 | 4 | 3.20E-05 | 1.58E-05 | 0.013018 | 5.75E-05 | 0.000317649 | HAUS3 |
| chr9 | 114610443 | 114610959 | 517 | 4 | 3.43E-05 | 0.0003105 | 0.002106 | 5.75E-05 | -0.00618507 | NA |
| chr12 | 127455117 | 127455541 | 425 | 2 | 2.11E-06 | 0.0096569 | 2.64E-05 | 5.75E-05 | -0.01949987 | AC068787.4 |
| chr11 | 128811351 | 128811387 | 37 | 2 | 6.28E-05 | 4.92E-05 | 0.000491 | 5.75E-05 | -0.00038594 | FLI1 |
| chr16 | 28950579 | 28951204 | 626 | 9 | 2.97E-05 | 0.0005745 | 0.001189 | 5.75E-05 | -0.00100277 | AC009093.11, NFATC2IP |
| chr12 | 3491050 | 3491063 | 14 | 3 | 0.000165375 | 2.33E-05 | 0.005664 | 5.75E-05 | 0.001445676 | PRMT8, THCAT155 |
| chr14 | 100155511 | 100156476 | 966 | 6 | 4.05E-05 | 2.82E-05 | 0.005314 | 5.76E-05 | -0.00359498 | DEGS2 |
| chr1 | 32393852 | 32394004 | 153 | 3 | 6.36E-06 | 0.0066061 | 1.61E-05 | 5.76E-05 | -0.00828897 | BSDC1 |
| chr1 | 172639426 | 172639564 | 139 | 3 | 7.02E-05 | 0.0019076 | 9.80E-05 | 5.76E-05 | -0.00617416 | NA |
| chr11 | 117951353 | 117951379 | 27 | 2 | 8.14E-05 | 3.48E-05 | 0.001867 | 5.76E-05 | 0.006005964 | NA |
| chr13 | 40220691 | 40220707 | 17 | 2 | 7.62E-05 | 0.0002092 | 7.89E-05 | 5.78E-05 | -0.00987973 | LINC00598 |
| chr6 | 27374281 | 27374533 | 253 | 3 | 2.66E-05 | 0.0139545 | 1.37E-05 | 5.78E-05 | 0.003072691 | NA |
| chr8 | 56320116 | 56320339 | 224 | 5 | 6.82E-05 | 0.0001754 | 0.000372 | 5.78E-05 | -0.00618421 | SDR16C5 |
| chr9 | 126704759 | 126704904 | 146 | 3 | 5.59E-05 | 0.0001361 | 0.000289 | 5.78E-05 | -0.00537379 | NA |
| chr12 | 6905124 | 6905524 | 401 | 2 | 1.86E-06 | 6.63E-05 | 0.000265 | 5.78E-05 | -0.00814446 | LRRC23 |
| chr14 | 61263805 | 61264222 | 418 | 4 | 2.05E-05 | 0.0050674 | 1.25E-05 | 5.78E-05 | -0.00394923 | PRKCH |
| chr11 | 102268661 | 102269050 | 390 | 3 | 3.54E-06 | 0.0001066 | 0.000899 | 5.78E-05 | -0.01150949 | NA |
| chr1 | 5867357 | 5867496 | 140 | 2 | 0.000102099 | 3.46E-05 | 0.001993 | 5.78E-05 | -0.00383072 | NPHP4 |
| chr6 | 170380061 | 170380155 | 95 | 2 | 0.00011536 | 0.0001548 | 9.72E-05 | 5.79E-05 | -0.00847143 | FAM120B |
| chr6 | 50844884 | 50845282 | 399 | 3 | 3.79E-06 | 0.0001884 | 0.00019 | 5.79E-05 | -0.00707508 | TFAP2B |
| chr6 | 158475385 | 158475758 | 374 | 2 | 5.42E-05 | 4.52E-05 | 0.000639 | 5.79E-05 | -0.01004836 | TULP4 |
| chr12 | 126742567 | 126742596 | 30 | 3 | 7.15E-05 | 0.0001168 | 0.00091 | 5.79E-05 | -0.01044883 | LINC00943, LINC00944 |
| chr13 | 28584200 | 28584254 | 55 | 3 | 0.000125639 | 2.70E-05 | 0.003907 | 5.79E-05 | -0.00051566 | NA |
| chr14 | 104264747 | 104265215 | 469 | 2 | 1.74E-05 | 7.79E-05 | 0.000204 | 5.80E-05 | -0.0013259 | LINC02691 |
| chr2 | 10123092 | 10123569 | 478 | 3 | 7.57E-06 | 5.03E-05 | 0.000683 | 5.80E-05 | -0.00649526 | RRM2 |
| chr1 | 1528909 | 1529129 | 221 | 2 | 3.05E-05 | 0.0040178 | 2.96E-05 | 5.81E-05 | -0.00278658 | ATAD3A |
| chr10 | 117196619 | 117197587 | 969 | 10 | 3.50E-05 | 0.0001507 | 0.012461 | 5.81E-05 | -0.00071359 | KCNK18, RPL12P26 |
| chr6 | 167290700 | 167291454 | 755 | 5 | 1.92E-05 | 6.72E-05 | 0.000954 | 5.81E-05 | 0.000110528 | UNC93A |
| chr16 | 85028815 | 85029275 | 461 | 2 | 4.81E-05 | 3.52E-05 | 0.001865 | 5.81E-05 | -0.00894823 | KIAA0513 |
| chr4 | 610614 | 610658 | 45 | 2 | 0.000139709 | 3.48E-05 | 0.002004 | 5.81E-05 | -0.01107433 | AC116565.1 |
| chr19 | 55385081 | 55385597 | 517 | 4 | 3.42E-06 | 0.0001102 | 0.00131 | 5.82E-05 | 0.000157124 | NA |
| chr17 | 35141814 | 35142469 | 656 | 10 | 8.36E-05 | 0.0012476 | 0.001132 | 5.82E-05 | -0.00223659 | NLE1 |
| chr8 | 29080916 | 29081091 | 176 | 3 | 9.62E-05 | 0.000364 | 0.000101 | 5.82E-05 | -0.00152639 | KIF13B |
| chr11 | 70148046 | 70148191 | 146 | 2 | 0.000122029 | 0.000139 | 0.000107 | 5.83E-05 | -0.00975544 | ANO1, AP000879.2 |
| chr1 | 37883599 | 37883815 | 217 | 3 | 0.000161213 | 0.0001594 | 0.000713 | 5.83E-05 | -0.02119395 | INPP5B |
| chr1 | 32322736 | 32322890 | 155 | 3 | 0.00012186 | 6.08E-05 | 0.00079 | 5.83E-05 | -0.00480476 | HDAC1 |
| chr1 | 3823486 | 3823556 | 71 | 2 | 0.000117163 | 3.55E-05 | 0.001773 | 5.83E-05 | -0.00757497 | CEP104 |
| chr6 | 32859425 | 32859816 | 392 | 2 | 3.16E-05 | 0.0001881 | 8.58E-05 | 5.83E-05 | -0.01249124 | PSMB9 |
| chr3 | 74267587 | 74267594 | 8 | 2 | 0.0001496 | 0.0057945 | 2.83E-05 | 5.83E-05 | -0.00844008 | CNTN3 |
| chr12 | 10723275 | 10723927 | 653 | 9 | 3.41E-05 | 0.0045114 | 6.37E-05 | 5.83E-05 | 0.000352305 | YBX3 |
| chr12 | 109206841 | 109206880 | 40 | 2 | 7.23E-05 | 0.0022568 | 3.28E-05 | 5.84E-05 | -0.00187409 | ACACB |
| chr1 | 119811536 | 119812149 | 614 | 7 | 8.63E-05 | 0.0004531 | 0.001115 | 5.84E-05 | -0.00476342 | REG4 |
| chr2 | 240587103 | 240587545 | 443 | 3 | 7.51E-06 | 0.002679 | 2.28E-05 | 5.84E-05 | -0.00146294 | CAPN10 |
| chr4 | 55395360 | 55395598 | 239 | 5 | 0.000116235 | 1.69E-05 | 0.007169 | 5.85E-05 | 0.000532052 | SRD5A3-AS1 |
| chr13 | 35471476 | 35471919 | 444 | 3 | 2.51E-06 | 0.0073645 | 5.97E-05 | 5.85E-05 | -0.00626491 | NBEA |
| chr10 | 125823258 | 125824027 | 770 | 12 | 1.16E-05 | 0.0050466 | 0.000496 | 5.85E-05 | 0.000439095 | BCCIP, UROS |
| chr3 | 159273317 | 159273665 | 349 | 3 | 9.37E-05 | 6.64E-05 | 0.000383 | 5.86E-05 | -0.0091169 | IQCJ-SCHIP1 |
| chr8 | 28428096 | 28428440 | 345 | 2 | 4.77E-05 | 0.000172 | 9.18E-05 | 5.86E-05 | -0.00798635 | FBXO16 |
| chr8 | 40885013 | 40885187 | 175 | 4 | 6.20E-05 | 0.0055431 | 0.000595 | 5.86E-05 | -0.00214398 | ZMAT4 |
| chr19 | 10252579 | 10252652 | 74 | 2 | 8.85E-05 | 5.27E-05 | 0.000442 | 5.86E-05 | -0.00120441 | MRPL4, AC011511.2 |
| chr13 | 49539543 | 49539643 | 101 | 3 | 6.24E-05 | 0.0004337 | 0.001211 | 5.86E-05 | 0.013229963 | RCBTB1 |
| chr7 | 80923222 | 80923367 | 146 | 2 | 9.14E-05 | 3.63E-05 | 0.001618 | 5.86E-05 | 0.011477252 | NA |
| chr5 | 61245603 | 61246038 | 436 | 3 | 0.000108958 | 0.0004038 | 0.000451 | 5.86E-05 | -0.00025659 | LINC02057 |
| chr14 | 100285921 | 100286439 | 519 | 4 | 1.92E-05 | 0.0016136 | 3.70E-05 | 5.86E-05 | 0.006299028 | AL157871.6 |
| chr15 | 88614584 | 88615165 | 582 | 3 | 3.48E-05 | 9.71E-05 | 0.000593 | 5.87E-05 | -0.01843944 | NA |
| chr12 | 21941527 | 21942543 | 1017 | 10 | 2.74E-06 | 0.0004289 | 0.000238 | 5.87E-05 | -0.00168365 | ABCC9 |
| chr6 | 157146688 | 157146854 | 167 | 2 | 1.08E-05 | 5.94E-05 | 0.000342 | 5.88E-05 | -0.00071463 | ARID1B |

| | | | | | | | | | | |
|---|---|---|---|---|---|---|---|---|---|---|
| chr18 | 76495482 | 76495595 | 114 | 2 | 7.01E-05 | 5.41E-05 | 0.000419 | 5.88E-05 | -0.00103697 | ZNF516-DT |
| chr9 | 130580012 | 130580108 | 97 | 2 | 0.000111031 | 5.33E-05 | 0.000433 | 5.88E-05 | -0.00026706 | FUBP3 |
| chr1 | 244835125 | 244835839 | 715 | 10 | 3.30E-05 | 0.0004059 | 0.004724 | 5.88E-05 | -0.00026511 | COX20 |
| chr10 | 28761362 | 28761982 | 621 | 3 | 1.46E-05 | 8.25E-05 | 0.000273 | 5.88E-05 | -0.01177521 | LINC01517 |
| chr12 | 53096350 | 53096848 | 499 | 4 | 6.94E-05 | 9.13E-05 | 0.000248 | 5.88E-05 | -0.00612295 | NA |
| chr11 | 72780547 | 72781416 | 870 | 4 | 6.44E-06 | 0.0001127 | 0.000819 | 5.88E-05 | -0.00479381 | ARAP1, STARD10 |
| chr22 | 17983501 | 17983672 | 172 | 2 | 5.91E-05 | 4.65E-05 | 0.000625 | 5.89E-05 | -0.00632134 | MICAL3, AC016027.3 |
| chr9 | 130393260 | 130393438 | 179 | 2 | 0.00016994 | 8.30E-05 | 0.000193 | 5.89E-05 | -0.0038251 | HMCN2 |
| chr14 | 21924397 | 21924601 | 205 | 3 | 2.09E-05 | 0.0037829 | 2.75E-05 | 5.89E-05 | 0.009890219 | TRAV14DV4 |
| chr4 | 985291 | 985543 | 253 | 4 | 4.76E-05 | 0.0012448 | 0.003588 | 5.89E-05 | -0.00089617 | DGKQ, SLC26A1 |
| chr15 | 98647072 | 98647566 | 495 | 4 | 8.83E-05 | 5.13E-05 | 0.001256 | 5.89E-05 | -0.00038675 | IRAIN |
| chr7 | 98948368 | 98948395 | 28 | 2 | 9.26E-05 | 9.54E-05 | 0.000162 | 5.89E-05 | -0.00260256 | TRRAP |
| chr5 | 77844719 | 77844984 | 266 | 2 | 5.11E-05 | 3.99E-05 | 0.001064 | 5.89E-05 | 0.005614347 | TBCA |
| chr22 | 37281793 | 37281871 | 79 | 2 | 4.01E-05 | 3.54E-05 | 0.001991 | 5.90E-05 | -0.0282041 | NA |
| chr1 | 119648366 | 119648743 | 378 | 2 | 1.40E-05 | 0.0006433 | 4.70E-05 | 5.90E-05 | 0.001468477 | PHGDH |
| chr17 | 66458852 | 66458968 | 117 | 2 | 0.000144045 | 5.78E-05 | 0.000367 | 5.90E-05 | -0.00866313 | PRKCA |
| chr17 | 50536685 | 50537075 | 391 | 3 | 1.80E-05 | 0.0001198 | 0.000192 | 5.90E-05 | 0.002672764 | EPN3, AC021491.2 |
| chr9 | 137547402 | 137547805 | 404 | 2 | 0.000132964 | 5.34E-05 | 0.000436 | 5.90E-05 | -0.00544218 | PNPLA7 |
| chr2 | 43225422 | 43225699 | 278 | 2 | 7.14E-07 | 8.09E-05 | 0.000201 | 5.90E-05 | -0.00100314 | ZFP36L2 |
| chr12 | 5741161 | 5741233 | 73 | 2 | 6.45E-05 | 4.40E-05 | 0.000748 | 5.90E-05 | -0.00764369 | ANO2 |
| chr1 | 203007323 | 203007975 | 653 | 9 | 3.67E-06 | 0.0043846 | 0.000302 | 5.91E-05 | -5.45E-05 | TMEM183A |
| chr15 | 52558445 | 52558531 | 87 | 2 | 1.54E-05 | 0.000579 | 4.90E-05 | 5.91E-05 | -0.00883228 | ARPP19 |
| chr15 | 58760733 | 58760811 | 79 | 2 | 0.00011327 | 4.01E-05 | 0.001061 | 5.91E-05 | -0.0015664 | AC090515.6 |
| chr3 | 157152290 | 157152572 | 283 | 2 | 8.52E-06 | 0.0246699 | 2.56E-05 | 5.91E-05 | -0.00508537 | CCNL1 |
| chr1 | 29182999 | 29183513 | 515 | 2 | 1.69E-05 | 3.67E-05 | 0.001609 | 5.91E-05 | -0.00393508 | NA |
| chr3 | 70251770 | 70251981 | 212 | 2 | 8.28E-05 | 0.000344 | 6.18E-05 | 5.91E-05 | 0.014783286 | SAMMSON, MDFIC2 |
| chr14 | 55024291 | 55024385 | 95 | 2 | 6.05E-05 | 0.002347 | 3.30E-05 | 5.92E-05 | -0.00255985 | WDHD1 |
| chr5 | 148312335 | 148312610 | 276 | 4 | 2.59E-05 | 0.0099139 | 0.000174 | 5.92E-05 | -0.00188387 | SPINK7, AC011346.1 |
| chr14 | 103209149 | 103209411 | 263 | 2 | 3.67E-05 | 3.55E-05 | 0.001984 | 5.92E-05 | -0.01550201 | NA |
| chr5 | 43515703 | 43516380 | 678 | 2 | 9.33E-06 | 3.84E-05 | 0.001292 | 5.92E-05 | -0.03168163 | C5orf34-AS1, AC114956.1 |
| chr11 | 35200933 | 35201096 | 164 | 2 | 6.43E-05 | 0.0014493 | 3.67E-05 | 5.92E-05 | -0.00476576 | CD44 |
| chr1 | 3082632 | 3082712 | 81 | 2 | 6.38E-05 | 5.65E-05 | 0.000389 | 5.92E-05 | -0.00795045 | PRDM16 |
| chr7 | 2103287 | 2103872 | 586 | 3 | 4.73E-05 | 2.67E-05 | 0.003933 | 5.93E-05 | -0.00484319 | MAD1L1, AC069288.1 |
| chr2 | 85758485 | 85759063 | 579 | 3 | 1.51E-05 | 0.000313 | 0.000739 | 5.93E-05 | -0.00556524 | ATOH8 |
| chr7 | 6412991 | 6413241 | 251 | 2 | 1.39E-05 | 0.0272686 | 2.55E-05 | 5.93E-05 | -0.00352176 | DAGLB |
| chr5 | 53035762 | 53036042 | 281 | 2 | 8.30E-05 | 3.68E-05 | 0.001635 | 5.93E-05 | -0.0134233 | ITGA2, AC008966.3 |
| chr14 | 23007217 | 23007777 | 561 | 3 | 1.23E-05 | 9.53E-05 | 0.000291 | 5.93E-05 | 0.007837852 | C14orf93 |
| chr11 | 64613504 | 64613813 | 310 | 2 | 3.08E-05 | 0.0009088 | 4.20E-05 | 5.94E-05 | -0.00205256 | NRXN2 |
| chr7 | 158400273 | 158400298 | 26 | 2 | 0.000130056 | 4.24E-05 | 0.00087 | 5.94E-05 | -0.00533316 | PTPRN2 |
| chr6 | 94045800 | 94045821 | 22 | 2 | 0.000126185 | 0.0002259 | 7.83E-05 | 5.94E-05 | -0.00119259 | NA |
| chr22 | 33165985 | 33166188 | 204 | 3 | 6.69E-05 | 0.0002269 | 0.000114 | 5.94E-05 | 0.003328513 | Z82198.2, LARGE1 |
| chr2 | 232419951 | 232420224 | 274 | 2 | 6.00E-05 | 7.14E-05 | 0.000251 | 5.95E-05 | -0.01545776 | NA |
| chr16 | 85921297 | 85921476 | 180 | 2 | 9.65E-05 | 0.0002602 | 7.21E-05 | 5.95E-05 | 0.003549204 | IRF8 |
| chr6 | 136270956 | 136271101 | 146 | 2 | 4.84E-05 | 0.0012193 | 3.87E-05 | 5.95E-05 | 0.005057906 | BCLAF1 |
| chr16 | 89733526 | 89733951 | 426 | 3 | 1.78E-05 | 0.0001693 | 0.000197 | 5.95E-05 | -0.0022423 | ZNF276 |
| chr19 | 49818038 | 49818466 | 429 | 9 | 8.07E-05 | 0.0001629 | 0.000126 | 5.95E-05 | 0.000451965 | MED25 |
| chr9 | 114806184 | 114806495 | 312 | 3 | 8.70E-05 | 0.0038161 | 0.000888 | 5.96E-05 | -0.00351141 | NA |
| chr1 | 15362660 | 15362711 | 52 | 2 | 0.000133915 | 5.68E-05 | 0.000394 | 5.97E-05 | 0.001453709 | FHAD1 |
| chr11 | 76659466 | 76659752 | 287 | 3 | 0.000121642 | 0.0001782 | 0.00149 | 5.98E-05 | -0.00616712 | AP001189.1, LRRC32 |
| chr2 | 38469538 | 38469882 | 345 | 4 | 7.70E-05 | 0.0009386 | 0.001306 | 5.99E-05 | -0.00574001 | LINC02613 |
| chr12 | 121391850 | 121392151 | 302 | 3 | 5.79E-05 | 0.00039 | 0.000141 | 5.99E-05 | 0.013824754 | AC048337.1, ANAPC5 |
| chr3 | 99394947 | 99395410 | 464 | 2 | 5.72E-05 | 6.48E-05 | 0.000304 | 5.99E-05 | -0.00628497 | NA |
| chr15 | 63599431 | 63599861 | 431 | 2 | 1.02E-05 | 0.000761 | 4.52E-05 | 6.00E-05 | -0.00782814 | FBXL22, USP3-AS1 |
| chr19 | 3671905 | 3672230 | 326 | 3 | 2.98E-05 | 7.73E-05 | 0.001233 | 6.00E-05 | 0.001174967 | PIP5K1C |
| chr18 | 3874883 | 3875619 | 737 | 6 | 8.11E-05 | 0.0001482 | 0.007038 | 6.01E-05 | -0.00451518 | DLGAP1 |
| chr2 | 109501507 | 109501646 | 140 | 3 | 8.21E-05 | 0.0001403 | 0.000152 | 6.01E-05 | 0.002690857 | SH3RF3 |
| chr8 | 93234912 | 93235287 | 376 | 3 | 6.22E-05 | 0.0001289 | 0.000205 | 6.01E-05 | -0.00501303 | LINC00535 |
| chr7 | 44848091 | 44848455 | 365 | 10 | 0.000103054 | 0.0004771 | 0.001086 | 6.01E-05 | -0.00049064 | LINC01952 |
| chr11 | 78198887 | 78198921 | 35 | 3 | 0.000152414 | 3.85E-05 | 0.003127 | 6.02E-05 | 0.004320146 | USP35 |
| chr15 | 74443138 | 74443213 | 76 | 2 | 3.90E-05 | 0.0214028 | 2.61E-05 | 6.02E-05 | -0.00602082 | NA |
| chr4 | 68349587 | 68350393 | 807 | 16 | 2.21E-05 | 0.0011107 | 0.004152 | 6.02E-05 | 0.001493305 | YTHDC1 |
| chr13 | 78355814 | 78355903 | 90 | 2 | 0.000140186 | 4.64E-05 | 0.000686 | 6.03E-05 | -0.01402205 | OBI1-AS1 |
| chr17 | 76264611 | 76265055 | 445 | 9 | 6.47E-06 | 0.0019485 | 2.57E-05 | 6.03E-05 | -0.00352782 | NA |
| chr1 | 222547580 | 222548167 | 588 | 5 | 9.25E-06 | 0.0003365 | 0.003991 | 6.03E-05 | -0.0076271 | HHIPL2 |
| chr2 | 218992509 | 218993205 | 697 | 3 | 5.88E-05 | 9.65E-05 | 0.001786 | 6.03E-05 | 0.003750906 | CRYBA2 |
| chr17 | 6852609 | 6852951 | 343 | 2 | 0.000117519 | 3.64E-05 | 0.001945 | 6.03E-05 | 0.007046367 | NA |
| chr9 | 38424434 | 38424720 | 287 | 5 | 2.53E-05 | 0.0002078 | 0.00017 | 6.03E-05 | -0.00075186 | IGFBPL1 |
| chr19 | 1051080 | 1051161 | 82 | 3 | 8.07E-05 | 0.0005297 | 7.45E-05 | 6.03E-05 | -0.00329595 | ABCA7 |
| chr1 | 236551749 | 236551769 | 21 | 2 | 7.35E-05 | 0.0002363 | 7.78E-05 | 6.04E-05 | -0.0088808 | LGALS8, HEATR1 |
| chr7 | 149791125 | 149791865 | 741 | 2 | 1.53E-05 | 0.0001029 | 0.000155 | 6.04E-05 | -0.00284476 | SSPOP |
| chr9 | 114378930 | 114379055 | 126 | 2 | 4.84E-05 | 5.06E-05 | 0.000539 | 6.04E-05 | 0.002795049 | AKNA |
| chr5 | 157778244 | 157778251 | 8 | 2 | 7.69E-05 | 0.0022534 | 3.40E-05 | 6.04E-05 | -0.00954938 | NA |
| chr22 | 37659267 | 37659431 | 165 | 2 | 6.27E-06 | 0.000106 | 0.00015 | 6.04E-05 | 0.002820724 | Z83844.2, PDXP |
| chr1 | 167914060 | 167914844 | 785 | 6 | 2.80E-06 | 0.0158298 | 5.66E-06 | 6.04E-05 | 0.001794145 | ADCY10 |
| chr16 | 711108 | 711307 | 200 | 2 | 8.08E-05 | 5.16E-05 | 0.000514 | 6.05E-05 | -0.00500469 | Z97653.1 |
| chr8 | 10058683 | 10058715 | 33 | 2 | 0.000123038 | 0.0002708 | 7.20E-05 | 6.05E-05 | 0.002736506 | MSRA |
| chr9 | 131027079 | 131027749 | 671 | 5 | 1.15E-05 | 0.0034473 | 0.000349 | 6.05E-05 | -0.00135172 | LAMC3 |
| chr11 | 124758247 | 124759010 | 764 | 4 | 2.63E-05 | 0.0026437 | 0.000341 | 6.06E-05 | -0.00527139 | ESAM |
| chr3 | 52536232 | 52536800 | 569 | 7 | 3.75E-05 | 0.0006898 | 0.001063 | 6.06E-05 | -0.0004324 | SMIM4 |
| chr8 | 22574713 | 22574872 | 160 | 2 | 0.000111389 | 0.0002082 | 8.49E-05 | 6.07E-05 | -0.01130605 | SORBS3 |
| chr7 | 128879923 | 128880139 | 217 | 2 | 0.000135841 | 3.90E-05 | 0.001381 | 6.07E-05 | -0.02382364 | KCP |
| chr4 | 158801282 | 158801373 | 92 | 2 | 0.000126175 | 0.0002057 | 8.57E-05 | 6.07E-05 | 0.001811926 | FNIP2 |
| chr16 | 30051634 | 30051827 | 194 | 3 | 1.38E-05 | 0.000119 | 0.000234 | 6.08E-05 | -0.00457765 | TLCD3B |
| chr17 | 3514370 | 3514739 | 370 | 2 | 1.09E-05 | 0.0007967 | 4.52E-05 | 6.08E-05 | -0.00402065 | TRPV3 |
| chr21 | 39538872 | 39538896 | 25 | 2 | 7.05E-05 | 0.0048842 | 3.00E-05 | 6.09E-05 | -0.00394858 | NA |
| chr7 | 30164898 | 30165062 | 165 | 2 | 3.95E-05 | 0.0001998 | 8.77E-05 | 6.09E-05 | 0.002287514 | NA |
| chr16 | 49780684 | 49780705 | 22 | 3 | 4.26E-05 | 0.0001243 | 0.000495 | 6.09E-05 | -0.00348328 | ZNF423 |
| chr7 | 52325093 | 52325348 | 256 | 2 | 4.81E-05 | 0.0009699 | 4.23E-05 | 6.09E-05 | -0.01115426 | NA |
| chr4 | 184017138 | 184017317 | 180 | 4 | 9.11E-05 | 0.0017226 | 2.99E-05 | 6.09E-05 | -0.00277646 | STOX2 |
| chr17 | 35119918 | 35120841 | 924 | 12 | 7.09E-06 | 0.0073182 | 8.55E-06 | 6.09E-05 | -0.00197177 | AC004223.3, RAD51D |
| chr16 | 2462502 | 2462726 | 225 | 2 | 3.12E-05 | 0.0220048 | 2.64E-05 | 6.10E-05 | -0.00285479 | TEDC2 |
| chr17 | 8210159 | 8210714 | 556 | 11 | 6.07E-05 | 0.0021032 | 0.002321 | 6.10E-05 | -0.00017133 | AURKB |
| chr1 | 17424963 | 17425527 | 565 | 8 | 9.71E-05 | 0.0046531 | 0.00947 | 6.10E-05 | -0.00260878 | RCC2 |
| chr2 | 227633176 | 227633313 | 138 | 2 | 7.06E-05 | 6.02E-05 | 0.000369 | 6.10E-05 | -0.00951734 | C2orf83 |
| chr11 | 45921686 | 45922540 | 855 | 6 | 1.34E-05 | 4.91E-05 | 0.012622 | 6.10E-05 | -0.00602817 | LARGE2 |
| chr6 | 105994713 | 105995306 | 594 | 3 | 6.59E-06 | 0.0008501 | 0.000504 | 6.10E-05 | 0.002939732 | PRDM1 |
| chr3 | 81180892 | 81180927 | 36 | 2 | 0.000124068 | 0.0002744 | 7.23E-05 | 6.11E-05 | -0.01167832 | NA |
| chr18 | 46418863 | 46418930 | 68 | 2 | 0.000136997 | 0.000173 | 9.71E-05 | 6.11E-05 | -0.00654516 | RNF165 |
| chr22 | 36439123 | 36440009 | 887 | 3 | 2.02E-05 | 3.86E-05 | 0.003246 | 6.11E-05 | -0.00824399 | NA |
| chr10 | 119325483 | 119325717 | 235 | 4 | 6.34E-05 | 0.0012369 | 8.73E-05 | 6.11E-05 | -0.00046751 | GRK5 |
| chr1 | 63079718 | 63080018 | 301 | 2 | 1.20E-05 | 5.23E-05 | 0.000514 | 6.11E-05 | -0.00689995 | NA |
| chr16 | 84184591 | 84185333 | 743 | 6 | 1.41E-05 | 0.0178 | 0.000278 | 6.12E-05 | -0.00265378 | TAF1C |
| chr13 | 114278904 | 114279202 | 299 | 2 | 3.37E-05 | 0.0026058 | 3.34E-05 | 6.12E-05 | 0.00183326 | NA |
| chr3 | 183904312 | 183904477 | 166 | 5 | 0.000110291 | 6.76E-05 | 0.002106 | 6.12E-05 | -0.00641235 | CYP2AB1P |
| chr11 | 34309510 | 34310361 | 852 | 3 | 6.99E-06 | 0.0123798 | 2.31E-05 | 6.12E-05 | 0.007568171 | ABTB2 |
| chr3 | 160676738 | 160676952 | 215 | 2 | 6.66E-05 | 8.48E-05 | 0.000204 | 6.12E-05 | -0.01916699 | NA |
| chr11 | 2166191 | 2167014 | 824 | 5 | 7.52E-06 | 0.0039217 | 0.000346 | 6.13E-05 | 0.011067373 | TH |
| chr22 | 42640190 | 42640728 | 539 | 5 | 4.80E-05 | 0.0036455 | 0.000333 | 6.13E-05 | 0.005262253 | CYB5R3, ATP5MGL |
| chr11 | 45209624 | 45209994 | 371 | 3 | 7.95E-05 | 3.26E-05 | 0.003685 | 6.13E-05 | -0.00326364 | PRDM11 |
| chr17 | 9825655 | 9826107 | 453 | 9 | 4.25E-05 | 0.0009818 | 0.000802 | 6.13E-05 | -0.00665466 | GLP2R |
| chr7 | 22478913 | 22479394 | 482 | 4 | 2.04E-05 | 0.002884 | 6.74E-05 | 6.14E-05 | -0.00323651 | STEAP1B |
| chr17 | 60663448 | 60663617 | 170 | 2 | 0.000154036 | 6.53E-05 | 0.000319 | 6.14E-05 | -0.00599038 | PPM1D |
| chr2 | 219559537 | 219559559 | 23 | 2 | 8.65E-05 | 0.000119 | 0.000137 | 6.14E-05 | -0.01045339 | AC009955.4, OBSL1 |
| chr8 | 39912527 | 39913006 | 480 | 4 | 9.81E-05 | 0.0001634 | 0.001049 | 6.14E-05 | -0.0105336 | IDO1, AC007991.3 |
| chr10 | 132133170 | 132133377 | 208 | 3 | 2.09E-05 | 5.64E-05 | 0.001576 | 6.14E-05 | -0.01318879 | JAKMIP3 |
| chr12 | 13212209 | 13212523 | 315 | 3 | 0.000120949 | 9.51E-05 | 0.000274 | 6.15E-05 | -0.01539729 | EMP1 |
| chr5 | 42824943 | 42824955 | 13 | 2 | 7.02E-05 | 0.00443 | 3.08E-05 | 6.15E-05 | -0.00326478 | AC008945.2, SELENOP |
| chr3 | 62367637 | 62367829 | 193 | 2 | 2.75E-05 | 0.0083321 | 2.86E-05 | 6.15E-05 | -0.0069521 | PTPRG-AS1 |
| chr2 | 58046285 | 58046548 | 264 | 10 | 8.28E-05 | 0.0001998 | 0.002426 | 6.15E-05 | 0.00018494 | VRK2, AC068193.1 |
| chr10 | 1970921 | 1971074 | 154 | 2 | 7.27E-05 | 0.0001096 | 0.00015 | 6.15E-05 | 0.006305278 | NA |
| chr16 | 56194276 | 56194665 | 390 | 9 | 0.000137074 | 4.47E-05 | 0.007987 | 6.16E-05 | -0.00032146 | GNAO1, DKFZP434H168 |
| chr1 | 1891533 | 1891940 | 408 | 3 | 1.37E-06 | 0.0131645 | 1.33E-05 | 6.17E-05 | 0.008777579 | AL109917.1 |
| chr1 | 211258225 | 211258797 | 573 | 5 | 4.53E-06 | 0.0020723 | 0.00011 | 6.17E-05 | 0.002522952 | RCOR3 |
| chr10 | 3414926 | 3415249 | 324 | 3 | 5.61E-05 | 0.0037336 | 5.70E-05 | 6.17E-05 | 0.002636007 | NA |
| chr7 | 27868109 | 27868223 | 115 | 4 | 0.000125668 | 0.0002117 | 0.000207 | 6.17E-05 | -0.00089534 | JAZF1 |
| chr5 | 150318939 | 150319159 | 221 | 2 | 1.80E-05 | 0.0105898 | 2.80E-05 | 6.17E-05 | -0.00633423 | ARSI |
| chr19 | 1467009 | 1467033 | 25 | 3 | 2.68E-05 | 3.25E-05 | 0.003899 | 6.18E-05 | -0.0087025 | APC2, C19orf25 |
| chr16 | 81697644 | 81698433 | 790 | 3 | 1.20E-05 | 0.0001164 | 0.000196 | 6.18E-05 | -0.00307204 | CMIP |
| chr1 | 230279439 | 230280156 | 718 | 7 | 4.08E-05 | 0.0001428 | 0.005154 | 6.18E-05 | 0.008905595 | GALNT2 |
| chr14 | 102394811 | 102395191 | 381 | 3 | 6.21E-05 | 0.0004555 | 6.40E-05 | 6.18E-05 | -0.00549164 | TECPR2 |
| chr7 | 155581408 | 155581598 | 191 | 2 | 5.18E-05 | 0.0036488 | 3.19E-05 | 6.18E-05 | -0.00587963 | AC009403.2 |
| chr17 | 31563791 | 31564001 | 211 | 4 | 8.42E-05 | 0.0001419 | 0.000561 | 6.18E-05 | -0.00341259 | RNU6ATAC7P |
| chr2 | 178453127 | 178453395 | 269 | 3 | 3.28E-05 | 0.0008233 | 0.000147 | 6.18E-05 | -0.0045969 | PJVK |
| chr11 | 92883952 | 92884490 | 539 | 2 | 2.23E-05 | 5.62E-05 | 0.000448 | 6.19E-05 | -0.00075269 | FAT3 |
| chr11 | 69477266 | 69477906 | 641 | 3 | 6.00E-06 | 0.001257 | 0.000608 | 6.19E-05 | -0.00519853 | LINC02747 |
| chr11 | 77196438 | 77197113 | 676 | 2 | 3.41E-05 | 3.98E-05 | 0.001396 | 6.19E-05 | 0.006272864 | MYO7A |
| chr5 | 53672177 | 53672395 | 219 | 2 | 4.29E-05 | 0.0006278 | 5.01E-05 | 6.19E-05 | -0.00180074 | NDUFS4 |
| chr19 | 45363795 | 45364454 | 660 | 6 | 3.82E-05 | 1.77E-05 | 0.015805 | 6.19E-05 | -0.00283 | ERCC2 |
| chr5 | 43191585 | 43192326 | 742 | 9 | 4.00E-05 | 7.95E-05 | 0.004829 | 6.19E-05 | 0.000475623 | ZNF131, NIM1K |
| chr18 | 26470141 | 26470666 | 526 | 7 | 6.48E-05 | 0.0007121 | 5.79E-05 | 6.20E-05 | -0.00199153 | KCTD1 |
| chr14 | 65089963 | 65090124 | 162 | 2 | 0.000125625 | 0.000108 | 0.000155 | 6.20E-05 | 0.002399512 | AL139022.1, MAX |
| chr10 | 5502155 | 5502419 | 265 | 3 | 8.47E-05 | 0.0004867 | 0.000116 | 6.21E-05 | -0.00813249 | NA |
| chr2 | 127643965 | 127644840 | 876 | 5 | 3.78E-05 | 7.67E-05 | 0.000835 | 6.21E-05 | -0.00347402 | LIMS2 |
| chr16 | 71442986 | 71443149 | 164 | 2 | 4.01E-05 | 0.0038767 | 3.17E-05 | 6.21E-05 | -0.00388424 | AC010547.5 |
| chr17 | 8142297 | 8142299 | 3 | 2 | 0.000178483 | 0.0035382 | 3.22E-05 | 6.21E-05 | -0.00424588 | PER1 |
| chr18 | 9855610 | 9855836 | 227 | 2 | 4.67E-05 | 0.0034971 | 3.22E-05 | 6.22E-05 | -0.006338 | RAB31 |
| chr16 | 84186774 | 84187088 | 315 | 4 | 3.21E-05 | 0.000127 | 0.000161 | 6.22E-05 | -0.00149307 | TAF1C |
| chr16 | 68211811 | 68211891 | 81 | 3 | 0.000118476 | 0.0004144 | 9.70E-05 | 6.22E-05 | -0.00090422 | NFATC3 |
| chr1 | 150282028 | 150282118 | 91 | 3 | 0.000127458 | 0.0004919 | 0.000374 | 6.22E-05 | -0.00022114 | NA |
| chr8 | 47610487 | 47610936 | 450 | 2 | 6.79E-05 | 4.23E-05 | 0.001095 | 6.23E-05 | 0.006104984 | SPIDR |
| chr2 | 239026736 | 239027033 | 298 | 3 | 4.32E-05 | 0.0014592 | 3.88E-05 | 6.23E-05 | -0.00269303 | NA |
| chr10 | 87792812 | 87792927 | 116 | 2 | 5.39E-05 | 0.0024756 | 3.44E-05 | 6.23E-05 | 0.007011098 | ATAD1 |
| chr3 | 13792382 | 13792455 | 74 | 2 | 3.99E-05 | 4.65E-05 | 0.000776 | 6.23E-05 | -0.00341654 | NA |
| chr7 | 139722344 | 139722697 | 354 | 3 | 1.76E-05 | 0.0137473 | 1.72E-05 | 6.23E-05 | -0.00872079 | HIPK2 |
| chr10 | 650123 | 650416 | 294 | 4 | 1.04E-05 | 0.0020731 | 0.000777 | 6.24E-05 | -0.00082069 | PRR26, DIP2C |
| chr3 | 12759303 | 12759634 | 332 | 6 | 1.31E-05 | 0.0065685 | 0.000112 | 6.24E-05 | -0.00230281 | TMEM40 |
| chr7 | 158457019 | 158457777 | 759 | 4 | 2.08E-05 | 0.000939 | 4.30E-05 | 6.25E-05 | -0.00601796 | PTPRN2 |
| chr20 | 2871646 | 2871861 | 216 | 2 | 0.000111402 | 5.28E-05 | 0.000539 | 6.25E-05 | 0.007144573 | PTPRA |

| | | | | | | | | | | |
|---|---|---|---|---|---|---|---|---|---|---|
| chr12 | 1864956 | 1865648 | 693 | 6 | 1.09E-05 | 0.0038603 | 6.81E-05 | 6.25E-05 | -0.00811835 | CACNA2D4 |
| chr10 | 95390302 | 95390439 | 138 | 2 | 8.06E-05 | 6.71E-05 | 0.000318 | 6.26E-05 | 0.000610927 | SORBS1 |
| chr17 | 29566389 | 29566713 | 325 | 3 | 8.45E-05 | 0.0001759 | 0.000119 | 6.26E-05 | -0.00129172 | ABHD15-AS1, TP53I13, ABHD15 |
| chr5 | 73450516 | 73451372 | 857 | 9 | 2.73E-05 | 0.0046647 | 9.24E-05 | 6.26E-05 | 0.002678011 | NA |
| chr2 | 197675794 | 197676010 | 217 | 4 | 9.65E-05 | 3.80E-05 | 0.007516 | 6.26E-05 | -0.01994716 | RFTN2 |
| chr1 | 209585800 | 209585999 | 200 | 2 | 0.000106598 | 6.70E-05 | 0.000321 | 6.27E-05 | -0.00770737 | CAMK1G |
| chr1 | 26529435 | 26529929 | 495 | 3 | 2.47E-05 | 0.0003921 | 0.001088 | 6.27E-05 | -0.00331585 | RPS6KA1 |
| chr17 | 79797817 | 79797855 | 39 | 2 | 2.32E-05 | 6.00E-05 | 0.000404 | 6.27E-05 | -0.00388826 | CBX8 |
| chr1 | 119963634 | 119963949 | 316 | 3 | 9.62E-05 | 0.0002363 | 9.12E-05 | 6.27E-05 | 0.002677421 | NOTCH2 |
| chr11 | 58269525 | 58269703 | 179 | 3 | 0.000116011 | 0.0019824 | 0.000404 | 6.28E-05 | -0.01099534 | NA |
| chr11 | 45709762 | 45709992 | 231 | 4 | 0.000135069 | 7.89E-05 | 0.002438 | 6.28E-05 | 0.00636777 | NA |
| chr21 | 45319381 | 45319696 | 316 | 2 | 4.96E-05 | 3.87E-05 | 0.001798 | 6.30E-05 | 0.006312894 | NA |
| chr17 | 81912441 | 81912886 | 446 | 2 | 4.06E-06 | 7.78E-05 | 0.000249 | 6.30E-05 | -0.00279963 | SIRT7 |
| chr17 | 78186270 | 78186385 | 116 | 3 | 1.72E-05 | 2.46E-05 | 0.006731 | 6.30E-05 | -0.01289025 | TK1 |
| chr20 | 62914950 | 62915055 | 106 | 2 | 2.76E-05 | 5.32E-05 | 0.000543 | 6.30E-05 | 0.010427187 | DIDO1 |
| chr6 | 32828770 | 32828962 | 193 | 3 | 7.59E-05 | 0.0009169 | 0.000879 | 6.30E-05 | -0.00411293 | AL669918.1, TAP2 |
| chr16 | 17064811 | 17064953 | 143 | 2 | 0.00010201 | 0.0003966 | 6.25E-05 | 6.30E-05 | -0.00919357 | NA |
| chr18 | 10251331 | 10251343 | 13 | 2 | 0.000167306 | 0.0001004 | 0.000175 | 6.30E-05 | 0.002678338 | NA |
| chr9 | 97632214 | 97632372 | 159 | 2 | 2.27E-05 | 9.59E-05 | 0.000185 | 6.31E-05 | -0.00941463 | TSTD2 |
| chr11 | 118177497 | 118177831 | 335 | 4 | 7.12E-05 | 0.0049464 | 0.000531 | 6.31E-05 | -0.00558076 | NA |
| chr17 | 1498111 | 1498262 | 152 | 2 | 6.20E-05 | 3.88E-05 | 0.001795 | 6.31E-05 | -0.02101528 | INPP5K |
| chr9 | 122375101 | 122375315 | 215 | 4 | 7.01E-05 | 0.0003752 | 0.00011 | 6.32E-05 | -0.00587439 | PTGS1 |
| chr19 | 2425478 | 2425892 | 415 | 3 | 6.11E-05 | 0.000201 | 0.000184 | 6.32E-05 | -0.01122333 | TMPRSS9, TIMM13 |
| chr1 | 89121074 | 89121172 | 99 | 2 | 0.000121025 | 0.0002704 | 7.65E-05 | 6.32E-05 | -0.01064417 | GBP2 |
| chr9 | 131470926 | 131471030 | 105 | 2 | 7.34E-05 | 0.0015976 | 3.84E-05 | 6.33E-05 | -0.00339809 | PRRC2B |
| chr13 | 112928722 | 112929793 | 1072 | 6 | 1.89E-06 | 0.0001525 | 0.00051 | 6.33E-05 | 0.001067953 | MCF2L |
| chr4 | 26356915 | 26356930 | 16 | 2 | 0.000171005 | 6.53E-05 | 0.000346 | 6.33E-05 | -0.00411663 | RBPJ |
| chr7 | 5497306 | 5497495 | 190 | 3 | 4.09E-05 | 2.68E-05 | 0.005337 | 6.33E-05 | -0.01256985 | FBXL18 |
| chr10 | 132741617 | 132742217 | 601 | 3 | 4.72E-06 | 0.0088451 | 2.34E-05 | 6.33E-05 | 0.001076256 | INPP5A |
| chr7 | 5424867 | 5425244 | 378 | 3 | 7.30E-06 | 0.0003094 | 0.001186 | 6.33E-05 | 0.000391129 | TNRC18 |
| chr3 | 195909233 | 195909543 | 311 | 3 | 6.07E-05 | 0.0002942 | 0.000199 | 6.34E-05 | -0.00379867 | TNK2-AS1, TNK2 |
| chr2 | 10063692 | 10063890 | 199 | 2 | 5.74E-05 | 0.0022793 | 3.55E-05 | 6.34E-05 | -0.00415326 | CYS1 |
| chr17 | 63523408 | 63523596 | 189 | 2 | 3.17E-05 | 6.04E-05 | 0.00041 | 6.34E-05 | -0.01036122 | KCNH6 |
| chr10 | 70694219 | 70694270 | 52 | 3 | 0.000113971 | 0.0003624 | 0.000792 | 6.34E-05 | -0.00350958 | ADAMTS14 |
| chr3 | 69762494 | 69763647 | 1154 | 6 | 2.45E-06 | 0.0014563 | 9.84E-05 | 6.34E-05 | 0.002783521 | MITF |
| chr11 | 62538426 | 62538669 | 244 | 2 | 0.000139003 | 4.38E-05 | 0.001036 | 6.35E-05 | -0.00647385 | AP001363.1, AHNAK |
| chr7 | 152462488 | 152462732 | 245 | 2 | 3.66E-05 | 0.0002453 | 8.17E-05 | 6.35E-05 | -0.00222099 | NA |
| chr8 | 37069425 | 37069559 | 135 | 2 | 4.08E-05 | 0.0164613 | 2.79E-05 | 6.35E-05 | -0.01107669 | NA |
| chr11 | 118883571 | 118883884 | 314 | 8 | 2.19E-05 | 0.0093111 | 0.003466 | 6.36E-05 | -0.00315864 | NA |
| chr22 | 20642468 | 20642597 | 130 | 2 | 0.000136867 | 0.0001604 | 0.00011 | 6.36E-05 | -0.00082485 | SMPD4P1 |
| chr17 | 76505476 | 76505668 | 193 | 2 | 9.35E-05 | 4.13E-05 | 0.001361 | 6.36E-05 | -0.00277388 | NA |
| chr6 | 146543050 | 146543749 | 700 | 13 | 6.78E-05 | 0.000125 | 0.010201 | 6.36E-05 | 0.000587084 | NA |
| chr1 | 10785804 | 10786035 | 232 | 3 | 3.64E-05 | 0.0018068 | 2.74E-05 | 6.37E-05 | -0.00298581 | CASZ1 |
| chr2 | 234203172 | 234203356 | 185 | 3 | 0.000106558 | 3.37E-05 | 0.003599 | 6.37E-05 | -0.01941646 | NA |
| chr3 | 134485259 | 134486333 | 1075 | 21 | 1.39E-05 | 0.0040735 | 0.006916 | 6.39E-05 | 0.000912108 | CEP63, ANAPC13 |
| chr13 | 110906279 | 110906298 | 20 | 2 | 0.000132383 | 0.0002345 | 8.46E-05 | 6.39E-05 | -0.00947147 | ANKRD10 |
| chr14 | 36973392 | 36973532 | 141 | 2 | 9.85E-05 | 0.0004955 | 5.75E-05 | 6.39E-05 | -0.00844687 | SLC25A21 |
| chr16 | 10944451 | 10945028 | 578 | 5 | 8.30E-05 | 0.000173 | 0.002953 | 6.39E-05 | -0.00178058 | CLEC16A |
| chr2 | 48340854 | 48340979 | 126 | 2 | 3.35E-05 | 0.0003387 | 6.87E-05 | 6.40E-05 | -0.00228488 | FOXN2 |
| chr10 | 73247150 | 73247497 | 348 | 8 | 6.97E-05 | 0.0005236 | 0.000124 | 6.40E-05 | 0.000731895 | DNAJC9-AS1, DNAJC9, AC016394.2 |
| chr1 | 48182767 | 48182920 | 154 | 2 | 1.78E-05 | 0.0057647 | 3.09E-05 | 6.40E-05 | -0.00442467 | AL109659.1 |
| chr19 | 47741062 | 47741262 | 201 | 3 | 0.000108594 | 0.000967 | 0.000585 | 6.40E-05 | 0.001229467 | EHD2 |
| chr17 | 44664725 | 44664855 | 131 | 2 | 9.77E-05 | 0.0005644 | 5.45E-05 | 6.40E-05 | -0.01018643 | MEIOC |
| chr3 | 128576812 | 128577215 | 404 | 2 | 2.77E-06 | 0.0039193 | 3.26E-05 | 6.41E-05 | -0.00479164 | NA |
| chr12 | 119513076 | 119513625 | 550 | 3 | 6.94E-05 | 0.0002517 | 0.000126 | 6.41E-05 | 0.001540105 | CCDC60, AC002070.1 |
| chr13 | 26873060 | 26873159 | 100 | 3 | 0.000151087 | 0.0001833 | 0.000162 | 6.41E-05 | 0.010387215 | NA |
| chr1 | 2876072 | 2876179 | 108 | 2 | 6.42E-05 | 0.0001562 | 0.000114 | 6.42E-05 | 0.011734981 | NA |
| chr2 | 151507796 | 151508074 | 279 | 2 | 9.17E-06 | 0.0151592 | 2.83E-05 | 6.42E-05 | 0.011803229 | RIF1, NEB |
| chr3 | 169765107 | 169765507 | 401 | 8 | 5.05E-05 | 0.0010209 | 0.000279 | 6.42E-05 | -0.00465564 | NA |
| chr9 | 127186449 | 127186650 | 202 | 2 | 2.61E-05 | 0.0005085 | 5.73E-05 | 6.43E-05 | -0.00344745 | RALGPS1 |
| chr14 | 68979483 | 68979968 | 486 | 8 | 8.71E-05 | 0.0002897 | 0.017134 | 6.43E-05 | 0.001567043 | ACTN1-AS1 |
| chr12 | 105327320 | 105327440 | 121 | 2 | 0.000141584 | 7.95E-05 | 0.000253 | 6.44E-05 | -0.00356916 | C12orf75 |
| chr17 | 57044526 | 57044603 | 78 | 2 | 2.58E-05 | 5.06E-05 | 0.000674 | 6.44E-05 | 0.009198029 | NA |
| chr10 | 114239949 | 114240157 | 209 | 2 | 4.12E-05 | 4.38E-05 | 0.001104 | 6.44E-05 | -0.02210097 | VWA2 |
| chr6 | 19753398 | 19753996 | 599 | 2 | 0.00010678 | 3.85E-05 | 0.002161 | 6.44E-05 | -0.01443731 | LNC-LBCS |
| chr1 | 113072514 | 113073022 | 509 | 7 | 3.51E-05 | 0.0003444 | 0.001718 | 6.44E-05 | 0.000234136 | LRIG2-DT |
| chr10 | 7576511 | 7576849 | 339 | 3 | 7.01E-05 | 0.0003661 | 0.000715 | 6.44E-05 | -0.00188949 | ITIH5 |
| chr22 | 17221202 | 17222018 | 817 | 7 | 2.47E-05 | 0.0006519 | 0.003364 | 6.45E-05 | 0.005716263 | ADA2 |
| chr20 | 33299046 | 33299062 | 17 | 2 | 0.000169289 | 0.0002244 | 8.82E-05 | 6.45E-05 | -0.00580658 | BPIFB1 |
| chr1 | 30929243 | 30929395 | 153 | 2 | 0.000106489 | 6.33E-05 | 0.000389 | 6.45E-05 | -0.0015229 | NA |
| chr7 | 151807758 | 151808030 | 273 | 3 | 6.23E-05 | 0.0002289 | 0.000502 | 6.45E-05 | -0.01190528 | AC093583.1, PRKAG2 |
| chr17 | 36722299 | 36722750 | 452 | 5 | 5.82E-05 | 0.0005733 | 0.000253 | 6.45E-05 | 0.000925276 | AC243830.1 |
| chr12 | 199556 | 199986 | 431 | 4 | 7.81E-05 | 0.0007715 | 0.000887 | 6.46E-05 | 0.000735748 | SLC6A12 |
| chr15 | 75625822 | 75626772 | 951 | 15 | 7.62E-05 | 0.0010547 | 0.003864 | 6.46E-05 | 0.001028442 | SNUPN |
| chr20 | 1492765 | 1493077 | 313 | 3 | 3.72E-05 | 0.0004063 | 0.000669 | 6.46E-05 | -0.00211338 | NA |
| chr22 | 50245066 | 50245290 | 225 | 6 | 5.99E-05 | 0.0143153 | 8.35E-05 | 6.46E-05 | 0.000524659 | HDAC10 |
| chr16 | 67232137 | 67232178 | 42 | 3 | 0.000153717 | 7.56E-05 | 0.000367 | 6.46E-05 | -0.00403296 | FHOD1 |
| chr1 | 59816478 | 59816728 | 251 | 2 | 5.34E-05 | 0.0008566 | 4.71E-05 | 6.47E-05 | -0.00532466 | HOOK1 |
| chr2 | 96858982 | 96859272 | 291 | 4 | 1.64E-05 | 3.58E-05 | 0.004585 | 6.47E-05 | 0.005797095 | NA |
| chr5 | 1950587 | 1950728 | 142 | 3 | 2.84E-05 | 0.0018272 | 3.35E-05 | 6.47E-05 | -0.00150421 | AC124852.1 |
| chr19 | 57871181 | 57871679 | 499 | 3 | 1.75E-05 | 0.0197314 | 4.93E-05 | 6.47E-05 | 0.002144598 | ZNF814 |
| chr15 | 72473943 | 72474399 | 457 | 8 | 2.36E-05 | 3.01E-05 | 0.008956 | 6.47E-05 | -0.00044014 | ARIH1, TMEM202-AS1 |
| chr8 | 123416263 | 123416742 | 480 | 10 | 0.000113534 | 0.0004819 | 0.000808 | 6.47E-05 | 0.001359914 | NTAQ1, ATAD2 |
| chr16 | 5616304 | 5616498 | 195 | 4 | 8.62E-05 | 0.0032222 | 0.000397 | 6.47E-05 | -0.00112213 | RBFOX1, LINC01570 |
| chr16 | 84054209 | 84055072 | 864 | 4 | 1.36E-05 | 0.0008973 | 4.01E-05 | 6.47E-05 | -0.00225943 | MBTPS1 |
| chr10 | 5906324 | 5906545 | 222 | 4 | 9.81E-05 | 0.001758 | 0.00018 | 6.48E-05 | -0.00330646 | FBH1 |
| chr8 | 1195761 | 1195806 | 46 | 2 | 9.88E-05 | 0.0005404 | 5.64E-05 | 6.48E-05 | -0.0037195 | DLGAP2 |
| chr7 | 1097527 | 1097738 | 212 | 3 | 9.83E-05 | 0.0005497 | 5.58E-05 | 6.48E-05 | -0.0006484 | C7orf50 |
| chr3 | 195547852 | 195548117 | 266 | 2 | 2.48E-05 | 0.0035371 | 3.35E-05 | 6.48E-05 | -0.01623769 | AC069213.1 |
| chr19 | 53545815 | 53546031 | 217 | 3 | 9.55E-05 | 0.0789717 | 0.001035 | 6.48E-05 | -0.00072418 | ZNF331 |
| chr1 | 210185207 | 210185261 | 55 | 2 | 0.000122188 | 0.00039 | 6.53E-05 | 6.48E-05 | -0.01724377 | NA |
| chr12 | 125621158 | 125621443 | 286 | 3 | 5.27E-05 | 0.0013945 | 4.47E-05 | 6.49E-05 | -0.00496527 | TMEM132B |
| chr14 | 22979317 | 22979539 | 223 | 2 | 0.000137751 | 0.0002515 | 8.30E-05 | 6.49E-05 | 0.003238622 | AL132780.3, AJUBA |
| chr1 | 46432673 | 46432990 | 318 | 2 | 2.51E-05 | 0.000373 | 6.69E-05 | 6.50E-05 | 0.006846744 | FAAHP1 |
| chr13 | 114152052 | 114152165 | 114 | 4 | 0.000101145 | 0.0015274 | 0.000161 | 6.50E-05 | 0.003087256 | NA |
| chr9 | 121741346 | 121741516 | 171 | 2 | 0.000112662 | 4.61E-05 | 0.000951 | 6.50E-05 | -0.00758219 | DAB2IP |
| chr1 | 42035296 | 42035352 | 57 | 2 | 4.05E-05 | 0.0001149 | 0.000159 | 6.50E-05 | 0.002778576 | HIVEP3 |
| chr11 | 33491024 | 33491323 | 300 | 2 | 2.98E-05 | 0.0131262 | 2.89E-05 | 6.51E-05 | -0.01636581 | KIAA1549L |
| chr22 | 38272317 | 38273216 | 900 | 10 | 1.11E-06 | 0.0003553 | 0.002545 | 6.51E-05 | 6.96E-05 | TMEM184B |
| chr1 | 226388394 | 226388916 | 523 | 3 | 3.02E-05 | 0.0039634 | 3.15E-05 | 6.51E-05 | -0.00111167 | PARP1 |
| chr3 | 121730790 | 121731239 | 450 | 3 | 3.43E-05 | 0.0002956 | 8.63E-05 | 6.51E-05 | -0.00175014 | GOLGB1 |
| chr17 | 35578484 | 35579035 | 552 | 13 | 0.000117736 | 0.0025645 | 0.002818 | 6.51E-05 | 4.97E-05 | AP2B1, PEX12 |
| chr7 | 55107306 | 55107808 | 503 | 2 | 6.58E-06 | 0.0005317 | 5.72E-05 | 6.52E-05 | 0.003597471 | EGFR |
| chr22 | 26169215 | 26169307 | 93 | 5 | 0.000103044 | 0.0002296 | 0.003342 | 6.52E-05 | 0.002965326 | AL022337.1 |
| chr1 | 156420263 | 156420332 | 70 | 3 | 0.000143937 | 6.04E-05 | 0.002106 | 6.52E-05 | 0.004880446 | MIR9-1HG |
| chr3 | 58145819 | 58145940 | 122 | 2 | 7.78E-05 | 7.89E-05 | 0.000265 | 6.53E-05 | -0.00548492 | FLNB |
| chr11 | 32041334 | 32041468 | 135 | 5 | 0.000135214 | 0.0003337 | 0.000325 | 6.54E-05 | -0.00299856 | AL035078.4 |
| chr14 | 104229757 | 104230050 | 294 | 4 | 2.71E-05 | 0.0102545 | 9.16E-06 | 6.54E-05 | -0.00291654 | LINC02691 |
| chr2 | 11558249 | 11558420 | 172 | 2 | 6.95E-05 | 0.0002396 | 8.66E-05 | 6.55E-05 | -0.01762072 | GREB1 |
| chr12 | 56130714 | 56131046 | 333 | 2 | 8.36E-05 | 4.00E-05 | 0.00191 | 6.55E-05 | 0.002459128 | ESYT1 |
| chr4 | 3517872 | 3518662 | 791 | 5 | 1.01E-05 | 8.19E-05 | 0.002445 | 6.56E-05 | 0.001182023 | LRPAP1 |
| chr19 | 31279282 | 31279570 | 289 | 2 | 5.80E-05 | 0.0004959 | 5.93E-05 | 6.56E-05 | 0.00225903 | TSHZ3 |
| chr7 | 142864283 | 142864398 | 116 | 2 | 0.0001418 | 4.10E-05 | 0.001679 | 6.56E-05 | -0.00447708 | EPHB6 |
| chr22 | 50200333 | 50201056 | 724 | 7 | 7.20E-05 | 0.0001502 | 0.000718 | 6.57E-05 | -0.00014767 | SELENOO, AL022328.4 |
| chr11 | 118908308 | 118908522 | 215 | 2 | 8.92E-05 | 0.0001116 | 0.000168 | 6.57E-05 | -0.00385904 | BCL9L |
| chr1 | 167625954 | 167626269 | 316 | 2 | 3.66E-05 | 4.13E-05 | 0.001643 | 6.57E-05 | 0.011872557 | NA |
| chr2 | 233833195 | 233833277 | 83 | 2 | 0.000143093 | 0.0002073 | 9.57E-05 | 6.58E-05 | -0.00146435 | MROH2A |
| chr8 | 33722201 | 33722640 | 440 | 2 | 1.27E-05 | 0.0001491 | 0.000125 | 6.58E-05 | -0.00354929 | AF279873.3, VENTXP5 |
| chr17 | 15999187 | 15999879 | 693 | 11 | 5.07E-05 | 0.0299379 | 0.001989 | 6.59E-05 | 0.000179308 | TTC19, ZSWIM7 |
| chr6 | 11538295 | 11538534 | 240 | 3 | 5.38E-05 | 0.0003685 | 0.000404 | 6.59E-05 | 0.001715397 | TMEM170B |
| chr5 | 1345193 | 1345562 | 370 | 4 | 2.13E-05 | 0.0035973 | 6.88E-05 | 6.59E-05 | -0.0018784 | AC026748.7 |
| chr20 | 63361575 | 63362075 | 501 | 5 | 8.22E-05 | 6.47E-05 | 0.00121 | 6.60E-05 | -0.00131254 | AL121827.1, CHRNA4 |
| chr8 | 140578019 | 140578523 | 505 | 4 | 1.04E-05 | 0.0009372 | 4.18E-05 | 6.60E-05 | -0.00167554 | AGO2 |
| chr17 | 20009603 | 20010306 | 704 | 4 | 3.77E-05 | 0.0001987 | 0.003508 | 6.60E-05 | -0.00206629 | SPECC1 |
| chr3 | 45986073 | 45986147 | 75 | 2 | 3.60E-05 | 0.0055995 | 3.18E-05 | 6.60E-05 | -0.00312319 | FYCO1 |
| chr1 | 160102164 | 160102290 | 127 | 2 | 6.73E-05 | 9.97E-05 | 0.000195 | 6.60E-05 | -0.00479384 | NA |
| chr15 | 43382666 | 43382879 | 214 | 2 | 6.12E-05 | 0.0004587 | 6.19E-05 | 6.60E-05 | -0.00516222 | TUBGCP4 |
| chr12 | 47079405 | 47080212 | 808 | 17 | 5.36E-05 | 0.0013118 | 0.004121 | 6.60E-05 | 0.000336969 | PCED1B, AMIGO2 |
| chr12 | 98543330 | 98543614 | 285 | 3 | 3.99E-05 | 0.0003609 | 7.38E-05 | 6.61E-05 | -0.00367922 | TMPO |
| chr20 | 32762367 | 32762601 | 235 | 5 | 3.81E-05 | 0.0017796 | 0.001198 | 6.61E-05 | 0.001675817 | DNMT3B |
| chr19 | 35851823 | 35852628 | 806 | 6 | 2.82E-05 | 0.0175223 | 3.48E-05 | 6.62E-05 | -0.00021489 | NPHS1 |
| chr2 | 66440671 | 66440880 | 210 | 4 | 7.65E-05 | 0.0006465 | 8.98E-05 | 6.62E-05 | 0.003106532 | MEIS1, MEIS1-AS2 |
| chr6 | 146857633 | 146857881 | 249 | 3 | 0.000109741 | 5.83E-05 | 0.000659 | 6.62E-05 | -0.00847353 | AL138916.1, STXBP5-AS1 |
| chr1 | 202144464 | 202144933 | 470 | 6 | 3.97E-05 | 0.00064 | 0.000608 | 6.63E-05 | 0.000103541 | ARL8A |
| chr3 | 143341831 | 143342085 | 255 | 3 | 2.01E-05 | 0.0310807 | 1.28E-05 | 6.63E-05 | -0.00386894 | SLC9A9 |
| chr3 | 52527680 | 52527687 | 8 | 2 | 0.000117456 | 0.0001251 | 0.00015 | 6.63E-05 | 0.00220325 | NT5DC2 |
| chr16 | 87775011 | 87775067 | 57 | 2 | 0.000123715 | 0.0002575 | 8.42E-05 | 6.63E-05 | -0.01357204 | AC126696.3 |
| chr15 | 26866288 | 26866902 | 615 | 8 | 7.81E-06 | 0.0021627 | 0.000289 | 6.63E-05 | -0.00644891 | GABRB3 |
| chr11 | 118628546 | 118628818 | 273 | 2 | 2.20E-05 | 0.0010627 | 4.53E-05 | 6.63E-05 | 0.005214451 | PHLDB1 |
| chr22 | 26619421 | 26619755 | 335 | 2 | 0.000118662 | 4.07E-05 | 0.001903 | 6.64E-05 | -0.00555483 | NA |
| chr12 | 11486582 | 11487597 | 1016 | 5 | 5.41E-05 | 0.0001624 | 0.00382 | 6.64E-05 | 0.008292794 | PRB2, AC078950.1 |
| chr15 | 68874984 | 68875239 | 256 | 2 | 2.73E-05 | 0.0009814 | 4.65E-05 | 6.64E-05 | -0.00406004 | SPESP1 |
| chr14 | 72895117 | 72895506 | 390 | 3 | 2.20E-06 | 0.0524277 | 1.11E-05 | 6.65E-05 | -0.01250635 | NA |
| chr2 | 216659502 | 216659708 | 207 | 4 | 9.09E-05 | 0.0005458 | 0.001131 | 6.65E-05 | -0.0035644 | IGFBP2 |
| chr13 | 99956871 | 99956893 | 23 | 2 | 0.000154784 | 5.19E-05 | 0.000702 | 6.65E-05 | -0.00021411 | AL137139.2 |
| chr2 | 209579351 | 209579751 | 401 | 8 | 3.94E-05 | 0.0198764 | 6.64E-05 | 6.65E-05 | -0.00395335 | MAP2 |
| chr3 | 52452675 | 52452985 | 311 | 2 | 0.00010103 | 6.19E-05 | 0.000447 | 6.65E-05 | -0.00243565 | TNNC1 |
| chr17 | 57955454 | 57955768 | 315 | 2 | 6.79E-05 | 4.20E-05 | 0.001618 | 6.66E-05 | -0.00616179 | NA |
| chr1 | 116400947 | 116400994 | 48 | 2 | 5.76E-05 | 0.0042636 | 3.34E-05 | 6.67E-05 | -0.00296722 | ATP1A1, ATP1A1-AS1 |
| chr1 | 160742943 | 160742949 | 7 | 2 | 0.000115664 | 4.55E-05 | 0.001124 | 6.67E-05 | -0.00645967 | SLAMF7 |
| chr10 | 1505522 | 1505788 | 267 | 2 | 9.23E-06 | 0.0107905 | 3.00E-05 | 6.67E-05 | -0.00654881 | ADARB2 |
| chr11 | 120212643 | 120212662 | 20 | 2 | 0.000132803 | 4.07E-05 | 0.001973 | 6.68E-05 | -0.00893164 | OAF |
| chr11 | 35290827 | 35291014 | 188 | 2 | 3.62E-05 | 0.0043798 | 3.33E-05 | 6.68E-05 | 0.027705217 | SLC1A2 |
| chr4 | 26204458 | 26204508 | 51 | 2 | 4.95E-05 | 0.0001855 | 0.000106 | 6.68E-05 | 0.019644388 | RBPJ |
| chr17 | 5559275 | 5559555 | 281 | 3 | 6.16E-05 | 0.0015255 | 7.87E-05 | 6.68E-05 | -0.00245883 | NLRP1 |
| chr2 | 207334116 | 207334741 | 626 | 4 | 3.95E-05 | 7.53E-05 | 0.002456 | 6.69E-05 | -0.00846554 | AC009226.1 |
| chr21 | 41470389 | 41470657 | 269 | 2 | 2.12E-05 | 0.0258513 | 2.85E-05 | 6.69E-05 | -0.00650894 | TMPRSS2 |
| chr6 | 32067250 | 32067667 | 418 | 4 | 2.50E-05 | 0.0007346 | 0.000847 | 6.70E-05 | -0.00639479 | TNXB |

| | | | | | | | | | | |
|---|---|---|---|---|---|---|---|---|---|---|
| chr4 | 15939986 | 15940050 | 65 | 3 | 3.57E-05 | 0.0113897 | 2.29E-05 | 6.70E-05 | 0.003338063 | NA |
| chr10 | 30042050 | 30042272 | 223 | 2 | 1.59E-05 | 5.06E-05 | 0.000784 | 6.71E-05 | -0.00138419 | JCAD |
| chr22 | 45417724 | 45417942 | 219 | 4 | 0.00012791 | 0.0027185 | 5.27E-05 | 6.71E-05 | -0.0003648 | RIBC2 |
| chr1 | 29176428 | 29176434 | 7 | 2 | 0.000137549 | 5.01E-05 | 0.000814 | 6.71E-05 | -0.00469286 | SRSF4 |
| chr1 | 93846650 | 93847134 | 485 | 5 | 3.71E-05 | 0.0005172 | 7.08E-05 | 6.72E-05 | -0.00182389 | MIR760, BCAR3 |
| chr10 | 783393 | 783523 | 131 | 3 | 5.73E-05 | 0.0100411 | 3.03E-05 | 6.72E-05 | -0.00178418 | NA |
| chr9 | 117783112 | 117783375 | 264 | 2 | 2.15E-05 | 0.000106 | 0.000187 | 6.72E-05 | 0.00252664 | AL160272.2, AL160272.1 |
| chr15 | 99683069 | 99683105 | 37 | 2 | 0.000105016 | 0.0009801 | 4.71E-05 | 6.73E-05 | 0.010394669 | MEF2A |
| chr12 | 29791185 | 29791637 | 453 | 3 | 3.81E-05 | 0.0098355 | 3.68E-05 | 6.73E-05 | 0.015327749 | NA |
| chr16 | 19712735 | 19713045 | 311 | 2 | 3.83E-05 | 4.23E-05 | 0.001653 | 6.73E-05 | -0.00425637 | AC002550.1, KNOP1 |
| chr15 | 29968256 | 29968331 | 76 | 2 | 0.000109054 | 0.0001697 | 0.000116 | 6.74E-05 | 0.000467383 | TJP1 |
| chr11 | 58579615 | 58579705 | 91 | 2 | 9.18E-05 | 4.27E-05 | 0.001602 | 6.75E-05 | -0.0005915 | ZFP91, ZFP91-CNTF |
| chr10 | 72039966 | 72040209 | 244 | 2 | 0.000133742 | 5.98E-05 | 0.000508 | 6.75E-05 | -0.00363171 | NA |
| chr7 | 149882634 | 149883604 | 971 | 3 | 0.000110815 | 4.54E-05 | 0.001174 | 6.75E-05 | -0.00548032 | ATP6V0E2 |
| chr1 | 36306113 | 36306463 | 351 | 5 | 1.88E-05 | 0.0002342 | 0.000798 | 6.76E-05 | 0.000418075 | SH3D21 |
| chr11 | 3672179 | 3672788 | 610 | 3 | 8.10E-07 | 0.0242851 | 1.28E-05 | 6.76E-05 | -0.00170692 | NUP98 |
| chr16 | 4317590 | 4318123 | 534 | 2 | 9.21E-06 | 0.0006488 | 5.50E-05 | 6.76E-05 | -0.0030333 | GLIS2 |
| chr18 | 49476800 | 49476807 | 8 | 2 | 0.000133925 | 0.0003763 | 7.00E-05 | 6.77E-05 | 0.008467831 | NA |
| chr12 | 30677153 | 30677410 | 258 | 2 | 3.59E-05 | 0.0313189 | 2.86E-05 | 6.77E-05 | -0.00665506 | IPO8 |
| chr2 | 231483162 | 231483973 | 812 | 5 | 9.75E-06 | 0.0001138 | 0.00667 | 6.77E-05 | -0.01085607 | NCL |
| chr16 | 9160411 | 9160656 | 246 | 2 | 5.65E-05 | 0.0017552 | 4.04E-05 | 6.77E-05 | -0.01012885 | NA |
| chr20 | 57398610 | 57398629 | 20 | 2 | 7.72E-05 | 0.000811 | 5.06E-05 | 6.78E-05 | 0.008321624 | RBM38 |
| chr20 | 41087094 | 41087444 | 351 | 3 | 5.38E-05 | 0.0009249 | 0.001265 | 6.78E-05 | 0.004981091 | TOP1 |
| chr1 | 33048663 | 33049257 | 595 | 2 | 1.98E-05 | 0.0003614 | 7.16E-05 | 6.78E-05 | -0.00198355 | AK2 |
| chr11 | 65040609 | 65041166 | 558 | 10 | 5.76E-05 | 0.0048906 | 0.001028 | 6.78E-05 | 0.0019669 | SAC3D1 |
| chr2 | 187497074 | 187497309 | 236 | 2 | 5.31E-05 | 9.47E-05 | 0.000221 | 6.78E-05 | -0.00683723 | AC007319.1, TFPI |
| chr20 | 18467113 | 18467829 | 717 | 10 | 8.23E-05 | 2.22E-05 | 0.020455 | 6.79E-05 | 0.00017686 | POLR3F, DZANK1 |
| chr15 | 52161791 | 52161951 | 161 | 2 | 0.00010479 | 0.0003836 | 6.96E-05 | 6.79E-05 | 0.001557122 | GNB5 |
| chr1 | 3616419 | 3616464 | 46 | 2 | 3.71E-05 | 0.000911 | 4.88E-05 | 6.79E-05 | -0.00562099 | NA |
| chr7 | 155483402 | 155484185 | 784 | 8 | 9.10E-05 | 0.0006338 | 0.0023 | 6.80E-05 | 0.003829456 | CNPY1, AC009403.2 |
| chr16 | 12744480 | 12744546 | 67 | 2 | 6.96E-05 | 0.0095339 | 3.09E-05 | 6.80E-05 | -0.0139949 | CPPED1 |
| chr6 | 25223716 | 25223834 | 119 | 2 | 4.56E-05 | 0.0049777 | 3.33E-05 | 6.81E-05 | 0.01106163 | CMAHP |
| chr7 | 116861258 | 116861504 | 247 | 2 | 5.17E-06 | 0.0003948 | 6.90E-05 | 6.82E-05 | -0.00421884 | CAPZA2 |
| chr10 | 125776444 | 125776807 | 364 | 3 | 7.34E-05 | 3.05E-05 | 0.004196 | 6.82E-05 | -0.00692401 | NA |
| chr18 | 11851234 | 11851604 | 371 | 10 | 6.56E-05 | 0.0007613 | 0.001027 | 6.83E-05 | -0.00037788 | GNAL, CHMP1B, CHMP1B-AS1 |
| chr11 | 6474861 | 6475333 | 473 | 3 | 3.80E-05 | 0.000917 | 0.001207 | 6.83E-05 | 0.004401068 | ARFIP2 |
| chr8 | 38401707 | 38402049 | 343 | 4 | 2.06E-05 | 0.0007436 | 0.000183 | 6.83E-05 | 0.00133078 | LETM2, FGFR1 |
| chr2 | 159944946 | 159945025 | 80 | 2 | 7.56E-05 | 9.98E-05 | 0.000209 | 6.83E-05 | 0.002803652 | PLA2R1 |
| chr17 | 324163 | 324355 | 193 | 3 | 0.000152115 | 0.0001864 | 0.000137 | 6.83E-05 | -0.00268546 | RPH3AL |
| chr15 | 96046938 | 96047337 | 400 | 2 | 3.79E-05 | 4.24E-05 | 0.001777 | 6.83E-05 | -0.02047365 | AC012409.2, AC024337.1 |
| chr1 | 236155127 | 236155245 | 119 | 3 | 5.59E-05 | 0.0115478 | 2.93E-05 | 6.84E-05 | -4.94E-05 | GPR137B |
| chr12 | 111418021 | 111418125 | 105 | 5 | 4.76E-05 | 0.0003194 | 0.00029 | 6.84E-05 | -0.00091327 | SH2B3 |
| chr11 | 121534104 | 121534107 | 4 | 2 | 6.05E-05 | 0.0001334 | 0.000149 | 6.84E-05 | 0.000210543 | SORL1 |
| chr22 | 29816725 | 29817172 | 448 | 4 | 1.43E-05 | 0.0037643 | 3.11E-05 | 6.84E-05 | -0.0016364 | ASCC2 |
| chr10 | 132814541 | 132814648 | 108 | 2 | 6.92E-05 | 0.0001041 | 0.000198 | 6.84E-05 | -0.01128747 | CFAP46 |
| chr12 | 47210487 | 47211264 | 778 | 3 | 8.26E-07 | 0.1134772 | 9.81E-06 | 6.85E-05 | -0.01116658 | PCED1B, PCED1B-AS1 |
| chr2 | 33291614 | 33291724 | 111 | 2 | 4.35E-05 | 8.49E-05 | 0.000264 | 6.85E-05 | -0.009207 | LTBP1 |
| chr5 | 10758517 | 10758613 | 97 | 2 | 6.97E-05 | 4.10E-05 | 0.002244 | 6.85E-05 | -0.00547311 | DAP |
| chr17 | 82274220 | 82275127 | 908 | 4 | 1.55E-06 | 0.0018684 | 5.92E-05 | 6.85E-05 | -0.00409175 | AC132872.5 |
| chr2 | 233119930 | 233119979 | 50 | 2 | 0.000159797 | 0.0030556 | 3.62E-05 | 6.85E-05 | 0.00149259 | INPP5D |
| chr5 | 172766732 | 172766900 | 169 | 2 | 4.54E-05 | 0.0008744 | 5.00E-05 | 6.85E-05 | 0.000341217 | AC022217.3 |
| chr3 | 94028566 | 94029028 | 463 | 6 | 8.39E-05 | 0.0008498 | 0.000382 | 6.85E-05 | -0.00675476 | ARL13B, STX19 |
| chr5 | 14726308 | 14727053 | 746 | 3 | 4.46E-06 | 0.0010638 | 0.000677 | 6.86E-05 | -0.00295963 | ANKH |
| chr13 | 45391413 | 45391420 | 8 | 2 | 4.17E-05 | 0.0002286 | 9.51E-05 | 6.86E-05 | 0.00336126 | TPT1-AS1 |
| chr17 | 38606358 | 38607345 | 988 | 4 | 2.59E-05 | 0.0001144 | 0.000537 | 6.86E-05 | -0.00556828 | NA |
| chr12 | 130827428 | 130827651 | 224 | 2 | 0.000115418 | 0.0001241 | 0.000162 | 6.87E-05 | -0.01064194 | STX2 |
| chr2 | 241102601 | 241103226 | 626 | 5 | 3.36E-05 | 0.0001114 | 0.000332 | 6.87E-05 | 0.003066049 | NA |
| chr10 | 92834301 | 92834737 | 437 | 5 | 1.36E-05 | 0.002399 | 2.72E-05 | 6.87E-05 | -0.00096664 | EXOC6 |
| chr2 | 234082539 | 234082723 | 185 | 2 | 3.78E-05 | 0.0027328 | 3.71E-05 | 6.87E-05 | -0.00867969 | AC006037.2 |
| chr4 | 184826221 | 184826933 | 713 | 10 | 2.76E-05 | 0.0118515 | 6.75E-05 | 6.88E-05 | 0.00011869 | AC084871.3, ACSL1 |
| chr19 | 4471495 | 4471755 | 261 | 4 | 3.24E-05 | 0.0006144 | 0.00032 | 6.88E-05 | -0.00337306 | NA |
| chr1 | 54054441 | 54054445 | 5 | 2 | 4.18E-05 | 4.13E-05 | 0.002211 | 6.88E-05 | 0.004559548 | TCEANC2 |
| chr12 | 49950661 | 49950843 | 183 | 6 | 3.86E-05 | 0.0011282 | 0.002617 | 6.88E-05 | -0.00450827 | AQP2 |
| chr19 | 340965 | 341276 | 312 | 2 | 7.76E-05 | 4.60E-05 | 0.001244 | 6.88E-05 | 0.011186404 | MIER2 |
| chr5 | 135447385 | 135447583 | 199 | 3 | 7.31E-05 | 0.0018493 | 3.05E-05 | 6.89E-05 | -0.00378072 | TIFAB |
| chr20 | 13994633 | 13994857 | 225 | 3 | 9.11E-06 | 0.0005651 | 5.82E-05 | 6.89E-05 | -0.01077091 | SEL1L2 |
| chr16 | 30458911 | 30458967 | 57 | 2 | 5.64E-05 | 0.0001916 | 0.000109 | 6.90E-05 | 0.002338139 | NA |
| chr16 | 1038364 | 1038734 | 371 | 3 | 3.25E-05 | 0.0011495 | 0.000148 | 6.90E-05 | -0.01022829 | NA |
| chr1 | 21639817 | 21640007 | 191 | 3 | 0.000102727 | 0.0002621 | 0.001024 | 6.90E-05 | -0.0031186 | RAP1GAP |
| chr1 | 204962638 | 204962665 | 28 | 2 | 0.000146342 | 0.0003385 | 7.59E-05 | 6.91E-05 | 0.003305495 | NFASC |
| chr11 | 119206080 | 119206748 | 669 | 5 | 2.38E-05 | 5.31E-05 | 0.000767 | 6.91E-05 | -0.00017742 | CBL |
| chr12 | 132508311 | 132508676 | 366 | 3 | 2.53E-05 | 0.0053856 | 1.95E-05 | 6.91E-05 | -0.00048607 | FBRSL1 |
| chr9 | 21029853 | 21029866 | 14 | 2 | 0.000137436 | 0.0002396 | 9.31E-05 | 6.91E-05 | -0.00481948 | HACD4 |
| chr14 | 101052183 | 101052435 | 253 | 4 | 4.57E-05 | 0.0532983 | 9.04E-05 | 6.91E-05 | -0.00527226 | NA |
| chr18 | 24272154 | 24272369 | 216 | 5 | 0.000132574 | 0.0089617 | 0.000303 | 6.91E-05 | 0.00158375 | OSBPL1A |
| chr17 | 44503677 | 44504056 | 380 | 8 | 0.000118617 | 0.0015407 | 0.001628 | 6.92E-05 | -0.00010781 | NA |
| chr6 | 166505701 | 166505789 | 89 | 2 | 8.40E-05 | 5.61E-05 | 0.000647 | 6.92E-05 | -0.00444429 | RPS6KA2 |
| chr5 | 1293432 | 1294083 | 652 | 3 | 2.58E-06 | 0.0012516 | 5.10E-05 | 6.92E-05 | -0.0023135 | TERT |
| chr18 | 22176916 | 22177061 | 146 | 3 | 7.53E-05 | 8.05E-05 | 0.001819 | 6.93E-05 | -0.0071394 | GATA6 |
| chr11 | 122441911 | 122442041 | 131 | 2 | 2.99E-05 | 0.0087738 | 3.17E-05 | 6.93E-05 | -0.02068365 | MIR100HG |
| chr5 | 3324746 | 3325581 | 836 | 6 | 3.00E-05 | 0.003111 | 0.000569 | 6.93E-05 | -0.00054035 | NA |
| chr16 | 4239416 | 4239561 | 146 | 2 | 8.52E-05 | 4.38E-05 | 0.001637 | 6.94E-05 | -0.00801922 | SRL |
| chr13 | 21148253 | 21148262 | 10 | 2 | 6.85E-05 | 0.0003385 | 7.64E-05 | 6.94E-05 | -0.00117263 | SAP18 |
| chr15 | 40593978 | 40594637 | 660 | 10 | 7.48E-05 | 3.97E-05 | 0.016918 | 6.95E-05 | 0.000903927 | KNL1 |
| chr2 | 10451116 | 10451301 | 186 | 2 | 3.58E-05 | 0.0001639 | 0.000126 | 6.96E-05 | -0.00866363 | ODC1-DT |
| chr19 | 19063262 | 19063795 | 534 | 5 | 6.08E-06 | 0.0009891 | 0.00168 | 6.96E-05 | -0.00099345 | NA |
| chr11 | 13355405 | 13355441 | 37 | 2 | 0.000135217 | 5.82E-05 | 0.000599 | 6.96E-05 | -0.00040742 | ARNTL |
| chr1 | 31483423 | 31483428 | 6 | 2 | 0.000160294 | 4.28E-05 | 0.001913 | 6.96E-05 | -0.00852337 | NA |
| chr5 | 173888148 | 173888975 | 828 | 11 | 2.18E-05 | 0.0006207 | 0.000822 | 6.97E-05 | -0.00082993 | CPEB4 |
| chr12 | 93096782 | 93097223 | 442 | 2 | 9.60E-05 | 4.55E-05 | 0.001396 | 6.97E-05 | -0.00645847 | AC138123.2, AC138123.1 |
| chr1 | 33255633 | 33255858 | 226 | 3 | 4.31E-05 | 9.13E-05 | 0.001888 | 6.97E-05 | -0.00028729 | NA |
| chr17 | 39548097 | 39548417 | 321 | 3 | 2.49E-05 | 2.93E-05 | 0.005709 | 6.98E-05 | 0.003039788 | CDK12 |
| chr9 | 137739787 | 137739864 | 78 | 2 | 8.70E-05 | 0.0025718 | 3.81E-05 | 6.99E-05 | -0.004493 | EHMT1 |
| chr19 | 53756395 | 53756427 | 33 | 2 | 8.76E-05 | 0.0002713 | 8.77E-05 | 7.00E-05 | -0.01123423 | NA |
| chr5 | 96768922 | 96769004 | 83 | 2 | 0.000147172 | 0.0002577 | 9.06E-05 | 7.00E-05 | 0.001195739 | CAST, ERAP1 |
| chr1 | 201828070 | 201828594 | 525 | 4 | 4.79E-06 | 0.0116479 | 1.86E-05 | 7.00E-05 | -0.00344977 | IPO9-AS1 |
| chr17 | 16352246 | 16352570 | 325 | 4 | 9.49E-06 | 0.1014222 | 0.000138 | 7.00E-05 | -0.00404536 | CENPV |
| chr16 | 4686794 | 4686868 | 75 | 2 | 3.46E-05 | 4.19E-05 | 0.002274 | 7.00E-05 | -0.00252186 | MGRN1 |
| chr11 | 62922852 | 62923245 | 394 | 3 | 2.55E-05 | 5.95E-05 | 0.000705 | 7.00E-05 | -0.01247449 | NA |
| chr19 | 42197768 | 42197832 | 65 | 2 | 5.30E-05 | 0.0026124 | 3.81E-05 | 7.00E-05 | -0.00666125 | NA |
| chr1 | 227171012 | 227171026 | 15 | 2 | 0.00017604 | 0.000158 | 0.000132 | 7.01E-05 | 0.002336608 | CDC42BPA |
| chr9 | 89712411 | 89712483 | 73 | 2 | 9.84E-05 | 0.0004133 | 6.98E-05 | 7.01E-05 | 0.013653301 | UNQ6494 |
| chr7 | 149277872 | 149278408 | 537 | 5 | 3.29E-05 | 0.0032448 | 2.10E-05 | 7.01E-05 | -0.00158606 | ZNF783, AC004890.3 |
| chr2 | 10126822 | 10126859 | 38 | 2 | 0.000113724 | 0.0009084 | 5.06E-05 | 7.02E-05 | -0.01538193 | RRM2 |
| chr19 | 57252612 | 57252660 | 49 | 2 | 0.000145535 | 0.0001265 | 0.000166 | 7.02E-05 | -0.00042016 | ZNF805 |
| chr12 | 51863477 | 51864109 | 633 | 3 | 4.47E-06 | 0.0013711 | 0.000734 | 7.03E-05 | 0.009068488 | NA |
| chr1 | 66276222 | 66276662 | 441 | 2 | 4.94E-05 | 7.67E-05 | 0.000334 | 7.03E-05 | 0.014337722 | PDE4B |
| chr12 | 47808069 | 47808535 | 467 | 3 | 4.41E-05 | 0.0087926 | 0.000108 | 7.03E-05 | 0.002844763 | HDAC7 |
| chr12 | 54854079 | 54854778 | 700 | 5 | 3.78E-05 | 0.0001403 | 0.00038 | 7.04E-05 | -0.00498346 | MUCL1 |
| chr19 | 1957776 | 1957786 | 11 | 2 | 0.000102708 | 0.0001395 | 0.000151 | 7.05E-05 | 0.009588258 | CSNK1G2 |
| chr18 | 6881502 | 6881786 | 285 | 2 | 3.57E-05 | 0.0138566 | 3.09E-05 | 7.05E-05 | 0.003692419 | ARHGAP28 |
| chr21 | 42235809 | 42236297 | 489 | 2 | 1.18E-06 | 0.0291483 | 2.97E-05 | 7.06E-05 | 0.010210545 | ABCG1 |
| chr1 | 168135071 | 168135207 | 137 | 3 | 6.95E-05 | 0.0002578 | 0.001114 | 7.06E-05 | 0.00052491 | GPR161 |
| chr9 | 97039738 | 97040096 | 359 | 5 | 4.41E-06 | 0.0005695 | 0.000203 | 7.06E-05 | 0.008049277 | NA |
| chr12 | 28189865 | 28190585 | 721 | 8 | 4.37E-05 | 0.01479 | 0.000214 | 7.07E-05 | 0.001127045 | CCDC91, AC022364.1 |
| chr16 | 873475 | 873712 | 238 | 6 | 0.000158504 | 0.0007973 | 0.001415 | 7.07E-05 | -0.00366237 | LMF1 |
| chr21 | 40386043 | 40386167 | 125 | 2 | 0.000106726 | 0.0001086 | 0.000202 | 7.07E-05 | -0.00829365 | DSCAM |
| chr19 | 13830261 | 13830582 | 322 | 4 | 5.40E-05 | 0.0003272 | 0.001629 | 7.08E-05 | -0.00012092 | ZSWIM4, AC020916.1 |
| chr3 | 45274535 | 45274694 | 160 | 2 | 2.03E-05 | 0.0016361 | 4.29E-05 | 7.08E-05 | 0.003087979 | NA |
| chr21 | 20356761 | 20357004 | 244 | 2 | 5.14E-05 | 0.0037304 | 3.61E-05 | 7.08E-05 | 0.008952947 | RN7SKP147 |
| chr18 | 58864657 | 58865201 | 545 | 3 | 1.52E-06 | 0.0010356 | 0.000771 | 7.08E-05 | -0.01085277 | ZNF532 |
| chr16 | 88718414 | 88718524 | 111 | 3 | 0.000133673 | 5.19E-05 | 0.001467 | 7.09E-05 | 0.006372441 | PIEZO1 |
| chr9 | 108761756 | 108762031 | 276 | 2 | 0.000159607 | 0.0003615 | 7.59E-05 | 7.10E-05 | 0.011838845 | NA |
| chr12 | 131380265 | 131380349 | 85 | 2 | 2.84E-05 | 0.0001395 | 0.000153 | 7.10E-05 | 0.023606627 | NA |
| chr12 | 116458636 | 116458672 | 37 | 2 | 0.000115176 | 9.27E-05 | 0.000253 | 7.11E-05 | 0.001687548 | NA |
| chr6 | 15505718 | 15505854 | 137 | 2 | 0.000152053 | 0.0001741 | 0.000124 | 7.11E-05 | -0.01252137 | JARID2 |
| chr2 | 235834059 | 235834155 | 97 | 2 | 0.000143162 | 0.0002917 | 8.60E-05 | 7.12E-05 | 0.008001851 | AGAP1 |
| chr4 | 146224523 | 146224539 | 17 | 3 | 1.61E-05 | 0.0445852 | 1.17E-05 | 7.12E-05 | -0.00577931 | REELD1 |
| chr21 | 43718914 | 43719348 | 435 | 6 | 7.22E-05 | 6.61E-05 | 0.001969 | 7.12E-05 | -0.00096897 | PDXK |
| chr6 | 32234971 | 32235067 | 97 | 2 | 9.05E-05 | 7.07E-05 | 0.000407 | 7.12E-05 | -0.03703108 | NA |
| chr12 | 10243922 | 10243953 | 32 | 2 | 0.000168397 | 0.0001986 | 0.000111 | 7.13E-05 | -0.00224397 | KLRD1 |
| chr17 | 48908709 | 48909445 | 737 | 5 | 1.59E-05 | 0.0002861 | 0.004146 | 7.13E-05 | -0.00438243 | UBE2Z, SUMO2P17 |
| chr17 | 75270318 | 75270561 | 244 | 4 | 1.00E-04 | 0.0001488 | 0.000385 | 7.14E-05 | -0.00196038 | MIF4GD |
| chr6 | 34527199 | 34527531 | 333 | 4 | 1.53E-05 | 0.0014016 | 0.000158 | 7.15E-05 | 0.003674403 | PACSIN1 |
| chr1 | 226371862 | 226372321 | 460 | 2 | 2.39E-05 | 4.58E-05 | 0.001567 | 7.16E-05 | -0.00102092 | PARP1 |
| chr9 | 124467458 | 124467620 | 163 | 2 | 1.93E-05 | 0.0001167 | 0.00019 | 7.17E-05 | 0.004803147 | ADGRD2 |
| chr10 | 50553742 | 50554290 | 549 | 2 | 8.73E-05 | 6.50E-05 | 0.000501 | 7.18E-05 | -0.00404734 | SGMS1 |
| chr12 | 113232130 | 113232489 | 360 | 4 | 6.48E-05 | 0.0001177 | 0.003657 | 7.19E-05 | -0.00074882 | TPCN1 |
| chr11 | 96098638 | 96098785 | 148 | 2 | 2.69E-05 | 0.005566 | 3.44E-05 | 7.19E-05 | -0.00387937 | AP000779.1, MAML2 |
| chr6 | 34523132 | 34523190 | 59 | 2 | 9.49E-05 | 0.0001532 | 0.000142 | 7.19E-05 | -0.00377041 | PACSIN1 |
| chr1 | 156619156 | 156619239 | 84 | 3 | 4.83E-05 | 0.0032743 | 3.59E-05 | 7.19E-05 | -0.00561879 | NA |
| chr18 | 58695909 | 58695929 | 21 | 2 | 6.70E-05 | 4.56E-05 | 0.001619 | 7.19E-05 | -0.00174745 | MALT1, AC104365.1 |
| chr9 | 34637816 | 34638330 | 515 | 9 | 7.93E-05 | 0.0112218 | 0.005101 | 7.20E-05 | -0.00159363 | GALT |
| chr9 | 124888151 | 124888498 | 348 | 2 | 2.02E-05 | 4.60E-05 | 0.001566 | 7.20E-05 | -0.00506815 | GOLGA1 |
| chr3 | 180183340 | 180183524 | 185 | 3 | 3.31E-05 | 0.0402872 | 1.22E-05 | 7.20E-05 | 0.003482547 | NA |
| chr11 | 116838385 | 116838666 | 282 | 4 | 2.76E-05 | 0.0002594 | 0.00077 | 7.21E-05 | -0.00519097 | APOA1-AS |
| chr12 | 79689472 | 79690134 | 663 | 4 | 2.09E-05 | 0.0001292 | 0.002289 | 7.22E-05 | 0.00140445 | PAWR |
| chr11 | 7571539 | 7571790 | 252 | 2 | 8.39E-05 | 5.73E-05 | 0.000708 | 7.22E-05 | -0.01290201 | PPFIBP2, AC107884.2 |
| chr12 | 52316684 | 52316701 | 18 | 2 | 8.84E-05 | 6.97E-05 | 0.000436 | 7.23E-05 | -0.0094822 | KRT83 |
| chr2 | 231182660 | 231182811 | 152 | 2 | 5.45E-05 | 0.0066056 | 3.39E-05 | 7.23E-05 | -0.01129528 | NA |
| chr8 | 81721855 | 81722566 | 712 | 4 | 9.84E-06 | 0.0002398 | 0.000831 | 7.23E-05 | -0.0009233 | ZFAND1 |
| chr22 | 31104910 | 31104954 | 45 | 2 | 0.00010634 | 0.0046993 | 3.54E-05 | 7.23E-05 | -0.00720514 | SELENOM |
| chr3 | 62185754 | 62185968 | 215 | 2 | 3.02E-05 | 0.0095118 | 3.26E-05 | 7.23E-05 | -0.00636461 | PTPRG |
| chr17 | 69480208 | 69480300 | 93 | 2 | 0.000140995 | 0.000344 | 8.00E-05 | 7.24E-05 | 0.001084435 | MAP2K6, AC002546.1 |
| chr1 | 15410031 | 15410544 | 514 | 5 | 1.96E-06 | 0.1439745 | 1.77E-05 | 7.26E-05 | 0.001464655 | EFHD2 |
| chr7 | 465228 | 465244 | 17 | 2 | 0.000102205 | 4.52E-05 | 0.001809 | 7.27E-05 | -0.01194219 | NA |
| chr2 | 174345137 | 174345503 | 367 | 2 | 3.11E-05 | 4.35E-05 | 0.002297 | 7.28E-05 | -0.01291157 | NA |
| chr10 | 102424381 | 102424457 | 77 | 2 | 7.98E-05 | 0.0038095 | 3.68E-05 | 7.28E-05 | 0.001965665 | CUEDC2 |

| | | | | | | | | | | |
|---|---|---|---|---|---|---|---|---|---|---|
| chr1 | 20070025 | 20070257 | 233 | 5 | 2.53E-05 | 0.0016425 | 3.16E-05 | 7.29E-05 | -0.00548657 | PLA2G5 |
| chr1 | 43978578 | 43979530 | 953 | 7 | 1.94E-06 | 0.0008568 | 0.001283 | 7.30E-05 | -0.00331359 | B4GALT2 |
| chr18 | 57622093 | 57622593 | 501 | 9 | 4.86E-06 | 0.0004859 | 0.003628 | 7.30E-05 | -0.00459934 | NARS1 |
| chr14 | 74570390 | 74570506 | 117 | 2 | 8.81E-05 | 0.0016324 | 4.43E-05 | 7.30E-05 | 0.003286722 | LTBP2 |
| chr22 | 33568311 | 33568492 | 182 | 3 | 7.77E-05 | 0.0029222 | 7.26E-05 | 7.30E-05 | -0.0012882 | LARGE1 |
| chr19 | 5978403 | 5978779 | 377 | 8 | 6.39E-05 | 0.0009539 | 0.006098 | 7.31E-05 | 0.000889783 | AC011444.1 |
| chr19 | 1566840 | 1566951 | 112 | 2 | 2.71E-05 | 6.88E-05 | 0.000462 | 7.31E-05 | -0.01139681 | MEX3D |
| chr7 | 128201282 | 128201874 | 593 | 4 | 4.93E-05 | 0.0024258 | 0.000165 | 7.31E-05 | 0.00212069 | NA |
| chr19 | 8335970 | 8336173 | 204 | 2 | 5.69E-05 | 0.000699 | 5.84E-05 | 7.32E-05 | 0.016008147 | KANK3 |
| chr9 | 109968448 | 109968484 | 37 | 2 | 0.000144809 | 6.93E-05 | 0.000457 | 7.32E-05 | 0.013469916 | PALM2AKAP2 |
| chr16 | 77213115 | 77213229 | 115 | 2 | 0.000122924 | 0.000104 | 0.000229 | 7.32E-05 | -0.01232831 | SYCE1L, AC009139.2 |
| chr5 | 175878134 | 175878790 | 657 | 2 | 6.74E-05 | 4.38E-05 | 0.002303 | 7.32E-05 | -0.01980916 | CPLX2 |
| chr3 | 129161521 | 129161630 | 110 | 3 | 3.94E-05 | 0.0015431 | 3.79E-05 | 7.34E-05 | 0.001753925 | NA |
| chr15 | 64774032 | 64774511 | 480 | 2 | 0.000103908 | 5.08E-05 | 0.001108 | 7.34E-05 | -0.01073924 | RBPMS2 |
| chr17 | 29967540 | 29967773 | 234 | 3 | 0.000134173 | 3.27E-05 | 0.004201 | 7.34E-05 | -0.00093095 | EFCAB5 |
| chr9 | 131405188 | 131405531 | 344 | 3 | 0.000136978 | 0.0001297 | 0.001886 | 7.34E-05 | -0.01078007 | PRRC2B |
| chr5 | 154039415 | 154039417 | 3 | 2 | 0.000172456 | 4.86E-05 | 0.001336 | 7.35E-05 | -0.01080335 | MFAP3 |
| chr17 | 82215353 | 82215540 | 188 | 2 | 3.49E-05 | 0.003673 | 3.74E-05 | 7.35E-05 | -0.00886709 | AC132872.2 |
| chr8 | 142320923 | 142321087 | 165 | 2 | 6.84E-05 | 0.0047315 | 3.59E-05 | 7.35E-05 | -0.00349356 | TSNARE1 |
| chr14 | 51488806 | 51489326 | 521 | 5 | 2.89E-05 | 0.0013203 | 2.44E-05 | 7.35E-05 | -0.00489853 | FRMD6, FRMD6-AS2 |
| chr17 | 715793 | 715824 | 32 | 3 | 3.17E-05 | 5.67E-05 | 0.000866 | 7.35E-05 | -0.01738019 | VPS53 |
| chr17 | 28250537 | 28250687 | 151 | 3 | 0.000129446 | 0.0007543 | 0.000134 | 7.35E-05 | 0.00615015 | NA |
| chr17 | 67374708 | 67374723 | 16 | 2 | 7.41E-05 | 0.0024876 | 4.04E-05 | 7.36E-05 | -0.00225236 | NA |
| chr9 | 134768668 | 134768824 | 157 | 2 | 0.00013344 | 4.42E-05 | 0.002257 | 7.37E-05 | -0.020202 | COL5A1 |
| chr8 | 91247957 | 91248252 | 296 | 4 | 2.61E-05 | 0.0072403 | 2.03E-05 | 7.37E-05 | -0.006127 | SLC26A7 |
| chr13 | 27967005 | 27967340 | 336 | 3 | 1.79E-05 | 0.0022535 | 6.29E-05 | 7.38E-05 | -0.00124473 | CDX2 |
| chr19 | 1941706 | 1941929 | 224 | 2 | 0.000121564 | 5.50E-05 | 0.000861 | 7.38E-05 | 0.000958922 | CSNK1G2 |
| chr9 | 33468811 | 33469002 | 192 | 2 | 4.31E-05 | 4.88E-05 | 0.001359 | 7.40E-05 | -0.00425086 | NOL6 |
| chr12 | 65641938 | 65642757 | 820 | 6 | 5.83E-05 | 0.0002144 | 0.003752 | 7.41E-05 | 0.002467386 | AC090023.2, AC025419.1 |
| chr1 | 94238603 | 94238829 | 227 | 3 | 7.44E-05 | 0.0002872 | 0.000567 | 7.41E-05 | -0.01096755 | ARHGAP29 |
| chr1 | 210253013 | 210253214 | 202 | 2 | 2.39E-05 | 0.0004782 | 6.95E-05 | 7.41E-05 | -0.0065152 | NA |
| chr17 | 78852948 | 78853099 | 152 | 2 | 2.99E-05 | 0.0009824 | 5.22E-05 | 7.41E-05 | 0.002990313 | TIMP2 |
| chr17 | 35741405 | 35741552 | 148 | 2 | 0.000155564 | 4.70E-05 | 0.001642 | 7.42E-05 | 0.003595382 | TAF15, RASL10B |
| chr20 | 4023050 | 4023499 | 450 | 2 | 9.90E-05 | 4.44E-05 | 0.002301 | 7.42E-05 | 0.00103927 | NA |
| chr14 | 100063739 | 100063816 | 78 | 2 | 5.00E-05 | 0.0001243 | 0.000188 | 7.42E-05 | -0.00861581 | EVL, AL157912.1 |
| chr8 | 103020331 | 103020890 | 560 | 10 | 6.10E-05 | 0.0006398 | 0.000608 | 7.42E-05 | -0.00659155 | AP003550.1 |
| chr6 | 42423470 | 42423516 | 47 | 2 | 0.000169786 | 0.0002288 | 0.000106 | 7.42E-05 | 0.007489313 | TRERF1 |
| chr10 | 122290012 | 122290266 | 255 | 2 | 6.35E-05 | 0.0007112 | 5.89E-05 | 7.43E-05 | 0.004940328 | BTBD16 |
| chr4 | 141713327 | 141713750 | 424 | 2 | 4.77E-05 | 0.0001186 | 0.000199 | 7.43E-05 | -0.00750225 | IL15 |
| chr10 | 17226922 | 17227275 | 354 | 4 | 2.59E-05 | 0.0002661 | 0.004506 | 7.44E-05 | -0.00148058 | VIM-AS1 |
| chr19 | 10334139 | 10335232 | 1094 | 5 | 3.12E-05 | 3.60E-05 | 0.010349 | 7.44E-05 | -0.00731591 | AC114271.1, ICAM3 |
| chr13 | 44917724 | 44918194 | 471 | 6 | 5.03E-05 | 8.94E-05 | 0.000815 | 7.44E-05 | 0.00150265 | NA |
| chr15 | 73994074 | 73994838 | 765 | 12 | 2.82E-05 | 0.0014526 | 0.0002 | 7.45E-05 | 0.000368291 | PML, STOML1 |
| chr15 | 58137820 | 58138362 | 543 | 4 | 4.59E-05 | 0.000553 | 0.000634 | 7.45E-05 | -0.00101936 | AQP9, ALDH1A2 |
| chr17 | 50111726 | 50112197 | 472 | 2 | 7.48E-05 | 7.43E-05 | 0.000412 | 7.45E-05 | -0.00749875 | PDK2, SAMD14 |
| chr2 | 6309350 | 6309720 | 371 | 2 | 1.33E-05 | 0.0951444 | 3.03E-05 | 7.45E-05 | -0.00485273 | AC017053.1 |
| chr1 | 33159727 | 33159837 | 111 | 4 | 0.000145192 | 0.0013497 | 0.001447 | 7.46E-05 | 0.000435792 | TRIM62 |
| chr6 | 31537716 | 31537719 | 4 | 2 | 0.000122913 | 6.65E-05 | 0.00053 | 7.47E-05 | -0.00083229 | DDX39B, ATP6V1G2-DDX39B |
| chr7 | 152025901 | 152026144 | 244 | 2 | 1.40E-05 | 0.0001505 | 0.000155 | 7.47E-05 | -0.002455 | GALNT11 |
| chr18 | 78991616 | 78991952 | 337 | 2 | 0.000163443 | 5.07E-05 | 0.001209 | 7.47E-05 | 0.021323497 | SALL3 |
| chr3 | 196232934 | 196233116 | 183 | 2 | 1.82E-05 | 0.0104837 | 3.32E-05 | 7.48E-05 | -0.00683899 | SLC51A, PCYT1A |
| chr3 | 100994728 | 100994802 | 75 | 2 | 7.78E-05 | 0.0004397 | 7.32E-05 | 7.48E-05 | -0.00869452 | NA |
| chr1 | 204190370 | 204190601 | 232 | 3 | 6.54E-05 | 0.0006661 | 7.14E-05 | 7.49E-05 | 0.003833378 | KISS1 |
| chr15 | 39580623 | 39581352 | 730 | 13 | 3.91E-05 | 0.0004826 | 0.011213 | 7.49E-05 | -0.00075162 | THBS1 |
| chr22 | 22928662 | 22928738 | 77 | 2 | 0.000135565 | 4.48E-05 | 0.002302 | 7.49E-05 | 0.009491033 | NA |
| chr9 | 131270438 | 131270747 | 310 | 3 | 2.59E-05 | 0.0096022 | 2.12E-05 | 7.49E-05 | 0.002043613 | FAM78A |
| chr15 | 90932473 | 90932695 | 223 | 7 | 0.000105155 | 0.0002514 | 0.001558 | 7.50E-05 | 0.001874424 | UNC45A, HDDC3 |
| chr2 | 5673126 | 5673734 | 609 | 4 | 1.25E-05 | 0.0048852 | 1.89E-05 | 7.50E-05 | -0.00768514 | LINC01248, AC108025.1 |
| chr10 | 124791210 | 124791803 | 594 | 4 | 0.000102189 | 0.0017156 | 0.001044 | 7.50E-05 | -0.00229444 | AC068896.1, EEF1AKMT2 |
| chr17 | 82127206 | 82127456 | 251 | 2 | 3.47E-05 | 8.46E-05 | 0.000329 | 7.50E-05 | -0.00332644 | CCDC57 |
| chr9 | 973184 | 973198 | 15 | 2 | 1.80E-05 | 5.37E-05 | 0.000992 | 7.51E-05 | 0.00334668 | NA |
| chr5 | 94076132 | 94076180 | 49 | 3 | 0.000170674 | 0.0002486 | 0.000113 | 7.51E-05 | -0.00180237 | FAM172A |
| chr1 | 38138660 | 38138804 | 145 | 2 | 0.000106806 | 0.0008258 | 5.65E-05 | 7.51E-05 | 0.004440587 | LINC02786 |
| chr1 | 3186110 | 3186542 | 433 | 4 | 9.20E-05 | 0.0010379 | 0.000537 | 7.52E-05 | -0.00130059 | PRDM16 |
| chr8 | 98170194 | 98170884 | 691 | 5 | 9.08E-05 | 0.012428 | 7.21E-05 | 7.53E-05 | -0.00309621 | NA |
| chr6 | 2963434 | 2963624 | 191 | 3 | 8.17E-05 | 0.0008428 | 5.81E-05 | 7.53E-05 | 0.002032696 | SERPINB6 |
| chr16 | 695294 | 695765 | 472 | 6 | 3.66E-05 | 0.0001201 | 0.010233 | 7.54E-05 | -4.13E-05 | FBXL16 |
| chr4 | 110634284 | 110634347 | 64 | 2 | 0.000155086 | 0.0007533 | 5.86E-05 | 7.54E-05 | -0.0021682 | PITX2 |
| chr8 | 9573952 | 9574316 | 365 | 2 | 6.32E-05 | 5.54E-05 | 0.000911 | 7.55E-05 | -0.005181 | TNKS |
| chr11 | 65084882 | 65085522 | 641 | 5 | 6.92E-05 | 2.02E-05 | 0.015119 | 7.55E-05 | -0.00484945 | ZFPL1, TMEM262 |
| chr4 | 1225220 | 1225919 | 700 | 4 | 2.06E-05 | 0.0021414 | 4.69E-05 | 7.56E-05 | 0.000552413 | CTBP1 |
| chr11 | 57303657 | 57303787 | 131 | 2 | 5.18E-05 | 0.0011656 | 5.04E-05 | 7.56E-05 | -0.00726709 | TNKS1BP1 |
| chr14 | 105491405 | 105491859 | 455 | 2 | 2.60E-05 | 0.0001009 | 0.000256 | 7.57E-05 | -0.00173118 | AL928654.3, TEDC1 |
| chr16 | 54285492 | 54286318 | 827 | 4 | 4.01E-07 | 0.0010993 | 0.000358 | 7.57E-05 | 0.000790681 | AC018553.2, IRX3 |
| chr6 | 124962580 | 124963513 | 934 | 12 | 1.29E-05 | 0.0013589 | 0.000583 | 7.57E-05 | 0.001387588 | RNF217, RNF217-AS1 |
| chr17 | 74924081 | 74924503 | 423 | 10 | 1.44E-05 | 0.0123219 | 0.000138 | 7.58E-05 | -0.00084719 | OTOP2 |
| chr2 | 61877106 | 61877270 | 165 | 2 | 3.67E-05 | 0.0011643 | 5.06E-05 | 7.58E-05 | -0.00548023 | AC107081.1, CCT4 |
| chr6 | 41651811 | 41651928 | 118 | 2 | 6.49E-05 | 0.0093312 | 3.40E-05 | 7.58E-05 | -0.00671013 | MDFI |
| chr17 | 5514863 | 5514991 | 129 | 2 | 7.54E-05 | 6.89E-05 | 0.00051 | 7.59E-05 | 0.000423749 | NLRP1 |
| chr9 | 132161481 | 132161956 | 476 | 7 | 2.33E-05 | 0.000419 | 0.000183 | 7.60E-05 | -0.00126597 | NTNG2 |
| chr2 | 42567682 | 42568122 | 441 | 6 | 7.54E-05 | 0.0001889 | 0.002279 | 7.62E-05 | -0.00054907 | MTA3 |
| chr8 | 95133012 | 95133255 | 244 | 2 | 8.05E-05 | 4.69E-05 | 0.001946 | 7.63E-05 | -0.00182514 | NA |
| chr17 | 39635336 | 39635434 | 99 | 2 | 5.64E-06 | 0.020782 | 3.21E-05 | 7.63E-05 | -0.00542876 | PPP1R1B |
| chr9 | 128125066 | 128125445 | 380 | 2 | 8.32E-05 | 9.02E-05 | 0.000307 | 7.63E-05 | -0.00370408 | PTGES2 |
| chr12 | 75318248 | 75318295 | 48 | 2 | 0.000145529 | 8.55E-05 | 0.000338 | 7.65E-05 | 0.006077777 | CAPS2 |
| chr14 | 67285860 | 67285994 | 135 | 2 | 0.000167447 | 8.48E-05 | 0.000343 | 7.65E-05 | -0.0094714 | MPP5, AL135978.1 |
| chr21 | 36419998 | 36420198 | 201 | 3 | 0.00016372 | 0.0008159 | 0.001188 | 7.65E-05 | -0.0019994 | NA |
| chr5 | 73443941 | 73444706 | 766 | 4 | 1.26E-05 | 0.0003302 | 0.001589 | 7.65E-05 | -0.00161008 | NA |
| chr2 | 27129086 | 27129344 | 259 | 3 | 3.22E-05 | 0.0019421 | 3.46E-05 | 7.66E-05 | 0.005460771 | ABHD1 |
| chr13 | 99980900 | 99981638 | 739 | 8 | 4.73E-05 | 0.0003477 | 0.000147 | 7.67E-05 | 0.000574814 | NA |
| chr2 | 46915416 | 46916262 | 847 | 13 | 5.17E-05 | 0.0007107 | 0.004784 | 7.67E-05 | 0.000569543 | TTC7A, MCFD2 |
| chr20 | 47319709 | 47320109 | 401 | 3 | 1.39E-05 | 0.0004184 | 0.00011 | 7.67E-05 | -0.00957774 | AL031666.2, ZMYND8 |
| chr14 | 24334473 | 24334816 | 344 | 6 | 1.57E-05 | 0.0183758 | 1.45E-05 | 7.68E-05 | 0.000766408 | ADCY4, AL096870.1 |
| chr6 | 167978363 | 167978433 | 71 | 2 | 0.000151729 | 9.35E-05 | 0.000295 | 7.68E-05 | 3.04E-05 | HGC6.3 |
| chr2 | 240973290 | 240973686 | 397 | 2 | 4.91E-05 | 5.77E-05 | 0.000852 | 7.68E-05 | 0.003642204 | CROCC2 |
| chr2 | 60719430 | 60719562 | 133 | 2 | 5.98E-05 | 0.0001913 | 0.000129 | 7.69E-05 | -0.02033406 | NA |
| chr11 | 72227731 | 72227789 | 59 | 2 | 4.22E-05 | 0.0004505 | 7.47E-05 | 7.70E-05 | 0.005163692 | INPPL1 |
| chr7 | 128364318 | 128364385 | 68 | 2 | 7.37E-05 | 5.14E-05 | 0.001298 | 7.70E-05 | -0.00175884 | NA |
| chr4 | 109912728 | 109913434 | 707 | 5 | 2.07E-06 | 0.0476131 | 1.40E-05 | 7.70E-05 | -0.00174182 | EGF |
| chr2 | 26674474 | 26674650 | 177 | 3 | 6.15E-05 | 0.0079376 | 2.89E-05 | 7.71E-05 | -0.00570113 | NA |
| chr12 | 53756832 | 53757159 | 328 | 2 | 1.53E-05 | 0.0236287 | 3.22E-05 | 7.71E-05 | -0.00572945 | AC076968.2, AC076968.1, CISTR |
| chr4 | 163349680 | 163349808 | 129 | 2 | 4.60E-05 | 0.000512 | 7.05E-05 | 7.71E-05 | -0.01305462 | NPY5R |
| chr2 | 21650295 | 21650329 | 35 | 2 | 7.01E-05 | 0.003085 | 4.04E-05 | 7.71E-05 | 0.008125059 | AC018742.1 |
| chr14 | 37622439 | 37622970 | 532 | 5 | 2.99E-05 | 0.000778 | 0.001385 | 7.72E-05 | -0.00305449 | TTC6 |
| chr10 | 70363991 | 70364336 | 346 | 3 | 3.00E-05 | 0.0001071 | 0.001473 | 7.73E-05 | -0.00222947 | LRRC20 |
| chr12 | 132288247 | 132288626 | 380 | 4 | 7.91E-05 | 0.0001369 | 0.000971 | 7.73E-05 | 0.005607502 | GALNT9 |
| chr14 | 100977905 | 100978297 | 393 | 2 | 1.05E-05 | 5.77E-05 | 0.000875 | 7.74E-05 | -0.01365669 | MEG8 |
| chr17 | 75182149 | 75182372 | 224 | 2 | 1.84E-05 | 4.66E-05 | 0.002189 | 7.74E-05 | 0.00227362 | SUMO2 |
| chr8 | 53914598 | 53914704 | 107 | 3 | 4.38E-05 | 0.0069793 | 2.64E-05 | 7.74E-05 | -0.00310413 | RGS20 |
| chr19 | 1151961 | 1152359 | 399 | 5 | 8.29E-05 | 0.0003247 | 0.001856 | 7.74E-05 | -0.00061491 | SBNO2 |
| chr3 | 194580438 | 194580836 | 399 | 2 | 3.07E-05 | 0.0010403 | 5.37E-05 | 7.75E-05 | -0.00833997 | NA |
| chr15 | 62732707 | 62732785 | 79 | 2 | 7.95E-05 | 5.39E-05 | 0.001108 | 7.75E-05 | -0.00562535 | TLN2 |
| chr4 | 40631051 | 40631555 | 505 | 4 | 5.57E-05 | 0.0057298 | 0.000187 | 7.76E-05 | -0.00418188 | NA |
| chr5 | 179963655 | 179964548 | 894 | 3 | 5.88E-06 | 0.0009809 | 0.000125 | 7.77E-05 | -0.00354655 | AC010285.1, RNF130 |
| chr11 | 1743755 | 1743994 | 240 | 2 | 0.000107878 | 0.0001245 | 0.000204 | 7.77E-05 | -0.00476414 | IFITM10, AC068580.4 |
| chr1 | 31993034 | 31993092 | 59 | 2 | 0.000142621 | 0.0005543 | 6.86E-05 | 7.77E-05 | -0.00179182 | NA |
| chr12 | 132604984 | 132605014 | 31 | 2 | 3.43E-05 | 0.0003176 | 9.13E-05 | 7.77E-05 | -0.01336922 | LRCOL1 |
| chr12 | 121893453 | 121893627 | 175 | 2 | 5.04E-05 | 5.01E-05 | 0.001526 | 7.78E-05 | 0.001029773 | PSMD9, AC069503.2 |
| chr21 | 43911745 | 43911881 | 137 | 2 | 4.03E-05 | 0.0227977 | 3.25E-05 | 7.78E-05 | 0.002468904 | AGPAT3 |
| chr21 | 45086714 | 45086758 | 45 | 2 | 0.000155483 | 0.0003353 | 8.86E-05 | 7.78E-05 | -0.00650533 | ADARB1 |
| chr8 | 1823251 | 1823278 | 28 | 3 | 8.43E-05 | 3.96E-05 | 0.003227 | 7.78E-05 | -0.00069991 | AC019257.8 |
| chr4 | 146175633 | 146175757 | 125 | 5 | 3.96E-05 | 0.0004802 | 0.000371 | 7.78E-05 | -0.00125449 | LSM6 |
| chr2 | 6876236 | 6876378 | 143 | 2 | 6.62E-05 | 9.04E-05 | 0.000321 | 7.79E-05 | 0.007384997 | RSAD2 |
| chr2 | 48530338 | 48530583 | 246 | 4 | 1.70E-05 | 0.0008509 | 0.000108 | 7.80E-05 | -0.00131245 | STON1, STON1-GTF2A1L |
| chr10 | 77856082 | 77856479 | 398 | 2 | 0.000120811 | 4.74E-05 | 0.00209 | 7.80E-05 | 0.002972071 | DLG5 |
| chr19 | 50517040 | 50517277 | 238 | 2 | 5.01E-05 | 4.63E-05 | 0.002416 | 7.81E-05 | -0.02523336 | LRRC4B |
| chr2 | 203365918 | 203366548 | 631 | 4 | 1.38E-05 | 0.0067578 | 2.52E-05 | 7.81E-05 | 0.004197598 | ABI2 |
| chr13 | 41961063 | 41961468 | 406 | 10 | 4.75E-05 | 0.0001971 | 0.004157 | 7.82E-05 | 0.001646668 | VWA8-AS1, VWA8 |
| chr4 | 107990191 | 107990556 | 366 | 3 | 4.36E-05 | 0.0007263 | 0.000195 | 7.82E-05 | 0.000247202 | HADH |
| chr5 | 956406 | 957469 | 1064 | 6 | 2.52E-06 | 0.0006202 | 0.000329 | 7.83E-05 | -0.00549769 | AC122719.1 |
| chr7 | 1536758 | 1537380 | 623 | 5 | 2.82E-05 | 0.0013855 | 0.000659 | 7.83E-05 | -0.00078443 | MAFK |
| chr8 | 8893746 | 8894096 | 351 | 5 | 4.65E-06 | 0.0031795 | 6.41E-05 | 7.84E-05 | -0.00094776 | NA |
| chr12 | 113184757 | 113184973 | 217 | 3 | 1.27E-05 | 0.0002334 | 0.000359 | 7.84E-05 | 0.003575076 | DDX54 |
| chr9 | 125747450 | 125747591 | 142 | 4 | 4.44E-05 | 0.0003583 | 0.001523 | 7.85E-05 | -0.00014887 | PBX3 |
| chr4 | 186700645 | 186701010 | 366 | 4 | 3.08E-05 | 0.0013282 | 0.000605 | 7.85E-05 | -0.00245659 | FAT1 |
| chr2 | 202090836 | 202091062 | 227 | 2 | 0.000124234 | 4.88E-05 | 0.001826 | 7.85E-05 | 0.000265549 | KIAA2012, KIAA2012-AS1 |
| chr11 | 60455765 | 60455818 | 54 | 2 | 5.71E-05 | 0.0069475 | 3.61E-05 | 7.85E-05 | 0.00947965 | NA |
| chr3 | 56894855 | 56895407 | 553 | 2 | 5.62E-06 | 0.0131825 | 3.39E-05 | 7.85E-05 | 0.002019807 | ARHGEF3 |
| chr17 | 80800809 | 80800923 | 115 | 2 | 1.65E-05 | 0.000512 | 7.20E-05 | 7.85E-05 | 0.003565306 | RPTOR |
| chr1 | 39540825 | 39540999 | 175 | 2 | 8.89E-05 | 0.000324 | 9.14E-05 | 7.85E-05 | -0.01229144 | PPIEL |
| chr9 | 130714106 | 130714267 | 162 | 2 | 9.31E-05 | 0.0002085 | 0.000125 | 7.86E-05 | -0.01554609 | ABL1 |
| chr6 | 168081768 | 168082034 | 267 | 4 | 0.000161392 | 9.21E-05 | 0.000938 | 7.86E-05 | -0.00188099 | FRMD1 |
| chr1 | 226383016 | 226383069 | 54 | 2 | 0.000110542 | 0.0010356 | 5.46E-05 | 7.86E-05 | -0.00209839 | PARP1 |
| chr15 | 69731387 | 69731768 | 382 | 3 | 6.50E-05 | 5.74E-05 | 0.00182 | 7.86E-05 | 0.005444939 | DRAIC |
| chr7 | 12111595 | 12112012 | 418 | 4 | 9.83E-05 | 0.0002304 | 0.001697 | 7.86E-05 | -0.00424912 | NA |
| chr17 | 8392419 | 8392653 | 235 | 3 | 4.46E-05 | 0.0039988 | 3.57E-05 | 7.86E-05 | 0.007055069 | RNF222 |
| chr2 | 61019035 | 61019113 | 79 | 2 | 9.19E-06 | 5.51E-05 | 0.001086 | 7.87E-05 | 0.01246582 | PEX13 |
| chr10 | 22622608 | 22622700 | 93 | 2 | 0.000127474 | 0.0002143 | 0.000122 | 7.87E-05 | 0.003945684 | PIP4K2A |
| chr7 | 48663367 | 48663457 | 91 | 3 | 0.000119982 | 0.0001656 | 0.000231 | 7.88E-05 | -0.00173215 | AC091770.1 |
| chr5 | 95554460 | 95554707 | 248 | 2 | 1.95E-05 | 9.88E-05 | 0.000288 | 7.88E-05 | -0.00421075 | TTC37 |
| chr21 | 46635300 | 46635459 | 160 | 5 | 0.000138884 | 3.32E-05 | 0.006519 | 7.89E-05 | -0.00024961 | NA |
| chr3 | 126084414 | 126085016 | 603 | 5 | 4.89E-05 | 0.0001306 | 0.003098 | 7.89E-05 | 0.000782233 | AC079848.1, SLC41A3 |
| chr8 | 2032947 | 2033459 | 513 | 5 | 5.59E-05 | 0.0059819 | 8.33E-05 | 7.89E-05 | -0.00101222 | NA |
| chr5 | 178514878 | 178515469 | 592 | 3 | 6.78E-05 | 6.91E-05 | 0.000863 | 7.89E-05 | 0.003990649 | COL23A1 |
| chr16 | 85557845 | 85557908 | 64 | 2 | 6.11E-05 | 0.0004537 | 7.68E-05 | 7.90E-05 | -0.00171408 | GSE1 |
| chr11 | 15482857 | 15482975 | 119 | 2 | 2.37E-05 | 0.001575 | 4.83E-05 | 7.90E-05 | -0.01251905 | NA |
| chr11 | 1932431 | 1932616 | 186 | 2 | 1.00E-05 | 0.000121 | 0.000219 | 7.90E-05 | 0.007846725 | TNNT3 |
| chr17 | 15265166 | 15265341 | 176 | 3 | 3.33E-05 | 0.000525 | 7.14E-05 | 7.91E-05 | -0.01081131 | AC005703.2, PMP22 |

| | | | | | | | | | | |
|---|---|---|---|---|---|---|---|---|---|---|
| chr7 | 73739645 | 73740027 | 383 | 2 | 4.41E-05 | 0.0001109 | 0.000247 | 7.91E-05 | -0.007787 | NA |
| chr8 | 141172437 | 141173337 | 901 | 4 | 1.28E-06 | 0.0015281 | 8.82E-05 | 7.91E-05 | -0.00661935 | DENND3 |
| chr6 | 27867585 | 27867911 | 327 | 7 | 0.000151809 | 0.001643 | 0.002413 | 7.92E-05 | -0.00176879 | H1-5 |
| chr2 | 20177886 | 20178123 | 238 | 2 | 7.64E-05 | 0.0010618 | 5.46E-05 | 7.92E-05 | -0.0051154 | NA |
| chr1 | 6634271 | 6634332 | 62 | 2 | 0.000159398 | 5.29E-05 | 0.001315 | 7.92E-05 | -0.00764637 | THAP3, DNAJC11 |
| chr1 | 25340415 | 25340618 | 204 | 3 | 3.54E-05 | 0.0296082 | 1.40E-05 | 7.92E-05 | -0.00363211 | TMEM50A |
| chr15 | 34339163 | 34339561 | 399 | 2 | 1.39E-05 | 0.0001824 | 0.000142 | 7.93E-05 | -0.01132991 | NA |
| chr7 | 44570099 | 44570785 | 687 | 3 | 5.41E-05 | 4.07E-05 | 0.004518 | 7.93E-05 | 0.00385598 | DDX56 |
| chr3 | 149003740 | 149003881 | 142 | 2 | 0.000128226 | 5.16E-05 | 0.001474 | 7.93E-05 | -0.0007378 | GYG1 |
| chr13 | 95076198 | 95076261 | 64 | 2 | 7.60E-05 | 0.0004283 | 7.95E-05 | 7.93E-05 | 0.004164264 | ABCC4 |
| chr7 | 30134960 | 30135186 | 227 | 4 | 3.88E-05 | 0.0016053 | 6.74E-05 | 7.93E-05 | -0.00190631 | MTURN |
| chr2 | 32356061 | 32356526 | 466 | 2 | 3.50E-07 | 0.0004617 | 7.66E-05 | 7.94E-05 | -0.00524466 | NA |
| chr8 | 37699345 | 37699667 | 323 | 3 | 0.000131412 | 3.69E-05 | 0.004045 | 7.94E-05 | -0.01185697 | ZNF703 |
| chr9 | 86406089 | 86406681 | 593 | 3 | 0.000110151 | 4.61E-05 | 0.002393 | 7.94E-05 | -0.01275347 | NA |
| chr8 | 143570333 | 143570692 | 360 | 2 | 5.20E-05 | 0.0001692 | 0.000153 | 7.95E-05 | -0.00687521 | MROH6 |
| chr1 | 158203897 | 158204237 | 341 | 4 | 2.48E-05 | 9.90E-05 | 0.000491 | 7.95E-05 | -0.00018772 | NA |
| chr14 | 61528848 | 61529108 | 261 | 2 | 4.09E-05 | 0.0001401 | 0.000187 | 7.95E-05 | -0.00300507 | PRKCH |
| chr11 | 1084585 | 1084949 | 365 | 4 | 2.52E-05 | 0.002701 | 6.03E-05 | 7.95E-05 | -0.00801676 | MUC2 |
| chr14 | 77800365 | 77800504 | 140 | 3 | 0.000145972 | 0.0132849 | 2.51E-05 | 7.96E-05 | -0.00228611 | ADCK1 |
| chr6 | 55276956 | 55277213 | 258 | 2 | 4.60E-05 | 0.0179984 | 3.36E-05 | 7.97E-05 | -0.02392703 | HCRTR2 |
| chr6 | 2767598 | 2767883 | 286 | 2 | 5.35E-05 | 5.95E-05 | 0.000883 | 7.97E-05 | -0.01378134 | WRNIP1 |
| chr12 | 590127 | 590605 | 479 | 5 | 7.50E-05 | 0.0004527 | 0.000124 | 7.99E-05 | -0.003686 | NINJ2 |
| chr22 | 48547505 | 48547724 | 220 | 3 | 1.90E-05 | 0.0045528 | 2.47E-05 | 7.99E-05 | -0.00585591 | TAFA5 |
| chr2 | 28870572 | 28871039 | 468 | 6 | 7.03E-05 | 3.40E-05 | 0.005227 | 8.00E-05 | -0.00270463 | NA |
| chr15 | 92782047 | 92782055 | 9 | 2 | 8.95E-05 | 0.0073677 | 3.65E-05 | 8.01E-05 | -0.00493376 | FAM174B |
| chr22 | 41900078 | 41900937 | 860 | 7 | 2.63E-05 | 0.0015146 | 0.00082 | 8.01E-05 | -0.0005441 | SREBF2 |
| chr16 | 50684228 | 50684896 | 669 | 2 | 2.68E-06 | 0.0108307 | 3.50E-05 | 8.01E-05 | 0.003073002 | NA |
| chr1 | 234272534 | 234272561 | 28 | 2 | 0.000118555 | 0.0023665 | 4.43E-05 | 8.01E-05 | 0.00655707 | SLC35F3, AL122008.1 |
| chr12 | 57890336 | 57890796 | 461 | 3 | 5.50E-05 | 0.0004337 | 0.000331 | 8.02E-05 | -0.00513526 | AC083805.1 |
| chr4 | 3304950 | 3304978 | 29 | 2 | 4.57E-05 | 5.07E-05 | 0.001681 | 8.02E-05 | -0.0068341 | RGS12 |
| chr15 | 47438930 | 47439345 | 416 | 2 | 1.79E-05 | 0.0049311 | 3.85E-05 | 8.02E-05 | 0.003247072 | SEMA6D |
| chr14 | 72539434 | 72540059 | 626 | 3 | 2.69E-05 | 0.0087281 | 3.12E-05 | 8.02E-05 | 0.000476337 | RGS6, AC004828.2 |
| chr12 | 10153578 | 10153903 | 326 | 5 | 4.81E-05 | 0.0049433 | 4.56E-05 | 8.02E-05 | 0.000728975 | NA |
| chr15 | 40109519 | 40109684 | 166 | 2 | 2.08E-05 | 0.001264 | 5.23E-05 | 8.02E-05 | -0.00414483 | NA |
| chr6 | 32064503 | 32065112 | 610 | 6 | 2.37E-05 | 0.0002154 | 0.000689 | 8.02E-05 | -0.00622721 | TNXB |
| chr3 | 161371641 | 161373010 | 1370 | 14 | 5.77E-05 | 0.0014516 | 0.002244 | 8.03E-05 | -0.00014286 | SPTSSB |
| chr8 | 17921895 | 17922414 | 520 | 2 | 3.22E-07 | 0.000432 | 8.04E-05 | 8.03E-05 | -0.00695461 | NA |
| chr19 | 704346 | 704580 | 235 | 3 | 5.46E-05 | 0.0052677 | 2.30E-05 | 8.03E-05 | 0.001457136 | NA |
| chr20 | 57774346 | 57774537 | 192 | 2 | 9.61E-05 | 0.0011308 | 5.43E-05 | 8.03E-05 | 0.004100672 | NA |
| chr11 | 26309244 | 26309303 | 60 | 2 | 0.000132923 | 5.13E-05 | 0.001619 | 8.04E-05 | 0.016530826 | ANO3, ANO3-AS1 |
| chr22 | 43231142 | 43231407 | 266 | 3 | 0.000130065 | 0.0001734 | 0.000919 | 8.04E-05 | 0.003678238 | SCUBE1 |
| chr2 | 20609263 | 20609497 | 235 | 2 | 0.000131938 | 6.27E-05 | 0.000777 | 8.05E-05 | -0.00542504 | HS1BP3 |
| chr16 | 85979604 | 85979772 | 169 | 2 | 1.21E-05 | 0.0007322 | 6.36E-05 | 8.06E-05 | -0.00711136 | AC092723.4 |
| chr22 | 39532204 | 39532303 | 100 | 2 | 1.26E-05 | 0.0002646 | 0.000108 | 8.08E-05 | 0.001498443 | RPS19BP1 |
| chr17 | 81556616 | 81556858 | 243 | 2 | 4.23E-05 | 0.0010592 | 5.59E-05 | 8.08E-05 | -0.00332412 | NA |
| chr14 | 98819631 | 98819648 | 18 | 2 | 8.37E-05 | 0.0191604 | 3.39E-05 | 8.08E-05 | -0.01505455 | NA |
| chr4 | 56910497 | 56910560 | 64 | 2 | 0.000103799 | 5.32E-05 | 0.001399 | 8.08E-05 | 0.000102944 | REST |
| chr11 | 94876354 | 94876462 | 109 | 2 | 0.000134973 | 0.0048985 | 3.88E-05 | 8.08E-05 | 0.004558367 | AMOTL1, AP002383.2 |
| chr12 | 131755164 | 131755710 | 547 | 3 | 3.70E-05 | 0.0042384 | 9.06E-05 | 8.09E-05 | -0.00463324 | SFSWAP |
| chr20 | 2138077 | 2138137 | 61 | 2 | 0.000136861 | 0.0002037 | 0.000134 | 8.11E-05 | 0.00201418 | STK35 |
| chr2 | 61405981 | 61406286 | 306 | 3 | 1.08E-05 | 0.0177358 | 1.94E-05 | 8.11E-05 | -0.00834718 | USP34 |
| chr1 | 151721027 | 151721844 | 818 | 12 | 1.58E-05 | 0.0122493 | 0.003461 | 8.12E-05 | 0.000352767 | RIIAD1 |
| chr7 | 575194 | 576098 | 905 | 4 | 7.86E-06 | 0.0051895 | 1.97E-05 | 8.12E-05 | 0.002004047 | PRKAR1B |
| chr5 | 141753770 | 141754281 | 512 | 3 | 4.64E-05 | 6.34E-05 | 0.002066 | 8.12E-05 | 0.003145607 | AC005753.2 |
| chr15 | 58852518 | 58852646 | 129 | 2 | 0.000123915 | 0.0008173 | 6.16E-05 | 8.13E-05 | -0.02114591 | MINDY2 |
| chr5 | 154486043 | 154486335 | 293 | 4 | 4.19E-05 | 7.31E-05 | 0.003741 | 8.13E-05 | 0.006687403 | AC026688.2 |
| chr1 | 234531617 | 234531803 | 187 | 5 | 8.00E-05 | 0.0002527 | 0.0021 | 8.13E-05 | 0.000798936 | LINC01354 |
| chr2 | 144615508 | 144615684 | 177 | 2 | 7.68E-05 | 0.0022063 | 4.56E-05 | 8.13E-05 | -0.00494651 | NA |
| chr17 | 27245000 | 27245646 | 647 | 3 | 3.62E-06 | 0.0008468 | 6.56E-05 | 8.14E-05 | -0.00440081 | NA |
| chr19 | 1205013 | 1205324 | 312 | 3 | 5.45E-06 | 0.0010587 | 0.000178 | 8.14E-05 | 0.002061876 | STK11 |
| chr5 | 1809257 | 1809416 | 160 | 2 | 3.46E-05 | 0.0011297 | 5.51E-05 | 8.14E-05 | 0.011342718 | NDUFS6 |
| chr19 | 4185627 | 4185754 | 128 | 2 | 0.000111968 | 4.98E-05 | 0.002024 | 8.15E-05 | -0.00735542 | ANKRD24 |
| chr5 | 15111041 | 15111075 | 35 | 2 | 7.05E-05 | 5.92E-05 | 0.000974 | 8.15E-05 | -0.00987275 | NA |
| chr1 | 6701804 | 6702540 | 737 | 9 | 4.63E-05 | 0.0012152 | 0.005095 | 8.16E-05 | 0.001644873 | DNAJC11 |
| chr12 | 63898175 | 63898358 | 184 | 2 | 8.01E-05 | 0.0020489 | 4.65E-05 | 8.16E-05 | 0.012013242 | SRGAP1 |
| chr4 | 112647664 | 112647972 | 309 | 2 | 8.46E-05 | 0.005089 | 3.89E-05 | 8.17E-05 | -0.00345281 | LARP7, MIR302CHG, MIR367 |
| chr2 | 3559711 | 3559716 | 6 | 2 | 1.12E-05 | 0.0035112 | 4.15E-05 | 8.17E-05 | 0.002317107 | RNASEH1-AS1 |
| chr3 | 29644215 | 29644513 | 299 | 2 | 2.93E-05 | 0.0006295 | 6.87E-05 | 8.17E-05 | -0.00397003 | AC098650.1, RBMS3 |
| chr4 | 110643050 | 110643612 | 563 | 5 | 1.36E-05 | 0.021573 | 0.000324 | 8.18E-05 | 0.001330951 | NA |
| chr16 | 13244833 | 13244843 | 11 | 2 | 8.67E-05 | 0.0009984 | 5.78E-05 | 8.18E-05 | -0.00351482 | NA |
| chr12 | 49674771 | 49674827 | 57 | 2 | 0.000165956 | 0.0001123 | 0.00026 | 8.18E-05 | 0.010251244 | FMNL3 |
| chr16 | 31482039 | 31482051 | 13 | 2 | 3.12E-05 | 5.34E-05 | 0.001463 | 8.19E-05 | 0.000120492 | NA |
| chr7 | 99438670 | 99439079 | 410 | 11 | 0.000150031 | 0.0050556 | 0.003952 | 8.19E-05 | -0.00059463 | CPSF4, PTCD1, ATP5MF-PTCD1 |
| chr8 | 76673554 | 76674344 | 791 | 5 | 6.45E-06 | 0.023457 | 1.30E-05 | 8.19E-05 | -0.00283202 | ZFHX4-AS1 |
| chr18 | 3594175 | 3594398 | 224 | 4 | 0.000163322 | 0.0017932 | 0.000261 | 8.19E-05 | 0.002185063 | DLGAP1-AS1, DLGAP1 |
| chr7 | 2175049 | 2175155 | 107 | 4 | 0.000112403 | 0.003107 | 9.08E-05 | 8.19E-05 | 0.000212053 | MAD1L1, AC069288.1 |
| chr1 | 55215053 | 55215498 | 446 | 5 | 2.15E-05 | 0.0002269 | 0.000996 | 8.20E-05 | 0.001144885 | USP24 |
| chr10 | 89328294 | 89328393 | 100 | 2 | 4.74E-05 | 0.0001196 | 0.00024 | 8.20E-05 | 0.012045336 | IFIT3, LIPA |
| chr14 | 22957383 | 22957914 | 532 | 4 | 1.21E-05 | 0.0036687 | 3.49E-05 | 8.21E-05 | -0.00114001 | AL132780.3 |
| chr16 | 67648102 | 67648162 | 61 | 2 | 5.26E-05 | 8.33E-05 | 0.00042 | 8.21E-05 | -0.00354647 | CARMIL2 |
| chr17 | 82663760 | 82663982 | 223 | 4 | 4.64E-05 | 0.0070753 | 7.25E-05 | 8.21E-05 | 0.000374679 | RAB40B |
| chr1 | 19342665 | 19342790 | 126 | 2 | 0.000144516 | 0.0007215 | 6.54E-05 | 8.22E-05 | 0.002553286 | CAPZB |
| chr11 | 1937821 | 1938016 | 196 | 2 | 0.000105666 | 5.70E-05 | 0.001149 | 8.22E-05 | -0.00169323 | TNNT3 |
| chr12 | 76084675 | 76084888 | 214 | 5 | 6.51E-06 | 0.0207328 | 8.48E-06 | 8.23E-05 | -0.00128954 | NAP1L1 |
| chr1 | 53893459 | 53893473 | 15 | 3 | 5.63E-05 | 0.0002576 | 0.001524 | 8.23E-05 | 0.004874584 | DIO1 |
| chr8 | 48319095 | 48319226 | 132 | 3 | 7.77E-05 | 0.0010369 | 0.000818 | 8.24E-05 | -0.0055418 | NA |
| chr19 | 42095337 | 42095479 | 143 | 3 | 7.28E-05 | 3.12E-05 | 0.008282 | 8.24E-05 | -0.00826762 | POU2F2 |
| chr1 | 1907315 | 1907590 | 276 | 2 | 3.46E-05 | 0.0001289 | 0.00022 | 8.24E-05 | 0.008524902 | NA |
| chr17 | 1754154 | 1754971 | 818 | 5 | 2.86E-05 | 0.0138627 | 0.000768 | 8.24E-05 | -0.00616057 | SERPINF2 |
| chr18 | 74520227 | 74520241 | 15 | 2 | 0.000132027 | 0.0012089 | 5.46E-05 | 8.24E-05 | -0.00316207 | CNDP2 |
| chr11 | 63671090 | 63671862 | 773 | 12 | 4.75E-05 | 0.0001045 | 0.017594 | 8.24E-05 | -0.00017801 | ATL3 |
| chr20 | 62900946 | 62901290 | 345 | 3 | 2.36E-05 | 0.0008491 | 7.29E-05 | 8.25E-05 | 0.000499884 | DIDO1 |
| chr12 | 5565642 | 5565763 | 122 | 2 | 1.71E-05 | 4.87E-05 | 0.002498 | 8.25E-05 | 0.000782265 | ANO2 |
| chr3 | 50345258 | 50345937 | 680 | 11 | 1.83E-05 | 0.0003986 | 0.000729 | 8.26E-05 | -0.00210729 | ZMYND10-AS1, ZMYND10 |
| chr10 | 24255150 | 24255364 | 215 | 4 | 9.30E-05 | 0.0021845 | 0.000181 | 8.26E-05 | -0.0109751 | KIAA1217, AC063961.1 |
| chr17 | 79770125 | 79770285 | 161 | 2 | 5.63E-05 | 0.0091454 | 3.66E-05 | 8.28E-05 | -0.00643354 | NA |
| chr12 | 123905535 | 123905537 | 3 | 2 | 0.000128253 | 0.001436 | 5.19E-05 | 8.29E-05 | 0.007633399 | DNAH10 |
| chr4 | 7787815 | 7788434 | 620 | 3 | 2.19E-05 | 0.0003118 | 0.000113 | 8.29E-05 | -0.00416835 | AFAP1 |
| chr6 | 7107272 | 7108404 | 1133 | 12 | 1.37E-05 | 0.0022264 | 0.001796 | 8.29E-05 | 0.000220413 | RREB1 |
| chr11 | 65132304 | 65132692 | 389 | 3 | 7.69E-05 | 0.0007286 | 0.000218 | 8.30E-05 | 0.006347336 | SYVN1 |
| chr18 | 79378860 | 79379085 | 226 | 3 | 4.09E-05 | 0.0241936 | 1.52E-05 | 8.30E-05 | -0.00384997 | NA |
| chr11 | 61754706 | 61755335 | 630 | 7 | 5.71E-05 | 0.0010414 | 0.00214 | 8.30E-05 | -0.00135186 | MYRF, MYRF-AS1 |
| chr7 | 131908847 | 131908881 | 35 | 2 | 8.25E-05 | 4.91E-05 | 0.002473 | 8.30E-05 | 0.001230087 | AC009518.1 |
| chr19 | 34593497 | 34593671 | 175 | 2 | 9.38E-05 | 0.0001717 | 0.000163 | 8.30E-05 | 0.004344561 | SCGB2B2 |
| chr7 | 504888 | 505287 | 400 | 5 | 8.83E-05 | 0.0045605 | 0.000226 | 8.31E-05 | 0.002919958 | PDGFA |
| chr1 | 18481056 | 18481100 | 45 | 2 | 0.000120169 | 6.18E-05 | 0.000909 | 8.31E-05 | -0.01897168 | KLHDC7A |
| chr17 | 3801177 | 3801813 | 637 | 6 | 7.96E-05 | 0.001934 | 0.003405 | 8.32E-05 | -0.00605643 | ITGAE |
| chr8 | 108443025 | 108443435 | 411 | 7 | 9.48E-05 | 0.0002558 | 0.003609 | 8.32E-05 | 9.95E-05 | EIF3E |
| chr11 | 65572220 | 65572811 | 592 | 10 | 9.14E-05 | 0.0109367 | 0.005514 | 8.32E-05 | -0.00017326 | ZNRD2, FAM89B |
| chr19 | 50788977 | 50789208 | 232 | 2 | 2.64E-05 | 9.05E-05 | 0.000375 | 8.33E-05 | -0.00646956 | AC010325.1 |
| chr22 | 36870761 | 36871239 | 479 | 2 | 9.51E-06 | 0.0001859 | 0.000152 | 8.33E-05 | -0.00464334 | NCF4 |
| chr1 | 4656253 | 4656518 | 266 | 4 | 5.64E-06 | 0.0020527 | 3.51E-05 | 8.34E-05 | -0.00165992 | AJAP1 |
| chr4 | 122921870 | 122922428 | 559 | 4 | 2.08E-06 | 0.0001357 | 0.000566 | 8.34E-05 | -0.00604992 | NUDT6 |
| chr19 | 53193818 | 53194031 | 214 | 2 | 2.07E-05 | 5.33E-05 | 0.001619 | 8.34E-05 | -0.0039021 | NA |
| chr4 | 147485802 | 147486029 | 228 | 2 | 3.29E-05 | 0.001376 | 5.31E-05 | 8.35E-05 | -0.00864529 | EDNRA |
| chr4 | 25233747 | 25234143 | 397 | 10 | 5.45E-05 | 0.0895669 | 0.000115 | 8.35E-05 | -0.00103627 | PI4K2B |
| chr3 | 124585186 | 124585314 | 129 | 2 | 7.80E-05 | 0.0003257 | 9.86E-05 | 8.36E-05 | -0.00233325 | KALRN |
| chr14 | 50040811 | 50041760 | 950 | 6 | 3.96E-06 | 0.0068656 | 0.001416 | 8.37E-05 | 0.000483374 | LINC01588 |
| chr2 | 138100667 | 138100697 | 31 | 2 | 8.67E-05 | 0.0002715 | 0.000111 | 8.37E-05 | -0.00873785 | NA |
| chr6 | 144660000 | 144660067 | 68 | 4 | 2.58E-05 | 0.0258403 | 1.97E-05 | 8.37E-05 | -0.00585283 | UTRN |
| chr14 | 91804039 | 91804191 | 153 | 3 | 6.91E-05 | 0.0212667 | 2.90E-05 | 8.37E-05 | -0.00062405 | TC2N |
| chr1 | 110339198 | 110339526 | 329 | 9 | 0.000131036 | 0.0004971 | 0.00044 | 8.38E-05 | 4.07E-06 | RBM15 |
| chr12 | 88035766 | 88036360 | 595 | 4 | 4.07E-06 | 0.0001182 | 0.001568 | 8.38E-05 | -0.00340392 | C12orf29 |
| chr1 | 91891835 | 91891901 | 67 | 2 | 0.000101133 | 0.0043899 | 4.09E-05 | 8.38E-05 | 0.014108951 | TGFBR3 |
| chr2 | 170770648 | 170771211 | 564 | 7 | 5.80E-05 | 0.0010364 | 7.92E-05 | 8.38E-05 | -0.00020541 | AC007405.3, AC007405.1 |
| chr2 | 239246587 | 239246717 | 131 | 2 | 3.55E-05 | 7.02E-05 | 0.000652 | 8.39E-05 | -0.01322605 | HDAC4 |
| chr17 | 40921222 | 40921363 | 142 | 2 | 8.64E-05 | 7.01E-05 | 0.000655 | 8.39E-05 | 0.003013858 | NA |
| chr19 | 21750584 | 21750738 | 155 | 7 | 0.000100049 | 0.0108429 | 0.000461 | 8.40E-05 | 0.000501894 | ZNF100 |
| chr1 | 1247278 | 1247806 | 529 | 2 | 3.47E-05 | 4.96E-05 | 0.00251 | 8.41E-05 | -0.00407078 | NA |
| chr5 | 141848851 | 141849426 | 576 | 2 | 8.71E-06 | 0.0013771 | 5.35E-05 | 8.41E-05 | -0.00851145 | NA |
| chr11 | 80328582 | 80328730 | 149 | 2 | 7.23E-05 | 0.0001476 | 0.000195 | 8.42E-05 | -0.01388191 | NA |
| chr1 | 160204965 | 160205496 | 532 | 2 | 7.75E-05 | 0.0008387 | 6.33E-05 | 8.42E-05 | 0.003904181 | PEA15, AL121987.2 |
| chr7 | 1744951 | 1745661 | 711 | 3 | 2.11E-06 | 0.0029348 | 3.90E-05 | 8.42E-05 | 0.002190502 | ELFN1 |
| chr22 | 24554411 | 24555409 | 999 | 8 | 2.41E-05 | 0.0001363 | 0.005467 | 8.43E-05 | 0.000322294 | GUCD1 |
| chr11 | 7467975 | 7468246 | 272 | 2 | 7.61E-05 | 0.0001808 | 0.000159 | 8.43E-05 | -0.00438115 | SYT9, AC107884.1 |
| chr2 | 3576102 | 3576436 | 335 | 3 | 5.87E-05 | 0.0011009 | 0.000473 | 8.44E-05 | 0.009222526 | RPS7 |
| chr1 | 234372601 | 234373495 | 895 | 10 | 1.10E-06 | 0.0012883 | 0.001395 | 8.44E-05 | -0.0036869 | AL355472.3, COA6, COA6-AS1 |
| chr17 | 39953383 | 39953768 | 386 | 2 | 1.38E-05 | 0.0037609 | 4.22E-05 | 8.44E-05 | -0.00722224 | GSDMA |
| chr1 | 8335247 | 8335497 | 251 | 6 | 0.000154375 | 0.0002742 | 0.004964 | 8.45E-05 | -0.00292172 | SLC45A1 |
| chr9 | 127611353 | 127611480 | 128 | 3 | 0.000111018 | 0.0001657 | 0.00021 | 8.45E-05 | 0.006457305 | STXBP1 |
| chr4 | 41255674 | 41256357 | 684 | 4 | 6.69E-05 | 0.0001778 | 0.000827 | 8.45E-05 | -0.00940974 | UCHL1-AS1 |
| chr7 | 117322572 | 117323958 | 1387 | 11 | 2.37E-05 | 0.000496 | 0.006403 | 8.45E-05 | 0.002147508 | CFTR, WNT2 |
| chr8 | 22877875 | 22877965 | 91 | 2 | 0.000178831 | 7.23E-05 | 0.000621 | 8.46E-05 | -0.01267045 | PEBP4 |
| chr17 | 2695964 | 2696529 | 566 | 3 | 7.24E-05 | 3.30E-05 | 0.007581 | 8.47E-05 | -0.00053604 | CLUH |
| chr1 | 27391717 | 27392541 | 825 | 7 | 1.90E-05 | 0.0001262 | 0.001398 | 8.47E-05 | -0.0039989 | NA |
| chr1 | 177171067 | 177171676 | 610 | 9 | 2.74E-05 | 0.0027337 | 3.93E-05 | 8.48E-05 | 0.000936258 | BRINP2 |
| chr11 | 44607824 | 44608929 | 1106 | 5 | 8.37E-06 | 0.0093362 | 6.05E-05 | 8.48E-05 | -0.00230747 | CD82, AC010768.2 |
| chr11 | 16791775 | 16792134 | 360 | 4 | 4.77E-05 | 0.0005417 | 8.85E-05 | 8.49E-05 | -0.0079769 | PLEKHA7 |
| chr3 | 126703417 | 126703732 | 316 | 3 | 9.14E-05 | 0.001257 | 0.00064 | 8.49E-05 | 0.0213872 | NA |
| chr17 | 45169193 | 45169783 | 591 | 4 | 4.67E-05 | 0.0006474 | 6.16E-05 | 8.50E-05 | 0.00113639 | HEXIM2, AC138150.2 |
| chr11 | 44709947 | 44710143 | 197 | 2 | 7.03E-05 | 0.0048358 | 4.08E-05 | 8.50E-05 | -0.00952164 | NA |
| chr8 | 4514486 | 4515229 | 744 | 4 | 1.61E-05 | 9.07E-05 | 0.002788 | 8.51E-05 | -0.00277035 | CSMD1 |
| chr7 | 64902696 | 64903197 | 502 | 4 | 5.19E-06 | 3.27E-05 | 0.00604 | 8.51E-05 | -0.00564452 | ZNF273 |
| chr8 | 103110912 | 103111325 | 414 | 2 | 8.08E-05 | 6.78E-05 | 0.00075 | 8.52E-05 | 0.021020262 | NA |
| chr10 | 71793381 | 71793444 | 64 | 3 | 0.000118579 | 0.0001825 | 0.001023 | 8.52E-05 | -0.00350665 | CDH23 |
| chr17 | 46193950 | 46194193 | 244 | 2 | 1.09E-05 | 0.0003057 | 0.000105 | 8.53E-05 | 0.002031977 | KANSL1-AS1, KANSL1 |
| chr11 | 62339267 | 62339400 | 134 | 2 | 8.37E-05 | 7.64E-05 | 0.000556 | 8.53E-05 | 0.006772536 | ASRGL1 |
| chr17 | 75764214 | 75764493 | 280 | 3 | 8.45E-06 | 0.0298354 | 1.76E-05 | 8.53E-05 | -0.00546171 | GALK1 |

| | | | | | | | | | | |
|---|---|---|---|---|---|---|---|---|---|---|
| chr9 | 121370641 | 121371278 | 638 | 3 | 4.38E-06 | 4.64E-05 | 0.002199 | 8.55E-05 | -0.00784529 | AL359644.1 |
| chr19 | 50824100 | 50824221 | 122 | 2 | 4.65E-05 | 0.0026251 | 4.60E-05 | 8.56E-05 | 0.009188874 | NA |
| chr2 | 241725197 | 241725919 | 723 | 5 | 3.45E-05 | 0.0001995 | 0.001605 | 8.56E-05 | -0.00420927 | ING5, AC114730.3 |
| chr10 | 100534797 | 100535384 | 588 | 4 | 8.56E-06 | 0.0012213 | 0.00085 | 8.57E-05 | -0.00331655 | HIF1AN |
| chr7 | 32318452 | 32318719 | 268 | 2 | 0.000147713 | 5.50E-05 | 0.001602 | 8.58E-05 | 0.04050514 | PDE1C |
| chr6 | 165977418 | 165977430 | 13 | 2 | 0.000141216 | 0.0011236 | 5.83E-05 | 8.58E-05 | 0.001616534 | PDE10A |
| chr7 | 149881295 | 149881352 | 58 | 2 | 0.00013502 | 5.05E-05 | 0.002537 | 8.58E-05 | -0.01223056 | ATP6V0E2 |
| chr11 | 57459617 | 57460360 | 744 | 5 | 2.57E-05 | 8.08E-05 | 0.001433 | 8.58E-05 | 0.000489279 | NA |
| chr12 | 57628627 | 57628837 | 211 | 3 | 0.000138198 | 4.40E-05 | 0.004106 | 8.59E-05 | -0.01199328 | B4GALNT1 |
| chr14 | 69992946 | 69992955 | 10 | 2 | 0.000102324 | 0.0016345 | 5.19E-05 | 8.59E-05 | -0.00726987 | SMOC1 |
| chr17 | 39625406 | 39626432 | 1027 | 5 | 1.78E-05 | 0.0001416 | 0.004845 | 8.59E-05 | -0.00319691 | NA |
| chr1 | 183591778 | 183591788 | 11 | 2 | 8.55E-05 | 5.26E-05 | 0.002032 | 8.59E-05 | -0.0114005 | SMG7 |
| chr7 | 2167149 | 2167171 | 23 | 2 | 0.000102104 | 9.69E-05 | 0.000361 | 8.60E-05 | -0.00103617 | MAD1L1, AC069288.1 |
| chr10 | 124728443 | 124728535 | 93 | 3 | 0.000170241 | 8.02E-05 | 0.000787 | 8.61E-05 | 0.009128312 | AC068896.1, FAM53B |
| chr20 | 58514905 | 58515513 | 609 | 10 | 9.01E-05 | 0.0009131 | 0.007703 | 8.61E-05 | 0.001119615 | APCDD1L-DT, APCDD1L |
| chr16 | 69798786 | 69798928 | 143 | 2 | 0.000171804 | 0.0004939 | 8.16E-05 | 8.61E-05 | 0.002725211 | WWP2 |
| chr5 | 130386803 | 130386811 | 9 | 2 | 2.37E-05 | 0.0255667 | 3.51E-05 | 8.62E-05 | -0.01305418 | NA |
| chr13 | 110598414 | 110598604 | 191 | 3 | 3.21E-05 | 0.0134091 | 1.95E-05 | 8.63E-05 | -0.00176985 | NA |
| chr7 | 100827323 | 100828108 | 786 | 9 | 3.84E-05 | 0.0003341 | 0.00432 | 8.64E-05 | 0.000536975 | SLC12A9, EPHB4 |
| chr4 | 1640327 | 1640530 | 204 | 3 | 0.000110077 | 6.54E-05 | 0.000989 | 8.64E-05 | -0.00488093 | FAM53A |
| chr14 | 103563018 | 103563746 | 729 | 8 | 1.03E-05 | 0.0052376 | 2.83E-05 | 8.65E-05 | -0.00063278 | KLC1, COA8, AL139300.1 |
| chr6 | 145736995 | 145737079 | 85 | 2 | 7.88E-05 | 5.10E-05 | 0.002517 | 8.65E-05 | 0.003601434 | AL023806.1, AL023806.2 |
| chr1 | 1782237 | 1782832 | 596 | 6 | 6.31E-05 | 0.0001897 | 0.003272 | 8.66E-05 | 0.004568731 | NA |
| chr2 | 150485836 | 150487068 | 1233 | 6 | 2.54E-05 | 6.86E-05 | 0.004488 | 8.66E-05 | -0.00153528 | RND3 |
| chr1 | 46719564 | 46719905 | 342 | 2 | 3.77E-05 | 5.37E-05 | 0.001893 | 8.66E-05 | 0.015310705 | NA |
| chr2 | 161136804 | 161136853 | 50 | 2 | 5.31E-05 | 0.0014358 | 5.44E-05 | 8.67E-05 | -0.00270064 | TANK-AS1 |
| chr5 | 116449463 | 116449505 | 43 | 2 | 0.000112704 | 0.000102 | 0.000339 | 8.68E-05 | -0.01227039 | SEMA6A-AS1, SEMA6A |
| chr6 | 147406743 | 147407154 | 412 | 5 | 5.09E-06 | 0.0070152 | 1.22E-05 | 8.68E-05 | -0.00127483 | AL355365.1, YAP1P1 |
| chr20 | 54171995 | 54173107 | 1113 | 4 | 1.03E-05 | 0.004253 | 2.11E-05 | 8.69E-05 | -0.00397421 | CYP24A1 |
| chr13 | 83883203 | 83883205 | 3 | 2 | 9.64E-05 | 6.12E-05 | 0.001104 | 8.69E-05 | -0.01475788 | AL355481.1 |
| chr3 | 33797897 | 33798211 | 315 | 4 | 3.86E-05 | 9.00E-05 | 0.004262 | 8.70E-05 | 0.002731819 | AC112220.2 |
| chr7 | 123601698 | 123602545 | 848 | 5 | 1.67E-06 | 0.0031823 | 2.19E-05 | 8.70E-05 | -0.0032719 | ASB15, ASB15-AS1 |
| chr2 | 113848821 | 113848945 | 125 | 2 | 6.51E-05 | 0.0084384 | 3.85E-05 | 8.70E-05 | -0.00707138 | NA |
| chr9 | 29212691 | 29213103 | 413 | 4 | 4.84E-05 | 0.0002667 | 0.000427 | 8.70E-05 | 0.003980947 | NA |
| chr11 | 68914901 | 68914965 | 65 | 2 | 0.000103991 | 0.0050302 | 4.14E-05 | 8.72E-05 | -0.00275251 | IGHMBP2 |
| chr20 | 17929101 | 17929195 | 95 | 2 | 0.000125879 | 5.31E-05 | 0.002069 | 8.72E-05 | -0.0060784 | NA |
| chr2 | 176002117 | 176003158 | 1042 | 9 | 7.21E-06 | 0.0030142 | 0.000757 | 8.73E-05 | 0.000283148 | LNPK |
| chr17 | 792188 | 792421 | 234 | 2 | 7.42E-05 | 0.0015505 | 5.36E-05 | 8.73E-05 | -0.01188705 | MRM3 |
| chr9 | 110443581 | 110443672 | 92 | 2 | 0.000118953 | 5.92E-05 | 0.001278 | 8.73E-05 | 0.012478473 | SVEP1 |
| chr19 | 19660051 | 19660149 | 99 | 2 | 0.00010905 | 0.0020445 | 4.98E-05 | 8.75E-05 | -0.00639614 | ATP13A1 |
| chr11 | 120564171 | 120564347 | 177 | 9 | 0.000173445 | 1.52E-05 | 0.029724 | 8.75E-05 | -0.00902478 | GRIK4 |
| chr14 | 73489668 | 73490005 | 338 | 4 | 4.86E-05 | 0.0003572 | 0.003037 | 8.75E-05 | -0.00190088 | HEATR4 |
| chr11 | 597231 | 597597 | 367 | 4 | 3.62E-05 | 0.0034428 | 3.46E-05 | 8.76E-05 | -0.00062674 | PHRF1 |
| chr14 | 89932616 | 89932787 | 172 | 2 | 4.85E-05 | 0.0003311 | 0.000104 | 8.79E-05 | -0.01104494 | AL137230.2, EFCAB11 |
| chr17 | 80708985 | 80709472 | 488 | 4 | 6.55E-05 | 0.0002977 | 0.003158 | 8.81E-05 | -0.00545875 | RPTOR |
| chr9 | 68739713 | 68739874 | 162 | 2 | 5.02E-05 | 0.0146206 | 3.69E-05 | 8.81E-05 | 0.012774671 | PIP5K1B |
| chr20 | 10362393 | 10362480 | 88 | 2 | 0.000108249 | 0.003603 | 4.43E-05 | 8.81E-05 | -0.00928708 | SNAP25-AS1 |
| chr1 | 1752299 | 1752433 | 135 | 3 | 5.46E-05 | 0.0004288 | 0.000655 | 8.82E-05 | -0.00267301 | NADK |
| chr19 | 13962153 | 13962465 | 313 | 2 | 9.63E-05 | 5.50E-05 | 0.001834 | 8.82E-05 | -0.0086462 | RFX1 |
| chr12 | 14773829 | 14774417 | 589 | 11 | 5.17E-05 | 0.0010585 | 0.000189 | 8.82E-05 | -0.00419249 | H2AJ |
| chr14 | 99793464 | 99793523 | 60 | 3 | 2.08E-05 | 0.0002759 | 0.000144 | 8.84E-05 | 9.41E-05 | EML1 |
| chr10 | 89260710 | 89260824 | 115 | 2 | 0.000114318 | 0.0007272 | 7.05E-05 | 8.84E-05 | -0.00772752 | LIPA |
| chr17 | 30558947 | 30559170 | 224 | 2 | 5.93E-05 | 0.0213475 | 3.61E-05 | 8.84E-05 | -0.00811399 | TBC1D29P |
| chr3 | 184908006 | 184908017 | 12 | 2 | 0.000125769 | 7.72E-05 | 0.000601 | 8.85E-05 | -0.00634003 | VPS8 |
| chr13 | 113046504 | 113046680 | 177 | 3 | 2.65E-05 | 0.0004348 | 0.000114 | 8.85E-05 | -0.00319653 | MCF2L |
| chr2 | 106887713 | 106888067 | 355 | 2 | 6.97E-05 | 0.0186736 | 3.65E-05 | 8.85E-05 | -0.00736407 | NA |
| chr5 | 132813256 | 132813712 | 457 | 7 | 1.50E-05 | 0.0019592 | 0.000216 | 8.85E-05 | 0.000762016 | SOWAHA |
| chr14 | 70671314 | 70671350 | 37 | 2 | 0.000174923 | 0.0004728 | 8.63E-05 | 8.86E-05 | -0.00685344 | TTC9 |
| chr2 | 127081255 | 127081352 | 98 | 2 | 0.00013817 | 0.0001178 | 0.000287 | 8.87E-05 | 0.014243727 | BIN1 |
| chr21 | 42062519 | 42063293 | 775 | 8 | 4.50E-05 | 0.0009961 | 0.013185 | 8.88E-05 | 0.000114989 | UMODL1 |
| chr1 | 225652806 | 225653499 | 694 | 5 | 2.51E-05 | 0.0002164 | 0.000307 | 8.89E-05 | 0.001763505 | ENAH |
| chr12 | 132744189 | 132744389 | 201 | 2 | 5.10E-05 | 0.0099729 | 3.85E-05 | 8.89E-05 | -0.0029888 | ANKLE2 |
| chr17 | 7844789 | 7845126 | 338 | 4 | 5.41E-05 | 0.0001451 | 0.000356 | 8.89E-05 | -0.00133431 | KDM6B |
| chr3 | 46405507 | 46405763 | 257 | 2 | 8.12E-05 | 0.0001858 | 0.00017 | 8.91E-05 | -0.01083039 | CCR5AS |
| chr15 | 49155043 | 49155585 | 543 | 7 | 4.65E-05 | 0.0007083 | 0.002442 | 8.91E-05 | 0.001620874 | COPS2 |
| chr11 | 43922072 | 43922270 | 199 | 3 | 7.15E-05 | 0.0005677 | 0.000248 | 8.92E-05 | 0.002354895 | AC087521.3 |
| chr5 | 173824862 | 173825201 | 340 | 3 | 3.20E-05 | 0.0108184 | 2.54E-05 | 8.92E-05 | -0.00110618 | NA |
| chr12 | 74022075 | 74022230 | 156 | 2 | 0.000104164 | 0.0022942 | 4.93E-05 | 8.92E-05 | -0.01559759 | LINC02882 |
| chr5 | 35875345 | 35875524 | 180 | 2 | 6.34E-05 | 6.18E-05 | 0.001183 | 8.93E-05 | -0.01120598 | IL7R |
| chr5 | 141155309 | 141156178 | 870 | 6 | 2.15E-06 | 0.0001617 | 0.000385 | 8.93E-05 | -6.49E-05 | PCDHB17P, AC244517.2, AC244517.11 |
| chr3 | 128572695 | 128572760 | 66 | 2 | 9.71E-05 | 6.81E-05 | 0.000862 | 8.93E-05 | 0.001919459 | LINC01565 |
| chr11 | 3796462 | 3796619 | 158 | 4 | 4.62E-05 | 0.0026859 | 5.29E-05 | 8.93E-05 | 0.009201692 | NUP98 |
| chr5 | 132073466 | 132073715 | 250 | 4 | 4.79E-05 | 0.0001602 | 0.001824 | 8.94E-05 | -0.00781444 | NA |
| chr9 | 97500620 | 97501605 | 986 | 9 | 2.00E-05 | 0.000413 | 0.00058 | 8.96E-05 | 0.003834828 | TMOD1 |
| chr5 | 177503262 | 177503753 | 492 | 3 | 1.13E-05 | 8.25E-05 | 0.000649 | 8.97E-05 | 0.005453491 | AC145098.1, DOK3 |
| chr13 | 39678984 | 39679103 | 120 | 2 | 0.000140427 | 6.63E-05 | 0.000951 | 8.97E-05 | 0.005154727 | COG6 |
| chr3 | 62369644 | 62370209 | 566 | 5 | 1.87E-05 | 0.0022864 | 0.001108 | 8.97E-05 | 0.000117184 | FEZF2 |
| chr19 | 10780800 | 10780833 | 34 | 2 | 0.000130788 | 6.80E-05 | 0.000881 | 8.97E-05 | -0.008659 | DNM2 |
| chr8 | 143877587 | 143878048 | 462 | 2 | 1.54E-05 | 5.46E-05 | 0.002125 | 8.99E-05 | 0.017876511 | EPPK1 |
| chr11 | 65524714 | 65525195 | 482 | 6 | 2.98E-05 | 7.35E-05 | 0.004752 | 9.00E-05 | -0.00355044 | SCYL1 |
| chr8 | 27897427 | 27897505 | 79 | 2 | 0.000115395 | 0.0008509 | 6.75E-05 | 9.00E-05 | -0.00837522 | SCARA5 |
| chr7 | 99416150 | 99416226 | 77 | 2 | 0.00011224 | 0.0018809 | 5.24E-05 | 9.02E-05 | -0.00718488 | BUD31 |
| chr3 | 196966938 | 196967282 | 345 | 3 | 0.000100074 | 0.0004934 | 0.001191 | 9.03E-05 | 0.002903345 | PIGZ |
| chr8 | 24382858 | 24383084 | 227 | 2 | 0.000107722 | 7.21E-05 | 0.000767 | 9.06E-05 | -0.00992705 | AC120193.1 |
| chr17 | 35808732 | 35809008 | 277 | 4 | 0.000155757 | 7.10E-05 | 0.002946 | 9.06E-05 | -0.00405402 | TAF15, AC015849.2 |
| chr10 | 124540441 | 124540490 | 50 | 2 | 5.91E-05 | 0.0020776 | 5.12E-05 | 9.06E-05 | -0.01355655 | LHPP |
| chr1 | 151156822 | 151157368 | 547 | 2 | 7.55E-06 | 8.57E-05 | 0.000511 | 9.06E-05 | -0.0004857 | TNFAIP8L2, SCNM1 |
| chr19 | 37506965 | 37507269 | 305 | 3 | 9.80E-06 | 0.0028318 | 8.68E-05 | 9.08E-05 | 0.000185823 | ZNF793, ZNF793-AS1 |
| chr10 | 7943147 | 7943282 | 136 | 2 | 6.24E-05 | 6.35E-05 | 0.001155 | 9.10E-05 | 0.000307737 | TAF3 |
| chr8 | 141300911 | 141300973 | 63 | 3 | 0.000136888 | 0.0003043 | 0.000327 | 9.10E-05 | -0.00291717 | SLC45A4 |
| chr6 | 1553595 | 1553659 | 65 | 2 | 2.32E-05 | 0.000156 | 0.000213 | 9.12E-05 | 0.001034509 | NA |
| chr5 | 95682767 | 95683358 | 592 | 3 | 2.06E-05 | 0.0001201 | 0.000812 | 9.13E-05 | -0.00302768 | RFESD, SPATA9 |
| chr14 | 99197798 | 99197942 | 145 | 3 | 0.000120941 | 8.81E-05 | 0.000609 | 9.13E-05 | -0.00870649 | BCL11B |
| chr10 | 22475736 | 22476280 | 545 | 3 | 4.13E-05 | 0.0005048 | 9.00E-05 | 9.14E-05 | 0.003193122 | AL513128.3, AL513128.2 |
| chr1 | 1567034 | 1567226 | 193 | 2 | 0.000154155 | 0.0001206 | 0.000296 | 9.14E-05 | -0.01183764 | SSU72 |
| chr16 | 921556 | 922115 | 560 | 4 | 3.46E-05 | 4.03E-05 | 0.005058 | 9.14E-05 | -0.00087281 | LMF1-AS1, LMF1 |
| chr2 | 10303506 | 10303692 | 187 | 5 | 3.53E-05 | 0.0001522 | 0.001681 | 9.14E-05 | 0.001857912 | HPCAL1 |
| chr1 | 118920940 | 118921049 | 110 | 2 | 9.90E-05 | 0.0064779 | 4.16E-05 | 9.15E-05 | -0.00527361 | TBX15 |
| chr1 | 47713569 | 47713799 | 231 | 2 | 2.09E-05 | 0.0022609 | 5.07E-05 | 9.15E-05 | -0.0069822 | NA |
| chr10 | 6247459 | 6247550 | 92 | 2 | 5.82E-05 | 0.0002106 | 0.000158 | 9.16E-05 | 0.001144688 | PFKFB3 |
| chr15 | 64381256 | 64381673 | 418 | 9 | 3.63E-05 | 0.0100668 | 2.46E-05 | 9.16E-05 | 8.07E-05 | AC087632.2, PCLAF |
| chr21 | 46297950 | 46298166 | 217 | 3 | 5.12E-05 | 0.00039 | 0.000818 | 9.16E-05 | -0.01516734 | NA |
| chr22 | 36481504 | 36481690 | 187 | 6 | 4.43E-05 | 0.0011286 | 0.000247 | 9.17E-05 | -8.57E-05 | TXN2 |
| chr10 | 21147636 | 21147853 | 218 | 3 | 2.55E-05 | 0.0015156 | 7.92E-05 | 9.17E-05 | -0.00711939 | NEBL |
| chr1 | 204332150 | 204332222 | 73 | 2 | 0.000154494 | 7.05E-05 | 0.00085 | 9.17E-05 | -0.00065998 | PLEKHA6 |
| chr7 | 101154780 | 101155086 | 307 | 3 | 5.84E-05 | 0.001187 | 0.001386 | 9.17E-05 | 0.003346017 | AP1S1 |
| chr19 | 52190370 | 52190717 | 348 | 2 | 3.28E-06 | 6.57E-05 | 0.001063 | 9.18E-05 | 0.003833488 | PPP2R1A |
| chr3 | 195884058 | 195884108 | 51 | 2 | 8.27E-05 | 8.53E-05 | 0.000534 | 9.18E-05 | -0.00095746 | TNK2 |
| chr1 | 197180368 | 197180393 | 26 | 2 | 6.68E-05 | 0.0040676 | 4.50E-05 | 9.18E-05 | -0.01189346 | ZBTB41 |
| chr12 | 32755971 | 32756862 | 892 | 9 | 3.06E-06 | 0.0005853 | 0.000728 | 9.18E-05 | -0.00194055 | AC087588.2 |
| chr1 | 34885120 | 34885145 | 26 | 2 | 0.000137452 | 0.000936 | 6.66E-05 | 9.19E-05 | -0.01586196 | DLGAP3 |
| chr15 | 57438968 | 57439450 | 483 | 2 | 2.44E-05 | 6.08E-05 | 0.001426 | 9.19E-05 | -0.00124341 | CGNL1 |
| chr5 | 16464904 | 16465130 | 227 | 3 | 2.72E-05 | 7.42E-05 | 0.000993 | 9.19E-05 | -0.0011013 | ZNF622 |
| chr5 | 7325917 | 7325930 | 14 | 2 | 2.44E-05 | 0.0041542 | 4.48E-05 | 9.19E-05 | -0.00563113 | AC091951.3 |
| chr16 | 52229181 | 52229238 | 58 | 2 | 5.90E-05 | 0.0279645 | 3.68E-05 | 9.21E-05 | -0.01044359 | NA |
| chr17 | 38460399 | 38460582 | 184 | 2 | 2.57E-05 | 0.0073678 | 4.12E-05 | 9.21E-05 | -0.00229529 | ARHGAP23 |
| chr22 | 36323360 | 36323465 | 106 | 2 | 1.60E-05 | 0.0561241 | 3.61E-05 | 9.22E-05 | 0.002773275 | MYH9 |
| chr3 | 186979988 | 186980267 | 280 | 2 | 0.000153636 | 5.50E-05 | 0.002379 | 9.23E-05 | -0.00401872 | ST6GAL1 |
| chr8 | 15540128 | 15540390 | 263 | 5 | 5.42E-05 | 0.000105 | 0.003486 | 9.26E-05 | -0.00197824 | TUSC3 |
| chr12 | 57622573 | 57622843 | 271 | 2 | 7.08E-05 | 5.48E-05 | 0.002467 | 9.26E-05 | -0.01148693 | AC025165.6, SLC26A10 |
| chr11 | 103862932 | 103863015 | 84 | 3 | 0.00013207 | 7.17E-05 | 0.001077 | 9.27E-05 | -0.00457324 | AP002989.1 |
| chr3 | 42121131 | 42121140 | 10 | 2 | 0.000119351 | 0.0003347 | 0.000111 | 9.27E-05 | -0.00442292 | TRAK1 |
| chr16 | 166451 | 166758 | 308 | 4 | 1.95E-05 | 0.000101 | 0.001212 | 9.28E-05 | 0.002611221 | HBM |
| chr1 | 182953570 | 182954615 | 1046 | 4 | 2.69E-07 | 0.0077305 | 0.001072 | 9.28E-05 | -0.00095462 | NA |
| chr13 | 101917188 | 101917544 | 357 | 4 | 9.63E-05 | 4.13E-05 | 0.007143 | 9.29E-05 | 0.001243123 | FGF14 |
| chr3 | 52324654 | 52325044 | 391 | 3 | 9.42E-05 | 0.0001304 | 0.002201 | 9.29E-05 | -0.00210674 | DNAH1 |
| chr7 | 1957029 | 1957694 | 666 | 4 | 4.08E-05 | 0.000711 | 0.003138 | 9.29E-05 | -0.0045884 | MAD1L1, AC069288.1 |
| chr20 | 1965708 | 1965857 | 150 | 3 | 5.49E-05 | 0.0017216 | 4.66E-05 | 9.29E-05 | -0.00138121 | PDYN-AS1 |
| chr20 | 17646249 | 17646443 | 195 | 2 | 0.000142004 | 6.79E-05 | 0.001 | 9.30E-05 | -0.03102775 | RRBP1 |
| chr19 | 11449249 | 11449479 | 231 | 4 | 0.000123332 | 0.0008814 | 0.000457 | 9.30E-05 | -0.00405048 | PRKCSH |
| chr9 | 34701534 | 34701738 | 205 | 2 | 5.07E-05 | 7.54E-05 | 0.000735 | 9.30E-05 | -0.00337102 | NA |
| chr9 | 93127699 | 93127860 | 162 | 2 | 9.21E-05 | 0.0005723 | 8.30E-05 | 9.30E-05 | -0.00392297 | NINJ1 |
| chr19 | 43463678 | 43463981 | 304 | 2 | 4.78E-05 | 0.0002968 | 0.000122 | 9.30E-05 | 0.001830218 | LYPD3 |
| chr4 | 82871965 | 82872016 | 52 | 3 | 0.000159721 | 0.0006602 | 0.000146 | 9.30E-05 | -0.0038059 | SEC31A |
| chr11 | 63536255 | 63536865 | 611 | 3 | 5.62E-06 | 0.0005281 | 0.001682 | 9.31E-05 | -0.00471387 | PLAAT4 |
| chr11 | 123261622 | 123261856 | 235 | 3 | 0.000100154 | 0.000731 | 0.00014 | 9.33E-05 | -0.0114087 | NA |
| chr19 | 46745683 | 46745962 | 280 | 7 | 7.76E-05 | 0.0069611 | 2.41E-05 | 9.35E-05 | 0.001497975 | STRN4 |
| chr1 | 200831814 | 200832052 | 239 | 2 | 9.35E-05 | 0.0045744 | 4.48E-05 | 9.35E-05 | 0.010208751 | CAMSAP2 |
| chr1 | 3094332 | 3094740 | 409 | 3 | 0.000123553 | 0.0005998 | 9.17E-05 | 9.35E-05 | -0.00546703 | PRDM16 |
| chr1 | 2579726 | 2580205 | 480 | 2 | 2.76E-05 | 0.0002777 | 0.000129 | 9.35E-05 | -0.00051686 | NA |
| chr3 | 172039996 | 172040420 | 425 | 7 | 3.17E-05 | 6.46E-05 | 0.012975 | 9.36E-05 | 0.000352089 | FNDC3B |
| chr5 | 59587566 | 59587678 | 113 | 2 | 0.000106363 | 6.59E-05 | 0.001132 | 9.36E-05 | -0.02023201 | PDE4D |
| chr11 | 72103174 | 72103683 | 510 | 10 | 9.99E-05 | 6.94E-05 | 0.005009 | 9.36E-05 | -0.00114462 | AP000812.4, LAMTOR1 |
| chr2 | 108587774 | 108588231 | 458 | 4 | 2.79E-05 | 0.0002336 | 0.00055 | 9.37E-05 | -0.00275412 | LIMS1 |
| chr9 | 123930046 | 123930676 | 631 | 6 | 1.61E-05 | 0.0007919 | 0.000402 | 9.37E-05 | -0.00567337 | DENND1A |
| chr22 | 28796070 | 28796153 | 84 | 2 | 5.76E-05 | 0.0002085 | 0.000166 | 9.37E-05 | -0.00686112 | XBP1 |
| chr7 | 44103374 | 44103839 | 466 | 4 | 2.58E-05 | 0.0024171 | 0.00096 | 9.38E-05 | -0.00337332 | NA |
| chr1 | 205775258 | 205775620 | 363 | 8 | 9.92E-06 | 0.002565 | 3.12E-05 | 9.39E-05 | -0.00016914 | AC119673.1, RAB29 |
| chr1 | 156408222 | 156408610 | 389 | 2 | 7.23E-05 | 5.70E-05 | 0.002138 | 9.39E-05 | 0.001593403 | MIR9-1HG |
| chr8 | 143770400 | 143770783 | 384 | 7 | 2.37E-05 | 0.0024034 | 0.002002 | 9.39E-05 | -0.00250295 | IQANK1, AC105219.3 |
| chr5 | 1110934 | 1111309 | 376 | 2 | 1.66E-05 | 5.99E-05 | 0.001685 | 9.39E-05 | -0.01690764 | SLC12A7 |
| chr20 | 63523362 | 63523578 | 217 | 2 | 2.97E-05 | 0.0669223 | 3.65E-05 | 9.40E-05 | -0.00256564 | NA |
| chr2 | 70087879 | 70087959 | 81 | 2 | 5.94E-05 | 0.0014831 | 5.86E-05 | 9.40E-05 | 0.0025142 | PCBP1, PCBP1-AS1 |
| chr19 | 47754698 | 47755001 | 304 | 2 | 2.15E-05 | 0.002996 | 4.88E-05 | 9.41E-05 | 0.002499409 | NOP53 |
| chr12 | 114674384 | 114674821 | 438 | 3 | 8.73E-05 | 3.63E-05 | 0.007889 | 9.41E-05 | -0.01801555 | TBX3 |
| chr11 | 1469508 | 1470078 | 571 | 2 | 1.19E-06 | 0.0079873 | 4.15E-05 | 9.41E-05 | -0.00279744 | MOB2 |
| chr18 | 78313275 | 78313399 | 125 | 2 | 0.000121919 | 0.000312 | 0.00012 | 9.44E-05 | -0.00890554 | NA |
| chr3 | 13549468 | 13549584 | 117 | 2 | 7.40E-05 | 6.63E-05 | 0.001149 | 9.45E-05 | -0.010005 | FBLN2 |

| | | | | | | | | | | |
|---|---|---|---|---|---|---|---|---|---|---|
| chr4 | 76215178 | 76215395 | 218 | 2 | 1.24E-05 | 0.0101927 | 4.05E-05 | 9.45E-05 | -0.00229681 | FAM47E, SCARB2 |
| chr7 | 23531261 | 23531871 | 611 | 5 | 3.62E-05 | 0.0001032 | 0.003614 | 9.45E-05 | 0.000576378 | TRA2A |
| chr5 | 72508801 | 72508967 | 167 | 3 | 0.00015178 | 7.45E-05 | 0.000901 | 9.45E-05 | 0.000978703 | NA |
| chr2 | 176172903 | 176172982 | 80 | 2 | 8.98E-05 | 0.0057538 | 4.36E-05 | 9.47E-05 | -0.00512406 | HOXD3, HAGLR |
| chr2 | 240782505 | 240782864 | 360 | 4 | 2.40E-05 | 0.0044578 | 5.97E-05 | 9.48E-05 | 0.00289904 | KIF1A |
| chr7 | 97116868 | 97117976 | 1109 | 9 | 6.47E-05 | 0.0009681 | 0.000268 | 9.49E-05 | 0.001451033 | SDHAF3 |
| chr17 | 49389859 | 49390138 | 280 | 3 | 8.69E-05 | 0.0001322 | 0.002159 | 9.50E-05 | -0.01301196 | AC091180.5 |
| chr16 | 87407650 | 87407949 | 300 | 3 | 0.000112177 | 3.60E-05 | 0.008476 | 9.50E-05 | -0.01723823 | ZCCHC14 |
| chr2 | 45922795 | 45922951 | 157 | 2 | 0.000102383 | 0.00371 | 4.71E-05 | 9.51E-05 | -0.00488348 | PRKCE |
| chr20 | 64270518 | 64270594 | 77 | 2 | 8.43E-05 | 6.10E-05 | 0.001644 | 9.52E-05 | -0.01353572 | PCMTD2 |
| chr2 | 58107873 | 58107945 | 73 | 2 | 0.000146612 | 0.0010218 | 6.69E-05 | 9.52E-05 | 0.008203308 | VRK2 |
| chr17 | 8670654 | 8670820 | 167 | 3 | 9.67E-05 | 0.0013754 | 6.52E-05 | 9.53E-05 | -0.00727272 | NA |
| chr5 | 138622666 | 138623334 | 669 | 4 | 2.67E-05 | 0.0152421 | 2.38E-05 | 9.54E-05 | -0.00157069 | CTNNA1 |
| chr4 | 4872239 | 4872315 | 77 | 2 | 0.000120952 | 5.99E-05 | 0.001801 | 9.54E-05 | -0.0017048 | NA |
| chr20 | 53875670 | 53875709 | 40 | 2 | 7.82E-05 | 0.0030257 | 4.94E-05 | 9.56E-05 | 0.007994254 | NA |
| chr2 | 174026933 | 174027082 | 150 | 2 | 5.66E-05 | 6.84E-05 | 0.00108 | 9.56E-05 | 0.000178292 | LINC01960 |
| chr10 | 102868152 | 102868265 | 114 | 3 | 4.55E-05 | 6.94E-05 | 0.001582 | 9.57E-05 | 0.002628576 | BORCS7-ASMT |
| chr2 | 10689599 | 10690106 | 508 | 5 | 2.13E-05 | 0.0026955 | 0.004351 | 9.57E-05 | 0.000266173 | NOL10 |
| chr2 | 2732197 | 2732372 | 176 | 2 | 4.93E-05 | 0.0006561 | 8.05E-05 | 9.57E-05 | -0.01164801 | AC011995.2 |
| chr18 | 31381052 | 31381361 | 310 | 3 | 6.40E-05 | 0.0004064 | 0.000111 | 9.57E-05 | -0.00994846 | DSG4, DSG1-AS1 |
| chr17 | 39725571 | 39726621 | 1051 | 5 | 1.58E-05 | 0.0003932 | 0.001328 | 9.57E-05 | -0.00044302 | ERBB2, MIR4728 |
| chr16 | 72580369 | 72580392 | 24 | 2 | 0.000137575 | 0.0001404 | 0.000266 | 9.58E-05 | -0.00317972 | LINC01572 |
| chr14 | 105166380 | 105166420 | 41 | 2 | 0.000123978 | 0.0001398 | 0.000268 | 9.58E-05 | -0.00200831 | JAG2, AL512356.1 |
| chr11 | 126712288 | 126712422 | 135 | 3 | 0.000105105 | 0.0001091 | 0.000531 | 9.58E-05 | 0.026106883 | KIRREL3 |
| chr22 | 21632687 | 21632901 | 215 | 6 | 0.000156539 | 0.0006482 | 0.000729 | 9.58E-05 | -0.00693223 | CCDC116 |
| chr3 | 194617114 | 194617349 | 236 | 2 | 7.19E-05 | 0.0005613 | 8.69E-05 | 9.59E-05 | -0.00204943 | TMEM44 |
| chr11 | 117176598 | 117176602 | 5 | 2 | 4.27E-05 | 0.0017947 | 5.65E-05 | 9.60E-05 | 0.003170815 | PAFAH1B2 |
| chr2 | 46541311 | 46542172 | 862 | 8 | 5.00E-06 | 0.0016682 | 0.000948 | 9.60E-05 | -0.00066532 | RHOQ, ATP6V1E2 |
| chr10 | 5317725 | 5317795 | 71 | 2 | 2.97E-05 | 0.0004755 | 9.46E-05 | 9.61E-05 | 0.014276218 | NA |
| chr22 | 46148740 | 46149179 | 440 | 2 | 1.30E-05 | 8.35E-05 | 0.000632 | 9.61E-05 | -0.00965466 | NA |
| chr2 | 101435125 | 101435227 | 103 | 3 | 0.000112141 | 0.0064903 | 0.000117 | 9.61E-05 | -0.00504682 | RFX8 |
| chr12 | 104125001 | 104125195 | 195 | 2 | 8.99E-05 | 0.0044461 | 4.62E-05 | 9.63E-05 | -0.01609991 | NFYB |
| chr17 | 81684776 | 81684994 | 219 | 2 | 2.72E-05 | 5.78E-05 | 0.0023 | 9.64E-05 | -0.00911893 | HGS |
| chr12 | 116598898 | 116599167 | 270 | 6 | 4.23E-05 | 0.0004915 | 0.000403 | 9.66E-05 | -0.00136242 | NA |
| chr3 | 28348154 | 28348190 | 37 | 2 | 9.70E-05 | 0.0009536 | 6.97E-05 | 9.66E-05 | -0.00880563 | AZI2 |
| chr14 | 56513156 | 56513293 | 138 | 2 | 0.000121673 | 0.0021703 | 5.41E-05 | 9.67E-05 | -0.00719042 | TMEM260 |
| chr5 | 141487759 | 141488157 | 399 | 3 | 1.88E-05 | 0.0004463 | 0.000177 | 9.68E-05 | -0.00154337 | PCDHGA1, PCDHGA2, PCDHGA3, PCDHGB1, PCDHGA4, PCDHGB2, PCDHGA5, PCDHGB3, PCDHGA6, PCDHGA7, PCDHGB4, PCDHGA8, PCDHGB5, PCDHGA9, PCDHGB6, PCDHGA10, PCDHGB7, PCDHGA11, PCDHGA12, PCDHGC3, PCDHGC4 |
| chr17 | 64465169 | 64465484 | 316 | 5 | 8.80E-05 | 0.0004764 | 0.001371 | 9.68E-05 | -0.00265511 | MILR1 |
| chr14 | 32938128 | 32939054 | 927 | 5 | 2.69E-06 | 0.0051197 | 8.17E-05 | 9.69E-05 | -0.00346734 | NPAS3 |
| chr7 | 99558388 | 99558846 | 459 | 10 | 9.56E-05 | 0.0005515 | 0.001718 | 9.69E-05 | 0.001112509 | ZNF655, AC005020.2, FAM200A |
| chr10 | 116886373 | 116886862 | 490 | 3 | 2.24E-05 | 0.0300718 | 1.75E-05 | 9.70E-05 | 0.003006501 | ENO4, SHTN1 |
| chr4 | 138243569 | 138243636 | 68 | 2 | 6.04E-05 | 6.14E-05 | 0.001751 | 9.71E-05 | -0.01249574 | NA |
| chr12 | 7039501 | 7039517 | 17 | 2 | 7.29E-05 | 5.73E-05 | 0.002553 | 9.72E-05 | -0.00910071 | C1S |
| chr2 | 29242426 | 29242692 | 267 | 2 | 3.22E-05 | 0.0004291 | 0.000102 | 9.73E-05 | -0.00388039 | ALK |
| chr1 | 45523708 | 45523998 | 291 | 4 | 4.84E-05 | 0.0035477 | 2.42E-05 | 9.73E-05 | 0.002010125 | PRDX1 |
| chr19 | 45488481 | 45488620 | 140 | 3 | 7.66E-05 | 0.0058104 | 2.72E-05 | 9.73E-05 | -0.00610595 | RTN2 |
| chr13 | 26022637 | 26022858 | 222 | 3 | 3.08E-05 | 0.0328618 | 2.44E-05 | 9.74E-05 | -0.00284204 | ATP8A2 |
| chr5 | 114361633 | 114362967 | 1335 | 13 | 1.05E-05 | 0.0267437 | 0.000212 | 9.76E-05 | 0.000884566 | KCNN2 |
| chr7 | 66653338 | 66653413 | 76 | 2 | 0.000110719 | 0.0001224 | 0.000334 | 9.77E-05 | -0.00232282 | AC027644.4 |
| chr3 | 69865814 | 69866298 | 485 | 7 | 0.000112771 | 0.0013957 | 0.002605 | 9.77E-05 | -0.00478366 | MITF |
| chr19 | 35739436 | 35739445 | 10 | 2 | 4.47E-05 | 5.95E-05 | 0.002138 | 9.78E-05 | -0.00563954 | IGFLR1, AD000671.2 |
| chr17 | 16981514 | 16981985 | 472 | 3 | 3.99E-05 | 8.43E-05 | 0.002598 | 9.80E-05 | -0.00244006 | NA |
| chr5 | 151083399 | 151083603 | 205 | 2 | 5.67E-05 | 8.95E-05 | 0.000574 | 9.80E-05 | -0.00576023 | TNIP1 |
| chr18 | 36829375 | 36830004 | 630 | 7 | 1.61E-06 | 0.002994 | 0.001228 | 9.80E-05 | -0.00721683 | KIAA1328 |
| chr7 | 155951646 | 155951850 | 205 | 3 | 4.90E-05 | 0.0010291 | 6.91E-05 | 9.81E-05 | 0.002720582 | NA |
| chr1 | 109985403 | 109985455 | 53 | 2 | 8.03E-05 | 0.0009982 | 6.97E-05 | 9.81E-05 | 0.001740186 | AHCYL1 |
| chr6 | 1436029 | 1436036 | 8 | 2 | 6.31E-05 | 6.15E-05 | 0.001842 | 9.82E-05 | -0.00586987 | NA |
| chr1 | 110871021 | 110871112 | 92 | 2 | 5.02E-05 | 0.0001583 | 0.000241 | 9.82E-05 | 0.006664612 | NA |
| chr12 | 50201634 | 50202119 | 486 | 3 | 1.94E-05 | 0.0003152 | 0.00119 | 9.83E-05 | -0.0009042 | LIMA1 |
| chr9 | 114027599 | 114027756 | 158 | 2 | 0.000105069 | 0.000537 | 9.13E-05 | 9.83E-05 | -0.0057563 | ZNF618 |
| chr4 | 15427126 | 15427371 | 246 | 3 | 9.64E-05 | 0.0001837 | 0.000709 | 9.83E-05 | -0.00669292 | C1QTNF7, C1QTNF7-AS1 |
| chr11 | 114257202 | 114257595 | 394 | 3 | 0.000108569 | 0.0002004 | 0.000764 | 9.84E-05 | -0.00858664 | AP002518.2 |
| chr12 | 52082215 | 52082433 | 219 | 4 | 0.000130937 | 0.0002086 | 0.000487 | 9.86E-05 | -0.00355921 | SMIM41 |
| chr1 | 155857163 | 155857253 | 91 | 4 | 0.000166462 | 0.0002918 | 0.002949 | 9.86E-05 | -0.00617726 | GON4L |
| chr20 | 2006637 | 2007572 | 936 | 4 | 5.97E-06 | 0.001735 | 7.22E-05 | 9.87E-05 | -0.00155728 | PDYN-AS1 |
| chr1 | 167721489 | 167722473 | 985 | 8 | 4.23E-06 | 0.0003749 | 0.001763 | 9.87E-05 | 0.002236321 | MPZL1 |
| chr2 | 127068359 | 127068573 | 215 | 3 | 7.65E-05 | 0.0143239 | 2.67E-05 | 9.87E-05 | -0.00528458 | BIN1 |
| chr18 | 26657096 | 26657453 | 358 | 5 | 5.91E-05 | 0.0075258 | 1.34E-05 | 9.87E-05 | 0.010960257 | AQP4-AS1, KCTD1 |
| chr9 | 136941991 | 136942668 | 678 | 5 | 8.64E-06 | 0.0012558 | 7.09E-05 | 9.87E-05 | 8.16E-05 | FBXW5 |
| chr6 | 18265547 | 18265882 | 336 | 2 | 1.15E-06 | 0.0003849 | 0.000111 | 9.90E-05 | -0.00490841 | NA |
| chr6 | 149450426 | 149451210 | 785 | 5 | 2.35E-05 | 0.0008282 | 0.000183 | 9.90E-05 | -0.00600371 | ZC3H12D |
| chr7 | 149172 | 149795 | 624 | 8 | 2.02E-05 | 0.0004604 | 0.001037 | 9.90E-05 | -0.00082426 | AC093627.4, AC093627.5 |
| chr16 | 84309090 | 84309103 | 14 | 2 | 0.000105895 | 0.004666 | 4.69E-05 | 9.90E-05 | -0.0108641 | WFDC1 |
| chr17 | 81625346 | 81625365 | 20 | 2 | 9.96E-05 | 0.0790684 | 3.79E-05 | 9.90E-05 | 0.001645677 | NPLOC4 |
| chr14 | 90454297 | 90455324 | 1028 | 5 | 3.97E-05 | 0.0008428 | 0.002896 | 9.90E-05 | -0.00117156 | LINC00642, LINC02317 |
| chr3 | 123124053 | 123124279 | 227 | 3 | 8.56E-05 | 0.0004806 | 0.000144 | 9.90E-05 | -0.00064096 | PDIA5 |
| chr5 | 163437285 | 163437303 | 19 | 2 | 0.00015105 | 5.95E-05 | 0.002296 | 9.91E-05 | 0.005036247 | AC113414.1 |
| chr22 | 22921286 | 22921342 | 57 | 2 | 0.000108622 | 6.06E-05 | 0.002087 | 9.92E-05 | -0.00208318 | NA |
| chr1 | 20731734 | 20732473 | 740 | 4 | 2.19E-06 | 0.0002897 | 0.000788 | 9.92E-05 | 0.001164888 | SH2D5 |
| chr12 | 57243907 | 57244142 | 236 | 3 | 6.20E-05 | 7.79E-05 | 0.000929 | 9.92E-05 | -0.00529742 | STAC3 |
| chr10 | 122374066 | 122374728 | 663 | 11 | 1.76E-05 | 0.0364195 | 0.000134 | 9.92E-05 | 0.001724566 | PLEKHA1 |
| chr19 | 17304168 | 17304414 | 247 | 4 | 4.13E-05 | 0.0005336 | 0.000295 | 9.92E-05 | -0.00462219 | MRPL34, ABHD8 |
| chr5 | 178552175 | 178552294 | 120 | 2 | 8.97E-05 | 8.33E-05 | 0.0007 | 9.94E-05 | -0.00303778 | COL23A1 |
| chr1 | 147641727 | 147642036 | 310 | 2 | 4.21E-05 | 0.0022603 | 5.50E-05 | 9.94E-05 | -0.00684041 | ACP6 |
| chr12 | 132853801 | 132854121 | 321 | 3 | 3.94E-05 | 0.0002481 | 0.000194 | 9.94E-05 | 0.001216365 | CHFR |
| chr13 | 26253690 | 26253945 | 256 | 5 | 9.97E-05 | 0.000264 | 0.000499 | 9.95E-05 | 0.002741737 | NA |
| chr8 | 143715759 | 143715797 | 39 | 2 | 8.61E-05 | 7.07E-05 | 0.001122 | 9.96E-05 | -0.00096863 | NA |
| chr4 | 181365874 | 181366078 | 205 | 2 | 7.56E-05 | 0.0160764 | 4.07E-05 | 9.96E-05 | -0.00289329 | NA |
| chr7 | 1269472 | 1269553 | 82 | 2 | 5.64E-05 | 5.79E-05 | 0.002784 | 9.97E-05 | 0.020084706 | NA |
| chr7 | 156725154 | 156725224 | 71 | 2 | 7.59E-05 | 0.0004542 | 0.000102 | 9.98E-05 | -0.00355391 | LMBR1 |
| chr7 | 148809093 | 148809335 | 243 | 3 | 4.38E-05 | 0.0177029 | 6.00E-05 | 9.99E-05 | 0.001342652 | EZH2 |
| chr12 | 51238857 | 51239266 | 410 | 6 | 4.39E-06 | 0.0009927 | 0.000152 | 0.0001 | 0.00134779 | DAZAP2 |
| chr6 | 1494847 | 1494864 | 18 | 2 | 0.000143252 | 0.0007522 | 7.97E-05 | 0.0001 | -0.00536792 | NA |
| chr4 | 142596889 | 142596963 | 75 | 2 | 0.000157811 | 0.0006938 | 8.27E-05 | 0.0001 | 0.000897692 | AC139720.1, INPP4B |
| chr19 | 44341038 | 44341491 | 454 | 4 | 4.61E-05 | 0.0005952 | 0.00158 | 0.0001 | 0.004148914 | ZNF112, AC245748.1 |
| chr1 | 19814247 | 19814335 | 89 | 3 | 0.000104611 | 0.0006484 | 0.001657 | 0.0001 | -0.00264869 | RNF186 |
| chr9 | 135946921 | 135947554 | 634 | 2 | 2.00E-05 | 6.98E-05 | 0.001199 | 0.0001 | 0.003380604 | UBAC1 |
| chr6 | 79231236 | 79231293 | 58 | 2 | 0.000164991 | 0.000296 | 0.000137 | 0.0001 | -0.0069601 | HMGN3 |
| chr12 | 18737899 | 18738463 | 565 | 5 | 2.25E-05 | 7.22E-05 | 0.002084 | 0.0001 | -0.0018893 | CAPZA3, PLCZ1 |
| chr1 | 205257083 | 205257294 | 212 | 2 | 1.12E-05 | 0.0088548 | 4.33E-05 | 0.0001 | -0.00382981 | TMCC2 |
| chr17 | 80670446 | 80670966 | 521 | 5 | 3.90E-05 | 0.0072134 | 3.19E-05 | 0.0001 | 0.002063825 | RPTOR |
| chr13 | 110687812 | 110688062 | 251 | 5 | 0.00012399 | 0.0158687 | 0.000361 | 0.0001 | -0.00012077 | CARS2 |
| chr7 | 25326859 | 25326876 | 18 | 2 | 5.91E-05 | 0.0003081 | 0.000133 | 0.000101 | -0.00078549 | AC003985.1 |
| chr3 | 42659093 | 42659186 | 94 | 3 | 0.000125898 | 5.61E-05 | 0.004144 | 0.000101 | -0.01609374 | ZBTB47 |
| chr7 | 2593055 | 2593494 | 440 | 4 | 2.06E-05 | 0.0044554 | 4.71E-05 | 0.000101 | 0.000616792 | IQCE |
| chr11 | 65298568 | 65298897 | 330 | 2 | 2.37E-05 | 0.0001799 | 0.00022 | 0.000101 | -0.00500833 | POLA2, AP000944.5 |
| chr10 | 18339703 | 18339719 | 17 | 2 | 0.000177797 | 5.88E-05 | 0.002771 | 0.000101 | 0.007974972 | CACNB2 |
| chr20 | 56731134 | 56731632 | 499 | 4 | 1.22E-05 | 0.0001914 | 0.00045 | 0.000101 | -0.01335386 | AL133232.1 |
| chr7 | 41966485 | 41966522 | 38 | 2 | 0.000129213 | 0.0001774 | 0.000224 | 0.000101 | -0.00045675 | GLI3 |
| chr1 | 156115406 | 156116213 | 808 | 2 | 3.58E-05 | 0.0001316 | 0.000325 | 0.000101 | 0.010650033 | LMNA |
| chr17 | 51042385 | 51042462 | 78 | 3 | 0.000130436 | 3.99E-05 | 0.007151 | 0.000101 | -0.00787727 | SPAG9 |
| chr10 | 103772177 | 103772305 | 129 | 2 | 9.78E-05 | 0.0138292 | 4.17E-05 | 0.000101 | 0.00174566 | SH3PXD2A |
| chr16 | 66994502 | 66994504 | 3 | 2 | 0.000141486 | 0.0003788 | 0.000116 | 0.000101 | 0.00175125 | CES4A |
| chr9 | 75912260 | 75912293 | 34 | 2 | 6.39E-05 | 0.0001024 | 0.000486 | 0.000102 | -0.00056613 | PCSK5 |
| chr16 | 89620210 | 89620644 | 435 | 7 | 6.62E-05 | 0.0043991 | 5.59E-05 | 0.000102 | -0.00055747 | DPEP1 |
| chr19 | 46626080 | 46626380 | 301 | 4 | 8.03E-05 | 0.0001949 | 0.000907 | 0.000102 | -0.00304983 | NA |
| chr22 | 45976621 | 45976864 | 244 | 5 | 3.33E-05 | 0.0002526 | 0.000214 | 0.000102 | -0.00069115 | WNT7B |
| chr1 | 2883014 | 2883629 | 616 | 5 | 1.19E-05 | 0.0006283 | 0.001391 | 0.000102 | -0.00249163 | NA |
| chr17 | 82449155 | 82449503 | 349 | 2 | 6.67E-07 | 0.0042854 | 4.88E-05 | 0.000102 | 0.004108203 | CYBC1 |
| chr11 | 57656812 | 57657062 | 251 | 6 | 6.18E-05 | 0.0002885 | 0.002784 | 0.000102 | -0.00636741 | CLP1 |
| chr1 | 113758674 | 113759512 | 839 | 13 | 1.89E-05 | 0.0023305 | 0.000183 | 0.000102 | -0.00023499 | PHTF1 |
| chr1 | 51517824 | 51518378 | 555 | 6 | 5.04E-05 | 0.0033656 | 0.000972 | 0.000102 | -0.00504408 | EPS15-AS1, EPS15 |
| chr4 | 3027777 | 3027878 | 102 | 2 | 2.09E-05 | 0.0001896 | 0.000211 | 0.000102 | 0.001207842 | GRK4 |
| chr9 | 37515117 | 37515272 | 156 | 2 | 0.000112909 | 0.0036075 | 5.06E-05 | 0.000102 | -0.00218859 | FBXO10, AL513165.2 |
| chr2 | 11133572 | 11133775 | 204 | 2 | 8.39E-06 | 8.05E-05 | 0.000825 | 0.000102 | -0.01364938 | C2orf50 |
| chr16 | 1808876 | 1809329 | 454 | 4 | 0.000125878 | 0.0046653 | 0.000486 | 0.000102 | -0.00262765 | HAGH |
| chr22 | 39124641 | 39124660 | 20 | 2 | 0.000135247 | 0.0041474 | 4.94E-05 | 0.000102 | -0.00203438 | CBX7 |
| chr11 | 66971908 | 66972659 | 752 | 2 | 4.52E-05 | 0.0002132 | 0.000188 | 0.000102 | -0.00433587 | NA |
| chr7 | 149855693 | 149855921 | 229 | 4 | 0.000113693 | 0.0003658 | 0.004612 | 0.000102 | -0.00134236 | ZNF862 |
| chr12 | 6478510 | 6478837 | 328 | 3 | 5.36E-05 | 0.0010217 | 6.91E-05 | 0.000102 | -0.00423722 | AC005840.3 |
| chr9 | 90966023 | 90966175 | 153 | 2 | 3.04E-05 | 6.47E-05 | 0.001787 | 0.000102 | -0.0134502 | NA |
| chr5 | 163459490 | 163460102 | 613 | 7 | 4.14E-05 | 0.0015268 | 0.005034 | 0.000103 | 0.000235931 | NUDCD2 |
| chr11 | 27522256 | 27522259 | 4 | 2 | 0.000172218 | 5.96E-05 | 0.002814 | 0.000103 | 0.000766113 | BDNF-AS |
| chr6 | 166678595 | 166678669 | 75 | 2 | 0.000139166 | 0.0028737 | 5.36E-05 | 0.000103 | 0.011744982 | RPS6KA2 |
| chr1 | 87130822 | 87131251 | 430 | 4 | 1.83E-05 | 0.0002945 | 0.003892 | 0.000103 | -0.00454227 | AC093155.3, LINC01140 |
| chr6 | 87408347 | 87408856 | 510 | 4 | 5.16E-05 | 6.21E-05 | 0.006018 | 0.000103 | 0.006831208 | CFAP206, AL049697.1 |
| chr17 | 39696779 | 39696975 | 197 | 2 | 7.03E-05 | 0.0002584 | 0.000159 | 0.000103 | 0.000768975 | ERBB2, PGAP3 |
| chr8 | 51898306 | 51898814 | 509 | 3 | 2.81E-06 | 0.0066591 | 5.31E-05 | 0.000103 | 0.001772232 | PCMTD1 |
| chr13 | 95174676 | 95174775 | 100 | 3 | 0.000148513 | 0.001371 | 5.49E-05 | 0.000103 | 0.002007636 | ABCC4 |
| chr12 | 67989103 | 67989423 | 321 | 4 | 5.27E-05 | 0.0042843 | 3.64E-05 | 0.000103 | -0.00159167 | NA |
| chr1 | 33603526 | 33603556 | 31 | 2 | 8.71E-05 | 0.0005293 | 9.75E-05 | 0.000103 | -0.00449634 | CSMD2 |
| chr22 | 19722568 | 19723189 | 622 | 8 | 4.26E-05 | 0.0004621 | 0.000175 | 0.000103 | -0.00391967 | SEPTIN5, AC000093.1 |
| chr1 | 108711909 | 108712023 | 115 | 2 | 1.34E-05 | 7.40E-05 | 0.001107 | 0.000104 | -0.01110429 | NA |
| chr20 | 48680757 | 48680898 | 142 | 2 | 2.88E-05 | 0.1463777 | 3.91E-05 | 0.000104 | -0.00403364 | PREX1 |
| chr14 | 23375667 | 23375882 | 216 | 4 | 0.000149328 | 0.0009571 | 0.001676 | 0.000104 | -0.00262672 | IL25 |
| chr11 | 113147277 | 113147279 | 3 | 2 | 0.000107292 | 0.0407906 | 4.00E-05 | 0.000104 | 0.002304175 | NCAM1 |
| chr12 | 3105943 | 3106137 | 195 | 2 | 5.28E-05 | 0.0024808 | 5.61E-05 | 0.000104 | 0.00337753 | TSPAN9 |
| chr19 | 18318472 | 18318855 | 384 | 4 | 0.000100323 | 4.93E-05 | 0.007417 | 0.000104 | 0.011883307 | LSM4 |
| chr10 | 101007750 | 101008138 | 389 | 3 | 4.92E-05 | 5.63E-05 | 0.004333 | 0.000104 | -0.00219078 | LZTS2, PDZD7 |
| chr2 | 241314652 | 241315870 | 1219 | 19 | 3.16E-05 | 0.0040876 | 0.002917 | 0.000104 | 0.000846551 | SEPTIN2, HDLBP |
| chr11 | 26840555 | 26840574 | 20 | 2 | 0.000141534 | 0.0030299 | 5.36E-05 | 0.000104 | -0.00558707 | NA |
| chr22 | 49943192 | 49944024 | 833 | 4 | 2.97E-05 | 0.0002302 | 0.000293 | 0.000104 | 0.00144807 | NA |
| chr5 | 176613852 | 176613997 | 146 | 2 | 0.000127668 | 0.0011084 | 7.15E-05 | 0.000104 | 0.01404978 | NA |
| chr2 | 239976644 | 239976808 | 165 | 3 | 0.000152141 | 0.0024123 | 0.001381 | 0.000104 | -0.00209953 | NDUFA10 |
| chr5 | 176724770 | 176724777 | 8 | 2 | 0.000137804 | 6.24E-05 | 0.002384 | 0.000105 | -0.00417618 | AC113391.1 |
| chr16 | 88537354 | 88537479 | 126 | 2 | 3.12E-05 | 0.0133328 | 4.30E-05 | 0.000105 | -0.00374255 | NA |
| chr19 | 48392823 | 48392965 | 143 | 3 | 1.75E-05 | 5.77E-05 | 0.002831 | 0.000105 | -0.00930531 | NA |
| chr7 | 135438337 | 135438420 | 84 | 2 | 1.99E-05 | 0.0003246 | 0.000136 | 0.000105 | -0.00997629 | CNOT4 |

| | | | | | | | | | | |
|---|---|---|---|---|---|---|---|---|---|---|
| chr3 | 13650542 | 13650843 | 302 | 6 | 2.67E-05 | 0.0178682 | 9.63E-06 | 0.000105 | -0.00322335 | LINC00620 |
| chr5 | 171313543 | 171313933 | 391 | 6 | 4.95E-05 | 0.0004579 | 0.000222 | 0.000105 | 9.02E-05 | NA |
| chr6 | 123380388 | 123380446 | 59 | 2 | 0.000115704 | 0.0124567 | 4.34E-05 | 0.000105 | -0.01656145 | TRDN |
| chr13 | 111241001 | 111241190 | 190 | 3 | 8.05E-05 | 0.0006822 | 8.31E-05 | 0.000105 | 0.002047798 | ARHGEF7 |
| chr6 | 28207097 | 28207505 | 409 | 3 | 2.88E-05 | 0.0003035 | 0.00022 | 0.000105 | -0.00735132 | NA |
| chr11 | 61102129 | 61102649 | 521 | 7 | 2.36E-05 | 0.004501 | 0.000643 | 0.000105 | 0.004759835 | CD5 |
| chr12 | 93020731 | 93021474 | 744 | 5 | 6.29E-05 | 0.0013152 | 0.000302 | 0.000105 | -0.00131577 | AC138123.1 |
| chr11 | 56374353 | 56374384 | 32 | 3 | 0.000173525 | 0.0010014 | 0.0001 | 0.000105 | -0.00765282 | NA |
| chr3 | 10816819 | 10817267 | 449 | 3 | 1.04E-05 | 0.0028479 | 5.21E-05 | 0.000105 | -0.00975431 | SLC6A11 |
| chr4 | 83002064 | 83002315 | 252 | 3 | 6.95E-05 | 0.0021076 | 0.000237 | 0.000105 | -0.01100572 | LIN54 |
| chr12 | 47792026 | 47792084 | 59 | 3 | 0.000119724 | 0.0001139 | 0.000614 | 0.000105 | -0.00334593 | HDAC7 |
| chr20 | 32727792 | 32728325 | 534 | 2 | 6.66E-07 | 0.0068763 | 4.66E-05 | 0.000105 | 0.006250251 | FO393400.1, COMMD7 |
| chr17 | 59220122 | 59220689 | 568 | 12 | 2.21E-05 | 0.1390253 | 7.16E-05 | 0.000105 | -0.00181257 | GDPD1 |
| chr10 | 18533272 | 18533634 | 363 | 4 | 5.40E-05 | 0.0002461 | 0.000506 | 0.000105 | -0.00220847 | CACNB2, AL450384.2, AL450384.1 |
| chr18 | 22468072 | 22468121 | 50 | 2 | 8.43E-05 | 0.0005159 | 0.000101 | 0.000106 | -0.00557995 | NA |
| chr20 | 45348293 | 45348930 | 638 | 10 | 8.17E-05 | 0.0017718 | 0.000747 | 0.000106 | -0.00028678 | SDC4 |
| chr4 | 13248620 | 13248753 | 134 | 2 | 4.01E-05 | 0.0018384 | 6.18E-05 | 0.000106 | -0.00620837 | NA |
| chr2 | 156436252 | 156436443 | 192 | 5 | 5.83E-05 | 0.0011221 | 0.002028 | 0.000106 | 0.000763641 | GPD2 |
| chr3 | 14133037 | 14133265 | 229 | 2 | 2.54E-05 | 0.0300821 | 4.12E-05 | 0.000106 | -0.00246986 | TMEM43 |
| chr21 | 43768238 | 43768313 | 76 | 2 | 3.73E-05 | 0.029557 | 4.13E-05 | 0.000106 | -0.00808917 | NA |
| chr11 | 130868016 | 130868136 | 121 | 2 | 0.000100883 | 0.0012257 | 7.03E-05 | 0.000106 | -0.01073655 | SNX19, AP003486.1 |
| chr19 | 41945638 | 41945707 | 70 | 2 | 7.26E-05 | 0.0012599 | 6.97E-05 | 0.000106 | 0.010260248 | NA |
| chr15 | 96369726 | 96369954 | 229 | 2 | 0.000133345 | 7.97E-05 | 0.000966 | 0.000106 | 0.003899205 | AC087477.2 |
| chr1 | 153202023 | 153202178 | 156 | 2 | 5.98E-05 | 0.0004577 | 0.00011 | 0.000106 | 0.002381439 | NA |
| chr3 | 49930813 | 49931100 | 288 | 2 | 1.65E-06 | 6.93E-05 | 0.001587 | 0.000107 | -5.78E-05 | NA |
| chr10 | 60777766 | 60778525 | 760 | 14 | 8.31E-05 | 0.0002826 | 0.00319 | 0.000107 | -0.00029717 | CDK1 |
| chr6 | 117266400 | 117266425 | 26 | 2 | 0.000102392 | 7.73E-05 | 0.001071 | 0.000107 | -0.01045959 | VGLL2 |
| chr11 | 70670453 | 70670719 | 267 | 3 | 3.75E-05 | 0.0004807 | 0.000549 | 0.000107 | -0.01060663 | SHANK2 |
| chr17 | 8220430 | 8220693 | 264 | 2 | 0.000130657 | 0.000115 | 0.000449 | 0.000107 | -0.01046654 | LINC00324 |
| chr11 | 110429705 | 110430404 | 700 | 7 | 7.59E-05 | 0.0236314 | 0.000374 | 0.000107 | 0.002170786 | FDX1 |
| chr7 | 1322932 | 1323488 | 557 | 4 | 0.000113345 | 5.73E-05 | 0.005859 | 0.000107 | 0.009511076 | NA |
| chr2 | 232584554 | 232584573 | 20 | 2 | 0.000118561 | 0.0026084 | 5.70E-05 | 0.000107 | 0.000670058 | AC073254.1 |
| chr17 | 9645247 | 9646029 | 783 | 8 | 5.31E-05 | 0.0002506 | 0.006474 | 0.000107 | 0.001560453 | USP43, AC118755.2 |
| chr7 | 70792940 | 70793024 | 85 | 3 | 0.000140963 | 0.0012074 | 0.000355 | 0.000107 | -0.00547543 | AUTS2 |
| chr16 | 58507708 | 58507984 | 277 | 2 | 6.73E-05 | 8.78E-05 | 0.000768 | 0.000107 | -0.00636363 | NDRG4 |
| chr4 | 725102 | 725825 | 724 | 6 | 3.98E-05 | 0.0004163 | 0.00145 | 0.000107 | 0.004173279 | PCGF3 |
| chr9 | 2196089 | 2196156 | 68 | 2 | 0.00016647 | 0.0016053 | 6.50E-05 | 0.000107 | 0.001821645 | NA |
| chr1 | 230391832 | 230391895 | 64 | 2 | 9.78E-05 | 0.0080322 | 4.64E-05 | 0.000107 | 0.018216604 | PGBD5 |
| chr11 | 65918231 | 65919042 | 812 | 8 | 2.94E-06 | 0.0575621 | 0.000322 | 0.000107 | 0.002778656 | C11orf68 |
| chr10 | 13348286 | 13348394 | 109 | 5 | 0.000168496 | 8.75E-05 | 0.001298 | 0.000107 | 0.000381306 | SEPHS1 |
| chr9 | 128097175 | 128097685 | 511 | 4 | 1.07E-05 | 0.0318492 | 1.38E-05 | 0.000108 | -0.00338767 | SLC25A25 |
| chr14 | 64209166 | 64209444 | 279 | 3 | 5.63E-06 | 0.0022902 | 4.59E-05 | 0.000108 | 0.004301459 | SYNE2, ESR2 |
| chr5 | 72238871 | 72239092 | 222 | 2 | 0.000100357 | 0.0128616 | 4.42E-05 | 0.000108 | -0.00551762 | MRPS27 |
| chr10 | 1627022 | 1627134 | 113 | 3 | 8.95E-05 | 0.000223 | 0.000243 | 0.000108 | -0.00358338 | ADARB2 |
| chr5 | 113203526 | 113204165 | 640 | 2 | 2.45E-06 | 0.0008722 | 8.12E-05 | 0.000108 | 0.004562665 | MCC |
| chr17 | 82000716 | 82000809 | 94 | 2 | 0.00014912 | 0.0002003 | 0.000221 | 0.000108 | 0.010168828 | ASPSCR1 |
| chr12 | 16606001 | 16606300 | 300 | 6 | 7.83E-05 | 0.0005058 | 0.003881 | 0.000108 | 0.001736291 | MGST1, LMO3 |
| chr19 | 7787853 | 7788658 | 806 | 3 | 2.51E-05 | 0.0001029 | 0.002614 | 0.000108 | -0.00464024 | CLEC4GP1 |
| chr5 | 174724103 | 174724776 | 674 | 10 | 9.15E-05 | 0.0004559 | 0.003245 | 0.000108 | 0.000344439 | MSX2 |
| chr4 | 8108519 | 8108663 | 145 | 2 | 9.46E-05 | 0.0036205 | 5.34E-05 | 0.000108 | -0.00673203 | ABLIM2 |
| chr6 | 43046917 | 43046959 | 43 | 2 | 8.63E-05 | 0.0004105 | 0.000121 | 0.000108 | -0.00447869 | KLC4, CUL7 |
| chr10 | 627790 | 627951 | 162 | 2 | 0.000150526 | 8.88E-05 | 0.00077 | 0.000108 | -0.0037913 | DIP2C |
| chr7 | 106865243 | 106865516 | 274 | 7 | 0.000117507 | 0.0007226 | 6.85E-05 | 0.000108 | 0.004911637 | PIK3CG |
| chr21 | 40508607 | 40508626 | 20 | 2 | 0.000163262 | 0.0009679 | 7.84E-05 | 0.000108 | -0.01306087 | DSCAM |
| chr10 | 122270132 | 122270356 | 225 | 4 | 3.70E-05 | 0.0042163 | 2.43E-05 | 0.000108 | -0.00654301 | NA |
| chr6 | 47477936 | 47478462 | 527 | 3 | 1.08E-06 | 0.0017621 | 5.15E-05 | 0.000108 | 0.002373266 | CD2AP |
| chr3 | 14124735 | 14124981 | 247 | 7 | 0.000150672 | 0.0002382 | 0.007656 | 0.000108 | 0.000263414 | CHCHD4 |
| chr19 | 34255248 | 34255298 | 51 | 2 | 3.59E-06 | 0.0087442 | 4.64E-05 | 0.000109 | 0.001891554 | GARRE1 |
| chr7 | 47654845 | 47655147 | 303 | 5 | 7.26E-05 | 0.030987 | 0.000111 | 0.000109 | -0.00465495 | NA |
| chr1 | 16724581 | 16725134 | 554 | 5 | 3.09E-05 | 0.0015312 | 0.001754 | 0.000109 | 0.00137654 | NA |
| chr7 | 73578284 | 73578831 | 548 | 9 | 1.00E-05 | 0.0028046 | 0.000946 | 0.000109 | -3.68E-06 | TBL2 |
| chr9 | 130465515 | 130465852 | 338 | 4 | 8.92E-05 | 0.0001247 | 0.003773 | 0.000109 | 0.001322901 | ASS1 |
| chr17 | 50869429 | 50869441 | 13 | 2 | 0.000130476 | 0.0006459 | 9.38E-05 | 0.000109 | -0.00366523 | TOB1-AS1 |
| chr18 | 46560082 | 46560145 | 64 | 2 | 7.79E-05 | 0.002057 | 6.15E-05 | 0.000109 | 0.002362107 | LOXHD1 |
| chr7 | 130497256 | 130497289 | 34 | 2 | 8.54E-05 | 0.0003318 | 0.000142 | 0.000109 | 0.003228905 | MEST, AC007938.2 |
| chr5 | 168578902 | 168579301 | 400 | 2 | 5.95E-05 | 7.37E-05 | 0.001359 | 0.000109 | -0.00146387 | PANK3 |
| chr3 | 130345236 | 130345740 | 505 | 4 | 6.07E-05 | 0.0011493 | 0.000117 | 0.000109 | -0.00301024 | COL6A5 |
| chr5 | 124706892 | 124707416 | 525 | 3 | 8.55E-06 | 0.0001877 | 0.000462 | 0.000109 | -0.00351036 | ZNF608 |
| chr13 | 20717034 | 20717214 | 181 | 2 | 0.000145333 | 6.62E-05 | 0.002163 | 0.000109 | -0.00296188 | IL17D |
| chr20 | 63394910 | 63395110 | 201 | 3 | 0.000102935 | 0.0020047 | 4.90E-05 | 0.000109 | 0.004912201 | NA |
| chr2 | 105769269 | 105769948 | 680 | 14 | 6.32E-06 | 0.0091236 | 1.17E-05 | 0.000109 | 0.002144351 | NCK2 |
| chr6 | 38597023 | 38597478 | 456 | 2 | 4.83E-05 | 7.46E-05 | 0.001313 | 0.000109 | -0.00505374 | BTBD9 |
| chr8 | 19697522 | 19697858 | 337 | 2 | 1.27E-05 | 0.0075882 | 4.75E-05 | 0.000109 | -0.01751245 | CSGALNACT1 |
| chr14 | 74842994 | 74843039 | 46 | 2 | 0.000121108 | 0.0087598 | 4.66E-05 | 0.000109 | -0.00279744 | YLPM1 |
| chr1 | 46285970 | 46286303 | 334 | 2 | 4.55E-05 | 8.75E-05 | 0.000827 | 0.000109 | 0.003516907 | LRRC41 |
| chr15 | 26666484 | 26666723 | 240 | 3 | 0.000114272 | 0.0002718 | 0.000278 | 0.000109 | -0.01009335 | GABRB3 |
| chr11 | 111512190 | 111512563 | 374 | 3 | 4.23E-05 | 7.66E-05 | 0.001806 | 0.00011 | -0.00457791 | AP002008.4, BTG4 |
| chr14 | 102864439 | 102864832 | 394 | 3 | 2.38E-05 | 0.058855 | 1.63E-05 | 0.00011 | 0.002760721 | TRAF3 |
| chr5 | 126281270 | 126281290 | 21 | 2 | 0.000105252 | 0.0009037 | 8.19E-05 | 0.00011 | -0.01380634 | AC116362.1 |
| chr12 | 57112578 | 57112729 | 152 | 2 | 3.66E-05 | 0.0008009 | 8.62E-05 | 0.00011 | 0.005325144 | STAT6 |
| chr1 | 18937312 | 18937346 | 35 | 2 | 7.02E-05 | 0.012587 | 4.52E-05 | 0.00011 | -0.00281617 | IFFO2 |
| chr17 | 38924047 | 38924120 | 74 | 2 | 9.62E-05 | 6.60E-05 | 0.00232 | 0.00011 | -0.01296045 | NA |
| chr12 | 57467425 | 57467516 | 92 | 2 | 8.44E-05 | 0.0002192 | 0.000208 | 0.00011 | 0.000353734 | GLI1 |
| chr11 | 60912633 | 60912693 | 61 | 2 | 0.000127553 | 0.0075563 | 4.78E-05 | 0.00011 | -0.00917323 | NA |
| chr14 | 72742420 | 72742480 | 61 | 2 | 0.000144925 | 0.0046083 | 5.17E-05 | 0.00011 | -0.01178638 | DPF3 |
| chr7 | 4877676 | 4878230 | 555 | 4 | 6.41E-05 | 0.0054362 | 7.53E-05 | 0.00011 | -0.00185666 | RADIL |
| chr19 | 14081046 | 14081176 | 131 | 2 | 3.35E-05 | 0.0165767 | 4.42E-05 | 0.00011 | 0.001848671 | NA |
| chr20 | 63177339 | 63178489 | 1151 | 10 | 2.72E-05 | 0.0002137 | 0.012414 | 0.00011 | 0.00134021 | AL096828.1 |
| chr11 | 66011494 | 66011520 | 27 | 3 | 9.73E-05 | 0.1279504 | 1.46E-05 | 0.00011 | 0.00445671 | NA |
| chr2 | 176554177 | 176554503 | 327 | 3 | 1.82E-05 | 0.0005764 | 0.001273 | 0.00011 | 0.001221188 | LINC01117 |
| chr9 | 94909640 | 94909687 | 48 | 2 | 0.000104936 | 0.0018253 | 6.45E-05 | 0.00011 | -0.00388707 | AOPEP |
| chr10 | 112849539 | 112849961 | 423 | 2 | 0.000101966 | 7.93E-05 | 0.001111 | 0.00011 | 0.000569727 | NA |
| chr6 | 31529637 | 31529979 | 343 | 2 | 1.27E-05 | 0.0035794 | 5.47E-05 | 0.00011 | -0.00211883 | MCCD1 |
| chr11 | 74085735 | 74085864 | 130 | 2 | 0.000112098 | 0.0023519 | 6.02E-05 | 0.000111 | -0.01085428 | C2CD3 |
| chr6 | 43784785 | 43785384 | 600 | 3 | 4.61E-06 | 0.0073596 | 4.11E-05 | 0.000111 | -0.00339322 | VEGFA |
| chr6 | 30490050 | 30490824 | 775 | 16 | 3.98E-05 | 0.0019198 | 0.003788 | 0.000111 | 0.002608415 | HLA-E |
| chr19 | 11170146 | 11170237 | 92 | 3 | 0.000106806 | 0.0056161 | 6.55E-05 | 0.000111 | -0.00415616 | KANK2 |
| chr8 | 24387846 | 24388233 | 388 | 2 | 4.01E-05 | 0.0001667 | 0.000285 | 0.000111 | -0.00343066 | ADAMDEC1, AC120193.1 |
| chr3 | 141737452 | 141737463 | 12 | 2 | 9.20E-06 | 6.61E-05 | 0.002364 | 0.000111 | -0.01630137 | NA |
| chr8 | 42270980 | 42271835 | 856 | 9 | 2.69E-05 | 0.0005346 | 0.003435 | 0.000111 | 0.000802146 | IKBKB, AC083973.1 |
| chr8 | 73976242 | 73976833 | 592 | 5 | 3.02E-05 | 0.001274 | 0.002816 | 0.000111 | -0.0004616 | TMEM70 |
| chr4 | 686325 | 686867 | 543 | 4 | 3.65E-05 | 0.0001461 | 0.005486 | 0.000111 | 0.000580098 | SLC49A3 |
| chr11 | 44376117 | 44376191 | 75 | 2 | 0.000148693 | 0.0001138 | 0.000501 | 0.000111 | -0.00244419 | NA |
| chr10 | 78588256 | 78588259 | 4 | 2 | 0.000173327 | 8.75E-05 | 0.000869 | 0.000111 | -0.00362799 | AC012560.1 |
| chr4 | 188737253 | 188737272 | 20 | 2 | 4.64E-05 | 0.0001683 | 0.000285 | 0.000111 | -0.0110233 | NA |
| chr16 | 66933964 | 66934724 | 761 | 13 | 3.18E-05 | 0.0049372 | 0.000199 | 0.000111 | -0.00029411 | CES2, CIAO2B |
| chr1 | 116693139 | 116693631 | 493 | 2 | 4.73E-06 | 0.0007438 | 9.04E-05 | 0.000111 | -0.00546815 | NA |
| chr7 | 36642709 | 36642971 | 263 | 2 | 2.98E-05 | 0.0001889 | 0.00025 | 0.000112 | 0.014341772 | AOAH |
| chr17 | 37144243 | 37144371 | 129 | 2 | 0.000100351 | 0.0025253 | 5.97E-05 | 0.000112 | -0.00806432 | ACACA, HMGB1P24 |
| chr13 | 113670176 | 113670632 | 457 | 3 | 6.86E-05 | 0.0001765 | 0.001315 | 0.000112 | -0.00672526 | GRK1 |
| chr17 | 17203496 | 17203525 | 30 | 2 | 2.69E-05 | 0.0002106 | 0.000223 | 0.000112 | -0.00447358 | MPRIP, PLD6, AC055811.2 |
| chr11 | 1628717 | 1628885 | 169 | 3 | 0.000142765 | 0.0001134 | 0.000822 | 0.000112 | 0.008255143 | NA |
| chr5 | 173791547 | 173792202 | 656 | 3 | 1.56E-05 | 0.0011667 | 0.000102 | 0.000112 | 0.002776259 | LINC01485 |
| chr14 | 63728097 | 63728386 | 290 | 6 | 0.000112342 | 0.0006267 | 0.002972 | 0.000112 | -0.00087685 | NA |
| chr18 | 41954455 | 41954987 | 533 | 4 | 4.33E-06 | 0.0001088 | 0.001311 | 0.000112 | -0.00810113 | NA |
| chr20 | 49482609 | 49482822 | 214 | 5 | 0.000116141 | 3.90E-05 | 0.010724 | 0.000112 | 0.000343207 | KCNB1 |
| chr7 | 44490815 | 44491183 | 369 | 9 | 5.99E-05 | 0.0001236 | 0.030515 | 0.000112 | -1.63E-05 | NA |
| chr3 | 164832363 | 164832543 | 181 | 2 | 0.000119723 | 0.00016 | 0.00031 | 0.000112 | -0.00956351 | NA |
| chr9 | 83622878 | 83623229 | 352 | 4 | 8.03E-05 | 0.0077999 | 0.000966 | 0.000113 | -0.00357722 | IDNK |
| chr1 | 201540188 | 201540287 | 100 | 2 | 0.000140475 | 0.0001847 | 0.000261 | 0.000113 | -0.00552008 | RPS10P7 |
| chr7 | 97016677 | 97017286 | 610 | 4 | 1.87E-05 | 0.0032057 | 0.0018 | 0.000113 | -0.00330717 | NA |
| chr5 | 179195802 | 179196034 | 233 | 2 | 3.99E-05 | 0.001939 | 6.47E-05 | 0.000113 | -0.00490742 | ADAMTS2 |
| chr19 | 51759716 | 51759805 | 90 | 2 | 0.000113223 | 0.0002666 | 0.00018 | 0.000113 | 0.007466258 | FPR2, FPR1 |
| chr17 | 75132480 | 75133135 | 656 | 3 | 6.53E-06 | 0.0033452 | 4.48E-05 | 0.000113 | 0.001911219 | AC022211.4 |
| chr1 | 919666 | 920679 | 1014 | 7 | 7.83E-06 | 0.0003457 | 0.000828 | 0.000113 | 0.000617606 | LINC02593 |
| chr1 | 20634374 | 20634736 | 363 | 2 | 2.80E-05 | 6.49E-05 | 0.002979 | 0.000113 | -0.00643529 | PINK1 |
| chr10 | 117168999 | 117169055 | 57 | 2 | 0.00014401 | 7.16E-05 | 0.001784 | 0.000113 | -0.00544326 | MIR3663HG |
| chr1 | 150305593 | 150306013 | 421 | 3 | 8.52E-05 | 0.0011134 | 0.001753 | 0.000113 | -0.00647387 | MRPS21 |
| chr1 | 18373840 | 18374131 | 292 | 3 | 8.75E-05 | 4.72E-05 | 0.007124 | 0.000113 | -0.01303988 | IGSF21 |
| chr3 | 194582534 | 194583259 | 726 | 5 | 4.89E-05 | 0.0062653 | 0.000788 | 0.000113 | -5.02E-05 | NA |
| chr6 | 158628411 | 158628437 | 27 | 2 | 0.00016096 | 0.0002982 | 0.000165 | 0.000113 | -0.00176306 | TMEM181 |
| chr19 | 42313507 | 42313832 | 326 | 4 | 1.13E-05 | 0.0168616 | 0.000107 | 0.000114 | 0.005855543 | TMEM145 |
| chr13 | 40480765 | 40481124 | 360 | 2 | 3.58E-05 | 6.52E-05 | 0.002996 | 0.000114 | 0.003486486 | LINC00598, AL133318.1 |
| chr2 | 232696651 | 232697124 | 474 | 6 | 7.89E-06 | 0.0016743 | 0.000715 | 0.000114 | -0.00238954 | NA |
| chr16 | 28862986 | 28863750 | 765 | 10 | 3.04E-05 | 0.0059526 | 0.005097 | 0.000114 | -0.00206098 | AC009093.11, SH2B1, AC133550.3 |
| chr14 | 100375431 | 100376012 | 582 | 10 | 4.79E-05 | 0.0001823 | 0.006782 | 0.000114 | -0.00192713 | WARS1 |
| chr20 | 61023481 | 61023521 | 41 | 2 | 6.76E-05 | 0.0207467 | 4.46E-05 | 0.000114 | -0.00495544 | NA |
| chr1 | 12125378 | 12125788 | 411 | 3 | 0.000119803 | 0.0002989 | 0.000237 | 0.000114 | 0.003280319 | TNFRSF8 |
| chr14 | 60507666 | 60507807 | 142 | 5 | 3.54E-05 | 0.0006556 | 0.001069 | 0.000114 | -0.00607711 | C14orf39 |
| chr12 | 53208079 | 53208705 | 627 | 3 | 4.18E-05 | 4.28E-05 | 0.009078 | 0.000114 | 0.004325209 | NA |
| chr19 | 48670475 | 48670500 | 26 | 2 | 0.000100423 | 0.0102074 | 4.76E-05 | 0.000114 | -0.00258254 | SEC1P, NTN5 |
| chr13 | 51861215 | 51861588 | 374 | 2 | 2.61E-05 | 0.0001009 | 0.000684 | 0.000115 | 0.007507064 | NA |
| chr20 | 33490160 | 33490962 | 803 | 3 | 1.39E-05 | 0.0001321 | 0.002817 | 0.000115 | 0.005107338 | CBFA2T2 |
| chr10 | 987409 | 988090 | 682 | 5 | 5.90E-07 | 0.0036333 | 1.98E-05 | 0.000115 | -0.00450354 | AL359878.1 |
| chr16 | 88531118 | 88531165 | 48 | 2 | 6.96E-05 | 9.17E-05 | 0.000853 | 0.000115 | -0.00851163 | ZFPM1 |
| chr21 | 45405235 | 45405275 | 41 | 2 | 0.000170919 | 7.51E-05 | 0.001561 | 0.000115 | 0.000469857 | COL18A1 |
| chr2 | 2889370 | 2889837 | 468 | 4 | 4.24E-05 | 0.0006407 | 0.000198 | 0.000115 | -0.00607484 | NA |
| chr20 | 25032709 | 25033282 | 574 | 8 | 6.87E-05 | 0.002864 | 0.000601 | 0.000115 | -0.0012163 | ACSS1 |
| chr13 | 30159729 | 30159915 | 187 | 2 | 2.03E-05 | 0.0059483 | 5.15E-05 | 0.000115 | -0.00160262 | LINC00385 |
| chr11 | 64342351 | 64342706 | 356 | 2 | 1.13E-06 | 0.0003678 | 0.000143 | 0.000115 | -0.00732836 | CCDC88B |
| chr19 | 49664535 | 49664874 | 340 | 5 | 4.96E-05 | 0.0049533 | 0.001041 | 0.000115 | 0.0009094 | IRF3 |
| chr19 | 3135133 | 3135366 | 234 | 4 | 8.73E-05 | 0.0003512 | 0.000888 | 0.000115 | 0.004869011 | NA |
| chr8 | 26292872 | 26293115 | 244 | 4 | 3.13E-05 | 0.0018357 | 0.000117 | 0.000115 | 0.007256073 | PPP2R2A |
| chr19 | 16535517 | 16535532 | 16 | 2 | 0.000135007 | 0.0007151 | 9.55E-05 | 0.000115 | 0.001901247 | AC008764.1, CHERP |
| chr22 | 41688218 | 41689087 | 870 | 10 | 8.57E-05 | 0.0005149 | 0.012372 | 0.000116 | 6.74E-05 | C22orf46, SNU13 |
| chr5 | 1859650 | 1859735 | 86 | 3 | 0.000110946 | 6.17E-05 | 0.004197 | 0.000116 | -0.00480565 | AC025183.2 |
| chr5 | 136029281 | 136029712 | 432 | 4 | 4.39E-05 | 0.0003104 | 0.002296 | 0.000116 | -0.00219841 | TGFBI |
| chr7 | 55369447 | 55369636 | 190 | 2 | 7.25E-05 | 0.0026093 | 6.14E-05 | 0.000116 | -0.02045194 | LANCL2 |

| | | | | | | | | | | |
|---|---|---|---|---|---|---|---|---|---|---|
| chr3 | 69201708 | 69201734 | 27 | 2 | 1.33E-05 | 0.000503 | 0.000116 | 0.000116 | -0.00115856 | FRMD4B |
| chr12 | 65607542 | 65607697 | 156 | 2 | 8.19E-05 | 0.0181204 | 4.57E-05 | 0.000116 | -0.01418592 | LINC02454, AC025419.1 |
| chr2 | 70905116 | 70905388 | 273 | 2 | 1.49E-05 | 0.0003414 | 0.000153 | 0.000116 | 0.012825162 | VAX2 |
| chr1 | 107897609 | 107897700 | 92 | 2 | 0.000131939 | 0.003981 | 5.60E-05 | 0.000116 | 0.003581051 | VAV3 |
| chr17 | 42670924 | 42671554 | 631 | 4 | 3.50E-05 | 0.0001926 | 0.001193 | 0.000116 | -0.01564321 | PLEKHH3 |
| chr10 | 117536671 | 117537431 | 761 | 6 | 5.20E-06 | 0.0018398 | 0.0001 | 0.000116 | 0.003243304 | EMX2OS |
| chr8 | 141141042 | 141141055 | 14 | 2 | 0.000102568 | 6.84E-05 | 0.002614 | 0.000117 | -0.00328167 | DENND3 |
| chr4 | 6231807 | 6231925 | 119 | 3 | 0.00016014 | 0.0002335 | 0.000366 | 0.000117 | 0.00016399 | LINC02495 |
| chr3 | 128069614 | 128069730 | 117 | 3 | 0.000140632 | 0.0036393 | 6.69E-05 | 0.000117 | -0.00196969 | SEC61A1, RUVBL1 |
| chr6 | 150865033 | 150865768 | 736 | 8 | 0.000112042 | 4.18E-05 | 0.011168 | 0.000117 | -0.00275128 | MTHFD1L |
| chr16 | 73060280 | 73060444 | 165 | 3 | 5.42E-05 | 0.0075844 | 0.00049 | 0.000117 | 0.004341676 | AC116667.1, ZFHX3 |
| chr7 | 54833546 | 54833952 | 407 | 2 | 5.39E-05 | 8.89E-05 | 0.000972 | 0.000117 | 0.003144862 | NA |
| chr7 | 3043699 | 3044139 | 441 | 6 | 8.43E-06 | 0.0762222 | 3.35E-06 | 0.000117 | -0.00105427 | CARD11 |
| chr2 | 86195072 | 86195171 | 100 | 3 | 8.10E-05 | 0.0001784 | 0.002101 | 0.000117 | -0.00300319 | IMMT |
| chr7 | 151199922 | 151200538 | 617 | 2 | 3.71E-05 | 0.0002298 | 0.00022 | 0.000117 | -0.00733596 | IQCA1L |
| chr10 | 132794734 | 132794960 | 227 | 2 | 9.97E-05 | 0.0002345 | 0.000216 | 0.000117 | 0.013817701 | NA |
| chr12 | 100093120 | 100093178 | 59 | 2 | 7.95E-05 | 0.0003076 | 0.000168 | 0.000117 | 0.006966639 | UHRF1BP1L |
| chr8 | 33483748 | 33483843 | 96 | 2 | 4.55E-05 | 0.0024577 | 6.29E-05 | 0.000117 | 0.00742184 | TTI2 |
| chr20 | 50645240 | 50645406 | 167 | 5 | 0.00015297 | 0.0126904 | 2.58E-05 | 0.000117 | -0.00092191 | RIPOR3 |
| chr4 | 54276667 | 54277102 | 436 | 2 | 3.78E-05 | 0.0001397 | 0.000406 | 0.000117 | -0.00035841 | AC058822.1, PDGFRA |
| chr15 | 96171935 | 96172397 | 463 | 2 | 7.14E-05 | 7.35E-05 | 0.001858 | 0.000117 | 0.003548491 | AC012409.2, AC012409.1, NR2F2-AS1 |
| chr6 | 101398727 | 101399512 | 786 | 15 | 2.98E-05 | 0.0007782 | 0.000617 | 0.000117 | 0.001419851 | GRIK2 |
| chr1 | 42957925 | 42957976 | 52 | 2 | 2.95E-05 | 0.0119665 | 4.77E-05 | 0.000117 | 0.001637543 | SLC2A1 |
| chr6 | 33859919 | 33860553 | 635 | 2 | 1.49E-05 | 0.0001467 | 0.000379 | 0.000117 | 0.007139107 | NA |
| chr18 | 75428830 | 75429086 | 257 | 2 | 7.71E-05 | 0.0001422 | 0.000397 | 0.000117 | -0.01188001 | NA |
| chr6 | 35213132 | 35213336 | 205 | 2 | 2.47E-05 | 0.0206465 | 4.57E-05 | 0.000117 | -0.00294328 | NA |
| chr11 | 4044975 | 4045392 | 418 | 2 | 4.40E-05 | 9.41E-05 | 0.000858 | 0.000117 | 0.006581741 | STIM1 |
| chr1 | 193136259 | 193136577 | 319 | 3 | 0.000100093 | 0.0014773 | 6.16E-05 | 0.000118 | -0.00288869 | CDC73, MIR1278 |
| chr17 | 81026406 | 81026876 | 471 | 3 | 3.13E-05 | 6.99E-05 | 0.002032 | 0.000118 | -0.00526868 | AC127496.5 |
| chr19 | 5838724 | 5838891 | 168 | 4 | 0.000127734 | 0.0058493 | 8.57E-05 | 0.000118 | 0.006592693 | FUT6 |
| chr5 | 73565063 | 73566033 | 971 | 22 | 1.76E-05 | 0.0014197 | 0.000417 | 0.000118 | 0.000205142 | UTP15, ANKRA2 |
| chr22 | 46012123 | 46012157 | 35 | 2 | 0.00014926 | 7.13E-05 | 0.002232 | 0.000118 | -0.00567211 | NA |
| chr14 | 69208696 | 69208721 | 26 | 2 | 5.68E-05 | 0.0002773 | 0.000187 | 0.000118 | -0.00715797 | EXD2, AL359317.2 |
| chr11 | 134358898 | 134359257 | 360 | 2 | 6.34E-05 | 0.0130162 | 4.76E-05 | 0.000118 | -0.00561835 | GLB1L2 |
| chr6 | 33177746 | 33177942 | 197 | 4 | 8.42E-05 | 0.0028014 | 5.47E-05 | 0.000118 | -0.00308497 | COL11A2 |
| chr10 | 100912611 | 100913180 | 570 | 5 | 1.61E-05 | 0.0005044 | 0.0001 | 0.000118 | 0.001782082 | SLF2, AL138762.1 |
| chr5 | 146098589 | 146098661 | 73 | 2 | 5.78E-05 | 0.0053161 | 5.38E-05 | 0.000118 | 0.00852412 | PLAC8L1 |
| chr6 | 9199614 | 9199642 | 29 | 2 | 0.000163004 | 0.0002738 | 0.00019 | 0.000118 | -0.01882519 | HULC |
| chr2 | 3146716 | 3147307 | 592 | 3 | 1.64E-05 | 0.0009918 | 0.000348 | 0.000118 | -0.00607009 | NA |
| chr4 | 100518797 | 100519121 | 325 | 4 | 0.000125492 | 0.000103 | 0.003591 | 0.000118 | 0.007450114 | EMCN |
| chr8 | 1270276 | 1270477 | 202 | 2 | 3.87E-05 | 0.003676 | 5.81E-05 | 0.000119 | -0.01047141 | DLGAP2 |
| chr1 | 6211607 | 6211995 | 389 | 2 | 8.01E-06 | 0.0030203 | 6.06E-05 | 0.000119 | -0.0015144 | RNF207 |
| chr18 | 21956110 | 21956196 | 87 | 2 | 0.000130056 | 0.0001393 | 0.000421 | 0.000119 | 0.001447571 | NA |
| chr2 | 206159163 | 206159838 | 676 | 14 | 3.21E-05 | 0.0080708 | 0.000369 | 0.000119 | 0.001064141 | EEF1B2, NDUFS1 |
| chr1 | 205813744 | 205814261 | 518 | 3 | 1.92E-05 | 0.0008801 | 8.65E-05 | 0.000119 | 0.001188283 | AC119673.2, SLC41A1 |
| chr12 | 130452754 | 130452820 | 67 | 3 | 0.000111378 | 4.59E-05 | 0.008501 | 0.000119 | -0.01241811 | RIMBP2 |
| chr12 | 114669127 | 114669735 | 609 | 3 | 1.32E-05 | 0.005077 | 9.02E-05 | 0.000119 | -0.00408638 | NA |
| chr16 | 12436050 | 12436389 | 340 | 3 | 9.88E-05 | 0.0013766 | 6.65E-05 | 0.000119 | 0.0015195 | SNX29 |
| chr1 | 21887308 | 21887613 | 306 | 4 | 6.44E-05 | 0.0085213 | 0.0001 | 0.000119 | -0.00280274 | HSPG2 |
| chr14 | 93085783 | 93085999 | 217 | 2 | 6.25E-05 | 9.78E-05 | 0.000823 | 0.000119 | -0.00778277 | ITPK1 |
| chr2 | 163348522 | 163349312 | 791 | 5 | 6.90E-05 | 4.98E-05 | 0.006331 | 0.000119 | -0.01033574 | NA |
| chr11 | 1227127 | 1227462 | 336 | 4 | 0.000132288 | 0.0001631 | 0.002909 | 0.000119 | 0.003411872 | MUC5B |
| chr22 | 46622259 | 46622386 | 128 | 2 | 8.24E-05 | 0.0019456 | 6.84E-05 | 0.00012 | -0.00577875 | GRAMD4 |
| chr19 | 5903857 | 5904644 | 788 | 11 | 2.31E-05 | 0.0021947 | 0.010375 | 0.00012 | 0.000743878 | NDUFA11 |
| chr21 | 41226131 | 41226277 | 147 | 2 | 6.50E-05 | 0.0055995 | 5.40E-05 | 0.00012 | -0.00795137 | BACE2 |
| chr5 | 132826175 | 132826307 | 133 | 3 | 0.000103102 | 4.70E-05 | 0.008229 | 0.00012 | -0.0093501 | SHROOM1 |
| chr8 | 102124945 | 102125757 | 813 | 7 | 2.11E-05 | 0.0110787 | 0.000406 | 0.00012 | -8.30E-05 | MIR5680 |
| chr20 | 53753677 | 53753737 | 61 | 2 | 0.000123033 | 0.0004026 | 0.000142 | 0.00012 | -0.00301266 | NA |
| chr21 | 41821327 | 41821965 | 639 | 2 | 1.19E-07 | 0.1071193 | 4.43E-05 | 0.000121 | 0.0021414 | PRDM15 |
| chr9 | 89418247 | 89418330 | 84 | 2 | 7.97E-05 | 7.18E-05 | 0.002436 | 0.000121 | 0.01961715 | SEMA4D |
| chr6 | 6751504 | 6751780 | 277 | 4 | 0.00014143 | 0.0008455 | 0.0024 | 0.000121 | -0.0076625 | NA |
| chr7 | 45574701 | 45575249 | 549 | 3 | 7.55E-06 | 0.0002252 | 0.002337 | 0.000121 | 0.00291095 | ADCY1 |
| chr2 | 219538049 | 219538136 | 88 | 2 | 3.55E-05 | 9.87E-05 | 0.000844 | 0.000121 | -0.00806939 | ASIC4 |
| chr8 | 10833071 | 10833325 | 255 | 3 | 3.55E-05 | 0.2227479 | 1.59E-05 | 0.000121 | -0.00153954 | AC105001.2, PINX1 |
| chr13 | 77025642 | 77025745 | 104 | 2 | 5.41E-05 | 0.00014 | 0.000438 | 0.000121 | -0.01204519 | AC001226.2, FBXL3 |
| chr11 | 104898684 | 104899127 | 444 | 3 | 1.07E-05 | 0.0025582 | 0.00173 | 0.000122 | -0.01321381 | NA |
| chr20 | 5888834 | 5888859 | 26 | 2 | 0.000127656 | 0.0001542 | 0.000381 | 0.000122 | -0.00533743 | NA |
| chr1 | 229508530 | 229509346 | 817 | 7 | 2.51E-05 | 0.0001179 | 0.003895 | 0.000122 | 0.000706669 | AL121990.1 |
| chr1 | 44641883 | 44642020 | 138 | 2 | 6.11E-05 | 0.0007329 | 0.0001 | 0.000122 | -0.00290517 | RNF220, TMEM53 |
| chr17 | 9699995 | 9700023 | 29 | 2 | 3.63E-05 | 0.0003645 | 0.000156 | 0.000122 | -0.00683052 | USP43 |
| chr6 | 35496527 | 35496794 | 268 | 3 | 1.52E-05 | 0.0026961 | 0.00012 | 0.000122 | -0.00309436 | TEAD3 |
| chr17 | 29579467 | 29579525 | 59 | 2 | 0.000104483 | 8.29E-05 | 0.001375 | 0.000122 | -0.00752163 | ABHD15-AS1, GIT1 |
| chr2 | 241844912 | 241845091 | 180 | 2 | 2.19E-05 | 9.18E-05 | 0.001023 | 0.000122 | -0.00028643 | AC131097.3 |
| chr13 | 111530327 | 111530621 | 295 | 3 | 4.19E-05 | 0.0037169 | 4.47E-05 | 0.000122 | -0.00830382 | NA |
| chr8 | 1155035 | 1155087 | 53 | 2 | 0.000101306 | 0.00978 | 5.06E-05 | 0.000122 | -0.00227875 | DLGAP2 |
| chr7 | 96321817 | 96323042 | 1226 | 14 | 3.68E-05 | 0.0312624 | 0.001904 | 0.000122 | 0.001458812 | SLC25A13 |
| chr3 | 42684590 | 42684798 | 209 | 2 | 2.68E-05 | 0.0001002 | 0.000834 | 0.000122 | -0.01702795 | NA |
| chr3 | 29290731 | 29290843 | 113 | 2 | 9.24E-05 | 7.57E-05 | 0.001989 | 0.000122 | 0.003487394 | AC098650.1, RBMS3 |
| chr4 | 76204679 | 76204728 | 50 | 2 | 0.000112848 | 0.0012912 | 8.03E-05 | 0.000123 | 0.003548564 | SCARB2 |
| chr12 | 116835677 | 116835986 | 310 | 2 | 3.18E-05 | 0.0009414 | 9.08E-05 | 0.000123 | -0.00504604 | RNFT2 |
| chr11 | 790669 | 791310 | 642 | 4 | 1.68E-05 | 0.0009936 | 0.001143 | 0.000123 | -0.00145462 | SLC25A22 |
| chr6 | 70280484 | 70280658 | 175 | 4 | 5.68E-05 | 3.63E-05 | 0.013256 | 0.000123 | -0.01237227 | COL9A1 |
| chr10 | 71757615 | 71757640 | 26 | 3 | 6.91E-05 | 0.0002561 | 0.000272 | 0.000123 | -0.00975072 | CDH23, VSIR |
| chr1 | 212467246 | 212467553 | 308 | 4 | 5.27E-05 | 8.83E-05 | 0.002903 | 0.000123 | 0.004410654 | LINC02771 |
| chr19 | 48933461 | 48933710 | 250 | 9 | 6.16E-05 | 0.0012828 | 0.000343 | 0.000123 | 0.001039818 | DHDH |
| chr2 | 3076619 | 3077210 | 592 | 3 | 2.28E-05 | 0.0002778 | 0.000237 | 0.000123 | -0.00663793 | LINC01250 |
| chr11 | 60869888 | 60870387 | 500 | 2 | 1.03E-05 | 0.0002481 | 0.000224 | 0.000124 | -0.00453064 | ZP1 |
| chr4 | 84481244 | 84482070 | 827 | 5 | 6.42E-06 | 0.0609959 | 1.39E-05 | 0.000124 | -0.00094674 | NA |
| chr11 | 76115089 | 76115112 | 24 | 2 | 8.76E-05 | 0.0003105 | 0.000182 | 0.000124 | -0.00550081 | UVRAG |
| chr19 | 35890905 | 35890950 | 46 | 2 | 5.36E-05 | 0.0026148 | 6.53E-05 | 0.000124 | -0.00412307 | NFKBID |
| chr9 | 96219151 | 96219218 | 68 | 2 | 4.68E-05 | 7.05E-05 | 0.003145 | 0.000124 | 0.029113933 | NA |
| chr11 | 59172218 | 59172480 | 263 | 7 | 7.43E-05 | 0.0010618 | 0.000358 | 0.000124 | 0.001607698 | DTX4 |
| chr17 | 76735226 | 76735351 | 126 | 2 | 1.45E-05 | 0.0149381 | 4.91E-05 | 0.000124 | 0.005834728 | AC005837.2, SRSF2 |
| chr2 | 100143161 | 100143427 | 267 | 3 | 7.64E-05 | 7.54E-05 | 0.002581 | 0.000124 | -0.00423374 | AFF3 |
| chr2 | 27148669 | 27149023 | 355 | 8 | 7.45E-05 | 0.0009566 | 0.001485 | 0.000125 | -0.00200236 | TCF23 |
| chr1 | 23619517 | 23619884 | 368 | 4 | 0.00011443 | 0.0008747 | 9.70E-05 | 0.000125 | -0.00041728 | MDS2 |
| chr19 | 42397576 | 42397619 | 44 | 2 | 8.36E-05 | 0.0001579 | 0.000391 | 0.000125 | 0.003056811 | LIPE-AS1 |
| chr14 | 20382677 | 20383158 | 482 | 3 | 5.91E-06 | 0.0015082 | 6.39E-05 | 0.000125 | -0.00174757 | TEP1 |
| chr13 | 72755918 | 72755992 | 75 | 2 | 0.000149255 | 0.0002937 | 0.000195 | 0.000125 | 0.001337611 | BORA, DIS3 |
| chr2 | 161230639 | 161231025 | 387 | 2 | 0.000114074 | 8.94E-05 | 0.001207 | 0.000125 | -0.00644795 | TANK, AC009299.2 |
| chr2 | 231412032 | 231412303 | 272 | 2 | 9.83E-05 | 0.0070596 | 5.41E-05 | 0.000125 | -0.00197777 | NA |
| chr21 | 42838085 | 42838188 | 104 | 2 | 7.62E-05 | 0.000101 | 0.000883 | 0.000126 | 0.001926006 | NA |
| chr1 | 66774373 | 66774491 | 119 | 2 | 7.87E-05 | 7.13E-05 | 0.00317 | 0.000126 | 0.004657902 | TCTEX1D1 |
| chr3 | 195175798 | 195175997 | 200 | 3 | 2.86E-05 | 0.0019486 | 7.67E-05 | 0.000126 | -0.00340149 | XXYLT1 |
| chr5 | 113513459 | 113513745 | 287 | 8 | 7.89E-05 | 0.0021247 | 0.002167 | 0.000126 | 0.003716121 | YTHDC2 |
| chr10 | 97589105 | 97589246 | 142 | 2 | 8.98E-05 | 0.001255 | 8.31E-05 | 0.000126 | -0.00294021 | HOGA1, AL355315.1 |
| chr9 | 72592602 | 72592756 | 155 | 2 | 0.000114834 | 0.0186837 | 4.87E-05 | 0.000126 | -0.01502969 | TMC1 |
| chr11 | 61695955 | 61696013 | 59 | 2 | 6.41E-05 | 0.009422 | 5.22E-05 | 0.000126 | 0.002925405 | DAGLA |
| chr9 | 21996655 | 21996836 | 182 | 2 | 5.50E-05 | 0.0093875 | 5.23E-05 | 0.000126 | 0.010405102 | AL359922.1, CDKN2B-AS1 |
| chr17 | 41733941 | 41735206 | 1266 | 12 | 9.83E-05 | 0.0028273 | 0.01646 | 0.000126 | 0.000158892 | HAP1 |
| chr9 | 137254575 | 137255046 | 472 | 4 | 3.04E-05 | 0.0047882 | 0.000647 | 0.000126 | -0.00116446 | NA |
| chr10 | 93337529 | 93337850 | 322 | 2 | 4.43E-05 | 0.0002064 | 0.000284 | 0.000126 | -0.00343075 | MYOF |
| chr14 | 34540286 | 34540646 | 361 | 5 | 5.28E-06 | 0.0103493 | 1.66E-05 | 0.000126 | -0.00528787 | NA |
| chr5 | 1211545 | 1212526 | 982 | 5 | 2.69E-05 | 0.0040386 | 0.000716 | 0.000126 | -0.00359825 | SLC6A19 |
| chr17 | 1095192 | 1095471 | 280 | 3 | 5.04E-05 | 0.0330748 | 2.37E-05 | 0.000126 | -0.00316328 | ABR |
| chr12 | 122395017 | 122395285 | 269 | 2 | 3.79E-05 | 0.0002669 | 0.000217 | 0.000126 | -0.0057025 | CLIP1 |
| chr7 | 29990127 | 29990522 | 396 | 8 | 0.000105964 | 2.37E-05 | 0.031403 | 0.000126 | -0.00400181 | FKBP14-AS1, SCRN1 |
| chr8 | 118929158 | 118929172 | 15 | 2 | 0.000158633 | 0.0028495 | 6.52E-05 | 0.000126 | -0.00482998 | TNFRSF11B |
| chr7 | 75987530 | 75987710 | 181 | 2 | 5.49E-05 | 0.0099333 | 5.20E-05 | 0.000127 | 0.003797472 | TMEM120A |
| chr6 | 152696761 | 152696786 | 26 | 2 | 0.000130205 | 0.0002963 | 0.000197 | 0.000127 | -0.01587642 | NA |
| chr1 | 10639774 | 10639918 | 145 | 3 | 9.96E-05 | 0.0001021 | 0.001443 | 0.000127 | -0.00293051 | AL139423.1, CASZ1 |
| chr13 | 112572918 | 112572947 | 30 | 2 | 0.0001528 | 0.0005448 | 0.000125 | 0.000127 | -0.00974746 | TUBGCP3 |
| chr6 | 30167867 | 30168372 | 506 | 4 | 6.63E-05 | 0.0012709 | 0.000831 | 0.000127 | 0.001673912 | TRIM15 |
| chr13 | 112943899 | 112944244 | 346 | 2 | 2.62E-05 | 0.001631 | 7.67E-05 | 0.000127 | -0.01210294 | MCF2L |
| chr1 | 248238473 | 248238495 | 23 | 2 | 0.000110571 | 0.0004348 | 0.000146 | 0.000127 | -0.0070748 | OR2M4 |
| chr14 | 22875936 | 22876600 | 665 | 4 | 4.54E-06 | 0.007682 | 2.22E-05 | 0.000127 | -0.0024772 | LRP10 |
| chr19 | 49594490 | 49595185 | 696 | 4 | 8.62E-05 | 0.0010509 | 0.002423 | 0.000127 | -0.00262625 | PRR12 |
| chr19 | 2720849 | 2721065 | 217 | 2 | 1.95E-05 | 0.0004956 | 0.000134 | 0.000127 | -0.0052674 | DIRAS1, AC006538.2 |
| chr14 | 35534891 | 35534936 | 46 | 2 | 2.17E-05 | 0.0001914 | 0.000316 | 0.000128 | -0.00126518 | INSM2 |
| chr17 | 18857709 | 18858418 | 710 | 11 | 8.92E-05 | 0.0010954 | 0.000103 | 0.000128 | 0.001070378 | PRPSAP2 |
| chr9 | 130919132 | 130919338 | 207 | 3 | 4.86E-05 | 0.0315559 | 2.81E-05 | 0.000128 | -0.00472848 | FIBCD1 |
| chr7 | 48299627 | 48299921 | 295 | 2 | 0.000148576 | 8.25E-05 | 0.001708 | 0.000128 | -0.01178486 | ABCA13 |
| chr11 | 61488935 | 61489313 | 379 | 3 | 3.82E-05 | 0.0007786 | 0.001391 | 0.000128 | -0.01239351 | PPP1R32 |
| chr22 | 47060890 | 47061005 | 116 | 2 | 6.48E-05 | 0.0005721 | 0.000123 | 0.000128 | -0.00169023 | TBC1D22A |
| chr5 | 32613160 | 32613203 | 44 | 2 | 0.000143931 | 0.0002229 | 0.000268 | 0.000128 | -0.01597835 | NA |
| chr8 | 11701825 | 11702287 | 463 | 4 | 1.04E-05 | 0.0001207 | 0.002702 | 0.000128 | -0.00185187 | GATA4 |
| chr1 | 18948464 | 18948744 | 281 | 2 | 0.000111365 | 7.80E-05 | 0.002186 | 0.000128 | -0.01538987 | IFFO2 |
| chr16 | 85316038 | 85316370 | 333 | 2 | 9.46E-05 | 0.0001139 | 0.000726 | 0.000129 | -0.00702652 | GSE1 |
| chr9 | 137460675 | 137460866 | 192 | 4 | 0.000113874 | 0.0107272 | 0.000436 | 0.000129 | -0.00288291 | PNPLA7 |
| chr12 | 68610416 | 68610890 | 475 | 10 | 0.000113916 | 0.0001423 | 0.041278 | 0.000129 | 0.001146996 | RAP1B |
| chr1 | 27547747 | 27548063 | 317 | 4 | 7.29E-05 | 0.0003958 | 0.002045 | 0.000129 | 0.000254785 | AHDC1 |
| chr14 | 20873704 | 20873712 | 9 | 2 | 0.000141704 | 0.0051726 | 5.85E-05 | 0.000129 | -0.00450839 | AL133371.3 |
| chr8 | 144078954 | 144078996 | 43 | 2 | 8.80E-05 | 9.82E-05 | 0.001028 | 0.000129 | 0.003500518 | EXOSC4, AC109322.1 |
| chr5 | 179617051 | 179617692 | 642 | 2 | 1.68E-05 | 0.0003498 | 0.000176 | 0.000129 | -0.006129 | HNRNPH1 |
| chr19 | 34134287 | 34134338 | 52 | 2 | 0.00013081 | 0.0010253 | 9.23E-05 | 0.000129 | -0.00104351 | NA |
| chr22 | 23880412 | 23880513 | 102 | 2 | 0.00013036 | 0.0003907 | 0.000161 | 0.000129 | -0.00857415 | SLC2A11, AP000350.4 |
| chr20 | 44391518 | 44391647 | 130 | 2 | 6.44E-05 | 0.0001043 | 0.000891 | 0.000129 | -0.00607584 | HNF4A, HNF4A-AS1, AL117382.2 |
| chr16 | 1221741 | 1221998 | 258 | 2 | 0.000114611 | 0.0002131 | 0.000286 | 0.000129 | -0.00163204 | CACNA1H, TPSG1 |
| chr1 | 202160438 | 202160737 | 300 | 3 | 3.99E-05 | 0.0035239 | 0.001652 | 0.000129 | -0.00534672 | PTPN7 |
| chr8 | 28963823 | 28963853 | 31 | 2 | 0.000169704 | 0.0019206 | 7.42E-05 | 0.000129 | -0.00703244 | HMBOX1 |
| chr22 | 40008953 | 40009628 | 676 | 3 | 1.05E-05 | 0.0007028 | 0.000155 | 0.00013 | 0.009694526 | FAM83F |
| chr8 | 144316230 | 144316380 | 151 | 2 | 7.02E-05 | 0.0287498 | 4.87E-05 | 0.00013 | -0.00545436 | DGAT1 |
| chr10 | 78777847 | 78778141 | 295 | 2 | 1.26E-05 | 0.0050546 | 5.92E-05 | 0.00013 | -0.00748271 | NA |
| chr16 | 73092709 | 73093563 | 855 | 6 | 3.35E-05 | 0.0008471 | 0.000119 | 0.00013 | -0.00210656 | HCCAT5, ZFHX3 |
| chr21 | 36157212 | 36157602 | 391 | 4 | 8.28E-05 | 0.0023527 | 0.000208 | 0.00013 | 0.003266452 | DOP1B, CBR3-AS1 |
| chr1 | 1089033 | 1089228 | 196 | 3 | 8.14E-05 | 0.0005575 | 0.001875 | 0.00013 | -0.00365907 | C1orf159 |
| chr15 | 80060036 | 80060661 | 626 | 5 | 4.72E-05 | 0.000959 | 0.001394 | 0.000131 | 0.003790037 | ZFAND6 |
| chr1 | 237043338 | 237043956 | 619 | 4 | 1.20E-05 | 0.0382086 | 1.71E-05 | 0.000131 | 0.002466856 | RYR2 |

| | | | | | | | | | | |
|---|---|---|---|---|---|---|---|---|---|---|
| chr21 | 44682294 | 44682448 | 155 | 3 | 8.55E-05 | 0.0052755 | 0.000829 | 0.000131 | -0.01329717 | TSPEAR |
| chr13 | 108268453 | 108268704 | 252 | 3 | 5.48E-05 | 0.0045676 | 3.91E-05 | 0.000131 | 0.008871122 | TNFSF13B |
| chr3 | 159988561 | 159988667 | 107 | 3 | 8.96E-05 | 0.0010422 | 0.000111 | 0.000131 | 0.000152347 | IL12A-AS1 |
| chr14 | 104843844 | 104844584 | 741 | 3 | 1.33E-05 | 0.0003844 | 0.000282 | 0.000131 | 0.004753094 | AL583810.2 |
| chr16 | 28538850 | 28539298 | 449 | 6 | 0.000109319 | 0.0015434 | 0.000234 | 0.000131 | -0.00324809 | AC009093.11, NUPR1, AC020765.6 |
| chr9 | 35604318 | 35604512 | 195 | 2 | 1.52E-05 | 0.0058451 | 5.83E-05 | 0.000131 | -0.00222832 | AL357874.3 |
| chr12 | 45907963 | 45908160 | 198 | 2 | 6.90E-05 | 0.0046814 | 6.06E-05 | 0.000131 | 0.011174172 | ARID2 |
| chr19 | 1154277 | 1154486 | 210 | 2 | 6.63E-05 | 9.26E-05 | 0.00129 | 0.000132 | -0.01069329 | SBNO2 |
| chr16 | 77675161 | 77675375 | 215 | 2 | 5.25E-05 | 0.0009157 | 9.90E-05 | 0.000132 | -0.0103458 | NA |
| chr3 | 107882251 | 107882380 | 130 | 2 | 0.000124826 | 7.54E-05 | 0.002971 | 0.000132 | 4.46E-05 | LINC00636 |
| chr6 | 170282148 | 170283297 | 1150 | 7 | 2.87E-06 | 0.0343816 | 7.94E-05 | 0.000132 | -0.00046781 | DLL1 |
| chr6 | 34104946 | 34105021 | 76 | 2 | 0.000159604 | 0.0004288 | 0.000155 | 0.000132 | -0.00798474 | GRM4 |
| chr3 | 45916172 | 45917060 | 889 | 5 | 8.37E-05 | 0.0005554 | 0.000935 | 0.000132 | 0.00098918 | NA |
| chr1 | 34164552 | 34165202 | 651 | 5 | 8.03E-06 | 0.0006951 | 0.001084 | 0.000132 | 0.003506877 | CSMD2 |
| chr15 | 40289308 | 40289353 | 46 | 2 | 5.16E-05 | 0.0035595 | 6.46E-05 | 0.000133 | -0.00279939 | PLCB2 |
| chr2 | 15919167 | 15919417 | 251 | 2 | 5.05E-05 | 0.0037009 | 6.41E-05 | 0.000133 | -0.00545824 | MYCNOS |
| chr14 | 99319856 | 99320224 | 369 | 4 | 5.44E-05 | 0.0021133 | 0.000111 | 0.000133 | -0.00194934 | NA |
| chr9 | 93451844 | 93452652 | 809 | 5 | 2.21E-05 | 0.000264 | 0.000483 | 0.000133 | -0.00019368 | FAM120A, FAM120AOS |
| chr6 | 169700560 | 169701209 | 650 | 2 | 1.15E-07 | 0.003214 | 6.62E-05 | 0.000133 | -0.01153277 | AL031315.1, WDR27 |
| chr9 | 112717780 | 112718350 | 571 | 8 | 7.45E-05 | 0.0067766 | 0.000221 | 0.000133 | 0.002587625 | INIP |
| chr16 | 3654413 | 3655228 | 816 | 5 | 2.00E-05 | 0.0028716 | 0.000376 | 0.000133 | 0.000391267 | DNASE1, TRAP1 |
| chr17 | 61454836 | 61455465 | 630 | 4 | 2.09E-05 | 0.0064144 | 3.36E-05 | 0.000133 | -0.00297506 | TBX4 |
| chr5 | 168992428 | 168992964 | 537 | 4 | 4.88E-05 | 0.0006415 | 0.002417 | 0.000133 | 0.004972758 | SLIT3 |
| chr17 | 78723530 | 78723558 | 29 | 3 | 5.45E-05 | 0.009513 | 3.19E-05 | 0.000133 | -0.00168684 | CYTH1 |
| chr6 | 99606054 | 99606242 | 189 | 3 | 4.53E-06 | 0.0003392 | 0.000764 | 0.000133 | -0.00664786 | NA |
| chr22 | 50185847 | 50186200 | 354 | 2 | 2.65E-05 | 0.000137 | 0.000563 | 0.000133 | -0.00169079 | TRABD |
| chr15 | 91852293 | 91852606 | 314 | 3 | 1.95E-05 | 0.0021848 | 0.000311 | 0.000133 | 0.005574626 | NA |
| chr20 | 396273 | 396490 | 218 | 3 | 4.24E-05 | 0.0010175 | 8.92E-05 | 0.000134 | -0.00183399 | TRIB3 |
| chr19 | 43574953 | 43575735 | 783 | 7 | 1.32E-05 | 0.0106317 | 4.97E-05 | 0.000134 | 0.001100447 | L34079.3, XRCC1, L34079.1 |
| chr6 | 110646858 | 110646965 | 108 | 3 | 6.62E-05 | 0.0004465 | 0.000883 | 0.000134 | 0.001001009 | CDK19 |
| chr1 | 17623068 | 17623070 | 3 | 2 | 0.000140366 | 0.0006491 | 0.000121 | 0.000134 | -0.00661456 | ARHGEF10L |
| chr12 | 31502042 | 31502054 | 13 | 2 | 0.000163128 | 0.0008963 | 0.000102 | 0.000134 | 0.006198568 | DENND5B |
| chr13 | 99303877 | 99303929 | 53 | 2 | 0.000157813 | 0.0085469 | 5.59E-05 | 0.000134 | -0.00863476 | UBAC2, GPR183 |
| chr17 | 44268110 | 44268321 | 212 | 5 | 0.000104454 | 0.0468886 | 2.37E-05 | 0.000134 | 0.00229557 | SLC4A1 |
| chr10 | 48684158 | 48684388 | 231 | 3 | 4.00E-05 | 0.0002016 | 0.000483 | 0.000134 | -0.00654249 | NA |
| chr22 | 37745412 | 37745504 | 93 | 2 | 8.30E-05 | 0.005693 | 5.96E-05 | 0.000135 | -0.00718452 | TRIOBP |
| chr7 | 155787094 | 155787628 | 535 | 5 | 0.000103318 | 0.0003964 | 0.004083 | 0.000135 | -0.00341244 | NA |
| chr18 | 37393958 | 37394586 | 629 | 7 | 1.56E-05 | 0.1069787 | 1.95E-05 | 0.000135 | 0.001726469 | CELF4 |
| chr1 | 208244563 | 208244670 | 108 | 4 | 0.00012196 | 0.0002128 | 0.002972 | 0.000135 | 0.001495781 | NA |
| chr2 | 174682395 | 174682398 | 4 | 2 | 0.00015244 | 9.24E-05 | 0.001418 | 0.000135 | -0.00842544 | AC010894.2, WIPF1 |
| chr4 | 6524243 | 6524469 | 227 | 2 | 0.000125792 | 7.71E-05 | 0.00302 | 0.000135 | 0.000193482 | PPP2R2C |
| chr1 | 229024567 | 229024636 | 70 | 2 | 0.000112067 | 0.0022328 | 7.41E-05 | 0.000135 | 0.002480997 | LINC02815 |
| chr22 | 20981015 | 20981343 | 329 | 2 | 3.99E-05 | 0.0001825 | 0.000378 | 0.000135 | -0.00944186 | AIFM3, AC002470.2 |
| chr19 | 18682895 | 18682970 | 76 | 2 | 0.00010367 | 0.0003685 | 0.000182 | 0.000135 | -0.0184718 | NA |
| chr19 | 827429 | 827843 | 415 | 8 | 7.57E-05 | 0.0025121 | 0.002696 | 0.000136 | -0.00205781 | AZU1 |
| chr6 | 53347820 | 53348361 | 542 | 4 | 6.21E-05 | 0.0021206 | 0.000951 | 0.000136 | -0.00086286 | ELOVL5 |
| chr19 | 47675401 | 47675452 | 52 | 2 | 0.000151946 | 0.001375 | 8.72E-05 | 0.000136 | -0.00273857 | AC010519.1, BICRA |
| chr5 | 115545265 | 115545670 | 406 | 2 | 4.62E-05 | 8.79E-05 | 0.001727 | 0.000136 | -0.00633949 | NA |
| chr2 | 208163675 | 208163754 | 80 | 2 | 4.00E-05 | 0.0030251 | 6.87E-05 | 0.000136 | 0.007577439 | NA |
| chr11 | 8263199 | 8263539 | 341 | 4 | 3.16E-05 | 0.001192 | 0.000101 | 0.000136 | 0.000709375 | LMO1 |
| chr14 | 105528560 | 105529056 | 497 | 3 | 3.18E-05 | 0.0007369 | 0.000629 | 0.000136 | -0.01625246 | TMEM121 |
| chr6 | 149749856 | 149749933 | 78 | 4 | 6.21E-05 | 0.0054743 | 2.96E-05 | 0.000136 | 0.001812204 | PCMT1 |
| chr22 | 29581214 | 29581273 | 60 | 3 | 0.000140916 | 0.03676 | 4.57E-05 | 0.000136 | 0.001832607 | NIPSNAP1 |
| chr5 | 2169615 | 2169701 | 87 | 4 | 0.000124625 | 0.0001127 | 0.002897 | 0.000136 | 0.000415535 | NA |
| chr6 | 52244549 | 52245649 | 1101 | 6 | 2.29E-06 | 0.0075208 | 3.33E-05 | 0.000136 | -0.00503597 | NA |
| chr4 | 151175390 | 151176136 | 747 | 7 | 1.90E-05 | 0.0231429 | 0.00085 | 0.000136 | -0.00038559 | SH3D19 |
| chr2 | 6917022 | 6917814 | 793 | 7 | 3.59E-05 | 0.0005171 | 0.00037 | 0.000137 | 0.00186818 | RNF144A, GRASLND |
| chr20 | 45706110 | 45706230 | 121 | 2 | 0.000148184 | 0.0033325 | 6.74E-05 | 0.000137 | -0.00562747 | WFDC13 |
| chr7 | 158075003 | 158075120 | 118 | 2 | 9.74E-05 | 0.0004483 | 0.000159 | 0.000137 | -0.00932099 | PTPRN2 |
| chr5 | 42424502 | 42424991 | 490 | 9 | 8.41E-05 | 0.0009276 | 0.006089 | 0.000137 | 0.001075797 | GHR |
| chr22 | 43955309 | 43956371 | 1063 | 7 | 5.51E-07 | 0.0043029 | 0.000199 | 0.000137 | 0.000713726 | PNPLA3, SAMM50 |
| chr6 | 119149159 | 119149224 | 66 | 2 | 1.98E-05 | 0.000942 | 0.000102 | 0.000137 | -0.00821007 | AL137009.1, FAM184A |
| chr3 | 138592574 | 138592666 | 93 | 2 | 0.000117087 | 0.0005034 | 0.000147 | 0.000137 | -0.0195819 | CEP70 |
| chr19 | 56765549 | 56765920 | 372 | 2 | 8.54E-06 | 0.0114586 | 5.50E-05 | 0.000137 | -0.02144957 | ZIM2-AS1, AC006115.2 |
| chr11 | 65089015 | 65089675 | 661 | 7 | 1.56E-05 | 0.0710107 | 9.87E-06 | 0.000137 | -0.00233626 | VPS51, TMEM262 |
| chr9 | 130932787 | 130932975 | 189 | 2 | 0.000100402 | 0.000101 | 0.001163 | 0.000137 | -0.01173774 | FIBCD1 |
| chr6 | 109977948 | 109978706 | 759 | 8 | 3.28E-05 | 0.0004488 | 0.001725 | 0.000137 | 0.001126118 | GPR6 |
| chr5 | 180591562 | 180592265 | 704 | 7 | 4.37E-05 | 0.0013379 | 0.009981 | 0.000137 | 0.001461251 | NA |
| chr14 | 20437990 | 20438357 | 368 | 3 | 8.48E-05 | 0.0005049 | 0.001068 | 0.000138 | 0.000987206 | AL355075.5 |
| chr5 | 3854059 | 3854590 | 532 | 5 | 1.94E-05 | 0.0054188 | 0.000248 | 0.000138 | -0.00139039 | NA |
| chr19 | 34963987 | 34964485 | 499 | 9 | 1.01E-05 | 0.0774531 | 1.68E-06 | 0.000138 | 0.000642817 | ZNF792 |
| chr9 | 68715492 | 68715618 | 127 | 2 | 4.99E-05 | 0.0030605 | 6.93E-05 | 0.000138 | -0.00412557 | PIP5K1B |
| chr6 | 70667361 | 70667442 | 82 | 2 | 6.09E-05 | 0.0472195 | 4.99E-05 | 0.000138 | 0.005002165 | NA |
| chr10 | 88879008 | 88879049 | 42 | 2 | 2.01E-05 | 0.0002153 | 0.000318 | 0.000138 | -0.00180329 | NA |
| chr17 | 8309877 | 8310286 | 410 | 4 | 4.24E-05 | 0.003322 | 5.02E-05 | 0.000138 | 0.006138284 | ARHGEF15 |
| chr17 | 39766991 | 39767004 | 14 | 2 | 6.78E-05 | 0.000139 | 0.000594 | 0.000138 | -0.00572987 | IKZF3 |
| chr10 | 131088371 | 131088658 | 288 | 5 | 0.000107965 | 9.08E-05 | 0.005622 | 0.000138 | -0.00193936 | NA |
| chr6 | 71588539 | 71588721 | 183 | 3 | 5.32E-05 | 0.0002315 | 0.000983 | 0.000138 | -0.01199419 | NA |
| chr4 | 40056369 | 40057096 | 728 | 5 | 1.89E-05 | 4.23E-05 | 0.017068 | 0.000138 | -0.00030663 | N4BP2, AC098591.2 |
| chr20 | 45913102 | 45913268 | 167 | 4 | 8.69E-05 | 0.0003168 | 0.00031 | 0.000138 | -0.00606949 | NA |
| chr8 | 144496513 | 144496807 | 295 | 3 | 9.42E-05 | 0.000324 | 0.000417 | 0.000138 | 0.001661468 | PPP1R16A, AC084125.2 |
| chr1 | 101997034 | 101997321 | 288 | 8 | 0.000139156 | 0.002188 | 0.001072 | 0.000139 | 0.00290436 | NA |
| chr2 | 42763335 | 42764419 | 1085 | 7 | 6.05E-07 | 0.018776 | 0.000363 | 0.000139 | -0.00111331 | OXER1 |
| chr1 | 95318278 | 95318329 | 52 | 3 | 0.000146051 | 0.0039464 | 0.000221 | 0.000139 | -0.00219038 | NA |
| chr9 | 74497137 | 74497410 | 274 | 4 | 7.75E-05 | 5.10E-05 | 0.008429 | 0.000139 | 0.001725067 | RORB, RORB-AS1 |
| chr12 | 6958660 | 6959261 | 602 | 4 | 7.58E-05 | 0.0007767 | 0.00022 | 0.000139 | -0.00178998 | PTPN6 |
| chr13 | 94602025 | 94603004 | 980 | 4 | 6.42E-06 | 0.0002845 | 0.002654 | 0.000139 | 0.004907506 | GPR180 |
| chr11 | 60282454 | 60282670 | 217 | 2 | 2.49E-05 | 0.0003551 | 0.000197 | 0.00014 | 0.011809825 | MS4A4A |
| chr12 | 119988725 | 119988814 | 90 | 2 | 0.000100101 | 0.0001747 | 0.000428 | 0.00014 | 0.000269304 | NA |
| chr3 | 134375960 | 134376082 | 123 | 2 | 3.42E-05 | 0.0001004 | 0.001262 | 0.00014 | -0.00469253 | NA |
| chr11 | 64744301 | 64744851 | 551 | 8 | 1.02E-05 | 0.0110859 | 3.76E-05 | 0.00014 | 0.001473396 | RASGRP2 |
| chr6 | 44003184 | 44003500 | 317 | 3 | 6.20E-05 | 7.34E-05 | 0.002849 | 0.00014 | -0.00982179 | C6orf223, AL109615.3 |
| chr8 | 143161031 | 143161487 | 457 | 2 | 5.40E-06 | 0.0002327 | 0.0003 | 0.00014 | -0.00647799 | NA |
| chr10 | 13891996 | 13892438 | 443 | 4 | 3.67E-05 | 0.0025124 | 0.000138 | 0.00014 | -0.00528984 | FRMD4A |
| chr10 | 35335426 | 35335827 | 402 | 5 | 2.68E-05 | 0.0018262 | 0.001708 | 0.00014 | 0.009527677 | CCNY, AL117336.2 |
| chr22 | 26672753 | 26672945 | 193 | 5 | 0.000100556 | 0.0015243 | 0.000364 | 0.00014 | 0.000532364 | MIAT, MIATNB |
| chr15 | 75352865 | 75353254 | 390 | 2 | 2.30E-05 | 0.0002089 | 0.000341 | 0.00014 | -0.00309026 | NEIL1 |
| chr15 | 29821556 | 29822271 | 716 | 7 | 4.48E-05 | 0.0004905 | 0.008105 | 0.00014 | -0.00070712 | TJP1 |
| chr4 | 154781608 | 154782018 | 411 | 5 | 7.03E-05 | 6.30E-05 | 0.010026 | 0.00014 | -0.02065164 | RBM46, AC009567.1 |
| chr6 | 169255213 | 169255402 | 190 | 2 | 3.61E-05 | 0.0012 | 9.51E-05 | 0.000141 | -0.01714968 | NA |
| chr9 | 127450678 | 127451030 | 353 | 4 | 2.17E-05 | 0.014368 | 0.000119 | 0.000141 | -0.00067995 | RPL12 |
| chr17 | 79082917 | 79082949 | 33 | 2 | 0.000102415 | 0.0001201 | 0.000825 | 0.000141 | 0.015389606 | ENGASE |
| chr6 | 47029205 | 47029236 | 32 | 2 | 0.000124356 | 0.0001022 | 0.001227 | 0.000141 | -0.00822666 | ADGRF1 |
| chr2 | 207134870 | 207136042 | 1173 | 4 | 2.40E-06 | 0.0015087 | 0.000815 | 0.000141 | 0.00076249 | KLF7 |
| chr13 | 110493225 | 110493245 | 21 | 4 | 0.000169874 | 0.0001293 | 0.003125 | 0.000141 | -0.00160831 | COL4A2 |
| chr10 | 114697407 | 114697540 | 134 | 3 | 0.000132612 | 0.032441 | 4.93E-05 | 0.000142 | 0.00137937 | ABLIM1 |
| chr17 | 80958825 | 80958897 | 73 | 2 | 3.28E-05 | 0.0083533 | 5.88E-05 | 0.000142 | 0.002625828 | RPTOR |
| chr5 | 177510349 | 177510833 | 485 | 5 | 1.56E-05 | 8.16E-05 | 0.002218 | 0.000142 | 0.001798768 | DOK3 |
| chr19 | 44808448 | 44808934 | 487 | 8 | 7.59E-05 | 0.0017511 | 0.003385 | 0.000142 | -0.00062377 | NA |
| chr3 | 14807537 | 14807738 | 202 | 2 | 4.84E-05 | 0.0044347 | 6.56E-05 | 0.000142 | -0.00590343 | LINC02011 |
| chr4 | 10685495 | 10685851 | 357 | 3 | 2.34E-05 | 6.58E-05 | 0.004289 | 0.000142 | 0.006581709 | AC084048.1 |
| chr8 | 81036674 | 81036889 | 216 | 2 | 0.000116757 | 0.00305 | 7.14E-05 | 0.000142 | -0.01117073 | PAG1 |
| chr10 | 17398930 | 17399049 | 120 | 2 | 4.73E-05 | 0.0002063 | 0.000355 | 0.000142 | -0.01031849 | ST8SIA6-AS1, ST8SIA6 |
| chr15 | 55744040 | 55744257 | 218 | 2 | 3.87E-05 | 9.43E-05 | 0.001622 | 0.000142 | 0.008714463 | NA |
| chr18 | 57441715 | 57442022 | 308 | 4 | 4.59E-05 | 0.0026758 | 0.000326 | 0.000142 | 0.001810633 | ONECUT2 |
| chr15 | 68383970 | 68384108 | 139 | 2 | 3.07E-05 | 0.0062803 | 6.17E-05 | 0.000142 | -0.00754861 | ITGA11 |
| chr2 | 134125624 | 134126051 | 428 | 2 | 0.000123196 | 0.0001454 | 0.000594 | 0.000142 | -0.0062219 | MGAT5 |
| chr3 | 131026989 | 131027902 | 914 | 8 | 9.12E-06 | 0.0011251 | 0.002867 | 0.000143 | 0.000261448 | NEK11, ASTE1 |
| chr12 | 103672432 | 103672879 | 448 | 4 | 4.92E-05 | 0.0254308 | 1.73E-05 | 0.000143 | -0.00306037 | STAB2 |
| chr17 | 81492442 | 81492464 | 23 | 2 | 0.000141911 | 0.0003066 | 0.000233 | 0.000143 | -0.00901547 | NA |
| chr1 | 9264956 | 9265352 | 397 | 3 | 2.16E-05 | 0.0006684 | 0.000431 | 0.000143 | 0.007947719 | H6PD |
| chr10 | 15088624 | 15089037 | 414 | 9 | 5.81E-05 | 0.0041904 | 0.003112 | 0.000143 | 0.001134253 | ACBD7 |
| chr6 | 157322973 | 157323909 | 937 | 7 | 1.14E-05 | 0.0007004 | 0.001891 | 0.000143 | 0.000255149 | TMEM242 |
| chr17 | 80874733 | 80874879 | 147 | 3 | 0.000107297 | 0.0003875 | 0.000723 | 0.000143 | 0.000286031 | RPTOR |
| chr10 | 531773 | 532144 | 372 | 2 | 0.000123598 | 0.0002065 | 0.000362 | 0.000143 | -0.00630678 | DIP2C |
| chr19 | 49527760 | 49528016 | 257 | 3 | 7.58E-05 | 0.0024825 | 0.000597 | 0.000143 | 0.000747052 | RCN3 |
| chr8 | 22161383 | 22161668 | 286 | 3 | 0.000127222 | 9.04E-05 | 0.001957 | 0.000144 | 0.004261328 | SFTPC |
| chr9 | 136969068 | 136969207 | 140 | 2 | 5.66E-05 | 0.0112368 | 5.73E-05 | 0.000144 | -0.00638636 | NA |
| chr1 | 1275165 | 1275231 | 67 | 2 | 0.000112418 | 0.0001791 | 0.000438 | 0.000144 | 0.00251907 | LINC01786 |
| chr6 | 87015944 | 87015957 | 14 | 2 | 4.72E-05 | 0.0127532 | 5.66E-05 | 0.000144 | 0.001713727 | HTR1E |
| chr1 | 54298960 | 54298981 | 22 | 2 | 0.000156115 | 0.0214943 | 5.40E-05 | 0.000144 | -0.02358451 | SSBP3 |
| chr16 | 25014662 | 25015529 | 868 | 6 | 2.95E-05 | 0.0001418 | 0.008263 | 0.000144 | 0.000248303 | ARHGAP17 |
| chr19 | 55386792 | 55386957 | 166 | 2 | 7.82E-05 | 9.09E-05 | 0.001973 | 0.000145 | -0.00519909 | RPL28 |
| chr7 | 66921058 | 66921067 | 10 | 2 | 0.000124913 | 0.0003716 | 0.0002 | 0.000145 | -0.00636655 | NA |
| chr22 | 29311113 | 29311793 | 681 | 4 | 5.52E-05 | 0.0001123 | 0.005009 | 0.000145 | 0.006574733 | GAS2L1 |
| chr5 | 6714063 | 6714424 | 362 | 4 | 1.72E-05 | 0.000133 | 0.000946 | 0.000145 | 0.003395334 | TENT4A |
| chr1 | 7944146 | 7944269 | 124 | 2 | 4.71E-05 | 0.0155378 | 5.59E-05 | 0.000145 | -0.00254177 | NA |
| chr8 | 67343677 | 67343726 | 50 | 3 | 9.19E-05 | 0.0014358 | 8.65E-05 | 0.000145 | -0.0007165 | ARFGEF1 |
| chr4 | 172265192 | 172265245 | 54 | 2 | 3.81E-05 | 0.0002993 | 0.000246 | 0.000145 | -0.00734577 | GALNTL6 |
| chr7 | 140700068 | 140700418 | 351 | 3 | 9.93E-05 | 0.0002475 | 0.000989 | 0.000146 | -0.00203887 | NDUFB2 |
| chr8 | 91208157 | 91208324 | 168 | 3 | 0.000113667 | 0.0044838 | 5.84E-05 | 0.000146 | -0.00593627 | LRRC69 |
| chr8 | 25457438 | 25457771 | 334 | 5 | 8.10E-05 | 0.0018109 | 0.000293 | 0.000146 | -0.00102627 | KCTD9 |
| chr13 | 112219888 | 112219937 | 50 | 3 | 9.97E-05 | 7.81E-05 | 0.003109 | 0.000146 | -0.00508136 | NA |
| chr1 | 162381874 | 162382233 | 360 | 3 | 4.25E-05 | 0.0027086 | 7.47E-05 | 0.000146 | -0.00408031 | AL512785.2, C1orf226 |
| chr1 | 26791731 | 26792071 | 341 | 3 | 6.58E-05 | 0.0002808 | 0.00038 | 0.000146 | -0.00651056 | PIGV |
| chr17 | 62625409 | 62625567 | 159 | 2 | 7.75E-05 | 0.0006575 | 0.000135 | 0.000146 | -0.01064907 | NA |
| chr11 | 62983845 | 62984016 | 172 | 4 | 0.000104598 | 0.0112817 | 0.007091 | 0.000146 | -0.00260889 | SLC22A6 |
| chr7 | 101216965 | 101217496 | 532 | 4 | 2.71E-05 | 5.03E-05 | 0.008559 | 0.000146 | -0.00114356 | PLOD3 |
| chr8 | 67028694 | 67029003 | 310 | 7 | 9.27E-05 | 0.0009191 | 0.002266 | 0.000146 | -9.69E-05 | PPP1R42 |
| chr16 | 25690723 | 25691249 | 527 | 3 | 2.65E-06 | 0.1329188 | 1.84E-05 | 0.000146 | -0.00838462 | NA |
| chr19 | 1626510 | 1626758 | 249 | 4 | 9.31E-05 | 0.0004211 | 0.001854 | 0.000147 | -0.00312799 | TCF3 |
| chr19 | 1777393 | 1777680 | 288 | 2 | 8.56E-05 | 0.0011229 | 0.000102 | 0.000147 | -0.01368195 | ONECUT3 |
| chr2 | 130756590 | 130757015 | 426 | 2 | 5.57E-06 | 0.0001558 | 0.000572 | 0.000147 | -0.00974322 | AMER3 |
| chr7 | 4872914 | 4873224 | 311 | 2 | 1.92E-05 | 0.0058936 | 6.44E-05 | 0.000148 | 0.011357693 | RADIL |
| chr10 | 103917325 | 103918043 | 719 | 6 | 8.15E-05 | 0.0011126 | 0.009362 | 0.000148 | 0.002364014 | STN1 |
| chr4 | 69057349 | 69058047 | 699 | 4 | 5.34E-05 | 0.0015662 | 8.87E-05 | 0.000148 | -9.95E-05 | UGT2B7 |
| chr1 | 36124880 | 36125359 | 480 | 2 | 2.08E-05 | 0.0001934 | 0.00042 | 0.000148 | -0.00915355 | COL8A2 |
| chr14 | 37594275 | 37594608 | 334 | 4 | 6.32E-05 | 0.0096022 | 0.00033 | 0.000148 | -0.00091038 | FOXA1 |

| | | | | | | | | | | |
|---|---|---|---|---|---|---|---|---|---|---|
| chr8 | 1945627 | 1945740 | 114 | 2 | 9.28E-05 | 0.0002804 | 0.000271 | 0.000148 | -0.00103337 | AC019257.8, ARHGEF10 |
| chr9 | 126056826 | 126056920 | 95 | 2 | 0.000108763 | 0.0001196 | 0.000969 | 0.000148 | -0.01418314 | NA |
| chr7 | 157590594 | 157590867 | 274 | 2 | 7.95E-05 | 0.000387 | 0.000201 | 0.000148 | -0.01052638 | PTPRN2 |
| chr18 | 23649545 | 23649644 | 100 | 2 | 9.41E-05 | 0.0002426 | 0.000319 | 0.000149 | -0.00660245 | ANKRD29 |
| chr16 | 58039913 | 58040142 | 230 | 6 | 4.61E-05 | 0.1040402 | 6.94E-05 | 0.000149 | -0.00036168 | MMP15 |
| chr19 | 35491497 | 35491752 | 256 | 2 | 1.36E-05 | 0.000394 | 0.0002 | 0.000149 | 0.008716339 | KRTDAP |
| chr15 | 68300932 | 68301147 | 216 | 3 | 3.92E-05 | 0.0015992 | 7.53E-05 | 0.000149 | 0.003117208 | ITGA11 |
| chr12 | 67657899 | 67658539 | 641 | 4 | 2.48E-05 | 0.0034366 | 7.09E-05 | 0.000149 | -0.00176489 | DYRK2 |
| chr20 | 2393118 | 2393194 | 77 | 2 | 0.000140638 | 0.0016819 | 8.95E-05 | 0.000149 | 0.00068652 | TGM6 |
| chr6 | 5026090 | 5026325 | 236 | 3 | 8.15E-05 | 0.0001885 | 0.000532 | 0.000149 | 0.004249963 | LYRM4-AS1 |
| chr16 | 29901841 | 29901902 | 62 | 2 | 0.000106027 | 0.0030296 | 7.52E-05 | 0.00015 | -0.00801813 | ASPHD1 |
| chr5 | 43017249 | 43017900 | 652 | 5 | 1.60E-05 | 0.0082704 | 0.000324 | 0.00015 | -0.0023347 | AC025171.1 |
| chr22 | 20530699 | 20531036 | 338 | 2 | 2.44E-05 | 0.0001879 | 0.000447 | 0.00015 | -0.00062903 | MED15 |
| chr16 | 8620444 | 8620689 | 246 | 2 | 1.27E-05 | 0.0002305 | 0.000344 | 0.00015 | 0.001129268 | NA |
| chr9 | 135934161 | 135934235 | 75 | 2 | 7.93E-05 | 0.0003964 | 0.000201 | 0.00015 | 0.003898313 | UBAC1 |
| chr1 | 17053655 | 17054226 | 572 | 8 | 0.000134514 | 0.0011318 | 0.000856 | 0.00015 | 0.000162721 | SDHB |
| chr6 | 24748120 | 24748174 | 55 | 3 | 3.56E-05 | 0.1014859 | 2.32E-05 | 0.00015 | -0.00412364 | LINC02828 |
| chr8 | 60690855 | 60690858 | 4 | 2 | 0.000111524 | 0.0006065 | 0.000148 | 0.00015 | 0.003267303 | CHD7 |
| chr1 | 23437119 | 23437289 | 171 | 3 | 7.72E-05 | 8.95E-05 | 0.002059 | 0.000151 | -0.01265199 | ASAP3 |
| chr6 | 136064701 | 136065273 | 573 | 6 | 3.48E-05 | 0.0135065 | 2.18E-05 | 0.000151 | -0.00137766 | PDE7B, AL138828.1 |
| chr18 | 59035322 | 59035399 | 78 | 2 | 8.84E-05 | 0.0478378 | 5.37E-05 | 0.000151 | 0.003552027 | OACYLP |
| chr19 | 42303283 | 42303428 | 146 | 2 | 5.87E-05 | 8.82E-05 | 0.002728 | 0.000151 | -0.01092033 | PRR19, PAFAH1B3 |
| chr17 | 58540154 | 58540183 | 30 | 2 | 0.000120857 | 9.13E-05 | 0.002332 | 0.000151 | -0.00342402 | SEPTIN4-AS1, SEPTIN4 |
| chr3 | 85503678 | 85503939 | 262 | 3 | 6.32E-05 | 0.1026429 | 2.27E-05 | 0.000151 | -0.00630393 | CADM2 |
| chr3 | 143119840 | 143120180 | 341 | 4 | 0.000132923 | 0.0005308 | 0.000572 | 0.000151 | 0.00105901 | CHST2 |
| chr1 | 151070267 | 151070548 | 282 | 9 | 0.000132231 | 0.0002518 | 0.007479 | 0.000151 | 0.000270135 | CDC42SE1 |
| chr5 | 126775863 | 126776259 | 397 | 4 | 3.16E-05 | 0.0016694 | 0.000177 | 0.000151 | -0.0102462 | LMNB1-DT |
| chr22 | 31398536 | 31399317 | 782 | 5 | 6.06E-06 | 0.0107695 | 6.87E-05 | 0.000151 | -0.00240986 | NA |
| chr3 | 149657741 | 149658255 | 515 | 12 | 7.80E-05 | 0.0009132 | 0.011292 | 0.000151 | -0.00015127 | WWTR1-AS1, WWTR1 |
| chr8 | 144401629 | 144401846 | 218 | 2 | 1.61E-05 | 0.0021037 | 8.44E-05 | 0.000152 | 0.004378418 | CPSF1 |
| chr2 | 38536793 | 38536810 | 18 | 2 | 8.37E-05 | 0.0002385 | 0.000337 | 0.000152 | -0.00518801 | NA |
| chr12 | 53965928 | 53966015 | 88 | 4 | 0.00015346 | 6.57E-05 | 0.008328 | 0.000152 | 5.19E-05 | AC012531.7, HOTAIR |
| chr7 | 37433822 | 37433974 | 153 | 2 | 7.64E-05 | 0.0178896 | 5.73E-05 | 0.000152 | -0.00404048 | ELMO1 |
| chr19 | 12790667 | 12791152 | 486 | 3 | 4.21E-07 | 0.0037942 | 0.00012 | 0.000152 | 0.00250392 | HOOK2 |
| chr12 | 120737614 | 120738016 | 403 | 3 | 5.54E-05 | 0.0006354 | 0.000158 | 0.000152 | -0.00255463 | ACADS |
| chr16 | 1765967 | 1766289 | 323 | 3 | 2.37E-05 | 0.0011073 | 0.000266 | 0.000152 | 0.002245833 | MAPK8IP3 |
| chr10 | 104328451 | 104328997 | 547 | 5 | 8.30E-05 | 8.76E-05 | 0.006744 | 0.000152 | 0.003915279 | ITPRIP |
| chr22 | 46432514 | 46432570 | 57 | 2 | 0.000108557 | 0.0046088 | 6.94E-05 | 0.000152 | -0.00110628 | CELSR1 |
| chr13 | 114015296 | 114015518 | 223 | 4 | 0.000157332 | 0.0044578 | 0.000478 | 0.000152 | -0.00214208 | RASA3 |
| chr11 | 69200854 | 69200950 | 97 | 2 | 0.000129174 | 0.0134443 | 5.90E-05 | 0.000152 | -0.00596324 | NA |
| chr2 | 88627381 | 88627776 | 396 | 8 | 2.31E-05 | 0.0144065 | 1.70E-05 | 0.000152 | -0.0004336 | EIF2AK3-DT, EIF2AK3 |
| chr8 | 91041640 | 91041950 | 311 | 2 | 1.07E-05 | 0.0122198 | 5.96E-05 | 0.000153 | 0.003322868 | NA |
| chr18 | 5196875 | 5197072 | 198 | 3 | 0.000108943 | 0.0001449 | 0.000866 | 0.000153 | -0.00480067 | AKAIN1 |
| chr1 | 20290458 | 20290482 | 25 | 2 | 0.000143987 | 0.0012141 | 0.000104 | 0.000153 | 0.00857559 | NA |
| chr7 | 157815881 | 157815950 | 70 | 3 | 9.45E-05 | 0.0024807 | 8.92E-05 | 0.000153 | 0.00159014 | PTPRN2 |
| chr11 | 64804201 | 64804713 | 513 | 3 | 1.57E-05 | 0.001655 | 0.000122 | 0.000154 | -0.00287303 | MEN1 |
| chr12 | 50283677 | 50284224 | 548 | 7 | 6.79E-05 | 0.0001473 | 0.005151 | 0.000154 | 0.002719389 | NA |
| chr12 | 113104998 | 113105176 | 179 | 2 | 5.28E-05 | 0.0013418 | 0.0001 | 0.000154 | -0.00384374 | RASAL1 |
| chr20 | 20216262 | 20216266 | 5 | 2 | 7.95E-05 | 8.86E-05 | 0.00295 | 0.000154 | 0.020639702 | CFAP61 |
| chr5 | 1324953 | 1325878 | 926 | 5 | 3.44E-06 | 0.0920101 | 0.000546 | 0.000154 | 0.003350373 | CLPTM1L |
| chr11 | 95863981 | 95864313 | 333 | 2 | 0.00016224 | 0.0001861 | 0.00048 | 0.000154 | -0.00553577 | MTMR2 |
| chr21 | 34513057 | 34513337 | 281 | 2 | 1.34E-05 | 9.55E-05 | 0.002159 | 0.000154 | 0.000783375 | RCAN1 |
| chr4 | 55082030 | 55082612 | 583 | 3 | 3.19E-05 | 0.0964915 | 2.32E-05 | 0.000154 | -0.00773034 | AC111194.2, KDR |
| chr11 | 68039630 | 68040297 | 668 | 4 | 3.38E-05 | 0.0161739 | 0.000274 | 0.000154 | -0.0040393 | TCIRG1 |
| chr15 | 43794601 | 43794891 | 291 | 3 | 5.07E-06 | 0.1226002 | 2.19E-05 | 0.000154 | -0.00332708 | SERF2, AC018512.1, SERINC4 |
| chr20 | 58987889 | 58988116 | 228 | 2 | 6.20E-05 | 0.0001128 | 0.001266 | 0.000154 | 0.002056369 | NELFCD |
| chr1 | 95116936 | 95117517 | 582 | 6 | 2.33E-05 | 0.0013478 | 0.00015 | 0.000155 | 0.000485136 | TLCD4 |
| chr2 | 65309805 | 65309822 | 18 | 2 | 0.000136883 | 0.0001269 | 0.000955 | 0.000155 | 0.004591712 | NA |
| chr6 | 49949432 | 49949541 | 110 | 3 | 2.37E-05 | 0.0318912 | 2.71E-05 | 0.000155 | -0.00902634 | DEFB133 |
| chr11 | 65630238 | 65630406 | 169 | 2 | 1.35E-05 | 0.0120597 | 6.05E-05 | 0.000155 | -0.00490282 | PCNX3 |
| chr12 | 47759168 | 47759641 | 474 | 4 | 9.51E-05 | 0.0001996 | 0.00297 | 0.000155 | -0.0006582 | SLC48A1, RAPGEF3 |
| chr2 | 168901108 | 168901270 | 163 | 3 | 8.42E-05 | 6.92E-05 | 0.005341 | 0.000155 | 0.002054441 | SPC25 |
| chr4 | 152942456 | 152942960 | 505 | 4 | 1.41E-05 | 0.02586 | 1.83E-05 | 0.000156 | 0.001644083 | FHDC1 |
| chr4 | 181647581 | 181647885 | 305 | 3 | 4.35E-05 | 0.004055 | 8.62E-05 | 0.000156 | -0.01332397 | NA |
| chr20 | 57999550 | 57999618 | 69 | 2 | 0.000144038 | 0.0039001 | 7.34E-05 | 0.000156 | -0.00339475 | NA |
| chr1 | 155191993 | 155192042 | 50 | 2 | 0.000160118 | 0.0001784 | 0.000525 | 0.000156 | 0.008486799 | MUC1 |
| chr3 | 125140503 | 125141054 | 552 | 2 | 1.94E-06 | 0.0008428 | 0.000127 | 0.000156 | -0.00379855 | SLC12A8 |
| chr6 | 9399365 | 9399444 | 80 | 2 | 0.000104855 | 0.0002642 | 0.000317 | 0.000156 | -0.00714129 | NA |
| chr6 | 165827091 | 165827387 | 297 | 3 | 0.000108185 | 0.0001403 | 0.002461 | 0.000156 | -0.00446108 | PDE10A |
| chr6 | 166009138 | 166009253 | 116 | 3 | 0.000166073 | 5.62E-05 | 0.010783 | 0.000157 | -0.00805109 | NA |
| chr2 | 239008792 | 239009169 | 378 | 3 | 7.51E-05 | 0.0001191 | 0.002574 | 0.000157 | 0.000128776 | NA |
| chr12 | 123764878 | 123765059 | 182 | 2 | 9.13E-05 | 0.0032061 | 7.77E-05 | 0.000157 | -0.00571851 | DNAH10 |
| chr5 | 82304785 | 82304893 | 109 | 2 | 0.000144465 | 0.0043069 | 7.24E-05 | 0.000157 | 0.007664763 | ATP6AP1L |
| chr22 | 44281827 | 44281943 | 117 | 2 | 0.000157774 | 0.0012202 | 0.000107 | 0.000157 | -0.00413004 | SHISAL1 |
| chr4 | 99816664 | 99816982 | 319 | 4 | 8.26E-05 | 0.0051613 | 0.000719 | 0.000158 | 0.00180601 | DAPP1 |
| chr20 | 44162057 | 44162083 | 27 | 2 | 0.00011388 | 9.22E-05 | 0.002754 | 0.000158 | 0.006857955 | JPH2 |
| chr12 | 1923294 | 1923353 | 60 | 3 | 9.14E-05 | 0.0008512 | 0.000131 | 0.000158 | 6.00E-05 | NA |
| chr15 | 49877598 | 49877642 | 45 | 2 | 6.65E-05 | 9.16E-05 | 0.002899 | 0.000158 | -0.00908039 | AC025040.1, ATP8B4 |
| chr4 | 3317814 | 3317942 | 129 | 2 | 6.49E-05 | 0.000191 | 0.000489 | 0.000158 | 3.51E-05 | RGS12 |
| chr12 | 132859059 | 132859254 | 196 | 4 | 0.000148047 | 0.0023623 | 0.000114 | 0.000159 | 0.001735282 | CHFR |
| chr6 | 30465646 | 30466295 | 650 | 4 | 3.56E-05 | 0.0003025 | 0.000765 | 0.000159 | -0.01671043 | NA |
| chr12 | 14972973 | 14973086 | 114 | 2 | 4.94E-05 | 8.85E-05 | 0.003552 | 0.000159 | 0.007669402 | PDE6H |
| chr11 | 34495994 | 34496204 | 211 | 3 | 0.000108519 | 0.023039 | 3.36E-05 | 0.000159 | -0.00269061 | ELF5 |
| chr3 | 158800982 | 158801213 | 232 | 4 | 6.62E-05 | 5.09E-05 | 0.014719 | 0.000159 | -0.01518478 | MFSD1 |
| chr2 | 104746909 | 104747078 | 170 | 2 | 9.61E-05 | 8.87E-05 | 0.003533 | 0.000159 | -0.00336381 | LINC01114 |
| chr1 | 174022308 | 174022809 | 502 | 4 | 5.66E-05 | 0.0015276 | 0.001364 | 0.000159 | -0.00084555 | LINC02776, RC3H1 |
| chr16 | 87353490 | 87353623 | 134 | 2 | 3.10E-05 | 0.0010602 | 0.000116 | 0.000159 | -0.00241844 | FBXO31 |
| chr6 | 34249321 | 34249558 | 238 | 5 | 9.03E-05 | 0.0003707 | 0.003648 | 0.000159 | -0.0004856 | AL354740.1 |
| chr5 | 72847904 | 72848312 | 409 | 7 | 7.12E-05 | 0.0010121 | 0.004391 | 0.00016 | 0.000294054 | TNPO1 |
| chr10 | 12348908 | 12349240 | 333 | 3 | 1.99E-05 | 0.0006975 | 0.000221 | 0.00016 | 0.003295697 | NA |
| chr5 | 177996998 | 177997342 | 345 | 3 | 4.02E-05 | 0.0028863 | 7.40E-05 | 0.00016 | -0.00365844 | NA |
| chr6 | 37506995 | 37507263 | 269 | 4 | 0.000105026 | 0.0207942 | 9.45E-05 | 0.00016 | -0.00081214 | NA |
| chr1 | 15456663 | 15457018 | 356 | 3 | 8.32E-05 | 0.0056541 | 4.71E-05 | 0.00016 | -0.00043101 | CELA2A |
| chr7 | 99392642 | 99393003 | 362 | 2 | 2.15E-05 | 0.0007369 | 0.000142 | 0.00016 | -0.01592206 | AC004922.1, ARPC1B, PDAP1 |
| chr12 | 29148592 | 29149102 | 511 | 6 | 7.46E-05 | 5.60E-05 | 0.014663 | 0.00016 | 0.000789619 | AC012150.1 |
| chr14 | 57268854 | 57269489 | 636 | 13 | 6.83E-05 | 0.0039242 | 0.000388 | 0.00016 | 0.001005904 | AP5M1, EXOC5 |
| chr1 | 24491736 | 24491795 | 60 | 3 | 0.000145458 | 0.0514185 | 4.03E-05 | 0.00016 | -0.00654762 | NA |
| chr14 | 102362439 | 102363451 | 1013 | 13 | 2.94E-05 | 0.0260704 | 0.000256 | 0.00016 | 0.000469725 | TECPR2, CINP |
| chr17 | 57885161 | 57885741 | 581 | 5 | 4.29E-05 | 0.0001475 | 0.009993 | 0.000161 | 0.013444346 | CUEDC1 |
| chr11 | 75336067 | 75336136 | 70 | 2 | 5.45E-05 | 0.0005779 | 0.000167 | 0.000161 | 0.004251823 | ARRB1 |
| chr12 | 123273345 | 123273515 | 171 | 3 | 8.81E-05 | 0.0002882 | 0.000551 | 0.000161 | 6.39E-05 | NA |
| chr9 | 36135668 | 36135763 | 96 | 2 | 6.88E-05 | 0.0001371 | 0.000909 | 0.000161 | -0.01217408 | NA |
| chr22 | 23824875 | 23825362 | 488 | 2 | 4.75E-05 | 9.32E-05 | 0.00299 | 0.000162 | 0.00368344 | SMARCB1 |
| chr9 | 33024427 | 33024825 | 399 | 3 | 9.02E-05 | 0.0001016 | 0.004787 | 0.000162 | 0.003735778 | APTX |
| chr4 | 88456931 | 88458005 | 1075 | 10 | 5.89E-06 | 0.0004071 | 0.008096 | 0.000162 | -0.00257251 | HERC5 |
| chr1 | 37595550 | 37596275 | 726 | 10 | 1.40E-05 | 0.1299509 | 1.24E-05 | 0.000162 | 0.001881205 | AL513220.1, GNL2 |
| chr4 | 163344290 | 163344670 | 381 | 3 | 6.99E-05 | 0.0053613 | 0.001998 | 0.000162 | 0.004867041 | NPY5R, NPY1R |
| chr5 | 115297307 | 115297771 | 465 | 3 | 9.77E-06 | 0.0006457 | 0.002413 | 0.000162 | -0.00487383 | NA |
| chr11 | 2534267 | 2534346 | 80 | 2 | 7.79E-05 | 0.0001335 | 0.000978 | 0.000162 | 0.013666222 | KCNQ1 |
| chr2 | 95346167 | 95346580 | 414 | 3 | 1.30E-05 | 0.000433 | 0.001874 | 0.000163 | 0.002685221 | KCNIP3 |
| chr17 | 5434234 | 5434287 | 54 | 2 | 0.000102852 | 0.0020297 | 9.19E-05 | 0.000163 | 0.001097515 | C1QBP |
| chr8 | 142539270 | 142539341 | 72 | 3 | 5.49E-05 | 0.0018184 | 0.000462 | 0.000163 | -0.00543694 | ADGRB1 |
| chr1 | 45686822 | 45687524 | 703 | 6 | 5.06E-05 | 3.67E-05 | 0.027035 | 0.000164 | 0.001905634 | GPBP1L1 |
| chr12 | 52191027 | 52191466 | 440 | 2 | 1.28E-05 | 0.0020598 | 9.17E-05 | 0.000164 | -0.00747088 | LINC00592, KRT80 |
| chr2 | 145334427 | 145334480 | 54 | 2 | 0.000126508 | 0.0029521 | 8.26E-05 | 0.000164 | -0.0082494 | NA |
| chr1 | 193105047 | 193105647 | 601 | 10 | 4.42E-05 | 0.0004069 | 0.000966 | 0.000164 | -0.00085029 | GLRX2 |
| chr6 | 43427431 | 43427819 | 389 | 7 | 0.000107865 | 0.0023766 | 0.001142 | 0.000164 | 0.001077475 | ABCC10 |
| chr8 | 26051655 | 26052083 | 429 | 6 | 9.48E-05 | 0.0021404 | 0.000159 | 0.000164 | -0.00080771 | NA |
| chr9 | 129801746 | 129802161 | 416 | 4 | 4.78E-06 | 0.0419118 | 1.56E-05 | 0.000164 | -0.00288805 | NA |
| chr2 | 239196388 | 239196923 | 536 | 2 | 5.95E-05 | 0.0001931 | 0.000522 | 0.000164 | -0.00211435 | AC017028.2, HDAC4 |
| chr12 | 111032963 | 111033685 | 723 | 5 | 1.05E-05 | 0.0007022 | 0.002016 | 0.000164 | 0.003037403 | NA |
| chr2 | 96208990 | 96209235 | 246 | 7 | 2.87E-05 | 0.0087873 | 2.29E-05 | 0.000165 | 0.001007721 | STARD7-AS1 |
| chr10 | 80602886 | 80603123 | 238 | 2 | 1.71E-05 | 0.0001567 | 0.000736 | 0.000165 | -0.00657127 | SH2D4B |
| chr7 | 1498702 | 1498970 | 269 | 2 | 1.71E-05 | 0.0001624 | 0.000691 | 0.000165 | -0.0001981 | INTS1 |
| chr7 | 1952393 | 1952524 | 132 | 2 | 4.83E-05 | 0.0001474 | 0.000825 | 0.000165 | 0.001133762 | MAD1L1, AC069288.1 |
| chr19 | 4638921 | 4639496 | 576 | 9 | 8.41E-05 | 0.0003718 | 0.006639 | 0.000165 | -0.00024882 | NA |
| chr19 | 912831 | 913536 | 706 | 8 | 9.03E-06 | 0.0019458 | 0.000725 | 0.000165 | -6.06E-05 | R3HDM4 |
| chr5 | 141635998 | 141636092 | 95 | 2 | 5.55E-06 | 0.0023863 | 8.85E-05 | 0.000165 | 0.014960698 | HDAC3 |
| chr2 | 9770496 | 9771021 | 526 | 3 | 1.67E-05 | 0.0006306 | 0.000207 | 0.000166 | -0.00035714 | NA |
| chr10 | 52778745 | 52778939 | 195 | 2 | 5.65E-05 | 0.0001178 | 0.001419 | 0.000166 | 0.030147324 | NA |
| chr12 | 56688386 | 56688820 | 435 | 10 | 4.20E-05 | 0.0082181 | 0.000925 | 0.000166 | -0.00047307 | PTGES3 |
| chr17 | 5485814 | 5486780 | 967 | 14 | 3.71E-06 | 0.005053 | 0.005353 | 0.000166 | 0.001815888 | MIS12, DERL2 |
| chr8 | 138913945 | 138913993 | 49 | 2 | 0.0001433 | 0.000187 | 0.00056 | 0.000166 | -0.00791536 | COL22A1 |
| chr2 | 23346047 | 23346331 | 285 | 2 | 1.22E-05 | 0.0015421 | 0.000103 | 0.000166 | -0.00636962 | NA |
| chr1 | 31689542 | 31689638 | 97 | 2 | 0.000137923 | 9.23E-05 | 0.003707 | 0.000167 | -0.00845652 | COL16A1 |
| chr10 | 13707888 | 13708224 | 337 | 3 | 9.41E-05 | 0.0021365 | 7.19E-05 | 0.000167 | -0.00469399 | FRMD4A |
| chr21 | 41420375 | 41420776 | 402 | 8 | 8.00E-05 | 0.0041401 | 0.007004 | 0.000167 | -0.00442685 | MX1 |
| chr10 | 133373978 | 133374513 | 536 | 3 | 1.55E-05 | 9.13E-05 | 0.00293 | 0.000167 | -0.00092894 | NA |
| chr11 | 111540153 | 111541031 | 879 | 9 | 4.25E-05 | 0.011536 | 0.000308 | 0.000167 | -0.00154193 | LAYN |
| chr16 | 1542815 | 1543213 | 399 | 4 | 0.00010452 | 0.0002043 | 0.000823 | 0.000167 | -0.0057196 | TMEM204, IFT140 |
| chr15 | 91694219 | 91694417 | 199 | 2 | 6.93E-05 | 0.0026283 | 8.71E-05 | 0.000167 | 0.003477342 | NA |
| chr19 | 46415090 | 46415108 | 19 | 2 | 0.000123271 | 0.0002703 | 0.000351 | 0.000168 | -0.00223091 | NA |
| chr6 | 55173718 | 55174102 | 385 | 5 | 8.60E-05 | 0.0006107 | 0.00121 | 0.000168 | 0.001308497 | HCRTR2 |
| chr6 | 1515461 | 1515562 | 102 | 2 | 0.000121519 | 0.0044111 | 7.67E-05 | 0.000168 | -0.00389731 | NA |
| chr2 | 74369322 | 74369486 | 165 | 3 | 0.000119793 | 0.0145505 | 3.70E-05 | 0.000169 | 0.001101528 | DCTN1 |
| chr20 | 43457741 | 43457836 | 96 | 5 | 0.000136941 | 0.0002499 | 0.005415 | 0.000169 | 0.00034667 | NA |
| chr16 | 1507365 | 1507604 | 240 | 4 | 0.000125079 | 0.0043299 | 0.000341 | 0.000169 | -0.00189876 | TELO2 |
| chr12 | 3471023 | 3471058 | 36 | 2 | 7.78E-05 | 0.0011493 | 0.00012 | 0.00017 | -0.00528773 | PRMT8 |
| chr9 | 92669913 | 92670720 | 808 | 9 | 2.16E-05 | 0.0257141 | 1.82E-05 | 0.00017 | 0.001086262 | IPPK |
| chr4 | 112356710 | 112356831 | 122 | 2 | 4.05E-05 | 0.0005126 | 0.000198 | 0.00017 | -0.00337155 | ALPK1, RTEL1P1 |
| chr18 | 2912737 | 2913256 | 520 | 6 | 5.87E-05 | 0.0010558 | 0.000353 | 0.00017 | 0.000562876 | EMILIN2 |
| chr4 | 188412142 | 188412295 | 154 | 2 | 0.000117197 | 0.0002399 | 0.000417 | 0.00017 | -0.02240149 | LINC01060 |
| chr14 | 98135113 | 98135795 | 683 | 3 | 1.35E-05 | 0.0049636 | 0.000283 | 0.00017 | -0.00300432 | AL163932.1 |
| chr2 | 241557962 | 241558180 | 219 | 2 | 1.83E-05 | 0.0009187 | 0.000136 | 0.000171 | -0.00144238 | BOK, BOK-AS1 |
| chr1 | 76313772 | 76313862 | 91 | 2 | 0.000170838 | 0.000232 | 0.000437 | 0.000171 | 0.003466813 | ST6GALNAC3 |
| chr9 | 96049160 | 96049358 | 199 | 2 | 7.75E-05 | 0.000173 | 0.000677 | 0.000171 | -0.0044303 | ERCC6L2 |
| chr16 | 11742518 | 11743081 | 564 | 11 | 0.000107835 | 0.0034277 | 0.007563 | 0.000171 | 0.00026365 | AC007613.1, TXNDC11 |
| chr1 | 31703673 | 31703967 | 295 | 2 | 2.96E-05 | 0.0001323 | 0.001174 | 0.000172 | -0.01442788 | COL16A1 |

| | | | | | | | | | | |
|---|---|---|---|---|---|---|---|---|---|---|
| chr1 | 193689264 | 193689269 | 6 | 2 | 0.000145095 | 0.0003841 | 0.000256 | 0.000172 | 0.003048401 | AL136456.1, AL357793.1 |
| chr2 | 223051874 | 223052234 | 361 | 8 | 9.99E-05 | 0.0021822 | 0.006283 | 0.000172 | 0.00083152 | KCNE4 |
| chr6 | 165791712 | 165791752 | 41 | 2 | 0.00012712 | 0.001208 | 0.000119 | 0.000172 | -0.0019917 | PDE10A |
| chr17 | 81656988 | 81657434 | 447 | 5 | 2.88E-05 | 0.0026967 | 0.00127 | 0.000172 | -0.0045491 | PDE6G |
| chr18 | 54356771 | 54356791 | 21 | 2 | 0.000125434 | 0.0002149 | 0.000495 | 0.000173 | 0.018388248 | STARD6 |
| chr10 | 16933878 | 16934045 | 168 | 2 | 6.03E-05 | 0.0068469 | 7.19E-05 | 0.000173 | 0.003954786 | CUBN |
| chr1 | 94925999 | 94926288 | 290 | 3 | 0.000127867 | 0.0002754 | 0.002925 | 0.000173 | 0.003678985 | CNN3 |
| chr9 | 137429592 | 137430225 | 634 | 2 | 0.000111557 | 0.0001037 | 0.002581 | 0.000173 | -0.00599924 | NOXA1 |
| chr1 | 53839173 | 53839458 | 286 | 4 | 8.40E-06 | 0.0010324 | 0.000337 | 0.000173 | 0.009402903 | NA |
| chr17 | 12791361 | 12791526 | 166 | 3 | 6.92E-05 | 0.0161392 | 3.47E-05 | 0.000174 | -0.00755761 | ARHGAP44 |
| chr19 | 35249326 | 35249606 | 281 | 2 | 7.07E-05 | 0.0004402 | 0.000231 | 0.000174 | -0.00272174 | LSR |
| chr1 | 85576466 | 85576547 | 82 | 2 | 6.59E-05 | 9.63E-05 | 0.003771 | 0.000174 | 0.000374114 | DDAH1, AC092807.3 |
| chr15 | 42272436 | 42273453 | 1018 | 7 | 1.60E-05 | 0.0015773 | 0.007813 | 0.000174 | -0.00129245 | GANC, TMEM87A |
| chr10 | 49608808 | 49609260 | 453 | 5 | 1.40E-05 | 0.0165269 | 0.000959 | 0.000174 | 0.005190275 | CHAT |
| chr2 | 32063184 | 32063492 | 309 | 7 | 9.24E-05 | 0.0021939 | 0.00197 | 0.000174 | -0.00083231 | NA |
| chr8 | 144545401 | 144545537 | 137 | 2 | 6.64E-05 | 0.0722347 | 5.95E-05 | 0.000174 | -0.00172319 | ARHGAP39 |
| chr3 | 9743148 | 9743194 | 47 | 3 | 0.00015072 | 0.0020993 | 0.000101 | 0.000174 | -0.00103763 | BRPF1 |
| chr9 | 129666345 | 129666488 | 144 | 2 | 6.34E-05 | 0.0019682 | 9.89E-05 | 0.000174 | -0.00961009 | PRRX2 |
| chr15 | 78263836 | 78264192 | 357 | 5 | 7.63E-05 | 0.0346522 | 0.000609 | 0.000174 | 2.58E-05 | DNAJA4, AC090607.3 |
| chr1 | 5161297 | 5161461 | 165 | 2 | 9.33E-05 | 0.0063969 | 7.34E-05 | 0.000174 | 0.014126338 | NA |
| chr7 | 29884216 | 29884549 | 334 | 4 | 0.000102419 | 6.96E-05 | 0.009197 | 0.000174 | -0.00997392 | WIPF3 |
| chr8 | 143599676 | 143600230 | 555 | 3 | 5.35E-05 | 9.92E-05 | 0.003317 | 0.000175 | -0.0067678 | TIGD5 |
| chr12 | 91720236 | 91721256 | 1021 | 3 | 1.20E-05 | 0.0006544 | 0.002161 | 0.000175 | -0.01570774 | NA |
| chr22 | 24593970 | 24594295 | 326 | 2 | 2.21E-05 | 0.000816 | 0.00015 | 0.000175 | -0.01261747 | AP000356.5 |
| chr12 | 48328281 | 48328691 | 411 | 3 | 2.92E-05 | 0.0001015 | 0.00267 | 0.000175 | -0.00376946 | AC024257.5 |
| chr3 | 40765279 | 40765290 | 12 | 2 | 5.53E-05 | 0.0037309 | 8.29E-05 | 0.000175 | 0.005492586 | AC099560.1 |
| chr13 | 112753165 | 112753953 | 789 | 4 | 2.62E-05 | 0.0010441 | 0.001487 | 0.000175 | 0.001099762 | ATP11A, ATP11A-AS1 |
| chr7 | 96999123 | 96999651 | 529 | 2 | 1.65E-05 | 0.0001924 | 0.000603 | 0.000175 | -0.01243 | DLX6-AS1 |
| chr11 | 4848733 | 4849223 | 491 | 3 | 0.000146779 | 0.0041475 | 0.000772 | 0.000176 | -0.00855069 | MMP26, OR51S1 |
| chr3 | 35640135 | 35640391 | 257 | 4 | 5.25E-05 | 0.0004427 | 0.000289 | 0.000176 | 0.009893398 | ARPP21 |
| chr16 | 49854189 | 49854693 | 505 | 3 | 9.18E-05 | 0.0002099 | 0.002949 | 0.000176 | -0.00658523 | ZNF423 |
| chr10 | 13706133 | 13706597 | 465 | 2 | 1.49E-05 | 9.87E-05 | 0.003591 | 0.000176 | -0.00097884 | FRMD4A |
| chr11 | 116772995 | 116773482 | 488 | 9 | 0.000102214 | 0.0008945 | 0.001653 | 0.000177 | 0.00167853 | AP006216.1 |
| chr19 | 4502666 | 4502773 | 108 | 2 | 5.09E-05 | 0.0141104 | 6.65E-05 | 0.000177 | -0.00199127 | PLIN4 |
| chr19 | 45428430 | 45428650 | 221 | 2 | 1.47E-05 | 0.0003884 | 0.000267 | 0.000177 | 0.002549695 | ERCC1 |
| chr11 | 372999 | 373468 | 470 | 4 | 5.27E-05 | 0.0001885 | 0.001244 | 0.000178 | -0.00122049 | B4GALNT4 |
| chr19 | 796153 | 796257 | 105 | 2 | 1.55E-05 | 0.0001021 | 0.003147 | 0.000178 | -0.010685 | NA |
| chr16 | 10942838 | 10943265 | 428 | 2 | 5.28E-07 | 0.0002773 | 0.000379 | 0.000178 | -0.00260652 | CIITA |
| chr11 | 64710714 | 64710902 | 189 | 3 | 6.00E-05 | 0.0432839 | 2.63E-05 | 0.000178 | -0.00400364 | NRXN2 |
| chr17 | 39636485 | 39636524 | 40 | 3 | 5.56E-05 | 0.0001026 | 0.005413 | 0.000178 | 0.007774544 | PPP1R1B |
| chr7 | 44159300 | 44159667 | 368 | 4 | 5.26E-05 | 0.1351771 | 1.21E-05 | 0.000178 | -0.00271011 | GCK |
| chr8 | 23682426 | 23683433 | 1008 | 8 | 2.97E-05 | 0.0003565 | 0.002369 | 0.000178 | 9.64E-05 | NKX3-1 |
| chr10 | 131945335 | 131945383 | 49 | 2 | 0.0001569 | 0.0408859 | 6.19E-05 | 0.000178 | 0.002798861 | PPP2R2D |
| chr17 | 52984855 | 52984923 | 69 | 2 | 0.000149272 | 0.0018113 | 0.000105 | 0.000179 | -0.00590273 | NA |
| chr5 | 57172823 | 57173192 | 370 | 3 | 5.73E-05 | 0.0083879 | 4.23E-05 | 0.000179 | 0.002693443 | NA |
| chr9 | 137077957 | 137078035 | 79 | 2 | 3.38E-05 | 0.0045812 | 8.04E-05 | 0.000179 | 0.001970911 | UAP1L1 |
| chr4 | 3484312 | 3484653 | 342 | 5 | 4.89E-05 | 0.0005168 | 0.000744 | 0.000179 | -0.00313877 | DOK7 |
| chr9 | 137495174 | 137495298 | 125 | 2 | 3.03E-05 | 0.0001152 | 0.001999 | 0.000179 | -0.00333375 | PNPLA7 |
| chr13 | 113459688 | 113460010 | 323 | 5 | 2.36E-05 | 0.0668149 | 1.33E-05 | 0.000179 | -0.00465572 | DCUN1D2 |
| chr11 | 455955 | 456082 | 128 | 3 | 2.03E-05 | 0.0002047 | 0.003149 | 0.000179 | -0.00215266 | PTDSS2 |
| chr1 | 100351387 | 100351744 | 358 | 7 | 3.31E-05 | 0.0045982 | 0.000906 | 0.00018 | 0.001571769 | CDC14A |
| chr12 | 27778894 | 27779480 | 587 | 3 | 3.39E-05 | 0.00057 | 0.00142 | 0.00018 | -0.00468259 | MANSC4 |
| chr9 | 127898508 | 127898896 | 389 | 5 | 8.18E-05 | 0.0035827 | 7.74E-05 | 0.00018 | -0.00455997 | AL157935.2, ST6GALNAC6 |
| chr19 | 17448804 | 17448857 | 54 | 2 | 3.97E-05 | 0.0050942 | 7.90E-05 | 0.00018 | -0.00262691 | AC010319.2 |
| chr18 | 23827814 | 23828051 | 238 | 2 | 2.18E-05 | 0.0085305 | 7.20E-05 | 0.00018 | -0.01008022 | LAMA3 |
| chr6 | 133838165 | 133838274 | 110 | 2 | 0.000136522 | 0.0010915 | 0.000133 | 0.00018 | -0.00178401 | LINC01312, TARID |
| chr12 | 57236324 | 57236673 | 350 | 4 | 2.38E-05 | 0.0039473 | 4.66E-05 | 0.00018 | -0.00440506 | NDUFA4L2 |
| chr16 | 30483928 | 30484181 | 254 | 2 | 5.71E-05 | 0.0002064 | 0.000582 | 0.000181 | 0.002942569 | ITGAL, AC116348.3 |
| chr12 | 123437141 | 123437310 | 170 | 3 | 5.87E-05 | 0.0006901 | 0.002297 | 0.000181 | -0.01307238 | NA |
| chr4 | 1798427 | 1798721 | 295 | 4 | 7.02E-05 | 0.0001341 | 0.00354 | 0.000182 | -0.00343899 | FGFR3 |
| chr3 | 138936985 | 138937786 | 802 | 5 | 1.28E-05 | 0.0051291 | 3.59E-05 | 0.000182 | 0.001933691 | LINC01391 |
| chr16 | 58685011 | 58685296 | 286 | 4 | 4.86E-05 | 0.1363095 | 1.04E-05 | 0.000182 | -0.00532188 | NA |
| chr7 | 105961959 | 105962986 | 1028 | 5 | 4.99E-06 | 0.0200798 | 2.42E-05 | 0.000182 | -0.00228821 | CDHR3 |
| chr1 | 153509948 | 153509971 | 24 | 2 | 0.000109232 | 0.0001542 | 0.000985 | 0.000182 | -0.01015278 | NA |
| chr8 | 39989427 | 39989597 | 171 | 2 | 0.000147816 | 0.0001118 | 0.002413 | 0.000182 | -0.00719777 | IDO2, AC007991.3 |
| chr5 | 140965035 | 140965160 | 126 | 2 | 4.50E-05 | 0.000401 | 0.000272 | 0.000183 | 0.000200206 | PCDHA1, PCDHA2, PCDHA3, PCDHA4, PCDHA5, PCDHA6, PCDHA7, PCDHA8, PCDHA9, PCDHA10, PCDHA11, PCDHA12, PCDHA13, PCDHAC1 |
| chr8 | 143820411 | 143820666 | 256 | 2 | 9.81E-05 | 0.0001647 | 0.000869 | 0.000183 | -0.00095266 | PUF60 |
| chr2 | 160271673 | 160271915 | 243 | 4 | 9.27E-05 | 0.0003299 | 0.004591 | 0.000183 | 0.004622265 | AC092153.1, LINC02478 |
| chr2 | 212536145 | 212536609 | 465 | 3 | 6.60E-05 | 0.0004362 | 0.001031 | 0.000183 | 0.003232425 | ERBB4 |
| chr5 | 56862668 | 56863222 | 555 | 3 | 7.70E-05 | 0.0001206 | 0.005095 | 0.000184 | -0.00527589 | MAP3K1 |
| chr1 | 24293985 | 24294586 | 602 | 2 | 0.000105162 | 0.0001364 | 0.001338 | 0.000184 | -0.00578856 | NA |
| chr2 | 218958837 | 218959331 | 495 | 3 | 5.05E-06 | 0.0611025 | 2.64E-05 | 0.000184 | -0.00497976 | AC097468.2 |
| chr8 | 140302602 | 140302880 | 279 | 3 | 4.77E-05 | 0.0108554 | 5.87E-05 | 0.000184 | -0.00285499 | TRAPPC9 |
| chr12 | 123762370 | 123762676 | 307 | 7 | 0.000136894 | 0.0011231 | 0.001808 | 0.000185 | 6.70E-05 | DNAH10 |
| chr2 | 231132414 | 231132942 | 529 | 3 | 2.32E-05 | 0.0025991 | 0.000445 | 0.000185 | -0.00179534 | PSMD1 |
| chr17 | 77140167 | 77140244 | 78 | 3 | 8.03E-05 | 9.50E-05 | 0.005602 | 0.000185 | 0.001660782 | SEC14L1 |
| chr6 | 125953150 | 125953595 | 446 | 2 | 7.66E-05 | 0.0004359 | 0.000259 | 0.000186 | -0.01090678 | NA |
| chr19 | 40442790 | 40443545 | 756 | 6 | 3.65E-05 | 0.0003315 | 0.004654 | 0.000186 | -0.000274 | AC010271.1, SERTAD3 |
| chr17 | 81120615 | 81121016 | 402 | 2 | 2.58E-06 | 0.0092683 | 7.32E-05 | 0.000186 | -0.00220661 | AATK |
| chr10 | 4242027 | 4242061 | 35 | 2 | 6.97E-05 | 0.0062702 | 7.83E-05 | 0.000186 | -0.00780624 | LINC00702 |
| chr7 | 128911618 | 128911703 | 86 | 2 | 4.29E-05 | 0.0001882 | 0.000718 | 0.000186 | -0.01096039 | NA |
| chr7 | 763506 | 763816 | 311 | 2 | 1.43E-05 | 0.0307446 | 6.50E-05 | 0.000187 | 0.003390235 | DNAAF5 |
| chr1 | 9847014 | 9847217 | 204 | 2 | 2.81E-05 | 0.0132538 | 6.99E-05 | 0.000187 | 0.002098419 | NA |
| chr1 | 228277095 | 228278464 | 1370 | 5 | 9.11E-06 | 0.001149 | 0.000689 | 0.000187 | -0.00150871 | OBSCN |
| chr1 | 190475536 | 190476067 | 532 | 7 | 6.58E-05 | 0.0014912 | 0.002761 | 0.000188 | 0.000694778 | BRINP3 |
| chr7 | 98282200 | 98282837 | 638 | 3 | 0.000102755 | 0.0001577 | 0.002771 | 0.000188 | -0.01098497 | BRI3 |
| chr7 | 70615052 | 70615318 | 267 | 2 | 0.000139713 | 0.0001266 | 0.001789 | 0.000189 | -0.00071716 | AUTS2 |
| chr20 | 62656214 | 62656520 | 307 | 3 | 9.78E-05 | 0.000245 | 0.000664 | 0.000189 | -0.0051327 | SLCO4A1 |
| chr16 | 47460347 | 47461446 | 1100 | 10 | 1.42E-05 | 0.0003816 | 0.001951 | 0.000189 | -0.00128725 | PHKB, ITFG1 |
| chr2 | 174445604 | 174445625 | 22 | 2 | 0.000128476 | 0.0292643 | 6.60E-05 | 0.000189 | -0.00784225 | GPR155 |
| chr15 | 67165501 | 67166065 | 565 | 7 | 7.74E-05 | 4.53E-05 | 0.022067 | 0.000191 | -0.00203286 | SMAD3 |
| chr5 | 141234056 | 141234534 | 479 | 9 | 0.000111434 | 0.0089471 | 0.005252 | 0.000191 | 0.00059236 | AC244517.10, PCDHB18P, AC244517.11 |
| chr15 | 48897249 | 48897341 | 93 | 2 | 0.000128481 | 0.0025866 | 9.91E-05 | 0.000191 | -0.00476056 | SHC4 |
| chr16 | 67992971 | 67993172 | 202 | 2 | 0.000164556 | 0.0001168 | 0.002464 | 0.000191 | -0.00748904 | DUS2, DPEP2 |
| chr7 | 100477630 | 100477779 | 150 | 5 | 0.000145177 | 0.000848 | 0.000129 | 0.000191 | 0.000202776 | TSC22D4 |
| chr14 | 36520186 | 36520383 | 198 | 6 | 0.000156813 | 0.0004626 | 0.003303 | 0.000191 | 0.000832856 | AL132857.1, NKX2-1-AS1, SFTA3, NKX2-1 |
| chr19 | 35296029 | 35296134 | 106 | 3 | 0.000164644 | 7.68E-05 | 0.008419 | 0.000192 | 0.003813921 | MAG |
| chr2 | 130340896 | 130341933 | 1038 | 3 | 9.76E-05 | 0.0007932 | 0.000889 | 0.000192 | 0.001487798 | CCDC115 |
| chr20 | 51008064 | 51008078 | 15 | 2 | 0.000152589 | 0.0032905 | 9.30E-05 | 0.000192 | 0.006761083 | AL050404.1, KCNG1 |
| chr16 | 30659250 | 30660428 | 1179 | 6 | 0.000109983 | 0.000124 | 0.007735 | 0.000192 | -0.00043344 | FBRS |
| chr11 | 18588151 | 18588527 | 377 | 2 | 3.12E-07 | 0.0090226 | 7.54E-05 | 0.000192 | -0.00468239 | UEVLD |
| chr5 | 118562118 | 118562576 | 459 | 5 | 0.000108268 | 0.0018396 | 0.003097 | 0.000192 | -0.00292788 | LINC02208 |
| chr10 | 133278760 | 133279117 | 358 | 2 | 5.21E-05 | 0.0009861 | 0.000152 | 0.000192 | -0.01092842 | TUBGCP2 |
| chr4 | 337549 | 337896 | 348 | 8 | 1.65E-05 | 0.0666394 | 2.01E-05 | 0.000192 | 0.00056148 | ZNF141 |
| chr9 | 130835781 | 130835897 | 117 | 2 | 0.000115675 | 0.0003539 | 0.000335 | 0.000192 | 0.002152843 | ABL1 |
| chr12 | 10307313 | 10307373 | 61 | 3 | 0.000145409 | 0.0118876 | 4.25E-05 | 0.000192 | -0.00539799 | KLRD1 |
| chr12 | 14365457 | 14366040 | 584 | 8 | 0.0001236 | 0.0006898 | 0.005092 | 0.000193 | 0.001220607 | ATF7IP, RPL30P11 |
| chr6 | 32857263 | 32857924 | 662 | 9 | 3.19E-05 | 0.0035012 | 0.001259 | 0.000193 | -0.00357269 | PSMB9 |
| chr11 | 1156671 | 1157800 | 1130 | 4 | 9.97E-06 | 0.0041953 | 0.000396 | 0.000193 | 0.004238764 | NA |
| chr5 | 172833796 | 172834152 | 357 | 5 | 0.000106219 | 0.0214355 | 0.003722 | 0.000194 | 0.0059395 | NA |
| chr7 | 124032049 | 124032196 | 148 | 2 | 0.000127234 | 0.0001608 | 0.001063 | 0.000194 | -0.00242152 | AC006148.1, TMEM229A |
| chr7 | 145698893 | 145699352 | 460 | 2 | 0.00013232 | 0.000112 | 0.003184 | 0.000194 | -0.01530071 | NA |
| chr7 | 35695094 | 35695537 | 444 | 10 | 0.000122481 | 0.0004484 | 0.010034 | 0.000195 | 0.001285474 | AC018647.2, HERPUD2 |
| chr17 | 78466839 | 78467548 | 710 | 3 | 6.82E-06 | 0.0074421 | 8.83E-05 | 0.000195 | -0.00322212 | DNAH17 |
| chr19 | 13138309 | 13138387 | 79 | 2 | 0.000150555 | 0.0043379 | 8.81E-05 | 0.000195 | 0.008091084 | NACC1 |
| chr8 | 47961391 | 47961684 | 294 | 3 | 2.67E-05 | 0.0096467 | 0.000232 | 0.000195 | -0.00385295 | MCM4 |
| chr6 | 31057213 | 31057237 | 25 | 2 | 0.000125491 | 0.0001104 | 0.00355 | 0.000196 | -0.00950108 | HCG22 |
| chr2 | 186496406 | 186497008 | 603 | 2 | 2.63E-05 | 0.001865 | 0.000115 | 0.000196 | -0.0112294 | ZC3H15 |
| chr16 | 4693813 | 4693912 | 100 | 2 | 9.46E-05 | 0.0215942 | 6.93E-05 | 0.000196 | 0.002021151 | NUDT16L1 |
| chr1 | 16981537 | 16981808 | 272 | 3 | 4.20E-05 | 0.0395774 | 3.10E-05 | 0.000196 | -0.00293323 | AL049569.1 |
| chr12 | 124224812 | 124224967 | 156 | 2 | 0.000136443 | 0.0020704 | 0.000111 | 0.000197 | 0.00236972 | RFLNA |
| chr3 | 52802733 | 52802953 | 221 | 2 | 0.000126986 | 0.0013488 | 0.000133 | 0.000197 | -0.00077309 | ITIH3 |
| chr5 | 63959978 | 63961099 | 1122 | 5 | 8.71E-06 | 0.0004774 | 0.003016 | 0.000198 | 0.000491185 | HTR1A, AC122707.1 |
| chr6 | 159035875 | 159036363 | 489 | 2 | 4.10E-05 | 0.0001652 | 0.00106 | 0.000198 | -0.00364739 | AL035530.2, TAGAP |
| chr10 | 70478955 | 70479088 | 134 | 2 | 7.89E-05 | 0.0016535 | 0.000122 | 0.000198 | 0.001265637 | PALD1 |
| chr21 | 42120370 | 42120511 | 142 | 2 | 2.92E-05 | 0.0002726 | 0.000474 | 0.000198 | -0.02804258 | UMODL1 |
| chr17 | 17795899 | 17796147 | 249 | 3 | 0.000114646 | 0.0014327 | 0.000147 | 0.000198 | -0.00073801 | RAI1 |
| chr21 | 46318196 | 46319225 | 1030 | 8 | 0.00012573 | 0.0610265 | 8.77E-05 | 0.000198 | 0.001620827 | C21orf58 |
| chr3 | 47282065 | 47282819 | 755 | 13 | 2.22E-05 | 0.0021637 | 0.003675 | 0.000199 | -0.00093377 | KIF9 |
| chr15 | 29571444 | 29571965 | 522 | 4 | 9.79E-05 | 0.0089977 | 0.000241 | 0.0002 | -0.00085875 | NA |
| chr13 | 111623874 | 111623999 | 126 | 2 | 0.000102807 | 0.0003049 | 0.000422 | 0.0002 | -0.00204424 | AL359649.1, LINC02337 |
| chr3 | 43998017 | 43998449 | 433 | 6 | 7.08E-05 | 0.025254 | 2.85E-05 | 0.0002 | 0.000984958 | AC006058.3 |
| chr7 | 73647065 | 73647077 | 13 | 2 | 9.24E-05 | 0.0031578 | 9.81E-05 | 0.0002 | -0.00513142 | NA |
| chr12 | 81758450 | 81759511 | 1062 | 15 | 4.07E-06 | 0.0956473 | 2.75E-05 | 0.000201 | 0.001093228 | PPFIA2 |
| chr8 | 26624150 | 26624306 | 157 | 3 | 0.000124923 | 0.0045055 | 0.001182 | 0.000201 | -0.00559471 | DPYSL2 |
| chr10 | 683539 | 684016 | 478 | 5 | 5.66E-05 | 0.0017654 | 0.006043 | 0.000201 | 0.006582012 | DIP2C |
| chr11 | 44615658 | 44616096 | 439 | 4 | 6.89E-05 | 0.0255723 | 2.76E-05 | 0.000201 | -0.00087326 | CD82 |
| chr15 | 76341640 | 76342546 | 907 | 8 | 5.41E-05 | 0.0010799 | 0.000695 | 0.000201 | -0.00124749 | ISL2, AC027243.1 |
| chr19 | 2098101 | 2098125 | 25 | 2 | 0.000129016 | 0.001973 | 0.000116 | 0.000202 | -0.00566305 | IZUMO4 |
| chr10 | 43124323 | 43124445 | 123 | 2 | 0.000146702 | 0.0001915 | 0.00084 | 0.000202 | -0.00082917 | RET |
| chr8 | 38386726 | 38387161 | 436 | 5 | 1.86E-05 | 0.0016891 | 0.003105 | 0.000202 | -0.00182846 | LETM2 |
| chr11 | 2870172 | 2870265 | 94 | 2 | 3.17E-05 | 0.0001508 | 0.00139 | 0.000202 | -0.01155438 | KCNQ1DN |
| chr17 | 79940167 | 79940508 | 342 | 3 | 5.48E-05 | 0.0002678 | 0.000618 | 0.000202 | -0.00640572 | TBC1D16 |
| chr6 | 89672055 | 89672229 | 175 | 3 | 6.62E-05 | 0.0445756 | 2.92E-05 | 0.000202 | -0.00393768 | MDN1 |
| chr16 | 67933801 | 67933890 | 90 | 2 | 0.000116186 | 0.0231106 | 7.07E-05 | 0.000202 | 0.006380297 | AC040162.1 |
| chr17 | 82414617 | 82415036 | 420 | 6 | 9.34E-05 | 3.49E-05 | 0.028608 | 0.000202 | 0.007229425 | OGFOD3 |
| chr1 | 155255351 | 155256336 | 986 | 13 | 8.09E-06 | 0.0149207 | 0.000367 | 0.000203 | -0.00121836 | FAM189B, SCAMP3 |
| chr16 | 87215782 | 87215808 | 27 | 2 | 8.60E-05 | 0.0001152 | 0.003507 | 0.000203 | 0.000630197 | AC136285.1, C16orf95 |
| chr22 | 39519229 | 39519588 | 360 | 5 | 2.68E-05 | 0.0017542 | 0.00044 | 0.000203 | 0.001173988 | NA |
| chr1 | 111755479 | 111756007 | 529 | 8 | 1.83E-05 | 0.0155828 | 0.000133 | 0.000203 | 0.001593918 | DDX20, INKA2, AL049557.1 |
| chr1 | 93179394 | 93179921 | 528 | 5 | 4.68E-07 | 0.0296767 | 3.36E-05 | 0.000203 | 0.003746751 | CCDC18, TMED5 |
| chr7 | 30920610 | 30920803 | 194 | 2 | 2.88E-05 | 0.0001867 | 0.000896 | 0.000203 | -0.00844344 | AC004691.2, AQP1 |
| chr12 | 10130949 | 10131164 | 216 | 3 | 5.04E-05 | 0.000803 | 0.00024 | 0.000204 | 0.005679084 | NA |
| chr16 | 15055418 | 15055827 | 410 | 3 | 1.69E-05 | 0.0001769 | 0.00112 | 0.000204 | 0.000767344 | PDXDC1, NTAN1 |
| chr3 | 13497082 | 13497559 | 478 | 2 | 8.53E-06 | 0.0006241 | 0.000222 | 0.000204 | -0.00838342 | HDAC11 |
| chr4 | 8591780 | 8592206 | 427 | 2 | 4.09E-05 | 0.1810481 | 6.66E-05 | 0.000204 | 0.012584762 | GPR78 |
| chr14 | 21042155 | 21042225 | 71 | 3 | 0.000109809 | 0.004083 | 6.53E-05 | 0.000204 | 0.004053436 | NDRG2 |
| chr21 | 42681381 | 42681431 | 51 | 2 | 0.000129478 | 0.0074372 | 8.22E-05 | 0.000204 | -0.00568419 | PDE9A |

| | | | | | | | | | | |
|---|---|---|---|---|---|---|---|---|---|---|
| chr15 | 65423463 | 65424092 | 630 | 4 | 4.73E-05 | 6.80E-05 | 0.00997 | 0.000204 | -0.00454332 | NA |
| chr1 | 2339077 | 2339368 | 292 | 3 | 6.52E-05 | 0.002428 | 0.000155 | 0.000205 | -0.00276158 | MORN1 |
| chr20 | 25496706 | 25497268 | 563 | 3 | 4.43E-05 | 0.0008621 | 0.000196 | 0.000205 | -0.00375522 | NINL |
| chr13 | 113399922 | 113400640 | 719 | 4 | 9.54E-05 | 0.0001747 | 0.00831 | 0.000206 | -0.0038625 | ADPRHL1 |
| chr1 | 211675795 | 211676071 | 277 | 5 | 4.77E-05 | 0.000248 | 0.011368 | 0.000206 | 0.001195307 | AC096637.2 |
| chr2 | 216632820 | 216633226 | 407 | 6 | 8.19E-05 | 0.0016226 | 8.98E-05 | 0.000206 | -0.00095778 | IGFBP2 |
| chr19 | 8730244 | 8730423 | 180 | 3 | 0.00013722 | 0.0022573 | 8.88E-05 | 0.000206 | 0.006477066 | OR2Z1 |
| chr16 | 11798735 | 11798749 | 15 | 2 | 8.91E-05 | 0.0548452 | 6.87E-05 | 0.000207 | 0.003995038 | NA |
| chr13 | 110138006 | 110138433 | 428 | 5 | 8.17E-06 | 0.0452401 | 9.81E-06 | 0.000208 | 0.001614182 | NA |
| chr3 | 137997852 | 137997976 | 125 | 4 | 7.72E-05 | 0.0005437 | 0.000455 | 0.000208 | 0.000133851 | NA |
| chr5 | 891728 | 891871 | 144 | 3 | 0.000120287 | 0.0009179 | 0.002357 | 0.000208 | -0.00758735 | BRD9 |
| chr15 | 25070207 | 25070278 | 72 | 4 | 0.00012247 | 0.0007698 | 0.000218 | 0.000208 | 0.001357221 | SNHG14 |
| chr5 | 52989346 | 52989730 | 385 | 4 | 4.55E-06 | 0.0003667 | 0.001306 | 0.000209 | 0.002035717 | ITGA2, AC025180.1 |
| chr22 | 24423514 | 24423896 | 383 | 2 | 9.99E-06 | 0.0003953 | 0.000345 | 0.000209 | -0.00531119 | SPECC1L-ADORA2A, ADORA2A |
| chr1 | 59813880 | 59814831 | 952 | 7 | 7.45E-05 | 0.0001682 | 0.01498 | 0.000209 | 0.003764189 | HOOK1 |
| chr17 | 1560190 | 1560754 | 565 | 3 | 3.46E-05 | 0.0176678 | 0.002189 | 0.00021 | -0.00725573 | PITPNA |
| chr5 | 133252011 | 133252368 | 358 | 2 | 2.97E-05 | 0.0038345 | 9.72E-05 | 0.00021 | 0.003428093 | FSTL4 |
| chr19 | 33626117 | 33626336 | 220 | 2 | 6.62E-05 | 0.0044458 | 9.37E-05 | 0.00021 | -0.01161092 | CHST8 |
| chr11 | 47591906 | 47591944 | 39 | 3 | 9.19E-05 | 0.0012803 | 0.000134 | 0.00021 | 0.0014325 | C1QTNF4 |
| chr2 | 95207385 | 95207717 | 333 | 4 | 1.89E-05 | 0.0013568 | 0.000173 | 0.000211 | 0.001189984 | AC092835.1 |
| chr8 | 89900930 | 89901037 | 108 | 2 | 0.000100171 | 0.0001163 | 0.004074 | 0.000211 | 0.001667346 | NA |
| chr11 | 129694430 | 129694807 | 378 | 3 | 9.56E-05 | 0.0005584 | 0.001572 | 0.000211 | -0.00661535 | NA |
| chr17 | 45283417 | 45283964 | 548 | 2 | 9.97E-06 | 0.0026788 | 0.000109 | 0.000211 | 0.004774165 | MAP3K14 |
| chr20 | 5910461 | 5910572 | 112 | 2 | 6.94E-05 | 0.0005247 | 0.00027 | 0.000211 | 0.027073782 | NA |
| chr5 | 51382617 | 51382939 | 323 | 5 | 7.14E-05 | 0.0012279 | 0.000161 | 0.000211 | 0.001016509 | AC010478.1 |
| chr1 | 205275266 | 205275316 | 51 | 2 | 0.000142967 | 0.0006949 | 0.000217 | 0.000211 | -0.00721379 | TMCC2 |
| chr19 | 32252664 | 32253393 | 730 | 2 | 2.33E-05 | 0.0006817 | 0.00022 | 0.000212 | -0.01355028 | NA |
| chr12 | 26230761 | 26230821 | 61 | 3 | 3.97E-05 | 0.0213297 | 3.64E-05 | 0.000212 | -0.00418457 | SSPN, AC022509.2, AC022509.1 |
| chr16 | 30065159 | 30066066 | 908 | 7 | 8.88E-06 | 0.0029965 | 0.00025 | 0.000212 | 0.001670301 | AC093512.2, ALDOA |
| chr16 | 823426 | 823598 | 173 | 2 | 6.56E-05 | 0.0147185 | 7.69E-05 | 0.000212 | -0.0033495 | AL023882.1 |
| chr1 | 32395084 | 32395165 | 82 | 2 | 0.000121097 | 0.0001188 | 0.003864 | 0.000213 | -0.00523483 | NA |
| chr11 | 36377589 | 36377923 | 335 | 3 | 6.00E-05 | 0.0364492 | 5.93E-05 | 0.000213 | -0.00102472 | PRR5L |
| chr10 | 116803117 | 116803286 | 170 | 3 | 7.90E-05 | 0.0020631 | 0.000636 | 0.000214 | -0.00236385 | HSPA12A |
| chr19 | 10432334 | 10432468 | 135 | 4 | 0.000162184 | 0.0031244 | 0.000491 | 0.000214 | 0.003956344 | PDE4A |
| chr12 | 128266426 | 128266503 | 78 | 3 | 6.21E-05 | 0.0505934 | 3.00E-05 | 0.000214 | -0.00733352 | NA |
| chr1 | 213984593 | 213985030 | 438 | 3 | 4.41E-05 | 0.0005898 | 0.002064 | 0.000215 | 0.000173332 | PROX1, PROX1-AS1, AC011700.1 |
| chr1 | 53220225 | 53220336 | 112 | 2 | 7.41E-05 | 0.0003201 | 0.000455 | 0.000215 | -0.00076171 | CZIB |
| chr16 | 85286979 | 85287116 | 138 | 3 | 5.51E-05 | 0.0009646 | 0.000315 | 0.000215 | -0.00175465 | GSE1, AC092275.1 |
| chr17 | 297443 | 297855 | 413 | 2 | 5.00E-05 | 0.0004859 | 0.000297 | 0.000215 | -0.0040695 | RPH3AL |
| chr1 | 1909875 | 1909895 | 21 | 2 | 8.96E-05 | 0.0005329 | 0.000274 | 0.000215 | -0.01352553 | NA |
| chr10 | 101784029 | 101784076 | 48 | 2 | 8.43E-05 | 0.0008337 | 0.000196 | 0.000215 | -0.00209112 | NA |
| chr1 | 10399083 | 10399211 | 129 | 3 | 0.000112949 | 0.0005326 | 0.001749 | 0.000215 | -0.00212282 | PGD |
| chr4 | 69995732 | 69995825 | 94 | 2 | 8.17E-05 | 0.0001239 | 0.00337 | 0.000215 | -0.00354882 | NA |
| chr12 | 124381223 | 124381316 | 94 | 2 | 3.78E-05 | 0.0005487 | 0.000269 | 0.000216 | -0.01250049 | NCOR2 |
| chr3 | 52823596 | 52823915 | 320 | 5 | 4.38E-05 | 0.0074383 | 0.002041 | 0.000216 | -0.00192807 | ITIH4, AC006254.1 |
| chr1 | 201896083 | 201896349 | 267 | 2 | 1.67E-05 | 0.0084977 | 8.46E-05 | 0.000216 | 0.002667617 | AL513217.1 |
| chr18 | 23438253 | 23438760 | 508 | 3 | 7.40E-05 | 8.61E-05 | 0.008351 | 0.000216 | -0.00434113 | NA |
| chr2 | 127107186 | 127107388 | 203 | 7 | 0.00012567 | 0.0005244 | 0.004959 | 0.000217 | -0.0005856 | BIN1 |
| chr1 | 156591351 | 156592155 | 805 | 11 | 8.65E-05 | 0.0346127 | 0.000663 | 0.000217 | 0.001316445 | NAXE |
| chr9 | 36277718 | 36277750 | 33 | 2 | 0.000101732 | 0.0001958 | 0.000964 | 0.000217 | -0.00060957 | CLTA |
| chr6 | 113872049 | 113872181 | 133 | 2 | 0.000151179 | 0.0063212 | 8.96E-05 | 0.000217 | 0.014195207 | MROCKI |
| chr2 | 66577574 | 66577733 | 160 | 2 | 9.57E-05 | 0.0001198 | 0.004159 | 0.000217 | 0.002991185 | LINC01798 |
| chr1 | 2533965 | 2534105 | 141 | 3 | 0.000108926 | 0.0229925 | 4.18E-05 | 0.000217 | -0.00333833 | AL139246.5 |
| chr19 | 18397683 | 18397900 | 218 | 2 | 1.32E-05 | 0.0002918 | 0.000522 | 0.000217 | 0.007837549 | NA |
| chr7 | 156522969 | 156523033 | 65 | 2 | 8.06E-05 | 0.0001194 | 0.004283 | 0.000218 | -0.00476272 | LINC01006 |
| chr6 | 28526668 | 28527107 | 440 | 5 | 5.27E-05 | 0.0188821 | 6.78E-05 | 0.000218 | -0.00018541 | GPX5, GPX6 |
| chr1 | 10796886 | 10797379 | 494 | 4 | 3.81E-05 | 0.0001124 | 0.009859 | 0.000218 | -0.00600388 | NA |
| chr17 | 7205270 | 7205688 | 419 | 3 | 5.57E-05 | 0.0012919 | 0.000846 | 0.000218 | 0.000594262 | DLG4 |
| chr10 | 21069956 | 21070178 | 223 | 4 | 9.89E-05 | 0.0020936 | 0.000673 | 0.000218 | -0.01014226 | NEBL |
| chr10 | 129889828 | 129890481 | 654 | 4 | 0.00012181 | 0.0003897 | 0.008234 | 0.000219 | -0.00198513 | EBF3 |
| chr12 | 6312271 | 6312577 | 307 | 5 | 0.000112227 | 0.0193393 | 0.000913 | 0.000219 | -0.00385225 | PLEKHG6 |
| chr5 | 116470908 | 116470973 | 66 | 2 | 0.000154813 | 0.0006951 | 0.000229 | 0.000219 | -0.00896654 | SEMA6A-AS1, SEMA6A |
| chr5 | 73215585 | 73216000 | 416 | 3 | 3.42E-05 | 0.0001072 | 0.006629 | 0.00022 | -0.00133384 | AC116345.1 |
| chr10 | 70124791 | 70125044 | 254 | 2 | 9.03E-05 | 0.001023 | 0.000178 | 0.00022 | 0.004343142 | AIFM2 |
| chr22 | 19951393 | 19951538 | 146 | 5 | 2.81E-05 | 0.0075437 | 3.99E-05 | 0.00022 | -0.00134872 | COMT |
| chr2 | 28012206 | 28012688 | 483 | 2 | 2.45E-05 | 0.0002522 | 0.000656 | 0.00022 | 0.002242663 | BABAM2 |
| chr1 | 27367123 | 27367451 | 329 | 5 | 5.14E-05 | 0.0015386 | 0.000323 | 0.000221 | -0.0021219 | NA |
| chr12 | 50167080 | 50167535 | 456 | 7 | 2.72E-05 | 0.0098013 | 0.001664 | 0.000221 | 0.002218767 | CERS5 |
| chr14 | 58199544 | 58199889 | 346 | 4 | 8.59E-06 | 0.0035005 | 0.000121 | 0.000221 | -0.00472265 | ARMH4 |
| chr19 | 49119307 | 49119832 | 526 | 4 | 2.28E-05 | 6.86E-05 | 0.013867 | 0.000221 | -0.00318099 | PPFIA3 |
| chr9 | 132923480 | 132923749 | 270 | 2 | 2.87E-05 | 0.0998637 | 7.16E-05 | 0.000222 | -0.00801193 | TSC1 |
| chr5 | 158235059 | 158235436 | 378 | 3 | 5.91E-05 | 0.0014112 | 0.000138 | 0.000222 | 0.006199026 | NA |
| chr15 | 45586807 | 45587178 | 372 | 13 | 2.50E-05 | 0.0090119 | 0.001074 | 0.000222 | -0.0005924 | NA |
| chr8 | 143841024 | 143841434 | 411 | 5 | 6.39E-05 | 0.0006129 | 0.002251 | 0.000223 | 0.001900482 | NA |
| chr8 | 145002187 | 145002724 | 538 | 5 | 5.11E-05 | 0.0001094 | 0.011769 | 0.000223 | 0.003632001 | ZNF252P |
| chr6 | 28281390 | 28281768 | 379 | 5 | 0.000134542 | 0.0003259 | 0.001924 | 0.000223 | 0.00516522 | AL021997.2, PGBD1 |
| chr1 | 47434655 | 47434958 | 304 | 3 | 8.04E-06 | 0.0067093 | 9.83E-05 | 0.000224 | 0.029596554 | NA |
| chr7 | 94005246 | 94005673 | 428 | 2 | 3.41E-05 | 0.0002688 | 0.000618 | 0.000224 | -0.02243376 | BET1-AS1 |
| chr20 | 23365595 | 23366096 | 502 | 4 | 3.80E-05 | 0.0159333 | 3.46E-05 | 0.000224 | -0.00707487 | GZF1 |
| chr1 | 171257198 | 171257885 | 688 | 6 | 1.22E-05 | 0.0019238 | 0.001002 | 0.000224 | -0.00066386 | FMO1 |
| chr17 | 43212102 | 43213075 | 974 | 3 | 3.69E-05 | 0.0004911 | 0.002926 | 0.000224 | -0.01804732 | TMEM106A |
| chr1 | 229303659 | 229304087 | 429 | 3 | 4.06E-06 | 0.0005259 | 0.00324 | 0.000225 | 0.008141832 | RAB4A |
| chr12 | 6271530 | 6271732 | 203 | 2 | 9.54E-06 | 0.0010118 | 0.000184 | 0.000225 | -0.00264781 | NA |
| chr3 | 5095864 | 5095945 | 82 | 2 | 0.000131687 | 0.0001243 | 0.004128 | 0.000225 | -0.00293053 | NA |
| chr6 | 43370663 | 43370776 | 114 | 2 | 9.43E-05 | 0.0006224 | 0.000258 | 0.000225 | -0.00176851 | AL359813.1 |
| chr10 | 103368705 | 103368861 | 157 | 2 | 4.39E-05 | 0.000277 | 0.000597 | 0.000225 | 0.003297668 | TAF5 |
| chr5 | 173229578 | 173229710 | 133 | 2 | 8.01E-05 | 0.0001246 | 0.004076 | 0.000225 | -0.00026772 | NA |
| chr2 | 20350770 | 20350988 | 219 | 6 | 0.000149469 | 0.0005369 | 0.006892 | 0.000225 | 0.001793568 | PUM2 |
| chr7 | 122305094 | 122306159 | 1066 | 10 | 6.10E-05 | 0.0003582 | 0.009962 | 0.000225 | -0.0013337 | FEZF1-AS1, FEZF1 |
| chr6 | 43456019 | 43456453 | 435 | 3 | 5.85E-05 | 0.0002393 | 0.003071 | 0.000226 | 0.000476713 | DLK2 |
| chr9 | 137276645 | 137276890 | 246 | 3 | 6.87E-05 | 0.0002416 | 0.00412 | 0.000226 | -0.00332001 | NA |
| chr1 | 247373857 | 247373922 | 66 | 2 | 0.000124465 | 0.000156 | 0.001831 | 0.000226 | -0.03606665 | NA |
| chr22 | 46531636 | 46531639 | 4 | 2 | 0.000118286 | 0.000858 | 0.000207 | 0.000227 | -0.00136498 | CELSR1 |
| chr2 | 218880925 | 218881026 | 102 | 2 | 1.11E-05 | 0.0001285 | 0.003732 | 0.000227 | -0.0088865 | WNT10A |
| chr9 | 128420217 | 128420450 | 234 | 3 | 3.75E-05 | 0.0009515 | 0.00099 | 0.000228 | 0.000109929 | CERCAM |
| chr7 | 143954744 | 143954784 | 41 | 2 | 0.000169611 | 0.0023806 | 0.000124 | 0.000228 | 0.0052805 | NA |
| chr20 | 25059071 | 25059978 | 908 | 7 | 9.35E-06 | 0.0086651 | 0.000397 | 0.000228 | -0.00015813 | NA |
| chr11 | 92408814 | 92408822 | 9 | 2 | 9.54E-05 | 0.0021331 | 0.000129 | 0.000228 | 0.003572096 | FAT3 |
| chr12 | 12717756 | 12718398 | 643 | 6 | 3.75E-05 | 5.45E-05 | 0.016754 | 0.000229 | 0.001556454 | CDKN1B |
| chr22 | 48536503 | 48536512 | 10 | 2 | 0.000164042 | 0.0004362 | 0.000365 | 0.000229 | 0.001366213 | TAFA5 |
| chr17 | 73232131 | 73232244 | 114 | 4 | 7.11E-05 | 0.0006808 | 0.000268 | 0.000229 | 0.001362768 | C17orf80, FAM104A |
| chr19 | 55166297 | 55166847 | 551 | 9 | 4.28E-05 | 0.0040419 | 0.003286 | 0.000229 | 0.00052572 | AC010327.4, DNAAF3 |
| chr1 | 19468758 | 19468875 | 118 | 2 | 0.000146976 | 0.0012379 | 0.000169 | 0.00023 | 0.015685218 | CAPZB |
| chr9 | 36189768 | 36190874 | 1107 | 9 | 1.42E-07 | 0.0073457 | 7.19E-05 | 0.00023 | -0.00199556 | CLTA |
| chr2 | 101477990 | 101478094 | 105 | 2 | 5.51E-05 | 0.0019219 | 0.000137 | 0.000231 | -0.00760379 | NA |
| chr1 | 154609531 | 154609680 | 150 | 2 | 7.94E-05 | 0.0001683 | 0.001624 | 0.000232 | -0.02793414 | ADAR |
| chr4 | 121932309 | 121932538 | 230 | 2 | 0.000101531 | 0.0001732 | 0.001527 | 0.000233 | -0.02302661 | TRPC3 |
| chr11 | 120266048 | 120266256 | 209 | 4 | 0.000144997 | 7.66E-05 | 0.013351 | 0.000233 | 0.00315103 | POU2F3 |
| chr11 | 105046033 | 105046046 | 14 | 2 | 0.000169936 | 0.001258 | 0.000171 | 0.000233 | 0.003004073 | CARD16 |
| chr11 | 128481473 | 128481484 | 12 | 2 | 7.27E-05 | 0.0001383 | 0.003083 | 0.000233 | 0.015218167 | ETS1 |
| chr17 | 81413826 | 81413925 | 100 | 2 | 9.41E-05 | 0.013153 | 8.47E-05 | 0.000233 | 0.001932965 | BAHCC1 |
| chr16 | 72008692 | 72009086 | 395 | 6 | 8.54E-05 | 0.000772 | 0.00487 | 0.000234 | 0.000129464 | DHODH |
| chr12 | 77324477 | 77324522 | 46 | 3 | 0.000159355 | 0.0205868 | 0.000101 | 0.000234 | -0.0046944 | NA |
| chr5 | 152005127 | 152005168 | 42 | 2 | 0.000119637 | 0.0002105 | 0.00103 | 0.000235 | -0.00891469 | LINC01933 |
| chr1 | 180233649 | 180234008 | 360 | 3 | 2.73E-05 | 0.0002186 | 0.002573 | 0.000235 | 0.002135592 | LHX4 |
| chr12 | 6309982 | 6310618 | 637 | 10 | 0.000102713 | 0.0002425 | 0.02107 | 0.000235 | -0.00227759 | PLEKHG6 |
| chr18 | 50375265 | 50375649 | 385 | 3 | 1.11E-05 | 0.0002484 | 0.001942 | 0.000236 | -0.0010203 | SKA1 |
| chr1 | 201982807 | 201982974 | 168 | 3 | 0.000158201 | 0.0228664 | 4.15E-05 | 0.000236 | 0.001912851 | RNPEP |
| chr1 | 92837351 | 92837368 | 18 | 2 | 5.37E-05 | 0.0082942 | 9.17E-05 | 0.000236 | -0.0032552 | RPL5, SNORD21, DIPK1A |
| chr22 | 41366298 | 41366495 | 198 | 3 | 4.94E-05 | 0.0003042 | 0.00358 | 0.000237 | 0.003826314 | NA |
| chr6 | 138691636 | 138692009 | 374 | 5 | 6.76E-05 | 5.77E-05 | 0.020627 | 0.000237 | -0.05290673 | NHSL1 |
| chr12 | 68831793 | 68832570 | 778 | 2 | 0.000100446 | 0.0002059 | 0.001101 | 0.000237 | -0.0113909 | MDM2 |
| chr2 | 133951808 | 133951846 | 39 | 2 | 0.000161499 | 0.0002209 | 0.000965 | 0.000238 | -0.00925983 | NA |
| chr14 | 77454701 | 77454980 | 280 | 2 | 0.000111278 | 0.0016698 | 0.000152 | 0.000238 | 0.004384729 | VIPAS39 |
| chr16 | 66919529 | 66920079 | 551 | 3 | 1.26E-05 | 0.0004619 | 0.003354 | 0.000238 | 0.002211368 | NA |
| chr1 | 211579553 | 211580088 | 536 | 3 | 2.20E-05 | 9.19E-05 | 0.00977 | 0.000239 | -0.00764005 | NA |
| chr2 | 26641366 | 26641383 | 18 | 2 | 0.00011262 | 0.0013027 | 0.000173 | 0.000239 | -0.00896101 | CIB4 |
| chr14 | 31207001 | 31207122 | 122 | 2 | 9.62E-06 | 0.0003958 | 0.000436 | 0.000239 | -0.0018627 | HECTD1 |
| chr10 | 7170853 | 7171342 | 490 | 4 | 0.000141466 | 0.0019183 | 0.000172 | 0.000241 | -0.0009549 | SFMBT2 |
| chr9 | 4804014 | 4804137 | 124 | 2 | 0.000141371 | 0.0005013 | 0.000347 | 0.000241 | -0.00354837 | RCL1, KLF4P1 |
| chr2 | 73981653 | 73982033 | 381 | 4 | 0.00012791 | 0.0006247 | 0.004905 | 0.000241 | 0.001607493 | NA |
| chr16 | 54287348 | 54288582 | 1235 | 5 | 8.12E-05 | 0.0003928 | 0.001446 | 0.000241 | -0.00439035 | AC018553.2 |
| chr8 | 118111468 | 118112223 | 756 | 6 | 1.42E-05 | 0.0843193 | 4.94E-06 | 0.000241 | -0.00032049 | EXT1 |
| chr8 | 46954561 | 46955162 | 602 | 3 | 1.04E-05 | 0.0175317 | 0.0007 | 0.000241 | 0.000307503 | NA |
| chr11 | 888316 | 888570 | 255 | 2 | 5.77E-05 | 0.0002347 | 0.000904 | 0.000242 | -0.00105807 | CHID1 |
| chr16 | 4335297 | 4335434 | 138 | 4 | 0.000114931 | 0.0013252 | 0.000485 | 0.000242 | -0.00209313 | GLIS2, PAM16 |
| chr6 | 167443313 | 167443622 | 310 | 3 | 0.000141705 | 0.0143549 | 8.60E-05 | 0.000242 | 0.002496199 | NA |
| chr10 | 132848046 | 132848136 | 91 | 2 | 0.000134251 | 0.00026 | 0.000769 | 0.000242 | 0.010913285 | CFAP46 |
| chr1 | 53961949 | 53962307 | 359 | 2 | 8.79E-05 | 0.0002009 | 0.001238 | 0.000244 | -0.00991195 | LRRC42 |
| chr15 | 100602931 | 100603238 | 308 | 3 | 0.000101821 | 0.0001258 | 0.003772 | 0.000244 | 0.00141071 | ASB7, LINS1 |
| chr2 | 9288535 | 9288597 | 63 | 2 | 3.45E-05 | 0.0266713 | 8.19E-05 | 0.000245 | -0.00688163 | ASAP2 |
| chr4 | 3745950 | 3746427 | 478 | 3 | 3.33E-05 | 0.0009874 | 0.002356 | 0.000245 | 0.016499769 | NA |
| chr1 | 212701210 | 212701400 | 191 | 3 | 6.85E-05 | 0.001987 | 0.000487 | 0.000245 | -0.004999 | NA |
| chr19 | 7496522 | 7496853 | 332 | 3 | 2.83E-05 | 0.0083574 | 5.81E-05 | 0.000247 | -0.01614706 | TEX45, PEX11G |
| chr20 | 44745598 | 44745765 | 168 | 7 | 0.00014748 | 0.0006276 | 0.000757 | 0.000247 | -0.00124179 | KCNK15-AS1 |
| chr22 | 47859710 | 47859841 | 132 | 2 | 7.45E-05 | 0.0267623 | 8.28E-05 | 0.000248 | -0.01683791 | AL117329.1 |
| chr8 | 38856150 | 38856158 | 9 | 2 | 0.000126432 | 0.0001357 | 0.004618 | 0.000249 | 0.022575047 | NA |
| chr6 | 27288596 | 27288627 | 32 | 2 | 0.000111557 | 0.0002712 | 0.000767 | 0.000249 | -0.00057281 | POM121L2 |
| chr1 | 112707352 | 112707764 | 413 | 6 | 0.000113267 | 0.0010576 | 0.000667 | 0.00025 | -0.00061869 | RHOC, AL603832.3 |
| chr2 | 176162712 | 176163434 | 723 | 7 | 1.49E-05 | 0.0136546 | 0.000905 | 0.00025 | -0.00331971 | HOXD3 |
| chr11 | 94020928 | 94021241 | 314 | 6 | 7.81E-05 | 0.0011131 | 0.000894 | 0.00025 | 0.008607621 | NA |
| chr8 | 73879408 | 73879796 | 389 | 3 | 2.44E-05 | 0.0110045 | 0.000135 | 0.000251 | -0.00465719 | AC022868.1 |
| chr10 | 121595940 | 121596527 | 588 | 4 | 2.66E-05 | 7.13E-05 | 0.019151 | 0.000251 | -0.04131523 | FGFR2 |
| chr1 | 39025787 | 39026521 | 735 | 6 | 1.21E-05 | 0.0013077 | 0.000202 | 0.000251 | 0.000151086 | NDUFS5 |
| chr6 | 166562072 | 166562269 | 198 | 2 | 6.81E-05 | 0.020171 | 8.59E-05 | 0.000252 | -0.00498286 | RPS6KA2 |
| chr10 | 99330714 | 99330801 | 88 | 2 | 8.93E-05 | 0.0002291 | 0.001044 | 0.000252 | -0.00538306 | CNNM1 |
| chr7 | 99919513 | 99919755 | 243 | 10 | 0.000145025 | 0.0001067 | 0.018411 | 0.000252 | -0.00482481 | TRIM4 |
| chr12 | 6656875 | 6656891 | 17 | 2 | 9.61E-05 | 0.000185 | 0.001653 | 0.000252 | 0.005474495 | ING4 |

| | | | | | | | | | | |
|---|---|---|---|---|---|---|---|---|---|---|
| chr20 | 62895124 | 62895533 | 410 | 4 | 6.54E-05 | 0.0001126 | 0.009657 | 0.000252 | -0.00165634 | DIDO1 |
| chr10 | 119537519 | 119537755 | 237 | 2 | 2.31E-05 | 0.0029191 | 0.000129 | 0.000252 | 0.006242166 | RGS10 |
| chr11 | 119361031 | 119361219 | 189 | 2 | 1.59E-05 | 0.0017967 | 0.000157 | 0.000252 | -0.00491381 | USP2 |
| chr6 | 1602880 | 1603297 | 418 | 2 | 7.86E-05 | 0.0001381 | 0.004639 | 0.000253 | 0.003560344 | FOXCUT |
| chr12 | 107580057 | 107580676 | 620 | 8 | 1.36E-05 | 0.0004719 | 0.004251 | 0.000253 | -0.00185716 | BTBD11 |
| chr17 | 82310704 | 82310947 | 244 | 2 | 1.42E-05 | 0.0035895 | 0.000121 | 0.000254 | -0.0053463 | NA |
| chr6 | 31271011 | 31271409 | 399 | 3 | 6.78E-06 | 0.0022722 | 0.001059 | 0.000254 | -0.03272467 | HLA-C |
| chr1 | 236280912 | 236281041 | 130 | 2 | 8.67E-05 | 0.0001395 | 0.004573 | 0.000254 | -0.01136176 | ERO1B |
| chr14 | 50821369 | 50821803 | 435 | 2 | 5.73E-05 | 0.0002086 | 0.001296 | 0.000255 | 0.005016254 | NIN |
| chr3 | 168287103 | 168287111 | 9 | 2 | 0.000156498 | 0.0021564 | 0.000147 | 0.000255 | -0.00301472 | EGFEM1P |
| chr13 | 52967861 | 52968128 | 268 | 2 | 8.30E-05 | 0.0003847 | 0.00051 | 0.000256 | -0.000181 | NA |
| chr1 | 231026981 | 231027113 | 133 | 3 | 0.000105515 | 0.0004306 | 0.000802 | 0.000257 | 0.002254055 | FAM89A |
| chr10 | 102064303 | 102064696 | 394 | 2 | 9.62E-06 | 0.0069023 | 0.000103 | 0.000257 | -0.0074731 | NA |
| chr2 | 233869235 | 233869523 | 289 | 2 | 4.75E-05 | 0.0377781 | 8.33E-05 | 0.000257 | -0.01046464 | NA |
| chr3 | 110529595 | 110529667 | 73 | 2 | 0.000108467 | 0.0009577 | 0.00023 | 0.000258 | -0.00293324 | NA |
| chr11 | 85627727 | 85628214 | 488 | 6 | 2.36E-05 | 0.0021052 | 0.001166 | 0.000258 | -0.00050971 | DLG2 |
| chr14 | 91399360 | 91399575 | 216 | 2 | 5.32E-05 | 0.000215 | 0.001256 | 0.000258 | 0.001415992 | CCDC88C |
| chr16 | 3946294 | 3946415 | 122 | 4 | 5.31E-05 | 0.1024975 | 1.97E-05 | 0.000258 | -0.00617711 | LINC02861 |
| chr2 | 70553772 | 70554193 | 422 | 9 | 7.57E-05 | 0.0039682 | 0.003622 | 0.000258 | -0.00061409 | TGFA |
| chr15 | 68205913 | 68206519 | 607 | 4 | 8.04E-06 | 0.0109628 | 0.000256 | 0.000259 | 0.001768841 | AC107871.1, CALML4 |
| chr1 | 181312460 | 181312564 | 105 | 2 | 7.59E-05 | 0.00941 | 9.71E-05 | 0.000259 | -0.00807019 | NA |
| chr5 | 151225416 | 151225583 | 168 | 2 | 0.000123973 | 0.0005932 | 0.000335 | 0.000259 | 0.01252625 | GM2A |
| chr19 | 48918013 | 48918591 | 579 | 3 | 3.49E-05 | 0.0010342 | 0.001106 | 0.000259 | -0.00671472 | NUCB1, NUCB1-AS1 |
| chr13 | 112119228 | 112119636 | 409 | 2 | 5.36E-05 | 0.0034758 | 0.000125 | 0.000259 | -0.00630222 | NA |
| chr15 | 44536692 | 44536739 | 48 | 4 | 0.000127397 | 0.0010164 | 0.007565 | 0.000259 | -0.00063918 | EIF3J-DT |
| chr20 | 48621669 | 48621708 | 40 | 2 | 0.000109944 | 0.0032906 | 0.000127 | 0.00026 | 0.007306927 | NA |
| chr14 | 39177632 | 39177638 | 7 | 2 | 0.000132589 | 0.0001437 | 0.004376 | 0.00026 | -0.00585487 | PNN |
| chr4 | 174217226 | 174217535 | 310 | 3 | 9.57E-05 | 0.0002121 | 0.002907 | 0.00026 | -0.00278314 | LINC02268 |
| chr12 | 52904203 | 52905104 | 902 | 5 | 2.15E-06 | 0.0048788 | 0.001637 | 0.00026 | -0.00091822 | KRT8 |
| chr3 | 184711846 | 184712689 | 844 | 12 | 6.02E-05 | 0.0064589 | 0.002317 | 0.000261 | 0.002141993 | AC107294.2, MAGEF1 |
| chr14 | 76370415 | 76370595 | 181 | 3 | 2.27E-05 | 0.0002189 | 0.004849 | 0.000261 | -0.00858843 | ESRRB |
| chr8 | 101126783 | 101126879 | 97 | 2 | 0.000114666 | 0.0002991 | 0.000735 | 0.000261 | 0.001477791 | NA |
| chr1 | 6142117 | 6142349 | 233 | 2 | 5.88E-05 | 0.0015818 | 0.000175 | 0.000262 | -0.00342059 | CHD5 |
| chr3 | 114900635 | 114900967 | 333 | 2 | 7.11E-05 | 0.0102248 | 9.70E-05 | 0.000262 | -0.00296162 | ZBTB20 |
| chr1 | 154558942 | 154559417 | 476 | 7 | 3.14E-05 | 0.0914525 | 1.86E-05 | 0.000262 | 0.001800707 | UBE2Q1 |
| chr6 | 44387804 | 44388213 | 410 | 3 | 1.97E-05 | 9.41E-05 | 0.011906 | 0.000262 | 0.00327993 | CDC5L |
| chr12 | 46371271 | 46371832 | 562 | 5 | 8.19E-05 | 0.0006464 | 0.008858 | 0.000263 | -0.00016276 | AC025031.2, SLC38A2 |
| chr3 | 52410601 | 52410817 | 217 | 5 | 4.60E-05 | 0.0005007 | 0.004409 | 0.000263 | 0.001761425 | PHF7 |
| chr8 | 141179950 | 141180541 | 592 | 5 | 2.73E-05 | 0.0007046 | 0.005324 | 0.000264 | -0.00125521 | DENND3 |
| chr9 | 128094903 | 128095134 | 232 | 2 | 0.000100034 | 0.0001891 | 0.001802 | 0.000264 | 0.007415254 | SLC25A25 |
| chr15 | 55408696 | 55409548 | 853 | 7 | 4.25E-07 | 0.0189502 | 0.000475 | 0.000265 | 0.000442283 | C15orf65, DNAAF4-CCPG1 |
| chr12 | 53079008 | 53079623 | 616 | 9 | 3.72E-05 | 0.0019543 | 0.005351 | 0.000265 | 0.001502952 | SPRYD3 |
| chr4 | 110622860 | 110623231 | 372 | 7 | 0.000107418 | 0.0162169 | 0.000161 | 0.000265 | -8.65E-05 | PITX2 |
| chr15 | 65068793 | 65068997 | 205 | 4 | 3.66E-05 | 0.0063848 | 0.001266 | 0.000266 | -0.0055505 | RASL12 |
| chr15 | 25990015 | 25990215 | 201 | 2 | 0.000164098 | 0.0001647 | 0.002786 | 0.000266 | -0.00013314 | LINC02346 |
| chr15 | 39250492 | 39250676 | 185 | 2 | 0.000100567 | 0.0002542 | 0.000988 | 0.000266 | -0.00247466 | AC013652.1 |
| chr3 | 48371400 | 48371434 | 35 | 2 | 0.000102939 | 0.0361895 | 8.59E-05 | 0.000266 | 0.003718941 | NA |
| chr3 | 49786677 | 49787042 | 366 | 7 | 5.18E-05 | 0.018929 | 0.000114 | 0.000267 | -0.00049687 | NA |
| chr7 | 155372300 | 155373028 | 729 | 5 | 4.39E-05 | 0.0145938 | 0.003885 | 0.000267 | -0.00107232 | NA |
| chr2 | 74374304 | 74375161 | 858 | 6 | 3.83E-06 | 0.0655886 | 5.08E-05 | 0.000267 | -0.00250564 | DCTN1 |
| chr4 | 87421377 | 87421533 | 157 | 2 | 5.11E-05 | 0.0004901 | 0.000423 | 0.000268 | -0.0105379 | NA |
| chr4 | 48907022 | 48907129 | 108 | 4 | 8.16E-05 | 0.0012016 | 0.002719 | 0.000268 | -0.00468342 | NA |
| chr9 | 94508149 | 94508544 | 396 | 2 | 5.05E-05 | 0.0018049 | 0.00017 | 0.000269 | -0.00288233 | AL358232.1, AL358232.2 |
| chr12 | 32485735 | 32485905 | 171 | 4 | 8.63E-05 | 0.0095594 | 5.13E-05 | 0.000269 | -0.00319768 | FGD4 |
| chr7 | 96996643 | 96997738 | 1096 | 6 | 1.40E-05 | 0.0181301 | 0.000385 | 0.000269 | -0.00260539 | DLX6-AS1 |
| chr13 | 30133128 | 30133448 | 321 | 2 | 2.86E-05 | 0.0002306 | 0.001223 | 0.000269 | -0.00490008 | NA |
| chr10 | 1056895 | 1057962 | 1068 | 5 | 2.06E-05 | 0.0001792 | 0.00315 | 0.00027 | -0.00311279 | WDR37 |
| chr16 | 2252392 | 2252947 | 556 | 3 | 5.03E-05 | 0.0019598 | 0.001418 | 0.00027 | 0.002045484 | NA |
| chr16 | 15948910 | 15949099 | 190 | 2 | 2.76E-05 | 0.0001789 | 0.002269 | 0.00027 | 0.016002922 | NA |
| chr16 | 74700353 | 74700987 | 635 | 4 | 2.00E-05 | 0.1390224 | 2.03E-05 | 0.00027 | -0.00062285 | MLKL |
| chr10 | 104642284 | 104642410 | 127 | 2 | 9.41E-05 | 0.001046 | 0.000233 | 0.000271 | 0.004410995 | SORCS3 |
| chr16 | 87391402 | 87391689 | 288 | 4 | 7.54E-05 | 0.000472 | 0.000929 | 0.000271 | 0.0050517 | MAP1LC3B, FBXO31 |
| chr8 | 123395483 | 123395953 | 471 | 3 | 1.57E-05 | 0.0009136 | 0.000958 | 0.000271 | 0.002201387 | ATAD2 |
| chr8 | 81720104 | 81720739 | 636 | 2 | 1.30E-05 | 0.0019156 | 0.000167 | 0.000272 | 0.003674023 | ZFAND1 |
| chr10 | 132960349 | 132960392 | 44 | 2 | 0.000100906 | 0.0002148 | 0.001456 | 0.000272 | -0.00473867 | LINC01166 |
| chr22 | 31631475 | 31631619 | 145 | 5 | 4.34E-05 | 0.0023448 | 0.003627 | 0.000272 | 0.00208494 | PISD |
| chr10 | 102120070 | 102120372 | 303 | 3 | 1.01E-05 | 0.0019311 | 0.000202 | 0.000272 | -0.00315862 | LDB1 |
| chr1 | 200890874 | 200891554 | 681 | 8 | 1.52E-05 | 0.0104633 | 4.53E-05 | 0.000273 | 0.000695979 | INAVA |
| chr12 | 106683978 | 106684875 | 898 | 6 | 8.17E-05 | 0.0046838 | 0.002107 | 0.000274 | -0.00134176 | RFX4, AC079385.1, AC079385.2 |
| chr17 | 81846237 | 81846546 | 310 | 3 | 2.93E-05 | 0.0002307 | 0.004425 | 0.000275 | -0.00470563 | P4HB |
| chr8 | 123073051 | 123073429 | 379 | 5 | 4.11E-05 | 0.0008751 | 0.000447 | 0.000275 | -0.00123725 | TBC1D31 |
| chr19 | 42267691 | 42268294 | 604 | 3 | 7.94E-06 | 0.0026321 | 0.000169 | 0.000276 | -0.00517577 | NA |
| chr5 | 160008260 | 160008426 | 167 | 3 | 1.62E-05 | 0.0015955 | 0.002806 | 0.000276 | -0.00382714 | NA |
| chr5 | 152599256 | 152599677 | 422 | 2 | 0.000126624 | 0.0001509 | 0.004812 | 0.000276 | -0.00231486 | AC008571.2 |
| chr15 | 60509500 | 60509544 | 45 | 2 | 0.000119019 | 0.0008681 | 0.000273 | 0.000277 | -0.00334173 | RORA-AS1, RORA |
| chr12 | 131799267 | 131799550 | 284 | 4 | 8.32E-05 | 0.0004479 | 0.001265 | 0.000277 | -0.00032784 | SFSWAP |
| chr5 | 181200716 | 181200784 | 69 | 3 | 0.000154688 | 9.63E-05 | 0.012935 | 0.000277 | -0.00339234 | TRIM7-AS1, TRIM7 |
| chr9 | 137010168 | 137010346 | 179 | 3 | 0.000135469 | 0.0001031 | 0.010082 | 0.000277 | 0.00107932 | ABCA2 |
| chr6 | 28266747 | 28267249 | 503 | 8 | 3.24E-05 | 0.0088338 | 0.000315 | 0.000278 | 0.006032455 | ZSCAN26, AL021997.2 |
| chr2 | 74466205 | 74466352 | 148 | 3 | 0.000110571 | 0.0059333 | 0.000361 | 0.000279 | 0.004811626 | AC005041.5 |
| chr6 | 28988800 | 28989027 | 228 | 6 | 9.49E-05 | 0.0006609 | 0.003321 | 0.000281 | 0.004225039 | NA |
| chr1 | 155308485 | 155308553 | 69 | 2 | 0.000145287 | 0.0010098 | 0.000252 | 0.000282 | 0.002455904 | NA |
| chr1 | 109934023 | 109934046 | 24 | 2 | 0.000150505 | 0.0001552 | 0.004662 | 0.000282 | -0.00679866 | NA |
| chr11 | 774038 | 774070 | 33 | 2 | 0.000150876 | 0.0124561 | 0.0001 | 0.000282 | -0.00279688 | GATD1 |
| chr2 | 143935953 | 143936126 | 174 | 2 | 4.60E-05 | 0.0004375 | 0.000524 | 0.000282 | 0.001482268 | NA |
| chr8 | 64586151 | 64587144 | 994 | 6 | 5.93E-06 | 0.0015736 | 0.000113 | 0.000282 | -0.00356351 | NA |
| chr6 | 12033892 | 12034041 | 150 | 2 | 8.06E-05 | 0.000213 | 0.001654 | 0.000283 | -0.00070964 | HIVEP1 |
| chr8 | 102655825 | 102656300 | 476 | 10 | 6.32E-05 | 0.0014209 | 0.008434 | 0.000283 | 0.000582122 | NA |
| chr6 | 15245851 | 15245928 | 78 | 5 | 0.000130734 | 0.0009552 | 0.002869 | 0.000283 | 0.001182325 | NA |
| chr17 | 75286819 | 75286932 | 114 | 3 | 9.29E-05 | 0.000433 | 0.000867 | 0.000283 | -0.00177944 | SLC25A19 |
| chr16 | 3646401 | 3646411 | 11 | 2 | 0.000162197 | 0.0001627 | 0.003841 | 0.000283 | 0.001388476 | DNASE1 |
| chr2 | 208358855 | 208359501 | 647 | 3 | 3.48E-05 | 0.0009357 | 0.000293 | 0.000284 | 0.002056029 | NA |
| chr3 | 119279985 | 119280063 | 79 | 2 | 0.000158158 | 0.0048107 | 0.000125 | 0.000284 | -0.00633334 | B4GALT4-AS1 |
| chr6 | 2415047 | 2415171 | 125 | 3 | 0.000146351 | 0.0002948 | 0.00172 | 0.000284 | 0.001289072 | GMDS-DT |
| chr2 | 85133929 | 85134318 | 390 | 3 | 4.12E-05 | 0.0025531 | 0.000153 | 0.000285 | -0.00024624 | TCF7L1 |
| chr2 | 74198622 | 74198636 | 15 | 2 | 0.000160118 | 0.000984 | 0.000261 | 0.000285 | 0.001110185 | MTHFD2 |
| chr4 | 6781545 | 6782448 | 904 | 5 | 1.18E-05 | 0.0001765 | 0.01081 | 0.000285 | 0.003475407 | KIAA0232 |
| chr19 | 49680567 | 49680796 | 230 | 3 | 7.21E-05 | 0.0066602 | 0.000492 | 0.000286 | -0.00212412 | PRMT1 |
| chr3 | 184134541 | 184134968 | 428 | 7 | 7.76E-05 | 0.0012265 | 0.010055 | 0.000286 | 0.000409463 | AC131235.2 |
| chr19 | 7068662 | 7068948 | 287 | 2 | 4.13E-05 | 0.0001867 | 0.002465 | 0.000287 | -0.00919165 | NA |
| chr15 | 100601433 | 100601750 | 318 | 3 | 3.03E-05 | 0.0020298 | 0.000229 | 0.000287 | 0.001764916 | LINS1 |
| chr8 | 142451374 | 142451471 | 98 | 2 | 0.000159477 | 0.0002088 | 0.001831 | 0.000288 | 0.005030982 | ADGRB1 |
| chr6 | 40378560 | 40378770 | 211 | 2 | 5.04E-05 | 0.0003268 | 0.00079 | 0.000288 | -0.00201499 | TDRG1 |
| chr2 | 191677822 | 191677900 | 79 | 6 | 0.000164931 | 0.0003637 | 0.002721 | 0.000288 | -0.00017008 | NA |
| chr1 | 3509039 | 3509197 | 159 | 2 | 4.84E-05 | 0.0001583 | 0.004752 | 0.000288 | -0.00044786 | MEGF6 |
| chr2 | 240465355 | 240465779 | 425 | 3 | 4.98E-05 | 0.0100474 | 7.91E-05 | 0.000288 | -0.00163312 | GPC1 |
| chr2 | 234171653 | 234171738 | 86 | 2 | 0.000161142 | 0.000969 | 0.000268 | 0.000288 | -0.00548892 | NA |
| chr6 | 34520926 | 34521338 | 413 | 2 | 7.95E-05 | 0.0003947 | 0.000618 | 0.000288 | 0.003254531 | PACSIN1 |
| chr9 | 127754416 | 127754537 | 122 | 2 | 2.66E-05 | 0.0005429 | 0.000435 | 0.000289 | -0.00348991 | AL162586.2, SH2D3C |
| chr11 | 47152877 | 47153099 | 223 | 2 | 8.27E-05 | 0.0020293 | 0.000175 | 0.000289 | -0.00449654 | C11orf49 |
| chr4 | 6690583 | 6690693 | 111 | 3 | 0.000101767 | 0.0098911 | 6.46E-05 | 0.000289 | -0.00047039 | NA |
| chr10 | 84138743 | 84139135 | 393 | 4 | 7.99E-06 | 0.011215 | 0.000112 | 0.000289 | -0.00600642 | AL603756.1 |
| chr14 | 99604849 | 99605107 | 259 | 3 | 5.45E-05 | 0.0001931 | 0.006135 | 0.000291 | -0.00213928 | AL160313.1 |
| chr17 | 43399903 | 43399982 | 80 | 3 | 2.79E-05 | 0.0239464 | 5.79E-05 | 0.000292 | -0.00184733 | ARL4D |
| chr1 | 209674650 | 209675134 | 485 | 3 | 3.66E-05 | 0.000732 | 0.000415 | 0.000292 | 0.008031512 | HSD11B1-AS1 |
| chr17 | 17681191 | 17682331 | 1141 | 6 | 9.69E-06 | 0.0007873 | 0.009024 | 0.000292 | 0.000675657 | RAI1 |
| chr1 | 50104940 | 50104959 | 20 | 2 | 0.000112455 | 0.0001605 | 0.00479 | 0.000292 | -0.00747815 | ELAVL4 |
| chr20 | 31849481 | 31849757 | 277 | 2 | 5.80E-05 | 0.0001893 | 0.002533 | 0.000292 | -0.01903762 | DUSP15 |
| chr7 | 157568348 | 157569000 | 653 | 7 | 2.13E-05 | 0.0034022 | 0.007679 | 0.000292 | 0.00471831 | PTPRN2 |
| chr9 | 105447424 | 105447503 | 80 | 2 | 0.000122497 | 0.0004724 | 0.000513 | 0.000292 | -0.00654611 | NA |
| chr16 | 7899629 | 7900243 | 615 | 3 | 1.35E-05 | 0.0003915 | 0.001056 | 0.000292 | -0.00903428 | AC093515.1 |
| chr4 | 3307286 | 3307677 | 392 | 3 | 9.06E-05 | 0.0039711 | 0.000247 | 0.000293 | 0.003784791 | RGS12 |
| chr20 | 54157302 | 54157469 | 168 | 2 | 6.21E-05 | 0.0038746 | 0.000138 | 0.000293 | -0.00726773 | CYP24A1 |
| chr1 | 30716766 | 30717314 | 549 | 2 | 3.21E-06 | 0.0063025 | 0.00012 | 0.000294 | -0.00511451 | MATN1 |
| chr22 | 31495343 | 31496088 | 746 | 8 | 3.28E-05 | 0.0114195 | 0.012271 | 0.000294 | 0.004377696 | DRG1, SFI1, EIF4ENIF1 |
| chr1 | 74198315 | 74198772 | 458 | 7 | 2.57E-05 | 0.0002494 | 0.008549 | 0.000294 | 0.002569341 | FPGT, FPGT-TNNI3K |
| chr3 | 197734762 | 197735313 | 552 | 2 | 1.41E-05 | 0.0039186 | 0.000138 | 0.000294 | 0.002311781 | RUBCN |
| chr16 | 56932563 | 56933088 | 526 | 2 | 1.68E-05 | 0.0007265 | 0.000345 | 0.000294 | -0.00063363 | HERPUD1 |
| chr1 | 204411821 | 204412124 | 304 | 9 | 0.000104591 | 0.0043372 | 0.006084 | 0.000295 | -0.00229183 | AL606489.1 |
| chr11 | 93741293 | 93742060 | 768 | 17 | 3.50E-05 | 0.017475 | 0.000488 | 0.000295 | 0.000292929 | C11orf54, AP001273.2, TAF1D |
| chr8 | 26049474 | 26049798 | 325 | 3 | 5.70E-05 | 0.0005612 | 0.000607 | 0.000296 | -0.00238802 | NA |
| chr12 | 109439461 | 109439475 | 15 | 2 | 0.000114564 | 0.0004253 | 0.000594 | 0.000296 | 0.003691396 | MYO1H |
| chr6 | 57089184 | 57089193 | 10 | 3 | 4.70E-05 | 0.000423 | 0.003912 | 0.000296 | 0.009436256 | ZNF451 |
| chr17 | 28365406 | 28365823 | 418 | 3 | 5.68E-05 | 0.000431 | 0.000821 | 0.000297 | 0.000101652 | AC002094.1, SARM1, AC002094.3 |
| chr7 | 42911260 | 42911837 | 578 | 5 | 1.75E-05 | 0.0016378 | 0.004179 | 0.000297 | 3.23E-05 | C7orf25, AC010132.3 |
| chr20 | 45779815 | 45779898 | 84 | 2 | 6.97E-05 | 0.0223107 | 9.77E-05 | 0.000297 | -0.0067415 | WFDC3 |
| chr19 | 662740 | 663394 | 655 | 6 | 7.96E-05 | 0.0002011 | 0.015947 | 0.000297 | 0.001187449 | RNF126 |
| chr11 | 5507084 | 5507439 | 356 | 2 | 4.62E-05 | 0.0010153 | 0.000272 | 0.000298 | -0.00720116 | AC104389.5, UBQLN3 |
| chr5 | 178589594 | 178589795 | 202 | 2 | 2.93E-05 | 0.0025023 | 0.000166 | 0.000298 | -0.00376509 | COL23A1 |
| chr17 | 4641669 | 4642058 | 390 | 6 | 4.65E-05 | 0.0028848 | 0.000724 | 0.000298 | -0.00108079 | ALOX15 |
| chr17 | 64007876 | 64008172 | 297 | 4 | 0.000155582 | 0.0076603 | 0.000168 | 0.000299 | -0.00466975 | ICAM2 |
| chr7 | 65983157 | 65983628 | 472 | 2 | 8.05E-05 | 0.0004491 | 0.000568 | 0.0003 | 0.00125334 | NA |
| chr16 | 85399365 | 85399703 | 339 | 3 | 2.97E-05 | 0.0056893 | 0.002194 | 0.0003 | -0.00931423 | GSE1 |
| chr17 | 1769708 | 1770398 | 691 | 3 | 1.35E-05 | 0.0041837 | 0.000119 | 0.0003 | 0.007192185 | SERPINF1 |
| chr12 | 53221245 | 53221695 | 451 | 4 | 3.73E-05 | 0.000954 | 0.000456 | 0.000301 | 0.003520201 | RARG |
| chr12 | 106140125 | 106140996 | 872 | 5 | 2.73E-05 | 0.000525 | 0.011355 | 0.000302 | -0.00805894 | NA |
| chr15 | 82711236 | 82711311 | 76 | 3 | 0.000148813 | 0.0009302 | 0.002755 | 0.000303 | -0.00082492 | AC105339.1 |
| chr12 | 9234663 | 9235003 | 341 | 3 | 5.68E-06 | 0.046893 | 5.42E-05 | 0.000303 | 0.001437947 | A2MP1 |
| chr20 | 5445853 | 5446442 | 590 | 5 | 1.30E-05 | 0.0142208 | 0.00047 | 0.000303 | -0.00492306 | AL121757.2, AL121757.1, LINC00658 |
| chr15 | 55270205 | 55271096 | 892 | 7 | 4.67E-05 | 0.0076918 | 4.67E-05 | 0.000305 | 0.001387056 | RAB27A |
| chr20 | 50730834 | 50731651 | 818 | 7 | 5.01E-05 | 0.0001745 | 0.00883 | 0.000306 | 0.000673073 | PARD6B |
| chr20 | 23350534 | 23350705 | 172 | 6 | 0.000149227 | 0.0022241 | 0.004323 | 0.000306 | -0.00068487 | NXT1-AS1 |
| chr11 | 65786189 | 65786290 | 102 | 2 | 7.34E-05 | 0.0603126 | 9.38E-05 | 0.000306 | -0.01005346 | NA |
| chr16 | 2531731 | 2532141 | 411 | 3 | 1.40E-05 | 0.0006517 | 0.000435 | 0.000307 | 0.000565381 | MIR3178 |
| chr4 | 25030850 | 25030942 | 93 | 3 | 0.000137867 | 0.000715 | 0.000581 | 0.000309 | 0.002395754 | LGI2 |
| chr3 | 32502434 | 32502670 | 237 | 4 | 0.000130101 | 0.0183421 | 4.77E-05 | 0.000309 | -0.00127593 | CMTM6 |
| chr17 | 79024785 | 79024956 | 172 | 2 | 1.57E-05 | 0.000541 | 0.000491 | 0.000309 | 0.009533122 | C1QTNF1, C1QTNF1-AS1 |
| chr10 | 13586544 | 13587255 | 712 | 10 | 3.48E-05 | 0.0179135 | 0.000416 | 0.000309 | 0.000283449 | PRPF18 |

| | | | | | | | | | | |
|---|---|---|---|---|---|---|---|---|---|---|
| chr3 | 119293078 | 119293202 | 125 | 2 | 3.42E-05 | 0.0019038 | 0.000197 | 0.00031 | 0.007933035 | NA |
| chr8 | 77001026 | 77001105 | 80 | 5 | 6.96E-05 | 0.0002912 | 0.005632 | 0.00031 | -0.01493119 | PEX2 |
| chr2 | 3479826 | 3480196 | 371 | 3 | 8.68E-05 | 0.0030284 | 0.003065 | 0.00031 | 0.005239662 | TRAPPC12 |
| chr6 | 122995843 | 122996067 | 225 | 7 | 0.000123591 | 0.0025453 | 0.000142 | 0.00031 | 0.0026327 | AL591428.1 |
| chr16 | 88535038 | 88535121 | 84 | 2 | 7.52E-05 | 0.0005813 | 0.00046 | 0.000311 | -0.01012743 | ZFPM1 |
| chr6 | 75927498 | 75927582 | 85 | 2 | 0.000122653 | 0.0002893 | 0.001144 | 0.000311 | 0.007060586 | IMPG1 |
| chr14 | 50559173 | 50559308 | 136 | 3 | 6.47E-05 | 0.0032282 | 0.000121 | 0.000311 | -0.00195689 | ATL1, MAP4K5 |
| chr17 | 82795361 | 82795487 | 127 | 2 | 1.13E-05 | 0.0060963 | 0.000129 | 0.000312 | -0.00224028 | TBCD, AC068014.1 |
| chr9 | 136848127 | 136848773 | 647 | 7 | 5.06E-06 | 0.0007635 | 0.009937 | 0.000313 | 0.00161138 | AJM1, PHPT1 |
| chr3 | 148990304 | 148990454 | 151 | 2 | 4.64E-05 | 0.0001778 | 0.004331 | 0.000314 | 0.009189552 | NA |
| chr20 | 11918203 | 11918610 | 408 | 2 | 1.81E-06 | 0.0001872 | 0.003525 | 0.000314 | 0.000784503 | BTBD3, AL035448.1 |
| chr5 | 177491919 | 177492581 | 663 | 4 | 3.27E-05 | 0.0103576 | 0.003341 | 0.000315 | -0.00059708 | PDLIM7 |
| chr20 | 36051402 | 36051764 | 363 | 2 | 1.13E-05 | 0.0009789 | 0.000302 | 0.000315 | -0.00811784 | AL035420.1 |
| chr7 | 27129589 | 27130055 | 467 | 6 | 9.72E-05 | 0.0080434 | 6.69E-05 | 0.000315 | -0.00164002 | HOXA-AS2, HOXA-AS3, HOXA3, HOXA4, AC004080.6 |
| chr17 | 6556819 | 6557252 | 434 | 3 | 3.92E-05 | 0.0044468 | 0.00024 | 0.000316 | -0.00658766 | NA |
| chr1 | 23483940 | 23484378 | 439 | 4 | 8.69E-05 | 0.0016763 | 0.000289 | 0.000318 | 0.001969907 | ASAP3 |
| chr6 | 82248034 | 82248607 | 574 | 7 | 7.52E-06 | 0.0070251 | 0.000637 | 0.000318 | 0.005047263 | NA |
| chr19 | 10929336 | 10929918 | 583 | 5 | 3.10E-05 | 0.0014247 | 0.004682 | 0.000319 | -0.00585338 | TIMM29 |
| chr15 | 22880225 | 22880471 | 247 | 2 | 0.000152706 | 0.0015801 | 0.000226 | 0.000319 | 0.002519466 | CYFIP1 |
| chr5 | 54310809 | 54311323 | 515 | 4 | 1.70E-05 | 0.0004368 | 0.001345 | 0.000321 | 0.000936886 | NA |
| chr12 | 21817385 | 21817468 | 84 | 2 | 0.000171109 | 0.0001825 | 0.00433 | 0.000322 | -0.0049825 | ABCC9 |
| chr2 | 189179365 | 189179778 | 414 | 3 | 2.34E-05 | 0.0060594 | 0.000415 | 0.000322 | -0.00711552 | COL5A2 |
| chr5 | 141401612 | 141401626 | 15 | 2 | 0.000152839 | 0.0016821 | 0.000221 | 0.000322 | -0.02543582 | PCDHGA1, PCDHGA2, PCDHGA3, PCDHGB1, PCDHGA4, PCDHGB2, PCDHGA5, PCDHGB3, PCDHGA6, PCDHGA7, PCDHGB4, PCDHGA8, PCDHGB5 |
| chr15 | 88255100 | 88255435 | 336 | 4 | 0.000138651 | 0.0004618 | 0.002104 | 0.000323 | -0.00181849 | NTRK3-AS1, NTRK3 |
| chr16 | 6017746 | 6017800 | 55 | 2 | 8.00E-05 | 0.003787 | 0.000155 | 0.000323 | 0.007346644 | RBFOX1 |
| chr22 | 18078342 | 18078643 | 302 | 3 | 8.30E-05 | 0.0007021 | 0.000527 | 0.000324 | 0.000112476 | PEX26, AC016027.1 |
| chr5 | 160312694 | 160312844 | 151 | 5 | 9.28E-05 | 0.0010441 | 0.001398 | 0.000324 | -0.00105559 | CCNJL |
| chr14 | 100734095 | 100734380 | 286 | 2 | 0.000159664 | 0.0002142 | 0.002538 | 0.000324 | 0.002580568 | DLK1 |
| chr10 | 129872686 | 129873352 | 667 | 4 | 3.26E-05 | 0.1494789 | 3.39E-05 | 0.000326 | 0.00305612 | EBF3 |
| chr2 | 73779538 | 73780197 | 660 | 10 | 3.10E-05 | 0.0008588 | 0.00713 | 0.000327 | 0.00058647 | DUSP11 |
| chr15 | 78338837 | 78339284 | 448 | 6 | 6.64E-05 | 0.0022448 | 0.000386 | 0.000327 | 0.003085337 | AC011270.2 |
| chr10 | 96043561 | 96043768 | 208 | 2 | 9.05E-05 | 0.0001791 | 0.005121 | 0.000327 | -6.61E-05 | CCNJ, ENTPD1-AS1 |
| chr11 | 2889568 | 2889970 | 403 | 4 | 4.90E-05 | 0.0418529 | 6.29E-05 | 0.000327 | -0.00173001 | SLC22A18AS |
| chr11 | 71240969 | 71241129 | 161 | 2 | 0.000124198 | 0.0001979 | 0.003426 | 0.000328 | -0.00139244 | SHANK2 |
| chr2 | 241618662 | 241618929 | 268 | 2 | 8.76E-06 | 0.0002492 | 0.001788 | 0.000329 | -0.00522803 | THAP4 |
| chr14 | 22883279 | 22883876 | 598 | 3 | 1.38E-06 | 0.0054037 | 0.000152 | 0.000329 | -0.00778961 | REM2 |
| chr12 | 57754105 | 57754606 | 502 | 2 | 8.88E-05 | 0.0001788 | 0.005387 | 0.000329 | 0.005529187 | CDK4 |
| chr19 | 15380021 | 15380997 | 977 | 3 | 6.40E-06 | 0.0046401 | 0.001764 | 0.00033 | -0.00407318 | AC005785.1, AKAP8L |
| chr16 | 21952844 | 21953471 | 628 | 10 | 8.24E-05 | 0.0015164 | 0.018488 | 0.00033 | 0.000392827 | UQCRC2 |
| chr7 | 159092252 | 159092890 | 639 | 2 | 2.84E-05 | 0.0006283 | 0.000472 | 0.00033 | -0.00316823 | VIPR2 |
| chr3 | 47578200 | 47578474 | 275 | 4 | 5.79E-05 | 0.0038564 | 0.00156 | 0.000331 | -0.00055536 | CSPG5 |
| chr15 | 66386626 | 66386836 | 211 | 5 | 8.54E-05 | 0.0709805 | 0.00028 | 0.000331 | 0.000173904 | TIPIN |
| chr20 | 32581762 | 32581837 | 76 | 2 | 8.52E-05 | 0.0180029 | 0.00011 | 0.000331 | 0.006622641 | NOL4L-DT, NOL4L |
| chr12 | 4446096 | 4446554 | 459 | 2 | 9.32E-05 | 0.0001891 | 0.004301 | 0.000332 | -0.00101041 | NA |
| chr2 | 121736638 | 121736803 | 166 | 3 | 0.00012009 | 0.000182 | 0.00384 | 0.000332 | -0.00142648 | NIFK |
| chr19 | 13506357 | 13507149 | 793 | 8 | 3.45E-05 | 0.0088314 | 0.001367 | 0.000332 | -0.0053897 | CACNA1A |
| chr18 | 63970460 | 63970921 | 462 | 3 | 4.53E-05 | 0.002875 | 0.001045 | 0.000332 | -0.00176088 | HMSD, SERPINB8 |
| chr1 | 157046051 | 157046367 | 317 | 3 | 0.000158381 | 0.0004308 | 0.003935 | 0.000334 | 0.004156431 | NA |
| chr11 | 121653242 | 121653408 | 167 | 2 | 5.69E-05 | 0.0124872 | 0.000118 | 0.000336 | -0.00657286 | NA |
| chr11 | 45681495 | 45681554 | 60 | 2 | 0.000103524 | 0.0695424 | 0.000101 | 0.000336 | -0.00244711 | NA |
| chr9 | 21396658 | 21396675 | 18 | 2 | 0.000106213 | 0.00023 | 0.002376 | 0.000338 | -0.01312335 | NA |
| chr5 | 131974978 | 131975231 | 254 | 2 | 2.48E-05 | 0.0088191 | 0.000127 | 0.000338 | -0.00496712 | AC034228.3, ACSL6 |
| chr5 | 141374747 | 141374884 | 138 | 2 | 0.000156533 | 0.0002333 | 0.002295 | 0.000338 | -0.02636007 | PCDHGA1, PCDHGA2, PCDHGA3, PCDHGB1, PCDHGA4, PCDHGB2, PCDHGA5, PCDHGB3, PCDHGA6 |
| chr14 | 77707408 | 77708009 | 602 | 9 | 3.14E-05 | 0.0012664 | 0.011844 | 0.000338 | 0.000249253 | ALKBH1 |
| chr17 | 1125942 | 1126409 | 468 | 3 | 4.72E-05 | 0.0017003 | 0.001488 | 0.000339 | -0.00369589 | ABR |
| chr1 | 2212351 | 2212620 | 270 | 3 | 9.94E-05 | 0.0002312 | 0.003491 | 0.000339 | -0.00302556 | AL590822.1, FAAP20 |
| chr10 | 100987948 | 100988391 | 444 | 3 | 1.12E-05 | 0.0002154 | 0.002997 | 0.00034 | -0.0073723 | TWNK |
| chr3 | 160566505 | 160566594 | 90 | 2 | 0.000103994 | 0.0007125 | 0.00044 | 0.00034 | -0.02180963 | KRT8P12 |
| chr14 | 21070866 | 21071553 | 688 | 8 | 3.72E-05 | 0.0011626 | 0.001558 | 0.000341 | 0.000749631 | ARHGEF40, NDRG2 |
| chr2 | 190882497 | 190882998 | 502 | 3 | 2.33E-05 | 0.0009962 | 0.000375 | 0.000342 | 0.008318319 | GLS |
| chr4 | 75672775 | 75673069 | 295 | 2 | 2.51E-05 | 0.0002013 | 0.003938 | 0.000344 | 0.000807928 | G3BP2 |
| chr18 | 51029989 | 51030594 | 606 | 8 | 3.60E-05 | 0.0040545 | 0.001592 | 0.000344 | 0.001363518 | AC091551.1, SMAD4 |
| chr15 | 64912171 | 64912229 | 59 | 3 | 0.00015811 | 0.0046145 | 0.000218 | 0.000345 | -0.00887774 | AC069368.1, ANKDD1A |
| chr11 | 3028538 | 3028666 | 129 | 2 | 0.000119277 | 0.000493 | 0.000666 | 0.000346 | 0.002640744 | CARS1 |
| chr10 | 8058292 | 8058798 | 507 | 5 | 1.34E-05 | 0.0007062 | 0.004538 | 0.000347 | -0.00304151 | GATA3 |
| chr20 | 3889025 | 3889312 | 288 | 6 | 8.72E-05 | 0.0003285 | 0.000961 | 0.000348 | 0.000121145 | PANK2 |
| chr15 | 40238753 | 40239329 | 577 | 12 | 8.67E-05 | 0.0017063 | 0.003656 | 0.000349 | -4.50E-05 | PAK6, BUB1B-PAK6 |
| chr17 | 83012558 | 83013016 | 459 | 4 | 0.000135602 | 0.0006894 | 0.000554 | 0.00035 | -0.00388038 | B3GNTL1 |
| chr1 | 161726049 | 161726512 | 464 | 5 | 2.10E-05 | 0.0007616 | 0.005244 | 0.00035 | 0.001717734 | FCRLB |
| chr20 | 17967599 | 17968557 | 959 | 5 | 2.55E-06 | 0.0008778 | 0.003704 | 0.00035 | -0.00189104 | SNX5, OVOL2 |
| chr17 | 38825777 | 38825820 | 44 | 2 | 0.000114611 | 0.0003179 | 0.001294 | 0.000351 | -0.00799875 | NA |
| chr19 | 58154276 | 58154842 | 567 | 3 | 6.62E-06 | 0.0050783 | 0.000142 | 0.000351 | -0.00718643 | ZNF329 |
| chr17 | 74211379 | 74211456 | 78 | 2 | 9.14E-05 | 0.0001919 | 0.005405 | 0.000352 | 0.000435369 | MGC16275 |
| chr6 | 49636729 | 49637009 | 281 | 2 | 9.01E-05 | 0.0001923 | 0.005404 | 0.000352 | -0.00247699 | RHAG |
| chr12 | 118775288 | 118775355 | 68 | 3 | 2.05E-05 | 0.0158909 | 0.000157 | 0.000353 | 0.001470536 | NA |
| chr7 | 99551568 | 99551792 | 225 | 3 | 6.15E-05 | 0.0437497 | 0.00018 | 0.000353 | -0.00280581 | FAM200A |
| chr5 | 59216868 | 59216980 | 113 | 2 | 2.58E-05 | 0.0115097 | 0.000126 | 0.000354 | -0.0052409 | PDE4D |
| chr15 | 43825790 | 43825867 | 78 | 2 | 3.66E-05 | 0.0001965 | 0.004993 | 0.000354 | -0.00514492 | NA |
| chr2 | 98730172 | 98730959 | 788 | 4 | 3.47E-05 | 0.0142846 | 4.79E-05 | 0.000354 | 0.001045812 | MGAT4A |
| chr6 | 168520267 | 168520557 | 291 | 2 | 7.98E-05 | 0.0005447 | 0.000621 | 0.000355 | -0.00620228 | SMOC2 |
| chr3 | 4977015 | 4977809 | 795 | 7 | 2.96E-05 | 0.0097015 | 0.002144 | 0.000355 | -0.00061875 | BHLHE40-AS1 |
| chr17 | 48054221 | 48054367 | 147 | 2 | 0.000120885 | 0.0008656 | 0.0004 | 0.000357 | -0.00249731 | NFE2L1 |
| chr1 | 2906141 | 2906257 | 117 | 2 | 0.000125719 | 0.0001992 | 0.004917 | 0.000357 | 0.011025554 | NA |
| chr7 | 144835840 | 144836378 | 539 | 10 | 9.69E-05 | 0.0020486 | 0.003447 | 0.000358 | 0.000211659 | TPK1 |
| chr2 | 19346869 | 19346943 | 75 | 3 | 0.000147799 | 0.0042582 | 0.001055 | 0.000359 | 0.006807487 | LINC01376 |
| chr7 | 102395603 | 102395908 | 306 | 4 | 6.06E-05 | 0.0003844 | 0.001102 | 0.00036 | -0.00610467 | PRKRIP1 |
| chr7 | 157558173 | 157558348 | 176 | 3 | 6.56E-05 | 0.0104743 | 0.002746 | 0.000362 | 0.000771298 | PTPRN2 |
| chr15 | 60398468 | 60398775 | 308 | 3 | 9.30E-05 | 0.0001694 | 0.008885 | 0.000363 | 0.002043646 | ANXA2 |
| chr7 | 4811313 | 4811381 | 69 | 3 | 0.000101714 | 0.0017883 | 0.000332 | 0.000364 | 0.007678034 | RADIL |
| chr7 | 159144802 | 159144918 | 117 | 2 | 2.97E-05 | 0.0002075 | 0.004531 | 0.000364 | -0.00046878 | VIPR2 |
| chr12 | 3365897 | 3366346 | 450 | 4 | 7.09E-05 | 0.0007614 | 0.000754 | 0.000365 | 0.00151135 | AC005865.1 |
| chr7 | 47762743 | 47763297 | 555 | 4 | 2.79E-05 | 0.012639 | 8.78E-05 | 0.000366 | -0.00030928 | LINC00525, PKD1L1 |
| chr3 | 52518670 | 52519417 | 748 | 4 | 1.46E-05 | 0.0714177 | 2.68E-05 | 0.000366 | 0.002177573 | STAB1 |
| chr16 | 35746543 | 35747023 | 481 | 3 | 9.65E-05 | 0.0003333 | 0.005473 | 0.000366 | -0.00715874 | LINC02167, RNA5SP411, RNA5SP412 |
| chr19 | 12956740 | 12956869 | 130 | 4 | 0.0001567 | 0.0018 | 0.003488 | 0.000367 | -0.00282118 | GADD45GIP1 |
| chr1 | 219928356 | 219928443 | 88 | 4 | 0.000120843 | 0.0001984 | 0.013956 | 0.000369 | 0.000801128 | SLC30A10 |
| chr17 | 4710345 | 4711127 | 783 | 8 | 4.51E-05 | 0.0014534 | 0.010004 | 0.000369 | 0.002436479 | ARRB2 |
| chr14 | 22835614 | 22835944 | 331 | 2 | 5.97E-05 | 0.0007919 | 0.00046 | 0.000371 | -0.00365082 | NA |
| chr16 | 1310922 | 1311062 | 141 | 2 | 4.59E-05 | 0.0023434 | 0.000223 | 0.000371 | 0.002448188 | UBE2I |
| chr2 | 30820553 | 30820895 | 343 | 3 | 0.000108258 | 0.0002234 | 0.007536 | 0.000372 | 0.003515322 | NA |
| chr20 | 62860852 | 62861317 | 466 | 2 | 2.75E-05 | 0.0031131 | 0.000197 | 0.000373 | -0.00840431 | TCFL5 |
| chr10 | 104268761 | 104269181 | 421 | 9 | 5.24E-05 | 0.0057549 | 0.000413 | 0.000373 | 0.000688177 | GSTO2 |
| chr1 | 89632289 | 89632422 | 134 | 4 | 0.000100949 | 0.0004304 | 0.00443 | 0.000373 | -0.00566012 | AC093423.2, LRRC8C-DT |
| chr19 | 40391344 | 40391350 | 7 | 2 | 9.43E-05 | 0.0006228 | 0.000588 | 0.000374 | -0.00190474 | NA |
| chr1 | 147598729 | 147598943 | 215 | 2 | 9.95E-05 | 0.0008573 | 0.000434 | 0.000374 | -0.00810695 | BCL9 |
| chr19 | 51084688 | 51084753 | 66 | 2 | 7.05E-05 | 0.0062536 | 0.000155 | 0.000375 | -0.00523241 | NA |
| chr1 | 3288979 | 3289369 | 391 | 2 | 4.08E-05 | 0.009132 | 0.000141 | 0.000375 | 0.005843282 | PRDM16 |
| chr12 | 106986485 | 106986721 | 237 | 2 | 4.96E-05 | 0.0011947 | 0.000338 | 0.000376 | 0.003669338 | MTERF2 |
| chr7 | 135977268 | 135977814 | 547 | 6 | 9.69E-05 | 0.0033471 | 0.000618 | 0.000376 | 0.001074125 | MTPN |
| chr12 | 98647347 | 98647866 | 520 | 3 | 9.75E-05 | 0.0725876 | 6.29E-05 | 0.000377 | -0.00156239 | APAF1 |
| chr4 | 7042208 | 7042868 | 661 | 2 | 8.45E-07 | 0.0040033 | 0.000181 | 0.000378 | -0.00694695 | TADA2B, AC097382.2, CCDC96 |
| chr2 | 160270290 | 160270346 | 57 | 2 | 0.000167229 | 0.0005849 | 0.000643 | 0.000379 | 0.004944714 | AC092153.1, LINC02478 |
| chr16 | 58629031 | 58629084 | 54 | 2 | 3.46E-05 | 0.0441717 | 0.000114 | 0.000379 | 0.004706057 | CNOT1 |
| chr19 | 3062810 | 3062885 | 76 | 2 | 5.01E-05 | 0.000513 | 0.000759 | 0.000381 | 0.001712138 | TLE5 |
| chr1 | 174447908 | 174448237 | 330 | 4 | 1.58E-05 | 0.0979082 | 2.12E-05 | 0.000381 | -0.00170979 | RABGAP1L, GPR52 |
| chr10 | 89645520 | 89645676 | 157 | 4 | 0.000114394 | 0.0001773 | 0.010934 | 0.000382 | -0.0002323 | AL157400.2, PANK1 |
| chr6 | 5996424 | 5996795 | 372 | 3 | 0.000111235 | 0.0004314 | 0.001084 | 0.000384 | -4.35E-06 | NA |
| chr14 | 64914311 | 64914890 | 580 | 10 | 1.05E-05 | 0.0010257 | 0.000991 | 0.000384 | 0.000556035 | CHURC1, CHURC1-FNTB |
| chr14 | 74816973 | 74817317 | 345 | 2 | 0.000114668 | 0.0002219 | 0.004403 | 0.000384 | -0.00060829 | YLPM1 |
| chr16 | 30182743 | 30183095 | 353 | 3 | 9.78E-06 | 0.005181 | 0.000291 | 0.000385 | -0.00822523 | CORO1A |
| chr8 | 100908771 | 100909030 | 260 | 2 | 6.55E-05 | 0.0008161 | 0.000476 | 0.000386 | 0.011530039 | NA |
| chr13 | 113759277 | 113759819 | 543 | 8 | 5.98E-05 | 0.0134436 | 0.000656 | 0.000386 | -0.00017958 | TMEM255B |
| chr1 | 156096807 | 156097195 | 389 | 3 | 6.01E-06 | 0.1020871 | 8.42E-05 | 0.000387 | 0.000687791 | LMNA |
| chr12 | 53058197 | 53058360 | 164 | 2 | 0.000124024 | 0.0210976 | 0.000125 | 0.000387 | -0.00344115 | TNS2 |
| chr19 | 6772359 | 6772919 | 561 | 6 | 3.12E-05 | 0.0282784 | 0.001227 | 0.000388 | 0.000734818 | VAV1 |
| chr11 | 528653 | 528924 | 272 | 4 | 9.90E-05 | 0.0086212 | 0.002261 | 0.000388 | 8.95E-05 | AC137894.1 |
| chr3 | 137770830 | 137770925 | 96 | 2 | 9.74E-05 | 0.0063628 | 0.00016 | 0.000388 | 0.003379813 | NA |
| chr2 | 241547355 | 241547454 | 100 | 2 | 0.000144235 | 0.0070635 | 0.000156 | 0.000388 | -0.01240944 | BOK-AS1 |
| chr6 | 84774082 | 84774569 | 488 | 3 | 7.21E-05 | 0.0019526 | 0.000565 | 0.000389 | 0.002850861 | NA |
| chr7 | 29566008 | 29566281 | 274 | 3 | 1.10E-05 | 0.0026527 | 0.000197 | 0.000389 | 0.009892412 | PRR15 |
| chr6 | 44263568 | 44263919 | 352 | 2 | 1.83E-05 | 0.0235167 | 0.000124 | 0.00039 | -0.01478934 | NFKBIE |
| chr9 | 137435043 | 137435118 | 76 | 2 | 4.73E-05 | 0.0195417 | 0.000127 | 0.00039 | 0.007940462 | ENTPD8 |
| chr19 | 1132538 | 1133210 | 673 | 5 | 2.70E-05 | 0.0013568 | 0.00064 | 0.000391 | 0.005069126 | SBNO2 |
| chr14 | 58244353 | 58244404 | 52 | 3 | 7.63E-05 | 0.0016733 | 0.000475 | 0.000391 | 0.004823414 | ARMH4 |
| chr19 | 48414559 | 48414859 | 301 | 2 | 7.45E-05 | 0.0005734 | 0.000701 | 0.000392 | -0.01045988 | GRIN2D |
| chr11 | 44564822 | 44565053 | 232 | 3 | 7.65E-05 | 0.0017984 | 0.000388 | 0.000392 | -0.00372121 | CD82 |
| chr10 | 131454069 | 131454256 | 188 | 4 | 7.73E-05 | 0.0378035 | 5.71E-05 | 0.000395 | -0.00060646 | NA |
| chr11 | 61553824 | 61553894 | 71 | 2 | 9.94E-05 | 0.098921 | 0.000115 | 0.000397 | 0.010575311 | SYT7 |
| chr20 | 37315177 | 37315736 | 560 | 4 | 7.79E-05 | 0.0029582 | 0.000201 | 0.000398 | 0.005268406 | MANBAL |
| chr12 | 132147462 | 132147588 | 127 | 2 | 0.000134034 | 0.0005419 | 0.000772 | 0.000398 | -0.00185821 | NOC4L |
| chr1 | 11980085 | 11980820 | 736 | 7 | 1.66E-05 | 0.0028745 | 0.000822 | 0.0004 | 0.000842692 | MFN2, AL096840.2 |
| chr11 | 96389794 | 96389871 | 78 | 3 | 0.00011437 | 0.0005545 | 0.000881 | 0.0004 | 0.000113027 | CCDC82 |
| chr2 | 170817330 | 170817669 | 340 | 4 | 3.86E-05 | 0.0052866 | 0.000301 | 0.000402 | 0.002407369 | GAD1, AC007405.2 |
| chr4 | 122378492 | 122379503 | 1012 | 17 | 6.08E-05 | 0.0266246 | 0.001495 | 0.000402 | -0.00116737 | ADAD1 |
| chr17 | 67825017 | 67826117 | 1101 | 4 | 0.000119255 | 0.0041457 | 0.001968 | 0.000404 | -0.00154597 | BPTF |
| chr21 | 44353686 | 44354114 | 429 | 3 | 5.82E-05 | 0.0001711 | 0.007825 | 0.000405 | -0.00554236 | TRPM2 |
| chr6 | 30206413 | 30206927 | 515 | 4 | 0.000124047 | 0.0002028 | 0.012845 | 0.000406 | 0.000807196 | TRIM26 |
| chr2 | 70978136 | 70978390 | 255 | 5 | 0.000158907 | 0.0001834 | 0.01476 | 0.000406 | 0.001993714 | ANKRD53, AC007040.2 |
| chr11 | 64035886 | 64035904 | 19 | 3 | 0.000113723 | 0.0015395 | 0.000561 | 0.000409 | -0.00171941 | MACROD1 |
| chr9 | 137479454 | 137479833 | 380 | 3 | 0.000158885 | 0.0031461 | 0.002943 | 0.000409 | -0.00374871 | PNPLA7 |
| chr2 | 177553132 | 177553325 | 194 | 6 | 6.64E-05 | 0.0012295 | 0.003262 | 0.00041 | -0.0007344 | AGPS |
| chr19 | 19653164 | 19653930 | 767 | 4 | 2.59E-05 | 0.0085816 | 0.00156 | 0.00041 | -8.40E-05 | ATP13A1 |
| chr8 | 57143317 | 57143467 | 151 | 2 | 0.000102754 | 0.0002304 | 0.005095 | 0.00041 | -0.0589588 | LINC01606 |
| chr7 | 143284983 | 143285370 | 388 | 4 | 0.00010532 | 0.0007613 | 0.001195 | 0.000411 | 0.000352 | TMEM139, TMEM139-AS1 |
| chr13 | 112588425 | 112588827 | 403 | 6 | 8.55E-05 | 0.0052685 | 0.003972 | 0.000411 | 0.010004509 | NA |
| chr16 | 88616283 | 88616288 | 6 | 2 | 0.000157255 | 0.0004249 | 0.001152 | 0.000411 | 0.000788041 | ZC3H18 |
| chr12 | 55966132 | 55966445 | 314 | 5 | 0.000116009 | 0.0006551 | 0.004819 | 0.000411 | -0.00132671 | PMEL |
| chr19 | 2799964 | 2799975 | 12 | 2 | 0.000177996 | 0.0002412 | 0.004381 | 0.000413 | -0.00098548 | THOP1 |

| | | | | | | | | | | |
|---|---|---|---|---|---|---|---|---|---|---|
| chr21 | 32392591 | 32392939 | 349 | 4 | 7.24E-05 | 0.0241146 | 8.61E-05 | 0.000413 | 0.001139404 | URB1 |
| chr17 | 48574817 | 48575188 | 372 | 5 | 4.68E-05 | 0.0044019 | 0.000219 | 0.000415 | -0.00236448 | HOXB-AS3, HOXB3 |
| chr16 | 152369 | 152483 | 115 | 2 | 0.000155805 | 0.0003767 | 0.001449 | 0.000416 | -0.00451345 | HBZ |
| chr3 | 161161899 | 161162195 | 297 | 3 | 0.000149117 | 0.0003293 | 0.002389 | 0.000417 | -0.00406237 | NMD3 |
| chr1 | 170075512 | 170075569 | 58 | 2 | 5.36E-05 | 0.0003801 | 0.001434 | 0.000417 | -0.00318257 | KIFAP3 |
| chr3 | 139538871 | 139539453 | 583 | 7 | 3.66E-05 | 0.0204735 | 6.65E-05 | 0.000419 | -0.00340549 | AC046134.2, RBP1 |
| chr14 | 85621780 | 85622236 | 457 | 3 | 2.25E-05 | 0.0001609 | 0.012343 | 0.000419 | 0.000574258 | FLRT2 |
| chr17 | 82231349 | 82231505 | 157 | 2 | 6.90E-06 | 0.0053922 | 0.000184 | 0.00042 | -0.00437341 | SLC16A3 |
| chr16 | 28259169 | 28259352 | 184 | 2 | 7.86E-05 | 0.0098953 | 0.000156 | 0.000422 | -0.00558153 | AC138904.3, SBK1 |
| chr1 | 97921894 | 97922049 | 156 | 2 | 8.53E-05 | 0.0006779 | 0.000669 | 0.000423 | -0.01706431 | DPYD |
| chr12 | 57771473 | 57771609 | 137 | 2 | 8.70E-05 | 0.0014035 | 0.000359 | 0.000424 | -0.00486057 | EEF1AKMT3, METTL1 |
| chr4 | 82042926 | 82043159 | 234 | 2 | 0.000111915 | 0.0016478 | 0.000324 | 0.000424 | -0.01324737 | RASGEF1B |
| chr17 | 7307336 | 7308605 | 1270 | 13 | 5.26E-07 | 0.0112473 | 0.000174 | 0.000424 | -0.00042328 | EIF5A |
| chr1 | 248905313 | 248905840 | 528 | 5 | 1.67E-05 | 0.026096 | 0.003143 | 0.000425 | -0.00273508 | NA |
| chr20 | 48921046 | 48921392 | 347 | 2 | 9.74E-06 | 0.0012649 | 0.00039 | 0.000427 | 0.004963413 | NA |
| chr2 | 20670602 | 20671435 | 834 | 4 | 3.57E-05 | 0.0001279 | 0.019053 | 0.000428 | -0.03932852 | GDF7 |
| chr3 | 9732381 | 9732617 | 237 | 2 | 2.05E-05 | 0.0002396 | 0.005325 | 0.000429 | 0.003036556 | BRPF1 |
| chr1 | 90850666 | 90850761 | 96 | 3 | 0.000119055 | 0.0007262 | 0.003615 | 0.000432 | -0.00235551 | LINC02609 |
| chr1 | 161168307 | 161168486 | 180 | 2 | 0.000165361 | 0.0015975 | 0.000338 | 0.000432 | -0.00123445 | PPOX |
| chr15 | 45114733 | 45114741 | 9 | 2 | 0.000173051 | 0.0002341 | 0.006163 | 0.000432 | 0.006389086 | DUOXA2 |
| chr2 | 24841648 | 24841790 | 143 | 3 | 5.19E-05 | 0.0009165 | 0.001119 | 0.000434 | -0.00805305 | ADCY3 |
| chr5 | 4971299 | 4971528 | 230 | 2 | 6.19E-05 | 0.0007548 | 0.000632 | 0.000437 | -0.00250386 | NA |
| chr8 | 143974599 | 143975021 | 423 | 3 | 5.19E-05 | 0.0004821 | 0.002981 | 0.000437 | -4.15E-05 | PLEC |
| chr19 | 38255898 | 38256448 | 551 | 5 | 2.42E-05 | 0.0005359 | 0.006235 | 0.000439 | -0.00325232 | SPINT2, PPP1R14A |
| chr22 | 20117930 | 20118838 | 909 | 4 | 9.51E-06 | 0.0029199 | 0.000912 | 0.00044 | -0.00670361 | RANBP1 |
| chr1 | 970983 | 971585 | 603 | 3 | 3.48E-05 | 0.0008057 | 0.002637 | 0.00044 | 0.013586164 | PLEKHN1 |
| chr17 | 8295309 | 8295407 | 99 | 3 | 0.00015339 | 0.0008633 | 0.000923 | 0.000443 | -0.00258546 | SLC25A35 |
| chr2 | 74472085 | 74472140 | 56 | 2 | 8.89E-05 | 0.0059196 | 0.00019 | 0.000443 | 0.002981798 | AC005041.5, MRPL53, CCDC142 |
| chr16 | 84505279 | 84505504 | 226 | 3 | 0.000129961 | 0.0004682 | 0.004845 | 0.000443 | 0.016271812 | MEAK7 |
| chr11 | 47337466 | 47337564 | 99 | 2 | 5.13E-05 | 0.0002458 | 0.005667 | 0.000444 | -0.00047034 | MYBPC3 |
| chr17 | 8177244 | 8177335 | 92 | 3 | 8.62E-06 | 0.0237178 | 7.10E-05 | 0.000445 | -0.00347385 | AC129492.4 |
| chr14 | 23351940 | 23352387 | 448 | 6 | 0.000139594 | 0.0006686 | 0.002765 | 0.000445 | -0.00250791 | SLC22A17 |
| chr12 | 3874599 | 3874655 | 57 | 2 | 0.000123622 | 0.0009284 | 0.000537 | 0.000446 | 0.003702069 | PARP11-AS1 |
| chr1 | 231160894 | 231161511 | 618 | 6 | 4.55E-05 | 0.0327166 | 3.65E-05 | 0.000447 | 0.00240904 | NA |
| chr6 | 100463870 | 100464292 | 423 | 5 | 5.41E-05 | 0.0007341 | 0.003592 | 0.000447 | -0.00298893 | SIM1 |
| chr15 | 52788543 | 52789329 | 787 | 4 | 1.55E-05 | 0.0364943 | 0.000161 | 0.000448 | -0.00186579 | ONECUT1 |
| chr3 | 10395004 | 10395941 | 938 | 3 | 2.18E-06 | 0.0768666 | 5.29E-05 | 0.000449 | 0.004513113 | ATP2B2 |
| chr22 | 50272060 | 50272159 | 100 | 2 | 7.51E-05 | 0.0002528 | 0.005327 | 0.00045 | 0.004382711 | NA |
| chr16 | 3734090 | 3734244 | 155 | 2 | 0.000116751 | 0.0002493 | 0.005753 | 0.000451 | 0.00221492 | CREBBP |
| chr17 | 75843868 | 75843891 | 24 | 2 | 9.09E-05 | 0.0004039 | 0.001559 | 0.000451 | -0.00129737 | UNC13D |
| chr15 | 96323454 | 96323921 | 468 | 3 | 0.000121167 | 0.0001814 | 0.012097 | 0.000452 | 0.00230473 | NR2F2-AS1 |
| chr14 | 49853270 | 49854090 | 821 | 4 | 4.80E-05 | 0.0004718 | 0.003234 | 0.000452 | -0.00680012 | RN7SL3 |
| chr2 | 113172753 | 113172780 | 28 | 2 | 8.26E-05 | 0.0041868 | 0.00022 | 0.000453 | 0.004426863 | PSD4, AC016683.1 |
| chr7 | 98399954 | 98400786 | 833 | 3 | 5.63E-05 | 0.0007434 | 0.002506 | 0.000453 | -0.02182569 | BAIAP2L1 |
| chr7 | 95772000 | 95772955 | 956 | 9 | 5.35E-05 | 0.0144845 | 0.000594 | 0.000455 | 0.001275858 | DYNC1I1 |
| chr6 | 133895782 | 133896104 | 323 | 3 | 5.52E-05 | 0.0036166 | 0.000441 | 0.000455 | -0.00398393 | NA |
| chr8 | 17722098 | 17722250 | 153 | 2 | 0.000102827 | 0.0054139 | 0.000203 | 0.000458 | 0.012077389 | MTUS1 |
| chr2 | 8645132 | 8645598 | 467 | 2 | 4.15E-05 | 0.0021388 | 0.000307 | 0.000458 | -0.01439512 | NA |
| chr11 | 45286900 | 45287125 | 226 | 3 | 9.90E-05 | 0.0002646 | 0.007046 | 0.000458 | -0.00371602 | NA |
| chr16 | 49679067 | 49679217 | 151 | 2 | 7.39E-05 | 0.0011279 | 0.000475 | 0.000459 | -0.00476294 | ZNF423 |
| chr6 | 105179371 | 105179537 | 167 | 2 | 1.49E-05 | 0.041242 | 0.000138 | 0.000462 | 0.001576173 | POPDC3 |
| chr1 | 88891955 | 88892360 | 406 | 6 | 3.91E-05 | 0.0029659 | 0.002688 | 0.000463 | -0.00687785 | NA |
| chr1 | 226739516 | 226739626 | 111 | 2 | 4.80E-05 | 0.0010826 | 0.000503 | 0.000466 | -0.00329463 | NA |
| chr12 | 56079021 | 56079090 | 70 | 2 | 0.000150931 | 0.0261401 | 0.000146 | 0.000466 | -0.00583189 | ERBB3 |
| chr9 | 122264550 | 122264794 | 245 | 10 | 0.000116781 | 0.1009554 | 6.20E-05 | 0.000467 | 0.001530688 | MRRF, RBM18 |
| chr11 | 113876574 | 113876705 | 132 | 3 | 5.21E-05 | 0.0662798 | 5.72E-05 | 0.000468 | 0.001430328 | NA |
| chr8 | 760997 | 761085 | 89 | 3 | 9.48E-05 | 0.0167339 | 8.33E-05 | 0.000469 | -0.00187466 | DLGAP2 |
| chr17 | 38558567 | 38558838 | 272 | 2 | 8.75E-05 | 0.0031455 | 0.00026 | 0.000469 | 0.005921899 | SRCIN1 |
| chr12 | 6328742 | 6329336 | 595 | 4 | 8.75E-05 | 0.0018476 | 0.003546 | 0.00047 | -0.00402752 | TNFRSF1A |
| chr22 | 31753747 | 31754132 | 386 | 6 | 9.69E-05 | 0.0063909 | 0.007142 | 0.00047 | 0.001296248 | DEPDC5 |
| chr16 | 88856045 | 88856726 | 682 | 4 | 2.81E-05 | 0.0002152 | 0.01711 | 0.000471 | -0.00186627 | TRAPPC2L, GALNS |
| chr6 | 69866631 | 69866979 | 349 | 5 | 6.93E-05 | 0.0002399 | 0.013825 | 0.000472 | -0.00106559 | COL19A1, LMBRD1 |
| chr10 | 97185543 | 97186192 | 650 | 10 | 6.29E-05 | 0.0423518 | 0.001276 | 0.000472 | -0.00165713 | SLIT1, ARHGAP19-SLIT1 |
| chr7 | 133081639 | 133082115 | 477 | 4 | 7.52E-06 | 0.0128386 | 0.000256 | 0.000472 | -0.00191934 | CHCHD3 |
| chr11 | 70952191 | 70952921 | 731 | 3 | 3.12E-05 | 0.0008124 | 0.004506 | 0.000473 | -0.00555973 | SHANK2 |
| chr12 | 6200295 | 6200351 | 57 | 4 | 7.24E-05 | 0.0002114 | 0.011035 | 0.000474 | 0.002389888 | CD9 |
| chr15 | 62164937 | 62165010 | 74 | 2 | 9.20E-05 | 0.0021027 | 0.000328 | 0.000478 | -0.00067497 | C2CD4B |
| chr16 | 83486491 | 83486598 | 108 | 2 | 6.36E-05 | 0.0153589 | 0.000162 | 0.000478 | -0.00214978 | CDH13 |
| chr6 | 26223471 | 26223785 | 315 | 2 | 3.68E-05 | 0.0133772 | 0.000166 | 0.000478 | 0.004283042 | NA |
| chr12 | 93442877 | 93443471 | 595 | 2 | 5.31E-05 | 0.0004229 | 0.001683 | 0.000482 | 0.007420844 | NA |
| chr10 | 100970265 | 100970373 | 109 | 2 | 3.34E-05 | 0.0005755 | 0.001039 | 0.000482 | -0.01227205 | SEMA4G, MRPL43 |
| chr1 | 160631010 | 160631413 | 404 | 2 | 9.72E-06 | 0.0022566 | 0.00032 | 0.000484 | 0.002437515 | SLAMF1 |
| chr20 | 23049896 | 23050926 | 1031 | 4 | 3.47E-05 | 0.0001538 | 0.020188 | 0.000485 | 0.001440291 | NA |
| chr4 | 105473234 | 105473962 | 729 | 5 | 5.19E-06 | 0.0443884 | 3.31E-05 | 0.000488 | 0.001671829 | PPA2 |
| chr12 | 121096030 | 121096628 | 599 | 2 | 1.17E-05 | 0.0010993 | 0.000535 | 0.000489 | -0.00554046 | AC079602.3 |
| chr16 | 66879624 | 66879913 | 290 | 3 | 1.45E-05 | 0.0003702 | 0.002387 | 0.000489 | -0.0046192 | PDP2 |
| chr10 | 122112438 | 122112494 | 57 | 3 | 9.32E-05 | 0.005562 | 0.000148 | 0.00049 | -0.01085512 | TACC2 |
| chr12 | 106356414 | 106356426 | 13 | 2 | 0.000136602 | 0.0005593 | 0.001117 | 0.00049 | -0.00391706 | NA |
| chr7 | 43582833 | 43583270 | 438 | 6 | 8.62E-05 | 0.002766 | 0.005348 | 0.00049 | 0.000906943 | STK17A |
| chr5 | 1855785 | 1855846 | 62 | 3 | 6.33E-05 | 0.004099 | 0.000182 | 0.000491 | 0.007189471 | NA |
| chr11 | 1443255 | 1443562 | 308 | 2 | 4.59E-05 | 0.0098812 | 0.000183 | 0.000491 | -0.00207029 | BRSK2 |
| chr16 | 527442 | 527628 | 187 | 3 | 5.26E-05 | 0.0056324 | 0.001023 | 0.000491 | -8.85E-05 | NA |
| chr22 | 38039196 | 38039669 | 474 | 4 | 0.000122604 | 0.0033061 | 0.010958 | 0.000495 | -0.00336124 | POLR2F |
| chr17 | 74973182 | 74973472 | 291 | 3 | 2.58E-05 | 0.0074319 | 0.000588 | 0.000497 | -0.01148508 | HID1-AS1 |
| chr9 | 92735341 | 92735740 | 400 | 3 | 2.56E-05 | 0.0562329 | 7.01E-05 | 0.000498 | -0.00281599 | BICD2 |
| chr1 | 241640378 | 241640518 | 141 | 5 | 0.000143945 | 0.0010314 | 0.000779 | 0.000499 | 0.00107625 | OPN3 |
| chr7 | 1495130 | 1495505 | 376 | 5 | 0.000113174 | 0.0005428 | 0.007834 | 0.000501 | -2.00E-05 | INTS1 |
| chr19 | 1011667 | 1011796 | 130 | 2 | 8.62E-05 | 0.0126075 | 0.000177 | 0.000501 | -0.00343882 | TMEM259 |
| chr3 | 2099015 | 2099293 | 279 | 2 | 3.58E-05 | 0.0025849 | 0.000312 | 0.000503 | -0.00220583 | CNTN4 |
| chr1 | 213982860 | 213983499 | 640 | 6 | 3.20E-05 | 0.0314556 | 6.62E-05 | 0.000503 | -0.00104177 | PROX1, PROX1-AS1 |
| chr1 | 8872120 | 8872377 | 258 | 3 | 3.32E-05 | 0.0003512 | 0.002938 | 0.000503 | -0.01200129 | ENO1 |
| chr8 | 37520758 | 37520837 | 80 | 2 | 0.000165236 | 0.0002721 | 0.006802 | 0.000505 | -0.01074179 | LINC01605, AC124067.2 |
| chr19 | 55407796 | 55408015 | 220 | 3 | 9.40E-05 | 0.0073617 | 0.000257 | 0.000505 | 0.002136868 | NA |
| chr6 | 36129782 | 36130204 | 423 | 4 | 6.63E-05 | 0.0067811 | 0.000192 | 0.000506 | -0.00415748 | MAPK13 |
| chr1 | 6393464 | 6393468 | 5 | 2 | 0.000172424 | 0.0170715 | 0.000168 | 0.000506 | -0.00858163 | ACOT7 |
| chr7 | 155072219 | 155072227 | 9 | 2 | 1.49E-05 | 0.0194142 | 0.000165 | 0.000507 | 0.00999665 | HTR5A, HTR5A-AS1 |
| chr14 | 100948633 | 100948986 | 354 | 2 | 4.95E-05 | 0.0009765 | 0.000633 | 0.000508 | -0.0141456 | MEG8 |
| chr20 | 19212413 | 19212416 | 4 | 2 | 0.000156504 | 0.0003303 | 0.003524 | 0.000511 | -0.00312545 | NA |
| chr16 | 1960794 | 1961116 | 323 | 2 | 2.25E-05 | 0.0011129 | 0.00057 | 0.000512 | -0.01074043 | NDUFB10 |
| chr19 | 19501497 | 19501929 | 433 | 3 | 6.97E-05 | 0.0004435 | 0.00318 | 0.000513 | -2.00E-05 | GATAD2A |
| chr2 | 27628951 | 27629000 | 50 | 3 | 0.000153338 | 0.000201 | 0.011055 | 0.000514 | 0.001125359 | AC074091.2, GPN1, CCDC121 |
| chr10 | 133308222 | 133308516 | 295 | 6 | 7.65E-05 | 0.0030665 | 0.000213 | 0.000514 | 0.000606801 | TUBGCP2 |
| chr3 | 9917002 | 9917444 | 443 | 5 | 6.66E-05 | 0.0020701 | 0.007922 | 0.000514 | -0.00452328 | IL17RC |
| chr6 | 72182894 | 72183198 | 305 | 2 | 9.21E-05 | 0.0002799 | 0.006585 | 0.000515 | 0.006367607 | RIMS1 |
| chr18 | 75427478 | 75427772 | 295 | 3 | 0.00014581 | 0.0042784 | 0.002018 | 0.000515 | -0.00207075 | SMIM21 |
| chr22 | 47113172 | 47113730 | 559 | 2 | 5.55E-05 | 0.0003159 | 0.004187 | 0.000516 | 0.001654596 | TBC1D22A |
| chr12 | 47716902 | 47717574 | 673 | 3 | 4.51E-05 | 0.0002379 | 0.009589 | 0.000517 | -0.00395869 | AC004241.1, ENDOU |
| chr1 | 32200434 | 32201134 | 701 | 8 | 3.28E-05 | 0.0028487 | 0.00247 | 0.000518 | -0.00114018 | CCDC28B |
| chr10 | 70169741 | 70170135 | 395 | 4 | 3.71E-05 | 0.0556418 | 8.63E-05 | 0.000523 | -0.0023334 | SAR1A |
| chr9 | 114377635 | 114377744 | 110 | 2 | 0.000162003 | 0.0003061 | 0.005018 | 0.000525 | 5.89E-05 | AKNA |
| chr21 | 42530367 | 42530558 | 192 | 3 | 6.78E-05 | 0.0191618 | 9.17E-05 | 0.000527 | -0.00392914 | SLC37A1 |
| chr4 | 4487647 | 4488162 | 516 | 2 | 4.15E-05 | 0.0037509 | 0.000276 | 0.000527 | -0.00214797 | STX18 |
| chr17 | 2940465 | 2940742 | 278 | 2 | 4.07E-05 | 0.0554817 | 0.000153 | 0.000529 | -0.00408651 | RAP1GAP2 |
| chr14 | 64388263 | 64388635 | 373 | 4 | 3.82E-05 | 0.0019454 | 0.001875 | 0.000529 | 0.000518096 | MTHFD1 |
| chr8 | 12755302 | 12755781 | 480 | 4 | 7.92E-05 | 0.0081312 | 0.00528 | 0.000529 | -0.0012435 | LONRF1 |
| chr21 | 31873595 | 31873903 | 309 | 2 | 4.45E-05 | 0.0004121 | 0.002273 | 0.00053 | 0.00456291 | HUNK |
| chr7 | 73719068 | 73719340 | 273 | 2 | 5.75E-05 | 0.0003176 | 0.004605 | 0.000531 | 0.002835624 | STX1A |
| chr4 | 165874108 | 165874621 | 514 | 8 | 7.70E-05 | 0.0165106 | 0.002431 | 0.000532 | 0.000907713 | TLL1 |
| chr3 | 14304768 | 14304931 | 164 | 2 | 1.46E-05 | 0.0069632 | 0.000221 | 0.000532 | -0.00339583 | NA |
| chr12 | 103930238 | 103930792 | 555 | 6 | 4.82E-05 | 0.0020689 | 0.000909 | 0.000534 | 0.001514423 | HSP90B1, MIR3652 |
| chr10 | 124469801 | 124470046 | 246 | 2 | 6.27E-05 | 0.0167672 | 0.000179 | 0.000536 | -0.00333995 | LHPP |
| chr3 | 184561101 | 184561740 | 640 | 9 | 6.81E-05 | 0.0005865 | 0.020307 | 0.000538 | 0.000616233 | NA |
| chr14 | 23056511 | 23057365 | 855 | 3 | 8.40E-05 | 0.001156 | 0.003379 | 0.000539 | 0.001962745 | CDH24 |
| chr16 | 89185228 | 89185567 | 340 | 2 | 4.93E-05 | 0.0031084 | 0.000312 | 0.000541 | 0.010241143 | CDH15 |
| chr6 | 168319796 | 168319999 | 204 | 6 | 0.000165371 | 0.0561358 | 2.08E-05 | 0.000542 | -0.000429 | NA |
| chr9 | 4850209 | 4850517 | 309 | 2 | 4.20E-05 | 0.0057124 | 0.000243 | 0.000546 | -0.00694887 | RCL1, MIR101-2, AL158147.1 |
| chr13 | 19634111 | 19634273 | 163 | 2 | 6.54E-05 | 0.0010531 | 0.000665 | 0.000547 | -0.00182345 | MPHOSPH8 |
| chr6 | 111793412 | 111794006 | 595 | 4 | 2.40E-06 | 0.0088489 | 0.000276 | 0.000548 | -0.00027712 | FYN |
| chr19 | 56314275 | 56314817 | 543 | 6 | 6.23E-05 | 0.000623 | 0.00496 | 0.000549 | -0.00147054 | AC006116.7, ZSCAN5A |
| chr6 | 6001609 | 6001799 | 191 | 3 | 5.41E-05 | 0.0083972 | 0.00014 | 0.000551 | -0.00976828 | NRN1 |
| chr8 | 32594533 | 32595094 | 562 | 4 | 7.83E-05 | 0.0031225 | 0.000285 | 0.000551 | -0.00334779 | NRG1 |
| chr20 | 44886225 | 44886310 | 86 | 2 | 0.000103625 | 0.0010496 | 0.00068 | 0.000553 | 0.001611278 | YWHAB |
| chr7 | 157003451 | 157004235 | 785 | 3 | 1.13E-06 | 0.0322763 | 8.22E-05 | 0.000555 | -0.00399411 | MNX1 |
| chr10 | 27998778 | 27998950 | 173 | 2 | 6.57E-05 | 0.002724 | 0.000347 | 0.000557 | -0.002956 | ARMC4 |
| chr12 | 6567981 | 6568485 | 505 | 5 | 3.20E-05 | 0.436972 | 6.31E-05 | 0.000558 | -0.00083321 | NOP2, AC006064.6 |
| chr1 | 3650739 | 3651681 | 943 | 19 | 7.59E-06 | 0.0510552 | 5.54E-05 | 0.000559 | -0.00106101 | WRAP73 |
| chr2 | 218661910 | 218662295 | 386 | 3 | 1.46E-05 | 0.1121313 | 6.14E-05 | 0.000561 | -0.00350038 | BCS1L |
| chr11 | 114060248 | 114060428 | 181 | 4 | 8.90E-05 | 0.0001887 | 0.016593 | 0.000562 | -0.00104128 | ZBTB16 |
| chr13 | 27821134 | 27821646 | 513 | 4 | 0.000109856 | 0.0017121 | 0.005279 | 0.000569 | -0.00018406 | PLUT |
| chr15 | 40694504 | 40694811 | 308 | 3 | 3.22E-06 | 0.0284169 | 8.36E-05 | 0.00057 | 0.001384387 | RAD51, RAD51-AS1 |
| chr2 | 3102450 | 3102523 | 74 | 2 | 0.000150677 | 0.0003652 | 0.003873 | 0.000571 | -0.00134356 | LINC01250 |
| chr11 | 66690099 | 66690150 | 52 | 2 | 0.000163381 | 0.0003456 | 0.004623 | 0.000572 | -0.00528996 | SPTBN2 |
| chr19 | 11179788 | 11180326 | 539 | 2 | 9.37E-06 | 0.0054341 | 0.000261 | 0.000572 | -0.00795393 | KANK2 |
| chr7 | 157812877 | 157813278 | 402 | 3 | 6.34E-05 | 0.0159527 | 0.000112 | 0.000576 | -0.00348114 | PTPRN2 |
| chr6 | 31739242 | 31739524 | 283 | 5 | 2.67E-05 | 0.0180029 | 0.000706 | 0.000576 | -0.00404333 | CLIC1 |
| chr12 | 13530074 | 13530465 | 392 | 3 | 4.13E-05 | 0.0068442 | 0.000191 | 0.000577 | -0.00582235 | AC007527.1, GRIN2B |
| chr2 | 85693751 | 85694129 | 379 | 4 | 4.88E-05 | 0.0033908 | 0.003607 | 0.000578 | 0.003982584 | GNLY |
| chr6 | 168706544 | 168706680 | 137 | 3 | 0.000126224 | 0.0003695 | 0.007794 | 0.000579 | 0.002239577 | NA |
| chr3 | 184381308 | 184381471 | 164 | 2 | 0.000101815 | 0.0003734 | 0.003973 | 0.000586 | 0.002913514 | CHRD, THPO |
| chr3 | 40525182 | 40525486 | 305 | 3 | 4.92E-05 | 0.0002605 | 0.010419 | 0.000589 | -0.0037049 | ZNF621 |
| chr16 | 2451146 | 2451225 | 80 | 2 | 0.000106699 | 0.0133208 | 0.000208 | 0.000591 | -0.00198993 | CCNF |
| chr1 | 108660988 | 108661301 | 314 | 9 | 5.65E-05 | 0.0260986 | 0.000142 | 0.000594 | -0.00068299 | HENMT1 |
| chr1 | 154822443 | 154822479 | 37 | 2 | 0.000148883 | 0.0357026 | 0.000178 | 0.000595 | -0.00587564 | KCNN3 |
| chr12 | 57773700 | 57773767 | 68 | 2 | 8.62E-05 | 0.0046892 | 0.000292 | 0.000597 | 0.004508435 | EEF1AKMT3, AC025165.3 |
| chr14 | 95714489 | 95715280 | 792 | 4 | 3.52E-05 | 0.0015664 | 0.005338 | 0.000597 | -0.00378486 | AL139020.1 |
| chr8 | 98116904 | 98117091 | 188 | 3 | 9.82E-05 | 0.0007431 | 0.001254 | 0.0006 | 0.000800633 | RIDA |
| chr12 | 114241906 | 114241941 | 36 | 2 | 7.42E-05 | 0.0077674 | 0.000244 | 0.0006 | 0.010408265 | NA |
| chr12 | 132275197 | 132275381 | 185 | 4 | 0.000145647 | 0.009787 | 0.000734 | 0.0006 | -0.00753722 | GALNT9 |

| | | | | | | | | | | |
|---|---|---|---|---|---|---|---|---|---|---|
| chr4 | 1406903 | 1407081 | 179 | 2 | 1.72E-05 | 0.0018146 | 0.000501 | 0.000606 | 0.00761767 | NA |
| chr9 | 34663029 | 34663051 | 23 | 2 | 0.000144515 | 0.0004687 | 0.002508 | 0.000607 | -0.00675321 | CCL27, AL162231.4 |
| chr2 | 181684023 | 181684681 | 659 | 6 | 4.94E-05 | 0.0033767 | 0.011438 | 0.000607 | 0.001726438 | AC013733.2 |
| chr6 | 28999547 | 28999989 | 443 | 4 | 2.17E-05 | 0.0086791 | 0.000142 | 0.000608 | 0.000851438 | ZNF311 |
| chr7 | 128862319 | 128862406 | 88 | 2 | 5.57E-05 | 0.0956115 | 0.000171 | 0.000609 | -0.00308402 | FLNC-AS1, KCP |
| chr7 | 124765083 | 124765706 | 624 | 7 | 7.33E-05 | 0.001223 | 0.003732 | 0.000613 | -0.00051659 | GPR37 |
| chr17 | 3916566 | 3916624 | 59 | 3 | 9.71E-05 | 0.0053827 | 0.000598 | 0.000613 | -0.00052902 | NA |
| chr14 | 22546420 | 22546715 | 296 | 2 | 0.000169912 | 0.0009477 | 0.0009 | 0.000614 | -0.00654064 | NA |
| chr19 | 1276281 | 1276642 | 362 | 2 | 3.36E-05 | 0.0277864 | 0.00019 | 0.000614 | 0.001944587 | FAM174C |
| chr9 | 6646386 | 6647012 | 627 | 3 | 5.49E-05 | 0.0358433 | 0.000172 | 0.000615 | -0.0037242 | AL162411.1 |
| chr12 | 38905699 | 38906434 | 736 | 10 | 5.08E-05 | 0.0004317 | 0.008285 | 0.000616 | -2.41E-05 | CPNE8 |
| chr19 | 51065003 | 51065266 | 264 | 5 | 0.000132087 | 0.0061804 | 0.003025 | 0.000617 | -0.00053585 | KLK13 |
| chr17 | 29618194 | 29618369 | 176 | 4 | 0.000124959 | 0.0081192 | 0.000535 | 0.000617 | -0.00109688 | ABHD15-AS1, CORO6 |
| chr8 | 81279340 | 81279603 | 264 | 3 | 0.000137375 | 0.0014211 | 0.004961 | 0.000619 | 0.00695644 | NA |
| chr19 | 50873383 | 50873455 | 73 | 2 | 7.20E-05 | 0.1632072 | 0.000171 | 0.000622 | -0.00905687 | KLK2 |
| chr1 | 68497773 | 68497859 | 87 | 2 | 8.81E-05 | 0.004413 | 0.000315 | 0.000622 | 0.002320507 | DEPDC1-AS1 |
| chr1 | 47724909 | 47725359 | 451 | 3 | 6.25E-06 | 0.0124745 | 0.000307 | 0.000623 | -0.00555305 | NA |
| chr2 | 44087257 | 44087523 | 267 | 3 | 9.35E-05 | 0.0002829 | 0.009241 | 0.000625 | -0.00074666 | NA |
| chr7 | 134316804 | 134316921 | 118 | 2 | 0.000138528 | 0.0011786 | 0.000747 | 0.000625 | 0.000971456 | SLC35B4 |
| chr2 | 88857749 | 88858416 | 668 | 4 | 5.33E-05 | 0.0215451 | 0.001574 | 0.000626 | -0.00408235 | AC244205.1 |
| chr1 | 90719471 | 90719884 | 414 | 6 | 1.30E-05 | 0.0141638 | 0.001496 | 0.000626 | -0.00492001 | AC092805.1 |
| chr19 | 18099207 | 18099632 | 426 | 5 | 2.60E-05 | 0.0029297 | 0.002249 | 0.000627 | 0.005387824 | MAST3 |
| chr10 | 132350288 | 132350321 | 34 | 3 | 0.000111454 | 0.0003149 | 0.009642 | 0.000629 | 0.002996297 | LRRC27 |
| chr6 | 122788246 | 122788758 | 513 | 4 | 7.28E-05 | 0.000619 | 0.00725 | 0.000629 | 0.010580342 | NA |
| chr11 | 119380980 | 119381500 | 521 | 3 | 2.41E-05 | 0.0004125 | 0.006983 | 0.00063 | 0.000225183 | USP2-AS1, AP003396.3, USP2 |
| chr10 | 125162096 | 125162254 | 159 | 2 | 0.000109996 | 0.0005086 | 0.002357 | 0.000631 | 0.024580893 | NA |
| chr1 | 209370635 | 209370775 | 141 | 2 | 6.63E-05 | 0.0127269 | 0.000226 | 0.000633 | -0.00143144 | LINC01698 |
| chr7 | 22082994 | 22083579 | 586 | 4 | 6.12E-05 | 0.0164231 | 8.61E-05 | 0.000634 | 0.000614746 | NA |
| chr1 | 3166846 | 3166971 | 126 | 2 | 0.000145204 | 0.0003631 | 0.006084 | 0.000637 | -0.00431632 | PRDM16 |
| chr14 | 60724044 | 60724357 | 314 | 8 | 0.000131824 | 0.0010704 | 0.020865 | 0.000638 | 0.000794921 | SIX4 |
| chr5 | 113294528 | 113295360 | 833 | 11 | 4.75E-05 | 0.0539412 | 0.005134 | 0.00064 | 0.000589018 | MCC |
| chr4 | 121765164 | 121765537 | 374 | 10 | 5.96E-05 | 0.0224251 | 0.000873 | 0.000642 | -0.00182637 | PP12613, TMEM155 |
| chr3 | 126476739 | 126476874 | 136 | 2 | 8.83E-05 | 0.0087809 | 0.000256 | 0.000646 | -0.01208502 | NA |
| chr19 | 38321135 | 38321143 | 9 | 2 | 0.000108079 | 0.0220719 | 0.000208 | 0.000651 | 0.003080199 | KCNK6 |
| chr20 | 56005014 | 56005140 | 127 | 4 | 4.74E-05 | 0.0055563 | 0.000165 | 0.000651 | 0.005584694 | CBLN4 |
| chr13 | 52739394 | 52739793 | 400 | 4 | 7.73E-05 | 0.0001757 | 0.027208 | 0.000653 | -0.00420536 | CNMD |
| chr4 | 151099788 | 151100562 | 775 | 4 | 5.68E-05 | 0.0007358 | 0.002362 | 0.000653 | 0.002981059 | RPS3A |
| chr2 | 240140755 | 240141047 | 293 | 5 | 0.000136358 | 0.0046881 | 0.004058 | 0.000654 | -0.00417096 | OTOS |
| chr19 | 33303650 | 33303840 | 191 | 3 | 4.27E-05 | 0.1043014 | 7.51E-05 | 0.000655 | 0.001339947 | CEBPA-DT |
| chr8 | 75538846 | 75539151 | 306 | 2 | 0.000127097 | 0.003671 | 0.000369 | 0.000658 | -0.00944597 | HNF4G |
| chr1 | 39883231 | 39883305 | 75 | 2 | 0.000145448 | 0.0049353 | 0.000323 | 0.00066 | 0.005035733 | TRIT1 |
| chr2 | 209808015 | 209808587 | 573 | 3 | 0.000103373 | 0.0003396 | 0.008951 | 0.000667 | -0.00223271 | UNC80 |
| chr6 | 31884999 | 31885741 | 743 | 6 | 1.91E-05 | 0.0053603 | 0.003622 | 0.000675 | -0.00201581 | EHMT2 |
| chr6 | 44342361 | 44342903 | 543 | 7 | 0.000135512 | 0.0089558 | 0.001081 | 0.000675 | -0.00090553 | AL353588.1, SPATS1 |
| chr4 | 3374190 | 3374417 | 228 | 3 | 0.000142586 | 0.0005772 | 0.004882 | 0.000678 | -0.00212497 | RGS12 |
| chr19 | 48954958 | 48955339 | 382 | 5 | 6.03E-05 | 0.0134253 | 0.009519 | 0.000678 | -0.00245883 | BAX |
| chr18 | 50825334 | 50825663 | 330 | 4 | 0.000128107 | 0.0033017 | 0.000446 | 0.000679 | -0.00378612 | MRO |
| chr3 | 58667170 | 58667469 | 300 | 2 | 7.15E-05 | 0.0015386 | 0.000677 | 0.00068 | -0.00243391 | NA |
| chr11 | 65048654 | 65048855 | 202 | 3 | 0.000125424 | 0.3409537 | 5.95E-05 | 0.000682 | -0.00500469 | NAALADL1 |
| chr11 | 18589668 | 18590002 | 335 | 2 | 5.42E-05 | 0.007057 | 0.000295 | 0.000687 | -0.00528877 | SPTY2D1OS |
| chr1 | 52842672 | 52842982 | 311 | 3 | 0.000103229 | 0.0004452 | 0.00536 | 0.000687 | 0.002493151 | ZYG11A |
| chr15 | 31435666 | 31435940 | 275 | 2 | 7.10E-05 | 0.0003764 | 0.007721 | 0.000692 | -0.00464139 | NA |
| chr5 | 139903766 | 139904451 | 686 | 5 | 4.20E-06 | 0.0371192 | 0.000294 | 0.000695 | 0.000504529 | NRG2 |
| chr11 | 72238318 | 72238503 | 186 | 2 | 8.60E-05 | 0.0004574 | 0.004056 | 0.000697 | -0.00822997 | INPPL1 |
| chr22 | 21938709 | 21939421 | 713 | 5 | 1.72E-05 | 0.0538108 | 4.45E-05 | 0.000699 | -0.00299992 | PPM1F-AS1, PPM1F |
| chr1 | 84635136 | 84635647 | 512 | 3 | 4.26E-05 | 0.0302035 | 0.0001 | 0.0007 | -0.00361318 | NA |
| chr6 | 75207752 | 75207844 | 93 | 2 | 4.47E-05 | 0.0012454 | 0.000857 | 0.000701 | -0.00902784 | NA |
| chr16 | 3035866 | 3036395 | 530 | 4 | 9.91E-05 | 0.0002062 | 0.021451 | 0.000701 | 0.00846564 | AC108134.1, BICDL2 |
| chr2 | 438095 | 438323 | 229 | 3 | 3.83E-05 | 0.0823113 | 8.55E-05 | 0.000702 | 0.002264165 | NA |
| chr3 | 45841194 | 45841757 | 564 | 3 | 7.57E-05 | 0.0005475 | 0.004879 | 0.000703 | -0.00287938 | LZTFL1 |
| chr10 | 122979100 | 122979234 | 135 | 2 | 3.92E-05 | 0.0014926 | 0.000732 | 0.000704 | -0.00030973 | PSTK |
| chr2 | 61904843 | 61905401 | 559 | 4 | 3.30E-05 | 0.0030359 | 0.000757 | 0.000709 | -0.00319547 | COMMD1 |
| chr3 | 1092181 | 1092321 | 141 | 2 | 0.000100612 | 0.0009121 | 0.001233 | 0.00071 | 0.002249829 | NA |
| chr14 | 24231746 | 24232182 | 437 | 4 | 4.21E-05 | 0.0049753 | 0.000275 | 0.00071 | 0.001563508 | NEDD8-MDP1, NEDD8 |
| chr5 | 150302387 | 150303075 | 689 | 5 | 7.65E-05 | 0.0018108 | 0.002553 | 0.000712 | 0.001848288 | ARSI |
| chr2 | 97645489 | 97645558 | 70 | 3 | 9.57E-05 | 0.004748 | 0.000485 | 0.000713 | 0.006443891 | NA |
| chr14 | 45135458 | 45135969 | 512 | 7 | 0.000112185 | 0.0009041 | 0.014211 | 0.000713 | 0.001130369 | FANCM |
| chr1 | 152514133 | 152515223 | 1091 | 6 | 2.16E-05 | 0.0013156 | 0.014904 | 0.000714 | 0.005502952 | CRCT1 |
| chr3 | 147388112 | 147388421 | 310 | 3 | 0.000110945 | 0.0298 | 0.001095 | 0.000716 | -0.00178244 | ZIC4 |
| chr4 | 1510521 | 1510542 | 22 | 2 | 0.000107081 | 0.0136466 | 0.000255 | 0.00072 | 0.00451544 | NA |
| chr9 | 77178398 | 77178904 | 507 | 3 | 6.09E-05 | 0.0011064 | 0.002425 | 0.00072 | -0.00481662 | VPS13A |
| chr11 | 63813647 | 63814159 | 513 | 3 | 3.39E-05 | 0.0031701 | 0.004568 | 0.000721 | -0.00032568 | SPINDOC |
| chr8 | 58033833 | 58034132 | 300 | 2 | 0.000115808 | 0.0006055 | 0.0024 | 0.000724 | 0.011755962 | FAM110B |
| chr10 | 132187742 | 132187951 | 210 | 2 | 0.000131741 | 0.0004408 | 0.005383 | 0.000731 | 0.011087096 | DPYSL4 |
| chr15 | 39595513 | 39595848 | 336 | 3 | 0.000134856 | 0.0069791 | 0.001301 | 0.000732 | 0.002138058 | THBS1, FSIP1 |
| chr3 | 138009241 | 138009284 | 44 | 2 | 0.000119484 | 0.0036012 | 0.000426 | 0.000732 | -0.00353504 | CLDN18 |
| chr6 | 33431129 | 33431374 | 246 | 4 | 0.000106558 | 0.0022249 | 0.00594 | 0.000732 | 0.004908198 | SYNGAP1 |
| chr20 | 31713169 | 31713376 | 208 | 2 | 8.89E-05 | 0.0008276 | 0.001493 | 0.000733 | -0.00771256 | BCL2L1-AS1, BCL2L1 |
| chr2 | 73925959 | 73926673 | 715 | 6 | 6.51E-06 | 0.0092966 | 0.003438 | 0.000736 | 0.000323924 | NA |
| chr1 | 23558875 | 23559278 | 404 | 4 | 4.89E-05 | 0.0012703 | 0.009289 | 0.000736 | 0.000672697 | ID3 |
| chr17 | 50197963 | 50198558 | 596 | 2 | 0.000113634 | 0.0004468 | 0.005394 | 0.00074 | -0.01045214 | COL1A1 |
| chr2 | 236508183 | 236508384 | 202 | 3 | 9.35E-05 | 0.0030642 | 0.004316 | 0.000743 | 0.004542907 | NA |
| chr12 | 3200098 | 3200329 | 232 | 2 | 7.38E-05 | 0.0004186 | 0.006965 | 0.000743 | 0.000812375 | TSPAN9 |
| chr10 | 26217079 | 26217424 | 346 | 2 | 2.58E-06 | 0.0733853 | 0.000209 | 0.000744 | -0.00465848 | GAD2 |
| chr8 | 22668146 | 22668802 | 657 | 3 | 4.62E-05 | 0.0011275 | 0.002252 | 0.000745 | -0.00934485 | BIN3 |
| chr7 | 950063 | 950514 | 452 | 2 | 7.26E-05 | 0.0020982 | 0.000612 | 0.000746 | -0.00666693 | ADAP1, COX19 |
| chr6 | 168227029 | 168227745 | 717 | 3 | 8.39E-05 | 0.0018139 | 0.000753 | 0.00075 | 0.008460043 | NA |
| chr1 | 43933668 | 43933756 | 89 | 4 | 0.000151773 | 0.0009361 | 0.004658 | 0.000758 | -0.00026836 | ARTN |
| chr7 | 5231297 | 5232024 | 728 | 5 | 8.55E-05 | 0.0005156 | 0.007588 | 0.00076 | 0.022003644 | WIPI2 |
| chr7 | 4707535 | 4707630 | 96 | 3 | 0.000150134 | 0.0009234 | 0.002571 | 0.000761 | 0.003079862 | FOXK1 |
| chr6 | 16761437 | 16761837 | 401 | 7 | 0.000114934 | 0.0349984 | 0.000824 | 0.000763 | 0.001928066 | ATXN1-AS1, ATXN1 |
| chr19 | 9538866 | 9538954 | 89 | 2 | 0.00013722 | 0.0010775 | 0.001162 | 0.000764 | -0.0095009 | ZNF426-DT |
| chr10 | 49612232 | 49612558 | 327 | 2 | 6.95E-05 | 0.0183775 | 0.000257 | 0.000771 | 0.008528924 | CHAT, SLC18A3 |
| chr3 | 49423088 | 49423133 | 46 | 2 | 3.24E-05 | 0.0495434 | 0.000223 | 0.000774 | -0.00224493 | AC104452.1, NICN1 |
| chr17 | 79046373 | 79047462 | 1090 | 7 | 1.15E-05 | 0.0103042 | 0.00088 | 0.000774 | -0.00193404 | C1QTNF1 |
| chr10 | 119209227 | 119209332 | 106 | 2 | 4.65E-05 | 0.0020739 | 0.000651 | 0.000774 | -0.01560089 | GRK5, GRK5-IT1 |
| chr3 | 33218397 | 33218428 | 32 | 2 | 3.67E-05 | 0.0070993 | 0.000341 | 0.00078 | 0.003268272 | SUSD5 |
| chr17 | 80837736 | 80837881 | 146 | 2 | 0.000130833 | 0.0024533 | 0.000588 | 0.000783 | -0.00304541 | RPTOR |
| chr6 | 56247014 | 56247226 | 213 | 3 | 0.000133909 | 0.0002788 | 0.018838 | 0.000785 | -0.0111151 | COL21A1 |
| chr7 | 20778307 | 20779037 | 731 | 4 | 2.95E-05 | 0.0030211 | 0.004757 | 0.000787 | -0.00509013 | NA |
| chr2 | 33133992 | 33134222 | 231 | 4 | 6.92E-05 | 0.0058451 | 0.002405 | 0.000788 | -0.00396315 | LTBP1 |
| chr12 | 55707865 | 55708061 | 197 | 4 | 0.000128817 | 0.0024584 | 0.00045 | 0.000793 | 0.001465501 | ITGA7 |
| chr6 | 30683734 | 30684048 | 315 | 3 | 1.62E-05 | 0.0004314 | 0.007 | 0.000793 | 0.004249338 | PPP1R18 |
| chr9 | 131277927 | 131278403 | 477 | 3 | 6.38E-05 | 0.0027172 | 0.005785 | 0.000796 | -0.00522093 | NA |
| chr11 | 93521354 | 93521423 | 70 | 2 | 0.000122743 | 0.0222221 | 0.000257 | 0.000798 | 0.008612116 | SMCO4 |
| chr2 | 203328604 | 203329767 | 1164 | 6 | 3.33E-06 | 0.1684721 | 1.58E-05 | 0.0008 | -0.0062388 | ABI2, AC080075.1 |
| chr20 | 46684965 | 46685169 | 205 | 2 | 0.000176652 | 0.0054461 | 0.000391 | 0.000801 | -0.00324158 | TP53RK |
| chr11 | 7576583 | 7577071 | 489 | 5 | 7.04E-05 | 0.009208 | 0.00363 | 0.000814 | -0.00796024 | PPFIBP2 |
| chr10 | 5499460 | 5499860 | 401 | 5 | 0.00014166 | 0.0019111 | 0.014336 | 0.000815 | 0.000527482 | CALML5 |
| chr16 | 3769293 | 3769795 | 503 | 2 | 1.42E-05 | 0.0117933 | 0.000306 | 0.00082 | -0.00184528 | CREBBP |
| chr1 | 3644401 | 3644512 | 112 | 2 | 0.000137768 | 0.0031157 | 0.000547 | 0.00083 | -0.00564759 | WRAP73 |
| chr22 | 42078652 | 42079131 | 480 | 4 | 3.95E-06 | 0.0171781 | 0.00031 | 0.000835 | -0.00224038 | PHETA2 |
| chr6 | 33264405 | 33264867 | 463 | 7 | 4.90E-05 | 0.002311 | 0.006479 | 0.000845 | -0.00103104 | VPS52 |
| chr12 | 102917061 | 102917360 | 300 | 4 | 0.00010326 | 0.0212963 | 0.000499 | 0.000852 | 0.005639945 | PAH |
| chr1 | 15151710 | 15152465 | 756 | 5 | 4.10E-05 | 0.0376487 | 0.00184 | 0.000852 | -0.0052629 | TMEM51-AS1 |
| chr3 | 49870233 | 49870399 | 167 | 3 | 0.00013715 | 0.0097221 | 0.002277 | 0.000854 | -0.00797071 | NA |
| chr21 | 38661230 | 38661312 | 83 | 3 | 0.000137345 | 0.0003828 | 0.013348 | 0.000854 | -0.00403695 | ERG |
| chr12 | 50057895 | 50058124 | 230 | 2 | 7.85E-05 | 0.0010733 | 0.00148 | 0.000866 | 0.001183296 | ASIC1 |
| chr1 | 8816954 | 8817198 | 245 | 3 | 3.04E-05 | 0.1976073 | 7.73E-05 | 0.000868 | -0.0013707 | RERE |
| chr13 | 36999203 | 36999899 | 697 | 6 | 1.71E-05 | 0.0057176 | 0.006086 | 0.000869 | -6.85E-05 | EXOSC8, ALG5 |
| chr16 | 85574618 | 85575053 | 436 | 4 | 6.51E-05 | 0.005355 | 0.000823 | 0.00087 | 0.000994795 | GSE1 |
| chr2 | 146587346 | 146587769 | 424 | 4 | 0.000104862 | 0.1300664 | 4.43E-05 | 0.00087 | -0.00252125 | PABPC1P2 |
| chr16 | 578357 | 579464 | 1108 | 3 | 2.53E-06 | 0.0148319 | 0.000175 | 0.000872 | -0.00756952 | PIGQ |
| chr4 | 3525363 | 3525850 | 488 | 2 | 3.77E-05 | 0.0007883 | 0.00238 | 0.000875 | 0.000449197 | LRPAP1 |
| chr1 | 9365692 | 9365879 | 188 | 2 | 7.68E-05 | 0.0061787 | 0.000414 | 0.000878 | -0.01653418 | SPSB1 |
| chr17 | 60525501 | 60526259 | 759 | 5 | 1.40E-05 | 0.0045496 | 0.001235 | 0.000879 | 0.002290223 | APPBP2 |
| chr6 | 30893089 | 30893183 | 95 | 3 | 6.91E-05 | 0.0065145 | 0.000974 | 0.000885 | -0.003863 | DDR1 |
| chr2 | 9207163 | 9207447 | 285 | 2 | 0.0001133 | 0.0006447 | 0.003658 | 0.000887 | 0.003080187 | ASAP2 |
| chr17 | 39451355 | 39451410 | 56 | 5 | 0.000145327 | 0.0022284 | 0.017904 | 0.000897 | 0.001229841 | NA |
| chr2 | 240915962 | 240916037 | 76 | 2 | 0.000141925 | 0.0006117 | 0.004312 | 0.000898 | 0.000809479 | CROCC2 |
| chr11 | 62880369 | 62880507 | 139 | 2 | 7.27E-05 | 0.002077 | 0.000816 | 0.0009 | 0.002580874 | SLC3A2 |
| chr17 | 42457808 | 42457958 | 151 | 2 | 0.000117629 | 0.0008717 | 0.002181 | 0.000905 | 0.008184127 | NA |
| chr16 | 58391749 | 58391807 | 59 | 2 | 9.87E-05 | 0.0006196 | 0.004321 | 0.000908 | -0.00081329 | GINS3 |
| chr5 | 172919892 | 172920574 | 683 | 3 | 2.21E-05 | 0.0021293 | 0.00136 | 0.000912 | -0.00161026 | ERGIC1 |
| chr2 | 176113045 | 176113561 | 517 | 2 | 7.19E-05 | 0.0005596 | 0.006075 | 0.000923 | 0.006446509 | HOXD10 |
| chr18 | 69400485 | 69401032 | 548 | 4 | 0.000151658 | 0.000297 | 0.024275 | 0.000924 | 0.000425746 | DOK6 |
| chr17 | 4704303 | 4704445 | 143 | 7 | 0.000130208 | 0.0517395 | 0.00096 | 0.000928 | -0.00144787 | AC091153.3, PELP1 |
| chr8 | 17246382 | 17247343 | 962 | 11 | 1.43E-05 | 0.188998 | 2.09E-05 | 0.000929 | -0.00026238 | VPS37A, CNOT7 |
| chr12 | 79934383 | 79934745 | 363 | 3 | 2.14E-05 | 0.053563 | 0.000128 | 0.00093 | -0.0017726 | PPP1R12A |
| chr6 | 70282338 | 70283120 | 783 | 3 | 4.63E-05 | 0.0026924 | 0.000648 | 0.000939 | -0.00195847 | COL9A1 |
| chr10 | 128069489 | 128069666 | 178 | 2 | 9.18E-05 | 0.0037644 | 0.000584 | 0.000946 | 0.00037556 | PTPRE |
| chr3 | 108090848 | 108091514 | 667 | 7 | 3.24E-05 | 0.0018384 | 0.001951 | 0.000949 | 0.000116553 | CD47 |
| chr15 | 67522467 | 67522542 | 76 | 2 | 0.000108144 | 0.0009913 | 0.00199 | 0.00095 | -0.01005373 | C15orf61 |
| chr20 | 21102301 | 21102456 | 156 | 2 | 9.55E-05 | 0.0782977 | 0.000264 | 0.00095 | -0.00234891 | LINC00237 |
| chr7 | 94663577 | 94664073 | 497 | 4 | 4.37E-05 | 0.0141985 | 0.000469 | 0.000951 | 0.001534081 | PEG10 |
| chr14 | 105489408 | 105489542 | 135 | 2 | 6.88E-05 | 0.0055765 | 0.00048 | 0.000952 | -0.01053812 | AL928654.3 |
| chr1 | 23344284 | 23344399 | 116 | 4 | 0.00017542 | 0.0020607 | 0.013905 | 0.000953 | 0.00092995 | HNRNPR |
| chr13 | 31161968 | 31162421 | 454 | 10 | 4.94E-05 | 0.0079687 | 0.001873 | 0.000953 | -0.00014889 | HSPH1 |
| chr7 | 36725537 | 36725744 | 208 | 2 | 3.67E-05 | 0.005121 | 0.000503 | 0.000957 | 0.007250499 | NA |
| chr12 | 52974941 | 52975385 | 445 | 3 | 0.000106196 | 0.0036997 | 0.000932 | 0.000971 | -0.00555624 | NA |
| chr10 | 49540382 | 49540692 | 311 | 2 | 3.77E-05 | 0.0599301 | 0.000276 | 0.000973 | 0.008444787 | NA |
| chr9 | 136364486 | 136364622 | 137 | 2 | 5.81E-05 | 0.0006031 | 0.005947 | 0.000976 | -0.02065338 | CARD9 |
| chr20 | 6767237 | 6767474 | 238 | 3 | 5.48E-05 | 0.0088676 | 0.000878 | 0.000976 | -0.00025536 | NA |
| chr16 | 19303900 | 19304125 | 226 | 2 | 0.00010215 | 0.004519 | 0.000556 | 0.000984 | -0.00731456 | CLEC19A |
| chr20 | 3045322 | 3045704 | 383 | 4 | 0.000127103 | 0.0037843 | 0.004834 | 0.000993 | 0.004314525 | GNRH2 |
| chr8 | 143804913 | 143805177 | 265 | 3 | 8.62E-05 | 0.004548 | 0.000525 | 0.000995 | 0.002057438 | SCRIB |
| chr5 | 108381523 | 108381929 | 407 | 3 | 6.54E-05 | 0.0096034 | 0.000377 | 0.001 | 0.002224212 | FBXL17 |
| chr4 | 107824890 | 107825495 | 606 | 5 | 8.12E-06 | 0.7770461 | 1.44E-05 | 0.001006 | 0.001003539 | SGMS2 |

| | | | | | | | | | | |
|---|---|---|---|---|---|---|---|---|---|---|
| chr12 | 53180026 | 53180801 | 776 | 10 | 7.44E-05 | 0.0137456 | 0.001626 | 0.001007 | 0.000718695 | ZNF740, CSAD |
| chr5 | 171308182 | 171308854 | 673 | 5 | 3.33E-05 | 0.0012149 | 0.013991 | 0.001008 | -0.00024276 | AC091980.2 |
| chr17 | 82905452 | 82906006 | 555 | 2 | 4.45E-05 | 0.0006761 | 0.004915 | 0.001014 | 0.000859432 | TBCD |
| chr1 | 201496649 | 201496661 | 13 | 2 | 0.000130256 | 0.1159168 | 0.000277 | 0.001017 | 0.001736113 | CSRP1 |
| chr14 | 94769065 | 94769152 | 88 | 2 | 0.000107519 | 0.0052134 | 0.000546 | 0.001028 | 0.001944339 | GSC |
| chr22 | 38283372 | 38283919 | 548 | 2 | 1.69E-05 | 0.0176768 | 0.000354 | 0.001031 | -0.00688286 | NA |
| chr1 | 6360653 | 6360766 | 114 | 3 | 4.57E-05 | 0.0413836 | 0.000278 | 0.001035 | -0.00604762 | ACOT7 |
| chr4 | 5050891 | 5051777 | 887 | 9 | 1.61E-05 | 0.002316 | 0.005596 | 0.001037 | -0.00019706 | STK32B |
| chr18 | 12657693 | 12657750 | 58 | 2 | 0.000156845 | 0.0005961 | 0.007831 | 0.001038 | 0.002108493 | SPIRE1 |
| chr19 | 43533543 | 43533714 | 172 | 3 | 8.36E-05 | 0.2119432 | 9.58E-05 | 0.001042 | -0.00263213 | ZNF575 |
| chr4 | 36281966 | 36282368 | 403 | 2 | 0.000115085 | 0.0006103 | 0.007553 | 0.001049 | -0.00120882 | DTHD1 |
| chr6 | 169851571 | 169851664 | 94 | 2 | 4.40E-05 | 0.0140177 | 0.000383 | 0.001049 | -0.00380403 | NA |
| chr2 | 78993755 | 78994140 | 386 | 2 | 4.25E-05 | 0.0015427 | 0.001398 | 0.001052 | 0.016609313 | NA |
| chr11 | 3378878 | 3379157 | 280 | 3 | 6.59E-05 | 0.001352 | 0.001871 | 0.001054 | 0.003013503 | ZNF195 |
| chr2 | 9638557 | 9638683 | 127 | 2 | 0.00014021 | 0.0094225 | 0.000442 | 0.001064 | -0.00561274 | NA |
| chr8 | 13132704 | 13133235 | 532 | 5 | 4.28E-05 | 0.0034386 | 0.00217 | 0.001065 | 7.05E-05 | DLC1 |
| chr11 | 62082389 | 62082487 | 99 | 2 | 0.000106044 | 0.0010298 | 0.002467 | 0.001074 | 0.003418601 | NA |
| chr10 | 99431007 | 99431167 | 161 | 3 | 0.000145861 | 0.000672 | 0.012098 | 0.001083 | 0.001668159 | AL391684.1 |
| chr7 | 21427445 | 21427787 | 343 | 3 | 1.58E-05 | 0.0114574 | 0.000433 | 0.001096 | 0.001591759 | NA |
| chr16 | 89695253 | 89696313 | 1061 | 4 | 7.32E-05 | 0.0065532 | 0.000604 | 0.001104 | -0.00183088 | CDK10 |
| chr8 | 9905649 | 9906042 | 394 | 3 | 0.000135669 | 0.0063614 | 0.006165 | 0.001109 | -0.00321132 | MIR124-1HG |
| chr16 | 88655110 | 88655557 | 448 | 2 | 3.35E-05 | 0.0006711 | 0.006853 | 0.001109 | -0.00433533 | MVD |
| chr7 | 151816155 | 151816242 | 88 | 2 | 0.000108289 | 0.0075691 | 0.000507 | 0.00111 | -0.00358615 | PRKAG2 |
| chr7 | 151411715 | 151411725 | 11 | 2 | 0.000116664 | 0.0103893 | 0.000452 | 0.001118 | 0.016178417 | WDR86-AS1 |
| chr17 | 48998396 | 48998518 | 123 | 3 | 4.10E-05 | 0.0385952 | 0.000171 | 0.001121 | -0.00256172 | IGF2BP1 |
| chr5 | 150778164 | 150778496 | 333 | 2 | 8.76E-05 | 0.0044342 | 0.000683 | 0.00114 | -0.00934693 | SMIM3 |
| chr13 | 87671938 | 87672624 | 687 | 5 | 9.61E-05 | 0.0057933 | 0.000735 | 0.00114 | 0.00210947 | SLITRK5, MIR4500HG |
| chr1 | 41022651 | 41023260 | 610 | 3 | 4.54E-05 | 0.0593173 | 0.001471 | 0.001142 | -0.00107418 | SLFNL1-AS1, SLFNL1 |
| chr8 | 18683936 | 18684275 | 340 | 6 | 6.07E-05 | 0.0202626 | 0.000359 | 0.001143 | -0.00367984 | PSD3 |
| chr17 | 44199257 | 44199563 | 307 | 2 | 7.77E-05 | 0.0015372 | 0.001641 | 0.001146 | 0.001914984 | AC004596.1, ATXN7L3 |
| chr4 | 53558309 | 53558620 | 312 | 5 | 0.000130789 | 0.0020037 | 0.007105 | 0.001146 | -0.00042022 | AC058822.1, LNX1 |
| chr17 | 27293570 | 27294204 | 635 | 10 | 8.58E-05 | 0.0595462 | 0.005645 | 0.001158 | -0.00069037 | WSB1, MIR4522 |
| chr15 | 39435082 | 39435722 | 641 | 3 | 9.50E-05 | 0.0126221 | 0.00032 | 0.001175 | 0.004682206 | NA |
| chr5 | 88269652 | 88269735 | 84 | 2 | 2.42E-05 | 0.0247017 | 0.000379 | 0.001178 | -0.01283594 | TMEM161B-AS1 |
| chr12 | 98893351 | 98893691 | 341 | 2 | 4.61E-05 | 0.0014291 | 0.001938 | 0.001194 | 0.021155986 | ANKS1B |
| chr1 | 113811820 | 113812371 | 552 | 6 | 5.69E-05 | 0.0173398 | 0.000749 | 0.001195 | 0.000466953 | RSBN1 |
| chr4 | 76895097 | 76895362 | 266 | 3 | 9.71E-05 | 0.0835578 | 0.00013 | 0.001201 | -0.00319061 | SOWAHB |
| chr12 | 104216202 | 104216286 | 85 | 2 | 7.40E-05 | 0.0066194 | 0.000597 | 0.001207 | 0.001110487 | TXNRD1 |
| chr16 | 73048636 | 73049115 | 480 | 2 | 8.68E-05 | 0.0264653 | 0.000389 | 0.001225 | -0.00305062 | ZFHX3 |
| chr4 | 88696774 | 88697516 | 743 | 6 | 3.85E-05 | 0.0319115 | 0.00383 | 0.001228 | -0.00262638 | AC098582.1, HERC3, NAP1L5 |
| chr19 | 36214020 | 36214193 | 174 | 2 | 0.000115049 | 0.0032342 | 0.000931 | 0.001229 | -0.00955845 | ZNF565 |
| chr17 | 62670032 | 62670235 | 204 | 2 | 7.56E-05 | 0.0007421 | 0.007375 | 0.00123 | -0.00578368 | MRC2 |
| chr13 | 113635744 | 113635941 | 198 | 3 | 8.73E-05 | 0.0019748 | 0.004848 | 0.001235 | -0.00168756 | TFDP1 |
| chr20 | 31513982 | 31514020 | 39 | 2 | 0.000116422 | 0.0008275 | 0.005587 | 0.001241 | -0.0091656 | NA |
| chr22 | 41149526 | 41149786 | 261 | 2 | 0.000157596 | 0.0124754 | 0.000481 | 0.001242 | -0.00286026 | EP300 |
| chr10 | 133350742 | 133350779 | 38 | 3 | 0.000132499 | 0.001072 | 0.003598 | 0.001242 | -0.00709141 | ZNF511-PRAP1, PRAP1 |
| chr5 | 1466828 | 1467202 | 375 | 2 | 0.000133436 | 0.0007886 | 0.006415 | 0.001244 | -0.00616803 | LPCAT1 |
| chr2 | 56091303 | 56091479 | 177 | 2 | 0.00012039 | 0.0013771 | 0.002229 | 0.001248 | -0.0011368 | AC011306.1 |
| chr11 | 65721453 | 65721669 | 217 | 2 | 0.000123493 | 0.0007682 | 0.007108 | 0.001254 | -0.00072806 | NA |
| chr5 | 65925544 | 65925936 | 393 | 3 | 3.06E-05 | 0.003111 | 0.002259 | 0.001261 | 0.000444115 | NA |
| chr2 | 238081444 | 238081649 | 206 | 4 | 7.51E-05 | 0.0042301 | 0.005948 | 0.00127 | -0.00174256 | UBE2F-SCLY, SCLY |
| chr14 | 102922425 | 102922730 | 306 | 6 | 7.82E-05 | 0.0023028 | 0.008428 | 0.001276 | -0.01060004 | AMN |
| chr18 | 46072206 | 46072289 | 84 | 3 | 0.000165837 | 0.0008136 | 0.012291 | 0.001277 | 0.000259177 | PSTPIP2 |
| chr14 | 80211504 | 80211722 | 219 | 2 | 0.000138725 | 0.0017406 | 0.001743 | 0.001278 | 0.017969864 | DIO2-AS1, DIO2 |
| chr20 | 63979854 | 63979946 | 93 | 2 | 6.65E-05 | 0.0010873 | 0.003481 | 0.001291 | -0.00187725 | SAMD10 |
| chr19 | 8592808 | 8593032 | 225 | 3 | 9.27E-05 | 0.0036384 | 0.006652 | 0.001295 | -0.00403399 | ADAMTS10 |
| chr8 | 30082482 | 30083068 | 587 | 5 | 8.61E-05 | 0.0059333 | 0.004946 | 0.001303 | -0.0007248 | AC044849.1, SARAF |
| chr10 | 88583528 | 88584263 | 736 | 4 | 1.85E-05 | 0.0005368 | 0.019553 | 0.001306 | -0.00771393 | RNLS |
| chr2 | 11235677 | 11235720 | 44 | 2 | 0.000111987 | 0.0187605 | 0.000452 | 0.001308 | -0.00467182 | ROCK2 |
| chr12 | 49707026 | 49707244 | 219 | 2 | 7.49E-05 | 0.1010235 | 0.000359 | 0.001318 | 0.002578064 | FMNL3 |
| chr8 | 22691590 | 22692055 | 466 | 3 | 4.52E-05 | 0.0011315 | 0.007832 | 0.001325 | -0.00136924 | AC055854.1, EGR3 |
| chr17 | 45983751 | 45983989 | 239 | 3 | 6.98E-05 | 0.1352228 | 0.000125 | 0.001336 | -0.00722341 | MAPT |
| chr3 | 5188124 | 5188249 | 126 | 4 | 0.000114364 | 0.0211923 | 0.000238 | 0.001374 | 0.001349124 | EDEM1, AC026202.3 |
| chr5 | 139276044 | 139276077 | 34 | 2 | 0.000112354 | 0.1385663 | 0.000369 | 0.001375 | -0.01125591 | MATR3, SNHG4, SIL1 |
| chr19 | 19663125 | 19663295 | 171 | 3 | 0.00015348 | 0.0009074 | 0.012779 | 0.001387 | -0.00297987 | ATP13A1 |
| chr11 | 46239067 | 46239298 | 232 | 4 | 0.000128183 | 0.0008252 | 0.00742 | 0.00142 | 0.000903366 | AC116021.1 |
| chr4 | 1211571 | 1211659 | 89 | 2 | 0.000141949 | 0.0011745 | 0.003914 | 0.001431 | -0.00127175 | CTBP1-AS, CTBP1 |
| chr6 | 83095698 | 83096502 | 805 | 2 | 8.92E-05 | 0.001575 | 0.002518 | 0.001447 | 0.003191084 | DOP1A |
| chr17 | 3929315 | 3929342 | 28 | 2 | 0.000146703 | 0.0191411 | 0.000506 | 0.001453 | -0.00548374 | ATP2A3 |
| chr16 | 69335167 | 69335329 | 163 | 2 | 0.000113393 | 0.0077672 | 0.000705 | 0.00146 | -0.00276789 | COG8, AC026464.3, AC026464.1 |
| chr15 | 29103997 | 29104226 | 230 | 3 | 0.000130974 | 0.0142016 | 0.000349 | 0.001462 | -0.00194041 | APBA2 |
| chr11 | 34641658 | 34641681 | 24 | 2 | 0.000169677 | 0.0128879 | 0.000576 | 0.001464 | -0.00956469 | EHF |
| chr11 | 76656133 | 76656321 | 189 | 2 | 3.86E-05 | 0.0018285 | 0.002166 | 0.001477 | -0.00477664 | AP001189.3 |
| chr22 | 45329041 | 45329067 | 27 | 2 | 0.000107817 | 0.0023434 | 0.001669 | 0.00149 | -0.02152438 | FAM118A |
| chr1 | 182392123 | 182392525 | 403 | 5 | 6.61E-05 | 0.0019361 | 0.004858 | 0.001494 | -0.00076242 | GLUL |
| chr4 | 73836904 | 73837064 | 161 | 5 | 0.00011074 | 0.0018309 | 0.024773 | 0.001498 | -2.54E-05 | CXCL6 |
| chr1 | 85464771 | 85465182 | 412 | 4 | 0.000112679 | 0.0061373 | 0.001053 | 0.001507 | -0.00055075 | DDAH1 |
| chr7 | 151167874 | 151168574 | 701 | 8 | 5.72E-05 | 0.0017125 | 0.021931 | 0.001515 | 0.001840188 | GBX1 |
| chr5 | 88678552 | 88678730 | 179 | 7 | 0.000142144 | 0.0077259 | 0.009892 | 0.001519 | -0.00154833 | MEF2C-AS2, LINC00461 |
| chr19 | 35628297 | 35628764 | 468 | 4 | 1.16E-05 | 0.0140103 | 0.006615 | 0.00152 | -0.00802438 | NA |
| chr1 | 48052731 | 48053322 | 592 | 2 | 5.48E-05 | 0.0048885 | 0.000959 | 0.001524 | -0.00978301 | LINC02794 |
| chr10 | 91219583 | 91220484 | 902 | 9 | 9.49E-06 | 0.015413 | 0.003621 | 0.001542 | 0.000656494 | PCGF5 |
| chr2 | 149038848 | 149039363 | 516 | 4 | 4.39E-05 | 0.0016209 | 0.002949 | 0.001545 | -0.00319059 | LYPD6B |
| chr8 | 32646591 | 32647400 | 810 | 6 | 7.56E-05 | 0.0020929 | 0.011163 | 0.001564 | 0.008113938 | NRG1, AC083977.1 |
| chr1 | 118979565 | 118979763 | 199 | 2 | 0.000106593 | 0.005069 | 0.001 | 0.001595 | -0.00753703 | TBX15 |
| chr6 | 109095032 | 109095417 | 386 | 8 | 0.000158211 | 0.0209615 | 0.008211 | 0.001613 | 0.000916838 | CEP57L1 |
| chr5 | 66828339 | 66828735 | 397 | 7 | 0.000141263 | 0.0123566 | 0.006656 | 0.001614 | -0.00298537 | MAST4 |
| chr19 | 43619675 | 43619880 | 206 | 4 | 0.000104991 | 0.0013425 | 0.0069 | 0.001621 | 0.002066114 | NA |
| chr2 | 85603323 | 85603361 | 39 | 2 | 6.42E-05 | 0.0013955 | 0.003949 | 0.001626 | -0.00232359 | USP39 |
| chr6 | 34055641 | 34056031 | 391 | 2 | 7.34E-05 | 0.0149939 | 0.000621 | 0.001631 | -0.00352718 | GRM4 |
| chr5 | 1839301 | 1840067 | 767 | 4 | 1.50E-05 | 0.0540763 | 0.000874 | 0.001636 | -0.00277134 | NA |
| chr1 | 24744173 | 24744181 | 9 | 2 | 2.55E-05 | 0.0028343 | 0.001656 | 0.001653 | 0.002020518 | NA |
| chr12 | 10172622 | 10172982 | 361 | 3 | 0.000129069 | 0.0097582 | 0.004445 | 0.001655 | -0.00573199 | TMEM52B |
| chr16 | 2075674 | 2075874 | 201 | 4 | 8.84E-05 | 0.0180516 | 0.000481 | 0.001664 | 0.000152252 | TSC2 |
| chr17 | 35731949 | 35732241 | 293 | 2 | 6.30E-05 | 0.0027005 | 0.001763 | 0.001669 | -0.00540417 | TAF15, RASL10B |
| chr16 | 714262 | 714420 | 159 | 2 | 3.99E-05 | 0.001317 | 0.004787 | 0.001682 | 0.008643088 | NA |
| chr6 | 35511851 | 35512352 | 502 | 5 | 0.000107372 | 0.0059667 | 0.009196 | 0.001686 | 0.001794502 | TULP1 |
| chr12 | 27161360 | 27161526 | 167 | 3 | 0.000108734 | 0.0050663 | 0.008868 | 0.001756 | -0.00055841 | AC092747.1 |
| chr22 | 50342765 | 50343101 | 337 | 3 | 0.00014216 | 0.0051766 | 0.00872 | 0.001782 | -0.00039177 | NA |
| chr7 | 150824535 | 150824582 | 48 | 2 | 0.000158947 | 0.0037831 | 0.00148 | 0.001802 | -0.00381275 | NA |
| chr2 | 54330358 | 54330837 | 480 | 7 | 0.000111711 | 0.0088994 | 0.008625 | 0.00184 | -7.50E-05 | C2orf73 |
| chr21 | 36479549 | 36479643 | 95 | 2 | 7.56E-05 | 0.0016586 | 0.004037 | 0.001855 | 0.006675048 | AP000695.1, AP000695.2, CLDN14 |
| chr16 | 86929141 | 86929473 | 333 | 2 | 6.85E-05 | 0.0324829 | 0.000592 | 0.001876 | -0.00124037 | AC093519.2 |
| chr10 | 72354405 | 72354855 | 451 | 3 | 7.42E-05 | 0.1247472 | 0.000187 | 0.001878 | -0.00131427 | DNAJB12 |
| chr18 | 21600401 | 21600709 | 309 | 2 | 0.000161579 | 0.0012489 | 0.007413 | 0.001881 | 0.000399446 | ESCO1 |
| chr2 | 240573863 | 240574272 | 410 | 3 | 5.05E-05 | 0.0021827 | 0.003992 | 0.001923 | -0.00260716 | RNPEPL1 |
| chr13 | 70107672 | 70107966 | 295 | 7 | 9.57E-05 | 0.0024195 | 0.038684 | 0.001924 | 0.002100558 | ATXN8OS, KLHL1 |
| chr3 | 134796039 | 134796301 | 263 | 2 | 7.01E-05 | 0.002429 | 0.002614 | 0.001946 | 0.00196407 | EPHB1, AC016931.1 |
| chr22 | 46366944 | 46367046 | 103 | 2 | 0.000158152 | 0.0012115 | 0.009057 | 0.00195 | -0.02840997 | CELSR1 |
| chr3 | 181718855 | 181719357 | 503 | 2 | 3.85E-05 | 0.003873 | 0.001651 | 0.001954 | -0.01370078 | SOX2-OT |
| chr5 | 72815922 | 72816536 | 615 | 11 | 7.13E-05 | 0.0029475 | 0.034186 | 0.001994 | 0.001991132 | TNPO1, AC035140.1 |
| chr19 | 49930005 | 49930692 | 688 | 4 | 4.82E-06 | 0.134153 | 9.67E-05 | 0.002001 | -0.00343907 | ATF5, AC011452.1 |
| chr1 | 12063678 | 12063879 | 202 | 3 | 0.000114936 | 0.0243147 | 0.006494 | 0.002026 | 0.004463786 | TNFRSF8 |
| chr2 | 10961277 | 10961466 | 190 | 3 | 0.000108453 | 0.0009538 | 0.01656 | 0.002029 | -0.0134645 | NA |
| chr17 | 3723466 | 3723946 | 481 | 10 | 0.00010595 | 0.0689323 | 0.000721 | 0.002031 | 0.000466902 | HASPIN, ITGAE |
| chr9 | 135700962 | 135701165 | 204 | 2 | 7.49E-06 | 0.0191345 | 0.000743 | 0.002036 | 0.005617844 | SOHLH1 |
| chr9 | 124852720 | 124852739 | 20 | 2 | 0.00012151 | 0.001503 | 0.006157 | 0.002037 | -0.00686313 | NA |
| chr12 | 53454497 | 53455326 | 830 | 2 | 5.28E-05 | 0.0404399 | 0.000624 | 0.002049 | -0.01135352 | AC023509.1, PCBP2 |
| chr20 | 42981438 | 42981943 | 506 | 2 | 7.92E-05 | 0.0032414 | 0.002142 | 0.002071 | -0.00282099 | PTPRT |
| chr14 | 101060148 | 101061351 | 1204 | 10 | 5.09E-05 | 0.1659194 | 8.90E-05 | 0.002076 | 0.000199714 | MIR496 |
| chr13 | 113086912 | 113087291 | 380 | 3 | 7.78E-05 | 0.0041032 | 0.002689 | 0.002079 | -0.00206131 | MCF2L |
| chr14 | 101986350 | 101986502 | 153 | 2 | 9.84E-05 | 0.0257149 | 0.000703 | 0.002086 | -0.0033884 | DYNC1H1 |
| chr17 | 56592867 | 56593514 | 648 | 4 | 8.60E-05 | 0.0225608 | 0.000467 | 0.002129 | -0.00124801 | NA |
| chr8 | 104589387 | 104589413 | 27 | 2 | 0.000170446 | 0.0040118 | 0.001844 | 0.002133 | 0.003332103 | NA |
| chr16 | 30677640 | 30678142 | 503 | 3 | 8.66E-05 | 0.000758 | 0.031044 | 0.002165 | -0.0022987 | NA |
| chr1 | 110211635 | 110212084 | 450 | 2 | 2.45E-05 | 0.0158603 | 0.000852 | 0.002166 | -0.00746144 | KCNC4 |
| chr12 | 132625426 | 132625548 | 123 | 2 | 6.67E-05 | 0.0013345 | 0.010186 | 0.002181 | 0.0254798 | POLE |
| chr2 | 97995131 | 97995232 | 102 | 2 | 8.47E-05 | 0.0034704 | 0.002243 | 0.002211 | 0.001747826 | TMEM131 |
| chr15 | 45130651 | 45131345 | 695 | 2 | 8.99E-05 | 0.0288699 | 0.00073 | 0.002213 | -0.00199876 | DUOX1 |
| chr2 | 32010171 | 32010701 | 531 | 3 | 9.55E-05 | 0.0010004 | 0.01777 | 0.002279 | -0.00188635 | MEMO1, DPY30 |
| chr2 | 218568856 | 218569279 | 424 | 5 | 0.000118417 | 0.0171875 | 0.00779 | 0.002295 | 0.000389044 | CNOT9 |
| chr7 | 1747099 | 1747558 | 460 | 2 | 5.28E-05 | 0.0101995 | 0.001119 | 0.002317 | -0.00049466 | ELFN1 |
| chr8 | 43139946 | 43140421 | 476 | 4 | 3.46E-05 | 0.0059862 | 0.006784 | 0.002344 | 0.000398063 | NA |
| chr10 | 14536264 | 14536794 | 531 | 3 | 0.000133361 | 0.0020947 | 0.00477 | 0.002367 | -0.0069812 | FAM107B |
| chr20 | 64061757 | 64062076 | 320 | 2 | 0.000132406 | 0.0126585 | 0.001058 | 0.002397 | -0.00515895 | TCEA2 |
| chr1 | 169104313 | 169104607 | 295 | 2 | 0.00014539 | 0.0038619 | 0.002339 | 0.002406 | 0.001681935 | AL031726.1 |
| chr11 | 64917411 | 64917733 | 323 | 6 | 0.000117374 | 0.0019443 | 0.011163 | 0.002428 | -0.0026774 | PPP2R5B |
| chr11 | 133064882 | 133065225 | 344 | 4 | 0.000101727 | 0.0031036 | 0.021146 | 0.002436 | 0.005293183 | AP004782.1, OPCML |
| chr4 | 47031378 | 47031617 | 240 | 6 | 9.55E-05 | 0.0078276 | 0.023121 | 0.002436 | -0.00217254 | GABRB1 |
| chr1 | 26913987 | 26914023 | 37 | 2 | 0.000176692 | 0.0014919 | 0.012816 | 0.002543 | -0.00313267 | NUDC |
| chr1 | 201023421 | 201023660 | 240 | 4 | 0.000159839 | 0.1995573 | 0.000409 | 0.002543 | 0.001792329 | KIF21B |
| chr7 | 150802405 | 150802649 | 245 | 2 | 0.000110348 | 0.0035743 | 0.002821 | 0.002558 | -0.00177718 | TMEM176A |
| chr16 | 29815433 | 29815605 | 173 | 3 | 6.21E-05 | 0.0089619 | 0.001595 | 0.00256 | -0.00397279 | PRRT2, AC009133.6, AC009133.1 |
| chr8 | 141220134 | 141220322 | 189 | 3 | 9.21E-05 | 0.1756929 | 0.000237 | 0.002575 | 0.001382499 | SLC45A4 |
| chr1 | 38876979 | 38877641 | 663 | 2 | 0.000105866 | 0.0170666 | 0.001039 | 0.002608 | -0.00719284 | AL139260.1, AL139260.3, GJA9 |
| chr14 | 50831652 | 50832019 | 368 | 2 | 7.46E-05 | 0.0286514 | 0.000889 | 0.002633 | 0.006484173 | AL133485.3 |
| chr10 | 5512077 | 5512584 | 508 | 2 | 0.000130954 | 0.0017416 | 0.009828 | 0.002669 | -0.00491173 | CALML3-AS1 |
| chr6 | 158559945 | 158559949 | 5 | 2 | 0.000101448 | 0.0028478 | 0.00416 | 0.002711 | -0.00253803 | TMEM181 |
| chr3 | 62318580 | 62319610 | 1031 | 18 | 0.000155345 | 0.0153613 | 0.017632 | 0.002714 | -0.00026339 | C3orf14, PTPRG-AS1 |
| chr14 | 24216135 | 24216779 | 645 | 6 | 6.75E-05 | 0.0087 | 0.005732 | 0.002725 | 4.54E-05 | NEDD8-MDP1 |
| chr12 | 55729174 | 55729199 | 26 | 3 | 0.000160139 | 0.0020287 | 0.01186 | 0.00274 | 0.001812848 | AC009779.2, CD63 |
| chr15 | 39829163 | 39829356 | 194 | 2 | 6.01E-05 | 0.0596782 | 0.00081 | 0.002773 | -0.0030471 | GPR176 |
| chr1 | 153773735 | 153774112 | 378 | 3 | 9.85E-05 | 0.038558 | 0.000701 | 0.002783 | -0.00450695 | INTS3 |
| chr12 | 123115326 | 123115590 | 265 | 2 | 6.65E-05 | 0.0083513 | 0.001623 | 0.002799 | -0.00652967 | PITPNM2 |
| chr13 | 23428076 | 23428580 | 505 | 2 | 0.000130128 | 0.0109594 | 0.00139 | 0.002799 | -0.00607565 | SACS-AS1, SACS |
| chr8 | 102560204 | 102560277 | 74 | 3 | 0.000144623 | 0.0027709 | 0.004789 | 0.002802 | 0.001314931 | ODF1 |
| chr1 | 16440831 | 16441346 | 516 | 5 | 5.26E-05 | 0.0015958 | 0.027539 | 0.002811 | -0.00053373 | NECAP2 |
| chr10 | 92353894 | 92354712 | 819 | 2 | 0.000128055 | 0.019509 | 0.001094 | 0.002832 | -0.01303309 | MARCHF5 |

| | | | | | | | | | | |
|---|---|---|---|---|---|---|---|---|---|---|
| chr12 | 49351453 | 49351623 | 171 | 3 | 0.00016689 | 0.0077238 | 0.00778 | 0.00284 | -0.00630309 | DNAJC22 |
| chr3 | 171899612 | 171900446 | 835 | 4 | 6.33E-05 | 0.0035526 | 0.003679 | 0.002848 | -0.00050726 | TMEM212, TMEM212-AS1 |
| chr1 | 153967006 | 153967375 | 370 | 5 | 5.88E-05 | 0.0265265 | 0.000424 | 0.002942 | -0.00230413 | SLC39A1, AL358472.7 |
| chr1 | 100038210 | 100038330 | 121 | 4 | 0.000104817 | 0.0510105 | 0.002172 | 0.002952 | 0.001217783 | AC118553.2, MFSD14A |
| chr10 | 68105353 | 68105730 | 378 | 2 | 8.08E-05 | 0.0166075 | 0.001232 | 0.002962 | -0.00779357 | NA |
| chr2 | 46941301 | 46941483 | 183 | 3 | 6.87E-05 | 0.0024963 | 0.017175 | 0.002963 | 0.001823102 | TTC7A, MCFD2 |
| chr20 | 20017660 | 20017914 | 255 | 8 | 0.000156086 | 0.0091417 | 0.00406 | 0.003012 | 0.000601015 | NAA20 |
| chr1 | 169486291 | 169486661 | 371 | 4 | 5.44E-05 | 0.0167341 | 0.000723 | 0.003019 | 0.003739114 | Z99572.1 |
| chr15 | 57591194 | 57591313 | 120 | 2 | 2.93E-05 | 0.0106689 | 0.001572 | 0.003034 | 0.009888654 | NA |
| chr19 | 52139758 | 52139954 | 197 | 5 | 0.000151257 | 0.0039588 | 0.022637 | 0.00304 | 0.000618938 | ZNF616, AC011468.2 |
| chr1 | 6045264 | 6045799 | 536 | 4 | 9.66E-05 | 0.0046384 | 0.009815 | 0.003057 | -0.00259309 | KCNAB2 |
| chr6 | 10420463 | 10420836 | 374 | 5 | 0.000128187 | 0.0051424 | 0.015353 | 0.00307 | 0.00086946 | NA |
| chr17 | 75510377 | 75510449 | 73 | 2 | 0.00011231 | 0.0045676 | 0.003091 | 0.003101 | -0.00183866 | CASKIN2 |
| chr1 | 19882927 | 19882968 | 42 | 2 | 4.64E-05 | 0.0060162 | 0.002429 | 0.003124 | 0.001952707 | OTUD3 |
| chr18 | 48949927 | 48950452 | 526 | 3 | 5.47E-05 | 0.2055768 | 0.000732 | 0.003184 | 0.002422255 | SMAD7 |
| chr2 | 5932007 | 5932021 | 15 | 2 | 0.000104763 | 0.0752755 | 0.000916 | 0.003209 | -0.00306817 | NA |
| chr1 | 43172266 | 43172804 | 539 | 12 | 0.000151057 | 0.0144686 | 0.022567 | 0.003215 | -0.00030599 | CFAP57, EBNA1BP2 |
| chr11 | 122881532 | 122882228 | 697 | 3 | 6.56E-05 | 0.0059104 | 0.005343 | 0.003226 | 0.002084608 | NA |
| chr16 | 28846568 | 28847121 | 554 | 7 | 9.58E-05 | 0.1253855 | 0.000932 | 0.003262 | -0.00194093 | AC009093.11, SH2B1 |
| chr16 | 57619180 | 57619701 | 522 | 5 | 0.000101739 | 0.0046666 | 0.027813 | 0.003265 | -0.00335789 | ADGRG1 |
| chr2 | 118774465 | 118774738 | 274 | 4 | 0.000108346 | 0.0511044 | 0.000587 | 0.00331 | -4.73E-05 | LINC01956 |
| chr17 | 48817587 | 48817657 | 71 | 2 | 0.000151842 | 0.0035039 | 0.004909 | 0.003354 | -0.00645023 | NA |
| chr12 | 56257618 | 56258128 | 511 | 2 | 3.30E-05 | 0.0018624 | 0.01865 | 0.00336 | -0.00075145 | ANKRD52 |
| chr13 | 26051954 | 26051996 | 43 | 2 | 0.000162223 | 0.016759 | 0.001458 | 0.00339 | -0.00168279 | SHISA2 |
| chr12 | 6235158 | 6235213 | 56 | 2 | 0.000124062 | 0.0019224 | 0.018555 | 0.003446 | 0.004178829 | CD9 |
| chr14 | 73491568 | 73491576 | 9 | 2 | 4.35E-05 | 0.0061652 | 0.002819 | 0.003466 | 0.00238607 | RIOX1, HEATR4 |
| chr3 | 123415953 | 123415993 | 41 | 2 | 0.00015607 | 0.0019598 | 0.018629 | 0.003507 | -0.00282202 | ADCY5 |
| chr16 | 66853912 | 66853945 | 34 | 2 | 0.000156036 | 0.0105532 | 0.001942 | 0.003512 | 0.001503495 | CA7, NAE1 |
| chr11 | 65899881 | 65900453 | 573 | 2 | 0.000138884 | 0.0126689 | 0.001756 | 0.003524 | -0.00601811 | FOSL1 |
| chr3 | 184244296 | 184244831 | 536 | 2 | 3.68E-05 | 0.0045783 | 0.003969 | 0.00356 | 0.000421189 | ALG3 |
| chr11 | 10694617 | 10694908 | 292 | 2 | 4.14E-05 | 0.0139849 | 0.00171 | 0.00359 | -0.00435696 | NA |
| chr5 | 40841391 | 40841488 | 98 | 3 | 0.000130216 | 0.0050526 | 0.011504 | 0.003606 | -0.00396779 | CARD6 |
| chr8 | 127735102 | 127735449 | 348 | 4 | 9.36E-05 | 0.0460365 | 0.00039 | 0.003637 | 0.001176328 | MYC, CASC11 |
| chr1 | 112714661 | 112714980 | 320 | 2 | 0.000131243 | 0.0338952 | 0.00124 | 0.003646 | -0.00150906 | PPM1J |
| chr6 | 32395344 | 32395548 | 205 | 3 | 4.55E-05 | 0.0036996 | 0.005762 | 0.003669 | -0.0008827 | TSBP1-AS1, BTNL2 |
| chr16 | 69319941 | 69320388 | 448 | 2 | 0.00010905 | 0.0020759 | 0.01904 | 0.003699 | 0.002810588 | AC026464.4, VPS4A, COG8 |
| chr4 | 26583223 | 26583596 | 374 | 4 | 1.99E-05 | 0.0682719 | 0.0004 | 0.003751 | 0.007398997 | TBC1D19 |
| chr3 | 84959436 | 84959527 | 92 | 2 | 0.000108659 | 0.1062666 | 0.001052 | 0.003786 | 0.005548873 | CADM2 |
| chr16 | 85780325 | 85780560 | 236 | 3 | 3.12E-05 | 0.055177 | 0.000576 | 0.003794 | -0.00584297 | EMC8 |
| chr9 | 124777663 | 124777801 | 139 | 2 | 8.52E-05 | 0.0030983 | 0.007898 | 0.0038 | -0.00670313 | OLFML2A |
| chr3 | 13282459 | 13282632 | 174 | 2 | 9.20E-05 | 0.0083541 | 0.002673 | 0.003911 | 0.001325844 | IQSEC1 |
| chr3 | 52685559 | 52685905 | 347 | 3 | 9.22E-05 | 0.0016756 | 0.029525 | 0.003949 | -0.00132605 | GNL3, PBRM1 |
| chr7 | 209447 | 209587 | 141 | 2 | 0.000116408 | 0.0547029 | 0.001213 | 0.00395 | 0.009439862 | FAM20C |
| chr14 | 77457347 | 77457358 | 12 | 2 | 0.000143726 | 0.0048105 | 0.00462 | 0.003971 | 0.002337141 | VIPAS39 |
| chr12 | 132847980 | 132848750 | 771 | 7 | 1.66E-05 | 0.3377201 | 0.002106 | 0.003988 | -0.00216577 | CHFR |
| chr6 | 30987935 | 30988215 | 281 | 4 | 0.000121142 | 0.0042522 | 0.010501 | 0.004023 | 0.000641866 | MUC21 |
| chr2 | 99141999 | 99142341 | 343 | 2 | 6.97E-05 | 0.0038906 | 0.006701 | 0.004147 | -0.00990999 | AC079447.1, C2orf15, TSGA10 |
| chr20 | 62407355 | 62407466 | 112 | 2 | 0.000163892 | 0.0050641 | 0.004735 | 0.004149 | 0.002836952 | NA |
| chr14 | 71932836 | 71932874 | 39 | 2 | 6.53E-05 | 0.0077678 | 0.003137 | 0.004168 | 0.003746356 | RGS6 |
| chr1 | 46550442 | 46550729 | 288 | 2 | 5.78E-05 | 0.008351 | 0.002977 | 0.004185 | -0.0104621 | MKNK1-AS1, KNCN |
| chr1 | 235328048 | 235328796 | 749 | 11 | 5.84E-05 | 0.0148108 | 0.002384 | 0.004218 | 0.000195353 | GGPS1, AL357556.3, ARID4B |
| chr10 | 95560516 | 95561110 | 595 | 3 | 2.68E-05 | 0.029191 | 0.00272 | 0.004234 | -0.00171903 | SORBS1 |
| chr10 | 27746702 | 27746802 | 101 | 4 | 9.24E-05 | 0.0091193 | 0.006697 | 0.004248 | -0.00338491 | MKX-AS1 |
| chr12 | 123934331 | 123934549 | 219 | 2 | 8.39E-05 | 0.0048339 | 0.005333 | 0.004284 | -0.0066104 | DNAH10, CCDC92, DNAH10OS |
| chr3 | 128123779 | 128123861 | 83 | 4 | 0.000112451 | 0.0156938 | 0.001106 | 0.004298 | 0.001967463 | RUVBL1 |
| chr3 | 47781577 | 47781801 | 225 | 3 | 0.000150654 | 0.0227038 | 0.002539 | 0.004371 | 0.000959977 | SMARCC1 |
| chr14 | 22589718 | 22589873 | 156 | 3 | 0.000144803 | 0.0069791 | 0.003675 | 0.004372 | -0.00263594 | NA |
| chr10 | 110210877 | 110210964 | 88 | 2 | 8.66E-05 | 0.0067148 | 0.004071 | 0.004499 | -0.00229695 | MXI1 |
| chr2 | 108942139 | 108942556 | 418 | 3 | 6.09E-05 | 0.1243981 | 0.000932 | 0.004501 | 0.002173366 | EDAR |
| chr14 | 22501519 | 22501587 | 69 | 2 | 0.000130468 | 0.0155717 | 0.002201 | 0.004501 | -0.00875167 | NA |
| chr20 | 49811781 | 49812370 | 590 | 3 | 7.30E-05 | 0.0057427 | 0.007164 | 0.004533 | -0.00492279 | NA |
| chr16 | 50497645 | 50498240 | 596 | 2 | 0.000154357 | 0.0155671 | 0.002242 | 0.004559 | -0.00415137 | AC007493.1 |
| chr1 | 2983390 | 2983599 | 210 | 2 | 0.000105155 | 0.0256797 | 0.001782 | 0.004563 | -0.0045146 | NA |
| chr7 | 32956533 | 32956702 | 170 | 3 | 0.000105815 | 0.068708 | 0.000811 | 0.00462 | -0.0046734 | NA |
| chr4 | 185143040 | 185143096 | 57 | 3 | 0.000157138 | 0.0114741 | 0.003948 | 0.004629 | 0.002002695 | NA |
| chr17 | 2401852 | 2401866 | 15 | 2 | 0.000161274 | 0.02262 | 0.001943 | 0.00469 | 0.00090601 | NA |
| chr14 | 104513035 | 104513041 | 7 | 2 | 0.000170485 | 0.0277858 | 0.001816 | 0.004743 | 0.009063567 | TMEM179 |
| chr6 | 33248989 | 33249526 | 538 | 12 | 7.06E-05 | 0.0376936 | 0.012612 | 0.004773 | 0.000224412 | NA |
| chr16 | 1774040 | 1774176 | 137 | 2 | 7.24E-05 | 0.0036379 | 0.011185 | 0.004877 | -0.01001413 | EME2 |
| chr7 | 92589965 | 92590024 | 60 | 2 | 0.000122121 | 0.0031517 | 0.016111 | 0.004954 | -0.00040803 | FAM133B |
| chr17 | 21125876 | 21126383 | 508 | 3 | 4.67E-05 | 0.0214187 | 0.015029 | 0.004988 | -0.00343942 | DHRS7B |
| chr17 | 76969532 | 76969626 | 95 | 2 | 0.00013092 | 0.0129803 | 0.002883 | 0.005043 | 0.001904253 | NA |
| chr14 | 20802248 | 20802458 | 211 | 3 | 0.000132513 | 0.1340487 | 0.003597 | 0.005078 | 0.004507062 | RNASE1 |
| chr2 | 55952372 | 55952566 | 195 | 2 | 0.000141157 | 0.0091713 | 0.003797 | 0.0051 | 0.008522648 | AC011306.1 |
| chr22 | 20106215 | 20106563 | 349 | 2 | 0.000172103 | 0.0108468 | 0.003327 | 0.005101 | 0.002921849 | DGCR8 |
| chr7 | 40135244 | 40135437 | 194 | 2 | 9.57E-05 | 0.0070889 | 0.005192 | 0.005312 | 0.006878292 | SUGCT |
| chr16 | 23713580 | 23713622 | 43 | 2 | 0.000157113 | 0.0036474 | 0.014525 | 0.00536 | -0.01450829 | NA |
| chr16 | 89584096 | 89584123 | 28 | 2 | 0.000139634 | 0.0093169 | 0.004108 | 0.005391 | -0.00184772 | CPNE7 |
| chr8 | 85177237 | 85178334 | 1098 | 10 | 2.80E-05 | 0.0265224 | 0.011979 | 0.005391 | 0.000503027 | E2F5, AC011773.1 |
| chr21 | 46228731 | 46228764 | 34 | 2 | 0.000159182 | 0.0032067 | 0.022389 | 0.005522 | -0.00039505 | LSS |
| chr12 | 53074758 | 53075071 | 314 | 2 | 0.000115574 | 0.0242566 | 0.002371 | 0.005589 | -0.00384522 | SPRYD3 |
| chr12 | 10674795 | 10674984 | 190 | 3 | 4.44E-05 | 0.0033712 | 0.016982 | 0.005654 | 0.009092277 | NA |
| chr5 | 138213702 | 138213848 | 147 | 2 | 0.000144478 | 0.0287481 | 0.002247 | 0.005667 | 0.002305299 | NA |
| chr16 | 85611965 | 85612583 | 619 | 3 | 9.56E-05 | 0.0230057 | 0.017066 | 0.00587 | -0.00023155 | GSE1 |
| chr2 | 176083868 | 176084289 | 422 | 4 | 1.00E-04 | 0.0044213 | 0.009639 | 0.005937 | -0.00361524 | EVX2 |
| chr19 | 57560399 | 57560811 | 413 | 2 | 5.03E-05 | 0.0043302 | 0.013478 | 0.005937 | -0.00153121 | NA |
| chr22 | 44765775 | 44765919 | 145 | 2 | 5.20E-05 | 0.0042641 | 0.013996 | 0.00595 | -0.00701992 | PRR5-ARHGAP8, ARHGAP8 |
| chr17 | 58219552 | 58219832 | 281 | 5 | 0.000143937 | 0.0284095 | 0.005528 | 0.006025 | 0.00452159 | LPO, AC005962.2, MKS1 |
| chr18 | 55590109 | 55590370 | 262 | 3 | 0.000127883 | 0.0083051 | 0.004931 | 0.00606 | -0.00309554 | AC022031.2, TCF4 |
| chr3 | 143972593 | 143973164 | 572 | 6 | 8.69E-05 | 0.0808634 | 0.000508 | 0.006165 | 0.000431539 | DIPK2A |
| chr2 | 219171518 | 219171938 | 421 | 2 | 5.96E-05 | 0.0094325 | 0.005388 | 0.006362 | -0.00121641 | CNPPD1 |
| chr10 | 62818371 | 62818716 | 346 | 4 | 8.88E-05 | 0.0614277 | 0.022432 | 0.006369 | 0.00320762 | EGR2 |
| chr19 | 44234964 | 44235236 | 273 | 2 | 3.01E-05 | 0.0072814 | 0.0075 | 0.006534 | -0.00114442 | ZNF227, ZNF235 |
| chr20 | 1118811 | 1118911 | 101 | 2 | 0.000170284 | 0.003769 | 0.027057 | 0.006624 | 8.80E-06 | PSMF1 |
| chr1 | 2194537 | 2194540 | 4 | 2 | 0.000176431 | 0.0081777 | 0.006781 | 0.006651 | 0.010304876 | FAAP20 |
| chr7 | 4010946 | 4011015 | 70 | 2 | 9.77E-05 | 0.0041425 | 0.021072 | 0.006661 | -0.00546814 | SDK1 |
| chr8 | 144046281 | 144046423 | 143 | 2 | 9.62E-05 | 0.0139745 | 0.004247 | 0.006722 | -0.00547698 | SPATC1 |
| chr17 | 36544130 | 36544384 | 255 | 3 | 8.72E-05 | 0.2461968 | 0.000866 | 0.00676 | 0.001210586 | NA |
| chr11 | 47848253 | 47848562 | 310 | 5 | 0.000138456 | 0.0039387 | 0.020293 | 0.006833 | -0.00036975 | NUP160 |
| chr11 | 85647973 | 85648516 | 544 | 4 | 2.84E-05 | 0.2440407 | 0.000542 | 0.006871 | 0.008200571 | TMEM126A |
| chr2 | 68734363 | 68734465 | 103 | 2 | 0.000107974 | 0.0299327 | 0.002908 | 0.006995 | 0.003823204 | ARHGAP25 |
| chr11 | 74988008 | 74988425 | 418 | 2 | 1.09E-05 | 0.0141104 | 0.004548 | 0.007055 | -0.00903754 | NEU3 |
| chr19 | 1848351 | 1848441 | 91 | 2 | 0.00016731 | 0.0169202 | 0.004451 | 0.007567 | -0.00059114 | REXO1 |
| chr19 | 7596039 | 7596731 | 693 | 5 | 4.70E-05 | 0.1784475 | 0.00023 | 0.007627 | -0.00434899 | CAMSAP3 |
| chr12 | 122865405 | 122865530 | 126 | 2 | 0.000125784 | 0.0073245 | 0.010286 | 0.00763 | -0.00357642 | AC027290.1, VPS37B |
| chr22 | 36365907 | 36365926 | 20 | 2 | 0.00014569 | 0.0044538 | 0.027597 | 0.00763 | 0.002480958 | MYH9 |
| chr6 | 169501376 | 169501408 | 33 | 2 | 0.000106196 | 0.0215072 | 0.003937 | 0.00772 | -0.00129135 | AL031315.1, WDR27 |
| chr10 | 486469 | 486968 | 500 | 3 | 7.63E-05 | 0.0164135 | 0.004698 | 0.007759 | -0.00123094 | DIP2C |
| chr6 | 30883136 | 30883861 | 726 | 9 | 6.89E-05 | 0.0558282 | 0.009695 | 0.007959 | -0.00071478 | DDR1 |
| chr1 | 905687 | 905909 | 223 | 2 | 0.00012379 | 0.0353097 | 0.003435 | 0.008368 | 0.004448246 | AL645608.6 |
| chr19 | 29664683 | 29664959 | 277 | 2 | 1.82E-05 | 0.0407776 | 0.003274 | 0.008435 | 0.003403405 | NA |
| chr1 | 50418872 | 50419460 | 589 | 2 | 0.000123313 | 0.0462673 | 0.003212 | 0.008618 | -0.00400935 | DMRTA2 |
| chr20 | 62219812 | 62220100 | 289 | 2 | 0.0001113 | 0.0174827 | 0.005479 | 0.008783 | 0.001662034 | HRH3 |
| chr11 | 27363049 | 27363261 | 213 | 2 | 6.37E-05 | 0.008465 | 0.011918 | 0.00896 | 0.003636526 | CCDC34 |
| chr9 | 134409897 | 134410300 | 404 | 2 | 0.00011247 | 0.0068105 | 0.018119 | 0.0092 | -0.00786316 | RXRA |
| chr17 | 50867003 | 50867301 | 299 | 3 | 0.000129787 | 0.0041211 | 0.04637 | 0.009204 | 0.000515287 | TOB1-AS1, TOB1 |
| chr1 | 67307445 | 67307757 | 313 | 5 | 0.000131993 | 0.0179901 | 0.038228 | 0.009356 | -0.00019101 | IL12RB2 |
| chr1 | 167455427 | 167455897 | 471 | 2 | 9.49E-05 | 0.0064025 | 0.021829 | 0.009405 | -0.00188937 | AL359962.3, CD247 |
| chr7 | 158141313 | 158141808 | 496 | 3 | 5.79E-05 | 0.0197002 | 0.01081 | 0.009571 | 0.001341736 | PTPRN2 |
| chr9 | 136510752 | 136511215 | 464 | 5 | 0.000171005 | 0.0116848 | 0.021536 | 0.009712 | 0.000416146 | NOTCH1 |
| chr14 | 103628442 | 103628581 | 140 | 2 | 0.000128831 | 0.0107756 | 0.010841 | 0.009964 | 0.002795474 | KLC1, AL139300.1 |
| chr2 | 148882043 | 148882253 | 211 | 2 | 0.00012163 | 0.0193906 | 0.00636 | 0.010124 | -0.00307324 | KIF5C, KIF5C-AS1 |
| chr12 | 108696812 | 108697245 | 434 | 2 | 0.000115033 | 0.0128994 | 0.009424 | 0.010302 | -0.0008659 | CORO1C |
| chr18 | 12702186 | 12702229 | 44 | 2 | 0.00015082 | 0.0097043 | 0.014374 | 0.010657 | 0.003403795 | PSMG2, CEP76 |
| chr1 | 36323787 | 36323950 | 164 | 4 | 4.89E-05 | 0.0101151 | 0.033892 | 0.010846 | 0.00073106 | SH3D21, EVA1B |
| chr1 | 18109996 | 18110240 | 245 | 2 | 7.79E-05 | 0.1489439 | 0.00331 | 0.011072 | -0.00132708 | IGSF21, AL365209.1 |
| chr10 | 75210363 | 75210738 | 376 | 4 | 0.000110513 | 0.0534708 | 0.008133 | 0.011515 | 0.000426478 | VDAC2 |
| chr18 | 10032281 | 10032702 | 422 | 3 | 0.000101889 | 0.1362745 | 0.001598 | 0.011666 | 0.002524395 | NA |
| chr11 | 46120044 | 46120449 | 406 | 2 | 5.79E-05 | 0.0094248 | 0.018582 | 0.011668 | -5.32E-05 | PHF21A |
| chr8 | 11770303 | 11770465 | 163 | 2 | 8.10E-05 | 0.0127114 | 0.012669 | 0.011896 | 0.008239294 | NEIL2 |
| chr6 | 30730807 | 30731003 | 197 | 6 | 0.000134397 | 0.0410465 | 0.040669 | 0.012606 | -0.00331348 | FLOT1 |
| chr4 | 139301704 | 139301909 | 206 | 2 | 9.60E-05 | 0.0073296 | 0.040572 | 0.012668 | 0.000830048 | NAA15, NDUFC1 |
| chr11 | 65109146 | 65109304 | 159 | 2 | 0.000145401 | 0.0096691 | 0.021978 | 0.012703 | -0.00056203 | VPS51 |
| chr1 | 31617362 | 31617934 | 573 | 4 | 6.61E-05 | 0.0095486 | 0.016145 | 0.012955 | -0.0040028 | HCRTR1 |
| chr22 | 40636663 | 40636688 | 26 | 2 | 0.000163778 | 0.1529347 | 0.004074 | 0.013296 | -0.00172877 | MRTFA |
| chr3 | 16884040 | 16884215 | 176 | 2 | 0.000124223 | 0.012074 | 0.01856 | 0.013767 | -0.00647138 | PLCL2 |
| chr2 | 69013063 | 69013622 | 560 | 6 | 9.52E-05 | 0.3654679 | 0.001988 | 0.013915 | 0.000778543 | ANTXR1 |
| chr6 | 95576616 | 95577060 | 445 | 3 | 0.000140246 | 0.0095646 | 0.026826 | 0.014216 | -0.00109456 | MANEA-DT |
| chr15 | 50182557 | 50182842 | 286 | 2 | 5.29E-05 | 0.0467653 | 0.006259 | 0.014302 | 0.001819344 | SLC27A2, ATP8B4 |
| chr2 | 32947375 | 32947377 | 3 | 2 | 9.26E-05 | 0.1048632 | 0.004854 | 0.014535 | 0.001220453 | LTBP1 |
| chr10 | 133114551 | 133114591 | 41 | 2 | 0.00015687 | 0.011739 | 0.0239 | 0.015032 | -0.00251403 | ADGRA1 |
| chr5 | 50963899 | 50963941 | 43 | 2 | 9.79E-05 | 0.0091308 | 0.040877 | 0.015103 | -0.00868123 | NA |
| chr5 | 176743703 | 176743794 | 92 | 2 | 0.000136162 | 0.1324541 | 0.004944 | 0.015365 | 0.006163186 | LINC01574 |
| chr6 | 31764739 | 31765657 | 919 | 2 | 6.31E-05 | 0.0154395 | 0.016898 | 0.015403 | 0.002110468 | MSH5-SAPCD1, SAPCD1, SAPCD1-AS1, VWA7 |
| chr2 | 219552800 | 219553131 | 332 | 2 | 7.03E-05 | 0.0105182 | 0.035796 | 0.016133 | -0.00313457 | OBSL1 |
| chr12 | 54005406 | 54005710 | 305 | 4 | 8.37E-05 | 0.0170301 | 0.034377 | 0.017156 | -0.00107532 | AC012531.3, HOXC6 |
| chr7 | 134890773 | 134891321 | 549 | 4 | 6.55E-05 | 0.0192573 | 0.037689 | 0.017367 | 0.001041166 | CALD1, AC083870.1 |
| chr17 | 7688246 | 7688353 | 108 | 4 | 0.000116702 | 0.1259141 | 0.004329 | 0.017371 | 0.001226842 | WRAP53 |
| chr11 | 33257538 | 33257590 | 53 | 4 | 0.000134943 | 0.0265412 | 0.028791 | 0.017413 | 0.000600257 | HIPK3 |
| chr7 | 102249881 | 102250133 | 253 | 2 | 0.000126254 | 0.0219719 | 0.014841 | 0.017691 | -0.00752671 | CUX1 |
| chr20 | 2814231 | 2814955 | 725 | 4 | 0.00014095 | 0.0637509 | 0.005705 | 0.018437 | -0.00374314 | NA |
| chr13 | 41311159 | 41311521 | 363 | 4 | 6.92E-05 | 0.0105011 | 0.040804 | 0.018866 | -0.00031299 | NAA16 |
| chr10 | 69577348 | 69577830 | 483 | 2 | 0.000117959 | 0.099928 | 0.00694 | 0.019167 | -0.0043576 | NA |
| chr11 | 2868372 | 2868399 | 28 | 3 | 0.000149711 | 0.0208196 | 0.02503 | 0.019204 | 0.003059901 | NA |
| chr5 | 147454374 | 147454398 | 25 | 2 | 0.0001629 | 0.0374012 | 0.014759 | 0.023116 | -0.00083559 | DPYSL3 |
| chr12 | 94149201 | 94149673 | 473 | 2 | 0.000137775 | 0.0143597 | 0.057744 | 0.023863 | 0.001634222 | PLXNC1 |
| chr12 | 132341719 | 132341859 | 141 | 2 | 0.00016677 | 0.0143906 | 0.060369 | 0.024231 | -0.00743306 | NA |
| chr6 | 6008148 | 6008409 | 262 | 2 | 7.28E-05 | 0.0162348 | 0.047476 | 0.024527 | -0.00026322 | NA |

| | | | | | | | | | | |
|---|---|---|---|---|---|---|---|---|---|---|
| chr21 | 42338322 | 42338734 | 413 | 2 | 0.000163231 | 0.029181 | 0.022315 | 0.025712 | -0.00364939 | NA |
| chr14 | 59188141 | 59188203 | 63 | 2 | 5.17E-05 | 0.0209498 | 0.035192 | 0.026087 | 0.001585383 | NA |
| chr3 | 129278212 | 129278452 | 241 | 2 | 0.000110785 | 0.0308183 | 0.022177 | 0.026413 | 0.001248066 | AC137695.3 |
| chr3 | 45500342 | 45500463 | 122 | 2 | 0.000140863 | 0.0194465 | 0.042238 | 0.026718 | -0.00450069 | LARS2, LARS2-AS1 |
| chr19 | 38703866 | 38704716 | 851 | 2 | 0.000114606 | 0.1030789 | 0.010635 | 0.02681 | -0.00204103 | ACTN4 |
| chr2 | 202265870 | 202265922 | 53 | 2 | 0.000139361 | 0.0293786 | 0.024178 | 0.026908 | 0.001975039 | NOP58 |
| chr17 | 50479099 | 50479147 | 49 | 2 | 0.000141799 | 0.167796 | 0.00985 | 0.028054 | 0.000927939 | RSAD1 |
| chr14 | 21056507 | 21056714 | 208 | 2 | 0.000165998 | 0.0190786 | 0.064775 | 0.030724 | -0.00318723 | NDRG2 |
| chr12 | 56158020 | 56158348 | 329 | 8 | 0.00015495 | 0.2411562 | 0.002929 | 0.032827 | 0.00147391 | MYL6B, MYL6, AC034102.5 |
| chr7 | 107661348 | 107661397 | 50 | 2 | 9.20E-05 | 0.0484926 | 0.02427 | 0.035118 | 0.000484342 | SLC26A4, SLC26A4-AS1 |
| chr3 | 52220826 | 52221389 | 564 | 2 | 0.000169207 | 0.0221196 | 0.07706 | 0.03636 | -0.00120271 | TLR9, AC097637.1 |
| chr15 | 62823066 | 62823141 | 76 | 2 | 0.000160924 | 0.1115632 | 0.017014 | 0.038675 | -0.00315061 | TLN2 |
| chr22 | 40819820 | 40819935 | 116 | 2 | 0.000158179 | 0.1757292 | 0.014764 | 0.038906 | -0.00449201 | NA |
| chr17 | 47323270 | 47323426 | 157 | 3 | 0.000100509 | 0.0417191 | 0.068372 | 0.041404 | 0.008458646 | AC068234.1, EFCAB13, THCAT158 |
| chr5 | 35048672 | 35048840 | 169 | 2 | 0.000151239 | 0.0528127 | 0.030352 | 0.041632 | -0.00504205 | PRLR |
| chr7 | 150450448 | 150450465 | 18 | 2 | 0.000162061 | 0.0672655 | 0.027126 | 0.043715 | 0.002122172 | NA |
| chr4 | 10117311 | 10117360 | 50 | 2 | 0.000142327 | 0.0290533 | 0.086987 | 0.046408 | -0.00016385 | NA |
| chr22 | 22559639 | 22559831 | 193 | 4 | 0.000135941 | 0.0365016 | 0.05647 | 0.046673 | -0.00526831 | LL22NC03-63E9.3 |
| chr19 | 23070514 | 23070995 | 482 | 3 | 0.000111453 | 0.0507026 | 0.041887 | 0.047164 | 0.000751433 | AC074135.2 |
| chr3 | 42590349 | 42590799 | 451 | 5 | 0.000116383 | 0.1416116 | 0.066936 | 0.047709 | 0.000177478 | SS18L2, SEC22C |
| chr11 | 70435862 | 70435946 | 85 | 2 | 2.78E-05 | 0.171341 | 0.022178 | 0.052668 | -0.00182913 | CTTN |
| chr12 | 21501353 | 21501730 | 378 | 6 | 0.000120896 | 0.1160527 | 0.026015 | 0.052725 | 0.000852659 | RECQL |
| chr18 | 9136290 | 9136386 | 97 | 3 | 0.000153237 | 0.0885324 | 0.071514 | 0.053145 | -0.0015728 | AP005263.1, ANKRD12, NDUFV2-AS1 |
| chr10 | 46023498 | 46023708 | 211 | 6 | 0.000135726 | 0.0203601 | 0.150653 | 0.054303 | -0.00171702 | NCOA4 |
| chr14 | 96269295 | 96269917 | 623 | 2 | 4.54E-05 | 0.1378638 | 0.026383 | 0.056259 | 0.004996303 | NA |
| chr17 | 40863293 | 40863304 | 12 | 2 | 0.000174793 | 0.085757 | 0.037221 | 0.059015 | -0.01007487 | KRT12 |
| chr13 | 113208201 | 113208913 | 713 | 9 | 0.000123465 | 0.3097924 | 0.021779 | 0.060183 | -0.00123667 | CUL4A, PCID2 |
| chr2 | 70888550 | 70889058 | 509 | 4 | 0.000154547 | 0.0368114 | 0.104012 | 0.063552 | 0.001532433 | LINC01143 |
| chr22 | 37952867 | 37953038 | 172 | 2 | 0.000142144 | 0.0545429 | 0.067931 | 0.064272 | -0.00239099 | POLR2F, C22orf23 |
| chr5 | 126594325 | 126594975 | 651 | 3 | 7.88E-05 | 0.0938737 | 0.084779 | 0.066981 | -0.00059333 | ALDH7A1 |
| chr8 | 1844047 | 1844083 | 37 | 2 | 0.000155495 | 0.1170758 | 0.037401 | 0.06732 | -0.0016763 | AC019257.8, ARHGEF10 |
| chr15 | 89751240 | 89751353 | 114 | 5 | 0.00015236 | 0.0355054 | 0.08602 | 0.068406 | 0.000838656 | MESP1 |
| chr10 | 15719499 | 15719658 | 160 | 2 | 0.000152962 | 0.0461059 | 0.111036 | 0.070718 | 0.000289589 | ITGA8 |
| chr17 | 75845114 | 75845511 | 398 | 2 | 0.000109988 | 0.0644275 | 0.072461 | 0.073243 | -0.00156869 | NA |
| chr17 | 63476577 | 63476593 | 17 | 2 | 0.000168263 | 0.0561075 | 0.092159 | 0.075011 | 0.001776539 | NA |
| chr10 | 96981224 | 96981592 | 369 | 3 | 0.0001008 | 0.1067879 | 0.07858 | 0.07625 | 0.001300721 | LCOR |
| chr2 | 196927145 | 196927213 | 69 | 2 | 0.00012968 | 0.0745058 | 0.072682 | 0.07978 | 0.001830302 | PGAP1 |
| chr12 | 53695150 | 53695188 | 39 | 2 | 0.00015348 | 0.2173675 | 0.04028 | 0.08637 | 0.002110304 | NA |
| chr15 | 41659807 | 41660037 | 231 | 2 | 0.000122938 | 0.0847805 | 0.076063 | 0.087741 | -0.00511986 | MGA |
| chr19 | 13747791 | 13748056 | 266 | 2 | 9.37E-05 | 0.059989 | 0.130687 | 0.090475 | 0.000579378 | CCDC130 |
| chr3 | 188901731 | 188901866 | 136 | 2 | 0.000135215 | 0.0983125 | 0.072551 | 0.092555 | 0.001372451 | NA |
| chr9 | 36400913 | 36401085 | 173 | 5 | 0.000144812 | 0.204683 | 0.038213 | 0.093561 | -0.00057257 | RNF38 |
| chr10 | 126905055 | 126905231 | 177 | 3 | 0.000152937 | 0.5163977 | 0.017303 | 0.097859 | 0.001360813 | NA |
| chr15 | 73972826 | 73973055 | 230 | 2 | 0.000148133 | 0.0725603 | 0.117328 | 0.098284 | -0.00123848 | NA |
| chr19 | 12550918 | 12550991 | 74 | 2 | 0.000121942 | 0.2737269 | 0.045031 | 0.098908 | -0.00064221 | AC008758.1, ZNF564, AC010422.6 |
| chr4 | 112636890 | 112637027 | 138 | 5 | 0.000168307 | 0.0958314 | 0.110721 | 0.104662 | 0.001355549 | ZGRF1 |
| chr1 | 54172753 | 54172817 | 65 | 2 | 0.000165879 | 0.1271013 | 0.073585 | 0.105753 | -0.00228926 | AL357673.1, CYB5RL |
| chr10 | 101130539 | 101130757 | 219 | 2 | 0.000111306 | 0.1481495 | 0.067207 | 0.107095 | 0.003929944 | TLX1NB |
| chr2 | 54456181 | 54456185 | 5 | 3 | 0.000168377 | 0.2040024 | 0.041289 | 0.114012 | 0.000916187 | NA |
| chr3 | 50338114 | 50338243 | 130 | 4 | 0.00014498 | 0.114655 | 0.117472 | 0.124651 | 0.000635007 | RASSF1-AS1, RASSF1 |
| chr14 | 94774107 | 94774223 | 117 | 3 | 0.000160525 | 0.1608151 | 0.134946 | 0.143017 | -0.0034495 | AL121612.2 |
| chr8 | 142448345 | 142448460 | 116 | 2 | 0.000149606 | 0.1262472 | 0.14322 | 0.15112 | -0.00637512 | NA |
| chr19 | 49581026 | 49581062 | 37 | 2 | 0.000128683 | 0.1142846 | 0.193388 | 0.163617 | 0.009926948 | PRRG2, NOSIP |
| chr12 | 56042420 | 56042656 | 237 | 2 | 0.000107239 | 0.1228405 | 0.203715 | 0.175316 | 0.001363323 | RPS26 |
| chr3 | 194071281 | 194071476 | 196 | 2 | 0.000137235 | 0.389816 | 0.094458 | 0.183268 | 0.001220654 | AC024559.2 |
| chr5 | 62404221 | 62404908 | 688 | 3 | 8.20E-05 | 0.1214077 | 0.219489 | 0.201113 | 0.001166342 | AC114982.2, IPO11 |
| chr16 | 4327892 | 4328160 | 269 | 2 | 0.000107747 | 0.4878074 | 0.127932 | 0.238728 | 0.001045935 | GLIS2, GLIS2-AS1 |
| chr8 | 23068253 | 23068292 | 40 | 2 | 0.000136364 | 0.2396317 | 0.183138 | 0.2404 | -0.00133946 | AC107959.2, TNFRSF10B |
| chr14 | 57390580 | 57390652 | 73 | 3 | 0.000171028 | 0.239679 | 0.159463 | 0.242963 | -0.00123451 | NAA30 |
| chr7 | 128937872 | 128937969 | 98 | 2 | 0.000174732 | 0.2864286 | 0.232448 | 0.299366 | 0.000476746 | IRF5 |
| chr10 | 112446675 | 112446753 | 79 | 2 | 0.000173838 | 0.3227202 | 0.227249 | 0.311053 | 0.000635323 | ZDHHC6 |
| chr8 | 10524394 | 10524408 | 15 | 2 | 7.57E-05 | 0.2932716 | 0.347303 | 0.378268 | -0.00144866 | PRSS51 |
| chr1 | 206865663 | 206865695 | 33 | 2 | 0.000172635 | 0.3230621 | 0.369524 | 0.411093 | 0.000226492 | IL20 |
| chr19 | 53133145 | 53133697 | 553 | 2 | 0.000116058 | 0.4454126 | 0.453796 | 0.536982 | 0.001021213 | ZNF347 |
| chr8 | 70104344 | 70104451 | 108 | 2 | 0.000141218 | 0.4728089 | 0.460175 | 0.555353 | 0.000758698 | NA |
| chr2 | 53767904 | 53768274 | 371 | 4 | 0.000122014 | 0.5084371 | 0.420115 | 0.640538 | 0.00054187 | CHAC2, ASB3 |
| chr1 | 245685626 | 245685800 | 175 | 2 | 0.000134043 | 0.6789077 | 0.615099 | 0.752315 | -0.00051161 | KIF26B |
| chr16 | 71358470 | 71358692 | 223 | 5 | 0.000171114 | 0.8161179 | 0.500869 | 0.843398 | -0.00047772 | NA |

Supplementary Table S5.3. Significant Differentially methylated regions between perinatal SI v NC at 8w post-partum ranked by Fisher's p-value.

| Chr | Start | End | Width | *n* CpGs | Minimum smoothed FDR | Stouffer | HMFDR | Fisher | Mean differential | Overlapping genes |
|---|---|---|---|---|---|---|---|---|---|---|
| chr6 | 30879679 | 30887405 | 7727 | 60 | 9.85E-29 | 9.82E-52 | 8.42E-05 | 8.58E-59 | 0.008269414 | DDR1 |
| chr1 | 19642391 | 19646283 | 3893 | 30 | 2.88E-32 | 4.19E-39 | 1.03E-05 | 8.12E-50 | -0.00686316 | NBL1, MICOS10-NBL1 |
| chr10 | 93059802 | 93062445 | 2644 | 19 | 1.92E-41 | 2.40E-46 | 2.15E-05 | 7.09E-46 | 0.020856412 | CYP26C1, AL358613.1 |
| chr13 | 35474755 | 35479271 | 4517 | 24 | 9.57E-28 | 1.94E-42 | 2.33E-05 | 1.44E-44 | 0.014345239 | NBEA, MAB21L1 |
| chr17 | 48601357 | 48606047 | 4691 | 27 | 1.18E-23 | 4.11E-41 | 9.94E-05 | 4.44E-44 | 0.02256986 | HOXB-AS3, HOXB3, HOXB6 |
| chr6 | 32149064 | 32154523 | 5460 | 93 | 3.00E-28 | 9.98E-37 | 0.001038 | 9.90E-42 | 0.005334617 | PPT2, PPT2-EGFL8, PRRT1, AL662884.4, AL662884.3 |
| chr2 | 156326127 | 156330829 | 4703 | 24 | 2.57E-26 | 2.08E-41 | 1.85E-05 | 2.71E-40 | 0.018251599 | NR4A2 |
| chr1 | 153627003 | 153631863 | 4861 | 29 | 1.22E-33 | 3.19E-39 | 0.000372 | 9.70E-40 | -0.02944501 | S100A1, S100A13, AL162258.1, AL162258.2 |
| chr10 | 132407463 | 132409077 | 1615 | 11 | 1.31E-39 | 2.86E-41 | 5.19E-06 | 1.75E-39 | 0.025855335 | PWWP2B |
| chr19 | 13023440 | 13024994 | 1555 | 16 | 2.14E-41 | 5.82E-38 | 1.42E-05 | 5.69E-39 | 0.018992422 | NFIX |
| chr1 | 203348120 | 203351604 | 3485 | 20 | 5.47E-31 | 6.92E-37 | 7.77E-05 | 6.38E-37 | 0.027826308 | FMOD |
| chr17 | 81832829 | 81836060 | 3232 | 26 | 7.05E-38 | 2.95E-27 | 4.82E-06 | 7.00E-37 | 0.006281546 | MCRIP1, PPP1R27 |
| chr6 | 31571762 | 31575909 | 4148 | 38 | 1.07E-18 | 3.13E-28 | 0.000393 | 1.71E-32 | 0.008122044 | LTA, TNF |
| chr20 | 767789 | 768976 | 1188 | 11 | 2.11E-31 | 3.73E-35 | 2.24E-05 | 5.46E-32 | 0.026885323 | SLC52A3 |
| chr19 | 13009741 | 13013401 | 3661 | 12 | 5.18E-18 | 7.71E-28 | 1.21E-05 | 3.93E-28 | 0.020613968 | NFIX |
| chr6 | 37648634 | 37650738 | 2105 | 14 | 7.69E-23 | 5.21E-30 | 8.61E-05 | 6.52E-28 | 0.055321135 | MDGA1 |
| chr12 | 6960695 | 6964101 | 3407 | 21 | 3.16E-18 | 2.96E-24 | 1.97E-05 | 7.70E-28 | 0.006222423 | PTPN6, MIR200CHG, MIR200C, MIR141 |
| chr8 | 42376725 | 42378737 | 2013 | 13 | 1.95E-27 | 5.00E-26 | 5.54E-05 | 1.22E-27 | 0.012536455 | DKK4 |
| chr2 | 238098555 | 238100861 | 2307 | 15 | 1.30E-25 | 7.83E-29 | 5.92E-05 | 4.06E-27 | 0.02730166 | UBE2F-SCLY, SCLY, ESPNL |
| chr8 | 17001400 | 17002612 | 1213 | 15 | 7.44E-28 | 1.33E-25 | 9.58E-05 | 2.89E-26 | 0.018913049 | FGF20 |
| chr15 | 93036102 | 93037616 | 1515 | 11 | 8.91E-26 | 1.46E-25 | 2.40E-05 | 7.78E-25 | 0.021484335 | RGMA |
| chr11 | 70433261 | 70436683 | 3423 | 19 | 9.79E-15 | 9.94E-17 | 9.20E-07 | 6.29E-24 | 0.004309153 | CTTN |
| chr1 | 84996800 | 84999073 | 2274 | 14 | 4.91E-19 | 7.07E-22 | 3.42E-05 | 9.41E-24 | 0.009123352 | MCOLN2 |
| chr9 | 88989558 | 88991535 | 1978 | 16 | 9.94E-25 | 1.21E-18 | 9.01E-06 | 9.99E-24 | 0.011250412 | S1PR3 |
| chr6 | 29630214 | 29632685 | 2472 | 27 | 1.15E-21 | 3.60E-22 | 0.000438 | 1.03E-23 | 0.008336032 | GABBR1 |
| chr14 | 22508023 | 22510179 | 2157 | 12 | 1.44E-24 | 1.39E-20 | 1.21E-06 | 2.73E-23 | 0.012277613 | TRAJ33, TRAJ32 |
| chr13 | 87675754 | 87677945 | 2192 | 11 | 6.98E-19 | 3.36E-25 | 9.02E-05 | 2.76E-23 | 0.031970953 | SLITRK5 |
| chr1 | 180953493 | 180954961 | 1469 | 11 | 6.18E-20 | 2.14E-24 | 0.000156 | 2.93E-22 | -0.0245966 | AL162431.1, KIAA1614-AS1 |
| chr6 | 32107986 | 32109326 | 1341 | 16 | 1.99E-26 | 1.28E-18 | 3.24E-05 | 4.93E-22 | 0.011914483 | AL662884.5, TNXB, AL662884.2 |
| chr7 | 27101178 | 27104187 | 3010 | 30 | 5.31E-19 | 5.87E-22 | 0.00044 | 1.67E-21 | 0.012382523 | HOXA2 |
| chr3 | 147407500 | 147414073 | 6574 | 50 | 9.63E-10 | 5.01E-20 | 0.000254 | 1.73E-21 | 0.004883233 | ZIC1 |
| chr17 | 8338447 | 8339839 | 1393 | 6 | 4.80E-24 | 5.16E-21 | 8.51E-07 | 1.87E-21 | 0.018575386 | AC135178.5 |
| chr2 | 3376194 | 3380204 | 4011 | 27 | 7.68E-17 | 1.38E-13 | 5.38E-06 | 2.00E-21 | 0.007093723 | TRAPPC12, EIPR1 |
| chr6 | 31883581 | 31888840 | 5260 | 63 | 1.47E-09 | 2.37E-15 | 0.000293 | 4.10E-21 | 0.000677042 | EHMT2-AS1, EHMT2 |
| chr6 | 31627870 | 31634962 | 7093 | 69 | 1.05E-09 | 9.74E-17 | 9.49E-05 | 4.12E-21 | 0.000526197 | PRRC2A, MIR6832 |
| chr2 | 161413942 | 161419414 | 5473 | 38 | 1.74E-11 | 9.24E-19 | 0.000494 | 5.77E-21 | 0.004547159 | TBR1 |
| chr10 | 100518882 | 100520794 | 1913 | 17 | 3.47E-22 | 2.13E-20 | 0.000181 | 7.22E-21 | 0.01324155 | SEC31B, AL133352.1 |
| chr3 | 154120337 | 154122929 | 2593 | 25 | 4.65E-18 | 3.07E-20 | 0.000119 | 7.32E-21 | 0.00420467 | ARHGEF26, ARHGEF26-AS1 |
| chr6 | 31307371 | 31308892 | 1522 | 18 | 3.41E-25 | 2.17E-17 | 7.54E-05 | 1.33E-20 | 0.029372078 | AL671883.1 |
| chr8 | 22050511 | 22054963 | 4453 | 17 | 1.39E-09 | 2.05E-21 | 0.000204 | 1.34E-20 | 0.00845187 | DMTN |
| chr1 | 204213829 | 204214527 | 699 | 13 | 2.37E-24 | 1.43E-16 | 2.49E-06 | 1.46E-20 | 0.010885821 | GOLT1A |
| chr17 | 44003279 | 44005279 | 2001 | 13 | 8.97E-23 | 3.65E-16 | 4.37E-05 | 1.95E-20 | 0.009981666 | NAGS, PYY |
| chr14 | 22481406 | 22482431 | 1026 | 8 | 2.55E-22 | 2.38E-20 | 8.58E-06 | 2.67E-20 | 0.027629701 | TRAJ55, TRAJ54, AC244502.1 |
| chr7 | 100892361 | 100896190 | 3830 | 24 | 6.17E-16 | 2.35E-17 | 5.96E-05 | 3.24E-20 | 0.003755793 | ACHE |
| chr2 | 218872670 | 218874010 | 1341 | 6 | 1.62E-15 | 3.78E-21 | 4.14E-06 | 3.71E-20 | 0.028282694 | WNT6 |
| chr16 | 89320390 | 89321988 | 1599 | 16 | 5.66E-20 | 7.34E-17 | 4.01E-05 | 7.95E-20 | 0.008558647 | AC137932.2, ANKRD11 |
| chr7 | 142796464 | 142798162 | 1699 | 18 | 2.14E-21 | 1.93E-17 | 0.000295 | 8.65E-20 | 0.01675454 | TRBJ2-2, TRBJ2-2P, TRBJ2-3, TRBJ2-4, TRBJ2-5, TRBJ2-6, TRBJ2-7 |
| chr8 | 22040984 | 22044562 | 3579 | 18 | 3.49E-12 | 2.28E-16 | 0.000116 | 8.65E-20 | 0.01037734 | FGF17 |
| chr4 | 1047662 | 1051009 | 3348 | 14 | 5.82E-12 | 1.52E-15 | 8.80E-05 | 9.88E-20 | 0.014496151 | NA |
| chr6 | 101398197 | 101400360 | 2164 | 21 | 1.06E-18 | 1.07E-16 | 0.000308 | 1.12E-19 | 0.008190717 | GRIK2 |
| chr19 | 10430531 | 10432877 | 2347 | 18 | 3.59E-15 | 4.15E-13 | 2.20E-05 | 1.26E-19 | 0.00708528 | PDE4A |
| chr13 | 114108813 | 114110915 | 2103 | 11 | 8.89E-20 | 1.23E-18 | 1.09E-05 | 1.28E-19 | 0.016432415 | RASA3, RASA3-IT1 |
| chr15 | 40767429 | 40770026 | 2598 | 18 | 1.02E-13 | 9.85E-16 | 4.14E-05 | 1.49E-19 | 0.006995347 | GCHFR, C15orf62, DNAJC17 |
| chr9 | 22004917 | 22006483 | 1567 | 18 | 9.56E-19 | 3.47E-21 | 0.00065 | 1.61E-19 | 0.00876889 | AL359922.1, CDKN2B-AS1, CDKN2B |
| chr1 | 43933214 | 43935761 | 2548 | 21 | 2.44E-16 | 5.16E-19 | 9.18E-05 | 1.67E-19 | 0.00995762 | ARTN |
| chr17 | 81821273 | 81823119 | 1847 | 27 | 8.26E-21 | 1.55E-18 | 0.001836 | 3.75E-19 | 0.009195987 | MCRIP1 |
| chr3 | 181723247 | 181728186 | 4940 | 26 | 1.64E-09 | 2.01E-17 | 0.00032 | 4.20E-19 | 0.005866665 | SOX2-OT |
| chr15 | 74725573 | 74727861 | 2289 | 30 | 1.47E-21 | 1.46E-14 | 0.00076 | 4.69E-19 | 0.008581424 | NA |
| chr3 | 138948933 | 138950592 | 1660 | 8 | 9.48E-15 | 7.43E-18 | 7.59E-06 | 6.79E-19 | 0.013811873 | FOXL2NB |
| chr17 | 79791193 | 79794888 | 3696 | 15 | 8.56E-12 | 2.30E-15 | 9.01E-06 | 6.79E-19 | 0.010622517 | CBX8 |
| chr5 | 173244523 | 173247510 | 2988 | 14 | 1.79E-09 | 1.89E-17 | 1.16E-05 | 6.91E-19 | 0.009145773 | NA |
| chr2 | 544788 | 545736 | 949 | 6 | 6.41E-21 | 8.05E-15 | 1.46E-05 | 1.30E-18 | 0.01532685 | NA |
| chr2 | 223050804 | 223052872 | 2069 | 15 | 1.05E-20 | 4.93E-17 | 0.000144 | 1.44E-18 | -0.01304483 | KCNE4 |
| chr11 | 14971232 | 14973209 | 1978 | 26 | 2.38E-20 | 1.58E-15 | 0.000152 | 2.51E-18 | 0.007529446 | CALCB, CALCA |
| chr6 | 30912780 | 30918023 | 5244 | 65 | 8.53E-13 | 9.57E-16 | 0.001288 | 2.84E-18 | 0.001446393 | GTF2H4, AL669830.1, VARS2 |
| chr17 | 78979862 | 78980975 | 1114 | 10 | 3.17E-18 | 2.14E-19 | 0.000185 | 3.88E-18 | 0.016643681 | LGALS3BP |
| chr12 | 49292836 | 49295902 | 3067 | 16 | 3.45E-13 | 2.07E-17 | 9.68E-05 | 4.15E-18 | 0.009856849 | PRPH, AC125611.4 |
| chr22 | 19763395 | 19765841 | 2447 | 9 | 1.65E-15 | 1.36E-18 | 6.02E-05 | 4.56E-18 | 0.02113127 | TBX1 |
| chr16 | 66959808 | 66962125 | 2318 | 12 | 3.97E-18 | 7.79E-17 | 2.76E-05 | 4.85E-18 | 0.008868507 | AC009084.1, CES3 |
| chr6 | 100455815 | 100459019 | 3205 | 19 | 2.28E-13 | 1.62E-17 | 0.000116 | 4.85E-18 | 0.007562505 | SIM1 |
| chr17 | 45143526 | 45145153 | 1628 | 11 | 7.14E-19 | 4.00E-19 | 0.000103 | 6.35E-18 | 0.01570855 | ACBD4 |
| chr8 | 22131211 | 22134847 | 3637 | 15 | 3.07E-10 | 4.47E-15 | 7.28E-05 | 6.48E-18 | 0.007894743 | HR |
| chr7 | 374529 | 377179 | 2651 | 22 | 4.41E-19 | 4.72E-10 | 9.79E-05 | 6.76E-18 | -0.0016416 | NA |
| chr2 | 44932006 | 44934194 | 2189 | 20 | 6.19E-16 | 4.10E-16 | 0.000262 | 7.60E-18 | 0.007448315 | AC012354.2, AC012354.7, LINC01833 |
| chr3 | 73623247 | 73625664 | 2418 | 21 | 3.55E-15 | 3.76E-17 | 0.000532 | 7.87E-18 | 0.009671021 | PDZRN3-AS1, AC117489.1, PDZRN3 |
| chr6 | 30162332 | 30164449 | 2118 | 32 | 7.41E-20 | 1.82E-15 | 0.001833 | 8.17E-18 | 0.009964775 | TRIM15 |
| chr6 | 28921644 | 28925315 | 3672 | 62 | 7.60E-11 | 2.75E-14 | 0.000264 | 9.40E-18 | 0.001164086 | TRIM27 |
| chr2 | 70903753 | 70907806 | 4054 | 15 | 1.20E-09 | 2.96E-18 | 0.000179 | 1.08E-17 | 0.010287636 | VAX2 |
| chr6 | 34145516 | 34147280 | 1765 | 14 | 1.90E-19 | 3.48E-14 | 8.85E-05 | 1.63E-17 | 0.010709877 | GRM4 |
| chr2 | 63048374 | 63052558 | 4185 | 50 | 5.57E-10 | 3.79E-15 | 0.000478 | 1.70E-17 | 0.002304315 | OTX1, AC007098.1 |
| chr16 | 54937039 | 54940054 | 3016 | 20 | 6.18E-13 | 9.85E-16 | 0.000126 | 1.79E-17 | 0.005042993 | AC106738.1, AC106738.2 |
| chr11 | 2459976 | 2461505 | 1530 | 22 | 8.88E-19 | 1.68E-09 | 7.05E-06 | 2.02E-17 | 0.004026448 | KCNQ1 |
| chr4 | 153788226 | 153791270 | 3045 | 44 | 3.44E-18 | 4.84E-12 | 0.001515 | 2.40E-17 | 0.003736395 | SFRP2 |
| chr3 | 21750576 | 21752269 | 1694 | 13 | 8.22E-17 | 6.16E-19 | 0.000448 | 3.57E-17 | 0.013138054 | ZNF385D |
| chr17 | 4785870 | 4789495 | 3626 | 19 | 4.03E-10 | 5.94E-17 | 0.000133 | 3.57E-17 | 0.010433498 | AC233723.2, GLTPD2, VMO1 |
| chr2 | 219433825 | 219435520 | 1696 | 14 | 1.32E-21 | 4.67E-13 | 2.18E-06 | 3.96E-17 | 0.009879688 | SPEG |
| chr14 | 56809339 | 56813045 | 3707 | 23 | 1.87E-11 | 2.38E-17 | 0.00058 | 4.00E-17 | 0.005625493 | OTX2 |
| chr11 | 31796262 | 31799024 | 2763 | 12 | 1.22E-12 | 7.25E-17 | 0.000213 | 5.09E-17 | 0.008369073 | PAX6 |
| chr9 | 98942879 | 98945251 | 2373 | 15 | 5.45E-14 | 7.43E-18 | 6.35E-05 | 5.55E-17 | 0.006753157 | COL15A1, AL136084.2 |
| chr16 | 68233664 | 68237274 | 3611 | 20 | 2.91E-13 | 1.06E-11 | 1.89E-05 | 7.01E-17 | 0.006847275 | ESRP2, AC020978.4 |
| chr19 | 3369480 | 3370474 | 995 | 6 | 2.50E-17 | 5.43E-18 | 2.92E-05 | 7.80E-17 | 0.021885511 | NFIC |
| chr17 | 81458851 | 81462064 | 3214 | 14 | 2.54E-10 | 3.85E-15 | 4.11E-05 | 8.21E-17 | 0.0097897 | BAHCC1, AC110285.4 |
| chr6 | 32194918 | 32198875 | 3958 | 63 | 3.31E-12 | 2.86E-11 | 0.000307 | 1.10E-16 | 0.000570553 | GPSM3, NOTCH4 |
| chr18 | 79679074 | 79681810 | 2737 | 16 | 1.17E-11 | 1.59E-14 | 0.000114 | 1.18E-16 | 0.006250569 | CTDP1, AC068473.1, AC068473.5 |
| chr10 | 70494558 | 70495313 | 756 | 4 | 3.00E-15 | 8.16E-18 | 2.05E-05 | 1.19E-16 | 0.060656724 | PALD1 |
| chr6 | 31579927 | 31583648 | 3722 | 36 | 2.25E-12 | 2.19E-10 | 0.001533 | 1.42E-16 | 0.003850364 | LTB |
| chr16 | 85168234 | 85169026 | 793 | 10 | 1.98E-17 | 8.98E-16 | 0.000108 | 1.56E-16 | 0.016343249 | NA |
| chr15 | 93046982 | 93050060 | 3079 | 8 | 2.26E-13 | 8.16E-18 | 0.000123 | 1.84E-16 | 0.018242627 | RGMA |
| chr16 | 3011834 | 3012974 | 1141 | 13 | 8.21E-17 | 2.53E-17 | 0.000479 | 1.92E-16 | 0.015147653 | CLDN9 |
| chr3 | 45035107 | 45036883 | 1777 | 11 | 3.38E-18 | 1.15E-13 | 9.13E-05 | 1.96E-16 | 0.024445474 | EXOSC7, CLEC3B |
| chr9 | 34651277 | 34655259 | 3983 | 16 | 5.28E-09 | 1.62E-13 | 0.000291 | 1.96E-16 | 0.007122107 | AL162231.3, IL11RA |
| chr7 | 27156936 | 27159699 | 2764 | 18 | 5.45E-13 | 2.88E-17 | 0.000305 | 1.99E-16 | 0.008454042 | AC004080.4, HOXA7 |
| chr19 | 17334264 | 17338393 | 4130 | 27 | 1.25E-09 | 2.91E-13 | 0.00027 | 2.01E-16 | 0.0045595 | GTPBP3, ANO8 |
| chr2 | 207256294 | 207258757 | 2464 | 5 | 1.18E-13 | 9.94E-17 | 2.68E-06 | 2.21E-16 | 0.011187429 | AC009226.1 |
| chr1 | 247517800 | 247518629 | 830 | 11 | 1.25E-15 | 1.05E-18 | 0.001146 | 2.65E-16 | -0.02490744 | GCSAML |
| chr8 | 109973624 | 109976887 | 3264 | 25 | 2.98E-10 | 3.85E-16 | 0.000997 | 2.65E-16 | 0.007057279 | AC027451.1, KCNV1 |
| chr1 | 9714684 | 9717034 | 2351 | 14 | 1.35E-12 | 1.83E-17 | 0.000851 | 3.41E-16 | 0.009509237 | PIK3CD |
| chr11 | 503270 | 504933 | 1664 | 13 | 1.55E-15 | 8.73E-15 | 9.79E-05 | 3.53E-16 | 0.009310242 | RNH1 |
| chr2 | 222320330 | 222321129 | 800 | 11 | 2.72E-17 | 5.34E-17 | 0.000168 | 3.69E-16 | 0.011899294 | CT75 |
| chr9 | 121283998 | 121286880 | 2883 | 14 | 1.34E-09 | 1.84E-14 | 3.88E-05 | 4.49E-16 | 0.009514634 | GSN, GSN-AS1 |
| chr18 | 49613149 | 49614189 | 1041 | 5 | 1.68E-15 | 3.74E-17 | 2.12E-05 | 4.77E-16 | 0.020273447 | NA |
| chr9 | 21967470 | 21969692 | 2223 | 20 | 1.39E-14 | 1.89E-17 | 0.000534 | 5.15E-16 | 0.006589316 | AL359922.1, CDKN2A-DT, CDKN2A |
| chr19 | 35755050 | 35758553 | 3504 | 29 | 8.28E-09 | 4.79E-14 | 0.00054 | 6.04E-16 | 0.005583692 | AC002398.2, PROSER3, HSPB6 |
| chr1 | 211258486 | 211260791 | 2306 | 21 | 1.42E-18 | 4.20E-13 | 5.66E-05 | 6.19E-16 | 0.000657454 | RCOR3 |
| chr2 | 176148343 | 176151891 | 3549 | 26 | 3.50E-11 | 1.45E-12 | 0.001034 | 6.36E-16 | 0.010906301 | HOXD3, MIR10B, HOXD4 |
| chr1 | 110674572 | 110675932 | 1361 | 14 | 1.14E-21 | 1.36E-12 | 1.83E-05 | 6.57E-16 | 0.008465627 | KCNA3 |
| chr20 | 54070468 | 54071385 | 918 | 6 | 1.07E-18 | 3.64E-15 | 3.44E-06 | 6.95E-16 | 0.023255775 | BCAS1 |
| chr22 | 36817360 | 36820023 | 2664 | 12 | 4.24E-10 | 5.95E-15 | 0.000399 | 7.45E-16 | 0.012221485 | Z82185.1, PVALB |
| chr12 | 49961524 | 49963569 | 2046 | 11 | 6.98E-16 | 1.38E-14 | 7.24E-05 | 7.48E-16 | 0.011852235 | AQP5, AC025154.2 |
| chr6 | 31775992 | 31777404 | 1413 | 24 | 5.69E-17 | 3.52E-14 | 0.000599 | 7.99E-16 | 0.004936731 | VWA7 |
| chr17 | 76076876 | 76079255 | 2380 | 14 | 8.83E-13 | 1.46E-11 | 1.76E-05 | 7.99E-16 | 0.013830941 | ZACN, GALR2 |
| chr11 | 32433172 | 32436332 | 3161 | 31 | 1.32E-12 | 1.10E-15 | 6.01E-05 | 8.24E-16 | 0.004430458 | WT1-AS, AL049692.6, AL049692.1, WT1 |
| chr14 | 24365800 | 24368630 | 2831 | 24 | 2.77E-14 | 2.02E-12 | 5.52E-05 | 8.95E-16 | 0.005226536 | NFATC4 |
| chr12 | 52903429 | 52908426 | 4998 | 23 | 9.95E-10 | 1.42E-12 | 0.000109 | 8.95E-16 | 0.007636629 | KRT8 |
| chr16 | 69932738 | 69933863 | 1126 | 8 | 1.79E-15 | 1.70E-17 | 0.000298 | 9.01E-16 | 0.02894677 | WWP2, MIR140 |
| chr11 | 7019849 | 7020790 | 942 | 17 | 1.60E-15 | 1.07E-15 | 8.85E-05 | 1.17E-15 | 0.007219828 | NLRP14, ZNF214 |
| chr6 | 31869662 | 31871552 | 1891 | 15 | 1.96E-16 | 2.69E-15 | 0.000139 | 1.25E-15 | 0.004869295 | SLC44A4 |
| chr11 | 78022477 | 78024006 | 1530 | 10 | 1.83E-20 | 6.42E-11 | 1.77E-05 | 1.42E-15 | 0.013491943 | AP003032.1, KCTD14, NDUFC2-KCTD14 |
| chr14 | 68363583 | 68364493 | 911 | 6 | 2.24E-18 | 6.04E-11 | 4.56E-07 | 1.42E-15 | 0.01878355 | RAD51B |
| chr1 | 153615834 | 153617767 | 1934 | 11 | 2.60E-14 | 3.56E-15 | 1.84E-05 | 1.54E-15 | -0.01336454 | S100A14 |
| chr8 | 22056495 | 22057999 | 1505 | 7 | 5.59E-16 | 2.55E-16 | 1.47E-05 | 1.62E-15 | 0.012618787 | DMTN |
| chr7 | 100600759 | 100604107 | 3349 | 19 | 1.99E-09 | 2.68E-15 | 0.000447 | 1.66E-15 | 0.005878055 | FBXO24, PCOLCE, PCOLCE-AS1 |
| chr20 | 25147493 | 25149300 | 1808 | 15 | 1.05E-18 | 2.08E-11 | 2.22E-05 | 1.70E-15 | 0.007244838 | AL035252.2 |
| chr7 | 128714808 | 128715663 | 856 | 9 | 6.81E-16 | 2.30E-16 | 9.06E-05 | 1.85E-15 | 0.025555295 | FAM71F1 |
| chr4 | 1308393 | 1312065 | 3673 | 16 | 3.08E-11 | 6.11E-16 | 0.000229 | 1.86E-15 | 0.007508913 | MAEA |
| chr16 | 75237974 | 75239692 | 1719 | 9 | 9.79E-17 | 9.51E-14 | 1.64E-05 | 2.14E-15 | 0.00927794 | BCAR1 |
| chr1 | 3649740 | 3652822 | 3083 | 45 | 1.03E-12 | 3.31E-12 | 0.002475 | 2.14E-15 | 0.002628099 | TP73, WRAP73 |
| chr7 | 25858287 | 25861148 | 2862 | 16 | 1.95E-11 | 5.30E-15 | 0.000564 | 2.88E-15 | 0.009177395 | NA |
| chr19 | 32719260 | 32719945 | 686 | 11 | 6.19E-16 | 3.14E-15 | 0.001018 | 2.93E-15 | -0.0155251 | TDRD12, AC008736.2 |
| chr17 | 80921264 | 80924569 | 3306 | 15 | 7.05E-14 | 1.93E-11 | 1.41E-05 | 2.93E-15 | 0.00274877 | RPTOR |
| chr5 | 177509562 | 177511888 | 2327 | 14 | 1.96E-14 | 9.61E-11 | 3.72E-06 | 3.22E-15 | 0.008643671 | DOK3, DDX41 |
| chr6 | 31576917 | 31578308 | 1392 | 11 | 7.24E-12 | 9.42E-13 | 4.22E-06 | 3.22E-15 | 0.008640992 | TNF |
| chr4 | 41878302 | 41881155 | 2854 | 20 | 1.55E-10 | 1.38E-12 | 1.78E-05 | 3.30E-15 | 0.007268848 | LINC00682 |
| chr15 | 89376762 | 89379758 | 2997 | 21 | 9.29E-12 | 4.79E-13 | 0.000143 | 3.80E-15 | 0.007218455 | MIR9-3HG |
| chr6 | 3848009 | 3849584 | 1576 | 29 | 4.93E-14 | 6.93E-14 | 9.80E-05 | 4.15E-15 | 0.006692473 | AL391422.3, AL391422.2, FAM50B |
| chr1 | 3657943 | 3659353 | 1411 | 6 | 5.09E-12 | 1.29E-15 | 1.88E-05 | 4.79E-15 | 0.020471375 | TP73, TP73-AS3 |
| chr16 | 30759907 | 30763232 | 3326 | 32 | 3.73E-10 | 8.98E-14 | 0.001116 | 5.25E-15 | 0.002505284 | PHKG2, RNF40, CCDC189 |
| chr22 | 48499846 | 48501388 | 1543 | 5 | 3.25E-17 | 6.96E-11 | 8.68E-06 | 5.36E-15 | 0.01033832 | TAFA5 |
| chr14 | 21722899 | 21723641 | 743 | 7 | 2.11E-15 | 6.77E-15 | 0.000112 | 5.61E-15 | 0.014331039 | NA |
| chr10 | 70602536 | 70603516 | 981 | 11 | 4.69E-14 | 1.94E-15 | 7.01E-05 | 6.26E-15 | 0.008803402 | PRF1 |
| chr6 | 31877333 | 31879251 | 1919 | 19 | 2.90E-12 | 7.76E-14 | 0.000182 | 6.97E-15 | 0.009813525 | EHMT2-AS1, SLC44A4 |
| chr1 | 16977684 | 16981808 | 4125 | 15 | 8.51E-09 | 4.47E-11 | 1.99E-05 | 6.99E-15 | 0.004188649 | AL049569.2, AL049569.1, MFAP2 |
| chr8 | 79766448 | 79769229 | 2782 | 15 | 5.32E-11 | 7.00E-15 | 0.000524 | 8.66E-15 | 0.009211237 | LINC01607, HEY1 |
| chr6 | 100446753 | 100449569 | 2817 | 14 | 5.53E-11 | 6.89E-14 | 0.000288 | 9.44E-15 | 0.007695903 | SIM1 |
| chr10 | 46019369 | 46021045 | 1677 | 9 | 6.39E-13 | 6.16E-13 | 1.61E-05 | 9.71E-15 | 0.012189736 | NCOA4 |
| chr1 | 202631352 | 202631369 | 18 | 2 | 1.03E-15 | 4.92E-15 | 1.74E-07 | 1.10E-14 | 0.04716645 | SYT2 |
| chr6 | 32068753 | 32072658 | 3906 | 45 | 1.94E-08 | 3.23E-10 | 0.000123 | 1.13E-14 | 0.002795661 | TNXB |

| | | | | | | | | | | |
|---|---|---|---|---|---|---|---|---|---|---|
| chr2 | 25168173 | 25170072 | 1900 | 18 | 2.12E-13 | 2.26E-12 | 0.000274 | 1.25E-14 | 0.006203809 | POMC |
| chr1 | 236141041 | 236143011 | 1971 | 16 | 2.67E-10 | 3.57E-12 | 0.000103 | 1.29E-14 | 0.005198559 | GPR137B |
| chr7 | 97001777 | 97005348 | 3572 | 17 | 2.43E-09 | 1.03E-11 | 0.000298 | 1.29E-14 | 0.006729404 | DLX6-AS1 |
| chr17 | 48579328 | 48582735 | 3408 | 21 | 1.14E-07 | 2.16E-13 | 0.000397 | 1.79E-14 | 0.008987956 | HOXB-AS3, HOXB3, AC103702.1, MIR10A |
| chr2 | 240895335 | 240897087 | 1753 | 10 | 7.37E-13 | 4.51E-14 | 0.000258 | 1.93E-14 | 0.01222412 | MAB21L4 |
| chr2 | 130915112 | 130917321 | 2210 | 18 | 1.23E-15 | 6.79E-12 | 0.00013 | 2.07E-14 | -0.00850084 | ARHGEF4 |
| chr1 | 182587958 | 182589740 | 1783 | 11 | 3.28E-17 | 2.43E-12 | 2.46E-07 | 2.10E-14 | 0.007429059 | RNASEL |
| chr1 | 111199893 | 111201881 | 1989 | 15 | 2.30E-12 | 5.06E-14 | 0.00036 | 2.14E-14 | 0.007934923 | CHI3L2, DENND2D |
| chr12 | 51027442 | 51029044 | 1603 | 12 | 6.56E-17 | 7.64E-11 | 5.14E-05 | 2.28E-14 | 0.009639335 | SLC11A2 |
| chr3 | 111998687 | 111999610 | 924 | 14 | 1.41E-15 | 1.61E-13 | 0.000186 | 2.38E-14 | 0.009189913 | TAGLN3 |
| chr11 | 76669877 | 76670927 | 1051 | 15 | 2.58E-17 | 9.69E-12 | 0.00017 | 2.53E-14 | 0.007974609 | LRRC32 |
| chr16 | 979745 | 981205 | 1461 | 11 | 1.91E-16 | 2.33E-12 | 4.19E-05 | 2.53E-14 | 0.008474832 | LMF1, CEROX1 |
| chr1 | 7388997 | 7390574 | 1578 | 6 | 2.14E-15 | 7.76E-14 | 8.20E-06 | 2.53E-14 | 0.014153135 | CAMTA1, CAMTA1-AS2 |
| chr2 | 205763364 | 205764590 | 1227 | 12 | 2.07E-13 | 1.21E-16 | 0.002588 | 2.55E-14 | 0.029229829 | NRP2, AC007362.1 |
| chr4 | 56466030 | 56467997 | 1968 | 17 | 1.51E-12 | 3.62E-09 | 7.13E-05 | 2.55E-14 | 0.005288461 | SRP72 |
| chr22 | 37696636 | 37697200 | 565 | 12 | 5.50E-13 | 1.53E-16 | 0.001054 | 2.67E-14 | 0.011952777 | Z83844.1, TRIOBP |
| chr5 | 1856145 | 1858987 | 2843 | 10 | 1.03E-10 | 1.59E-14 | 0.000121 | 2.77E-14 | -0.03447489 | LINC02116, AC025183.2 |
| chr1 | 15764223 | 15765769 | 1547 | 8 | 1.95E-13 | 7.91E-14 | 4.70E-05 | 3.11E-14 | 0.010621198 | FBLIM1 |
| chr20 | 37343094 | 37346619 | 3526 | 17 | 3.82E-09 | 2.94E-12 | 0.000141 | 3.17E-14 | 0.004011773 | SRC, AL034422.1 |
| chr7 | 129208845 | 129211966 | 3122 | 26 | 5.47E-10 | 4.95E-10 | 8.85E-05 | 3.36E-14 | 0.003337185 | SMO, AC011005.4 |
| chr9 | 128392068 | 128394502 | 2435 | 9 | 2.43E-11 | 2.55E-13 | 6.60E-05 | 4.58E-14 | 0.006931455 | MIR219B, AL359091.1, MIR219A2 |
| chr13 | 20191963 | 20192927 | 965 | 5 | 5.05E-20 | 3.36E-11 | 7.19E-06 | 4.60E-14 | 0.020648986 | GJB2 |
| chr15 | 40279796 | 40282343 | 2548 | 8 | 1.03E-10 | 1.62E-13 | 3.51E-05 | 4.85E-14 | 0.011691822 | PLCB2, ANKRD63 |
| chr5 | 103562449 | 103563299 | 851 | 12 | 3.18E-14 | 6.55E-15 | 0.001065 | 4.96E-14 | -0.01781848 | NUDT12 |
| chr1 | 109398132 | 109399212 | 1081 | 9 | 3.94E-17 | 1.27E-10 | 2.82E-05 | 5.36E-14 | 0.014837095 | PSMA5 |
| chr17 | 49992990 | 49997054 | 4065 | 27 | 5.03E-08 | 4.56E-10 | 0.001472 | 5.47E-14 | 0.005457431 | AC009720.1, DLX3 |
| chr13 | 36430399 | 36432474 | 2076 | 32 | 1.70E-11 | 2.92E-10 | 0.00058 | 5.48E-14 | 0.003598905 | CCNA1 |
| chr14 | 105862948 | 105865308 | 2361 | 14 | 2.72E-11 | 4.60E-14 | 0.000168 | 5.65E-14 | 0.015770486 | IGHJ6, IGHJ3P, IGHJ5, IGHJ4, IGHJ3, IGHJ2P, IGHJ2 |
| chr8 | 143715759 | 143717914 | 2156 | 15 | 4.91E-16 | 2.53E-10 | 3.39E-05 | 5.85E-14 | 0.008628189 | MAPK15 |
| chr17 | 48310865 | 48311103 | 239 | 4 | 2.41E-14 | 1.40E-14 | 5.57E-06 | 6.07E-14 | 0.016049969 | SKAP1 |
| chr5 | 177607316 | 177608575 | 1260 | 7 | 4.16E-13 | 2.45E-12 | 5.89E-07 | 6.24E-14 | 0.006700914 | B4GALT7 |
| chr19 | 48501266 | 48501577 | 312 | 4 | 5.55E-14 | 7.28E-15 | 1.69E-05 | 6.54E-14 | 0.024598681 | LMTK3 |
| chr17 | 8124068 | 8127461 | 3394 | 21 | 2.00E-08 | 1.87E-13 | 8.18E-05 | 6.65E-14 | 0.001067629 | HES7 |
| chr13 | 50843333 | 50844478 | 1146 | 13 | 2.02E-14 | 1.12E-14 | 0.000245 | 6.66E-14 | 0.010118893 | DLEU1, DLEU7 |
| chr7 | 2523550 | 2528259 | 4710 | 21 | 1.65E-06 | 4.72E-15 | 0.002228 | 6.66E-14 | 0.005580433 | LFNG, MIR4648 |
| chr7 | 151186274 | 151188567 | 2294 | 16 | 6.04E-13 | 5.33E-13 | 0.000424 | 7.22E-14 | 0.00598491 | ASB10 |
| chr13 | 21670817 | 21675384 | 4568 | 26 | 5.14E-06 | 9.58E-13 | 0.002105 | 9.69E-14 | 0.000795736 | FGF9 |
| chr10 | 97195875 | 97196955 | 1081 | 7 | 3.69E-17 | 4.88E-10 | 1.67E-06 | 9.97E-14 | 0.011280451 | AL512424.1, ARHGAP19-SLIT1 |
| chr6 | 32080686 | 32084080 | 3395 | 24 | 7.00E-14 | 3.49E-08 | 0.000163 | 1.00E-13 | 0.003262357 | TNXB |
| chr1 | 202809315 | 202810369 | 1055 | 8 | 5.16E-13 | 8.43E-14 | 4.99E-05 | 1.01E-13 | 0.018874011 | AC098934.2 |
| chr15 | 74432221 | 74434565 | 2345 | 20 | 7.75E-13 | 5.60E-08 | 9.25E-07 | 1.05E-13 | 0.002799179 | SEMA7A |
| chr14 | 37171042 | 37174060 | 3019 | 23 | 2.23E-10 | 5.46E-11 | 0.000189 | 1.10E-13 | 0.005461958 | SLC25A21-AS1, SLC25A21 |
| chr7 | 16465469 | 16466651 | 1183 | 7 | 1.91E-15 | 4.79E-13 | 4.10E-05 | 1.19E-13 | 0.02400576 | CRPPA, SOSTDC1 |
| chr5 | 136027315 | 136029712 | 2398 | 17 | 2.37E-08 | 4.89E-14 | 8.33E-05 | 1.24E-13 | 0.007057817 | TGFBI |
| chr19 | 17404677 | 17406953 | 2277 | 15 | 3.97E-11 | 1.94E-12 | 0.000743 | 1.26E-13 | 0.006656216 | BISPR, MVB12A, BST2 |
| chr17 | 54899194 | 54901495 | 2302 | 18 | 3.03E-13 | 1.40E-13 | 0.0017 | 1.30E-13 | 0.008335066 | TOM1L1 |
| chr16 | 769064 | 770155 | 1092 | 6 | 1.68E-12 | 5.81E-15 | 0.000172 | 1.34E-13 | -0.01380369 | AL031258.1, MSLNL |
| chr6 | 37691836 | 37692627 | 792 | 4 | 5.58E-14 | 2.17E-13 | 2.15E-06 | 1.53E-13 | 0.025015009 | MDGA1 |
| chr11 | 126302787 | 126304795 | 2009 | 17 | 7.88E-15 | 1.30E-11 | 4.38E-05 | 1.62E-13 | -0.00870406 | DCPS, AP001318.2 |
| chr11 | 95149665 | 95152233 | 2569 | 12 | 4.33E-11 | 1.70E-12 | 0.000138 | 1.79E-13 | 0.006960574 | AP000787.1 |
| chr2 | 222297175 | 222301375 | 4201 | 34 | 5.43E-08 | 2.72E-11 | 0.005343 | 1.81E-13 | 0.004075633 | CCDC140, PAX3 |
| chr7 | 55188190 | 55189719 | 1530 | 8 | 4.09E-15 | 1.65E-12 | 3.12E-06 | 1.91E-13 | 0.00865575 | EGFR, EGFR-AS1 |
| chr12 | 122226692 | 122228378 | 1687 | 11 | 4.36E-16 | 1.09E-11 | 6.09E-05 | 1.94E-13 | 0.00684836 | AC048338.2, DIABLO, AC048338.1 |
| chr1 | 47332797 | 47334495 | 1699 | 11 | 2.89E-16 | 2.84E-11 | 9.55E-06 | 2.03E-13 | 0.007212244 | CMPK1 |
| chr21 | 43778621 | 43778711 | 91 | 2 | 7.11E-15 | 1.19E-13 | 2.28E-07 | 2.15E-13 | 0.031310157 | NA |
| chr1 | 46448856 | 46450586 | 1731 | 7 | 1.31E-13 | 6.19E-13 | 1.56E-05 | 2.15E-13 | 0.010127662 | FAAHP1, LINC01398 |
| chr20 | 2535610 | 2536967 | 1358 | 11 | 9.36E-12 | 6.04E-13 | 1.75E-05 | 2.22E-13 | 0.009924418 | TMC2 |
| chr13 | 50132447 | 50134284 | 1838 | 13 | 5.68E-12 | 5.30E-11 | 6.14E-05 | 2.41E-13 | 0.006508482 | DLEU1 |
| chr16 | 823426 | 826216 | 2791 | 9 | 5.21E-09 | 6.04E-14 | 0.000508 | 2.61E-13 | 0.019779802 | AL023882.1 |
| chr21 | 44338160 | 44340476 | 2317 | 18 | 3.76E-13 | 3.22E-11 | 0.00018 | 2.62E-13 | 0.001541813 | AP001062.2, CFAP410 |
| chr19 | 35281754 | 35282815 | 1062 | 12 | 2.96E-12 | 4.34E-13 | 0.00084 | 2.83E-13 | 0.009642668 | HAMP |
| chr6 | 13923681 | 13924811 | 1131 | 9 | 3.90E-15 | 1.75E-12 | 9.57E-05 | 2.99E-13 | 0.011112407 | RNF182 |
| chr17 | 49217270 | 49220150 | 2881 | 12 | 2.56E-07 | 2.08E-14 | 0.000188 | 3.03E-13 | 0.006965374 | ABI3 |
| chr7 | 102912564 | 102913171 | 608 | 8 | 1.74E-14 | 3.47E-10 | 0.000105 | 3.06E-13 | 0.017500923 | LRRC17, FBXL13 |
| chr4 | 6730661 | 6731667 | 1007 | 5 | 5.41E-13 | 6.52E-14 | 4.97E-05 | 3.30E-13 | 0.016398134 | NA |
| chr13 | 113816942 | 113818367 | 1426 | 16 | 1.22E-12 | 8.82E-14 | 0.000839 | 3.30E-13 | 0.012095097 | TMEM255B, GAS6-AS1 |
| chr6 | 31111333 | 31112752 | 1420 | 9 | 1.90E-11 | 1.21E-14 | 0.000342 | 3.31E-13 | 0.002489705 | C6orf15 |
| chr17 | 10197693 | 10199241 | 1549 | 7 | 9.31E-11 | 1.87E-07 | 8.20E-06 | 3.33E-13 | 0.00238753 | GAS7 |
| chr1 | 200042145 | 200043398 | 1254 | 16 | 1.32E-12 | 2.65E-12 | 1.53E-05 | 3.37E-13 | 0.00238936 | NR5A2 |
| chr19 | 15010387 | 15011412 | 1026 | 14 | 1.95E-14 | 4.58E-11 | 0.000845 | 3.38E-13 | 0.010743636 | CCDC105, SLC1A6 |
| chr7 | 130491077 | 130493269 | 2193 | 34 | 1.69E-13 | 2.29E-10 | 0.000513 | 3.99E-13 | 0.005826158 | MEST |
| chr2 | 170714497 | 170717381 | 2885 | 23 | 1.01E-09 | 5.80E-11 | 0.000234 | 4.10E-13 | 0.001103023 | SP5, AC007405.1, LINC01124 |
| chr17 | 43907706 | 43910432 | 2727 | 17 | 1.10E-12 | 4.03E-12 | 0.000201 | 4.12E-13 | 0.009628854 | MPP2 |
| chr2 | 161623337 | 161625640 | 2304 | 12 | 8.76E-10 | 1.25E-08 | 9.06E-05 | 4.12E-13 | 0.007904105 | SLC4A10, AC062022.2 |
| chr17 | 49963937 | 49967101 | 3165 | 17 | 1.21E-08 | 1.44E-12 | 0.001249 | 4.31E-13 | 0.005881268 | RNU6-1313P |
| chr17 | 82316900 | 82318258 | 1359 | 8 | 1.38E-10 | 1.81E-13 | 3.48E-05 | 4.44E-13 | 0.008200088 | CD7 |
| chr13 | 113837046 | 113838183 | 1138 | 14 | 7.04E-12 | 1.04E-14 | 0.00297 | 4.61E-13 | 0.016744181 | GAS6-AS1, GAS6 |
| chr17 | 74970911 | 74972594 | 1684 | 7 | 4.20E-19 | 8.73E-10 | 4.33E-06 | 4.74E-13 | 0.007485425 | HID1-AS1, HID1 |
| chr12 | 50102423 | 50105125 | 2703 | 13 | 1.35E-11 | 8.85E-11 | 2.31E-05 | 4.92E-13 | 0.007134696 | GPD1 |
| chr4 | 676558 | 677754 | 1197 | 6 | 3.05E-16 | 8.39E-12 | 1.48E-05 | 5.03E-13 | 0.009124584 | MYL5 |
| chr7 | 41693235 | 41694202 | 968 | 7 | 2.88E-14 | 5.11E-11 | 8.59E-05 | 5.16E-13 | 0.013566214 | INHBA-AS1, INHBA |
| chr7 | 44188162 | 44190315 | 2154 | 8 | 4.98E-10 | 1.47E-12 | 3.50E-05 | 5.16E-13 | 0.007650019 | GCK |
| chr12 | 48004495 | 48005710 | 1216 | 13 | 1.06E-13 | 1.04E-10 | 0.000297 | 5.37E-13 | 0.006651043 | COL2A1, AC004801.5 |
| chr14 | 95360169 | 95362275 | 2107 | 9 | 1.75E-11 | 1.41E-10 | 0.000152 | 5.50E-13 | 0.016905778 | NA |
| chr5 | 979820 | 981715 | 1896 | 8 | 1.09E-14 | 5.07E-11 | 1.31E-05 | 5.64E-13 | 0.011596886 | AC122719.2 |
| chr5 | 135546280 | 135547230 | 951 | 4 | 1.04E-11 | 3.28E-12 | 1.03E-06 | 5.80E-13 | 0.02530613 | NA |
| chr4 | 47835758 | 47838716 | 2959 | 20 | 7.99E-09 | 2.00E-12 | 0.003057 | 5.80E-13 | 0.00506574 | AC107068.2, CORIN |
| chr13 | 52450021 | 52452103 | 2083 | 13 | 4.86E-12 | 9.54E-10 | 5.04E-05 | 6.13E-13 | 0.000471135 | VPS36 |
| chr9 | 121650308 | 121651887 | 1580 | 8 | 3.47E-15 | 1.66E-10 | 1.20E-06 | 6.19E-13 | 0.005412601 | DAB2IP |
| chr3 | 50154511 | 50157185 | 2675 | 17 | 1.76E-09 | 7.59E-12 | 0.000281 | 6.47E-13 | 0.007401873 | SEMA3F, SEMA3F-AS1 |
| chr3 | 46923615 | 46924042 | 428 | 6 | 3.02E-12 | 1.40E-13 | 8.80E-05 | 7.67E-13 | 0.010765821 | CCDC12 |
| chr2 | 10329115 | 10331262 | 2148 | 12 | 3.48E-10 | 2.41E-13 | 0.000431 | 7.67E-13 | 0.007328812 | HPCAL1 |
| chr1 | 2143583 | 2145057 | 1475 | 7 | 1.13E-11 | 2.66E-13 | 9.40E-05 | 8.17E-13 | 0.007224577 | PRKCZ, AL590822.2 |
| chr10 | 127735467 | 127736349 | 883 | 6 | 1.62E-13 | 6.57E-13 | 8.39E-05 | 8.22E-13 | 0.024053956 | AL391005.1 |
| chr1 | 35120071 | 35120987 | 917 | 5 | 2.58E-13 | 5.97E-13 | 6.09E-05 | 8.22E-13 | -0.02320473 | NA |
| chr12 | 6548578 | 6549779 | 1202 | 10 | 3.38E-13 | 1.26E-10 | 0.000193 | 8.63E-13 | 0.007784315 | IFFO1 |
| chr4 | 41215545 | 41217788 | 2244 | 10 | 1.31E-10 | 8.63E-10 | 3.45E-05 | 9.46E-13 | 0.005913778 | APBB2 |
| chr1 | 203764786 | 203765431 | 646 | 10 | 1.18E-12 | 1.95E-12 | 0.000551 | 9.84E-13 | 0.012639429 | LAX1 |
| chr19 | 12294872 | 12295608 | 737 | 8 | 8.74E-17 | 2.77E-08 | 2.18E-07 | 9.95E-13 | 0.006750736 | ZNF44 |
| chr11 | 68041804 | 68044254 | 2451 | 17 | 8.52E-09 | 2.57E-11 | 0.000246 | 1.02E-12 | 0.003685243 | TCIRG1 |
| chr7 | 50560457 | 50562386 | 1930 | 11 | 1.08E-12 | 2.08E-12 | 0.000575 | 1.04E-12 | 0.007810769 | DDC |
| chr12 | 57521593 | 57522839 | 1247 | 11 | 5.88E-14 | 1.23E-10 | 7.41E-05 | 1.08E-12 | 0.007786489 | MBD6, DDIT3 |
| chr6 | 30073950 | 30076482 | 2533 | 41 | 2.46E-09 | 7.48E-10 | 0.00082 | 1.17E-12 | 0.004542368 | RNF39 |
| chr2 | 230661369 | 230662068 | 700 | 2 | 4.05E-12 | 6.83E-13 | 5.61E-07 | 1.33E-12 | 0.044932361 | NA |
| chr1 | 25566342 | 25568983 | 2642 | 6 | 1.27E-08 | 5.50E-12 | 6.80E-06 | 1.34E-12 | 0.006739494 | LDLRAP1 |
| chr8 | 141356240 | 141358216 | 1977 | 13 | 2.47E-13 | 2.88E-11 | 8.60E-05 | 1.34E-12 | 0.003897391 | GPR20 |
| chr9 | 93346037 | 93347617 | 1581 | 8 | 1.03E-10 | 1.75E-13 | 0.00014 | 1.60E-12 | 0.016102787 | C9orf129 |
| chr11 | 118620603 | 118623384 | 2782 | 9 | 3.27E-09 | 2.93E-11 | 0.00015 | 1.67E-12 | 0.008090764 | PHLDB1 |
| chr14 | 100824219 | 100826753 | 2535 | 29 | 3.02E-10 | 1.12E-10 | 0.000695 | 1.72E-12 | 0.005826252 | MEG3, AL117190.3 |
| chr6 | 54845601 | 54846678 | 1078 | 10 | 1.51E-11 | 6.48E-11 | 0.000124 | 1.74E-12 | 0.00612743 | NA |
| chr3 | 149656279 | 149658885 | 2607 | 26 | 5.03E-11 | 2.99E-09 | 8.44E-05 | 1.80E-12 | 0.003355813 | WWTR1-AS1, WWTR1 |
| chr19 | 58395237 | 58397083 | 1847 | 12 | 3.97E-12 | 1.61E-10 | 3.30E-05 | 1.82E-12 | 0.009456126 | RNF225 |
| chr2 | 74501496 | 74503821 | 2326 | 20 | 2.95E-12 | 1.42E-09 | 0.000123 | 1.83E-12 | 0.004713605 | AC005041.3, LBX2-AS1, LBX2 |
| chr10 | 61453057 | 61454730 | 1674 | 13 | 2.00E-15 | 5.91E-10 | 6.14E-05 | 1.84E-12 | 0.006540264 | TMEM26-AS1, TMEM26 |
| chr6 | 33276395 | 33279732 | 3338 | 61 | 6.17E-10 | 1.81E-09 | 0.010258 | 1.85E-12 | 0.003325004 | RPS18, B3GALT4, WDR46 |
| chr2 | 11495905 | 11496168 | 264 | 4 | 1.32E-12 | 1.10E-12 | 2.90E-06 | 1.86E-12 | -0.02422054 | GREB1 |
| chr1 | 11847267 | 11848889 | 1623 | 10 | 4.43E-13 | 1.10E-12 | 0.000459 | 1.88E-12 | -0.01736215 | CLCN6, AL021155.1, NPPA |
| chr6 | 32046197 | 32049271 | 3075 | 40 | 8.63E-09 | 9.54E-10 | 0.001114 | 1.89E-12 | 0.004669143 | TNXB |
| chr16 | 67165772 | 67167816 | 2045 | 10 | 1.59E-11 | 3.05E-10 | 7.01E-05 | 1.97E-12 | 0.008197024 | AC074143.1, HSF4 |
| chr19 | 38912183 | 38913156 | 974 | 8 | 4.94E-13 | 9.81E-13 | 9.87E-05 | 2.00E-12 | 0.016793128 | NA |
| chr14 | 101945328 | 101945399 | 72 | 2 | 8.22E-13 | 8.52E-13 | 2.18E-06 | 2.03E-12 | 0.050476518 | NA |
| chr1 | 110067351 | 110069090 | 1740 | 8 | 4.40E-14 | 6.43E-08 | 1.20E-06 | 2.04E-12 | 0.006427433 | STRIP1, ALX3 |
| chr15 | 52790938 | 52792482 | 1545 | 8 | 3.18E-14 | 2.76E-09 | 1.89E-05 | 2.21E-12 | 0.008313473 | ONECUT1 |
| chr17 | 38672201 | 38673505 | 1305 | 5 | 2.87E-12 | 6.46E-12 | 3.80E-05 | 2.21E-12 | 0.015715 | EPOP |
| chr16 | 89052430 | 89053700 | 1271 | 12 | 1.40E-11 | 6.00E-14 | 0.000484 | 2.21E-12 | 0.014845408 | AC135782.1 |
| chr6 | 32852437 | 32854575 | 2139 | 50 | 2.15E-10 | 5.50E-11 | 0.0012 | 2.21E-12 | 0.000686566 | PSMB9, TAP1 |
| chr1 | 2256018 | 2257455 | 1438 | 5 | 2.96E-14 | 3.84E-10 | 3.88E-06 | 2.23E-12 | 0.003296754 | SKI |
| chr2 | 178108292 | 178109485 | 1194 | 12 | 8.64E-15 | 2.34E-09 | 3.27E-06 | 2.27E-12 | 0.000780204 | PDE11A |
| chr3 | 53495147 | 53496820 | 1674 | 10 | 3.47E-19 | 2.49E-06 | 1.21E-06 | 2.33E-12 | 0.008171483 | CACNA1D |
| chr1 | 109392289 | 109393572 | 1284 | 3 | 2.74E-15 | 6.17E-11 | 2.39E-07 | 2.33E-12 | 0.025735573 | SORT1 |
| chr15 | 90062096 | 90064172 | 2077 | 6 | 3.71E-09 | 1.54E-11 | 2.19E-05 | 2.47E-12 | 0.009413201 | ZNF710 |
| chr5 | 170388289 | 170390243 | 1955 | 10 | 2.92E-12 | 1.05E-09 | 6.10E-05 | 2.56E-12 | 0.01050247 | KCNIP1, KCNIP1-OT1, KCNMB1 |
| chr7 | 4682205 | 4684109 | 1905 | 10 | 2.99E-10 | 2.99E-10 | 6.86E-05 | 2.59E-12 | 0.000724407 | FOXK1 |
| chr10 | 133236543 | 133238500 | 1958 | 25 | 4.26E-12 | 4.42E-12 | 0.000534 | 2.65E-12 | 0.006808306 | VENTX |
| chr11 | 64290299 | 64292465 | 2167 | 18 | 1.79E-10 | 1.24E-09 | 8.65E-05 | 2.71E-12 | 0.003249712 | KCNK4, KCNK4-TEX40 |
| chr6 | 28943422 | 28944389 | 968 | 15 | 9.07E-12 | 2.65E-12 | 0.000591 | 2.72E-12 | 0.010639567 | LINC01556 |
| chr2 | 240455111 | 240456870 | 1760 | 9 | 2.74E-10 | 3.71E-13 | 0.00055 | 2.72E-12 | 0.009645452 | GPC1, MIR149, AC110619.1 |
| chr20 | 23187038 | 23188020 | 983 | 3 | 2.31E-09 | 5.65E-13 | 2.13E-05 | 2.72E-12 | 0.046774243 | AL118508.1 |
| chr20 | 37527523 | 37529362 | 1840 | 48 | 9.23E-09 | 6.18E-10 | 0.002095 | 2.72E-12 | -0.00097978 | BLCAP |
| chr6 | 31161552 | 31162739 | 1188 | 20 | 1.03E-10 | 7.78E-11 | 0.000547 | 2.85E-12 | 0.000461085 | TCF19 |
| chr6 | 28587547 | 28590336 | 2790 | 37 | 1.03E-09 | 8.06E-10 | 0.003145 | 2.87E-12 | 0.005157425 | AL049543.1, ZBED9 |
| chr10 | 122460048 | 122461550 | 1503 | 15 | 3.67E-18 | 4.07E-07 | 6.44E-05 | 2.90E-12 | -0.00767459 | HTRA1, BX842242.1 |
| chr17 | 73231560 | 73233448 | 1889 | 23 | 1.09E-12 | 1.64E-12 | 0.001248 | 2.92E-12 | 0.001521497 | C17orf80, FAM104A |
| chr15 | 93029379 | 93030805 | 1427 | 6 | 1.41E-10 | 5.00E-13 | 0.000105 | 3.14E-12 | 0.009715502 | NA |
| chr1 | 54996601 | 54998894 | 2294 | 11 | 5.70E-13 | 5.66E-10 | 1.29E-05 | 3.15E-12 | 0.002055309 | NA |
| chr8 | 142784755 | 142788057 | 3303 | 24 | 2.12E-07 | 1.66E-09 | 0.000529 | 3.17E-12 | 0.002859528 | AC083841.2, LY6D |
| chr1 | 26189160 | 26191095 | 1936 | 10 | 9.28E-12 | 3.01E-10 | 9.06E-05 | 3.18E-12 | 0.007171152 | CNKSR1, CATSPER4 |
| chr11 | 66267362 | 66269461 | 2100 | 45 | 3.01E-11 | 3.73E-11 | 0.000211 | 3.33E-12 | 9.88E-05 | KLC2, RAB1B, AP001107.7 |
| chr19 | 38647380 | 38648187 | 808 | 11 | 2.53E-12 | 1.28E-12 | 0.000639 | 3.56E-12 | 0.000549926 | ACTN4 |
| chr9 | 135505343 | 135507793 | 2451 | 8 | 4.96E-10 | 1.35E-11 | 7.75E-06 | 4.00E-12 | 0.005613196 | AL161452.1 |
| chr6 | 33163992 | 33166971 | 2980 | 34 | 3.15E-11 | 3.18E-09 | 0.000746 | 4.01E-12 | 0.003829376 | COL11A2 |
| chr20 | 62815610 | 62816973 | 1364 | 34 | 2.85E-15 | 1.33E-09 | 5.78E-05 | 4.35E-12 | 0.005851629 | COL9A3 |
| chr19 | 45071223 | 45072503 | 1281 | 6 | 7.67E-10 | 1.36E-12 | 8.17E-05 | 4.40E-12 | 0.003668647 | ZNF296 |
| chr6 | 30328912 | 30330164 | 1253 | 13 | 9.44E-12 | 3.26E-12 | 0.000271 | 4.45E-12 | 0.007161658 | TRIM39, TRIM39-RPP21 |
| chr2 | 208358855 | 208361147 | 2293 | 14 | 3.72E-10 | 8.77E-10 | 5.39E-05 | 4.81E-12 | 0.0081083 | PTH2R, ARPC1BP1 |
| chr1 | 151720723 | 151722106 | 1384 | 18 | 6.38E-13 | 8.38E-08 | 4.23E-05 | 5.14E-12 | 0.006035259 | RIIAD1 |
| chr1 | 33180772 | 33183218 | 2447 | 18 | 1.42E-09 | 3.96E-10 | 0.000124 | 5.14E-12 | 0.004733404 | TRIM62 |
| chr14 | 23407921 | 23410105 | 2185 | 8 | 5.44E-09 | 1.94E-11 | 0.000129 | 5.19E-12 | 0.007091347 | MYH6 |
| chr17 | 74670279 | 74671826 | 1548 | 16 | 2.18E-12 | 5.30E-11 | 0.000581 | 5.32E-12 | 0.00965761 | RAB37, AC064805.2 |
| chr7 | 4739088 | 4739711 | 624 | 8 | 8.85E-12 | 2.50E-12 | 0.000281 | 5.32E-12 | 0.012732103 | FOXK1 |
| chr1 | 24964456 | 24965921 | 1466 | 19 | 1.42E-13 | 1.74E-11 | 0.001851 | 5.45E-12 | 0.009020665 | RUNX3 |
| chr20 | 45812185 | 45813380 | 1196 | 18 | 3.76E-15 | 1.32E-08 | 0.000156 | 5.80E-12 | 0.003989306 | UBE2C |

| | | | | | | | | | | |
|---|---|---|---|---|---|---|---|---|---|---|
| chr3 | 46692903 | 46694372 | 1470 | 13 | 6.87E-11 | 9.18E-07 | 4.24E-05 | 5.86E-12 | 0.005920974 | ALS2CL |
| chr6 | 82361716 | 82363902 | 2187 | 14 | 9.73E-11 | 9.96E-11 | 0.000712 | 6.12E-12 | 0.007831814 | TPBG, AL121977.2 |
| chr1 | 11653483 | 11655190 | 1708 | 14 | 1.61E-10 | 4.86E-12 | 0.00254 | 6.56E-12 | 0.007208303 | FBXO44, FBXO2 |
| chr13 | 114115286 | 114116493 | 1208 | 11 | 3.22E-13 | 1.82E-09 | 3.95E-05 | 6.61E-12 | 0.008142082 | RASA3 |
| chr1 | 232949386 | 232951963 | 2578 | 16 | 3.42E-10 | 2.14E-07 | 1.20E-05 | 6.61E-12 | 0.003516517 | NTPCR |
| chr10 | 26214888 | 26217784 | 2897 | 16 | 2.00E-07 | 2.26E-12 | 0.004424 | 6.61E-12 | 0.003386115 | GAD2 |
| chr11 | 64240097 | 64241611 | 1515 | 15 | 5.37E-12 | 5.42E-07 | 5.88E-05 | 6.68E-12 | 0.002220392 | FKBP2, AP001453.3 |
| chr7 | 48089027 | 48090600 | 1574 | 15 | 7.07E-12 | 2.11E-10 | 9.67E-05 | 6.68E-12 | 0.005912329 | UPP1 |
| chr2 | 169360697 | 169363070 | 2374 | 15 | 6.23E-11 | 1.65E-07 | 1.79E-05 | 6.68E-12 | 0.004516274 | LRP2 |
| chr1 | 90724448 | 90727636 | 3189 | 25 | 8.09E-10 | 7.94E-08 | 8.70E-05 | 6.84E-12 | 0.004080502 | AC092805.1 |
| chr21 | 37366563 | 37368592 | 2030 | 7 | 3.36E-10 | 3.88E-08 | 1.23E-06 | 6.87E-12 | -0.00013174 | DYRK1A, AP001437.2 |
| chr11 | 66546350 | 66548399 | 2050 | 14 | 4.11E-14 | 3.32E-10 | 5.89E-07 | 7.25E-12 | 0.004530492 | ACTN3 |
| chr14 | 97457980 | 97459339 | 1360 | 10 | 9.91E-13 | 3.02E-10 | 0.000593 | 7.33E-12 | 0.015048246 | LINC02325 |
| chr1 | 85579664 | 85580253 | 590 | 6 | 1.27E-13 | 2.05E-12 | 0.000177 | 7.50E-12 | 0.014158805 | NA |
| chr1 | 27634823 | 27635901 | 1079 | 12 | 3.54E-13 | 3.34E-11 | 4.60E-05 | 8.38E-12 | 0.008441624 | FGR |
| chr1 | 62194262 | 62194952 | 691 | 11 | 1.33E-11 | 8.67E-12 | 0.000614 | 8.47E-12 | 0.013905275 | L1TD1 |
| chr13 | 110409433 | 110409940 | 508 | 6 | 1.73E-12 | 6.28E-12 | 0.000148 | 8.77E-12 | 0.022926458 | COL4A2 |
| chr2 | 68773954 | 68775272 | 1319 | 20 | 3.06E-10 | 1.33E-09 | 0.000114 | 8.87E-12 | -5.85E-05 | ARHGAP25 |
| chr2 | 118847865 | 118852537 | 4673 | 27 | 4.71E-06 | 3.54E-09 | 0.001132 | 8.87E-12 | 0.004162153 | NA |
| chr12 | 110743222 | 110744288 | 1067 | 6 | 1.64E-13 | 1.51E-11 | 2.31E-05 | 9.05E-12 | 0.012426867 | NA |
| chr14 | 105855010 | 105856881 | 1872 | 14 | 1.46E-09 | 5.91E-10 | 0.000239 | 9.05E-12 | 0.006594067 | IGHM |
| chr19 | 55536176 | 55537890 | 1715 | 10 | 1.59E-14 | 2.42E-09 | 0.000167 | 9.08E-12 | 0.01237404 | SBK2 |
| chr12 | 62739007 | 62739798 | 792 | 3 | 1.26E-12 | 3.86E-11 | 2.18E-07 | 9.39E-12 | 0.012744383 | PPM1H |
| chr22 | 42142905 | 42142987 | 83 | 2 | 4.94E-12 | 4.35E-12 | 1.92E-06 | 9.48E-12 | -0.02744099 | CYP2D7 |
| chr14 | 23538645 | 23540837 | 2193 | 6 | 6.51E-08 | 4.01E-10 | 1.10E-05 | 9.56E-12 | 0.003091008 | ZFHX2-AS1, ZFHX2 |
| chr12 | 49041686 | 49045674 | 3989 | 27 | 1.28E-07 | 1.70E-09 | 0.000278 | 9.62E-12 | 0.002517344 | KMT2D |
| chr17 | 47777899 | 47778513 | 615 | 4 | 7.20E-12 | 2.52E-12 | 4.03E-05 | 9.71E-12 | -0.02562584 | NA |
| chr1 | 225941781 | 225942857 | 1077 | 5 | 7.00E-10 | 1.51E-11 | 6.28E-05 | 9.71E-12 | 0.009289403 | NA |
| chr22 | 38317603 | 38318766 | 1164 | 13 | 1.84E-11 | 4.10E-13 | 0.004526 | 9.95E-12 | 0.008911571 | CSNK1E, TPTEP2-CSNK1E |
| chr11 | 77127503 | 77129195 | 1693 | 10 | 3.32E-09 | 5.63E-13 | 0.001348 | 9.99E-12 | 0.011334683 | MYO7A |
| chr17 | 3695461 | 3696341 | 881 | 9 | 1.94E-14 | 2.15E-08 | 1.46E-05 | 1.02E-11 | 0.005147198 | P2RX5-TAX1BP3, P2RX5 |
| chr4 | 99518798 | 99519143 | 346 | 4 | 3.43E-11 | 1.07E-12 | 0.000129 | 1.05E-11 | 0.017633523 | C4orf17 |
| chr5 | 139377438 | 139378965 | 1528 | 9 | 3.23E-11 | 1.10E-12 | 0.000847 | 1.05E-11 | -0.01840984 | SLC23A1 |
| chr14 | 73829631 | 73830721 | 1091 | 6 | 7.13E-14 | 9.61E-09 | 2.12E-05 | 1.06E-11 | 0.013273401 | LINC02274 |
| chr13 | 100533346 | 100534283 | 938 | 8 | 3.00E-12 | 1.45E-10 | 4.76E-05 | 1.07E-11 | 0.004027731 | GGACT |
| chr3 | 64267858 | 64268436 | 579 | 7 | 4.52E-12 | 2.02E-10 | 3.18E-05 | 1.08E-11 | 0.011563921 | PRICKLE2 |
| chr17 | 60421510 | 60422825 | 1316 | 13 | 1.35E-12 | 5.46E-11 | 0.000175 | 1.14E-11 | 0.005720899 | C17orf64, USP32 |
| chr19 | 906759 | 907816 | 1058 | 3 | 1.21E-09 | 3.73E-12 | 1.17E-05 | 1.14E-11 | 0.017805203 | R3HDM4 |
| chr12 | 132273679 | 132275381 | 1703 | 9 | 1.08E-14 | 1.44E-07 | 2.12E-06 | 1.16E-11 | 0.009850948 | GALNT9 |
| chr16 | 28877426 | 28879165 | 1740 | 9 | 1.36E-09 | 9.56E-12 | 0.000333 | 1.17E-11 | 0.00871594 | AC009093.11, ATP2A1, ATP2A1-AS1 |
| chr10 | 131981407 | 131983823 | 2417 | 16 | 8.61E-10 | 3.49E-10 | 8.74E-05 | 1.22E-11 | -0.00355514 | BNIP3 |
| chr17 | 44123154 | 44124476 | 1323 | 14 | 1.95E-12 | 1.11E-08 | 1.22E-06 | 1.22E-11 | 0.000208831 | HDAC5 |
| chr7 | 2632766 | 2634547 | 1782 | 9 | 4.44E-10 | 1.18E-11 | 7.87E-05 | 1.22E-11 | 0.007981991 | TTYH3 |
| chr9 | 136996556 | 136997419 | 864 | 9 | 2.36E-11 | 2.79E-12 | 0.000671 | 1.24E-11 | 0.008891316 | CLIC3 |
| chr10 | 31927103 | 31929040 | 1938 | 19 | 2.94E-09 | 1.97E-08 | 0.000162 | 1.25E-11 | 0.003572419 | ARHGAP12 |
| chr17 | 28742653 | 28745233 | 2581 | 17 | 5.15E-08 | 5.91E-10 | 0.000304 | 1.26E-11 | 0.002938048 | NEK8, TRAF4, AC010761.2 |
| chr11 | 10454695 | 10455914 | 1220 | 9 | 1.02E-12 | 1.68E-11 | 4.20E-05 | 1.27E-11 | 0.006835269 | AMPD3 |
| chr13 | 36998186 | 37002080 | 3895 | 30 | 4.47E-08 | 3.14E-08 | 0.000922 | 1.28E-11 | 0.000950974 | EXOSC8, ALG5 |
| chr17 | 79831381 | 79833605 | 2225 | 8 | 1.06E-09 | 9.23E-12 | 0.000218 | 1.29E-11 | 0.015857096 | CBX4 |
| chr2 | 113278042 | 113279547 | 1506 | 11 | 2.71E-10 | 9.23E-12 | 4.83E-05 | 1.33E-11 | 0.008883716 | PAX8 |
| chr10 | 4825581 | 4826320 | 740 | 13 | 2.64E-13 | 3.51E-09 | 0.001012 | 1.34E-11 | 0.014610765 | AKR1E2 |
| chr10 | 133565066 | 133566343 | 1278 | 6 | 3.35E-12 | 1.66E-11 | 9.05E-05 | 1.40E-11 | -0.01730522 | AL731769.2, SYCE1 |
| chr1 | 160098251 | 160100148 | 1898 | 18 | 1.69E-10 | 1.99E-07 | 1.65E-05 | 1.40E-11 | 0.002697161 | IGSF8 |
| chr6 | 31765657 | 31766803 | 1147 | 19 | 1.63E-10 | 3.10E-12 | 0.00464 | 1.41E-11 | 0.010783638 | VWA7 |
| chr13 | 47904223 | 47904456 | 234 | 3 | 1.12E-11 | 8.57E-12 | 9.50E-06 | 1.47E-11 | 0.020900412 | SUCLA2 |
| chr12 | 107580057 | 107581522 | 1466 | 15 | 1.18E-11 | 4.72E-10 | 0.001296 | 1.47E-11 | 0.006511787 | BTBD11 |
| chr11 | 3218172 | 3219781 | 1610 | 12 | 9.42E-12 | 2.40E-12 | 0.000601 | 1.50E-11 | 0.007073875 | MRGPRG-AS1, MRGPRG |
| chr20 | 63940311 | 63943165 | 2855 | 16 | 2.91E-08 | 3.01E-11 | 5.52E-05 | 1.52E-11 | 0.012040098 | UCKL1, MIR1914, MIR647 |
| chr5 | 77957554 | 77958454 | 901 | 8 | 2.54E-11 | 1.52E-12 | 9.94E-05 | 1.54E-11 | 0.011678492 | AC108482.2 |
| chr1 | 2496216 | 2497639 | 1424 | 11 | 1.76E-09 | 1.61E-10 | 0.000304 | 1.54E-11 | 0.004173811 | PLCH2 |
| chr5 | 132776757 | 132778931 | 2175 | 25 | 8.89E-11 | 1.33E-09 | 7.26E-06 | 1.57E-11 | 0.004540409 | SEPTIN8 |
| chr9 | 128097685 | 128099140 | 1456 | 10 | 6.17E-12 | 2.71E-11 | 0.000312 | 1.57E-11 | 0.01085401 | SLC25A25 |
| chr6 | 30126523 | 30128025 | 1503 | 26 | 9.41E-10 | 8.67E-12 | 0.007516 | 1.58E-11 | 0.010015187 | NA |
| chr7 | 95771174 | 95773421 | 2248 | 14 | 2.53E-10 | 1.37E-08 | 0.000385 | 1.58E-11 | 0.006551688 | DYNC1I1 |
| chr16 | 21194438 | 21195050 | 613 | 6 | 7.83E-14 | 1.80E-07 | 1.87E-07 | 1.60E-11 | 0.009786573 | AF001550.1 |
| chr19 | 4908669 | 4910786 | 2118 | 23 | 1.25E-10 | 2.21E-09 | 0.00185 | 1.64E-11 | 0.00111804 | UHRF1 |
| chr6 | 33319727 | 33323334 | 3608 | 67 | 1.47E-07 | 4.86E-10 | 0.006072 | 1.65E-11 | 0.000211864 | DAXX, SMIM40 |
| chr6 | 170262055 | 170262928 | 874 | 10 | 1.82E-11 | 1.84E-09 | 0.000144 | 1.68E-11 | 0.015618992 | AL109910.1 |
| chr20 | 53736427 | 53738625 | 2199 | 8 | 1.81E-08 | 2.35E-11 | 3.78E-05 | 1.70E-11 | 0.008165522 | NA |
| chr12 | 49338638 | 49339552 | 915 | 3 | 4.31E-13 | 1.46E-10 | 4.97E-06 | 1.70E-11 | 0.012951957 | NA |
| chr9 | 127927919 | 127931322 | 3404 | 18 | 1.16E-06 | 2.23E-10 | 0.001065 | 1.70E-11 | 0.005443027 | PIP5KL1 |
| chr17 | 39174208 | 39176102 | 1895 | 9 | 1.39E-09 | 2.76E-12 | 0.000266 | 1.78E-11 | 0.009032217 | AC004408.1, CACNB1 |
| chr15 | 67831149 | 67833261 | 2113 | 10 | 1.25E-10 | 1.25E-11 | 0.000563 | 1.79E-11 | 0.012346819 | SKOR1, AC009292.2 |
| chr21 | 43757657 | 43758450 | 794 | 5 | 1.76E-12 | 1.57E-10 | 6.46E-06 | 1.80E-11 | 0.009088455 | PDXK |
| chr8 | 38029437 | 38031246 | 1810 | 13 | 1.10E-10 | 1.33E-07 | 0.000605 | 1.82E-11 | 0.002755068 | EIF4EBP1 |
| chr2 | 64858109 | 64860235 | 2127 | 10 | 1.01E-06 | 6.95E-13 | 0.000421 | 1.94E-11 | 0.011379625 | LINC01800 |
| chr22 | 33564906 | 33565840 | 935 | 5 | 9.55E-11 | 2.01E-12 | 9.18E-05 | 1.98E-11 | 0.01049192 | LARGE1 |
| chr6 | 143449475 | 143451591 | 2117 | 23 | 1.92E-09 | 1.72E-11 | 0.00177 | 2.02E-11 | 0.000862768 | RNA5SP221, PEX3, ADAT2 |
| chr10 | 132883538 | 132885433 | 1896 | 10 | 1.43E-08 | 1.40E-10 | 3.11E-05 | 2.07E-11 | 0.004741506 | CFAP46 |
| chr15 | 93071529 | 93072489 | 961 | 8 | 1.32E-12 | 1.28E-11 | 0.000245 | 2.15E-11 | 0.024498144 | RGMA |
| chr5 | 131944031 | 131946239 | 2209 | 13 | 3.26E-07 | 2.09E-07 | 4.74E-05 | 2.20E-11 | 4.86E-06 | AC034228.2, MEIKIN, AC034228.3 |
| chr6 | 170244045 | 170246188 | 2144 | 8 | 3.05E-08 | 1.08E-12 | 0.000496 | 2.23E-11 | 0.023672978 | NA |
| chr6 | 10414117 | 10415857 | 1741 | 23 | 1.11E-10 | 6.98E-09 | 0.000108 | 2.28E-11 | 0.003786136 | TFAP2A-AS1, TFAP2A |
| chr8 | 11703329 | 11704858 | 1530 | 14 | 3.89E-11 | 1.24E-08 | 0.000539 | 2.33E-11 | 0.00410151 | GATA4 |
| chr1 | 56818601 | 56820449 | 1849 | 9 | 1.24E-10 | 6.28E-11 | 7.96E-05 | 2.49E-11 | 0.007888443 | FYB2 |
| chr22 | 50301869 | 50305425 | 3557 | 11 | 2.05E-06 | 1.62E-09 | 0.000106 | 2.65E-11 | 0.00637436 | PLXNB2 |
| chr17 | 9646179 | 9647742 | 1564 | 10 | 2.62E-12 | 1.61E-10 | 0.000305 | 2.70E-11 | 0.01352926 | USP43, AC118755.1 |
| chr1 | 201421078 | 201422974 | 1897 | 12 | 6.26E-11 | 4.24E-08 | 4.27E-05 | 2.71E-11 | 0.006280582 | TNNI1 |
| chr16 | 66578387 | 66580067 | 1681 | 14 | 4.23E-09 | 5.03E-12 | 0.004322 | 2.77E-11 | -0.00342689 | CKLF-CMTM1, CMTM1, CMTM2, AC010542.1 |
| chr14 | 104691295 | 104692771 | 1477 | 6 | 7.49E-09 | 8.57E-12 | 0.000119 | 2.82E-11 | 0.00890526 | INF2 |
| chr6 | 33417459 | 33420306 | 2848 | 57 | 2.40E-12 | 2.66E-06 | 3.80E-05 | 2.90E-11 | 0.000665952 | SYNGAP1, CUTA |
| chr11 | 68007831 | 68009106 | 1276 | 8 | 1.15E-12 | 2.08E-09 | 6.43E-05 | 2.90E-11 | 0.011479028 | ALDH3B1 |
| chr11 | 45846479 | 45848786 | 2308 | 17 | 3.77E-09 | 6.27E-08 | 2.72E-06 | 2.91E-11 | -0.00065661 | CRY2 |
| chr22 | 23144460 | 23145782 | 1323 | 15 | 4.95E-11 | 2.68E-07 | 0.00116 | 2.92E-11 | 0.006693021 | RAB36, RSPH14 |
| chr1 | 231019703 | 231020613 | 911 | 12 | 5.97E-10 | 1.69E-12 | 0.002294 | 3.03E-11 | 0.007263509 | FAM89A, MIR1182 |
| chr1 | 26862014 | 26863839 | 1826 | 12 | 5.71E-11 | 2.12E-12 | 0.000661 | 3.12E-11 | 0.007355279 | SFN |
| chr13 | 113001056 | 113002110 | 1055 | 15 | 2.38E-10 | 2.55E-13 | 0.002776 | 3.14E-11 | 0.008148086 | MCF2L |
| chr10 | 68559544 | 68563117 | 3574 | 28 | 1.89E-05 | 4.93E-10 | 0.001205 | 3.15E-11 | 0.004359227 | TET1 |
| chr13 | 27921771 | 27924908 | 3138 | 17 | 8.77E-08 | 3.84E-10 | 6.86E-05 | 3.15E-11 | 0.003838927 | PDX1 |
| chr1 | 41484566 | 41486550 | 1985 | 9 | 1.76E-07 | 3.12E-11 | 0.000403 | 3.16E-11 | 0.008799803 | EDN2 |
| chr13 | 113839818 | 113841102 | 1285 | 15 | 2.93E-12 | 7.83E-09 | 0.000232 | 3.20E-11 | 0.006109107 | GAS6-AS1, GAS6 |
| chr11 | 8810133 | 8812426 | 2294 | 17 | 1.60E-08 | 1.97E-09 | 0.000135 | 3.23E-11 | 0.006666218 | AC026894.1, DENND2B |
| chr5 | 43040072 | 43042186 | 2115 | 17 | 3.69E-09 | 4.32E-10 | 4.65E-05 | 3.28E-11 | -0.00113624 | AC025171.2, AC025171.1, ANXA2R |
| chr9 | 88281211 | 88281296 | 86 | 3 | 4.87E-11 | 1.21E-11 | 2.06E-05 | 3.34E-11 | 0.027962114 | NA |
| chr6 | 97669013 | 97669665 | 653 | 3 | 4.55E-11 | 1.01E-11 | 2.60E-05 | 3.60E-11 | 0.018392151 | AL589740.1 |
| chr6 | 108165183 | 108168088 | 2906 | 24 | 1.64E-06 | 8.65E-10 | 0.00082 | 3.60E-11 | 0.003913611 | NR2E1, OSTM1 |
| chr14 | 94770202 | 94774003 | 3802 | 21 | 8.84E-06 | 1.05E-07 | 6.43E-05 | 3.63E-11 | 0.004025282 | AL121612.2 |
| chr18 | 3623487 | 3625022 | 1536 | 7 | 1.69E-09 | 3.20E-12 | 0.00019 | 3.68E-11 | 0.015001271 | DLGAP1 |
| chr21 | 46160496 | 46162437 | 1942 | 7 | 3.62E-10 | 2.08E-10 | 0.000445 | 3.72E-11 | 0.043572552 | SPATC1L |
| chr21 | 43811142 | 43812720 | 1579 | 7 | 7.63E-08 | 3.70E-12 | 0.000477 | 3.84E-11 | 0.011185924 | AATBC |
| chr2 | 127205754 | 127206798 | 1045 | 10 | 1.00E-10 | 5.36E-12 | 0.000569 | 3.96E-11 | -0.00929545 | CYP27C1 |
| chr22 | 50217724 | 50219169 | 1446 | 6 | 2.32E-12 | 2.51E-08 | 3.89E-05 | 4.16E-11 | 0.004952067 | TUBGCP6 |
| chr3 | 42935464 | 42936912 | 1449 | 15 | 7.88E-09 | 4.52E-13 | 0.005962 | 4.25E-11 | -0.01246812 | AC092042.3, KRBOX1, KRBOX1-AS1 |
| chr10 | 102403134 | 102404678 | 1545 | 5 | 8.10E-13 | 1.35E-09 | 1.64E-05 | 4.36E-11 | 0.014716992 | PSD |
| chr19 | 39154294 | 39156387 | 2094 | 7 | 1.07E-09 | 1.50E-08 | 0.000104 | 4.40E-11 | 0.008979423 | PAK4 |
| chr8 | 144427346 | 144429471 | 2126 | 15 | 1.92E-09 | 3.33E-10 | 0.003931 | 4.42E-11 | -0.00536219 | VPS28, TONSL |
| chr1 | 89762212 | 89764072 | 1861 | 11 | 3.26E-10 | 6.81E-06 | 4.23E-06 | 4.46E-11 | 0.003881344 | LRRC8C, AC093423.3 |
| chr2 | 84515778 | 84516811 | 1034 | 14 | 3.33E-11 | 3.06E-11 | 0.002393 | 4.50E-11 | 0.007615609 | DNAH6 |
| chr14 | 49692250 | 49693589 | 1340 | 11 | 1.10E-10 | 7.21E-11 | 0.0004 | 4.54E-11 | 0.002969869 | KLHDC1 |
| chr19 | 35513331 | 35515004 | 1674 | 8 | 2.07E-11 | 9.67E-10 | 0.000266 | 4.91E-11 | 0.012774805 | DMKN |
| chr3 | 11601793 | 11602357 | 565 | 7 | 5.36E-11 | 1.13E-11 | 0.000345 | 5.11E-11 | 0.013348867 | VGLL4 |
| chr18 | 31101158 | 31103350 | 2193 | 12 | 3.62E-09 | 2.02E-09 | 6.25E-05 | 5.23E-11 | 0.007168337 | DSCAS, DSC2 |
| chr16 | 66999824 | 67001485 | 1662 | 12 | 1.05E-10 | 4.07E-11 | 0.000442 | 5.51E-11 | 0.008090546 | CES4A |
| chr7 | 4837083 | 4838952 | 1870 | 9 | 2.36E-13 | 4.72E-09 | 4.27E-05 | 5.69E-11 | 0.00812411 | RADIL |
| chr1 | 10230214 | 10232137 | 1924 | 9 | 5.61E-10 | 3.22E-08 | 3.92E-07 | 5.75E-11 | 0.002417404 | KIF1B |
| chr2 | 218879423 | 218880575 | 1153 | 13 | 9.43E-14 | 5.50E-09 | 0.000217 | 5.76E-11 | 0.006500478 | NA |
| chr11 | 1862314 | 1864573 | 2260 | 8 | 2.84E-08 | 1.06E-11 | 0.00085 | 6.01E-11 | 0.008924176 | LSP1, AC051649.1 |
| chr1 | 26959890 | 26961628 | 1739 | 12 | 3.53E-11 | 6.71E-10 | 0.000752 | 6.04E-11 | 0.005892348 | KDF1 |
| chr1 | 163202859 | 163204354 | 1496 | 8 | 7.10E-09 | 1.65E-11 | 0.000521 | 6.24E-11 | 0.01355091 | RGS5-AS1, RGS5 |
| chr7 | 100641127 | 100643615 | 2489 | 13 | 2.18E-08 | 1.53E-09 | 0.000739 | 6.36E-11 | -0.00154152 | TFR2, ACTL6B |
| chr3 | 12158458 | 12160715 | 2258 | 13 | 2.85E-09 | 1.66E-11 | 0.002413 | 6.46E-11 | 0.010878011 | SYN2, TIMP4 |
| chr19 | 4277325 | 4278058 | 734 | 5 | 6.07E-13 | 5.67E-08 | 3.14E-06 | 6.50E-11 | 0.014984582 | NA |
| chr9 | 137493086 | 137494152 | 1067 | 6 | 3.76E-13 | 6.80E-08 | 1.10E-05 | 6.50E-11 | 0.005840431 | PNPLA7 |
| chr15 | 48190791 | 48192272 | 1482 | 21 | 5.33E-10 | 5.21E-09 | 0.000712 | 6.56E-11 | 0.003217563 | CTXN2, SLC12A1, AC066612.1, AC066612.2 |
| chr19 | 54802595 | 54803628 | 1034 | 9 | 1.08E-10 | 2.51E-10 | 0.000866 | 6.57E-11 | 0.010856895 | KIR2DL4 |
| chr11 | 61656794 | 61657750 | 957 | 4 | 1.72E-09 | 3.46E-11 | 3.18E-05 | 6.60E-11 | 0.020611444 | NA |
| chr4 | 76757999 | 76758997 | 999 | 4 | 1.53E-10 | 2.71E-11 | 1.10E-05 | 6.80E-11 | 0.018933957 | SHROOM3, SHROOM3-AS1, AC107072.1 |
| chr16 | 66800938 | 66802317 | 1380 | 15 | 8.79E-10 | 3.66E-10 | 0.00046 | 7.16E-11 | 0.00225224 | TERB1 |
| chr9 | 137127664 | 137128964 | 1301 | 4 | 1.59E-10 | 2.71E-11 | 3.30E-05 | 7.54E-11 | 0.013122558 | NA |
| chr4 | 742540 | 744411 | 1872 | 13 | 1.91E-08 | 3.84E-10 | 0.000375 | 7.59E-11 | 0.002498588 | PCGF3 |
| chr6 | 166097394 | 166098426 | 1033 | 6 | 5.85E-12 | 1.33E-09 | 5.73E-05 | 7.65E-11 | 0.009774288 | AL627443.3 |
| chr22 | 26671443 | 26671758 | 316 | 3 | 7.09E-12 | 1.31E-10 | 1.67E-06 | 7.74E-11 | 0.009113502 | MIAT, Z99774.4, Z99774.1 |
| chr18 | 32469353 | 32471840 | 2488 | 30 | 2.79E-07 | 4.01E-09 | 0.002506 | 7.80E-11 | 0.002934234 | AC025887.2, GAREM1 |
| chr6 | 32058370 | 32061375 | 3006 | 23 | 1.17E-06 | 3.12E-11 | 0.005733 | 7.89E-11 | 0.005296456 | TNXB |
| chr17 | 79926725 | 79927693 | 969 | 8 | 7.13E-11 | 5.45E-09 | 0.000101 | 7.96E-11 | 0.013562747 | LINC01979 |
| chr17 | 19744660 | 19746756 | 2097 | 14 | 3.46E-09 | 2.35E-08 | 3.60E-05 | 8.00E-11 | 0.000298909 | ALDH3A1 |
| chr10 | 101235683 | 101238324 | 2642 | 20 | 6.12E-07 | 3.84E-10 | 0.001507 | 8.32E-11 | 0.00496516 | LBX1-AS1 |
| chr11 | 2700106 | 2701483 | 1378 | 34 | 3.95E-12 | 4.62E-09 | 0.006737 | 8.41E-11 | 0.003513127 | KCNQ1 |
| chr12 | 117140962 | 117142202 | 1241 | 4 | 1.66E-12 | 9.52E-10 | 1.22E-06 | 8.53E-11 | -0.00119353 | TESC-AS1, FBXO21 |
| chr10 | 79243900 | 79246309 | 2410 | 7 | 1.70E-08 | 2.88E-11 | 0.000164 | 8.85E-11 | 0.013338066 | ZMIZ1 |
| chr5 | 88677621 | 88678730 | 1110 | 13 | 2.29E-10 | 2.06E-08 | 0.000214 | 8.89E-11 | 0.005970873 | MEF2C-AS2, LINC00461 |
| chr21 | 34885249 | 34887500 | 2252 | 17 | 5.66E-07 | 2.26E-12 | 0.00722 | 8.90E-11 | 0.025224325 | RUNX1 |
| chr6 | 4806071 | 4806092 | 22 | 2 | 6.19E-11 | 6.42E-11 | 1.23E-06 | 9.01E-11 | 0.020546951 | CDYL |
| chr6 | 29602730 | 29605312 | 2583 | 22 | 1.68E-10 | 2.14E-09 | 0.000406 | 9.26E-11 | 0.001448061 | GABBR1 |
| chr6 | 32085323 | 32088472 | 3150 | 33 | 1.14E-12 | 0.000153 | 0.000274 | 9.47E-11 | 0.004439102 | TNXB |
| chr22 | 37122046 | 37123735 | 1690 | 6 | 4.80E-10 | 7.38E-08 | 4.77E-06 | 9.47E-11 | 0.0035362 | NA |
| chr11 | 116787812 | 116789798 | 1987 | 12 | 6.12E-07 | 3.91E-12 | 0.000656 | 9.67E-11 | 0.001992421 | ZPR1, APOA5 |
| chr15 | 74994282 | 74995948 | 1667 | 9 | 1.04E-13 | 8.21E-08 | 9.18E-05 | 9.78E-11 | 0.009819866 | SCAMP5 |
| chr17 | 42121797 | 42123341 | 1545 | 13 | 1.51E-11 | 2.43E-08 | 0.000168 | 9.78E-11 | 0.006350132 | HSPB9, AC099811.2 |
| chr11 | 126212639 | 126214979 | 2341 | 4 | 4.05E-09 | 6.18E-11 | 2.13E-05 | 1.01E-10 | 0.012805262 | FAM118B, RNU4-86P |
| chr9 | 137074174 | 137074798 | 625 | 4 | 8.50E-11 | 6.62E-07 | 2.28E-05 | 1.01E-10 | 0.009719878 | NA |
| chr22 | 41445815 | 41449210 | 3396 | 18 | 8.33E-09 | 1.30E-08 | 0.000336 | 1.03E-10 | 0.001887377 | ACO2, TOB2 |

| | | | | | | | | | | |
|---|---|---|---|---|---|---|---|---|---|---|
| chr17 | 15942043 | 15943919 | 1877 | 6 | 5.05E-14 | 1.75E-06 | 3.02E-06 | 1.04E-10 | 0.011105105 | NA |
| chr1 | 182952525 | 182953570 | 1046 | 12 | 5.96E-12 | 2.34E-09 | 0.000132 | 1.04E-10 | -0.01358055 | SHCBP1L |
| chr12 | 112005978 | 112007487 | 1510 | 6 | 1.34E-09 | 2.83E-11 | 5.79E-05 | 1.04E-10 | 0.008493185 | TMEM116 |
| chr6 | 27247015 | 27247896 | 882 | 7 | 1.34E-12 | 2.69E-06 | 5.81E-05 | 1.16E-10 | 0.01181573 | PRSS16 |
| chr16 | 3064846 | 3065808 | 963 | 11 | 7.70E-11 | 4.20E-10 | 2.56E-05 | 1.16E-10 | 0.008933202 | IL32 |
| chr21 | 33229381 | 33230564 | 1184 | 16 | 2.42E-10 | 4.37E-11 | 0.000711 | 1.16E-10 | -0.00231686 | IFNAR2, AP000295.1 |
| chr15 | 96340958 | 96343908 | 2951 | 15 | 4.10E-07 | 4.90E-07 | 0.000558 | 1.16E-10 | 0.003413724 | AC103746.1 |
| chr6 | 31120411 | 31122503 | 2093 | 11 | 7.46E-07 | 1.53E-12 | 0.004965 | 1.17E-10 | 0.012159086 | PSORS1C1, CDSN |
| chr6 | 32219490 | 32222779 | 3290 | 23 | 1.29E-06 | 1.45E-08 | 0.001098 | 1.28E-10 | 0.003488913 | NOTCH4 |
| chr14 | 21022430 | 21024313 | 1884 | 15 | 6.32E-13 | 4.91E-07 | 0.00038 | 1.29E-10 | 0.004641349 | TPPP2, AL161668.4, NDRG2, MIR6717 |
| chr19 | 55062204 | 55063998 | 1795 | 12 | 2.67E-09 | 6.85E-11 | 0.000542 | 1.29E-10 | 0.008964384 | RDH13 |
| chr17 | 81683489 | 81685748 | 2260 | 21 | 1.31E-11 | 9.85E-07 | 0.000574 | 1.29E-10 | 0.001862923 | HGS, ARL16 |
| chr11 | 129439600 | 129440968 | 1369 | 5 | 1.03E-10 | 3.23E-10 | 8.99E-05 | 1.29E-10 | 0.011803336 | BARX2 |
| chr1 | 153773735 | 153775644 | 1910 | 16 | 5.00E-09 | 1.05E-09 | 0.000521 | 1.32E-10 | 0.006792119 | INTS3, SLC27A3 |
| chr5 | 135034623 | 135036708 | 2086 | 8 | 6.78E-11 | 2.90E-07 | 0.000115 | 1.34E-10 | 0.013159361 | C5orf66, AC008406.3, PITX1 |
| chr7 | 123624646 | 123625790 | 1145 | 10 | 2.68E-10 | 1.25E-09 | 6.15E-05 | 1.34E-10 | 0.010934645 | ASB15, ASB15-AS1 |
| chr8 | 42895767 | 42898344 | 2578 | 13 | 6.70E-08 | 8.59E-10 | 0.000938 | 1.34E-10 | 0.002305063 | HOOK3, RNF170, MIR4469 |
| chr14 | 104967446 | 104967961 | 516 | 4 | 5.56E-12 | 1.29E-08 | 1.09E-05 | 1.35E-10 | 0.010396578 | AHNAK2 |
| chr11 | 60923500 | 60924857 | 1358 | 11 | 2.66E-11 | 8.50E-07 | 1.35E-05 | 1.35E-10 | 0.003377962 | TMEM132A, AP003721.1 |
| chr5 | 132746402 | 132748376 | 1975 | 13 | 8.36E-10 | 8.82E-10 | 0.002015 | 1.35E-10 | 0.007477407 | CCNI2 |
| chr11 | 3807587 | 3809141 | 1555 | 16 | 7.81E-10 | 7.22E-08 | 0.000259 | 1.36E-10 | 0.003394888 | PGAP2 |
| chr19 | 6729138 | 6732032 | 2895 | 9 | 1.23E-07 | 9.49E-09 | 1.51E-05 | 1.43E-10 | 0.001283527 | C3, GPR108 |
| chr18 | 49723780 | 49724526 | 747 | 3 | 5.01E-11 | 1.36E-10 | 1.45E-05 | 1.45E-10 | 0.052991912 | NA |
| chr10 | 61662115 | 61663477 | 1363 | 13 | 4.72E-11 | 2.78E-09 | 0.000255 | 1.46E-10 | 0.007749673 | CABCOCO1 |
| chr7 | 151367412 | 151368738 | 1327 | 4 | 5.60E-09 | 1.10E-10 | 1.45E-05 | 1.48E-10 | 0.007756272 | NUB1 |
| chr1 | 24535114 | 24535891 | 778 | 7 | 7.99E-10 | 1.52E-11 | 0.000539 | 1.50E-10 | 0.011725898 | RCAN3, RCAN3AS |
| chr17 | 28981711 | 28983009 | 1299 | 8 | 1.66E-08 | 1.50E-10 | 0.000193 | 1.53E-10 | 0.008520756 | PIPOX, SEZ6 |
| chr18 | 14747037 | 14748299 | 1263 | 11 | 5.04E-09 | 1.69E-09 | 0.000844 | 1.53E-10 | 0.016022518 | ANKRD30B |
| chr4 | 8747531 | 8747869 | 339 | 5 | 3.66E-10 | 8.75E-11 | 7.98E-05 | 1.56E-10 | 0.009832675 | AC209005.1 |
| chr2 | 227870847 | 227872527 | 1681 | 15 | 3.21E-12 | 1.28E-07 | 0.000505 | 1.56E-10 | 0.004210861 | DAW1 |
| chr1 | 32246239 | 32247674 | 1436 | 6 | 2.65E-11 | 2.74E-07 | 2.22E-05 | 1.57E-10 | 0.008073565 | FAM167B |
| chr14 | 93745472 | 93745794 | 323 | 2 | 1.21E-12 | 1.53E-09 | 9.40E-08 | 1.58E-10 | 0.022918105 | PRIMA1 |
| chr22 | 19524705 | 19525653 | 949 | 11 | 1.99E-10 | 2.07E-10 | 0.000377 | 1.59E-10 | 0.0071854 | CLDN5 |
| chr6 | 33174809 | 33176294 | 1486 | 8 | 9.31E-11 | 8.17E-09 | 1.82E-05 | 1.59E-10 | 0.004718327 | COL11A2 |
| chr1 | 43284969 | 43286514 | 1546 | 12 | 1.88E-08 | 1.17E-10 | 0.001455 | 1.60E-10 | 0.005550875 | C1orf210 |
| chr7 | 150406024 | 150408625 | 2602 | 9 | 5.88E-09 | 1.09E-07 | 9.49E-05 | 1.61E-10 | 0.00604737 | AC073111.2, AC073111.3 |
| chr3 | 156290338 | 156291891 | 1554 | 12 | 1.49E-08 | 3.37E-11 | 0.002108 | 1.61E-10 | 0.005636108 | KCNAB1, AC091607.2 |
| chr22 | 37674994 | 37676094 | 1101 | 14 | 2.11E-11 | 2.41E-09 | 0.000735 | 1.63E-10 | 0.007867194 | LGALS1 |
| chr2 | 120465958 | 120466751 | 794 | 8 | 1.10E-10 | 7.28E-10 | 0.000782 | 1.64E-10 | 0.020629519 | LINC01101 |
| chr12 | 110920384 | 110921158 | 775 | 12 | 7.95E-11 | 2.35E-09 | 8.36E-05 | 1.65E-10 | 0.006878513 | MYL2 |
| chr15 | 65421138 | 65424092 | 2955 | 11 | 4.06E-08 | 6.61E-09 | 0.000368 | 1.69E-10 | 0.006761849 | IGDCC4 |
| chr7 | 27243983 | 27245170 | 1188 | 6 | 3.73E-12 | 3.45E-10 | 0.000158 | 1.70E-10 | 0.007180552 | EVX1, EVX1-AS |
| chr7 | 151061132 | 151063035 | 1904 | 16 | 3.75E-09 | 6.47E-08 | 0.000639 | 1.70E-10 | 0.002826665 | SLC4A2, AC010973.3 |
| chr10 | 54801185 | 54802398 | 1214 | 7 | 1.84E-10 | 1.84E-09 | 5.67E-05 | 1.76E-10 | 0.014523872 | PCDH15 |
| chr4 | 110628724 | 110631197 | 2474 | 14 | 1.75E-07 | 3.84E-10 | 0.001156 | 1.80E-10 | 0.005253817 | PITX2 |
| chr7 | 55448574 | 55449845 | 1272 | 8 | 3.80E-10 | 2.02E-11 | 0.000262 | 1.83E-10 | -0.0174201 | VOPP1 |
| chr10 | 110005915 | 110007621 | 1707 | 8 | 8.06E-11 | 4.82E-09 | 3.51E-05 | 1.86E-10 | 0.005728909 | ADD3, ADD3-AS1 |
| chr6 | 36336679 | 36338047 | 1369 | 6 | 1.09E-09 | 5.80E-11 | 9.58E-05 | 1.87E-10 | 0.007366673 | BNIP5 |
| chr10 | 43044124 | 43044775 | 652 | 5 | 1.70E-10 | 1.08E-09 | 8.29E-05 | 1.89E-10 | 0.012101029 | NA |
| chr16 | 31088774 | 31090058 | 1285 | 8 | 1.44E-09 | 9.49E-10 | 2.77E-05 | 1.89E-10 | 0.00544572 | PRSS53, AC135050.2 |
| chr1 | 1419855 | 1421981 | 2127 | 12 | 1.65E-08 | 5.46E-10 | 0.00262 | 1.92E-10 | 0.005838022 | AL391244.1, ANKRD65 |
| chr7 | 51326987 | 51327749 | 763 | 6 | 2.41E-10 | 5.95E-11 | 0.00013 | 1.94E-10 | 0.013959267 | NA |
| chr10 | 43322533 | 43323983 | 1451 | 11 | 2.36E-09 | 4.94E-11 | 0.000514 | 1.94E-10 | 0.006960038 | LINC02633 |
| chr1 | 9720848 | 9722781 | 1934 | 12 | 2.76E-11 | 2.06E-09 | 0.000247 | 1.95E-10 | 0.004439564 | PIK3CD |
| chr3 | 58260497 | 58261032 | 536 | 3 | 1.04E-12 | 4.06E-07 | 1.77E-07 | 1.99E-10 | 0.013950525 | ABHD6 |
| chr7 | 128791224 | 128793207 | 1984 | 11 | 1.98E-08 | 7.45E-08 | 0.000182 | 2.02E-10 | 0.006617212 | CCDC136 |
| chr6 | 135181656 | 135183851 | 2196 | 11 | 1.30E-08 | 4.63E-10 | 0.000856 | 2.02E-10 | 0.000797413 | MYB |
| chr3 | 195837877 | 195839815 | 1939 | 6 | 1.56E-08 | 3.74E-08 | 7.91E-06 | 2.04E-10 | 0.004676235 | LINC01983 |
| chr5 | 1874899 | 1877195 | 2297 | 14 | 2.32E-07 | 2.04E-11 | 0.004244 | 2.06E-10 | 0.003184025 | NA |
| chr6 | 47476401 | 47477785 | 1385 | 9 | 1.52E-10 | 7.73E-10 | 0.000401 | 2.09E-10 | 0.010980027 | AL355353.1 |
| chr19 | 39872645 | 39873511 | 867 | 3 | 2.49E-12 | 1.32E-08 | 7.37E-06 | 2.11E-10 | 0.007924273 | FCGBP |
| chr20 | 52128176 | 52128817 | 642 | 4 | 6.30E-11 | 4.11E-10 | 1.53E-05 | 2.12E-10 | 0.023359956 | ZFP64 |
| chr6 | 30891399 | 30893183 | 1785 | 19 | 8.72E-09 | 6.58E-10 | 0.001328 | 2.12E-10 | 0.006125933 | DDR1 |
| chr4 | 47485064 | 47486603 | 1540 | 12 | 1.13E-08 | 1.83E-09 | 3.06E-05 | 2.13E-10 | 0.002350253 | ATP10D, AC107398.5 |
| chr12 | 124737208 | 124737869 | 662 | 3 | 1.43E-11 | 9.78E-10 | 6.22E-07 | 2.16E-10 | 0.007535618 | NA |
| chr22 | 50484532 | 50486684 | 2153 | 9 | 4.83E-07 | 4.95E-08 | 0.000118 | 2.18E-10 | 0.007508702 | ADM2 |
| chr3 | 185497744 | 185499507 | 1764 | 14 | 8.12E-12 | 2.60E-07 | 8.39E-05 | 2.19E-10 | 0.000808842 | AC099661.1, TMEM41A |
| chr2 | 9271443 | 9271511 | 69 | 3 | 4.57E-10 | 6.04E-11 | 3.20E-05 | 2.21E-10 | 0.022745213 | ASAP2, EIF1P7 |
| chr2 | 17754304 | 17755263 | 960 | 6 | 1.38E-11 | 1.16E-10 | 0.000133 | 2.22E-10 | 0.010178762 | GEN1, SMC6 |
| chr6 | 41704563 | 41706091 | 1529 | 4 | 2.77E-08 | 3.25E-08 | 1.90E-05 | 2.22E-10 | 0.009702187 | TFEB |
| chr1 | 940226 | 941613 | 1388 | 8 | 2.44E-10 | 1.60E-10 | 0.000422 | 2.27E-10 | 0.002550553 | SAMD11 |
| chr13 | 43878348 | 43880519 | 2172 | 19 | 2.89E-08 | 2.25E-09 | 0.000154 | 2.29E-10 | 0.005016768 | LACC1, CCDC122 |
| chr11 | 72079014 | 72081160 | 2147 | 21 | 3.26E-07 | 1.59E-11 | 0.001493 | 2.31E-10 | 0.001659478 | AP000812.4, LRTOMT, NUMA1 |
| chr8 | 109642766 | 109644213 | 1448 | 8 | 1.30E-09 | 2.09E-09 | 3.51E-05 | 2.33E-10 | 0.011018823 | AC079061.1, SYBU |
| chr1 | 114854008 | 114855502 | 1495 | 11 | 1.18E-11 | 7.14E-10 | 0.000123 | 2.35E-10 | -0.01524449 | SYCP1 |
| chr12 | 57751967 | 57754105 | 2139 | 22 | 5.62E-11 | 6.07E-06 | 0.000113 | 2.35E-10 | 0.002773214 | CDK4 |
| chr10 | 90163682 | 90165370 | 1689 | 4 | 9.10E-12 | 1.77E-08 | 7.41E-06 | 2.36E-10 | 0.012606982 | LINC01374, RN7SKP143 |
| chr10 | 97102563 | 97103924 | 1362 | 7 | 2.82E-10 | 3.21E-08 | 3.85E-05 | 2.37E-10 | 0.007326718 | SLIT1-AS1, SLIT1 |
| chr15 | 49421780 | 49424448 | 2669 | 10 | 4.93E-06 | 2.71E-11 | 0.002745 | 2.37E-10 | 0.021733078 | FGF7, FAM227B |
| chr19 | 1913401 | 1915312 | 1912 | 8 | 1.78E-06 | 9.52E-10 | 0.000546 | 2.43E-10 | 0.003283218 | SCAMP4, ADAT3 |
| chr5 | 135491446 | 135492346 | 901 | 7 | 2.26E-11 | 2.41E-07 | 4.62E-05 | 2.49E-10 | 0.010090033 | NA |
| chr6 | 138545728 | 138545988 | 261 | 7 | 2.51E-11 | 2.59E-06 | 1.42E-05 | 2.49E-10 | 0.00469058 | NHSL1 |
| chr12 | 10190044 | 10191165 | 1122 | 8 | 2.89E-13 | 9.98E-08 | 2.05E-05 | 2.53E-10 | 0.009864558 | TMEM52B |
| chr6 | 1606642 | 1608432 | 1791 | 11 | 8.75E-10 | 2.67E-09 | 5.31E-05 | 2.54E-10 | 0.004917904 | FOXCUT |
| chr2 | 218882357 | 218884483 | 2127 | 6 | 7.04E-09 | 4.35E-09 | 7.31E-05 | 2.57E-10 | 0.010505909 | WNT10A |
| chr8 | 41646559 | 41648177 | 1619 | 15 | 1.17E-12 | 4.69E-08 | 3.21E-05 | 2.57E-10 | 0.005660843 | NKX6-3 |
| chr15 | 62886494 | 62886980 | 487 | 3 | 7.45E-14 | 4.55E-06 | 2.08E-08 | 2.60E-10 | 0.009723385 | NA |
| chr18 | 3447018 | 3449545 | 2528 | 26 | 6.00E-07 | 4.28E-08 | 7.84E-05 | 2.61E-10 | 0.00138239 | TGIF1 |
| chr16 | 2234124 | 2235774 | 1651 | 12 | 3.53E-11 | 6.96E-06 | 1.69E-05 | 2.71E-10 | 0.002864579 | E4F1, AC009065.1 |
| chr21 | 46184138 | 46185260 | 1123 | 7 | 8.26E-08 | 9.73E-12 | 0.001934 | 2.73E-10 | -0.05351584 | AP001468.1, SPATC1L |
| chr6 | 31137934 | 31140331 | 2398 | 20 | 1.90E-09 | 1.25E-08 | 0.001083 | 2.75E-10 | 0.004336702 | PSORS1C1, PSORS1C2 |
| chr11 | 88103463 | 88103945 | 483 | 2 | 7.19E-11 | 3.07E-10 | 1.11E-06 | 2.82E-10 | 0.025072361 | AP005436.2 |
| chr16 | 53433469 | 53435613 | 2145 | 17 | 6.58E-09 | 1.77E-07 | 0.000121 | 2.82E-10 | 0.002649545 | RBL2 |
| chr11 | 108221360 | 108224052 | 2693 | 46 | 2.46E-07 | 1.26E-06 | 9.87E-05 | 2.85E-10 | 0.001247406 | ATM, NPAT |
| chr6 | 41501398 | 41503935 | 2538 | 10 | 3.77E-09 | 2.13E-08 | 0.000468 | 2.91E-10 | 0.009164849 | LINC01276, FOXP4-AS1 |
| chr12 | 46825843 | 46826414 | 572 | 12 | 1.26E-09 | 6.35E-11 | 0.003888 | 2.91E-10 | 0.025974798 | AC008014.1, SLC38A4 |
| chr11 | 61480457 | 61482721 | 2265 | 11 | 2.56E-07 | 1.67E-09 | 0.000288 | 2.92E-10 | 0.003079974 | AP003108.2, PPP1R32 |
| chr1 | 9151777 | 9152695 | 919 | 5 | 9.73E-13 | 2.33E-08 | 6.46E-07 | 2.92E-10 | 0.005738705 | MIR34AHG, MIR34A |
| chr1 | 156906276 | 156908781 | 2506 | 9 | 1.24E-08 | 4.31E-08 | 9.55E-06 | 2.92E-10 | 0.005715203 | PEAR1 |
| chr17 | 77318520 | 77320398 | 1879 | 16 | 2.19E-09 | 2.93E-08 | 0.000587 | 2.95E-10 | 0.007381312 | SEPTIN9 |
| chr17 | 1477737 | 1479645 | 1909 | 9 | 5.88E-08 | 1.04E-08 | 6.96E-06 | 2.97E-10 | 0.003324074 | MYO1C |
| chr6 | 30942043 | 30943164 | 1122 | 9 | 7.86E-11 | 8.88E-08 | 1.59E-05 | 2.99E-10 | 0.00762453 | MUCL3, SFTA2 |
| chr20 | 31473019 | 31474893 | 1875 | 8 | 2.04E-08 | 1.96E-09 | 7.57E-05 | 3.00E-10 | 0.010096007 | DEFB124 |
| chr16 | 474300 | 474912 | 613 | 6 | 1.88E-10 | 1.75E-10 | 0.000406 | 3.02E-10 | 0.016668814 | RAB11FIP3 |
| chr11 | 2532968 | 2534346 | 1379 | 10 | 3.87E-10 | 6.06E-10 | 0.000135 | 3.02E-10 | 0.008548828 | KCNQ1 |
| chr10 | 76114270 | 76115918 | 1649 | 4 | 1.70E-08 | 4.12E-10 | 1.72E-05 | 3.04E-10 | 0.008210943 | LRMDA |
| chr15 | 42272954 | 42274464 | 1511 | 24 | 2.03E-10 | 1.33E-07 | 0.001013 | 3.10E-10 | 0.002042076 | GANC, TMEM87A |
| chr3 | 147359399 | 147360232 | 834 | 9 | 8.96E-10 | 7.37E-10 | 0.000141 | 3.15E-10 | 0.009374687 | AC092957.1 |
| chr4 | 187063258 | 187063352 | 95 | 2 | 9.10E-12 | 2.96E-10 | 1.49E-06 | 3.15E-10 | -0.02343571 | NA |
| chr9 | 125840364 | 125841060 | 697 | 2 | 6.76E-09 | 1.57E-10 | 1.04E-05 | 3.25E-10 | 0.019113333 | PBX3 |
| chr20 | 36173176 | 36173888 | 713 | 5 | 2.18E-10 | 4.35E-10 | 0.000124 | 3.36E-10 | 0.008613566 | EPB41L1 |
| chr11 | 57656451 | 57658259 | 1809 | 18 | 3.54E-07 | 4.89E-09 | 0.003482 | 3.41E-10 | -0.00017722 | CLP1 |
| chr13 | 113120627 | 113122846 | 2220 | 15 | 5.94E-07 | 7.22E-11 | 0.000994 | 3.49E-10 | 0.007018368 | F7, F10 |
| chr10 | 4024174 | 4024930 | 757 | 6 | 3.99E-10 | 6.87E-08 | 3.88E-05 | 3.53E-10 | 0.004400675 | AC025822.1 |
| chr10 | 86967302 | 86970104 | 2803 | 19 | 6.06E-07 | 2.85E-09 | 0.001039 | 3.53E-10 | 0.003047146 | ADIRF, MMRN2, ADIRF-AS1 |
| chr19 | 289762 | 291851 | 2090 | 10 | 8.06E-09 | 2.18E-10 | 0.000934 | 3.54E-10 | -0.00236814 | PLPP2 |
| chr6 | 163309336 | 163310957 | 1622 | 11 | 1.03E-09 | 2.47E-08 | 0.00035 | 3.57E-10 | 0.006664826 | PACRG, PACRG-AS1 |
| chr1 | 205565860 | 205566104 | 245 | 3 | 1.70E-10 | 1.71E-08 | 1.96E-06 | 3.58E-10 | 0.007804706 | NA |
| chr14 | 75126382 | 75128072 | 1691 | 18 | 6.46E-09 | 6.26E-11 | 0.000508 | 3.60E-10 | 0.001820145 | AL049780.2, NEK9 |
| chr2 | 86791259 | 86792489 | 1231 | 10 | 5.36E-13 | 2.22E-08 | 0.000101 | 3.64E-10 | 0.009661749 | CD8A |
| chr7 | 1501377 | 1503133 | 1757 | 7 | 4.82E-09 | 2.34E-06 | 1.34E-05 | 3.74E-10 | 0.005368389 | INTS1 |
| chr1 | 31735859 | 31738117 | 2259 | 6 | 1.55E-05 | 4.65E-11 | 0.000168 | 3.76E-10 | 0.006642073 | ADGRB2 |
| chr18 | 24731551 | 24732142 | 592 | 2 | 1.31E-11 | 1.64E-09 | 2.38E-07 | 3.81E-10 | 0.015709458 | AC018697.1 |
| chr11 | 128548656 | 128549617 | 962 | 6 | 7.28E-09 | 1.33E-10 | 0.00066 | 3.82E-10 | 0.009546907 | ETS1 |
| chr1 | 116167148 | 116168456 | 1309 | 8 | 2.68E-11 | 1.84E-09 | 4.88E-05 | 3.84E-10 | 0.012859879 | NA |
| chr7 | 151850884 | 151851829 | 946 | 6 | 1.58E-11 | 6.53E-08 | 4.47E-05 | 3.85E-10 | 0.012910902 | PRKAG2 |
| chr9 | 35538215 | 35539512 | 1298 | 7 | 1.67E-12 | 2.53E-07 | 1.63E-05 | 3.87E-10 | 0.009475979 | RUSC2 |
| chr17 | 2054202 | 2056326 | 2125 | 37 | 3.08E-09 | 2.01E-08 | 0.005648 | 3.90E-10 | 0.001660686 | HIC1 |
| chr3 | 48563684 | 48565361 | 1678 | 10 | 1.43E-11 | 5.26E-07 | 0.00012 | 4.15E-10 | 0.005272033 | UCN2, COL7A1 |
| chr5 | 154485407 | 154486656 | 1250 | 8 | 7.65E-08 | 4.55E-10 | 0.000267 | 4.22E-10 | 0.008147697 | AC026688.2 |
| chr14 | 24070126 | 24071564 | 1439 | 15 | 4.16E-08 | 3.18E-10 | 0.000556 | 4.37E-10 | 0.005337906 | CPNE6 |
| chr17 | 82871152 | 82873658 | 2507 | 12 | 1.01E-06 | 1.98E-09 | 0.000562 | 4.42E-10 | 0.003864438 | TBCD |
| chr11 | 2928733 | 2930751 | 2019 | 30 | 2.07E-08 | 4.12E-09 | 0.002199 | 4.48E-10 | -0.0012738 | PHLDA2 |
| chr7 | 47634971 | 47635469 | 499 | 5 | 8.03E-10 | 1.22E-10 | 0.000351 | 4.48E-10 | -0.03366722 | NA |
| chr21 | 42403495 | 42404153 | 659 | 10 | 3.03E-10 | 6.71E-10 | 0.000302 | 4.55E-10 | 0.006421353 | UBASH3A |
| chr8 | 29459116 | 29459388 | 273 | 3 | 1.18E-10 | 6.55E-09 | 1.23E-06 | 4.56E-10 | 0.018899382 | NA |
| chr1 | 40708587 | 40709527 | 941 | 9 | 8.23E-10 | 1.61E-10 | 0.000882 | 4.59E-10 | 0.009768015 | NFYC |
| chr11 | 68750061 | 68752267 | 2207 | 15 | 1.21E-07 | 2.75E-10 | 0.000297 | 4.67E-10 | -0.00436738 | TESMIN |
| chr18 | 47035347 | 47036547 | 1201 | 11 | 4.89E-09 | 1.35E-11 | 0.003487 | 4.81E-10 | 0.01058414 | KATNAL2, ELOA2 |
| chr7 | 1566630 | 1568151 | 1522 | 8 | 3.88E-09 | 8.73E-09 | 6.21E-05 | 4.84E-10 | -0.00117092 | PSMG3 |
| chr3 | 147388421 | 147390113 | 1693 | 8 | 2.28E-11 | 8.35E-08 | 9.12E-05 | 4.96E-10 | 0.00595549 | ZIC4 |
| chr7 | 27213118 | 27214452 | 1335 | 7 | 4.54E-11 | 1.16E-08 | 0.000767 | 5.00E-10 | 0.020820731 | NA |
| chr16 | 1781467 | 1784322 | 2856 | 27 | 1.48E-06 | 2.18E-08 | 0.001384 | 5.04E-10 | 0.00042549 | EME2, NUBP2, SPSB3 |
| chr14 | 24171686 | 24172497 | 812 | 12 | 3.40E-10 | 1.39E-10 | 0.001434 | 5.06E-10 | 0.006377476 | REC8 |
| chr20 | 47317527 | 47319709 | 2183 | 10 | 2.28E-09 | 5.62E-11 | 9.31E-05 | 5.07E-10 | 0.00894998 | AL031666.2, ZMYND8 |
| chr1 | 108591141 | 108592634 | 1494 | 3 | 1.38E-09 | 1.05E-07 | 5.42E-06 | 5.29E-10 | 0.007767023 | FAM102B |
| chr9 | 105655279 | 105656135 | 857 | 2 | 3.94E-14 | 1.66E-08 | 8.20E-08 | 5.37E-10 | 0.008409387 | NA |
| chr6 | 30102235 | 30104607 | 2373 | 36 | 6.18E-10 | 2.16E-07 | 0.00075 | 5.39E-10 | 0.004042464 | TRIM31 |
| chr11 | 44310027 | 44311642 | 1616 | 40 | 6.13E-09 | 4.08E-09 | 0.001436 | 5.39E-10 | 0.003054635 | ALX4 |
| chr11 | 118887226 | 118888692 | 1467 | 8 | 4.45E-11 | 1.87E-07 | 7.91E-05 | 5.41E-10 | 0.006871787 | CXCR5, AP004609.1 |
| chr7 | 27240966 | 27242825 | 1860 | 23 | 1.63E-07 | 5.50E-09 | 0.001849 | 5.52E-10 | 0.004946659 | EVX1, AC004080.1, EVX1-AS |
| chr3 | 39106807 | 39109759 | 2953 | 22 | 4.21E-06 | 2.76E-09 | 0.000129 | 5.55E-10 | 0.001016747 | TTC21A, GORASP1 |
| chr12 | 124294592 | 124296492 | 1901 | 14 | 6.31E-08 | 8.65E-09 | 0.000465 | 5.56E-10 | 0.00351048 | RFLNA |
| chr5 | 140145164 | 140146641 | 1478 | 8 | 7.58E-07 | 1.14E-08 | 0.000163 | 5.69E-10 | 0.007628647 | NA |
| chr3 | 27723792 | 27724891 | 1100 | 12 | 2.73E-11 | 1.27E-06 | 2.49E-05 | 5.87E-10 | 0.006979233 | NA |
| chr20 | 59006951 | 59008654 | 1704 | 26 | 2.34E-10 | 5.50E-09 | 0.000938 | 5.90E-10 | 0.005267085 | CTSZ |
| chr3 | 48608942 | 48610909 | 1968 | 14 | 5.38E-10 | 8.55E-07 | 9.35E-06 | 6.01E-10 | 0.001986754 | UQCRC1 |
| chr20 | 63685907 | 63687235 | 1329 | 10 | 4.61E-08 | 1.56E-10 | 0.000988 | 6.01E-10 | 0.009404281 | RTEL1, RTEL1-TNFRSF6B |
| chr2 | 241723828 | 241726651 | 2824 | 19 | 1.25E-06 | 3.51E-10 | 0.001492 | 6.01E-10 | -0.00106574 | ING5, AC114730.3 |
| chr10 | 13706597 | 13709379 | 2783 | 18 | 1.87E-07 | 2.12E-06 | 0.000165 | 6.10E-10 | 0.003073196 | FRMD4A |
| chr15 | 68365319 | 68367420 | 2102 | 5 | 2.75E-09 | 3.19E-08 | 6.30E-05 | 6.17E-10 | 0.00470422 | ITGA11 |
| chr6 | 89638290 | 89640141 | 1852 | 18 | 2.96E-10 | 1.23E-06 | 1.72E-05 | 6.18E-10 | 0.0052848 | LYRM2 |

| | | | | | | | | | | |
|---|---|---|---|---|---|---|---|---|---|---|
| chr5 | 95861167 | 95861772 | 606 | 5 | 5.23E-10 | 1.09E-09 | 1.20E-05 | 6.19E-10 | 0.006956947 | NA |
| chr20 | 38164920 | 38165926 | 1007 | 12 | 1.87E-10 | 1.48E-08 | 0.000227 | 6.22E-10 | 0.004980544 | TGM2 |
| chr16 | 30984306 | 30985501 | 1196 | 11 | 1.12E-08 | 2.07E-10 | 0.000693 | 6.32E-10 | 0.004552665 | SETD1A, HSD3B7, AC135048.4 |
| chr5 | 159021285 | 159022752 | 1468 | 5 | 6.31E-08 | 7.34E-11 | 0.000291 | 6.40E-10 | 0.013956674 | EBF1 |
| chr9 | 133114295 | 133115910 | 1616 | 4 | 1.65E-07 | 1.10E-08 | 1.96E-05 | 6.42E-10 | 0.013798913 | RALGDS, AL162417.1 |
| chr4 | 139279911 | 139281173 | 1263 | 13 | 7.55E-09 | 1.13E-08 | 0.000395 | 6.50E-10 | 0.007514432 | MGARP, NDUFC1 |
| chr6 | 83852607 | 83854191 | 1585 | 18 | 6.12E-09 | 2.92E-09 | 0.000287 | 6.79E-10 | 0.002357043 | RIPPLY2, AL139232.1 |
| chr12 | 10098937 | 10100497 | 1561 | 7 | 2.42E-06 | 1.11E-09 | 0.000127 | 6.81E-10 | 0.007065157 | CLEC1A |
| chr17 | 19707441 | 19707870 | 430 | 3 | 8.10E-12 | 3.63E-06 | 2.46E-07 | 6.85E-10 | 0.008986456 | SLC47A2 |
| chr19 | 53865704 | 53869371 | 3668 | 33 | 3.28E-08 | 1.13E-05 | 0.005172 | 6.99E-10 | 0.003843935 | MYADM, MYADM-AS1 |
| chr1 | 155070937 | 155071869 | 933 | 7 | 8.29E-12 | 8.19E-08 | 5.58E-05 | 7.16E-10 | 0.009920128 | AL691442.1 |
| chr19 | 32869693 | 32871154 | 1462 | 9 | 7.93E-09 | 3.47E-08 | 0.000166 | 7.26E-10 | 0.008987926 | SLC7A9 |
| chr12 | 52368221 | 52369914 | 1694 | 4 | 1.33E-11 | 1.05E-07 | 3.25E-07 | 7.34E-10 | 0.007638174 | NA |
| chr6 | 128882363 | 128883395 | 1033 | 9 | 2.74E-09 | 5.78E-09 | 0.000692 | 7.35E-10 | 0.007558131 | LAMA2 |
| chr1 | 33626037 | 33626780 | 744 | 4 | 2.28E-12 | 2.28E-07 | 8.32E-06 | 7.41E-10 | 0.01122492 | CSMD2 |
| chr11 | 61675069 | 61675659 | 591 | 4 | 3.16E-09 | 2.76E-10 | 8.15E-05 | 7.52E-10 | -0.02417993 | NA |
| chr7 | 150955697 | 150956361 | 665 | 11 | 2.62E-09 | 1.10E-09 | 0.001461 | 7.53E-10 | -0.01261693 | KCNH2 |
| chr1 | 162366192 | 162367585 | 1394 | 10 | 6.04E-09 | 8.65E-10 | 0.000603 | 7.56E-10 | 0.005899884 | NOS1AP, AL512785.2 |
| chr12 | 101695769 | 101697995 | 2227 | 17 | 1.27E-07 | 3.85E-10 | 0.001756 | 7.81E-10 | 0.002673073 | CHPT1, AC010205.1 |
| chr11 | 6457780 | 6458233 | 454 | 5 | 5.68E-12 | 5.57E-07 | 2.03E-07 | 7.82E-10 | 0.002202293 | TRIM3 |
| chr16 | 84503923 | 84505504 | 1582 | 11 | 1.60E-07 | 1.16E-08 | 0.000312 | 8.00E-10 | 0.001288923 | MEAK7 |
| chr5 | 179622815 | 179625006 | 2192 | 22 | 2.14E-07 | 5.60E-09 | 0.00291 | 8.18E-10 | 0.001824627 | HNRNPH1 |
| chr14 | 101456413 | 101457849 | 1437 | 8 | 7.02E-09 | 2.65E-08 | 1.98E-05 | 8.19E-10 | 0.00934153 | AL049836.2 |
| chr8 | 143549354 | 143550305 | 952 | 8 | 3.27E-08 | 1.27E-10 | 0.000249 | 8.36E-10 | 0.019658728 | AC067930.3 |
| chr3 | 179251607 | 179252619 | 1013 | 8 | 2.04E-09 | 1.99E-08 | 8.67E-05 | 8.38E-10 | 0.003421405 | KCNMB3 |
| chr1 | 26201379 | 26202592 | 1214 | 5 | 8.95E-08 | 6.25E-11 | 0.000905 | 8.44E-10 | 0.038000694 | CATSPER4 |
| chr13 | 110307830 | 110309477 | 1648 | 13 | 5.42E-08 | 4.11E-09 | 0.002624 | 8.58E-10 | 0.006635978 | COL4A2 |
| chr3 | 147422337 | 147424628 | 2292 | 18 | 1.05E-07 | 1.72E-08 | 0.000655 | 8.63E-10 | 0.004893042 | ZIC1 |
| chr19 | 45914771 | 45916603 | 1833 | 7 | 3.48E-07 | 5.36E-09 | 7.90E-05 | 8.63E-10 | -9.49E-05 | NANOS2 |
| chr21 | 39312343 | 39314871 | 2529 | 16 | 2.19E-06 | 8.56E-10 | 0.000187 | 8.82E-10 | 6.01E-05 | BRWD1-AS2, BRWD1 |
| chr11 | 128684691 | 128686685 | 1995 | 14 | 3.88E-08 | 1.13E-08 | 0.001548 | 9.06E-10 | 0.006584795 | FLI1, AP001122.1 |
| chr11 | 1785984 | 1786103 | 120 | 3 | 1.78E-09 | 3.84E-10 | 1.90E-05 | 9.10E-10 | 0.017364464 | NA |
| chr9 | 126293785 | 126294753 | 969 | 2 | 2.14E-08 | 7.18E-10 | 2.99E-06 | 9.15E-10 | 0.018228244 | AL162391.1 |
| chr16 | 85061929 | 85063627 | 1699 | 11 | 2.71E-08 | 3.09E-10 | 0.000493 | 9.30E-10 | 0.007349295 | KIAA0513 |
| chr9 | 70042630 | 70043921 | 1292 | 9 | 8.31E-11 | 2.86E-07 | 1.14E-05 | 9.45E-10 | 0.007142134 | MAMDC2, MAMDC2-AS1 |
| chr11 | 3670957 | 3672788 | 1832 | 8 | 4.91E-07 | 1.44E-10 | 0.00132 | 9.55E-10 | 0.002051617 | CHRNA10, NUP98 |
| chr19 | 45643321 | 45644290 | 970 | 11 | 1.03E-09 | 8.65E-10 | 0.000614 | 9.70E-10 | 0.007926842 | EML2 |
| chr8 | 47259691 | 47260723 | 1033 | 12 | 4.04E-10 | 2.86E-07 | 3.59E-05 | 9.71E-10 | -0.00128887 | NA |
| chr13 | 108214101 | 108215673 | 1573 | 6 | 5.93E-11 | 2.90E-10 | 0.000398 | 9.77E-10 | 0.031338356 | LIG4 |
| chr5 | 135544519 | 135545277 | 759 | 6 | 1.27E-16 | 0.000159 | 4.61E-06 | 9.92E-10 | 0.008200477 | NA |
| chr1 | 44355054 | 44356140 | 1087 | 16 | 9.36E-11 | 2.45E-08 | 0.000392 | 9.92E-10 | 0.002970472 | ERI3 |
| chr2 | 86561751 | 86563362 | 1612 | 7 | 3.49E-09 | 4.39E-08 | 7.80E-06 | 9.94E-10 | 0.003774525 | AC015971.1, CHMP3, RNF103-CHMP3 |
| chr4 | 109849892 | 109850098 | 207 | 2 | 5.04E-11 | 5.95E-10 | 5.75E-06 | 9.95E-10 | 0.025263431 | LRIT3 |
| chr14 | 54691119 | 54691936 | 818 | 4 | 2.26E-09 | 3.36E-10 | 3.11E-05 | 9.95E-10 | 0.02947996 | SAMD4A |
| chr2 | 73985604 | 73986635 | 1032 | 9 | 2.35E-10 | 4.24E-09 | 0.000223 | 1.01E-09 | 0.005892817 | AC073046.1, TET3 |
| chr17 | 80187164 | 80188146 | 983 | 9 | 7.81E-10 | 6.26E-07 | 0.000105 | 1.01E-09 | 0.004628262 | CARD14 |
| chr3 | 52409287 | 52410817 | 1531 | 16 | 5.03E-11 | 7.64E-08 | 7.70E-05 | 1.02E-09 | 0.000458578 | PHF7, BAP1 |
| chr13 | 114146091 | 114146852 | 762 | 8 | 8.60E-10 | 1.17E-07 | 3.44E-05 | 1.02E-09 | 0.004130538 | NA |
| chr16 | 683709 | 686175 | 2467 | 17 | 1.19E-07 | 1.65E-09 | 0.000222 | 1.04E-09 | 0.004176253 | JMJD8, WDR24 |
| chr15 | 99250673 | 99252604 | 1932 | 16 | 1.10E-10 | 7.45E-08 | 1.42E-05 | 1.06E-09 | 0.00708231 | LRRC28, TTC23 |
| chr10 | 120980893 | 120982250 | 1358 | 6 | 3.65E-09 | 9.66E-10 | 3.45E-05 | 1.06E-09 | 0.013320515 | NA |
| chr11 | 68032084 | 68033757 | 1674 | 6 | 7.53E-08 | 2.85E-09 | 7.90E-05 | 1.06E-09 | 0.003530473 | NDUFS8, MIR7113 |
| chr21 | 46110263 | 46113030 | 2768 | 11 | 3.60E-07 | 1.65E-07 | 8.37E-05 | 1.06E-09 | -0.00087297 | COL6A2 |
| chr1 | 32336132 | 32338484 | 2353 | 13 | 1.52E-06 | 5.00E-09 | 7.03E-05 | 1.06E-09 | 0.004710904 | MARCKSL1 |
| chr8 | 143741908 | 143743677 | 1770 | 8 | 1.03E-07 | 1.59E-08 | 4.88E-05 | 1.06E-09 | 0.003223162 | IQANK1 |
| chr22 | 38727054 | 38728072 | 1019 | 3 | 5.06E-09 | 6.37E-10 | 1.18E-05 | 1.07E-09 | 0.00368676 | GTPBP1 |
| chr11 | 57646772 | 57648609 | 1838 | 10 | 1.51E-09 | 8.18E-06 | 0.000274 | 1.08E-09 | 0.008302953 | AP000662.1, YPEL4 |
| chr15 | 82043606 | 82044877 | 1272 | 8 | 4.08E-10 | 1.99E-08 | 6.25E-05 | 1.09E-09 | 0.011300194 | MEX3B |
| chr6 | 27837180 | 27839064 | 1885 | 13 | 2.85E-08 | 1.44E-09 | 0.000725 | 1.10E-09 | -6.45E-05 | H2BC15, H2AC15 |
| chr6 | 30465972 | 30466775 | 804 | 18 | 3.90E-09 | 1.96E-09 | 0.004381 | 1.11E-09 | 0.004254888 | TMPOP1 |
| chr1 | 24875126 | 24876126 | 1001 | 3 | 9.17E-10 | 2.16E-09 | 3.85E-06 | 1.12E-09 | 0.010696079 | NA |
| chr17 | 19533296 | 19534940 | 1645 | 14 | 4.05E-08 | 2.52E-08 | 0.00054 | 1.14E-09 | 0.007270334 | SLC47A1 |
| chr5 | 149728434 | 149730561 | 2128 | 12 | 4.64E-08 | 3.43E-08 | 0.000313 | 1.15E-09 | -0.00387914 | PPARGC1B |
| chr19 | 36849740 | 36851683 | 1944 | 28 | 3.01E-08 | 1.31E-08 | 0.000658 | 1.15E-09 | -0.0003618 | ZNF345, ZNF790 |
| chr1 | 203463860 | 203465133 | 1274 | 3 | 2.99E-08 | 1.37E-09 | 6.38E-06 | 1.16E-09 | 0.012550151 | NA |
| chr22 | 43127010 | 43127778 | 769 | 4 | 1.11E-09 | 7.48E-10 | 6.68E-05 | 1.17E-09 | 0.007505331 | BIK |
| chr1 | 26020184 | 26021993 | 1810 | 8 | 5.39E-07 | 2.74E-08 | 8.35E-05 | 1.18E-09 | 0.006701907 | EXTL1 |
| chr9 | 69671472 | 69673263 | 1792 | 14 | 1.97E-07 | 6.24E-09 | 0.000601 | 1.19E-09 | 0.001776626 | AL162412.1, APBA1 |
| chr11 | 64890754 | 64891593 | 840 | 11 | 6.87E-10 | 1.62E-09 | 3.29E-05 | 1.19E-09 | 0.002266907 | MIR194-2HG, MIR192, MIR194-2 |
| chr19 | 10717669 | 10719127 | 1459 | 23 | 3.73E-09 | 1.96E-09 | 0.010373 | 1.20E-09 | 0.000640115 | DNM2, MIR638 |
| chr9 | 137253557 | 137255090 | 1534 | 7 | 1.27E-09 | 2.02E-07 | 2.09E-05 | 1.20E-09 | -0.0001344 | NA |
| chr4 | 169659513 | 169660555 | 1043 | 7 | 1.36E-09 | 5.14E-10 | 0.000209 | 1.20E-09 | 0.014210875 | CLCN3 |
| chr2 | 176091668 | 176093180 | 1513 | 12 | 4.16E-11 | 1.60E-06 | 0.000139 | 1.21E-09 | 0.004733393 | HOXD13 |
| chr10 | 70597979 | 70599185 | 1207 | 8 | 7.05E-13 | 7.11E-06 | 2.52E-05 | 1.23E-09 | 0.004916382 | PRF1 |
| chr2 | 164955294 | 164955968 | 675 | 12 | 8.90E-09 | 1.61E-09 | 0.000229 | 1.23E-09 | -0.0094826 | AC019197.1, SLC38A11 |
| chr7 | 88795671 | 88796711 | 1041 | 9 | 2.50E-08 | 2.60E-08 | 9.71E-05 | 1.23E-09 | -0.00356781 | ZNF804B, TEX47 |
| chr12 | 57477910 | 57479393 | 1484 | 7 | 9.47E-08 | 1.96E-09 | 0.000104 | 1.24E-09 | 0.008568178 | MARS1, ARHGAP9 |
| chr22 | 42692720 | 42695747 | 3028 | 17 | 5.60E-06 | 5.55E-09 | 0.000797 | 1.25E-09 | 0.002465524 | A4GALT |
| chr1 | 1205704 | 1207512 | 1809 | 14 | 6.56E-08 | 4.21E-09 | 0.001492 | 1.25E-09 | 0.008431191 | TNFRSF18 |
| chr16 | 546670 | 548833 | 2164 | 9 | 2.03E-06 | 1.01E-09 | 0.000149 | 1.25E-09 | 0.011731451 | CAPN15, Z97986.1 |
| chr2 | 237858583 | 237860776 | 2194 | 19 | 2.69E-06 | 2.04E-11 | 0.003184 | 1.26E-09 | 0.013483005 | RAMP1 |
| chr11 | 64778159 | 64779281 | 1123 | 14 | 1.14E-08 | 7.09E-09 | 0.00154 | 1.27E-09 | 0.003152008 | AP001462.1, SF1 |
| chr17 | 49209083 | 49211187 | 2105 | 18 | 3.65E-06 | 8.45E-09 | 0.006717 | 1.30E-09 | 0.006422521 | ABI3, GNGT2 |
| chr16 | 1748740 | 1749706 | 967 | 3 | 1.36E-10 | 1.24E-08 | 1.26E-05 | 1.31E-09 | -0.00104356 | MAPK8IP3 |
| chr1 | 53837934 | 53839821 | 1888 | 17 | 4.74E-10 | 1.64E-06 | 8.93E-05 | 1.31E-09 | 0.001973527 | NDC1 |
| chr8 | 133249614 | 133251549 | 1936 | 7 | 4.89E-08 | 4.20E-10 | 0.000539 | 1.31E-09 | 0.010556615 | NDRG1 |
| chr10 | 87861662 | 87863314 | 1653 | 44 | 7.43E-11 | 1.36E-07 | 0.004789 | 1.32E-09 | 0.000675267 | KLLN |
| chr5 | 176548439 | 176549401 | 963 | 9 | 3.66E-09 | 7.87E-09 | 0.000257 | 1.33E-09 | 0.005962382 | CDHR2 |
| chr13 | 113845847 | 113846724 | 878 | 6 | 9.51E-12 | 2.72E-08 | 7.30E-05 | 1.34E-09 | 0.013937036 | GAS6 |
| chr10 | 117005010 | 117006086 | 1077 | 15 | 9.73E-10 | 9.34E-08 | 0.001178 | 1.34E-09 | 0.006171026 | AC023283.1, SHTN1 |
| chr4 | 185369990 | 185370708 | 719 | 5 | 1.84E-10 | 1.16E-08 | 1.60E-05 | 1.35E-09 | 0.011296386 | SNX25, LRP2BP |
| chr16 | 67198277 | 67200264 | 1988 | 15 | 5.22E-07 | 5.32E-10 | 0.002433 | 1.35E-09 | 0.01067182 | E2F4, ELMO3 |
| chr14 | 105528279 | 105530176 | 1898 | 8 | 1.88E-07 | 2.84E-08 | 9.08E-05 | 1.39E-09 | -0.02264053 | TMEM121 |
| chr7 | 149790250 | 149791865 | 1616 | 5 | 1.13E-08 | 2.23E-10 | 0.000271 | 1.40E-09 | 0.009009022 | SSPOP |
| chr22 | 19716875 | 19718842 | 1968 | 11 | 9.16E-11 | 2.71E-08 | 1.22E-05 | 1.41E-09 | 0.001698773 | SEPTIN5, AC000093.1 |
| chr8 | 23403604 | 23405135 | 1532 | 12 | 3.16E-10 | 6.74E-08 | 8.80E-05 | 1.46E-09 | 0.005633891 | LOXL2, ENTPD4 |
| chr16 | 75248153 | 75249108 | 956 | 11 | 1.42E-09 | 4.77E-08 | 0.000165 | 1.46E-09 | 0.004389898 | BCAR1, AC009078.3 |
| chr11 | 1275239 | 1276328 | 1090 | 8 | 3.90E-08 | 2.35E-10 | 0.001263 | 1.48E-09 | 0.013481917 | TOLLIP |
| chr15 | 101206962 | 101208768 | 1807 | 4 | 4.50E-06 | 5.45E-10 | 8.96E-05 | 1.50E-09 | 0.013155409 | CHSY1 |
| chr13 | 98044576 | 98044606 | 31 | 2 | 2.26E-09 | 7.28E-10 | 1.83E-05 | 1.51E-09 | 0.020796234 | NA |
| chr17 | 9904600 | 9906797 | 2198 | 12 | 9.08E-07 | 1.28E-09 | 0.000709 | 1.51E-09 | 0.004392442 | RCVRN |
| chr4 | 4761532 | 4762026 | 495 | 7 | 6.17E-10 | 9.61E-09 | 0.00049 | 1.51E-09 | 0.016620728 | STX18-AS1 |
| chr1 | 170661256 | 170663282 | 2027 | 10 | 1.60E-08 | 8.35E-09 | 0.000472 | 1.53E-09 | 0.006554354 | PRRX1 |
| chr10 | 101587178 | 101588813 | 1636 | 17 | 6.29E-08 | 7.74E-06 | 5.39E-05 | 1.55E-09 | 0.00019071 | DPCD, POLL |
| chr18 | 11850908 | 11852568 | 1661 | 14 | 1.54E-08 | 1.34E-07 | 0.000297 | 1.55E-09 | 0.003247313 | GNAL, CHMP1B, CHMP1B-AS1 |
| chr1 | 206555706 | 206556926 | 1221 | 9 | 1.44E-10 | 1.28E-09 | 0.001188 | 1.56E-09 | 0.007776637 | RASSF5 |
| chr10 | 93495774 | 93497002 | 1229 | 13 | 3.09E-09 | 1.79E-06 | 5.39E-05 | 1.56E-09 | 0.000777682 | CEP55 |
| chr7 | 75611756 | 75612377 | 622 | 3 | 5.32E-09 | 6.37E-10 | 3.80E-05 | 1.60E-09 | 0.019921447 | HIP1 |
| chr1 | 61043160 | 61043656 | 497 | 6 | 8.67E-11 | 1.54E-06 | 6.19E-05 | 1.62E-09 | 0.007610929 | NFIA, NFIA-AS2 |
| chr3 | 136818327 | 136820148 | 1822 | 17 | 6.51E-07 | 2.60E-10 | 0.006662 | 1.62E-09 | -0.00029015 | SLC35G2 |
| chr12 | 1837791 | 1838313 | 523 | 3 | 1.52E-10 | 6.71E-10 | 3.80E-05 | 1.63E-09 | 0.01525637 | CACNA2D4 |
| chr17 | 50273107 | 50274834 | 1728 | 10 | 1.14E-07 | 2.35E-06 | 7.90E-06 | 1.67E-09 | 0.000472857 | TMEM92 |
| chr9 | 34692054 | 34692581 | 528 | 3 | 9.46E-12 | 7.71E-08 | 9.55E-06 | 1.68E-09 | 0.013582933 | NA |
| chr15 | 25862937 | 25864467 | 1531 | 30 | 4.92E-09 | 3.44E-07 | 0.003153 | 1.68E-09 | 0.002429315 | ATP10A |
| chr11 | 118892150 | 118893782 | 1633 | 11 | 5.92E-09 | 1.38E-08 | 0.00045 | 1.68E-09 | 0.007302283 | CXCR5 |
| chr2 | 285936 | 287760 | 1825 | 13 | 2.88E-08 | 9.13E-09 | 0.002361 | 1.70E-09 | 0.010000501 | AC079779.2, ALKAL2 |
| chr6 | 29649543 | 29650605 | 1063 | 18 | 2.40E-09 | 5.21E-08 | 0.000322 | 1.70E-09 | 0.001821962 | NA |
| chr17 | 74247642 | 74248800 | 1159 | 12 | 6.44E-09 | 2.19E-07 | 0.000103 | 1.71E-09 | 0.003949682 | TTYH2 |
| chr11 | 6298744 | 6298761 | 18 | 2 | 6.66E-10 | 8.63E-10 | 1.56E-05 | 1.72E-09 | 0.029858643 | NA |
| chr19 | 48628853 | 48630588 | 1736 | 8 | 7.34E-10 | 3.27E-09 | 0.000205 | 1.76E-09 | 0.005312971 | SPHK2, DBP |
| chr4 | 3369509 | 3371279 | 1771 | 20 | 7.30E-08 | 4.04E-07 | 0.000405 | 1.77E-09 | 0.007556254 | RGS12 |
| chr1 | 182301347 | 182301803 | 457 | 3 | 4.33E-09 | 5.14E-10 | 6.46E-05 | 1.78E-09 | 0.031996822 | LINC01344 |
| chr7 | 100560090 | 100560720 | 631 | 3 | 2.76E-09 | 2.76E-08 | 1.65E-05 | 1.78E-09 | 0.005681475 | AGFG2 |
| chr7 | 157387079 | 157387432 | 354 | 6 | 3.30E-09 | 1.85E-08 | 0.000234 | 1.81E-09 | 0.011238725 | DNAJB6 |
| chr19 | 4784928 | 4785593 | 666 | 3 | 5.95E-09 | 6.00E-09 | 2.21E-05 | 1.84E-09 | -0.04315529 | MIR7-3HG, AC005523.1 |
| chr7 | 128696565 | 128697671 | 1107 | 6 | 6.95E-10 | 5.00E-09 | 3.47E-05 | 1.85E-09 | 0.010742123 | RNA5SP242 |
| chr16 | 56576895 | 56577499 | 605 | 4 | 1.57E-08 | 4.95E-10 | 0.000162 | 1.91E-09 | 0.010362868 | NA |
| chr1 | 6465989 | 6468201 | 2213 | 12 | 4.68E-08 | 1.03E-08 | 0.000311 | 1.92E-09 | 0.003526265 | TNFRSF25, PLEKHG5 |
| chr10 | 12194834 | 12196716 | 1883 | 18 | 6.03E-09 | 1.05E-08 | 0.000222 | 1.94E-09 | 0.000865215 | CDC123, NUDT5 |
| chr4 | 40630282 | 40631555 | 1274 | 12 | 1.33E-10 | 1.83E-07 | 5.83E-05 | 1.95E-09 | 0.008057611 | RBM47 |
| chr20 | 63235148 | 63236717 | 1570 | 11 | 3.70E-10 | 8.20E-08 | 0.000275 | 1.96E-09 | 0.010682544 | BIRC7 |
| chr3 | 128425458 | 128427221 | 1764 | 10 | 1.05E-07 | 7.38E-08 | 0.000165 | 1.97E-09 | 0.003134141 | NA |
| chr1 | 15731534 | 15732009 | 476 | 5 | 9.82E-10 | 3.26E-09 | 0.00013 | 1.98E-09 | 0.001358215 | PLEKHM2, AL450998.2 |
| chr11 | 72232143 | 72233444 | 1302 | 3 | 9.97E-10 | 1.23E-09 | 3.05E-05 | 1.98E-09 | 0.005375552 | INPPL1 |
| chr19 | 38318566 | 38321143 | 2578 | 15 | 3.37E-06 | 7.85E-08 | 0.000335 | 1.99E-09 | 0.00376851 | KCNK6 |
| chr6 | 31127499 | 31128136 | 638 | 6 | 3.48E-09 | 1.03E-08 | 9.68E-05 | 1.99E-09 | 0.0017791 | PSORS1C1 |
| chr8 | 47600196 | 47600954 | 759 | 2 | 5.97E-12 | 2.51E-08 | 2.24E-07 | 2.04E-09 | 0.012119835 | SPIDR |
| chr15 | 22950766 | 22951883 | 1118 | 7 | 7.15E-09 | 4.81E-07 | 2.80E-06 | 2.05E-09 | 0.002023093 | CYFIP1 |
| chr13 | 106917020 | 106918163 | 1144 | 9 | 1.77E-08 | 1.34E-06 | 0.000169 | 2.06E-09 | 0.002227101 | NA |
| chr6 | 31862944 | 31865675 | 2732 | 42 | 2.82E-08 | 2.71E-08 | 0.003139 | 2.06E-09 | 0.000271058 | SLC44A4 |
| chr13 | 99085110 | 99085531 | 422 | 4 | 4.14E-10 | 3.45E-09 | 1.29E-05 | 2.07E-09 | 0.019059718 | DOCK9 |
| chr9 | 35859093 | 35860915 | 1823 | 9 | 6.79E-07 | 2.20E-08 | 0.000228 | 2.07E-09 | 0.004125738 | TMEM8B, OR13E1P |
| chr17 | 81888329 | 81889810 | 1482 | 10 | 1.17E-10 | 7.80E-07 | 4.70E-05 | 2.08E-09 | 0.000730877 | ALYREF |
| chr17 | 1231612 | 1231617 | 6 | 3 | 2.87E-09 | 9.54E-10 | 5.34E-05 | 2.08E-09 | 0.027373731 | NA |
| chr12 | 53215618 | 53217864 | 2247 | 5 | 7.26E-08 | 6.84E-09 | 2.58E-05 | 2.08E-09 | 0.009179484 | RARG |
| chr20 | 577324 | 577513 | 190 | 2 | 2.36E-09 | 1.41E-09 | 5.57E-06 | 2.08E-09 | 0.018884125 | NA |
| chr9 | 129110641 | 129111907 | 1267 | 13 | 6.02E-09 | 3.69E-09 | 0.000286 | 2.15E-09 | 0.000551504 | PTPA, CRAT |
| chr19 | 49155275 | 49156042 | 768 | 8 | 1.59E-11 | 5.39E-08 | 0.000116 | 2.17E-09 | -0.00476029 | HRC |
| chr15 | 24764270 | 24765007 | 738 | 2 | 6.73E-09 | 1.64E-09 | 4.61E-06 | 2.18E-09 | -0.00998546 | PWRN1, AC090983.2 |
| chr10 | 132640657 | 132642832 | 2176 | 10 | 6.25E-07 | 3.14E-08 | 0.000243 | 2.18E-09 | 0.000530674 | INPP5A |
| chr2 | 28979946 | 28981829 | 1884 | 9 | 1.49E-07 | 5.57E-07 | 0.000146 | 2.19E-09 | 0.003636821 | TOGARAM2 |
| chr5 | 94784920 | 94785566 | 647 | 5 | 6.35E-11 | 2.28E-07 | 1.08E-05 | 2.19E-09 | 0.013971483 | MCTP1 |
| chr1 | 17123141 | 17124087 | 947 | 4 | 3.83E-08 | 1.09E-09 | 0.000117 | 2.20E-09 | 0.003050376 | NA |
| chr19 | 35532506 | 35533974 | 1469 | 13 | 5.15E-11 | 2.27E-06 | 1.49E-05 | 2.23E-09 | 0.012091143 | GAPDHS |
| chr1 | 14996690 | 14997988 | 1299 | 3 | 1.91E-06 | 7.10E-09 | 2.17E-05 | 2.27E-09 | 0.011840365 | KAZN |
| chr17 | 4196606 | 4197527 | 922 | 2 | 2.16E-08 | 1.84E-09 | 3.94E-06 | 2.30E-09 | 0.000870762 | ANKFY1 |
| chr1 | 35719034 | 35720338 | 1305 | 10 | 3.83E-11 | 7.91E-08 | 4.63E-05 | 2.30E-09 | 0.00173343 | CLSPN |
| chr1 | 15735621 | 15738653 | 3033 | 17 | 6.58E-06 | 1.94E-09 | 0.007746 | 2.31E-09 | 0.003692586 | SLC25A34, AL450998.2 |
| chr7 | 1950358 | 1952393 | 2036 | 10 | 1.41E-08 | 5.83E-08 | 0.000584 | 2.34E-09 | 0.004538246 | MAD1L1, AC069288.1 |
| chr11 | 84923082 | 84924252 | 1171 | 11 | 2.42E-08 | 2.55E-08 | 0.001047 | 2.35E-09 | 0.014695594 | DLG2 |
| chr1 | 23867008 | 23869169 | 2162 | 19 | 4.10E-07 | 1.77E-08 | 0.001842 | 2.41E-09 | 0.005730145 | FUCA1 |
| chr12 | 108598001 | 108598906 | 906 | 8 | 4.50E-10 | 6.30E-08 | 5.25E-05 | 2.42E-09 | 0.007628653 | TMEM119 |

| | | | | | | | | | | |
|---|---|---|---|---|---|---|---|---|---|---|
| chr6 | 13709129 | 13710862 | 1734 | 6 | 1.76E-09 | 4.43E-09 | 7.49E-06 | 2.49E-09 | 0.000312584 | RANBP9 |
| chr1 | 202018030 | 202018855 | 826 | 3 | 1.26E-09 | 1.80E-09 | 3.89E-05 | 2.49E-09 | 0.01653252 | NA |
| chr10 | 70671986 | 70673462 | 1477 | 10 | 6.66E-09 | 2.62E-08 | 9.36E-05 | 2.56E-09 | 0.002503766 | ADAMTS14 |
| chr14 | 22505965 | 22506536 | 572 | 4 | 1.07E-09 | 5.32E-08 | 4.26E-05 | 2.58E-09 | 0.017224364 | NA |
| chr6 | 6003513 | 6005116 | 1604 | 12 | 4.41E-09 | 2.15E-08 | 0.000474 | 2.58E-09 | 0.004422192 | NRN1 |
| chr16 | 88938646 | 88940469 | 1824 | 14 | 9.39E-08 | 1.73E-09 | 0.005351 | 2.58E-09 | 0.006186469 | AC092384.1, CBFA2T3, AC092384.3 |
| chr17 | 78230041 | 78232152 | 2112 | 15 | 8.96E-09 | 7.54E-07 | 0.000142 | 2.58E-09 | 0.00539493 | TMEM235 |
| chr10 | 13271037 | 13271693 | 657 | 4 | 9.52E-11 | 1.05E-07 | 3.30E-05 | 2.59E-09 | 0.013157191 | NA |
| chr16 | 86984750 | 86986598 | 1849 | 4 | 2.14E-07 | 5.83E-09 | 2.18E-05 | 2.61E-09 | 0.00949501 | NA |
| chr10 | 47325205 | 47326492 | 1288 | 9 | 1.23E-12 | 9.88E-05 | 1.26E-05 | 2.62E-09 | 0.001605666 | GDF2 |
| chr6 | 30617109 | 30618691 | 1583 | 34 | 1.44E-11 | 1.39E-06 | 0.002854 | 2.63E-09 | 0.000763561 | MRPS18B, PPP1R10 |
| chr20 | 41361287 | 41362821 | 1535 | 6 | 2.05E-08 | 6.39E-10 | 0.000424 | 2.65E-09 | 0.004508006 | EMILIN3 |
| chr20 | 33653647 | 33654544 | 898 | 4 | 3.93E-09 | 5.13E-09 | 6.48E-06 | 2.65E-09 | 0.008371033 | NA |
| chr17 | 76420469 | 76421138 | 670 | 3 | 1.22E-09 | 1.36E-09 | 3.51E-05 | 2.65E-09 | 0.006694567 | UBE2O |
| chr12 | 122965669 | 122967327 | 1659 | 14 | 3.03E-07 | 6.84E-09 | 0.002755 | 2.65E-09 | 0.004315871 | ABCB9 |
| chr5 | 135535944 | 135537267 | 1324 | 21 | 1.45E-11 | 1.13E-07 | 0.000713 | 2.73E-09 | 0.004565977 | NEUROG1 |
| chr6 | 137217447 | 137220250 | 2804 | 13 | 3.62E-05 | 5.65E-08 | 0.000531 | 2.78E-09 | 0.001766571 | IFNGR1 |
| chr12 | 53750596 | 53752349 | 1754 | 11 | 2.88E-08 | 5.00E-09 | 0.00035 | 2.82E-09 | 0.007872527 | AC076968.2, CISTR |
| chr16 | 4681717 | 4683252 | 1536 | 9 | 5.89E-08 | 3.79E-10 | 0.002784 | 2.82E-09 | 0.005332521 | MGRN1 |
| chr1 | 980966 | 982512 | 1547 | 13 | 1.30E-10 | 1.43E-07 | 5.77E-06 | 2.83E-09 | 0.007399387 | PERM1 |
| chr8 | 144852521 | 144853980 | 1460 | 17 | 1.02E-09 | 4.23E-09 | 0.000804 | 2.87E-09 | -0.00073379 | COMMD5 |
| chr11 | 71998510 | 71999975 | 1466 | 12 | 1.52E-09 | 1.85E-05 | 1.79E-05 | 2.87E-09 | 0.004835273 | IL18BP |
| chr8 | 141287022 | 141287485 | 464 | 5 | 4.64E-10 | 1.34E-08 | 4.49E-05 | 2.89E-09 | 0.015024584 | AC011676.3, SLC45A4 |
| chr11 | 45693222 | 45694895 | 1674 | 9 | 1.45E-10 | 4.17E-07 | 0.000288 | 2.91E-09 | 0.006884692 | NA |
| chr1 | 156149629 | 156150667 | 1039 | 6 | 1.22E-08 | 1.48E-09 | 0.000334 | 2.92E-09 | 0.007752247 | SEMA4A |
| chr17 | 49952853 | 49953440 | 588 | 4 | 6.81E-11 | 4.35E-08 | 1.48E-05 | 2.93E-09 | 0.010554705 | NA |
| chr11 | 2129457 | 2130495 | 1039 | 7 | 3.52E-10 | 3.50E-09 | 0.000114 | 2.95E-09 | 0.000967408 | IGF2, AC132217.2, AC132217.1 |
| chr10 | 132336615 | 132337256 | 642 | 12 | 3.29E-09 | 7.70E-10 | 0.005544 | 2.96E-09 | -0.01933097 | LRRC27 |
| chr14 | 104590686 | 104591687 | 1002 | 4 | 6.85E-11 | 5.20E-08 | 6.57E-06 | 2.96E-09 | 0.012252504 | TMEM179 |
| chr11 | 119148217 | 119149135 | 919 | 6 | 1.49E-10 | 1.46E-08 | 0.000233 | 2.96E-09 | 0.011536061 | ABCG4 |
| chr17 | 80023399 | 80024198 | 800 | 10 | 1.20E-09 | 1.03E-07 | 2.09E-05 | 2.96E-09 | 0.00276471 | AC116025.1, TBC1D16 |
| chr1 | 74732527 | 74733951 | 1425 | 17 | 5.83E-09 | 4.05E-10 | 0.008489 | 2.98E-09 | -0.00730716 | TYW3, CRYZ |
| chr4 | 39637387 | 39638421 | 1035 | 10 | 3.34E-12 | 7.01E-06 | 1.20E-05 | 3.00E-09 | 0.003586112 | SMIM14 |
| chr2 | 53859717 | 53860604 | 888 | 13 | 2.28E-08 | 9.52E-10 | 0.001269 | 3.03E-09 | 0.009823583 | ASB3, GPR75 |
| chr9 | 130936429 | 130937336 | 908 | 4 | 1.26E-09 | 2.97E-09 | 3.70E-05 | 3.03E-09 | 0.009102588 | AL161733.1, FIBCD1 |
| chr1 | 61052135 | 61053225 | 1091 | 4 | 6.03E-09 | 1.96E-09 | 9.36E-05 | 3.04E-09 | 0.025230405 | NFIA, NFIA-AS2 |
| chr20 | 58837890 | 58840122 | 2233 | 16 | 1.21E-06 | 4.39E-08 | 0.000975 | 3.06E-09 | 0.00487614 | GNAS, GNAS-AS1 |
| chr6 | 31528294 | 31529979 | 1686 | 8 | 4.47E-06 | 1.30E-08 | 0.000402 | 3.06E-09 | 0.003264264 | RPL15P4, MCCD1 |
| chr11 | 45696020 | 45698276 | 2257 | 13 | 3.00E-06 | 8.11E-07 | 0.000322 | 3.06E-09 | 0.005818578 | NA |
| chr3 | 193869701 | 193871362 | 1662 | 5 | 4.47E-08 | 7.82E-08 | 5.51E-05 | 3.07E-09 | 0.013499366 | NA |
| chr20 | 44909359 | 44910832 | 1474 | 14 | 7.64E-09 | 3.13E-08 | 0.000889 | 3.12E-09 | 0.004092816 | PABPC1L |
| chr6 | 170175978 | 170177623 | 1646 | 4 | 4.11E-08 | 1.26E-09 | 6.57E-05 | 3.12E-09 | 0.007595221 | NA |
| chr22 | 49449610 | 49450453 | 844 | 5 | 9.96E-09 | 4.58E-10 | 0.000438 | 3.16E-09 | 0.044745298 | C22orf34 |
| chr12 | 108827631 | 108829019 | 1389 | 11 | 8.92E-08 | 6.92E-08 | 2.17E-05 | 3.20E-09 | 0.002104898 | SSH1 |
| chr20 | 34544523 | 34545461 | 939 | 4 | 5.83E-10 | 1.33E-09 | 0.000134 | 3.22E-09 | 0.016277151 | NA |
| chr1 | 9823408 | 9824981 | 1574 | 16 | 6.79E-09 | 1.74E-08 | 0.000935 | 3.22E-09 | 0.003138818 | CLSTN1 |
| chr20 | 1993823 | 1994800 | 978 | 12 | 1.59E-08 | 2.60E-08 | 0.000224 | 3.22E-09 | 0.005013895 | PDYN-AS1, PDYN |
| chr10 | 125773607 | 125774827 | 1221 | 5 | 1.18E-10 | 5.43E-07 | 2.52E-06 | 3.25E-09 | -0.01448224 | MMP21 |
| chr4 | 106315113 | 106318004 | 2892 | 22 | 9.27E-08 | 3.18E-05 | 0.000555 | 3.25E-09 | 0.003726933 | AIMP1, TBCK |
| chr5 | 56908578 | 56909443 | 866 | 8 | 2.66E-09 | 3.50E-09 | 0.000365 | 3.28E-09 | 0.013034188 | SETD9, AC008937.1 |
| chr5 | 139389275 | 139391152 | 1878 | 11 | 7.00E-09 | 2.90E-08 | 0.000466 | 3.28E-09 | 0.007617938 | MZB1, PROB1 |
| chr4 | 3483599 | 3484653 | 1055 | 8 | 1.84E-08 | 2.23E-10 | 0.001793 | 3.28E-09 | 0.022439335 | DOK7 |
| chr4 | 41873176 | 41873858 | 683 | 6 | 5.33E-09 | 3.32E-09 | 0.000411 | 3.29E-09 | 0.011062082 | LINC00682 |
| chr7 | 97024305 | 97025793 | 1489 | 20 | 1.09E-09 | 1.96E-07 | 0.000276 | 3.29E-09 | 0.0043152 | DLX5 |
| chr12 | 4379583 | 4380613 | 1031 | 9 | 2.55E-09 | 1.63E-08 | 0.000342 | 3.30E-09 | 0.010613615 | AC008012.1, FGF23 |
| chr6 | 132813259 | 132814876 | 1618 | 13 | 1.39E-07 | 1.84E-08 | 0.00206 | 3.35E-09 | 0.002408139 | RPS12, SLC18B1 |
| chr16 | 1175588 | 1176302 | 715 | 4 | 8.37E-11 | 2.71E-08 | 3.37E-05 | 3.35E-09 | 0.010312817 | CACNA1H |
| chr13 | 67226773 | 67228752 | 1980 | 8 | 3.42E-07 | 3.65E-10 | 0.000902 | 3.38E-09 | -0.01837457 | PCDH9 |
| chr16 | 2087579 | 2089661 | 2083 | 12 | 6.77E-08 | 1.65E-08 | 0.000459 | 3.42E-09 | 0.003415641 | TSC2, PKD1 |
| chr15 | 40251973 | 40253310 | 1338 | 16 | 8.94E-11 | 3.63E-07 | 5.84E-05 | 3.44E-09 | 0.003039949 | PAK6, BUB1B-PAK6, C15orf56 |
| chr8 | 22023334 | 22024341 | 1008 | 12 | 1.39E-09 | 1.33E-07 | 0.00086 | 3.45E-09 | 0.004333478 | NPM2 |
| chr15 | 65377225 | 65378719 | 1495 | 10 | 1.59E-07 | 6.79E-09 | 0.000501 | 3.56E-09 | 0.003494675 | IGDCC3 |
| chr1 | 17247683 | 17249112 | 1430 | 8 | 4.77E-08 | 1.61E-08 | 2.18E-05 | 3.56E-09 | 0.006953674 | PADI3 |
| chr13 | 111534818 | 111535049 | 232 | 3 | 6.95E-09 | 1.96E-09 | 5.65E-05 | 3.57E-09 | 0.0261095 | NA |
| chr22 | 36481504 | 36482822 | 1319 | 11 | 3.27E-08 | 3.52E-06 | 0.000117 | 3.58E-09 | 0.002106672 | TXN2 |
| chr7 | 77797925 | 77799532 | 1608 | 29 | 1.33E-08 | 4.23E-08 | 0.000697 | 3.64E-09 | -3.48E-05 | PHTF2, TMEM60 |
| chr22 | 49966260 | 49967926 | 1667 | 6 | 7.86E-07 | 2.18E-09 | 0.000125 | 3.66E-09 | 0.0120613 | NA |
| chr6 | 33851292 | 33852196 | 905 | 4 | 3.95E-08 | 5.59E-10 | 0.000394 | 3.66E-09 | 0.017229087 | NA |
| chr5 | 138138669 | 138139843 | 1175 | 8 | 1.20E-09 | 4.57E-08 | 3.08E-05 | 3.67E-09 | 0.005914132 | NME5, BRD8 |
| chr3 | 64225315 | 64226318 | 1004 | 9 | 4.46E-09 | 7.16E-07 | 2.94E-05 | 3.67E-09 | 0.006600812 | PRICKLE2 |
| chr17 | 43843047 | 43843151 | 105 | 3 | 3.47E-09 | 1.90E-09 | 4.60E-05 | 3.68E-09 | 0.02675219 | NA |
| chr15 | 62835517 | 62836526 | 1010 | 6 | 8.16E-10 | 3.30E-09 | 8.90E-05 | 3.72E-09 | 0.003282077 | TLN2, AC103740.1 |
| chr4 | 186723466 | 186724531 | 1066 | 9 | 3.81E-09 | 2.51E-09 | 0.002783 | 3.72E-09 | 0.005745368 | FAT1 |
| chr18 | 8721209 | 8721742 | 534 | 3 | 2.12E-09 | 2.52E-09 | 3.51E-05 | 3.79E-09 | 0.004563343 | MTCL1 |
| chr20 | 33282363 | 33283546 | 1184 | 11 | 6.88E-11 | 1.63E-06 | 0.000295 | 3.82E-09 | 0.006106099 | BPIFB1 |
| chr11 | 62921276 | 62922571 | 1296 | 9 | 1.72E-10 | 9.72E-07 | 4.31E-05 | 3.84E-09 | 0.009839001 | CHRM1 |
| chr19 | 42204233 | 42204412 | 180 | 2 | 5.80E-09 | 1.97E-09 | 1.92E-05 | 3.89E-09 | 0.010946676 | DEDD2 |
| chr18 | 5196398 | 5197633 | 1236 | 14 | 1.35E-07 | 3.79E-09 | 0.000273 | 3.89E-09 | 0.004973733 | AP001496.4, AKAIN1 |
| chr4 | 89308337 | 89309247 | 911 | 5 | 3.07E-10 | 2.32E-08 | 6.86E-06 | 3.89E-09 | 0.007341901 | NA |
| chr12 | 9115783 | 9117075 | 1293 | 8 | 4.18E-09 | 1.07E-07 | 1.17E-05 | 3.93E-09 | -0.00074305 | A2M |
| chr3 | 187669401 | 187670493 | 1093 | 14 | 9.99E-10 | 4.66E-09 | 0.00276 | 3.98E-09 | 0.009418739 | SST |
| chr1 | 31208201 | 31208849 | 649 | 2 | 6.44E-08 | 1.96E-09 | 2.51E-05 | 3.98E-09 | 0.057442333 | NKAIN1 |
| chr12 | 52870756 | 52871306 | 551 | 4 | 2.33E-08 | 6.12E-10 | 0.000449 | 4.00E-09 | 0.019854491 | NA |
| chr9 | 128090865 | 128092434 | 1570 | 9 | 5.71E-07 | 6.18E-09 | 0.001337 | 4.03E-09 | 0.00544456 | SLC25A25, AL360268.1 |
| chr15 | 68205392 | 68206842 | 1451 | 10 | 5.71E-11 | 1.28E-05 | 0.000258 | 4.04E-09 | 0.007924651 | AC107871.1, CALML4 |
| chr8 | 101491638 | 101493758 | 2121 | 14 | 1.93E-07 | 1.16E-08 | 0.005409 | 4.04E-09 | 0.004759295 | GRHL2, AP001207.3 |
| chr1 | 202212741 | 202214316 | 1576 | 10 | 1.69E-10 | 7.24E-07 | 7.03E-05 | 4.04E-09 | 0.010393357 | LGR6 |
| chr12 | 109814791 | 109816037 | 1247 | 12 | 1.10E-07 | 5.77E-11 | 0.00899 | 4.08E-09 | 0.00819997 | TRPV4 |
| chr19 | 15469199 | 15469910 | 712 | 7 | 5.49E-09 | 4.23E-09 | 0.000227 | 4.11E-09 | 0.005125338 | PGLYRP2 |
| chr2 | 70086815 | 70088357 | 1543 | 13 | 7.61E-08 | 7.48E-07 | 0.000317 | 4.12E-09 | 0.000566861 | PCBP1, PCBP1-AS1 |
| chr1 | 153609018 | 153610340 | 1323 | 10 | 1.87E-10 | 7.85E-07 | 3.84E-05 | 4.12E-09 | 0.024031613 | S100A16 |
| chr1 | 153698311 | 153699019 | 709 | 4 | 4.65E-08 | 8.73E-10 | 0.000216 | 4.16E-09 | 0.015059422 | NA |
| chr17 | 49890357 | 49890989 | 633 | 4 | 1.25E-08 | 8.20E-10 | 0.000235 | 4.17E-09 | 0.022278363 | AC027801.2 |
| chr5 | 132011022 | 132012306 | 1285 | 18 | 8.75E-08 | 1.76E-08 | 0.009811 | 4.24E-09 | 0.002536555 | AC034228.1, AC034228.3, ACSL6 |
| chr14 | 105525761 | 105526282 | 522 | 3 | 6.89E-10 | 5.65E-09 | 2.93E-05 | 4.25E-09 | 0.019512393 | NA |
| chr1 | 231929445 | 231930043 | 599 | 5 | 7.17E-09 | 1.40E-09 | 0.000448 | 4.31E-09 | 0.015154324 | DISC1, DISC1-IT1 |
| chr1 | 171185162 | 171185663 | 502 | 4 | 3.53E-10 | 2.01E-06 | 1.08E-05 | 4.36E-09 | 0.017921181 | FMO2, BX284613.2 |
| chr5 | 112825864 | 112827386 | 1523 | 5 | 2.43E-07 | 3.24E-07 | 1.93E-05 | 4.40E-09 | -6.94E-05 | APC, AC008575.1 |
| chr15 | 59657137 | 59658578 | 1442 | 12 | 4.62E-11 | 1.08E-05 | 9.05E-05 | 4.41E-09 | -0.00528704 | GTF2A2, AC092755.1 |
| chr2 | 178450350 | 178451979 | 1630 | 25 | 6.39E-09 | 1.91E-07 | 9.52E-05 | 4.41E-09 | 0.000731225 | PJVK, PRKRA |
| chr19 | 19514952 | 19516562 | 1611 | 19 | 1.39E-09 | 1.69E-08 | 0.000948 | 4.44E-09 | 0.004453667 | NDUFA13, AC011448.1, TSSK6 |
| chr17 | 48996607 | 48998518 | 1912 | 18 | 5.21E-09 | 4.04E-08 | 6.43E-05 | 4.49E-09 | 0.002451944 | IGF2BP1, AC091133.3 |
| chr19 | 6261266 | 6261831 | 566 | 2 | 9.86E-10 | 3.67E-09 | 5.37E-06 | 4.51E-09 | 0.038111839 | MLLT1 |
| chr2 | 8574118 | 8574677 | 560 | 4 | 4.65E-10 | 2.48E-08 | 1.94E-05 | 4.55E-09 | 0.006641404 | LINC01814 |
| chr19 | 18397305 | 18398548 | 1244 | 4 | 4.35E-09 | 1.44E-09 | 0.000103 | 4.55E-09 | 0.011397457 | LRRC25 |
| chr16 | 28507570 | 28508245 | 676 | 3 | 2.71E-11 | 7.28E-09 | 5.27E-06 | 4.59E-09 | 0.010510028 | AC009093.11, IL27 |
| chr17 | 64505362 | 64507078 | 1717 | 16 | 8.94E-10 | 7.40E-06 | 1.42E-05 | 4.61E-09 | -0.0004311 | CEP95, DDX5 |
| chr2 | 199955095 | 199956303 | 1209 | 19 | 2.55E-09 | 5.13E-09 | 0.000106 | 4.65E-09 | 0.000699432 | C2orf69, MAIP1, TYW5 |
| chr14 | 92255039 | 92255161 | 123 | 2 | 4.62E-09 | 2.48E-09 | 1.76E-05 | 4.68E-09 | 0.03398167 | AL133240.1 |
| chr21 | 42495815 | 42497303 | 1489 | 15 | 5.21E-09 | 1.73E-05 | 1.90E-05 | 4.70E-09 | 0.003252997 | SLC37A1, AP001625.3, RSPH1 |
| chr1 | 75785773 | 75786965 | 1193 | 12 | 1.87E-08 | 8.45E-09 | 0.000105 | 4.73E-09 | 0.004130816 | ACADM, RABGGTB |
| chr14 | 75279526 | 75281549 | 2024 | 10 | 5.77E-08 | 6.43E-09 | 0.000604 | 4.76E-09 | 0.005066161 | FOS |
| chr1 | 118989002 | 118990227 | 1226 | 17 | 2.58E-09 | 2.64E-07 | 0.000994 | 4.76E-09 | 0.003869245 | TBX15 |
| chr9 | 131358171 | 131358949 | 779 | 4 | 8.53E-09 | 2.23E-09 | 4.09E-05 | 4.82E-09 | 0.004934903 | PLPP7 |
| chr1 | 156878952 | 156880026 | 1075 | 6 | 8.40E-08 | 5.57E-08 | 6.75E-05 | 4.84E-09 | -0.00069927 | NTRK1 |
| chr9 | 116686999 | 116688585 | 1587 | 13 | 6.09E-09 | 8.84E-06 | 1.85E-05 | 4.88E-09 | 0.001394713 | TRIM32, ASTN2 |
| chr11 | 66509584 | 66510887 | 1304 | 12 | 1.30E-08 | 2.96E-08 | 0.000198 | 4.89E-09 | 0.002294425 | DPP3, AP002748.5, BBS1 |
| chr1 | 208090101 | 208091164 | 1064 | 3 | 5.60E-10 | 1.21E-08 | 6.21E-06 | 4.89E-09 | 0.010339606 | PLXNA2 |
| chr7 | 44103374 | 44104761 | 1388 | 8 | 1.04E-07 | 3.00E-07 | 0.000372 | 4.91E-09 | 0.004337928 | AEBP1 |
| chr1 | 234659843 | 234660296 | 454 | 6 | 1.52E-08 | 5.77E-10 | 0.000821 | 4.95E-09 | 0.01089696 | AL160408.1, AL160408.4 |
| chr7 | 99503577 | 99505420 | 1844 | 18 | 3.14E-08 | 5.71E-07 | 0.00019 | 4.97E-09 | 0.001582047 | ZNF789, ZKSCAN5, ZNF394 |
| chr9 | 8858259 | 8859139 | 881 | 4 | 5.09E-09 | 3.75E-09 | 0.000161 | 5.00E-09 | 0.035601038 | PTPRD-AS1, PTPRD |
| chr11 | 75524957 | 75526481 | 1525 | 10 | 1.04E-08 | 8.57E-09 | 0.000303 | 5.00E-09 | 0.000889106 | GDPD5 |
| chr8 | 33472776 | 33474082 | 1307 | 16 | 1.45E-09 | 4.45E-07 | 1.75E-05 | 5.18E-09 | 0.000458853 | FUT10, TTI2 |
| chr3 | 16604868 | 16606204 | 1337 | 13 | 3.85E-08 | 2.89E-08 | 0.002939 | 5.18E-09 | 0.000584982 | DAZL |
| chr8 | 12755630 | 12757165 | 1536 | 11 | 1.10E-09 | 1.50E-06 | 1.95E-05 | 5.25E-09 | 0.005610068 | LONRF1 |
| chr7 | 53811510 | 53812096 | 587 | 7 | 1.36E-09 | 2.49E-08 | 0.000134 | 5.25E-09 | 0.00889586 | LINC01446 |
| chr22 | 38120142 | 38121549 | 1408 | 5 | 6.62E-08 | 2.53E-09 | 0.000124 | 5.28E-09 | 0.004637019 | PLA2G6 |
| chr16 | 85286253 | 85287276 | 1024 | 9 | 1.31E-11 | 5.39E-06 | 2.03E-07 | 5.32E-09 | 0.003338445 | GSE1, AC092275.1 |
| chr6 | 10514627 | 10515578 | 952 | 3 | 5.22E-10 | 1.58E-06 | 3.55E-06 | 5.37E-09 | 0.00780251 | GCNT2, AL139039.1 |
| chr1 | 3266056 | 3267267 | 1212 | 6 | 3.23E-08 | 3.21E-06 | 2.08E-05 | 5.48E-09 | 0.000945947 | PRDM16 |
| chr12 | 1812892 | 1813174 | 283 | 4 | 4.80E-09 | 1.05E-09 | 0.000392 | 5.51E-09 | 0.014577203 | CACNA2D4 |
| chr16 | 774262 | 775176 | 915 | 3 | 1.08E-07 | 2.24E-07 | 2.01E-05 | 5.53E-09 | 0.006472084 | MSLNL |
| chr8 | 64577786 | 64580379 | 2594 | 21 | 3.85E-06 | 1.10E-06 | 0.000227 | 5.54E-09 | 0.002100115 | BHLHE22, AC090136.3 |
| chr1 | 1614651 | 1616145 | 1495 | 13 | 4.33E-09 | 1.34E-06 | 0.000352 | 5.57E-09 | 6.79E-05 | MIB2, AL691432.2 |
| chr13 | 51584246 | 51585196 | 951 | 12 | 6.64E-12 | 9.91E-06 | 0.000142 | 5.57E-09 | 0.002464385 | WDFY2 |
| chr16 | 1524268 | 1526772 | 2505 | 12 | 5.26E-06 | 7.61E-08 | 0.00018 | 5.59E-09 | 0.003149995 | IFT140 |
| chr7 | 157682856 | 157684238 | 1383 | 8 | 1.67E-09 | 2.58E-08 | 0.000141 | 5.59E-09 | 0.008596171 | PTPRN2 |
| chr17 | 82496043 | 82497183 | 1141 | 4 | 2.43E-09 | 4.93E-08 | 0.000103 | 5.61E-09 | 0.008252361 | NA |
| chr15 | 45113591 | 45115620 | 2030 | 16 | 1.15E-09 | 7.36E-07 | 9.67E-05 | 5.63E-09 | 0.003998793 | DUOXA2, DUOX2 |
| chr11 | 16605987 | 16607742 | 1756 | 15 | 1.18E-08 | 1.17E-07 | 6.91E-05 | 5.65E-09 | 0.00405567 | SOX6 |
| chr6 | 30645938 | 30648363 | 2426 | 36 | 1.73E-07 | 1.96E-07 | 0.002503 | 5.65E-09 | 0.002174059 | ATAT1, C6orf136 |
| chr2 | 74552672 | 74553490 | 819 | 9 | 8.32E-09 | 4.20E-08 | 0.00012 | 5.66E-09 | 0.007447641 | DOK1, LOXL3 |
| chr1 | 3496189 | 3496727 | 539 | 3 | 3.77E-09 | 4.06E-09 | 3.93E-05 | 5.81E-09 | 0.011424168 | MEGF6 |
| chr16 | 16121308 | 16122251 | 944 | 4 | 7.03E-08 | 4.23E-09 | 6.19E-05 | 5.82E-09 | 0.013140666 | ABCC1 |
| chr14 | 74493044 | 74494187 | 1144 | 16 | 1.57E-10 | 7.49E-06 | 6.08E-05 | 5.82E-09 | -0.00078571 | ISCA2, NPC2 |
| chr20 | 44335785 | 44337113 | 1329 | 9 | 2.18E-11 | 2.42E-07 | 9.33E-06 | 5.87E-09 | 0.009596232 | R3HDML |
| chr16 | 1443858 | 1445121 | 1264 | 9 | 3.40E-10 | 3.45E-08 | 0.000405 | 5.92E-09 | 0.006550095 | CCDC154, CLCN7 |
| chr22 | 37817783 | 37819197 | 1415 | 6 | 3.30E-07 | 8.81E-09 | 7.14E-05 | 5.94E-09 | 0.004942141 | NA |
| chr15 | 41513332 | 41514390 | 1059 | 12 | 6.51E-09 | 9.20E-08 | 0.000785 | 5.99E-09 | 0.00653442 | LTK |
| chr1 | 234214517 | 234215707 | 1191 | 8 | 1.94E-10 | 6.45E-08 | 0.000247 | 6.10E-09 | 0.005590993 | SLC35F3, AL122008.3 |
| chr2 | 10846367 | 10847816 | 1450 | 6 | 7.79E-07 | 1.43E-08 | 6.08E-05 | 6.11E-09 | 0.005723354 | LINC01954, AC092687.1 |
| chr19 | 46745233 | 46747350 | 2118 | 17 | 9.01E-08 | 2.43E-08 | 0.002396 | 6.14E-09 | 0.001427974 | FKRP, STRN4 |
| chr18 | 2845400 | 2846239 | 840 | 5 | 7.80E-09 | 6.14E-09 | 7.59E-05 | 6.28E-09 | 0.014345784 | NA |
| chr19 | 2513001 | 2514624 | 1624 | 5 | 2.05E-06 | 1.47E-09 | 0.000292 | 6.28E-09 | 0.002771739 | GNG7 |
| chr2 | 127674950 | 127676995 | 2046 | 12 | 4.75E-06 | 2.13E-07 | 0.000213 | 6.30E-09 | 0.002809619 | LIMS2 |
| chr17 | 79251707 | 79252409 | 703 | 2 | 1.13E-07 | 3.18E-09 | 2.69E-05 | 6.30E-09 | 0.018319532 | RBFOX3 |
| chr7 | 5177117 | 5177789 | 673 | 3 | 2.73E-09 | 6.30E-08 | 8.87E-06 | 6.38E-09 | 0.019172558 | NA |
| chr19 | 50383458 | 50384506 | 1049 | 7 | 3.79E-10 | 4.73E-06 | 2.91E-05 | 6.40E-09 | 0.004873648 | POLD1 |
| chr6 | 31810230 | 31811805 | 1576 | 16 | 1.76E-10 | 0.000225 | 5.59E-05 | 6.53E-09 | 0.000671675 | HSPA1L |
| chr16 | 79262559 | 79263261 | 703 | 3 | 6.75E-08 | 1.87E-09 | 0.00012 | 6.53E-09 | 0.007780978 | NA |
| chr4 | 7646213 | 7647879 | 1667 | 6 | 5.06E-07 | 7.38E-08 | 0.000203 | 6.59E-09 | 0.009882463 | SORCS2 |

| | | | | | | | | | | |
|---|---|---|---|---|---|---|---|---|---|---|
| chr1 | 150604651 | 150604946 | 296 | 3 | 1.40E-08 | 2.51E-09 | 8.65E-05 | 6.64E-09 | 0.019292055 | ENSA |
| chr2 | 11105281 | 11105388 | 108 | 3 | 1.59E-08 | 3.67E-09 | 2.76E-05 | 6.64E-09 | 0.007392376 | AC062028.1 |
| chr7 | 155806332 | 155808003 | 1672 | 8 | 1.80E-07 | 7.73E-08 | 0.001258 | 6.71E-09 | 0.008058337 | SHH |
| chr7 | 106111887 | 106113169 | 1283 | 17 | 3.51E-07 | 3.50E-07 | 0.001641 | 6.71E-09 | 0.001134606 | SYPL1 |
| chr11 | 1358250 | 1359125 | 876 | 4 | 7.64E-09 | 3.82E-07 | 3.88E-06 | 6.73E-09 | 0.002443947 | LINC02689 |
| chr20 | 63864435 | 63866004 | 1570 | 13 | 1.28E-08 | 5.60E-07 | 0.000846 | 6.78E-09 | 0.000448687 | TPD52L2 |
| chr22 | 50626061 | 50627966 | 1906 | 4 | 1.10E-05 | 3.27E-09 | 0.000115 | 6.82E-09 | 0.022041518 | ARSA |
| chr2 | 42567220 | 42569001 | 1782 | 15 | 1.65E-08 | 1.20E-07 | 0.000196 | 6.86E-09 | 0.000763152 | MTA3 |
| chr16 | 67931569 | 67932718 | 1150 | 12 | 6.54E-09 | 4.80E-08 | 0.00089 | 6.86E-09 | 0.000775517 | CTRL, AC040162.1 |
| chr6 | 3356150 | 3357388 | 1239 | 4 | 4.15E-09 | 4.31E-09 | 4.20E-05 | 6.89E-09 | 0.010449002 | SLC22A23 |
| chr9 | 129178300 | 129179185 | 886 | 8 | 6.29E-09 | 1.64E-06 | 0.000347 | 6.98E-09 | 0.005718411 | AL158151.3, AL158151.1 |
| chr20 | 23363036 | 23365595 | 2560 | 12 | 7.00E-07 | 5.77E-08 | 0.000182 | 7.03E-09 | 0.002934967 | GZF1 |
| chr1 | 209747069 | 209748757 | 1689 | 8 | 2.16E-06 | 2.04E-09 | 0.001698 | 7.04E-09 | 0.006971199 | NA |
| chr6 | 31157201 | 31159750 | 2550 | 44 | 1.58E-07 | 5.07E-06 | 0.002483 | 7.06E-09 | 0.001175927 | TCF19, CCHCR1 |
| chr20 | 37295573 | 37296833 | 1261 | 6 | 1.15E-06 | 4.13E-09 | 0.000259 | 7.08E-09 | 0.007414196 | MANBAL |
| chr9 | 114396905 | 114397936 | 1032 | 6 | 5.81E-10 | 4.64E-07 | 1.89E-05 | 7.22E-09 | 0.008903521 | AL138895.1 |
| chr17 | 48049414 | 48050216 | 803 | 3 | 1.13E-09 | 3.89E-09 | 2.66E-05 | 7.24E-09 | 0.008733345 | NFE2L1 |
| chr4 | 115113715 | 115114860 | 1146 | 8 | 7.93E-09 | 1.80E-06 | 3.55E-05 | 7.24E-09 | 0.007997054 | NDST4 |
| chr2 | 8118462 | 8119343 | 882 | 6 | 1.36E-09 | 9.19E-08 | 2.33E-05 | 7.40E-09 | 0.003745543 | LINC00298, LINC00299 |
| chr19 | 50417915 | 50419227 | 1313 | 11 | 9.32E-08 | 1.00E-08 | 0.001873 | 7.48E-09 | 0.00755179 | POLD1, AC020909.1, SPIB |
| chr4 | 54093252 | 54094805 | 1554 | 8 | 5.48E-07 | 4.33E-10 | 0.002035 | 7.53E-09 | 0.008020384 | AC058822.1 |
| chr17 | 17205532 | 17206622 | 1091 | 10 | 1.02E-08 | 1.96E-09 | 0.004306 | 7.59E-09 | -0.02065816 | MPRIP, PLD6, AC055811.2 |
| chr11 | 69702171 | 69705206 | 3036 | 21 | 6.22E-07 | 3.54E-06 | 7.59E-05 | 7.60E-09 | 0.001097433 | FGF19 |
| chr1 | 156423669 | 156424935 | 1267 | 3 | 5.14E-11 | 3.38E-07 | 4.86E-07 | 7.61E-09 | 0.019779803 | MIR9-1HG |
| chr7 | 98251251 | 98252964 | 1714 | 14 | 1.82E-07 | 5.57E-07 | 0.000227 | 7.67E-09 | 0.002711593 | BRI3, TECPR1 |
| chr6 | 157510759 | 157511148 | 390 | 6 | 2.96E-08 | 4.57E-09 | 0.000392 | 7.69E-09 | 0.017857638 | ZDHHC14 |
| chr11 | 126282344 | 126283088 | 745 | 12 | 3.84E-12 | 5.88E-07 | 0.000638 | 7.70E-09 | 0.004241407 | TIRAP |
| chr2 | 232928378 | 232929624 | 1247 | 5 | 2.20E-10 | 5.08E-08 | 8.87E-06 | 7.70E-09 | 0.010459221 | NGEF |
| chr21 | 34613874 | 34614577 | 704 | 8 | 1.43E-10 | 2.00E-07 | 1.52E-05 | 7.81E-09 | 0.002922534 | RCAN1 |
| chr12 | 92702205 | 92703068 | 864 | 6 | 7.08E-11 | 0.000113 | 8.69E-08 | 7.83E-09 | 0.002827919 | PLEKHG7 |
| chr15 | 85768923 | 85770742 | 1820 | 12 | 7.00E-07 | 7.11E-07 | 0.000258 | 7.92E-09 | 0.000957176 | KLHL25, MIR1276 |
| chr1 | 18668279 | 18668967 | 689 | 2 | 1.44E-08 | 6.54E-09 | 6.65E-06 | 7.95E-09 | 0.019053082 | PAX7 |
| chr12 | 98644068 | 98645800 | 1733 | 25 | 1.75E-08 | 2.64E-06 | 0.000202 | 7.95E-09 | 0.000360984 | APAF1, IKBIP |
| chr19 | 38930175 | 38931890 | 1716 | 21 | 4.25E-08 | 7.28E-09 | 0.002039 | 7.95E-09 | 0.001460945 | MRPS12, SARS2, AC011455.2 |
| chr16 | 89114179 | 89114435 | 257 | 3 | 3.54E-09 | 3.51E-09 | 3.60E-05 | 7.98E-09 | 0.01002737 | ACSF3, AC135782.3 |
| chr9 | 132902693 | 132904640 | 1948 | 6 | 3.31E-07 | 2.63E-07 | 4.37E-05 | 7.98E-09 | 0.002969454 | TSC1 |
| chr2 | 241248482 | 241250065 | 1584 | 4 | 2.32E-07 | 1.72E-09 | 0.000183 | 8.05E-09 | -0.00057036 | HDLBP |
| chr6 | 41733828 | 41735875 | 2048 | 19 | 5.72E-06 | 1.24E-06 | 9.35E-05 | 8.08E-09 | 0.001374916 | AL035588.1, TFEB |
| chr19 | 8430787 | 8430986 | 200 | 4 | 7.46E-09 | 2.03E-07 | 3.74E-06 | 8.12E-09 | 0.004084086 | MARCHF2 |
| chr10 | 33290921 | 33292184 | 1264 | 5 | 8.87E-09 | 6.67E-09 | 0.000124 | 8.13E-09 | 0.004876804 | NRP1 |
| chr12 | 51269700 | 51271359 | 1660 | 14 | 1.06E-08 | 1.13E-07 | 6.84E-05 | 8.25E-09 | 0.001691208 | DAZAP2, SMAGP |
| chr15 | 92595915 | 92596837 | 923 | 10 | 2.01E-08 | 1.75E-08 | 0.000693 | 8.45E-09 | -0.00169249 | AC091544.2, AC091544.4 |
| chr16 | 85251061 | 85252118 | 1058 | 4 | 2.82E-11 | 2.38E-05 | 2.08E-05 | 8.46E-09 | 0.005202936 | GSE1, AC092275.1 |
| chr16 | 15649631 | 15650540 | 910 | 11 | 2.68E-11 | 0.00019 | 2.18E-07 | 8.54E-09 | -8.30E-05 | NDE1 |
| chr2 | 240621590 | 240623233 | 1644 | 7 | 5.99E-06 | 1.77E-08 | 3.37E-05 | 8.62E-09 | -0.01686417 | GPR35 |
| chr8 | 133101882 | 133103417 | 1536 | 11 | 1.05E-08 | 6.68E-07 | 2.34E-05 | 8.67E-09 | 0.003448695 | TG, SLA |
| chr6 | 153002228 | 153003717 | 1490 | 15 | 8.87E-09 | 6.06E-07 | 5.42E-06 | 8.72E-09 | 0.000744094 | MTRF1L |
| chr12 | 57628134 | 57628837 | 704 | 5 | 1.29E-09 | 1.11E-08 | 3.87E-05 | 8.77E-09 | 0.013001655 | B4GALNT1 |
| chr6 | 33204423 | 33205804 | 1382 | 26 | 2.08E-08 | 1.77E-07 | 0.006714 | 8.78E-09 | 0.003513681 | SLC39A7, HSD17B8 |
| chr12 | 12656119 | 12656286 | 168 | 2 | 4.76E-09 | 7.13E-09 | 7.12E-06 | 8.81E-09 | 0.016753194 | NA |
| chr12 | 93570809 | 93572432 | 1624 | 26 | 1.50E-08 | 1.32E-05 | 0.002238 | 8.81E-09 | -0.00087207 | SOCS2, SOCS2-AS1 |
| chr3 | 147356202 | 147357434 | 1233 | 8 | 8.21E-08 | 2.74E-07 | 4.91E-05 | 8.86E-09 | 0.005742086 | AC092957.1 |
| chr10 | 91908181 | 91909490 | 1310 | 8 | 6.55E-10 | 6.87E-07 | 1.29E-05 | 8.88E-09 | 0.00062995 | AL359198.1, FGFBP3 |
| chr1 | 85276851 | 85278049 | 1199 | 10 | 4.50E-07 | 4.10E-09 | 0.001848 | 8.96E-09 | -0.00298685 | AL078459.1 |
| chr14 | 24295816 | 24297878 | 2063 | 7 | 4.54E-06 | 7.21E-09 | 0.000223 | 8.96E-09 | 0.004559065 | AL096870.12, DHRS1 |
| chr13 | 113666574 | 113668736 | 2163 | 15 | 1.18E-05 | 3.29E-08 | 0.001244 | 8.96E-09 | 0.00507724 | GRK1 |
| chr1 | 222814156 | 222815524 | 1369 | 10 | 1.85E-10 | 1.96E-06 | 0.000144 | 8.98E-09 | 0.008692471 | DISP1 |
| chr19 | 10602190 | 10602867 | 678 | 7 | 1.52E-08 | 9.64E-09 | 0.000103 | 8.98E-09 | 0.008625949 | SLC44A2 |
| chr11 | 65213348 | 65214125 | 778 | 9 | 1.18E-07 | 3.96E-10 | 0.00353 | 8.99E-09 | 0.012515269 | SLC22A20P |
| chr2 | 8837202 | 8838820 | 1619 | 11 | 5.92E-09 | 3.80E-08 | 0.000283 | 8.99E-09 | -0.00192337 | KIDINS220 |
| chr22 | 19961925 | 19962643 | 719 | 10 | 4.19E-08 | 3.36E-10 | 0.006745 | 9.03E-09 | 0.009580909 | COMT |
| chr7 | 77416570 | 77417246 | 677 | 6 | 2.29E-09 | 2.50E-07 | 6.94E-05 | 9.06E-09 | 0.01201342 | AC004921.1 |
| chr12 | 57236324 | 57237904 | 1581 | 9 | 1.84E-09 | 2.99E-08 | 0.000202 | 9.07E-09 | 0.007160892 | NDUFA4L2 |
| chr8 | 38873850 | 38874512 | 663 | 2 | 9.79E-09 | 8.75E-09 | 5.37E-06 | 9.07E-09 | -0.00475096 | NA |
| chr10 | 132558077 | 132560428 | 2352 | 10 | 5.37E-06 | 9.22E-08 | 0.000357 | 9.23E-09 | 0.007392503 | INPP5A |
| chr19 | 48600996 | 48601151 | 156 | 2 | 1.24E-08 | 7.10E-09 | 8.19E-06 | 9.25E-09 | 0.011819425 | FAM83E |
| chr6 | 41200447 | 41202073 | 1627 | 9 | 1.02E-07 | 2.20E-07 | 0.000123 | 9.25E-09 | 0.009779171 | TREML2 |
| chr5 | 64164440 | 64166103 | 1664 | 10 | 3.33E-09 | 3.28E-07 | 0.000331 | 9.37E-09 | 0.009559575 | RNF180 |
| chr17 | 7018146 | 7020207 | 2062 | 7 | 1.01E-06 | 4.28E-07 | 0.000214 | 9.37E-09 | 0.007398532 | AC040977.2, MIR497HG |
| chr22 | 27311933 | 27312062 | 130 | 2 | 6.99E-09 | 5.13E-09 | 2.15E-05 | 9.38E-09 | 0.029286328 | LINC02554, AL020994.1 |
| chr20 | 45114660 | 45116237 | 1578 | 11 | 2.71E-08 | 5.57E-07 | 0.00016 | 9.38E-09 | 0.003432488 | WFDC5 |
| chr1 | 116467008 | 116467622 | 615 | 3 | 1.57E-09 | 1.21E-06 | 2.07E-05 | 9.45E-09 | 0.006700776 | LINC01762 |
| chr17 | 82267105 | 82267803 | 699 | 5 | 3.41E-11 | 3.42E-05 | 2.39E-06 | 9.48E-09 | 0.009757838 | CSNK1D |
| chr8 | 17412838 | 17414026 | 1189 | 13 | 5.04E-08 | 2.91E-09 | 0.002999 | 9.48E-09 | 0.006032803 | MTMR7 |
| chr9 | 100472659 | 100473254 | 596 | 6 | 7.84E-10 | 7.57E-08 | 6.60E-05 | 9.53E-09 | 0.007417466 | MSANTD3-TMEFF1, TMEFF1 |
| chr5 | 150256732 | 150258405 | 1674 | 6 | 2.20E-06 | 5.41E-09 | 0.000364 | 9.67E-09 | 0.009241221 | CAMK2A |
| chr11 | 18455606 | 18456588 | 983 | 7 | 1.09E-07 | 7.20E-10 | 0.002937 | 1.00E-08 | 0.025148351 | LDHAL6A |
| chr12 | 14938024 | 14939500 | 1477 | 8 | 1.50E-07 | 1.03E-08 | 0.000689 | 1.00E-08 | 0.007610493 | ERP27 |
| chr13 | 113734071 | 113735493 | 1423 | 5 | 8.08E-08 | 4.83E-08 | 3.05E-05 | 1.02E-08 | 0.010607835 | GRK1 |
| chr3 | 23751178 | 23751515 | 338 | 3 | 2.08E-09 | 8.50E-08 | 3.34E-06 | 1.02E-08 | 0.010383295 | NA |
| chr3 | 177358436 | 177359541 | 1106 | 6 | 8.42E-09 | 6.30E-09 | 0.000425 | 1.02E-08 | 0.009404501 | NA |
| chr6 | 168885862 | 168886325 | 464 | 3 | 4.94E-10 | 6.25E-08 | 8.23E-06 | 1.02E-08 | 0.009867718 | NA |
| chr12 | 1864706 | 1865648 | 943 | 7 | 3.08E-08 | 1.96E-09 | 0.000653 | 1.02E-08 | -0.01334779 | CACNA2D4 |
| chr6 | 33247872 | 33249851 | 1980 | 43 | 1.31E-06 | 3.11E-05 | 0.006835 | 1.03E-08 | 0.001082026 | HCG25 |
| chr18 | 12911360 | 12912419 | 1060 | 5 | 9.79E-08 | 3.16E-08 | 5.70E-05 | 1.04E-08 | 0.013910017 | STK25P1, PTPN2 |
| chr13 | 113364549 | 113366266 | 1718 | 6 | 9.73E-06 | 1.48E-08 | 0.000363 | 1.04E-08 | 0.005442393 | NA |
| chr11 | 73664759 | 73666046 | 1288 | 4 | 6.15E-11 | 1.70E-06 | 3.16E-07 | 1.04E-08 | 0.006729009 | NA |
| chr19 | 4803870 | 4804437 | 568 | 3 | 1.54E-08 | 1.06E-08 | 1.12E-05 | 1.05E-08 | 0.012192809 | NA |
| chr3 | 69012424 | 69014184 | 1761 | 17 | 3.39E-07 | 4.54E-06 | 0.000673 | 1.05E-08 | -0.00125783 | AC109587.1, EOGT |
| chr7 | 97021095 | 97022603 | 1509 | 8 | 7.00E-08 | 1.21E-08 | 0.001347 | 1.05E-08 | 0.007642716 | DLX5 |
| chr8 | 42540963 | 42543211 | 2249 | 23 | 6.27E-08 | 1.24E-06 | 0.002252 | 1.05E-08 | 0.000712615 | SMIM19, SLC20A2 |
| chr4 | 1568019 | 1569064 | 1046 | 6 | 1.70E-07 | 1.71E-09 | 0.000966 | 1.05E-08 | -0.02011389 | NA |
| chr16 | 85781899 | 85782245 | 347 | 5 | 1.46E-08 | 1.03E-08 | 9.12E-05 | 1.06E-08 | 0.009602571 | EMC8, RNU1-103P |
| chr19 | 48111455 | 48112136 | 682 | 5 | 8.51E-10 | 6.74E-08 | 1.60E-05 | 1.06E-08 | 0.007038936 | NA |
| chr6 | 33593176 | 33593672 | 497 | 9 | 4.30E-08 | 4.86E-10 | 0.005005 | 1.06E-08 | 0.01330504 | LINC00336 |
| chr11 | 8932659 | 8933590 | 932 | 7 | 2.09E-08 | 9.52E-09 | 0.000552 | 1.08E-08 | 0.00679871 | C11orf16 |
| chr22 | 35840227 | 35840713 | 487 | 8 | 1.07E-08 | 8.61E-08 | 0.000263 | 1.09E-08 | 0.010661632 | RBFOX2 |
| chr5 | 1933488 | 1934051 | 564 | 4 | 1.19E-09 | 2.21E-07 | 1.97E-05 | 1.09E-08 | -0.01555463 | AC124852.1, AC126768.3 |
| chr14 | 97410664 | 97410960 | 297 | 2 | 4.66E-09 | 6.29E-09 | 1.89E-05 | 1.09E-08 | 0.031444684 | NA |
| chr10 | 47321962 | 47322970 | 1009 | 12 | 1.59E-08 | 3.69E-09 | 0.000963 | 1.09E-08 | 0.006351602 | GDF2 |
| chr17 | 5418637 | 5420595 | 1959 | 28 | 3.87E-07 | 1.50E-07 | 0.008745 | 1.09E-08 | -0.00010841 | RPAIN, NUP88 |
| chr3 | 46833407 | 46834864 | 1458 | 11 | 3.49E-08 | 4.62E-07 | 0.000106 | 1.10E-08 | 0.002227811 | PRSS42P |
| chr17 | 76140677 | 76142315 | 1639 | 12 | 8.21E-07 | 4.08E-09 | 0.001832 | 1.10E-08 | 0.002919908 | RNF157-AS1, FOXJ1 |
| chr1 | 182061618 | 182061683 | 66 | 2 | 2.44E-09 | 5.73E-09 | 3.18E-05 | 1.11E-08 | 0.018598826 | ZNF648 |
| chr11 | 63670612 | 63672506 | 1895 | 22 | 2.10E-08 | 1.22E-06 | 0.000147 | 1.12E-08 | -0.00037886 | ATL3 |
| chr1 | 109983536 | 109984627 | 1092 | 8 | 1.30E-10 | 5.16E-07 | 5.97E-06 | 1.12E-08 | 0.003618636 | NA |
| chr9 | 134370639 | 134371885 | 1247 | 6 | 3.26E-09 | 2.05E-08 | 2.99E-05 | 1.14E-08 | 0.005781198 | RXRA |
| chr17 | 60163224 | 60163743 | 520 | 2 | 5.27E-09 | 8.82E-09 | 9.11E-06 | 1.15E-08 | 0.014898402 | CA4 |
| chr3 | 105367147 | 105369702 | 2556 | 15 | 2.25E-07 | 1.86E-06 | 3.62E-05 | 1.17E-08 | 0.001997341 | ALCAM |
| chr2 | 32277157 | 32278473 | 1317 | 14 | 3.57E-10 | 1.34E-05 | 0.000231 | 1.18E-08 | 0.000123429 | YIPF4 |
| chr11 | 95788980 | 95790878 | 1899 | 19 | 7.71E-09 | 6.89E-06 | 0.000172 | 1.20E-08 | 0.000406475 | CEP57, FAM76B |
| chr1 | 66862426 | 66863226 | 801 | 3 | 3.47E-09 | 1.33E-07 | 1.20E-06 | 1.20E-08 | 0.018156018 | DNAI4 |
| chr15 | 77671462 | 77671661 | 200 | 2 | 2.10E-08 | 7.03E-09 | 1.89E-05 | 1.21E-08 | 0.013249914 | LINGO1 |
| chr7 | 21169304 | 21170162 | 859 | 7 | 8.70E-08 | 1.77E-08 | 0.000657 | 1.21E-08 | 0.015314999 | NA |
| chr13 | 113044094 | 113045713 | 1620 | 16 | 1.26E-07 | 2.96E-06 | 0.000233 | 1.21E-08 | 0.000835278 | MCF2L |
| chr6 | 88169679 | 88170880 | 1202 | 4 | 5.30E-11 | 1.86E-07 | 1.23E-06 | 1.23E-08 | 0.00779035 | NA |
| chr20 | 20366851 | 20368597 | 1747 | 9 | 3.46E-06 | 3.04E-06 | 6.15E-05 | 1.23E-08 | 0.002415469 | INSM1 |
| chr4 | 40516007 | 40516940 | 934 | 8 | 1.83E-09 | 2.20E-08 | 0.000384 | 1.25E-08 | 0.006029098 | RBM47 |
| chr7 | 138662785 | 138664698 | 1914 | 14 | 3.99E-07 | 1.82E-06 | 0.000418 | 1.26E-08 | 0.009627006 | SVOPL |
| chr1 | 155940732 | 155942778 | 2047 | 9 | 4.12E-06 | 1.07E-07 | 0.000854 | 1.27E-08 | 0.005922296 | RXFP4 |
| chr10 | 17228432 | 17230617 | 2186 | 19 | 2.51E-09 | 6.89E-07 | 0.000194 | 1.28E-08 | 0.002099982 | VIM, VIM-AS1 |
| chr19 | 38733730 | 38734945 | 1216 | 4 | 6.53E-08 | 1.06E-08 | 0.000101 | 1.28E-08 | 0.012279447 | CAPN12 |
| chr19 | 6463938 | 6464874 | 937 | 11 | 1.47E-07 | 1.16E-08 | 0.002121 | 1.28E-08 | 0.007206176 | CRB3, SLC25A23 |
| chr8 | 81279340 | 81280428 | 1089 | 12 | 1.68E-09 | 3.86E-06 | 4.78E-05 | 1.28E-08 | 0.000292912 | AC009902.2 |
| chr6 | 15065884 | 15067136 | 1253 | 5 | 8.24E-08 | 1.63E-07 | 0.000199 | 1.28E-08 | 0.009198646 | AL138720.1 |
| chr1 | 156579408 | 156580363 | 956 | 5 | 1.46E-07 | 2.23E-09 | 0.000269 | 1.28E-08 | 0.007248027 | TTC24 |
| chr10 | 6220907 | 6222813 | 1907 | 10 | 1.73E-07 | 1.09E-05 | 0.000175 | 1.29E-08 | 0.00257407 | PFKFB3 |
| chr2 | 224944527 | 224945091 | 565 | 4 | 5.66E-09 | 1.10E-05 | 1.15E-05 | 1.31E-08 | 0.017530644 | DOCK10 |
| chr17 | 83186958 | 83187207 | 250 | 2 | 4.88E-09 | 1.01E-08 | 9.35E-06 | 1.32E-08 | 0.000306172 | AC139099.2 |
| chr8 | 26047105 | 26049273 | 2169 | 12 | 6.02E-07 | 1.33E-09 | 0.001553 | 1.33E-08 | 0.0062264 | NA |
| chr22 | 43109886 | 43111659 | 1774 | 30 | 5.95E-09 | 8.01E-07 | 0.000163 | 1.33E-08 | 0.000942585 | BIK |
| chr19 | 53603339 | 53604025 | 687 | 5 | 4.41E-09 | 8.04E-09 | 0.000161 | 1.33E-08 | 0.032416149 | AC011487.2 |
| chr2 | 152717369 | 152719203 | 1835 | 18 | 2.12E-05 | 4.03E-07 | 0.000958 | 1.33E-08 | 0.001176058 | ARL6IP6, PRPF40A |
| chr12 | 9064483 | 9065263 | 781 | 8 | 1.97E-08 | 2.03E-09 | 0.00245 | 1.34E-08 | 0.027822903 | A2M-AS1, LINC00612 |
| chr17 | 61450641 | 61452262 | 1622 | 9 | 4.88E-07 | 9.54E-09 | 0.000351 | 1.35E-08 | 0.006096712 | NA |
| chr18 | 23498395 | 23499109 | 715 | 4 | 9.49E-10 | 2.65E-08 | 9.35E-06 | 1.35E-08 | 0.014377281 | NA |
| chr8 | 143695739 | 143697788 | 2050 | 10 | 1.73E-06 | 1.91E-07 | 0.000404 | 1.35E-08 | 0.002118532 | ZNF707, LINC02878 |
| chr19 | 36604430 | 36605813 | 1384 | 19 | 4.51E-09 | 2.72E-07 | 0.000507 | 1.37E-08 | -0.00310019 | ZNF382, ZNF529 |
| chr6 | 69865438 | 69867437 | 2000 | 19 | 2.05E-09 | 1.90E-07 | 0.000539 | 1.38E-08 | 0.0029026 | COL19A1, LMBRD1 |
| chr2 | 216694297 | 216696223 | 1927 | 14 | 6.21E-08 | 4.08E-06 | 1.42E-05 | 1.38E-08 | 0.002099469 | AC007563.2, IGFBP5 |
| chr5 | 136080004 | 136080924 | 921 | 15 | 4.83E-07 | 5.21E-11 | 0.026114 | 1.40E-08 | -0.04175881 | VTRNA2-1, Vault |
| chr19 | 33432401 | 33432836 | 436 | 2 | 1.44E-08 | 9.80E-09 | 1.20E-05 | 1.41E-08 | 0.023641742 | PEPD |
| chr8 | 17632912 | 17634059 | 1148 | 6 | 2.53E-08 | 2.06E-07 | 0.000266 | 1.41E-08 | 0.005018924 | PDGFRL |
| chr14 | 37602763 | 37604306 | 1544 | 6 | 1.71E-08 | 9.94E-09 | 0.000671 | 1.42E-08 | 0.015591171 | TTC6 |
| chr1 | 47191181 | 47192298 | 1118 | 5 | 2.00E-07 | 5.21E-08 | 5.92E-05 | 1.42E-08 | 0.003850683 | NA |
| chr4 | 182800030 | 182801357 | 1328 | 6 | 8.68E-08 | 2.88E-08 | 0.00034 | 1.44E-08 | -0.01381501 | TENM3 |
| chr3 | 49123896 | 49124581 | 686 | 4 | 2.54E-09 | 1.96E-07 | 3.19E-06 | 1.44E-08 | 0.004280848 | LAMB2 |
| chr17 | 40106837 | 40107842 | 1006 | 4 | 8.43E-08 | 5.57E-09 | 9.27E-05 | 1.44E-08 | 0.011616478 | NA |
| chr21 | 39095259 | 39095430 | 172 | 2 | 8.92E-09 | 5.87E-08 | 1.23E-06 | 1.45E-08 | 0.013703302 | NA |
| chr7 | 39085598 | 39085710 | 113 | 3 | 8.91E-09 | 1.48E-08 | 1.85E-05 | 1.45E-08 | 0.031646538 | POU6F2 |
| chr5 | 132830636 | 132831790 | 1155 | 9 | 2.82E-10 | 1.46E-05 | 7.28E-05 | 1.46E-08 | 0.005376953 | SHROOM1 |
| chr2 | 42924798 | 42926687 | 1890 | 6 | 1.12E-05 | 6.22E-09 | 0.000462 | 1.46E-08 | 0.009673644 | NA |
| chr16 | 1280146 | 1280747 | 602 | 4 | 3.27E-09 | 9.47E-09 | 7.36E-05 | 1.46E-08 | 0.008987437 | AC120498.5 |
| chr16 | 661611 | 663101 | 1491 | 12 | 6.18E-07 | 1.07E-07 | 0.002333 | 1.46E-08 | -0.00011248 | WDR90 |
| chr6 | 41071900 | 41073450 | 1551 | 21 | 2.12E-09 | 1.40E-06 | 0.001024 | 1.46E-08 | -0.00037145 | NFYA, OARD1 |
| chr15 | 68342363 | 68343151 | 789 | 3 | 3.31E-11 | 1.38E-06 | 2.78E-07 | 1.47E-08 | 0.010022649 | ITGA11 |
| chr12 | 118015695 | 118017280 | 1586 | 15 | 2.24E-08 | 3.41E-07 | 0.000355 | 1.47E-08 | 0.003880756 | RFC5 |
| chr10 | 80699541 | 80700448 | 908 | 4 | 1.27E-07 | 2.78E-09 | 0.000422 | 1.47E-08 | 0.005973674 | NA |
| chr8 | 141149893 | 141150973 | 1081 | 6 | 2.21E-06 | 2.46E-09 | 0.000904 | 1.47E-08 | 0.003327727 | DENND3 |
| chr2 | 241035500 | 241036827 | 1328 | 7 | 6.10E-07 | 7.28E-10 | 0.004452 | 1.48E-08 | 0.019717912 | SNED1, SNED1-AS1 |
| chr19 | 10587014 | 10588041 | 1028 | 8 | 3.89E-08 | 4.23E-08 | 0.00034 | 1.48E-08 | 0.011924233 | AP1M2 |
| chr14 | 70187247 | 70189794 | 2548 | 14 | 1.31E-06 | 8.31E-07 | 0.000285 | 1.48E-08 | 0.000360073 | AL160191.1, SLC8A3 |

| | | | | | | | | | | |
|---|---|---|---|---|---|---|---|---|---|---|
| chr16 | 68370894 | 68371828 | 935 | 6 | 3.23E-07 | 3.82E-07 | 0.000321 | 1.49E-08 | 0.007464172 | SMPD3 |
| chr2 | 152421712 | 152422331 | 620 | 4 | 1.73E-09 | 2.80E-07 | 1.09E-05 | 1.52E-08 | 0.004387883 | FMNL2 |
| chr19 | 10835987 | 10837219 | 1233 | 13 | 2.95E-11 | 1.24E-05 | 4.94E-05 | 1.53E-08 | 0.000797383 | C19orf38, TMED1 |
| chr17 | 35448276 | 35450200 | 1925 | 15 | 2.70E-08 | 8.41E-07 | 0.001412 | 1.53E-08 | 0.007552312 | SLFN13 |
| chr14 | 21017741 | 21017883 | 143 | 2 | 6.53E-10 | 1.26E-08 | 8.95E-06 | 1.55E-08 | 0.017524888 | NDRG2 |
| chr14 | 100158879 | 100160675 | 1797 | 14 | 1.19E-07 | 1.99E-07 | 0.000193 | 1.56E-08 | 0.003900623 | DEGS2 |
| chr6 | 29975432 | 29977951 | 2520 | 24 | 3.35E-05 | 1.24E-07 | 0.008995 | 1.57E-08 | 0.004275797 | HCG9 |
| chr11 | 597231 | 597651 | 421 | 5 | 3.62E-09 | 2.20E-07 | 0.000171 | 1.57E-08 | 0.000797768 | PHRF1 |
| chr7 | 122303651 | 122305982 | 2332 | 23 | 2.05E-07 | 2.18E-06 | 0.00044 | 1.58E-08 | 0.002827316 | FEZF1-AS1, FEZF1 |
| chr1 | 41394969 | 41395436 | 468 | 2 | 9.40E-10 | 1.73E-07 | 6.39E-07 | 1.58E-08 | 0.009212285 | NA |
| chr8 | 73669176 | 73670425 | 1250 | 5 | 5.78E-08 | 3.37E-07 | 3.34E-05 | 1.58E-08 | 0.005549553 | STAU2 |
| chr6 | 30195165 | 30195877 | 713 | 5 | 6.71E-12 | 1.34E-07 | 6.38E-06 | 1.59E-08 | 0.004416884 | TRIM26 |
| chr11 | 18725768 | 18726620 | 853 | 6 | 4.21E-10 | 2.74E-06 | 4.17E-05 | 1.59E-08 | 0.004969557 | AC103974.1, IGSF22 |
| chr19 | 43925475 | 43926886 | 1412 | 5 | 3.01E-06 | 2.40E-07 | 0.00014 | 1.59E-08 | 0.003118348 | AC006213.3, ZNF45 |
| chr7 | 129500536 | 129502140 | 1605 | 6 | 1.19E-07 | 1.89E-08 | 0.000449 | 1.59E-08 | 0.008394247 | NA |
| chr1 | 38475144 | 38477304 | 2161 | 11 | 9.41E-07 | 1.13E-06 | 0.000479 | 1.59E-08 | 0.007747423 | LINC01685 |
| chr10 | 30433042 | 30434171 | 1130 | 16 | 1.55E-08 | 3.62E-07 | 0.000208 | 1.59E-08 | 0.003255996 | MAP3K8 |
| chr14 | 73238749 | 73239968 | 1220 | 9 | 8.34E-10 | 8.79E-08 | 6.73E-05 | 1.60E-08 | 0.006617477 | PAPLN |
| chr1 | 27383085 | 27383780 | 696 | 15 | 2.45E-08 | 8.43E-07 | 0.000686 | 1.60E-08 | 0.006739517 | CD164L2 |
| chr7 | 44146920 | 44147711 | 792 | 4 | 1.74E-08 | 3.81E-08 | 5.69E-05 | 1.61E-08 | 0.006651121 | GCK |
| chr21 | 36966131 | 36967999 | 1869 | 11 | 2.17E-06 | 4.73E-08 | 0.001314 | 1.61E-08 | -0.00024514 | AP000704.1, HLCS |
| chr5 | 153497128 | 153497487 | 360 | 2 | 1.71E-09 | 2.58E-07 | 5.10E-07 | 1.61E-08 | 0.028467483 | GRIA1 |
| chr12 | 24562316 | 24564244 | 1929 | 16 | 2.33E-05 | 6.65E-08 | 0.002844 | 1.62E-08 | 0.001789259 | SOX5 |
| chr1 | 36581996 | 36582201 | 206 | 3 | 1.25E-08 | 1.58E-07 | 4.82E-06 | 1.62E-08 | 0.011490947 | NA |
| chr3 | 127575763 | 127576899 | 1137 | 7 | 4.48E-07 | 1.50E-07 | 6.32E-05 | 1.62E-08 | 0.001966095 | TPRA1 |
| chr7 | 128910357 | 128911703 | 1347 | 12 | 4.12E-09 | 9.22E-08 | 0.000255 | 1.62E-08 | 0.005997761 | KCP |
| chr12 | 121672181 | 121673274 | 1094 | 8 | 5.23E-08 | 3.79E-07 | 0.000337 | 1.63E-08 | 0.003529729 | MORN3 |
| chr5 | 43556589 | 43558349 | 1761 | 12 | 2.17E-08 | 2.76E-08 | 0.000268 | 1.66E-08 | 0.002700394 | PAIP1 |
| chr6 | 31656065 | 31657382 | 1318 | 7 | 1.37E-08 | 2.29E-07 | 0.000261 | 1.66E-08 | 0.001195856 | APOM |
| chr19 | 49511457 | 49513299 | 1843 | 15 | 2.73E-07 | 2.10E-06 | 7.38E-05 | 1.66E-08 | 0.004650768 | FCGRT |
| chr1 | 26176033 | 26177442 | 1410 | 6 | 4.90E-08 | 4.13E-06 | 2.91E-06 | 1.67E-08 | -0.00069884 | NA |
| chr18 | 22559020 | 22560282 | 1263 | 7 | 3.60E-07 | 1.26E-08 | 0.000321 | 1.67E-08 | 5.12E-05 | NA |
| chr22 | 23118314 | 23118490 | 177 | 2 | 3.49E-09 | 7.10E-07 | 3.11E-07 | 1.69E-08 | 0.014006816 | GNAZ, RSPH14 |
| chr1 | 170666819 | 170667428 | 610 | 4 | 2.95E-08 | 2.19E-08 | 4.23E-05 | 1.69E-08 | 0.017615447 | PRRX1, Z97200.1 |
| chr11 | 66256111 | 66257589 | 1479 | 15 | 4.30E-09 | 5.26E-06 | 0.000459 | 1.70E-08 | 0.002392702 | KLC2 |
| chr6 | 143606733 | 143608163 | 1431 | 6 | 5.69E-09 | 3.53E-08 | 0.000286 | 1.70E-08 | 0.014781023 | PHACTR2 |
| chr12 | 4975901 | 4975997 | 97 | 2 | 3.65E-08 | 9.01E-09 | 3.51E-05 | 1.70E-08 | 0.046191649 | AC005906.2 |
| chr7 | 116672050 | 116672850 | 801 | 8 | 2.67E-09 | 6.43E-09 | 0.000158 | 1.70E-08 | 0.007025125 | MET |
| chr9 | 37034936 | 37036497 | 1562 | 9 | 4.11E-08 | 1.58E-08 | 0.000468 | 1.70E-08 | 0.007278743 | NA |
| chr4 | 1009906 | 1011617 | 1712 | 27 | 3.07E-09 | 2.07E-06 | 0.000143 | 1.71E-08 | 7.85E-05 | FGFRL1 |
| chr7 | 63925848 | 63926769 | 922 | 7 | 1.82E-07 | 1.01E-09 | 0.003486 | 1.71E-08 | 0.023644957 | AC092634.8, AC092634.3 |
| chr19 | 17441314 | 17441673 | 360 | 3 | 1.74E-08 | 2.36E-08 | 2.83E-05 | 1.71E-08 | 0.009841299 | TMEM221, AC010319.2 |
| chr20 | 2508401 | 2509784 | 1384 | 17 | 6.37E-08 | 6.92E-07 | 0.000794 | 1.71E-08 | 0.002039132 | AL049650.1, ZNF343 |
| chr8 | 142777727 | 142778674 | 948 | 13 | 9.20E-08 | 9.52E-09 | 0.005753 | 1.71E-08 | 0.012283059 | LYNX1-SLURP2, LYNX1 |
| chr6 | 36947722 | 36948347 | 626 | 5 | 3.21E-08 | 5.99E-08 | 5.08E-05 | 1.72E-08 | 0.012757061 | PI16 |
| chr16 | 30429243 | 30430732 | 1490 | 17 | 9.03E-07 | 1.76E-07 | 0.000984 | 1.73E-08 | 0.001968365 | ZNF771, DCTPP1 |
| chr2 | 9638524 | 9639327 | 804 | 5 | 6.55E-08 | 2.85E-09 | 0.000464 | 1.77E-08 | 0.007726608 | AC082651.1 |
| chr6 | 43221394 | 43221785 | 392 | 4 | 2.75E-08 | 1.10E-07 | 2.82E-05 | 1.77E-08 | 0.006897354 | CUL9, AL133375.1 |
| chr22 | 39642697 | 39643315 | 619 | 3 | 1.61E-08 | 3.07E-07 | 3.02E-05 | 1.78E-08 | 0.012108759 | CACNA1I |
| chr13 | 57630087 | 57631660 | 1574 | 10 | 9.03E-09 | 5.96E-08 | 0.000709 | 1.78E-08 | 0.005286144 | NA |
| chr22 | 19712588 | 19714348 | 1761 | 9 | 1.28E-07 | 2.26E-07 | 5.44E-05 | 1.78E-08 | 0.003648049 | NA |
| chr19 | 10102595 | 10103141 | 547 | 6 | 1.16E-07 | 1.37E-09 | 0.002417 | 1.79E-08 | 0.012183808 | ANGPTL6 |
| chr4 | 80206665 | 80207818 | 1154 | 7 | 2.66E-08 | 2.79E-08 | 0.000499 | 1.79E-08 | 0.011354758 | NA |
| chr18 | 54356771 | 54359425 | 2655 | 15 | 1.71E-05 | 5.77E-06 | 0.000969 | 1.79E-08 | 0.005117686 | C18orf54, STARD6 |
| chr6 | 1600994 | 1601902 | 909 | 6 | 3.84E-08 | 8.54E-09 | 0.000465 | 1.81E-08 | 0.014432213 | FOXCUT |
| chr1 | 241956586 | 241958761 | 2176 | 10 | 1.08E-06 | 1.16E-07 | 0.003029 | 1.81E-08 | 0.004662341 | BECN2 |
| chr14 | 100857013 | 100857517 | 505 | 5 | 4.99E-08 | 1.16E-07 | 0.000142 | 1.81E-08 | 0.003690415 | MEG3, AL117190.1, AL117190.2 |
| chr4 | 134934262 | 134934622 | 361 | 5 | 1.46E-08 | 4.56E-08 | 0.000101 | 1.82E-08 | -0.00411461 | NA |
| chr1 | 150282028 | 150283470 | 1443 | 15 | 5.51E-09 | 5.89E-08 | 0.002613 | 1.83E-08 | 0.006876405 | CIART |
| chr14 | 24113847 | 24114773 | 927 | 11 | 3.77E-09 | 1.01E-05 | 0.00011 | 1.83E-08 | 0.002558743 | DCAF11, NRL |
| chr2 | 3322190 | 3323565 | 1376 | 8 | 1.20E-07 | 2.11E-08 | 0.000235 | 1.83E-08 | 0.000913237 | EIPR1 |
| chr2 | 236217592 | 236217730 | 139 | 2 | 3.20E-09 | 7.87E-07 | 3.16E-07 | 1.84E-08 | 0.000319525 | AC019068.1, ASB18 |
| chr16 | 84593901 | 84594413 | 513 | 6 | 4.96E-09 | 3.43E-08 | 0.00015 | 1.84E-08 | 0.010822829 | AC092145.1, COTL1 |
| chr4 | 52751512 | 52752964 | 1453 | 11 | 3.79E-07 | 3.50E-07 | 9.68E-05 | 1.84E-08 | 0.007387696 | LINC01618, ERVMER34-1 |
| chr9 | 34635019 | 34636073 | 1055 | 3 | 4.97E-08 | 1.12E-08 | 8.46E-05 | 1.84E-08 | -0.00032416 | SIGMAR1 |
| chr12 | 106581893 | 106583784 | 1892 | 18 | 6.52E-07 | 1.44E-06 | 0.000934 | 1.85E-08 | 0.003346095 | RFX4, AC079385.1 |
| chr19 | 14114137 | 14114835 | 699 | 7 | 2.63E-08 | 3.00E-08 | 0.000111 | 1.87E-08 | 0.004002837 | PRKACA |
| chr13 | 72772040 | 72772866 | 827 | 4 | 3.35E-08 | 4.20E-08 | 4.55E-05 | 1.91E-08 | -0.00391533 | DIS3 |
| chr8 | 100336131 | 100337471 | 1341 | 11 | 5.35E-07 | 2.91E-08 | 0.001408 | 1.91E-08 | 0.010523528 | RNF19A |
| chr8 | 22156181 | 22156616 | 436 | 3 | 5.80E-11 | 5.59E-08 | 1.29E-05 | 1.91E-08 | 0.011320154 | LGI3 |
| chr17 | 50397203 | 50398399 | 1197 | 13 | 1.41E-07 | 3.51E-08 | 0.000625 | 1.92E-08 | 0.006024246 | AC004707.1, LRRC59 |
| chr3 | 154430164 | 154430871 | 708 | 5 | 4.94E-09 | 2.69E-08 | 9.70E-05 | 1.93E-08 | 0.017181089 | GPR149 |
| chr8 | 142128948 | 142129624 | 677 | 5 | 2.18E-07 | 6.14E-09 | 0.000678 | 1.93E-08 | 0.011906027 | AC103758.1 |
| chr17 | 57994384 | 57995718 | 1335 | 5 | 4.51E-07 | 2.35E-08 | 4.19E-05 | 1.95E-08 | 0.009394585 | AC015813.2, AC015813.1 |
| chr12 | 48970311 | 48971340 | 1030 | 7 | 2.70E-10 | 2.46E-07 | 5.17E-06 | 1.96E-08 | 0.007494358 | WNT10B |
| chr11 | 859532 | 860157 | 626 | 5 | 2.26E-09 | 2.84E-07 | 1.18E-05 | 1.96E-08 | 0.007617227 | TSPAN4, AP006623.1 |
| chr22 | 30386815 | 30388006 | 1192 | 11 | 6.67E-09 | 2.09E-07 | 0.001475 | 1.96E-08 | 0.004523225 | RNF215 |
| chr11 | 115760030 | 115761137 | 1108 | 14 | 2.01E-08 | 9.47E-07 | 0.001064 | 1.96E-08 | 0.00242592 | LINC02698, LINC00900 |
| chr19 | 18939141 | 18940162 | 1022 | 4 | 2.51E-08 | 1.53E-08 | 8.51E-05 | 1.96E-08 | 0.009758393 | HOMER3 |
| chr3 | 45688997 | 45689913 | 917 | 19 | 2.87E-08 | 2.96E-07 | 0.002845 | 1.96E-08 | -0.00116438 | SACM1L, LIMD1-AS1 |
| chr6 | 42048129 | 42049757 | 1629 | 17 | 6.09E-07 | 6.67E-07 | 0.000825 | 1.96E-08 | 0.001365719 | CCND3 |
| chr1 | 6490111 | 6492343 | 2233 | 17 | 6.95E-06 | 6.40E-06 | 5.28E-05 | 1.96E-08 | 0.003172617 | PLEKHG5 |
| chr7 | 45921690 | 45922977 | 1288 | 26 | 8.22E-09 | 3.28E-06 | 0.00196 | 1.99E-08 | 0.003467669 | IGFBP3 |
| chr20 | 5688958 | 5689650 | 693 | 4 | 3.14E-08 | 1.24E-08 | 9.74E-05 | 1.99E-08 | 0.021068625 | NA |
| chr19 | 39307326 | 39308739 | 1414 | 5 | 1.51E-10 | 2.80E-05 | 4.06E-05 | 2.01E-08 | 0.009320595 | LRFN1 |
| chr2 | 241562997 | 241563437 | 441 | 2 | 7.68E-09 | 2.17E-08 | 6.49E-06 | 2.01E-08 | 0.010505232 | BOK |
| chr1 | 33219981 | 33220834 | 854 | 2 | 4.45E-10 | 1.65E-07 | 9.20E-07 | 2.04E-08 | 0.012118555 | NA |
| chr14 | 103532607 | 103533441 | 835 | 6 | 4.27E-09 | 1.91E-08 | 0.000342 | 2.04E-08 | 0.003084949 | TRMT61A |
| chr1 | 178058297 | 178058898 | 602 | 2 | 2.37E-09 | 6.93E-08 | 1.82E-06 | 2.05E-08 | 0.015129932 | NA |
| chr11 | 2271131 | 2272435 | 1305 | 38 | 5.42E-09 | 1.89E-07 | 0.013633 | 2.05E-08 | 0.006160492 | NA |
| chr18 | 37243082 | 37244775 | 1694 | 5 | 1.83E-08 | 6.37E-07 | 1.56E-05 | 2.05E-08 | 0.005494427 | AC015961.2, CELF4 |
| chr6 | 16322885 | 16323054 | 170 | 2 | 3.33E-08 | 1.62E-08 | 1.13E-05 | 2.06E-08 | 0.024819448 | ATXN1 |
| chr11 | 74009286 | 74010329 | 1044 | 5 | 1.01E-07 | 4.75E-08 | 5.69E-05 | 2.06E-08 | 0.008642898 | NA |
| chr6 | 24910965 | 24912668 | 1704 | 15 | 3.56E-08 | 6.99E-07 | 6.89E-05 | 2.07E-08 | 0.002136189 | RIPOR2 |
| chr1 | 153545812 | 153547374 | 1563 | 9 | 1.87E-09 | 2.89E-06 | 1.93E-05 | 2.07E-08 | 0.002453317 | S100A4, S100A3 |
| chr1 | 15343956 | 15344778 | 823 | 6 | 2.30E-08 | 6.34E-09 | 0.00038 | 2.09E-08 | 0.008392634 | FHAD1 |
| chr20 | 41348567 | 41349288 | 722 | 2 | 1.19E-09 | 5.90E-08 | 2.16E-06 | 2.09E-08 | 0.009551581 | LPIN3 |
| chr11 | 69638235 | 69641429 | 3195 | 33 | 7.86E-06 | 6.50E-08 | 0.001617 | 2.09E-08 | 0.001946896 | CCND1 |
| chr2 | 127650510 | 127651694 | 1185 | 7 | 7.56E-07 | 1.18E-07 | 0.000146 | 2.11E-08 | 0.003017154 | GPR17, LIMS2 |
| chr1 | 111426897 | 111427909 | 1013 | 5 | 1.33E-09 | 8.93E-08 | 3.73E-05 | 2.13E-08 | 0.010373641 | OVGP1 |
| chr16 | 1450566 | 1452072 | 1507 | 8 | 2.09E-09 | 1.91E-05 | 7.07E-06 | 2.14E-08 | 0.002638805 | AL031600.1, CLCN7 |
| chr11 | 36266100 | 36266778 | 679 | 4 | 2.15E-10 | 2.58E-07 | 5.27E-06 | 2.14E-08 | 0.007294238 | NA |
| chr17 | 19411006 | 19411305 | 300 | 4 | 1.42E-08 | 1.00E-08 | 0.000227 | 2.18E-08 | 0.015777565 | RNF112 |
| chr15 | 75723185 | 75723994 | 810 | 7 | 2.73E-08 | 6.67E-09 | 0.0008 | 2.19E-08 | 0.009186432 | NA |
| chr19 | 40749985 | 40751328 | 1344 | 19 | 3.96E-08 | 2.62E-08 | 0.003178 | 2.20E-08 | -0.00197919 | SNRPA, C19orf54 |
| chr17 | 80108536 | 80110429 | 1894 | 6 | 6.54E-06 | 5.40E-09 | 0.000285 | 2.22E-08 | 0.000798527 | GAA |
| chr4 | 70721723 | 70722597 | 875 | 7 | 6.37E-07 | 5.35E-09 | 0.001004 | 2.22E-08 | 0.007858766 | RUFY3 |
| chr17 | 82583673 | 82584242 | 570 | 8 | 4.01E-08 | 1.27E-09 | 0.004016 | 2.24E-08 | 0.016725119 | FOXK2 |
| chr20 | 38259798 | 38260962 | 1165 | 9 | 2.51E-09 | 1.42E-06 | 7.59E-05 | 2.27E-08 | 0.012368865 | AL359555.3, AL359555.4, KIAA1755 |
| chr14 | 33949577 | 33951966 | 2390 | 19 | 1.75E-07 | 3.38E-08 | 0.000201 | 2.27E-08 | 0.003570747 | EGLN3 |
| chr17 | 80068780 | 80069258 | 479 | 3 | 4.21E-08 | 9.62E-09 | 8.98E-05 | 2.28E-08 | 0.011441659 | CCDC40 |
| chr13 | 113654304 | 113654434 | 131 | 3 | 5.96E-08 | 2.88E-08 | 5.81E-05 | 2.28E-08 | 0.010297108 | ATP4B |
| chr13 | 21377874 | 21379204 | 1331 | 5 | 2.81E-06 | 8.46E-08 | 6.47E-05 | 2.28E-08 | 0.022826204 | ZDHHC20, ZDHHC20-IT1 |
| chr14 | 96698830 | 96698960 | 131 | 2 | 4.89E-08 | 1.30E-08 | 2.82E-05 | 2.29E-08 | 0.019431273 | NA |
| chr9 | 130171291 | 130172420 | 1130 | 8 | 9.01E-10 | 3.02E-06 | 5.66E-05 | 2.29E-08 | 0.007230927 | NCS1 |
| chr5 | 153490183 | 153491235 | 1053 | 11 | 6.66E-11 | 2.18E-06 | 0.000798 | 2.29E-08 | 0.009748681 | GRIA1 |
| chr3 | 120560673 | 120561279 | 607 | 2 | 1.02E-07 | 1.20E-08 | 4.38E-05 | 2.29E-08 | 0.014396016 | NA |
| chr11 | 86418217 | 86420025 | 1809 | 3 | 2.42E-05 | 1.21E-08 | 9.36E-05 | 2.30E-08 | 0.005199972 | CCDC81 |
| chr14 | 93231707 | 93233674 | 1968 | 9 | 4.76E-07 | 1.47E-08 | 0.003593 | 2.30E-08 | -0.01389619 | NA |
| chr15 | 89003871 | 89004342 | 472 | 2 | 1.69E-08 | 2.38E-08 | 7.45E-06 | 2.32E-08 | 0.009874637 | NA |
| chr3 | 10423693 | 10423728 | 36 | 2 | 4.98E-08 | 1.48E-08 | 1.96E-05 | 2.32E-08 | 0.025769914 | ATP2B2 |
| chr5 | 116447492 | 116449505 | 2014 | 9 | 6.70E-07 | 1.07E-06 | 6.56E-05 | 2.33E-08 | 0.002917626 | SEMA6A-AS1, SEMA6A |
| chr1 | 28682066 | 28683524 | 1459 | 5 | 3.40E-06 | 7.35E-08 | 0.000179 | 2.33E-08 | -0.00353321 | GMEB1 |
| chr4 | 8376449 | 8377511 | 1063 | 3 | 2.91E-08 | 4.03E-08 | 4.03E-05 | 2.33E-08 | 0.004742944 | ACOX3 |
| chr2 | 207505015 | 207505308 | 294 | 3 | 8.63E-08 | 6.49E-09 | 0.000238 | 2.35E-08 | 0.030102811 | AC009226.1 |
| chr21 | 36702178 | 36702871 | 694 | 4 | 3.22E-09 | 2.14E-07 | 4.39E-05 | 2.35E-08 | 0.012543176 | SIM2 |
| chr14 | 59187195 | 59188711 | 1517 | 13 | 5.73E-08 | 1.68E-05 | 2.31E-05 | 2.36E-08 | 0.003425979 | DAAM1 |
| chr16 | 57946240 | 57947072 | 833 | 7 | 9.76E-08 | 1.05E-08 | 8.34E-05 | 2.37E-08 | 0.005920808 | CNGB1 |
| chr1 | 960603 | 963017 | 2415 | 12 | 3.59E-06 | 2.16E-07 | 0.001628 | 2.37E-08 | 0.00329294 | KLHL17 |
| chr10 | 63720166 | 63720872 | 707 | 2 | 2.82E-09 | 9.39E-08 | 1.67E-06 | 2.37E-08 | 0.011280472 | AC013287.1 |
| chr20 | 1619891 | 1620059 | 169 | 3 | 4.45E-08 | 9.52E-09 | 8.64E-05 | 2.39E-08 | 0.01259555 | AL049634.2, SIRPB1 |
| chr10 | 126521411 | 126522825 | 1415 | 8 | 1.54E-06 | 3.43E-08 | 0.00191 | 2.40E-08 | 0.003490048 | C10orf90 |
| chr3 | 47421290 | 47423009 | 1720 | 7 | 4.18E-09 | 1.02E-07 | 4.59E-05 | 2.41E-08 | 0.002092115 | SCAP |
| chr20 | 63536589 | 63537981 | 1393 | 14 | 7.70E-08 | 1.56E-07 | 0.000848 | 2.46E-08 | 0.006752582 | PTK6 |
| chr14 | 57493746 | 57494735 | 990 | 8 | 1.35E-09 | 0.00025 | 3.16E-06 | 2.46E-08 | 0.006709562 | CCDC198 |
| chr22 | 30634534 | 30635698 | 1165 | 10 | 6.87E-10 | 1.73E-06 | 4.70E-05 | 2.47E-08 | 0.004035356 | NA |
| chr13 | 77919068 | 77920752 | 1685 | 36 | 1.51E-08 | 1.13E-06 | 0.004582 | 2.49E-08 | -0.00169475 | OBI1-AS1, EDNRB |
| chr8 | 143944654 | 143946213 | 1560 | 13 | 7.96E-08 | 1.07E-06 | 0.004226 | 2.49E-08 | 0.002602853 | PLEC, MIR661 |
| chr3 | 30893651 | 30895065 | 1415 | 11 | 6.20E-07 | 1.74E-07 | 0.000935 | 2.49E-08 | -0.00443468 | GADL1 |
| chr5 | 141959521 | 141960627 | 1107 | 6 | 3.39E-09 | 1.06E-07 | 4.10E-05 | 2.49E-08 | 0.004038617 | RNF14, PCDH12 |
| chr7 | 22823594 | 22823884 | 291 | 4 | 4.96E-10 | 1.23E-06 | 8.17E-07 | 2.51E-08 | 0.006215598 | AC005682.2 |
| chr19 | 4284364 | 4285052 | 689 | 2 | 1.31E-07 | 1.43E-08 | 3.10E-05 | 2.52E-08 | 0.032913768 | SHD |
| chr2 | 27494842 | 27496677 | 1836 | 11 | 5.07E-06 | 4.42E-07 | 0.000509 | 2.53E-08 | 0.002557329 | FNDC4 |
| chr7 | 22726104 | 22728819 | 2716 | 14 | 3.70E-05 | 4.41E-07 | 0.000955 | 2.53E-08 | 0.002692792 | IL6, STEAP1B, IL6-AS1 |
| chr16 | 89629730 | 89630160 | 431 | 5 | 7.57E-08 | 5.06E-08 | 0.000245 | 2.54E-08 | 0.006782017 | DPEP1 |
| chr19 | 2545988 | 2546940 | 953 | 5 | 2.00E-07 | 3.18E-09 | 0.00125 | 2.55E-08 | 0.023314158 | GNG7 |
| chr17 | 79797817 | 79798917 | 1101 | 8 | 7.02E-09 | 6.92E-08 | 0.000278 | 2.57E-08 | 0.001847606 | CBX8 |
| chr20 | 41699877 | 41701105 | 1229 | 5 | 1.39E-07 | 1.24E-07 | 6.99E-05 | 2.58E-08 | 0.004056775 | NA |
| chr17 | 75557489 | 75559230 | 1742 | 9 | 1.56E-08 | 0.000108 | 0.000115 | 2.58E-08 | 0.004455929 | LLGL2 |
| chr6 | 31623566 | 31625638 | 2073 | 24 | 6.56E-06 | 1.24E-07 | 0.003225 | 2.60E-08 | 0.000249614 | PRRC2A |
| chr17 | 81309093 | 81309938 | 846 | 12 | 1.71E-07 | 1.14E-09 | 0.010101 | 2.61E-08 | 0.007793585 | LINC00482 |
| chr6 | 33404990 | 33405164 | 175 | 5 | 1.49E-09 | 3.58E-05 | 1.34E-05 | 2.62E-08 | 0.001799751 | KIFC1 |
| chr7 | 102170091 | 102170829 | 739 | 6 | 2.26E-08 | 1.48E-07 | 0.000262 | 2.64E-08 | 0.004213165 | CUX1 |
| chr3 | 133745203 | 133747091 | 1889 | 15 | 3.24E-06 | 4.01E-09 | 0.003617 | 2.67E-08 | 0.007849455 | INHCAP, TF |
| chr1 | 37454921 | 37456006 | 1086 | 17 | 4.43E-09 | 8.44E-06 | 0.000475 | 2.68E-08 | 0.005342129 | LINC01137 |
| chr4 | 48906452 | 48907129 | 678 | 12 | 6.90E-09 | 2.24E-05 | 0.000144 | 2.68E-08 | 0.003934622 | OCIAD2 |
| chr19 | 1437370 | 1438597 | 1228 | 12 | 3.30E-09 | 2.18E-05 | 2.21E-05 | 2.71E-08 | -5.74E-05 | RPS15 |
| chr1 | 40508048 | 40509247 | 1200 | 17 | 7.14E-08 | 4.02E-08 | 0.004139 | 2.74E-08 | -0.00098587 | EXO5, AL603839.3 |
| chr6 | 33312275 | 33314165 | 1891 | 41 | 2.23E-07 | 4.16E-06 | 0.00886 | 2.78E-08 | 0.003767511 | TAPBP |
| chr7 | 13980715 | 13981490 | 776 | 2 | 9.01E-08 | 2.50E-08 | 1.00E-05 | 2.78E-08 | 0.004332361 | ETV1 |
| chr1 | 240313162 | 240313292 | 131 | 2 | 6.10E-08 | 1.62E-08 | 2.88E-05 | 2.79E-08 | 0.047824901 | FMN2 |
| chr9 | 125885630 | 125886543 | 914 | 2 | 1.15E-06 | 1.66E-08 | 2.60E-05 | 2.79E-08 | 0.036493906 | PBX3 |

| | | | | | | | | | | |
|---|---|---|---|---|---|---|---|---|---|---|
| chr12 | 121989947 | 121991478 | 1532 | 4 | 1.48E-09 | 1.20E-06 | 2.27E-05 | 2.80E-08 | 0.006968716 | CFAP251 |
| chr1 | 3078693 | 3080086 | 1394 | 4 | 1.41E-08 | 2.10E-07 | 4.30E-05 | 2.80E-08 | -0.01767347 | PRDM16 |
| chr14 | 77265940 | 77266298 | 359 | 5 | 1.07E-08 | 1.78E-06 | 5.55E-05 | 2.82E-08 | -9.01E-05 | MIR1260A, NGB |
| chr2 | 105037449 | 105038821 | 1373 | 15 | 1.53E-08 | 1.23E-07 | 0.002407 | 2.83E-08 | -0.00109521 | MRPS9, MRPS9-AS2 |
| chr19 | 42219757 | 42221246 | 1490 | 16 | 4.37E-07 | 1.32E-05 | 0.000203 | 2.85E-08 | 0.002718997 | ZNF526, DEDD2, AC006486.3 |
| chr15 | 40338590 | 40340198 | 1609 | 7 | 9.74E-06 | 6.17E-08 | 9.84E-05 | 2.85E-08 | -0.00025991 | CCDC9B |
| chr8 | 138083589 | 138083967 | 379 | 4 | 5.69E-09 | 4.13E-07 | 6.63E-06 | 2.87E-08 | 0.008279764 | AC046195.1 |
| chr14 | 100486120 | 100486783 | 664 | 4 | 9.54E-08 | 7.25E-09 | 0.000356 | 2.87E-08 | 0.014478851 | WDR25 |
| chr3 | 44423710 | 44425160 | 1451 | 7 | 3.50E-06 | 7.37E-09 | 0.000487 | 2.87E-08 | 0.003386589 | LINC01988, C3orf86 |
| chr19 | 40358920 | 40360366 | 1447 | 8 | 3.95E-08 | 1.39E-07 | 0.000507 | 2.88E-08 | 0.004327897 | PLD3, AC118344.2 |
| chr3 | 127544329 | 127544804 | 476 | 3 | 5.67E-09 | 1.22E-07 | 8.87E-06 | 2.89E-08 | 0.010989498 | NA |
| chr8 | 6660763 | 6661589 | 827 | 2 | 2.66E-08 | 5.11E-08 | 3.81E-06 | 2.90E-08 | 0.027832862 | MCPH1-AS1 |
| chr10 | 45425037 | 45425744 | 708 | 2 | 4.46E-10 | 1.86E-07 | 1.22E-06 | 2.90E-08 | 0.013492414 | ALOX5 |
| chr6 | 166764125 | 166765467 | 1343 | 8 | 7.05E-08 | 8.60E-08 | 0.001306 | 2.91E-08 | 0.01389382 | RPS6KA2 |
| chr16 | 4257905 | 4258819 | 915 | 3 | 2.53E-09 | 1.24E-06 | 3.01E-06 | 2.92E-08 | 0.004190496 | TFAP4 |
| chr20 | 41357595 | 41358825 | 1231 | 5 | 3.04E-09 | 2.51E-06 | 8.13E-07 | 2.92E-08 | 0.004085518 | LPIN3 |
| chr9 | 86353716 | 86355013 | 1298 | 12 | 4.60E-08 | 6.31E-08 | 0.003521 | 2.92E-08 | 0.000534587 | TUT7 |
| chr17 | 5234597 | 5235401 | 805 | 8 | 4.79E-08 | 1.75E-07 | 0.000562 | 2.92E-08 | 0.008377834 | AC087500.1, SCIMP |
| chr5 | 181200171 | 181201689 | 1519 | 9 | 5.20E-08 | 4.33E-07 | 0.00078 | 2.92E-08 | 0.008387598 | TRIM7-AS1, TRIM7 |
| chr14 | 100949791 | 100950999 | 1209 | 4 | 1.97E-06 | 9.81E-09 | 0.000181 | 2.95E-08 | 0.013237739 | MEG8, SNORD114-1 |
| chr6 | 34555057 | 34556560 | 1504 | 9 | 4.50E-08 | 1.65E-07 | 0.001607 | 2.95E-08 | 0.016258006 | SPDEF |
| chr8 | 144103888 | 144105048 | 1161 | 12 | 2.09E-09 | 5.37E-07 | 2.26E-05 | 2.97E-08 | -0.00102509 | MAF1, SHARPIN |
| chr3 | 30631339 | 30631967 | 629 | 2 | 5.50E-08 | 1.93E-08 | 2.11E-05 | 3.00E-08 | 0.020073463 | TGFBR2 |
| chr8 | 101791009 | 101792688 | 1680 | 9 | 1.34E-05 | 3.41E-08 | 0.003285 | 3.01E-08 | 0.0107126 | NCALD |
| chr3 | 195883213 | 195884108 | 896 | 7 | 8.92E-08 | 1.11E-07 | 0.000205 | 3.02E-08 | 0.0014891 | TNK2 |
| chr1 | 2406124 | 2407299 | 1176 | 5 | 9.22E-15 | 0.000472 | 5.55E-08 | 3.03E-08 | 0.001904379 | PEX10 |
| chr11 | 120324757 | 120325796 | 1040 | 11 | 2.49E-08 | 1.21E-06 | 0.000114 | 3.05E-08 | 0.001592886 | TLCD5 |
| chr7 | 151080767 | 151083423 | 2657 | 33 | 1.65E-05 | 5.61E-07 | 0.009814 | 3.06E-08 | -2.42E-05 | FASTK, TMUB1 |
| chr6 | 119348949 | 119350733 | 1785 | 8 | 2.06E-08 | 1.02E-05 | 9.24E-06 | 3.07E-08 | 0.000682025 | AL078600.1, MAN1A1 |
| chr4 | 41128604 | 41129584 | 981 | 2 | 2.28E-06 | 1.85E-08 | 2.61E-05 | 3.07E-08 | -0.00125362 | APBB2 |
| chr13 | 52159157 | 52160297 | 1141 | 17 | 5.50E-08 | 9.60E-07 | 0.000489 | 3.12E-08 | 0.003538411 | NEK3 |
| chr5 | 135488198 | 135488926 | 729 | 5 | 2.79E-11 | 6.32E-07 | 6.29E-05 | 3.12E-08 | 0.009047881 | NA |
| chr10 | 124691076 | 124691972 | 897 | 2 | 2.06E-09 | 1.00E-07 | 2.30E-06 | 3.12E-08 | 0.010112946 | AC068896.1, FAM53B |
| chr10 | 35214948 | 35215265 | 318 | 4 | 6.11E-08 | 2.86E-08 | 3.57E-05 | 3.12E-08 | -0.00162432 | NA |
| chr19 | 18192759 | 18194204 | 1446 | 15 | 1.91E-10 | 1.59E-06 | 2.99E-05 | 3.13E-08 | 0.001307136 | MPV17L2 |
| chr1 | 83860502 | 83861173 | 672 | 10 | 2.71E-08 | 1.20E-08 | 0.001826 | 3.16E-08 | 0.005275486 | LINC01725 |
| chr19 | 55209369 | 55210453 | 1085 | 8 | 9.26E-08 | 3.26E-07 | 0.000227 | 3.16E-08 | 0.007123201 | PTPRH |
| chr17 | 82860233 | 82861542 | 1310 | 9 | 8.17E-10 | 4.87E-06 | 0.000381 | 3.17E-08 | 0.003309459 | TBCD |
| chr11 | 68437867 | 68438657 | 791 | 6 | 2.14E-07 | 2.82E-09 | 0.002387 | 3.18E-08 | 0.00294379 | LRP5 |
| chr11 | 85854516 | 85855698 | 1183 | 13 | 4.87E-08 | 1.34E-07 | 0.000776 | 3.20E-08 | 0.003815646 | CCDC83, AP000974.1 |
| chr8 | 69941752 | 69942822 | 1071 | 4 | 1.27E-06 | 1.14E-08 | 0.000338 | 3.22E-08 | -0.00787614 | NA |
| chr7 | 2216020 | 2216699 | 680 | 6 | 2.94E-09 | 4.94E-06 | 1.15E-05 | 3.24E-08 | 0.002961428 | MAD1L1, AC069288.1 |
| chr16 | 24495627 | 24496897 | 1271 | 4 | 5.26E-07 | 5.89E-08 | 0.000145 | 3.27E-08 | 0.008408995 | NA |
| chr17 | 82064942 | 82066450 | 1509 | 13 | 1.37E-08 | 7.30E-08 | 0.002977 | 3.30E-08 | -0.0015575 | DUS1L |
| chr15 | 93051555 | 93053254 | 1700 | 6 | 2.37E-08 | 1.42E-06 | 4.43E-05 | 3.31E-08 | 0.012515801 | RGMA |
| chr10 | 117196512 | 117197748 | 1237 | 12 | 2.63E-07 | 1.03E-07 | 0.001609 | 3.31E-08 | 0.004132112 | KCNK18, RPL12P26 |
| chr1 | 36150244 | 36151244 | 1001 | 5 | 3.22E-09 | 9.42E-08 | 2.69E-05 | 3.31E-08 | 0.00949766 | TRAPPC3 |
| chr6 | 154038423 | 154039597 | 1175 | 13 | 2.08E-09 | 2.19E-06 | 0.000106 | 3.32E-08 | 0.007678023 | OPRM1 |
| chr8 | 140598217 | 140598547 | 331 | 4 | 8.70E-08 | 5.30E-09 | 0.001031 | 3.32E-08 | 0.013746193 | AGO2 |
| chr11 | 69210678 | 69210705 | 28 | 2 | 7.95E-08 | 1.84E-08 | 4.01E-05 | 3.33E-08 | 0.020423013 | NA |
| chr1 | 161367610 | 161368597 | 988 | 5 | 1.74E-09 | 2.92E-07 | 8.62E-06 | 3.34E-08 | 0.011382206 | AL592295.6, CFAP126 |
| chr1 | 9320025 | 9320741 | 717 | 4 | 2.30E-09 | 1.44E-06 | 1.02E-05 | 3.37E-08 | 0.009884225 | SPSB1 |
| chr1 | 3247146 | 3248627 | 1482 | 8 | 5.37E-10 | 2.54E-06 | 0.00016 | 3.37E-08 | 0.010616469 | PRDM16 |
| chr11 | 22828569 | 22830359 | 1791 | 17 | 4.02E-06 | 2.00E-07 | 0.003419 | 3.38E-08 | 0.002976346 | AC006299.1, SVIP |
| chr11 | 134358333 | 134358898 | 566 | 3 | 3.90E-09 | 1.09E-07 | 1.33E-05 | 3.38E-08 | 0.001578082 | GLB1L2 |
| chr21 | 39611682 | 39612853 | 1172 | 11 | 8.28E-09 | 1.57E-07 | 0.001648 | 3.38E-08 | 0.005962489 | B3GALT5, B3GALT5-AS1 |
| chr6 | 154731670 | 154733945 | 2276 | 11 | 2.13E-06 | 4.37E-07 | 0.001437 | 3.40E-08 | 0.002235846 | SCAF8 |
| chr2 | 231900464 | 231901044 | 581 | 5 | 2.16E-08 | 5.92E-07 | 9.77E-05 | 3.40E-08 | 0.01066221 | NA |
| chr6 | 132763150 | 132764066 | 917 | 4 | 1.77E-07 | 9.96E-09 | 0.000252 | 3.42E-08 | 0.014169547 | VNN2 |
| chr15 | 73767841 | 73767974 | 134 | 3 | 2.30E-08 | 1.06E-06 | 1.60E-06 | 3.42E-08 | 0.016446211 | INSYN1-AS1 |
| chr1 | 150558055 | 150559869 | 1815 | 8 | 1.15E-06 | 3.88E-05 | 0.000468 | 3.43E-08 | 0.003017047 | ADAMTSL4 |
| chr11 | 67371401 | 67372436 | 1036 | 5 | 1.33E-08 | 4.31E-08 | 0.000145 | 3.43E-08 | 0.008666318 | RAD9A, AP003419.1, CLCF1 |
| chr16 | 1379050 | 1380366 | 1317 | 10 | 1.41E-07 | 6.15E-07 | 8.43E-05 | 3.43E-08 | 0.005873612 | UNKL |
| chr4 | 54226717 | 54228413 | 1697 | 12 | 4.05E-08 | 1.46E-05 | 0.000165 | 3.44E-08 | 0.001507573 | AC058822.1 |
| chr8 | 81111172 | 81112826 | 1655 | 11 | 7.25E-07 | 2.30E-07 | 0.000482 | 3.44E-08 | 0.003428627 | PAG1 |
| chr5 | 141507735 | 141509267 | 1533 | 5 | 9.14E-07 | 7.95E-09 | 0.000767 | 3.44E-08 | 0.017460936 | PCDHGA1, PCDHGA2, PCDHGA3, PCDHGB1, PCDHGA4, PCDHGB2, PCDHGA5, PCDHGB3, PCDHGA6, PCDHGA7, PCDHGB4, PCDHGA8, PCDHGB5, PCDHGA9, PCDHGB6, PCDHGA10, PCDHGB7, PCDHGA11, PCDHGA12, PCDHGC3, PCDHGC4, PCDHGC5 |
| chr13 | 27822985 | 27823222 | 238 | 4 | 1.25E-08 | 4.30E-07 | 4.88E-05 | 3.46E-08 | 0.010919507 | PLUT |
| chr17 | 74361853 | 74363051 | 1199 | 8 | 1.27E-08 | 3.18E-08 | 0.000615 | 3.46E-08 | 0.009381207 | BTBD17 |
| chr8 | 1899833 | 1900448 | 616 | 6 | 2.83E-08 | 1.54E-07 | 6.02E-05 | 3.46E-08 | 0.011829196 | AC019257.8, ARHGEF10 |
| chr1 | 203303675 | 203304565 | 891 | 4 | 3.33E-09 | 2.91E-08 | 3.39E-05 | 3.47E-08 | 0.008452948 | LINC01136 |
| chr6 | 11805591 | 11807190 | 1600 | 6 | 2.00E-09 | 2.55E-07 | 0.000127 | 3.50E-08 | 0.006686758 | ADTRP |
| chr7 | 2039015 | 2040365 | 1351 | 8 | 7.31E-08 | 6.50E-06 | 6.21E-05 | 3.51E-08 | 0.003398948 | MAD1L1, AC069288.1 |
| chr6 | 169765589 | 169766421 | 833 | 3 | 1.37E-07 | 1.86E-08 | 8.04E-05 | 3.53E-08 | 0.006811144 | ERMARD |
| chr3 | 47513610 | 47514389 | 780 | 9 | 9.06E-10 | 1.65E-05 | 0.000197 | 3.56E-08 | 0.000599372 | ELP6 |
| chr3 | 129121776 | 129123207 | 1432 | 19 | 6.85E-08 | 3.28E-07 | 0.001128 | 3.56E-08 | 0.000397247 | RAB43, ISY1-RAB43 |
| chr20 | 8305544 | 8306589 | 1046 | 4 | 1.54E-06 | 7.77E-09 | 0.000651 | 3.56E-08 | 0.003645067 | PLCB1 |
| chr22 | 19722025 | 19723189 | 1165 | 11 | 7.09E-10 | 3.34E-07 | 0.000135 | 3.56E-08 | 0.008562025 | SEPTIN5, AC000093.1 |
| chr7 | 121312954 | 121314495 | 1542 | 10 | 5.41E-07 | 2.29E-08 | 0.001079 | 3.56E-08 | 0.009160655 | NA |
| chr16 | 70646201 | 70646914 | 714 | 8 | 2.03E-08 | 1.60E-06 | 8.30E-05 | 3.57E-08 | 0.00376971 | IL34 |
| chr1 | 161177188 | 161178569 | 1382 | 17 | 7.65E-08 | 6.53E-07 | 0.000879 | 3.59E-08 | 0.001510375 | PPOX, B4GALT3 |
| chr17 | 66374433 | 66375182 | 750 | 3 | 8.46E-09 | 8.74E-08 | 4.48E-05 | 3.59E-08 | 0.029265074 | PRKCA |
| chr10 | 34739970 | 34740109 | 140 | 3 | 7.03E-08 | 5.84E-08 | 1.71E-05 | 3.61E-08 | 0.018286792 | PARD3 |
| chr6 | 6006474 | 6008148 | 1675 | 19 | 8.59E-07 | 2.96E-07 | 0.004551 | 3.61E-08 | 0.002242629 | NRN1 |
| chr20 | 46689532 | 46690711 | 1180 | 12 | 1.94E-07 | 2.19E-08 | 0.001694 | 3.62E-08 | 0.003961906 | AL031055.1 |
| chr10 | 132776335 | 132777268 | 934 | 6 | 4.19E-09 | 1.36E-07 | 3.39E-05 | 3.63E-08 | 0.003582319 | INPP5A |
| chr1 | 145691388 | 145692988 | 1601 | 5 | 8.60E-09 | 2.15E-07 | 0.000159 | 3.64E-08 | 0.003347877 | PDZK1 |
| chr9 | 136944911 | 136946377 | 1467 | 15 | 1.45E-09 | 8.68E-06 | 1.99E-05 | 3.69E-08 | 0.003137695 | C8G |
| chr19 | 34327904 | 34328150 | 247 | 2 | 3.74E-08 | 5.65E-08 | 5.21E-06 | 3.73E-08 | 0.003502238 | GARRE1 |
| chr10 | 86264772 | 86265885 | 1114 | 8 | 5.37E-07 | 4.61E-08 | 0.000255 | 3.73E-08 | 0.006457561 | GRID1, MIR346 |
| chr2 | 16664144 | 16665301 | 1158 | 4 | 2.03E-06 | 5.60E-08 | 0.000141 | 3.73E-08 | 0.015695282 | CYRIA |
| chr3 | 119240332 | 119241493 | 1162 | 12 | 5.39E-07 | 4.10E-09 | 0.001934 | 3.74E-08 | 0.00321275 | B4GALT4-AS1, B4GALT4 |
| chr7 | 27179725 | 27181364 | 1640 | 11 | 2.12E-06 | 2.51E-09 | 0.002738 | 3.74E-08 | 0.003901647 | AC004080.3, HOXA10, HOXA11 |
| chr7 | 5607196 | 5608491 | 1296 | 8 | 9.10E-08 | 2.48E-07 | 4.18E-05 | 3.75E-08 | 0.003029793 | NA |
| chr5 | 154472994 | 154474238 | 1245 | 10 | 1.44E-08 | 4.31E-08 | 0.000602 | 3.76E-08 | 0.006085169 | NA |
| chr2 | 11784799 | 11785249 | 451 | 5 | 5.74E-08 | 4.74E-08 | 0.00018 | 3.76E-08 | 0.005285712 | LPIN1 |
| chr21 | 42684840 | 42685364 | 525 | 3 | 1.04E-09 | 1.41E-07 | 5.30E-05 | 3.78E-08 | -0.03377322 | PDE9A |
| chr16 | 57357743 | 57358706 | 964 | 8 | 1.52E-07 | 1.11E-08 | 0.003133 | 3.81E-08 | 0.008996119 | NA |
| chr14 | 93995129 | 93996261 | 1133 | 7 | 2.83E-08 | 1.10E-07 | 0.000138 | 3.82E-08 | 0.012226324 | CCDC197 |
| chr3 | 39807865 | 39809293 | 1429 | 14 | 4.32E-08 | 1.00E-07 | 0.004031 | 3.82E-08 | 0.006906877 | MYRIP |
| chr6 | 31682958 | 31683899 | 942 | 18 | 3.51E-07 | 2.08E-09 | 0.031399 | 3.83E-08 | 0.024938881 | LY6G5C |
| chr1 | 44244295 | 44245401 | 1107 | 4 | 1.24E-07 | 1.32E-07 | 0.000219 | 3.83E-08 | 0.005214135 | ERI3 |
| chr5 | 134524021 | 134525751 | 1731 | 16 | 3.95E-07 | 2.71E-08 | 0.003575 | 3.83E-08 | 0.001223925 | JADE2 |
| chr11 | 113875000 | 113875743 | 744 | 6 | 9.25E-11 | 1.34E-06 | 4.58E-06 | 3.83E-08 | -0.00208937 | USP28 |
| chr7 | 369667 | 372038 | 2372 | 15 | 4.41E-06 | 6.79E-06 | 0.000496 | 3.86E-08 | 0.001739929 | NA |
| chr6 | 39791077 | 39792640 | 1564 | 11 | 1.85E-08 | 5.83E-07 | 1.24E-05 | 3.87E-08 | 0.004915048 | DAAM2 |
| chr8 | 85106666 | 85108072 | 1407 | 16 | 7.92E-07 | 3.31E-06 | 0.000184 | 3.87E-08 | -0.00140053 | LRRCC1 |
| chr14 | 103099530 | 103100251 | 722 | 8 | 7.84E-09 | 3.26E-06 | 3.48E-05 | 3.90E-08 | 0.004387452 | EXOC3L4 |
| chr11 | 368351 | 369810 | 1460 | 21 | 8.40E-08 | 3.32E-06 | 0.001906 | 3.90E-08 | 0.004532312 | B4GALNT4 |
| chr1 | 99849534 | 99851944 | 2411 | 26 | 2.93E-05 | 8.56E-07 | 0.002299 | 3.92E-08 | 0.000909359 | AGL |
| chr11 | 117262705 | 117263737 | 1033 | 4 | 1.03E-06 | 2.47E-08 | 0.000116 | 3.95E-08 | 0.007153768 | RNF214 |
| chr22 | 49425449 | 49425620 | 172 | 2 | 9.94E-08 | 2.13E-08 | 5.26E-05 | 3.95E-08 | 0.020360613 | C22orf34 |
| chr9 | 123851660 | 123851911 | 252 | 3 | 6.46E-09 | 0.00053 | 1.06E-06 | 3.97E-08 | 0.004940347 | DENND1A |
| chr9 | 94639223 | 94640847 | 1625 | 12 | 1.21E-07 | 3.89E-07 | 0.001119 | 3.98E-08 | 0.004077645 | FBP1 |
| chr9 | 77013572 | 77015389 | 1818 | 14 | 7.72E-07 | 3.52E-07 | 0.000459 | 3.99E-08 | 0.005901449 | NA |
| chr8 | 143633776 | 143636355 | 2580 | 17 | 3.66E-05 | 1.21E-06 | 0.000777 | 4.03E-08 | -0.00086757 | ZNF623 |
| chr1 | 201739291 | 201740262 | 972 | 13 | 1.94E-08 | 3.74E-07 | 0.00227 | 4.05E-08 | 0.009074579 | NAV1, IPO9-AS1 |
| chr10 | 28531438 | 28533153 | 1716 | 15 | 1.73E-06 | 6.37E-07 | 0.002684 | 4.09E-08 | -0.00204571 | WAC, WAC-AS1 |
| chr12 | 123930713 | 123932119 | 1407 | 7 | 5.26E-06 | 7.83E-08 | 0.000581 | 4.09E-08 | 0.005869185 | DNAH10, CCDC92, DNAH10OS |
| chr21 | 46037824 | 46039419 | 1596 | 4 | 9.89E-06 | 5.06E-08 | 7.44E-05 | 4.09E-08 | 0.004928627 | AP001476.1 |
| chr2 | 162768556 | 162768824 | 269 | 4 | 1.23E-08 | 3.91E-05 | 1.99E-05 | 4.10E-08 | 0.016691261 | KCNH7 |
| chr10 | 683286 | 684016 | 731 | 7 | 2.02E-08 | 1.11E-07 | 0.000373 | 4.11E-08 | 0.010109855 | DIP2C |
| chr7 | 158165879 | 158166861 | 983 | 4 | 1.42E-10 | 1.12E-05 | 7.32E-06 | 4.13E-08 | 0.004904286 | PTPRN2 |
| chr6 | 56247014 | 56248236 | 1223 | 12 | 3.72E-09 | 1.88E-06 | 6.86E-05 | 4.13E-08 | 0.005518788 | COL21A1 |
| chr6 | 83430331 | 83431828 | 1498 | 13 | 1.63E-07 | 1.84E-07 | 0.000427 | 4.16E-08 | 0.005954616 | ME1 |
| chr20 | 56466001 | 56466489 | 489 | 3 | 6.27E-08 | 2.60E-08 | 3.88E-05 | 4.17E-08 | 0.020775586 | NA |
| chr6 | 46921887 | 46922675 | 789 | 7 | 1.87E-08 | 7.81E-07 | 0.000505 | 4.22E-08 | 0.0034017 | ADGRF5 |
| chr1 | 183185643 | 183186928 | 1286 | 11 | 5.41E-08 | 1.03E-06 | 0.000588 | 4.22E-08 | 0.004895698 | LAMC2 |
| chr12 | 7095651 | 7097614 | 1964 | 7 | 1.80E-06 | 1.91E-06 | 0.000399 | 4.22E-08 | 0.005965451 | C1RL |
| chr5 | 180807518 | 180807984 | 467 | 2 | 6.52E-09 | 1.87E-07 | 2.03E-06 | 4.25E-08 | 0.008444894 | MGAT1 |
| chr9 | 121822182 | 121822195 | 14 | 2 | 4.05E-08 | 2.41E-08 | 4.24E-05 | 4.25E-08 | 0.015338235 | TTLL11 |
| chr12 | 67729521 | 67730541 | 1021 | 5 | 1.60E-09 | 8.94E-07 | 1.74E-05 | 4.26E-08 | 0.001431589 | NA |
| chr5 | 177609765 | 177609886 | 122 | 2 | 5.58E-09 | 3.29E-08 | 1.59E-05 | 4.26E-08 | 0.022595074 | B4GALT7 |
| chr17 | 74882914 | 74883762 | 849 | 5 | 3.98E-08 | 4.01E-08 | 0.00027 | 4.26E-08 | 0.009355067 | FADS6 |
| chr11 | 113386896 | 113388186 | 1291 | 10 | 8.08E-08 | 1.34E-06 | 0.000476 | 4.31E-08 | 0.006792325 | ANKK1 |
| chr1 | 151006883 | 151008615 | 1733 | 16 | 3.10E-09 | 3.04E-08 | 0.007241 | 4.33E-08 | 0.002749869 | PRUNE1, MINDY1 |
| chr16 | 14308300 | 14309568 | 1269 | 9 | 3.39E-06 | 7.82E-08 | 0.003573 | 4.34E-08 | -0.00992843 | MIR193BHG, MIR365A |
| chr11 | 120475847 | 120476802 | 956 | 2 | 3.37E-06 | 2.58E-08 | 3.51E-05 | 4.37E-08 | 0.015246459 | ARHGEF12 |
| chr1 | 7062481 | 7063255 | 775 | 5 | 2.20E-09 | 2.93E-05 | 1.67E-05 | 4.39E-08 | -0.00772243 | CAMTA1 |
| chr17 | 10858083 | 10858873 | 791 | 4 | 4.06E-08 | 3.78E-08 | 0.000152 | 4.41E-08 | 0.039719656 | AC015908.7, AC015908.2 |
| chr3 | 42702252 | 42703170 | 919 | 7 | 6.75E-09 | 2.14E-07 | 0.000785 | 4.42E-08 | 0.009516457 | HHATL-AS1, HHATL, AC006059.3 |
| chr12 | 94147689 | 94150171 | 2483 | 11 | 4.74E-06 | 3.82E-07 | 0.000225 | 4.42E-08 | 0.001167971 | PLXNC1 |
| chr1 | 22051919 | 22053900 | 1982 | 12 | 1.37E-05 | 8.62E-06 | 0.001083 | 4.44E-08 | 0.001012358 | CDC42, CDC42-AS1 |
| chr15 | 88836177 | 88836458 | 282 | 2 | 3.30E-09 | 4.17E-06 | 3.86E-07 | 4.45E-08 | 0.005982436 | ACAN |
| chr19 | 35509615 | 35511151 | 1537 | 16 | 5.18E-08 | 1.66E-07 | 7.45E-05 | 4.45E-08 | 0.004821736 | DMKN |
| chr6 | 29658214 | 29660068 | 1855 | 10 | 1.78E-08 | 9.44E-06 | 4.52E-05 | 4.48E-08 | 0.003933506 | MOG |
| chr1 | 107055604 | 107057469 | 1866 | 10 | 2.46E-05 | 1.77E-07 | 0.000142 | 4.49E-08 | 0.00186083 | PRMT6 |
| chr7 | 1603662 | 1604807 | 1146 | 4 | 1.30E-07 | 2.91E-08 | 0.000121 | 4.50E-08 | 0.008051176 | NA |
| chr11 | 58575845 | 58578244 | 2400 | 12 | 9.62E-08 | 3.52E-06 | 4.53E-05 | 4.50E-08 | 0.003338894 | LPXN |
| chr14 | 89493230 | 89493321 | 92 | 2 | 8.25E-08 | 3.32E-08 | 1.78E-05 | 4.51E-08 | 0.013270688 | FOXN3, AL137230.2 |
| chr5 | 179797562 | 179799186 | 1625 | 10 | 1.94E-06 | 9.83E-06 | 0.000293 | 4.51E-08 | 0.002522559 | MGAT4B, MIR1229 |
| chr20 | 32744008 | 32745262 | 1255 | 9 | 1.30E-06 | 2.77E-08 | 0.004201 | 4.51E-08 | 0.006202299 | NA |
| chr20 | 21390909 | 21392225 | 1317 | 8 | 1.54E-08 | 1.34E-07 | 0.001033 | 4.51E-08 | 0.009569015 | NA |
| chr17 | 57744405 | 57745490 | 1086 | 7 | 1.14E-08 | 6.03E-07 | 4.81E-05 | 4.52E-08 | 0.006222985 | CCDC182 |
| chr17 | 39626049 | 39627771 | 1723 | 13 | 1.04E-05 | 1.65E-06 | 0.001881 | 4.53E-08 | 0.006749699 | PPP1R1B |
| chr3 | 13647641 | 13649720 | 2080 | 7 | 8.88E-07 | 3.23E-06 | 8.29E-05 | 4.54E-08 | 0.00451328 | NA |
| chr16 | 1970989 | 1972306 | 1318 | 11 | 1.96E-06 | 1.23E-09 | 0.011228 | 4.54E-08 | 0.000640467 | TBL3, AC005363.2 |
| chr21 | 45607206 | 45607381 | 176 | 4 | 5.88E-08 | 6.03E-07 | 2.83E-05 | 4.58E-08 | 0.01341119 | AL133493.1 |
| chr3 | 50322152 | 50323259 | 1108 | 9 | 1.60E-08 | 1.05E-06 | 0.000989 | 4.61E-08 | 0.007005867 | HYAL2, TUSC2 |
| chr17 | 10064052 | 10064182 | 131 | 4 | 1.60E-07 | 3.16E-08 | 7.67E-05 | 4.62E-08 | 0.01662088 | GAS7 |
| chr10 | 133445075 | 133446850 | 1776 | 8 | 1.35E-07 | 2.77E-08 | 0.000607 | 4.64E-08 | -0.01797831 | NA |
| chr8 | 92877683 | 92878038 | 356 | 3 | 3.57E-10 | 0.008786 | 6.79E-08 | 4.64E-08 | 0.004210502 | FLJ46284 |
| chr20 | 36064034 | 36064994 | 961 | 6 | 1.42E-08 | 5.67E-08 | 0.000288 | 4.66E-08 | 0.005561129 | AL035420.1, AL035420.3 |

| | | | | | | | | | | |
|---|---|---|---|---|---|---|---|---|---|---|
| chr5 | 138273992 | 138275807 | 1816 | 14 | 7.07E-08 | 1.61E-07 | 0.003144 | 4.70E-08 | 0.002947941 | GFRA3 |
| chr8 | 102124471 | 102125757 | 1287 | 12 | 2.07E-07 | 9.99E-08 | 0.001614 | 4.70E-08 | 0.005033018 | MIR5680, NCALD |
| chr1 | 152718649 | 152719282 | 634 | 5 | 2.18E-08 | 2.10E-07 | 1.04E-05 | 4.72E-08 | 0.004456585 | NA |
| chr17 | 7444194 | 7445878 | 1685 | 14 | 1.08E-08 | 8.98E-07 | 9.99E-05 | 4.74E-08 | 0.004059934 | FGF11, AC113189.4, CHRNB1 |
| chr22 | 38951302 | 38952634 | 1333 | 5 | 1.85E-07 | 3.36E-07 | 0.000134 | 4.74E-08 | 0.007003305 | NA |
| chr4 | 153257112 | 153258249 | 1138 | 4 | 1.62E-06 | 2.85E-08 | 4.80E-05 | 4.74E-08 | 0.012959321 | TRIM2, AC013477.1 |
| chr15 | 75200641 | 75202069 | 1429 | 11 | 3.32E-07 | 3.57E-06 | 0.000626 | 4.77E-08 | 0.001180659 | C15orf39 |
| chr5 | 88883193 | 88884278 | 1086 | 18 | 3.95E-09 | 1.41E-05 | 3.12E-05 | 4.78E-08 | -0.00085524 | MEF2C-AS1, MEF2C |
| chr15 | 69730650 | 69731577 | 928 | 5 | 4.87E-08 | 1.65E-07 | 3.81E-05 | 4.79E-08 | 0.004421852 | DRAIC |
| chr10 | 48305986 | 48307331 | 1346 | 9 | 4.65E-08 | 9.59E-06 | 4.14E-05 | 4.79E-08 | 0.002177432 | MAPK8 |
| chr14 | 22518119 | 22518764 | 646 | 5 | 3.11E-09 | 2.17E-06 | 3.98E-05 | 4.82E-08 | 0.00724119 | TRAJ26 |
| chr1 | 235650709 | 235651646 | 938 | 13 | 2.09E-12 | 1.21E-05 | 1.54E-05 | 4.83E-08 | 0.008860658 | GNG4 |
| chr9 | 135106359 | 135107124 | 766 | 3 | 8.30E-09 | 1.74E-07 | 5.65E-05 | 4.83E-08 | 0.020410661 | OLFM1 |
| chr17 | 18107399 | 18108820 | 1422 | 11 | 1.37E-05 | 1.18E-07 | 0.000821 | 4.83E-08 | 0.00233895 | DRG2, MYO15A, AC087164.2 |
| chr17 | 864140 | 864902 | 763 | 6 | 9.99E-08 | 2.19E-07 | 0.000121 | 4.83E-08 | -0.00604407 | NXN |
| chr3 | 16932143 | 16933189 | 1047 | 5 | 9.87E-08 | 1.16E-08 | 0.000479 | 4.88E-08 | 0.02100445 | PLCL2 |
| chr15 | 79412100 | 79413314 | 1215 | 5 | 4.59E-09 | 1.40E-06 | 1.31E-05 | 4.88E-08 | 0.00620779 | TMED3 |
| chr21 | 36746548 | 36748523 | 1976 | 8 | 1.85E-06 | 2.84E-07 | 0.00017 | 4.88E-08 | 0.003086647 | SIM2 |
| chr19 | 49073698 | 49074046 | 349 | 4 | 5.76E-08 | 1.53E-08 | 0.000467 | 4.89E-08 | 0.017307337 | NA |
| chr1 | 25544331 | 25545323 | 993 | 4 | 1.05E-08 | 5.60E-08 | 2.94E-05 | 4.89E-08 | 0.00971502 | LDLRAP1 |
| chr11 | 96343105 | 96343751 | 647 | 12 | 1.10E-07 | 2.85E-07 | 6.06E-05 | 4.93E-08 | 0.002547721 | MAML2 |
| chr9 | 31304136 | 31304806 | 671 | 3 | 1.70E-07 | 2.05E-08 | 8.46E-05 | 4.93E-08 | 0.038053258 | NA |
| chr6 | 168055916 | 168057621 | 1706 | 4 | 1.87E-06 | 1.13E-08 | 0.000315 | 4.94E-08 | 0.004036027 | FRMD1 |
| chr1 | 117210691 | 117211871 | 1181 | 11 | 1.25E-06 | 3.65E-07 | 0.001144 | 4.94E-08 | 0.002362232 | VTCN1 |
| chr22 | 23803123 | 23803331 | 209 | 2 | 6.91E-08 | 6.10E-08 | 7.78E-06 | 4.94E-08 | -0.00085026 | SMARCB1 |
| chr20 | 3237499 | 3238259 | 761 | 9 | 4.52E-08 | 3.58E-07 | 5.73E-05 | 4.95E-08 | 0.010482257 | SLC4A11 |
| chr15 | 29135455 | 29137022 | 1568 | 5 | 2.00E-05 | 9.00E-07 | 0.000345 | 4.98E-08 | 0.004092465 | FAM189A1 |
| chr3 | 50204670 | 50205321 | 652 | 7 | 1.58E-09 | 9.34E-08 | 0.000308 | 4.99E-08 | 0.010271691 | SLC38A3 |
| chr3 | 13088179 | 13088319 | 141 | 3 | 2.00E-07 | 1.66E-08 | 0.000172 | 5.05E-08 | 0.017431655 | IQSEC1 |
| chr8 | 29341299 | 29342069 | 771 | 4 | 2.68E-08 | 8.53E-08 | 6.74E-05 | 5.08E-08 | 0.014418376 | DUSP4 |
| chr11 | 72821846 | 72822619 | 774 | 5 | 1.90E-07 | 6.41E-09 | 0.001776 | 5.09E-08 | -0.02930959 | ATG16L2 |
| chr12 | 3456510 | 3457010 | 501 | 5 | 1.19E-08 | 9.72E-07 | 0.000132 | 5.11E-08 | 0.011833932 | PRMT8 |
| chr17 | 81039911 | 81040410 | 500 | 3 | 5.01E-08 | 4.97E-08 | 8.74E-05 | 5.14E-08 | 0.01306699 | BAIAP2 |
| chr8 | 142742316 | 142742825 | 510 | 7 | 2.11E-08 | 1.90E-07 | 0.000104 | 5.16E-08 | 0.004828231 | SLURP1 |
| chr1 | 38093699 | 38094020 | 322 | 3 | 1.22E-08 | 1.48E-07 | 6.62E-06 | 5.20E-08 | 0.020709768 | MIR3659HG |
| chr5 | 173237992 | 173238900 | 909 | 8 | 1.13E-07 | 3.13E-07 | 0.000345 | 5.21E-08 | 0.007594996 | NA |
| chr2 | 106269470 | 106271109 | 1640 | 6 | 1.09E-06 | 8.73E-07 | 0.00027 | 5.24E-08 | 0.0193523 | NA |
| chr17 | 1188121 | 1188994 | 874 | 5 | 1.37E-07 | 3.47E-08 | 0.000525 | 5.25E-08 | 0.007540385 | ABR |
| chr15 | 60678775 | 60679721 | 947 | 2 | 2.92E-06 | 2.82E-08 | 5.85E-05 | 5.26E-08 | 0.011605342 | RORA |
| chr7 | 99011977 | 99013545 | 1569 | 10 | 1.94E-07 | 4.95E-06 | 5.40E-05 | 5.29E-08 | 0.000906085 | TRRAP |
| chr6 | 91691768 | 91691827 | 60 | 3 | 1.34E-07 | 3.54E-08 | 0.000109 | 5.29E-08 | 0.007466709 | NA |
| chr13 | 30932133 | 30933441 | 1309 | 14 | 1.32E-07 | 1.43E-08 | 0.005499 | 5.31E-08 | 0.013523273 | TEX26, TEX26-AS1 |
| chr15 | 72756003 | 72756805 | 803 | 3 | 1.72E-07 | 4.78E-06 | 3.74E-06 | 5.31E-08 | 0.006072029 | ADPGK |
| chr12 | 117155454 | 117155923 | 470 | 6 | 1.73E-07 | 3.25E-08 | 0.000111 | 5.31E-08 | 0.006907779 | FBXO21 |
| chr2 | 104843805 | 104845338 | 1534 | 7 | 1.49E-07 | 1.41E-06 | 0.000376 | 5.31E-08 | 0.000370896 | PANTR1 |
| chr2 | 239065864 | 239067436 | 1573 | 10 | 1.39E-08 | 7.01E-06 | 8.16E-05 | 5.32E-08 | 0.001242771 | HDAC4 |
| chr11 | 31509231 | 31510212 | 982 | 15 | 1.04E-08 | 3.29E-07 | 0.00031 | 5.32E-08 | 0.000914726 | ELP4, IMMP1L |
| chr10 | 43208975 | 43210556 | 1582 | 6 | 1.02E-07 | 2.16E-07 | 0.000186 | 5.35E-08 | 0.006150567 | RASGEF1A |
| chr1 | 206612062 | 206612827 | 766 | 6 | 9.98E-08 | 1.28E-07 | 0.000425 | 5.36E-08 | -0.01729294 | EIF2D |
| chr8 | 23705405 | 23707661 | 2257 | 21 | 2.74E-07 | 2.62E-07 | 0.001694 | 5.37E-08 | 0.003057568 | AC012574.1, NKX2-6 |
| chr1 | 119896660 | 119897607 | 948 | 7 | 9.05E-09 | 7.78E-07 | 1.67E-05 | 5.38E-08 | -0.00046092 | NA |
| chr6 | 157892146 | 157893454 | 1309 | 6 | 2.06E-06 | 8.41E-09 | 0.001705 | 5.39E-08 | 0.001329339 | SNX9, SNX9-AS1 |
| chr20 | 63894130 | 63894935 | 806 | 5 | 5.36E-08 | 2.64E-08 | 0.000798 | 5.41E-08 | 0.00843546 | NA |
| chr5 | 77643508 | 77644624 | 1117 | 7 | 5.02E-07 | 2.71E-06 | 0.000262 | 5.41E-08 | 0.006366939 | NA |
| chr7 | 529781 | 531160 | 1380 | 7 | 1.05E-07 | 1.30E-06 | 8.33E-06 | 5.44E-08 | 0.005209208 | NA |
| chr4 | 56387098 | 56388027 | 930 | 14 | 4.22E-10 | 2.40E-05 | 2.51E-05 | 5.49E-08 | -0.00075794 | AC068620.1, AASDH |
| chr8 | 123041544 | 123042906 | 1363 | 15 | 1.07E-07 | 6.40E-08 | 0.001872 | 5.51E-08 | 0.000754017 | TBC1D31, DERL1 |
| chr19 | 535548 | 536244 | 697 | 2 | 1.61E-09 | 1.18E-07 | 4.19E-06 | 5.53E-08 | 0.008104515 | CDC34 |
| chr8 | 143722570 | 143723570 | 1001 | 4 | 4.48E-09 | 1.10E-06 | 1.76E-05 | 5.53E-08 | 0.009015665 | NA |
| chr1 | 16119443 | 16120207 | 765 | 3 | 3.61E-07 | 5.61E-07 | 1.73E-05 | 5.54E-08 | 0.005281269 | NA |
| chr16 | 67881335 | 67882044 | 710 | 3 | 8.86E-08 | 4.21E-07 | 3.80E-05 | 5.55E-08 | 0.005729214 | EDC4 |
| chr14 | 22815309 | 22816156 | 848 | 5 | 1.12E-06 | 5.51E-08 | 0.000171 | 5.59E-08 | 0.012044905 | SLC7A7 |
| chr14 | 81220437 | 81222140 | 1704 | 22 | 1.37E-05 | 3.72E-08 | 0.009322 | 5.59E-08 | 0.002741771 | AL136040.1, GTF2A1 |
| chr1 | 109306923 | 109308215 | 1293 | 8 | 1.93E-07 | 1.26E-06 | 0.000314 | 5.65E-08 | 0.00515414 | MYBPHL |
| chr20 | 11001812 | 11002260 | 449 | 2 | 5.85E-09 | 5.92E-07 | 1.21E-06 | 5.65E-08 | 0.019897652 | LINC02871 |
| chr1 | 147517645 | 147518295 | 651 | 3 | 7.55E-09 | 4.65E-07 | 2.90E-06 | 5.69E-08 | 0.008459395 | LINC00624 |
| chr9 | 109167267 | 109167917 | 651 | 8 | 5.38E-09 | 4.24E-06 | 0.00011 | 5.69E-08 | 0.004427796 | NA |
| chr17 | 28380457 | 28381754 | 1298 | 4 | 7.32E-06 | 2.88E-08 | 0.000415 | 5.72E-08 | 0.016270969 | AC002094.1, SARM1 |
| chr17 | 82238843 | 82240022 | 1180 | 9 | 1.02E-06 | 1.03E-07 | 0.000811 | 5.72E-08 | 0.007465047 | SLC16A3, CSNK1D |
| chr3 | 184261303 | 184262307 | 1005 | 6 | 2.63E-08 | 2.76E-07 | 6.29E-05 | 5.73E-08 | 0.002775513 | EEF1AKMT4-ECE2, CAMK2N2 |
| chr1 | 151837715 | 151839144 | 1430 | 9 | 9.23E-07 | 1.04E-06 | 0.001141 | 5.75E-08 | 0.003978018 | C2CD4D |
| chr11 | 64566745 | 64568423 | 1679 | 6 | 6.00E-07 | 5.37E-06 | 6.63E-05 | 5.78E-08 | 0.011137484 | SLC22A11 |
| chr12 | 57736072 | 57738331 | 2260 | 12 | 1.88E-06 | 2.49E-05 | 1.61E-05 | 5.79E-08 | 0.000249785 | TSPAN31, AGAP2 |
| chr1 | 29181348 | 29182999 | 1652 | 16 | 6.06E-07 | 9.80E-09 | 0.000279 | 5.81E-08 | -0.00106996 | SRSF4 |
| chr19 | 5787454 | 5787855 | 402 | 2 | 5.23E-09 | 8.53E-07 | 1.05E-06 | 5.87E-08 | 0.003675853 | DUS3L |
| chr6 | 33297057 | 33300021 | 2965 | 54 | 4.38E-05 | 1.33E-06 | 0.011228 | 5.92E-08 | 0.000754563 | PFDN6, RGL2, TAPBP |
| chr3 | 52063562 | 52065275 | 1714 | 6 | 2.46E-09 | 3.32E-06 | 4.19E-05 | 5.92E-08 | 0.005465151 | NA |
| chr11 | 119491957 | 119493370 | 1414 | 4 | 5.05E-07 | 5.14E-08 | 0.000153 | 5.93E-08 | 0.00743596 | USP2-AS1 |
| chr22 | 46107566 | 46110305 | 2740 | 13 | 4.53E-07 | 6.15E-07 | 3.88E-05 | 5.96E-08 | 0.004406101 | MIRLET7BHG |
| chr12 | 126425097 | 126425942 | 846 | 3 | 1.01E-06 | 2.93E-08 | 0.000124 | 5.97E-08 | -0.00893652 | AC007368.1 |
| chr1 | 2526787 | 2527785 | 999 | 10 | 1.08E-08 | 6.82E-07 | 0.001087 | 5.99E-08 | 0.003636173 | NA |
| chr1 | 37989664 | 37990859 | 1196 | 15 | 6.87E-08 | 9.20E-06 | 0.001791 | 5.99E-08 | -0.0009312 | SF3A3 |
| chr6 | 24495036 | 24495761 | 726 | 6 | 1.27E-08 | 3.07E-07 | 2.11E-05 | 6.01E-08 | 0.000306942 | ALDH5A1, GPLD1 |
| chr3 | 196503786 | 196505235 | 1450 | 8 | 2.19E-05 | 3.40E-08 | 0.002479 | 6.09E-08 | 0.002075517 | NA |
| chr14 | 23435469 | 23436607 | 1139 | 8 | 8.30E-07 | 1.03E-08 | 0.003358 | 6.09E-08 | 0.00196031 | MYH7 |
| chr14 | 94639464 | 94640682 | 1219 | 4 | 2.53E-08 | 6.81E-07 | 4.19E-05 | 6.10E-08 | 0.007976629 | NA |
| chr16 | 70700256 | 70701516 | 1261 | 7 | 1.15E-09 | 2.94E-07 | 7.20E-05 | 6.10E-08 | 0.007119723 | VAC14 |
| chr2 | 237315493 | 237315614 | 122 | 2 | 1.55E-07 | 3.50E-08 | 4.32E-05 | 6.10E-08 | 0.037688079 | NA |
| chr5 | 115961281 | 115963023 | 1743 | 16 | 4.26E-06 | 9.84E-09 | 0.008132 | 6.11E-08 | 0.009536102 | LVRN |
| chr4 | 89836478 | 89838037 | 1560 | 15 | 2.46E-08 | 5.60E-07 | 0.000633 | 6.14E-08 | 0.002554187 | SNCA-AS1, SNCA |
| chr17 | 39199708 | 39200814 | 1107 | 11 | 8.53E-08 | 3.46E-06 | 0.00151 | 6.14E-08 | 0.001813416 | RPL19 |
| chr16 | 21728357 | 21728440 | 84 | 2 | 1.56E-07 | 3.74E-08 | 3.54E-05 | 6.17E-08 | 0.014782688 | OTOA |
| chr1 | 159900112 | 159901125 | 1014 | 8 | 9.55E-07 | 2.50E-08 | 0.0022 | 6.19E-08 | 0.012458321 | AL590560.5, CFAP45 |
| chr15 | 44710399 | 44712379 | 1981 | 13 | 1.11E-06 | 4.51E-07 | 0.001219 | 6.19E-08 | 0.000545176 | B2M, PATL2 |
| chr3 | 181718855 | 181719924 | 1070 | 6 | 1.01E-07 | 2.80E-08 | 0.000875 | 6.24E-08 | 0.009802709 | SOX2-OT |
| chr3 | 42159174 | 42160406 | 1233 | 8 | 2.75E-07 | 1.59E-08 | 0.000746 | 6.24E-08 | 0.017134988 | TRAK1 |
| chr7 | 35258088 | 35259606 | 1519 | 7 | 5.37E-06 | 5.85E-08 | 0.001441 | 6.25E-08 | 0.006864368 | AC009531.1 |
| chr7 | 56447817 | 56448811 | 995 | 9 | 6.57E-07 | 1.72E-08 | 0.00367 | 6.26E-08 | 0.020670732 | AC092447.7 |
| chr5 | 178462944 | 178464150 | 1207 | 7 | 1.94E-06 | 5.39E-06 | 0.000358 | 6.28E-08 | 0.006995629 | COL23A1 |
| chr1 | 12198265 | 12199482 | 1218 | 3 | 6.09E-06 | 3.01E-08 | 9.14E-05 | 6.28E-08 | 0.000668516 | TNFRSF1B |
| chr10 | 73413340 | 73415378 | 2039 | 16 | 3.10E-06 | 2.48E-06 | 0.000153 | 6.28E-08 | 0.000763691 | ANXA7 |
| chr4 | 175748041 | 175748394 | 354 | 2 | 1.22E-08 | 7.35E-07 | 1.20E-06 | 6.30E-08 | -0.00965796 | GPM6A |
| chr10 | 44384788 | 44386892 | 2105 | 14 | 2.37E-06 | 1.16E-06 | 0.000436 | 6.31E-08 | 0.003230366 | CXCL12 |
| chr12 | 8066326 | 8067406 | 1081 | 5 | 3.59E-08 | 1.45E-07 | 4.17E-05 | 6.31E-08 | 0.005811331 | C3AR1 |
| chr1 | 234661511 | 234662095 | 585 | 2 | 1.20E-07 | 4.76E-08 | 1.99E-05 | 6.32E-08 | 0.007204267 | AL160408.1, AL160408.4 |
| chr6 | 32166152 | 32168275 | 2124 | 43 | 4.25E-06 | 1.44E-05 | 0.00224 | 6.35E-08 | 0.00043336 | PPT2-EGFL8, EGFL8, AGPAT1 |
| chr17 | 61454836 | 61456483 | 1648 | 7 | 1.35E-06 | 1.46E-05 | 0.00018 | 6.42E-08 | 0.008672766 | TBX4 |
| chr5 | 66958091 | 66959192 | 1102 | 8 | 2.26E-07 | 3.61E-07 | 0.000694 | 6.44E-08 | 0.008058766 | MAST4 |
| chr7 | 72337589 | 72338579 | 991 | 6 | 9.46E-08 | 1.26E-07 | 0.000839 | 6.44E-08 | -0.0140848 | CALN1 |
| chr17 | 31878410 | 31878531 | 122 | 2 | 1.05E-07 | 1.02E-07 | 6.46E-06 | 6.47E-08 | 0.006864493 | AC004253.1, UTP6 |
| chr17 | 28661589 | 28663184 | 1596 | 17 | 3.97E-06 | 2.15E-07 | 0.001354 | 6.47E-08 | -0.00015974 | SUPT6H, SDF2 |
| chr2 | 70687007 | 70687507 | 501 | 4 | 1.44E-08 | 6.32E-08 | 0.000324 | 6.49E-08 | 0.009344549 | AC005234.1, ADD2 |
| chr4 | 792456 | 792930 | 475 | 3 | 1.10E-08 | 4.85E-07 | 3.45E-05 | 6.51E-08 | 0.019784203 | CPLX1 |
| chr17 | 36850023 | 36850121 | 99 | 2 | 1.13E-07 | 8.95E-08 | 7.75E-06 | 6.60E-08 | 0.018899742 | NA |
| chr16 | 89595996 | 89597204 | 1209 | 4 | 5.21E-08 | 1.33E-07 | 4.19E-05 | 6.62E-08 | 0.009523759 | CPNE7 |
| chr17 | 44758320 | 44758807 | 488 | 4 | 3.03E-09 | 3.11E-06 | 3.09E-05 | 6.63E-08 | 0.008199005 | NA |
| chr20 | 290259 | 291100 | 842 | 12 | 1.78E-08 | 1.20E-05 | 0.00035 | 6.73E-08 | 0.001889931 | C20orf96 |
| chr12 | 107845693 | 107846108 | 416 | 4 | 2.54E-07 | 1.49E-08 | 0.00085 | 6.73E-08 | 0.026067678 | AC126177.3 |
| chr1 | 31751028 | 31751997 | 970 | 3 | 2.63E-06 | 2.49E-08 | 0.000151 | 6.73E-08 | 0.011737863 | ADGRB2 |
| chr15 | 93235683 | 93236605 | 923 | 2 | 3.46E-06 | 4.04E-08 | 3.92E-05 | 6.78E-08 | 0.017785477 | AC091078.1 |
| chr22 | 41891363 | 41891605 | 243 | 2 | 1.32E-07 | 4.65E-08 | 2.55E-05 | 6.79E-08 | -0.00045725 | SREBF2 |
| chr15 | 101578201 | 101578674 | 474 | 3 | 9.19E-09 | 4.22E-05 | 4.01E-05 | 6.81E-08 | 0.036215426 | NA |
| chr1 | 3182669 | 3183353 | 685 | 4 | 3.44E-09 | 8.38E-08 | 8.56E-05 | 6.82E-08 | -0.01559194 | PRDM16 |
| chr9 | 127711317 | 127712306 | 990 | 8 | 7.99E-07 | 1.67E-08 | 0.001756 | 6.82E-08 | 0.009491045 | CFAP157, PTRH1 |
| chr1 | 7247952 | 7248306 | 355 | 4 | 7.91E-08 | 9.91E-07 | 0.000204 | 6.87E-08 | 0.022954683 | CAMTA1 |
| chr22 | 50479932 | 50480518 | 587 | 3 | 8.61E-08 | 2.07E-08 | 0.000338 | 6.99E-08 | 0.021609234 | NA |
| chr10 | 71313742 | 71314508 | 767 | 4 | 1.10E-06 | 5.47E-08 | 0.000216 | 6.99E-08 | 0.002571022 | NA |
| chr1 | 110209425 | 110210067 | 643 | 6 | 4.99E-08 | 1.73E-07 | 0.001771 | 7.01E-08 | -0.02192401 | LINC02586 |
| chr2 | 19901378 | 19902305 | 928 | 15 | 1.37E-11 | 1.71E-05 | 5.10E-06 | 7.05E-08 | -0.00011629 | AC013400.1, TTC32 |
| chr4 | 133148515 | 133149664 | 1150 | 11 | 1.19E-07 | 2.75E-07 | 0.000602 | 7.07E-08 | 0.001560526 | PCDH10, AC105383.1 |
| chr6 | 2952793 | 2953208 | 416 | 3 | 1.07E-07 | 2.69E-08 | 0.000136 | 7.08E-08 | 0.058247703 | SERPINB6 |
| chr7 | 16644696 | 16646686 | 1991 | 20 | 1.13E-06 | 6.48E-06 | 0.00079 | 7.10E-08 | 0.000644579 | BZW2, ANKMY2 |
| chr10 | 79285074 | 79285409 | 336 | 5 | 1.19E-07 | 3.67E-08 | 0.000604 | 7.10E-08 | 0.012879281 | ZMIZ1 |
| chr7 | 73682882 | 73684288 | 1407 | 18 | 1.07E-08 | 1.35E-05 | 1.46E-05 | 7.17E-08 | 0.001898731 | BUD23, DNAJC30 |
| chr3 | 57142497 | 57143875 | 1379 | 7 | 6.13E-07 | 1.24E-08 | 0.001601 | 7.19E-08 | 0.010247165 | IL17RD |
| chr11 | 82718386 | 82719449 | 1064 | 5 | 1.14E-06 | 4.38E-08 | 0.000141 | 7.19E-08 | 0.003376754 | NA |
| chr5 | 158984647 | 158985182 | 536 | 3 | 7.66E-08 | 6.21E-08 | 8.80E-05 | 7.19E-08 | 0.014253399 | AC136424.2, EBF1 |
| chr19 | 51501376 | 51502736 | 1361 | 7 | 1.08E-06 | 2.29E-07 | 0.001059 | 7.19E-08 | 0.005310867 | SIGLEC12 |
| chr19 | 17931804 | 17933323 | 1520 | 13 | 3.52E-06 | 2.83E-07 | 0.001685 | 7.23E-08 | 0.003950111 | CCDC124 |
| chr11 | 73216787 | 73218128 | 1342 | 8 | 6.62E-09 | 7.57E-07 | 0.000495 | 7.26E-08 | 0.005303169 | NA |
| chr17 | 75320199 | 75320880 | 682 | 12 | 6.45E-08 | 1.75E-06 | 5.71E-05 | 7.35E-08 | 0.000425042 | GRB2 |
| chr19 | 1400852 | 1402420 | 1569 | 13 | 1.68E-06 | 1.35E-07 | 0.001384 | 7.37E-08 | 0.001966174 | GAMT |
| chr6 | 45423628 | 45424725 | 1098 | 6 | 3.54E-07 | 2.52E-08 | 0.001197 | 7.39E-08 | 0.009604494 | RUNX2 |
| chr3 | 193878564 | 193879776 | 1213 | 3 | 5.85E-10 | 8.33E-06 | 3.12E-06 | 7.41E-08 | 0.016394354 | NA |
| chr6 | 45576857 | 45577841 | 985 | 5 | 1.27E-06 | 6.10E-07 | 0.000158 | 7.43E-08 | 0.00346497 | RUNX2 |
| chr5 | 177515041 | 177517606 | 2566 | 19 | 3.95E-06 | 1.19E-05 | 0.000108 | 7.47E-08 | -0.00106289 | DDX41 |
| chr20 | 63336623 | 63338225 | 1603 | 8 | 2.44E-06 | 1.39E-07 | 0.000919 | 7.52E-08 | 0.006224354 | RNU6-994P |
| chr6 | 28863523 | 28864895 | 1373 | 34 | 1.61E-06 | 2.00E-06 | 0.020187 | 7.54E-08 | 0.000695019 | LINC01623 |
| chr12 | 114395860 | 114396908 | 1049 | 5 | 9.51E-08 | 4.97E-08 | 7.28E-05 | 7.65E-08 | 0.001781398 | TBX5 |
| chr10 | 34723686 | 34724090 | 405 | 3 | 5.49E-08 | 1.12E-07 | 3.95E-05 | 7.67E-08 | 0.017776068 | PARD3 |
| chr11 | 1927703 | 1927900 | 198 | 6 | 2.77E-07 | 6.03E-09 | 0.004386 | 7.67E-08 | 0.032363874 | TNNT3 |
| chr11 | 18394094 | 18395220 | 1127 | 15 | 2.82E-09 | 1.02E-06 | 0.000238 | 7.68E-08 | 0.002260305 | LDHA |
| chr16 | 80892573 | 80893139 | 567 | 5 | 3.17E-07 | 9.81E-08 | 0.000292 | 7.69E-08 | 0.005664491 | ARLNC1, AC009070.1 |
| chr12 | 56157606 | 56158695 | 1090 | 15 | 2.69E-07 | 2.54E-07 | 0.001267 | 7.76E-08 | 0.000754844 | MYL6B, MYL6, AC034102.5 |
| chr1 | 112505323 | 112505609 | 287 | 2 | 6.80E-09 | 6.69E-06 | 5.39E-07 | 7.77E-08 | 0.008567999 | WNT2B |
| chr16 | 2062787 | 2064117 | 1331 | 10 | 3.00E-09 | 4.56E-06 | 2.19E-05 | 7.83E-08 | 0.00303083 | TSC2 |
| chr4 | 18021478 | 18022822 | 1345 | 11 | 3.41E-08 | 4.77E-07 | 0.00044 | 7.89E-08 | 0.000843097 | LCORL |
| chr8 | 98167923 | 98168512 | 590 | 5 | 1.31E-07 | 1.84E-07 | 0.000105 | 7.92E-08 | 0.003676051 | NA |
| chr16 | 88062958 | 88064260 | 1303 | 7 | 2.23E-07 | 2.11E-06 | 0.000266 | 7.92E-08 | 0.002351987 | BANP |
| chr9 | 130692400 | 130693971 | 1572 | 12 | 8.77E-06 | 1.01E-06 | 7.80E-05 | 7.94E-08 | 0.002120484 | EXOSC2 |
| chr14 | 53957089 | 53958222 | 1134 | 6 | 6.10E-07 | 7.57E-08 | 0.000435 | 8.01E-08 | 0.008144337 | AL138479.2, BMP4 |
| chr16 | 72989828 | 72990303 | 476 | 3 | 3.24E-07 | 2.40E-07 | 6.72E-05 | 8.04E-08 | 0.016526472 | ZFHX3 |

| | | | | | | | | | | |
|---|---|---|---|---|---|---|---|---|---|---|
| chr2 | 3422198 | 3423715 | 1518 | 5 | 1.41E-07 | 1.89E-06 | 3.53E-05 | 8.07E-08 | -0.00066688 | TRAPPC12 |
| chr12 | 132144840 | 132146416 | 1577 | 7 | 6.00E-13 | 2.36E-05 | 1.78E-05 | 8.08E-08 | -6.63E-05 | NOC4L |
| chr7 | 4805863 | 4805994 | 132 | 3 | 1.08E-08 | 8.55E-05 | 2.18E-07 | 8.17E-08 | 0.001485925 | RADIL |
| chr7 | 151852945 | 151853011 | 67 | 2 | 1.74E-08 | 1.13E-05 | 4.86E-07 | 8.17E-08 | 0.005109957 | PRKAG2 |
| chr7 | 151410551 | 151411725 | 1175 | 12 | 3.64E-07 | 5.37E-07 | 0.000382 | 8.17E-08 | 0.008529492 | WDR86-AS1, WDR86 |
| chr1 | 108830482 | 108830876 | 395 | 3 | 3.94E-07 | 2.77E-08 | 0.000252 | 8.17E-08 | 0.01631492 | AKNAD1 |
| chr4 | 5710699 | 5711373 | 675 | 11 | 1.82E-07 | 1.61E-07 | 0.000839 | 8.19E-08 | 0.004704164 | EVC |
| chr20 | 20055988 | 20056702 | 715 | 8 | 6.13E-07 | 4.83E-09 | 0.006817 | 8.21E-08 | 0.018753325 | CFAP61, CRNKL1 |
| chr11 | 113690345 | 113691289 | 945 | 3 | 1.09E-07 | 4.40E-08 | 5.88E-05 | 8.22E-08 | 0.009266908 | TMPRSS5 |
| chr2 | 68643922 | 68644722 | 801 | 5 | 9.19E-09 | 1.26E-07 | 0.000113 | 8.27E-08 | 0.008439069 | APLF, PROKR1 |
| chr6 | 143513124 | 143514004 | 881 | 4 | 1.86E-09 | 9.65E-07 | 1.33E-05 | 8.29E-08 | 0.004948078 | NA |
| chr5 | 123511072 | 123512720 | 1649 | 16 | 3.98E-08 | 2.76E-07 | 0.002088 | 8.29E-08 | -0.00271272 | CSNK1G3 |
| chr9 | 136497156 | 136498169 | 1014 | 4 | 1.28E-07 | 3.72E-08 | 0.000225 | 8.29E-08 | 0.002254013 | NOTCH1 |
| chr12 | 2295694 | 2296146 | 453 | 3 | 2.44E-09 | 7.94E-07 | 2.79E-06 | 8.29E-08 | 0.010449591 | CACNA1C |
| chr13 | 43785279 | 43785860 | 582 | 4 | 6.03E-09 | 3.84E-06 | 6.33E-05 | 8.29E-08 | 0.010798037 | ENOX1 |
| chr2 | 197704943 | 197706002 | 1060 | 17 | 1.87E-07 | 5.31E-07 | 0.000361 | 8.30E-08 | -9.87E-05 | AC011997.1, MARS2 |
| chr16 | 67482259 | 67484276 | 2018 | 11 | 4.87E-05 | 3.74E-07 | 0.004437 | 8.31E-08 | 0.005868983 | AC009061.2, AGRP |
| chr4 | 13527478 | 13529589 | 2112 | 14 | 5.10E-05 | 1.65E-07 | 0.004203 | 8.33E-08 | 0.002285535 | LINC01097 |
| chr9 | 136665445 | 136666552 | 1108 | 6 | 1.70E-07 | 1.01E-05 | 7.84E-05 | 8.33E-08 | 0.002445642 | EGFL7 |
| chr8 | 143613523 | 143614701 | 1179 | 9 | 1.44E-08 | 1.00E-05 | 0.000184 | 8.34E-08 | 0.002594666 | GFUS |
| chr17 | 63972678 | 63973359 | 682 | 8 | 2.31E-08 | 5.81E-07 | 9.86E-06 | 8.40E-08 | 0.001932085 | SCN4A, AC127029.2 |
| chr8 | 2561751 | 2562352 | 602 | 4 | 1.37E-07 | 3.70E-08 | 0.000396 | 8.42E-08 | 0.021808048 | NA |
| chr16 | 31529238 | 31529977 | 740 | 3 | 6.46E-08 | 1.12E-07 | 5.91E-05 | 8.45E-08 | 0.012638187 | NA |
| chr8 | 84920506 | 84920922 | 417 | 3 | 2.30E-07 | 4.31E-08 | 6.57E-05 | 8.46E-08 | -0.00682564 | RALYL |
| chr7 | 50590684 | 50591284 | 601 | 4 | 3.16E-10 | 3.99E-05 | 2.46E-07 | 8.46E-08 | 0.003576095 | GRB10 |
| chr5 | 55994356 | 55995718 | 1363 | 12 | 6.02E-08 | 1.24E-06 | 0.000352 | 8.52E-08 | 6.05E-05 | AC008892.1, FLJ31104, IL6ST |
| chr7 | 151375908 | 151377433 | 1526 | 6 | 4.83E-07 | 2.49E-06 | 0.000107 | 8.56E-08 | 0.000507968 | NUB1, WDR86 |
| chr12 | 120264672 | 120266231 | 1560 | 13 | 1.02E-05 | 1.96E-07 | 0.003644 | 8.58E-08 | 0.003560013 | PXN |
| chr5 | 66143481 | 66144988 | 1508 | 19 | 1.15E-08 | 5.61E-05 | 0.000252 | 8.60E-08 | -8.20E-05 | SREK1, AC025442.1 |
| chr4 | 107719633 | 107720678 | 1046 | 11 | 8.01E-08 | 3.97E-06 | 0.000571 | 8.61E-08 | 0.001024492 | PAPSS1 |
| chr10 | 116849187 | 116850676 | 1490 | 16 | 1.58E-06 | 2.58E-06 | 0.001659 | 8.62E-08 | 0.00196252 | ENO4, HSPA12A |
| chr8 | 106617905 | 106618031 | 127 | 3 | 2.30E-07 | 5.03E-08 | 0.000138 | 8.66E-08 | 0.032846391 | OXR1, AC090579.1 |
| chr5 | 139439096 | 139440581 | 1486 | 17 | 2.24E-09 | 0.000363 | 7.33E-06 | 8.71E-08 | -0.00248096 | DNAJC18 |
| chr5 | 171437203 | 171437688 | 486 | 2 | 3.04E-09 | 7.85E-08 | 1.58E-05 | 8.71E-08 | 0.000668791 | FGF18 |
| chr14 | 101031246 | 101032006 | 761 | 8 | 5.80E-08 | 2.94E-07 | 2.83E-05 | 8.72E-08 | 0.004205952 | MEG8, MIR543 |
| chr1 | 39025646 | 39026898 | 1253 | 12 | 3.96E-07 | 1.50E-07 | 0.00229 | 8.78E-08 | -0.00016953 | NDUFS5 |
| chr2 | 24048916 | 24050223 | 1308 | 14 | 1.24E-07 | 4.14E-07 | 0.000459 | 8.84E-08 | 0.00588222 | MFSD2B, FKBP1B, WDCP |
| chr12 | 56096382 | 56097177 | 796 | 5 | 2.01E-08 | 6.79E-07 | 7.00E-05 | 8.86E-08 | 0.006422956 | ERBB3 |
| chr6 | 87589474 | 87591250 | 1777 | 18 | 1.03E-08 | 3.21E-05 | 5.55E-05 | 8.86E-08 | 0.00122813 | ORC3, RARS2 |
| chr1 | 37514301 | 37515688 | 1388 | 15 | 1.14E-07 | 3.77E-07 | 0.000844 | 8.89E-08 | 0.000181456 | MEAF6 |
| chr15 | 45198748 | 45200908 | 2161 | 15 | 2.28E-07 | 0.000188 | 0.000161 | 8.93E-08 | 0.000678752 | AC051619.3, SHF, AC051619.6, AC051619.7 |
| chr4 | 122378492 | 122380230 | 1739 | 23 | 4.15E-06 | 8.30E-07 | 0.007012 | 8.95E-08 | 0.001679517 | ADAD1 |
| chr7 | 41940495 | 41940776 | 282 | 3 | 9.62E-08 | 1.11E-07 | 1.99E-05 | 9.02E-08 | 0.008004732 | NA |
| chr4 | 41748705 | 41750435 | 1731 | 11 | 5.16E-12 | 4.44E-05 | 1.11E-05 | 9.04E-08 | 0.005201107 | PHOX2B-AS1, AC105389.1, PHOX2B |
| chr4 | 1718084 | 1718712 | 629 | 5 | 5.12E-07 | 1.26E-08 | 0.001341 | 9.07E-08 | 0.008190171 | TACC3, TMEM129 |
| chr17 | 76948693 | 76949137 | 445 | 5 | 1.17E-07 | 2.75E-07 | 0.000108 | 9.10E-08 | 0.006087223 | MGAT5B |
| chr15 | 50181657 | 50182842 | 1186 | 12 | 3.58E-06 | 5.77E-08 | 0.003704 | 9.11E-08 | 0.006319954 | SLC27A2, ATP8B4 |
| chr8 | 17536929 | 17538675 | 1747 | 7 | 6.85E-08 | 8.57E-07 | 1.30E-05 | 9.13E-08 | 0.003939267 | SLC7A2 |
| chr7 | 151394516 | 151396790 | 2275 | 13 | 3.64E-06 | 3.38E-06 | 0.000325 | 9.14E-08 | 0.002093266 | WDR86 |
| chr4 | 3430110 | 3432046 | 1937 | 7 | 5.23E-05 | 7.97E-08 | 0.000191 | 9.15E-08 | 0.005108651 | RGS12 |
| chr12 | 56448908 | 56450555 | 1648 | 13 | 1.62E-07 | 1.38E-07 | 0.000794 | 9.16E-08 | 0.000992841 | TIMELESS, MIP, AC097104.1 |
| chr5 | 22853025 | 22854531 | 1507 | 10 | 6.51E-09 | 1.72E-07 | 0.000461 | 9.17E-08 | 0.002114411 | CDH12 |
| chr5 | 466414 | 467037 | 624 | 6 | 7.47E-10 | 5.88E-05 | 8.00E-07 | 9.18E-08 | 0.001083879 | EXOC3, PP7080 |
| chr7 | 13989065 | 13991182 | 2118 | 22 | 5.28E-08 | 2.54E-06 | 0.006356 | 9.18E-08 | 0.000794212 | ETV1 |
| chr11 | 2140084 | 2141700 | 1617 | 34 | 1.16E-08 | 3.49E-06 | 0.000823 | 9.22E-08 | 0.003298263 | IGF2-AS, IGF2, AC132217.2, INS-IGF2 |
| chr2 | 135984716 | 135986351 | 1636 | 20 | 3.56E-07 | 1.21E-05 | 0.001088 | 9.22E-08 | 0.001399245 | DARS-AS1, DARS1 |
| chr15 | 31223547 | 31224278 | 732 | 9 | 1.01E-06 | 7.35E-09 | 0.004083 | 9.24E-08 | 0.021568765 | LINC02352, AC012236.1 |
| chr13 | 94703560 | 94705578 | 2019 | 10 | 1.57E-05 | 1.14E-08 | 0.002667 | 9.29E-08 | 0.005149772 | SOX21-AS1, LINC00391 |
| chr3 | 12286585 | 12288091 | 1507 | 16 | 6.02E-08 | 8.98E-07 | 0.001076 | 9.30E-08 | 0.006299553 | PPARG |
| chr10 | 86253453 | 86253531 | 79 | 2 | 2.41E-07 | 5.21E-08 | 5.83E-05 | 9.30E-08 | 0.017475088 | GRID1 |
| chr2 | 218381579 | 218382332 | 754 | 8 | 5.26E-07 | 1.19E-06 | 0.001062 | 9.31E-08 | 0.007342737 | SLC11A1 |
| chr5 | 127705416 | 127705812 | 397 | 3 | 2.25E-07 | 4.79E-08 | 8.25E-05 | 9.32E-08 | 0.013327126 | CCDC192 |
| chr5 | 115295848 | 115297096 | 1249 | 17 | 1.34E-07 | 2.60E-07 | 0.001018 | 9.32E-08 | -0.00079642 | CCDC112 |
| chr2 | 263112 | 265084 | 1973 | 23 | 1.75E-05 | 1.49E-06 | 0.007846 | 9.35E-08 | -0.00289378 | ACP1, SH3YL1 |
| chr19 | 10120025 | 10121035 | 1011 | 7 | 4.27E-09 | 2.04E-05 | 2.14E-06 | 9.39E-08 | -0.00373097 | NA |
| chr13 | 99307944 | 99308703 | 760 | 4 | 9.80E-07 | 1.22E-07 | 0.000121 | 9.39E-08 | 0.012413485 | UBAC2 |
| chr7 | 1022292 | 1023721 | 1430 | 13 | 1.25E-08 | 3.32E-06 | 0.000447 | 9.42E-08 | 0.004430929 | C7orf50, MIR339 |
| chr1 | 244969480 | 244970565 | 1086 | 18 | 9.97E-08 | 2.43E-06 | 0.000153 | 9.42E-08 | -0.00049243 | EFCAB2, AL356512.1 |
| chr7 | 76047589 | 76049078 | 1490 | 18 | 1.05E-06 | 1.08E-06 | 0.003041 | 9.42E-08 | 0.000804841 | MDH2, STYXL1 |
| chr8 | 100906934 | 100908771 | 1838 | 6 | 3.05E-07 | 3.51E-07 | 0.000765 | 9.44E-08 | 0.00469456 | NA |
| chr9 | 114162451 | 114162660 | 210 | 5 | 1.76E-07 | 7.65E-08 | 7.10E-05 | 9.49E-08 | 0.000808552 | COL27A1 |
| chr12 | 49332717 | 49333327 | 611 | 5 | 1.57E-07 | 5.89E-08 | 0.000404 | 9.52E-08 | 0.018205987 | C1QL4 |
| chr2 | 218853808 | 218854125 | 318 | 2 | 5.66E-08 | 5.90E-08 | 3.97E-05 | 9.53E-08 | 0.028668388 | NA |
| chr6 | 31759932 | 31761054 | 1123 | 5 | 6.57E-08 | 4.93E-08 | 0.000448 | 9.53E-08 | 0.001978549 | MSH5, MSH5-SAPCD1 |
| chr17 | 381637 | 382816 | 1180 | 4 | 4.90E-06 | 2.15E-08 | 0.000607 | 9.53E-08 | 0.014876514 | RPH3AL |
| chr14 | 69263028 | 69263761 | 734 | 4 | 2.26E-08 | 3.94E-06 | 2.57E-05 | 9.56E-08 | 0.007034276 | GALNT16 |
| chr15 | 73741184 | 73742042 | 859 | 3 | 1.85E-08 | 1.90E-06 | 8.20E-06 | 9.56E-08 | -0.0043645 | INSYN1 |
| chr5 | 92687665 | 92688839 | 1175 | 5 | 4.74E-08 | 9.04E-07 | 0.000338 | 9.56E-08 | 0.013341063 | AC026780.2 |
| chr6 | 30555536 | 30557335 | 1800 | 51 | 6.21E-07 | 1.03E-07 | 0.005514 | 9.56E-08 | 0.000408412 | PRR3, GNL1 |
| chr5 | 73565063 | 73566283 | 1221 | 23 | 7.43E-08 | 3.71E-07 | 0.000498 | 9.65E-08 | -0.00034761 | UTP15, ANKRA2 |
| chr15 | 65185082 | 65185963 | 882 | 15 | 1.45E-08 | 1.89E-06 | 0.000513 | 9.68E-08 | -0.00047297 | CLPX |
| chr4 | 140523742 | 140525495 | 1754 | 13 | 5.92E-07 | 0.000423 | 0.000218 | 9.69E-08 | -0.00158206 | ELMOD2 |
| chr1 | 6485754 | 6486672 | 919 | 6 | 2.51E-09 | 5.49E-05 | 1.83E-05 | 9.74E-08 | 0.00155845 | PLEKHG5 |
| chr12 | 53499758 | 53501528 | 1771 | 23 | 2.04E-05 | 9.80E-08 | 0.008839 | 9.75E-08 | 0.000327405 | TARBP2, MAP3K12, AC023509.3 |
| chr15 | 84883680 | 84884665 | 986 | 6 | 2.34E-07 | 3.37E-07 | 0.000381 | 9.79E-08 | 0.004039769 | SLC28A1 |
| chr16 | 73058018 | 73059571 | 1554 | 8 | 4.40E-07 | 2.25E-06 | 0.000107 | 9.80E-08 | -0.00362462 | ZFHX3 |
| chr21 | 36134136 | 36135092 | 957 | 9 | 1.08E-08 | 2.27E-06 | 0.000936 | 9.85E-08 | 0.004655787 | CBR3, CBR3-AS1 |
| chr2 | 112720749 | 112721573 | 825 | 6 | 3.02E-08 | 3.58E-07 | 9.38E-05 | 9.93E-08 | 0.005076134 | NT5DC4 |
| chr7 | 141119531 | 141119582 | 52 | 2 | 2.16E-07 | 1.16E-07 | 1.13E-05 | 1.00E-07 | 0.007069548 | TMEM178B |
| chr19 | 49160228 | 49161208 | 981 | 2 | 3.74E-06 | 6.30E-08 | 3.89E-05 | 1.00E-07 | 0.014782405 | TRPM4 |
| chr22 | 41543977 | 41545445 | 1469 | 11 | 3.28E-08 | 3.59E-06 | 0.000299 | 1.01E-07 | 0.003165501 | POLR3H |
| chr10 | 133276877 | 133279117 | 2241 | 10 | 3.33E-06 | 9.42E-07 | 0.00188 | 1.01E-07 | 0.006112027 | TUBGCP2 |
| chr7 | 38630812 | 38632261 | 1450 | 15 | 4.39E-07 | 7.26E-07 | 0.001409 | 1.01E-07 | 0.000218392 | AMPH |
| chr17 | 81498285 | 81499412 | 1128 | 5 | 1.12E-06 | 3.59E-08 | 0.000969 | 1.01E-07 | -0.01363834 | NA |
| chr11 | 57597368 | 57598252 | 885 | 10 | 4.61E-08 | 5.54E-07 | 0.002796 | 1.02E-07 | 0.003013282 | SERPING1 |
| chr22 | 49657958 | 49658952 | 995 | 5 | 1.41E-07 | 6.36E-08 | 0.000679 | 1.02E-07 | -0.01632905 | NA |
| chr17 | 75729643 | 75731632 | 1990 | 12 | 6.22E-06 | 1.09E-05 | 0.000229 | 1.02E-07 | 0.000156198 | ITGB4 |
| chr1 | 14969325 | 14969487 | 163 | 2 | 9.21E-08 | 8.39E-08 | 2.01E-05 | 1.02E-07 | 0.019268028 | KAZN |
| chr2 | 88055249 | 88056265 | 1017 | 13 | 9.23E-08 | 1.53E-05 | 0.000445 | 1.02E-07 | -0.00230641 | KRCC1 |
| chr8 | 98293882 | 98295292 | 1411 | 10 | 8.42E-09 | 3.18E-06 | 9.27E-05 | 1.02E-07 | 0.006818198 | NIPAL2 |
| chr2 | 224401546 | 224402402 | 857 | 13 | 4.33E-08 | 9.29E-07 | 0.000177 | 1.02E-07 | 0.00362661 | FAM124B |
| chr1 | 66923838 | 66924860 | 1023 | 9 | 1.00E-07 | 8.67E-08 | 0.004045 | 1.03E-07 | 0.006107828 | DNAI4 |
| chr7 | 55552663 | 55553546 | 884 | 7 | 3.56E-09 | 3.02E-06 | 2.03E-05 | 1.03E-07 | 0.008474081 | VOPP1 |
| chr19 | 44718312 | 44718584 | 273 | 3 | 2.72E-07 | 6.91E-08 | 0.00012 | 1.03E-07 | 0.001684302 | AC243964.3 |
| chr7 | 1668478 | 1670314 | 1837 | 9 | 1.18E-07 | 2.95E-06 | 0.00094 | 1.04E-07 | 0.005960936 | NA |
| chr11 | 61495972 | 61496310 | 339 | 5 | 2.89E-07 | 7.38E-08 | 0.000315 | 1.04E-07 | 0.01061253 | NA |
| chr15 | 30925204 | 30925214 | 11 | 2 | 1.10E-07 | 9.65E-07 | 1.82E-06 | 1.05E-07 | -0.0013397 | FAN1 |
| chr5 | 1962197 | 1962440 | 244 | 3 | 1.82E-07 | 3.03E-08 | 0.000448 | 1.05E-07 | 0.033105326 | AC124852.1 |
| chr11 | 65824126 | 65824663 | 538 | 3 | 1.94E-07 | 5.67E-08 | 7.33E-05 | 1.05E-07 | 0.001319894 | CFL1 |
| chr8 | 144059671 | 144060357 | 687 | 4 | 6.94E-08 | 3.03E-08 | 0.000247 | 1.06E-07 | 0.008226407 | OPLAH |
| chr3 | 127002288 | 127002743 | 456 | 3 | 2.30E-08 | 3.35E-07 | 1.14E-05 | 1.06E-07 | 0.01032773 | PLXNA1 |
| chr5 | 178323823 | 178324793 | 971 | 3 | 3.89E-07 | 6.37E-07 | 1.52E-05 | 1.06E-07 | 0.007956258 | COL23A1 |
| chr16 | 67109401 | 67110516 | 1116 | 10 | 2.13E-08 | 6.85E-07 | 0.000115 | 1.07E-07 | 0.002518138 | C16orf70 |
| chr16 | 912080 | 913219 | 1140 | 6 | 5.78E-07 | 4.83E-08 | 0.000298 | 1.07E-07 | 0.00231844 | LMF1 |
| chr11 | 72684463 | 72685331 | 869 | 5 | 1.58E-09 | 2.27E-05 | 1.98E-05 | 1.07E-07 | 0.004374384 | ARAP1-AS1, ARAP1 |
| chr11 | 1353621 | 1354704 | 1084 | 6 | 3.93E-08 | 3.92E-07 | 0.000451 | 1.07E-07 | 0.004442125 | LINC02689 |
| chr12 | 5433989 | 5434103 | 115 | 2 | 6.93E-09 | 5.84E-08 | 7.47E-05 | 1.07E-07 | 0.03249384 | NTF3 |
| chr20 | 36090463 | 36092730 | 2268 | 9 | 2.00E-07 | 8.42E-08 | 0.000751 | 1.07E-07 | 0.005290204 | EPB41L1 |
| chr21 | 38296552 | 38297032 | 481 | 3 | 2.66E-07 | 4.10E-08 | 0.000147 | 1.07E-07 | 0.014684908 | KCNJ15 |
| chr1 | 14300619 | 14302359 | 1741 | 7 | 1.41E-06 | 2.71E-08 | 0.002873 | 1.07E-07 | -0.00495318 | KAZN, AL357873.1 |
| chr6 | 106509124 | 106510513 | 1390 | 10 | 2.59E-06 | 3.42E-07 | 0.003677 | 1.08E-07 | 0.008005589 | CRYBG1 |
| chr10 | 71986831 | 71987028 | 198 | 3 | 9.58E-08 | 1.08E-06 | 3.19E-06 | 1.08E-07 | 0.007996459 | CHST3 |
| chr17 | 82272784 | 82274564 | 1781 | 14 | 5.02E-07 | 5.86E-06 | 0.00077 | 1.08E-07 | 0.000580509 | AC132872.5, CSNK1D |
| chr12 | 54584230 | 54584958 | 729 | 3 | 2.34E-07 | 2.76E-07 | 1.25E-05 | 1.08E-07 | 0.011446488 | PPP1R1A |
| chr1 | 12549278 | 12550074 | 797 | 6 | 1.08E-07 | 1.45E-06 | 0.000243 | 1.08E-07 | 0.005755965 | NA |
| chr4 | 113979275 | 113980883 | 1609 | 13 | 9.28E-06 | 1.86E-07 | 0.007012 | 1.08E-07 | 0.002754701 | ARSJ |
| chr13 | 105705519 | 105706225 | 707 | 4 | 2.15E-07 | 2.65E-07 | 0.000337 | 1.09E-07 | 0.002101509 | LINC00343 |
| chr16 | 85282317 | 85283005 | 689 | 7 | 3.46E-08 | 5.96E-07 | 5.73E-05 | 1.09E-07 | 0.005639085 | GSE1, AC092275.1, LINC00311 |
| chr13 | 78355814 | 78355903 | 90 | 2 | 6.24E-08 | 2.96E-07 | 4.53E-06 | 1.10E-07 | 0.00734192 | OBI1-AS1 |
| chr1 | 198160506 | 198160535 | 30 | 2 | 1.32E-07 | 1.44E-07 | 1.01E-05 | 1.10E-07 | 0.007543435 | NEK7 |
| chr19 | 13233929 | 13234785 | 857 | 5 | 9.51E-07 | 1.70E-08 | 0.001646 | 1.10E-07 | 0.017781241 | CACNA1A |
| chr14 | 102944179 | 102945791 | 1613 | 11 | 1.43E-06 | 3.85E-07 | 0.000354 | 1.10E-07 | 0.002772302 | CDC42BPB |
| chr5 | 147508882 | 147511018 | 2137 | 17 | 4.98E-07 | 1.35E-05 | 0.000177 | 1.10E-07 | 0.00380023 | DPYSL3 |
| chr1 | 155236558 | 155237161 | 604 | 2 | 7.64E-10 | 4.39E-05 | 4.60E-07 | 1.11E-07 | 0.001872327 | GBA |
| chr14 | 104119611 | 104120256 | 646 | 3 | 1.72E-07 | 1.57E-07 | 9.68E-05 | 1.11E-07 | 0.012937105 | NA |
| chr17 | 35146464 | 35147651 | 1188 | 6 | 6.40E-08 | 5.89E-06 | 0.000206 | 1.12E-07 | 0.005314898 | NA |
| chr11 | 130166647 | 130166665 | 19 | 2 | 1.36E-07 | 7.35E-07 | 2.35E-06 | 1.12E-07 | 0.010105591 | ST14 |
| chr7 | 19772704 | 19773961 | 1258 | 18 | 2.63E-07 | 1.42E-06 | 0.004167 | 1.12E-07 | 0.00147675 | TMEM196 |
| chr8 | 37674409 | 37675305 | 897 | 6 | 1.65E-06 | 1.05E-07 | 0.000277 | 1.12E-07 | 0.002919173 | NA |
| chr12 | 132362579 | 132363406 | 828 | 7 | 2.94E-07 | 2.37E-07 | 0.000295 | 1.12E-07 | 0.00496287 | AC148477.9 |
| chr17 | 81845617 | 81847334 | 1718 | 10 | 5.59E-06 | 1.54E-06 | 0.000245 | 1.12E-07 | 0.002474592 | P4HB |
| chr15 | 85408341 | 85408581 | 241 | 3 | 2.32E-07 | 1.47E-07 | 7.65E-05 | 1.13E-07 | 0.051862287 | AKAP13 |
| chr10 | 107915200 | 107915721 | 522 | 5 | 5.30E-08 | 2.26E-07 | 9.26E-05 | 1.14E-07 | -0.00600609 | LINC01435 |
| chr2 | 100322822 | 100323763 | 942 | 5 | 4.01E-09 | 6.22E-06 | 1.69E-05 | 1.14E-07 | 0.013391791 | NA |
| chr13 | 111588223 | 111588730 | 508 | 3 | 3.23E-08 | 4.21E-06 | 4.59E-06 | 1.14E-07 | 0.013792904 | AL359649.1 |
| chr12 | 132120480 | 132122062 | 1583 | 4 | 1.28E-08 | 6.56E-07 | 3.87E-05 | 1.14E-07 | 0.006418499 | EP400P1 |
| chr7 | 44979059 | 44980487 | 1429 | 10 | 4.89E-07 | 7.53E-07 | 0.001293 | 1.14E-07 | -0.00214785 | MYO1G |
| chr9 | 116157142 | 116157636 | 495 | 2 | 2.83E-07 | 6.66E-08 | 5.55E-05 | 1.15E-07 | 0.021089586 | PAPPA |
| chr18 | 3593463 | 3594398 | 936 | 10 | 1.53E-07 | 7.04E-08 | 0.000816 | 1.15E-07 | -0.00140756 | DLGAP1-AS1, DLGAP1 |
| chr12 | 53753977 | 53754433 | 457 | 4 | 4.12E-08 | 1.24E-06 | 0.000341 | 1.15E-07 | 0.035923426 | AC076968.2, AC076968.1, CISTR |
| chr7 | 92417842 | 92418074 | 233 | 3 | 2.82E-07 | 9.46E-08 | 7.37E-05 | 1.15E-07 | 0.007938415 | TMBIM7P |
| chr9 | 5449663 | 5451023 | 1361 | 12 | 7.10E-07 | 3.56E-06 | 0.00053 | 1.15E-07 | 0.004974552 | CD274 |
| chr16 | 46883584 | 46885282 | 1699 | 18 | 8.10E-06 | 3.29E-07 | 0.00263 | 1.15E-07 | 0.003512962 | GPT2 |
| chr22 | 39900403 | 39901285 | 883 | 5 | 1.29E-07 | 1.79E-06 | 4.70E-05 | 1.16E-07 | 0.010244434 | GRAP2 |
| chr2 | 112092382 | 112093328 | 947 | 2 | 1.78E-06 | 9.34E-08 | 2.22E-05 | 1.16E-07 | 0.00554073 | TMEM87B |
| chr7 | 155252313 | 155253079 | 767 | 3 | 8.57E-09 | 5.79E-06 | 9.26E-06 | 1.16E-07 | 0.005022005 | NA |
| chr12 | 131035338 | 131036642 | 1305 | 9 | 3.20E-07 | 2.58E-08 | 0.001363 | 1.16E-07 | 0.021621847 | ADGRD1, AC078925.1 |
| chr6 | 25652019 | 25652587 | 569 | 12 | 9.16E-10 | 6.72E-06 | 4.81E-06 | 1.18E-07 | 0.00668462 | SCGN |
| chr4 | 6732847 | 6732995 | 149 | 2 | 3.16E-08 | 4.80E-07 | 3.25E-06 | 1.18E-07 | 0.018460548 | NA |
| chr8 | 26190386 | 26191601 | 1216 | 5 | 2.28E-06 | 3.60E-07 | 0.000296 | 1.18E-07 | -0.01171136 | NA |
| chr19 | 39833547 | 39834765 | 1219 | 17 | 1.18E-07 | 2.99E-06 | 0.000209 | 1.19E-07 | -0.00057604 | DYRK1B, FBL |
| chr17 | 81994266 | 81996032 | 1767 | 8 | 1.08E-05 | 3.59E-08 | 0.002757 | 1.19E-07 | 0.000733637 | ASPSCR1 |

| | | | | | | | | | | |
|---|---|---|---|---|---|---|---|---|---|---|
| chr2 | 219039106 | 219039379 | 274 | 3 | 5.85E-09 | 9.92E-06 | 9.20E-07 | 1.21E-07 | 0.02177425 | CFAP65 |
| chr2 | 144519419 | 144521241 | 1823 | 22 | 3.26E-06 | 4.19E-06 | 0.002301 | 1.21E-07 | -0.00106183 | ZEB2-AS1, AC009951.1, AC009951.3, ZEB2 |
| chr8 | 141391273 | 141392716 | 1444 | 5 | 2.06E-05 | 7.11E-06 | 0.000218 | 1.21E-07 | 0.017767874 | PTP4A3, AC100803.2 |
| chr2 | 96146528 | 96147369 | 842 | 8 | 3.90E-07 | 1.87E-07 | 0.001716 | 1.21E-07 | 0.005654626 | AC012307.1 |
| chr11 | 94171470 | 94172001 | 532 | 3 | 3.21E-08 | 1.11E-06 | 3.21E-05 | 1.22E-07 | 0.01740315 | PANX1 |
| chr10 | 102151668 | 102152822 | 1155 | 15 | 3.68E-08 | 1.55E-06 | 0.001547 | 1.22E-07 | -0.00087068 | NOLC1 |
| chr17 | 43543054 | 43544257 | 1204 | 3 | 2.06E-07 | 9.36E-08 | 0.00013 | 1.22E-07 | 0.008826488 | DHX8, ETV4 |
| chr1 | 203127555 | 203128500 | 946 | 8 | 3.42E-08 | 1.52E-06 | 0.000275 | 1.22E-07 | 0.005140403 | ADORA1, AC105940.2 |
| chr2 | 241773994 | 241775607 | 1614 | 6 | 1.73E-06 | 7.73E-06 | 0.000245 | 1.23E-07 | 0.011394555 | NA |
| chr11 | 3827273 | 3827993 | 721 | 4 | 1.50E-07 | 1.94E-07 | 6.59E-05 | 1.23E-07 | 0.003132188 | RHOG |
| chr9 | 136545309 | 136546339 | 1031 | 7 | 9.01E-10 | 1.84E-05 | 1.60E-05 | 1.23E-07 | 0.001070967 | NALT1, NOTCH1, MIR4674 |
| chr10 | 127737629 | 127739044 | 1416 | 7 | 6.78E-08 | 1.44E-07 | 0.000793 | 1.23E-07 | 0.018065988 | FOXI2 |
| chr8 | 141265550 | 141266505 | 956 | 5 | 1.49E-07 | 7.82E-08 | 0.000147 | 1.25E-07 | 0.010355336 | SLC45A4 |
| chr6 | 52575631 | 52577666 | 2036 | 12 | 1.22E-05 | 8.15E-05 | 8.90E-05 | 1.26E-07 | 0.004637595 | TRAM2-AS1, TRAM2 |
| chr20 | 57310790 | 57311304 | 515 | 3 | 2.85E-09 | 1.16E-06 | 3.12E-06 | 1.26E-07 | 0.001788651 | NA |
| chr19 | 13241460 | 13242706 | 1247 | 3 | 9.79E-06 | 1.54E-07 | 9.51E-05 | 1.26E-07 | 0.0016318 | CACNA1A |
| chr22 | 39388459 | 39388977 | 519 | 6 | 8.51E-07 | 1.87E-08 | 0.002367 | 1.27E-07 | -0.03120907 | NA |
| chr11 | 73645271 | 73646696 | 1426 | 13 | 2.21E-07 | 3.52E-07 | 0.000683 | 1.28E-07 | 0.006492138 | PLEKHB1 |
| chr6 | 30153997 | 30156522 | 2526 | 24 | 5.05E-06 | 6.79E-06 | 0.000835 | 1.28E-07 | 0.003723204 | TRIM10 |
| chr8 | 56438176 | 56438508 | 333 | 6 | 5.63E-07 | 6.74E-08 | 0.000497 | 1.28E-07 | -0.02748783 | PENK |
| chr16 | 83402637 | 83403294 | 658 | 3 | 4.33E-08 | 6.96E-07 | 1.45E-05 | 1.29E-07 | 6.82E-05 | CDH13 |
| chr20 | 51013909 | 51014601 | 693 | 4 | 4.92E-08 | 8.59E-06 | 0.000288 | 1.29E-07 | 0.01791449 | KCNG1 |
| chr12 | 121223035 | 121223959 | 925 | 2 | 1.89E-06 | 7.06E-08 | 7.93E-05 | 1.29E-07 | 0.018718244 | P2RX4 |
| chr4 | 96310358 | 96310642 | 285 | 4 | 2.32E-07 | 6.06E-07 | 2.61E-05 | 1.29E-07 | 0.011482118 | NA |
| chr10 | 97413136 | 97414075 | 940 | 3 | 7.84E-07 | 6.18E-08 | 0.000102 | 1.30E-07 | 0.014725194 | AL355490.2, RRP12 |
| chr17 | 47941288 | 47941808 | 521 | 13 | 2.27E-07 | 1.60E-06 | 0.000392 | 1.30E-07 | 0.001195401 | PNPO, SP2-AS1 |
| chr7 | 29989770 | 29991342 | 1573 | 14 | 2.05E-07 | 1.11E-06 | 0.000579 | 1.30E-07 | 0.003785955 | FKBP14-AS1, SCRN1 |
| chr11 | 728374 | 729135 | 762 | 3 | 9.10E-08 | 1.10E-07 | 0.000101 | 1.31E-07 | 0.016370602 | NA |
| chr2 | 84458921 | 84460649 | 1729 | 14 | 2.52E-07 | 1.08E-05 | 2.56E-05 | 1.32E-07 | 0.000931082 | SUCLG1 |
| chr17 | 3690657 | 3692460 | 1804 | 8 | 1.77E-07 | 1.04E-06 | 0.000154 | 1.34E-07 | 0.003564953 | P2RX5-TAX1BP3, P2RX5 |
| chr1 | 167118572 | 167119421 | 850 | 3 | 8.49E-08 | 5.26E-06 | 2.23E-06 | 1.34E-07 | 0.003177412 | STYXL2, GPA33 |
| chr12 | 27161360 | 27161997 | 638 | 6 | 3.53E-07 | 1.06E-06 | 0.000286 | 1.34E-07 | 0.001555679 | AC092747.1 |
| chr1 | 110488509 | 110490417 | 1909 | 5 | 7.86E-05 | 3.82E-06 | 0.000683 | 1.34E-07 | 0.003562523 | AL358215.3, CYMP, CYMP-AS1 |
| chr17 | 40355340 | 40356510 | 1171 | 8 | 9.09E-09 | 1.15E-05 | 5.37E-06 | 1.34E-07 | 0.003279325 | RARA |
| chr8 | 93846769 | 93847591 | 823 | 3 | 4.21E-07 | 3.25E-07 | 4.86E-05 | 1.34E-07 | 0.00219478 | NA |
| chr15 | 31159577 | 31159811 | 235 | 2 | 1.13E-08 | 1.03E-05 | 7.98E-07 | 1.34E-07 | 0.004527504 | TRPM1 |
| chr6 | 31702619 | 31704706 | 2088 | 37 | 1.86E-05 | 2.51E-05 | 0.001811 | 1.34E-07 | -0.00028358 | ABHD16A, AL662899.1 |
| chr20 | 3174253 | 3174843 | 591 | 5 | 8.44E-08 | 5.01E-08 | 0.000251 | 1.34E-07 | 0.015988891 | NA |
| chr16 | 28193060 | 28194776 | 1717 | 4 | 7.92E-05 | 2.86E-08 | 0.000962 | 1.34E-07 | 0.012052602 | XPO6 |
| chr15 | 89774798 | 89776285 | 1488 | 10 | 5.73E-10 | 3.03E-06 | 1.12E-05 | 1.35E-07 | 0.00222709 | MESP2 |
| chr10 | 77781892 | 77782850 | 959 | 3 | 1.08E-06 | 4.27E-07 | 6.40E-05 | 1.35E-07 | 0.00615834 | NA |
| chr8 | 141299553 | 141300973 | 1421 | 10 | 1.12E-07 | 6.28E-08 | 0.000376 | 1.35E-07 | 0.002687184 | SLC45A4 |
| chr1 | 227235270 | 227235735 | 466 | 2 | 3.14E-07 | 8.80E-08 | 3.95E-05 | 1.35E-07 | 0.016281287 | AL627308.1, CDC42BPA |
| chr9 | 127976971 | 127978320 | 1350 | 6 | 5.41E-07 | 1.38E-07 | 0.000195 | 1.35E-07 | 0.008101405 | FAM102A |
| chr8 | 144354481 | 144355762 | 1282 | 8 | 1.43E-08 | 7.08E-06 | 2.23E-05 | 1.35E-07 | 0.002445524 | SLC52A2, TMEM249, AC233992.2, FBXL6 |
| chr19 | 696832 | 696946 | 115 | 4 | 2.04E-07 | 5.82E-08 | 0.000108 | 1.36E-07 | 0.008704733 | NA |
| chr8 | 133273077 | 133273116 | 40 | 2 | 4.13E-08 | 7.97E-06 | 8.60E-07 | 1.36E-07 | 0.002136386 | NDRG1 |
| chr19 | 694837 | 695642 | 806 | 7 | 2.38E-07 | 3.01E-08 | 0.003814 | 1.36E-07 | 0.005046075 | PRSS57 |
| chr19 | 19191536 | 19193170 | 1635 | 18 | 8.40E-08 | 3.16E-06 | 0.000596 | 1.36E-07 | 0.000235656 | RFXANK, MEF2B, BORCS8-MEF2B, BORCS8 |
| chr15 | 77620482 | 77620801 | 320 | 3 | 1.51E-07 | 2.95E-07 | 3.60E-05 | 1.36E-07 | 0.004814459 | LINGO1 |
| chr5 | 52709841 | 52710238 | 398 | 2 | 5.23E-09 | 5.83E-06 | 9.79E-07 | 1.38E-07 | 0.008055905 | AC022126.1 |
| chr12 | 92927317 | 92927997 | 681 | 2 | 8.24E-07 | 8.61E-08 | 4.60E-05 | 1.39E-07 | 0.013938087 | EEA1 |
| chr15 | 39587430 | 39588708 | 1279 | 8 | 2.56E-05 | 1.24E-07 | 0.000525 | 1.39E-07 | 0.002624033 | THBS1, FSIP1, AC037198.2 |
| chr22 | 45239719 | 45241541 | 1823 | 10 | 1.48E-05 | 3.92E-06 | 0.001176 | 1.40E-07 | 0.005664274 | KIAA0930 |
| chr2 | 231085031 | 231085203 | 173 | 3 | 5.98E-08 | 1.33E-06 | 6.86E-06 | 1.40E-07 | 0.001268316 | PSMD1 |
| chr5 | 124310360 | 124310875 | 516 | 2 | 1.04E-07 | 1.78E-07 | 1.15E-05 | 1.40E-07 | 0.014232313 | LINC01170 |
| chr20 | 36431491 | 36432303 | 813 | 7 | 2.86E-09 | 2.14E-06 | 0.00019 | 1.41E-07 | 0.00150147 | DLGAP4 |
| chr1 | 173205511 | 173206189 | 679 | 8 | 5.18E-07 | 5.21E-08 | 0.000533 | 1.41E-07 | 0.007356366 | TNFSF4 |
| chr10 | 614187 | 615448 | 1262 | 6 | 4.10E-07 | 2.87E-07 | 0.00041 | 1.43E-07 | -0.00335778 | DIP2C |
| chr6 | 144007780 | 144009208 | 1429 | 21 | 4.02E-06 | 3.56E-07 | 0.002424 | 1.43E-07 | 0.003369279 | PLAGL1, HYMAI |
| chr4 | 141133055 | 141134531 | 1477 | 12 | 2.44E-09 | 0.000182 | 1.06E-06 | 1.44E-07 | 0.000235524 | RNF150 |
| chr1 | 67054509 | 67055804 | 1296 | 7 | 2.55E-07 | 4.56E-06 | 0.000134 | 1.44E-07 | 0.003025077 | NA |
| chr14 | 101947667 | 101948392 | 726 | 6 | 1.08E-08 | 5.47E-06 | 3.63E-05 | 1.44E-07 | 0.011943584 | AL118558.3 |
| chr12 | 122835347 | 122836707 | 1361 | 9 | 3.04E-08 | 2.21E-06 | 0.000548 | 1.44E-07 | 0.0023294 | HIP1R |
| chr2 | 113977644 | 113978562 | 919 | 5 | 5.76E-08 | 2.34E-06 | 0.000291 | 1.44E-07 | 0.003468246 | LINC01191 |
| chr3 | 139534996 | 139535692 | 697 | 3 | 1.20E-07 | 1.53E-07 | 4.40E-05 | 1.44E-07 | 0.008806174 | AC046134.2, RBP1 |
| chr1 | 233115354 | 233115371 | 18 | 2 | 1.51E-07 | 2.62E-07 | 7.55E-06 | 1.44E-07 | 0.015261579 | PCNX2 |
| chr8 | 144394273 | 144395548 | 1276 | 6 | 3.23E-08 | 3.85E-07 | 0.000281 | 1.44E-07 | 0.003784014 | CPSF1 |
| chr10 | 117532255 | 117533076 | 822 | 8 | 6.59E-08 | 2.52E-07 | 0.001377 | 1.45E-07 | 0.006031905 | EMX2OS |
| chr1 | 44806961 | 44808020 | 1060 | 11 | 7.27E-08 | 1.15E-06 | 0.00059 | 1.45E-07 | 0.005437027 | TCTEX1D4 |
| chr3 | 151460939 | 151461116 | 178 | 3 | 7.56E-08 | 4.24E-08 | 0.00052 | 1.45E-07 | -0.04191397 | NA |
| chr3 | 188946843 | 188948335 | 1493 | 13 | 1.69E-07 | 4.04E-08 | 0.002573 | 1.45E-07 | 0.003408503 | TPRG1, TPRG1-AS1 |
| chr8 | 22047951 | 22048838 | 888 | 5 | 3.55E-07 | 5.39E-08 | 0.000566 | 1.45E-07 | 0.015927873 | FGF17 |
| chr2 | 197514353 | 197516776 | 2424 | 29 | 4.92E-06 | 7.93E-05 | 0.000124 | 1.45E-07 | 0.000340609 | HSPE1-MOB4, MOB4, HSPD1 |
| chr17 | 10033468 | 10033830 | 363 | 3 | 2.97E-07 | 8.45E-08 | 9.04E-05 | 1.45E-07 | 0.013160404 | GAS7 |
| chr4 | 110611523 | 110612795 | 1273 | 10 | 6.14E-07 | 6.51E-07 | 0.002122 | 1.46E-07 | 0.008002176 | PANCR |
| chr10 | 25061307 | 25062598 | 1292 | 8 | 5.48E-06 | 1.24E-07 | 0.003833 | 1.47E-07 | 0.006691038 | AL512598.2, ENKUR |
| chr3 | 49421710 | 49422729 | 1020 | 11 | 2.87E-08 | 1.48E-06 | 8.42E-05 | 1.47E-07 | 0.006782876 | AC104452.1, AMT, NICN1 |
| chr14 | 75727223 | 75727541 | 319 | 2 | 6.31E-08 | 8.80E-07 | 2.79E-06 | 1.47E-07 | 0.008635901 | TTLL5 |
| chr1 | 42849299 | 42850167 | 869 | 4 | 3.32E-09 | 5.53E-07 | 1.82E-05 | 1.47E-07 | 0.002841494 | ZNF691 |
| chr22 | 50313688 | 50314639 | 952 | 5 | 5.83E-08 | 2.33E-07 | 0.000188 | 1.47E-07 | 0.030955641 | CR559946.1, DENND6B |
| chr1 | 37634583 | 37635980 | 1398 | 11 | 1.61E-07 | 3.15E-07 | 0.004631 | 1.47E-07 | 0.005003897 | RSPO1 |
| chr18 | 54260801 | 54260940 | 140 | 2 | 2.86E-07 | 1.26E-07 | 2.16E-05 | 1.48E-07 | 0.010971525 | NA |
| chr8 | 118073892 | 118074523 | 632 | 4 | 1.18E-06 | 3.89E-08 | 0.000955 | 1.48E-07 | 0.009917375 | EXT1 |
| chr14 | 28765684 | 28767692 | 2009 | 21 | 2.43E-06 | 1.30E-06 | 0.00107 | 1.49E-07 | 0.004018376 | AL049777.1, FOXG1 |
| chr15 | 89416555 | 89417512 | 958 | 8 | 6.45E-08 | 3.35E-06 | 0.001211 | 1.49E-07 | -0.01540213 | NA |
| chr6 | 152380490 | 152381195 | 706 | 2 | 5.55E-14 | 0.000266 | 4.72E-07 | 1.50E-07 | 0.021978306 | SYNE1-AS1, SYNE1 |
| chr18 | 49669269 | 49669367 | 99 | 2 | 3.54E-07 | 1.41E-07 | 1.82E-05 | 1.50E-07 | 0.03622285 | NA |
| chr3 | 9249049 | 9250341 | 1293 | 17 | 4.69E-06 | 9.80E-08 | 0.010021 | 1.50E-07 | 0.001445099 | SRGAP3-AS4, SRGAP3 |
| chr2 | 73932757 | 73932815 | 59 | 2 | 3.08E-07 | 2.35E-07 | 9.23E-06 | 1.51E-07 | 0.010278196 | DGUOK |
| chr17 | 80855485 | 80856708 | 1224 | 6 | 8.14E-10 | 0.000223 | 4.27E-05 | 1.51E-07 | 0.005861709 | RPTOR |
| chr8 | 37330917 | 37331159 | 243 | 2 | 7.01E-08 | 8.50E-08 | 7.37E-05 | 1.51E-07 | 0.008215937 | AC091182.1 |
| chr1 | 2207318 | 2208773 | 1456 | 13 | 1.73E-07 | 9.14E-06 | 0.000187 | 1.52E-07 | 0.003180795 | FAAP20 |
| chr4 | 849530 | 850040 | 511 | 4 | 3.11E-08 | 9.60E-07 | 7.77E-05 | 1.52E-07 | 0.002855899 | GAK |
| chr2 | 232487729 | 232488494 | 766 | 5 | 8.79E-08 | 4.08E-06 | 0.000269 | 1.52E-07 | 0.007961943 | ECEL1 |
| chr18 | 79399684 | 79400758 | 1075 | 11 | 8.77E-08 | 7.33E-06 | 0.000194 | 1.53E-07 | 0.002433957 | NFATC1 |
| chr11 | 68118138 | 68119318 | 1181 | 4 | 8.27E-07 | 5.98E-06 | 3.90E-05 | 1.53E-07 | 0.009552592 | CHKA |
| chr11 | 2825789 | 2827262 | 1474 | 8 | 5.41E-08 | 3.60E-07 | 1.64E-05 | 1.54E-07 | 0.00575586 | KCNQ1 |
| chr12 | 120725220 | 120726843 | 1624 | 15 | 3.02E-07 | 1.46E-06 | 0.000229 | 1.55E-07 | 0.004874654 | ACADS |
| chr6 | 159799487 | 159801084 | 1598 | 12 | 1.25E-07 | 1.36E-05 | 0.000745 | 1.56E-07 | 0.001407147 | PNLDC1 |
| chr3 | 138186363 | 138186532 | 170 | 2 | 4.36E-10 | 1.37E-07 | 2.11E-05 | 1.57E-07 | 0.015893186 | NA |
| chr6 | 41783861 | 41785031 | 1171 | 5 | 4.10E-08 | 1.45E-06 | 2.32E-05 | 1.57E-07 | 0.006608478 | AL365205.1, PRICKLE4, FRS3 |
| chr15 | 96326641 | 96327730 | 1090 | 11 | 3.24E-08 | 2.14E-06 | 0.000438 | 1.57E-07 | 0.006263042 | NR2F2, NR2F2-AS1 |
| chr12 | 109713523 | 109714781 | 1259 | 9 | 4.48E-08 | 9.12E-07 | 0.000236 | 1.57E-07 | 0.000820815 | FAM222A |
| chr5 | 142685974 | 142687093 | 1120 | 11 | 5.45E-07 | 3.67E-08 | 0.002043 | 1.58E-07 | 0.00181342 | FGF1 |
| chr1 | 204462486 | 204463243 | 758 | 2 | 2.45E-07 | 1.22E-07 | 2.72E-05 | 1.58E-07 | 0.011695757 | PIK3C2B |
| chr1 | 73749688 | 73750472 | 785 | 5 | 4.04E-08 | 4.25E-05 | 4.77E-06 | 1.58E-07 | 0.007481591 | NA |
| chr7 | 157299988 | 157300840 | 853 | 6 | 9.83E-08 | 3.44E-07 | 0.000121 | 1.59E-07 | 0.010483735 | NA |
| chr11 | 60931852 | 60932094 | 243 | 2 | 4.85E-07 | 8.72E-08 | 8.43E-05 | 1.59E-07 | 0.009407085 | TMEM132A |
| chr12 | 53998209 | 54000063 | 1855 | 19 | 4.41E-09 | 6.05E-05 | 0.000179 | 1.59E-07 | 0.001632843 | AC012531.3, HOXC6, HOXC9, HOXC-AS1 |
| chr20 | 45377423 | 45378707 | 1285 | 5 | 9.67E-09 | 3.08E-06 | 0.000197 | 1.59E-07 | 0.001877242 | SYS1-DBNDD2, TP53TG5 |
| chr12 | 6470482 | 6471525 | 1044 | 13 | 2.96E-07 | 7.33E-06 | 0.000146 | 1.59E-07 | 0.001047796 | VAMP1 |
| chr5 | 140699508 | 140700990 | 1483 | 15 | 4.19E-07 | 5.49E-07 | 0.000643 | 1.59E-07 | 0.005141119 | ZMAT2 |
| chr6 | 56890768 | 56891021 | 254 | 4 | 4.62E-07 | 1.54E-07 | 4.20E-05 | 1.59E-07 | 0.012092832 | DST |
| chr19 | 50639028 | 50640202 | 1175 | 14 | 2.05E-06 | 5.40E-06 | 0.005225 | 1.59E-07 | -0.00044958 | SYT3 |
| chr6 | 167093959 | 167095049 | 1091 | 10 | 1.97E-07 | 4.33E-07 | 0.000445 | 1.59E-07 | 0.006271821 | CEP43, Z94721.2 |
| chr2 | 218998726 | 218999424 | 699 | 3 | 1.33E-07 | 1.99E-07 | 0.000113 | 1.60E-07 | 0.013256786 | NA |
| chr1 | 26346122 | 26346818 | 697 | 3 | 4.72E-08 | 5.20E-07 | 1.36E-05 | 1.60E-07 | 0.00338472 | CRYBG2 |
| chr9 | 114485067 | 114485108 | 42 | 2 | 2.05E-07 | 9.98E-08 | 4.92E-05 | 1.60E-07 | 0.019249701 | WHRN |
| chr1 | 221741429 | 221742454 | 1026 | 13 | 1.27E-07 | 3.68E-06 | 0.000321 | 1.60E-07 | -0.00146532 | DUSP10 |
| chr17 | 7041840 | 7043208 | 1369 | 6 | 1.38E-07 | 1.47E-06 | 6.84E-05 | 1.61E-07 | 0.009456722 | SLC16A11 |
| chr1 | 151791390 | 151792027 | 638 | 7 | 2.42E-11 | 0.000136 | 3.16E-07 | 1.61E-07 | 0.008109512 | TDRKH-AS1, TDRKH |
| chr14 | 105485631 | 105486963 | 1333 | 12 | 8.01E-07 | 3.00E-07 | 0.0021 | 1.61E-07 | 0.005915047 | CRIP1 |
| chr8 | 27611167 | 27612708 | 1542 | 12 | 8.68E-07 | 2.20E-07 | 0.003902 | 1.61E-07 | 0.007191648 | CLU |
| chr9 | 33447034 | 33448384 | 1351 | 8 | 1.98E-05 | 1.65E-07 | 0.000538 | 1.61E-07 | 0.005707155 | AQP3 |
| chr21 | 45228290 | 45228729 | 440 | 2 | 1.53E-07 | 1.87E-07 | 1.42E-05 | 1.62E-07 | 0.01390778 | NA |
| chr7 | 143391379 | 143391624 | 246 | 5 | 5.36E-07 | 8.61E-08 | 0.00066 | 1.63E-07 | 0.002845465 | EPHA1 |
| chr20 | 52493357 | 52494554 | 1198 | 5 | 2.29E-05 | 2.12E-08 | 0.002888 | 1.63E-07 | 0.005906804 | LINC01524, AL049765.1 |
| chr7 | 41965268 | 41967078 | 1811 | 9 | 3.86E-05 | 8.56E-08 | 0.002065 | 1.63E-07 | 0.000313068 | GLI3 |
| chr2 | 206158434 | 206159838 | 1405 | 16 | 1.41E-07 | 6.95E-07 | 0.001924 | 1.64E-07 | 0.000191479 | EEF1B2, NDUFS1 |
| chr16 | 3086856 | 3087689 | 834 | 6 | 8.62E-07 | 2.62E-08 | 0.003151 | 1.64E-07 | 0.015233123 | AC108134.3 |
| chr11 | 111912624 | 111914218 | 1595 | 13 | 5.75E-08 | 8.28E-07 | 0.00145 | 1.64E-07 | 0.002519351 | HSPB2, HSPB2-C11orf52, CRYAB |
| chr12 | 56115819 | 56117369 | 1551 | 16 | 2.06E-07 | 4.67E-07 | 0.001084 | 1.64E-07 | -0.00170733 | RPL41 |
| chr4 | 6268953 | 6270177 | 1225 | 13 | 1.63E-06 | 5.47E-06 | 0.001684 | 1.64E-07 | 1.77E-05 | WFS1 |
| chr11 | 15650045 | 15651310 | 1266 | 9 | 6.84E-06 | 1.63E-06 | 0.000333 | 1.64E-07 | -0.00259902 | AC087379.2 |
| chr10 | 132897700 | 132897942 | 243 | 2 | 2.62E-07 | 1.96E-07 | 1.36E-05 | 1.64E-07 | 0.006257985 | CFAP46 |
| chr1 | 53367720 | 53368779 | 1060 | 4 | 1.92E-07 | 6.17E-07 | 0.0001 | 1.65E-07 | 0.011805952 | LINC02812 |
| chr6 | 29666718 | 29668234 | 1517 | 10 | 3.46E-07 | 1.85E-08 | 0.00449 | 1.66E-07 | 0.009390925 | MOG |
| chr12 | 42482288 | 42483083 | 796 | 5 | 1.07E-08 | 1.05E-06 | 4.23E-05 | 1.66E-07 | 0.007102094 | PRICKLE1 |
| chr19 | 49119104 | 49120377 | 1274 | 8 | 3.54E-06 | 2.53E-07 | 0.000642 | 1.67E-07 | -0.00044564 | PPFIA3, C19orf73 |
| chr19 | 19636736 | 19636925 | 190 | 2 | 6.60E-08 | 5.60E-06 | 1.20E-06 | 1.67E-07 | 0.003014316 | GMIP |
| chr10 | 6103087 | 6103714 | 628 | 2 | 6.58E-11 | 0.000632 | 4.86E-07 | 1.67E-07 | -0.00864799 | RBM17 |
| chr3 | 197748649 | 197750974 | 2326 | 25 | 4.56E-05 | 1.13E-05 | 0.002729 | 1.67E-07 | 0.001052084 | FYTTD1, RUBCN |
| chr2 | 73779538 | 73780931 | 1394 | 16 | 1.58E-07 | 3.10E-05 | 0.001183 | 1.68E-07 | 0.000994327 | DUSP11 |
| chr22 | 38872503 | 38874134 | 1632 | 7 | 3.53E-05 | 1.18E-07 | 0.000514 | 1.68E-07 | 0.003358151 | NA |
| chr8 | 113173829 | 113174475 | 647 | 5 | 4.61E-08 | 5.40E-07 | 2.76E-05 | 1.69E-07 | 0.01471469 | CSMD3 |
| chr2 | 240737685 | 240737915 | 231 | 2 | 1.19E-07 | 1.03E-07 | 5.71E-05 | 1.71E-07 | 0.021027535 | KIF1A |
| chr3 | 137768928 | 137769855 | 928 | 6 | 2.29E-07 | 6.33E-07 | 0.000446 | 1.71E-07 | 0.007423719 | NA |
| chr7 | 139798242 | 139798513 | 272 | 3 | 4.17E-07 | 1.44E-07 | 5.23E-05 | 1.72E-07 | 0.020228582 | TBXAS1 |
| chr14 | 91396469 | 91396563 | 95 | 4 | 2.27E-07 | 2.54E-06 | 8.89E-06 | 1.73E-07 | 0.015044715 | CCDC88C |
| chr2 | 181682674 | 181684838 | 2165 | 14 | 1.76E-05 | 3.16E-06 | 0.00118 | 1.73E-07 | 0.004502999 | AC013733.2 |
| chr4 | 1328607 | 1330030 | 1424 | 8 | 1.64E-07 | 5.39E-06 | 0.000318 | 1.73E-07 | 0.001239872 | MAEA |
| chr11 | 63505292 | 63506558 | 1267 | 8 | 5.44E-08 | 8.29E-07 | 6.66E-05 | 1.73E-07 | 0.005651211 | LGALS12 |
| chr12 | 76158611 | 76158956 | 346 | 3 | 2.92E-07 | 2.19E-07 | 6.47E-05 | 1.74E-07 | 0.017757301 | NA |
| chr13 | 112984558 | 112985855 | 1298 | 7 | 3.48E-07 | 3.13E-08 | 0.002277 | 1.74E-07 | 0.008888847 | MCF2L |
| chr11 | 134026569 | 134026981 | 413 | 5 | 8.61E-08 | 1.41E-07 | 7.57E-05 | 1.74E-07 | 0.012357334 | LINC02730 |
| chr6 | 34121573 | 34122237 | 665 | 2 | 1.23E-08 | 1.58E-06 | 2.35E-06 | 1.74E-07 | 0.015159352 | GRM4 |
| chr12 | 56258128 | 56259161 | 1034 | 12 | 2.07E-08 | 7.88E-06 | 1.45E-05 | 1.76E-07 | -9.37E-05 | ANKRD52 |
| chr2 | 219177729 | 219178808 | 1080 | 12 | 2.21E-07 | 1.10E-05 | 0.000508 | 1.76E-07 | 0.001116163 | RETREG2, CNPPD1 |
| chr9 | 99824716 | 99825950 | 1235 | 7 | 1.24E-07 | 7.07E-08 | 0.001398 | 1.76E-07 | 0.008437093 | NR4A3, STX17-AS1 |
| chr3 | 101676076 | 101677692 | 1617 | 19 | 2.65E-05 | 7.01E-07 | 0.000557 | 1.77E-07 | -0.00134829 | ZBTB11-AS1, ZBTB11 |
| chr7 | 28958165 | 28959608 | 1444 | 14 | 1.22E-08 | 4.52E-07 | 0.000406 | 1.77E-07 | 0.005603683 | AC005013.1, TRIL |

| | | | | | | | | | | |
|---|---|---|---|---|---|---|---|---|---|---|
| chr3 | 50348958 | 50351493 | 2536 | 27 | 4.63E-05 | 2.97E-05 | 0.000115 | 1.77E-07 | 0.001237587 | CYB561D2, Z84492.2, NPRL2 |
| chr17 | 80586678 | 80587116 | 439 | 3 | 1.08E-07 | 1.06E-06 | 1.29E-05 | 1.79E-07 | 0.01614885 | RPTOR |
| chr1 | 65264103 | 65265749 | 1647 | 11 | 3.14E-07 | 5.53E-07 | 0.001144 | 1.79E-07 | 0.005911238 | DNAJC6 |
| chr2 | 1707631 | 1708755 | 1125 | 6 | 2.68E-07 | 7.79E-08 | 0.000695 | 1.79E-07 | 0.015386506 | PXDN |
| chr22 | 24133125 | 24133290 | 166 | 2 | 1.85E-07 | 9.52E-07 | 3.18E-06 | 1.79E-07 | 0.007435329 | CABIN1 |
| chr16 | 14015993 | 14016498 | 506 | 3 | 7.81E-07 | 1.07E-07 | 0.000155 | 1.79E-07 | 0.001828424 | LINC02185 |
| chr12 | 120998899 | 120999453 | 555 | 3 | 1.80E-08 | 4.87E-06 | 2.18E-06 | 1.79E-07 | -0.00114047 | HNF1A |
| chr11 | 63574247 | 63575322 | 1076 | 5 | 7.63E-07 | 1.58E-06 | 0.000106 | 1.80E-07 | 0.004432599 | PLAAT3 |
| chr6 | 14001402 | 14002239 | 838 | 4 | 6.98E-07 | 2.10E-07 | 9.62E-05 | 1.81E-07 | 0.005126274 | NA |
| chr10 | 133180296 | 133181441 | 1146 | 7 | 1.15E-06 | 1.96E-07 | 0.00037 | 1.81E-07 | 0.001158311 | KNDC1 |
| chr12 | 57534476 | 57535533 | 1058 | 3 | 2.33E-07 | 3.63E-07 | 8.31E-05 | 1.81E-07 | 0.003527461 | DCTN2 |
| chr15 | 63049015 | 63050133 | 1119 | 8 | 6.77E-07 | 1.40E-06 | 0.000697 | 1.82E-07 | 0.002530416 | TPM1, TPM1-AS |
| chr17 | 14303255 | 14304384 | 1130 | 9 | 3.05E-08 | 1.11E-07 | 0.004525 | 1.84E-07 | 0.014417223 | HS3ST3B1, AC005224.3 |
| chr1 | 154860059 | 154861246 | 1188 | 5 | 1.68E-07 | 5.39E-08 | 0.000546 | 1.85E-07 | 0.012044863 | KCNN3 |
| chr6 | 37699281 | 37699742 | 462 | 3 | 1.70E-08 | 8.77E-07 | 2.10E-05 | 1.85E-07 | 0.021061829 | MDGA1 |
| chr2 | 85243497 | 85243764 | 268 | 2 | 3.54E-07 | 1.54E-07 | 2.50E-05 | 1.85E-07 | 0.015711461 | TCF7L1 |
| chr12 | 108820805 | 108821533 | 729 | 3 | 1.03E-06 | 6.10E-08 | 0.000371 | 1.85E-07 | 0.014730734 | SSH1 |
| chr15 | 74044473 | 74044516 | 44 | 2 | 3.10E-07 | 1.11E-07 | 5.97E-05 | 1.86E-07 | 0.007398681 | PML |
| chr13 | 113842669 | 113844288 | 1620 | 11 | 9.23E-06 | 3.77E-07 | 0.000646 | 1.86E-07 | 0.002791763 | GAS6-AS1, GAS6 |
| chr9 | 95551666 | 95553436 | 1771 | 6 | 1.74E-05 | 3.58E-07 | 0.000704 | 1.86E-07 | 0.008467798 | NA |
| chr16 | 4316838 | 4318123 | 1286 | 4 | 2.62E-07 | 1.06E-07 | 0.000159 | 1.86E-07 | 0.000251414 | GLIS2 |
| chr2 | 110772708 | 110773384 | 677 | 3 | 9.82E-07 | 6.50E-08 | 0.000338 | 1.87E-07 | 0.009126807 | ACOXL |
| chr15 | 82572550 | 82573763 | 1214 | 5 | 1.13E-07 | 7.16E-08 | 0.000657 | 1.87E-07 | 0.006486228 | AC245033.1, CPEB1 |
| chr21 | 44284903 | 44286218 | 1316 | 13 | 1.61E-06 | 1.69E-08 | 0.007806 | 1.87E-07 | 0.006570442 | AIRE |
| chr7 | 30970844 | 30971187 | 344 | 3 | 2.86E-08 | 4.69E-06 | 5.83E-06 | 1.88E-07 | 0.016802944 | GHRHR |
| chr1 | 160400050 | 160401954 | 1905 | 16 | 1.18E-06 | 3.72E-05 | 0.000231 | 1.88E-07 | 0.002421808 | VANGL2 |
| chr16 | 78770654 | 78771146 | 493 | 3 | 8.04E-08 | 1.89E-06 | 7.14E-05 | 1.88E-07 | 0.012307913 | WWOX |
| chr2 | 218858338 | 218859816 | 1479 | 8 | 1.03E-07 | 1.08E-05 | 0.001397 | 1.88E-07 | 0.006250796 | WNT6 |
| chr5 | 36875435 | 36876765 | 1331 | 9 | 6.15E-09 | 6.56E-07 | 2.62E-05 | 1.89E-07 | -0.0005319 | NIPBL-DT |
| chr12 | 124869427 | 124869612 | 186 | 3 | 4.54E-07 | 5.85E-08 | 0.000494 | 1.89E-07 | 0.012343648 | SCARB1 |
| chr7 | 143409444 | 143410189 | 746 | 4 | 2.62E-10 | 8.28E-06 | 2.14E-06 | 1.89E-07 | 0.012052755 | EPHA1-AS1 |
| chr17 | 77099290 | 77100300 | 1011 | 4 | 3.06E-06 | 4.40E-08 | 0.000756 | 1.89E-07 | 0.017363683 | SNHG20, SEC14L1 |
| chr21 | 37418501 | 37419933 | 1433 | 6 | 1.73E-06 | 6.07E-06 | 8.14E-05 | 1.90E-07 | 0.005291556 | DYRK1A |
| chr2 | 112913779 | 112914819 | 1041 | 4 | 4.33E-08 | 7.82E-08 | 0.000151 | 1.91E-07 | 0.005434249 | IL37 |
| chr1 | 167473375 | 167474633 | 1259 | 3 | 4.19E-05 | 8.95E-08 | 0.000159 | 1.91E-07 | 0.013184682 | CD247 |
| chr7 | 83648625 | 83649278 | 654 | 11 | 1.01E-07 | 2.42E-06 | 0.002827 | 1.92E-07 | 0.006908313 | SEMA3E |
| chr5 | 141626200 | 141626599 | 400 | 6 | 2.26E-07 | 3.31E-05 | 3.35E-05 | 1.96E-07 | 0.002529609 | AC008781.1, HDAC3 |
| chr6 | 159820073 | 159820524 | 452 | 5 | 5.07E-07 | 2.65E-08 | 0.002796 | 1.98E-07 | 0.038252818 | PNLDC1 |
| chr17 | 39650320 | 39651050 | 731 | 3 | 6.22E-08 | 1.90E-07 | 9.27E-05 | 1.98E-07 | 0.007987609 | STARD3 |
| chr11 | 66367798 | 66368156 | 359 | 5 | 3.00E-07 | 6.20E-07 | 3.81E-05 | 1.98E-07 | 0.004995789 | SLC29A2 |
| chr10 | 129867319 | 129868744 | 1426 | 8 | 6.23E-08 | 2.86E-07 | 0.00056 | 1.99E-07 | -0.00039817 | AL354950.3, EBF3 |
| chr6 | 130148463 | 130148473 | 11 | 2 | 6.25E-07 | 1.12E-07 | 8.09E-05 | 2.00E-07 | 0.036574737 | SAMD3 |
| chr19 | 10551029 | 10552106 | 1078 | 8 | 6.07E-07 | 0.000154 | 0.000199 | 2.00E-07 | 0.004283704 | ATG4D, RNU7-140P |
| chr11 | 60838245 | 60838900 | 656 | 3 | 1.92E-06 | 9.52E-08 | 0.000126 | 2.01E-07 | 0.007880276 | AP000777.2 |
| chr4 | 931024 | 932742 | 1719 | 17 | 1.22E-05 | 7.81E-06 | 0.002143 | 2.01E-07 | 0.00211719 | TMEM175, GAK |
| chr17 | 42116300 | 42116363 | 64 | 3 | 5.93E-08 | 4.26E-05 | 2.15E-05 | 2.01E-07 | 0.003018523 | KAT2A |
| chr7 | 42690887 | 42691257 | 371 | 2 | 5.72E-07 | 1.18E-07 | 6.47E-05 | 2.02E-07 | 0.01812474 | LINC01448 |
| chr12 | 6632483 | 6632716 | 234 | 4 | 1.39E-08 | 0.000537 | 7.98E-07 | 2.02E-07 | 0.001689262 | LPAR5 |
| chr3 | 196270614 | 196271017 | 404 | 5 | 8.84E-07 | 8.18E-08 | 0.000769 | 2.02E-07 | 0.004093555 | PCYT1A, AC069257.3 |
| chr3 | 10247168 | 10248822 | 1655 | 14 | 1.44E-05 | 8.35E-06 | 0.001565 | 2.02E-07 | 0.002879341 | TATDN2 |
| chr18 | 8717325 | 8717928 | 604 | 6 | 9.32E-08 | 2.06E-07 | 0.00017 | 2.03E-07 | 0.004632765 | MTCL1 |
| chr2 | 174343416 | 174345137 | 1722 | 8 | 5.16E-06 | 8.67E-07 | 0.000788 | 2.03E-07 | 0.004674762 | NA |
| chr19 | 42293796 | 42294907 | 1112 | 6 | 4.80E-07 | 3.48E-08 | 0.001417 | 2.04E-07 | 0.003193269 | CIC |
| chr8 | 144359875 | 144360344 | 470 | 2 | 2.01E-09 | 3.40E-07 | 9.66E-06 | 2.04E-07 | 0.008080491 | SLC52A2 |
| chr2 | 120735830 | 120737775 | 1946 | 15 | 5.92E-05 | 2.02E-06 | 0.000424 | 2.04E-07 | 0.001948446 | GLI2 |
| chr17 | 48719929 | 48720936 | 1008 | 4 | 4.40E-07 | 1.98E-07 | 0.000219 | 2.05E-07 | -0.00153746 | NA |
| chr8 | 123657418 | 123658733 | 1316 | 14 | 1.85E-06 | 2.06E-05 | 0.001674 | 2.06E-07 | 0.004010697 | AC135166.1 |
| chr5 | 179119866 | 179121699 | 1834 | 8 | 1.77E-05 | 3.32E-06 | 0.000106 | 2.06E-07 | -0.01288624 | ADAMTS2 |
| chr10 | 7334186 | 7334764 | 579 | 4 | 1.04E-07 | 2.14E-06 | 7.08E-05 | 2.07E-07 | 0.003265416 | SFMBT2 |
| chr15 | 84914562 | 84915596 | 1035 | 3 | 4.35E-06 | 7.85E-08 | 0.000215 | 2.07E-07 | 0.002233349 | SLC28A1, AC087468.2 |
| chr10 | 102460618 | 102461260 | 643 | 9 | 1.06E-06 | 2.76E-07 | 0.004509 | 2.08E-07 | 0.002003057 | RPARP-AS1 |
| chr1 | 26354099 | 26355488 | 1390 | 10 | 6.77E-06 | 1.14E-05 | 0.000659 | 2.08E-07 | 0.001037014 | CRYBG2 |
| chr2 | 27377021 | 27379010 | 1990 | 10 | 8.41E-05 | 5.60E-07 | 0.00126 | 2.08E-07 | -0.00079637 | SNX17, ZNF513 |
| chr20 | 19993509 | 19994188 | 680 | 4 | 2.81E-08 | 4.74E-06 | 2.80E-06 | 2.08E-07 | 0.001554318 | RIN2 |
| chr10 | 71512543 | 71513258 | 716 | 2 | 1.54E-06 | 1.16E-07 | 8.29E-05 | 2.08E-07 | 0.007400194 | CDH23 |
| chr17 | 50507855 | 50509208 | 1354 | 15 | 1.20E-07 | 6.04E-06 | 0.000113 | 2.08E-07 | -0.01400049 | MYCBPAP, AC021491.1 |
| chr1 | 120176325 | 120177638 | 1314 | 12 | 4.13E-08 | 1.41E-06 | 0.000123 | 2.11E-07 | -0.00018945 | SEC22B |
| chr14 | 103889492 | 103889835 | 344 | 3 | 6.15E-07 | 6.92E-08 | 0.000374 | 2.11E-07 | 0.008975809 | NA |
| chr1 | 21279558 | 21280274 | 717 | 7 | 1.23E-07 | 8.35E-06 | 0.000134 | 2.13E-07 | 0.004149932 | ECE1 |
| chr17 | 42670406 | 42671554 | 1149 | 7 | 1.81E-07 | 8.38E-08 | 0.000145 | 2.13E-07 | -0.01948365 | PLEKHH3 |
| chr1 | 155934293 | 155935125 | 833 | 14 | 1.23E-06 | 6.05E-07 | 0.004334 | 2.15E-07 | 0.002819003 | KHDC4 |
| chr9 | 98056141 | 98057260 | 1120 | 13 | 4.86E-08 | 1.38E-05 | 0.000132 | 2.15E-07 | 0.001387981 | NANS |
| chr5 | 83076967 | 83078125 | 1159 | 20 | 3.93E-08 | 9.65E-05 | 0.000425 | 2.15E-07 | 0.001887652 | XRCC4, TMEM167A |
| chr1 | 200979644 | 200979771 | 128 | 2 | 2.10E-07 | 1.41E-06 | 3.07E-06 | 2.15E-07 | 0.003104569 | KIF21B |
| chr4 | 25863556 | 25864774 | 1219 | 9 | 2.31E-06 | 1.44E-07 | 0.00541 | 2.16E-07 | 0.007033271 | SMIM20, SEL1L3 |
| chr1 | 178724460 | 178725346 | 887 | 10 | 4.90E-09 | 0.001038 | 1.29E-05 | 2.16E-07 | 0.001418482 | RALGPS2, AL449106.1 |
| chr16 | 89039578 | 89041250 | 1673 | 4 | 1.18E-06 | 3.04E-06 | 0.000172 | 2.16E-07 | 0.006171991 | NA |
| chr17 | 17577015 | 17578078 | 1064 | 12 | 3.67E-09 | 0.005895 | 2.31E-05 | 2.17E-07 | 0.003877461 | PEMT |
| chr7 | 2221266 | 2221871 | 606 | 5 | 5.90E-08 | 2.42E-07 | 0.000214 | 2.17E-07 | 0.005476004 | MAD1L1, AC069288.1 |
| chr11 | 1309363 | 1310601 | 1239 | 13 | 1.11E-06 | 4.06E-07 | 0.001833 | 2.17E-07 | -0.00502674 | TOLLIP-AS1, TOLLIP |
| chr6 | 15245436 | 15246382 | 947 | 14 | 1.47E-06 | 4.92E-07 | 0.003289 | 2.17E-07 | 0.000763243 | JARID2 |
| chr16 | 35506149 | 35507452 | 1304 | 7 | 6.09E-06 | 5.03E-07 | 0.001138 | 2.17E-07 | 0.015702806 | AC018558.7, AC018558.1, TP53TG3HP |
| chr7 | 76423907 | 76425438 | 1532 | 6 | 0.000135172 | 2.96E-07 | 0.000339 | 2.17E-07 | 0.005328945 | ZP3 |
| chr7 | 156177607 | 156178704 | 1098 | 7 | 1.48E-08 | 7.23E-06 | 4.62E-05 | 2.18E-07 | 0.007937401 | NA |
| chr12 | 53211607 | 53211803 | 197 | 2 | 4.42E-07 | 2.06E-07 | 2.11E-05 | 2.18E-07 | 0.009227424 | RARG |
| chr21 | 16587624 | 16588751 | 1128 | 4 | 1.52E-06 | 4.01E-08 | 0.001802 | 2.19E-07 | 0.012777466 | MIR99AHG |
| chr5 | 6378484 | 6380068 | 1585 | 13 | 4.78E-06 | 1.56E-07 | 0.003771 | 2.20E-07 | -0.00099144 | AC093307.1, MED10 |
| chr1 | 211578847 | 211580240 | 1394 | 10 | 3.96E-06 | 0.000116 | 3.88E-05 | 2.21E-07 | -0.00076471 | SLC30A1 |
| chr2 | 8525264 | 8526090 | 827 | 2 | 2.43E-07 | 3.76E-07 | 9.69E-06 | 2.21E-07 | 0.00747968 | LINC01814 |
| chr8 | 28709203 | 28710184 | 982 | 3 | 4.53E-07 | 9.76E-08 | 0.000239 | 2.21E-07 | 0.009173101 | EXTL3 |
| chr14 | 99578820 | 99579273 | 454 | 2 | 3.53E-09 | 1.04E-05 | 1.20E-06 | 2.22E-07 | 0.008789793 | CCDC85C |
| chr16 | 50644993 | 50645738 | 746 | 4 | 6.22E-07 | 0.0001 | 4.43E-05 | 2.22E-07 | 0.008441468 | NKD1 |
| chr4 | 3415122 | 3417333 | 2212 | 9 | 3.35E-05 | 3.60E-06 | 0.00011 | 2.23E-07 | 0.003296459 | RGS12 |
| chr14 | 92321561 | 92322258 | 698 | 6 | 2.40E-08 | 6.64E-06 | 0.000154 | 2.23E-07 | 0.009560519 | NA |
| chr12 | 54015423 | 54016539 | 1117 | 15 | 1.58E-08 | 4.80E-07 | 0.000176 | 2.23E-07 | 0.006926577 | AC012531.3, HOXC6 |
| chr1 | 58576603 | 58578201 | 1599 | 11 | 2.55E-06 | 1.89E-08 | 0.004431 | 2.24E-07 | 0.020316155 | TACSTD2 |
| chr9 | 128947882 | 128949074 | 1193 | 9 | 2.75E-07 | 7.36E-07 | 0.000593 | 2.24E-07 | 0.001314345 | AL672142.1, NUP188 |
| chr7 | 55964133 | 55964911 | 779 | 12 | 4.00E-08 | 5.81E-06 | 9.15E-05 | 2.25E-07 | 0.002948825 | NIPSNAP2 |
| chr6 | 29436890 | 29436956 | 67 | 4 | 3.88E-07 | 1.03E-07 | 0.000258 | 2.25E-07 | -0.02267388 | OR11A1 |
| chr19 | 11738213 | 11739789 | 1577 | 11 | 3.01E-07 | 1.42E-06 | 2.05E-05 | 2.25E-07 | 0.000168798 | ZNF823 |
| chr10 | 111079225 | 111079688 | 464 | 3 | 4.52E-08 | 9.47E-07 | 8.80E-05 | 2.26E-07 | 0.010786742 | ADRA2A |
| chr2 | 239308757 | 239309360 | 604 | 6 | 7.50E-07 | 3.81E-07 | 0.000708 | 2.26E-07 | 0.010019912 | HDAC4 |
| chr7 | 2520332 | 2521482 | 1151 | 5 | 2.95E-05 | 3.78E-08 | 0.002453 | 2.26E-07 | 0.009471867 | LFNG |
| chr5 | 173866226 | 173867006 | 781 | 4 | 4.36E-07 | 3.34E-07 | 5.40E-05 | 2.27E-07 | 0.01156094 | NA |
| chr13 | 77326044 | 77327343 | 1300 | 12 | 4.25E-06 | 1.00E-05 | 0.002065 | 2.29E-07 | -5.95E-05 | MYCBP2 |
| chr20 | 915706 | 916902 | 1197 | 7 | 6.24E-07 | 3.94E-07 | 0.000846 | 2.29E-07 | 0.006301316 | ANGPT4 |
| chr7 | 100435949 | 100437437 | 1489 | 16 | 1.07E-07 | 1.19E-06 | 0.001933 | 2.29E-07 | -3.50E-05 | AC092849.1, PPP1R35-AS1, PPP1R35 |
| chr22 | 39987719 | 39988895 | 1177 | 3 | 1.63E-05 | 1.10E-07 | 0.000164 | 2.29E-07 | 0.014461384 | NA |
| chr17 | 10696703 | 10698684 | 1982 | 14 | 8.58E-06 | 1.63E-05 | 0.002164 | 2.29E-07 | 0.00061273 | ADPRM, SCO1 |
| chr16 | 53377531 | 53378529 | 999 | 3 | 5.32E-06 | 7.90E-07 | 2.81E-05 | 2.29E-07 | 0.010190169 | AC007342.4 |
| chr20 | 31554738 | 31555348 | 611 | 3 | 1.31E-08 | 1.14E-05 | 1.67E-06 | 2.29E-07 | 0.004389661 | HM13 |
| chr3 | 66139788 | 66139797 | 10 | 2 | 2.06E-07 | 9.22E-07 | 4.42E-06 | 2.30E-07 | 0.008854738 | SLC25A26 |
| chr19 | 4944461 | 4945205 | 745 | 3 | 2.53E-09 | 3.11E-06 | 2.08E-05 | 2.31E-07 | 0.002925452 | UHRF1 |
| chr13 | 46183260 | 46183520 | 261 | 5 | 2.67E-07 | 8.41E-07 | 0.000468 | 2.31E-07 | 0.020899922 | LCP1 |
| chr1 | 210236317 | 210237259 | 943 | 6 | 9.07E-07 | 6.74E-08 | 0.000946 | 2.31E-07 | 0.008779439 | SERTAD4 |
| chr16 | 11781950 | 11783056 | 1107 | 9 | 7.88E-08 | 1.81E-06 | 0.000245 | 2.32E-07 | 0.005888769 | ZC3H7A |
| chr17 | 74779755 | 74780147 | 393 | 3 | 2.89E-07 | 2.29E-07 | 5.35E-05 | 2.32E-07 | 0.014882308 | TMEM104 |
| chr12 | 45050025 | 45051119 | 1095 | 10 | 1.09E-05 | 5.43E-07 | 0.002428 | 2.32E-07 | 0.002274652 | AC008127.1, DBX2 |
| chr15 | 64147323 | 64147696 | 374 | 3 | 1.99E-07 | 2.72E-06 | 4.76E-05 | 2.34E-07 | 0.003374827 | NA |
| chr18 | 3493955 | 3494437 | 483 | 3 | 2.92E-07 | 4.90E-07 | 1.45E-05 | 2.34E-07 | 0.014013316 | NA |
| chr16 | 86120668 | 86121250 | 583 | 2 | 2.58E-09 | 2.70E-05 | 9.35E-07 | 2.34E-07 | 0.023479832 | NA |
| chr6 | 32839129 | 32840118 | 990 | 14 | 3.57E-09 | 4.54E-06 | 0.000316 | 2.35E-07 | 0.003005854 | NA |
| chr11 | 63534350 | 63535479 | 1130 | 3 | 3.37E-06 | 1.82E-07 | 5.40E-05 | 2.35E-07 | 0.014103981 | NA |
| chr12 | 106138713 | 106140842 | 2130 | 15 | 2.07E-05 | 1.23E-05 | 0.000957 | 2.35E-07 | 0.003910676 | NUAK1 |
| chr17 | 18246010 | 18247640 | 1631 | 8 | 2.12E-05 | 1.71E-07 | 0.000811 | 2.35E-07 | 0.009658994 | FLII |
| chr5 | 77639894 | 77640968 | 1075 | 6 | 3.12E-07 | 5.57E-06 | 0.000461 | 2.36E-07 | 0.008156004 | NA |
| chr22 | 37641172 | 37642342 | 1171 | 7 | 3.42E-06 | 3.92E-08 | 0.004034 | 2.36E-07 | 0.007782857 | SH3BP1, Z83844.2, PDXP-DT |
| chr13 | 113927317 | 113928125 | 809 | 5 | 3.50E-06 | 3.47E-08 | 0.002413 | 2.37E-07 | 0.022743749 | LINC00565 |
| chr9 | 37602574 | 37603218 | 645 | 3 | 7.61E-08 | 8.12E-06 | 2.39E-06 | 2.37E-07 | 0.005565871 | AL138752.2 |
| chr8 | 30555426 | 30556790 | 1365 | 7 | 6.22E-07 | 1.56E-05 | 0.00027 | 2.38E-07 | 0.003148757 | RBPMS |
| chr10 | 133247755 | 133248651 | 897 | 8 | 1.18E-08 | 7.17E-06 | 2.73E-05 | 2.39E-07 | 0.002560096 | MIR202HG |
| chr20 | 63700580 | 63701558 | 979 | 8 | 8.27E-08 | 3.85E-05 | 0.000163 | 2.39E-07 | 0.002558435 | ARFRP1 |
| chr12 | 41412507 | 41412726 | 220 | 2 | 5.09E-07 | 2.14E-07 | 2.42E-05 | 2.39E-07 | -0.00056708 | PDZRN4, AC090531.1 |
| chr11 | 706526 | 707166 | 641 | 7 | 4.63E-08 | 1.18E-06 | 8.29E-05 | 2.39E-07 | 0.010169052 | EPS8L2, DEAF1 |
| chr4 | 2320230 | 2320980 | 751 | 6 | 4.46E-07 | 4.36E-06 | 0.001233 | 2.39E-07 | 0.006859086 | ZFYVE28 |
| chr19 | 18756691 | 18757059 | 369 | 3 | 5.85E-07 | 1.53E-07 | 7.37E-05 | 2.39E-07 | 0.004348335 | CRTC1 |
| chr6 | 152750266 | 152750956 | 691 | 6 | 1.12E-06 | 6.56E-07 | 0.001717 | 2.39E-07 | 0.018907327 | VIP |
| chr3 | 49132567 | 49133752 | 1186 | 10 | 8.40E-07 | 1.64E-08 | 0.010001 | 2.40E-07 | 0.020497644 | LAMB2 |
| chr7 | 136869984 | 136871810 | 1827 | 8 | 1.64E-05 | 2.74E-07 | 0.001436 | 2.40E-07 | 0.008736318 | CHRM2, AC009264.1 |
| chr20 | 57262427 | 57263422 | 996 | 6 | 7.74E-07 | 1.30E-06 | 3.94E-05 | 2.41E-07 | 0.016893372 | BMP7 |
| chr11 | 102390800 | 102391404 | 605 | 3 | 9.77E-07 | 4.22E-07 | 9.75E-05 | 2.41E-07 | 0.007798523 | NA |
| chr1 | 93908712 | 93910042 | 1331 | 15 | 3.68E-07 | 1.29E-05 | 0.000437 | 2.41E-07 | 0.000824166 | GCLM |
| chr12 | 52543848 | 52544636 | 789 | 3 | 1.57E-07 | 3.17E-07 | 2.58E-05 | 2.44E-07 | 0.008373214 | KRT71 |
| chr6 | 31945238 | 31946528 | 1291 | 13 | 4.30E-07 | 1.09E-07 | 0.006531 | 2.45E-07 | 0.00618102 | C2, AL645922.1, CFB |
| chr1 | 38570262 | 38570358 | 97 | 3 | 7.53E-07 | 1.48E-07 | 0.00011 | 2.45E-07 | 0.017011537 | NA |
| chr12 | 106357282 | 106358626 | 1345 | 17 | 6.29E-08 | 2.61E-05 | 9.94E-05 | 2.46E-07 | 0.000946254 | POLR3B |
| chr8 | 26576021 | 26577176 | 1156 | 7 | 1.06E-09 | 0.000131 | 8.79E-05 | 2.47E-07 | 0.011017987 | DPYSL2 |
| chr7 | 152145107 | 152146605 | 1499 | 5 | 5.69E-06 | 4.04E-08 | 0.001864 | 2.47E-07 | 0.007259039 | KMT2C |
| chr17 | 2883701 | 2884201 | 501 | 3 | 7.92E-07 | 1.98E-07 | 5.23E-05 | 2.47E-07 | 0.010046385 | RAP1GAP2 |
| chr1 | 62906632 | 62906792 | 161 | 2 | 5.22E-07 | 2.87E-07 | 1.64E-05 | 2.47E-07 | 0.029820145 | NA |
| chr2 | 219441199 | 219442485 | 1287 | 7 | 2.94E-05 | 2.14E-07 | 0.002295 | 2.48E-07 | 0.003136464 | SPEG |
| chr22 | 30615656 | 30615832 | 177 | 2 | 6.86E-07 | 1.66E-07 | 4.67E-05 | 2.49E-07 | 0.02816711 | TCN2 |
| chr10 | 132547620 | 132548666 | 1047 | 6 | 1.92E-06 | 5.57E-07 | 0.002421 | 2.50E-07 | 0.009526731 | INPP5A |
| chr5 | 91279324 | 91279642 | 319 | 5 | 3.83E-07 | 2.74E-07 | 0.001287 | 2.50E-07 | 0.025930429 | LUCAT1 |
| chr3 | 159586563 | 159587014 | 452 | 3 | 4.34E-07 | 2.76E-07 | 3.14E-05 | 2.50E-07 | 0.016303191 | IQCJ-SCHIP1 |
| chr1 | 153921987 | 153923485 | 1499 | 15 | 2.69E-06 | 6.04E-08 | 0.005842 | 2.50E-07 | -0.00014043 | AL358472.5, GATAD2B |
| chr5 | 2702224 | 2703281 | 1058 | 9 | 5.46E-08 | 3.90E-05 | 6.21E-05 | 2.51E-07 | 0.001393429 | NA |
| chr1 | 205068833 | 205069955 | 1123 | 5 | 7.70E-06 | 5.50E-07 | 0.000345 | 2.51E-07 | 0.007094696 | CNTN2 |
| chr12 | 56599171 | 56599760 | 590 | 2 | 1.03E-06 | 1.57E-07 | 5.71E-05 | 2.51E-07 | -0.00016148 | BAZ2A |
| chr16 | 85056171 | 85057234 | 1064 | 5 | 2.96E-07 | 2.27E-05 | 1.01E-05 | 2.52E-07 | -0.00505752 | KIAA0513 |
| chr7 | 95096860 | 95096993 | 134 | 2 | 1.56E-07 | 2.59E-06 | 2.57E-06 | 2.52E-07 | 0.004056195 | PPP1R9A, PPP1R9A-AS1 |
| chr8 | 142612238 | 142613259 | 1022 | 6 | 2.75E-07 | 1.63E-06 | 0.00046 | 2.53E-07 | 0.004739733 | ARC |

| | | | | | | | | | | |
|---|---|---|---|---|---|---|---|---|---|---|
| chr19 | 1361623 | 1362386 | 764 | 2 | 3.25E-09 | 1.05E-06 | 4.56E-06 | 2.53E-07 | 0.005744784 | PWWP3A |
| chr6 | 33788305 | 33789300 | 996 | 10 | 9.91E-07 | 1.27E-06 | 0.001531 | 2.53E-07 | -9.03E-05 | LEMD2 |
| chr10 | 132850339 | 132851629 | 1291 | 10 | 6.13E-09 | 1.01E-05 | 1.42E-05 | 2.55E-07 | 0.002258102 | CFAP46 |
| chr2 | 24885466 | 24885850 | 385 | 3 | 1.92E-07 | 2.95E-06 | 5.42E-06 | 2.55E-07 | 0.005507361 | ADCY3 |
| chr4 | 1644223 | 1645434 | 1212 | 6 | 3.81E-06 | 5.98E-08 | 0.00256 | 2.55E-07 | 0.001919916 | FAM53A |
| chr12 | 6752313 | 6753802 | 1490 | 14 | 5.05E-06 | 8.11E-07 | 0.001348 | 2.55E-07 | 0.001744042 | MLF2 |
| chr19 | 35476368 | 35476650 | 283 | 2 | 1.41E-07 | 1.82E-06 | 3.12E-06 | 2.56E-07 | 0.011180739 | NA |
| chr17 | 34960573 | 34961913 | 1341 | 16 | 2.83E-07 | 8.26E-06 | 0.000222 | 2.56E-07 | 0.000936395 | ZNF830, CCT6B |
| chr10 | 78302258 | 78304034 | 1777 | 8 | 5.65E-06 | 2.41E-06 | 5.76E-05 | 2.58E-07 | 0.005474012 | AC012560.1, LINC00595 |
| chr5 | 149427604 | 149428924 | 1321 | 5 | 1.09E-05 | 5.36E-07 | 0.000113 | 2.59E-07 | 0.002044081 | CARMN, MIR143, AC131025.2 |
| chr19 | 48739696 | 48740869 | 1174 | 6 | 6.15E-07 | 2.86E-08 | 0.004452 | 2.59E-07 | 0.011987438 | RASIP1, IZUMO1 |
| chr19 | 1617800 | 1619402 | 1603 | 6 | 1.36E-08 | 2.14E-06 | 0.000286 | 2.59E-07 | 0.003556982 | TCF3 |
| chr17 | 78985930 | 78986569 | 640 | 3 | 7.86E-07 | 1.24E-07 | 0.000123 | 2.60E-07 | 0.015039076 | NA |
| chr11 | 77112872 | 77113479 | 608 | 4 | 9.45E-08 | 1.62E-06 | 1.65E-05 | 2.60E-07 | 0.003926056 | CAPN5 |
| chr6 | 34053739 | 34053787 | 49 | 3 | 1.11E-06 | 1.22E-07 | 0.000151 | 2.60E-07 | 0.004406427 | GRM4 |
| chr11 | 69891517 | 69892616 | 1100 | 9 | 1.96E-06 | 9.86E-08 | 0.0029 | 2.60E-07 | 0.01201886 | NA |
| chr11 | 104074132 | 104074294 | 163 | 3 | 6.25E-07 | 5.46E-07 | 0.000102 | 2.61E-07 | 0.01701218 | PDGFD |
| chr2 | 1715389 | 1715551 | 163 | 3 | 9.39E-07 | 1.13E-07 | 0.000296 | 2.63E-07 | 0.03041678 | PXDN |
| chr15 | 41558414 | 41559771 | 1358 | 7 | 3.15E-06 | 0.000287 | 0.000143 | 2.63E-07 | 0.002772218 | TYRO3 |
| chr11 | 1298418 | 1299983 | 1566 | 11 | 1.33E-05 | 2.64E-05 | 0.00078 | 2.63E-07 | 0.002808988 | TOLLIP |
| chr3 | 38645267 | 38645893 | 627 | 3 | 3.64E-07 | 1.91E-07 | 5.72E-05 | 2.63E-07 | 0.010907807 | SCN5A |
| chr17 | 74955145 | 74955275 | 131 | 2 | 6.04E-07 | 1.48E-07 | 8.93E-05 | 2.63E-07 | 0.017440968 | HID1 |
| chr3 | 184811374 | 184812466 | 1093 | 11 | 4.20E-07 | 1.47E-05 | 0.001702 | 2.64E-07 | 0.001166497 | VPS8 |
| chr14 | 50396523 | 50397697 | 1175 | 13 | 2.59E-07 | 7.15E-05 | 0.000589 | 2.64E-07 | 0.004745964 | AL359397.1, CDKL1 |
| chr13 | 112342032 | 112343329 | 1298 | 7 | 4.16E-06 | 2.07E-07 | 0.004766 | 2.66E-07 | -0.01646125 | NA |
| chr9 | 109012615 | 109013883 | 1269 | 11 | 5.34E-06 | 7.09E-06 | 0.000188 | 2.68E-07 | 0.001743344 | CTNNAL1 |
| chr1 | 30723565 | 30724371 | 807 | 8 | 1.38E-07 | 4.99E-06 | 0.000907 | 2.68E-07 | 0.005634521 | MATN1-AS1, MATN1 |
| chr14 | 101125721 | 101126143 | 423 | 3 | 5.57E-07 | 4.17E-07 | 3.97E-05 | 2.68E-07 | 0.003168947 | NA |
| chr1 | 154551806 | 154552855 | 1050 | 7 | 2.44E-07 | 2.21E-07 | 7.24E-05 | 2.68E-07 | 0.0016279 | UBE2Q1 |
| chr11 | 57321383 | 57322801 | 1419 | 8 | 1.33E-07 | 1.30E-06 | 0.000127 | 2.69E-07 | 0.005556787 | TNKS1BP1 |
| chr21 | 45836506 | 45837464 | 959 | 6 | 1.73E-06 | 3.06E-07 | 0.001458 | 2.69E-07 | 0.00881717 | PCBP3, PCBP3-AS1 |
| chr16 | 2999876 | 3000152 | 277 | 3 | 2.22E-07 | 4.95E-07 | 2.79E-05 | 2.71E-07 | -0.00250726 | LINC00514 |
| chr6 | 137495764 | 137497190 | 1427 | 8 | 1.74E-07 | 9.92E-07 | 0.00043 | 2.71E-07 | 0.004275191 | NA |
| chr8 | 94217297 | 94217644 | 348 | 7 | 5.79E-07 | 5.10E-07 | 0.001206 | 2.72E-07 | 0.01008042 | CDH17 |
| chr1 | 159945584 | 159946682 | 1099 | 9 | 2.29E-06 | 5.14E-07 | 0.005488 | 2.72E-07 | 0.003664974 | IGSF9 |
| chr22 | 43409959 | 43411115 | 1157 | 6 | 5.50E-08 | 1.68E-05 | 0.000398 | 2.73E-07 | 0.008307911 | NA |
| chr8 | 1361488 | 1363410 | 1923 | 10 | 1.21E-05 | 2.91E-05 | 0.001652 | 2.73E-07 | -0.00121425 | DLGAP2 |
| chr10 | 71712558 | 71713478 | 921 | 4 | 3.26E-07 | 1.34E-07 | 0.000122 | 2.75E-07 | 0.010991624 | CDH23, C10orf105 |
| chr7 | 32955902 | 32957261 | 1360 | 10 | 2.15E-06 | 8.28E-06 | 0.000388 | 2.75E-07 | 0.004029846 | NA |
| chr6 | 31658615 | 31660663 | 2049 | 29 | 9.72E-06 | 4.09E-07 | 0.010608 | 2.75E-07 | 0.001584217 | C6orf47-AS1, C6orf47 |
| chr18 | 80033812 | 80034836 | 1025 | 9 | 2.73E-07 | 9.52E-08 | 0.005425 | 2.75E-07 | -0.00139399 | AC090360.1, RBFA, TXNL4A |
| chr3 | 69250204 | 69250341 | 138 | 2 | 7.64E-07 | 1.55E-07 | 9.19E-05 | 2.76E-07 | 0.02313547 | FRMD4B |
| chr3 | 101561139 | 101562107 | 969 | 13 | 2.62E-07 | 1.52E-06 | 0.001129 | 2.77E-07 | -0.0009595 | TRMT10C |
| chr1 | 17826255 | 17826564 | 310 | 2 | 2.11E-08 | 2.24E-06 | 3.07E-06 | 2.78E-07 | 0.011784257 | ACTL8 |
| chr1 | 205211399 | 205212453 | 1055 | 12 | 7.25E-08 | 2.56E-06 | 0.001794 | 2.78E-07 | 0.004007588 | DSTYK |
| chr14 | 93918265 | 93918903 | 639 | 4 | 1.10E-07 | 2.54E-06 | 6.48E-05 | 2.78E-07 | 0.000465662 | FAM181A, FAM181A-AS1 |
| chr7 | 25949904 | 25950406 | 503 | 6 | 4.21E-07 | 6.94E-07 | 0.000108 | 2.78E-07 | 0.007272925 | MIR148A |
| chr11 | 2883794 | 2886524 | 2731 | 26 | 1.61E-05 | 1.39E-05 | 0.001741 | 2.79E-07 | 0.000853232 | CDKN1C |
| chr16 | 89281316 | 89283058 | 1743 | 6 | 3.42E-08 | 2.60E-06 | 0.00033 | 2.80E-07 | 0.001834391 | ANKRD11 |
| chr11 | 31813986 | 31815125 | 1140 | 6 | 2.76E-07 | 3.01E-07 | 0.000505 | 2.80E-07 | 0.004633677 | PAUPAR, AL035078.4, PAX6 |
| chr8 | 141284722 | 141285530 | 809 | 4 | 1.04E-06 | 7.49E-08 | 0.000871 | 2.80E-07 | 7.09E-05 | AC011676.3, SLC45A4 |
| chr2 | 142877562 | 142878047 | 486 | 3 | 1.05E-06 | 1.51E-07 | 0.000209 | 2.82E-07 | 0.016837446 | KYNU |
| chr1 | 161165821 | 161166874 | 1054 | 13 | 7.78E-08 | 1.31E-05 | 0.000131 | 2.82E-07 | -0.00111381 | PPOX |
| chr3 | 50191438 | 50192593 | 1156 | 4 | 6.08E-06 | 8.87E-06 | 2.49E-05 | 2.82E-07 | 0.001202259 | GNAT1 |
| chr2 | 229920981 | 229923131 | 2151 | 17 | 7.79E-06 | 2.06E-06 | 0.00093 | 2.82E-07 | 0.002816271 | FBXO36, TRIP12 |
| chr19 | 37218805 | 37219192 | 388 | 2 | 2.60E-10 | 1.94E-05 | 1.20E-06 | 2.83E-07 | 0.016760081 | ZNF383 |
| chr11 | 64316849 | 64318548 | 1700 | 22 | 2.10E-05 | 1.11E-06 | 0.005871 | 2.83E-07 | 0.000573804 | PRDX5, TRMT112 |
| chr14 | 103002245 | 103003542 | 1298 | 5 | 1.75E-05 | 8.32E-07 | 0.000737 | 2.83E-07 | 0.005394433 | CDC42BPB |
| chr17 | 4519039 | 4519463 | 425 | 3 | 1.02E-06 | 3.90E-07 | 9.54E-05 | 2.84E-07 | 0.008682194 | SPNS2 |
| chr8 | 133571488 | 133572449 | 962 | 8 | 4.23E-08 | 1.49E-05 | 0.00016 | 2.84E-07 | 0.002170473 | ST3GAL1 |
| chr2 | 241665798 | 241667748 | 1951 | 10 | 1.91E-05 | 2.74E-07 | 0.001253 | 2.84E-07 | -0.00458913 | ATG4B |
| chr17 | 81447708 | 81449524 | 1817 | 6 | 2.55E-05 | 3.09E-06 | 0.00011 | 2.84E-07 | 0.007113954 | BAHCC1 |
| chr22 | 41752836 | 41753821 | 986 | 3 | 1.58E-09 | 2.23E-05 | 1.20E-06 | 2.85E-07 | 0.008848148 | MEI1 |
| chr7 | 1580401 | 1580685 | 285 | 2 | 1.32E-07 | 3.53E-07 | 1.59E-05 | 2.85E-07 | 0.007979411 | PSMG3-AS1 |
| chr17 | 68456202 | 68457341 | 1140 | 5 | 8.01E-09 | 3.53E-07 | 0.000282 | 2.85E-07 | 0.010918842 | AC007780.1, WIPI1 |
| chr2 | 238912746 | 238913435 | 690 | 2 | 1.81E-09 | 6.12E-05 | 9.28E-07 | 2.86E-07 | 0.011174694 | NA |
| chr2 | 226995390 | 226996264 | 875 | 3 | 1.48E-09 | 5.67E-05 | 8.02E-07 | 2.88E-07 | 0.012612114 | RHBDD1 |
| chr7 | 4960382 | 4960588 | 207 | 4 | 7.95E-07 | 2.37E-07 | 0.000514 | 2.88E-07 | 0.037858229 | NA |
| chr7 | 130441609 | 130442518 | 910 | 5 | 7.32E-10 | 1.48E-05 | 1.67E-06 | 2.88E-07 | 0.006881187 | CEP41 |
| chr18 | 36309241 | 36309824 | 584 | 2 | 2.03E-07 | 4.16E-07 | 1.34E-05 | 2.88E-07 | 0.016788409 | FHOD3 |
| chr7 | 24256034 | 24256316 | 283 | 3 | 9.83E-07 | 1.86E-07 | 0.000104 | 2.88E-07 | 0.009774182 | NA |
| chr5 | 76379293 | 76379424 | 132 | 2 | 9.07E-07 | 1.75E-07 | 7.10E-05 | 2.90E-07 | 0.03630525 | NA |
| chr5 | 179792499 | 179794089 | 1591 | 10 | 1.18E-05 | 1.76E-06 | 0.00321 | 2.91E-07 | 0.006121717 | LTC4S |
| chr17 | 7425052 | 7426400 | 1349 | 10 | 3.18E-07 | 1.16E-05 | 0.000345 | 2.93E-07 | 0.00425159 | AC113189.2, SPEM2 |
| chr17 | 76385703 | 76387193 | 1491 | 8 | 1.33E-05 | 1.00E-05 | 0.000803 | 2.93E-07 | 0.004332144 | SPHK1 |
| chr15 | 48201542 | 48202132 | 591 | 3 | 3.41E-08 | 1.92E-06 | 5.57E-06 | 2.93E-07 | 0.01357565 | CTXN2, SLC12A1, AC066612.2 |
| chr1 | 25027370 | 25027875 | 506 | 2 | 2.33E-10 | 0.000228 | 8.13E-07 | 2.94E-07 | 0.002670925 | NA |
| chr1 | 160672587 | 160673185 | 599 | 5 | 3.65E-07 | 5.50E-07 | 0.000199 | 2.94E-07 | 0.021299673 | AL121985.1 |
| chr3 | 190717859 | 190719021 | 1163 | 3 | 2.67E-06 | 1.02E-07 | 0.000457 | 2.94E-07 | 0.001908692 | NA |
| chr20 | 63950038 | 63951347 | 1310 | 8 | 3.87E-06 | 5.31E-06 | 0.00047 | 2.94E-07 | 0.003439096 | UCKL1 |
| chr11 | 67317708 | 67319200 | 1493 | 14 | 3.41E-05 | 2.64E-07 | 0.002338 | 2.94E-07 | 0.001108663 | RAD9A |
| chr11 | 134046065 | 134047825 | 1761 | 6 | 3.36E-05 | 5.75E-07 | 0.000523 | 2.94E-07 | 0.009242414 | LINC02731 |
| chr20 | 63745355 | 63745703 | 349 | 4 | 2.73E-07 | 1.05E-07 | 0.000561 | 2.96E-07 | 0.002984262 | AL121845.1, ZBTB46 |
| chr11 | 44605158 | 44606106 | 949 | 6 | 1.92E-07 | 1.96E-06 | 0.001845 | 2.96E-07 | 0.00617866 | CD82, AC010768.1 |
| chr15 | 66504297 | 66505460 | 1164 | 12 | 2.60E-07 | 1.50E-05 | 8.10E-05 | 2.96E-07 | 0.000363123 | ZWILCH, RPL4 |
| chr17 | 40500625 | 40502887 | 2263 | 13 | 7.45E-06 | 2.31E-05 | 0.000574 | 2.97E-07 | 0.003053857 | TNS4 |
| chr22 | 50506929 | 50508217 | 1289 | 13 | 1.57E-07 | 1.89E-06 | 0.003957 | 2.97E-07 | -0.00021217 | LMF2 |
| chr7 | 102249353 | 102250451 | 1099 | 5 | 5.01E-08 | 1.17E-06 | 0.000114 | 2.98E-07 | 0.000880584 | CUX1 |
| chr7 | 100588209 | 100589110 | 902 | 4 | 1.73E-08 | 0.001854 | 1.56E-05 | 2.99E-07 | 0.008010462 | FBXO24 |
| chr2 | 199324741 | 199325206 | 466 | 3 | 1.68E-07 | 7.32E-07 | 1.39E-05 | 2.99E-07 | 0.006109367 | SATB2 |
| chr13 | 80654739 | 80655406 | 668 | 6 | 5.32E-07 | 2.43E-07 | 0.002003 | 3.00E-07 | -0.02129392 | PWWP2AP1 |
| chr3 | 141051351 | 141052530 | 1180 | 19 | 1.09E-06 | 6.79E-07 | 0.005743 | 3.00E-07 | 0.001844018 | SPSB4 |
| chr1 | 214753397 | 214753603 | 207 | 3 | 1.39E-06 | 9.86E-08 | 0.000492 | 3.00E-07 | 0.017935174 | NA |
| chr3 | 3798237 | 3799773 | 1537 | 12 | 8.24E-07 | 2.26E-06 | 0.004215 | 3.00E-07 | 0.001926547 | LRRN1, SUMF1 |
| chr2 | 119159423 | 119160179 | 757 | 3 | 1.67E-08 | 3.40E-07 | 3.28E-05 | 3.02E-07 | -0.01325715 | NA |
| chr2 | 239280285 | 239280665 | 381 | 3 | 2.66E-07 | 6.10E-07 | 1.66E-05 | 3.02E-07 | 0.020733955 | HDAC4 |
| chr3 | 119676995 | 119678107 | 1113 | 15 | 7.21E-07 | 4.08E-06 | 0.003605 | 3.02E-07 | -0.00023618 | COX17 |
| chr19 | 47698541 | 47699339 | 799 | 6 | 2.17E-06 | 3.43E-08 | 0.00369 | 3.02E-07 | 0.000884749 | AC010519.1, BICRA |
| chr12 | 123364370 | 123365879 | 1510 | 7 | 1.07E-05 | 1.54E-06 | 9.80E-05 | 3.02E-07 | 0.000809737 | SBNO1-AS1, SBNO1, MIR8072 |
| chr9 | 91722416 | 91722906 | 491 | 2 | 3.56E-07 | 2.13E-07 | 4.53E-05 | 3.02E-07 | 0.00941129 | ROR2 |
| chr15 | 70460200 | 70460397 | 198 | 3 | 3.64E-07 | 1.12E-06 | 7.14E-05 | 3.03E-07 | 0.014029666 | NA |
| chr5 | 40840411 | 40841488 | 1078 | 8 | 1.00E-07 | 3.88E-06 | 3.80E-05 | 3.04E-07 | 0.011030951 | CARD6 |
| chr6 | 43687230 | 43689137 | 1908 | 14 | 2.54E-06 | 3.65E-05 | 0.000452 | 3.04E-07 | 0.002788252 | MRPS18A |
| chr20 | 64188788 | 64189601 | 814 | 3 | 1.90E-07 | 3.23E-07 | 4.68E-05 | 3.05E-07 | 0.015350868 | MYT1 |
| chr10 | 98029820 | 98031153 | 1334 | 9 | 8.12E-06 | 1.15E-06 | 0.001875 | 3.06E-07 | 0.002318245 | CRTAC1 |
| chr5 | 170055264 | 170056277 | 1014 | 3 | 3.67E-05 | 1.05E-07 | 0.000406 | 3.06E-07 | -0.00146442 | DOCK2 |
| chr14 | 93043739 | 93044446 | 708 | 3 | 2.00E-08 | 1.95E-05 | 6.44E-05 | 3.06E-07 | 0.0104793 | ITPK1 |
| chr6 | 108114371 | 108116480 | 2110 | 12 | 3.03E-05 | 9.10E-07 | 0.000143 | 3.07E-07 | 0.004640272 | OSTM1 |
| chr9 | 137230222 | 137231244 | 1023 | 9 | 1.76E-07 | 2.55E-06 | 0.00015 | 3.08E-07 | 0.005640695 | SLC34A3 |
| chr7 | 27361329 | 27361921 | 593 | 5 | 2.05E-09 | 2.31E-05 | 8.20E-06 | 3.08E-07 | 0.010262551 | AC004009.1 |
| chr15 | 32640740 | 32641984 | 1245 | 9 | 8.85E-07 | 1.13E-06 | 9.49E-05 | 3.08E-07 | 0.008337078 | SCG5, AC123768.4 |
| chr8 | 27992658 | 27993080 | 423 | 9 | 2.17E-06 | 1.56E-07 | 0.003004 | 3.09E-07 | 0.008162213 | SCARA5 |
| chr1 | 42958956 | 42959666 | 711 | 13 | 4.62E-08 | 1.33E-06 | 0.003827 | 3.09E-07 | 0.005395959 | SLC2A1-AS1, AC099795.1 |
| chr6 | 37547683 | 37548530 | 848 | 3 | 1.00E-06 | 1.16E-07 | 0.000338 | 3.10E-07 | 0.007173275 | AL353597.1 |
| chr16 | 23826277 | 23826701 | 425 | 2 | 6.91E-08 | 2.55E-06 | 3.14E-06 | 3.10E-07 | 0.014752217 | NA |
| chr6 | 157887365 | 157888102 | 738 | 5 | 4.32E-08 | 4.05E-05 | 8.87E-05 | 3.10E-07 | 0.009121987 | SNX9, SNX9-AS1 |
| chr6 | 151239442 | 151240532 | 1091 | 14 | 7.79E-07 | 9.74E-07 | 0.002393 | 3.10E-07 | 0.004855504 | AKAP12 |
| chr1 | 53078467 | 53078942 | 476 | 5 | 8.78E-07 | 4.43E-07 | 0.000829 | 3.11E-07 | 0.003094253 | PODN, AL445183.1 |
| chr19 | 44746906 | 44747497 | 592 | 4 | 1.24E-08 | 1.60E-05 | 8.39E-05 | 3.11E-07 | 0.00575832 | NA |
| chr19 | 11155361 | 11156353 | 993 | 12 | 3.17E-07 | 1.03E-06 | 0.000147 | 3.11E-07 | 0.002157744 | SPC24 |
| chr10 | 110496260 | 110496971 | 712 | 5 | 8.93E-07 | 1.67E-07 | 0.00192 | 3.11E-07 | 0.011973509 | AL355512.1 |
| chr6 | 30719680 | 30722067 | 2388 | 38 | 0.000104615 | 2.67E-05 | 0.010363 | 3.12E-07 | 0.000554679 | TUBB |
| chr11 | 68010148 | 68011031 | 884 | 8 | 3.51E-07 | 1.11E-06 | 0.000135 | 3.12E-07 | 0.003275202 | ALDH3B1 |
| chr19 | 10624798 | 10625772 | 975 | 10 | 8.82E-07 | 1.82E-06 | 0.007926 | 3.12E-07 | 0.012707419 | SLC44A2 |
| chr17 | 73901888 | 73902973 | 1086 | 8 | 2.22E-06 | 4.86E-07 | 0.000471 | 3.13E-07 | 0.007910294 | AC137735.1 |
| chr10 | 97633491 | 97634383 | 893 | 13 | 8.47E-07 | 1.83E-07 | 0.001642 | 3.13E-07 | 0.003867754 | AL355315.1, MORN4 |
| chr2 | 223603177 | 223603854 | 678 | 2 | 1.17E-07 | 4.28E-07 | 1.49E-05 | 3.14E-07 | 0.028951572 | NA |
| chr13 | 42969056 | 42969871 | 816 | 5 | 2.69E-06 | 1.11E-07 | 0.001616 | 3.14E-07 | 0.002524405 | EPSTI1 |
| chr20 | 62302011 | 62303211 | 1201 | 15 | 1.83E-07 | 6.40E-07 | 6.76E-05 | 3.14E-07 | 0.001606768 | ADRM1 |
| chr14 | 24271081 | 24271933 | 853 | 11 | 8.95E-07 | 5.16E-07 | 0.000777 | 3.15E-07 | 0.002783801 | AL096870.12, AL096870.10, RABGGTA |
| chr5 | 66897401 | 66898065 | 665 | 3 | 1.16E-09 | 0.000316 | 6.92E-07 | 3.16E-07 | 0.002846102 | MAST4 |
| chr9 | 98848293 | 98849081 | 789 | 6 | 8.72E-07 | 1.82E-07 | 0.000335 | 3.16E-07 | 0.001212631 | GALNT12, AL136084.1 |
| chr16 | 87836151 | 87837613 | 1463 | 8 | 2.06E-05 | 1.55E-06 | 0.001673 | 3.16E-07 | 0.003671223 | AC126696.2, SLC7A5 |
| chr1 | 156152631 | 156154188 | 1558 | 12 | 2.38E-07 | 8.91E-06 | 4.97E-05 | 3.16E-07 | 0.004487421 | SEMA4A |
| chr20 | 58850460 | 58851991 | 1532 | 30 | 6.66E-09 | 3.38E-05 | 0.000694 | 3.16E-07 | 0.002179713 | GNAS, AL132655.4, AL132655.5, GNAS-AS1 |
| chr1 | 181087514 | 181088710 | 1197 | 14 | 3.85E-07 | 2.72E-06 | 0.008057 | 3.17E-07 | 0.003773683 | IER5 |
| chr1 | 39738043 | 39739175 | 1133 | 10 | 6.56E-08 | 0.000233 | 3.52E-05 | 3.17E-07 | 0.001313755 | PPIE |
| chr3 | 187701502 | 187702920 | 1419 | 8 | 2.44E-07 | 2.00E-07 | 0.000497 | 3.18E-07 | 0.002223536 | AC072022.1, RTP2 |
| chr10 | 63976587 | 63977288 | 702 | 4 | 6.89E-08 | 7.69E-07 | 0.000156 | 3.18E-07 | 0.008205323 | AC013287.1 |
| chr19 | 11174452 | 11174655 | 204 | 3 | 1.49E-06 | 1.23E-07 | 0.00039 | 3.19E-07 | 0.007437984 | KANK2 |
| chr20 | 46132874 | 46133157 | 284 | 3 | 1.47E-07 | 8.90E-05 | 7.07E-06 | 3.20E-07 | 0.011345162 | NA |
| chr2 | 130341933 | 130343022 | 1090 | 16 | 2.25E-06 | 3.61E-07 | 0.003842 | 3.20E-07 | 0.000473451 | IMP4, CCDC115 |
| chr20 | 62427539 | 62428247 | 709 | 8 | 7.36E-07 | 2.74E-07 | 0.000536 | 3.20E-07 | 0.008454535 | AL121832.1, RBBP8NL |
| chr2 | 104659143 | 104660200 | 1058 | 7 | 1.88E-06 | 1.96E-07 | 0.002731 | 3.20E-07 | -0.01435088 | AC013402.1, AC068057.3 |
| chr11 | 70179052 | 70179242 | 191 | 2 | 1.27E-07 | 1.45E-06 | 4.77E-06 | 3.21E-07 | 0.006727056 | ANO1 |
| chr5 | 52786552 | 52788579 | 2028 | 18 | 2.41E-05 | 4.70E-05 | 0.000789 | 3.21E-07 | 0.00233279 | ITGA1, PELO, AC022126.1 |
| chr5 | 43010409 | 43011003 | 595 | 2 | 1.23E-07 | 3.36E-07 | 2.18E-05 | 3.21E-07 | 0.028449034 | NA |
| chr16 | 18983279 | 18984806 | 1528 | 11 | 1.25E-07 | 4.09E-06 | 0.000234 | 3.21E-07 | 0.001003258 | TMC7 |
| chr22 | 46609887 | 46610445 | 559 | 3 | 3.15E-09 | 0.000229 | 2.80E-06 | 3.21E-07 | 0.014120541 | GRAMD4 |
| chr9 | 129141306 | 129142762 | 1457 | 5 | 1.99E-06 | 9.68E-07 | 0.00109 | 3.21E-07 | 0.015477544 | PTPA, AL158151.3 |
| chr3 | 181709692 | 181710909 | 1218 | 15 | 2.29E-06 | 1.92E-07 | 0.003582 | 3.21E-07 | 0.004377666 | SOX2-OT |
| chr3 | 139020880 | 139021512 | 633 | 7 | 7.58E-07 | 1.44E-07 | 0.001208 | 3.22E-07 | -0.01660393 | MRPS22, PRR23B |
| chr15 | 78262868 | 78263836 | 969 | 5 | 1.69E-07 | 9.54E-07 | 0.000209 | 3.23E-07 | 0.005033386 | AC090607.3 |
| chr14 | 23366003 | 23366838 | 836 | 5 | 2.55E-08 | 1.75E-06 | 0.00076 | 3.23E-07 | 0.009814451 | NA |
| chr11 | 77589718 | 77590732 | 1015 | 5 | 7.14E-08 | 3.26E-07 | 9.78E-05 | 3.23E-07 | 0.007972678 | AQP11, CLNS1A |
| chr19 | 54162194 | 54163024 | 831 | 6 | 9.35E-08 | 2.33E-05 | 0.000115 | 3.23E-07 | -0.00696603 | TMC4 |
| chr11 | 120659184 | 120660325 | 1142 | 7 | 6.91E-07 | 5.95E-07 | 0.000959 | 3.23E-07 | 0.012726996 | GRIK4 |
| chr12 | 113774954 | 113775067 | 114 | 2 | 2.43E-07 | 1.95E-06 | 3.88E-06 | 3.23E-07 | 0.007157612 | NA |
| chr15 | 43776048 | 43777090 | 1043 | 9 | 1.09E-08 | 3.22E-06 | 0.001986 | 3.24E-07 | 0.005837456 | SERF2, ELL3, AC018512.1 |
| chr1 | 1965572 | 1966346 | 775 | 4 | 2.76E-07 | 1.96E-07 | 0.000674 | 3.24E-07 | 0.014345393 | CFAP74 |
| chr14 | 94864543 | 94865129 | 587 | 5 | 3.02E-06 | 4.16E-08 | 0.004124 | 3.24E-07 | -0.0197509 | NA |
| chr1 | 223392785 | 223393831 | 1047 | 12 | 2.83E-08 | 4.61E-06 | 0.004182 | 3.25E-07 | -0.01867621 | CCDC185 |

| | | | | | | | | | | |
|---|---|---|---|---|---|---|---|---|---|---|
| chr1 | 89719790 | 89720237 | 448 | 2 | 8.11E-07 | 2.20E-07 | 5.26E-05 | 3.25E-07 | 0.008135822 | LRRC8C, AC093423.3 |
| chr22 | 23204771 | 23205957 | 1187 | 4 | 3.01E-06 | 9.27E-05 | 3.08E-05 | 3.26E-07 | 0.004221602 | BCR |
| chr1 | 10449625 | 10450442 | 818 | 12 | 3.12E-07 | 2.04E-05 | 0.000129 | 3.27E-07 | 0.003115982 | CENPS-CORT, CORT |
| chr20 | 32364988 | 32365789 | 802 | 2 | 1.02E-07 | 4.03E-07 | 1.75E-05 | 3.27E-07 | 0.010283007 | ASXL1 |
| chr11 | 119754393 | 119755084 | 692 | 2 | 2.52E-08 | 3.31E-06 | 3.00E-06 | 3.28E-07 | 0.014924909 | NA |
| chr6 | 148745982 | 148747080 | 1099 | 8 | 7.99E-07 | 4.70E-08 | 0.002343 | 3.28E-07 | -0.00094109 | UST |
| chr12 | 47753251 | 47753692 | 442 | 3 | 2.92E-07 | 3.07E-07 | 4.30E-05 | 3.28E-07 | 0.012768423 | RAPGEF3 |
| chr5 | 150544436 | 150544824 | 389 | 4 | 5.44E-07 | 1.18E-07 | 0.000255 | 3.28E-07 | 0.006441662 | NDST1 |
| chr7 | 1483933 | 1485602 | 1670 | 9 | 3.85E-05 | 5.18E-06 | 0.001751 | 3.28E-07 | -0.00119422 | INTS1 |
| chr11 | 117797022 | 117798099 | 1078 | 12 | 8.24E-07 | 7.62E-07 | 0.000753 | 3.28E-07 | 0.005072626 | DSCAML1 |
| chr14 | 24135502 | 24136573 | 1072 | 14 | 1.45E-05 | 5.75E-08 | 0.005242 | 3.28E-07 | 0.001495988 | PSME1 |
| chr10 | 124671111 | 124671318 | 208 | 3 | 5.95E-07 | 9.46E-07 | 1.35E-05 | 3.28E-07 | 0.01233668 | AC068896.1, FAM53B |
| chr1 | 154869120 | 154870838 | 1719 | 11 | 1.58E-05 | 1.75E-05 | 0.000119 | 3.29E-07 | 0.004441681 | KCNN3 |
| chr7 | 158133252 | 158133850 | 599 | 5 | 3.21E-08 | 5.20E-05 | 4.43E-05 | 3.29E-07 | 0.003539172 | PTPRN2 |
| chr20 | 58692120 | 58693363 | 1244 | 8 | 2.71E-06 | 7.34E-08 | 0.004286 | 3.31E-07 | 0.010008577 | STX16-NPEPL1, NPEPL1 |
| chr5 | 133256394 | 133256463 | 70 | 3 | 3.31E-07 | 1.61E-07 | 0.000233 | 3.33E-07 | 0.004744241 | FSTL4 |
| chr17 | 49887301 | 49887516 | 216 | 4 | 6.74E-07 | 4.36E-07 | 0.000258 | 3.33E-07 | 0.012742774 | NA |
| chr16 | 89759922 | 89760132 | 211 | 2 | 8.00E-07 | 2.34E-07 | 4.87E-05 | 3.34E-07 | 0.002777435 | FANCA |
| chr2 | 238107292 | 238107635 | 344 | 2 | 4.63E-07 | 2.03E-07 | 7.80E-05 | 3.35E-07 | 0.018745291 | ESPNL |
| chr22 | 19597656 | 19597696 | 41 | 2 | 7.29E-07 | 2.20E-07 | 5.97E-05 | 3.37E-07 | 0.024181749 | NA |
| chr17 | 75740023 | 75740850 | 828 | 4 | 1.29E-06 | 7.15E-07 | 0.000137 | 3.37E-07 | 0.006432105 | ITGB4 |
| chr17 | 6654401 | 6655496 | 1096 | 5 | 2.37E-06 | 9.28E-08 | 0.00139 | 3.37E-07 | 0.077518396 | C17orf100, MIR4520-2, AC004706.1, MIR4520-1 |
| chr14 | 31457570 | 31458309 | 740 | 8 | 2.02E-07 | 1.74E-06 | 0.000121 | 3.41E-07 | 0.00724856 | NA |
| chr10 | 100286072 | 100287011 | 940 | 23 | 4.00E-07 | 1.01E-06 | 0.004393 | 3.41E-07 | 0.00194865 | BLOC1S2 |
| chr14 | 23300256 | 23302047 | 1792 | 9 | 3.76E-06 | 1.69E-05 | 0.000158 | 3.41E-07 | 0.002491558 | BCL2L2, PPP1R3E |
| chr11 | 4186501 | 4187493 | 993 | 7 | 1.11E-06 | 1.01E-07 | 0.002728 | 3.41E-07 | 0.007145977 | LINC02749 |
| chr7 | 1713760 | 1714273 | 514 | 6 | 9.51E-07 | 2.51E-07 | 0.000245 | 3.41E-07 | 0.013804987 | ELFN1 |
| chr14 | 22471725 | 22472631 | 907 | 4 | 9.64E-08 | 1.61E-06 | 0.000106 | 3.43E-07 | 0.010967792 | AC244502.1 |
| chr6 | 32204009 | 32204920 | 912 | 11 | 1.60E-07 | 1.60E-05 | 0.002613 | 3.45E-07 | 0.000719365 | NOTCH4 |
| chr11 | 65497210 | 65498500 | 1291 | 12 | 3.14E-06 | 8.60E-08 | 0.005696 | 3.46E-07 | 0.002051267 | MALAT1, AP000769.2 |
| chr16 | 57984976 | 57986080 | 1105 | 7 | 2.85E-07 | 2.10E-07 | 0.000229 | 3.46E-07 | 0.005285315 | TEPP |
| chr6 | 13272106 | 13272915 | 810 | 6 | 6.40E-07 | 4.73E-07 | 0.00023 | 3.47E-07 | 0.007812921 | PHACTR1, AL008729.1, TBC1D7 |
| chr17 | 28953623 | 28953653 | 31 | 2 | 2.18E-07 | 1.96E-07 | 0.000111 | 3.48E-07 | 0.011024004 | PIPOX |
| chr13 | 114064838 | 114066075 | 1238 | 6 | 2.51E-07 | 3.38E-06 | 0.000343 | 3.48E-07 | 0.009509413 | RASA3 |
| chr7 | 107659842 | 107660843 | 1002 | 11 | 8.36E-07 | 3.61E-07 | 0.000894 | 3.48E-07 | 0.002966557 | SLC26A4, SLC26A4-AS1 |
| chr13 | 41020336 | 41020768 | 433 | 4 | 2.50E-07 | 1.99E-05 | 3.88E-05 | 3.49E-07 | 0.006433578 | ELF1 |
| chr10 | 19817858 | 19818856 | 999 | 5 | 3.03E-07 | 6.95E-07 | 0.000478 | 3.49E-07 | 0.017303572 | PLXDC2 |
| chr7 | 151113894 | 151115195 | 1302 | 7 | 3.42E-07 | 1.71E-06 | 9.31E-05 | 3.49E-07 | 0.005876703 | AGAP3 |
| chr16 | 52547493 | 52548552 | 1060 | 11 | 1.02E-05 | 1.01E-07 | 0.004175 | 3.50E-07 | 0.009143223 | TOX3 |
| chr11 | 44936682 | 44937573 | 892 | 4 | 9.63E-07 | 1.38E-07 | 0.000441 | 3.51E-07 | 0.001979322 | TP53I11 |
| chr17 | 8309877 | 8310763 | 887 | 6 | 2.00E-07 | 1.75E-06 | 0.001535 | 3.51E-07 | 0.012228322 | ARHGEF15 |
| chr7 | 143510752 | 143511574 | 823 | 4 | 3.58E-06 | 8.79E-08 | 0.001345 | 3.52E-07 | 0.007858238 | EPHA1-AS1, OR10AC1 |
| chr4 | 112516645 | 112517399 | 755 | 5 | 1.05E-06 | 1.21E-06 | 0.000192 | 3.53E-07 | -0.0018361 | NEUROG2-AS1 |
| chr4 | 147616398 | 147617803 | 1406 | 13 | 6.27E-08 | 4.74E-05 | 0.000709 | 3.53E-07 | 0.003680472 | TMEM184C, AC093835.1 |
| chr5 | 42889335 | 42890039 | 705 | 3 | 3.58E-06 | 2.87E-06 | 6.73E-05 | 3.54E-07 | 0.024493751 | AC008945.2 |
| chr3 | 33883825 | 33884674 | 850 | 3 | 6.14E-08 | 6.10E-06 | 1.26E-05 | 3.55E-07 | 0.015900827 | NA |
| chr12 | 52675408 | 52676149 | 742 | 3 | 1.35E-08 | 7.08E-06 | 3.37E-06 | 3.58E-07 | 0.010320125 | KRT1 |
| chr16 | 86495130 | 86496447 | 1318 | 6 | 5.04E-08 | 8.76E-06 | 6.50E-05 | 3.58E-07 | 0.006853054 | FENDRR |
| chr21 | 44294818 | 44295645 | 828 | 4 | 4.20E-07 | 6.16E-06 | 0.000138 | 3.58E-07 | 0.009642704 | AIRE |
| chr6 | 32105258 | 32105813 | 556 | 4 | 1.27E-07 | 2.47E-07 | 0.000259 | 3.59E-07 | 0.006767573 | TNXB, AL662884.2 |
| chr14 | 36513910 | 36514927 | 1018 | 13 | 5.17E-08 | 2.21E-06 | 0.000136 | 3.59E-07 | 0.004940504 | AL132857.1, SFTA3 |
| chr16 | 88322277 | 88323083 | 807 | 5 | 1.05E-07 | 4.67E-06 | 8.39E-05 | 3.60E-07 | 0.007482555 | NA |
| chr4 | 7852486 | 7852956 | 471 | 2 | 7.98E-07 | 2.62E-07 | 4.54E-05 | 3.60E-07 | 0.010080911 | AFAP1 |
| chr19 | 34772580 | 34773523 | 944 | 15 | 1.60E-06 | 3.10E-07 | 0.010039 | 3.60E-07 | 0.00190911 | ZNF599 |
| chr14 | 37202481 | 37202657 | 177 | 2 | 5.87E-07 | 2.48E-07 | 5.25E-05 | 3.60E-07 | -0.00010291 | MIPOL1 |
| chr5 | 139393379 | 139394230 | 852 | 4 | 2.02E-06 | 1.18E-07 | 0.00038 | 3.60E-07 | 0.015987451 | PROB1 |
| chr14 | 92684737 | 92685103 | 367 | 2 | 1.40E-07 | 2.31E-06 | 3.94E-06 | 3.61E-07 | 0.0025272 | RIN3 |
| chr1 | 156814515 | 156815766 | 1252 | 6 | 3.08E-05 | 7.14E-07 | 0.000476 | 3.61E-07 | 0.006931726 | NTRK1, SH2D2A |
| chr13 | 46752336 | 46752712 | 377 | 2 | 1.68E-07 | 4.92E-07 | 1.58E-05 | 3.61E-07 | 0.011584838 | LRCH1 |
| chr3 | 141077171 | 141077341 | 171 | 4 | 1.76E-06 | 1.05E-07 | 0.001261 | 3.62E-07 | 0.013213651 | SPSB4 |
| chr5 | 179061693 | 179062200 | 508 | 2 | 1.48E-08 | 1.02E-06 | 7.48E-06 | 3.63E-07 | -0.00120351 | ZNF354C |
| chr18 | 62610375 | 62611215 | 841 | 3 | 3.77E-07 | 3.60E-07 | 4.12E-05 | 3.64E-07 | 0.015772997 | NA |
| chr15 | 80591587 | 80591656 | 70 | 2 | 2.79E-07 | 3.19E-07 | 3.16E-05 | 3.65E-07 | 7.49E-06 | ARNT2 |
| chr18 | 50606883 | 50607497 | 615 | 3 | 1.43E-06 | 2.06E-07 | 0.000118 | 3.65E-07 | 0.006723489 | MAPK4 |
| chr16 | 66517405 | 66518006 | 602 | 3 | 1.47E-07 | 1.87E-06 | 1.20E-05 | 3.65E-07 | 0.001933454 | TK2 |
| chr17 | 58525012 | 58525390 | 379 | 3 | 2.48E-07 | 7.09E-06 | 7.96E-05 | 3.65E-07 | 0.008140043 | SEPTIN4-AS1, SEPTIN4 |
| chr12 | 13098361 | 13098561 | 201 | 2 | 3.38E-07 | 3.03E-07 | 3.48E-05 | 3.65E-07 | 0.016877869 | FAM234B, GSG1 |
| chr2 | 213282985 | 213284311 | 1327 | 10 | 3.45E-06 | 2.54E-05 | 0.0002 | 3.66E-07 | -0.00116759 | SPAG16-DT |
| chr3 | 13850809 | 13851327 | 519 | 2 | 2.55E-07 | 4.05E-07 | 2.12E-05 | 3.66E-07 | 0.008916378 | WNT7A |
| chr14 | 68625885 | 68626487 | 603 | 2 | 1.31E-07 | 5.03E-07 | 1.59E-05 | 3.67E-07 | 0.003091965 | RAD51B |
| chr14 | 103131720 | 103132560 | 841 | 4 | 1.62E-07 | 4.95E-07 | 3.73E-05 | 3.68E-07 | 0.005076234 | TNFAIP2 |
| chr9 | 136415506 | 136416376 | 871 | 7 | 4.86E-07 | 3.08E-06 | 8.20E-05 | 3.68E-07 | 0.001855607 | PMPCA |
| chr18 | 3246934 | 3248150 | 1217 | 13 | 5.40E-07 | 6.77E-06 | 0.001728 | 3.68E-07 | -8.86E-05 | AP005329.2, MYL12A, AP005329.3 |
| chr14 | 103016179 | 103016945 | 767 | 5 | 2.96E-07 | 1.08E-06 | 0.00015 | 3.68E-07 | 0.007484773 | CDC42BPB |
| chr16 | 87345908 | 87346874 | 967 | 7 | 1.60E-07 | 2.83E-06 | 0.000541 | 3.70E-07 | 0.000757673 | FBXO31 |
| chr14 | 67617473 | 67618289 | 817 | 4 | 7.21E-07 | 1.04E-07 | 0.000642 | 3.71E-07 | 0.014506146 | NA |
| chr7 | 47304981 | 47305152 | 172 | 4 | 1.66E-06 | 1.39E-07 | 0.000871 | 3.71E-07 | 0.009830773 | TNS3 |
| chr19 | 10333341 | 10334917 | 1577 | 9 | 6.06E-07 | 7.52E-06 | 0.000562 | 3.71E-07 | 0.002197152 | AC114271.1, RAVER1, ICAM3 |
| chr3 | 50327891 | 50329910 | 2020 | 13 | 3.51E-05 | 3.01E-06 | 0.000357 | 3.72E-07 | -0.00109161 | TUSC2, RASSF1 |
| chr22 | 18064734 | 18065436 | 703 | 3 | 7.76E-07 | 1.71E-07 | 0.000176 | 3.73E-07 | 0.009586902 | NA |
| chr10 | 79320647 | 79321642 | 996 | 3 | 1.61E-05 | 1.32E-07 | 0.000441 | 3.74E-07 | 0.00849303 | NA |
| chr7 | 1139276 | 1139559 | 284 | 3 | 2.33E-08 | 1.89E-06 | 1.32E-05 | 3.74E-07 | 0.012354963 | NA |
| chr1 | 87151004 | 87152289 | 1286 | 11 | 8.56E-07 | 4.66E-06 | 0.000158 | 3.74E-07 | 0.006225117 | AC093155.3, LINC01140 |
| chr10 | 1725288 | 1726471 | 1184 | 7 | 1.37E-06 | 7.14E-06 | 0.000159 | 3.74E-07 | 0.000420822 | ADARB2 |
| chr2 | 199467905 | 199469006 | 1102 | 6 | 3.75E-05 | 5.57E-07 | 0.001541 | 3.74E-07 | 0.009583058 | SATB2-AS1, SATB2 |
| chr3 | 78607963 | 78608589 | 627 | 3 | 6.89E-07 | 3.47E-07 | 5.10E-05 | 3.75E-07 | 0.006697768 | ROBO1 |
| chr15 | 86261948 | 86262455 | 508 | 3 | 1.60E-07 | 9.90E-06 | 3.12E-06 | 3.76E-07 | 0.002766204 | AGBL1 |
| chr5 | 146515960 | 146516875 | 916 | 5 | 3.18E-07 | 8.19E-07 | 0.000108 | 3.77E-07 | 0.014433086 | GPR151 |
| chr6 | 43254417 | 43254772 | 356 | 2 | 1.12E-07 | 4.82E-06 | 2.86E-06 | 3.79E-07 | 0.006688099 | TTBK1 |
| chr12 | 130115048 | 130115184 | 137 | 3 | 1.92E-06 | 1.24E-07 | 0.000654 | 3.81E-07 | 0.013819671 | NA |
| chr7 | 88306664 | 88307608 | 945 | 12 | 8.09E-08 | 0.000763 | 0.000139 | 3.83E-07 | 0.001929371 | STEAP4 |
| chr12 | 113018252 | 113019070 | 819 | 3 | 2.78E-06 | 2.13E-07 | 0.000106 | 3.83E-07 | 0.006486907 | NA |
| chr6 | 564040 | 564609 | 570 | 6 | 1.29E-08 | 4.21E-05 | 0.000201 | 3.83E-07 | 0.005183617 | EXOC2 |
| chr3 | 136078591 | 136078738 | 148 | 2 | 4.41E-07 | 2.90E-06 | 3.73E-06 | 3.84E-07 | 0.012914633 | PPP2R3A |
| chr12 | 121602873 | 121603004 | 132 | 2 | 5.14E-07 | 3.28E-07 | 3.41E-05 | 3.85E-07 | 0.000511034 | NA |
| chr16 | 22346505 | 22346923 | 419 | 3 | 1.17E-06 | 7.41E-07 | 6.01E-05 | 3.85E-07 | 0.002713767 | CDR2 |
| chr14 | 105141811 | 105142230 | 420 | 4 | 2.44E-07 | 6.37E-07 | 0.000161 | 3.85E-07 | 0.001204411 | JAG2 |
| chr19 | 45711716 | 45712921 | 1206 | 7 | 1.33E-07 | 1.89E-06 | 0.000229 | 3.85E-07 | 0.003848367 | FBXO46 |
| chr8 | 1946648 | 1947392 | 745 | 4 | 4.01E-08 | 2.32E-06 | 9.30E-05 | 3.86E-07 | 0.005632947 | AC019257.8, ARHGEF10 |
| chr16 | 86479642 | 86479694 | 53 | 2 | 5.29E-07 | 1.07E-06 | 7.84E-06 | 3.86E-07 | 0.014793784 | FENDRR |
| chr10 | 125626028 | 125626396 | 369 | 2 | 2.12E-08 | 0.000136 | 1.06E-06 | 3.86E-07 | 0.01638348 | TEX36 |
| chr3 | 126105111 | 126105889 | 779 | 4 | 4.34E-08 | 6.37E-07 | 3.24E-05 | 3.87E-07 | -0.00090612 | ALDH1L1-AS1, ALDH1L1 |
| chr2 | 23501260 | 23501361 | 102 | 2 | 1.87E-07 | 3.12E-06 | 3.62E-06 | 3.87E-07 | 0.006260683 | KLHL29 |
| chr12 | 309946 | 311009 | 1064 | 3 | 2.66E-09 | 2.58E-05 | 1.53E-06 | 3.87E-07 | 0.002643607 | KDM5A |
| chr17 | 29074459 | 29075788 | 1330 | 6 | 8.44E-08 | 7.47E-05 | 1.52E-05 | 3.87E-07 | 0.000536178 | MYO18A, TIAF1 |
| chr7 | 150857332 | 150858214 | 883 | 7 | 6.10E-07 | 3.32E-06 | 0.000403 | 3.87E-07 | 0.002560187 | AOC1 |
| chr4 | 7787254 | 7788564 | 1311 | 5 | 1.17E-07 | 6.96E-06 | 2.63E-05 | 3.87E-07 | 0.004899651 | AFAP1 |
| chr16 | 2210189 | 2211236 | 1048 | 12 | 1.65E-06 | 4.07E-06 | 0.001557 | 3.89E-07 | 0.000513461 | BRICD5 |
| chr5 | 176898835 | 176899923 | 1089 | 8 | 3.02E-07 | 7.48E-07 | 0.001652 | 3.90E-07 | 0.006145231 | HK3 |
| chr2 | 231197940 | 231199133 | 1194 | 13 | 2.82E-06 | 1.77E-05 | 0.000944 | 3.91E-07 | 0.002669224 | ARMC9 |
| chr2 | 74834714 | 74835677 | 964 | 7 | 1.89E-07 | 1.02E-06 | 0.000201 | 3.93E-07 | 0.000289509 | HK2 |
| chr8 | 103980265 | 103980408 | 144 | 2 | 1.40E-06 | 2.21E-07 | 0.00012 | 3.93E-07 | 0.018472114 | RIMS2 |
| chr3 | 138942998 | 138944098 | 1101 | 5 | 9.76E-08 | 8.56E-07 | 2.69E-05 | 3.93E-07 | 0.009434737 | LINC01391 |
| chr7 | 18086836 | 18087880 | 1045 | 8 | 1.86E-06 | 1.02E-07 | 0.008301 | 3.94E-07 | 0.005516282 | HDAC9 |
| chr12 | 69584598 | 69585515 | 918 | 13 | 2.81E-08 | 3.96E-05 | 4.37E-05 | 3.95E-07 | 0.004682584 | MIR3913-2, CCT2, MIR3913-1 |
| chr13 | 99983929 | 99985306 | 1378 | 15 | 6.17E-08 | 5.29E-06 | 0.001043 | 3.96E-07 | 0.002914329 | ZIC2 |
| chr11 | 69381637 | 69382556 | 920 | 6 | 3.38E-07 | 1.60E-05 | 0.000237 | 3.96E-07 | -0.00567629 | NA |
| chr1 | 201114991 | 201116266 | 1276 | 11 | 1.02E-05 | 1.17E-07 | 0.008685 | 3.97E-07 | 0.007056644 | ASCL5 |
| chr22 | 20437013 | 20438248 | 1236 | 12 | 1.39E-06 | 4.79E-06 | 0.002008 | 3.97E-07 | 0.002391417 | SCARF2, AC007731.4 |
| chr20 | 32006959 | 32008450 | 1492 | 6 | 7.00E-05 | 2.30E-07 | 0.001365 | 3.97E-07 | 0.00130117 | RNA5SP481, AL031658.1 |
| chr1 | 32894848 | 32895776 | 929 | 3 | 4.03E-08 | 6.79E-06 | 8.28E-05 | 3.98E-07 | 0.00849504 | HPCA, TMEM54 |
| chr12 | 57167367 | 57167406 | 40 | 2 | 9.06E-07 | 2.95E-07 | 4.64E-05 | 4.00E-07 | 0.018120927 | LRP1 |
| chr18 | 74082112 | 74082748 | 637 | 4 | 9.37E-07 | 1.07E-06 | 0.000124 | 4.00E-07 | 0.005526823 | FBXO15 |
| chr6 | 32094616 | 32095626 | 1011 | 10 | 5.25E-07 | 3.40E-06 | 0.002987 | 4.00E-07 | 0.004536605 | TNXB |
| chr4 | 152679790 | 152681099 | 1310 | 11 | 9.32E-07 | 1.09E-05 | 2.07E-05 | 4.00E-07 | 0.00308427 | TMEM154 |
| chr16 | 57970556 | 57971630 | 1075 | 7 | 3.11E-06 | 2.61E-05 | 9.91E-05 | 4.00E-07 | 0.004010635 | CNGB1 |
| chr6 | 35370271 | 35371147 | 877 | 3 | 1.76E-06 | 3.40E-07 | 5.42E-05 | 4.01E-07 | 0.000727448 | PPARD |
| chr6 | 31794853 | 31796096 | 1244 | 28 | 4.10E-08 | 5.39E-06 | 0.002095 | 4.03E-07 | 0.000622196 | VARS1 |
| chr12 | 105083748 | 105085541 | 1794 | 16 | 8.79E-06 | 6.15E-07 | 0.003531 | 4.03E-07 | 0.004524256 | ALDH1L2 |
| chr10 | 361781 | 362229 | 449 | 4 | 5.64E-07 | 1.25E-05 | 0.000109 | 4.04E-07 | 0.0114064 | DIP2C |
| chr16 | 89657550 | 89658861 | 1312 | 16 | 3.06E-06 | 6.16E-05 | 0.001759 | 4.04E-07 | -0.00171749 | SPATA33, CHMP1A |
| chr2 | 232194597 | 232195210 | 614 | 2 | 2.92E-06 | 2.29E-07 | 0.000119 | 4.05E-07 | -0.00484837 | DIS3L2 |
| chr2 | 23524327 | 23525373 | 1047 | 6 | 5.37E-07 | 9.40E-07 | 8.79E-05 | 4.06E-07 | 0.004684839 | KLHL29, AC011239.1 |
| chr4 | 169612124 | 169612796 | 673 | 13 | 4.40E-08 | 6.29E-05 | 0.000276 | 4.06E-07 | -0.00088272 | CLCN3, NEK1 |
| chr11 | 36015342 | 36015938 | 597 | 3 | 5.41E-08 | 2.07E-05 | 4.94E-05 | 4.07E-07 | 0.009982524 | LDLRAD3 |
| chr19 | 45444530 | 45445349 | 820 | 5 | 1.73E-07 | 9.52E-07 | 0.00018 | 4.08E-07 | 0.006147961 | ERCC1 |
| chr5 | 93594346 | 93596288 | 1943 | 10 | 4.60E-08 | 8.24E-06 | 8.89E-05 | 4.08E-07 | 0.004106591 | NR2F1 |
| chr3 | 73383301 | 73384172 | 872 | 5 | 5.43E-06 | 8.50E-07 | 0.000312 | 4.09E-07 | 0.008215273 | PDZRN3 |
| chr19 | 50478000 | 50479052 | 1053 | 7 | 7.72E-07 | 2.78E-07 | 0.001178 | 4.09E-07 | -0.00063819 | EMC10 |
| chr11 | 121575493 | 121576567 | 1075 | 3 | 1.51E-09 | 3.04E-06 | 6.48E-06 | 4.09E-07 | 0.010675235 | SORL1 |
| chr6 | 33607942 | 33608000 | 59 | 2 | 1.01E-06 | 2.65E-07 | 6.78E-05 | 4.09E-07 | 0.013639042 | NA |
| chr5 | 171365204 | 171365551 | 348 | 3 | 1.97E-07 | 2.07E-05 | 1.66E-05 | 4.11E-07 | 0.012510317 | NA |
| chr10 | 48604641 | 48605889 | 1249 | 10 | 1.98E-06 | 1.46E-07 | 0.000955 | 4.12E-07 | 0.007512429 | ARHGAP22 |
| chr1 | 228148773 | 228150075 | 1303 | 9 | 4.68E-07 | 2.42E-06 | 0.002854 | 4.12E-07 | 0.000277067 | GUK1, GJC2 |
| chr1 | 16066876 | 16067296 | 421 | 2 | 5.46E-07 | 4.44E-07 | 2.32E-05 | 4.12E-07 | 0.011247736 | FAM131C |
| chr6 | 13281828 | 13282538 | 711 | 3 | 1.59E-06 | 1.62E-07 | 0.000342 | 4.12E-07 | 0.003596833 | PHACTR1, AL008729.1, TBC1D7 |
| chr12 | 124685836 | 124686597 | 762 | 3 | 1.65E-06 | 2.57E-07 | 0.000232 | 4.13E-07 | 0.007870012 | NA |
| chr10 | 43039939 | 43040440 | 502 | 3 | 5.70E-07 | 4.07E-07 | 9.77E-05 | 4.14E-07 | 0.00632493 | NA |
| chr4 | 121763748 | 121765537 | 1790 | 18 | 3.84E-05 | 7.54E-06 | 0.000206 | 4.14E-07 | 0.005798476 | PP12613, TMEM155 |
| chr1 | 42738492 | 42738514 | 23 | 2 | 5.33E-07 | 3.51E-07 | 3.54E-05 | 4.15E-07 | 0.008534822 | CLDN19 |
| chr5 | 4970540 | 4971528 | 989 | 5 | 7.21E-06 | 2.01E-06 | 0.000283 | 4.15E-07 | -0.00765913 | NA |
| chr10 | 71850809 | 71851788 | 980 | 13 | 4.09E-06 | 2.74E-07 | 0.005511 | 4.15E-07 | 0.002935313 | PSAP |
| chr7 | 552828 | 554013 | 1186 | 4 | 1.86E-07 | 1.62E-06 | 0.000344 | 4.16E-07 | 0.004545099 | PRKAR1B |
| chr1 | 171511015 | 171511142 | 128 | 2 | 1.89E-07 | 3.50E-06 | 3.69E-06 | 4.16E-07 | 0.000580538 | PRRC2C |
| chr2 | 105398622 | 105400005 | 1384 | 16 | 3.18E-07 | 3.37E-05 | 0.001452 | 4.16E-07 | 0.003991022 | FHL2 |
| chr6 | 8435498 | 8436919 | 1422 | 13 | 4.67E-07 | 1.55E-06 | 0.006521 | 4.16E-07 | 0.00606714 | AL355499.2, HULC, SLC35B3 |
| chr9 | 108889844 | 108890790 | 947 | 4 | 1.31E-06 | 5.83E-06 | 0.00022 | 4.17E-07 | 0.003444826 | ELP1 |
| chr15 | 74245173 | 74245689 | 517 | 5 | 1.75E-07 | 2.33E-05 | 1.41E-05 | 4.17E-07 | 0.006458777 | CCDC33 |
| chr8 | 126300441 | 126301165 | 725 | 4 | 1.51E-06 | 3.54E-06 | 0.000162 | 4.19E-07 | 0.010019847 | NA |
| chr20 | 64155378 | 64156250 | 873 | 4 | 6.37E-06 | 3.36E-07 | 0.000225 | 4.19E-07 | 0.007234555 | MYT1 |
| chr1 | 235993287 | 235994192 | 906 | 7 | 1.43E-05 | 6.05E-07 | 0.000238 | 4.20E-07 | 0.002534142 | NID1 |
| chr6 | 154847532 | 154848066 | 535 | 2 | 2.64E-07 | 6.87E-07 | 1.42E-05 | 4.27E-07 | 0.009179979 | TIAM2 |

| | | | | | | | | | | |
|---|---|---|---|---|---|---|---|---|---|---|
| chr7 | 150448286 | 150449438 | 1153 | 6 | 2.54E-06 | 1.66E-07 | 0.002873 | 4.28E-07 | 0.006612069 | NA |
| chr9 | 136492916 | 136493412 | 497 | 3 | 2.52E-07 | 2.83E-07 | 0.000123 | 4.29E-07 | 0.004870161 | NA |
| chr3 | 4976222 | 4977532 | 1311 | 8 | 5.08E-06 | 6.69E-06 | 0.000476 | 4.30E-07 | 0.006155919 | BHLHE40-AS1 |
| chr11 | 30012413 | 30012943 | 531 | 4 | 7.77E-08 | 1.25E-05 | 5.89E-05 | 4.31E-07 | 0.002856704 | KCNA4 |
| chr17 | 67559148 | 67559566 | 419 | 2 | 5.02E-08 | 2.06E-06 | 5.37E-06 | 4.32E-07 | 0.007550675 | PITPNC1 |
| chr10 | 5315208 | 5315409 | 202 | 2 | 5.01E-07 | 1.96E-06 | 5.55E-06 | 4.32E-07 | 0.003616343 | NA |
| chr2 | 27148088 | 27149264 | 1177 | 11 | 7.89E-07 | 5.07E-05 | 9.79E-05 | 4.33E-07 | 0.00298693 | TCF23 |
| chr4 | 48489748 | 48491829 | 2082 | 14 | 0.000104446 | 4.22E-06 | 0.001629 | 4.34E-07 | -0.00198188 | ZAR1 |
| chr16 | 89737867 | 89739404 | 1538 | 7 | 2.01E-07 | 1.03E-05 | 0.000225 | 4.36E-07 | 0.00182857 | ZNF276, FANCA |
| chr22 | 23913824 | 23914832 | 1009 | 6 | 5.51E-06 | 1.46E-07 | 0.00282 | 4.36E-07 | 0.003482409 | NA |
| chr3 | 16263783 | 16265316 | 1534 | 14 | 4.04E-05 | 1.99E-05 | 0.00061 | 4.36E-07 | -6.96E-05 | OXNAD1, DPH3 |
| chr2 | 88885092 | 88885898 | 807 | 5 | 2.61E-06 | 6.31E-05 | 0.000141 | 4.39E-07 | 0.011512817 | IGKV4-1 |
| chr11 | 128718199 | 128719095 | 897 | 2 | 1.16E-05 | 2.54E-07 | 0.000108 | 4.39E-07 | 0.011729218 | FLI1 |
| chr2 | 70886438 | 70887802 | 1365 | 5 | 5.22E-05 | 1.59E-07 | 0.001242 | 4.40E-07 | 0.006321932 | NA |
| chr10 | 98134223 | 98135568 | 1346 | 10 | 3.31E-07 | 4.23E-06 | 0.003881 | 4.40E-07 | 0.001596657 | R3HCC1L |
| chr5 | 146339034 | 146340461 | 1428 | 6 | 4.47E-05 | 4.06E-07 | 0.002157 | 4.40E-07 | 0.006063306 | AC091959.1, POU4F3, AC011396.2 |
| chr15 | 78899491 | 78899876 | 386 | 3 | 8.42E-07 | 4.65E-06 | 7.08E-05 | 4.41E-07 | 0.003506374 | NA |
| chr9 | 98087418 | 98088236 | 819 | 5 | 3.43E-06 | 7.52E-08 | 0.002577 | 4.41E-07 | 0.018863547 | TRIM14 |
| chr20 | 50625532 | 50626441 | 910 | 4 | 2.96E-06 | 1.65E-07 | 0.000607 | 4.42E-07 | 0.006622964 | RIPOR3 |
| chr15 | 40044434 | 40044603 | 170 | 2 | 2.26E-07 | 1.31E-06 | 7.93E-06 | 4.45E-07 | 0.010423713 | SRP14-AS1 |
| chr5 | 150309015 | 150309998 | 984 | 4 | 6.59E-07 | 8.30E-07 | 0.000252 | 4.45E-07 | 0.006561431 | ARSI |
| chr17 | 3531897 | 3532930 | 1034 | 5 | 5.53E-07 | 4.16E-07 | 0.000333 | 4.45E-07 | 0.000992335 | TRPV3 |
| chr8 | 102961571 | 102962054 | 484 | 3 | 1.38E-06 | 2.17E-07 | 0.000243 | 4.45E-07 | 0.017847455 | MAILR |
| chr3 | 130345011 | 130346092 | 1082 | 7 | 6.52E-07 | 5.25E-07 | 0.002101 | 4.46E-07 | 0.003637286 | COL6A5 |
| chr14 | 24188107 | 24189086 | 980 | 14 | 1.45E-06 | 8.22E-07 | 0.00927 | 4.46E-07 | 0.000391237 | AL136295.22, IPO4, AL136295.4 |
| chr16 | 89403133 | 89403437 | 305 | 4 | 3.61E-07 | 4.69E-06 | 0.00021 | 4.47E-07 | 0.009311526 | ANKRD11 |
| chr11 | 32333196 | 32334353 | 1158 | 6 | 2.14E-07 | 8.80E-07 | 9.04E-05 | 4.48E-07 | 0.006277681 | THEM7P |
| chr1 | 15438373 | 15439573 | 1201 | 7 | 1.40E-06 | 9.16E-06 | 0.000311 | 4.51E-07 | -0.0011405 | CTRC |
| chr20 | 32206808 | 32208368 | 1561 | 14 | 1.81E-06 | 0.000241 | 0.000133 | 4.52E-07 | 0.002231475 | POFUT1, PLAGL2 |
| chr9 | 130128855 | 130129884 | 1030 | 5 | 4.93E-07 | 5.60E-07 | 0.000639 | 4.53E-07 | 0.011605797 | GPR107 |
| chr5 | 141392456 | 141393555 | 1100 | 9 | 6.03E-06 | 4.40E-08 | 0.008338 | 4.53E-07 | 0.010164659 | PCDHGA1, PCDHGA2, PCDHGA3, PCDHGB1, PCDHGA4, PCDHGB2, PCDHGA5, PCDHGB3, PCDHGA6, PCDHGA7, PCDHGB4, PCDHGA8 |
| chr13 | 50128778 | 50130234 | 1457 | 7 | 3.23E-08 | 9.16E-06 | 0.002596 | 4.54E-07 | 0.016328903 | DLEU1 |
| chr9 | 84258378 | 84258517 | 140 | 2 | 3.50E-07 | 1.41E-05 | 2.15E-06 | 4.54E-07 | 0.012858046 | NA |
| chr2 | 1591962 | 1592282 | 321 | 4 | 1.17E-06 | 1.71E-07 | 0.001022 | 4.54E-07 | 0.019803633 | AC141930.1 |
| chr17 | 8162866 | 8164005 | 1140 | 8 | 2.51E-06 | 2.81E-07 | 0.008113 | 4.54E-07 | 0.015472016 | AC129492.3, VAMP2 |
| chr10 | 97756491 | 97757214 | 724 | 4 | 2.42E-07 | 1.97E-06 | 1.43E-05 | 4.55E-07 | -0.00146865 | ZFYVE27 |
| chr12 | 12785148 | 12785531 | 384 | 7 | 2.68E-06 | 3.50E-07 | 0.004159 | 4.55E-07 | 0.007414533 | APOLD1 |
| chr14 | 96264145 | 96265750 | 1606 | 6 | 7.49E-05 | 2.94E-07 | 0.000971 | 4.55E-07 | 0.006098607 | BDKRB1, AL355102.4 |
| chr16 | 69166348 | 69166943 | 596 | 3 | 1.39E-06 | 1.01E-06 | 4.17E-05 | 4.56E-07 | -0.0112682 | UTP4 |
| chr11 | 16806861 | 16807633 | 773 | 3 | 1.04E-07 | 1.27E-06 | 2.04E-05 | 4.56E-07 | 0.010047171 | PLEKHA7 |
| chr5 | 151757869 | 151759236 | 1368 | 11 | 9.95E-06 | 2.87E-06 | 0.002368 | 4.56E-07 | 0.001813596 | AC091982.1, ATOX1 |
| chr10 | 96719666 | 96721141 | 1476 | 13 | 1.53E-05 | 1.38E-06 | 0.008719 | 4.57E-07 | 0.002068917 | PIK3AP1 |
| chr7 | 27139242 | 27140300 | 1059 | 8 | 3.91E-07 | 6.59E-07 | 0.000108 | 4.58E-07 | 0.001850163 | HOXA-AS3, HOXA3, AC004080.6 |
| chr22 | 27275035 | 27276168 | 1134 | 5 | 7.05E-07 | 1.96E-06 | 0.0006 | 4.58E-07 | 0.013470572 | NA |
| chr4 | 105145042 | 105146875 | 1834 | 19 | 4.16E-06 | 4.20E-06 | 0.000566 | 4.58E-07 | 0.000631773 | TET2 |
| chr13 | 111530327 | 111531065 | 739 | 4 | 2.66E-07 | 4.07E-07 | 0.000153 | 4.59E-07 | 0.01246865 | NA |
| chr3 | 147377434 | 147378343 | 910 | 4 | 2.66E-05 | 1.17E-07 | 0.001541 | 4.59E-07 | 0.010938335 | NA |
| chr13 | 112252953 | 112254315 | 1363 | 5 | 4.57E-05 | 0.000195 | 5.58E-05 | 4.60E-07 | -0.00649791 | NA |
| chr5 | 3599572 | 3600458 | 887 | 7 | 8.78E-07 | 1.14E-06 | 0.000797 | 4.60E-07 | 0.007133769 | IRX1, AC016595.1 |
| chr1 | 31296400 | 31297945 | 1546 | 15 | 5.21E-06 | 1.90E-05 | 0.00263 | 4.62E-07 | 0.000965369 | ZCCHC17, SNRNP40 |
| chr3 | 167156326 | 167156440 | 115 | 3 | 2.27E-06 | 1.72E-07 | 0.000431 | 4.62E-07 | -0.00465885 | NA |
| chr1 | 109041560 | 109042865 | 1306 | 14 | 4.61E-06 | 1.81E-06 | 0.000832 | 4.62E-07 | -0.0013138 | WDR47 |
| chr6 | 84764055 | 84765085 | 1031 | 14 | 1.72E-06 | 4.09E-07 | 0.009434 | 4.64E-07 | 0.002705183 | TBX18 |
| chr13 | 52739710 | 52740091 | 382 | 6 | 3.15E-07 | 8.23E-07 | 0.000515 | 4.65E-07 | 0.009942732 | CNMD |
| chr10 | 104339278 | 104339889 | 612 | 4 | 6.59E-07 | 1.24E-07 | 0.001174 | 4.65E-07 | 0.017036172 | NA |
| chr1 | 25055407 | 25055708 | 302 | 2 | 2.69E-07 | 8.11E-07 | 1.37E-05 | 4.69E-07 | 0.011764099 | AL050344.1 |
| chr3 | 124986301 | 124986657 | 357 | 2 | 2.01E-07 | 3.24E-06 | 4.39E-06 | 4.70E-07 | 0.013033938 | HEG1 |
| chr8 | 142638248 | 142638564 | 317 | 4 | 1.28E-06 | 8.50E-07 | 6.50E-05 | 4.70E-07 | 0.004007178 | NA |
| chr17 | 82229843 | 82230798 | 956 | 7 | 1.88E-06 | 1.08E-07 | 0.003855 | 4.70E-07 | 0.00889776 | SLC16A3 |
| chr2 | 201619076 | 201619860 | 785 | 10 | 3.63E-09 | 1.87E-05 | 0.000268 | 4.71E-07 | 0.007697857 | C2CD6 |
| chr7 | 2144045 | 2144393 | 349 | 3 | 9.51E-08 | 7.27E-06 | 8.61E-05 | 4.71E-07 | 0.000161505 | MAD1L1, AC069288.1 |
| chr5 | 95852741 | 95852804 | 64 | 2 | 2.26E-07 | 2.85E-07 | 9.26E-05 | 4.71E-07 | 0.010419378 | LINC01554 |
| chr1 | 248739466 | 248740632 | 1167 | 8 | 4.65E-06 | 2.25E-07 | 0.002462 | 4.71E-07 | 0.007025234 | CR589904.2, LYPD8 |
| chr9 | 36140809 | 36141431 | 623 | 3 | 1.24E-07 | 6.42E-06 | 4.15E-05 | 4.73E-07 | 0.008304949 | GLIPR2 |
| chr6 | 134175731 | 134176733 | 1003 | 10 | 1.90E-07 | 5.06E-07 | 0.0011 | 4.73E-07 | 0.006580796 | SGK1 |
| chr9 | 136200475 | 136202006 | 1532 | 8 | 8.22E-08 | 2.78E-06 | 0.001649 | 4.74E-07 | 0.010555436 | LHX3 |
| chr10 | 103492963 | 103493585 | 623 | 5 | 8.14E-07 | 1.83E-07 | 0.000891 | 4.74E-07 | 0.008242976 | NEURL1-AS1 |
| chr5 | 141484453 | 141485267 | 815 | 10 | 2.75E-06 | 2.47E-07 | 0.005782 | 4.74E-07 | 0.002879269 | PCDHGA1, PCDHGA2, PCDHGA3, PCDHGB1, PCDHGA4, PCDHGB2, PCDHGA5, PCDHGB3, PCDHGA6, PCDHGA7, PCDHGB4, PCDHGA8, PCDHGB5, PCDHGA9, PCDHGB6, PCDHGA10, PCDHGB7, PCDHGA11, PCDHGA12, PCDHGC3, PCDHGC4 |
| chr5 | 17436262 | 17436632 | 371 | 5 | 4.66E-07 | 5.66E-06 | 0.000255 | 4.77E-07 | 0.003422673 | LINC02217 |
| chr11 | 67611099 | 67612250 | 1152 | 6 | 4.45E-06 | 9.74E-07 | 8.60E-05 | 4.78E-07 | 0.003388496 | NDUFV1 |
| chr1 | 110604939 | 110605717 | 779 | 3 | 3.27E-08 | 1.45E-06 | 8.68E-05 | 4.78E-07 | 0.009178186 | KCNA2 |
| chr4 | 25160731 | 25161724 | 994 | 5 | 7.94E-10 | 1.18E-05 | 4.56E-05 | 4.79E-07 | -0.0198138 | SEPSECS-AS1, PI4K2B |
| chr16 | 25118904 | 25119300 | 397 | 2 | 8.90E-07 | 3.11E-07 | 7.23E-05 | 4.79E-07 | 0.010754274 | LCMT1 |
| chr14 | 94560551 | 94561768 | 1218 | 8 | 1.18E-06 | 6.73E-06 | 0.000195 | 4.79E-07 | 0.006591771 | SERPINA4, SERPINA5 |
| chr5 | 140042940 | 140043768 | 829 | 13 | 7.84E-07 | 1.39E-06 | 0.00078 | 4.81E-07 | 0.000112551 | NRG2 |
| chr15 | 65862881 | 65863315 | 435 | 2 | 1.91E-06 | 2.84E-07 | 0.000106 | 4.81E-07 | 0.006397138 | RAB11A |
| chr7 | 3951038 | 3951927 | 890 | 6 | 2.66E-06 | 1.91E-07 | 0.001128 | 4.83E-07 | -0.00038053 | SDK1 |
| chr17 | 49494601 | 49495435 | 835 | 7 | 4.19E-07 | 1.92E-05 | 0.000381 | 4.85E-07 | 0.007072233 | NGFR, AC015656.1 |
| chr2 | 72886838 | 72887481 | 644 | 12 | 8.50E-09 | 0.000124 | 7.28E-05 | 4.85E-07 | 0.003790962 | SPR |
| chr16 | 81956794 | 81958035 | 1242 | 3 | 1.72E-05 | 1.27E-06 | 8.42E-05 | 4.85E-07 | -0.00024568 | PLCG2 |
| chr12 | 14365385 | 14366519 | 1135 | 10 | 6.34E-09 | 7.98E-05 | 1.11E-05 | 4.85E-07 | 0.00030506 | ATF7IP, RPL30P11 |
| chr6 | 56852304 | 56852935 | 632 | 4 | 6.02E-08 | 5.99E-06 | 9.66E-05 | 4.87E-07 | 0.010786835 | DST-AS1, DST |
| chr17 | 5438708 | 5439891 | 1184 | 12 | 5.79E-06 | 4.08E-06 | 0.000721 | 4.88E-07 | 0.000196654 | C1QBP |
| chr11 | 125167330 | 125167452 | 123 | 3 | 3.15E-07 | 1.89E-06 | 1.36E-05 | 4.89E-07 | 0.013153893 | PKNOX2 |
| chr15 | 96366226 | 96367311 | 1086 | 7 | 5.09E-07 | 2.56E-06 | 5.99E-05 | 4.89E-07 | 0.002634862 | AC087477.2 |
| chr15 | 28918185 | 28919274 | 1090 | 6 | 7.28E-06 | 8.02E-08 | 0.002948 | 4.89E-07 | -0.0103113 | APBA2 |
| chr1 | 119965514 | 119966584 | 1071 | 5 | 3.00E-07 | 4.02E-06 | 1.94E-05 | 4.89E-07 | 0.001051225 | NOTCH2 |
| chr13 | 45344067 | 45344201 | 135 | 3 | 1.34E-06 | 2.27E-06 | 0.000105 | 4.91E-07 | 0.008424758 | TPT1-AS1 |
| chr20 | 2864013 | 2864572 | 560 | 7 | 2.76E-07 | 5.00E-06 | 0.000248 | 4.92E-07 | 0.000632716 | VPS16, PTPRA |
| chr12 | 121466210 | 121466677 | 468 | 4 | 2.33E-06 | 1.31E-07 | 0.001504 | 4.93E-07 | 0.004780219 | KDM2B |
| chr15 | 48149314 | 48149391 | 78 | 2 | 3.34E-07 | 9.10E-07 | 1.30E-05 | 4.94E-07 | 0.013511633 | MYEF2 |
| chr3 | 52226074 | 52226949 | 876 | 8 | 8.88E-06 | 1.16E-07 | 0.003131 | 4.95E-07 | 0.003128826 | TLR9, AC097637.1 |
| chr8 | 54457611 | 54458019 | 409 | 10 | 5.41E-07 | 3.96E-06 | 0.001234 | 4.96E-07 | -0.00049289 | SOX17 |
| chr12 | 132395670 | 132396354 | 685 | 4 | 3.99E-07 | 0.000118 | 3.73E-05 | 4.96E-07 | 0.011848339 | NA |
| chr3 | 122993176 | 122993870 | 695 | 5 | 2.16E-07 | 1.21E-06 | 8.46E-05 | 4.98E-07 | 0.01364084 | SEMA5B |
| chr19 | 38692171 | 38692928 | 758 | 4 | 5.09E-07 | 1.11E-06 | 4.55E-05 | 4.98E-07 | 0.008869534 | ACTN4, AC008649.2 |
| chr17 | 81986494 | 81986738 | 245 | 3 | 3.28E-07 | 3.76E-06 | 1.82E-05 | 4.98E-07 | 0.006475865 | ASPSCR1 |
| chr19 | 49403818 | 49404255 | 438 | 3 | 1.03E-06 | 5.92E-07 | 0.000144 | 5.00E-07 | 0.007479108 | KASH5 |
| chr1 | 210343750 | 210343795 | 46 | 2 | 1.26E-06 | 3.84E-07 | 4.74E-05 | 5.04E-07 | 0.02559645 | HHAT |
| chr6 | 98842408 | 98843467 | 1060 | 8 | 4.40E-07 | 3.09E-07 | 0.002131 | 5.06E-07 | 0.007447574 | NA |
| chr11 | 30322284 | 30323498 | 1215 | 10 | 1.53E-06 | 1.25E-05 | 0.000379 | 5.08E-07 | 0.005798005 | ARL14EP, AL358944.1 |
| chr3 | 197949125 | 197950087 | 963 | 9 | 9.36E-10 | 0.000775 | 3.18E-05 | 5.08E-07 | 0.004530477 | IQCG |
| chr14 | 22766069 | 22767516 | 1448 | 18 | 1.86E-06 | 1.71E-05 | 0.00215 | 5.08E-07 | 0.000529895 | OXA1L, AL160314.2 |
| chr16 | 78925216 | 78925498 | 283 | 2 | 1.47E-06 | 3.41E-07 | 6.67E-05 | 5.09E-07 | 0.016051397 | WWOX |
| chr6 | 27553262 | 27553698 | 437 | 6 | 3.21E-07 | 2.46E-06 | 3.14E-05 | 5.09E-07 | 0.015546336 | NA |
| chr1 | 156858410 | 156859585 | 1176 | 10 | 2.19E-06 | 1.96E-07 | 0.009944 | 5.11E-07 | 0.007455679 | NTRK1, INSRR |
| chr11 | 62393322 | 62393621 | 300 | 4 | 1.98E-06 | 1.72E-07 | 0.00078 | 5.11E-07 | 0.035253153 | ASRGL1, AP003064.1 |
| chr15 | 89912100 | 89913155 | 1056 | 10 | 5.66E-06 | 1.90E-07 | 0.001119 | 5.12E-07 | 0.00493808 | ARPIN-AP3S2, ARPIN |
| chr16 | 11071348 | 11072041 | 694 | 3 | 2.35E-06 | 2.49E-07 | 0.000156 | 5.12E-07 | 0.004195486 | CLEC16A |
| chr1 | 46485917 | 46486794 | 878 | 3 | 3.69E-08 | 4.85E-06 | 2.72E-05 | 5.12E-07 | 0.010939591 | NA |
| chr10 | 119178237 | 119179176 | 940 | 14 | 3.84E-06 | 4.63E-06 | 0.000297 | 5.12E-07 | 0.000826645 | PRDX3 |
| chr10 | 121973878 | 121975374 | 1497 | 13 | 3.54E-06 | 3.61E-07 | 0.006744 | 5.13E-07 | 0.002187814 | NSMCE4A |
| chr16 | 24255004 | 24256205 | 1202 | 8 | 1.19E-07 | 1.87E-06 | 9.43E-05 | 5.14E-07 | 0.008532022 | NA |
| chr5 | 157108797 | 157109385 | 589 | 7 | 3.68E-06 | 6.46E-07 | 0.001315 | 5.15E-07 | 0.007114932 | AC011377.1, HAVCR2 |
| chr2 | 240624752 | 240625526 | 775 | 8 | 1.57E-06 | 4.45E-07 | 0.005834 | 5.15E-07 | -0.01090318 | GPR35 |
| chr2 | 222311964 | 222313023 | 1060 | 11 | 4.95E-06 | 3.88E-08 | 0.004554 | 5.15E-07 | 0.005288915 | NA |
| chr2 | 152334517 | 152336424 | 1908 | 13 | 3.07E-05 | 1.18E-06 | 0.001992 | 5.15E-07 | 0.001735301 | FMNL2 |
| chr18 | 26459604 | 26460191 | 588 | 2 | 6.61E-09 | 5.89E-06 | 3.42E-06 | 5.16E-07 | 0.005367672 | KCTD1 |
| chr12 | 112170205 | 112171170 | 966 | 6 | 1.69E-06 | 6.19E-06 | 0.000185 | 5.16E-07 | 0.00094299 | HECTD4 |
| chr19 | 42151938 | 42152513 | 576 | 4 | 7.77E-07 | 2.45E-06 | 5.84E-05 | 5.17E-07 | 0.007137695 | POU2F2 |
| chr8 | 140451001 | 140451142 | 142 | 3 | 4.25E-07 | 3.89E-05 | 3.88E-06 | 5.17E-07 | -0.0016396 | TRAPPC9 |
| chr5 | 131974768 | 131975777 | 1010 | 4 | 1.95E-06 | 1.39E-07 | 0.001388 | 5.19E-07 | 0.007972915 | AC034228.3, ACSL6 |
| chr17 | 46170867 | 46171275 | 409 | 5 | 1.75E-07 | 1.19E-05 | 0.000594 | 5.20E-07 | 0.006927487 | KANSL1 |
| chr19 | 15672915 | 15672981 | 67 | 2 | 1.94E-06 | 2.96E-07 | 0.000135 | 5.21E-07 | 0.018587027 | NA |
| chr1 | 151729373 | 151729778 | 406 | 3 | 6.47E-07 | 5.54E-07 | 7.24E-05 | 5.22E-07 | 0.011021596 | RIIAD1 |
| chr1 | 153549183 | 153550522 | 1340 | 11 | 1.03E-06 | 9.34E-07 | 0.000443 | 5.22E-07 | 0.004474249 | S100A4, S100A3 |
| chr10 | 80603123 | 80603678 | 556 | 6 | 4.06E-07 | 3.52E-06 | 5.34E-05 | 5.22E-07 | 0.005094954 | SH2D4B |
| chr2 | 27274887 | 27275995 | 1109 | 14 | 7.28E-07 | 1.76E-06 | 0.001954 | 5.22E-07 | 0.001835344 | DNAJC5G, SLC30A3 |
| chr17 | 30476540 | 30477858 | 1319 | 11 | 3.44E-06 | 6.63E-06 | 0.000572 | 5.22E-07 | 0.004795073 | GOSR1 |
| chr5 | 1278676 | 1279467 | 792 | 6 | 4.45E-08 | 0.000177 | 1.25E-06 | 5.23E-07 | -0.00057049 | TERT |
| chr12 | 111752654 | 111753718 | 1065 | 6 | 9.05E-06 | 4.23E-07 | 0.002386 | 5.24E-07 | 0.003601288 | ACAD10 |
| chr16 | 3105858 | 3107367 | 1510 | 9 | 7.91E-05 | 9.78E-06 | 0.001757 | 5.25E-07 | 0.007692761 | AC108134.4, ZNF213-AS1 |
| chr12 | 57434397 | 57435064 | 668 | 5 | 4.32E-07 | 6.21E-06 | 3.30E-05 | 5.26E-07 | 0.005564117 | INHBC |
| chr6 | 31538827 | 31540580 | 1754 | 17 | 1.83E-06 | 2.24E-07 | 0.00353 | 5.26E-07 | -0.00602775 | DDX39B, ATP6V1G2-DDX39B |
| chr3 | 190586171 | 190586877 | 707 | 4 | 6.11E-07 | 4.60E-07 | 0.000137 | 5.28E-07 | 0.012634393 | IL1RAP |
| chr4 | 40514623 | 40514874 | 252 | 4 | 2.73E-07 | 4.52E-07 | 0.000209 | 5.28E-07 | 0.015272202 | RBM47 |
| chr6 | 31647608 | 31647764 | 157 | 4 | 5.71E-07 | 1.24E-06 | 0.000336 | 5.29E-07 | 0.00325164 | BAG6 |
| chr14 | 96087467 | 96087780 | 314 | 2 | 5.83E-07 | 3.10E-07 | 0.000115 | 5.29E-07 | 0.000338067 | C14orf132 |
| chr2 | 95181128 | 95181553 | 426 | 2 | 4.04E-07 | 3.60E-07 | 6.54E-05 | 5.31E-07 | 0.001659701 | ZNF2 |
| chr4 | 88591721 | 88593090 | 1370 | 14 | 3.53E-07 | 2.72E-05 | 0.001263 | 5.32E-07 | -0.00023718 | AC098582.1, HERC3 |
| chr19 | 16330562 | 16330646 | 85 | 2 | 1.21E-06 | 3.04E-07 | 0.000137 | 5.33E-07 | 0.014760538 | NA |
| chr17 | 48293805 | 48294302 | 498 | 5 | 9.54E-07 | 4.55E-07 | 0.00079 | 5.33E-07 | 0.01416117 | SKAP1 |
| chr12 | 52926452 | 52927804 | 1353 | 7 | 8.89E-06 | 8.75E-06 | 0.000191 | 5.33E-07 | 0.002369972 | KRT8 |
| chr1 | 6519524 | 6520465 | 942 | 9 | 1.05E-06 | 4.88E-06 | 0.002121 | 5.33E-07 | 0.005375754 | PLEKHG5 |
| chr15 | 99408593 | 99409718 | 1126 | 5 | 2.34E-07 | 1.89E-06 | 0.000148 | 5.35E-07 | 0.010669997 | NA |
| chr13 | 61415070 | 61415980 | 911 | 15 | 1.09E-06 | 1.71E-07 | 0.004998 | 5.35E-07 | 0.00557893 | PCDH20, AL592490.1 |
| chr6 | 168372978 | 168373561 | 584 | 5 | 2.30E-07 | 1.05E-05 | 0.000508 | 5.37E-07 | 0.012895724 | NA |
| chr6 | 32169928 | 32171343 | 1416 | 16 | 3.10E-07 | 2.74E-05 | 0.00066 | 5.37E-07 | 0.001107025 | PPT2-EGFL8, AGPAT1, MIR6721 |
| chr17 | 45118010 | 45119018 | 1009 | 4 | 1.40E-05 | 2.86E-07 | 0.000383 | 5.37E-07 | 0.001220773 | PLCD3 |
| chr18 | 9913437 | 9915101 | 1665 | 15 | 2.83E-05 | 3.19E-06 | 0.001714 | 5.38E-07 | 0.000736628 | VAPA |
| chr4 | 146400422 | 146400764 | 343 | 3 | 2.11E-07 | 2.67E-05 | 1.26E-05 | 5.38E-07 | 0.005263132 | SLC10A7 |
| chr12 | 131558827 | 131559679 | 853 | 2 | 4.88E-06 | 3.43E-07 | 8.21E-05 | 5.40E-07 | 0.010215218 | NA |
| chr10 | 101902566 | 101904029 | 1464 | 3 | 5.58E-05 | 1.84E-07 | 0.000657 | 5.41E-07 | 0.008837882 | ARMH3 |
| chr11 | 115941630 | 115941703 | 74 | 3 | 2.62E-06 | 2.34E-07 | 0.00023 | 5.42E-07 | 0.019556189 | LINC02703 |
| chr16 | 30874643 | 30875538 | 896 | 11 | 2.93E-06 | 2.21E-06 | 0.002163 | 5.44E-07 | 0.001427909 | MIR762HG, BCL7C, MIR4519 |
| chr19 | 45681586 | 45682090 | 505 | 5 | 4.74E-08 | 7.82E-06 | 9.29E-06 | 5.44E-07 | 0.006359933 | GIPR |
| chr15 | 90528815 | 90529873 | 1059 | 7 | 7.85E-07 | 0.000155 | 0.000249 | 5.44E-07 | 0.003655186 | NA |
| chr19 | 4677218 | 4678674 | 1457 | 7 | 1.29E-05 | 8.53E-07 | 0.000782 | 5.44E-07 | 0.003721564 | DPP9 |
| chr2 | 112784514 | 112784816 | 303 | 7 | 4.91E-06 | 4.97E-08 | 0.008354 | 5.46E-07 | 0.014573009 | NA |
| chr9 | 19032780 | 19034019 | 1240 | 11 | 1.51E-07 | 9.07E-05 | 3.32E-05 | 5.47E-07 | 0.001953941 | SAXO1 |
| chr5 | 6819361 | 6819788 | 428 | 2 | 1.71E-07 | 3.20E-06 | 5.29E-06 | 5.47E-07 | 0.021404279 | LINC02236 |
| chr19 | 47231859 | 47232896 | 1038 | 6 | 2.04E-06 | 2.30E-06 | 0.000851 | 5.47E-07 | 0.009393199 | BBC3 |

| | | | | | | | | | | |
|---|---|---|---|---|---|---|---|---|---|---|
| chr1 | 7334964 | 7335395 | 432 | 5 | 6.56E-07 | 1.15E-06 | 0.000293 | 5.47E-07 | 0.013680145 | CAMTA1 |
| chr22 | 30289717 | 30291364 | 1648 | 11 | 6.52E-07 | 0.000288 | 0.000116 | 5.48E-07 | 0.004854445 | AC004997.1 |
| chr22 | 30286807 | 30287954 | 1148 | 8 | 9.09E-06 | 2.62E-07 | 0.003297 | 5.51E-07 | 0.006756855 | CASTOR1, AC004997.1 |
| chr7 | 151302481 | 151302505 | 25 | 2 | 2.69E-07 | 8.59E-07 | 1.64E-05 | 5.52E-07 | 0.016735354 | NA |
| chr18 | 74533783 | 74534550 | 768 | 11 | 3.71E-07 | 2.94E-05 | 0.002043 | 5.53E-07 | 0.000408858 | CNDP1 |
| chr11 | 1826776 | 1827512 | 737 | 6 | 2.50E-08 | 4.20E-07 | 0.000231 | 5.54E-07 | 0.004750675 | NA |
| chr15 | 68817786 | 68819172 | 1387 | 9 | 4.06E-05 | 6.26E-07 | 0.001812 | 5.54E-07 | 0.003553917 | SPESP1, ANP32A |
| chr1 | 204196981 | 204197618 | 638 | 2 | 4.81E-08 | 6.12E-07 | 2.54E-05 | 5.54E-07 | 0.008021374 | NA |
| chr1 | 62435528 | 62436666 | 1139 | 11 | 1.38E-08 | 5.30E-06 | 5.67E-05 | 5.55E-07 | -0.00058741 | USP1 |
| chr11 | 9661094 | 9661312 | 219 | 2 | 1.36E-06 | 4.97E-07 | 3.61E-05 | 5.55E-07 | 0.021612478 | NA |
| chr15 | 100860421 | 100861832 | 1412 | 4 | 9.53E-06 | 5.77E-05 | 0.000562 | 5.55E-07 | 0.007362865 | AC015712.1, AC015712.6 |
| chr17 | 40308427 | 40309258 | 832 | 12 | 5.18E-06 | 4.40E-08 | 0.012384 | 5.55E-07 | 0.006875904 | RARA |
| chr14 | 22821589 | 22822883 | 1295 | 9 | 1.46E-06 | 7.17E-07 | 0.001944 | 5.56E-07 | 0.005667277 | SLC7A7 |
| chr10 | 49761951 | 49763075 | 1125 | 14 | 6.46E-08 | 3.70E-05 | 2.04E-05 | 5.57E-07 | -0.00287406 | OGDHL |
| chr2 | 237485562 | 237486798 | 1237 | 5 | 2.01E-07 | 3.17E-06 | 0.000126 | 5.57E-07 | 0.006548202 | MLPH |
| chr1 | 112673760 | 112675474 | 1715 | 19 | 1.34E-06 | 1.82E-05 | 0.003111 | 5.58E-07 | 0.00237434 | MOV10 |
| chr10 | 5317725 | 5318333 | 609 | 4 | 4.11E-07 | 5.54E-07 | 0.000287 | 5.58E-07 | 0.017246948 | NA |
| chr2 | 174847261 | 174848461 | 1201 | 4 | 8.74E-06 | 1.45E-07 | 0.001035 | 5.58E-07 | 0.016981338 | CHN1 |
| chr17 | 8393001 | 8393538 | 538 | 4 | 2.52E-07 | 5.26E-07 | 0.000511 | 5.58E-07 | 0.006975886 | RNF222 |
| chr9 | 133667853 | 133667942 | 90 | 2 | 5.40E-08 | 6.36E-07 | 2.45E-05 | 5.58E-07 | 0.013075565 | SARDH |
| chr17 | 44220589 | 44222560 | 1972 | 11 | 1.60E-05 | 6.41E-06 | 0.00233 | 5.59E-07 | 0.001869152 | AC003102.1, UBTF |
| chr17 | 75048163 | 75049334 | 1172 | 4 | 1.56E-05 | 1.67E-06 | 0.000167 | 5.59E-07 | -0.00508158 | KCTD2 |
| chr21 | 25911744 | 25911924 | 181 | 7 | 4.43E-07 | 3.25E-05 | 1.06E-05 | 5.60E-07 | 0.004900205 | APP |
| chr20 | 3798771 | 3800004 | 1234 | 9 | 5.30E-06 | 5.68E-06 | 0.000805 | 5.60E-07 | 0.006013693 | CDC25B |
| chr5 | 150520171 | 150521400 | 1230 | 5 | 1.64E-08 | 7.09E-05 | 2.42E-06 | 5.60E-07 | 0.003797691 | NDST1 |
| chr10 | 46285537 | 46287449 | 1913 | 5 | 2.21E-05 | 2.13E-07 | 0.00091 | 5.60E-07 | 0.009575109 | ANTXRL |
| chr19 | 30373636 | 30374579 | 944 | 4 | 1.73E-06 | 1.28E-07 | 0.00184 | 5.60E-07 | 0.031434654 | ZNF536 |
| chr17 | 82300758 | 82301757 | 1000 | 4 | 5.08E-06 | 1.01E-05 | 0.000198 | 5.60E-07 | 0.008306216 | NA |
| chr12 | 114440089 | 114441349 | 1261 | 9 | 4.09E-07 | 1.52E-06 | 0.001219 | 5.61E-07 | 0.004167783 | NA |
| chr1 | 113390548 | 113391437 | 890 | 11 | 3.05E-07 | 8.39E-07 | 0.00402 | 5.62E-07 | 0.000543521 | MAGI3 |
| chr3 | 153161062 | 153162312 | 1251 | 10 | 7.85E-07 | 0.003106 | 1.85E-05 | 5.62E-07 | -2.49E-05 | RAP2B, AC117394.2 |
| chr4 | 624325 | 626076 | 1752 | 16 | 3.62E-05 | 4.62E-05 | 6.75E-05 | 5.63E-07 | 0.003492753 | PDE6B |
| chr16 | 2973755 | 2974778 | 1024 | 4 | 1.48E-06 | 2.02E-07 | 0.001013 | 5.63E-07 | 0.007292126 | PKMYT1 |
| chr11 | 112174918 | 112176240 | 1323 | 10 | 2.60E-07 | 9.57E-06 | 0.000174 | 5.64E-07 | 0.003080624 | AP002884.2, BCO2 |
| chr17 | 65055754 | 65056508 | 755 | 7 | 8.96E-07 | 3.06E-05 | 0.000231 | 5.65E-07 | 0.000409893 | GNA13 |
| chr1 | 151783563 | 151784498 | 936 | 3 | 5.91E-06 | 1.43E-06 | 7.23E-05 | 5.66E-07 | 0.010429164 | TDRKH |
| chr8 | 1858279 | 1859538 | 1260 | 5 | 3.11E-05 | 1.08E-07 | 0.002361 | 5.67E-07 | 0.016222647 | AC019257.8, ARHGEF10 |
| chr15 | 64162628 | 64164166 | 1539 | 12 | 1.76E-05 | 8.14E-06 | 0.000312 | 5.69E-07 | 0.001900691 | PPIB |
| chr19 | 45815181 | 45816235 | 1055 | 11 | 6.60E-06 | 4.05E-08 | 0.014093 | 5.72E-07 | 0.008970573 | RSPH6A, SYMPK |
| chr10 | 114100521 | 114101200 | 680 | 9 | 1.72E-06 | 8.94E-07 | 0.000678 | 5.72E-07 | 0.009787661 | NA |
| chr19 | 15550896 | 15551804 | 909 | 5 | 1.08E-05 | 1.76E-05 | 0.000223 | 5.72E-07 | 0.010353132 | CYP4F22 |
| chr15 | 77584185 | 77584475 | 291 | 3 | 4.17E-07 | 2.06E-05 | 2.39E-05 | 5.74E-07 | 0.013395476 | AC046168.2 |
| chr4 | 8506235 | 8506877 | 643 | 6 | 3.22E-06 | 3.02E-05 | 0.000108 | 5.75E-07 | 0.006134751 | AC105345.1 |
| chr1 | 1781382 | 1782832 | 1451 | 8 | 6.79E-05 | 5.48E-07 | 0.001091 | 5.75E-07 | 0.007938587 | NA |
| chr16 | 56607461 | 56608849 | 1389 | 12 | 4.26E-07 | 2.42E-06 | 0.000776 | 5.75E-07 | 0.00569671 | MT2A |
| chr3 | 42539914 | 42540544 | 631 | 10 | 9.74E-07 | 1.19E-05 | 4.67E-05 | 5.79E-07 | 0.004480309 | NA |
| chr20 | 49188015 | 49188617 | 603 | 9 | 5.25E-07 | 8.63E-06 | 0.0009 | 5.80E-07 | 0.000181755 | STAU1 |
| chr1 | 153347938 | 153349535 | 1598 | 9 | 7.03E-05 | 9.10E-07 | 0.002834 | 5.80E-07 | 0.002515202 | PGLYRP4 |
| chr4 | 6088088 | 6088203 | 116 | 2 | 1.89E-06 | 4.49E-07 | 4.99E-05 | 5.81E-07 | 0.021777651 | C4orf50, JAKMIP1 |
| chr12 | 48715997 | 48717070 | 1074 | 9 | 6.58E-07 | 1.10E-05 | 0.000274 | 5.82E-07 | -0.00246759 | CCNT1 |
| chr2 | 25156938 | 25157773 | 836 | 5 | 9.83E-07 | 4.89E-07 | 0.000682 | 5.82E-07 | 0.003060696 | EFR3B |
| chr8 | 71356597 | 71357508 | 912 | 7 | 6.73E-06 | 1.24E-07 | 0.002594 | 5.83E-07 | 0.020217742 | EYA1 |
| chr22 | 25741557 | 25742462 | 906 | 12 | 3.47E-07 | 8.01E-06 | 0.002547 | 5.84E-07 | 0.004318794 | MYO18B |
| chr5 | 141303629 | 141304583 | 955 | 10 | 4.07E-07 | 1.13E-05 | 0.002747 | 5.84E-07 | -0.00949067 | TAF7, SLC25A2 |
| chr1 | 149936206 | 149937231 | 1026 | 11 | 4.60E-06 | 3.09E-07 | 0.005791 | 5.85E-07 | 0.008106407 | MTMR11 |
| chr4 | 17632473 | 17633849 | 1377 | 6 | 2.68E-07 | 6.90E-06 | 4.24E-05 | 5.85E-07 | 0.004990867 | MED28, FAM184B |
| chr6 | 30742859 | 30744903 | 2045 | 52 | 4.18E-07 | 1.25E-05 | 0.024678 | 5.85E-07 | 0.000817582 | IER3-AS1, HCG20, IER3 |
| chr1 | 205247857 | 205248423 | 567 | 2 | 2.85E-07 | 5.96E-07 | 3.00E-05 | 5.85E-07 | 0.0041711 | TMCC2 |
| chr10 | 100330125 | 100331164 | 1040 | 9 | 2.28E-07 | 1.18E-05 | 0.000104 | 5.85E-07 | 0.00784048 | PKD2L1 |
| chr3 | 52286816 | 52287916 | 1101 | 13 | 1.36E-06 | 1.42E-06 | 0.000654 | 5.87E-07 | 0.001096331 | GLYCTK, WDR82 |
| chr4 | 6420636 | 6421644 | 1009 | 7 | 1.51E-07 | 4.82E-06 | 5.37E-05 | 5.88E-07 | 0.003586426 | PPP2R2C |
| chr10 | 119726126 | 119726694 | 569 | 3 | 1.68E-07 | 9.01E-06 | 1.50E-05 | 5.88E-07 | 0.006699013 | INPP5F |
| chr18 | 48942676 | 48943569 | 894 | 6 | 6.86E-06 | 8.08E-08 | 0.004051 | 5.89E-07 | 0.005708355 | SMAD7 |
| chr8 | 133202597 | 133203347 | 751 | 4 | 1.75E-07 | 3.76E-05 | 5.04E-05 | 5.90E-07 | 0.008082447 | CCN4 |
| chr17 | 44218153 | 44218947 | 795 | 12 | 1.58E-07 | 0.000384 | 0.000381 | 5.93E-07 | -0.00151426 | UBTF |
| chr20 | 46406104 | 46407228 | 1125 | 10 | 9.17E-07 | 1.06E-06 | 0.001553 | 5.97E-07 | 0.000813404 | ELMO2 |
| chr15 | 67819345 | 67820550 | 1206 | 9 | 2.31E-06 | 1.31E-06 | 0.002522 | 5.98E-07 | 0.009269262 | SKOR1 |
| chr11 | 93543426 | 93544025 | 600 | 7 | 3.33E-07 | 1.35E-06 | 5.49E-05 | 5.99E-07 | 0.002637982 | NA |
| chr15 | 55318376 | 55319409 | 1034 | 12 | 5.69E-06 | 4.16E-07 | 0.006197 | 5.99E-07 | 0.00174471 | PIGB, RAB27A, PIGBOS1 |
| chr16 | 67847059 | 67848172 | 1114 | 9 | 2.03E-07 | 3.58E-05 | 5.59E-05 | 6.04E-07 | -0.0005093 | NUTF2, CENPT |
| chr1 | 212226698 | 212227996 | 1299 | 4 | 1.15E-05 | 1.87E-06 | 0.000287 | 6.05E-07 | 0.002459715 | LINC02608 |
| chr5 | 161846833 | 161848948 | 2116 | 25 | 2.62E-05 | 9.17E-06 | 0.000199 | 6.05E-07 | 0.002429895 | GABRA1 |
| chr8 | 26394802 | 26395461 | 660 | 3 | 1.48E-07 | 1.29E-06 | 2.05E-05 | 6.06E-07 | 0.002237659 | BNIP3L |
| chr3 | 57352415 | 57352483 | 69 | 3 | 2.24E-06 | 5.92E-07 | 5.07E-05 | 6.06E-07 | -0.00556287 | DNAH12 |
| chr16 | 30113482 | 30114082 | 601 | 10 | 6.74E-07 | 4.38E-06 | 0.000161 | 6.06E-07 | 0.003521018 | GDPD3 |
| chr2 | 176138552 | 176139287 | 736 | 6 | 6.19E-07 | 1.13E-06 | 0.000308 | 6.06E-07 | 0.00716612 | HOXD3 |
| chr11 | 1851523 | 1851526 | 4 | 2 | 3.12E-07 | 4.13E-07 | 7.04E-05 | 6.08E-07 | 0.015622632 | LSP1 |
| chr14 | 76958146 | 76959487 | 1342 | 4 | 4.85E-06 | 1.99E-07 | 0.000997 | 6.09E-07 | 0.005354264 | AC007686.4 |
| chr6 | 41636652 | 41638256 | 1605 | 10 | 8.67E-05 | 3.62E-07 | 0.005264 | 6.09E-07 | 0.002934522 | MDFI |
| chr11 | 47268263 | 47269950 | 1688 | 17 | 7.09E-08 | 2.97E-05 | 2.13E-05 | 6.12E-07 | 0.002533601 | NR1H3, MADD |
| chr6 | 143443864 | 143444356 | 493 | 4 | 7.62E-07 | 4.67E-07 | 8.17E-05 | 6.12E-07 | 0.005034968 | ADAT2 |
| chr1 | 2605903 | 2606335 | 433 | 6 | 1.48E-06 | 1.57E-05 | 0.000179 | 6.13E-07 | 0.00255738 | MMEL1 |
| chr11 | 315102 | 316247 | 1146 | 7 | 4.85E-06 | 4.26E-06 | 0.002561 | 6.14E-07 | 0.010158141 | IFITM2, IFITM1 |
| chr2 | 65344735 | 65344814 | 80 | 2 | 1.15E-06 | 2.46E-06 | 7.37E-06 | 6.15E-07 | 0.012927766 | SPRED2 |
| chr1 | 116499094 | 116499451 | 358 | 4 | 4.86E-07 | 2.42E-06 | 2.39E-05 | 6.15E-07 | 0.007019458 | AL390066.2, AL390066.1 |
| chr3 | 44975805 | 44977045 | 1241 | 11 | 2.59E-05 | 1.01E-06 | 0.00259 | 6.15E-07 | -0.00140423 | EXOSC7, ZDHHC3 |
| chr9 | 129587135 | 129587705 | 571 | 2 | 2.41E-07 | 1.64E-06 | 1.02E-05 | 6.18E-07 | 0.026765709 | NA |
| chr9 | 137578732 | 137579423 | 692 | 9 | 1.71E-06 | 8.74E-06 | 0.000722 | 6.18E-07 | -0.00273374 | DPH7 |
| chr12 | 54161859 | 54163129 | 1271 | 4 | 2.16E-05 | 3.53E-06 | 0.000121 | 6.19E-07 | 0.003845613 | SMUG1, AC023794.4 |
| chr5 | 135477523 | 135477639 | 117 | 2 | 1.32E-06 | 3.55E-07 | 0.000146 | 6.20E-07 | 0.0234088 | NA |
| chr10 | 117968729 | 117968803 | 75 | 2 | 2.27E-06 | 3.89E-07 | 9.13E-05 | 6.21E-07 | 0.014675319 | NA |
| chr3 | 150384998 | 150385956 | 959 | 4 | 4.20E-05 | 1.07E-06 | 0.000326 | 6.21E-07 | 0.006478002 | NA |
| chr10 | 124589177 | 124589933 | 757 | 6 | 8.00E-07 | 6.07E-06 | 0.000377 | 6.22E-07 | 0.002320594 | LHPP |
| chr3 | 113745969 | 113747346 | 1378 | 20 | 7.65E-06 | 3.59E-05 | 0.006719 | 6.23E-07 | 0.002095891 | ATP6V1A, NAA50, AC108693.2 |
| chr10 | 127983591 | 127983952 | 362 | 3 | 2.81E-06 | 2.31E-07 | 0.000543 | 6.25E-07 | 0.03273035 | PTPRE |
| chr11 | 20386990 | 20388101 | 1112 | 15 | 1.52E-06 | 1.06E-05 | 0.000609 | 6.25E-07 | 0.001831604 | PRMT3 |
| chr8 | 143804289 | 143805177 | 889 | 5 | 2.34E-06 | 1.03E-06 | 0.001036 | 6.25E-07 | 0.003997028 | SCRIB |
| chr7 | 101630747 | 101631531 | 785 | 3 | 3.27E-06 | 2.48E-07 | 0.000521 | 6.25E-07 | 0.034395363 | NA |
| chr1 | 11132725 | 11133112 | 388 | 3 | 1.65E-06 | 5.45E-07 | 9.36E-05 | 6.26E-07 | 0.005101633 | MTOR |
| chr6 | 150229982 | 150230510 | 529 | 2 | 4.33E-07 | 1.08E-06 | 1.56E-05 | 6.27E-07 | 0.0178773 | PPP1R14C |
| chr1 | 63321423 | 63322257 | 835 | 6 | 9.88E-08 | 9.15E-07 | 0.000313 | 6.27E-07 | 0.006670518 | FOXD3-AS1 |
| chr17 | 40061190 | 40063339 | 2150 | 13 | 4.40E-05 | 3.74E-05 | 0.001143 | 6.28E-07 | 0.003288478 | THRA, MED24 |
| chr10 | 121595940 | 121597000 | 1061 | 10 | 7.40E-07 | 7.81E-08 | 0.006123 | 6.28E-07 | 0.028310974 | FGFR2 |
| chr11 | 65027138 | 65027977 | 840 | 11 | 1.88E-07 | 9.29E-05 | 4.24E-05 | 6.29E-07 | 0.000621915 | ARL2-SNX15, SNX15 |
| chr1 | 32773175 | 32773937 | 763 | 2 | 1.72E-06 | 6.30E-07 | 3.18E-05 | 6.29E-07 | 0.008054715 | KIAA1522 |
| chr4 | 108166156 | 108167773 | 1618 | 16 | 1.15E-06 | 0.000166 | 5.51E-05 | 6.34E-07 | 0.004156976 | LEF1-AS1, LEF1 |
| chr4 | 143512693 | 143513818 | 1126 | 15 | 8.67E-07 | 1.46E-06 | 0.001652 | 6.37E-07 | 0.000293359 | SMARCA5, SMARCA5-AS1 |
| chr18 | 32525041 | 32526024 | 984 | 5 | 2.49E-05 | 7.14E-07 | 0.001097 | 6.38E-07 | 0.00293554 | AC025887.2 |
| chr16 | 31442085 | 31442627 | 543 | 4 | 1.09E-07 | 3.73E-06 | 0.000161 | 6.39E-07 | -0.01205904 | ZNF843 |
| chr14 | 37406467 | 37406674 | 208 | 2 | 1.33E-06 | 6.59E-07 | 3.06E-05 | 6.43E-07 | 0.005545385 | MIPOL1 |
| chr20 | 59719810 | 59720678 | 869 | 4 | 1.39E-07 | 7.54E-06 | 1.34E-05 | 6.43E-07 | 0.003326008 | PHACTR3 |
| chr5 | 93583629 | 93584792 | 1164 | 5 | 5.52E-06 | 7.26E-07 | 0.000947 | 6.43E-07 | 0.002727623 | NR2F1, NR2F1-AS1 |
| chr22 | 45201846 | 45203178 | 1333 | 9 | 7.30E-07 | 6.60E-06 | 0.001807 | 6.46E-07 | 0.001988539 | KIAA0930 |
| chr19 | 2607728 | 2607905 | 178 | 4 | 3.31E-06 | 2.21E-07 | 0.00044 | 6.48E-07 | 0.009592358 | GNG7 |
| chr1 | 220690120 | 220690979 | 860 | 10 | 2.41E-08 | 1.57E-06 | 0.00379 | 6.50E-07 | 0.003841016 | C1orf115 |
| chr7 | 2639433 | 2640337 | 905 | 5 | 8.78E-07 | 3.25E-07 | 0.000769 | 6.51E-07 | 0.007198325 | TTYH3 |
| chr1 | 183635654 | 183636825 | 1172 | 15 | 1.54E-06 | 4.41E-07 | 0.012097 | 6.51E-07 | 0.001991952 | RGL1, ARPC5 |
| chr16 | 782110 | 783190 | 1081 | 8 | 1.19E-08 | 5.03E-05 | 5.52E-05 | 6.52E-07 | 0.005948675 | MSLNL |
| chr12 | 130870795 | 130872052 | 1258 | 11 | 9.22E-07 | 3.50E-06 | 0.003644 | 6.52E-07 | 0.003716551 | RAN |
| chr2 | 31138238 | 31139588 | 1351 | 13 | 1.95E-06 | 1.36E-05 | 0.005905 | 6.53E-07 | 0.001362493 | GALNT14 |
| chr9 | 18776342 | 18777685 | 1344 | 4 | 6.96E-05 | 1.03E-05 | 0.000163 | 6.53E-07 | 0.004194021 | ADAMTSL1 |
| chr18 | 70180947 | 70181289 | 343 | 2 | 1.55E-06 | 5.16E-07 | 4.98E-05 | 6.56E-07 | 0.001804694 | RTTN |
| chr11 | 17546565 | 17547460 | 896 | 10 | 3.43E-07 | 4.49E-06 | 8.41E-05 | 6.58E-07 | 0.007427302 | OTOG |
| chr15 | 67106861 | 67108235 | 1375 | 6 | 2.14E-06 | 4.25E-07 | 0.000835 | 6.58E-07 | 0.007204363 | SMAD3 |
| chr4 | 3243476 | 3243827 | 352 | 3 | 3.98E-08 | 0.001234 | 8.60E-07 | 6.59E-07 | 0.004612445 | HTT |
| chr12 | 126874585 | 126874870 | 286 | 5 | 9.70E-07 | 3.44E-05 | 0.000348 | 6.59E-07 | 0.005221055 | AC078878.2, LINC02372 |
| chr10 | 49558402 | 49558471 | 70 | 2 | 2.30E-06 | 4.74E-07 | 6.15E-05 | 6.59E-07 | -0.01916921 | NA |
| chr17 | 48617830 | 48618715 | 886 | 6 | 3.72E-07 | 7.83E-07 | 0.000519 | 6.59E-07 | 0.006373431 | HOXB7 |
| chr7 | 155456324 | 155457248 | 925 | 9 | 4.95E-08 | 3.82E-06 | 0.001598 | 6.60E-07 | 0.003902988 | AC008060.3 |
| chr3 | 183228241 | 183228876 | 636 | 2 | 1.76E-07 | 2.39E-06 | 8.20E-06 | 6.63E-07 | 0.001209628 | MCF2L2 |
| chr15 | 90903281 | 90905105 | 1825 | 11 | 6.72E-06 | 1.14E-05 | 0.001834 | 6.63E-07 | 0.002757213 | MAN2A2 |
| chr10 | 69811023 | 69811191 | 169 | 2 | 4.72E-07 | 6.06E-07 | 3.80E-05 | 6.66E-07 | 0.010940824 | COL13A1 |
| chr6 | 35394049 | 35394269 | 221 | 2 | 1.30E-06 | 6.12E-07 | 3.75E-05 | 6.67E-07 | 0.018747966 | PPARD |
| chr4 | 184504107 | 184504552 | 446 | 2 | 2.69E-06 | 4.01E-07 | 0.000114 | 6.68E-07 | 0.014038775 | LINC02427 |
| chr20 | 5001550 | 5001645 | 96 | 4 | 1.64E-06 | 1.15E-06 | 0.000126 | 6.68E-07 | 0.007369165 | SLC23A2 |
| chr1 | 33336876 | 33337634 | 759 | 3 | 1.93E-06 | 4.11E-07 | 0.000205 | 6.69E-07 | -0.00466014 | AL513327.1, PHC2 |
| chr2 | 42931470 | 42931787 | 318 | 2 | 6.99E-07 | 1.63E-06 | 1.14E-05 | 6.71E-07 | -0.03030596 | NA |
| chr10 | 110205646 | 110206336 | 691 | 5 | 4.70E-07 | 4.51E-07 | 0.000559 | 6.71E-07 | 0.017103363 | NA |
| chr14 | 32938559 | 32939742 | 1184 | 8 | 5.42E-07 | 3.08E-06 | 0.000214 | 6.73E-07 | 0.001877596 | NPAS3 |
| chr2 | 110869980 | 110870662 | 683 | 3 | 2.86E-06 | 8.43E-06 | 8.60E-05 | 6.74E-07 | 0.007201978 | ACOXL |
| chr22 | 49490747 | 49491233 | 487 | 2 | 3.76E-06 | 3.86E-07 | 0.000146 | 6.74E-07 | 0.014655346 | C22orf34 |
| chr9 | 83707449 | 83708757 | 1309 | 13 | 1.99E-08 | 0.000102 | 6.99E-06 | 6.74E-07 | -0.00055014 | AL354920.1, UBQLN1 |
| chr17 | 81540654 | 81541116 | 463 | 2 | 5.49E-08 | 2.32E-06 | 8.59E-06 | 6.75E-07 | 0.003878616 | FAAP100 |
| chr16 | 67647162 | 67648162 | 1001 | 3 | 1.69E-06 | 7.92E-05 | 1.77E-05 | 6.77E-07 | 0.000149887 | CARMIL2 |
| chr17 | 82418996 | 82419996 | 1001 | 6 | 6.04E-07 | 4.78E-07 | 0.000414 | 6.83E-07 | 0.006784859 | HEXD |
| chr11 | 86301335 | 86302557 | 1223 | 13 | 5.65E-06 | 5.77E-06 | 0.001707 | 6.86E-07 | 0.00014975 | HIKESHI |
| chr3 | 197366872 | 197367330 | 459 | 2 | 3.48E-08 | 5.09E-06 | 5.10E-06 | 6.88E-07 | 0.005721698 | NA |
| chr19 | 10489300 | 10489770 | 471 | 5 | 2.49E-07 | 9.07E-05 | 6.51E-05 | 6.90E-07 | 0.003105273 | KEAP1 |
| chr4 | 766140 | 766251 | 112 | 2 | 9.64E-07 | 9.09E-07 | 2.18E-05 | 6.90E-07 | 0.008343779 | PCGF3, AC139887.2 |
| chr2 | 241880852 | 241881818 | 967 | 8 | 5.80E-07 | 3.91E-06 | 0.000672 | 6.91E-07 | 0.003773038 | LINC01237 |
| chr6 | 166495323 | 166495704 | 382 | 2 | 5.84E-08 | 2.18E-06 | 9.29E-06 | 6.91E-07 | 0.011708198 | RPS6KA2 |
| chr17 | 81917582 | 81918247 | 666 | 6 | 2.22E-07 | 4.66E-06 | 1.93E-05 | 6.91E-07 | -0.00536138 | SIRT7 |
| chr10 | 44291977 | 44293179 | 1203 | 6 | 2.39E-07 | 1.91E-05 | 0.000365 | 6.91E-07 | 0.01143527 | LINC02881, AL137026.1 |
| chr22 | 19207523 | 19208829 | 1307 | 4 | 0.000107931 | 3.37E-07 | 0.00047 | 6.91E-07 | 0.001103043 | CLTCL1 |
| chr11 | 22666036 | 22666928 | 893 | 8 | 3.00E-07 | 2.80E-06 | 0.000588 | 6.91E-07 | 0.010381838 | GAS2 |
| chr2 | 18560425 | 18561648 | 1224 | 11 | 1.92E-07 | 1.03E-06 | 0.000194 | 6.92E-07 | 0.004757471 | RDH14, NT5C1B-RDH14 |
| chr6 | 44126623 | 44128471 | 1849 | 13 | 5.85E-05 | 8.80E-07 | 0.003487 | 6.92E-07 | -0.00024378 | TMEM63B, MRPL14 |
| chr15 | 41283223 | 41284549 | 1327 | 14 | 7.49E-07 | 7.73E-06 | 0.000513 | 6.92E-07 | 0.000820506 | OIP5-AS1, AC087721.2 |
| chr11 | 72237104 | 72238503 | 1400 | 6 | 1.29E-05 | 9.88E-06 | 0.00011 | 6.94E-07 | 0.005522245 | INPPL1 |
| chr5 | 5330371 | 5330996 | 626 | 2 | 1.38E-07 | 3.19E-06 | 7.06E-06 | 6.97E-07 | -0.02024223 | NA |
| chr1 | 47993564 | 47994289 | 726 | 4 | 5.74E-07 | 3.00E-07 | 0.000718 | 7.00E-07 | 0.005859058 | TRABD2B |
| chr10 | 132827266 | 132827557 | 292 | 4 | 3.52E-06 | 2.14E-07 | 0.000934 | 7.03E-07 | 0.00268802 | CFAP46 |

| | | | | | | | | | | |
|---|---|---|---|---|---|---|---|---|---|---|
| chr1 | 245997338 | 245997524 | 187 | 2 | 2.40E-06 | 4.55E-07 | 8.56E-05 | 7.06E-07 | 0.013020919 | SMYD3 |
| chr6 | 39114826 | 39115429 | 604 | 8 | 4.05E-08 | 0.000218 | 3.06E-06 | 7.06E-07 | 0.001696802 | SAYSD1 |
| chr1 | 201530795 | 201531265 | 471 | 3 | 3.20E-08 | 9.88E-06 | 5.48E-06 | 7.08E-07 | 0.017561366 | AC096677.1, RPS10P7 |
| chr17 | 289501 | 290464 | 964 | 5 | 5.72E-07 | 6.20E-06 | 4.03E-05 | 7.10E-07 | 0.004224627 | RPH3AL |
| chr3 | 186716380 | 186717736 | 1357 | 9 | 2.95E-05 | 5.19E-06 | 0.002295 | 7.10E-07 | 0.006958371 | KNG1, AC068631.1 |
| chr11 | 83034242 | 83035603 | 1362 | 6 | 1.65E-05 | 5.33E-07 | 0.00146 | 7.12E-07 | 0.006522493 | AP000893.2, RAB30 |
| chr2 | 237566670 | 237566773 | 104 | 2 | 1.94E-06 | 4.31E-07 | 0.00011 | 7.14E-07 | 0.010528258 | PRLH |
| chr1 | 114509213 | 114510027 | 815 | 2 | 2.41E-06 | 4.11E-07 | 0.000145 | 7.15E-07 | 0.011640054 | TRIM33 |
| chr11 | 118921359 | 118922073 | 715 | 2 | 2.07E-07 | 5.76E-07 | 4.81E-05 | 7.16E-07 | 0.014143196 | BCL9L |
| chr17 | 81940856 | 81942065 | 1210 | 7 | 1.94E-06 | 1.39E-07 | 0.002178 | 7.16E-07 | 0.00814153 | AC145207.1, PYCR1, MYADML2 |
| chr9 | 137515624 | 137516896 | 1273 | 5 | 5.17E-06 | 1.16E-07 | 0.003564 | 7.16E-07 | 0.000950252 | PNPLA7 |
| chr6 | 166633822 | 166634389 | 568 | 3 | 6.19E-08 | 7.54E-06 | 2.45E-05 | 7.17E-07 | 0.015399666 | RPS6KA2 |
| chr1 | 81498522 | 81498825 | 304 | 2 | 2.31E-06 | 4.55E-07 | 9.15E-05 | 7.19E-07 | 0.025346755 | ADGRL2 |
| chr3 | 179261061 | 179261554 | 494 | 4 | 3.34E-06 | 2.02E-07 | 0.000786 | 7.19E-07 | -0.01313466 | KCNMB3, LRRFIP1P1 |
| chr2 | 172889004 | 172889702 | 699 | 5 | 4.60E-07 | 3.39E-06 | 8.05E-05 | 7.19E-07 | 0.005513007 | RAPGEF4 |
| chr1 | 10298564 | 10299147 | 584 | 6 | 8.96E-07 | 7.14E-06 | 0.000102 | 7.21E-07 | 0.007398694 | KIF1B, RNU6-37P |
| chr16 | 86431896 | 86432821 | 926 | 5 | 5.82E-06 | 3.69E-07 | 0.001724 | 7.22E-07 | 0.006302293 | NA |
| chr16 | 1133462 | 1133656 | 195 | 4 | 7.03E-07 | 6.82E-05 | 3.95E-05 | 7.23E-07 | 0.00425653 | NA |
| chr12 | 113130355 | 113131241 | 887 | 3 | 2.77E-06 | 4.31E-07 | 0.000164 | 7.23E-07 | 0.011603366 | RASAL1 |
| chr18 | 41312313 | 41312833 | 521 | 2 | 2.62E-06 | 4.13E-07 | 0.000155 | 7.24E-07 | 0.018343659 | NA |
| chr3 | 57544146 | 57545072 | 927 | 8 | 3.43E-06 | 6.58E-07 | 0.001567 | 7.24E-07 | 0.008310917 | DNAH12 |
| chr7 | 12210535 | 12211716 | 1182 | 8 | 3.16E-07 | 1.82E-06 | 0.000821 | 7.26E-07 | 0.000104475 | TMEM106B |
| chr13 | 110438086 | 110438896 | 811 | 8 | 8.38E-07 | 1.10E-06 | 0.000117 | 7.26E-07 | 0.007248336 | COL4A2 |
| chr13 | 114164782 | 114164962 | 181 | 3 | 8.45E-07 | 2.54E-06 | 6.66E-05 | 7.26E-07 | 0.001235355 | CFAP97D2 |
| chr18 | 63367091 | 63368000 | 910 | 12 | 7.21E-07 | 5.27E-06 | 0.001113 | 7.26E-07 | -0.000316 | AC036176.1, KDSR |
| chr10 | 46024210 | 46024459 | 250 | 3 | 3.10E-08 | 5.07E-06 | 7.86E-06 | 7.27E-07 | 0.012477508 | NCOA4 |
| chr12 | 6904293 | 6905524 | 1232 | 14 | 7.04E-09 | 5.31E-05 | 1.23E-06 | 7.27E-07 | -0.00140261 | LRRC23 |
| chr3 | 129439340 | 129440347 | 1008 | 20 | 1.53E-07 | 3.99E-06 | 0.002087 | 7.27E-07 | -0.00076471 | IFT122, MBD4 |
| chr14 | 104513035 | 104513449 | 415 | 5 | 1.93E-06 | 3.78E-07 | 0.000689 | 7.27E-07 | 0.011402675 | TMEM179 |
| chr6 | 32761341 | 32762522 | 1182 | 24 | 2.65E-07 | 9.87E-06 | 0.001584 | 7.27E-07 | -0.00927147 | HLA-DQB2 |
| chr5 | 73299285 | 73301299 | 2015 | 15 | 1.38E-05 | 8.08E-05 | 0.00132 | 7.28E-07 | 0.002661689 | NA |
| chr17 | 35981520 | 35982541 | 1022 | 8 | 2.41E-06 | 3.92E-06 | 0.001328 | 7.30E-07 | 0.005709571 | NA |
| chr6 | 3013765 | 3014382 | 618 | 3 | 8.30E-07 | 4.07E-05 | 8.62E-05 | 7.31E-07 | 0.009884107 | NQO2 |
| chr11 | 134383715 | 134384983 | 1269 | 9 | 5.34E-07 | 2.79E-06 | 0.004226 | 7.33E-07 | -0.00216546 | B3GAT1 |
| chr19 | 2050860 | 2052565 | 1706 | 11 | 3.77E-06 | 4.82E-05 | 0.000292 | 7.36E-07 | 0.003611785 | MKNK2 |
| chr16 | 5065303 | 5066673 | 1371 | 8 | 5.51E-06 | 1.17E-05 | 0.000199 | 7.39E-07 | 0.005919375 | ALG1, C16orf89 |
| chr21 | 44994612 | 44995489 | 878 | 6 | 2.32E-07 | 6.19E-06 | 4.37E-05 | 7.39E-07 | 0.001949493 | LINC00165 |
| chr10 | 32357151 | 32357317 | 167 | 3 | 3.36E-06 | 3.43E-07 | 0.000298 | 7.39E-07 | 0.012371467 | AL391839.1, EPC1 |
| chr2 | 232549967 | 232551715 | 1749 | 23 | 3.10E-05 | 5.18E-06 | 0.003379 | 7.40E-07 | 0.00297343 | EIF4E2, TIGD1, MIR5001 |
| chr9 | 127825809 | 127826401 | 593 | 2 | 4.69E-06 | 4.51E-07 | 0.00011 | 7.41E-07 | 0.011843195 | ENG |
| chr17 | 47957089 | 47957977 | 889 | 5 | 8.65E-07 | 4.17E-06 | 0.000212 | 7.41E-07 | 0.003704859 | AC018521.1, PRR15L |
| chr8 | 141131356 | 141132773 | 1418 | 5 | 9.16E-05 | 7.23E-07 | 0.0005 | 7.42E-07 | 0.007698469 | DENND3 |
| chr19 | 44737334 | 44737892 | 559 | 3 | 3.85E-06 | 6.28E-07 | 0.000175 | 7.42E-07 | 0.01250423 | NA |
| chr17 | 10026211 | 10027563 | 1353 | 10 | 9.07E-06 | 1.14E-05 | 0.000372 | 7.42E-07 | 0.004868407 | GAS7 |
| chr8 | 22668709 | 22670358 | 1650 | 13 | 1.24E-06 | 3.41E-06 | 0.000193 | 7.43E-07 | 0.002468535 | BIN3 |
| chr14 | 104428555 | 104428789 | 235 | 2 | 3.30E-07 | 6.18E-07 | 4.65E-05 | 7.45E-07 | 0.015416103 | NA |
| chr7 | 1542365 | 1543493 | 1129 | 8 | 2.45E-06 | 4.33E-07 | 0.003652 | 7.45E-07 | 0.000663237 | MAFK, TMEM184A |
| chr6 | 166656808 | 166657755 | 948 | 4 | 3.92E-06 | 2.13E-06 | 0.000708 | 7.45E-07 | 0.021349584 | RPS6KA2 |
| chr6 | 41371157 | 41372555 | 1399 | 9 | 3.97E-05 | 7.97E-07 | 0.000618 | 7.45E-07 | 0.003177146 | NA |
| chr1 | 116374190 | 116374997 | 808 | 7 | 2.29E-06 | 3.18E-06 | 0.0007 | 7.45E-07 | 0.008227431 | ATP1A1 |
| chr14 | 64464943 | 64465997 | 1055 | 11 | 9.36E-06 | 4.45E-07 | 0.00401 | 7.48E-07 | 0.002267011 | AKAP5, ZBTB25 |
| chr12 | 76031798 | 76033055 | 1258 | 12 | 1.82E-07 | 3.43E-07 | 0.007527 | 7.48E-07 | 0.005850574 | AC011611.4, PHLDA1 |
| chr6 | 137044163 | 137045408 | 1246 | 17 | 2.39E-06 | 1.41E-05 | 0.001612 | 7.48E-07 | 0.002904149 | IL20RA |
| chr17 | 50520715 | 50520964 | 250 | 2 | 2.07E-06 | 5.60E-07 | 5.81E-05 | 7.49E-07 | 0.014689535 | MYCBPAP |
| chr4 | 8440363 | 8441362 | 1000 | 11 | 2.66E-06 | 4.22E-06 | 0.002433 | 7.49E-07 | 0.000953381 | TRMT44, ACOX3 |
| chr20 | 63643763 | 63643884 | 122 | 2 | 2.54E-07 | 1.00E-06 | 2.22E-05 | 7.50E-07 | 0.008238414 | STMN3 |
| chr10 | 131221724 | 131221964 | 241 | 3 | 1.12E-06 | 4.39E-05 | 2.52E-05 | 7.50E-07 | -0.005167 | TCERG1L |
| chr17 | 7845003 | 7846323 | 1321 | 6 | 4.61E-08 | 3.57E-06 | 4.42E-05 | 7.52E-07 | 0.004793208 | KDM6B |
| chr4 | 2746201 | 2746663 | 463 | 5 | 1.01E-06 | 1.34E-06 | 0.000618 | 7.52E-07 | 0.015416696 | TNIP2 |
| chr6 | 27684447 | 27684575 | 129 | 2 | 1.81E-06 | 6.03E-07 | 5.03E-05 | 7.52E-07 | 0.019781897 | NA |
| chr3 | 49869864 | 49871273 | 1410 | 10 | 1.60E-05 | 7.72E-06 | 0.000339 | 7.52E-07 | 0.00426207 | CAMKV |
| chr3 | 169966181 | 169967530 | 1350 | 20 | 2.05E-05 | 1.49E-05 | 0.004925 | 7.52E-07 | -0.00492175 | SEC62, AC008040.1 |
| chr9 | 93295288 | 93295522 | 235 | 2 | 7.72E-07 | 4.31E-06 | 6.38E-06 | 7.52E-07 | 0.001393417 | WNK2 |
| chr9 | 104928204 | 104929349 | 1146 | 8 | 8.84E-08 | 1.46E-06 | 0.000141 | 7.52E-07 | -0.00925776 | AL359182.1 |
| chr11 | 71548205 | 71549277 | 1073 | 6 | 1.44E-08 | 7.21E-06 | 3.24E-05 | 7.53E-07 | 0.009402772 | KRTAP5-9 |
| chr6 | 147407020 | 147408059 | 1040 | 5 | 8.39E-07 | 3.40E-06 | 9.17E-05 | 7.53E-07 | 0.003216968 | YAP1P1 |
| chr10 | 26642802 | 26643062 | 261 | 6 | 2.27E-06 | 1.89E-06 | 5.61E-05 | 7.53E-07 | 0.010960534 | NA |
| chr2 | 46645864 | 46646787 | 924 | 2 | 3.78E-06 | 5.28E-07 | 7.02E-05 | 7.53E-07 | 0.050355945 | NA |
| chr19 | 38223452 | 38224744 | 1293 | 13 | 5.47E-06 | 1.07E-05 | 0.000592 | 7.54E-07 | 0.002808738 | DPF1 |
| chr8 | 144058254 | 144058633 | 380 | 5 | 9.70E-07 | 5.00E-06 | 0.000404 | 7.54E-07 | 0.002738818 | OPLAH |
| chr1 | 230084119 | 230084780 | 662 | 2 | 3.63E-09 | 1.22E-05 | 3.59E-06 | 7.55E-07 | 0.007327027 | GALNT2 |
| chr2 | 107985905 | 107986788 | 884 | 13 | 6.19E-07 | 6.48E-06 | 0.000287 | 7.57E-07 | 0.00555089 | SLC5A7 |
| chr14 | 95292058 | 95292731 | 674 | 3 | 8.60E-07 | 5.35E-07 | 0.000119 | 7.57E-07 | 0.014026564 | CLMN |
| chr17 | 75778734 | 75779883 | 1150 | 10 | 6.54E-06 | 1.48E-05 | 0.000883 | 7.59E-07 | -0.00013484 | H3-3B |
| chr2 | 1503752 | 1504060 | 309 | 4 | 1.80E-06 | 1.03E-06 | 0.000187 | 7.59E-07 | 0.005649172 | TPO, AC105450.1 |
| chr9 | 125820593 | 125821092 | 500 | 2 | 8.08E-07 | 9.77E-07 | 2.37E-05 | 7.60E-07 | 0.031823361 | PBX3 |
| chr1 | 9163944 | 9164317 | 374 | 3 | 1.65E-06 | 5.73E-07 | 7.96E-05 | 7.60E-07 | 0.020140878 | MIR34AHG |
| chr19 | 35138452 | 35140269 | 1818 | 18 | 1.43E-05 | 1.15E-05 | 0.00056 | 7.60E-07 | 0.001622803 | FXYD1, AC020907.2, LGI4, AC020907.5 |
| chr21 | 41166957 | 41168034 | 1078 | 11 | 2.44E-07 | 1.69E-05 | 0.000887 | 7.62E-07 | 0.000977993 | MIR3197, BACE2 |
| chr11 | 68992793 | 68992977 | 185 | 3 | 1.87E-06 | 7.21E-07 | 7.45E-05 | 7.63E-07 | 0.013944637 | NA |
| chr20 | 62549937 | 62551477 | 1541 | 13 | 8.79E-07 | 9.58E-06 | 0.002621 | 7.64E-07 | 0.004105176 | MIR1-1HG, MIR1-1HG-AS1 |
| chr11 | 62553664 | 62554805 | 1142 | 4 | 2.10E-07 | 0.000883 | 4.77E-05 | 7.66E-07 | 0.008721483 | AHNAK |
| chr3 | 47380310 | 47381650 | 1341 | 12 | 1.66E-06 | 8.52E-06 | 0.002078 | 7.66E-07 | 0.000899886 | PTPN23, AC099778.1 |
| chr15 | 62165257 | 62166016 | 760 | 9 | 2.31E-06 | 1.75E-07 | 0.004463 | 7.67E-07 | 0.007822092 | C2CD4B |
| chr4 | 175812258 | 175813151 | 894 | 8 | 1.29E-06 | 6.79E-07 | 0.001436 | 7.67E-07 | 0.018194653 | GPM6A |
| chr8 | 141581169 | 141581827 | 659 | 2 | 3.01E-07 | 1.97E-06 | 1.16E-05 | 7.68E-07 | 0.017307048 | AC025839.1 |
| chr18 | 26470141 | 26470666 | 526 | 7 | 2.14E-06 | 8.20E-06 | 0.000542 | 7.69E-07 | 0.002503455 | KCTD1 |
| chr12 | 50924084 | 50924998 | 915 | 8 | 3.36E-07 | 9.44E-06 | 0.000124 | 7.69E-07 | 0.004073049 | METTL7A |
| chr11 | 70569083 | 70570204 | 1122 | 7 | 8.95E-06 | 4.17E-07 | 0.001476 | 7.70E-07 | 0.0029481 | SHANK2 |
| chr15 | 78977815 | 78979174 | 1360 | 5 | 2.55E-05 | 3.75E-05 | 0.000453 | 7.71E-07 | 0.005827879 | AC011944.1, RASGRF1 |
| chr14 | 74294411 | 74295287 | 877 | 6 | 2.93E-06 | 2.26E-06 | 0.000101 | 7.72E-07 | 0.00687805 | ABCD4, AC005519.1 |
| chr5 | 68434244 | 68435816 | 1573 | 6 | 3.36E-05 | 4.71E-06 | 0.001021 | 7.74E-07 | 0.008979666 | AC010280.2 |
| chr12 | 121580272 | 121582059 | 1788 | 21 | 5.94E-06 | 2.58E-06 | 0.009639 | 7.74E-07 | 0.001860894 | KDM2B-DT, KDM2B |
| chr6 | 157275536 | 157276741 | 1206 | 5 | 1.01E-07 | 1.62E-06 | 4.32E-05 | 7.75E-07 | -0.00047122 | NA |
| chr4 | 138688811 | 138689034 | 224 | 4 | 1.14E-06 | 1.38E-05 | 2.96E-05 | 7.75E-07 | -0.0014438 | NA |
| chr1 | 63317704 | 63318306 | 603 | 6 | 6.84E-07 | 8.57E-07 | 0.000236 | 7.77E-07 | 0.004697017 | NA |
| chr11 | 69666966 | 69668466 | 1501 | 7 | 0.000112805 | 9.85E-07 | 0.004737 | 7.77E-07 | 0.002786783 | LTO1 |
| chr15 | 73753897 | 73754367 | 471 | 2 | 1.42E-06 | 4.56E-07 | 0.000142 | 7.77E-07 | 0.017061203 | INSYN1-AS1 |
| chr10 | 12898223 | 12898696 | 474 | 3 | 2.57E-07 | 8.15E-06 | 1.59E-05 | 7.79E-07 | 0.005455381 | AL353586.1, CCDC3 |
| chr3 | 186193419 | 186194464 | 1046 | 10 | 1.80E-07 | 6.69E-06 | 0.000206 | 7.80E-07 | -0.00657319 | DGKG |
| chr1 | 20289947 | 20290963 | 1017 | 9 | 9.73E-06 | 5.20E-05 | 0.001402 | 7.81E-07 | 0.008113303 | VWA5B1 |
| chr5 | 80406851 | 80408290 | 1440 | 13 | 3.92E-06 | 9.33E-07 | 0.008318 | 7.81E-07 | 0.000292047 | ZFYVE16 |
| chr6 | 37653456 | 37654354 | 899 | 3 | 4.27E-06 | 3.42E-07 | 0.000289 | 7.82E-07 | 0.017735504 | MDGA1 |
| chr20 | 63690633 | 63691914 | 1282 | 5 | 2.15E-06 | 1.20E-06 | 0.000489 | 7.83E-07 | 0.006106133 | RTEL1, RTEL1-TNFRSF6B |
| chr19 | 11339257 | 11340065 | 809 | 11 | 3.25E-07 | 2.00E-05 | 0.001041 | 7.83E-07 | 0.004564074 | RAB3D |
| chr19 | 16032434 | 16032559 | 126 | 2 | 2.01E-06 | 1.08E-06 | 2.20E-05 | 7.84E-07 | 0.000254418 | LINC00905 |
| chr9 | 21989777 | 21990278 | 502 | 6 | 2.51E-06 | 5.57E-07 | 0.001202 | 7.84E-07 | 0.01137683 | AL359922.1, CDKN2A |
| chr19 | 41993727 | 41995467 | 1741 | 10 | 3.39E-06 | 5.05E-07 | 0.000922 | 7.85E-07 | 0.003559145 | AC010616.1, ATP1A3 |
| chr17 | 7740480 | 7740923 | 444 | 5 | 1.87E-06 | 3.50E-07 | 0.00084 | 7.86E-07 | -0.02964753 | DNAH2 |
| chr12 | 57631202 | 57631813 | 612 | 3 | 6.87E-07 | 4.29E-07 | 0.000348 | 7.86E-07 | 0.010617031 | B4GALNT1 |
| chr9 | 34372091 | 34372238 | 148 | 2 | 7.00E-08 | 5.50E-07 | 7.24E-05 | 7.87E-07 | 0.002526753 | MYORG |
| chr15 | 36897098 | 36897206 | 109 | 2 | 1.32E-06 | 4.63E-07 | 0.000147 | 7.93E-07 | 0.023055149 | MEIS2 |
| chr11 | 17389007 | 17390405 | 1399 | 12 | 2.52E-06 | 2.42E-06 | 0.001303 | 7.93E-07 | 0.00015023 | KCNJ11 |
| chr5 | 171629819 | 171630571 | 753 | 6 | 7.93E-07 | 8.48E-06 | 2.20E-05 | 7.95E-07 | 0.019159031 | NA |
| chr3 | 126987875 | 126988911 | 1037 | 6 | 5.29E-07 | 5.78E-07 | 0.000723 | 8.00E-07 | 0.002948637 | PLXNA1 |
| chr5 | 14726927 | 14727933 | 1007 | 5 | 1.07E-06 | 7.22E-05 | 6.94E-05 | 8.03E-07 | 0.008822785 | ANKH |
| chr14 | 91417211 | 91418930 | 1720 | 12 | 9.36E-06 | 1.96E-06 | 0.000169 | 8.03E-07 | -0.00034763 | AL133153.2, CCDC88C |
| chr17 | 47831600 | 47832162 | 563 | 9 | 1.40E-07 | 4.24E-06 | 0.000626 | 8.04E-07 | 0.003402126 | LRRC46 |
| chr9 | 136975842 | 136977709 | 1868 | 11 | 8.12E-05 | 4.85E-06 | 0.001204 | 8.04E-07 | 0.007270134 | PTGDS, AL807752.7 |
| chr5 | 67605158 | 67606704 | 1547 | 4 | 0.000223034 | 8.12E-07 | 0.000626 | 8.04E-07 | -0.0044669 | AC112206.2 |
| chr4 | 182142041 | 182142882 | 842 | 7 | 5.96E-07 | 4.70E-07 | 0.000893 | 8.05E-07 | 0.003856057 | TENM3-AS1 |
| chr11 | 57391311 | 57391701 | 391 | 4 | 1.49E-06 | 2.62E-07 | 0.001253 | 8.06E-07 | -0.04108243 | AP000781.2 |
| chr17 | 76871336 | 76872138 | 803 | 3 | 3.38E-09 | 2.56E-05 | 4.42E-06 | 8.07E-07 | 0.0054773 | MGAT5B |
| chr8 | 140464694 | 140465024 | 331 | 4 | 1.24E-07 | 5.16E-05 | 2.74E-06 | 8.07E-07 | -0.00064493 | NA |
| chr11 | 59142454 | 59143110 | 657 | 18 | 1.01E-06 | 4.13E-05 | 0.001816 | 8.07E-07 | 0.000918078 | FAM111A, FAM111A-DT, AP001258.1 |
| chr13 | 94595993 | 94596836 | 844 | 11 | 2.73E-07 | 8.23E-06 | 0.000146 | 8.07E-07 | 0.000961987 | TGDS |
| chr11 | 121459639 | 121459692 | 54 | 3 | 3.05E-06 | 2.92E-07 | 0.000628 | 8.10E-07 | 0.012332386 | SORL1 |
| chr9 | 79572294 | 79573226 | 933 | 8 | 1.00E-05 | 1.15E-06 | 0.00073 | 8.10E-07 | 0.000506951 | TLE4 |
| chr4 | 38761818 | 38761863 | 46 | 2 | 2.84E-06 | 5.87E-07 | 6.62E-05 | 8.12E-07 | 0.01233268 | NA |
| chr9 | 136777721 | 136777732 | 12 | 2 | 2.92E-06 | 6.29E-07 | 5.69E-05 | 8.12E-07 | 0.006701527 | NA |
| chr1 | 110071311 | 110072068 | 758 | 4 | 2.19E-07 | 2.38E-06 | 0.000337 | 8.15E-07 | 0.009831676 | STRIP1 |
| chr20 | 45963343 | 45963925 | 583 | 2 | 7.71E-08 | 1.29E-06 | 1.92E-05 | 8.17E-07 | 0.008974109 | ZNF335 |
| chr22 | 50262013 | 50262804 | 792 | 7 | 8.66E-07 | 3.32E-06 | 8.36E-05 | 8.21E-07 | 0.002812523 | NA |
| chr3 | 183287271 | 183287331 | 61 | 2 | 2.27E-06 | 4.74E-07 | 0.000163 | 8.24E-07 | 0.017530993 | B3GNT5, MCF2L2 |
| chr10 | 5524297 | 5525561 | 1265 | 12 | 1.56E-06 | 1.61E-05 | 0.003851 | 8.25E-07 | 0.00247466 | CALML3, CALML3-AS1, AL732437.1 |
| chr11 | 65121721 | 65123068 | 1348 | 21 | 2.51E-06 | 1.64E-06 | 0.007101 | 8.25E-07 | -0.00029118 | MRPL49, FAU, SYVN1 |
| chr11 | 43969460 | 43969629 | 170 | 2 | 2.48E-06 | 7.14E-07 | 4.53E-05 | 8.25E-07 | 0.02204086 | AC087521.3 |
| chr5 | 173988627 | 173989424 | 798 | 9 | 8.81E-06 | 6.02E-07 | 0.002408 | 8.26E-07 | 0.002122344 | C5orf47 |
| chr6 | 49333975 | 49334963 | 989 | 3 | 3.91E-05 | 1.78E-06 | 0.000152 | 8.26E-07 | 0.004584219 | NA |
| chr1 | 107480627 | 107480864 | 238 | 5 | 4.80E-06 | 2.54E-07 | 0.002256 | 8.28E-07 | -0.01990222 | NTNG1 |
| chr12 | 52146994 | 52148083 | 1090 | 5 | 5.99E-08 | 1.60E-05 | 7.02E-06 | 8.28E-07 | 0.005690489 | NA |
| chr9 | 91857402 | 91858129 | 728 | 2 | 5.13E-06 | 4.82E-07 | 0.000149 | 8.28E-07 | 0.044855943 | ROR2 |
| chr17 | 82151673 | 82152863 | 1191 | 6 | 1.17E-08 | 0.000297 | 1.54E-05 | 8.28E-07 | 0.002267879 | CCDC57 |
| chr6 | 31592664 | 31593094 | 431 | 5 | 1.55E-07 | 4.40E-05 | 5.86E-05 | 8.28E-07 | 0.004138999 | NCR3 |
| chr7 | 151008270 | 151009267 | 998 | 7 | 6.36E-07 | 4.42E-07 | 0.001532 | 8.29E-07 | -0.01634332 | NOS3 |
| chr14 | 56117767 | 56118768 | 1002 | 9 | 8.11E-07 | 1.87E-05 | 4.41E-05 | 8.29E-07 | 0.000378665 | PELI2, AL138995.1 |
| chr12 | 132471171 | 132471360 | 190 | 2 | 2.43E-06 | 4.79E-07 | 0.000164 | 8.30E-07 | 0.013871599 | NA |
| chr18 | 26752116 | 26752427 | 312 | 2 | 3.52E-06 | 4.79E-07 | 0.000163 | 8.30E-07 | 0.007327619 | AQP4-AS1 |
| chr12 | 11855732 | 11856233 | 502 | 2 | 6.89E-07 | 6.28E-07 | 6.04E-05 | 8.31E-07 | 0.014483617 | ETV6 |
| chr7 | 112484924 | 112484929 | 6 | 2 | 3.54E-07 | 6.73E-07 | 5.19E-05 | 8.31E-07 | 0.006869065 | AC005192.1, LSMEM1 |
| chr8 | 125568886 | 125569837 | 952 | 4 | 4.88E-06 | 1.50E-06 | 0.000341 | 8.32E-07 | 0.008422699 | NA |
| chr21 | 42836922 | 42837246 | 325 | 4 | 8.10E-08 | 0.000254 | 1.23E-06 | 8.36E-07 | 0.00480446 | NA |
| chr14 | 74501506 | 74502147 | 642 | 2 | 1.96E-06 | 6.92E-07 | 4.97E-05 | 8.36E-07 | 0.009520311 | LTBP2 |
| chr17 | 66425780 | 66426691 | 912 | 3 | 1.36E-06 | 5.10E-07 | 0.000156 | 8.37E-07 | 0.017920575 | PRKCA |
| chr17 | 48968479 | 48968942 | 464 | 4 | 4.79E-07 | 9.29E-05 | 9.29E-06 | 8.40E-07 | 0.007472319 | GIP |
| chr20 | 35284605 | 35285860 | 1256 | 16 | 1.41E-05 | 2.15E-06 | 0.006995 | 8.41E-07 | 0.001032916 | FAM83C-AS1, EIF6, FAM83C |
| chr17 | 60154433 | 60154980 | 548 | 3 | 2.38E-07 | 4.08E-06 | 1.26E-05 | 8.43E-07 | 0.003712328 | CA4 |
| chr19 | 5114403 | 5115613 | 1211 | 3 | 2.08E-10 | 0.000935 | 1.20E-06 | 8.45E-07 | -0.00403563 | KDM4B |
| chr12 | 124422559 | 124423566 | 1008 | 5 | 2.37E-06 | 6.82E-06 | 2.17E-05 | 8.47E-07 | 0.000919778 | NCOR2 |
| chr19 | 39970516 | 39971580 | 1065 | 12 | 7.14E-07 | 0.000282 | 0.000143 | 8.50E-07 | 0.0014054 | PSMC4 |
| chr20 | 18056395 | 18057652 | 1258 | 3 | 6.36E-06 | 2.92E-07 | 0.000849 | 8.51E-07 | 0.000878189 | OVOL2 |
| chr10 | 27192492 | 27192558 | 67 | 2 | 1.05E-06 | 4.90E-07 | 0.000163 | 8.51E-07 | 0.010064869 | ACBD5 |

| | | | | | | | | | | |
|---|---|---|---|---|---|---|---|---|---|---|
| chr16 | 67399281 | 67399733 | 453 | 4 | 3.66E-06 | 2.52E-07 | 0.001357 | 8.53E-07 | 0.010696848 | ZDHHC1 |
| chr7 | 4774523 | 4776078 | 1556 | 13 | 1.39E-05 | 4.80E-05 | 0.001472 | 8.54E-07 | 0.001464452 | AP5Z1 |
| chr10 | 8083636 | 8083808 | 173 | 3 | 1.59E-06 | 2.57E-06 | 1.96E-05 | 8.57E-07 | 0.006331697 | NA |
| chr6 | 36677220 | 36678350 | 1131 | 8 | 1.32E-07 | 1.33E-05 | 0.000283 | 8.60E-07 | 0.011694412 | CDKN1A, DINOL |
| chr3 | 185473308 | 185473950 | 643 | 6 | 4.02E-07 | 2.11E-05 | 0.000269 | 8.60E-07 | 0.000336351 | MAP3K13 |
| chr6 | 158853421 | 158854354 | 934 | 3 | 2.26E-05 | 2.86E-07 | 0.00099 | 8.61E-07 | 0.005240976 | AL627422.2, OSTCP1 |
| chr1 | 95233757 | 95234716 | 960 | 12 | 7.47E-07 | 5.56E-06 | 2.09E-05 | 8.62E-07 | 0.002021452 | TLCD4-RWDD3, RWDD3, AC092802.1 |
| chr3 | 155745220 | 155745930 | 711 | 5 | 2.01E-07 | 7.01E-07 | 0.00026 | 8.64E-07 | 0.009430054 | NA |
| chr4 | 7704820 | 7704938 | 119 | 2 | 1.85E-06 | 2.24E-06 | 1.22E-05 | 8.64E-07 | 0.005728106 | SORCS2 |
| chr2 | 63587769 | 63589294 | 1526 | 23 | 2.97E-06 | 2.66E-05 | 0.007931 | 8.64E-07 | 0.000415653 | MDH1, WDPCP |
| chr17 | 50835499 | 50836063 | 565 | 5 | 1.34E-08 | 3.34E-06 | 6.16E-05 | 8.66E-07 | 0.007192256 | WFIKKN2 |
| chr19 | 51387332 | 51388305 | 974 | 5 | 3.59E-06 | 2.29E-07 | 0.001743 | 8.66E-07 | 0.007842335 | LIM2, C19orf84 |
| chr5 | 119913174 | 119913927 | 754 | 3 | 8.48E-06 | 4.30E-07 | 0.000321 | 8.66E-07 | 0.001597664 | NA |
| chr11 | 9572903 | 9573997 | 1095 | 10 | 4.61E-07 | 8.31E-06 | 0.002316 | 8.68E-07 | 0.001292723 | WEE1 |
| chr4 | 186919457 | 186919513 | 57 | 2 | 3.48E-06 | 5.00E-07 | 0.000163 | 8.68E-07 | 0.012530703 | NA |
| chr1 | 184754390 | 184755666 | 1277 | 15 | 2.35E-05 | 3.30E-07 | 0.005871 | 8.70E-07 | 0.000932599 | EDEM3 |
| chr19 | 4517286 | 4517792 | 507 | 8 | 1.75E-06 | 2.51E-06 | 0.00053 | 8.72E-07 | 0.005400908 | PLIN4 |
| chr10 | 103966835 | 103968014 | 1180 | 11 | 1.60E-10 | 0.006404 | 5.39E-07 | 8.72E-07 | 0.000535821 | SLK |
| chr11 | 65013229 | 65013831 | 603 | 5 | 1.57E-07 | 3.12E-06 | 0.000126 | 8.72E-07 | 0.005293446 | NA |
| chr22 | 38769004 | 38769966 | 963 | 4 | 2.76E-05 | 3.11E-07 | 0.001029 | 8.72E-07 | 0.005709865 | SUN2 |
| chr22 | 24100593 | 24101667 | 1075 | 6 | 5.72E-06 | 1.18E-06 | 0.00059 | 8.72E-07 | 0.001760846 | CABIN1 |
| chr1 | 15829797 | 15830119 | 323 | 3 | 1.68E-06 | 8.03E-07 | 9.49E-05 | 8.76E-07 | 0.012998785 | NA |
| chr10 | 102419267 | 102420722 | 1456 | 19 | 5.75E-08 | 3.43E-05 | 0.000525 | 8.80E-07 | 0.002462674 | FBXL15, PSD |
| chr9 | 95126989 | 95127785 | 797 | 7 | 1.90E-06 | 1.48E-06 | 0.003108 | 8.81E-07 | -0.01101857 | FANCC |
| chr15 | 42457138 | 42457687 | 550 | 6 | 5.66E-06 | 2.73E-07 | 0.001512 | 8.82E-07 | 0.004720929 | ZNF106 |
| chr10 | 3166248 | 3166293 | 46 | 2 | 4.33E-07 | 8.87E-06 | 4.85E-06 | 8.82E-07 | 0.004503717 | PITRM1-AS1, PITRM1 |
| chr2 | 120588909 | 120589634 | 726 | 3 | 1.61E-06 | 1.09E-05 | 3.20E-05 | 8.83E-07 | 0.009535644 | NA |
| chr5 | 132657922 | 132658369 | 448 | 5 | 1.76E-06 | 4.92E-07 | 0.000494 | 8.84E-07 | 0.00551833 | IL13, TH2LCRR |
| chr20 | 19751108 | 19751408 | 301 | 2 | 1.75E-07 | 0.000328 | 1.88E-06 | 8.84E-07 | 0.004001811 | NA |
| chr6 | 33899170 | 33900661 | 1492 | 13 | 6.42E-06 | 1.28E-05 | 6.79E-05 | 8.84E-07 | 0.001723656 | AL138889.1, MIR7159 |
| chr3 | 195209441 | 195210906 | 1466 | 8 | 8.07E-05 | 0.00024 | 0.000258 | 8.85E-07 | 0.003075004 | XXYLT1 |
| chr10 | 104037832 | 104038388 | 557 | 2 | 7.04E-08 | 2.03E-05 | 3.33E-06 | 8.85E-07 | 0.004414973 | COL17A1 |
| chr12 | 6542743 | 6543435 | 693 | 4 | 1.69E-06 | 1.30E-06 | 6.70E-05 | 8.86E-07 | 0.007227168 | IFFO1 |
| chr2 | 239239923 | 239240627 | 705 | 5 | 7.78E-07 | 6.24E-07 | 0.000542 | 8.86E-07 | 0.009064846 | HDAC4 |
| chr7 | 1268529 | 1269472 | 944 | 6 | 3.97E-06 | 1.38E-05 | 0.00037 | 8.86E-07 | 0.003217803 | AC073094.1 |
| chr2 | 1769493 | 1770118 | 626 | 3 | 8.30E-07 | 1.78E-06 | 3.57E-05 | 8.86E-07 | 0.008739725 | NA |
| chr22 | 29073731 | 29074359 | 629 | 4 | 4.90E-08 | 4.28E-05 | 1.71E-05 | 8.88E-07 | 0.009326711 | KREMEN1 |
| chr8 | 124474345 | 124475563 | 1219 | 11 | 1.08E-05 | 1.94E-05 | 0.0007 | 8.90E-07 | -0.00035184 | RNF139, RNF139-AS1 |
| chr16 | 15948910 | 15950000 | 1091 | 10 | 8.63E-06 | 1.24E-06 | 0.007163 | 8.91E-07 | 0.004309412 | ABCC1 |
| chr1 | 29148157 | 29149173 | 1017 | 5 | 1.86E-06 | 9.76E-06 | 0.000125 | 8.93E-07 | 0.002471695 | SRSF4 |
| chr1 | 169894353 | 169895114 | 762 | 4 | 1.48E-09 | 0.000128 | 3.73E-05 | 8.94E-07 | -0.00127578 | NA |
| chr11 | 66336352 | 66337677 | 1326 | 11 | 8.35E-07 | 6.85E-06 | 0.000153 | 8.94E-07 | 0.005276287 | AP001107.6, RIN1, BRMS1 |
| chr2 | 203781086 | 203781268 | 183 | 3 | 2.61E-07 | 3.66E-05 | 2.79E-06 | 8.95E-07 | 0.003945847 | NA |
| chr6 | 3252784 | 3253296 | 513 | 2 | 1.53E-08 | 0.000147 | 2.12E-06 | 8.96E-07 | 0.002597115 | PSMG4 |
| chr7 | 98961442 | 98961523 | 82 | 3 | 4.79E-06 | 3.63E-07 | 0.000473 | 8.99E-07 | 0.000264271 | TRRAP |
| chr14 | 89469282 | 89469840 | 559 | 2 | 6.06E-06 | 5.31E-07 | 0.000147 | 9.01E-07 | 0.009806522 | FOXN3, AL137230.2 |
| chr7 | 73624529 | 73625223 | 695 | 9 | 3.69E-07 | 3.40E-05 | 0.000132 | 9.03E-07 | -0.00113699 | MLXIPL |
| chr3 | 37992279 | 37993419 | 1141 | 6 | 1.52E-06 | 8.79E-05 | 0.000305 | 9.04E-07 | 0.003341133 | VILL |
| chr12 | 122279183 | 122280493 | 1311 | 4 | 1.49E-07 | 1.20E-06 | 0.00036 | 9.05E-07 | 0.008981821 | CLIP1 |
| chr1 | 111619113 | 111620310 | 1198 | 12 | 2.80E-05 | 6.21E-07 | 0.005549 | 9.07E-07 | -0.00438054 | RAP1A |
| chr20 | 36154860 | 36155992 | 1133 | 6 | 4.24E-05 | 1.94E-07 | 0.001601 | 9.07E-07 | 0.000498887 | EPB41L1, AL121895.1 |
| chr12 | 30925856 | 30927489 | 1634 | 17 | 1.64E-05 | 1.24E-05 | 0.005314 | 9.07E-07 | 0.002384383 | TSPAN11 |
| chr2 | 120985686 | 120986989 | 1304 | 6 | 6.74E-07 | 3.31E-05 | 0.000163 | 9.10E-07 | 0.000772645 | GLI2 |
| chr13 | 44435915 | 44437335 | 1421 | 18 | 3.57E-09 | 0.000833 | 0.000506 | 9.11E-07 | 0.000836896 | AL139184.1, TSC22D1 |
| chr3 | 58586931 | 58587593 | 663 | 9 | 2.98E-06 | 9.84E-07 | 0.002715 | 9.11E-07 | -0.00896401 | FAM107A |
| chr6 | 1436029 | 1436932 | 904 | 5 | 2.14E-05 | 2.36E-07 | 0.00232 | 9.11E-07 | 0.006579474 | NA |
| chr19 | 17943551 | 17944725 | 1175 | 5 | 1.43E-07 | 0.000358 | 3.10E-05 | 9.12E-07 | 0.000626578 | CCDC124 |
| chr1 | 47443923 | 47445347 | 1425 | 9 | 6.48E-06 | 1.45E-05 | 0.000755 | 9.13E-07 | 0.00255342 | NA |
| chr11 | 6563442 | 6564423 | 982 | 9 | 1.37E-06 | 2.46E-06 | 0.00017 | 9.16E-07 | 0.002783924 | DNHD1 |
| chr17 | 4591443 | 4592440 | 998 | 4 | 2.15E-06 | 1.18E-06 | 0.000474 | 9.16E-07 | 0.007884491 | SMTNL2 |
| chr11 | 111299665 | 111300605 | 941 | 10 | 2.05E-06 | 3.13E-05 | 0.000345 | 9.17E-07 | 0.00260587 | COLCA2, COLCA1 |
| chr14 | 76918823 | 76919712 | 890 | 5 | 3.00E-06 | 6.60E-07 | 0.000429 | 9.17E-07 | 0.010816697 | NA |
| chr11 | 107345778 | 107346570 | 793 | 2 | 3.33E-06 | 8.77E-07 | 4.03E-05 | 9.17E-07 | 0.007211154 | CWF19L2 |
| chr10 | 14953626 | 14954928 | 1303 | 19 | 1.78E-06 | 1.26E-06 | 0.001538 | 9.17E-07 | -0.00085872 | DCLRE1C |
| chr9 | 34380892 | 34381701 | 810 | 7 | 3.79E-07 | 5.94E-07 | 0.002277 | 9.18E-07 | 0.007029116 | C9orf24 |
| chr12 | 72271502 | 72272534 | 1033 | 8 | 4.64E-06 | 7.29E-06 | 4.74E-05 | 9.21E-07 | 0.002815245 | TRHDE, TRHDE-AS1 |
| chr14 | 91253829 | 91254474 | 646 | 11 | 2.63E-07 | 7.53E-06 | 0.000193 | 9.25E-07 | 0.005657425 | GPR68 |
| chr13 | 114067732 | 114068417 | 686 | 3 | 3.77E-07 | 1.05E-06 | 7.16E-05 | 9.27E-07 | 0.002387662 | RASA3 |
| chr3 | 160384743 | 160385138 | 396 | 4 | 7.34E-07 | 1.52E-05 | 0.000276 | 9.27E-07 | 0.003814524 | AC079594.1, IFT80 |
| chr1 | 18373840 | 18374477 | 638 | 6 | 1.19E-06 | 3.10E-06 | 7.19E-05 | 9.30E-07 | 0.006713868 | IGSF21 |
| chr1 | 248028642 | 248028937 | 296 | 3 | 1.68E-06 | 1.24E-05 | 2.05E-05 | 9.30E-07 | 0.01198502 | NA |
| chr21 | 46254290 | 46255245 | 956 | 3 | 3.65E-05 | 5.33E-07 | 0.000266 | 9.30E-07 | 0.010430675 | MCM3AP-AS1, MCM3AP |
| chr12 | 89525271 | 89526873 | 1603 | 21 | 4.78E-06 | 4.62E-06 | 0.001073 | 9.32E-07 | 0.001751643 | POC1B-AS1, POC1B, POC1B-GALNT4 |
| chr18 | 903976 | 905449 | 1474 | 19 | 8.37E-07 | 8.24E-06 | 0.005452 | 9.33E-07 | 0.00402413 | ADCYAP1, AP000894.3, AP000894.2 |
| chr16 | 3067861 | 3068748 | 888 | 4 | 4.86E-06 | 5.21E-07 | 0.000609 | 9.34E-07 | 0.010592884 | IL32 |
| chr4 | 3363172 | 3363715 | 544 | 5 | 3.81E-06 | 7.00E-07 | 0.000491 | 9.35E-07 | 0.011344086 | RGS12 |
| chr15 | 77027607 | 77028043 | 437 | 5 | 8.36E-07 | 3.89E-07 | 0.000821 | 9.37E-07 | 0.004109471 | PSTPIP1 |
| chr15 | 82647889 | 82648881 | 993 | 13 | 2.02E-06 | 1.17E-05 | 0.004104 | 9.37E-07 | 0.005680774 | CPEB1-AS1, CPEB1 |
| chr8 | 143868995 | 143869641 | 647 | 4 | 2.69E-07 | 7.34E-06 | 6.03E-05 | 9.40E-07 | 0.001742893 | EPPK1 |
| chr11 | 123062318 | 123063406 | 1089 | 10 | 3.83E-06 | 7.88E-06 | 0.002121 | 9.41E-07 | 0.001461269 | AP000926.2, HSPA8 |
| chr21 | 45618519 | 45618991 | 473 | 3 | 5.67E-06 | 3.24E-07 | 0.000898 | 9.41E-07 | 0.021347961 | AL133493.1 |
| chr16 | 67672726 | 67673193 | 468 | 4 | 8.46E-07 | 1.06E-06 | 5.69E-05 | 9.44E-07 | 0.010305658 | NA |
| chr2 | 234289059 | 234289352 | 294 | 3 | 1.01E-06 | 1.06E-06 | 6.27E-05 | 9.44E-07 | 0.013722529 | NA |
| chr22 | 30470216 | 30470578 | 363 | 2 | 1.17E-06 | 6.85E-07 | 7.04E-05 | 9.45E-07 | 0.01240489 | SEC14L3 |
| chr19 | 13835956 | 13837100 | 1145 | 10 | 1.93E-06 | 7.96E-07 | 0.005667 | 9.45E-07 | 0.006459888 | AC020916.1, MIR24-2, MIR27A, MIR23A |
| chr16 | 47299140 | 47299396 | 257 | 5 | 3.08E-06 | 1.62E-06 | 0.000216 | 9.45E-07 | 0.00349722 | ITFG1 |
| chr12 | 55727032 | 55727967 | 936 | 9 | 1.51E-06 | 2.30E-06 | 0.002289 | 9.47E-07 | 0.007539368 | CD63 |
| chr12 | 542145 | 542958 | 814 | 4 | 6.97E-07 | 6.26E-06 | 1.17E-05 | 9.48E-07 | 0.001537679 | B4GALNT3 |
| chr8 | 120529758 | 120530325 | 568 | 3 | 6.25E-07 | 1.61E-06 | 3.43E-05 | 9.48E-07 | 0.006218851 | MTBP |
| chr12 | 16605178 | 16606657 | 1480 | 23 | 2.76E-06 | 6.93E-06 | 0.004447 | 9.50E-07 | 0.000333077 | MGST1, LMO3 |
| chr3 | 127366700 | 127367151 | 452 | 2 | 3.81E-06 | 5.52E-07 | 0.000165 | 9.52E-07 | 0.016958614 | LINC02016 |
| chr1 | 6554281 | 6555544 | 1264 | 18 | 2.49E-06 | 7.78E-06 | 0.000546 | 9.52E-07 | 0.000783133 | TAS1R1, NOL9 |
| chr4 | 3246523 | 3247454 | 932 | 6 | 3.38E-06 | 4.69E-06 | 0.00079 | 9.52E-07 | 0.004118435 | MSANTD1 |
| chr2 | 169507648 | 169508702 | 1055 | 3 | 2.57E-05 | 1.11E-06 | 0.000162 | 9.54E-07 | 0.015131667 | AC093899.2 |
| chr3 | 125958022 | 125959147 | 1126 | 5 | 3.48E-06 | 2.90E-07 | 0.002012 | 9.55E-07 | 0.016645985 | AC092903.1 |
| chr12 | 2223542 | 2223995 | 454 | 4 | 1.86E-06 | 1.35E-05 | 4.65E-05 | 9.55E-07 | 0.007838317 | CACNA1C |
| chr1 | 98749003 | 98749343 | 341 | 2 | 8.30E-07 | 3.04E-06 | 1.09E-05 | 9.56E-07 | -0.01209238 | SNX7 |
| chr9 | 113011458 | 113012261 | 804 | 3 | 7.05E-07 | 2.57E-06 | 1.00E-04 | 9.56E-07 | 0.007779417 | ZNF883, ZNF883 |
| chr4 | 110198141 | 110199298 | 1158 | 17 | 5.54E-06 | 1.63E-06 | 0.005774 | 9.57E-07 | 0.000889608 | ELOVL6 |
| chr16 | 66731145 | 66731624 | 480 | 3 | 2.44E-07 | 2.17E-05 | 3.86E-06 | 9.59E-07 | 0.001845144 | AC018557.1, AC018557.3, DYNC1LI2 |
| chr1 | 42420333 | 42420755 | 423 | 2 | 1.51E-06 | 5.87E-07 | 0.000123 | 9.59E-07 | 0.005894133 | RIMKLA |
| chr9 | 132409104 | 132409820 | 717 | 4 | 3.13E-06 | 4.88E-07 | 0.000684 | 9.64E-07 | 0.00466253 | NA |
| chr11 | 35200933 | 35201096 | 164 | 2 | 3.93E-06 | 5.54E-07 | 0.000176 | 9.64E-07 | 0.008026095 | CD44 |
| chr13 | 106287012 | 106287123 | 112 | 2 | 3.29E-06 | 8.04E-07 | 5.30E-05 | 9.66E-07 | 0.024467415 | AL139379.1 |
| chr8 | 143367746 | 143368439 | 694 | 4 | 1.10E-05 | 1.21E-06 | 0.000139 | 9.68E-07 | 0.005847073 | RHPN1-AS1 |
| chr16 | 31371497 | 31372592 | 1096 | 9 | 1.63E-06 | 3.09E-05 | 0.00131 | 9.68E-07 | 0.002497481 | ITGAX |
| chr8 | 11853402 | 11854354 | 953 | 6 | 8.40E-06 | 3.31E-06 | 8.06E-05 | 9.68E-07 | 0.000823555 | CTSB |
| chr11 | 126331015 | 126331227 | 213 | 3 | 1.20E-06 | 1.79E-06 | 2.99E-05 | 9.69E-07 | 0.001090001 | DCPS |
| chr2 | 148873622 | 148874399 | 778 | 5 | 1.04E-07 | 3.08E-06 | 0.000101 | 9.70E-07 | 0.005243115 | KIF5C-AS1 |
| chr22 | 31972502 | 31973299 | 798 | 2 | 5.23E-06 | 7.59E-07 | 5.97E-05 | 9.70E-07 | 0.008781218 | LINC02558 |
| chr9 | 132201300 | 132201894 | 595 | 4 | 6.01E-07 | 1.20E-05 | 6.17E-05 | 9.71E-07 | 0.006372103 | NTNG2 |
| chr1 | 150009705 | 150009785 | 81 | 3 | 4.82E-07 | 7.09E-07 | 0.00032 | 9.72E-07 | 0.0044539 | OTUD7B |
| chr10 | 114544827 | 114545038 | 212 | 2 | 9.75E-07 | 6.47E-06 | 6.62E-06 | 9.72E-07 | 0.011941807 | ABLIM1 |
| chr19 | 14086519 | 14087074 | 556 | 4 | 1.01E-06 | 4.79E-06 | 0.000401 | 9.72E-07 | 0.002349533 | NA |
| chr17 | 80430226 | 80430263 | 38 | 2 | 2.02E-06 | 1.40E-06 | 2.29E-05 | 9.72E-07 | 0.004225855 | ENDOV |
| chr10 | 101047114 | 101047830 | 717 | 5 | 4.26E-06 | 2.43E-07 | 0.002576 | 9.72E-07 | 0.016484555 | NA |
| chr11 | 62504623 | 62505644 | 1022 | 5 | 5.07E-06 | 1.38E-06 | 0.000509 | 9.72E-07 | 0.009592699 | AHNAK |
| chr20 | 44343386 | 44343556 | 171 | 3 | 3.14E-06 | 5.29E-07 | 0.000163 | 9.74E-07 | 0.000269485 | R3HDML |
| chr4 | 7810315 | 7811748 | 1434 | 3 | 5.18E-06 | 1.24E-06 | 0.000102 | 9.77E-07 | 0.007033938 | AFAP1 |
| chr8 | 70770801 | 70771355 | 555 | 2 | 4.45E-07 | 2.57E-06 | 1.29E-05 | 9.79E-07 | 0.007348782 | XKR9, AC015687.1 |
| chr16 | 1182084 | 1183262 | 1179 | 4 | 4.98E-05 | 8.96E-07 | 0.000627 | 9.79E-07 | 0.013991193 | CACNA1H |
| chr17 | 47970268 | 47971580 | 1313 | 21 | 1.29E-05 | 1.27E-05 | 0.010492 | 9.80E-07 | 0.002603819 | AC018521.1, CDK5RAP3 |
| chr20 | 48734796 | 48734987 | 192 | 3 | 1.44E-06 | 5.55E-06 | 1.29E-05 | 9.88E-07 | 0.002814022 | PREX1 |
| chr8 | 109086008 | 109087353 | 1346 | 9 | 2.43E-06 | 9.05E-07 | 0.002107 | 9.89E-07 | 0.008321379 | TRHR |
| chr10 | 14838333 | 14839097 | 765 | 6 | 3.73E-08 | 1.48E-05 | 4.70E-05 | 9.90E-07 | -0.00017552 | AC069544.2, HSPA14, CDNF |
| chr11 | 2393789 | 2394657 | 869 | 5 | 1.69E-05 | 7.62E-07 | 0.00119 | 9.90E-07 | 0.007348852 | CD81 |
| chr3 | 50299444 | 50300822 | 1379 | 11 | 5.45E-06 | 5.75E-07 | 0.00042 | 9.91E-07 | 0.003220038 | HYAL1 |
| chr9 | 135817898 | 135819132 | 1235 | 7 | 3.41E-06 | 2.60E-05 | 3.90E-05 | 9.93E-07 | 0.002839722 | CAMSAP1 |
| chr10 | 71812295 | 71812780 | 486 | 2 | 1.28E-06 | 9.83E-07 | 3.96E-05 | 9.95E-07 | 0.013873251 | CDH23 |
| chr6 | 20117519 | 20118087 | 569 | 3 | 9.91E-08 | 7.28E-05 | 6.11E-06 | 9.97E-07 | 0.0066933 | MBOAT1 |
| chr1 | 9009195 | 9010068 | 874 | 3 | 3.89E-06 | 4.63E-07 | 0.00026 | 9.97E-07 | 0.00890984 | SLC2A7 |
| chr6 | 40586640 | 40587622 | 983 | 9 | 3.48E-06 | 5.98E-05 | 0.000269 | 9.98E-07 | 0.002067158 | LRFN2 |
| chr7 | 2019515 | 2020544 | 1030 | 9 | 1.80E-05 | 2.46E-07 | 0.003351 | 9.98E-07 | 0.009218016 | MAD1L1, AC069288.1 |
| chr22 | 39363952 | 39364262 | 311 | 7 | 1.22E-06 | 1.04E-05 | 0.000165 | 1.00E-06 | 0.007067443 | SYNGR1 |
| chr5 | 174503585 | 174503866 | 282 | 3 | 1.43E-06 | 2.07E-05 | 5.70E-05 | 1.00E-06 | 0.000485523 | LINC01411 |
| chr4 | 6673582 | 6674869 | 1288 | 11 | 7.84E-05 | 8.77E-06 | 0.002404 | 1.00E-06 | 0.003883419 | AC093323.1, LINC02482 |
| chr1 | 160954507 | 160954786 | 280 | 3 | 8.78E-07 | 7.79E-06 | 3.30E-05 | 1.00E-06 | 0.006301018 | ITLN2 |
| chr11 | 64659944 | 64661756 | 1813 | 10 | 0.000330666 | 2.27E-05 | 0.002244 | 1.00E-06 | 0.004411928 | NRXN2 |
| chr14 | 24231930 | 24233378 | 1449 | 20 | 9.06E-06 | 2.56E-06 | 0.013956 | 1.00E-06 | 0.000776814 | GMPR2, NEDD8-MDP1, NEDD8 |
| chr5 | 172167749 | 172168483 | 735 | 5 | 1.47E-07 | 3.76E-05 | 0.000137 | 1.01E-06 | 0.004460314 | STK10 |
| chr16 | 19717794 | 19718976 | 1183 | 15 | 1.96E-06 | 4.24E-06 | 0.001815 | 1.02E-06 | 0.001170391 | IQCK, KNOP1 |
| chr2 | 135875400 | 135876638 | 1239 | 10 | 5.50E-08 | 1.96E-05 | 6.86E-05 | 1.02E-06 | -0.00075531 | MCM6 |
| chr2 | 27770980 | 27771673 | 694 | 8 | 5.40E-06 | 2.65E-07 | 0.004065 | 1.02E-06 | 0.00755544 | NA |
| chr16 | 759910 | 761347 | 1438 | 16 | 3.67E-06 | 1.05E-06 | 0.013471 | 1.02E-06 | 0.002503442 | MSLN |
| chr17 | 49972588 | 49973725 | 1138 | 11 | 1.53E-05 | 1.47E-05 | 0.000821 | 1.02E-06 | 0.004898871 | DLX4 |
| chr11 | 61598950 | 61599468 | 519 | 3 | 1.57E-07 | 0.000205 | 1.48E-06 | 1.03E-06 | 0.007175821 | AP002754.1 |
| chr20 | 58066196 | 58066322 | 127 | 3 | 1.24E-06 | 8.84E-06 | 0.000121 | 1.03E-06 | 0.013500926 | NA |
| chr4 | 81031499 | 81032048 | 550 | 3 | 3.42E-08 | 1.98E-05 | 5.70E-05 | 1.03E-06 | 0.011557204 | BMP3 |
| chr3 | 122793323 | 122794218 | 896 | 10 | 7.96E-07 | 3.00E-06 | 0.001256 | 1.03E-06 | -0.00019601 | HSPBAP1 |
| chr8 | 11067123 | 11067904 | 782 | 5 | 3.38E-06 | 8.45E-07 | 0.000869 | 1.03E-06 | 0.010012199 | XKR6 |
| chr4 | 70838784 | 70839967 | 1184 | 17 | 1.14E-05 | 1.99E-05 | 0.000917 | 1.03E-06 | 0.001065819 | GRSF1 |
| chr8 | 363712 | 364269 | 558 | 3 | 2.20E-06 | 2.11E-06 | 6.82E-05 | 1.03E-06 | 0.006107018 | NA |
| chr16 | 3443133 | 3444155 | 1023 | 15 | 1.15E-05 | 3.59E-07 | 0.009805 | 1.03E-06 | 0.007753794 | AC025283.2, NAA60, ZNF597 |
| chr10 | 79270487 | 79270970 | 484 | 2 | 1.72E-06 | 8.90E-07 | 5.03E-05 | 1.03E-06 | 0.013929416 | ZMIZ1 |
| chr20 | 44210101 | 44211285 | 1185 | 14 | 1.68E-06 | 5.44E-05 | 9.28E-05 | 1.03E-06 | -0.00015086 | OSER1-DT, OSER1 |
| chr15 | 52799910 | 52801208 | 1299 | 4 | 1.94E-07 | 1.07E-06 | 9.72E-05 | 1.03E-06 | 0.013544527 | AC016044.1 |
| chr20 | 461737 | 463035 | 1299 | 14 | 1.25E-05 | 3.35E-05 | 0.000329 | 1.03E-06 | 0.001301219 | TBC1D20 |
| chr9 | 38227285 | 38228012 | 728 | 3 | 1.84E-08 | 0.00148 | 1.42E-05 | 1.04E-06 | 0.008983333 | NA |
| chr6 | 11043873 | 11045137 | 1265 | 9 | 3.16E-06 | 6.09E-06 | 0.000839 | 1.04E-06 | 0.001097283 | ELOVL2-AS1, ELOVL2 |
| chr14 | 36583463 | 36584484 | 1022 | 6 | 1.16E-05 | 4.03E-07 | 0.002879 | 1.04E-06 | 0.007168323 | AL132857.1 |
| chr3 | 133573194 | 133574616 | 1423 | 20 | 2.11E-06 | 1.78E-05 | 0.002843 | 1.04E-06 | 0.000284639 | CDV3 |
| chr6 | 163971523 | 163972828 | 1306 | 8 | 0.000130471 | 4.17E-07 | 0.002335 | 1.04E-06 | 0.004084843 | AL078602.1 |

| | | | | | | | | | | |
|---|---|---|---|---|---|---|---|---|---|---|
| chr21 | 34204170 | 34205022 | 853 | 5 | 1.01E-07 | 6.45E-05 | 1.99E-06 | 1.04E-06 | 0.00377598 | AP000317.2 |
| chr1 | 202130549 | 202131174 | 626 | 2 | 6.79E-07 | 1.50E-06 | 2.35E-05 | 1.04E-06 | 0.003908266 | GPR37L1 |
| chr2 | 199189759 | 199190345 | 587 | 2 | 7.06E-06 | 6.52E-07 | 0.000114 | 1.04E-06 | 0.015032419 | NA |
| chr12 | 1924594 | 1924683 | 90 | 2 | 1.67E-07 | 0.000103 | 2.51E-06 | 1.04E-06 | 0.014898077 | NA |
| chr17 | 48816829 | 48817657 | 829 | 13 | 2.23E-06 | 4.58E-05 | 0.000396 | 1.04E-06 | 0.002232467 | TTLL6 |
| chr12 | 31728533 | 31729692 | 1160 | 13 | 3.85E-07 | 3.83E-06 | 0.001388 | 1.04E-06 | 0.001459151 | AC023157.2, AMN1 |
| chr7 | 101915854 | 101916971 | 1118 | 12 | 6.04E-08 | 0.00163 | 8.50E-05 | 1.04E-06 | -0.00133092 | CUX1 |
| chr11 | 61485824 | 61487249 | 1426 | 5 | 1.22E-05 | 6.29E-07 | 0.001186 | 1.04E-06 | 0.004518753 | PPP1R32 |
| chr13 | 48318415 | 48319038 | 624 | 2 | 3.43E-06 | 6.02E-07 | 0.000185 | 1.04E-06 | 0.021394234 | RB1, PPP1R26P1 |
| chr14 | 103135934 | 103137007 | 1074 | 7 | 3.30E-05 | 1.23E-05 | 0.000129 | 1.04E-06 | 0.008073144 | TNFAIP2 |
| chr16 | 58503323 | 58503726 | 404 | 3 | 1.12E-08 | 0.00399 | 2.90E-06 | 1.05E-06 | 0.006867514 | NDRG4 |
| chr11 | 19524586 | 19524609 | 24 | 2 | 2.93E-06 | 6.61E-07 | 0.000112 | 1.05E-06 | 0.021437488 | NAV2 |
| chr11 | 111450752 | 111451293 | 542 | 5 | 4.01E-08 | 0.000674 | 9.20E-07 | 1.05E-06 | 0.001656732 | AP002008.2, POU2AF1 |
| chr20 | 32858638 | 32859275 | 638 | 5 | 1.31E-06 | 1.66E-06 | 0.000585 | 1.05E-06 | 0.004825677 | AL035071.1, EFCAB8 |
| chr13 | 78586750 | 78587484 | 735 | 4 | 6.51E-07 | 2.94E-05 | 0.000318 | 1.06E-06 | 0.011340249 | OBI1-AS1 |
| chr22 | 19478808 | 19480287 | 1480 | 18 | 1.41E-05 | 4.80E-07 | 0.006426 | 1.06E-06 | 0.000982684 | CDC45, UFD1 |
| chr11 | 14364477 | 14365312 | 836 | 7 | 2.03E-06 | 1.31E-05 | 0.000446 | 1.06E-06 | -0.00077445 | RRAS2 |
| chr17 | 50055350 | 50057069 | 1720 | 17 | 2.80E-05 | 2.28E-05 | 0.001215 | 1.06E-06 | 0.001783006 | ITGA3, PICART1 |
| chr15 | 68053542 | 68055069 | 1528 | 13 | 4.31E-05 | 1.19E-05 | 0.000956 | 1.07E-06 | 0.002396296 | PIAS1 |
| chr2 | 40450460 | 40451946 | 1487 | 8 | 1.39E-06 | 2.59E-06 | 0.001173 | 1.07E-06 | 0.005843278 | SLC8A1 |
| chr7 | 99551792 | 99552641 | 850 | 9 | 6.84E-07 | 4.76E-05 | 0.002214 | 1.07E-06 | 0.005511985 | FAM200A |
| chr18 | 23982646 | 23982913 | 268 | 3 | 3.38E-06 | 1.12E-06 | 5.71E-05 | 1.07E-06 | 0.006497517 | NA |
| chr6 | 27873179 | 27874320 | 1142 | 14 | 8.76E-07 | 4.03E-06 | 0.007271 | 1.07E-06 | 0.001790127 | H4C13 |
| chr7 | 1529433 | 1530771 | 1339 | 8 | 9.95E-06 | 4.49E-07 | 0.007441 | 1.07E-06 | 0.004364541 | MAFK |
| chr12 | 33440242 | 33440978 | 737 | 3 | 1.13E-06 | 5.69E-06 | 0.000107 | 1.08E-06 | 0.009889447 | NA |
| chr8 | 6706925 | 6708801 | 1877 | 14 | 1.36E-05 | 0.000598 | 0.000143 | 1.08E-06 | 0.000745841 | AGPAT5, MCPH1-AS1 |
| chr2 | 3805422 | 3805658 | 237 | 3 | 5.08E-06 | 4.80E-07 | 0.000567 | 1.08E-06 | 0.018013556 | DCDC2C |
| chr10 | 102235809 | 102236025 | 217 | 2 | 3.13E-06 | 6.59E-07 | 0.000128 | 1.08E-06 | 0.008291662 | PITX3 |
| chr5 | 172452606 | 172453852 | 1247 | 4 | 3.32E-07 | 1.01E-05 | 0.000153 | 1.08E-06 | 0.009539536 | SH3PXD2B |
| chr18 | 45828805 | 45830056 | 1252 | 6 | 1.45E-06 | 1.68E-06 | 0.000238 | 1.09E-06 | 0.008306551 | SIGLEC15 |
| chr11 | 507006 | 507970 | 965 | 11 | 8.22E-06 | 3.55E-07 | 0.003787 | 1.09E-06 | -0.00072248 | RNH1 |
| chr6 | 31735362 | 31736997 | 1636 | 11 | 3.12E-05 | 0.000183 | 0.000614 | 1.09E-06 | 0.001629576 | CLIC1 |
| chr1 | 38482257 | 38482450 | 194 | 2 | 3.48E-06 | 8.76E-07 | 5.99E-05 | 1.10E-06 | 0.01480469 | LINC01685 |
| chr3 | 46936042 | 46937120 | 1079 | 4 | 4.84E-05 | 6.81E-07 | 0.000854 | 1.10E-06 | 0.006981488 | CCDC12 |
| chr14 | 101039186 | 101040242 | 1057 | 12 | 8.83E-07 | 5.54E-05 | 0.000545 | 1.10E-06 | 0.00293017 | MIR376C, MIR376A2, MIR654 |
| chr6 | 36340612 | 36341170 | 559 | 3 | 4.86E-06 | 1.43E-06 | 7.02E-05 | 1.10E-06 | 0.009508773 | NA |
| chr6 | 32051265 | 32052189 | 925 | 7 | 9.89E-06 | 2.37E-05 | 0.000401 | 1.10E-06 | 0.006169065 | TNXB |
| chr12 | 7188984 | 7190049 | 1066 | 16 | 9.34E-07 | 2.76E-06 | 0.002618 | 1.10E-06 | -0.00066742 | PEX5, AC018653.1 |
| chr3 | 23809763 | 23810665 | 903 | 15 | 1.95E-06 | 5.26E-06 | 0.000184 | 1.10E-06 | 0.002948726 | UBE2E1 |
| chr6 | 33116212 | 33117286 | 1075 | 19 | 6.61E-07 | 8.39E-07 | 0.010273 | 1.10E-06 | 0.01090279 | HLA-DPB2 |
| chr12 | 118874855 | 118874970 | 116 | 2 | 2.27E-06 | 6.42E-07 | 0.000167 | 1.10E-06 | 0.022488179 | AC087863.2 |
| chr18 | 57930193 | 57930921 | 729 | 3 | 7.48E-06 | 5.53E-07 | 0.000222 | 1.10E-06 | 0.014842452 | NA |
| chr8 | 71557653 | 71558918 | 1266 | 8 | 6.67E-05 | 4.58E-06 | 0.000935 | 1.10E-06 | 0.002621306 | EYA1 |
| chr12 | 54036900 | 54037207 | 308 | 3 | 2.33E-06 | 4.88E-07 | 0.000396 | 1.10E-06 | 0.013579238 | HOXC4 |
| chr19 | 7628573 | 7629608 | 1036 | 7 | 1.04E-08 | 0.00044 | 6.77E-05 | 1.11E-06 | 0.00408786 | XAB2 |
| chr17 | 39604106 | 39605063 | 958 | 5 | 3.85E-07 | 6.55E-05 | 0.000131 | 1.11E-06 | 0.006978384 | NEUROD2 |
| chr2 | 114460450 | 114460987 | 538 | 2 | 4.12E-09 | 0.000168 | 2.42E-06 | 1.11E-06 | 0.011770533 | DPP10 |
| chr8 | 57492743 | 57492791 | 49 | 2 | 1.46E-06 | 1.16E-06 | 3.78E-05 | 1.11E-06 | 0.015254448 | NA |
| chr5 | 611581 | 612846 | 1266 | 13 | 1.29E-05 | 5.62E-06 | 0.004472 | 1.11E-06 | 0.002570937 | CEP72, AC106772.1 |
| chr14 | 77376087 | 77377245 | 1159 | 9 | 2.31E-07 | 5.79E-06 | 0.000548 | 1.11E-06 | -0.00074484 | SAMD15, TMED8 |
| chr16 | 14364614 | 14364705 | 92 | 3 | 1.45E-06 | 1.73E-05 | 5.24E-05 | 1.11E-06 | 0.005066136 | LINC02130 |
| chr6 | 33271425 | 33272611 | 1187 | 32 | 1.51E-05 | 1.33E-06 | 0.02326 | 1.11E-06 | 0.000208395 | RPS18, VPS52 |
| chr14 | 103401494 | 103402014 | 521 | 2 | 3.52E-06 | 7.85E-07 | 8.18E-05 | 1.12E-06 | 0.021462548 | MARK3 |
| chr2 | 242038234 | 242038937 | 704 | 3 | 4.81E-06 | 8.52E-07 | 9.00E-05 | 1.12E-06 | 0.006125312 | LINC01237, AC093642.6 |
| chr3 | 56467552 | 56468719 | 1168 | 10 | 1.49E-07 | 3.53E-05 | 0.00114 | 1.12E-06 | 0.00277926 | ERC2 |
| chr5 | 177383554 | 177384304 | 751 | 6 | 2.00E-06 | 6.85E-07 | 0.001205 | 1.13E-06 | 0.003320942 | SLC34A1 |
| chr13 | 42883221 | 42883858 | 638 | 2 | 3.72E-08 | 0.000101 | 2.69E-06 | 1.13E-06 | 0.024183987 | NA |
| chr12 | 48184293 | 48185462 | 1170 | 5 | 8.65E-08 | 7.61E-06 | 0.000153 | 1.13E-06 | 0.010551498 | CCDC184 |
| chr1 | 173824292 | 173825948 | 1657 | 18 | 7.61E-12 | 4.15E-05 | 7.14E-06 | 1.14E-06 | 0.001310535 | DARS2, CENPL |
| chr3 | 40924954 | 40925474 | 521 | 2 | 1.27E-06 | 1.42E-06 | 2.98E-05 | 1.14E-06 | 0.009718391 | NA |
| chr10 | 132202763 | 132203411 | 649 | 4 | 3.53E-07 | 5.89E-06 | 2.73E-05 | 1.14E-06 | 0.007095062 | DPYSL4 |
| chr15 | 31351739 | 31352398 | 660 | 3 | 1.07E-05 | 4.64E-07 | 0.000632 | 1.14E-06 | 0.008546362 | KLF13 |
| chr7 | 155509039 | 155510213 | 1175 | 8 | 4.20E-05 | 4.55E-06 | 0.000785 | 1.14E-06 | 0.002531255 | CNPY1, AC009403.2 |
| chr19 | 3227774 | 3227984 | 211 | 2 | 7.62E-07 | 1.67E-06 | 2.41E-05 | 1.14E-06 | 0.018004199 | CELF5 |
| chr2 | 143435234 | 143435275 | 42 | 2 | 3.44E-06 | 1.66E-06 | 2.42E-05 | 1.14E-06 | 0.015666498 | ARHGAP15, AC079793.1 |
| chr9 | 131289193 | 131289881 | 689 | 7 | 6.23E-06 | 2.24E-07 | 0.002918 | 1.14E-06 | -0.00932326 | PLPP7 |
| chr16 | 693925 | 695296 | 1372 | 7 | 5.85E-06 | 3.12E-06 | 0.001053 | 1.15E-06 | 0.006342799 | FBXL16 |
| chr17 | 75404857 | 75406686 | 1830 | 18 | 5.33E-05 | 1.28E-06 | 0.006965 | 1.15E-06 | 0.002024476 | MIR3678, GRB2 |
| chr19 | 18248113 | 18248762 | 650 | 8 | 2.01E-06 | 7.74E-06 | 0.000318 | 1.15E-06 | 0.00413162 | AC008397.2, PDE4C |
| chr12 | 53251210 | 53252191 | 982 | 11 | 1.03E-06 | 5.16E-07 | 0.006957 | 1.15E-06 | 0.002443493 | MFSD5 |
| chr2 | 135530329 | 135532081 | 1753 | 19 | 2.39E-06 | 5.30E-06 | 0.003331 | 1.15E-06 | -0.00033538 | R3HDM1, ZRANB3 |
| chr6 | 33170331 | 33170892 | 562 | 4 | 7.67E-07 | 3.56E-07 | 0.001392 | 1.15E-06 | 0.008050536 | COL11A2 |
| chr15 | 85788826 | 85789351 | 526 | 2 | 1.90E-06 | 1.14E-06 | 4.20E-05 | 1.15E-06 | 0.008906418 | KLHL25 |
| chr16 | 86558782 | 86559046 | 265 | 3 | 1.98E-06 | 1.41E-06 | 5.28E-05 | 1.15E-06 | 0.009986928 | NA |
| chr17 | 82082095 | 82083363 | 1269 | 8 | 2.30E-05 | 5.47E-06 | 0.001933 | 1.15E-06 | 0.002179203 | FASN |
| chr19 | 48551651 | 48552989 | 1339 | 12 | 5.30E-07 | 3.43E-05 | 0.000705 | 1.15E-06 | 0.004919958 | SULT2B1, AC008403.3 |
| chr8 | 141095306 | 141096112 | 807 | 4 | 5.37E-06 | 4.96E-06 | 9.79E-05 | 1.16E-06 | 0.007560338 | NA |
| chr8 | 108462952 | 108463507 | 556 | 2 | 2.18E-07 | 3.39E-06 | 1.27E-05 | 1.16E-06 | -9.03E-06 | EMC2 |
| chr19 | 16096610 | 16096748 | 139 | 2 | 4.30E-06 | 6.96E-07 | 0.000143 | 1.16E-06 | -0.02464074 | TPM4 |
| chr11 | 118701527 | 118702247 | 721 | 3 | 6.09E-06 | 5.10E-07 | 0.000513 | 1.16E-06 | -0.00475453 | AP002954.1 |
| chr1 | 209427581 | 209428657 | 1077 | 4 | 6.72E-05 | 9.90E-07 | 0.000261 | 1.16E-06 | 0.00923265 | NA |
| chr15 | 22911431 | 22912429 | 999 | 4 | 3.48E-05 | 3.01E-06 | 0.000152 | 1.16E-06 | -0.00114914 | CYFIP1 |
| chr2 | 43595856 | 43596988 | 1133 | 15 | 1.05E-06 | 4.97E-05 | 0.003032 | 1.16E-06 | 0.003237047 | THADA |
| chr1 | 153789725 | 153790513 | 789 | 8 | 2.47E-06 | 6.60E-07 | 0.004306 | 1.16E-06 | 0.009882877 | GATAD2B |
| chr16 | 57536145 | 57537187 | 1043 | 10 | 3.20E-06 | 6.41E-06 | 0.000178 | 1.16E-06 | -0.00104107 | CCDC102A |
| chr2 | 84022824 | 84022851 | 28 | 2 | 9.23E-07 | 0.000264 | 2.39E-06 | 1.16E-06 | 0.021611462 | NA |
| chr8 | 132049967 | 132050727 | 761 | 3 | 2.09E-06 | 2.11E-06 | 3.21E-05 | 1.16E-06 | -0.00385196 | OC90, AC100868.1 |
| chr7 | 128454656 | 128454973 | 318 | 3 | 1.45E-07 | 1.54E-06 | 4.56E-05 | 1.17E-06 | 0.011048415 | NA |
| chr5 | 78575123 | 78575862 | 740 | 2 | 3.74E-06 | 1.06E-06 | 4.87E-05 | 1.17E-06 | 0.009732309 | LHFPL2 |
| chr9 | 93201220 | 93202202 | 983 | 5 | 5.28E-06 | 1.89E-05 | 0.000818 | 1.17E-06 | 0.005099997 | WNK2 |
| chr17 | 19286824 | 19287163 | 340 | 2 | 2.92E-07 | 5.28E-05 | 3.12E-06 | 1.17E-06 | 0.006053212 | EPN2 |
| chr1 | 119327274 | 119328179 | 906 | 11 | 1.77E-06 | 8.60E-05 | 0.000428 | 1.17E-06 | 0.002841891 | LINC01780, AL359915.1 |
| chr17 | 7786176 | 7787211 | 1036 | 8 | 4.48E-05 | 3.43E-06 | 0.001682 | 1.17E-06 | 0.001399945 | DNAH2 |
| chr2 | 70978136 | 70979681 | 1546 | 11 | 3.92E-08 | 9.36E-05 | 0.000254 | 1.17E-06 | 0.012292809 | ANKRD53, AC007040.2 |
| chr19 | 8403305 | 8404397 | 1093 | 7 | 1.95E-06 | 4.58E-06 | 0.000129 | 1.17E-06 | 0.002717447 | RAB11B |
| chr1 | 231626492 | 231626918 | 427 | 6 | 7.06E-06 | 5.28E-07 | 0.00224 | 1.17E-06 | 0.009067346 | TSNAX-DISC1, DISC1 |
| chr5 | 142357060 | 142357701 | 642 | 4 | 6.10E-08 | 5.18E-05 | 1.09E-05 | 1.17E-06 | -4.58E-05 | SPRY4-AS1 |
| chr13 | 29526709 | 29527025 | 317 | 4 | 5.32E-06 | 7.39E-07 | 0.000152 | 1.17E-06 | 0.001217478 | SLC7A1 |
| chr11 | 67997183 | 67998250 | 1068 | 6 | 1.78E-05 | 9.47E-07 | 0.001698 | 1.17E-06 | 0.009503537 | UNC93B1 |
| chr17 | 58328093 | 58329906 | 1814 | 13 | 4.64E-05 | 0.00055 | 0.000264 | 1.18E-06 | 0.002441083 | TSPOAP1-AS1, TSPOAP1 |
| chr11 | 69593095 | 69593264 | 170 | 2 | 1.20E-06 | 3.12E-06 | 1.41E-05 | 1.18E-06 | 0.005210244 | NA |
| chr21 | 29155692 | 29155723 | 32 | 2 | 4.97E-06 | 6.79E-07 | 0.000196 | 1.18E-06 | 0.038463342 | MAP3K7CL |
| chr3 | 195804427 | 195804762 | 336 | 2 | 8.86E-07 | 7.76E-07 | 0.000104 | 1.18E-06 | 0.014665558 | MUC4 |
| chr20 | 17613426 | 17613885 | 460 | 2 | 3.57E-07 | 7.90E-07 | 9.80E-05 | 1.18E-06 | 0.009024146 | RRBP1 |
| chr22 | 50473507 | 50474367 | 861 | 4 | 4.97E-07 | 1.02E-05 | 0.00019 | 1.18E-06 | 0.008805789 | SBF1 |
| chr8 | 108785918 | 108786596 | 679 | 2 | 1.13E-07 | 2.15E-06 | 1.96E-05 | 1.18E-06 | 0.011257522 | TMEM74 |
| chr8 | 27258350 | 27258869 | 520 | 4 | 8.07E-06 | 3.52E-07 | 0.001164 | 1.19E-06 | 0.013658475 | STMN4 |
| chr7 | 156616364 | 156617094 | 731 | 5 | 2.27E-06 | 1.61E-06 | 0.00028 | 1.19E-06 | 0.008226234 | LINC01006 |
| chr14 | 80955256 | 80955816 | 561 | 8 | 1.72E-06 | 9.61E-07 | 0.002736 | 1.19E-06 | 0.009099786 | TSHR, CEP128 |
| chr11 | 61533103 | 61534224 | 1122 | 6 | 6.68E-06 | 1.54E-05 | 0.000126 | 1.20E-06 | 0.004399012 | SYT7 |
| chr12 | 55966132 | 55967454 | 1323 | 20 | 7.45E-07 | 0.000602 | 2.21E-05 | 1.20E-06 | 0.001414314 | CDK2, PMEL, AC025162.1 |
| chr9 | 113650001 | 113650940 | 940 | 5 | 3.27E-06 | 1.08E-05 | 0.000393 | 1.20E-06 | 0.005381941 | NA |
| chr10 | 43689420 | 43690726 | 1307 | 7 | 2.07E-05 | 2.04E-05 | 0.001415 | 1.20E-06 | 0.007481505 | NA |
| chr16 | 48384007 | 48385220 | 1214 | 4 | 6.60E-05 | 1.36E-05 | 0.000446 | 1.20E-06 | 0.006003609 | SIAH1 |
| chr6 | 155216461 | 155217021 | 561 | 8 | 6.56E-07 | 0.000245 | 5.96E-05 | 1.20E-06 | 0.009140617 | TIAM2 |
| chr5 | 9547057 | 9547713 | 657 | 4 | 1.10E-06 | 8.45E-07 | 0.000258 | 1.20E-06 | -0.02680071 | SNHG18 |
| chr2 | 10762529 | 10762782 | 254 | 3 | 3.72E-06 | 7.35E-07 | 0.000311 | 1.20E-06 | 0.009239257 | ATP6V1C2 |
| chr20 | 61498295 | 61499464 | 1170 | 7 | 6.20E-06 | 6.25E-06 | 0.00063 | 1.20E-06 | 0.000864605 | CDH4 |
| chr14 | 68962186 | 68962214 | 29 | 2 | 2.17E-06 | 7.35E-07 | 0.000137 | 1.20E-06 | 0.008591446 | ACTN1 |
| chr2 | 127591324 | 127591590 | 267 | 2 | 2.18E-06 | 7.15E-07 | 0.000156 | 1.20E-06 | 0.014081917 | MYO7B |
| chr6 | 28634766 | 28635515 | 750 | 24 | 3.65E-06 | 1.16E-06 | 0.004204 | 1.20E-06 | 0.00290916 | AL121932.2 |
| chr20 | 4699662 | 4700197 | 536 | 4 | 5.16E-06 | 1.67E-05 | 0.000175 | 1.20E-06 | 0.001304854 | PRNP |
| chr5 | 149041573 | 149042132 | 560 | 4 | 4.12E-07 | 4.66E-06 | 7.50E-05 | 1.21E-06 | -0.00755395 | SH3TC2 |
| chr3 | 128608346 | 128609316 | 971 | 5 | 2.12E-07 | 1.09E-05 | 0.000197 | 1.21E-06 | -0.00382705 | NA |
| chr18 | 45836995 | 45837576 | 582 | 4 | 2.36E-06 | 3.38E-07 | 0.001476 | 1.21E-06 | 0.014793573 | SIGLEC15 |
| chr17 | 81131759 | 81132693 | 935 | 6 | 3.06E-07 | 0.00019 | 7.14E-05 | 1.21E-06 | 0.001887847 | AATK |
| chr17 | 69414327 | 69415286 | 960 | 12 | 1.66E-06 | 2.48E-06 | 8.73E-05 | 1.21E-06 | 0.001327507 | MAP2K6 |
| chr20 | 21397407 | 21398788 | 1382 | 17 | 5.58E-06 | 9.02E-05 | 0.001723 | 1.21E-06 | 0.006176239 | AL158013.1, NKX2-4 |
| chr16 | 19766088 | 19767161 | 1074 | 5 | 1.24E-06 | 2.65E-06 | 0.000128 | 1.21E-06 | 0.020506925 | IQCK, AC027130.1 |
| chr18 | 23872302 | 23872931 | 630 | 8 | 7.72E-07 | 7.37E-05 | 0.000196 | 1.21E-06 | 0.005119847 | LAMA3 |
| chr2 | 27071230 | 27072723 | 1494 | 12 | 4.34E-06 | 5.19E-06 | 0.001659 | 1.22E-06 | 0.000876691 | OST4 |
| chr17 | 76517712 | 76518030 | 319 | 2 | 6.00E-07 | 1.75E-05 | 4.85E-06 | 1.22E-06 | 0.004014554 | NA |
| chr7 | 378639 | 380502 | 1864 | 10 | 3.90E-05 | 3.07E-06 | 0.000741 | 1.22E-06 | 0.014848413 | AC226118.1 |
| chr12 | 5222539 | 5223064 | 526 | 3 | 4.52E-06 | 7.62E-07 | 0.000132 | 1.22E-06 | 0.003964954 | NA |
| chr6 | 108123302 | 108124330 | 1029 | 9 | 1.98E-06 | 1.76E-05 | 0.00213 | 1.22E-06 | 0.012370211 | OSTM1-AS1, OSTM1 |
| chr16 | 3611764 | 3613071 | 1308 | 8 | 1.45E-05 | 5.32E-06 | 0.0052 | 1.22E-06 | 0.006454252 | DNASE1 |
| chr6 | 30570746 | 30571864 | 1119 | 27 | 1.12E-06 | 4.96E-06 | 0.000113 | 1.22E-06 | 0.000652043 | ABCF1 |
| chr10 | 104085962 | 104086546 | 585 | 4 | 1.76E-06 | 3.11E-06 | 4.42E-05 | 1.22E-06 | 0.003109978 | NA |
| chr19 | 29831350 | 29832042 | 693 | 2 | 2.17E-06 | 1.29E-06 | 3.93E-05 | 1.22E-06 | 0.000968857 | NA |
| chr8 | 102388713 | 102389345 | 633 | 5 | 5.27E-06 | 9.51E-07 | 0.000904 | 1.22E-06 | -0.00147896 | UBR5 |
| chr3 | 196967109 | 196968061 | 953 | 4 | 2.85E-05 | 4.16E-07 | 0.000858 | 1.22E-06 | 0.017869489 | PIGZ |
| chr10 | 84179307 | 84179695 | 389 | 3 | 7.36E-06 | 8.80E-07 | 0.000363 | 1.23E-06 | 0.010908356 | C10orf99 |
| chr1 | 37690379 | 37691267 | 889 | 12 | 1.62E-06 | 6.76E-06 | 0.000819 | 1.23E-06 | 0.014026955 | C1orf109 |
| chr7 | 122300162 | 122300938 | 777 | 7 | 1.04E-05 | 4.92E-07 | 0.004777 | 1.23E-06 | 0.007245448 | NA |
| chr8 | 47377777 | 47377779 | 3 | 2 | 2.67E-06 | 5.61E-06 | 9.49E-06 | 1.23E-06 | 0.015307088 | SPIDR |
| chr13 | 111975982 | 111976728 | 747 | 6 | 2.86E-07 | 1.41E-05 | 8.40E-05 | 1.23E-06 | -0.00700246 | NA |
| chr20 | 37521348 | 37522936 | 1589 | 7 | 7.37E-06 | 6.14E-06 | 3.70E-05 | 1.23E-06 | 0.008155821 | NNAT, BLCAP |
| chr10 | 5977563 | 5978622 | 1060 | 11 | 8.95E-06 | 4.98E-06 | 0.003342 | 1.23E-06 | 0.001915556 | IL15RA |
| chr22 | 25026922 | 25027096 | 175 | 2 | 1.58E-07 | 1.28E-06 | 4.05E-05 | 1.23E-06 | 0.007266402 | KIAA1671 |
| chr7 | 29798635 | 29798646 | 12 | 2 | 5.13E-06 | 7.62E-07 | 0.000135 | 1.23E-06 | 0.014929168 | NA |
| chr1 | 156875602 | 156876544 | 943 | 6 | 2.00E-06 | 6.48E-06 | 0.000395 | 1.24E-06 | 0.010971155 | NTRK1 |
| chr19 | 5578315 | 5578744 | 430 | 4 | 1.35E-07 | 5.28E-05 | 2.42E-05 | 1.24E-06 | 0.009468414 | TINCR |
| chr8 | 19602586 | 19603557 | 972 | 9 | 3.94E-06 | 5.98E-06 | 0.001037 | 1.24E-06 | -0.00380904 | CSGALNACT1 |
| chr4 | 74157502 | 74158482 | 981 | 12 | 5.61E-07 | 1.53E-05 | 3.00E-05 | 1.24E-06 | -0.00110783 | MTHFD2L, AC093677.2 |
| chr7 | 140024899 | 140024975 | 77 | 3 | 3.57E-06 | 7.55E-07 | 0.000166 | 1.24E-06 | 0.005229987 | PARP12 |
| chr3 | 46566109 | 46567272 | 1164 | 11 | 1.22E-06 | 4.41E-06 | 0.001412 | 1.24E-06 | 0.002533615 | LRRC2 |
| chr9 | 130576116 | 130576529 | 414 | 2 | 2.40E-06 | 1.17E-06 | 4.70E-05 | 1.24E-06 | 0.024959838 | NA |
| chr20 | 50292629 | 50293614 | 986 | 5 | 2.04E-05 | 1.55E-06 | 0.000942 | 1.24E-06 | 0.010401046 | LINC01270 |
| chr14 | 54698885 | 54699286 | 402 | 2 | 5.00E-07 | 1.06E-06 | 5.69E-05 | 1.24E-06 | 0.01238593 | SAMD4A |
| chr12 | 12449038 | 12449320 | 283 | 3 | 3.33E-06 | 1.71E-06 | 6.30E-05 | 1.24E-06 | 0.005603908 | BORCS5 |
| chr20 | 407338 | 408259 | 922 | 13 | 4.02E-07 | 0.000233 | 0.001433 | 1.24E-06 | 0.002653512 | RBCK1 |
| chr10 | 56537955 | 56538499 | 545 | 3 | 1.30E-06 | 2.57E-06 | 0.000202 | 1.24E-06 | -0.00440837 | NA |

| | | | | | | | | | | |
|---|---|---|---|---|---|---|---|---|---|---|
| chr12 | 93814241 | 93815107 | 867 | 3 | 1.59E-05 | 1.51E-06 | 0.000154 | 1.25E-06 | 0.005349519 | CRADD |
| chr11 | 45666340 | 45667527 | 1188 | 3 | 3.18E-07 | 4.92E-06 | 1.78E-05 | 1.25E-06 | 0.012896178 | NA |
| chr15 | 40843234 | 40844039 | 806 | 6 | 1.26E-06 | 5.74E-07 | 0.000366 | 1.25E-06 | 0.006781634 | SPINT1, SPINT1-AS1 |
| chr20 | 50138063 | 50138862 | 800 | 3 | 1.56E-05 | 2.66E-06 | 7.26E-05 | 1.25E-06 | 0.003549915 | PEDS1-UBE2V1, PEDS1 |
| chr16 | 31428351 | 31429545 | 1195 | 6 | 1.12E-05 | 5.07E-07 | 0.003619 | 1.25E-06 | 0.001731795 | AC026471.5, COX6A2 |
| chr16 | 90009082 | 90010800 | 1719 | 4 | 0.000154573 | 3.74E-07 | 0.001074 | 1.26E-06 | 0.005923728 | DBNDD1 |
| chr1 | 54237139 | 54237899 | 761 | 5 | 3.21E-08 | 0.000123 | 5.57E-05 | 1.26E-06 | 0.006669935 | SSBP3-AS1, SSBP3 |
| chr17 | 6190686 | 6190902 | 217 | 3 | 1.74E-06 | 4.23E-06 | 2.12E-05 | 1.26E-06 | 0.013036111 | NA |
| chr21 | 17738394 | 17738813 | 420 | 4 | 4.66E-07 | 0.000111 | 2.98E-05 | 1.26E-06 | 0.004813063 | NA |
| chr2 | 1891419 | 1892189 | 771 | 7 | 1.17E-06 | 1.48E-06 | 0.001721 | 1.26E-06 | 0.003397106 | MYT1L |
| chr2 | 231514617 | 231515006 | 390 | 4 | 3.08E-06 | 5.28E-07 | 0.001128 | 1.26E-06 | 0.011536066 | NA |
| chr9 | 137197393 | 137198412 | 1020 | 5 | 5.55E-07 | 1.57E-05 | 3.51E-05 | 1.27E-06 | 0.002229616 | TPRN |
| chr17 | 5483849 | 5484012 | 164 | 2 | 2.69E-06 | 8.03E-07 | 0.000124 | 1.27E-06 | -0.00061899 | DERL2 |
| chr20 | 62358667 | 62359544 | 878 | 3 | 9.33E-06 | 5.77E-06 | 7.85E-05 | 1.27E-06 | 0.008433051 | LAMA5 |
| chr11 | 33927193 | 33927467 | 275 | 2 | 5.65E-06 | 7.35E-07 | 0.000204 | 1.27E-06 | 0.005100988 | NA |
| chr17 | 66304611 | 66305209 | 599 | 3 | 7.13E-06 | 5.26E-07 | 0.000719 | 1.27E-06 | 0.013424275 | PRKCA |
| chr17 | 77884187 | 77885150 | 964 | 5 | 2.26E-05 | 1.88E-06 | 0.000412 | 1.27E-06 | 0.007491387 | NA |
| chr19 | 45501876 | 45502547 | 672 | 2 | 1.96E-05 | 7.38E-07 | 0.000197 | 1.27E-06 | 0.013259468 | PPM1N |
| chr11 | 118264086 | 118265062 | 977 | 15 | 1.11E-06 | 5.98E-05 | 0.004413 | 1.27E-06 | 0.002839104 | AP002800.1, MPZL2 |
| chr5 | 176676446 | 176676505 | 60 | 2 | 4.11E-06 | 7.36E-07 | 0.000205 | 1.27E-06 | 0.012435541 | NA |
| chr4 | 159018195 | 159018416 | 222 | 2 | 1.85E-06 | 4.81E-06 | 1.10E-05 | 1.28E-06 | 0.013657458 | C4orf45 |
| chr6 | 41040360 | 41041574 | 1215 | 3 | 3.57E-06 | 9.68E-07 | 0.000312 | 1.28E-06 | 0.013756462 | OARD1 |
| chr10 | 26557091 | 26557322 | 232 | 2 | 1.31E-06 | 1.05E-05 | 6.80E-06 | 1.28E-06 | 0.013318952 | APBB1IP |
| chr17 | 1936175 | 1937252 | 1078 | 5 | 8.36E-06 | 5.54E-07 | 0.000409 | 1.28E-06 | 0.006165012 | RTN4RL1 |
| chr2 | 44949412 | 44950493 | 1082 | 5 | 1.08E-05 | 1.84E-06 | 0.000294 | 1.28E-06 | 0.0093974 | NA |
| chr5 | 146877213 | 146878949 | 1737 | 17 | 5.45E-07 | 4.12E-06 | 0.00194 | 1.28E-06 | 4.97E-05 | PPP2R2B |
| chr2 | 110731357 | 110731812 | 456 | 2 | 1.18E-09 | 7.15E-06 | 8.47E-06 | 1.29E-06 | 0.005586536 | NA |
| chr20 | 32465772 | 32466587 | 816 | 4 | 7.22E-06 | 3.79E-07 | 0.001082 | 1.29E-06 | 0.007837994 | NOL4L |
| chr1 | 152662750 | 152662763 | 14 | 2 | 5.16E-07 | 1.65E-06 | 3.06E-05 | 1.29E-06 | 0.016560335 | NA |
| chr14 | 105818502 | 105818742 | 241 | 2 | 2.92E-06 | 7.72E-07 | 0.000162 | 1.29E-06 | 0.017231516 | NA |
| chr15 | 31339037 | 31339887 | 851 | 6 | 8.85E-06 | 1.49E-06 | 0.002578 | 1.29E-06 | 0.006881664 | KLF13 |
| chr6 | 105608765 | 105609830 | 1066 | 3 | 9.78E-05 | 5.38E-07 | 0.000448 | 1.29E-06 | 0.009240966 | NA |
| chr10 | 1159918 | 1160750 | 833 | 6 | 2.39E-07 | 0.000445 | 0.000339 | 1.29E-06 | 0.003573112 | LINC00200 |
| chr10 | 22713585 | 22714850 | 1266 | 8 | 5.09E-06 | 1.38E-05 | 0.000162 | 1.29E-06 | 0.000396349 | PIP4K2A |
| chr5 | 132579144 | 132579672 | 529 | 3 | 6.27E-06 | 4.50E-07 | 0.001107 | 1.29E-06 | 0.010027591 | AC116366.2, RAD50 |
| chr8 | 48659004 | 48659864 | 861 | 7 | 2.07E-07 | 0.000162 | 4.58E-05 | 1.29E-06 | 0.002627833 | AC022915.2 |
| chr19 | 58479708 | 58480518 | 811 | 4 | 7.53E-06 | 3.00E-06 | 9.80E-05 | 1.30E-06 | 0.003502272 | ZNF446, SLC27A5 |
| chr13 | 98173738 | 98173943 | 206 | 2 | 1.03E-06 | 3.60E-06 | 1.42E-05 | 1.30E-06 | 0.011848841 | FARP1 |
| chr10 | 104260880 | 104261172 | 293 | 3 | 3.30E-06 | 5.85E-06 | 3.56E-05 | 1.30E-06 | 0.010095381 | GSTO1 |
| chr12 | 116605049 | 116605826 | 778 | 4 | 4.87E-06 | 1.10E-06 | 0.00049 | 1.30E-06 | 0.025296608 | NA |
| chr11 | 67054194 | 67055433 | 1240 | 6 | 3.16E-05 | 4.33E-07 | 0.002349 | 1.30E-06 | 0.008014912 | NA |
| chr6 | 30687420 | 30688800 | 1381 | 20 | 1.40E-06 | 4.72E-06 | 0.004755 | 1.30E-06 | -1.46E-05 | PPP1R18, NRM |
| chr1 | 34791874 | 34793016 | 1143 | 7 | 1.81E-07 | 6.80E-06 | 7.59E-05 | 1.30E-06 | 0.004151308 | GJA4, SMIM12 |
| chr10 | 13234306 | 13234816 | 511 | 5 | 7.16E-07 | 6.08E-06 | 0.000169 | 1.30E-06 | 0.007505751 | UCMA |
| chr4 | 64726517 | 64726889 | 373 | 4 | 2.30E-06 | 4.66E-05 | 0.000127 | 1.30E-06 | 0.005485203 | NA |
| chr6 | 83708940 | 83710108 | 1169 | 22 | 1.42E-05 | 2.09E-06 | 0.008835 | 1.30E-06 | 0.002420911 | SNAP91 |
| chr17 | 40800110 | 40801255 | 1146 | 3 | 4.22E-07 | 2.92E-06 | 4.30E-05 | 1.31E-06 | 0.018743708 | NA |
| chr17 | 896844 | 897521 | 678 | 8 | 1.67E-06 | 8.17E-05 | 0.000368 | 1.31E-06 | 0.005819854 | NXN |
| chr6 | 53794068 | 53795079 | 1012 | 9 | 5.05E-06 | 9.26E-06 | 0.0001 | 1.31E-06 | 0.000414065 | LRRC1, AL590652.1 |
| chr5 | 68287770 | 68288826 | 1057 | 14 | 6.66E-06 | 4.63E-07 | 0.001743 | 1.31E-06 | 0.001810255 | PIK3R1 |
| chr5 | 150507446 | 150508225 | 780 | 7 | 5.71E-06 | 3.26E-07 | 0.002879 | 1.31E-06 | 0.005004401 | NDST1 |
| chr6 | 169838993 | 169839615 | 623 | 4 | 4.93E-06 | 1.62E-06 | 0.000164 | 1.31E-06 | 0.003918288 | NA |
| chr4 | 186298076 | 186299308 | 1233 | 6 | 7.52E-07 | 1.06E-06 | 0.000141 | 1.32E-06 | 0.013764455 | F11-AS1 |
| chr11 | 122672720 | 122673105 | 386 | 3 | 1.03E-06 | 5.98E-05 | 3.03E-05 | 1.32E-06 | 0.007309468 | UBASH3B |
| chr18 | 46103289 | 46105140 | 1852 | 18 | 9.22E-06 | 0.000196 | 0.000607 | 1.32E-06 | -0.0004474 | HAUS1, ATP5F1A |
| chr1 | 233395127 | 233395262 | 136 | 2 | 7.22E-07 | 0.000718 | 2.42E-06 | 1.32E-06 | 0.009232697 | NA |
| chr17 | 1190735 | 1191261 | 527 | 7 | 4.86E-06 | 1.37E-06 | 0.001869 | 1.32E-06 | 0.003720141 | ABR |
| chr13 | 26253690 | 26254743 | 1054 | 10 | 8.40E-07 | 1.42E-06 | 0.001643 | 1.32E-06 | 0.000407292 | CDK8 |
| chr13 | 108219661 | 108220104 | 444 | 3 | 1.75E-06 | 5.39E-07 | 0.000756 | 1.33E-06 | 0.00201702 | ABHD13 |
| chr16 | 70746256 | 70747044 | 789 | 5 | 5.55E-06 | 4.40E-07 | 0.000806 | 1.33E-06 | 0.014298814 | VAC14 |
| chr15 | 25159888 | 25160687 | 800 | 2 | 8.73E-06 | 8.20E-07 | 0.000143 | 1.33E-06 | 0.031249391 | SNHG14 |
| chr17 | 10145224 | 10145672 | 449 | 4 | 7.90E-06 | 4.10E-07 | 0.000893 | 1.33E-06 | 0.007721125 | GAS7 |
| chr18 | 48050110 | 48050480 | 371 | 3 | 2.28E-06 | 3.07E-06 | 2.61E-05 | 1.33E-06 | 0.005432141 | ZBTB7C |
| chr15 | 25903578 | 25903617 | 40 | 2 | 4.61E-07 | 4.45E-05 | 3.73E-06 | 1.33E-06 | 0.004565633 | LINC02346 |
| chr22 | 30648973 | 30649523 | 551 | 3 | 7.87E-07 | 7.94E-06 | 0.000144 | 1.33E-06 | 0.007680221 | SLC35E4 |
| chr19 | 38316087 | 38317447 | 1361 | 11 | 1.69E-05 | 3.72E-06 | 0.002521 | 1.33E-06 | 0.001198745 | YIF1B |
| chr1 | 53470245 | 53470515 | 271 | 2 | 4.42E-06 | 9.44E-07 | 8.85E-05 | 1.34E-06 | 0.018691624 | NA |
| chr19 | 4586766 | 4586789 | 24 | 2 | 3.33E-06 | 2.39E-06 | 2.12E-05 | 1.34E-06 | 0.013306267 | NA |
| chr20 | 6122982 | 6124239 | 1258 | 11 | 3.23E-08 | 2.93E-05 | 0.000675 | 1.35E-06 | 0.006987244 | FERMT1 |
| chr19 | 42065793 | 42066254 | 462 | 6 | 2.95E-06 | 2.75E-06 | 0.001705 | 1.35E-06 | 0.002789029 | GRIK5 |
| chr12 | 54380460 | 54381523 | 1064 | 3 | 5.94E-05 | 4.92E-07 | 0.000849 | 1.35E-06 | 0.0077755 | AC079313.2, AC079313.1, ZNF385A |
| chr19 | 3659349 | 3660028 | 680 | 7 | 1.68E-06 | 7.17E-06 | 0.000412 | 1.35E-06 | 0.002882669 | PIP5K1C |
| chr16 | 69565330 | 69566630 | 1301 | 12 | 9.77E-07 | 4.58E-06 | 0.003682 | 1.35E-06 | 0.000414393 | NFAT5, MIR1538 |
| chr5 | 134758282 | 134759182 | 901 | 12 | 7.31E-06 | 7.89E-06 | 0.001652 | 1.35E-06 | 0.000159541 | DDX46 |
| chr6 | 168909085 | 168909723 | 639 | 3 | 6.32E-07 | 1.20E-06 | 0.000166 | 1.36E-06 | 0.011121942 | NA |
| chr1 | 225961918 | 225962685 | 768 | 5 | 1.82E-06 | 5.83E-06 | 0.000203 | 1.36E-06 | 0.005492551 | NA |
| chr16 | 19871934 | 19873057 | 1124 | 6 | 3.41E-08 | 0.000181 | 7.98E-06 | 1.36E-06 | 0.002147634 | GPRC5B |
| chr13 | 40350347 | 40351950 | 1604 | 6 | 1.75E-05 | 1.67E-06 | 0.001065 | 1.36E-06 | -0.00164242 | LINC00598 |
| chr4 | 8319569 | 8320824 | 1256 | 6 | 4.21E-05 | 2.44E-06 | 0.001367 | 1.36E-06 | 0.002785554 | LINC02517 |
| chr5 | 132295061 | 132295917 | 857 | 9 | 6.17E-07 | 4.95E-06 | 0.001472 | 1.37E-06 | 0.003944959 | SLC22A4, P4HA2 |
| chr3 | 13565286 | 13565731 | 446 | 2 | 6.52E-07 | 3.69E-06 | 1.48E-05 | 1.37E-06 | 0.005556629 | FBLN2 |
| chr17 | 64007876 | 64008172 | 297 | 4 | 8.63E-08 | 5.52E-05 | 4.39E-06 | 1.37E-06 | 0.002773984 | ICAM2 |
| chr9 | 124446688 | 124446888 | 201 | 2 | 4.45E-06 | 1.07E-06 | 7.04E-05 | 1.38E-06 | 0.010263232 | NA |
| chr1 | 161089477 | 161089864 | 388 | 7 | 4.92E-06 | 9.94E-07 | 0.003326 | 1.38E-06 | 0.007208832 | NECTIN4 |
| chr17 | 15999031 | 15999976 | 946 | 14 | 1.67E-06 | 4.75E-06 | 0.000108 | 1.38E-06 | 0.001338389 | TTC19, ZSWIM7 |
| chr7 | 87875744 | 87876796 | 1053 | 13 | 6.40E-06 | 4.09E-06 | 0.005995 | 1.38E-06 | 0.001010786 | DBF4, SLC25A40 |
| chr8 | 103498526 | 103499063 | 538 | 5 | 6.81E-09 | 0.001052 | 3.58E-05 | 1.38E-06 | 0.009226466 | AC012213.5, AC012213.1 |
| chr20 | 19260758 | 19260932 | 175 | 2 | 5.82E-06 | 8.15E-07 | 0.000191 | 1.39E-06 | 0.076115606 | SLC24A3, SLC24A3-AS1 |
| chr2 | 230713589 | 230714082 | 494 | 4 | 2.19E-06 | 5.35E-07 | 0.000911 | 1.39E-06 | -0.00207884 | CAB39 |
| chr7 | 102455300 | 102455875 | 576 | 2 | 6.91E-08 | 1.82E-05 | 5.51E-06 | 1.39E-06 | 0.005500956 | ORAI2 |
| chr22 | 21952825 | 21953901 | 1077 | 10 | 1.54E-06 | 9.93E-05 | 0.000384 | 1.39E-06 | 0.003504785 | PPM1F-AS1, PPM1F |
| chr8 | 23102200 | 23103106 | 907 | 8 | 1.77E-07 | 3.74E-05 | 2.37E-05 | 1.40E-06 | 0.003011898 | AC107959.4, AC107959.5, TNFRSF10C |
| chr19 | 45615765 | 45616378 | 614 | 3 | 7.14E-07 | 1.71E-06 | 5.50E-05 | 1.40E-06 | 0.011096388 | EML2 |
| chr1 | 24083979 | 24084525 | 547 | 6 | 1.71E-06 | 2.76E-05 | 0.000115 | 1.40E-06 | 0.004648978 | AL591178.1, MYOM3 |
| chr1 | 156646762 | 156647282 | 521 | 6 | 1.92E-07 | 0.000149 | 0.000137 | 1.40E-06 | 0.009612521 | BCAN, AL590666.2 |
| chr1 | 33268787 | 33269973 | 1187 | 3 | 7.84E-07 | 3.00E-06 | 9.52E-05 | 1.40E-06 | 0.004320307 | ZNF362 |
| chr10 | 124592569 | 124593263 | 695 | 5 | 2.39E-07 | 8.43E-05 | 1.28E-05 | 1.40E-06 | 0.002285055 | LHPP |
| chr6 | 4078709 | 4079820 | 1112 | 14 | 1.12E-05 | 7.40E-05 | 0.002025 | 1.40E-06 | -0.00307273 | C6orf201, FAM217A |
| chr3 | 49557507 | 49557844 | 338 | 2 | 5.11E-07 | 2.28E-05 | 5.03E-06 | 1.41E-06 | -0.01230804 | BSN |
| chr17 | 46819382 | 46820146 | 765 | 7 | 3.20E-06 | 1.36E-06 | 0.001935 | 1.41E-06 | 0.002306179 | WNT3 |
| chr19 | 39320232 | 39320718 | 487 | 4 | 1.52E-07 | 0.000263 | 2.80E-06 | 1.41E-06 | -0.00033291 | LRFN1 |
| chr10 | 132134703 | 132135936 | 1234 | 7 | 1.20E-07 | 0.000285 | 7.32E-05 | 1.41E-06 | 0.00064042 | JAKMIP3 |
| chr11 | 120137512 | 120138427 | 916 | 11 | 1.66E-06 | 1.45E-05 | 0.000907 | 1.41E-06 | 0.007064351 | TRIM29 |
| chr14 | 95307544 | 95307896 | 353 | 3 | 3.26E-06 | 4.49E-06 | 4.10E-05 | 1.41E-06 | 0.014765356 | CLMN |
| chr15 | 94167763 | 94168743 | 981 | 3 | 1.14E-05 | 1.90E-06 | 0.000148 | 1.41E-06 | -0.00444179 | H3P40 |
| chr1 | 7518956 | 7519347 | 392 | 2 | 8.00E-07 | 4.13E-06 | 1.42E-05 | 1.41E-06 | 0.009473463 | CAMTA1 |
| chr19 | 11577431 | 11577896 | 466 | 5 | 2.94E-07 | 5.78E-07 | 0.00123 | 1.41E-06 | 0.01027072 | ZNF627, ACP5 |
| chr1 | 12622028 | 12622386 | 359 | 2 | 3.62E-06 | 8.53E-07 | 0.000165 | 1.42E-06 | 0.011399444 | NA |
| chr15 | 66707371 | 66708617 | 1247 | 8 | 1.02E-05 | 9.07E-07 | 0.000675 | 1.42E-06 | 0.003469285 | SMAD6 |
| chr1 | 209684994 | 209686005 | 1012 | 7 | 2.52E-06 | 7.17E-06 | 5.79E-05 | 1.42E-06 | 0.000692299 | HSD11B1-AS1 |
| chr6 | 149897543 | 149898539 | 997 | 4 | 5.62E-05 | 9.02E-07 | 0.000513 | 1.43E-06 | 0.007502726 | RAET1E-AS1, RAET1E-AS1, RAET1E |
| chr20 | 50224631 | 50225140 | 510 | 3 | 7.71E-07 | 3.20E-05 | 6.04E-06 | 1.43E-06 | 0.004973673 | NA |
| chr8 | 130465790 | 130466695 | 906 | 2 | 4.58E-05 | 8.93E-07 | 0.000137 | 1.43E-06 | 0.01255379 | NA |
| chr11 | 112073696 | 112074560 | 865 | 13 | 1.48E-06 | 5.15E-05 | 2.93E-05 | 1.43E-06 | 0.001086746 | NKAPD1, PIH1D2 |
| chr8 | 58594834 | 58595009 | 176 | 2 | 1.37E-06 | 1.89E-06 | 3.11E-05 | 1.43E-06 | 0.018317613 | NSMAF |
| chr5 | 180139792 | 180140375 | 584 | 2 | 4.05E-08 | 0.000421 | 2.69E-06 | 1.43E-06 | 0.007531309 | RASGEF1C |
| chr19 | 50716718 | 50717526 | 809 | 12 | 1.85E-06 | 5.55E-05 | 0.000146 | 1.44E-06 | 0.005608124 | SHANK1 |
| chr1 | 150882910 | 150882913 | 4 | 2 | 3.48E-06 | 3.37E-06 | 1.71E-05 | 1.44E-06 | -0.01095918 | NA |
| chr17 | 48728118 | 48729442 | 1325 | 13 | 2.27E-07 | 2.42E-05 | 4.28E-05 | 1.44E-06 | -0.00092179 | HOXB13 |
| chr6 | 38702661 | 38704567 | 1907 | 10 | 3.95E-05 | 0.00024 | 0.000448 | 1.44E-06 | 0.001274126 | GLO1 |
| chr5 | 115179644 | 115181157 | 1514 | 16 | 4.41E-06 | 5.44E-05 | 0.002443 | 1.45E-06 | 0.002252939 | TRIM36 |
| chr19 | 12659146 | 12659426 | 281 | 3 | 4.59E-06 | 7.90E-07 | 0.000505 | 1.45E-06 | -0.00081469 | MAN2B1 |
| chr6 | 29374115 | 29375038 | 924 | 10 | 5.13E-05 | 1.16E-07 | 0.01544 | 1.45E-06 | 0.004697494 | OR5V1, OR12D3 |
| chr7 | 23471335 | 23471824 | 490 | 5 | 4.66E-07 | 4.27E-06 | 8.94E-05 | 1.45E-06 | 0.006186706 | NA |
| chr7 | 34069932 | 34069934 | 3 | 2 | 6.27E-06 | 8.50E-07 | 0.000199 | 1.45E-06 | -0.02594581 | BMPER |
| chr1 | 206690259 | 206690773 | 515 | 2 | 1.64E-06 | 1.95E-06 | 3.05E-05 | 1.45E-06 | 0.013204772 | MAPKAPK2 |
| chr6 | 137492447 | 137493219 | 773 | 4 | 6.16E-06 | 3.51E-07 | 0.002551 | 1.46E-06 | 0.012224309 | OLIG3 |
| chr3 | 69122584 | 69123052 | 469 | 3 | 4.04E-06 | 5.50E-07 | 0.000859 | 1.46E-06 | -0.00681964 | LMOD3 |
| chr4 | 40476648 | 40477245 | 598 | 2 | 8.10E-07 | 1.70E-06 | 3.65E-05 | 1.46E-06 | 0.006184273 | RBM47 |
| chr19 | 12642424 | 12642515 | 92 | 2 | 2.33E-06 | 8.68E-07 | 0.00018 | 1.46E-06 | 0.006960838 | NA |
| chr16 | 89994364 | 89994760 | 397 | 5 | 2.48E-06 | 7.19E-06 | 7.90E-05 | 1.46E-06 | 0.004433404 | AFG3L1P |
| chr1 | 161039673 | 161040281 | 609 | 6 | 2.26E-07 | 4.20E-05 | 6.24E-05 | 1.46E-06 | 0.002063272 | USF1 |
| chr14 | 103519890 | 103520813 | 924 | 5 | 6.65E-05 | 4.35E-06 | 0.00045 | 1.47E-06 | 0.016758795 | CKB |
| chr3 | 69200125 | 69200567 | 443 | 2 | 1.12E-06 | 8.70E-07 | 0.000185 | 1.47E-06 | 0.017537425 | FRMD4B |
| chr4 | 109702818 | 109704025 | 1208 | 14 | 1.92E-05 | 4.96E-05 | 0.001231 | 1.47E-06 | 0.001939879 | CASP6 |
| chr2 | 157628797 | 157629719 | 923 | 10 | 4.59E-06 | 4.73E-06 | 0.001179 | 1.47E-06 | 0.002026763 | ACVR1C |
| chr2 | 118844636 | 118845913 | 1278 | 11 | 1.49E-05 | 2.00E-07 | 0.011317 | 1.47E-06 | 0.00424731 | EN1 |
| chr1 | 225824252 | 225825511 | 1260 | 9 | 8.21E-06 | 1.06E-05 | 0.000136 | 1.47E-06 | 0.002672307 | EPHX1 |
| chr17 | 48584813 | 48585555 | 743 | 5 | 1.33E-06 | 1.36E-05 | 0.000932 | 1.47E-06 | 0.010619484 | HOXB-AS3, HOXB3 |
| chr5 | 37837361 | 37838188 | 828 | 4 | 6.46E-07 | 8.31E-07 | 0.000651 | 1.48E-06 | 0.005973877 | GDNF-AS1, GDNF |
| chr16 | 888049 | 889156 | 1108 | 4 | 2.45E-06 | 8.86E-07 | 0.000601 | 1.48E-06 | -0.00067691 | LMF1 |
| chr8 | 41684574 | 41684654 | 81 | 3 | 4.62E-07 | 0.000111 | 2.90E-06 | 1.48E-06 | -0.01912699 | ANK1 |
| chr19 | 48754874 | 48755659 | 786 | 5 | 1.01E-07 | 5.15E-05 | 0.000115 | 1.48E-06 | 0.002293675 | FGF21, FUT1 |
| chr3 | 179450972 | 179452312 | 1341 | 12 | 2.47E-06 | 1.65E-05 | 3.15E-05 | 1.48E-06 | 0.000834447 | GNB4 |
| chr17 | 82206012 | 82206658 | 647 | 5 | 1.66E-06 | 4.90E-06 | 0.000366 | 1.48E-06 | 0.00278659 | CCDC57 |
| chr12 | 46382573 | 46383407 | 835 | 4 | 4.24E-06 | 4.23E-07 | 0.00119 | 1.49E-06 | 0.004020648 | NA |
| chr3 | 165837315 | 165837474 | 160 | 4 | 5.75E-06 | 6.74E-07 | 0.001614 | 1.49E-06 | 0.021122865 | LINC01322, BCHE |
| chr2 | 138603984 | 138604118 | 135 | 2 | 1.10E-06 | 1.89E-06 | 3.31E-05 | 1.49E-06 | -0.00088461 | LINC02631 |
| chr22 | 46334738 | 46335853 | 1116 | 14 | 4.77E-07 | 9.71E-05 | 3.57E-05 | 1.49E-06 | -0.00120779 | TRMU |
| chr7 | 4882196 | 4883077 | 882 | 5 | 2.48E-06 | 7.13E-07 | 0.001286 | 1.49E-06 | 0.010418464 | RADIL |
| chr16 | 58027316 | 58028088 | 773 | 4 | 3.91E-06 | 2.23E-06 | 0.000243 | 1.50E-06 | 0.008515512 | MMP15 |
| chr12 | 111736747 | 111737058 | 312 | 2 | 3.55E-06 | 1.46E-06 | 4.82E-05 | 1.50E-06 | 0.005510133 | ACAD10 |
| chr2 | 237056277 | 237056884 | 608 | 5 | 3.56E-06 | 6.37E-06 | 0.000551 | 1.50E-06 | -0.00116478 | AC012063.1 |
| chr17 | 3916045 | 3917389 | 1345 | 11 | 6.38E-06 | 2.80E-05 | 0.001328 | 1.50E-06 | 0.00571088 | P2RX1 |
| chr17 | 7177219 | 7178575 | 1357 | 10 | 2.43E-06 | 1.44E-05 | 0.000133 | 1.51E-06 | 0.001144232 | ASGR1 |
| chr5 | 96814182 | 96814751 | 570 | 4 | 1.84E-06 | 0.000135 | 7.53E-05 | 1.51E-06 | -0.00138551 | AC009126.1 |
| chr12 | 101743180 | 101743613 | 434 | 2 | 4.03E-06 | 8.77E-07 | 0.000217 | 1.51E-06 | 0.000319748 | CHPT1 |
| chr2 | 112651013 | 112652208 | 1196 | 4 | 2.90E-05 | 3.80E-06 | 0.000247 | 1.51E-06 | 0.006324333 | SLC20A1 |
| chr2 | 177392240 | 177393697 | 1458 | 14 | 1.17E-06 | 4.20E-06 | 0.007589 | 1.51E-06 | 0.000134465 | AGPS, NFE2L2, AC019080.1 |
| chr3 | 51384188 | 51385599 | 1412 | 14 | 2.42E-06 | 4.88E-05 | 0.000961 | 1.51E-06 | -0.00034967 | DOCK3, MANF |
| chr11 | 27506378 | 27507156 | 779 | 11 | 6.95E-06 | 1.46E-05 | 0.00468 | 1.51E-06 | 0.001515181 | BDNF-AS, LIN7C |
| chr12 | 105233010 | 105233668 | 659 | 5 | 1.32E-06 | 1.56E-06 | 0.000241 | 1.52E-06 | 0.001894977 | APPL2 |

| | | | | | | | | | | |
|---|---|---|---|---|---|---|---|---|---|---|
| chr1 | 12496514 | 12497169 | 656 | 2 | 2.16E-07 | 2.36E-06 | 2.61E-05 | 1.52E-06 | 0.014015636 | VPS13D |
| chr8 | 54615539 | 54616108 | 570 | 3 | 8.79E-08 | 0.000124 | 3.43E-05 | 1.52E-06 | 0.013298168 | RP1 |
| chr5 | 163437285 | 163438230 | 946 | 13 | 2.18E-05 | 1.66E-07 | 0.022738 | 1.52E-06 | 0.001894444 | CCNG1, AC113414.1 |
| chr6 | 31729944 | 31731122 | 1179 | 22 | 1.52E-06 | 6.77E-05 | 0.00125 | 1.52E-06 | 0.001830258 | DDAH2, CLIC1 |
| chr9 | 134590067 | 134590838 | 772 | 3 | 3.20E-05 | 5.26E-07 | 0.001243 | 1.53E-06 | 0.007546601 | NA |
| chr1 | 200038275 | 200040319 | 2045 | 9 | 0.00014444 | 1.63E-06 | 0.001241 | 1.53E-06 | 0.004883846 | NR5A2 |
| chr17 | 81344670 | 81346062 | 1393 | 6 | 2.43E-06 | 0.000119 | 8.83E-05 | 1.53E-06 | 0.004174083 | AC027601.5 |
| chr3 | 39100468 | 39100769 | 302 | 4 | 1.80E-06 | 2.59E-05 | 1.86E-05 | 1.53E-06 | 0.003392411 | GORASP1 |
| chr4 | 127843927 | 127844689 | 763 | 5 | 1.20E-06 | 1.96E-06 | 0.000945 | 1.53E-06 | -0.00104821 | AC093591.2 |
| chr19 | 43438650 | 43439401 | 752 | 2 | 3.48E-05 | 9.00E-07 | 0.000206 | 1.54E-06 | 0.007439859 | NA |
| chr11 | 110429157 | 110429908 | 752 | 8 | 9.19E-07 | 8.09E-05 | 9.18E-05 | 1.54E-06 | 0.007396691 | NA |
| chr19 | 4151821 | 4153448 | 1628 | 7 | 3.12E-06 | 1.17E-06 | 0.001761 | 1.54E-06 | 0.005923458 | NA |
| chr11 | 8016496 | 8016976 | 481 | 4 | 7.05E-07 | 4.39E-06 | 0.000129 | 1.54E-06 | 0.011221695 | CASC23 |
| chr22 | 43386992 | 43388330 | 1339 | 3 | 5.85E-05 | 6.59E-07 | 0.00064 | 1.54E-06 | 0.010468189 | NA |
| chr16 | 67718974 | 67719651 | 678 | 11 | 1.35E-06 | 1.99E-06 | 0.001932 | 1.54E-06 | 0.000392898 | GFOD2 |
| chr6 | 73523477 | 73523917 | 441 | 7 | 6.50E-06 | 1.89E-06 | 0.00137 | 1.55E-06 | -0.00028816 | AL121972.1, EEF1A1 |
| chr10 | 103671785 | 103672966 | 1182 | 4 | 4.57E-05 | 9.66E-06 | 0.000505 | 1.55E-06 | 0.00100471 | SH3PXD2A |
| chr8 | 140556633 | 140557680 | 1048 | 4 | 1.23E-06 | 1.47E-06 | 0.000121 | 1.55E-06 | 0.004024488 | AGO2 |
| chr20 | 54058812 | 54059148 | 337 | 2 | 2.67E-06 | 2.06E-06 | 3.18E-05 | 1.55E-06 | 0.011661968 | BCAS1 |
| chr3 | 50229961 | 50230827 | 867 | 7 | 2.49E-06 | 2.70E-06 | 0.000672 | 1.55E-06 | 0.005911054 | GNAI2 |
| chr10 | 3105904 | 3106134 | 231 | 2 | 3.51E-08 | 0.00066 | 2.75E-06 | 1.56E-06 | -0.00541568 | PFKP |
| chr11 | 27701175 | 27702242 | 1068 | 15 | 5.63E-07 | 4.20E-05 | 0.000128 | 1.57E-06 | 0.001885072 | BDNF, AC103796.1 |
| chr11 | 134731217 | 134731923 | 707 | 3 | 1.16E-05 | 5.61E-07 | 0.000986 | 1.57E-06 | 0.023167005 | NA |
| chr19 | 12919282 | 12920294 | 1013 | 6 | 2.19E-05 | 1.52E-05 | 8.56E-05 | 1.57E-06 | 0.002976731 | SYCE2 |
| chr8 | 139914476 | 139915082 | 607 | 6 | 5.42E-06 | 2.09E-05 | 0.000133 | 1.57E-06 | 0.002931163 | AC021744.1, TRAPPC9 |
| chr6 | 32118648 | 32119413 | 766 | 11 | 1.66E-06 | 1.04E-05 | 0.001143 | 1.58E-06 | 0.005986506 | ATF6B |
| chr17 | 1709723 | 1710703 | 981 | 10 | 6.28E-07 | 3.39E-05 | 7.54E-05 | 1.58E-06 | -0.00140314 | TLCD2 |
| chr17 | 48930763 | 48931659 | 897 | 5 | 0.000122865 | 4.98E-06 | 0.000244 | 1.58E-06 | -0.00050708 | SNF8 |
| chr2 | 6876236 | 6877440 | 1205 | 5 | 2.28E-08 | 2.75E-05 | 8.82E-06 | 1.59E-06 | 0.013992505 | RSAD2 |
| chr14 | 92323675 | 92324454 | 780 | 8 | 3.60E-07 | 2.84E-05 | 6.50E-05 | 1.59E-06 | 0.001825669 | SLC24A4 |
| chr17 | 66229228 | 66229985 | 758 | 5 | 1.78E-06 | 1.21E-05 | 3.30E-05 | 1.59E-06 | 0.003880981 | APOH |
| chr11 | 74417483 | 74418555 | 1073 | 3 | 6.33E-05 | 1.42E-05 | 0.000119 | 1.59E-06 | 0.012344518 | AP001372.1 |
| chr16 | 51150077 | 51151090 | 1014 | 12 | 7.26E-07 | 2.91E-05 | 0.000264 | 1.59E-06 | 0.003562983 | AC087564.1, SALL1 |
| chr20 | 21511314 | 21512111 | 798 | 5 | 3.12E-06 | 9.30E-06 | 0.000413 | 1.59E-06 | 0.011526126 | NKX2-2-AS1, NKX2-2 |
| chr21 | 37257933 | 37258427 | 495 | 4 | 7.61E-06 | 3.85E-07 | 0.00263 | 1.59E-06 | 0.024718069 | VPS26C |
| chr1 | 92029341 | 92030603 | 1263 | 14 | 9.17E-06 | 7.91E-06 | 0.005904 | 1.59E-06 | 0.000308217 | EPHX4 |
| chr17 | 82832209 | 82833201 | 993 | 6 | 1.12E-05 | 5.49E-05 | 0.000606 | 1.59E-06 | 0.003164183 | TBCD, ZNF750 |
| chr5 | 135039737 | 135041368 | 1632 | 8 | 1.66E-05 | 6.03E-06 | 0.000165 | 1.60E-06 | 0.007218433 | C5orf66, C5orf66-AS1 |
| chr17 | 59707081 | 59708182 | 1102 | 18 | 1.36E-06 | 4.45E-05 | 0.000133 | 1.60E-06 | -0.00032428 | VMP1, PTRH2 |
| chr19 | 48321365 | 48322363 | 999 | 7 | 1.36E-06 | 4.66E-05 | 0.000563 | 1.60E-06 | 0.000846982 | EMP3, CCDC114 |
| chr22 | 27460150 | 27460979 | 830 | 4 | 7.90E-06 | 1.06E-06 | 0.000448 | 1.60E-06 | 0.013968154 | NA |
| chr6 | 43628015 | 43629725 | 1711 | 17 | 9.94E-06 | 7.28E-05 | 0.000302 | 1.60E-06 | 0.002877988 | MAD2L1BP, GTPBP2 |
| chr7 | 44220314 | 44220992 | 679 | 2 | 2.75E-07 | 1.84E-06 | 3.88E-05 | 1.60E-06 | 0.01175126 | CAMK2B |
| chr9 | 128202095 | 128202639 | 545 | 5 | 3.36E-07 | 1.60E-05 | 7.33E-05 | 1.60E-06 | 0.001766457 | DNM1, CIZ1 |
| chr9 | 95880173 | 95880551 | 379 | 3 | 1.58E-06 | 6.03E-05 | 1.04E-05 | 1.60E-06 | 0.00189345 | ERCC6L2 |
| chr20 | 50099589 | 50099758 | 170 | 2 | 5.16E-06 | 1.52E-06 | 5.14E-05 | 1.60E-06 | 0.001633731 | UBE2V1, PEDS1-UBE2V1 |
| chr17 | 1716157 | 1716750 | 594 | 13 | 6.54E-07 | 4.32E-05 | 2.82E-05 | 1.60E-06 | -0.00071859 | WDR81, MIR22HG |
| chr13 | 114059466 | 114060133 | 668 | 5 | 1.82E-08 | 0.056323 | 4.50E-06 | 1.61E-06 | 0.003614572 | RASA3 |
| chr22 | 42431635 | 42432380 | 746 | 4 | 6.95E-08 | 1.72E-06 | 0.000292 | 1.61E-06 | 0.008504499 | NFAM1 |
| chr12 | 27971020 | 27971937 | 918 | 7 | 5.88E-06 | 1.02E-06 | 0.001196 | 1.61E-06 | 0.009832675 | PTHLH |
| chr4 | 1556724 | 1557517 | 794 | 5 | 4.36E-06 | 6.73E-07 | 0.001124 | 1.61E-06 | -0.01564873 | NA |
| chr7 | 5393189 | 5393655 | 467 | 2 | 1.98E-06 | 2.13E-06 | 3.26E-05 | 1.61E-06 | -0.02128028 | TNRC18 |
| chr1 | 198776357 | 198776991 | 635 | 4 | 3.74E-06 | 1.08E-06 | 0.001084 | 1.61E-06 | 0.00379834 | NA |
| chr9 | 2024342 | 2024728 | 387 | 3 | 3.24E-06 | 8.62E-07 | 0.000386 | 1.61E-06 | 0.011372116 | SMARCA2 |
| chr19 | 46296248 | 46297345 | 1098 | 8 | 1.69E-05 | 8.02E-07 | 0.001534 | 1.61E-06 | 0.006073751 | HIF3A |
| chr17 | 79836291 | 79837113 | 823 | 7 | 1.69E-06 | 2.70E-05 | 0.000345 | 1.62E-06 | 0.003497611 | CBX4 |
| chr12 | 3214855 | 3215264 | 410 | 3 | 2.63E-06 | 1.67E-06 | 0.000283 | 1.62E-06 | 0.022752446 | TSPAN9 |
| chr19 | 2217801 | 2218068 | 268 | 3 | 1.74E-06 | 5.51E-06 | 0.000183 | 1.62E-06 | -0.00114972 | DOT1L |
| chr6 | 169564883 | 169565159 | 277 | 3 | 3.34E-06 | 8.36E-06 | 4.65E-05 | 1.62E-06 | 0.003069468 | AL031315.1, WDR27 |
| chr5 | 146620730 | 146621295 | 566 | 2 | 2.40E-08 | 7.36E-05 | 3.80E-06 | 1.62E-06 | 0.003853311 | PPP2R2B |
| chr9 | 70258429 | 70259335 | 907 | 11 | 3.94E-07 | 0.000271 | 6.32E-05 | 1.62E-06 | -0.0006508 | SMC5, SMC5-AS1 |
| chr9 | 99807502 | 99807669 | 168 | 2 | 3.83E-06 | 1.06E-06 | 0.000122 | 1.62E-06 | -0.01329128 | AL359710.1, STX17-AS1 |
| chr19 | 3112650 | 3113261 | 612 | 2 | 4.87E-06 | 1.28E-06 | 7.32E-05 | 1.62E-06 | 0.007931778 | GNA11 |
| chr20 | 35568813 | 35568847 | 35 | 2 | 5.52E-06 | 2.00E-06 | 3.54E-05 | 1.62E-06 | 0.016203302 | FER1L4 |
| chr16 | 50671351 | 50671851 | 501 | 3 | 1.57E-06 | 1.82E-06 | 6.76E-05 | 1.63E-06 | -0.00136822 | AC007608.3, SNX20 |
| chr11 | 47577668 | 47579420 | 1753 | 20 | 5.03E-05 | 3.99E-05 | 0.002562 | 1.63E-06 | -3.65E-06 | NDUFS3, KF459542.1, KBTBD4 |
| chr16 | 4788142 | 4788905 | 764 | 8 | 5.72E-07 | 7.09E-06 | 0.000103 | 1.63E-06 | 0.002706265 | SMIM22, SEPTIN12, AC020663.4 |
| chr11 | 128809989 | 128810587 | 599 | 4 | 1.15E-06 | 6.26E-06 | 0.000473 | 1.63E-06 | -0.00027136 | FLI1 |
| chr7 | 20798904 | 20799012 | 109 | 4 | 4.63E-06 | 4.57E-07 | 0.00134 | 1.63E-06 | 0.007749773 | NA |
| chr3 | 127672092 | 127673560 | 1469 | 9 | 6.11E-07 | 6.26E-05 | 0.000121 | 1.64E-06 | -0.00050651 | PODXL2, ABTB1 |
| chr19 | 15448308 | 15449472 | 1165 | 10 | 1.62E-05 | 1.39E-05 | 0.001722 | 1.64E-06 | -0.00057607 | WIZ |
| chr19 | 5821780 | 5822619 | 840 | 2 | 3.87E-05 | 1.01E-06 | 0.000154 | 1.64E-06 | 0.022731233 | NA |
| chr10 | 73998669 | 73999392 | 724 | 4 | 7.86E-07 | 1.61E-06 | 0.000259 | 1.66E-06 | 0.00903817 | VCL |
| chr22 | 22594141 | 22594420 | 280 | 4 | 8.58E-07 | 3.16E-05 | 1.12E-05 | 1.66E-06 | 0.013226269 | NA |
| chr1 | 2165143 | 2165719 | 577 | 4 | 1.03E-05 | 1.16E-06 | 0.00061 | 1.66E-06 | 0.003614345 | PRKCZ |
| chr12 | 57613839 | 57614519 | 681 | 4 | 4.59E-07 | 1.65E-05 | 0.00026 | 1.66E-06 | 0.00912268 | ARHGEF25, AC025165.1 |
| chr21 | 44873757 | 44874700 | 944 | 10 | 9.82E-06 | 1.42E-05 | 0.000236 | 1.66E-06 | -0.00078891 | PTTG1IP |
| chr17 | 58403025 | 58403618 | 594 | 3 | 1.36E-06 | 3.51E-06 | 7.13E-05 | 1.67E-06 | 0.005282292 | TSPOAP1-AS1, AC004687.2, RNF43 |
| chr20 | 18813453 | 18814331 | 879 | 4 | 4.04E-05 | 1.15E-06 | 0.000649 | 1.67E-06 | 0.009201283 | SCP2D1, SCP2D1-AS1 |
| chr4 | 55853195 | 55854061 | 867 | 11 | 8.67E-07 | 1.59E-05 | 0.000588 | 1.67E-06 | 0.00160459 | EXOC1 |
| chr9 | 19228801 | 19229369 | 569 | 3 | 5.36E-08 | 4.37E-06 | 2.79E-05 | 1.67E-06 | 0.008535173 | NA |
| chr15 | 40756985 | 40756999 | 15 | 2 | 2.97E-06 | 1.34E-06 | 7.10E-05 | 1.67E-06 | 0.019390695 | NA |
| chr2 | 100976385 | 100977859 | 1475 | 10 | 6.25E-05 | 3.34E-06 | 0.004598 | 1.67E-06 | 0.003706389 | NPAS2, NPAS2-AS1 |
| chr1 | 18632329 | 18633891 | 1563 | 7 | 1.13E-07 | 3.24E-05 | 5.69E-05 | 1.67E-06 | 0.00722316 | PAX7 |
| chr2 | 3648275 | 3649158 | 884 | 4 | 1.52E-05 | 5.26E-07 | 0.001523 | 1.67E-06 | 0.007263491 | NA |
| chr19 | 4556970 | 4558107 | 1138 | 4 | 2.00E-07 | 4.52E-06 | 0.000657 | 1.68E-06 | 0.008073567 | SEMA6B |
| chr10 | 1736335 | 1737238 | 904 | 9 | 2.31E-06 | 1.26E-06 | 0.004192 | 1.68E-06 | 0.004018845 | ADARB2 |
| chr11 | 67674596 | 67675660 | 1065 | 10 | 9.07E-08 | 0.000116 | 1.06E-05 | 1.68E-06 | 0.001623778 | ALDH3B2 |
| chr1 | 117686144 | 117686179 | 36 | 2 | 3.84E-06 | 2.30E-06 | 3.19E-05 | 1.68E-06 | 0.008100815 | NA |
| chr2 | 10475626 | 10476111 | 486 | 4 | 2.68E-06 | 1.78E-06 | 8.65E-05 | 1.69E-06 | 0.001563959 | NA |
| chr20 | 57329233 | 57330020 | 788 | 11 | 1.62E-06 | 2.71E-06 | 0.000825 | 1.69E-06 | 0.008762401 | SPO11 |
| chr7 | 101551346 | 101551966 | 621 | 3 | 4.23E-06 | 7.62E-07 | 0.000637 | 1.69E-06 | 0.008573338 | COL26A1 |
| chr11 | 119674296 | 119674398 | 103 | 2 | 1.37E-06 | 4.62E-06 | 1.60E-05 | 1.69E-06 | 0.011238181 | NECTIN1 |
| chr10 | 77661982 | 77662634 | 653 | 3 | 2.32E-05 | 8.20E-07 | 0.000615 | 1.69E-06 | 0.004581068 | NA |
| chr19 | 54173053 | 54173380 | 328 | 6 | 3.86E-06 | 3.02E-05 | 0.000173 | 1.69E-06 | 0.006484098 | TMC4 |
| chr7 | 44150329 | 44151712 | 1384 | 4 | 2.05E-05 | 2.91E-05 | 6.34E-05 | 1.69E-06 | 0.001180538 | GCK |
| chr15 | 80059756 | 80060930 | 1175 | 7 | 3.50E-07 | 7.05E-05 | 7.52E-05 | 1.69E-06 | -0.00247801 | ZFAND6 |
| chr13 | 50909612 | 50909924 | 313 | 9 | 3.64E-06 | 8.04E-06 | 0.000219 | 1.69E-06 | -0.00104025 | RNASEH2B, RNASEH2B-AS1 |
| chr8 | 37827531 | 37827544 | 14 | 2 | 4.35E-06 | 9.78E-07 | 0.000236 | 1.69E-06 | 0.00401236 | ADGRA2 |
| chr19 | 36009542 | 36010677 | 1136 | 6 | 9.38E-09 | 0.001055 | 5.24E-05 | 1.70E-06 | 0.00429379 | AC002116.1, ALKBH6 |
| chr1 | 151993628 | 151995221 | 1594 | 14 | 2.37E-07 | 6.96E-05 | 0.002238 | 1.70E-06 | 0.006403876 | AL450992.1, S100A10 |
| chr17 | 7317936 | 7319263 | 1328 | 7 | 6.97E-07 | 5.11E-05 | 1.77E-05 | 1.70E-06 | 0.001254027 | AC026954.2, NEURL4 |
| chr12 | 34205654 | 34206367 | 714 | 3 | 2.06E-08 | 4.56E-06 | 2.46E-05 | 1.70E-06 | -0.00339283 | RNA5SP357, AC140847.2 |
| chr17 | 1050602 | 1051250 | 649 | 3 | 3.00E-06 | 2.29E-06 | 7.55E-05 | 1.71E-06 | 0.009997386 | ABR |
| chr1 | 234724869 | 234725662 | 794 | 6 | 4.57E-06 | 4.79E-07 | 0.004861 | 1.71E-06 | 0.007430534 | LINC01132 |
| chr12 | 7936222 | 7937362 | 1141 | 7 | 3.27E-05 | 1.50E-06 | 0.00329 | 1.72E-06 | 0.00088356 | NA |
| chr20 | 33009704 | 33010247 | 544 | 4 | 2.15E-06 | 1.59E-06 | 0.000302 | 1.72E-06 | 0.007814811 | BPIFB2 |
| chr11 | 72003305 | 72004122 | 818 | 4 | 9.73E-06 | 3.59E-05 | 0.000124 | 1.73E-06 | 0.000943117 | IL18BP, NUMA1 |
| chr3 | 52016379 | 52016531 | 153 | 2 | 9.36E-07 | 4.52E-05 | 4.69E-06 | 1.73E-06 | 0.004993688 | NA |
| chr15 | 85750751 | 85751413 | 663 | 3 | 3.52E-07 | 7.27E-05 | 7.20E-05 | 1.73E-06 | 0.015451266 | AC021739.4 |
| chr1 | 1540561 | 1542236 | 1676 | 9 | 2.38E-06 | 3.06E-05 | 0.000588 | 1.73E-06 | 0.002776777 | TMEM240, SSU72 |
| chr12 | 52059649 | 52060562 | 914 | 5 | 3.40E-07 | 1.95E-05 | 0.000185 | 1.73E-06 | 0.007098267 | NA |
| chr8 | 63167962 | 63169192 | 1231 | 14 | 3.97E-06 | 1.43E-05 | 0.000468 | 1.73E-06 | -0.00066582 | YTHDF3, YTHDF3-AS1 |
| chr20 | 2840435 | 2841356 | 922 | 17 | 2.99E-05 | 8.61E-07 | 0.006482 | 1.73E-06 | 0.001137322 | VPS16, PCED1A |
| chr2 | 19989447 | 19990750 | 1304 | 17 | 2.62E-06 | 1.82E-05 | 0.001022 | 1.73E-06 | 0.0009147 | AC079145.1, WDR35 |
| chr6 | 31180555 | 31180880 | 326 | 9 | 1.95E-05 | 1.48E-07 | 0.016928 | 1.73E-06 | -0.02212462 | POU5F1 |
| chr6 | 30954266 | 30955973 | 1708 | 18 | 5.53E-06 | 6.38E-06 | 0.001083 | 1.74E-06 | 0.000955842 | SFTA2, HCG21 |
| chr8 | 94949591 | 94950235 | 645 | 9 | 1.89E-06 | 2.02E-06 | 0.00811 | 1.74E-06 | 0.018535915 | NDUFAF6, AC087752.3 |
| chr1 | 84300841 | 84301641 | 801 | 6 | 2.24E-06 | 2.57E-06 | 0.001692 | 1.74E-06 | -0.00716915 | SAMD13 |
| chr8 | 47610487 | 47610936 | 450 | 2 | 6.79E-07 | 9.43E-06 | 1.00E-05 | 1.74E-06 | 0.0097191 | SPIDR |
| chr17 | 42676879 | 42678239 | 1361 | 11 | 0.000106255 | 1.79E-05 | 0.002118 | 1.74E-06 | -0.0005137 | PLEKHH3 |
| chr15 | 74128930 | 74130420 | 1491 | 24 | 1.02E-05 | 3.91E-05 | 0.001657 | 1.75E-06 | 0.002037815 | ISLR2, AC010931.1 |
| chr7 | 2277001 | 2277940 | 940 | 10 | 1.10E-05 | 1.13E-06 | 0.002866 | 1.75E-06 | 0.005222391 | SNX8 |
| chr20 | 1423591 | 1424068 | 478 | 3 | 1.14E-05 | 1.74E-06 | 0.000272 | 1.75E-06 | 0.00722906 | NA |
| chr1 | 224615047 | 224616135 | 1089 | 5 | 5.12E-07 | 1.08E-06 | 0.001878 | 1.75E-06 | -0.00901167 | CNIH3, CNIH3-AS2 |
| chr1 | 10074203 | 10074562 | 360 | 2 | 4.22E-06 | 1.40E-06 | 7.24E-05 | 1.75E-06 | 0.005560101 | UBE4B |
| chr6 | 138107430 | 138108174 | 745 | 10 | 5.98E-06 | 3.01E-05 | 0.00014 | 1.75E-06 | 0.002526775 | NA |
| chr1 | 223142424 | 223144206 | 1783 | 17 | 4.87E-05 | 1.02E-05 | 0.002262 | 1.75E-06 | 0.002891396 | AL359979.2, TLR5 |
| chr6 | 6724609 | 6725262 | 654 | 6 | 2.74E-07 | 4.57E-06 | 0.000724 | 1.75E-06 | 0.007580307 | AL031123.5, AL031123.1 |
| chr2 | 191847804 | 191848317 | 514 | 6 | 3.60E-07 | 6.47E-05 | 1.65E-05 | 1.75E-06 | -0.00569702 | CAVIN2-AS1 |
| chr4 | 73708998 | 73709611 | 614 | 3 | 4.28E-06 | 9.47E-07 | 0.000274 | 1.76E-06 | 0.002863077 | NA |
| chr16 | 2204665 | 2205399 | 735 | 9 | 1.00E-08 | 0.000107 | 4.03E-05 | 1.76E-06 | 0.001124576 | MLST8 |
| chr10 | 3103134 | 3104357 | 1224 | 5 | 1.02E-07 | 2.19E-05 | 4.45E-05 | 1.76E-06 | -0.00055312 | PFKP |
| chr7 | 152077332 | 152077801 | 470 | 2 | 3.58E-08 | 0.001076 | 2.90E-06 | 1.77E-06 | 0.003202657 | GALNT11 |
| chr3 | 127923512 | 127924009 | 498 | 4 | 9.74E-07 | 1.54E-05 | 1.60E-05 | 1.77E-06 | 0.004687144 | KBTBD12, MGLL |
| chr19 | 39685136 | 39685219 | 84 | 2 | 1.27E-06 | 1.04E-06 | 0.000212 | 1.77E-06 | 0.006777805 | LGALS17A |
| chr19 | 10361589 | 10362290 | 702 | 5 | 2.10E-06 | 2.72E-05 | 6.60E-05 | 1.77E-06 | 0.003813677 | TYK2 |
| chr14 | 24141118 | 24142019 | 902 | 13 | 2.42E-05 | 1.70E-06 | 0.014227 | 1.77E-06 | -0.00034574 | EMC9 |
| chr8 | 10524282 | 10524408 | 127 | 4 | 8.51E-07 | 4.44E-05 | 6.79E-06 | 1.78E-06 | 0.004422507 | PRSS51 |
| chr8 | 129938998 | 129940087 | 1090 | 10 | 2.85E-06 | 3.63E-06 | 0.000903 | 1.78E-06 | 0.000272918 | AC022973.3, CYRIB |
| chr17 | 21028564 | 21029294 | 731 | 3 | 8.15E-06 | 9.77E-07 | 0.000234 | 1.78E-06 | 0.00456816 | USP22 |
| chr17 | 39669910 | 39670291 | 382 | 3 | 9.79E-07 | 1.13E-06 | 0.00015 | 1.78E-06 | 0.004646747 | PNMT |
| chr12 | 124617442 | 124618069 | 628 | 4 | 7.64E-06 | 5.61E-07 | 0.001813 | 1.78E-06 | 0.007312161 | NA |
| chr15 | 25346238 | 25346540 | 303 | 2 | 1.60E-07 | 0.00011 | 3.74E-06 | 1.78E-06 | 0.006297131 | SNHG14, UBE3A, AC100774.1 |
| chr19 | 48619037 | 48619756 | 720 | 21 | 3.08E-06 | 6.56E-06 | 0.021361 | 1.79E-06 | 0.000634039 | SPHK2, RPL18, AC022154.1 |
| chr1 | 53176562 | 53176939 | 378 | 2 | 3.19E-06 | 1.52E-06 | 6.50E-05 | 1.79E-06 | 0.003303066 | NA |
| chr20 | 62060027 | 62060577 | 551 | 3 | 4.37E-06 | 8.27E-07 | 0.000691 | 1.79E-06 | -0.00035893 | TAF4 |
| chr1 | 93584425 | 93585549 | 1125 | 7 | 2.38E-05 | 2.32E-05 | 0.000586 | 1.79E-06 | 0.001039771 | BCAR3 |
| chr7 | 128754295 | 128754508 | 214 | 2 | 3.76E-06 | 3.35E-06 | 2.30E-05 | 1.79E-06 | 0.002441545 | CALU |
| chr16 | 67160964 | 67161837 | 874 | 7 | 4.24E-07 | 0.000109 | 3.30E-05 | 1.79E-06 | 0.002388437 | FBXL8 |
| chr7 | 346851 | 346946 | 96 | 5 | 6.11E-06 | 4.69E-06 | 5.53E-05 | 1.79E-06 | 0.003761281 | NA |
| chr19 | 56104526 | 56105411 | 886 | 6 | 1.24E-05 | 1.81E-06 | 0.00014 | 1.79E-06 | 0.005504984 | ZNF787 |
| chr20 | 31259599 | 31260401 | 803 | 2 | 1.51E-05 | 1.16E-06 | 0.000134 | 1.80E-06 | 0.029879744 | DEFB115 |
| chr14 | 101053741 | 101054652 | 912 | 8 | 4.29E-06 | 3.47E-05 | 0.000139 | 1.80E-06 | 0.000429075 | MIR382 |
| chr10 | 95168319 | 95169336 | 1018 | 4 | 3.15E-05 | 4.30E-05 | 0.00028 | 1.80E-06 | 0.008316841 | AL157834.4 |
| chr18 | 78585144 | 78585570 | 427 | 2 | 6.72E-06 | 1.16E-06 | 0.000133 | 1.80E-06 | 0.005908026 | NA |
| chr22 | 30591562 | 30592488 | 927 | 11 | 2.11E-05 | 4.62E-06 | 0.001573 | 1.80E-06 | 0.001281809 | PES1 |
| chr8 | 144417646 | 144418739 | 1094 | 6 | 2.04E-05 | 3.42E-07 | 0.007073 | 1.80E-06 | 0.009369471 | NA |
| chr8 | 126843889 | 126845352 | 1464 | 3 | 0.000213456 | 6.31E-07 | 0.001219 | 1.80E-06 | 0.004791195 | PCAT1, AC024382.1 |
| chr16 | 68247018 | 68248076 | 1059 | 5 | 4.16E-05 | 3.37E-06 | 0.001276 | 1.81E-06 | 0.005449281 | PLA2G15 |
| chr7 | 139713779 | 139713849 | 71 | 2 | 3.22E-06 | 1.05E-06 | 0.000245 | 1.81E-06 | -0.00065183 | HIPK2 |
| chr2 | 37816338 | 37817281 | 944 | 3 | 3.70E-06 | 1.15E-06 | 0.000172 | 1.81E-06 | 0.01528044 | AC010878.1 |

| | | | | | | | | | | |
|---|---|---|---|---|---|---|---|---|---|---|
| chr13 | 48303298 | 48304503 | 1206 | 19 | 2.03E-06 | 1.85E-05 | 0.000413 | 1.81E-06 | 0.000298795 | RB1, RB1-DT |
| chr2 | 216415439 | 216415908 | 470 | 2 | 5.24E-07 | 2.09E-06 | 4.13E-05 | 1.82E-06 | 0.004391666 | SMARCAL1 |
| chr17 | 74842227 | 74842899 | 673 | 3 | 5.39E-06 | 6.37E-07 | 0.001242 | 1.82E-06 | -0.0032801 | GRIN2C |
| chr21 | 41612250 | 41612477 | 228 | 3 | 8.85E-06 | 8.17E-07 | 0.00043 | 1.82E-06 | 0.015615121 | NA |
| chr1 | 160085463 | 160085906 | 444 | 3 | 1.44E-06 | 1.80E-06 | 0.000152 | 1.82E-06 | 0.013446355 | KCNJ9 |
| chr1 | 98920244 | 98920508 | 265 | 2 | 1.86E-06 | 1.21E-05 | 9.16E-06 | 1.82E-06 | 0.044711786 | PLPPR5 |
| chr14 | 60641673 | 60642827 | 1155 | 5 | 1.93E-07 | 3.31E-05 | 0.000152 | 1.82E-06 | 0.005620745 | SALRNA1 |
| chr3 | 32239474 | 32240379 | 906 | 3 | 4.27E-06 | 2.49E-06 | 0.00017 | 1.82E-06 | 0.013933447 | CMTM8 |
| chr11 | 58927447 | 58928069 | 623 | 6 | 6.74E-06 | 1.46E-06 | 0.002304 | 1.82E-06 | 0.010718488 | GLYATL1, AP001652.1 |
| chr6 | 29800442 | 29800845 | 404 | 6 | 4.59E-06 | 5.34E-06 | 8.87E-05 | 1.83E-06 | 0.00499532 | HLA-P |
| chr2 | 3503633 | 3504221 | 589 | 3 | 6.90E-06 | 9.47E-07 | 0.000297 | 1.83E-06 | 0.005190497 | ADI1 |
| chr20 | 58888210 | 58889687 | 1478 | 32 | 1.86E-06 | 7.78E-06 | 0.011115 | 1.83E-06 | 0.002555778 | GNAS, AL121917.1 |
| chr1 | 3746121 | 3746871 | 751 | 6 | 3.18E-07 | 1.09E-05 | 0.000134 | 1.83E-06 | 0.00469631 | CCDC27, TP73-AS1 |
| chr11 | 133930790 | 133931330 | 541 | 4 | 7.69E-06 | 8.98E-07 | 0.000668 | 1.83E-06 | 0.011218056 | IGSF9B |
| chr8 | 117520386 | 117521015 | 630 | 11 | 9.54E-06 | 2.89E-06 | 0.00382 | 1.83E-06 | -0.00249601 | MED30 |
| chr2 | 382090 | 382356 | 267 | 3 | 4.49E-06 | 7.66E-07 | 0.00078 | 1.84E-06 | 0.019378057 | NA |
| chr10 | 1210399 | 1210711 | 313 | 4 | 2.18E-06 | 6.26E-05 | 6.29E-05 | 1.84E-06 | 0.007635837 | LINC00200, ADARB2 |
| chr17 | 4967006 | 4968531 | 1526 | 15 | 1.89E-06 | 3.31E-05 | 0.0025 | 1.84E-06 | 3.63E-05 | AC004771.3, SPAG7, CAMTA2 |
| chr7 | 94655692 | 94657050 | 1359 | 42 | 9.78E-07 | 5.15E-06 | 0.002732 | 1.84E-06 | 0.002449191 | PEG10, SGCE |
| chr12 | 131840526 | 131841673 | 1148 | 5 | 2.00E-05 | 5.95E-05 | 0.000253 | 1.84E-06 | 0.003068673 | MMP17 |
| chr7 | 65429785 | 65430919 | 1135 | 5 | 2.23E-07 | 0.000332 | 9.34E-06 | 1.84E-06 | 0.007363768 | NA |
| chr3 | 141777017 | 141778156 | 1140 | 7 | 0.000165196 | 1.32E-05 | 0.001917 | 1.84E-06 | 0.004011461 | AC112504.2, GRK7 |
| chr19 | 9584256 | 9584879 | 624 | 10 | 1.61E-06 | 1.74E-05 | 8.89E-05 | 1.84E-06 | 0.00046592 | ZNF121 |
| chr9 | 126913805 | 126914752 | 948 | 10 | 1.41E-05 | 1.40E-06 | 0.004726 | 1.85E-06 | 0.003033018 | NA |
| chr5 | 140635180 | 140635608 | 429 | 2 | 2.40E-08 | 4.32E-05 | 5.10E-06 | 1.85E-06 | 0.009421404 | NA |
| chr7 | 92165626 | 92166089 | 464 | 2 | 3.94E-06 | 1.64E-06 | 6.10E-05 | 1.85E-06 | 0.011295932 | CYP51A1-AS1, AC000120.2, AC000120.3, LRRD1 |
| chr5 | 172648456 | 172649161 | 706 | 3 | 1.13E-06 | 2.22E-06 | 7.99E-05 | 1.85E-06 | 0.009778008 | NEURL1B |
| chr5 | 4511441 | 4512441 | 1001 | 8 | 1.39E-05 | 9.98E-06 | 0.00046 | 1.86E-06 | 0.012217892 | AC106799.1, AC106799.2 |
| chr1 | 2503648 | 2504775 | 1128 | 5 | 3.13E-06 | 9.50E-07 | 0.000849 | 1.86E-06 | 0.009589087 | PLCH2 |
| chr15 | 23642139 | 23642592 | 454 | 3 | 8.03E-06 | 1.48E-06 | 0.000163 | 1.86E-06 | 0.004912768 | NA |
| chr1 | 20322889 | 20323929 | 1041 | 6 | 6.91E-06 | 9.87E-07 | 0.001514 | 1.87E-06 | 0.009139357 | VWA5B1, AL020998.1 |
| chr4 | 8240659 | 8240732 | 74 | 2 | 5.12E-06 | 1.26E-06 | 0.000117 | 1.87E-06 | 0.012394 | SH3TC1 |
| chr10 | 68900073 | 68900791 | 719 | 3 | 2.13E-07 | 1.29E-06 | 0.000132 | 1.88E-06 | -0.00024713 | AL359844.1 |
| chr8 | 103375616 | 103376526 | 911 | 4 | 1.06E-06 | 0.038714 | 5.42E-06 | 1.88E-06 | 0.00210626 | CTHRC1 |
| chr12 | 121494463 | 121494647 | 185 | 2 | 6.22E-06 | 1.67E-06 | 6.09E-05 | 1.88E-06 | 0.012191341 | KDM2B |
| chr13 | 106530191 | 106530744 | 554 | 4 | 3.84E-07 | 2.71E-06 | 5.53E-05 | 1.88E-06 | 0.010897942 | EFNB2 |
| chr19 | 1854024 | 1854820 | 797 | 6 | 1.14E-06 | 9.98E-06 | 0.001716 | 1.88E-06 | 0.008472096 | KLF16 |
| chr17 | 77376958 | 77377375 | 418 | 6 | 6.45E-06 | 1.82E-05 | 0.001194 | 1.88E-06 | 0.004718931 | SEPTIN9, AC111182.1 |
| chr3 | 52786701 | 52787685 | 985 | 5 | 5.04E-07 | 6.61E-05 | 1.18E-05 | 1.89E-06 | 6.52E-06 | ITIH1 |
| chr20 | 47351460 | 47352846 | 1387 | 9 | 1.13E-05 | 8.28E-06 | 0.003881 | 1.89E-06 | 0.008135154 | AL031666.1, ZMYND8 |
| chr21 | 29129774 | 29131154 | 1381 | 9 | 8.19E-07 | 2.98E-05 | 7.81E-05 | 1.89E-06 | 0.003728305 | MAP3K7CL |
| chr8 | 142462665 | 142463052 | 388 | 4 | 5.95E-07 | 1.21E-05 | 4.59E-05 | 1.89E-06 | 0.009043254 | ADGRB1 |
| chr11 | 65864524 | 65864744 | 221 | 2 | 3.50E-06 | 1.99E-06 | 4.70E-05 | 1.89E-06 | 0.004780515 | MUS81 |
| chr11 | 61638490 | 61639036 | 547 | 4 | 4.08E-06 | 0.001926 | 4.03E-05 | 1.89E-06 | -0.00602635 | RPLP0P2 |
| chr6 | 84032427 | 84033314 | 888 | 5 | 1.53E-05 | 3.45E-07 | 0.004478 | 1.89E-06 | 0.009255383 | NA |
| chr16 | 25111510 | 25112542 | 1033 | 10 | 3.67E-06 | 2.57E-06 | 0.007816 | 1.90E-06 | 0.000694929 | LCMT1, LCMT1-AS1 |
| chr19 | 18098774 | 18099362 | 589 | 10 | 7.20E-07 | 3.39E-06 | 0.003681 | 1.90E-06 | 0.005885931 | MAST3, IL12RB1 |
| chr11 | 35040217 | 35041118 | 902 | 4 | 1.24E-07 | 0.000217 | 1.12E-05 | 1.90E-06 | 0.007938837 | NA |
| chr11 | 117816913 | 117817941 | 1029 | 7 | 8.87E-06 | 1.02E-06 | 0.003823 | 1.90E-06 | 0.010071124 | DSCAML1, FXYD2 |
| chr3 | 184389211 | 184389746 | 536 | 2 | 2.08E-06 | 2.17E-06 | 4.24E-05 | 1.90E-06 | 0.013222329 | CHRD |
| chr19 | 32343631 | 32344143 | 513 | 2 | 1.65E-06 | 1.11E-06 | 0.000239 | 1.91E-06 | 0.010961941 | NA |
| chr20 | 50294625 | 50295142 | 518 | 4 | 6.80E-08 | 0.000329 | 1.93E-06 | 1.91E-06 | 0.003770333 | LINC01270 |
| chr16 | 18800472 | 18801655 | 1184 | 13 | 2.22E-06 | 9.57E-06 | 0.001201 | 1.91E-06 | 0.001443561 | AC138811.2, ARL6IP1 |
| chr3 | 141805307 | 141805491 | 185 | 2 | 7.89E-06 | 1.19E-06 | 0.000157 | 1.91E-06 | 0.008550481 | AC112504.2, GRK7 |
| chr15 | 27421547 | 27422358 | 812 | 2 | 4.25E-05 | 1.16E-06 | 0.000182 | 1.91E-06 | 0.013778219 | GABRG3 |
| chr8 | 143973811 | 143975112 | 1302 | 8 | 8.10E-07 | 5.87E-05 | 0.00055 | 1.91E-06 | 0.00935006 | PLEC |
| chr4 | 8415960 | 8416405 | 446 | 4 | 6.93E-06 | 2.90E-06 | 0.000369 | 1.91E-06 | 2.47E-05 | ACOX3 |
| chr10 | 101030627 | 101031709 | 1083 | 12 | 1.73E-05 | 1.47E-06 | 0.007956 | 1.92E-06 | 0.001320631 | SFXN3, PDZD7 |
| chr17 | 16981424 | 16981985 | 562 | 5 | 2.08E-06 | 1.46E-05 | 4.07E-05 | 1.92E-06 | 0.004580537 | NA |
| chr6 | 33315993 | 33317807 | 1815 | 25 | 4.11E-06 | 7.12E-06 | 0.013081 | 1.92E-06 | 0.001594807 | ZBTB22 |
| chr6 | 150069758 | 150069827 | 70 | 3 | 7.44E-07 | 8.14E-07 | 0.000795 | 1.92E-06 | 0.018715795 | NA |
| chr8 | 43206389 | 43206920 | 532 | 3 | 7.61E-06 | 1.25E-06 | 0.000229 | 1.92E-06 | 0.005890007 | NA |
| chr2 | 79521813 | 79522509 | 697 | 5 | 1.55E-06 | 1.55E-05 | 0.000102 | 1.92E-06 | 0.001042593 | CTNNA2 |
| chr5 | 389648 | 390748 | 1101 | 3 | 8.60E-06 | 1.39E-06 | 0.000366 | 1.92E-06 | 0.014837151 | PDCD6-AHRR, AHRR |
| chr10 | 89535288 | 89536554 | 1267 | 14 | 1.52E-06 | 0.000168 | 0.000443 | 1.92E-06 | 0.002041297 | SLC16A12 |
| chr2 | 112728921 | 112729590 | 670 | 2 | 1.39E-05 | 1.25E-06 | 0.000137 | 1.92E-06 | 0.010534562 | NT5DC4 |
| chr2 | 218975494 | 218976240 | 747 | 4 | 1.98E-06 | 6.33E-07 | 0.000845 | 1.92E-06 | 0.017032442 | LINC00608 |
| chr22 | 42067255 | 42068231 | 977 | 4 | 5.36E-05 | 7.34E-06 | 0.000592 | 1.92E-06 | 0.003001045 | NAGA |
| chr15 | 90922195 | 90923096 | 902 | 2 | 7.90E-05 | 1.14E-06 | 0.000216 | 1.92E-06 | 0.00939924 | MAN2A2 |
| chr1 | 107140565 | 107142383 | 1819 | 18 | 9.70E-05 | 1.43E-05 | 0.001509 | 1.92E-06 | 0.001372667 | NTNG1 |
| chr15 | 74367063 | 74368225 | 1163 | 11 | 3.48E-07 | 0.000204 | 7.61E-05 | 1.92E-06 | 0.002677938 | AC090826.1, CYP11A1 |
| chr5 | 77776041 | 77776965 | 925 | 6 | 1.31E-06 | 2.90E-06 | 0.000165 | 1.92E-06 | -9.84E-05 | TBCA |
| chr6 | 93254747 | 93254932 | 186 | 2 | 2.98E-06 | 5.26E-06 | 1.72E-05 | 1.92E-06 | 0.008614691 | EPHA7 |
| chr7 | 75823445 | 75824388 | 944 | 4 | 3.74E-05 | 1.68E-06 | 0.000437 | 1.92E-06 | 0.007315041 | NA |
| chr12 | 48985121 | 48985422 | 302 | 2 | 4.84E-06 | 1.89E-06 | 5.32E-05 | 1.93E-06 | 0.009576114 | NA |
| chr11 | 1349099 | 1349748 | 650 | 4 | 1.14E-05 | 9.22E-07 | 0.000506 | 1.93E-06 | 0.007602029 | NA |
| chr17 | 50908659 | 50909608 | 950 | 3 | 7.66E-06 | 2.81E-05 | 7.49E-05 | 1.94E-06 | 0.007741812 | TOB1-AS1 |
| chr17 | 82489059 | 82489612 | 554 | 3 | 7.27E-07 | 0.000128 | 0.000137 | 1.94E-06 | 0.004845959 | NARF |
| chr1 | 26598865 | 26599309 | 445 | 2 | 4.03E-06 | 1.75E-06 | 6.08E-05 | 1.94E-06 | 0.015163665 | NA |
| chr10 | 19629381 | 19629805 | 425 | 3 | 1.62E-06 | 7.23E-06 | 2.95E-05 | 1.94E-06 | 0.010979038 | MALRD1 |
| chr5 | 160228418 | 160229359 | 942 | 5 | 8.25E-06 | 2.60E-05 | 0.000691 | 1.94E-06 | 0.00737922 | FABP6 |
| chr9 | 134135630 | 134135941 | 312 | 7 | 6.63E-06 | 5.54E-07 | 0.003766 | 1.94E-06 | 0.001322777 | WDR5, BX649632.1 |
| chr1 | 153958034 | 153959745 | 1712 | 16 | 2.76E-05 | 0.002362 | 0.000618 | 1.95E-06 | 0.000905295 | CRTC2, SLC39A1, AL358472.7 |
| chr12 | 4443863 | 4444404 | 542 | 2 | 1.80E-06 | 2.31E-06 | 4.12E-05 | 1.95E-06 | 0.003922804 | FGF6 |
| chr19 | 20814050 | 20814516 | 467 | 2 | 1.92E-06 | 3.46E-06 | 2.55E-05 | 1.96E-06 | 0.020505347 | NA |
| chr11 | 34512299 | 34514032 | 1734 | 16 | 2.02E-06 | 7.12E-05 | 0.000552 | 1.96E-06 | 0.002838515 | ELF5 |
| chr6 | 170294594 | 170295915 | 1322 | 10 | 1.03E-05 | 1.26E-06 | 0.001207 | 1.96E-06 | 0.005054392 | FAM120B, DLL1 |
| chr6 | 40713190 | 40713913 | 724 | 2 | 5.23E-06 | 1.91E-06 | 5.39E-05 | 1.96E-06 | 0.015821293 | AL583854.1 |
| chr4 | 163330790 | 163331266 | 477 | 3 | 1.75E-06 | 9.83E-06 | 5.01E-05 | 1.96E-06 | 0.017817366 | NPY1R |
| chr16 | 3058040 | 3059205 | 1166 | 9 | 2.32E-05 | 1.69E-05 | 0.000119 | 1.96E-06 | 0.000925669 | MMP25, MMP25-AS1 |
| chr6 | 118710759 | 118711039 | 281 | 5 | 4.53E-06 | 1.37E-06 | 0.000738 | 1.96E-06 | 0.010143967 | NA |
| chr6 | 17817348 | 17817862 | 515 | 3 | 5.59E-06 | 3.92E-06 | 0.000237 | 1.96E-06 | -0.00505702 | KIF13A |
| chr4 | 156970935 | 156972079 | 1145 | 10 | 6.54E-06 | 3.69E-06 | 0.002033 | 1.96E-06 | 0.002557386 | PDGFC |
| chr7 | 134295024 | 134295368 | 345 | 3 | 5.70E-07 | 0.000151 | 9.51E-06 | 1.96E-06 | 0.000854691 | SLC35B4 |
| chr5 | 124660143 | 124661303 | 1161 | 3 | 3.18E-05 | 3.80E-06 | 0.000129 | 1.97E-06 | 0.001226633 | ZNF608 |
| chr11 | 75800518 | 75800985 | 468 | 2 | 2.49E-06 | 2.13E-06 | 4.65E-05 | 1.97E-06 | 0.007463828 | DGAT2, AP003031.1 |
| chr13 | 112837452 | 112838462 | 1011 | 8 | 5.67E-05 | 2.43E-06 | 0.001083 | 1.97E-06 | -0.00022265 | ATP11A |
| chr21 | 39666845 | 39667282 | 438 | 3 | 2.13E-07 | 0.000283 | 3.12E-06 | 1.97E-06 | 0.005178977 | B3GALT5, AF064860.1 |
| chr5 | 113294528 | 113295829 | 1302 | 12 | 3.55E-06 | 6.33E-05 | 0.000283 | 1.97E-06 | 0.001300227 | MCC |
| chr16 | 81266289 | 81267066 | 778 | 4 | 5.74E-07 | 5.92E-05 | 1.94E-05 | 1.98E-06 | 0.008558947 | BCO1 |
| chr19 | 48363449 | 48364555 | 1107 | 13 | 1.92E-06 | 4.55E-05 | 0.000664 | 1.99E-06 | 0.001275709 | SYNGR4, TMEM143 |
| chr2 | 176136528 | 176137181 | 654 | 8 | 2.02E-06 | 5.77E-06 | 0.001271 | 1.99E-06 | 0.006200532 | HOXD3, HOXD-AS2 |
| chr6 | 1615608 | 1616195 | 588 | 4 | 3.13E-06 | 1.31E-06 | 0.000755 | 1.99E-06 | 0.01775618 | NA |
| chr7 | 39184597 | 39184632 | 36 | 2 | 8.43E-06 | 1.34E-06 | 0.000121 | 1.99E-06 | 0.013318419 | POU6F2 |
| chr4 | 1502927 | 1504111 | 1185 | 8 | 4.07E-06 | 9.72E-05 | 0.000171 | 1.99E-06 | 0.005484705 | NA |
| chr16 | 803160 | 803533 | 374 | 3 | 1.17E-06 | 7.00E-07 | 0.001268 | 1.99E-06 | 0.014039009 | NA |
| chr19 | 6056512 | 6057192 | 681 | 3 | 1.89E-06 | 3.63E-06 | 0.000258 | 2.00E-06 | 0.007058095 | RFX2 |
| chr15 | 74906919 | 74907690 | 772 | 12 | 7.20E-07 | 3.82E-05 | 0.004795 | 2.00E-06 | 0.004874537 | NA |
| chr12 | 559387 | 560289 | 903 | 2 | 2.48E-05 | 1.16E-06 | 0.000252 | 2.00E-06 | 0.008551957 | B4GALNT3 |
| chr8 | 126840767 | 126840818 | 52 | 2 | 4.26E-06 | 1.06E-05 | 1.10E-05 | 2.00E-06 | 0.006040228 | PCAT1, AC024382.1 |
| chr16 | 48609394 | 48610695 | 1302 | 14 | 9.00E-06 | 8.17E-05 | 0.001306 | 2.00E-06 | -0.00467244 | N4BP1 |
| chr2 | 176066275 | 176067421 | 1147 | 5 | 1.99E-05 | 4.63E-07 | 0.003617 | 2.00E-06 | 0.007819518 | NA |
| chr8 | 101109170 | 101109908 | 739 | 5 | 2.07E-05 | 1.24E-06 | 0.001202 | 2.00E-06 | 0.009121808 | NA |
| chr6 | 156630966 | 156631454 | 489 | 5 | 2.68E-06 | 3.69E-06 | 0.000391 | 2.01E-06 | 0.003544625 | NA |
| chr12 | 131785018 | 131785673 | 656 | 6 | 1.15E-06 | 1.82E-05 | 0.000789 | 2.01E-06 | 0.001966581 | SFSWAP |
| chr20 | 62053333 | 62054218 | 886 | 3 | 1.19E-05 | 1.24E-06 | 0.000288 | 2.01E-06 | 0.011461466 | TAF4 |
| chr16 | 29666648 | 29667883 | 1236 | 4 | 3.91E-05 | 5.43E-07 | 0.00222 | 2.01E-06 | 0.008525758 | SPN, QPRT |
| chr11 | 67652148 | 67652672 | 525 | 2 | 1.47E-08 | 0.000105 | 4.23E-06 | 2.01E-06 | 0.002644862 | AP003385.1 |
| chr8 | 55878408 | 55878440 | 33 | 2 | 7.93E-07 | 2.13E-06 | 4.85E-05 | 2.01E-06 | -0.01434308 | NA |
| chr9 | 35650529 | 35651146 | 618 | 5 | 1.17E-06 | 2.99E-06 | 0.000736 | 2.01E-06 | 0.007491773 | SIT1 |
| chr19 | 2799759 | 2800912 | 1154 | 5 | 5.62E-06 | 4.81E-05 | 0.000445 | 2.01E-06 | 0.003401013 | THOP1 |
| chr1 | 196897048 | 196897382 | 335 | 4 | 6.83E-06 | 1.37E-06 | 0.000446 | 2.01E-06 | 0.023798451 | CFHR4 |
| chr20 | 59933021 | 59934504 | 1484 | 13 | 6.76E-05 | 3.17E-07 | 0.022298 | 2.02E-06 | -0.00146916 | FAM217B, SYCP2 |
| chr15 | 45429391 | 45430148 | 758 | 3 | 9.02E-05 | 2.21E-06 | 0.000194 | 2.02E-06 | 0.008107931 | NA |
| chr5 | 150245118 | 150245681 | 564 | 5 | 6.20E-06 | 1.49E-06 | 0.000776 | 2.02E-06 | 0.005002775 | CAMK2A |
| chr1 | 157598801 | 157599013 | 213 | 4 | 3.58E-06 | 4.51E-05 | 0.000138 | 2.03E-06 | 0.012452305 | NA |
| chr10 | 97408629 | 97408894 | 266 | 3 | 5.59E-06 | 7.90E-07 | 0.000815 | 2.03E-06 | 0.010608867 | AL355490.2, RRP12 |
| chr3 | 10505146 | 10506208 | 1063 | 10 | 1.53E-05 | 5.57E-06 | 0.001538 | 2.03E-06 | 0.000165147 | ATP2B2 |
| chr14 | 100831879 | 100832583 | 705 | 6 | 1.16E-06 | 2.26E-05 | 0.000751 | 2.03E-06 | -0.00064169 | MEG3 |
| chr5 | 136132661 | 136133426 | 766 | 8 | 3.81E-06 | 9.75E-06 | 0.002783 | 2.03E-06 | 0.000580767 | SMAD5, SMAD5-AS1 |
| chr3 | 183542432 | 183542587 | 156 | 2 | 7.96E-06 | 1.38E-06 | 0.000118 | 2.03E-06 | 0.011191073 | KLHL6 |
| chr20 | 64151362 | 64152224 | 863 | 5 | 9.81E-07 | 3.91E-05 | 0.000254 | 2.04E-06 | 0.006415133 | MYT1 |
| chr6 | 170270244 | 170270386 | 143 | 2 | 1.79E-06 | 1.33E-06 | 0.000138 | 2.04E-06 | -0.00138357 | AL109910.2 |
| chr20 | 50320663 | 50321538 | 876 | 4 | 8.54E-06 | 1.35E-05 | 8.41E-05 | 2.04E-06 | 0.006321368 | LINC01271 |
| chr15 | 85716534 | 85717512 | 979 | 3 | 3.85E-06 | 4.22E-06 | 9.33E-05 | 2.05E-06 | 0.006120051 | AKAP13 |
| chr22 | 38864330 | 38865084 | 755 | 4 | 1.24E-05 | 1.14E-06 | 0.000906 | 2.05E-06 | 0.000373276 | CBX6 |
| chr3 | 196978758 | 196979027 | 270 | 6 | 1.83E-05 | 2.50E-07 | 0.009539 | 2.05E-06 | -0.05239709 | AC011322.1 |
| chr7 | 107748390 | 107749101 | 712 | 2 | 4.67E-06 | 1.85E-06 | 6.32E-05 | 2.06E-06 | -0.01741145 | CBLL1 |
| chr9 | 132079911 | 132080401 | 491 | 7 | 8.94E-06 | 1.22E-06 | 0.000977 | 2.06E-06 | -0.00209878 | NA |
| chr1 | 15128630 | 15129226 | 597 | 3 | 2.07E-06 | 4.23E-06 | 0.000142 | 2.06E-06 | 0.008970893 | TMEM51-AS1 |
| chr1 | 156491806 | 156492410 | 605 | 4 | 1.14E-06 | 4.46E-06 | 4.65E-05 | 2.07E-06 | 0.008409019 | MEF2D |
| chr10 | 101121084 | 101121788 | 705 | 7 | 1.53E-06 | 7.68E-05 | 4.71E-05 | 2.07E-06 | 0.001714794 | TLX1NB |
| chr6 | 35017059 | 35017176 | 118 | 4 | 9.79E-06 | 7.55E-07 | 0.00099 | 2.07E-06 | 0.015027633 | ANKS1A |
| chr5 | 39073599 | 39075155 | 1557 | 17 | 0.000165231 | 7.94E-06 | 0.010194 | 2.07E-06 | -0.00084633 | RICTOR |
| chr2 | 175257190 | 175257199 | 10 | 3 | 7.45E-06 | 9.24E-07 | 0.000773 | 2.07E-06 | -0.01684042 | NA |
| chr13 | 33728376 | 33728907 | 532 | 2 | 8.16E-07 | 8.15E-06 | 1.36E-05 | 2.07E-06 | 0.007705178 | NA |
| chr11 | 61804698 | 61805004 | 307 | 3 | 4.27E-06 | 7.91E-06 | 8.50E-05 | 2.07E-06 | 0.003364066 | FADS2, FADS1 |
| chr16 | 4495138 | 4495538 | 401 | 3 | 2.59E-07 | 0.001421 | 1.90E-06 | 2.07E-06 | 0.004294072 | HMOX2, NMRAL1 |
| chr13 | 112619992 | 112620221 | 230 | 4 | 1.54E-05 | 4.82E-07 | 0.00351 | 2.08E-06 | 0.031590863 | NA |
| chr11 | 7656763 | 7657429 | 667 | 5 | 1.84E-06 | 1.55E-05 | 0.000138 | 2.08E-06 | 0.007518506 | PPFIBP2 |
| chr15 | 72175604 | 72176322 | 719 | 3 | 8.49E-06 | 1.63E-06 | 0.00011 | 2.08E-06 | 0.007293808 | GRAMD2A |
| chr3 | 194604339 | 194604948 | 610 | 2 | 1.71E-06 | 2.83E-06 | 3.57E-05 | 2.09E-06 | 0.003946858 | TMEM44 |
| chr3 | 188036300 | 188036908 | 609 | 3 | 8.79E-06 | 8.24E-07 | 0.000834 | 2.09E-06 | -0.04579114 | NA |
| chr2 | 190859704 | 190859856 | 153 | 3 | 2.19E-06 | 4.66E-05 | 3.67E-05 | 2.09E-06 | 0.005389399 | NA |
| chr21 | 45513460 | 45514152 | 693 | 3 | 8.50E-07 | 8.63E-06 | 3.44E-05 | 2.09E-06 | 0.001872014 | COL18A1, SLC19A1 |
| chr12 | 6199488 | 6200125 | 638 | 6 | 3.16E-06 | 1.03E-06 | 0.001246 | 2.09E-06 | 0.009478598 | CD9 |
| chr3 | 50473090 | 50473793 | 704 | 3 | 1.89E-07 | 0.000103 | 4.42E-06 | 2.09E-06 | 0.00622548 | CACNA2D2 |
| chr2 | 203857844 | 203858620 | 777 | 4 | 8.54E-06 | 1.64E-06 | 0.000147 | 2.09E-06 | 0.001864448 | NA |
| chr1 | 210231867 | 210233393 | 1527 | 10 | 1.21E-06 | 5.20E-05 | 0.000433 | 2.10E-06 | 0.001957727 | SERTAD4, SERTAD4-AS1 |
| chr2 | 18584177 | 18585028 | 852 | 6 | 3.00E-06 | 5.63E-06 | 0.001715 | 2.10E-06 | -0.01591716 | NT5C1B-RDH14, NT5C1B |
| chr16 | 1426898 | 1427110 | 213 | 2 | 6.39E-06 | 1.25E-06 | 0.000227 | 2.10E-06 | 0.004031043 | NA |
| chr1 | 153390451 | 153391385 | 935 | 7 | 3.33E-05 | 8.04E-07 | 0.002889 | 2.10E-06 | 0.006123274 | S100A8 |

| | | | | | | | | | | |
|---|---|---|---|---|---|---|---|---|---|---|
| chr10 | 103616998 | 103617081 | 84 | 3 | 1.15E-05 | 8.19E-07 | 0.000984 | 2.10E-06 | 0.032339874 | SH3PXD2A |
| chr9 | 93952765 | 93953809 | 1045 | 8 | 2.09E-05 | 1.19E-06 | 0.004768 | 2.10E-06 | 0.003711088 | BARX1 |
| chr3 | 119660082 | 119660683 | 602 | 4 | 5.26E-08 | 0.001742 | 4.67E-06 | 2.10E-06 | 0.004981351 | POPDC2, COX17 |
| chr1 | 116667019 | 116667998 | 980 | 12 | 3.77E-06 | 6.15E-06 | 0.001282 | 2.11E-06 | 0.001093259 | IGSF3 |
| chr11 | 69491038 | 69491694 | 657 | 2 | 2.74E-06 | 1.39E-06 | 0.000136 | 2.11E-06 | 0.003806504 | LINC01488 |
| chr4 | 107823719 | 107824741 | 1023 | 12 | 5.25E-06 | 2.41E-05 | 0.0022 | 2.11E-06 | 0.003099195 | SGMS2 |
| chr6 | 151681724 | 151681988 | 265 | 4 | 1.18E-05 | 1.10E-06 | 0.000822 | 2.11E-06 | 0.008918831 | ESR1 |
| chr4 | 684494 | 685962 | 1469 | 4 | 1.26E-05 | 9.45E-07 | 0.00038 | 2.11E-06 | 0.006164466 | SLC49A3 |
| chr20 | 5470577 | 5471254 | 678 | 8 | 1.95E-07 | 0.008566 | 4.57E-05 | 2.11E-06 | 0.001533722 | AL121757.2, AL121757.1, LINC00658 |
| chr3 | 114290758 | 114291079 | 322 | 6 | 4.58E-06 | 6.07E-06 | 0.001032 | 2.12E-06 | 0.007692027 | TIGIT |
| chr19 | 38565072 | 38565954 | 883 | 5 | 5.92E-06 | 6.89E-07 | 0.002388 | 2.12E-06 | 0.017834057 | RYR1 |
| chr19 | 9538200 | 9539564 | 1365 | 13 | 2.27E-06 | 0.000188 | 0.000647 | 2.12E-06 | 0.002201232 | ZNF426-DT, ZNF426 |
| chr4 | 182925404 | 182925407 | 4 | 2 | 4.22E-06 | 1.28E-06 | 0.000206 | 2.13E-06 | 0.025687956 | NA |
| chr19 | 58117368 | 58119245 | 1878 | 11 | 3.50E-05 | 0.000163 | 0.000825 | 2.13E-06 | -0.00758988 | ZSCAN18 |
| chr3 | 114383718 | 114384498 | 781 | 9 | 1.96E-06 | 4.35E-05 | 0.000764 | 2.14E-06 | 0.006333585 | ZBTB20-AS1, ZBTB20 |
| chr4 | 83485335 | 83486741 | 1407 | 8 | 4.06E-05 | 1.31E-06 | 0.00225 | 2.14E-06 | 0.001184388 | ABRAXAS1 |
| chr6 | 52360298 | 52361195 | 898 | 3 | 2.89E-06 | 2.73E-05 | 0.000162 | 2.14E-06 | 0.007807173 | NA |
| chr3 | 149812143 | 149812864 | 722 | 12 | 9.92E-06 | 4.85E-07 | 0.008048 | 2.14E-06 | 0.006164959 | RNF13, ANKUB1 |
| chr20 | 32182116 | 32182217 | 102 | 2 | 9.65E-06 | 1.28E-06 | 0.000225 | 2.14E-06 | 0.010234436 | NA |
| chr16 | 66365843 | 66366059 | 217 | 3 | 6.94E-07 | 9.89E-07 | 0.000773 | 2.15E-06 | 0.005897551 | NA |
| chr7 | 97006804 | 97007873 | 1070 | 8 | 1.60E-06 | 2.35E-06 | 0.001074 | 2.15E-06 | 0.005757794 | DLX6, DLX6-AS1 |
| chr17 | 62670032 | 62670496 | 465 | 6 | 2.01E-06 | 0.000295 | 0.000153 | 2.15E-06 | 0.004366655 | MRC2 |
| chr3 | 184005573 | 184006227 | 655 | 6 | 2.65E-07 | 2.48E-05 | 0.000111 | 2.15E-06 | 0.009997097 | ABCC5 |
| chr19 | 48898464 | 48899167 | 704 | 9 | 5.72E-07 | 1.96E-06 | 0.002158 | 2.15E-06 | 0.001337029 | TULP2 |
| chr6 | 145686459 | 145687283 | 825 | 5 | 7.05E-06 | 2.45E-05 | 0.000155 | 2.16E-06 | 0.004629636 | EPM2A |
| chr19 | 38307448 | 38307892 | 445 | 2 | 7.70E-06 | 1.27E-06 | 0.000247 | 2.16E-06 | 0.00506687 | YIF1B |
| chr14 | 71574550 | 71575238 | 689 | 2 | 1.68E-06 | 3.90E-06 | 2.63E-05 | 2.16E-06 | 0.016548724 | SIPA1L1 |
| chr5 | 141968637 | 141969538 | 902 | 15 | 1.05E-05 | 7.28E-06 | 0.009571 | 2.17E-06 | 0.005494283 | RNF14, PCDH12 |
| chr3 | 11566855 | 11568289 | 1435 | 9 | 1.56E-05 | 9.84E-06 | 0.005886 | 2.17E-06 | 0.001907416 | VGLL4 |
| chr8 | 143792671 | 143793724 | 1054 | 3 | 2.41E-05 | 9.85E-07 | 0.00066 | 2.17E-06 | 0.006490208 | SCRIB |
| chr1 | 2425399 | 2425860 | 462 | 5 | 2.20E-06 | 4.03E-05 | 5.90E-05 | 2.17E-06 | 0.014990831 | NA |
| chr20 | 51855194 | 51855771 | 578 | 2 | 4.13E-06 | 2.14E-06 | 5.58E-05 | 2.17E-06 | 0.009385958 | LINC01429 |
| chr19 | 42080260 | 42081305 | 1046 | 9 | 1.70E-06 | 3.83E-06 | 0.001087 | 2.18E-06 | 0.000554243 | ZNF574 |
| chr3 | 50502790 | 50504121 | 1332 | 15 | 2.61E-06 | 4.73E-06 | 0.003125 | 2.18E-06 | -0.00118141 | CACNA2D2 |
| chr5 | 114055642 | 114056420 | 779 | 7 | 2.88E-05 | 4.06E-07 | 0.010591 | 2.18E-06 | 0.005423375 | KCNN2 |
| chr7 | 128030517 | 128031208 | 692 | 9 | 4.81E-06 | 5.95E-06 | 0.001659 | 2.18E-06 | 0.005606167 | SND1, LRRC4 |
| chr12 | 1510194 | 1510440 | 247 | 2 | 7.12E-06 | 1.76E-06 | 7.87E-05 | 2.18E-06 | 0.014880802 | NA |
| chr7 | 231371 | 232095 | 725 | 2 | 2.32E-05 | 1.35E-06 | 0.000178 | 2.18E-06 | 0.008588188 | FAM20C |
| chr17 | 72119690 | 72120984 | 1295 | 11 | 1.75E-06 | 3.61E-05 | 0.004568 | 2.18E-06 | 0.004183849 | AC007461.1, SOX9-AS1 |
| chr4 | 85748259 | 85748568 | 310 | 2 | 4.19E-06 | 2.92E-06 | 3.73E-05 | 2.18E-06 | 0.023464545 | ARHGAP24 |
| chr2 | 9720676 | 9721243 | 568 | 3 | 9.65E-07 | 0.000343 | 7.35E-05 | 2.18E-06 | 0.002848861 | NA |
| chr8 | 80565937 | 80566099 | 163 | 3 | 4.34E-06 | 4.68E-05 | 0.000147 | 2.18E-06 | 0.00773901 | NA |
| chr20 | 44450102 | 44450455 | 354 | 4 | 5.09E-06 | 2.05E-06 | 0.000942 | 2.18E-06 | -0.00215077 | LINC01620 |
| chr7 | 7254946 | 7254983 | 38 | 3 | 1.06E-05 | 1.13E-06 | 0.000386 | 2.18E-06 | 0.000312838 | NA |
| chr11 | 105609440 | 105610782 | 1343 | 18 | 6.18E-07 | 7.44E-05 | 0.001236 | 2.18E-06 | 0.003885968 | GRIA4 |
| chr19 | 3375658 | 3376155 | 498 | 4 | 4.70E-06 | 8.97E-06 | 0.000223 | 2.18E-06 | 0.006304392 | NFIC |
| chr4 | 186708078 | 186709362 | 1285 | 13 | 1.29E-05 | 1.76E-05 | 0.002332 | 2.18E-06 | -0.00028734 | FAT1 |
| chr5 | 169302263 | 169302843 | 581 | 4 | 4.53E-06 | 1.31E-06 | 0.000939 | 2.19E-06 | 0.008953384 | NA |
| chr14 | 77180720 | 77181697 | 978 | 8 | 2.57E-08 | 3.13E-05 | 0.0003 | 2.19E-06 | 0.009105131 | AC007375.2, TMEM63C |
| chr6 | 2767883 | 2768613 | 731 | 3 | 8.71E-07 | 1.08E-05 | 1.78E-05 | 2.19E-06 | -0.02380087 | WRNIP1 |
| chr8 | 37590679 | 37591213 | 535 | 2 | 5.15E-07 | 1.28E-05 | 1.09E-05 | 2.19E-06 | 0.007580038 | NA |
| chr19 | 13134876 | 13135394 | 519 | 2 | 3.50E-06 | 2.32E-06 | 5.06E-05 | 2.19E-06 | -0.00282086 | NACC1 |
| chr14 | 105907252 | 105907281 | 30 | 2 | 9.78E-06 | 1.39E-06 | 0.00016 | 2.19E-06 | 0.029629515 | NA |
| chr3 | 23945444 | 23946600 | 1157 | 11 | 1.26E-05 | 2.99E-06 | 0.009127 | 2.20E-06 | -0.00083709 | NR1D2, NKIRAS1 |
| chr5 | 93407686 | 93408410 | 725 | 2 | 1.65E-05 | 1.50E-06 | 0.000121 | 2.20E-06 | 0.015612943 | NR2F1-AS1 |
| chr7 | 748424 | 749210 | 787 | 5 | 1.32E-06 | 2.71E-05 | 2.34E-05 | 2.20E-06 | 0.00122255 | DNAAF5 |
| chr1 | 96863278 | 96863799 | 522 | 3 | 2.88E-06 | 4.33E-06 | 5.10E-05 | 2.21E-06 | 0.010439356 | NA |
| chr5 | 168747684 | 168748339 | 656 | 5 | 4.71E-06 | 2.57E-05 | 0.000361 | 2.21E-06 | 0.009066641 | SLIT3 |
| chr11 | 10634322 | 10635190 | 869 | 2 | 4.34E-05 | 1.33E-06 | 0.000215 | 2.21E-06 | 0.002316688 | IRAG1 |
| chr14 | 103084115 | 103084581 | 467 | 2 | 9.54E-06 | 1.37E-06 | 0.000179 | 2.21E-06 | 0.011676474 | LBHD2 |
| chr21 | 14658977 | 14659230 | 254 | 3 | 5.04E-06 | 9.63E-06 | 0.000209 | 2.21E-06 | 0.004143354 | NA |
| chr15 | 93044471 | 93044907 | 437 | 3 | 1.64E-06 | 3.18E-05 | 5.73E-05 | 2.21E-06 | 0.001846368 | RGMA |
| chr9 | 125888222 | 125889663 | 1442 | 5 | 0.000119288 | 2.88E-06 | 0.001474 | 2.21E-06 | 0.009692622 | PBX3 |
| chr10 | 128366781 | 128367115 | 335 | 4 | 4.17E-06 | 6.81E-06 | 0.00042 | 2.22E-06 | -0.00189757 | NA |
| chr1 | 210252455 | 210253214 | 760 | 5 | 3.93E-06 | 2.07E-06 | 0.001025 | 2.22E-06 | 0.008409781 | NA |
| chr5 | 135139682 | 135139916 | 235 | 2 | 1.80E-06 | 6.23E-06 | 1.79E-05 | 2.22E-06 | 0.009859653 | C5orf66 |
| chr22 | 19122312 | 19123441 | 1130 | 8 | 5.29E-06 | 0.000805 | 0.001462 | 2.23E-06 | 0.000859105 | AC004471.1, DGCR2 |
| chr20 | 31712129 | 31713169 | 1041 | 4 | 1.82E-07 | 0.000135 | 6.21E-05 | 2.24E-06 | 0.008344013 | BCL2L1-AS1, BCL2L1 |
| chr12 | 57782005 | 57782911 | 907 | 9 | 8.69E-07 | 2.66E-05 | 0.000517 | 2.24E-06 | 0.0037866 | EEF1AKMT3, AC025165.3, TSFM |
| chr3 | 46574019 | 46574522 | 504 | 6 | 6.66E-06 | 4.06E-06 | 0.00031 | 2.24E-06 | 0.008147648 | LRRC2 |
| chr1 | 8563414 | 8564088 | 675 | 2 | 7.82E-06 | 1.82E-06 | 7.90E-05 | 2.24E-06 | 0.000427362 | RERE |
| chr10 | 130021589 | 130022461 | 873 | 4 | 3.62E-06 | 4.21E-06 | 0.000116 | 2.25E-06 | 0.008197296 | C10orf143 |
| chr19 | 35637201 | 35637591 | 391 | 4 | 4.93E-06 | 1.91E-05 | 0.000317 | 2.27E-06 | 0.002523529 | RBM42 |
| chr9 | 126830903 | 126831259 | 357 | 3 | 1.26E-05 | 8.77E-07 | 0.000862 | 2.27E-06 | 0.009647333 | ZBTB43 |
| chr9 | 136053980 | 136055266 | 1287 | 12 | 2.11E-05 | 3.02E-06 | 0.000592 | 2.27E-06 | 0.001724792 | NACC2 |
| chr13 | 21603536 | 21604461 | 926 | 10 | 2.49E-06 | 1.16E-05 | 0.000515 | 2.27E-06 | 0.000998317 | MICU2 |
| chr21 | 36990330 | 36990860 | 531 | 9 | 5.99E-06 | 2.73E-05 | 0.003249 | 2.27E-06 | 0.009305743 | NA |
| chr19 | 41844264 | 41845409 | 1146 | 17 | 2.99E-05 | 4.80E-07 | 0.011262 | 2.27E-06 | 0.003638933 | DMRTC2, LYPD4 |
| chr12 | 114241906 | 114241941 | 36 | 2 | 1.48E-06 | 1.98E-06 | 7.00E-05 | 2.28E-06 | 0.014734878 | NA |
| chr7 | 746228 | 747045 | 818 | 4 | 3.61E-06 | 5.47E-06 | 8.69E-05 | 2.28E-06 | 0.001941728 | DNAAF5 |
| chr2 | 232419951 | 232420744 | 794 | 3 | 2.21E-06 | 1.13E-06 | 0.000422 | 2.28E-06 | 0.012446504 | AC068134.2 |
| chr16 | 11190920 | 11191851 | 932 | 2 | 0.000184156 | 1.33E-06 | 0.000275 | 2.28E-06 | 0.009273356 | NA |
| chr11 | 9363276 | 9364433 | 1158 | 5 | 1.21E-08 | 0.001326 | 2.69E-05 | 2.28E-06 | 0.008457453 | NA |
| chr7 | 155369372 | 155369383 | 12 | 2 | 8.16E-06 | 2.91E-06 | 4.03E-05 | 2.28E-06 | 0.005457686 | NA |
| chr14 | 97789264 | 97790106 | 843 | 4 | 5.52E-06 | 9.35E-07 | 0.001301 | 2.29E-06 | 0.012893492 | NA |
| chr12 | 123670750 | 123671623 | 874 | 10 | 3.00E-06 | 9.88E-06 | 0.000244 | 2.29E-06 | -0.00027897 | TCTN2 |
| chr3 | 53196094 | 53197162 | 1069 | 4 | 0.000101687 | 6.42E-06 | 0.000887 | 2.30E-06 | 0.00574023 | AC097015.1 |
| chr13 | 45218447 | 45218490 | 44 | 2 | 2.19E-07 | 0.000905 | 3.50E-06 | 2.30E-06 | 0.015241067 | GTF2F2 |
| chr2 | 43391697 | 43392113 | 417 | 2 | 1.67E-06 | 7.74E-06 | 1.59E-05 | 2.30E-06 | 0.014093546 | THADA |
| chr3 | 194628308 | 194628418 | 111 | 2 | 7.89E-06 | 2.65E-06 | 4.56E-05 | 2.30E-06 | 0.00139658 | TMEM44 |
| chr1 | 167517741 | 167518634 | 894 | 7 | 9.08E-06 | 3.31E-06 | 0.00217 | 2.30E-06 | 0.007428179 | CD247 |
| chr20 | 52670799 | 52671764 | 966 | 3 | 0.000156342 | 2.94E-06 | 0.000331 | 2.30E-06 | -0.00350855 | AL031674.1 |
| chr14 | 53571718 | 53572222 | 505 | 2 | 4.63E-06 | 1.55E-06 | 0.000128 | 2.30E-06 | 0.014149398 | AL365295.1 |
| chr19 | 55362423 | 55363332 | 910 | 7 | 6.78E-05 | 6.22E-06 | 0.000293 | 2.30E-06 | 0.003799315 | FAM71E2 |
| chr8 | 22097304 | 22097752 | 449 | 2 | 1.94E-06 | 5.26E-06 | 2.15E-05 | 2.30E-06 | 0.004114233 | FAM160B2 |
| chr2 | 200585783 | 200586580 | 798 | 12 | 7.79E-08 | 6.63E-05 | 0.000759 | 2.30E-06 | 0.002807093 | AOX1 |
| chr8 | 144529609 | 144530590 | 982 | 5 | 2.36E-06 | 1.07E-06 | 0.000539 | 2.31E-06 | -0.01628159 | ARHGAP39 |
| chr8 | 109639767 | 109639771 | 5 | 2 | 9.92E-06 | 1.34E-06 | 0.000279 | 2.31E-06 | 0.015617878 | SYBU |
| chr3 | 126025826 | 126026438 | 613 | 3 | 1.60E-05 | 1.24E-06 | 0.000292 | 2.31E-06 | 0.009559137 | SLC41A3 |
| chr6 | 119149159 | 119150116 | 958 | 6 | 1.70E-05 | 5.87E-06 | 0.000129 | 2.31E-06 | 0.002977401 | AL137009.1, FAM184A |
| chr19 | 30444270 | 30445198 | 929 | 4 | 1.12E-07 | 7.34E-05 | 4.09E-05 | 2.31E-06 | 0.000239381 | ZNF536 |
| chr11 | 64502752 | 64502866 | 115 | 2 | 7.04E-06 | 2.05E-06 | 6.84E-05 | 2.31E-06 | 0.009262039 | AP005273.1 |
| chr6 | 93259304 | 93259637 | 334 | 3 | 4.15E-06 | 4.03E-06 | 0.000176 | 2.31E-06 | -0.00650119 | EPHA7 |
| chr11 | 70336624 | 70336907 | 284 | 2 | 1.30E-06 | 3.25E-06 | 3.57E-05 | 2.31E-06 | 0.002783692 | PPFIA1, AP002336.2 |
| chr9 | 129881845 | 129881901 | 57 | 2 | 4.04E-06 | 1.44E-06 | 0.000177 | 2.31E-06 | 0.009276293 | NA |
| chr2 | 96895407 | 96895992 | 586 | 5 | 1.16E-05 | 9.02E-07 | 0.000798 | 2.31E-06 | 0.005950076 | FAM178B |
| chr21 | 31659079 | 31659855 | 777 | 12 | 4.27E-06 | 2.57E-06 | 0.000103 | 2.32E-06 | 0.000801349 | SOD1, AP000253.1 |
| chr10 | 95513218 | 95513237 | 20 | 2 | 1.07E-05 | 1.35E-06 | 0.000277 | 2.32E-06 | -0.00722949 | SORBS1 |
| chr21 | 44932530 | 44932902 | 373 | 2 | 5.14E-08 | 0.000177 | 4.33E-06 | 2.32E-06 | 0.006079235 | LINC01547 |
| chr1 | 13892986 | 13893770 | 785 | 7 | 9.17E-07 | 1.34E-05 | 0.001256 | 2.32E-06 | 0.004155191 | KAZN |
| chr22 | 25942639 | 25942884 | 246 | 2 | 1.91E-06 | 1.19E-05 | 1.21E-05 | 2.32E-06 | 0.012978407 | MYO18B |
| chr4 | 171812880 | 171813692 | 813 | 8 | 5.86E-06 | 4.48E-06 | 0.001124 | 2.32E-06 | 0.005704644 | GALNTL6 |
| chr13 | 112451583 | 112452159 | 577 | 6 | 9.68E-07 | 2.00E-06 | 0.000806 | 2.32E-06 | 0.002756164 | NA |
| chr17 | 41351765 | 41353048 | 1284 | 4 | 1.52E-05 | 0.00013 | 0.00018 | 2.33E-06 | 0.009769368 | NA |
| chr6 | 503773 | 504774 | 1002 | 7 | 2.68E-06 | 5.70E-06 | 0.000521 | 2.33E-06 | 0.000779981 | EXOC2 |
| chr15 | 77069831 | 77070511 | 681 | 4 | 3.98E-07 | 7.06E-05 | 2.17E-05 | 2.33E-06 | 0.004898671 | TSPAN3 |
| chr9 | 4155366 | 4155739 | 374 | 3 | 3.57E-06 | 3.82E-06 | 4.74E-05 | 2.33E-06 | -0.00046632 | GLIS3 |
| chr2 | 76446384 | 76447332 | 949 | 2 | 4.21E-05 | 1.61E-06 | 0.00012 | 2.33E-06 | 0.017790923 | NA |
| chr15 | 26666484 | 26666723 | 240 | 3 | 7.49E-06 | 8.07E-07 | 0.001569 | 2.33E-06 | 0.012726809 | GABRB3 |
| chr1 | 32217519 | 32217889 | 371 | 5 | 6.54E-06 | 2.06E-06 | 0.001252 | 2.33E-06 | -0.00027812 | TMEM234 |
| chr7 | 150340899 | 150342016 | 1118 | 5 | 1.65E-06 | 3.46E-05 | 0.002605 | 2.33E-06 | 0.02746019 | AC005586.1, RARRES2 |
| chr9 | 37040360 | 37040789 | 430 | 3 | 1.23E-05 | 1.04E-06 | 0.000699 | 2.33E-06 | 0.015998209 | NA |
| chr2 | 200525571 | 200526627 | 1057 | 12 | 2.42E-06 | 2.88E-05 | 0.002572 | 2.34E-06 | -0.00409605 | SGO2 |
| chr1 | 94541670 | 94542561 | 892 | 13 | 2.44E-06 | 8.69E-06 | 0.000292 | 2.35E-06 | -1.76E-05 | F3 |
| chr12 | 26402902 | 26403281 | 380 | 3 | 3.52E-06 | 3.00E-06 | 7.16E-05 | 2.35E-06 | 0.000100526 | AC024145.1, ITPR2 |
| chr12 | 4909063 | 4909549 | 487 | 4 | 3.74E-06 | 4.78E-06 | 0.000454 | 2.35E-06 | 0.005591252 | AC006063.4 |
| chr1 | 230332422 | 230333584 | 1163 | 7 | 9.84E-06 | 1.40E-06 | 0.006075 | 2.35E-06 | -0.02313951 | PGBD5 |
| chr11 | 10856981 | 10857786 | 806 | 5 | 1.34E-06 | 3.62E-06 | 0.000179 | 2.36E-06 | -0.0026337 | ZBED5 |
| chr3 | 35678990 | 35679906 | 917 | 11 | 2.07E-06 | 4.74E-06 | 0.000963 | 2.36E-06 | -0.00049843 | ARPP21 |
| chr2 | 173306857 | 173307367 | 511 | 3 | 1.34E-05 | 8.80E-07 | 0.001114 | 2.36E-06 | 0.004550695 | NA |
| chr7 | 151992521 | 151993665 | 1145 | 3 | 3.76E-05 | 1.30E-06 | 0.000511 | 2.37E-06 | 0.014170904 | GALNTL5 |
| chr17 | 36535024 | 36536188 | 1165 | 18 | 4.09E-06 | 4.18E-06 | 0.004761 | 2.37E-06 | 0.00057339 | PIGW, MYO19 |
| chr3 | 32503050 | 32503981 | 932 | 9 | 9.23E-07 | 8.73E-06 | 0.006012 | 2.37E-06 | 0.000630649 | NA |
| chr17 | 3968009 | 3969275 | 1267 | 4 | 0.000119634 | 2.25E-06 | 0.000681 | 2.37E-06 | 0.006143383 | NA |
| chr1 | 204370358 | 204371178 | 821 | 3 | 4.23E-05 | 8.76E-07 | 0.00124 | 2.37E-06 | 0.001254486 | PLEKHA6 |
| chr1 | 201260058 | 201260119 | 62 | 2 | 8.20E-06 | 1.40E-06 | 0.000248 | 2.37E-06 | 0.009617771 | NA |
| chr15 | 40238753 | 40239979 | 1227 | 18 | 7.05E-05 | 9.47E-07 | 0.011415 | 2.37E-06 | 0.002367054 | PAK6, BUB1B-PAK6 |
| chr9 | 129674159 | 129674211 | 53 | 2 | 5.57E-06 | 1.89E-06 | 8.50E-05 | 2.37E-06 | 0.007509367 | PRRX2 |
| chr4 | 2436500 | 2437058 | 559 | 4 | 1.36E-07 | 3.41E-05 | 2.96E-05 | 2.38E-06 | 0.007752784 | CFAP99 |
| chr2 | 26483285 | 26483562 | 278 | 2 | 7.74E-07 | 2.89E-05 | 7.74E-06 | 2.38E-06 | 0.003656811 | OTOF |
| chr16 | 23175338 | 23175345 | 8 | 2 | 6.05E-06 | 9.44E-06 | 1.46E-05 | 2.39E-06 | 0.023551249 | AC099482.2 |
| chr11 | 63016372 | 63016967 | 596 | 2 | 9.38E-08 | 1.98E-06 | 7.90E-05 | 2.39E-06 | 0.016924839 | NA |
| chr9 | 107122121 | 107122545 | 425 | 2 | 5.35E-07 | 1.84E-05 | 9.69E-06 | 2.40E-06 | 0.004167681 | NA |
| chr18 | 76689261 | 76689714 | 454 | 2 | 7.41E-06 | 1.43E-06 | 0.000235 | 2.40E-06 | 0.022369688 | LINC01879 |
| chr6 | 125154504 | 125155261 | 758 | 5 | 3.28E-07 | 1.15E-05 | 0.000728 | 2.40E-06 | 0.010417678 | TPD52L1 |
| chr3 | 124032070 | 124033170 | 1101 | 4 | 4.77E-07 | 1.54E-05 | 9.28E-05 | 2.40E-06 | 0.006730363 | NA |
| chr1 | 58784139 | 58785294 | 1156 | 14 | 1.40E-05 | 0.000104 | 0.000787 | 2.40E-06 | 0.001727384 | LINC01135 |
| chr16 | 85011512 | 85012549 | 1038 | 11 | 7.29E-06 | 1.20E-05 | 0.000643 | 2.41E-06 | 7.72E-05 | ZDHHC7 |
| chr6 | 22220222 | 22220596 | 375 | 5 | 3.99E-06 | 2.20E-06 | 0.00083 | 2.42E-06 | -0.00236137 | CASC15 |
| chr6 | 44155877 | 44156597 | 721 | 2 | 7.62E-06 | 1.43E-06 | 0.000249 | 2.42E-06 | 0.008344733 | NA |
| chr1 | 22799720 | 22800652 | 933 | 4 | 2.81E-05 | 2.14E-06 | 0.001477 | 2.42E-06 | 0.007451132 | EPHB2 |
| chr6 | 41545428 | 41546694 | 1267 | 9 | 7.67E-07 | 2.38E-05 | 0.000881 | 2.42E-06 | 0.001711519 | FOXP4, FOXP4-AS1 |
| chr17 | 75105839 | 75106050 | 212 | 3 | 6.53E-06 | 5.29E-06 | 4.44E-05 | 2.42E-06 | 0.007794358 | SLC16A5 |
| chr10 | 80355284 | 80356313 | 1030 | 5 | 1.41E-05 | 3.35E-05 | 0.000286 | 2.43E-06 | -0.0008942 | DYDC2, DYDC1 |
| chr7 | 45174902 | 45175172 | 271 | 2 | 2.20E-06 | 1.70E-06 | 0.000119 | 2.44E-06 | 0.008536372 | RAMP3 |
| chr6 | 14397099 | 14397609 | 511 | 3 | 3.14E-06 | 1.22E-05 | 2.07E-05 | 2.44E-06 | 0.021552516 | AL080313.1 |
| chr11 | 2861643 | 2862682 | 1040 | 5 | 3.01E-05 | 4.90E-06 | 0.000136 | 2.44E-06 | 0.006922585 | NA |
| chr1 | 30689513 | 30689839 | 327 | 2 | 4.43E-06 | 2.42E-06 | 5.87E-05 | 2.44E-06 | 0.0122399 | NA |
| chr18 | 47772826 | 47773204 | 379 | 4 | 7.46E-06 | 4.00E-06 | 0.000395 | 2.44E-06 | 0.013626588 | NA |
| chr10 | 27251884 | 27252688 | 805 | 6 | 2.99E-05 | 2.72E-05 | 0.001481 | 2.44E-06 | -0.0122132 | LRRC37A6P |
| chr22 | 41512896 | 41513275 | 380 | 2 | 1.82E-06 | 2.63E-06 | 5.20E-05 | 2.44E-06 | 0.006690841 | ACO2 |
| chr18 | 13219223 | 13219730 | 508 | 4 | 2.52E-07 | 8.59E-06 | 3.91E-05 | 2.45E-06 | 0.014925664 | LDLRAD4 |

| | | | | | | | | | | |
|---|---|---|---|---|---|---|---|---|---|---|
| chr6 | 135851415 | 135851947 | 533 | 6 | 2.56E-06 | 4.85E-06 | 0.000107 | 2.45E-06 | 0.006276232 | PDE7B |
| chr7 | 130149818 | 130150069 | 252 | 2 | 6.59E-06 | 1.45E-06 | 0.000255 | 2.45E-06 | 0.011338937 | NA |
| chr7 | 47380555 | 47381364 | 810 | 3 | 4.46E-05 | 5.25E-05 | 0.000166 | 2.45E-06 | 0.008773417 | TNS3 |
| chr2 | 201781379 | 201781726 | 348 | 5 | 6.89E-07 | 9.17E-06 | 7.30E-05 | 2.45E-06 | 0.002695173 | NA |
| chr7 | 50565297 | 50566076 | 780 | 6 | 1.95E-05 | 8.70E-07 | 0.002243 | 2.45E-06 | 0.010774654 | DDC |
| chr20 | 11687479 | 11688256 | 778 | 4 | 2.78E-05 | 1.04E-06 | 0.001233 | 2.45E-06 | 0.005505606 | NA |
| chr16 | 67517922 | 67518810 | 889 | 3 | 3.66E-07 | 0.000111 | 1.31E-05 | 2.45E-06 | 0.008689391 | RIPOR1, AC027682.6 |
| chr6 | 31664394 | 31665968 | 1575 | 46 | 2.35E-06 | 3.65E-06 | 0.015273 | 2.47E-06 | 0.000548929 | CSNK2B, GPANK1 |
| chr14 | 23635532 | 23635808 | 277 | 2 | 6.64E-06 | 2.54E-06 | 5.59E-05 | 2.47E-06 | 0.011443114 | DHRS2 |
| chr15 | 90872302 | 90872888 | 587 | 8 | 1.69E-06 | 4.38E-06 | 0.003974 | 2.47E-06 | 0.009009084 | FURIN |
| chr22 | 37822677 | 37823393 | 717 | 7 | 1.90E-06 | 3.35E-05 | 0.000119 | 2.47E-06 | 0.005775121 | GALR3 |
| chr15 | 88799615 | 88800495 | 881 | 2 | 1.79E-05 | 1.95E-06 | 8.79E-05 | 2.47E-06 | 0.007859184 | NA |
| chr2 | 135814067 | 135814269 | 203 | 3 | 5.77E-06 | 1.19E-05 | 0.000105 | 2.47E-06 | 0.002957516 | LCT |
| chr1 | 221487932 | 221488341 | 410 | 3 | 4.42E-07 | 0.000189 | 4.03E-06 | 2.47E-06 | 0.006130739 | NA |
| chr18 | 32090365 | 32090433 | 69 | 3 | 2.27E-06 | 0.000238 | 3.21E-05 | 2.47E-06 | 0.00472178 | AC011825.3 |
| chr14 | 105477534 | 105478950 | 1417 | 7 | 6.88E-05 | 2.53E-07 | 0.011557 | 2.47E-06 | 0.022837232 | CRIP2 |
| chr1 | 243421366 | 243421622 | 257 | 4 | 5.46E-06 | 3.77E-06 | 9.07E-05 | 2.47E-06 | 0.010726362 | SDCCAG8 |
| chr2 | 151049548 | 151049864 | 317 | 2 | 7.52E-07 | 9.74E-06 | 1.48E-05 | 2.47E-06 | 0.010418538 | NA |
| chr7 | 150993430 | 150993881 | 452 | 4 | 1.60E-07 | 0.003294 | 1.61E-05 | 2.48E-06 | 0.0037929 | NOS3 |
| chr13 | 52377978 | 52378184 | 207 | 2 | 4.48E-06 | 8.77E-06 | 1.59E-05 | 2.48E-06 | 0.001459729 | THSD1 |
| chr11 | 128691780 | 128692513 | 734 | 8 | 5.04E-06 | 1.35E-06 | 0.001836 | 2.48E-06 | 0.006272326 | FLI1, SENCR |
| chr3 | 10766422 | 10767196 | 775 | 3 | 1.72E-05 | 1.62E-06 | 0.000214 | 2.48E-06 | 0.010059423 | NA |
| chr1 | 28886313 | 28887351 | 1039 | 12 | 6.17E-07 | 6.74E-05 | 0.001357 | 2.49E-06 | 0.004280741 | EPB41 |
| chr22 | 46245841 | 46246237 | 397 | 2 | 7.00E-07 | 2.61E-05 | 8.47E-06 | 2.49E-06 | 0.006990565 | CDPF1 |
| chr11 | 70269939 | 70270954 | 1016 | 7 | 8.20E-06 | 5.87E-06 | 0.000421 | 2.50E-06 | -7.76E-05 | PPFIA1 |
| chr7 | 98870415 | 98870963 | 549 | 5 | 2.08E-06 | 1.24E-05 | 0.000311 | 2.51E-06 | 0.000624942 | TMEM130 |
| chr11 | 85757994 | 85758876 | 883 | 6 | 1.16E-06 | 2.97E-05 | 0.000959 | 2.51E-06 | 0.006297237 | SYTL2 |
| chr1 | 39899827 | 39900438 | 612 | 2 | 4.28E-08 | 5.37E-05 | 6.41E-06 | 2.51E-06 | 0.011655358 | MYCL |
| chr6 | 28906925 | 28907593 | 669 | 4 | 2.70E-06 | 2.04E-06 | 0.001196 | 2.51E-06 | 0.011423087 | TRIM27 |
| chr10 | 122454011 | 122454635 | 625 | 3 | 3.88E-06 | 3.21E-05 | 6.62E-05 | 2.52E-06 | 0.007771067 | BX842242.1 |
| chr1 | 52365608 | 52367322 | 1715 | 11 | 1.47E-06 | 2.76E-06 | 0.000178 | 2.52E-06 | 3.67E-05 | AL513218.2, CC2D1B |
| chr1 | 18100640 | 18100658 | 19 | 2 | 9.92E-06 | 1.61E-06 | 0.000167 | 2.52E-06 | 0.003035105 | NA |
| chr7 | 123635712 | 123636103 | 392 | 2 | 4.91E-07 | 0.000194 | 4.59E-06 | 2.52E-06 | 0.020112982 | ASB15 |
| chr19 | 55433497 | 55433866 | 370 | 3 | 1.08E-06 | 8.77E-06 | 4.70E-05 | 2.52E-06 | -0.01719532 | SHISA7 |
| chr15 | 65337415 | 65337418 | 4 | 2 | 7.74E-06 | 5.52E-06 | 2.36E-05 | 2.52E-06 | 0.006846815 | IGDCC3 |
| chr11 | 61688599 | 61688915 | 317 | 3 | 1.03E-05 | 3.69E-06 | 0.000269 | 2.52E-06 | 0.01318455 | DAGLA |
| chr1 | 91765793 | 91766592 | 800 | 2 | 1.00E-07 | 1.97E-05 | 9.88E-06 | 2.52E-06 | 0.044339898 | TGFBR3 |
| chr16 | 524452 | 524506 | 55 | 2 | 4.40E-06 | 1.49E-06 | 0.000267 | 2.53E-06 | 0.012396077 | NA |
| chr2 | 176637546 | 176638310 | 765 | 7 | 1.92E-08 | 0.000153 | 4.01E-06 | 2.53E-06 | 0.002225652 | LINC01117, LINC01116 |
| chr6 | 33620354 | 33621641 | 1288 | 14 | 8.25E-06 | 5.88E-05 | 0.006427 | 2.53E-06 | 0.000802395 | ITPR3 |
| chr11 | 62706187 | 62706646 | 460 | 10 | 2.45E-06 | 3.61E-05 | 0.002216 | 2.54E-06 | -0.00065844 | BSCL2, HNRNPUL2-BSCL2 |
| chr2 | 43223826 | 43224729 | 904 | 7 | 1.68E-06 | 7.16E-06 | 0.00018 | 2.54E-06 | 0.001931657 | ZFP36L2 |
| chr5 | 41509974 | 41511048 | 1075 | 11 | 3.02E-06 | 3.73E-06 | 0.002969 | 2.54E-06 | 0.008042501 | PLCXD3 |
| chr10 | 509605 | 510752 | 1148 | 7 | 1.08E-05 | 3.19E-06 | 0.000859 | 2.54E-06 | -0.00060556 | DIP2C |
| chr10 | 94030759 | 94031117 | 359 | 2 | 7.35E-07 | 4.34E-05 | 7.00E-06 | 2.55E-06 | 0.000745444 | PLCE1 |
| chr13 | 113069981 | 113070234 | 254 | 3 | 2.88E-06 | 2.06E-05 | 1.21E-05 | 2.55E-06 | 0.002504494 | MCF2L |
| chr12 | 43728925 | 43729766 | 842 | 2 | 3.37E-05 | 1.75E-06 | 0.000128 | 2.55E-06 | 0.013789938 | PUS7L |
| chr2 | 69914573 | 69915391 | 819 | 10 | 2.99E-06 | 4.63E-05 | 8.87E-05 | 2.55E-06 | 0.000542792 | MXD1 |
| chr11 | 111549545 | 111549660 | 116 | 2 | 6.06E-06 | 8.72E-06 | 1.66E-05 | 2.55E-06 | 0.007153079 | LAYN |
| chr12 | 132769170 | 132769720 | 551 | 6 | 1.83E-06 | 3.11E-05 | 0.000179 | 2.55E-06 | 0.006445066 | GOLGA3 |
| chr2 | 6445207 | 6445489 | 283 | 2 | 4.41E-06 | 5.24E-06 | 2.52E-05 | 2.55E-06 | 0.00388156 | NA |
| chr5 | 141806289 | 141806345 | 57 | 2 | 9.84E-06 | 2.64E-06 | 5.66E-05 | 2.56E-06 | 0.02105127 | NA |
| chr10 | 97450238 | 97450839 | 602 | 4 | 1.07E-06 | 6.49E-06 | 0.000119 | 2.56E-06 | 0.001573071 | ZDHHC16 |
| chr19 | 42243675 | 42244410 | 736 | 2 | 4.23E-09 | 1.94E-06 | 9.87E-05 | 2.56E-06 | 0.012919404 | AC006486.2, AC006486.1 |
| chr1 | 174964341 | 174964795 | 455 | 4 | 5.61E-06 | 1.20E-06 | 0.00057 | 2.56E-06 | 0.007845298 | RABGAP1L |
| chr19 | 54159481 | 54160839 | 1359 | 15 | 5.70E-06 | 3.83E-05 | 0.002 | 2.56E-06 | 0.000132515 | LENG1, TMC4 |
| chr22 | 20224991 | 20225337 | 347 | 2 | 6.28E-06 | 2.49E-06 | 6.21E-05 | 2.56E-06 | 0.019166755 | NA |
| chr17 | 82521463 | 82522151 | 689 | 4 | 7.18E-06 | 8.10E-07 | 0.002058 | 2.56E-06 | 0.004091551 | FOXK2 |
| chr13 | 111586326 | 111586999 | 674 | 5 | 1.82E-05 | 1.26E-06 | 0.001683 | 2.56E-06 | 0.001892211 | NA |
| chr8 | 103212957 | 103213162 | 206 | 3 | 1.07E-05 | 1.48E-06 | 0.000507 | 2.56E-06 | 0.002662299 | BAALC, BAALC-AS1 |
| chr11 | 94767446 | 94768470 | 1025 | 12 | 1.68E-06 | 1.11E-05 | 0.001043 | 2.56E-06 | 0.00331935 | AMOTL1 |
| chr6 | 166497521 | 166498279 | 759 | 3 | 1.21E-05 | 1.22E-06 | 0.000462 | 2.56E-06 | 0.004339248 | RPS6KA2 |
| chr19 | 8363260 | 8364014 | 755 | 8 | 3.23E-06 | 1.80E-06 | 0.001129 | 2.57E-06 | 0.007211733 | ANGPTL4 |
| chr5 | 171446091 | 171446151 | 61 | 3 | 8.82E-06 | 7.03E-06 | 5.95E-05 | 2.57E-06 | 0.003448879 | FGF18 |
| chr17 | 80991447 | 80992395 | 949 | 13 | 1.63E-06 | 0.00019 | 0.000461 | 2.57E-06 | 0.001596427 | CHMP6 |
| chr1 | 64895206 | 64896745 | 1540 | 5 | 0.000102744 | 4.16E-05 | 0.000118 | 2.57E-06 | 7.44E-05 | JAK1 |
| chr4 | 189035170 | 189035783 | 614 | 4 | 5.91E-07 | 3.89E-05 | 1.95E-05 | 2.58E-06 | -0.00364223 | NA |
| chr4 | 186500851 | 186501189 | 339 | 7 | 1.39E-05 | 3.69E-07 | 0.01153 | 2.58E-06 | -0.02385488 | RNU6-1055P, F11-AS1, AC018709.1 |
| chr11 | 11243800 | 11244293 | 494 | 5 | 1.22E-05 | 1.14E-06 | 0.001355 | 2.58E-06 | 0.009944123 | NA |
| chr14 | 100834926 | 100836170 | 1245 | 6 | 4.21E-07 | 1.42E-05 | 5.76E-05 | 2.58E-06 | 0.004268201 | MEG3, AL117190.2 |
| chr11 | 75774782 | 75775503 | 722 | 4 | 6.45E-07 | 5.00E-06 | 0.000102 | 2.58E-06 | 0.004960538 | DGAT2 |
| chr17 | 68971982 | 68972214 | 233 | 2 | 1.38E-06 | 0.00041 | 4.19E-06 | 2.58E-06 | 0.018615354 | ABCA9-AS1 |
| chr18 | 46466355 | 46466746 | 392 | 3 | 1.76E-06 | 4.39E-05 | 7.21E-05 | 2.59E-06 | 0.017641071 | NA |
| chr11 | 101912852 | 101913674 | 823 | 2 | 1.75E-05 | 1.54E-06 | 0.000263 | 2.59E-06 | 0.012380938 | ANGPTL5 |
| chr11 | 45805076 | 45806212 | 1137 | 8 | 1.07E-06 | 1.89E-06 | 0.003061 | 2.60E-06 | 0.007044975 | SLC35C1 |
| chr5 | 1076051 | 1077116 | 1066 | 8 | 1.17E-05 | 6.52E-06 | 0.000258 | 2.60E-06 | 0.002275786 | SLC12A7 |
| chr7 | 31513335 | 31513752 | 418 | 2 | 3.73E-05 | 1.59E-06 | 0.000219 | 2.61E-06 | 0.004268631 | NA |
| chr1 | 226363999 | 226364453 | 455 | 3 | 3.01E-06 | 5.57E-06 | 0.000282 | 2.61E-06 | 0.003692511 | PARP1 |
| chr14 | 102910673 | 102911806 | 1134 | 4 | 0.000234751 | 0.000206 | 0.000106 | 2.61E-06 | -0.00257879 | TRAF3 |
| chr10 | 112686823 | 112687017 | 195 | 2 | 2.96E-06 | 6.30E-06 | 2.20E-05 | 2.62E-06 | 0.005859816 | VTI1A |
| chr4 | 88697173 | 88698488 | 1316 | 18 | 1.14E-06 | 0.000284 | 0.000251 | 2.62E-06 | 0.003759243 | AC098582.1, HERC3, NAP1L5 |
| chr17 | 58540154 | 58541044 | 891 | 5 | 4.16E-06 | 2.40E-06 | 0.00035 | 2.63E-06 | -0.00180022 | SEPTIN4-AS1, SEPTIN4 |
| chr17 | 63445910 | 63447352 | 1443 | 8 | 0.000109143 | 7.92E-06 | 0.001635 | 2.64E-06 | 0.00147191 | CYB561 |
| chr10 | 98251812 | 98251967 | 156 | 4 | 3.31E-06 | 8.27E-06 | 0.0003 | 2.64E-06 | 0.002341505 | LOXL4 |
| chr16 | 67992971 | 67993936 | 966 | 8 | 8.19E-06 | 2.47E-05 | 0.001545 | 2.64E-06 | 0.002013282 | DUS2, DPEP2 |
| chr8 | 43122231 | 43123006 | 776 | 2 | 2.52E-05 | 1.82E-06 | 0.000131 | 2.64E-06 | 0.003848276 | POMK |
| chr15 | 64151101 | 64152938 | 1838 | 14 | 0.000110243 | 2.62E-06 | 0.012558 | 2.65E-06 | 0.001265426 | SNX22 |
| chr1 | 43198892 | 43199460 | 569 | 2 | 4.61E-06 | 2.62E-06 | 6.09E-05 | 2.65E-06 | 0.008113642 | CFAP57, EBNA1BP2 |
| chr4 | 102431997 | 102432343 | 347 | 2 | 7.38E-06 | 1.58E-06 | 0.000263 | 2.65E-06 | -0.0023749 | AC098487.1 |
| chr16 | 70429262 | 70429315 | 54 | 2 | 3.89E-06 | 1.86E-06 | 0.000123 | 2.66E-06 | 0.012129823 | ST3GAL2 |
| chr1 | 217275472 | 217275520 | 49 | 2 | 8.95E-06 | 1.56E-06 | 0.000279 | 2.66E-06 | 0.001011446 | NA |
| chr20 | 31998159 | 31999096 | 938 | 4 | 0.000104252 | 3.57E-06 | 0.000315 | 2.66E-06 | 0.007397346 | XKR7 |
| chr17 | 48722278 | 48723676 | 1399 | 19 | 2.41E-05 | 1.93E-05 | 0.005092 | 2.66E-06 | 0.003847286 | PRAC2, AC091179.3, PRAC1 |
| chr4 | 54315345 | 54315871 | 527 | 3 | 1.68E-05 | 1.06E-06 | 0.000864 | 2.67E-06 | 0.012965944 | NA |
| chr14 | 31106760 | 31107566 | 807 | 4 | 2.85E-05 | 6.36E-07 | 0.003527 | 2.67E-06 | 0.008192822 | AP4S1, HECTD1 |
| chr7 | 6614791 | 6615601 | 811 | 6 | 2.02E-05 | 4.71E-07 | 0.008163 | 2.67E-06 | 0.00762525 | NA |
| chr6 | 149456848 | 149457639 | 792 | 3 | 8.62E-06 | 5.10E-05 | 2.85E-05 | 2.67E-06 | -0.00119292 | ZC3H12D |
| chr10 | 97845671 | 97846138 | 468 | 2 | 1.53E-05 | 1.59E-06 | 0.000265 | 2.67E-06 | 0.003105547 | LINC00866 |
| chr22 | 46625327 | 46626270 | 944 | 6 | 2.01E-06 | 1.14E-05 | 0.000102 | 2.67E-06 | 0.005985373 | GRAMD4 |
| chr10 | 71246889 | 71246978 | 90 | 2 | 8.05E-06 | 1.56E-06 | 0.000301 | 2.67E-06 | 0.018645603 | UNC5B |
| chr12 | 54018722 | 54019774 | 1053 | 9 | 1.94E-07 | 4.57E-05 | 4.72E-05 | 2.67E-06 | 0.009880377 | AC012531.3, HOXC6, HOXC4, AC012531.2 |
| chr3 | 123322038 | 123322109 | 72 | 2 | 3.45E-06 | 3.08E-05 | 8.42E-06 | 2.67E-06 | 0.006251672 | ADCY5 |
| chr10 | 78543725 | 78544085 | 361 | 2 | 1.18E-06 | 4.39E-05 | 7.31E-06 | 2.68E-06 | 0.001242924 | AC012560.1, LINC00595, AL583852.1 |
| chr3 | 169764926 | 169765507 | 582 | 9 | 1.65E-06 | 1.89E-05 | 0.000858 | 2.68E-06 | 0.005308759 | TERC, Telomerase-vert |
| chr15 | 90191244 | 90192190 | 947 | 5 | 1.07E-05 | 7.27E-07 | 0.001849 | 2.68E-06 | 0.012192038 | SEMA4B |
| chr20 | 1326608 | 1327252 | 645 | 4 | 7.66E-07 | 2.01E-05 | 1.49E-05 | 2.69E-06 | -0.00311835 | SDCBP2-AS1, SDCBP2, AL136531.2 |
| chr1 | 94010907 | 94011383 | 477 | 3 | 3.28E-06 | 2.33E-05 | 1.93E-05 | 2.69E-06 | 0.006010923 | ABCA4 |
| chr8 | 1964920 | 1965258 | 339 | 5 | 3.09E-06 | 2.67E-05 | 0.000193 | 2.69E-06 | 0.0052332 | NA |
| chr7 | 137454352 | 137455132 | 781 | 5 | 1.70E-05 | 5.15E-06 | 0.00059 | 2.70E-06 | 0.009381633 | DGKI |
| chr8 | 30227647 | 30228028 | 382 | 3 | 1.06E-05 | 1.62E-06 | 0.000212 | 2.70E-06 | -0.00311784 | NA |
| chr2 | 70341082 | 70341215 | 134 | 2 | 1.25E-05 | 1.62E-06 | 0.000252 | 2.71E-06 | -0.00404867 | NA |
| chr18 | 46173507 | 46174083 | 577 | 11 | 1.62E-05 | 5.19E-06 | 0.005053 | 2.71E-06 | 0.001650831 | C18orf25 |
| chr17 | 17515377 | 17515929 | 553 | 3 | 1.84E-05 | 9.56E-07 | 0.001551 | 2.71E-06 | 0.003865405 | PEMT |
| chr3 | 49459320 | 49459600 | 281 | 2 | 2.00E-06 | 2.23E-05 | 1.00E-05 | 2.72E-06 | 0.008615935 | NA |
| chr11 | 66958349 | 66958996 | 648 | 9 | 2.77E-06 | 6.55E-06 | 0.00103 | 2.72E-06 | -0.00068336 | PC |
| chr7 | 128920334 | 128920770 | 437 | 2 | 7.78E-06 | 1.89E-06 | 0.000128 | 2.72E-06 | 0.007421711 | NA |
| chr10 | 100997220 | 100998646 | 1427 | 6 | 1.25E-07 | 2.93E-06 | 0.000341 | 2.72E-06 | 0.005233408 | LZTS2 |
| chr10 | 128958959 | 128960121 | 1163 | 7 | 4.15E-06 | 1.05E-05 | 0.000195 | 2.72E-06 | 0.000750599 | AL355537.1 |
| chr4 | 4407043 | 4407751 | 709 | 2 | 2.63E-05 | 1.71E-06 | 0.000189 | 2.72E-06 | 0.016654263 | NSG1 |
| chr3 | 44916002 | 44916063 | 62 | 3 | 4.12E-06 | 4.89E-05 | 2.41E-05 | 2.72E-06 | 0.008145757 | ZDHHC3 |
| chr7 | 90212121 | 90212729 | 609 | 5 | 1.75E-06 | 1.26E-06 | 0.001262 | 2.73E-06 | 0.012784848 | STEAP2 |
| chr12 | 89350011 | 89351200 | 1190 | 11 | 2.74E-07 | 7.97E-06 | 0.001542 | 2.73E-06 | 0.007234822 | AC024909.1, DUSP6 |
| chr6 | 152544773 | 152544821 | 49 | 2 | 1.20E-05 | 1.99E-06 | 0.000112 | 2.73E-06 | -0.00084916 | SYNE1 |
| chr19 | 46353927 | 46354058 | 132 | 2 | 6.38E-06 | 9.21E-06 | 1.74E-05 | 2.74E-06 | 0.004793405 | PPP5C |
| chr4 | 3834686 | 3835492 | 807 | 2 | 5.34E-05 | 1.63E-06 | 0.000266 | 2.74E-06 | 0.013841024 | NA |
| chr22 | 37538871 | 37539273 | 403 | 3 | 1.01E-05 | 4.93E-06 | 0.000119 | 2.74E-06 | 0.004108184 | NA |
| chr15 | 36887069 | 36888242 | 1174 | 6 | 8.09E-06 | 1.89E-05 | 0.000147 | 2.74E-06 | 0.009593597 | NA |
| chr12 | 121452360 | 121452857 | 498 | 5 | 1.46E-06 | 1.86E-05 | 8.79E-05 | 2.75E-06 | 0.006251599 | KDM2B |
| chr15 | 81426922 | 81427891 | 970 | 2 | 3.77E-05 | 1.94E-06 | 0.000125 | 2.76E-06 | -0.01232467 | TMC3-AS1, AC060809.1 |
| chr4 | 151227058 | 151228044 | 987 | 5 | 6.86E-08 | 0.035778 | 1.36E-06 | 2.76E-06 | 0.008883313 | SH3D19 |
| chr15 | 91854239 | 91855496 | 1258 | 8 | 4.63E-06 | 1.82E-05 | 0.003683 | 2.76E-06 | 0.002846763 | SLCO3A1 |
| chr8 | 144436503 | 144436951 | 449 | 7 | 1.45E-05 | 6.53E-07 | 0.007585 | 2.76E-06 | 0.001484931 | TONSL |
| chr1 | 2082491 | 2083189 | 699 | 5 | 2.96E-06 | 2.80E-05 | 0.000143 | 2.77E-06 | 0.003337768 | PRKCZ |
| chr2 | 53959351 | 53959874 | 524 | 2 | 3.86E-06 | 3.48E-06 | 4.48E-05 | 2.77E-06 | 0.007793577 | PSME4 |
| chr8 | 10474381 | 10474758 | 378 | 2 | 7.18E-06 | 1.66E-06 | 0.000248 | 2.78E-06 | -0.00348966 | LINCR-0001 |
| chr9 | 128418877 | 128419814 | 938 | 7 | 2.98E-06 | 9.30E-05 | 0.000176 | 2.78E-06 | 0.004891156 | CERCAM |
| chr5 | 892900 | 894108 | 1209 | 8 | 1.02E-05 | 8.71E-06 | 0.002644 | 2.78E-06 | -0.00029884 | TRIP13 |
| chr1 | 39691085 | 39692259 | 1175 | 16 | 1.27E-06 | 3.05E-05 | 8.48E-05 | 2.80E-06 | -0.00049306 | PPIE, HPCAL4 |
| chr12 | 52692448 | 52692508 | 61 | 3 | 8.25E-06 | 1.50E-06 | 0.000353 | 2.80E-06 | 0.007603453 | KRT77 |
| chr11 | 74170749 | 74171691 | 943 | 22 | 9.09E-06 | 2.44E-05 | 0.001375 | 2.80E-06 | 0.000449045 | PPME1, C2CD3 |
| chr18 | 77598172 | 77599062 | 891 | 4 | 7.36E-05 | 2.65E-06 | 0.001151 | 2.80E-06 | 0.001533832 | NA |
| chr20 | 45934900 | 45936005 | 1106 | 5 | 2.66E-05 | 8.01E-07 | 0.003895 | 2.80E-06 | 0.001607855 | PCIF1, AL162458.1 |
| chr13 | 113086609 | 113087841 | 1233 | 6 | 2.21E-06 | 1.31E-06 | 0.000652 | 2.82E-06 | 0.001761671 | MCF2L |
| chr17 | 45393605 | 45394173 | 569 | 3 | 1.42E-06 | 1.97E-05 | 1.59E-05 | 2.82E-06 | -0.01166222 | ARHGAP27 |
| chr6 | 168308387 | 168308913 | 527 | 2 | 2.75E-06 | 1.66E-06 | 0.000282 | 2.82E-06 | 0.008726937 | DACT2 |
| chr1 | 179954379 | 179955596 | 1218 | 14 | 3.12E-05 | 1.77E-05 | 0.004444 | 2.82E-06 | 0.000307778 | CEP350, AL353708.1 |
| chr12 | 130704835 | 130705500 | 666 | 5 | 5.12E-06 | 2.64E-06 | 0.000693 | 2.82E-06 | 0.007021072 | RIMBP2 |
| chr20 | 31721153 | 31721952 | 800 | 5 | 2.89E-06 | 2.56E-05 | 0.000264 | 2.82E-06 | 0.009165271 | ABALON, BCL2L1 |
| chr19 | 47607656 | 47608586 | 931 | 10 | 3.49E-06 | 2.38E-05 | 0.000205 | 2.82E-06 | -4.47E-05 | AC010519.1, BICRA, AC010331.1 |
| chr7 | 42911260 | 42912490 | 1231 | 15 | 8.63E-06 | 2.48E-06 | 0.004494 | 2.82E-06 | 9.92E-05 | C7orf25, AC010132.3 |
| chr10 | 86681309 | 86681782 | 474 | 3 | 7.63E-07 | 0.000286 | 5.80E-05 | 2.82E-06 | 0.004486659 | LDB3 |
| chr1 | 151346787 | 151347905 | 1119 | 14 | 1.50E-06 | 6.78E-05 | 0.00063 | 2.82E-06 | 0.001033138 | RFX5-AS1, RFX5 |
| chr3 | 45841876 | 45842912 | 1037 | 14 | 1.69E-05 | 7.29E-07 | 0.021446 | 2.84E-06 | 0.001314594 | AC099782.1, LZTFL1 |
| chr4 | 145761225 | 145761603 | 379 | 4 | 5.24E-06 | 6.03E-06 | 0.001033 | 2.84E-06 | 0.002803366 | C4orf51, ZNF827 |
| chr20 | 64066190 | 64066961 | 772 | 4 | 5.72E-06 | 4.46E-06 | 0.000107 | 2.84E-06 | 0.00354172 | TCEA2 |
| chr10 | 131519809 | 131520492 | 684 | 3 | 4.86E-06 | 1.97E-06 | 0.000263 | 2.85E-06 | 0.00706116 | NA |
| chr10 | 22003083 | 22003947 | 865 | 11 | 5.70E-06 | 2.98E-05 | 0.001675 | 2.85E-06 | -0.00050845 | DNAJC1 |
| chr1 | 54017680 | 54018549 | 870 | 7 | 1.49E-06 | 3.41E-05 | 0.000271 | 2.85E-06 | 0.006313576 | LDLRAD1 |
| chr15 | 89783580 | 89784009 | 430 | 3 | 2.45E-06 | 1.67E-05 | 0.000164 | 2.85E-06 | 0.004233657 | NA |
| chr5 | 83673711 | 83674157 | 447 | 2 | 1.17E-06 | 1.99E-05 | 1.11E-05 | 2.85E-06 | 0.002937825 | HAPLN1 |
| chr10 | 86957607 | 86958636 | 1030 | 10 | 1.41E-07 | 8.65E-05 | 0.000136 | 2.85E-06 | -0.00282262 | SNCG, MMRN2 |
| chr17 | 78501304 | 78502755 | 1452 | 8 | 1.66E-06 | 6.12E-05 | 0.002038 | 2.85E-06 | 0.00177901 | DNAH17-AS1, DNAH17 |

| | | | | | | | | | | |
|---|---|---|---|---|---|---|---|---|---|---|
| chr9 | 113298634 | 113299574 | 941 | 7 | 6.97E-06 | 2.07E-05 | 0.000235 | 2.85E-06 | 0.002657631 | RNF183 |
| chr12 | 4537866 | 4539108 | 1243 | 12 | 2.92E-05 | 1.42E-06 | 0.002729 | 2.86E-06 | 0.001975299 | RAD51AP1, C12orf4 |
| chr6 | 42960237 | 42961182 | 946 | 25 | 3.17E-06 | 7.94E-06 | 0.021213 | 2.86E-06 | 0.006157185 | GNMT |
| chr10 | 4188221 | 4188560 | 340 | 5 | 1.70E-05 | 9.24E-07 | 0.003025 | 2.86E-06 | 0.00678803 | NA |
| chr9 | 129738150 | 129738213 | 64 | 3 | 1.71E-05 | 1.20E-06 | 0.001036 | 2.87E-06 | 0.008823788 | NA |
| chr1 | 147758953 | 147759416 | 464 | 2 | 1.10E-07 | 0.000158 | 5.36E-06 | 2.87E-06 | 0.006463926 | GJA5 |
| chr19 | 44613396 | 44614147 | 752 | 8 | 8.23E-06 | 1.40E-06 | 0.001765 | 2.87E-06 | 0.001060607 | IGSF23, CEACAM22P |
| chr1 | 155972294 | 155972551 | 258 | 4 | 1.78E-05 | 8.53E-07 | 0.002262 | 2.87E-06 | 0.008347704 | ARHGEF2 |
| chr6 | 166251864 | 166252410 | 547 | 2 | 1.49E-07 | 5.84E-06 | 2.69E-05 | 2.88E-06 | 0.010309415 | AL121956.1 |
| chr10 | 95289853 | 95291206 | 1354 | 5 | 1.64E-06 | 1.82E-06 | 0.002403 | 2.88E-06 | 0.001272093 | PDLIM1 |
| chr13 | 44475143 | 44475597 | 455 | 2 | 6.38E-06 | 1.81E-06 | 0.000191 | 2.88E-06 | 0.018262379 | TSC22D1 |
| chr16 | 58391213 | 58392580 | 1368 | 11 | 0.000157518 | 1.10E-05 | 0.000629 | 2.88E-06 | 0.000641146 | GINS3, AC009118.2 |
| chr19 | 57119260 | 57120110 | 851 | 11 | 3.22E-06 | 3.49E-05 | 0.002337 | 2.88E-06 | -0.00255159 | USP29 |
| chr4 | 54571487 | 54571850 | 364 | 4 | 1.08E-05 | 8.54E-06 | 0.000162 | 2.88E-06 | 7.41E-05 | NA |
| chr11 | 117181060 | 117181681 | 622 | 6 | 6.86E-06 | 4.63E-06 | 8.56E-05 | 2.88E-06 | 0.009746555 | SIDT2 |
| chr5 | 141604490 | 141604493 | 4 | 2 | 8.30E-06 | 8.28E-06 | 1.99E-05 | 2.88E-06 | 0.007426321 | DIAPH1 |
| chr11 | 7088575 | 7088917 | 343 | 6 | 3.82E-06 | 3.80E-05 | 0.002519 | 2.89E-06 | -0.03400243 | NA |
| chr16 | 89920720 | 89921376 | 657 | 4 | 4.25E-07 | 2.75E-06 | 0.00058 | 2.89E-06 | 0.005292454 | MC1R, AC092143.1, AC092143.2 |
| chr8 | 47188787 | 47189334 | 548 | 5 | 6.86E-06 | 9.63E-07 | 0.003484 | 2.89E-06 | 0.007991112 | AC120036.3, AC120036.1 |
| chr19 | 14061612 | 14062274 | 663 | 2 | 8.57E-06 | 2.52E-06 | 7.93E-05 | 2.90E-06 | 0.012533754 | PALM3 |
| chr1 | 91845173 | 91846072 | 900 | 2 | 0.000173028 | 1.69E-06 | 0.000308 | 2.90E-06 | -0.01624122 | TGFBR3 |
| chr11 | 9433507 | 9434262 | 756 | 4 | 4.53E-05 | 9.41E-07 | 0.001777 | 2.90E-06 | 0.000903123 | IPO7 |
| chr15 | 80468854 | 80468953 | 100 | 3 | 6.38E-06 | 6.44E-06 | 3.98E-05 | 2.90E-06 | 0.009506256 | ARNT2 |
| chr22 | 22201479 | 22202146 | 668 | 3 | 4.01E-05 | 1.30E-06 | 0.000924 | 2.90E-06 | -0.00305725 | IGLV10-54, IGLV11-55 |
| chr2 | 227471293 | 227472212 | 920 | 10 | 7.09E-06 | 4.62E-06 | 0.008222 | 2.91E-06 | 0.000802143 | MIR5703, AGFG1 |
| chr10 | 110492808 | 110493538 | 731 | 2 | 2.61E-05 | 1.92E-06 | 0.000156 | 2.91E-06 | 0.00693941 | AL355512.1 |
| chr7 | 159010029 | 159010226 | 198 | 3 | 8.60E-06 | 8.36E-06 | 8.10E-05 | 2.91E-06 | 0.003136394 | LINC00689 |
| chr1 | 178122116 | 178122289 | 174 | 2 | 1.15E-05 | 2.19E-06 | 0.000106 | 2.91E-06 | 0.021240422 | RASAL2 |
| chr10 | 14586302 | 14587396 | 1095 | 3 | 8.33E-06 | 8.84E-06 | 0.000116 | 2.91E-06 | 0.006602897 | FAM107B |
| chr17 | 58527586 | 58528658 | 1073 | 6 | 2.82E-05 | 6.48E-06 | 0.001294 | 2.93E-06 | 0.006250188 | SEPTIN4-AS1, SEPTIN4 |
| chr19 | 48695772 | 48696923 | 1152 | 7 | 1.02E-05 | 0.000151 | 0.000121 | 2.93E-06 | 0.003894842 | FUT2 |
| chr6 | 26016446 | 26017052 | 607 | 3 | 1.20E-06 | 1.38E-06 | 0.000864 | 2.94E-06 | 0.01039084 | H3P26, H1-1 |
| chr17 | 28572194 | 28572668 | 475 | 3 | 7.41E-08 | 1.17E-05 | 3.49E-05 | 2.94E-06 | 0.009544221 | AC005726.1 |
| chr1 | 18879496 | 18880065 | 570 | 2 | 7.22E-06 | 2.58E-06 | 7.83E-05 | 2.94E-06 | 0.003694455 | ALDH4A1 |
| chr8 | 121641286 | 121642326 | 1041 | 8 | 1.00E-05 | 3.11E-06 | 0.002031 | 2.94E-06 | 0.000313229 | HAS2-AS1, HAS2 |
| chr17 | 16064895 | 16065489 | 595 | 3 | 1.10E-06 | 0.000188 | 7.07E-05 | 2.94E-06 | 0.002855226 | NCOR1 |
| chr9 | 131842017 | 131842307 | 291 | 4 | 7.36E-06 | 1.48E-05 | 0.000271 | 2.95E-06 | 0.000747389 | NA |
| chr3 | 125154924 | 125155004 | 81 | 2 | 1.13E-05 | 3.11E-06 | 5.83E-05 | 2.95E-06 | 0.012882886 | SLC12A8 |
| chr21 | 40923423 | 40923530 | 108 | 2 | 1.24E-05 | 2.30E-06 | 9.94E-05 | 2.96E-06 | 0.019520288 | NA |
| chr12 | 55935512 | 55936119 | 608 | 5 | 2.84E-06 | 6.03E-05 | 0.000537 | 2.96E-06 | 0.007833924 | DGKA |
| chr11 | 117703609 | 117703714 | 106 | 2 | 7.00E-06 | 9.59E-06 | 1.85E-05 | 2.97E-06 | 0.004892267 | DSCAML1 |
| chr11 | 1195217 | 1196302 | 1086 | 5 | 9.08E-06 | 2.06E-05 | 7.43E-05 | 2.98E-06 | 0.005337317 | MUC5AC |
| chr1 | 245677366 | 245677965 | 600 | 3 | 1.73E-05 | 9.76E-06 | 0.000209 | 2.98E-06 | 0.001167079 | KIF26B |
| chr7 | 101153663 | 101154502 | 840 | 7 | 7.88E-05 | 2.42E-06 | 0.002156 | 2.98E-06 | 0.002643717 | AP1S1 |
| chr1 | 156703503 | 156704356 | 854 | 3 | 1.07E-06 | 2.82E-05 | 8.75E-05 | 2.98E-06 | 0.009455429 | AL590666.4, CRABP2 |
| chr14 | 21796735 | 21797197 | 463 | 2 | 7.40E-06 | 2.55E-06 | 8.25E-05 | 2.98E-06 | 0.012424777 | NA |
| chr8 | 22067508 | 22068137 | 630 | 4 | 8.79E-06 | 1.61E-05 | 5.95E-05 | 2.99E-06 | 0.004284983 | DMTN |
| chr10 | 6701522 | 6701669 | 148 | 2 | 6.40E-06 | 1.19E-05 | 1.60E-05 | 2.99E-06 | 0.007859438 | AL158210.1 |
| chr3 | 49411438 | 49412593 | 1156 | 12 | 6.29E-07 | 2.38E-05 | 0.000614 | 2.99E-06 | -0.00033953 | TCTA, RHOA |
| chr17 | 40648514 | 40648905 | 392 | 6 | 4.82E-06 | 8.58E-06 | 0.001527 | 3.00E-06 | 0.010758478 | AC073508.3, SMARCE1, AC073508.2 |
| chr17 | 81385206 | 81387226 | 2021 | 14 | 2.70E-05 | 0.000109 | 0.004021 | 3.00E-06 | 0.002152731 | AC110285.1 |
| chr2 | 106085490 | 106085841 | 352 | 2 | 1.35E-06 | 5.91E-05 | 7.29E-06 | 3.00E-06 | 0.014080844 | NA |
| chr6 | 34155438 | 34156106 | 669 | 7 | 1.53E-05 | 3.49E-06 | 0.00132 | 3.00E-06 | 0.005847786 | GRM4 |
| chr6 | 32971047 | 32973019 | 1973 | 47 | 7.75E-07 | 0.000275 | 0.003956 | 3.01E-06 | 0.000629339 | BRD2, AL645941.3 |
| chr1 | 8211858 | 8212217 | 360 | 5 | 9.89E-06 | 1.39E-06 | 0.001568 | 3.01E-06 | 0.006558136 | LINC01714 |
| chr2 | 127702362 | 127703213 | 852 | 3 | 2.36E-05 | 1.68E-06 | 0.000279 | 3.01E-06 | -0.00250276 | SFT2D3, WDR33 |
| chr16 | 1408205 | 1408756 | 552 | 5 | 3.12E-06 | 7.92E-06 | 0.000156 | 3.02E-06 | 0.010213174 | UNKL |
| chr1 | 159953618 | 159954449 | 832 | 5 | 5.38E-06 | 5.89E-06 | 0.000372 | 3.02E-06 | 0.007963512 | SLAMF9 |
| chr1 | 24931014 | 24932188 | 1175 | 19 | 7.27E-06 | 5.83E-05 | 0.002718 | 3.02E-06 | 0.001984834 | RUNX3 |
| chr14 | 49620375 | 49621542 | 1168 | 14 | 2.82E-06 | 6.79E-05 | 0.000536 | 3.03E-06 | 0.000910452 | MGAT2, RPL36AL, AL139099.1 |
| chr4 | 182443488 | 182443672 | 185 | 2 | 4.90E-06 | 1.68E-05 | 1.31E-05 | 3.03E-06 | 0.009823215 | TENM3 |
| chr17 | 49901574 | 49902366 | 793 | 5 | 3.67E-07 | 7.90E-05 | 0.000128 | 3.03E-06 | 0.009343384 | NA |
| chr16 | 88983848 | 88984323 | 476 | 6 | 1.08E-05 | 1.10E-06 | 0.002904 | 3.03E-06 | 0.01342872 | AC135782.2 |
| chr6 | 35048672 | 35048804 | 133 | 2 | 1.17E-05 | 2.02E-06 | 0.000152 | 3.03E-06 | 0.020113624 | ANKS1A, AL591002.1 |
| chr12 | 54364638 | 54365512 | 875 | 6 | 3.03E-06 | 2.50E-05 | 0.000542 | 3.03E-06 | 0.003678216 | AC079313.2, AC079313.1 |
| chr15 | 99706285 | 99706497 | 213 | 2 | 1.09E-05 | 2.42E-06 | 9.49E-05 | 3.03E-06 | 0.01373417 | MEF2A |
| chr2 | 59249618 | 59250113 | 496 | 4 | 9.68E-07 | 0.000145 | 9.03E-06 | 3.04E-06 | 0.010028978 | AC007092.1, LINC01793, AC007100.1, AC007179.2 |
| chr19 | 57362380 | 57363620 | 1241 | 10 | 1.39E-06 | 3.88E-05 | 0.000462 | 3.04E-06 | 0.002832479 | ZNF547, TRAPPC2B, AC003002.1 |
| chr12 | 51238074 | 51238882 | 809 | 16 | 4.46E-07 | 5.67E-05 | 4.31E-05 | 3.04E-06 | -0.00102793 | DAZAP2 |
| chr14 | 22559804 | 22561112 | 1309 | 3 | 0.000373163 | 2.02E-06 | 0.000596 | 3.04E-06 | 0.012182323 | NA |
| chr7 | 36153209 | 36154245 | 1037 | 7 | 1.27E-05 | 4.56E-05 | 0.000303 | 3.05E-06 | 0.004190575 | EEPD1 |
| chr1 | 156435173 | 156436006 | 834 | 6 | 2.23E-06 | 2.46E-05 | 3.89E-05 | 3.05E-06 | 0.004016109 | MIR9-1HG |
| chr3 | 186241807 | 186242651 | 845 | 4 | 5.41E-06 | 3.00E-06 | 0.000775 | 3.05E-06 | 0.018524059 | DGKG |
| chr4 | 69930682 | 69931124 | 443 | 3 | 3.46E-06 | 4.89E-06 | 0.000192 | 3.06E-06 | 0.012957571 | CSN1S1 |
| chr5 | 141477625 | 141478540 | 916 | 9 | 4.66E-06 | 1.08E-05 | 0.000149 | 3.08E-06 | 0.011283352 | PCDHGA1, PCDHGA2, PCDHGA3, PCDHGB1, PCDHGA4, PCDHGB2, PCDHGA5, PCDHGB3, PCDHGA6, PCDHGA7, PCDHGB4, PCDHGA8, PCDHGB5, PCDHGA9, PCDHGB6, PCDHGA10, PCDHGB7, PCDHGA11, PCDHGA12, PCDHGC3 |
| chr1 | 20717538 | 20718930 | 1393 | 15 | 3.82E-05 | 2.74E-05 | 0.001923 | 3.08E-06 | 0.002565107 | KIF17 |
| chr11 | 19519903 | 19520946 | 1044 | 4 | 4.20E-05 | 2.52E-05 | 0.000225 | 3.08E-06 | 0.005440314 | NAV2 |
| chr16 | 67947939 | 67948357 | 419 | 3 | 2.34E-06 | 5.29E-06 | 0.000146 | 3.09E-06 | 0.003934403 | SLC12A4 |
| chr10 | 84715689 | 84716026 | 338 | 2 | 5.40E-06 | 4.96E-06 | 3.53E-05 | 3.10E-06 | 0.024177597 | NA |
| chr5 | 645092 | 646024 | 933 | 7 | 4.38E-05 | 5.45E-06 | 0.002188 | 3.10E-06 | -0.00185903 | CEP72 |
| chr11 | 1758907 | 1759094 | 188 | 5 | 4.89E-06 | 5.36E-06 | 0.000459 | 3.10E-06 | 0.005171874 | AC068580.4, CTSD |
| chr5 | 159331131 | 159331895 | 765 | 7 | 2.20E-05 | 2.14E-06 | 0.008619 | 3.10E-06 | 0.005410736 | AC008691.1 |
| chr19 | 12936842 | 12937914 | 1073 | 5 | 7.87E-06 | 6.33E-07 | 0.003472 | 3.10E-06 | 0.004684448 | NA |
| chr10 | 13437287 | 13437844 | 558 | 4 | 1.04E-05 | 2.69E-06 | 0.000427 | 3.10E-06 | 0.007124071 | NA |
| chr4 | 143336532 | 143338348 | 1817 | 13 | 5.13E-05 | 0.000789 | 0.00021 | 3.10E-06 | -0.00058794 | GAB1 |
| chr12 | 93848339 | 93849158 | 820 | 5 | 1.02E-05 | 5.56E-06 | 0.000588 | 3.11E-06 | 0.006771566 | CRADD |
| chr20 | 38291667 | 38291690 | 24 | 2 | 1.30E-05 | 2.73E-06 | 7.98E-05 | 3.11E-06 | 0.021156062 | AL359555.3, AL359555.4, AL359555.1 |
| chr5 | 171858551 | 171859089 | 539 | 3 | 5.15E-08 | 0.000271 | 3.86E-06 | 3.11E-06 | 0.004081733 | NA |
| chr7 | 138090846 | 138091136 | 291 | 3 | 1.11E-05 | 1.09E-06 | 0.00173 | 3.11E-06 | 0.013121553 | AKR1D1 |
| chr9 | 131809832 | 131810689 | 858 | 4 | 1.62E-05 | 3.18E-06 | 0.000824 | 3.11E-06 | -0.00544844 | NA |
| chr12 | 49885275 | 49886289 | 1015 | 3 | 5.55E-06 | 2.92E-06 | 0.000151 | 3.11E-06 | 0.014084833 | FAIM2 |
| chr12 | 130839381 | 130840179 | 799 | 7 | 3.49E-05 | 1.97E-06 | 0.001915 | 3.11E-06 | 0.002399758 | NA |
| chr8 | 55772793 | 55773838 | 1046 | 19 | 7.99E-07 | 3.93E-05 | 0.002467 | 3.12E-06 | 0.000204239 | TGS1, TMEM68 |
| chr10 | 132333453 | 132334022 | 570 | 3 | 3.04E-06 | 7.26E-06 | 7.65E-05 | 3.12E-06 | -0.00473366 | LRRC27 |
| chr13 | 110714824 | 110715568 | 745 | 12 | 5.43E-06 | 3.32E-06 | 0.00047 | 3.12E-06 | 6.83E-05 | ING1 |
| chr12 | 106585948 | 106586768 | 821 | 5 | 7.66E-06 | 2.01E-06 | 0.00046 | 3.12E-06 | 0.00298794 | RFX4, AC079385.1 |
| chr15 | 75138401 | 75139061 | 661 | 3 | 1.33E-05 | 1.55E-06 | 0.000546 | 3.12E-06 | 0.008419751 | NA |
| chr21 | 45470935 | 45470944 | 10 | 2 | 8.84E-06 | 2.84E-06 | 7.59E-05 | 3.12E-06 | 0.021217589 | COL18A1 |
| chr17 | 54860783 | 54860802 | 20 | 2 | 2.70E-06 | 0.000286 | 5.18E-06 | 3.13E-06 | 0.003608285 | NA |
| chr2 | 233011550 | 233013305 | 1756 | 11 | 2.49E-06 | 3.56E-06 | 0.001524 | 3.14E-06 | 0.004543516 | AC106876.1, NGEF |
| chr20 | 63208640 | 63209100 | 461 | 5 | 6.18E-06 | 1.60E-05 | 0.00019 | 3.14E-06 | 0.002195079 | YTHDF1 |
| chr6 | 158066509 | 158066576 | 68 | 3 | 9.82E-06 | 4.17E-06 | 9.25E-05 | 3.14E-06 | 0.001241214 | SYNJ2 |
| chr3 | 154135794 | 154136007 | 214 | 2 | 8.77E-06 | 4.24E-06 | 4.32E-05 | 3.14E-06 | 0.001911958 | ARHGEF26 |
| chr10 | 43121742 | 43122004 | 263 | 2 | 9.10E-06 | 1.90E-06 | 0.000251 | 3.15E-06 | 0.014971624 | RET |
| chr20 | 45313575 | 45313925 | 351 | 3 | 7.01E-06 | 1.37E-05 | 7.32E-05 | 3.15E-06 | 0.005902488 | RBPJL |
| chr12 | 113057761 | 113058363 | 603 | 6 | 6.50E-06 | 4.71E-06 | 0.003947 | 3.15E-06 | 0.006538111 | DTX1 |
| chr7 | 139339654 | 139340533 | 880 | 12 | 5.30E-06 | 2.30E-05 | 0.002113 | 3.16E-06 | -0.00051101 | FMC1, LUC7L2 |
| chr12 | 105107176 | 105108126 | 951 | 17 | 1.09E-05 | 4.46E-06 | 0.000296 | 3.16E-06 | -0.00025331 | WASHC4, ALDH1L2 |
| chr5 | 73444327 | 73444921 | 595 | 6 | 5.04E-06 | 1.15E-05 | 0.000873 | 3.16E-06 | 0.006322839 | FOXD1 |
| chr22 | 49623780 | 49624744 | 965 | 2 | 0.000226793 | 1.85E-06 | 0.000316 | 3.16E-06 | 0.005477583 | C22orf34 |
| chr14 | 35333878 | 35334375 | 498 | 5 | 6.16E-06 | 2.21E-06 | 0.001623 | 3.17E-06 | 0.002623295 | NA |
| chr1 | 206023513 | 206024691 | 1179 | 10 | 2.34E-05 | 4.12E-06 | 0.00084 | 3.17E-06 | 0.002436863 | CTSE |
| chr7 | 76358895 | 76359988 | 1094 | 13 | 1.14E-05 | 3.31E-05 | 6.67E-05 | 3.17E-06 | 0.001903003 | YWHAG |
| chr3 | 195815682 | 195815709 | 28 | 2 | 1.11E-05 | 2.69E-06 | 8.60E-05 | 3.17E-06 | 0.016699564 | NA |
| chr15 | 28026768 | 28027510 | 743 | 5 | 1.41E-05 | 1.48E-06 | 0.001671 | 3.17E-06 | -0.00087621 | OCA2 |
| chr19 | 41883178 | 41884655 | 1478 | 13 | 4.28E-06 | 5.09E-05 | 3.19E-05 | 3.18E-06 | 0.001054779 | ARHGEF1 |
| chr2 | 19746675 | 19747106 | 432 | 2 | 1.21E-06 | 4.09E-06 | 4.64E-05 | 3.18E-06 | 0.011692192 | NA |
| chr1 | 112502830 | 112503528 | 699 | 2 | 2.09E-05 | 1.89E-06 | 0.000285 | 3.18E-06 | 0.00921173 | WNT2B |
| chr2 | 9941403 | 9942363 | 961 | 5 | 1.45E-06 | 2.69E-05 | 0.000191 | 3.18E-06 | 0.009073811 | AC010969.3 |
| chr1 | 23553990 | 23554245 | 256 | 2 | 1.33E-06 | 1.89E-06 | 0.000282 | 3.18E-06 | 0.027387313 | NA |
| chr7 | 65955038 | 65955957 | 920 | 7 | 3.60E-06 | 2.58E-06 | 0.001289 | 3.18E-06 | 0.002490115 | VKORC1L1 |
| chr12 | 116662596 | 116663095 | 500 | 3 | 1.10E-06 | 1.70E-05 | 9.65E-05 | 3.18E-06 | 0.014425623 | AC125603.2 |
| chr20 | 3795525 | 3796608 | 1084 | 14 | 1.90E-05 | 1.70E-05 | 0.000472 | 3.18E-06 | 0.001960161 | CDC25B |
| chr13 | 114020975 | 114021428 | 454 | 5 | 6.32E-06 | 1.07E-05 | 0.000326 | 3.18E-06 | 0.002245018 | RASA3 |
| chr19 | 51391265 | 51391645 | 381 | 5 | 9.58E-06 | 2.04E-06 | 0.002093 | 3.19E-06 | 0.00804659 | NA |
| chr16 | 23670686 | 23671177 | 492 | 3 | 1.81E-07 | 0.000206 | 7.68E-06 | 3.19E-06 | 0.000808988 | DCTN5, AC008870.1 |
| chr11 | 44535023 | 44535616 | 594 | 3 | 6.86E-06 | 9.86E-06 | 0.000102 | 3.19E-06 | 0.0096714 | NA |
| chr3 | 52238480 | 52239465 | 986 | 12 | 1.41E-05 | 9.52E-05 | 0.0058 | 3.20E-06 | 0.002094896 | AC006252.1, TWF2 |
| chr1 | 155125959 | 155127602 | 1644 | 10 | 3.99E-09 | 0.002472 | 5.39E-05 | 3.20E-06 | 0.004939818 | NA |
| chr5 | 79299571 | 79300189 | 619 | 4 | 3.36E-05 | 3.81E-06 | 0.000247 | 3.20E-06 | 0.000401907 | JMY |
| chr11 | 129432934 | 129433513 | 580 | 2 | 7.53E-08 | 0.002736 | 4.26E-06 | 3.21E-06 | 0.003729434 | BARX2 |
| chr1 | 31776572 | 31776948 | 377 | 2 | 2.40E-06 | 1.59E-05 | 1.46E-05 | 3.21E-06 | 0.012095304 | NA |
| chr22 | 43197575 | 43197979 | 405 | 3 | 1.03E-05 | 1.15E-06 | 0.001568 | 3.22E-06 | 0.010948354 | SCUBE1 |
| chr16 | 30604690 | 30605328 | 639 | 3 | 2.73E-06 | 4.54E-06 | 0.000369 | 3.23E-06 | 0.010080565 | AC093249.2, ZNF689 |
| chr4 | 73222844 | 73223452 | 609 | 4 | 7.68E-06 | 8.79E-06 | 0.000649 | 3.23E-06 | 0.00907961 | ANKRD17 |
| chr8 | 38717047 | 38717831 | 785 | 3 | 2.42E-05 | 9.34E-06 | 0.000142 | 3.23E-06 | 0.007460173 | NA |
| chr3 | 28315053 | 28316145 | 1093 | 3 | 0.000137285 | 1.29E-05 | 0.000245 | 3.23E-06 | 0.003845474 | CMC1, AZI2 |
| chr12 | 12017501 | 12018320 | 820 | 3 | 2.28E-06 | 7.47E-06 | 3.88E-05 | 3.23E-06 | -0.00526402 | NA |
| chr12 | 62631710 | 62632705 | 996 | 8 | 1.40E-05 | 3.30E-06 | 0.006032 | 3.23E-06 | 0.00323708 | NA |
| chr2 | 192193446 | 192193902 | 457 | 3 | 8.12E-06 | 1.58E-06 | 0.000419 | 3.23E-06 | 0.02641742 | TMEFF2 |
| chr5 | 72105883 | 72106814 | 932 | 4 | 3.02E-05 | 3.40E-06 | 0.000371 | 3.23E-06 | 0.001958167 | AC025774.1 |
| chr12 | 93574960 | 93575438 | 479 | 2 | 1.04E-06 | 4.00E-06 | 4.88E-05 | 3.24E-06 | 0.011913134 | SOCS2 |
| chr8 | 53945099 | 53945332 | 234 | 4 | 3.71E-06 | 1.81E-05 | 0.000359 | 3.25E-06 | 0.018050655 | RGS20 |
| chr3 | 101827239 | 101828452 | 1214 | 13 | 1.01E-05 | 5.59E-05 | 0.001627 | 3.25E-06 | 0.001776445 | NXPE3, NFKBIZ |
| chr17 | 1493004 | 1493659 | 656 | 5 | 2.25E-07 | 7.48E-06 | 0.000151 | 3.25E-06 | 0.006971372 | NA |
| chr1 | 153655725 | 153656507 | 783 | 3 | 3.86E-05 | 2.56E-06 | 0.000202 | 3.25E-06 | 0.000483862 | NA |
| chr11 | 31810805 | 31812405 | 1601 | 22 | 1.76E-05 | 1.18E-05 | 0.002701 | 3.26E-06 | 0.001467282 | PAUPAR, AL035078.4, PAX6 |
| chr10 | 182255 | 182861 | 607 | 2 | 2.63E-05 | 1.97E-06 | 0.00026 | 3.26E-06 | -0.00775963 | ZMYND11 |
| chr16 | 67661064 | 67662278 | 1215 | 19 | 5.11E-06 | 0.001018 | 0.000176 | 3.26E-06 | 0.001778485 | PARD6A |
| chr20 | 52105778 | 52106688 | 911 | 4 | 6.19E-06 | 1.81E-05 | 0.000233 | 3.27E-06 | 0.010761668 | ZFP64 |
| chr17 | 4931746 | 4932876 | 1131 | 8 | 1.53E-06 | 9.12E-06 | 0.00044 | 3.27E-06 | -0.00894737 | GP1BA, CHRNE |
| chr2 | 236402067 | 236402130 | 64 | 2 | 1.56E-05 | 1.96E-06 | 0.000271 | 3.27E-06 | 0.010785092 | IQCA1 |
| chr15 | 30991419 | 30992210 | 792 | 5 | 2.70E-05 | 8.69E-07 | 0.004521 | 3.28E-06 | -0.00142271 | AC009562.1, MTMR10 |
| chr7 | 50732281 | 50732910 | 630 | 6 | 8.50E-06 | 1.40E-05 | 0.001636 | 3.29E-06 | 0.003141975 | GRB10 |
| chr14 | 104985713 | 104986684 | 972 | 10 | 1.31E-05 | 5.99E-06 | 0.006028 | 3.29E-06 | 0.003397006 | CLBA1 |
| chr16 | 89100610 | 89101582 | 973 | 6 | 5.63E-06 | 1.90E-05 | 0.000283 | 3.29E-06 | 0.007457491 | ACSF3 |
| chr20 | 52456581 | 52456881 | 301 | 3 | 7.10E-06 | 8.19E-06 | 6.85E-05 | 3.29E-06 | 0.013138421 | LINC01524 |
| chr14 | 88163687 | 88164427 | 741 | 3 | 1.55E-05 | 0.000234 | 0.000171 | 3.29E-06 | 0.005092347 | NA |
| chr10 | 11337113 | 11337740 | 628 | 2 | 7.45E-06 | 2.21E-06 | 0.000159 | 3.30E-06 | 0.02145654 | NA |
| chr20 | 5609532 | 5610665 | 1134 | 3 | 4.12E-06 | 9.94E-05 | 4.53E-05 | 3.31E-06 | 0.008610543 | GPCPD1 |
| chr12 | 56725000 | 56726006 | 1007 | 16 | 6.56E-06 | 1.71E-05 | 0.006273 | 3.31E-06 | 0.003193344 | NACA, AC117378.1 |
| chr8 | 9555413 | 9556605 | 1193 | 15 | 2.60E-05 | 8.14E-06 | 0.002154 | 3.32E-06 | 0.001542148 | TNKS, AC021242.3 |

| | | | | | | | | | | |
|---|---|---|---|---|---|---|---|---|---|---|
| chr10 | 30431646 | 30431888 | 243 | 4 | 9.05E-06 | 4.09E-06 | 0.000369 | 3.33E-06 | -0.00339878 | NA |
| chr10 | 12271132 | 12271168 | 37 | 2 | 1.34E-05 | 3.28E-06 | 6.78E-05 | 3.33E-06 | 0.018705292 | NA |
| chr3 | 121749180 | 121750321 | 1142 | 13 | 4.86E-05 | 3.06E-06 | 0.000513 | 3.33E-06 | 0.001408511 | GOLGB1 |
| chr15 | 45134004 | 45134188 | 185 | 2 | 6.74E-06 | 2.23E-06 | 0.00016 | 3.34E-06 | 0.008124699 | DUOX1 |
| chr2 | 3172377 | 3173122 | 746 | 3 | 3.65E-05 | 2.56E-06 | 0.000487 | 3.34E-06 | -0.00369042 | NA |
| chr17 | 81572067 | 81572623 | 557 | 4 | 7.30E-06 | 4.37E-06 | 0.000526 | 3.35E-06 | -0.00015595 | NPLOC4 |
| chr8 | 11701478 | 11702287 | 810 | 7 | 1.33E-05 | 3.86E-06 | 0.003107 | 3.35E-06 | 0.005708962 | GATA4 |
| chr15 | 64774032 | 64774511 | 480 | 2 | 1.99E-06 | 2.79E-06 | 9.17E-05 | 3.36E-06 | 0.012866595 | RBPMS2 |
| chr12 | 32597218 | 32597572 | 355 | 3 | 1.55E-05 | 1.19E-06 | 0.001736 | 3.36E-06 | -0.00359394 | FGD4 |
| chr18 | 63421921 | 63422860 | 940 | 12 | 2.19E-06 | 2.29E-05 | 0.000598 | 3.36E-06 | -0.00110377 | VPS4B |
| chr21 | 41316264 | 41316959 | 696 | 5 | 3.97E-07 | 2.38E-05 | 0.000141 | 3.36E-06 | 0.003875815 | FAM3B |
| chr8 | 9573952 | 9574702 | 751 | 3 | 1.31E-06 | 0.000513 | 2.27E-05 | 3.36E-06 | 0.004950684 | TNKS |
| chr11 | 122979432 | 122980414 | 983 | 8 | 3.40E-05 | 1.54E-06 | 0.0046 | 3.37E-06 | 0.00407453 | BSX |
| chr2 | 236107102 | 236107828 | 727 | 3 | 9.74E-07 | 2.07E-05 | 1.63E-05 | 3.37E-06 | 0.007686597 | AGAP1 |
| chr7 | 74505559 | 74506271 | 713 | 3 | 5.08E-07 | 5.59E-05 | 0.000134 | 3.37E-06 | 0.005229457 | GTF2IRD1 |
| chr9 | 128245635 | 128245701 | 67 | 2 | 7.30E-06 | 1.99E-06 | 0.000302 | 3.37E-06 | 0.00977211 | DNM1 |
| chr2 | 240351027 | 240351653 | 627 | 7 | 3.58E-06 | 1.43E-05 | 0.001934 | 3.37E-06 | 0.011991235 | NA |
| chr7 | 324003 | 324024 | 22 | 2 | 9.73E-06 | 6.50E-06 | 3.06E-05 | 3.37E-06 | -0.00089368 | NA |
| chr9 | 121156876 | 121158020 | 1145 | 4 | 0.000141786 | 3.89E-05 | 0.000176 | 3.37E-06 | 0.002519737 | CNTRL |
| chr10 | 720101 | 720561 | 461 | 4 | 2.68E-06 | 5.08E-05 | 1.22E-05 | 3.38E-06 | -0.01007009 | NA |
| chr12 | 50351079 | 50351824 | 746 | 4 | 2.13E-05 | 1.51E-06 | 0.001989 | 3.38E-06 | 0.006126201 | FAM186A |
| chr8 | 11867000 | 11867983 | 984 | 4 | 1.54E-05 | 1.95E-05 | 0.000239 | 3.38E-06 | 0.003003538 | CTSB |
| chr5 | 36744787 | 36745516 | 730 | 5 | 1.45E-05 | 1.73E-06 | 0.00183 | 3.39E-06 | 0.003338182 | NA |
| chr3 | 52339376 | 52339731 | 356 | 2 | 2.05E-06 | 3.26E-05 | 1.04E-05 | 3.39E-06 | 0.003210221 | DNAH1 |
| chr7 | 128904972 | 128905299 | 328 | 2 | 9.73E-06 | 2.87E-06 | 8.85E-05 | 3.39E-06 | 0.004860614 | KCP |
| chr14 | 26595548 | 26596540 | 993 | 5 | 4.07E-05 | 7.76E-07 | 0.002545 | 3.39E-06 | 0.012361631 | NOVA1 |
| chr20 | 17314576 | 17315672 | 1097 | 5 | 1.09E-05 | 5.05E-05 | 8.83E-05 | 3.40E-06 | 0.007623956 | PCSK2 |
| chr8 | 67952288 | 67952952 | 665 | 7 | 4.18E-06 | 4.23E-06 | 0.001126 | 3.40E-06 | 0.001777639 | PREX2 |
| chr1 | 21337216 | 21338317 | 1102 | 4 | 2.03E-06 | 4.78E-06 | 0.000378 | 3.41E-06 | 0.010207509 | ECE1 |
| chr7 | 11831909 | 11833119 | 1211 | 12 | 2.65E-05 | 1.02E-06 | 0.009299 | 3.41E-06 | 0.011675582 | THSD7A |
| chr20 | 51311895 | 51312195 | 301 | 2 | 1.31E-05 | 2.31E-06 | 0.000153 | 3.41E-06 | 0.017663155 | NA |
| chr19 | 1368816 | 1369516 | 701 | 3 | 2.76E-06 | 6.58E-06 | 0.000231 | 3.42E-06 | 0.002631699 | PWWP3A |
| chr7 | 100119858 | 100120641 | 784 | 12 | 3.88E-07 | 2.28E-05 | 0.000614 | 3.42E-06 | 0.005156839 | CNPY4, AC073842.2 |
| chr5 | 177473277 | 177473841 | 565 | 8 | 1.04E-05 | 1.56E-05 | 0.00128 | 3.42E-06 | -0.00087669 | DBN1 |
| chr6 | 12715629 | 12715905 | 277 | 2 | 5.74E-07 | 5.77E-06 | 3.51E-05 | 3.42E-06 | 0.017484284 | NA |
| chr11 | 116216469 | 116216913 | 445 | 2 | 1.58E-05 | 2.13E-06 | 0.000225 | 3.43E-06 | 0.030244916 | NA |
| chr3 | 44873179 | 44873402 | 224 | 2 | 6.53E-07 | 6.03E-05 | 8.15E-06 | 3.43E-06 | 0.0020155 | KIF15 |
| chr12 | 14259084 | 14259801 | 718 | 2 | 4.88E-05 | 2.54E-06 | 0.00012 | 3.43E-06 | -0.00093913 | NA |
| chr11 | 69647892 | 69648555 | 664 | 7 | 2.83E-06 | 1.95E-05 | 0.002836 | 3.43E-06 | -0.01120822 | CCND1 |
| chr7 | 28410228 | 28411392 | 1165 | 8 | 1.11E-05 | 5.10E-06 | 0.001517 | 3.43E-06 | 0.002634083 | CREB5 |
| chr13 | 51803594 | 51804608 | 1015 | 13 | 4.63E-06 | 0.000125 | 0.001777 | 3.43E-06 | 0.000609021 | AL162377.1, AL162377.3, DHRS12 |
| chr15 | 37093699 | 37094385 | 687 | 3 | 4.27E-07 | 3.82E-06 | 0.000339 | 3.43E-06 | 0.005914108 | MEIS2 |
| chr6 | 41191870 | 41192476 | 607 | 2 | 2.58E-05 | 2.10E-06 | 0.000248 | 3.43E-06 | 0.020394992 | TREML2 |
| chr3 | 63912447 | 63913421 | 975 | 7 | 1.92E-05 | 1.01E-05 | 0.000417 | 3.43E-06 | 0.000544653 | ATXN7 |
| chr9 | 137216275 | 137216736 | 462 | 2 | 2.50E-06 | 4.69E-06 | 4.44E-05 | 3.44E-06 | 0.004950908 | NDOR1 |
| chr13 | 46142858 | 46142933 | 76 | 2 | 9.63E-06 | 2.51E-06 | 0.000125 | 3.44E-06 | 0.009905124 | CPB2-AS1, LCP1 |
| chr19 | 7732145 | 7732469 | 325 | 6 | 1.66E-05 | 3.63E-06 | 0.000671 | 3.44E-06 | 0.011038946 | CLEC4G, AC008763.4 |
| chr1 | 27686472 | 27686830 | 359 | 2 | 2.78E-06 | 1.80E-05 | 1.46E-05 | 3.45E-06 | 0.005167456 | AL445490.1, CHMP1AP1 |
| chr6 | 38179764 | 38179918 | 155 | 2 | 1.11E-05 | 3.97E-06 | 5.53E-05 | 3.45E-06 | 0.009346893 | BTBD9 |
| chr2 | 178193298 | 178194009 | 712 | 5 | 4.16E-06 | 3.17E-06 | 0.001229 | 3.45E-06 | 0.004893042 | NA |
| chr1 | 1954509 | 1955832 | 1324 | 7 | 2.62E-05 | 0.00044 | 0.000609 | 3.46E-06 | 0.00900275 | CFAP74 |
| chr2 | 201451959 | 201452879 | 921 | 7 | 5.98E-06 | 5.57E-05 | 0.001242 | 3.46E-06 | 0.006416515 | STRADB |
| chr6 | 127344107 | 127344736 | 630 | 5 | 8.30E-06 | 3.44E-06 | 0.000704 | 3.46E-06 | -0.00501295 | NA |
| chr16 | 84360363 | 84360436 | 74 | 3 | 8.69E-06 | 6.61E-06 | 6.20E-05 | 3.46E-06 | 0.005316779 | NA |
| chr10 | 43093769 | 43093978 | 210 | 2 | 1.12E-05 | 3.61E-06 | 6.32E-05 | 3.47E-06 | 0.011650449 | RET |
| chr5 | 34944848 | 34944879 | 32 | 2 | 1.68E-05 | 2.08E-06 | 0.000288 | 3.47E-06 | 0.000812443 | DNAJC21 |
| chr12 | 114683410 | 114685614 | 2205 | 14 | 0.000135225 | 5.36E-06 | 0.005443 | 3.48E-06 | 0.001924794 | AC026765.3, TBX3 |
| chr11 | 124863142 | 124863661 | 520 | 4 | 8.25E-07 | 1.82E-05 | 4.15E-05 | 3.48E-06 | 0.007736396 | NA |
| chr9 | 125970117 | 125970537 | 421 | 2 | 1.21E-06 | 4.89E-05 | 9.01E-06 | 3.49E-06 | 0.005847039 | NA |
| chr2 | 169733980 | 169734729 | 750 | 3 | 8.33E-05 | 1.80E-06 | 0.000449 | 3.49E-06 | 0.007306643 | KLHL23 |
| chr17 | 44186438 | 44187503 | 1066 | 18 | 3.40E-06 | 1.60E-05 | 0.002851 | 3.50E-06 | -0.00044242 | TMUB2, ASB16-AS1 |
| chr7 | 15685026 | 15686541 | 1516 | 9 | 2.65E-05 | 5.54E-06 | 0.002018 | 3.50E-06 | 0.002990603 | MEOX2 |
| chr5 | 157743405 | 157744312 | 908 | 12 | 2.55E-05 | 9.79E-06 | 0.001153 | 3.50E-06 | 0.000534322 | LSM11 |
| chr2 | 218890039 | 218890118 | 80 | 3 | 1.72E-05 | 3.30E-06 | 0.000289 | 3.50E-06 | 0.00690643 | WNT10A |
| chr9 | 134052598 | 134054022 | 1425 | 7 | 4.52E-06 | 4.99E-05 | 0.00059 | 3.51E-06 | 0.004553622 | BRD3 |
| chr3 | 142723692 | 142725360 | 1669 | 15 | 0.000184034 | 0.000108 | 0.001961 | 3.52E-06 | -7.94E-05 | TRPC1, AC072028.1 |
| chr11 | 67505165 | 67506341 | 1177 | 7 | 3.33E-06 | 1.27E-05 | 0.00245 | 3.53E-06 | 0.004515465 | PITPNM1 |
| chr3 | 125024335 | 125024758 | 424 | 2 | 6.96E-06 | 2.07E-06 | 0.000338 | 3.53E-06 | 0.014979854 | HEG1 |
| chr17 | 78442799 | 78443222 | 424 | 2 | 2.80E-06 | 9.31E-06 | 2.33E-05 | 3.53E-06 | 0.015119374 | DNAH17 |
| chr2 | 235811223 | 235812270 | 1048 | 8 | 2.00E-05 | 7.12E-06 | 0.002665 | 3.54E-06 | 0.001165421 | AGAP1 |
| chr2 | 213226063 | 213226393 | 331 | 3 | 8.89E-06 | 4.99E-06 | 0.000167 | 3.54E-06 | 0.010160745 | NA |
| chr4 | 83219568 | 83219861 | 294 | 3 | 1.22E-05 | 4.11E-06 | 8.52E-05 | 3.54E-06 | 0.009678371 | NA |
| chr17 | 7014674 | 7015633 | 960 | 10 | 1.67E-05 | 6.21E-06 | 0.00275 | 3.55E-06 | -0.00057417 | RNASEK-C17orf49, C17orf49, AC040977.2 |
| chr11 | 67021268 | 67021830 | 563 | 4 | 2.79E-06 | 4.92E-06 | 0.000713 | 3.57E-06 | 0.012071536 | SYT12 |
| chr11 | 22466899 | 22467066 | 168 | 2 | 1.75E-05 | 2.08E-06 | 0.000354 | 3.57E-06 | 0.00198122 | LINC01495 |
| chr19 | 2041347 | 2042392 | 1046 | 7 | 2.30E-05 | 7.00E-06 | 0.001533 | 3.57E-06 | -0.00545219 | MKNK2 |
| chr11 | 44591805 | 44592430 | 626 | 2 | 4.29E-06 | 2.66E-06 | 0.00012 | 3.58E-06 | 0.014625309 | CD82 |
| chr16 | 85799314 | 85799914 | 601 | 9 | 2.98E-06 | 0.000102 | 0.000172 | 3.60E-06 | -0.00037284 | COX4I1, EMC8 |
| chr10 | 119818459 | 119819347 | 889 | 9 | 5.74E-06 | 4.63E-05 | 0.002861 | 3.60E-06 | 0.011433767 | INPP5F, AL133461.1 |
| chr16 | 86929690 | 86930196 | 507 | 2 | 3.75E-08 | 0.007711 | 4.54E-06 | 3.60E-06 | 0.018307532 | AC093519.2 |
| chr11 | 1103351 | 1103694 | 344 | 2 | 3.26E-07 | 1.51E-05 | 1.70E-05 | 3.60E-06 | 0.010454397 | MUC2 |
| chr5 | 10246886 | 10247214 | 329 | 2 | 1.07E-06 | 0.000193 | 6.27E-06 | 3.60E-06 | 0.002958647 | ATPSCKMT |
| chr12 | 104108957 | 104109346 | 390 | 2 | 1.76E-05 | 2.16E-06 | 0.000289 | 3.61E-06 | 0.017769947 | NA |
| chr16 | 27176785 | 27177384 | 600 | 3 | 7.12E-06 | 3.12E-06 | 0.000118 | 3.61E-06 | 1.54E-05 | NA |
| chr20 | 21125191 | 21126046 | 856 | 9 | 7.58E-06 | 1.46E-05 | 0.001052 | 3.61E-06 | 0.002749639 | KIZ |
| chr2 | 206086922 | 206087435 | 514 | 2 | 5.55E-08 | 6.69E-05 | 8.20E-06 | 3.61E-06 | -0.02014573 | NA |
| chr20 | 2688609 | 2689767 | 1159 | 4 | 2.04E-05 | 5.98E-06 | 0.000488 | 3.62E-06 | 0.002079938 | NA |
| chr10 | 45377748 | 45378425 | 678 | 2 | 6.07E-05 | 2.17E-06 | 0.000286 | 3.62E-06 | 0.007432141 | ALOX5 |
| chr13 | 30388836 | 30389242 | 407 | 2 | 6.15E-06 | 3.21E-06 | 8.34E-05 | 3.63E-06 | 0.014970337 | AL161893.1 |
| chr5 | 78907022 | 78907488 | 467 | 3 | 2.44E-06 | 2.94E-05 | 4.63E-05 | 3.63E-06 | 0.011677835 | ARSB |
| chr17 | 7893112 | 7893644 | 533 | 2 | 6.57E-06 | 3.61E-06 | 6.91E-05 | 3.63E-06 | 0.001759291 | CHD3 |
| chr11 | 65107634 | 65108278 | 645 | 4 | 7.22E-06 | 1.64E-06 | 0.000643 | 3.63E-06 | -0.000553 | VPS51 |
| chr4 | 41120432 | 41121268 | 837 | 3 | 6.26E-05 | 1.30E-06 | 0.001698 | 3.64E-06 | 0.024019908 | APBB2 |
| chr19 | 42287312 | 42288206 | 895 | 6 | 1.57E-05 | 1.31E-06 | 0.00269 | 3.64E-06 | 0.000812837 | CIC |
| chr22 | 33477084 | 33477375 | 292 | 2 | 5.46E-06 | 1.07E-05 | 2.18E-05 | 3.64E-06 | 0.006982388 | LARGE1 |
| chr10 | 132370281 | 132371043 | 763 | 3 | 2.27E-05 | 5.35E-06 | 0.00034 | 3.64E-06 | 0.003652844 | LRRC27 |
| chr14 | 35044412 | 35045127 | 716 | 4 | 5.68E-07 | 3.63E-06 | 0.000606 | 3.64E-06 | 0.005212532 | FAM177A1 |
| chr2 | 1741858 | 1743074 | 1217 | 7 | 1.65E-05 | 0.000198 | 0.000131 | 3.65E-06 | 0.004430219 | PXDN |
| chr4 | 122826292 | 122826894 | 603 | 10 | 6.68E-06 | 0.000171 | 0.000366 | 3.65E-06 | 0.004045417 | FGF2 |
| chr13 | 48896284 | 48896303 | 20 | 2 | 1.79E-05 | 2.13E-06 | 0.000354 | 3.65E-06 | 0.000851881 | NA |
| chr19 | 459731 | 460135 | 405 | 3 | 1.17E-06 | 1.11E-05 | 4.95E-05 | 3.65E-06 | 0.022878529 | SHC2 |
| chr16 | 81740282 | 81740423 | 142 | 2 | 9.65E-07 | 2.22E-06 | 0.00027 | 3.65E-06 | 0.020215271 | AC099524.1, PLCG2 |
| chr17 | 36837362 | 36837874 | 513 | 3 | 1.61E-05 | 1.90E-06 | 0.000368 | 3.65E-06 | 0.017627374 | NA |
| chr1 | 25112777 | 25113940 | 1164 | 4 | 0.000108203 | 2.19E-05 | 0.000388 | 3.65E-06 | 0.008784878 | AL050344.1 |
| chr20 | 10419682 | 10419772 | 91 | 2 | 7.43E-06 | 2.76E-05 | 1.21E-05 | 3.66E-06 | 0.005919267 | MKKS, AL034430.1 |
| chr20 | 59041842 | 59042376 | 535 | 6 | 4.90E-06 | 2.07E-06 | 0.005681 | 3.66E-06 | 0.016643647 | PRELID3B |
| chr18 | 62186683 | 62187889 | 1207 | 10 | 2.37E-07 | 0.000454 | 9.30E-05 | 3.66E-06 | -5.17E-05 | RELCH, PIGN |
| chr1 | 118985261 | 118986737 | 1477 | 8 | 3.94E-06 | 8.33E-06 | 0.002107 | 3.66E-06 | 0.006274217 | TBX15 |
| chr2 | 741268 | 741887 | 620 | 4 | 2.69E-05 | 1.02E-05 | 0.000227 | 3.66E-06 | 0.012947211 | NA |
| chr15 | 85840168 | 85840810 | 643 | 4 | 7.06E-06 | 2.15E-05 | 0.000136 | 3.66E-06 | 0.000255444 | NA |
| chr17 | 43545439 | 43546866 | 1428 | 18 | 1.75E-06 | 0.00042 | 0.003186 | 3.66E-06 | 0.002233216 | DHX8, ETV4 |
| chr10 | 100993434 | 100994071 | 638 | 2 | 2.35E-05 | 2.23E-06 | 0.00027 | 3.66E-06 | -0.00355021 | TWNK |
| chr2 | 72922615 | 72923107 | 493 | 3 | 1.03E-06 | 0.000561 | 3.89E-05 | 3.67E-06 | 0.005241364 | EMX1 |
| chr4 | 88158861 | 88159549 | 689 | 10 | 2.23E-06 | 1.12E-05 | 0.000556 | 3.67E-06 | 0.004047789 | ABCG2 |
| chr9 | 107638145 | 107638990 | 846 | 3 | 5.41E-05 | 5.55E-05 | 0.000124 | 3.67E-06 | 0.005002746 | NA |
| chr16 | 1672992 | 1674288 | 1297 | 5 | 1.72E-05 | 5.36E-05 | 8.93E-05 | 3.67E-06 | 0.00124752 | CRAMP1, AL031708.1 |
| chr15 | 73929499 | 73930613 | 1115 | 4 | 0.000120933 | 0.000203 | 0.000173 | 3.67E-06 | 0.002608778 | LOXL1 |
| chr11 | 26993926 | 26994444 | 519 | 8 | 3.79E-05 | 1.20E-06 | 0.007322 | 3.67E-06 | -0.01146147 | FIBIN |
| chr6 | 166478728 | 166479594 | 867 | 4 | 4.61E-06 | 2.36E-05 | 0.000286 | 3.68E-06 | 0.001242181 | RPS6KA2 |
| chr21 | 42668310 | 42669233 | 924 | 6 | 1.39E-05 | 2.23E-05 | 0.000176 | 3.68E-06 | -0.00098131 | PDE9A |
| chr19 | 45443014 | 45443527 | 514 | 4 | 3.86E-06 | 6.78E-05 | 0.000176 | 3.68E-06 | 0.003255194 | ERCC1 |
| chr6 | 127514044 | 127515014 | 971 | 6 | 1.61E-06 | 1.79E-05 | 7.85E-05 | 3.68E-06 | 0.006461769 | AL096711.2, SOGA3 |
| chr20 | 48236734 | 48236946 | 213 | 2 | 8.54E-06 | 7.40E-06 | 3.06E-05 | 3.68E-06 | 0.013702696 | NA |
| chr15 | 58933614 | 58934210 | 597 | 9 | 2.18E-06 | 0.000337 | 0.000688 | 3.68E-06 | 0.001634612 | RNF111, SLTM |
| chr1 | 2150545 | 2151409 | 865 | 6 | 9.87E-06 | 7.71E-06 | 0.001568 | 3.68E-06 | 0.008306917 | PRKCZ |
| chr9 | 122861212 | 122861218 | 7 | 2 | 8.96E-06 | 2.07E-05 | 1.45E-05 | 3.69E-06 | 0.009018764 | RC3H2 |
| chr1 | 87327681 | 87327945 | 265 | 5 | 1.78E-06 | 1.48E-05 | 0.000967 | 3.71E-06 | 0.008767477 | NA |
| chr14 | 75579320 | 75580374 | 1055 | 4 | 4.19E-07 | 0.000122 | 0.000344 | 3.73E-06 | 0.008659108 | FLVCR2, AC007182.1 |
| chr3 | 42978232 | 42979301 | 1070 | 10 | 6.64E-06 | 2.73E-05 | 0.002596 | 3.75E-06 | 0.003033878 | AC092042.3, GASK1A |
| chr20 | 48002603 | 48002618 | 16 | 2 | 9.74E-06 | 5.70E-06 | 4.12E-05 | 3.76E-06 | -0.01820717 | NA |
| chr12 | 56151644 | 56152626 | 983 | 16 | 9.54E-06 | 3.69E-06 | 0.012425 | 3.77E-06 | 0.002388626 | MYL6B, AC034102.5 |
| chr12 | 9647262 | 9648420 | 1159 | 11 | 3.78E-06 | 8.52E-06 | 7.00E-05 | 3.77E-06 | 0.000951503 | AC092821.3, AC010186.4, AC010186.2, AC010186.1 |
| chr6 | 137874891 | 137875676 | 786 | 3 | 1.37E-05 | 1.63E-06 | 0.00099 | 3.77E-06 | 0.002534832 | TNFAIP3 |
| chr15 | 76333957 | 76334157 | 201 | 2 | 3.08E-06 | 6.12E-06 | 3.82E-05 | 3.77E-06 | 0.015064036 | NA |
| chr10 | 45426774 | 45427926 | 1153 | 4 | 4.33E-06 | 7.91E-06 | 6.65E-05 | 3.78E-06 | -0.01065252 | ALOX5 |
| chr3 | 196947886 | 196948658 | 773 | 5 | 2.29E-06 | 7.68E-05 | 2.11E-05 | 3.78E-06 | 0.003102975 | PIGZ |
| chr3 | 72348196 | 72348340 | 145 | 2 | 1.55E-05 | 2.97E-06 | 0.000109 | 3.79E-06 | 0.029862208 | NA |
| chr12 | 130787473 | 130787728 | 256 | 3 | 1.87E-06 | 4.25E-05 | 2.46E-05 | 3.79E-06 | 9.35E-05 | NA |
| chr6 | 44257911 | 44258783 | 873 | 9 | 1.63E-06 | 0.00025 | 0.00034 | 3.79E-06 | 0.004150919 | NFKBIE |
| chr12 | 66169636 | 66170486 | 851 | 9 | 1.57E-05 | 2.62E-06 | 0.001463 | 3.79E-06 | -0.00138842 | AC078927.1, TMBIM4 |
| chr2 | 74106554 | 74107427 | 874 | 3 | 5.89E-05 | 7.73E-06 | 9.68E-05 | 3.79E-06 | 0.002105651 | TET3 |
| chr19 | 31149288 | 31149736 | 449 | 3 | 1.24E-06 | 6.91E-05 | 0.0002 | 3.81E-06 | 0.009059588 | NA |
| chr11 | 14905458 | 14906003 | 546 | 4 | 1.64E-06 | 1.22E-05 | 9.40E-05 | 3.81E-06 | 0.00946796 | CALCB |
| chr11 | 61361677 | 61362661 | 985 | 18 | 5.05E-06 | 0.000209 | 0.003049 | 3.81E-06 | 0.00039534 | TMEM138, CYB561A3 |
| chr4 | 78056069 | 78056536 | 468 | 3 | 5.70E-06 | 1.61E-06 | 0.001036 | 3.81E-06 | 0.00760158 | NA |
| chr9 | 137247754 | 137248145 | 392 | 5 | 1.09E-05 | 1.78E-05 | 0.000715 | 3.82E-06 | 0.003637684 | FAM166A |
| chr2 | 231190794 | 231190867 | 74 | 2 | 1.01E-05 | 4.20E-06 | 6.12E-05 | 3.82E-06 | 0.028181876 | NA |
| chr7 | 129830890 | 129831164 | 275 | 2 | 4.43E-06 | 3.65E-06 | 7.59E-05 | 3.83E-06 | -0.00391555 | UBE2H |
| chr10 | 85786396 | 85786862 | 467 | 2 | 1.49E-05 | 2.58E-06 | 0.000165 | 3.83E-06 | 0.012441374 | GRID1 |
| chr11 | 3165319 | 3166786 | 1468 | 29 | 2.36E-05 | 0.000441 | 0.002016 | 3.83E-06 | 0.000665566 | OSBPL5 |
| chr18 | 50972772 | 50973475 | 704 | 3 | 4.04E-05 | 0.000105 | 9.00E-05 | 3.83E-06 | 0.010648766 | ELAC1, AC091551.1 |
| chr8 | 102813989 | 102814350 | 362 | 2 | 1.46E-05 | 2.37E-06 | 0.000249 | 3.83E-06 | 0.010379629 | NA |
| chr2 | 114661960 | 114662683 | 724 | 7 | 6.07E-06 | 1.17E-05 | 0.003626 | 3.84E-06 | -0.01758058 | DPP10 |
| chr11 | 118500416 | 118501021 | 606 | 3 | 8.62E-06 | 1.59E-06 | 0.001061 | 3.84E-06 | 0.00361966 | KMT2A |
| chr12 | 57617981 | 57618818 | 838 | 5 | 5.23E-07 | 3.37E-05 | 1.59E-05 | 3.84E-06 | 0.011821143 | AC025165.6, AC025165.1 |
| chr20 | 43569004 | 43569426 | 423 | 4 | 1.41E-06 | 0.000621 | 0.000107 | 3.84E-06 | 0.004597378 | Z98752.3, SGK2 |
| chr1 | 76866255 | 76867144 | 890 | 3 | 9.50E-08 | 1.34E-05 | 7.54E-05 | 3.85E-06 | 0.006245236 | NA |
| chr11 | 76368693 | 76369363 | 671 | 2 | 2.16E-06 | 5.13E-06 | 4.75E-05 | 3.85E-06 | 0.004196563 | THAP12 |
| chr11 | 6418576 | 6419049 | 474 | 4 | 5.40E-06 | 3.01E-06 | 0.000449 | 3.85E-06 | 0.001441021 | APBB1 |
| chr11 | 72722014 | 72722574 | 561 | 7 | 1.00E-06 | 2.72E-05 | 0.000297 | 3.85E-06 | 0.001266845 | ARAP1 |
| chr6 | 35342045 | 35342664 | 620 | 10 | 2.27E-06 | 1.27E-05 | 0.000369 | 3.85E-06 | 0.00226749 | PPARD |
| chr11 | 93783971 | 93784963 | 993 | 12 | 4.21E-06 | 6.79E-05 | 0.001177 | 3.85E-06 | -0.00043061 | AP001273.2, MED17, TAF1D |

| | | | | | | | | | | |
|---|---|---|---|---|---|---|---|---|---|---|
| chr11 | 43576682 | 43577805 | 1124 | 7 | 2.12E-06 | 0.000207 | 0.000104 | 3.86E-06 | 0.006615339 | AC068205.1 |
| chr11 | 64629861 | 64630665 | 805 | 5 | 5.32E-06 | 1.44E-05 | 0.000192 | 3.86E-06 | 0.000773806 | NRXN2 |
| chr10 | 129887839 | 129888621 | 783 | 8 | 2.48E-05 | 2.11E-05 | 0.000238 | 3.86E-06 | 0.004353824 | EBF3 |
| chr20 | 37524411 | 37524823 | 413 | 4 | 1.39E-06 | 1.67E-05 | 3.22E-05 | 3.87E-06 | 0.006023226 | BLCAP |
| chr4 | 114599775 | 114600345 | 571 | 3 | 1.41E-05 | 1.45E-06 | 0.001513 | 3.87E-06 | 0.009640422 | UGT8 |
| chr19 | 8589505 | 8589924 | 420 | 3 | 7.84E-06 | 2.04E-05 | 8.21E-05 | 3.87E-06 | 0.005292172 | ADAMTS10 |
| chr1 | 16073154 | 16074259 | 1106 | 7 | 1.83E-05 | 1.79E-06 | 0.00075 | 3.88E-06 | 0.003634649 | FAM131C |
| chr3 | 15049790 | 15050258 | 469 | 2 | 7.14E-07 | 6.55E-06 | 3.73E-05 | 3.88E-06 | 0.007566313 | NR2C2, MRPS25 |
| chr11 | 103108678 | 103109737 | 1060 | 13 | 5.52E-06 | 1.94E-05 | 0.001071 | 3.89E-06 | -0.00059435 | DYNC2H1 |
| chr6 | 33305683 | 33306704 | 1022 | 6 | 2.54E-05 | 2.52E-06 | 0.000963 | 3.89E-06 | 0.004639332 | TAPBP |
| chr1 | 172347653 | 172348441 | 789 | 3 | 4.05E-05 | 1.38E-06 | 0.001817 | 3.89E-06 | 0.009461007 | DNM3 |
| chr9 | 26892474 | 26893345 | 872 | 12 | 2.50E-06 | 6.57E-05 | 0.000373 | 3.91E-06 | -0.00070927 | CAAP1 |
| chr1 | 109438788 | 109438957 | 170 | 2 | 4.23E-06 | 2.54E-05 | 1.37E-05 | 3.91E-06 | 0.007282595 | NA |
| chr19 | 14505910 | 14506410 | 501 | 4 | 5.60E-06 | 1.81E-05 | 0.000185 | 3.91E-06 | 0.002371743 | NA |
| chr10 | 114685366 | 114685932 | 567 | 4 | 4.85E-08 | 0.00064 | 8.68E-06 | 3.91E-06 | 0.00515951 | ABLIM1 |
| chr4 | 38523228 | 38523254 | 27 | 3 | 4.61E-06 | 1.94E-06 | 0.000717 | 3.91E-06 | 0.012223108 | NA |
| chr7 | 88759689 | 88760326 | 638 | 5 | 2.24E-05 | 7.29E-07 | 0.006368 | 3.91E-06 | 0.007285609 | ZNF804B |
| chr11 | 112164078 | 112164956 | 879 | 8 | 1.74E-05 | 2.45E-06 | 0.003082 | 3.91E-06 | 0.001277652 | AP002884.2, IL18 |
| chr7 | 22589429 | 22589855 | 427 | 3 | 1.82E-07 | 0.000119 | 7.05E-06 | 3.92E-06 | 0.004539849 | AC002480.2, STEAP1B |
| chr11 | 75814836 | 75815244 | 409 | 8 | 4.92E-05 | 1.55E-06 | 0.004668 | 3.92E-06 | 0.001401084 | UVRAG, UVRAG-DT |
| chr12 | 132441522 | 132442452 | 931 | 3 | 1.74E-05 | 1.80E-06 | 0.00058 | 3.92E-06 | 0.005243883 | NA |
| chr9 | 104991881 | 104992255 | 375 | 4 | 2.65E-07 | 0.002292 | 3.10E-06 | 3.92E-06 | 0.003447629 | NA |
| chr2 | 241987888 | 241988253 | 366 | 3 | 1.15E-05 | 1.78E-06 | 0.000817 | 3.92E-06 | 0.001915776 | LINC01237 |
| chr11 | 9784868 | 9785496 | 629 | 3 | 5.05E-06 | 5.17E-06 | 0.000106 | 3.93E-06 | 0.010086978 | SBF2-AS1, SBF2 |
| chr7 | 142856370 | 142856777 | 408 | 3 | 4.46E-06 | 5.08E-06 | 0.000271 | 3.93E-06 | 0.009015091 | EPHB6 |
| chr7 | 97012725 | 97014142 | 1418 | 9 | 3.16E-06 | 1.95E-05 | 0.000267 | 3.93E-06 | 0.004806954 | DLX6-AS1, AC004774.2 |
| chr2 | 47040274 | 47040673 | 400 | 3 | 5.21E-06 | 0.000355 | 6.06E-05 | 3.93E-06 | -0.00077228 | TTC7A, AC093732.1 |
| chr9 | 14372712 | 14373046 | 335 | 2 | 1.58E-05 | 2.60E-06 | 0.000185 | 3.94E-06 | 0.037479968 | NFIB |
| chr19 | 2044541 | 2045013 | 473 | 2 | 9.38E-07 | 9.64E-06 | 2.63E-05 | 3.94E-06 | 0.006950438 | MKNK2 |
| chr10 | 119536498 | 119536530 | 33 | 2 | 3.75E-06 | 1.58E-05 | 1.82E-05 | 3.94E-06 | 0.011260435 | RGS10 |
| chr6 | 30060961 | 30062233 | 1273 | 29 | 7.01E-06 | 1.84E-05 | 0.003674 | 3.94E-06 | -0.00123036 | POLR1H, AL669914.4, ZNRD1ASP |
| chr20 | 58130882 | 58131686 | 805 | 2 | 3.95E-05 | 2.37E-06 | 0.00031 | 3.94E-06 | -0.00992534 | NA |
| chr6 | 125749572 | 125750392 | 821 | 8 | 5.39E-06 | 1.31E-06 | 0.003393 | 3.94E-06 | 0.002572374 | HEY2 |
| chr6 | 167678092 | 167678747 | 656 | 6 | 4.14E-06 | 2.36E-05 | 0.000138 | 3.94E-06 | 0.004908854 | AL356123.2, LINC02538 |
| chr1 | 211680931 | 211681801 | 871 | 6 | 1.45E-05 | 2.38E-05 | 0.000271 | 3.94E-06 | 0.009411342 | AC096637.2 |
| chr4 | 7054240 | 7054853 | 614 | 4 | 2.18E-07 | 0.00031 | 0.000155 | 3.94E-06 | 0.0035609 | TADA2B |
| chr6 | 168826071 | 168826602 | 532 | 4 | 1.58E-07 | 0.000325 | 7.29E-06 | 3.95E-06 | 0.002018668 | NA |
| chr14 | 20305250 | 20306060 | 811 | 13 | 1.55E-05 | 1.03E-05 | 0.000138 | 3.95E-06 | -0.00107832 | AL356019.1, TTC5 |
| chr13 | 20526052 | 20526759 | 708 | 6 | 2.14E-06 | 9.90E-06 | 0.000386 | 3.95E-06 | 0.000953874 | NA |
| chr12 | 25957585 | 25958888 | 1304 | 11 | 1.20E-05 | 4.39E-06 | 0.005613 | 3.95E-06 | -0.00039485 | RASSF8-AS1 |
| chr3 | 111069891 | 111070625 | 735 | 6 | 2.66E-05 | 5.26E-07 | 0.01122 | 3.95E-06 | 0.012360925 | NECTIN3, NECTIN3-AS1 |
| chr15 | 74540442 | 74541497 | 1056 | 8 | 1.30E-05 | 4.87E-06 | 0.003491 | 3.95E-06 | 0.001389385 | ARID3B |
| chr19 | 11179788 | 11180326 | 539 | 2 | 4.73E-07 | 4.63E-06 | 5.74E-05 | 3.96E-06 | 0.002338619 | KANK2 |
| chr3 | 119145618 | 119146631 | 1014 | 14 | 1.64E-05 | 2.56E-06 | 0.012808 | 3.96E-06 | 0.002380352 | TEX55, IGSF11 |
| chr5 | 179362650 | 179363409 | 760 | 2 | 3.56E-05 | 2.71E-06 | 0.000163 | 3.96E-06 | 0.001793961 | NA |
| chr3 | 123450574 | 123450721 | 148 | 3 | 3.24E-06 | 0.000111 | 0.000114 | 3.96E-06 | 0.006765517 | NA |
| chr15 | 86141795 | 86142734 | 940 | 5 | 3.44E-06 | 0.000444 | 3.74E-05 | 3.96E-06 | 0.001696939 | AGBL1 |
| chr20 | 25213365 | 25214276 | 912 | 3 | 7.93E-05 | 1.35E-05 | 0.000112 | 3.97E-06 | 0.003361109 | ENTPD6 |
| chr8 | 19661406 | 19661562 | 157 | 2 | 7.21E-06 | 2.57E-05 | 1.40E-05 | 3.97E-06 | 0.005354076 | CSGALNACT1 |
| chr8 | 141339675 | 141340434 | 760 | 3 | 1.02E-05 | 2.87E-06 | 0.000172 | 3.97E-06 | 0.00549693 | NA |
| chr10 | 133086721 | 133087402 | 682 | 4 | 2.34E-07 | 1.24E-05 | 5.92E-05 | 3.98E-06 | 0.002721568 | ADGRA1, ADGRA1-AS1 |
| chr13 | 66994341 | 66994386 | 46 | 2 | 1.11E-05 | 2.35E-06 | 0.000363 | 3.99E-06 | 0.02185961 | PCDH9-AS4, PCDH9 |
| chr5 | 139903275 | 139903766 | 492 | 7 | 3.21E-06 | 1.44E-05 | 0.003823 | 3.99E-06 | 0.008334225 | NRG2 |
| chr18 | 48700785 | 48700879 | 95 | 2 | 1.49E-05 | 2.95E-06 | 0.000132 | 4.01E-06 | 0.011227847 | CTIF |
| chr1 | 22637112 | 22638108 | 997 | 4 | 2.19E-05 | 1.08E-06 | 0.003584 | 4.01E-06 | 0.005577566 | C1QA |
| chr11 | 65781789 | 65781970 | 182 | 3 | 4.89E-06 | 2.13E-06 | 0.000386 | 4.03E-06 | 0.006791759 | NA |
| chr6 | 36387711 | 36388467 | 757 | 10 | 1.19E-06 | 0.000141 | 0.00268 | 4.03E-06 | 0.010239952 | ETV7-AS1, ETV7 |
| chr13 | 113361442 | 113361825 | 384 | 2 | 7.15E-06 | 5.26E-06 | 5.00E-05 | 4.03E-06 | 0.017171449 | GRTP1-AS1, GRTP1 |
| chr3 | 188212419 | 188213008 | 590 | 6 | 1.66E-05 | 2.51E-06 | 0.00223 | 4.03E-06 | 0.003136518 | LPP |
| chr7 | 36287475 | 36287947 | 473 | 4 | 2.13E-05 | 4.15E-06 | 0.000864 | 4.03E-06 | 0.007082165 | EEPD1 |
| chr1 | 239387877 | 239388828 | 952 | 3 | 9.68E-05 | 7.28E-05 | 0.000136 | 4.03E-06 | -0.01130336 | CHRM3 |
| chr3 | 13019415 | 13020167 | 753 | 4 | 4.05E-05 | 1.27E-06 | 0.001564 | 4.04E-06 | 0.009059381 | IQSEC1 |
| chr5 | 138177864 | 138178998 | 1135 | 20 | 1.37E-05 | 2.56E-05 | 0.001687 | 4.04E-06 | 0.002371814 | KIF20A, BRD8 |
| chr17 | 35257534 | 35257736 | 203 | 2 | 2.03E-05 | 2.37E-06 | 0.00038 | 4.04E-06 | 0.058540703 | SLFN5 |
| chr1 | 150579449 | 150580341 | 893 | 15 | 5.63E-05 | 1.30E-06 | 0.008966 | 4.04E-06 | -0.0016711 | MCL1 |
| chr1 | 6154712 | 6155565 | 854 | 4 | 2.43E-05 | 6.35E-06 | 0.000127 | 4.04E-06 | 0.011922192 | CHD5 |
| chr11 | 33694073 | 33694488 | 416 | 2 | 1.64E-06 | 3.32E-05 | 1.24E-05 | 4.05E-06 | 0.008080833 | AL049629.1 |
| chr8 | 66522287 | 66523098 | 812 | 3 | 1.84E-06 | 4.31E-05 | 2.12E-05 | 4.05E-06 | 0.007623081 | NA |
| chr16 | 70749841 | 70750616 | 776 | 2 | 1.71E-05 | 2.87E-06 | 0.000148 | 4.05E-06 | 0.009157675 | VAC14 |
| chr1 | 203768430 | 203768844 | 415 | 4 | 4.85E-06 | 7.37E-06 | 0.000256 | 4.06E-06 | 0.006278315 | LAX1 |
| chr6 | 27809871 | 27810469 | 599 | 8 | 3.88E-06 | 6.45E-05 | 0.000386 | 4.06E-06 | 0.000962202 | H3C10 |
| chr7 | 27048288 | 27048517 | 230 | 2 | 1.36E-05 | 3.48E-06 | 9.36E-05 | 4.06E-06 | 0.005071632 | NHP2P2 |
| chr1 | 211832068 | 211832113 | 46 | 2 | 1.33E-07 | 9.90E-05 | 8.15E-06 | 4.06E-06 | 0.003668818 | LPGAT1-AS1 |
| chr2 | 39664589 | 39665472 | 884 | 6 | 4.88E-06 | 5.61E-06 | 0.001317 | 4.07E-06 | 0.007857927 | MAP4K3-DT, TMEM178A |
| chr14 | 70810131 | 70811120 | 990 | 8 | 1.89E-05 | 1.03E-05 | 0.002938 | 4.07E-06 | 0.011480388 | AC004816.1 |
| chr20 | 49046006 | 49046699 | 694 | 11 | 1.90E-05 | 3.52E-05 | 0.000819 | 4.07E-06 | -0.00035967 | CSE1L, CSE1L-AS1 |
| chr13 | 112886086 | 112886317 | 232 | 4 | 8.66E-06 | 1.07E-06 | 0.003497 | 4.10E-06 | 0.039391455 | ATP11A |
| chr17 | 36601157 | 36602394 | 1238 | 11 | 1.85E-07 | 0.001548 | 1.10E-05 | 4.10E-06 | -0.00075412 | MRM1 |
| chr7 | 70790224 | 70791017 | 794 | 6 | 2.16E-05 | 4.14E-06 | 0.002105 | 4.10E-06 | 0.000315002 | AUTS2 |
| chr3 | 58151872 | 58152793 | 922 | 3 | 7.22E-06 | 7.79E-06 | 7.76E-05 | 4.10E-06 | -0.00192715 | FLNB |
| chr2 | 240559494 | 240560674 | 1181 | 7 | 3.14E-07 | 0.00011 | 4.85E-05 | 4.11E-06 | 0.000307965 | DUSP28, ANKMY1 |
| chr1 | 165356832 | 165357768 | 937 | 8 | 2.20E-06 | 5.18E-06 | 0.00015 | 4.11E-06 | 0.006890876 | NA |
| chr2 | 65431449 | 65432225 | 777 | 3 | 6.30E-07 | 8.20E-06 | 0.000342 | 4.11E-06 | 0.001407407 | SPRED2 |
| chr11 | 130197911 | 130198553 | 643 | 5 | 1.02E-05 | 5.54E-05 | 0.00018 | 4.11E-06 | 0.003641184 | ST14 |
| chr1 | 117591393 | 117591915 | 523 | 2 | 7.35E-07 | 2.23E-05 | 1.55E-05 | 4.11E-06 | 0.008815484 | NA |
| chr8 | 9884450 | 9884514 | 65 | 2 | 9.08E-06 | 2.93E-05 | 1.35E-05 | 4.11E-06 | 0.029198628 | NA |
| chr9 | 82272050 | 82272054 | 5 | 2 | 9.11E-06 | 1.29E-05 | 2.21E-05 | 4.11E-06 | -0.00124618 | AL162726.3 |
| chr7 | 1980247 | 1980948 | 702 | 5 | 1.01E-06 | 4.26E-05 | 0.000599 | 4.11E-06 | 0.002467223 | MAD1L1, AC069288.1 |
| chr10 | 73359306 | 73360495 | 1190 | 7 | 2.58E-05 | 1.64E-05 | 0.003347 | 4.11E-06 | 0.002100408 | NA |
| chr14 | 63203993 | 63205019 | 1027 | 13 | 4.20E-05 | 1.48E-06 | 0.013222 | 4.12E-06 | 0.022003284 | RHOJ |
| chr19 | 56507245 | 56507564 | 320 | 3 | 2.59E-06 | 0.000558 | 4.06E-06 | 4.12E-06 | 0.015267436 | NA |
| chr18 | 76923806 | 76924447 | 642 | 2 | 3.25E-06 | 5.25E-06 | 5.25E-05 | 4.12E-06 | 0.002896651 | ZNF236 |
| chr2 | 43226419 | 43227833 | 1415 | 23 | 1.39E-05 | 9.21E-06 | 0.007182 | 4.12E-06 | 0.000723589 | LINC01126, ZFP36L2 |
| chr19 | 12887183 | 12887950 | 768 | 7 | 6.23E-05 | 1.23E-05 | 0.000924 | 4.13E-06 | 0.004314334 | KLF1 |
| chr9 | 87725302 | 87725865 | 564 | 9 | 7.90E-06 | 1.90E-05 | 0.001755 | 4.14E-06 | 0.003805969 | CTSL |
| chr2 | 70108901 | 70108986 | 86 | 3 | 2.58E-05 | 1.74E-06 | 0.00093 | 4.14E-06 | 0.009772396 | NA |
| chr3 | 186825689 | 186825985 | 297 | 3 | 2.04E-05 | 2.15E-06 | 0.00074 | 4.14E-06 | 0.013777399 | NA |
| chr17 | 28497405 | 28498277 | 873 | 2 | 6.30E-05 | 2.50E-06 | 0.000318 | 4.14E-06 | 0.006737722 | SLC13A2, AC005726.1 |
| chr11 | 395625 | 396114 | 490 | 4 | 7.07E-06 | 6.16E-06 | 0.000149 | 4.15E-06 | 0.006519423 | PKP3 |
| chr1 | 118184908 | 118185603 | 696 | 12 | 2.56E-05 | 1.14E-05 | 0.008647 | 4.15E-06 | 0.005165302 | AL391557.1, SPAG17 |
| chr19 | 48867822 | 48868844 | 1023 | 3 | 1.14E-06 | 1.11E-05 | 6.29E-05 | 4.16E-06 | 0.001359882 | PLEKHA4 |
| chr10 | 23694337 | 23696048 | 1712 | 11 | 0.000253834 | 0.000129 | 0.001693 | 4.16E-06 | 0.003166862 | KIAA1217 |
| chr14 | 60510180 | 60511138 | 959 | 5 | 1.09E-05 | 1.55E-06 | 0.001546 | 4.16E-06 | 0.004093031 | SIX6, C14orf39 |
| chr21 | 43694092 | 43695059 | 968 | 6 | 1.94E-05 | 8.04E-06 | 0.00125 | 4.16E-06 | 0.000951316 | RRP1B |
| chr2 | 70790079 | 70791691 | 1613 | 12 | 0.000230289 | 7.11E-06 | 0.003171 | 4.17E-06 | 0.006355099 | FIGLA |
| chr17 | 31916809 | 31917611 | 803 | 6 | 1.09E-05 | 3.38E-06 | 0.002723 | 4.17E-06 | -0.02104226 | NA |
| chr6 | 31645088 | 31646435 | 1348 | 5 | 1.22E-05 | 5.67E-06 | 0.00207 | 4.18E-06 | 0.000827995 | BAG6 |
| chr3 | 188573450 | 188573845 | 396 | 2 | 1.78E-05 | 2.66E-06 | 0.000229 | 4.18E-06 | 0.012754118 | LPP |
| chr2 | 241652263 | 241652989 | 727 | 3 | 5.91E-07 | 0.000349 | 6.84E-06 | 4.18E-06 | 0.001585455 | ATG4B |
| chr7 | 114086048 | 114086818 | 771 | 4 | 1.21E-06 | 3.39E-05 | 8.08E-05 | 4.18E-06 | 0.000379757 | FOXP2 |
| chr5 | 62412341 | 62413307 | 967 | 12 | 1.55E-05 | 1.40E-05 | 0.007097 | 4.20E-06 | 0.000490233 | AC114982.2, IPO11 |
| chr7 | 149003732 | 149004312 | 581 | 4 | 6.41E-06 | 1.50E-05 | 0.00072 | 4.20E-06 | 0.003776444 | PDIA4 |
| chr8 | 23726585 | 23727673 | 1089 | 7 | 2.90E-06 | 5.71E-05 | 0.000798 | 4.21E-06 | 0.004886477 | AC012574.1, AC012574.2 |
| chr10 | 112951056 | 112952173 | 1118 | 5 | 0.000114905 | 1.85E-05 | 0.000788 | 4.22E-06 | -0.00035873 | TCF7L2, AL158212.2 |
| chr1 | 3633168 | 3633696 | 529 | 5 | 1.98E-05 | 3.53E-06 | 0.000974 | 4.22E-06 | 0.004009657 | WRAP73 |
| chr11 | 67423804 | 67424302 | 499 | 3 | 2.99E-06 | 3.08E-05 | 0.000231 | 4.22E-06 | 0.003640453 | CARNS1 |
| chr11 | 27721711 | 27722816 | 1106 | 12 | 1.29E-05 | 5.30E-06 | 0.017504 | 4.22E-06 | 0.003374002 | BDNF, AC103796.1 |
| chr12 | 64171181 | 64171797 | 617 | 2 | 3.08E-05 | 2.64E-06 | 0.000256 | 4.23E-06 | -0.00940692 | NA |
| chr7 | 8442364 | 8443247 | 884 | 8 | 3.97E-06 | 1.18E-05 | 0.001132 | 4.24E-06 | 0.00258032 | NXPH1 |
| chr5 | 97142791 | 97143649 | 859 | 4 | 2.79E-05 | 0.000311 | 9.41E-05 | 4.24E-06 | 0.003587273 | LIX1-AS1 |
| chr12 | 122096880 | 122097358 | 479 | 6 | 1.58E-05 | 2.69E-06 | 0.003466 | 4.24E-06 | 0.000955847 | MLXIP |
| chr16 | 4329698 | 4330752 | 1055 | 5 | 1.71E-05 | 1.31E-05 | 0.000522 | 4.24E-06 | 0.006867607 | GLIS2 |
| chr7 | 100463988 | 100465198 | 1211 | 4 | 6.86E-05 | 4.81E-06 | 0.000411 | 4.24E-06 | 0.002463301 | C7orf61, TSC22D4 |
| chr11 | 70109979 | 70110104 | 126 | 2 | 2.23E-06 | 1.71E-05 | 1.88E-05 | 4.25E-06 | 0.005377547 | ANO1 |
| chr3 | 107598153 | 107598907 | 755 | 3 | 1.21E-06 | 0.000226 | 0.000132 | 4.25E-06 | 0.008872072 | BBX |
| chr3 | 114450849 | 114451621 | 773 | 2 | 1.65E-05 | 3.13E-06 | 0.000134 | 4.25E-06 | 0.006703735 | ZBTB20-AS5, ZBTB20 |
| chr17 | 45146354 | 45148432 | 2079 | 15 | 0.000121549 | 1.72E-05 | 0.004504 | 4.25E-06 | -0.00024953 | AC142472.1 |
| chr6 | 10694312 | 10695604 | 1293 | 17 | 3.68E-05 | 0.000144 | 0.004417 | 4.25E-06 | -0.00026806 | PAK1IP1, C6orf52 |
| chr10 | 28302306 | 28303170 | 865 | 7 | 1.57E-05 | 1.69E-05 | 0.002279 | 4.25E-06 | 0.001430429 | MPP7 |
| chr7 | 100088707 | 100089333 | 627 | 9 | 2.93E-06 | 2.92E-05 | 0.000597 | 4.26E-06 | -0.00030652 | COPS6 |
| chr5 | 146446962 | 146448174 | 1213 | 11 | 2.25E-06 | 0.000108 | 0.0001 | 4.26E-06 | 0.000754816 | TCERG1 |
| chr16 | 89905007 | 89905662 | 656 | 3 | 2.61E-05 | 5.28E-05 | 8.41E-05 | 4.26E-06 | 0.003593435 | TCF25 |
| chr1 | 13779317 | 13780492 | 1176 | 5 | 1.21E-05 | 4.00E-05 | 0.000199 | 4.26E-06 | -0.00013632 | PRDM2 |
| chr15 | 85389922 | 85390703 | 782 | 5 | 4.47E-06 | 7.52E-05 | 0.000146 | 4.26E-06 | -0.00028564 | AKAP13 |
| chr9 | 121733556 | 121734036 | 481 | 2 | 2.05E-06 | 6.25E-06 | 4.51E-05 | 4.27E-06 | 0.01021567 | DAB2IP |
| chr15 | 61226775 | 61227287 | 513 | 3 | 3.33E-06 | 3.26E-06 | 0.000161 | 4.28E-06 | 0.011173946 | RORA |
| chr1 | 151146318 | 151147051 | 734 | 8 | 7.11E-05 | 2.68E-06 | 0.005885 | 4.29E-06 | 0.000635216 | SEMA6C |
| chr22 | 49305962 | 49306466 | 505 | 2 | 3.15E-06 | 7.99E-06 | 3.53E-05 | 4.29E-06 | 0.006974365 | NA |
| chr3 | 48174833 | 48175403 | 571 | 4 | 9.16E-06 | 5.68E-06 | 8.23E-05 | 4.29E-06 | 0.002291135 | CDC25A |
| chr12 | 7060009 | 7061144 | 1136 | 10 | 2.66E-06 | 0.000108 | 0.000188 | 4.29E-06 | 0.006449225 | C1S |
| chr11 | 108406483 | 108406800 | 318 | 2 | 9.01E-06 | 2.77E-06 | 0.000222 | 4.29E-06 | 0.014936553 | C11orf65 |
| chr11 | 101915928 | 101916679 | 752 | 3 | 7.81E-06 | 2.76E-06 | 0.000485 | 4.29E-06 | 0.008401295 | CEP126, ANGPTL5 |
| chr6 | 106092798 | 106092836 | 39 | 2 | 1.20E-05 | 1.49E-05 | 2.09E-05 | 4.29E-06 | 0.003001188 | PRDM1, ATG5 |
| chr3 | 133805162 | 133805814 | 653 | 7 | 3.34E-05 | 8.50E-07 | 0.007618 | 4.29E-06 | 0.003895679 | SRPRB, AC080128.2 |
| chr15 | 41834554 | 41835023 | 470 | 4 | 3.88E-06 | 1.91E-05 | 2.56E-05 | 4.29E-06 | 0.002173749 | JMJD7, JMJD7-PLA2G4B |
| chr9 | 92617647 | 92618318 | 672 | 2 | 5.99E-06 | 3.23E-06 | 0.000128 | 4.29E-06 | 0.007180071 | CENPP, IPPK |
| chr16 | 80039543 | 80039968 | 426 | 2 | 1.88E-06 | 3.11E-05 | 1.37E-05 | 4.30E-06 | 0.008338522 | NA |
| chr3 | 14178180 | 14179335 | 1156 | 20 | 4.49E-05 | 9.36E-06 | 0.003562 | 4.30E-06 | 0.000829217 | LSM3, XPC |
| chr14 | 22829263 | 22830347 | 1085 | 14 | 4.18E-06 | 2.23E-05 | 0.002192 | 4.30E-06 | 0.000805497 | MRPL52, SLC7A7 |
| chr1 | 207891255 | 207891896 | 642 | 3 | 8.45E-07 | 5.70E-06 | 0.000431 | 4.31E-06 | -0.00353296 | CD34 |
| chr5 | 176643734 | 176644467 | 734 | 4 | 1.29E-05 | 2.90E-06 | 0.000777 | 4.32E-06 | 0.005156172 | EIF4E1B |
| chr3 | 56675810 | 56676348 | 539 | 2 | 9.37E-06 | 4.04E-06 | 8.35E-05 | 4.32E-06 | 0.007636755 | TASOR |
| chr6 | 30626409 | 30627595 | 1187 | 17 | 8.07E-06 | 4.35E-05 | 0.002423 | 4.32E-06 | 0.000793079 | ATAT1 |
| chr11 | 43881236 | 43881871 | 636 | 4 | 8.55E-07 | 1.02E-05 | 0.000107 | 4.32E-06 | -0.00953758 | ALKBH3 |
| chr4 | 2364758 | 2365104 | 347 | 3 | 1.34E-05 | 2.36E-06 | 0.000361 | 4.33E-06 | -0.03353112 | ZFYVE28 |
| chr1 | 234980890 | 234981821 | 932 | 6 | 3.06E-05 | 2.32E-06 | 0.002166 | 4.33E-06 | 0.006412774 | NA |
| chr5 | 177085895 | 177086703 | 809 | 5 | 2.05E-06 | 5.23E-06 | 0.000168 | 4.33E-06 | 0.008944126 | NA |
| chr1 | 2413559 | 2414455 | 897 | 11 | 6.32E-06 | 3.25E-06 | 0.001139 | 4.35E-06 | 0.006056371 | PEX10 |
| chr17 | 78278059 | 78279052 | 994 | 5 | 3.29E-05 | 1.87E-06 | 0.002137 | 4.37E-06 | 0.013382825 | LINC01993 |
| chr2 | 100285667 | 100285783 | 117 | 2 | 1.95E-05 | 2.72E-06 | 0.000266 | 4.37E-06 | 0.037377102 | LONRF2 |
| chr6 | 28479017 | 28479338 | 322 | 5 | 3.96E-05 | 8.21E-07 | 0.007153 | 4.37E-06 | 0.026497305 | Z98745.2 |
| chr16 | 1964766 | 1965898 | 1133 | 12 | 5.51E-05 | 1.83E-06 | 0.004 | 4.38E-06 | -0.00188159 | SNHG9, SNORA78, RPS2 |

| | | | | | | | | | | |
|---|---|---|---|---|---|---|---|---|---|---|
| chr19 | 11295438 | 11295499 | 62 | 3 | 2.96E-06 | 1.01E-05 | 0.000103 | 4.38E-06 | 0.003579688 | NA |
| chr11 | 67451770 | 67452847 | 1078 | 10 | 6.53E-05 | 4.93E-06 | 0.00464 | 4.38E-06 | 0.003161342 | CABP4, GPR152 |
| chr16 | 76263505 | 76263551 | 47 | 2 | 4.12E-06 | 8.05E-05 | 9.31E-06 | 4.38E-06 | -0.01609061 | NA |
| chr5 | 167754452 | 167754840 | 389 | 6 | 1.28E-06 | 0.000588 | 1.56E-05 | 4.38E-06 | 0.004391594 | TENM2 |
| chr16 | 72921262 | 72921396 | 135 | 3 | 2.70E-05 | 1.79E-06 | 0.001333 | 4.38E-06 | 0.012386441 | ZFHX3 |
| chr8 | 55991700 | 55992251 | 552 | 2 | 1.73E-07 | 0.000971 | 5.90E-06 | 4.39E-06 | 0.013096785 | LYN |
| chr6 | 33630175 | 33630226 | 52 | 2 | 1.94E-05 | 3.70E-06 | 0.0001 | 4.39E-06 | 0.012311567 | ITPR3 |
| chr19 | 40613372 | 40614423 | 1052 | 10 | 1.34E-06 | 0.000598 | 0.000396 | 4.39E-06 | 9.13E-05 | LTBP4 |
| chr22 | 22118370 | 22118374 | 5 | 2 | 2.21E-05 | 2.58E-06 | 0.0004 | 4.40E-06 | -0.05647065 | IGLV10-54 |
| chr3 | 136070506 | 136071258 | 753 | 3 | 4.29E-06 | 3.66E-05 | 2.72E-05 | 4.40E-06 | 0.010349661 | PPP2R3A |
| chr12 | 124818071 | 124818209 | 139 | 2 | 2.97E-06 | 1.81E-05 | 1.89E-05 | 4.40E-06 | 0.001014485 | SCARB1 |
| chr1 | 30807296 | 30807897 | 602 | 5 | 4.20E-06 | 1.96E-05 | 0.000294 | 4.41E-06 | -0.00915573 | NA |
| chr11 | 102767701 | 102768047 | 347 | 6 | 2.54E-05 | 2.18E-06 | 0.003641 | 4.41E-06 | -0.01864949 | WTAPP1, AP000619.3 |
| chr14 | 60654994 | 60655609 | 616 | 4 | 1.48E-06 | 1.22E-05 | 0.000158 | 4.41E-06 | 0.008991004 | SIX1 |
| chr8 | 6374528 | 6374630 | 103 | 2 | 1.73E-05 | 4.70E-06 | 6.87E-05 | 4.42E-06 | 0.022877689 | NA |
| chr4 | 881490 | 882668 | 1179 | 6 | 3.78E-06 | 9.38E-06 | 0.001255 | 4.42E-06 | 0.001418535 | GAK |
| chr4 | 185534855 | 185535858 | 1004 | 11 | 3.94E-06 | 0.00014 | 5.81E-05 | 4.42E-06 | 0.000995369 | PDLIM3 |
| chr20 | 38727322 | 38728201 | 880 | 4 | 7.78E-06 | 0.000106 | 0.000269 | 4.42E-06 | 0.006524872 | SLC32A1 |
| chr5 | 95391070 | 95391111 | 42 | 2 | 8.82E-06 | 3.45E-06 | 0.000119 | 4.42E-06 | 0.010323276 | NA |
| chr16 | 4537747 | 4538951 | 1205 | 14 | 1.35E-05 | 0.000123 | 0.001088 | 4.42E-06 | 0.001908907 | CDIP1 |
| chr12 | 131872217 | 131872824 | 608 | 4 | 4.07E-05 | 7.54E-06 | 0.000848 | 4.42E-06 | 0.0081023 | NA |
| chr19 | 56134274 | 56134751 | 478 | 3 | 4.28E-06 | 0.000825 | 0.00013 | 4.42E-06 | 0.009029441 | ZNF444 |
| chr10 | 133019393 | 133020321 | 929 | 8 | 8.91E-06 | 0.000416 | 0.002578 | 4.43E-06 | 0.003411189 | NA |
| chr19 | 50432048 | 50432915 | 868 | 9 | 1.17E-05 | 6.33E-07 | 0.014296 | 4.43E-06 | 0.004895077 | MYBPC2 |
| chr5 | 173195860 | 173196823 | 964 | 3 | 9.42E-05 | 4.15E-05 | 0.000252 | 4.44E-06 | 0.008999319 | NA |
| chr1 | 63773720 | 63774531 | 812 | 10 | 3.99E-06 | 0.000162 | 0.000121 | 4.44E-06 | 0.000110243 | ROR1 |
| chr11 | 18526663 | 18527742 | 1080 | 15 | 1.59E-05 | 0.000109 | 0.006756 | 4.45E-06 | 0.003672197 | TSG101 |
| chr6 | 39901204 | 39902064 | 861 | 6 | 5.74E-05 | 5.53E-06 | 0.000662 | 4.45E-06 | 0.006292186 | DAAM2, MOCS1 |
| chr10 | 118754145 | 118755076 | 932 | 4 | 1.30E-06 | 1.68E-06 | 0.002788 | 4.45E-06 | -0.00054168 | CACUL1 |
| chr6 | 128504118 | 128504938 | 821 | 4 | 1.33E-05 | 3.66E-06 | 0.000239 | 4.45E-06 | 0.005663478 | PTPRK |
| chr11 | 64231619 | 64231970 | 352 | 3 | 2.69E-06 | 1.62E-05 | 3.49E-05 | 4.45E-06 | 0.009641803 | DNAJC4, AP001453.1 |
| chr14 | 103548948 | 103549700 | 753 | 5 | 5.26E-06 | 6.84E-06 | 0.000347 | 4.45E-06 | 0.001722693 | NA |
| chr4 | 3179168 | 3179203 | 36 | 2 | 1.07E-05 | 4.65E-06 | 7.10E-05 | 4.46E-06 | 0.004163126 | HTT |
| chr12 | 109798038 | 109798896 | 859 | 4 | 1.31E-05 | 0.000197 | 0.000847 | 4.46E-06 | 0.005403511 | TRPV4 |
| chr16 | 15501997 | 15502566 | 570 | 10 | 1.03E-05 | 8.97E-05 | 0.000675 | 4.47E-06 | 0.000578641 | AC140504.1, BMERB1 |
| chr5 | 178591185 | 178592598 | 1414 | 4 | 1.52E-06 | 3.74E-05 | 5.12E-05 | 4.47E-06 | 0.006891057 | NA |
| chr15 | 29131994 | 29132733 | 740 | 5 | 2.62E-05 | 9.84E-07 | 0.006175 | 4.47E-06 | 0.010707923 | FAM189A1 |
| chr8 | 119982346 | 119982891 | 546 | 2 | 2.31E-06 | 8.84E-06 | 3.40E-05 | 4.47E-06 | -0.00401237 | DEPTOR |
| chr3 | 33426320 | 33426643 | 324 | 2 | 7.13E-06 | 9.51E-06 | 3.19E-05 | 4.48E-06 | 0.001305051 | UBP1 |
| chr12 | 123787719 | 123787886 | 168 | 2 | 1.11E-05 | 3.49E-06 | 0.000121 | 4.48E-06 | 0.005771709 | DNAH10 |
| chr16 | 29661847 | 29662359 | 513 | 3 | 2.14E-06 | 3.68E-06 | 0.000151 | 4.48E-06 | 0.007885837 | NA |
| chr13 | 111521662 | 111521839 | 178 | 2 | 1.97E-05 | 2.87E-06 | 0.000236 | 4.48E-06 | 0.057757738 | NA |
| chr14 | 69637230 | 69638350 | 1121 | 3 | 1.49E-06 | 9.30E-06 | 4.70E-05 | 4.49E-06 | 0.006125047 | SUSD6 |
| chr10 | 3484531 | 3484882 | 352 | 2 | 6.26E-06 | 3.74E-06 | 0.000104 | 4.49E-06 | 0.004672233 | LINC02669 |
| chr5 | 1494865 | 1495241 | 377 | 4 | 3.11E-05 | 1.49E-06 | 0.001352 | 4.49E-06 | 0.012044574 | LPCAT1 |
| chr15 | 77710896 | 77711639 | 744 | 2 | 8.44E-08 | 2.00E-05 | 1.82E-05 | 4.51E-06 | 0.005456676 | LINGO1 |
| chr16 | 88922661 | 88922979 | 319 | 3 | 3.99E-06 | 1.80E-06 | 0.001249 | 4.53E-06 | 0.013526982 | CBFA2T3 |
| chr9 | 134170267 | 134170934 | 668 | 4 | 1.92E-05 | 1.48E-05 | 9.35E-05 | 4.53E-06 | 0.003227892 | NA |
| chr5 | 139794944 | 139795814 | 871 | 4 | 1.52E-05 | 3.48E-06 | 0.000573 | 4.55E-06 | 0.008514986 | PSD2 |
| chr17 | 48156352 | 48156822 | 471 | 5 | 1.80E-05 | 1.53E-06 | 0.004171 | 4.56E-06 | 0.009427053 | SKAP1, MIR1203 |
| chr8 | 70604264 | 70604449 | 186 | 3 | 2.11E-05 | 1.74E-06 | 0.001456 | 4.57E-06 | 0.011893628 | TRAM1 |
| chr6 | 70302688 | 70303686 | 999 | 7 | 2.79E-05 | 2.55E-06 | 0.004497 | 4.57E-06 | 0.007201699 | COL9A1 |
| chr6 | 152981431 | 152983053 | 1623 | 5 | 5.14E-08 | 3.07E-05 | 0.000221 | 4.58E-06 | 0.013010321 | FBXO5 |
| chr2 | 63056220 | 63057633 | 1414 | 10 | 1.49E-06 | 0.000124 | 0.000132 | 4.58E-06 | 0.003500032 | OTX1 |
| chr20 | 46220568 | 46221373 | 806 | 4 | 7.79E-06 | 2.23E-06 | 0.001225 | 4.58E-06 | 0.002258983 | CDH22 |
| chr14 | 76310127 | 76310833 | 707 | 4 | 3.00E-06 | 3.66E-05 | 0.000234 | 4.58E-06 | 0.013310623 | ESRRB |
| chr17 | 31570426 | 31571243 | 818 | 3 | 2.79E-06 | 3.02E-06 | 0.000225 | 4.59E-06 | 0.004916166 | AC003101.1 |
| chr5 | 171308854 | 171310392 | 1539 | 13 | 0.000125384 | 3.05E-05 | 0.001893 | 4.59E-06 | 0.002402521 | TLX3, AC091980.2 |
| chr7 | 849656 | 849927 | 272 | 4 | 1.20E-06 | 0.005212 | 3.12E-06 | 4.59E-06 | 0.001531056 | SUN1 |
| chr9 | 70390043 | 70390288 | 246 | 3 | 5.07E-06 | 0.001256 | 3.37E-05 | 4.59E-06 | 0.00242687 | KLF9 |
| chr11 | 76719471 | 76720168 | 698 | 6 | 8.00E-06 | 8.54E-05 | 0.000382 | 4.60E-06 | 0.00189244 | GUCY2EP |
| chr16 | 89220343 | 89221017 | 675 | 5 | 8.78E-07 | 0.010853 | 1.85E-05 | 4.60E-06 | -0.00161492 | ZNF778 |
| chr20 | 34557652 | 34559213 | 1562 | 11 | 8.29E-05 | 8.63E-06 | 0.014515 | 4.60E-06 | -0.00295496 | MAP1LC3A |
| chr14 | 73727297 | 73727813 | 517 | 3 | 4.90E-07 | 4.07E-05 | 1.59E-05 | 4.61E-06 | 0.004561687 | MIDEAS |
| chr10 | 17398427 | 17398930 | 504 | 3 | 1.34E-06 | 4.00E-05 | 1.46E-05 | 4.61E-06 | 0.003549838 | ST8SIA6-AS1, ST8SIA6 |
| chr11 | 10568263 | 10569453 | 1191 | 10 | 5.14E-06 | 9.21E-06 | 0.00029 | 4.61E-06 | 0.000909137 | IRAG1-AS1, LYVE1 |
| chr4 | 82223513 | 82223948 | 436 | 2 | 1.08E-05 | 4.41E-06 | 8.25E-05 | 4.61E-06 | 0.007944943 | NA |
| chr2 | 191245616 | 191246454 | 839 | 7 | 4.30E-06 | 6.38E-06 | 0.001902 | 4.61E-06 | 0.005356151 | MYO1B, AC092614.1 |
| chr20 | 56735268 | 56735271 | 4 | 2 | 1.62E-05 | 2.71E-06 | 0.000411 | 4.61E-06 | 0.025384951 | NA |
| chr10 | 132577963 | 132578435 | 473 | 2 | 3.24E-05 | 2.71E-06 | 0.000408 | 4.61E-06 | 0.008353922 | INPP5A |
| chr2 | 101742436 | 101742565 | 130 | 2 | 3.61E-06 | 0.010905 | 5.42E-06 | 4.62E-06 | 0.011960573 | MAP4K4 |
| chr17 | 61410348 | 61411082 | 735 | 6 | 2.85E-05 | 5.66E-07 | 0.013752 | 4.62E-06 | 0.005848097 | TBX2-AS1 |
| chr8 | 143025505 | 143025814 | 310 | 2 | 7.06E-06 | 2.83E-06 | 0.000306 | 4.62E-06 | 0.008763739 | NA |
| chr10 | 92045219 | 92046113 | 895 | 7 | 2.42E-05 | 9.94E-07 | 0.008164 | 4.63E-06 | 0.009140698 | NA |
| chr1 | 9365692 | 9366491 | 800 | 4 | 5.63E-05 | 2.08E-06 | 0.002695 | 4.63E-06 | -0.00949388 | SPSB1 |
| chr5 | 139805069 | 139805475 | 407 | 2 | 2.25E-05 | 2.81E-06 | 0.000326 | 4.63E-06 | 0.011357258 | PSD2 |
| chr19 | 46786153 | 46786354 | 202 | 3 | 2.60E-06 | 3.90E-06 | 0.000272 | 4.65E-06 | 0.00995186 | SLC1A5 |
| chr2 | 64227963 | 64228446 | 484 | 6 | 1.97E-05 | 8.77E-06 | 0.000546 | 4.65E-06 | 0.005189991 | AC012368.1 |
| chr9 | 94913133 | 94913648 | 516 | 2 | 6.64E-06 | 5.15E-06 | 6.59E-05 | 4.65E-06 | -0.00028891 | AOPEP |
| chr14 | 88326961 | 88327638 | 678 | 9 | 1.29E-05 | 5.06E-06 | 0.005268 | 4.65E-06 | 0.011378179 | NA |
| chr19 | 45280400 | 45280716 | 317 | 3 | 2.28E-05 | 3.17E-06 | 0.000231 | 4.65E-06 | 0.010540697 | MARK4 |
| chr12 | 109131254 | 109132002 | 749 | 7 | 8.17E-05 | 6.55E-07 | 0.013068 | 4.65E-06 | 0.011565401 | ACACB |
| chr1 | 23562473 | 23562608 | 136 | 3 | 1.83E-05 | 1.82E-06 | 0.001367 | 4.65E-06 | 0.012710528 | NA |
| chr10 | 654113 | 654515 | 403 | 2 | 1.34E-05 | 3.86E-06 | 0.000107 | 4.66E-06 | 0.019683603 | PRR26, DIP2C |
| chr1 | 32374874 | 32374880 | 7 | 2 | 3.34E-06 | 9.47E-06 | 3.39E-05 | 4.66E-06 | 0.006615445 | BSDC1 |
| chr10 | 119222030 | 119222251 | 222 | 2 | 4.15E-06 | 4.03E-06 | 9.91E-05 | 4.66E-06 | 0.00695218 | GRK5 |
| chr10 | 67696234 | 67696549 | 316 | 3 | 4.91E-06 | 4.35E-05 | 0.000257 | 4.68E-06 | 0.018186162 | NA |
| chr6 | 33454042 | 33455621 | 1580 | 40 | 1.13E-07 | 0.000357 | 0.000101 | 4.68E-06 | 0.00049286 | ZBTB9, SYNGAP1-AS1 |
| chr19 | 1227853 | 1228386 | 534 | 4 | 2.30E-06 | 0.000366 | 1.18E-05 | 4.68E-06 | 0.003257982 | STK11, CBARP |
| chr2 | 240687795 | 240688138 | 344 | 3 | 1.10E-05 | 4.15E-06 | 0.000554 | 4.69E-06 | 0.012073113 | AC011298.1 |
| chr16 | 1045865 | 1046754 | 890 | 4 | 4.48E-06 | 3.17E-06 | 0.000298 | 4.70E-06 | 0.00796885 | NA |
| chr14 | 104096938 | 104097734 | 797 | 3 | 2.01E-05 | 2.62E-06 | 0.000762 | 4.71E-06 | 0.007948399 | ASPG |
| chr9 | 16679658 | 16679749 | 92 | 3 | 1.31E-05 | 1.82E-05 | 2.69E-05 | 4.71E-06 | 0.015402726 | BNC2 |
| chr20 | 34080620 | 34081020 | 401 | 3 | 1.25E-05 | 3.46E-06 | 0.000455 | 4.71E-06 | 0.005191953 | RALY |
| chr18 | 13824073 | 13824397 | 325 | 3 | 8.16E-07 | 0.000115 | 8.02E-05 | 4.72E-06 | -0.00819669 | MC5R |
| chr18 | 35972239 | 35973154 | 916 | 9 | 5.39E-06 | 1.92E-05 | 0.001873 | 4.72E-06 | 2.27E-05 | AC091060.1, C18orf21 |
| chr11 | 45751631 | 45751982 | 352 | 4 | 8.66E-06 | 6.34E-05 | 2.25E-05 | 4.72E-06 | 0.015404263 | LINC02690 |
| chr10 | 80458192 | 80459202 | 1011 | 5 | 8.57E-06 | 5.93E-05 | 0.000108 | 4.72E-06 | 0.00475523 | TSPAN14 |
| chr17 | 17591859 | 17593169 | 1311 | 7 | 2.33E-05 | 3.61E-05 | 0.000896 | 4.73E-06 | 0.002601848 | AC020558.1 |
| chr3 | 179604407 | 179605183 | 777 | 15 | 2.28E-06 | 0.00013 | 0.002145 | 4.74E-06 | 0.000853151 | NDUFB5, MRPL47 |
| chr19 | 56175828 | 56176019 | 192 | 4 | 7.48E-06 | 0.000331 | 0.000524 | 4.74E-06 | 0.012470934 | GALP |
| chr14 | 104110691 | 104111276 | 586 | 2 | 1.66E-05 | 3.04E-06 | 0.000236 | 4.74E-06 | 0.009356368 | ASPG |
| chr12 | 94152425 | 94153157 | 733 | 2 | 1.98E-05 | 2.83E-06 | 0.000371 | 4.74E-06 | 0.001873912 | PLXNC1 |
| chr14 | 50943971 | 50945069 | 1099 | 13 | 6.97E-05 | 9.11E-05 | 0.002122 | 4.74E-06 | 0.001036701 | PYGL |
| chr8 | 22046164 | 22046568 | 405 | 2 | 2.16E-05 | 2.81E-06 | 0.000389 | 4.74E-06 | 0.024174691 | FGF17 |
| chr20 | 37396139 | 37396266 | 128 | 5 | 8.43E-06 | 6.85E-05 | 0.000114 | 4.74E-06 | 0.006001738 | SRC |
| chr21 | 36319449 | 36320542 | 1094 | 15 | 1.70E-05 | 2.39E-05 | 0.001149 | 4.75E-06 | 0.000349328 | MORC3, AP000692.2 |
| chr7 | 152594885 | 152595894 | 1010 | 6 | 4.60E-05 | 5.42E-06 | 0.001203 | 4.75E-06 | 0.005191088 | NA |
| chr2 | 41733808 | 41734367 | 560 | 4 | 2.80E-05 | 2.40E-06 | 0.002372 | 4.75E-06 | -0.00518525 | NA |
| chr2 | 30148503 | 30149124 | 622 | 3 | 1.48E-05 | 4.24E-06 | 0.000211 | 4.75E-06 | 0.023491278 | YPEL5 |
| chr7 | 74062007 | 74062345 | 339 | 3 | 1.17E-05 | 5.09E-06 | 0.000241 | 4.76E-06 | 0.015106498 | ELN, ELN-AS1 |
| chr14 | 100068152 | 100068243 | 92 | 2 | 1.96E-05 | 2.85E-06 | 0.000368 | 4.76E-06 | 0.013867864 | EVL, AL157912.1 |
| chr6 | 151764112 | 151764997 | 886 | 6 | 3.09E-05 | 2.81E-06 | 0.004909 | 4.76E-06 | 0.013859459 | ESR1 |
| chr8 | 103413610 | 103415491 | 1882 | 20 | 4.01E-05 | 0.001115 | 0.000167 | 4.76E-06 | -8.54E-05 | DCAF13, SLC25A32, AC012213.5 |
| chr12 | 114407375 | 114409044 | 1670 | 16 | 8.74E-06 | 3.53E-06 | 0.006528 | 4.76E-06 | 0.005121465 | TBX5-AS1, TBX5 |
| chr9 | 134405766 | 134406967 | 1202 | 10 | 4.73E-06 | 7.47E-06 | 0.003399 | 4.77E-06 | 0.004694387 | RXRA |
| chr3 | 13635731 | 13636280 | 550 | 2 | 6.91E-06 | 5.92E-06 | 5.71E-05 | 4.77E-06 | -0.00604487 | FBLN2 |
| chr1 | 4711065 | 4711925 | 861 | 5 | 2.22E-05 | 5.37E-06 | 0.000256 | 4.77E-06 | -0.00296353 | AJAP1 |
| chr12 | 52360863 | 52360951 | 89 | 2 | 9.80E-06 | 3.46E-06 | 0.000149 | 4.77E-06 | 0.004872914 | KRT85 |
| chr7 | 30910598 | 30911471 | 874 | 4 | 1.93E-05 | 1.26E-06 | 0.003426 | 4.77E-06 | 0.006326884 | AC004691.2 |
| chr11 | 18105745 | 18106579 | 835 | 7 | 1.11E-05 | 2.69E-05 | 0.002297 | 4.78E-06 | -9.96E-05 | SAAL1 |
| chr3 | 191330519 | 191331051 | 533 | 8 | 2.18E-05 | 1.50E-06 | 0.004888 | 4.79E-06 | 0.008090093 | CCDC50, UTS2B |
| chr6 | 24125365 | 24126096 | 732 | 7 | 3.30E-05 | 7.04E-05 | 0.001616 | 4.79E-06 | -0.00061516 | NA |
| chr1 | 28143235 | 28144235 | 1001 | 4 | 1.42E-06 | 7.11E-06 | 0.000127 | 4.79E-06 | 0.006610249 | NA |
| chr1 | 43951896 | 43951992 | 97 | 2 | 6.89E-07 | 6.26E-05 | 1.11E-05 | 4.80E-06 | 0.003332733 | IPO13 |
| chr9 | 98502417 | 98502973 | 557 | 2 | 4.25E-06 | 4.80E-06 | 7.78E-05 | 4.80E-06 | 0.0148209 | GABBR2 |
| chr8 | 28483546 | 28484007 | 462 | 3 | 2.52E-06 | 4.98E-05 | 0.000111 | 4.80E-06 | 0.003999942 | FBXO16 |
| chr7 | 157574241 | 157575132 | 892 | 9 | 3.69E-06 | 2.27E-05 | 0.001552 | 4.81E-06 | 0.005185527 | PTPRN2, MIR153-2 |
| chr5 | 891092 | 891728 | 637 | 4 | 1.54E-08 | 0.017728 | 1.23E-06 | 4.81E-06 | 0.00281977 | BRD9 |
| chr6 | 36781803 | 36782207 | 405 | 2 | 4.14E-07 | 0.000283 | 7.48E-06 | 4.81E-06 | 0.010213946 | CPNE5 |
| chr14 | 75620671 | 75621094 | 424 | 3 | 1.33E-05 | 1.19E-05 | 0.000188 | 4.81E-06 | 0.004049724 | FLVCR2 |
| chr1 | 43312497 | 43313289 | 793 | 4 | 4.53E-05 | 3.32E-06 | 0.001148 | 4.82E-06 | 0.000347454 | TIE1 |
| chr12 | 6361988 | 6362185 | 198 | 3 | 2.20E-05 | 2.27E-06 | 0.00071 | 4.82E-06 | 0.01419877 | SCNN1A |
| chr4 | 22692051 | 22692116 | 66 | 2 | 5.29E-06 | 1.84E-05 | 2.07E-05 | 4.82E-06 | -0.01347553 | NA |
| chr7 | 150388582 | 150389081 | 500 | 3 | 6.11E-06 | 9.76E-06 | 0.000387 | 4.83E-06 | 0.007431842 | ZNF775, AC073111.2 |
| chr19 | 5977955 | 5978779 | 825 | 11 | 1.12E-05 | 0.000157 | 0.000903 | 4.83E-06 | -0.00185317 | AC011444.1, RANBP3 |
| chr4 | 6901003 | 6901205 | 203 | 3 | 1.81E-05 | 5.90E-06 | 0.000121 | 4.83E-06 | 0.005158106 | NA |
| chr5 | 180328324 | 180329082 | 759 | 5 | 1.99E-05 | 4.99E-06 | 0.000547 | 4.83E-06 | 0.003873544 | GFPT2 |
| chr22 | 20564343 | 20564522 | 180 | 3 | 9.86E-06 | 4.90E-06 | 0.000108 | 4.84E-06 | 0.016081513 | MED15 |
| chr4 | 1826241 | 1827067 | 827 | 5 | 3.14E-05 | 1.57E-05 | 0.001278 | 4.85E-06 | 0.001008593 | LETM1 |
| chr6 | 144186690 | 144187548 | 859 | 4 | 1.39E-06 | 3.15E-06 | 0.000906 | 4.86E-06 | -0.00019948 | STX11 |
| chr14 | 75069858 | 75070967 | 1110 | 3 | 8.23E-06 | 2.51E-06 | 0.000617 | 4.86E-06 | 0.0045574 | ZC2HC1C |
| chr17 | 57988666 | 57989124 | 459 | 2 | 1.75E-05 | 2.89E-06 | 0.000395 | 4.86E-06 | 0.014518495 | AC015813.7, AC015813.2, AC015813.1 |
| chr11 | 15623634 | 15623971 | 338 | 4 | 1.19E-05 | 5.57E-06 | 0.001931 | 4.86E-06 | 0.015798845 | AC087379.2 |
| chr5 | 75336287 | 75337197 | 911 | 11 | 2.29E-06 | 2.42E-05 | 0.001538 | 4.87E-06 | 2.49E-05 | HMGCR, AC008897.2 |
| chr3 | 42490449 | 42491101 | 653 | 3 | 1.47E-05 | 3.02E-06 | 0.000398 | 4.87E-06 | -0.02270743 | VIPR1 |
| chr19 | 2231869 | 2232619 | 751 | 3 | 4.21E-05 | 1.79E-06 | 0.001851 | 4.87E-06 | 0.003994257 | DOT1L, PLEKHJ1 |
| chr19 | 54102843 | 54103275 | 433 | 4 | 0.000120525 | 2.14E-06 | 0.001495 | 4.87E-06 | 0.005677479 | NDUFA3 |
| chr12 | 118136463 | 118136952 | 490 | 4 | 1.51E-06 | 2.94E-06 | 0.000532 | 4.87E-06 | 0.002218387 | PEBP1 |
| chr16 | 69941417 | 69942325 | 909 | 5 | 2.00E-05 | 8.39E-05 | 0.000294 | 4.87E-06 | 0.010679831 | WWP2 |
| chr1 | 247409991 | 247410387 | 397 | 2 | 1.54E-05 | 3.81E-06 | 0.000125 | 4.87E-06 | 0.01843049 | NA |
| chr3 | 160449089 | 160450202 | 1114 | 14 | 4.09E-06 | 2.06E-06 | 0.008064 | 4.89E-06 | 0.000771685 | AC079594.1, TRIM59 |
| chr11 | 12862739 | 12863158 | 420 | 2 | 1.29E-05 | 4.88E-06 | 7.86E-05 | 4.89E-06 | 0.008852409 | TEAD1 |
| chr3 | 196632331 | 196633118 | 788 | 7 | 1.58E-06 | 0.001206 | 1.33E-05 | 4.89E-06 | 0.000541756 | LINC01063 |
| chr14 | 61697315 | 61698035 | 721 | 5 | 9.30E-06 | 7.21E-06 | 0.002576 | 4.89E-06 | 0.013856151 | HIF1A |
| chr12 | 112937756 | 112938876 | 1121 | 10 | 3.16E-06 | 5.24E-05 | 0.000128 | 4.89E-06 | 0.00190339 | OAS3, AC004551.1 |
| chr9 | 125706991 | 125707504 | 514 | 7 | 1.18E-05 | 2.75E-06 | 0.002643 | 4.89E-06 | 0.002065941 | MAPKAP1 |
| chr2 | 96500264 | 96500685 | 422 | 3 | 1.57E-05 | 2.55E-06 | 0.000501 | 4.89E-06 | 0.018238544 | NEURL3 |
| chr21 | 46155616 | 46156596 | 981 | 5 | 2.26E-05 | 2.25E-06 | 0.001046 | 4.89E-06 | 0.005647339 | NA |
| chr17 | 75455567 | 75456521 | 955 | 10 | 6.36E-05 | 6.89E-05 | 0.000928 | 4.89E-06 | 0.001208459 | TMEM94 |
| chr17 | 77825763 | 77826587 | 825 | 2 | 0.000158145 | 2.90E-06 | 0.000398 | 4.89E-06 | 0.006180495 | NA |
| chr18 | 49812615 | 49814131 | 1517 | 12 | 0.000173169 | 3.02E-05 | 0.003711 | 4.89E-06 | 6.80E-06 | SNHG22, ACAA2, AC090227.2 |

| | | | | | | | | | | |
|---|---|---|---|---|---|---|---|---|---|---|
| chr16 | 68377424 | 68378066 | 643 | 2 | 1.06E-05 | 3.87E-06 | 0.000122 | 4.89E-06 | 0.00770296 | SMPD3 |
| chr19 | 58362784 | 58363196 | 413 | 9 | 1.46E-05 | 1.80E-05 | 0.012873 | 4.90E-06 | -0.000729 | AC012313.6, ZNF497 |
| chr14 | 100533849 | 100534276 | 428 | 3 | 3.40E-06 | 1.55E-05 | 4.14E-05 | 4.90E-06 | 0.00858308 | NA |
| chr11 | 48101293 | 48101364 | 72 | 2 | 9.51E-06 | 5.98E-05 | 1.15E-05 | 4.91E-06 | 0.011116835 | PTPRJ |
| chr3 | 71573511 | 71574578 | 1068 | 5 | 3.69E-07 | 3.64E-05 | 6.60E-05 | 4.91E-06 | -0.00302433 | FOXP1, AC097634.4, FOXP1-IT1 |
| chr6 | 168318193 | 168319274 | 1082 | 5 | 1.78E-05 | 1.89E-06 | 0.003187 | 4.92E-06 | 0.009246963 | DACT2 |
| chr16 | 84085062 | 84085185 | 124 | 3 | 9.21E-06 | 3.41E-05 | 3.22E-05 | 4.92E-06 | 0.013602238 | MBTPS1 |
| chr6 | 170115859 | 170115926 | 68 | 2 | 1.77E-05 | 3.82E-06 | 0.000129 | 4.92E-06 | 0.000189449 | NA |
| chr19 | 55255485 | 55256442 | 958 | 3 | 1.53E-05 | 6.77E-05 | 0.000137 | 4.93E-06 | 0.006891777 | PPP6R1 |
| chr11 | 118435084 | 118435638 | 555 | 4 | 4.87E-07 | 9.46E-05 | 0.000112 | 4.93E-06 | 0.000474279 | AP001267.5, AP001267.2 |
| chr14 | 91317275 | 91317856 | 582 | 2 | 1.52E-06 | 1.47E-05 | 2.52E-05 | 4.93E-06 | 0.004101306 | CCDC88C |
| chr18 | 79816950 | 79817401 | 452 | 4 | 3.76E-06 | 9.50E-05 | 1.49E-05 | 4.93E-06 | -0.0066401 | NA |
| chr2 | 128090754 | 128091906 | 1153 | 11 | 6.47E-05 | 3.41E-05 | 0.005461 | 4.93E-06 | 0.000781709 | UGGT1 |
| chr14 | 24263028 | 24263345 | 318 | 3 | 1.68E-06 | 1.63E-05 | 0.000212 | 4.94E-06 | 0.00394542 | TGM1 |
| chr8 | 128048468 | 128048808 | 341 | 4 | 1.07E-05 | 4.23E-06 | 0.000213 | 4.95E-06 | 0.012288579 | PVT1 |
| chr7 | 97116868 | 97117751 | 884 | 8 | 2.91E-06 | 5.94E-06 | 0.002966 | 4.95E-06 | 0.000963403 | SDHAF3 |
| chr15 | 75728282 | 75728746 | 465 | 3 | 3.30E-06 | 4.73E-05 | 3.48E-05 | 4.95E-06 | 0.012061999 | DNM1P35 |
| chr12 | 48781450 | 48782737 | 1288 | 4 | 0.000294062 | 2.37E-06 | 0.001436 | 4.95E-06 | 0.004537235 | ADCY6 |
| chr1 | 220894320 | 220896230 | 1911 | 12 | 7.53E-05 | 3.83E-06 | 0.001004 | 4.95E-06 | 0.003524036 | NA |
| chr8 | 42731780 | 42731988 | 209 | 2 | 1.61E-05 | 5.83E-06 | 6.26E-05 | 4.95E-06 | 0.004447383 | CHRNB3 |
| chr17 | 60435464 | 60436161 | 698 | 4 | 5.31E-05 | 2.00E-05 | 0.000777 | 4.95E-06 | 0.004433443 | RPL12P38 |
| chr17 | 79119391 | 79120090 | 700 | 4 | 4.73E-05 | 3.30E-06 | 0.000975 | 4.95E-06 | 0.005779487 | RBFOX3 |
| chr4 | 48986222 | 48986784 | 563 | 8 | 4.74E-06 | 5.68E-06 | 0.003851 | 4.96E-06 | 0.006783878 | CWH43 |
| chr5 | 121961339 | 121962683 | 1345 | 13 | 1.22E-05 | 7.96E-05 | 0.001242 | 4.96E-06 | -0.00065692 | SRFBP1 |
| chr9 | 77458166 | 77458226 | 61 | 2 | 1.15E-05 | 5.65E-06 | 6.64E-05 | 4.96E-06 | 0.008930649 | GNA14-AS1, GNA14 |
| chr7 | 118214226 | 118214711 | 486 | 6 | 2.91E-05 | 1.03E-06 | 0.006343 | 4.96E-06 | 0.009854265 | ANKRD7 |
| chr3 | 115693164 | 115693416 | 253 | 2 | 2.08E-05 | 3.12E-06 | 0.000283 | 4.97E-06 | 0.014824879 | GAP43 |
| chr12 | 6551757 | 6552578 | 822 | 6 | 4.18E-05 | 4.38E-06 | 0.002466 | 4.98E-06 | 0.009030444 | IFFO1 |
| chr11 | 133833199 | 133833812 | 614 | 2 | 5.95E-06 | 6.17E-06 | 5.90E-05 | 4.98E-06 | -0.02858708 | NA |
| chr3 | 66990433 | 66990480 | 48 | 2 | 1.03E-05 | 3.34E-06 | 0.000207 | 4.98E-06 | 0.015580289 | NA |
| chr7 | 150369567 | 150370036 | 470 | 4 | 1.28E-06 | 9.11E-05 | 4.37E-05 | 4.98E-06 | 0.011574135 | REPIN1, ZNF775, REPIN1-AS1 |
| chr4 | 139665242 | 139665834 | 593 | 6 | 3.17E-05 | 1.42E-05 | 0.00051 | 4.98E-06 | 0.004848771 | MGST2 |
| chr16 | 30977389 | 30977757 | 369 | 3 | 1.94E-05 | 1.97E-06 | 0.001532 | 4.99E-06 | 0.000864017 | SETD1A |
| chr15 | 89120354 | 89120764 | 411 | 2 | 5.50E-06 | 3.95E-06 | 0.000124 | 4.99E-06 | 0.019523989 | ABHD2 |
| chr8 | 22746080 | 22746459 | 380 | 3 | 5.98E-06 | 2.45E-05 | 0.000279 | 4.99E-06 | 0.009569994 | AC055854.1, PEBP4 |
| chr4 | 139302715 | 139302994 | 280 | 5 | 1.15E-05 | 9.08E-05 | 5.40E-05 | 4.99E-06 | 0.003350242 | NAA15 |
| chr9 | 137456052 | 137456633 | 582 | 2 | 1.20E-05 | 2.96E-06 | 0.000426 | 5.00E-06 | 0.008463206 | NSMF |
| chr1 | 211829157 | 211830160 | 1004 | 6 | 3.19E-06 | 7.52E-06 | 0.001136 | 5.00E-06 | 0.009938432 | LPGAT1-AS1, LPGAT1 |
| chr18 | 46946373 | 46947063 | 691 | 10 | 1.89E-05 | 4.30E-05 | 0.000351 | 5.01E-06 | 0.006632132 | KATNAL2 |
| chr11 | 892568 | 892898 | 331 | 2 | 7.45E-07 | 0.000103 | 9.74E-06 | 5.01E-06 | 0.005607075 | CHID1 |
| chr15 | 47301736 | 47301741 | 6 | 2 | 1.53E-05 | 1.47E-05 | 2.57E-05 | 5.02E-06 | 0.012968164 | SEMA6D |
| chr6 | 89311133 | 89311147 | 15 | 2 | 1.09E-05 | 6.19E-06 | 5.98E-05 | 5.03E-06 | 0.00969932 | GABRR2 |
| chr3 | 108102537 | 108102708 | 172 | 2 | 1.16E-05 | 3.16E-06 | 0.000284 | 5.04E-06 | 0.014036768 | NA |
| chr11 | 118076499 | 118077282 | 784 | 8 | 8.47E-06 | 1.68E-05 | 0.001039 | 5.05E-06 | 0.002383947 | TMPRSS4, SMIM35 |
| chr11 | 75404332 | 75404957 | 626 | 5 | 3.27E-05 | 2.22E-06 | 0.001204 | 5.05E-06 | -0.00453007 | RPS3, SNORD15B |
| chr9 | 81559123 | 81559585 | 463 | 2 | 8.16E-06 | 1.40E-05 | 2.72E-05 | 5.05E-06 | 0.007390554 | NA |
| chr2 | 3313698 | 3314268 | 571 | 5 | 2.54E-06 | 4.49E-05 | 0.000262 | 5.05E-06 | -2.37E-05 | EIPR1 |
| chr4 | 140255930 | 140256602 | 673 | 3 | 7.11E-06 | 7.10E-06 | 8.28E-05 | 5.05E-06 | 0.006920333 | NA |
| chr6 | 33413903 | 33414543 | 641 | 6 | 7.18E-06 | 6.76E-06 | 0.000422 | 5.05E-06 | 0.00230428 | PHF1 |
| chr3 | 48592012 | 48592585 | 574 | 3 | 1.68E-05 | 3.01E-06 | 0.000821 | 5.05E-06 | 0.005087395 | COL7A1 |
| chr19 | 13794770 | 13795543 | 774 | 9 | 3.43E-05 | 1.96E-06 | 0.008109 | 5.06E-06 | -0.0032712 | ZSWIM4 |
| chr7 | 726686 | 728256 | 1571 | 11 | 6.97E-06 | 3.05E-05 | 0.005521 | 5.07E-06 | 0.002617825 | DNAAF5, PRKAR1B |
| chr11 | 116580202 | 116581071 | 870 | 7 | 4.73E-06 | 1.66E-05 | 0.000111 | 5.08E-06 | 0.002583054 | NA |
| chr15 | 90934794 | 90935752 | 959 | 10 | 5.06E-05 | 5.27E-05 | 0.005263 | 5.08E-06 | 0.00107146 | UNC45A, HDDC3 |
| chr12 | 9760799 | 9760949 | 151 | 2 | 9.24E-06 | 4.19E-06 | 0.000115 | 5.08E-06 | -0.01264638 | CD69 |
| chr6 | 75368315 | 75368471 | 157 | 2 | 9.41E-06 | 3.85E-05 | 1.47E-05 | 5.11E-06 | 0.005136605 | AL445465.1, FILIP1 |
| chr2 | 219712931 | 219713338 | 408 | 2 | 1.44E-05 | 4.41E-06 | 0.000104 | 5.11E-06 | 0.006847807 | LINC02832 |
| chr6 | 105726869 | 105727052 | 184 | 2 | 2.42E-05 | 3.25E-06 | 0.000268 | 5.11E-06 | 0.015168869 | AL591518.1 |
| chr2 | 238499867 | 238500214 | 348 | 2 | 1.29E-05 | 3.12E-06 | 0.000339 | 5.11E-06 | 0.011483627 | NA |
| chr2 | 89841876 | 89842694 | 819 | 2 | 0.000152608 | 3.04E-06 | 0.000422 | 5.11E-06 | 0.012391639 | NA |
| chr16 | 74632588 | 74632771 | 184 | 3 | 1.15E-05 | 5.84E-05 | 0.000114 | 5.11E-06 | -0.00634225 | RFWD3 |
| chr19 | 47804847 | 47805352 | 506 | 2 | 1.86E-05 | 3.73E-06 | 0.000155 | 5.12E-06 | 0.009590961 | TPRX1 |
| chr14 | 74881870 | 74882452 | 583 | 4 | 1.94E-06 | 1.11E-05 | 0.000612 | 5.13E-06 | 0.001614294 | DLST |
| chr12 | 80053173 | 80053258 | 86 | 2 | 1.14E-05 | 1.40E-05 | 2.77E-05 | 5.13E-06 | 0.014176029 | NA |
| chr19 | 38433405 | 38433796 | 392 | 8 | 1.04E-05 | 7.99E-05 | 0.001899 | 5.13E-06 | 0.003550167 | RYR1 |
| chr22 | 40651738 | 40652825 | 1088 | 5 | 1.10E-05 | 3.71E-06 | 0.001534 | 5.15E-06 | 0.008347908 | NA |
| chr3 | 143119478 | 143120180 | 703 | 5 | 4.07E-06 | 1.13E-05 | 0.000369 | 5.16E-06 | -0.00108091 | CHST2 |
| chr9 | 138046857 | 138046985 | 129 | 5 | 1.26E-05 | 1.36E-05 | 7.22E-05 | 5.16E-06 | 0.003082371 | CACNA1B |
| chr11 | 32090545 | 32090911 | 367 | 5 | 1.56E-05 | 2.59E-06 | 0.00175 | 5.16E-06 | 0.007601279 | AL035078.4 |
| chr2 | 23656267 | 23656589 | 323 | 2 | 6.14E-06 | 2.13E-05 | 2.05E-05 | 5.16E-06 | 0.000591006 | KLHL29 |
| chr11 | 102109624 | 102109763 | 140 | 3 | 1.84E-06 | 5.01E-05 | 6.30E-05 | 5.16E-06 | 0.006599243 | AP001527.1 |
| chr12 | 56643873 | 56644333 | 461 | 2 | 2.80E-06 | 5.55E-06 | 7.33E-05 | 5.16E-06 | 0.005350174 | ATP5F1B |
| chr9 | 121735484 | 121736503 | 1020 | 7 | 6.89E-06 | 8.42E-06 | 0.002597 | 5.16E-06 | 0.007093546 | DAB2IP |
| chr19 | 3958346 | 3958509 | 164 | 2 | 1.26E-05 | 3.36E-06 | 0.000238 | 5.16E-06 | 0.004078953 | DAPK3 |
| chr16 | 50246584 | 50246812 | 229 | 3 | 1.86E-05 | 2.12E-06 | 0.001316 | 5.16E-06 | 0.010355781 | ADCY7 |
| chr16 | 86761792 | 86762552 | 761 | 5 | 1.45E-05 | 2.80E-06 | 0.001055 | 5.17E-06 | 0.02060926 | NA |
| chr1 | 10660212 | 10660936 | 725 | 6 | 2.90E-08 | 0.00037 | 1.63E-05 | 5.17E-06 | 0.003003254 | CASZ1 |
| chr16 | 23302936 | 23304055 | 1120 | 4 | 9.69E-05 | 1.38E-05 | 0.000337 | 5.17E-06 | 0.005778871 | SCNN1B |
| chr18 | 50339978 | 50340461 | 484 | 6 | 7.51E-06 | 3.69E-05 | 0.000192 | 5.18E-06 | 0.002814154 | NA |
| chr22 | 31122721 | 31122977 | 257 | 5 | 1.69E-05 | 7.70E-06 | 0.001101 | 5.18E-06 | 0.011520104 | INPP5J |
| chr14 | 61608078 | 61608392 | 315 | 2 | 8.34E-06 | 5.04E-06 | 8.49E-05 | 5.19E-06 | 0.005032385 | AL355916.3, AL355916.1 |
| chr6 | 170276576 | 170276883 | 308 | 4 | 5.12E-06 | 0.000935 | 0.000181 | 5.20E-06 | -0.00292187 | LINC01624, AL109910.2 |
| chr12 | 44876007 | 44877284 | 1278 | 25 | 2.26E-05 | 9.42E-05 | 0.000146 | 5.20E-06 | 0.002298817 | NELL2 |
| chr17 | 33667018 | 33667177 | 160 | 2 | 6.76E-06 | 0.000104 | 1.01E-05 | 5.20E-06 | 0.01202108 | ASIC2 |
| chr1 | 230112975 | 230113545 | 571 | 2 | 5.01E-08 | 0.00025 | 8.20E-06 | 5.21E-06 | 0.007961162 | GALNT2 |
| chr1 | 2205406 | 2206226 | 821 | 6 | 3.40E-05 | 2.72E-05 | 0.000618 | 5.21E-06 | 0.004919269 | FAAP20 |
| chr5 | 135337018 | 135337813 | 796 | 4 | 3.89E-05 | 5.67E-06 | 0.000506 | 5.21E-06 | -0.00156059 | C5orf66, MACROH2A1 |
| chr1 | 109467148 | 109467762 | 615 | 4 | 2.05E-06 | 5.50E-06 | 0.000133 | 5.21E-06 | 0.00709374 | SYPL2 |
| chr9 | 129664417 | 129664942 | 526 | 3 | 5.26E-06 | 3.13E-06 | 0.000395 | 5.21E-06 | 0.011234664 | NA |
| chr6 | 159041932 | 159042355 | 424 | 5 | 8.30E-06 | 7.01E-06 | 0.001824 | 5.22E-06 | 0.005965647 | AL035530.2, TAGAP, AL035530.1 |
| chr8 | 104353375 | 104354429 | 1055 | 4 | 1.68E-05 | 1.04E-05 | 7.87E-05 | 5.22E-06 | 0.001458052 | DCSTAMP, DPYS |
| chr20 | 3785615 | 3786314 | 700 | 5 | 2.72E-06 | 3.31E-05 | 0.000211 | 5.22E-06 | 0.005749382 | CENPB |
| chr18 | 13392189 | 13392227 | 39 | 2 | 2.39E-05 | 4.33E-06 | 0.000115 | 5.22E-06 | 0.010507999 | LDLRAD4 |
| chr2 | 162073944 | 162074887 | 944 | 7 | 5.41E-06 | 1.83E-05 | 0.000218 | 5.23E-06 | 0.002245186 | DPP4-DT, DPP4 |
| chr14 | 93788240 | 93789167 | 928 | 11 | 6.45E-06 | 9.30E-05 | 0.00303 | 5.23E-06 | 0.001193068 | PRIMA1 |
| chr12 | 52209516 | 52210119 | 604 | 3 | 3.05E-07 | 3.87E-05 | 1.82E-05 | 5.23E-06 | 0.007607371 | LINC00592, LINC02874 |
| chr7 | 23000620 | 23001200 | 581 | 2 | 2.79E-05 | 3.38E-06 | 0.000251 | 5.23E-06 | 0.010377315 | FAM126A |
| chr4 | 3422476 | 3422965 | 490 | 3 | 1.40E-06 | 7.61E-05 | 1.81E-05 | 5.23E-06 | 0.004115448 | RGS12 |
| chr1 | 161083061 | 161083768 | 708 | 3 | 1.27E-06 | 1.15E-05 | 0.000158 | 5.24E-06 | 0.009196544 | NECTIN4 |
| chr2 | 44943183 | 44943586 | 404 | 3 | 1.71E-06 | 2.69E-06 | 0.000495 | 5.24E-06 | 0.009884031 | SIX3 |
| chr3 | 121545997 | 121546911 | 915 | 12 | 7.35E-06 | 0.000222 | 0.000303 | 5.24E-06 | 0.002140598 | POLQ |
| chr16 | 58652193 | 58652916 | 724 | 2 | 0.000108571 | 3.11E-06 | 0.000435 | 5.24E-06 | 0.015127094 | NA |
| chr1 | 30842688 | 30843310 | 623 | 3 | 4.32E-06 | 6.52E-06 | 9.53E-05 | 5.25E-06 | 0.015272671 | NA |
| chr1 | 36858850 | 36860008 | 1159 | 7 | 3.73E-05 | 5.12E-06 | 0.001774 | 5.25E-06 | 0.002748459 | GRIK3 |
| chr19 | 7658339 | 7658460 | 122 | 2 | 7.10E-06 | 3.97E-06 | 0.000144 | 5.25E-06 | 0.016657017 | NA |
| chr17 | 440346 | 440715 | 370 | 4 | 2.84E-06 | 1.07E-05 | 0.000122 | 5.26E-06 | 0.00110934 | RFLNB |
| chr12 | 113368567 | 113368731 | 165 | 2 | 1.45E-05 | 1.20E-05 | 3.28E-05 | 5.26E-06 | 0.00851627 | PLBD2 |
| chr19 | 40276094 | 40276405 | 312 | 4 | 3.80E-06 | 5.73E-05 | 1.65E-05 | 5.27E-06 | -0.00285289 | AKT2 |
| chr2 | 48768749 | 48768902 | 154 | 2 | 2.55E-05 | 3.36E-06 | 0.00027 | 5.27E-06 | 0.030861477 | STON1-GTF2A1L |
| chr17 | 45792261 | 45792357 | 97 | 3 | 1.47E-05 | 4.13E-06 | 0.000633 | 5.29E-06 | 0.009125971 | LINC02210-CRHR1, CRHR1 |
| chr10 | 62376272 | 62376463 | 192 | 3 | 9.46E-06 | 2.42E-05 | 6.16E-05 | 5.29E-06 | 0.001823552 | ZNF365, AC024598.1 |
| chr1 | 150148837 | 150149826 | 990 | 9 | 1.80E-06 | 6.10E-05 | 0.00114 | 5.30E-06 | 0.005057827 | PLEKHO1 |
| chr1 | 160376142 | 160376806 | 665 | 3 | 2.11E-05 | 2.58E-06 | 0.000835 | 5.30E-06 | 0.010140161 | NA |
| chr22 | 20114307 | 20115057 | 751 | 4 | 2.94E-06 | 6.41E-06 | 0.000115 | 5.31E-06 | 0.003258676 | TRMT2A, MIR6816 |
| chr19 | 16017696 | 16018327 | 632 | 2 | 1.09E-05 | 3.95E-06 | 0.000149 | 5.31E-06 | 0.008300657 | LINC00661 |
| chr6 | 41375472 | 41376092 | 621 | 6 | 3.11E-05 | 1.01E-05 | 0.002216 | 5.31E-06 | 0.007209498 | NA |
| chr1 | 234093846 | 234093979 | 134 | 2 | 1.14E-05 | 3.66E-06 | 0.000193 | 5.32E-06 | 0.014694468 | SLC35F3 |
| chr4 | 107868435 | 107868454 | 20 | 2 | 9.88E-06 | 2.05E-05 | 2.17E-05 | 5.32E-06 | 0.011742093 | SGMS2, CYP2U1-AS1 |
| chr6 | 28837525 | 28837631 | 107 | 2 | 1.45E-06 | 3.36E-06 | 0.000288 | 5.33E-06 | -0.00077135 | NA |
| chr17 | 4997812 | 4998986 | 1175 | 7 | 1.63E-06 | 3.91E-05 | 0.000196 | 5.33E-06 | 0.004087578 | KIF1C |
| chr12 | 94831710 | 94832183 | 474 | 3 | 2.06E-05 | 2.01E-06 | 0.001765 | 5.34E-06 | 0.009490258 | NA |
| chr6 | 52049878 | 52050452 | 575 | 2 | 1.36E-05 | 4.81E-06 | 9.74E-05 | 5.34E-06 | 0.015889843 | PKHD1 |
| chr8 | 58066713 | 58067302 | 590 | 2 | 5.02E-06 | 9.74E-06 | 4.05E-05 | 5.35E-06 | 0.015904604 | FAM110B |
| chr12 | 8049379 | 8049892 | 514 | 4 | 4.68E-06 | 2.95E-05 | 3.67E-05 | 5.36E-06 | 0.005076233 | FOXJ2 |
| chr12 | 117407783 | 117408396 | 614 | 2 | 5.11E-06 | 4.43E-06 | 0.000117 | 5.36E-06 | 0.017432759 | NOS1 |
| chr10 | 100081442 | 100081755 | 314 | 4 | 3.70E-06 | 0.000289 | 5.19E-05 | 5.36E-06 | 0.004425202 | CPN1 |
| chr17 | 42185005 | 42185874 | 870 | 11 | 1.44E-05 | 2.10E-05 | 0.003125 | 5.37E-06 | 0.002708295 | HCRT |
| chr16 | 31471866 | 31472711 | 846 | 10 | 1.28E-05 | 5.66E-06 | 0.005349 | 5.37E-06 | 0.004986641 | TGFB1I1, AC026471.6 |
| chr8 | 11343741 | 11343749 | 9 | 2 | 2.42E-05 | 4.91E-06 | 9.54E-05 | 5.38E-06 | 0.007382228 | TDH |
| chr14 | 72714565 | 72715066 | 502 | 2 | 6.34E-06 | 8.93E-06 | 4.41E-05 | 5.38E-06 | 0.013546693 | DPF3 |
| chr19 | 14408309 | 14408444 | 136 | 2 | 8.93E-06 | 3.51E-06 | 0.000243 | 5.38E-06 | 0.008545227 | ADGRE5, AC008569.1 |
| chr16 | 85360392 | 85360571 | 180 | 2 | 1.69E-05 | 4.95E-06 | 9.47E-05 | 5.39E-06 | 0.012070418 | GSE1 |
| chr2 | 96761184 | 96762238 | 1055 | 7 | 8.56E-09 | 0.000796 | 9.48E-06 | 5.39E-06 | 0.001790241 | CNNM4 |
| chr15 | 98519038 | 98519527 | 490 | 3 | 3.34E-07 | 0.006055 | 3.07E-06 | 5.39E-06 | 0.008746984 | FAM169B |
| chr10 | 31827126 | 31827529 | 404 | 3 | 2.66E-06 | 4.86E-05 | 2.98E-05 | 5.39E-06 | 0.002056602 | ARHGAP12 |
| chr7 | 90244873 | 90246426 | 1554 | 13 | 0.000142605 | 2.52E-05 | 0.001601 | 5.39E-06 | 0.001591914 | CFAP69 |
| chr3 | 183513457 | 183513470 | 14 | 2 | 2.33E-05 | 4.54E-06 | 0.000113 | 5.39E-06 | 0.012048988 | KLHL6 |
| chr10 | 22700462 | 22701199 | 738 | 2 | 4.99E-06 | 6.94E-06 | 5.87E-05 | 5.40E-06 | 0.010606998 | PIP4K2A |
| chr6 | 165987448 | 165988212 | 765 | 10 | 4.35E-05 | 1.19E-05 | 0.002545 | 5.41E-06 | 0.000615924 | LINC00602, PDE10A |
| chr17 | 72609802 | 72609934 | 133 | 2 | 1.14E-05 | 2.94E-05 | 1.78E-05 | 5.41E-06 | -0.00040401 | LINC00511 |
| chr7 | 135509451 | 135510403 | 953 | 10 | 1.73E-08 | 0.006576 | 2.04E-06 | 5.41E-06 | -0.00071096 | CNOT4 |
| chr2 | 85692873 | 85692877 | 5 | 2 | 3.74E-06 | 1.25E-05 | 3.29E-05 | 5.42E-06 | 0.004325839 | GNLY |
| chr21 | 43882048 | 43882386 | 339 | 5 | 2.04E-05 | 3.85E-05 | 0.000501 | 5.42E-06 | 0.003337928 | AGPAT3 |
| chr14 | 75660483 | 75661506 | 1024 | 11 | 2.85E-05 | 2.99E-05 | 0.000682 | 5.45E-06 | 0.002464812 | FLVCR2, TTLL5, ERG28 |
| chr7 | 56092716 | 56093327 | 612 | 10 | 2.52E-05 | 4.66E-06 | 0.003368 | 5.46E-06 | 0.008752106 | PHKG1 |
| chr7 | 155482918 | 155483958 | 1041 | 7 | 1.17E-06 | 1.97E-05 | 0.000146 | 5.46E-06 | 0.003979266 | CNPY1, AC009403.2 |
| chr19 | 51108559 | 51109027 | 469 | 4 | 2.00E-06 | 0.003701 | 3.12E-06 | 5.46E-06 | -0.00051125 | NA |
| chr16 | 1986529 | 1986960 | 432 | 4 | 2.19E-06 | 5.81E-06 | 0.00115 | 5.47E-06 | 0.002979908 | GFER, AC005606.2 |
| chr12 | 16236887 | 16237337 | 451 | 3 | 1.88E-05 | 3.35E-06 | 0.000789 | 5.48E-06 | -0.00286731 | SLC15A5 |
| chr11 | 65859388 | 65859933 | 546 | 10 | 1.07E-05 | 0.000101 | 0.000118 | 5.48E-06 | -0.00024337 | MUS81, CFL1 |
| chr17 | 17711552 | 17711875 | 324 | 2 | 1.39E-05 | 3.30E-06 | 0.000395 | 5.48E-06 | 0.007574759 | RAI1 |
| chr14 | 63852527 | 63853094 | 568 | 7 | 6.23E-05 | 6.67E-06 | 0.003978 | 5.48E-06 | 0.002000934 | SYNE2 |
| chr3 | 49702801 | 49703505 | 705 | 3 | 8.27E-05 | 2.25E-06 | 0.001205 | 5.48E-06 | 0.009855016 | RNF123 |
| chr1 | 44655713 | 44656230 | 518 | 3 | 1.53E-06 | 1.01E-05 | 7.03E-05 | 5.49E-06 | 0.002622474 | TMEM53 |
| chr21 | 17784364 | 17785313 | 950 | 3 | 1.44E-06 | 1.05E-05 | 0.000333 | 5.49E-06 | 0.020032914 | C21orf91-OT1 |
| chr17 | 36629358 | 36629637 | 280 | 3 | 1.44E-05 | 1.15E-05 | 0.000147 | 5.49E-06 | 0.002264658 | NA |
| chr8 | 144626600 | 144626612 | 13 | 2 | 1.68E-05 | 1.37E-05 | 3.10E-05 | 5.49E-06 | 0.001574343 | NA |
| chr5 | 180071943 | 180072824 | 882 | 6 | 1.18E-05 | 4.77E-06 | 0.001703 | 5.50E-06 | 0.000146093 | RNF130 |
| chr5 | 123099158 | 123100267 | 1110 | 8 | 3.93E-05 | 1.54E-06 | 0.002686 | 5.50E-06 | 0.003154272 | PRDM6 |
| chr5 | 173055669 | 173056445 | 777 | 13 | 6.24E-05 | 1.21E-05 | 0.004143 | 5.50E-06 | 0.000309314 | CREBRF |
| chr4 | 4387715 | 4388191 | 477 | 5 | 4.27E-06 | 5.69E-05 | 5.10E-05 | 5.50E-06 | 0.002700238 | NSG1 |
| chr17 | 51461784 | 51461831 | 48 | 3 | 3.63E-05 | 2.26E-06 | 0.001164 | 5.50E-06 | 0.025214444 | LINC02073 |

| | | | | | | | | | | |
|---|---|---|---|---|---|---|---|---|---|---|
| chr11 | 45021646 | 45022009 | 364 | 3 | 1.98E-05 | 2.26E-06 | 0.001505 | 5.51E-06 | 0.011052783 | AC068858.1 |
| chr13 | 73113131 | 73113588 | 458 | 4 | 4.34E-05 | 1.36E-06 | 0.005056 | 5.51E-06 | 0.003740422 | NA |
| chr20 | 33402223 | 33402740 | 518 | 2 | 1.69E-07 | 3.85E-05 | 1.58E-05 | 5.51E-06 | 0.005367815 | NA |
| chr14 | 69397562 | 69398739 | 1178 | 16 | 1.72E-05 | 5.42E-05 | 0.013222 | 5.51E-06 | 0.002189116 | SLC39A9, ERH |
| chr2 | 171574162 | 171574306 | 145 | 4 | 3.51E-05 | 3.01E-06 | 0.001326 | 5.53E-06 | 0.011044618 | NA |
| chr7 | 150433620 | 150434424 | 805 | 2 | 2.31E-05 | 3.84E-06 | 0.000191 | 5.53E-06 | 0.010910679 | LINC00996 |
| chr11 | 69293777 | 69294891 | 1115 | 8 | 1.19E-06 | 0.000132 | 0.000207 | 5.53E-06 | 0.003821482 | MYEOV |
| chr16 | 85329357 | 85329603 | 247 | 4 | 2.73E-05 | 1.49E-06 | 0.004 | 5.54E-06 | 0.022793565 | GSE1 |
| chr21 | 42614462 | 42615111 | 650 | 3 | 7.28E-06 | 5.70E-06 | 0.000526 | 5.54E-06 | 0.00671362 | LINC01671 |
| chr4 | 131975111 | 131975424 | 314 | 2 | 2.74E-05 | 3.37E-06 | 0.00037 | 5.54E-06 | -0.04491474 | AC096711.2 |
| chr7 | 8433649 | 8434914 | 1266 | 11 | 1.30E-05 | 1.02E-05 | 0.004529 | 5.54E-06 | 0.003688164 | NXPH1 |
| chr7 | 55133624 | 55133852 | 229 | 2 | 6.46E-06 | 4.82E-06 | 0.000106 | 5.55E-06 | 0.011789902 | EGFR |
| chr6 | 3230953 | 3232012 | 1060 | 10 | 9.33E-06 | 1.19E-05 | 0.002834 | 5.55E-06 | 0.004583514 | PSMG4, TUBB2B |
| chr6 | 68653278 | 68653388 | 111 | 2 | 9.76E-06 | 3.29E-06 | 0.000453 | 5.55E-06 | 0.017125086 | ADGRB3 |
| chr16 | 89769813 | 89770659 | 847 | 7 | 9.16E-07 | 0.001232 | 4.95E-05 | 5.55E-06 | 0.004316048 | FANCA |
| chr20 | 61914058 | 61914540 | 483 | 3 | 9.76E-06 | 7.08E-06 | 9.26E-05 | 5.57E-06 | 0.003795871 | CDH4 |
| chr1 | 160285968 | 160286554 | 587 | 7 | 3.66E-05 | 8.37E-07 | 0.012167 | 5.57E-06 | 0.002617408 | PEX19 |
| chr11 | 114062791 | 114063726 | 936 | 7 | 2.20E-05 | 2.99E-06 | 0.001569 | 5.58E-06 | 0.006449692 | ZBTB16 |
| chr2 | 100818149 | 100818705 | 557 | 3 | 1.35E-05 | 2.42E-05 | 0.000323 | 5.58E-06 | 0.005753152 | NA |
| chr5 | 62443960 | 62444528 | 569 | 2 | 2.11E-05 | 3.34E-06 | 0.000421 | 5.58E-06 | -0.00159162 | AC114982.2, IPO11 |
| chr7 | 144403537 | 144403926 | 390 | 5 | 2.08E-06 | 0.000793 | 1.92E-05 | 5.58E-06 | 0.002457524 | NOBOX |
| chr8 | 142004389 | 142005098 | 710 | 6 | 9.92E-06 | 0.0001 | 0.000314 | 5.59E-06 | 0.009430168 | NA |
| chr19 | 40596342 | 40597262 | 921 | 6 | 1.51E-05 | 3.37E-06 | 0.000461 | 5.60E-06 | 0.006992233 | LTBP4 |
| chr15 | 49169435 | 49170463 | 1029 | 14 | 7.72E-06 | 9.33E-06 | 0.001091 | 5.63E-06 | 0.002234754 | GALK2 |
| chr20 | 59994510 | 59995468 | 959 | 4 | 1.12E-05 | 2.39E-06 | 0.000995 | 5.64E-06 | 0.011107182 | CDH26 |
| chr18 | 159265 | 159793 | 529 | 4 | 2.19E-06 | 1.72E-05 | 0.00068 | 5.65E-06 | 0.005760759 | USP14 |
| chr9 | 83538643 | 83539337 | 695 | 4 | 5.75E-06 | 1.86E-05 | 4.60E-05 | 5.65E-06 | 0.008488496 | NA |
| chr19 | 1592511 | 1593201 | 691 | 6 | 1.76E-06 | 1.43E-05 | 0.000856 | 5.65E-06 | -0.00090873 | MBD3, AC005943.1 |
| chr2 | 241670081 | 241670910 | 830 | 4 | 2.67E-05 | 1.45E-05 | 0.001776 | 5.65E-06 | 0.0005174 | ATG4B |
| chr19 | 53537520 | 53538144 | 625 | 8 | 5.10E-05 | 2.35E-06 | 0.007376 | 5.66E-06 | 0.003780347 | ZNF331 |
| chr11 | 60976751 | 60977039 | 289 | 3 | 1.43E-05 | 1.10E-05 | 0.00014 | 5.66E-06 | 0.009070552 | CD6 |
| chr1 | 205592678 | 205592778 | 101 | 2 | 1.25E-06 | 7.23E-06 | 6.02E-05 | 5.66E-06 | 0.005673428 | MFSD4A |
| chr19 | 43407530 | 43408008 | 479 | 2 | 5.97E-07 | 1.19E-05 | 3.62E-05 | 5.67E-06 | 0.014664637 | TEX101 |
| chr7 | 57449049 | 57449560 | 512 | 3 | 1.25E-05 | 3.81E-06 | 0.000786 | 5.67E-06 | 0.015560676 | NA |
| chr4 | 6574858 | 6575357 | 500 | 7 | 1.15E-05 | 5.68E-05 | 0.004153 | 5.67E-06 | -0.00019234 | MAN2B2 |
| chr18 | 50281657 | 50282435 | 779 | 9 | 5.64E-05 | 3.49E-06 | 0.003133 | 5.67E-06 | -9.74E-05 | MBD1, CXXC1 |
| chr1 | 148309719 | 148310364 | 646 | 5 | 7.08E-05 | 1.28E-06 | 0.00589 | 5.68E-06 | 0.035208129 | AC245100.1, LINC01138 |
| chr9 | 113220769 | 113221530 | 762 | 6 | 1.18E-05 | 5.09E-06 | 0.000274 | 5.69E-06 | 0.002337624 | FKBP15 |
| chr2 | 70768217 | 70768889 | 673 | 10 | 1.51E-05 | 1.68E-05 | 0.002141 | 5.71E-06 | 0.002619633 | ADD2 |
| chr17 | 48570425 | 48571220 | 796 | 5 | 1.83E-05 | 8.56E-05 | 0.001233 | 5.71E-06 | 0.007078913 | HOXB-AS3, HOXB3 |
| chr21 | 46439560 | 46440548 | 989 | 4 | 0.00012359 | 1.67E-06 | 0.003425 | 5.71E-06 | 0.006613742 | PCNT |
| chr9 | 127979602 | 127980164 | 563 | 3 | 2.45E-07 | 1.42E-05 | 0.000131 | 5.71E-06 | 0.007811042 | FAM102A |
| chr11 | 1834621 | 1835534 | 914 | 9 | 3.81E-06 | 1.83E-05 | 0.001554 | 5.71E-06 | 0.003973129 | SYT8 |
| chr3 | 148990966 | 148992015 | 1050 | 13 | 1.32E-05 | 6.88E-05 | 0.00459 | 5.71E-06 | 0.001274181 | GYG1 |
| chr6 | 36198818 | 36199120 | 303 | 2 | 2.94E-06 | 5.59E-05 | 1.40E-05 | 5.72E-06 | 0.013550063 | BRPF3 |
| chr16 | 49814270 | 49815078 | 809 | 2 | 0.000107609 | 3.53E-06 | 0.000339 | 5.72E-06 | 0.011977876 | ZNF423 |
| chr11 | 250601 | 250989 | 389 | 4 | 3.62E-07 | 0.001609 | 4.23E-06 | 5.73E-06 | 0.004490496 | PSMD13 |
| chr8 | 120444294 | 120445695 | 1402 | 16 | 3.93E-07 | 0.000335 | 0.001848 | 5.73E-06 | -0.00076403 | MTBP, MRPL13 |
| chr6 | 52087037 | 52087563 | 527 | 2 | 1.18E-05 | 3.77E-06 | 0.000245 | 5.73E-06 | 0.009299321 | PKHD1 |
| chr14 | 76509459 | 76509794 | 336 | 2 | 1.60E-05 | 5.70E-06 | 8.54E-05 | 5.73E-06 | 0.009920839 | AC008050.1 |
| chr2 | 44937356 | 44938388 | 1033 | 8 | 0.000105704 | 7.11E-06 | 0.00263 | 5.73E-06 | 0.006642735 | AC012354.4, LINC01833 |
| chr9 | 123372890 | 123373129 | 240 | 3 | 8.11E-06 | 2.31E-06 | 0.001286 | 5.73E-06 | 0.031032782 | CRB2 |
| chr6 | 31792270 | 31793278 | 1009 | 15 | 5.23E-07 | 1.60E-05 | 0.000548 | 5.73E-06 | -0.00494615 | VARS1 |
| chr6 | 34001334 | 34002389 | 1056 | 6 | 9.70E-07 | 0.000298 | 0.000106 | 5.73E-06 | 0.004344385 | NA |
| chr1 | 234523601 | 234523801 | 201 | 2 | 7.71E-06 | 1.31E-05 | 3.40E-05 | 5.73E-06 | 0.012022645 | NA |
| chr17 | 8189860 | 8190993 | 1134 | 12 | 2.97E-06 | 4.43E-05 | 0.003263 | 5.74E-06 | 0.000620271 | BORCS6 |
| chr9 | 137025621 | 137026329 | 709 | 3 | 4.18E-06 | 4.15E-06 | 0.000226 | 5.74E-06 | 0.006156264 | ABCA2 |
| chr6 | 65838049 | 65838702 | 654 | 4 | 8.73E-06 | 1.35E-05 | 0.000334 | 5.74E-06 | 0.003593357 | NA |
| chr1 | 220527454 | 220528427 | 974 | 9 | 0.000159691 | 1.75E-05 | 0.001493 | 5.74E-06 | 0.000842622 | MARK1 |
| chr7 | 75953563 | 75954103 | 541 | 4 | 3.70E-05 | 3.09E-06 | 0.002042 | 5.75E-06 | 0.009479258 | POR |
| chr10 | 92848182 | 92848836 | 655 | 10 | 1.19E-05 | 2.56E-05 | 0.001686 | 5.75E-06 | -0.00011367 | EXOC6 |
| chr19 | 38251069 | 38251288 | 220 | 2 | 1.63E-05 | 8.91E-06 | 4.87E-05 | 5.75E-06 | 0.006775957 | SPINT2, PPP1R14A |
| chr11 | 120242377 | 120243215 | 839 | 2 | 6.62E-05 | 4.06E-06 | 0.000188 | 5.75E-06 | 0.00934734 | POU2F3 |
| chr19 | 1815426 | 1816514 | 1089 | 13 | 4.07E-05 | 1.40E-05 | 0.003254 | 5.76E-06 | 0.00139976 | AC012615.2, REXO1, MIR1909 |
| chr2 | 94993209 | 94993774 | 566 | 3 | 6.82E-06 | 3.72E-05 | 0.000198 | 5.76E-06 | 0.004478356 | AC103563.8 |
| chr17 | 36544714 | 36545515 | 802 | 8 | 7.62E-06 | 3.18E-05 | 0.001213 | 5.76E-06 | -0.00045118 | GGNBP2 |
| chr1 | 153566488 | 153567563 | 1076 | 5 | 1.17E-06 | 8.83E-05 | 0.000176 | 5.76E-06 | 0.006498685 | S100A2 |
| chr8 | 56320042 | 56320832 | 791 | 11 | 3.20E-05 | 4.12E-06 | 0.010592 | 5.77E-06 | 0.006148947 | SDR16C5 |
| chr16 | 1351194 | 1352334 | 1141 | 12 | 2.35E-08 | 0.003025 | 4.60E-06 | 5.77E-06 | 9.41E-05 | GNPTG, TSR3 |
| chr12 | 3264500 | 3264598 | 99 | 2 | 1.24E-05 | 4.27E-05 | 1.58E-05 | 5.77E-06 | 0.004249327 | TSPAN9 |
| chr6 | 27726886 | 27726982 | 97 | 2 | 2.99E-05 | 3.46E-06 | 0.00043 | 5.77E-06 | 0.025751408 | NA |
| chr1 | 175385499 | 175386226 | 728 | 4 | 4.68E-05 | 2.36E-06 | 0.000929 | 5.77E-06 | -0.00038657 | TNR |
| chr15 | 88257773 | 88258243 | 471 | 4 | 1.53E-05 | 2.55E-06 | 0.001646 | 5.78E-06 | 0.008753009 | NTRK3-AS1 |
| chr6 | 151347305 | 151347706 | 402 | 3 | 7.08E-06 | 0.000105 | 1.77E-05 | 5.79E-06 | 0.011080234 | AKAP12 |
| chr1 | 10032241 | 10033422 | 1182 | 15 | 2.21E-05 | 5.07E-06 | 0.016907 | 5.79E-06 | 0.00031738 | UBE4B |
| chr5 | 103556944 | 103557511 | 568 | 2 | 1.52E-05 | 5.17E-06 | 0.000103 | 5.79E-06 | 0.017245188 | NUDT12 |
| chr3 | 32423180 | 32423585 | 406 | 2 | 2.20E-06 | 3.43E-06 | 0.00047 | 5.80E-06 | 0.013801413 | CMTM7 |
| chr14 | 99971614 | 99972400 | 787 | 6 | 2.65E-06 | 7.20E-05 | 0.001154 | 5.80E-06 | 0.000524845 | EVL |
| chr4 | 6555693 | 6556501 | 809 | 7 | 8.88E-06 | 5.20E-05 | 0.000563 | 5.80E-06 | 0.006944894 | PPP2R2C |
| chr3 | 186652719 | 186653087 | 369 | 2 | 1.45E-06 | 9.69E-05 | 1.14E-05 | 5.81E-06 | 0.004174757 | FETUB, AC068631.1 |
| chr3 | 12003975 | 12004249 | 275 | 6 | 1.60E-05 | 7.36E-05 | 0.002358 | 5.82E-06 | 0.011222879 | NA |
| chr11 | 8963830 | 8964217 | 388 | 3 | 2.25E-06 | 2.44E-06 | 0.001114 | 5.82E-06 | 0.006847152 | TMEM9B |
| chr19 | 7230977 | 7231260 | 284 | 2 | 1.19E-05 | 7.12E-06 | 6.49E-05 | 5.83E-06 | 0.006083783 | INSR |
| chr6 | 109466157 | 109467425 | 1269 | 5 | 1.85E-05 | 5.83E-05 | 0.000231 | 5.83E-06 | -0.0016073 | ZBTB24 |
| chr17 | 57635267 | 57635868 | 602 | 2 | 1.92E-05 | 3.65E-06 | 0.00032 | 5.83E-06 | 0.002896345 | MSI2 |
| chr2 | 232352369 | 232352907 | 539 | 2 | 1.15E-05 | 4.92E-06 | 0.000117 | 5.84E-06 | -0.01047842 | NA |
| chr14 | 100073954 | 100074113 | 160 | 2 | 1.04E-05 | 4.79E-06 | 0.000124 | 5.84E-06 | 0.014933257 | EVL, AL157912.1 |
| chr17 | 80168515 | 80168955 | 441 | 2 | 4.90E-06 | 8.12E-06 | 5.59E-05 | 5.85E-06 | 0.00603449 | NA |
| chr5 | 150758968 | 150760150 | 1183 | 11 | 4.64E-05 | 5.65E-06 | 0.003447 | 5.85E-06 | 0.003030693 | DCTN4 |
| chr20 | 18567118 | 18567375 | 258 | 8 | 6.54E-05 | 1.81E-05 | 0.000867 | 5.85E-06 | 0.001329809 | SMIM26 |
| chr21 | 43691129 | 43691558 | 430 | 3 | 6.00E-06 | 2.42E-05 | 0.000287 | 5.86E-06 | 0.001952912 | RRP1B |
| chr1 | 46533539 | 46534317 | 779 | 4 | 1.18E-05 | 1.23E-05 | 0.000146 | 5.86E-06 | 0.007082396 | TMEM275 |
| chr16 | 51131684 | 51131765 | 82 | 3 | 2.56E-05 | 5.08E-06 | 0.000171 | 5.86E-06 | 0.00804825 | NA |
| chr8 | 143986631 | 143987618 | 988 | 6 | 5.43E-05 | 2.04E-06 | 0.004146 | 5.86E-06 | 0.006150678 | PARP10 |
| chr15 | 40232015 | 40232706 | 692 | 4 | 5.67E-05 | 1.49E-06 | 0.004418 | 5.86E-06 | 0.002132382 | PAK6, BUB1B-PAK6, AC020658.3 |
| chr10 | 133459059 | 133460180 | 1122 | 4 | 5.44E-06 | 2.44E-05 | 0.000363 | 5.87E-06 | -0.00865495 | SCART1 |
| chr5 | 97087822 | 97088033 | 212 | 2 | 2.98E-05 | 3.54E-06 | 0.000409 | 5.88E-06 | -0.00025263 | NA |
| chr3 | 196211177 | 196211565 | 389 | 6 | 2.03E-05 | 2.09E-05 | 0.000129 | 5.88E-06 | 0.007475914 | SLC51A, ZDHHC19 |
| chr22 | 50538016 | 50538510 | 495 | 3 | 4.06E-06 | 0.000174 | 1.44E-05 | 5.88E-06 | 0.009576854 | NA |
| chr3 | 119322357 | 119323108 | 752 | 6 | 1.52E-05 | 3.45E-05 | 0.000686 | 5.90E-06 | -0.00128346 | ARHGAP31, ARHGAP31-AS1 |
| chr6 | 107115292 | 107116026 | 735 | 6 | 9.28E-07 | 4.17E-05 | 5.97E-05 | 5.90E-06 | 0.000933825 | BEND3 |
| chr19 | 3907915 | 3908399 | 485 | 4 | 3.72E-06 | 0.000684 | 0.000173 | 5.90E-06 | 0.010558744 | ATCAY |
| chr11 | 3797465 | 3798309 | 845 | 16 | 6.46E-05 | 8.11E-07 | 0.027942 | 5.91E-06 | 0.000552128 | PGAP2, NUP98 |
| chr8 | 30095548 | 30096085 | 538 | 3 | 1.34E-06 | 6.75E-06 | 0.000294 | 5.91E-06 | 0.002402615 | LEPROTL1 |
| chr18 | 54959896 | 54960396 | 501 | 5 | 1.48E-06 | 9.90E-05 | 2.18E-05 | 5.91E-06 | 0.004794511 | NA |
| chr12 | 6643715 | 6643851 | 137 | 3 | 9.07E-06 | 8.68E-05 | 0.000149 | 5.91E-06 | 0.009698023 | ACRBP |
| chr11 | 113908394 | 113909214 | 821 | 6 | 1.60E-05 | 1.25E-05 | 7.82E-05 | 5.91E-06 | 0.005040752 | HTR3B |
| chr5 | 135063101 | 135063685 | 585 | 3 | 2.71E-05 | 2.78E-06 | 0.000959 | 5.91E-06 | 0.00755503 | C5orf66 |
| chr15 | 78437327 | 78437992 | 666 | 7 | 1.97E-06 | 1.71E-05 | 0.002019 | 5.92E-06 | 0.006312096 | IREB2 |
| chr12 | 120262558 | 120263362 | 805 | 3 | 2.21E-05 | 3.05E-06 | 0.000659 | 5.92E-06 | 0.012218178 | PXN |
| chr19 | 45076120 | 45077369 | 1250 | 16 | 4.83E-05 | 1.45E-05 | 0.022383 | 5.92E-06 | 0.003104842 | ZNF296, GEMIN7-AS1 |
| chr20 | 48722089 | 48722300 | 212 | 2 | 1.68E-05 | 9.57E-06 | 4.74E-05 | 5.92E-06 | 0.013469841 | PREX1 |
| chr3 | 12502799 | 12503351 | 553 | 3 | 1.75E-06 | 6.81E-05 | 4.06E-05 | 5.93E-06 | 0.001704982 | TSEN2 |
| chr19 | 38305090 | 38305454 | 365 | 3 | 3.23E-06 | 3.05E-05 | 5.13E-05 | 5.93E-06 | 0.001684238 | YIF1B |
| chr15 | 37358287 | 37358477 | 191 | 2 | 2.59E-05 | 4.45E-06 | 0.000157 | 5.93E-06 | -0.00189323 | NA |
| chr16 | 50741565 | 50742740 | 1176 | 12 | 3.58E-05 | 7.16E-05 | 0.001773 | 5.93E-06 | 0.001147076 | CYLD, MIR3181, AC007728.2 |
| chr7 | 12570192 | 12571611 | 1420 | 17 | 1.89E-06 | 0.000178 | 0.000975 | 5.94E-06 | 0.001122303 | SCIN, AC005281.1 |
| chr5 | 29906005 | 29906594 | 590 | 2 | 1.80E-05 | 4.99E-06 | 0.000118 | 5.94E-06 | 0.00339117 | NA |
| chr7 | 25179537 | 25180793 | 1257 | 15 | 1.98E-06 | 8.44E-05 | 0.000617 | 5.94E-06 | 0.001000122 | C7orf31 |
| chr12 | 52778374 | 52778816 | 443 | 5 | 7.65E-06 | 1.81E-05 | 6.69E-05 | 5.94E-06 | 0.007365096 | NA |
| chr1 | 42152370 | 42153492 | 1123 | 11 | 3.08E-05 | 5.06E-05 | 0.006051 | 5.94E-06 | 0.003723217 | GUCA2B |
| chr17 | 7323862 | 7324418 | 557 | 7 | 3.49E-05 | 4.92E-06 | 0.006935 | 5.94E-06 | 0.00594172 | NEURL4 |
| chr17 | 77474485 | 77475409 | 925 | 11 | 9.39E-05 | 1.48E-06 | 0.006886 | 5.94E-06 | 0.003940934 | SEPTIN9 |
| chr1 | 219928356 | 219929186 | 831 | 16 | 8.21E-06 | 3.82E-05 | 0.001256 | 5.94E-06 | 0.003654798 | SLC30A10 |
| chr7 | 2008862 | 2009125 | 264 | 3 | 1.77E-05 | 2.81E-06 | 0.001133 | 5.95E-06 | 0.006745072 | MAD1L1, AC069288.1 |
| chr7 | 1495130 | 1495835 | 706 | 6 | 3.45E-05 | 1.06E-06 | 0.005394 | 5.95E-06 | 0.00175234 | INTS1 |
| chr11 | 47177936 | 47178371 | 436 | 2 | 6.30E-08 | 0.00057 | 8.12E-06 | 5.96E-06 | 0.007649182 | PACSIN3 |
| chr18 | 46357953 | 46358444 | 492 | 3 | 1.65E-05 | 4.80E-06 | 0.00072 | 5.97E-06 | 0.005378482 | RNF165 |
| chr1 | 220093712 | 220094753 | 1042 | 6 | 5.45E-06 | 3.91E-05 | 0.00019 | 5.98E-06 | -0.00274219 | IARS2 |
| chr7 | 3809708 | 3809970 | 263 | 4 | 1.61E-05 | 1.87E-05 | 3.81E-05 | 5.98E-06 | -0.00048054 | SDK1 |
| chr17 | 59565056 | 59565932 | 877 | 14 | 4.79E-05 | 4.66E-06 | 0.008215 | 5.99E-06 | 0.003408785 | DHX40 |
| chr4 | 183443934 | 183444467 | 534 | 4 | 1.39E-06 | 3.50E-06 | 0.00136 | 5.99E-06 | 0.003825001 | NA |
| chr11 | 118659703 | 118660367 | 665 | 4 | 2.39E-06 | 9.62E-05 | 4.07E-05 | 5.99E-06 | 0.00097545 | TREH |
| chr3 | 48570591 | 48571472 | 882 | 3 | 4.77E-06 | 4.73E-06 | 0.000205 | 6.00E-06 | -0.00049726 | COL7A1 |
| chr20 | 2691717 | 2692556 | 840 | 5 | 4.54E-06 | 4.39E-06 | 0.000505 | 6.00E-06 | 0.006815758 | NA |
| chr19 | 55038046 | 55038980 | 935 | 11 | 3.53E-05 | 3.81E-06 | 0.015935 | 6.00E-06 | 0.011260168 | AC011476.3, GP6, AC011476.2 |
| chr2 | 219612025 | 219612371 | 347 | 2 | 2.72E-06 | 7.06E-06 | 6.98E-05 | 6.01E-06 | 0.007506323 | STK11IP |
| chr22 | 18990566 | 18990598 | 33 | 2 | 2.84E-05 | 3.57E-06 | 0.000481 | 6.02E-06 | 0.012526047 | DGCR5, DGCR5 |
| chr16 | 2268025 | 2268987 | 963 | 16 | 3.16E-05 | 6.58E-06 | 0.005889 | 6.02E-06 | 0.000569067 | AC009065.4, RNPS1 |
| chr18 | 37537896 | 37538740 | 845 | 4 | 3.51E-05 | 3.84E-05 | 0.000369 | 6.03E-06 | 0.010974741 | CELF4 |
| chr17 | 50096898 | 50097482 | 585 | 2 | 1.73E-05 | 4.15E-06 | 0.000216 | 6.04E-06 | 0.005465641 | PDK2 |
| chr2 | 69948460 | 69949161 | 702 | 5 | 2.07E-05 | 4.13E-06 | 0.000733 | 6.05E-06 | -0.01067454 | NA |
| chr13 | 114042631 | 114043616 | 986 | 4 | 7.36E-06 | 0.000122 | 2.46E-05 | 6.05E-06 | 0.003436527 | RASA3 |
| chr12 | 39619710 | 39620169 | 460 | 5 | 1.92E-05 | 5.61E-06 | 0.002421 | 6.06E-06 | 0.004612727 | ABCD2 |
| chr14 | 105326973 | 105327542 | 570 | 4 | 5.76E-06 | 1.68E-06 | 0.003241 | 6.06E-06 | 0.001924631 | PACS2 |
| chr14 | 21301081 | 21301220 | 140 | 3 | 6.23E-06 | 6.75E-05 | 7.66E-05 | 6.06E-06 | 0.004654822 | RPGRIP1 |
| chr15 | 64762591 | 64763106 | 516 | 5 | 5.75E-05 | 1.90E-06 | 0.002072 | 6.06E-06 | 0.0048542 | RBPMS2 |
| chr1 | 4410896 | 4412007 | 1112 | 6 | 5.96E-06 | 0.000199 | 0.000673 | 6.07E-06 | 0.002605606 | NA |
| chr15 | 45522613 | 45523338 | 726 | 10 | 3.23E-05 | 1.85E-05 | 0.007961 | 6.07E-06 | 0.002016527 | HMGN2P46, SLC30A4 |
| chr17 | 7477969 | 7479529 | 1561 | 10 | 6.12E-07 | 6.36E-05 | 3.98E-05 | 6.07E-06 | 0.001840828 | ZBTB4 |
| chr2 | 206084924 | 206085699 | 776 | 6 | 6.62E-06 | 4.76E-06 | 0.000454 | 6.07E-06 | -0.00231503 | AC007383.1, INO80D |
| chr3 | 65381681 | 65382020 | 340 | 2 | 8.47E-06 | 1.69E-05 | 2.98E-05 | 6.07E-06 | 0.002243017 | MAGI1 |
| chr10 | 133024829 | 133025152 | 324 | 4 | 8.71E-06 | 5.57E-06 | 0.000826 | 6.07E-06 | 0.007929029 | NA |
| chr5 | 139724 | 140452 | 729 | 9 | 1.53E-05 | 1.57E-05 | 0.003856 | 6.07E-06 | 0.001442291 | PLEKHG4B |
| chr5 | 54517015 | 54518085 | 1071 | 10 | 1.49E-05 | 0.000366 | 0.001303 | 6.07E-06 | 0.000251179 | SNX18 |
| chr8 | 97640219 | 97640863 | 645 | 2 | 9.38E-05 | 3.61E-06 | 0.000484 | 6.08E-06 | 0.009500663 | NA |
| chr22 | 49481495 | 49481646 | 152 | 2 | 2.92E-05 | 4.11E-06 | 0.000234 | 6.08E-06 | 0.052249761 | C22orf34 |
| chr20 | 61818844 | 61819373 | 530 | 2 | 1.99E-06 | 1.62E-05 | 3.11E-05 | 6.08E-06 | -0.00288765 | CDH4 |
| chr4 | 112650985 | 112651265 | 281 | 2 | 5.03E-06 | 8.97E-06 | 5.33E-05 | 6.09E-06 | 0.010467616 | LARP7 |
| chr1 | 154219686 | 154220650 | 965 | 11 | 6.84E-06 | 1.53E-05 | 9.02E-05 | 6.09E-06 | 0.002590946 | UBAP2L, C1orf43 |
| chr1 | 160767146 | 160767768 | 623 | 2 | 4.08E-05 | 3.63E-06 | 0.000468 | 6.09E-06 | 0.020457961 | NA |
| chr20 | 31836461 | 31837303 | 843 | 3 | 0.00010405 | 1.16E-05 | 0.000225 | 6.09E-06 | 0.007943375 | NA |

| | | | | | | | | | | |
|---|---|---|---|---|---|---|---|---|---|---|
| chr16 | 71524865 | 71525635 | 771 | 4 | 4.32E-07 | 0.000137 | 1.09E-05 | 6.09E-06 | -0.00480416 | CHST4, ZNF19 |
| chr9 | 128684925 | 128686197 | 1273 | 6 | 1.04E-06 | 0.000613 | 4.31E-05 | 6.09E-06 | 0.00358812 | SET |
| chr19 | 41442748 | 41443213 | 466 | 2 | 1.05E-05 | 4.49E-06 | 0.000168 | 6.10E-06 | 0.011795476 | ERICH4 |
| chr19 | 30046471 | 30046929 | 459 | 2 | 2.27E-05 | 4.40E-06 | 0.00018 | 6.10E-06 | 0.021812566 | NA |
| chr10 | 93993136 | 93994071 | 936 | 6 | 4.67E-06 | 2.98E-05 | 0.000805 | 6.10E-06 | 0.008622549 | PLCE1 |
| chr3 | 47007111 | 47007643 | 533 | 3 | 1.20E-05 | 3.41E-06 | 0.000506 | 6.10E-06 | -0.00139096 | NBEAL2 |
| chr20 | 6769959 | 6770788 | 830 | 6 | 3.77E-05 | 1.21E-06 | 0.011337 | 6.11E-06 | 0.006157684 | BMP2 |
| chr3 | 49786151 | 49786792 | 642 | 16 | 6.92E-06 | 2.21E-05 | 0.006054 | 6.11E-06 | -4.55E-05 | IP6K1 |
| chr5 | 9631005 | 9631686 | 682 | 4 | 2.06E-05 | 4.80E-06 | 0.000323 | 6.11E-06 | -0.00194422 | AC026787.1, TAS2R1 |
| chr15 | 22795029 | 22795304 | 276 | 2 | 1.65E-05 | 4.39E-06 | 0.000184 | 6.11E-06 | 0.000532056 | NIPA1 |
| chr7 | 157018340 | 157018961 | 622 | 7 | 7.52E-06 | 1.99E-05 | 0.000566 | 6.11E-06 | -0.00532662 | NA |
| chr4 | 1262020 | 1262484 | 465 | 4 | 2.78E-05 | 3.89E-06 | 0.000907 | 6.11E-06 | 0.003661966 | CTBP1-DT |
| chr19 | 1275761 | 1276642 | 882 | 6 | 5.29E-06 | 4.53E-06 | 0.004202 | 6.11E-06 | 0.004867442 | FAM174C |
| chr7 | 1545816 | 1546620 | 805 | 8 | 1.81E-05 | 8.34E-06 | 0.000412 | 6.11E-06 | 0.005394815 | TMEM184A |
| chr15 | 91098432 | 91098489 | 58 | 2 | 2.90E-06 | 6.08E-05 | 1.45E-05 | 6.11E-06 | 0.00693261 | AC127520.1 |
| chr3 | 62483007 | 62483038 | 32 | 2 | 2.18E-06 | 0.012604 | 6.86E-06 | 6.12E-06 | 0.006730738 | CADPS |
| chr10 | 73784638 | 73785749 | 1112 | 11 | 1.75E-05 | 9.79E-05 | 0.008044 | 6.14E-06 | 0.000871673 | ZSWIM8 |
| chr7 | 47627745 | 47628333 | 589 | 2 | 3.47E-05 | 4.09E-06 | 0.000252 | 6.14E-06 | 0.002795308 | LINC01447 |
| chr2 | 203014055 | 203015302 | 1248 | 14 | 2.55E-06 | 0.000506 | 0.001575 | 6.15E-06 | -0.00063757 | NBEAL1, WDR12 |
| chr9 | 125877316 | 125878179 | 864 | 2 | 0.000284321 | 3.66E-06 | 0.000483 | 6.15E-06 | 0.009491856 | PBX3 |
| chr2 | 168455532 | 168456423 | 892 | 11 | 7.51E-07 | 0.000227 | 7.15E-06 | 6.15E-06 | 0.002048442 | CERS6 |
| chr8 | 140560746 | 140560997 | 252 | 2 | 6.14E-06 | 3.69E-06 | 0.000456 | 6.16E-06 | -0.00464106 | AGO2 |
| chr2 | 100397540 | 100397636 | 97 | 2 | 2.01E-05 | 1.15E-05 | 4.24E-05 | 6.16E-06 | 0.00662257 | CHST10 |
| chr10 | 78763410 | 78763959 | 550 | 3 | 1.12E-05 | 1.41E-05 | 4.76E-05 | 6.17E-06 | 0.006008735 | NA |
| chr3 | 112034750 | 112034837 | 88 | 2 | 4.90E-06 | 6.83E-06 | 7.62E-05 | 6.17E-06 | 0.006243583 | NA |
| chr14 | 73590950 | 73591782 | 833 | 4 | 1.22E-05 | 7.22E-06 | 0.000679 | 6.17E-06 | 0.004691522 | ACOT4, AC005225.2 |
| chr18 | 8615247 | 8615607 | 361 | 3 | 3.43E-06 | 1.97E-05 | 3.80E-05 | 6.17E-06 | 0.006356548 | RAB12 |
| chr12 | 132207311 | 132208060 | 750 | 5 | 1.65E-05 | 3.47E-06 | 0.001413 | 6.17E-06 | -0.00199962 | GALNT9 |
| chr6 | 1577424 | 1578344 | 921 | 6 | 1.81E-06 | 0.001176 | 9.11E-05 | 6.17E-06 | 0.003731867 | NA |
| chr6 | 43225783 | 43226084 | 302 | 2 | 3.86E-06 | 1.05E-05 | 4.63E-05 | 6.17E-06 | 0.005939771 | DNPH1 |
| chr9 | 137607630 | 137608474 | 845 | 3 | 6.33E-05 | 2.24E-06 | 0.002328 | 6.17E-06 | 0.011215213 | ARRDC1, AL590627.2 |
| chr20 | 38440566 | 38440844 | 279 | 2 | 5.23E-06 | 1.50E-05 | 3.37E-05 | 6.17E-06 | 0.009553085 | NA |
| chr18 | 78797159 | 78797541 | 383 | 3 | 3.61E-05 | 3.49E-06 | 0.001006 | 6.19E-06 | 0.004267411 | NA |
| chr3 | 194132194 | 194132792 | 599 | 3 | 4.20E-07 | 0.000811 | 5.26E-06 | 6.19E-06 | 0.003910618 | NA |
| chr7 | 149238970 | 149240020 | 1051 | 11 | 2.98E-05 | 4.41E-05 | 0.001324 | 6.20E-06 | 0.002150792 | ZNF212 |
| chr16 | 89096446 | 89097392 | 947 | 6 | 1.88E-06 | 2.56E-05 | 0.000429 | 6.20E-06 | 0.011109063 | ACSF3 |
| chr20 | 31951741 | 31952501 | 761 | 9 | 3.20E-06 | 0.000186 | 0.000139 | 6.20E-06 | 0.001441076 | PDRG1 |
| chr12 | 10454765 | 10455183 | 419 | 3 | 5.09E-06 | 6.80E-05 | 0.00012 | 6.20E-06 | 0.007688277 | NA |
| chr19 | 41707112 | 41707560 | 449 | 5 | 7.55E-06 | 1.29E-05 | 0.00011 | 6.20E-06 | 0.010924218 | NA |
| chr16 | 68239687 | 68240180 | 494 | 4 | 2.21E-05 | 1.78E-06 | 0.003384 | 6.20E-06 | 0.005311461 | NA |
| chr19 | 40366379 | 40366820 | 442 | 2 | 6.49E-07 | 2.63E-05 | 2.22E-05 | 6.21E-06 | 0.012177625 | PLD3 |
| chr11 | 65134408 | 65135196 | 789 | 7 | 7.47E-06 | 0.000181 | 0.00026 | 6.21E-06 | 0.00207425 | SYVN1 |
| chr6 | 57313815 | 57313954 | 140 | 3 | 3.50E-05 | 8.36E-06 | 0.000101 | 6.21E-06 | 0.003902202 | NA |
| chr4 | 74531717 | 74532050 | 334 | 3 | 2.46E-06 | 0.000871 | 8.31E-06 | 6.21E-06 | 0.01383776 | NA |
| chr11 | 86431365 | 86431789 | 425 | 5 | 5.83E-05 | 1.34E-06 | 0.007927 | 6.22E-06 | -0.02035386 | AP001831.1 |
| chr20 | 47262796 | 47262845 | 50 | 2 | 3.26E-05 | 3.81E-06 | 0.000401 | 6.22E-06 | 0.008763957 | ZMYND8 |
| chr5 | 141353829 | 141354034 | 206 | 3 | 8.24E-06 | 3.46E-06 | 0.000967 | 6.22E-06 | 0.023157286 | PCDHGA1, PCDHGA2, PCDHGA3, PCDHGB1 |
| chr10 | 132677701 | 132678089 | 389 | 4 | 2.51E-05 | 1.35E-05 | 0.000205 | 6.22E-06 | 0.001671296 | INPP5A |
| chr4 | 84503042 | 84503316 | 275 | 3 | 2.39E-05 | 2.75E-06 | 0.001234 | 6.22E-06 | 0.010691098 | NA |
| chr12 | 53139158 | 53139695 | 538 | 3 | 1.42E-06 | 3.11E-05 | 2.75E-05 | 6.23E-06 | 0.006719938 | NA |
| chr17 | 43846577 | 43847209 | 633 | 13 | 2.62E-05 | 0.000125 | 0.000458 | 6.23E-06 | 0.004930355 | CD300LG |
| chr15 | 51105866 | 51106362 | 497 | 4 | 3.77E-06 | 5.44E-05 | 4.30E-05 | 6.23E-06 | 0.010453844 | MIR4713HG |
| chr7 | 151806183 | 151806522 | 340 | 5 | 1.34E-05 | 1.81E-05 | 0.000456 | 6.24E-06 | 0.003874101 | AC093583.1, PRKAG2 |
| chr4 | 94406824 | 94407212 | 389 | 3 | 9.17E-06 | 1.81E-05 | 0.00012 | 6.25E-06 | 0.014699767 | NA |
| chr14 | 91947613 | 91948558 | 946 | 10 | 6.49E-05 | 3.04E-06 | 0.008304 | 6.25E-06 | 0.003684526 | FBLN5 |
| chr8 | 139842001 | 139843059 | 1059 | 3 | 3.21E-07 | 0.000155 | 1.04E-05 | 6.25E-06 | 0.007155683 | TRAPPC9 |
| chr20 | 41340291 | 41341228 | 938 | 10 | 1.38E-05 | 4.73E-06 | 0.002401 | 6.26E-06 | 0.007655787 | LPIN3 |
| chr20 | 48668129 | 48668361 | 233 | 2 | 4.57E-06 | 0.000865 | 8.05E-06 | 6.26E-06 | 0.001957541 | PREX1 |
| chr17 | 78705861 | 78706388 | 528 | 3 | 4.23E-06 | 4.96E-06 | 0.000273 | 6.27E-06 | 0.000291995 | CYTH1 |
| chr11 | 47260374 | 47260543 | 170 | 3 | 2.25E-06 | 6.32E-05 | 1.82E-05 | 6.27E-06 | 0.000960748 | NR1H3 |
| chr6 | 29729923 | 29730172 | 250 | 3 | 9.64E-06 | 1.27E-05 | 0.000117 | 6.28E-06 | 0.004949239 | HLA-F, HLA-F-AS1 |
| chr17 | 82917617 | 82918521 | 905 | 8 | 1.14E-05 | 3.50E-05 | 0.00127 | 6.29E-06 | 0.006522088 | TBCD, AC087222.1 |
| chr8 | 42391184 | 42391563 | 380 | 5 | 2.42E-05 | 1.59E-05 | 0.000977 | 6.30E-06 | 0.006183973 | NA |
| chr1 | 31396934 | 31397661 | 728 | 2 | 9.56E-05 | 3.86E-06 | 0.000404 | 6.30E-06 | 0.019820791 | NA |
| chr21 | 41919119 | 41919702 | 584 | 2 | 7.00E-06 | 1.02E-05 | 4.92E-05 | 6.31E-06 | 0.005984405 | C2CD2 |
| chr22 | 46421464 | 46421755 | 292 | 3 | 1.76E-05 | 1.08E-05 | 7.68E-05 | 6.31E-06 | 0.007078487 | CELSR1 |
| chr3 | 53134295 | 53134649 | 355 | 2 | 5.91E-06 | 1.68E-05 | 3.18E-05 | 6.31E-06 | -0.00127577 | NA |
| chr10 | 70683064 | 70683121 | 58 | 2 | 1.79E-05 | 3.79E-06 | 0.000487 | 6.34E-06 | 0.008984088 | ADAMTS14 |
| chr4 | 123397368 | 123398807 | 1440 | 8 | 4.57E-06 | 4.03E-05 | 0.000132 | 6.34E-06 | -0.00110302 | SPRY1 |
| chr16 | 57185844 | 57186750 | 907 | 10 | 1.82E-05 | 7.32E-06 | 0.000961 | 6.34E-06 | -0.00014945 | RSPRY1, PSME3IP1 |
| chr7 | 100675080 | 100676320 | 1241 | 6 | 2.13E-05 | 2.47E-05 | 0.001075 | 6.34E-06 | -2.98E-05 | GNB2 |
| chr12 | 128800809 | 128800857 | 49 | 2 | 5.50E-06 | 4.24E-06 | 0.000253 | 6.35E-06 | 0.005169371 | SLC15A4 |
| chr1 | 152534366 | 152534446 | 81 | 2 | 1.24E-05 | 1.18E-05 | 4.32E-05 | 6.35E-06 | -0.00103983 | NA |
| chr14 | 101048305 | 101049048 | 744 | 5 | 3.43E-05 | 1.28E-06 | 0.007216 | 6.36E-06 | 0.004781906 | MIR381HG, MIR544A |
| chr19 | 47377417 | 47377688 | 272 | 2 | 1.26E-06 | 5.14E-05 | 1.60E-05 | 6.36E-06 | 0.00368535 | DHX34 |
| chr2 | 69642830 | 69643394 | 565 | 4 | 9.08E-06 | 3.07E-06 | 0.00082 | 6.36E-06 | -0.00110798 | AAK1 |
| chr20 | 63588494 | 63588615 | 122 | 2 | 4.50E-06 | 4.81E-06 | 0.00016 | 6.37E-06 | 0.008631898 | GMEB2 |
| chr15 | 58979728 | 58980606 | 879 | 8 | 5.57E-06 | 0.000119 | 0.000179 | 6.39E-06 | 0.007043588 | RNF111 |
| chr9 | 90914553 | 90915010 | 458 | 2 | 5.94E-07 | 7.21E-05 | 1.42E-05 | 6.40E-06 | 0.007297026 | NA |
| chr10 | 45071600 | 45071712 | 113 | 2 | 2.69E-05 | 6.52E-06 | 8.73E-05 | 6.40E-06 | -0.00087191 | CEP164P1, AL358394.3 |
| chr3 | 3171919 | 3171950 | 32 | 2 | 1.94E-05 | 2.26E-05 | 2.55E-05 | 6.42E-06 | 0.002286339 | CRBN |
| chr12 | 114735201 | 114736246 | 1046 | 3 | 8.72E-08 | 1.62E-05 | 4.60E-05 | 6.42E-06 | 0.010902025 | AC026765.3, AC026765.4 |
| chr12 | 26523846 | 26523976 | 131 | 3 | 3.24E-05 | 5.99E-06 | 0.000578 | 6.42E-06 | 0.015937282 | ITPR2 |
| chr19 | 48445607 | 48446206 | 600 | 9 | 2.64E-05 | 5.71E-06 | 0.001928 | 6.42E-06 | 0.001850188 | GRWD1 |
| chr11 | 68980085 | 68980167 | 83 | 2 | 2.93E-06 | 1.15E-05 | 4.49E-05 | 6.43E-06 | 0.011301637 | MRGPRD |
| chr8 | 86154299 | 86154914 | 616 | 2 | 1.33E-05 | 7.51E-06 | 7.25E-05 | 6.43E-06 | -0.01235703 | NA |
| chr4 | 3768027 | 3768083 | 57 | 2 | 3.49E-06 | 2.82E-05 | 2.21E-05 | 6.43E-06 | -0.01623549 | ADRA2C |
| chr1 | 184035929 | 184036400 | 472 | 3 | 5.42E-06 | 3.32E-06 | 0.000555 | 6.43E-06 | 0.013814314 | COLGALT2 |
| chr6 | 34055641 | 34056639 | 999 | 5 | 6.31E-05 | 1.63E-06 | 0.004779 | 6.43E-06 | 0.011079973 | GRM4 |
| chr19 | 44724872 | 44725591 | 720 | 3 | 2.52E-05 | 2.89E-06 | 0.001018 | 6.45E-06 | 0.011284471 | AC243964.3 |
| chr6 | 39567227 | 39567247 | 21 | 2 | 3.06E-05 | 5.35E-06 | 0.000128 | 6.45E-06 | 0.007771959 | KIF6 |
| chr11 | 116276690 | 116277212 | 523 | 6 | 5.89E-06 | 3.79E-05 | 0.000496 | 6.45E-06 | 0.004738996 | NA |
| chr17 | 78974775 | 78974851 | 77 | 3 | 1.08E-05 | 7.50E-05 | 3.44E-05 | 6.46E-06 | 0.001600774 | LGALS3BP |
| chr9 | 136642820 | 136644083 | 1264 | 4 | 0.000456677 | 3.29E-06 | 0.001268 | 6.46E-06 | 0.008140675 | NA |
| chr7 | 157738205 | 157738264 | 60 | 2 | 1.45E-06 | 7.56E-06 | 7.29E-05 | 6.48E-06 | 0.02765919 | PTPRN2 |
| chr12 | 120361529 | 120362263 | 735 | 7 | 7.51E-05 | 2.27E-06 | 0.003302 | 6.48E-06 | 0.005654846 | MSI1 |
| chr5 | 172114823 | 172115323 | 501 | 3 | 3.07E-05 | 3.92E-06 | 0.000539 | 6.49E-06 | 0.002072633 | STK10 |
| chr9 | 16575762 | 16576173 | 412 | 3 | 1.22E-05 | 1.14E-05 | 0.000127 | 6.50E-06 | 0.013118376 | BNC2 |
| chr6 | 31652747 | 31653592 | 846 | 26 | 2.43E-05 | 1.45E-05 | 0.020233 | 6.50E-06 | 0.000797732 | APOM |
| chr11 | 67206829 | 67207584 | 756 | 5 | 9.12E-06 | 2.49E-05 | 0.000208 | 6.50E-06 | 0.002456197 | KDM2A |
| chr19 | 9323594 | 9324182 | 589 | 7 | 1.46E-05 | 2.13E-06 | 0.002189 | 6.50E-06 | -0.0051278 | ZNF559, ZNF559-ZNF177 |
| chr16 | 88662444 | 88663560 | 1117 | 14 | 4.10E-06 | 6.29E-05 | 0.005891 | 6.51E-06 | -0.00021668 | SNAI3-AS1, MVD |
| chr3 | 196247106 | 196247478 | 373 | 5 | 2.01E-05 | 1.11E-05 | 0.002682 | 6.52E-06 | 0.010480616 | PCYT1A |
| chr10 | 133785 | 134248 | 464 | 3 | 1.81E-05 | 6.87E-06 | 0.000141 | 6.52E-06 | 0.016986993 | NA |
| chr1 | 24925239 | 24925672 | 434 | 2 | 3.24E-05 | 3.89E-06 | 0.000504 | 6.52E-06 | 0.014257989 | RUNX3 |
| chr3 | 13301899 | 13302398 | 500 | 2 | 7.35E-07 | 2.82E-05 | 2.24E-05 | 6.52E-06 | 0.004124155 | NA |
| chr15 | 51731502 | 51731942 | 441 | 2 | 1.80E-05 | 5.78E-06 | 0.000112 | 6.53E-06 | 0.010958743 | LYSMD2 |
| chr1 | 202712996 | 202713498 | 503 | 2 | 3.42E-07 | 0.00088 | 8.28E-06 | 6.53E-06 | 0.014570703 | NA |
| chr8 | 1326512 | 1326520 | 9 | 2 | 2.82E-06 | 5.87E-06 | 0.000108 | 6.54E-06 | 0.005051593 | DLGAP2 |
| chr1 | 247451425 | 247452381 | 957 | 9 | 3.79E-05 | 2.91E-06 | 0.006187 | 6.54E-06 | 0.000518515 | OR2B11 |
| chr10 | 10057050 | 10057542 | 493 | 2 | 1.04E-05 | 5.54E-06 | 0.000124 | 6.54E-06 | 0.013109886 | NA |
| chr11 | 65572220 | 65573372 | 1153 | 12 | 6.95E-07 | 8.73E-05 | 9.33E-05 | 6.55E-06 | 0.000663713 | ZNRD2, FAM89B |
| chr4 | 3447573 | 3448177 | 605 | 5 | 3.82E-05 | 2.56E-06 | 0.00159 | 6.55E-06 | 0.003332434 | HGFAC |
| chr3 | 37974323 | 37975232 | 910 | 4 | 8.24E-06 | 8.77E-06 | 0.001023 | 6.56E-06 | 0.00485513 | CTDSPL |
| chr5 | 78887063 | 78887198 | 136 | 2 | 2.36E-05 | 4.75E-06 | 0.000185 | 6.56E-06 | 0.007724616 | ARSB |
| chr3 | 14409842 | 14410663 | 822 | 3 | 9.13E-05 | 7.99E-06 | 0.00046 | 6.57E-06 | 0.00120052 | SLC6A6 |
| chr1 | 227747986 | 227748505 | 520 | 2 | 1.88E-06 | 3.51E-05 | 1.99E-05 | 6.57E-06 | 0.002750608 | SNAP47 |
| chr12 | 121858774 | 121859502 | 729 | 6 | 6.51E-05 | 1.63E-05 | 0.006755 | 6.57E-06 | 0.00161521 | HPD |
| chr2 | 158969321 | 158969960 | 640 | 4 | 2.49E-06 | 1.20E-05 | 0.00029 | 6.57E-06 | 0.002898874 | TANC1 |
| chr17 | 75979403 | 75980702 | 1300 | 8 | 4.99E-06 | 0.000117 | 0.000367 | 6.59E-06 | 0.004166887 | TEN1-CDK3, TEN1 |
| chr7 | 30635938 | 30636223 | 286 | 2 | 1.85E-05 | 8.00E-06 | 6.97E-05 | 6.59E-06 | 0.000857013 | NA |
| chr1 | 2203276 | 2204150 | 875 | 3 | 6.01E-06 | 0.000165 | 0.00028 | 6.61E-06 | 0.006224076 | FAAP20 |
| chr8 | 40199492 | 40199773 | 282 | 3 | 2.95E-05 | 5.03E-06 | 0.000777 | 6.61E-06 | 0.016794975 | NA |
| chr16 | 4364674 | 4365278 | 605 | 2 | 4.13E-05 | 4.35E-06 | 0.000281 | 6.61E-06 | 0.005100139 | CORO7-PAM16, CORO7 |
| chr3 | 98086696 | 98087250 | 555 | 3 | 3.68E-06 | 0.000122 | 8.62E-05 | 6.62E-06 | 0.003001651 | OR5AC2 |
| chr1 | 181189865 | 181190538 | 674 | 2 | 7.04E-08 | 0.000232 | 1.04E-05 | 6.62E-06 | 0.013308371 | AL358393.1 |
| chr15 | 67174255 | 67174650 | 396 | 3 | 1.18E-05 | 4.62E-05 | 3.50E-05 | 6.62E-06 | 0.004823654 | SMAD3 |
| chr5 | 175478126 | 175478615 | 490 | 9 | 1.40E-05 | 3.75E-05 | 0.000438 | 6.62E-06 | 0.002474922 | SFXN1 |
| chr6 | 154832358 | 154833195 | 838 | 4 | 7.92E-05 | 1.01E-05 | 0.000182 | 6.66E-06 | 0.001172796 | SCAF8, TIAM2 |
| chr14 | 64539233 | 64540609 | 1377 | 26 | 0.000125735 | 6.49E-05 | 0.003513 | 6.66E-06 | 0.001124061 | HSPA2, AL049869.2 |
| chr1 | 63074534 | 63074546 | 13 | 2 | 4.77E-05 | 5.69E-06 | 0.000121 | 6.66E-06 | -0.01368586 | NA |
| chr8 | 143360259 | 143361047 | 789 | 4 | 2.03E-05 | 1.73E-06 | 0.004831 | 6.66E-06 | 0.005875204 | NA |
| chr16 | 88300633 | 88300666 | 34 | 2 | 3.41E-05 | 4.49E-06 | 0.000248 | 6.66E-06 | 0.014816678 | AC138512.1 |
| chr17 | 5111828 | 5113237 | 1410 | 8 | 6.78E-05 | 0.000401 | 0.000927 | 6.67E-06 | 0.001288513 | ZNF232-AS1, ZNF232 |
| chr17 | 39726621 | 39727516 | 896 | 5 | 1.17E-06 | 9.41E-05 | 0.000385 | 6.68E-06 | -0.00101044 | ERBB2 |
| chr11 | 114040419 | 114040890 | 472 | 2 | 1.43E-05 | 7.10E-06 | 8.40E-05 | 6.69E-06 | 0.008844485 | NA |
| chr13 | 112888866 | 112889979 | 1114 | 3 | 4.90E-06 | 9.06E-06 | 0.000102 | 6.69E-06 | 0.002267855 | NA |
| chr1 | 18902122 | 18903139 | 1018 | 16 | 3.08E-05 | 1.03E-05 | 0.009421 | 6.69E-06 | -0.00036728 | ALDH4A1 |
| chr11 | 111454347 | 111455029 | 683 | 5 | 3.15E-05 | 1.60E-05 | 0.000386 | 6.69E-06 | 0.007588856 | AP002008.2, POU2AF1 |
| chr2 | 241771409 | 241772184 | 776 | 5 | 6.02E-05 | 1.91E-06 | 0.004049 | 6.69E-06 | -0.01819941 | NA |
| chr7 | 108001045 | 108001351 | 307 | 3 | 6.35E-06 | 3.23E-06 | 0.000822 | 6.69E-06 | 0.000451139 | LAMB1 |
| chr1 | 173669551 | 173670241 | 691 | 7 | 6.27E-06 | 8.62E-05 | 0.00474 | 6.69E-06 | 0.006465082 | AL139142.2, ANKRD45 |
| chr3 | 58577370 | 58578180 | 811 | 10 | 7.43E-06 | 6.18E-05 | 0.000506 | 6.70E-06 | 0.006474742 | FAM107A |
| chr4 | 1035622 | 1036266 | 645 | 5 | 1.16E-05 | 5.64E-05 | 6.25E-05 | 6.70E-06 | 0.003379452 | NA |
| chr1 | 235104159 | 235105306 | 1148 | 8 | 2.57E-05 | 1.05E-05 | 0.002691 | 6.70E-06 | 0.003831442 | AL732292.2 |
| chr6 | 121619013 | 121619841 | 829 | 3 | 0.000187374 | 9.89E-06 | 0.000406 | 6.71E-06 | 0.011110976 | NA |
| chr1 | 3164514 | 3164917 | 404 | 3 | 6.10E-08 | 0.01105 | 4.60E-06 | 6.71E-06 | 0.003872778 | PRDM16 |
| chr14 | 100304759 | 100305585 | 827 | 3 | 6.65E-06 | 2.90E-06 | 0.001533 | 6.71E-06 | 0.017108351 | SLC25A29 |
| chr16 | 11597373 | 11597814 | 442 | 3 | 2.91E-06 | 3.34E-05 | 0.000275 | 6.72E-06 | 0.020249954 | LITAF |
| chr3 | 45167252 | 45167704 | 453 | 5 | 1.71E-05 | 4.81E-06 | 0.000622 | 6.72E-06 | 0.01034388 | NA |
| chr7 | 16526476 | 16526926 | 451 | 9 | 1.69E-05 | 1.01E-05 | 0.002065 | 6.72E-06 | 0.001758312 | LRRC72, SOSTDC1 |
| chr5 | 181052236 | 181052829 | 594 | 4 | 1.12E-05 | 1.49E-05 | 0.000106 | 6.72E-06 | 0.012504898 | BTNL9 |
| chr15 | 82108476 | 82108953 | 478 | 3 | 3.29E-05 | 3.02E-06 | 0.001445 | 6.73E-06 | 0.010731693 | NA |
| chr8 | 66036434 | 66036493 | 60 | 2 | 1.13E-05 | 0.000162 | 1.14E-05 | 6.74E-06 | 0.011656455 | DNAJC5B |
| chr7 | 149431080 | 149431845 | 766 | 5 | 3.38E-06 | 7.95E-05 | 6.85E-05 | 6.74E-06 | 0.001247878 | ZNF777 |
| chr1 | 1081090 | 1082003 | 914 | 5 | 1.79E-06 | 0.001129 | 8.29E-05 | 6.76E-06 | 0.001884339 | C1orf159 |
| chr22 | 49993341 | 49993489 | 149 | 3 | 4.27E-05 | 3.01E-06 | 0.001303 | 6.76E-06 | 0.016595026 | NA |
| chr14 | 64756023 | 64756242 | 220 | 2 | 3.16E-05 | 4.56E-06 | 0.000251 | 6.76E-06 | 0.019104087 | SPTB |
| chr8 | 144711057 | 144711562 | 506 | 2 | 2.59E-06 | 1.61E-05 | 3.57E-05 | 6.76E-06 | 0.021467272 | NA |
| chr20 | 38806307 | 38807073 | 767 | 5 | 6.30E-07 | 0.000443 | 2.11E-05 | 6.76E-06 | 0.001224323 | PPP1R16B |
| chr1 | 101376364 | 101376869 | 506 | 2 | 2.44E-06 | 5.61E-05 | 1.64E-05 | 6.76E-06 | 0.000390943 | LINC01307 |
| chr4 | 73102531 | 73102856 | 326 | 3 | 3.51E-05 | 4.51E-06 | 0.000321 | 6.76E-06 | 0.00678551 | ANKRD17 |

| | | | | | | | | | | |
|---|---|---|---|---|---|---|---|---|---|---|
| chr1 | 200971899 | 200972578 | 680 | 5 | 3.69E-05 | 1.89E-06 | 0.005288 | 6.76E-06 | 0.0075569 | KIF21B |
| chr2 | 240955738 | 240956317 | 580 | 2 | 4.66E-05 | 4.08E-06 | 0.000499 | 6.77E-06 | 0.011755048 | CROCC2, UICLM |
| chr16 | 21202973 | 21203187 | 215 | 3 | 4.14E-05 | 3.53E-06 | 0.001036 | 6.77E-06 | 0.010508077 | AF001550.1, ZP2 |
| chr11 | 810820 | 811954 | 1135 | 8 | 3.58E-05 | 0.00079 | 0.000643 | 6.78E-06 | 0.000658263 | RPLP2, SNORA52 |
| chr16 | 67326353 | 67327251 | 899 | 11 | 3.88E-06 | 0.000107 | 0.001064 | 6.78E-06 | 0.000566803 | LRRC36, KCTD19 |
| chr18 | 74741283 | 74741818 | 536 | 2 | 1.84E-05 | 5.98E-06 | 0.000115 | 6.78E-06 | 0.001890647 | ZNF407 |
| chr5 | 93578330 | 93579122 | 793 | 6 | 1.09E-05 | 1.52E-05 | 0.00351 | 6.79E-06 | 0.00663152 | NR2F1-AS1 |
| chr11 | 2904364 | 2905371 | 1008 | 14 | 1.71E-05 | 3.62E-05 | 0.004753 | 6.79E-06 | 0.000426305 | SLC22A18 |
| chr8 | 80767772 | 80767908 | 137 | 2 | 2.00E-05 | 1.74E-05 | 3.39E-05 | 6.80E-06 | 0.007871756 | ZNF704 |
| chr21 | 41660258 | 41660496 | 239 | 2 | 1.40E-05 | 8.20E-06 | 7.18E-05 | 6.81E-06 | 0.008132044 | NA |
| chr20 | 32778631 | 32778680 | 50 | 3 | 3.76E-05 | 3.78E-06 | 0.000711 | 6.81E-06 | 0.017798972 | DNMT3B |
| chr2 | 240782505 | 240782864 | 360 | 4 | 1.94E-05 | 8.24E-06 | 0.000217 | 6.82E-06 | 0.003677167 | KIF1A |
| chr9 | 127463973 | 127464310 | 338 | 2 | 1.35E-05 | 1.15E-05 | 4.90E-05 | 6.83E-06 | -0.00494187 | LRSAM1 |
| chr12 | 110501427 | 110502028 | 602 | 10 | 2.41E-06 | 1.71E-05 | 0.003374 | 6.83E-06 | 0.000602234 | AC002350.2, RAD9B, VPS29 |
| chr17 | 82009146 | 82009166 | 21 | 3 | 5.46E-06 | 1.59E-05 | 7.24E-05 | 6.83E-06 | 0.010474054 | ASPSCR1 |
| chr9 | 99303630 | 99303953 | 324 | 2 | 2.14E-05 | 4.22E-06 | 0.000408 | 6.83E-06 | -0.00166397 | NA |
| chr17 | 78782359 | 78782810 | 452 | 7 | 7.25E-06 | 0.001029 | 2.88E-05 | 6.85E-06 | 0.002184965 | NA |
| chr16 | 88657393 | 88657579 | 187 | 2 | 9.47E-06 | 1.62E-05 | 3.60E-05 | 6.85E-06 | 0.004144021 | MVD |
| chr19 | 14006270 | 14006729 | 460 | 9 | 6.84E-06 | 0.000125 | 5.14E-05 | 6.85E-06 | -0.00056756 | AC022098.4, RFX1 |
| chr2 | 189783852 | 189784648 | 797 | 14 | 4.21E-05 | 5.63E-06 | 0.023669 | 6.85E-06 | 0.00073376 | PMS1, ORMDL1 |
| chr1 | 17010851 | 17011888 | 1038 | 6 | 4.53E-06 | 6.18E-06 | 0.002314 | 6.85E-06 | -0.00194516 | ATP13A2, AL049569.3 |
| chr2 | 8570912 | 8571126 | 215 | 5 | 1.53E-05 | 4.75E-05 | 9.74E-05 | 6.85E-06 | 0.002669608 | LINC01814 |
| chr20 | 63917948 | 63918032 | 85 | 3 | 1.08E-05 | 1.85E-05 | 6.24E-05 | 6.85E-06 | 0.006497356 | DNAJC5 |
| chr6 | 13448131 | 13448271 | 141 | 2 | 3.58E-05 | 4.21E-06 | 0.000437 | 6.87E-06 | 0.013768458 | GFOD1 |
| chr7 | 982835 | 983520 | 686 | 11 | 2.13E-05 | 2.60E-06 | 0.001454 | 6.88E-06 | 0.007819406 | CYP2W1 |
| chr14 | 37598760 | 37600001 | 1242 | 10 | 6.92E-06 | 1.22E-05 | 0.003608 | 6.89E-06 | 0.004213761 | TTC6 |
| chr15 | 70853423 | 70854968 | 1546 | 11 | 2.46E-05 | 1.51E-05 | 0.001796 | 6.89E-06 | 0.004585661 | LRRC49, LARP6 |
| chr10 | 129709921 | 129710326 | 406 | 2 | 9.15E-06 | 4.38E-06 | 0.000345 | 6.89E-06 | 0.003797316 | MGMT |
| chr19 | 7926220 | 7927230 | 1011 | 10 | 4.83E-05 | 7.11E-05 | 0.000705 | 6.89E-06 | 0.002429379 | AC010336.5, TIMM44 |
| chr17 | 77036719 | 77037257 | 539 | 4 | 5.18E-06 | 0.000102 | 7.35E-05 | 6.91E-06 | 0.008847323 | NA |
| chr1 | 63326251 | 63327192 | 942 | 8 | 4.36E-06 | 0.000187 | 3.45E-05 | 6.91E-06 | 0.005805048 | MIR6068 |
| chr4 | 2393978 | 2394207 | 230 | 3 | 6.68E-06 | 2.12E-05 | 0.000362 | 6.91E-06 | 0.010528018 | ZFYVE28 |
| chr4 | 8020920 | 8022163 | 1244 | 5 | 6.56E-05 | 6.58E-06 | 0.000502 | 6.91E-06 | -0.00381846 | ABLIM2 |
| chr11 | 64130203 | 64130802 | 600 | 2 | 8.10E-07 | 5.27E-05 | 1.73E-05 | 6.91E-06 | 0.013141656 | MACROD1 |
| chr7 | 25301275 | 25301861 | 587 | 2 | 1.40E-05 | 7.40E-06 | 8.43E-05 | 6.92E-06 | 0.009093451 | NA |
| chr1 | 41737846 | 41738647 | 802 | 3 | 6.82E-05 | 3.09E-06 | 0.001209 | 6.92E-06 | 0.005806588 | HIVEP3 |
| chr17 | 36594202 | 36594318 | 117 | 2 | 9.24E-06 | 3.99E-05 | 1.98E-05 | 6.93E-06 | 0.002927474 | DHRS11 |
| chr12 | 3747678 | 3748080 | 403 | 2 | 1.10E-05 | 9.16E-06 | 6.40E-05 | 6.93E-06 | 0.012856443 | CRACR2A |
| chr2 | 10033550 | 10033572 | 23 | 2 | 2.33E-05 | 1.83E-05 | 3.32E-05 | 6.93E-06 | 0.006221043 | NA |
| chr3 | 170062343 | 170062950 | 608 | 4 | 3.33E-05 | 1.68E-05 | 0.000331 | 6.93E-06 | 0.002097388 | GPR160, KMT5AP3 |
| chr10 | 80289216 | 80290102 | 887 | 6 | 6.89E-05 | 9.78E-07 | 0.012804 | 6.93E-06 | 0.012834801 | MAT1A |
| chr20 | 54637168 | 54637586 | 419 | 2 | 6.89E-06 | 2.10E-05 | 2.99E-05 | 6.93E-06 | 0.00784614 | DOK5 |
| chr19 | 7520959 | 7521412 | 454 | 2 | 8.73E-07 | 5.42E-06 | 0.000153 | 6.94E-06 | 0.004137838 | ZNF358 |
| chr17 | 7133515 | 7134087 | 573 | 7 | 2.17E-06 | 0.000963 | 2.88E-05 | 6.94E-06 | 0.008165828 | NA |
| chr6 | 158858604 | 158859050 | 447 | 3 | 1.21E-06 | 0.000509 | 8.57E-06 | 6.94E-06 | 0.002088181 | AL627422.2 |
| chr1 | 201551735 | 201551878 | 144 | 2 | 1.43E-05 | 5.05E-05 | 1.77E-05 | 6.94E-06 | 0.016727629 | RPS10P7 |
| chr1 | 116909682 | 116910283 | 602 | 11 | 3.86E-05 | 1.85E-05 | 0.003931 | 6.94E-06 | 0.000336579 | PTGFRN |
| chr16 | 68283576 | 68284349 | 774 | 5 | 3.15E-05 | 1.30E-06 | 0.009188 | 6.95E-06 | 0.012769687 | SLC7A6, RNU6-1262P |
| chr18 | 50195371 | 50195529 | 159 | 3 | 4.13E-06 | 8.48E-05 | 2.07E-05 | 6.97E-06 | -0.00561111 | NA |
| chr9 | 128454415 | 128455201 | 787 | 5 | 3.32E-06 | 3.03E-05 | 4.59E-05 | 6.97E-06 | 0.001227474 | ODF2 |
| chr10 | 124646348 | 124646508 | 161 | 2 | 9.25E-06 | 1.67E-05 | 3.60E-05 | 6.97E-06 | 0.009412831 | AC068896.1, FAM53B |
| chr12 | 57549224 | 57549952 | 729 | 7 | 2.42E-07 | 0.000366 | 5.97E-05 | 6.98E-06 | 0.00510456 | KIF5A |
| chr20 | 22283569 | 22283902 | 334 | 2 | 1.68E-05 | 4.40E-06 | 0.000366 | 6.98E-06 | 0.003380145 | LINC01427 |
| chr1 | 172322587 | 172323017 | 431 | 2 | 2.20E-05 | 5.68E-06 | 0.00014 | 6.98E-06 | 0.002559218 | DNM3 |
| chr13 | 34155443 | 34156193 | 751 | 2 | 0.000135491 | 4.19E-06 | 0.000522 | 6.98E-06 | 0.031148987 | NA |
| chr10 | 103367858 | 103368256 | 399 | 7 | 1.10E-05 | 0.000327 | 0.000249 | 6.99E-06 | 0.00423107 | TAF5 |
| chr11 | 86459402 | 86459980 | 579 | 3 | 1.42E-05 | 6.67E-06 | 0.000193 | 6.99E-06 | 0.007104468 | AP001831.1, ME3 |
| chr10 | 91554938 | 91555004 | 67 | 2 | 1.30E-05 | 0.000135 | 1.24E-05 | 7.00E-06 | 0.006937215 | NA |
| chr6 | 127517748 | 127518962 | 1215 | 5 | 8.52E-06 | 0.000305 | 0.000193 | 7.01E-06 | 0.00650089 | AL096711.2, SOGA3 |
| chr15 | 101497090 | 101498178 | 1089 | 5 | 1.63E-05 | 4.33E-06 | 0.001585 | 7.01E-06 | 0.007987693 | PCSK6 |
| chr7 | 4203654 | 4204102 | 449 | 5 | 2.66E-06 | 0.000242 | 1.24E-05 | 7.02E-06 | 0.002951808 | SDK1 |
| chr7 | 23473420 | 23473970 | 551 | 5 | 1.17E-06 | 0.000211 | 2.50E-05 | 7.03E-06 | 0.007186446 | NA |
| chr3 | 184030909 | 184031439 | 531 | 4 | 1.97E-05 | 5.25E-06 | 0.000598 | 7.04E-06 | 0.010320642 | NA |
| chr11 | 33135060 | 33135291 | 232 | 2 | 2.37E-05 | 7.94E-06 | 7.93E-05 | 7.04E-06 | -1.65E-05 | CSTF3 |
| chr10 | 26810257 | 26810742 | 486 | 2 | 8.69E-06 | 1.07E-05 | 5.57E-05 | 7.04E-06 | 0.006757185 | ABI1 |
| chr16 | 31073103 | 31073963 | 861 | 9 | 3.75E-06 | 0.000599 | 8.15E-05 | 7.06E-06 | 0.00012004 | ZNF668 |
| chr1 | 154193964 | 154194624 | 661 | 4 | 3.46E-05 | 1.75E-05 | 0.001751 | 7.06E-06 | 0.009580518 | TPM3 |
| chr5 | 100891046 | 100891471 | 426 | 3 | 1.49E-05 | 6.96E-06 | 0.000666 | 7.06E-06 | 0.019399151 | ST8SIA4 |
| chr12 | 124500726 | 124501109 | 384 | 3 | 2.98E-05 | 5.05E-06 | 0.000829 | 7.06E-06 | -0.01610115 | NCOR2 |
| chr19 | 5288203 | 5288620 | 418 | 2 | 3.41E-05 | 4.54E-06 | 0.00033 | 7.06E-06 | 0.01402839 | PTPRS |
| chr16 | 75409632 | 75409670 | 39 | 2 | 3.06E-05 | 8.20E-06 | 7.65E-05 | 7.06E-06 | 3.29E-05 | CFDP1 |
| chr16 | 87765528 | 87766357 | 830 | 8 | 1.96E-05 | 4.02E-05 | 0.002272 | 7.07E-06 | 0.002262565 | KLHDC4 |
| chr12 | 64625549 | 64625678 | 130 | 3 | 4.10E-05 | 3.77E-06 | 0.000548 | 7.08E-06 | 0.009283165 | RASSF3 |
| chr2 | 20093832 | 20094248 | 417 | 3 | 3.18E-05 | 2.69E-05 | 0.000165 | 7.08E-06 | 0.008820546 | AC098828.1 |
| chr1 | 205664557 | 205664631 | 75 | 3 | 5.72E-06 | 0.000683 | 6.38E-06 | 7.08E-06 | 0.004030334 | SLC45A3 |
| chr1 | 9305435 | 9305456 | 22 | 2 | 1.67E-05 | 5.18E-06 | 0.000189 | 7.09E-06 | 0.007353619 | SPSB1 |
| chr15 | 92011861 | 92012158 | 298 | 3 | 3.46E-05 | 3.80E-06 | 0.000635 | 7.09E-06 | 0.014570031 | SLCO3A1 |
| chr3 | 52892306 | 52892859 | 554 | 2 | 1.58E-07 | 0.006097 | 7.78E-06 | 7.09E-06 | 0.011253669 | STIMATE-MUSTN1, STIMATE |
| chr3 | 9216349 | 9216889 | 541 | 5 | 3.01E-06 | 0.000191 | 0.000193 | 7.11E-06 | 0.003153042 | SRGAP3 |
| chr5 | 93741044 | 93741275 | 232 | 2 | 5.14E-06 | 4.73E-06 | 0.000274 | 7.11E-06 | 0.004466175 | FAM172A, POU5F2 |
| chr5 | 145835882 | 145836221 | 340 | 4 | 6.60E-06 | 3.72E-06 | 0.000558 | 7.12E-06 | 0.012397548 | NA |
| chr9 | 37693182 | 37693400 | 219 | 2 | 2.50E-05 | 7.83E-06 | 8.29E-05 | 7.13E-06 | 0.009276871 | FRMPD1, AL138752.2 |
| chr22 | 40176840 | 40177623 | 784 | 5 | 2.42E-06 | 0.000208 | 0.000118 | 7.14E-06 | 0.007472009 | TNRC6B |
| chr16 | 8866976 | 8868389 | 1414 | 13 | 1.15E-05 | 1.92E-05 | 0.012706 | 7.14E-06 | 0.009308581 | CARHSP1 |
| chr12 | 121961852 | 121962194 | 343 | 2 | 1.26E-05 | 4.91E-06 | 0.000238 | 7.15E-06 | 0.006399403 | CFAP251 |
| chr16 | 31149684 | 31150050 | 367 | 2 | 9.34E-06 | 4.30E-06 | 0.000516 | 7.15E-06 | 0.009124501 | PRSS36 |
| chr1 | 143691956 | 143692498 | 543 | 2 | 4.92E-05 | 4.43E-06 | 0.000418 | 7.15E-06 | 0.028663926 | NA |
| chr7 | 157291933 | 157292862 | 930 | 4 | 5.44E-05 | 5.42E-06 | 0.000848 | 7.15E-06 | 0.001466298 | NA |
| chr19 | 33193194 | 33193940 | 747 | 3 | 1.96E-05 | 3.77E-06 | 0.000566 | 7.16E-06 | 0.010014018 | LRP3 |
| chr17 | 14299439 | 14299872 | 434 | 3 | 5.59E-06 | 3.08E-06 | 0.001642 | 7.17E-06 | 0.000975229 | NA |
| chr6 | 87122600 | 87122948 | 349 | 3 | 2.17E-05 | 5.79E-06 | 0.000196 | 7.17E-06 | -0.02584304 | RCN1P1 |
| chr7 | 66377450 | 66377500 | 51 | 2 | 1.54E-05 | 4.77E-06 | 0.000277 | 7.17E-06 | 0.007341393 | TPST1, LINC00174 |
| chr1 | 94063173 | 94063707 | 535 | 4 | 3.33E-05 | 2.48E-06 | 0.002328 | 7.17E-06 | 0.005452385 | ABCA4 |
| chr14 | 21526651 | 21527154 | 504 | 4 | 1.77E-05 | 1.39E-05 | 0.000415 | 7.17E-06 | 0.003473275 | SALL2 |
| chr20 | 51121040 | 51121506 | 467 | 2 | 5.30E-05 | 4.30E-06 | 0.000532 | 7.17E-06 | 0.024195823 | NA |
| chr8 | 41796886 | 41797636 | 751 | 7 | 4.53E-06 | 1.45E-05 | 0.000602 | 7.17E-06 | 0.004260776 | ANK1 |
| chr3 | 49339986 | 49340838 | 853 | 9 | 1.23E-05 | 2.72E-05 | 0.001264 | 7.18E-06 | 0.002180333 | USP4 |
| chr4 | 56655928 | 56656889 | 962 | 10 | 4.25E-07 | 0.001915 | 0.000346 | 7.18E-06 | 8.80E-05 | HOPX |
| chr7 | 149281413 | 149282176 | 764 | 4 | 1.23E-05 | 5.81E-05 | 4.70E-05 | 7.18E-06 | 0.002731389 | ZNF783 |
| chr9 | 98172910 | 98173410 | 501 | 5 | 4.41E-05 | 1.38E-06 | 0.009163 | 7.18E-06 | 0.003620117 | CORO2A |
| chr15 | 77599493 | 77600159 | 667 | 4 | 3.85E-07 | 0.007137 | 8.87E-06 | 7.18E-06 | 0.008106519 | AC046168.2 |
| chr19 | 47788635 | 47789015 | 381 | 3 | 2.63E-05 | 4.85E-06 | 0.000339 | 7.19E-06 | 0.014795337 | NA |
| chr7 | 100168580 | 100169511 | 932 | 10 | 6.50E-06 | 2.39E-05 | 0.000168 | 7.20E-06 | 0.006126763 | GAL3ST4 |
| chr15 | 65217684 | 65218238 | 555 | 3 | 4.67E-06 | 7.55E-05 | 8.51E-05 | 7.20E-06 | 0.007504768 | NA |
| chr12 | 94872934 | 94873531 | 598 | 3 | 5.14E-06 | 0.000121 | 2.98E-05 | 7.22E-06 | -0.00441285 | NA |
| chr1 | 111139433 | 111140331 | 899 | 12 | 1.27E-05 | 9.61E-05 | 0.000614 | 7.22E-06 | -0.00062547 | CEPT1, DRAM2 |
| chr16 | 57058601 | 57059686 | 1086 | 6 | 5.84E-06 | 4.17E-05 | 0.000833 | 7.23E-06 | 0.003839272 | NLRC5 |
| chr11 | 13668575 | 13669163 | 589 | 7 | 9.90E-06 | 2.30E-05 | 0.000158 | 7.23E-06 | 0.00172138 | FAR1 |
| chr16 | 27400815 | 27401175 | 361 | 3 | 8.36E-06 | 4.28E-06 | 0.000407 | 7.24E-06 | -0.00343873 | NA |
| chr7 | 100745460 | 100745487 | 28 | 2 | 3.78E-05 | 4.84E-06 | 0.000272 | 7.24E-06 | -0.04733624 | ZAN |
| chr4 | 185425593 | 185426612 | 1020 | 12 | 8.30E-06 | 5.61E-05 | 0.000104 | 7.25E-06 | 0.000559759 | C4orf47, UFSP2 |
| chr3 | 45943093 | 45943676 | 584 | 6 | 1.67E-05 | 9.05E-06 | 0.003919 | 7.25E-06 | 0.005118732 | CXCR6, FYCO1 |
| chr3 | 12180193 | 12180748 | 556 | 3 | 1.42E-05 | 6.39E-06 | 0.000221 | 7.26E-06 | 0.006016439 | SYN2 |
| chr17 | 75509751 | 75510449 | 699 | 9 | 1.29E-05 | 4.50E-05 | 0.003493 | 7.26E-06 | 0.003991514 | CASKIN2 |
| chr14 | 23376661 | 23377697 | 1037 | 7 | 8.65E-06 | 5.05E-05 | 0.002935 | 7.26E-06 | 0.00502415 | CMTM5 |
| chr17 | 1730157 | 1731041 | 885 | 6 | 6.79E-05 | 5.54E-05 | 0.002613 | 7.26E-06 | 0.005681083 | WDR81, AC130343.1 |
| chr8 | 37830778 | 37831462 | 685 | 3 | 5.56E-05 | 2.89E-06 | 0.001713 | 7.27E-06 | 0.005856948 | ADGRA2 |
| chr6 | 31560377 | 31561138 | 762 | 3 | 2.88E-07 | 0.004169 | 0.000149 | 7.27E-06 | 0.008609126 | NA |
| chr12 | 6329764 | 6330282 | 519 | 5 | 6.44E-05 | 1.63E-06 | 0.007083 | 7.27E-06 | -0.00415958 | TNFRSF1A |
| chr5 | 72109705 | 72110361 | 657 | 5 | 6.60E-06 | 4.41E-05 | 4.98E-05 | 7.27E-06 | 0.001079217 | MAP1B |
| chr17 | 21377705 | 21378195 | 491 | 5 | 6.94E-06 | 1.05E-05 | 0.000244 | 7.27E-06 | 0.016212428 | KCNJ12 |
| chr21 | 34957184 | 34957353 | 170 | 2 | 3.58E-05 | 4.88E-06 | 0.00027 | 7.28E-06 | 0.009503583 | RUNX1 |
| chr12 | 96219989 | 96220133 | 145 | 2 | 5.50E-06 | 5.68E-05 | 1.77E-05 | 7.29E-06 | 0.00590411 | ELK3 |
| chr17 | 55963158 | 55963188 | 31 | 2 | 3.94E-05 | 4.42E-06 | 0.00049 | 7.29E-06 | 0.006867811 | ANKFN1 |
| chr10 | 45472036 | 45472050 | 15 | 3 | 3.98E-05 | 5.69E-06 | 0.000458 | 7.32E-06 | 0.003972763 | MARCHF8 |
| chr3 | 158643793 | 158644429 | 637 | 7 | 2.11E-06 | 4.54E-05 | 0.000129 | 7.34E-06 | 0.001695886 | NA |
| chr12 | 53748164 | 53748219 | 56 | 2 | 2.57E-05 | 4.42E-06 | 0.00052 | 7.34E-06 | 0.01553923 | AC076968.2, CISTR |
| chr4 | 13537183 | 13537974 | 792 | 8 | 1.20E-05 | 0.000311 | 0.001129 | 7.34E-06 | 0.006096916 | NA |
| chr15 | 38252782 | 38253330 | 549 | 6 | 1.33E-05 | 2.05E-05 | 0.003796 | 7.35E-06 | -0.00171527 | SPRED1 |
| chr7 | 116315407 | 116315901 | 495 | 2 | 3.36E-05 | 4.55E-06 | 0.000425 | 7.36E-06 | -0.00249061 | CAV2, AC073130.2, AC002066.1 |
| chr20 | 58854222 | 58855258 | 1037 | 4 | 3.90E-05 | 2.23E-06 | 0.003831 | 7.36E-06 | 0.008021844 | GNAS |
| chr3 | 31981502 | 31982119 | 618 | 10 | 3.95E-05 | 1.71E-05 | 0.003207 | 7.36E-06 | 0.003881079 | ZNF860, OSBPL10 |
| chr6 | 29656637 | 29657179 | 543 | 5 | 1.04E-06 | 7.41E-05 | 5.39E-05 | 7.37E-06 | 0.004389907 | MOG |
| chr11 | 65857104 | 65857682 | 579 | 5 | 3.42E-05 | 9.28E-06 | 0.003086 | 7.37E-06 | 0.005744593 | MUS81, CFL1 |
| chr14 | 59871042 | 59871500 | 459 | 5 | 2.32E-06 | 1.16E-05 | 0.000639 | 7.37E-06 | 0.008140624 | NA |
| chr6 | 30110855 | 30111885 | 1031 | 17 | 6.87E-06 | 2.52E-06 | 0.020618 | 7.38E-06 | -0.01262836 | TRIM31-AS1, TRIM31 |
| chr4 | 1172482 | 1173451 | 970 | 14 | 2.17E-05 | 2.65E-05 | 0.008194 | 7.39E-06 | 0.003680357 | SPON2 |
| chr7 | 30478011 | 30478696 | 686 | 4 | 3.59E-08 | 8.35E-05 | 6.23E-05 | 7.39E-06 | 0.004080099 | NOD1 |
| chr11 | 67417565 | 67417832 | 268 | 2 | 1.78E-06 | 1.94E-05 | 3.45E-05 | 7.39E-06 | 0.00823926 | CARNS1, PPP1CA |
| chr17 | 38898459 | 38898926 | 468 | 3 | 1.22E-05 | 3.35E-06 | 0.000969 | 7.40E-06 | 0.00147992 | LASP1 |
| chr9 | 28671204 | 28671714 | 511 | 2 | 1.66E-05 | 7.48E-06 | 9.51E-05 | 7.40E-06 | 0.016914677 | NA |
| chr22 | 24805697 | 24806644 | 948 | 12 | 3.60E-05 | 8.97E-06 | 0.000948 | 7.40E-06 | 0.005150203 | SGSM1 |
| chr3 | 184135338 | 184136188 | 851 | 7 | 3.77E-07 | 5.75E-05 | 0.000216 | 7.40E-06 | 0.001079985 | EIF2B5 |
| chr8 | 125091439 | 125092298 | 860 | 8 | 6.26E-06 | 0.004809 | 1.85E-05 | 7.41E-06 | 0.000652194 | NSMCE2, WASHC5 |
| chr3 | 50319740 | 50320891 | 1152 | 8 | 3.03E-06 | 0.000167 | 0.002163 | 7.41E-06 | -0.00048175 | HYAL2, TUSC2 |
| chr1 | 45750285 | 45751205 | 921 | 11 | 5.97E-06 | 9.90E-05 | 0.000937 | 7.41E-06 | 0.0002186 | IPP |
| chr19 | 35908283 | 35909147 | 865 | 5 | 9.75E-05 | 1.42E-06 | 0.008938 | 7.42E-06 | 0.009788378 | TYROBP |
| chr6 | 69961077 | 69961397 | 321 | 2 | 1.65E-05 | 7.44E-06 | 9.63E-05 | 7.42E-06 | 0.005728717 | COL19A1 |
| chr11 | 8868917 | 8869823 | 907 | 2 | 0.000218363 | 4.49E-06 | 0.000505 | 7.42E-06 | 0.012025994 | DENND2B |
| chr1 | 50422788 | 50423838 | 1051 | 12 | 1.85E-05 | 0.000456 | 9.27E-05 | 7.42E-06 | 0.000557481 | AL049637.1, DMRTA2 |
| chr21 | 29297781 | 29298095 | 315 | 3 | 4.09E-06 | 0.000623 | 0.000255 | 7.42E-06 | 0.010382265 | BACH1 |
| chr11 | 66708181 | 66708250 | 70 | 2 | 1.35E-05 | 7.89E-05 | 1.56E-05 | 7.43E-06 | 0.001424859 | SPTBN2 |
| chr3 | 55481140 | 55482568 | 1429 | 11 | 1.82E-05 | 2.12E-05 | 0.00263 | 7.43E-06 | 0.004486053 | WNT5A |
| chr14 | 24316104 | 24316727 | 624 | 4 | 3.04E-05 | 6.15E-06 | 0.002196 | 7.43E-06 | 0.003010424 | LTB4R |
| chr1 | 181180187 | 181180273 | 87 | 3 | 5.32E-05 | 2.74E-06 | 0.002565 | 7.45E-06 | 0.021503252 | LINC01732 |
| chr12 | 109458690 | 109459761 | 1072 | 7 | 3.09E-05 | 8.75E-06 | 0.000962 | 7.46E-06 | 0.000232461 | KCTD10 |
| chr9 | 37369885 | 37370305 | 421 | 2 | 2.20E-05 | 4.84E-06 | 0.000322 | 7.46E-06 | 0.005413442 | NA |
| chr4 | 4482280 | 4482354 | 75 | 2 | 8.18E-06 | 7.13E-06 | 0.000105 | 7.46E-06 | 0.004229629 | STX18 |
| chr3 | 182755260 | 182755716 | 457 | 2 | 3.22E-06 | 3.26E-05 | 2.41E-05 | 7.47E-06 | 0.013424331 | NA |
| chr11 | 2443592 | 2444261 | 670 | 15 | 1.51E-05 | 1.65E-05 | 0.002261 | 7.47E-06 | 0.007252127 | NA |
| chr17 | 42325853 | 42326151 | 299 | 2 | 1.78E-05 | 4.92E-06 | 0.000304 | 7.50E-06 | 0.010037984 | STAT3 |
| chr6 | 7301479 | 7301721 | 243 | 2 | 3.18E-06 | 5.59E-05 | 1.82E-05 | 7.50E-06 | 0.0056485 | SSR1 |

| | | | | | | | | | | |
|---|---|---|---|---|---|---|---|---|---|---|
| chr12 | 122206280 | 122206648 | 369 | 3 | 1.04E-05 | 5.82E-06 | 0.000804 | 7.50E-06 | 0.010817686 | B3GNT4 |
| chr15 | 63998608 | 63999317 | 710 | 4 | 4.67E-06 | 2.95E-05 | 5.01E-05 | 7.51E-06 | 0.008769874 | DAPK2 |
| chr15 | 45714651 | 45715270 | 620 | 3 | 1.12E-05 | 9.20E-05 | 0.000312 | 7.53E-06 | 0.011454964 | AC068722.1 |
| chr4 | 133154319 | 133154743 | 425 | 2 | 2.46E-05 | 5.57E-06 | 0.000189 | 7.53E-06 | 0.007084401 | PCDH10 |
| chr9 | 34567204 | 34567947 | 744 | 3 | 1.23E-06 | 0.00047 | 8.34E-06 | 7.54E-06 | 0.004428304 | CNTFR |
| chr7 | 63054295 | 63054550 | 256 | 3 | 1.69E-05 | 5.07E-05 | 0.000345 | 7.54E-06 | 0.025435005 | AC069285.2 |
| chr19 | 19647708 | 19647711 | 4 | 2 | 2.42E-05 | 1.08E-05 | 6.10E-05 | 7.54E-06 | 0.005657798 | ATP13A1 |
| chr8 | 946697 | 947033 | 337 | 2 | 3.06E-05 | 4.69E-06 | 0.000422 | 7.54E-06 | 0.030231723 | DLGAP2 |
| chr1 | 23800499 | 23801576 | 1078 | 16 | 5.14E-06 | 0.000234 | 0.001565 | 7.54E-06 | 0.001439579 | GALE |
| chr19 | 43768674 | 43769441 | 768 | 3 | 2.26E-05 | 6.97E-05 | 0.00012 | 7.56E-06 | 0.004481498 | KCNN4 |
| chr2 | 208255026 | 208255584 | 559 | 9 | 7.34E-06 | 2.54E-05 | 0.000201 | 7.56E-06 | 0.001295786 | IDH1-AS1, IDH1 |
| chr14 | 30375373 | 30375874 | 502 | 2 | 1.58E-05 | 8.14E-06 | 8.74E-05 | 7.56E-06 | 0.012373127 | NA |
| chr14 | 77761222 | 77762035 | 814 | 11 | 3.20E-05 | 3.33E-05 | 0.0009 | 7.57E-06 | -0.00048568 | C14orf178 |
| chr16 | 54120984 | 54122081 | 1098 | 7 | 2.50E-06 | 5.10E-05 | 6.32E-05 | 7.57E-06 | 0.001295759 | FTO |
| chr19 | 42422360 | 42423524 | 1165 | 5 | 5.84E-05 | 3.57E-06 | 0.003589 | 7.57E-06 | 0.003982994 | LIPE-AS1, LIPE |
| chr7 | 158061240 | 158062236 | 997 | 7 | 5.13E-06 | 0.000504 | 0.000468 | 7.60E-06 | 0.000502019 | PTPRN2 |
| chr5 | 567142 | 567723 | 582 | 3 | 5.73E-05 | 4.90E-06 | 0.000945 | 7.60E-06 | 0.002961741 | NA |
| chr2 | 15664470 | 15664888 | 419 | 5 | 7.67E-06 | 5.25E-05 | 0.00024 | 7.60E-06 | 0.001260535 | NA |
| chr3 | 10332383 | 10332731 | 349 | 2 | 2.77E-05 | 4.56E-06 | 0.00055 | 7.62E-06 | -0.00122488 | ATP2B2 |
| chr11 | 13008175 | 13009382 | 1208 | 14 | 0.000105272 | 9.54E-06 | 0.010892 | 7.62E-06 | 0.001621242 | RASSF10, RASSF10-DT |
| chr7 | 2005489 | 2006192 | 704 | 3 | 1.12E-06 | 1.72E-05 | 0.000244 | 7.62E-06 | 0.00360531 | MAD1L1, AC069288.1 |
| chr1 | 160366610 | 160367248 | 639 | 7 | 2.42E-05 | 6.39E-06 | 0.000178 | 7.62E-06 | 0.004050702 | NHLH1 |
| chr5 | 169583263 | 169583833 | 571 | 12 | 3.50E-05 | 3.11E-06 | 0.005658 | 7.62E-06 | 0.002911088 | SPDL1 |
| chr2 | 70553252 | 70554193 | 942 | 14 | 1.98E-05 | 4.31E-05 | 0.005014 | 7.64E-06 | 0.000647756 | TGFA |
| chr15 | 77024235 | 77024443 | 209 | 2 | 1.38E-05 | 1.32E-05 | 5.07E-05 | 7.64E-06 | 0.00161583 | PSTPIP1 |
| chr16 | 66521956 | 66522230 | 275 | 2 | 1.56E-05 | 6.52E-06 | 0.000132 | 7.64E-06 | 0.014126074 | TK2 |
| chr9 | 98877221 | 98877627 | 407 | 2 | 1.63E-05 | 9.27E-06 | 7.50E-05 | 7.64E-06 | 0.009566707 | NA |
| chr14 | 104752893 | 104753689 | 797 | 9 | 3.64E-05 | 2.08E-05 | 0.003813 | 7.64E-06 | 0.001908074 | SIVA1 |
| chr11 | 117201564 | 117202199 | 636 | 6 | 1.24E-05 | 1.36E-05 | 0.006374 | 7.65E-06 | 0.006204059 | TAGLN |
| chr5 | 180291685 | 180292756 | 1072 | 9 | 3.29E-06 | 0.001321 | 0.001366 | 7.65E-06 | -0.00049823 | MAPK9 |
| chr6 | 28903958 | 28904877 | 920 | 11 | 1.64E-06 | 0.000173 | 7.83E-05 | 7.66E-06 | 0.001180839 | TRIM27 |
| chr11 | 59363202 | 59363530 | 329 | 2 | 2.40E-06 | 0.005451 | 8.28E-06 | 7.66E-06 | 0.009826169 | OR5AN1 |
| chr1 | 113197924 | 113198106 | 183 | 2 | 7.48E-06 | 3.68E-05 | 2.31E-05 | 7.66E-06 | 0.027290605 | NA |
| chr2 | 54708668 | 54708890 | 223 | 3 | 2.26E-05 | 5.07E-06 | 0.00065 | 7.66E-06 | 0.016232411 | NA |
| chr16 | 2028823 | 2029440 | 618 | 4 | 5.12E-05 | 4.03E-06 | 0.000526 | 7.67E-06 | 0.003555878 | SLC9A3R2 |
| chr5 | 191012 | 191691 | 680 | 12 | 9.49E-05 | 8.30E-07 | 0.026829 | 7.67E-06 | -0.00259318 | LRRC14B |
| chr5 | 78540994 | 78541510 | 517 | 4 | 1.99E-06 | 0.001532 | 6.20E-05 | 7.67E-06 | 0.009941073 | LHFPL2 |
| chr12 | 109477277 | 109478191 | 915 | 14 | 5.79E-06 | 0.000946 | 0.001502 | 7.67E-06 | 0.000882418 | UBE3B, KCTD10 |
| chr17 | 76390736 | 76391500 | 765 | 7 | 3.12E-05 | 3.10E-05 | 0.002449 | 7.67E-06 | 0.000681533 | UBE2O |
| chr14 | 22492269 | 22492844 | 576 | 3 | 3.37E-05 | 6.42E-06 | 0.000269 | 7.67E-06 | 0.011437564 | NA |
| chr7 | 101693940 | 101694014 | 75 | 2 | 1.58E-05 | 0.000102 | 1.47E-05 | 7.69E-06 | 0.01032985 | NA |
| chr17 | 73091717 | 73093484 | 1768 | 12 | 3.51E-05 | 0.032944 | 0.000271 | 7.69E-06 | -0.00408784 | SLC39A11 |
| chr14 | 19874894 | 19875225 | 332 | 2 | 5.27E-06 | 2.19E-05 | 3.32E-05 | 7.69E-06 | 0.023560375 | OR4K2 |
| chr14 | 34539544 | 34540317 | 774 | 8 | 4.23E-05 | 6.90E-06 | 0.000287 | 7.70E-06 | -0.00276747 | EAPP |
| chr22 | 29267519 | 29268450 | 932 | 15 | 1.43E-05 | 0.000105 | 0.006138 | 7.70E-06 | 6.09E-05 | EWSR1, RHBDD3 |
| chr20 | 45112282 | 45112599 | 318 | 2 | 1.53E-05 | 1.50E-05 | 4.52E-05 | 7.70E-06 | 0.006364071 | WFDC5 |
| chr2 | 15940379 | 15941057 | 679 | 9 | 3.67E-05 | 2.42E-05 | 0.000541 | 7.70E-06 | -0.0014447 | MYCN, MYCNOS |
| chr3 | 49356276 | 49357189 | 914 | 2 | 0.000109052 | 4.73E-06 | 0.000462 | 7.70E-06 | 0.011649781 | GPX1 |
| chr14 | 75837198 | 75837616 | 419 | 3 | 3.51E-05 | 7.17E-06 | 0.000177 | 7.72E-06 | 0.004301524 | TTLL5, AF107885.2 |
| chr3 | 123799788 | 123800555 | 768 | 4 | 5.96E-06 | 3.67E-05 | 4.81E-05 | 7.73E-06 | 0.001760567 | MYLK |
| chr7 | 6576792 | 6577663 | 872 | 8 | 3.35E-06 | 6.65E-05 | 0.000249 | 7.73E-06 | 0.007777229 | ZDHHC4 |
| chr5 | 41057879 | 41058227 | 349 | 3 | 1.79E-05 | 1.02E-05 | 0.000292 | 7.73E-06 | 0.004332259 | MROH2B |
| chr4 | 8152271 | 8152678 | 408 | 5 | 4.10E-06 | 0.000133 | 2.80E-05 | 7.73E-06 | 0.002193162 | ABLIM2 |
| chr6 | 41894252 | 41894466 | 215 | 3 | 3.82E-05 | 3.09E-06 | 0.001803 | 7.74E-06 | 0.009731987 | USP49 |
| chr12 | 52010892 | 52011096 | 205 | 4 | 8.27E-06 | 0.000131 | 3.13E-05 | 7.75E-06 | 0.004370355 | TAMALIN |
| chr21 | 34521358 | 34521915 | 558 | 3 | 1.82E-05 | 3.21E-05 | 4.81E-05 | 7.75E-06 | 0.001398336 | RCAN1 |
| chr16 | 81500967 | 81501476 | 510 | 4 | 5.05E-05 | 2.39E-06 | 0.003282 | 7.75E-06 | 0.009899432 | CMIP |
| chr9 | 98989377 | 98989992 | 616 | 3 | 7.88E-05 | 5.13E-06 | 0.000958 | 7.76E-06 | 0.008349164 | COL15A1 |
| chr5 | 141876570 | 141877083 | 514 | 2 | 6.08E-07 | 5.03E-05 | 1.99E-05 | 7.76E-06 | -0.00596789 | PCDH1 |
| chr19 | 29548137 | 29548201 | 65 | 2 | 3.30E-05 | 8.27E-06 | 8.98E-05 | 7.76E-06 | 0.014630476 | VSTM2B |
| chr2 | 95346838 | 95348173 | 1336 | 7 | 5.53E-06 | 7.03E-05 | 0.00128 | 7.76E-06 | 0.00355159 | KCNIP3 |
| chr5 | 73450516 | 73451372 | 857 | 9 | 9.88E-06 | 6.88E-05 | 0.000415 | 7.76E-06 | 0.002594035 | NA |
| chr1 | 147245226 | 147245249 | 24 | 2 | 2.01E-05 | 1.06E-05 | 6.57E-05 | 7.76E-06 | 0.012658819 | AC242426.2, CHD1L |
| chr9 | 19408002 | 19408798 | 797 | 7 | 2.95E-05 | 0.001299 | 0.000235 | 7.76E-06 | 0.00066513 | AL391834.3 |
| chr5 | 154466388 | 154466504 | 117 | 2 | 1.62E-05 | 1.59E-05 | 4.37E-05 | 7.77E-06 | -0.00821969 | NA |
| chr17 | 50550098 | 50550586 | 489 | 3 | 8.25E-06 | 7.29E-06 | 0.000558 | 7.77E-06 | 0.006475404 | SPATA20 |
| chr5 | 179823371 | 179824299 | 929 | 6 | 3.83E-05 | 3.12E-06 | 0.00512 | 7.77E-06 | 0.002477232 | SQSTM1 |
| chr1 | 151265822 | 151266031 | 210 | 2 | 1.02E-05 | 5.77E-06 | 0.00019 | 7.77E-06 | -0.00013893 | PSMD4 |
| chr10 | 47350824 | 47351074 | 251 | 3 | 1.33E-05 | 4.81E-06 | 0.000375 | 7.77E-06 | -0.00368237 | RBP3 |
| chr15 | 41852068 | 41852382 | 315 | 4 | 6.05E-06 | 0.000212 | 3.12E-05 | 7.78E-06 | 0.005365869 | SPTBN5 |
| chr11 | 120332503 | 120332836 | 334 | 3 | 3.09E-05 | 5.10E-06 | 0.00049 | 7.78E-06 | 0.005503256 | TLCD5 |
| chr7 | 49771596 | 49772169 | 574 | 3 | 9.96E-06 | 1.54E-05 | 7.14E-05 | 7.78E-06 | 0.003395796 | NA |
| chr13 | 75226171 | 75226337 | 167 | 2 | 1.43E-05 | 1.92E-05 | 3.74E-05 | 7.78E-06 | 0.005879796 | NA |
| chr19 | 5270204 | 5271035 | 832 | 2 | 1.70E-05 | 5.05E-06 | 0.000332 | 7.78E-06 | 0.008427021 | PTPRS |
| chr22 | 25112709 | 25112871 | 163 | 3 | 6.07E-06 | 0.000114 | 0.000129 | 7.79E-06 | 0.008044717 | KIAA1671 |
| chr4 | 70728297 | 70728352 | 56 | 2 | 1.07E-05 | 0.001192 | 9.34E-06 | 7.79E-06 | 0.009385953 | RUFY3 |
| chr1 | 19468758 | 19468875 | 118 | 2 | 3.56E-06 | 0.000185 | 1.27E-05 | 7.79E-06 | 0.013416258 | CAPZB |
| chr2 | 42339045 | 42339416 | 372 | 4 | 4.70E-06 | 0.000482 | 0.000203 | 7.79E-06 | 0.005288127 | COX7A2L |
| chr13 | 45975999 | 45976068 | 70 | 2 | 1.02E-05 | 5.86E-06 | 0.000183 | 7.79E-06 | -0.00088715 | ZC3H13 |
| chr19 | 7482202 | 7482500 | 299 | 2 | 1.17E-05 | 6.93E-06 | 0.000123 | 7.79E-06 | 0.006401939 | PEX11G |
| chr2 | 73981475 | 73982033 | 559 | 6 | 7.66E-06 | 4.16E-05 | 0.001241 | 7.80E-06 | 0.007724179 | NA |
| chr14 | 22531191 | 22531840 | 650 | 4 | 4.92E-06 | 9.74E-06 | 0.000761 | 7.83E-06 | 0.008468599 | NA |
| chr10 | 85643950 | 85643972 | 23 | 2 | 4.26E-05 | 4.88E-06 | 0.000428 | 7.85E-06 | 0.01023236 | GRID1 |
| chr12 | 14802873 | 14803968 | 1096 | 15 | 1.92E-06 | 0.000313 | 0.000538 | 7.85E-06 | -9.60E-05 | C12orf60, WBP11 |
| chr7 | 27196114 | 27196372 | 259 | 2 | 4.98E-06 | 6.54E-06 | 0.000141 | 7.86E-06 | 0.004873239 | HOXA13 |
| chr4 | 165872637 | 165873319 | 683 | 6 | 2.98E-06 | 0.000602 | 0.000298 | 7.86E-06 | 0.006082646 | TLL1 |
| chr2 | 169583436 | 169584830 | 1395 | 11 | 1.22E-05 | 0.001264 | 0.000488 | 7.86E-06 | 0.002662876 | PPIG |
| chr6 | 31733234 | 31734031 | 798 | 5 | 1.58E-05 | 3.08E-05 | 0.000126 | 7.86E-06 | 0.0019279 | CLIC1 |
| chr19 | 36527864 | 36528712 | 849 | 12 | 7.88E-05 | 1.51E-05 | 0.01033 | 7.87E-06 | 0.000448964 | AC092295.1, ZNF260 |
| chr19 | 913261 | 913714 | 454 | 6 | 9.00E-06 | 4.55E-05 | 0.000884 | 7.91E-06 | 0.003250196 | NA |
| chr7 | 50308578 | 50309274 | 697 | 4 | 2.44E-05 | 2.75E-05 | 0.001303 | 7.91E-06 | 0.005530338 | IKZF1 |
| chr2 | 98444503 | 98445283 | 781 | 13 | 5.92E-06 | 0.000205 | 0.003626 | 7.91E-06 | 0.001578668 | INPP4A |
| chr13 | 113018789 | 113019970 | 1182 | 5 | 3.13E-05 | 1.63E-05 | 0.000419 | 7.92E-06 | 0.000959272 | MCF2L |
| chr13 | 77740660 | 77741471 | 812 | 7 | 8.24E-05 | 3.22E-06 | 0.005771 | 7.92E-06 | 0.008186202 | SLAIN1 |
| chr12 | 91111420 | 91111897 | 478 | 5 | 1.09E-05 | 0.000462 | 0.000262 | 7.92E-06 | 0.010394704 | LUM |
| chr6 | 132400906 | 132402025 | 1120 | 9 | 7.28E-06 | 0.000626 | 0.001748 | 7.94E-06 | 0.002178019 | MOXD1 |
| chr1 | 2632476 | 2633145 | 670 | 6 | 2.20E-05 | 2.46E-06 | 0.006332 | 7.96E-06 | 0.006449266 | MMEL1-AS1, MMEL1 |
| chr20 | 32359030 | 32360408 | 1379 | 12 | 0.000148232 | 1.42E-05 | 0.000826 | 7.96E-06 | 0.000536925 | ASXL1 |
| chr21 | 46146362 | 46147441 | 1080 | 6 | 4.36E-07 | 0.000171 | 7.49E-05 | 7.96E-06 | 0.008246965 | FTCD |
| chr9 | 105558834 | 105559052 | 219 | 2 | 4.54E-07 | 4.61E-05 | 2.13E-05 | 7.96E-06 | -0.00553478 | FKTN |
| chr17 | 48559101 | 48559171 | 71 | 2 | 7.61E-06 | 8.17E-05 | 1.66E-05 | 7.96E-06 | 0.005256321 | HOXB-AS3, HOXB-AS2, HOXB3 |
| chr11 | 441365 | 441456 | 92 | 2 | 5.39E-07 | 7.76E-06 | 0.000105 | 7.97E-06 | 0.007558264 | ANO9 |
| chr5 | 42922866 | 42923911 | 1046 | 4 | 5.86E-05 | 4.55E-05 | 0.000489 | 7.98E-06 | 0.012054272 | AC008945.2 |
| chr2 | 101696607 | 101697526 | 920 | 5 | 5.21E-06 | 1.06E-05 | 0.000164 | 7.99E-06 | 0.003744223 | MAP4K4 |
| chr1 | 23178645 | 23179335 | 691 | 3 | 7.89E-06 | 1.30E-05 | 0.000162 | 7.99E-06 | 0.00887882 | NA |
| chr7 | 30172660 | 30173226 | 567 | 2 | 1.17E-05 | 8.70E-06 | 8.81E-05 | 7.99E-06 | 0.008606907 | NA |
| chr8 | 38414599 | 38414802 | 204 | 3 | 1.18E-05 | 1.25E-05 | 0.000524 | 7.99E-06 | 0.003318681 | FGFR1 |
| chr11 | 62984647 | 62984792 | 146 | 3 | 1.64E-06 | 1.71E-05 | 8.85E-05 | 7.99E-06 | 0.011727727 | SLC22A6 |
| chr9 | 138104531 | 138104925 | 395 | 2 | 1.18E-05 | 1.18E-05 | 6.09E-05 | 7.99E-06 | 0.012974593 | CACNA1B |
| chr1 | 33080550 | 33081624 | 1075 | 15 | 5.90E-05 | 8.17E-05 | 0.005922 | 7.99E-06 | 0.001449375 | AZIN2, AK2 |
| chr13 | 25171268 | 25172395 | 1128 | 14 | 9.32E-06 | 0.001488 | 0.000789 | 8.00E-06 | 0.001382376 | LINC01053, AMER2 |
| chr18 | 5420952 | 5421106 | 155 | 2 | 9.51E-06 | 8.09E-06 | 9.94E-05 | 8.02E-06 | 0.019311779 | EPB41L3 |
| chr18 | 55422661 | 55423699 | 1039 | 6 | 2.21E-05 | 7.22E-06 | 0.002285 | 8.02E-06 | 0.005388082 | TCF4 |
| chr1 | 108559428 | 108560473 | 1046 | 7 | 0.000104805 | 4.52E-06 | 0.006506 | 8.02E-06 | 0.000815112 | FAM102B |
| chr16 | 798997 | 799062 | 66 | 2 | 1.37E-05 | 7.35E-06 | 0.000118 | 8.04E-06 | 0.011674957 | CHTF18, GNG13 |
| chr2 | 65129729 | 65130724 | 996 | 12 | 7.33E-06 | 0.000343 | 0.001072 | 8.04E-06 | 0.000603588 | RAB1A |
| chr2 | 148885440 | 148885762 | 323 | 4 | 4.12E-06 | 0.000748 | 7.52E-06 | 8.04E-06 | 0.003444772 | KIF5C, KIF5C-AS1 |
| chr1 | 155966193 | 155966673 | 481 | 2 | 9.78E-06 | 1.44E-05 | 5.01E-05 | 8.04E-06 | 0.00495003 | ARHGEF2 |
| chr1 | 6329993 | 6330770 | 778 | 5 | 1.63E-05 | 1.94E-05 | 0.001052 | 8.04E-06 | 0.00306875 | ACOT7 |
| chr14 | 104229757 | 104230050 | 294 | 4 | 3.30E-05 | 8.83E-05 | 0.000612 | 8.06E-06 | 0.00368228 | LINC02691 |
| chr14 | 60647758 | 60648037 | 280 | 3 | 0.000100301 | 4.20E-06 | 0.000727 | 8.06E-06 | 0.007479897 | SIX1 |
| chr16 | 84407827 | 84408376 | 550 | 2 | 7.27E-07 | 0.001599 | 9.34E-06 | 8.07E-06 | 0.014347128 | ATP2C2 |
| chr22 | 24548585 | 24548975 | 391 | 3 | 3.00E-05 | 7.45E-06 | 0.000206 | 8.08E-06 | 0.001081627 | GUCD1 |
| chr1 | 2025634 | 2026366 | 733 | 5 | 1.95E-05 | 2.04E-05 | 0.002174 | 8.08E-06 | 0.010788234 | GABRD |
| chr1 | 227563144 | 227563911 | 768 | 13 | 4.45E-05 | 1.34E-05 | 0.006502 | 8.08E-06 | 0.001521808 | ZNF678 |
| chr6 | 30263862 | 30264238 | 377 | 3 | 2.42E-05 | 5.82E-06 | 0.000361 | 8.08E-06 | 0.007482504 | HLA-L, HCG17 |
| chr6 | 6856587 | 6857200 | 614 | 3 | 3.03E-05 | 5.52E-06 | 0.00061 | 8.08E-06 | 0.014041334 | NA |
| chr1 | 233629371 | 233629604 | 234 | 4 | 3.00E-05 | 4.34E-05 | 0.00184 | 8.09E-06 | 0.008251949 | KCNK1 |
| chr9 | 136825792 | 136826520 | 729 | 5 | 8.49E-06 | 2.31E-05 | 0.000216 | 8.09E-06 | 0.002526579 | AL355987.2, RABL6 |
| chr6 | 36834692 | 36835307 | 616 | 4 | 8.15E-06 | 1.09E-05 | 0.000492 | 8.10E-06 | 0.014272437 | CPNE5 |
| chr8 | 20231958 | 20232714 | 757 | 3 | 9.44E-06 | 2.37E-05 | 6.06E-05 | 8.10E-06 | 0.00242126 | NA |
| chr2 | 132415766 | 132416145 | 380 | 5 | 1.81E-05 | 2.66E-05 | 0.001078 | 8.10E-06 | 0.007037985 | NA |
| chr11 | 3838076 | 3838403 | 328 | 2 | 2.23E-05 | 4.95E-06 | 0.000493 | 8.10E-06 | 0.015653825 | RHOG |
| chr17 | 45327559 | 45327856 | 298 | 3 | 2.63E-05 | 8.12E-06 | 0.00059 | 8.10E-06 | -0.00303179 | NA |
| chr16 | 19211134 | 19211154 | 21 | 3 | 0.000165689 | 3.82E-06 | 0.0014 | 8.10E-06 | 0.016485811 | SYT17 |
| chr11 | 125496525 | 125497216 | 692 | 6 | 1.10E-06 | 0.000151 | 0.001316 | 8.11E-06 | 0.004927151 | AP000708.1, FEZ1 |
| chr1 | 54558822 | 54559171 | 350 | 2 | 1.61E-05 | 7.64E-06 | 0.000112 | 8.11E-06 | 0.004480226 | ACOT11 |
| chr19 | 50518420 | 50519345 | 926 | 5 | 1.69E-05 | 3.74E-05 | 0.000191 | 8.12E-06 | 0.000629496 | LRRC4B |
| chr2 | 39778648 | 39779421 | 774 | 5 | 5.36E-05 | 2.48E-06 | 0.003879 | 8.12E-06 | -0.00016381 | THUMPD2 |
| chr17 | 66834877 | 66835259 | 383 | 4 | 8.68E-06 | 8.90E-05 | 4.65E-05 | 8.14E-06 | 0.003183789 | CACNG5 |
| chr1 | 8323986 | 8324599 | 614 | 9 | 1.22E-05 | 0.001411 | 0.000926 | 8.14E-06 | 0.003037391 | SLC45A1 |
| chr22 | 38273489 | 38273545 | 57 | 3 | 0.000314058 | 2.33E-05 | 0.000396 | 8.14E-06 | 0.011041403 | NA |
| chr20 | 45791094 | 45791892 | 799 | 11 | 1.42E-05 | 2.94E-05 | 0.012537 | 8.14E-06 | -0.00352559 | WFDC3 |
| chr8 | 133040096 | 133040531 | 436 | 3 | 8.07E-06 | 3.09E-05 | 3.72E-05 | 8.15E-06 | 0.003389054 | TG, SLA |
| chr1 | 167533985 | 167534176 | 192 | 3 | 8.03E-06 | 0.006178 | 1.99E-05 | 8.16E-06 | 0.008926311 | CREG1 |
| chr8 | 139830430 | 139830956 | 527 | 2 | 3.16E-06 | 1.63E-05 | 4.57E-05 | 8.16E-06 | 0.006948555 | TRAPPC9 |
| chr12 | 94459569 | 94460239 | 671 | 14 | 6.09E-06 | 6.34E-05 | 4.90E-05 | 8.16E-06 | 0.00112068 | CEP83-DT, CEP83 |
| chr6 | 155381153 | 155381385 | 233 | 2 | 3.33E-05 | 5.34E-06 | 0.000335 | 8.17E-06 | 0.019770802 | AL031773.1 |
| chr14 | 22421479 | 22422392 | 914 | 5 | 6.09E-06 | 4.20E-05 | 0.000423 | 8.18E-06 | 0.008952596 | TRDV2, AC244502.1 |
| chr1 | 180585047 | 180585697 | 651 | 2 | 2.84E-05 | 6.80E-06 | 0.000145 | 8.18E-06 | 0.008993586 | NA |
| chr12 | 130709413 | 130709634 | 222 | 4 | 2.71E-05 | 1.38E-05 | 0.00016 | 8.18E-06 | 0.015617595 | RIMBP2 |
| chr2 | 130819852 | 130820389 | 538 | 3 | 6.66E-05 | 3.18E-06 | 0.002129 | 8.18E-06 | 0.01214346 | NA |
| chr22 | 23837169 | 23838305 | 1137 | 3 | 2.68E-07 | 5.53E-05 | 0.000221 | 8.19E-06 | 0.006547532 | SMARCB1, DERL3 |
| chr6 | 3283860 | 3284189 | 330 | 3 | 9.44E-05 | 4.59E-06 | 0.000893 | 8.20E-06 | 0.00200932 | PSMG4, SLC22A23 |
| chr8 | 2071504 | 2072427 | 924 | 4 | 3.97E-05 | 1.35E-05 | 0.002198 | 8.21E-06 | 0.000437386 | MYOM2 |
| chr11 | 34366442 | 34367066 | 625 | 3 | 5.90E-05 | 5.00E-06 | 0.001079 | 8.21E-06 | 0.008702108 | NA |
| chr16 | 833622 | 833720 | 99 | 2 | 1.67E-06 | 5.06E-06 | 0.000472 | 8.22E-06 | 0.015465733 | AL023882.1 |
| chr19 | 19554702 | 19555164 | 463 | 2 | 2.18E-05 | 7.68E-06 | 0.000115 | 8.22E-06 | 0.017345136 | NA |
| chr17 | 36934180 | 36935391 | 1212 | 8 | 1.94E-06 | 0.001624 | 4.65E-05 | 8.22E-06 | 0.006635297 | LHX1-DT |
| chr19 | 42320048 | 42320428 | 381 | 3 | 2.14E-05 | 5.28E-05 | 7.47E-05 | 8.22E-06 | 0.006840933 | TMEM145 |
| chr6 | 33287103 | 33288664 | 1562 | 23 | 9.71E-07 | 3.44E-05 | 0.007748 | 8.24E-06 | 0.000662408 | WDR46, MIR6873 |
| chr4 | 54275975 | 54276667 | 693 | 3 | 4.50E-05 | 5.70E-06 | 0.000379 | 8.24E-06 | 0.001383108 | AC058822.1, PDGFRA |

| | | | | | | | | | | |
|---|---|---|---|---|---|---|---|---|---|---|
| chr17 | 7455155 | 7455626 | 472 | 5 | 1.61E-05 | 8.80E-06 | 0.001329 | 8.24E-06 | 0.00424329 | CHRNB1 |
| chr6 | 117341596 | 117341870 | 275 | 2 | 3.19E-05 | 6.86E-06 | 0.000145 | 8.24E-06 | 0.005289316 | ROS1, AL132671.2 |
| chr12 | 116276434 | 116277409 | 976 | 8 | 2.42E-06 | 0.000306 | 0.000707 | 8.24E-06 | 0.000297631 | MED13L |
| chr18 | 77248673 | 77249538 | 866 | 16 | 1.48E-06 | 0.000103 | 0.001783 | 8.25E-06 | -0.00562413 | NA |
| chr10 | 5041851 | 5041878 | 28 | 2 | 4.07E-05 | 5.67E-06 | 0.00027 | 8.26E-06 | 0.0094762 | AKR1C3 |
| chr14 | 49598338 | 49598853 | 516 | 9 | 1.16E-05 | 2.60E-05 | 0.000164 | 8.28E-06 | 0.001189257 | LRR1, RPS29 |
| chr9 | 136024705 | 136024818 | 114 | 2 | 1.68E-05 | 2.41E-05 | 3.40E-05 | 8.28E-06 | -2.33E-06 | NACC2 |
| chr1 | 161505180 | 161506079 | 900 | 6 | 5.60E-05 | 4.45E-06 | 0.001752 | 8.28E-06 | 0.007794477 | FCGR2A |
| chr18 | 75208922 | 75209491 | 570 | 8 | 1.38E-05 | 8.37E-05 | 0.00035 | 8.28E-06 | 0.000841176 | ZADH2 |
| chr1 | 38818705 | 38818937 | 233 | 3 | 2.32E-05 | 4.26E-06 | 0.00133 | 8.29E-06 | 0.008859864 | NA |
| chr6 | 163707394 | 163707954 | 561 | 2 | 1.76E-05 | 8.70E-06 | 9.49E-05 | 8.29E-06 | 0.029533534 | AL078602.1, AL137005.1 |
| chr4 | 23889267 | 23890212 | 946 | 6 | 5.33E-06 | 1.65E-05 | 0.000236 | 8.30E-06 | 0.00465265 | PPARGC1A |
| chr3 | 155854231 | 155855163 | 933 | 15 | 3.77E-05 | 2.13E-06 | 0.006548 | 8.30E-06 | 0.001977525 | AC104472.4, AC104472.3, SLC33A1 |
| chr20 | 53610043 | 53610785 | 743 | 4 | 1.07E-05 | 1.17E-05 | 0.00042 | 8.30E-06 | 0.010942067 | AC005808.1 |
| chr3 | 16536035 | 16536280 | 246 | 4 | 1.63E-05 | 5.72E-06 | 0.00061 | 8.30E-06 | -0.00776492 | LINC00690 |
| chr9 | 128001146 | 128001637 | 492 | 3 | 1.35E-05 | 1.58E-05 | 0.00053 | 8.31E-06 | 0.009915678 | NA |
| chr1 | 26005662 | 26006059 | 398 | 3 | 3.29E-05 | 7.54E-06 | 0.000465 | 8.31E-06 | 0.009335514 | AL592064.1 |
| chr3 | 171459120 | 171460387 | 1268 | 7 | 1.63E-06 | 1.00E-04 | 7.95E-05 | 8.31E-06 | -0.00083141 | AC092919.1, TNIK |
| chr5 | 150000282 | 150001458 | 1177 | 15 | 2.51E-05 | 0.000168 | 0.000274 | 8.32E-06 | 0.000690872 | HMGXB3, TIGD6 |
| chr3 | 64030777 | 64030878 | 102 | 3 | 4.46E-05 | 6.83E-06 | 0.000233 | 8.32E-06 | -0.0033586 | NA |
| chr4 | 33104295 | 33104716 | 422 | 2 | 4.83E-06 | 7.16E-05 | 1.82E-05 | 8.33E-06 | -0.00897321 | NA |
| chr13 | 110914455 | 110915260 | 806 | 9 | 4.72E-05 | 4.93E-05 | 0.003079 | 8.33E-06 | 0.000600354 | ANKRD10 |
| chr2 | 65063052 | 65063604 | 553 | 2 | 3.41E-05 | 5.83E-06 | 0.000246 | 8.34E-06 | 0.011980185 | CEP68 |
| chr17 | 75286927 | 75287872 | 946 | 7 | 9.14E-05 | 0.000182 | 0.000267 | 8.34E-06 | 0.001172654 | SLC25A19 |
| chr19 | 48613166 | 48613582 | 417 | 3 | 1.06E-05 | 1.03E-05 | 0.000639 | 8.34E-06 | 0.006602945 | FAM83E |
| chr1 | 209354003 | 209354634 | 632 | 5 | 4.25E-05 | 1.64E-05 | 0.000791 | 8.35E-06 | 0.013960611 | NA |
| chr22 | 30204700 | 30204718 | 19 | 2 | 2.56E-05 | 1.27E-05 | 6.06E-05 | 8.37E-06 | 0.009385072 | AC002378.1 |
| chr17 | 45395359 | 45396142 | 784 | 3 | 4.82E-05 | 3.52E-05 | 0.000238 | 8.37E-06 | 0.005090262 | ARHGAP27 |
| chr11 | 732768 | 733384 | 617 | 5 | 9.22E-06 | 4.31E-05 | 9.36E-05 | 8.39E-06 | 0.006990417 | NA |
| chr1 | 9505471 | 9505590 | 120 | 2 | 1.49E-05 | 6.20E-06 | 0.000203 | 8.39E-06 | 0.008970987 | NA |
| chr6 | 168384640 | 168384952 | 313 | 3 | 1.77E-05 | 2.99E-05 | 0.000431 | 8.40E-06 | 0.017678086 | NA |
| chr14 | 61108751 | 61109398 | 648 | 4 | 2.61E-05 | 6.52E-06 | 0.001263 | 8.41E-06 | 0.007493448 | NA |
| chr1 | 16212518 | 16212846 | 329 | 4 | 5.41E-05 | 2.69E-06 | 0.003118 | 8.41E-06 | 0.011140863 | ARHGEF19 |
| chr4 | 112511071 | 112511614 | 544 | 4 | 1.24E-05 | 1.17E-05 | 0.000853 | 8.42E-06 | 0.008068266 | NA |
| chr17 | 8397658 | 8397874 | 217 | 5 | 5.89E-05 | 2.40E-06 | 0.006525 | 8.43E-06 | 0.016631639 | RNF222 |
| chr9 | 129513423 | 129513780 | 358 | 2 | 4.38E-05 | 5.17E-06 | 0.000503 | 8.44E-06 | 0.006626577 | LINC00963 |
| chr17 | 82392315 | 82393396 | 1082 | 7 | 5.22E-05 | 5.65E-06 | 0.005141 | 8.44E-06 | 0.002496883 | OGFOD3 |
| chr1 | 17634220 | 17634937 | 718 | 2 | 0.000149969 | 5.96E-06 | 0.000239 | 8.44E-06 | 0.004218659 | ARHGEF10L |
| chr1 | 243853757 | 243854094 | 338 | 2 | 4.55E-05 | 5.18E-06 | 0.000495 | 8.45E-06 | 0.01186236 | NA |
| chr10 | 70588664 | 70589410 | 747 | 2 | 0.000124148 | 5.28E-06 | 0.000441 | 8.45E-06 | 0.014954533 | NA |
| chr10 | 86354802 | 86354958 | 157 | 3 | 2.09E-05 | 1.32E-05 | 0.000137 | 8.45E-06 | 0.018749728 | GRID1 |
| chr19 | 21081325 | 21081549 | 225 | 3 | 3.02E-05 | 9.05E-06 | 0.000196 | 8.46E-06 | 0.008846925 | NA |
| chr1 | 24108607 | 24109092 | 486 | 5 | 8.73E-06 | 0.000905 | 3.37E-05 | 8.46E-06 | 0.003933607 | MYOM3 |
| chr18 | 31446338 | 31446565 | 228 | 4 | 2.03E-05 | 4.08E-05 | 0.000709 | 8.46E-06 | 0.011534178 | NA |
| chr11 | 44951023 | 44952099 | 1077 | 15 | 8.18E-06 | 0.00012 | 0.000276 | 8.46E-06 | 0.002895551 | TP53I11 |
| chr1 | 155016245 | 155017181 | 937 | 4 | 1.50E-06 | 6.31E-05 | 4.22E-05 | 8.46E-06 | 0.004033802 | ZBTB7B |
| chr22 | 20956439 | 20956926 | 488 | 3 | 6.10E-06 | 4.99E-06 | 0.000535 | 8.46E-06 | 0.017248757 | NA |
| chr4 | 184347973 | 184348532 | 560 | 3 | 2.05E-05 | 4.06E-06 | 0.001505 | 8.47E-06 | 0.011973604 | LINC02363 |
| chr2 | 11398751 | 11399163 | 413 | 5 | 3.13E-05 | 1.01E-05 | 0.000808 | 8.47E-06 | 0.000775211 | LINC00570 |
| chr17 | 44936931 | 44937290 | 360 | 3 | 6.48E-06 | 0.000184 | 1.51E-05 | 8.47E-06 | 0.000673361 | KIF18B |
| chr17 | 35730187 | 35730769 | 583 | 4 | 1.96E-06 | 1.62E-05 | 0.000223 | 8.48E-06 | 0.005685951 | TAF15 |
| chr6 | 117602109 | 117602482 | 374 | 4 | 1.38E-05 | 9.43E-06 | 0.000401 | 8.48E-06 | -0.00065167 | GOPC |
| chr11 | 110165041 | 110165372 | 332 | 3 | 4.79E-05 | 4.14E-06 | 0.00122 | 8.48E-06 | 0.002845759 | ZC3H12C, RDX |
| chr22 | 42289728 | 42290049 | 322 | 4 | 5.51E-05 | 4.05E-06 | 0.002421 | 8.49E-06 | 0.002273319 | TCF20 |
| chr20 | 723997 | 724275 | 279 | 2 | 5.40E-06 | 3.82E-05 | 2.55E-05 | 8.49E-06 | 0.004547271 | NA |
| chr3 | 52834966 | 52835792 | 827 | 8 | 6.25E-06 | 2.70E-05 | 0.000195 | 8.49E-06 | 0.002132237 | AC006254.1, MUSTN1, STIMATE-MUSTN1 |
| chr8 | 143609392 | 143610098 | 707 | 13 | 1.48E-05 | 3.40E-05 | 0.008943 | 8.49E-06 | -0.00051568 | PYCR3 |
| chr2 | 62305886 | 62306005 | 120 | 3 | 1.77E-05 | 8.32E-06 | 0.00024 | 8.49E-06 | 0.003424576 | NA |
| chr5 | 150527904 | 150528275 | 372 | 4 | 2.83E-05 | 0.000145 | 0.000212 | 8.50E-06 | -0.00034564 | NDST1 |
| chr8 | 22688727 | 22689906 | 1180 | 5 | 8.67E-06 | 6.46E-06 | 0.001322 | 8.50E-06 | 0.002651731 | EGR3 |
| chr11 | 3854309 | 3854593 | 285 | 6 | 6.44E-06 | 4.99E-05 | 0.000263 | 8.50E-06 | 0.00678433 | STIM1, STIM1-AS1 |
| chr5 | 132409829 | 132410990 | 1162 | 12 | 6.99E-06 | 0.000147 | 0.001545 | 8.50E-06 | 3.34E-05 | IRF1-AS1, AC116366.2 |
| chr4 | 109302442 | 109303132 | 691 | 16 | 3.82E-05 | 5.06E-06 | 0.024144 | 8.50E-06 | 0.000949903 | COL25A1-DT, COL25A1 |
| chr19 | 47778169 | 47779093 | 925 | 14 | 9.04E-06 | 0.000155 | 0.002568 | 8.50E-06 | 0.0002178 | SELENOW |
| chr6 | 106731986 | 106732794 | 809 | 2 | 0.000156511 | 5.11E-06 | 0.000588 | 8.51E-06 | 0.003251828 | LINC02532 |
| chr11 | 67104145 | 67104627 | 483 | 2 | 4.00E-05 | 5.72E-06 | 0.000299 | 8.52E-06 | 0.003487532 | NA |
| chr12 | 100266676 | 100267867 | 1192 | 18 | 1.90E-05 | 0.001882 | 0.000652 | 8.52E-06 | 0.000232169 | SCYL2, DEPDC4 |
| chr19 | 35491497 | 35492483 | 987 | 3 | 8.01E-05 | 9.10E-05 | 0.000286 | 8.55E-06 | 0.005641916 | KRTDAP |
| chr3 | 72086529 | 72086765 | 237 | 4 | 2.20E-05 | 3.14E-05 | 4.47E-05 | 8.56E-06 | 0.008524245 | LINC00877 |
| chr12 | 121632526 | 121632767 | 242 | 3 | 2.98E-05 | 1.26E-05 | 9.80E-05 | 8.56E-06 | 0.012973354 | ORAI1 |
| chr13 | 28018434 | 28018568 | 135 | 2 | 3.91E-05 | 6.92E-06 | 0.000158 | 8.56E-06 | -0.00567277 | FLT3 |
| chr11 | 72668448 | 72669682 | 1235 | 5 | 5.53E-06 | 6.83E-05 | 5.78E-05 | 8.57E-06 | 0.007410481 | PDE2A |
| chr10 | 129353271 | 129353389 | 119 | 3 | 5.02E-05 | 5.72E-06 | 0.001019 | 8.57E-06 | 0.004884891 | NA |
| chr8 | 37966669 | 37967247 | 579 | 8 | 9.28E-05 | 7.71E-06 | 0.008056 | 8.57E-06 | 0.008654686 | NA |
| chr8 | 28494567 | 28495126 | 560 | 4 | 9.16E-06 | 3.48E-05 | 0.000179 | 8.57E-06 | 0.001536861 | FZD3 |
| chr2 | 112138574 | 112138637 | 64 | 2 | 6.69E-05 | 2.71E-05 | 3.26E-05 | 8.57E-06 | 0.004481898 | FBLN7 |
| chr12 | 125113676 | 125113723 | 48 | 2 | 1.98E-05 | 9.07E-06 | 9.55E-05 | 8.58E-06 | 0.003824713 | AACS |
| chr11 | 62299061 | 62299243 | 183 | 4 | 2.61E-05 | 1.66E-05 | 0.000128 | 8.58E-06 | 0.007411681 | SCGB1D4 |
| chr7 | 105678990 | 105679372 | 383 | 8 | 5.92E-05 | 1.66E-06 | 0.010892 | 8.58E-06 | 0.00721513 | ATXN7L1 |
| chr5 | 33935691 | 33936157 | 467 | 5 | 8.54E-05 | 6.82E-05 | 4.94E-05 | 8.58E-06 | 0.000268568 | NA |
| chr11 | 34570363 | 34570736 | 374 | 5 | 6.56E-05 | 2.06E-06 | 0.005325 | 8.58E-06 | 0.012533447 | NA |
| chr14 | 73715684 | 73715857 | 174 | 3 | 2.34E-05 | 4.92E-06 | 0.000487 | 8.58E-06 | 0.002351141 | MIDEAS |
| chr14 | 23161126 | 23161993 | 868 | 4 | 3.87E-05 | 2.84E-06 | 0.00309 | 8.58E-06 | 0.011355724 | SLC7A8 |
| chr17 | 8186235 | 8186639 | 405 | 4 | 0.00013264 | 1.49E-05 | 0.000285 | 8.58E-06 | 0.001311762 | NA |
| chr16 | 88484553 | 88485081 | 529 | 6 | 5.29E-06 | 0.000366 | 0.000144 | 8.59E-06 | 0.001869015 | ZFPM1 |
| chr1 | 182673051 | 182673613 | 563 | 3 | 1.99E-05 | 3.04E-05 | 0.000287 | 8.60E-06 | 0.008536931 | RGS8 |
| chr1 | 18108750 | 18109403 | 654 | 5 | 1.22E-05 | 8.62E-06 | 0.001659 | 8.61E-06 | 0.007426865 | IGSF21, AL365209.1 |
| chr18 | 35442016 | 35442088 | 73 | 2 | 1.09E-05 | 0.002638 | 9.49E-06 | 8.61E-06 | 0.005407479 | NA |
| chr5 | 126776360 | 126777191 | 832 | 12 | 2.92E-05 | 6.17E-06 | 0.000552 | 8.62E-06 | -0.00051288 | LMNB1, LMNB1-DT |
| chr6 | 30982211 | 30983002 | 792 | 5 | 3.42E-05 | 2.00E-05 | 0.005 | 8.62E-06 | 0.008810322 | NA |
| chr5 | 93591442 | 93592090 | 649 | 3 | 6.43E-06 | 5.75E-05 | 2.82E-05 | 8.62E-06 | 0.012362404 | NR2F1 |
| chr8 | 21787566 | 21788797 | 1232 | 11 | 1.04E-05 | 1.64E-05 | 0.004702 | 8.62E-06 | 0.002104833 | GFRA2 |
| chr19 | 13774389 | 13775124 | 736 | 5 | 7.06E-05 | 5.63E-06 | 0.00502 | 8.62E-06 | -0.0001574 | C19orf53 |
| chr8 | 102910243 | 102910439 | 197 | 2 | 2.27E-05 | 1.77E-05 | 4.57E-05 | 8.62E-06 | 0.012121932 | MAILR |
| chr12 | 129710812 | 129711232 | 421 | 2 | 1.80E-06 | 0.018748 | 8.88E-06 | 8.62E-06 | 0.006415719 | TMEM132D |
| chr1 | 920679 | 921534 | 856 | 4 | 6.30E-06 | 2.14E-05 | 6.03E-05 | 8.63E-06 | 0.000913671 | LINC02593 |
| chr2 | 98236241 | 98236475 | 235 | 5 | 7.18E-06 | 0.002248 | 1.57E-05 | 8.65E-06 | 0.008143773 | VWA3B |
| chr17 | 82813465 | 82813841 | 377 | 6 | 9.23E-06 | 9.21E-05 | 6.29E-05 | 8.65E-06 | -4.52E-05 | TBCD |
| chr10 | 127619160 | 127619513 | 354 | 2 | 2.77E-05 | 7.88E-06 | 0.000125 | 8.66E-06 | 0.022502365 | NA |
| chr1 | 164562792 | 164562859 | 68 | 2 | 1.99E-05 | 8.55E-06 | 0.000108 | 8.67E-06 | -0.01348887 | PBX1 |
| chr6 | 6850634 | 6851134 | 501 | 2 | 2.36E-05 | 5.96E-06 | 0.000272 | 8.67E-06 | 0.008409544 | NA |
| chr10 | 126066181 | 126067128 | 948 | 4 | 0.000102973 | 2.46E-05 | 0.001345 | 8.68E-06 | -0.00283559 | ADAM12 |
| chr22 | 23827223 | 23827829 | 607 | 3 | 8.49E-05 | 8.84E-06 | 0.000445 | 8.68E-06 | 0.001452514 | SMARCB1 |
| chr19 | 45493114 | 45493643 | 530 | 8 | 1.65E-05 | 1.74E-05 | 0.000602 | 8.69E-06 | 0.003080771 | PPM1N, RTN2 |
| chr7 | 37919880 | 37920364 | 485 | 4 | 1.51E-05 | 1.16E-05 | 0.000908 | 8.70E-06 | -0.00163037 | EPDR1, SFRP4 |
| chr17 | 16286240 | 16287253 | 1014 | 5 | 1.83E-05 | 4.11E-06 | 0.004335 | 8.70E-06 | 0.007248088 | PIGL |
| chr6 | 144095083 | 144095853 | 771 | 10 | 1.21E-05 | 2.60E-05 | 0.000847 | 8.70E-06 | 0.000161828 | SF3B5 |
| chr18 | 12423850 | 12423977 | 128 | 2 | 2.72E-06 | 2.38E-05 | 3.64E-05 | 8.70E-06 | -0.00147402 | PRELID3A |
| chr1 | 236170455 | 236170514 | 60 | 2 | 1.53E-05 | 6.08E-05 | 2.05E-05 | 8.71E-06 | 0.00068169 | GPR137B |
| chr1 | 173442177 | 173442281 | 105 | 2 | 1.98E-05 | 1.44E-05 | 5.69E-05 | 8.71E-06 | 0.005340827 | PRDX6-AS1 |
| chr19 | 39934509 | 39935056 | 548 | 3 | 1.04E-05 | 4.33E-06 | 0.001112 | 8.71E-06 | 0.006347847 | FCGBP |
| chr1 | 241531634 | 241532316 | 683 | 5 | 4.00E-06 | 0.000581 | 0.001197 | 8.71E-06 | 0.002817036 | KMO |
| chr2 | 189674684 | 189675096 | 413 | 4 | 1.44E-05 | 3.21E-06 | 0.002704 | 8.71E-06 | -0.00082477 | ANKAR |
| chr16 | 67233468 | 67234177 | 710 | 4 | 6.94E-06 | 2.31E-05 | 0.000105 | 8.71E-06 | -0.00069068 | FHOD1 |
| chr11 | 69312203 | 69312593 | 391 | 4 | 6.60E-06 | 1.24E-05 | 0.00032 | 8.71E-06 | 0.005853225 | MYEOV |
| chr1 | 27100461 | 27101258 | 798 | 3 | 4.49E-05 | 1.22E-05 | 0.000601 | 8.72E-06 | 0.003937379 | SLC9A1 |
| chr1 | 172444209 | 172445012 | 804 | 5 | 1.99E-05 | 3.99E-05 | 0.001546 | 8.72E-06 | 0.005876634 | C1orf105 |
| chr6 | 32950529 | 32951190 | 662 | 6 | 9.97E-07 | 4.10E-05 | 0.000245 | 8.72E-06 | 0.002904168 | AL645941.2, HLA-DMA |
| chr6 | 30335039 | 30335907 | 869 | 10 | 8.10E-06 | 0.000566 | 0.000129 | 8.72E-06 | -0.00042557 | TRIM39, TRIM39-RPP21 |
| chr1 | 243451619 | 243452107 | 489 | 3 | 3.11E-05 | 3.59E-06 | 0.001674 | 8.73E-06 | 0.014634793 | SDCCAG8 |
| chr11 | 76630439 | 76631276 | 838 | 3 | 4.34E-05 | 1.94E-05 | 0.00039 | 8.73E-06 | -0.00246984 | LINC02757 |
| chr11 | 112555819 | 112556545 | 727 | 5 | 4.50E-05 | 1.99E-05 | 0.000781 | 8.73E-06 | 0.004006312 | LINC02763 |
| chr3 | 176814349 | 176814886 | 538 | 3 | 6.88E-06 | 9.72E-05 | 7.24E-05 | 8.74E-06 | 0.01075105 | LINC01208, LINC01209 |
| chr22 | 36783868 | 36784241 | 374 | 2 | 1.51E-05 | 1.65E-05 | 4.99E-05 | 8.76E-06 | 0.004224481 | NA |
| chr4 | 56907134 | 56908036 | 903 | 11 | 1.29E-05 | 1.40E-05 | 0.005862 | 8.77E-06 | 0.00292549 | REST |
| chr5 | 76716766 | 76717002 | 237 | 3 | 1.82E-05 | 4.48E-06 | 0.001145 | 8.77E-06 | -0.00204305 | F2R |
| chr12 | 93377527 | 93377882 | 356 | 6 | 1.21E-05 | 8.35E-05 | 0.000673 | 8.78E-06 | 0.003133493 | AC124947.1 |
| chr10 | 133045954 | 133046706 | 753 | 8 | 3.21E-06 | 0.00133 | 7.65E-05 | 8.79E-06 | 0.00284426 | NA |
| chr9 | 36135045 | 36135763 | 719 | 4 | 2.20E-05 | 6.89E-06 | 0.000571 | 8.79E-06 | 0.008598546 | NA |
| chr5 | 177307342 | 177308072 | 731 | 5 | 5.68E-05 | 2.32E-06 | 0.005433 | 8.79E-06 | 0.008913036 | MXD3 |
| chr2 | 174394841 | 174395886 | 1046 | 16 | 1.61E-05 | 6.44E-05 | 0.000341 | 8.80E-06 | 0.001276101 | SCRN3, CIR1 |
| chr7 | 22587757 | 22588363 | 607 | 3 | 4.26E-05 | 3.83E-06 | 0.001381 | 8.81E-06 | -0.00106764 | STEAP1B |
| chr1 | 174990579 | 174990612 | 34 | 2 | 1.51E-05 | 5.30E-06 | 0.000597 | 8.81E-06 | 0.020936597 | RABGAP1L |
| chr1 | 7491516 | 7492176 | 661 | 3 | 3.53E-05 | 5.25E-06 | 0.000459 | 8.81E-06 | 0.004292977 | CAMTA1 |
| chr17 | 48614646 | 48615497 | 852 | 14 | 5.81E-06 | 0.000213 | 0.000981 | 8.81E-06 | 0.003842562 | HOXB7, HOXB8 |
| chr1 | 185015164 | 185015618 | 455 | 5 | 1.73E-05 | 1.78E-05 | 0.001021 | 8.81E-06 | 0.005080876 | NA |
| chr4 | 70049510 | 70049615 | 106 | 2 | 1.04E-05 | 0.001561 | 1.01E-05 | 8.82E-06 | 0.017491504 | NA |
| chr7 | 128081742 | 128082254 | 513 | 4 | 3.62E-07 | 0.000866 | 8.05E-06 | 8.82E-06 | 0.001232098 | SND1, MIR593 |
| chr4 | 89129747 | 89130184 | 438 | 2 | 1.61E-05 | 6.92E-06 | 0.000178 | 8.84E-06 | 0.011291544 | NA |
| chr10 | 123380983 | 123381283 | 301 | 2 | 2.81E-05 | 9.05E-06 | 0.000102 | 8.85E-06 | 0.010704176 | LINC02641 |
| chr6 | 1522845 | 1523588 | 744 | 3 | 1.72E-05 | 8.64E-05 | 0.000366 | 8.85E-06 | 0.00953824 | NA |
| chr6 | 44151841 | 44152688 | 848 | 6 | 0.000136807 | 6.39E-05 | 0.001048 | 8.85E-06 | 0.004043911 | TMEM63B |
| chr5 | 3030857 | 3030874 | 18 | 2 | 1.04E-05 | 0.000631 | 1.12E-05 | 8.86E-06 | 0.002867941 | NA |
| chr16 | 70473191 | 70473212 | 22 | 3 | 2.13E-05 | 2.38E-05 | 5.21E-05 | 8.86E-06 | 0.002635681 | FCSK |
| chr19 | 1732553 | 1732906 | 354 | 2 | 9.06E-06 | 4.74E-05 | 2.36E-05 | 8.87E-06 | -0.00593969 | NA |
| chr20 | 36149390 | 36149692 | 303 | 3 | 9.96E-06 | 3.15E-05 | 0.000114 | 8.87E-06 | 0.008228876 | EPB41L1, AL121895.1 |
| chr10 | 112446675 | 112447251 | 577 | 13 | 5.07E-06 | 0.003532 | 0.000143 | 8.88E-06 | 0.000123882 | VTI1A, ZDHHC6 |
| chr1 | 165630214 | 165631336 | 1123 | 13 | 3.70E-05 | 3.93E-05 | 0.002065 | 8.88E-06 | 0.003521015 | MGST3 |
| chr12 | 120363995 | 120364449 | 455 | 2 | 2.82E-06 | 0.000229 | 1.37E-05 | 8.88E-06 | 0.00400516 | MSI1 |
| chr3 | 130929798 | 130930422 | 625 | 5 | 3.73E-05 | 5.77E-06 | 0.001702 | 8.89E-06 | -0.00055708 | ATP2C1, AC055733.2 |
| chr12 | 117099779 | 117100522 | 744 | 3 | 7.22E-06 | 6.82E-06 | 0.000497 | 8.91E-06 | 0.012273449 | TESC-AS1 |
| chr17 | 66673621 | 66673954 | 334 | 2 | 1.83E-05 | 1.58E-05 | 5.36E-05 | 8.91E-06 | 0.013431431 | PRKCA |
| chr11 | 1921174 | 1921784 | 611 | 5 | 2.16E-05 | 3.99E-06 | 0.002982 | 8.91E-06 | 0.013069003 | TNNT3 |
| chr17 | 49764918 | 49764996 | 79 | 2 | 4.34E-06 | 0.002281 | 9.86E-06 | 8.91E-06 | 0.011686121 | FAM117A |
| chr11 | 31172897 | 31173323 | 427 | 2 | 9.50E-06 | 1.54E-05 | 5.50E-05 | 8.91E-06 | 0.013008944 | DCDC1 |
| chr3 | 46097302 | 46097813 | 512 | 2 | 1.13E-05 | 1.40E-05 | 6.02E-05 | 8.91E-06 | 0.01107665 | NA |
| chr16 | 2271167 | 2271695 | 529 | 5 | 2.25E-05 | 6.23E-05 | 0.001201 | 8.91E-06 | 0.00176652 | AC009065.4 |
| chr11 | 130389713 | 130390424 | 712 | 3 | 5.08E-05 | 5.56E-06 | 0.001124 | 8.91E-06 | 0.00910547 | ZBTB44-DT |
| chr12 | 28583764 | 28584180 | 417 | 2 | 2.14E-06 | 0.003529 | 9.55E-06 | 8.92E-06 | 0.006078817 | NA |
| chr19 | 42060485 | 42060981 | 497 | 2 | 6.93E-06 | 2.80E-05 | 3.37E-05 | 8.92E-06 | -0.00893016 | GRIK5 |
| chr1 | 25922406 | 25923234 | 829 | 6 | 2.90E-06 | 0.000424 | 6.62E-05 | 8.93E-06 | -0.00272532 | NA |
| chr10 | 100866419 | 100866895 | 477 | 3 | 2.46E-05 | 7.74E-06 | 0.000187 | 8.94E-06 | 0.00758585 | NA |

| | | | | | | | | | | |
|---|---|---|---|---|---|---|---|---|---|---|
| chr6 | 38721894 | 38722804 | 911 | 4 | 2.05E-05 | 0.000159 | 0.000545 | 8.95E-06 | -0.0029557 | DNAH8 |
| chr1 | 41898401 | 41898664 | 264 | 3 | 4.54E-05 | 5.66E-06 | 0.000425 | 8.95E-06 | 0.008018898 | HIVEP3 |
| chr16 | 87185312 | 87185578 | 267 | 2 | 2.26E-05 | 5.56E-06 | 0.000492 | 8.95E-06 | 0.008564832 | C16orf95 |
| chr2 | 176172484 | 176172982 | 499 | 4 | 1.23E-05 | 4.60E-05 | 0.001257 | 8.96E-06 | 0.008496518 | HOXD3, HAGLR |
| chr5 | 119629065 | 119629652 | 588 | 7 | 3.19E-05 | 2.53E-05 | 0.002453 | 8.96E-06 | -0.00033338 | HSD17B4, FAM170A |
| chr12 | 125117668 | 125118215 | 548 | 4 | 1.02E-05 | 0.000136 | 4.76E-05 | 8.97E-06 | -0.00215922 | AACS |
| chr1 | 204928916 | 204929295 | 380 | 2 | 1.34E-05 | 8.12E-06 | 0.000129 | 8.98E-06 | 0.01089853 | NFASC |
| chr22 | 25199385 | 25199873 | 489 | 8 | 1.61E-05 | 1.45E-05 | 0.000187 | 8.98E-06 | 0.004189814 | CRYBB3 |
| chr1 | 14959060 | 14959285 | 226 | 2 | 2.36E-05 | 1.19E-05 | 7.35E-05 | 8.98E-06 | 0.019071936 | KAZN |
| chr3 | 66582912 | 66583357 | 446 | 4 | 4.21E-05 | 4.42E-06 | 0.001351 | 8.98E-06 | 0.009055281 | NA |
| chr16 | 81625550 | 81625923 | 374 | 2 | 2.76E-05 | 8.28E-06 | 0.000124 | 8.99E-06 | 0.008371215 | CMIP |
| chr1 | 205630860 | 205631439 | 580 | 4 | 5.41E-06 | 4.10E-05 | 0.00043 | 9.00E-06 | 0.004186378 | ELK4 |
| chr9 | 136964447 | 136964644 | 198 | 2 | 3.10E-05 | 5.42E-06 | 0.000611 | 9.00E-06 | 0.009188176 | NA |
| chr5 | 1067996 | 1069120 | 1125 | 5 | 2.22E-06 | 1.81E-05 | 0.000112 | 9.01E-06 | 0.00282451 | SLC12A7 |
| chr1 | 212497878 | 212498565 | 688 | 3 | 6.11E-05 | 4.53E-06 | 0.000797 | 9.04E-06 | 0.005877843 | NA |
| chr7 | 23347746 | 23348243 | 498 | 5 | 2.39E-05 | 0.00013 | 0.000582 | 9.05E-06 | 0.006513386 | IGF2BP3 |
| chr5 | 43602014 | 43603044 | 1031 | 10 | 8.16E-06 | 1.36E-05 | 0.004378 | 9.05E-06 | 0.002089912 | NNT, NNT-AS1 |
| chr15 | 33968232 | 33968971 | 740 | 8 | 6.77E-05 | 3.21E-06 | 0.008207 | 9.05E-06 | 0.000853002 | CHRM5, AVEN |
| chr6 | 6001609 | 6002310 | 702 | 4 | 7.82E-06 | 1.10E-05 | 0.00032 | 9.06E-06 | 0.011192658 | NRN1 |
| chr1 | 22735071 | 22735797 | 727 | 2 | 0.000114769 | 5.72E-06 | 0.000439 | 9.06E-06 | 0.006380292 | EPHB2 |
| chr6 | 52595125 | 52595466 | 342 | 2 | 3.03E-05 | 7.76E-06 | 0.000144 | 9.07E-06 | 0.008271992 | TRAM2-AS1 |
| chr2 | 2767890 | 2768022 | 133 | 3 | 5.42E-05 | 5.23E-06 | 0.000546 | 9.11E-06 | 0.005965755 | AC011995.2 |
| chr1 | 212515075 | 212515655 | 581 | 5 | 5.65E-05 | 6.37E-06 | 0.003398 | 9.11E-06 | 0.014155559 | AC092803.2 |
| chr3 | 150226310 | 150226625 | 316 | 2 | 3.49E-06 | 6.89E-05 | 2.03E-05 | 9.11E-06 | 0.005651821 | NA |
| chr15 | 92878439 | 92878961 | 523 | 3 | 5.93E-06 | 4.09E-05 | 9.13E-05 | 9.11E-06 | 0.008086206 | CHASERR |
| chr11 | 64371934 | 64372551 | 618 | 3 | 6.92E-07 | 0.000567 | 0.000118 | 9.13E-06 | 0.002686438 | RPS6KA4 |
| chr6 | 36270051 | 36270633 | 583 | 5 | 2.46E-05 | 2.95E-05 | 0.000498 | 9.13E-06 | 0.005847768 | PNPLA1 |
| chr2 | 241553747 | 241554391 | 645 | 2 | 1.29E-05 | 6.95E-06 | 0.000196 | 9.14E-06 | 0.010741875 | BOK, BOK-AS1 |
| chr4 | 183241128 | 183241445 | 318 | 3 | 4.76E-05 | 2.08E-05 | 0.000129 | 9.14E-06 | 0.004207198 | WWC2 |
| chr14 | 95516533 | 95517046 | 514 | 5 | 6.16E-05 | 1.99E-05 | 0.000861 | 9.14E-06 | 0.000492495 | AL133467.1 |
| chr17 | 40818635 | 40819197 | 563 | 8 | 1.69E-06 | 0.002955 | 6.88E-06 | 9.15E-06 | -0.00076879 | TMEM99, KRT10 |
| chr9 | 108862100 | 108862659 | 560 | 4 | 1.02E-05 | 3.83E-05 | 0.000358 | 9.15E-06 | 0.001661439 | ACTL7A |
| chr22 | 43155302 | 43155947 | 646 | 4 | 4.82E-05 | 7.71E-06 | 0.001352 | 9.15E-06 | 0.009841184 | TSPO |
| chr22 | 38143377 | 38143622 | 246 | 2 | 4.82E-05 | 5.66E-06 | 0.000509 | 9.15E-06 | 0.012383271 | AL022322.2, PLA2G6 |
| chr16 | 67211526 | 67212211 | 686 | 2 | 0.000155199 | 5.54E-06 | 0.000609 | 9.16E-06 | 0.008953151 | LRRC29 |
| chr2 | 219243125 | 219243788 | 664 | 9 | 3.85E-05 | 9.47E-07 | 0.013241 | 9.16E-06 | -0.02946055 | GLB1L |
| chr15 | 48877568 | 48878124 | 557 | 13 | 5.10E-06 | 0.000158 | 0.000956 | 9.17E-06 | 0.003178391 | SHC4 |
| chr1 | 26542670 | 26543146 | 477 | 5 | 4.84E-05 | 3.04E-06 | 0.006179 | 9.19E-06 | 0.008576169 | RPS6KA1 |
| chr2 | 9991873 | 9992113 | 241 | 2 | 1.32E-05 | 3.50E-05 | 2.99E-05 | 9.20E-06 | 0.008382897 | GRHL1 |
| chr11 | 118565808 | 118566853 | 1046 | 17 | 1.77E-05 | 4.01E-05 | 8.44E-05 | 9.21E-06 | 0.001869035 | IFT46 |
| chr8 | 53876765 | 53877812 | 1048 | 4 | 9.95E-06 | 8.74E-05 | 0.00014 | 9.21E-06 | 0.005370252 | RGS20 |
| chr6 | 37657253 | 37658259 | 1007 | 3 | 2.90E-07 | 2.95E-05 | 6.94E-05 | 9.21E-06 | 0.022435413 | MDGA1 |
| chr6 | 168315390 | 168315652 | 263 | 3 | 7.01E-06 | 0.001473 | 7.87E-06 | 9.21E-06 | 0.005714676 | DACT2 |
| chr3 | 189316612 | 189316846 | 235 | 2 | 1.84E-05 | 3.64E-05 | 2.93E-05 | 9.21E-06 | 0.010105177 | TPRG1 |
| chr3 | 196318249 | 196319222 | 974 | 8 | 8.19E-05 | 2.24E-05 | 0.000179 | 9.21E-06 | 0.001837689 | TM4SF19-AS1, TCTEX1D2, TM4SF19-TCTEX1D2 |
| chr20 | 62831078 | 62831687 | 610 | 3 | 9.47E-06 | 7.28E-06 | 0.000276 | 9.21E-06 | 0.015016834 | COL9A3 |
| chr2 | 193105 | 194026 | 922 | 4 | 2.99E-05 | 1.21E-05 | 0.00021 | 9.22E-06 | 0.003991929 | NA |
| chr2 | 30206697 | 30206902 | 206 | 2 | 5.17E-05 | 5.56E-06 | 0.000619 | 9.22E-06 | 0.012260971 | NA |
| chr2 | 45278453 | 45279019 | 567 | 2 | 7.53E-05 | 5.64E-06 | 0.000556 | 9.22E-06 | 0.006293265 | LINC01121 |
| chr12 | 99647263 | 99648206 | 944 | 6 | 1.03E-05 | 9.07E-05 | 0.000831 | 9.22E-06 | 0.001587341 | FAM71C, ANKS1B |
| chr3 | 35743885 | 35744988 | 1104 | 7 | 2.00E-06 | 2.84E-05 | 0.000221 | 9.24E-06 | 0.00245017 | ARPP21, MIR128-2 |
| chr5 | 132496620 | 132496806 | 187 | 2 | 6.12E-05 | 7.25E-06 | 0.00018 | 9.24E-06 | 0.002236748 | AC116366.2 |
| chr10 | 132183655 | 132184390 | 736 | 2 | 1.77E-05 | 5.68E-06 | 0.000538 | 9.26E-06 | 0.000147835 | JAKMIP3, AL512622.1 |
| chr5 | 88381966 | 88382327 | 362 | 3 | 6.37E-05 | 5.57E-06 | 0.001142 | 9.26E-06 | 0.006447974 | TMEM161B-AS1, RPS3AP22 |
| chr18 | 76351900 | 76352352 | 453 | 3 | 3.65E-05 | 4.39E-06 | 0.001413 | 9.26E-06 | 0.016583073 | NA |
| chr4 | 158120626 | 158121356 | 731 | 2 | 0.000168965 | 5.65E-06 | 0.000578 | 9.28E-06 | 0.021476643 | NA |
| chr17 | 38512264 | 38512506 | 243 | 2 | 8.57E-06 | 4.52E-05 | 2.56E-05 | 9.28E-06 | 0.007146741 | ARHGAP23 |
| chr10 | 113854075 | 113854953 | 879 | 19 | 6.18E-05 | 1.35E-06 | 0.036057 | 9.28E-06 | 0.000297652 | NHLRC2, DCLRE1A |
| chr10 | 71397465 | 71398315 | 851 | 6 | 4.45E-06 | 0.001079 | 0.0004 | 9.28E-06 | 0.006718967 | CDH23 |
| chr7 | 19116463 | 19117797 | 1335 | 8 | 4.76E-06 | 0.000347 | 0.000172 | 9.31E-06 | 0.000500471 | TWIST1 |
| chr14 | 22478753 | 22479509 | 757 | 4 | 4.18E-06 | 0.000274 | 1.82E-05 | 9.31E-06 | -0.00836927 | TRAJ57, AC244502.1 |
| chr10 | 69232469 | 69233279 | 811 | 5 | 4.38E-05 | 8.72E-06 | 0.002876 | 9.31E-06 | 0.006772051 | HKDC1, AL596223.1 |
| chr1 | 24952535 | 24952719 | 185 | 2 | 1.10E-05 | 1.81E-05 | 4.98E-05 | 9.32E-06 | 0.015642421 | RUNX3 |
| chr3 | 122969792 | 122970014 | 223 | 2 | 3.08E-06 | 0.000196 | 1.47E-05 | 9.33E-06 | 0.013078369 | SEMA5B |
| chr10 | 101818498 | 101819554 | 1057 | 10 | 6.25E-05 | 6.63E-05 | 0.00842 | 9.33E-06 | 0.001449396 | KCNIP2-AS1 |
| chr12 | 108779094 | 108779292 | 199 | 2 | 4.59E-06 | 0.000452 | 1.24E-05 | 9.34E-06 | 0.003400572 | AC087893.3, SSH1 |
| chr5 | 172932523 | 172932677 | 155 | 4 | 1.05E-05 | 6.37E-05 | 0.000512 | 9.34E-06 | 0.007361103 | ERGIC1 |
| chr17 | 67217256 | 67217446 | 191 | 2 | 3.17E-05 | 1.40E-05 | 6.47E-05 | 9.34E-06 | 0.005218471 | HELZ |
| chr11 | 65711810 | 65712930 | 1121 | 15 | 3.79E-06 | 2.96E-05 | 0.000578 | 9.34E-06 | 0.000221978 | KAT5 |
| chr15 | 78619482 | 78620400 | 919 | 4 | 3.16E-05 | 3.51E-06 | 0.002937 | 9.34E-06 | 0.004213122 | CHRNA3 |
| chr6 | 24933687 | 24934041 | 355 | 4 | 3.87E-05 | 7.94E-06 | 0.000565 | 9.34E-06 | 0.006671697 | RIPOR2 |
| chr11 | 113399121 | 113399715 | 595 | 5 | 1.06E-05 | 0.000488 | 9.48E-05 | 9.34E-06 | -0.01121385 | ANKK1 |
| chr11 | 47414739 | 47415124 | 386 | 3 | 1.92E-05 | 1.35E-05 | 0.000107 | 9.34E-06 | 0.002824849 | SLC39A13 |
| chr15 | 42094528 | 42095092 | 565 | 6 | 1.98E-05 | 1.15E-05 | 0.001632 | 9.35E-06 | 0.004328588 | PLA2G4D |
| chr9 | 33164439 | 33164687 | 249 | 2 | 5.07E-06 | 2.88E-05 | 3.48E-05 | 9.35E-06 | 0.008078753 | B4GALT1 |
| chr19 | 45449293 | 45450040 | 748 | 4 | 9.24E-06 | 3.74E-06 | 0.00209 | 9.35E-06 | 0.007963543 | ERCC1 |
| chr1 | 46132603 | 46133456 | 854 | 16 | 0.000164961 | 1.81E-05 | 0.021584 | 9.35E-06 | 0.001154476 | PIK3R3, P3R3URF-PIK3R3 |
| chr16 | 87680563 | 87680995 | 433 | 4 | 3.47E-05 | 6.79E-06 | 0.001417 | 9.37E-06 | 0.00564382 | JPH3 |
| chr6 | 70957948 | 70958326 | 379 | 3 | 2.67E-06 | 0.000446 | 7.13E-05 | 9.38E-06 | 0.007794836 | NA |
| chr20 | 37872030 | 37872328 | 299 | 2 | 5.41E-05 | 5.68E-06 | 0.00061 | 9.38E-06 | 0.003493567 | CTNNBL1 |
| chr18 | 9922738 | 9923435 | 698 | 3 | 9.00E-06 | 3.25E-05 | 4.60E-05 | 9.40E-06 | 0.02028002 | VAPA |
| chr1 | 245352836 | 245353594 | 759 | 5 | 0.00013581 | 6.36E-06 | 0.002918 | 9.40E-06 | 0.000604929 | KIF26B |
| chr20 | 63186675 | 63187055 | 381 | 2 | 3.29E-06 | 0.000581 | 1.20E-05 | 9.41E-06 | 0.004561488 | NA |
| chr4 | 183320261 | 183321135 | 875 | 5 | 1.44E-06 | 0.002279 | 1.01E-05 | 9.41E-06 | 0.007133325 | WWC2, CLDN22 |
| chr9 | 3270389 | 3271163 | 775 | 3 | 1.00E-05 | 0.001161 | 9.11E-05 | 9.41E-06 | -0.0033473 | RFX3 |
| chr21 | 36698224 | 36699507 | 1284 | 15 | 2.94E-07 | 0.000166 | 7.13E-05 | 9.41E-06 | 0.001392983 | SIM2, AP000697.1 |
| chr16 | 2740807 | 2741086 | 280 | 4 | 4.11E-05 | 2.96E-06 | 0.003951 | 9.44E-06 | 0.00339521 | SRRM2-AS1 |
| chr7 | 130128392 | 130128538 | 147 | 2 | 5.18E-05 | 5.69E-06 | 0.000629 | 9.46E-06 | 0.001657983 | KLHDC10 |
| chr1 | 70410684 | 70411698 | 1015 | 13 | 1.79E-05 | 1.80E-05 | 0.001165 | 9.46E-06 | -0.00048282 | CTH |
| chr10 | 117134558 | 117135104 | 547 | 3 | 1.72E-06 | 4.21E-05 | 0.000157 | 9.47E-06 | 0.008329864 | VAX1 |
| chr18 | 63192404 | 63193056 | 653 | 4 | 1.84E-05 | 1.93E-05 | 0.001413 | 9.47E-06 | 0.011122318 | BCL2 |
| chr7 | 98228044 | 98228980 | 937 | 9 | 1.78E-05 | 7.26E-05 | 0.000668 | 9.47E-06 | -0.00070711 | TECPR1 |
| chr4 | 787068 | 787484 | 417 | 4 | 3.02E-05 | 6.09E-06 | 0.000424 | 9.48E-06 | 0.01953113 | CPLX1 |
| chr3 | 23629269 | 23629354 | 86 | 2 | 4.76E-05 | 7.56E-06 | 0.000175 | 9.48E-06 | 0.010454528 | NA |
| chr8 | 139629268 | 139629540 | 273 | 3 | 2.06E-05 | 5.99E-06 | 0.000779 | 9.48E-06 | 0.004775904 | KCNK9 |
| chr12 | 107349047 | 107349325 | 279 | 2 | 3.95E-05 | 7.38E-06 | 0.000188 | 9.49E-06 | 0.008437144 | BTBD11 |
| chr15 | 50118765 | 50119338 | 574 | 2 | 1.67E-06 | 8.59E-06 | 0.000132 | 9.50E-06 | 0.019697552 | ATP8B4 |
| chr4 | 57194175 | 57194607 | 433 | 3 | 6.81E-06 | 1.11E-05 | 0.000715 | 9.50E-06 | -0.04294763 | IGFBP7-AS1 |
| chr1 | 241423799 | 241424483 | 685 | 8 | 1.68E-05 | 4.30E-05 | 0.002479 | 9.50E-06 | -7.66E-05 | AL359764.2 |
| chr7 | 17947122 | 17947354 | 233 | 3 | 4.25E-05 | 8.20E-06 | 0.000561 | 9.51E-06 | 0.013067544 | NA |
| chr19 | 17247936 | 17248508 | 573 | 3 | 3.12E-06 | 9.34E-05 | 5.17E-05 | 9.52E-06 | 0.004731546 | AC010646.1 |
| chr13 | 78605147 | 78605238 | 92 | 2 | 2.30E-05 | 6.49E-06 | 0.000307 | 9.53E-06 | 0.008505642 | OBI1-AS1 |
| chr9 | 98034960 | 98035619 | 660 | 4 | 3.38E-05 | 1.71E-05 | 0.000171 | 9.53E-06 | 0.006864815 | NA |
| chr4 | 155665331 | 155665558 | 228 | 4 | 1.35E-05 | 4.76E-05 | 0.000377 | 9.54E-06 | 0.00823343 | NA |
| chr9 | 88998964 | 88999258 | 295 | 3 | 1.92E-05 | 1.71E-05 | 7.76E-05 | 9.54E-06 | 0.001309872 | S1PR3 |
| chr7 | 159015288 | 159015933 | 646 | 4 | 2.13E-06 | 0.000298 | 5.30E-05 | 9.54E-06 | 0.008115931 | LINC00689 |
| chr12 | 6829654 | 6830461 | 808 | 5 | 1.54E-05 | 1.82E-05 | 0.000544 | 9.55E-06 | 0.003534644 | GPR162, P3H3 |
| chr1 | 53594310 | 53594541 | 232 | 3 | 3.87E-05 | 1.91E-05 | 0.000377 | 9.55E-06 | 0.007124288 | GLIS1 |
| chr11 | 64314073 | 64314402 | 330 | 4 | 4.24E-05 | 9.15E-06 | 0.000266 | 9.55E-06 | 0.000746907 | ESRRA |
| chr1 | 39268138 | 39268668 | 531 | 5 | 2.56E-05 | 1.41E-05 | 0.001188 | 9.56E-06 | 0.009243057 | MACF1 |
| chr2 | 177754190 | 177754635 | 446 | 2 | 1.57E-06 | 0.000708 | 1.18E-05 | 9.56E-06 | 0.016410163 | PDE11A, PDE11A |
| chr17 | 80191293 | 80191890 | 598 | 5 | 1.12E-05 | 0.000119 | 0.000765 | 9.56E-06 | 0.00396716 | CARD14 |
| chr2 | 207770193 | 207771308 | 1116 | 8 | 2.02E-05 | 8.16E-06 | 0.005746 | 9.57E-06 | 0.002940336 | NA |
| chr2 | 231388353 | 231388908 | 556 | 5 | 5.25E-05 | 5.85E-05 | 0.000992 | 9.57E-06 | 0.008842203 | NA |
| chr13 | 23153007 | 23153806 | 800 | 2 | 0.000285333 | 5.89E-06 | 0.000533 | 9.57E-06 | -0.02059167 | NA |
| chr6 | 474297 | 474629 | 333 | 3 | 1.22E-05 | 5.00E-06 | 0.000723 | 9.58E-06 | 0.011512549 | AL512308.1 |
| chr12 | 31589714 | 31590530 | 817 | 9 | 7.19E-05 | 6.76E-06 | 0.005767 | 9.58E-06 | 0.002175413 | DENND5B-AS1, DENND5B |
| chr22 | 39465107 | 39465485 | 379 | 2 | 3.45E-06 | 4.31E-05 | 2.76E-05 | 9.58E-06 | 0.008607934 | MGAT3 |
| chr1 | 173635575 | 173635785 | 211 | 3 | 5.33E-05 | 7.07E-06 | 0.000431 | 9.58E-06 | 0.01361738 | TEX50, AL139142.2, ANKRD45 |
| chr13 | 20161177 | 20162203 | 1027 | 9 | 2.63E-05 | 1.82E-05 | 0.000374 | 9.58E-06 | 0.004778586 | NA |
| chr22 | 41575231 | 41575959 | 729 | 3 | 0.00012809 | 4.09E-06 | 0.001968 | 9.58E-06 | 0.002162743 | CSDC2 |
| chr6 | 149429444 | 149430279 | 836 | 4 | 0.000196177 | 3.16E-06 | 0.0045 | 9.58E-06 | 0.001353618 | NA |
| chr2 | 127811489 | 127811939 | 451 | 6 | 5.33E-06 | 4.26E-05 | 0.000638 | 9.59E-06 | 0.012066202 | NA |
| chr10 | 124999084 | 124999143 | 60 | 2 | 1.94E-05 | 6.01E-06 | 0.000471 | 9.59E-06 | 0.007302382 | AL731571.1, CTBP2 |
| chr16 | 2816833 | 2817682 | 850 | 6 | 3.51E-05 | 3.35E-06 | 0.005722 | 9.59E-06 | -0.01935746 | PRSS21 |
| chr6 | 77464290 | 77464569 | 280 | 3 | 1.01E-06 | 3.35E-05 | 4.07E-05 | 9.60E-06 | 0.016431998 | NA |
| chr16 | 89909879 | 89910380 | 502 | 4 | 5.89E-06 | 1.57E-05 | 0.000837 | 9.60E-06 | 0.005906122 | TCF25, AC092143.3 |
| chr15 | 100973951 | 100974278 | 328 | 4 | 4.59E-05 | 8.35E-06 | 0.000397 | 9.62E-06 | 0.005960014 | LRRK1 |
| chr19 | 34963575 | 34964103 | 529 | 5 | 0.000114918 | 1.54E-05 | 0.000604 | 9.63E-06 | 0.007110107 | ZNF792 |
| chr17 | 825465 | 826021 | 557 | 5 | 1.77E-05 | 5.79E-06 | 0.001029 | 9.63E-06 | 0.001610965 | NXN |
| chr5 | 109688859 | 109689289 | 431 | 9 | 6.21E-05 | 4.73E-06 | 0.009879 | 9.63E-06 | 0.0017124 | NA |
| chr4 | 154742537 | 154743159 | 623 | 7 | 1.30E-05 | 1.18E-05 | 0.000142 | 9.64E-06 | 0.001677104 | LRAT |
| chr20 | 25696248 | 25697114 | 867 | 11 | 2.65E-05 | 0.000141 | 0.005542 | 9.64E-06 | 0.000449546 | AL390198.1, ZNF337 |
| chr8 | 133076274 | 133076455 | 182 | 3 | 1.37E-05 | 0.000115 | 5.15E-05 | 9.65E-06 | 0.004234254 | TG, SLA |
| chr12 | 53930707 | 53931111 | 405 | 4 | 2.45E-05 | 1.85E-05 | 0.000688 | 9.66E-06 | 0.00507467 | NA |
| chr2 | 66438296 | 66439969 | 1674 | 11 | 0.000107147 | 1.19E-05 | 0.011481 | 9.66E-06 | 0.004291288 | MEIS1, MEIS1-AS2 |
| chr8 | 41907124 | 41907652 | 529 | 2 | 4.00E-06 | 3.85E-05 | 2.99E-05 | 9.66E-06 | 0.007686959 | NA |
| chr13 | 43390031 | 43390089 | 59 | 2 | 1.18E-05 | 1.67E-05 | 5.70E-05 | 9.66E-06 | 0.025100068 | ENOX1 |
| chr6 | 99515373 | 99516238 | 866 | 11 | 5.96E-05 | 2.32E-05 | 0.000291 | 9.66E-06 | 0.001481113 | USP45 |
| chr7 | 2425620 | 2426060 | 441 | 2 | 2.65E-05 | 9.17E-06 | 0.000122 | 9.66E-06 | 0.007565545 | CHST12 |
| chr18 | 615471 | 615496 | 26 | 2 | 6.14E-05 | 5.82E-06 | 0.000634 | 9.67E-06 | 0.020725262 | CLUL1 |
| chr5 | 141682296 | 141683004 | 709 | 10 | 9.20E-06 | 1.47E-05 | 0.006297 | 9.67E-06 | 0.004779092 | NA |
| chr7 | 155711318 | 155711444 | 127 | 2 | 1.98E-05 | 1.26E-05 | 7.74E-05 | 9.67E-06 | -0.00076386 | RBM33 |
| chr12 | 132523747 | 132524128 | 382 | 6 | 2.09E-05 | 4.23E-05 | 0.000882 | 9.67E-06 | 0.003883768 | FBRSL1 |
| chr13 | 45713584 | 45714378 | 795 | 5 | 5.02E-05 | 3.76E-06 | 0.003141 | 9.68E-06 | 0.001967968 | CBY2 |
| chr11 | 48043542 | 48044299 | 758 | 3 | 9.20E-05 | 2.09E-05 | 0.00018 | 9.68E-06 | 0.008281084 | PTPRJ |
| chr14 | 75270108 | 75270723 | 616 | 2 | 1.04E-05 | 1.23E-05 | 8.03E-05 | 9.68E-06 | 0.010651271 | AF111167.1, U2 |
| chr12 | 51906086 | 51907138 | 1053 | 5 | 2.61E-07 | 0.00332 | 3.15E-05 | 9.69E-06 | 0.006656598 | ACVRL1 |
| chr11 | 1277921 | 1278561 | 641 | 5 | 2.44E-06 | 0.000133 | 0.00014 | 9.69E-06 | 0.00568546 | TOLLIP |
| chr19 | 43414254 | 43414887 | 634 | 9 | 5.10E-05 | 0.000152 | 0.002964 | 9.69E-06 | -0.00040498 | TEX101 |
| chr2 | 9102747 | 9103317 | 571 | 4 | 1.01E-05 | 0.000434 | 0.000144 | 9.70E-06 | 0.003168129 | NA |
| chr7 | 46536666 | 46536693 | 28 | 2 | 2.17E-05 | 0.00014 | 1.67E-05 | 9.70E-06 | 0.004025563 | NA |
| chr4 | 134327699 | 134328258 | 560 | 3 | 4.03E-05 | 1.55E-05 | 0.000476 | 9.70E-06 | -0.0029936 | PES1P1, AC015631.1 |
| chr5 | 119478708 | 119478994 | 287 | 2 | 1.34E-05 | 6.07E-05 | 2.31E-05 | 9.73E-06 | -0.00249453 | HSD17B4 |
| chr1 | 170669666 | 170670148 | 483 | 3 | 3.14E-05 | 4.26E-06 | 0.001669 | 9.74E-06 | 0.018970793 | PRRX1 |
| chr5 | 172771711 | 172772664 | 954 | 21 | 1.41E-06 | 0.000496 | 1.56E-05 | 9.74E-06 | -0.0001286 | AC022217.3 |
| chr20 | 57593904 | 57594552 | 649 | 4 | 8.69E-07 | 0.000125 | 7.24E-05 | 9.74E-06 | 0.004990708 | NA |
| chr22 | 25219120 | 25219951 | 832 | 11 | 5.29E-05 | 2.76E-05 | 0.010327 | 9.74E-06 | 0.001354114 | CRYBB2 |
| chr20 | 32031719 | 32032043 | 325 | 3 | 1.83E-05 | 9.38E-06 | 0.00017 | 9.77E-06 | 0.008652403 | CCM2L |
| chr2 | 11160233 | 11160966 | 734 | 6 | 9.28E-05 | 7.01E-06 | 0.004828 | 9.77E-06 | 0.001494186 | SLC66A3 |
| chr7 | 135798172 | 135798600 | 429 | 4 | 4.19E-05 | 1.02E-05 | 0.000332 | 9.78E-06 | 0.007226793 | AC015987.1 |
| chr8 | 97643617 | 97644339 | 723 | 8 | 2.45E-05 | 1.43E-05 | 0.001426 | 9.79E-06 | -0.00019005 | MTDH, AP002982.2 |

| | | | | | | | | | | |
|---|---|---|---|---|---|---|---|---|---|---|
| chr17 | 20153180 | 20153422 | 243 | 2 | 3.31E-05 | 8.37E-06 | 0.000151 | 9.80E-06 | 0.006797127 | SPECC1 |
| chr5 | 95646457 | 95647069 | 613 | 12 | 2.64E-05 | 0.000197 | 0.001656 | 9.82E-06 | 0.000969062 | RFESD |
| chr17 | 39711533 | 39712433 | 901 | 6 | 2.52E-05 | 6.09E-06 | 0.003788 | 9.82E-06 | 0.002969428 | ERBB2 |
| chr3 | 137764221 | 137764971 | 751 | 12 | 2.62E-05 | 5.09E-05 | 0.005084 | 9.84E-06 | 0.002310557 | SOX14 |
| chr1 | 24705012 | 24705456 | 445 | 2 | 2.05E-05 | 1.20E-05 | 8.45E-05 | 9.84E-06 | -0.0380768 | AL445648.1 |
| chr5 | 132822654 | 132823311 | 658 | 4 | 5.10E-05 | 3.08E-06 | 0.004001 | 9.84E-06 | 0.010511605 | SHROOM1 |
| chr22 | 31143315 | 31143842 | 528 | 2 | 7.00E-05 | 6.07E-06 | 0.000539 | 9.84E-06 | 0.006913617 | NA |
| chr16 | 21158010 | 21159606 | 1597 | 16 | 8.20E-05 | 0.000755 | 0.006054 | 9.84E-06 | 0.002638233 | TMEM159, DNAH3 |
| chr17 | 44967619 | 44967646 | 28 | 2 | 0.000447246 | 5.97E-06 | 0.000608 | 9.84E-06 | -0.01288621 | NMT1, C1QL1 |
| chr19 | 17485880 | 17486725 | 846 | 6 | 8.32E-06 | 7.43E-05 | 0.000104 | 9.85E-06 | 0.003703939 | SLC27A1 |
| chr5 | 146334044 | 146334534 | 491 | 7 | 4.26E-05 | 1.11E-05 | 0.005834 | 9.87E-06 | 0.006802267 | AC091959.1, AC011396.2 |
| chr6 | 108294257 | 108295084 | 828 | 12 | 3.29E-06 | 0.000417 | 0.000295 | 9.88E-06 | 0.00117115 | AFG1L |
| chr2 | 103656138 | 103656566 | 429 | 2 | 3.13E-05 | 8.49E-06 | 0.00015 | 9.88E-06 | 0.001595417 | NA |
| chr5 | 146104641 | 146105684 | 1044 | 5 | 1.01E-05 | 5.54E-05 | 7.44E-05 | 9.89E-06 | 0.006074773 | AC091887.1, PLAC8L1 |
| chr17 | 40175807 | 40176451 | 645 | 3 | 6.24E-08 | 0.000252 | 1.46E-05 | 9.90E-06 | 0.005899419 | NA |
| chr12 | 109573138 | 109574263 | 1126 | 22 | 3.22E-05 | 1.47E-05 | 0.00265 | 9.90E-06 | -0.00029681 | MVK, MMAB |
| chr17 | 48526873 | 48527412 | 540 | 4 | 3.34E-05 | 1.02E-05 | 0.000342 | 9.90E-06 | 0.014017135 | NA |
| chr5 | 179697977 | 179698945 | 969 | 12 | 2.29E-05 | 1.60E-05 | 0.001507 | 9.91E-06 | 0.001292389 | CANX |
| chr9 | 2621037 | 2621094 | 58 | 3 | 0.000202042 | 6.42E-06 | 0.000544 | 9.91E-06 | 0.017064867 | VLDLR-AS1 |
| chr19 | 3961639 | 3962196 | 558 | 8 | 1.50E-06 | 7.53E-05 | 0.00096 | 9.92E-06 | -0.00130451 | DAPK3 |
| chr11 | 60842990 | 60843404 | 415 | 5 | 1.07E-05 | 1.67E-05 | 0.00062 | 9.92E-06 | 0.00847369 | CCDC86, AP000777.3 |
| chr11 | 14262278 | 14262879 | 602 | 2 | 9.05E-05 | 5.99E-06 | 0.00063 | 9.92E-06 | -0.00050527 | SPON1, SPON1-AS1 |
| chr10 | 70132537 | 70133403 | 867 | 12 | 7.46E-06 | 0.000212 | 0.003338 | 9.92E-06 | 0.005015741 | AIFM2 |
| chr3 | 179321633 | 179322077 | 445 | 2 | 3.10E-05 | 6.37E-06 | 0.00043 | 9.92E-06 | 0.022951113 | NA |
| chr18 | 6561762 | 6561806 | 45 | 2 | 5.41E-05 | 6.75E-06 | 0.00032 | 9.92E-06 | 0.028857556 | LINC01387 |
| chr1 | 17272867 | 17273314 | 448 | 2 | 6.01E-05 | 5.98E-06 | 0.000643 | 9.92E-06 | 0.007204781 | PADI3 |
| chr18 | 49976819 | 49977056 | 238 | 2 | 2.76E-05 | 1.82E-05 | 5.48E-05 | 9.92E-06 | 0.006858771 | MYO5B |
| chr17 | 82374135 | 82374402 | 268 | 6 | 2.85E-05 | 2.67E-05 | 0.001402 | 9.93E-06 | 0.007755958 | UTS2R |
| chr14 | 64772769 | 64773225 | 457 | 3 | 1.13E-05 | 7.79E-05 | 9.05E-05 | 9.93E-06 | 0.006150365 | SPTB |
| chr12 | 109273336 | 109273488 | 153 | 2 | 5.66E-05 | 5.98E-06 | 0.000648 | 9.93E-06 | 0.01965093 | NA |
| chr3 | 195900460 | 195900712 | 253 | 3 | 6.01E-05 | 4.65E-06 | 0.001588 | 9.93E-06 | 0.012087182 | TNK2 |
| chr17 | 17025789 | 17026073 | 285 | 3 | 1.13E-05 | 0.001071 | 4.68E-05 | 9.93E-06 | 0.00387905 | NA |
| chr9 | 128205541 | 128205793 | 253 | 3 | 3.94E-05 | 1.95E-05 | 0.000429 | 9.93E-06 | 0.006831809 | DNM1 |
| chr11 | 19553715 | 19554515 | 801 | 3 | 0.000155442 | 1.17E-05 | 0.000648 | 9.93E-06 | 0.009174145 | NAV2 |
| chr15 | 58770535 | 58771686 | 1152 | 16 | 0.00011388 | 7.40E-05 | 0.002612 | 9.93E-06 | 0.000495818 | MINDY2, AC090515.2 |
| chr20 | 46709145 | 46710114 | 970 | 12 | 4.83E-05 | 3.76E-06 | 0.002311 | 9.94E-06 | 0.001158207 | SLC2A10 |
| chr7 | 155918468 | 155919119 | 652 | 2 | 5.06E-05 | 7.20E-06 | 0.00025 | 9.94E-06 | 0.012453277 | NA |
| chr1 | 225866614 | 225867153 | 540 | 2 | 2.99E-06 | 7.74E-06 | 0.000197 | 9.94E-06 | 0.021325341 | TMEM63A |
| chr22 | 42906361 | 42906798 | 438 | 2 | 5.39E-05 | 6.21E-06 | 0.000505 | 9.94E-06 | 0.000835176 | PACSIN2 |
| chr10 | 104322000 | 104322486 | 487 | 2 | 3.92E-05 | 7.39E-06 | 0.000228 | 9.96E-06 | 0.011345429 | ITPRIP |
| chr2 | 206274342 | 206275579 | 1238 | 13 | 8.19E-05 | 2.61E-05 | 0.020233 | 9.96E-06 | 0.001782169 | ZDBF2 |
| chr17 | 50905259 | 50905856 | 598 | 4 | 3.98E-05 | 0.000165 | 0.0003 | 9.96E-06 | 0.003626798 | TOB1-AS1 |
| chr1 | 3595146 | 3595679 | 534 | 4 | 6.91E-06 | 0.000209 | 1.58E-05 | 9.96E-06 | 0.004686847 | MEGF6 |
| chr1 | 211975416 | 211975514 | 99 | 2 | 3.09E-05 | 1.03E-05 | 0.000108 | 9.96E-06 | 0.014200962 | INTS7 |
| chr1 | 58805536 | 58805825 | 290 | 3 | 5.71E-05 | 4.15E-06 | 0.001895 | 9.98E-06 | 0.003965035 | LINC01135 |
| chr8 | 20249944 | 20250247 | 304 | 4 | 9.07E-06 | 0.001559 | 1.01E-05 | 1.00E-05 | 0.005035718 | LZTS1 |
| chr6 | 7051070 | 7051521 | 452 | 4 | 5.12E-05 | 3.37E-06 | 0.00406 | 1.00E-05 | 0.017787552 | NA |
| chr21 | 45051440 | 45051540 | 101 | 2 | 3.38E-05 | 8.37E-06 | 0.000162 | 1.00E-05 | 0.009864865 | NA |
| chr12 | 116441580 | 116441591 | 12 | 2 | 4.47E-05 | 1.33E-05 | 7.75E-05 | 1.00E-05 | 0.009634843 | NA |
| chr1 | 7549350 | 7549866 | 517 | 6 | 1.40E-05 | 2.55E-05 | 0.000112 | 1.00E-05 | 0.004239479 | CAMTA1 |
| chr16 | 1938448 | 1939085 | 638 | 3 | 0.000105845 | 1.10E-05 | 0.000747 | 1.00E-05 | 0.008225932 | MSRB1 |
| chr5 | 87412842 | 87413206 | 365 | 9 | 0.000210601 | 2.38E-06 | 0.008583 | 1.01E-05 | 0.000581105 | AC018754.1, CCNH |
| chr12 | 101766798 | 101766958 | 161 | 3 | 2.28E-05 | 0.000124 | 5.36E-05 | 1.01E-05 | 0.004252914 | GNPTAB |
| chr3 | 150147406 | 150147772 | 367 | 2 | 2.40E-05 | 1.31E-05 | 7.96E-05 | 1.01E-05 | 0.013280064 | AC117386.2 |
| chr9 | 3180374 | 3181007 | 634 | 3 | 1.46E-05 | 4.55E-06 | 0.001694 | 1.01E-05 | 0.021308863 | NA |
| chr12 | 44914275 | 44914776 | 502 | 3 | 3.43E-05 | 1.64E-05 | 0.000131 | 1.01E-05 | 0.011317351 | NELL2 |
| chr12 | 27969005 | 27969693 | 689 | 6 | 2.76E-05 | 2.30E-06 | 0.003664 | 1.01E-05 | 0.005406452 | AC008011.2, PTHLH |
| chr19 | 42202201 | 42202648 | 448 | 2 | 1.37E-05 | 6.90E-06 | 0.000327 | 1.02E-05 | 0.0115067 | DEDD2 |
| chr18 | 23252490 | 23252777 | 288 | 2 | 2.90E-05 | 6.24E-06 | 0.000567 | 1.02E-05 | 0.005681623 | CABLES1, TMEM241 |
| chr12 | 11543930 | 11543960 | 31 | 2 | 3.14E-05 | 6.89E-06 | 0.00033 | 1.02E-05 | -0.00134645 | AC007450.1 |
| chr2 | 241891343 | 241891923 | 581 | 3 | 1.06E-06 | 6.49E-05 | 0.000141 | 1.02E-05 | 0.002787785 | LINC01237 |
| chr17 | 47802069 | 47802613 | 545 | 2 | 1.89E-05 | 9.05E-06 | 0.000142 | 1.02E-05 | 0.010394286 | NA |
| chr2 | 58275748 | 58275751 | 4 | 2 | 2.05E-05 | 4.42E-05 | 2.93E-05 | 1.02E-05 | -0.00523925 | LINC01795 |
| chr14 | 59525485 | 59525707 | 223 | 2 | 4.01E-05 | 1.01E-05 | 0.000116 | 1.02E-05 | 0.014536199 | CCDC175 |
| chr12 | 49912332 | 49913103 | 772 | 5 | 3.35E-05 | 5.07E-05 | 0.000413 | 1.02E-05 | 0.004123679 | LINC02395 |
| chr1 | 32983290 | 32983561 | 272 | 2 | 1.87E-05 | 1.52E-05 | 6.86E-05 | 1.02E-05 | 0.007667833 | NA |
| chr17 | 58006396 | 58007146 | 751 | 6 | 3.08E-05 | 1.60E-05 | 0.00608 | 1.02E-05 | -0.00148804 | AC015813.5, AC015813.2, SRSF1 |
| chr12 | 132438934 | 132440082 | 1149 | 7 | 4.80E-05 | 1.87E-06 | 0.008441 | 1.02E-05 | 0.004719362 | NA |
| chr20 | 35559330 | 35560146 | 817 | 3 | 2.92E-07 | 0.007066 | 5.21E-06 | 1.02E-05 | -0.00133259 | FER1L4 |
| chr9 | 98778945 | 98779404 | 460 | 2 | 1.49E-06 | 0.018651 | 1.01E-05 | 1.03E-05 | 0.005715361 | ANKS6 |
| chr6 | 28281329 | 28281927 | 599 | 7 | 1.33E-05 | 2.67E-05 | 0.003273 | 1.03E-05 | 0.000746039 | AL021997.2, PGBD1 |
| chr12 | 52878034 | 52878218 | 185 | 2 | 3.77E-05 | 8.73E-06 | 0.000155 | 1.03E-05 | 0.014361565 | NA |
| chr5 | 10441127 | 10441791 | 665 | 11 | 3.15E-05 | 8.24E-05 | 0.00492 | 1.03E-05 | 0.003337673 | ROPN1L, ROPN1L-AS1 |
| chr9 | 83980236 | 83981017 | 782 | 14 | 4.51E-05 | 2.23E-05 | 0.006024 | 1.03E-05 | 0.001508402 | RMI1, HNRNPK |
| chr11 | 86671858 | 86672898 | 1041 | 17 | 4.60E-05 | 2.95E-05 | 0.004649 | 1.03E-05 | 0.002641277 | ME3 |
| chr3 | 9397705 | 9398075 | 371 | 5 | 9.09E-05 | 3.95E-05 | 0.000833 | 1.03E-05 | 0.004007116 | SETD5, THUMPD3-AS1 |
| chr9 | 137055125 | 137055968 | 844 | 3 | 2.49E-06 | 5.72E-05 | 3.07E-05 | 1.03E-05 | 0.008621481 | NA |
| chr3 | 169660887 | 169661766 | 880 | 8 | 6.90E-06 | 5.61E-05 | 0.001604 | 1.03E-05 | 0.000662163 | MECOM |
| chr8 | 128150739 | 128151130 | 392 | 2 | 5.66E-06 | 3.46E-05 | 3.45E-05 | 1.03E-05 | 0.006173154 | PVT1 |
| chr19 | 13804769 | 13805271 | 503 | 9 | 9.89E-06 | 4.05E-05 | 0.000265 | 1.03E-05 | 0.002716297 | ZSWIM4 |
| chr2 | 232381858 | 232382212 | 355 | 2 | 9.97E-06 | 7.02E-05 | 2.28E-05 | 1.03E-05 | 0.014203401 | ALPP |
| chr6 | 132537299 | 132537622 | 324 | 4 | 4.09E-05 | 4.52E-06 | 0.002683 | 1.03E-05 | 0.004911717 | NA |
| chr19 | 50835567 | 50836232 | 666 | 5 | 6.21E-05 | 5.89E-06 | 0.000719 | 1.03E-05 | 0.002354239 | AC011523.1, KLK15 |
| chr12 | 117188972 | 117189226 | 255 | 3 | 2.36E-05 | 1.77E-05 | 0.000272 | 1.03E-05 | -0.00851005 | FBXO21 |
| chr1 | 29256727 | 29256860 | 134 | 2 | 1.76E-05 | 9.85E-06 | 0.000125 | 1.03E-05 | 0.011735759 | PTPRU |
| chr6 | 170225531 | 170226064 | 534 | 6 | 5.85E-05 | 3.17E-05 | 0.001089 | 1.03E-05 | 0.005877799 | NA |
| chr17 | 29005541 | 29006497 | 957 | 15 | 2.11E-05 | 4.42E-05 | 0.001818 | 1.03E-05 | 0.001079285 | PIPOX, SEZ6 |
| chr4 | 20699952 | 20700754 | 803 | 11 | 2.83E-05 | 6.31E-05 | 0.00548 | 1.03E-05 | 0.000212792 | PACRGL |
| chr7 | 77537028 | 77537630 | 603 | 12 | 5.02E-05 | 7.60E-06 | 0.007301 | 1.03E-05 | -0.00037275 | PTPN12 |
| chr6 | 32076607 | 32077182 | 576 | 4 | 2.14E-05 | 3.09E-06 | 0.00446 | 1.03E-05 | 0.007771275 | TNXB |
| chr17 | 65538501 | 65539105 | 605 | 4 | 0.000303269 | 2.32E-05 | 0.000535 | 1.03E-05 | -0.00775259 | AXIN2, AC004805.1 |
| chr2 | 101752906 | 101752932 | 27 | 2 | 3.15E-05 | 1.31E-05 | 8.28E-05 | 1.04E-05 | 0.003604121 | MAP4K4 |
| chr20 | 25507905 | 25508514 | 610 | 4 | 9.49E-05 | 3.91E-06 | 0.004221 | 1.04E-05 | 0.00248508 | NINL |
| chr6 | 31834822 | 31836301 | 1480 | 23 | 2.49E-06 | 0.000215 | 0.001136 | 1.04E-05 | 0.001894171 | AL671762.1, SNHG32, SNORD48 |
| chr3 | 66501106 | 66501378 | 273 | 5 | 2.65E-05 | 0.000443 | 0.000235 | 1.04E-05 | -0.00045249 | LRIG1 |
| chr19 | 18007991 | 18008260 | 270 | 6 | 4.00E-05 | 1.76E-05 | 0.000437 | 1.04E-05 | -0.00273427 | ARRDC2 |
| chr1 | 23558481 | 23560239 | 1759 | 19 | 5.48E-05 | 4.05E-05 | 0.007085 | 1.04E-05 | 0.001869873 | ID3 |
| chr16 | 399685 | 400051 | 367 | 2 | 2.38E-06 | 6.48E-06 | 0.000536 | 1.04E-05 | 0.010236505 | NME4 |
| chr11 | 34052594 | 34052980 | 387 | 4 | 2.47E-05 | 3.36E-06 | 0.003331 | 1.04E-05 | 0.00088482 | CAPRIN1 |
| chr18 | 31342211 | 31342227 | 17 | 3 | 3.44E-05 | 2.80E-05 | 0.000149 | 1.04E-05 | 0.004863263 | DSG1 |
| chr21 | 44711184 | 44711963 | 780 | 10 | 5.90E-06 | 0.000192 | 0.000273 | 1.04E-05 | 0.001659597 | TSPEAR |
| chr14 | 102592114 | 102592723 | 610 | 11 | 6.94E-05 | 1.72E-05 | 0.003542 | 1.04E-05 | 0.000651376 | RCOR1 |
| chr11 | 134036798 | 134036866 | 69 | 2 | 9.83E-06 | 9.23E-06 | 0.000145 | 1.04E-05 | -0.00065562 | LINC02731, AP000911.1 |
| chr2 | 24359784 | 24360741 | 958 | 10 | 1.39E-05 | 0.000157 | 0.002021 | 1.04E-05 | 0.000682734 | ITSN2 |
| chr15 | 62065511 | 62065581 | 71 | 2 | 1.60E-06 | 0.000372 | 1.42E-05 | 1.04E-05 | 0.000108369 | NA |
| chr10 | 11441365 | 11441528 | 164 | 2 | 6.91E-05 | 6.31E-06 | 0.000654 | 1.04E-05 | -0.0035624 | NA |
| chr17 | 62689218 | 62689953 | 736 | 5 | 1.88E-05 | 2.09E-05 | 0.000227 | 1.05E-05 | 0.000922604 | MRC2 |
| chr2 | 37659891 | 37659985 | 95 | 2 | 5.75E-05 | 6.81E-06 | 0.000407 | 1.05E-05 | 0.006120631 | CDC42EP3 |
| chr17 | 66944628 | 66945035 | 408 | 2 | 3.71E-06 | 7.73E-06 | 0.000239 | 1.05E-05 | 0.027768607 | NA |
| chr17 | 66728130 | 66728664 | 535 | 3 | 1.11E-05 | 4.52E-05 | 4.51E-05 | 1.05E-05 | -0.00223697 | PRKCA |
| chr1 | 216423752 | 216424365 | 614 | 3 | 7.16E-05 | 4.47E-06 | 0.002018 | 1.05E-05 | 0.012522421 | NA |
| chr9 | 33452951 | 33453245 | 295 | 2 | 4.07E-05 | 8.83E-06 | 0.000161 | 1.05E-05 | 0.013274348 | NA |
| chr1 | 230746641 | 230747806 | 1166 | 10 | 1.90E-05 | 0.000313 | 3.46E-05 | 1.05E-05 | 0.002899027 | CAPN9, AL512328.1 |
| chr2 | 70891162 | 70891693 | 532 | 2 | 1.21E-05 | 1.94E-05 | 5.56E-05 | 1.05E-05 | 0.016603861 | NA |
| chr19 | 54710713 | 54710744 | 32 | 2 | 5.40E-05 | 8.81E-06 | 0.000163 | 1.05E-05 | 0.020819342 | LILRP2 |
| chr20 | 38222013 | 38222961 | 949 | 3 | 2.24E-05 | 4.49E-06 | 0.001951 | 1.05E-05 | 0.009948383 | KIAA1755 |
| chr1 | 3914235 | 3914750 | 516 | 4 | 4.30E-05 | 7.54E-06 | 0.000329 | 1.05E-05 | 0.001980547 | LINC01134 |
| chr16 | 88768437 | 88768907 | 471 | 2 | 3.27E-06 | 2.05E-05 | 5.32E-05 | 1.05E-05 | 0.007894538 | PIEZO1 |
| chr17 | 39980361 | 39981236 | 876 | 15 | 6.20E-05 | 9.42E-06 | 0.019565 | 1.05E-05 | 0.000220559 | PSMD3 |
| chr19 | 55528819 | 55529009 | 191 | 2 | 2.12E-05 | 3.02E-05 | 3.90E-05 | 1.05E-05 | 0.010369396 | SBK2 |
| chr11 | 76597717 | 76598161 | 445 | 2 | 4.44E-06 | 1.30E-05 | 8.64E-05 | 1.06E-05 | 0.00534914 | NA |
| chr1 | 43581756 | 43582297 | 542 | 6 | 1.05E-05 | 2.79E-05 | 0.000202 | 1.06E-05 | 0.005114209 | PTPRF |
| chr16 | 89153007 | 89154108 | 1102 | 5 | 3.02E-05 | 4.54E-06 | 0.005424 | 1.06E-05 | 0.001178741 | ACSF3 |
| chr10 | 33210920 | 33211303 | 384 | 6 | 6.13E-05 | 4.73E-06 | 0.001349 | 1.06E-05 | 0.005435689 | AL121748.1, NRP1 |
| chr2 | 146586662 | 146586850 | 189 | 2 | 1.58E-06 | 0.001894 | 1.14E-05 | 1.06E-05 | -0.00302878 | NA |
| chr12 | 109718551 | 109719459 | 909 | 4 | 0.000248348 | 5.87E-06 | 0.002859 | 1.06E-05 | 0.009594828 | FAM222A |
| chr7 | 74033717 | 74034443 | 727 | 3 | 5.28E-05 | 6.83E-05 | 0.000292 | 1.06E-05 | 0.005935742 | ELN |
| chr1 | 41021421 | 41022337 | 917 | 7 | 2.77E-05 | 1.22E-05 | 0.006429 | 1.06E-05 | 0.007324973 | SLFNL1-AS1, SLFNL1 |
| chr3 | 5016750 | 5016865 | 116 | 2 | 1.63E-05 | 0.000279 | 1.52E-05 | 1.06E-05 | 0.009229164 | NA |
| chr5 | 1536376 | 1536647 | 272 | 2 | 3.44E-06 | 5.50E-05 | 2.69E-05 | 1.06E-05 | 0.013970461 | NA |
| chr11 | 1514816 | 1515697 | 882 | 6 | 9.35E-06 | 8.40E-05 | 4.54E-05 | 1.06E-05 | 0.006592344 | NA |
| chr10 | 365859 | 366385 | 527 | 4 | 3.73E-05 | 9.18E-06 | 0.002315 | 1.06E-05 | 0.002661997 | DIP2C |
| chr6 | 28919364 | 28920252 | 889 | 7 | 3.55E-05 | 2.82E-05 | 0.002306 | 1.06E-05 | 0.004861853 | TRIM27 |
| chr4 | 95620719 | 95621187 | 469 | 2 | 3.56E-06 | 0.000249 | 1.56E-05 | 1.06E-05 | 0.00686662 | NA |
| chr6 | 107732198 | 107732759 | 562 | 3 | 2.61E-06 | 0.000175 | 8.32E-05 | 1.06E-05 | 0.010682276 | SCML4 |
| chr2 | 25702125 | 25702181 | 57 | 2 | 2.81E-05 | 7.68E-05 | 2.26E-05 | 1.06E-05 | -0.0052705 | NA |
| chr1 | 4775238 | 4775951 | 714 | 3 | 0.000162749 | 6.00E-06 | 0.000803 | 1.06E-05 | -0.00282203 | AJAP1 |
| chr17 | 81400381 | 81401454 | 1074 | 6 | 2.33E-05 | 4.80E-06 | 0.007237 | 1.06E-05 | 0.013557016 | BAHCC1, MIR4740 |
| chr14 | 95476156 | 95476490 | 335 | 5 | 2.22E-05 | 2.63E-05 | 7.61E-05 | 1.07E-05 | 0.003810572 | NA |
| chr12 | 53298861 | 53300018 | 1158 | 13 | 1.10E-05 | 1.28E-05 | 0.003036 | 1.07E-05 | 0.00384897 | PFDN5, AC073611.1, MYG1, MYG1-AS1 |
| chr8 | 41650325 | 41651099 | 775 | 7 | 0.000149506 | 1.01E-05 | 0.006192 | 1.07E-05 | 0.003266937 | NKX6-3 |
| chr14 | 55270878 | 55272065 | 1188 | 14 | 1.15E-05 | 0.000148 | 0.00193 | 1.07E-05 | 0.000856127 | FBXO34, AL158801.2 |
| chr15 | 29110472 | 29111130 | 659 | 2 | 4.51E-07 | 4.55E-05 | 3.05E-05 | 1.07E-05 | 0.007860788 | APBA2 |
| chr22 | 21773539 | 21773727 | 189 | 2 | 2.88E-06 | 0.000495 | 1.39E-05 | 1.07E-05 | 0.002308424 | MAPK1 |
| chr3 | 126982844 | 126983462 | 619 | 2 | 8.81E-05 | 6.50E-06 | 0.00064 | 1.07E-05 | -0.00325262 | NA |
| chr4 | 39480753 | 39481161 | 409 | 4 | 3.72E-05 | 8.51E-06 | 0.000892 | 1.07E-05 | 0.008035166 | LIAS, AC021148.1 |
| chr16 | 2174552 | 2175775 | 1224 | 7 | 5.55E-07 | 3.98E-05 | 0.000102 | 1.07E-05 | 0.002385321 | TRAF7 |
| chr3 | 196969640 | 196970316 | 677 | 4 | 3.97E-05 | 2.36E-05 | 0.000914 | 1.07E-05 | -0.00746065 | NA |
| chr5 | 34684022 | 34684523 | 502 | 5 | 7.18E-06 | 0.000436 | 2.60E-05 | 1.07E-05 | 0.002405848 | RAI14 |
| chr13 | 75332883 | 75333458 | 576 | 2 | 6.70E-05 | 6.78E-06 | 0.000499 | 1.07E-05 | 0.012587779 | TBC1D4 |
| chr5 | 62143866 | 62143963 | 98 | 2 | 3.79E-05 | 1.98E-05 | 5.64E-05 | 1.07E-05 | 0.01406498 | NA |
| chr4 | 141053742 | 141054836 | 1095 | 4 | 2.42E-06 | 0.002734 | 9.29E-06 | 1.07E-05 | 0.007404129 | RNF150 |
| chr19 | 46529829 | 46530347 | 519 | 3 | 6.08E-05 | 6.33E-05 | 0.000184 | 1.07E-05 | 0.014864245 | PPP5D1 |
| chr5 | 181234392 | 181234554 | 163 | 3 | 6.30E-05 | 4.63E-06 | 0.002106 | 1.07E-05 | 0.009082961 | TRIM41 |
| chr21 | 44065319 | 44065829 | 511 | 2 | 6.74E-05 | 6.61E-06 | 0.000592 | 1.08E-05 | 0.025620339 | TRAPPC10 |
| chr4 | 73836358 | 73837155 | 798 | 9 | 0.000111057 | 2.29E-05 | 0.007519 | 1.08E-05 | 0.005875334 | CXCL6 |
| chr10 | 46550360 | 46550713 | 354 | 3 | 1.30E-05 | 9.26E-05 | 0.000169 | 1.08E-05 | 0.00477877 | GPRIN2 |
| chr7 | 92424284 | 92424903 | 620 | 3 | 5.77E-06 | 0.00068 | 0.000128 | 1.08E-05 | 0.002922806 | TMBIM7P |
| chr9 | 120713089 | 120713708 | 620 | 3 | 1.28E-06 | 5.13E-05 | 5.35E-05 | 1.08E-05 | 0.007979339 | MEGF9 |
| chr1 | 28552635 | 28553464 | 830 | 11 | 1.26E-06 | 0.001953 | 6.41E-05 | 1.08E-05 | 0.001929857 | TRNAU1AP |
| chr15 | 75648214 | 75649235 | 1022 | 10 | 1.18E-06 | 0.000135 | 4.30E-05 | 1.08E-05 | 0.003575807 | SNX33, IMP3 |
| chr1 | 2882670 | 2883629 | 960 | 7 | 1.63E-05 | 2.81E-05 | 0.000182 | 1.08E-05 | 0.005416509 | NA |

| | | | | | | | | | | |
|---|---|---|---|---|---|---|---|---|---|---|
| chr20 | 34949342 | 34949713 | 372 | 3 | 5.91E-05 | 5.10E-06 | 0.001494 | 1.08E-05 | -9.73E-05 | GSS |
| chr7 | 965063 | 965453 | 391 | 4 | 8.67E-06 | 4.87E-05 | 0.000465 | 1.08E-05 | 0.006680004 | COX19 |
| chr2 | 172102484 | 172103251 | 768 | 11 | 1.82E-05 | 7.01E-05 | 0.001938 | 1.08E-05 | 0.000974562 | DLX2-DT, DLX2 |
| chr20 | 47413219 | 47413457 | 239 | 3 | 6.73E-05 | 7.76E-06 | 0.000603 | 1.08E-05 | 0.005027033 | NA |
| chr14 | 104216543 | 104216978 | 436 | 3 | 2.73E-05 | 1.02E-05 | 0.000304 | 1.08E-05 | 0.001698876 | NA |
| chr10 | 531147 | 532407 | 1261 | 6 | 3.37E-05 | 3.16E-06 | 0.008985 | 1.08E-05 | 0.00378617 | DIP2C |
| chr4 | 8069276 | 8069475 | 200 | 2 | 2.20E-05 | 6.39E-05 | 2.53E-05 | 1.08E-05 | 0.006058722 | ABLIM2 |
| chr16 | 87636724 | 87636962 | 239 | 4 | 7.82E-05 | 8.35E-06 | 0.000685 | 1.08E-05 | 0.002873027 | JPH3 |
| chr4 | 6301966 | 6302315 | 350 | 4 | 1.55E-05 | 1.92E-05 | 0.000137 | 1.08E-05 | 0.001471745 | WFS1, AC116317.1 |
| chr5 | 61636754 | 61636961 | 208 | 2 | 2.33E-05 | 4.95E-05 | 2.94E-05 | 1.08E-05 | 0.001555069 | C5orf64 |
| chr3 | 37694459 | 37694554 | 96 | 3 | 4.72E-05 | 6.21E-06 | 0.000632 | 1.08E-05 | 0.023235166 | ITGA9, ITGA9-AS1 |
| chr8 | 143246330 | 143247374 | 1045 | 14 | 1.11E-05 | 0.000157 | 0.011635 | 1.08E-05 | 0.000938898 | ZFP41, AC138696.1 |
| chr10 | 117544976 | 117545824 | 849 | 9 | 5.03E-05 | 9.23E-05 | 0.001313 | 1.09E-05 | 0.003029637 | EMX2, EMX2OS |
| chr19 | 54141259 | 54141289 | 31 | 3 | 7.41E-06 | 0.000152 | 1.74E-05 | 1.09E-05 | -0.00285048 | CNOT3 |
| chr8 | 22147084 | 22148089 | 1006 | 4 | 6.80E-06 | 7.97E-06 | 0.001131 | 1.09E-05 | 0.007740255 | LGI3 |
| chr15 | 64824056 | 64824514 | 459 | 3 | 9.97E-06 | 3.89E-05 | 0.000105 | 1.09E-05 | -0.00736021 | PIF1 |
| chr7 | 907020 | 907514 | 495 | 3 | 1.07E-05 | 3.02E-05 | 5.98E-05 | 1.09E-05 | 0.013073023 | ADAP1, COX19 |
| chr1 | 229102661 | 229102719 | 59 | 2 | 3.50E-05 | 3.98E-05 | 3.37E-05 | 1.09E-05 | 0.006913679 | LINC02814 |
| chr9 | 122947457 | 122947577 | 121 | 2 | 3.03E-05 | 8.35E-06 | 0.000215 | 1.09E-05 | 0.015352808 | RABGAP1 |
| chr9 | 137293535 | 137294573 | 1039 | 4 | 1.43E-06 | 0.001951 | 8.37E-06 | 1.09E-05 | 0.00415986 | BX255925.1 |
| chr5 | 173235738 | 173236216 | 479 | 3 | 1.05E-05 | 1.45E-05 | 0.000175 | 1.09E-05 | 0.013674285 | NA |
| chr12 | 53005309 | 53005760 | 452 | 5 | 1.97E-05 | 0.000116 | 0.000189 | 1.09E-05 | 0.010126539 | NA |
| chr9 | 128771033 | 128772192 | 1160 | 13 | 2.80E-05 | 3.52E-06 | 0.005843 | 1.09E-05 | 4.00E-05 | ZER1 |
| chr1 | 6025450 | 6026296 | 847 | 10 | 3.19E-05 | 7.55E-06 | 0.00886 | 1.09E-05 | -0.00810493 | KCNAB2 |
| chr6 | 98825342 | 98825954 | 613 | 8 | 1.50E-05 | 3.53E-06 | 0.006029 | 1.09E-05 | 0.004567858 | NA |
| chr19 | 45136729 | 45137147 | 419 | 2 | 2.57E-05 | 9.81E-06 | 0.000144 | 1.09E-05 | 0.010854999 | MARK4, PPP1R37, AC005757.1 |
| chr2 | 174340988 | 174341773 | 786 | 7 | 5.04E-05 | 1.82E-05 | 0.000532 | 1.09E-05 | -0.00096864 | NA |
| chr22 | 48503886 | 48504237 | 352 | 3 | 5.83E-06 | 0.000195 | 1.55E-05 | 1.09E-05 | 0.005479025 | TAFA5 |
| chr1 | 201283072 | 201283696 | 625 | 8 | 2.27E-05 | 1.62E-05 | 0.001219 | 1.09E-05 | -0.00102488 | PKP1 |
| chr12 | 47078534 | 47079908 | 1375 | 12 | 1.98E-05 | 1.09E-06 | 0.013014 | 1.09E-05 | 0.001058223 | PCED1B, AMIGO2 |
| chr19 | 2702865 | 2703117 | 253 | 9 | 3.94E-05 | 6.19E-05 | 0.000741 | 1.09E-05 | 0.000126713 | NA |
| chr14 | 103330100 | 103330403 | 304 | 2 | 4.43E-05 | 8.68E-06 | 0.000194 | 1.09E-05 | 0.010521641 | NA |
| chr2 | 10526459 | 10526736 | 278 | 4 | 1.76E-05 | 0.00026 | 7.96E-05 | 1.09E-05 | 0.005903012 | NA |
| chr13 | 112183838 | 112184656 | 819 | 6 | 1.85E-05 | 4.45E-05 | 0.001478 | 1.09E-05 | 0.005904122 | NA |
| chr3 | 49006127 | 49007370 | 1244 | 15 | 2.83E-05 | 0.00014 | 0.004506 | 1.09E-05 | 0.001272942 | P4HTM, WDR6 |
| chr3 | 186262050 | 186262277 | 228 | 2 | 7.31E-06 | 0.000219 | 1.66E-05 | 1.09E-05 | 0.004023761 | DGKG |
| chr17 | 5587366 | 5587567 | 202 | 2 | 1.42E-05 | 3.25E-05 | 3.89E-05 | 1.09E-05 | 0.003496088 | NLRP1 |
| chr17 | 49402929 | 49403756 | 828 | 4 | 3.36E-05 | 2.21E-05 | 0.000792 | 1.09E-05 | 0.009249543 | AC091180.5 |
| chr1 | 10754967 | 10755465 | 499 | 3 | 6.76E-05 | 4.26E-06 | 0.0027 | 1.09E-05 | 0.006251916 | CASZ1 |
| chr12 | 92649659 | 92650003 | 345 | 3 | 2.40E-05 | 8.76E-06 | 0.000362 | 1.09E-05 | 0.009273986 | NA |
| chr5 | 158954061 | 158954260 | 200 | 3 | 2.69E-05 | 3.25E-05 | 0.000506 | 1.09E-05 | 0.013357131 | EBF1 |
| chr8 | 6454641 | 6454895 | 255 | 2 | 5.85E-05 | 6.66E-06 | 0.000657 | 1.09E-05 | 0.009933398 | MCPH1 |
| chr7 | 99398073 | 99398223 | 151 | 3 | 3.37E-05 | 1.13E-05 | 0.000784 | 1.09E-05 | 0.005278931 | PDAP1 |
| chr5 | 65925936 | 65926763 | 828 | 12 | 1.48E-05 | 0.000344 | 0.002367 | 1.09E-05 | -0.00085671 | ERBIN |
| chr2 | 177212241 | 177212969 | 729 | 6 | 3.41E-06 | 0.001488 | 6.57E-05 | 1.09E-05 | 0.000523279 | HNRNPA3, MIR4444-1, AC079305.2 |
| chr1 | 17717582 | 17717676 | 95 | 3 | 4.76E-05 | 1.28E-05 | 0.000132 | 1.09E-05 | 0.016262907 | LINC02810 |
| chr21 | 46634985 | 46635803 | 819 | 14 | 6.44E-05 | 7.74E-05 | 0.00506 | 1.09E-05 | 0.001811315 | PRMT2 |
| chr11 | 57349188 | 57350192 | 1005 | 5 | 8.11E-05 | 3.71E-05 | 0.00054 | 1.09E-05 | 0.006780869 | P2RX3 |
| chr6 | 30492467 | 30493021 | 555 | 7 | 2.22E-05 | 5.45E-05 | 0.000342 | 1.09E-05 | -0.00184192 | HLA-E |
| chr3 | 49104324 | 49105382 | 1059 | 13 | 7.70E-05 | 3.34E-05 | 0.015728 | 1.09E-05 | 0.00278323 | QARS1 |
| chr6 | 89819419 | 89820474 | 1056 | 11 | 8.74E-06 | 0.000108 | 0.000807 | 1.09E-05 | 0.002085478 | MDN1 |
| chr17 | 21397423 | 21397479 | 57 | 3 | 1.78E-05 | 4.12E-05 | 4.08E-05 | 1.09E-05 | 0.003506585 | KCNJ12 |
| chr5 | 152505513 | 152505607 | 95 | 3 | 8.10E-05 | 4.23E-06 | 0.002801 | 1.09E-05 | 0.023322203 | AC008571.2 |
| chr17 | 80837736 | 80838028 | 293 | 4 | 4.02E-05 | 1.97E-05 | 0.0002 | 1.10E-05 | -0.00041893 | RPTOR |
| chr4 | 1951944 | 1952077 | 134 | 3 | 2.51E-05 | 1.07E-05 | 0.000169 | 1.10E-05 | 0.002285184 | NSD2 |
| chr16 | 765553 | 766874 | 1322 | 9 | 2.67E-06 | 0.000146 | 0.000909 | 1.10E-05 | 0.002116904 | MSLN |
| chr6 | 29620391 | 29621279 | 889 | 6 | 3.73E-06 | 2.15E-05 | 0.002256 | 1.10E-05 | 0.00293337 | GABBR1 |
| chr15 | 74208976 | 74209859 | 884 | 7 | 1.31E-05 | 1.31E-05 | 0.002981 | 1.10E-05 | 0.007456681 | CCDC33, STRA6 |
| chr9 | 106717790 | 106718201 | 412 | 2 | 3.13E-05 | 1.08E-05 | 0.000125 | 1.10E-05 | 0.012181389 | LINC01505 |
| chr6 | 31419462 | 31419960 | 499 | 2 | 7.44E-05 | 6.82E-06 | 0.000577 | 1.10E-05 | 0.027114922 | AL645933.3 |
| chr17 | 10657302 | 10658033 | 732 | 5 | 1.44E-05 | 3.07E-05 | 0.000635 | 1.10E-05 | 0.00251101 | MYH3 |
| chr17 | 10518872 | 10519740 | 869 | 4 | 2.36E-06 | 1.95E-05 | 0.000244 | 1.10E-05 | 0.002242529 | AC005323.2, MYHAS |
| chr5 | 37834069 | 37836144 | 2076 | 20 | 6.24E-08 | 0.002122 | 0.000169 | 1.10E-05 | 0.003628642 | GDNF-AS1, GDNF |
| chr1 | 26043510 | 26043916 | 407 | 2 | 4.33E-06 | 1.30E-05 | 9.36E-05 | 1.10E-05 | 0.004661076 | SLC30A2 |
| chr3 | 69084608 | 69085351 | 744 | 8 | 2.63E-05 | 6.00E-05 | 0.000302 | 1.10E-05 | -0.00015075 | ARL6IP5 |
| chr19 | 49704511 | 49704887 | 377 | 2 | 5.04E-05 | 7.61E-06 | 0.000328 | 1.10E-05 | 0.005909061 | CPT1C |
| chr13 | 49350886 | 49351402 | 517 | 4 | 2.23E-05 | 2.14E-05 | 0.001129 | 1.11E-05 | 0.000855639 | CAB39L |
| chr1 | 27351369 | 27351647 | 279 | 4 | 5.66E-06 | 0.001763 | 9.27E-05 | 1.11E-05 | 0.00356535 | SYTL1 |
| chr16 | 29863187 | 29863662 | 476 | 9 | 0.000110984 | 8.93E-06 | 0.017471 | 1.11E-05 | 0.000516268 | AC120114.2, CDIPTOSP, CDIPT |
| chr2 | 173353639 | 173354481 | 843 | 3 | 7.85E-06 | 3.34E-05 | 5.18E-05 | 1.11E-05 | -0.00264799 | AC092573.1 |
| chr3 | 194397884 | 194398459 | 576 | 6 | 2.21E-05 | 3.40E-05 | 0.000258 | 1.11E-05 | -0.00014993 | GP5 |
| chr22 | 39299898 | 39300101 | 204 | 3 | 7.15E-05 | 7.38E-06 | 0.000629 | 1.11E-05 | 0.013503721 | NA |
| chr20 | 44395801 | 44396108 | 308 | 3 | 6.21E-06 | 0.000232 | 5.83E-05 | 1.11E-05 | 0.01240648 | HNF4A |
| chr14 | 102139264 | 102139910 | 647 | 10 | 1.24E-05 | 2.82E-05 | 0.000537 | 1.11E-05 | 0.001052884 | WDR20, HSP90AA1 |
| chr7 | 44911911 | 44912156 | 246 | 2 | 3.17E-05 | 1.71E-05 | 6.90E-05 | 1.11E-05 | 0.007969962 | NA |
| chr4 | 169620658 | 169621020 | 363 | 5 | 3.87E-05 | 2.92E-05 | 0.002559 | 1.11E-05 | 0.001557726 | CLCN3 |
| chr19 | 17206200 | 17206252 | 53 | 2 | 3.42E-05 | 6.97E-06 | 0.000542 | 1.11E-05 | 0.008483648 | MYO9B |
| chr2 | 71276116 | 71276959 | 844 | 11 | 1.48E-05 | 0.000417 | 0.000186 | 1.11E-05 | 0.001436328 | ZNF638 |
| chr5 | 151638015 | 151638115 | 101 | 2 | 5.57E-05 | 6.78E-06 | 0.000677 | 1.11E-05 | 0.018044542 | NA |
| chr15 | 77568696 | 77569331 | 636 | 3 | 5.02E-05 | 8.31E-06 | 0.001013 | 1.12E-05 | 0.004887198 | AC046168.2 |
| chr2 | 207269914 | 207270378 | 465 | 2 | 3.89E-05 | 8.99E-06 | 0.000191 | 1.12E-05 | 0.00641926 | LINC01802, AC009226.1 |
| chr11 | 63901731 | 63902659 | 929 | 3 | 5.24E-06 | 4.65E-05 | 3.84E-05 | 1.12E-05 | 0.000422218 | MARK2 |
| chr1 | 201023257 | 201024183 | 927 | 14 | 1.94E-05 | 0.000148 | 0.000845 | 1.12E-05 | -0.00018166 | AL358473.1, KIF21B |
| chr10 | 7619659 | 7619819 | 161 | 2 | 2.12E-05 | 9.76E-06 | 0.000157 | 1.12E-05 | 0.008500001 | ITIH5 |
| chr18 | 78914210 | 78915070 | 861 | 6 | 4.29E-05 | 6.83E-06 | 0.00137 | 1.13E-05 | -0.01378576 | AC091027.2 |
| chr7 | 27663817 | 27664446 | 630 | 4 | 5.58E-06 | 4.46E-06 | 0.004028 | 1.13E-05 | -0.00472301 | NA |
| chr5 | 9265730 | 9266074 | 345 | 2 | 2.50E-05 | 1.81E-05 | 6.61E-05 | 1.13E-05 | 0.021601883 | SEMA5A |
| chr6 | 25930666 | 25931431 | 766 | 8 | 7.53E-05 | 4.84E-05 | 0.002894 | 1.13E-05 | 0.004330883 | SLC17A2 |
| chr16 | 12804573 | 12804798 | 226 | 5 | 8.19E-06 | 8.84E-05 | 0.000212 | 1.13E-05 | 0.003818569 | NA |
| chr1 | 150272362 | 150273134 | 773 | 5 | 2.25E-05 | 2.36E-05 | 0.001601 | 1.13E-05 | 0.013155328 | C1orf54 |
| chr2 | 160493620 | 160494881 | 1262 | 14 | 4.21E-05 | 7.47E-05 | 0.001552 | 1.13E-05 | 0.000518263 | RBMS1 |
| chr16 | 19776433 | 19776442 | 10 | 2 | 6.05E-06 | 2.31E-05 | 5.26E-05 | 1.13E-05 | 0.009883431 | IQCK |
| chr6 | 33209720 | 33210696 | 977 | 12 | 3.70E-06 | 0.000273 | 6.05E-05 | 1.13E-05 | 0.001023534 | RING1 |
| chr12 | 121274442 | 121274577 | 136 | 4 | 0.000180394 | 1.01E-05 | 0.000653 | 1.13E-05 | -0.00036593 | CAMKK2 |
| chr2 | 172109489 | 172110232 | 744 | 5 | 1.25E-05 | 1.07E-05 | 0.00154 | 1.13E-05 | 0.015163101 | DLX2-DT |
| chr17 | 82232086 | 82232999 | 914 | 10 | 0.000106339 | 4.55E-05 | 0.004433 | 1.13E-05 | 0.006982469 | SLC16A3 |
| chr15 | 53759042 | 53759704 | 663 | 8 | 8.37E-06 | 7.99E-05 | 0.002827 | 1.13E-05 | 0.005112753 | WDR72 |
| chr10 | 43229827 | 43230694 | 868 | 8 | 2.09E-05 | 0.000254 | 0.000994 | 1.13E-05 | 0.004247106 | RASGEF1A |
| chr13 | 23793903 | 23794067 | 165 | 3 | 2.29E-05 | 0.000117 | 2.07E-05 | 1.13E-05 | 0.004303945 | MIPEP |
| chr17 | 5526928 | 5527650 | 723 | 2 | 0.000138648 | 7.21E-06 | 0.000498 | 1.13E-05 | 0.011036825 | NLRP1 |
| chr16 | 7805303 | 7805620 | 318 | 3 | 6.72E-05 | 5.26E-06 | 0.001708 | 1.14E-05 | 0.025509592 | NA |
| chr12 | 1505555 | 1505715 | 161 | 2 | 1.72E-05 | 2.14E-05 | 5.68E-05 | 1.14E-05 | 0.003763643 | NA |
| chr17 | 17396062 | 17396322 | 261 | 2 | 7.15E-06 | 0.021123 | 1.10E-05 | 1.14E-05 | 0.003517821 | NA |
| chr1 | 2790395 | 2791008 | 614 | 6 | 3.84E-05 | 4.12E-05 | 0.000843 | 1.14E-05 | -0.0074164 | TTC34 |
| chr8 | 19312730 | 19313379 | 650 | 4 | 5.52E-05 | 7.09E-05 | 0.001535 | 1.14E-05 | 0.00688759 | NA |
| chr7 | 39622868 | 39623591 | 724 | 8 | 3.04E-05 | 2.81E-05 | 0.001381 | 1.14E-05 | 0.00214972 | RALA, AC004837.4 |
| chr6 | 16409913 | 16410075 | 163 | 3 | 6.30E-05 | 1.10E-05 | 0.000451 | 1.14E-05 | 0.015013953 | ATXN1 |
| chr11 | 46694904 | 46695141 | 238 | 2 | 6.59E-06 | 1.27E-05 | 0.000103 | 1.14E-05 | -0.00313031 | ARHGAP1 |
| chr5 | 1019994 | 1020603 | 610 | 2 | 2.49E-05 | 7.65E-06 | 0.000377 | 1.14E-05 | 0.014503162 | NKD2 |
| chr2 | 96507976 | 96508654 | 679 | 12 | 4.09E-05 | 2.14E-05 | 0.006474 | 1.14E-05 | -0.00311454 | NEURL3 |
| chr14 | 60649330 | 60650182 | 853 | 9 | 2.15E-05 | 0.000345 | 0.004203 | 1.14E-05 | 0.000552692 | SIX1 |
| chr7 | 122080960 | 122081539 | 580 | 4 | 4.60E-05 | 1.42E-05 | 0.00045 | 1.14E-05 | 0.001156883 | AASS |
| chr19 | 49182589 | 49182997 | 409 | 5 | 3.29E-05 | 0.00048 | 0.000874 | 1.14E-05 | 0.004727202 | TRPM4 |
| chr22 | 37559920 | 37560422 | 503 | 4 | 5.36E-05 | 3.46E-06 | 0.005284 | 1.14E-05 | 0.007605817 | NA |
| chr11 | 424617 | 424667 | 51 | 2 | 1.90E-05 | 1.05E-05 | 0.000143 | 1.14E-05 | 0.01235001 | ANO9 |
| chr3 | 9952047 | 9952632 | 586 | 9 | 5.02E-05 | 0.000103 | 0.004344 | 1.14E-05 | 0.001232374 | PRRT3-AS1, PRRT3 |
| chr11 | 72226471 | 72227127 | 657 | 3 | 1.78E-05 | 6.91E-05 | 9.25E-05 | 1.14E-05 | 0.000637308 | INPPL1 |
| chr6 | 166956003 | 166956788 | 786 | 9 | 1.04E-05 | 3.66E-05 | 0.000498 | 1.15E-05 | 0.000633678 | AL159163.1, RNASET2 |
| chr9 | 109907580 | 109907965 | 386 | 2 | 2.62E-05 | 1.25E-05 | 0.000106 | 1.15E-05 | 0.01005855 | PALM2AKAP2 |
| chr1 | 154627418 | 154628312 | 895 | 12 | 0.00016122 | 3.17E-06 | 0.032909 | 1.15E-05 | 0.002931488 | ADAR |
| chr17 | 49131153 | 49131413 | 261 | 3 | 4.82E-06 | 0.000182 | 3.79E-05 | 1.15E-05 | 0.007234826 | NA |
| chr17 | 40909497 | 40910163 | 667 | 3 | 6.81E-06 | 5.04E-05 | 4.14E-05 | 1.15E-05 | 0.008874526 | NA |
| chr5 | 50966343 | 50967161 | 819 | 5 | 1.58E-05 | 2.71E-05 | 0.000813 | 1.15E-05 | 0.00707424 | AC008808.2 |
| chr15 | 39594102 | 39594940 | 839 | 4 | 3.51E-05 | 1.67E-05 | 0.000237 | 1.15E-05 | 0.006355009 | THBS1, FSIP1 |
| chr11 | 2423014 | 2424212 | 1199 | 16 | 1.70E-06 | 0.00046 | 0.000179 | 1.15E-05 | 0.00212594 | TRPM5 |
| chr13 | 110629037 | 110629132 | 96 | 2 | 0.00015178 | 7.90E-05 | 2.45E-05 | 1.15E-05 | 0.006501326 | NAXD |
| chr10 | 11907839 | 11908052 | 214 | 3 | 4.05E-05 | 4.77E-06 | 0.002257 | 1.15E-05 | 0.004451932 | NA |
| chr17 | 82793996 | 82794311 | 316 | 3 | 1.94E-05 | 8.63E-05 | 9.07E-05 | 1.15E-05 | 0.003286707 | TBCD |
| chr4 | 2229184 | 2229933 | 750 | 4 | 0.00011497 | 1.34E-05 | 0.000818 | 1.15E-05 | 0.002247695 | POLN, HAUS3 |
| chr1 | 33471832 | 33472706 | 875 | 11 | 7.11E-06 | 0.000897 | 2.68E-05 | 1.15E-05 | 0.001040327 | ZSCAN20 |
| chr19 | 13003707 | 13004427 | 721 | 2 | 7.51E-06 | 8.73E-06 | 0.000234 | 1.15E-05 | 0.011333576 | NFIX |
| chr8 | 36784153 | 36784406 | 254 | 6 | 1.70E-05 | 0.00084 | 0.000158 | 1.15E-05 | 0.000168748 | KCNU1 |
| chr19 | 11539529 | 11539984 | 456 | 4 | 2.15E-05 | 2.22E-05 | 0.00064 | 1.15E-05 | -0.01382165 | CNN1 |
| chr3 | 42658793 | 42659550 | 758 | 6 | 4.44E-06 | 2.93E-05 | 0.002523 | 1.15E-05 | -0.00664945 | ZBTB47 |
| chr10 | 128721560 | 128721621 | 62 | 3 | 7.89E-05 | 5.82E-06 | 0.001352 | 1.15E-05 | 0.000351022 | NA |
| chr1 | 22285648 | 22286759 | 1112 | 5 | 0.000153038 | 4.79E-05 | 0.000476 | 1.16E-05 | 0.004325653 | NA |
| chr5 | 170752539 | 170752821 | 283 | 2 | 6.40E-06 | 0.000361 | 1.56E-05 | 1.16E-05 | 0.007786255 | AC008514.1 |
| chr2 | 56182639 | 56183698 | 1060 | 14 | 2.68E-05 | 1.42E-05 | 0.001456 | 1.16E-05 | 0.00158176 | AC007744.1, AC007743.1 |
| chr11 | 36274646 | 36275079 | 434 | 5 | 1.57E-05 | 4.87E-05 | 0.000818 | 1.16E-05 | 0.002382352 | COMMD9 |
| chr1 | 3461305 | 3461814 | 510 | 4 | 6.01E-07 | 0.001051 | 1.16E-05 | 1.16E-05 | 0.002066395 | ARHGEF16 |
| chr9 | 20624152 | 20624968 | 817 | 6 | 0.000136796 | 0.000118 | 0.000162 | 1.16E-05 | 0.003816825 | NA |
| chr16 | 30123024 | 30123561 | 538 | 8 | 6.31E-05 | 0.000161 | 0.001805 | 1.16E-05 | 0.002227671 | MAPK3 |
| chr3 | 9428907 | 9429429 | 523 | 3 | 0.000105369 | 7.26E-06 | 0.001166 | 1.17E-05 | 0.002494716 | SETD5 |
| chr8 | 144619241 | 144619469 | 229 | 3 | 1.79E-05 | 2.81E-05 | 8.06E-05 | 1.17E-05 | 0.002807078 | NA |
| chr2 | 117814440 | 117815092 | 653 | 12 | 2.21E-05 | 0.000173 | 0.001671 | 1.17E-05 | 0.001937225 | DDX18 |
| chr11 | 10807587 | 10808686 | 1100 | 12 | 2.27E-05 | 1.32E-05 | 0.014185 | 1.17E-05 | 0.001228177 | EIF4G2 |
| chr19 | 57745924 | 57746957 | 1034 | 9 | 9.76E-06 | 0.004953 | 0.000544 | 1.17E-05 | 0.00066082 | AC003006.1, ZNF776 |
| chr12 | 5017603 | 5018056 | 454 | 2 | 1.12E-05 | 2.37E-05 | 5.40E-05 | 1.17E-05 | 0.005449407 | AC005906.2 |
| chr17 | 75502046 | 75502696 | 651 | 5 | 1.84E-05 | 7.52E-05 | 0.000705 | 1.17E-05 | 0.002843525 | CASKIN2 |
| chr7 | 1533656 | 1533815 | 160 | 2 | 1.10E-05 | 8.06E-06 | 0.000338 | 1.17E-05 | 0.013236819 | MAFK |
| chr5 | 180351527 | 180352664 | 1138 | 5 | 0.000163857 | 1.43E-05 | 0.00464 | 1.17E-05 | 0.008776664 | GFPT2 |
| chr17 | 73156192 | 73156405 | 214 | 2 | 4.85E-05 | 7.10E-06 | 0.0007 | 1.17E-05 | 0.009161964 | NA |
| chr4 | 4289538 | 4290560 | 1023 | 16 | 5.19E-05 | 1.06E-05 | 0.005791 | 1.17E-05 | 0.000424417 | ZBTB49, LYAR |
| chr4 | 730493 | 730956 | 464 | 5 | 6.11E-05 | 1.58E-05 | 0.000331 | 1.17E-05 | 0.002329004 | PCGF3 |
| chr3 | 78497179 | 78497631 | 453 | 4 | 1.22E-05 | 0.000127 | 0.000333 | 1.17E-05 | 0.000940492 | NA |
| chr7 | 129776128 | 129776154 | 27 | 2 | 3.62E-06 | 7.38E-06 | 0.000545 | 1.18E-05 | 0.015870793 | NA |
| chr17 | 17570356 | 17570579 | 224 | 2 | 2.11E-05 | 2.93E-05 | 4.57E-05 | 1.18E-05 | 0.007970449 | PEMT |
| chr6 | 134909949 | 134910453 | 505 | 2 | 7.71E-05 | 7.29E-06 | 0.000596 | 1.18E-05 | 0.018295774 | NA |
| chr17 | 78894041 | 78894694 | 654 | 2 | 7.54E-05 | 7.94E-06 | 0.000372 | 1.18E-05 | 0.012029266 | TIMP2, CEP295NL |
| chr13 | 102694461 | 102695041 | 581 | 7 | 5.24E-05 | 2.76E-05 | 0.001149 | 1.18E-05 | 0.007271062 | METTL21C |
| chr15 | 63871019 | 63871700 | 682 | 5 | 2.51E-06 | 0.000245 | 1.78E-05 | 1.18E-05 | 0.001096842 | MIR422A |
| chr19 | 14181886 | 14182414 | 529 | 3 | 4.54E-05 | 6.67E-06 | 0.000945 | 1.18E-05 | 0.000466842 | ADGRL1 |

| | | | | | | | | | | |
|---|---|---|---|---|---|---|---|---|---|---|
| chr1 | 203007201 | 203007798 | 598 | 11 | 1.16E-05 | 0.000174 | 0.003263 | 1.18E-05 | 0.000816848 | TMEM183A |
| chr22 | 37608869 | 37610052 | 1184 | 9 | 2.92E-05 | 2.89E-05 | 0.006207 | 1.18E-05 | 0.004125477 | GGA1 |
| chr22 | 47060890 | 47061005 | 116 | 2 | 1.17E-05 | 6.84E-05 | 2.70E-05 | 1.18E-05 | 0.002881918 | TBC1D22A |
| chr20 | 38083490 | 38083840 | 351 | 6 | 5.82E-05 | 6.89E-06 | 0.002223 | 1.18E-05 | 0.003714269 | RPRD1B |
| chr15 | 88531086 | 88531356 | 271 | 3 | 7.83E-05 | 4.89E-06 | 0.002423 | 1.18E-05 | -0.00078558 | AC013489.1, DET1 |
| chr11 | 6603179 | 6604100 | 922 | 16 | 1.11E-05 | 0.000365 | 0.003139 | 1.18E-05 | 0.000321198 | ILK, RRP8, AC091564.2 |
| chr4 | 8865151 | 8865499 | 349 | 5 | 2.78E-05 | 4.38E-06 | 0.00477 | 1.18E-05 | 0.015050818 | HMX1 |
| chr6 | 159762069 | 159763009 | 941 | 13 | 1.04E-06 | 0.000289 | 7.54E-05 | 1.18E-05 | -0.00024744 | ACAT2, SOD2, SOD2-OT1 |
| chr2 | 11124563 | 11124640 | 78 | 2 | 5.99E-05 | 1.05E-05 | 0.000157 | 1.19E-05 | 0.01236072 | AC062028.1 |
| chr16 | 85182037 | 85182563 | 527 | 3 | 3.78E-05 | 6.21E-06 | 0.001375 | 1.19E-05 | 0.022818233 | GSE1 |
| chr6 | 39431090 | 39431797 | 708 | 4 | 3.72E-05 | 1.91E-05 | 0.001483 | 1.19E-05 | 0.007186758 | KIF6 |
| chr3 | 40524287 | 40525486 | 1200 | 15 | 6.87E-05 | 6.18E-05 | 0.001163 | 1.19E-05 | 0.002932109 | ZNF621 |
| chr11 | 111309869 | 111310171 | 303 | 2 | 6.70E-05 | 7.20E-06 | 0.000714 | 1.19E-05 | 0.010115297 | NA |
| chr20 | 20969286 | 20970203 | 918 | 3 | 6.66E-06 | 3.51E-05 | 0.00011 | 1.19E-05 | 0.003565258 | NA |
| chr7 | 32489928 | 32490724 | 797 | 15 | 3.32E-06 | 0.024312 | 0.000492 | 1.19E-05 | 0.00140066 | LSM5 |
| chr8 | 48620315 | 48620797 | 483 | 5 | 7.24E-06 | 4.01E-05 | 0.000194 | 1.19E-05 | 0.004835001 | AC022915.2, AC022915.1, LINC02847 |
| chr17 | 29291330 | 29291826 | 497 | 2 | 3.19E-05 | 7.94E-06 | 0.000405 | 1.19E-05 | 0.000985459 | NUFIP2 |
| chr12 | 105330495 | 105331334 | 840 | 8 | 4.29E-05 | 3.02E-05 | 0.003827 | 1.19E-05 | 0.001087513 | C12orf75 |
| chr3 | 127620632 | 127620802 | 171 | 3 | 6.40E-05 | 8.71E-06 | 0.000341 | 1.19E-05 | 0.004607449 | MCM2, AC023593.1 |
| chr6 | 30341347 | 30342538 | 1192 | 22 | 0.000140913 | 3.35E-05 | 0.001285 | 1.20E-05 | -0.00094848 | TRIM39, TRIM39-RPP21 |
| chr17 | 40364797 | 40365230 | 434 | 8 | 4.55E-05 | 2.52E-05 | 0.001258 | 1.20E-05 | 0.002004132 | NA |
| chr11 | 112280109 | 112280582 | 474 | 6 | 3.52E-06 | 0.002156 | 1.99E-05 | 1.20E-05 | 0.002947073 | LINC02762 |
| chr2 | 85887553 | 85887760 | 208 | 3 | 2.71E-05 | 5.55E-06 | 0.001706 | 1.20E-05 | 0.007011927 | ST3GAL5 |
| chr18 | 76619510 | 76619863 | 354 | 2 | 2.32E-05 | 2.62E-05 | 5.12E-05 | 1.20E-05 | 0.012572622 | LINC00683 |
| chr5 | 149853193 | 149853794 | 602 | 2 | 2.48E-05 | 1.17E-05 | 0.000131 | 1.20E-05 | 0.013126164 | PPARGC1B |
| chr5 | 138337445 | 138338082 | 638 | 9 | 0.000113099 | 2.89E-05 | 0.001995 | 1.20E-05 | 0.001670637 | FAM53C, CDC25C |
| chr11 | 3036669 | 3037248 | 580 | 3 | 3.10E-05 | 8.58E-06 | 0.000343 | 1.20E-05 | 0.011837886 | CARS1-AS1, CARS1 |
| chr5 | 44389180 | 44390113 | 934 | 5 | 1.21E-05 | 2.88E-05 | 0.001085 | 1.20E-05 | 0.001524871 | FGF10-AS1, FGF10 |
| chr10 | 44999987 | 45000789 | 803 | 11 | 3.06E-05 | 5.11E-05 | 0.001539 | 1.20E-05 | 0.005210286 | ZNF22-AS1 |
| chr15 | 89398492 | 89398925 | 434 | 2 | 9.50E-07 | 0.000939 | 1.40E-05 | 1.20E-05 | 0.010979435 | NA |
| chr2 | 30807413 | 30807866 | 454 | 3 | 5.49E-06 | 9.89E-06 | 0.000882 | 1.20E-05 | 0.001830344 | CAPN13 |
| chr2 | 96692700 | 96692846 | 147 | 2 | 4.83E-05 | 1.08E-05 | 0.000155 | 1.20E-05 | 0.00626948 | FER1L5 |
| chr11 | 12008541 | 12009888 | 1348 | 20 | 3.01E-05 | 1.31E-05 | 0.007046 | 1.20E-05 | 0.001494146 | DKK3 |
| chr2 | 188292147 | 188292839 | 693 | 12 | 2.54E-05 | 1.95E-05 | 0.000589 | 1.21E-05 | 0.00348985 | GULP1, LINC01090 |
| chr19 | 45529596 | 45530027 | 432 | 2 | 8.81E-07 | 8.86E-05 | 2.43E-05 | 1.21E-05 | 0.007183209 | OPA3 |
| chr12 | 21946186 | 21946256 | 71 | 2 | 1.95E-05 | 0.001092 | 1.37E-05 | 1.21E-05 | 0.004148606 | NA |
| chr13 | 114024258 | 114024873 | 616 | 3 | 1.52E-05 | 6.79E-05 | 0.000156 | 1.21E-05 | 0.005072034 | RASA3 |
| chr3 | 151330549 | 151330618 | 70 | 2 | 1.65E-05 | 7.34E-06 | 0.000704 | 1.21E-05 | 0.013964459 | MED12L |
| chr10 | 92699426 | 92699908 | 483 | 6 | 4.03E-05 | 1.34E-05 | 0.001868 | 1.21E-05 | 5.44E-05 | NA |
| chr17 | 7313462 | 7314048 | 587 | 3 | 5.32E-05 | 2.26E-05 | 0.000522 | 1.21E-05 | -0.0052105 | GPS2, AC026954.2 |
| chr1 | 24150042 | 24150371 | 330 | 4 | 3.19E-05 | 1.39E-05 | 0.000202 | 1.21E-05 | 0.010425017 | NA |
| chr6 | 129672188 | 129672522 | 335 | 2 | 8.54E-06 | 0.000373 | 1.61E-05 | 1.21E-05 | 0.019786424 | ARHGAP18 |
| chr15 | 89628962 | 89630095 | 1134 | 5 | 8.21E-05 | 7.46E-05 | 0.000781 | 1.21E-05 | 0.001122361 | TICRR, KIF7 |
| chr6 | 116003133 | 116003799 | 667 | 3 | 2.12E-06 | 8.14E-05 | 0.000465 | 1.21E-05 | 0.012304865 | FRK |
| chr4 | 107931297 | 107932191 | 895 | 10 | 3.95E-06 | 7.28E-05 | 0.001378 | 1.21E-05 | -6.06E-05 | CYP2U1, CYP2U1-AS1 |
| chr11 | 17386890 | 17387122 | 233 | 2 | 4.55E-06 | 9.09E-05 | 2.42E-05 | 1.21E-05 | 0.011068391 | KCNJ11 |
| chr15 | 41604382 | 41604482 | 101 | 2 | 2.97E-05 | 7.53E-06 | 0.000605 | 1.21E-05 | 0.013910323 | NA |
| chr3 | 111541909 | 111542456 | 548 | 8 | 5.20E-05 | 7.33E-06 | 0.004661 | 1.21E-05 | 0.002632213 | CD96 |
| chr7 | 41882493 | 41882930 | 438 | 4 | 2.60E-05 | 3.06E-05 | 0.000261 | 1.21E-05 | -0.00203166 | NA |
| chr20 | 51578310 | 51578951 | 642 | 2 | 0.000174632 | 7.35E-06 | 0.00073 | 1.21E-05 | 0.008141294 | NA |
| chr8 | 37899424 | 37900028 | 605 | 9 | 1.60E-06 | 0.002015 | 9.01E-06 | 1.22E-05 | 0.000570614 | RAB11FIP1 |
| chr8 | 119870447 | 119871004 | 558 | 2 | 4.49E-05 | 8.21E-06 | 0.000383 | 1.22E-05 | -0.00593933 | AP005717.1 |
| chr1 | 16164296 | 16165130 | 835 | 5 | 4.12E-05 | 8.13E-06 | 0.001031 | 1.22E-05 | 0.009223879 | NA |
| chr6 | 151807730 | 151809072 | 1343 | 11 | 2.90E-06 | 8.35E-05 | 0.002256 | 1.22E-05 | 0.006495115 | ESR1 |
| chr5 | 17918250 | 17918341 | 92 | 2 | 5.31E-05 | 7.59E-06 | 0.000601 | 1.22E-05 | 0.02679421 | LINC02223 |
| chr16 | 30445440 | 30445994 | 555 | 8 | 1.78E-05 | 0.000594 | 0.001724 | 1.22E-05 | -0.00196536 | SEPHS2 |
| chr5 | 179963115 | 179963259 | 145 | 2 | 3.47E-05 | 1.58E-05 | 8.69E-05 | 1.22E-05 | -0.0064645 | RNF130 |
| chr1 | 208252235 | 208252455 | 221 | 3 | 8.68E-05 | 4.95E-06 | 0.002446 | 1.22E-05 | 0.024590867 | NA |
| chr12 | 57625864 | 57626114 | 251 | 3 | 7.26E-05 | 6.51E-06 | 0.001461 | 1.22E-05 | 0.015123265 | AC025165.6, SLC26A10, B4GALNT1 |
| chr1 | 201946861 | 201947517 | 657 | 3 | 3.11E-05 | 4.18E-05 | 0.000495 | 1.22E-05 | 0.006796121 | NA |
| chr7 | 44606467 | 44606672 | 206 | 7 | 0.00031503 | 4.12E-05 | 0.001142 | 1.22E-05 | 0.000775858 | OGDH |
| chr3 | 122563810 | 122564837 | 1028 | 16 | 3.30E-06 | 0.000728 | 0.000127 | 1.22E-05 | 0.001279083 | DTX3L, PARP9 |
| chr6 | 142838510 | 142838629 | 120 | 3 | 5.67E-05 | 1.49E-05 | 0.000376 | 1.22E-05 | -0.00365653 | HIVEP2 |
| chr10 | 594798 | 595701 | 904 | 7 | 1.80E-05 | 0.000162 | 0.002658 | 1.22E-05 | 0.00400382 | DIP2C |
| chr1 | 208107167 | 208107509 | 343 | 2 | 2.68E-05 | 9.89E-06 | 0.000201 | 1.22E-05 | 0.005916711 | AL590138.1, PLXNA2 |
| chr8 | 23287371 | 23287947 | 577 | 6 | 7.47E-06 | 2.10E-05 | 0.000964 | 1.23E-05 | 0.007736653 | R3HCC1 |
| chr8 | 131309452 | 131309670 | 219 | 2 | 2.85E-05 | 2.80E-05 | 4.98E-05 | 1.23E-05 | -0.00251864 | AC104257.1 |
| chr20 | 22578667 | 22579229 | 563 | 7 | 1.84E-05 | 2.61E-05 | 0.001128 | 1.23E-05 | 0.006382354 | NA |
| chr22 | 31159684 | 31160581 | 898 | 12 | 1.10E-05 | 0.002849 | 0.000363 | 1.23E-05 | 0.000174002 | RNF185, MIR3928 |
| chr22 | 22380570 | 22380591 | 22 | 3 | 3.21E-05 | 1.26E-05 | 0.000205 | 1.23E-05 | 0.01501627 | NA |
| chr16 | 85272120 | 85272285 | 166 | 3 | 4.43E-05 | 2.35E-05 | 0.000342 | 1.23E-05 | 0.003096202 | GSE1, AC092275.1 |
| chr11 | 72779826 | 72780547 | 722 | 4 | 5.95E-06 | 0.000595 | 0.000289 | 1.23E-05 | 0.005022337 | ARAP1, STARD10 |
| chr1 | 180198974 | 180199032 | 59 | 2 | 3.49E-05 | 1.15E-05 | 0.000144 | 1.23E-05 | 0.005858879 | QSOX1 |
| chr3 | 33797897 | 33798821 | 925 | 16 | 1.80E-05 | 7.01E-05 | 0.005838 | 1.23E-05 | -0.00099031 | PDCD6IP, AC112220.2 |
| chr8 | 8821940 | 8822325 | 386 | 3 | 9.81E-06 | 2.63E-05 | 0.000549 | 1.23E-05 | 0.009442231 | MFHAS1 |
| chr1 | 6634271 | 6634766 | 496 | 4 | 6.10E-05 | 6.60E-06 | 0.001392 | 1.23E-05 | 0.006114242 | THAP3, DNAJC11 |
| chr8 | 948407 | 949171 | 765 | 3 | 2.13E-05 | 5.17E-06 | 0.002405 | 1.24E-05 | 0.009864321 | DLGAP2 |
| chr3 | 134527288 | 134527837 | 550 | 2 | 4.97E-05 | 9.18E-06 | 0.000261 | 1.24E-05 | 0.00654692 | CEP63 |
| chr1 | 15337686 | 15337889 | 204 | 3 | 5.32E-05 | 4.91E-06 | 0.00262 | 1.24E-05 | -0.00420588 | FHAD1, FHAD1-AS1 |
| chr19 | 5250365 | 5250644 | 280 | 4 | 5.55E-05 | 1.81E-05 | 0.002486 | 1.24E-05 | 0.006476422 | PTPRS |
| chr7 | 97731889 | 97732096 | 208 | 5 | 0.000324534 | 3.91E-06 | 0.003459 | 1.24E-05 | 0.005972055 | TAC1 |
| chr7 | 151035587 | 151035927 | 341 | 3 | 6.01E-05 | 8.23E-06 | 0.000601 | 1.24E-05 | 0.003436177 | ABCB8 |
| chr14 | 51651115 | 51651970 | 856 | 8 | 2.00E-05 | 0.00012 | 0.004746 | 1.24E-05 | 0.00066563 | FRMD6, FRMD6-AS1 |
| chr11 | 47988026 | 47988101 | 76 | 2 | 5.68E-05 | 1.45E-05 | 0.0001 | 1.24E-05 | 0.012176686 | PTPRJ |
| chr17 | 31095345 | 31095864 | 520 | 5 | 5.85E-06 | 0.000223 | 8.87E-05 | 1.24E-05 | 0.001175381 | NF1, AC138207.2 |
| chr11 | 8092758 | 8092849 | 92 | 2 | 5.45E-05 | 7.91E-06 | 0.00053 | 1.24E-05 | 0.014915329 | TUB |
| chr11 | 8283624 | 8283755 | 132 | 2 | 4.98E-05 | 1.82E-05 | 7.62E-05 | 1.25E-05 | -0.00088383 | NA |
| chr1 | 66929482 | 66930052 | 571 | 6 | 7.19E-05 | 2.74E-05 | 0.000577 | 1.25E-05 | -0.00414663 | MIER1 |
| chr6 | 28861169 | 28861969 | 801 | 12 | 9.36E-06 | 0.000257 | 0.003752 | 1.25E-05 | 0.001867216 | RPL13P, LINC01623 |
| chr16 | 3348367 | 3348766 | 400 | 2 | 1.15E-05 | 6.54E-05 | 2.95E-05 | 1.25E-05 | 0.01900369 | NA |
| chr2 | 232538353 | 232538671 | 319 | 3 | 8.45E-07 | 0.003575 | 7.19E-06 | 1.25E-05 | 0.003051417 | NA |
| chr7 | 116771521 | 116771874 | 354 | 3 | 5.20E-05 | 6.61E-06 | 0.00115 | 1.25E-05 | 0.000167278 | MET |
| chr5 | 135030687 | 135031704 | 1018 | 8 | 5.78E-05 | 3.12E-06 | 0.016203 | 1.25E-05 | 0.00333634 | PITX1 |
| chr10 | 119458220 | 119458516 | 297 | 2 | 3.46E-05 | 1.75E-05 | 8.01E-05 | 1.25E-05 | 0.005007877 | GRK5 |
| chr2 | 68366114 | 68366681 | 568 | 3 | 1.67E-05 | 0.00204 | 0.000101 | 1.25E-05 | 0.008984287 | PLEK |
| chr17 | 13602434 | 13602967 | 534 | 5 | 6.41E-05 | 2.36E-05 | 0.00027 | 1.25E-05 | 0.012738966 | NA |
| chr9 | 132154640 | 132154657 | 18 | 2 | 3.11E-05 | 1.68E-05 | 8.38E-05 | 1.25E-05 | 0.005172919 | NA |
| chr8 | 90082830 | 90083678 | 849 | 6 | 3.14E-05 | 1.84E-05 | 0.004703 | 1.25E-05 | 0.005601046 | CALB1 |
| chr14 | 22982118 | 22982902 | 785 | 8 | 7.87E-05 | 9.64E-05 | 0.004508 | 1.25E-05 | 0.000221128 | AL132780.2, AL132780.3, AJUBA |
| chr17 | 39721093 | 39721572 | 480 | 2 | 3.20E-06 | 1.82E-05 | 7.70E-05 | 1.25E-05 | -0.00404754 | ERBB2 |
| chr1 | 3400788 | 3401356 | 569 | 3 | 4.88E-05 | 8.58E-06 | 0.001217 | 1.25E-05 | -0.00524565 | PRDM16 |
| chr17 | 8408276 | 8408518 | 243 | 4 | 6.34E-05 | 1.43E-05 | 0.000234 | 1.25E-05 | 0.006997924 | NA |
| chr22 | 40407124 | 40407325 | 202 | 2 | 4.15E-06 | 1.32E-05 | 0.000118 | 1.25E-05 | 0.004904106 | AL022238.3, SGSM3 |
| chr3 | 126315605 | 126315997 | 393 | 2 | 6.25E-06 | 0.000424 | 1.61E-05 | 1.25E-05 | -0.00486621 | AC063919.2 |
| chr1 | 1939839 | 1940594 | 756 | 5 | 5.30E-05 | 0.000124 | 0.000258 | 1.25E-05 | 0.004808867 | CFAP74 |
| chr19 | 4867229 | 4867885 | 657 | 7 | 1.65E-07 | 0.00185 | 1.83E-05 | 1.25E-05 | -4.12E-05 | PLIN3 |
| chr21 | 43920003 | 43920032 | 30 | 3 | 1.51E-05 | 0.000659 | 1.16E-05 | 1.25E-05 | 0.002619121 | AGPAT3 |
| chr1 | 26282437 | 26282657 | 221 | 2 | 3.87E-05 | 2.19E-05 | 6.37E-05 | 1.25E-05 | 0.004759106 | UBXN11 |
| chr10 | 89476844 | 89477290 | 447 | 3 | 2.34E-06 | 0.00761 | 7.68E-06 | 1.25E-05 | 0.00175933 | SLC16A12 |
| chr1 | 230120647 | 230121320 | 674 | 2 | 8.11E-06 | 1.76E-05 | 8.05E-05 | 1.25E-05 | 0.005706049 | GALNT2 |
| chr15 | 89690362 | 89691001 | 640 | 16 | 1.57E-05 | 0.000422 | 0.000603 | 1.25E-05 | 0.001980027 | WDR93, PEX11A |
| chr16 | 86059967 | 86060720 | 754 | 4 | 8.92E-05 | 1.72E-05 | 0.001659 | 1.25E-05 | 0.007861262 | NA |
| chr10 | 132236523 | 132236793 | 271 | 2 | 4.41E-05 | 1.62E-05 | 8.87E-05 | 1.25E-05 | 0.007303335 | STK32C |
| chr22 | 20192862 | 20193062 | 201 | 2 | 1.72E-05 | 1.57E-05 | 9.26E-05 | 1.25E-05 | 0.006553258 | NA |
| chr10 | 103838756 | 103839268 | 513 | 3 | 1.85E-06 | 0.002954 | 9.01E-06 | 1.26E-05 | -0.00465475 | SH3PXD2A |
| chr8 | 42471449 | 42471806 | 358 | 2 | 5.45E-05 | 7.62E-06 | 0.000749 | 1.26E-05 | -0.0048881 | SLC20A2 |
| chr20 | 36236035 | 36236164 | 130 | 4 | 7.39E-05 | 0.000101 | 7.66E-05 | 1.26E-05 | 0.005856049 | NA |
| chr17 | 45034158 | 45034264 | 107 | 2 | 5.21E-05 | 1.13E-05 | 0.000156 | 1.26E-05 | 0.005137909 | NMT1, DCAKD |
| chr4 | 73435392 | 73435590 | 199 | 2 | 4.38E-06 | 1.66E-05 | 8.66E-05 | 1.26E-05 | 0.010584229 | AFP |
| chr11 | 117478633 | 117478963 | 331 | 2 | 2.89E-05 | 2.10E-05 | 6.72E-05 | 1.26E-05 | 6.06E-05 | DSCAML1 |
| chr6 | 40805220 | 40805645 | 426 | 2 | 3.09E-05 | 9.09E-06 | 0.000295 | 1.26E-05 | 0.008935014 | NA |
| chr6 | 16064692 | 16064913 | 222 | 3 | 6.96E-05 | 8.24E-06 | 0.001263 | 1.26E-05 | 0.006929519 | NA |
| chr2 | 113243084 | 113243100 | 17 | 2 | 2.07E-05 | 0.000102 | 2.39E-05 | 1.26E-05 | 0.005444999 | PAX8-AS1, PAX8 |
| chr1 | 12018764 | 12019954 | 1191 | 17 | 1.23E-05 | 0.000563 | 0.00213 | 1.26E-05 | -0.00110516 | MIIP |
| chr12 | 81269677 | 81269793 | 117 | 3 | 4.04E-05 | 1.45E-05 | 0.000204 | 1.26E-05 | 0.011443048 | PPFIA2 |
| chr2 | 119655629 | 119655869 | 241 | 5 | 3.08E-05 | 7.87E-05 | 0.000316 | 1.26E-05 | 0.008653038 | CFAP221 |
| chr1 | 26850958 | 26851561 | 604 | 2 | 7.78E-06 | 1.32E-05 | 0.00012 | 1.26E-05 | 0.002442536 | ZDHHC18 |
| chr17 | 4984229 | 4984534 | 306 | 2 | 3.12E-05 | 1.11E-05 | 0.000167 | 1.27E-05 | 0.011325556 | CAMTA2 |
| chr15 | 59668962 | 59668987 | 26 | 2 | 4.87E-05 | 1.35E-05 | 0.000117 | 1.27E-05 | 0.014878544 | AC092755.1, BNIP2 |
| chr11 | 123607079 | 123607646 | 568 | 3 | 1.83E-06 | 0.006441 | 6.28E-06 | 1.27E-05 | 0.001306131 | GRAMD1B |
| chr2 | 240026503 | 240026837 | 335 | 2 | 6.56E-06 | 7.99E-06 | 0.000573 | 1.27E-05 | 0.015865392 | NA |
| chr13 | 37105187 | 37105841 | 655 | 6 | 1.02E-05 | 5.48E-05 | 0.000369 | 1.27E-05 | 0.002650464 | CSNK1A1L |
| chr4 | 3784045 | 3784200 | 156 | 2 | 6.87E-05 | 8.57E-06 | 0.000394 | 1.27E-05 | 0.002587626 | NA |
| chr5 | 1266259 | 1267058 | 800 | 4 | 8.68E-05 | 1.42E-05 | 0.00241 | 1.27E-05 | 0.001180831 | TERT |
| chr14 | 74069478 | 74069484 | 7 | 2 | 6.81E-05 | 1.03E-05 | 0.000204 | 1.27E-05 | 0.000949361 | BBOF1, ALDH6A1 |
| chr19 | 39599000 | 39599322 | 323 | 2 | 1.73E-05 | 6.27E-05 | 3.09E-05 | 1.27E-05 | 0.011337396 | NA |
| chr15 | 98485430 | 98486140 | 711 | 2 | 5.85E-05 | 7.83E-06 | 0.00066 | 1.27E-05 | 0.008486165 | FAM169B |
| chr22 | 36365907 | 36366464 | 558 | 4 | 1.15E-05 | 1.67E-05 | 0.000134 | 1.27E-05 | 0.006319971 | MYH9 |
| chr15 | 52528816 | 52529720 | 905 | 9 | 1.86E-05 | 0.000148 | 0.000515 | 1.27E-05 | 0.001102852 | MYO5A |
| chr19 | 1419465 | 1419753 | 289 | 2 | 2.03E-05 | 1.46E-05 | 0.000104 | 1.27E-05 | 0.00201798 | DAZAP1 |
| chr10 | 113045106 | 113045247 | 142 | 2 | 6.18E-05 | 1.10E-05 | 0.000175 | 1.27E-05 | 0.005307832 | TCF7L2 |
| chr14 | 103848397 | 103848886 | 490 | 4 | 9.89E-05 | 4.97E-05 | 0.000385 | 1.27E-05 | 0.007149638 | LINC00637 |
| chr19 | 49468408 | 49468953 | 546 | 2 | 2.90E-05 | 1.35E-05 | 0.000118 | 1.27E-05 | 0.007263773 | ALDH16A1, AC010619.1 |
| chr10 | 4457801 | 4458242 | 442 | 3 | 4.38E-05 | 9.77E-06 | 0.000742 | 1.27E-05 | 0.000661613 | NA |
| chr14 | 101480831 | 101481192 | 362 | 3 | 5.25E-05 | 1.57E-05 | 0.000149 | 1.27E-05 | 0.014551538 | NA |
| chr1 | 174998049 | 174999268 | 1220 | 10 | 1.14E-06 | 3.56E-05 | 0.001406 | 1.27E-05 | 0.007200076 | CACYBP, Z99127.3 |
| chr2 | 237967482 | 237968523 | 1042 | 5 | 7.33E-05 | 7.13E-06 | 0.002701 | 1.27E-05 | 0.005998489 | UBE2F, UBE2F-SCLY |
| chr16 | 56675591 | 56676186 | 596 | 3 | 6.97E-05 | 7.85E-06 | 0.000561 | 1.27E-05 | 0.009458657 | MT1IP |
| chr3 | 46368680 | 46368931 | 252 | 4 | 6.19E-05 | 4.46E-06 | 0.002927 | 1.28E-05 | 0.012194502 | CCR5AS |
| chr2 | 70257482 | 70258624 | 1143 | 17 | 2.25E-05 | 2.69E-05 | 0.006055 | 1.28E-05 | 0.000390417 | PCYOX1 |
| chr11 | 76257001 | 76257147 | 147 | 2 | 1.79E-05 | 0.001055 | 1.45E-05 | 1.28E-05 | -0.0052719 | NA |
| chr3 | 150237052 | 150237523 | 472 | 5 | 9.45E-05 | 3.51E-06 | 0.00887 | 1.28E-05 | 0.011629575 | NA |
| chr19 | 32336848 | 32337248 | 401 | 3 | 2.36E-05 | 6.76E-05 | 3.54E-05 | 1.28E-05 | 0.00585553 | NA |
| chr2 | 720516 | 720852 | 337 | 2 | 6.82E-05 | 9.75E-06 | 0.000248 | 1.28E-05 | 0.004851706 | NA |
| chr20 | 3781269 | 3781873 | 605 | 8 | 8.67E-06 | 0.000111 | 0.001189 | 1.28E-05 | 0.003257969 | SPEF1 |
| chr7 | 105690863 | 105691465 | 603 | 5 | 9.04E-06 | 2.82E-05 | 0.000218 | 1.28E-05 | 0.004121621 | ATXN7L1 |
| chr10 | 70499169 | 70499947 | 779 | 2 | 9.53E-06 | 1.72E-05 | 8.54E-05 | 1.28E-05 | 0.01409413 | PALD1 |
| chr1 | 28260222 | 28260737 | 516 | 5 | 2.37E-05 | 6.08E-05 | 0.000546 | 1.28E-05 | 0.005717069 | SESN2 |
| chr16 | 85436879 | 85437068 | 190 | 3 | 4.29E-05 | 6.95E-05 | 6.50E-05 | 1.28E-05 | 0.011009963 | GSE1 |
| chr19 | 4096127 | 4096187 | 61 | 2 | 6.14E-05 | 1.40E-05 | 0.000113 | 1.28E-05 | 0.007828959 | MAP2K2 |
| chr12 | 106736564 | 106736590 | 27 | 2 | 7.17E-05 | 9.16E-06 | 0.000311 | 1.28E-05 | 0.020762152 | RFX4, AC079385.1 |
| chr3 | 50340384 | 50341643 | 1260 | 16 | 6.93E-05 | 0.000188 | 0.000432 | 1.28E-05 | 0.000467724 | ZMYND10-AS1, RASSF1, ZMYND10 |

| | | | | | | | | | | |
|---|---|---|---|---|---|---|---|---|---|---|
| chr11 | 70816452 | 70817424 | 973 | 3 | 3.08E-05 | 9.20E-06 | 0.000348 | 1.28E-05 | 0.003342399 | SHANK2 |
| chr5 | 65198507 | 65198933 | 427 | 3 | 2.17E-05 | 9.10E-05 | 0.000445 | 1.28E-05 | -0.01192439 | ADAMTS6 |
| chr6 | 34027640 | 34028334 | 695 | 6 | 3.39E-05 | 4.33E-05 | 0.000629 | 1.28E-05 | 0.002591413 | GRM4 |
| chr7 | 51315422 | 51315964 | 543 | 3 | 2.94E-05 | 8.06E-06 | 0.001051 | 1.28E-05 | 0.006224018 | COBL |
| chr5 | 141869534 | 141870255 | 722 | 5 | 1.07E-05 | 0.000331 | 0.000408 | 1.28E-05 | 0.005891021 | PCDH1 |
| chr15 | 22917910 | 22918780 | 871 | 6 | 6.12E-06 | 8.12E-05 | 0.000166 | 1.28E-05 | 0.002500401 | CYFIP1 |
| chr9 | 35790474 | 35791193 | 720 | 6 | 1.56E-05 | 9.51E-06 | 0.000759 | 1.28E-05 | -0.00648604 | NA |
| chr2 | 2185007 | 2185429 | 423 | 2 | 1.41E-06 | 0.000122 | 2.29E-05 | 1.28E-05 | 0.006302276 | MYT1L |
| chr4 | 183658670 | 183659355 | 686 | 10 | 2.51E-05 | 0.000297 | 0.004053 | 1.28E-05 | -0.00085853 | TRAPPC11, RWDD4 |
| chr17 | 17872707 | 17873308 | 602 | 3 | 3.38E-05 | 9.32E-06 | 0.000766 | 1.29E-05 | 0.00781193 | TOM1L2 |
| chr6 | 3748122 | 3748421 | 300 | 2 | 4.35E-05 | 1.11E-05 | 0.000176 | 1.29E-05 | 0.008356225 | PXDC1 |
| chr8 | 90939339 | 90940069 | 731 | 4 | 1.35E-05 | 1.58E-05 | 0.001008 | 1.29E-05 | 0.004632438 | NECAB1 |
| chr22 | 40521439 | 40521793 | 355 | 2 | 5.25E-06 | 0.008122 | 1.24E-05 | 1.29E-05 | -0.00285235 | MRTFA |
| chr7 | 50794053 | 50794569 | 517 | 9 | 5.14E-05 | 5.23E-05 | 0.003238 | 1.29E-05 | 0.000862825 | NA |
| chr6 | 70221851 | 70221967 | 117 | 3 | 3.48E-05 | 8.35E-05 | 0.000244 | 1.29E-05 | 0.013302528 | COL9A1 |
| chr22 | 41828777 | 41828788 | 12 | 2 | 4.74E-05 | 1.03E-05 | 0.000216 | 1.29E-05 | 0.006474474 | CCDC134 |
| chr8 | 97564467 | 97564692 | 226 | 3 | 2.63E-05 | 1.55E-05 | 0.000164 | 1.29E-05 | -0.00343596 | NA |
| chr8 | 116912865 | 116913538 | 674 | 2 | 0.000112391 | 8.50E-06 | 0.000462 | 1.29E-05 | 0.010052374 | NA |
| chr4 | 186889068 | 186889459 | 392 | 3 | 1.40E-05 | 9.48E-05 | 5.21E-05 | 1.30E-05 | -0.00405348 | NA |
| chr18 | 48952785 | 48953167 | 383 | 3 | 3.20E-05 | 1.84E-05 | 0.000132 | 1.30E-05 | 0.006832554 | NA |
| chr3 | 9808874 | 9809488 | 615 | 3 | 0.000138745 | 2.73E-05 | 0.000166 | 1.30E-05 | 0.006275466 | ARPC4-TTLL3, TTLL3 |
| chr1 | 201497609 | 201498043 | 435 | 2 | 1.94E-05 | 1.36E-05 | 0.000121 | 1.30E-05 | 0.005533293 | CSRP1 |
| chr1 | 6494631 | 6494817 | 187 | 2 | 4.72E-05 | 7.92E-06 | 0.00074 | 1.30E-05 | 0.006834259 | PLEKHG5 |
| chr5 | 141753770 | 141754281 | 512 | 3 | 2.63E-05 | 1.51E-05 | 0.000788 | 1.30E-05 | 0.006992782 | AC005753.2 |
| chr17 | 19716592 | 19717282 | 691 | 5 | 5.76E-05 | 6.59E-06 | 0.003522 | 1.30E-05 | 0.006779434 | SLC47A2 |
| chr5 | 35230064 | 35231170 | 1107 | 11 | 2.70E-05 | 0.000132 | 0.003002 | 1.30E-05 | -0.00655443 | PRLR |
| chr17 | 80087320 | 80087933 | 614 | 7 | 3.13E-05 | 1.84E-05 | 0.004028 | 1.30E-05 | -0.00050759 | CCDC40 |
| chr1 | 98046381 | 98047314 | 934 | 4 | 8.90E-05 | 1.44E-05 | 0.000186 | 1.30E-05 | 0.017445223 | MIR137HG |
| chr12 | 46219962 | 46220724 | 763 | 2 | 2.09E-05 | 1.38E-05 | 0.00012 | 1.30E-05 | 0.021226242 | SLC38A1 |
| chr15 | 75234040 | 75234379 | 340 | 3 | 3.21E-05 | 6.64E-06 | 0.000965 | 1.30E-05 | 0.008558897 | AC113208.3 |
| chr8 | 22636418 | 22636969 | 552 | 2 | 4.31E-06 | 1.04E-05 | 0.000218 | 1.30E-05 | 0.003825828 | BIN3 |
| chr16 | 89000179 | 89000555 | 377 | 4 | 2.77E-05 | 0.000268 | 0.0002 | 1.30E-05 | 0.009202155 | NA |
| chr6 | 166308271 | 166308677 | 407 | 4 | 3.83E-05 | 5.10E-06 | 0.003761 | 1.30E-05 | 0.005122225 | PRR18 |
| chr4 | 103019554 | 103020143 | 590 | 12 | 0.000123153 | 2.87E-06 | 0.025633 | 1.31E-05 | -0.00339148 | SLC9B1, SLC9B2 |
| chr22 | 42719931 | 42720632 | 702 | 5 | 8.15E-06 | 0.000724 | 0.000167 | 1.31E-05 | 0.004131826 | A4GALT |
| chr22 | 29049382 | 29049915 | 534 | 4 | 6.71E-05 | 1.81E-05 | 0.000473 | 1.31E-05 | -0.00205399 | ZNRF3 |
| chr10 | 49609167 | 49609764 | 598 | 5 | 3.52E-05 | 8.39E-05 | 0.000573 | 1.31E-05 | 0.004823468 | CHAT |
| chr16 | 13205847 | 13206143 | 297 | 2 | 4.60E-05 | 1.33E-05 | 0.000128 | 1.31E-05 | -0.01444103 | SHISA9 |
| chr18 | 48993417 | 48994101 | 685 | 3 | 1.39E-05 | 2.16E-05 | 0.00019 | 1.31E-05 | 0.005154223 | NA |
| chr11 | 7848691 | 7848857 | 167 | 2 | 0.000279909 | 1.11E-05 | 0.000185 | 1.31E-05 | 0.013434741 | OR5E1P, AC044810.3, AC044810.2 |
| chr16 | 1370040 | 1370536 | 497 | 5 | 0.000128877 | 3.52E-06 | 0.0078 | 1.31E-05 | 0.002690538 | UNKL |
| chr19 | 48716845 | 48717228 | 384 | 6 | 2.99E-05 | 4.78E-05 | 0.001671 | 1.31E-05 | 0.006437958 | MAMSTR |
| chr1 | 67165729 | 67166345 | 617 | 4 | 0.000103201 | 9.67E-06 | 0.002229 | 1.31E-05 | -0.00054425 | IL23R, C1orf141 |
| chr22 | 35551562 | 35551771 | 210 | 3 | 0.000102594 | 5.55E-06 | 0.002116 | 1.31E-05 | 0.003428161 | RASD2 |
| chr17 | 1011526 | 1012025 | 500 | 2 | 1.00E-05 | 8.67E-06 | 0.000453 | 1.31E-05 | 0.004281884 | ABR |
| chr20 | 64079420 | 64080180 | 761 | 9 | 2.41E-05 | 0.00042 | 0.001469 | 1.31E-05 | -0.00177676 | OPRL1, RGS19 |
| chr16 | 47143692 | 47144812 | 1121 | 10 | 1.41E-06 | 0.017339 | 3.12E-05 | 1.32E-05 | 0.00113453 | ITFG1-AS1, NETO2 |
| chr1 | 27604773 | 27604992 | 220 | 3 | 3.25E-05 | 8.44E-06 | 0.000595 | 1.32E-05 | 0.008014645 | NA |
| chr4 | 89894142 | 89894822 | 681 | 3 | 5.01E-05 | 6.00E-05 | 0.000412 | 1.32E-05 | 0.005569728 | MMRN1 |
| chr9 | 128687827 | 128688696 | 870 | 7 | 0.00019175 | 8.59E-06 | 0.007384 | 1.32E-05 | 0.00496777 | SET |
| chr17 | 7903071 | 7903803 | 733 | 6 | 4.24E-05 | 5.56E-05 | 0.000525 | 1.32E-05 | -0.00149852 | CHD3 |
| chr1 | 19532346 | 19533099 | 754 | 4 | 4.88E-05 | 1.63E-05 | 0.000215 | 1.32E-05 | 0.001937617 | MICOS10 |
| chr19 | 17727583 | 17728006 | 424 | 3 | 6.00E-06 | 2.13E-05 | 0.00023 | 1.32E-05 | 0.003588047 | MAP1S, AC008761.1 |
| chr1 | 110337830 | 110338957 | 1128 | 8 | 8.52E-08 | 0.018805 | 1.36E-05 | 1.32E-05 | -9.78E-05 | RBM15, RBM15-AS1 |
| chr2 | 169071412 | 169071741 | 330 | 3 | 3.12E-05 | 4.27E-05 | 0.00011 | 1.32E-05 | 0.01320714 | DHRS9 |
| chr21 | 44682294 | 44682651 | 358 | 4 | 9.52E-06 | 7.74E-05 | 9.99E-05 | 1.32E-05 | 0.007272413 | TSPEAR |
| chr9 | 33044613 | 33045035 | 423 | 3 | 8.69E-06 | 2.00E-05 | 0.00034 | 1.32E-05 | 0.014502987 | SMU1 |
| chr2 | 159712178 | 159712646 | 469 | 9 | 1.94E-05 | 0.000313 | 0.000145 | 1.32E-05 | 0.000224094 | MARCHF7, AC009961.1 |
| chr2 | 238644242 | 238644765 | 524 | 3 | 7.26E-05 | 6.17E-06 | 0.001897 | 1.32E-05 | -0.06687555 | LINC01937 |
| chr12 | 118151124 | 118151282 | 159 | 3 | 5.15E-05 | 2.76E-05 | 9.33E-05 | 1.32E-05 | -0.00161831 | TAOK3 |
| chr7 | 5488038 | 5489070 | 1033 | 6 | 1.42E-05 | 8.59E-05 | 0.000719 | 1.32E-05 | -0.01323647 | FBXL18 |
| chr6 | 2770276 | 2770381 | 106 | 2 | 7.35E-05 | 8.14E-06 | 0.000708 | 1.32E-05 | -0.02917765 | WRNIP1 |
| chr12 | 53205556 | 53206022 | 467 | 3 | 3.71E-05 | 8.59E-06 | 0.00114 | 1.32E-05 | 0.010799541 | ITGB7 |
| chr19 | 15614644 | 15615063 | 420 | 2 | 3.71E-05 | 8.70E-06 | 0.000467 | 1.33E-05 | 0.019719385 | NA |
| chr8 | 98423997 | 98424562 | 566 | 2 | 1.65E-05 | 1.47E-05 | 0.000111 | 1.33E-05 | -0.01102851 | STK3 |
| chr15 | 101333584 | 101333715 | 132 | 2 | 2.68E-05 | 0.00013 | 2.31E-05 | 1.33E-05 | -0.00280832 | PCSK6 |
| chr11 | 78495524 | 78495667 | 144 | 3 | 2.90E-06 | 0.211044 | 5.57E-06 | 1.33E-05 | -0.00178735 | NARS2 |
| chr21 | 17799024 | 17799717 | 694 | 3 | 7.63E-05 | 6.88E-06 | 0.001158 | 1.33E-05 | 0.016940191 | AL109761.1, C21orf91 |
| chr3 | 151279583 | 151279900 | 318 | 2 | 1.19E-06 | 0.005121 | 1.29E-05 | 1.33E-05 | 0.013066075 | MED12L |
| chr8 | 346808 | 347436 | 629 | 3 | 7.41E-05 | 2.97E-05 | 0.000639 | 1.33E-05 | 0.01322149 | NA |
| chr12 | 121400188 | 121401221 | 1034 | 5 | 2.75E-05 | 1.52E-05 | 0.003304 | 1.33E-05 | 0.002972303 | RNF34 |
| chr6 | 12585831 | 12586300 | 470 | 3 | 7.54E-06 | 3.16E-05 | 0.000259 | 1.33E-05 | -0.00620516 | NA |
| chr14 | 64387300 | 64388305 | 1006 | 12 | 6.66E-05 | 7.46E-05 | 0.004356 | 1.33E-05 | 0.002511129 | MTHFD1, TEX21P |
| chr22 | 24420713 | 24421629 | 917 | 3 | 2.18E-05 | 1.37E-05 | 0.000333 | 1.33E-05 | 0.008407306 | SPECC1L-ADORA2A, ADORA2A |
| chr4 | 689042 | 689587 | 546 | 10 | 7.83E-05 | 2.66E-06 | 0.019315 | 1.33E-05 | 0.007511816 | SLC49A3 |
| chr2 | 219626708 | 219627462 | 755 | 9 | 6.99E-05 | 5.59E-05 | 0.001196 | 1.33E-05 | 0.00439939 | SLC4A3 |
| chr1 | 63426835 | 63427304 | 470 | 3 | 1.65E-05 | 7.23E-05 | 3.51E-05 | 1.33E-05 | 0.004896069 | ALG6 |
| chr5 | 172640909 | 172641689 | 781 | 6 | 1.64E-05 | 6.64E-05 | 0.000159 | 1.33E-05 | 0.002755078 | NEURL1B |
| chr19 | 36121469 | 36122009 | 541 | 4 | 2.51E-05 | 3.50E-05 | 0.000876 | 1.33E-05 | 0.006893177 | TBCB |
| chr1 | 200213298 | 200213815 | 518 | 4 | 4.13E-05 | 0.000102 | 0.00024 | 1.34E-05 | -0.00235522 | CCNQP1 |
| chr10 | 101231098 | 101231427 | 330 | 2 | 5.44E-06 | 1.60E-05 | 0.000101 | 1.34E-05 | 0.01265984 | LBX1-AS1 |
| chr19 | 10085238 | 10085914 | 677 | 5 | 6.97E-05 | 1.13E-05 | 0.000808 | 1.34E-05 | -0.00044849 | NA |
| chr8 | 26589997 | 26590070 | 74 | 2 | 2.00E-05 | 0.001403 | 1.45E-05 | 1.34E-05 | 0.007084954 | DPYSL2 |
| chr12 | 2902348 | 2902575 | 228 | 2 | 7.93E-05 | 8.16E-06 | 0.000761 | 1.34E-05 | 0.022533696 | TULP3 |
| chr1 | 213996474 | 213997650 | 1177 | 10 | 1.50E-06 | 0.000702 | 7.23E-05 | 1.34E-05 | -0.00143475 | PROX1 |
| chr5 | 60628709 | 60629176 | 468 | 3 | 4.62E-05 | 3.41E-05 | 0.000174 | 1.34E-05 | -0.00255465 | DEPDC1B |
| chr19 | 15297533 | 15297590 | 58 | 2 | 2.84E-05 | 5.92E-05 | 3.39E-05 | 1.34E-05 | 0.003700642 | BRD4 |
| chr16 | 86618359 | 86618896 | 538 | 3 | 5.62E-05 | 6.56E-06 | 0.001727 | 1.34E-05 | 0.004495209 | NA |
| chr19 | 35856374 | 35857513 | 1140 | 14 | 8.36E-06 | 0.000437 | 0.000344 | 1.34E-05 | -0.00017926 | KIRREL2, NPHS1 |
| chr19 | 9796573 | 9796893 | 321 | 3 | 7.97E-05 | 7.40E-06 | 0.000747 | 1.34E-05 | 0.01113723 | AC008752.1 |
| chr2 | 27280573 | 27281827 | 1255 | 5 | 9.82E-05 | 8.69E-06 | 0.002395 | 1.34E-05 | 0.005953613 | DNAJC5G |
| chr19 | 43352922 | 43353722 | 801 | 5 | 0.000136913 | 4.75E-06 | 0.006219 | 1.34E-05 | 0.017296606 | CD177 |
| chr14 | 67533035 | 67533915 | 881 | 11 | 2.85E-05 | 0.000435 | 0.001213 | 1.35E-05 | 0.002766498 | PLEKHH1, TMEM229B |
| chr5 | 141618777 | 141619361 | 585 | 8 | 0.000110947 | 1.69E-05 | 0.000681 | 1.35E-05 | 0.001283206 | AC008781.1, DIAPH1 |
| chr19 | 11142751 | 11143369 | 619 | 4 | 7.22E-05 | 1.31E-05 | 0.0005 | 1.35E-05 | 0.012402906 | SPC24 |
| chr11 | 64760856 | 64761726 | 871 | 6 | 1.35E-05 | 0.001104 | 0.00042 | 1.35E-05 | -0.00235328 | NA |
| chr2 | 191677012 | 191677222 | 211 | 2 | 3.48E-06 | 0.000143 | 2.27E-05 | 1.35E-05 | 0.013428152 | NA |
| chr6 | 89950043 | 89950379 | 337 | 4 | 1.41E-05 | 2.56E-05 | 0.000291 | 1.35E-05 | -4.59E-05 | AL121787.1, BACH2 |
| chr8 | 47132993 | 47133378 | 386 | 3 | 3.87E-05 | 4.69E-05 | 0.00023 | 1.35E-05 | 0.007049977 | AC120036.1 |
| chr4 | 1805855 | 1806859 | 1005 | 12 | 2.15E-05 | 5.55E-05 | 0.000451 | 1.35E-05 | 0.002780672 | FGFR3 |
| chr8 | 133883257 | 133883691 | 435 | 2 | 5.51E-06 | 0.000397 | 1.76E-05 | 1.35E-05 | 0.012679324 | NA |
| chr5 | 59768409 | 59769263 | 855 | 13 | 5.25E-05 | 3.71E-05 | 0.00283 | 1.35E-05 | 0.003627839 | PDE4D |
| chr2 | 160416189 | 160416265 | 77 | 2 | 2.27E-05 | 1.73E-05 | 9.25E-05 | 1.35E-05 | 0.008969499 | RBMS1 |
| chr15 | 55087769 | 55087784 | 16 | 2 | 4.85E-05 | 4.15E-05 | 4.24E-05 | 1.35E-05 | 0.005714098 | AC025272.1 |
| chr9 | 90773987 | 90774624 | 638 | 2 | 0.000224759 | 8.39E-06 | 0.00065 | 1.35E-05 | 0.015859043 | NA |
| chr2 | 197453306 | 197453622 | 317 | 12 | 0.000338336 | 3.73E-05 | 0.010366 | 1.35E-05 | 0.000740833 | COQ10B |
| chr1 | 19647636 | 19647682 | 47 | 2 | 4.75E-05 | 1.05E-05 | 0.000238 | 1.35E-05 | -0.03516566 | NBL1, MICOS10-NBL1 |
| chr18 | 23951182 | 23951724 | 543 | 5 | 1.74E-05 | 6.87E-05 | 0.000809 | 1.35E-05 | 0.001695472 | LAMA3 |
| chr12 | 49599484 | 49600501 | 1018 | 7 | 8.10E-05 | 2.37E-05 | 0.009126 | 1.35E-05 | 0.002658053 | PRPF40B, FAM186B |
| chr2 | 238170511 | 238170574 | 64 | 3 | 4.24E-05 | 2.68E-05 | 0.000132 | 1.35E-05 | 0.004995665 | ILKAP |
| chr10 | 13302468 | 13302622 | 155 | 3 | 0.000393249 | 1.17E-05 | 0.000341 | 1.35E-05 | 0.001043409 | NA |
| chr10 | 45651093 | 45651237 | 145 | 2 | 2.65E-05 | 0.000208 | 2.05E-05 | 1.35E-05 | 0.006376356 | ZFAND4 |
| chr16 | 67390448 | 67391316 | 869 | 6 | 0.000109845 | 7.17E-06 | 0.006108 | 1.35E-05 | 0.007115133 | TPPP3, RNU1-123P |
| chr15 | 84657808 | 84658578 | 771 | 6 | 1.83E-06 | 0.000161 | 0.000535 | 1.35E-05 | 0.011083904 | NMB |
| chr2 | 37083848 | 37084851 | 1004 | 12 | 2.09E-05 | 7.19E-05 | 0.002396 | 1.35E-05 | 0.000931553 | GPATCH11, HEATR5B |
| chr7 | 131284582 | 131285366 | 785 | 5 | 1.45E-05 | 0.000153 | 5.73E-05 | 1.36E-05 | 0.005898768 | MKLN1 |
| chr5 | 141811864 | 141811992 | 129 | 3 | 6.90E-05 | 1.59E-05 | 0.000153 | 1.36E-05 | 0.008278948 | NA |
| chr16 | 85444974 | 85445695 | 722 | 8 | 0.00013676 | 7.74E-06 | 0.01051 | 1.36E-05 | 0.005617776 | GSE1 |
| chr6 | 31961805 | 31962546 | 742 | 6 | 5.54E-06 | 3.01E-05 | 0.001067 | 1.36E-05 | 0.003169747 | SKIV2L |
| chr9 | 12814398 | 12814644 | 247 | 3 | 4.81E-05 | 2.36E-05 | 0.00013 | 1.36E-05 | 0.010733171 | LURAP1L |
| chr12 | 132784084 | 132784254 | 171 | 4 | 5.60E-05 | 3.87E-05 | 0.000674 | 1.36E-05 | 0.007510442 | GOLGA3 |
| chr19 | 5507263 | 5507529 | 267 | 5 | 2.21E-05 | 9.75E-05 | 3.53E-05 | 1.36E-05 | 0.003086952 | NA |
| chr20 | 4822345 | 4823406 | 1062 | 6 | 6.42E-05 | 9.03E-05 | 0.002288 | 1.36E-05 | 0.001957617 | RASSF2 |
| chr9 | 94232703 | 94232761 | 59 | 2 | 5.10E-05 | 3.38E-05 | 4.94E-05 | 1.36E-05 | 0.004027525 | LINC02603 |
| chr19 | 42010357 | 42010475 | 119 | 2 | 5.90E-05 | 9.58E-06 | 0.000345 | 1.36E-05 | -0.0063876 | GRIK5 |
| chr9 | 126474117 | 126474221 | 105 | 2 | 3.64E-05 | 8.58E-06 | 0.00061 | 1.36E-05 | 0.005968649 | MVB12B |
| chr12 | 116592349 | 116592666 | 318 | 2 | 1.07E-05 | 0.000417 | 1.76E-05 | 1.36E-05 | 0.011936722 | NA |
| chr16 | 2977538 | 2978108 | 571 | 4 | 9.14E-06 | 7.95E-05 | 3.51E-05 | 1.36E-05 | 0.002818204 | PKMYT1 |
| chr6 | 47203822 | 47204079 | 258 | 3 | 2.99E-05 | 1.68E-05 | 0.000841 | 1.37E-05 | 0.010264983 | NA |
| chr15 | 73633583 | 73634423 | 841 | 6 | 1.08E-05 | 2.82E-05 | 0.00028 | 1.37E-05 | 0.000344129 | NPTN |
| chr12 | 120719786 | 120720499 | 714 | 3 | 0.00012138 | 2.42E-05 | 0.000674 | 1.37E-05 | -0.00346096 | UNC119B |
| chr1 | 153541532 | 153542006 | 475 | 5 | 4.35E-05 | 3.98E-06 | 0.006176 | 1.37E-05 | 0.006166621 | S100A5 |
| chr14 | 77139992 | 77140647 | 656 | 6 | 4.35E-05 | 0.00013 | 0.000225 | 1.37E-05 | 0.00516773 | AC007375.2, TMEM63C, ZDHHC22 |
| chr1 | 212035174 | 212036157 | 984 | 19 | 2.50E-05 | 0.000492 | 0.002582 | 1.37E-05 | 0.001102404 | DTL, INTS7 |
| chr17 | 7437872 | 7438617 | 746 | 6 | 5.78E-05 | 4.98E-06 | 0.004342 | 1.37E-05 | 0.009043492 | AC113189.2, FGF11 |
| chr11 | 64887642 | 64888352 | 711 | 5 | 6.57E-05 | 1.28E-05 | 0.000632 | 1.37E-05 | 0.003643794 | EHD1 |
| chr18 | 23689116 | 23689938 | 823 | 8 | 7.08E-06 | 0.000498 | 9.49E-05 | 1.37E-05 | -0.00066948 | LAMA3 |
| chr13 | 51111059 | 51111522 | 464 | 5 | 1.94E-05 | 8.50E-05 | 0.000432 | 1.37E-05 | -0.00180983 | C13orf42 |
| chr17 | 3737621 | 3738591 | 971 | 5 | 1.23E-06 | 0.014863 | 6.23E-05 | 1.37E-05 | 0.004925054 | ITGAE |
| chr21 | 44965054 | 44965521 | 468 | 2 | 0.00010569 | 8.33E-06 | 0.00079 | 1.37E-05 | 0.004616458 | FAM207A |
| chr13 | 98727076 | 98727487 | 412 | 3 | 1.38E-05 | 0.00014 | 7.61E-05 | 1.37E-05 | -0.0024224 | SLC15A1 |
| chr7 | 1071148 | 1071793 | 646 | 3 | 2.55E-05 | 0.000182 | 5.84E-05 | 1.37E-05 | 0.004462513 | C7orf50 |
| chr15 | 60518674 | 60518920 | 247 | 3 | 2.81E-05 | 1.75E-05 | 0.000159 | 1.37E-05 | 0.001859869 | RORA-AS1, RORA |
| chr11 | 12377951 | 12378523 | 573 | 4 | 1.73E-07 | 0.000829 | 1.03E-05 | 1.37E-05 | -0.00746198 | PARVA |
| chr17 | 32265014 | 32265387 | 374 | 3 | 1.70E-05 | 7.83E-06 | 0.000968 | 1.37E-05 | 0.014952838 | AC026620.2 |
| chr2 | 28338718 | 28339190 | 473 | 2 | 1.33E-05 | 3.64E-05 | 4.74E-05 | 1.38E-05 | 0.007087783 | BABAM2 |
| chr21 | 46304747 | 46305227 | 481 | 3 | 7.28E-05 | 6.50E-06 | 0.001622 | 1.38E-05 | 0.001124889 | C21orf58 |
| chr22 | 45947873 | 45948124 | 252 | 2 | 2.46E-05 | 7.97E-05 | 3.00E-05 | 1.38E-05 | 0.008389853 | WNT7B |
| chr2 | 36516278 | 36516849 | 572 | 2 | 3.62E-05 | 9.56E-06 | 0.000373 | 1.38E-05 | 0.004450817 | CRIM1 |
| chr22 | 46026665 | 46026872 | 208 | 2 | 2.29E-05 | 8.40E-06 | 0.00078 | 1.38E-05 | 0.011799101 | NA |
| chr10 | 114633174 | 114633664 | 491 | 2 | 2.23E-06 | 2.48E-05 | 6.56E-05 | 1.38E-05 | 0.019624896 | ABLIM1 |
| chr18 | 50308335 | 50308734 | 400 | 2 | 7.30E-06 | 8.62E-05 | 2.89E-05 | 1.38E-05 | 0.01108009 | AC105227.1 |
| chr7 | 101090131 | 101090770 | 640 | 2 | 3.20E-05 | 8.57E-06 | 0.000699 | 1.38E-05 | -0.00102344 | TRIM56 |
| chr9 | 87034340 | 87034433 | 94 | 2 | 4.92E-05 | 3.64E-05 | 4.77E-05 | 1.38E-05 | 0.012610741 | AL513318.1 |
| chr4 | 76306376 | 76306726 | 351 | 5 | 7.35E-05 | 0.000165 | 8.69E-05 | 1.38E-05 | -0.00090275 | FAM47E-STBD1, STBD1, AC034139.1 |
| chr6 | 159786138 | 159786453 | 316 | 2 | 3.98E-05 | 1.77E-05 | 9.39E-05 | 1.38E-05 | 0.008078675 | TCP1 |
| chr21 | 45266993 | 45267744 | 752 | 4 | 9.94E-06 | 9.21E-05 | 6.99E-05 | 1.38E-05 | -0.00233393 | POFUT2 |
| chr2 | 46815303 | 46815564 | 262 | 3 | 4.67E-05 | 8.83E-06 | 0.000601 | 1.38E-05 | 0.005747425 | LINC01118 |
| chr11 | 131246347 | 131246765 | 419 | 3 | 1.85E-05 | 0.000132 | 0.000229 | 1.39E-05 | 0.007548383 | AP002856.4 |
| chr9 | 98210840 | 98211323 | 484 | 2 | 5.56E-05 | 1.04E-05 | 0.000278 | 1.39E-05 | 0.001243259 | TBC1D2 |
| chr13 | 29884781 | 29885168 | 388 | 3 | 5.81E-05 | 1.12E-05 | 0.000322 | 1.39E-05 | 0.005206433 | LINC00297 |
| chr3 | 155492284 | 155492286 | 3 | 2 | 5.81E-05 | 2.63E-05 | 6.25E-05 | 1.39E-05 | 0.003312365 | PLCH1 |

| | | | | | | | | | | |
|---|---|---|---|---|---|---|---|---|---|---|
| chr14 | 104735650 | 104736350 | 701 | 4 | 0.000102798 | 5.39E-06 | 0.004647 | 1.39E-05 | 0.00811665 | ADSS1 |
| chr11 | 86473220 | 86473877 | 658 | 2 | 0.000178522 | 8.44E-06 | 0.000794 | 1.39E-05 | 0.004664622 | AP001831.1, ME3 |
| chr12 | 28190318 | 28191233 | 916 | 10 | 4.42E-05 | 6.15E-06 | 0.01443 | 1.39E-05 | 0.001007768 | CCDC91, AC022364.1 |
| chr6 | 134170025 | 134170393 | 369 | 4 | 7.85E-06 | 0.006395 | 5.83E-06 | 1.39E-05 | 0.001836958 | SGK1 |
| chr2 | 212832070 | 212832325 | 256 | 3 | 2.93E-05 | 2.00E-05 | 0.000194 | 1.39E-05 | -0.00138198 | PCED1CP, AC093865.1 |
| chr2 | 162018238 | 162018953 | 716 | 3 | 1.55E-05 | 0.000811 | 0.000372 | 1.39E-05 | 0.006919357 | DPP4 |
| chr22 | 48396812 | 48396890 | 79 | 2 | 7.37E-05 | 1.10E-05 | 0.000239 | 1.40E-05 | 0.018800783 | NA |
| chr1 | 14945334 | 14946071 | 738 | 9 | 9.85E-06 | 0.000258 | 0.000224 | 1.40E-05 | 0.001812619 | KAZN |
| chr2 | 68252488 | 68253090 | 603 | 4 | 2.43E-05 | 0.000122 | 4.80E-05 | 1.40E-05 | 0.001660834 | AC017083.1, AC017083.3, PPP3R1 |
| chr7 | 140479484 | 140479823 | 340 | 8 | 0.000137093 | 1.78E-05 | 0.010921 | 1.40E-05 | 0.000677377 | MKRN1 |
| chr10 | 24623087 | 24624262 | 1176 | 4 | 4.22E-05 | 6.09E-05 | 0.000858 | 1.40E-05 | 0.010207329 | ARHGAP21 |
| chr11 | 60056520 | 60057068 | 549 | 6 | 8.87E-05 | 1.13E-05 | 0.008546 | 1.40E-05 | 0.007737872 | MS4A3 |
| chr17 | 81721654 | 81722302 | 649 | 2 | 4.28E-05 | 9.64E-06 | 0.000386 | 1.40E-05 | 0.002620656 | NA |
| chr6 | 158560583 | 158561220 | 638 | 6 | 3.98E-05 | 2.54E-05 | 0.000221 | 1.40E-05 | 0.000577796 | TMEM181 |
| chr11 | 72398611 | 72398623 | 13 | 2 | 2.03E-05 | 0.01148 | 1.31E-05 | 1.40E-05 | -0.00329908 | CLPB |
| chr8 | 138914328 | 138914758 | 431 | 4 | 2.88E-05 | 0.000492 | 0.000785 | 1.40E-05 | 0.007495507 | NA |
| chr14 | 68824907 | 68825139 | 233 | 3 | 8.42E-05 | 8.35E-06 | 0.001287 | 1.41E-05 | 0.008410112 | NA |
| chr19 | 45638954 | 45640173 | 1220 | 16 | 2.21E-06 | 0.001004 | 0.001247 | 1.41E-05 | 0.000300417 | EML2, MIR330 |
| chr3 | 4995051 | 4995539 | 489 | 7 | 7.56E-05 | 1.55E-05 | 0.002628 | 1.41E-05 | 0.003761907 | NA |
| chr13 | 100314014 | 100314055 | 42 | 2 | 8.12E-05 | 9.46E-06 | 0.000433 | 1.41E-05 | 0.005504519 | PCCA |
| chr8 | 19823342 | 19823734 | 393 | 2 | 5.25E-06 | 0.000259 | 2.00E-05 | 1.41E-05 | -0.0055413 | INTS10 |
| chr14 | 57068618 | 57069129 | 512 | 3 | 2.79E-05 | 2.31E-05 | 9.77E-05 | 1.41E-05 | 0.02140139 | OTX2-AS1, AL391152.1 |
| chr10 | 132307909 | 132308787 | 879 | 8 | 2.05E-06 | 0.002751 | 5.10E-05 | 1.41E-05 | -0.00092252 | STK32C |
| chr1 | 31244254 | 31244412 | 159 | 3 | 7.84E-05 | 1.09E-05 | 0.000312 | 1.41E-05 | 0.011178965 | NA |
| chr2 | 97734188 | 97735085 | 898 | 9 | 5.77E-06 | 8.38E-05 | 0.000134 | 1.41E-05 | 0.001610754 | ZAP70 |
| chr13 | 99217212 | 99217643 | 432 | 2 | 7.58E-06 | 0.000192 | 2.17E-05 | 1.41E-05 | 0.018130084 | UBAC2 |
| chr1 | 85581544 | 85582228 | 685 | 6 | 1.73E-05 | 1.94E-05 | 0.000663 | 1.41E-05 | 0.006268477 | CCN1 |
| chr16 | 20805493 | 20806639 | 1147 | 18 | 3.01E-05 | 1.21E-05 | 0.008653 | 1.41E-05 | 0.00136132 | REXO5, ERI2 |
| chr10 | 125448100 | 125448110 | 11 | 2 | 7.67E-05 | 1.16E-05 | 0.000212 | 1.41E-05 | 0.010496146 | NA |
| chr7 | 56537632 | 56538142 | 511 | 6 | 8.79E-05 | 9.26E-06 | 0.00342 | 1.41E-05 | 0.015887565 | AC092447.5 |
| chr17 | 78645331 | 78646013 | 683 | 7 | 1.57E-05 | 3.83E-05 | 0.004459 | 1.41E-05 | 0.005755688 | NA |
| chr17 | 79850835 | 79851590 | 756 | 3 | 6.77E-05 | 3.52E-05 | 0.000472 | 1.41E-05 | 0.000212579 | NA |
| chr8 | 31497217 | 31497502 | 286 | 5 | 8.78E-05 | 7.28E-06 | 0.001621 | 1.41E-05 | 0.023669706 | AC068672.1 |
| chr15 | 82680668 | 82681112 | 445 | 4 | 1.26E-05 | 5.24E-05 | 0.00015 | 1.41E-05 | 0.006654909 | CPEB1-AS1, AP3B2 |
| chr11 | 61767291 | 61767332 | 42 | 2 | 5.77E-05 | 8.59E-06 | 0.000799 | 1.41E-05 | 0.00850499 | MYRF |
| chr17 | 70128785 | 70129553 | 769 | 4 | 2.12E-05 | 0.000717 | 0.000329 | 1.41E-05 | -2.42E-05 | KCNJ16, AC005208.1 |
| chr19 | 19033163 | 19034219 | 1057 | 12 | 3.98E-05 | 2.44E-05 | 0.00724 | 1.41E-05 | -0.00041535 | ARMC6, SUGP2 |
| chr7 | 39609097 | 39610300 | 1204 | 7 | 0.000129074 | 1.65E-05 | 0.0041 | 1.42E-05 | 0.005371729 | YAE1, AC004837.1, AC004837.3 |
| chr14 | 36661839 | 36663285 | 1447 | 8 | 1.97E-05 | 1.83E-05 | 0.008942 | 1.42E-05 | 0.002642755 | PAX9 |
| chr22 | 49983267 | 49983934 | 668 | 12 | 4.24E-05 | 0.000234 | 0.000106 | 1.42E-05 | -0.00663701 | NA |
| chr13 | 98543565 | 98543780 | 216 | 2 | 4.00E-05 | 9.68E-06 | 0.000412 | 1.42E-05 | 0.012810678 | STK24 |
| chr17 | 8858656 | 8858823 | 168 | 3 | 1.34E-05 | 0.001883 | 2.54E-05 | 1.42E-05 | 0.008847582 | PIK3R6 |
| chr22 | 37259574 | 37259734 | 161 | 2 | 4.29E-05 | 3.61E-05 | 4.98E-05 | 1.42E-05 | 0.010374168 | NA |
| chr16 | 81644892 | 81645363 | 472 | 7 | 4.68E-05 | 1.67E-05 | 0.00055 | 1.42E-05 | 0.001181082 | CMIP |
| chr4 | 186167015 | 186167037 | 23 | 2 | 3.06E-05 | 0.00038 | 1.85E-05 | 1.42E-05 | 0.007589695 | FAM149A |
| chr9 | 34478166 | 34478553 | 388 | 3 | 1.32E-05 | 4.89E-05 | 4.88E-05 | 1.43E-05 | 0.010232851 | DNAI1 |
| chr19 | 4390017 | 4390307 | 291 | 3 | 5.52E-05 | 1.40E-05 | 0.000238 | 1.43E-05 | 0.007075893 | SH3GL1 |
| chr6 | 30672377 | 30673443 | 1067 | 34 | 1.88E-05 | 0.000247 | 0.00014 | 1.43E-05 | 0.001114927 | DHX16 |
| chr5 | 141813943 | 141814271 | 329 | 2 | 8.24E-05 | 8.84E-06 | 0.000696 | 1.43E-05 | 0.016880906 | NA |
| chr3 | 101778631 | 101779439 | 809 | 13 | 5.42E-06 | 0.00043 | 0.004023 | 1.43E-05 | 0.002462826 | NXPE3 |
| chr2 | 68721858 | 68721957 | 100 | 2 | 5.32E-05 | 1.04E-05 | 0.000315 | 1.43E-05 | 0.01225657 | ARHGAP25 |
| chr17 | 75896800 | 75897936 | 1137 | 11 | 0.000160891 | 1.10E-05 | 0.018107 | 1.43E-05 | 0.000824939 | AC087289.2, TRIM65 |
| chr1 | 192788257 | 192788468 | 212 | 2 | 2.52E-05 | 0.000145 | 2.41E-05 | 1.43E-05 | 0.014107587 | AL390957.1 |
| chr6 | 41548347 | 41548765 | 419 | 5 | 8.11E-05 | 5.42E-06 | 0.007063 | 1.43E-05 | 0.005411522 | FOXP4, FOXP4-AS1 |
| chr1 | 26921521 | 26922127 | 607 | 13 | 0.000132282 | 0.000217 | 0.000831 | 1.43E-05 | -0.00022665 | NUDC |
| chr13 | 83057549 | 83057632 | 84 | 2 | 7.75E-05 | 1.13E-05 | 0.000242 | 1.43E-05 | 0.012939397 | AL445255.1 |
| chr9 | 136203917 | 136204411 | 495 | 4 | 4.88E-06 | 0.000216 | 0.000151 | 1.44E-05 | 0.014388129 | LHX3 |
| chr1 | 156456225 | 156457153 | 929 | 7 | 1.76E-06 | 0.001117 | 0.000628 | 1.44E-05 | 0.00117641 | MIR9-1HG |
| chr22 | 50519071 | 50519468 | 398 | 3 | 2.72E-05 | 2.13E-05 | 0.000238 | 1.44E-05 | 0.006521565 | NCAPH2 |
| chr22 | 22085930 | 22086285 | 356 | 2 | 2.97E-05 | 2.64E-05 | 6.56E-05 | 1.44E-05 | 0.00606238 | IGLV10-54 |
| chr14 | 103225024 | 103225767 | 744 | 10 | 1.52E-05 | 0.000103 | 0.00062 | 1.44E-05 | 0.006273578 | NA |
| chr4 | 38109311 | 38110165 | 855 | 8 | 5.25E-05 | 9.97E-05 | 0.001794 | 1.44E-05 | -0.00036171 | TBC1D1 |
| chr11 | 61792490 | 61792654 | 165 | 7 | 0.000435425 | 8.19E-06 | 0.001197 | 1.44E-05 | 0.000897754 | TMEM258, MIR611 |
| chr19 | 3548801 | 3548979 | 179 | 2 | 1.18E-05 | 1.05E-05 | 0.000315 | 1.44E-05 | 0.010517906 | AC005786.3, MFSD12 |
| chr12 | 107092710 | 107093707 | 998 | 9 | 1.93E-06 | 0.000195 | 0.000392 | 1.44E-05 | -0.00126219 | AC078929.1, CRY1 |
| chr6 | 42563783 | 42564406 | 624 | 13 | 5.02E-05 | 0.000161 | 0.007362 | 1.44E-05 | 0.001312145 | UBR2 |
| chr14 | 103071182 | 103071806 | 625 | 3 | 9.15E-05 | 6.31E-06 | 0.00184 | 1.44E-05 | 0.005846612 | NA |
| chr9 | 22147984 | 22148136 | 153 | 2 | 8.66E-05 | 8.76E-06 | 0.000812 | 1.44E-05 | -0.00781661 | NA |
| chr11 | 47524050 | 47524753 | 704 | 8 | 8.63E-05 | 4.26E-06 | 0.004937 | 1.44E-05 | 0.008464346 | CELF1 |
| chr5 | 168683684 | 168683686 | 3 | 2 | 8.48E-05 | 8.78E-06 | 0.000797 | 1.44E-05 | 0.019765906 | SLIT3 |
| chr15 | 25802235 | 25802506 | 272 | 3 | 8.01E-05 | 9.16E-06 | 0.000549 | 1.44E-05 | 0.001359218 | ATP10A |
| chr3 | 131361639 | 131361911 | 273 | 6 | 8.75E-05 | 5.93E-06 | 0.007349 | 1.45E-05 | 0.011843428 | NUDT16P1, AC010210.1 |
| chr19 | 4536309 | 4536888 | 580 | 2 | 9.87E-06 | 1.78E-05 | 0.000101 | 1.45E-05 | 0.003177632 | AC011498.4, LRG1 |
| chr6 | 29486846 | 29486978 | 133 | 3 | 8.05E-05 | 2.67E-05 | 8.54E-05 | 1.45E-05 | -0.0130447 | MAS1L |
| chr2 | 85758062 | 85759063 | 1002 | 4 | 8.58E-06 | 0.000178 | 0.000313 | 1.45E-05 | 0.004220036 | ATOH8 |
| chr5 | 7825088 | 7825374 | 287 | 3 | 4.80E-05 | 2.56E-05 | 0.000142 | 1.45E-05 | 0.005002739 | ADCY2 |
| chr12 | 108339476 | 108340269 | 794 | 10 | 0.000102865 | 1.96E-05 | 0.001604 | 1.45E-05 | 0.004030024 | NA |
| chr5 | 55159873 | 55160228 | 356 | 6 | 6.03E-05 | 1.68E-05 | 0.003941 | 1.45E-05 | 0.010250403 | GPX8, CDC20B |
| chr5 | 4880575 | 4880597 | 23 | 2 | 3.28E-05 | 1.73E-05 | 0.000105 | 1.45E-05 | 0.012229961 | NA |
| chr14 | 67189312 | 67189524 | 213 | 3 | 4.11E-05 | 1.19E-05 | 0.000958 | 1.45E-05 | 0.016183401 | FAM71D |
| chr4 | 39366079 | 39366592 | 514 | 10 | 2.11E-05 | 4.80E-05 | 0.00385 | 1.45E-05 | 0.000356721 | RFC1 |
| chr2 | 11539458 | 11540018 | 561 | 8 | 3.57E-05 | 8.81E-05 | 0.001777 | 1.45E-05 | -0.0084712 | GREB1 |
| chr12 | 49544214 | 49544235 | 22 | 2 | 8.41E-05 | 8.84E-06 | 0.000816 | 1.45E-05 | 0.015683883 | KCNH3 |
| chr8 | 1200488 | 1201175 | 688 | 4 | 7.54E-05 | 6.33E-06 | 0.002835 | 1.45E-05 | -0.00352705 | DLGAP2 |
| chr14 | 51095248 | 51096716 | 1469 | 18 | 1.13E-05 | 4.76E-06 | 0.006953 | 1.46E-05 | 0.004000744 | TRIM9 |
| chr6 | 26021540 | 26022045 | 506 | 6 | 1.90E-05 | 0.000212 | 0.000566 | 1.46E-05 | 8.33E-05 | H4C1 |
| chr13 | 110870304 | 110870638 | 335 | 5 | 6.05E-05 | 8.57E-06 | 0.001969 | 1.46E-05 | 0.009435623 | NA |
| chr5 | 177351471 | 177352088 | 618 | 12 | 1.27E-06 | 6.07E-05 | 0.005049 | 1.46E-05 | -0.00124087 | LMAN2 |
| chr8 | 1160520 | 1160781 | 262 | 3 | 6.82E-06 | 0.00014 | 3.21E-05 | 1.46E-05 | 0.001604124 | DLGAP2 |
| chr5 | 1337211 | 1337632 | 422 | 3 | 8.10E-05 | 9.92E-05 | 0.000331 | 1.46E-05 | -0.00033516 | CLPTM1L |
| chr2 | 26502596 | 26502848 | 253 | 2 | 9.65E-06 | 2.27E-05 | 7.77E-05 | 1.46E-05 | -0.00132948 | OTOF |
| chr9 | 75028054 | 75028724 | 671 | 9 | 1.44E-05 | 8.46E-05 | 0.000318 | 1.46E-05 | 0.001558585 | CARNMT1 |
| chr17 | 7701485 | 7701926 | 442 | 5 | 6.74E-06 | 0.000763 | 1.73E-05 | 1.47E-05 | 0.001148098 | WRAP53 |
| chr7 | 16397326 | 16397822 | 497 | 4 | 2.97E-06 | 3.38E-05 | 0.000528 | 1.47E-05 | 0.002854326 | CRPPA |
| chr20 | 35680746 | 35680868 | 123 | 2 | 3.40E-05 | 0.000151 | 2.43E-05 | 1.47E-05 | 0.017896156 | NFS1 |
| chr8 | 94896580 | 94897181 | 602 | 6 | 5.10E-05 | 2.37E-05 | 0.002143 | 1.47E-05 | 0.001089963 | NDUFAF6, CCNE2 |
| chr6 | 33804010 | 33804589 | 580 | 8 | 2.27E-06 | 0.006 | 5.25E-05 | 1.47E-05 | 0.002283068 | NA |
| chr21 | 17820140 | 17820627 | 488 | 3 | 1.61E-05 | 0.000157 | 0.000198 | 1.47E-05 | 0.004727777 | NA |
| chr2 | 70936510 | 70936823 | 314 | 2 | 1.43E-05 | 4.26E-05 | 4.60E-05 | 1.47E-05 | 0.008467449 | VAX2, ATP6V1B1 |
| chr16 | 21653681 | 21653917 | 237 | 4 | 7.29E-05 | 1.49E-05 | 0.000597 | 1.47E-05 | 0.006459311 | METTL9 |
| chr2 | 113062447 | 113063021 | 575 | 4 | 3.32E-06 | 6.72E-05 | 0.000403 | 1.47E-05 | 0.007078946 | IL36RN |
| chr16 | 233070 | 233464 | 395 | 2 | 2.04E-05 | 1.34E-05 | 0.000169 | 1.47E-05 | -0.00658349 | NA |
| chr5 | 139670574 | 139670830 | 257 | 3 | 4.56E-05 | 1.56E-05 | 0.000897 | 1.47E-05 | 0.013492764 | CXXC5 |
| chr6 | 33870531 | 33870731 | 201 | 2 | 5.47E-05 | 2.02E-05 | 8.87E-05 | 1.47E-05 | 0.009279643 | LINC01016 |
| chr1 | 92398545 | 92398805 | 261 | 2 | 2.36E-05 | 0.000121 | 2.65E-05 | 1.47E-05 | 0.008334054 | RPAP2 |
| chr11 | 113045383 | 113045528 | 146 | 3 | 8.98E-05 | 9.93E-06 | 0.000512 | 1.47E-05 | 0.011626245 | NCAM1 |
| chr4 | 579830 | 580221 | 392 | 2 | 7.15E-05 | 9.06E-06 | 0.000761 | 1.47E-05 | 0.018099583 | AC116565.1 |
| chr11 | 2708323 | 2708899 | 577 | 4 | 7.40E-05 | 7.12E-06 | 0.00265 | 1.47E-05 | 0.00458701 | KCNQ1 |
| chr12 | 29604249 | 29604613 | 365 | 2 | 2.99E-05 | 2.68E-05 | 6.73E-05 | 1.47E-05 | 0.002703043 | TMTC1 |
| chr6 | 31951923 | 31952368 | 446 | 7 | 2.04E-05 | 0.001643 | 0.000164 | 1.48E-05 | 0.002048303 | AL645922.1, CFB, NELFE |
| chr7 | 1647128 | 1647608 | 481 | 5 | 2.18E-05 | 1.95E-05 | 0.001081 | 1.48E-05 | 0.010840796 | NA |
| chr12 | 64099004 | 64099032 | 29 | 2 | 6.76E-05 | 1.20E-05 | 0.000222 | 1.48E-05 | 0.003925436 | SRGAP1 |
| chr6 | 99604757 | 99605562 | 806 | 5 | 9.92E-06 | 0.000158 | 0.00038 | 1.48E-05 | 0.008511839 | NA |
| chr4 | 2443100 | 2443594 | 495 | 2 | 5.57E-05 | 1.11E-05 | 0.000292 | 1.48E-05 | 0.004791873 | CFAP99 |
| chr3 | 72318683 | 72318920 | 238 | 2 | 1.24E-05 | 0.000112 | 2.76E-05 | 1.48E-05 | 0.006864196 | AC134508.2 |
| chr20 | 62775434 | 62775648 | 215 | 4 | 3.28E-05 | 2.94E-05 | 0.000242 | 1.48E-05 | 0.004612249 | LINC00659 |
| chr5 | 134592297 | 134592350 | 54 | 2 | 5.02E-05 | 1.31E-05 | 0.000179 | 1.48E-05 | 0.010853711 | NA |
| chr5 | 65611543 | 65611871 | 329 | 3 | 1.26E-05 | 0.000133 | 5.02E-05 | 1.48E-05 | 0.004431033 | TRIM23 |
| chr1 | 182739798 | 182739807 | 10 | 2 | 8.27E-05 | 1.14E-05 | 0.000266 | 1.48E-05 | 0.016306123 | NA |
| chr6 | 105587404 | 105587653 | 250 | 4 | 8.00E-05 | 1.47E-05 | 0.001807 | 1.48E-05 | 0.009390951 | AL096794.1 |
| chr15 | 85700422 | 85700936 | 515 | 2 | 8.15E-05 | 9.24E-06 | 0.000686 | 1.48E-05 | 0.00945485 | AKAP13 |
| chr21 | 45945234 | 45945651 | 418 | 2 | 3.30E-05 | 9.05E-06 | 0.000798 | 1.48E-05 | 0.005580907 | NA |
| chr18 | 62455078 | 62455216 | 139 | 2 | 3.63E-05 | 3.27E-05 | 5.70E-05 | 1.48E-05 | 0.008441038 | NA |
| chr19 | 41262098 | 41262597 | 500 | 11 | 6.32E-05 | 2.31E-05 | 0.017222 | 1.48E-05 | 0.001069142 | HNRNPUL1 |
| chr3 | 127536709 | 127536844 | 136 | 2 | 7.59E-05 | 9.23E-06 | 0.0007 | 1.48E-05 | 0.017225121 | LINC01471 |
| chr4 | 10021360 | 10021577 | 218 | 4 | 7.96E-05 | 8.61E-06 | 0.003372 | 1.48E-05 | 0.00857634 | SLC2A9 |
| chr12 | 57810514 | 57810526 | 13 | 2 | 8.93E-05 | 1.02E-05 | 0.000417 | 1.48E-05 | 0.00446001 | AVIL |
| chr15 | 43483574 | 43483882 | 309 | 2 | 9.10E-06 | 0.003846 | 1.45E-05 | 1.48E-05 | 0.001749676 | TP53BP1 |
| chr13 | 99024483 | 99025118 | 636 | 2 | 1.47E-05 | 1.17E-05 | 0.000249 | 1.48E-05 | 0.018760676 | DOCK9 |
| chr12 | 4648551 | 4649337 | 787 | 13 | 8.52E-05 | 4.76E-05 | 0.009234 | 1.48E-05 | 0.001396824 | AC005832.4, NDUFA9, AC005833.1, AKAP3 |
| chr19 | 17194723 | 17195050 | 328 | 2 | 4.66E-05 | 1.15E-05 | 0.000261 | 1.49E-05 | 0.00306735 | MYO9B |
| chr19 | 10111142 | 10111499 | 358 | 6 | 2.85E-05 | 2.50E-05 | 0.000994 | 1.49E-05 | 0.010593621 | PPAN-P2RY11, PPAN |
| chr19 | 53786757 | 53787180 | 424 | 2 | 1.41E-05 | 1.17E-05 | 0.000252 | 1.49E-05 | 0.00753617 | NA |
| chr5 | 141673450 | 141673782 | 333 | 3 | 6.31E-05 | 8.59E-06 | 0.001559 | 1.49E-05 | 0.010545795 | ARAP3 |
| chr9 | 136100608 | 136100969 | 362 | 4 | 8.50E-05 | 6.64E-06 | 0.002811 | 1.49E-05 | 0.011957246 | NA |
| chr17 | 55458602 | 55459187 | 586 | 3 | 4.01E-05 | 5.51E-05 | 0.000273 | 1.49E-05 | 0.006270087 | SMIM36 |
| chr7 | 1504300 | 1504724 | 425 | 8 | 1.89E-05 | 0.000205 | 0.000215 | 1.49E-05 | 0.000286797 | INTS1 |
| chr3 | 196216403 | 196216765 | 363 | 6 | 2.92E-05 | 6.09E-05 | 0.001952 | 1.49E-05 | 0.006196142 | SLC51A, PCYT1A |
| chr17 | 15029757 | 15030219 | 463 | 3 | 1.37E-05 | 7.09E-06 | 0.001756 | 1.49E-05 | 0.010800337 | AC005772.1 |
| chr22 | 35538817 | 35538905 | 89 | 2 | 2.51E-05 | 0.000119 | 2.73E-05 | 1.50E-05 | 0.016770058 | NA |
| chr5 | 56585778 | 56586031 | 254 | 2 | 1.54E-05 | 0.000821 | 1.70E-05 | 1.50E-05 | -0.01027269 | C5orf67 |
| chr4 | 36158838 | 36159285 | 448 | 2 | 1.17E-05 | 1.35E-05 | 0.000174 | 1.50E-05 | 0.010796805 | ARAP2 |
| chr19 | 3691015 | 3691091 | 77 | 2 | 1.35E-05 | 1.44E-05 | 0.000152 | 1.50E-05 | 0.018664955 | PIP5K1C |
| chr8 | 125428798 | 125429151 | 354 | 3 | 2.83E-05 | 6.01E-06 | 0.002991 | 1.50E-05 | 0.004653832 | NA |
| chr1 | 24928958 | 24929886 | 929 | 8 | 4.89E-05 | 1.14E-05 | 0.005291 | 1.50E-05 | 0.001586838 | RUNX3 |
| chr12 | 122845318 | 122845979 | 662 | 3 | 3.67E-05 | 3.58E-05 | 0.000225 | 1.50E-05 | 0.0034439 | HIP1R |
| chr15 | 87974028 | 87974246 | 219 | 2 | 3.06E-05 | 7.89E-05 | 3.31E-05 | 1.50E-05 | 0.008287743 | NTRK3 |
| chr8 | 26422115 | 26422437 | 323 | 4 | 4.71E-05 | 9.64E-06 | 0.001418 | 1.50E-05 | -0.00169151 | BNIP3L |
| chr19 | 42595330 | 42596079 | 750 | 4 | 9.84E-06 | 0.00024 | 4.30E-05 | 1.50E-05 | 0.006394666 | LIPE-AS1 |
| chr12 | 69157050 | 69157415 | 366 | 2 | 5.88E-05 | 9.37E-06 | 0.000708 | 1.50E-05 | 0.009260482 | NA |
| chr1 | 180913140 | 180913545 | 406 | 4 | 3.00E-05 | 3.50E-05 | 0.000571 | 1.51E-05 | 0.011028725 | KIAA1614 |
| chr3 | 138329259 | 138330169 | 911 | 12 | 3.57E-05 | 6.03E-05 | 0.001366 | 1.51E-05 | 0.000571928 | NME9 |
| chr1 | 19081877 | 19082385 | 509 | 4 | 1.59E-05 | 8.28E-05 | 0.000569 | 1.51E-05 | 0.005675691 | UBR4 |
| chr7 | 148589903 | 148590628 | 726 | 5 | 3.81E-05 | 1.63E-05 | 0.001249 | 1.51E-05 | 0.00463134 | AC005229.5 |
| chr9 | 126587356 | 126587681 | 326 | 3 | 0.000105438 | 1.12E-05 | 0.001252 | 1.51E-05 | -0.00341533 | NA |
| chr1 | 150269541 | 150269725 | 185 | 2 | 0.000178541 | 8.58E-05 | 3.18E-05 | 1.51E-05 | -0.01384055 | C1orf54, APH1A |
| chr7 | 134317994 | 134318372 | 379 | 2 | 1.00E-05 | 9.19E-06 | 0.000826 | 1.51E-05 | -2.70E-05 | NA |
| chr1 | 202122737 | 202123168 | 432 | 9 | 4.77E-05 | 7.24E-05 | 0.000744 | 1.51E-05 | 0.001749051 | GPR37L1 |
| chr14 | 60169892 | 60170526 | 635 | 3 | 6.22E-05 | 4.91E-05 | 0.00011 | 1.51E-05 | 0.004701704 | NA |
| chr12 | 124390935 | 124391555 | 621 | 3 | 0.000114177 | 1.15E-05 | 0.000666 | 1.51E-05 | 0.010309585 | NCOR2 |
| chr6 | 123636782 | 123637281 | 500 | 5 | 0.000125151 | 5.54E-06 | 0.004091 | 1.51E-05 | 0.001146189 | AL133257.1, TRDN |
| chr6 | 170064083 | 170064153 | 71 | 2 | 6.06E-05 | 2.67E-05 | 7.02E-05 | 1.51E-05 | 0.009905132 | NA |

| | | | | | | | | | | |
|---|---|---|---|---|---|---|---|---|---|---|
| chr1 | 37889686 | 37890504 | 819 | 4 | 9.23E-06 | 3.01E-05 | 9.15E-05 | 1.51E-05 | 0.002130954 | INPP5B |
| chr19 | 15401233 | 15401739 | 507 | 4 | 4.84E-05 | 5.72E-06 | 0.004017 | 1.52E-05 | -0.00019028 | AKAP8L |
| chr1 | 43389326 | 43390515 | 1190 | 15 | 1.30E-05 | 0.000173 | 0.004389 | 1.52E-05 | -0.00055682 | SZT2, MED8 |
| chr10 | 101139505 | 101139893 | 389 | 5 | 2.18E-05 | 2.88E-05 | 0.002231 | 1.52E-05 | 0.007202623 | NA |
| chr12 | 105485558 | 105485856 | 299 | 2 | 7.24E-05 | 1.09E-05 | 0.000343 | 1.52E-05 | 0.005672164 | NA |
| chr15 | 28112301 | 28112572 | 272 | 3 | 7.29E-06 | 2.49E-05 | 0.000522 | 1.52E-05 | -0.00416409 | HERC2 |
| chr16 | 30011900 | 30012753 | 854 | 10 | 4.03E-05 | 3.31E-05 | 0.001802 | 1.52E-05 | 0.010051184 | DOC2A |
| chr10 | 103602028 | 103602631 | 604 | 4 | 0.000103388 | 9.64E-05 | 0.00041 | 1.52E-05 | 0.003625422 | SH3PXD2A |
| chr12 | 29381158 | 29382049 | 892 | 10 | 1.37E-06 | 0.002828 | 1.67E-05 | 1.52E-05 | 0.002125401 | ERGIC2 |
| chr1 | 18294253 | 18294705 | 453 | 2 | 1.61E-05 | 1.19E-05 | 0.000255 | 1.52E-05 | 0.013975233 | IGSF21 |
| chr3 | 49346621 | 49347059 | 439 | 2 | 5.16E-05 | 1.15E-05 | 0.000282 | 1.52E-05 | 0.012095294 | NA |
| chr15 | 32866788 | 32867315 | 528 | 2 | 0.00011786 | 9.42E-06 | 0.000741 | 1.52E-05 | 0.022841846 | FMN1 |
| chr8 | 124537940 | 124539108 | 1169 | 12 | 2.17E-07 | 0.000241 | 0.001946 | 1.52E-05 | 0.001536002 | NDUFB9, TATDN1 |
| chr16 | 88952995 | 88953630 | 636 | 4 | 6.06E-05 | 5.30E-06 | 0.004273 | 1.52E-05 | 0.002217424 | CBFA2T3 |
| chr20 | 34362408 | 34363604 | 1197 | 10 | 3.48E-05 | 5.34E-05 | 0.00134 | 1.52E-05 | 0.001380578 | ITCH |
| chr6 | 30909962 | 30910622 | 661 | 4 | 0.000113981 | 1.39E-05 | 0.000366 | 1.52E-05 | 0.003566178 | GTF2H4, AL669830.1 |
| chr7 | 77008156 | 77008176 | 21 | 3 | 0.000112784 | 6.89E-06 | 0.002336 | 1.53E-05 | 0.018930289 | DTX2P1-UPK3BP1-PMS2P11 |
| chr6 | 3724288 | 3724456 | 169 | 2 | 5.95E-06 | 0.00018 | 2.39E-05 | 1.53E-05 | 0.001079422 | PXDC1 |
| chr15 | 75359231 | 75360169 | 939 | 5 | 6.36E-05 | 0.000658 | 0.000169 | 1.53E-05 | 0.002661324 | MAN2C1 |
| chr3 | 43080657 | 43081202 | 546 | 4 | 3.61E-06 | 0.000848 | 4.04E-05 | 1.53E-05 | 0.002142479 | POMGNT2 |
| chr17 | 44830569 | 44831042 | 474 | 6 | 6.74E-05 | 2.89E-05 | 0.005506 | 1.53E-05 | 0.009734724 | GJC1 |
| chr10 | 59138226 | 59138327 | 102 | 3 | 8.54E-05 | 1.66E-05 | 0.000297 | 1.53E-05 | -0.01132257 | NA |
| chr8 | 143391280 | 143391778 | 499 | 2 | 9.61E-05 | 9.77E-06 | 0.000611 | 1.53E-05 | 0.015532458 | NA |
| chr2 | 176099220 | 176099860 | 641 | 10 | 3.21E-05 | 0.000276 | 0.003714 | 1.53E-05 | 0.005742874 | HOXD12 |
| chr1 | 165900432 | 165900853 | 422 | 2 | 1.42E-05 | 1.92E-05 | 0.000101 | 1.53E-05 | -0.01618235 | UCK2, AL358115.1 |
| chr11 | 74002958 | 74003091 | 134 | 2 | 3.79E-06 | 0.00047 | 1.90E-05 | 1.53E-05 | 0.003322867 | UCP3 |
| chr2 | 241144053 | 241144451 | 399 | 3 | 1.91E-05 | 2.72E-05 | 0.000236 | 1.53E-05 | 0.003724313 | PASK |
| chr8 | 136794750 | 136795214 | 465 | 2 | 7.25E-05 | 9.65E-06 | 0.000666 | 1.53E-05 | 0.00043974 | LINC02055 |
| chr19 | 35874405 | 35874898 | 494 | 5 | 3.65E-05 | 2.73E-05 | 0.001144 | 1.53E-05 | 0.005356773 | APLP1 |
| chr19 | 18435909 | 18436548 | 640 | 3 | 6.89E-06 | 2.11E-05 | 0.000157 | 1.53E-05 | 0.00677213 | ISYNA1 |
| chr7 | 33814585 | 33815050 | 466 | 2 | 4.41E-06 | 0.000283 | 2.12E-05 | 1.54E-05 | 0.014720072 | BBS9 |
| chr4 | 184433823 | 184434155 | 333 | 4 | 7.70E-05 | 9.28E-06 | 0.001222 | 1.54E-05 | 0.002022493 | IRF2 |
| chr16 | 80563299 | 80563354 | 56 | 2 | 3.36E-05 | 1.24E-05 | 0.000235 | 1.54E-05 | 0.004152316 | AC105411.1, LINC01227 |
| chr8 | 91208324 | 91208829 | 506 | 3 | 5.48E-05 | 5.49E-05 | 0.000597 | 1.54E-05 | 0.002992695 | LRRC69 |
| chr15 | 85718760 | 85719105 | 346 | 3 | 2.72E-05 | 0.000116 | 0.000465 | 1.54E-05 | 0.006227513 | AKAP13 |
| chr10 | 72260670 | 72261264 | 595 | 2 | 1.46E-05 | 1.58E-05 | 0.000138 | 1.54E-05 | 0.009912096 | NA |
| chr8 | 100396273 | 100396340 | 68 | 2 | 1.16E-05 | 0.003343 | 1.49E-05 | 1.54E-05 | 0.007027827 | AP003472.1 |
| chr7 | 2621189 | 2621328 | 140 | 2 | 6.04E-05 | 9.39E-06 | 0.000842 | 1.54E-05 | 0.016856703 | NA |
| chr17 | 19364810 | 19365156 | 347 | 3 | 8.19E-05 | 7.76E-06 | 0.001266 | 1.54E-05 | 0.005013462 | AC124066.1, B9D1 |
| chr19 | 3200079 | 3200968 | 890 | 3 | 0.000342266 | 0.000144 | 0.000126 | 1.55E-05 | 0.0035058 | NCLN |
| chr14 | 21852149 | 21852454 | 306 | 3 | 1.92E-05 | 0.000271 | 2.68E-05 | 1.55E-05 | -0.00574359 | NA |
| chr3 | 23916473 | 23917583 | 1111 | 20 | 1.16E-05 | 0.000473 | 0.000103 | 1.55E-05 | 0.00100047 | RPL15, NKIRAS1 |
| chr6 | 37539221 | 37539347 | 127 | 2 | 8.37E-06 | 0.001024 | 1.69E-05 | 1.55E-05 | 0.004384622 | NA |
| chr3 | 9920277 | 9920568 | 292 | 3 | 9.21E-06 | 0.005503 | 7.94E-06 | 1.55E-05 | 0.001684279 | IL17RC |
| chr7 | 74655550 | 74656163 | 614 | 2 | 2.78E-05 | 9.83E-06 | 0.000648 | 1.55E-05 | 0.013687986 | GTF2I |
| chr22 | 48531895 | 48531927 | 33 | 2 | 4.24E-05 | 3.10E-05 | 6.29E-05 | 1.55E-05 | 0.016115803 | TAFA5 |
| chr16 | 88925770 | 88927033 | 1264 | 3 | 0.000151458 | 6.13E-06 | 0.003249 | 1.55E-05 | 0.006888987 | CBFA2T3 |
| chr20 | 43514111 | 43514862 | 752 | 17 | 1.11E-06 | 0.000172 | 0.000688 | 1.55E-05 | 0.006093266 | AL031681.2, L3MBTL1 |
| chr17 | 17685683 | 17686157 | 475 | 2 | 1.47E-06 | 5.03E-05 | 4.41E-05 | 1.55E-05 | 0.00631195 | RAI1 |
| chr4 | 83212231 | 83212682 | 452 | 2 | 9.45E-05 | 9.83E-06 | 0.000651 | 1.55E-05 | 0.001971998 | NA |
| chr12 | 215145 | 215581 | 437 | 3 | 5.48E-05 | 5.97E-06 | 0.003582 | 1.55E-05 | 0.00344375 | NA |
| chr16 | 10983007 | 10983597 | 591 | 3 | 1.32E-05 | 7.76E-05 | 3.88E-05 | 1.55E-05 | 0.00419983 | CLEC16A |
| chr12 | 1616622 | 1617126 | 505 | 8 | 2.79E-05 | 9.67E-05 | 0.000848 | 1.56E-05 | 0.006964374 | WNT5B |
| chr15 | 26629584 | 26629938 | 355 | 5 | 1.51E-05 | 6.93E-06 | 0.004494 | 1.56E-05 | -0.00265256 | GABRB3 |
| chr7 | 105521851 | 105522675 | 825 | 14 | 1.72E-05 | 0.000341 | 0.000419 | 1.56E-05 | 0.001573105 | PUS7 |
| chr6 | 32406385 | 32407309 | 925 | 7 | 2.26E-05 | 0.003785 | 0.000651 | 1.56E-05 | 0.003345695 | TSBP1-AS1, BTNL2 |
| chr5 | 132864316 | 132865678 | 1363 | 10 | 3.81E-07 | 0.002068 | 0.000141 | 1.56E-05 | 5.30E-05 | GDF9 |
| chr8 | 85229073 | 85229461 | 389 | 3 | 3.73E-05 | 3.94E-05 | 7.59E-05 | 1.56E-05 | 0.004211823 | CA13, AC011773.3 |
| chr3 | 118479932 | 118480575 | 644 | 2 | 0.000123921 | 1.03E-05 | 0.000526 | 1.56E-05 | 0.022623992 | AC068633.1 |
| chr11 | 68800174 | 68800231 | 58 | 2 | 9.34E-06 | 8.09E-05 | 3.41E-05 | 1.56E-05 | -0.00419648 | CPT1A |
| chr6 | 28775650 | 28775923 | 274 | 5 | 0.000121063 | 7.66E-06 | 0.004012 | 1.56E-05 | 0.007334501 | NA |
| chr1 | 23423608 | 23424641 | 1034 | 4 | 1.39E-07 | 0.00675 | 0.000146 | 1.56E-05 | 0.008318442 | TCEA3 |
| chr4 | 165377811 | 165379536 | 1726 | 14 | 1.88E-06 | 0.07892 | 3.06E-05 | 1.56E-05 | 0.001810515 | CPE |
| chr8 | 142599458 | 142599920 | 463 | 3 | 1.19E-05 | 0.00043 | 2.33E-05 | 1.56E-05 | 0.007382663 | NA |
| chr7 | 106758273 | 106758413 | 141 | 2 | 1.45E-05 | 0.000948 | 1.73E-05 | 1.56E-05 | 0.005728785 | AC004917.1 |
| chr2 | 230797597 | 230797867 | 271 | 2 | 2.37E-05 | 2.63E-05 | 7.45E-05 | 1.56E-05 | 0.010038769 | CAB39 |
| chr6 | 46658173 | 46658442 | 270 | 2 | 6.14E-05 | 4.96E-05 | 4.50E-05 | 1.56E-05 | 0.009509823 | SLC25A27 |
| chr19 | 46860045 | 46860692 | 648 | 6 | 6.43E-05 | 7.16E-06 | 0.004993 | 1.56E-05 | 0.00061209 | NA |
| chr7 | 461573 | 461921 | 349 | 3 | 0.000114896 | 8.59E-06 | 0.001675 | 1.56E-05 | 0.009535387 | NA |
| chr15 | 89383714 | 89384519 | 806 | 2 | 0.000401028 | 9.55E-06 | 0.000837 | 1.56E-05 | 0.012547002 | MIR9-3HG |
| chr11 | 113477467 | 113477515 | 49 | 2 | 7.81E-05 | 1.20E-05 | 0.000279 | 1.56E-05 | 0.017294756 | NA |
| chr1 | 179877241 | 179878214 | 974 | 13 | 1.62E-05 | 0.001679 | 0.001076 | 1.56E-05 | 0.002054972 | TOR1AIP2 |
| chr15 | 62822698 | 62823141 | 444 | 3 | 2.43E-05 | 1.82E-05 | 0.000168 | 1.56E-05 | 0.005563032 | TLN2 |
| chr4 | 184476853 | 184477334 | 482 | 2 | 1.09E-05 | 5.86E-05 | 4.10E-05 | 1.56E-05 | 0.010271349 | IRF2-DT |
| chr15 | 73965145 | 73965301 | 157 | 2 | 3.84E-05 | 2.72E-05 | 7.24E-05 | 1.57E-05 | 0.013559204 | NA |
| chr7 | 106365251 | 106365382 | 132 | 2 | 8.87E-05 | 1.03E-05 | 0.000536 | 1.57E-05 | 0.00682435 | NA |
| chr20 | 59604359 | 59605186 | 828 | 8 | 1.92E-05 | 6.10E-05 | 0.003097 | 1.57E-05 | 0.000903658 | PHACTR3 |
| chr7 | 44325326 | 44327053 | 1728 | 17 | 8.01E-05 | 0.020901 | 0.000519 | 1.57E-05 | 0.002745557 | CAMK2B |
| chr5 | 1003782 | 1004664 | 883 | 6 | 3.28E-05 | 2.23E-05 | 0.003866 | 1.57E-05 | 0.016701163 | NA |
| chr6 | 28511607 | 28511935 | 329 | 4 | 7.86E-06 | 0.000288 | 2.65E-05 | 1.57E-05 | 0.004945916 | GPX6 |
| chr3 | 159260463 | 159260594 | 132 | 2 | 4.69E-05 | 4.59E-05 | 4.74E-05 | 1.57E-05 | 0.028210872 | IQCJ-SCHIP1, IQCJ |
| chr14 | 29928202 | 29928557 | 356 | 5 | 3.41E-06 | 0.000387 | 2.20E-05 | 1.57E-05 | 0.008764495 | AL355053.1, PRKD1 |
| chr5 | 150177582 | 150178249 | 668 | 4 | 0.000108322 | 7.51E-06 | 0.003482 | 1.57E-05 | 4.67E-05 | CDX1 |
| chr9 | 134669527 | 134670144 | 618 | 3 | 4.00E-05 | 1.97E-05 | 0.00014 | 1.57E-05 | 0.012041314 | COL5A1 |
| chr16 | 11640051 | 11640799 | 749 | 15 | 2.20E-05 | 0.000213 | 0.001087 | 1.57E-05 | 0.001453568 | NA |
| chr15 | 41920106 | 41921261 | 1156 | 4 | 4.03E-06 | 0.000118 | 0.000166 | 1.57E-05 | 0.007966593 | EHD4 |
| chr17 | 55732066 | 55732203 | 138 | 3 | 6.41E-05 | 7.34E-06 | 0.002155 | 1.58E-05 | 0.014998369 | TMEM100 |
| chr3 | 2156335 | 2156405 | 71 | 2 | 8.54E-05 | 1.32E-05 | 0.000218 | 1.58E-05 | 0.02867914 | CNTN4 |
| chr1 | 25239840 | 25240749 | 910 | 6 | 3.44E-05 | 3.42E-05 | 0.001316 | 1.58E-05 | 0.000473762 | AL031432.5 |
| chr9 | 114414319 | 114414861 | 543 | 2 | 0.00015388 | 9.79E-06 | 0.000777 | 1.58E-05 | 0.000765945 | WHRN |
| chr7 | 94663577 | 94664307 | 731 | 5 | 1.17E-05 | 0.000507 | 0.000249 | 1.58E-05 | -0.00051588 | PEG10 |
| chr12 | 3381048 | 3381516 | 469 | 3 | 9.99E-05 | 1.32E-05 | 0.00031 | 1.58E-05 | 0.009324367 | PRMT8 |
| chr1 | 74673897 | 74674605 | 709 | 5 | 4.76E-06 | 2.82E-05 | 0.00047 | 1.58E-05 | 0.009490347 | NA |
| chr14 | 52244779 | 52245284 | 506 | 2 | 4.50E-05 | 1.51E-05 | 0.00016 | 1.58E-05 | 0.005943236 | NA |
| chr19 | 50968108 | 50968299 | 192 | 2 | 2.96E-05 | 9.89E-06 | 0.000733 | 1.58E-05 | 0.017519809 | AC011483.1, KLK6 |
| chr5 | 159207809 | 159208665 | 857 | 13 | 3.55E-05 | 0.001191 | 0.000918 | 1.58E-05 | -0.00116522 | RNF145 |
| chr6 | 16128695 | 16129176 | 482 | 11 | 5.78E-05 | 3.67E-05 | 0.002899 | 1.58E-05 | -0.00022306 | MYLIP |
| chr1 | 48202124 | 48202313 | 190 | 2 | 0.000102015 | 9.72E-06 | 0.000836 | 1.58E-05 | -0.01408448 | AL109659.1 |
| chr3 | 12967300 | 12967831 | 532 | 11 | 8.27E-05 | 0.000346 | 0.000353 | 1.58E-05 | -0.00124687 | IQSEC1 |
| chr7 | 87151838 | 87152257 | 420 | 8 | 0.000109231 | 1.18E-05 | 0.000748 | 1.59E-05 | -0.00031002 | AC005076.1 |
| chr9 | 131402499 | 131403005 | 507 | 3 | 3.33E-05 | 2.56E-05 | 0.000166 | 1.59E-05 | 0.006199519 | PRRC2B |
| chr9 | 87713796 | 87714161 | 366 | 2 | 2.38E-05 | 2.30E-05 | 8.71E-05 | 1.59E-05 | 0.006895354 | NA |
| chr5 | 1753186 | 1753235 | 50 | 2 | 2.12E-05 | 1.06E-05 | 0.000517 | 1.59E-05 | -0.01215298 | NA |
| chr19 | 6393761 | 6394129 | 369 | 5 | 0.000140948 | 2.35E-05 | 0.000841 | 1.59E-05 | -0.00021319 | AC011491.1, GTF2F1 |
| chr13 | 99501112 | 99502161 | 1050 | 15 | 0.000173713 | 0.000144 | 0.01007 | 1.59E-05 | 4.20E-05 | TM9SF2, AL139035.1 |
| chr22 | 30605779 | 30606375 | 597 | 4 | 3.27E-05 | 1.13E-05 | 0.001715 | 1.59E-05 | 0.011663318 | AC005006.1, PES1 |
| chr2 | 106144729 | 106145349 | 621 | 3 | 7.91E-05 | 2.34E-05 | 0.000339 | 1.59E-05 | 0.011937528 | UXS1 |
| chr2 | 217366669 | 217367205 | 537 | 3 | 0.000116524 | 6.17E-06 | 0.003549 | 1.59E-05 | 0.002892621 | DIRC3 |
| chr7 | 569338 | 570700 | 1363 | 6 | 4.13E-06 | 0.000122 | 9.09E-05 | 1.59E-05 | 0.001842508 | PRKAR1B |
| chr1 | 248691586 | 248692244 | 659 | 7 | 6.38E-05 | 3.06E-05 | 0.001069 | 1.59E-05 | -0.01065441 | LYPD9P, CR589904.2 |
| chr12 | 98773 | 99116 | 344 | 2 | 2.94E-06 | 0.000144 | 2.72E-05 | 1.59E-05 | 0.010242056 | IQSEC3 |
| chr4 | 6587184 | 6587561 | 378 | 2 | 1.45E-05 | 4.00E-05 | 5.39E-05 | 1.59E-05 | 0.003404597 | MAN2B2 |
| chr13 | 112992379 | 112992585 | 207 | 3 | 6.79E-05 | 6.21E-06 | 0.003501 | 1.59E-05 | 0.014840758 | MCF2L |
| chr17 | 74744346 | 74744385 | 40 | 2 | 3.36E-05 | 1.06E-05 | 0.000525 | 1.59E-05 | 0.008447141 | RAB37 |
| chr12 | 3318297 | 3318647 | 351 | 5 | 2.21E-05 | 0.000269 | 0.00154 | 1.60E-05 | 0.006556436 | AC005865.2, AC005865.1 |
| chr14 | 100851283 | 100851482 | 200 | 3 | 8.98E-05 | 8.68E-06 | 0.001623 | 1.60E-05 | 0.018696789 | MEG3, AL117190.1, AL117190.2 |
| chr3 | 50218875 | 50219561 | 687 | 2 | 0.000170213 | 9.84E-06 | 0.00082 | 1.60E-05 | -0.00105273 | SLC38A3 |
| chr16 | 56389632 | 56389948 | 317 | 2 | 5.07E-05 | 1.18E-05 | 0.000332 | 1.60E-05 | 0.003631814 | AMFR |
| chr20 | 543961 | 544958 | 998 | 6 | 1.58E-05 | 8.64E-05 | 0.001162 | 1.60E-05 | -0.0002094 | NA |
| chr1 | 35269201 | 35269566 | 366 | 4 | 4.81E-07 | 0.006049 | 2.53E-05 | 1.60E-05 | -0.00161758 | ZMYM4 |
| chr2 | 80525460 | 80525842 | 383 | 5 | 2.36E-05 | 1.71E-05 | 0.001068 | 1.60E-05 | 0.005827601 | CTNNA2 |
| chr4 | 146057879 | 146058106 | 228 | 2 | 5.70E-05 | 1.08E-05 | 0.000496 | 1.60E-05 | -0.00707079 | NA |
| chr19 | 44754048 | 44755008 | 961 | 4 | 8.77E-06 | 0.000424 | 0.000647 | 1.60E-05 | 0.001649182 | BCL3 |
| chr6 | 168192969 | 168193489 | 521 | 3 | 5.68E-06 | 2.36E-05 | 0.00012 | 1.60E-05 | -0.00511465 | NA |
| chr5 | 138609727 | 138610910 | 1184 | 3 | 3.22E-05 | 8.93E-06 | 0.000943 | 1.60E-05 | 0.010132848 | NA |
| chr8 | 109652746 | 109652754 | 9 | 2 | 9.74E-05 | 1.01E-05 | 0.000736 | 1.60E-05 | 0.018310016 | SYBU |
| chr10 | 1505264 | 1505522 | 259 | 3 | 5.64E-05 | 1.82E-05 | 0.000167 | 1.61E-05 | 0.002192299 | ADARB2 |
| chr15 | 90931226 | 90931659 | 434 | 2 | 1.04E-06 | 1.18E-05 | 0.000332 | 1.61E-05 | 0.020174565 | UNC45A, HDDC3 |
| chr1 | 150256129 | 150257048 | 920 | 4 | 6.67E-06 | 0.000339 | 0.000159 | 1.61E-05 | 0.008000601 | AC242988.1 |
| chr21 | 39006285 | 39006953 | 669 | 4 | 2.13E-05 | 3.80E-05 | 9.32E-05 | 1.61E-05 | 0.004844188 | AF064858.1 |
| chr5 | 127652133 | 127652955 | 823 | 6 | 6.66E-05 | 5.27E-06 | 0.006804 | 1.61E-05 | 0.00108086 | CTXN3, AC022118.1 |
| chr1 | 207818248 | 207818739 | 492 | 7 | 3.77E-05 | 0.001355 | 0.001092 | 1.61E-05 | 0.001940839 | MIR29B2CHG |
| chr1 | 83999317 | 84000005 | 689 | 5 | 3.52E-05 | 6.19E-05 | 7.87E-05 | 1.61E-05 | 0.00707388 | NA |
| chr2 | 102419155 | 102419665 | 511 | 4 | 2.55E-05 | 0.000224 | 0.000419 | 1.61E-05 | -0.00048086 | IL18RAP |
| chr13 | 23427828 | 23428076 | 249 | 2 | 3.20E-05 | 1.81E-05 | 0.000122 | 1.61E-05 | 0.005284299 | SACS-AS1, SACS |
| chr18 | 13726492 | 13726785 | 294 | 10 | 0.000150074 | 7.94E-06 | 0.002749 | 1.61E-05 | 0.001248719 | RNMT, FAM210A |
| chr6 | 105981082 | 105981568 | 487 | 7 | 3.12E-05 | 1.80E-05 | 0.007166 | 1.61E-05 | 0.004756891 | NA |
| chr11 | 57551853 | 57552399 | 547 | 3 | 9.79E-05 | 7.20E-06 | 0.002502 | 1.61E-05 | 0.003134374 | UBE2L6 |
| chr9 | 95858910 | 95859052 | 143 | 3 | 4.28E-05 | 5.40E-05 | 0.000119 | 1.61E-05 | 0.000904343 | LINC00476 |
| chr2 | 106194205 | 106195004 | 800 | 11 | 2.42E-05 | 0.00044 | 0.003387 | 1.62E-05 | 0.000756417 | UXS1 |
| chr6 | 43297601 | 43298108 | 508 | 4 | 2.16E-05 | 2.17E-05 | 0.001225 | 1.62E-05 | 0.007544636 | SLC22A7 |
| chr2 | 1593351 | 1593830 | 480 | 2 | 2.47E-06 | 0.000192 | 2.50E-05 | 1.62E-05 | 0.007502416 | AC141930.1 |
| chr7 | 30157123 | 30157514 | 392 | 6 | 5.12E-05 | 3.13E-05 | 0.000609 | 1.62E-05 | 0.014023361 | MTURN |
| chr6 | 31144891 | 31145481 | 591 | 6 | 2.68E-05 | 4.44E-05 | 0.000456 | 1.62E-05 | 0.002345696 | CCHCR1 |
| chr8 | 120376920 | 120377106 | 187 | 2 | 4.60E-05 | 9.91E-06 | 0.000874 | 1.62E-05 | 0.00319855 | NA |
| chr5 | 2133844 | 2134126 | 283 | 2 | 1.17E-05 | 3.64E-05 | 5.91E-05 | 1.62E-05 | 0.00129163 | NA |
| chr19 | 53535576 | 53535733 | 158 | 2 | 8.37E-05 | 1.20E-05 | 0.00033 | 1.62E-05 | 0.009801855 | ZNF331 |
| chr1 | 31800045 | 31800063 | 19 | 2 | 2.32E-05 | 4.20E-05 | 5.32E-05 | 1.62E-05 | 0.006636623 | SPOCD1 |
| chr14 | 21868153 | 21868222 | 70 | 2 | 5.48E-05 | 1.34E-05 | 0.000233 | 1.62E-05 | 0.00761802 | NA |
| chr8 | 1652885 | 1653148 | 264 | 2 | 2.33E-05 | 0.000173 | 2.60E-05 | 1.63E-05 | -0.01014132 | DLGAP2 |
| chr15 | 60118862 | 60119356 | 495 | 3 | 9.40E-05 | 2.42E-05 | 0.000848 | 1.63E-05 | 0.005592547 | NA |
| chr16 | 276064 | 276728 | 665 | 9 | 2.58E-05 | 7.23E-06 | 0.007936 | 1.63E-05 | 0.003851981 | Z69667.1 |
| chr5 | 172097468 | 172097521 | 54 | 2 | 5.50E-05 | 4.79E-05 | 4.84E-05 | 1.63E-05 | -0.00477701 | STK10 |
| chr14 | 103542912 | 103543390 | 479 | 2 | 1.78E-05 | 2.06E-05 | 0.000104 | 1.63E-05 | 0.01235492 | NA |
| chr17 | 74641833 | 74642166 | 334 | 2 | 5.93E-05 | 1.05E-05 | 0.000629 | 1.63E-05 | -0.00273694 | NA |
| chr19 | 2774522 | 2774551 | 30 | 3 | 0.000211961 | 1.71E-05 | 0.000222 | 1.63E-05 | 0.000991719 | SGTA |
| chr1 | 94238492 | 94238829 | 338 | 4 | 6.57E-06 | 0.000506 | 0.00019 | 1.63E-05 | -0.00959726 | ARHGAP29 |
| chr3 | 112409238 | 112409293 | 56 | 2 | 1.28E-05 | 1.68E-05 | 0.000141 | 1.63E-05 | 0.009943858 | NA |
| chr10 | 110642990 | 110643340 | 351 | 4 | 8.65E-05 | 1.33E-05 | 0.001299 | 1.63E-05 | 0.007140851 | NA |
| chr13 | 51325247 | 51325843 | 597 | 2 | 3.04E-05 | 1.40E-05 | 0.000213 | 1.63E-05 | 0.008228253 | NA |
| chr14 | 24147204 | 24148323 | 1120 | 13 | 3.18E-06 | 0.00038 | 6.72E-05 | 1.63E-05 | 0.00125187 | RNF31, AL136295.5, PSME2 |
| chr13 | 23497720 | 23498006 | 287 | 2 | 4.46E-05 | 1.24E-05 | 0.000299 | 1.64E-05 | 5.75E-05 | NA |

| | | | | | | | | | | |
|---|---|---|---|---|---|---|---|---|---|---|
| chr1 | 54199760 | 54200728 | 969 | 13 | 3.26E-05 | 0.001319 | 0.000639 | 1.64E-05 | 0.000868874 | MRPL37, AL357673.1, CYB5RL |
| chr3 | 18424691 | 18424989 | 299 | 3 | 1.50E-05 | 1.14E-05 | 0.00108 | 1.64E-05 | -0.00062827 | TBC1D5, SATB1 |
| chr2 | 9740821 | 9740892 | 72 | 2 | 2.50E-05 | 1.45E-05 | 0.000194 | 1.64E-05 | 0.009008278 | NA |
| chr10 | 75782006 | 75782824 | 819 | 8 | 0.000107929 | 1.19E-05 | 0.003555 | 1.64E-05 | -0.01058616 | LRMDA |
| chr11 | 108592758 | 108594009 | 1252 | 12 | 2.94E-05 | 0.000182 | 0.005712 | 1.64E-05 | 0.000225162 | EXPH5 |
| chr17 | 45785544 | 45786032 | 489 | 6 | 4.73E-05 | 2.34E-05 | 0.002106 | 1.64E-05 | 0.009310009 | LINC02210-CRHR1, CRHR1 |
| chr11 | 65814194 | 65814291 | 98 | 2 | 5.84E-05 | 2.65E-05 | 7.99E-05 | 1.65E-05 | 0.006222566 | NA |
| chr9 | 113378064 | 113378128 | 65 | 2 | 3.51E-06 | 0.000283 | 2.27E-05 | 1.65E-05 | -0.01250026 | NA |
| chr22 | 37419122 | 37419421 | 300 | 3 | 3.06E-05 | 1.37E-05 | 0.000315 | 1.65E-05 | 0.01569243 | ELFN2, ELFN2 |
| chr22 | 43435085 | 43435241 | 157 | 3 | 9.60E-05 | 7.68E-06 | 0.001584 | 1.65E-05 | 0.006044448 | MPPED1 |
| chr2 | 119428646 | 119429227 | 582 | 3 | 9.68E-05 | 8.35E-06 | 0.001234 | 1.65E-05 | 0.007334068 | NA |
| chr12 | 51291926 | 51292333 | 408 | 2 | 1.17E-05 | 0.00019 | 2.55E-05 | 1.65E-05 | 0.01241839 | BIN2 |
| chr19 | 998979 | 999689 | 711 | 2 | 0.000326468 | 1.01E-05 | 0.000862 | 1.65E-05 | -0.01587578 | NA |
| chr15 | 66650236 | 66650481 | 246 | 2 | 2.85E-05 | 0.000129 | 2.94E-05 | 1.65E-05 | 0.003985455 | LINC01169 |
| chr14 | 105300641 | 105301545 | 905 | 13 | 2.77E-05 | 0.00011 | 0.002614 | 1.65E-05 | 0.001038663 | PACS2, BRF1 |
| chr17 | 79910791 | 79910832 | 42 | 2 | 0.00014864 | 1.02E-05 | 0.000851 | 1.65E-05 | 0.005983339 | NA |
| chr11 | 6319053 | 6319660 | 608 | 3 | 7.34E-05 | 1.00E-05 | 0.001551 | 1.65E-05 | 0.00753396 | AC068733.3, CAVIN3 |
| chr6 | 34091863 | 34091941 | 79 | 2 | 4.13E-05 | 0.000199 | 2.52E-05 | 1.65E-05 | 0.003931738 | GRM4 |
| chr5 | 148646747 | 148647101 | 355 | 2 | 1.70E-05 | 6.94E-05 | 3.96E-05 | 1.65E-05 | 0.008490824 | HTR4 |
| chr19 | 51083845 | 51084152 | 308 | 3 | 4.43E-06 | 0.000239 | 2.31E-05 | 1.66E-05 | 0.000302449 | KLK14 |
| chr2 | 127858052 | 127858593 | 542 | 12 | 2.76E-05 | 5.88E-05 | 0.003557 | 1.66E-05 | 0.001941764 | POLR2D |
| chr8 | 58248712 | 58249077 | 366 | 2 | 6.76E-05 | 1.32E-05 | 0.000259 | 1.66E-05 | -0.00017859 | NA |
| chr19 | 1095522 | 1096085 | 564 | 9 | 7.41E-05 | 0.000132 | 0.006727 | 1.66E-05 | 0.00428524 | NA |
| chr11 | 66729019 | 66729144 | 126 | 2 | 7.17E-05 | 1.93E-05 | 0.000117 | 1.66E-05 | 0.017094176 | SPTBN2 |
| chr17 | 37642869 | 37643571 | 703 | 9 | 7.33E-05 | 3.13E-05 | 0.006469 | 1.66E-05 | 0.002321414 | AC243571.2, DDX52 |
| chr15 | 47716950 | 47717459 | 510 | 5 | 4.52E-05 | 0.000927 | 0.001119 | 1.66E-05 | 0.002082347 | SEMA6D |
| chr6 | 34756671 | 34757339 | 669 | 6 | 1.00E-05 | 0.000226 | 0.001429 | 1.66E-05 | 0.005262173 | NA |
| chr5 | 116573847 | 116574904 | 1058 | 10 | 2.08E-05 | 0.003939 | 8.53E-05 | 1.66E-05 | 0.001620222 | SEMA6A-AS2, SEMA6A |
| chr22 | 50176197 | 50176448 | 252 | 2 | 3.31E-05 | 2.30E-05 | 9.45E-05 | 1.66E-05 | 0.010364839 | PANX2 |
| chr12 | 52192851 | 52193325 | 475 | 2 | 5.08E-07 | 0.000759 | 1.88E-05 | 1.66E-05 | 0.003263506 | LINC00592 |
| chr3 | 31132433 | 31132740 | 308 | 2 | 1.50E-05 | 0.000557 | 1.98E-05 | 1.66E-05 | 0.014974096 | NA |
| chr12 | 130953732 | 130954243 | 512 | 9 | 7.17E-05 | 1.71E-05 | 0.005189 | 1.67E-05 | -0.00397412 | ADGRD1 |
| chr20 | 56624437 | 56625389 | 953 | 5 | 1.98E-06 | 0.000119 | 3.51E-05 | 1.67E-05 | 0.007214699 | NA |
| chr10 | 8052130 | 8052897 | 768 | 8 | 2.51E-05 | 6.05E-06 | 0.012189 | 1.67E-05 | 0.006304728 | GATA3, AL390294.1, GATA3-AS1 |
| chr11 | 20598741 | 20599368 | 628 | 6 | 4.33E-05 | 4.81E-05 | 0.001032 | 1.67E-05 | 0.007397352 | NA |
| chr3 | 155382520 | 155382609 | 90 | 2 | 8.24E-05 | 1.82E-05 | 0.000131 | 1.67E-05 | 0.016781958 | PLCH1 |
| chr14 | 104653046 | 104653225 | 180 | 2 | 2.19E-05 | 0.000101 | 3.31E-05 | 1.67E-05 | -0.00510225 | NA |
| chr3 | 128399346 | 128399378 | 33 | 2 | 2.25E-05 | 1.87E-05 | 0.000125 | 1.67E-05 | 0.003995859 | EEFSEC |
| chr1 | 37556229 | 37557393 | 1165 | 13 | 0.000181359 | 2.58E-06 | 0.033982 | 1.67E-05 | 0.006328528 | AL034379.1, DNALI1 |
| chr5 | 180851031 | 180851633 | 603 | 4 | 2.27E-05 | 0.000104 | 8.79E-05 | 1.68E-05 | -0.00190904 | ZFP62 |
| chr1 | 1943049 | 1943459 | 411 | 3 | 8.35E-05 | 9.98E-06 | 0.00145 | 1.68E-05 | 0.004900797 | CFAP74 |
| chr17 | 4276700 | 4277275 | 576 | 2 | 8.87E-05 | 1.16E-05 | 0.00044 | 1.68E-05 | -0.00950233 | UBE2G1 |
| chr19 | 3030092 | 3031029 | 938 | 5 | 8.63E-05 | 1.82E-05 | 0.002826 | 1.68E-05 | 0.006679979 | TLE2 |
| chr17 | 82094878 | 82095329 | 452 | 4 | 9.19E-06 | 0.00019 | 3.40E-05 | 1.68E-05 | 0.001562772 | FASN |
| chr11 | 64249865 | 64250147 | 283 | 2 | 4.50E-06 | 8.97E-05 | 3.51E-05 | 1.68E-05 | 0.00258373 | NA |
| chr14 | 99382915 | 99383850 | 936 | 6 | 2.25E-06 | 0.001176 | 0.000111 | 1.68E-05 | 0.001717079 | NA |
| chr1 | 147756774 | 147757243 | 470 | 3 | 1.82E-05 | 0.000115 | 3.35E-05 | 1.68E-05 | 0.000626792 | AC241644.3, GJA5 |
| chr2 | 1234541 | 1234654 | 114 | 4 | 1.45E-05 | 0.000136 | 0.000493 | 1.68E-05 | 0.004253954 | SNTG2 |
| chr5 | 132737589 | 132738344 | 756 | 9 | 0.000128232 | 5.93E-06 | 0.015907 | 1.69E-05 | 0.00428855 | KIF3A |
| chr6 | 89561620 | 89562773 | 1154 | 11 | 3.22E-06 | 0.011923 | 7.28E-05 | 1.69E-05 | -0.00036512 | ANKRD6, AL159174.1 |
| chr1 | 235504048 | 235505121 | 1074 | 10 | 2.99E-06 | 0.000742 | 0.000171 | 1.69E-05 | 0.000526829 | B3GALNT2 |
| chr19 | 5669463 | 5669898 | 436 | 2 | 4.58E-05 | 1.78E-05 | 0.000138 | 1.69E-05 | 0.012473386 | NA |
| chr2 | 54749771 | 54749805 | 35 | 2 | 9.68E-05 | 1.28E-05 | 0.000311 | 1.69E-05 | 0.00927456 | EML6, EML6-AS1 |
| chr4 | 121380852 | 121381525 | 674 | 9 | 3.79E-05 | 3.23E-05 | 0.000192 | 1.69E-05 | 0.004078347 | QRFPR |
| chr9 | 29213783 | 29214451 | 669 | 5 | 1.86E-05 | 0.000436 | 0.000125 | 1.69E-05 | 0.00316719 | AL162388.2 |
| chr3 | 55486806 | 55487833 | 1028 | 15 | 2.82E-05 | 4.05E-05 | 0.000706 | 1.69E-05 | 0.001583348 | WNT5A-AS1, WNT5A |
| chr8 | 81522233 | 81522308 | 76 | 2 | 5.73E-05 | 2.15E-05 | 0.000105 | 1.69E-05 | 0.006038264 | AC023644.1 |
| chr5 | 141330419 | 141331121 | 703 | 6 | 3.18E-05 | 9.40E-06 | 0.002309 | 1.69E-05 | 0.00277906 | PCDHGA1 |
| chr8 | 95305240 | 95305377 | 138 | 2 | 9.71E-05 | 1.07E-05 | 0.000725 | 1.69E-05 | 0.021637804 | C8orf37-AS1 |
| chr4 | 184779506 | 184780170 | 665 | 2 | 0.000109889 | 1.09E-05 | 0.000654 | 1.69E-05 | -0.00701947 | ACSL1 |
| chr22 | 26902872 | 26903337 | 466 | 6 | 5.21E-06 | 0.000155 | 5.64E-05 | 1.69E-05 | 0.00783688 | LINC01422, Z97353.2 |
| chr5 | 1088089 | 1088455 | 367 | 4 | 4.25E-05 | 2.83E-05 | 0.000862 | 1.70E-05 | 0.002792563 | SLC12A7 |
| chr1 | 1894422 | 1894788 | 367 | 2 | 7.10E-05 | 1.38E-05 | 0.000248 | 1.70E-05 | 0.011180908 | NA |
| chr18 | 24014057 | 24014286 | 230 | 8 | 6.60E-05 | 1.19E-05 | 0.002599 | 1.70E-05 | -5.48E-05 | TTC39C, TTC39C-AS1 |
| chr19 | 15524816 | 15525292 | 477 | 5 | 0.000156687 | 3.61E-06 | 0.011284 | 1.70E-05 | 0.004314011 | CYP4F22 |
| chr8 | 22161383 | 22161762 | 380 | 4 | 2.98E-05 | 6.07E-05 | 7.93E-05 | 1.70E-05 | 0.006504522 | SFTPC |
| chr11 | 73970236 | 73970292 | 57 | 2 | 1.85E-05 | 0.00042 | 2.13E-05 | 1.70E-05 | 0.012524915 | DNAJB13, AP003717.1 |
| chr19 | 55385081 | 55386191 | 1111 | 16 | 2.22E-05 | 9.37E-05 | 0.008954 | 1.70E-05 | -0.00031305 | RPL28 |
| chr16 | 30980406 | 30980807 | 402 | 2 | 2.35E-05 | 1.12E-05 | 0.000573 | 1.70E-05 | 0.005087712 | SETD1A |
| chr3 | 54967093 | 54967623 | 531 | 3 | 3.93E-05 | 5.95E-05 | 8.80E-05 | 1.70E-05 | 0.008415251 | CACNA2D3 |
| chr11 | 36374962 | 36375577 | 616 | 4 | 2.32E-05 | 3.17E-05 | 0.001644 | 1.70E-05 | 0.009394323 | PRR5L |
| chr7 | 76305097 | 76305167 | 71 | 2 | 5.40E-05 | 2.94E-05 | 7.60E-05 | 1.70E-05 | 0.011122377 | NA |
| chr1 | 202057041 | 202057196 | 156 | 2 | 4.65E-05 | 1.10E-05 | 0.000629 | 1.71E-05 | 0.004509154 | NA |
| chr12 | 53960149 | 53960806 | 658 | 4 | 5.78E-05 | 4.07E-05 | 0.000289 | 1.71E-05 | 0.010428551 | NA |
| chr5 | 9066513 | 9066553 | 41 | 2 | 4.04E-05 | 0.000349 | 2.22E-05 | 1.71E-05 | 0.004159055 | SEMA5A |
| chr8 | 54101706 | 54102483 | 778 | 8 | 5.31E-05 | 3.12E-05 | 0.000333 | 1.71E-05 | 0.000534409 | LYPLA1 |
| chr1 | 197915975 | 197917288 | 1314 | 7 | 1.63E-07 | 0.000306 | 3.18E-05 | 1.71E-05 | 0.001776728 | LHX9 |
| chr15 | 41877927 | 41878435 | 509 | 2 | 4.87E-05 | 2.02E-05 | 0.000116 | 1.71E-05 | -0.00408373 | SPTBN5 |
| chr15 | 25053706 | 25054064 | 359 | 4 | 7.08E-05 | 2.28E-05 | 0.000771 | 1.71E-05 | -0.00279227 | SNHG14 |
| chr16 | 19086410 | 19086876 | 467 | 6 | 3.34E-05 | 1.14E-05 | 0.006398 | 1.71E-05 | -0.00019652 | AC099518.1 |
| chr20 | 44710677 | 44710999 | 323 | 2 | 1.65E-05 | 6.60E-05 | 4.23E-05 | 1.71E-05 | 0.004960964 | KCNK15-AS1 |
| chr15 | 75903326 | 75904221 | 896 | 8 | 5.92E-06 | 0.000131 | 0.000365 | 1.71E-05 | 0.001411192 | FBXO22 |
| chr11 | 118928529 | 118929145 | 617 | 8 | 1.76E-05 | 0.000143 | 5.69E-05 | 1.71E-05 | 0.001837737 | UPK2 |
| chr3 | 42042759 | 42043562 | 804 | 3 | 5.80E-05 | 0.000593 | 0.000111 | 1.71E-05 | 0.002872828 | TRAK1 |
| chr8 | 53807871 | 53808164 | 294 | 2 | 6.68E-05 | 1.48E-05 | 0.000214 | 1.72E-05 | 0.006324099 | ATP6V1H |
| chr19 | 12858390 | 12858797 | 408 | 3 | 3.87E-05 | 4.80E-05 | 0.000475 | 1.72E-05 | 0.000720266 | MAST1, HOOK2 |
| chr8 | 66428513 | 66429125 | 613 | 10 | 5.75E-05 | 0.006433 | 0.000296 | 1.72E-05 | 0.000863524 | RRS1, AC009879.4, RRS1-AS1 |
| chr8 | 143888763 | 143888813 | 51 | 2 | 7.77E-05 | 1.15E-05 | 0.000515 | 1.72E-05 | 0.007300354 | AC109322.2 |
| chr16 | 67206301 | 67206735 | 435 | 2 | 1.00E-05 | 5.48E-05 | 4.72E-05 | 1.72E-05 | 0.005381505 | AC040160.2 |
| chr9 | 136484363 | 136485204 | 842 | 5 | 1.20E-05 | 1.91E-05 | 0.001115 | 1.72E-05 | 0.006807347 | C9orf163 |
| chr5 | 177996597 | 177996998 | 402 | 2 | 3.04E-05 | 1.08E-05 | 0.000761 | 1.72E-05 | 0.014238517 | NA |
| chr19 | 7846234 | 7846665 | 432 | 6 | 6.88E-05 | 7.88E-05 | 0.000552 | 1.72E-05 | -0.00011019 | EVI5L |
| chr17 | 45135375 | 45135903 | 529 | 11 | 0.000181447 | 2.87E-05 | 0.002857 | 1.72E-05 | 0.000665232 | ACBD4 |
| chr11 | 126269112 | 126270215 | 1104 | 7 | 4.01E-05 | 0.000166 | 0.000161 | 1.73E-05 | 0.000511134 | FOXRED1, SRPRA |
| chr7 | 2127681 | 2127868 | 188 | 3 | 4.33E-05 | 0.000859 | 0.000461 | 1.73E-05 | -0.00054296 | MAD1L1, AC069288.1 |
| chr6 | 16681274 | 16681609 | 336 | 3 | 4.24E-05 | 1.48E-05 | 0.000686 | 1.73E-05 | 0.008823591 | ATXN1 |
| chr12 | 7132314 | 7132884 | 571 | 3 | 8.56E-05 | 6.78E-06 | 0.003542 | 1.73E-05 | 0.001415598 | CLSTN3 |
| chr18 | 70157959 | 70158546 | 588 | 2 | 0.000164683 | 1.08E-05 | 0.000819 | 1.73E-05 | 0.011707109 | RTTN |
| chr5 | 58378690 | 58378840 | 151 | 2 | 3.59E-05 | 0.000239 | 2.50E-05 | 1.73E-05 | 0.002311823 | NA |
| chr19 | 51337188 | 51337300 | 113 | 2 | 7.52E-05 | 1.41E-05 | 0.000248 | 1.73E-05 | 0.003894525 | VSIG10L |
| chr6 | 48837740 | 48837992 | 253 | 3 | 0.000116202 | 7.70E-06 | 0.002618 | 1.73E-05 | 0.001847579 | NA |
| chr6 | 2377126 | 2377466 | 341 | 2 | 4.67E-05 | 2.25E-05 | 0.000103 | 1.73E-05 | 0.001752291 | GMDS-DT, AL031768.2 |
| chr12 | 123606449 | 123606501 | 53 | 2 | 9.16E-05 | 1.66E-05 | 0.000167 | 1.73E-05 | -0.00354684 | DDX55 |
| chr15 | 40931468 | 40932037 | 570 | 2 | 3.94E-07 | 0.000364 | 2.23E-05 | 1.73E-05 | 0.00565856 | DLL4 |
| chr19 | 54533925 | 54534035 | 111 | 3 | 8.70E-05 | 8.83E-06 | 0.002071 | 1.73E-05 | -0.00400381 | KIR3DX1 |
| chr3 | 51963369 | 51964035 | 667 | 5 | 2.12E-06 | 0.000249 | 0.001264 | 1.73E-05 | 0.005032655 | PCBP4, AC115284.1 |
| chr14 | 74713102 | 74713831 | 730 | 10 | 3.59E-05 | 0.000164 | 0.002336 | 1.73E-05 | 0.005131321 | FCF1, AREL1 |
| chr8 | 143328531 | 143328802 | 272 | 3 | 6.49E-05 | 7.55E-06 | 0.002243 | 1.73E-05 | 0.008598099 | TOP1MT |
| chr13 | 43787488 | 43788120 | 633 | 5 | 1.79E-05 | 2.52E-05 | 0.000858 | 1.73E-05 | 0.010272715 | AL162713.2, AL162713.1 |
| chr2 | 7012103 | 7012814 | 712 | 4 | 1.78E-05 | 0.000601 | 3.03E-05 | 1.73E-05 | 0.002160043 | RNF144A |
| chr12 | 12717185 | 12718176 | 992 | 11 | 6.77E-06 | 0.000131 | 0.000233 | 1.74E-05 | 0.002140259 | CDKN1B |
| chr5 | 143243881 | 143244227 | 347 | 2 | 4.37E-05 | 2.24E-05 | 0.000104 | 1.74E-05 | 0.007800006 | NA |
| chr12 | 69611263 | 69611829 | 567 | 3 | 3.49E-05 | 2.36E-05 | 0.00029 | 1.74E-05 | 0.0047573 | NA |
| chr17 | 42609120 | 42609354 | 235 | 9 | 0.000223947 | 0.000435 | 5.71E-05 | 1.74E-05 | -0.00059224 | RETREG3 |
| chr1 | 202603501 | 202604067 | 567 | 2 | 7.06E-07 | 0.000834 | 1.92E-05 | 1.74E-05 | 0.005761888 | SYT2 |
| chr2 | 98508407 | 98508829 | 423 | 4 | 2.75E-05 | 6.20E-05 | 0.000648 | 1.74E-05 | 0.01208015 | INPP4A |
| chr19 | 12757713 | 12758133 | 421 | 6 | 3.06E-05 | 0.000159 | 0.000151 | 1.74E-05 | -0.00088087 | BEST2 |
| chr15 | 79035160 | 79035469 | 310 | 2 | 7.04E-05 | 1.09E-05 | 0.000764 | 1.74E-05 | 0.015168347 | RASGRF1 |
| chr7 | 158941939 | 158942439 | 501 | 6 | 7.14E-05 | 3.25E-05 | 0.00224 | 1.74E-05 | -0.00132196 | DYNC2I1 |
| chr1 | 167795935 | 167795974 | 40 | 2 | 4.92E-05 | 1.54E-05 | 0.000203 | 1.74E-05 | 0.010800295 | NA |
| chr1 | 8283211 | 8283515 | 305 | 2 | 7.93E-06 | 0.000354 | 2.26E-05 | 1.74E-05 | 0.011094362 | NA |
| chr10 | 103396209 | 103396933 | 725 | 20 | 4.40E-07 | 0.003928 | 0.001112 | 1.74E-05 | 5.28E-05 | PDCD11, ATP5MD |
| chr11 | 74921277 | 74921442 | 166 | 2 | 7.95E-05 | 1.85E-05 | 0.000138 | 1.74E-05 | 0.00046886 | XRRA1 |
| chr22 | 27299457 | 27300242 | 786 | 3 | 1.59E-05 | 0.000112 | 4.73E-05 | 1.74E-05 | 0.001620171 | NA |
| chr7 | 1909996 | 1910842 | 847 | 6 | 1.92E-05 | 4.28E-05 | 0.002241 | 1.74E-05 | 0.003218047 | MAD1L1, AC069288.1 |
| chr19 | 38374558 | 38375013 | 456 | 6 | 5.61E-05 | 3.72E-05 | 0.001247 | 1.74E-05 | 0.000794798 | PSMD8 |
| chr4 | 6692294 | 6692463 | 170 | 2 | 7.89E-06 | 1.39E-05 | 0.000271 | 1.74E-05 | 0.008710853 | AC093323.2 |
| chr16 | 5071458 | 5071972 | 515 | 8 | 1.95E-05 | 0.000142 | 0.000831 | 1.74E-05 | 0.006399453 | ALG1 |
| chr15 | 72289228 | 72289381 | 154 | 2 | 1.77E-05 | 1.21E-05 | 0.000454 | 1.74E-05 | -0.00036922 | AC009690.2, AC009690.3, CELF6, AC009690.1 |
| chr11 | 86395122 | 86395431 | 310 | 2 | 2.32E-05 | 1.14E-05 | 0.00062 | 1.74E-05 | 0.010117141 | CCDC81 |
| chr1 | 31181219 | 31181606 | 388 | 2 | 1.34E-06 | 4.46E-05 | 5.58E-05 | 1.74E-05 | 0.013503706 | NKAIN1 |
| chr11 | 13964364 | 13964801 | 438 | 3 | 7.58E-05 | 1.10E-05 | 0.000621 | 1.75E-05 | 0.010634554 | SPON1 |
| chr6 | 26330989 | 26332024 | 1036 | 6 | 9.21E-06 | 3.09E-05 | 0.004046 | 1.75E-05 | 0.002144486 | AL021917.1 |
| chr5 | 103413430 | 103413713 | 284 | 3 | 2.15E-05 | 0.00119 | 1.50E-05 | 1.75E-05 | -0.00016857 | NA |
| chr9 | 706082 | 706836 | 755 | 7 | 4.90E-05 | 1.23E-05 | 0.003664 | 1.75E-05 | 0.013313575 | KANK1 |
| chr19 | 51342605 | 51343549 | 945 | 4 | 2.74E-06 | 0.000532 | 5.99E-05 | 1.75E-05 | 0.00323121 | AC008750.2 |
| chr11 | 72776518 | 72777235 | 718 | 3 | 0.000112995 | 1.04E-05 | 0.001357 | 1.75E-05 | 0.008176229 | ARAP1, STARD10 |
| chr5 | 1171855 | 1172426 | 572 | 2 | 8.21E-05 | 1.36E-05 | 0.000296 | 1.75E-05 | 0.007786493 | NA |
| chr16 | 2712568 | 2712898 | 331 | 6 | 3.93E-05 | 0.000156 | 0.000523 | 1.75E-05 | 0.000565417 | PRSS27 |
| chr17 | 41401112 | 41401337 | 226 | 2 | 1.71E-05 | 1.34E-05 | 0.00031 | 1.75E-05 | 0.001095193 | NA |
| chr19 | 813735 | 814520 | 786 | 2 | 5.94E-07 | 0.000125 | 3.18E-05 | 1.75E-05 | 0.003194353 | PLPPR3 |
| chr1 | 165543743 | 165544213 | 471 | 7 | 1.67E-05 | 3.33E-05 | 0.002266 | 1.75E-05 | 0.007556533 | LRRC52, LRRC52-AS1 |
| chr1 | 74752995 | 74753237 | 243 | 2 | 2.08E-05 | 0.000205 | 2.66E-05 | 1.75E-05 | 0.00268537 | TYW3 |
| chr10 | 4121439 | 4121689 | 251 | 2 | 5.51E-05 | 1.27E-05 | 0.000376 | 1.75E-05 | 0.016775139 | NA |
| chr16 | 3731319 | 3731829 | 511 | 5 | 3.26E-06 | 0.00075 | 4.50E-05 | 1.75E-05 | -0.00173284 | CREBBP |
| chr9 | 37984947 | 37985120 | 174 | 2 | 8.78E-05 | 1.11E-05 | 0.00076 | 1.76E-05 | 0.010148371 | AL138752.2, SHB |
| chr1 | 212791391 | 212792406 | 1016 | 17 | 1.11E-05 | 0.00014 | 6.86E-05 | 1.76E-05 | 0.000941045 | TATDN3, NSL1 |
| chr18 | 58220531 | 58221163 | 633 | 2 | 3.66E-08 | 0.000746 | 1.98E-05 | 1.76E-05 | -0.00286645 | NEDD4L |
| chr11 | 45907818 | 45908269 | 452 | 3 | 4.82E-06 | 2.32E-05 | 0.000263 | 1.76E-05 | 0.004933032 | NA |
| chr20 | 56575968 | 56576451 | 484 | 2 | 2.80E-05 | 1.54E-05 | 0.000209 | 1.76E-05 | 0.010961372 | NA |
| chr1 | 17527447 | 17527779 | 333 | 2 | 4.00E-05 | 3.39E-05 | 7.02E-05 | 1.76E-05 | 0.014202823 | NA |
| chr16 | 87722012 | 87722148 | 137 | 2 | 0.000192378 | 1.50E-05 | 0.000222 | 1.76E-05 | 0.005103164 | KLHDC4 |
| chr19 | 48452683 | 48453140 | 458 | 5 | 3.35E-05 | 2.83E-05 | 0.000219 | 1.77E-05 | 0.002130638 | GRWD1 |
| chr22 | 19756166 | 19756716 | 551 | 8 | 8.94E-05 | 2.25E-05 | 0.0002 | 1.77E-05 | 0.003516721 | TBX1 |
| chr4 | 5888562 | 5889024 | 463 | 9 | 0.000101793 | 6.42E-05 | 0.003358 | 1.77E-05 | 0.000451365 | CRMP1 |
| chr5 | 1765197 | 1765636 | 440 | 2 | 2.27E-06 | 0.000397 | 2.23E-05 | 1.77E-05 | 0.007740988 | NA |
| chr1 | 180252693 | 180253160 | 468 | 2 | 4.34E-06 | 0.008675 | 1.58E-05 | 1.77E-05 | 0.011173421 | LHX4 |
| chr2 | 219685529 | 219685641 | 113 | 3 | 0.000120264 | 1.15E-05 | 0.001561 | 1.77E-05 | 0.022072259 | LINC02832 |
| chr19 | 436541 | 437061 | 521 | 2 | 0.000143794 | 1.09E-05 | 0.000864 | 1.77E-05 | 0.012293029 | SHC2 |
| chr15 | 63596894 | 63597699 | 806 | 9 | 5.32E-05 | 2.05E-05 | 0.003147 | 1.77E-05 | 0.003386449 | FBXL22, USP3-AS1 |
| chr5 | 180809652 | 180810544 | 893 | 11 | 2.44E-05 | 6.96E-05 | 0.004325 | 1.77E-05 | 0.001062973 | AC022413.1, MGAT1 |
| chr9 | 108997383 | 108997570 | 188 | 2 | 0.000103029 | 1.14E-05 | 0.000651 | 1.77E-05 | 0.009653298 | CTNNAL1 |

| | | | | | | | | | | |
|---|---|---|---|---|---|---|---|---|---|---|
| chr14 | 24334918 | 24336183 | 1266 | 8 | 6.64E-06 | 0.000106 | 0.000813 | 1.77E-05 | 0.004959047 | ADCY4, AL096870.1, RIPK3 |
| chr1 | 37859723 | 37860302 | 580 | 6 | 7.78E-05 | 1.08E-05 | 0.001068 | 1.77E-05 | -0.0002075 | NA |
| chr4 | 141635375 | 141635959 | 585 | 2 | 0.000119333 | 1.09E-05 | 0.00092 | 1.77E-05 | 0.010501062 | NA |
| chr12 | 132956294 | 132956509 | 216 | 5 | 0.000239421 | 3.14E-05 | 0.001316 | 1.77E-05 | -0.00126099 | CHFR, ZNF605 |
| chr12 | 56193302 | 56193742 | 441 | 2 | 4.19E-05 | 2.15E-05 | 0.000114 | 1.77E-05 | 0.017180413 | NA |
| chr10 | 31756419 | 31756939 | 521 | 3 | 9.68E-05 | 7.11E-06 | 0.003272 | 1.77E-05 | 0.00569385 | NA |
| chr19 | 17969683 | 17969986 | 304 | 2 | 4.91E-05 | 1.55E-05 | 0.000211 | 1.77E-05 | 0.009270562 | KCNN1 |
| chr8 | 26451660 | 26451998 | 339 | 2 | 2.94E-05 | 4.27E-05 | 5.87E-05 | 1.77E-05 | 0.009882293 | BNIP3L |
| chr1 | 1163520 | 1164271 | 752 | 8 | 2.41E-05 | 0.000162 | 0.000115 | 1.77E-05 | -0.00031572 | NA |
| chr21 | 42395950 | 42396246 | 297 | 6 | 2.80E-05 | 0.000166 | 0.000688 | 1.78E-05 | 0.002469857 | TMPRSS3 |
| chr3 | 50612563 | 50613332 | 770 | 4 | 1.41E-05 | 1.05E-05 | 0.003094 | 1.78E-05 | 0.009922315 | MAPKAPK3 |
| chr15 | 93476998 | 93477109 | 112 | 2 | 8.92E-05 | 1.12E-05 | 0.00078 | 1.78E-05 | 0.006669775 | AC091078.1 |
| chr10 | 62000113 | 62000150 | 38 | 2 | 6.53E-05 | 2.06E-05 | 0.000123 | 1.78E-05 | 0.001469559 | ARID5B |
| chr2 | 111899052 | 111899372 | 321 | 3 | 3.24E-05 | 4.96E-05 | 0.000104 | 1.78E-05 | 0.000165744 | MERTK |
| chr3 | 184275292 | 184275938 | 647 | 9 | 4.08E-05 | 3.67E-05 | 0.00069 | 1.78E-05 | 0.006131142 | EEF1AKMT4-ECE2 |
| chr19 | 57279898 | 57281026 | 1129 | 12 | 1.39E-06 | 0.000513 | 0.000926 | 1.78E-05 | -0.00152526 | ZNF460, ZNF460-AS1 |
| chr8 | 29885976 | 29886071 | 96 | 2 | 8.37E-05 | 2.24E-05 | 0.00011 | 1.78E-05 | 0.015322975 | NA |
| chr10 | 99236313 | 99237266 | 954 | 7 | 4.50E-06 | 8.62E-05 | 0.001634 | 1.78E-05 | 0.005855511 | NA |
| chr8 | 123206333 | 123206837 | 505 | 4 | 1.02E-05 | 2.07E-05 | 0.000388 | 1.78E-05 | -0.00894254 | FAM83A |
| chr19 | 5967062 | 5967152 | 91 | 2 | 2.13E-05 | 7.02E-05 | 4.27E-05 | 1.78E-05 | 0.00733592 | RANBP3 |
| chr22 | 38033766 | 38033886 | 121 | 2 | 3.74E-05 | 1.37E-05 | 0.000307 | 1.78E-05 | 0.009932466 | POLR2F, AL031587.1 |
| chr11 | 109424564 | 109425041 | 478 | 3 | 1.00E-05 | 4.42E-05 | 0.000255 | 1.78E-05 | 0.014725496 | C11orf87, AP003049.2 |
| chr20 | 1657911 | 1659170 | 1260 | 6 | 3.28E-05 | 0.001072 | 6.50E-05 | 1.78E-05 | 0.002538098 | NA |
| chr14 | 85622150 | 85622919 | 770 | 5 | 8.53E-06 | 0.000157 | 8.54E-05 | 1.78E-05 | 0.004224976 | FLRT2 |
| chr7 | 73616098 | 73616109 | 12 | 2 | 7.45E-05 | 3.97E-05 | 6.25E-05 | 1.79E-05 | 0.01201138 | MLXIPL |
| chr19 | 6039815 | 6040069 | 255 | 3 | 5.17E-05 | 2.47E-05 | 0.000307 | 1.79E-05 | 0.005430241 | RFX2 |
| chr20 | 3712511 | 3712574 | 64 | 3 | 7.47E-05 | 9.01E-06 | 0.001611 | 1.79E-05 | 0.009840991 | NA |
| chr18 | 12237045 | 12237370 | 326 | 3 | 1.23E-05 | 0.001603 | 3.47E-05 | 1.79E-05 | 0.009745065 | NA |
| chr18 | 58257443 | 58257656 | 214 | 2 | 1.37E-05 | 0.000167 | 2.93E-05 | 1.79E-05 | 0.01371555 | NEDD4L |
| chr17 | 7393475 | 7394017 | 543 | 6 | 0.000103115 | 1.64E-05 | 0.000872 | 1.79E-05 | 0.002089394 | PLSCR3, TMEM256-PLSCR3 |
| chr22 | 40646383 | 40646470 | 88 | 2 | 3.87E-05 | 0.000239 | 2.59E-05 | 1.79E-05 | -0.00223341 | NA |
| chr3 | 153388359 | 153388692 | 334 | 2 | 0.000103518 | 1.14E-05 | 0.000731 | 1.79E-05 | 0.003297525 | LINC02006 |
| chr16 | 88316118 | 88316176 | 59 | 2 | 2.34E-05 | 0.000732 | 2.01E-05 | 1.79E-05 | 0.00957508 | NA |
| chr17 | 7579138 | 7579424 | 287 | 5 | 6.96E-05 | 1.97E-05 | 0.000267 | 1.79E-05 | 0.008175075 | AC016876.2 |
| chr18 | 79714974 | 79715340 | 367 | 3 | 1.26E-05 | 0.000698 | 5.46E-05 | 1.79E-05 | -0.00249176 | CTDP1 |
| chr15 | 101611714 | 101612314 | 601 | 3 | 0.000111527 | 8.37E-06 | 0.001633 | 1.79E-05 | 0.006411502 | LINC02348 |
| chr19 | 10751187 | 10751424 | 238 | 2 | 4.39E-05 | 3.58E-05 | 6.88E-05 | 1.79E-05 | 0.010427869 | DNM2 |
| chr19 | 42428694 | 42428872 | 179 | 2 | 4.45E-05 | 3.21E-05 | 7.57E-05 | 1.79E-05 | 0.004408343 | LIPE-AS1, CXCL17 |
| chr7 | 140041756 | 140042285 | 530 | 4 | 2.49E-05 | 6.75E-05 | 0.000238 | 1.79E-05 | 0.001693263 | PARP12 |
| chr7 | 155469795 | 155470654 | 860 | 3 | 7.22E-06 | 0.000141 | 3.94E-05 | 1.79E-05 | 0.011135059 | NA |
| chr2 | 150611941 | 150612210 | 270 | 2 | 3.66E-05 | 1.35E-05 | 0.000341 | 1.80E-05 | 0.007394441 | AC104777.1 |
| chr2 | 74924912 | 74925213 | 302 | 4 | 2.17E-05 | 0.000144 | 0.000295 | 1.80E-05 | -0.0037027 | LINC01291 |
| chr17 | 79747208 | 79747460 | 253 | 2 | 3.00E-05 | 1.11E-05 | 0.000882 | 1.80E-05 | 0.01386061 | NA |
| chr3 | 169665243 | 169665504 | 262 | 2 | 1.31E-05 | 2.12E-05 | 0.000121 | 1.80E-05 | 0.0088018 | NA |
| chr3 | 10949282 | 10949392 | 111 | 2 | 5.10E-05 | 1.53E-05 | 0.00023 | 1.80E-05 | 0.010606868 | AC027128.1 |
| chr1 | 84505328 | 84505830 | 503 | 6 | 5.17E-06 | 0.000804 | 0.001337 | 1.80E-05 | -0.00318893 | GNG5, AL359762.3 |
| chr4 | 186147903 | 186147997 | 95 | 3 | 4.95E-05 | 0.000149 | 0.000367 | 1.80E-05 | 0.00294887 | FAM149A |
| chr11 | 56029714 | 56030730 | 1017 | 4 | 7.26E-07 | 0.028944 | 1.40E-05 | 1.80E-05 | 0.003722583 | OR5AS1 |
| chr9 | 37159137 | 37159665 | 529 | 3 | 3.16E-05 | 0.0003 | 0.000537 | 1.80E-05 | 0.012905255 | ZCCHC7 |
| chr5 | 111782206 | 111782372 | 167 | 2 | 8.81E-05 | 1.67E-05 | 0.000185 | 1.80E-05 | 0.003918019 | NREP |
| chr3 | 157174100 | 157174240 | 141 | 2 | 6.40E-06 | 7.02E-05 | 4.32E-05 | 1.80E-05 | -0.00773256 | AC092944.1 |
| chr1 | 225412419 | 225412589 | 171 | 4 | 6.12E-05 | 6.76E-06 | 0.004055 | 1.80E-05 | 0.007069467 | LBR |
| chr19 | 46600861 | 46601606 | 746 | 10 | 3.58E-05 | 0.000248 | 0.000274 | 1.80E-05 | 0.000108815 | CALM3, PPP5D1 |
| chr5 | 154376687 | 154376778 | 92 | 2 | 1.90E-05 | 9.00E-05 | 3.82E-05 | 1.80E-05 | 0.004058058 | GALNT10, SAP30L-AS1 |
| chr16 | 6019008 | 6019730 | 723 | 11 | 3.06E-05 | 6.31E-05 | 0.001715 | 1.80E-05 | 0.006096093 | RBFOX1 |
| chr8 | 98045183 | 98045702 | 520 | 7 | 0.000200575 | 2.54E-05 | 0.000439 | 1.80E-05 | -2.43E-08 | RPL30 |
| chr15 | 76311316 | 76311772 | 457 | 12 | 5.10E-05 | 3.04E-05 | 0.012689 | 1.80E-05 | 0.004151739 | ETFA |
| chr5 | 73813395 | 73813772 | 378 | 5 | 0.000108583 | 1.28E-05 | 0.003326 | 1.81E-05 | 0.007500253 | ARHGEF28 |
| chr17 | 39912454 | 39912621 | 168 | 2 | 2.47E-05 | 0.000195 | 2.81E-05 | 1.81E-05 | 0.004481366 | GSDMB |
| chr1 | 224498767 | 224499099 | 333 | 3 | 5.49E-05 | 3.46E-05 | 0.000269 | 1.81E-05 | 0.01091457 | CNIH3 |
| chr16 | 14078991 | 14079088 | 98 | 2 | 0.000120936 | 1.12E-05 | 0.000914 | 1.81E-05 | -0.01020875 | MRTFB |
| chr3 | 47578200 | 47578474 | 275 | 4 | 3.53E-05 | 4.72E-05 | 0.000766 | 1.81E-05 | 0.002323867 | CSPG5 |
| chr12 | 111320082 | 111320288 | 207 | 2 | 6.18E-05 | 1.51E-05 | 0.000246 | 1.81E-05 | -0.01025367 | CUX2 |
| chr19 | 39125656 | 39126178 | 523 | 8 | 6.68E-06 | 0.000206 | 0.000689 | 1.81E-05 | 0.003180589 | PAK4 |
| chr13 | 99845199 | 99845535 | 337 | 4 | 1.56E-05 | 0.000268 | 8.30E-05 | 1.81E-05 | 0.000867799 | CLYBL, AL137139.2 |
| chr12 | 109184352 | 109184392 | 41 | 2 | 2.04E-05 | 1.71E-05 | 0.000178 | 1.81E-05 | 0.003916451 | ACACB |
| chr3 | 72848389 | 72848918 | 530 | 13 | 9.74E-05 | 1.48E-05 | 0.018699 | 1.81E-05 | 0.000111189 | SHQ1 |
| chr17 | 8294977 | 8295771 | 795 | 7 | 9.31E-05 | 0.001368 | 0.001567 | 1.81E-05 | 0.001387326 | SLC25A35 |
| chr16 | 67115360 | 67115709 | 350 | 2 | 8.25E-06 | 0.000359 | 2.35E-05 | 1.81E-05 | 0.012381185 | C16orf70 |
| chr13 | 68713720 | 68713912 | 193 | 2 | 1.05E-05 | 0.002937 | 1.73E-05 | 1.81E-05 | 0.011151002 | NA |
| chr1 | 104072214 | 104072261 | 48 | 3 | 7.35E-05 | 1.20E-05 | 0.000567 | 1.82E-05 | -0.00236402 | NA |
| chr20 | 62474721 | 62474828 | 108 | 3 | 1.46E-05 | 0.000198 | 2.69E-05 | 1.82E-05 | 0.011513246 | GATA5 |
| chr2 | 232519779 | 232520058 | 280 | 5 | 3.49E-05 | 1.03E-05 | 0.003003 | 1.82E-05 | 0.008024604 | NA |
| chr17 | 20009904 | 20010306 | 403 | 3 | 1.29E-05 | 5.23E-05 | 6.31E-05 | 1.82E-05 | 0.000596631 | SPECC1 |
| chr3 | 117789329 | 117789826 | 498 | 3 | 3.17E-05 | 5.44E-05 | 6.19E-05 | 1.82E-05 | 0.005167736 | AC092691.3, AC092691.1 |
| chr17 | 11884160 | 11884314 | 155 | 3 | 7.10E-05 | 4.62E-05 | 0.000254 | 1.82E-05 | -0.00169074 | DNAH9 |
| chr16 | 88069429 | 88070023 | 595 | 4 | 3.11E-07 | 0.000145 | 4.27E-05 | 1.82E-05 | 0.007712862 | BANP |
| chr2 | 40480811 | 40480814 | 4 | 2 | 2.26E-05 | 0.000201 | 2.81E-05 | 1.82E-05 | 0.009748307 | SLC8A1 |
| chr9 | 34206366 | 34206680 | 315 | 2 | 1.65E-05 | 6.34E-05 | 4.65E-05 | 1.83E-05 | 0.016365596 | UBAP1 |
| chr17 | 76073209 | 76073626 | 418 | 2 | 1.14E-05 | 1.75E-05 | 0.000174 | 1.83E-05 | 0.013190288 | ZACN |
| chr17 | 73312055 | 73312667 | 613 | 10 | 4.11E-05 | 9.43E-06 | 0.011706 | 1.83E-05 | 0.002204139 | NA |
| chr2 | 121755334 | 121755993 | 660 | 5 | 6.83E-05 | 1.40E-05 | 0.001445 | 1.83E-05 | 0.004145746 | TSN |
| chr12 | 32750743 | 32750779 | 37 | 2 | 0.000113754 | 1.14E-05 | 0.000886 | 1.83E-05 | 0.006514791 | YARS2 |
| chr3 | 149618342 | 149618938 | 597 | 2 | 0.000359483 | 1.15E-05 | 0.000791 | 1.83E-05 | 0.006356151 | WWTR1 |
| chr11 | 1074682 | 1074869 | 188 | 5 | 6.77E-05 | 8.67E-06 | 0.004304 | 1.83E-05 | 0.004221593 | NA |
| chr4 | 25312307 | 25313027 | 721 | 10 | 1.80E-06 | 7.99E-05 | 3.91E-05 | 1.83E-05 | 0.00220028 | ZCCHC4 |
| chr7 | 33725125 | 33725797 | 673 | 4 | 7.36E-05 | 1.90E-05 | 0.000279 | 1.83E-05 | 0.006222002 | BBS9 |
| chr6 | 155389487 | 155389507 | 21 | 2 | 0.000107563 | 1.37E-05 | 0.000355 | 1.83E-05 | 0.028530508 | NA |
| chr1 | 163321985 | 163322450 | 466 | 6 | 2.66E-05 | 1.33E-05 | 0.003946 | 1.83E-05 | -0.00026236 | NUF2 |
| chr22 | 30373448 | 30373584 | 137 | 2 | 4.62E-05 | 2.54E-05 | 9.98E-05 | 1.84E-05 | 0.009131698 | CCDC157, RNF215 |
| chr7 | 31192918 | 31193486 | 569 | 8 | 2.93E-05 | 0.000312 | 0.000658 | 1.84E-05 | 0.002266431 | NA |
| chr11 | 101129216 | 101129986 | 771 | 11 | 7.59E-05 | 0.00103 | 0.000135 | 1.84E-05 | 0.002731985 | PGR-AS1, PGR |
| chr3 | 108821307 | 108821939 | 633 | 5 | 1.64E-05 | 0.000342 | 9.58E-05 | 1.84E-05 | -0.00292604 | NA |
| chr1 | 17460268 | 17460538 | 271 | 2 | 2.44E-05 | 0.000294 | 2.51E-05 | 1.84E-05 | 0.003970583 | NA |
| chr22 | 29031500 | 29031863 | 364 | 3 | 2.75E-05 | 1.54E-05 | 0.000895 | 1.84E-05 | 0.016744494 | ZNRF3 |
| chr14 | 104199365 | 104199710 | 346 | 3 | 5.72E-05 | 2.93E-05 | 0.000132 | 1.84E-05 | 0.012009593 | NA |
| chr16 | 89358385 | 89358703 | 319 | 2 | 9.58E-06 | 0.000133 | 3.28E-05 | 1.84E-05 | 0.018526737 | ANKRD11 |
| chr20 | 63766866 | 63767422 | 557 | 3 | 1.81E-05 | 3.83E-05 | 0.000132 | 1.84E-05 | 0.005464116 | ZBTB46 |
| chr13 | 113158210 | 113158368 | 159 | 3 | 0.000226222 | 9.40E-05 | 5.02E-05 | 1.84E-05 | 0.00407121 | NA |
| chr4 | 75673069 | 75673633 | 565 | 11 | 0.000155498 | 9.20E-05 | 0.003096 | 1.84E-05 | 0.001060389 | G3BP2 |
| chr9 | 92483523 | 92483683 | 161 | 2 | 5.17E-05 | 5.73E-05 | 5.00E-05 | 1.84E-05 | 0.00312341 | CENPP |
| chr16 | 24978301 | 24978604 | 304 | 4 | 2.84E-05 | 0.000373 | 4.28E-05 | 1.84E-05 | 0.000828934 | ARHGAP17 |
| chr21 | 26405878 | 26406090 | 213 | 2 | 3.32E-05 | 2.97E-05 | 8.50E-05 | 1.84E-05 | 0.024282543 | AP001596.1, CYYR1-AS1 |
| chr16 | 56194202 | 56194989 | 788 | 12 | 0.000127336 | 4.73E-05 | 0.008619 | 1.84E-05 | 0.001069872 | GNAO1, DKFZP434H168 |
| chr14 | 100145995 | 100146005 | 11 | 2 | 3.36E-05 | 1.21E-05 | 0.000629 | 1.84E-05 | 0.008521105 | DEGS2 |
| chr17 | 81842892 | 81843702 | 811 | 9 | 2.05E-05 | 0.000353 | 0.000474 | 1.84E-05 | 0.004827581 | AC145207.3, P4HB |
| chr12 | 12725445 | 12725953 | 509 | 9 | 2.19E-05 | 0.000428 | 0.000765 | 1.84E-05 | -0.00032407 | APOLD1 |
| chr16 | 26322057 | 26322215 | 159 | 3 | 3.85E-05 | 0.000386 | 4.67E-05 | 1.84E-05 | 0.006952866 | AC009158.1 |
| chr15 | 60478872 | 60479118 | 247 | 4 | 0.000122521 | 0.000185 | 5.23E-05 | 1.84E-05 | -0.00235713 | ICE2 |
| chr22 | 18109499 | 18110209 | 711 | 5 | 8.22E-06 | 2.25E-05 | 0.000468 | 1.84E-05 | 0.007714207 | PEX26 |
| chr14 | 81533119 | 81533159 | 41 | 2 | 2.56E-05 | 2.50E-05 | 0.000103 | 1.85E-05 | 0.024982566 | SEL1L |
| chr19 | 40425790 | 40426369 | 580 | 11 | 2.82E-05 | 0.000248 | 0.000578 | 1.85E-05 | -0.00045867 | AC010271.2, SERTAD1 |
| chr6 | 168377545 | 168377632 | 88 | 3 | 0.000146037 | 7.24E-06 | 0.003853 | 1.85E-05 | 0.021184623 | NA |
| chr14 | 50590316 | 50590849 | 534 | 3 | 1.11E-05 | 0.000991 | 1.62E-05 | 1.85E-05 | 0.001897996 | ATL1 |
| chr8 | 38936736 | 38937184 | 449 | 2 | 1.33E-05 | 2.31E-05 | 0.000114 | 1.85E-05 | 0.011222783 | PLEKHA2 |
| chr1 | 222618919 | 222619245 | 327 | 3 | 4.85E-05 | 1.08E-05 | 0.000799 | 1.85E-05 | 0.011978983 | MIA3 |
| chr1 | 15412100 | 15412306 | 207 | 3 | 7.71E-05 | 8.70E-06 | 0.002085 | 1.85E-05 | 0.012988714 | EFHD2 |
| chr4 | 105552076 | 105552649 | 574 | 6 | 4.01E-05 | 7.64E-05 | 0.002149 | 1.85E-05 | 0.005588704 | ARHGEF38, AC004066.1 |
| chr6 | 28573259 | 28573685 | 427 | 2 | 4.11E-06 | 1.99E-05 | 0.000141 | 1.86E-05 | 0.007260573 | ZBED9 |
| chr19 | 4371347 | 4371937 | 591 | 5 | 5.11E-05 | 0.000134 | 0.000902 | 1.86E-05 | -0.00321859 | SH3GL1 |
| chr11 | 121570140 | 121570183 | 44 | 3 | 9.45E-05 | 3.53E-05 | 0.000271 | 1.86E-05 | -0.00065675 | SORL1 |
| chr19 | 42095337 | 42095479 | 143 | 3 | 9.73E-05 | 1.35E-05 | 0.00044 | 1.86E-05 | 0.009463757 | POU2F2 |
| chr2 | 209772024 | 209772370 | 347 | 3 | 5.02E-05 | 2.42E-05 | 0.000906 | 1.86E-05 | 0.008570217 | UNC80 |
| chr3 | 49176279 | 49176670 | 392 | 2 | 4.91E-05 | 1.57E-05 | 0.000238 | 1.86E-05 | 0.004503911 | KLHDC8B |
| chr7 | 2912291 | 2912330 | 40 | 2 | 5.17E-05 | 1.27E-05 | 0.000508 | 1.86E-05 | -0.00045194 | CARD11 |
| chr1 | 193106061 | 193106501 | 441 | 4 | 8.05E-06 | 1.95E-05 | 0.000867 | 1.86E-05 | 0.008413574 | GLRX2 |
| chr2 | 24025514 | 24025593 | 80 | 2 | 0.000112269 | 1.18E-05 | 0.000783 | 1.86E-05 | 0.005968598 | MFSD2B |
| chr10 | 31697445 | 31697796 | 352 | 2 | 4.20E-05 | 1.40E-05 | 0.000351 | 1.86E-05 | 0.014195257 | MACORIS |
| chr12 | 41437684 | 41437793 | 110 | 4 | 0.000136447 | 7.94E-06 | 0.005073 | 1.86E-05 | -0.03495641 | PDZRN4, AC090531.1 |
| chr12 | 116558554 | 116559380 | 827 | 10 | 4.01E-05 | 4.80E-05 | 0.000901 | 1.86E-05 | 0.001176262 | MAP1LC3B2 |
| chr3 | 182740852 | 182740987 | 136 | 3 | 5.80E-05 | 1.04E-05 | 0.000921 | 1.86E-05 | 0.014719835 | NA |
| chr11 | 128890198 | 128890472 | 275 | 2 | 9.02E-06 | 1.43E-05 | 0.000324 | 1.86E-05 | 0.01106377 | NA |
| chr1 | 213609965 | 213610477 | 513 | 3 | 2.98E-05 | 0.000255 | 0.000321 | 1.86E-05 | 0.013125133 | NA |
| chr10 | 26391800 | 26392418 | 619 | 4 | 0.000171528 | 7.20E-06 | 0.003265 | 1.86E-05 | 0.013049876 | NA |
| chr16 | 85898388 | 85899247 | 860 | 9 | 5.05E-05 | 0.0003 | 0.000796 | 1.86E-05 | 0.001398159 | IRF8 |
| chr10 | 70600500 | 70600692 | 193 | 4 | 0.000123291 | 8.23E-06 | 0.004717 | 1.87E-05 | 0.007603259 | PRF1 |
| chr4 | 42152823 | 42154165 | 1343 | 10 | 7.92E-05 | 4.10E-05 | 0.009837 | 1.87E-05 | 0.001561551 | AC111006.1, BEND4 |
| chr5 | 157142676 | 157143346 | 671 | 12 | 0.000167999 | 1.78E-05 | 0.027899 | 1.87E-05 | -0.00108339 | ITK, HAVCR2, MED7 |
| chr2 | 96897996 | 96898764 | 769 | 7 | 0.00015897 | 1.07E-05 | 0.00908 | 1.87E-05 | 0.002504868 | FAM178B |
| chr16 | 35352466 | 35352749 | 284 | 3 | 0.000130507 | 8.35E-06 | 0.002291 | 1.87E-05 | 0.023671554 | C2orf69P4 |
| chr7 | 1592629 | 1593231 | 603 | 4 | 5.97E-05 | 2.12E-05 | 0.000894 | 1.87E-05 | 0.010968967 | NA |
| chr12 | 132423442 | 132423797 | 356 | 5 | 0.000154522 | 3.79E-06 | 0.01405 | 1.87E-05 | 0.010891663 | NA |
| chr9 | 127502873 | 127502908 | 36 | 2 | 7.05E-05 | 1.35E-05 | 0.000419 | 1.88E-05 | 0.005388515 | LRSAM1 |
| chr1 | 216722395 | 216723660 | 1266 | 6 | 4.55E-05 | 0.000122 | 0.001265 | 1.88E-05 | 0.005884162 | ESRRG |
| chr17 | 79089733 | 79089780 | 48 | 2 | 5.03E-05 | 1.48E-05 | 0.000293 | 1.88E-05 | 0.010620741 | RBFOX3 |
| chr17 | 81487898 | 81488796 | 899 | 7 | 5.78E-05 | 1.71E-05 | 0.004417 | 1.88E-05 | -0.00485803 | NA |
| chr17 | 1160601 | 1160916 | 316 | 4 | 0.000154635 | 5.34E-06 | 0.007677 | 1.88E-05 | 0.006077653 | ABR |
| chr11 | 535387 | 535740 | 354 | 4 | 6.39E-05 | 9.06E-06 | 0.00295 | 1.88E-05 | 0.003388455 | HRAS |
| chr8 | 144756560 | 144756674 | 115 | 2 | 6.70E-07 | 0.001192 | 1.95E-05 | 1.88E-05 | -0.00395774 | NA |
| chr5 | 40904936 | 40905153 | 218 | 2 | 5.70E-05 | 3.34E-05 | 7.84E-05 | 1.88E-05 | 0.010529593 | NA |
| chr6 | 166313416 | 166313749 | 334 | 2 | 6.29E-05 | 2.02E-05 | 0.000142 | 1.88E-05 | 0.019849189 | AL121956.7 |
| chr1 | 25564047 | 25564593 | 547 | 4 | 0.000133769 | 8.10E-06 | 0.001951 | 1.88E-05 | -0.01123298 | LDLRAP1 |
| chr22 | 24145443 | 24145842 | 400 | 2 | 3.80E-05 | 2.65E-05 | 9.92E-05 | 1.88E-05 | -0.0068116 | CABIN1 |
| chr21 | 45287866 | 45288263 | 398 | 10 | 4.18E-05 | 1.37E-05 | 0.014182 | 1.88E-05 | 0.000395645 | LINC00205, POFUT2 |
| chr20 | 4244113 | 4244297 | 185 | 2 | 6.02E-05 | 1.90E-05 | 0.000158 | 1.88E-05 | 0.007756119 | ADRA1D |
| chr3 | 73621612 | 73622044 | 433 | 2 | 0.000129128 | 1.16E-05 | 0.000961 | 1.88E-05 | 0.023841612 | PDZRN3-AS1, PDZRN3 |
| chr7 | 79453101 | 79454193 | 1093 | 10 | 1.52E-06 | 1.46E-05 | 0.001912 | 1.88E-05 | 0.003180189 | MAGI2-AS3, MAGI2 |
| chr6 | 137739056 | 137739321 | 266 | 4 | 9.13E-05 | 8.63E-06 | 0.002019 | 1.88E-05 | 0.000958275 | AL356234.2, LINC02539 |
| chr17 | 39750114 | 39750633 | 520 | 2 | 3.60E-05 | 1.55E-05 | 0.00026 | 1.88E-05 | 0.00901238 | NA |
| chr15 | 65286492 | 65287715 | 1224 | 10 | 0.000127442 | 0.000727 | 0.00029 | 1.88E-05 | 0.001958733 | PARP16 |
| chr6 | 130191127 | 130191657 | 531 | 3 | 0.000167208 | 7.59E-06 | 0.003512 | 1.88E-05 | 0.014460379 | SAMD3 |
| chr20 | 33662229 | 33662603 | 375 | 6 | 1.22E-05 | 0.00201 | 0.000281 | 1.88E-05 | 0.002013215 | C20orf144, NECAB3 |

| | | | | | | | | | | |
|---|---|---|---|---|---|---|---|---|---|---|
| chr7 | 76317944 | 76318582 | 639 | 6 | 5.89E-05 | 2.49E-05 | 0.00077 | 1.89E-05 | -0.00071111 | NA |
| chr21 | 26079432 | 26079861 | 430 | 3 | 3.43E-05 | 0.000917 | 5.86E-05 | 1.89E-05 | 0.009113465 | APP |
| chr16 | 87409295 | 87409934 | 640 | 4 | 3.69E-05 | 5.60E-05 | 0.000168 | 1.89E-05 | 0.002892462 | ZCCHC14 |
| chr11 | 84317519 | 84318084 | 566 | 4 | 4.28E-06 | 0.000135 | 5.81E-05 | 1.89E-05 | 0.005214356 | DLG2 |
| chr10 | 60572594 | 60573554 | 961 | 10 | 6.26E-06 | 0.000335 | 0.004417 | 1.89E-05 | 0.008799286 | ANK3 |
| chr12 | 123616542 | 123616745 | 204 | 3 | 6.54E-05 | 2.44E-05 | 0.000898 | 1.89E-05 | -0.00122437 | DDX55, SNORA9B |
| chr1 | 202988862 | 202989038 | 177 | 2 | 7.29E-05 | 3.07E-05 | 8.56E-05 | 1.89E-05 | 0.007177034 | NA |
| chr11 | 112166950 | 112167752 | 803 | 8 | 5.32E-05 | 1.46E-05 | 0.001825 | 1.89E-05 | -0.00558034 | AP002884.2, TEX12, AP002884.1 |
| chr8 | 38348211 | 38348950 | 740 | 3 | 5.93E-05 | 1.27E-05 | 0.000856 | 1.89E-05 | 0.010340456 | NSD3 |
| chr21 | 37914212 | 37914353 | 142 | 3 | 1.29E-05 | 0.001661 | 3.09E-05 | 1.89E-05 | 0.004858287 | KCNJ6 |
| chr9 | 125824393 | 125824848 | 456 | 3 | 4.91E-05 | 1.19E-05 | 0.000652 | 1.90E-05 | 0.021839489 | PBX3 |
| chr22 | 44568860 | 44569098 | 239 | 2 | 5.29E-05 | 3.93E-05 | 6.91E-05 | 1.90E-05 | 0.009405623 | NA |
| chr1 | 21293578 | 21294319 | 742 | 3 | 5.44E-05 | 9.38E-06 | 0.002254 | 1.90E-05 | 0.009964236 | ECE1-AS1, ECE1 |
| chr2 | 24174352 | 24175558 | 1207 | 10 | 9.17E-05 | 6.14E-05 | 0.002658 | 1.90E-05 | -0.00582622 | AC008073.3, FAM228A, AC008073.2 |
| chr12 | 24133344 | 24133603 | 260 | 2 | 7.54E-05 | 2.00E-05 | 0.000148 | 1.90E-05 | 0.009095613 | SOX5 |
| chr16 | 75350748 | 75351128 | 381 | 2 | 0.000107998 | 1.24E-05 | 0.000676 | 1.90E-05 | 0.003985642 | CFDP1 |
| chr5 | 166087757 | 166088345 | 589 | 2 | 0.000146514 | 1.22E-05 | 0.000711 | 1.90E-05 | -0.00099342 | NA |
| chr11 | 35345174 | 35345489 | 316 | 2 | 3.61E-05 | 6.11E-05 | 4.99E-05 | 1.90E-05 | -0.03353328 | SLC1A2 |
| chr12 | 19306534 | 19307453 | 920 | 3 | 0.000179943 | 0.000187 | 0.000394 | 1.90E-05 | 0.003999327 | PLEKHA5 |
| chr15 | 99443887 | 99443937 | 51 | 2 | 0.000101965 | 1.85E-05 | 0.000171 | 1.90E-05 | 0.009564365 | NA |
| chr16 | 30021385 | 30021921 | 537 | 5 | 0.000118072 | 1.47E-05 | 0.001415 | 1.90E-05 | 0.002759881 | DOC2A |
| chr14 | 75147483 | 75148141 | 659 | 4 | 3.75E-05 | 0.000398 | 0.000736 | 1.90E-05 | 0.012602868 | TMED10 |
| chr5 | 180313743 | 180314343 | 601 | 5 | 9.11E-05 | 4.23E-06 | 0.011176 | 1.90E-05 | 0.060779936 | GFPT2 |
| chr16 | 55876582 | 55876821 | 240 | 3 | 4.47E-05 | 3.50E-05 | 0.000117 | 1.90E-05 | 0.005545861 | CES5A |
| chr1 | 18854546 | 18854965 | 420 | 6 | 6.91E-05 | 3.58E-05 | 0.0038 | 1.90E-05 | 0.011392957 | TAS1R2 |
| chr7 | 117427229 | 117427924 | 696 | 10 | 0.00012106 | 2.25E-05 | 0.016237 | 1.90E-05 | 0.002455318 | CFTR, ASZ1 |
| chr17 | 48646776 | 48647413 | 638 | 5 | 0.000131425 | 9.44E-05 | 0.004466 | 1.90E-05 | 0.002435951 | LINC02086, AC103702.2 |
| chr2 | 64987406 | 64987491 | 86 | 2 | 1.52E-05 | 1.20E-05 | 0.000826 | 1.90E-05 | 0.027828138 | LINC02245 |
| chr17 | 4771548 | 4771997 | 450 | 5 | 6.00E-05 | 2.64E-05 | 0.00285 | 1.90E-05 | 0.007428552 | TM4SF5 |
| chr5 | 72480917 | 72481022 | 106 | 2 | 9.06E-05 | 1.26E-05 | 0.000628 | 1.90E-05 | 0.018038728 | ZNF366 |
| chr11 | 76129538 | 76129589 | 52 | 2 | 4.78E-05 | 1.64E-05 | 0.000234 | 1.91E-05 | 0.011192275 | UVRAG |
| chr2 | 130370578 | 130370590 | 13 | 2 | 3.59E-05 | 7.84E-05 | 4.37E-05 | 1.91E-05 | 0.011630701 | PTPN18 |
| chr19 | 55590260 | 55590550 | 291 | 2 | 3.59E-05 | 4.96E-05 | 5.81E-05 | 1.91E-05 | 0.009631098 | NA |
| chr10 | 47340553 | 47340949 | 397 | 2 | 5.62E-05 | 1.82E-05 | 0.00018 | 1.91E-05 | 0.017721109 | NA |
| chr2 | 48601219 | 48601499 | 281 | 2 | 9.08E-05 | 1.24E-05 | 0.000709 | 1.91E-05 | 0.025610955 | STON1-GTF2A1L |
| chr3 | 159273067 | 159273849 | 783 | 7 | 2.50E-05 | 6.85E-05 | 0.001432 | 1.91E-05 | 0.012781069 | IQCJ-SCHIP1 |
| chr17 | 43982268 | 43982628 | 361 | 2 | 8.17E-05 | 1.27E-05 | 0.000607 | 1.91E-05 | 0.009559638 | PYY |
| chr7 | 6170300 | 6171036 | 737 | 3 | 1.42E-05 | 2.09E-05 | 0.000244 | 1.91E-05 | -0.01820835 | CYTH3 |
| chr2 | 71129747 | 71130604 | 858 | 19 | 8.56E-05 | 0.000261 | 0.021044 | 1.91E-05 | 0.000582356 | MCEE |
| chr14 | 91285827 | 91287044 | 1218 | 6 | 2.81E-05 | 3.18E-05 | 0.003565 | 1.92E-05 | 0.002087884 | CCDC88C |
| chr12 | 14962455 | 14963008 | 554 | 4 | 1.57E-05 | 4.99E-05 | 0.000491 | 1.92E-05 | 0.013931966 | NA |
| chr17 | 50668294 | 50668955 | 662 | 2 | 2.60E-05 | 1.85E-05 | 0.000177 | 1.92E-05 | 0.012951334 | ABCC3 |
| chr10 | 101130539 | 101131523 | 985 | 12 | 3.01E-05 | 7.18E-05 | 0.009478 | 1.92E-05 | 0.002935842 | TLX1, TLX1NB |
| chr11 | 13463707 | 13464009 | 303 | 3 | 8.52E-05 | 2.15E-05 | 0.000194 | 1.92E-05 | 0.008313881 | AC021269.3 |
| chr6 | 169254031 | 169254624 | 594 | 5 | 1.34E-06 | 0.000122 | 0.000214 | 1.92E-05 | 0.003437239 | THBS2 |
| chr1 | 37756639 | 37756709 | 71 | 2 | 3.80E-05 | 2.31E-05 | 0.000123 | 1.92E-05 | -0.00264752 | EPHA10 |
| chr12 | 48104986 | 48106077 | 1092 | 15 | 5.15E-05 | 9.13E-06 | 0.035505 | 1.92E-05 | -0.0008213 | PFKM, SENP1 |
| chr4 | 186251799 | 186252297 | 499 | 6 | 0.000178172 | 8.86E-06 | 0.004438 | 1.92E-05 | 0.003737007 | KLKB1 |
| chr10 | 59283390 | 59283828 | 439 | 4 | 0.000196171 | 5.23E-06 | 0.008729 | 1.92E-05 | 0.033933585 | FAM13C |
| chr7 | 130485549 | 130486347 | 799 | 15 | 8.85E-05 | 1.28E-05 | 0.001895 | 1.92E-05 | 0.005671978 | MEST, MESTIT1 |
| chr8 | 121164677 | 121164902 | 226 | 3 | 3.08E-05 | 0.000376 | 7.28E-05 | 1.93E-05 | 0.000703835 | NA |
| chr17 | 44947655 | 44948287 | 633 | 13 | 7.73E-05 | 9.54E-05 | 0.00635 | 1.93E-05 | 0.002141261 | AC015936.1, KIF18B |
| chr11 | 20861304 | 20861772 | 469 | 2 | 7.74E-05 | 1.52E-05 | 0.000302 | 1.93E-05 | 0.006715229 | NELL1 |
| chr16 | 31066064 | 31066429 | 366 | 2 | 3.71E-06 | 9.66E-05 | 3.97E-05 | 1.93E-05 | -0.00132401 | AC135050.4, ZNF668 |
| chr6 | 160348327 | 160348722 | 396 | 2 | 2.67E-05 | 3.62E-05 | 7.60E-05 | 1.93E-05 | -0.0044257 | SLC22A3 |
| chr7 | 102177792 | 102177844 | 53 | 2 | 3.87E-05 | 0.000858 | 2.10E-05 | 1.93E-05 | 0.003250704 | CUX1 |
| chr10 | 26211792 | 26212136 | 345 | 3 | 2.01E-05 | 0.000447 | 0.000341 | 1.93E-05 | 0.007227546 | MYO3A |
| chr19 | 1834986 | 1835071 | 86 | 3 | 9.65E-05 | 2.02E-05 | 0.000927 | 1.93E-05 | 0.006680757 | REXO1 |
| chr3 | 107774724 | 107775157 | 434 | 3 | 0.000138224 | 8.30E-06 | 0.002684 | 1.93E-05 | 0.005519799 | BBX |
| chr13 | 109667215 | 109667260 | 46 | 3 | 0.000150343 | 7.60E-06 | 0.00397 | 1.93E-05 | 0.026858685 | AL163541.1 |
| chr17 | 50892973 | 50892996 | 24 | 2 | 1.95E-05 | 1.27E-05 | 0.000657 | 1.93E-05 | 0.011003784 | TOB1-AS1 |
| chr3 | 32109567 | 32110209 | 643 | 3 | 2.54E-05 | 0.000739 | 5.58E-05 | 1.93E-05 | 0.003754946 | GPD1L |
| chr2 | 25484098 | 25484348 | 251 | 2 | 0.000111443 | 1.26E-05 | 0.000676 | 1.93E-05 | 0.012640158 | DTNB |
| chr5 | 167587703 | 167587900 | 198 | 2 | 0.00012082 | 1.20E-05 | 0.000946 | 1.93E-05 | -0.00308526 | TENM2 |
| chr8 | 139991007 | 139991360 | 354 | 3 | 1.65E-05 | 0.000141 | 7.29E-05 | 1.93E-05 | 0.006199379 | TRAPPC9 |
| chr1 | 186450016 | 186450493 | 478 | 2 | 4.83E-06 | 3.67E-05 | 7.57E-05 | 1.94E-05 | 0.003815703 | PDC-AS1, PDC |
| chr19 | 3207504 | 3208478 | 975 | 5 | 0.000142589 | 6.03E-06 | 0.009983 | 1.94E-05 | 0.003111155 | NCLN |
| chr11 | 12221394 | 12221674 | 281 | 2 | 8.13E-05 | 1.62E-05 | 0.000258 | 1.94E-05 | 0.003217784 | MICAL2 |
| chr8 | 29408453 | 29408690 | 238 | 3 | 7.70E-05 | 2.95E-05 | 0.000576 | 1.94E-05 | 0.013370623 | NA |
| chr7 | 151248521 | 151249270 | 750 | 11 | 2.03E-05 | 0.000619 | 0.000348 | 1.94E-05 | 0.001410911 | SMARCD3 |
| chr5 | 1182922 | 1183309 | 388 | 5 | 4.55E-05 | 0.001885 | 0.00134 | 1.94E-05 | 0.005314786 | NA |
| chr19 | 43639990 | 43640842 | 853 | 6 | 4.61E-05 | 7.17E-05 | 0.000345 | 1.94E-05 | 0.001005189 | NA |
| chr22 | 18048262 | 18048681 | 420 | 4 | 0.000104406 | 6.92E-06 | 0.006028 | 1.94E-05 | -0.0108198 | NA |
| chr12 | 3083410 | 3083762 | 353 | 3 | 0.000108484 | 8.57E-06 | 0.002669 | 1.95E-05 | 0.01368402 | TSPAN9 |
| chr12 | 121209797 | 121210290 | 494 | 9 | 3.39E-05 | 9.08E-05 | 0.006173 | 1.95E-05 | 0.000940815 | P2RX4 |
| chr4 | 3441881 | 3442420 | 540 | 6 | 3.44E-05 | 1.63E-05 | 0.002541 | 1.95E-05 | 0.004132822 | HGFAC |
| chr18 | 25077321 | 25077604 | 284 | 2 | 7.69E-05 | 1.96E-05 | 0.000165 | 1.95E-05 | -0.00469171 | ZNF521 |
| chr10 | 1465701 | 1465866 | 166 | 2 | 2.23E-05 | 2.16E-05 | 0.00014 | 1.95E-05 | 0.010032208 | ADARB2 |
| chr17 | 3964500 | 3964698 | 199 | 4 | 9.19E-05 | 7.35E-05 | 0.000289 | 1.95E-05 | -0.00090903 | NA |
| chr11 | 95024100 | 95024594 | 495 | 2 | 0.000116425 | 1.23E-05 | 0.000843 | 1.95E-05 | 0.007877583 | NA |
| chr14 | 88562454 | 88563581 | 1128 | 16 | 1.65E-05 | 0.000946 | 0.002038 | 1.95E-05 | 0.001318317 | ZC3H14 |
| chr2 | 207827535 | 207828372 | 838 | 4 | 6.31E-06 | 0.004155 | 1.03E-05 | 1.96E-05 | 0.001862585 | PLEKHM3 |
| chr4 | 8177158 | 8177625 | 468 | 2 | 3.50E-06 | 8.66E-05 | 4.26E-05 | 1.96E-05 | 0.007439181 | NA |
| chr1 | 175505301 | 175505397 | 97 | 3 | 0.000125388 | 1.44E-05 | 0.000505 | 1.96E-05 | -0.02164599 | TNR |
| chr8 | 143574827 | 143575466 | 640 | 3 | 9.02E-05 | 9.46E-06 | 0.002246 | 1.96E-05 | 0.008437139 | AC067930.4, NAPRT |
| chr7 | 157137870 | 157138688 | 819 | 3 | 7.42E-06 | 4.33E-05 | 0.000316 | 1.96E-05 | 0.00943678 | NA |
| chr5 | 34686395 | 34687428 | 1034 | 7 | 1.24E-05 | 3.75E-05 | 0.001043 | 1.96E-05 | 0.007485737 | RAI14 |
| chr8 | 26044353 | 26045042 | 690 | 5 | 1.90E-05 | 0.000254 | 0.00016 | 1.96E-05 | 0.006511669 | EBF2 |
| chr17 | 15917702 | 15917816 | 115 | 2 | 0.000259541 | 1.24E-05 | 0.000868 | 1.96E-05 | 0.014937643 | NA |
| chr2 | 240573863 | 240574927 | 1065 | 9 | 1.08E-05 | 0.000766 | 0.001333 | 1.97E-05 | 0.002165842 | RNPEPL1 |
| chr5 | 10563246 | 10564093 | 848 | 4 | 1.05E-05 | 2.20E-05 | 0.001431 | 1.97E-05 | 0.000880715 | ANKRD33B |
| chr11 | 87037619 | 87038195 | 577 | 8 | 4.34E-05 | 6.19E-05 | 0.000493 | 1.97E-05 | 0.000119448 | TMEM135 |
| chr11 | 82733904 | 82734342 | 439 | 7 | 0.000108614 | 8.25E-06 | 0.004075 | 1.97E-05 | 0.00609367 | NA |
| chr20 | 50794875 | 50795684 | 810 | 10 | 2.39E-05 | 9.89E-06 | 0.003081 | 1.97E-05 | 0.00154534 | BCAS4 |
| chr10 | 15814371 | 15814705 | 335 | 4 | 7.39E-05 | 1.67E-05 | 0.000452 | 1.97E-05 | -0.00320554 | MINDY3 |
| chr20 | 31440420 | 31441090 | 671 | 6 | 0.00011661 | 4.13E-05 | 0.001249 | 1.97E-05 | 0.003026394 | DEFB123 |
| chr17 | 75755731 | 75755794 | 64 | 2 | 3.38E-05 | 4.32E-05 | 6.73E-05 | 1.97E-05 | 0.009460572 | ITGB4, GALK1 |
| chr2 | 157327624 | 157328129 | 506 | 6 | 0.000105536 | 1.15E-05 | 0.00124 | 1.97E-05 | 0.007698372 | ERMN |
| chr16 | 58384501 | 58385617 | 1117 | 6 | 2.90E-05 | 0.004123 | 0.00049 | 1.97E-05 | 0.001590876 | GINS3, RNU6-1110P |
| chr22 | 30207119 | 30207329 | 211 | 4 | 0.000127975 | 1.01E-05 | 0.001998 | 1.97E-05 | 0.001027528 | AC002378.1 |
| chr7 | 2589666 | 2589773 | 108 | 3 | 0.0002002 | 1.08E-05 | 0.001077 | 1.97E-05 | 0.003607182 | IQCE |
| chr3 | 172324103 | 172324492 | 390 | 2 | 2.51E-05 | 3.58E-05 | 7.92E-05 | 1.98E-05 | 0.010955552 | FNDC3B |
| chr14 | 74409597 | 74409965 | 369 | 2 | 7.43E-05 | 1.32E-05 | 0.00061 | 1.98E-05 | 0.000429537 | SYNDIG1L |
| chr1 | 46757501 | 46757896 | 396 | 2 | 0.000140313 | 1.22E-05 | 0.000981 | 1.98E-05 | 0.01618312 | CYP4B1 |
| chr12 | 2855881 | 2856345 | 465 | 3 | 3.20E-05 | 3.28E-05 | 0.000384 | 1.98E-05 | -0.00011828 | ITFG2, TEX52 |
| chr6 | 30699848 | 30700432 | 585 | 2 | 4.81E-05 | 1.33E-05 | 0.000599 | 1.98E-05 | 0.012704053 | MDC1 |
| chr3 | 71130881 | 71131708 | 828 | 12 | 3.56E-06 | 3.32E-05 | 0.013162 | 1.98E-05 | 0.004420397 | FOXP1, AC097634.4 |
| chr1 | 155046240 | 155046453 | 214 | 3 | 9.36E-05 | 5.98E-05 | 0.00018 | 1.98E-05 | 0.004524732 | DCST1 |
| chr10 | 77636826 | 77638182 | 1357 | 11 | 3.68E-05 | 0.004706 | 0.000345 | 1.98E-05 | 0.002601559 | KCNMA1 |
| chr15 | 51861934 | 51862141 | 208 | 2 | 0.000124625 | 1.23E-05 | 0.00097 | 1.98E-05 | 0.033574454 | TMOD3 |
| chr11 | 72674654 | 72675496 | 843 | 5 | 5.68E-05 | 6.58E-06 | 0.005304 | 1.98E-05 | 0.009246691 | NA |
| chr3 | 8659116 | 8659138 | 23 | 2 | 2.65E-05 | 0.000152 | 3.39E-05 | 1.99E-05 | 0.004530956 | SSUH2 |
| chr12 | 27460734 | 27460921 | 188 | 2 | 9.55E-05 | 1.72E-05 | 0.000236 | 1.99E-05 | 0.008049287 | NA |
| chr1 | 96845556 | 96845777 | 222 | 2 | 7.31E-05 | 2.94E-05 | 9.73E-05 | 1.99E-05 | 0.018197609 | NA |
| chr10 | 8037242 | 8037774 | 533 | 5 | 2.28E-05 | 1.63E-05 | 0.001833 | 2.00E-05 | 0.006116149 | NA |
| chr6 | 29887333 | 29887859 | 527 | 6 | 1.63E-05 | 0.000386 | 0.000137 | 2.00E-05 | 0.014653092 | HLA-H, AL645929.1 |
| chr3 | 139388101 | 139388271 | 171 | 3 | 2.18E-05 | 6.23E-05 | 6.62E-05 | 2.00E-05 | 0.002280856 | COPB2 |
| chr11 | 365961 | 366487 | 527 | 5 | 3.65E-05 | 2.31E-05 | 0.003126 | 2.00E-05 | 0.007535559 | NA |
| chr9 | 133478721 | 133479534 | 814 | 10 | 6.37E-05 | 3.79E-05 | 0.004378 | 2.00E-05 | 0.001465651 | SLC2A6 |
| chr15 | 70462795 | 70463320 | 526 | 2 | 0.00011393 | 1.32E-05 | 0.000646 | 2.00E-05 | 0.021051958 | NA |
| chr12 | 89862937 | 89863452 | 516 | 2 | 8.43E-06 | 2.22E-05 | 0.000141 | 2.00E-05 | -0.00156542 | AC009522.1 |
| chr22 | 50424527 | 50424537 | 11 | 2 | 2.16E-05 | 9.59E-05 | 4.15E-05 | 2.00E-05 | -0.00814669 | PPP6R2 |
| chr20 | 58833675 | 58833740 | 66 | 2 | 2.55E-05 | 0.000443 | 2.47E-05 | 2.00E-05 | 0.015717317 | GNAS-AS1 |
| chr15 | 90571110 | 90571174 | 65 | 2 | 0.000125428 | 1.26E-05 | 0.000863 | 2.00E-05 | 0.018261263 | CRTC3 |
| chr7 | 1664184 | 1664948 | 765 | 4 | 1.69E-05 | 3.91E-05 | 0.00021 | 2.00E-05 | 0.000797709 | NA |
| chr17 | 39904672 | 39905064 | 393 | 2 | 2.72E-05 | 1.82E-05 | 0.00021 | 2.00E-05 | 0.003598764 | GSDMB |
| chr19 | 8333706 | 8334679 | 974 | 6 | 4.98E-05 | 1.82E-05 | 0.00241 | 2.00E-05 | 0.01537591 | KANK3 |
| chr5 | 177457683 | 177457707 | 25 | 2 | 7.26E-05 | 6.59E-05 | 5.09E-05 | 2.00E-05 | 0.010559389 | DBN1 |
| chr1 | 33633242 | 33633937 | 696 | 4 | 0.000135669 | 4.45E-05 | 0.000248 | 2.00E-05 | 0.008886253 | CSMD2 |
| chr1 | 53459985 | 53460554 | 570 | 5 | 9.88E-06 | 0.000254 | 0.000692 | 2.01E-05 | 0.000290388 | DMRTB1 |
| chr7 | 151254396 | 151255013 | 618 | 7 | 3.20E-06 | 0.017214 | 9.69E-06 | 2.01E-05 | 0.001096271 | SMARCD3 |
| chr19 | 8151693 | 8152048 | 356 | 2 | 9.76E-05 | 1.31E-05 | 0.000695 | 2.01E-05 | 0.016305457 | NA |
| chr5 | 586428 | 586983 | 556 | 3 | 3.25E-05 | 8.60E-05 | 0.000122 | 2.01E-05 | 0.012833542 | NA |
| chr20 | 2807377 | 2807986 | 610 | 3 | 2.02E-05 | 0.000134 | 3.76E-05 | 2.01E-05 | -0.00853632 | NA |
| chr12 | 131790547 | 131790737 | 191 | 2 | 4.22E-05 | 1.67E-05 | 0.000265 | 2.01E-05 | 0.009462183 | SFSWAP |
| chr1 | 236894246 | 236894302 | 57 | 2 | 2.59E-05 | 0.001011 | 2.12E-05 | 2.01E-05 | -0.00611598 | MTR |
| chr17 | 28854234 | 28855119 | 886 | 11 | 3.68E-05 | 0.00012 | 0.001066 | 2.01E-05 | -0.00046065 | ERAL1, FAM222B |
| chr3 | 119749799 | 119750244 | 446 | 3 | 0.000138667 | 5.11E-05 | 0.000349 | 2.01E-05 | 0.009294737 | CFAP91, AC069444.2, AC069444.1 |
| chr7 | 102465306 | 102465964 | 659 | 4 | 1.47E-06 | 5.44E-05 | 0.000178 | 2.01E-05 | 0.003171292 | LRWD1, MIR5090 |
| chr20 | 41318302 | 41318810 | 509 | 3 | 1.70E-05 | 2.85E-05 | 0.000174 | 2.02E-05 | 0.008275531 | NA |
| chr21 | 41507731 | 41508521 | 791 | 8 | 0.000130792 | 9.55E-05 | 0.006325 | 2.02E-05 | -0.00178102 | TMPRSS2 |
| chr22 | 37488514 | 37488623 | 110 | 2 | 8.77E-05 | 1.46E-05 | 0.00043 | 2.02E-05 | 0.011404152 | NA |
| chr16 | 893000 | 893632 | 633 | 6 | 3.93E-05 | 3.00E-05 | 0.000804 | 2.02E-05 | -0.00353285 | LMF1 |
| chr6 | 16302726 | 16302938 | 213 | 4 | 8.23E-05 | 3.74E-05 | 0.000717 | 2.02E-05 | -0.0002942 | ATXN1 |
| chr2 | 32926490 | 32926682 | 193 | 2 | 0.000114211 | 1.41E-05 | 0.000498 | 2.02E-05 | 0.000522967 | LINC00486 |
| chr19 | 16238136 | 16238534 | 399 | 3 | 9.48E-05 | 1.29E-05 | 0.001076 | 2.02E-05 | 0.012517176 | AP1M1 |
| chr12 | 123949341 | 123949905 | 565 | 4 | 1.42E-05 | 0.000157 | 3.78E-05 | 2.02E-05 | -0.00998992 | CCDC92 |
| chr13 | 98444215 | 98444593 | 379 | 5 | 0.000108371 | 2.37E-05 | 0.000871 | 2.02E-05 | 0.002304328 | FARP1 |
| chr8 | 133150430 | 133150597 | 168 | 2 | 0.000130505 | 1.67E-05 | 0.00027 | 2.02E-05 | 0.010279279 | NA |
| chr8 | 134659099 | 134659154 | 56 | 2 | 4.02E-05 | 0.001357 | 2.05E-05 | 2.02E-05 | 0.002503934 | ZFAT |
| chr12 | 51820974 | 51821432 | 459 | 4 | 4.83E-05 | 6.76E-06 | 0.005723 | 2.02E-05 | 0.016348499 | AC068987.2, FIGNL2 |
| chr6 | 32442079 | 32442682 | 604 | 6 | 1.30E-05 | 0.000597 | 2.68E-05 | 2.03E-05 | 0.002781518 | HLA-DRA |
| chr8 | 23748030 | 23748088 | 59 | 2 | 7.27E-05 | 1.96E-05 | 0.00018 | 2.03E-05 | 0.026686754 | AC012574.1 |
| chr16 | 89306887 | 89307180 | 294 | 3 | 9.36E-05 | 1.69E-05 | 0.000934 | 2.03E-05 | 0.003089442 | ANKRD11 |
| chr19 | 53981081 | 53981622 | 542 | 4 | 7.06E-06 | 1.60E-05 | 0.00096 | 2.03E-05 | 0.004026457 | CACNG8 |
| chr9 | 98193328 | 98193975 | 648 | 4 | 1.45E-05 | 0.000144 | 4.60E-05 | 2.03E-05 | 0.001744537 | NA |
| chr15 | 88638194 | 88638918 | 725 | 7 | 1.65E-05 | 0.000542 | 0.000327 | 2.03E-05 | -0.00646203 | ISG20 |
| chr1 | 91505165 | 91505188 | 24 | 2 | 0.000127137 | 1.31E-05 | 0.000758 | 2.03E-05 | 0.014846574 | CDC7 |
| chr1 | 3706538 | 3706936 | 399 | 4 | 6.56E-05 | 1.19E-05 | 0.001213 | 2.04E-05 | 0.007352052 | TP73 |
| chr12 | 30989842 | 30990030 | 189 | 2 | 9.03E-05 | 1.32E-05 | 0.00075 | 2.04E-05 | 0.01516314 | TSPAN11, AC008013.1 |
| chr14 | 93926516 | 93927445 | 930 | 9 | 2.96E-05 | 9.62E-05 | 0.001213 | 2.04E-05 | 0.003893637 | FAM181A, FAM181A-AS1 |
| chr13 | 110536251 | 110536507 | 257 | 4 | 6.21E-05 | 9.48E-05 | 0.000158 | 2.04E-05 | 0.004578156 | RAB20 |

| | | | | | | | | | | |
|---|---|---|---|---|---|---|---|---|---|---|
| chr11 | 889148 | 889231 | 84 | 3 | 6.75E-05 | 1.23E-05 | 0.000916 | 2.04E-05 | 0.004753954 | CHID1 |
| chr12 | 55973426 | 55974442 | 1017 | 16 | 0.000113812 | 4.56E-05 | 0.007881 | 2.04E-05 | 0.000392109 | RAB5B |
| chr20 | 33664000 | 33664523 | 524 | 3 | 1.09E-05 | 1.59E-05 | 0.001392 | 2.04E-05 | -0.00048478 | C20orf144, NECAB3 |
| chr20 | 3664879 | 3664992 | 114 | 2 | 3.90E-05 | 1.64E-05 | 0.000292 | 2.04E-05 | 0.004717692 | NA |
| chr10 | 73766512 | 73766695 | 184 | 3 | 6.58E-05 | 5.09E-05 | 0.000774 | 2.04E-05 | 0.003345198 | SEC24C |
| chr11 | 6474017 | 6474861 | 845 | 14 | 1.96E-05 | 0.000686 | 0.004184 | 2.04E-05 | 0.004482154 | TRIM3, ARFIP2 |
| chr17 | 73423507 | 73424315 | 809 | 7 | 6.98E-05 | 0.000551 | 0.002124 | 2.04E-05 | 0.002490803 | SDK2 |
| chr22 | 50562901 | 50563418 | 518 | 8 | 8.10E-05 | 2.12E-05 | 0.011025 | 2.04E-05 | 0.002296887 | SYCE3 |
| chr16 | 4160850 | 4161239 | 390 | 2 | 4.91E-05 | 2.94E-05 | 0.000102 | 2.04E-05 | 0.026899963 | NA |
| chr12 | 52005616 | 52006385 | 770 | 4 | 4.05E-05 | 1.77E-05 | 0.002247 | 2.04E-05 | 0.006854886 | NA |
| chr11 | 46895670 | 46896209 | 540 | 2 | 5.40E-05 | 1.65E-05 | 0.000291 | 2.04E-05 | 0.009924136 | LRP4 |
| chr16 | 1568461 | 1568845 | 385 | 2 | 6.76E-05 | 1.71E-05 | 0.000265 | 2.04E-05 | 0.003806682 | IFT140 |
| chr2 | 105299369 | 105299486 | 118 | 3 | 1.83E-05 | 0.000115 | 4.23E-05 | 2.05E-05 | 0.004438831 | TGFBRAP1 |
| chr2 | 3641766 | 3642730 | 965 | 5 | 0.000391117 | 0.000135 | 0.000321 | 2.05E-05 | 0.004871099 | COLEC11 |
| chr17 | 73643176 | 73644114 | 939 | 5 | 1.56E-07 | 0.002893 | 1.82E-05 | 2.05E-05 | 0.00219486 | SDK2 |
| chr14 | 104724102 | 104724781 | 680 | 7 | 2.63E-05 | 0.000229 | 0.000292 | 2.05E-05 | -0.00156426 | ADSS1 |
| chr16 | 65571813 | 65571907 | 95 | 2 | 6.91E-05 | 2.30E-05 | 0.000141 | 2.05E-05 | 0.008653406 | LINC00922 |
| chr13 | 97363102 | 97363308 | 207 | 3 | 0.000121218 | 1.25E-05 | 0.00079 | 2.05E-05 | -0.00130975 | MBNL2 |
| chr3 | 193782537 | 193782975 | 439 | 2 | 7.48E-05 | 1.38E-05 | 0.000603 | 2.05E-05 | 0.008618297 | AC069421.3 |
| chr3 | 58121045 | 58121481 | 437 | 2 | 8.40E-06 | 6.21E-05 | 5.48E-05 | 2.05E-05 | 0.003841882 | FLNB |
| chr7 | 2607148 | 2607955 | 808 | 6 | 2.44E-05 | 1.16E-05 | 0.006849 | 2.05E-05 | 0.011078471 | IQCE |
| chr1 | 86553077 | 86553672 | 596 | 2 | 0.000164527 | 1.29E-05 | 0.00089 | 2.05E-05 | -0.01199499 | CLCA4, CDCA4P2 |
| chr15 | 98960884 | 98961026 | 143 | 2 | 0.000323815 | 1.40E-05 | 0.000571 | 2.06E-05 | 0.004835222 | IGF1R, AC036108.1 |
| chr14 | 50714116 | 50714570 | 455 | 2 | 5.04E-05 | 2.45E-05 | 0.000129 | 2.06E-05 | 0.008614206 | NA |
| chr6 | 53440536 | 53440782 | 247 | 2 | 0.000122079 | 1.33E-05 | 0.000769 | 2.06E-05 | 0.011309603 | NA |
| chr13 | 20415506 | 20416104 | 599 | 4 | 2.51E-05 | 0.000571 | 7.54E-05 | 2.06E-05 | 0.000295046 | CRYL1 |
| chr3 | 127617700 | 127617706 | 7 | 2 | 9.21E-05 | 1.87E-05 | 0.000212 | 2.06E-05 | -0.00166127 | MCM2 |
| chr12 | 52009108 | 52009238 | 131 | 4 | 2.01E-05 | 0.000249 | 3.81E-05 | 2.06E-05 | 0.002645385 | TAMALIN |
| chr17 | 442430 | 443025 | 596 | 4 | 0.000119288 | 3.03E-05 | 0.002158 | 2.06E-05 | 0.002179316 | RFLNB |
| chr17 | 34156541 | 34157240 | 700 | 13 | 0.000189196 | 9.51E-06 | 0.01704 | 2.06E-05 | 0.003789231 | AC004147.2, ASIC2 |
| chr22 | 20656921 | 20657505 | 585 | 4 | 0.000100177 | 1.50E-05 | 0.001957 | 2.06E-05 | 0.006758197 | NA |
| chr5 | 73626611 | 73627132 | 522 | 3 | 3.56E-05 | 1.52E-05 | 0.000464 | 2.06E-05 | 0.00660413 | ARHGEF28 |
| chr12 | 57806323 | 57806616 | 294 | 4 | 0.00011171 | 1.30E-05 | 0.002667 | 2.07E-05 | 0.000748831 | TSFM, AVIL |
| chr2 | 227327376 | 227328065 | 690 | 6 | 1.97E-05 | 0.000397 | 5.74E-05 | 2.07E-05 | 0.003961171 | MFF |
| chr21 | 39343444 | 39343719 | 276 | 3 | 0.000124123 | 1.12E-05 | 0.001594 | 2.07E-05 | -0.00702308 | HMGN1 |
| chr10 | 97081328 | 97081831 | 504 | 3 | 4.20E-05 | 6.35E-05 | 0.000106 | 2.07E-05 | 0.011110386 | SLIT1 |
| chr1 | 233361796 | 233362170 | 375 | 4 | 1.79E-05 | 0.000112 | 0.00014 | 2.07E-05 | 0.002757688 | MAP3K21 |
| chr19 | 22602351 | 22603022 | 672 | 3 | 7.70E-06 | 0.000996 | 2.12E-05 | 2.07E-05 | 0.00781675 | GOLGA2P9 |
| chr8 | 139806053 | 139806264 | 212 | 2 | 0.00012793 | 1.49E-05 | 0.000435 | 2.07E-05 | 0.006384788 | TRAPPC9 |
| chr16 | 89922502 | 89923078 | 577 | 8 | 4.88E-06 | 0.000728 | 0.000295 | 2.07E-05 | 0.000869389 | AC092143.1, TUBB3, AC092143.2 |
| chr17 | 48733325 | 48733572 | 248 | 5 | 4.47E-05 | 0.000434 | 8.33E-05 | 2.07E-05 | 0.004921976 | AC091179.1 |
| chr19 | 57943006 | 57943650 | 645 | 2 | 9.34E-05 | 1.38E-05 | 0.000648 | 2.07E-05 | -0.00973901 | ZNF256 |
| chr13 | 112867218 | 112867352 | 135 | 2 | 6.21E-05 | 1.28E-05 | 0.001022 | 2.07E-05 | 0.006210379 | ATP11A |
| chr22 | 44962827 | 44962956 | 130 | 2 | 0.000126284 | 1.34E-05 | 0.000767 | 2.07E-05 | -0.00490188 | PHF21B |
| chr13 | 28433919 | 28434140 | 222 | 3 | 5.73E-05 | 0.000179 | 0.000145 | 2.07E-05 | -0.00251838 | FLT1 |
| chr1 | 27357318 | 27357750 | 433 | 6 | 0.000189286 | 1.18E-05 | 0.008376 | 2.08E-05 | -0.00855669 | MAP3K6 |
| chr9 | 69468770 | 69468853 | 84 | 3 | 8.69E-05 | 1.18E-05 | 0.000956 | 2.08E-05 | 0.001883054 | APBA1 |
| chr10 | 3451832 | 3452335 | 504 | 3 | 0.000130289 | 8.87E-05 | 0.000369 | 2.08E-05 | 0.002857849 | LINC02669 |
| chr11 | 1370658 | 1370869 | 212 | 2 | 0.000121278 | 4.34E-05 | 7.24E-05 | 2.08E-05 | 0.006657764 | NA |
| chr13 | 30307390 | 30308136 | 747 | 9 | 2.35E-05 | 8.50E-05 | 0.001811 | 2.09E-05 | 0.005294081 | KATNAL1 |
| chr17 | 75703291 | 75703869 | 579 | 3 | 0.00011885 | 3.07E-05 | 0.000953 | 2.09E-05 | 0.000486805 | SAP30BP |
| chr21 | 38753656 | 38753903 | 248 | 2 | 1.66E-05 | 1.61E-05 | 0.00034 | 2.09E-05 | 0.002371833 | NA |
| chr19 | 1818745 | 1818887 | 143 | 2 | 0.000258462 | 1.68E-05 | 0.000294 | 2.09E-05 | 0.003376673 | REXO1 |
| chr16 | 11205456 | 11205909 | 454 | 3 | 3.24E-06 | 0.000341 | 3.72E-05 | 2.09E-05 | 0.007611728 | AC007220.1 |
| chr8 | 27361995 | 27362295 | 301 | 3 | 6.25E-05 | 4.60E-05 | 0.00063 | 2.09E-05 | 0.008735247 | PTK2B |
| chr10 | 112887375 | 112887519 | 145 | 2 | 6.14E-05 | 4.93E-05 | 6.55E-05 | 2.09E-05 | 0.010324894 | NA |
| chr6 | 43181303 | 43181891 | 589 | 6 | 1.64E-05 | 0.002639 | 0.000511 | 2.09E-05 | 0.006053382 | SRF |
| chr17 | 1341688 | 1341723 | 36 | 2 | 7.51E-05 | 2.63E-05 | 0.000121 | 2.09E-05 | 0.005426576 | NA |
| chr12 | 54982075 | 54982329 | 255 | 2 | 1.22E-05 | 0.00025 | 3.03E-05 | 2.09E-05 | 0.007312614 | TESPA1 |
| chr1 | 167430884 | 167431328 | 445 | 2 | 2.58E-05 | 5.92E-05 | 5.75E-05 | 2.09E-05 | 0.00901373 | CD247 |
| chr15 | 40927040 | 40927543 | 504 | 3 | 1.21E-05 | 1.45E-05 | 0.000564 | 2.09E-05 | 0.010829859 | NA |
| chr1 | 6056951 | 6057099 | 149 | 2 | 6.56E-05 | 6.85E-05 | 5.25E-05 | 2.09E-05 | 0.008440441 | KCNAB2 |
| chr12 | 53739638 | 53739869 | 232 | 4 | 5.41E-05 | 1.57E-05 | 0.000439 | 2.09E-05 | 0.001809002 | AC076968.2 |
| chr8 | 60877168 | 60877334 | 167 | 3 | 9.73E-06 | 0.004443 | 1.29E-05 | 2.09E-05 | 0.004067084 | AC113143.2 |
| chr16 | 55654418 | 55654484 | 67 | 2 | 5.31E-06 | 0.001729 | 2.05E-05 | 2.10E-05 | 0.006194049 | NA |
| chr20 | 41415253 | 41415416 | 164 | 2 | 0.000208777 | 5.33E-05 | 6.20E-05 | 2.10E-05 | -0.00137401 | CHD6 |
| chr10 | 133286165 | 133286616 | 452 | 7 | 6.19E-05 | 0.000176 | 0.001782 | 2.10E-05 | 0.003512293 | TUBGCP2 |
| chr9 | 126482459 | 126482967 | 509 | 2 | 7.35E-05 | 1.39E-05 | 0.000666 | 2.10E-05 | 0.045239079 | MVB12B |
| chr17 | 2379121 | 2379977 | 857 | 6 | 4.39E-05 | 0.000624 | 8.93E-05 | 2.10E-05 | 0.00307131 | SGSM2, AC006435.2 |
| chr16 | 50094114 | 50094161 | 48 | 2 | 4.58E-05 | 0.000892 | 2.24E-05 | 2.10E-05 | 0.007422783 | HEATR3 |
| chr12 | 257320 | 257707 | 388 | 5 | 0.000183281 | 8.63E-06 | 0.007204 | 2.10E-05 | 0.003407591 | SLC6A13 |
| chr12 | 57078291 | 57079184 | 894 | 14 | 0.000270727 | 5.71E-05 | 0.002883 | 2.10E-05 | 0.001101799 | NEMP1 |
| chr15 | 23878085 | 23878370 | 286 | 5 | 0.000302296 | 5.06E-06 | 0.01224 | 2.11E-05 | 0.011509017 | NA |
| chr1 | 27376160 | 27376201 | 42 | 3 | 7.27E-05 | 0.000202 | 0.00011 | 2.11E-05 | 0.000158837 | NA |
| chr15 | 80202789 | 80203261 | 473 | 2 | 6.37E-05 | 2.04E-05 | 0.000187 | 2.11E-05 | 0.011971297 | CTXND1 |
| chr2 | 16522997 | 16523180 | 184 | 2 | 9.65E-05 | 1.46E-05 | 0.000539 | 2.11E-05 | 0.002324177 | AC104623.1 |
| chr5 | 1062972 | 1064018 | 1047 | 4 | 0.000141456 | 4.04E-05 | 0.002258 | 2.11E-05 | 0.0034635 | SLC12A7, MIR4635 |
| chr21 | 36004573 | 36004596 | 24 | 2 | 4.42E-05 | 0.000303 | 2.89E-05 | 2.11E-05 | 0.002327394 | RUNX1 |
| chr17 | 63928997 | 63929351 | 355 | 2 | 2.67E-05 | 1.89E-05 | 0.000224 | 2.11E-05 | 0.006672317 | AC127029.3, CD79B |
| chr3 | 32467526 | 32468051 | 526 | 8 | 0.000127522 | 1.73E-05 | 0.00235 | 2.12E-05 | -0.01422871 | CMTM7 |
| chr2 | 113275783 | 113276253 | 471 | 4 | 1.13E-05 | 0.00026 | 9.94E-05 | 2.12E-05 | 0.00783698 | PAX8-AS1, PAX8 |
| chr13 | 113405207 | 113405456 | 250 | 3 | 7.18E-05 | 1.62E-05 | 0.000491 | 2.12E-05 | 0.004706953 | ADPRHL1 |
| chr5 | 35622193 | 35622726 | 534 | 2 | 0.000174028 | 1.33E-05 | 0.000924 | 2.12E-05 | 0.001908227 | SPEF2 |
| chr10 | 132666151 | 132666380 | 230 | 2 | 5.09E-05 | 2.19E-05 | 0.000163 | 2.12E-05 | 0.004251 | INPP5A |
| chr20 | 63975146 | 63975727 | 582 | 3 | 0.000151765 | 1.29E-05 | 0.001128 | 2.12E-05 | 0.001795912 | SAMD10 |
| chr16 | 88943521 | 88943739 | 219 | 2 | 2.24E-05 | 0.000129 | 3.87E-05 | 2.12E-05 | -0.00198593 | AC092384.1, CBFA2T3 |
| chr19 | 45766986 | 45767442 | 457 | 3 | 1.35E-05 | 2.98E-05 | 0.000388 | 2.12E-05 | 0.004084687 | AC074212.1, SIX5 |
| chr18 | 3874222 | 3874363 | 142 | 4 | 3.49E-05 | 0.000162 | 0.000214 | 2.12E-05 | 0.004867676 | DLGAP1 |
| chr17 | 40442681 | 40443114 | 434 | 4 | 4.90E-06 | 9.54E-05 | 0.000572 | 2.12E-05 | 0.006517044 | NA |
| chr15 | 85330298 | 85330390 | 93 | 2 | 2.11E-05 | 1.33E-05 | 0.000965 | 2.12E-05 | 0.017081264 | ADAMTS7P4 |
| chr5 | 73436078 | 73436659 | 582 | 5 | 6.11E-05 | 2.25E-05 | 0.001385 | 2.12E-05 | 0.001233663 | NA |
| chr1 | 37475212 | 37475662 | 451 | 2 | 9.52E-06 | 2.20E-05 | 0.000163 | 2.12E-05 | 0.011099598 | ZC3H12A |
| chr5 | 112976328 | 112977254 | 927 | 12 | 3.89E-05 | 7.01E-05 | 0.00436 | 2.12E-05 | 4.92E-05 | DCP2 |
| chr9 | 98118973 | 98120094 | 1122 | 11 | 8.30E-05 | 8.38E-05 | 0.002003 | 2.12E-05 | -0.00220704 | TRIM14 |
| chr4 | 8641890 | 8642242 | 353 | 4 | 4.49E-05 | 0.000206 | 0.001753 | 2.13E-05 | 0.004579037 | NA |
| chr2 | 135052123 | 135052455 | 333 | 7 | 0.000115062 | 1.76E-05 | 0.002605 | 2.13E-05 | -0.00067914 | RAB3GAP1 |
| chr11 | 78209046 | 78210228 | 1183 | 6 | 1.20E-05 | 2.13E-05 | 0.001528 | 2.13E-05 | 0.001505325 | USP35 |
| chr7 | 101874472 | 101874655 | 184 | 3 | 3.15E-05 | 1.47E-05 | 0.000546 | 2.14E-05 | 0.004484006 | CUX1 |
| chr5 | 163505294 | 163505598 | 305 | 6 | 8.68E-05 | 3.41E-05 | 0.001461 | 2.14E-05 | 0.00146606 | MAT2B |
| chr5 | 7755687 | 7756161 | 475 | 3 | 2.57E-06 | 0.030389 | 1.08E-05 | 2.14E-05 | 0.00616534 | ADCY2 |
| chr20 | 62236909 | 62237288 | 380 | 2 | 2.75E-05 | 1.34E-05 | 0.000941 | 2.14E-05 | 0.009747426 | OSBPL2 |
| chr16 | 4601731 | 4602236 | 506 | 2 | 6.58E-05 | 1.99E-05 | 0.000202 | 2.14E-05 | 0.011849035 | NA |
| chr8 | 18209822 | 18210261 | 440 | 6 | 7.61E-05 | 0.00015 | 0.000258 | 2.14E-05 | 0.002892014 | NAT1 |
| chr19 | 50786303 | 50786395 | 93 | 4 | 0.000129866 | 5.19E-05 | 9.91E-05 | 2.14E-05 | 0.002299584 | AC010325.1 |
| chr1 | 24355042 | 24355679 | 638 | 2 | 0.000189568 | 1.32E-05 | 0.001044 | 2.14E-05 | 0.004933968 | GRHL3 |
| chr7 | 6059102 | 6059619 | 518 | 11 | 6.46E-05 | 0.000207 | 0.007965 | 2.14E-05 | 0.003678099 | EIF2AK1 |
| chr17 | 48607388 | 48608484 | 1097 | 5 | 0.000155699 | 5.27E-06 | 0.011665 | 2.14E-05 | 0.016598157 | HOXB7 |
| chr11 | 17442778 | 17443276 | 499 | 5 | 8.83E-05 | 1.56E-05 | 0.000921 | 2.14E-05 | 0.002019682 | ABCC8 |
| chr6 | 87343587 | 87344018 | 432 | 2 | 4.89E-06 | 4.77E-05 | 6.96E-05 | 2.14E-05 | 0.007665625 | SMIM8 |
| chr17 | 60148588 | 60148827 | 240 | 2 | 1.63E-05 | 6.76E-05 | 5.46E-05 | 2.14E-05 | 0.015810117 | NA |
| chr7 | 4079053 | 4079236 | 184 | 3 | 6.14E-05 | 7.96E-05 | 9.33E-05 | 2.15E-05 | 0.003034753 | SDK1 |
| chr14 | 104775035 | 104775855 | 821 | 5 | 7.81E-05 | 6.93E-06 | 0.008145 | 2.15E-05 | 0.003244239 | AKT1 |
| chr18 | 49564043 | 49564440 | 398 | 3 | 0.000112242 | 1.03E-05 | 0.001704 | 2.15E-05 | 0.011207018 | LIPG |
| chr2 | 73999973 | 74000347 | 375 | 2 | 0.000122884 | 1.34E-05 | 0.001009 | 2.15E-05 | 0.002338527 | TET3 |
| chr19 | 4338772 | 4339220 | 449 | 8 | 0.000203593 | 8.26E-06 | 0.017304 | 2.15E-05 | -0.00178106 | STAP2 |
| chr2 | 98505365 | 98505464 | 100 | 2 | 4.51E-05 | 9.61E-05 | 4.47E-05 | 2.15E-05 | 0.0133164 | INPP4A |
| chr20 | 32052013 | 32052453 | 441 | 12 | 0.000164888 | 4.24E-05 | 0.003791 | 2.15E-05 | 0.001670848 | HCK |
| chr10 | 100830411 | 100830644 | 234 | 4 | 7.25E-05 | 1.92E-05 | 0.000608 | 2.15E-05 | 0.0050073 | NA |
| chr6 | 33746854 | 33747333 | 480 | 4 | 0.000139087 | 2.53E-05 | 0.000401 | 2.15E-05 | 0.008770853 | IP6K3 |
| chr11 | 2273547 | 2273961 | 415 | 5 | 6.88E-06 | 0.000256 | 7.20E-05 | 2.15E-05 | 0.002602232 | NA |
| chr5 | 151447388 | 151447886 | 499 | 9 | 5.63E-05 | 0.000119 | 0.001313 | 2.15E-05 | 0.00065389 | SLC36A1 |
| chr1 | 37721800 | 37721913 | 114 | 2 | 0.00012779 | 1.36E-05 | 0.000895 | 2.16E-05 | 0.014768877 | EPHA10 |
| chr17 | 76943944 | 76944355 | 412 | 4 | 0.000121113 | 7.52E-06 | 0.006617 | 2.16E-05 | -0.00068321 | MGAT5B |
| chr7 | 128352262 | 128352580 | 319 | 3 | 0.000110579 | 1.13E-05 | 0.001375 | 2.16E-05 | 0.011599643 | PRRT4 |
| chr1 | 24322494 | 24323004 | 511 | 5 | 0.000280467 | 1.54E-05 | 0.000783 | 2.16E-05 | 0.006812713 | GRHL3 |
| chr16 | 51544994 | 51545085 | 92 | 2 | 0.000130453 | 2.29E-05 | 0.000158 | 2.16E-05 | 0.023803218 | NA |
| chr1 | 228171052 | 228171085 | 34 | 3 | 5.22E-05 | 0.001256 | 8.17E-05 | 2.17E-05 | -0.0005215 | IBA57 |
| chr16 | 442818 | 442969 | 152 | 2 | 5.78E-05 | 3.95E-05 | 8.29E-05 | 2.17E-05 | 0.013019005 | RAB11FIP3 |
| chr17 | 48554548 | 48554792 | 245 | 3 | 8.52E-05 | 2.52E-05 | 0.000903 | 2.17E-05 | 0.005375865 | HOXB-AS3, HOXB3 |
| chr11 | 117876315 | 117877159 | 845 | 11 | 3.34E-06 | 0.002647 | 0.000377 | 2.17E-05 | 0.002347761 | FXYD6-FXYD2, FXYD6 |
| chr7 | 98300178 | 98300436 | 259 | 2 | 0.000104248 | 1.34E-05 | 0.001053 | 2.17E-05 | -0.00212404 | BRI3, BAIAP2L1 |
| chr11 | 7485424 | 7485709 | 286 | 3 | 3.37E-05 | 2.48E-05 | 0.000239 | 2.17E-05 | 0.006873441 | OLFML1, AC107884.1 |
| chr14 | 105364269 | 105364522 | 254 | 3 | 0.000105967 | 1.52E-05 | 0.001558 | 2.17E-05 | 0.009954557 | PACS2 |
| chr14 | 100286126 | 100286439 | 314 | 2 | 0.000182171 | 1.62E-05 | 0.000395 | 2.17E-05 | 0.011795989 | AL157871.6 |
| chr15 | 74873465 | 74873959 | 495 | 8 | 0.000185496 | 2.36E-05 | 0.004661 | 2.17E-05 | -0.00408027 | NA |
| chr7 | 6536337 | 6536927 | 591 | 6 | 3.85E-05 | 0.000602 | 0.000273 | 2.18E-05 | -0.00081902 | GRID2IP |
| chr12 | 109289906 | 109290545 | 640 | 6 | 9.15E-05 | 0.000168 | 0.001297 | 2.18E-05 | 0.003523986 | FOXN4 |
| chr9 | 32384121 | 32384423 | 303 | 4 | 1.54E-05 | 0.000134 | 0.000106 | 2.18E-05 | -0.00338319 | NA |
| chr5 | 7868774 | 7869315 | 542 | 13 | 0.000169733 | 3.97E-05 | 0.001818 | 2.18E-05 | -0.00014616 | MTRR, FASTKD3 |
| chr9 | 127568466 | 127568888 | 423 | 2 | 0.000222535 | 3.24E-05 | 0.000101 | 2.18E-05 | 0.004234542 | NIBAN2 |
| chr2 | 235868433 | 235868478 | 46 | 2 | 4.44E-05 | 0.001502 | 2.14E-05 | 2.18E-05 | 0.005590187 | AGAP1 |
| chr15 | 92391489 | 92391913 | 425 | 3 | 7.66E-05 | 1.02E-05 | 0.002139 | 2.18E-05 | 0.008968363 | NA |
| chr2 | 66872067 | 66872667 | 601 | 3 | 0.000220001 | 1.02E-05 | 0.002445 | 2.18E-05 | -0.00012807 | NA |
| chr2 | 241148088 | 241148889 | 802 | 5 | 2.39E-07 | 0.005121 | 1.60E-05 | 2.18E-05 | 0.005709993 | PASK |
| chr11 | 72435212 | 72435969 | 758 | 4 | 1.62E-05 | 0.000682 | 0.000594 | 2.18E-05 | 0.003412031 | NA |
| chr16 | 89620210 | 89620644 | 435 | 7 | 5.21E-05 | 0.000189 | 0.000192 | 2.18E-05 | 0.003997578 | DPEP1 |
| chr4 | 73403808 | 73404394 | 587 | 4 | 0.00010064 | 0.000116 | 0.000169 | 2.18E-05 | 0.003581316 | ALB |
| chr15 | 64045484 | 64046558 | 1075 | 5 | 4.14E-06 | 2.85E-05 | 0.000264 | 2.18E-05 | 0.004507368 | DAPK2 |
| chr16 | 62034033 | 62034543 | 511 | 2 | 5.89E-06 | 6.86E-05 | 5.52E-05 | 2.18E-05 | 0.021280865 | CDH8 |
| chr6 | 165826574 | 165826708 | 135 | 3 | 3.70E-05 | 0.00027 | 3.01E-05 | 2.18E-05 | 0.00445967 | PDE10A |
| chr3 | 115658513 | 115658926 | 414 | 3 | 0.000155117 | 0.000425 | 0.00013 | 2.18E-05 | 0.001054152 | GAP43, AC092468.1 |
| chr2 | 200735186 | 200735197 | 12 | 2 | 2.53E-05 | 2.57E-05 | 0.000136 | 2.18E-05 | 0.008229552 | AOX3P-AOX2P, LINC01792 |
| chr5 | 75713603 | 75713874 | 272 | 2 | 5.21E-06 | 0.000246 | 3.18E-05 | 2.18E-05 | -0.00219808 | POC5 |
| chr12 | 31354021 | 31354179 | 159 | 2 | 0.000105168 | 2.33E-05 | 0.000158 | 2.19E-05 | 0.011123262 | NA |
| chr7 | 128813089 | 128813332 | 244 | 2 | 4.38E-05 | 0.000121 | 4.13E-05 | 2.20E-05 | 0.009791971 | CCDC136 |
| chr6 | 33085815 | 33086014 | 200 | 2 | 6.36E-05 | 4.87E-05 | 7.07E-05 | 2.20E-05 | -0.00210535 | HLA-DPB1 |
| chr3 | 170869905 | 170870631 | 727 | 12 | 4.35E-05 | 0.000145 | 0.010382 | 2.20E-05 | 2.23E-05 | RPL22L1 |
| chr19 | 2227430 | 2227700 | 271 | 2 | 4.40E-05 | 1.44E-05 | 0.000727 | 2.20E-05 | 0.009206 | DOT1L |
| chr21 | 45555891 | 45556261 | 371 | 3 | 7.82E-05 | 9.35E-06 | 0.003408 | 2.20E-05 | 0.062148936 | SLC19A1 |
| chr20 | 61916849 | 61917562 | 714 | 3 | 0.000144083 | 0.000111 | 0.000672 | 2.20E-05 | 0.007730998 | CDH4 |
| chr5 | 181039679 | 181040519 | 841 | 8 | 4.53E-05 | 0.000101 | 0.000484 | 2.20E-05 | 0.007387381 | BTNL9 |
| chr20 | 47484420 | 47484702 | 283 | 6 | 0.000128931 | 1.92E-05 | 0.006176 | 2.21E-05 | -0.00069772 | NA |

| | | | | | | | | | | |
|---|---|---|---|---|---|---|---|---|---|---|
| chr14 | 60117653 | 60118341 | 689 | 4 | 4.53E-06 | 4.74E-05 | 0.000789 | 2.21E-05 | 0.005113854 | PCNX4 |
| chr1 | 167786988 | 167787338 | 351 | 2 | 6.62E-05 | 2.73E-05 | 0.000127 | 2.21E-05 | 0.008112307 | MPZL1 |
| chr19 | 41192643 | 41193459 | 817 | 12 | 0.000322568 | 1.06E-05 | 0.01686 | 2.21E-05 | 0.004168285 | CYP2S1 |
| chr6 | 43488904 | 43489536 | 633 | 6 | 0.000152765 | 6.35E-05 | 0.001635 | 2.21E-05 | 0.004425827 | TJAP1 |
| chr3 | 10449050 | 10450025 | 976 | 6 | 0.000209438 | 9.21E-05 | 0.001884 | 2.21E-05 | 0.003535115 | ATP2B2 |
| chr15 | 63503931 | 63504709 | 779 | 14 | 4.33E-05 | 0.000259 | 0.000191 | 2.21E-05 | -0.00010803 | USP3 |
| chr3 | 174056238 | 174056511 | 274 | 2 | 6.17E-05 | 1.67E-05 | 0.000387 | 2.21E-05 | -0.00996548 | NLGN1 |
| chr12 | 123585102 | 123585211 | 110 | 3 | 0.000292844 | 3.62E-05 | 0.000156 | 2.21E-05 | 0.000145451 | TMED2, TMED2-DT |
| chr1 | 214438194 | 214438270 | 77 | 2 | 0.000119474 | 1.52E-05 | 0.000571 | 2.22E-05 | 0.009583776 | PTPN14 |
| chr11 | 44957093 | 44957775 | 683 | 5 | 9.04E-05 | 1.85E-05 | 0.003153 | 2.22E-05 | 0.012156926 | NA |
| chr10 | 49299169 | 49299759 | 591 | 3 | 1.87E-08 | 0.000539 | 2.13E-05 | 2.22E-05 | 0.000726757 | C10orf71 |
| chr13 | 46639212 | 46639217 | 6 | 2 | 0.000129779 | 1.82E-05 | 0.000292 | 2.22E-05 | 0.014913459 | LRCH1 |
| chr7 | 1047181 | 1047212 | 32 | 2 | 3.17E-05 | 7.20E-05 | 5.46E-05 | 2.22E-05 | 0.004326039 | GPR146, C7orf50 |
| chr9 | 21031679 | 21031932 | 254 | 6 | 6.26E-05 | 0.001685 | 0.000396 | 2.22E-05 | 0.006003393 | NA |
| chr3 | 186640373 | 186640683 | 311 | 3 | 7.04E-05 | 4.37E-05 | 0.000386 | 2.22E-05 | 0.006291569 | FETUB, AC068631.1 |
| chr3 | 52221389 | 52222170 | 782 | 5 | 1.01E-05 | 2.17E-05 | 0.001135 | 2.22E-05 | 0.001801588 | TLR9, AC097637.1 |
| chr6 | 55040433 | 55040580 | 148 | 2 | 5.96E-05 | 4.51E-05 | 7.61E-05 | 2.22E-05 | -0.00562887 | NA |
| chr20 | 52190409 | 52190889 | 481 | 2 | 2.83E-05 | 1.65E-05 | 0.000405 | 2.22E-05 | 0.012395718 | ZFP64 |
| chr16 | 11586219 | 11586756 | 538 | 6 | 8.85E-05 | 1.01E-05 | 0.005058 | 2.23E-05 | 0.00525157 | LITAF |
| chr10 | 16519298 | 16519606 | 309 | 3 | 0.000225665 | 9.91E-06 | 0.002679 | 2.23E-05 | 0.018161461 | C1QL3 |
| chr19 | 45764054 | 45765251 | 1198 | 5 | 0.000102774 | 2.02E-05 | 0.0015 | 2.23E-05 | 0.010038635 | BHMG1, AC074212.1, SIX5 |
| chr4 | 13543008 | 13543667 | 660 | 3 | 1.84E-05 | 1.88E-05 | 0.001408 | 2.23E-05 | 0.004468098 | NKX3-2 |
| chr10 | 5500365 | 5500978 | 614 | 4 | 4.84E-05 | 3.44E-05 | 0.001808 | 2.23E-05 | 0.002231292 | NA |
| chr5 | 141369791 | 141370593 | 803 | 9 | 0.000105536 | 1.68E-05 | 0.00774 | 2.23E-05 | 0.012961826 | PCDHGA1, PCDHGA2, PCDHGA3, PCDHGB1, PCDHGA4, PCDHGB2, PCDHGA5, PCDHGB3 |
| chr22 | 29370806 | 29371153 | 348 | 2 | 0.000118903 | 1.40E-05 | 0.000935 | 2.23E-05 | 0.013061287 | AP1B1 |
| chr5 | 180324165 | 180324817 | 653 | 4 | 6.07E-05 | 0.000178 | 0.000797 | 2.23E-05 | -0.00067985 | GFPT2 |
| chr1 | 36374197 | 36374427 | 231 | 2 | 4.88E-05 | 1.83E-05 | 0.000292 | 2.23E-05 | 0.008847703 | STK40 |
| chr20 | 62352866 | 62352950 | 85 | 3 | 0.00011379 | 1.53E-05 | 0.000587 | 2.23E-05 | 0.006412552 | LAMA5 |
| chr11 | 124777348 | 124777451 | 104 | 3 | 6.11E-05 | 1.60E-05 | 0.000689 | 2.23E-05 | 0.012222746 | MSANTD2 |
| chr2 | 10816629 | 10816843 | 215 | 3 | 6.22E-05 | 0.00028 | 0.000274 | 2.23E-05 | 0.003478145 | PDIA6 |
| chr8 | 22392151 | 22392395 | 245 | 2 | 8.47E-05 | 8.20E-05 | 5.08E-05 | 2.23E-05 | 0.004282048 | SLC39A14 |
| chr4 | 186604109 | 186604550 | 442 | 3 | 6.00E-05 | 1.90E-05 | 0.000922 | 2.23E-05 | 0.001980055 | FAT1 |
| chr1 | 11176372 | 11176590 | 219 | 2 | 5.95E-05 | 0.007585 | 1.91E-05 | 2.23E-05 | 0.002402464 | MTOR |
| chr12 | 124681070 | 124681473 | 404 | 2 | 9.19E-05 | 1.47E-05 | 0.000708 | 2.23E-05 | 0.014418177 | NA |
| chr3 | 171811038 | 171811820 | 783 | 4 | 6.75E-05 | 0.00025 | 4.24E-05 | 2.24E-05 | 0.008054581 | NA |
| chr5 | 180016379 | 180016887 | 509 | 3 | 6.52E-05 | 1.24E-05 | 0.001243 | 2.24E-05 | -0.00076381 | RNF130 |
| chr1 | 67528820 | 67528964 | 145 | 2 | 0.000106366 | 2.61E-05 | 0.00014 | 2.24E-05 | 0.005727451 | LINC01702 |
| chr9 | 125545299 | 125545386 | 88 | 2 | 3.33E-05 | 0.059416 | 1.82E-05 | 2.24E-05 | 0.008870816 | MAPKAP1 |
| chr6 | 56642872 | 56643073 | 202 | 3 | 4.41E-05 | 6.21E-05 | 7.37E-05 | 2.24E-05 | -0.00033529 | DST |
| chr4 | 16083557 | 16084191 | 635 | 11 | 0.000189209 | 0.000229 | 0.000549 | 2.25E-05 | 0.001345256 | PROM1 |
| chr1 | 54096367 | 54096870 | 504 | 5 | 0.000136537 | 1.02E-05 | 0.002857 | 2.25E-05 | -0.00782843 | TCEANC2 |
| chr5 | 1827557 | 1827984 | 428 | 2 | 9.59E-06 | 2.19E-05 | 0.00019 | 2.25E-05 | -0.01417677 | NA |
| chr12 | 610325 | 610583 | 259 | 3 | 8.58E-05 | 1.99E-05 | 0.000487 | 2.25E-05 | 0.006728478 | NINJ2 |
| chr6 | 166861907 | 166862871 | 965 | 12 | 3.78E-05 | 0.000187 | 0.001376 | 2.25E-05 | 0.000757818 | RPS6KA2, AL159163.1 |
| chr3 | 183508237 | 183509009 | 773 | 2 | 3.28E-06 | 6.51E-05 | 5.91E-05 | 2.25E-05 | 0.012682036 | KLHL6 |
| chr7 | 344824 | 344989 | 166 | 2 | 5.62E-05 | 1.42E-05 | 0.000945 | 2.25E-05 | 0.001713397 | NA |
| chr19 | 38213859 | 38214293 | 435 | 4 | 3.57E-05 | 0.000125 | 0.000126 | 2.26E-05 | 0.01234396 | DPF1 |
| chr17 | 81229147 | 81229306 | 160 | 3 | 4.34E-05 | 8.76E-05 | 5.73E-05 | 2.26E-05 | 0.004353853 | AC027601.1, TEPSIN |
| chr2 | 231056579 | 231057238 | 660 | 10 | 9.21E-05 | 0.000221 | 0.000182 | 2.26E-05 | 6.70E-05 | PSMD1 |
| chr16 | 74303952 | 74304079 | 128 | 2 | 8.84E-05 | 4.91E-05 | 7.28E-05 | 2.26E-05 | -0.0003258 | PSMD7 |
| chr1 | 31531210 | 31531819 | 610 | 4 | 0.000166423 | 1.72E-05 | 0.001079 | 2.26E-05 | -0.003366 | LINC01226 |
| chr7 | 56483058 | 56483947 | 890 | 8 | 3.41E-05 | 0.00021 | 0.002487 | 2.26E-05 | 0.01370723 | AC092447.4 |
| chr15 | 24848097 | 24848838 | 742 | 5 | 2.37E-05 | 0.002137 | 0.000318 | 2.26E-05 | -0.01116796 | AC090983.2, SNRPN, AC090983.1 |
| chr10 | 104614421 | 104614603 | 183 | 2 | 2.62E-05 | 6.03E-05 | 6.24E-05 | 2.26E-05 | 0.019487731 | AL161646.1 |
| chr13 | 50140897 | 50141568 | 672 | 5 | 5.54E-05 | 0.000436 | 0.001006 | 2.26E-05 | 0.006648297 | DLEU1 |
| chr17 | 79950470 | 79951648 | 1179 | 13 | 8.40E-06 | 0.000167 | 0.000198 | 2.26E-05 | 0.007765223 | TBC1D16 |
| chr12 | 56751574 | 56752575 | 1002 | 17 | 2.19E-05 | 0.001791 | 0.002107 | 2.26E-05 | 0.000993315 | HSD17B6, PRIM1 |
| chr8 | 66922748 | 66923618 | 871 | 3 | 1.40E-06 | 0.00258 | 1.42E-05 | 2.27E-05 | 0.012568101 | SNHG6 |
| chr5 | 42912418 | 42912638 | 221 | 2 | 8.44E-05 | 3.40E-05 | 0.000102 | 2.27E-05 | 0.010682124 | AC008945.2 |
| chr3 | 193551771 | 193551807 | 37 | 2 | 4.03E-05 | 4.64E-05 | 7.65E-05 | 2.27E-05 | -0.00138069 | ATP13A4 |
| chr3 | 133949390 | 133949719 | 330 | 2 | 9.70E-06 | 0.000849 | 2.42E-05 | 2.27E-05 | 0.003176177 | SLCO2A1 |
| chr5 | 124644390 | 124645043 | 654 | 5 | 0.0001104 | 2.88E-05 | 0.000988 | 2.27E-05 | -0.0003736 | ZNF608 |
| chr12 | 67590020 | 67590583 | 564 | 4 | 2.15E-06 | 0.010876 | 1.56E-05 | 2.27E-05 | -0.00459991 | LINC02408 |
| chr3 | 98594554 | 98595149 | 596 | 3 | 1.57E-05 | 1.08E-05 | 0.002031 | 2.27E-05 | 0.005407078 | NA |
| chr12 | 47504858 | 47505007 | 150 | 3 | 3.61E-05 | 0.000249 | 0.000114 | 2.27E-05 | 0.010653696 | NA |
| chr1 | 112360235 | 112360607 | 373 | 3 | 9.56E-05 | 1.29E-05 | 0.001908 | 2.27E-05 | 0.012111026 | LINC02884 |
| chr12 | 114090979 | 114091197 | 219 | 2 | 0.000112336 | 1.92E-05 | 0.000268 | 2.27E-05 | 0.002896058 | AC010183.2 |
| chr22 | 24950387 | 24950787 | 401 | 2 | 0.000140238 | 1.41E-05 | 0.001064 | 2.27E-05 | 0.009080693 | NA |
| chr16 | 80553644 | 80553663 | 20 | 2 | 3.35E-05 | 1.43E-05 | 0.000936 | 2.27E-05 | 0.007942548 | AC105411.1, LINC01227 |
| chr9 | 89010491 | 89010857 | 367 | 3 | 1.86E-05 | 0.000544 | 0.000115 | 2.27E-05 | 0.003964341 | SHC3 |
| chr7 | 50782867 | 50783173 | 307 | 5 | 1.09E-05 | 9.38E-06 | 0.005193 | 2.27E-05 | 0.009591506 | GRB10 |
| chr19 | 36572866 | 36573478 | 613 | 13 | 0.000106409 | 2.91E-05 | 0.010845 | 2.27E-05 | 0.001429831 | ZNF529-AS1, ZNF529 |
| chr1 | 54463903 | 54464483 | 581 | 2 | 0.000216822 | 1.41E-05 | 0.001078 | 2.27E-05 | 0.012764653 | NA |
| chr19 | 40691144 | 40691359 | 216 | 4 | 2.62E-05 | 5.47E-05 | 0.001146 | 2.27E-05 | -0.00077492 | NA |
| chr14 | 76812974 | 76813385 | 412 | 7 | 3.87E-05 | 1.59E-05 | 0.001951 | 2.27E-05 | 0.005639909 | ANGEL1 |
| chr8 | 37691684 | 37691750 | 67 | 2 | 7.49E-05 | 4.18E-05 | 8.42E-05 | 2.27E-05 | 0.014416159 | NA |
| chr12 | 6941787 | 6942343 | 557 | 2 | 2.88E-05 | 1.49E-05 | 0.00073 | 2.27E-05 | -0.00421588 | ATN1 |
| chr12 | 121823509 | 121823922 | 414 | 4 | 0.000139263 | 3.79E-05 | 0.000154 | 2.27E-05 | 0.000771093 | SETD1B |
| chr8 | 38303076 | 38303186 | 111 | 2 | 3.49E-05 | 0.007515 | 1.94E-05 | 2.27E-05 | 0.005423606 | NSD3 |
| chr17 | 50163708 | 50164709 | 1002 | 4 | 1.66E-05 | 0.000477 | 0.000111 | 2.27E-05 | 0.006472726 | SGCA, AC015909.3 |
| chr22 | 38412810 | 38413251 | 442 | 4 | 0.000116513 | 2.55E-05 | 0.000905 | 2.27E-05 | 0.005493255 | NA |
| chr2 | 143128757 | 143129371 | 615 | 6 | 6.53E-05 | 0.000112 | 0.000846 | 2.27E-05 | 0.000405752 | ARHGAP15 |
| chr6 | 14721236 | 14721506 | 271 | 2 | 6.82E-05 | 1.98E-05 | 0.00025 | 2.27E-05 | 0.01511796 | AL138720.1 |
| chr8 | 11802905 | 11803459 | 555 | 8 | 2.19E-07 | 0.002788 | 2.05E-05 | 2.28E-05 | -0.00052722 | FDFT1 |
| chr19 | 5806153 | 5806293 | 141 | 2 | 1.82E-05 | 4.86E-05 | 7.45E-05 | 2.28E-05 | 0.012892933 | NA |
| chr3 | 156586909 | 156586937 | 29 | 2 | 0.000143052 | 1.51E-05 | 0.00069 | 2.28E-05 | 0.016862303 | NA |
| chr15 | 98982024 | 98982583 | 560 | 4 | 2.41E-05 | 9.81E-05 | 8.13E-05 | 2.28E-05 | 0.004176997 | AC036108.1, PGPEP1L |
| chr5 | 66209353 | 66209574 | 222 | 2 | 6.27E-05 | 1.50E-05 | 0.00071 | 2.28E-05 | 0.010127538 | LINC02065 |
| chr17 | 59220396 | 59221314 | 919 | 10 | 4.41E-06 | 0.000635 | 0.000239 | 2.28E-05 | 0.002325145 | GDPD1 |
| chr1 | 216601246 | 216601614 | 369 | 4 | 5.95E-05 | 1.87E-05 | 0.00159 | 2.28E-05 | 0.009496271 | ESRRG |
| chr2 | 101991159 | 101991887 | 729 | 9 | 0.000146938 | 4.13E-05 | 0.004213 | 2.28E-05 | 0.004781211 | NA |
| chr15 | 95286808 | 95287456 | 649 | 2 | 0.000362133 | 1.45E-05 | 0.000896 | 2.28E-05 | 0.015071211 | LINC01197 |
| chr16 | 56989279 | 56989923 | 645 | 10 | 8.08E-05 | 0.000334 | 0.002176 | 2.28E-05 | 0.000467913 | NLRC5 |
| chr2 | 173965725 | 173966328 | 604 | 7 | 8.49E-05 | 1.53E-05 | 0.008111 | 2.28E-05 | 0.001245199 | NA |
| chr8 | 1061358 | 1061835 | 478 | 3 | 1.26E-05 | 0.000394 | 0.000589 | 2.28E-05 | -0.00074668 | DLGAP2 |
| chr7 | 8261805 | 8263101 | 1297 | 11 | 2.29E-05 | 0.000359 | 0.000597 | 2.28E-05 | 0.001736789 | AC007009.1, AC007128.1, ICA1 |
| chr3 | 64023420 | 64023731 | 312 | 7 | 0.000340537 | 0.000502 | 9.47E-05 | 2.28E-05 | 0.000115514 | PSMD6 |
| chr15 | 90906808 | 90907434 | 627 | 5 | 4.91E-05 | 2.90E-05 | 0.00383 | 2.28E-05 | 0.003920215 | MAN2A2 |
| chr8 | 23304251 | 23304870 | 620 | 4 | 0.000231503 | 9.70E-06 | 0.004576 | 2.28E-05 | 0.014723993 | LOXL2 |
| chr7 | 98413463 | 98413674 | 212 | 2 | 0.000141954 | 1.45E-05 | 0.00093 | 2.28E-05 | -0.00057146 | NA |
| chr14 | 22575170 | 22575196 | 27 | 2 | 9.36E-05 | 1.85E-05 | 0.000306 | 2.28E-05 | 0.01603891 | DAD1 |
| chr8 | 29067101 | 29067162 | 62 | 3 | 0.000105048 | 3.07E-05 | 0.00036 | 2.29E-05 | 0.003069128 | NA |
| chr4 | 8387740 | 8388421 | 682 | 5 | 1.33E-05 | 6.15E-05 | 0.001602 | 2.29E-05 | 0.003040749 | ACOX3 |
| chr1 | 169539119 | 169539436 | 318 | 2 | 2.19E-05 | 0.000783 | 2.49E-05 | 2.29E-05 | 0.002534018 | F5 |
| chr9 | 78030400 | 78030640 | 241 | 2 | 3.87E-05 | 2.67E-05 | 0.000143 | 2.29E-05 | 0.005640399 | GNAQ |
| chr7 | 44218112 | 44218455 | 344 | 2 | 6.24E-05 | 1.44E-05 | 0.000971 | 2.30E-05 | 0.004848921 | CAMK2B |
| chr5 | 168579707 | 168580334 | 628 | 8 | 0.000134372 | 0.000106 | 0.00582 | 2.30E-05 | 0.002749675 | NA |
| chr2 | 205776364 | 205776906 | 543 | 2 | 0.00018055 | 1.46E-05 | 0.000898 | 2.30E-05 | 0.010591945 | NRP2 |
| chr19 | 8158423 | 8158772 | 350 | 4 | 1.89E-05 | 0.002665 | 1.87E-05 | 2.30E-05 | 0.008018274 | NA |
| chr14 | 104378989 | 104379595 | 607 | 5 | 7.13E-05 | 0.00032 | 0.000204 | 2.30E-05 | 0.005390129 | NA |
| chr7 | 44044460 | 44045287 | 828 | 7 | 4.36E-05 | 0.000147 | 0.001136 | 2.30E-05 | 0.000495994 | DBNL |
| chr10 | 74103827 | 74104372 | 546 | 2 | 1.37E-05 | 3.41E-05 | 0.000105 | 2.30E-05 | 0.006669697 | VCL |
| chr19 | 36110141 | 36110198 | 58 | 2 | 4.61E-05 | 0.000209 | 3.51E-05 | 2.30E-05 | -0.00376001 | NA |
| chr16 | 87912567 | 87912938 | 372 | 2 | 5.63E-05 | 3.00E-05 | 0.000123 | 2.30E-05 | 0.011804428 | CA5A |
| chr5 | 140111310 | 140111645 | 336 | 2 | 9.38E-05 | 1.85E-05 | 0.000315 | 2.30E-05 | 0.020885948 | PURA |
| chr1 | 247210208 | 247211055 | 848 | 5 | 7.97E-05 | 6.53E-05 | 0.000175 | 2.30E-05 | -0.00129194 | AL390728.4, AL390728.5 |
| chr7 | 151878192 | 151878592 | 401 | 2 | 6.87E-06 | 2.89E-05 | 0.000129 | 2.31E-05 | 0.012646087 | PRKAG2-AS1 |
| chr9 | 126616766 | 126617040 | 275 | 2 | 2.52E-05 | 0.000225 | 3.44E-05 | 2.31E-05 | 0.017513816 | LMX1B |
| chr1 | 15761257 | 15761507 | 251 | 3 | 9.81E-05 | 3.11E-05 | 0.000508 | 2.31E-05 | 0.010875022 | FBLIM1 |
| chr6 | 26246561 | 26247288 | 728 | 11 | 0.000179156 | 5.72E-05 | 0.009554 | 2.31E-05 | 0.000571033 | H4C7 |
| chr9 | 5465479 | 5465685 | 207 | 3 | 4.13E-05 | 0.000995 | 4.26E-05 | 2.31E-05 | -0.00143822 | CD274, AL162253.2 |
| chr6 | 149863780 | 149864718 | 939 | 12 | 1.36E-05 | 0.000311 | 0.000332 | 2.31E-05 | -0.00074873 | RAET1E-AS1, AL355312.5, LRP11 |
| chr17 | 81155027 | 81155278 | 252 | 5 | 6.18E-05 | 0.000147 | 0.00064 | 2.31E-05 | 0.003812819 | AATK |
| chr19 | 45423932 | 45424438 | 507 | 10 | 8.81E-05 | 0.001465 | 0.001304 | 2.31E-05 | 0.001799595 | ERCC1 |
| chr16 | 17155198 | 17155729 | 532 | 2 | 0.000136495 | 1.58E-05 | 0.000613 | 2.31E-05 | 0.011592809 | XYLT1 |
| chr7 | 27252526 | 27252935 | 410 | 3 | 6.15E-05 | 1.63E-05 | 0.001086 | 2.31E-05 | 0.005645823 | NA |
| chr20 | 43557981 | 43559010 | 1030 | 7 | 1.91E-06 | 0.000496 | 0.001242 | 2.31E-05 | 0.01500605 | Z98752.3, SGK2 |
| chr6 | 106997800 | 106998137 | 338 | 3 | 4.37E-05 | 0.000105 | 5.71E-05 | 2.32E-05 | 0.005795746 | NA |
| chr22 | 31749615 | 31750719 | 1105 | 11 | 1.01E-05 | 0.000301 | 0.000585 | 2.32E-05 | 0.000346062 | PRR14L |
| chr10 | 123568062 | 123568142 | 81 | 3 | 0.000138252 | 2.45E-05 | 0.00032 | 2.32E-05 | 0.006638311 | NA |
| chr18 | 74315595 | 74316387 | 793 | 9 | 4.60E-05 | 0.00013 | 0.005867 | 2.32E-05 | -0.00078626 | C18orf63 |
| chr7 | 134683782 | 134684029 | 248 | 2 | 4.61E-05 | 6.04E-05 | 6.47E-05 | 2.32E-05 | 0.008264987 | NA |
| chr15 | 82739183 | 82739282 | 100 | 2 | 8.54E-05 | 6.66E-05 | 6.06E-05 | 2.32E-05 | 0.01536481 | NA |
| chr1 | 93847583 | 93848234 | 652 | 7 | 3.47E-05 | 0.000151 | 0.000488 | 2.32E-05 | 0.003086929 | AL049796.1 |
| chr17 | 81027168 | 81027619 | 452 | 3 | 0.000161355 | 1.55E-05 | 0.000635 | 2.32E-05 | 0.00439349 | AC127496.5 |
| chr9 | 84261777 | 84261982 | 206 | 2 | 2.32E-06 | 0.000526 | 2.76E-05 | 2.32E-05 | 0.008135608 | NA |
| chr17 | 39940080 | 39940395 | 316 | 4 | 9.36E-05 | 5.41E-05 | 0.001364 | 2.33E-05 | 0.00461608 | NA |
| chr2 | 200778471 | 200778773 | 303 | 2 | 0.000124647 | 1.62E-05 | 0.000553 | 2.33E-05 | -0.00354504 | AOX2P |
| chr17 | 42107706 | 42108530 | 825 | 6 | 6.76E-07 | 0.008961 | 1.47E-05 | 2.33E-05 | 0.003873542 | DHX58 |
| chr7 | 3244762 | 3244817 | 56 | 2 | 0.000179047 | 1.44E-05 | 0.001089 | 2.33E-05 | -0.01703256 | NA |
| chr11 | 64742420 | 64742674 | 255 | 3 | 0.000149395 | 6.68E-05 | 0.000339 | 2.33E-05 | -0.00097631 | RASGRP2 |
| chr13 | 51962441 | 51962514 | 74 | 3 | 0.000169544 | 1.29E-05 | 0.001142 | 2.33E-05 | 0.000973366 | ATP7B |
| chr16 | 1487774 | 1488663 | 890 | 6 | 1.49E-06 | 3.00E-05 | 0.003583 | 2.33E-05 | 0.007449935 | PTX4 |
| chr19 | 1304575 | 1305253 | 679 | 5 | 2.71E-05 | 0.000489 | 4.98E-05 | 2.33E-05 | 0.002475858 | NA |
| chr1 | 100423918 | 100424116 | 199 | 2 | 4.05E-05 | 4.16E-05 | 8.77E-05 | 2.33E-05 | 0.010037181 | CDC14A |
| chr1 | 225454429 | 225455044 | 616 | 2 | 0.000284636 | 1.45E-05 | 0.001077 | 2.33E-05 | 0.014002102 | LINC02765 |
| chr2 | 30231280 | 30231777 | 498 | 8 | 8.82E-06 | 0.012134 | 1.78E-05 | 2.33E-05 | -0.00106579 | LBH |
| chr16 | 3965230 | 3965760 | 531 | 2 | 0.000201736 | 1.72E-05 | 0.000438 | 2.33E-05 | -0.00582981 | ADCY9 |
| chr15 | 42841674 | 42841984 | 311 | 3 | 8.15E-05 | 8.97E-05 | 0.000722 | 2.33E-05 | 0.002280746 | TTBK2 |
| chr16 | 57627387 | 57627605 | 219 | 3 | 2.97E-05 | 4.45E-05 | 0.000165 | 2.33E-05 | 0.005251332 | ADGRG1, AC018552.3 |
| chr19 | 14031235 | 14031773 | 539 | 10 | 0.000191712 | 2.64E-05 | 0.000937 | 2.33E-05 | 0.003104107 | RLN3, IL27RA, AC022098.2 |
| chr1 | 151282024 | 151282595 | 572 | 9 | 0.000155916 | 5.46E-05 | 0.002773 | 2.33E-05 | 0.002094454 | ZNF687 |
| chr7 | 129206171 | 129206750 | 580 | 4 | 4.36E-05 | 0.00033 | 0.00222 | 2.34E-05 | 0.004965534 | SMO |
| chr16 | 47450745 | 47450983 | 239 | 2 | 7.78E-05 | 8.53E-05 | 5.27E-05 | 2.34E-05 | 0.006427716 | ITFG1 |
| chr17 | 40054220 | 40054744 | 525 | 10 | 0.000296695 | 7.84E-05 | 0.003931 | 2.34E-05 | -0.00164952 | MED24 |
| chr15 | 34345366 | 34345668 | 303 | 3 | 0.000100572 | 1.70E-05 | 0.000651 | 2.34E-05 | 0.00637719 | NUTM1 |
| chr15 | 77035157 | 77035546 | 390 | 2 | 1.93E-05 | 7.77E-05 | 5.57E-05 | 2.34E-05 | 0.00545828 | PSTPIP1 |
| chr19 | 40603532 | 40603964 | 433 | 4 | 7.25E-05 | 5.67E-05 | 0.000721 | 2.34E-05 | 0.006379181 | LTBP4 |
| chr11 | 60986181 | 60986856 | 676 | 4 | 0.000134383 | 0.000116 | 0.001423 | 2.34E-05 | 0.007165749 | CD6 |
| chr17 | 48430392 | 48430743 | 352 | 8 | 0.000218014 | 6.01E-05 | 0.000454 | 2.34E-05 | -0.00662436 | NA |
| chr17 | 81919438 | 81919631 | 194 | 3 | 4.94E-05 | 1.09E-05 | 0.002108 | 2.34E-05 | 0.010483014 | SIRT7, MAFG |
| chr12 | 15001247 | 15002297 | 1051 | 4 | 3.16E-06 | 0.000507 | 5.34E-05 | 2.34E-05 | 0.01005111 | LINC01489 |

| | | | | | | | | | | |
|---|---|---|---|---|---|---|---|---|---|---|
| chr3 | 111977426 | 111977984 | 559 | 3 | 6.88E-06 | 7.36E-05 | 6.85E-05 | 2.34E-05 | -0.00706409 | NA |
| chr3 | 149969073 | 149969528 | 456 | 3 | 5.22E-06 | 0.000757 | 2.72E-05 | 2.34E-05 | 0.007580429 | PFN2 |
| chr18 | 73461685 | 73462277 | 593 | 2 | 0.00021571 | 1.48E-05 | 0.000935 | 2.34E-05 | 0.024228173 | NA |
| chr21 | 43003419 | 43003719 | 301 | 2 | 3.11E-05 | 0.000305 | 3.19E-05 | 2.34E-05 | 0.002447452 | PKNOX1 |
| chr1 | 107449255 | 107449558 | 304 | 3 | 4.64E-05 | 9.62E-05 | 5.53E-05 | 2.35E-05 | 0.004244024 | NTNG1 |
| chr11 | 18680733 | 18680902 | 170 | 2 | 6.56E-05 | 0.000118 | 4.49E-05 | 2.35E-05 | -0.00208958 | NA |
| chr7 | 158016291 | 158016904 | 614 | 7 | 0.000135656 | 3.37E-05 | 0.011272 | 2.35E-05 | 0.01682724 | PTPRN2 |
| chr8 | 140540561 | 140541554 | 994 | 4 | 8.56E-07 | 0.000549 | 2.82E-05 | 2.35E-05 | 0.001139093 | AGO2 |
| chr7 | 92626181 | 92626410 | 230 | 3 | 0.000175195 | 9.72E-06 | 0.003651 | 2.35E-05 | 0.012149847 | CDK6 |
| chr16 | 85509143 | 85509691 | 549 | 5 | 2.17E-05 | 6.18E-05 | 0.00084 | 2.35E-05 | 0.006389762 | GSE1 |
| chr8 | 69833501 | 69834431 | 931 | 14 | 0.000179114 | 4.31E-05 | 0.003566 | 2.35E-05 | -8.43E-05 | AC079089.1, SLCO5A1 |
| chr11 | 829035 | 829182 | 148 | 3 | 0.000247023 | 1.46E-05 | 0.000794 | 2.36E-05 | 0.010398844 | CRACR2B, AP006621.1 |
| chr2 | 176083868 | 176084289 | 422 | 4 | 1.23E-05 | 0.000416 | 6.09E-05 | 2.36E-05 | 0.008104943 | EVX2 |
| chr17 | 48440279 | 48440486 | 208 | 2 | 0.000128054 | 1.46E-05 | 0.001099 | 2.36E-05 | 0.007359471 | NA |
| chr20 | 63133155 | 63133211 | 57 | 2 | 0.000152216 | 1.47E-05 | 0.001058 | 2.36E-05 | 0.012377174 | NA |
| chr20 | 64074076 | 64074581 | 506 | 2 | 1.57E-06 | 6.52E-05 | 6.25E-05 | 2.36E-05 | 0.008775928 | RGS19 |
| chr14 | 77604112 | 77604127 | 16 | 2 | 0.000127795 | 2.30E-05 | 0.000199 | 2.36E-05 | 0.016974323 | SPTLC2 |
| chr3 | 129881640 | 129881909 | 270 | 2 | 1.93E-05 | 0.000442 | 2.93E-05 | 2.36E-05 | 0.008984929 | TMCC1 |
| chr12 | 53371294 | 53371403 | 110 | 2 | 0.000111539 | 1.47E-05 | 0.001086 | 2.36E-05 | 0.002343856 | NA |
| chr5 | 134249715 | 134249800 | 86 | 2 | 0.000142232 | 1.64E-05 | 0.000572 | 2.36E-05 | 0.010313516 | AC104109.4 |
| chr14 | 24204954 | 24205272 | 319 | 3 | 1.43E-05 | 2.27E-05 | 0.000305 | 2.36E-05 | -0.00089446 | AL136295.1 |
| chr16 | 51154461 | 51155933 | 1473 | 6 | 0.000164162 | 2.23E-05 | 0.00234 | 2.36E-05 | 0.005382907 | AC087564.1 |
| chr19 | 55474857 | 55475107 | 251 | 2 | 8.41E-06 | 1.78E-05 | 0.000412 | 2.37E-05 | 0.004060631 | NA |
| chr11 | 62841356 | 62842100 | 745 | 6 | 2.08E-05 | 1.97E-05 | 0.001392 | 2.37E-05 | -0.00099785 | WDR74, RNU2-2P |
| chr1 | 58816250 | 58816810 | 561 | 3 | 3.14E-05 | 0.000982 | 0.000149 | 2.37E-05 | 0.006387998 | LINC01135 |
| chr17 | 31394709 | 31394920 | 212 | 3 | 0.000178623 | 1.14E-05 | 0.002383 | 2.37E-05 | 0.012663238 | RAB11FIP4 |
| chr13 | 30154397 | 30154682 | 286 | 3 | 1.42E-05 | 4.86E-05 | 0.000387 | 2.37E-05 | 0.000890671 | LINC00385, LINC00384 |
| chr19 | 39514711 | 39515552 | 842 | 11 | 1.97E-05 | 0.003045 | 0.002979 | 2.37E-05 | 0.000937864 | SELENOV |
| chr12 | 95956485 | 95957017 | 533 | 6 | 8.27E-05 | 5.68E-05 | 0.00351 | 2.37E-05 | -0.01788863 | AMDHD1 |
| chr6 | 30159588 | 30160477 | 890 | 6 | 2.83E-05 | 3.72E-05 | 0.002511 | 2.37E-05 | 0.003405065 | TRIM10 |
| chr17 | 80189469 | 80189991 | 523 | 4 | 2.01E-05 | 0.003931 | 0.001068 | 2.37E-05 | 0.007062042 | CARD14 |
| chr8 | 53842859 | 53843664 | 806 | 17 | 5.98E-05 | 0.000324 | 0.000701 | 2.37E-05 | 0.000374606 | ATP6V1H |
| chr6 | 108844152 | 108844595 | 444 | 2 | 6.92E-05 | 2.51E-05 | 0.000171 | 2.37E-05 | 0.004405529 | NA |
| chr6 | 31170643 | 31170834 | 192 | 2 | 0.000103106 | 1.49E-05 | 0.000994 | 2.37E-05 | 0.017002308 | POU5F1 |
| chr7 | 129781073 | 129781688 | 616 | 3 | 0.000102198 | 1.11E-05 | 0.002838 | 2.38E-05 | -0.00154616 | NA |
| chr10 | 28681605 | 28682069 | 465 | 2 | 0.000103864 | 1.91E-05 | 0.00033 | 2.38E-05 | 0.006122201 | BAMBI |
| chr9 | 137076923 | 137077388 | 466 | 7 | 1.63E-05 | 9.70E-05 | 0.000529 | 2.39E-05 | 0.005260444 | NA |
| chr4 | 7800465 | 7800902 | 438 | 6 | 0.000128432 | 5.18E-05 | 0.002189 | 2.39E-05 | 0.004197626 | AC112254.1, AFAP1 |
| chr19 | 12064292 | 12064904 | 613 | 10 | 4.86E-05 | 0.000673 | 0.003044 | 2.39E-05 | 0.001896175 | AC008770.4, ZNF844 |
| chr8 | 132775140 | 132775444 | 305 | 8 | 2.13E-05 | 0.000208 | 8.42E-05 | 2.39E-05 | 0.000184937 | PHF20L1 |
| chr19 | 50512447 | 50512573 | 127 | 3 | 1.88E-05 | 0.000148 | 0.000127 | 2.39E-05 | 0.003714441 | ASPDH |
| chr18 | 3585486 | 3585935 | 450 | 2 | 1.98E-05 | 2.85E-05 | 0.000141 | 2.39E-05 | 0.01090163 | DLGAP1 |
| chr8 | 142614466 | 142615170 | 705 | 9 | 8.99E-05 | 0.000109 | 0.005262 | 2.39E-05 | 0.005606338 | ARC |
| chr14 | 81617105 | 81617137 | 33 | 2 | 0.000104462 | 1.93E-05 | 0.00032 | 2.39E-05 | 0.007890998 | LINC01467 |
| chr5 | 108747746 | 108748527 | 782 | 8 | 2.37E-05 | 0.000406 | 0.000651 | 2.39E-05 | 0.00412601 | FER |
| chr12 | 131919804 | 131920311 | 508 | 4 | 2.59E-05 | 0.000114 | 0.000705 | 2.39E-05 | 0.004420458 | ULK1 |
| chr1 | 168609022 | 168609496 | 475 | 3 | 8.34E-05 | 1.61E-05 | 0.001644 | 2.39E-05 | 0.00519754 | NA |
| chr2 | 127024659 | 127025506 | 848 | 7 | 5.44E-05 | 1.37E-05 | 0.008654 | 2.40E-05 | 0.010456488 | AC012508.3, AC012508.1, AC012508.2 |
| chr11 | 47979746 | 47980283 | 538 | 4 | 4.19E-05 | 0.000185 | 0.001328 | 2.40E-05 | 0.008862955 | NA |
| chr1 | 230336310 | 230336959 | 650 | 4 | 0.000196898 | 7.01E-06 | 0.007662 | 2.40E-05 | 0.007291858 | PGBD5 |
| chr2 | 240584641 | 240585445 | 805 | 3 | 1.77E-06 | 0.00039 | 2.69E-05 | 2.40E-05 | 0.00281659 | CAPN10-DT |
| chr4 | 144646096 | 144646579 | 484 | 7 | 9.17E-07 | 0.003842 | 1.46E-05 | 2.40E-05 | 0.001740957 | HHIP, AC098588.3, AC098588.2, HHIP-AS1 |
| chr18 | 31942471 | 31943115 | 645 | 5 | 7.76E-05 | 2.76E-05 | 0.00111 | 2.41E-05 | 0.001117028 | AC009831.1, TRAPPC8 |
| chr1 | 16115884 | 16116135 | 252 | 2 | 7.70E-05 | 2.73E-05 | 0.000152 | 2.41E-05 | 0.010644671 | NA |
| chr9 | 133561977 | 133562264 | 288 | 3 | 6.80E-05 | 2.93E-05 | 0.000208 | 2.41E-05 | 0.005863249 | ADAMTSL2 |
| chr4 | 55819679 | 55819951 | 273 | 4 | 7.25E-05 | 3.67E-05 | 0.000461 | 2.41E-05 | 0.006522662 | EXOC1L |
| chr2 | 119678854 | 119679433 | 580 | 9 | 9.78E-05 | 1.48E-05 | 0.016015 | 2.41E-05 | -0.00171536 | TMEM177 |
| chr6 | 34022067 | 34022404 | 338 | 2 | 5.16E-05 | 3.74E-05 | 0.000103 | 2.41E-05 | 0.011366541 | GRM4 |
| chr16 | 71483383 | 71484329 | 947 | 7 | 4.21E-05 | 0.00196 | 0.00117 | 2.42E-05 | 0.001320605 | AC010547.4, ZNF19 |
| chr17 | 77388808 | 77389004 | 197 | 2 | 0.000226623 | 1.65E-05 | 0.000629 | 2.42E-05 | 0.013859797 | SEPTIN9 |
| chr1 | 223129368 | 223129768 | 401 | 4 | 0.000211674 | 1.13E-05 | 0.003778 | 2.42E-05 | 0.008022566 | TLR5 |
| chr6 | 155154426 | 155154519 | 94 | 3 | 5.85E-05 | 0.000359 | 0.00011 | 2.42E-05 | 0.01020777 | TIAM2 |
| chr8 | 58272605 | 58273262 | 658 | 3 | 1.84E-05 | 0.000464 | 3.80E-05 | 2.42E-05 | 0.002424785 | NA |
| chr20 | 25045129 | 25045551 | 423 | 2 | 0.000100998 | 1.52E-05 | 0.001017 | 2.42E-05 | 0.008386874 | ACSS1 |
| chr5 | 178995414 | 178995430 | 17 | 2 | 0.00036296 | 1.53E-05 | 0.000994 | 2.42E-05 | 0.018578494 | GRM6 |
| chr13 | 112874061 | 112874687 | 627 | 3 | 4.14E-05 | 9.76E-06 | 0.004215 | 2.43E-05 | 0.011069236 | ATP11A |
| chr14 | 20295176 | 20295480 | 305 | 2 | 9.65E-05 | 2.39E-05 | 0.000197 | 2.43E-05 | -0.01121665 | TTC5 |
| chr21 | 27954825 | 27954850 | 26 | 2 | 0.000179558 | 1.54E-05 | 0.000945 | 2.43E-05 | 0.010206929 | NA |
| chr22 | 26756669 | 26757052 | 384 | 4 | 8.88E-05 | 3.56E-05 | 0.002804 | 2.43E-05 | 0.012849388 | MIATNB |
| chr1 | 27027037 | 27027628 | 592 | 4 | 0.000243762 | 1.08E-05 | 0.005732 | 2.43E-05 | 0.006293559 | NA |
| chr20 | 4720273 | 4721342 | 1070 | 5 | 0.000181569 | 1.51E-05 | 0.004332 | 2.43E-05 | 0.002043917 | NA |
| chr7 | 44184401 | 44184533 | 133 | 2 | 0.000254864 | 1.66E-05 | 0.000632 | 2.44E-05 | 0.001185042 | GCK |
| chr22 | 30881259 | 30882102 | 844 | 7 | 6.69E-05 | 1.69E-05 | 0.000917 | 2.44E-05 | 0.005288293 | OSBP2 |
| chr12 | 39332192 | 39332404 | 213 | 2 | 7.30E-05 | 2.73E-05 | 0.000155 | 2.44E-05 | 0.007134581 | KIF21A |
| chr2 | 197499805 | 197500947 | 1143 | 19 | 5.62E-06 | 0.028888 | 7.03E-05 | 2.44E-05 | 0.002734761 | HSPE1, HSPE1-MOB4, HSPD1 |
| chr15 | 92399347 | 92399578 | 232 | 2 | 1.04E-05 | 0.002431 | 2.22E-05 | 2.44E-05 | 0.009112418 | ST8SIA2 |
| chr14 | 56797921 | 56798550 | 630 | 3 | 1.59E-06 | 0.000331 | 3.09E-05 | 2.44E-05 | 0.005375819 | NA |
| chr3 | 66972378 | 66972885 | 508 | 4 | 0.00021029 | 8.76E-06 | 0.003489 | 2.44E-05 | 0.016472173 | NA |
| chr1 | 17766303 | 17766435 | 133 | 3 | 7.12E-05 | 0.000154 | 4.49E-05 | 2.44E-05 | 0.010377368 | ACTL8 |
| chr16 | 87391402 | 87392281 | 880 | 12 | 0.000134562 | 1.82E-05 | 0.010389 | 2.45E-05 | 0.002340768 | MAP1LC3B, FBXO31 |
| chr19 | 53912237 | 53912911 | 675 | 7 | 9.25E-05 | 0.000996 | 0.002248 | 2.45E-05 | 0.008995687 | CACNG7 |
| chr1 | 229944550 | 229944583 | 34 | 2 | 0.000158754 | 1.53E-05 | 0.001061 | 2.45E-05 | 0.004347928 | NA |
| chr16 | 57069860 | 57070225 | 366 | 2 | 0.000121498 | 1.52E-05 | 0.001129 | 2.45E-05 | 0.012694398 | NLRC5 |
| chr16 | 1460711 | 1460889 | 179 | 2 | 1.60E-05 | 1.58E-05 | 0.000866 | 2.45E-05 | -0.00062824 | CLCN7 |
| chr16 | 71461596 | 71462482 | 887 | 14 | 0.000152064 | 4.86E-05 | 0.007004 | 2.45E-05 | 0.000570175 | AC010547.1, ZNF23, AC010547.4 |
| chr3 | 129528080 | 129528709 | 630 | 4 | 5.81E-05 | 7.87E-05 | 0.000397 | 2.45E-05 | 0.000251198 | RHO |
| chr20 | 59514942 | 59515131 | 190 | 4 | 2.18E-05 | 0.000702 | 3.06E-05 | 2.45E-05 | 0.006898016 | NA |
| chr13 | 77027524 | 77028365 | 842 | 7 | 5.67E-05 | 3.36E-05 | 0.006315 | 2.45E-05 | 0.004123543 | AC001226.2, MYCBP2-AS1, AC001226.1 |
| chr12 | 84867427 | 84868028 | 602 | 2 | 4.46E-06 | 8.24E-05 | 5.69E-05 | 2.45E-05 | 0.018250614 | SLC6A15 |
| chr1 | 7561678 | 7561789 | 112 | 2 | 4.10E-05 | 2.86E-05 | 0.000147 | 2.45E-05 | 0.013227408 | CAMTA1 |
| chr9 | 19114204 | 19114270 | 67 | 2 | 6.36E-05 | 0.000436 | 3.05E-05 | 2.45E-05 | -0.01514651 | PLIN2 |
| chr11 | 43269124 | 43269660 | 537 | 5 | 0.000101213 | 2.37E-05 | 0.000361 | 2.45E-05 | -0.02243263 | HNRNPKP3 |
| chr2 | 127218775 | 127218824 | 50 | 2 | 0.0001312 | 1.53E-05 | 0.001095 | 2.45E-05 | 0.00771602 | CYP27C1 |
| chr4 | 76585949 | 76586955 | 1007 | 7 | 1.86E-05 | 0.000167 | 0.000228 | 2.46E-05 | 0.005059208 | SHROOM3 |
| chr10 | 43574527 | 43575267 | 741 | 8 | 2.57E-05 | 0.002344 | 0.001532 | 2.46E-05 | 0.001638806 | ZNF239 |
| chr12 | 105288869 | 105289232 | 364 | 2 | 0.000106535 | 1.54E-05 | 0.001031 | 2.46E-05 | 0.000220842 | C12orf75 |
| chr8 | 78665383 | 78665561 | 179 | 3 | 7.43E-05 | 9.70E-06 | 0.004608 | 2.46E-05 | 0.040643543 | AC068700.2 |
| chr10 | 123146905 | 123147414 | 510 | 7 | 4.24E-05 | 0.000199 | 0.002454 | 2.46E-05 | 0.007217166 | NA |
| chr16 | 67393703 | 67394594 | 892 | 10 | 2.03E-05 | 0.000287 | 0.003589 | 2.46E-05 | 0.005097188 | ZDHHC1 |
| chr19 | 38613936 | 38614403 | 468 | 2 | 6.87E-05 | 2.49E-05 | 0.000189 | 2.46E-05 | -0.00099962 | MAP4K1 |
| chr15 | 99015198 | 99015792 | 595 | 4 | 3.74E-05 | 2.31E-05 | 0.000564 | 2.46E-05 | 0.005646099 | LUNAR1, AC036108.1 |
| chr9 | 131485058 | 131485312 | 255 | 2 | 1.05E-05 | 5.80E-05 | 7.23E-05 | 2.46E-05 | 0.001730811 | PRRC2B |
| chr19 | 40923818 | 40924284 | 467 | 4 | 0.000105079 | 1.72E-05 | 0.001114 | 2.46E-05 | 0.010271976 | CYP2B7P |
| chr2 | 97995868 | 97996213 | 346 | 9 | 8.74E-05 | 3.35E-05 | 0.002697 | 2.46E-05 | -0.00094772 | TMEM131 |
| chr17 | 82623825 | 82624049 | 225 | 3 | 0.00017615 | 1.25E-05 | 0.001625 | 2.46E-05 | 0.009404415 | FOXK2, WDR45B |
| chr2 | 98454331 | 98454566 | 236 | 2 | 6.59E-05 | 1.54E-05 | 0.00105 | 2.46E-05 | 0.011444605 | INPP4A |
| chr6 | 42915846 | 42916024 | 179 | 4 | 0.000303657 | 6.03E-05 | 0.000159 | 2.46E-05 | 0.008266078 | PTCRA |
| chr6 | 82248219 | 82248607 | 389 | 5 | 2.04E-05 | 9.64E-05 | 0.000463 | 2.46E-05 | -0.01239283 | NA |
| chr2 | 199380434 | 199380460 | 27 | 2 | 0.000142464 | 2.21E-05 | 0.000248 | 2.46E-05 | 0.003545161 | SATB2 |
| chr2 | 186599341 | 186599344 | 4 | 2 | 7.28E-05 | 1.55E-05 | 0.001027 | 2.46E-05 | -0.00791766 | ITGAV |
| chr6 | 108666632 | 108667051 | 420 | 2 | 0.000101087 | 1.71E-05 | 0.00059 | 2.47E-05 | 0.003641314 | FOXO3 |
| chr9 | 136469905 | 136470059 | 155 | 3 | 2.31E-05 | 2.47E-05 | 0.000268 | 2.47E-05 | 0.001441343 | SEC16A |
| chr8 | 22069174 | 22069438 | 265 | 2 | 1.26E-05 | 0.001487 | 2.40E-05 | 2.47E-05 | 0.003381212 | DMTN |
| chr20 | 63737484 | 63738252 | 769 | 6 | 0.000121408 | 2.02E-05 | 0.00904 | 2.47E-05 | 0.005442041 | AL121845.3, LIME1 |
| chr11 | 65294003 | 65294156 | 154 | 3 | 7.07E-05 | 0.000106 | 5.54E-05 | 2.47E-05 | 0.004101306 | POLA2, AP000944.5 |
| chr2 | 158481547 | 158482170 | 624 | 2 | 0.000291785 | 1.54E-05 | 0.001084 | 2.47E-05 | 0.010527299 | PKP4 |
| chr1 | 2318178 | 2318971 | 794 | 4 | 6.75E-05 | 7.12E-06 | 0.008821 | 2.47E-05 | 0.00359542 | AL590822.3 |
| chr1 | 203089826 | 203090050 | 225 | 3 | 8.04E-05 | 0.000104 | 0.000138 | 2.47E-05 | 0.007187049 | NA |
| chr11 | 3156392 | 3157092 | 701 | 6 | 3.60E-05 | 5.92E-05 | 0.001521 | 2.48E-05 | 0.000937922 | OSBPL5 |
| chr2 | 228181242 | 228181874 | 633 | 9 | 7.64E-05 | 9.34E-05 | 0.000816 | 2.48E-05 | 0.006609125 | SPHKAP |
| chr1 | 47766247 | 47766758 | 512 | 2 | 5.35E-05 | 2.16E-05 | 0.000267 | 2.48E-05 | 0.009428311 | TRABD2B |
| chr9 | 131709919 | 131710205 | 287 | 5 | 0.000325772 | 1.01E-05 | 0.005026 | 2.48E-05 | 0.011952437 | RAPGEF1 |
| chr10 | 124947091 | 124947478 | 388 | 2 | 3.01E-05 | 3.18E-05 | 0.000131 | 2.48E-05 | 0.01129619 | ZRANB1 |
| chr3 | 36379739 | 36380248 | 510 | 4 | 1.56E-05 | 0.001172 | 0.000198 | 2.48E-05 | -0.00021846 | NA |
| chr1 | 66802172 | 66802632 | 461 | 3 | 3.60E-05 | 0.000295 | 0.00049 | 2.49E-05 | -0.00814981 | NA |
| chr16 | 381255 | 381628 | 374 | 3 | 6.93E-05 | 1.99E-05 | 0.001004 | 2.49E-05 | -0.00267427 | PGAP6 |
| chr16 | 68529 | 68538 | 10 | 2 | 3.85E-05 | 3.02E-05 | 0.000141 | 2.49E-05 | 0.008071964 | RHBDF1 |
| chr3 | 46866354 | 46866931 | 578 | 3 | 1.71E-05 | 2.36E-05 | 0.000296 | 2.49E-05 | 0.002208118 | MYL3 |
| chr3 | 48464927 | 48465514 | 588 | 7 | 2.93E-05 | 8.28E-05 | 0.001158 | 2.49E-05 | 0.001933656 | ATRIP |
| chr1 | 9311347 | 9311747 | 401 | 2 | 9.35E-05 | 1.56E-05 | 0.001078 | 2.49E-05 | 0.006117569 | SPSB1 |
| chr8 | 124372486 | 124373018 | 533 | 6 | 0.00010101 | 1.53E-05 | 0.005386 | 2.49E-05 | -0.00107056 | TMEM65 |
| chr1 | 243931567 | 243932113 | 547 | 5 | 5.80E-06 | 0.000113 | 0.000137 | 2.49E-05 | 0.007663432 | LINC02774 |
| chr3 | 146314032 | 146314799 | 768 | 3 | 5.07E-05 | 3.51E-05 | 0.000155 | 2.49E-05 | 0.012952222 | NA |
| chr19 | 15372920 | 15373318 | 399 | 4 | 7.15E-05 | 5.81E-05 | 0.001928 | 2.49E-05 | 0.002698359 | AKAP8 |
| chr6 | 87649806 | 87650202 | 397 | 2 | 0.000122939 | 1.80E-05 | 0.000503 | 2.49E-05 | 0.013386737 | ORC3 |
| chr2 | 203935763 | 203936167 | 405 | 3 | 0.000270883 | 2.04E-05 | 0.000714 | 2.50E-05 | 0.008937036 | NA |
| chr15 | 78002168 | 78003174 | 1007 | 3 | 3.38E-05 | 1.84E-05 | 0.000515 | 2.50E-05 | 0.005007255 | TBC1D2B |
| chr15 | 51737804 | 51738549 | 746 | 9 | 0.000100036 | 4.15E-05 | 0.003826 | 2.50E-05 | 0.001351901 | LYSMD2 |
| chr7 | 151745265 | 151745881 | 617 | 4 | 0.000129826 | 4.27E-05 | 0.000634 | 2.50E-05 | 0.007968953 | PRKAG2 |
| chr8 | 103403234 | 103403719 | 486 | 3 | 0.000139657 | 2.42E-05 | 0.000372 | 2.50E-05 | 0.009174009 | SLC25A32, AC012213.5 |
| chr10 | 86374268 | 86374696 | 429 | 3 | 3.07E-05 | 0.000293 | 8.95E-05 | 2.50E-05 | 0.011338217 | NA |
| chr7 | 73543768 | 73544107 | 340 | 3 | 0.000118114 | 2.54E-05 | 0.000278 | 2.50E-05 | 0.003979372 | BCL7B |
| chr7 | 101987185 | 101987668 | 484 | 7 | 0.00012184 | 2.75E-05 | 0.002111 | 2.50E-05 | 0.002685766 | CUX1 |
| chr20 | 62307591 | 62307745 | 155 | 3 | 5.88E-05 | 1.37E-05 | 0.001785 | 2.50E-05 | 0.003122215 | ADRM1 |
| chr11 | 64225886 | 64226443 | 558 | 11 | 9.80E-06 | 0.00406 | 0.00212 | 2.50E-05 | -0.00054515 | NUDT22, TRPT1 |
| chr8 | 113436430 | 113437597 | 1168 | 13 | 0.000101881 | 0.000229 | 0.01 | 2.50E-05 | 0.000461624 | CSMD3 |
| chr5 | 136992921 | 136993431 | 511 | 2 | 0.000112067 | 2.48E-05 | 0.000202 | 2.51E-05 | -0.00451067 | SPOCK1 |
| chr3 | 108806322 | 108806663 | 342 | 4 | 0.00016274 | 1.59E-05 | 0.000704 | 2.51E-05 | 0.015408805 | NA |
| chr1 | 151398800 | 151399496 | 697 | 6 | 4.08E-05 | 8.00E-05 | 0.003231 | 2.51E-05 | -0.00213698 | NA |
| chr12 | 132796020 | 132796966 | 947 | 5 | 6.32E-06 | 0.0012 | 2.98E-05 | 2.51E-05 | 0.001621567 | GOLGA3 |
| chr1 | 39560473 | 39560848 | 376 | 4 | 7.50E-05 | 3.11E-05 | 0.000227 | 2.51E-05 | 0.001377684 | PABPC4 |
| chr2 | 3785537 | 3785800 | 264 | 3 | 0.000103591 | 4.82E-05 | 0.000691 | 2.51E-05 | 0.006563133 | DCDC2C |
| chr6 | 131610236 | 131610802 | 567 | 4 | 0.00013782 | 2.79E-05 | 0.000565 | 2.51E-05 | 0.006150881 | MED23 |
| chr7 | 129774258 | 129775054 | 797 | 13 | 0.000215271 | 2.25E-05 | 0.012808 | 2.51E-05 | 0.002207875 | MIR96, MIR183 |
| chr9 | 127450271 | 127451030 | 760 | 7 | 3.31E-05 | 2.06E-05 | 0.011235 | 2.51E-05 | 0.003553623 | RPL12 |
| chr2 | 174599101 | 174599525 | 425 | 2 | 0.000139684 | 1.71E-05 | 0.000647 | 2.51E-05 | 0.01114299 | AC010894.2, WIPF1 |
| chr1 | 3014567 | 3014993 | 427 | 4 | 2.91E-05 | 6.84E-05 | 0.000208 | 2.52E-05 | 0.002907891 | NA |
| chr2 | 241893031 | 241893297 | 267 | 2 | 2.86E-05 | 1.78E-05 | 0.000544 | 2.52E-05 | 0.007658669 | LINC01237 |
| chr8 | 48977626 | 48978227 | 602 | 5 | 0.000187951 | 0.000102 | 0.00119 | 2.52E-05 | 0.002560276 | NA |
| chr16 | 938973 | 939298 | 326 | 2 | 7.18E-05 | 2.07E-05 | 0.000317 | 2.52E-05 | -0.00164997 | LMF1 |
| chr14 | 104159131 | 104159583 | 453 | 2 | 3.78E-06 | 4.73E-05 | 8.74E-05 | 2.52E-05 | 0.000968171 | KIF26A |
| chr12 | 53053998 | 53054645 | 648 | 9 | 3.44E-05 | 0.001144 | 0.002366 | 2.52E-05 | 0.002055132 | TNS2, AC068888.1 |

| | | | | | | | | | | |
|---|---|---|---|---|---|---|---|---|---|---|
| chr18 | 9960639 | 9961038 | 400 | 2 | 0.00011528 | 1.75E-05 | 0.000583 | 2.52E-05 | 0.018710294 | NA |
| chr7 | 5045621 | 5045832 | 212 | 5 | 2.66E-05 | 0.002753 | 2.47E-05 | 2.52E-05 | 0.00827878 | RBAK-RBAKDN, RBAK |
| chr16 | 23375370 | 23375548 | 179 | 2 | 2.66E-05 | 1.59E-05 | 0.001028 | 2.52E-05 | 0.012869595 | SCNN1B |
| chr19 | 38593392 | 38593668 | 277 | 2 | 2.05E-05 | 0.054007 | 2.00E-05 | 2.52E-05 | 0.009818173 | MAP4K1 |
| chr2 | 7671793 | 7672127 | 335 | 2 | 2.11E-05 | 0.000161 | 4.25E-05 | 2.52E-05 | 0.010811356 | AC092580.4, AC092580.2 |
| chr17 | 49016836 | 49016890 | 55 | 2 | 5.82E-05 | 1.60E-05 | 0.00099 | 2.52E-05 | -0.02276509 | IGF2BP1 |
| chr10 | 122699795 | 122700195 | 401 | 5 | 5.72E-05 | 0.000168 | 0.00185 | 2.52E-05 | 0.010616464 | C10orf120 |
| chr1 | 48530624 | 48530955 | 332 | 2 | 0.0001711 | 1.57E-05 | 0.001132 | 2.52E-05 | 0.01203223 | NA |
| chr11 | 134032155 | 134032615 | 461 | 5 | 0.000144083 | 1.13E-05 | 0.00374 | 2.52E-05 | 0.009941281 | LINC02731 |
| chr10 | 27743441 | 27744648 | 1208 | 9 | 1.08E-05 | 5.42E-05 | 0.001296 | 2.52E-05 | -0.00042864 | MKX |
| chr11 | 67442562 | 67443546 | 985 | 7 | 2.20E-05 | 0.000128 | 0.002759 | 2.52E-05 | -9.50E-05 | CORO1B |
| chr5 | 135330196 | 135330266 | 71 | 2 | 0.000101066 | 1.73E-05 | 0.00063 | 2.52E-05 | 0.012199674 | C5orf66 |
| chr14 | 93764434 | 93764880 | 447 | 2 | 0.00019992 | 1.57E-05 | 0.001156 | 2.52E-05 | 0.015206001 | PRIMA1 |
| chr10 | 117541272 | 117541527 | 256 | 4 | 7.53E-06 | 0.00103 | 0.000436 | 2.53E-05 | 0.005175219 | EMX2OS |
| chr11 | 36377466 | 36377923 | 458 | 4 | 1.01E-05 | 0.003379 | 3.07E-05 | 2.53E-05 | 0.000272358 | PRR5L |
| chr17 | 32943336 | 32943467 | 132 | 2 | 6.08E-05 | 0.000287 | 3.51E-05 | 2.53E-05 | 0.003181764 | TMEM98 |
| chr13 | 41354640 | 41355193 | 554 | 3 | 2.03E-05 | 9.80E-05 | 0.000381 | 2.53E-05 | 0.008265625 | NAA16, RNU6-57P |
| chr12 | 131827561 | 131828330 | 770 | 10 | 6.48E-05 | 0.000631 | 0.003298 | 2.53E-05 | 0.001939163 | NA |
| chr8 | 41661197 | 41661508 | 312 | 3 | 9.38E-05 | 5.32E-05 | 0.000145 | 2.53E-05 | 0.007866227 | AC113133.1, ANK1 |
| chr12 | 98593014 | 98593881 | 868 | 11 | 0.000119825 | 0.000275 | 0.004193 | 2.53E-05 | 0.001681989 | SLC25A3 |
| chr13 | 113657057 | 113657790 | 734 | 3 | 1.37E-05 | 0.005039 | 8.26E-05 | 2.53E-05 | 0.0086042 | ATP4B |
| chr19 | 50607928 | 50608131 | 204 | 2 | 0.000116806 | 2.70E-05 | 0.000176 | 2.53E-05 | 0.018000079 | NA |
| chr1 | 32716771 | 32717526 | 756 | 3 | 3.58E-06 | 0.002376 | 2.31E-05 | 2.53E-05 | 0.004182578 | NA |
| chr4 | 54099517 | 54100169 | 653 | 10 | 2.14E-05 | 0.005316 | 0.000206 | 2.53E-05 | -0.00095885 | AC058822.1, GSX2 |
| chr17 | 2058838 | 2059333 | 496 | 3 | 6.48E-05 | 1.37E-05 | 0.002164 | 2.53E-05 | 0.013209238 | HIC1 |
| chr2 | 9288535 | 9288597 | 63 | 2 | 7.48E-05 | 1.82E-05 | 0.000517 | 2.53E-05 | 0.010363251 | ASAP2 |
| chr12 | 125326142 | 125326618 | 477 | 5 | 6.00E-05 | 1.80E-05 | 0.001408 | 2.53E-05 | 0.00691095 | TMEM132B |
| chr2 | 238140748 | 238140909 | 162 | 4 | 0.000105794 | 4.70E-05 | 0.000798 | 2.53E-05 | 0.001975438 | KLHL30 |
| chr3 | 128077272 | 128077664 | 393 | 2 | 1.70E-05 | 2.01E-05 | 0.000355 | 2.54E-05 | 0.012254066 | RUVBL1-AS1, RUVBL1 |
| chr20 | 54475061 | 54475501 | 441 | 5 | 2.17E-05 | 5.68E-05 | 0.000776 | 2.54E-05 | 0.004479756 | NA |
| chr2 | 222292182 | 222292546 | 365 | 3 | 5.51E-05 | 1.49E-05 | 0.000966 | 2.54E-05 | 0.014345259 | PAX3 |
| chr6 | 7140698 | 7141013 | 316 | 3 | 8.09E-05 | 4.71E-05 | 0.000747 | 2.54E-05 | 0.005557079 | RREB1 |
| chr11 | 772787 | 773106 | 320 | 3 | 9.45E-05 | 8.13E-05 | 0.000736 | 2.54E-05 | 0.003643458 | GATD1 |
| chr17 | 39653396 | 39653982 | 587 | 5 | 0.000124079 | 6.67E-06 | 0.010532 | 2.54E-05 | 0.007226316 | STARD3 |
| chr9 | 133939315 | 133940076 | 762 | 6 | 2.64E-05 | 0.00027 | 0.000606 | 2.54E-05 | 0.006536856 | VAV2 |
| chr17 | 62625409 | 62625567 | 159 | 2 | 0.000104235 | 1.68E-05 | 0.00075 | 2.54E-05 | 0.015647286 | NA |
| chr13 | 113857595 | 113857886 | 292 | 6 | 7.97E-05 | 5.11E-05 | 0.001978 | 2.54E-05 | -0.00681137 | GAS6 |
| chr1 | 177970090 | 177970538 | 449 | 8 | 7.00E-05 | 1.81E-05 | 0.00778 | 2.54E-05 | 0.006950535 | SEC16B, CRYZL2P-SEC16B |
| chr10 | 113679498 | 113680387 | 890 | 9 | 3.92E-05 | 0.000503 | 0.006322 | 2.54E-05 | -6.16E-05 | CASP7 |
| chr15 | 72319784 | 72320129 | 346 | 7 | 4.45E-05 | 4.40E-05 | 0.005116 | 2.54E-05 | 0.002774465 | AC009690.2, AC009690.3, CELF6, AC009690.1 |
| chr12 | 51970915 | 51971438 | 524 | 4 | 2.35E-05 | 0.000313 | 0.000534 | 2.54E-05 | 0.007753386 | ACVR1B |
| chr20 | 643012 | 643215 | 204 | 2 | 0.000189925 | 2.34E-05 | 0.00024 | 2.54E-05 | 0.020247187 | NA |
| chr4 | 138308426 | 138308860 | 435 | 3 | 0.000121083 | 2.06E-05 | 0.000452 | 2.54E-05 | 0.003241571 | LINC00498 |
| chr20 | 23019505 | 23019808 | 304 | 3 | 0.000101736 | 3.95E-05 | 0.000214 | 2.55E-05 | 0.005709679 | AL049651.1 |
| chr10 | 121611539 | 121611642 | 104 | 2 | 0.000102143 | 1.65E-05 | 0.00086 | 2.55E-05 | 0.011818827 | NA |
| chr3 | 49020900 | 49021776 | 877 | 15 | 1.71E-05 | 0.000117 | 0.001834 | 2.55E-05 | 0.000224313 | NDUFAF3, DALRD3 |
| chr9 | 95086454 | 95086491 | 38 | 2 | 0.000157096 | 4.07E-05 | 0.000103 | 2.55E-05 | 0.003930127 | AOPEP |
| chr8 | 143023638 | 143023842 | 205 | 3 | 5.10E-05 | 1.74E-05 | 0.001581 | 2.55E-05 | 0.002749765 | LY6E |
| chr11 | 64072007 | 64072056 | 50 | 3 | 0.00017603 | 1.84E-05 | 0.000866 | 2.55E-05 | -0.00043318 | AP000721.2, MACROD1 |
| chr6 | 98833194 | 98833752 | 559 | 5 | 1.24E-05 | 0.000197 | 0.000154 | 2.55E-05 | 0.001302132 | NA |
| chr17 | 5486405 | 5486981 | 577 | 9 | 0.000109 | 4.63E-05 | 0.003585 | 2.55E-05 | 0.001920139 | MIS12, DERL2 |
| chr10 | 48945554 | 48945998 | 445 | 3 | 4.76E-05 | 9.96E-05 | 6.00E-05 | 2.55E-05 | -0.00286372 | WDFY4 |
| chr6 | 122609892 | 122610460 | 569 | 13 | 0.000150758 | 1.80E-05 | 0.015882 | 2.56E-05 | 0.001088437 | PKIB, AL512283.2 |
| chr18 | 63915110 | 63915313 | 204 | 2 | 4.53E-05 | 0.000362 | 3.34E-05 | 2.56E-05 | 0.005681212 | SERPINB10 |
| chr20 | 49483353 | 49484425 | 1073 | 4 | 9.92E-05 | 1.48E-05 | 0.00101 | 2.56E-05 | 0.001314488 | KCNB1 |
| chr16 | 841260 | 841400 | 141 | 4 | 0.000230649 | 2.62E-05 | 0.002238 | 2.56E-05 | 0.006154418 | AL023882.1 |
| chr14 | 104897498 | 104897775 | 278 | 2 | 2.26E-05 | 0.000899 | 2.71E-05 | 2.56E-05 | 0.003550603 | NA |
| chr10 | 49394944 | 49395566 | 623 | 2 | 4.07E-05 | 2.00E-05 | 0.000376 | 2.56E-05 | 0.013604405 | DRGX |
| chr1 | 178294392 | 178294635 | 244 | 2 | 5.01E-05 | 0.000195 | 4.05E-05 | 2.56E-05 | 0.00384393 | RASAL2 |
| chr10 | 48424740 | 48425072 | 333 | 3 | 3.61E-05 | 0.000137 | 4.98E-05 | 2.56E-05 | 0.00234718 | MAPK8 |
| chr16 | 88005343 | 88006228 | 886 | 6 | 5.25E-05 | 0.000622 | 0.000332 | 2.56E-05 | 0.00316525 | BANP |
| chr5 | 14508823 | 14508836 | 14 | 2 | 0.000179309 | 2.31E-05 | 0.000252 | 2.56E-05 | -0.00455372 | TRIO |
| chr1 | 65058401 | 65059202 | 802 | 6 | 8.77E-05 | 2.97E-05 | 0.003076 | 2.56E-05 | -0.0007722 | JAK1, MIR101-1 |
| chr6 | 30715784 | 30716629 | 846 | 6 | 1.67E-05 | 7.77E-05 | 0.000368 | 2.56E-05 | 0.001769664 | MDC1 |
| chr17 | 80160344 | 80160992 | 649 | 3 | 6.33E-06 | 0.012422 | 1.09E-05 | 2.56E-05 | 0.002416354 | NA |
| chr3 | 41238567 | 41239060 | 494 | 4 | 3.93E-05 | 0.006115 | 0.000395 | 2.57E-05 | 0.004255768 | CTNNB1 |
| chr17 | 37232530 | 37233371 | 842 | 3 | 0.000145314 | 7.36E-05 | 0.000249 | 2.57E-05 | 0.002085417 | ACACA |
| chr20 | 64205357 | 64205835 | 479 | 3 | 4.58E-05 | 0.000267 | 0.000146 | 2.57E-05 | 0.010738104 | MYT1 |
| chr1 | 68050418 | 68051030 | 613 | 10 | 1.94E-05 | 1.34E-05 | 0.005322 | 2.57E-05 | 0.006627632 | GNG12-AS1, DIRAS3 |
| chr20 | 48783640 | 48783963 | 324 | 2 | 0.000103997 | 1.62E-05 | 0.001067 | 2.57E-05 | -0.00310648 | PREX1 |
| chr6 | 42879229 | 42880172 | 944 | 10 | 4.63E-05 | 0.000252 | 0.000135 | 2.57E-05 | 0.000809803 | RPL7L1 |
| chr4 | 1301179 | 1301290 | 112 | 4 | 0.000275208 | 8.70E-06 | 0.006638 | 2.57E-05 | 0.011445385 | MAEA |
| chr8 | 41895830 | 41895922 | 93 | 2 | 2.03E-05 | 1.72E-05 | 0.000726 | 2.57E-05 | -0.01153436 | ANK1 |
| chr12 | 18810961 | 18811363 | 403 | 3 | 0.000390171 | 1.32E-05 | 0.001758 | 2.57E-05 | -0.01256726 | NA |
| chr4 | 76741266 | 76741835 | 570 | 3 | 3.38E-05 | 1.67E-05 | 0.000733 | 2.57E-05 | 0.000992164 | SHROOM3, SHROOM3-AS1 |
| chr21 | 33429305 | 33429584 | 280 | 2 | 2.65E-05 | 7.08E-05 | 6.64E-05 | 2.57E-05 | -0.00831681 | IFNGR2 |
| chr9 | 38391435 | 38391802 | 368 | 2 | 4.96E-05 | 1.82E-05 | 0.000563 | 2.57E-05 | 0.007256626 | NA |
| chr2 | 192402864 | 192402915 | 52 | 2 | 5.51E-05 | 0.001665 | 2.46E-05 | 2.57E-05 | -0.0064111 | NA |
| chr7 | 99002999 | 99003176 | 178 | 3 | 7.17E-05 | 9.93E-05 | 0.000297 | 2.57E-05 | 0.005749558 | TRRAP |
| chr19 | 46661885 | 46662138 | 254 | 3 | 3.38E-06 | 0.000428 | 5.76E-05 | 2.57E-05 | 0.00248229 | DACT3-AS1 |
| chr12 | 131835113 | 131835359 | 247 | 2 | 1.20E-05 | 0.001193 | 2.59E-05 | 2.57E-05 | 0.00721691 | MMP17 |
| chr14 | 31337559 | 31337637 | 79 | 2 | 3.77E-05 | 0.000135 | 4.66E-05 | 2.57E-05 | 0.000800203 | HEATR5A, AL139353.1 |
| chr12 | 40143623 | 40144035 | 413 | 2 | 8.05E-05 | 2.72E-05 | 0.00018 | 2.57E-05 | 0.015914063 | NA |
| chr9 | 112861830 | 112862322 | 493 | 2 | 0.000185462 | 1.65E-05 | 0.000967 | 2.58E-05 | 0.008326199 | SNX30 |
| chr1 | 3703654 | 3704074 | 421 | 2 | 2.44E-06 | 0.000169 | 4.27E-05 | 2.58E-05 | 0.004991732 | TP73 |
| chr2 | 105336923 | 105337873 | 951 | 15 | 2.17E-05 | 0.006052 | 8.85E-05 | 2.58E-05 | 0.001210259 | C2orf49, AC012360.3 |
| chr2 | 96869587 | 96870550 | 964 | 9 | 4.00E-05 | 6.99E-05 | 0.000835 | 2.58E-05 | 0.000585663 | SEMA4C |
| chr17 | 74429944 | 74430794 | 851 | 7 | 4.46E-05 | 3.96E-05 | 0.005761 | 2.58E-05 | 0.004892799 | GPRC5C |
| chr6 | 41164085 | 41164317 | 233 | 3 | 0.000279689 | 1.46E-05 | 0.002266 | 2.58E-05 | 0.004941163 | NA |
| chr22 | 44675460 | 44676004 | 545 | 3 | 0.000102913 | 0.000116 | 0.000366 | 2.58E-05 | 0.00584127 | PRR5 |
| chr19 | 53693595 | 53693813 | 219 | 2 | 0.000118739 | 1.61E-05 | 0.001175 | 2.59E-05 | 0.009558721 | NA |
| chr19 | 7617952 | 7618590 | 639 | 4 | 0.00018388 | 1.08E-05 | 0.005689 | 2.59E-05 | 0.005267721 | CAMSAP3 |
| chr18 | 69949082 | 69949294 | 213 | 2 | 4.13E-05 | 0.000128 | 4.80E-05 | 2.59E-05 | 0.011537346 | CD226 |
| chr5 | 138725235 | 138725659 | 425 | 2 | 0.000107455 | 2.09E-05 | 0.000345 | 2.59E-05 | -0.00215997 | CTNNA1 |
| chr1 | 181483691 | 181483915 | 225 | 3 | 0.000169857 | 1.12E-05 | 0.003165 | 2.59E-05 | 0.012449044 | CACNA1E |
| chr5 | 147234456 | 147235156 | 701 | 10 | 0.000111159 | 3.81E-05 | 0.011359 | 2.59E-05 | 0.001604525 | STK32A, STK32A-AS1 |
| chr4 | 87391912 | 87392514 | 603 | 3 | 5.73E-05 | 1.20E-05 | 0.003111 | 2.59E-05 | 0.004873339 | NA |
| chr11 | 4906390 | 4906685 | 296 | 2 | 3.99E-06 | 0.000846 | 2.79E-05 | 2.59E-05 | 0.00958629 | MMP26, OR51A7 |
| chr1 | 151035935 | 151036495 | 561 | 4 | 3.30E-05 | 7.68E-05 | 8.59E-05 | 2.59E-05 | 0.001185391 | BNIPL |
| chr2 | 29247110 | 29247183 | 74 | 2 | 8.63E-05 | 0.000151 | 4.50E-05 | 2.59E-05 | 0.001817846 | ALK |
| chr16 | 67616502 | 67616833 | 332 | 3 | 2.75E-05 | 0.001957 | 2.54E-05 | 2.59E-05 | -0.00242745 | CTCF |
| chr9 | 135639443 | 135639795 | 353 | 7 | 6.51E-05 | 4.08E-05 | 0.001221 | 2.59E-05 | -0.00226845 | GLT6D1 |
| chr6 | 30898112 | 30898248 | 137 | 5 | 7.27E-05 | 0.000269 | 0.001798 | 2.59E-05 | 0.001694534 | DDR1 |
| chr10 | 125009944 | 125010078 | 135 | 2 | 0.000166659 | 1.64E-05 | 0.001052 | 2.59E-05 | 0.005919982 | CTBP2 |
| chr12 | 124607825 | 124607970 | 146 | 3 | 6.89E-05 | 2.69E-05 | 0.000424 | 2.59E-05 | 0.003484105 | NA |
| chr1 | 160646814 | 160647334 | 521 | 5 | 2.37E-06 | 0.001189 | 0.000144 | 2.60E-05 | 0.004657212 | SLAMF1 |
| chr1 | 72773838 | 72774579 | 742 | 3 | 7.46E-05 | 2.15E-05 | 0.000915 | 2.60E-05 | 0.008883824 | LINC02796, AL583808.1 |
| chr3 | 109136286 | 109136581 | 296 | 4 | 4.77E-05 | 7.20E-05 | 0.001084 | 2.60E-05 | 0.006138487 | C3orf85 |
| chr17 | 41185736 | 41186254 | 519 | 4 | 0.000202391 | 1.23E-05 | 0.0045 | 2.60E-05 | 0.013706853 | NA |
| chr8 | 22864611 | 22864847 | 237 | 2 | 1.88E-05 | 4.08E-05 | 0.000106 | 2.60E-05 | 0.012065537 | PEBP4 |
| chr15 | 25204470 | 25204984 | 515 | 5 | 6.50E-05 | 0.001334 | 0.000594 | 2.60E-05 | 0.004900219 | SNHG14 |
| chr1 | 22836859 | 22837581 | 723 | 3 | 6.36E-05 | 1.75E-05 | 0.000678 | 2.60E-05 | 0.004171088 | EPHB2 |
| chr16 | 48537363 | 48537444 | 82 | 2 | 7.16E-05 | 1.62E-05 | 0.00115 | 2.60E-05 | 0.013282297 | NA |
| chr7 | 1989442 | 1990117 | 676 | 5 | 7.78E-05 | 0.000201 | 0.002074 | 2.60E-05 | -0.00053052 | MAD1L1, AC069288.1 |
| chr7 | 2084931 | 2085394 | 464 | 4 | 6.42E-05 | 1.11E-05 | 0.00254 | 2.60E-05 | 0.008511862 | MAD1L1, AC069288.1 |
| chr11 | 67582682 | 67583040 | 359 | 5 | 7.93E-05 | 2.10E-05 | 0.000815 | 2.60E-05 | 0.013716112 | NA |
| chr7 | 27152568 | 27153037 | 470 | 5 | 3.06E-05 | 9.31E-05 | 0.00183 | 2.60E-05 | 0.006105032 | HOXA-AS3, HOXA3, AC004080.5 |
| chr14 | 103518641 | 103518764 | 124 | 2 | 5.99E-05 | 2.06E-05 | 0.000372 | 2.60E-05 | 0.010921477 | NA |
| chr22 | 49996984 | 49997422 | 439 | 3 | 4.40E-05 | 1.22E-05 | 0.002763 | 2.60E-05 | 0.008053376 | IL17REL |
| chr12 | 52221931 | 52222313 | 383 | 3 | 5.30E-05 | 0.000134 | 0.000312 | 2.60E-05 | 0.014868436 | LINC00592 |
| chr9 | 87724069 | 87724135 | 67 | 2 | 4.17E-05 | 0.000365 | 3.40E-05 | 2.61E-05 | 0.00934504 | CTSL |
| chr16 | 70421000 | 70421531 | 532 | 2 | 3.34E-05 | 2.70E-05 | 0.00019 | 2.61E-05 | 0.006037257 | ST3GAL2 |
| chr14 | 96391230 | 96391553 | 324 | 4 | 0.000113188 | 9.20E-05 | 0.000417 | 2.61E-05 | -0.00081729 | NA |
| chr8 | 96853255 | 96853644 | 390 | 2 | 0.000150995 | 1.88E-05 | 0.000518 | 2.61E-05 | 0.011507652 | CPQ |
| chr8 | 51898814 | 51899751 | 938 | 15 | 0.000167104 | 0.000107 | 0.005232 | 2.61E-05 | 0.000818967 | AC064807.1, PCMTD1 |
| chr1 | 20413266 | 20413707 | 442 | 5 | 1.28E-05 | 0.010607 | 0.000125 | 2.61E-05 | 0.002001557 | LINC01141 |
| chr19 | 38724741 | 38724753 | 13 | 2 | 5.94E-05 | 1.64E-05 | 0.001115 | 2.61E-05 | 0.003794551 | ACTN4 |
| chr17 | 81390971 | 81391457 | 487 | 3 | 7.03E-05 | 2.07E-05 | 0.000481 | 2.61E-05 | 0.008178424 | NA |
| chr18 | 35497875 | 35498755 | 881 | 11 | 4.51E-05 | 0.000288 | 0.000111 | 2.61E-05 | 0.002885232 | AC007998.2, INO80C |
| chr21 | 44324950 | 44325222 | 273 | 2 | 0.000168358 | 1.67E-05 | 0.000976 | 2.61E-05 | 0.003910218 | PFKL |
| chr2 | 242011867 | 242012279 | 413 | 4 | 8.04E-05 | 1.76E-05 | 0.002107 | 2.62E-05 | 0.013550311 | LINC01237 |
| chr12 | 120495391 | 120495590 | 200 | 4 | 8.29E-05 | 7.28E-05 | 7.90E-05 | 2.62E-05 | 0.004299218 | DYNLL1, NRAV |
| chr20 | 63406150 | 63406758 | 609 | 5 | 4.16E-05 | 0.00131 | 0.000189 | 2.62E-05 | 0.002714226 | KCNQ2 |
| chr16 | 89697408 | 89697906 | 499 | 5 | 0.000215022 | 4.60E-05 | 0.001494 | 2.62E-05 | 0.001397983 | SPATA2L |
| chr3 | 62374742 | 62375774 | 1033 | 4 | 3.32E-05 | 0.000182 | 0.000342 | 2.62E-05 | 0.01035451 | NA |
| chr11 | 64294724 | 64294859 | 136 | 2 | 3.62E-05 | 8.64E-05 | 5.99E-05 | 2.62E-05 | 0.01322799 | KCNK4, KCNK4-TEX40 |
| chr5 | 114361790 | 114362594 | 805 | 10 | 7.17E-05 | 8.47E-06 | 0.009641 | 2.62E-05 | 0.002540323 | KCNN2 |
| chr12 | 3172354 | 3172492 | 139 | 3 | 0.000238946 | 1.49E-05 | 0.001794 | 2.63E-05 | 0.016088408 | TSPAN9, AC005912.2 |
| chr6 | 71288607 | 71288695 | 89 | 3 | 0.000126765 | 4.26E-05 | 0.00015 | 2.63E-05 | 0.00123623 | AL589935.1 |
| chr6 | 398505 | 398883 | 379 | 2 | 7.00E-05 | 2.39E-05 | 0.000252 | 2.63E-05 | 0.006488748 | IRF4 |
| chr9 | 122246942 | 122247120 | 179 | 2 | 0.00012502 | 2.72E-05 | 0.000192 | 2.63E-05 | -0.00029623 | RBM18 |
| chr15 | 89749252 | 89749499 | 248 | 2 | 5.28E-06 | 1.64E-05 | 0.001193 | 2.63E-05 | 0.00402974 | MESP1 |
| chr11 | 66852416 | 66853398 | 983 | 6 | 2.27E-05 | 0.00091 | 0.000487 | 2.64E-05 | 0.00337075 | PC |
| chr7 | 45111449 | 45111990 | 542 | 8 | 0.000251204 | 0.000107 | 0.000782 | 2.64E-05 | 0.001215506 | TBRG4 |
| chr22 | 21001571 | 21002226 | 656 | 12 | 0.000118722 | 8.38E-05 | 0.009276 | 2.64E-05 | -1.65E-05 | THAP7-AS1, THAP7 |
| chr12 | 85273575 | 85273838 | 264 | 3 | 8.80E-05 | 0.000135 | 8.61E-05 | 2.64E-05 | 0.006556332 | NA |
| chr17 | 17680565 | 17680987 | 423 | 2 | 0.000164001 | 1.66E-05 | 0.001102 | 2.64E-05 | 0.00675027 | NA |
| chr17 | 50559987 | 50560829 | 843 | 20 | 0.000114928 | 5.60E-05 | 0.007664 | 2.64E-05 | 7.53E-05 | CACNA1G, CACNA1G-AS1 |
| chr20 | 50233346 | 50233520 | 175 | 2 | 1.90E-05 | 1.65E-05 | 0.001146 | 2.64E-05 | 0.007755836 | NA |
| chr14 | 96378705 | 96378845 | 141 | 3 | 0.000101884 | 6.61E-05 | 0.000381 | 2.65E-05 | 0.001131399 | GSKIP |
| chr1 | 93680939 | 93681545 | 607 | 4 | 0.000115476 | 5.27E-05 | 0.000931 | 2.65E-05 | 0.00283934 | BCAR3 |
| chr16 | 70689551 | 70689698 | 148 | 2 | 0.000197036 | 1.74E-05 | 0.000834 | 2.65E-05 | 0.008307506 | VAC14 |
| chr9 | 91037488 | 91037516 | 29 | 2 | 0.000103591 | 1.99E-05 | 0.000443 | 2.65E-05 | 0.00827246 | NA |
| chr12 | 48893183 | 48893532 | 350 | 3 | 0.000153525 | 1.78E-05 | 0.000684 | 2.65E-05 | 0.004905923 | NA |
| chr9 | 99183159 | 99183343 | 185 | 3 | 5.62E-05 | 3.17E-05 | 0.000203 | 2.65E-05 | 0.007196402 | NA |
| chr10 | 11742191 | 11742474 | 284 | 5 | 7.88E-05 | 0.000292 | 0.000108 | 2.65E-05 | 0.003089723 | ECHDC3 |
| chr6 | 168370142 | 168370959 | 818 | 6 | 0.000301457 | 0.000719 | 0.002067 | 2.65E-05 | 0.003879268 | NA |
| chr2 | 61471272 | 61472030 | 759 | 5 | 0.0001147 | 8.63E-05 | 0.001899 | 2.65E-05 | -0.00068095 | AC016727.1 |
| chr2 | 165839832 | 165839994 | 163 | 2 | 9.80E-05 | 1.70E-05 | 0.000968 | 2.66E-05 | 0.006303583 | AC009495.1 |
| chr2 | 25279171 | 25279271 | 101 | 2 | 6.75E-05 | 0.000189 | 4.24E-05 | 2.66E-05 | 0.011087825 | DNMT3A |
| chr19 | 8147107 | 8147836 | 730 | 6 | 0.000134677 | 1.97E-05 | 0.005384 | 2.66E-05 | 0.007835316 | FBN3 |
| chr6 | 35569222 | 35569376 | 155 | 2 | 4.92E-05 | 0.001276 | 2.64E-05 | 2.66E-05 | 0.009521235 | NA |

| | | | | | | | | | | |
|---|---|---|---|---|---|---|---|---|---|---|
| chr1 | 58896308 | 58896880 | 573 | 6 | 0.000196848 | 1.30E-05 | 0.011456 | 2.66E-05 | 0.007356469 | LINC01135, LINC02777 |
| chr16 | 74997049 | 74997258 | 210 | 2 | 8.74E-05 | 2.14E-05 | 0.000359 | 2.66E-05 | 0.015246243 | WDR59 |
| chr1 | 46299992 | 46300475 | 484 | 2 | 9.36E-06 | 3.13E-05 | 0.000153 | 2.66E-05 | -0.00490164 | LRRC41 |
| chr20 | 17692663 | 17693092 | 430 | 3 | 6.00E-05 | 4.60E-05 | 0.000304 | 2.66E-05 | 0.004206955 | NA |
| chr21 | 43277879 | 43278382 | 504 | 3 | 8.39E-05 | 1.61E-05 | 0.002121 | 2.66E-05 | 0.015960286 | NA |
| chr21 | 21179759 | 21179835 | 77 | 2 | 9.72E-05 | 0.000108 | 5.39E-05 | 2.66E-05 | -0.00722456 | NCAM2 |
| chr14 | 23346589 | 23346900 | 312 | 5 | 0.000167001 | 1.20E-05 | 0.003136 | 2.67E-05 | 0.00466684 | SLC22A17 |
| chr8 | 6877440 | 6878106 | 667 | 4 | 9.31E-05 | 1.46E-05 | 0.001421 | 2.67E-05 | 0.012495838 | GS1-24F4.2, DEFB1 |
| chr1 | 202236619 | 202237294 | 676 | 4 | 7.51E-05 | 3.76E-05 | 0.001481 | 2.67E-05 | 0.003508133 | LGR6 |
| chr3 | 190512843 | 190513235 | 393 | 2 | 2.71E-05 | 1.66E-05 | 0.001203 | 2.67E-05 | 0.008454537 | NA |
| chr19 | 41414929 | 41415235 | 307 | 4 | 6.51E-05 | 5.40E-05 | 0.000147 | 2.67E-05 | 0.005721718 | AC011462.1, BCKDHA |
| chr16 | 4769303 | 4769631 | 329 | 2 | 3.56E-06 | 4.93E-05 | 9.23E-05 | 2.67E-05 | 0.010471048 | NA |
| chr2 | 239938409 | 239939124 | 716 | 4 | 0.00017869 | 8.28E-06 | 0.007768 | 2.67E-05 | 0.026798567 | NDUFA10 |
| chr7 | 157620002 | 157620365 | 364 | 4 | 4.58E-05 | 0.000408 | 9.17E-05 | 2.67E-05 | 0.003844064 | PTPRN2 |
| chr12 | 112892565 | 112892996 | 432 | 2 | 1.79E-05 | 0.000223 | 4.05E-05 | 2.68E-05 | 0.00972329 | RPH3A |
| chr6 | 36977317 | 36977657 | 341 | 4 | 5.43E-05 | 0.000102 | 0.000196 | 2.68E-05 | 0.001832388 | MTCH1 |
| chr3 | 25789604 | 25790458 | 855 | 15 | 8.80E-05 | 3.70E-05 | 0.023422 | 2.68E-05 | 1.62E-05 | OXSM, NGLY1 |
| chr16 | 58463491 | 58464681 | 1191 | 15 | 8.03E-05 | 5.95E-05 | 0.014831 | 2.68E-05 | 0.000404635 | NDRG4 |
| chr21 | 32270714 | 32270967 | 254 | 2 | 7.53E-05 | 2.80E-05 | 0.000191 | 2.68E-05 | 0.005120637 | MIS18A |
| chr19 | 6502480 | 6502752 | 273 | 7 | 0.000448746 | 2.52E-05 | 0.007873 | 2.68E-05 | -0.00283249 | TUBB4A |
| chr16 | 57123730 | 57124107 | 378 | 3 | 9.80E-05 | 2.57E-05 | 0.000355 | 2.69E-05 | 0.013268181 | CPNE2 |
| chr3 | 31831933 | 31832628 | 696 | 2 | 0.000135987 | 1.98E-05 | 0.000489 | 2.69E-05 | -0.00121597 | OSBPL10 |
| chr7 | 76302289 | 76302615 | 327 | 9 | 7.54E-05 | 2.45E-05 | 0.001483 | 2.69E-05 | 0.0021443 | NA |
| chr8 | 127083137 | 127083739 | 603 | 4 | 6.25E-05 | 0.000128 | 0.000134 | 2.69E-05 | -0.00113641 | PCAT1, PRNCR1, CASC19 |
| chr11 | 118498003 | 118498309 | 307 | 2 | 2.96E-05 | 0.000271 | 3.82E-05 | 2.69E-05 | 0.004009898 | KMT2A |
| chr10 | 97627824 | 97628021 | 198 | 2 | 0.000153127 | 1.99E-05 | 0.000478 | 2.69E-05 | 0.001552843 | AL355315.1, MORN4 |
| chr10 | 92833511 | 92833801 | 291 | 2 | 1.32E-05 | 0.000137 | 4.87E-05 | 2.69E-05 | 0.008457829 | EXOC6 |
| chr1 | 226701795 | 226702586 | 792 | 3 | 2.44E-06 | 0.001738 | 2.09E-05 | 2.69E-05 | 0.0061836 | ITPKB |
| chr12 | 46373506 | 46374233 | 728 | 6 | 6.52E-05 | 9.77E-06 | 0.010124 | 2.69E-05 | 0.005028653 | AC025031.2 |
| chr17 | 28861419 | 28862267 | 849 | 7 | 1.16E-05 | 0.001978 | 9.14E-05 | 2.70E-05 | 0.004785267 | MIR451B, AC024267.1, MIR451A, MIR144, MIR4732 |
| chr12 | 49033980 | 49034633 | 654 | 5 | 4.00E-05 | 2.42E-05 | 0.002526 | 2.70E-05 | 0.00161451 | KMT2D |
| chr21 | 25822626 | 25823021 | 396 | 2 | 0.000157015 | 1.72E-05 | 0.000999 | 2.70E-05 | 0.004725558 | NA |
| chr16 | 67470444 | 67470692 | 249 | 2 | 8.56E-05 | 5.50E-05 | 8.55E-05 | 2.70E-05 | 0.011973367 | ATP6V0D1 |
| chr6 | 33068687 | 33068770 | 84 | 4 | 6.34E-05 | 0.000103 | 0.00031 | 2.70E-05 | 0.003874479 | HLA-DPA1 |
| chr19 | 4544083 | 4544562 | 480 | 3 | 9.98E-05 | 1.37E-05 | 0.002118 | 2.70E-05 | 0.014893766 | SEMA6B |
| chr5 | 8766683 | 8766929 | 247 | 2 | 3.80E-05 | 0.00099 | 2.82E-05 | 2.70E-05 | 0.004691951 | NA |
| chr22 | 21567539 | 21568123 | 585 | 7 | 7.13E-05 | 4.93E-05 | 0.007818 | 2.70E-05 | 0.000737322 | UBE2L3 |
| chr1 | 8944996 | 8945854 | 859 | 8 | 6.58E-05 | 2.84E-05 | 0.004508 | 2.70E-05 | 0.002883278 | NA |
| chr6 | 3511118 | 3511636 | 519 | 2 | 1.46E-05 | 6.56E-05 | 7.51E-05 | 2.71E-05 | 0.009723116 | NA |
| chr10 | 89698199 | 89698272 | 74 | 2 | 5.18E-05 | 0.0036 | 2.35E-05 | 2.71E-05 | 0.009708232 | AL157400.3 |
| chr9 | 108552479 | 108552877 | 399 | 2 | 1.81E-05 | 3.51E-05 | 0.000137 | 2.71E-05 | -0.00485182 | NA |
| chr16 | 2025239 | 2025475 | 237 | 3 | 3.48E-05 | 0.000197 | 0.000204 | 2.71E-05 | 0.002437134 | SLC9A3R2 |
| chr9 | 76459150 | 76459644 | 495 | 5 | 0.000205561 | 5.89E-05 | 0.001121 | 2.71E-05 | -0.00196043 | GCNT1 |
| chr22 | 46674354 | 46674595 | 242 | 4 | 0.000120953 | 2.45E-05 | 0.001096 | 2.71E-05 | -0.0066688 | GRAMD4 |
| chr9 | 136567229 | 136567268 | 40 | 2 | 4.96E-05 | 2.16E-05 | 0.000372 | 2.71E-05 | 1.51E-05 | NA |
| chr13 | 113664276 | 113664761 | 486 | 3 | 0.000102449 | 0.000166 | 6.67E-05 | 2.71E-05 | 0.007104781 | NA |
| chr9 | 136094096 | 136094533 | 438 | 4 | 4.77E-05 | 7.94E-06 | 0.009047 | 2.71E-05 | 0.006660259 | NACC2 |
| chr5 | 129126534 | 129126841 | 308 | 2 | 9.06E-05 | 3.64E-05 | 0.000131 | 2.71E-05 | 0.007598225 | NA |
| chr19 | 38310699 | 38311013 | 315 | 4 | 0.000126555 | 4.93E-05 | 0.001283 | 2.71E-05 | 0.001372682 | YIF1B |
| chr17 | 18166531 | 18167092 | 562 | 2 | 0.000265753 | 1.70E-05 | 0.001156 | 2.71E-05 | 0.009347049 | MYO15A |
| chr1 | 113887193 | 113887658 | 466 | 5 | 8.11E-05 | 4.89E-05 | 0.001439 | 2.72E-05 | -0.00558433 | AP4B1-AS1, BCL2L15 |
| chr15 | 23564798 | 23565187 | 390 | 5 | 4.56E-05 | 0.000676 | 0.002056 | 2.72E-05 | 0.011463778 | NA |
| chr9 | 129761645 | 129761859 | 215 | 2 | 9.68E-05 | 1.69E-05 | 0.001192 | 2.72E-05 | 0.01222399 | NA |
| chr1 | 161379105 | 161379326 | 222 | 3 | 8.97E-05 | 0.000198 | 0.000752 | 2.72E-05 | -0.03545998 | RRM2P2 |
| chr10 | 11242585 | 11242639 | 55 | 2 | 0.000109095 | 2.58E-05 | 0.000238 | 2.72E-05 | 0.012819761 | CELF2 |
| chr14 | 100848018 | 100848439 | 422 | 2 | 9.32E-05 | 1.82E-05 | 0.000751 | 2.72E-05 | 0.009398198 | MEG3, AL117190.1, AL117190.2 |
| chr8 | 66566782 | 66566958 | 177 | 2 | 0.00010672 | 4.23E-05 | 0.00011 | 2.72E-05 | 0.010407751 | MYBL1 |
| chr17 | 50866227 | 50867301 | 1075 | 14 | 2.01E-05 | 0.000111 | 0.000679 | 2.73E-05 | -0.00048354 | TOB1-AS1, TOB1 |
| chr1 | 234918059 | 234918149 | 91 | 2 | 5.44E-05 | 4.09E-05 | 0.000115 | 2.73E-05 | 0.006750624 | AL391832.4 |
| chr13 | 51339588 | 51340065 | 478 | 4 | 1.39E-05 | 6.42E-05 | 0.001019 | 2.73E-05 | 0.00385255 | SERPINE3 |
| chr2 | 73202411 | 73202663 | 253 | 2 | 3.47E-05 | 1.91E-05 | 0.00061 | 2.73E-05 | 0.009936345 | NOTO |
| chr16 | 729218 | 729861 | 644 | 3 | 3.45E-05 | 2.68E-05 | 0.000294 | 2.73E-05 | 0.007473106 | HAGHL, CIAO3 |
| chr15 | 52112035 | 52112829 | 795 | 7 | 7.42E-05 | 1.08E-05 | 0.004519 | 2.73E-05 | -0.01140257 | BCL2L10 |
| chr8 | 26039685 | 26040018 | 334 | 3 | 4.18E-05 | 7.79E-05 | 0.000283 | 2.73E-05 | 0.00651965 | EBF2 |
| chr6 | 44301426 | 44301433 | 8 | 2 | 5.98E-05 | 2.36E-05 | 0.000295 | 2.73E-05 | 0.005572521 | TMEM151B, AL353588.1, AARS2 |
| chr1 | 245091895 | 245092074 | 180 | 2 | 4.19E-05 | 2.08E-05 | 0.000438 | 2.73E-05 | 0.010961393 | EFCAB2 |
| chr6 | 33078805 | 33079408 | 604 | 3 | 0.000149374 | 2.18E-05 | 0.001634 | 2.74E-05 | 0.008957367 | HLA-DPB1, HLA-DPA1 |
| chr10 | 69269888 | 69270123 | 236 | 2 | 0.000159806 | 1.71E-05 | 0.001185 | 2.74E-05 | 0.001939313 | HK1 |
| chr18 | 80191067 | 80191416 | 350 | 2 | 0.000176322 | 1.74E-05 | 0.001053 | 2.74E-05 | 0.009361045 | AC139100.1, PARD6G |
| chr22 | 19617849 | 19617930 | 82 | 2 | 8.31E-05 | 2.44E-05 | 0.000275 | 2.74E-05 | 0.001297131 | NA |
| chr15 | 89574943 | 89575625 | 683 | 9 | 6.82E-05 | 0.00015 | 0.001069 | 2.74E-05 | -0.00046709 | AC013391.2, TICRR |
| chr17 | 36936899 | 36937927 | 1029 | 11 | 2.79E-05 | 0.000153 | 0.009982 | 2.74E-05 | 0.000178236 | LHX1 |
| chr20 | 50168700 | 50169499 | 800 | 6 | 4.87E-05 | 8.87E-05 | 0.000222 | 2.74E-05 | 0.002069407 | AL161937.1 |
| chr8 | 124082577 | 124082646 | 70 | 2 | 0.000176693 | 1.83E-05 | 0.000763 | 2.74E-05 | 0.005694058 | FER1L6, FER1L6-AS2 |
| chr13 | 111167033 | 111167134 | 102 | 2 | 0.000114999 | 1.71E-05 | 0.001225 | 2.75E-05 | 0.019902209 | ARHGEF7 |
| chr6 | 165804749 | 165805379 | 631 | 4 | 0.000176205 | 8.59E-06 | 0.008121 | 2.75E-05 | 0.000333628 | PDE10A |
| chr4 | 7046332 | 7046743 | 412 | 6 | 0.000219528 | 1.30E-05 | 0.007702 | 2.75E-05 | 0.002001066 | TADA2B |
| chr22 | 41589638 | 41590341 | 704 | 12 | 1.40E-05 | 0.001288 | 0.001194 | 2.75E-05 | 0.000690489 | PMM1 |
| chr3 | 23202850 | 23203333 | 484 | 8 | 0.000113354 | 9.53E-05 | 0.002555 | 2.75E-05 | -0.00049533 | UBE2E2 |
| chr6 | 84226264 | 84226840 | 577 | 2 | 2.10E-05 | 2.21E-05 | 0.000374 | 2.75E-05 | 0.008400062 | CEP162 |
| chr2 | 129337182 | 129337486 | 305 | 2 | 4.79E-05 | 0.000165 | 4.64E-05 | 2.76E-05 | 0.002301221 | NA |
| chr1 | 12079149 | 12079772 | 624 | 3 | 7.10E-05 | 9.43E-05 | 0.000234 | 2.76E-05 | 0.00809493 | TNFRSF8 |
| chr12 | 120468359 | 120468855 | 497 | 3 | 5.43E-05 | 3.78E-05 | 0.000168 | 2.76E-05 | -0.00117824 | SRSF9 |
| chr1 | 32291508 | 32292233 | 726 | 10 | 4.16E-05 | 0.000795 | 0.005628 | 2.76E-05 | 0.0018173 | HDAC1 |
| chr6 | 30198184 | 30199129 | 946 | 22 | 4.53E-05 | 7.76E-05 | 0.00177 | 2.76E-05 | 0.000673581 | TRIM26 |
| chr12 | 130640606 | 130640655 | 50 | 2 | 3.51E-05 | 0.000123 | 5.30E-05 | 2.77E-05 | 0.000636341 | RIMBP2 |
| chr10 | 120979143 | 120979877 | 735 | 3 | 2.16E-05 | 7.93E-05 | 0.000872 | 2.77E-05 | -0.00642121 | AC010998.1 |
| chr13 | 96994087 | 96994643 | 557 | 9 | 0.000194954 | 1.03E-05 | 0.01844 | 2.77E-05 | -0.00496863 | OXGR1 |
| chr2 | 16202014 | 16202408 | 395 | 3 | 7.88E-05 | 1.12E-05 | 0.004693 | 2.77E-05 | 0.004221713 | NA |
| chr21 | 32392591 | 32392990 | 400 | 6 | 5.74E-05 | 2.09E-05 | 0.005649 | 2.77E-05 | -0.00205822 | URB1 |
| chr13 | 99316862 | 99317019 | 158 | 3 | 0.000114232 | 9.67E-05 | 0.000677 | 2.77E-05 | -0.00319279 | UBAC2 |
| chr6 | 132824227 | 132824497 | 271 | 2 | 1.75E-05 | 1.74E-05 | 0.00116 | 2.77E-05 | 0.014304993 | NA |
| chr12 | 49882729 | 49883201 | 473 | 2 | 0.000183747 | 1.80E-05 | 0.000922 | 2.77E-05 | 0.011033892 | FAIM2 |
| chr1 | 3623513 | 3624385 | 873 | 4 | 7.93E-06 | 1.60E-05 | 0.004773 | 2.77E-05 | 0.011433661 | AL513320.1 |
| chr7 | 148804740 | 148805022 | 283 | 2 | 0.000111746 | 1.98E-05 | 0.000566 | 2.77E-05 | -0.00406195 | NA |
| chr1 | 116763847 | 116764047 | 201 | 2 | 0.000178624 | 1.82E-05 | 0.000861 | 2.77E-05 | 0.01024846 | CD2 |
| chr10 | 72088857 | 72089409 | 553 | 8 | 0.000114245 | 2.89E-05 | 0.017356 | 2.78E-05 | 0.006159083 | SPOCK2 |
| chr8 | 33512494 | 33513230 | 737 | 11 | 0.000178862 | 1.96E-05 | 0.003579 | 2.78E-05 | -0.00034472 | TTI2 |
| chr4 | 146272750 | 146273136 | 387 | 2 | 8.12E-05 | 3.44E-05 | 0.000147 | 2.78E-05 | 0.003229441 | SLC10A7 |
| chr2 | 218290525 | 218291067 | 543 | 4 | 6.09E-05 | 4.86E-05 | 0.000473 | 2.78E-05 | 0.005867174 | PNKD, TMBIM1 |
| chr19 | 9185462 | 9186012 | 551 | 4 | 6.73E-05 | 2.18E-05 | 0.002556 | 2.78E-05 | 0.004187821 | OR7D2 |
| chr16 | 12078977 | 12079105 | 129 | 2 | 0.000162643 | 1.73E-05 | 0.001234 | 2.78E-05 | 0.019866391 | SNX29 |
| chr4 | 664902 | 665492 | 591 | 5 | 1.66E-05 | 0.003383 | 0.000114 | 2.78E-05 | 0.002013276 | PDE6B |
| chr16 | 10743417 | 10744303 | 887 | 9 | 4.59E-05 | 0.000303 | 0.000442 | 2.78E-05 | 0.001383345 | NUBP1 |
| chr15 | 69919829 | 69920083 | 255 | 2 | 9.77E-05 | 1.73E-05 | 0.001232 | 2.78E-05 | 0.006415432 | NA |
| chr11 | 11501944 | 11502175 | 232 | 3 | 0.000116996 | 4.06E-05 | 0.000222 | 2.78E-05 | 0.007099335 | GALNT18 |
| chr10 | 119723411 | 119723685 | 275 | 2 | 0.000130128 | 2.03E-05 | 0.00052 | 2.78E-05 | 0.000690993 | NA |
| chr17 | 81924753 | 81925000 | 248 | 4 | 0.000167322 | 1.32E-05 | 0.003798 | 2.78E-05 | 0.007397653 | MAFG |
| chr8 | 23163812 | 23164676 | 865 | 8 | 2.31E-05 | 8.18E-05 | 0.00046 | 2.78E-05 | 0.000771603 | TNFRSF10D |
| chr1 | 244590211 | 244590782 | 572 | 3 | 4.66E-05 | 1.14E-05 | 0.004568 | 2.78E-05 | -0.00529874 | CATSPERE |
| chr14 | 95363572 | 95363838 | 267 | 5 | 0.000231348 | 1.12E-05 | 0.004904 | 2.79E-05 | 0.010563809 | NA |
| chr12 | 45729770 | 45730125 | 356 | 7 | 9.86E-05 | 5.94E-05 | 0.003615 | 2.79E-05 | 0.004189344 | ARID2 |
| chr22 | 22341846 | 22342093 | 248 | 3 | 8.56E-05 | 8.01E-05 | 0.000179 | 2.79E-05 | 0.006497496 | NA |
| chr11 | 9138373 | 9138774 | 402 | 2 | 4.98E-05 | 1.92E-05 | 0.000676 | 2.79E-05 | 0.008763722 | NA |
| chr16 | 627526 | 627664 | 139 | 2 | 0.000147633 | 2.79E-05 | 0.000216 | 2.79E-05 | 0.008524861 | RAB40C |
| chr15 | 68229843 | 68230449 | 607 | 7 | 0.000204066 | 5.41E-06 | 0.016478 | 2.80E-05 | 0.004824727 | CLN6 |
| chr19 | 55408272 | 55408688 | 417 | 6 | 4.62E-05 | 8.32E-05 | 0.0009 | 2.80E-05 | 0.000893674 | NA |
| chr4 | 83455668 | 83456369 | 702 | 14 | 7.59E-05 | 0.000208 | 0.00199 | 2.80E-05 | 0.000121133 | MRPS18C, HELQ |
| chr17 | 50201417 | 50201904 | 488 | 8 | 0.00011315 | 7.80E-06 | 0.011433 | 2.80E-05 | 0.006164118 | AC015909.1, COL1A1 |
| chr6 | 30847967 | 30848670 | 704 | 5 | 0.000133539 | 2.63E-05 | 0.001788 | 2.80E-05 | 0.007970458 | LINC02570 |
| chr19 | 4302998 | 4304449 | 1452 | 7 | 0.00020901 | 4.35E-05 | 0.004458 | 2.80E-05 | 0.005942481 | NA |
| chr12 | 130777293 | 130777352 | 60 | 2 | 6.15E-05 | 4.04E-05 | 0.000123 | 2.80E-05 | 0.005162183 | NA |
| chr8 | 68095747 | 68095978 | 232 | 2 | 0.000155356 | 1.85E-05 | 0.000817 | 2.80E-05 | 0.006230996 | PREX2 |
| chr5 | 177112027 | 177112325 | 299 | 3 | 0.000199749 | 1.42E-05 | 0.001733 | 2.80E-05 | 0.010718893 | NA |
| chr20 | 64051746 | 64051756 | 11 | 2 | 4.30E-05 | 3.52E-05 | 0.000146 | 2.81E-05 | -0.00320099 | TCEA2 |
| chr12 | 481079 | 481648 | 570 | 3 | 0.000183775 | 1.28E-05 | 0.002757 | 2.81E-05 | 0.008480221 | B4GALNT3 |
| chr19 | 54115273 | 54115786 | 514 | 11 | 8.04E-05 | 0.00032 | 0.000266 | 2.81E-05 | 0.000415477 | PRPF31, TFPT |
| chr4 | 110639306 | 110639651 | 346 | 7 | 1.62E-05 | 0.000191 | 0.000224 | 2.81E-05 | 0.005371104 | PITX2 |
| chr10 | 101585086 | 101585590 | 505 | 3 | 0.000110063 | 0.000136 | 0.000201 | 2.81E-05 | -0.00157469 | DPCD, POLL |
| chr1 | 151044350 | 151044908 | 559 | 2 | 0.000149986 | 1.97E-05 | 0.000615 | 2.81E-05 | 0.005944149 | BNIPL |
| chr3 | 123143871 | 123144365 | 495 | 6 | 0.000155916 | 4.37E-05 | 0.001128 | 2.81E-05 | -0.00149348 | PDIA5 |
| chr16 | 4334966 | 4335434 | 469 | 8 | 0.000292332 | 1.39E-05 | 0.009607 | 2.82E-05 | 0.002653288 | GLIS2, PAM16 |
| chr6 | 423875 | 424126 | 252 | 3 | 0.000103878 | 7.68E-05 | 0.000244 | 2.82E-05 | 0.002327788 | NA |
| chr5 | 124394559 | 124394925 | 367 | 2 | 2.45E-05 | 2.02E-05 | 0.000562 | 2.82E-05 | 0.012575995 | LINC01170 |
| chr19 | 8412824 | 8413285 | 462 | 7 | 0.000212306 | 1.76E-05 | 0.011658 | 2.82E-05 | 0.005245422 | MARCHF2 |
| chr1 | 186447444 | 186447533 | 90 | 2 | 0.000104154 | 6.79E-05 | 7.73E-05 | 2.83E-05 | 0.017860392 | PDC-AS1, PDC |
| chr11 | 121466675 | 121466799 | 125 | 2 | 0.000119258 | 5.42E-05 | 9.23E-05 | 2.83E-05 | 0.012312236 | SORL1 |
| chr5 | 16701085 | 16701466 | 382 | 5 | 0.000128416 | 1.41E-05 | 0.001552 | 2.83E-05 | 0.000491171 | MYO10 |
| chr11 | 132332083 | 132332695 | 613 | 3 | 2.09E-05 | 0.000272 | 4.28E-05 | 2.83E-05 | 0.00080542 | NTM |
| chr7 | 156893294 | 156893484 | 191 | 6 | 0.000290466 | 2.07E-05 | 0.007664 | 2.83E-05 | -0.00062781 | NA |
| chr19 | 415722 | 416188 | 467 | 2 | 0.000172362 | 1.88E-05 | 0.000809 | 2.83E-05 | 0.012692999 | NA |
| chr14 | 47626291 | 47626838 | 548 | 3 | 0.000207413 | 1.25E-05 | 0.003656 | 2.83E-05 | -0.0022083 | MDGA2 |
| chr5 | 123701003 | 123701405 | 403 | 2 | 6.88E-06 | 0.00063 | 3.21E-05 | 2.83E-05 | -0.00888206 | NA |
| chr6 | 108678222 | 108678383 | 162 | 2 | 0.000164456 | 2.19E-05 | 0.000431 | 2.84E-05 | 0.001124322 | FOXO3 |
| chr12 | 120583870 | 120583983 | 114 | 2 | 0.000182028 | 8.72E-05 | 6.58E-05 | 2.84E-05 | 0.015443201 | NA |
| chr18 | 24425803 | 24426771 | 969 | 13 | 1.66E-05 | 0.000366 | 0.000239 | 2.84E-05 | 0.002592225 | IMPACT |
| chr5 | 10595548 | 10595947 | 400 | 2 | 4.16E-05 | 2.92E-05 | 0.000207 | 2.84E-05 | 0.011181523 | ANKRD33B |
| chr2 | 217850170 | 217850362 | 193 | 3 | 0.000118471 | 5.19E-05 | 0.000809 | 2.84E-05 | 0.001300305 | TNS1 |
| chr17 | 57261095 | 57261808 | 714 | 2 | 0.000202383 | 1.96E-05 | 0.000669 | 2.84E-05 | 0.000281219 | MSI2 |
| chr2 | 70889281 | 70889635 | 355 | 4 | 7.37E-05 | 3.19E-05 | 0.002437 | 2.84E-05 | 0.007171748 | LINC01143 |
| chr22 | 24939546 | 24940319 | 774 | 6 | 4.49E-05 | 0.000146 | 0.000111 | 2.84E-05 | 0.001525265 | TMEM211 |
| chr11 | 23730114 | 23730676 | 563 | 6 | 9.05E-05 | 7.33E-05 | 0.00046 | 2.85E-05 | -0.00429098 | LINC02726 |
| chr14 | 72893883 | 72894614 | 732 | 9 | 2.57E-05 | 0.000519 | 0.005257 | 2.85E-05 | 0.003363369 | DPF3 |
| chr17 | 8120320 | 8121228 | 909 | 7 | 2.01E-05 | 0.002918 | 7.96E-05 | 2.85E-05 | 0.001351902 | HES7 |
| chr17 | 21043731 | 21044337 | 607 | 8 | 0.000149692 | 4.29E-05 | 0.005512 | 2.85E-05 | 0.004378474 | USP22 |
| chr16 | 1999383 | 1999658 | 276 | 3 | 7.66E-05 | 0.000361 | 0.00021 | 2.85E-05 | 0.006606807 | ZNF598 |
| chr17 | 1723215 | 1723281 | 67 | 2 | 2.72E-05 | 0.000113 | 5.70E-05 | 2.85E-05 | 0.005915978 | WDR81 |
| chr17 | 40552147 | 40552386 | 240 | 4 | 6.32E-05 | 1.62E-05 | 0.001398 | 2.85E-05 | 0.001321742 | NA |
| chr7 | 64562665 | 64563446 | 782 | 10 | 9.66E-05 | 4.16E-05 | 0.004705 | 2.85E-05 | 0.002955776 | ZNF680 |
| chr16 | 346393 | 346560 | 168 | 3 | 0.000194812 | 4.48E-05 | 0.000144 | 2.85E-05 | 0.001413952 | AXIN1 |
| chr2 | 33728128 | 33728136 | 9 | 2 | 0.000465661 | 0.000109 | 5.81E-05 | 2.85E-05 | 0.004533808 | LINC01320, MYADML |
| chr22 | 19290726 | 19291149 | 424 | 2 | 3.08E-06 | 0.007177 | 2.34E-05 | 2.86E-05 | 0.005059524 | CLTCL1 |

| | | | | | | | | | | |
|---|---|---|---|---|---|---|---|---|---|---|
| chr5 | 55853051 | 55853146 | 96 | 2 | 0.00012357 | 2.79E-05 | 0.00023 | 2.86E-05 | -0.01973213 | IL31RA |
| chr12 | 132887359 | 132888306 | 948 | 26 | 6.30E-05 | 8.63E-05 | 0.013559 | 2.86E-05 | 0.000358737 | AC127070.1, CHFR |
| chr11 | 72697141 | 72697467 | 327 | 4 | 0.000104852 | 3.41E-05 | 0.00235 | 2.86E-05 | 0.002895119 | ARAP1 |
| chr3 | 129086556 | 129086669 | 114 | 2 | 0.000171262 | 1.85E-05 | 0.000928 | 2.86E-05 | 0.006640483 | NA |
| chr11 | 69894327 | 69894456 | 130 | 2 | 0.000110519 | 7.40E-05 | 7.41E-05 | 2.86E-05 | 0.001235766 | NA |
| chr8 | 142326292 | 142326686 | 395 | 2 | 0.000118791 | 1.90E-05 | 0.000825 | 2.86E-05 | 0.026263543 | TSNARE1 |
| chr22 | 37329180 | 37329212 | 33 | 3 | 0.000210914 | 1.15E-05 | 0.004998 | 2.86E-05 | -0.01754716 | NA |
| chr13 | 102879809 | 102880085 | 277 | 8 | 1.29E-05 | 0.002034 | 1.99E-05 | 2.86E-05 | 0.000156289 | NA |
| chr6 | 158039694 | 158040377 | 684 | 3 | 1.65E-05 | 0.00073 | 2.52E-05 | 2.86E-05 | 0.005488044 | SYNJ2 |
| chr1 | 109909994 | 109910618 | 625 | 8 | 7.40E-05 | 0.00014 | 0.002412 | 2.87E-05 | 0.000923695 | CSF1 |
| chr2 | 55498647 | 55499053 | 407 | 2 | 3.34E-05 | 5.02E-05 | 0.000101 | 2.87E-05 | 0.008591588 | NA |
| chr6 | 29429219 | 29429525 | 307 | 3 | 4.54E-05 | 2.26E-05 | 0.000657 | 2.87E-05 | 0.00373739 | OR5V1, OR11A1 |
| chr12 | 119676798 | 119677205 | 408 | 2 | 8.27E-05 | 1.92E-05 | 0.000793 | 2.87E-05 | 0.007002895 | PRKAB1 |
| chr17 | 1004403 | 1004646 | 244 | 4 | 6.11E-05 | 7.04E-05 | 0.00039 | 2.87E-05 | 0.005385407 | ABR |
| chr3 | 160038861 | 160039142 | 282 | 3 | 0.000134452 | 4.39E-05 | 0.000928 | 2.87E-05 | 0.005116041 | IL12A-AS1 |
| chr5 | 2224897 | 2225368 | 472 | 5 | 6.10E-05 | 8.51E-05 | 0.000465 | 2.87E-05 | 0.001814072 | NA |
| chr9 | 112717780 | 112718350 | 571 | 8 | 0.000250077 | 1.43E-05 | 0.004192 | 2.88E-05 | 0.0027079 | INIP |
| chr16 | 17633315 | 17633464 | 150 | 2 | 0.000167731 | 2.26E-05 | 0.000412 | 2.88E-05 | 0.01810912 | NA |
| chr16 | 89267569 | 89267612 | 44 | 2 | 0.000124575 | 2.72E-05 | 0.00025 | 2.88E-05 | 0.006635986 | NA |
| chr3 | 184572806 | 184573006 | 201 | 4 | 6.28E-05 | 0.001257 | 3.03E-05 | 2.88E-05 | 0.00304385 | EPHB3 |
| chr8 | 10511142 | 10511167 | 26 | 2 | 6.35E-05 | 2.32E-05 | 0.000378 | 2.88E-05 | 0.015922016 | PRSS51 |
| chr2 | 146413052 | 146413239 | 188 | 2 | 0.00017212 | 2.04E-05 | 0.00061 | 2.88E-05 | -0.01056436 | NA |
| chr2 | 70948409 | 70949157 | 749 | 6 | 6.28E-05 | 8.71E-05 | 0.000291 | 2.88E-05 | 0.009462353 | VAX2, ATP6V1B1, AC007040.2, ATP6V1B1-AS1 |
| chr18 | 69957610 | 69958187 | 578 | 5 | 3.32E-05 | 0.000104 | 0.000138 | 2.88E-05 | -0.00017735 | CD226 |
| chr2 | 217335796 | 217335932 | 137 | 2 | 7.91E-05 | 0.000271 | 4.12E-05 | 2.88E-05 | -0.00689021 | DIRC3-AS1, DIRC3 |
| chr6 | 127342055 | 127342499 | 445 | 4 | 2.93E-05 | 0.000133 | 0.00093 | 2.88E-05 | -0.01006997 | ECHDC1 |
| chr12 | 95942343 | 95943054 | 712 | 6 | 9.89E-06 | 0.000421 | 0.000624 | 2.89E-05 | 0.003140588 | CCDC38 |
| chr15 | 101121240 | 101121629 | 390 | 6 | 0.000122546 | 4.23E-05 | 0.001252 | 2.89E-05 | 0.006549326 | NA |
| chr2 | 200305968 | 200306681 | 714 | 13 | 1.78E-05 | 0.000161 | 5.76E-05 | 2.89E-05 | 0.002668864 | SPATS2L |
| chr4 | 184165509 | 184165847 | 339 | 2 | 3.29E-05 | 0.000165 | 4.86E-05 | 2.89E-05 | 0.007135935 | ENPP6 |
| chr14 | 93100381 | 93100784 | 404 | 2 | 0.00016566 | 1.80E-05 | 0.001258 | 2.89E-05 | 0.006911273 | ITPK1 |
| chr6 | 134725115 | 134725338 | 224 | 2 | 0.000132687 | 1.83E-05 | 0.001102 | 2.89E-05 | 0.012750294 | NA |
| chr3 | 88149832 | 88150685 | 854 | 9 | 4.35E-05 | 8.72E-05 | 0.000104 | 2.89E-05 | -0.00041285 | C3orf38, CGGBP1 |
| chr13 | 48533557 | 48534491 | 935 | 4 | 8.62E-07 | 0.001723 | 3.66E-05 | 2.89E-05 | 0.009429106 | RB1, AL157813.2 |
| chr16 | 88059738 | 88060400 | 663 | 5 | 8.52E-05 | 0.000103 | 0.00022 | 2.89E-05 | -0.00174195 | BANP |
| chr20 | 63065411 | 63066057 | 647 | 4 | 0.000120933 | 2.12E-05 | 0.003312 | 2.89E-05 | 0.007950192 | LINC01749 |
| chr5 | 142913973 | 142914281 | 309 | 3 | 0.000127232 | 6.36E-05 | 0.000155 | 2.89E-05 | 0.00450492 | ARHGAP26 |
| chr15 | 44735885 | 44736072 | 188 | 4 | 0.00012739 | 9.88E-06 | 0.006127 | 2.89E-05 | 0.012947187 | TRIM69 |
| chr18 | 79513440 | 79513814 | 375 | 3 | 0.000133454 | 1.49E-05 | 0.001687 | 2.89E-05 | 0.005152627 | NFATC1 |
| chr4 | 2067956 | 2068023 | 68 | 2 | 4.68E-05 | 2.37E-05 | 0.00036 | 2.89E-05 | 0.00563546 | NAT8L |
| chr14 | 54902494 | 54903513 | 1020 | 8 | 1.65E-05 | 0.000437 | 0.000149 | 2.89E-05 | 0.000477795 | GCH1 |
| chr8 | 140544092 | 140544281 | 190 | 3 | 0.000124525 | 2.40E-05 | 0.000999 | 2.89E-05 | 0.009269377 | AGO2 |
| chr8 | 38996335 | 38996910 | 576 | 13 | 0.000241607 | 4.85E-05 | 0.010682 | 2.89E-05 | 0.000719969 | ADAM9, TM2D2 |
| chr17 | 78923058 | 78923786 | 729 | 2 | 6.87E-06 | 2.07E-05 | 0.000585 | 2.89E-05 | 0.003986194 | TIMP2 |
| chr10 | 89207050 | 89207943 | 894 | 9 | 1.51E-05 | 5.46E-05 | 0.002506 | 2.89E-05 | 0.000994695 | CH25H |
| chr20 | 3001260 | 3001478 | 219 | 2 | 0.000176572 | 1.94E-05 | 0.000784 | 2.90E-05 | -0.00873658 | PTPRA |
| chr7 | 74092970 | 74093130 | 161 | 5 | 0.000297683 | 9.38E-06 | 0.007181 | 2.90E-05 | 0.007207333 | LIMK1 |
| chr11 | 63918305 | 63918433 | 129 | 2 | 3.73E-05 | 2.65E-05 | 0.000272 | 2.90E-05 | 0.01324288 | NA |
| chr1 | 77292999 | 77293533 | 535 | 3 | 6.47E-05 | 2.47E-05 | 0.000474 | 2.90E-05 | 0.014375233 | AK5 |
| chr13 | 113053157 | 113053265 | 109 | 3 | 0.000132492 | 0.004103 | 7.54E-05 | 2.90E-05 | 0.008351454 | MCF2L |
| chr14 | 100332991 | 100332994 | 4 | 2 | 0.000185047 | 2.36E-05 | 0.000369 | 2.90E-05 | 0.010255999 | NA |
| chr4 | 57195693 | 57195825 | 133 | 2 | 6.98E-05 | 1.84E-05 | 0.001093 | 2.90E-05 | -0.04220927 | IGFBP7-AS1 |
| chr15 | 74302364 | 74302933 | 570 | 4 | 0.000153465 | 0.000226 | 0.00026 | 2.90E-05 | 0.008450737 | CCDC33, AC023300.3 |
| chr4 | 25925921 | 25926238 | 318 | 2 | 0.000176637 | 1.90E-05 | 0.000908 | 2.90E-05 | 0.020118004 | SMIM20 |
| chr5 | 157390585 | 157390602 | 18 | 2 | 6.81E-05 | 0.001469 | 2.82E-05 | 2.90E-05 | 0.006750812 | CYFIP2, AC008676.3 |
| chr18 | 37273726 | 37273932 | 207 | 2 | 4.75E-05 | 1.84E-05 | 0.001089 | 2.90E-05 | -0.02048746 | AC090386.1, CELF4 |
| chr2 | 238231391 | 238231728 | 338 | 8 | 0.000339793 | 6.41E-06 | 0.020904 | 2.90E-05 | 0.009462555 | AC012485.1, LINC02610 |
| chr22 | 46737874 | 46739274 | 1401 | 7 | 6.50E-06 | 0.000551 | 0.000858 | 2.91E-05 | 0.003855152 | CERK |
| chr17 | 4476497 | 4476712 | 216 | 2 | 4.49E-05 | 1.98E-05 | 0.000723 | 2.91E-05 | 0.017846427 | SPNS3 |
| chr14 | 22485879 | 22486178 | 300 | 3 | 4.55E-05 | 0.000102 | 8.54E-05 | 2.91E-05 | 0.009779731 | NA |
| chr5 | 39202905 | 39203653 | 749 | 4 | 0.000121649 | 0.0002 | 0.000569 | 2.91E-05 | 0.001827384 | FYB1 |
| chr16 | 1135985 | 1136286 | 302 | 6 | 0.000151294 | 2.99E-05 | 0.001312 | 2.91E-05 | -0.00279036 | NA |
| chr12 | 113421622 | 113422450 | 829 | 6 | 4.24E-05 | 0.000224 | 0.00125 | 2.91E-05 | 0.001331638 | SDSL, SDS |
| chr5 | 3197616 | 3198061 | 446 | 3 | 8.38E-05 | 0.000125 | 0.000355 | 2.91E-05 | 0.010467741 | NA |
| chr3 | 125211492 | 125211978 | 487 | 4 | 0.00013115 | 5.01E-05 | 0.001321 | 2.91E-05 | 0.007471969 | SLC12A8 |
| chr11 | 62791867 | 62792241 | 375 | 8 | 0.000259667 | 9.53E-05 | 0.004641 | 2.91E-05 | 0.000190888 | TMEM223, NXF1 |
| chr2 | 172427549 | 172428250 | 702 | 6 | 3.32E-05 | 0.000368 | 0.001643 | 2.91E-05 | -0.00134678 | ITGA6, AC078883.2 |
| chr13 | 87129491 | 87129852 | 362 | 2 | 0.000131622 | 2.36E-05 | 0.000377 | 2.91E-05 | -0.0129648 | NA |
| chr7 | 116731206 | 116731455 | 250 | 2 | 0.000245667 | 2.69E-05 | 0.000266 | 2.91E-05 | 0.001458886 | MET |
| chr10 | 48608207 | 48608639 | 433 | 2 | 9.17E-05 | 2.98E-05 | 0.000212 | 2.92E-05 | 0.011406385 | ARHGAP22 |
| chr8 | 4991425 | 4992000 | 576 | 3 | 7.34E-05 | 1.58E-05 | 0.002639 | 2.92E-05 | 0.003131532 | CSMD1 |
| chr17 | 77141050 | 77141791 | 742 | 5 | 6.02E-05 | 2.25E-05 | 0.003073 | 2.92E-05 | 0.003748905 | SEC14L1 |
| chr14 | 31015262 | 31015290 | 29 | 2 | 0.000195463 | 1.83E-05 | 0.001205 | 2.92E-05 | 0.003405642 | STRN3 |
| chr3 | 42778782 | 42779058 | 277 | 2 | 0.00012357 | 3.03E-05 | 0.000207 | 2.92E-05 | 0.004911312 | AC006059.2 |
| chr16 | 73483165 | 73483594 | 430 | 4 | 7.71E-06 | 0.000282 | 7.27E-05 | 2.92E-05 | 0.005926335 | LINC01568, ZFHX3 |
| chr20 | 31988659 | 31988710 | 52 | 2 | 0.000131095 | 6.59E-05 | 8.29E-05 | 2.92E-05 | 0.006767613 | XKR7 |
| chr9 | 121990638 | 121990908 | 271 | 2 | 3.42E-05 | 0.001626 | 2.80E-05 | 2.92E-05 | 0.002876442 | TTLL11 |
| chr11 | 107591447 | 107592328 | 882 | 5 | 7.13E-05 | 1.25E-05 | 0.008356 | 2.92E-05 | 0.003715373 | ELMOD1 |
| chr14 | 60658732 | 60659202 | 471 | 2 | 4.66E-07 | 0.000317 | 3.97E-05 | 2.92E-05 | 0.011263509 | NA |
| chr13 | 105139539 | 105139995 | 457 | 3 | 0.000207474 | 1.82E-05 | 0.001045 | 2.92E-05 | 0.005457781 | NA |
| chr2 | 238408324 | 238409187 | 864 | 2 | 0.000166407 | 1.87E-05 | 0.001037 | 2.93E-05 | 0.004458294 | NA |
| chr7 | 24285357 | 24285752 | 396 | 4 | 3.87E-05 | 0.000363 | 4.18E-05 | 2.93E-05 | 0.008489891 | NPY |
| chr4 | 186930179 | 186930570 | 392 | 5 | 0.000195582 | 4.52E-05 | 0.001968 | 2.93E-05 | 0.000556868 | NA |
| chr13 | 23324225 | 23324725 | 501 | 4 | 0.000127271 | 1.36E-05 | 0.00204 | 2.94E-05 | 0.012504017 | SGCG |
| chr8 | 30442500 | 30442790 | 291 | 4 | 0.000139957 | 4.43E-05 | 0.000773 | 2.94E-05 | -0.00022018 | RBPMS |
| chr21 | 45430535 | 45431198 | 664 | 6 | 0.000182516 | 2.60E-05 | 0.006867 | 2.94E-05 | -0.01466393 | COL18A1 |
| chr7 | 154599093 | 154599219 | 127 | 2 | 0.000184103 | 2.05E-05 | 0.000659 | 2.94E-05 | -0.00334961 | DPP6 |
| chr17 | 17352113 | 17352457 | 345 | 4 | 0.000100844 | 6.35E-05 | 0.000167 | 2.94E-05 | 0.007000818 | NA |
| chr16 | 67516608 | 67516670 | 63 | 2 | 0.000138556 | 2.61E-05 | 0.000293 | 2.94E-05 | 0.014951599 | NA |
| chr6 | 164105801 | 164106404 | 604 | 5 | 3.60E-05 | 0.000622 | 8.56E-05 | 2.94E-05 | 0.001424055 | NA |
| chr10 | 104056002 | 104056148 | 147 | 3 | 0.000139151 | 1.50E-05 | 0.002272 | 2.94E-05 | 0.002460654 | COL17A1 |
| chr13 | 52128709 | 52129704 | 996 | 10 | 3.25E-05 | 0.000229 | 0.00196 | 2.94E-05 | 0.002068914 | AL139082.1, NEK5 |
| chr1 | 228103785 | 228104361 | 577 | 6 | 9.49E-05 | 4.81E-05 | 0.003968 | 2.95E-05 | -0.01584271 | C1orf35 |
| chr9 | 108562243 | 108562393 | 151 | 2 | 5.71E-05 | 0.001384 | 2.88E-05 | 2.95E-05 | 0.004612478 | NA |
| chr17 | 41467824 | 41467836 | 13 | 2 | 0.000345336 | 3.56E-05 | 0.000159 | 2.95E-05 | 0.015815309 | NA |
| chr2 | 69960608 | 69960787 | 180 | 2 | 8.08E-06 | 0.000112 | 5.97E-05 | 2.95E-05 | 0.002908324 | ASPRV1 |
| chr1 | 167434090 | 167434373 | 284 | 2 | 0.000164636 | 1.93E-05 | 0.000908 | 2.95E-05 | 0.002725947 | CD247 |
| chr1 | 36464148 | 36464781 | 634 | 9 | 0.000166644 | 8.13E-05 | 0.00365 | 2.95E-05 | 0.001658545 | MRPS15 |
| chr1 | 155196719 | 155197652 | 934 | 5 | 1.80E-06 | 0.000801 | 0.000163 | 2.95E-05 | 0.003110819 | THBS3-AS1, THBS3 |
| chr19 | 48354732 | 48354893 | 162 | 2 | 4.64E-05 | 1.84E-05 | 0.001277 | 2.95E-05 | 0.003338631 | TMEM143 |
| chr1 | 19377868 | 19378509 | 642 | 3 | 6.54E-05 | 3.72E-05 | 0.000276 | 2.95E-05 | -0.0037028 | CAPZB |
| chr5 | 55976285 | 55976963 | 679 | 2 | 9.01E-05 | 2.45E-05 | 0.000354 | 2.95E-05 | 0.006346469 | IL6ST |
| chr3 | 71070036 | 71070264 | 229 | 2 | 0.000122869 | 3.05E-05 | 0.000211 | 2.95E-05 | 0.005880624 | FOXP1, AC097634.4 |
| chr14 | 35122027 | 35123162 | 1136 | 19 | 0.000123769 | 0.000572 | 0.004114 | 2.95E-05 | 0.000463659 | PRORP, AL121594.1, PPP2R3C |
| chr14 | 102085724 | 102086446 | 723 | 3 | 0.000135537 | 0.000296 | 0.000676 | 2.95E-05 | -0.0079139 | HSP90AA1 |
| chr12 | 40554338 | 40554957 | 620 | 2 | 0.000203297 | 1.84E-05 | 0.001261 | 2.95E-05 | 0.01269321 | MUC19 |
| chr11 | 64091348 | 64091408 | 61 | 2 | 0.000189672 | 1.84E-05 | 0.001253 | 2.95E-05 | 0.012597155 | AP000721.2, MACROD1 |
| chr7 | 106178519 | 106179128 | 610 | 3 | 3.20E-05 | 0.000436 | 4.22E-05 | 2.96E-05 | 0.004855996 | NA |
| chr12 | 15221369 | 15222363 | 995 | 11 | 3.98E-05 | 0.000456 | 0.000299 | 2.96E-05 | 0.001399242 | RERG |
| chr6 | 44192365 | 44192485 | 121 | 2 | 9.46E-05 | 0.000149 | 5.25E-05 | 2.96E-05 | 0.029685399 | NA |
| chr4 | 105057495 | 105057863 | 369 | 2 | 4.40E-05 | 0.000115 | 5.90E-05 | 2.96E-05 | 0.014658239 | AC096577.1 |
| chr2 | 65436498 | 65436705 | 208 | 4 | 7.58E-05 | 0.000192 | 6.92E-05 | 2.96E-05 | 9.45E-05 | NA |
| chr14 | 91257680 | 91258105 | 426 | 3 | 0.000334226 | 1.37E-05 | 0.002618 | 2.96E-05 | 0.009892694 | NA |
| chr14 | 37902081 | 37902179 | 99 | 2 | 0.000347512 | 2.61E-05 | 0.000304 | 2.96E-05 | 0.011729035 | TTC6, LINC00517 |
| chr8 | 41828806 | 41828949 | 144 | 3 | 0.000302551 | 6.62E-05 | 0.000127 | 2.96E-05 | -0.00238575 | ANK1, AC027702.1 |
| chr2 | 23422911 | 23423305 | 395 | 2 | 3.15E-05 | 0.000252 | 4.30E-05 | 2.96E-05 | 0.009435688 | KLHL29 |
| chr1 | 244863071 | 244863833 | 763 | 7 | 2.19E-05 | 9.01E-05 | 7.80E-05 | 2.96E-05 | 0.000752429 | HNRNPU |
| chr5 | 3608029 | 3608637 | 609 | 3 | 2.74E-05 | 0.000335 | 0.000178 | 2.97E-05 | -0.00412751 | NA |
| chr1 | 147699832 | 147699993 | 162 | 2 | 0.000182288 | 2.08E-05 | 0.000649 | 2.97E-05 | 0.015408649 | NA |
| chr9 | 128920978 | 128922317 | 1340 | 9 | 3.36E-05 | 2.99E-05 | 0.020273 | 2.97E-05 | 0.001283954 | PHYHD1 |
| chr7 | 116526116 | 116526770 | 655 | 12 | 0.000155202 | 6.47E-05 | 0.008672 | 2.97E-05 | 0.00054127 | CAV1 |
| chr1 | 197413641 | 197413830 | 190 | 2 | 9.80E-05 | 7.16E-05 | 7.96E-05 | 2.97E-05 | -0.00257146 | CRB1 |
| chr8 | 143475036 | 143475349 | 314 | 3 | 8.94E-05 | 0.000112 | 0.000787 | 2.97E-05 | 0.001843482 | ZC3H3 |
| chr6 | 168519256 | 168519651 | 396 | 5 | 5.06E-05 | 8.83E-05 | 0.000499 | 2.97E-05 | 0.001094119 | SMOC2 |
| chr17 | 1482265 | 1482528 | 264 | 3 | 3.19E-05 | 3.10E-05 | 0.000318 | 2.98E-05 | -0.00108661 | MYO1C |
| chr6 | 11709666 | 11709695 | 30 | 2 | 3.35E-05 | 2.24E-05 | 0.000493 | 2.98E-05 | 0.004646681 | NA |
| chr7 | 40196640 | 40196689 | 50 | 2 | 0.000124977 | 8.62E-05 | 7.05E-05 | 2.98E-05 | 0.004735937 | SUGCT |
| chr8 | 142528148 | 142528556 | 409 | 4 | 0.000266934 | 4.32E-05 | 0.000531 | 2.98E-05 | -0.00327091 | ADGRB1 |
| chr2 | 174328087 | 174329026 | 940 | 6 | 0.000108525 | 0.000186 | 0.006958 | 2.98E-05 | 0.004776323 | LINC01305 |
| chr10 | 132164800 | 132164946 | 147 | 2 | 0.000174922 | 1.90E-05 | 0.001099 | 2.98E-05 | 0.026633443 | JAKMIP3 |
| chr8 | 133375765 | 133376372 | 608 | 4 | 1.48E-05 | 0.00046 | 3.01E-05 | 2.98E-05 | 0.008669274 | NA |
| chr5 | 88683188 | 88683687 | 500 | 3 | 0.000242269 | 1.47E-05 | 0.002062 | 2.98E-05 | 0.008121349 | MEF2C-AS2, LINC00461 |
| chr15 | 75625822 | 75626466 | 645 | 14 | 0.000137055 | 0.000341 | 0.005195 | 2.99E-05 | 0.001583038 | SNUPN |
| chr22 | 35646974 | 35647023 | 50 | 2 | 7.72E-05 | 2.56E-05 | 0.00033 | 2.99E-05 | -0.00067297 | NA |
| chr2 | 231791487 | 231791737 | 251 | 2 | 6.55E-05 | 8.03E-05 | 7.45E-05 | 2.99E-05 | 0.000556549 | COPS7B |
| chr22 | 42090495 | 42091355 | 861 | 12 | 0.000125648 | 0.00067 | 0.000696 | 3.00E-05 | 6.46E-05 | NDUFA6-DT, NDUFA6 |
| chr11 | 47767025 | 47767641 | 617 | 9 | 6.87E-05 | 2.35E-05 | 0.017135 | 3.00E-05 | -0.00018599 | FNBP4 |
| chr22 | 49119805 | 49120162 | 358 | 3 | 3.00E-05 | 0.000122 | 0.000662 | 3.00E-05 | 0.007987701 | NA |
| chr3 | 183386055 | 183386486 | 432 | 2 | 0.00018089 | 1.99E-05 | 0.00085 | 3.00E-05 | 0.010117088 | MCF2L2 |
| chr14 | 64823010 | 64823409 | 400 | 9 | 5.53E-05 | 0.000969 | 0.004272 | 3.00E-05 | 0.001737542 | SPTB |
| chr17 | 16653245 | 16654101 | 857 | 11 | 8.63E-05 | 9.49E-05 | 0.003812 | 3.00E-05 | 0.000643051 | AC098850.2, ZNF624 |
| chr12 | 27003397 | 27003805 | 409 | 2 | 0.00010701 | 2.89E-05 | 0.000247 | 3.00E-05 | 0.008353339 | TM7SF3 |
| chr8 | 123653883 | 123654389 | 507 | 2 | 3.41E-05 | 2.78E-05 | 0.00027 | 3.00E-05 | 0.00545948 | AC135166.1 |
| chr5 | 151224705 | 151225583 | 879 | 4 | 0.000109494 | 0.000203 | 0.000468 | 3.00E-05 | 0.00347073 | GM2A |
| chr12 | 124574294 | 124574338 | 45 | 2 | 0.000191466 | 2.19E-05 | 0.000557 | 3.00E-05 | 0.012768294 | NA |
| chr8 | 20542793 | 20543117 | 325 | 2 | 0.000167999 | 2.08E-05 | 0.000685 | 3.00E-05 | 0.009775108 | NA |
| chr2 | 241053074 | 241053295 | 222 | 3 | 5.01E-05 | 0.000728 | 2.85E-05 | 3.00E-05 | 0.004500633 | SNED1, SNED1-AS1 |
| chr11 | 1031856 | 1032259 | 404 | 4 | 0.00015494 | 6.49E-05 | 0.001358 | 3.01E-05 | -0.00264309 | MUC6 |
| chr1 | 156591542 | 156592155 | 614 | 9 | 3.08E-05 | 0.000155 | 0.002024 | 3.01E-05 | 0.00097387 | NAXE |
| chr10 | 11563833 | 11564113 | 281 | 2 | 6.20E-05 | 9.16E-05 | 6.87E-05 | 3.01E-05 | 0.008523708 | USP6NL |
| chr14 | 37099285 | 37099746 | 462 | 3 | 6.23E-05 | 9.97E-05 | 7.44E-05 | 3.01E-05 | -0.00552356 | SLC25A21 |
| chr14 | 29390100 | 29390311 | 212 | 4 | 0.000140647 | 3.44E-05 | 0.001209 | 3.01E-05 | 0.020515193 | AL158058.1, AL133166.1 |
| chr7 | 2094921 | 2095526 | 606 | 2 | 2.44E-05 | 2.16E-05 | 0.000594 | 3.01E-05 | 0.022005396 | MAD1L1, AC069288.1 |
| chr19 | 12752481 | 12753347 | 867 | 5 | 0.000141217 | 0.000144 | 0.001602 | 3.01E-05 | 0.007379242 | BEST2 |
| chr17 | 19724422 | 19724774 | 353 | 3 | 6.66E-05 | 1.38E-05 | 0.002841 | 3.01E-05 | -0.01866115 | NA |
| chr15 | 40933164 | 40933539 | 376 | 2 | 1.18E-05 | 7.57E-05 | 7.81E-05 | 3.01E-05 | 0.006228643 | DLL4 |
| chr2 | 120440404 | 120440464 | 61 | 2 | 1.28E-05 | 2.08E-05 | 0.000711 | 3.02E-05 | 0.004686319 | NA |
| chr17 | 57893805 | 57893833 | 29 | 2 | 7.73E-05 | 0.000568 | 3.51E-05 | 3.02E-05 | 0.009148442 | CUEDC1 |
| chr9 | 106862898 | 106863338 | 441 | 6 | 0.000164063 | 7.32E-05 | 0.005316 | 3.03E-05 | 0.001776051 | ZNF462 |
| chr15 | 66329018 | 66329101 | 84 | 3 | 0.000425481 | 0.000276 | 0.000282 | 3.03E-05 | 0.000133703 | DIS3L, AC055855.1 |
| chr10 | 12169479 | 12169797 | 319 | 3 | 0.000108895 | 0.000129 | 0.00011 | 3.03E-05 | -0.00159069 | SEC61A2, NUDT5 |
| chr6 | 33781666 | 33782302 | 637 | 3 | 0.000111303 | 1.85E-05 | 0.001548 | 3.03E-05 | 0.002628295 | LEMD2 |

| | | | | | | | | | | |
|---|---|---|---|---|---|---|---|---|---|---|
| chr12 | 52231702 | 52232113 | 412 | 2 | 0.000187874 | 5.47E-05 | 0.000101 | 3.03E-05 | -0.00071098 | NA |
| chr8 | 38151958 | 38152030 | 73 | 2 | 6.09E-05 | 7.11E-05 | 8.23E-05 | 3.03E-05 | 0.012622147 | AC084024.3 |
| chr7 | 25856626 | 25857230 | 605 | 5 | 7.66E-05 | 1.24E-05 | 0.007655 | 3.03E-05 | 0.00038718 | NA |
| chr22 | 21686915 | 21686948 | 34 | 2 | 0.000197398 | 2.09E-05 | 0.000711 | 3.03E-05 | 0.00412879 | PPIL2 |
| chr1 | 201222807 | 201223158 | 352 | 3 | 4.67E-05 | 0.000519 | 0.000318 | 3.04E-05 | 0.007088093 | IGFN1, AC103925.1 |
| chr7 | 986124 | 986727 | 604 | 7 | 8.89E-05 | 3.93E-05 | 0.007946 | 3.04E-05 | 0.009509728 | CYP2W1 |
| chr9 | 132601745 | 132602092 | 348 | 2 | 5.70E-05 | 0.000101 | 6.52E-05 | 3.04E-05 | -0.00582229 | DDX31 |
| chr14 | 79325090 | 79325506 | 417 | 2 | 0.000115476 | 2.10E-05 | 0.000699 | 3.04E-05 | 0.014202075 | NRXN3 |
| chr11 | 36510364 | 36510875 | 512 | 13 | 5.60E-06 | 0.008089 | 2.32E-05 | 3.04E-05 | 0.002113604 | AC061999.1, RAG1 |
| chr2 | 60880951 | 60882020 | 1070 | 13 | 6.58E-06 | 0.003898 | 3.89E-05 | 3.04E-05 | 1.43E-05 | REL, LINC01185 |
| chr20 | 23656482 | 23656832 | 351 | 3 | 2.83E-05 | 0.000266 | 5.10E-05 | 3.04E-05 | 0.006310563 | AL121894.3 |
| chr15 | 79091043 | 79091638 | 596 | 15 | 9.40E-06 | 0.005887 | 0.000822 | 3.05E-05 | 0.001209637 | NA |
| chr9 | 129989310 | 129989342 | 33 | 2 | 7.19E-05 | 0.000799 | 3.29E-05 | 3.05E-05 | 0.006964148 | FNBP1 |
| chr19 | 2976791 | 2977619 | 829 | 11 | 7.21E-06 | 0.030801 | 0.00028 | 3.05E-05 | -0.00180898 | TLE6 |
| chr1 | 2247345 | 2247919 | 575 | 5 | 7.92E-06 | 0.000433 | 0.000333 | 3.05E-05 | 0.00238647 | SKI |
| chr16 | 89587427 | 89587677 | 251 | 2 | 6.78E-05 | 1.90E-05 | 0.001304 | 3.06E-05 | 0.017212705 | CPNE7 |
| chr12 | 111312344 | 111312937 | 594 | 2 | 8.08E-05 | 2.01E-05 | 0.000907 | 3.06E-05 | 0.000566259 | CUX2 |
| chr1 | 212857680 | 212858766 | 1087 | 13 | 4.66E-05 | 9.57E-05 | 0.004456 | 3.06E-05 | 0.000288231 | FLVCR1, FLVCR1-DT |
| chr15 | 32718965 | 32719240 | 276 | 7 | 4.04E-05 | 0.000556 | 2.69E-05 | 3.06E-05 | -0.00117538 | GREM1, AC090877.2 |
| chr11 | 125682192 | 125682291 | 100 | 2 | 4.67E-05 | 0.002531 | 2.74E-05 | 3.06E-05 | -0.00631249 | NA |
| chr7 | 82506907 | 82506961 | 55 | 2 | 0.000136535 | 2.24E-05 | 0.000551 | 3.06E-05 | 0.00972807 | NA |
| chr9 | 133734083 | 133734157 | 75 | 3 | 9.94E-05 | 1.83E-05 | 0.001685 | 3.06E-05 | 0.004333371 | SARDH |
| chr4 | 2904964 | 2905143 | 180 | 3 | 0.000123061 | 0.000121 | 0.000114 | 3.06E-05 | 0.001917677 | ADD1 |
| chr19 | 17027241 | 17028048 | 808 | 5 | 0.000286528 | 1.15E-05 | 0.005459 | 3.07E-05 | 0.006438301 | NA |
| chr17 | 68942599 | 68942690 | 92 | 3 | 8.39E-05 | 1.44E-05 | 0.002826 | 3.07E-05 | 0.012343837 | ABCA8 |
| chr7 | 19144804 | 19145784 | 981 | 11 | 5.12E-05 | 5.05E-05 | 0.008917 | 3.07E-05 | 0.003626563 | AC003986.1, FERD3L |
| chr5 | 181188984 | 181190100 | 1117 | 6 | 1.49E-05 | 0.043778 | 5.63E-05 | 3.07E-05 | 0.001153325 | LINC01962 |
| chr6 | 26103859 | 26104332 | 474 | 8 | 0.000236644 | 0.001218 | 0.000853 | 3.07E-05 | 0.001628095 | H4C3 |
| chr12 | 1908483 | 1908580 | 98 | 2 | 0.000296186 | 3.85E-05 | 0.000154 | 3.07E-05 | -0.01368036 | CACNA2D4 |
| chr17 | 8087387 | 8087851 | 465 | 4 | 4.20E-05 | 3.96E-05 | 0.00053 | 3.07E-05 | 0.007120868 | ALOX12B |
| chr5 | 176450836 | 176451399 | 564 | 3 | 0.000156977 | 1.79E-05 | 0.001277 | 3.07E-05 | 0.006519681 | FAF2 |
| chr20 | 35771310 | 35771646 | 337 | 5 | 2.74E-05 | 8.84E-05 | 0.00112 | 3.08E-05 | 0.00324917 | NA |
| chr11 | 62622298 | 62622564 | 267 | 5 | 0.000105893 | 3.89E-05 | 0.002438 | 3.08E-05 | 0.003938587 | NA |
| chr22 | 43152690 | 43153010 | 321 | 3 | 5.39E-05 | 3.23E-05 | 0.000279 | 3.08E-05 | 0.015755582 | TSPO |
| chr7 | 127593580 | 127593975 | 396 | 6 | 6.87E-05 | 3.21E-05 | 0.000885 | 3.08E-05 | 0.004346049 | FSCN3, GCC1 |
| chr12 | 124545604 | 124546198 | 595 | 3 | 0.000365815 | 1.41E-05 | 0.003562 | 3.08E-05 | 0.0099789 | NCOR2 |
| chr17 | 51041520 | 51041906 | 387 | 2 | 2.68E-05 | 3.64E-05 | 0.000169 | 3.08E-05 | 0.010667614 | SPAG9 |
| chr21 | 41740201 | 41741152 | 952 | 5 | 2.82E-05 | 4.28E-05 | 0.003108 | 3.08E-05 | 0.004063289 | AP001615.1, RIPK4 |
| chr9 | 114633754 | 114633825 | 72 | 2 | 0.000112397 | 0.000146 | 5.54E-05 | 3.08E-05 | 0.002477681 | TMEM268 |
| chr9 | 34899593 | 34899916 | 324 | 2 | 0.000100197 | 4.22E-05 | 0.000138 | 3.08E-05 | 0.014960786 | NA |
| chr10 | 3208582 | 3208957 | 376 | 2 | 0.000179035 | 2.07E-05 | 0.000811 | 3.08E-05 | 0.021133942 | AL451164.3 |
| chr15 | 59615113 | 59615228 | 116 | 2 | 0.000116838 | 2.79E-05 | 0.000293 | 3.09E-05 | 0.01360684 | GCNT3 |
| chr13 | 45419277 | 45419614 | 338 | 2 | 1.83E-05 | 2.13E-05 | 0.000716 | 3.09E-05 | -0.01224024 | SLC25A30-AS1 |
| chr1 | 226708655 | 226709416 | 762 | 4 | 0.000118654 | 1.05E-05 | 0.006946 | 3.09E-05 | 0.006923675 | ITPKB |
| chr12 | 104050147 | 104050657 | 511 | 10 | 2.29E-05 | 0.000285 | 0.001977 | 3.09E-05 | 0.006241124 | GLT8D2 |
| chr11 | 114058392 | 114058565 | 174 | 5 | 0.000219869 | 1.14E-05 | 0.005926 | 3.09E-05 | 0.016167757 | NA |
| chr14 | 97686288 | 97686694 | 407 | 3 | 1.31E-05 | 0.000953 | 2.85E-05 | 3.09E-05 | 0.005254564 | AL163872.1, LINC02291 |
| chr2 | 219245500 | 219246382 | 883 | 10 | 2.99E-05 | 4.63E-05 | 0.005398 | 3.09E-05 | 0.001928381 | STK16 |
| chr2 | 145021993 | 145022443 | 451 | 2 | 3.50E-05 | 2.72E-05 | 0.000312 | 3.09E-05 | 0.01354575 | TEX41 |
| chr2 | 98296623 | 98296659 | 37 | 2 | 0.000130557 | 1.92E-05 | 0.001321 | 3.09E-05 | -0.00627892 | VWA3B |
| chr1 | 244368416 | 244368618 | 203 | 3 | 0.000110654 | 1.66E-05 | 0.001575 | 3.09E-05 | 0.011933176 | C1orf100 |
| chr11 | 125618399 | 125618480 | 82 | 3 | 0.000117813 | 8.43E-05 | 0.000102 | 3.10E-05 | -0.00320914 | STT3A |
| chr2 | 48440668 | 48441112 | 445 | 5 | 0.000198276 | 1.44E-05 | 0.007384 | 3.10E-05 | -0.0010967 | PPP1R21 |
| chr18 | 35939339 | 35939617 | 279 | 3 | 7.97E-05 | 0.000212 | 0.000833 | 3.10E-05 | 0.010370801 | NA |
| chr20 | 43766410 | 43766618 | 209 | 2 | 3.02E-05 | 0.000627 | 3.51E-05 | 3.10E-05 | 0.009327825 | NA |
| chr11 | 44306319 | 44306873 | 555 | 3 | 0.000121465 | 1.63E-05 | 0.001642 | 3.11E-05 | 0.012005672 | ALX4 |
| chr1 | 109484079 | 109484163 | 85 | 2 | 0.000355159 | 3.73E-05 | 0.000167 | 3.11E-05 | 0.00101095 | ATXN7L2 |
| chr17 | 17714025 | 17714031 | 7 | 2 | 0.000134498 | 3.32E-05 | 0.000205 | 3.12E-05 | 0.013697484 | RAI1 |
| chr9 | 114225801 | 114225834 | 34 | 2 | 0.000152678 | 1.94E-05 | 0.001302 | 3.12E-05 | 0.011420442 | COL27A1 |
| chr10 | 13972419 | 13972651 | 233 | 5 | 9.51E-06 | 0.033676 | 4.61E-06 | 3.12E-05 | 0.008366304 | FRMD4A |
| chr6 | 163047218 | 163047522 | 305 | 2 | 0.000110118 | 2.19E-05 | 0.000674 | 3.12E-05 | 0.003975252 | PACRG, PACRG-AS2 |
| chr8 | 28622119 | 28622122 | 4 | 2 | 0.000386342 | 0.00011 | 6.40E-05 | 3.12E-05 | 0.011270937 | EXTL3 |
| chr9 | 92718296 | 92718839 | 544 | 3 | 6.07E-07 | 0.007416 | 0.000134 | 3.12E-05 | 0.003560931 | BICD2 |
| chr8 | 17754727 | 17755160 | 434 | 2 | 9.39E-06 | 0.000884 | 3.30E-05 | 3.12E-05 | 0.007342069 | MTUS1 |
| chr20 | 16551950 | 16551952 | 3 | 2 | 0.000136653 | 8.51E-05 | 7.55E-05 | 3.12E-05 | 0.006358259 | KIF16B |
| chr17 | 57772988 | 57773023 | 36 | 2 | 0.000205808 | 2.09E-05 | 0.000833 | 3.12E-05 | 0.017200749 | AC015845.2 |
| chr5 | 76564872 | 76565087 | 216 | 3 | 0.000148173 | 0.000102 | 0.000262 | 3.12E-05 | 0.00755461 | IQGAP2 |
| chr9 | 122905259 | 122905665 | 407 | 8 | 0.000104291 | 0.000383 | 0.000909 | 3.13E-05 | 0.000961056 | RC3H2 |
| chr3 | 10291066 | 10291971 | 906 | 4 | 2.04E-05 | 0.00017 | 0.000155 | 3.14E-05 | 0.001413339 | GHRLOS, GHRL |
| chr3 | 128787321 | 128787676 | 356 | 2 | 3.85E-05 | 5.23E-05 | 0.000111 | 3.14E-05 | 0.011937278 | RAB7A |
| chr17 | 42019443 | 42019952 | 510 | 9 | 0.000159655 | 0.00064 | 0.000826 | 3.14E-05 | 0.001966288 | NKIRAS2, DNAJC7 |
| chr11 | 3228254 | 3228841 | 588 | 3 | 0.000239487 | 1.30E-05 | 0.004491 | 3.14E-05 | 0.008513242 | AC109309.1, MRGPRE |
| chr7 | 116030741 | 116030750 | 10 | 2 | 0.000183541 | 2.06E-05 | 0.000927 | 3.14E-05 | 0.0002571 | TFEC |
| chr6 | 169810836 | 169811293 | 458 | 2 | 0.000171158 | 2.08E-05 | 0.000877 | 3.14E-05 | 0.004983159 | NA |
| chr1 | 185054339 | 185054609 | 271 | 2 | 7.24E-05 | 0.000134 | 5.87E-05 | 3.14E-05 | 0.002803711 | RNF2 |
| chr13 | 113350839 | 113351712 | 874 | 8 | 4.69E-05 | 0.000562 | 0.000195 | 3.15E-05 | 0.003709763 | GRTP1-AS1, GRTP1 |
| chr8 | 74244804 | 74245164 | 361 | 4 | 0.000129193 | 6.61E-05 | 0.000684 | 3.15E-05 | 0.005801069 | JPH1 |
| chr11 | 111878829 | 111879553 | 725 | 15 | 0.000165992 | 0.001996 | 0.000855 | 3.15E-05 | 0.000583842 | C11orf1, AP001781.2, FDXACB1 |
| chr19 | 13236271 | 13236523 | 253 | 3 | 0.000154442 | 4.17E-05 | 0.000652 | 3.15E-05 | 0.000331408 | CACNA1A |
| chr10 | 79146764 | 79147246 | 483 | 3 | 4.36E-05 | 0.000157 | 5.84E-05 | 3.16E-05 | 0.005926782 | ZMIZ1 |
| chr3 | 58335622 | 58335968 | 347 | 2 | 7.57E-05 | 6.60E-05 | 9.17E-05 | 3.16E-05 | 0.014198996 | PXK |
| chr14 | 100292527 | 100292550 | 24 | 2 | 4.25E-05 | 0.000101 | 6.85E-05 | 3.16E-05 | 0.002902366 | AL157871.1, SLC25A29 |
| chr17 | 40016601 | 40016748 | 148 | 2 | 7.13E-05 | 2.59E-05 | 0.00039 | 3.17E-05 | 0.010198789 | CSF3 |
| chr11 | 26572009 | 26572545 | 537 | 6 | 9.38E-05 | 0.000155 | 0.001089 | 3.18E-05 | 0.007109882 | ANO3, MUC15 |
| chr1 | 225840752 | 225841141 | 390 | 2 | 4.08E-05 | 0.000162 | 5.48E-05 | 3.18E-05 | 0.012718681 | EPHX1, AL591895.1 |
| chr2 | 7031355 | 7031832 | 478 | 2 | 4.83E-06 | 4.76E-05 | 0.000126 | 3.18E-05 | 0.0034469 | RNF144A |
| chr9 | 70311843 | 70312356 | 514 | 2 | 0.000195382 | 2.15E-05 | 0.000796 | 3.18E-05 | 0.01158058 | SMC5, AL162390.1 |
| chr16 | 30184675 | 30185228 | 554 | 4 | 2.39E-05 | 0.000176 | 0.000646 | 3.18E-05 | 0.003542072 | CORO1A, AC012645.3 |
| chr18 | 23259989 | 23260500 | 512 | 3 | 0.000197193 | 1.41E-05 | 0.003854 | 3.18E-05 | 0.005908123 | CABLES1, TMEM241, AC011731.1 |
| chr15 | 25171625 | 25171814 | 190 | 2 | 8.69E-05 | 3.03E-05 | 0.000261 | 3.18E-05 | 0.004068105 | SNHG14 |
| chr5 | 179975240 | 179975497 | 258 | 2 | 0.000173402 | 2.09E-05 | 0.000924 | 3.19E-05 | 0.004596919 | AC010285.3, RNF130 |
| chr10 | 50623678 | 50624575 | 898 | 11 | 1.23E-05 | 0.002867 | 0.000273 | 3.19E-05 | 0.00103334 | SGMS1-AS1, SGMS1 |
| chr21 | 41304162 | 41304855 | 694 | 6 | 3.70E-05 | 0.002745 | 0.000401 | 3.19E-05 | 0.001081746 | FAM3B |
| chr4 | 15777338 | 15778463 | 1126 | 9 | 1.54E-05 | 0.000169 | 0.000542 | 3.19E-05 | 0.003109202 | CD38 |
| chr12 | 123075541 | 123075932 | 392 | 3 | 8.33E-05 | 8.43E-05 | 0.000152 | 3.19E-05 | 0.0087491 | PITPNM2 |
| chr1 | 43569293 | 43569658 | 366 | 3 | 9.76E-05 | 1.63E-05 | 0.002541 | 3.19E-05 | 0.011412589 | PTPRF |
| chr11 | 2168117 | 2168904 | 788 | 8 | 9.94E-05 | 0.00131 | 0.000802 | 3.19E-05 | 0.001853513 | TH |
| chr5 | 42952633 | 42953441 | 809 | 3 | 4.46E-05 | 2.19E-05 | 0.000762 | 3.19E-05 | 0.016699963 | NA |
| chr22 | 18970165 | 18970367 | 203 | 7 | 0.000115283 | 0.000447 | 0.002102 | 3.19E-05 | 0.00112427 | NA |
| chr21 | 30337739 | 30338345 | 607 | 4 | 1.30E-06 | 0.007339 | 1.51E-05 | 3.20E-05 | 0.000850149 | NA |
| chr13 | 113148572 | 113149265 | 694 | 7 | 6.19E-05 | 0.000406 | 8.07E-05 | 3.20E-05 | 0.002597763 | F10 |
| chr2 | 85437590 | 85438049 | 460 | 5 | 4.36E-05 | 5.73E-05 | 0.000207 | 3.20E-05 | -0.00113817 | NA |
| chr11 | 67627447 | 67627701 | 255 | 4 | 7.17E-05 | 7.86E-05 | 0.000794 | 3.20E-05 | -0.00040719 | NA |
| chr4 | 11414334 | 11414648 | 315 | 5 | 0.00014127 | 0.000172 | 0.002961 | 3.21E-05 | 0.002104876 | HS3ST1 |
| chr12 | 101359960 | 101360302 | 343 | 3 | 4.88E-05 | 0.001186 | 0.000151 | 3.21E-05 | 0.007916113 | UTP20 |
| chr2 | 70469628 | 70469815 | 188 | 3 | 0.000203078 | 2.33E-05 | 0.000755 | 3.21E-05 | 0.013848183 | TGFA |
| chr12 | 81758690 | 81759685 | 996 | 13 | 3.52E-05 | 0.000311 | 0.000195 | 3.21E-05 | 0.001761734 | PPFIA2 |
| chr11 | 17439434 | 17439619 | 186 | 3 | 0.000271933 | 1.68E-05 | 0.002692 | 3.21E-05 | 0.013399836 | ABCC8 |
| chr19 | 39390753 | 39391487 | 735 | 12 | 5.65E-05 | 0.000564 | 0.002572 | 3.21E-05 | 0.001245909 | MED29, PAF1 |
| chr19 | 53329324 | 53329417 | 94 | 2 | 0.000354384 | 2.33E-05 | 0.000587 | 3.21E-05 | 0.026876521 | NA |
| chr10 | 70052171 | 70053200 | 1030 | 14 | 5.32E-05 | 0.000612 | 0.002727 | 3.21E-05 | -0.00106614 | MACROH2A2 |
| chr2 | 181679580 | 181679686 | 107 | 3 | 2.03E-05 | 2.80E-05 | 0.000452 | 3.21E-05 | 0.00867369 | CERKL, NEUROD1 |
| chr16 | 2970785 | 2971098 | 314 | 2 | 3.01E-06 | 0.000208 | 4.98E-05 | 3.22E-05 | 0.013526182 | PAQR4, PKMYT1 |
| chr1 | 2315025 | 2315425 | 401 | 3 | 3.28E-05 | 0.001509 | 4.19E-05 | 3.23E-05 | -0.0019444 | AL590822.3 |
| chr4 | 148441657 | 148442670 | 1014 | 10 | 0.000252166 | 2.63E-05 | 0.011144 | 3.23E-05 | 9.46E-05 | NR3C2 |
| chr13 | 40188013 | 40188332 | 320 | 6 | 7.03E-05 | 0.000782 | 0.000799 | 3.23E-05 | -0.00270919 | LINC00332, LINC00598 |
| chr17 | 77460005 | 77460235 | 231 | 2 | 0.000240784 | 2.84E-05 | 0.000323 | 3.23E-05 | 0.011262066 | SEPTIN9 |
| chr19 | 50794907 | 50795248 | 342 | 2 | 0.000105157 | 5.92E-05 | 0.000104 | 3.23E-05 | 0.010747529 | ACP4 |
| chr19 | 8142197 | 8142513 | 317 | 2 | 2.77E-05 | 0.002613 | 2.87E-05 | 3.24E-05 | 0.003957249 | FBN3 |
| chr21 | 33542424 | 33543286 | 863 | 16 | 6.69E-05 | 0.000131 | 0.004053 | 3.24E-05 | 0.000586703 | SON, GART |
| chr20 | 32437885 | 32438063 | 179 | 2 | 0.000187899 | 2.50E-05 | 0.00047 | 3.24E-05 | 0.012230493 | ASXL1 |
| chr19 | 1632633 | 1633074 | 442 | 2 | 3.24E-05 | 2.32E-05 | 0.000618 | 3.24E-05 | 0.003710959 | TCF3 |
| chr12 | 56080189 | 56081280 | 1092 | 6 | 2.53E-05 | 3.07E-05 | 0.000362 | 3.24E-05 | 0.004885401 | ERBB3 |
| chr2 | 45010550 | 45011539 | 990 | 6 | 5.65E-05 | 2.93E-05 | 0.004374 | 3.24E-05 | 0.002039097 | NA |
| chr7 | 39987358 | 39987886 | 529 | 7 | 1.15E-05 | 0.017274 | 1.14E-05 | 3.24E-05 | 0.0054797 | CDK13 |
| chr2 | 241755834 | 241756274 | 441 | 5 | 7.44E-05 | 0.000707 | 5.00E-05 | 3.24E-05 | 0.001810242 | D2HGDH |
| chr8 | 24911289 | 24911988 | 700 | 3 | 0.000108158 | 1.39E-05 | 0.003925 | 3.24E-05 | 0.023846145 | AC120193.1 |
| chr19 | 36916560 | 36917208 | 649 | 5 | 0.000146143 | 1.02E-05 | 0.009449 | 3.24E-05 | 0.006763 | ZNF568 |
| chr1 | 230377531 | 230377820 | 290 | 5 | 0.000228413 | 8.70E-06 | 0.012604 | 3.24E-05 | 0.00121703 | PGBD5 |
| chr3 | 154970363 | 154970913 | 551 | 3 | 1.72E-05 | 0.000459 | 3.51E-05 | 3.25E-05 | 0.005763827 | NA |
| chr16 | 680898 | 681069 | 172 | 4 | 5.11E-05 | 0.000618 | 3.13E-05 | 3.25E-05 | -0.00167358 | STUB1 |
| chr1 | 248347833 | 248348020 | 188 | 2 | 1.62E-05 | 0.000296 | 4.48E-05 | 3.25E-05 | 0.013393576 | NA |
| chr3 | 97961740 | 97961914 | 175 | 2 | 0.000100776 | 0.000143 | 5.91E-05 | 3.25E-05 | 0.005928722 | RIOX2 |
| chr22 | 49414978 | 49415707 | 730 | 2 | 0.00022583 | 2.22E-05 | 0.000784 | 3.25E-05 | 0.020467347 | C22orf34 |
| chr7 | 80644791 | 80645371 | 581 | 2 | 9.01E-05 | 2.08E-05 | 0.001095 | 3.25E-05 | 0.007965074 | CD36 |
| chr1 | 33519511 | 33519806 | 296 | 2 | 0.000136663 | 3.32E-05 | 0.00023 | 3.25E-05 | -0.00908755 | CSMD2 |
| chr17 | 65558418 | 65559346 | 929 | 7 | 4.97E-05 | 1.73E-05 | 0.008961 | 3.26E-05 | 0.008575167 | AXIN2, AC004805.1 |
| chr7 | 87476082 | 87476735 | 654 | 4 | 9.53E-05 | 8.04E-05 | 0.001646 | 3.26E-05 | 0.013035633 | ABCB4 |
| chr16 | 47699457 | 47699497 | 41 | 2 | 0.000149805 | 7.25E-05 | 8.86E-05 | 3.26E-05 | -0.00032646 | PHKB |
| chr10 | 112421491 | 112421517 | 27 | 2 | 0.000100252 | 2.73E-05 | 0.000372 | 3.26E-05 | 0.001863302 | ACSL5, AL157786.1 |
| chr6 | 15533185 | 15533589 | 405 | 2 | 0.000217076 | 2.05E-05 | 0.001238 | 3.26E-05 | 0.004636935 | DTNBP1 |
| chr19 | 8485540 | 8486156 | 617 | 5 | 9.53E-05 | 0.000123 | 0.00116 | 3.26E-05 | 0.001351557 | HNRNPM |
| chr19 | 49473181 | 49473240 | 60 | 2 | 1.74E-06 | 2.17E-05 | 0.00089 | 3.26E-05 | 0.006431291 | AC010619.1 |
| chr15 | 81295728 | 81295967 | 240 | 4 | 0.000106459 | 5.34E-05 | 0.001427 | 3.26E-05 | 0.008632561 | IL16 |
| chr21 | 42415184 | 42415483 | 300 | 2 | 0.000126555 | 2.10E-05 | 0.001052 | 3.26E-05 | 0.01876775 | UBASH3A |
| chr7 | 1916102 | 1916910 | 809 | 4 | 5.43E-05 | 9.48E-05 | 0.00033 | 3.26E-05 | 0.003426961 | MAD1L1, AC069288.1 |
| chr12 | 118152647 | 118153545 | 899 | 4 | 3.38E-06 | 0.004445 | 1.94E-05 | 3.26E-05 | 0.001685572 | TAOK3 |
| chr11 | 118491016 | 118491276 | 261 | 2 | 3.25E-05 | 0.000291 | 4.52E-05 | 3.26E-05 | -0.00219278 | KMT2A |
| chr6 | 38640029 | 38640452 | 424 | 10 | 0.000180262 | 6.13E-05 | 0.004337 | 3.26E-05 | 0.00063985 | BTBD9 |
| chr22 | 30560583 | 30560967 | 385 | 4 | 2.79E-05 | 0.001146 | 0.000173 | 3.27E-05 | 0.006138477 | GAL3ST1 |
| chr16 | 57760899 | 57761402 | 504 | 4 | 0.00010636 | 8.98E-05 | 0.000184 | 3.27E-05 | -0.00427371 | KIFC3 |
| chr5 | 10306835 | 10307576 | 742 | 4 | 0.00029147 | 1.39E-05 | 0.002796 | 3.27E-05 | 0.001385809 | CMBL |
| chr2 | 113756626 | 113757174 | 549 | 14 | 0.000118364 | 8.54E-05 | 0.000654 | 3.27E-05 | -0.00037296 | SLC35F5 |
| chr8 | 139790316 | 139790374 | 59 | 2 | 0.000355814 | 5.20E-05 | 0.00012 | 3.27E-05 | 0.003942612 | TRAPPC9 |
| chr8 | 141209866 | 141210463 | 598 | 3 | 2.08E-05 | 3.21E-05 | 0.000715 | 3.27E-05 | 0.008348403 | SLC45A4 |
| chr1 | 197222236 | 197222256 | 21 | 2 | 7.13E-05 | 7.79E-05 | 8.48E-05 | 3.27E-05 | 0.000814815 | AL513325.1 |
| chr17 | 56914037 | 56914632 | 596 | 12 | 9.53E-06 | 0.002549 | 0.001059 | 3.27E-05 | -0.0004885 | AC015912.3, TRIM25 |
| chr6 | 10850802 | 10850914 | 113 | 3 | 0.000116022 | 0.000257 | 0.000103 | 3.27E-05 | 0.012893185 | TMEM14B, AL024498.2 |
| chr19 | 15244093 | 15244393 | 301 | 2 | 9.56E-06 | 7.64E-05 | 8.61E-05 | 3.27E-05 | 0.001767258 | BRD4 |
| chr12 | 122749674 | 122750037 | 364 | 3 | 0.000179698 | 7.15E-05 | 0.000242 | 3.27E-05 | 0.007220109 | NA |
| chr8 | 8325102 | 8325546 | 445 | 3 | 2.12E-05 | 4.09E-05 | 0.000609 | 3.27E-05 | 0.003497054 | PRAG1 |
| chr14 | 52272538 | 52272902 | 365 | 2 | 4.42E-05 | 0.000228 | 4.92E-05 | 3.28E-05 | 0.010343014 | PTGDR |

| | | | | | | | | | | |
|---|---|---|---|---|---|---|---|---|---|---|
| chr18 | 499393 | 499887 | 495 | 3 | 7.87E-05 | 6.12E-05 | 0.001089 | 3.28E-05 | 0.004509667 | COLEC12 |
| chr7 | 74209546 | 74210043 | 498 | 5 | 0.000122661 | 0.000118 | 0.00032 | 3.28E-05 | 0.007036666 | LAT2 |
| chr17 | 81135713 | 81136223 | 511 | 6 | 5.73E-05 | 0.000102 | 0.005021 | 3.28E-05 | 0.00703274 | AC115099.1, AATK |
| chr8 | 27948497 | 27948757 | 261 | 3 | 9.18E-05 | 0.00036 | 0.000748 | 3.28E-05 | 0.008461426 | SCARA5 |
| chr2 | 196799584 | 196799831 | 248 | 8 | 4.75E-05 | 0.000642 | 4.33E-05 | 3.28E-05 | -0.00177644 | GTF3C3 |
| chr3 | 52794377 | 52794802 | 426 | 7 | 5.12E-05 | 0.000382 | 0.000302 | 3.28E-05 | 0.002927428 | ITIH3 |
| chr17 | 78240046 | 78240321 | 276 | 2 | 5.13E-05 | 0.00052 | 3.88E-05 | 3.29E-05 | 0.005304297 | TMEM235 |
| chr10 | 12709738 | 12710270 | 533 | 2 | 0.000185271 | 2.25E-05 | 0.000782 | 3.29E-05 | 0.008918638 | CAMK1D |
| chr3 | 141261701 | 141261925 | 225 | 3 | 6.78E-05 | 0.00023 | 4.91E-05 | 3.29E-05 | 0.001864014 | PXYLP1, AC022215.2 |
| chr4 | 92304782 | 92305344 | 563 | 4 | 0.000107247 | 1.54E-05 | 0.006242 | 3.29E-05 | 0.001746245 | GRID2 |
| chr5 | 81232879 | 81233521 | 643 | 9 | 9.43E-05 | 4.62E-05 | 0.002559 | 3.29E-05 | 0.006452613 | CKMT2, CKMT2-AS1 |
| chr11 | 12111468 | 12111786 | 319 | 3 | 0.000149438 | 1.42E-05 | 0.004145 | 3.29E-05 | 0.003419773 | MICAL2 |
| chr7 | 150400349 | 150400356 | 8 | 2 | 0.000239322 | 4.35E-05 | 0.000149 | 3.29E-05 | 0.005459798 | AC073111.2 |
| chr16 | 4313831 | 4314149 | 319 | 4 | 7.74E-05 | 2.07E-05 | 0.00241 | 3.29E-05 | 0.000391947 | NA |
| chr8 | 101087892 | 101087942 | 51 | 2 | 0.000185586 | 2.21E-05 | 0.000854 | 3.29E-05 | 0.000573196 | NA |
| chr2 | 96861181 | 96861786 | 606 | 3 | 0.000101508 | 0.000108 | 0.000337 | 3.29E-05 | 0.003683767 | SEMA4C |
| chr5 | 148826661 | 148827104 | 444 | 2 | 7.93E-07 | 0.000763 | 3.54E-05 | 3.29E-05 | 0.011015888 | ADRB2 |
| chr6 | 89925109 | 89925522 | 414 | 4 | 5.81E-05 | 0.000433 | 7.99E-05 | 3.30E-05 | 0.002594501 | NA |
| chr3 | 142578382 | 142579176 | 795 | 12 | 0.000118351 | 4.02E-05 | 0.015216 | 3.30E-05 | -0.00060196 | ATR |
| chr20 | 22220246 | 22220473 | 228 | 4 | 0.00024345 | 1.27E-05 | 0.006683 | 3.30E-05 | 0.003801476 | NA |
| chr11 | 35111254 | 35111727 | 474 | 2 | 3.30E-05 | 0.000115 | 6.73E-05 | 3.31E-05 | 0.010293382 | NA |
| chr15 | 94120146 | 94120432 | 287 | 2 | 3.25E-05 | 0.005882 | 2.70E-05 | 3.31E-05 | 0.011402307 | NA |
| chr2 | 201185302 | 201185409 | 108 | 2 | 0.000136495 | 3.41E-05 | 0.000229 | 3.31E-05 | 0.013271197 | CASP10 |
| chr1 | 52842193 | 52842672 | 480 | 3 | 2.27E-05 | 2.46E-05 | 0.001272 | 3.31E-05 | 0.019703503 | ZYG11A |
| chr10 | 48511259 | 48511879 | 621 | 4 | 4.85E-05 | 0.002005 | 0.001013 | 3.32E-05 | 0.001168425 | ARHGAP22, ARHGAP22-IT1 |
| chr8 | 142324302 | 142324831 | 530 | 4 | 9.66E-05 | 9.21E-05 | 0.000126 | 3.32E-05 | 0.002137691 | TSNARE1 |
| chr8 | 101166280 | 101166595 | 316 | 2 | 6.76E-05 | 2.65E-05 | 0.000436 | 3.32E-05 | 0.013875602 | NA |
| chr2 | 95053237 | 95053647 | 411 | 5 | 6.72E-05 | 5.22E-05 | 0.007518 | 3.32E-05 | 0.007999058 | MAL |
| chr14 | 23321016 | 23321425 | 410 | 8 | 0.000124729 | 3.98E-05 | 0.002087 | 3.32E-05 | 0.000176322 | BCL2L2-PABPN1, PABPN1 |
| chr22 | 31055577 | 31055901 | 325 | 3 | 0.000156236 | 4.59E-05 | 0.001326 | 3.32E-05 | -0.01418292 | AC005005.4 |
| chr19 | 5699234 | 5699440 | 207 | 3 | 5.14E-05 | 0.012981 | 4.58E-05 | 3.32E-05 | 0.002179954 | LONP1 |
| chr2 | 142131798 | 142132114 | 317 | 6 | 0.000152781 | 5.45E-05 | 0.00499 | 3.32E-05 | -0.00975476 | AC078882.1 |
| chr20 | 57694923 | 57695268 | 346 | 2 | 0.000111572 | 4.42E-05 | 0.000149 | 3.32E-05 | 0.007679722 | PMEPA1 |
| chr8 | 42468679 | 42468971 | 293 | 2 | 0.000158622 | 2.29E-05 | 0.000759 | 3.32E-05 | 0.011780204 | SLC20A2 |
| chr8 | 93917000 | 93917802 | 803 | 6 | 5.99E-07 | 0.000567 | 5.52E-05 | 3.33E-05 | -0.00198792 | PDP1 |
| chr16 | 68793051 | 68793120 | 70 | 2 | 0.000133538 | 2.50E-05 | 0.000544 | 3.34E-05 | 0.012661872 | CDH1 |
| chr5 | 179873085 | 179873200 | 116 | 2 | 6.37E-05 | 9.02E-05 | 7.88E-05 | 3.34E-05 | 0.001723227 | TBC1D9B |
| chr18 | 53417100 | 53417274 | 175 | 3 | 0.000204164 | 3.09E-05 | 0.001089 | 3.34E-05 | -0.0191653 | DCC |
| chr2 | 20632301 | 20632337 | 37 | 2 | 0.00013379 | 2.76E-05 | 0.00039 | 3.34E-05 | -0.00542199 | HS1BP3 |
| chr8 | 120761000 | 120761177 | 178 | 3 | 4.87E-05 | 4.64E-05 | 0.001311 | 3.34E-05 | 0.002034782 | SNTB1 |
| chr1 | 161421915 | 161422017 | 103 | 2 | 0.000137525 | 2.15E-05 | 0.001089 | 3.34E-05 | 0.033120881 | AL592295.4, AL592295.3 |
| chr16 | 27229349 | 27229484 | 136 | 5 | 0.000142593 | 0.000432 | 0.000649 | 3.34E-05 | 0.005875347 | NSMCE1 |
| chr17 | 44916526 | 44916901 | 376 | 5 | 9.07E-05 | 3.21E-05 | 0.000846 | 3.34E-05 | 0.002573957 | GFAP |
| chr17 | 41775588 | 41775678 | 91 | 2 | 0.000308306 | 2.19E-05 | 0.000978 | 3.34E-05 | 0.006238504 | JUP |
| chr10 | 104333901 | 104333960 | 60 | 3 | 0.000331021 | 0.000415 | 4.26E-05 | 3.35E-05 | 0.007519546 | ITPRIP |
| chr10 | 125763436 | 125763517 | 82 | 3 | 0.00013211 | 0.000226 | 0.000521 | 3.35E-05 | -0.00056333 | EDRF1, MMP21 |
| chr2 | 232632939 | 232633308 | 370 | 6 | 3.20E-05 | 0.000156 | 0.001604 | 3.35E-05 | 0.00593661 | EFHD1 |
| chr20 | 46249735 | 46249851 | 117 | 2 | 0.00010034 | 0.00018 | 5.54E-05 | 3.35E-05 | 0.002915286 | CDH22 |
| chr11 | 117930617 | 117930777 | 161 | 2 | 6.00E-05 | 4.50E-05 | 0.000148 | 3.35E-05 | 0.005915178 | NA |
| chr15 | 63290761 | 63290887 | 127 | 2 | 0.000173426 | 2.09E-05 | 0.001336 | 3.35E-05 | -0.00477047 | APH1B |
| chr17 | 77241088 | 77241888 | 801 | 7 | 0.000229495 | 0.000194 | 0.001186 | 3.35E-05 | 0.0012621 | NA |
| chr1 | 206062562 | 206062869 | 308 | 2 | 9.72E-05 | 4.10E-05 | 0.000169 | 3.36E-05 | -0.00306621 | RHEX |
| chr13 | 113548368 | 113548546 | 179 | 3 | 5.98E-05 | 0.000104 | 0.000162 | 3.36E-05 | 0.002938953 | TMCO3 |
| chr7 | 44754744 | 44755163 | 420 | 4 | 2.65E-05 | 0.000213 | 5.04E-05 | 3.36E-05 | 0.002552369 | ZMIZ2 |
| chr21 | 43344654 | 43345164 | 511 | 4 | 3.82E-05 | 3.58E-05 | 0.000835 | 3.36E-05 | 0.007568733 | NA |
| chr2 | 177806953 | 177807235 | 283 | 3 | 0.000155557 | 2.30E-05 | 0.000773 | 3.36E-05 | 0.007431861 | PDE11A, PDE11A |
| chr18 | 12658234 | 12659101 | 868 | 10 | 3.24E-05 | 0.000464 | 0.000628 | 3.36E-05 | 0.002626196 | PSMG2 |
| chr9 | 92658075 | 92658162 | 88 | 2 | 6.79E-05 | 0.001219 | 3.32E-05 | 3.37E-05 | -0.00446645 | IPPK |
| chr14 | 77029329 | 77029372 | 44 | 3 | 0.00013294 | 5.15E-05 | 0.000253 | 3.37E-05 | 0.012114946 | AC007686.3 |
| chr8 | 127989230 | 127989408 | 179 | 2 | 0.000149818 | 2.47E-05 | 0.000591 | 3.37E-05 | -0.00397207 | PVT1 |
| chr10 | 4767386 | 4767747 | 362 | 4 | 0.000136976 | 6.24E-05 | 0.001794 | 3.37E-05 | 0.009071234 | AC018978.1 |
| chr12 | 1503498 | 1503506 | 9 | 2 | 6.32E-05 | 9.75E-05 | 7.59E-05 | 3.37E-05 | 0.006549266 | LINC00942 |
| chr19 | 2056487 | 2056558 | 72 | 2 | 0.000140811 | 9.60E-05 | 7.65E-05 | 3.37E-05 | 0.000500977 | NA |
| chr2 | 8425851 | 8425859 | 9 | 2 | 0.000206631 | 2.97E-05 | 0.000332 | 3.37E-05 | 0.014452182 | LINC00299 |
| chr22 | 38767422 | 38767488 | 67 | 2 | 4.43E-05 | 2.97E-05 | 0.000332 | 3.37E-05 | 0.008724702 | SUN2 |
| chr6 | 5994002 | 5994510 | 509 | 4 | 0.00011529 | 1.31E-05 | 0.006484 | 3.37E-05 | 0.013760553 | NA |
| chr19 | 1423754 | 1423968 | 215 | 3 | 7.89E-05 | 2.07E-05 | 0.002397 | 3.37E-05 | 0.008962165 | DAZAP1 |
| chr6 | 10425320 | 10425866 | 547 | 4 | 3.35E-05 | 1.16E-05 | 0.007815 | 3.37E-05 | 0.006861705 | AL138885.2 |
| chr11 | 364623 | 364809 | 187 | 3 | 0.000147913 | 2.12E-05 | 0.001131 | 3.37E-05 | 0.013121769 | NA |
| chr19 | 12881256 | 12881665 | 410 | 10 | 0.000113885 | 0.001278 | 0.000791 | 3.38E-05 | -0.00154688 | AC020934.2, DNASE2 |
| chr16 | 87705771 | 87705847 | 77 | 4 | 7.45E-05 | 0.000318 | 5.36E-05 | 3.38E-05 | 0.001606292 | KLHDC4 |
| chr7 | 66087239 | 66087243 | 5 | 2 | 8.37E-05 | 0.000352 | 4.41E-05 | 3.38E-05 | 0.003522068 | ASL |
| chr5 | 19039995 | 19040127 | 133 | 2 | 7.66E-05 | 0.000585 | 3.88E-05 | 3.38E-05 | 0.012223437 | AC106744.1 |
| chr5 | 159104124 | 159104639 | 516 | 4 | 0.000107753 | 9.79E-06 | 0.010545 | 3.38E-05 | 0.015741901 | LINC02202 |
| chr22 | 31293176 | 31293345 | 170 | 2 | 6.44E-05 | 2.10E-05 | 0.001397 | 3.38E-05 | 0.015032819 | PIK3IP1-DT |
| chr13 | 29851832 | 29852123 | 292 | 2 | 6.58E-05 | 2.14E-05 | 0.001241 | 3.38E-05 | -0.00323545 | NA |
| chr4 | 7984524 | 7985603 | 1080 | 4 | 2.15E-05 | 0.001665 | 0.001034 | 3.38E-05 | 0.004960137 | ABLIM2 |
| chr11 | 69257994 | 69258183 | 190 | 2 | 0.000144284 | 5.21E-05 | 0.000127 | 3.38E-05 | 0.007442176 | NA |
| chr1 | 16018041 | 16018482 | 442 | 6 | 1.81E-05 | 0.000517 | 9.68E-05 | 3.39E-05 | 0.002650383 | HSPB7 |
| chr1 | 31188069 | 31188541 | 473 | 5 | 0.000125276 | 2.08E-05 | 0.004053 | 3.39E-05 | 0.00385306 | NKAIN1 |
| chr19 | 2484582 | 2484940 | 359 | 2 | 0.00020304 | 2.25E-05 | 0.000923 | 3.39E-05 | 0.009479568 | NA |
| chr11 | 129852559 | 129852617 | 59 | 3 | 0.000148173 | 4.84E-05 | 0.001299 | 3.39E-05 | 0.002188684 | TMEM45B |
| chr8 | 142453275 | 142454076 | 802 | 4 | 1.00E-06 | 0.002309 | 0.000178 | 3.39E-05 | 0.003917704 | ADGRB1 |
| chr2 | 68239186 | 68239203 | 18 | 2 | 8.25E-05 | 6.87E-05 | 9.79E-05 | 3.39E-05 | 0.019045209 | AC017083.3, PPP3R1 |
| chr13 | 20107061 | 20107390 | 330 | 2 | 9.22E-05 | 6.37E-05 | 0.000104 | 3.39E-05 | 0.008377056 | NA |
| chr4 | 7664043 | 7664532 | 490 | 6 | 0.000138957 | 0.000228 | 0.002728 | 3.39E-05 | 0.000150763 | SORCS2 |
| chr6 | 168060434 | 168060586 | 153 | 2 | 0.000254597 | 2.67E-05 | 0.000463 | 3.39E-05 | 0.00712854 | FRMD1 |
| chr2 | 97710017 | 97710098 | 82 | 2 | 0.000144474 | 2.15E-05 | 0.00121 | 3.39E-05 | 0.010504206 | NA |
| chr12 | 123974943 | 123975155 | 213 | 2 | 7.88E-05 | 2.36E-05 | 0.000743 | 3.39E-05 | -0.01056756 | ZNF664, AC068790.8, RFLNA |
| chr15 | 73946177 | 73946403 | 227 | 2 | 9.59E-05 | 2.37E-05 | 0.000725 | 3.40E-05 | 0.011753281 | LOXL1 |
| chr13 | 36346507 | 36347037 | 531 | 13 | 0.000151274 | 8.99E-05 | 0.021288 | 3.40E-05 | 4.15E-05 | SPART-AS1, SPART |
| chr14 | 59483645 | 59484347 | 703 | 8 | 0.000143797 | 0.001873 | 0.001036 | 3.40E-05 | 0.002428156 | L3HYPDH, AL121694.1 |
| chr15 | 70220189 | 70220524 | 336 | 2 | 0.000247055 | 2.11E-05 | 0.001397 | 3.40E-05 | 0.003985524 | NA |
| chr12 | 7162566 | 7162969 | 404 | 5 | 4.99E-05 | 0.000349 | 0.000368 | 3.40E-05 | 0.010073515 | NA |
| chr19 | 18927975 | 18928185 | 211 | 2 | 4.96E-05 | 0.000388 | 4.32E-05 | 3.41E-05 | 0.00286471 | DDX49, AC002985.1 |
| chr10 | 73946091 | 73946110 | 20 | 2 | 5.74E-05 | 0.007817 | 2.72E-05 | 3.41E-05 | 0.008874505 | NA |
| chr1 | 179142942 | 179143242 | 301 | 2 | 6.67E-05 | 2.32E-05 | 0.000821 | 3.41E-05 | 0.013711001 | ABL2 |
| chr10 | 113567246 | 113567435 | 190 | 4 | 0.00010636 | 0.000143 | 0.000639 | 3.41E-05 | 0.008832306 | HABP2 |
| chr1 | 3638443 | 3639479 | 1037 | 5 | 1.19E-05 | 0.001268 | 0.000812 | 3.41E-05 | -0.00078039 | WRAP73 |
| chr4 | 11429629 | 11429913 | 285 | 5 | 9.11E-05 | 6.82E-05 | 0.000225 | 3.42E-05 | 0.008295182 | AC006230.1 |
| chr20 | 31407420 | 31407562 | 143 | 3 | 7.73E-05 | 8.23E-05 | 0.001028 | 3.42E-05 | -0.01107842 | DEFB121 |
| chr3 | 53891804 | 53893006 | 1203 | 12 | 8.71E-06 | 0.000271 | 0.000972 | 3.42E-05 | 0.005728642 | SELENOK |
| chr3 | 18745237 | 18745731 | 495 | 3 | 1.98E-05 | 9.15E-05 | 0.000836 | 3.42E-05 | 0.01353529 | SATB1-AS1 |
| chr10 | 118309488 | 118309652 | 165 | 2 | 6.38E-05 | 7.38E-05 | 9.36E-05 | 3.42E-05 | -0.01199153 | FAM204A |
| chr12 | 111599582 | 111600064 | 483 | 11 | 0.000177954 | 3.10E-05 | 0.007965 | 3.42E-05 | -0.00052276 | ATXN2-AS, ATXN2 |
| chr12 | 116737831 | 116738308 | 478 | 9 | 5.11E-05 | 0.00016 | 0.004056 | 3.42E-05 | 0.00103593 | RNFT2, SPRING1 |
| chr8 | 127743024 | 127743048 | 25 | 2 | 0.000166159 | 4.07E-05 | 0.00018 | 3.43E-05 | 0.002335853 | NA |
| chr6 | 10763104 | 10763190 | 87 | 2 | 9.68E-05 | 0.000536 | 4.02E-05 | 3.43E-05 | 0.005592461 | TMEM14B, AL024498.2, MAK |
| chr20 | 43636786 | 43636955 | 170 | 2 | 9.89E-05 | 0.00022 | 5.26E-05 | 3.43E-05 | 0.001384497 | IFT52 |
| chr17 | 50802161 | 50802521 | 361 | 2 | 0.000108224 | 2.16E-05 | 0.001298 | 3.43E-05 | 0.003851363 | NA |
| chr6 | 104214378 | 104214507 | 130 | 2 | 4.97E-05 | 0.000185 | 5.63E-05 | 3.43E-05 | 0.004573047 | NA |
| chr10 | 1182953 | 1183359 | 407 | 5 | 0.000119168 | 4.34E-05 | 0.000832 | 3.43E-05 | 0.006169874 | LINC00200, ADARB2 |
| chr14 | 20955610 | 20955678 | 69 | 2 | 0.000199169 | 3.42E-05 | 0.00025 | 3.43E-05 | -0.01683807 | RNASE2 |
| chr10 | 558092 | 558717 | 626 | 4 | 5.74E-05 | 2.11E-05 | 0.002507 | 3.43E-05 | 0.001312707 | DIP2C |
| chr16 | 8621669 | 8622073 | 405 | 5 | 5.89E-05 | 0.000132 | 0.000591 | 3.43E-05 | 0.000936037 | METTL22 |
| chr16 | 16190158 | 16190229 | 72 | 2 | 0.00014251 | 2.14E-05 | 0.001397 | 3.43E-05 | 0.006920041 | ABCC6 |
| chr11 | 66188731 | 66189035 | 305 | 2 | 5.62E-05 | 4.76E-05 | 0.000145 | 3.44E-05 | 0.010580241 | PACS1 |
| chr11 | 123654954 | 123655483 | 530 | 6 | 6.46E-06 | 0.004704 | 0.000333 | 3.44E-05 | 0.009613648 | SCN3B |
| chr1 | 172774747 | 172775139 | 393 | 3 | 6.52E-05 | 1.54E-05 | 0.003414 | 3.44E-05 | -0.00283802 | NA |
| chr2 | 178478531 | 178479144 | 614 | 8 | 0.000114013 | 0.000419 | 0.006006 | 3.44E-05 | 0.001002462 | FKBP7 |
| chr11 | 27363362 | 27363957 | 596 | 9 | 0.000125133 | 9.36E-06 | 0.011972 | 3.44E-05 | 0.004066271 | NA |
| chr16 | 796463 | 797279 | 817 | 3 | 3.35E-05 | 3.16E-05 | 0.000767 | 3.44E-05 | 0.003624268 | CHTF18 |
| chr5 | 102753480 | 102754056 | 577 | 4 | 1.27E-05 | 0.000658 | 3.98E-05 | 3.44E-05 | 0.009163677 | PAM |
| chr1 | 1561030 | 1561556 | 527 | 5 | 0.000196693 | 8.78E-05 | 0.00382 | 3.45E-05 | 0.003496712 | SSU72 |
| chr3 | 160720185 | 160720671 | 487 | 2 | 0.000280486 | 2.16E-05 | 0.001319 | 3.45E-05 | 0.00841009 | NA |
| chr5 | 176542166 | 176542805 | 640 | 8 | 0.000116034 | 3.86E-05 | 0.005653 | 3.45E-05 | 0.003206157 | CDHR2 |
| chr17 | 63697434 | 63697436 | 3 | 2 | 6.76E-05 | 2.48E-05 | 0.000651 | 3.45E-05 | 0.00919748 | STRADA, LIMD2 |
| chr15 | 82082497 | 82083210 | 714 | 2 | 8.44E-05 | 2.32E-05 | 0.00088 | 3.45E-05 | 0.010462857 | NA |
| chr2 | 85538948 | 85539685 | 738 | 9 | 0.000102192 | 0.001063 | 0.000442 | 3.45E-05 | -0.0014315 | MAT2A |
| chr6 | 30326698 | 30327551 | 854 | 13 | 5.19E-05 | 4.70E-05 | 0.005118 | 3.45E-05 | 0.00080968 | TRIM39, HCG18 |
| chr12 | 56987739 | 56987742 | 4 | 2 | 0.000277266 | 3.35E-05 | 0.000269 | 3.46E-05 | 0.003609188 | AC026120.3 |
| chr10 | 59906663 | 59907329 | 667 | 8 | 4.07E-05 | 0.000448 | 0.000341 | 3.46E-05 | 0.000574889 | NA |
| chr2 | 9758099 | 9758470 | 372 | 2 | 0.000140721 | 3.15E-05 | 0.000308 | 3.46E-05 | 0.01466072 | AC082651.3 |
| chr14 | 100377605 | 100378015 | 411 | 3 | 1.67E-05 | 0.000222 | 0.000185 | 3.46E-05 | 0.009233613 | WDR25 |
| chr12 | 132724964 | 132725680 | 717 | 8 | 5.57E-05 | 0.006181 | 0.003207 | 3.46E-05 | 0.001153996 | ANKLE2 |
| chr5 | 19886305 | 19886766 | 462 | 4 | 6.12E-05 | 2.95E-05 | 0.000572 | 3.46E-05 | 0.00501033 | CDH18 |
| chr11 | 520862 | 521046 | 185 | 3 | 0.000265024 | 1.71E-05 | 0.002606 | 3.47E-05 | 0.002586388 | NA |
| chr19 | 37468202 | 37469059 | 858 | 7 | 8.52E-05 | 0.000119 | 0.001508 | 3.47E-05 | 0.000329095 | ZNF570, ZNF569 |
| chr8 | 142834945 | 142836298 | 1354 | 6 | 1.26E-05 | 0.000918 | 0.000486 | 3.47E-05 | 0.005174826 | GML |
| chr19 | 1230769 | 1231293 | 525 | 4 | 6.74E-05 | 0.000128 | 0.000228 | 3.47E-05 | 0.004416102 | CBARP |
| chr2 | 99554304 | 99554674 | 371 | 5 | 0.000284321 | 1.22E-05 | 0.006241 | 3.47E-05 | 0.000279364 | AFF3 |
| chr20 | 31608238 | 31608911 | 674 | 4 | 6.84E-05 | 8.62E-05 | 0.002488 | 3.47E-05 | 0.00793641 | NA |
| chr1 | 59813880 | 59814831 | 952 | 7 | 0.000104682 | 1.68E-05 | 0.011277 | 3.47E-05 | -0.00331821 | HOOK1 |
| chr8 | 1948854 | 1949531 | 678 | 5 | 3.18E-05 | 5.22E-05 | 0.002134 | 3.47E-05 | 0.008399273 | AC019257.8, ARHGEF10 |
| chr22 | 31945205 | 31945794 | 590 | 11 | 4.22E-05 | 0.001699 | 9.94E-05 | 3.48E-05 | 0.002032476 | Z82190.2, YWHAH, C22orf24 |
| chr16 | 56519618 | 56520359 | 742 | 11 | 9.10E-05 | 0.000593 | 0.000158 | 3.48E-05 | 0.000237224 | BBS2 |
| chr10 | 32022179 | 32022502 | 324 | 2 | 0.00020771 | 2.19E-05 | 0.001336 | 3.48E-05 | 0.012199482 | KIF5B |
| chr2 | 42493184 | 42493186 | 3 | 2 | 2.27E-05 | 2.34E-05 | 0.000885 | 3.48E-05 | 0.010938504 | KCNG3 |
| chr10 | 102713430 | 102714194 | 765 | 3 | 3.41E-05 | 1.60E-05 | 0.003634 | 3.48E-05 | 0.000295682 | ARL3 |
| chr12 | 49567470 | 49568718 | 1249 | 12 | 2.15E-05 | 0.000414 | 0.000518 | 3.48E-05 | -0.00092703 | PRPF40B, MCRS1 |
| chr7 | 112080753 | 112081178 | 426 | 2 | 2.27E-05 | 0.000461 | 4.24E-05 | 3.48E-05 | 0.008623907 | DOCK4 |
| chr17 | 62965357 | 62965711 | 355 | 5 | 3.53E-05 | 0.004261 | 0.000123 | 3.49E-05 | 0.00654878 | AC005972.3 |
| chr15 | 63509404 | 63509431 | 28 | 2 | 0.000105007 | 0.000538 | 4.09E-05 | 3.49E-05 | 0.01002534 | USP3 |
| chr10 | 69337571 | 69337979 | 409 | 6 | 0.000185359 | 7.92E-05 | 0.00137 | 3.49E-05 | 0.007098471 | HK1 |
| chr6 | 169861640 | 169862129 | 490 | 2 | 6.87E-06 | 0.002331 | 3.12E-05 | 3.49E-05 | 0.010043343 | NA |
| chr2 | 74526564 | 74527110 | 547 | 5 | 6.02E-05 | 0.000237 | 0.001009 | 3.49E-05 | 0.004216993 | AUP1 |
| chr17 | 28022944 | 28023361 | 418 | 3 | 4.40E-05 | 0.001912 | 2.70E-05 | 3.49E-05 | 0.005846815 | NA |
| chr11 | 19564159 | 19564253 | 95 | 2 | 0.000248314 | 2.85E-05 | 0.000423 | 3.49E-05 | 0.00456938 | NAV2 |
| chr10 | 124008279 | 124008449 | 171 | 2 | 0.000312827 | 6.91E-05 | 0.000102 | 3.50E-05 | 0.003914813 | CHST15 |
| chr6 | 32074322 | 32075252 | 931 | 9 | 2.08E-05 | 0.000175 | 0.001869 | 3.50E-05 | 0.003255781 | TNXB |
| chr19 | 56229594 | 56229650 | 57 | 2 | 0.00021354 | 2.25E-05 | 0.001132 | 3.50E-05 | -0.00209246 | ZSCAN5A |
| chr19 | 5585637 | 5586084 | 448 | 3 | 0.000128696 | 2.02E-05 | 0.001452 | 3.50E-05 | 0.008168118 | NA |

| | | | | | | | | | | |
|---|---|---|---|---|---|---|---|---|---|---|
| chr11 | 1760275 | 1760480 | 206 | 5 | 0.000102821 | 1.27E-05 | 0.003564 | 3.50E-05 | 0.004040515 | AC068580.3, AC068580.4, CTSD |
| chr5 | 82640224 | 82640698 | 475 | 3 | 4.21E-05 | 0.000109 | 0.000809 | 3.50E-05 | 0.009358082 | NA |
| chr14 | 104422574 | 104422779 | 206 | 3 | 6.39E-05 | 0.001096 | 3.30E-05 | 3.50E-05 | 0.009892104 | NA |
| chr10 | 5093259 | 5093261 | 3 | 2 | 3.64E-05 | 0.000646 | 3.91E-05 | 3.50E-05 | 0.010618299 | AKR1C3 |
| chr15 | 75229713 | 75230346 | 634 | 4 | 4.36E-05 | 2.61E-05 | 0.000842 | 3.50E-05 | 0.006451181 | AC113208.3 |
| chr7 | 99470484 | 99470735 | 252 | 2 | 7.91E-05 | 2.19E-05 | 0.001376 | 3.50E-05 | 0.011219205 | NA |
| chr1 | 115314764 | 115314917 | 154 | 3 | 0.000182989 | 8.28E-05 | 0.000195 | 3.50E-05 | 0.004367569 | NGF-AS1, NGF |
| chr7 | 130272553 | 130273418 | 866 | 6 | 9.52E-05 | 6.99E-05 | 0.000985 | 3.50E-05 | -0.00226836 | CPA2 |
| chr11 | 3921490 | 3921806 | 317 | 2 | 1.78E-05 | 0.001297 | 3.41E-05 | 3.50E-05 | 0.002531781 | STIM1 |
| chr2 | 189094826 | 189095031 | 206 | 2 | 0.000222844 | 2.45E-05 | 0.000744 | 3.50E-05 | 0.009947489 | COL5A2 |
| chr10 | 27944180 | 27944259 | 80 | 3 | 0.000300957 | 1.44E-05 | 0.005244 | 3.51E-05 | 0.003119839 | ARMC4 |
| chr18 | 10525450 | 10526141 | 692 | 8 | 7.35E-05 | 0.000193 | 0.000693 | 3.51E-05 | -0.00012652 | NAPG |
| chr9 | 112750250 | 112750765 | 516 | 10 | 0.000142786 | 5.10E-05 | 0.002901 | 3.51E-05 | 0.002240135 | SNX30, AL390067.1 |
| chr3 | 195180628 | 195181065 | 438 | 2 | 0.000227642 | 2.24E-05 | 0.001199 | 3.51E-05 | 0.01121131 | XXYLT1 |
| chr6 | 70864349 | 70864702 | 354 | 3 | 0.000180427 | 4.60E-05 | 0.000241 | 3.51E-05 | 0.011563704 | B3GAT2 |
| chr20 | 64069647 | 64070259 | 613 | 3 | 0.000129122 | 4.90E-05 | 0.001343 | 3.51E-05 | 0.006943832 | TCEA2 |
| chr11 | 64892604 | 64892914 | 311 | 2 | 3.00E-07 | 2.46E-05 | 0.00074 | 3.51E-05 | 0.010581878 | MIR194-2HG |
| chr4 | 740514 | 740958 | 445 | 4 | 1.22E-05 | 0.00204 | 0.000144 | 3.52E-05 | -0.00034363 | PCGF3 |
| chr16 | 68222569 | 68222986 | 418 | 3 | 3.75E-05 | 6.67E-05 | 0.000149 | 3.52E-05 | 0.001192354 | NFATC3 |
| chr19 | 16070159 | 16070724 | 566 | 4 | 0.000360681 | 3.16E-05 | 0.002854 | 3.52E-05 | 0.000910892 | TPM4 |
| chr3 | 52232039 | 52232057 | 19 | 2 | 0.000498768 | 3.91E-05 | 0.000206 | 3.52E-05 | 0.007946901 | TWF2 |
| chr6 | 30651020 | 30651465 | 446 | 8 | 8.32E-05 | 0.000111 | 0.000178 | 3.52E-05 | 0.003321904 | C6orf136 |
| chr17 | 45894526 | 45895207 | 682 | 8 | 1.21E-05 | 0.000698 | 8.85E-05 | 3.53E-05 | 0.000896217 | MAPT, MAPT-AS1 |
| chr5 | 141703293 | 141703549 | 257 | 2 | 9.98E-05 | 2.96E-05 | 0.000392 | 3.53E-05 | 0.012244613 | NA |
| chr2 | 86037113 | 86037265 | 153 | 2 | 0.000295638 | 3.24E-05 | 0.000306 | 3.53E-05 | 0.011213482 | POLR1A |
| chr11 | 106076530 | 106077779 | 1250 | 17 | 3.34E-06 | 0.00015 | 0.001973 | 3.53E-05 | -0.00014292 | AASDHPPT, KBTBD3 |
| chr16 | 24540088 | 24540758 | 671 | 10 | 4.30E-05 | 0.00032 | 0.003955 | 3.53E-05 | 0.000828756 | RBBP6 |
| chr21 | 41425541 | 41425661 | 121 | 2 | 0.000151857 | 2.61E-05 | 0.000595 | 3.53E-05 | 0.012922809 | MX1 |
| chr16 | 89719278 | 89720092 | 815 | 3 | 8.97E-06 | 4.85E-05 | 0.00068 | 3.53E-05 | 0.007966165 | VPS9D1 |
| chr3 | 66394046 | 66394799 | 754 | 3 | 5.02E-05 | 2.05E-05 | 0.002199 | 3.53E-05 | 0.008036407 | LRIG1 |
| chr1 | 237043182 | 237043343 | 162 | 4 | 0.000187185 | 1.31E-05 | 0.005793 | 3.53E-05 | 0.003578833 | RYR2 |
| chr5 | 16936349 | 16937270 | 922 | 8 | 3.26E-05 | 9.43E-05 | 0.001121 | 3.53E-05 | -0.000682 | NA |
| chr9 | 134000136 | 134000482 | 347 | 2 | 0.000217257 | 2.21E-05 | 0.001425 | 3.54E-05 | 0.014097822 | NA |
| chr19 | 1904289 | 1905253 | 965 | 5 | 1.39E-05 | 0.004855 | 0.000744 | 3.54E-05 | 0.004952783 | NA |
| chr8 | 28984160 | 28984599 | 440 | 3 | 0.0001602 | 9.61E-05 | 0.000316 | 3.54E-05 | 0.000823052 | HMBOX1 |
| chr20 | 47183182 | 47183260 | 79 | 2 | 8.73E-05 | 0.001124 | 3.51E-05 | 3.54E-05 | -0.00052496 | EYA2 |
| chr17 | 50633143 | 50633817 | 675 | 2 | 4.71E-05 | 2.21E-05 | 0.00144 | 3.55E-05 | 0.008175356 | NA |
| chr17 | 76137673 | 76137978 | 306 | 2 | 6.00E-05 | 7.39E-05 | 9.85E-05 | 3.55E-05 | 0.009547411 | FOXJ1 |
| chr15 | 65571247 | 65571568 | 322 | 3 | 0.000145084 | 4.55E-05 | 0.000436 | 3.55E-05 | 0.002681414 | HACD3 |
| chr11 | 72216537 | 72216894 | 358 | 3 | 6.06E-05 | 7.73E-05 | 0.000258 | 3.55E-05 | 0.001752974 | FOLR2 |
| chr1 | 1359249 | 1359938 | 690 | 4 | 3.37E-05 | 2.01E-05 | 0.002159 | 3.56E-05 | 0.007057579 | MXRA8 |
| chr18 | 74457992 | 74458722 | 731 | 5 | 0.000110867 | 1.09E-05 | 0.009132 | 3.56E-05 | 0.015228807 | NA |
| chr4 | 3021563 | 3021773 | 211 | 2 | 8.11E-05 | 4.89E-05 | 0.000149 | 3.56E-05 | -0.00123621 | GRK4 |
| chr3 | 138174722 | 138174965 | 244 | 8 | 0.000189869 | 8.02E-05 | 0.000208 | 3.56E-05 | 0.001138191 | DBR1 |
| chr15 | 68572338 | 68573248 | 911 | 2 | 2.30E-06 | 0.000501 | 4.23E-05 | 3.56E-05 | 0.004733742 | NA |
| chr6 | 11767603 | 11767862 | 260 | 2 | 0.000120718 | 4.39E-05 | 0.000175 | 3.56E-05 | -0.01888369 | ADTRP |
| chr1 | 156299257 | 156299377 | 121 | 3 | 0.000188001 | 2.30E-05 | 0.001492 | 3.57E-05 | -0.0024723 | VHLL |
| chr10 | 95755562 | 95755996 | 435 | 4 | 3.39E-05 | 3.23E-05 | 0.000529 | 3.57E-05 | 0.008894409 | ENTPD1, ENTPD1-AS1 |
| chr2 | 73233212 | 73234052 | 841 | 9 | 6.58E-05 | 1.05E-05 | 0.017558 | 3.57E-05 | -0.00100968 | CCT7, PRADC1 |
| chr19 | 11177713 | 11178700 | 988 | 9 | 0.000112519 | 0.000426 | 0.000725 | 3.57E-05 | 0.002229516 | KANK2 |
| chr8 | 139799688 | 139799980 | 293 | 2 | 0.000233185 | 2.23E-05 | 0.001426 | 3.57E-05 | 0.005169004 | TRAPPC9 |
| chr12 | 121512712 | 121513013 | 302 | 2 | 8.55E-05 | 2.72E-05 | 0.00054 | 3.58E-05 | 0.006419108 | KDM2B |
| chr20 | 33103400 | 33103543 | 144 | 2 | 6.42E-05 | 0.000252 | 5.24E-05 | 3.58E-05 | 0.010179877 | BPIFB4 |
| chr16 | 85812020 | 85812578 | 559 | 5 | 9.22E-05 | 0.0002 | 0.000497 | 3.58E-05 | -0.01658419 | NA |
| chr17 | 46832621 | 46833249 | 629 | 5 | 8.65E-05 | 0.000103 | 0.002557 | 3.58E-05 | 0.004836399 | WNT9B, WNT3 |
| chr22 | 35352546 | 35352902 | 357 | 2 | 1.86E-05 | 3.57E-05 | 0.00026 | 3.58E-05 | 0.011119884 | NA |
| chr10 | 18710216 | 18710496 | 281 | 3 | 8.48E-05 | 0.005058 | 0.000525 | 3.58E-05 | -0.02026966 | AL512641.1 |
| chr1 | 234607942 | 234608582 | 641 | 9 | 4.88E-05 | 0.001617 | 0.00022 | 3.58E-05 | 0.00106459 | AL160408.2, IRF2BP2 |
| chr5 | 139395752 | 139396133 | 382 | 4 | 0.000171989 | 0.000185 | 7.99E-05 | 3.58E-05 | 0.005282196 | NA |
| chr8 | 19820341 | 19820502 | 162 | 2 | 0.000139637 | 2.29E-05 | 0.001207 | 3.58E-05 | 0.007597696 | INTS10 |
| chr16 | 3960965 | 3961160 | 196 | 2 | 0.000220912 | 2.50E-05 | 0.000766 | 3.58E-05 | 0.025136296 | ADCY9 |
| chr7 | 92834851 | 92835167 | 317 | 3 | 1.17E-05 | 3.02E-05 | 0.001342 | 3.58E-05 | -0.00200106 | CDK6 |
| chr3 | 143341690 | 143342085 | 396 | 4 | 3.80E-05 | 0.001647 | 0.000182 | 3.59E-05 | 0.003749806 | SLC9A9 |
| chr1 | 12636327 | 12636549 | 223 | 3 | 0.000138364 | 0.000713 | 0.000269 | 3.59E-05 | 0.006607111 | NA |
| chr22 | 27331359 | 27331696 | 338 | 5 | 0.00012213 | 8.28E-05 | 0.000417 | 3.59E-05 | 0.009164331 | AL020994.2 |
| chr6 | 16306146 | 16306626 | 481 | 4 | 0.000211912 | 3.32E-05 | 0.001791 | 3.59E-05 | 0.00566567 | ATXN1 |
| chr17 | 77438188 | 77438530 | 343 | 2 | 9.63E-05 | 2.24E-05 | 0.001418 | 3.59E-05 | 0.00607994 | SEPTIN9 |
| chr14 | 69008657 | 69008908 | 252 | 2 | 0.000183568 | 3.11E-05 | 0.000366 | 3.59E-05 | -6.85E-05 | NA |
| chr3 | 32171878 | 32172102 | 225 | 3 | 0.000154727 | 4.60E-05 | 0.00022 | 3.59E-05 | 0.002514632 | NA |
| chr3 | 105869692 | 105870209 | 518 | 3 | 3.12E-06 | 0.000112 | 0.000214 | 3.59E-05 | 0.006844714 | NA |
| chr19 | 47702504 | 47702862 | 359 | 2 | 0.000172548 | 2.26E-05 | 0.001361 | 3.60E-05 | 0.013988532 | AC010519.1, BICRA |
| chr19 | 55312459 | 55313197 | 739 | 4 | 3.69E-05 | 0.000293 | 6.43E-05 | 3.60E-05 | 0.000634545 | BRSK1, AC020922.2, TMEM150B |
| chr13 | 41310889 | 41311521 | 633 | 10 | 1.75E-05 | 0.000116 | 0.000389 | 3.60E-05 | 0.000230517 | NAA16 |
| chr2 | 9692972 | 9693293 | 322 | 2 | 0.00022544 | 2.34E-05 | 0.0011 | 3.61E-05 | 0.018847717 | AC082651.1, AC082651.4 |
| chr6 | 83065768 | 83066180 | 413 | 8 | 5.41E-05 | 0.002239 | 0.00012 | 3.61E-05 | -0.00139087 | UBE3D |
| chr17 | 50663915 | 50664028 | 114 | 2 | 6.93E-05 | 6.85E-05 | 0.000107 | 3.61E-05 | 0.007156004 | ABCC3 |
| chr1 | 201693753 | 201694098 | 346 | 2 | 2.34E-05 | 0.001195 | 3.53E-05 | 3.61E-05 | 0.007168693 | NAV1, IPO9-AS1 |
| chr8 | 144820712 | 144820950 | 239 | 4 | 0.000113083 | 4.37E-05 | 0.001082 | 3.61E-05 | 0.003040613 | NA |
| chr1 | 202166643 | 202166769 | 127 | 2 | 0.000235807 | 2.29E-05 | 0.001292 | 3.61E-05 | 0.010335675 | NA |
| chr1 | 118983437 | 118984259 | 823 | 6 | 1.28E-05 | 9.52E-05 | 8.98E-05 | 3.61E-05 | 0.005321893 | TBX15 |
| chr17 | 3754999 | 3755468 | 470 | 4 | 9.74E-05 | 3.61E-05 | 0.000956 | 3.61E-05 | 0.00432609 | ITGAE |
| chr11 | 695873 | 696675 | 803 | 11 | 3.03E-05 | 0.000176 | 0.000451 | 3.61E-05 | 0.000532564 | EPS8L2, TMEM80, DEAF1 |
| chr12 | 40195237 | 40195311 | 75 | 2 | 0.000247128 | 2.30E-05 | 0.001266 | 3.62E-05 | 0.012023041 | LINC02471, LRRK2-DT |
| chr17 | 6120541 | 6120697 | 157 | 3 | 0.000224759 | 7.89E-05 | 0.001127 | 3.62E-05 | -0.00048718 | WSCD1 |
| chr5 | 134436592 | 134437106 | 515 | 4 | 0.000263963 | 1.24E-05 | 0.006775 | 3.62E-05 | 0.011147337 | AC005355.1, AC109454.3 |
| chr1 | 55285142 | 55285239 | 98 | 2 | 0.000129955 | 5.14E-05 | 0.000145 | 3.62E-05 | 0.018789834 | MIR4422HG |
| chr19 | 3286179 | 3286246 | 68 | 2 | 0.000339835 | 3.39E-05 | 0.0003 | 3.62E-05 | 0.011321233 | CELF5 |
| chr1 | 19924520 | 19924787 | 268 | 2 | 3.92E-05 | 3.84E-05 | 0.000231 | 3.62E-05 | 0.006058813 | NA |
| chr17 | 75153387 | 75153909 | 523 | 6 | 9.40E-05 | 5.94E-05 | 0.000186 | 3.62E-05 | 0.002245687 | JPT1 |
| chr2 | 38073742 | 38074613 | 872 | 4 | 1.75E-05 | 9.28E-05 | 0.000472 | 3.62E-05 | 0.006962669 | CYP1B1-AS1, CYP1B1 |
| chr17 | 48999542 | 48999803 | 262 | 2 | 9.86E-05 | 5.46E-05 | 0.000136 | 3.62E-05 | 0.019864279 | IGF2BP1 |
| chr14 | 57247731 | 57248202 | 472 | 2 | 8.70E-05 | 2.74E-05 | 0.000554 | 3.62E-05 | 0.0176881 | EXOC5 |
| chr18 | 21611757 | 21612541 | 785 | 11 | 0.000132553 | 0.000524 | 0.004339 | 3.62E-05 | -0.00272029 | SNRPD1 |
| chr19 | 40390157 | 40390676 | 520 | 4 | 1.08E-05 | 4.88E-05 | 0.001713 | 3.62E-05 | 0.003370462 | HIPK4 |
| chr12 | 63904150 | 63904341 | 192 | 2 | 0.000184098 | 3.11E-05 | 0.000376 | 3.62E-05 | 0.004744474 | SRGAP1 |
| chr4 | 2297445 | 2297507 | 63 | 2 | 0.000219051 | 2.87E-05 | 0.000479 | 3.62E-05 | 0.002609505 | AL158068.2, ZFYVE28 |
| chr6 | 54847860 | 54848511 | 652 | 2 | 0.000179253 | 2.51E-05 | 0.000809 | 3.62E-05 | 0.009051807 | FAM83B |
| chr11 | 47184972 | 47185284 | 313 | 3 | 1.47E-05 | 2.01E-05 | 0.001643 | 3.63E-05 | 0.007406114 | PACSIN3 |
| chr14 | 24819540 | 24820029 | 490 | 4 | 5.80E-05 | 0.000526 | 0.000227 | 3.63E-05 | -0.00737924 | AL161663.2, STXBP6 |
| chr17 | 82848259 | 82848512 | 254 | 3 | 0.000130296 | 2.48E-05 | 0.001215 | 3.63E-05 | 0.010938404 | TBCD |
| chr6 | 166759769 | 166759855 | 87 | 2 | 0.000225834 | 2.85E-05 | 0.000497 | 3.63E-05 | 0.018989844 | RPS6KA2 |
| chr14 | 22867271 | 22867379 | 109 | 2 | 0.000181033 | 2.30E-05 | 0.001313 | 3.63E-05 | -0.00014259 | NA |
| chr16 | 74998611 | 74999402 | 792 | 7 | 9.46E-05 | 4.28E-05 | 0.001846 | 3.64E-05 | 0.001418669 | ZNRF1, WDR59 |
| chr2 | 240512028 | 240512150 | 123 | 2 | 0.000240268 | 2.53E-05 | 0.000793 | 3.64E-05 | 0.0034112 | ANKMY1 |
| chr1 | 20186124 | 20186909 | 786 | 5 | 2.35E-05 | 3.77E-05 | 0.002341 | 3.64E-05 | 0.003183998 | UBXN10, PLA2G2C |
| chr3 | 122576749 | 122577562 | 814 | 7 | 5.01E-06 | 0.006599 | 3.42E-05 | 3.64E-05 | 0.002072182 | NA |
| chr2 | 176112074 | 176112492 | 419 | 2 | 0.000432764 | 9.68E-05 | 8.41E-05 | 3.64E-05 | 0.009750908 | HOXD10 |
| chr13 | 50927595 | 50927718 | 124 | 2 | 9.57E-05 | 0.000159 | 6.40E-05 | 3.64E-05 | 0.017314779 | RNASEH2B |
| chr19 | 55114740 | 55115096 | 357 | 2 | 5.55E-06 | 0.00317 | 3.12E-05 | 3.65E-05 | 0.006166317 | PPP1R12C |
| chr6 | 44065874 | 44066243 | 370 | 2 | 0.000138414 | 3.06E-05 | 0.000404 | 3.65E-05 | 0.006840565 | AL109615.2, AL109615.3 |
| chr6 | 166908178 | 166908180 | 3 | 2 | 7.81E-05 | 0.004643 | 3.01E-05 | 3.65E-05 | 0.005461846 | AL159163.1 |
| chr13 | 27971158 | 27971429 | 272 | 4 | 5.27E-05 | 0.001281 | 0.000941 | 3.65E-05 | 0.009810271 | NA |
| chr15 | 90620500 | 90620961 | 462 | 3 | 9.67E-05 | 0.000142 | 9.55E-05 | 3.65E-05 | 0.007282779 | CRTC3, CRTC3-AS1 |
| chr22 | 43171459 | 43171884 | 426 | 3 | 3.07E-05 | 0.000394 | 4.37E-05 | 3.65E-05 | 0.003522229 | TTLL12 |
| chr1 | 99969318 | 99970299 | 982 | 17 | 0.000281788 | 4.68E-05 | 0.030937 | 3.66E-05 | 0.003521739 | SLC35A3, AC118553.2, AC118553.1 |
| chr13 | 54133836 | 54134209 | 374 | 2 | 6.71E-05 | 0.000116 | 7.59E-05 | 3.66E-05 | 0.014394482 | LINC00458 |
| chr16 | 56701699 | 56701809 | 111 | 2 | 0.000103979 | 0.001679 | 3.40E-05 | 3.66E-05 | 0.005629045 | NA |
| chr6 | 138376642 | 138376939 | 298 | 3 | 9.83E-05 | 0.000417 | 0.000174 | 3.66E-05 | -0.01008152 | NA |
| chr10 | 61410487 | 61410614 | 128 | 3 | 0.000145538 | 0.000476 | 0.000848 | 3.66E-05 | 0.000926446 | TMEM26 |
| chr17 | 82863690 | 82864304 | 615 | 3 | 9.07E-05 | 2.61E-05 | 0.002127 | 3.67E-05 | 0.004950701 | TBCD |
| chr19 | 19447640 | 19447789 | 150 | 2 | 0.000230465 | 2.66E-05 | 0.00067 | 3.67E-05 | 0.010392379 | GATAD2A |
| chr20 | 25281098 | 25281596 | 499 | 4 | 4.21E-06 | 0.010156 | 1.31E-05 | 3.67E-05 | 0.002256694 | PYGB |
| chr1 | 3814737 | 3815271 | 535 | 6 | 0.000116078 | 0.000218 | 0.001436 | 3.67E-05 | 0.000536444 | CEP104 |
| chr13 | 111095590 | 111095827 | 238 | 4 | 0.000276887 | 1.49E-05 | 0.004051 | 3.67E-05 | 0.007981758 | NA |
| chr1 | 2397409 | 2398150 | 742 | 4 | 0.000108864 | 0.000102 | 0.001341 | 3.67E-05 | -0.0040696 | RER1 |
| chr1 | 17610127 | 17610147 | 21 | 2 | 0.000197924 | 2.78E-05 | 0.000569 | 3.67E-05 | 0.010421456 | ARHGEF10L |
| chr1 | 247639437 | 247639864 | 428 | 7 | 0.000122207 | 3.68E-05 | 0.002997 | 3.67E-05 | -0.02035595 | AL390860.1, AL606804.1 |
| chr19 | 51065606 | 51066365 | 760 | 5 | 1.43E-05 | 0.000669 | 0.000123 | 3.68E-05 | 0.008074752 | NA |
| chr19 | 39342870 | 39342969 | 100 | 3 | 3.97E-05 | 9.08E-05 | 0.000676 | 3.68E-05 | 0.004916876 | SAMD4B |
| chr20 | 3092655 | 3092915 | 261 | 2 | 2.71E-05 | 3.42E-05 | 0.00031 | 3.68E-05 | -0.02615366 | NA |
| chr19 | 12766032 | 12766374 | 343 | 4 | 0.000349339 | 1.10E-05 | 0.010382 | 3.68E-05 | 0.12985053 | HOOK2 |
| chr1 | 239804898 | 239805253 | 356 | 2 | 8.37E-05 | 2.35E-05 | 0.001264 | 3.68E-05 | 0.01386671 | CHRM3 |
| chr1 | 9315568 | 9316433 | 866 | 4 | 3.19E-06 | 0.002727 | 0.000315 | 3.68E-05 | 0.009275037 | SPSB1 |
| chr19 | 14127336 | 14127492 | 157 | 2 | 2.09E-05 | 0.003727 | 3.10E-05 | 3.68E-05 | 0.007067446 | ASF1B |
| chr12 | 95549338 | 95549355 | 18 | 2 | 0.000413877 | 3.35E-05 | 0.000328 | 3.68E-05 | -0.02070406 | USP44 |
| chr14 | 60507781 | 60508819 | 1039 | 7 | 4.94E-06 | 2.33E-05 | 0.003104 | 3.69E-05 | 0.005054647 | C14orf39 |
| chr3 | 195850998 | 195851409 | 412 | 8 | 0.000439495 | 5.90E-06 | 0.01614 | 3.69E-05 | -0.01131942 | LINC01983 |
| chr7 | 43869445 | 43869709 | 265 | 7 | 0.000181935 | 4.32E-05 | 0.006629 | 3.69E-05 | 0.001241811 | MRPS24, URGCP-MRPS24 |
| chr6 | 170584857 | 170585841 | 985 | 6 | 7.73E-06 | 0.001548 | 0.000835 | 3.69E-05 | 0.001806362 | NA |
| chr16 | 72093916 | 72094847 | 932 | 16 | 2.77E-05 | 0.000115 | 0.001885 | 3.69E-05 | 0.001400792 | DHX38, TXNL4B |
| chr6 | 348772 | 348879 | 108 | 2 | 8.30E-05 | 5.89E-05 | 0.00013 | 3.70E-05 | 0.002063622 | DUSP22 |
| chr10 | 132296251 | 132296415 | 165 | 2 | 0.000122065 | 0.000102 | 8.28E-05 | 3.70E-05 | 0.017407675 | STK32C |
| chr1 | 50368384 | 50368631 | 248 | 3 | 0.000110845 | 0.000107 | 9.47E-05 | 3.70E-05 | 0.007433385 | NA |
| chr11 | 27521814 | 27522259 | 446 | 3 | 3.05E-05 | 0.000251 | 0.000238 | 3.70E-05 | 0.002428605 | BDNF-AS |
| chr3 | 69387640 | 69387721 | 82 | 2 | 2.93E-05 | 0.001347 | 3.54E-05 | 3.70E-05 | 0.009129977 | FRMD4B |
| chr12 | 131083896 | 131084219 | 324 | 7 | 2.40E-05 | 0.002684 | 0.000832 | 3.70E-05 | 0.002134007 | ADGRD1 |
| chr1 | 16724581 | 16725279 | 699 | 6 | 8.30E-05 | 0.001706 | 0.000178 | 3.70E-05 | 0.003105481 | NA |
| chr16 | 1954675 | 1955061 | 387 | 7 | 4.62E-05 | 0.001222 | 0.000158 | 3.71E-05 | 0.003657018 | RPL3L |
| chr6 | 168244641 | 168245078 | 438 | 3 | 4.62E-05 | 0.00016 | 0.000522 | 3.71E-05 | 0.003167561 | AL606970.3 |
| chr1 | 6006958 | 6007069 | 112 | 2 | 0.000159685 | 2.41E-05 | 0.001142 | 3.71E-05 | 0.004359335 | KCNAB2 |
| chr14 | 55663025 | 55663521 | 497 | 4 | 5.22E-05 | 0.000995 | 6.29E-05 | 3.71E-05 | 0.012141098 | KTN1 |
| chr20 | 46177767 | 46178141 | 375 | 4 | 5.90E-05 | 8.09E-05 | 0.001255 | 3.71E-05 | 0.005390145 | CDH22 |
| chr1 | 107245709 | 107245775 | 67 | 2 | 0.000191965 | 5.66E-05 | 0.000137 | 3.71E-05 | 0.005202747 | NTNG1 |
| chr7 | 342905 | 343274 | 370 | 2 | 9.77E-05 | 4.05E-05 | 0.000223 | 3.71E-05 | -0.00022374 | NA |
| chr1 | 53438646 | 53439496 | 851 | 5 | 9.36E-06 | 1.79E-05 | 0.003879 | 3.71E-05 | 0.002074327 | SLC25A3P1 |
| chr3 | 51501181 | 51501492 | 312 | 3 | 3.15E-05 | 1.85E-05 | 0.002441 | 3.72E-05 | 0.001182602 | NA |
| chr1 | 167714258 | 167714660 | 403 | 4 | 2.82E-06 | 0.000222 | 5.71E-05 | 3.72E-05 | 0.002357843 | NA |
| chr14 | 24440788 | 24440931 | 144 | 2 | 0.000107258 | 2.95E-05 | 0.000489 | 3.73E-05 | 0.003002476 | KHNYN, SDR39U1 |
| chr2 | 115163036 | 115163682 | 647 | 4 | 1.76E-05 | 0.000379 | 0.00014 | 3.73E-05 | 0.012965727 | DPP10 |
| chr12 | 131944766 | 131945127 | 362 | 4 | 1.92E-05 | 0.002025 | 2.13E-05 | 3.73E-05 | 0.003292745 | PUS1 |
| chr11 | 3614373 | 3614551 | 179 | 2 | 0.000214334 | 2.36E-05 | 0.001355 | 3.73E-05 | 0.00516001 | TRPC2 |
| chr16 | 3500026 | 3500926 | 901 | 7 | 2.00E-05 | 0.000198 | 0.000149 | 3.73E-05 | 0.00511141 | AC004224.2 |

| | | | | | | | | | | |
|---|---|---|---|---|---|---|---|---|---|---|
| chr16 | 68842656 | 68842895 | 240 | 3 | 1.18E-05 | 1.98E-05 | 0.001861 | 3.73E-05 | 0.003314669 | NA |
| chr6 | 25926784 | 25926788 | 5 | 2 | 4.74E-05 | 0.00047 | 4.49E-05 | 3.73E-05 | 0.007895796 | SLC17A2 |
| chr19 | 2700765 | 2700929 | 165 | 5 | 0.000184368 | 8.46E-05 | 0.000556 | 3.73E-05 | 0.004266347 | GNG7, AC006538.3 |
| chr2 | 75570996 | 75571106 | 111 | 2 | 0.000165564 | 7.31E-05 | 0.000107 | 3.74E-05 | 0.009503864 | NA |
| chr6 | 7238820 | 7238976 | 157 | 2 | 0.000110163 | 8.98E-05 | 9.15E-05 | 3.74E-05 | 0.005357394 | RREB1 |
| chr14 | 89490348 | 89490473 | 126 | 2 | 0.000211963 | 3.46E-05 | 0.000317 | 3.74E-05 | 0.000291601 | FOXN3, AL137230.2 |
| chr1 | 18362190 | 18362569 | 380 | 2 | 0.000200245 | 2.62E-05 | 0.000781 | 3.74E-05 | 0.004366253 | IGSF21 |
| chr17 | 17416690 | 17416715 | 26 | 2 | 0.000225293 | 2.57E-05 | 0.000859 | 3.74E-05 | 0.00764833 | NA |
| chr16 | 67935464 | 67936081 | 618 | 5 | 1.46E-05 | 0.000378 | 0.000174 | 3.75E-05 | 0.001398086 | AC040162.1, PSMB10 |
| chr9 | 76926343 | 76926695 | 353 | 2 | 9.94E-05 | 6.55E-05 | 0.00012 | 3.75E-05 | 0.009275012 | NA |
| chr17 | 1516755 | 1517255 | 501 | 8 | 0.000160576 | 3.99E-05 | 0.000249 | 3.75E-05 | -0.00021854 | PITPNA-AS1 |
| chr15 | 79317077 | 79317431 | 355 | 2 | 5.05E-05 | 0.000292 | 5.24E-05 | 3.75E-05 | 0.006132417 | TMED3 |
| chr6 | 118923792 | 118924004 | 213 | 2 | 0.000118019 | 0.000124 | 7.53E-05 | 3.75E-05 | 0.002648707 | MCM9 |
| chr1 | 75133847 | 75135435 | 1589 | 13 | 2.41E-06 | 0.005165 | 0.000352 | 3.75E-05 | 0.002061614 | LHX8 |
| chr10 | 92101628 | 92102067 | 440 | 2 | 4.86E-06 | 0.047838 | 2.79E-05 | 3.76E-05 | 0.010028706 | CPEB3 |
| chr10 | 132022318 | 132022423 | 106 | 2 | 5.74E-05 | 2.52E-05 | 0.000969 | 3.76E-05 | 0.008546541 | NA |
| chr17 | 82186036 | 82186377 | 342 | 3 | 5.85E-05 | 0.000309 | 0.000894 | 3.76E-05 | 0.003042516 | CCDC57 |
| chr1 | 231528390 | 231528925 | 536 | 12 | 0.000243625 | 1.67E-05 | 0.020769 | 3.76E-05 | -0.00058022 | TSNAX, TSNAX-DISC1, AL445524.2, AL445524.1 |
| chr9 | 100597378 | 100597586 | 209 | 2 | 7.01E-05 | 0.000242 | 5.59E-05 | 3.76E-05 | 0.012785822 | NA |
| chr16 | 81096386 | 81097197 | 812 | 9 | 0.00017157 | 4.49E-05 | 0.008014 | 3.76E-05 | -0.00017747 | GCSH |
| chr15 | 69325450 | 69325452 | 3 | 2 | 0.000102774 | 2.37E-05 | 0.001379 | 3.76E-05 | 0.004240323 | PAQR5 |
| chr17 | 28334348 | 28335363 | 1016 | 7 | 1.90E-05 | 0.012139 | 0.00014 | 3.76E-05 | 0.00113707 | IFT20 |
| chr6 | 32396098 | 32396298 | 201 | 4 | 3.95E-05 | 0.00022 | 0.000837 | 3.77E-05 | 0.002400503 | TSBP1-AS1, BTNL2 |
| chr3 | 125520183 | 125520468 | 286 | 7 | 0.000114745 | 0.000146 | 0.000647 | 3.77E-05 | -0.00047594 | SNX4 |
| chr20 | 35715014 | 35715018 | 5 | 2 | 0.000158644 | 5.55E-05 | 0.000143 | 3.77E-05 | 0.001947824 | RBM39 |
| chr17 | 6435156 | 6436199 | 1044 | 10 | 1.11E-05 | 4.16E-05 | 0.002585 | 3.77E-05 | 0.001658891 | AIPL1 |
| chr12 | 53321217 | 53321808 | 592 | 8 | 8.96E-05 | 0.000191 | 0.000357 | 3.77E-05 | 0.002536835 | AAAS |
| chr16 | 89817580 | 89818151 | 572 | 7 | 0.000125177 | 0.000174 | 0.000755 | 3.77E-05 | 0.001459601 | NA |
| chr19 | 17999751 | 17999867 | 117 | 2 | 5.43E-05 | 2.39E-05 | 0.001341 | 3.77E-05 | 0.005067451 | KCNN1 |
| chr11 | 69005242 | 69005493 | 252 | 3 | 5.85E-05 | 1.98E-05 | 0.002895 | 3.77E-05 | 0.00405473 | MRGPRF |
| chr12 | 132186517 | 132187342 | 826 | 5 | 5.46E-05 | 9.63E-06 | 0.013978 | 3.77E-05 | 0.022514897 | LINC02361 |
| chr6 | 1424369 | 1424442 | 74 | 2 | 7.80E-05 | 0.005725 | 3.05E-05 | 3.77E-05 | 0.011461149 | NA |
| chr2 | 98518204 | 98519008 | 805 | 5 | 7.23E-05 | 7.81E-05 | 0.001222 | 3.77E-05 | 0.002662228 | INPP4A |
| chr15 | 76190940 | 76191296 | 357 | 3 | 7.89E-05 | 0.000526 | 0.000728 | 3.77E-05 | 0.011578838 | TMEM266 |
| chr17 | 82235576 | 82236182 | 607 | 7 | 6.04E-05 | 2.31E-05 | 0.016423 | 3.77E-05 | 0.002531281 | SLC16A3 |
| chr3 | 101732584 | 101732595 | 12 | 3 | 0.000167405 | 6.67E-05 | 0.000147 | 3.78E-05 | -0.00169697 | CEP97 |
| chr1 | 203722566 | 203722793 | 228 | 3 | 0.000145099 | 7.38E-05 | 0.001227 | 3.78E-05 | -0.00089708 | ATP2B4 |
| chr7 | 113086341 | 113087159 | 819 | 11 | 4.20E-05 | 0.001275 | 0.000307 | 3.78E-05 | 0.003962246 | GPR85 |
| chr6 | 33423130 | 33423564 | 435 | 9 | 2.45E-05 | 0.006284 | 0.000111 | 3.78E-05 | -0.0002784 | SYNGAP1 |
| chr5 | 168162687 | 168163021 | 335 | 2 | 0.000188935 | 2.99E-05 | 0.000499 | 3.78E-05 | 0.00683961 | TENM2, AC011369.1 |
| chr13 | 36447521 | 36447735 | 215 | 3 | 1.99E-05 | 0.176186 | 1.11E-05 | 3.79E-05 | 0.00514229 | NA |
| chr10 | 132771156 | 132771617 | 462 | 2 | 0.000206079 | 2.63E-05 | 0.00083 | 3.79E-05 | -0.00340322 | INPP5A |
| chr10 | 131945335 | 131945383 | 49 | 2 | 0.000209231 | 2.47E-05 | 0.001151 | 3.79E-05 | 0.003727052 | PPP2R2D |
| chr3 | 87547040 | 87547225 | 186 | 2 | 0.00011078 | 6.49E-05 | 0.000123 | 3.79E-05 | 0.01524531 | NA |
| chr17 | 15310368 | 15310845 | 478 | 2 | 0.000114947 | 2.46E-05 | 0.001179 | 3.79E-05 | 0.01060183 | AC005703.6, TEKT3 |
| chr2 | 105881839 | 105882057 | 219 | 2 | 0.000124287 | 0.000211 | 5.96E-05 | 3.79E-05 | 0.004590023 | NCK2, AC009505.1 |
| chr1 | 6045704 | 6046479 | 776 | 12 | 0.000201536 | 6.31E-05 | 0.00308 | 3.79E-05 | 0.000268241 | KCNAB2 |
| chr10 | 67885168 | 67885254 | 87 | 3 | 2.85E-05 | 6.69E-05 | 0.000254 | 3.79E-05 | 0.000156526 | SIRT1 |
| chr10 | 45453128 | 45453280 | 153 | 3 | 0.00023515 | 5.10E-05 | 0.000213 | 3.80E-05 | 0.009466832 | NA |
| chr10 | 27414318 | 27414618 | 301 | 5 | 0.000311511 | 2.56E-05 | 0.008226 | 3.80E-05 | -0.01869932 | PTCHD3 |
| chr6 | 33167976 | 33168734 | 759 | 7 | 5.35E-05 | 5.22E-05 | 0.003086 | 3.80E-05 | -0.00100273 | COL11A2 |
| chr16 | 28931269 | 28932356 | 1088 | 8 | 3.79E-05 | 0.000296 | 0.002231 | 3.80E-05 | 0.00282889 | AC009093.11, CD19, RABEP2 |
| chr19 | 55659141 | 55659251 | 111 | 3 | 0.000239671 | 2.36E-05 | 0.001307 | 3.80E-05 | 0.003764496 | U2AF2 |
| chr8 | 144515923 | 144516607 | 685 | 4 | 0.00025495 | 3.20E-05 | 0.003928 | 3.80E-05 | 0.000977998 | RECQL4 |
| chr15 | 75451412 | 75451921 | 510 | 9 | 0.00011711 | 2.42E-05 | 0.007099 | 3.80E-05 | -3.61E-05 | SIN3A |
| chr11 | 82901501 | 82902108 | 608 | 11 | 0.000154913 | 0.000273 | 0.003837 | 3.81E-05 | -0.00029717 | DDIAS, PRCP |
| chr11 | 131371391 | 131371893 | 503 | 2 | 3.34E-05 | 8.12E-05 | 0.000101 | 3.81E-05 | 0.021166667 | NTM |
| chr5 | 141186288 | 141186465 | 178 | 2 | 6.47E-05 | 2.51E-05 | 0.001079 | 3.81E-05 | 0.022995697 | PCDHB16, AC244517.6, AC244517.11, AC244517.4 |
| chr1 | 160032137 | 160032908 | 772 | 7 | 1.73E-05 | 3.44E-05 | 0.001543 | 3.81E-05 | 0.000328352 | KCNJ10 |
| chr2 | 219027639 | 219027794 | 156 | 2 | 0.000151057 | 9.50E-05 | 9.01E-05 | 3.81E-05 | 0.00727252 | CFAP65 |
| chr4 | 56505202 | 56505761 | 560 | 7 | 0.000129141 | 3.34E-05 | 0.003501 | 3.82E-05 | -0.00604549 | ARL9 |
| chr7 | 132830602 | 132830805 | 204 | 2 | 4.67E-05 | 0.001428 | 3.61E-05 | 3.82E-05 | 0.007786482 | AC009365.3, CHCHD3 |
| chr8 | 1500584 | 1501331 | 748 | 6 | 7.60E-05 | 5.84E-05 | 0.003374 | 3.82E-05 | 0.002619071 | DLGAP2 |
| chr10 | 124523033 | 124523445 | 413 | 5 | 0.000214212 | 2.62E-05 | 0.006625 | 3.82E-05 | 0.007439246 | LHPP |
| chr21 | 17611688 | 17611976 | 289 | 3 | 6.46E-05 | 2.37E-05 | 0.002583 | 3.82E-05 | 0.000781536 | BTG3-AS1, BTG3 |
| chr9 | 130768594 | 130768644 | 51 | 2 | 0.00022905 | 2.53E-05 | 0.001048 | 3.82E-05 | 0.011810848 | ABL1 |
| chr1 | 1373781 | 1374097 | 317 | 3 | 1.01E-05 | 9.20E-05 | 0.000147 | 3.82E-05 | 0.00603087 | AURKAIP1 |
| chr8 | 81281931 | 81282488 | 558 | 3 | 7.28E-05 | 4.08E-05 | 0.000321 | 3.83E-05 | -0.00929425 | FABP5 |
| chr1 | 77888697 | 77889654 | 958 | 8 | 4.73E-06 | 4.07E-05 | 0.001216 | 3.83E-05 | 0.00170847 | NEXN, NEXN-AS1 |
| chr20 | 3081303 | 3081698 | 396 | 2 | 0.000225037 | 2.43E-05 | 0.001367 | 3.83E-05 | 0.011605118 | NA |
| chr5 | 177302761 | 177302811 | 51 | 2 | 0.000104171 | 6.43E-05 | 0.000126 | 3.83E-05 | 0.013456706 | RAB24, MXD3 |
| chr12 | 50134833 | 50134971 | 139 | 2 | 2.76E-05 | 8.13E-05 | 0.000102 | 3.83E-05 | 0.007842052 | AC074032.1, CERS5 |
| chr12 | 57622843 | 57623091 | 249 | 2 | 3.00E-05 | 4.03E-05 | 0.000247 | 3.84E-05 | 0.009369801 | AC025165.6, SLC26A10 |
| chr11 | 4638359 | 4638581 | 223 | 2 | 5.87E-05 | 0.003623 | 3.21E-05 | 3.84E-05 | 0.004152915 | OR51D1 |
| chr1 | 11067837 | 11068316 | 480 | 5 | 6.23E-07 | 0.042831 | 5.21E-06 | 3.84E-05 | 0.000816767 | EXOSC10 |
| chr5 | 180136497 | 180137268 | 772 | 6 | 0.000129342 | 0.000105 | 0.000233 | 3.84E-05 | 0.000612749 | RASGEF1C |
| chr16 | 67884392 | 67884783 | 392 | 4 | 0.000338941 | 0.000109 | 0.001626 | 3.84E-05 | 0.003000476 | EDC4, AC040162.3 |
| chr5 | 94111249 | 94112121 | 873 | 12 | 7.83E-05 | 0.000158 | 0.008223 | 3.85E-05 | 0.001058268 | AC117528.1, FAM172A |
| chr10 | 114571383 | 114571390 | 8 | 2 | 0.000246718 | 3.55E-05 | 0.000328 | 3.85E-05 | 0.004743571 | ABLIM1 |
| chr17 | 35241579 | 35241772 | 194 | 2 | 2.22E-05 | 6.43E-05 | 0.000127 | 3.85E-05 | 0.012609671 | AC022916.1 |
| chr11 | 70428854 | 70429416 | 563 | 3 | 9.00E-05 | 6.26E-05 | 0.000179 | 3.85E-05 | 0.002666615 | CTTN |
| chr3 | 38109619 | 38109634 | 16 | 2 | 0.000100912 | 3.67E-05 | 0.000305 | 3.85E-05 | 0.007279667 | DLEC1, ACAA1 |
| chr2 | 32948096 | 32948234 | 139 | 2 | 0.000157932 | 3.23E-05 | 0.000421 | 3.85E-05 | 0.013579997 | LTBP1 |
| chr20 | 46153400 | 46153487 | 88 | 2 | 0.000196159 | 2.48E-05 | 0.001249 | 3.85E-05 | -0.04704976 | NA |
| chr22 | 35481221 | 35481544 | 324 | 2 | 7.03E-05 | 0.00018 | 6.48E-05 | 3.85E-05 | 0.011243959 | NA |
| chr13 | 99303877 | 99303929 | 53 | 2 | 0.000223806 | 3.59E-05 | 0.000323 | 3.86E-05 | 0.017577083 | UBAC2, GPR183 |
| chr16 | 56300325 | 56300572 | 248 | 4 | 0.000147202 | 8.32E-05 | 0.00014 | 3.86E-05 | 0.0067223 | GNAO1 |
| chr4 | 170273808 | 170273863 | 56 | 2 | 0.000393683 | 2.77E-05 | 0.000726 | 3.86E-05 | 0.00131272 | LINC01612 |
| chr2 | 148644799 | 148644879 | 81 | 2 | 0.000379351 | 0.000397 | 4.87E-05 | 3.86E-05 | 0.002817303 | EPC2 |
| chr6 | 45421798 | 45422547 | 750 | 13 | 0.000136077 | 5.45E-05 | 0.003793 | 3.86E-05 | 0.000425714 | RUNX2, AL096865.1 |
| chr6 | 73461214 | 73462019 | 806 | 12 | 7.40E-05 | 0.007118 | 0.001532 | 3.87E-05 | -0.00115572 | MTO1 |
| chr12 | 25195039 | 25195892 | 854 | 12 | 2.45E-05 | 0.003677 | 0.000117 | 3.87E-05 | -0.00190732 | ETFRF1, CFAP94 |
| chr10 | 102433606 | 102433632 | 27 | 2 | 1.99E-05 | 3.16E-05 | 0.000458 | 3.87E-05 | 0.014008989 | NA |
| chr20 | 56358465 | 56359054 | 590 | 8 | 9.69E-05 | 0.001703 | 0.000848 | 3.87E-05 | -0.00095878 | FAM210B |
| chr13 | 113623878 | 113624111 | 234 | 2 | 9.93E-05 | 3.67E-05 | 0.00031 | 3.87E-05 | 0.007170294 | TFDP1 |
| chr10 | 71773547 | 71774529 | 983 | 9 | 4.06E-05 | 0.000466 | 0.004163 | 3.88E-05 | 0.001224837 | CDH23 |
| chr8 | 11691664 | 11692492 | 829 | 5 | 1.01E-05 | 0.006983 | 4.37E-05 | 3.88E-05 | 0.006381585 | GATA4 |
| chr4 | 1640327 | 1640530 | 204 | 3 | 5.19E-05 | 0.003357 | 2.47E-05 | 3.88E-05 | 0.005331884 | FAM53A |
| chr1 | 36482712 | 36483380 | 669 | 4 | 0.000161119 | 1.82E-05 | 0.004311 | 3.88E-05 | 0.010225074 | CSF3R |
| chr17 | 73340251 | 73340638 | 388 | 3 | 0.000208126 | 2.11E-05 | 0.002346 | 3.88E-05 | 0.007397039 | SDK2 |
| chr20 | 48638645 | 48638716 | 72 | 2 | 0.000282334 | 3.82E-05 | 0.000285 | 3.88E-05 | 0.01212476 | PREX1 |
| chr6 | 151615991 | 151616518 | 528 | 4 | 7.66E-05 | 0.000305 | 6.03E-05 | 3.88E-05 | 0.008623684 | CCDC170 |
| chr6 | 136784087 | 136784111 | 25 | 2 | 0.000193244 | 3.91E-05 | 0.000272 | 3.88E-05 | 0.009300194 | AL121933.2, MAP3K5 |
| chr7 | 108180903 | 108181034 | 132 | 2 | 0.000230576 | 3.39E-05 | 0.000382 | 3.88E-05 | 0.018497952 | NRCAM |
| chr20 | 47236391 | 47236479 | 89 | 2 | 8.25E-05 | 3.84E-05 | 0.000283 | 3.88E-05 | 0.000799108 | ZMYND8 |
| chr15 | 83208110 | 83208562 | 453 | 7 | 3.06E-05 | 0.003199 | 5.60E-05 | 3.89E-05 | -0.00255328 | AC103876.1 |
| chr11 | 44574204 | 44574305 | 102 | 2 | 0.000117379 | 2.73E-05 | 0.000805 | 3.89E-05 | 0.018554388 | CD82 |
| chr17 | 82983596 | 82983682 | 87 | 3 | 9.82E-05 | 0.000864 | 3.80E-05 | 3.89E-05 | 0.004959705 | B3GNTL1 |
| chr17 | 81688133 | 81688439 | 307 | 3 | 4.64E-05 | 0.003187 | 0.000106 | 3.89E-05 | 0.000282019 | HGS |
| chr12 | 123115326 | 123115866 | 541 | 3 | 0.000141253 | 1.83E-05 | 0.003749 | 3.89E-05 | 0.011860799 | PITPNM2 |
| chr19 | 1218483 | 1219345 | 863 | 3 | 9.22E-06 | 0.002359 | 3.39E-05 | 3.89E-05 | 0.004166275 | STK11 |
| chr12 | 10723275 | 10723927 | 653 | 9 | 0.000125 | 0.000203 | 0.004033 | 3.89E-05 | 0.000688704 | YBX3 |
| chr20 | 46308578 | 46309070 | 493 | 6 | 0.000197471 | 1.92E-05 | 0.001767 | 3.89E-05 | -0.00032902 | NA |
| chr4 | 984935 | 985543 | 609 | 6 | 6.56E-05 | 0.00074 | 0.001087 | 3.89E-05 | 0.011217224 | DGKQ, SLC26A1 |
| chr1 | 203076800 | 203076842 | 43 | 2 | 7.59E-05 | 7.46E-05 | 0.000113 | 3.90E-05 | 0.009669706 | PPFIA4 |
| chr11 | 2991080 | 2991649 | 570 | 6 | 1.37E-05 | 0.000282 | 0.001876 | 3.90E-05 | 0.001691207 | AC131971.1, NAP1L4 |
| chr1 | 201000545 | 201000778 | 234 | 4 | 0.000161856 | 9.02E-05 | 0.001308 | 3.90E-05 | 0.000834785 | KIF21B |
| chr11 | 114082528 | 114083116 | 589 | 5 | 0.000162158 | 4.27E-05 | 0.001387 | 3.91E-05 | 0.008012148 | ZBTB16 |
| chr16 | 29744367 | 29744414 | 48 | 2 | 0.000185879 | 4.16E-05 | 0.000244 | 3.91E-05 | 0.009295509 | C16orf54 |
| chr12 | 53421633 | 53421901 | 269 | 3 | 0.000215374 | 1.99E-05 | 0.002825 | 3.91E-05 | 0.004476241 | NA |
| chr14 | 104173243 | 104173862 | 620 | 4 | 4.18E-05 | 0.000826 | 5.99E-05 | 3.91E-05 | 0.002682594 | KIF26A |
| chr11 | 2916173 | 2916804 | 632 | 5 | 6.85E-05 | 6.74E-05 | 0.000825 | 3.92E-05 | 0.001676276 | SLC22A18 |
| chr1 | 240765507 | 240765948 | 442 | 2 | 8.69E-05 | 3.96E-05 | 0.00027 | 3.92E-05 | 0.007582899 | AL365184.1 |
| chr5 | 79512371 | 79513187 | 817 | 6 | 0.000110845 | 0.000599 | 0.001355 | 3.92E-05 | 0.005673345 | HOMER1 |
| chr7 | 23897352 | 23897714 | 363 | 3 | 0.000115638 | 8.18E-05 | 0.000129 | 3.92E-05 | 0.003279635 | NA |
| chr13 | 94468872 | 94468986 | 115 | 2 | 0.000269626 | 2.50E-05 | 0.001322 | 3.92E-05 | -0.00466201 | DCT |
| chr16 | 25692509 | 25692579 | 71 | 2 | 2.56E-05 | 3.51E-05 | 0.000358 | 3.92E-05 | -0.00010683 | HS3ST4 |
| chr2 | 45611890 | 45611996 | 107 | 2 | 0.000155299 | 2.98E-05 | 0.000586 | 3.92E-05 | 0.003980721 | SRBD1 |
| chr19 | 11381163 | 11381969 | 807 | 5 | 3.19E-05 | 0.000116 | 0.000178 | 3.92E-05 | 0.003460533 | EPOR |
| chr17 | 7383781 | 7384682 | 902 | 8 | 5.73E-05 | 0.000352 | 0.000908 | 3.92E-05 | 0.001218734 | TNK1 |
| chr10 | 123144610 | 123144674 | 65 | 2 | 8.84E-05 | 2.59E-05 | 0.001107 | 3.92E-05 | 0.013173066 | NA |
| chr22 | 46963744 | 46963965 | 222 | 2 | 0.000106875 | 9.15E-05 | 9.63E-05 | 3.92E-05 | 0.010938804 | TBC1D22A |
| chr14 | 101917671 | 101917740 | 70 | 2 | 0.000198583 | 3.49E-05 | 0.000367 | 3.92E-05 | 0.001371602 | PPP2R5C |
| chr3 | 32816788 | 32817168 | 381 | 3 | 2.06E-05 | 0.000119 | 0.000166 | 3.93E-05 | -0.00172548 | NA |
| chr15 | 79310531 | 79310937 | 407 | 5 | 0.000226834 | 3.52E-05 | 0.00309 | 3.93E-05 | 0.003806167 | NA |
| chr7 | 129214044 | 129214284 | 241 | 2 | 0.000209728 | 2.53E-05 | 0.001268 | 3.93E-05 | 0.012481455 | NA |
| chr1 | 28315476 | 28315813 | 338 | 2 | 5.35E-05 | 5.84E-05 | 0.000146 | 3.93E-05 | 0.01787461 | NA |
| chr1 | 220878681 | 220879786 | 1106 | 11 | 7.33E-05 | 3.79E-05 | 0.021082 | 3.93E-05 | -0.00096516 | AL445423.3, HLX, HLX-AS1 |
| chr3 | 141381110 | 141381631 | 522 | 3 | 0.00011656 | 0.00013 | 0.000109 | 3.93E-05 | 0.002405305 | ZBTB38 |
| chr13 | 50082218 | 50082821 | 604 | 7 | 0.000102204 | 0.000171 | 0.007967 | 3.93E-05 | 0.000709772 | DLEU1, AL137060.6, DLEU2 |
| chr17 | 40015202 | 40015444 | 243 | 5 | 8.48E-05 | 1.46E-05 | 0.007167 | 3.94E-05 | 0.008767967 | CSF3 |
| chr19 | 5680603 | 5680964 | 362 | 11 | 0.000343602 | 0.000212 | 0.003135 | 3.94E-05 | 0.000396924 | RPL36, HSD11B1L |
| chr10 | 507517 | 507544 | 28 | 2 | 0.000191024 | 2.67E-05 | 0.000957 | 3.94E-05 | -0.00511144 | DIP2C |
| chr1 | 207751759 | 207752655 | 897 | 11 | 4.01E-05 | 0.000418 | 0.000389 | 3.95E-05 | 0.000488437 | CD46 |
| chr1 | 167601474 | 167601725 | 252 | 3 | 0.000204501 | 2.31E-05 | 0.002682 | 3.95E-05 | 0.007251626 | NA |
| chr15 | 89524813 | 89525265 | 453 | 3 | 6.34E-05 | 0.00024 | 0.000815 | 3.95E-05 | 0.006950002 | AC013391.2 |
| chr7 | 38178012 | 38178939 | 928 | 12 | 4.26E-05 | 0.003226 | 0.001359 | 3.96E-05 | -0.00039174 | STARD3NL |
| chr4 | 128287570 | 128287940 | 371 | 4 | 0.000184856 | 3.35E-05 | 0.003962 | 3.96E-05 | 0.003692607 | PGRMC2 |
| chr19 | 10976041 | 10976279 | 239 | 4 | 0.000140351 | 8.06E-05 | 0.000166 | 3.96E-05 | 0.003081302 | SMARCA4 |
| chr11 | 74694223 | 74694451 | 229 | 3 | 0.000107988 | 0.000269 | 0.000102 | 3.96E-05 | 0.0103318 | NA |
| chr15 | 43220857 | 43221365 | 509 | 4 | 0.000333019 | 0.00037 | 0.00271 | 3.96E-05 | 0.007650239 | EPB42 |
| chr6 | 1516378 | 1516408 | 31 | 2 | 5.61E-05 | 0.000233 | 5.99E-05 | 3.96E-05 | 0.004844262 | NA |
| chr4 | 154332791 | 154333174 | 384 | 5 | 0.000346891 | 2.74E-05 | 0.000831 | 3.97E-05 | -9.96E-05 | DCHS2 |
| chr16 | 87957516 | 87957989 | 474 | 5 | 1.70E-05 | 0.001314 | 7.70E-05 | 3.97E-05 | 0.003176512 | BANP |
| chr3 | 11633233 | 11634186 | 954 | 5 | 7.36E-05 | 0.000146 | 0.000463 | 3.97E-05 | 0.007908388 | VGLL4 |
| chr3 | 13022418 | 13022501 | 84 | 2 | 0.000187816 | 3.09E-05 | 0.000548 | 3.97E-05 | 0.007156146 | IQSEC1 |
| chr12 | 103659006 | 103659401 | 396 | 2 | 3.18E-05 | 0.000142 | 7.54E-05 | 3.97E-05 | 0.019276954 | STAB2 |
| chr16 | 75064648 | 75064678 | 31 | 2 | 0.000113368 | 0.000168 | 6.94E-05 | 3.98E-05 | 0.003810991 | ZNRF1 |
| chr6 | 137878934 | 137879542 | 609 | 9 | 0.000157206 | 3.23E-05 | 0.011259 | 3.98E-05 | 0.002708304 | TNFAIP3 |
| chr15 | 80777996 | 80778533 | 538 | 4 | 0.000110208 | 6.02E-05 | 0.001393 | 3.99E-05 | 0.006482121 | NA |
| chr4 | 185761541 | 185761657 | 117 | 2 | 4.10E-05 | 0.006416 | 3.16E-05 | 3.99E-05 | 0.00279077 | SORBS2 |
| chr20 | 62927269 | 62928404 | 1136 | 5 | 0.00013915 | 0.002482 | 0.001225 | 3.99E-05 | 0.005725841 | DIDO1 |

| | | | | | | | | | | |
|---|---|---|---|---|---|---|---|---|---|---|
| chr13 | 102799869 | 102800439 | 571 | 7 | 3.47E-06 | 0.001331 | 0.000434 | 4.00E-05 | 0.000373838 | BIVM-ERCC5, BIVM |
| chr9 | 5832985 | 5833864 | 880 | 10 | 7.17E-05 | 0.000595 | 0.000285 | 4.00E-05 | 0.002412398 | ERMP1 |
| chr9 | 34457131 | 34457900 | 770 | 5 | 3.99E-05 | 0.000228 | 0.000721 | 4.00E-05 | 0.005248079 | DNAI1, FAM219A |
| chr12 | 132403703 | 132404100 | 398 | 4 | 4.39E-05 | 9.06E-05 | 0.00027 | 4.00E-05 | 0.007310781 | NA |
| chr3 | 128710077 | 128710162 | 86 | 2 | 0.000253026 | 2.80E-05 | 0.000836 | 4.00E-05 | 0.013372995 | RAB7A |
| chr20 | 62310214 | 62311021 | 808 | 4 | 2.13E-06 | 0.000219 | 6.91E-05 | 4.00E-05 | 0.005328012 | LAMA5 |
| chr22 | 46125692 | 46126201 | 510 | 3 | 0.000181452 | 3.60E-05 | 0.001493 | 4.01E-05 | 0.005009713 | NA |
| chr11 | 19299736 | 19299885 | 150 | 2 | 0.000111473 | 2.66E-05 | 0.001089 | 4.01E-05 | 0.0039022 | AC009652.1 |
| chr4 | 16075897 | 16076362 | 466 | 6 | 0.000354019 | 0.000419 | 0.002737 | 4.01E-05 | 0.00467643 | PROM1 |
| chr7 | 100093504 | 100094319 | 816 | 14 | 6.90E-05 | 0.001124 | 0.004305 | 4.01E-05 | 0.00040857 | MCM7, MIR25, MIR93, MIR106B |
| chr6 | 169158801 | 169159621 | 821 | 7 | 9.85E-05 | 4.27E-05 | 0.003949 | 4.01E-05 | -0.00154996 | LINC01615 |
| chr5 | 141155309 | 141155811 | 503 | 3 | 9.81E-05 | 0.000131 | 9.03E-05 | 4.01E-05 | -0.01874706 | AC244517.2, AC244517.11 |
| chr14 | 96739615 | 96739912 | 298 | 4 | 0.000138957 | 9.49E-05 | 0.000207 | 4.01E-05 | 0.0134104 | LINC02299 |
| chr8 | 85438660 | 85438966 | 307 | 6 | 3.88E-05 | 0.000741 | 0.000436 | 4.01E-05 | 0.006116548 | CA3 |
| chr17 | 49414418 | 49415358 | 941 | 14 | 5.47E-05 | 2.45E-05 | 0.003431 | 4.01E-05 | 0.001076987 | AC091180.5, PHB |
| chr18 | 26226225 | 26226467 | 243 | 7 | 5.67E-05 | 0.000569 | 0.000153 | 4.02E-05 | 0.002485296 | TAF4B |
| chr6 | 15595556 | 15595766 | 211 | 3 | 0.000127933 | 0.000525 | 8.82E-05 | 4.02E-05 | -0.0042189 | DTNBP1 |
| chr11 | 119680700 | 119681418 | 719 | 4 | 0.000164961 | 1.52E-05 | 0.00612 | 4.02E-05 | 0.008959463 | NECTIN1 |
| chr6 | 37169295 | 37169939 | 645 | 4 | 2.07E-05 | 9.02E-05 | 0.000248 | 4.02E-05 | -0.00029349 | NA |
| chr11 | 22192840 | 22194155 | 1316 | 10 | 6.32E-05 | 0.050367 | 6.91E-05 | 4.02E-05 | 0.001084487 | ANO5 |
| chr16 | 24844783 | 24844841 | 59 | 2 | 2.63E-05 | 6.58E-05 | 0.000134 | 4.02E-05 | 0.003043707 | NA |
| chr6 | 30041403 | 30041968 | 566 | 2 | 4.29E-05 | 2.64E-05 | 0.001163 | 4.03E-05 | -0.01129936 | ZNRD1ASP |
| chr18 | 22169415 | 22170078 | 664 | 6 | 0.000104344 | 0.000673 | 5.41E-05 | 4.03E-05 | 0.002323182 | GATA6, GATA6-AS1 |
| chr16 | 86700570 | 86700617 | 48 | 2 | 5.20E-05 | 0.003785 | 3.35E-05 | 4.04E-05 | 0.003116643 | NA |
| chr11 | 77027786 | 77027845 | 60 | 2 | 0.00021455 | 4.02E-05 | 0.000285 | 4.04E-05 | -0.00831451 | NA |
| chr12 | 66495875 | 66495950 | 76 | 2 | 0.000224869 | 2.53E-05 | 0.001569 | 4.04E-05 | 0.005101516 | GRIP1 |
| chr6 | 34236682 | 34237136 | 455 | 8 | 0.000220983 | 3.78E-05 | 0.008216 | 4.04E-05 | -0.00053721 | HMGA1 |
| chr2 | 26730899 | 26731188 | 290 | 2 | 0.00012661 | 3.39E-05 | 0.00044 | 4.04E-05 | 0.012907237 | KCNK3 |
| chr16 | 10180207 | 10180775 | 569 | 4 | 0.000176049 | 9.31E-05 | 0.001165 | 4.04E-05 | -0.00764934 | GRIN2A |
| chr10 | 43606048 | 43606779 | 732 | 9 | 4.61E-05 | 0.001267 | 2.53E-05 | 4.05E-05 | -0.0009807 | ZNF485, CAP1P2 |
| chr14 | 77962064 | 77962488 | 425 | 2 | 0.000260049 | 2.58E-05 | 0.001377 | 4.05E-05 | 0.006961331 | NA |
| chr16 | 2072295 | 2072884 | 590 | 4 | 6.08E-05 | 8.01E-05 | 0.001294 | 4.05E-05 | -0.00132869 | TSC2 |
| chr2 | 236264321 | 236264478 | 158 | 5 | 0.000208744 | 3.96E-05 | 0.004608 | 4.05E-05 | 0.008647952 | AC019068.1, ASB18 |
| chr6 | 38599216 | 38599565 | 350 | 3 | 0.000139849 | 5.79E-05 | 0.000604 | 4.05E-05 | 0.003097987 | BTBD9 |
| chr1 | 27872201 | 27872598 | 398 | 7 | 0.000283196 | 7.68E-05 | 0.005252 | 4.05E-05 | 0.000707677 | THEMIS2 |
| chr8 | 27885762 | 27885957 | 196 | 2 | 0.000144634 | 0.000113 | 8.69E-05 | 4.05E-05 | 0.003192525 | SCARA5 |
| chr3 | 191283531 | 191283624 | 94 | 2 | 0.000228063 | 2.56E-05 | 0.001474 | 4.05E-05 | 0.016568384 | UTS2B |
| chr5 | 134370966 | 134371489 | 524 | 10 | 0.000113818 | 6.88E-05 | 0.001636 | 4.05E-05 | -0.00100699 | UBE2B, AC104109.4, CDKL3 |
| chr17 | 2266228 | 2266695 | 468 | 7 | 0.000340222 | 6.62E-05 | 0.00496 | 4.05E-05 | 0.009042054 | SMG6 |
| chr11 | 102268611 | 102269050 | 440 | 4 | 7.45E-05 | 0.000763 | 0.000476 | 4.06E-05 | 0.008555937 | NA |
| chr2 | 1237918 | 1237980 | 63 | 2 | 8.67E-05 | 0.006201 | 3.20E-05 | 4.06E-05 | 0.00426346 | SNTG2 |
| chr20 | 62413946 | 62414302 | 357 | 3 | 0.000127421 | 6.93E-05 | 0.000595 | 4.06E-05 | 0.006161403 | RBBP8NL |
| chr1 | 2456634 | 2457212 | 579 | 2 | 2.38E-05 | 2.86E-05 | 0.000815 | 4.06E-05 | 0.008562345 | PLCH2 |
| chr16 | 977022 | 977604 | 583 | 3 | 5.93E-05 | 5.03E-05 | 0.000387 | 4.06E-05 | 0.01296909 | LMF1, CEROX1 |
| chr16 | 57775095 | 57775341 | 247 | 4 | 0.000106934 | 0.000195 | 7.75E-05 | 4.06E-05 | 0.005667674 | KIFC3 |
| chr11 | 72699080 | 72699320 | 241 | 2 | 3.33E-05 | 0.000831 | 4.26E-05 | 4.07E-05 | 0.003841912 | ARAP1 |
| chr7 | 70646194 | 70646427 | 234 | 2 | 0.00022271 | 3.35E-05 | 0.000465 | 4.07E-05 | -0.00437692 | AUTS2 |
| chr2 | 237414037 | 237414784 | 748 | 8 | 6.45E-05 | 0.001237 | 0.000252 | 4.07E-05 | 0.004002056 | COL6A3 |
| chr18 | 31071671 | 31071710 | 40 | 3 | 5.99E-05 | 0.005023 | 3.82E-05 | 4.08E-05 | 0.007014271 | DSC2 |
| chr3 | 134484546 | 134484947 | 402 | 4 | 2.92E-05 | 4.05E-05 | 0.002201 | 4.08E-05 | 0.000225056 | ANAPC13 |
| chr13 | 112062309 | 112062414 | 106 | 2 | 0.000209596 | 5.58E-05 | 0.000166 | 4.08E-05 | 0.011283074 | SOX1-OT |
| chr10 | 123891854 | 123892210 | 357 | 8 | 0.000127537 | 0.000228 | 0.007337 | 4.09E-05 | -0.00754812 | CPXM2 |
| chr3 | 4740709 | 4741101 | 393 | 2 | 9.73E-05 | 3.15E-05 | 0.000577 | 4.09E-05 | 0.014728934 | ITPR1 |
| chr6 | 43038880 | 43039089 | 210 | 2 | 0.000129321 | 0.000124 | 8.38E-05 | 4.09E-05 | 0.003712496 | CUL7 |
| chr6 | 26224164 | 26224440 | 277 | 3 | 0.000101 | 0.004395 | 6.48E-05 | 4.09E-05 | -0.00674912 | H3C6 |
| chr3 | 113403816 | 113403946 | 131 | 3 | 0.000217822 | 4.16E-05 | 0.000863 | 4.09E-05 | 0.004327441 | CFAP44, AC112128.1 |
| chr5 | 53035762 | 53036387 | 626 | 3 | 2.01E-05 | 0.010385 | 3.19E-05 | 4.10E-05 | 0.016080003 | ITGA2, AC008966.3 |
| chr15 | 74826086 | 74826677 | 592 | 10 | 0.000153625 | 0.00028 | 0.000599 | 4.10E-05 | 0.00374803 | AC091230.1, CPLX3 |
| chr6 | 28765810 | 28765918 | 109 | 4 | 0.000262243 | 6.34E-05 | 0.001915 | 4.10E-05 | 0.010819666 | NA |
| chr9 | 88578260 | 88578553 | 294 | 3 | 3.75E-05 | 0.001073 | 0.000332 | 4.10E-05 | 5.20E-05 | NXNL2 |
| chr16 | 68023045 | 68023563 | 519 | 10 | 4.45E-05 | 0.002283 | 0.00023 | 4.10E-05 | 0.000319059 | DUS2, DDX28 |
| chr5 | 128201432 | 128201603 | 172 | 3 | 0.000162344 | 8.00E-05 | 0.000169 | 4.10E-05 | -0.00299269 | NA |
| chr11 | 128905618 | 128906321 | 704 | 10 | 1.51E-05 | 0.005263 | 5.29E-05 | 4.10E-05 | 0.000825305 | KCNJ5, C11orf45 |
| chr15 | 99717818 | 99718014 | 197 | 3 | 0.000372668 | 3.50E-05 | 0.000548 | 4.10E-05 | 0.002487671 | LYSMD4 |
| chr2 | 31318484 | 31318866 | 383 | 2 | 5.46E-05 | 0.000126 | 8.32E-05 | 4.11E-05 | 0.006773265 | NA |
| chr2 | 48566734 | 48566798 | 65 | 2 | 0.000170218 | 3.57E-05 | 0.000404 | 4.11E-05 | 0.016977975 | STON1, STON1-GTF2A1L |
| chr1 | 206469958 | 206470498 | 541 | 7 | 0.000194358 | 4.00E-05 | 0.004196 | 4.11E-05 | 0.004400156 | IKBKE |
| chr12 | 27589391 | 27589433 | 43 | 2 | 0.000203144 | 3.40E-05 | 0.000464 | 4.11E-05 | 0.000380946 | PPFIBP1 |
| chr8 | 138767574 | 138767795 | 222 | 2 | 0.000171497 | 0.000233 | 6.24E-05 | 4.11E-05 | 0.014335281 | COL22A1 |
| chr22 | 19129444 | 19129802 | 359 | 4 | 0.000126069 | 0.000134 | 0.001366 | 4.11E-05 | 0.006053217 | NA |
| chr20 | 46058815 | 46059091 | 277 | 2 | 9.52E-05 | 3.77E-05 | 0.00035 | 4.12E-05 | 0.014635507 | SLC12A5 |
| chr4 | 150256868 | 150257053 | 186 | 3 | 0.000187381 | 2.22E-05 | 0.002853 | 4.12E-05 | 0.009128039 | DCLK2 |
| chr16 | 71624252 | 71624760 | 509 | 2 | 8.73E-05 | 2.98E-05 | 0.000734 | 4.12E-05 | 0.0127456 | AC009097.2 |
| chr18 | 78933954 | 78934174 | 221 | 3 | 2.08E-05 | 0.052012 | 1.45E-05 | 4.13E-05 | 0.001715516 | NA |
| chr17 | 29593916 | 29594647 | 732 | 4 | 2.77E-05 | 6.28E-05 | 0.003226 | 4.13E-05 | 0.00333644 | ABHD15-AS1, ANKRD13B, GIT1 |
| chr4 | 6138516 | 6138736 | 221 | 3 | 0.000166619 | 0.001592 | 0.000441 | 4.13E-05 | 0.008290221 | C4orf50, JAKMIP1 |
| chr14 | 103522852 | 103523746 | 895 | 12 | 0.000114798 | 0.001289 | 0.000903 | 4.13E-05 | 0.002833326 | NA |
| chr2 | 218983611 | 218983907 | 297 | 5 | 0.000178993 | 5.08E-05 | 0.00785 | 4.13E-05 | 0.004837563 | LINC00608, FEV |
| chr1 | 46602964 | 46603981 | 1018 | 4 | 2.77E-07 | 0.003004 | 2.23E-05 | 4.13E-05 | 0.003534919 | MKNK1 |
| chr20 | 20713467 | 20713493 | 27 | 2 | 0.000142966 | 2.71E-05 | 0.001179 | 4.13E-05 | 0.012207422 | NA |
| chr17 | 41536015 | 41536109 | 95 | 2 | 0.000213456 | 3.39E-05 | 0.000476 | 4.13E-05 | 0.008481392 | NA |
| chr12 | 54428047 | 54428289 | 243 | 2 | 0.000112414 | 3.60E-05 | 0.000405 | 4.14E-05 | 0.012786055 | AC079313.2, AC079313.1 |
| chr10 | 118502131 | 118502204 | 74 | 2 | 0.000125143 | 6.58E-05 | 0.000141 | 4.15E-05 | -0.0096133 | NA |
| chr8 | 20196861 | 20197204 | 344 | 11 | 0.000144894 | 0.000282 | 0.00399 | 4.15E-05 | 0.00072222 | NA |
| chr2 | 101474329 | 101474859 | 531 | 11 | 0.000195182 | 0.000246 | 0.013126 | 4.15E-05 | 0.001029867 | RFX8 |
| chr20 | 62666716 | 62667325 | 610 | 4 | 5.70E-05 | 0.000281 | 0.001635 | 4.15E-05 | 0.006070593 | SLCO4A1, SLCO4A1-AS1 |
| chr1 | 181128423 | 181128761 | 339 | 3 | 7.68E-05 | 7.05E-05 | 0.000852 | 4.15E-05 | 0.011802135 | NA |
| chr7 | 4724024 | 4724114 | 91 | 2 | 8.13E-05 | 0.000116 | 8.85E-05 | 4.15E-05 | 0.004725784 | FOXK1 |
| chr12 | 115576524 | 115576654 | 131 | 2 | 0.000115862 | 3.60E-05 | 0.00041 | 4.15E-05 | 0.012227386 | AC009803.2 |
| chr1 | 151133701 | 151134295 | 595 | 4 | 1.35E-05 | 0.003489 | 9.23E-05 | 4.16E-05 | 0.003609286 | SEMA6C |
| chr18 | 79848384 | 79849087 | 704 | 5 | 7.41E-05 | 0.000103 | 0.000438 | 4.16E-05 | 0.003175425 | NA |
| chr4 | 2337987 | 2338163 | 177 | 2 | 0.000257686 | 2.68E-05 | 0.001328 | 4.16E-05 | 5.17E-05 | ZFYVE28 |
| chr15 | 25093643 | 25093921 | 279 | 3 | 9.41E-05 | 0.000148 | 8.63E-05 | 4.17E-05 | 0.008642029 | SNHG14, AC124312.5 |
| chr12 | 132643881 | 132644050 | 170 | 3 | 0.000183088 | 1.76E-05 | 0.005292 | 4.17E-05 | 0.000840841 | POLE |
| chr22 | 17232542 | 17232552 | 11 | 2 | 0.000272034 | 3.09E-05 | 0.000673 | 4.17E-05 | 0.009421298 | ADA2 |
| chr1 | 30252542 | 30252617 | 76 | 2 | 0.00029411 | 2.62E-05 | 0.001596 | 4.17E-05 | 0.008096794 | NA |
| chr10 | 8014699 | 8015043 | 345 | 4 | 0.000324242 | 1.30E-05 | 0.010428 | 4.17E-05 | 0.002869615 | TAF3 |
| chr7 | 92738005 | 92738377 | 373 | 2 | 3.83E-05 | 0.001405 | 3.95E-05 | 4.18E-05 | 0.004408192 | CDK6 |
| chr10 | 71540485 | 71540684 | 200 | 2 | 5.95E-05 | 7.96E-05 | 0.000119 | 4.18E-05 | 0.007259852 | CDH23 |
| chr8 | 59593909 | 59594392 | 484 | 3 | 1.06E-05 | 0.000956 | 0.000824 | 4.18E-05 | 0.002616324 | AC087664.1 |
| chr6 | 44001399 | 44001840 | 442 | 2 | 9.36E-05 | 4.10E-05 | 0.000302 | 4.18E-05 | 0.010009252 | C6orf223, AL109615.3 |
| chr8 | 64806363 | 64806511 | 149 | 3 | 0.000141256 | 0.000462 | 8.94E-05 | 4.19E-05 | 0.006467742 | AC104232.3, AC104232.1 |
| chr6 | 170378487 | 170378789 | 303 | 3 | 5.24E-05 | 0.003935 | 2.59E-05 | 4.19E-05 | 0.004831709 | FAM120B |
| chr11 | 122180922 | 122181136 | 215 | 2 | 8.97E-05 | 0.000162 | 7.50E-05 | 4.19E-05 | 0.000711564 | MIR100HG |
| chr16 | 87350993 | 87351157 | 165 | 2 | 0.00027845 | 2.78E-05 | 0.001114 | 4.19E-05 | 0.004679259 | FBXO31 |
| chr8 | 123202713 | 123203134 | 422 | 6 | 0.000312196 | 2.30E-05 | 0.011745 | 4.19E-05 | 0.00877201 | FAM83A, FAM83A-AS1 |
| chr11 | 8832697 | 8832935 | 239 | 2 | 0.000224676 | 2.69E-05 | 0.001358 | 4.20E-05 | 0.01566999 | DENND2B |
| chr4 | 13546619 | 13547291 | 673 | 6 | 7.66E-05 | 1.10E-05 | 0.01187 | 4.20E-05 | 0.004984089 | LINC01096 |
| chr17 | 58417183 | 58417922 | 740 | 7 | 3.57E-05 | 0.002423 | 0.000126 | 4.20E-05 | 0.0034441 | AC004687.2, RNF43 |
| chr6 | 110875380 | 110875537 | 158 | 3 | 1.90E-05 | 0.000571 | 0.000687 | 4.20E-05 | 0.000316768 | AMD1 |
| chr11 | 61892663 | 61892718 | 56 | 2 | 4.48E-05 | 3.29E-05 | 0.000555 | 4.20E-05 | 0.006635694 | NA |
| chr11 | 78079750 | 78080252 | 503 | 9 | 0.000180268 | 1.91E-05 | 0.010783 | 4.20E-05 | 0.001593815 | NDUFC2-KCTD14, NDUFC2 |
| chr7 | 24716371 | 24716643 | 273 | 3 | 0.000224942 | 7.38E-05 | 0.000322 | 4.20E-05 | 0.004456215 | GSDME |
| chr3 | 158106420 | 158106721 | 302 | 3 | 0.000125384 | 0.000996 | 3.88E-05 | 4.20E-05 | 0.007685128 | RSRC1, SHOX2 |
| chr2 | 172985522 | 172985634 | 113 | 2 | 0.000296421 | 2.64E-05 | 0.001598 | 4.21E-05 | 0.010642613 | RAPGEF4 |
| chr17 | 48799560 | 48799694 | 135 | 2 | 0.000262715 | 2.66E-05 | 0.001541 | 4.21E-05 | 0.018328372 | TTLL6 |
| chr12 | 109736703 | 109736815 | 113 | 3 | 0.000169367 | 5.79E-05 | 0.001288 | 4.22E-05 | 0.008208368 | FAM222A, FAM222A-AS1 |
| chr1 | 226407971 | 226408277 | 307 | 7 | 0.000347748 | 0.000284 | 0.00053 | 4.22E-05 | 0.001158603 | PARP1 |
| chr7 | 142867142 | 142867161 | 20 | 2 | 0.00018067 | 0.000332 | 5.67E-05 | 4.22E-05 | 0.005780943 | EPHB6 |
| chr22 | 38943782 | 38943794 | 13 | 2 | 0.000285226 | 3.05E-05 | 0.000749 | 4.22E-05 | 0.018425261 | NA |
| chr4 | 55358029 | 55358123 | 95 | 2 | 9.29E-05 | 0.000131 | 8.44E-05 | 4.22E-05 | 0.004039054 | SRD5A3 |
| chr1 | 201448369 | 201448593 | 225 | 2 | 0.00013071 | 2.76E-05 | 0.001221 | 4.23E-05 | 0.01121289 | NA |
| chr16 | 657601 | 657712 | 112 | 2 | 1.40E-05 | 0.024417 | 3.11E-05 | 4.23E-05 | 0.002673445 | WDR90 |
| chr16 | 2452145 | 2452162 | 18 | 2 | 0.000103297 | 4.67E-05 | 0.00024 | 4.23E-05 | 0.002003717 | CCNF |
| chr8 | 93018544 | 93018948 | 405 | 2 | 0.000166885 | 2.66E-05 | 0.001616 | 4.24E-05 | 0.011215551 | NA |
| chr2 | 240946288 | 240946767 | 480 | 2 | 5.25E-05 | 3.77E-05 | 0.000387 | 4.24E-05 | 0.007712439 | CROCC2 |
| chr2 | 162092082 | 162092360 | 279 | 3 | 0.000197456 | 9.61E-05 | 0.001242 | 4.24E-05 | 0.003557282 | AC008063.1 |
| chr17 | 81527602 | 81527872 | 271 | 3 | 4.21E-05 | 0.001313 | 7.05E-05 | 4.24E-05 | 0.006299466 | AC139149.1 |
| chr11 | 7251422 | 7252147 | 726 | 7 | 4.54E-05 | 6.88E-05 | 0.004337 | 4.24E-05 | 0.004648378 | SYT9 |
| chr15 | 40739604 | 40739774 | 171 | 2 | 0.000140158 | 8.38E-05 | 0.000116 | 4.24E-05 | 0.002123895 | RMDN3 |
| chr1 | 16391577 | 16391634 | 58 | 2 | 0.000297594 | 2.70E-05 | 0.001436 | 4.24E-05 | 0.002279418 | SZRD1, SPATA21 |
| chr1 | 149927762 | 149928331 | 570 | 8 | 7.48E-05 | 0.00752 | 0.000425 | 4.25E-05 | 0.001960543 | SF3B4 |
| chr2 | 168244183 | 168244505 | 323 | 3 | 0.000243027 | 2.25E-05 | 0.001973 | 4.25E-05 | 0.014361864 | STK39 |
| chr8 | 11556082 | 11556495 | 414 | 2 | 4.75E-05 | 6.33E-05 | 0.000152 | 4.25E-05 | 0.007573593 | BLK, AC022239.1 |
| chr14 | 64197146 | 64197210 | 65 | 2 | 0.000135618 | 2.73E-05 | 0.001346 | 4.25E-05 | 0.015237403 | SYNE2, ESR2 |
| chr17 | 80959594 | 80960161 | 568 | 4 | 0.000224595 | 2.45E-05 | 0.005948 | 4.25E-05 | 0.002385756 | RPTOR |
| chr1 | 89812944 | 89813122 | 179 | 2 | 0.000181753 | 7.80E-05 | 0.000124 | 4.25E-05 | 0.010551509 | AC093423.3 |
| chr6 | 27478886 | 27478892 | 7 | 2 | 0.000503747 | 2.89E-05 | 0.001016 | 4.26E-05 | -0.00087184 | AL021918.5 |
| chr3 | 58666619 | 58667170 | 552 | 3 | 2.28E-05 | 0.002358 | 7.86E-05 | 4.26E-05 | 0.014660167 | FAM3D |
| chr5 | 119355007 | 119355762 | 756 | 13 | 0.000126064 | 0.000184 | 0.024198 | 4.27E-05 | 0.00134031 | TNFAIP8 |
| chr7 | 157904679 | 157904936 | 258 | 3 | 0.00012708 | 3.61E-05 | 0.001856 | 4.27E-05 | 0.015034698 | PTPRN2 |
| chr22 | 23117002 | 23117004 | 3 | 2 | 0.000162096 | 7.25E-05 | 0.000134 | 4.27E-05 | 0.005726955 | GNAZ, RSPH14 |
| chr4 | 911435 | 911653 | 219 | 3 | 0.000202854 | 2.21E-05 | 0.002282 | 4.27E-05 | 0.002651586 | GAK |
| chr4 | 6782441 | 6782669 | 229 | 6 | 0.000360018 | 4.09E-05 | 0.003326 | 4.27E-05 | -0.00074687 | KIAA0232 |
| chr12 | 6309982 | 6310409 | 428 | 7 | 7.22E-05 | 7.55E-05 | 0.000413 | 4.28E-05 | 0.004208 | NA |
| chr18 | 26549302 | 26550486 | 1185 | 12 | 5.06E-06 | 0.004039 | 6.11E-05 | 4.28E-05 | 0.000855296 | KCTD1 |
| chr16 | 57146142 | 57146263 | 122 | 3 | 0.00021904 | 2.18E-05 | 0.003286 | 4.28E-05 | 0.008911665 | CPNE2 |
| chr6 | 90296342 | 90296928 | 587 | 9 | 0.000287214 | 5.65E-05 | 0.008013 | 4.28E-05 | 0.001557327 | AL132996.1, BACH2 |
| chr14 | 99215373 | 99215932 | 560 | 6 | 0.000255393 | 4.98E-05 | 0.000913 | 4.29E-05 | 0.006703274 | BCL11B |
| chr12 | 109292316 | 109292964 | 649 | 2 | 0.000369758 | 2.93E-05 | 0.00098 | 4.29E-05 | 0.01018569 | FOXN4 |
| chr7 | 101171773 | 101172204 | 432 | 2 | 0.000114955 | 2.70E-05 | 0.001569 | 4.29E-05 | -0.00068461 | NAT16 |
| chr13 | 113520658 | 113520697 | 40 | 3 | 0.000348776 | 2.22E-05 | 0.002718 | 4.29E-05 | 0.001020537 | TMCO3 |
| chr16 | 87034165 | 87034499 | 335 | 2 | 0.000187075 | 4.07E-05 | 0.000334 | 4.30E-05 | 0.01042605 | NA |
| chr14 | 52853594 | 52853839 | 246 | 3 | 0.000148078 | 0.000211 | 0.000496 | 4.30E-05 | -0.00547006 | AL139317.5 |
| chr2 | 41896397 | 41896585 | 189 | 2 | 0.00025899 | 3.35E-05 | 0.000577 | 4.30E-05 | 0.00189369 | NA |
| chr11 | 8263958 | 8264470 | 513 | 11 | 4.63E-05 | 0.000105 | 0.001285 | 4.31E-05 | 0.003028674 | LMO1 |
| chr3 | 11583041 | 11583059 | 19 | 2 | 5.16E-06 | 0.000249 | 6.39E-05 | 4.31E-05 | 0.007619083 | VGLL4 |
| chr3 | 150728029 | 150728276 | 248 | 3 | 0.000119682 | 0.000436 | 6.25E-05 | 4.31E-05 | 0.008911539 | NA |
| chr1 | 44261271 | 44261565 | 295 | 3 | 0.000148982 | 0.000173 | 9.27E-05 | 4.32E-05 | 0.002165045 | ERI3 |
| chr19 | 7636212 | 7637077 | 866 | 9 | 2.15E-05 | 0.000353 | 0.000797 | 4.32E-05 | 0.000433558 | AC008763.2, STXBP2, AC008763.1 |
| chr17 | 1298798 | 1298958 | 161 | 2 | 0.000261436 | 8.99E-05 | 0.000111 | 4.32E-05 | 0.015650391 | TRARG1 |
| chr20 | 43564716 | 43565413 | 698 | 8 | 9.96E-05 | 7.34E-05 | 0.004368 | 4.32E-05 | 0.002293258 | Z98752.3, SGK2 |

| | | | | | | | | | | |
|---|---|---|---|---|---|---|---|---|---|---|
| chr17 | 38800436 | 38800536 | 101 | 2 | 0.000108187 | 3.80E-05 | 0.000402 | 4.32E-05 | 0.013526414 | CWC25 |
| chr14 | 32329181 | 32329556 | 376 | 4 | 6.01E-05 | 0.000865 | 0.00021 | 4.32E-05 | 7.56E-05 | AKAP6 |
| chr21 | 32351055 | 32351123 | 69 | 3 | 6.79E-05 | 0.013979 | 3.11E-05 | 4.32E-05 | 0.001869243 | URB1 |
| chr4 | 86594031 | 86594858 | 828 | 16 | 7.55E-05 | 0.000103 | 0.011131 | 4.32E-05 | 5.02E-05 | PTPN13, MAPK10 |
| chr6 | 44234388 | 44234962 | 575 | 2 | 0.000131435 | 2.72E-05 | 0.001564 | 4.32E-05 | 0.017899871 | NA |
| chr9 | 122016981 | 122017276 | 296 | 2 | 4.40E-05 | 0.004186 | 3.50E-05 | 4.33E-05 | 0.01319507 | TTLL11 |
| chr5 | 135466425 | 135466811 | 387 | 2 | 5.75E-05 | 4.89E-05 | 0.000233 | 4.33E-05 | 0.009668267 | NA |
| chr11 | 70210127 | 70210239 | 113 | 2 | 0.000154984 | 2.71E-05 | 0.001641 | 4.33E-05 | 0.008225467 | NA |
| chr12 | 101830597 | 101831335 | 739 | 6 | 9.23E-05 | 0.000602 | 0.000639 | 4.33E-05 | 0.000789921 | GNPTAB |
| chr16 | 55508418 | 55509254 | 837 | 10 | 9.71E-05 | 0.001197 | 0.001867 | 4.33E-05 | 0.001809681 | LPCAT2 |
| chr6 | 50513578 | 50513939 | 362 | 2 | 6.42E-05 | 7.01E-05 | 0.000141 | 4.33E-05 | 0.00962704 | NA |
| chr12 | 56332893 | 56333520 | 628 | 2 | 4.80E-07 | 0.00013 | 8.71E-05 | 4.33E-05 | 0.003683309 | PAN2 |
| chr5 | 88689044 | 88689466 | 423 | 6 | 3.73E-05 | 0.00134 | 0.000174 | 4.33E-05 | 0.003656178 | MEF2C-AS2, LINC00461 |
| chr2 | 151261686 | 151262214 | 529 | 10 | 6.29E-05 | 0.000231 | 0.000262 | 4.34E-05 | 0.000865348 | RBM43 |
| chr2 | 1604957 | 1605736 | 780 | 7 | 4.54E-05 | 0.000372 | 0.004432 | 4.34E-05 | 0.009718783 | AC141930.1 |
| chr1 | 42931200 | 42931380 | 181 | 2 | 0.000148243 | 2.82E-05 | 0.001273 | 4.34E-05 | 0.00491389 | SLC2A1 |
| chr19 | 14518381 | 14519105 | 725 | 9 | 4.71E-05 | 4.04E-05 | 0.001475 | 4.34E-05 | 0.000462735 | TECR, DNAJB1 |
| chr5 | 126463756 | 126464674 | 919 | 4 | 0.000102302 | 1.92E-05 | 0.003858 | 4.34E-05 | 0.00883485 | GRAMD2B, AC093535.1 |
| chr14 | 60606083 | 60606313 | 231 | 2 | 7.70E-05 | 0.00138 | 4.11E-05 | 4.34E-05 | 0.009246169 | NA |
| chr3 | 134406972 | 134407028 | 57 | 2 | 0.000376455 | 2.87E-05 | 0.001157 | 4.34E-05 | 0.0110363 | NA |
| chr9 | 136576178 | 136576216 | 39 | 2 | 0.000281954 | 3.58E-05 | 0.000489 | 4.35E-05 | 0.005152671 | NA |
| chr10 | 119526243 | 119526963 | 721 | 2 | 0.000271809 | 2.87E-05 | 0.001163 | 4.35E-05 | -0.00445273 | RGS10 |
| chr14 | 64942636 | 64943427 | 792 | 8 | 8.18E-05 | 0.014674 | 0.000412 | 4.35E-05 | 0.002873315 | CHURC1, CHURC1-FNTB, GPX2 |
| chr16 | 8644254 | 8644751 | 498 | 7 | 0.000142202 | 0.000279 | 0.003174 | 4.35E-05 | 0.00512839 | METTL22 |
| chr7 | 116524054 | 116524616 | 563 | 5 | 2.03E-05 | 1.14E-05 | 0.012922 | 4.35E-05 | 0.009126457 | NA |
| chr12 | 114677592 | 114677610 | 19 | 2 | 8.25E-05 | 0.000269 | 6.27E-05 | 4.35E-05 | 0.006669681 | TBX3 |
| chr2 | 930152 | 930324 | 173 | 3 | 0.000230289 | 4.07E-05 | 0.000466 | 4.35E-05 | 0.003043746 | NA |
| chr7 | 5349150 | 5349264 | 115 | 2 | 3.31E-05 | 0.000608 | 4.86E-05 | 4.35E-05 | 0.002311691 | TNRC18 |
| chr8 | 38148655 | 38148709 | 55 | 3 | 9.39E-05 | 0.001236 | 0.000848 | 4.35E-05 | 0.006345851 | STAR |
| chr14 | 93800103 | 93800372 | 270 | 2 | 0.000202187 | 2.87E-05 | 0.001204 | 4.36E-05 | 0.005503603 | NA |
| chr6 | 157120674 | 157120818 | 145 | 2 | 8.87E-05 | 5.09E-05 | 0.000222 | 4.36E-05 | 0.005441129 | ARID1B |
| chr5 | 1508971 | 1509184 | 214 | 4 | 0.000261122 | 1.81E-05 | 0.003486 | 4.36E-05 | 0.000665419 | LPCAT1 |
| chr5 | 71586895 | 71587620 | 726 | 11 | 0.000123028 | 9.78E-05 | 0.000289 | 4.36E-05 | 0.000436992 | MCCC2 |
| chr1 | 101432153 | 101432159 | 7 | 2 | 0.000253519 | 5.27E-05 | 0.00021 | 4.36E-05 | 0.002493365 | NA |
| chr4 | 52589104 | 52589383 | 280 | 2 | 5.70E-05 | 2.74E-05 | 0.001641 | 4.37E-05 | 0.001931527 | NA |
| chr17 | 6813049 | 6813481 | 433 | 2 | 4.99E-05 | 6.65E-05 | 0.000152 | 4.37E-05 | 0.006058978 | TEKT1 |
| chr19 | 4841204 | 4841344 | 141 | 3 | 0.000204887 | 3.40E-05 | 0.000776 | 4.37E-05 | 0.010461123 | PLIN3 |
| chr9 | 128911726 | 128912057 | 332 | 3 | 2.88E-05 | 0.010657 | 0.00089 | 4.38E-05 | 0.007748494 | LRRC8A |
| chr12 | 57184859 | 57184913 | 55 | 5 | 0.000254485 | 2.72E-05 | 0.007308 | 4.38E-05 | -0.00115513 | LRP1 |
| chr2 | 95275971 | 95276072 | 102 | 3 | 0.000102785 | 0.000176 | 8.85E-05 | 4.38E-05 | 0.003228857 | PROM2 |
| chr11 | 45145295 | 45146221 | 927 | 6 | 1.57E-05 | 0.000801 | 0.002697 | 4.38E-05 | 0.008643104 | PRDM11 |
| chr6 | 18823987 | 18823993 | 7 | 2 | 0.000307607 | 2.84E-05 | 0.00132 | 4.38E-05 | 0.016772586 | NA |
| chr17 | 16023383 | 16023636 | 254 | 4 | 0.000165045 | 0.000119 | 0.002555 | 4.38E-05 | 0.001157604 | TTC19, AC002553.2 |
| chr20 | 35604396 | 35604579 | 184 | 2 | 0.000149664 | 2.95E-05 | 0.001079 | 4.38E-05 | 0.010251154 | FER1L4 |
| chr16 | 67963955 | 67964563 | 609 | 9 | 0.000127791 | 1.46E-05 | 0.024537 | 4.39E-05 | 0.009794883 | SLC12A4 |
| chr4 | 9898894 | 9898901 | 8 | 2 | 0.000304697 | 2.95E-05 | 0.001082 | 4.39E-05 | 0.027458368 | SLC2A9 |
| chr16 | 67527939 | 67528769 | 831 | 9 | 2.80E-05 | 0.000438 | 0.00086 | 4.39E-05 | 0.000358051 | RIPOR1, AC027682.6 |
| chr19 | 43596197 | 43596814 | 618 | 14 | 6.85E-05 | 0.000203 | 0.003765 | 4.39E-05 | -0.00084273 | ZNF576, SRRM5 |
| chr16 | 87924675 | 87924887 | 213 | 3 | 3.93E-05 | 0.000122 | 0.000109 | 4.40E-05 | 0.005732487 | CA5A |
| chr1 | 147761002 | 147761198 | 197 | 2 | 0.000133578 | 0.027764 | 3.19E-05 | 4.40E-05 | 0.01700766 | GJA5 |
| chr1 | 2478567 | 2478949 | 383 | 2 | 4.11E-05 | 3.19E-05 | 0.00076 | 4.40E-05 | 0.007787859 | PLCH2 |
| chr2 | 219460900 | 219461319 | 420 | 4 | 3.06E-05 | 0.000339 | 0.001037 | 4.40E-05 | 0.010832386 | SPEG |
| chr5 | 181050662 | 181051185 | 524 | 2 | 0.000483296 | 3.50E-05 | 0.00055 | 4.40E-05 | -0.01962892 | BTNL9 |
| chr11 | 65649430 | 65649602 | 173 | 2 | 0.000115563 | 3.00E-05 | 0.001007 | 4.40E-05 | 0.000517486 | SIPA1 |
| chr3 | 142596005 | 142596724 | 720 | 10 | 3.62E-05 | 0.000597 | 0.000114 | 4.40E-05 | 0.000275677 | PLS1 |
| chr7 | 1897096 | 1897218 | 123 | 2 | 0.000265298 | 3.13E-05 | 0.000821 | 4.40E-05 | 0.006784313 | MAD1L1, AC069288.1 |
| chr19 | 1065377 | 1066350 | 974 | 9 | 3.84E-06 | 0.008352 | 7.10E-05 | 4.40E-05 | 0.00175793 | ABCA7, ARHGAP45 |
| chr1 | 228158480 | 228158574 | 95 | 2 | 0.00023839 | 8.43E-05 | 0.000122 | 4.41E-05 | -0.00027488 | GJC2 |
| chr19 | 17870883 | 17871790 | 908 | 5 | 3.62E-05 | 0.000257 | 0.001347 | 4.41E-05 | 0.006138894 | AC005796.1 |
| chr3 | 196287625 | 196287975 | 351 | 11 | 0.000281876 | 8.51E-05 | 0.003249 | 4.41E-05 | 0.000599523 | PCYT1A, AC069257.3 |
| chr22 | 49950726 | 49950744 | 19 | 2 | 0.000214334 | 2.79E-05 | 0.001568 | 4.41E-05 | 0.003430249 | NA |
| chr4 | 126817007 | 126817048 | 42 | 2 | 0.000241535 | 2.94E-05 | 0.001143 | 4.42E-05 | -0.00797519 | NA |
| chr6 | 93416423 | 93416925 | 503 | 2 | 8.90E-06 | 0.000822 | 4.59E-05 | 4.42E-05 | 0.013767147 | EPHA7 |
| chr11 | 64744851 | 64745817 | 967 | 8 | 9.04E-06 | 0.002776 | 7.82E-05 | 4.42E-05 | 0.000671639 | RASGRP2 |
| chr16 | 79088753 | 79089162 | 410 | 2 | 3.27E-05 | 8.27E-05 | 0.000125 | 4.42E-05 | 0.010516929 | WWOX |
| chr10 | 118568529 | 118568673 | 145 | 2 | 8.98E-05 | 0.000272 | 6.34E-05 | 4.42E-05 | 0.016070836 | NA |
| chr15 | 25230793 | 25230909 | 117 | 2 | 0.000209052 | 0.000103 | 0.000104 | 4.42E-05 | 0.002773171 | SNHG14, SNORD115-33 |
| chr8 | 56446641 | 56447667 | 1027 | 10 | 2.14E-05 | 0.000143 | 0.01297 | 4.42E-05 | -0.00744071 | AC012349.1, PENK |
| chr9 | 74951164 | 74951255 | 92 | 2 | 0.000143276 | 2.80E-05 | 0.00157 | 4.43E-05 | 0.000958236 | C9orf40 |
| chr7 | 151966956 | 151967105 | 150 | 3 | 0.000166705 | 0.000601 | 0.000935 | 4.43E-05 | 0.010210445 | GALNTL5 |
| chr5 | 178153210 | 178153923 | 714 | 7 | 4.20E-05 | 0.005322 | 0.000559 | 4.43E-05 | 0.002029547 | NHP2 |
| chr1 | 37765097 | 37765870 | 774 | 7 | 1.56E-05 | 0.000299 | 0.001531 | 4.43E-05 | 0.005115427 | EPHA10 |
| chr18 | 52339840 | 52340178 | 339 | 10 | 3.68E-05 | 0.00221 | 0.000388 | 4.44E-05 | 0.00256069 | NA |
| chr3 | 14304007 | 14304336 | 330 | 3 | 7.09E-05 | 3.21E-05 | 0.000815 | 4.44E-05 | -0.00726745 | NA |
| chr11 | 31823601 | 31824092 | 492 | 4 | 3.41E-05 | 6.10E-05 | 0.000477 | 4.44E-05 | 0.007589452 | PAUPAR, AL035078.4 |
| chr7 | 158188900 | 158189253 | 354 | 4 | 0.000227236 | 0.000165 | 0.000139 | 4.44E-05 | 7.43E-05 | PTPRN2 |
| chr6 | 73654772 | 73654977 | 206 | 4 | 0.000236561 | 1.68E-05 | 0.006917 | 4.44E-05 | 0.015917236 | NA |
| chr2 | 236577307 | 236577724 | 418 | 2 | 0.000241687 | 3.20E-05 | 0.000792 | 4.44E-05 | -0.001207 | ACKR3 |
| chr5 | 61330454 | 61331133 | 680 | 4 | 6.71E-05 | 0.006258 | 0.000488 | 4.44E-05 | 0.002081603 | NA |
| chr5 | 170254103 | 170254358 | 256 | 3 | 2.64E-05 | 0.023623 | 1.73E-05 | 4.45E-05 | 0.005788174 | LCP2 |
| chr11 | 67615442 | 67615906 | 465 | 2 | 1.58E-06 | 7.36E-05 | 0.000141 | 4.45E-05 | 0.016617356 | DOC2GP |
| chr8 | 30612366 | 30612415 | 50 | 2 | 0.000243052 | 4.78E-05 | 0.000263 | 4.45E-05 | 0.004652764 | GTF2E2 |
| chr1 | 205031635 | 205031782 | 148 | 2 | 0.000177186 | 2.82E-05 | 0.001541 | 4.45E-05 | 0.006138393 | NA |
| chr15 | 90888538 | 90889502 | 965 | 6 | 1.54E-05 | 0.0003 | 0.000238 | 4.45E-05 | 0.00309267 | FES |
| chr19 | 41859879 | 41859964 | 86 | 4 | 0.000357678 | 5.40E-05 | 0.003075 | 4.45E-05 | -0.00247261 | NA |
| chr11 | 64592186 | 64592221 | 36 | 2 | 7.78E-05 | 2.86E-05 | 0.001407 | 4.45E-05 | -0.0005565 | SLC22A12 |
| chr11 | 123580602 | 123580667 | 66 | 2 | 0.000144151 | 3.87E-05 | 0.000428 | 4.45E-05 | 0.014493535 | GRAMD1B |
| chr1 | 11907769 | 11908081 | 313 | 4 | 0.000161316 | 5.01E-05 | 0.001319 | 4.46E-05 | 0.002653662 | AL021155.4 |
| chr7 | 117322398 | 117322896 | 499 | 4 | 8.95E-05 | 0.000578 | 0.00093 | 4.46E-05 | 0.006239524 | CFTR, WNT2 |
| chr19 | 8307930 | 8308253 | 324 | 4 | 0.000195293 | 2.87E-05 | 0.005831 | 4.46E-05 | 0.002216414 | CD320 |
| chr7 | 1058989 | 1059440 | 452 | 6 | 0.00017276 | 0.000185 | 0.002685 | 4.47E-05 | 0.003734286 | GPR146, C7orf50, AC073957.2 |
| chr1 | 2242961 | 2243456 | 496 | 4 | 0.000186762 | 6.08E-05 | 0.000272 | 4.47E-05 | 0.004309171 | SKI |
| chr2 | 212764270 | 212764460 | 191 | 2 | 0.000285918 | 3.16E-05 | 0.000858 | 4.47E-05 | 0.00569517 | AC093865.1 |
| chr6 | 168648483 | 168648818 | 336 | 2 | 0.000276721 | 2.99E-05 | 0.001107 | 4.47E-05 | 0.021829854 | SMOC2 |
| chr19 | 46840249 | 46840677 | 429 | 2 | 3.69E-05 | 0.000545 | 5.16E-05 | 4.47E-05 | 0.002954225 | AP2S1 |
| chr11 | 3099794 | 3100360 | 567 | 5 | 0.000187289 | 0.000214 | 0.004249 | 4.47E-05 | -0.00109026 | OSBPL5 |
| chr1 | 201127161 | 201127263 | 103 | 3 | 0.000166338 | 1.84E-05 | 0.00581 | 4.47E-05 | 0.011601944 | ASCL5 |
| chr10 | 116803599 | 116803765 | 167 | 3 | 8.38E-05 | 0.002194 | 2.75E-05 | 4.48E-05 | 0.001476314 | HSPA12A |
| chr20 | 46608375 | 46608651 | 277 | 2 | 0.000114016 | 9.51E-05 | 0.000112 | 4.48E-05 | -0.04007569 | SLC13A3 |
| chr12 | 94282593 | 94282932 | 340 | 5 | 8.10E-05 | 0.000426 | 0.001313 | 4.48E-05 | 0.007235897 | PLXNC1, AC073655.2 |
| chr12 | 3227519 | 3227893 | 375 | 2 | 5.77E-05 | 9.76E-05 | 0.00011 | 4.48E-05 | 0.007441416 | TSPAN9 |
| chr3 | 167894738 | 167895041 | 304 | 3 | 0.000187615 | 4.85E-05 | 0.000743 | 4.48E-05 | 0.004400884 | LRRC77P, AC026353.1 |
| chr1 | 159850104 | 159850536 | 433 | 2 | 5.16E-05 | 3.58E-05 | 0.000555 | 4.48E-05 | 0.007201345 | AL590560.2, SNHG28 |
| chr5 | 173145864 | 173146291 | 428 | 2 | 3.10E-05 | 4.59E-05 | 0.000292 | 4.48E-05 | 0.006789326 | BNIP1 |
| chr21 | 14215287 | 14215799 | 513 | 4 | 3.72E-05 | 3.31E-05 | 0.001428 | 4.49E-05 | 0.009579143 | NA |
| chr8 | 68436651 | 68437090 | 440 | 2 | 9.73E-05 | 5.74E-05 | 0.000197 | 4.49E-05 | 0.009071206 | C8orf34 |
| chr3 | 43190792 | 43190957 | 166 | 2 | 0.000135933 | 4.46E-05 | 0.000311 | 4.49E-05 | 0.003327811 | NA |
| chr15 | 40987956 | 40988084 | 129 | 3 | 0.000326833 | 1.97E-05 | 0.004761 | 4.49E-05 | 0.001328225 | INO80 |
| chr1 | 182791765 | 182792127 | 363 | 2 | 4.41E-05 | 0.000136 | 8.89E-05 | 4.49E-05 | 0.00286784 | NPL |
| chr18 | 37484691 | 37484937 | 247 | 2 | 6.85E-05 | 0.000365 | 5.84E-05 | 4.49E-05 | 0.011687817 | CELF4 |
| chr2 | 3605893 | 3606880 | 988 | 5 | 0.00013375 | 0.000225 | 0.001973 | 4.50E-05 | -0.00132352 | COLEC11 |
| chr22 | 30693986 | 30694760 | 775 | 6 | 0.000219229 | 2.67E-05 | 0.009641 | 4.50E-05 | -0.00473825 | OSBP2 |
| chr22 | 43213853 | 43214148 | 296 | 2 | 0.00014219 | 3.97E-05 | 0.000417 | 4.50E-05 | 0.004362862 | SCUBE1 |
| chr15 | 52295969 | 52296361 | 393 | 5 | 0.000323303 | 0.000171 | 0.000163 | 4.50E-05 | 0.005661176 | NA |
| chr5 | 77620690 | 77620766 | 77 | 2 | 0.000286142 | 3.86E-05 | 0.00045 | 4.50E-05 | 0.016788816 | NA |
| chr14 | 77705281 | 77705560 | 280 | 3 | 0.000135999 | 0.000109 | 0.000167 | 4.51E-05 | 7.14E-05 | ALKBH1 |
| chr11 | 44927042 | 44927188 | 147 | 2 | 0.000192561 | 2.87E-05 | 0.00154 | 4.51E-05 | 0.00394959 | TSPAN18, TP53I11 |
| chr2 | 228678093 | 228678164 | 72 | 2 | 0.000223671 | 8.69E-05 | 0.000123 | 4.51E-05 | -0.00934296 | LINC01807 |
| chr2 | 10043493 | 10044330 | 838 | 7 | 4.12E-05 | 5.95E-05 | 0.000517 | 4.52E-05 | 0.003371712 | KLF11 |
| chr21 | 41927886 | 41928020 | 135 | 3 | 0.000181592 | 2.75E-05 | 0.002742 | 4.52E-05 | 0.012421948 | C2CD2 |
| chr11 | 61973497 | 61973507 | 11 | 2 | 0.000202161 | 2.88E-05 | 0.001505 | 4.52E-05 | 0.013462635 | NA |
| chr1 | 54517500 | 54517830 | 331 | 3 | 0.000295956 | 2.18E-05 | 0.003148 | 4.52E-05 | 0.00901061 | NA |
| chr8 | 860491 | 861003 | 513 | 6 | 0.000209381 | 0.00064 | 0.001026 | 4.53E-05 | 0.001870506 | DLGAP2 |
| chr7 | 1005521 | 1005952 | 432 | 4 | 3.65E-05 | 0.003912 | 0.000596 | 4.53E-05 | 0.003130596 | C7orf50 |
| chr17 | 7512819 | 7513051 | 233 | 4 | 5.43E-05 | 0.002992 | 3.54E-05 | 4.54E-05 | 0.003527174 | POLR2A |
| chr8 | 73879560 | 73879796 | 237 | 2 | 6.29E-05 | 4.07E-05 | 0.000401 | 4.54E-05 | 0.017748904 | AC022868.1 |
| chr3 | 11770598 | 11771124 | 527 | 3 | 0.000195878 | 8.55E-05 | 0.000536 | 4.54E-05 | 0.001828752 | VGLL4 |
| chr7 | 139035829 | 139036319 | 491 | 9 | 8.36E-05 | 0.000449 | 0.003137 | 4.54E-05 | -0.0011701 | ZC3HAV1L |
| chr4 | 182145071 | 182145184 | 114 | 3 | 0.000106576 | 0.000131 | 0.000355 | 4.54E-05 | 0.003539706 | TENM3, AC108142.1, TENM3-AS1 |
| chr17 | 1871751 | 1871830 | 80 | 2 | 0.000380191 | 4.12E-05 | 0.000388 | 4.54E-05 | 0.012042772 | RPA1 |
| chr3 | 13885597 | 13885904 | 308 | 3 | 2.69E-05 | 0.012365 | 1.78E-05 | 4.54E-05 | 0.011512704 | NA |
| chr13 | 20713098 | 20713143 | 46 | 2 | 0.000190948 | 5.41E-05 | 0.000222 | 4.54E-05 | 0.007578493 | IL17D, AL512652.2 |
| chr14 | 70730611 | 70730958 | 348 | 2 | 0.000140086 | 3.02E-05 | 0.001177 | 4.54E-05 | -0.00772844 | MAP3K9 |
| chr11 | 79393554 | 79393725 | 172 | 3 | 0.000299842 | 3.55E-05 | 0.00073 | 4.55E-05 | 0.018054815 | TENM4 |
| chr3 | 160014959 | 160015241 | 283 | 2 | 0.000139492 | 3.83E-05 | 0.000477 | 4.55E-05 | -0.00350081 | IL12A-AS1 |
| chr9 | 136215964 | 136216071 | 108 | 3 | 0.000307661 | 4.16E-05 | 0.000752 | 4.55E-05 | 0.002505328 | QSOX2 |
| chr1 | 150574861 | 150575201 | 341 | 2 | 9.91E-05 | 0.00012 | 9.77E-05 | 4.55E-05 | 0.017087998 | MCL1 |
| chr8 | 140532063 | 140532734 | 672 | 4 | 8.79E-05 | 0.000287 | 0.000272 | 4.55E-05 | 0.00459991 | AGO2 |
| chr2 | 46698614 | 46698930 | 317 | 3 | 0.00017106 | 5.67E-05 | 0.000289 | 4.55E-05 | 0.002551367 | NA |
| chr2 | 236718317 | 236718391 | 75 | 3 | 0.000182018 | 0.000554 | 0.000242 | 4.55E-05 | 0.003591756 | NA |
| chr3 | 119557839 | 119558070 | 232 | 3 | 0.000194381 | 2.85E-05 | 0.001293 | 4.55E-05 | 0.00885376 | CD80 |
| chr15 | 52643141 | 52643249 | 109 | 2 | 0.000127409 | 0.000872 | 4.65E-05 | 4.55E-05 | -0.00331265 | FAM214A |
| chr17 | 80126127 | 80126390 | 264 | 2 | 0.000259952 | 3.41E-05 | 0.000696 | 4.55E-05 | 0.01112206 | NA |
| chr14 | 93042234 | 93042271 | 38 | 2 | 0.000107458 | 3.75E-05 | 0.000509 | 4.56E-05 | 0.009652057 | ITPK1 |
| chr16 | 87849172 | 87850046 | 875 | 4 | 0.000112451 | 0.000224 | 0.000581 | 4.56E-05 | 0.006881008 | SLC7A5 |
| chr17 | 67379014 | 67379058 | 45 | 2 | 0.00016212 | 3.10E-05 | 0.001052 | 4.57E-05 | 0.002614369 | PITPNC1 |
| chr2 | 233461141 | 233461424 | 284 | 4 | 9.67E-05 | 0.000375 | 8.83E-05 | 4.57E-05 | 0.001619596 | DGKD |
| chr14 | 104799116 | 104799572 | 457 | 3 | 0.000263298 | 1.91E-05 | 0.005496 | 4.57E-05 | 0.011039274 | NA |
| chr14 | 105466475 | 105466946 | 472 | 4 | 0.000136495 | 1.54E-05 | 0.008947 | 4.57E-05 | 0.003208147 | MTA1 |
| chr1 | 57755650 | 57755961 | 312 | 3 | 0.000391896 | 2.28E-05 | 0.003412 | 4.57E-05 | -0.00536204 | DAB1 |
| chr6 | 152301959 | 152302344 | 386 | 5 | 6.30E-05 | 0.000161 | 0.000157 | 4.57E-05 | 0.003569272 | SYNE1 |
| chr20 | 298941 | 299332 | 392 | 2 | 5.52E-05 | 3.23E-05 | 0.000873 | 4.58E-05 | 0.009868444 | ZCCHC3 |
| chr3 | 197960121 | 197960462 | 342 | 7 | 0.000397423 | 1.91E-05 | 0.007901 | 4.58E-05 | 0.000389916 | LMLN, IQCG |
| chr7 | 134890214 | 134891060 | 847 | 9 | 4.15E-05 | 0.000654 | 0.000201 | 4.58E-05 | -0.00069119 | CALD1, AC083870.1 |
| chr9 | 109556346 | 109556842 | 497 | 3 | 0.000311692 | 2.50E-05 | 0.001897 | 4.58E-05 | -0.00752675 | PALM2AKAP2 |
| chr4 | 173526933 | 173527690 | 758 | 8 | 0.000116941 | 5.03E-05 | 0.003418 | 4.58E-05 | 0.004457638 | HAND2-AS1, HAND2 |
| chr1 | 61973195 | 61973349 | 155 | 3 | 8.15E-05 | 0.001147 | 0.000141 | 4.58E-05 | 0.007398361 | PATJ |
| chr22 | 20117832 | 20118294 | 463 | 6 | 8.71E-05 | 0.000244 | 0.002664 | 4.58E-05 | 0.000233849 | RANBP1 |
| chr2 | 132669854 | 132670309 | 456 | 5 | 0.000260527 | 0.000138 | 0.000399 | 4.58E-05 | -8.38E-05 | LYPD1 |
| chr2 | 109335809 | 109335811 | 3 | 2 | 0.000125102 | 0.000168 | 8.20E-05 | 4.60E-05 | -0.00038181 | SH3RF3 |
| chr1 | 12166086 | 12167360 | 1275 | 10 | 0.000154468 | 0.001773 | 0.001056 | 4.60E-05 | -0.0004968 | MIR7846, TNFRSF1B |
| chr19 | 16828893 | 16829329 | 437 | 9 | 0.000240172 | 5.72E-05 | 0.003195 | 4.61E-05 | -0.00145442 | NA |

| | | | | | | | | | | |
|---|---|---|---|---|---|---|---|---|---|---|
| chr19 | 35300289 | 35300506 | 218 | 3 | 0.000192744 | 0.000148 | 0.000355 | 4.61E-05 | 0.003761776 | MAG |
| chr3 | 39180854 | 39181263 | 410 | 4 | 0.000142827 | 5.39E-05 | 0.00135 | 4.61E-05 | 0.003333694 | NA |
| chr14 | 95132638 | 95133330 | 693 | 3 | 6.35E-05 | 0.000102 | 0.000248 | 4.61E-05 | 0.012407009 | DICER1 |
| chr19 | 50475900 | 50476908 | 1009 | 13 | 0.000335262 | 0.000169 | 0.011278 | 4.62E-05 | 0.000809562 | EMC10, FAM71E1 |
| chr2 | 164842839 | 164843173 | 335 | 3 | 2.82E-05 | 0.010657 | 0.000222 | 4.62E-05 | 0.008227769 | AC019197.1, COBLL1 |
| chr1 | 115839475 | 115839742 | 268 | 2 | 1.04E-05 | 0.000356 | 6.06E-05 | 4.62E-05 | 0.00272154 | NHLH2 |
| chr4 | 1208721 | 1209865 | 1145 | 17 | 1.32E-05 | 0.002457 | 0.001547 | 4.62E-05 | 0.001352639 | SPON2 |
| chr22 | 50542496 | 50542977 | 482 | 3 | 0.000249266 | 3.39E-05 | 0.000796 | 4.62E-05 | 0.003808084 | U62317.3, KLHDC7B-DT |
| chr7 | 3001445 | 3001476 | 32 | 3 | 0.000188211 | 0.0001 | 0.000247 | 4.63E-05 | 0.000838364 | CARD11 |
| chr3 | 162536789 | 162537020 | 232 | 2 | 5.30E-05 | 0.005245 | 3.62E-05 | 4.63E-05 | 0.01112842 | NA |
| chr18 | 58998848 | 58999007 | 160 | 2 | 0.000209902 | 3.21E-05 | 0.000953 | 4.63E-05 | 0.008797068 | OACYLP |
| chr9 | 127816567 | 127817387 | 821 | 3 | 0.00011872 | 0.00012 | 0.000118 | 4.64E-05 | 0.005626041 | AL162586.1, ENG |
| chr20 | 38645540 | 38646049 | 510 | 2 | 4.45E-06 | 0.001476 | 4.27E-05 | 4.64E-05 | 0.009820531 | ARHGAP40 |
| chr2 | 175006143 | 175006366 | 224 | 2 | 4.25E-05 | 5.57E-05 | 0.000222 | 4.64E-05 | 0.004509383 | NA |
| chr20 | 52156164 | 52156417 | 254 | 2 | 9.18E-05 | 0.000644 | 5.10E-05 | 4.64E-05 | 0.004399709 | ZFP64 |
| chr6 | 30490050 | 30490372 | 323 | 10 | 0.00019551 | 0.00016 | 0.00166 | 4.64E-05 | 0.001280382 | HLA-E |
| chr9 | 101398177 | 101399009 | 833 | 12 | 3.23E-05 | 0.0055 | 0.000297 | 4.65E-05 | 0.000213778 | ZNF189, MRPL50 |
| chr2 | 48616506 | 48617056 | 551 | 2 | 0.00017758 | 3.55E-05 | 0.000662 | 4.65E-05 | 0.011618912 | STON1-GTF2A1L |
| chr1 | 178512672 | 178512895 | 224 | 5 | 0.000186942 | 0.000372 | 0.001195 | 4.66E-05 | 0.001589961 | AL513013.1 |
| chr17 | 18243603 | 18243999 | 397 | 2 | 0.000214866 | 3.50E-05 | 0.000698 | 4.66E-05 | -8.07E-05 | LLGL1 |
| chr16 | 879729 | 880077 | 349 | 3 | 1.40E-05 | 0.01191 | 2.32E-05 | 4.66E-05 | 0.003663808 | LMF1 |
| chr17 | 74261824 | 74262092 | 269 | 4 | 0.000166436 | 0.000129 | 0.000565 | 4.66E-05 | 0.004656859 | TTYH2, AC100786.1 |
| chr17 | 76239554 | 76239678 | 125 | 2 | 0.000203763 | 3.03E-05 | 0.001342 | 4.66E-05 | 0.015011913 | RNF157 |
| chr5 | 149116497 | 149116515 | 19 | 2 | 0.000268222 | 2.93E-05 | 0.001709 | 4.66E-05 | 0.006724412 | SH3TC2-DT |
| chr13 | 112164624 | 112165106 | 483 | 2 | 0.000340443 | 2.93E-05 | 0.00172 | 4.66E-05 | 0.010001424 | NA |
| chr17 | 40316709 | 40317130 | 422 | 5 | 7.91E-05 | 0.003598 | 8.46E-05 | 4.67E-05 | 0.00651374 | RARA |
| chr2 | 175606061 | 175606129 | 69 | 2 | 0.000314642 | 3.40E-05 | 0.00078 | 4.67E-05 | 0.002175273 | NA |
| chr4 | 139085235 | 139085865 | 631 | 3 | 1.04E-05 | 0.006591 | 2.12E-05 | 4.67E-05 | 0.00474594 | ELF2 |
| chr12 | 103841185 | 103841776 | 592 | 7 | 0.000177095 | 0.000137 | 0.001541 | 4.67E-05 | 0.003564029 | AC012555.1, NT5DC3 |
| chr13 | 113022720 | 113023578 | 859 | 6 | 6.05E-05 | 0.000111 | 0.000606 | 4.67E-05 | 0.009069319 | MCF2L |
| chr1 | 200894431 | 200895095 | 665 | 5 | 7.68E-05 | 3.76E-05 | 0.005584 | 4.67E-05 | 0.003809606 | INAVA |
| chr19 | 38717723 | 38718058 | 336 | 4 | 0.000101437 | 8.60E-05 | 0.000331 | 4.68E-05 | 0.003181215 | ACTN4 |
| chr11 | 92829327 | 92829561 | 235 | 2 | 0.000231625 | 3.77E-05 | 0.000553 | 4.69E-05 | -0.00251041 | FAT3 |
| chr4 | 181159236 | 181159498 | 263 | 3 | 0.000198383 | 0.000102 | 0.000487 | 4.69E-05 | 0.008564936 | NA |
| chr2 | 25877738 | 25878252 | 515 | 4 | 8.85E-05 | 2.73E-05 | 0.002435 | 4.69E-05 | -0.00235701 | ASXL2 |
| chr7 | 129126970 | 129127421 | 452 | 3 | 5.81E-05 | 0.00063 | 6.27E-05 | 4.69E-05 | -0.00303062 | AC011005.1 |
| chr3 | 156793640 | 156794025 | 386 | 2 | 0.000371834 | 2.94E-05 | 0.001724 | 4.69E-05 | 0.012127449 | LINC00886 |
| chr16 | 4953248 | 4953279 | 32 | 2 | 0.000251121 | 2.94E-05 | 0.001724 | 4.69E-05 | 0.014441816 | PPL |
| chr16 | 69361592 | 69361614 | 23 | 2 | 0.000317769 | 3.36E-05 | 0.000835 | 4.69E-05 | 0.007946974 | TERF2 |
| chr19 | 17859423 | 17859935 | 513 | 12 | 0.000287282 | 0.000817 | 0.00061 | 4.70E-05 | 0.001391109 | RPL18A |
| chr2 | 38215321 | 38215456 | 136 | 2 | 0.00029651 | 3.74E-05 | 0.000577 | 4.70E-05 | 0.014289855 | CYP1B1-AS1, AC009229.1 |
| chr1 | 27153615 | 27154177 | 563 | 3 | 1.95E-06 | 0.000246 | 8.85E-05 | 4.70E-05 | 0.005642705 | SLC9A1 |
| chr19 | 39195295 | 39195478 | 184 | 2 | 0.000101014 | 2.96E-05 | 0.001656 | 4.70E-05 | 0.017219908 | NA |
| chr1 | 206559387 | 206559761 | 375 | 2 | 5.69E-05 | 7.95E-05 | 0.000143 | 4.71E-05 | 5.59E-05 | RASSF5 |
| chr17 | 19380838 | 19381314 | 477 | 3 | 0.00028123 | 3.97E-05 | 0.002189 | 4.71E-05 | 0.000603008 | MAPK7 |
| chr16 | 29897929 | 29898113 | 185 | 2 | 0.000123002 | 4.06E-05 | 0.000458 | 4.71E-05 | 0.015279709 | SEZ6L2 |
| chr10 | 132909987 | 132910193 | 207 | 3 | 6.58E-05 | 0.000942 | 3.95E-05 | 4.72E-05 | 0.00341757 | CFAP46 |
| chr7 | 78771067 | 78771453 | 387 | 8 | 0.000138632 | 0.001506 | 0.001303 | 4.72E-05 | 0.007474426 | MAGI2 |
| chr17 | 6755579 | 6756243 | 665 | 8 | 0.00014997 | 2.29E-05 | 0.003242 | 4.72E-05 | 0.002401859 | XAF1 |
| chr5 | 150700649 | 150701140 | 492 | 6 | 0.000197533 | 0.000135 | 0.000403 | 4.72E-05 | -0.00161138 | RBM22 |
| chr1 | 39214565 | 39215066 | 502 | 3 | 0.000278928 | 0.000104 | 0.000421 | 4.72E-05 | 0.007288063 | MACF1 |
| chr10 | 79137587 | 79137985 | 399 | 3 | 7.01E-05 | 0.000514 | 5.76E-05 | 4.72E-05 | 0.003963208 | ZMIZ1 |
| chr11 | 61948723 | 61949055 | 333 | 2 | 0.000111558 | 6.47E-05 | 0.000183 | 4.72E-05 | 0.0107817 | NA |
| chr4 | 185838902 | 185839224 | 323 | 3 | 3.40E-05 | 0.001938 | 0.000561 | 4.73E-05 | -0.00531869 | SORBS2 |
| chr12 | 67648665 | 67649019 | 355 | 5 | 3.05E-05 | 0.000838 | 5.49E-05 | 4.73E-05 | -3.59E-05 | DYRK2 |
| chr6 | 30212911 | 30213936 | 1026 | 34 | 2.24E-05 | 0.000153 | 0.007834 | 4.74E-05 | -7.06E-05 | TRIM26 |
| chr12 | 101594740 | 101595025 | 286 | 4 | 0.000325091 | 3.01E-05 | 0.002838 | 4.74E-05 | 0.013328746 | MYBPC1 |
| chr17 | 76716967 | 76717199 | 233 | 2 | 0.00023739 | 2.99E-05 | 0.001631 | 4.74E-05 | 0.011418265 | JMJD6 |
| chr12 | 131103995 | 131104150 | 156 | 2 | 0.000272235 | 9.61E-05 | 0.000121 | 4.74E-05 | 0.008341489 | ADGRD1 |
| chr11 | 120013196 | 120013407 | 212 | 3 | 7.88E-05 | 0.000241 | 7.45E-05 | 4.74E-05 | 0.006278957 | AP001360.1 |
| chr14 | 52266401 | 52267094 | 694 | 4 | 2.15E-06 | 0.000523 | 0.00013 | 4.74E-05 | 0.011652028 | NA |
| chr14 | 22846657 | 22847130 | 474 | 2 | 1.04E-05 | 0.000129 | 9.84E-05 | 4.75E-05 | 0.005234416 | MMP14 |
| chr1 | 26761063 | 26761616 | 554 | 4 | 2.36E-05 | 0.000632 | 0.000166 | 4.75E-05 | 0.003369259 | ARID1A |
| chr16 | 69744775 | 69745011 | 237 | 2 | 0.000190394 | 4.08E-05 | 0.000465 | 4.76E-05 | 0.000574357 | NOB1 |
| chr22 | 17167606 | 17167613 | 8 | 2 | 0.000167109 | 3.39E-05 | 0.000867 | 4.76E-05 | 0.00702472 | NA |
| chr16 | 73064004 | 73064457 | 454 | 3 | 1.36E-05 | 0.000357 | 0.000643 | 4.76E-05 | -0.00506732 | ZFHX3 |
| chr16 | 285547 | 285830 | 284 | 2 | 7.20E-05 | 3.08E-05 | 0.001396 | 4.76E-05 | 0.008147227 | PDIA2 |
| chr1 | 1543850 | 1543858 | 9 | 2 | 0.000112086 | 4.39E-05 | 0.000383 | 4.76E-05 | 0.015778672 | SSU72 |
| chr5 | 180646137 | 180646780 | 644 | 3 | 0.000120037 | 3.41E-05 | 0.000992 | 4.76E-05 | 0.000501665 | FLT4 |
| chr11 | 134168157 | 134168594 | 438 | 2 | 0.000392206 | 2.99E-05 | 0.001717 | 4.77E-05 | -0.0009172 | NCAPD3 |
| chr16 | 56972228 | 56972580 | 353 | 2 | 0.000104075 | 5.27E-05 | 0.000261 | 4.77E-05 | 0.006988061 | CETP |
| chr5 | 150781737 | 150781989 | 253 | 2 | 0.000121879 | 8.36E-05 | 0.000139 | 4.77E-05 | 0.013572318 | SMIM3 |
| chr3 | 131072793 | 131073091 | 299 | 3 | 3.90E-05 | 0.000517 | 0.000743 | 4.77E-05 | 0.005760989 | NEK11 |
| chr9 | 91786603 | 91786893 | 291 | 3 | 3.22E-05 | 0.019399 | 1.73E-05 | 4.77E-05 | -0.00528262 | ROR2 |
| chr6 | 30010537 | 30010860 | 324 | 2 | 0.000323009 | 5.75E-05 | 0.000225 | 4.77E-05 | 0.007908349 | ZNRD1ASP |
| chr8 | 144409833 | 144409980 | 148 | 4 | 5.58E-05 | 1.65E-05 | 0.008818 | 4.77E-05 | 0.005294498 | AC233992.3, SLC39A4 |
| chr11 | 18697761 | 18698081 | 321 | 3 | 0.000188527 | 3.06E-05 | 0.001992 | 4.78E-05 | 0.003753519 | TMEM86A |
| chr17 | 36638601 | 36638628 | 28 | 2 | 0.000217288 | 3.14E-05 | 0.001282 | 4.78E-05 | 0.015259151 | NA |
| chr22 | 38125813 | 38126118 | 306 | 2 | 8.54E-05 | 8.62E-05 | 0.000136 | 4.78E-05 | 0.001081791 | PLA2G6 |
| chr11 | 12249875 | 12250154 | 280 | 3 | 0.000117134 | 0.000427 | 0.000217 | 4.78E-05 | 0.005130625 | MICAL2 |
| chr6 | 17392030 | 17392751 | 722 | 3 | 3.54E-06 | 0.000134 | 0.00011 | 4.79E-05 | 0.005406471 | NA |
| chr1 | 75723507 | 75724743 | 1237 | 15 | 3.38E-06 | 0.000824 | 0.005852 | 4.79E-05 | 0.010645361 | ACADM, AL137803.1 |
| chr18 | 59219770 | 59220553 | 784 | 9 | 0.000133481 | 0.000322 | 0.001807 | 4.79E-05 | 0.003786493 | GRP |
| chr11 | 1871162 | 1871658 | 497 | 3 | 0.00016905 | 0.000859 | 0.000361 | 4.79E-05 | 0.005322194 | LSP1 |
| chr7 | 92538009 | 92538296 | 288 | 3 | 0.000124136 | 0.00062 | 8.25E-05 | 4.79E-05 | 0.00637697 | RBM48 |
| chr15 | 51341184 | 51342053 | 870 | 6 | 2.84E-05 | 0.000366 | 0.000708 | 4.79E-05 | -0.01056217 | GLDN |
| chr19 | 35936272 | 35936877 | 606 | 6 | 6.20E-05 | 0.000514 | 0.000272 | 4.80E-05 | 0.002656648 | LRFN3 |
| chr7 | 158689928 | 158689944 | 17 | 3 | 0.000159444 | 3.17E-05 | 0.001132 | 4.80E-05 | 0.000439273 | NCAPG2 |
| chr16 | 12010875 | 12010902 | 28 | 3 | 0.000259401 | 3.21E-05 | 0.001359 | 4.80E-05 | 0.016281668 | SNX29 |
| chr3 | 98343830 | 98344094 | 265 | 2 | 0.000284747 | 3.53E-05 | 0.000773 | 4.80E-05 | 0.006011904 | AC117473.1 |
| chr11 | 1757294 | 1757397 | 104 | 3 | 0.000135246 | 2.07E-05 | 0.005026 | 4.80E-05 | 0.010075604 | AC068580.4, CTSD |
| chr10 | 132591369 | 132591978 | 610 | 6 | 1.18E-05 | 0.002251 | 0.000194 | 4.81E-05 | 0.006217401 | INPP5A |
| chr15 | 75354198 | 75354442 | 245 | 2 | 8.07E-05 | 3.70E-05 | 0.000654 | 4.81E-05 | 0.003293245 | NEIL1 |
| chr4 | 110637308 | 110638199 | 892 | 6 | 7.35E-06 | 0.000162 | 0.000232 | 4.81E-05 | 0.006911099 | PITX2 |
| chr9 | 137311533 | 137312205 | 673 | 3 | 7.48E-05 | 0.000242 | 0.000186 | 4.81E-05 | 0.006098283 | EXD3 |
| chr22 | 42757713 | 42757892 | 180 | 2 | 0.000284258 | 3.90E-05 | 0.000557 | 4.82E-05 | 0.010000642 | NA |
| chr17 | 75064303 | 75064485 | 183 | 2 | 2.57E-05 | 0.000269 | 7.03E-05 | 4.82E-05 | 0.001029215 | KCTD2 |
| chr1 | 34866048 | 34866231 | 184 | 2 | 5.65E-05 | 0.000216 | 7.70E-05 | 4.82E-05 | 0.002493141 | DLGAP3 |
| chr14 | 76565175 | 76565397 | 223 | 2 | 0.00018108 | 3.58E-05 | 0.00075 | 4.82E-05 | 0.00542922 | AC008050.1 |
| chr15 | 32721771 | 32721820 | 50 | 2 | 0.000154957 | 0.000383 | 6.17E-05 | 4.82E-05 | 0.009150958 | GREM1 |
| chr16 | 30396240 | 30397104 | 865 | 3 | 7.41E-06 | 0.000431 | 5.69E-05 | 4.82E-05 | 0.006277345 | ZNF48 |
| chr14 | 24373768 | 24374151 | 384 | 3 | 0.000235997 | 2.52E-05 | 0.002276 | 4.82E-05 | 0.005384624 | NFATC4 |
| chr4 | 75756201 | 75756284 | 84 | 2 | 0.000236357 | 7.52E-05 | 0.000158 | 4.82E-05 | 0.011226519 | USO1 |
| chr11 | 2401155 | 2402356 | 1202 | 24 | 6.93E-06 | 0.001094 | 0.001815 | 4.82E-05 | 0.00209643 | TSSC4 |
| chr16 | 2297282 | 2297566 | 285 | 5 | 0.000218816 | 6.63E-05 | 0.002977 | 4.83E-05 | 1.44E-05 | ABCA3 |
| chr18 | 67516508 | 67517161 | 654 | 4 | 0.00014859 | 0.001107 | 8.68E-05 | 4.83E-05 | 0.002925549 | AC114689.3, DSEL |
| chr4 | 158393971 | 158394288 | 318 | 2 | 0.00013071 | 5.95E-05 | 0.000219 | 4.83E-05 | 0.01116997 | RXFP1 |
| chr14 | 61154370 | 61154629 | 260 | 2 | 0.000148211 | 5.72E-05 | 0.000235 | 4.83E-05 | -0.00584399 | NA |
| chr2 | 111219546 | 111219980 | 435 | 2 | 0.000340141 | 3.11E-05 | 0.001446 | 4.83E-05 | 0.013661137 | MIR4435-2HG |
| chr1 | 26359224 | 26359336 | 113 | 2 | 0.000140324 | 6.31E-05 | 0.000201 | 4.83E-05 | 0.019769855 | CRYBG2 |
| chr12 | 59113938 | 59114366 | 429 | 3 | 0.000346258 | 5.79E-05 | 0.000821 | 4.84E-05 | -0.00604612 | NA |
| chr5 | 157058619 | 157059175 | 557 | 4 | 4.06E-05 | 0.011997 | 4.26E-05 | 4.84E-05 | 0.008231263 | HAVCR1 |
| chr9 | 126409213 | 126409264 | 52 | 2 | 6.86E-05 | 0.001039 | 4.73E-05 | 4.84E-05 | 0.004868895 | MVB12B |
| chr16 | 30356339 | 30356679 | 341 | 3 | 0.000183192 | 5.94E-05 | 0.000336 | 4.84E-05 | 0.007313142 | CD2BP2-DT |
| chr19 | 5929831 | 5929892 | 62 | 2 | 0.000250527 | 8.54E-05 | 0.00014 | 4.84E-05 | -5.13E-05 | RANBP3 |
| chr22 | 38680605 | 38680722 | 118 | 2 | 5.02E-05 | 0.000143 | 9.48E-05 | 4.84E-05 | -0.00038226 | AL021707.2 |
| chr15 | 66564795 | 66565670 | 876 | 4 | 7.10E-06 | 9.12E-05 | 0.00068 | 4.84E-05 | 0.007015212 | LCTL |
| chr9 | 110579744 | 110580381 | 638 | 8 | 7.01E-05 | 0.001055 | 0.001259 | 4.84E-05 | 0.003808941 | SVEP1 |
| chr9 | 136746950 | 136746956 | 7 | 2 | 9.21E-05 | 3.18E-05 | 0.001299 | 4.84E-05 | 0.004685761 | AL355987.1, LCN6, MIR6722 |
| chr7 | 151704726 | 151704937 | 212 | 4 | 0.000127094 | 0.000229 | 0.000257 | 4.84E-05 | 0.001343239 | PRKAG2 |
| chr1 | 33176572 | 33176593 | 22 | 2 | 0.000150755 | 3.75E-05 | 0.000643 | 4.84E-05 | 0.0039075 | TRIM62 |
| chr16 | 21663884 | 21664134 | 251 | 4 | 0.000296581 | 5.55E-05 | 0.000733 | 4.84E-05 | 0.004865668 | OTOA |
| chr1 | 243615736 | 243616045 | 310 | 2 | 0.000336582 | 3.08E-05 | 0.001597 | 4.84E-05 | 0.014082254 | AKT3 |
| chr7 | 138002021 | 138002514 | 494 | 13 | 4.47E-05 | 0.000821 | 0.002176 | 4.84E-05 | 0.000496202 | AKR1D1, CREB3L2 |
| chr11 | 10382652 | 10382978 | 327 | 2 | 0.000354115 | 3.05E-05 | 0.00171 | 4.85E-05 | 0.015772736 | AMPD3 |
| chr1 | 157524124 | 157524308 | 185 | 2 | 0.000185881 | 4.79E-05 | 0.000335 | 4.85E-05 | 0.013497437 | FCRL5 |
| chr17 | 7704453 | 7704750 | 298 | 3 | 0.000105749 | 2.34E-05 | 0.003608 | 4.86E-05 | 0.007773976 | NA |
| chr3 | 128075376 | 128075988 | 613 | 7 | 0.000107605 | 7.37E-05 | 0.011079 | 4.86E-05 | 0.006882105 | RUVBL1-AS1, RUVBL1 |
| chr1 | 41017906 | 41018368 | 463 | 3 | 3.88E-05 | 4.56E-05 | 0.002023 | 4.86E-05 | 0.004317587 | SLFNL1-AS1, SLFNL1 |
| chr12 | 93096782 | 93097223 | 442 | 2 | 0.000201606 | 3.05E-05 | 0.001748 | 4.86E-05 | 0.006494659 | AC138123.2, AC138123.1 |
| chr1 | 119140107 | 119140727 | 621 | 10 | 0.000167491 | 7.20E-05 | 0.007326 | 4.86E-05 | 0.000668852 | WARS2-AS1, WARS2 |
| chr5 | 1362585 | 1362634 | 50 | 2 | 4.25E-05 | 0.000972 | 4.82E-05 | 4.86E-05 | -0.00582146 | LINC01511 |
| chr17 | 81937130 | 81937708 | 579 | 17 | 0.00016427 | 0.000245 | 0.000264 | 4.86E-05 | 0.000373214 | PYCR1 |
| chr3 | 55610593 | 55611308 | 716 | 3 | 3.10E-05 | 0.000134 | 0.000421 | 4.86E-05 | 0.005513542 | AC025572.1, ERC2 |
| chr11 | 13357386 | 13357482 | 97 | 2 | 0.000110708 | 3.13E-05 | 0.001478 | 4.86E-05 | 0.004865725 | ARNTL |
| chr1 | 205257083 | 205257294 | 212 | 2 | 3.56E-05 | 0.000124 | 0.000104 | 4.86E-05 | 0.004143774 | TMCC2 |
| chr22 | 38467226 | 38467600 | 375 | 3 | 9.64E-05 | 7.64E-05 | 0.000905 | 4.87E-05 | 0.008057915 | NA |
| chr9 | 129549822 | 129549829 | 8 | 2 | 0.000255136 | 3.15E-05 | 0.001415 | 4.87E-05 | 0.013410332 | NA |
| chr6 | 3494779 | 3494804 | 26 | 2 | 0.000273352 | 6.96E-05 | 0.000177 | 4.87E-05 | 0.005879623 | NA |
| chr12 | 49539756 | 49540246 | 491 | 3 | 8.33E-05 | 2.11E-05 | 0.004956 | 4.87E-05 | 0.003880856 | KCNH3 |
| chr18 | 63953882 | 63954190 | 309 | 2 | 0.000245408 | 3.09E-05 | 0.001643 | 4.88E-05 | 0.015262696 | HMSD |
| chr19 | 10491911 | 10492284 | 374 | 5 | 0.000342161 | 1.90E-05 | 0.008381 | 4.88E-05 | 0.003498837 | KEAP1 |
| chr19 | 48661715 | 48662137 | 423 | 2 | 9.88E-06 | 0.000237 | 7.53E-05 | 4.88E-05 | -0.00533639 | SEC1P, NTN5 |
| chr1 | 213983963 | 213984593 | 631 | 4 | 4.88E-05 | 9.00E-05 | 0.001515 | 4.88E-05 | 0.009987496 | PROX1, PROX1-AS1, AC011700.1 |
| chr11 | 1293629 | 1293941 | 313 | 2 | 7.34E-05 | 8.33E-05 | 0.000146 | 4.88E-05 | -0.00065694 | TOLLIP |
| chr9 | 83956899 | 83957351 | 453 | 6 | 0.00015015 | 5.68E-05 | 0.010594 | 4.88E-05 | 0.004612186 | C9orf64 |
| chr19 | 2540431 | 2541106 | 676 | 3 | 0.000191715 | 8.87E-05 | 0.00031 | 4.89E-05 | 0.003087066 | GNG7 |
| chr20 | 58103788 | 58104173 | 386 | 3 | 0.000215689 | 0.000164 | 0.00024 | 4.89E-05 | 0.004439156 | NA |
| chr6 | 149745700 | 149745843 | 144 | 2 | 1.23E-05 | 0.001285 | 4.59E-05 | 4.89E-05 | 0.021929766 | NUP43 |
| chr17 | 45161803 | 45162377 | 575 | 4 | 1.98E-05 | 0.001643 | 0.000132 | 4.89E-05 | 0.003425362 | HEXIM2 |
| chr6 | 47781978 | 47782306 | 329 | 5 | 0.000221165 | 0.001289 | 0.00042 | 4.89E-05 | 0.002769658 | OPN5 |
| chr14 | 93999373 | 93999482 | 110 | 2 | 6.58E-05 | 0.006225 | 3.76E-05 | 4.90E-05 | 0.005416754 | CCDC197 |
| chr15 | 101294889 | 101294918 | 30 | 2 | 2.46E-05 | 3.07E-05 | 0.001769 | 4.90E-05 | 0.000451846 | SNRPA1, PCSK6 |
| chr10 | 114993600 | 114993814 | 215 | 3 | 0.00039327 | 2.21E-05 | 0.004819 | 4.90E-05 | 0.000808825 | NA |
| chr9 | 135590546 | 135590795 | 250 | 2 | 0.00035183 | 3.07E-05 | 0.001756 | 4.90E-05 | 0.002501509 | PAEPP1 |
| chr16 | 86531186 | 86531322 | 137 | 2 | 3.31E-05 | 3.10E-05 | 0.00166 | 4.90E-05 | 0.005731412 | MTHFSD |
| chr19 | 29714865 | 29715466 | 602 | 4 | 7.34E-05 | 0.000264 | 0.000183 | 4.90E-05 | -0.00279593 | C19orf12 |
| chr5 | 141629226 | 141629295 | 70 | 2 | 0.000225954 | 3.84E-05 | 0.000625 | 4.90E-05 | 0.000965804 | HDAC3 |
| chr16 | 1344642 | 1345357 | 716 | 4 | 4.55E-05 | 4.96E-05 | 0.003092 | 4.90E-05 | -0.00235379 | BAIAP3 |
| chr22 | 42314070 | 42314435 | 366 | 3 | 0.000453342 | 2.01E-05 | 0.006372 | 4.90E-05 | 0.02478823 | TCF20 |
| chr19 | 15361785 | 15361922 | 138 | 2 | 0.000154144 | 0.000435 | 6.03E-05 | 4.90E-05 | 0.00260047 | AKAP8 |
| chr21 | 36480036 | 36480681 | 646 | 7 | 1.85E-05 | 0.007385 | 2.46E-05 | 4.91E-05 | 0.003077592 | AP000695.1, AP000695.2, CLDN14 |

| | | | | | | | | | | |
|---|---|---|---|---|---|---|---|---|---|---|
| chr14 | 103685649 | 103686619 | 971 | 7 | 0.000206809 | 0.00028 | 0.001981 | 4.91E-05 | -0.00135423 | KLC1, AL139300.1 |
| chr10 | 122190434 | 122190688 | 255 | 2 | 0.000309591 | 3.11E-05 | 0.001636 | 4.91E-05 | 0.003108704 | TACC2 |
| chr3 | 184298200 | 184298293 | 94 | 3 | 0.000123468 | 4.63E-05 | 0.000473 | 4.91E-05 | 0.000777526 | NA |
| chr1 | 57877329 | 57877342 | 14 | 2 | 0.000167305 | 0.0002 | 8.15E-05 | 4.91E-05 | -0.00690072 | DAB1-AS1, DAB1 |
| chr15 | 27771291 | 27771809 | 519 | 5 | 9.76E-05 | 6.30E-05 | 0.002957 | 4.91E-05 | 0.001887925 | OCA2 |
| chr19 | 18408182 | 18408201 | 20 | 2 | 0.000343383 | 3.30E-05 | 0.001178 | 4.91E-05 | 0.011961798 | NA |
| chr11 | 2219743 | 2220338 | 596 | 5 | 6.23E-05 | 0.000358 | 0.000132 | 4.92E-05 | 0.007947656 | NA |
| chr6 | 44254909 | 44255108 | 200 | 2 | 0.000241177 | 6.03E-05 | 0.000225 | 4.92E-05 | 0.009584414 | SLC35B2 |
| chr2 | 171434190 | 171435173 | 984 | 12 | 1.06E-05 | 0.000179 | 0.002908 | 4.92E-05 | 0.003037464 | DCAF17, METTL8 |
| chr15 | 66622087 | 66622545 | 459 | 3 | 0.000216133 | 2.62E-05 | 0.003652 | 4.92E-05 | 0.015431132 | LINC01169 |
| chr6 | 41051998 | 41052537 | 540 | 3 | 1.79E-05 | 0.009617 | 0.000157 | 4.92E-05 | 0.004453002 | OARD1 |
| chr2 | 9424119 | 9424913 | 795 | 4 | 2.65E-05 | 0.000239 | 0.000183 | 4.92E-05 | 0.009369933 | CPSF3 |
| chr12 | 121298575 | 121298776 | 202 | 3 | 2.96E-05 | 0.005455 | 2.72E-05 | 4.92E-05 | -0.00701965 | NA |
| chr17 | 76014487 | 76014554 | 68 | 2 | 0.000186748 | 4.13E-05 | 0.000511 | 4.93E-05 | 0.011481779 | EVPL |
| chr15 | 34038997 | 34039687 | 691 | 6 | 3.00E-05 | 0.003121 | 7.50E-05 | 4.93E-05 | -0.00331298 | CHRM5, AVEN |
| chr3 | 138879091 | 138879662 | 572 | 2 | 8.77E-05 | 3.17E-05 | 0.001489 | 4.93E-05 | 0.009235855 | NA |
| chr10 | 132260316 | 132260730 | 415 | 3 | 5.82E-05 | 0.000186 | 9.13E-05 | 4.93E-05 | 0.003358205 | STK32C |
| chr15 | 78951084 | 78951677 | 594 | 4 | 0.000166764 | 5.28E-05 | 0.003801 | 4.93E-05 | 0.007188365 | NA |
| chr1 | 228805059 | 228805315 | 257 | 3 | 0.000370011 | 2.41E-05 | 0.003171 | 4.93E-05 | 0.011391247 | NA |
| chr19 | 7294076 | 7294576 | 501 | 4 | 0.000131796 | 0.00153 | 0.000375 | 4.94E-05 | 0.001883035 | INSR |
| chr5 | 125575946 | 125576161 | 216 | 2 | 0.000331709 | 3.26E-05 | 0.001284 | 4.94E-05 | 0.020424976 | LINC02240, AC116362.2, AC116362.1 |
| chr11 | 57236383 | 57237054 | 672 | 3 | 2.23E-05 | 0.000129 | 0.00018 | 4.94E-05 | -0.0049184 | APLNR |
| chr3 | 185254037 | 185254383 | 347 | 6 | 0.000314345 | 2.23E-05 | 0.009239 | 4.94E-05 | -0.00705188 | EHHADH |
| chr19 | 38832017 | 38832518 | 502 | 5 | 8.53E-05 | 0.000153 | 0.001535 | 4.94E-05 | 0.00467718 | AC008982.1 |
| chr20 | 50152149 | 50152537 | 389 | 3 | 0.000283087 | 3.64E-05 | 0.000925 | 4.94E-05 | 0.002754984 | PEDS1-UBE2V1, PEDS1 |
| chr22 | 41781014 | 41781258 | 245 | 3 | 0.000178964 | 5.84E-05 | 0.000498 | 4.95E-05 | 0.005674761 | MEI1 |
| chr8 | 96644844 | 96645848 | 1005 | 14 | 4.07E-06 | 0.025473 | 0.000167 | 4.95E-05 | 0.000805401 | CPQ |
| chr11 | 76130853 | 76131343 | 491 | 5 | 0.000105564 | 0.000365 | 0.000187 | 4.95E-05 | 0.00362053 | UVRAG |
| chr5 | 622799 | 623229 | 431 | 3 | 6.68E-05 | 0.001618 | 0.000949 | 4.95E-05 | 0.005023568 | CEP72 |
| chr7 | 4179111 | 4179542 | 432 | 4 | 3.44E-05 | 0.005867 | 6.68E-05 | 4.95E-05 | 0.000408839 | SDK1 |
| chr11 | 126210667 | 126211033 | 367 | 3 | 8.63E-06 | 0.00011 | 0.000137 | 4.95E-05 | 0.003688889 | RPUSD4 |
| chr2 | 169827111 | 169827687 | 577 | 8 | 8.68E-05 | 0.000218 | 4.60E-05 | 4.96E-05 | -0.00258724 | UBR3 |
| chr16 | 84460527 | 84460681 | 155 | 2 | 0.00014234 | 0.000117 | 0.000111 | 4.96E-05 | 0.010122387 | ATP2C2, ATP2C2-AS1 |
| chr18 | 8704168 | 8704724 | 557 | 4 | 8.26E-05 | 0.00035 | 0.000272 | 4.97E-05 | 0.010291236 | GACAT2 |
| chr1 | 180946221 | 180946496 | 276 | 3 | 0.000380298 | 2.08E-05 | 0.00583 | 4.97E-05 | 0.01120062 | KIAA1614, AL162431.1 |
| chr17 | 74584658 | 74584942 | 285 | 5 | 0.000372176 | 0.000441 | 0.003707 | 4.97E-05 | 0.005370166 | C17orf77, CD300LD |
| chr1 | 35841418 | 35841643 | 226 | 3 | 0.000133151 | 0.000143 | 0.000111 | 4.97E-05 | 0.004098244 | AGO4 |
| chr11 | 5631078 | 5631633 | 556 | 3 | 0.000123073 | 4.91E-05 | 0.000449 | 4.97E-05 | 0.002952313 | TRIM6-TRIM34, TRIM34, AC104389.5 |
| chr20 | 35191656 | 35191857 | 202 | 2 | 0.000127955 | 0.000521 | 5.81E-05 | 4.97E-05 | 0.003093284 | PROCR |
| chr9 | 134380209 | 134380920 | 712 | 3 | 0.000153412 | 0.000558 | 0.000301 | 4.97E-05 | 0.008186652 | RXRA |
| chr1 | 52054921 | 52055374 | 454 | 7 | 3.35E-05 | 0.000544 | 6.84E-05 | 4.98E-05 | -0.00044789 | TXNDC12 |
| chr1 | 218324344 | 218324464 | 121 | 3 | 0.000164026 | 0.000522 | 6.67E-05 | 4.98E-05 | 0.003142242 | RRP15 |
| chr11 | 120353319 | 120353365 | 47 | 2 | 0.000199752 | 0.000264 | 7.36E-05 | 4.98E-05 | 0.00410487 | ARHGEF12 |
| chr11 | 61627992 | 61628317 | 326 | 2 | 0.000108712 | 3.13E-05 | 0.001786 | 4.98E-05 | 0.007310224 | RPLP0P2 |
| chr8 | 92726124 | 92726224 | 101 | 2 | 0.000223671 | 8.52E-05 | 0.000147 | 4.98E-05 | 0.009860412 | FLJ46284 |
| chr14 | 22541540 | 22541879 | 340 | 2 | 6.57E-05 | 0.000106 | 0.000121 | 4.98E-05 | -0.00737804 | NA |
| chr12 | 121520960 | 121521029 | 70 | 3 | 0.000397117 | 3.45E-05 | 0.001206 | 4.98E-05 | 0.004706704 | KDM2B |
| chr5 | 180207383 | 180207789 | 407 | 3 | 9.18E-05 | 4.35E-05 | 0.001076 | 4.99E-05 | -0.00915172 | RASGEF1C |
| chr17 | 81839162 | 81839497 | 336 | 4 | 4.96E-05 | 0.001246 | 0.000881 | 4.99E-05 | 0.007457739 | NA |
| chr1 | 111610020 | 111610042 | 23 | 2 | 0.000184631 | 0.00018 | 8.74E-05 | 4.99E-05 | -0.00327643 | RAP1A |
| chr12 | 452899 | 453682 | 784 | 2 | 2.11E-05 | 9.05E-05 | 0.00014 | 4.99E-05 | 0.007095496 | NA |
| chr22 | 17266863 | 17266927 | 65 | 3 | 0.000154723 | 5.00E-05 | 0.00176 | 4.99E-05 | 0.01209813 | NA |
| chr1 | 116665716 | 116665874 | 159 | 2 | 6.49E-05 | 3.20E-05 | 0.001566 | 5.00E-05 | 0.00013621 | IGSF3 |
| chr15 | 52013395 | 52013747 | 353 | 2 | 0.000368845 | 3.16E-05 | 0.001722 | 5.01E-05 | 0.016871235 | MAPK6, MAPK6-DT |
| chr1 | 13661668 | 13662005 | 338 | 3 | 0.000368073 | 2.79E-05 | 0.001848 | 5.01E-05 | 0.001876589 | NA |
| chr5 | 132375939 | 132376145 | 207 | 2 | 0.000322708 | 3.33E-05 | 0.001253 | 5.01E-05 | 0.006003105 | SLC22A5 |
| chr11 | 61554635 | 61554783 | 149 | 2 | 5.37E-06 | 5.89E-05 | 0.000245 | 5.01E-05 | 0.016882936 | SYT7 |
| chr18 | 2979976 | 2980159 | 184 | 2 | 2.26E-05 | 0.00061 | 5.59E-05 | 5.01E-05 | 0.015772368 | AP000919.2, LPIN2 |
| chr7 | 139829056 | 139829592 | 537 | 4 | 0.000134679 | 1.63E-05 | 0.010776 | 5.01E-05 | 0.009015359 | TBXAS1 |
| chr17 | 81492442 | 81492464 | 23 | 2 | 0.000251954 | 3.30E-05 | 0.001342 | 5.02E-05 | 0.011985519 | NA |
| chr8 | 134691460 | 134691541 | 82 | 2 | 0.00024318 | 0.000106 | 0.000122 | 5.02E-05 | 0.011213743 | ZFAT |
| chr12 | 55996943 | 55997293 | 351 | 11 | 0.000375051 | 0.000322 | 0.006933 | 5.02E-05 | 0.004091658 | SUOX |
| chr5 | 147877656 | 147877914 | 259 | 5 | 0.000164206 | 0.000172 | 0.001384 | 5.03E-05 | 0.002941697 | SCGB3A2 |
| chr1 | 225924104 | 225924834 | 731 | 11 | 0.000149895 | 0.00022 | 0.00112 | 5.03E-05 | 0.001526554 | AL117348.2, PYCR2 |
| chr1 | 6098583 | 6098631 | 49 | 3 | 0.000330685 | 2.98E-05 | 0.002284 | 5.03E-05 | 0.001924077 | KCNAB2 |
| chr11 | 3642261 | 3642858 | 598 | 9 | 0.000161668 | 0.000175 | 0.002101 | 5.03E-05 | -0.00141425 | ART5 |
| chr6 | 166505701 | 166505789 | 89 | 2 | 0.000164892 | 0.000486 | 5.98E-05 | 5.03E-05 | 0.003790482 | RPS6KA2 |
| chr16 | 79229445 | 79229842 | 398 | 6 | 0.000108525 | 0.000161 | 0.000345 | 5.03E-05 | 0.00130304 | AC092376.1 |
| chr6 | 168842386 | 168843315 | 930 | 4 | 5.70E-05 | 0.000213 | 0.000902 | 5.03E-05 | -0.00015396 | NA |
| chr5 | 134967882 | 134968013 | 132 | 2 | 0.000275515 | 6.12E-05 | 0.000233 | 5.03E-05 | 0.003953925 | PCBD2, CATSPER3 |
| chr4 | 186609711 | 186610140 | 430 | 3 | 0.000244429 | 3.71E-05 | 0.001132 | 5.04E-05 | 0.009497198 | FAT1 |
| chr9 | 134662908 | 134663133 | 226 | 2 | 3.57E-05 | 0.009523 | 3.73E-05 | 5.04E-05 | 0.00504675 | COL5A1 |
| chr9 | 109745202 | 109745470 | 269 | 2 | 0.000112121 | 3.41E-05 | 0.001158 | 5.05E-05 | 0.005239191 | PALM2AKAP2 |
| chr15 | 62245672 | 62245960 | 289 | 5 | 0.000209639 | 6.59E-05 | 0.002999 | 5.05E-05 | 0.000318262 | AC126323.1 |
| chr17 | 39617595 | 39618096 | 502 | 4 | 0.000100452 | 0.000138 | 0.000724 | 5.06E-05 | 0.003970052 | NA |
| chr6 | 7389429 | 7389946 | 518 | 13 | 0.00014859 | 0.000566 | 0.008679 | 5.06E-05 | 0.000443911 | RIOK1, CAGE1 |
| chr19 | 58519295 | 58520439 | 1145 | 13 | 1.63E-05 | 0.00719 | 0.000288 | 5.06E-05 | 0.001293318 | ZBTB45 |
| chr2 | 24971504 | 24972344 | 841 | 8 | 9.12E-05 | 0.002084 | 0.000497 | 5.06E-05 | 0.002141258 | DNAJC27-AS1, DNAJC27 |
| chr18 | 23703096 | 23703313 | 218 | 3 | 0.000248473 | 8.25E-05 | 0.000229 | 5.06E-05 | 0.010984938 | LAMA3 |
| chr1 | 109185906 | 109186544 | 639 | 5 | 7.37E-06 | 0.011168 | 0.000328 | 5.06E-05 | 0.005125866 | ELAPOR1 |
| chr1 | 235011997 | 235012059 | 63 | 2 | 0.000135189 | 0.00073 | 5.40E-05 | 5.06E-05 | 0.002403547 | NA |
| chr1 | 2305211 | 2305761 | 551 | 4 | 0.000124107 | 9.69E-05 | 0.001127 | 5.07E-05 | 0.002040967 | SKI |
| chr12 | 1844851 | 1845155 | 305 | 2 | 0.000278154 | 3.61E-05 | 0.000924 | 5.07E-05 | 0.01439608 | CACNA2D4 |
| chr19 | 40541045 | 40541537 | 493 | 3 | 0.000134065 | 6.37E-05 | 0.001156 | 5.07E-05 | 0.00316926 | SPTBN4 |
| chr10 | 86672553 | 86672568 | 16 | 2 | 0.000205768 | 6.88E-05 | 0.000197 | 5.07E-05 | 0.004198841 | LDB3 |
| chr16 | 85077977 | 85078350 | 374 | 2 | 2.00E-05 | 4.13E-05 | 0.000571 | 5.07E-05 | 0.014635489 | KIAA0513 |
| chr10 | 7410988 | 7411909 | 922 | 9 | 5.03E-05 | 0.000368 | 0.000354 | 5.07E-05 | 0.002967155 | AL139125.1, SFMBT2 |
| chr13 | 21663146 | 21663414 | 269 | 2 | 0.000314719 | 3.58E-05 | 0.000969 | 5.07E-05 | -0.00329206 | NA |
| chr16 | 57701873 | 57702003 | 131 | 4 | 0.000101353 | 0.005487 | 4.61E-05 | 5.08E-05 | 0.00365657 | DRC7 |
| chr10 | 112563574 | 112563600 | 27 | 2 | 0.000337083 | 3.41E-05 | 0.001207 | 5.08E-05 | 0.005457676 | VTI1A |
| chr2 | 236856120 | 236856405 | 286 | 2 | 0.000118952 | 3.19E-05 | 0.001811 | 5.08E-05 | 0.009243237 | NA |
| chr17 | 70074989 | 70075256 | 268 | 3 | 7.03E-05 | 7.40E-05 | 0.001382 | 5.08E-05 | 0.006688167 | KCNJ16, AC005208.1 |
| chr17 | 38463749 | 38464189 | 441 | 3 | 2.49E-05 | 0.00088 | 4.57E-05 | 5.08E-05 | 0.006319461 | ARHGAP23 |
| chr16 | 55668373 | 55668553 | 181 | 2 | 0.000351506 | 3.30E-05 | 0.001454 | 5.08E-05 | 0.011333808 | SLC6A2 |
| chr15 | 92809344 | 92810721 | 1378 | 8 | 0.000144052 | 0.000225 | 0.005798 | 5.09E-05 | 0.004351693 | FAM174B |
| chr9 | 130998655 | 130998734 | 80 | 2 | 0.000169585 | 3.24E-05 | 0.001632 | 5.09E-05 | 0.010331282 | NA |
| chr7 | 4107339 | 4107421 | 83 | 2 | 6.33E-05 | 0.002246 | 4.33E-05 | 5.09E-05 | 0.006374277 | SDK1 |
| chr20 | 45254905 | 45255105 | 201 | 3 | 8.56E-05 | 2.75E-05 | 0.002836 | 5.09E-05 | 0.009716884 | NA |
| chr15 | 74802665 | 74803168 | 504 | 6 | 0.000128626 | 4.60E-05 | 0.001796 | 5.09E-05 | 0.002006257 | CSK |
| chr12 | 95789688 | 95790802 | 1115 | 8 | 7.15E-05 | 0.000119 | 0.009697 | 5.09E-05 | 0.002167808 | NTN4 |
| chr14 | 100363010 | 100363254 | 245 | 2 | 0.000213032 | 3.34E-05 | 0.001383 | 5.09E-05 | 0.003222492 | WARS1 |
| chr11 | 131910904 | 131911366 | 463 | 6 | 6.81E-05 | 1.79E-05 | 0.003574 | 5.09E-05 | 0.002920312 | NTM |
| chr9 | 110125096 | 110125623 | 528 | 9 | 0.000296365 | 2.87E-05 | 0.009981 | 5.09E-05 | 0.005845494 | PALM2AKAP2 |
| chr18 | 59308779 | 59308954 | 176 | 4 | 0.00018797 | 0.000652 | 0.000222 | 5.10E-05 | 0.000606882 | CPLX4 |
| chr8 | 123878074 | 123878440 | 367 | 3 | 0.000272918 | 4.72E-05 | 0.002157 | 5.10E-05 | 0.010051148 | FER1L6 |
| chr6 | 40339814 | 40339883 | 70 | 2 | 0.000193361 | 4.83E-05 | 0.000384 | 5.11E-05 | 0.011392384 | TDRG1 |
| chr13 | 81988873 | 81988974 | 102 | 2 | 0.00034692 | 3.20E-05 | 0.001824 | 5.11E-05 | 0.017769357 | NA |
| chr7 | 55344858 | 55345302 | 445 | 3 | 6.32E-05 | 0.000103 | 0.001129 | 5.11E-05 | -0.02910735 | AC073347.1 |
| chr8 | 121677838 | 121678107 | 270 | 3 | 0.000200243 | 0.000631 | 0.000319 | 5.11E-05 | -0.00040636 | HAS2-AS1, LINC02855 |
| chr2 | 239401008 | 239402125 | 1118 | 15 | 9.59E-05 | 0.000775 | 0.000954 | 5.11E-05 | 0.000281279 | HDAC4-AS1, HDAC4 |
| chr11 | 118342557 | 118342936 | 380 | 4 | 0.000135006 | 5.13E-05 | 0.004522 | 5.11E-05 | 0.00771693 | CD3D |
| chr1 | 71068984 | 71069175 | 192 | 3 | 0.000124287 | 0.00069 | 8.00E-05 | 5.12E-05 | 0.005834636 | ZRANB2 |
| chr1 | 6109953 | 6110057 | 105 | 2 | 0.000148046 | 3.35E-05 | 0.001396 | 5.12E-05 | -0.01318422 | CHD5 |
| chr2 | 44802146 | 44802838 | 693 | 4 | 1.55E-06 | 8.37E-05 | 0.001071 | 5.12E-05 | 0.006058473 | NA |
| chr2 | 8608768 | 8608864 | 97 | 2 | 0.000125922 | 5.18E-05 | 0.000331 | 5.12E-05 | 0.019911996 | AC011747.1 |
| chr1 | 108746346 | 108746974 | 629 | 11 | 0.000132794 | 0.003761 | 0.000457 | 5.12E-05 | 0.001432625 | STXBP3 |
| chr16 | 2289259 | 2289535 | 277 | 4 | 0.000310049 | 1.98E-05 | 0.006146 | 5.12E-05 | 0.003459196 | ABCA3 |
| chr12 | 54854423 | 54854778 | 356 | 4 | 7.21E-05 | 9.82E-05 | 0.000433 | 5.13E-05 | 0.008074242 | MUCL1 |
| chr6 | 10838051 | 10838681 | 631 | 8 | 0.000151983 | 0.000703 | 0.002769 | 5.13E-05 | 0.000607061 | TMEM14B, AL024498.2, MAK |
| chr11 | 134781708 | 134781925 | 218 | 2 | 0.000296116 | 4.31E-05 | 0.000528 | 5.13E-05 | 0.016719157 | NA |
| chr12 | 6509972 | 6510637 | 666 | 4 | 8.75E-05 | 0.000442 | 9.03E-05 | 5.13E-05 | 0.000611306 | NCAPD2, SCARNA10, AC006064.5 |
| chr11 | 804761 | 804976 | 216 | 3 | 0.000254414 | 3.62E-05 | 0.002284 | 5.13E-05 | 0.002118298 | PIDD1 |
| chr3 | 50337733 | 50338243 | 511 | 15 | 0.000187291 | 0.003206 | 0.002397 | 5.14E-05 | 0.000775726 | RASSF1-AS1, RASSF1 |
| chr11 | 75430967 | 75431606 | 640 | 3 | 4.80E-05 | 9.55E-05 | 0.000962 | 5.14E-05 | 0.015660852 | KLHL35 |
| chr18 | 47264143 | 47264470 | 328 | 2 | 1.93E-05 | 0.014496 | 3.71E-05 | 5.14E-05 | -0.01684506 | AC051635.1 |
| chr1 | 46492803 | 46493184 | 382 | 4 | 0.000196725 | 4.61E-05 | 0.001366 | 5.15E-05 | 0.008240239 | DMBX1 |
| chr14 | 23372284 | 23372613 | 330 | 3 | 0.00024955 | 0.000305 | 0.00031 | 5.15E-05 | 0.005337563 | NA |
| chr18 | 21682615 | 21682858 | 244 | 2 | 0.00038721 | 4.15E-05 | 0.000599 | 5.15E-05 | 0.00994826 | ABHD3 |
| chr15 | 52019183 | 52019434 | 252 | 4 | 0.000271182 | 3.06E-05 | 0.004201 | 5.15E-05 | -0.0007097 | MAPK6 |
| chr3 | 126464821 | 126464888 | 68 | 2 | 0.000319019 | 5.12E-05 | 0.000345 | 5.15E-05 | -0.00745227 | ZXDC |
| chr1 | 212623980 | 212624624 | 645 | 4 | 6.01E-06 | 0.000252 | 0.000231 | 5.15E-05 | 0.004798349 | FAM71A, AL590648.2 |
| chr2 | 218962938 | 218963448 | 511 | 5 | 6.42E-05 | 0.000507 | 0.001017 | 5.16E-05 | 0.005470361 | NA |
| chr2 | 165813219 | 165814109 | 891 | 4 | 2.60E-05 | 0.000178 | 0.000209 | 5.16E-05 | 0.004209696 | AC009495.1 |
| chr2 | 215436618 | 215437043 | 426 | 4 | 8.63E-05 | 0.000272 | 8.06E-05 | 5.17E-05 | 0.007542628 | AC012462.3 |
| chr1 | 27725635 | 27725887 | 253 | 7 | 0.000384992 | 3.61E-05 | 0.007527 | 5.17E-05 | -7.60E-05 | AL020997.5 |
| chr6 | 73225402 | 73226162 | 761 | 6 | 0.000127773 | 0.000228 | 0.002167 | 5.17E-05 | 0.002131233 | AL365232.1, KHDC1L |
| chr10 | 34252052 | 34252164 | 113 | 2 | 0.00019736 | 0.000253 | 7.81E-05 | 5.17E-05 | 0.003872527 | PARD3 |
| chr22 | 50085257 | 50085695 | 439 | 11 | 0.000266181 | 0.000432 | 0.019508 | 5.17E-05 | 0.00209951 | MLC1 |
| chr7 | 100585599 | 100586371 | 773 | 10 | 5.25E-05 | 6.23E-05 | 0.00961 | 5.17E-05 | -0.00075526 | FBXO24, LRCH4 |
| chr4 | 101348325 | 101348622 | 298 | 2 | 8.77E-05 | 4.47E-05 | 0.000487 | 5.17E-05 | 0.00688827 | FLJ20021 |
| chr2 | 74938994 | 74939565 | 572 | 2 | 1.77E-05 | 0.000259 | 7.73E-05 | 5.17E-05 | 0.006038218 | NA |
| chr8 | 28083888 | 28084035 | 148 | 3 | 8.84E-05 | 0.000565 | 5.99E-05 | 5.17E-05 | 0.006801179 | NUGGC |
| chr12 | 49950428 | 49950843 | 416 | 10 | 0.000112035 | 0.000436 | 0.00099 | 5.18E-05 | 0.007228805 | AQP2 |
| chr16 | 75377886 | 75378163 | 278 | 2 | 0.000178271 | 8.26E-05 | 0.000163 | 5.18E-05 | 0.014709968 | CFDP1 |
| chr21 | 46269711 | 46269772 | 62 | 3 | 0.00040017 | 2.73E-05 | 0.002411 | 5.18E-05 | 0.001641479 | MCM3AP |
| chr12 | 123036027 | 123036152 | 126 | 2 | 0.000229879 | 6.59E-05 | 0.000222 | 5.18E-05 | -0.00243701 | PITPNM2 |
| chr5 | 154189337 | 154189686 | 350 | 2 | 0.000164818 | 3.84E-05 | 0.000798 | 5.18E-05 | 0.009105257 | MFAP3 |
| chr13 | 32310103 | 32310320 | 218 | 2 | 5.06E-05 | 0.000498 | 6.12E-05 | 5.18E-05 | 0.010244101 | ZAR1L |
| chr13 | 98441502 | 98441565 | 64 | 2 | 0.000175228 | 3.74E-05 | 0.00089 | 5.18E-05 | 0.006051069 | FARP1 |
| chr13 | 41454396 | 41454464 | 69 | 2 | 0.000355274 | 3.32E-05 | 0.001616 | 5.18E-05 | 0.011040333 | NA |
| chr12 | 14756433 | 14756622 | 190 | 2 | 0.000321892 | 4.03E-05 | 0.00068 | 5.19E-05 | 0.025034375 | AC010168.1 |
| chr19 | 7535531 | 7536121 | 591 | 4 | 3.87E-05 | 0.000372 | 0.000146 | 5.19E-05 | -0.00178169 | AC008878.2, PNPLA6 |
| chr7 | 140647207 | 140647317 | 111 | 2 | 0.000330538 | 4.21E-05 | 0.000589 | 5.19E-05 | 0.008602803 | AC006452.1, DENND2A |
| chr20 | 18507352 | 18508001 | 650 | 15 | 0.000153283 | 0.001104 | 0.002086 | 5.19E-05 | 3.53E-06 | SEC23B |
| chr7 | 74217971 | 74218114 | 144 | 3 | 0.000201327 | 6.89E-05 | 0.000351 | 5.19E-05 | 0.007664279 | LAT2 |
| chr16 | 85535749 | 85536278 | 530 | 3 | 0.000350102 | 2.44E-05 | 0.004083 | 5.19E-05 | 0.010298638 | GSE1 |
| chr3 | 184500708 | 184500737 | 30 | 2 | 0.000369152 | 3.45E-05 | 0.001299 | 5.19E-05 | 0.014135857 | NA |
| chr2 | 169694274 | 169694589 | 316 | 10 | 0.000353171 | 0.00112 | 0.003619 | 5.19E-05 | 0.000981229 | PHOSPHO2, KLHL23, CCDC173 |
| chr16 | 87668929 | 87669097 | 169 | 3 | 0.000192077 | 4.06E-05 | 0.001784 | 5.19E-05 | 0.003808093 | JPH3 |
| chr6 | 151493757 | 151494106 | 350 | 9 | 0.000196449 | 8.91E-05 | 0.007046 | 5.19E-05 | 0.005940222 | CCDC170 |
| chr3 | 15315372 | 15315374 | 3 | 2 | 0.000369422 | 3.44E-05 | 0.001333 | 5.19E-05 | 0.013367969 | SH3BP5 |
| chr16 | 85112482 | 85112572 | 91 | 3 | 4.56E-05 | 0.001575 | 0.000777 | 5.20E-05 | 0.004290324 | NA |

| | | | | | | | | | | |
|---|---|---|---|---|---|---|---|---|---|---|
| chr17 | 7404078 | 7404382 | 305 | 9 | 0.000276366 | 0.000561 | 0.002099 | 5.20E-05 | -0.0009282 | TMEM256-PLSCR3, TMEM256 |
| chr4 | 76074020 | 76074341 | 322 | 4 | 0.000275293 | 3.85E-05 | 0.004053 | 5.20E-05 | 0.003515217 | ART3 |
| chr5 | 150318939 | 150319159 | 221 | 2 | 4.58E-05 | 0.000227 | 8.25E-05 | 5.20E-05 | 0.005441966 | ARSI |
| chr10 | 79311648 | 79311887 | 240 | 3 | 0.000253039 | 0.000149 | 0.001237 | 5.20E-05 | -0.00495606 | ZMIZ1 |
| chr2 | 25375895 | 25375996 | 102 | 3 | 0.000242893 | 0.000105 | 0.000482 | 5.21E-05 | 0.009511785 | NA |
| chr3 | 99532492 | 99532692 | 201 | 2 | 0.000104634 | 0.001983 | 4.52E-05 | 5.21E-05 | 0.013069331 | NA |
| chr19 | 39406796 | 39407375 | 580 | 6 | 0.000418932 | 9.20E-05 | 0.010286 | 5.21E-05 | -6.16E-05 | ZFP36 |
| chr7 | 6571670 | 6571787 | 118 | 2 | 0.000224723 | 0.000151 | 0.000101 | 5.21E-05 | 0.002902086 | NA |
| chr2 | 96547825 | 96547905 | 81 | 2 | 0.000337862 | 3.30E-05 | 0.001776 | 5.21E-05 | 0.00402896 | ARID5A |
| chr20 | 63565034 | 63565288 | 255 | 2 | 1.07E-05 | 0.003059 | 4.26E-05 | 5.22E-05 | 0.003129813 | HELZ2 |
| chr8 | 100106615 | 100106964 | 350 | 3 | 0.000269319 | 2.94E-05 | 0.003338 | 5.22E-05 | -0.0188122 | RGS22 |
| chr6 | 41806286 | 41807018 | 733 | 7 | 2.83E-05 | 0.013904 | 9.34E-05 | 5.22E-05 | 0.000268352 | USP49 |
| chr8 | 87873823 | 87874296 | 474 | 10 | 0.000119163 | 0.000247 | 0.000929 | 5.23E-05 | 0.001412765 | DCAF4L2 |
| chr1 | 113140899 | 113141393 | 495 | 3 | 0.000242382 | 0.004434 | 0.000914 | 5.23E-05 | 0.012103837 | NA |
| chr6 | 35512649 | 35513139 | 491 | 6 | 7.37E-05 | 0.000237 | 0.000331 | 5.23E-05 | 0.003605709 | TULP1 |
| chr9 | 136760446 | 136760809 | 364 | 5 | 0.00018047 | 0.000127 | 0.000809 | 5.23E-05 | 0.005969793 | LCN15 |
| chr12 | 124793106 | 124793334 | 229 | 4 | 0.000261505 | 0.000149 | 0.000289 | 5.24E-05 | 0.000689722 | AC073593.2, SCARB1 |
| chr5 | 142769825 | 142770450 | 626 | 7 | 0.000212443 | 0.00025 | 0.000913 | 5.24E-05 | -0.00021559 | ARHGAP26 |
| chr6 | 7730704 | 7731047 | 344 | 3 | 0.000251438 | 0.000165 | 0.000709 | 5.24E-05 | 0.017784334 | BMP6 |
| chr17 | 50527508 | 50527548 | 41 | 3 | 0.000343326 | 4.38E-05 | 0.000649 | 5.24E-05 | 0.012915634 | MYCBPAP, AC021491.2 |
| chr12 | 131817341 | 131817573 | 233 | 3 | 9.61E-05 | 0.00013 | 0.000386 | 5.24E-05 | 0.007336972 | NA |
| chr1 | 150516329 | 150516851 | 523 | 3 | 3.70E-05 | 0.000158 | 0.000117 | 5.24E-05 | 0.006436784 | FALEC |
| chr3 | 185943835 | 185943874 | 40 | 2 | 0.000257904 | 4.86E-05 | 0.000416 | 5.24E-05 | 0.015098903 | NA |
| chr9 | 119690064 | 119690076 | 13 | 2 | 0.000380292 | 3.31E-05 | 0.001827 | 5.24E-05 | 0.025314034 | NA |
| chr20 | 61998188 | 61998467 | 280 | 2 | 7.23E-05 | 0.003144 | 4.26E-05 | 5.25E-05 | -0.00248905 | TAF4 |
| chr4 | 170090244 | 170090609 | 366 | 11 | 0.000127721 | 0.00077 | 0.003826 | 5.25E-05 | 0.002924759 | AADAT |
| chr16 | 87543667 | 87543933 | 267 | 3 | 0.000368833 | 2.45E-05 | 0.003775 | 5.25E-05 | 0.00764117 | NA |
| chr18 | 74520227 | 74520241 | 15 | 2 | 0.000286126 | 8.83E-05 | 0.000155 | 5.25E-05 | 0.004809909 | CNDP2 |
| chr2 | 24792728 | 24793469 | 742 | 15 | 1.77E-05 | 0.002302 | 0.000857 | 5.25E-05 | 0.000651108 | CENPO, PTRHD1 |
| chr4 | 2935246 | 2935654 | 409 | 5 | 1.70E-05 | 0.002038 | 0.001491 | 5.25E-05 | -0.00091432 | NOP14-AS1 |
| chr19 | 2307593 | 2307902 | 310 | 3 | 3.35E-05 | 4.17E-05 | 0.000968 | 5.26E-05 | 0.017574128 | AC005258.1 |
| chr9 | 113402166 | 113402516 | 351 | 2 | 5.11E-05 | 6.11E-05 | 0.000261 | 5.26E-05 | 0.007061214 | NA |
| chr8 | 11681745 | 11681896 | 152 | 3 | 0.000117339 | 0.000665 | 0.000458 | 5.27E-05 | 0.008694823 | GATA4 |
| chr11 | 123188231 | 123188751 | 521 | 3 | 0.000275116 | 2.66E-05 | 0.003931 | 5.27E-05 | 0.017859196 | CLMP |
| chr1 | 156161422 | 156161588 | 167 | 2 | 0.000461959 | 3.33E-05 | 0.001819 | 5.27E-05 | -0.01401283 | SEMA4A |
| chr20 | 2559149 | 2559373 | 225 | 4 | 0.000105848 | 0.000585 | 0.000184 | 5.28E-05 | 0.000317767 | TMC2 |
| chr22 | 50056732 | 50056945 | 214 | 2 | 0.000189569 | 9.73E-05 | 0.000142 | 5.28E-05 | 0.009586761 | TTLL8 |
| chr2 | 196926219 | 196927282 | 1064 | 9 | 4.63E-05 | 2.15E-05 | 0.008034 | 5.28E-05 | -0.0002154 | PGAP1 |
| chr6 | 27473787 | 27474327 | 541 | 4 | 4.69E-05 | 0.000403 | 0.001569 | 5.28E-05 | -0.00065454 | AL021918.5 |
| chr7 | 128883460 | 128883766 | 307 | 3 | 0.000258093 | 0.000302 | 0.000582 | 5.28E-05 | 0.009201231 | KCP |
| chr3 | 11878983 | 11879140 | 158 | 2 | 0.000199495 | 0.000197 | 8.94E-05 | 5.28E-05 | 0.01268137 | FANCD2P2, CYCSP12 |
| chr17 | 76881011 | 76881331 | 321 | 2 | 7.98E-05 | 4.17E-05 | 0.000653 | 5.29E-05 | 0.013152355 | MGAT5B |
| chr7 | 157910528 | 157910733 | 206 | 4 | 0.000394802 | 2.81E-05 | 0.005365 | 5.29E-05 | 0.000974505 | PTPRN2 |
| chr1 | 109208604 | 109208843 | 240 | 3 | 0.000103514 | 0.001504 | 6.08E-05 | 5.29E-05 | 0.005867168 | NA |
| chr12 | 51977919 | 51978041 | 123 | 2 | 0.00023715 | 3.67E-05 | 0.001083 | 5.29E-05 | 0.009464892 | ACVR1B |
| chr6 | 166626475 | 166626952 | 478 | 2 | 2.17E-06 | 0.000101 | 0.00014 | 5.31E-05 | -0.00378186 | RPS6KA2 |
| chr10 | 29538112 | 29538813 | 702 | 7 | 0.000125319 | 0.000562 | 0.011011 | 5.31E-05 | 0.00118958 | SVIL |
| chr4 | 152846870 | 152847098 | 229 | 2 | 0.000388392 | 3.35E-05 | 0.001868 | 5.31E-05 | 0.029194865 | ARFIP1 |
| chr7 | 46776630 | 46776993 | 364 | 2 | 0.000253762 | 3.71E-05 | 0.001052 | 5.31E-05 | 0.015274069 | NA |
| chr18 | 48925024 | 48925211 | 188 | 2 | 0.000301415 | 3.62E-05 | 0.001174 | 5.31E-05 | 0.01798752 | SMAD7 |
| chr2 | 11723707 | 11724674 | 968 | 6 | 8.43E-06 | 0.108851 | 0.000211 | 5.31E-05 | 0.002620899 | LPIN1, AC012456.1 |
| chr16 | 58683903 | 58684159 | 257 | 3 | 9.79E-05 | 0.000164 | 0.00041 | 5.31E-05 | 0.005267925 | SLC38A7 |
| chr3 | 72339293 | 72339461 | 169 | 2 | 0.000279818 | 3.35E-05 | 0.001833 | 5.31E-05 | 0.00674709 | NA |
| chr11 | 35420195 | 35420678 | 484 | 8 | 0.000152827 | 0.000121 | 0.000588 | 5.32E-05 | 0.001894166 | AC090625.2 |
| chr19 | 15237992 | 15238924 | 933 | 10 | 0.000186638 | 0.000189 | 0.01727 | 5.32E-05 | 0.001855567 | BRD4 |
| chr11 | 75597455 | 75597838 | 384 | 3 | 0.000349859 | 4.07E-05 | 0.000811 | 5.32E-05 | 0.006890956 | MAP6 |
| chr22 | 30870907 | 30871249 | 343 | 2 | 0.000201055 | 6.56E-05 | 0.000238 | 5.32E-05 | 0.006673936 | OSBP2 |
| chr16 | 84076240 | 84076344 | 105 | 2 | 0.000155368 | 3.40E-05 | 0.001705 | 5.32E-05 | 0.013808223 | MBTPS1 |
| chr1 | 159008678 | 159009167 | 490 | 4 | 2.23E-05 | 0.004637 | 4.49E-05 | 5.32E-05 | 0.004385051 | IFI16 |
| chr1 | 84277300 | 84277531 | 232 | 2 | 0.000200297 | 0.000325 | 7.32E-05 | 5.33E-05 | 0.002257175 | AL359273.1 |
| chr8 | 38425706 | 38426241 | 536 | 3 | 0.000180526 | 0.000107 | 0.000559 | 5.33E-05 | 0.003223992 | AC087623.1, FGFR1 |
| chr11 | 14643052 | 14643508 | 457 | 7 | 0.000300146 | 0.000304 | 0.004988 | 5.33E-05 | 0.006744245 | PSMA1 |
| chr20 | 36305565 | 36305859 | 295 | 2 | 6.91E-05 | 3.79E-05 | 0.00098 | 5.34E-05 | -0.00789122 | NA |
| chr21 | 35244671 | 35244697 | 27 | 2 | 0.000336287 | 5.39E-05 | 0.000346 | 5.34E-05 | -0.00318158 | RUNX1 |
| chr15 | 29269845 | 29270338 | 494 | 8 | 0.000154405 | 0.00041 | 0.003572 | 5.34E-05 | 9.00E-05 | FAM189A1 |
| chr11 | 123084382 | 123084745 | 364 | 4 | 0.000110603 | 0.000753 | 7.34E-05 | 5.34E-05 | 0.000866485 | AP000926.2, CLMP |
| chr7 | 103345057 | 103345140 | 84 | 6 | 0.000382429 | 1.99E-05 | 0.009613 | 5.34E-05 | -0.00346573 | PSMC2 |
| chr2 | 105857278 | 105857717 | 440 | 3 | 1.93E-05 | 0.000211 | 0.00014 | 5.35E-05 | 0.008655981 | NCK2 |
| chr12 | 55844109 | 55844141 | 33 | 2 | 6.72E-05 | 0.000101 | 0.00014 | 5.35E-05 | 0.01117227 | NA |
| chr10 | 79263442 | 79263481 | 40 | 2 | 0.000125054 | 3.43E-05 | 0.001674 | 5.35E-05 | 0.011532671 | ZMIZ1 |
| chr7 | 157643814 | 157644290 | 477 | 3 | 4.15E-05 | 0.000609 | 0.001012 | 5.35E-05 | 0.003255972 | PTPRN2 |
| chr19 | 40192644 | 40192977 | 334 | 2 | 0.000184688 | 4.50E-05 | 0.000542 | 5.35E-05 | 0.00301592 | MAP3K10 |
| chr10 | 99899725 | 99899971 | 247 | 2 | 5.12E-05 | 0.000848 | 5.51E-05 | 5.35E-05 | 0.00393566 | DNMBP |
| chr17 | 63932028 | 63932475 | 448 | 6 | 3.61E-05 | 3.23E-05 | 0.005549 | 5.35E-05 | 0.005525398 | CD79B |
| chr14 | 90777877 | 90778077 | 201 | 2 | 0.000147553 | 4.50E-05 | 0.000546 | 5.36E-05 | 0.01535956 | TTC7B |
| chr19 | 24004392 | 24004756 | 365 | 4 | 0.000448663 | 2.28E-05 | 0.003599 | 5.36E-05 | 0.008059097 | RNA5-8SP4 |
| chr18 | 50507189 | 50507463 | 275 | 2 | 0.000385256 | 3.40E-05 | 0.001786 | 5.36E-05 | 0.017243365 | NA |
| chr16 | 77787890 | 77788874 | 985 | 10 | 7.00E-05 | 0.002846 | 0.001161 | 5.36E-05 | -0.00243863 | VAT1L |
| chr3 | 126101529 | 126102081 | 553 | 9 | 9.98E-05 | 0.004994 | 0.000306 | 5.36E-05 | 0.002260583 | SLC41A3 |
| chr21 | 24953040 | 24953301 | 262 | 3 | 0.000300673 | 0.000103 | 0.001436 | 5.36E-05 | 0.008147252 | LINC01692, AP000146.1 |
| chr15 | 72229634 | 72230025 | 392 | 8 | 0.000290885 | 8.61E-05 | 0.016333 | 5.36E-05 | 0.000685417 | PKM |
| chr15 | 27685952 | 27685967 | 16 | 2 | 9.48E-05 | 0.000821 | 5.55E-05 | 5.36E-05 | 0.00455296 | NA |
| chr8 | 10526579 | 10526968 | 390 | 2 | 0.000178828 | 3.62E-05 | 0.001252 | 5.36E-05 | 0.015043159 | PRSS55, PRSS51 |
| chr7 | 108003148 | 108004125 | 978 | 12 | 8.79E-05 | 0.000276 | 0.001792 | 5.37E-05 | 0.000104444 | LAMB1 |
| chr7 | 136874674 | 136874915 | 242 | 2 | 0.00010121 | 0.000543 | 6.19E-05 | 5.37E-05 | 0.020389238 | CHRM2, AC009264.1 |
| chr5 | 1232980 | 1233005 | 26 | 2 | 0.000388874 | 4.28E-05 | 0.000645 | 5.38E-05 | 0.003243004 | SLC6A18 |
| chr1 | 103525471 | 103526489 | 1019 | 17 | 1.84E-05 | 0.010274 | 0.000307 | 5.38E-05 | 0.0002808 | RNPC3, AC095032.1 |
| chr15 | 97959653 | 97960214 | 562 | 3 | 0.000120616 | 2.77E-05 | 0.003126 | 5.38E-05 | 0.003359062 | NA |
| chr17 | 9575518 | 9575901 | 384 | 5 | 0.000287341 | 5.06E-05 | 0.001695 | 5.39E-05 | 0.001112681 | STX8 |
| chr10 | 50603372 | 50603589 | 218 | 2 | 0.000120745 | 0.000225 | 8.61E-05 | 5.40E-05 | 0.012667228 | SGMS1 |
| chr18 | 44778894 | 44778949 | 56 | 2 | 9.53E-05 | 9.15E-05 | 0.000157 | 5.40E-05 | 0.021184826 | SETBP1 |
| chr9 | 95475995 | 95476236 | 242 | 3 | 0.000197465 | 0.000905 | 0.000796 | 5.40E-05 | -0.00492839 | PTCH1 |
| chr2 | 232838410 | 232838478 | 69 | 2 | 0.000370815 | 3.62E-05 | 0.001304 | 5.40E-05 | 0.012514495 | GIGYF2 |
| chr9 | 137435624 | 137436234 | 611 | 5 | 1.39E-05 | 0.001102 | 9.96E-05 | 5.40E-05 | 0.007085931 | ENTPD8 |
| chr19 | 11087880 | 11088288 | 409 | 2 | 0.000241612 | 4.73E-05 | 0.000494 | 5.41E-05 | 0.01053115 | NA |
| chr14 | 73136010 | 73136621 | 612 | 13 | 9.78E-05 | 0.000471 | 9.00E-05 | 5.41E-05 | 0.00279048 | PSEN1 |
| chr11 | 73194285 | 73194779 | 495 | 3 | 0.000141529 | 2.81E-05 | 0.00313 | 5.41E-05 | 0.003562971 | NA |
| chr7 | 157265405 | 157265699 | 295 | 5 | 0.000355094 | 9.75E-05 | 0.004372 | 5.41E-05 | 7.60E-05 | UBE3C |
| chr1 | 75357257 | 75357336 | 80 | 2 | 4.49E-05 | 5.18E-05 | 0.000392 | 5.41E-05 | 0.001653755 | SLC44A5 |
| chr2 | 86808155 | 86808859 | 705 | 10 | 5.43E-05 | 0.000893 | 0.004544 | 5.41E-05 | 0.002088507 | CD8A |
| chr2 | 1768127 | 1768303 | 177 | 2 | 6.11E-05 | 0.005153 | 4.16E-05 | 5.41E-05 | 0.003687512 | NA |
| chr5 | 38845498 | 38846000 | 503 | 7 | 7.30E-05 | 0.001218 | 0.008856 | 5.41E-05 | -0.00394017 | OSMR, OSMR-AS1 |
| chr8 | 144623536 | 144624222 | 687 | 6 | 6.09E-05 | 0.000289 | 0.000603 | 5.41E-05 | 0.002635633 | NA |
| chr17 | 78639171 | 78639519 | 349 | 3 | 0.000110477 | 0.000922 | 0.000184 | 5.42E-05 | 0.007558055 | NA |
| chr2 | 237563393 | 237563762 | 370 | 3 | 5.03E-05 | 0.001118 | 4.85E-05 | 5.42E-05 | 0.004618407 | NA |
| chr6 | 1317327 | 1317512 | 186 | 4 | 0.000212574 | 1.58E-05 | 0.01391 | 5.42E-05 | -0.02575824 | NA |
| chr10 | 99330714 | 99330801 | 88 | 2 | 0.000137099 | 4.95E-05 | 0.000445 | 5.43E-05 | 0.007764182 | CNNM1 |
| chr3 | 194685721 | 194685843 | 123 | 6 | 0.000406204 | 2.35E-05 | 0.004603 | 5.43E-05 | 0.002558523 | NA |
| chr8 | 23854076 | 23854539 | 464 | 6 | 0.000251236 | 0.00055 | 0.001909 | 5.43E-05 | 0.00345378 | STC1 |
| chr10 | 123142833 | 123143328 | 496 | 5 | 0.000174007 | 0.001445 | 0.000443 | 5.44E-05 | 0.004721489 | NA |
| chr1 | 153962310 | 153962966 | 657 | 2 | 2.55E-06 | 0.00015 | 0.000108 | 5.45E-05 | 0.00728364 | SLC39A1, AL358472.7, MIR6737 |
| chr9 | 126488581 | 126488682 | 102 | 2 | 0.000296993 | 7.39E-05 | 0.00021 | 5.45E-05 | 0.016730371 | MVB12B |
| chr19 | 45008535 | 45008864 | 330 | 2 | 0.000346429 | 3.71E-05 | 0.001216 | 5.45E-05 | 0.009435239 | RELB |
| chr1 | 24066396 | 24067094 | 699 | 6 | 8.56E-05 | 0.000281 | 0.002216 | 5.45E-05 | 0.004577114 | AL591178.1, MYOM3-AS1, MYOM3 |
| chr10 | 132502896 | 132503082 | 187 | 2 | 0.000326951 | 4.17E-05 | 0.000748 | 5.46E-05 | 0.006357357 | NA |
| chr12 | 26938530 | 26938972 | 443 | 4 | 0.000127883 | 5.33E-05 | 0.002509 | 5.46E-05 | -0.00063397 | FGFR1OP2 |
| chr9 | 36092671 | 36092683 | 13 | 2 | 0.000232131 | 3.45E-05 | 0.001887 | 5.47E-05 | 0.020216173 | RECK |
| chr4 | 8616011 | 8616183 | 173 | 3 | 0.000349075 | 5.19E-05 | 0.001256 | 5.48E-05 | 0.002800856 | GPR78, CPZ |
| chr11 | 9219576 | 9219613 | 38 | 2 | 0.000235249 | 0.000446 | 6.73E-05 | 5.48E-05 | 0.011066737 | DENND5A |
| chr8 | 56074731 | 56075439 | 709 | 6 | 0.000162835 | 4.00E-05 | 0.012432 | 5.49E-05 | -0.0016792 | CERNA3 |
| chr19 | 10566815 | 10567339 | 525 | 5 | 3.28E-05 | 0.001118 | 0.002387 | 5.49E-05 | 0.006518109 | CDKN2D |
| chr5 | 914363 | 914414 | 52 | 2 | 0.000108319 | 0.000139 | 0.000115 | 5.50E-05 | 0.00157577 | TRIP13 |
| chr1 | 155858046 | 155858566 | 521 | 3 | 1.64E-05 | 0.000282 | 0.000107 | 5.50E-05 | 0.000643392 | GON4L |
| chr12 | 118060149 | 118060519 | 371 | 3 | 0.000156459 | 3.15E-05 | 0.001783 | 5.50E-05 | 0.008984051 | WSB2 |
| chr12 | 54378904 | 54379020 | 117 | 2 | 0.000164823 | 6.05E-05 | 0.000298 | 5.50E-05 | -0.023873 | AC079313.2, AC079313.1, ZNF385A |
| chr12 | 124289122 | 124289573 | 452 | 6 | 0.000250453 | 0.000165 | 0.00833 | 5.51E-05 | 0.002182995 | RFLNA |
| chr16 | 89098545 | 89099197 | 653 | 5 | 0.000120235 | 7.50E-05 | 0.002133 | 5.51E-05 | 0.01057206 | ACSF3 |
| chr19 | 1925797 | 1926144 | 348 | 2 | 7.37E-05 | 0.000598 | 6.18E-05 | 5.52E-05 | 0.007446738 | SCAMP4 |
| chr16 | 31401638 | 31402155 | 518 | 3 | 9.44E-05 | 0.000253 | 0.0001 | 5.52E-05 | 0.006647439 | ITGAD |
| chr12 | 1915245 | 1915503 | 259 | 4 | 0.000365117 | 3.53E-05 | 0.002918 | 5.52E-05 | 0.005237649 | CACNA2D4 |
| chr13 | 113724499 | 113725197 | 699 | 19 | 0.000157832 | 0.001944 | 0.000773 | 5.52E-05 | 0.000468146 | GRK1 |
| chr3 | 49171081 | 49171727 | 647 | 9 | 8.54E-05 | 0.000398 | 0.000408 | 5.52E-05 | 0.002054897 | KLHDC8B |
| chr12 | 31645931 | 31646184 | 254 | 4 | 5.64E-05 | 0.00258 | 0.000121 | 5.52E-05 | 0.011927566 | AC068774.1 |
| chr1 | 8232518 | 8232597 | 80 | 2 | 0.000312987 | 6.37E-05 | 0.000274 | 5.53E-05 | -0.01559945 | NA |
| chr10 | 102399528 | 102399886 | 359 | 5 | 0.00019873 | 0.000183 | 0.000539 | 5.53E-05 | 0.003099262 | NFKB2 |
| chr17 | 969522 | 969530 | 9 | 2 | 0.000297594 | 3.66E-05 | 0.001425 | 5.53E-05 | -0.00531854 | NXN |
| chr2 | 44941654 | 44941785 | 132 | 5 | 0.000177693 | 0.000173 | 0.000218 | 5.54E-05 | 0.002607703 | SIX3, SIX3-AS1 |
| chr8 | 143829574 | 143830316 | 743 | 6 | 2.05E-05 | 0.006652 | 1.61E-05 | 5.54E-05 | 0.00011023 | AC234917.3 |
| chr16 | 80629107 | 80629286 | 180 | 2 | 3.08E-05 | 0.000261 | 8.33E-05 | 5.54E-05 | 0.001905948 | CDYL2 |
| chr1 | 43529629 | 43530730 | 1102 | 6 | 3.23E-05 | 0.00148 | 0.002049 | 5.54E-05 | 0.007841271 | PTPRF |
| chr7 | 149714286 | 149715059 | 774 | 4 | 5.34E-05 | 0.000731 | 0.001205 | 5.54E-05 | -0.00789417 | KRBA1 |
| chr16 | 1884668 | 1885057 | 390 | 2 | 9.65E-05 | 3.49E-05 | 0.001928 | 5.54E-05 | 0.010313101 | NA |
| chr4 | 146241611 | 146242015 | 405 | 3 | 0.000101884 | 4.43E-05 | 0.001263 | 5.54E-05 | 0.011337061 | AC097372.1 |
| chr2 | 241603046 | 241603139 | 94 | 3 | 6.64E-05 | 7.48E-05 | 0.000593 | 5.54E-05 | 0.002177237 | THAP4 |
| chr3 | 48528033 | 48528264 | 232 | 2 | 0.00029932 | 3.68E-05 | 0.001405 | 5.55E-05 | 0.00944206 | PFKFB4 |
| chr1 | 20553885 | 20553968 | 84 | 2 | 0.000383594 | 0.000101 | 0.000149 | 5.55E-05 | 0.007738565 | FAM43B |
| chr5 | 1169258 | 1169517 | 260 | 2 | 4.63E-05 | 3.54E-05 | 0.00177 | 5.55E-05 | 0.005374382 | NA |
| chr6 | 116280500 | 116281004 | 505 | 3 | 2.53E-05 | 0.00029 | 0.001261 | 5.55E-05 | 0.006423655 | DSE |
| chr16 | 35145844 | 35146086 | 243 | 2 | 0.000140158 | 0.000402 | 7.08E-05 | 5.56E-05 | 0.000992248 | AC023824.5 |
| chr10 | 101354176 | 101354977 | 802 | 3 | 1.02E-05 | 0.000399 | 0.0001 | 5.56E-05 | 0.004365987 | BTRC |
| chr7 | 16398840 | 16399909 | 1070 | 4 | 2.01E-06 | 0.001217 | 9.13E-05 | 5.56E-05 | 0.00230266 | CRPPA |
| chr5 | 180117122 | 180117404 | 283 | 2 | 0.000264765 | 3.62E-05 | 0.001577 | 5.56E-05 | 0.005719497 | RASGEF1C |
| chr6 | 166006281 | 166006724 | 444 | 4 | 0.000205035 | 0.006267 | 0.000453 | 5.57E-05 | 0.002719632 | NA |
| chr5 | 177356737 | 177357549 | 813 | 5 | 1.07E-05 | 0.012371 | 0.000246 | 5.58E-05 | 0.006110468 | NA |
| chr17 | 59209948 | 59210430 | 483 | 10 | 0.000290417 | 0.000759 | 0.002737 | 5.58E-05 | 9.23E-06 | AC099850.2, SMG8 |
| chr7 | 43614157 | 43614192 | 36 | 2 | 0.000254673 | 3.58E-05 | 0.001722 | 5.59E-05 | 0.010464481 | STK17A, COA1 |
| chr19 | 44890128 | 44890668 | 541 | 4 | 0.000278393 | 1.90E-05 | 0.01009 | 5.59E-05 | -0.00541152 | TOMM40, AC011481.2 |
| chr3 | 169812239 | 169813132 | 894 | 16 | 0.000387177 | 4.97E-05 | 0.03083 | 5.60E-05 | 0.009536306 | LRRC34 |
| chr8 | 29767204 | 29767367 | 164 | 2 | 0.00023446 | 0.000125 | 0.000127 | 5.61E-05 | 0.00958974 | LINC02099 |
| chr16 | 9134844 | 9134925 | 82 | 2 | 0.000295154 | 3.61E-05 | 0.001675 | 5.61E-05 | 0.008063092 | NA |
| chr11 | 44775320 | 44775762 | 443 | 3 | 0.000340024 | 2.36E-05 | 0.006187 | 5.61E-05 | 0.01243614 | TSPAN18 |
| chr10 | 6124824 | 6124914 | 91 | 2 | 0.000257591 | 3.93E-05 | 0.001074 | 5.61E-05 | 0.018416358 | NA |

| | | | | | | | | | | |
|---|---|---|---|---|---|---|---|---|---|---|
| chr18 | 12540174 | 12540328 | 155 | 2 | 0.000409969 | 3.54E-05 | 0.001937 | 5.62E-05 | 0.011943351 | SPIRE1 |
| chr11 | 31801593 | 31801734 | 142 | 5 | 0.000157892 | 6.28E-05 | 0.000606 | 5.62E-05 | 0.008169713 | PAX6 |
| chr5 | 177396319 | 177396423 | 105 | 2 | 0.000387747 | 3.96E-05 | 0.001047 | 5.62E-05 | 0.015810752 | SLC34A1 |
| chr11 | 108219403 | 108219432 | 30 | 2 | 0.000408348 | 3.56E-05 | 0.00188 | 5.62E-05 | 0.013362167 | NPAT |
| chr14 | 100821121 | 100821339 | 219 | 3 | 0.000103023 | 0.001262 | 7.83E-05 | 5.62E-05 | -0.0096004 | MEG3 |
| chr6 | 44222512 | 44222992 | 481 | 4 | 2.43E-05 | 0.000996 | 5.82E-05 | 5.63E-05 | 0.006407927 | SLC29A1 |
| chr16 | 56363943 | 56364193 | 251 | 5 | 0.000144583 | 0.002444 | 0.00103 | 5.64E-05 | 0.001582375 | AMFR |
| chr9 | 127897391 | 127898122 | 732 | 5 | 0.000147683 | 1.58E-05 | 0.012099 | 5.64E-05 | 0.001652211 | AL157935.2, ST6GALNAC6 |
| chr16 | 2531121 | 2532141 | 1021 | 10 | 0.000142635 | 5.39E-05 | 0.013245 | 5.64E-05 | 0.003097941 | AMDHD2, CEMP1, MIR3178 |
| chr2 | 219294873 | 219295254 | 382 | 3 | 0.000187787 | 4.56E-05 | 0.000728 | 5.64E-05 | 0.005885752 | PTPRN |
| chr6 | 132275016 | 132275154 | 139 | 3 | 0.000120985 | 0.005193 | 4.24E-05 | 5.64E-05 | -0.00172846 | NA |
| chr1 | 23975664 | 23975730 | 67 | 2 | 0.000154964 | 3.92E-05 | 0.001116 | 5.64E-05 | 0.010190334 | SRSF10 |
| chr10 | 5498458 | 5498861 | 404 | 3 | 0.000225858 | 6.41E-05 | 0.001826 | 5.64E-05 | 0.002618716 | CALML5 |
| chr17 | 17380356 | 17380397 | 42 | 2 | 0.000107603 | 0.000268 | 8.40E-05 | 5.64E-05 | 0.00358685 | NA |
| chr6 | 13815015 | 13815336 | 322 | 2 | 6.11E-05 | 3.68E-05 | 0.00156 | 5.65E-05 | 0.009856552 | NA |
| chr6 | 88046636 | 88046658 | 23 | 2 | 6.81E-05 | 3.56E-05 | 0.001944 | 5.65E-05 | 0.000952621 | NA |
| chr19 | 4101813 | 4102603 | 791 | 5 | 0.000206615 | 0.004328 | 0.001623 | 5.65E-05 | 0.003294695 | MAP2K2 |
| chr14 | 91375611 | 91375805 | 195 | 2 | 0.000284294 | 3.64E-05 | 0.001666 | 5.65E-05 | 0.006288812 | CCDC88C |
| chr1 | 246566270 | 246566390 | 121 | 5 | 0.000484274 | 0.000285 | 0.0028 | 5.66E-05 | -0.0005005 | NA |
| chr16 | 74107767 | 74107905 | 139 | 2 | 0.000213826 | 0.000258 | 8.56E-05 | 5.67E-05 | 0.003986856 | AC138627.1 |
| chr3 | 27728568 | 27728900 | 333 | 3 | 0.000119885 | 3.71E-05 | 0.00124 | 5.67E-05 | 0.006531443 | NA |
| chr2 | 27119930 | 27120009 | 80 | 2 | 0.000342428 | 3.74E-05 | 0.001464 | 5.67E-05 | -0.00056405 | NA |
| chr19 | 14575808 | 14575902 | 95 | 2 | 0.000377301 | 4.25E-05 | 0.00082 | 5.67E-05 | 0.016189177 | NA |
| chr12 | 63779762 | 63780404 | 643 | 10 | 0.000277261 | 7.52E-05 | 0.012635 | 5.67E-05 | 0.000349964 | RXYLT1, AC084357.2 |
| chr17 | 77402208 | 77402501 | 294 | 4 | 0.000124708 | 0.000264 | 8.69E-05 | 5.67E-05 | 0.000652427 | SEPTIN9 |
| chr2 | 241570078 | 241570294 | 217 | 2 | 0.000337504 | 4.26E-05 | 0.000816 | 5.67E-05 | 0.025594776 | BOK |
| chr8 | 144294918 | 144295106 | 189 | 2 | 0.000193178 | 0.000131 | 0.000125 | 5.69E-05 | 0.006689691 | HSF1 |
| chr12 | 129831470 | 129831513 | 44 | 3 | 0.000392747 | 5.66E-05 | 0.00209 | 5.69E-05 | 0.008668523 | TMEM132D |
| chr7 | 117648388 | 117648656 | 269 | 3 | 8.10E-05 | 0.00119 | 0.000206 | 5.69E-05 | 0.006497243 | CFTR |
| chr21 | 43340962 | 43341600 | 639 | 4 | 0.000115129 | 0.005448 | 7.02E-05 | 5.70E-05 | 0.004620088 | NA |
| chr16 | 974642 | 974838 | 197 | 2 | 0.000207482 | 0.000139 | 0.000121 | 5.70E-05 | 0.007757843 | LMF1 |
| chr7 | 44565255 | 44565935 | 681 | 3 | 0.000169298 | 8.33E-05 | 0.000261 | 5.70E-05 | 0.004911973 | DDX56 |
| chr11 | 62817243 | 62817345 | 103 | 2 | 0.000195056 | 3.93E-05 | 0.001163 | 5.71E-05 | 0.008655349 | STX5 |
| chr4 | 48681368 | 48681370 | 3 | 2 | 0.00041686 | 3.61E-05 | 0.00191 | 5.71E-05 | 0.004911288 | FRYL |
| chr9 | 135352706 | 135353618 | 913 | 2 | 0.000435705 | 3.86E-05 | 0.001277 | 5.71E-05 | 0.002713116 | AL353615.1 |
| chr22 | 24986152 | 24986212 | 61 | 2 | 0.000355598 | 5.86E-05 | 0.000352 | 5.71E-05 | 0.023297317 | KIAA1671 |
| chr12 | 34350241 | 34350659 | 419 | 3 | 0.000141867 | 4.57E-05 | 0.000848 | 5.71E-05 | 0.016164637 | NA |
| chr2 | 148020221 | 148020612 | 392 | 4 | 0.000219297 | 0.000107 | 0.001546 | 5.71E-05 | -0.00074651 | ORC4 |
| chr7 | 101019904 | 101020232 | 329 | 4 | 0.000357022 | 2.92E-05 | 0.00294 | 5.71E-05 | 0.008816403 | MUC17 |
| chr19 | 542548 | 542796 | 249 | 3 | 0.00017817 | 4.87E-05 | 0.002324 | 5.71E-05 | 0.003659416 | NA |
| chr20 | 19886099 | 19886382 | 284 | 5 | 0.000118396 | 3.71E-05 | 0.004617 | 5.71E-05 | 0.002830392 | RIN2 |
| chr2 | 218216662 | 218217217 | 556 | 10 | 0.000115787 | 0.000278 | 0.001436 | 5.72E-05 | 0.002214224 | ARPC2 |
| chr4 | 37243892 | 37244658 | 767 | 8 | 3.94E-05 | 0.000635 | 0.000321 | 5.72E-05 | 0.001784189 | NA |
| chr14 | 34649766 | 34649815 | 50 | 2 | 0.000290395 | 0.000125 | 0.000131 | 5.72E-05 | 0.013206914 | NA |
| chr1 | 9129448 | 9129615 | 168 | 5 | 0.000119976 | 0.00039 | 0.000144 | 5.72E-05 | 0.001803256 | NA |
| chr13 | 64078755 | 64078767 | 13 | 2 | 0.00037376 | 5.25E-05 | 0.000452 | 5.73E-05 | -0.00760035 | NA |
| chr12 | 122062791 | 122063554 | 764 | 4 | 0.000184368 | 3.30E-05 | 0.003174 | 5.73E-05 | 0.007515062 | AC156455.1 |
| chr1 | 3732185 | 3732271 | 87 | 2 | 0.000176533 | 5.01E-05 | 0.000513 | 5.74E-05 | 0.010265769 | TP73 |
| chr12 | 125092251 | 125092314 | 64 | 2 | 0.000325749 | 4.03E-05 | 0.001077 | 5.74E-05 | 0.000187878 | AACS |
| chr20 | 50824561 | 50824573 | 13 | 2 | 0.000159032 | 0.002779 | 4.65E-05 | 5.75E-05 | 0.00306368 | BCAS4 |
| chr11 | 65487027 | 65487065 | 39 | 2 | 0.000258422 | 3.68E-05 | 0.001773 | 5.75E-05 | 0.005661712 | NA |
| chr6 | 33456893 | 33457486 | 594 | 3 | 3.69E-05 | 4.13E-05 | 0.002015 | 5.75E-05 | -0.00085681 | ZBTB9 |
| chr14 | 93017653 | 93017858 | 206 | 4 | 0.000133092 | 0.000162 | 0.001218 | 5.76E-05 | 9.95E-05 | ITPK1 |
| chr8 | 120051241 | 120051306 | 66 | 2 | 6.09E-05 | 0.007926 | 4.21E-05 | 5.76E-05 | 0.009026919 | NA |
| chr11 | 78017080 | 78017134 | 55 | 3 | 0.00018321 | 0.00132 | 0.000167 | 5.76E-05 | -0.00058444 | KCTD14, NDUFC2-KCTD14 |
| chr1 | 206793100 | 206793117 | 18 | 2 | 0.000265461 | 5.38E-05 | 0.000437 | 5.76E-05 | 0.00386981 | IL19 |
| chr22 | 44669457 | 44669874 | 418 | 2 | 0.000298565 | 3.77E-05 | 0.001544 | 5.76E-05 | 0.016350477 | PRR5 |
| chr12 | 51174049 | 51174320 | 272 | 3 | 0.000107522 | 5.25E-05 | 0.000609 | 5.76E-05 | 0.00102526 | NA |
| chr1 | 202274570 | 202274763 | 194 | 2 | 6.65E-05 | 4.95E-05 | 0.000541 | 5.76E-05 | 0.010905605 | LGR6 |
| chr22 | 46849203 | 46849259 | 57 | 2 | 0.000193576 | 0.000114 | 0.000142 | 5.76E-05 | 0.00318546 | TBC1D22A |
| chr9 | 113471715 | 113471844 | 130 | 3 | 0.000369202 | 7.75E-05 | 0.000807 | 5.76E-05 | 0.000747536 | RGS3 |
| chr3 | 97447781 | 97448141 | 361 | 3 | 3.12E-05 | 0.081139 | 1.66E-05 | 5.76E-05 | -0.00214678 | EPHA6 |
| chr8 | 139959090 | 139959526 | 437 | 3 | 0.000135878 | 0.006596 | 0.000111 | 5.77E-05 | 0.003611701 | TRAPPC9 |
| chr9 | 125747450 | 125747893 | 444 | 5 | 0.000211897 | 1.71E-05 | 0.014762 | 5.77E-05 | 0.003106795 | PBX3 |
| chr6 | 163834986 | 163835073 | 88 | 2 | 0.000189133 | 8.36E-05 | 0.000201 | 5.78E-05 | -0.00471774 | AL078602.1 |
| chr11 | 64284693 | 64285359 | 667 | 9 | 4.73E-05 | 0.000306 | 0.008602 | 5.78E-05 | -0.0013769 | GPR137, BAD |
| chr17 | 29931364 | 29931629 | 266 | 3 | 0.00012788 | 0.002103 | 0.000601 | 5.78E-05 | -0.00516803 | EFCAB5 |
| chr15 | 75023383 | 75023778 | 396 | 8 | 0.000198285 | 0.001058 | 7.92E-05 | 5.79E-05 | 0.000448697 | PPCDC |
| chr7 | 45684859 | 45685178 | 320 | 2 | 0.000209193 | 4.31E-05 | 0.000853 | 5.79E-05 | 0.010549006 | ADCY1 |
| chr8 | 28412447 | 28412516 | 70 | 2 | 0.000190994 | 3.74E-05 | 0.001692 | 5.80E-05 | -0.0036436 | FBXO16 |
| chr10 | 16484151 | 16484573 | 423 | 7 | 0.000117564 | 0.00208 | 7.37E-05 | 5.80E-05 | 0.001793775 | PTER |
| chr2 | 218417691 | 218417805 | 115 | 2 | 0.000409097 | 3.69E-05 | 0.001852 | 5.80E-05 | 0.007540904 | NA |
| chr1 | 27104966 | 27105024 | 59 | 2 | 0.000158126 | 0.000124 | 0.000135 | 5.80E-05 | 0.003372549 | SLC9A1 |
| chr8 | 65643685 | 65643912 | 228 | 5 | 7.62E-05 | 4.07E-05 | 0.004966 | 5.81E-05 | 0.007289622 | NA |
| chr16 | 3984239 | 3984503 | 265 | 4 | 6.84E-05 | 0.003129 | 4.52E-05 | 5.81E-05 | 0.002660703 | ADCY9 |
| chr15 | 45631570 | 45631942 | 373 | 2 | 0.00030723 | 3.76E-05 | 0.001659 | 5.82E-05 | 0.008648067 | AC090527.2, SQOR |
| chr16 | 22091126 | 22091293 | 168 | 2 | 8.05E-05 | 5.76E-05 | 0.000385 | 5.82E-05 | 0.005450893 | AC009019.1 |
| chr7 | 157831624 | 157832149 | 526 | 3 | 1.64E-05 | 9.71E-05 | 0.000817 | 5.82E-05 | 0.00555997 | PTPRN2 |
| chr1 | 212626747 | 212626962 | 216 | 2 | 3.18E-05 | 3.72E-05 | 0.001816 | 5.82E-05 | 0.004476922 | FAM71A, AL590648.2 |
| chr19 | 45000984 | 45001258 | 275 | 4 | 0.000100524 | 0.0001 | 0.00057 | 5.82E-05 | 0.00882331 | NA |
| chr13 | 114097318 | 114097640 | 323 | 3 | 7.54E-05 | 0.000268 | 0.000143 | 5.82E-05 | 0.004411749 | RASA3 |
| chr22 | 39436915 | 39437279 | 365 | 3 | 0.000277576 | 3.41E-05 | 0.001882 | 5.82E-05 | 0.008196869 | TAB1 |
| chr5 | 633868 | 633928 | 61 | 3 | 0.000466255 | 3.53E-05 | 0.001707 | 5.83E-05 | -0.00162323 | CEP72 |
| chr14 | 20469062 | 20469713 | 652 | 10 | 0.00017492 | 0.000309 | 0.004133 | 5.83E-05 | -0.00065079 | PNP |
| chr20 | 51366749 | 51366987 | 239 | 2 | 8.72E-05 | 6.40E-05 | 0.000312 | 5.83E-05 | 0.008402302 | NA |
| chr2 | 182426835 | 182427200 | 366 | 3 | 0.000117015 | 7.03E-05 | 0.000499 | 5.83E-05 | 0.008297233 | PDE1A |
| chr8 | 123100736 | 123100930 | 195 | 3 | 0.00020987 | 0.000305 | 8.71E-05 | 5.83E-05 | 0.000953311 | TBC1D31 |
| chr12 | 124385566 | 124385848 | 283 | 4 | 8.82E-05 | 0.004811 | 8.24E-05 | 5.84E-05 | 0.002395794 | NCOR2 |
| chr19 | 53394647 | 53395027 | 381 | 3 | 0.000258373 | 0.000189 | 0.000288 | 5.84E-05 | 0.006010281 | ZNF765 |
| chr16 | 81077613 | 81077621 | 9 | 2 | 0.000321843 | 0.000236 | 9.23E-05 | 5.84E-05 | -0.00136647 | AC092718.1, AC092718.8, AC092718.3 |
| chr17 | 75843868 | 75843891 | 24 | 2 | 1.59E-05 | 4.86E-05 | 0.000601 | 5.84E-05 | 0.002882063 | UNC13D |
| chr14 | 69881116 | 69881387 | 272 | 2 | 8.74E-05 | 0.000124 | 0.000136 | 5.84E-05 | 0.020339641 | SMOC1 |
| chr12 | 8564674 | 8564795 | 122 | 2 | 0.000403104 | 0.000136 | 0.000127 | 5.85E-05 | -0.00838879 | AC092746.1 |
| chr20 | 57712996 | 57713727 | 732 | 3 | 2.01E-05 | 0.000873 | 0.000155 | 5.85E-05 | 0.007840868 | NA |
| chr20 | 24987963 | 24988213 | 251 | 2 | 6.50E-05 | 8.03E-05 | 0.000218 | 5.85E-05 | -0.00337247 | APMAP |
| chr5 | 178940114 | 178940422 | 309 | 3 | 9.60E-05 | 0.000187 | 0.000128 | 5.86E-05 | 0.009253849 | NA |
| chr2 | 1738957 | 1739237 | 281 | 2 | 0.000111715 | 0.000322 | 8.13E-05 | 5.86E-05 | 0.002423147 | PXDN |
| chr6 | 136593950 | 136594418 | 469 | 3 | 0.000161142 | 3.68E-05 | 0.003276 | 5.86E-05 | 0.007468835 | MAP3K5 |
| chr4 | 6008844 | 6009415 | 572 | 4 | 0.000115228 | 8.95E-05 | 0.002047 | 5.87E-05 | 0.007052219 | C4orf50 |
| chr1 | 8408031 | 8408251 | 221 | 2 | 0.000177481 | 9.65E-05 | 0.000173 | 5.88E-05 | 0.007411735 | RERE |
| chr7 | 27225194 | 27225690 | 497 | 4 | 0.000171015 | 0.000114 | 0.000696 | 5.89E-05 | 0.006645636 | NA |
| chr22 | 49367507 | 49367627 | 121 | 2 | 5.36E-05 | 0.000105 | 0.000157 | 5.89E-05 | 0.01669085 | NA |
| chr5 | 56233380 | 56233938 | 559 | 5 | 0.000198523 | 9.37E-05 | 0.002247 | 5.89E-05 | -9.59E-05 | NA |
| chr16 | 2715619 | 2715899 | 281 | 5 | 0.00011385 | 0.000489 | 0.000108 | 5.90E-05 | -0.00316497 | PRSS27 |
| chr2 | 69584642 | 69584715 | 74 | 2 | 0.000197662 | 0.001222 | 5.53E-05 | 5.90E-05 | 0.016372853 | AAK1 |
| chr3 | 195825470 | 195825558 | 89 | 2 | 0.000330115 | 4.08E-05 | 0.001165 | 5.90E-05 | 0.007705626 | NA |
| chr11 | 44621318 | 44621382 | 65 | 3 | 0.000490989 | 2.50E-05 | 0.006495 | 5.90E-05 | 0.019657446 | NA |
| chr17 | 3825042 | 3825343 | 302 | 3 | 0.00026192 | 9.35E-05 | 0.000391 | 5.92E-05 | 0.005955491 | NCBP3 |
| chr16 | 2194614 | 2195032 | 419 | 2 | 0.000280497 | 3.78E-05 | 0.001803 | 5.92E-05 | -0.00189988 | CASKIN1 |
| chr2 | 20166682 | 20166739 | 58 | 2 | 0.000418002 | 4.13E-05 | 0.001123 | 5.92E-05 | 0.011149228 | NA |
| chr10 | 116771134 | 116771447 | 314 | 4 | 0.000219011 | 0.000122 | 0.000285 | 5.93E-05 | 0.012925837 | HSPA12A |
| chr16 | 71383780 | 71384122 | 343 | 2 | 0.000283228 | 3.87E-05 | 0.001589 | 5.93E-05 | 0.017184275 | CALB2, LINC02136 |
| chr7 | 16687154 | 16687190 | 37 | 2 | 0.000124996 | 0.001198 | 5.58E-05 | 5.93E-05 | 0.005958294 | BZW2 |
| chr16 | 64735705 | 64735839 | 135 | 3 | 0.000187189 | 8.27E-05 | 0.001143 | 5.94E-05 | -0.00156753 | NA |
| chr9 | 126425424 | 126425747 | 324 | 2 | 0.000114579 | 0.000246 | 9.22E-05 | 5.94E-05 | 0.003994163 | MVB12B |
| chr2 | 178112254 | 178112367 | 114 | 5 | 0.000309089 | 2.86E-05 | 0.010377 | 5.94E-05 | 0.003987135 | NA |
| chr1 | 200890874 | 200891554 | 681 | 8 | 0.000118634 | 0.000366 | 0.009504 | 5.94E-05 | 0.005648864 | INAVA |
| chr18 | 35344129 | 35344688 | 560 | 11 | 0.000152701 | 0.001043 | 0.001855 | 5.94E-05 | 0.000780348 | ZNF24 |
| chr6 | 157273876 | 157274030 | 155 | 4 | 0.000215623 | 0.010302 | 0.000876 | 5.94E-05 | 0.008701504 | NA |
| chr10 | 103551599 | 103551934 | 336 | 2 | 5.50E-05 | 0.015105 | 4.15E-05 | 5.94E-05 | 0.004495387 | NEURL1 |
| chr11 | 64939681 | 64939868 | 188 | 2 | 0.000107198 | 7.80E-05 | 0.000236 | 5.95E-05 | 0.004299405 | MAJIN |
| chr13 | 99124821 | 99124847 | 27 | 2 | 0.000263824 | 4.07E-05 | 0.00125 | 5.96E-05 | 0.0102618 | NA |
| chr8 | 81731432 | 81731445 | 14 | 2 | 0.000177914 | 3.81E-05 | 0.001797 | 5.96E-05 | -0.01041496 | ZFAND1 |
| chr4 | 83010560 | 83010910 | 351 | 5 | 0.000424681 | 1.82E-05 | 0.009645 | 5.96E-05 | 0.001031092 | LIN54 |
| chr3 | 134356692 | 134356741 | 50 | 3 | 0.00015062 | 0.000171 | 0.00116 | 5.96E-05 | -0.00408809 | AMOTL2 |
| chr1 | 40531389 | 40531879 | 491 | 10 | 0.000199928 | 0.000188 | 0.005033 | 5.96E-05 | 0.000752985 | ZNF684 |
| chr16 | 53509772 | 53510186 | 415 | 4 | 0.000227956 | 4.32E-05 | 0.002724 | 5.96E-05 | -0.01678038 | NA |
| chr17 | 78159291 | 78159985 | 695 | 6 | 6.49E-05 | 0.000108 | 0.001085 | 5.97E-05 | 0.00674835 | C17orf99 |
| chr20 | 62643490 | 62643681 | 192 | 3 | 0.000145884 | 5.38E-05 | 0.000963 | 5.97E-05 | 0.008088025 | SLCO4A1 |
| chr17 | 76044690 | 76044799 | 110 | 2 | 0.000170701 | 0.001365 | 5.48E-05 | 5.97E-05 | -0.00567034 | SRP68 |
| chr5 | 1968036 | 1968173 | 138 | 3 | 0.000212055 | 7.62E-05 | 0.001164 | 5.97E-05 | 0.025663396 | AC124852.1, AC126768.2 |
| chr1 | 2274331 | 2274463 | 133 | 3 | 9.27E-05 | 0.000154 | 0.000143 | 5.97E-05 | 0.003547765 | SKI |
| chr6 | 92126345 | 92126553 | 209 | 2 | 0.000276491 | 8.82E-05 | 0.0002 | 5.97E-05 | 9.60E-05 | AL590814.1 |
| chr12 | 1925742 | 1925989 | 248 | 2 | 4.53E-05 | 3.76E-05 | 0.002016 | 5.97E-05 | 0.008001303 | NA |
| chr11 | 57337771 | 57338340 | 570 | 6 | 0.000194897 | 0.000826 | 0.001671 | 5.97E-05 | 0.002899207 | NA |
| chr6 | 35259140 | 35259614 | 475 | 8 | 0.000177366 | 0.00413 | 0.000277 | 5.97E-05 | 0.00058898 | ZNF76 |
| chr17 | 28356528 | 28357235 | 708 | 5 | 2.37E-05 | 8.52E-05 | 0.001131 | 5.97E-05 | 0.001533266 | POLDIP2 |
| chr1 | 246775269 | 246775302 | 34 | 2 | 3.02E-05 | 0.006269 | 4.40E-05 | 5.97E-05 | -0.00242088 | AL591848.3, KIF28P |
| chr4 | 89227237 | 89227442 | 206 | 3 | 0.000313215 | 8.88E-05 | 0.000306 | 5.98E-05 | 0.006314914 | NA |
| chr8 | 36764082 | 36764470 | 389 | 3 | 0.00020742 | 0.000142 | 0.000147 | 5.98E-05 | 0.014839913 | RNA5SP264, AC090809.1 |
| chr1 | 1286990 | 1287562 | 573 | 3 | 2.23E-05 | 9.51E-05 | 0.000377 | 5.98E-05 | 0.005567802 | SCNN1D |
| chr17 | 21287669 | 21287984 | 316 | 5 | 5.36E-05 | 0.018646 | 0.000122 | 5.98E-05 | 0.00205626 | MAP2K3 |
| chr22 | 33648922 | 33649097 | 176 | 2 | 0.000186654 | 0.000336 | 8.17E-05 | 5.98E-05 | -0.00517825 | LARGE1 |
| chr22 | 30506628 | 30506909 | 282 | 5 | 8.84E-05 | 6.05E-05 | 0.007765 | 5.99E-05 | 0.00514814 | NA |
| chr11 | 128937165 | 128938008 | 844 | 4 | 0.000104179 | 3.95E-05 | 0.002085 | 5.99E-05 | 0.001089349 | TP53AIP1 |
| chr1 | 68174158 | 68174330 | 173 | 2 | 0.000251841 | 3.84E-05 | 0.001804 | 5.99E-05 | 0.010944798 | GNG12-AS1, WLS |
| chr6 | 5139193 | 5139329 | 137 | 2 | 0.000348885 | 6.39E-05 | 0.000337 | 5.99E-05 | 0.005670183 | LYRM4 |
| chr15 | 98281426 | 98281478 | 53 | 2 | 0.000315988 | 0.000114 | 0.00015 | 5.99E-05 | 0.010238929 | AC022523.1 |
| chr13 | 20177118 | 20177806 | 689 | 4 | 0.000115024 | 6.46E-05 | 0.001882 | 6.00E-05 | 0.01836398 | NA |
| chr1 | 236559779 | 236559854 | 76 | 2 | 0.000244969 | 0.00022 | 9.87E-05 | 6.00E-05 | 0.003204129 | HEATR1 |
| chr13 | 26557993 | 26558636 | 644 | 4 | 1.40E-05 | 0.006888 | 0.000188 | 6.00E-05 | -0.00140462 | WASF3 |
| chr2 | 236708547 | 236708584 | 38 | 2 | 0.000249117 | 0.000333 | 8.23E-05 | 6.00E-05 | 0.001462218 | NA |
| chr10 | 2266757 | 2266778 | 22 | 2 | 0.00026699 | 0.000252 | 9.25E-05 | 6.00E-05 | 0.0060331 | NA |
| chr1 | 4602088 | 4602245 | 158 | 2 | 0.000202697 | 0.000145 | 0.000127 | 6.01E-05 | 0.007510349 | NA |
| chr2 | 2657347 | 2657463 | 117 | 2 | 0.000141084 | 3.79E-05 | 0.001995 | 6.01E-05 | 0.011559817 | AC018685.2 |
| chr17 | 31859043 | 31859751 | 709 | 10 | 0.000178412 | 0.000215 | 0.00297 | 6.01E-05 | -0.00214721 | COPRS |
| chr5 | 1065660 | 1065807 | 148 | 2 | 0.000242169 | 0.000119 | 0.000146 | 6.01E-05 | 0.006371938 | SLC12A7 |
| chr14 | 102900969 | 102901254 | 286 | 3 | 0.00030566 | 2.73E-05 | 0.005342 | 6.01E-05 | 0.024087783 | TRAF3 |
| chr9 | 137089127 | 137089141 | 15 | 2 | 0.000452392 | 4.04E-05 | 0.001372 | 6.01E-05 | -0.00755992 | MAN1B1 |
| chr5 | 9117163 | 9117187 | 25 | 2 | 0.00015232 | 0.005217 | 4.50E-05 | 6.01E-05 | -0.00395556 | SEMA5A |
| chr9 | 89448622 | 89448694 | 73 | 3 | 0.000430723 | 5.35E-05 | 0.00105 | 6.01E-05 | 0.010055489 | SEMA4D |
| chr17 | 28598834 | 28599329 | 496 | 8 | 0.000184613 | 0.000171 | 0.000571 | 6.01E-05 | 0.000200102 | SPAG5-AS1, AC005726.1, SPAG5 |

| | | | | | | | | | | |
|---|---|---|---|---|---|---|---|---|---|---|
| chr6 | 39322383 | 39322948 | 566 | 6 | 5.79E-05 | 0.00348 | 0.001339 | 6.02E-05 | 0.002414161 | KCNK16 |
| chr7 | 100485461 | 100485505 | 45 | 2 | 5.13E-05 | 0.001157 | 5.70E-05 | 6.02E-05 | 0.003931062 | NYAP1 |
| chr3 | 161372684 | 161373387 | 704 | 5 | 2.47E-05 | 0.000118 | 0.002324 | 6.02E-05 | 0.005610505 | SPTSSB |
| chr3 | 184400318 | 184401402 | 1085 | 5 | 5.11E-06 | 0.000204 | 0.000222 | 6.02E-05 | 0.000946951 | LINC02054 |
| chr19 | 11085561 | 11085791 | 231 | 2 | 0.000245667 | 9.53E-05 | 0.000184 | 6.02E-05 | 0.004194726 | NA |
| chr9 | 78440976 | 78441420 | 445 | 4 | 0.000162835 | 4.83E-05 | 0.001276 | 6.04E-05 | 0.004112465 | NA |
| chr12 | 6568459 | 6568861 | 403 | 7 | 7.08E-05 | 0.003859 | 5.10E-05 | 6.04E-05 | 0.002403677 | NOP2, AC006064.6 |
| chr3 | 184560162 | 184560480 | 319 | 2 | 1.67E-05 | 4.16E-05 | 0.001213 | 6.04E-05 | 0.006902525 | NA |
| chr7 | 103073765 | 103074698 | 934 | 6 | 2.06E-06 | 0.000953 | 0.000104 | 6.04E-05 | 0.010184848 | FBXL13 |
| chr20 | 47361723 | 47361735 | 13 | 2 | 0.000278268 | 9.12E-05 | 0.000197 | 6.05E-05 | 0.010328418 | NA |
| chr4 | 37245964 | 37246332 | 369 | 2 | 3.19E-05 | 0.000162 | 0.000119 | 6.05E-05 | 0.012623142 | NWD2 |
| chr6 | 106103157 | 106103952 | 796 | 4 | 7.37E-05 | 0.001285 | 0.000125 | 6.05E-05 | 0.008075382 | PRDM1, ATG5 |
| chr3 | 197459066 | 197459285 | 220 | 3 | 0.000146288 | 9.25E-05 | 0.001756 | 6.05E-05 | 0.004566956 | AC128709.3 |
| chr1 | 165662823 | 165663017 | 195 | 2 | 0.00015404 | 3.98E-05 | 0.001551 | 6.05E-05 | 0.003967454 | ALDH9A1 |
| chr15 | 78131188 | 78131222 | 35 | 2 | 6.61E-05 | 3.82E-05 | 0.002044 | 6.06E-05 | 0.006746166 | CIB2 |
| chr12 | 52582311 | 52583215 | 905 | 3 | 0.000116669 | 4.07E-05 | 0.002338 | 6.06E-05 | 0.004272561 | NA |
| chr7 | 105040949 | 105040993 | 45 | 2 | 0.000111513 | 0.001213 | 5.69E-05 | 6.06E-05 | 0.008901024 | KMT2E |
| chr3 | 13332255 | 13332471 | 217 | 2 | 0.000152732 | 0.000701 | 6.48E-05 | 6.06E-05 | 0.007548674 | NUP210 |
| chr13 | 43391200 | 43391205 | 6 | 2 | 0.000151163 | 3.89E-05 | 0.001806 | 6.06E-05 | 0.010957096 | ENOX1 |
| chr1 | 234272940 | 234272942 | 3 | 2 | 0.000124719 | 4.31E-05 | 0.001061 | 6.06E-05 | 0.00467298 | SLC35F3, AL122008.1 |
| chr15 | 92639262 | 92639686 | 425 | 4 | 9.41E-05 | 0.000384 | 0.000292 | 6.07E-05 | 0.008429434 | FAM174B |
| chr6 | 29670798 | 29671141 | 344 | 2 | 0.000340509 | 4.75E-05 | 0.000745 | 6.07E-05 | 0.014167466 | MOG |
| chr8 | 104423941 | 104423967 | 27 | 2 | 0.00042984 | 0.000156 | 0.000123 | 6.07E-05 | 0.000132687 | DPYS |
| chr10 | 14605078 | 14605754 | 677 | 5 | 8.68E-05 | 0.00142 | 0.000189 | 6.08E-05 | 0.009926697 | FAM107B |
| chr19 | 7463752 | 7463899 | 148 | 3 | 8.27E-05 | 0.001505 | 4.32E-05 | 6.08E-05 | 0.002554366 | AC008878.3, ARHGEF18 |
| chr1 | 154259621 | 154259702 | 82 | 2 | 0.000178993 | 5.01E-05 | 0.000634 | 6.08E-05 | 0.004706186 | UBAP2L |
| chr17 | 41655660 | 41656015 | 356 | 2 | 3.68E-05 | 9.30E-05 | 0.000194 | 6.08E-05 | 0.0076713 | NA |
| chr6 | 32211790 | 32212864 | 1075 | 7 | 3.38E-05 | 0.01502 | 4.82E-05 | 6.08E-05 | 0.00151501 | NOTCH4 |
| chr15 | 76341995 | 76343016 | 1022 | 8 | 6.00E-05 | 3.23E-05 | 0.014925 | 6.09E-05 | 0.004798737 | ISL2, AC027243.1 |
| chr8 | 42193421 | 42193455 | 35 | 2 | 0.000231625 | 0.000507 | 7.22E-05 | 6.10E-05 | 0.004060801 | PLAT |
| chr19 | 11964957 | 11965067 | 111 | 4 | 0.000348007 | 0.002146 | 0.000226 | 6.10E-05 | 0.007308601 | AC008770.2, ZNF763 |
| chr14 | 68507803 | 68507972 | 170 | 2 | 0.000286743 | 6.04E-05 | 0.000399 | 6.10E-05 | 0.008821368 | RAD51B |
| chr17 | 39612530 | 39612551 | 22 | 2 | 0.000273278 | 3.89E-05 | 0.001889 | 6.10E-05 | 0.009346943 | NA |
| chr13 | 31139656 | 31139882 | 227 | 3 | 0.000135332 | 0.000846 | 0.000131 | 6.10E-05 | 0.003642966 | HSPH1 |
| chr11 | 78109980 | 78110481 | 502 | 3 | 0.000200231 | 7.24E-05 | 0.001852 | 6.11E-05 | -5.34E-05 | ALG8 |
| chr2 | 240593972 | 240594277 | 306 | 3 | 0.000105947 | 3.80E-05 | 0.003372 | 6.12E-05 | -0.00027828 | CAPN10 |
| chr7 | 2609103 | 2609461 | 359 | 3 | 5.78E-06 | 7.02E-05 | 0.000449 | 6.12E-05 | 0.008214567 | IQCE |
| chr9 | 93300305 | 93300331 | 27 | 2 | 0.000426097 | 0.000361 | 8.15E-05 | 6.12E-05 | 0.000691298 | WNK2 |
| chr16 | 55331903 | 55332218 | 316 | 5 | 0.000274323 | 5.74E-05 | 0.003549 | 6.13E-05 | 0.005426568 | AC109462.1 |
| chr5 | 176969004 | 176969115 | 112 | 3 | 0.000352779 | 2.74E-05 | 0.005703 | 6.14E-05 | 0.012098281 | UIMC1 |
| chr7 | 73593677 | 73593965 | 289 | 3 | 0.000159814 | 0.003318 | 0.000598 | 6.14E-05 | 0.010404785 | MLXIPL |
| chr11 | 20669082 | 20669580 | 499 | 9 | 0.000347378 | 0.000129 | 0.005478 | 6.14E-05 | 0.003080223 | NELL1 |
| chr10 | 119003905 | 119004001 | 97 | 2 | 0.000359822 | 4.07E-05 | 0.001493 | 6.14E-05 | 0.011695938 | AL157388.1 |
| chr18 | 69841430 | 69841746 | 317 | 2 | 3.19E-05 | 0.01797 | 4.22E-05 | 6.14E-05 | -0.00369766 | DOK6, CD226 |
| chr11 | 68633008 | 68633023 | 16 | 2 | 0.000402281 | 5.72E-05 | 0.000463 | 6.15E-05 | 0.014689143 | NA |
| chr19 | 6273833 | 6274197 | 365 | 3 | 7.98E-05 | 0.000694 | 0.000116 | 6.15E-05 | 0.009424563 | MLLT1 |
| chr7 | 116716684 | 116717038 | 355 | 2 | 4.99E-05 | 0.037319 | 4.14E-05 | 6.15E-05 | 0.012964196 | MET |
| chr3 | 128361975 | 128362244 | 270 | 4 | 0.000180738 | 6.49E-05 | 0.001261 | 6.15E-05 | 0.001911802 | EEFSEC |
| chr6 | 28349616 | 28350019 | 404 | 8 | 0.000194333 | 0.003855 | 0.000212 | 6.15E-05 | 0.000134357 | ZKSCAN3, ZSCAN31 |
| chr18 | 77111698 | 77112198 | 501 | 2 | 0.000313008 | 6.43E-05 | 0.000358 | 6.15E-05 | 0.02194138 | MBP |
| chr17 | 41498283 | 41498631 | 349 | 4 | 0.000108804 | 0.000793 | 9.15E-05 | 6.16E-05 | 0.006003063 | NA |
| chr8 | 48913490 | 48913841 | 352 | 4 | 0.000105664 | 0.000782 | 9.68E-05 | 6.16E-05 | -0.00209812 | NA |
| chr11 | 44949220 | 44949498 | 279 | 2 | 9.92E-05 | 3.88E-05 | 0.002066 | 6.16E-05 | 0.013660804 | TP53I11 |
| chr6 | 166998832 | 166999561 | 730 | 13 | 7.86E-06 | 0.08808 | 3.88E-05 | 6.17E-05 | -0.0003457 | CEP43, Z94721.2, Z94721.1, Z94721.3 |
| chr2 | 241936345 | 241936660 | 316 | 4 | 0.000100638 | 0.001102 | 0.000326 | 6.17E-05 | -0.00433052 | LINC01237 |
| chr1 | 10662120 | 10662134 | 15 | 2 | 8.80E-05 | 3.96E-05 | 0.001818 | 6.18E-05 | 0.008844491 | CASZ1 |
| chr19 | 17797003 | 17797437 | 435 | 2 | 0.000216039 | 4.69E-05 | 0.00083 | 6.18E-05 | 0.004799158 | B3GNT3 |
| chr6 | 132735515 | 132735820 | 306 | 3 | 0.000112628 | 0.000167 | 0.001486 | 6.18E-05 | 0.008889344 | VNN3 |
| chr8 | 96328042 | 96328392 | 351 | 3 | 0.000114025 | 5.20E-05 | 0.000737 | 6.18E-05 | 0.007512756 | PTDSS1 |
| chr12 | 119178388 | 119178877 | 490 | 8 | 0.000209944 | 4.42E-05 | 0.009747 | 6.18E-05 | 0.004774152 | HSPB8 |
| chr10 | 132586936 | 132587002 | 67 | 2 | 0.000110654 | 0.000105 | 0.000173 | 6.19E-05 | 0.013818719 | INPP5A |
| chr14 | 23469209 | 23469787 | 579 | 10 | 0.000181607 | 8.49E-05 | 0.005596 | 6.19E-05 | 0.001027127 | NGDN |
| chr2 | 36705557 | 36705829 | 273 | 2 | 0.000254693 | 0.000148 | 0.00013 | 6.20E-05 | 0.00027097 | VIT |
| chr11 | 66593299 | 66593610 | 312 | 4 | 0.000221233 | 0.000103 | 0.000868 | 6.20E-05 | 0.00371134 | CCS |
| chr6 | 42771901 | 42772049 | 149 | 2 | 0.000199793 | 0.00012 | 0.000152 | 6.20E-05 | 0.009409613 | BICRAL |
| chr16 | 4783705 | 4784031 | 327 | 4 | 0.000253206 | 0.007877 | 0.000785 | 6.20E-05 | 0.003799175 | SEPTIN12, AC020663.4 |
| chr12 | 96043311 | 96043713 | 403 | 3 | 0.000219037 | 0.000116 | 0.000629 | 6.21E-05 | 0.007507268 | LTA4H |
| chr17 | 81257888 | 81258529 | 642 | 3 | 9.39E-05 | 0.000357 | 0.00012 | 6.21E-05 | 0.002971972 | SLC38A10 |
| chr16 | 74453287 | 74453519 | 233 | 2 | 0.000371226 | 4.92E-05 | 0.000724 | 6.21E-05 | 0.004033639 | GLG1 |
| chr7 | 4758200 | 4758387 | 188 | 3 | 0.000283317 | 9.88E-05 | 0.000254 | 6.21E-05 | 0.001875794 | FOXK1 |
| chr2 | 2107851 | 2107876 | 26 | 2 | 0.000428269 | 4.86E-05 | 0.000754 | 6.22E-05 | 0.010206866 | MYT1L |
| chr12 | 124632561 | 124632812 | 252 | 2 | 0.000353488 | 3.91E-05 | 0.002076 | 6.22E-05 | 0.008912372 | NA |
| chr9 | 122095621 | 122095698 | 78 | 2 | 9.57E-05 | 0.001995 | 5.29E-05 | 6.22E-05 | 0.010646786 | NA |
| chr8 | 23681875 | 23682426 | 552 | 4 | 0.000122163 | 4.35E-05 | 0.005415 | 6.23E-05 | 0.004523984 | NKX3-1 |
| chr22 | 40714850 | 40714956 | 107 | 2 | 0.000430229 | 4.50E-05 | 0.00101 | 6.23E-05 | 0.016182536 | NA |
| chr1 | 94218489 | 94218552 | 64 | 2 | 0.000182902 | 0.001879 | 5.35E-05 | 6.24E-05 | 0.004348313 | ARHGAP29 |
| chr19 | 5057444 | 5057532 | 89 | 2 | 0.000220405 | 0.000481 | 7.51E-05 | 6.24E-05 | 0.004326255 | KDM4B |
| chr5 | 88786358 | 88786496 | 139 | 2 | 0.000243734 | 3.95E-05 | 0.002023 | 6.24E-05 | 0.013411817 | MEF2C |
| chr17 | 82022865 | 82023416 | 552 | 14 | 0.000196341 | 0.001614 | 0.000426 | 6.24E-05 | -0.00017808 | LRRC45, CENPX |
| chr5 | 660065 | 660135 | 71 | 2 | 0.000168496 | 0.000439 | 7.74E-05 | 6.24E-05 | 0.002363812 | CEP72, TPPP |
| chr12 | 53224253 | 53224623 | 371 | 2 | 6.65E-05 | 6.10E-05 | 0.000419 | 6.25E-05 | 0.009313254 | RARG |
| chr20 | 62070141 | 62070363 | 223 | 3 | 0.000163135 | 0.000419 | 7.86E-05 | 6.25E-05 | -0.00248366 | NA |
| chr12 | 29563825 | 29563858 | 34 | 2 | 0.000451906 | 3.94E-05 | 0.002085 | 6.25E-05 | 0.021356781 | TMTC1 |
| chr2 | 207260584 | 207261045 | 462 | 3 | 8.83E-05 | 0.000572 | 0.000183 | 6.25E-05 | 0.009059139 | LINC01802, AC009226.1 |
| chr9 | 89598201 | 89598610 | 410 | 4 | 0.000173344 | 0.000183 | 0.000168 | 6.26E-05 | 0.011387088 | AL161910.1 |
| chr1 | 223665443 | 223665650 | 208 | 3 | 0.000307521 | 3.20E-05 | 0.002857 | 6.26E-05 | 0.006597736 | CAPN8 |
| chr11 | 64300213 | 64300674 | 462 | 8 | 0.000335122 | 0.00028 | 0.004569 | 6.27E-05 | -0.00016925 | KCNK4-TEX40, CATSPERZ |
| chr13 | 49476547 | 49476753 | 207 | 2 | 0.000124463 | 0.002931 | 4.98E-05 | 6.27E-05 | -0.00917642 | SETDB2 |
| chr2 | 219010021 | 219010185 | 165 | 2 | 0.000198398 | 0.000518 | 7.36E-05 | 6.27E-05 | 0.014397044 | AC097468.1, CFAP65 |
| chr14 | 103909670 | 103909862 | 193 | 4 | 0.000113092 | 0.08888 | 8.12E-05 | 6.27E-05 | 0.003757534 | NA |
| chr7 | 157702789 | 157702840 | 52 | 2 | 0.000225028 | 4.99E-05 | 0.000716 | 6.27E-05 | 0.014197829 | PTPRN2 |
| chr17 | 18855942 | 18856790 | 849 | 11 | 0.00013223 | 0.000272 | 0.002016 | 6.28E-05 | -0.00022039 | PRPSAP2 |
| chr6 | 162443429 | 162443554 | 126 | 3 | 0.000376109 | 5.90E-05 | 0.001535 | 6.28E-05 | 0.012168889 | PRKN |
| chr8 | 143159528 | 143160017 | 490 | 5 | 0.000142995 | 0.000255 | 0.000453 | 6.28E-05 | 0.004384904 | LY6H |
| chr7 | 32870713 | 32870813 | 101 | 3 | 0.000165118 | 3.19E-05 | 0.004197 | 6.29E-05 | 0.003047556 | KBTBD2 |
| chr5 | 4942356 | 4942499 | 144 | 2 | 0.000334855 | 4.02E-05 | 0.001893 | 6.30E-05 | 0.020475048 | NA |
| chr12 | 122381651 | 122381774 | 124 | 3 | 0.000235903 | 0.00079 | 0.000457 | 6.30E-05 | 0.009090047 | CLIP1 |
| chr11 | 124936016 | 124936612 | 597 | 5 | 6.06E-05 | 0.00041 | 0.000253 | 6.30E-05 | 0.000513136 | HEPACAM |
| chr1 | 35181234 | 35181410 | 177 | 3 | 0.000305004 | 4.86E-05 | 0.002704 | 6.30E-05 | -0.00057999 | SFPQ |
| chr5 | 1813658 | 1813959 | 302 | 4 | 0.000376216 | 5.16E-05 | 0.003789 | 6.30E-05 | -0.00123807 | NDUFS6 |
| chr17 | 79458196 | 79458355 | 160 | 2 | 0.000286966 | 5.27E-05 | 0.000616 | 6.30E-05 | 0.011879883 | RBFOX3 |
| chr7 | 576414 | 576554 | 141 | 2 | 0.000376613 | 3.97E-05 | 0.00208 | 6.30E-05 | -0.0005407 | PRKAR1B |
| chr2 | 8103236 | 8103246 | 11 | 2 | 0.000133237 | 4.71E-05 | 0.000896 | 6.31E-05 | -0.00135539 | LINC00298, LINC00299 |
| chr5 | 154939789 | 154940152 | 364 | 3 | 5.74E-05 | 0.000135 | 0.000313 | 6.31E-05 | 0.004523353 | NA |
| chr20 | 45856847 | 45857123 | 277 | 4 | 4.91E-05 | 0.000148 | 0.000993 | 6.31E-05 | 0.005299455 | ACOT8 |
| chr2 | 134774996 | 134775057 | 62 | 2 | 0.000189453 | 4.52E-05 | 0.001047 | 6.31E-05 | 0.015459154 | CCNT2-AS1 |
| chr8 | 51962132 | 51962616 | 485 | 2 | 0.000363797 | 4.02E-05 | 0.001923 | 6.31E-05 | 0.009071281 | AC064807.2 |
| chr10 | 23195096 | 23195627 | 532 | 5 | 0.000126044 | 0.002291 | 0.000347 | 6.31E-05 | 0.002793354 | NA |
| chr3 | 96610246 | 96610336 | 91 | 2 | 0.000113793 | 0.005841 | 4.63E-05 | 6.32E-05 | 0.00870234 | NA |
| chr22 | 38222967 | 38223084 | 118 | 2 | 0.000288763 | 0.001334 | 5.76E-05 | 6.32E-05 | -0.00654072 | TMEM184B |
| chr21 | 40692859 | 40692976 | 118 | 2 | 0.000424897 | 4.83E-05 | 0.000825 | 6.32E-05 | -0.00817115 | DSCAM |
| chr5 | 154583798 | 154584014 | 217 | 2 | 0.000254983 | 0.000125 | 0.000152 | 6.32E-05 | 0.009575416 | NA |
| chr16 | 31354797 | 31355108 | 312 | 6 | 0.000123789 | 0.001364 | 0.000111 | 6.32E-05 | 0.007177329 | NA |
| chr1 | 206051268 | 206051904 | 637 | 4 | 3.59E-05 | 0.000265 | 0.000101 | 6.33E-05 | 0.003575885 | BX571818.1 |
| chr10 | 6298944 | 6299418 | 475 | 3 | 0.000227357 | 2.61E-05 | 0.007345 | 6.33E-05 | 0.001214793 | LINC02649 |
| chr16 | 2687219 | 2687644 | 426 | 4 | 0.000202981 | 0.002906 | 0.000498 | 6.34E-05 | 0.005524929 | KCTD5 |
| chr1 | 113031055 | 113031175 | 121 | 2 | 0.000224196 | 0.000482 | 7.62E-05 | 6.36E-05 | 0.005549139 | SLC16A1-AS1, LRIG2-DT |
| chr1 | 162855927 | 162856325 | 399 | 3 | 0.000249386 | 7.37E-05 | 0.001384 | 6.36E-05 | -0.0039262 | CCDC190 |
| chr6 | 168128972 | 168129747 | 776 | 5 | 0.000152187 | 0.000174 | 0.000218 | 6.36E-05 | 0.003014893 | NA |
| chr16 | 52652182 | 52652697 | 516 | 3 | 3.93E-05 | 0.004302 | 0.001152 | 6.37E-05 | -5.90E-05 | AC026462.4 |
| chr10 | 112153308 | 112153322 | 15 | 2 | 0.000395793 | 5.86E-05 | 0.000488 | 6.37E-05 | 0.003283145 | GPAM |
| chr7 | 128410061 | 128410562 | 502 | 9 | 0.000274448 | 0.000807 | 0.000807 | 6.37E-05 | -0.0001089 | IMPDH1 |
| chr16 | 87654873 | 87655083 | 211 | 3 | 0.000340247 | 7.89E-05 | 0.000461 | 6.37E-05 | 0.003114198 | JPH3 |
| chr5 | 170491615 | 170491860 | 246 | 2 | 0.000211011 | 0.000195 | 0.000113 | 6.37E-05 | 0.012803681 | KCNIP1 |
| chr5 | 76114088 | 76114322 | 235 | 2 | 0.000166407 | 0.00052 | 7.48E-05 | 6.38E-05 | 0.006120874 | SV2C |
| chr1 | 24143074 | 24143738 | 665 | 7 | 0.000271211 | 0.001824 | 0.007237 | 6.38E-05 | 0.001387412 | IL22RA1 |
| chr11 | 61815662 | 61816484 | 823 | 12 | 7.61E-05 | 5.16E-05 | 0.01812 | 6.38E-05 | 1.39E-05 | FADS2, FADS1 |
| chr16 | 1050756 | 1050847 | 92 | 2 | 0.000283995 | 7.68E-05 | 0.000285 | 6.38E-05 | 0.00759756 | NA |
| chr12 | 120220211 | 120220724 | 514 | 3 | 0.000218216 | 3.75E-05 | 0.001809 | 6.38E-05 | 0.001761159 | AC004263.2, PXN |
| chr4 | 116926254 | 116926389 | 136 | 3 | 0.000289971 | 0.000415 | 8.23E-05 | 6.38E-05 | 0.009052332 | AC093765.3 |
| chr17 | 45287399 | 45287782 | 384 | 2 | 9.42E-05 | 4.13E-05 | 0.001758 | 6.38E-05 | 0.006550818 | MAP3K14 |
| chr12 | 6355352 | 6355620 | 269 | 2 | 5.69E-05 | 0.000462 | 7.76E-05 | 6.38E-05 | 0.005552916 | SCNN1A |
| chr12 | 97567946 | 97568203 | 258 | 2 | 0.000174296 | 4.21E-05 | 0.001565 | 6.38E-05 | 0.009677635 | RMST |
| chr2 | 11657870 | 11658179 | 310 | 2 | 0.000161442 | 5.09E-05 | 0.000719 | 6.39E-05 | 0.010088965 | NTSR2 |
| chr9 | 112890544 | 112891263 | 720 | 9 | 0.000138854 | 0.000103 | 0.003947 | 6.39E-05 | -0.00148037 | SLC46A2 |
| chr19 | 45498292 | 45498500 | 209 | 5 | 0.000136845 | 0.000645 | 0.000349 | 6.40E-05 | 0.005451012 | PPM1N |
| chr9 | 96567118 | 96567519 | 402 | 7 | 0.000168279 | 0.000185 | 0.007104 | 6.40E-05 | 0.000798507 | CDC14B |
| chr21 | 43921367 | 43921521 | 155 | 2 | 1.00E-04 | 0.002033 | 5.40E-05 | 6.40E-05 | 0.002434091 | AGPAT3 |
| chr13 | 28049444 | 28050019 | 576 | 4 | 0.000134372 | 0.000685 | 0.000167 | 6.40E-05 | 0.001722785 | FLT3 |
| chr2 | 240449764 | 240450726 | 963 | 4 | 1.54E-06 | 0.000556 | 6.85E-05 | 6.41E-05 | 0.010121293 | GPC1, AC110619.1 |
| chr4 | 154529292 | 154529706 | 415 | 2 | 2.47E-05 | 0.000284 | 9.40E-05 | 6.41E-05 | 0.014975694 | NA |
| chr22 | 46061216 | 46061708 | 493 | 3 | 0.000211286 | 3.07E-05 | 0.004888 | 6.41E-05 | 0.034078018 | MIRLET7BHG |
| chr17 | 50707648 | 50708631 | 984 | 9 | 1.21E-05 | 0.013233 | 0.000458 | 6.42E-05 | 0.001675713 | ANKRD40 |
| chr16 | 83813944 | 83814260 | 317 | 3 | 0.00020609 | 5.41E-05 | 0.000906 | 6.42E-05 | 0.008467333 | HSBP1 |
| chr14 | 76531746 | 76531892 | 147 | 2 | 0.000404547 | 5.51E-05 | 0.000588 | 6.42E-05 | 0.003150578 | AC008050.1 |
| chr11 | 57751810 | 57752187 | 378 | 8 | 8.31E-05 | 0.000406 | 0.002024 | 6.43E-05 | 0.004146339 | AP001931.2, AP001931.1 |
| chr12 | 53966347 | 53966696 | 350 | 4 | 0.000229557 | 3.62E-05 | 0.005395 | 6.43E-05 | 0.007205352 | HOTAIR |
| chr2 | 31276096 | 31276271 | 176 | 2 | 0.000182019 | 4.86E-05 | 0.000868 | 6.43E-05 | 0.00486742 | NA |
| chr17 | 28577845 | 28578176 | 332 | 3 | 0.000137613 | 0.000117 | 0.001758 | 6.43E-05 | 0.00257818 | AC005726.1, SPAG5 |
| chr4 | 155666732 | 155666740 | 9 | 2 | 0.000215122 | 0.000238 | 0.000103 | 6.44E-05 | 0.010954997 | GUCY1A1 |
| chr19 | 3801550 | 3802560 | 1011 | 9 | 4.39E-05 | 0.000675 | 0.004869 | 6.44E-05 | 0.004151588 | MATK |
| chr22 | 45863763 | 45864120 | 358 | 2 | 0.000267091 | 4.08E-05 | 0.002006 | 6.44E-05 | 0.012324543 | NA |
| chr5 | 88274543 | 88274838 | 296 | 3 | 0.000237173 | 0.001014 | 0.000203 | 6.45E-05 | 0.006302729 | TMEM161B-AS1 |
| chr1 | 211382935 | 211383234 | 300 | 4 | 8.92E-05 | 5.11E-05 | 0.002534 | 6.45E-05 | 0.012252251 | LINC00467 |
| chr13 | 32421848 | 32421902 | 55 | 2 | 9.78E-05 | 0.000307 | 9.17E-05 | 6.45E-05 | 0.007882239 | N4BP2L1 |
| chr18 | 57351419 | 57352480 | 1062 | 8 | 2.06E-06 | 0.002077 | 3.51E-05 | 6.45E-05 | 0.006665499 | NA |
| chr12 | 104958589 | 104958781 | 193 | 3 | 3.31E-05 | 0.013227 | 2.41E-05 | 6.45E-05 | 0.002033314 | SLC41A2 |
| chr8 | 141326023 | 141326319 | 297 | 4 | 4.37E-05 | 0.001818 | 0.00013 | 6.45E-05 | 0.002843348 | AC011676.4 |
| chr1 | 15876221 | 15876607 | 387 | 6 | 0.000233259 | 0.008964 | 0.000264 | 6.46E-05 | 0.00175387 | SPEN |
| chr11 | 126101889 | 126102567 | 679 | 4 | 0.000149808 | 3.02E-05 | 0.006487 | 6.46E-05 | 0.013324833 | AP000821.1 |
| chr19 | 49148887 | 49149187 | 301 | 2 | 2.03E-05 | 0.001647 | 5.66E-05 | 6.47E-05 | -0.00368282 | PPFIA3 |
| chr20 | 46013731 | 46014149 | 419 | 3 | 0.000287451 | 0.00014 | 0.000275 | 6.47E-05 | 0.00158548 | MMP9, SLC12A5-AS1 |

| | | | | | | | | | | |
|---|---|---|---|---|---|---|---|---|---|---|
| chr11 | 68037783 | 68037989 | 207 | 3 | 9.62E-05 | 3.96E-05 | 0.003402 | 6.48E-05 | 0.007477357 | NA |
| chr8 | 141001570 | 141002418 | 849 | 4 | 0.000102433 | 2.52E-05 | 0.008217 | 6.48E-05 | -0.00114529 | PTK2 |
| chr16 | 89064523 | 89064609 | 87 | 2 | 0.000388236 | 6.59E-05 | 0.000394 | 6.48E-05 | 0.000433264 | NA |
| chr19 | 36246276 | 36246667 | 392 | 4 | 0.000107285 | 0.000321 | 0.002383 | 6.48E-05 | 0.011938303 | NA |
| chr16 | 30929944 | 30930612 | 669 | 5 | 0.000201552 | 0.000126 | 0.000593 | 6.48E-05 | -0.00030482 | FBXL19 |
| chr9 | 78807582 | 78808059 | 478 | 4 | 4.66E-05 | 0.003072 | 3.44E-05 | 6.48E-05 | -0.00101072 | NA |
| chr5 | 129459365 | 129459912 | 548 | 4 | 8.33E-05 | 0.000158 | 0.001076 | 6.49E-05 | 0.009151689 | ADAMTS19-AS1 |
| chr3 | 173395320 | 173395786 | 467 | 5 | 0.000217291 | 0.003255 | 0.003807 | 6.49E-05 | 0.000551201 | NA |
| chr11 | 70116252 | 70116359 | 108 | 3 | 0.000288039 | 0.001132 | 7.47E-05 | 6.49E-05 | 0.006799528 | ANO1 |
| chr12 | 57619904 | 57620159 | 256 | 4 | 9.70E-05 | 0.000223 | 0.0034 | 6.49E-05 | 0.009570866 | AC025165.6, SLC26A10 |
| chr14 | 76761202 | 76761816 | 615 | 8 | 0.000113185 | 0.000799 | 0.001041 | 6.49E-05 | -0.00019424 | VASH1, AC007376.2 |
| chr3 | 184229544 | 184229857 | 314 | 4 | 0.00017783 | 3.25E-05 | 0.002879 | 6.49E-05 | 0.001658259 | NA |
| chr9 | 136510752 | 136511413 | 662 | 9 | 0.000235126 | 0.001553 | 0.009677 | 6.49E-05 | 0.000295254 | NOTCH1 |
| chr20 | 42494305 | 42494556 | 252 | 2 | 8.95E-05 | 0.017683 | 4.39E-05 | 6.49E-05 | 0.010660777 | PTPRT |
| chr5 | 31825182 | 31825219 | 38 | 2 | 0.000265833 | 4.14E-05 | 0.001976 | 6.50E-05 | -0.01513639 | PDZD2 |
| chr1 | 156063660 | 156063810 | 151 | 2 | 0.000335062 | 4.86E-05 | 0.00091 | 6.50E-05 | 0.007476844 | RAB25 |
| chr8 | 58147385 | 58147587 | 203 | 3 | 0.000124446 | 2.76E-05 | 0.006532 | 6.50E-05 | 0.007338525 | FAM110B |
| chr7 | 44159126 | 44159667 | 542 | 6 | 8.61E-05 | 0.001263 | 0.000228 | 6.50E-05 | 0.00257217 | GCK |
| chr6 | 3160268 | 3160400 | 133 | 4 | 0.000222455 | 9.89E-05 | 0.000603 | 6.52E-05 | 0.012113442 | NA |
| chr12 | 88034848 | 88035117 | 270 | 3 | 5.71E-05 | 0.001414 | 6.55E-05 | 6.52E-05 | 0.00817488 | C12orf29 |
| chr1 | 116493341 | 116493710 | 370 | 4 | 4.39E-05 | 0.001239 | 7.66E-05 | 6.52E-05 | 0.002729272 | AL390066.2, AL390066.1 |
| chr12 | 121509443 | 121510200 | 758 | 7 | 0.000121274 | 0.000178 | 0.001061 | 6.53E-05 | 0.002463024 | KDM2B |
| chr3 | 4981365 | 4981709 | 345 | 3 | 2.01E-05 | 0.014017 | 2.84E-05 | 6.53E-05 | 0.011286743 | BHLHE40 |
| chr15 | 41496151 | 41496170 | 20 | 2 | 0.000230222 | 7.26E-05 | 0.000334 | 6.54E-05 | 0.013850073 | ITPKA |
| chr6 | 72215068 | 72215719 | 652 | 5 | 5.18E-05 | 0.007378 | 0.000315 | 6.54E-05 | 0.003018771 | RIMS1 |
| chr4 | 699532 | 699544 | 13 | 2 | 0.00028295 | 8.73E-05 | 0.000248 | 6.55E-05 | 0.00841686 | NA |
| chr22 | 30825866 | 30825965 | 100 | 2 | 0.000382157 | 4.95E-05 | 0.000876 | 6.55E-05 | 0.008021075 | OSBP2 |
| chr11 | 563860 | 563990 | 131 | 2 | 0.000334479 | 4.15E-05 | 0.002045 | 6.56E-05 | 0.006991373 | RASSF7 |
| chr12 | 47988523 | 47988751 | 229 | 4 | 0.000369051 | 4.62E-05 | 0.00114 | 6.56E-05 | -0.00122144 | COL2A1 |
| chr6 | 159165661 | 159165778 | 118 | 3 | 0.000289779 | 0.000553 | 0.000422 | 6.57E-05 | 0.002492521 | NA |
| chr19 | 1025999 | 1026492 | 494 | 5 | 2.55E-05 | 0.000261 | 0.000321 | 6.57E-05 | 0.00334475 | NA |
| chr10 | 95860398 | 95860629 | 232 | 3 | 0.000213036 | 5.43E-05 | 0.001435 | 6.57E-05 | 0.006318677 | ENTPD1, AL365273.1, AL365273.2, ENTPD1-AS1 |
| chr15 | 29674623 | 29675416 | 794 | 7 | 3.92E-05 | 0.002178 | 5.00E-05 | 6.57E-05 | 3.58E-06 | AC022613.1 |
| chr12 | 121794290 | 121795088 | 799 | 5 | 3.32E-05 | 7.68E-05 | 0.003361 | 6.57E-05 | 0.001981497 | RHOF |
| chr12 | 99253346 | 99253544 | 199 | 2 | 0.00020648 | 0.0004 | 8.43E-05 | 6.57E-05 | 0.006601999 | ANKS1B |
| chr11 | 115504702 | 115505464 | 763 | 6 | 4.33E-05 | 0.000251 | 0.000929 | 6.58E-05 | 0.000511551 | CADM1 |
| chr8 | 144198985 | 144199108 | 124 | 3 | 0.000160255 | 0.000154 | 0.001148 | 6.58E-05 | 0.004341381 | MROH1 |
| chr7 | 7271268 | 7271361 | 94 | 2 | 0.000431172 | 5.01E-05 | 0.000855 | 6.58E-05 | 0.01135738 | AC005532.1 |
| chr19 | 15808195 | 15808306 | 112 | 3 | 0.00017169 | 0.000105 | 0.00039 | 6.59E-05 | 0.005621424 | OR10H1 |
| chr19 | 3851246 | 3851518 | 273 | 2 | 6.31E-05 | 0.002015 | 5.54E-05 | 6.59E-05 | 0.001188467 | ZFR2 |
| chr1 | 156892222 | 156892507 | 286 | 5 | 0.000196909 | 3.16E-05 | 0.010744 | 6.59E-05 | 0.001398478 | NA |
| chr9 | 68779886 | 68780570 | 685 | 8 | 0.000175341 | 0.000132 | 0.004336 | 6.59E-05 | 0.002365993 | PIP5K1B, FAM122A |
| chr16 | 31038466 | 31038588 | 123 | 2 | 0.000105389 | 0.003562 | 5.06E-05 | 6.59E-05 | 0.003656024 | STX4 |
| chr2 | 66445421 | 66446184 | 764 | 4 | 3.69E-05 | 4.10E-05 | 0.007765 | 6.60E-05 | 0.001870467 | MEIS1 |
| chr14 | 73272629 | 73272643 | 15 | 2 | 0.000287598 | 0.000215 | 0.000112 | 6.60E-05 | -0.0034491 | PAPLN, AC004846.2 |
| chr7 | 156122679 | 156122778 | 100 | 2 | 0.000140156 | 0.000633 | 7.28E-05 | 6.61E-05 | -0.00174447 | NA |
| chr16 | 47013393 | 47013777 | 385 | 2 | 3.45E-06 | 0.00052 | 7.73E-05 | 6.61E-05 | 0.005325716 | AC018845.1 |
| chr11 | 124321352 | 124321493 | 142 | 2 | 6.51E-05 | 0.000343 | 8.99E-05 | 6.62E-05 | 0.002388037 | NA |
| chr2 | 54556636 | 54557265 | 630 | 4 | 4.14E-05 | 0.00033 | 0.001179 | 6.62E-05 | 0.016799711 | SPTBN1 |
| chr10 | 86668056 | 86668672 | 617 | 9 | 0.000145741 | 0.000166 | 0.003993 | 6.63E-05 | 0.005311456 | LDB3 |
| chr3 | 177516307 | 177516385 | 79 | 2 | 0.000247468 | 0.000549 | 7.62E-05 | 6.63E-05 | 0.001659191 | LINC00578 |
| chr14 | 95257622 | 95257687 | 66 | 2 | 0.000382639 | 9.14E-05 | 0.000238 | 6.64E-05 | 0.001107076 | CLMN |
| chr21 | 34110689 | 34110919 | 231 | 2 | 0.000182543 | 0.000391 | 8.59E-05 | 6.64E-05 | 0.002623055 | MRPS6, AP000317.2 |
| chr9 | 133706157 | 133706418 | 262 | 2 | 9.76E-05 | 0.000742 | 6.99E-05 | 6.64E-05 | -0.00265259 | SARDH |
| chr16 | 70579619 | 70579822 | 204 | 4 | 0.0004029 | 0.000444 | 0.001893 | 6.64E-05 | 0.003122673 | NA |
| chr1 | 38047573 | 38047969 | 397 | 4 | 2.32E-05 | 0.001552 | 0.000994 | 6.65E-05 | 0.00993572 | MIR3659HG |
| chr17 | 73507584 | 73507722 | 139 | 3 | 0.000125398 | 0.012004 | 4.51E-05 | 6.65E-05 | 0.005696536 | SDK2 |
| chr22 | 19758633 | 19758845 | 213 | 3 | 0.000106584 | 4.71E-05 | 0.001308 | 6.65E-05 | 0.01520467 | TBX1 |
| chr6 | 31927213 | 31927821 | 609 | 8 | 4.17E-05 | 0.00163 | 0.000674 | 6.66E-05 | 0.005836589 | C2, AL645922.1 |
| chr4 | 179624895 | 179624904 | 10 | 2 | 0.000407641 | 8.08E-05 | 0.000292 | 6.66E-05 | 0.014464653 | NA |
| chr13 | 21014416 | 21014456 | 41 | 2 | 0.00049384 | 4.18E-05 | 0.00217 | 6.66E-05 | -0.01076599 | LATS2-AS1, LATS2 |
| chr11 | 20363115 | 20364093 | 979 | 16 | 5.03E-05 | 0.003317 | 0.005091 | 6.67E-05 | -0.00312206 | HTATIP2 |
| chr11 | 123260548 | 123260703 | 156 | 2 | 0.000118023 | 0.000589 | 7.53E-05 | 6.68E-05 | 0.006728414 | NA |
| chr5 | 14900903 | 14901063 | 161 | 2 | 6.44E-05 | 0.000136 | 0.000154 | 6.68E-05 | 0.007557878 | NA |
| chr1 | 119632178 | 119632406 | 229 | 3 | 0.000190933 | 6.70E-05 | 0.000778 | 6.71E-05 | -0.00221919 | ZNF697 |
| chr10 | 131954452 | 131954823 | 372 | 5 | 0.000344786 | 0.000102 | 0.0029 | 6.71E-05 | -0.00111879 | PPP2R2D |
| chr8 | 38386078 | 38386427 | 350 | 10 | 0.000249076 | 5.79E-05 | 0.024802 | 6.71E-05 | 0.003182186 | LETM2 |
| chr2 | 8685243 | 8685777 | 535 | 3 | 7.02E-05 | 0.001119 | 5.67E-05 | 6.71E-05 | 0.001210481 | NA |
| chr16 | 46712712 | 46712975 | 264 | 3 | 0.000246276 | 7.03E-05 | 0.001753 | 6.71E-05 | 0.013496945 | MYLK3 |
| chr1 | 63457710 | 63457817 | 108 | 2 | 0.000278858 | 0.000817 | 6.87E-05 | 6.72E-05 | 0.004490441 | ITGB3BP |
| chr5 | 162067430 | 162068043 | 614 | 12 | 0.000192734 | 6.30E-05 | 0.008724 | 6.73E-05 | 0.004193671 | GABRG2 |
| chr6 | 32310876 | 32311239 | 364 | 2 | 0.000467331 | 4.25E-05 | 0.00209 | 6.73E-05 | 0.007586445 | TSBP1-AS1, TSBP1 |
| chr12 | 74022075 | 74022230 | 156 | 2 | 0.000239421 | 0.000387 | 8.74E-05 | 6.73E-05 | 0.016250441 | LINC02882 |
| chr3 | 71784379 | 71784756 | 378 | 6 | 6.89E-05 | 0.000197 | 0.003342 | 6.73E-05 | 0.004232468 | PROK2 |
| chr6 | 31720651 | 31720925 | 275 | 2 | 6.41E-05 | 0.000143 | 0.00015 | 6.74E-05 | -0.00083504 | MPIG6B, LY6G6C |
| chr19 | 37719783 | 37720128 | 346 | 10 | 0.00024308 | 0.000194 | 0.001901 | 6.74E-05 | -0.00135015 | NA |
| chr5 | 1335020 | 1335164 | 145 | 3 | 7.54E-05 | 0.000197 | 0.000141 | 6.74E-05 | 0.003242641 | CLPTM1L |
| chr1 | 33013380 | 33013928 | 549 | 2 | 8.16E-06 | 0.117869 | 4.37E-05 | 6.75E-05 | 0.003045826 | AL020995.1, AK2 |
| chr8 | 96161998 | 96162154 | 157 | 2 | 0.000203907 | 9.28E-05 | 0.000242 | 6.76E-05 | 0.020683662 | NA |
| chr4 | 99946135 | 99946776 | 642 | 4 | 3.50E-05 | 0.000255 | 0.000141 | 6.76E-05 | 0.000561858 | DNAJB14 |
| chr19 | 11237763 | 11238187 | 425 | 5 | 0.00038084 | 0.001466 | 0.000242 | 6.76E-05 | 0.00342238 | ANGPTL8, DOCK6 |
| chr17 | 45039159 | 45039208 | 50 | 2 | 0.000181359 | 4.64E-05 | 0.001306 | 6.76E-05 | 0.012328779 | NMT1, DCAKD |
| chr11 | 46346175 | 46346495 | 321 | 3 | 2.87E-05 | 0.000512 | 8.00E-05 | 6.76E-05 | 0.007077664 | DGKZ |
| chr19 | 37210491 | 37210836 | 346 | 8 | 0.000240736 | 0.000792 | 0.000121 | 6.76E-05 | 0.001631999 | AC012309.1, ZNF585B |
| chr1 | 35470626 | 35470943 | 318 | 2 | 0.000100189 | 0.000384 | 8.80E-05 | 6.77E-05 | 0.004468842 | KIAA0319L |
| chr22 | 21632671 | 21632901 | 231 | 7 | 0.000219147 | 8.36E-05 | 0.009415 | 6.77E-05 | 0.010073351 | CCDC116 |
| chr17 | 16325725 | 16326005 | 281 | 3 | 7.92E-05 | 0.002795 | 7.33E-05 | 6.78E-05 | 0.001568687 | PIGL |
| chr17 | 43530167 | 43530890 | 724 | 6 | 0.000180738 | 3.46E-05 | 0.00961 | 6.78E-05 | 0.007363254 | DHX8, ETV4 |
| chr17 | 80008601 | 80009107 | 507 | 3 | 0.00028146 | 0.000387 | 0.000808 | 6.78E-05 | 0.001630168 | TBC1D16 |
| chr8 | 37874451 | 37874706 | 256 | 2 | 0.000312406 | 6.42E-05 | 0.000473 | 6.78E-05 | 0.010121909 | RAB11FIP1 |
| chr19 | 6712310 | 6712582 | 273 | 6 | 8.80E-05 | 0.00271 | 0.001254 | 6.78E-05 | 0.003204188 | C3 |
| chr19 | 55685301 | 55685605 | 305 | 3 | 5.87E-05 | 3.10E-05 | 0.005205 | 6.79E-05 | -0.00260726 | EPN1 |
| chr19 | 2328392 | 2329404 | 1013 | 16 | 8.58E-05 | 0.000315 | 0.002911 | 6.79E-05 | 0.000239036 | AC005258.1, SPPL2B, LSM7 |
| chr10 | 72319545 | 72319601 | 57 | 2 | 0.00011984 | 0.001656 | 5.87E-05 | 6.79E-05 | -0.00209847 | NA |
| chr12 | 92593991 | 92594084 | 94 | 2 | 0.000503892 | 4.26E-05 | 0.002197 | 6.79E-05 | 0.015328948 | NA |
| chr8 | 136046132 | 136046174 | 43 | 2 | 0.00035709 | 4.40E-05 | 0.001784 | 6.80E-05 | 0.000372955 | LINC02055 |
| chr6 | 11771045 | 11771447 | 403 | 3 | 0.000156878 | 7.36E-05 | 0.001638 | 6.80E-05 | 0.008569383 | ADTRP |
| chr7 | 151059305 | 151059686 | 382 | 9 | 4.23E-05 | 0.001619 | 0.000231 | 6.80E-05 | 0.000180729 | SLC4A2 |
| chr20 | 1299511 | 1299874 | 364 | 3 | 0.000301246 | 0.000545 | 0.000839 | 6.80E-05 | 0.005342155 | SNPH |
| chr7 | 128209282 | 128209304 | 23 | 2 | 0.000223792 | 4.39E-05 | 0.00182 | 6.80E-05 | 0.012222786 | NA |
| chr7 | 151889781 | 151889984 | 204 | 2 | 0.000146673 | 9.97E-05 | 0.000223 | 6.82E-05 | 0.002462597 | NA |
| chr12 | 10130022 | 10130203 | 182 | 2 | 8.10E-05 | 0.000417 | 8.62E-05 | 6.83E-05 | 0.007862122 | CLEC7A |
| chr1 | 8027361 | 8027530 | 170 | 3 | 0.00028349 | 4.14E-05 | 0.001711 | 6.83E-05 | 0.009596259 | AL034417.2 |
| chr13 | 113183949 | 113184378 | 430 | 3 | 0.000205619 | 0.000166 | 0.000587 | 6.84E-05 | 0.01162761 | PCID2 |
| chr10 | 84245365 | 84245956 | 592 | 2 | 0.000131337 | 6.50E-05 | 0.000473 | 6.84E-05 | 0.012482614 | RGR |
| chr2 | 223700455 | 223700720 | 266 | 2 | 0.00016592 | 0.000262 | 0.000105 | 6.84E-05 | -0.00340388 | NA |
| chr7 | 45721830 | 45722295 | 466 | 2 | 0.000119927 | 4.58E-05 | 0.001501 | 6.85E-05 | 0.000157226 | ADCY1 |
| chr14 | 56648709 | 56649089 | 381 | 3 | 0.000135732 | 7.68E-05 | 0.001204 | 6.85E-05 | -0.00393597 | TMEM260, AL161757.5 |
| chr6 | 34695098 | 34695500 | 403 | 3 | 0.00022898 | 4.95E-05 | 0.001622 | 6.85E-05 | 0.010253133 | ILRUN |
| chr5 | 140966614 | 140966818 | 205 | 6 | 0.000190901 | 0.000311 | 0.000373 | 6.85E-05 | 7.35E-05 | PCDHA1, PCDHA2, PCDHA3, PCDHA4, PCDHA5, PCDHA6, PCDHA7, PCDHA8, PCDHA9, PCDHA10, PCDHA11, PCDHA12, PCDHA13, PCDHAC1, PCDHAC2 |
| chr16 | 84123344 | 84123487 | 144 | 3 | 0.000344234 | 0.000149 | 0.001375 | 6.86E-05 | -0.00062041 | HSDL1 |
| chr11 | 77096001 | 77096038 | 38 | 2 | 0.000298673 | 4.31E-05 | 0.002189 | 6.87E-05 | 0.003481228 | CAPN5 |
| chr5 | 88667588 | 88668230 | 643 | 4 | 0.000250997 | 0.000129 | 0.002535 | 6.87E-05 | 0.007486569 | LINC00461 |
| chr16 | 554188 | 555144 | 957 | 5 | 3.42E-05 | 0.000148 | 0.003085 | 6.88E-05 | 0.0040434 | CAPN15 |
| chr15 | 66957158 | 66957298 | 141 | 3 | 8.89E-05 | 0.052413 | 2.01E-05 | 6.88E-05 | 0.002784893 | NA |
| chr17 | 2695095 | 2695505 | 411 | 3 | 0.00013852 | 2.97E-05 | 0.006553 | 6.88E-05 | 0.004788227 | CLUH |
| chr17 | 39656563 | 39657116 | 554 | 3 | 0.000256884 | 0.007058 | 0.001029 | 6.89E-05 | 0.0040801 | STARD3 |
| chr5 | 1834301 | 1834893 | 593 | 5 | 0.00010735 | 0.000187 | 0.000446 | 6.90E-05 | 0.010964785 | NA |
| chr19 | 38362695 | 38363476 | 782 | 2 | 7.61E-06 | 0.001773 | 5.87E-05 | 6.90E-05 | 0.007783168 | CATSPERG |
| chr3 | 53795773 | 53796447 | 675 | 5 | 0.000102906 | 0.020061 | 0.001183 | 6.90E-05 | 0.005705793 | CACNA1D |
| chr15 | 75114274 | 75114389 | 116 | 3 | 0.000186391 | 0.00179 | 5.43E-05 | 6.91E-05 | 0.003518652 | PPCDC, AC113208.2 |
| chr13 | 110468424 | 110468511 | 88 | 3 | 0.00035847 | 8.40E-05 | 0.000396 | 6.91E-05 | 0.000299586 | COL4A2 |
| chr7 | 3979299 | 3979551 | 253 | 2 | 7.84E-05 | 0.002907 | 5.44E-05 | 6.91E-05 | -0.00812756 | SDK1 |
| chr16 | 73050981 | 73051193 | 213 | 3 | 0.000409502 | 4.60E-05 | 0.001618 | 6.92E-05 | 0.002542705 | ZFHX3 |
| chr2 | 131040129 | 131040514 | 386 | 3 | 0.000255739 | 2.87E-05 | 0.007393 | 6.92E-05 | -0.03316158 | ARHGEF4 |
| chr19 | 17497301 | 17497585 | 285 | 3 | 0.000164374 | 9.43E-05 | 0.000448 | 6.92E-05 | 0.003397522 | SLC27A1, AC010618.3 |
| chr16 | 4699246 | 4699555 | 310 | 4 | 0.000269531 | 8.62E-05 | 0.000742 | 6.92E-05 | -0.00384025 | ANKS3 |
| chr16 | 2428780 | 2429627 | 848 | 8 | 6.90E-05 | 5.74E-05 | 0.007382 | 6.92E-05 | 0.002964144 | CCNF |
| chr6 | 133892854 | 133893169 | 316 | 3 | 0.000299207 | 3.87E-05 | 0.002235 | 6.92E-05 | 0.007631007 | TCF21 |
| chr14 | 22839085 | 22839162 | 78 | 2 | 0.000209625 | 5.84E-05 | 0.000649 | 6.92E-05 | 0.010180356 | MMP14 |
| chr1 | 246681088 | 246681729 | 642 | 3 | 0.000106161 | 0.000125 | 0.000601 | 6.93E-05 | 0.001957909 | NA |
| chr1 | 24201981 | 24202343 | 363 | 2 | 4.07E-05 | 0.022789 | 4.56E-05 | 6.93E-05 | 0.007171595 | LINC02800 |
| chr16 | 73231483 | 73232148 | 666 | 6 | 0.000173426 | 0.000286 | 0.000933 | 6.93E-05 | 0.003363012 | AC140912.1, ZFHX3 |
| chr7 | 101165201 | 101165768 | 568 | 6 | 0.000122699 | 0.018021 | 0.003117 | 6.94E-05 | 0.00234617 | VGF |
| chr17 | 38739310 | 38739604 | 295 | 2 | 9.73E-05 | 4.64E-05 | 0.0015 | 6.94E-05 | 0.007966432 | PCGF2 |
| chr3 | 197505531 | 197505741 | 211 | 3 | 0.000250638 | 0.00022 | 0.000503 | 6.94E-05 | 0.002943856 | LINC02012 |
| chr11 | 6568531 | 6568862 | 332 | 2 | 0.000462882 | 4.38E-05 | 0.002146 | 6.94E-05 | 0.012211664 | DNHD1 |
| chr19 | 14419502 | 14419858 | 357 | 8 | 0.000285639 | 0.000142 | 0.003555 | 6.95E-05 | 0.002378899 | NA |
| chr21 | 37564807 | 37565327 | 521 | 3 | 8.71E-05 | 3.40E-05 | 0.004179 | 6.95E-05 | 0.015939048 | NA |
| chr17 | 8175994 | 8176308 | 315 | 3 | 3.38E-05 | 0.000572 | 0.001364 | 6.95E-05 | 0.000536987 | TMEM107 |
| chr1 | 1307617 | 1308445 | 829 | 11 | 4.33E-05 | 9.96E-05 | 0.031233 | 6.96E-05 | -0.00130379 | ACAP3 |
| chr1 | 28591649 | 28592014 | 366 | 4 | 0.000334843 | 0.000127 | 0.005818 | 6.96E-05 | 0.004813719 | TAF12 |
| chr1 | 245153583 | 245153872 | 290 | 4 | 0.000199643 | 2.38E-05 | 0.011243 | 6.98E-05 | 0.004206298 | NA |
| chr16 | 46843919 | 46844252 | 334 | 2 | 0.000169986 | 4.51E-05 | 0.001833 | 6.98E-05 | -0.02310391 | CKBP1 |
| chr9 | 117413778 | 117413842 | 65 | 2 | 0.000288138 | 4.54E-05 | 0.001778 | 6.99E-05 | 0.005148285 | ASTN2 |
| chr11 | 44316803 | 44317418 | 616 | 5 | 1.90E-05 | 0.001129 | 0.001332 | 6.99E-05 | -0.00057531 | NA |
| chr12 | 132642314 | 132642719 | 406 | 6 | 0.000116236 | 0.002734 | 0.000907 | 6.99E-05 | 0.00112307 | POLE |
| chr6 | 102849739 | 102850035 | 297 | 2 | 0.000109005 | 0.002118 | 5.75E-05 | 6.99E-05 | 0.014439058 | NA |
| chr8 | 116765126 | 116765830 | 705 | 4 | 2.74E-05 | 0.000296 | 0.000859 | 6.99E-05 | 0.005293053 | EIF3H |
| chr17 | 42180402 | 42180991 | 590 | 5 | 4.65E-05 | 8.90E-05 | 0.000451 | 7.00E-05 | 0.000549244 | KCNH4 |
| chr22 | 24408030 | 24408059 | 30 | 2 | 0.000143458 | 7.78E-05 | 0.00035 | 7.00E-05 | -0.01567049 | SPECC1L, SPECC1L-ADORA2A |
| chr15 | 24954566 | 24955106 | 541 | 6 | 7.08E-05 | 0.00015 | 0.001609 | 7.00E-05 | 0.002811541 | SNRPN, SNURF, AC124312.1 |
| chr7 | 107959319 | 107959433 | 115 | 2 | 0.000277485 | 0.001104 | 6.59E-05 | 7.01E-05 | 0.001850486 | LAMB1 |
| chr1 | 161119167 | 161119549 | 383 | 4 | 3.49E-05 | 0.00039 | 0.000166 | 7.03E-05 | 0.005154524 | NIT1 |
| chr7 | 2048208 | 2048270 | 63 | 2 | 0.000290518 | 4.77E-05 | 0.001413 | 7.03E-05 | 0.005803869 | MAD1L1, AC069288.1 |
| chr14 | 105394617 | 105394923 | 307 | 3 | 0.000321222 | 5.39E-05 | 0.001074 | 7.03E-05 | 0.0015067 | PACS2 |
| chr21 | 26573389 | 26573672 | 284 | 3 | 0.000104976 | 0.00017 | 0.001694 | 7.03E-05 | 0.002230268 | NA |
| chr9 | 97307012 | 97308141 | 1130 | 8 | 0.000268951 | 2.41E-05 | 0.025042 | 7.04E-05 | 0.015048049 | STRA6LP, CCDC180 |
| chr22 | 44929909 | 44930110 | 202 | 2 | 0.000163112 | 0.000731 | 7.41E-05 | 7.04E-05 | 0.006992801 | PHF21B |
| chr19 | 2567918 | 2567960 | 43 | 2 | 0.000495091 | 4.75E-05 | 0.001457 | 7.04E-05 | 0.027450732 | GNG7 |
| chr19 | 41431529 | 41432133 | 605 | 3 | 0.000339039 | 7.61E-05 | 0.002359 | 7.05E-05 | 0.006280842 | DMAC2 |
| chr1 | 203086534 | 203087069 | 536 | 3 | 4.81E-05 | 0.000215 | 0.000148 | 7.05E-05 | 0.006957507 | NA |

| | | | | | | | | | | |
|---|---|---|---|---|---|---|---|---|---|---|
| chr15 | 45448642 | 45449162 | 521 | 4 | 0.000103657 | 0.00028 | 0.00016 | 7.05E-05 | 0.004191342 | C15orf48, AC025580.2 |
| chr12 | 102958916 | 102959584 | 669 | 4 | 0.000207489 | 0.000116 | 0.004083 | 7.05E-05 | 0.005541121 | ASCL1 |
| chr22 | 42947078 | 42947269 | 192 | 2 | 3.91E-05 | 4.72E-05 | 0.001521 | 7.06E-05 | 0.012974946 | PACSIN2 |
| chr19 | 47331745 | 47332083 | 339 | 6 | 0.000146543 | 7.02E-05 | 0.003571 | 7.06E-05 | 0.005640212 | NA |
| chr2 | 129071839 | 129071885 | 47 | 3 | 0.000237104 | 0.000667 | 0.000209 | 7.06E-05 | 0.002107208 | NA |
| chr8 | 46039713 | 46040275 | 563 | 2 | 0.000178928 | 5.01E-05 | 0.001156 | 7.06E-05 | -0.00381678 | NA |
| chr17 | 7838808 | 7839714 | 907 | 6 | 0.000248961 | 1.86E-05 | 0.017747 | 7.07E-05 | 0.008280029 | NA |
| chr7 | 151927220 | 151927596 | 377 | 2 | 0.000127185 | 0.000199 | 0.000128 | 7.07E-05 | 0.004416447 | NA |
| chr19 | 1907973 | 1908389 | 417 | 4 | 0.000265806 | 0.000339 | 0.001452 | 7.08E-05 | 0.004557435 | SCAMP4, ADAT3 |
| chr19 | 7508075 | 7508080 | 6 | 2 | 0.000334262 | 6.15E-05 | 0.0006 | 7.08E-05 | 0.021695798 | TEX45, AC008878.1 |
| chr7 | 155003541 | 155003964 | 424 | 4 | 0.00010771 | 9.30E-05 | 0.000427 | 7.08E-05 | -0.00109111 | PAXIP1-AS1 |
| chr12 | 93089147 | 93089390 | 244 | 2 | 0.000198728 | 7.87E-05 | 0.000354 | 7.08E-05 | -0.00858736 | AC138123.1 |
| chr6 | 44245251 | 44245640 | 390 | 4 | 2.85E-05 | 0.000973 | 5.71E-05 | 7.09E-05 | 0.001221324 | NA |
| chr9 | 137031298 | 137031404 | 107 | 2 | 0.000122956 | 4.44E-05 | 0.002251 | 7.09E-05 | 0.011414651 | C9orf139, FUT7, AL807752.6 |
| chr7 | 218121 | 218310 | 190 | 2 | 0.000132913 | 4.44E-05 | 0.002256 | 7.09E-05 | 0.006570414 | FAM20C |
| chr8 | 2030763 | 2030774 | 12 | 2 | 0.000297904 | 9.39E-05 | 0.000264 | 7.09E-05 | -0.00068751 | NA |
| chr19 | 18097125 | 18097695 | 571 | 9 | 7.23E-05 | 0.001717 | 0.001493 | 7.09E-05 | 0.000800927 | IL12RB1 |
| chr6 | 144582103 | 144582128 | 26 | 2 | 7.28E-05 | 0.001239 | 6.49E-05 | 7.10E-05 | 0.006206997 | UTRN |
| chr1 | 8096258 | 8096606 | 349 | 3 | 0.000205227 | 0.000586 | 0.000377 | 7.10E-05 | 0.001087918 | AL034417.2 |
| chr14 | 36666230 | 36666466 | 237 | 3 | 0.000344962 | 0.000519 | 0.001051 | 7.10E-05 | 0.006199061 | PAX9 |
| chr15 | 26117868 | 26118694 | 827 | 5 | 4.27E-05 | 0.003554 | 0.000104 | 7.10E-05 | 0.003579722 | LINC00929 |
| chr3 | 46406593 | 46407472 | 880 | 11 | 9.00E-05 | 3.00E-05 | 0.023488 | 7.10E-05 | 0.003109393 | CCRL2, CCR5AS |
| chr16 | 81797337 | 81797757 | 421 | 2 | 0.000124742 | 4.82E-05 | 0.001442 | 7.11E-05 | 0.005255578 | PLCG2 |
| chr1 | 15849377 | 15849930 | 554 | 6 | 0.000259074 | 0.000273 | 0.000663 | 7.11E-05 | -0.00080765 | SPEN |
| chr7 | 21917835 | 21917909 | 75 | 2 | 0.000278659 | 0.000155 | 0.000153 | 7.11E-05 | -0.00330199 | CDCA7L |
| chr16 | 1334310 | 1335157 | 848 | 11 | 5.37E-05 | 0.002761 | 0.001143 | 7.12E-05 | -0.00324373 | BAIAP3 |
| chr10 | 79273968 | 79274748 | 781 | 4 | 2.60E-05 | 0.000132 | 0.001058 | 7.12E-05 | 0.006969661 | ZMIZ1 |
| chr17 | 77088089 | 77088636 | 548 | 8 | 0.000129837 | 0.000258 | 0.004781 | 7.12E-05 | 0.000934899 | SNHG20 |
| chr10 | 3750730 | 3751055 | 326 | 4 | 0.000351033 | 0.000128 | 0.000336 | 7.13E-05 | 0.006115396 | AL450322.2 |
| chr7 | 2543768 | 2543907 | 140 | 2 | 0.0001824 | 4.47E-05 | 0.002263 | 7.14E-05 | 0.004980008 | BRAT1 |
| chr8 | 1632832 | 1633481 | 650 | 5 | 0.000159315 | 0.009043 | 0.000275 | 7.14E-05 | 0.00187101 | DLGAP2 |
| chr3 | 127915345 | 127915616 | 272 | 6 | 0.000207362 | 6.27E-05 | 0.008226 | 7.14E-05 | 0.001237985 | KBTBD12, MGLL |
| chr11 | 20963785 | 20963975 | 191 | 2 | 0.000292933 | 0.000181 | 0.000138 | 7.14E-05 | -0.00499512 | NELL1 |
| chr21 | 42351011 | 42351246 | 236 | 5 | 0.000188999 | 0.001465 | 0.000403 | 7.14E-05 | 0.004454518 | NA |
| chr10 | 125718700 | 125719199 | 500 | 4 | 2.58E-05 | 4.42E-05 | 0.007559 | 7.14E-05 | -0.01532245 | EDRF1-DT, AL158835.3, AL158835.2 |
| chr7 | 131507979 | 131508030 | 52 | 2 | 0.000201647 | 5.63E-05 | 0.00081 | 7.14E-05 | 0.020341165 | PODXL |
| chr5 | 157291431 | 157291584 | 154 | 4 | 0.000420387 | 3.47E-05 | 0.008647 | 7.14E-05 | 0.006957476 | CYFIP2 |
| chr9 | 136686030 | 136686347 | 318 | 2 | 2.30E-05 | 0.000311 | 0.000102 | 7.15E-05 | 0.011193438 | AGPAT2 |
| chr10 | 73909097 | 73909337 | 241 | 2 | 0.000130638 | 4.50E-05 | 0.002218 | 7.16E-05 | 0.003653944 | PLAU |
| chr9 | 127726271 | 127726437 | 167 | 4 | 0.000443247 | 8.33E-05 | 0.002827 | 7.17E-05 | 7.08E-05 | TTC16 |
| chr10 | 124448038 | 124448360 | 323 | 3 | 5.19E-05 | 0.007937 | 0.000111 | 7.17E-05 | 0.005421147 | AL445237.1, NKX1-2 |
| chr5 | 149606381 | 149606780 | 400 | 3 | 0.000125643 | 0.001191 | 0.000209 | 7.18E-05 | 0.009142255 | ARHGEF37, RNU6-588P |
| chr2 | 3766949 | 3767060 | 112 | 2 | 0.000298197 | 0.000312 | 0.000102 | 7.18E-05 | 0.004795254 | DCDC2C |
| chr19 | 2462044 | 2462067 | 24 | 2 | 0.000248133 | 0.000479 | 8.64E-05 | 7.19E-05 | -0.00197867 | LINC01775 |
| chr2 | 216222836 | 216223018 | 183 | 2 | 0.000128593 | 0.000614 | 7.96E-05 | 7.20E-05 | 0.011565824 | NA |
| chr2 | 75654694 | 75654804 | 111 | 4 | 0.000259191 | 0.000141 | 0.000792 | 7.20E-05 | 0.001429179 | MRPL19, GCFC2 |
| chr2 | 94947863 | 94947885 | 23 | 2 | 0.000115812 | 4.89E-05 | 0.001421 | 7.21E-05 | 0.003282227 | NA |
| chr2 | 230615000 | 230615071 | 72 | 2 | 0.000487818 | 4.80E-05 | 0.001571 | 7.21E-05 | 0.013587072 | NA |
| chr13 | 30422432 | 30422835 | 404 | 3 | 2.42E-05 | 0.129973 | 1.93E-05 | 7.21E-05 | 0.002127738 | UBE2L5 |
| chr6 | 40276244 | 40276456 | 213 | 4 | 0.000116236 | 0.001345 | 0.000993 | 7.22E-05 | 0.003709901 | NA |
| chr8 | 133217858 | 133217914 | 57 | 2 | 0.000330035 | 0.000175 | 0.000143 | 7.22E-05 | 0.011504511 | CCN4 |
| chr20 | 58989911 | 58990122 | 212 | 2 | 0.000116806 | 0.000442 | 8.93E-05 | 7.23E-05 | 0.004899289 | NELFCD |
| chr11 | 68505793 | 68505839 | 47 | 2 | 0.000418303 | 4.58E-05 | 0.002105 | 7.23E-05 | 0.014346671 | PPP6R3 |
| chr2 | 218266391 | 218266760 | 370 | 2 | 2.63E-05 | 0.000256 | 0.000114 | 7.23E-05 | 0.001171564 | AAMP |
| chr13 | 36672911 | 36673023 | 113 | 2 | 0.000375754 | 5.08E-05 | 0.001229 | 7.23E-05 | 0.001317612 | NA |
| chr6 | 166235312 | 166235878 | 567 | 2 | 7.35E-06 | 0.003394 | 5.52E-05 | 7.23E-05 | 0.011221537 | NA |
| chr13 | 110712500 | 110713205 | 706 | 8 | 0.000194812 | 2.21E-05 | 0.017222 | 7.23E-05 | 0.001042601 | ING1, CARS2 |
| chr2 | 10434533 | 10435090 | 558 | 2 | 8.70E-05 | 6.17E-05 | 0.000642 | 7.23E-05 | 0.000723649 | NA |
| chr7 | 881455 | 881519 | 65 | 2 | 0.000146833 | 4.53E-05 | 0.002276 | 7.23E-05 | 0.003349858 | SUN1, GET4, AC073957.3 |
| chr10 | 130485925 | 130486129 | 205 | 4 | 0.000117486 | 0.000255 | 0.004185 | 7.24E-05 | 0.005433777 | NA |
| chr6 | 169225319 | 169225509 | 191 | 2 | 0.000373144 | 4.57E-05 | 0.002162 | 7.24E-05 | 0.005309694 | BX322234.1, THBS2 |
| chr21 | 26170431 | 26171403 | 973 | 12 | 8.25E-05 | 0.00118 | 0.000316 | 7.24E-05 | 0.000176747 | AP001439.1, AP000229.1, APP |
| chr16 | 654976 | 655079 | 104 | 3 | 0.000214596 | 7.83E-05 | 0.00098 | 7.24E-05 | 0.001007654 | WDR90, AL022341.2 |
| chr10 | 104352452 | 104352681 | 230 | 4 | 0.000324077 | 6.24E-05 | 0.002115 | 7.25E-05 | 0.003696174 | CFAP58-DT |
| chr6 | 53153943 | 53154188 | 246 | 3 | 0.000223 | 8.35E-05 | 0.00197 | 7.25E-05 | 0.010940644 | NA |
| chr10 | 71793381 | 71793444 | 64 | 3 | 0.000400874 | 3.58E-05 | 0.003672 | 7.25E-05 | 0.004112097 | CDH23 |
| chr5 | 34044399 | 34044670 | 272 | 3 | 0.000185271 | 0.000245 | 0.00013 | 7.25E-05 | -0.00447318 | C1QTNF3-AMACR |
| chr1 | 109539832 | 109540302 | 471 | 5 | 0.000376162 | 1.82E-05 | 0.019551 | 7.25E-05 | 0.003018619 | GPR61, AL355310.3 |
| chr11 | 66134693 | 66134701 | 9 | 2 | 0.000157188 | 5.42E-05 | 0.000973 | 7.26E-05 | 0.017886211 | PACS1 |
| chr11 | 64590278 | 64590603 | 326 | 4 | 0.000334957 | 0.000391 | 0.00154 | 7.26E-05 | 0.002976766 | NA |
| chr6 | 26056312 | 26056569 | 258 | 7 | 0.000324696 | 3.05E-05 | 0.012044 | 7.26E-05 | -0.00101149 | H1-2 |
| chr20 | 61866733 | 61866896 | 164 | 2 | 0.000454784 | 4.64E-05 | 0.00202 | 7.27E-05 | -0.00058268 | CDH4 |
| chr1 | 152784981 | 152785223 | 243 | 3 | 0.000230032 | 6.21E-05 | 0.000997 | 7.27E-05 | 0.003650128 | NA |
| chr12 | 74283675 | 74283882 | 208 | 2 | 0.000172076 | 0.00012 | 0.000203 | 7.27E-05 | 0.019888453 | LINC02882 |
| chr12 | 1574194 | 1574283 | 90 | 2 | 8.49E-05 | 0.000498 | 8.62E-05 | 7.27E-05 | 0.005665753 | WNT5B, FBXL14 |
| chr19 | 54143691 | 54144033 | 343 | 4 | 0.000328911 | 0.000418 | 0.002706 | 7.27E-05 | -0.00024249 | CNOT3 |
| chr2 | 235678066 | 235678100 | 35 | 2 | 7.77E-05 | 0.000994 | 7.03E-05 | 7.27E-05 | 0.002811089 | AGAP1 |
| chr5 | 171387450 | 171387546 | 97 | 6 | 0.000396466 | 0.000424 | 0.000398 | 7.27E-05 | 0.001915519 | NPM1 |
| chr16 | 46689136 | 46689727 | 592 | 13 | 0.000155852 | 5.23E-05 | 0.015749 | 7.28E-05 | -0.00053886 | ORC6, VPS35 |
| chr4 | 5892964 | 5893371 | 408 | 4 | 0.000217068 | 2.66E-05 | 0.009142 | 7.28E-05 | -0.00925949 | CRMP1 |
| chr4 | 8158568 | 8159458 | 891 | 12 | 0.000171187 | 8.35E-05 | 0.016194 | 7.28E-05 | 0.000299997 | ABLIM2 |
| chr12 | 114403731 | 114404226 | 496 | 6 | 0.000102872 | 0.000571 | 0.001716 | 7.29E-05 | 0.005473158 | TBX5 |
| chr10 | 99791727 | 99792162 | 436 | 5 | 0.000249444 | 0.000844 | 0.003423 | 7.29E-05 | 0.000277341 | ABCC2 |
| chr16 | 30947994 | 30948341 | 348 | 6 | 7.49E-05 | 0.000322 | 0.000637 | 7.30E-05 | 0.003632334 | FBXL19 |
| chr6 | 157822160 | 157822354 | 195 | 2 | 0.000140371 | 0.000139 | 0.000176 | 7.30E-05 | 0.011150541 | SNX9 |
| chr2 | 239776472 | 239776795 | 324 | 2 | 4.69E-05 | 0.00018 | 0.000144 | 7.31E-05 | -0.0001694 | AC093802.1 |
| chr2 | 174325542 | 174325545 | 4 | 2 | 0.00022472 | 4.58E-05 | 0.002299 | 7.31E-05 | 0.003812482 | NA |
| chr11 | 74455149 | 74455300 | 152 | 2 | 0.000141741 | 0.02411 | 4.73E-05 | 7.32E-05 | 0.008870546 | AP001372.1, KCNE3 |
| chr1 | 3021435 | 3022082 | 648 | 5 | 3.98E-06 | 0.012404 | 0.00021 | 7.33E-05 | 0.004757906 | ACTRT2 |
| chr10 | 118046949 | 118047461 | 513 | 6 | 1.10E-05 | 0.000953 | 8.02E-05 | 7.33E-05 | 0.006608928 | CASC2 |
| chr2 | 141810420 | 141810887 | 468 | 3 | 0.000351001 | 6.33E-05 | 0.002769 | 7.33E-05 | 0.014648481 | LRP1B |
| chr17 | 81837930 | 81837967 | 38 | 2 | 0.000202309 | 0.000147 | 0.000169 | 7.33E-05 | 0.008804754 | NA |
| chr21 | 42245499 | 42245804 | 306 | 2 | 0.000153777 | 6.88E-05 | 0.000513 | 7.33E-05 | -0.00507136 | ABCG1 |
| chr20 | 3240073 | 3240441 | 369 | 5 | 8.14E-05 | 0.008168 | 0.003193 | 7.34E-05 | 0.006449119 | AL109976.1 |
| chr12 | 131583520 | 131583570 | 51 | 2 | 0.000228792 | 0.000124 | 0.000201 | 7.35E-05 | -0.0048421 | NA |
| chr19 | 46345846 | 46346679 | 834 | 4 | 0.000103296 | 9.80E-05 | 0.003488 | 7.35E-05 | -0.00488354 | NA |
| chr17 | 77287349 | 77288111 | 763 | 6 | 5.73E-05 | 0.000616 | 0.000487 | 7.35E-05 | -6.77E-05 | SEPTIN9 |
| chr10 | 63522096 | 63522506 | 411 | 4 | 7.09E-05 | 0.000713 | 0.00032 | 7.35E-05 | 0.000512325 | REEP3 |
| chr19 | 49423329 | 49423819 | 491 | 7 | 0.000193867 | 3.05E-05 | 0.005013 | 7.37E-05 | 0.001940305 | GFY, PTH2 |
| chr5 | 151332488 | 151332571 | 84 | 3 | 0.000279599 | 0.000418 | 9.26E-05 | 7.37E-05 | 0.006128674 | SLC36A2 |
| chr3 | 116051861 | 116052568 | 708 | 2 | 3.24E-05 | 9.47E-05 | 0.000287 | 7.39E-05 | 0.00831054 | LSAMP |
| chr4 | 4858824 | 4858971 | 148 | 4 | 0.000412946 | 4.69E-05 | 0.004617 | 7.39E-05 | 0.011124646 | NA |
| chr11 | 74397318 | 74398176 | 859 | 6 | 6.80E-05 | 0.000289 | 0.004127 | 7.39E-05 | 0.006200338 | AP001372.1, PGM2L1 |
| chr4 | 1712292 | 1712672 | 381 | 7 | 0.000102535 | 0.000628 | 0.001943 | 7.39E-05 | 0.002154842 | SLBP |
| chr20 | 63908086 | 63908522 | 437 | 4 | 0.000288278 | 2.43E-05 | 0.012288 | 7.39E-05 | 0.012080253 | DNAJC5 |
| chr11 | 67482575 | 67482693 | 119 | 4 | 0.000332668 | 0.000983 | 0.003614 | 7.39E-05 | 0.005981279 | NA |
| chr14 | 21385311 | 21386021 | 711 | 2 | 0.000404237 | 5.21E-05 | 0.001223 | 7.39E-05 | 0.008872292 | CHD8 |
| chr6 | 162728135 | 162728642 | 508 | 5 | 0.00010714 | 2.09E-05 | 0.01329 | 7.39E-05 | -0.01486113 | PACRG |
| chr4 | 3343647 | 3343768 | 122 | 2 | 0.000149773 | 0.001691 | 6.29E-05 | 7.39E-05 | 0.002474497 | RGS12 |
| chr10 | 69571772 | 69571902 | 131 | 2 | 0.000120201 | 0.003218 | 5.68E-05 | 7.39E-05 | -0.00607734 | NEUROG3 |
| chr21 | 43361304 | 43361806 | 503 | 3 | 9.22E-05 | 8.36E-05 | 0.001523 | 7.41E-05 | 0.014024091 | LINC01679 |
| chr14 | 61261074 | 61261181 | 108 | 2 | 0.000415302 | 7.87E-05 | 0.0004 | 7.41E-05 | 0.009687938 | PRKCH |
| chr8 | 11493344 | 11494337 | 994 | 8 | 0.000164162 | 0.000122 | 0.00247 | 7.41E-05 | 0.003752927 | BLK |
| chr13 | 112465057 | 112465550 | 494 | 5 | 0.000179746 | 0.000165 | 0.002882 | 7.42E-05 | -0.00568965 | NA |
| chr10 | 132186505 | 132187233 | 729 | 10 | 4.08E-05 | 0.00516 | 0.000159 | 7.42E-05 | 0.000487447 | DPYSL4 |
| chr3 | 177730340 | 177730364 | 25 | 2 | 0.00029678 | 0.000229 | 0.000126 | 7.42E-05 | 0.010313287 | LINC00578 |
| chr17 | 58692196 | 58693063 | 868 | 14 | 4.31E-05 | 0.000648 | 0.008063 | 7.43E-05 | 0.00177866 | RAD51C |
| chr9 | 72061496 | 72061580 | 85 | 2 | 0.000337337 | 4.71E-05 | 0.002165 | 7.44E-05 | 0.014350206 | AL583829.1, C9orf57 |
| chr8 | 142197832 | 142198229 | 398 | 4 | 0.00010704 | 0.000197 | 0.00154 | 7.44E-05 | 0.011943057 | NA |
| chr17 | 15265166 | 15265341 | 176 | 3 | 0.000281379 | 6.24E-05 | 0.00248 | 7.44E-05 | 0.008568084 | AC005703.2, PMP22 |
| chr16 | 57846638 | 57846754 | 117 | 2 | 0.000261579 | 0.0001 | 0.000267 | 7.45E-05 | 0.013079715 | KIFC3 |
| chr6 | 36131346 | 36131418 | 73 | 2 | 6.98E-05 | 8.10E-05 | 0.000387 | 7.46E-05 | 0.007334092 | MAPK13 |
| chr11 | 611390 | 611692 | 303 | 2 | 0.000343589 | 5.46E-05 | 0.001082 | 7.47E-05 | 0.005036733 | PHRF1 |
| chr12 | 20694220 | 20694424 | 205 | 3 | 0.00021384 | 3.12E-05 | 0.007513 | 7.47E-05 | 0.001549665 | NA |
| chr2 | 238370873 | 238370900 | 28 | 2 | 0.000495919 | 6.90E-05 | 0.000539 | 7.47E-05 | 0.011403488 | TRAF3IP1 |
| chr12 | 48814244 | 48814336 | 93 | 4 | 0.000355572 | 3.93E-05 | 0.002255 | 7.49E-05 | 0.001255695 | CACNB3 |
| chr21 | 32950845 | 32950944 | 100 | 2 | 0.000390597 | 0.000141 | 0.000182 | 7.50E-05 | 0.004756445 | C21orf62-AS1, AP000282.1 |
| chr2 | 55900807 | 55900867 | 61 | 2 | 0.000338212 | 0.000255 | 0.00012 | 7.50E-05 | 0.007447134 | EFEMP1 |
| chr3 | 197555732 | 197556280 | 549 | 6 | 0.000166862 | 0.001124 | 0.002023 | 7.52E-05 | 8.81E-06 | BDH1 |
| chr1 | 198983244 | 198983525 | 282 | 5 | 8.79E-05 | 0.004711 | 9.89E-05 | 7.53E-05 | 0.001834087 | AC096631.1 |
| chr15 | 89334892 | 89335296 | 405 | 7 | 0.00022898 | 0.000803 | 0.000919 | 7.54E-05 | 0.001856379 | AC124068.2 |
| chr16 | 26594854 | 26594888 | 35 | 2 | 0.000470151 | 0.000795 | 7.71E-05 | 7.54E-05 | -0.00965531 | AC002331.1 |
| chr1 | 29251659 | 29251663 | 5 | 2 | 0.000274706 | 5.08E-05 | 0.001533 | 7.55E-05 | 0.011932596 | PTPRU |
| chr17 | 45241368 | 45242015 | 648 | 6 | 0.00018706 | 0.00012 | 0.003186 | 7.55E-05 | -0.00019022 | FMNL1, AC008105.1 |
| chr11 | 4585358 | 4585471 | 114 | 3 | 0.000200611 | 0.001421 | 0.000188 | 7.55E-05 | -0.00333639 | OR52I2 |
| chr15 | 92904547 | 92904651 | 105 | 5 | 0.000299927 | 0.016646 | 4.41E-05 | 7.56E-05 | -0.00302176 | AC013394.1, CHD2 |
| chr2 | 46915416 | 46915746 | 331 | 3 | 2.43E-05 | 0.001049 | 0.00012 | 7.56E-05 | -0.0005128 | MCFD2 |
| chr19 | 3428953 | 3429022 | 70 | 2 | 0.0001204 | 0.002281 | 6.08E-05 | 7.56E-05 | 0.005018122 | NFIC |
| chr3 | 49278889 | 49279352 | 464 | 2 | 0.000334747 | 4.88E-05 | 0.001935 | 7.56E-05 | 0.005829501 | USP4 |
| chr19 | 1460247 | 1460527 | 281 | 4 | 5.11E-05 | 0.01465 | 3.18E-05 | 7.56E-05 | 0.000849365 | APC2 |
| chr21 | 39079445 | 39079520 | 76 | 2 | 0.000259854 | 0.000474 | 9.15E-05 | 7.57E-05 | 0.011500546 | NA |
| chr17 | 38839984 | 38840297 | 314 | 2 | 0.000120459 | 4.76E-05 | 0.002294 | 7.57E-05 | -0.00662735 | C17orf98 |
| chr11 | 45651486 | 45651860 | 375 | 3 | 0.000187546 | 0.00015 | 0.000601 | 7.57E-05 | 0.003838591 | AC087442.1, CHST1 |
| chr21 | 42314294 | 42314368 | 75 | 2 | 1.12E-05 | 0.001209 | 6.97E-05 | 7.58E-05 | 0.003983652 | TFF3 |
| chr15 | 72293458 | 72293596 | 139 | 3 | 0.000202207 | 0.001306 | 5.77E-05 | 7.58E-05 | 0.002575304 | AC009690.2, AC009690.3, CELF6, AC009690.1 |
| chr10 | 133022020 | 133022625 | 606 | 3 | 6.98E-05 | 0.00152 | 0.000776 | 7.59E-05 | 0.007671365 | NA |
| chr19 | 3025667 | 3025831 | 165 | 4 | 0.00022583 | 4.36E-05 | 0.007656 | 7.59E-05 | 0.012575326 | TLE2 |
| chr19 | 2475132 | 2476058 | 927 | 7 | 4.90E-05 | 0.000226 | 0.004057 | 7.59E-05 | 0.001172172 | NA |
| chr9 | 127919371 | 127919528 | 158 | 2 | 0.00014457 | 4.82E-05 | 0.002164 | 7.59E-05 | 0.025030819 | NA |
| chr22 | 35311018 | 35311295 | 278 | 2 | 0.000207073 | 5.20E-05 | 0.001421 | 7.60E-05 | 0.00979219 | TOM1 |
| chr9 | 37428824 | 37428922 | 99 | 3 | 0.000316504 | 9.12E-05 | 0.000423 | 7.60E-05 | 0.003339027 | GRHPR |
| chr7 | 17298192 | 17298754 | 563 | 7 | 2.67E-05 | 0.001232 | 3.04E-05 | 7.61E-05 | 0.004943852 | AHR, AC073332.1 |
| chr2 | 164109211 | 164109481 | 271 | 3 | 0.000319328 | 0.000597 | 0.000563 | 7.61E-05 | 0.000252339 | AC016766.1 |
| chr10 | 101224636 | 101224924 | 289 | 3 | 6.30E-05 | 7.92E-05 | 0.000536 | 7.62E-05 | 0.008005131 | NA |
| chr8 | 1202941 | 1203004 | 64 | 2 | 0.000172967 | 0.020216 | 4.91E-05 | 7.63E-05 | 0.002600826 | DLGAP2 |
| chr8 | 68329511 | 68329688 | 178 | 2 | 1.53E-05 | 0.039665 | 4.81E-05 | 7.63E-05 | 0.004327094 | C8orf34-AS1 |
| chr14 | 92867535 | 92867776 | 242 | 2 | 0.000200349 | 0.000629 | 8.39E-05 | 7.63E-05 | -0.00399108 | NA |
| chr6 | 33294545 | 33295516 | 972 | 11 | 0.000122502 | 0.000221 | 0.003026 | 7.64E-05 | 0.000976635 | PFDN6, RGL2 |
| chr7 | 130291889 | 130292193 | 305 | 4 | 0.000261395 | 0.00235 | 0.005437 | 7.64E-05 | 0.0018001 | NA |
| chr9 | 133459253 | 133459868 | 616 | 5 | 0.000144817 | 5.71E-05 | 0.004222 | 7.65E-05 | 0.00716643 | ADAMTS13 |
| chr8 | 141227884 | 141228939 | 1056 | 9 | 6.17E-05 | 0.007731 | 0.000173 | 7.66E-05 | 0.004404967 | SLC45A4 |
| chr16 | 56403063 | 56403287 | 225 | 4 | 0.000369079 | 0.000199 | 0.001231 | 7.66E-05 | 0.003498111 | AMFR |

| | | | | | | | | | | |
|---|---|---|---|---|---|---|---|---|---|---|
| chr7 | 43649214 | 43649539 | 326 | 2 | 5.30E-05 | 0.000116 | 0.000234 | 7.67E-05 | 0.013153474 | STK17A, COA1 |
| chr11 | 43680435 | 43680749 | 315 | 6 | 0.000105576 | 0.017802 | 0.000777 | 7.68E-05 | 0.000225624 | AC068205.2, HSD17B12 |
| chr16 | 2033150 | 2033392 | 243 | 4 | 0.000371043 | 5.86E-05 | 0.006916 | 7.68E-05 | 0.013274785 | SLC9A3R2 |
| chr2 | 20052493 | 20053191 | 699 | 3 | 1.16E-05 | 0.000193 | 0.000875 | 7.69E-05 | -0.00474355 | LAPTM4A-DT |
| chr1 | 216750086 | 216750247 | 162 | 2 | 0.000267837 | 9.24E-05 | 0.000327 | 7.69E-05 | 0.013281284 | ESRRG |
| chr12 | 110348925 | 110349302 | 378 | 2 | 0.000185068 | 5.49E-05 | 0.001196 | 7.69E-05 | 0.00271882 | ATP2A2 |
| chr7 | 159144415 | 159144918 | 504 | 3 | 0.00017063 | 4.07E-05 | 0.002842 | 7.69E-05 | -0.00212589 | VIPR2 |
| chr1 | 84753582 | 84753716 | 135 | 2 | 7.78E-05 | 5.01E-05 | 0.001843 | 7.70E-05 | 0.023232648 | NA |
| chr6 | 157647423 | 157647465 | 43 | 2 | 0.000383486 | 0.000278 | 0.000118 | 7.70E-05 | 0.009432718 | ZDHHC14 |
| chr11 | 61161089 | 61161773 | 685 | 10 | 5.17E-05 | 0.00452 | 0.001937 | 7.70E-05 | -0.00022478 | VPS37C |
| chr14 | 89955484 | 89956282 | 799 | 8 | 0.000288611 | 4.56E-05 | 0.004824 | 7.71E-05 | 0.000524534 | TDP1 |
| chr10 | 43137153 | 43137810 | 658 | 3 | 1.59E-05 | 0.000678 | 7.70E-05 | 7.71E-05 | 0.017597608 | AC010864.1 |
| chr17 | 38725400 | 38725533 | 134 | 2 | 0.000261353 | 0.000127 | 0.000214 | 7.72E-05 | 0.007664717 | MLLT6 |
| chr7 | 1428676 | 1428746 | 71 | 3 | 9.79E-05 | 0.0009 | 0.000249 | 7.74E-05 | 0.003114833 | MICALL2 |
| chr12 | 25151634 | 25151748 | 115 | 2 | 0.000408102 | 0.000131 | 0.000208 | 7.74E-05 | 0.006458039 | CFAP94 |
| chr12 | 104792676 | 104792707 | 32 | 2 | 0.000139593 | 6.65E-05 | 0.000659 | 7.74E-05 | 0.003371787 | NA |
| chr3 | 197298527 | 197299536 | 1010 | 14 | 0.000201303 | 0.000274 | 0.007001 | 7.74E-05 | 0.001631147 | DLG1-AS1, DLG1 |
| chr4 | 173517779 | 173518327 | 549 | 3 | 0.00010234 | 3.35E-05 | 0.006775 | 7.74E-05 | 0.009299185 | NA |
| chr8 | 143272698 | 143273187 | 490 | 3 | 0.000240487 | 3.96E-05 | 0.003826 | 7.75E-05 | 0.007488894 | AC138696.1, GLI4 |
| chr15 | 72272327 | 72272698 | 372 | 3 | 0.000251858 | 0.000148 | 0.001119 | 7.75E-05 | -0.00047907 | PARP6, AC009690.3 |
| chr15 | 48402841 | 48402956 | 116 | 2 | 0.000350118 | 0.000239 | 0.00013 | 7.75E-05 | 0.006965025 | NA |
| chr15 | 90762219 | 90762948 | 730 | 6 | 3.87E-05 | 0.000291 | 0.000725 | 7.75E-05 | 0.0055555 | BLM |
| chr13 | 28750877 | 28751097 | 221 | 2 | 6.29E-05 | 0.000598 | 8.63E-05 | 7.76E-05 | 0.005259937 | NA |
| chr9 | 137551308 | 137551310 | 3 | 2 | 2.41E-05 | 9.59E-05 | 0.000314 | 7.76E-05 | 0.003725039 | NA |
| chr12 | 120368814 | 120369077 | 264 | 2 | 0.000110845 | 0.000257 | 0.000125 | 7.76E-05 | 0.005595842 | MSI1 |
| chr15 | 74815069 | 74815313 | 245 | 2 | 0.000273502 | 4.86E-05 | 0.002372 | 7.76E-05 | 0.009613524 | LMAN1L |
| chr12 | 113135013 | 113136137 | 1125 | 14 | 4.51E-05 | 0.000906 | 0.004405 | 7.77E-05 | 0.002509605 | RASAL1 |
| chr7 | 2998420 | 2999033 | 614 | 4 | 4.43E-05 | 0.000753 | 9.13E-05 | 7.78E-05 | 0.004306158 | CARD11 |
| chr7 | 157005139 | 157005294 | 156 | 2 | 0.000109292 | 6.84E-05 | 0.000628 | 7.80E-05 | 0.005800207 | MNX1 |
| chr5 | 141530145 | 141530540 | 396 | 3 | 0.000290608 | 0.000468 | 0.00022 | 7.80E-05 | 0.004079418 | DIAPH1 |
| chr3 | 184378152 | 184378188 | 37 | 2 | 0.000139736 | 7.27E-05 | 0.000545 | 7.81E-05 | 0.015812275 | THPO |
| chr2 | 108614651 | 108614700 | 50 | 2 | 0.00034977 | 5.73E-05 | 0.00109 | 7.82E-05 | 0.007627659 | LIMS1 |
| chr19 | 649039 | 649464 | 426 | 3 | 0.000355719 | 5.75E-05 | 0.001103 | 7.82E-05 | 0.004759166 | RNF126 |
| chr14 | 28876790 | 28876803 | 14 | 2 | 0.000284788 | 4.96E-05 | 0.002174 | 7.82E-05 | -0.01591669 | LINC02327, LINC02326 |
| chr20 | 50172401 | 50172639 | 239 | 4 | 0.000109921 | 0.000869 | 0.000148 | 7.83E-05 | 0.008908333 | LINC01273 |
| chr12 | 70242702 | 70242879 | 178 | 9 | 0.000380191 | 0.000113 | 0.00572 | 7.83E-05 | 0.001702589 | PRANCR |
| chr2 | 128899943 | 128900153 | 211 | 2 | 0.000133794 | 5.69E-05 | 0.00113 | 7.83E-05 | 0.003675166 | NA |
| chr6 | 33111372 | 33111915 | 544 | 2 | 4.42E-05 | 0.000148 | 0.000188 | 7.84E-05 | 0.008940674 | NA |
| chr4 | 2449915 | 2449957 | 43 | 2 | 2.47E-05 | 0.029715 | 4.95E-05 | 7.84E-05 | -0.00355959 | CFAP99 |
| chr2 | 77787276 | 77787492 | 217 | 2 | 0.000120439 | 0.000122 | 0.000231 | 7.85E-05 | 0.02056722 | AC012494.1 |
| chr15 | 40256983 | 40256990 | 8 | 2 | 0.000218731 | 0.002364 | 6.22E-05 | 7.85E-05 | 0.006085328 | PAK6, BUB1B-PAK6 |
| chr5 | 139462875 | 139463622 | 748 | 3 | 2.12E-05 | 0.011743 | 5.09E-05 | 7.86E-05 | 0.003083724 | NA |
| chr18 | 76274636 | 76274916 | 281 | 2 | 0.000348823 | 6.19E-05 | 0.000852 | 7.86E-05 | 0.006817075 | NA |
| chr18 | 44771989 | 44772106 | 118 | 2 | 0.000314962 | 5.30E-05 | 0.001558 | 7.87E-05 | 0.016698721 | SETBP1 |
| chr4 | 8006660 | 8006760 | 101 | 2 | 0.000211655 | 7.06E-05 | 0.000597 | 7.87E-05 | 0.011612844 | ABLIM2 |
| chr19 | 30612758 | 30612942 | 185 | 2 | 0.000310854 | 0.000794 | 8.05E-05 | 7.88E-05 | 0.009213386 | ZNF536 |
| chr1 | 245685335 | 245685800 | 466 | 3 | 0.000163891 | 3.71E-05 | 0.004532 | 7.89E-05 | 0.000547255 | KIF26B |
| chr11 | 558659 | 558938 | 280 | 5 | 0.00017847 | 4.87E-05 | 0.004019 | 7.89E-05 | 0.003745605 | LMNTD2-AS1, LMNTD2 |
| chr9 | 111599434 | 111600427 | 994 | 5 | 5.53E-06 | 0.00274 | 0.000672 | 7.90E-05 | 0.007597577 | PTGR1 |
| chr19 | 11435071 | 11435331 | 261 | 9 | 0.000411028 | 0.009412 | 7.82E-06 | 7.90E-05 | 0.00189334 | PRKCSH, CCDC151 |
| chr17 | 38869021 | 38869791 | 771 | 6 | 7.37E-05 | 0.004081 | 7.37E-05 | 7.90E-05 | -0.00121907 | NA |
| chr17 | 10350947 | 10350959 | 13 | 3 | 0.000183653 | 0.00039 | 0.000111 | 7.91E-05 | 0.008544585 | MYH13 |
| chr17 | 2262107 | 2262137 | 31 | 2 | 0.000240925 | 0.001516 | 6.86E-05 | 7.91E-05 | 0.003240277 | SMG6 |
| chr16 | 85542680 | 85542949 | 270 | 2 | 0.000340986 | 5.69E-05 | 0.001185 | 7.91E-05 | 0.011964269 | GSE1 |
| chr10 | 113043248 | 113043331 | 84 | 2 | 0.000235677 | 0.002207 | 6.34E-05 | 7.92E-05 | 0.010778862 | TCF7L2 |
| chr18 | 79461937 | 79462022 | 86 | 2 | 0.000387425 | 0.000448 | 9.80E-05 | 7.92E-05 | 0.002605715 | NFATC1, AC018445.5 |
| chr17 | 68423872 | 68423937 | 66 | 2 | 0.000105858 | 0.0012 | 7.25E-05 | 7.92E-05 | 0.002777267 | AC007780.1, WIPI1 |
| chr8 | 37761637 | 37762126 | 490 | 4 | 0.000258654 | 0.000209 | 0.001803 | 7.92E-05 | 0.002772475 | NA |
| chr10 | 28431077 | 28431195 | 119 | 2 | 0.000383494 | 0.000133 | 0.000214 | 7.92E-05 | 0.004227404 | NA |
| chr1 | 32884781 | 32885108 | 328 | 3 | 9.83E-05 | 0.001459 | 5.84E-05 | 7.92E-05 | 0.007692422 | NA |
| chr5 | 964081 | 964115 | 35 | 2 | 0.000347035 | 5.92E-05 | 0.001029 | 7.92E-05 | 0.010530123 | NA |
| chr1 | 3646761 | 3646851 | 91 | 2 | 0.000187933 | 6.12E-05 | 0.000917 | 7.93E-05 | 0.002445898 | WRAP73 |
| chr6 | 28079235 | 28079622 | 388 | 2 | 4.10E-05 | 6.56E-05 | 0.000746 | 7.93E-05 | 0.008914084 | ZSCAN16-AS1, AL121944.1 |
| chr2 | 61405981 | 61406286 | 306 | 3 | 0.000226086 | 0.001631 | 0.001151 | 7.94E-05 | 0.009828572 | USP34 |
| chr14 | 93009629 | 93009726 | 98 | 2 | 0.000281123 | 4.98E-05 | 0.002363 | 7.94E-05 | 0.024231983 | ITPK1 |
| chr8 | 37716147 | 37716195 | 49 | 2 | 0.000165636 | 5.02E-05 | 0.002243 | 7.94E-05 | 0.003683233 | NA |
| chr2 | 196638749 | 196639322 | 574 | 4 | 0.00021699 | 0.000397 | 0.001711 | 7.94E-05 | 0.006606633 | AC068544.2 |
| chr7 | 43095246 | 43095339 | 94 | 2 | 0.000425194 | 0.000141 | 0.000203 | 7.95E-05 | 0.011779341 | AC005537.1 |
| chr20 | 20095715 | 20095906 | 192 | 3 | 8.44E-05 | 0.000119 | 0.000307 | 7.95E-05 | 4.39E-05 | CFAP61 |
| chr5 | 88722664 | 88722788 | 125 | 3 | 0.000399713 | 5.94E-05 | 0.003052 | 7.96E-05 | 0.003852245 | MEF2C-AS2, MEF2C |
| chr9 | 26956116 | 26956771 | 656 | 6 | 0.000159046 | 0.001096 | 0.003357 | 7.97E-05 | 0.001488925 | IFT74, IFT74-AS1 |
| chr1 | 40737636 | 40737728 | 93 | 2 | 0.000187545 | 0.000452 | 9.85E-05 | 7.97E-05 | 0.001633625 | NFYC |
| chr8 | 141278117 | 141278747 | 631 | 3 | 0.000189651 | 7.99E-05 | 0.000771 | 7.97E-05 | 0.008812319 | AC011676.3, SLC45A4 |
| chr8 | 33805956 | 33806108 | 153 | 2 | 0.000277376 | 0.000566 | 9.05E-05 | 7.98E-05 | 0.011997206 | AF279873.3 |
| chr14 | 101061351 | 101062242 | 892 | 10 | 2.82E-05 | 0.009408 | 0.001202 | 7.99E-05 | 0.000880313 | MIR377 |
| chr9 | 134234268 | 134234270 | 3 | 2 | 0.00026609 | 0.001718 | 6.74E-05 | 7.99E-05 | 0.002162275 | NA |
| chr3 | 46108504 | 46108605 | 102 | 2 | 0.000249961 | 0.001549 | 6.90E-05 | 7.99E-05 | 0.004000947 | NA |
| chr12 | 109501281 | 109501377 | 97 | 2 | 0.000310834 | 0.0001 | 0.000315 | 8.00E-05 | -0.00081199 | UBE3B |
| chr20 | 63829806 | 63830186 | 381 | 4 | 0.000248502 | 0.000633 | 0.001296 | 8.01E-05 | 0.002842008 | ZBTB46 |
| chr17 | 49515246 | 49515318 | 73 | 2 | 0.000232333 | 5.65E-05 | 0.001297 | 8.01E-05 | 0.011202693 | AC006487.1 |
| chr15 | 92820334 | 92820534 | 201 | 3 | 0.00022586 | 0.000537 | 0.00154 | 8.03E-05 | -0.00161417 | CHASERR |
| chr3 | 160399604 | 160401055 | 1452 | 15 | 6.31E-05 | 9.55E-05 | 0.002633 | 8.03E-05 | -0.00051695 | SMC4, AC079594.1, IFT80 |
| chr19 | 36060373 | 36060576 | 204 | 3 | 0.000178977 | 0.000313 | 0.000128 | 8.03E-05 | 0.000581488 | WDR62 |
| chr8 | 64585883 | 64586151 | 269 | 4 | 0.000208693 | 7.48E-05 | 0.005577 | 8.03E-05 | 0.006938473 | NA |
| chr14 | 61752648 | 61752882 | 235 | 2 | 0.000305373 | 8.30E-05 | 0.000448 | 8.03E-05 | 0.014752941 | AL137129.1 |
| chr1 | 228082180 | 228082782 | 603 | 14 | 0.000307479 | 0.000797 | 0.002374 | 8.04E-05 | 0.000704562 | ARF1 |
| chr6 | 151349313 | 151349489 | 177 | 2 | 8.16E-05 | 9.27E-05 | 0.000363 | 8.04E-05 | 0.01173465 | AKAP12 |
| chr11 | 14381010 | 14381397 | 388 | 4 | 8.11E-05 | 0.000484 | 0.000837 | 8.05E-05 | 0.00462993 | NA |
| chr22 | 17628511 | 17629145 | 635 | 9 | 0.000195369 | 0.000209 | 0.002769 | 8.06E-05 | 0.000437404 | BCL2L13, ATP6V1E1 |
| chr16 | 58421481 | 58421744 | 264 | 2 | 0.000253197 | 0.000178 | 0.000167 | 8.07E-05 | 0.007593564 | LINC02137 |
| chr6 | 30481232 | 30481304 | 73 | 4 | 0.000460479 | 0.000755 | 0.000405 | 8.07E-05 | 0.004700044 | NA |
| chr11 | 81970474 | 81970609 | 136 | 3 | 0.000193584 | 0.002308 | 4.81E-05 | 8.08E-05 | 0.005558051 | MIR4300HG |
| chr17 | 7242200 | 7242770 | 571 | 13 | 0.000163726 | 0.001875 | 0.006744 | 8.08E-05 | 0.000461827 | AC120057.3, GABARAP, AC120057.2 |
| chr12 | 119714085 | 119714322 | 238 | 3 | 0.000309939 | 6.05E-05 | 0.002349 | 8.08E-05 | 0.005990188 | CIT |
| chr1 | 17579810 | 17579847 | 38 | 2 | 0.000160477 | 0.007481 | 5.52E-05 | 8.09E-05 | 0.007130196 | ARHGEF10L |
| chr14 | 95209254 | 95209320 | 67 | 3 | 0.000221642 | 0.000389 | 0.000184 | 8.10E-05 | 0.004907787 | CLMN |
| chr10 | 113709169 | 113709335 | 167 | 5 | 0.000493619 | 0.000532 | 0.001013 | 8.11E-05 | 0.004240531 | CASP7 |
| chr11 | 128876319 | 128876543 | 225 | 3 | 0.000279773 | 0.003164 | 0.000311 | 8.12E-05 | 0.001804461 | NA |
| chr6 | 169725877 | 169726929 | 1053 | 4 | 1.61E-05 | 0.000722 | 8.48E-05 | 8.12E-05 | 0.001558067 | AL354892.1 |
| chr19 | 10983899 | 10984255 | 357 | 5 | 0.000403184 | 2.91E-05 | 0.013749 | 8.13E-05 | 0.001460946 | SMARCA4 |
| chr17 | 18104331 | 18104411 | 81 | 2 | 0.000332003 | 0.000143 | 0.000208 | 8.14E-05 | 0.006364481 | DRG2 |
| chr17 | 35488432 | 35489129 | 698 | 4 | 2.67E-05 | 5.35E-05 | 0.001741 | 8.15E-05 | 0.007307044 | SLFN12L |
| chr17 | 6852951 | 6853536 | 586 | 2 | 0.000342941 | 5.33E-05 | 0.001845 | 8.15E-05 | 0.003994243 | NA |
| chr16 | 1129514 | 1129941 | 428 | 5 | 5.90E-05 | 0.022463 | 4.64E-05 | 8.15E-05 | 0.003215487 | NA |
| chr8 | 26865344 | 26865848 | 505 | 7 | 0.000203035 | 0.00015 | 0.006061 | 8.15E-05 | 0.001780463 | ADRA1A |
| chr15 | 77420396 | 77420771 | 376 | 6 | 0.000212207 | 0.002815 | 8.85E-05 | 8.15E-05 | -0.00060475 | HMG20A |
| chr16 | 85372746 | 85373199 | 454 | 2 | 0.000190952 | 6.01E-05 | 0.001098 | 8.16E-05 | 0.006957181 | GSE1 |
| chr4 | 152064204 | 152064246 | 43 | 2 | 0.000420993 | 0.000188 | 0.000163 | 8.16E-05 | 0.016092919 | NA |
| chr6 | 32858120 | 32858504 | 385 | 4 | 7.89E-05 | 0.000204 | 0.000473 | 8.18E-05 | 0.004359839 | PSMB9 |
| chr22 | 44765775 | 44765919 | 145 | 2 | 8.01E-05 | 0.000492 | 9.79E-05 | 8.18E-05 | 0.007238954 | PRR5-ARHGAP8, ARHGAP8 |
| chr19 | 54850230 | 54850306 | 77 | 2 | 0.000319048 | 0.000134 | 0.000226 | 8.19E-05 | 0.009229663 | NA |
| chr1 | 50104940 | 50104959 | 20 | 2 | 7.48E-05 | 7.50E-05 | 0.000589 | 8.19E-05 | 0.007227148 | ELAVL4 |
| chr2 | 796347 | 796524 | 178 | 2 | 0.000176604 | 9.95E-05 | 0.000337 | 8.20E-05 | 0.004199347 | LINC01115 |
| chr21 | 42860234 | 42860600 | 367 | 2 | 0.000246841 | 8.64E-05 | 0.000436 | 8.21E-05 | -0.00754354 | WDR4 |
| chr12 | 64534791 | 64534989 | 199 | 3 | 0.000308763 | 0.000218 | 0.001013 | 8.21E-05 | 0.006375753 | RASSF3 |
| chr8 | 18720741 | 18720762 | 22 | 3 | 0.000184466 | 0.000118 | 0.000399 | 8.22E-05 | 0.001467951 | PSD3 |
| chr16 | 47037775 | 47038132 | 358 | 2 | 0.000379439 | 5.25E-05 | 0.002139 | 8.23E-05 | -0.00310418 | NA |
| chr8 | 8892409 | 8892428 | 20 | 2 | 0.000245684 | 5.13E-05 | 0.002472 | 8.24E-05 | 0.024230409 | MFHAS1 |
| chr5 | 95942934 | 95943033 | 100 | 2 | 0.000202718 | 0.007769 | 5.58E-05 | 8.25E-05 | 0.009519059 | ELL2 |
| chr6 | 111568510 | 111568736 | 227 | 2 | 0.00015927 | 5.27E-05 | 0.002106 | 8.25E-05 | 0.00480854 | TRAF3IP2-AS1, TRAF3IP2 |
| chr8 | 17885668 | 17885898 | 231 | 3 | 0.00048208 | 9.63E-05 | 0.000494 | 8.26E-05 | 0.002337252 | FGL1 |
| chr19 | 4818347 | 4818603 | 257 | 2 | 0.00015999 | 0.00123 | 7.49E-05 | 8.26E-05 | 0.00287069 | TICAM1 |
| chr5 | 141388864 | 141389373 | 510 | 2 | 4.02E-06 | 0.001826 | 6.86E-05 | 8.26E-05 | 0.014556961 | PCDHGA1, PCDHGA2, PCDHGA3, PCDHGB1, PCDHGA4, PCDHGB2, PCDHGA5, PCDHGB3, PCDHGA6, PCDHGA7, PCDHGB4 |
| chr10 | 1083788 | 1084465 | 678 | 8 | 0.000202232 | 0.001638 | 0.000544 | 8.26E-05 | 0.001623616 | WDR37 |
| chr15 | 82750312 | 82750653 | 342 | 7 | 0.000302876 | 0.000346 | 0.000189 | 8.26E-05 | 0.001361233 | SNHG21, AC105339.6 |
| chr6 | 122399507 | 122400378 | 872 | 8 | 1.70E-06 | 0.001255 | 5.52E-05 | 8.27E-05 | -0.00100709 | HSF2 |
| chr20 | 21706053 | 21706090 | 38 | 3 | 0.000227064 | 0.000248 | 0.000196 | 8.28E-05 | 0.00472926 | PAX1 |
| chr1 | 184821595 | 184821648 | 54 | 2 | 0.000369211 | 6.09E-05 | 0.001111 | 8.28E-05 | 0.006258021 | NIBAN1 |
| chr1 | 16493164 | 16493198 | 35 | 2 | 0.000266116 | 0.002108 | 6.67E-05 | 8.28E-05 | 0.005063127 | CROCCP3 |
| chr11 | 3824394 | 3825023 | 630 | 3 | 8.45E-05 | 0.00061 | 0.000247 | 8.28E-05 | 0.004961416 | PGAP2 |
| chr1 | 61726720 | 61726876 | 157 | 2 | 0.000180313 | 5.19E-05 | 0.002433 | 8.29E-05 | 0.001249952 | NA |
| chr1 | 2492334 | 2492564 | 231 | 2 | 0.000442333 | 5.27E-05 | 0.002207 | 8.30E-05 | 0.017482312 | PLCH2, AL139246.1 |
| chr2 | 11154612 | 11155100 | 489 | 4 | 0.000127007 | 0.002393 | 0.000673 | 8.30E-05 | 0.002620843 | NA |
| chr19 | 1507933 | 1508043 | 111 | 2 | 5.38E-05 | 0.004133 | 5.99E-05 | 8.30E-05 | 0.002274926 | PLK5, ADAMTSL5 |
| chr7 | 101527864 | 101528049 | 186 | 2 | 0.000289623 | 0.000443 | 0.000104 | 8.31E-05 | -0.00368547 | COL26A1 |
| chr10 | 23345992 | 23346271 | 280 | 3 | 0.000103007 | 0.00031 | 0.000289 | 8.31E-05 | 0.008388653 | NA |
| chr2 | 229893323 | 229893436 | 114 | 2 | 0.000165873 | 5.36E-05 | 0.00202 | 8.31E-05 | 0.009864939 | TRIP12 |
| chr9 | 123232721 | 123232734 | 14 | 2 | 0.000379873 | 0.000364 | 0.000113 | 8.31E-05 | 0.002276297 | STRBP |
| chr12 | 3591965 | 3592184 | 220 | 2 | 0.000126434 | 0.000132 | 0.000237 | 8.32E-05 | 0.00595849 | PRMT8 |
| chr3 | 111858755 | 111858810 | 56 | 3 | 0.000388289 | 5.08E-05 | 0.001853 | 8.32E-05 | -0.00936739 | PHLDB2, AC117509.1 |
| chr1 | 1331320 | 1333089 | 1770 | 10 | 0.000184473 | 0.000764 | 0.001037 | 8.32E-05 | 0.000339021 | TAS1R3 |
| chr10 | 73874034 | 73874594 | 561 | 3 | 0.000115801 | 7.40E-05 | 0.001346 | 8.33E-05 | -0.00108637 | CAMK2G |
| chr17 | 78168030 | 78168063 | 34 | 3 | 0.000351414 | 0.000225 | 0.000595 | 8.34E-05 | 0.010616376 | NA |
| chr10 | 22334531 | 22334981 | 451 | 4 | 0.00025192 | 6.75E-05 | 0.005358 | 8.35E-05 | 0.009564785 | NA |
| chr14 | 102773370 | 102773653 | 284 | 2 | 0.000268396 | 0.000154 | 0.000203 | 8.35E-05 | 0.016096496 | AL132801.1 |
| chr15 | 43516210 | 43516242 | 33 | 2 | 0.000368643 | 0.000112 | 0.000294 | 8.35E-05 | 0.003997383 | MAP1A |
| chr9 | 69976346 | 69976525 | 180 | 2 | 0.00016045 | 6.38E-05 | 0.000975 | 8.35E-05 | 0.013004711 | NA |
| chr22 | 44042526 | 44042687 | 162 | 3 | 0.000290806 | 0.00127 | 0.000133 | 8.36E-05 | 0.006374941 | PARVB |
| chr1 | 233160418 | 233160539 | 122 | 3 | 0.000305076 | 9.10E-05 | 0.002411 | 8.36E-05 | -0.0030277 | PCNX2 |
| chr1 | 63023430 | 63023445 | 16 | 2 | 0.000324696 | 0.000805 | 8.47E-05 | 8.36E-05 | 0.004033755 | NA |
| chr10 | 75235800 | 75236069 | 270 | 5 | 0.000171567 | 0.000759 | 0.002405 | 8.36E-05 | -0.00077366 | COMTD1 |
| chr4 | 183097374 | 183097972 | 599 | 2 | 5.67E-06 | 0.000105 | 0.00032 | 8.36E-05 | 0.018856385 | WWC2-AS2 |
| chr18 | 76802181 | 76802335 | 155 | 2 | 0.000434706 | 6.12E-05 | 0.001141 | 8.37E-05 | -0.01274507 | ZNF236-DT |
| chr22 | 38083701 | 38084581 | 881 | 6 | 4.23E-05 | 8.11E-05 | 0.001081 | 8.38E-05 | -0.00033174 | SLC16A8 |
| chr17 | 81486549 | 81486627 | 79 | 3 | 0.00027369 | 0.000721 | 0.000206 | 8.38E-05 | 0.011514385 | NA |
| chr19 | 55069484 | 55069746 | 263 | 6 | 0.000315507 | 0.00014 | 0.004823 | 8.40E-05 | 0.005836535 | RDH13 |
| chr6 | 35148959 | 35149401 | 443 | 2 | 1.18E-05 | 5.38E-05 | 0.002121 | 8.40E-05 | 0.008266253 | NA |
| chr16 | 57802701 | 57803397 | 697 | 11 | 0.000101793 | 0.005083 | 0.015051 | 8.40E-05 | 0.004548993 | AC092118.1, KIFC3 |
| chr1 | 169105057 | 169105696 | 640 | 5 | 0.000233209 | 0.00019 | 0.002257 | 8.40E-05 | 0.002798622 | NA |
| chr9 | 130263768 | 130264131 | 364 | 2 | 0.000319519 | 5.24E-05 | 0.002481 | 8.40E-05 | 0.026592485 | NA |
| chr16 | 11396333 | 11396582 | 250 | 3 | 0.000321496 | 0.000881 | 0.000919 | 8.40E-05 | 0.005708124 | AC099489.1 |
| chr1 | 43268982 | 43269503 | 522 | 5 | 0.000285956 | 0.004568 | 0.001557 | 8.41E-05 | -0.00175386 | EBNA1BP2 |

| | | | | | | | | | | |
|---|---|---|---|---|---|---|---|---|---|---|
| chr4 | 8466452 | 8466749 | 298 | 3 | 0.000161891 | 0.000211 | 0.000559 | 8.42E-05 | 0.000112356 | TRMT44 |
| chr2 | 109006025 | 109006110 | 86 | 3 | 0.000471014 | 0.00017 | 0.000258 | 8.42E-05 | 0.004247717 | NA |
| chr12 | 56235280 | 56235686 | 407 | 3 | 0.000192598 | 0.000447 | 0.000106 | 8.43E-05 | 0.004405631 | SLC39A5 |
| chr11 | 93523345 | 93523376 | 32 | 2 | 0.000288011 | 8.36E-05 | 0.000505 | 8.44E-05 | 0.005133112 | SMCO4 |
| chr7 | 95434862 | 95435609 | 748 | 12 | 0.000205653 | 0.000309 | 0.017426 | 8.44E-05 | 0.000731523 | PON2 |
| chr10 | 43129040 | 43129190 | 151 | 2 | 0.000451127 | 7.21E-05 | 0.000711 | 8.45E-05 | 0.000204066 | RET |
| chr9 | 97877002 | 97877231 | 230 | 3 | 0.000154906 | 0.011217 | 4.32E-05 | 8.45E-05 | 0.005710717 | NA |
| chr16 | 66692377 | 66692440 | 64 | 2 | 0.000425194 | 0.000136 | 0.000236 | 8.46E-05 | 0.004344826 | CMTM4 |
| chr4 | 7024614 | 7024783 | 170 | 4 | 7.81E-05 | 0.000537 | 0.000693 | 8.46E-05 | 0.001617456 | TBC1D14 |
| chr21 | 44258420 | 44258660 | 241 | 3 | 0.00018817 | 0.001452 | 0.000109 | 8.47E-05 | 0.000717209 | DNMT3L |
| chr19 | 58469484 | 58470502 | 1019 | 5 | 9.47E-05 | 0.000482 | 0.00193 | 8.48E-05 | -0.00192683 | ZNF324 |
| chr3 | 98901421 | 98902247 | 827 | 11 | 7.28E-05 | 0.013012 | 0.002085 | 8.48E-05 | 0.000517351 | DCBLD2 |
| chr2 | 11670334 | 11670557 | 224 | 6 | 0.00026596 | 0.000166 | 0.000499 | 8.48E-05 | -0.0060397 | NA |
| chr10 | 124354952 | 124354980 | 29 | 2 | 0.000500219 | 0.000103 | 0.000346 | 8.49E-05 | 0.0111944 | NA |
| chr14 | 105493222 | 105493779 | 558 | 3 | 0.000190008 | 6.18E-05 | 0.001719 | 8.49E-05 | 0.000537748 | TEDC1 |
| chr7 | 31106514 | 31106646 | 133 | 2 | 0.000233873 | 0.000527 | 9.91E-05 | 8.49E-05 | 0.00671504 | ADCYAP1R1 |
| chr4 | 16256356 | 16256603 | 248 | 3 | 0.000170477 | 0.000889 | 0.000352 | 8.49E-05 | 0.00510835 | TAPT1-AS1, RPS21P4 |
| chr2 | 187123825 | 187124042 | 218 | 2 | 0.000432302 | 0.000164 | 0.000197 | 8.50E-05 | 0.017666953 | AC007319.1 |
| chr10 | 126459273 | 126459435 | 163 | 2 | 0.000148483 | 8.38E-05 | 0.000513 | 8.51E-05 | 0.005540721 | AL589787.1, C10orf90 |
| chr2 | 225400416 | 225401083 | 668 | 6 | 2.84E-05 | 0.002246 | 0.000127 | 8.52E-05 | 0.000788624 | NYAP2 |
| chr7 | 150378463 | 150378759 | 297 | 2 | 0.000336119 | 8.72E-05 | 0.000474 | 8.53E-05 | 0.010828169 | ZNF775 |
| chr6 | 125789644 | 125789751 | 108 | 3 | 0.000260696 | 0.001513 | 0.001455 | 8.53E-05 | 0.020340824 | NCOA7 |
| chr16 | 83912019 | 83912373 | 355 | 4 | 0.000208579 | 4.69E-05 | 0.005386 | 8.54E-05 | -0.00110757 | MLYCD |
| chr17 | 79944546 | 79945106 | 561 | 3 | 2.51E-05 | 0.00125 | 0.000139 | 8.54E-05 | 0.002987456 | TBC1D16 |
| chr15 | 84715957 | 84716362 | 406 | 5 | 0.000364838 | 0.002081 | 6.49E-05 | 8.55E-05 | -0.00015735 | SEC11A |
| chr11 | 68029341 | 68029385 | 45 | 2 | 3.28E-05 | 0.00033 | 0.000123 | 8.56E-05 | 0.010582018 | AC004923.4 |
| chr7 | 101923457 | 101923794 | 338 | 4 | 0.000234609 | 0.000161 | 0.000844 | 8.56E-05 | 0.005310291 | CUX1 |
| chr4 | 2843944 | 2844428 | 485 | 7 | 0.000357152 | 0.0002 | 0.003133 | 8.56E-05 | 0.000915056 | ADD1 |
| chr7 | 13986120 | 13986282 | 163 | 3 | 9.18E-05 | 0.03126 | 3.56E-05 | 8.57E-05 | 0.008803303 | ETV1 |
| chr9 | 69012700 | 69012751 | 52 | 2 | 0.000222745 | 0.000322 | 0.000125 | 8.58E-05 | -0.01996452 | PRKACG |
| chr11 | 68855271 | 68855435 | 165 | 2 | 0.000224138 | 9.39E-05 | 0.000419 | 8.59E-05 | -0.00090007 | NA |
| chr8 | 26088860 | 26089178 | 319 | 3 | 0.000208555 | 0.001311 | 6.34E-05 | 8.59E-05 | 0.002776218 | NA |
| chr3 | 98522714 | 98523134 | 421 | 7 | 0.000181054 | 8.33E-05 | 0.016774 | 8.59E-05 | 0.000786994 | AC021660.2, CLDND1, AC021660.3 |
| chr3 | 112778504 | 112778643 | 140 | 2 | 0.000279072 | 0.001026 | 8.15E-05 | 8.61E-05 | 0.012530913 | NA |
| chr16 | 66512896 | 66513395 | 500 | 4 | 6.09E-05 | 0.000954 | 7.10E-05 | 8.62E-05 | 0.000991214 | AC010542.3, TK2 |
| chr8 | 144824067 | 144824437 | 371 | 2 | 6.80E-05 | 0.000129 | 0.000263 | 8.62E-05 | 0.006173652 | NA |
| chr4 | 57002308 | 57002318 | 11 | 2 | 0.000191872 | 5.48E-05 | 0.00225 | 8.63E-05 | 0.007152658 | POLR2B |
| chr19 | 577938 | 578440 | 503 | 3 | 0.000109845 | 0.000452 | 0.000148 | 8.63E-05 | 0.001114815 | BSG |
| chr14 | 100345898 | 100346138 | 241 | 4 | 0.000334079 | 0.000939 | 0.001436 | 8.63E-05 | -0.00019364 | AL157871.4, WARS1 |
| chr7 | 41732841 | 41733219 | 379 | 2 | 0.000340122 | 5.81E-05 | 0.001656 | 8.63E-05 | 0.020590669 | INHBA-AS1 |
| chr16 | 7895753 | 7895773 | 21 | 2 | 0.000456463 | 8.41E-05 | 0.00053 | 8.63E-05 | 0.00361951 | AC093515.1 |
| chr17 | 4545236 | 4545539 | 304 | 2 | 2.92E-05 | 0.001041 | 8.13E-05 | 8.63E-05 | 0.002898784 | MYBBP1A |
| chr12 | 48682265 | 48682517 | 253 | 10 | 0.000339944 | 0.000674 | 7.29E-05 | 8.63E-05 | -0.00079985 | NA |
| chr14 | 104827241 | 104827423 | 183 | 2 | 0.000223253 | 0.000345 | 0.000122 | 8.64E-05 | 0.003312261 | NA |
| chr12 | 8081418 | 8081525 | 108 | 2 | 0.000136421 | 5.78E-05 | 0.001722 | 8.65E-05 | 0.008713934 | NECAP1, AC006511.3 |
| chr17 | 63440805 | 63440993 | 189 | 3 | 0.000234587 | 0.000415 | 0.000326 | 8.65E-05 | 0.005107124 | CYB561 |
| chr9 | 89381014 | 89381310 | 297 | 5 | 0.000406212 | 0.000973 | 0.001889 | 8.66E-05 | 0.001520507 | SEMA4D |
| chr20 | 63239339 | 63239715 | 377 | 4 | 0.000275126 | 6.04E-05 | 0.005221 | 8.66E-05 | 0.00163099 | BIRC7 |
| chr9 | 137423334 | 137423361 | 28 | 4 | 0.000305479 | 0.001401 | 0.000226 | 8.66E-05 | 0.000928963 | NOXA1 |
| chr6 | 19143903 | 19143935 | 33 | 2 | 0.000343272 | 0.000747 | 8.95E-05 | 8.66E-05 | -0.00463468 | AL589647.1 |
| chr16 | 81710230 | 81710406 | 177 | 5 | 0.000381131 | 0.001306 | 0.000415 | 8.67E-05 | -0.00232399 | CMIP |
| chr2 | 199958185 | 199958226 | 42 | 2 | 8.46E-05 | 0.009904 | 5.70E-05 | 8.69E-05 | 0.002478394 | MAIP1 |
| chr6 | 41646219 | 41646400 | 182 | 2 | 0.000305292 | 5.69E-05 | 0.00191 | 8.69E-05 | 0.00375392 | MDFI |
| chr2 | 40431202 | 40431225 | 24 | 2 | 7.20E-05 | 0.000192 | 0.000178 | 8.70E-05 | 0.002280162 | SLC8A1 |
| chr19 | 35136256 | 35136316 | 61 | 2 | 0.000218343 | 7.14E-05 | 0.000811 | 8.70E-05 | 0.004761416 | LGI4 |
| chr2 | 98822077 | 98822440 | 364 | 2 | 0.000211655 | 9.33E-05 | 0.00044 | 8.70E-05 | -0.014701 | CRACDL |
| chr2 | 85615570 | 85616257 | 688 | 12 | 0.000178561 | 0.005994 | 0.00011 | 8.70E-05 | -3.60E-05 | USP39 |
| chr11 | 121102488 | 121102565 | 78 | 3 | 0.000365724 | 0.000208 | 0.001107 | 8.70E-05 | -0.00279284 | TBCEL-TECTA, TECTA |
| chr7 | 605065 | 605764 | 700 | 3 | 1.83E-05 | 6.73E-05 | 0.001043 | 8.71E-05 | 0.006929461 | PRKAR1B-AS1, PRKAR1B |
| chr1 | 31122376 | 31122442 | 67 | 2 | 0.000294832 | 0.001496 | 7.51E-05 | 8.71E-05 | 0.003000521 | NA |
| chr12 | 120213943 | 120214139 | 197 | 5 | 0.000268692 | 0.000213 | 0.001722 | 8.71E-05 | 0.003048096 | PXN |
| chr11 | 15938469 | 15938964 | 496 | 3 | 1.82E-06 | 0.000683 | 8.74E-05 | 8.71E-05 | -0.00331422 | NA |
| chr14 | 102768440 | 102768720 | 281 | 2 | 9.40E-05 | 0.000299 | 0.000133 | 8.73E-05 | 0.010774888 | NA |
| chr11 | 44468027 | 44468254 | 228 | 2 | 0.00020679 | 5.83E-05 | 0.001753 | 8.74E-05 | 0.010548333 | NA |
| chr8 | 97775561 | 97776008 | 448 | 7 | 7.15E-05 | 0.001899 | 0.000156 | 8.74E-05 | 0.001382777 | LAPTM4B |
| chr13 | 97433693 | 97433695 | 3 | 2 | 0.000261905 | 0.000351 | 0.000123 | 8.75E-05 | 0.003317879 | AL442067.3 |
| chr5 | 1106983 | 1107462 | 480 | 5 | 0.000307288 | 4.07E-05 | 0.009554 | 8.75E-05 | -0.016731 | SLC12A7 |
| chr1 | 99645831 | 99646393 | 563 | 6 | 0.000135327 | 5.17E-05 | 0.00163 | 8.75E-05 | 0.011436057 | PALMD |
| chr19 | 39204353 | 39204597 | 245 | 5 | 8.30E-05 | 0.007483 | 0.000858 | 8.76E-05 | 0.009265823 | NA |
| chr11 | 66950247 | 66950268 | 22 | 2 | 0.000313398 | 7.18E-05 | 0.00082 | 8.76E-05 | 0.010215882 | PC |
| chr17 | 74212074 | 74212649 | 576 | 5 | 1.55E-05 | 0.000124 | 0.000845 | 8.78E-05 | 0.006199866 | MGC16275 |
| chr10 | 49326391 | 49326658 | 268 | 2 | 0.000342157 | 0.000159 | 0.000216 | 8.78E-05 | 0.000828789 | C10orf71 |
| chr1 | 77682919 | 77683881 | 963 | 9 | 3.88E-05 | 0.000225 | 0.014102 | 8.78E-05 | 0.000858823 | ZZZ3 |
| chr2 | 176162712 | 176163986 | 1275 | 12 | 6.84E-06 | 0.003839 | 0.000109 | 8.80E-05 | 0.002366556 | HOXD3 |
| chr4 | 37438896 | 37439111 | 216 | 4 | 0.00024478 | 0.000221 | 0.000718 | 8.80E-05 | 0.004691254 | NWD2 |
| chr13 | 26221631 | 26222111 | 481 | 13 | 0.000297007 | 5.18E-05 | 0.005202 | 8.83E-05 | -0.00049658 | RNF6 |
| chr20 | 62711870 | 62712020 | 151 | 2 | 0.000388855 | 0.000107 | 0.000361 | 8.85E-05 | 0.009567715 | NTSR1 |
| chr15 | 47241225 | 47241352 | 128 | 3 | 0.000428611 | 6.88E-05 | 0.001047 | 8.85E-05 | -0.00675355 | SEMA6D |
| chr15 | 74131557 | 74131746 | 190 | 3 | 0.000179209 | 0.000126 | 0.000884 | 8.87E-05 | 0.008254546 | ISLR2 |
| chr12 | 132632479 | 132632750 | 272 | 2 | 0.000113622 | 0.00449 | 6.26E-05 | 8.89E-05 | 0.00281929 | POLE |
| chr11 | 72676989 | 72677238 | 250 | 3 | 0.00024924 | 9.09E-05 | 0.000659 | 8.89E-05 | 0.00299304 | NA |
| chr12 | 52119837 | 52119839 | 3 | 2 | 0.000335346 | 6.93E-05 | 0.00096 | 8.91E-05 | 0.010294697 | NA |
| chr18 | 47918185 | 47918398 | 214 | 2 | 0.000205405 | 0.002227 | 7.04E-05 | 8.91E-05 | 0.009211177 | SMAD2 |
| chr15 | 99059137 | 99059789 | 653 | 6 | 0.000114499 | 0.002743 | 0.000724 | 8.91E-05 | -0.00030201 | AC036108.1 |
| chr11 | 6926321 | 6926542 | 222 | 8 | 0.00044633 | 0.00026 | 0.004006 | 8.92E-05 | 0.003353186 | ZNF215, AC087280.2 |
| chr16 | 1436383 | 1436535 | 153 | 2 | 0.000106161 | 0.028642 | 5.50E-05 | 8.92E-05 | -0.00247544 | CCDC154 |
| chr14 | 74650100 | 74650147 | 48 | 2 | 0.000285397 | 0.00063 | 9.79E-05 | 8.93E-05 | 0.001776903 | NA |
| chr14 | 101761159 | 101761884 | 726 | 8 | 0.000190771 | 0.000143 | 0.01924 | 8.94E-05 | -0.00116704 | PPP2R5C |
| chr11 | 2383154 | 2383829 | 676 | 4 | 8.30E-05 | 0.000311 | 0.001097 | 8.94E-05 | 0.005360095 | CD81 |
| chr3 | 114153103 | 114153319 | 217 | 2 | 8.20E-05 | 7.89E-05 | 0.000688 | 8.95E-05 | 0.007840762 | DRD3 |
| chr11 | 70082836 | 70082996 | 161 | 2 | 0.000260617 | 6.17E-05 | 0.001514 | 8.96E-05 | 0.017090703 | ANO1 |
| chr1 | 21176820 | 21177145 | 326 | 11 | 0.00031939 | 0.001031 | 0.010365 | 8.96E-05 | 0.000431841 | AL031005.2, EIF4G3 |
| chr1 | 44031865 | 44032503 | 639 | 4 | 9.95E-05 | 0.012692 | 0.000126 | 8.96E-05 | 0.002594975 | AL139220.2 |
| chr17 | 79805627 | 79806201 | 575 | 2 | 0.000254505 | 6.65E-05 | 0.001133 | 8.97E-05 | -7.56E-05 | NA |
| chr7 | 42156276 | 42156577 | 302 | 3 | 0.000219286 | 0.002608 | 8.61E-05 | 8.98E-05 | 0.007087669 | GLI3 |
| chr8 | 142274752 | 142274829 | 78 | 2 | 0.000200614 | 0.000464 | 0.000111 | 8.98E-05 | 0.003346346 | TSNARE1 |
| chr16 | 89246986 | 89247056 | 71 | 2 | 7.50E-05 | 0.004509 | 6.31E-05 | 8.98E-05 | 0.014632517 | NA |
| chr20 | 38581247 | 38581356 | 110 | 2 | 0.000166359 | 5.95E-05 | 0.001842 | 8.98E-05 | 0.00137311 | ADIG |
| chr4 | 912703 | 913072 | 370 | 3 | 9.87E-05 | 0.000313 | 0.000514 | 9.01E-05 | 0.002871974 | GAK |
| chr6 | 22569652 | 22570319 | 668 | 3 | 7.48E-05 | 3.77E-05 | 0.008027 | 9.01E-05 | -0.0041731 | CASC15, HDGFL1 |
| chr7 | 139777675 | 139778042 | 368 | 4 | 0.000316465 | 0.000298 | 0.000208 | 9.01E-05 | 0.001855512 | TBXAS1, HIPK2 |
| chr7 | 6017756 | 6017846 | 91 | 2 | 0.000324077 | 7.44E-05 | 0.000817 | 9.01E-05 | 0.001202627 | AIMP2 |
| chr11 | 64714010 | 64714027 | 18 | 2 | 0.000214312 | 0.000144 | 0.000254 | 9.02E-05 | 0.005317443 | NRXN2 |
| chr10 | 24208784 | 24209166 | 383 | 6 | 0.00034156 | 0.000129 | 0.00137 | 9.02E-05 | 0.002285236 | KIAA1217 |
| chr12 | 30197723 | 30198206 | 484 | 4 | 4.74E-05 | 0.029975 | 1.83E-05 | 9.03E-05 | -0.00549671 | NA |
| chr3 | 28241029 | 28241682 | 654 | 9 | 0.000145778 | 0.015996 | 6.18E-05 | 9.04E-05 | -0.00080938 | CMC1 |
| chr5 | 147442275 | 147442386 | 112 | 3 | 0.000436692 | 0.000441 | 0.000581 | 9.05E-05 | -0.00541106 | DPYSL3 |
| chr5 | 111071177 | 111071840 | 664 | 6 | 4.61E-05 | 0.000494 | 0.008949 | 9.06E-05 | 0.009152709 | TSLP |
| chr22 | 37925825 | 37926032 | 208 | 2 | 0.000198181 | 0.000622 | 9.99E-05 | 9.07E-05 | 0.002304991 | MICALL1 |
| chr7 | 20785926 | 20786726 | 801 | 7 | 9.56E-05 | 0.001223 | 0.00052 | 9.08E-05 | 0.004236839 | SP8 |
| chr6 | 32110744 | 32110847 | 104 | 5 | 0.000317545 | 0.000984 | 0.000598 | 9.09E-05 | 0.00626154 | AL662884.5, TNXB, AL662884.2 |
| chr10 | 110885115 | 110885365 | 251 | 2 | 5.65E-05 | 0.003419 | 6.66E-05 | 9.09E-05 | 0.003160941 | PDCD4 |
| chr7 | 128201676 | 128201874 | 199 | 3 | 0.000239327 | 0.001068 | 7.65E-05 | 9.10E-05 | 0.004925405 | NA |
| chr14 | 20323098 | 20323359 | 262 | 4 | 0.000323681 | 0.000662 | 0.002166 | 9.10E-05 | 0.001437428 | CCNB1IP1, SNORA79B |
| chr17 | 30219103 | 30219228 | 126 | 2 | 0.000341835 | 0.000496 | 0.00011 | 9.11E-05 | 0.009376446 | SLC6A4 |
| chr16 | 56332857 | 56332993 | 137 | 3 | 0.000157076 | 0.000245 | 0.000282 | 9.11E-05 | 0.007477991 | GNAO1 |
| chr11 | 57530608 | 57531121 | 514 | 8 | 0.000255859 | 0.000647 | 0.00031 | 9.11E-05 | -0.0008235 | TIMM10 |
| chr3 | 197317545 | 197317706 | 162 | 2 | 0.00029444 | 0.000908 | 8.85E-05 | 9.11E-05 | 0.003916074 | NA |
| chr9 | 129236218 | 129236255 | 38 | 2 | 0.000483014 | 8.32E-05 | 0.000642 | 9.12E-05 | 0.008137946 | AL158151.3, AL158151.1 |
| chr9 | 137382320 | 137382768 | 449 | 2 | 3.88E-05 | 0.001193 | 8.26E-05 | 9.12E-05 | 0.00474525 | EXD3 |
| chr15 | 23036126 | 23036248 | 123 | 2 | 0.000262996 | 5.70E-05 | 0.002593 | 9.12E-05 | 0.013610712 | TUBGCP5 |
| chr7 | 3974438 | 3974633 | 196 | 3 | 7.89E-05 | 0.047668 | 2.90E-05 | 9.13E-05 | -0.00362085 | SDK1 |
| chr1 | 94593228 | 94593760 | 533 | 2 | 0.000104604 | 6.76E-05 | 0.001154 | 9.13E-05 | 0.006622763 | NA |
| chr7 | 1064240 | 1064399 | 160 | 2 | 0.000320969 | 0.000107 | 0.00039 | 9.16E-05 | 0.003258869 | C7orf50 |
| chr10 | 5551423 | 5551596 | 174 | 3 | 0.000238068 | 6.06E-05 | 0.001573 | 9.16E-05 | 0.005872948 | AL732437.3 |
| chr1 | 6171941 | 6172210 | 270 | 2 | 0.000182341 | 6.87E-05 | 0.001101 | 9.16E-05 | 0.009795101 | CHD5, AL031847.1 |
| chr20 | 57265360 | 57265577 | 218 | 3 | 0.000288763 | 0.000179 | 0.002006 | 9.16E-05 | 0.005873476 | BMP7 |
| chr7 | 74234557 | 74234574 | 18 | 2 | 0.000340882 | 0.000778 | 9.35E-05 | 9.17E-05 | 0.002884139 | RFC2 |
| chr4 | 183904291 | 183904689 | 399 | 2 | 5.93E-06 | 0.000144 | 0.000263 | 9.18E-05 | 0.007478083 | STOX2 |
| chr8 | 123180696 | 123182121 | 1426 | 9 | 0.000141867 | 5.08E-05 | 0.012029 | 9.20E-05 | 0.007947273 | FAM83A, AC068228.1 |
| chr7 | 151023415 | 151023675 | 261 | 2 | 0.00021165 | 0.000342 | 0.000132 | 9.20E-05 | 0.007414102 | ATG9B |
| chr1 | 11501851 | 11502077 | 227 | 3 | 9.29E-05 | 0.000284 | 0.00016 | 9.21E-05 | 0.012185993 | DISP3, AL590989.1 |
| chr10 | 1242098 | 1242418 | 321 | 4 | 8.52E-05 | 0.000532 | 0.000898 | 9.22E-05 | -0.00121712 | LINC00200, ADARB2 |
| chr15 | 40466817 | 40466828 | 12 | 2 | 0.00044277 | 0.00139 | 8.01E-05 | 9.22E-05 | -0.00234967 | BAHD1 |
| chr20 | 62242209 | 62242377 | 169 | 2 | 0.000165652 | 0.000552 | 0.000106 | 9.22E-05 | -0.00769588 | OSBPL2 |
| chr11 | 67382368 | 67382628 | 261 | 3 | 0.000310298 | 4.68E-05 | 0.003746 | 9.23E-05 | 0.003925855 | RAD9A |
| chr14 | 39265760 | 39266007 | 248 | 3 | 0.000304534 | 4.65E-05 | 0.005183 | 9.24E-05 | 0.001185773 | MIA2, AL132639.2 |
| chr9 | 137087208 | 137087697 | 490 | 2 | 4.95E-05 | 8.09E-05 | 0.000718 | 9.25E-05 | -0.00288671 | MAN1B1 |
| chr2 | 130728923 | 130729033 | 111 | 2 | 0.000470537 | 0.000206 | 0.000186 | 9.26E-05 | 0.010964792 | NA |
| chr16 | 23277044 | 23277081 | 38 | 2 | 0.000463045 | 6.94E-05 | 0.001109 | 9.26E-05 | -0.02044231 | NA |
| chr11 | 119340730 | 119341537 | 808 | 7 | 0.000219924 | 0.000119 | 0.007695 | 9.27E-05 | 0.005108228 | C1QTNF5, MFRP |
| chr4 | 182824945 | 182824971 | 27 | 2 | 0.000373418 | 0.00019 | 0.0002 | 9.27E-05 | 0.005114923 | NA |
| chr18 | 63320784 | 63321232 | 449 | 6 | 0.000243385 | 0.000304 | 0.000414 | 9.27E-05 | 0.005385593 | NA |
| chr4 | 3317184 | 3317400 | 217 | 4 | 0.000391733 | 0.00068 | 0.000137 | 9.29E-05 | 0.003331956 | RGS12 |
| chr11 | 65477417 | 65477658 | 242 | 3 | 8.12E-05 | 0.000816 | 9.29E-05 | 9.30E-05 | -0.00123698 | NA |
| chr17 | 29166186 | 29166780 | 595 | 5 | 0.000286806 | 0.000602 | 0.008901 | 9.30E-05 | 0.001130896 | MYO18A |
| chr11 | 120299990 | 120300390 | 401 | 4 | 9.99E-05 | 0.001151 | 0.000894 | 9.31E-05 | 0.010680292 | POU2F3 |
| chr1 | 242524842 | 242525379 | 538 | 4 | 0.000196177 | 0.000828 | 8.32E-05 | 9.32E-05 | 0.003671636 | NA |
| chr1 | 161070254 | 161070933 | 680 | 4 | 0.000109845 | 8.78E-05 | 0.003509 | 9.32E-05 | 0.003268746 | NA |
| chr8 | 144336887 | 144337056 | 170 | 3 | 0.000332029 | 4.48E-05 | 0.005962 | 9.33E-05 | -0.00079274 | SLC52A2 |
| chr12 | 121625679 | 121626062 | 384 | 3 | 2.62E-05 | 9.44E-05 | 0.001269 | 9.33E-05 | 0.005689412 | NA |
| chr18 | 56651093 | 56651709 | 617 | 12 | 0.0002697 | 0.000643 | 0.00791 | 9.34E-05 | 0.000280945 | WDR7, TXNL1 |
| chr2 | 190880923 | 190881347 | 425 | 3 | 2.39E-05 | 0.004117 | 0.000819 | 9.35E-05 | -9.76E-05 | GLS, AC005540.1 |
| chr7 | 108178096 | 108178194 | 99 | 2 | 0.000424115 | 0.000292 | 0.000147 | 9.35E-05 | -0.00129839 | NRCAM |
| chr16 | 4647773 | 4648219 | 447 | 4 | 0.000284821 | 0.000168 | 0.000285 | 9.36E-05 | 0.000926789 | MGRN1 |
| chr11 | 5046697 | 5046927 | 231 | 2 | 0.000116988 | 0.003268 | 6.87E-05 | 9.36E-05 | -0.00323221 | OR52J3 |
| chr17 | 49914297 | 49914442 | 146 | 2 | 0.000157655 | 0.000178 | 0.000217 | 9.36E-05 | 0.0144552 | AC027801.4 |
| chr16 | 81692294 | 81692648 | 355 | 3 | 0.000143396 | 0.000606 | 0.000377 | 9.36E-05 | -0.0003816 | CMIP |
| chr17 | 4675184 | 4675865 | 682 | 4 | 0.000173083 | 3.79E-05 | 0.009334 | 9.36E-05 | 0.00156111 | AC091153.2, PELP1 |
| chr14 | 100880707 | 100881021 | 315 | 5 | 0.000442651 | 9.89E-05 | 0.004226 | 9.37E-05 | 0.002832497 | AL117190.1, MIR431, RTL1 |
| chr18 | 70382239 | 70382510 | 272 | 2 | 7.27E-05 | 0.001249 | 8.36E-05 | 9.37E-05 | 0.00565784 | LINC01910 |
| chr8 | 1872748 | 1873238 | 491 | 3 | 2.78E-05 | 0.006758 | 0.000108 | 9.37E-05 | -0.00071328 | AC019257.8, ARHGEF10 |

| | | | | | | | | | | |
|---|---|---|---|---|---|---|---|---|---|---|
| chr8 | 113378150 | 113378597 | 448 | 3 | 0.000224893 | 5.83E-05 | 0.00226 | 9.39E-05 | 0.005595314 | CSMD3 |
| chr14 | 90273663 | 90273693 | 31 | 3 | 0.000407997 | 0.001389 | 0.000476 | 9.39E-05 | 0.000212087 | PSMC1, NRDE2 |
| chr13 | 52903502 | 52903606 | 105 | 3 | 0.000419318 | 0.000807 | 0.00061 | 9.39E-05 | -0.00457887 | NA |
| chr7 | 4225348 | 4225916 | 569 | 5 | 0.00020975 | 0.001015 | 0.000771 | 9.39E-05 | 0.002207938 | SDK1 |
| chr7 | 27203657 | 27203844 | 188 | 2 | 0.000167253 | 5.85E-05 | 0.002692 | 9.39E-05 | 0.012248175 | HOTTIP |
| chr4 | 151686860 | 151687259 | 400 | 4 | 0.000370989 | 0.000482 | 0.003224 | 9.39E-05 | 0.00774988 | GATB |
| chr5 | 90529496 | 90529986 | 491 | 6 | 0.000216118 | 0.000107 | 0.004148 | 9.39E-05 | 0.00425974 | ADGRV1, LYSMD3 |
| chr12 | 106773765 | 106774855 | 1091 | 12 | 0.000185286 | 0.000322 | 0.010346 | 9.39E-05 | 0.001529756 | RIC8B, AC079385.1 |
| chr22 | 44442875 | 44443242 | 368 | 4 | 0.000295179 | 0.001478 | 0.000253 | 9.40E-05 | 0.012790439 | NA |
| chr16 | 74449796 | 74450058 | 263 | 5 | 0.000299742 | 2.72E-05 | 0.015117 | 9.42E-05 | 0.000786075 | GLG1 |
| chr2 | 238025152 | 238025213 | 62 | 3 | 0.000392369 | 0.000301 | 0.001383 | 9.42E-05 | 0.004434055 | UBE2F, UBE2F-SCLY |
| chr8 | 131923876 | 131923972 | 97 | 2 | 0.000444384 | 0.000148 | 0.000267 | 9.43E-05 | 0.011989455 | EFR3A |
| chr12 | 109760796 | 109761323 | 528 | 3 | 0.000278558 | 0.000325 | 0.000153 | 9.44E-05 | 0.001506554 | FAM222A, FAM222A-AS1 |
| chr6 | 113855418 | 113856320 | 903 | 5 | 1.89E-05 | 0.000137 | 0.001745 | 9.44E-05 | 0.003655849 | NA |
| chr11 | 16610319 | 16610764 | 446 | 5 | 0.000377869 | 4.57E-05 | 0.005566 | 9.47E-05 | 0.00559879 | SOX6 |
| chr7 | 151227270 | 151227588 | 319 | 11 | 0.000368691 | 0.000317 | 0.002722 | 9.48E-05 | -0.00023629 | NA |
| chr12 | 132808648 | 132808732 | 85 | 3 | 0.000362411 | 0.001697 | 0.000264 | 9.49E-05 | 0.00231698 | GOLGA3 |
| chr8 | 141435668 | 141435686 | 19 | 2 | 0.000492427 | 0.000242 | 0.00017 | 9.49E-05 | 0.002446787 | AC100803.4, MROH5 |
| chr9 | 136430475 | 136430500 | 26 | 2 | 0.000145514 | 5.94E-05 | 0.002603 | 9.50E-05 | 0.004069961 | INPP5E |
| chr12 | 65741747 | 65741954 | 208 | 2 | 0.000169691 | 6.62E-05 | 0.001485 | 9.50E-05 | -0.0004391 | NA |
| chr4 | 858005 | 858372 | 368 | 3 | 8.59E-05 | 0.000815 | 0.000111 | 9.50E-05 | 0.003663864 | GAK |
| chr16 | 78241740 | 78242109 | 370 | 2 | 1.90E-05 | 0.000212 | 0.00019 | 9.50E-05 | -0.00400848 | WWOX, WWOX-AS1 |
| chr1 | 111503849 | 111504293 | 445 | 4 | 7.61E-05 | 0.00177 | 0.00194 | 9.50E-05 | 0.005811198 | TMIGD3 |
| chr17 | 62807454 | 62807889 | 436 | 3 | 0.000140238 | 0.000181 | 0.000264 | 9.50E-05 | 0.00696275 | MARCHF10 |
| chr12 | 120978698 | 120978993 | 296 | 6 | 0.000183702 | 0.00019 | 0.000184 | 9.50E-05 | 0.003763025 | HNF1A, HNF1A-AS1 |
| chr19 | 418899 | 419190 | 292 | 6 | 0.000225866 | 0.00033 | 0.001418 | 9.50E-05 | 0.003448377 | SHC2 |
| chr16 | 2856069 | 2856918 | 850 | 3 | 0.000349157 | 0.000113 | 0.000506 | 9.50E-05 | 0.007832909 | PRSS22 |
| chr3 | 133852045 | 133852160 | 116 | 2 | 0.000432154 | 0.000149 | 0.00027 | 9.50E-05 | 0.005861686 | RAB6B |
| chr8 | 141058525 | 141059236 | 712 | 4 | 2.51E-05 | 0.004022 | 0.000122 | 9.51E-05 | 0.005315316 | AC100860.1 |
| chr17 | 40704233 | 40704843 | 611 | 4 | 0.000118665 | 7.45E-05 | 0.005095 | 9.51E-05 | 0.009646483 | NA |
| chr20 | 2816657 | 2817075 | 419 | 2 | 0.000276851 | 0.000124 | 0.00034 | 9.52E-05 | 0.006887607 | AL035460.1, TMEM239 |
| chr19 | 296480 | 296698 | 219 | 2 | 0.000180824 | 0.000269 | 0.000158 | 9.52E-05 | 0.007812561 | NA |
| chr1 | 8424097 | 8424643 | 547 | 9 | 1.29E-05 | 0.001046 | 0.007482 | 9.53E-05 | 0.017556633 | RERE |
| chr16 | 89534356 | 89534806 | 451 | 3 | 3.30E-05 | 0.00011 | 0.002297 | 9.53E-05 | 0.007867025 | SPG7 |
| chr8 | 125578796 | 125579069 | 274 | 2 | 3.88E-05 | 0.001202 | 8.56E-05 | 9.53E-05 | 0.009125221 | NA |
| chr12 | 57152094 | 57152314 | 221 | 2 | 0.000160667 | 0.000284 | 0.000153 | 9.53E-05 | 0.00215572 | LRP1 |
| chr2 | 216301924 | 216302045 | 122 | 2 | 0.00034522 | 0.00049 | 0.000116 | 9.53E-05 | 0.021246868 | MARCHF4 |
| chr10 | 133288210 | 133288471 | 262 | 2 | 0.000416372 | 6.54E-05 | 0.001591 | 9.54E-05 | 0.005010095 | TUBGCP2 |
| chr5 | 31193387 | 31193901 | 515 | 6 | 0.000162227 | 0.000235 | 0.003172 | 9.54E-05 | 0.00456795 | CDH6, AC113386.1 |
| chr4 | 2829260 | 2829632 | 373 | 3 | 0.000103042 | 0.029795 | 3.58E-05 | 9.54E-05 | 0.002699219 | SH3BP2 |
| chr13 | 49935766 | 49936015 | 250 | 4 | 0.000238329 | 3.32E-05 | 0.011681 | 9.55E-05 | -0.00181299 | SPRYD7 |
| chr19 | 1490415 | 1490526 | 112 | 5 | 0.000429233 | 0.000875 | 0.000786 | 9.55E-05 | 0.002509097 | PCSK4 |
| chr11 | 63837947 | 63838643 | 697 | 5 | 8.52E-05 | 0.000365 | 0.003261 | 9.57E-05 | 0.004196949 | NA |
| chr14 | 71321843 | 71322090 | 248 | 5 | 0.000287814 | 0.000206 | 0.000392 | 9.57E-05 | 0.006063236 | SIPA1L1, AC005476.2 |
| chr13 | 75549335 | 75549771 | 437 | 11 | 0.000207367 | 0.001054 | 0.000258 | 9.58E-05 | 0.000528016 | UCHL3, COMMD6 |
| chr4 | 183653772 | 183653955 | 184 | 3 | 0.000400723 | 6.21E-05 | 0.00224 | 9.60E-05 | 0.00593002 | RWDD4 |
| chr1 | 36087976 | 36089024 | 1049 | 6 | 7.05E-05 | 0.008942 | 0.000229 | 9.60E-05 | 0.000529371 | TEKT2, ADPRS |
| chr12 | 133129542 | 133129836 | 295 | 3 | 0.000165062 | 0.00038 | 0.000835 | 9.61E-05 | 0.023286239 | ZNF891 |
| chr2 | 77523249 | 77523740 | 492 | 3 | 0.000132157 | 0.001113 | 0.001283 | 9.61E-05 | -0.0038961 | LRRTM4 |
| chr17 | 31498329 | 31498594 | 266 | 2 | 0.000123914 | 0.000719 | 0.000101 | 9.62E-05 | 0.005676753 | RAB11FIP4 |
| chr16 | 68995544 | 68995555 | 12 | 2 | 0.000418093 | 0.000568 | 0.00011 | 9.62E-05 | 0.003303672 | TANGO6 |
| chr11 | 66279367 | 66279758 | 392 | 2 | 9.07E-06 | 6.96E-05 | 0.001288 | 9.63E-05 | 0.015063085 | CNIH2 |
| chr1 | 63592852 | 63593626 | 775 | 13 | 0.000104235 | 0.000268 | 0.01839 | 9.64E-05 | 0.00407235 | PGM1, ITGB3BP |
| chr19 | 58278308 | 58279073 | 766 | 12 | 2.13E-06 | 0.009965 | 1.52E-05 | 9.64E-05 | 0.000269486 | AC020915.4, ZNF8, ZNF8-ERVK3-1, AC020915.2 |
| chr9 | 4305355 | 4305588 | 234 | 2 | 6.36E-05 | 0.011029 | 6.09E-05 | 9.64E-05 | 0.004691564 | AL162419.1, GLIS3 |
| chr7 | 128135578 | 128135765 | 188 | 2 | 2.20E-05 | 0.002158 | 7.59E-05 | 9.64E-05 | -0.01651988 | NA |
| chr20 | 52103792 | 52104369 | 578 | 3 | 0.000106455 | 0.001458 | 0.000155 | 9.67E-05 | 0.003261494 | ZFP64 |
| chr19 | 6333536 | 6333690 | 155 | 5 | 0.000387943 | 0.000673 | 0.000231 | 9.68E-05 | 0.003209743 | ACER1 |
| chr10 | 101776945 | 101776974 | 30 | 2 | 0.000286171 | 0.000119 | 0.000377 | 9.69E-05 | 0.00718088 | FGF8 |
| chr10 | 45463311 | 45463861 | 551 | 4 | 0.000218181 | 7.77E-05 | 0.00192 | 9.70E-05 | 0.006876868 | MARCHF8 |
| chr17 | 44251255 | 44251319 | 65 | 2 | 0.000191662 | 0.000277 | 0.000159 | 9.70E-05 | 0.00359693 | SLC4A1 |
| chr12 | 69639619 | 69639684 | 66 | 2 | 0.000439083 | 6.65E-05 | 0.001607 | 9.70E-05 | 0.003366079 | NA |
| chr16 | 67929735 | 67930205 | 471 | 3 | 0.000281659 | 0.000246 | 0.000248 | 9.70E-05 | 0.003387525 | CTRL, AC040162.1 |
| chr22 | 46592627 | 46592747 | 121 | 3 | 0.000141888 | 4.24E-05 | 0.006716 | 9.71E-05 | 0.006177078 | GRAMD4 |
| chr2 | 72142758 | 72143203 | 446 | 3 | 0.00039647 | 8.55E-05 | 0.000865 | 9.71E-05 | -0.00097672 | CYP26B1 |
| chr17 | 78694877 | 78694892 | 16 | 2 | 0.000392382 | 6.62E-05 | 0.001637 | 9.71E-05 | -0.00499937 | CYTH1 |
| chr3 | 52403630 | 52403660 | 31 | 2 | 0.000366989 | 9.60E-05 | 0.000555 | 9.71E-05 | 0.000451821 | BAP1 |
| chr1 | 40317562 | 40317592 | 31 | 2 | 0.00046574 | 6.05E-05 | 0.002693 | 9.71E-05 | 0.021872188 | COL9A2 |
| chr16 | 14371323 | 14371423 | 101 | 2 | 9.45E-05 | 0.000229 | 0.000185 | 9.72E-05 | 0.004422438 | NA |
| chr11 | 94566608 | 94566935 | 328 | 3 | 0.000283543 | 0.006153 | 0.000143 | 9.73E-05 | 0.005329807 | PIWIL4, AP000943.2 |
| chr14 | 103058073 | 103058652 | 580 | 6 | 0.000236157 | 4.70E-05 | 0.011459 | 9.74E-05 | 0.00801255 | NA |
| chr8 | 40898345 | 40898715 | 371 | 4 | 0.000108224 | 5.88E-05 | 0.001777 | 9.76E-05 | 0.00584471 | NA |
| chr6 | 30134398 | 30134729 | 332 | 2 | 9.75E-05 | 0.000742 | 0.000101 | 9.78E-05 | 0.001505218 | NA |
| chr5 | 180789985 | 180790071 | 87 | 2 | 0.000330704 | 6.27E-05 | 0.002246 | 9.80E-05 | 0.000802133 | MGAT1 |
| chr22 | 50077556 | 50077818 | 263 | 2 | 0.000295059 | 6.25E-05 | 0.002298 | 9.80E-05 | 0.002821538 | MLC1 |
| chr19 | 55635940 | 55636754 | 815 | 4 | 0.000313072 | 0.000782 | 0.000185 | 9.81E-05 | 0.001295185 | ZNF580, ZNF581 |
| chr22 | 42647751 | 42648381 | 631 | 4 | 0.00016636 | 0.00124 | 0.000635 | 9.81E-05 | 0.005519591 | CYB5R3 |
| chr9 | 114691179 | 114691403 | 225 | 3 | 0.000389734 | 5.82E-05 | 0.002456 | 9.81E-05 | 0.010923231 | NA |
| chr19 | 53962303 | 53962342 | 40 | 2 | 0.000145695 | 0.029918 | 5.86E-05 | 9.82E-05 | -0.00978724 | NA |
| chr2 | 75710006 | 75710721 | 716 | 4 | 7.03E-06 | 0.000276 | 0.002263 | 9.82E-05 | 0.007167523 | GCFC2 |
| chr12 | 123934331 | 123934549 | 219 | 2 | 9.82E-05 | 6.38E-05 | 0.002071 | 9.83E-05 | 0.006879804 | DNAH10, CCDC92, DNAH10OS |
| chr21 | 18115789 | 18115934 | 146 | 2 | 0.000335994 | 7.45E-05 | 0.001103 | 9.83E-05 | -0.00760757 | CHODL |
| chr7 | 1092709 | 1093217 | 509 | 5 | 0.000173076 | 0.003538 | 0.000147 | 9.83E-05 | 0.002553107 | GPER1, C7orf50 |
| chr2 | 239155688 | 239155861 | 174 | 2 | 0.000252995 | 6.13E-05 | 0.002651 | 9.83E-05 | 0.005256472 | HDAC4 |
| chr8 | 28339355 | 28339415 | 61 | 3 | 0.000245052 | 0.00015 | 0.000368 | 9.85E-05 | 0.007859762 | PNOC |
| chr4 | 47463659 | 47463882 | 224 | 7 | 0.000214359 | 0.000698 | 0.000608 | 9.86E-05 | 0.002412353 | AC107398.4, COMMD8 |
| chr13 | 98843124 | 98843596 | 473 | 3 | 0.000344755 | 0.000526 | 0.000216 | 9.86E-05 | 0.005196794 | DOCK9 |
| chr13 | 33015648 | 33016205 | 558 | 7 | 0.000185092 | 0.004221 | 0.001662 | 9.87E-05 | -0.0017822 | NA |
| chr1 | 55429201 | 55429280 | 80 | 2 | 0.000171344 | 0.000568 | 0.000113 | 9.88E-05 | 0.0021434 | AL603840.1 |
| chr14 | 99481850 | 99481952 | 103 | 2 | 1.82E-05 | 0.000728 | 0.000103 | 9.88E-05 | -0.00180524 | CCNK |
| chr3 | 137766424 | 137766527 | 104 | 2 | 0.000101882 | 6.39E-05 | 0.002124 | 9.89E-05 | 0.010775755 | NA |
| chr9 | 91425432 | 91426348 | 917 | 6 | 4.48E-05 | 0.001001 | 0.000114 | 9.89E-05 | 0.003362334 | AL353764.1 |
| chr3 | 64025109 | 64025228 | 120 | 2 | 0.000173629 | 0.000954 | 9.41E-05 | 9.89E-05 | 0.002312962 | NA |
| chr11 | 1043852 | 1043951 | 100 | 3 | 0.000454047 | 0.000938 | 0.000647 | 9.89E-05 | -0.00327175 | NA |
| chr19 | 14487529 | 14487616 | 88 | 3 | 0.000208904 | 0.001954 | 7.67E-05 | 9.90E-05 | 0.006007211 | GIPC1 |
| chr14 | 102555305 | 102555768 | 464 | 3 | 0.000186582 | 5.65E-05 | 0.003731 | 9.90E-05 | 0.002886094 | LINC02323 |
| chr1 | 117122354 | 117122932 | 579 | 7 | 0.00012074 | 0.001077 | 0.001996 | 9.91E-05 | 0.002417595 | TRIM45 |
| chr12 | 611829 | 611949 | 121 | 3 | 0.000362418 | 0.000467 | 0.000635 | 9.91E-05 | -0.01558729 | NINJ2 |
| chr14 | 24057196 | 24057336 | 141 | 2 | 0.000351124 | 0.000123 | 0.000379 | 9.92E-05 | 0.012485433 | CARMIL3 |
| chr1 | 21257634 | 21257887 | 254 | 2 | 2.33E-05 | 0.001989 | 7.89E-05 | 9.94E-05 | 0.0023681 | ECE1 |
| chr11 | 13495937 | 13496247 | 311 | 3 | 9.73E-05 | 0.000877 | 0.000102 | 9.94E-05 | -9.29E-05 | PTH |
| chr2 | 218710726 | 218710784 | 59 | 6 | 0.000338822 | 0.000994 | 0.000644 | 9.96E-05 | 0.000118627 | NA |
| chr16 | 57026579 | 57026836 | 258 | 2 | 0.000219477 | 0.000179 | 0.00024 | 9.97E-05 | 0.013071902 | NLRC5 |
| chr20 | 12316523 | 12316651 | 129 | 2 | 0.000342838 | 9.30E-05 | 0.000638 | 9.97E-05 | -0.00943575 | AL109838.1 |
| chr9 | 128127331 | 128127634 | 304 | 3 | 0.000219202 | 0.000863 | 0.000542 | 9.98E-05 | 0.000778468 | PTGES2 |
| chr3 | 152835272 | 152835300 | 29 | 2 | 0.000256417 | 0.000297 | 0.000158 | 9.99E-05 | 0.002183086 | P2RY1 |
| chr11 | 7596951 | 7597480 | 530 | 5 | 0.000200575 | 0.000468 | 0.001565 | 9.99E-05 | -0.00119896 | PPFIBP2 |
| chr19 | 1822215 | 1822221 | 7 | 2 | 0.00045444 | 0.00103 | 9.30E-05 | 9.99E-05 | 0.003852196 | AC012615.6, REXO1 |
| chr10 | 27155558 | 27155969 | 412 | 3 | 0.000291813 | 4.41E-05 | 0.006693 | 9.99E-05 | 0.002571381 | MASTL |
| chr8 | 30432894 | 30432973 | 80 | 2 | 0.000409097 | 0.000638 | 0.000109 | 9.99E-05 | 0.009227196 | RBPMS |
| chr2 | 230067770 | 230068351 | 582 | 3 | 0.000287651 | 5.54E-05 | 0.002812 | 0.0001 | 0.001140131 | SLC16A14 |
| chr6 | 1379459 | 1379560 | 102 | 2 | 0.000102073 | 6.59E-05 | 0.001948 | 0.0001001 | 0.006118801 | NA |
| chr1 | 223190014 | 223190137 | 124 | 2 | 0.000451185 | 6.29E-05 | 0.002507 | 0.0001001 | 0.007285816 | NA |
| chr2 | 215755948 | 215755957 | 10 | 2 | 0.000452584 | 0.000492 | 0.000122 | 0.0001001 | -0.01347267 | LINC00607 |
| chr4 | 41256543 | 41257266 | 724 | 8 | 0.000172398 | 0.006665 | 0.003881 | 0.0001002 | 0.00403293 | UCHL1, UCHL1-AS1 |
| chr11 | 95232305 | 95232760 | 456 | 9 | 0.000302551 | 8.09E-05 | 0.009508 | 0.0001003 | 0.002471045 | AP000787.1, SESN3 |
| chr21 | 46605778 | 46606385 | 608 | 5 | 0.000136765 | 0.000263 | 0.00076 | 0.0001003 | 0.003957625 | NA |
| chr5 | 181198089 | 181198506 | 418 | 4 | 4.03E-05 | 0.000306 | 0.001972 | 0.0001004 | 0.006427448 | TRIM7-AS1, TRIM7 |
| chr16 | 15395749 | 15395762 | 14 | 3 | 0.000325087 | 0.002135 | 0.000234 | 0.0001005 | -0.00117983 | MPV17L, AC140504.1 |
| chr15 | 34343251 | 34343675 | 425 | 9 | 0.000105956 | 0.004574 | 0.000259 | 0.0001006 | 0.002164236 | NUTM1 |
| chr5 | 14405523 | 14405912 | 390 | 6 | 0.000253949 | 0.000256 | 0.000196 | 0.000101 | 0.001910455 | TRIO |
| chr6 | 30679784 | 30680243 | 460 | 4 | 3.58E-05 | 0.000641 | 0.001328 | 0.000101 | 0.007687777 | PPP1R18 |
| chr16 | 29962973 | 29963070 | 98 | 3 | 0.00043959 | 9.63E-05 | 0.000795 | 0.000101 | -0.00101774 | TMEM219 |
| chr22 | 25067899 | 25068332 | 434 | 2 | 0.000142782 | 0.000137 | 0.000337 | 0.000101 | 0.013543803 | KIAA1671 |
| chr7 | 39949872 | 39950455 | 584 | 10 | 6.34E-05 | 0.004675 | 0.00136 | 0.0001014 | 0.000628945 | CDK13 |
| chr6 | 167122558 | 167122598 | 41 | 5 | 0.000426103 | 6.94E-05 | 0.001309 | 0.0001014 | 0.00965601 | Z94721.2, CCR6 |
| chr2 | 46331990 | 46332060 | 71 | 2 | 0.000291234 | 0.000284 | 0.000167 | 0.0001015 | 0.00415488 | EPAS1 |
| chr8 | 38559535 | 38559596 | 62 | 2 | 0.000326405 | 0.015904 | 6.16E-05 | 0.0001015 | 0.002261933 | C8orf86, AC069120.1 |
| chr14 | 65103033 | 65103535 | 503 | 5 | 0.000128232 | 0.000207 | 0.002328 | 0.0001016 | 0.002042902 | NA |
| chr19 | 6738751 | 6739613 | 863 | 6 | 4.64E-05 | 6.77E-05 | 0.014093 | 0.0001016 | 0.002695309 | TRIP10 |
| chr4 | 55793977 | 55794294 | 318 | 4 | 0.000190786 | 3.00E-05 | 0.016971 | 0.0001016 | 0.014938885 | NA |
| chr15 | 88476678 | 88476828 | 151 | 3 | 0.000397721 | 0.000803 | 0.000126 | 0.0001016 | 0.007807303 | MRPS11, AC013489.1 |
| chr22 | 46103857 | 46103943 | 87 | 3 | 0.000256095 | 4.97E-05 | 0.005994 | 0.0001016 | 0.005023913 | MIRLET7BHG |
| chr2 | 25203282 | 25203353 | 72 | 2 | 0.000278225 | 6.69E-05 | 0.001942 | 0.0001017 | 0.005134623 | NA |
| chr1 | 156061018 | 156061704 | 687 | 5 | 5.61E-05 | 9.32E-05 | 0.004619 | 0.0001018 | 0.002928713 | RAB25 |
| chr1 | 234899807 | 234899940 | 134 | 2 | 0.000411855 | 0.000194 | 0.00023 | 0.0001018 | -0.00109332 | AL391832.4 |
| chr12 | 22334285 | 22335242 | 958 | 10 | 8.51E-06 | 0.000248 | 0.002284 | 0.0001018 | -0.00020682 | ST8SIA1 |
| chr15 | 48120348 | 48121147 | 800 | 8 | 1.15E-05 | 0.030348 | 1.17E-05 | 0.0001018 | 0.003261467 | SLC24A5 |
| chr14 | 60485478 | 60486227 | 750 | 8 | 0.00011288 | 0.000717 | 0.002136 | 0.0001019 | -0.00038892 | C14orf39 |
| chr11 | 62612858 | 62613884 | 1027 | 9 | 1.79E-05 | 0.000517 | 0.002155 | 0.0001019 | 0.00069993 | ROM1 |
| chr9 | 133126867 | 133126940 | 74 | 2 | 7.23E-05 | 8.13E-05 | 0.000967 | 0.000102 | 0.012104426 | RALGDS, AL162417.1 |
| chr15 | 31306832 | 31306887 | 56 | 2 | 0.000435024 | 0.000119 | 0.000422 | 0.000102 | 0.008334106 | NA |
| chr1 | 84573029 | 84573338 | 310 | 2 | 0.000135315 | 0.000102 | 0.000553 | 0.0001021 | 0.028178034 | CTBS |
| chr7 | 158565203 | 158565602 | 400 | 3 | 6.38E-05 | 0.019467 | 4.74E-05 | 0.0001021 | 0.011509098 | PTPRN2 |
| chr19 | 8390054 | 8390407 | 354 | 7 | 0.000395188 | 0.000528 | 0.001941 | 0.0001024 | 6.02E-05 | RAB11B, RAB11B-AS1 |
| chr12 | 52407286 | 52407710 | 425 | 3 | 0.000204269 | 9.98E-05 | 0.00292 | 0.0001025 | 0.006407346 | AC055736.1 |
| chr19 | 57578836 | 57579097 | 262 | 10 | 0.000289707 | 5.23E-05 | 0.01924 | 0.0001025 | 9.80E-05 | ZIK1, ZNF416 |
| chr11 | 67261474 | 67262130 | 657 | 4 | 0.000118476 | 0.001209 | 0.000167 | 0.0001025 | 0.003719172 | AP001885.3 |
| chr14 | 88498596 | 88499102 | 507 | 2 | 7.32E-06 | 0.000795 | 0.000103 | 0.0001025 | 0.004636695 | PTPN21 |
| chr3 | 158672540 | 158673251 | 712 | 12 | 4.35E-05 | 0.000653 | 0.003859 | 0.0001025 | 0.003872333 | GFM1, LXN |
| chr10 | 17169240 | 17170032 | 793 | 4 | 3.06E-05 | 0.040323 | 4.24E-05 | 0.0001025 | 0.007082834 | TRDMT1 |
| chr17 | 28494864 | 28495102 | 239 | 2 | 0.00013906 | 0.010313 | 6.40E-05 | 0.0001025 | 0.004334507 | SLC13A2, AC005726.1 |
| chr10 | 82813744 | 82813998 | 255 | 2 | 0.000213036 | 0.000124 | 0.000402 | 0.0001025 | 0.011826236 | NRG3 |
| chr17 | 15398587 | 15399401 | 815 | 4 | 4.00E-05 | 0.009191 | 4.24E-05 | 0.0001026 | 0.001418031 | AC005703.3 |
| chr2 | 239762668 | 239762819 | 152 | 3 | 0.00039498 | 0.00184 | 0.000911 | 0.0001027 | 0.002696364 | NA |
| chr4 | 139890083 | 139890091 | 9 | 2 | 0.000475668 | 0.000122 | 0.000416 | 0.0001027 | 0.004876293 | MAML3 |
| chr19 | 38484310 | 38484605 | 296 | 2 | 7.25E-05 | 0.000709 | 0.000108 | 0.0001027 | 0.006652485 | RYR1 |
| chr3 | 48424658 | 48425342 | 685 | 4 | 0.000137182 | 0.001969 | 0.001508 | 0.0001028 | 0.002152157 | PLXNB1 |
| chr17 | 8002318 | 8003203 | 886 | 9 | 9.61E-05 | 0.000178 | 0.004229 | 0.0001028 | 0.000874551 | GUCY2D |
| chr5 | 1806357 | 1806511 | 155 | 2 | 0.000360229 | 6.79E-05 | 0.001928 | 0.0001029 | 0.006586226 | NDUFS6 |
| chr3 | 157159658 | 157160112 | 455 | 2 | 0.000116307 | 6.59E-05 | 0.002257 | 0.0001029 | 0.002042756 | CCNL1 |
| chr13 | 112074262 | 112074650 | 389 | 5 | 0.000137856 | 0.000351 | 0.000925 | 0.000103 | 0.004534267 | SOX1-OT |

| | | | | | | | | | | |
|---|---|---|---|---|---|---|---|---|---|---|
| chr2 | 236049102 | 236049310 | 209 | 2 | 0.000364022 | 0.000102 | 0.000563 | 0.000103 | 0.004718104 | AGAP1 |
| chr1 | 3157062 | 3157242 | 181 | 3 | 0.000254038 | 0.00039 | 0.000178 | 0.000103 | 7.92E-05 | PRDM16 |
| chr16 | 89894936 | 89895064 | 129 | 2 | 2.33E-05 | 0.002295 | 7.90E-05 | 0.0001032 | -0.00198985 | TCF25 |
| chr19 | 45972546 | 45972788 | 243 | 2 | 4.15E-05 | 0.003901 | 7.23E-05 | 0.0001033 | -0.01422354 | NOVA2 |
| chr19 | 3808584 | 3808864 | 281 | 3 | 9.60E-05 | 0.00029 | 0.000191 | 0.0001033 | 0.002011214 | ZFR2 |
| chr11 | 71094488 | 71094854 | 367 | 3 | 0.000116616 | 0.000168 | 0.001848 | 0.0001034 | 0.003268637 | SHANK2 |
| chr7 | 44623933 | 44624007 | 75 | 2 | 0.00014075 | 0.000162 | 0.000289 | 0.0001035 | 0.008391028 | OGDH |
| chr5 | 181195072 | 181195276 | 205 | 3 | 0.000274545 | 0.000341 | 0.001091 | 0.0001037 | 0.003716549 | TRIM7 |
| chr6 | 34659020 | 34659164 | 145 | 2 | 0.000252162 | 0.005193 | 6.98E-05 | 0.0001037 | 0.003133769 | ILRUN |
| chr19 | 16541074 | 16541869 | 796 | 4 | 0.000101298 | 0.001205 | 0.000436 | 0.0001042 | -0.00677332 | AC008764.1, CHERP |
| chr21 | 10417729 | 10417768 | 40 | 2 | 0.000465742 | 0.000513 | 0.000125 | 0.0001042 | 0.005234042 | BAGE2 |
| chr15 | 38565057 | 38565807 | 751 | 8 | 5.67E-05 | 0.00185 | 0.004402 | 0.0001043 | 0.002326297 | RASGRP1 |
| chr17 | 819597 | 820102 | 506 | 4 | 0.000272271 | 9.03E-05 | 0.003107 | 0.0001045 | 0.001919492 | NXN |
| chr17 | 2689782 | 2690316 | 535 | 3 | 0.000212517 | 5.50E-05 | 0.003383 | 0.0001045 | 0.005057502 | CLUH |
| chr7 | 100672859 | 100673208 | 350 | 2 | 1.02E-05 | 0.000705 | 0.00011 | 0.0001049 | -0.00800161 | NA |
| chr16 | 19289882 | 19289991 | 110 | 2 | 0.000398263 | 0.000141 | 0.000353 | 0.0001052 | 0.005543531 | CLEC19A |
| chr10 | 92572745 | 92572949 | 205 | 2 | 0.000231158 | 7.29E-05 | 0.001556 | 0.0001052 | 0.007834909 | IDE |
| chr16 | 30483928 | 30484181 | 254 | 2 | 0.000401535 | 6.94E-05 | 0.001917 | 0.0001052 | -5.20E-05 | ITGAL, AC116348.3 |
| chr7 | 100657325 | 100657420 | 96 | 2 | 6.41E-05 | 0.010187 | 6.54E-05 | 0.0001052 | 0.006925594 | NA |
| chr19 | 48793427 | 48793600 | 174 | 4 | 0.00038692 | 0.00195 | 0.001044 | 0.0001052 | 0.002652568 | NA |
| chr21 | 46411038 | 46412013 | 976 | 3 | 7.93E-06 | 0.000886 | 0.000102 | 0.0001057 | 0.002920166 | PCNT |
| chr16 | 84817424 | 84817464 | 41 | 2 | 0.000283156 | 0.00759 | 6.76E-05 | 0.0001059 | -0.00583996 | AC025280.3 |
| chr9 | 35757161 | 35757643 | 483 | 3 | 1.60E-05 | 0.017951 | 4.44E-05 | 0.0001061 | 0.001111065 | RGP1, AL133410.2 |
| chr6 | 166423155 | 166423303 | 149 | 3 | 0.000142998 | 0.000199 | 0.00048 | 0.0001061 | 0.002812605 | RPS6KA2 |
| chr4 | 91112960 | 91113451 | 492 | 3 | 0.000196983 | 0.011857 | 6.58E-05 | 0.0001061 | 0.00994056 | CCSER1 |
| chr11 | 92843979 | 92844699 | 721 | 7 | 0.000118739 | 0.001033 | 0.007188 | 0.0001061 | 0.000583534 | FAT3 |
| chr16 | 88906058 | 88906457 | 400 | 4 | 0.000260191 | 8.28E-05 | 0.001263 | 0.0001063 | 0.006692558 | CBFA2T3 |
| chr6 | 154330024 | 154330146 | 123 | 2 | 0.000340923 | 0.00012 | 0.000457 | 0.0001063 | 0.008928666 | IPCEF1, AL357075.4 |
| chr13 | 74133901 | 74134336 | 436 | 9 | 0.000138957 | 0.008873 | 0.000203 | 0.0001063 | 0.00132233 | NA |
| chr3 | 186439292 | 186439357 | 66 | 2 | 0.000382877 | 0.000502 | 0.000129 | 0.0001064 | 0.007839568 | LINC02020 |
| chr1 | 36475385 | 36475508 | 124 | 2 | 0.000443078 | 0.000139 | 0.000368 | 0.0001064 | 0.004755551 | CSF3R |
| chr2 | 86106044 | 86106385 | 342 | 11 | 0.000197026 | 0.006033 | 0.000298 | 0.0001065 | 0.000433416 | PTCD3, POLR1A |
| chr19 | 2808072 | 2808387 | 316 | 2 | 0.000177335 | 9.63E-05 | 0.000705 | 0.0001065 | 0.005502722 | THOP1 |
| chr21 | 46228621 | 46229019 | 399 | 9 | 8.72E-05 | 0.00486 | 0.001394 | 0.0001067 | -0.0008873 | LSS |
| chr2 | 27527200 | 27527267 | 68 | 2 | 0.000267336 | 0.017323 | 6.34E-05 | 0.0001068 | 0.005817665 | NA |
| chr1 | 225461013 | 225461626 | 614 | 2 | 5.77E-05 | 0.000556 | 0.000124 | 0.0001068 | 0.008551114 | LINC02765 |
| chr17 | 73823861 | 73823883 | 23 | 2 | 0.000415784 | 7.16E-05 | 0.001807 | 0.0001069 | 0.0090338 | LINC00469 |
| chr12 | 2640887 | 2641341 | 455 | 3 | 0.000399753 | 0.000189 | 0.000505 | 0.0001069 | 0.007309485 | CACNA1C |
| chr2 | 121009159 | 121009174 | 16 | 2 | 0.000394698 | 0.001417 | 9.04E-05 | 0.000107 | 0.009578415 | NA |
| chr1 | 159898452 | 159898772 | 321 | 3 | 0.000196332 | 0.000327 | 0.00097 | 0.0001071 | 0.007811167 | CFAP45 |
| chr16 | 88722482 | 88722686 | 205 | 2 | 0.000411704 | 6.67E-05 | 0.002648 | 0.0001071 | -0.00365655 | PIEZO1 |
| chr9 | 136728225 | 136728249 | 25 | 2 | 0.000200353 | 0.00123 | 9.40E-05 | 0.0001072 | -0.00187743 | NA |
| chr17 | 13017734 | 13018192 | 459 | 9 | 0.000278915 | 0.020672 | 0.001576 | 0.0001072 | 0.000281553 | ELAC2 |
| chr18 | 59402163 | 59402458 | 296 | 2 | 0.000221969 | 8.11E-05 | 0.001164 | 0.0001075 | 0.007443549 | NA |
| chr1 | 54800157 | 54801064 | 908 | 4 | 1.79E-07 | 0.026442 | 3.18E-05 | 0.0001076 | 0.001664707 | TTC22 |
| chr2 | 200812313 | 200812433 | 121 | 2 | 0.000468577 | 7.37E-05 | 0.001651 | 0.0001077 | 0.000167658 | BZW1 |
| chr6 | 127459207 | 127459430 | 224 | 2 | 0.000130685 | 0.003581 | 7.56E-05 | 0.0001077 | 0.005163122 | KIAA0408, AL096711.2 |
| chr8 | 117712191 | 117712216 | 26 | 2 | 0.000435167 | 0.000143 | 0.000365 | 0.0001077 | 0.009793857 | NA |
| chr5 | 179150497 | 179150541 | 45 | 2 | 0.000475184 | 0.000194 | 0.000254 | 0.0001077 | 0.003754447 | ADAMTS2 |
| chr1 | 13469506 | 13469604 | 99 | 2 | 0.000194812 | 9.85E-05 | 0.000694 | 0.0001079 | 0.021162641 | NA |
| chr18 | 10453972 | 10454088 | 117 | 3 | 0.000111165 | 7.01E-05 | 0.002325 | 0.0001079 | 0.012458019 | NA |
| chr8 | 1180775 | 1180858 | 84 | 2 | 0.000331006 | 0.000557 | 0.000125 | 0.0001079 | -0.01280261 | DLGAP2 |
| chr2 | 96867898 | 96868320 | 423 | 2 | 8.66E-05 | 7.43E-05 | 0.001635 | 0.0001083 | 0.00858207 | SEMA4C |
| chr6 | 111048470 | 111048575 | 106 | 2 | 0.000381704 | 0.000238 | 0.000213 | 0.0001084 | 0.004767371 | NA |
| chr2 | 219568093 | 219568150 | 58 | 3 | 0.000421764 | 8.17E-05 | 0.003657 | 0.0001084 | 0.003201127 | OBSL1 |
| chr14 | 101224913 | 101225344 | 432 | 2 | 7.58E-05 | 6.98E-05 | 0.002195 | 0.0001085 | 0.008295958 | NA |
| chr17 | 1917151 | 1917239 | 89 | 2 | 0.000344181 | 7.16E-05 | 0.001949 | 0.0001085 | 0.008869327 | NA |
| chr9 | 121809110 | 121809346 | 237 | 2 | 0.000427959 | 0.000214 | 0.000234 | 0.0001085 | 0.000561158 | NA |
| chr11 | 57335491 | 57335860 | 370 | 3 | 5.12E-05 | 0.012147 | 4.52E-05 | 0.0001086 | 0.001447361 | SSRP1 |
| chr10 | 11435531 | 11436184 | 654 | 3 | 0.000251248 | 0.000128 | 0.00115 | 0.0001086 | 0.004660904 | NA |
| chr16 | 89506904 | 89507547 | 644 | 7 | 0.000145228 | 3.25E-05 | 0.023713 | 0.0001087 | -0.00849939 | SPG7 |
| chr2 | 100902977 | 100903332 | 356 | 2 | 0.000194346 | 0.000242 | 0.00021 | 0.0001088 | 0.008206181 | NPAS2 |
| chr8 | 134457795 | 134458026 | 232 | 2 | 0.00014864 | 6.74E-05 | 0.002748 | 0.0001089 | 0.009824663 | NA |
| chr13 | 36132096 | 36132296 | 201 | 3 | 0.000181231 | 0.00071 | 0.001452 | 0.0001089 | 0.00375839 | NA |
| chr2 | 21636348 | 21636471 | 124 | 3 | 0.000323179 | 0.003432 | 0.000329 | 0.0001089 | -0.0041535 | AC018742.1, AC009411.2 |
| chr9 | 92426552 | 92426608 | 57 | 2 | 0.000427834 | 0.000249 | 0.000206 | 0.0001089 | 0.001723964 | CENPP |
| chr1 | 40782537 | 40783209 | 673 | 3 | 2.62E-05 | 0.007292 | 0.000557 | 0.000109 | 0.013066412 | NA |
| chr8 | 56558603 | 56558732 | 130 | 2 | 3.85E-05 | 0.000425 | 0.000144 | 0.000109 | 0.009681286 | LINC00968 |
| chr10 | 48266746 | 48266883 | 138 | 2 | 0.000323138 | 0.002252 | 8.30E-05 | 0.0001092 | -0.00204005 | FRMPD2 |
| chr7 | 150716476 | 150716599 | 124 | 3 | 0.000449806 | 0.049521 | 3.06E-05 | 0.0001092 | -0.00123033 | NA |
| chr3 | 167474049 | 167474183 | 135 | 3 | 0.000342161 | 0.000932 | 9.41E-05 | 0.0001093 | 0.006906511 | SERPINI2 |
| chr6 | 36874431 | 36874475 | 45 | 2 | 0.000193983 | 7.67E-05 | 0.001517 | 0.0001093 | 0.00431963 | C6orf89, PPIL1 |
| chr7 | 73734841 | 73734860 | 20 | 2 | 0.000343089 | 0.00027 | 0.000194 | 0.0001093 | 0.007458335 | NA |
| chr6 | 31677900 | 31678191 | 292 | 2 | 0.000397984 | 0.000101 | 0.000689 | 0.0001097 | -0.00520035 | LY6G5C |
| chr7 | 2130708 | 2130995 | 288 | 3 | 0.000220582 | 0.000492 | 0.000441 | 0.0001098 | -0.00341411 | MAD1L1, AC069288.1 |
| chr14 | 20197226 | 20197396 | 171 | 3 | 0.000359942 | 0.001102 | 9.18E-05 | 0.0001098 | 0.009186498 | OR11G2 |
| chr2 | 230886017 | 230886765 | 749 | 6 | 3.07E-05 | 0.000392 | 0.006845 | 0.0001098 | 0.005805012 | GCSIR |
| chr2 | 202033293 | 202033626 | 334 | 3 | 8.30E-05 | 6.90E-05 | 0.002901 | 0.0001098 | -0.00084008 | AC069148.1 |
| chr17 | 34253850 | 34254201 | 352 | 3 | 0.000207344 | 0.00041 | 0.000402 | 0.0001098 | -0.00385329 | NA |
| chr1 | 18897627 | 18898610 | 984 | 3 | 3.16E-05 | 0.000404 | 0.000219 | 0.0001099 | 0.005106577 | ALDH4A1 |
| chr17 | 17770845 | 17770974 | 130 | 3 | 0.000174642 | 0.002156 | 8.17E-05 | 0.0001099 | 0.004278251 | RAI1 |
| chr2 | 227634225 | 227634670 | 446 | 4 | 0.000291307 | 0.0006 | 0.001233 | 0.0001102 | 0.003020709 | C2orf83 |
| chr16 | 73171514 | 73172095 | 582 | 3 | 0.000368643 | 0.000536 | 0.000368 | 0.0001103 | 0.00471197 | ZFHX3 |
| chr6 | 136526557 | 136527175 | 619 | 5 | 6.55E-05 | 0.001807 | 0.000324 | 0.0001104 | 0.004065922 | MAP7 |
| chr7 | 2010312 | 2010535 | 224 | 4 | 0.000494612 | 0.002303 | 0.000307 | 0.0001106 | 0.002609174 | MAD1L1, AC069288.1 |
| chr10 | 3265567 | 3265571 | 5 | 2 | 0.000185115 | 0.006828 | 7.06E-05 | 0.0001107 | 0.008803569 | AL139280.1 |
| chr7 | 108526278 | 108527071 | 794 | 9 | 1.28E-05 | 0.007623 | 1.92E-05 | 0.0001107 | 0.000871146 | PNPLA8 |
| chr3 | 187734030 | 187734333 | 304 | 2 | 5.53E-05 | 0.000722 | 0.000116 | 0.0001108 | -8.76E-06 | BCL6 |
| chr9 | 131797329 | 131797448 | 120 | 2 | 0.000238803 | 0.106374 | 6.20E-05 | 0.0001108 | 0.003543995 | NA |
| chr7 | 178613 | 179032 | 420 | 4 | 0.000138531 | 0.004191 | 7.01E-05 | 0.0001109 | 0.001323281 | NA |
| chr6 | 151803947 | 151804042 | 96 | 2 | 0.000273417 | 8.33E-05 | 0.001197 | 0.0001109 | 0.000146837 | ESR1 |
| chr12 | 102917061 | 102917360 | 300 | 4 | 0.000108571 | 0.00011 | 0.002057 | 0.000111 | 0.009780022 | PAH |
| chr13 | 114234012 | 114234267 | 256 | 3 | 7.52E-05 | 5.98E-05 | 0.00446 | 0.0001111 | 0.003585767 | NA |
| chr2 | 104857927 | 104858014 | 88 | 2 | 0.000112269 | 0.000653 | 0.000121 | 0.0001111 | 0.010237482 | AC018730.1, POU3F3 |
| chr15 | 78565229 | 78565235 | 7 | 2 | 0.000451783 | 8.04E-05 | 0.001363 | 0.0001111 | -0.00351245 | NA |
| chr7 | 312601 | 312677 | 77 | 2 | 0.000181109 | 8.42E-05 | 0.001164 | 0.0001112 | 0.010576462 | NA |
| chr15 | 49619978 | 49620408 | 431 | 2 | 2.74E-06 | 0.015785 | 6.59E-05 | 0.0001114 | -0.00592601 | FAM227B |
| chr17 | 35930932 | 35931399 | 468 | 3 | 2.68E-05 | 0.010356 | 4.24E-05 | 0.0001114 | 0.01112576 | NA |
| chr3 | 122614117 | 122614210 | 94 | 2 | 0.000292591 | 0.000276 | 0.000196 | 0.0001115 | 0.009629391 | PARP15 |
| chr12 | 131623802 | 131623963 | 162 | 2 | 0.000303697 | 0.002706 | 8.15E-05 | 0.0001115 | 0.005597613 | NA |
| chr11 | 67634984 | 67635012 | 29 | 2 | 0.000421801 | 0.000392 | 0.000154 | 0.0001115 | 0.005908653 | TBX10 |
| chr2 | 68062840 | 68063188 | 349 | 6 | 0.000138579 | 0.019651 | 0.000123 | 0.0001118 | -0.00040238 | C1D |
| chr4 | 77075091 | 77075687 | 597 | 3 | 2.72E-05 | 8.76E-05 | 0.003273 | 0.0001119 | 0.00262986 | CCNI |
| chr10 | 125756628 | 125756708 | 81 | 3 | 0.000427113 | 0.008592 | 5.35E-05 | 0.000112 | -0.00012392 | EDRF1, MMP21 |
| chr18 | 11856967 | 11857619 | 653 | 5 | 9.82E-05 | 0.000831 | 0.000453 | 0.0001122 | 0.008333155 | GNAL, AP005137.2 |
| chr7 | 129144043 | 129144658 | 616 | 9 | 0.000163945 | 0.002518 | 0.002177 | 0.0001122 | -0.00010342 | NA |
| chr1 | 161021188 | 161021759 | 572 | 9 | 0.000102827 | 0.004548 | 0.000182 | 0.0001122 | -0.00177254 | F11R, AL591806.2 |
| chr5 | 131796731 | 131797521 | 791 | 11 | 0.000105286 | 0.034044 | 0.000174 | 0.0001122 | -2.21E-05 | AC005593.1, AC008695.1, FNIP1 |
| chr4 | 38313274 | 38313342 | 69 | 2 | 0.000262481 | 0.006418 | 7.21E-05 | 0.0001123 | 0.009513958 | NA |
| chr16 | 1152468 | 1153382 | 915 | 5 | 1.33E-05 | 0.005689 | 0.00013 | 0.0001124 | -6.49E-05 | CACNA1H |
| chr1 | 230424572 | 230425047 | 476 | 2 | 3.99E-05 | 0.001332 | 9.59E-05 | 0.0001124 | 0.004895112 | PGBD5 |
| chr7 | 150947543 | 150948004 | 462 | 4 | 8.15E-05 | 9.20E-05 | 0.001127 | 0.0001126 | 0.01009745 | KCNH2 |
| chr11 | 65117490 | 65118396 | 907 | 9 | 5.41E-05 | 0.001469 | 0.007997 | 0.0001127 | 0.000828845 | AP003068.4, ZNHIT2 |
| chr21 | 43767818 | 43768313 | 496 | 3 | 0.000141492 | 0.000109 | 0.003211 | 0.0001127 | 0.007862474 | NA |
| chr13 | 101915995 | 101916205 | 211 | 3 | 0.000226418 | 0.000291 | 0.000924 | 0.0001128 | 0.007260124 | FGF14 |
| chr3 | 49120925 | 49121281 | 357 | 8 | 0.000148626 | 0.000285 | 0.007078 | 0.0001129 | 0.002137177 | USP19, LAMB2 |
| chr22 | 23106280 | 23106713 | 434 | 2 | 3.32E-05 | 0.000567 | 0.000131 | 0.0001134 | 0.006466676 | GNAZ, RSPH14 |
| chr14 | 88793207 | 88793404 | 198 | 5 | 0.000236329 | 0.000953 | 0.00046 | 0.0001134 | 0.004187292 | NA |
| chr19 | 35812978 | 35813177 | 200 | 2 | 0.00024936 | 9.86E-05 | 0.0008 | 0.0001136 | -0.00450537 | PRODH2 |
| chr5 | 88672840 | 88673568 | 729 | 7 | 0.000169109 | 0.00012 | 0.023576 | 0.0001139 | 0.005459118 | LINC00461 |
| chr22 | 42993790 | 42993812 | 23 | 2 | 8.33E-05 | 0.000472 | 0.000143 | 0.0001139 | 0.005360505 | PACSIN2 |
| chr1 | 3282847 | 3283086 | 240 | 2 | 0.000186845 | 0.00098 | 0.000106 | 0.0001139 | 0.00188375 | PRDM16 |
| chr4 | 185813263 | 185813551 | 289 | 4 | 0.000206825 | 0.002369 | 0.000172 | 0.000114 | 3.86E-05 | SORBS2 |
| chr19 | 57240743 | 57241387 | 645 | 4 | 0.000126562 | 0.002056 | 0.000104 | 0.000114 | -0.00278574 | ZNF805 |
| chr18 | 75161752 | 75161806 | 55 | 2 | 0.000401563 | 9.03E-05 | 0.001026 | 0.000114 | 0.011999182 | NA |
| chr5 | 154851607 | 154851610 | 4 | 2 | 0.000211408 | 8.27E-05 | 0.00136 | 0.000114 | 0.004317252 | FAXDC2 |
| chr21 | 45476041 | 45476260 | 220 | 2 | 0.000349434 | 6.96E-05 | 0.003002 | 0.0001141 | 0.006605061 | COL18A1 |
| chr13 | 114314017 | 114314766 | 750 | 5 | 2.12E-05 | 0.000634 | 0.000175 | 0.0001142 | -0.00138467 | CHAMP1 |
| chr2 | 20650763 | 20651392 | 630 | 9 | 0.000148237 | 0.007124 | 0.000475 | 0.0001142 | -0.00048867 | HS1BP3 |
| chr20 | 61251082 | 61251151 | 70 | 2 | 0.0003895 | 0.000177 | 0.000313 | 0.0001144 | 0.013669367 | NA |
| chr3 | 48635544 | 48635704 | 161 | 6 | 0.0001964 | 0.000622 | 0.000334 | 0.0001145 | 0.000186793 | NA |
| chr2 | 55694432 | 55694740 | 309 | 4 | 0.00015133 | 9.96E-05 | 0.002396 | 0.0001147 | -0.00394715 | NA |
| chr13 | 100545073 | 100545416 | 344 | 3 | 0.000208035 | 0.000549 | 0.001763 | 0.0001147 | 0.004847761 | GGACT, AL356966.1 |
| chr9 | 113461756 | 113462376 | 621 | 2 | 0.000196121 | 0.000151 | 0.000385 | 0.0001147 | 0.005237824 | RGS3 |
| chr5 | 127648126 | 127648301 | 176 | 2 | 5.66E-05 | 7.40E-05 | 0.002209 | 0.0001148 | 0.008237698 | NA |
| chr1 | 2403346 | 2403535 | 190 | 4 | 0.000218699 | 0.00185 | 0.000982 | 0.0001151 | 0.002751036 | RER1 |
| chr17 | 1629562 | 1629882 | 321 | 3 | 7.97E-05 | 0.000285 | 0.00051 | 0.0001155 | -0.0025846 | NA |
| chr16 | 66502357 | 66502817 | 461 | 3 | 6.45E-05 | 0.000331 | 0.001726 | 0.0001155 | 0.014758007 | AC010542.3 |
| chr2 | 240257439 | 240257470 | 32 | 2 | 0.000267478 | 0.00012 | 0.000559 | 0.0001156 | 0.003297032 | NA |
| chr3 | 185152613 | 185152917 | 305 | 6 | 0.000296892 | 0.000107 | 0.009846 | 0.0001156 | 0.001467487 | C3orf70 |
| chr15 | 43206561 | 43206575 | 15 | 2 | 0.000501696 | 0.000199 | 0.00028 | 0.0001156 | 0.003270531 | EPB42 |
| chr11 | 9384249 | 9384568 | 320 | 7 | 0.000377208 | 8.88E-05 | 0.010814 | 0.0001157 | 0.002118035 | NA |
| chr14 | 69046069 | 69046161 | 93 | 2 | 0.000290232 | 0.001289 | 9.93E-05 | 0.0001157 | 0.007097229 | NA |
| chr14 | 23568455 | 23568945 | 491 | 3 | 3.96E-06 | 0.000216 | 0.002385 | 0.0001157 | 0.000400885 | JPH4 |
| chr19 | 41908287 | 41908311 | 25 | 2 | 0.000221373 | 0.000173 | 0.000329 | 0.0001158 | 0.001877713 | ARHGEF1, ERFL |
| chr11 | 123573919 | 123573928 | 10 | 2 | 0.000479072 | 0.000841 | 0.000114 | 0.0001158 | 0.005514228 | GRAMD1B |
| chr9 | 77177731 | 77178112 | 382 | 5 | 0.000123399 | 0.00597 | 5.84E-05 | 0.0001159 | 0.002290123 | VPS13A, VPS13A-AS1 |
| chr1 | 25819514 | 25819717 | 204 | 5 | 0.000204433 | 0.000335 | 0.000265 | 0.000116 | 0.000412548 | AL020996.2, MTFR1L, AL020996.1 |
| chr1 | 29259341 | 29259787 | 447 | 3 | 0.000343272 | 6.37E-05 | 0.005192 | 0.0001162 | 0.000762083 | PTPRU |
| chr10 | 132248100 | 132248180 | 81 | 3 | 0.000226515 | 5.05E-05 | 0.007479 | 0.0001164 | 0.010991594 | STK32C |
| chr2 | 219056849 | 219057640 | 792 | 3 | 2.39E-05 | 0.003885 | 6.25E-05 | 0.0001165 | 0.006276418 | IHH |
| chr20 | 37652120 | 37652283 | 164 | 2 | 0.000257395 | 0.000974 | 0.000109 | 0.0001167 | 0.008660432 | NA |
| chr8 | 74350037 | 74350234 | 198 | 3 | 0.000228494 | 7.82E-05 | 0.003365 | 0.0001167 | 0.000924451 | GDAP1, AC103952.1 |
| chr12 | 26122378 | 26122450 | 73 | 2 | 0.00041922 | 9.11E-05 | 0.001077 | 0.0001168 | 0.001809473 | SSPN, BHLHE41 |
| chr2 | 95057708 | 95057778 | 71 | 2 | 0.000379505 | 0.000202 | 0.000281 | 0.0001168 | 0.005310215 | NA |
| chr8 | 77147466 | 77147631 | 166 | 2 | 0.000322865 | 0.002492 | 8.56E-05 | 0.0001169 | 0.018659077 | NA |
| chr1 | 24506126 | 24506305 | 180 | 2 | 0.000113259 | 0.003248 | 8.17E-05 | 0.0001169 | 0.011419874 | RCAN3, RCAN3AS |
| chr1 | 206971022 | 206971467 | 446 | 3 | 0.000102381 | 0.000517 | 0.00053 | 0.000117 | 0.007714948 | NA |
| chr4 | 6566322 | 6566393 | 72 | 2 | 0.000304141 | 0.011797 | 6.99E-05 | 0.0001171 | 0.013930607 | NA |
| chr14 | 22500712 | 22500773 | 62 | 2 | 0.000158532 | 0.001333 | 9.94E-05 | 0.0001172 | 0.008202959 | NA |
| chr18 | 76546701 | 76546975 | 275 | 2 | 9.39E-05 | 0.00134 | 9.94E-05 | 0.0001173 | 0.013498314 | LINC00683 |
| chr6 | 80004563 | 80005043 | 481 | 6 | 2.11E-05 | 0.000237 | 0.001352 | 0.0001175 | 0.002380721 | TTK |

| | | | | | | | | | | |
|---|---|---|---|---|---|---|---|---|---|---|
| chr1 | 11056282 | 11056822 | 541 | 3 | 7.73E-05 | 0.021164 | 3.98E-05 | 0.0001176 | 0.002213491 | SRM |
| chr4 | 185184138 | 185184237 | 100 | 2 | 0.000450338 | 0.000854 | 0.000115 | 0.0001176 | 0.0095032 | CFAP97 |
| chr1 | 1210238 | 1210693 | 456 | 3 | 0.000288649 | 0.000232 | 0.00049 | 0.0001178 | 0.005968209 | NA |
| chr1 | 1122697 | 1123059 | 363 | 2 | 6.03E-05 | 0.001412 | 9.84E-05 | 0.0001179 | 0.005411059 | NA |
| chr19 | 35743824 | 35743863 | 40 | 2 | 0.000349157 | 0.002174 | 8.85E-05 | 0.0001179 | -0.00221945 | AD000671.2, U2AF1L4 |
| chr3 | 42684028 | 42684590 | 563 | 3 | 0.000286126 | 8.54E-05 | 0.001515 | 0.0001183 | -0.00390154 | NA |
| chr15 | 63833892 | 63834226 | 335 | 3 | 0.000364291 | 0.000274 | 0.001793 | 0.0001184 | 0.000550842 | HERC1 |
| chr15 | 58065676 | 58066186 | 511 | 11 | 0.000106255 | 0.000104 | 0.004336 | 0.0001185 | 0.002568527 | AC025431.1, ALDH1A2 |
| chr10 | 133131397 | 133131595 | 199 | 2 | 0.000323681 | 0.006669 | 7.46E-05 | 0.0001186 | 0.009229401 | ADGRA1 |
| chr2 | 41857993 | 41858139 | 147 | 2 | 0.000353606 | 0.00013 | 0.000518 | 0.0001186 | 0.02452568 | NA |
| chr2 | 218607495 | 218608001 | 507 | 4 | 9.95E-05 | 0.000594 | 0.004761 | 0.0001188 | 0.012123699 | PLCD4 |
| chr2 | 8678283 | 8678428 | 146 | 4 | 0.000365426 | 5.40E-05 | 0.007056 | 0.0001188 | -0.00090289 | ID2-AS1 |
| chr10 | 104688803 | 104688830 | 28 | 2 | 0.000459695 | 0.001197 | 0.000104 | 0.0001189 | 0.019500415 | SORCS3 |
| chr2 | 170712281 | 170712871 | 591 | 6 | 0.000152378 | 0.000418 | 0.00288 | 0.0001191 | -0.00230855 | AC007405.1, LINC01124 |
| chr19 | 20237982 | 20238117 | 136 | 3 | 0.000495345 | 5.21E-05 | 0.007691 | 0.0001192 | 0.005063041 | AC011447.3 |
| chr1 | 243850606 | 243851163 | 558 | 5 | 0.00012417 | 0.000327 | 0.000934 | 0.0001192 | 0.000271909 | AKT3 |
| chr2 | 218994141 | 218994271 | 131 | 3 | 0.000284627 | 0.000186 | 0.000866 | 0.0001195 | 0.00633641 | NA |
| chr20 | 32581762 | 32581837 | 76 | 2 | 0.000316304 | 9.20E-05 | 0.001127 | 0.0001195 | 0.009821065 | NOL4L-DT, NOL4L |
| chr10 | 11209148 | 11209306 | 159 | 2 | 0.000331618 | 0.001952 | 9.18E-05 | 0.0001196 | 0.011949389 | CELF2 |
| chr13 | 100394134 | 100394192 | 59 | 2 | 0.000232935 | 7.35E-05 | 0.002881 | 0.0001196 | -0.01586946 | PCCA |
| chr7 | 101126324 | 101126622 | 299 | 2 | 0.000179342 | 7.75E-05 | 0.002181 | 0.0001197 | 0.020245687 | NA |
| chr16 | 27214435 | 27215149 | 715 | 8 | 2.59E-05 | 0.016916 | 3.11E-05 | 0.0001197 | 0.000655253 | KDM8, AC109449.1 |
| chr20 | 63573129 | 63573555 | 427 | 2 | 1.20E-05 | 7.98E-05 | 0.001916 | 0.0001197 | 0.009696212 | HELZ2 |
| chr10 | 70146201 | 70146765 | 565 | 5 | 2.24E-05 | 0.003318 | 0.001344 | 0.00012 | 0.001193354 | TYSND1 |
| chr12 | 7711417 | 7711462 | 46 | 3 | 0.000427088 | 0.010748 | 4.74E-05 | 0.00012 | 0.002185085 | DPPA3 |
| chr6 | 7129481 | 7129853 | 373 | 2 | 5.07E-05 | 0.001202 | 0.000105 | 0.0001202 | 0.014575062 | RREB1 |
| chr5 | 177093492 | 177093703 | 212 | 4 | 0.00014615 | 0.000194 | 0.000689 | 0.0001203 | 0.003489468 | FGFR4 |
| chr5 | 33229056 | 33229363 | 308 | 2 | 0.000174453 | 7.95E-05 | 0.001983 | 0.0001203 | -0.01377342 | AC010343.3 |
| chr17 | 5501001 | 5501261 | 261 | 6 | 0.000148919 | 0.000205 | 0.00563 | 0.0001204 | 0.001776823 | AC055839.2, NLRP1 |
| chr5 | 139272753 | 139272830 | 78 | 2 | 8.36E-05 | 0.000411 | 0.000165 | 0.0001205 | 0.005484256 | SIL1 |
| chr6 | 33292089 | 33292507 | 419 | 7 | 0.000141652 | 0.001633 | 0.000376 | 0.0001205 | 0.004402525 | PFDN6, RGL2 |
| chr20 | 22585651 | 22586338 | 688 | 4 | 0.00032351 | 0.003524 | 0.001105 | 0.0001207 | -0.00137802 | NA |
| chr3 | 36944206 | 36944663 | 458 | 3 | 0.000191136 | 0.000448 | 0.001002 | 0.0001207 | 0.002132219 | TRANK1 |
| chr11 | 57712522 | 57713113 | 592 | 7 | 6.75E-05 | 0.000708 | 0.003083 | 0.0001211 | 0.000424887 | AP001931.2, TMX2 |
| chr17 | 8152415 | 8152570 | 156 | 5 | 0.000196423 | 0.000724 | 0.002563 | 0.0001212 | 0.002235543 | PER1, AC129492.3 |
| chr17 | 2017802 | 2017815 | 14 | 3 | 0.000446679 | 0.001005 | 0.000168 | 0.0001212 | -0.00134641 | AC099684.1, RTN4RL1 |
| chr7 | 33062724 | 33063043 | 320 | 7 | 0.000194805 | 0.001902 | 0.000583 | 0.0001212 | 0.001563026 | NT5C3A |
| chr17 | 50124166 | 50124300 | 135 | 2 | 0.000274353 | 0.000963 | 0.000114 | 0.0001214 | 0.003396033 | SAMD14 |
| chr10 | 133158230 | 133158559 | 330 | 4 | 4.09E-05 | 0.051753 | 3.02E-05 | 0.0001214 | 0.002668513 | NA |
| chr19 | 38263889 | 38264290 | 402 | 6 | 0.000153004 | 0.000323 | 0.006486 | 0.0001215 | 0.005699709 | SPINT2 |
| chr19 | 4038853 | 4038958 | 106 | 3 | 0.000338055 | 0.000406 | 0.000198 | 0.0001215 | 0.003658676 | PIAS4 |
| chr1 | 50846419 | 50846447 | 29 | 2 | 0.000307327 | 0.026984 | 6.82E-05 | 0.0001215 | 0.003205234 | FAF1 |
| chr7 | 156063005 | 156063410 | 406 | 3 | 1.98E-05 | 0.002784 | 7.61E-05 | 0.0001216 | 0.002889092 | NA |
| chr4 | 865441 | 865683 | 243 | 3 | 0.000234239 | 0.000102 | 0.001912 | 0.0001216 | 0.000758742 | GAK |
| chr12 | 124380584 | 124381316 | 733 | 3 | 6.44E-05 | 0.00153 | 0.000221 | 0.0001218 | 0.007889926 | NCOR2 |
| chr17 | 2955916 | 2956074 | 159 | 2 | 7.21E-05 | 0.007104 | 7.55E-05 | 0.0001219 | -0.00703556 | RAP1GAP2 |
| chr4 | 153204056 | 153204568 | 513 | 9 | 0.000160164 | 0.004092 | 7.91E-05 | 0.000122 | 0.005821949 | TRIM2, AC013477.1 |
| chr6 | 158660547 | 158660693 | 147 | 2 | 0.00032773 | 0.000244 | 0.000251 | 0.000122 | 0.010126866 | SYTL3 |
| chr2 | 9572942 | 9572952 | 11 | 2 | 0.000422515 | 0.000239 | 0.000256 | 0.000122 | 0.003281375 | NA |
| chr11 | 67415859 | 67416145 | 287 | 4 | 7.22E-05 | 0.000423 | 0.000292 | 0.0001221 | 0.005109258 | CARNS1, PPP1CA |
| chr11 | 75399450 | 75399782 | 333 | 7 | 0.000314665 | 0.008615 | 0.000966 | 0.0001222 | 0.00155166 | RPS3 |
| chr16 | 85610777 | 85611112 | 336 | 2 | 0.000248728 | 0.000332 | 0.000195 | 0.0001222 | 0.001038686 | GSE1 |
| chr1 | 34784628 | 34785134 | 507 | 6 | 0.000258133 | 0.000366 | 0.000557 | 0.0001223 | 0.003891519 | GJB3, SMIM12, AL121988.1 |
| chr19 | 17243935 | 17244152 | 218 | 2 | 0.000333888 | 0.000154 | 0.000428 | 0.0001224 | 0.010137373 | NR2F6, AC010646.1 |
| chr17 | 47068667 | 47068673 | 7 | 2 | 0.000375843 | 0.001681 | 9.73E-05 | 0.0001226 | -0.00139246 | AC005670.3 |
| chr3 | 44477293 | 44477823 | 531 | 5 | 0.000102286 | 0.000782 | 0.002254 | 0.0001228 | 0.000782427 | ZNF445 |
| chr2 | 3318227 | 3318744 | 518 | 5 | 0.000124237 | 0.000905 | 0.000301 | 0.0001229 | -0.0001141 | EIPR1 |
| chr9 | 93958280 | 93958820 | 541 | 4 | 0.000131103 | 0.002753 | 6.26E-05 | 0.0001229 | 0.00795593 | BARX1-DT |
| chr11 | 494651 | 495198 | 548 | 5 | 6.02E-05 | 0.002004 | 0.002198 | 0.0001231 | 0.00259789 | RNH1 |
| chr19 | 1605619 | 1605845 | 227 | 8 | 0.000184701 | 0.002218 | 0.009369 | 0.0001231 | 0.001428295 | NA |
| chr14 | 24155467 | 24155627 | 161 | 2 | 0.000164527 | 0.001603 | 9.89E-05 | 0.0001233 | 0.005131583 | RNF31, AL136295.5 |
| chr2 | 238288498 | 238289141 | 644 | 5 | 0.000417355 | 0.00021 | 0.002809 | 0.0001233 | 0.001122 | AC012485.3, PER2 |
| chr3 | 12993569 | 12993851 | 283 | 2 | 0.000110281 | 0.001163 | 0.000108 | 0.0001233 | -0.00274175 | IQSEC1 |
| chr13 | 79480841 | 79481106 | 266 | 7 | 0.000294495 | 0.001565 | 0.003883 | 0.0001233 | -0.00362746 | NDFIP2-AS1 |
| chr6 | 137223937 | 137224161 | 225 | 2 | 0.000110804 | 0.000975 | 0.000115 | 0.0001235 | 0.011444513 | NA |
| chr2 | 231212136 | 231212508 | 373 | 2 | 4.98E-05 | 0.005004 | 7.96E-05 | 0.0001236 | 0.012134008 | ARMC9 |
| chr1 | 170145641 | 170145909 | 269 | 4 | 0.000339579 | 0.000159 | 0.001337 | 0.0001236 | 0.01109993 | NA |
| chr4 | 8619499 | 8619634 | 136 | 4 | 0.000278621 | 0.004932 | 4.87E-05 | 0.0001236 | 0.003909118 | GPR78, CPZ |
| chr22 | 25761007 | 25761072 | 66 | 2 | 0.000391671 | 0.00321 | 8.55E-05 | 0.0001239 | 0.005678776 | MYO18B |
| chr3 | 48631085 | 48631753 | 669 | 4 | 0.000486604 | 0.000104 | 0.00315 | 0.000124 | 0.00363761 | SLC26A6 |
| chr4 | 1355079 | 1355156 | 78 | 2 | 0.000346593 | 8.40E-05 | 0.001794 | 0.0001241 | 0.003706354 | UVSSA |
| chr3 | 20073413 | 20073957 | 545 | 2 | 1.26E-05 | 0.000331 | 0.0002 | 0.0001243 | 0.002009796 | KAT2B |
| chr2 | 16013332 | 16013925 | 594 | 3 | 2.13E-05 | 7.60E-05 | 0.002264 | 0.0001243 | 0.021452613 | AC010145.1 |
| chr15 | 90879687 | 90880344 | 658 | 6 | 0.000270157 | 0.000233 | 0.013937 | 0.0001243 | 0.002492504 | FURIN |
| chr4 | 155209054 | 155209936 | 883 | 5 | 7.14E-06 | 0.019999 | 0.000111 | 0.0001243 | 0.010814213 | AC097467.3, NPY2R |
| chr3 | 50153142 | 50153366 | 225 | 2 | 0.000172939 | 0.003199 | 8.57E-05 | 0.0001243 | 0.005252867 | SEMA3F-AS1 |
| chr9 | 27715150 | 27715218 | 69 | 2 | 0.000428178 | 0.001958 | 9.48E-05 | 0.0001243 | 0.010032312 | NA |
| chr1 | 109509578 | 109510060 | 483 | 9 | 0.000257573 | 0.000521 | 0.002571 | 0.0001245 | -0.00019546 | AMIGO1 |
| chr4 | 24980073 | 24980551 | 479 | 11 | 0.000338122 | 0.000159 | 0.004534 | 0.0001245 | 0.006248022 | CCDC149 |
| chr4 | 7050676 | 7050698 | 23 | 2 | 0.000453106 | 0.000208 | 0.000306 | 0.0001246 | 0.003282439 | TADA2B |
| chr10 | 79379483 | 79380151 | 669 | 3 | 4.58E-05 | 0.01154 | 8.55E-05 | 0.000125 | 0.004844265 | NA |
| chr10 | 110498568 | 110498922 | 355 | 5 | 3.96E-05 | 0.002969 | 0.00011 | 0.000125 | 0.004221072 | DUSP5 |
| chr15 | 78811653 | 78812139 | 487 | 5 | 0.000163694 | 4.32E-05 | 0.015141 | 0.000125 | -0.00254124 | MORF4L1 |
| chr1 | 2475356 | 2476279 | 924 | 4 | 5.28E-05 | 6.77E-05 | 0.004071 | 0.0001253 | 0.008345249 | PLCH2 |
| chr5 | 323679 | 323792 | 114 | 2 | 0.000137882 | 0.000783 | 0.000127 | 0.0001254 | 0.007772384 | PDCD6-AHRR, AHRR |
| chr19 | 35900540 | 35900686 | 147 | 3 | 0.000297842 | 0.000328 | 0.000418 | 0.0001255 | 0.006491686 | NFKBID |
| chr1 | 160061415 | 160061863 | 449 | 3 | 0.00010414 | 0.002914 | 7.89E-05 | 0.0001255 | 0.005712767 | KCNJ10 |
| chr2 | 235775924 | 235776643 | 720 | 4 | 5.66E-05 | 0.00389 | 5.94E-05 | 0.0001255 | 0.004424533 | AGAP1, AC064874.1 |
| chr1 | 175067381 | 175068223 | 843 | 6 | 5.66E-05 | 0.003923 | 7.05E-05 | 0.0001256 | 0.0023822 | TNN |
| chr14 | 104796854 | 104797363 | 510 | 3 | 0.000240601 | 0.00021 | 0.000424 | 0.0001258 | 0.006782491 | NA |
| chr7 | 157849045 | 157849133 | 89 | 2 | 0.000316546 | 0.003392 | 8.56E-05 | 0.0001259 | 0.003569171 | PTPRN2 |
| chr4 | 182793219 | 182793264 | 46 | 3 | 0.000418996 | 0.002678 | 0.000261 | 0.000126 | 0.000104409 | TENM3 |
| chr20 | 64163445 | 64163682 | 238 | 2 | 0.000274123 | 8.34E-05 | 0.002014 | 0.0001265 | 0.001767968 | MYT1 |
| chr1 | 53085627 | 53085755 | 129 | 3 | 0.000339438 | 0.004489 | 0.001741 | 0.0001267 | 0.003548841 | NA |
| chr5 | 472876 | 473337 | 462 | 5 | 0.000352814 | 0.000618 | 0.006565 | 0.0001267 | 2.24E-05 | SLC9A3-AS1, PP7080, SLC9A3 |
| chr22 | 24494077 | 24494362 | 286 | 3 | 4.23E-05 | 0.002353 | 0.000764 | 0.0001268 | 0.004856154 | UPB1, ADORA2A-AS1 |
| chr18 | 35240838 | 35241507 | 670 | 8 | 8.29E-05 | 0.000391 | 0.002436 | 0.0001269 | -0.00109571 | ZNF397 |
| chr8 | 89902628 | 89903111 | 484 | 3 | 0.000106771 | 0.035454 | 0.001022 | 0.000127 | 0.000611956 | OSGIN2 |
| chr7 | 23246123 | 23246184 | 62 | 3 | 0.000258421 | 7.36E-05 | 0.00477 | 0.0001274 | 0.005895477 | GPNMB |
| chr22 | 30821243 | 30821678 | 436 | 4 | 1.15E-05 | 0.010086 | 0.000358 | 0.0001276 | 0.003441297 | OSBP2 |
| chr5 | 139341098 | 139341338 | 241 | 3 | 3.03E-05 | 0.0105 | 5.59E-05 | 0.0001276 | 0.002777858 | NA |
| chr11 | 134714391 | 134714484 | 94 | 2 | 0.000194271 | 0.00012 | 0.000718 | 0.0001276 | 0.016871828 | LINC02706 |
| chr19 | 55100027 | 55100448 | 422 | 2 | 2.84E-05 | 0.000881 | 0.000124 | 0.0001278 | 0.009412038 | PPP1R12C |
| chr11 | 57238402 | 57238799 | 398 | 3 | 1.11E-05 | 0.021771 | 3.97E-05 | 0.0001278 | 0.00804432 | NA |
| chr1 | 34165950 | 34166517 | 568 | 6 | 5.41E-05 | 0.002368 | 0.002086 | 0.000128 | 0.003266676 | NA |
| chr20 | 36461435 | 36462099 | 665 | 7 | 0.000137748 | 5.86E-05 | 0.010788 | 0.0001285 | -0.00377074 | DLGAP4 |
| chr3 | 124094243 | 124094749 | 507 | 9 | 0.000172071 | 0.001037 | 0.000281 | 0.0001285 | 0.002690065 | KALRN |
| chr17 | 75182149 | 75182372 | 224 | 2 | 8.38E-05 | 0.001362 | 0.000107 | 0.0001288 | -0.0013826 | SUMO2 |
| chr8 | 51817088 | 51817139 | 52 | 2 | 0.000404957 | 0.027351 | 7.09E-05 | 0.000129 | -0.00248743 | NA |
| chr16 | 62036946 | 62037591 | 646 | 8 | 0.000343737 | 0.000236 | 0.009375 | 0.000129 | 0.00508458 | CDH8 |
| chr6 | 401311 | 401700 | 390 | 5 | 9.48E-05 | 0.032605 | 0.001371 | 0.0001291 | 0.001242086 | IRF4 |
| chr12 | 47716375 | 47716902 | 528 | 2 | 0.000156632 | 8.08E-05 | 0.002614 | 0.0001291 | 0.003879968 | AC004241.1, ENDOU |
| chr20 | 63283460 | 63283485 | 26 | 2 | 5.97E-05 | 0.0122 | 7.47E-05 | 0.0001291 | -0.00278181 | ARFGAP1 |
| chr15 | 95133495 | 95133556 | 62 | 2 | 0.000268586 | 0.005907 | 8.04E-05 | 0.0001293 | 0.003133792 | AC087633.2, AC087636.1 |
| chr1 | 151254980 | 151255060 | 81 | 2 | 0.000131056 | 0.000126 | 0.000676 | 0.0001293 | 0.004015689 | PSMD4 |
| chr12 | 115451704 | 115451793 | 90 | 2 | 0.000290877 | 0.000289 | 0.000238 | 0.0001296 | 0.006586517 | NA |
| chr6 | 163100339 | 163100576 | 238 | 3 | 0.000262336 | 8.89E-05 | 0.004378 | 0.0001296 | -6.74E-05 | PACRG |
| chr16 | 78567870 | 78568014 | 145 | 3 | 0.000434443 | 0.001694 | 0.000119 | 0.0001296 | -0.00192426 | WWOX |
| chr12 | 111597408 | 111597876 | 469 | 5 | 0.000227811 | 8.42E-05 | 0.005982 | 0.0001298 | 0.000963122 | ATXN2 |
| chr14 | 73246897 | 73247023 | 127 | 2 | 4.53E-05 | 0.000126 | 0.000679 | 0.0001298 | 0.007710826 | PAPLN |
| chr19 | 2035122 | 2035131 | 10 | 2 | 0.000250591 | 8.73E-05 | 0.001841 | 0.0001299 | 0.021735217 | NA |
| chr6 | 75102003 | 75102085 | 83 | 3 | 0.000452629 | 0.000805 | 0.000228 | 0.0001299 | 0.012088136 | COL12A1 |
| chr2 | 85411412 | 85411572 | 161 | 3 | 0.000202542 | 0.00092 | 0.001071 | 0.00013 | -0.00055447 | CAPG |
| chr19 | 42268081 | 42268294 | 214 | 2 | 0.000194449 | 7.87E-05 | 0.00319 | 0.00013 | -0.00330319 | NA |
| chr14 | 31488248 | 31488586 | 339 | 3 | 0.000165049 | 0.003986 | 6.25E-05 | 0.0001302 | 0.005322211 | NA |
| chr12 | 121641394 | 121641512 | 119 | 2 | 0.000376096 | 0.009712 | 7.65E-05 | 0.0001302 | 0.002197636 | ORAI1 |
| chr3 | 88137749 | 88137830 | 82 | 2 | 0.000119534 | 0.000394 | 0.000189 | 0.0001306 | 0.004607055 | ZNF654, CGGBP1 |
| chr16 | 71723337 | 71724135 | 799 | 5 | 7.51E-05 | 0.002675 | 0.00098 | 0.0001307 | -0.00121153 | AC009097.4, PHLPP2 |
| chr4 | 949774 | 949902 | 129 | 3 | 0.000187032 | 0.000323 | 0.000253 | 0.0001308 | 0.000986929 | TMEM175 |
| chr6 | 151025222 | 151025273 | 52 | 2 | 0.000212759 | 0.000274 | 0.000253 | 0.0001308 | 0.002888831 | MTHFD1L |
| chr5 | 917776 | 917785 | 10 | 2 | 0.000450392 | 0.002666 | 9.25E-05 | 0.0001311 | 0.0035772 | TRIP13 |
| chr18 | 31042019 | 31042234 | 216 | 3 | 7.91E-05 | 0.000671 | 0.00014 | 0.0001313 | 0.005544381 | DSC3 |
| chr17 | 40091060 | 40091148 | 89 | 2 | 0.000170493 | 0.000629 | 0.000146 | 0.0001316 | 0.003962348 | THRA |
| chr22 | 38283372 | 38283919 | 548 | 2 | 0.000329739 | 7.95E-05 | 0.003276 | 0.0001319 | 0.007362701 | NA |
| chr22 | 20032151 | 20032394 | 244 | 4 | 0.000218501 | 0.001864 | 6.94E-05 | 0.0001322 | 0.006852231 | TANGO2 |
| chr19 | 47675401 | 47675452 | 52 | 2 | 0.000323446 | 0.000397 | 0.000192 | 0.0001324 | 0.003789019 | AC010519.1, BICRA |
| chr20 | 1117856 | 1118503 | 648 | 8 | 9.81E-05 | 9.32E-05 | 0.008226 | 0.0001327 | 0.006773406 | PSMF1, AL110114.1 |
| chr3 | 44511753 | 44511924 | 172 | 3 | 0.00019146 | 0.001043 | 0.000109 | 0.0001328 | 0.001104199 | NA |
| chr13 | 113822231 | 113822677 | 447 | 4 | 9.00E-05 | 0.027863 | 4.91E-05 | 0.000133 | 0.003134018 | GAS6-AS1, GAS6 |
| chr8 | 141182619 | 141182889 | 271 | 2 | 0.000237399 | 8.09E-05 | 0.003081 | 0.000133 | 0.013008342 | DENND3 |
| chr4 | 17781020 | 17781436 | 417 | 4 | 0.000256446 | 0.000267 | 0.004034 | 0.000133 | 0.004938581 | FAM184B |
| chr19 | 54947518 | 54948109 | 592 | 6 | 0.000416243 | 0.000199 | 0.008379 | 0.0001331 | 0.000119064 | NLRP7 |
| chr2 | 231892270 | 231892704 | 435 | 5 | 0.000218809 | 0.008222 | 0.000102 | 0.0001331 | 0.005285938 | MIR1471 |
| chr11 | 126356046 | 126356259 | 214 | 6 | 0.00037423 | 0.000121 | 0.004464 | 0.0001336 | -0.0015451 | ST3GAL4 |
| chr17 | 44348643 | 44349138 | 496 | 3 | 0.000282694 | 0.000112 | 0.003119 | 0.0001336 | 0.003752695 | GRN |
| chr17 | 16492135 | 16492757 | 623 | 7 | 8.05E-05 | 0.003472 | 0.000672 | 0.0001339 | 0.003588253 | LRRC75A |
| chr4 | 84499846 | 84499897 | 52 | 2 | 0.000271497 | 0.000246 | 0.000292 | 0.000134 | 0.008344651 | NA |
| chr7 | 92528174 | 92529123 | 950 | 9 | 0.000256388 | 8.43E-05 | 0.012493 | 0.0001341 | -0.00080115 | RBM48, PEX1 |
| chr17 | 12998693 | 12998910 | 218 | 3 | 0.000321232 | 0.003543 | 6.61E-05 | 0.0001342 | -0.00072329 | ELAC2 |
| chr1 | 170085294 | 170085519 | 226 | 2 | 0.000188451 | 8.38E-05 | 0.00264 | 0.0001343 | 0.013997515 | NA |
| chr20 | 45404955 | 45405013 | 59 | 2 | 2.47E-05 | 8.26E-05 | 0.002865 | 0.0001343 | 0.019508891 | SYS1-DBNDD2, TP53TG5 |
| chr2 | 170455868 | 170456176 | 309 | 3 | 0.000276053 | 0.004307 | 6.18E-05 | 0.0001344 | 0.003937417 | MYO3B |
| chr17 | 77478968 | 77479158 | 191 | 2 | 0.000311081 | 0.001148 | 0.000118 | 0.0001346 | 0.011525073 | SEPTIN9 |
| chr12 | 118104222 | 118104601 | 380 | 5 | 0.000322096 | 6.87E-05 | 0.012934 | 0.0001351 | 0.004315613 | VSIG10 |
| chr5 | 1885253 | 1885602 | 350 | 3 | 0.000126658 | 0.0036 | 0.000179 | 0.0001353 | 0.004242627 | IRX4 |
| chr14 | 67527127 | 67527268 | 142 | 2 | 4.34E-05 | 0.004819 | 8.54E-05 | 0.0001353 | 0.007710047 | TMEM229B |
| chr3 | 69763283 | 69763822 | 540 | 6 | 0.000208617 | 0.003151 | 0.001786 | 0.0001353 | -0.00092431 | MITF |
| chr16 | 30030122 | 30030180 | 59 | 3 | 0.000369273 | 0.002092 | 0.000907 | 0.0001353 | 0.006933733 | TLCD3B |
| chr16 | 30090818 | 30091390 | 573 | 5 | 0.000219368 | 0.000286 | 0.007627 | 0.0001353 | 0.00608865 | TBX6 |
| chr3 | 113019935 | 113020987 | 1053 | 5 | 2.65E-06 | 0.001546 | 0.000418 | 0.0001354 | 0.001532468 | AC078785.1 |
| chr11 | 119174611 | 119174698 | 88 | 2 | 0.00019551 | 0.002629 | 9.52E-05 | 0.0001356 | 0.002984235 | NLRX1 |
| chr16 | 11345294 | 11345676 | 383 | 5 | 0.000289516 | 0.001828 | 0.000214 | 0.0001358 | 0.002294005 | RMI2 |

| | | | | | | | | | | |
|---|---|---|---|---|---|---|---|---|---|---|
| chr13 | 40535461 | 40535570 | 110 | 2 | 0.000458985 | 8.25E-05 | 0.00309 | 0.0001358 | 0.002110234 | LINC00598, AL133318.1 |
| chr2 | 219597455 | 219597853 | 399 | 9 | 9.98E-05 | 0.00734 | 0.000273 | 0.0001359 | -0.00119248 | NA |
| chr17 | 42016864 | 42017675 | 812 | 13 | 4.52E-05 | 0.010264 | 0.011526 | 0.0001359 | -0.00067772 | NKIRAS2, DNAJC7 |
| chr6 | 31171843 | 31172019 | 177 | 2 | 0.000308443 | 8.62E-05 | 0.002411 | 0.0001359 | 0.010702123 | POU5F1 |
| chr17 | 72339098 | 72339290 | 193 | 2 | 0.000195202 | 0.000387 | 0.000204 | 0.0001361 | 0.00693696 | LINC00511 |
| chr15 | 91866427 | 91866636 | 210 | 2 | 0.000337862 | 0.000132 | 0.000698 | 0.0001361 | 0.008721026 | SLCO3A1 |
| chr3 | 157087998 | 157088740 | 743 | 4 | 8.37E-05 | 0.000105 | 0.003384 | 0.0001365 | 0.008916627 | LINC02029, LINC00880 |
| chr2 | 119759052 | 119759547 | 496 | 9 | 0.000254198 | 0.007812 | 0.001321 | 0.0001365 | 0.001654748 | NA |
| chr7 | 1246387 | 1246688 | 302 | 3 | 3.07E-05 | 0.01492 | 9.36E-05 | 0.0001365 | 0.006490477 | NA |
| chr5 | 154630598 | 154631560 | 963 | 4 | 2.39E-05 | 0.007413 | 5.71E-05 | 0.0001367 | 0.002547652 | NA |
| chr7 | 752698 | 753081 | 384 | 3 | 0.000283473 | 0.027387 | 0.000349 | 0.0001367 | 0.002484205 | DNAAF5 |
| chr11 | 1488022 | 1488066 | 45 | 2 | 0.000442507 | 0.000162 | 0.000505 | 0.0001369 | 0.006279314 | MOB2 |
| chr3 | 150703280 | 150703553 | 274 | 5 | 0.000336345 | 6.11E-05 | 0.003473 | 0.0001374 | 0.006565872 | ERICH6 |
| chr3 | 133927221 | 133927251 | 31 | 2 | 0.000375336 | 8.25E-05 | 0.003359 | 0.0001376 | 0.011690022 | NA |
| chr17 | 6831621 | 6832220 | 600 | 9 | 0.000197754 | 0.001607 | 0.00136 | 0.0001378 | -0.00063304 | TEKT1 |
| chr9 | 81690029 | 81690172 | 144 | 2 | 0.000170679 | 0.017018 | 7.61E-05 | 0.0001379 | 0.002991345 | AL591368.1 |
| chr10 | 33706354 | 33706442 | 89 | 3 | 0.000347493 | 0.003115 | 0.000219 | 0.0001379 | -0.00690101 | AL450313.1 |
| chr17 | 27894141 | 27894862 | 722 | 3 | 9.73E-05 | 0.000125 | 0.003649 | 0.0001381 | 0.008429991 | LYRM9 |
| chr4 | 1979305 | 1979363 | 59 | 2 | 0.000384232 | 8.36E-05 | 0.003144 | 0.0001381 | -0.00200459 | NSD2 |
| chr20 | 34526369 | 34526636 | 268 | 2 | 0.000269896 | 0.000104 | 0.001245 | 0.0001381 | -0.00137076 | ITCH, DYNLRB1, Y RNA |
| chr15 | 92170918 | 92171115 | 198 | 2 | 0.000359609 | 0.000479 | 0.000179 | 0.0001382 | 0.00191194 | SLCO3A1, AC116903.2, AC116903.1 |
| chr6 | 100464946 | 100465716 | 771 | 7 | 3.61E-05 | 0.00488 | 0.000366 | 0.0001383 | 0.003126506 | NA |
| chr12 | 55931053 | 55931718 | 666 | 5 | 2.88E-05 | 0.000865 | 0.00235 | 0.0001384 | 0.006135705 | DGKA, PYM1 |
| chr10 | 124642276 | 124642611 | 336 | 5 | 5.00E-05 | 0.002354 | 7.24E-05 | 0.0001384 | 0.005808168 | AC068896.1, FAM53B |
| chr8 | 143835588 | 143836147 | 560 | 4 | 0.000146481 | 0.00356 | 0.000109 | 0.0001385 | -0.00050506 | NRBP2 |
| chr5 | 69369396 | 69369642 | 247 | 5 | 0.000356183 | 0.000261 | 0.012231 | 0.0001387 | 0.000587911 | RAD17, AK6, TAF9 |
| chr4 | 127880490 | 127881149 | 660 | 13 | 0.000195047 | 0.046099 | 0.000917 | 0.0001387 | 0.000924133 | PLK4 |
| chr8 | 76673213 | 76673554 | 342 | 3 | 6.90E-05 | 0.002317 | 0.000663 | 0.0001388 | 0.005656717 | ZFHX4-AS1 |
| chr22 | 37212571 | 37212779 | 209 | 3 | 4.28E-05 | 0.00221 | 0.000104 | 0.0001389 | 0.005111218 | NA |
| chr8 | 142089762 | 142089921 | 160 | 4 | 0.000160537 | 0.000471 | 0.002035 | 0.000139 | 0.001815451 | AC104417.2 |
| chr13 | 100513785 | 100513810 | 26 | 2 | 0.000429385 | 0.001438 | 0.000113 | 0.000139 | 0.002884403 | PCCA |
| chr6 | 165731168 | 165731280 | 113 | 2 | 0.000296755 | 0.00023 | 0.000334 | 0.0001391 | -0.00366828 | PDE10A |
| chr14 | 74816973 | 74817317 | 345 | 2 | 5.80E-05 | 9.90E-05 | 0.00152 | 0.0001393 | -0.00130253 | YLPM1 |
| chr2 | 97792729 | 97792929 | 201 | 5 | 0.000371983 | 0.000491 | 0.000762 | 0.0001397 | 0.002114985 | TMEM131 |
| chr20 | 3681865 | 3682311 | 447 | 4 | 0.000152368 | 6.07E-05 | 0.008132 | 0.0001397 | 0.002815572 | ADAM33 |
| chr18 | 79862587 | 79862593 | 7 | 2 | 7.13E-05 | 0.00055 | 0.000168 | 0.0001398 | 0.001445159 | NA |
| chr10 | 13502648 | 13503382 | 735 | 4 | 0.000266712 | 0.000216 | 0.002076 | 0.0001401 | 0.003880471 | BEND7 |
| chr2 | 11786754 | 11787123 | 370 | 5 | 0.000234566 | 0.004375 | 4.08E-05 | 0.0001402 | 0.001134207 | LPIN1 |
| chr6 | 215882 | 216043 | 162 | 2 | 0.000206197 | 0.002125 | 0.000103 | 0.0001402 | 0.013514753 | NA |
| chr11 | 1838280 | 1838608 | 329 | 2 | 0.00037878 | 8.48E-05 | 0.003179 | 0.0001405 | 0.010807949 | NA |
| chr3 | 138594169 | 138594324 | 156 | 3 | 0.000325124 | 0.000143 | 0.000921 | 0.0001405 | 0.001201095 | CEP70 |
| chr1 | 39683709 | 39683978 | 270 | 4 | 0.00029695 | 0.002823 | 0.000119 | 0.000141 | 0.008348946 | HPCAL4 |
| chr19 | 45716791 | 45716971 | 181 | 2 | 0.000298481 | 0.001194 | 0.000122 | 0.000141 | 0.002208989 | FBXO46, AC007191.1 |
| chr11 | 34685263 | 34685414 | 152 | 2 | 0.000339887 | 0.008434 | 8.21E-05 | 0.000141 | 0.005770334 | NA |
| chr21 | 41478838 | 41479498 | 661 | 3 | 0.000111333 | 5.73E-05 | 0.009341 | 0.000141 | 0.004903762 | TMPRSS2 |
| chr11 | 3834279 | 3834648 | 370 | 3 | 0.000289969 | 0.000325 | 0.000278 | 0.0001411 | 0.005324111 | RHOG |
| chr19 | 49670135 | 49670145 | 11 | 2 | 0.000408647 | 0.007525 | 8.32E-05 | 0.0001411 | -0.00386774 | BCL2L12 |
| chr9 | 131278806 | 131278813 | 8 | 2 | 0.000283854 | 8.44E-05 | 0.003408 | 0.0001413 | 0.009129655 | NA |
| chr11 | 82843179 | 82843515 | 337 | 2 | 0.000374024 | 0.000862 | 0.000138 | 0.0001414 | 0.004682242 | PRCP |
| chr6 | 31621588 | 31622012 | 425 | 4 | 0.000276905 | 5.02E-05 | 0.01027 | 0.0001414 | 0.009695794 | PRRC2A |
| chr12 | 53295555 | 53295818 | 264 | 3 | 0.000411875 | 0.000191 | 0.001583 | 0.0001414 | -0.0008636 | PFDN5, AC073611.1 |
| chr10 | 30042751 | 30042982 | 232 | 3 | 8.97E-05 | 0.000416 | 0.000452 | 0.0001414 | 0.003124756 | JCAD |
| chr16 | 75265372 | 75266145 | 774 | 7 | 0.000117743 | 7.74E-05 | 0.018004 | 0.0001414 | 0.003603714 | BCAR1 |
| chr9 | 129336032 | 129336286 | 255 | 2 | 0.000117286 | 0.000242 | 0.000327 | 0.0001416 | 0.00707932 | LINC01503 |
| chr6 | 108560667 | 108560677 | 11 | 3 | 0.000465517 | 0.000303 | 0.000367 | 0.000142 | 0.000105134 | FOXO3 |
| chr16 | 89161296 | 89161506 | 211 | 3 | 9.82E-05 | 0.004055 | 6.68E-05 | 0.0001421 | 0.006895655 | ACSF3, LINC00304 |
| chr6 | 43277501 | 43277833 | 333 | 2 | 0.000217274 | 0.001037 | 0.000129 | 0.0001422 | 0.00392849 | TTBK1 |
| chr3 | 63441973 | 63442190 | 218 | 2 | 4.22E-05 | 0.00272 | 9.85E-05 | 0.0001424 | 0.005359592 | SYNPR, SYNPR-AS1 |
| chr2 | 219543689 | 219544071 | 383 | 7 | 0.000312788 | 7.40E-05 | 0.024126 | 0.0001424 | -0.0004545 | TMEM198, CHPF |
| chr12 | 100573238 | 100573831 | 594 | 9 | 0.000304736 | 0.000814 | 0.007161 | 0.0001427 | -0.00062092 | GAS2L3 |
| chr3 | 128567811 | 128568127 | 317 | 2 | 0.000344996 | 9.16E-05 | 0.002307 | 0.0001429 | -0.00191432 | NA |
| chr12 | 112417386 | 112417467 | 82 | 2 | 0.00046118 | 0.000129 | 0.000834 | 0.000143 | 0.002353517 | RPL6 |
| chr14 | 102906732 | 102907265 | 534 | 4 | 0.00012843 | 5.54E-05 | 0.011147 | 0.0001433 | -0.00045346 | TRAF3 |
| chr14 | 21069177 | 21069759 | 583 | 3 | 6.16E-05 | 0.000365 | 0.000719 | 0.0001433 | 0.003133873 | NDRG2 |
| chr15 | 64990036 | 64990340 | 305 | 5 | 0.000283352 | 0.001193 | 9.25E-05 | 0.0001434 | 0.001649943 | SPG21 |
| chr18 | 74148286 | 74148534 | 249 | 7 | 0.000293874 | 0.001108 | 0.001893 | 0.0001434 | 0.000954226 | TIMM21 |
| chr14 | 24431605 | 24432228 | 624 | 3 | 0.000247215 | 0.000252 | 0.000688 | 0.0001437 | 0.00636092 | KHNYN |
| chr19 | 49064957 | 49065705 | 749 | 4 | 4.08E-05 | 0.000611 | 0.004067 | 0.0001437 | 0.010758127 | AC008687.8, NTF4 |
| chr12 | 131075273 | 131075463 | 191 | 2 | 0.000344165 | 8.85E-05 | 0.002851 | 0.0001439 | 0.000549039 | ADGRD1 |
| chr17 | 47322481 | 47323426 | 946 | 5 | 5.27E-06 | 0.001268 | 0.000101 | 0.0001443 | 0.011777752 | AC068234.1, EFCAB13, THCAT158 |
| chr3 | 113304241 | 113304409 | 169 | 2 | 0.000164886 | 0.00048 | 0.000191 | 0.0001446 | 0.005582659 | CFAP44 |
| chr3 | 48847756 | 48848270 | 515 | 10 | 0.000227974 | 0.001097 | 0.018777 | 0.000145 | 0.001617608 | PRKAR2A-AS1, PRKAR2A |
| chr5 | 158274824 | 158275059 | 236 | 2 | 0.000251333 | 0.001696 | 0.000112 | 0.0001451 | -0.00817856 | NA |
| chr10 | 42367428 | 42367620 | 193 | 3 | 0.000352852 | 6.43E-05 | 0.00676 | 0.0001451 | -0.02983712 | BX322639.1 |
| chr16 | 81715663 | 81715857 | 195 | 2 | 0.000165399 | 0.000203 | 0.00042 | 0.0001452 | 0.014846684 | NA |
| chr19 | 51702971 | 51703776 | 806 | 7 | 2.23E-05 | 0.001854 | 0.001538 | 0.0001454 | 0.007295359 | SPACA6 |
| chr6 | 166508272 | 166508402 | 131 | 3 | 0.000351 | 0.001044 | 0.000162 | 0.0001454 | 0.001350436 | RPS6KA2 |
| chr15 | 74173996 | 74175091 | 1096 | 8 | 0.000117728 | 0.000467 | 0.00516 | 0.0001459 | 0.003070377 | ISLR, AC010931.2 |
| chr5 | 154937995 | 154938369 | 375 | 7 | 0.000205542 | 0.001673 | 0.012185 | 0.0001459 | -0.0012394 | GEMIN5 |
| chr5 | 129903074 | 129903441 | 368 | 2 | 5.79E-05 | 0.000893 | 0.00014 | 0.000146 | -0.01294721 | AC008591.1 |
| chr7 | 157601645 | 157602278 | 634 | 5 | 0.000114929 | 0.041148 | 7.66E-05 | 0.0001461 | 0.00134198 | PTPRN2 |
| chr16 | 8861694 | 8861903 | 210 | 3 | 3.08E-05 | 0.000818 | 0.00016 | 0.0001462 | 0.006439057 | CARHSP1 |
| chr3 | 42600492 | 42601284 | 793 | 14 | 7.23E-05 | 0.001682 | 0.001856 | 0.0001463 | 0.000836948 | NKTR, SEC22C |
| chr19 | 49016056 | 49016356 | 301 | 2 | 7.65E-05 | 0.00084 | 0.000144 | 0.0001463 | 0.003796237 | LHB |
| chr3 | 49056985 | 49057185 | 201 | 2 | 7.46E-05 | 0.000343 | 0.000249 | 0.0001463 | 0.00161618 | QRICH1 |
| chr15 | 90966272 | 90966280 | 9 | 2 | 0.000393636 | 0.000934 | 0.000139 | 0.0001465 | 0.003704797 | PRC1, AC068831.6 |
| chr8 | 143982933 | 143983006 | 74 | 3 | 0.000357145 | 0.000381 | 0.000959 | 0.0001466 | 0.001046392 | PARP10 |
| chr7 | 27113280 | 27113593 | 314 | 2 | 0.000373853 | 9.67E-05 | 0.002032 | 0.0001467 | 0.004232542 | HOXA-AS2, HOXA3 |
| chr7 | 159026840 | 159026929 | 90 | 2 | 0.000181011 | 9.38E-05 | 0.002331 | 0.0001467 | 0.008959462 | LINC00689 |
| chr8 | 33485146 | 33485679 | 534 | 6 | 1.88E-05 | 0.001331 | 0.000605 | 0.0001467 | 0.000686047 | MAK16, TTI2 |
| chr9 | 136361280 | 136361353 | 74 | 3 | 0.000113923 | 0.000286 | 0.000982 | 0.0001467 | -0.00039451 | DNLZ |
| chr6 | 112362865 | 112363056 | 192 | 4 | 0.000238473 | 0.017808 | 3.26E-05 | 0.0001467 | 0.003267617 | NA |
| chr20 | 51523376 | 51524072 | 697 | 4 | 0.000313416 | 0.000209 | 0.005869 | 0.0001468 | 0.002771423 | NFATC2 |
| chr15 | 83011573 | 83011818 | 246 | 7 | 0.000369218 | 0.000207 | 0.002969 | 0.0001468 | -0.00018692 | AC022558.2, C15orf40 |
| chr7 | 2762198 | 2762439 | 242 | 3 | 0.000114048 | 0.006896 | 5.81E-05 | 0.000147 | 0.005620109 | AMZ1, GNA12 |
| chr14 | 105042727 | 105043039 | 313 | 3 | 0.00015264 | 0.001071 | 0.000123 | 0.0001471 | 0.00061391 | NA |
| chr9 | 129624085 | 129625040 | 956 | 4 | 1.11E-05 | 0.032956 | 0.001165 | 0.0001472 | 0.00694719 | NTMT1 |
| chr7 | 105760536 | 105760550 | 15 | 2 | 0.000422041 | 0.001416 | 0.00012 | 0.0001476 | 0.003347444 | ATXN7L1 |
| chr2 | 207134320 | 207134394 | 75 | 3 | 0.000312987 | 9.31E-05 | 0.003922 | 0.0001481 | 0.007341159 | KLF7 |
| chr1 | 20732473 | 20733252 | 780 | 12 | 0.000176667 | 0.011633 | 0.000748 | 0.0001481 | 0.000396293 | AL663074.1, SH2D5 |
| chr12 | 85279422 | 85279634 | 213 | 5 | 0.000223118 | 0.000388 | 0.015818 | 0.0001482 | 0.004806793 | NA |
| chr12 | 122006544 | 122006715 | 172 | 3 | 0.000409963 | 0.002223 | 0.000147 | 0.0001484 | 0.00659945 | NA |
| chr19 | 45352579 | 45353262 | 684 | 6 | 7.66E-05 | 0.000977 | 0.000807 | 0.0001485 | 0.001847922 | ERCC2 |
| chr6 | 165663995 | 165664181 | 187 | 2 | 6.33E-05 | 0.000244 | 0.000354 | 0.0001487 | 0.008949737 | PDE10A |
| chr2 | 2732197 | 2732372 | 176 | 2 | 2.80E-05 | 0.003812 | 9.53E-05 | 0.0001487 | 0.006112216 | AC011995.2 |
| chr14 | 20413906 | 20414269 | 364 | 5 | 0.00012159 | 0.002269 | 0.004107 | 0.0001488 | -1.97E-05 | NA |
| chr15 | 88620697 | 88621158 | 462 | 5 | 4.17E-05 | 0.000223 | 0.002048 | 0.0001491 | -0.00850844 | NA |
| chr14 | 95092603 | 95092885 | 283 | 2 | 0.000177121 | 8.99E-05 | 0.003208 | 0.0001492 | 0.007412749 | DICER1 |
| chr9 | 21812892 | 21813266 | 375 | 2 | 9.32E-05 | 0.000367 | 0.000243 | 0.0001493 | -0.00862869 | MTAP, AL359922.1 |
| chr1 | 31703967 | 31704465 | 499 | 9 | 0.000190893 | 0.000353 | 0.000306 | 0.0001494 | 0.005669361 | COL16A1 |
| chr22 | 19044611 | 19044742 | 132 | 2 | 9.43E-05 | 0.024744 | 7.87E-05 | 0.0001496 | 0.002380451 | DGCR2 |
| chr15 | 55587551 | 55588026 | 476 | 3 | 0.000154939 | 0.000176 | 0.000992 | 0.0001497 | 0.002720986 | PYGO1 |
| chr10 | 69961493 | 69961856 | 364 | 2 | 0.000249392 | 0.000146 | 0.000737 | 0.0001499 | 0.009722035 | COL13A1 |
| chr10 | 70364336 | 70364771 | 436 | 4 | 0.000137611 | 0.001347 | 0.00196 | 0.00015 | 0.00791106 | LRRC20 |
| chr1 | 248905313 | 248905840 | 528 | 5 | 0.000163518 | 0.000155 | 0.006533 | 0.00015 | 0.003233007 | NA |
| chr6 | 152628519 | 152628960 | 442 | 3 | 0.000316401 | 0.001945 | 0.000328 | 0.0001503 | -5.69E-05 | SYNE1 |
| chr11 | 9663752 | 9664221 | 470 | 8 | 0.000215477 | 0.001452 | 0.004068 | 0.0001504 | -0.00084332 | SWAP70 |
| chr3 | 50345878 | 50346600 | 723 | 6 | 5.52E-05 | 8.33E-05 | 0.004437 | 0.0001507 | 0.003650029 | NA |
| chr11 | 8038405 | 8038785 | 381 | 4 | 0.00038415 | 0.000381 | 0.000275 | 0.0001509 | 0.003692641 | TUB, AC116456.1 |
| chr11 | 12160688 | 12160830 | 143 | 4 | 4.33E-05 | 0.020949 | 2.99E-05 | 0.000151 | 0.003333186 | MICAL2 |
| chr17 | 57535078 | 57535096 | 19 | 2 | 0.000166891 | 0.006125 | 8.93E-05 | 0.000151 | 0.005097394 | MSI2 |
| chr14 | 75274946 | 75275030 | 85 | 2 | 0.000388263 | 0.000341 | 0.000263 | 0.0001511 | 0.011517451 | NA |
| chr17 | 75524111 | 75524678 | 568 | 3 | 7.29E-05 | 0.004359 | 0.000709 | 0.0001513 | 0.005802431 | TSEN54 |
| chr15 | 49651318 | 49651422 | 105 | 2 | 0.000373554 | 0.000734 | 0.000158 | 0.0001515 | 0.001514185 | DTWD1 |
| chr17 | 44314736 | 44315329 | 594 | 5 | 0.000226086 | 0.002459 | 0.001477 | 0.0001517 | 0.002358606 | RUNDC3A, RUNDC3A-AS1 |
| chr20 | 16747322 | 16747588 | 267 | 2 | 0.000185897 | 9.35E-05 | 0.0028 | 0.0001518 | 0.012688478 | NA |
| chr6 | 28895618 | 28895767 | 150 | 4 | 0.000189679 | 0.000148 | 0.006156 | 0.0001521 | -0.00547867 | NA |
| chr6 | 18155348 | 18155685 | 338 | 4 | 7.28E-05 | 0.000119 | 0.006172 | 0.0001522 | 7.69E-05 | KDM1B |
| chr11 | 72103971 | 72104162 | 192 | 4 | 4.14E-05 | 0.00086 | 0.000207 | 0.0001522 | -0.00032936 | AP000812.4 |
| chr13 | 23979404 | 23979622 | 219 | 6 | 0.000304632 | 0.000323 | 0.000449 | 0.0001524 | 0.003328114 | NA |
| chr1 | 111445966 | 111446063 | 98 | 3 | 0.000289403 | 0.026399 | 0.000176 | 0.0001524 | 0.002867177 | WDR77 |
| chr11 | 125887039 | 125887748 | 710 | 8 | 0.000443024 | 0.000481 | 0.008267 | 0.0001527 | -0.00445117 | HYLS1 |
| chr6 | 31742910 | 31743208 | 299 | 3 | 6.58E-05 | 0.000724 | 0.002113 | 0.0001528 | 0.000359928 | MSH5, MSH5-SAPCD1 |
| chr1 | 39405174 | 39405206 | 33 | 2 | 0.000472388 | 0.004996 | 9.30E-05 | 0.0001528 | 0.002670091 | MACF1 |
| chr5 | 150122272 | 150122343 | 72 | 2 | 0.000455562 | 0.000748 | 0.000158 | 0.0001529 | 0.012317968 | PDGFRB |
| chr3 | 158094974 | 158095628 | 655 | 4 | 0.000202127 | 0.003569 | 0.001672 | 0.0001531 | 0.007045105 | NA |
| chr19 | 4588338 | 4588421 | 84 | 2 | 0.000436765 | 0.001002 | 0.000141 | 0.0001534 | 0.008559269 | NA |
| chr6 | 46653762 | 46654119 | 358 | 2 | 1.74E-05 | 0.00029 | 0.000312 | 0.0001535 | 0.008941624 | SLC25A27 |
| chr17 | 41811839 | 41812167 | 329 | 7 | 0.000265344 | 0.000131 | 0.005717 | 0.0001537 | 0.000593618 | P3H4 |
| chr11 | 64562956 | 64563219 | 264 | 2 | 8.32E-05 | 0.000549 | 0.00019 | 0.0001538 | 0.004948243 | SLC22A11 |
| chr14 | 99674245 | 99674347 | 103 | 2 | 0.000449035 | 0.000495 | 0.000204 | 0.0001538 | 0.001831354 | HHIPL1 |
| chr6 | 32183569 | 32183954 | 386 | 6 | 2.08E-05 | 0.005326 | 6.16E-05 | 0.0001541 | 0.001224404 | AGER |
| chr17 | 21213604 | 21214765 | 1162 | 13 | 2.44E-05 | 0.007066 | 0.003867 | 0.0001543 | 0.001506621 | AC087294.1, TMEM11 |
| chr1 | 201507014 | 201507491 | 478 | 14 | 0.000314032 | 0.001061 | 0.003672 | 0.0001546 | 8.52E-06 | AC096677.1, CSRP1 |
| chr8 | 23458581 | 23458945 | 365 | 3 | 0.000207489 | 0.000148 | 0.002679 | 0.0001549 | 0.005376061 | AC104561.1 |
| chr8 | 81841442 | 81841545 | 104 | 3 | 0.000406281 | 0.000121 | 0.003146 | 0.0001549 | 0.006641402 | SNX16 |
| chr2 | 70009894 | 70009904 | 11 | 2 | 0.000364861 | 0.000116 | 0.001351 | 0.000155 | 0.003988654 | PCBP1-AS1 |
| chr20 | 11916667 | 11916916 | 250 | 4 | 5.39E-05 | 0.004996 | 5.49E-05 | 0.0001552 | 0.005650884 | BTBD3, AL035448.1 |
| chr15 | 59611769 | 59611810 | 42 | 2 | 0.000344279 | 9.51E-05 | 0.002877 | 0.0001553 | 0.008919968 | GCNT3 |
| chr5 | 157487533 | 157487611 | 79 | 2 | 0.000435317 | 0.007646 | 8.87E-05 | 0.0001555 | 0.005762042 | ADAM19 |
| chr19 | 18907821 | 18908097 | 277 | 3 | 0.000301992 | 0.01166 | 8.25E-05 | 0.0001556 | 0.003509053 | COPE, AC002985.1 |
| chr19 | 3204320 | 3204558 | 239 | 2 | 6.91E-05 | 0.00117 | 0.000135 | 0.0001557 | 0.00305678 | NCLN |
| chr21 | 45072719 | 45073254 | 536 | 2 | 2.88E-05 | 0.000168 | 0.00064 | 0.0001559 | 0.006850859 | SSR4P1 |
| chr1 | 145919723 | 145919761 | 39 | 2 | 0.000186939 | 9.20E-05 | 0.003591 | 0.0001559 | 0.015916699 | GNRHR2, RBM8A |
| chr10 | 87675099 | 87675640 | 542 | 3 | 0.000216778 | 0.031266 | 1.00E-04 | 0.0001559 | 0.011372517 | PAPSS2 |
| chr1 | 12510643 | 12510774 | 132 | 2 | 0.000285379 | 0.000102 | 0.00212 | 0.000156 | 0.013031675 | VPS13D |
| chr2 | 85612020 | 85612599 | 580 | 10 | 0.000135475 | 0.00132 | 0.001366 | 0.0001562 | 0.003022679 | USP39, C2orf68 |
| chr4 | 4541574 | 4542183 | 610 | 6 | 9.97E-05 | 8.80E-05 | 0.005797 | 0.0001563 | 0.000501213 | STX18-AS1, STX18 |
| chr1 | 241208752 | 241209256 | 505 | 6 | 0.000284644 | 0.004913 | 0.000135 | 0.0001565 | -0.00254705 | RGS7 |
| chr3 | 51983110 | 51983718 | 609 | 11 | 0.0001074 | 0.004222 | 0.001379 | 0.0001569 | 0.001536006 | ABHD14A-ACY1, ACY1, ABHD14B |
| chr1 | 201226085 | 201226208 | 124 | 2 | 0.000330609 | 0.000122 | 0.00122 | 0.000157 | 0.009431511 | IGFN1 |
| chr1 | 81803684 | 81803841 | 158 | 3 | 0.000231347 | 0.003558 | 0.001615 | 0.000157 | 0.004738581 | ADGRL2 |

| | | | | | | | | | | |
|---|---|---|---|---|---|---|---|---|---|---|
| chr11 | 60934446 | 60935133 | 688 | 3 | 6.06E-05 | 0.018808 | 4.91E-05 | 0.0001571 | 0.002222879 | TMEM132A |
| chr16 | 88607514 | 88607589 | 76 | 2 | 0.00045028 | 0.0001 | 0.002365 | 0.0001572 | 3.66E-05 | ZC3H18 |
| chr13 | 111343411 | 111343678 | 268 | 2 | 0.000190308 | 0.000184 | 0.000567 | 0.0001575 | 0.029472321 | TEX29 |
| chr1 | 25233054 | 25233252 | 199 | 3 | 7.59E-05 | 0.000579 | 0.001328 | 0.0001576 | -0.00200456 | AL031432.4 |
| chr10 | 124703542 | 124704065 | 524 | 6 | 0.000156614 | 0.004148 | 7.24E-05 | 0.0001579 | 0.003615353 | FAM53B-AS1, AC068896.1, FAM53B |
| chr10 | 97446437 | 97446545 | 109 | 2 | 8.54E-05 | 0.002354 | 0.000111 | 0.0001581 | 0.005763311 | ZDHHC16 |
| chr20 | 3663450 | 3663653 | 204 | 6 | 0.000308655 | 0.000167 | 0.00082 | 0.0001584 | 0.008477885 | NA |
| chr19 | 11346534 | 11347023 | 490 | 5 | 1.07E-05 | 0.237302 | 1.35E-05 | 0.0001584 | -0.00210605 | CCDC159 |
| chr6 | 31201166 | 31201437 | 272 | 3 | 0.00013063 | 0.00027 | 0.002857 | 0.0001585 | -0.00426812 | HCG27, AL662844.3 |
| chr8 | 120123775 | 120123901 | 127 | 2 | 0.000148843 | 0.007798 | 8.98E-05 | 0.0001587 | 0.006914415 | COL14A1 |
| chr19 | 2719891 | 2719992 | 102 | 2 | 0.000337862 | 9.37E-05 | 0.003555 | 0.0001587 | 0.016939224 | DIRAS1, AC006538.2 |
| chr13 | 49094017 | 49094241 | 225 | 2 | 0.00015997 | 0.018779 | 8.34E-05 | 0.0001592 | 0.003373623 | FNDC3A |
| chr12 | 50504628 | 50505235 | 608 | 11 | 0.000218824 | 0.003815 | 0.000823 | 0.0001592 | 0.000104611 | DIP2B |
| chr1 | 44775503 | 44776406 | 904 | 10 | 1.53E-05 | 0.000646 | 0.00309 | 0.0001593 | 0.002060372 | RPS8, SNORD55, AL592166.1 |
| chr11 | 69911383 | 69911389 | 7 | 2 | 0.000424741 | 9.37E-05 | 0.003626 | 0.0001593 | 0.008357727 | NA |
| chr6 | 158016934 | 158017387 | 454 | 7 | 0.000180224 | 0.001687 | 0.004673 | 0.0001596 | 0.00057229 | SYNJ2 |
| chr3 | 127029357 | 127029699 | 343 | 2 | 0.000276091 | 0.000489 | 0.000216 | 0.0001596 | -0.00480625 | PLXNA1 |
| chr2 | 240968030 | 240968226 | 197 | 2 | 8.95E-05 | 0.001639 | 0.000124 | 0.0001599 | 0.003524405 | CROCC2 |
| chr3 | 171900746 | 171900776 | 31 | 2 | 0.000354783 | 9.40E-05 | 0.003648 | 0.0001599 | 0.012680349 | TMEM212, TMEM212-AS1 |
| chr5 | 139709689 | 139710555 | 867 | 6 | 3.75E-05 | 0.001265 | 0.000606 | 0.0001599 | 0.003467293 | AC008667.1 |
| chr6 | 31649545 | 31649829 | 285 | 7 | 0.000170512 | 0.000901 | 0.000201 | 0.0001603 | 0.002996429 | BAG6 |
| chr19 | 55645064 | 55645176 | 113 | 2 | 0.00036296 | 9.78E-05 | 0.002956 | 0.0001605 | 0.004876915 | ZNF581, CCDC106 |
| chr4 | 2288160 | 2288456 | 297 | 2 | 0.000125177 | 0.000106 | 0.002024 | 0.0001605 | 0.003114886 | ZFYVE28 |
| chr11 | 70464431 | 70464878 | 448 | 2 | 0.000368529 | 9.76E-05 | 0.002998 | 0.0001605 | 0.009372273 | NA |
| chr1 | 235136516 | 235136625 | 110 | 2 | 0.000216003 | 0.000581 | 0.000195 | 0.0001606 | -0.00296957 | RBM34, ARID4B |
| chr5 | 171312849 | 171313543 | 695 | 4 | 2.09E-05 | 0.000999 | 0.000143 | 0.0001609 | 0.003231726 | NA |
| chr9 | 22009317 | 22009649 | 333 | 8 | 0.000180313 | 0.001887 | 0.000693 | 0.000161 | 0.002198883 | AL359922.1, CDKN2B-AS1 |
| chr10 | 69576485 | 69577017 | 533 | 4 | 1.80E-05 | 0.004081 | 0.00023 | 0.0001613 | 0.005100879 | AL450311.1 |
| chr19 | 9819548 | 9819817 | 270 | 4 | 0.000275129 | 0.006863 | 0.000238 | 0.0001614 | 0.002921568 | FBXL12 |
| chr22 | 33059429 | 33059521 | 93 | 2 | 4.42E-05 | 0.000117 | 0.001508 | 0.0001614 | 0.004520342 | NA |
| chr3 | 172315355 | 172315695 | 341 | 3 | 0.000291301 | 0.006047 | 6.57E-05 | 0.0001616 | 0.001664802 | FNDC3B |
| chr11 | 9758046 | 9758167 | 122 | 4 | 0.000187105 | 0.008305 | 0.000321 | 0.0001616 | -0.00115422 | LINC02709 |
| chr10 | 124091763 | 124092495 | 733 | 2 | 1.64E-05 | 0.163182 | 7.96E-05 | 0.0001618 | 0.003721325 | CHST15 |
| chr22 | 24431739 | 24432182 | 444 | 5 | 9.54E-05 | 0.062314 | 0.000519 | 0.000162 | 0.002056493 | SPECC1L-ADORA2A, ADORA2A, ADORA2A-AS1 |
| chr2 | 208124524 | 208124745 | 222 | 5 | 0.000246829 | 0.002244 | 0.001413 | 0.0001622 | 0.003981861 | CRYGD |
| chr14 | 76950856 | 76950900 | 45 | 2 | 0.000345116 | 0.010354 | 8.85E-05 | 0.0001623 | 0.006047326 | NA |
| chr19 | 4457268 | 4457983 | 716 | 7 | 9.83E-05 | 0.00199 | 0.007925 | 0.0001623 | -0.0005579 | UBXN6, AC011498.1 |
| chr20 | 50691509 | 50691811 | 303 | 7 | 0.000286214 | 0.001333 | 9.44E-05 | 0.0001624 | 0.003279293 | RIPOR3 |
| chr22 | 39499666 | 39499837 | 172 | 2 | 0.000354805 | 0.005584 | 9.57E-05 | 0.0001625 | 0.003615253 | MIEF1 |
| chr4 | 3574762 | 3574779 | 18 | 2 | 0.000409654 | 0.041372 | 8.16E-05 | 0.0001627 | 0.008037148 | NA |
| chr17 | 77467159 | 77467881 | 723 | 3 | 0.00028375 | 8.26E-05 | 0.004133 | 0.0001628 | -5.29E-05 | SEPTIN9 |
| chr10 | 131089289 | 131089455 | 167 | 2 | 0.000169458 | 0.029357 | 8.28E-05 | 0.000163 | 0.008014461 | NA |
| chr16 | 85555743 | 85556104 | 362 | 4 | 8.17E-05 | 0.003264 | 0.000423 | 0.0001631 | 0.001714965 | GSE1 |
| chr6 | 169702793 | 169702962 | 170 | 3 | 0.000262519 | 0.002908 | 0.000101 | 0.0001631 | -0.00051147 | C6orf120 |
| chr12 | 52211648 | 52212179 | 532 | 3 | 0.000255983 | 0.000148 | 0.001129 | 0.0001632 | 0.005690907 | LINC00592, LINC02874 |
| chr1 | 938346 | 938435 | 90 | 2 | 0.000302067 | 0.000256 | 0.000398 | 0.0001632 | 0.008507035 | SAMD11 |
| chr15 | 74812284 | 74812945 | 662 | 8 | 9.97E-05 | 0.010994 | 0.000431 | 0.0001633 | 0.002111952 | LMAN1L |
| chr17 | 82457544 | 82457942 | 399 | 4 | 0.000131796 | 0.006525 | 0.001438 | 0.0001636 | 0.002037647 | AC132938.3 |
| chr21 | 34822577 | 34822617 | 41 | 2 | 0.000376629 | 0.069618 | 8.09E-05 | 0.0001636 | 0.002578199 | RUNX1 |
| chr2 | 135742436 | 135742564 | 129 | 2 | 0.000122869 | 0.007002 | 9.33E-05 | 0.0001637 | 0.002295008 | UBXN4 |
| chr21 | 40508607 | 40508626 | 20 | 2 | 0.000138169 | 0.024409 | 8.38E-05 | 0.0001639 | 0.011650909 | DSCAM |
| chr1 | 197201761 | 197202192 | 432 | 2 | 1.40E-05 | 0.000127 | 0.001278 | 0.000164 | 0.009653736 | NA |
| chr6 | 85448173 | 85448640 | 468 | 3 | 5.52E-05 | 0.327542 | 4.74E-05 | 0.0001642 | 0.001659782 | NA |
| chr19 | 40799232 | 40799518 | 287 | 2 | 0.000162642 | 0.000105 | 0.002364 | 0.0001642 | 0.001383814 | RAB4B-EGLN2, EGLN2 |
| chr15 | 82806460 | 82806899 | 440 | 2 | 0.000245645 | 9.87E-05 | 0.003196 | 0.0001642 | 0.007097957 | NA |
| chr11 | 64015795 | 64015887 | 93 | 2 | 0.000221296 | 0.000584 | 0.000201 | 0.0001648 | 0.005866848 | MACROD1 |
| chr19 | 18203144 | 18203293 | 150 | 2 | 0.000242842 | 9.66E-05 | 0.003708 | 0.0001649 | 0.000574602 | RAB3A |
| chr4 | 128061237 | 128061307 | 71 | 4 | 0.000470492 | 0.000145 | 0.001221 | 0.0001651 | 0.002187579 | LARP1B |
| chr1 | 24186983 | 24187454 | 472 | 9 | 0.000239699 | 0.000183 | 0.001954 | 0.0001651 | 0.001914533 | IFNLR1 |
| chr9 | 135906757 | 135907150 | 394 | 2 | 0.000333144 | 0.0001 | 0.003013 | 0.0001652 | -0.00269872 | CAMSAP1 |
| chr20 | 35269126 | 35269374 | 249 | 3 | 0.000251565 | 0.001826 | 0.000138 | 0.0001657 | 0.00553658 | MMP24, MMP24OS, AL121753.1 |
| chr17 | 3602285 | 3602785 | 501 | 4 | 0.000359046 | 0.00032 | 0.002367 | 0.0001657 | 0.00913862 | TRPV1, AC027796.3 |
| chr11 | 6610055 | 6610309 | 255 | 4 | 0.000153905 | 0.001871 | 0.000129 | 0.0001678 | 0.002423338 | ILK, TAF10, AC091564.3 |
| chr12 | 93441922 | 93442279 | 358 | 5 | 0.000225858 | 0.000877 | 0.010317 | 0.0001678 | -0.00016367 | UBE2N |
| chr16 | 67944229 | 67944313 | 85 | 5 | 0.000473078 | 0.00024 | 0.002045 | 0.000168 | 0.004139389 | SLC12A4 |
| chr14 | 22497359 | 22497648 | 290 | 2 | 0.000143163 | 0.001119 | 0.000147 | 0.0001681 | -0.00740336 | NA |
| chr2 | 239349522 | 239349581 | 60 | 2 | 0.000410246 | 0.000161 | 0.000816 | 0.0001681 | 0.003683974 | HDAC4 |
| chr17 | 79022345 | 79023367 | 1023 | 9 | 6.91E-05 | 0.00133 | 0.000721 | 0.0001682 | 0.002534861 | C1QTNF1, C1QTNF1-AS1 |
| chr21 | 41465898 | 41466232 | 335 | 2 | 9.27E-05 | 0.00755 | 9.41E-05 | 0.0001684 | -0.0144291 | TMPRSS2 |
| chr20 | 58979738 | 58980068 | 331 | 4 | 0.000153506 | 0.007029 | 7.66E-05 | 0.0001684 | 0.000802676 | AL109840.2 |
| chr1 | 93579646 | 93579788 | 143 | 2 | 0.000303251 | 0.000112 | 0.002041 | 0.0001685 | 0.006837742 | BCAR3 |
| chr11 | 77103778 | 77104012 | 235 | 2 | 1.68E-05 | 0.008736 | 9.27E-05 | 0.0001685 | 0.002557831 | CAPN5 |
| chr22 | 50736936 | 50737087 | 152 | 2 | 0.00032214 | 0.000102 | 0.003049 | 0.0001687 | 5.53E-05 | AC000036.1 |
| chr2 | 223944338 | 223944357 | 20 | 2 | 0.00034197 | 0.000118 | 0.001715 | 0.0001687 | 0.007424632 | AC093884.1, WDFY1 |
| chr16 | 29746640 | 29747039 | 400 | 3 | 0.000144139 | 0.000204 | 0.001504 | 0.0001691 | 0.005670795 | AC009133.3 |
| chr12 | 54658732 | 54658915 | 184 | 2 | 0.000413872 | 9.99E-05 | 0.003472 | 0.0001691 | 0.018838855 | NA |
| chr1 | 206685409 | 206685722 | 314 | 4 | 4.57E-05 | 0.000902 | 0.002465 | 0.0001694 | 0.005099287 | MAPKAPK2 |
| chr19 | 41997246 | 41997311 | 66 | 2 | 0.000437683 | 0.007323 | 9.51E-05 | 0.0001696 | 0.002809109 | ATP1A3 |
| chr20 | 2749842 | 2750530 | 689 | 3 | 0.000224436 | 0.000166 | 0.001577 | 0.0001697 | 0.019652519 | EBF4 |
| chr19 | 2819643 | 2819810 | 168 | 4 | 0.000108383 | 0.00017 | 0.001339 | 0.00017 | 0.0051742 | NA |
| chr22 | 39292152 | 39292554 | 403 | 3 | 0.00011596 | 0.000147 | 0.004099 | 0.00017 | 0.007164048 | NA |
| chr2 | 101419490 | 101419653 | 164 | 2 | 0.000221206 | 0.004553 | 0.000102 | 0.00017 | -0.00150274 | RFX8 |
| chr2 | 219513100 | 219513989 | 890 | 4 | 0.000245675 | 0.000709 | 0.00024 | 0.00017 | 0.002198232 | ASIC4-AS1 |
| chr6 | 139375634 | 139376174 | 541 | 8 | 0.000137503 | 0.003271 | 0.000461 | 0.0001704 | 0.000930656 | AL592429.2 |
| chr1 | 185301307 | 185301765 | 459 | 3 | 6.04E-05 | 0.027974 | 4.70E-05 | 0.0001706 | 0.003340616 | IVNS1ABP |
| chr6 | 35140284 | 35140337 | 54 | 2 | 0.000215063 | 0.000109 | 0.002336 | 0.0001707 | 0.013180908 | TCP11 |
| chr8 | 11060642 | 11061062 | 421 | 2 | 0.000279583 | 0.000102 | 0.00319 | 0.0001707 | 0.006616155 | XKR6 |
| chr2 | 232014263 | 232014332 | 70 | 2 | 0.000116236 | 0.000102 | 0.003203 | 0.0001708 | 0.009691184 | DIS3L2, AC019130.1 |
| chr6 | 70183652 | 70183692 | 41 | 2 | 0.000450127 | 0.000372 | 0.000296 | 0.0001711 | 0.012026522 | COL19A1 |
| chr3 | 105867840 | 105868090 | 251 | 2 | 0.000195035 | 0.0001 | 0.003575 | 0.0001711 | -0.0034649 | CBLB |
| chr20 | 37384193 | 37384543 | 351 | 3 | 7.40E-05 | 0.000101 | 0.005547 | 0.0001711 | -0.01041924 | SRC |
| chr5 | 124736129 | 124736702 | 574 | 3 | 6.16E-05 | 0.0005 | 0.000304 | 0.0001715 | 0.005499057 | ZNF608 |
| chr2 | 240856766 | 240857136 | 371 | 2 | 0.000221865 | 0.000239 | 0.000475 | 0.0001716 | 0.003653099 | NA |
| chr21 | 33071939 | 33072278 | 340 | 4 | 2.37E-05 | 0.001406 | 0.000143 | 0.0001722 | 0.008124909 | OLIG1 |
| chr5 | 958693 | 958854 | 162 | 4 | 0.000481306 | 0.013407 | 9.61E-05 | 0.0001727 | 0.005468264 | NA |
| chr10 | 128030456 | 128030732 | 277 | 4 | 0.000447414 | 0.000319 | 0.005992 | 0.0001736 | 0.000681055 | PTPRE |
| chr6 | 155089857 | 155089943 | 87 | 2 | 0.000482006 | 0.000409 | 0.00028 | 0.0001737 | 0.004178845 | TIAM2 |
| chr22 | 28883215 | 28883609 | 395 | 3 | 0.000253801 | 0.00039 | 0.000826 | 0.0001742 | 0.000104919 | ZNRF3 |
| chr16 | 2172089 | 2172612 | 524 | 3 | 1.99E-05 | 0.000426 | 0.001565 | 0.0001749 | -0.00246781 | TRAF7 |
| chr9 | 34400749 | 34401184 | 436 | 3 | 0.000213395 | 0.000181 | 0.001045 | 0.000175 | 0.00366765 | FAM219A |
| chr7 | 97872105 | 97872776 | 672 | 10 | 0.00020969 | 0.012079 | 0.003086 | 0.000175 | 0.00221604 | ASNS, AC079781.5 |
| chr16 | 67278405 | 67278670 | 266 | 3 | 4.85E-05 | 7.21E-05 | 0.009335 | 0.0001752 | 0.009079372 | PLEKHG4 |
| chr17 | 42577774 | 42578549 | 776 | 12 | 0.000162017 | 0.000782 | 0.000591 | 0.0001756 | 4.58E-05 | AC067852.5, PSMC3IP |
| chr7 | 75073293 | 75074062 | 770 | 10 | 0.000313046 | 0.003352 | 0.002668 | 0.000176 | 0.001395862 | RCC1L |
| chr2 | 48315196 | 48315535 | 340 | 2 | 5.25E-05 | 0.000356 | 0.000323 | 0.0001761 | 0.002197164 | FOXN2 |
| chr12 | 48999625 | 49000052 | 428 | 3 | 1.24E-05 | 0.000905 | 0.001481 | 0.0001761 | 0.001928476 | DDN-AS1 |
| chr19 | 1629452 | 1629620 | 169 | 2 | 0.000267296 | 0.001742 | 0.000133 | 0.0001764 | 0.00362022 | TCF3 |
| chr16 | 304263 | 304374 | 112 | 3 | 0.000465861 | 0.093497 | 4.65E-05 | 0.0001764 | 0.002007368 | AXIN1 |
| chr15 | 34582916 | 34582937 | 22 | 2 | 0.000449296 | 0.000606 | 0.000215 | 0.0001765 | 0.002691693 | GOLGA8B |
| chr18 | 76442149 | 76442304 | 156 | 2 | 0.000465235 | 0.000137 | 0.001323 | 0.0001765 | 0.003799602 | ZNF516 |
| chr2 | 179264885 | 179265354 | 470 | 3 | 7.41E-05 | 0.003064 | 0.000104 | 0.0001766 | 0.004367582 | NA |
| chr4 | 1669773 | 1670242 | 470 | 4 | 0.000357488 | 0.000452 | 0.001088 | 0.0001766 | 0.001062821 | FAM53A |
| chr19 | 49127785 | 49127926 | 142 | 3 | 0.000218135 | 0.012057 | 0.000157 | 0.0001768 | 0.009539397 | PPFIA3 |
| chr1 | 184386085 | 184387421 | 1337 | 10 | 0.000282998 | 0.000555 | 0.010337 | 0.000177 | -0.0006234 | C1orf21, AL445228.2 |
| chr8 | 20304348 | 20304534 | 187 | 3 | 0.00038643 | 0.000187 | 0.001169 | 0.0001771 | 0.012109051 | NA |
| chr8 | 1475186 | 1475490 | 305 | 3 | 0.000235186 | 0.000149 | 0.001461 | 0.0001771 | -0.00326112 | DLGAP2 |
| chr17 | 37066105 | 37066164 | 60 | 3 | 0.000171167 | 0.000431 | 0.000305 | 0.0001772 | 0.011340923 | NA |
| chr1 | 229508024 | 229508530 | 507 | 10 | 0.000236166 | 0.000255 | 0.013001 | 0.0001772 | -0.00020361 | AL121990.1, NUP133 |
| chr15 | 101489745 | 101490012 | 268 | 3 | 0.000183096 | 0.002554 | 0.000772 | 0.0001772 | -3.03E-05 | PCSK6 |
| chr7 | 691617 | 691912 | 296 | 3 | 0.000218021 | 0.000127 | 0.001917 | 0.0001774 | 0.003288848 | PRKAR1B |
| chr5 | 37839305 | 37840056 | 752 | 11 | 0.000191961 | 0.009085 | 0.00044 | 0.0001774 | -0.00066714 | GDNF-AS1, GDNF |
| chr1 | 220887509 | 220887728 | 220 | 3 | 0.000254402 | 9.69E-05 | 0.003532 | 0.0001774 | 0.008431075 | NA |
| chr13 | 106376537 | 106377095 | 559 | 8 | 0.000191549 | 0.003298 | 0.00095 | 0.0001775 | 0.004060524 | LINC00460 |
| chr16 | 29808040 | 29808251 | 212 | 5 | 0.000158572 | 9.62E-05 | 0.013853 | 0.0001775 | -0.00093899 | MAZ |
| chr19 | 37078334 | 37078638 | 305 | 9 | 0.000381578 | 0.001916 | 0.001589 | 0.000178 | 0.000445999 | ZNF420, AC010632.1 |
| chr1 | 55217664 | 55217700 | 37 | 3 | 0.000312701 | 7.65E-05 | 0.008729 | 0.000178 | -0.00389775 | MIR4422HG |
| chr1 | 39154894 | 39154953 | 60 | 2 | 0.000375448 | 0.000175 | 0.000813 | 0.000178 | 0.015946157 | MACF1 |
| chr19 | 1409548 | 1410044 | 497 | 2 | 0.000218287 | 0.000105 | 0.003541 | 0.0001784 | 0.010297981 | DAZAP1 |
| chr5 | 91383571 | 91383726 | 156 | 4 | 0.000256807 | 0.00057 | 0.001544 | 0.0001786 | 0.003181373 | ARRDC3-AS1 |
| chr11 | 13870733 | 13870864 | 132 | 2 | 0.000157307 | 0.001273 | 0.000149 | 0.0001786 | -0.0038487 | AC027779.1 |
| chr6 | 36027652 | 36028594 | 943 | 11 | 0.00013212 | 0.000222 | 0.003881 | 0.0001789 | 0.000322394 | MAPK14 |
| chr16 | 48232368 | 48232394 | 27 | 2 | 0.000497282 | 0.000125 | 0.001769 | 0.0001793 | 0.003876542 | ABCC11 |
| chr7 | 1348944 | 1349081 | 138 | 2 | 0.00030851 | 0.005004 | 0.000105 | 0.0001793 | 0.006479797 | NA |
| chr15 | 51622549 | 51623216 | 668 | 9 | 0.00031472 | 0.000168 | 0.011254 | 0.0001793 | 8.21E-05 | DMXL2 |
| chr6 | 31587052 | 31587239 | 188 | 4 | 0.000376629 | 0.006018 | 7.05E-05 | 0.0001793 | 0.003659113 | LST1 |
| chr6 | 170178766 | 170179251 | 486 | 5 | 0.000355255 | 0.006244 | 0.00027 | 0.0001795 | 0.002796906 | AL596442.2 |
| chr3 | 46812683 | 46812821 | 139 | 5 | 0.00030356 | 0.035434 | 4.70E-05 | 0.0001802 | -0.0004445 | NA |
| chr4 | 1305354 | 1305726 | 373 | 4 | 9.62E-05 | 0.007918 | 8.95E-05 | 0.0001805 | 0.001767194 | MAEA |
| chr2 | 241844372 | 241845091 | 720 | 3 | 3.85E-05 | 0.000925 | 0.000165 | 0.0001811 | -0.00153432 | AC131097.3 |
| chr4 | 2274074 | 2274317 | 244 | 5 | 0.000215514 | 0.014251 | 0.000157 | 0.0001811 | 0.002410764 | ZFYVE28 |
| chr11 | 67523776 | 67524060 | 285 | 5 | 0.000134948 | 0.00162 | 0.004897 | 0.0001812 | 0.004959757 | CABP2 |
| chr12 | 57219390 | 57219710 | 321 | 3 | 0.000372914 | 0.000111 | 0.005294 | 0.0001812 | 0.01354364 | NXPH4 |
| chr1 | 202138627 | 202138748 | 122 | 2 | 0.000447734 | 0.000116 | 0.002411 | 0.0001814 | 0.007152198 | ARL8A |
| chr16 | 568794 | 569455 | 662 | 4 | 6.23E-05 | 0.000181 | 0.002428 | 0.0001818 | 0.002609699 | PIGQ, NHLRC4, Z98883.1 |
| chr6 | 75312731 | 75312840 | 110 | 3 | 0.000303798 | 0.000271 | 0.000583 | 0.0001824 | 0.005091473 | TMEM30A-DT, FILIP1 |
| chr12 | 93738068 | 93738181 | 114 | 2 | 0.000434139 | 0.000454 | 0.000277 | 0.0001824 | 0.01017861 | CRADD |
| chr17 | 58488647 | 58488824 | 178 | 2 | 9.98E-05 | 0.000646 | 0.000216 | 0.0001824 | 0.002310779 | NA |
| chr9 | 95517674 | 95518071 | 398 | 3 | 0.000199536 | 0.000117 | 0.003746 | 0.0001825 | 0.011709379 | NA |
| chr11 | 67303046 | 67303345 | 300 | 6 | 0.000346971 | 0.001933 | 0.00113 | 0.0001826 | 0.001363043 | NA |
| chr8 | 22603883 | 22604062 | 180 | 3 | 6.78E-05 | 0.002835 | 0.001039 | 0.0001827 | 0.001873701 | C8orf58 |
| chr11 | 66882870 | 66883044 | 175 | 2 | 0.00033498 | 0.00046 | 0.000276 | 0.0001833 | 0.012208303 | PC |
| chr2 | 86195072 | 86195456 | 385 | 4 | 0.000271294 | 0.003813 | 0.002259 | 0.0001838 | -0.00079591 | IMMT |
| chr14 | 20802765 | 20803332 | 568 | 11 | 0.000108514 | 0.00242 | 0.002149 | 0.0001842 | -8.26E-05 | RNASE1 |
| chr10 | 14554406 | 14554647 | 242 | 2 | 0.000151297 | 0.024449 | 9.02E-05 | 0.0001842 | 0.008612203 | FAM107B |
| chr21 | 31736364 | 31736999 | 636 | 2 | 1.56E-05 | 0.001124 | 0.000162 | 0.0001843 | 0.004105278 | AP000255.1 |
| chr15 | 90084811 | 90084979 | 169 | 3 | 0.000249526 | 0.00039 | 0.000463 | 0.0001844 | 0.00198943 | IDH2 |
| chr11 | 117842179 | 117842236 | 58 | 2 | 0.000210024 | 0.000781 | 0.000196 | 0.0001846 | 0.00898622 | FXYD6-FXYD2, FXYD6 |
| chr1 | 37478494 | 37478656 | 163 | 2 | 0.000170053 | 0.000191 | 0.000762 | 0.0001847 | 0.012506824 | ZC3H12A |
| chr12 | 81077148 | 81077415 | 268 | 3 | 6.93E-05 | 0.000268 | 0.001119 | 0.0001851 | 0.01070248 | ACSS3 |
| chr11 | 111540153 | 111540491 | 339 | 5 | 4.76E-05 | 0.00348 | 0.000282 | 0.0001857 | -0.00198288 | LAYN |
| chr1 | 24503497 | 24503564 | 68 | 2 | 0.000242438 | 0.001065 | 0.000167 | 0.0001857 | 0.004196553 | RCAN3, RCAN3AS |
| chr8 | 30657654 | 30657847 | 194 | 3 | 0.000261395 | 0.000168 | 0.001136 | 0.0001862 | 0.001359731 | GTF2E2 |

| | | | | | | | | | | |
|---|---|---|---|---|---|---|---|---|---|---|
| chr12 | 56221913 | 56222589 | 677 | 13 | 0.00035151 | 0.001759 | 0.005501 | 0.0001868 | -0.00013241 | NABP2, RNF41 |
| chr15 | 80695354 | 80695456 | 103 | 3 | 0.000391025 | 0.000137 | 0.001655 | 0.0001868 | 0.000808755 | ABHD17C |
| chr12 | 58086936 | 58087074 | 139 | 2 | 0.000118739 | 0.097355 | 8.78E-05 | 0.000188 | 0.008647089 | NA |
| chr12 | 107773953 | 107774785 | 833 | 8 | 0.000317306 | 0.000159 | 0.009105 | 0.0001881 | 0.000179071 | ASCL4 |
| chr16 | 73067170 | 73067365 | 196 | 2 | 0.000366829 | 0.000118 | 0.002668 | 0.0001883 | 0.01483941 | ZFHX3 |
| chr14 | 101819939 | 101820436 | 498 | 3 | 0.000252887 | 0.000195 | 0.003033 | 0.0001885 | 0.000516351 | PPP2R5C |
| chr7 | 100819782 | 100820170 | 389 | 2 | 0.000252741 | 0.000205 | 0.000709 | 0.0001885 | 0.003446035 | EPHB4 |
| chr17 | 6070046 | 6070225 | 180 | 3 | 1.42E-05 | 0.000247 | 0.001553 | 0.0001887 | 0.003054155 | AC007846.2, WSCD1 |
| chr21 | 41810179 | 41810650 | 472 | 7 | 0.000331709 | 0.001166 | 0.001331 | 0.0001893 | 0.001009284 | PRDM15 |
| chr11 | 77073151 | 77073370 | 220 | 2 | 0.000387593 | 0.000479 | 0.00028 | 0.0001893 | 0.006228793 | CAPN5 |
| chr1 | 8213292 | 8213800 | 509 | 2 | 3.64E-05 | 0.000114 | 0.003188 | 0.0001894 | 0.011845167 | LINC01714 |
| chr6 | 168090938 | 168090969 | 32 | 2 | 0.000252692 | 0.000144 | 0.001431 | 0.0001896 | 0.008838412 | FRMD1 |
| chr10 | 23171316 | 23171717 | 402 | 4 | 0.000135853 | 0.064406 | 0.00134 | 0.00019 | 0.009616793 | NA |
| chr1 | 119711554 | 119711599 | 46 | 2 | 0.000417755 | 0.000206 | 0.000719 | 0.00019 | 0.009121609 | PHGDH |
| chr4 | 105894932 | 105895226 | 295 | 4 | 0.000183954 | 0.00043 | 0.001913 | 0.0001903 | 0.006899138 | NPNT, INTS12 |
| chr20 | 44744429 | 44744465 | 37 | 3 | 0.000274997 | 0.00541 | 7.68E-05 | 0.0001905 | 0.001983934 | KCNK15-AS1 |
| chr9 | 128732813 | 128733028 | 216 | 9 | 0.000262519 | 0.00019 | 0.01894 | 0.0001907 | 0.002900983 | AL441992.1, ZER1 |
| chr1 | 201190371 | 201190860 | 490 | 5 | 4.98E-05 | 0.008835 | 0.000883 | 0.0001911 | 0.005641068 | IGFN1 |
| chr12 | 1445804 | 1445994 | 191 | 2 | 0.000352935 | 0.00693 | 0.000105 | 0.0001912 | 0.002294565 | ERC1 |
| chr19 | 14474397 | 14475037 | 641 | 3 | 0.000285148 | 0.000177 | 0.001537 | 0.0001916 | 0.009764007 | PTGER1 |
| chr8 | 38468816 | 38469436 | 621 | 9 | 0.000172617 | 0.001872 | 0.000386 | 0.0001917 | -0.00040973 | FGFR1 |
| chr2 | 112977800 | 112977874 | 75 | 4 | 0.000467828 | 0.000163 | 0.002399 | 0.0001917 | 0.004660643 | IL36G |
| chr2 | 159617389 | 159617836 | 448 | 2 | 9.53E-06 | 0.010088 | 9.99E-05 | 0.0001917 | 0.013240143 | NA |
| chr4 | 28402837 | 28402841 | 5 | 2 | 0.000500646 | 0.0038 | 0.000117 | 0.000192 | -0.00304098 | AC093791.1 |
| chr4 | 1767832 | 1768004 | 173 | 2 | 0.000230172 | 0.000113 | 0.00362 | 0.0001931 | 0.005881227 | NA |
| chr1 | 228486022 | 228486026 | 5 | 2 | 7.48E-05 | 0.02507 | 9.31E-05 | 0.0001933 | -0.00557679 | NA |
| chr1 | 35079413 | 35079751 | 339 | 6 | 0.000392933 | 0.00156 | 0.003188 | 0.0001935 | 0.000129966 | ZMYM1 |
| chr17 | 41492607 | 41492653 | 47 | 2 | 0.000458119 | 0.00019 | 0.000851 | 0.0001935 | 0.01204711 | NA |
| chr17 | 76915858 | 76916266 | 409 | 2 | 8.42E-05 | 0.001619 | 0.000148 | 0.0001941 | 0.004094735 | MGAT5B |
| chr1 | 18351792 | 18351881 | 90 | 2 | 0.0002431 | 0.007155 | 0.000105 | 0.0001944 | 0.013241619 | IGSF21 |
| chr18 | 79411705 | 79412117 | 413 | 3 | 0.000103856 | 0.004586 | 0.000168 | 0.0001948 | 0.005422471 | NFATC1 |
| chr6 | 32129643 | 32130392 | 750 | 20 | 0.000181054 | 0.001964 | 0.010345 | 0.0001957 | 0.001152129 | FKBPL |
| chr1 | 11139612 | 11139648 | 37 | 2 | 0.000450267 | 0.012056 | 9.94E-05 | 0.0001957 | 0.001935832 | MTOR |
| chr1 | 31616976 | 31617411 | 436 | 3 | 3.67E-05 | 0.000454 | 0.003016 | 0.0001968 | 0.007580503 | NA |
| chr1 | 223698374 | 223698594 | 221 | 2 | 0.000238325 | 0.000312 | 0.000447 | 0.0001971 | 0.006772864 | NA |
| chr11 | 126940199 | 126940504 | 306 | 2 | 0.000431544 | 0.000115 | 0.003811 | 0.0001974 | -0.00082702 | KIRREL3, AP002833.1 |
| chr12 | 118062561 | 118062634 | 74 | 2 | 0.000145666 | 0.000153 | 0.001413 | 0.0001977 | 0.009455497 | NA |
| chr1 | 197912611 | 197912873 | 263 | 3 | 0.000373803 | 0.007711 | 0.000291 | 0.0001982 | 0.004315589 | LHX9 |
| chr7 | 105531956 | 105532527 | 572 | 6 | 7.81E-05 | 0.146511 | 1.15E-05 | 0.0001985 | -0.0006925 | RINT1 |
| chr1 | 202012246 | 202012446 | 201 | 2 | 0.000161425 | 0.001067 | 0.000181 | 0.0001986 | 0.005796545 | ELF3, AL691482.3 |
| chr17 | 48627048 | 48627278 | 231 | 3 | 0.000323314 | 0.015498 | 6.09E-05 | 0.0001988 | 0.019346437 | HOXB7 |
| chr9 | 92679932 | 92680445 | 514 | 3 | 0.000102906 | 0.007237 | 7.48E-05 | 0.0001992 | 0.004122713 | NA |
| chr11 | 65224907 | 65225116 | 210 | 2 | 0.000260927 | 0.000205 | 0.000802 | 0.0001992 | 0.008310131 | SLC22A20P |
| chr1 | 153358282 | 153358300 | 19 | 2 | 6.72E-05 | 0.004123 | 0.000119 | 0.0001994 | 0.010895259 | S100A9 |
| chr9 | 109197097 | 109197295 | 199 | 2 | 0.000196341 | 0.005748 | 0.000111 | 0.0001994 | -0.00596937 | EPB41L4B |
| chr3 | 186571426 | 186571511 | 86 | 2 | 0.000147708 | 0.001628 | 0.000152 | 0.0002005 | 0.006450596 | DNAJB11 |
| chr7 | 128862319 | 128862597 | 279 | 5 | 0.000146277 | 0.000156 | 0.004731 | 0.0002006 | 0.00089724 | FLNC-AS1, KCP |
| chr20 | 6767237 | 6767474 | 238 | 3 | 0.000337966 | 0.003656 | 0.002373 | 0.0002014 | 0.005813765 | NA |
| chr2 | 74472455 | 74473119 | 665 | 7 | 0.000141038 | 0.002445 | 0.0072 | 0.0002017 | 0.003384437 | AC005041.5, MRPL53, CCDC142 |
| chr6 | 8102638 | 8103124 | 487 | 10 | 0.000153818 | 0.000116 | 0.012179 | 0.0002018 | 0.003641747 | AL355499.1 |
| chr3 | 195872234 | 195872946 | 713 | 4 | 0.000101793 | 0.0006 | 0.000917 | 0.0002023 | 0.003411273 | TNK2 |
| chr17 | 58331330 | 58331667 | 338 | 5 | 0.00023832 | 0.001996 | 0.000608 | 0.0002023 | 0.00091653 | TSPOAP1-AS1, AC004687.1 |
| chr7 | 90708753 | 90708776 | 24 | 2 | 0.000381938 | 0.000434 | 0.000338 | 0.0002026 | 0.01154539 | CDK14 |
| chr17 | 73323858 | 73324398 | 541 | 3 | 9.03E-06 | 0.003425 | 9.61E-05 | 0.0002031 | 0.004505452 | NA |
| chr2 | 25377582 | 25377883 | 302 | 2 | 0.000224395 | 0.000397 | 0.000371 | 0.000204 | 0.008176497 | DTNB |
| chr1 | 28232993 | 28233047 | 55 | 4 | 0.000485187 | 0.000644 | 0.001303 | 0.000204 | -0.00257309 | DNAJC8 |
| chr15 | 78092526 | 78092756 | 231 | 3 | 0.000466404 | 8.17E-05 | 0.009784 | 0.0002041 | 0.005170643 | SH2D7 |
| chr20 | 63101115 | 63101256 | 142 | 3 | 0.00047204 | 0.00085 | 0.00276 | 0.0002042 | 0.018448759 | HAR1B |
| chr1 | 109620536 | 109620908 | 373 | 8 | 5.66E-05 | 0.011812 | 0.000824 | 0.0002044 | 0.002260104 | AMPD2 |
| chr3 | 48429736 | 48430166 | 431 | 5 | 5.20E-05 | 0.000628 | 0.000756 | 0.0002049 | 0.001929525 | PLXNB1 |
| chr20 | 2694772 | 2695350 | 579 | 2 | 0.00027632 | 0.00018 | 0.001098 | 0.0002058 | 0.022088494 | EBF4 |
| chr14 | 88321999 | 88322587 | 589 | 7 | 3.67E-05 | 0.00923 | 0.00068 | 0.000206 | 0.008137773 | KCNK10 |
| chr2 | 191152660 | 191153014 | 355 | 2 | 0.000108791 | 0.007189 | 0.00011 | 0.0002062 | 0.018186986 | NA |
| chr19 | 8596197 | 8596380 | 184 | 2 | 0.000369713 | 0.000119 | 0.004125 | 0.0002066 | 0.00623695 | ADAMTS10 |
| chr13 | 30802962 | 30803487 | 526 | 6 | 7.10E-05 | 0.001323 | 0.000319 | 0.0002068 | 0.002553898 | LINC00398 |
| chr3 | 191329077 | 191329208 | 132 | 3 | 0.000342356 | 0.000221 | 0.002253 | 0.000207 | 0.004431941 | CCDC50, UTS2B |
| chr17 | 48944337 | 48944877 | 541 | 8 | 4.23E-05 | 0.012106 | 0.000841 | 0.0002073 | 0.001252113 | SNF8 |
| chr8 | 23224907 | 23225840 | 934 | 12 | 2.15E-05 | 0.015477 | 3.69E-05 | 0.0002081 | 0.001506238 | AC100861.1, TNFRSF10A, AC100861.2 |
| chr19 | 1103443 | 1103807 | 365 | 5 | 0.000268522 | 0.002941 | 0.002217 | 0.0002086 | 0.000695681 | NA |
| chr13 | 30847913 | 30847986 | 74 | 2 | 0.000351049 | 0.000171 | 0.001269 | 0.000209 | 0.013588151 | NA |
| chr9 | 136943207 | 136943305 | 99 | 2 | 0.000114051 | 0.000448 | 0.000345 | 0.0002093 | 0.004857461 | FBXW5 |
| chr20 | 58902501 | 58902714 | 214 | 2 | 0.000147316 | 0.000134 | 0.002507 | 0.0002093 | 0.003683685 | GNAS |
| chr15 | 93088349 | 93089501 | 1153 | 12 | 6.58E-05 | 0.002429 | 0.000915 | 0.0002095 | 0.000125967 | RGMA |
| chr9 | 127938163 | 127939233 | 1071 | 9 | 2.03E-05 | 0.030247 | 0.000336 | 0.0002102 | -0.00298137 | AL157935.1 |
| chr4 | 4865375 | 4866112 | 738 | 7 | 9.49E-05 | 0.007243 | 5.74E-05 | 0.0002102 | 0.003292299 | NA |
| chr13 | 57633516 | 57633725 | 210 | 2 | 0.000216339 | 0.000263 | 0.000622 | 0.0002102 | 0.007978871 | PCDH17, AL445288.1 |
| chr18 | 62569291 | 62569865 | 575 | 2 | 4.77E-05 | 0.002564 | 0.000139 | 0.0002102 | 0.010170225 | ZCCHC2 |
| chr13 | 113296073 | 113296397 | 325 | 3 | 7.27E-05 | 0.257723 | 4.24E-05 | 0.0002103 | 0.002253314 | NA |
| chr21 | 44558864 | 44559452 | 589 | 4 | 0.000138667 | 0.000158 | 0.002377 | 0.0002107 | 0.007613975 | TSPEAR |
| chr15 | 59994185 | 59994341 | 157 | 3 | 8.10E-05 | 0.000304 | 0.002545 | 0.000211 | 0.007724462 | NA |
| chr9 | 132546408 | 132546464 | 57 | 2 | 0.000229535 | 0.014406 | 0.000103 | 0.000211 | -0.00158455 | CFAP77 |
| chr2 | 24123345 | 24123397 | 53 | 5 | 0.000117886 | 0.022566 | 0.000685 | 0.0002113 | 0.001628308 | FAM228B, PFN4 |
| chr6 | 35319067 | 35319656 | 590 | 2 | 1.12E-05 | 0.001214 | 0.000183 | 0.0002117 | 0.001671904 | DEF6 |
| chr6 | 30210161 | 30210947 | 787 | 6 | 4.91E-05 | 0.027523 | 9.58E-05 | 0.0002119 | -0.00017433 | TRIM26 |
| chr2 | 26786073 | 26787031 | 959 | 5 | 1.94E-05 | 0.000162 | 0.00313 | 0.0002123 | 0.006015423 | CENPA |
| chr5 | 10308930 | 10309011 | 82 | 3 | 0.000326772 | 0.005022 | 8.63E-05 | 0.0002125 | -0.00100032 | NA |
| chr19 | 6271949 | 6272147 | 199 | 2 | 0.000126456 | 0.000122 | 0.004184 | 0.0002129 | 0.013778721 | MLLT1 |
| chr1 | 1669543 | 1669766 | 224 | 2 | 0.000154144 | 0.000487 | 0.000331 | 0.0002132 | 0.001863753 | SLC35E2B |
| chr9 | 124861561 | 124862429 | 869 | 10 | 0.000220919 | 0.008201 | 0.001564 | 0.0002134 | 0.001637878 | ARPC5L, RPL35 |
| chr10 | 132452167 | 132452559 | 393 | 3 | 0.000261786 | 0.000515 | 0.000718 | 0.0002137 | 0.00670236 | NA |
| chr10 | 11758113 | 11758294 | 182 | 2 | 0.000115294 | 0.002142 | 0.000148 | 0.0002139 | 0.003230709 | ECHDC3 |
| chr2 | 119543411 | 119543477 | 67 | 2 | 5.19E-05 | 0.000284 | 0.000585 | 0.0002143 | -0.0025125 | NA |
| chr8 | 143922229 | 143923056 | 828 | 3 | 5.73E-05 | 0.004848 | 0.00012 | 0.0002144 | -0.00140052 | PLEC |
| chr7 | 2979525 | 2979748 | 224 | 4 | 0.000225489 | 8.87E-05 | 0.013555 | 0.0002144 | 0.014541286 | CARD11 |
| chr5 | 147454374 | 147454694 | 321 | 3 | 2.12E-05 | 0.000963 | 0.001157 | 0.0002151 | 0.006799858 | DPYSL3 |
| chr17 | 2362098 | 2362581 | 484 | 3 | 2.58E-05 | 9.08E-05 | 0.008921 | 0.0002155 | 0.003245585 | SGSM2 |
| chr10 | 79444277 | 79444371 | 95 | 3 | 0.000173714 | 9.50E-05 | 0.007968 | 0.0002157 | 0.00854679 | ZCCHC24 |
| chr22 | 46457782 | 46457785 | 4 | 2 | 0.000420471 | 0.000343 | 0.000473 | 0.0002157 | 0.007646448 | CELSR1 |
| chr8 | 144508436 | 144508879 | 444 | 3 | 0.000323009 | 0.000795 | 0.000777 | 0.0002159 | 0.002222409 | NA |
| chr17 | 44067902 | 44067970 | 69 | 2 | 0.00029747 | 0.000361 | 0.000454 | 0.0002173 | 0.00749811 | NA |
| chr4 | 56909591 | 56909865 | 275 | 3 | 8.88E-05 | 0.001528 | 0.000398 | 0.0002176 | -0.000108 | REST |
| chr16 | 2799143 | 2799857 | 715 | 4 | 6.79E-06 | 0.053432 | 7.01E-05 | 0.0002179 | 0.003252813 | PRSS41 |
| chr1 | 46697377 | 46697494 | 118 | 3 | 0.000295769 | 0.002384 | 0.00012 | 0.0002183 | 0.001736754 | EFCAB14 |
| chr11 | 102453206 | 102454019 | 814 | 7 | 0.000208098 | 0.00115 | 0.000635 | 0.0002186 | 0.000890604 | AP001830.1, TMEM123 |
| chr19 | 48728930 | 48729347 | 418 | 3 | 3.78E-05 | 0.00192 | 0.000213 | 0.0002187 | 0.005669099 | RASIP1 |
| chr19 | 45368984 | 45369114 | 131 | 3 | 0.000151727 | 0.020284 | 7.61E-05 | 0.0002189 | 0.002968101 | ERCC2 |
| chr4 | 54103746 | 54104087 | 342 | 6 | 0.000348967 | 0.001119 | 0.001177 | 0.0002195 | 0.00444039 | AC058822.1 |
| chr12 | 10010431 | 10010933 | 503 | 3 | 0.000302664 | 0.000151 | 0.002005 | 0.0002206 | 0.008654735 | CLEC12B, CLEC1B |
| chr4 | 185020932 | 185021375 | 444 | 5 | 9.63E-05 | 0.000418 | 0.007387 | 0.0002209 | 0.00506665 | NA |
| chr9 | 133055214 | 133055249 | 36 | 3 | 0.000303732 | 0.002309 | 0.000254 | 0.0002215 | 0.003801931 | GTF3C5 |
| chr11 | 61586998 | 61587027 | 30 | 2 | 0.000439805 | 0.000131 | 0.003555 | 0.0002217 | 0.00970905 | NA |
| chr12 | 122980343 | 122980919 | 577 | 7 | 8.85E-05 | 0.000858 | 0.000453 | 0.0002218 | 0.001175481 | AC026362.1, ARL6IP4, ABCB9 |
| chr10 | 59176265 | 59177165 | 901 | 15 | 7.81E-05 | 0.003168 | 0.000499 | 0.0002222 | 0.002018725 | PHYHIPL |
| chr1 | 245330382 | 245331000 | 619 | 2 | 0.000139447 | 0.00014 | 0.002817 | 0.0002233 | 0.00510242 | KIF26B |
| chr19 | 39497583 | 39497619 | 37 | 2 | 0.000235172 | 0.003356 | 0.000137 | 0.0002239 | 0.010022958 | NA |
| chr1 | 226222697 | 226223558 | 862 | 8 | 0.000143375 | 0.001195 | 0.008697 | 0.0002239 | -0.00223012 | NA |
| chr17 | 75879336 | 75879422 | 87 | 2 | 6.12E-05 | 0.00014 | 0.002854 | 0.0002242 | 0.013053116 | AC087289.5 |
| chr6 | 170083441 | 170084086 | 646 | 4 | 0.000133641 | 0.003352 | 0.000266 | 0.0002246 | 3.57E-05 | NA |
| chr10 | 93065500 | 93065790 | 291 | 3 | 0.000178238 | 0.006066 | 8.98E-05 | 0.0002249 | 0.010014302 | CYP26C1, AL358613.3 |
| chr10 | 72216291 | 72216495 | 205 | 3 | 0.000401282 | 0.001642 | 0.002459 | 0.0002251 | 0.002342096 | ANAPC16, ASCC1 |
| chr20 | 17619830 | 17619920 | 91 | 2 | 0.00032176 | 0.001193 | 0.000199 | 0.0002256 | 0.003686268 | RRBP1 |
| chr16 | 68631189 | 68631234 | 46 | 2 | 0.000472055 | 0.0016 | 0.000174 | 0.0002257 | 0.003185551 | NA |
| chr19 | 3573545 | 3573842 | 298 | 4 | 0.000254696 | 0.000964 | 0.000212 | 0.0002258 | -0.0006186 | HMG20B, MFSD12 |
| chr17 | 79088147 | 79088536 | 390 | 2 | 8.50E-05 | 0.000311 | 0.000578 | 0.0002265 | 0.001317476 | ENGASE |
| chr15 | 52583823 | 52583899 | 77 | 2 | 0.000105937 | 0.01235 | 0.00011 | 0.0002266 | 0.003437336 | AC025917.1, FAM214A |
| chr20 | 18510176 | 18510387 | 212 | 3 | 0.000241365 | 0.000134 | 0.005044 | 0.0002266 | -0.00498825 | SEC23B |
| chr4 | 99894978 | 99895008 | 31 | 2 | 0.000443794 | 0.004302 | 0.00013 | 0.0002267 | 0.006515601 | NA |
| chr17 | 50919716 | 50919763 | 48 | 2 | 0.000346429 | 0.000448 | 0.000393 | 0.000227 | 0.008199216 | NA |
| chr20 | 36812789 | 36813285 | 497 | 3 | 0.000159231 | 0.065939 | 5.01E-05 | 0.0002283 | 0.001906973 | SOGA1 |
| chr9 | 85940948 | 85941343 | 396 | 5 | 0.000375012 | 0.00123 | 0.007524 | 0.000229 | 0.000171272 | NAA35 |
| chr1 | 113929190 | 113929581 | 392 | 10 | 0.000312651 | 0.037611 | 0.00053 | 0.000229 | -0.00081514 | HIPK1, HIPK1-AS1 |
| chr5 | 141338716 | 141339164 | 449 | 7 | 0.000211725 | 0.000229 | 0.011995 | 0.0002291 | 0.00793132 | PCDHGA1, PCDHGA2 |
| chr17 | 48101993 | 48102106 | 114 | 3 | 0.000333583 | 0.000446 | 0.000449 | 0.0002293 | -0.01609566 | NA |
| chr17 | 4365947 | 4366260 | 314 | 2 | 4.87E-05 | 0.010053 | 0.000114 | 0.0002297 | 0.003891671 | UBE2G1 |
| chr6 | 167796497 | 167797052 | 556 | 6 | 0.000329349 | 0.000564 | 0.005011 | 0.0002297 | 0.00679572 | AL009178.2, AL009178.3, LINC01558 |
| chr17 | 2336720 | 2337068 | 349 | 4 | 7.84E-05 | 0.003822 | 0.0003 | 0.00023 | -0.0001216 | NA |
| chr13 | 111186083 | 111186134 | 52 | 2 | 0.000209648 | 0.000676 | 0.000288 | 0.000231 | 0.006120092 | ARHGEF7 |
| chr5 | 6737665 | 6737735 | 71 | 2 | 0.000111031 | 0.000405 | 0.000448 | 0.0002311 | 0.002142955 | TENT4A |
| chr8 | 123232279 | 123232491 | 213 | 2 | 0.000169115 | 0.011133 | 0.000113 | 0.0002314 | 0.001599098 | C8orf76, ZHX1-C8orf76 |
| chr10 | 81873674 | 81873923 | 250 | 2 | 0.000409246 | 0.000262 | 0.000764 | 0.0002318 | -0.01607707 | AL096706.1 |
| chr18 | 45723812 | 45724114 | 303 | 4 | 0.000297022 | 0.000101 | 0.011144 | 0.0002326 | 0.008184098 | SLC14A1, AC023421.2 |
| chr6 | 157879104 | 157880172 | 1069 | 3 | 0.000185131 | 0.000576 | 0.00303 | 0.0002328 | 0.003302904 | SNX9 |
| chr2 | 176128592 | 176128915 | 324 | 3 | 0.000359587 | 0.000349 | 0.000743 | 0.0002336 | 0.005082012 | HOXD-AS2 |
| chr2 | 219538049 | 219538136 | 88 | 2 | 0.000177321 | 0.00015 | 0.00266 | 0.000234 | 0.007578226 | ASIC4 |
| chr9 | 124392841 | 124392908 | 68 | 2 | 0.000473254 | 0.000193 | 0.001338 | 0.0002353 | 0.008864793 | PSMB7 |
| chr20 | 64107191 | 64107198 | 8 | 2 | 0.000427385 | 0.005059 | 0.00013 | 0.000236 | 0.001757994 | MYT1 |
| chr16 | 31217106 | 31217399 | 294 | 6 | 0.00017265 | 0.000235 | 0.006848 | 0.0002362 | 0.007510313 | TRIM72, PYDC1 |
| chr20 | 34268941 | 34269092 | 152 | 2 | 0.000274755 | 0.000169 | 0.001876 | 0.0002364 | -0.02220661 | ASIP, AL035458.2 |
| chr2 | 11215527 | 11215829 | 303 | 3 | 0.000146769 | 0.002092 | 0.000146 | 0.0002366 | -0.00531321 | ROCK2 |
| chr6 | 28649384 | 28650014 | 631 | 3 | 3.96E-05 | 0.001638 | 0.000647 | 0.0002374 | -0.00185004 | AL121932.2 |
| chr11 | 118882656 | 118882961 | 306 | 4 | 0.00013258 | 0.000377 | 0.00155 | 0.0002376 | 0.005931613 | NA |
| chr17 | 17752772 | 17753001 | 230 | 2 | 0.000270431 | 0.00031 | 0.000637 | 0.000238 | 0.008496544 | RAI1 |
| chr5 | 172193979 | 172194694 | 716 | 3 | 6.87E-06 | 0.004424 | 0.000147 | 0.0002387 | 0.000539487 | EFCAB9 |
| chr6 | 32968958 | 32969637 | 680 | 12 | 0.000192873 | 0.000603 | 0.009894 | 0.0002391 | -0.00060026 | BRD2, HLA-DMA |
| chr11 | 3664329 | 3664503 | 175 | 2 | 0.00020026 | 0.000216 | 0.001119 | 0.0002392 | 0.001277912 | ART1 |
| chr8 | 11996296 | 11996411 | 116 | 3 | 0.000210444 | 0.000121 | 0.005566 | 0.0002398 | -0.01178444 | DEFB134 |
| chr5 | 68215561 | 68215827 | 267 | 6 | 0.000198537 | 0.003711 | 0.00374 | 0.00024 | -0.00121813 | PIK3R1 |
| chr20 | 55422115 | 55422899 | 785 | 4 | 8.71E-05 | 0.002254 | 0.000754 | 0.00024 | 0.005981185 | LINC01440, LINC01441 |
| chr3 | 194652926 | 194653161 | 236 | 2 | 0.000151984 | 0.002042 | 0.000168 | 0.0002405 | 0.003811501 | LSG1 |
| chr22 | 48258448 | 48258551 | 104 | 2 | 0.00045178 | 0.000512 | 0.000382 | 0.0002407 | 0.004580797 | AL008720.1 |
| chr1 | 18922024 | 18922458 | 435 | 4 | 0.000178862 | 0.030693 | 0.000279 | 0.000241 | 0.004103844 | IFFO2 |
| chr6 | 44067815 | 44068009 | 195 | 2 | 0.000310422 | 0.000139 | 0.004135 | 0.0002411 | 0.007177055 | AL109615.2, AL109615.3 |

| | | | | | | | | | | |
|---|---|---|---|---|---|---|---|---|---|---|
| chr7 | 739548 | 740099 | 552 | 3 | 0.000215681 | 9.23E-05 | 0.011477 | 0.0002418 | -0.00078565 | DNAAF5 |
| chr4 | 39526649 | 39526853 | 205 | 4 | 0.000317847 | 8.58E-05 | 0.013536 | 0.000242 | 0.003513268 | UGDH |
| chr11 | 69653569 | 69654107 | 539 | 5 | 0.000280467 | 0.002071 | 0.002437 | 0.0002421 | 0.003739976 | CCND1, LTO1 |
| chr11 | 64901003 | 64901032 | 30 | 2 | 0.000483353 | 0.074177 | 0.000104 | 0.0002427 | 0.003163622 | ATG2A |
| chr4 | 99088598 | 99089107 | 510 | 10 | 0.000319681 | 0.001965 | 0.00044 | 0.0002433 | 0.000369649 | AP002026.1, ADH5 |
| chr3 | 182986002 | 182986077 | 76 | 2 | 0.000335017 | 0.000186 | 0.001602 | 0.0002433 | 0.002948126 | NA |
| chr9 | 69121938 | 69122083 | 146 | 2 | 0.000237928 | 0.000161 | 0.002413 | 0.0002437 | 5.16E-05 | AL358113.1, TJP2 |
| chr10 | 97772381 | 97772879 | 499 | 6 | 0.000173921 | 0.000254 | 0.002292 | 0.0002438 | 0.003767893 | NA |
| chr7 | 151876507 | 151876963 | 457 | 7 | 0.00022819 | 0.000181 | 0.009011 | 0.0002439 | 0.001077712 | PRKAG2 |
| chr11 | 20208333 | 20208784 | 452 | 4 | 0.000323681 | 0.000385 | 0.001194 | 0.0002446 | 0.007262184 | NA |
| chr19 | 4181857 | 4182609 | 753 | 4 | 0.000238437 | 0.001649 | 0.000213 | 0.0002446 | 0.002731569 | SIRT6 |
| chr16 | 57798318 | 57798848 | 531 | 4 | 0.000350165 | 0.000862 | 0.001198 | 0.0002457 | 0.003482339 | AC092118.1, KIFC3 |
| chr6 | 75204990 | 75205087 | 98 | 2 | 8.57E-05 | 0.002076 | 0.000172 | 0.0002465 | 0.003350615 | COL12A1 |
| chr4 | 8105391 | 8106092 | 702 | 6 | 0.00017265 | 0.002187 | 0.000285 | 0.0002467 | -0.00278532 | ABLIM2 |
| chr7 | 157522394 | 157522604 | 211 | 2 | 0.000197375 | 0.001763 | 0.000184 | 0.0002468 | 0.003055499 | NA |
| chr18 | 51196711 | 51196813 | 103 | 2 | 9.75E-05 | 0.003665 | 0.000145 | 0.0002468 | -0.00203125 | MEX3C |
| chr15 | 88624414 | 88624505 | 92 | 2 | 0.000224514 | 0.001342 | 0.000209 | 0.0002471 | 0.004868564 | AEN |
| chr4 | 8579923 | 8580007 | 85 | 2 | 0.000341513 | 0.000556 | 0.000371 | 0.0002472 | 0.017503995 | GPR78 |
| chr11 | 10802143 | 10802367 | 225 | 3 | 9.78E-05 | 0.004788 | 0.000216 | 0.0002473 | 0.002328872 | EIF4G2 |
| chr17 | 48530737 | 48531100 | 364 | 9 | 0.000357289 | 0.00082 | 0.003493 | 0.0002475 | -0.00013413 | HOXB1 |
| chr5 | 123093483 | 123093822 | 340 | 4 | 0.000287397 | 0.00022 | 0.008701 | 0.0002476 | 0.005279369 | PRDM6 |
| chr3 | 4983867 | 4983885 | 19 | 2 | 0.000271423 | 0.002531 | 0.000161 | 0.0002477 | 0.009298225 | BHLHE40 |
| chr1 | 46670344 | 46670541 | 198 | 2 | 0.000481605 | 0.000229 | 0.001109 | 0.0002487 | 0.005783272 | TEX38, ATPAF1 |
| chr17 | 43994451 | 43995003 | 553 | 5 | 0.00017568 | 0.001266 | 0.000324 | 0.0002494 | 0.000516397 | PYY |
| chr2 | 70994950 | 70995209 | 260 | 8 | 0.000237458 | 0.003769 | 0.001081 | 0.0002496 | 0.001478471 | AC007040.1, AC007040.2 |
| chr20 | 10432776 | 10432897 | 122 | 2 | 0.000127969 | 0.000523 | 0.000398 | 0.00025 | 0.007996591 | MKKS |
| chr22 | 50170898 | 50171831 | 934 | 4 | 6.20E-05 | 0.001284 | 0.002273 | 0.0002503 | 0.005090195 | PANX2 |
| chr3 | 25427939 | 25428434 | 496 | 11 | 0.00026059 | 0.149474 | 0.000101 | 0.0002503 | 0.002502362 | RARB |
| chr4 | 1533946 | 1534002 | 57 | 2 | 0.000440701 | 0.006532 | 0.00013 | 0.0002503 | 0.009932111 | NA |
| chr4 | 76895857 | 76896239 | 383 | 2 | 0.000213826 | 0.000265 | 0.000883 | 0.0002506 | -0.00260803 | SOWAHB |
| chr6 | 167147239 | 167147313 | 75 | 2 | 0.000430952 | 0.000167 | 0.002395 | 0.0002512 | 0.009920357 | TCP10L2 |
| chr16 | 15726049 | 15726100 | 52 | 2 | 0.000194623 | 0.00015 | 0.003557 | 0.0002513 | -0.00426522 | NDE1, MYH11 |
| chr16 | 30372220 | 30372438 | 219 | 2 | 0.000257289 | 0.001336 | 0.000214 | 0.0002518 | 0.003182089 | MYLPF |
| chr12 | 1936644 | 1937064 | 421 | 4 | 0.000164583 | 0.002356 | 0.002029 | 0.0002521 | 0.002062959 | NA |
| chr12 | 101039753 | 101040351 | 599 | 3 | 0.000113782 | 0.099119 | 5.91E-05 | 0.0002525 | 0.006702018 | ANO4 |
| chr1 | 100266380 | 100266734 | 355 | 3 | 9.12E-05 | 0.014698 | 0.000819 | 0.0002526 | -4.36E-05 | RTCA |
| chr1 | 28058199 | 28058379 | 181 | 3 | 0.000343094 | 0.019559 | 0.000522 | 0.0002527 | 0.007296006 | EYA3 |
| chr2 | 222304633 | 222304890 | 258 | 4 | 0.000285891 | 0.001302 | 0.000184 | 0.0002529 | 0.005868358 | CCDC140 |
| chr14 | 105367683 | 105368135 | 453 | 3 | 0.000134245 | 0.002104 | 0.002838 | 0.000253 | 0.003698435 | PACS2 |
| chr2 | 101309005 | 101309692 | 688 | 4 | 8.37E-05 | 0.000137 | 0.003078 | 0.0002537 | 0.006600565 | MIR5696 |
| chr6 | 26295764 | 26296347 | 584 | 2 | 6.46E-05 | 0.000522 | 0.000409 | 0.000254 | 0.028558968 | NA |
| chr9 | 130446460 | 130446702 | 243 | 2 | 0.00014794 | 0.001197 | 0.000229 | 0.0002541 | 0.006369488 | ASS1 |
| chr19 | 7439683 | 7440458 | 776 | 6 | 2.03E-05 | 0.013268 | 0.000248 | 0.0002541 | 0.00107403 | AC008878.3, ARHGEF18 |
| chr10 | 786825 | 787350 | 526 | 3 | 0.000213983 | 0.039289 | 8.36E-05 | 0.0002541 | -0.00391638 | NA |
| chr5 | 56302533 | 56302611 | 79 | 2 | 0.000411494 | 0.005639 | 0.000136 | 0.0002545 | -0.00245712 | NA |
| chr2 | 1539449 | 1539555 | 107 | 2 | 0.000102357 | 0.001406 | 0.000212 | 0.0002549 | 0.001608497 | TPO, AC105450.1 |
| chr3 | 44241978 | 44242345 | 368 | 3 | 8.15E-05 | 0.001555 | 0.001089 | 0.0002549 | 0.001298608 | TOPAZ1 |
| chr2 | 620840 | 621426 | 587 | 3 | 0.00015618 | 0.002717 | 0.000146 | 0.000255 | 0.001391183 | NA |
| chr16 | 1990523 | 1991825 | 1303 | 10 | 3.20E-05 | 0.003003 | 0.000393 | 0.0002552 | 0.000573735 | SYNGR3 |
| chr11 | 67282765 | 67283040 | 276 | 2 | 0.000153936 | 0.000149 | 0.004082 | 0.0002559 | 0.007153433 | GRK2 |
| chr11 | 117145119 | 117145255 | 137 | 3 | 0.000184466 | 0.021754 | 9.14E-05 | 0.0002559 | 0.001512384 | PAFAH1B2 |
| chr8 | 1098486 | 1098844 | 359 | 2 | 0.000114574 | 0.000226 | 0.001229 | 0.0002568 | 0.004044923 | DLGAP2 |
| chr20 | 34709375 | 34710309 | 935 | 7 | 3.21E-05 | 0.002162 | 9.40E-05 | 0.0002578 | 0.001856879 | TP53INP2, NCOA6 |
| chr17 | 42050535 | 42050865 | 331 | 3 | 0.000259743 | 0.001383 | 0.001102 | 0.0002586 | 0.001521474 | ZNF385C, C17orf113 |
| chr16 | 86567904 | 86567914 | 11 | 2 | 0.000205322 | 0.000929 | 0.000272 | 0.0002591 | -0.00256017 | FOXC2 |
| chr21 | 45894154 | 45894851 | 698 | 5 | 5.98E-05 | 0.259605 | 0.000131 | 0.0002595 | 0.004413546 | PCBP3 |
| chr19 | 45496277 | 45496541 | 265 | 3 | 0.000163266 | 0.000172 | 0.005808 | 0.00026 | -0.00893221 | PPM1N, RTN2 |
| chr5 | 139648483 | 139649169 | 687 | 4 | 7.28E-05 | 0.000324 | 0.002322 | 0.0002601 | 0.000620965 | CXXC5, CXXC5-AS1 |
| chr17 | 45060780 | 45060991 | 212 | 4 | 0.00024933 | 0.002317 | 0.002158 | 0.0002607 | 0.001261405 | NMT1, DCAKD |
| chr1 | 218344077 | 218344436 | 360 | 2 | 6.03E-05 | 0.000148 | 0.004622 | 0.000261 | 0.009855326 | TGFB2-AS1 |
| chr7 | 510871 | 511003 | 133 | 3 | 0.000375102 | 0.038032 | 0.000632 | 0.0002616 | 0.007258414 | PDGFA |
| chr11 | 126265744 | 126265815 | 72 | 2 | 0.000199144 | 0.000271 | 0.000938 | 0.0002618 | 0.002453812 | SRPRA |
| chr7 | 150329465 | 150329642 | 178 | 2 | 7.13E-05 | 0.0034 | 0.000156 | 0.0002619 | 0.003170322 | ZBED6CL, LRRC61 |
| chr19 | 37595195 | 37595589 | 395 | 2 | 4.47E-05 | 0.00123 | 0.000235 | 0.0002622 | 0.010315538 | ZNF540 |
| chr8 | 123954822 | 123954871 | 50 | 2 | 0.000392369 | 0.000442 | 0.000511 | 0.0002631 | 0.003709029 | FER1L6 |
| chr10 | 131973209 | 131973419 | 211 | 2 | 0.000160434 | 0.038783 | 0.000112 | 0.0002631 | 0.007567312 | BNIP3 |
| chr4 | 67998459 | 67998833 | 375 | 2 | 0.000186542 | 0.005779 | 0.000139 | 0.0002636 | 0.001521561 | UBA6-AS1, TMPRSS11GP |
| chr1 | 159853304 | 159853311 | 8 | 2 | 0.000486603 | 0.004535 | 0.000146 | 0.0002645 | 0.003339845 | AL590560.2, SNHG28 |
| chr13 | 110651351 | 110651442 | 92 | 2 | 0.000300312 | 0.000209 | 0.00156 | 0.0002646 | -0.0046063 | CARS2 |
| chr1 | 153752563 | 153752842 | 280 | 3 | 0.000364514 | 0.00014 | 0.004232 | 0.0002651 | -0.00145801 | INTS3 |
| chr1 | 3510843 | 3510915 | 73 | 2 | 0.000432403 | 0.000282 | 0.000917 | 0.0002665 | 0.005130216 | MEGF6 |
| chr2 | 55268473 | 55269008 | 536 | 3 | 0.000185572 | 0.000671 | 0.000434 | 0.0002674 | 0.006254628 | MTIF2 |
| chr6 | 1385116 | 1385496 | 381 | 3 | 0.00039744 | 0.011106 | 0.001794 | 0.0002675 | 0.007257329 | NA |
| chr17 | 81002275 | 81002536 | 262 | 2 | 6.31E-05 | 0.004567 | 0.000147 | 0.0002676 | 0.005988574 | CHMP6 |
| chr11 | 10934454 | 10934528 | 75 | 2 | 0.000364444 | 0.000152 | 0.004861 | 0.0002692 | 0.00648149 | NA |
| chr5 | 177133593 | 177134446 | 854 | 6 | 0.000138008 | 0.001195 | 0.001363 | 0.0002697 | 0.00203781 | NSD1 |
| chr11 | 32426398 | 32426747 | 350 | 3 | 0.000295041 | 0.000139 | 0.006614 | 0.0002698 | 0.012750947 | WT1 |
| chr5 | 876612 | 876961 | 350 | 3 | 0.000232118 | 0.00217 | 0.000164 | 0.00027 | 0.002148453 | BRD9 |
| chr1 | 68819078 | 68819391 | 314 | 2 | 0.000172579 | 0.000376 | 0.000642 | 0.0002705 | 0.016604488 | AL033530.1 |
| chr9 | 134050505 | 134050568 | 64 | 2 | 0.000275398 | 0.000173 | 0.002817 | 0.0002707 | 0.003160611 | BRD3 |
| chr5 | 154480294 | 154480413 | 120 | 2 | 0.000450511 | 0.000323 | 0.00078 | 0.0002707 | 0.016878492 | NA |
| chr1 | 90716571 | 90716748 | 178 | 4 | 0.000287083 | 7.18E-05 | 0.023257 | 0.0002709 | 0.003238453 | BARHL2 |
| chr17 | 10128165 | 10128611 | 447 | 2 | 0.000242285 | 0.000214 | 0.001596 | 0.0002709 | -0.03435747 | GAS7 |
| chr5 | 55850782 | 55850816 | 35 | 2 | 0.000378251 | 0.000778 | 0.000327 | 0.0002709 | 0.012591035 | NA |
| chr19 | 45363616 | 45363827 | 212 | 4 | 0.000269451 | 0.001172 | 0.006393 | 0.0002713 | 0.003303083 | ERCC2 |
| chr11 | 119317758 | 119317954 | 197 | 2 | 0.000164537 | 0.000776 | 0.000329 | 0.0002716 | 0.007472933 | MCAM |
| chr15 | 59105311 | 59105596 | 286 | 2 | 3.38E-06 | 0.010959 | 0.000128 | 0.0002719 | -0.00238098 | CCNB2 |
| chr4 | 76422394 | 76422516 | 123 | 2 | 0.000295134 | 0.004445 | 0.00015 | 0.000272 | -0.01019447 | NA |
| chr5 | 139667982 | 139668154 | 173 | 2 | 5.76E-05 | 0.003937 | 0.000155 | 0.0002722 | 0.0046885 | CXXC5 |
| chr7 | 30853236 | 30853275 | 40 | 2 | 0.000134562 | 0.007802 | 0.000135 | 0.0002722 | -0.02587272 | INMT-MINDY4, MINDY4, AC004691.2 |
| chr5 | 157574553 | 157575167 | 615 | 4 | 0.000310223 | 0.000247 | 0.001339 | 0.0002725 | 0.008457115 | ADAM19 |
| chr10 | 72274886 | 72274909 | 24 | 3 | 0.000218412 | 0.000165 | 0.006427 | 0.0002725 | 0.011083634 | DDIT4 |
| chr11 | 561538 | 562547 | 1010 | 4 | 3.67E-05 | 0.001646 | 0.001433 | 0.000273 | 0.002156451 | RASSF7 |
| chr6 | 26533169 | 26533312 | 144 | 3 | 0.000434527 | 0.004688 | 0.00015 | 0.0002737 | -0.00172187 | NA |
| chr2 | 161136613 | 161136853 | 241 | 3 | 0.00032586 | 0.050604 | 0.002632 | 0.0002739 | 0.005331708 | TANK-AS1 |
| chr11 | 1485151 | 1485790 | 640 | 4 | 6.36E-06 | 0.014556 | 5.86E-05 | 0.0002748 | 0.001489661 | MOB2 |
| chr3 | 53845224 | 53845710 | 487 | 3 | 4.37E-05 | 0.000769 | 0.000366 | 0.0002751 | 0.00148972 | CHDH |
| chr3 | 128153249 | 128153584 | 336 | 7 | 0.00024703 | 0.001315 | 0.00108 | 0.0002752 | -0.00029029 | EEFSEC, RUVBL1 |
| chr15 | 55408064 | 55408983 | 920 | 17 | 0.000178045 | 0.028336 | 0.000473 | 0.0002757 | 5.33E-05 | C15orf65, CCPG1, DNAAF4-CCPG1 |
| chr1 | 78004428 | 78004502 | 75 | 2 | 0.000206197 | 0.000201 | 0.00194 | 0.0002761 | -0.00327126 | DNAJB4, GIPC2, AC103591.3 |
| chr3 | 28348154 | 28348885 | 732 | 7 | 2.87E-05 | 0.010959 | 4.26E-05 | 0.0002761 | -0.00092994 | ZCWPW2, AZI2 |
| chr17 | 30921586 | 30921830 | 245 | 6 | 0.000436983 | 0.00157 | 0.004473 | 0.0002767 | 0.005405616 | ADAP2 |
| chr2 | 3704003 | 3704763 | 761 | 3 | 3.42E-05 | 0.002671 | 0.000821 | 0.0002768 | -0.01508003 | DCDC2C, AC010907.1 |
| chr13 | 41457625 | 41458203 | 579 | 3 | 8.81E-05 | 0.001661 | 0.000259 | 0.0002768 | -0.00114932 | RGCC |
| chr5 | 57174073 | 57174693 | 621 | 4 | 3.47E-05 | 0.003067 | 0.000793 | 0.0002769 | 0.003274068 | GPBP1 |
| chr15 | 92581093 | 92581113 | 21 | 2 | 0.000224617 | 0.004532 | 0.000152 | 0.0002771 | -0.00184979 | AC091544.2 |
| chr6 | 170455501 | 170455676 | 176 | 4 | 0.000220896 | 0.035136 | 4.42E-05 | 0.0002775 | 0.000622065 | NA |
| chr19 | 7596576 | 7596731 | 156 | 2 | 0.000143506 | 0.000195 | 0.002182 | 0.0002783 | 0.016517433 | CAMSAP3 |
| chr1 | 47449197 | 47449336 | 140 | 2 | 0.000256526 | 0.001076 | 0.000274 | 0.0002792 | 0.012328836 | NA |
| chr13 | 113111083 | 113111320 | 238 | 2 | 4.52E-05 | 0.046021 | 0.000116 | 0.0002795 | 0.004732135 | F7 |
| chr3 | 53178823 | 53179684 | 862 | 3 | 7.98E-05 | 0.000194 | 0.006114 | 0.0002807 | 0.001729397 | PRKCD |
| chr7 | 149126514 | 149126873 | 360 | 7 | 7.60E-05 | 0.098812 | 2.53E-05 | 0.0002812 | 9.13E-05 | ZNF398 |
| chr8 | 145052040 | 145052617 | 578 | 6 | 0.000119533 | 0.003967 | 0.007631 | 0.0002816 | 0.001625859 | C8orf33 |
| chr3 | 35640135 | 35640213 | 79 | 2 | 0.000308239 | 0.00018 | 0.002892 | 0.0002817 | 0.005979172 | ARPP21 |
| chr3 | 46845959 | 46846161 | 203 | 2 | 0.0003633 | 0.000159 | 0.004918 | 0.0002817 | 0.005649293 | MYL3 |
| chr12 | 52520513 | 52521534 | 1022 | 8 | 0.000101985 | 0.007384 | 0.000344 | 0.0002822 | 0.003141496 | KRT5 |
| chr17 | 81565425 | 81565576 | 152 | 2 | 8.94E-05 | 0.015513 | 0.000127 | 0.0002828 | -0.00232179 | NPLOC4 |
| chr1 | 23792637 | 23792640 | 4 | 2 | 0.000177663 | 0.001998 | 0.000204 | 0.0002834 | -0.00390995 | LYPLA2 |
| chr1 | 1051180 | 1051514 | 335 | 2 | 0.00027911 | 0.008113 | 0.000139 | 0.000284 | 0.005346035 | AGRN |
| chr21 | 42366888 | 42367716 | 829 | 5 | 6.17E-05 | 0.006369 | 6.63E-05 | 0.0002849 | 0.003264283 | NA |
| chr10 | 88878534 | 88878712 | 179 | 2 | 0.000153604 | 0.004468 | 0.000156 | 0.0002849 | 0.006070647 | NA |
| chr2 | 85584348 | 85584900 | 553 | 3 | 0.00014444 | 0.000108 | 0.012205 | 0.0002851 | -0.01588601 | VAMP5 |
| chr10 | 73626327 | 73626534 | 208 | 3 | 0.000429607 | 0.043815 | 6.27E-05 | 0.0002851 | 0.00168586 | AC073389.1 |
| chr14 | 81170271 | 81170537 | 267 | 4 | 0.000158807 | 0.001389 | 0.002459 | 0.0002852 | 0.001757864 | AL136040.2, AC007262.2 |
| chr3 | 72324204 | 72324548 | 345 | 2 | 0.0004606 | 0.001813 | 0.000215 | 0.0002863 | 0.004946165 | NA |
| chr11 | 2149777 | 2150248 | 472 | 4 | 0.000127459 | 0.0039 | 0.001112 | 0.0002866 | 0.004754981 | AC132217.2, INS-IGF2 |
| chr1 | 151909309 | 151909498 | 190 | 2 | 9.62E-05 | 0.002378 | 0.000193 | 0.000287 | 0.002162863 | THEM4 |
| chr19 | 45338385 | 45338875 | 491 | 3 | 4.48E-05 | 0.002635 | 0.000287 | 0.000287 | 0.003234197 | KLC3 |
| chr2 | 176178419 | 176178773 | 355 | 4 | 5.53E-05 | 0.007061 | 0.000142 | 0.000287 | 0.003586675 | HAGLROS, HAGLR |
| chr17 | 28226541 | 28226661 | 121 | 2 | 0.000120144 | 0.000162 | 0.00501 | 0.0002886 | 0.007261611 | PYY2 |
| chr1 | 52707727 | 52707731 | 5 | 2 | 0.000402524 | 0.004181 | 0.000161 | 0.0002889 | 0.006466013 | NA |
| chr10 | 128210295 | 128210321 | 27 | 2 | 0.000131886 | 0.000818 | 0.000345 | 0.0002891 | -0.00510893 | LINC01163 |
| chr17 | 39224444 | 39224574 | 131 | 2 | 0.000297853 | 0.00022 | 0.001809 | 0.0002893 | 0.010662543 | STAC2 |
| chr15 | 76011293 | 76012209 | 917 | 4 | 0.000118691 | 0.000647 | 0.001942 | 0.0002901 | 0.000139083 | NRG4 |
| chr8 | 10839061 | 10839179 | 119 | 3 | 0.000199643 | 0.005322 | 0.000302 | 0.0002902 | 0.000349637 | AC105001.2, PINX1 |
| chr1 | 19664744 | 19665214 | 471 | 4 | 6.14E-05 | 0.002018 | 0.000927 | 0.0002907 | 0.001427531 | HTR6 |
| chr13 | 46053709 | 46054058 | 350 | 2 | 7.10E-05 | 0.004206 | 0.000162 | 0.0002911 | 0.009758516 | CPB2-AS1, CPB2 |
| chr16 | 20741535 | 20741580 | 46 | 2 | 0.000407367 | 0.000164 | 0.005086 | 0.000292 | 0.00691766 | ACSM3, THUMPD1 |
| chr11 | 837514 | 837944 | 431 | 2 | 0.000187786 | 0.000165 | 0.004908 | 0.0002926 | 0.008932096 | CD151, POLR2L |
| chr19 | 19528066 | 19528744 | 679 | 5 | 5.48E-05 | 0.003262 | 0.00041 | 0.000293 | 0.002523687 | NDUFA13, AC011448.1 |
| chr21 | 41538890 | 41539045 | 156 | 2 | 0.000420035 | 0.00019 | 0.002882 | 0.0002945 | -0.00012918 | NA |
| chr19 | 29673913 | 29674111 | 199 | 2 | 0.000111379 | 0.001278 | 0.000266 | 0.0002945 | 0.001960517 | PLEKHF1 |
| chr22 | 36498401 | 36498928 | 528 | 5 | 0.00015764 | 0.048651 | 0.001501 | 0.0002951 | -0.0001083 | FOXRED2 |
| chr8 | 71842927 | 71843549 | 623 | 9 | 0.000105152 | 0.002067 | 0.007129 | 0.0002959 | 0.001892264 | MSC-AS1, MSC |
| chr11 | 69443466 | 69443978 | 513 | 5 | 0.000332588 | 8.23E-05 | 0.023008 | 0.000296 | 0.004308737 | AP000439.1 |
| chr4 | 157074862 | 157075030 | 169 | 2 | 0.000279602 | 0.000255 | 0.001409 | 0.0002974 | 0.004393716 | NA |
| chr1 | 151848832 | 151848852 | 21 | 2 | 0.00049921 | 0.000194 | 0.002789 | 0.0002975 | -0.01000609 | C2CD4D-AS1, THEM5 |
| chr10 | 133013820 | 133013950 | 131 | 2 | 0.000402146 | 0.000279 | 0.001212 | 0.000299 | 0.015414116 | NA |
| chr7 | 18495249 | 18495709 | 461 | 5 | 0.000189956 | 0.001299 | 0.001923 | 0.000299 | 0.006752858 | HDAC9 |
| chr1 | 153533175 | 153533474 | 300 | 3 | 0.000391363 | 0.000192 | 0.003487 | 0.0002994 | 0.003699357 | NA |
| chr11 | 72308781 | 72308987 | 207 | 3 | 0.000170402 | 0.000932 | 0.000378 | 0.0002998 | 0.002733677 | CLPB |
| chr16 | 30473975 | 30474276 | 302 | 4 | 0.00024484 | 0.032528 | 0.000906 | 0.0003002 | 0.009671206 | ITGAL |
| chr17 | 28949238 | 28950030 | 793 | 6 | 0.000113686 | 0.001253 | 0.007293 | 0.0003008 | 0.000447894 | PHF12 |
| chr2 | 108942956 | 108943017 | 62 | 2 | 0.00048301 | 0.000811 | 0.000369 | 0.0003014 | 0.00091867 | EDAR |
| chr6 | 33009049 | 33010419 | 1371 | 16 | 0.000257572 | 0.00584 | 0.002063 | 0.0003023 | 0.003144931 | HLA-DOA |
| chr19 | 15232359 | 15232975 | 617 | 7 | 5.83E-05 | 0.148213 | 0.000111 | 0.0003024 | -0.00071782 | EPHX3 |
| chr11 | 65638289 | 65638840 | 552 | 6 | 0.000238625 | 0.010428 | 0.001111 | 0.0003037 | 0.001870464 | SIPA1 |
| chr6 | 29752742 | 29753238 | 497 | 7 | 0.000156326 | 0.012742 | 0.002453 | 0.0003037 | -0.0002762 | AL645939.5 |
| chr15 | 43824584 | 43825109 | 526 | 10 | 0.00037198 | 0.000693 | 0.01109 | 0.0003041 | 0.000911986 | MFAP1 |
| chr9 | 122228903 | 122229377 | 475 | 10 | 0.000201606 | 0.017395 | 6.38E-05 | 0.0003042 | -0.00273482 | LHX6 |
| chr18 | 63948330 | 63949628 | 1299 | 9 | 0.000275712 | 0.00348 | 0.010031 | 0.0003043 | 0.005487036 | HMSD |
| chr22 | 20017358 | 20017777 | 420 | 3 | 0.000134526 | 0.000168 | 0.004293 | 0.0003046 | 0.005665219 | TANGO2 |

| | | | | | | | | | | |
|---|---|---|---|---|---|---|---|---|---|---|
| chr15 | 70096009 | 70096223 | 215 | 5 | 0.00040203 | 0.000549 | 0.002964 | 0.0003047 | -0.00154359 | TLE3 |
| chr12 | 123436166 | 123436599 | 434 | 4 | 9.94E-05 | 0.011673 | 0.000158 | 0.0003052 | 0.002319317 | RILPL2 |
| chr20 | 33562157 | 33562505 | 349 | 3 | 8.21E-05 | 0.004187 | 0.000179 | 0.0003055 | 0.003251982 | CBFA2T2 |
| chr2 | 197784719 | 197785209 | 491 | 6 | 0.000304975 | 6.84E-05 | 0.032067 | 0.0003055 | 0.004911447 | BOLL |
| chr1 | 11151438 | 11151628 | 191 | 2 | 0.000126322 | 0.001226 | 0.000287 | 0.0003065 | 0.005276381 | MTOR |
| chr10 | 25821011 | 25821221 | 211 | 2 | 0.000392835 | 0.000187 | 0.003594 | 0.0003073 | -0.01273655 | NA |
| chr10 | 4384224 | 4384407 | 184 | 3 | 0.000264758 | 0.001097 | 0.000396 | 0.0003098 | 0.00166861 | LINC00703 |
| chr19 | 48955339 | 48955793 | 455 | 2 | 3.74E-05 | 0.01573 | 0.000136 | 0.0003107 | 0.002705649 | BAX |
| chr3 | 9902499 | 9902853 | 355 | 4 | 0.000378193 | 0.00015 | 0.008082 | 0.0003107 | 0.01113885 | IL17RE |
| chr7 | 27739029 | 27739320 | 292 | 3 | 1.61E-05 | 0.025374 | 7.33E-05 | 0.000311 | 0.008147818 | TAX1BP1-AS1 |
| chr22 | 37486798 | 37487218 | 421 | 4 | 9.76E-05 | 0.000308 | 0.003518 | 0.0003113 | 0.004770049 | NA |
| chr13 | 113413574 | 113413780 | 207 | 3 | 0.00014795 | 0.234831 | 5.35E-05 | 0.0003137 | 0.001380302 | ADPRHL1 |
| chr2 | 233250607 | 233250700 | 94 | 2 | 0.000452865 | 0.000294 | 0.001241 | 0.000314 | 0.001724882 | ATG16L1 |
| chr17 | 78723530 | 78723558 | 29 | 3 | 0.000360369 | 0.001568 | 0.000231 | 0.0003146 | 0.001795087 | CYTH1 |
| chr10 | 18141483 | 18141683 | 201 | 2 | 0.000335891 | 0.00021 | 0.002683 | 0.0003148 | 0.016731732 | CACNB2 |
| chr6 | 32829801 | 32830662 | 862 | 8 | 9.61E-05 | 8.66E-05 | 0.024286 | 0.0003149 | 0.001524288 | AL669918.1, TAP2 |
| chr12 | 48977845 | 48977968 | 124 | 2 | 0.000327826 | 0.000187 | 0.00401 | 0.0003151 | 0.001685193 | NA |
| chr16 | 58195619 | 58195766 | 148 | 2 | 0.000425587 | 0.000178 | 0.005083 | 0.0003151 | 0.003300553 | CSNK2A2 |
| chr3 | 49117202 | 49117774 | 573 | 3 | 0.00013437 | 0.000283 | 0.001595 | 0.000316 | 0.001871855 | USP19 |
| chr16 | 89547495 | 89548013 | 519 | 3 | 0.000114995 | 0.002378 | 0.000177 | 0.0003186 | 0.002130955 | SPG7 |
| chr16 | 57462633 | 57463258 | 626 | 5 | 0.00015408 | 0.002053 | 0.005152 | 0.0003193 | 0.001148818 | AC009052.1, POLR2C |
| chr8 | 84184445 | 84184821 | 377 | 6 | 0.00031252 | 0.000273 | 0.002543 | 0.0003193 | 0.004025797 | RALYL |
| chr11 | 69480105 | 69480399 | 295 | 4 | 0.000324053 | 0.002013 | 0.001088 | 0.0003199 | 0.002450585 | LINC02747 |
| chr19 | 50024839 | 50026031 | 1193 | 16 | 3.88E-05 | 0.006821 | 0.000341 | 0.000321 | 0.000372146 | ZNF473, VRK3 |
| chr5 | 181243701 | 181244165 | 465 | 9 | 0.000237673 | 0.000264 | 0.009913 | 0.000321 | 0.000115484 | RACK1 |
| chr1 | 160373814 | 160373945 | 132 | 2 | 0.000224248 | 0.00019 | 0.00411 | 0.0003216 | 0.01149309 | NA |
| chr19 | 49665822 | 49666244 | 423 | 10 | 0.00017169 | 0.0045 | 0.000583 | 0.0003217 | 0.00035877 | BCL2L12, IRF3 |
| chr16 | 86515631 | 86515670 | 40 | 2 | 0.0003545 | 0.000432 | 0.000753 | 0.0003222 | 0.002372829 | NA |
| chr1 | 227560440 | 227560741 | 302 | 3 | 0.000133633 | 0.000182 | 0.007618 | 0.0003225 | 0.01250425 | NA |
| chr2 | 1414256 | 1414522 | 267 | 4 | 0.000173534 | 0.002841 | 0.000209 | 0.0003226 | 0.003089413 | TPO, AC105450.1 |
| chr11 | 70076589 | 70076927 | 339 | 2 | 3.37E-05 | 0.00354 | 0.00019 | 0.0003229 | 0.007048343 | NA |
| chr19 | 49335221 | 49335520 | 300 | 3 | 0.000274316 | 0.002572 | 0.000187 | 0.000323 | 0.000593313 | CD37, AC011450.1 |
| chr7 | 155811098 | 155811397 | 300 | 4 | 0.000291164 | 0.011952 | 0.000128 | 0.0003239 | 0.001069951 | SHH |
| chr12 | 3236655 | 3237018 | 364 | 3 | 0.00033932 | 0.001014 | 0.00292 | 0.0003239 | 0.0120261 | TSPAN9 |
| chr8 | 98426396 | 98426740 | 345 | 3 | 0.000251479 | 0.228126 | 5.38E-05 | 0.0003242 | 0.004056608 | STK3 |
| chr4 | 80190548 | 80190831 | 284 | 2 | 0.000418881 | 0.00024 | 0.002063 | 0.0003255 | 0.007428966 | PRDM8, AC021127.1 |
| chr21 | 33481072 | 33481359 | 288 | 2 | 0.000124123 | 0.000183 | 0.005323 | 0.000326 | -0.00824573 | NA |
| chr10 | 132441341 | 132442101 | 761 | 6 | 0.000173169 | 0.001695 | 0.000181 | 0.0003266 | 0.002043321 | AL451069.3 |
| chr6 | 166008853 | 166009253 | 401 | 4 | 0.00014552 | 0.001121 | 0.000766 | 0.0003276 | 0.004412818 | NA |
| chr8 | 35236383 | 35236535 | 153 | 3 | 0.000190694 | 0.002886 | 0.00325 | 0.0003279 | 0.005006733 | UNC5D |
| chr12 | 132489885 | 132490176 | 292 | 12 | 0.000134062 | 0.003757 | 0.014491 | 0.0003281 | 0.000656184 | FBRSL1 |
| chr8 | 39989597 | 39990161 | 565 | 3 | 9.55E-05 | 0.005515 | 0.000129 | 0.000329 | 0.008372596 | IDO2, AC007991.3 |
| chr16 | 3953781 | 3954020 | 240 | 3 | 0.000449367 | 0.000125 | 0.012245 | 0.0003296 | 0.001456604 | ADCY9 |
| chr11 | 125904187 | 125904197 | 11 | 3 | 0.000374911 | 0.003558 | 0.000354 | 0.0003302 | -0.00262108 | DDX25, AP000842.2 |
| chr11 | 34107303 | 34107346 | 44 | 2 | 0.000352599 | 0.000509 | 0.000652 | 0.0003303 | 0.008244983 | NAT10 |
| chr11 | 57303657 | 57303787 | 131 | 2 | 0.000248008 | 0.039085 | 0.000132 | 0.0003306 | 0.006977408 | TNKS1BP1 |
| chr8 | 124728654 | 124728917 | 264 | 2 | 0.000322232 | 0.00027 | 0.001665 | 0.0003306 | 0.006748341 | NA |
| chr7 | 133252065 | 133252678 | 614 | 5 | 0.000182097 | 0.000351 | 0.011122 | 0.0003306 | 0.004330927 | NA |
| chr17 | 81711075 | 81711128 | 54 | 2 | 0.000460394 | 0.000406 | 0.000859 | 0.0003316 | 0.012723478 | AC139530.2 |
| chr19 | 1556285 | 1556415 | 131 | 2 | 0.000404717 | 0.000435 | 0.000794 | 0.0003328 | -0.00490612 | MEX3D |
| chr1 | 205271114 | 205271334 | 221 | 3 | 0.000392498 | 0.031676 | 0.001721 | 0.0003331 | 0.004611317 | TMCC2 |
| chr1 | 203487563 | 203487748 | 186 | 3 | 0.000241456 | 0.000181 | 0.005503 | 0.0003342 | -3.44E-05 | PRELP |
| chr16 | 87140354 | 87140597 | 244 | 2 | 4.77E-05 | 0.013307 | 0.000146 | 0.0003344 | 0.001816007 | C16orf95 |
| chr7 | 152759508 | 152759942 | 435 | 9 | 0.000142335 | 0.016028 | 0.001155 | 0.0003354 | 0.001156565 | ACTR3B |
| chr5 | 77850373 | 77850494 | 122 | 4 | 0.000318236 | 0.000208 | 0.008438 | 0.0003359 | 0.009156195 | TBCA |
| chr3 | 32818512 | 32818686 | 175 | 2 | 0.000121764 | 0.00382 | 0.000193 | 0.0003363 | -0.00281802 | TRIM71 |
| chr7 | 5189245 | 5189642 | 398 | 3 | 0.000164535 | 0.000247 | 0.006242 | 0.0003363 | 0.000306598 | NA |
| chr2 | 241762705 | 241763138 | 434 | 2 | 3.47E-05 | 0.000534 | 0.000639 | 0.0003364 | 0.012248968 | D2HGDH |
| chr22 | 40021756 | 40021865 | 110 | 2 | 0.000218532 | 0.004001 | 0.000191 | 0.0003381 | 0.003498153 | FAM83F |
| chr12 | 132367080 | 132367424 | 345 | 3 | 0.000229886 | 0.000702 | 0.00098 | 0.0003389 | 0.00815905 | NA |
| chr19 | 56840646 | 56840901 | 256 | 10 | 0.000321249 | 0.000532 | 0.015656 | 0.0003393 | 0.00363379 | MIMT1, ZIM2, PEG3 |
| chr5 | 55222308 | 55222380 | 73 | 2 | 0.000165458 | 0.000595 | 0.000582 | 0.0003397 | 0.00395111 | MCIDAS |
| chr6 | 30932420 | 30932783 | 364 | 4 | 0.000179947 | 0.007975 | 9.94E-05 | 0.0003406 | 0.004374839 | SFTA2, Y RNA |
| chr17 | 3048802 | 3049239 | 438 | 6 | 0.000255849 | 0.002791 | 0.003276 | 0.0003417 | 0.000308828 | NA |
| chr14 | 34875312 | 34875567 | 256 | 3 | 0.00040372 | 0.009574 | 0.000161 | 0.0003419 | -0.00163492 | AL121603.2, BAZ1A |
| chr17 | 2088229 | 2088907 | 679 | 3 | 0.000105638 | 0.001724 | 0.003561 | 0.0003428 | 0.004681991 | SMG6 |
| chr1 | 231041452 | 231041455 | 4 | 2 | 0.000490801 | 0.000415 | 0.000895 | 0.0003433 | 0.014210048 | NA |
| chr3 | 58333233 | 58333616 | 384 | 2 | 0.000127205 | 0.000263 | 0.001963 | 0.0003439 | 0.000848435 | PXK |
| chr16 | 2786680 | 2786815 | 136 | 3 | 0.000313934 | 0.002225 | 0.00024 | 0.0003442 | 0.004146087 | PRSS33 |
| chr8 | 26041323 | 26041943 | 621 | 5 | 0.000101892 | 0.000579 | 0.003657 | 0.0003444 | 0.005089672 | EBF2 |
| chr6 | 391680 | 392555 | 876 | 8 | 5.86E-05 | 0.020273 | 5.34E-05 | 0.0003451 | 0.001430497 | IRF4 |
| chr1 | 50047771 | 50048255 | 485 | 8 | 0.000268516 | 0.014698 | 0.000123 | 0.0003476 | -0.00363011 | ELAVL4 |
| chr14 | 73798075 | 73798196 | 122 | 2 | 0.000337171 | 0.002276 | 0.000244 | 0.0003487 | 0.001503332 | AC005520.2 |
| chr11 | 69708807 | 69709013 | 207 | 2 | 0.000341946 | 0.002527 | 0.000234 | 0.0003488 | 0.011184911 | NA |
| chr3 | 9703224 | 9703235 | 12 | 2 | 0.000457361 | 0.000254 | 0.002241 | 0.0003493 | 0.009109153 | AC022382.2 |
| chr16 | 69339312 | 69339368 | 57 | 5 | 0.000458718 | 0.001613 | 0.001971 | 0.0003504 | -8.24E-05 | NIP7, COG8, AC026464.3, AC026464.1 |
| chr9 | 129834520 | 129834563 | 44 | 2 | 0.000372095 | 0.007008 | 0.00017 | 0.0003512 | 0.008223005 | C9orf78 |
| chr9 | 136514631 | 136514757 | 127 | 2 | 0.000379093 | 0.000292 | 0.001663 | 0.000352 | 0.004115418 | NOTCH1 |
| chr3 | 113157088 | 113157370 | 283 | 2 | 0.000209691 | 0.00078 | 0.000482 | 0.0003523 | -0.00870854 | AC078785.1, LINC02044 |
| chr11 | 27471533 | 27471983 | 451 | 2 | 0.000164577 | 0.001776 | 0.00028 | 0.0003546 | 0.006685324 | LGR4-AS1, LGR4 |
| chr22 | 20964957 | 20965606 | 650 | 6 | 0.000208583 | 0.000129 | 0.016339 | 0.000356 | -0.00124326 | AIFM3 |
| chr19 | 19732647 | 19733125 | 479 | 6 | 2.20E-05 | 0.012587 | 0.003027 | 0.0003563 | -0.00033217 | ZNF14 |
| chr7 | 100538836 | 100539145 | 310 | 7 | 0.00029681 | 0.003735 | 0.003795 | 0.0003572 | -0.00100365 | NA |
| chr19 | 7496522 | 7497363 | 842 | 5 | 0.000217383 | 0.000294 | 0.006349 | 0.0003579 | -0.00901304 | TEX45, PEX11G |
| chr9 | 129458326 | 129458546 | 221 | 2 | 0.000483039 | 0.000531 | 0.000718 | 0.0003584 | 0.006024671 | NA |
| chr7 | 53035883 | 53036643 | 761 | 3 | 2.09E-05 | 0.020148 | 0.000126 | 0.0003591 | 0.0038916 | POM121L12 |
| chr15 | 37110287 | 37110522 | 236 | 4 | 9.78E-05 | 0.023325 | 6.94E-05 | 0.0003591 | 0.002144184 | NA |
| chr5 | 133935003 | 133935284 | 282 | 2 | 0.000111303 | 0.032203 | 0.000142 | 0.0003601 | 0.004562953 | NA |
| chr11 | 8718489 | 8718575 | 87 | 3 | 0.000213562 | 0.000164 | 0.009052 | 0.0003617 | 0.008017255 | DENND2B |
| chr5 | 51383746 | 51384131 | 386 | 6 | 0.000143019 | 0.006388 | 0.001886 | 0.0003619 | 0.004488681 | ISL1 |
| chr19 | 42290957 | 42291387 | 431 | 3 | 9.90E-05 | 0.000515 | 0.002312 | 0.0003621 | 0.006192171 | CIC |
| chr6 | 3089262 | 3089336 | 75 | 2 | 0.000314445 | 0.000204 | 0.005447 | 0.0003631 | -0.00268966 | RIPK1 |
| chr5 | 140794149 | 140794419 | 271 | 3 | 0.000176082 | 0.003348 | 0.000175 | 0.0003639 | 0.016072673 | PCDHA1, AC005609.5 |
| chr12 | 122856389 | 122857134 | 746 | 4 | 0.000293802 | 0.000317 | 0.005661 | 0.0003645 | -0.00068905 | HIP1R |
| chr15 | 93900380 | 93901012 | 633 | 7 | 6.08E-05 | 0.021703 | 4.05E-05 | 0.0003645 | 0.002282238 | LINC01580, LINC01579 |
| chr2 | 190407249 | 190407878 | 630 | 5 | 0.000280039 | 0.001054 | 0.00384 | 0.0003659 | 0.004054056 | NA |
| chr3 | 48918744 | 48918878 | 135 | 6 | 0.000313357 | 0.002796 | 0.000233 | 0.0003668 | 0.000423762 | ARIH2, ARIH2OS |
| chr14 | 95534840 | 95535032 | 193 | 6 | 0.000358667 | 0.001169 | 0.005456 | 0.000367 | 0.001296738 | GLRX5, SNHG10 |
| chr12 | 42691460 | 42691845 | 386 | 3 | 0.000257755 | 0.001899 | 0.000359 | 0.0003674 | -0.0075529 | LINC02450 |
| chr19 | 4066035 | 4066282 | 248 | 3 | 3.94E-05 | 0.000968 | 0.000444 | 0.0003675 | -0.00071918 | ZBTB7A |
| chr3 | 128994549 | 128995301 | 753 | 6 | 1.65E-05 | 0.01846 | 4.67E-05 | 0.0003677 | 0.001304736 | CFAP92 |
| chr17 | 79097324 | 79097449 | 126 | 3 | 0.000390691 | 0.011453 | 0.000118 | 0.0003701 | -0.00336181 | RBFOX3 |
| chr1 | 156601429 | 156601759 | 331 | 9 | 0.000449971 | 0.005069 | 0.00154 | 0.0003719 | 0.000544324 | GPATCH4 |
| chr2 | 102356277 | 102356332 | 56 | 3 | 0.000290423 | 0.004231 | 0.000413 | 0.0003726 | 0.001651977 | IL18R1 |
| chr12 | 113471913 | 113472143 | 231 | 6 | 0.000299048 | 0.031528 | 0.00295 | 0.0003732 | -0.0023206 | LHX5-AS1, LHX5 |
| chr5 | 129905388 | 129906307 | 920 | 8 | 0.000196741 | 0.009869 | 0.003976 | 0.0003733 | 0.003474656 | CHSY3, AC008591.1 |
| chr13 | 102596691 | 102597424 | 734 | 8 | 0.000138504 | 0.005872 | 0.003618 | 0.0003737 | 0.00028404 | TPP2 |
| chr5 | 43000688 | 43001108 | 421 | 3 | 0.000384753 | 0.001084 | 0.00191 | 0.0003745 | 0.009862091 | NA |
| chr14 | 92946023 | 92946423 | 401 | 4 | 8.59E-05 | 0.036725 | 8.86E-05 | 0.0003751 | 0.001146686 | ITPK1 |
| chr5 | 52990291 | 52990332 | 42 | 3 | 0.000198717 | 0.003994 | 0.000354 | 0.0003752 | 0.009618146 | ITGA2 |
| chr1 | 93880039 | 93880116 | 78 | 2 | 0.000396358 | 0.000211 | 0.005534 | 0.0003752 | 0.002496871 | NA |
| chr21 | 46249845 | 46250331 | 487 | 2 | 0.000306164 | 0.000283 | 0.002126 | 0.0003754 | 0.000677442 | MCM3AP-AS1, MCM3AP |
| chr5 | 80257603 | 80257787 | 185 | 2 | 0.000151329 | 0.000704 | 0.000584 | 0.0003756 | 0.010323733 | NA |
| chr8 | 1865797 | 1865987 | 191 | 4 | 0.000321942 | 0.010621 | 0.000475 | 0.0003765 | 0.000560205 | AC019257.8, ARHGEF10 |
| chr12 | 4289342 | 4289452 | 111 | 2 | 0.000218071 | 0.000307 | 0.001807 | 0.0003778 | 0.008095188 | CCND2, AC008012.1 |
| chr17 | 79082009 | 79082093 | 85 | 2 | 0.000214916 | 0.000361 | 0.001347 | 0.0003782 | 0.005253008 | ENGASE |
| chr16 | 10181910 | 10182828 | 919 | 14 | 6.30E-05 | 0.006922 | 0.016349 | 0.0003787 | 0.001193225 | GRIN2A |
| chr4 | 80264074 | 80264611 | 538 | 3 | 0.000257945 | 0.00016 | 0.009983 | 0.0003793 | 0.00983564 | NA |
| chr21 | 42513372 | 42513553 | 182 | 2 | 0.000335973 | 0.000229 | 0.004125 | 0.0003804 | 0.006008435 | SLC37A1 |
| chr17 | 39699566 | 39699977 | 412 | 5 | 3.77E-05 | 0.016985 | 0.000759 | 0.0003804 | 0.007341459 | ERBB2 |
| chr22 | 47632172 | 47632674 | 503 | 2 | 6.34E-05 | 0.000392 | 0.001205 | 0.0003814 | -0.00798185 | AL117329.1 |
| chr19 | 36417825 | 36417839 | 15 | 2 | 0.000422782 | 0.000217 | 0.005269 | 0.000382 | -0.02293365 | ZFP82 |
| chr1 | 2231243 | 2231492 | 250 | 3 | 0.000176588 | 0.000165 | 0.009642 | 0.0003824 | 0.014610485 | SKI |
| chr11 | 33891640 | 33891991 | 352 | 2 | 0.000128203 | 0.000332 | 0.001613 | 0.0003829 | 0.009681865 | LMO2 |
| chr3 | 98531769 | 98531879 | 111 | 2 | 0.000169574 | 0.00022 | 0.005115 | 0.0003831 | 0.010870657 | AC021660.3 |
| chr1 | 155323327 | 155323765 | 439 | 5 | 0.000247732 | 0.002959 | 0.000188 | 0.0003836 | 0.000571732 | RUSC1, RUSC1-AS1 |
| chr11 | 107457716 | 107458365 | 650 | 11 | 0.000281195 | 0.020298 | 2.05E-05 | 0.0003841 | -0.00034843 | CWF19L2 |
| chr13 | 27170825 | 27170882 | 58 | 3 | 0.000473615 | 0.003802 | 0.000185 | 0.0003856 | 0.001930159 | USP12 |
| chr6 | 100469040 | 100469330 | 291 | 3 | 4.74E-05 | 0.014829 | 0.000103 | 0.0003864 | 0.010802978 | NA |
| chr6 | 32190376 | 32191176 | 801 | 11 | 0.000146673 | 0.004789 | 0.005819 | 0.0003869 | 0.001379301 | GPSM3 |
| chr15 | 60509500 | 60510151 | 652 | 3 | 5.48E-05 | 0.001877 | 0.000499 | 0.0003882 | 0.005875613 | RORA-AS1, RORA |
| chr5 | 141363598 | 141363897 | 300 | 2 | 0.000180241 | 0.00035 | 0.001515 | 0.0003886 | 0.025782662 | PCDHGA1, PCDHGA2, PCDHGA3, PCDHGB1, PCDHGA4, PCDHGB2 |
| chr3 | 181712420 | 181712697 | 278 | 2 | 0.000205693 | 0.001119 | 0.000421 | 0.000389 | 0.006959594 | SOX2-OT, SOX2 |
| chr17 | 80803381 | 80803757 | 377 | 2 | 0.000260168 | 0.000253 | 0.003225 | 0.000389 | -0.00113135 | RPTOR, AC016245.1 |
| chr2 | 191792977 | 191793388 | 412 | 3 | 0.000270549 | 0.007195 | 0.000129 | 0.0003892 | -0.00230845 | AC098872.1 |
| chr1 | 46339906 | 46340703 | 798 | 10 | 0.000172705 | 0.000859 | 0.002747 | 0.0003913 | 0.000629803 | NSUN4 |
| chr5 | 160198450 | 160198583 | 134 | 2 | 0.00017935 | 0.000275 | 0.002607 | 0.0003918 | 0.010543232 | FABP6, FABP6-AS1 |
| chr5 | 123285235 | 123285256 | 22 | 2 | 0.000427731 | 0.000228 | 0.004889 | 0.0003924 | 0.009492791 | NA |
| chr7 | 120950414 | 120950661 | 248 | 7 | 0.000374664 | 0.002948 | 0.002161 | 0.0003939 | -0.00164656 | NA |
| chr15 | 90717154 | 90717687 | 534 | 10 | 0.000185267 | 0.007843 | 0.015225 | 0.0003962 | -0.00074826 | BLM |
| chr4 | 15374001 | 15374242 | 242 | 3 | 0.000370669 | 0.000781 | 0.00071 | 0.000397 | 0.00790293 | C1QTNF7, C1QTNF7-AS1, AC099550.1 |
| chr12 | 50400688 | 50400791 | 104 | 7 | 0.000453325 | 0.0607 | 4.03E-05 | 0.0003972 | 0.000358931 | LARP4 |
| chr9 | 91950011 | 91950627 | 617 | 9 | 7.50E-05 | 0.005296 | 0.001786 | 0.0003979 | -0.00048193 | ROR2 |
| chr15 | 56734601 | 56734835 | 235 | 3 | 0.00039575 | 0.000833 | 0.000864 | 0.0003982 | 0.002415678 | AC090517.4 |
| chr1 | 173866851 | 173867169 | 319 | 3 | 5.23E-05 | 0.149408 | 6.55E-05 | 0.0003983 | 0.006583464 | GAS5 |
| chr14 | 77799875 | 77799886 | 12 | 2 | 0.000498491 | 0.002069 | 0.0003 | 0.0003996 | 0.003788162 | NA |
| chr17 | 45844990 | 45845057 | 68 | 2 | 0.000496791 | 0.000226 | 0.005487 | 0.0003997 | 0.007600516 | SPPL2C, MAPT-AS1 |
| chr10 | 70114230 | 70114245 | 16 | 2 | 0.000467 | 0.001841 | 0.00032 | 0.0003999 | 0.00254761 | AIFM2 |
| chr16 | 49638706 | 49638776 | 71 | 2 | 0.000469027 | 0.007756 | 0.00019 | 0.0004027 | -0.00441701 | ZNF423 |
| chr5 | 536629 | 537579 | 951 | 5 | 5.03E-05 | 0.025084 | 4.50E-05 | 0.0004039 | 0.001792908 | NA |
| chr13 | 112753953 | 112754106 | 154 | 2 | 0.000402468 | 0.001345 | 0.000392 | 0.0004051 | 0.004718725 | ATP11A, ATP11A-AS1 |
| chr10 | 359422 | 359444 | 23 | 2 | 0.000472362 | 0.000228 | 0.005681 | 0.0004052 | 0.004228608 | DIP2C |
| chr14 | 36519334 | 36519649 | 316 | 4 | 0.000190482 | 0.011059 | 0.00062 | 0.0004065 | 0.003759002 | AL132857.1, NKX2-1-AS1, SFTA3, NKX2-1 |
| chr12 | 131710949 | 131711798 | 850 | 9 | 5.81E-06 | 0.046657 | 0.000755 | 0.0004072 | 0.002281995 | SFSWAP |
| chr7 | 158974323 | 158974444 | 122 | 2 | 0.000343435 | 0.029606 | 0.000157 | 0.000409 | -0.00533505 | NA |
| chr6 | 5998953 | 5999144 | 192 | 3 | 0.000320973 | 0.000817 | 0.001968 | 0.0004107 | 0.004661266 | NRN1 |
| chr16 | 71490009 | 71490232 | 224 | 2 | 0.000390349 | 0.00051 | 0.00097 | 0.0004108 | 0.011898941 | ZNF19 |
| chr17 | 13543821 | 13544348 | 528 | 3 | 0.000122883 | 0.000817 | 0.004682 | 0.0004108 | 0.007219718 | HS3ST3A1 |
| chr1 | 3530993 | 3531176 | 184 | 4 | 0.000227136 | 0.021244 | 0.000217 | 0.0004108 | 0.002294137 | MEGF6 |
| chr18 | 48138468 | 48138995 | 528 | 2 | 0.00010569 | 0.001931 | 0.000325 | 0.0004132 | 0.007524861 | ZBTB7C |
| chr1 | 6385572 | 6386174 | 603 | 11 | 0.000209917 | 0.041805 | 0.000136 | 0.000414 | 0.002616443 | ACOT7 |
| chr17 | 3891279 | 3891534 | 256 | 3 | 0.000386559 | 0.007359 | 0.002367 | 0.0004145 | 0.005976293 | CAMKK1 |

| | | | | | | | | | | |
|---|---|---|---|---|---|---|---|---|---|---|
| chr19 | 4852272 | 4852493 | 222 | 2 | 0.000341242 | 0.000332 | 0.001987 | 0.0004154 | -0.0005393 | PLIN3 |
| chr2 | 54329997 | 54330358 | 362 | 4 | 5.83E-05 | 0.005138 | 0.000176 | 0.0004157 | 0.004196019 | C2orf73 |
| chr17 | 44176968 | 44177294 | 327 | 5 | 0.000342271 | 0.007158 | 0.00068 | 0.0004166 | 0.001855722 | ASB16, ASB16-AS1 |
| chr18 | 47931650 | 47931872 | 223 | 2 | 6.45E-05 | 0.000699 | 0.000699 | 0.0004172 | 0.009171773 | NA |
| chr2 | 200963148 | 200963440 | 293 | 4 | 0.000412196 | 0.001966 | 0.000245 | 0.0004173 | 0.000757203 | AC005037.1, ORC2 |
| chr14 | 96592527 | 96592535 | 9 | 2 | 0.000324883 | 0.000246 | 0.004789 | 0.0004176 | -0.00989904 | AL137786.1 |
| chr14 | 24151142 | 24151215 | 74 | 3 | 0.000244857 | 0.000992 | 0.000558 | 0.0004186 | -0.00030405 | RNF31, AL136295.5 |
| chr19 | 18588516 | 18588821 | 306 | 6 | 6.16E-05 | 0.004615 | 0.000285 | 0.0004198 | -0.00576639 | REX1BD, CRLF1 |
| chr12 | 122865814 | 122866422 | 609 | 3 | 9.78E-05 | 0.000442 | 0.001674 | 0.000421 | 0.001507102 | AC027290.1, VPS37B |
| chr14 | 94025612 | 94026128 | 517 | 3 | 0.00030136 | 0.000223 | 0.008939 | 0.0004216 | 0.005085895 | NA |
| chr19 | 3786645 | 3787091 | 447 | 4 | 0.000200087 | 0.000155 | 0.017293 | 0.0004229 | 0.003575574 | MATK |
| chr2 | 45008102 | 45008373 | 272 | 3 | 0.000285956 | 0.001652 | 0.002496 | 0.0004233 | 0.010381375 | SIX2 |
| chr16 | 71809003 | 71809744 | 742 | 12 | 0.000216693 | 0.022671 | 0.004713 | 0.0004239 | -0.00077564 | AP1G1 |
| chr14 | 75299205 | 75299311 | 107 | 2 | 0.000319353 | 0.00035 | 0.001885 | 0.0004243 | -0.00249623 | NA |
| chr13 | 26760122 | 26760327 | 206 | 3 | 0.00019648 | 0.006178 | 0.001829 | 0.0004264 | 0.001058012 | GPR12 |
| chr21 | 45478696 | 45478875 | 180 | 2 | 0.000245858 | 0.001246 | 0.000446 | 0.0004276 | 0.018343914 | COL18A1 |
| chr2 | 230926062 | 230926098 | 37 | 2 | 0.000413566 | 0.000239 | 0.006047 | 0.0004279 | 0.011517301 | AC012507.3, GPR55 |
| chr20 | 64055718 | 64056697 | 980 | 4 | 4.25E-05 | 0.007916 | 0.000599 | 0.0004281 | 0.004396014 | TCEA2 |
| chr11 | 130868016 | 130868136 | 121 | 2 | 0.000300224 | 0.001271 | 0.000445 | 0.0004311 | 0.00574011 | SNX19, AP003486.1 |
| chr19 | 35666499 | 35666631 | 133 | 2 | 0.000371458 | 0.000317 | 0.002464 | 0.0004322 | 0.011527328 | UPK1A |
| chr11 | 111871567 | 111872066 | 500 | 7 | 0.000215503 | 0.000584 | 0.004487 | 0.0004329 | 0.003975849 | ALG9, AP001781.2 |
| chr1 | 155193751 | 155193870 | 120 | 2 | 0.00013386 | 0.006108 | 0.000218 | 0.0004344 | -0.00389283 | NA |
| chr6 | 111258691 | 111259180 | 490 | 9 | 0.000369273 | 0.000841 | 0.014333 | 0.0004347 | 0.003454112 | AL360227.1 |
| chr22 | 50307490 | 50307507 | 18 | 3 | 0.000215002 | 0.000713 | 0.000864 | 0.000435 | 0.001458109 | PLXNB2 |
| chr19 | 19501497 | 19502356 | 860 | 6 | 0.000226369 | 0.006319 | 0.001471 | 0.0004362 | 0.00187036 | GATAD2A |
| chr10 | 125024819 | 125025069 | 251 | 3 | 0.00022183 | 0.000673 | 0.002515 | 0.0004362 | 0.003373085 | CTBP2 |
| chr5 | 37248792 | 37248882 | 91 | 2 | 0.000422515 | 0.001964 | 0.000345 | 0.0004366 | -0.00218844 | CPLANE1 |
| chr11 | 73876415 | 73877226 | 812 | 13 | 0.000385898 | 0.000957 | 0.011674 | 0.0004394 | 0.000337318 | PAAF1, COA4 |
| chr19 | 12956740 | 12956869 | 130 | 4 | 0.000286309 | 0.073907 | 0.000229 | 0.0004396 | 0.00098717 | GADD45GIP1 |
| chr17 | 42183077 | 42183831 | 755 | 2 | 0.000196616 | 0.000594 | 0.000917 | 0.0004403 | 0.003699327 | NA |
| chr16 | 67830121 | 67830428 | 308 | 2 | 0.000465463 | 0.000251 | 0.005706 | 0.0004414 | 0.00062204 | TSNAXIP1, CENPT |
| chr19 | 11265529 | 11265832 | 304 | 2 | 0.000150058 | 0.000357 | 0.002039 | 0.0004438 | 0.004969955 | NA |
| chr3 | 123447714 | 123448675 | 962 | 4 | 0.000207869 | 0.000194 | 0.014858 | 0.0004476 | 0.002480313 | ADCY5 |
| chr6 | 30190217 | 30190923 | 707 | 10 | 0.000199639 | 0.033823 | 4.48E-05 | 0.0004492 | -0.00076902 | TRIM26 |
| chr11 | 64368403 | 64368791 | 389 | 7 | 0.000388619 | 0.001396 | 0.005757 | 0.00045 | 0.000442712 | RPS6KA4, MIR1237 |
| chr12 | 1635121 | 1635183 | 63 | 2 | 0.000383761 | 0.000302 | 0.003214 | 0.0004518 | 0.009653656 | WNT5B |
| chr20 | 2006637 | 2006874 | 238 | 2 | 0.000387942 | 0.001155 | 0.000513 | 0.0004536 | -0.01280765 | PDYN-AS1 |
| chr11 | 57567578 | 57567677 | 100 | 3 | 0.00046044 | 0.000375 | 0.002822 | 0.0004552 | -0.0005462 | UBE2L6 |
| chr19 | 35867711 | 35867881 | 171 | 2 | 0.000154293 | 0.002016 | 0.000359 | 0.0004554 | 0.005121344 | NPHS1 |
| chr3 | 141345970 | 141346403 | 434 | 4 | 6.07E-05 | 0.002025 | 0.000516 | 0.0004575 | 0.00105882 | PXYLP1, ZBTB38, AC117383.1 |
| chr10 | 133029398 | 133029680 | 283 | 3 | 0.000190762 | 0.004746 | 0.000342 | 0.0004585 | 0.008486946 | NA |
| chr1 | 109281818 | 109282465 | 648 | 2 | 0.000246639 | 0.000854 | 0.000674 | 0.0004593 | 0.007340542 | PSRC1 |
| chr15 | 91857700 | 91858143 | 444 | 2 | 0.00017991 | 0.000875 | 0.000661 | 0.0004603 | 0.010637242 | SLCO3A1 |
| chr11 | 59811566 | 59811814 | 249 | 2 | 0.000203545 | 0.000273 | 0.004849 | 0.000461 | 0.005869098 | NA |
| chr3 | 196338235 | 196338486 | 252 | 7 | 0.000155916 | 0.005052 | 0.000105 | 0.0004613 | 0.003936824 | TM4SF19-TCTEX1D2, TM4SF19 |
| chr3 | 129893519 | 129894272 | 754 | 8 | 0.00037642 | 0.000684 | 0.004107 | 0.0004613 | -0.00082796 | TMCC1-AS1, TMCC1 |
| chr16 | 58733784 | 58733819 | 36 | 3 | 0.000384419 | 0.000419 | 0.003884 | 0.0004621 | -0.00232489 | GOT2 |
| chr1 | 77759436 | 77759951 | 516 | 6 | 0.000312065 | 0.000963 | 0.006054 | 0.0004628 | 0.002090869 | USP33 |
| chr12 | 123634435 | 123634506 | 72 | 2 | 0.000184639 | 0.000269 | 0.005278 | 0.0004628 | 0.015884382 | GTF2H3 |
| chr11 | 68934562 | 68934823 | 262 | 2 | 0.000120997 | 0.036959 | 0.000171 | 0.0004632 | 0.001980484 | IGHMBP2 |
| chr6 | 149398970 | 149399123 | 154 | 2 | 0.000276774 | 0.000259 | 0.006274 | 0.0004639 | 0.011546956 | TAB2 |
| chr6 | 134636324 | 134636412 | 89 | 3 | 0.00030646 | 0.000281 | 0.005813 | 0.0004645 | 0.004467589 | AL078590.2 |
| chr11 | 126063658 | 126064016 | 359 | 5 | 0.000132642 | 0.00948 | 0.000235 | 0.0004647 | 0.000984377 | NA |
| chr3 | 53346776 | 53346846 | 71 | 2 | 0.000308443 | 0.008666 | 0.000213 | 0.000465 | 0.002886476 | CACNA1D, DCP1A |
| chr4 | 1251874 | 1251903 | 30 | 2 | 0.000382289 | 0.000432 | 0.001622 | 0.0004668 | -0.00299347 | CTBP1-DT |
| chr19 | 19489710 | 19489936 | 227 | 2 | 0.000196341 | 0.002446 | 0.000335 | 0.000467 | 0.006568512 | GATAD2A |
| chr3 | 71054462 | 71054703 | 242 | 2 | 0.000140724 | 0.009609 | 0.000209 | 0.0004676 | 0.002004438 | FOXP1, AC097634.4 |
| chr19 | 6238545 | 6238952 | 408 | 3 | 0.000423829 | 0.000609 | 0.001234 | 0.0004686 | 0.00403944 | MLLT1 |
| chr17 | 27559403 | 27559767 | 365 | 2 | 0.000131431 | 0.000953 | 0.000631 | 0.0004687 | 0.009936043 | KSR1 |
| chr10 | 117535545 | 117535889 | 345 | 2 | 0.000266284 | 0.004138 | 0.000269 | 0.0004699 | 0.011668491 | EMX2OS |
| chr22 | 26618451 | 26618929 | 479 | 3 | 0.00027948 | 0.000426 | 0.003976 | 0.0004722 | 0.005788364 | NA |
| chr17 | 5192154 | 5192950 | 797 | 5 | 5.08E-06 | 0.039536 | 6.47E-05 | 0.0004743 | 0.002322622 | AC087500.1 |
| chr10 | 124981291 | 124981944 | 654 | 3 | 9.77E-05 | 0.002104 | 0.00078 | 0.0004803 | 0.002472844 | ZRANB1 |
| chr16 | 31713149 | 31713253 | 105 | 8 | 0.000460946 | 0.00075 | 0.005051 | 0.0004807 | 0.001094849 | ZNF720 |
| chr10 | 99530411 | 99531201 | 791 | 4 | 0.000102598 | 0.000557 | 0.009376 | 0.0004836 | 0.004935945 | LINC01475 |
| chr13 | 25288300 | 25288677 | 378 | 3 | 8.03E-05 | 0.002488 | 0.001399 | 0.0004838 | 0.008836154 | NA |
| chr1 | 156660551 | 156660719 | 169 | 2 | 0.000258789 | 0.022681 | 0.000186 | 0.000484 | 0.007669881 | AL590666.2 |
| chr2 | 233760188 | 233760520 | 333 | 2 | 9.55E-05 | 0.000569 | 0.001158 | 0.0004844 | 0.007012195 | UGT1A8, UGT1A10, UGT1A9, UGT1A7, UGT1A6, UGT1A5, UGT1A4, UGT1A3, UGT1A1 |
| chr18 | 50559291 | 50559947 | 657 | 8 | 0.00014401 | 0.000265 | 0.017896 | 0.0004862 | 0.005537314 | NA |
| chr19 | 55599505 | 55599886 | 382 | 5 | 0.000327778 | 0.008807 | 0.00023 | 0.0004865 | -0.0014057 | FIZ1 |
| chr6 | 168964760 | 168964913 | 154 | 2 | 0.000267036 | 0.00066 | 0.000974 | 0.0004869 | 0.007338597 | NA |
| chr12 | 53442050 | 53442117 | 68 | 2 | 0.000173653 | 0.000289 | 0.004998 | 0.0004882 | 0.009365824 | PRR13, AC023509.1 |
| chr19 | 36378958 | 36378978 | 21 | 2 | 0.000435501 | 0.003518 | 0.000298 | 0.0004883 | -0.00089199 | ZFP14 |
| chr5 | 663524 | 663672 | 149 | 2 | 0.000430717 | 0.000668 | 0.000967 | 0.0004886 | 0.003560258 | CEP72, TPPP |
| chr6 | 50850972 | 50851341 | 370 | 3 | 0.000194913 | 0.000273 | 0.009244 | 0.000489 | 0.005028623 | NA |
| chr2 | 219252877 | 219253223 | 347 | 6 | 0.000327958 | 0.00956 | 0.00029 | 0.000493 | 0.000899783 | TUBA4A |
| chr14 | 102933663 | 102933900 | 238 | 2 | 0.000255 | 0.001641 | 0.000451 | 0.0004936 | 0.007293546 | AL117209.1, CDC42BPB |
| chr22 | 26464212 | 26464859 | 648 | 4 | 0.000147338 | 0.001211 | 0.000968 | 0.000494 | 0.004879942 | HPS4 |
| chr6 | 32089984 | 32090619 | 636 | 3 | 0.000318974 | 0.000726 | 0.004569 | 0.0004956 | -0.00528328 | TNXB |
| chr9 | 970297 | 970448 | 152 | 3 | 8.73E-05 | 0.023134 | 0.00044 | 0.0004985 | 0.006715301 | NA |
| chr11 | 67281202 | 67281391 | 190 | 2 | 0.000144219 | 0.016723 | 0.000202 | 0.0005024 | 0.00467458 | GRK2 |
| chr2 | 74515182 | 74515659 | 478 | 3 | 0.000261505 | 0.001415 | 0.00055 | 0.0005024 | 0.005777522 | TLX2 |
| chr7 | 115676326 | 115676364 | 39 | 2 | 0.000300999 | 0.000575 | 0.001234 | 0.0005034 | -0.00920529 | AC092590.1 |
| chr1 | 161196795 | 161197322 | 528 | 3 | 0.000302 | 0.000284 | 0.007452 | 0.0005041 | 0.00445712 | NDUFS2, ADAMTS4 |
| chr6 | 110358197 | 110358363 | 167 | 2 | 0.000214354 | 0.012264 | 0.000214 | 0.0005048 | 0.003929819 | METTL24 |
| chr16 | 19578813 | 19579232 | 420 | 4 | 5.96E-05 | 0.003941 | 0.0005 | 0.0005066 | 0.001015006 | VPS35L |
| chr12 | 53727522 | 53727930 | 409 | 8 | 0.000378364 | 0.007725 | 0.000812 | 0.0005086 | 0.000170072 | CALCOCO1 |
| chr3 | 126395506 | 126395863 | 358 | 3 | 2.99E-05 | 0.006027 | 0.001111 | 0.0005088 | -0.00121709 | CFAP100 |
| chr8 | 100682745 | 100683114 | 370 | 2 | 6.18E-05 | 0.012778 | 0.000214 | 0.0005092 | 0.002582277 | NA |
| chr2 | 163736711 | 163736754 | 44 | 2 | 0.000422966 | 0.003534 | 0.000313 | 0.0005093 | 0.002811931 | NA |
| chr7 | 155167197 | 155167331 | 135 | 4 | 0.000310268 | 0.002783 | 0.003764 | 0.0005107 | 0.00080119 | NA |
| chr17 | 81976978 | 81977864 | 887 | 8 | 7.30E-05 | 0.039459 | 0.000315 | 0.0005108 | 6.92E-05 | ASPSCR1 |
| chr15 | 45634852 | 45634928 | 77 | 4 | 0.000500215 | 0.00024 | 0.01005 | 0.0005135 | -0.00023228 | AC090527.2, SQOR |
| chr14 | 39431887 | 39432611 | 725 | 8 | 0.000116236 | 0.004172 | 0.000265 | 0.0005153 | -0.00049278 | AL049828.1, FBXO33 |
| chr8 | 1359967 | 1360114 | 148 | 3 | 0.000322585 | 0.005604 | 0.000193 | 0.0005156 | -0.00175375 | DLGAP2 |
| chr7 | 128240514 | 128240566 | 53 | 2 | 0.000150995 | 0.000297 | 0.005794 | 0.000517 | -0.00833619 | NA |
| chr7 | 1944283 | 1944349 | 67 | 2 | 0.000293181 | 0.000289 | 0.006625 | 0.0005176 | 0.02016831 | MAD1L1, AC069288.1 |
| chr5 | 93582362 | 93582366 | 5 | 2 | 0.000454468 | 0.000297 | 0.005916 | 0.000518 | 0.012425449 | NR2F1-AS1 |
| chr12 | 56996191 | 56996246 | 56 | 2 | 0.00038885 | 0.007429 | 0.000248 | 0.0005198 | 0.004197216 | GPR182 |
| chr3 | 128466847 | 128467026 | 180 | 2 | 7.60E-05 | 0.004222 | 0.000298 | 0.0005201 | 0.004869662 | DNAJB8-AS1, DNAJB8 |
| chr21 | 14382615 | 14382617 | 3 | 2 | 0.000352219 | 0.005066 | 0.00028 | 0.0005207 | 0.002276458 | HSPA13 |
| chr18 | 79899695 | 79899949 | 255 | 2 | 0.000318419 | 0.000301 | 0.005818 | 0.0005225 | -0.02379808 | KCNG2 |
| chr5 | 56872743 | 56872885 | 143 | 2 | 0.000162673 | 0.015981 | 0.000212 | 0.0005242 | -0.012646 | MAP3K1 |
| chr12 | 131844936 | 131845222 | 287 | 3 | 0.000350118 | 0.000528 | 0.00246 | 0.0005284 | 0.006642369 | MMP17 |
| chr11 | 832065 | 832357 | 293 | 3 | 0.000168013 | 0.002686 | 0.003042 | 0.0005311 | 0.006648669 | AP006621.1 |
| chr7 | 4144344 | 4144722 | 379 | 3 | 0.000176112 | 0.002319 | 0.000375 | 0.0005319 | -0.01253274 | SDK1 |
| chr1 | 197919764 | 197920033 | 270 | 2 | 0.000168989 | 0.000297 | 0.006897 | 0.0005348 | -0.00028042 | LHX9 |
| chr15 | 72197268 | 72197487 | 220 | 3 | 0.000274788 | 0.000507 | 0.004534 | 0.000536 | 0.005555567 | GRAMD2A |
| chr14 | 99939279 | 99939597 | 319 | 2 | 0.000254053 | 0.00093 | 0.000804 | 0.0005373 | 0.003475733 | EML1 |
| chr14 | 102039088 | 102039474 | 387 | 4 | 9.80E-05 | 0.011036 | 0.000319 | 0.0005376 | 0.001853747 | DYNC1H1, AL118558.1 |
| chr19 | 8592255 | 8592858 | 604 | 7 | 0.000307228 | 0.01073 | 0.00117 | 0.0005387 | -0.00087884 | ADAMTS10 |
| chr6 | 157672851 | 157673028 | 178 | 3 | 0.000165155 | 0.002803 | 0.003239 | 0.0005397 | -0.00187838 | ZDHHC14 |
| chr16 | 564645 | 564852 | 208 | 2 | 0.000178819 | 0.0004 | 0.002774 | 0.0005408 | 0.004676348 | PRR35 |
| chr2 | 24008887 | 24009733 | 847 | 3 | 4.51E-05 | 0.000307 | 0.00945 | 0.0005416 | 0.000702896 | NA |
| chr5 | 442270 | 442706 | 437 | 5 | 0.000299694 | 0.001714 | 0.018494 | 0.0005427 | -0.00342077 | EXOC3-AS1 |
| chr1 | 166876384 | 166876518 | 135 | 7 | 0.000484041 | 0.001225 | 0.005511 | 0.000543 | -0.00068233 | NA |
| chr9 | 136695630 | 136695641 | 12 | 2 | 0.000374396 | 0.000316 | 0.005663 | 0.0005435 | 0.006926967 | NA |
| chr14 | 24298916 | 24299657 | 742 | 8 | 1.76E-05 | 0.000931 | 0.005658 | 0.0005445 | 0.001694489 | DHRS1 |
| chr19 | 13014912 | 13015174 | 263 | 3 | 0.000316322 | 0.012451 | 0.001115 | 0.0005485 | 0.004995499 | NFIX |
| chr19 | 5689932 | 5690288 | 357 | 7 | 0.000286262 | 0.001566 | 0.007735 | 0.0005496 | 0.002996409 | RPL36 |
| chr16 | 30659759 | 30660428 | 670 | 4 | 7.59E-05 | 0.003781 | 0.000225 | 0.0005512 | 0.001670977 | FBRS |
| chr15 | 76336190 | 76336815 | 626 | 9 | 0.000467635 | 0.00213 | 0.007824 | 0.0005519 | -0.00047285 | ISL2 |
| chr7 | 100611162 | 100611327 | 166 | 2 | 5.16E-05 | 0.000589 | 0.001441 | 0.0005519 | 0.006523107 | NA |
| chr10 | 121736160 | 121736186 | 27 | 3 | 0.000303047 | 0.000786 | 0.005407 | 0.0005526 | 0.005558008 | NA |
| chr14 | 74762568 | 74762667 | 100 | 2 | 0.000124142 | 0.00046 | 0.002212 | 0.0005553 | 0.015112971 | NA |
| chr8 | 141270995 | 141271472 | 478 | 2 | 0.000294338 | 0.000488 | 0.002022 | 0.0005602 | 0.004799674 | SLC45A4 |
| chr13 | 45465197 | 45465636 | 440 | 3 | 0.000247762 | 0.000239 | 0.012847 | 0.0005634 | 0.003676602 | COG3 |
| chr15 | 73927304 | 73927607 | 304 | 3 | 0.000163533 | 0.009123 | 0.00142 | 0.0005649 | -0.00041973 | LOXL1, LOXL1-AS1 |
| chr9 | 33001638 | 33002023 | 386 | 9 | 0.000376806 | 0.00485 | 0.011053 | 0.0005657 | 0.003220179 | APTX |
| chr8 | 143732741 | 143733105 | 365 | 5 | 0.000216649 | 0.019146 | 0.000138 | 0.0005695 | 0.003215606 | FAM83H, MIR4664 |
| chr18 | 6966221 | 6966245 | 25 | 3 | 0.000378118 | 0.000566 | 0.001792 | 0.0005702 | -0.00231139 | LAMA1 |
| chr14 | 93957053 | 93957810 | 758 | 8 | 0.000281564 | 0.003703 | 0.001787 | 0.0005705 | 0.000215126 | ASB2 |
| chr17 | 80205512 | 80205961 | 450 | 3 | 0.000368048 | 0.002675 | 0.000505 | 0.0005744 | 0.003417658 | CARD14, AC087741.1 |
| chr10 | 129967115 | 129967514 | 400 | 2 | 0.000134094 | 0.000685 | 0.001271 | 0.0005749 | 0.004858281 | EBF3 |
| chr17 | 1486208 | 1486281 | 74 | 3 | 0.000317106 | 0.002001 | 0.000539 | 0.0005777 | -0.00153109 | MYO1C |
| chr1 | 219174545 | 219174897 | 353 | 3 | 0.000154144 | 0.008434 | 0.000183 | 0.0005778 | -0.00043783 | LYPLAL1 |
| chr10 | 12262571 | 12262878 | 308 | 2 | 0.000144043 | 0.001049 | 0.000808 | 0.0005783 | 0.008576945 | NA |
| chr1 | 1178792 | 1179199 | 408 | 2 | 0.000372461 | 0.003024 | 0.000393 | 0.0005794 | 0.003592402 | TTLL10, TTLL10-AS1 |
| chr12 | 12356373 | 12356791 | 419 | 5 | 0.000387861 | 0.000243 | 0.012875 | 0.0005804 | -0.00718224 | LOH12CR2 |
| chr22 | 22560259 | 22560340 | 82 | 2 | 0.000297302 | 0.002061 | 0.000487 | 0.0005808 | 0.002343604 | LL22NC03-63E9.3 |
| chr4 | 121150817 | 121150887 | 71 | 2 | 0.000307523 | 0.001101 | 0.00078 | 0.0005812 | 0.004183114 | TNIP3 |
| chr16 | 66887380 | 66887690 | 311 | 2 | 0.000207282 | 0.001035 | 0.000826 | 0.0005822 | 0.015318159 | PDP2 |
| chr21 | 45632583 | 45633158 | 576 | 2 | 0.000109192 | 0.000841 | 0.001029 | 0.0005841 | 0.01071698 | NA |
| chr1 | 31066084 | 31066767 | 684 | 6 | 0.000155926 | 0.000283 | 0.015143 | 0.000585 | 0.001116342 | NA |
| chr17 | 28711843 | 28712114 | 272 | 6 | 0.000254496 | 0.000113 | 0.037389 | 0.0005858 | 0.000564484 | PROCA1 |
| chr14 | 68793936 | 68794362 | 427 | 5 | 4.90E-05 | 0.025869 | 0.0001 | 0.0005866 | 0.003068399 | ZFP36L1 |
| chr17 | 3093777 | 3094111 | 335 | 2 | 0.000358635 | 0.000324 | 0.007356 | 0.0005872 | 0.018296125 | OR1D2 |
| chr6 | 151805650 | 151805803 | 154 | 3 | 0.000181605 | 0.000604 | 0.003532 | 0.0005887 | 0.01451949 | ESR1 |
| chr19 | 3924788 | 3924983 | 196 | 2 | 0.000187928 | 0.004856 | 0.000327 | 0.0005904 | 0.005534245 | ATCAY |
| chr11 | 20161029 | 20161462 | 434 | 4 | 0.00037331 | 0.000577 | 0.008152 | 0.0005916 | 0.004132155 | NA |
| chr3 | 179346673 | 179347049 | 377 | 3 | 0.000273945 | 0.001068 | 0.005514 | 0.0005928 | 0.011498298 | NA |
| chr12 | 49020370 | 49020678 | 309 | 2 | 7.93E-05 | 0.072565 | 0.000203 | 0.0005931 | 0.006041505 | KMT2D |
| chr19 | 15256248 | 15256254 | 7 | 2 | 0.000210699 | 0.005365 | 0.000317 | 0.000594 | -0.00237628 | BRD4 |
| chr3 | 48655630 | 48655686 | 57 | 2 | 0.000269001 | 0.000826 | 0.001088 | 0.0005962 | 0.005469887 | CELSR3 |
| chr2 | 79512737 | 79512792 | 56 | 2 | 0.000442944 | 0.007667 | 0.000285 | 0.0005997 | 0.002610909 | CTNNA2, CTNNA2-AS1 |
| chr1 | 151612886 | 151613229 | 344 | 2 | 3.26E-05 | 0.000339 | 0.006771 | 0.0006013 | -0.00062595 | SNX27, AL391335.1 |
| chr7 | 76397239 | 76397331 | 93 | 3 | 0.00039311 | 0.003823 | 0.000598 | 0.0006023 | 0.004850295 | SSC4D |
| chr6 | 41286019 | 41286733 | 715 | 4 | 0.000230817 | 0.008163 | 0.002158 | 0.0006032 | 0.001498351 | TREM1 |
| chr1 | 207323917 | 207324450 | 534 | 3 | 7.70E-05 | 0.003262 | 0.000386 | 0.0006036 | 0.002845382 | CD55 |
| chr6 | 3259503 | 3259727 | 225 | 3 | 9.61E-05 | 0.00491 | 0.002085 | 0.0006052 | 0.000838248 | PSMG4 |
| chr15 | 69463010 | 69463044 | 35 | 3 | 0.000472488 | 0.000344 | 0.007305 | 0.0006056 | -0.0011071 | DRAIC |
| chr11 | 64185419 | 64186040 | 622 | 9 | 0.000290405 | 0.00084 | 0.025514 | 0.0006089 | 0.001050628 | STIP1 |

| | | | | | | | | | | |
|---|---|---|---|---|---|---|---|---|---|---|
| chr8 | 140535423 | 140535649 | 227 | 5 | 0.000327916 | 0.063993 | 0.000121 | 0.0006137 | 0.000770222 | AGO2 |
| chr10 | 131927652 | 131928066 | 415 | 4 | 0.000314316 | 0.001156 | 0.000744 | 0.0006164 | 0.00010303 | PPP2R2D |
| chr19 | 13951885 | 13952219 | 335 | 4 | 0.000163221 | 0.000736 | 0.002285 | 0.000619 | 0.002980532 | PODNL1 |
| chr6 | 29723631 | 29723983 | 353 | 6 | 0.000297842 | 0.001919 | 0.009835 | 0.0006194 | 2.52E-05 | HLA-F, AL645939.1 |
| chr6 | 161273698 | 161273808 | 111 | 2 | 0.000328965 | 0.0019 | 0.00056 | 0.0006197 | -0.0034104 | AGPAT4 |
| chr4 | 183322484 | 183323323 | 840 | 5 | 0.000213713 | 0.002742 | 0.000499 | 0.0006207 | 0.003366845 | NA |
| chr19 | 42217308 | 42217726 | 419 | 5 | 9.84E-05 | 0.00054 | 0.006683 | 0.0006224 | -0.00055421 | DEDD2, AC006486.3 |
| chr5 | 1092951 | 1093623 | 673 | 4 | 9.36E-05 | 0.000558 | 0.003245 | 0.0006226 | 0.001945206 | SLC12A7 |
| chr6 | 158167731 | 158167879 | 149 | 2 | 0.000200471 | 0.006821 | 0.000309 | 0.0006266 | 0.002389108 | SERAC1 |
| chr14 | 21268844 | 21269539 | 696 | 6 | 7.88E-05 | 0.002199 | 0.001471 | 0.0006268 | -0.00308619 | HNRNPC |
| chr5 | 141234980 | 141235193 | 214 | 3 | 0.00027396 | 0.000846 | 0.001237 | 0.0006268 | -0.00132856 | AC244517.10, PCDHB18P, AC244517.11 |
| chr1 | 1074361 | 1075258 | 898 | 4 | 0.00024461 | 0.000215 | 0.015351 | 0.0006273 | 0.00661879 | NA |
| chr17 | 1605791 | 1606786 | 996 | 7 | 6.09E-05 | 0.003972 | 0.001734 | 0.0006282 | -0.0002184 | SLC43A2 |
| chr4 | 53653042 | 53653087 | 46 | 2 | 0.000376461 | 0.001217 | 0.000811 | 0.0006309 | 0.006676945 | AC058822.1, LNX1 |
| chr1 | 247604674 | 247604734 | 61 | 2 | 0.000269626 | 0.00106 | 0.00093 | 0.0006345 | 0.019511101 | AL606804.1 |
| chr12 | 57744021 | 57744412 | 392 | 2 | 0.000157194 | 0.000352 | 0.007747 | 0.0006386 | 0.012435948 | TSPAN31 |
| chr4 | 112145135 | 112145592 | 458 | 5 | 0.000142984 | 0.000772 | 0.002284 | 0.0006403 | 0.003930963 | FAM241A |
| chr3 | 194826789 | 194826795 | 7 | 2 | 0.000428471 | 0.000625 | 0.001814 | 0.0006404 | -0.00312037 | NA |
| chr8 | 127793836 | 127794025 | 190 | 2 | 0.000242632 | 0.000839 | 0.001217 | 0.0006406 | 0.010013811 | NA |
| chr1 | 54406435 | 54406440 | 6 | 2 | 0.000349415 | 0.001056 | 0.000954 | 0.0006426 | -0.00078814 | SSBP3 |
| chr6 | 36757510 | 36758683 | 1174 | 3 | 4.45E-06 | 0.007323 | 0.000274 | 0.0006441 | 0.004361632 | CPNE5 |
| chr12 | 2795465 | 2795746 | 282 | 2 | 0.000154008 | 0.00037 | 0.006566 | 0.0006448 | 0.00258047 | FKBP4, ITFG2-AS1 |
| chr12 | 49347744 | 49347840 | 97 | 2 | 0.000275801 | 0.0004 | 0.005097 | 0.0006488 | 0.005463498 | DNAJC22 |
| chr1 | 217136832 | 217137041 | 210 | 2 | 0.000378786 | 0.000539 | 0.00244 | 0.0006506 | 0.006919197 | ESRRG |
| chr6 | 33193070 | 33193434 | 365 | 11 | 0.000185089 | 0.008384 | 0.000173 | 0.0006525 | -7.30E-05 | NA |
| chr5 | 107389720 | 107390041 | 322 | 2 | 0.000416248 | 0.000414 | 0.004679 | 0.0006531 | 0.014785571 | EFNA5 |
| chr10 | 95561828 | 95561923 | 96 | 3 | 0.000245332 | 0.002336 | 0.000654 | 0.0006594 | 0.01178484 | NA |
| chr9 | 6646848 | 6647012 | 165 | 2 | 0.000270619 | 0.001311 | 0.000816 | 0.0006608 | -0.00579269 | AL162411.1 |
| chr17 | 9791676 | 9792099 | 424 | 3 | 0.000180822 | 0.000551 | 0.002618 | 0.000663 | 0.005734794 | AC027045.2, AC027045.1 |
| chr6 | 110813803 | 110813893 | 91 | 2 | 0.00016812 | 0.000432 | 0.004389 | 0.0006648 | 0.015297191 | CDK19 |
| chr12 | 124440897 | 124441206 | 310 | 3 | 0.000134536 | 0.002577 | 0.001089 | 0.0006659 | 0.000526615 | NCOR2 |
| chr12 | 9669390 | 9669691 | 302 | 3 | 0.000319496 | 0.015904 | 0.000216 | 0.0006663 | 0.012251955 | CLEC2D |
| chr16 | 29790281 | 29790655 | 375 | 8 | 0.000394802 | 0.002084 | 0.005216 | 0.0006668 | 0.002243948 | NA |
| chr3 | 18438842 | 18439572 | 731 | 5 | 0.000195463 | 0.001544 | 0.005011 | 0.0006739 | 0.007668272 | TBC1D5, SATB1 |
| chr2 | 55518879 | 55519151 | 273 | 4 | 0.000207329 | 0.000757 | 0.007042 | 0.0006872 | -0.00859148 | NA |
| chr6 | 33451989 | 33453028 | 1040 | 6 | 0.00015464 | 0.0011 | 0.018136 | 0.0006876 | 0.002667854 | SYNGAP1, SYNGAP1-AS1 |
| chr17 | 29139108 | 29139132 | 25 | 2 | 0.000385898 | 0.012772 | 0.000288 | 0.0006889 | -0.00101581 | MYO18A |
| chr21 | 44709477 | 44709705 | 229 | 2 | 0.000248348 | 0.000422 | 0.005444 | 0.0006907 | 0.006746991 | TSPEAR |
| chr8 | 144496177 | 144496513 | 337 | 3 | 0.00036705 | 0.010119 | 0.000252 | 0.000691 | 0.002985884 | PPP1R16A, AC084125.2 |
| chr8 | 143815942 | 143816155 | 214 | 3 | 0.000155663 | 0.005183 | 0.000337 | 0.0006921 | 0.002124045 | NA |
| chr17 | 59106245 | 59106826 | 582 | 8 | 0.000228171 | 0.007364 | 0.013515 | 0.0006941 | 0.000514129 | AC099850.1, TRIM37 |
| chr17 | 81238214 | 81238544 | 331 | 2 | 6.71E-05 | 0.160959 | 0.000228 | 0.000697 | 0.002472406 | TEPSIN |
| chr2 | 37323989 | 37324457 | 469 | 4 | 0.000217688 | 0.003343 | 0.003586 | 0.0007024 | 0.001898026 | PRKD3 |
| chr7 | 26199820 | 26200720 | 901 | 4 | 4.90E-05 | 0.000188 | 0.028983 | 0.0007093 | -0.00432453 | HNRNPA2B1 |
| chr11 | 86069804 | 86070002 | 199 | 3 | 0.000403965 | 0.00028 | 0.014753 | 0.000715 | 0.001369419 | PICALM |
| chr19 | 42283022 | 42283610 | 589 | 2 | 0.000280467 | 0.000398 | 0.007867 | 0.0007154 | 0.007842662 | CIC |
| chr14 | 100979650 | 100980000 | 351 | 2 | 0.0001677 | 0.012739 | 0.0003 | 0.0007194 | 0.008341438 | MEG8, AL132709.4 |
| chr10 | 28280716 | 28281368 | 653 | 3 | 9.98E-05 | 0.155726 | 9.53E-05 | 0.0007202 | 0.002259049 | MPP7 |
| chr1 | 201378351 | 201378789 | 439 | 5 | 0.00014355 | 0.0025 | 0.00055 | 0.0007203 | 0.001256546 | AC119427.2 |
| chr16 | 17158402 | 17158425 | 24 | 2 | 0.000412995 | 0.073298 | 0.00024 | 0.0007207 | -0.0029363 | XYLT1 |
| chr2 | 68466362 | 68467385 | 1024 | 9 | 4.79E-05 | 0.019646 | 0.003285 | 0.0007216 | -0.00107166 | FBXO48 |
| chr12 | 21765504 | 21765850 | 347 | 2 | 0.000258144 | 0.002661 | 0.000557 | 0.0007221 | 0.007135435 | AC010185.1, KCNJ8 |
| chr13 | 49792921 | 49793037 | 117 | 6 | 0.000334021 | 0.000472 | 0.014899 | 0.0007286 | 0.001200407 | AL136301.1 |
| chr1 | 247103763 | 247104023 | 261 | 2 | 0.000338599 | 0.000814 | 0.001621 | 0.0007286 | -0.00061311 | ZNF669 |
| chr10 | 35126584 | 35126678 | 95 | 7 | 0.00042475 | 0.001026 | 0.020722 | 0.00073 | 0.000420866 | AL392046.1 |
| chr9 | 114305856 | 114305879 | 24 | 2 | 0.000454602 | 0.019339 | 0.000281 | 0.0007325 | 0.008463999 | COL27A1 |
| chr9 | 33415280 | 33415603 | 324 | 3 | 0.000237585 | 0.052585 | 0.000304 | 0.0007352 | 0.006867639 | NA |
| chr1 | 155254978 | 155255072 | 95 | 3 | 0.000101729 | 0.017901 | 0.000193 | 0.0007357 | -0.0005001 | FAM189B |
| chr19 | 51021402 | 51021507 | 106 | 3 | 0.000438509 | 0.001862 | 0.000792 | 0.0007365 | 0.012374317 | NA |
| chr4 | 109433740 | 109434369 | 630 | 4 | 0.000314515 | 0.006779 | 0.002757 | 0.0007365 | 0.000550768 | SEC24B, SEC24B-AS1 |
| chr14 | 44896705 | 44897046 | 342 | 3 | 0.000208281 | 0.016227 | 0.00331 | 0.0007382 | -0.00370639 | AL049870.2 |
| chr14 | 45180017 | 45180405 | 389 | 2 | 0.000233454 | 0.000678 | 0.0022 | 0.0007391 | 0.002531402 | FANCM |
| chr4 | 37890195 | 37890213 | 19 | 2 | 0.000363858 | 0.000418 | 0.007546 | 0.0007409 | 0.009808577 | NA |
| chr6 | 34888108 | 34888440 | 333 | 7 | 0.000202832 | 0.007763 | 0.000507 | 0.0007413 | 0.000180931 | NA |
| chr9 | 131823190 | 131823759 | 570 | 3 | 0.000244037 | 0.001988 | 0.005928 | 0.0007466 | 0.004508739 | NA |
| chr5 | 93574262 | 93574346 | 85 | 2 | 0.000296402 | 0.003234 | 0.000528 | 0.000754 | 0.00645305 | NR2F1-AS1 |
| chr5 | 134367208 | 134367828 | 621 | 3 | 0.000390959 | 0.000451 | 0.010784 | 0.0007618 | 0.010857294 | AC104109.4, CDKL3 |
| chr10 | 92691237 | 92691868 | 632 | 7 | 8.68E-05 | 0.029716 | 0.003418 | 0.0007625 | 0.003156132 | HHEX |
| chr11 | 560244 | 560294 | 51 | 2 | 0.000432371 | 0.000682 | 0.002339 | 0.0007627 | 0.011077355 | LMNTD2 |
| chr11 | 73142279 | 73142361 | 83 | 3 | 0.000354476 | 0.009555 | 0.000336 | 0.0007628 | 0.002115247 | FCHSD2 |
| chr12 | 74537439 | 74537774 | 336 | 9 | 0.00033526 | 0.07261 | 0.006357 | 0.0007664 | 0.000383719 | NA |
| chr12 | 54261242 | 54261844 | 603 | 2 | 5.56E-05 | 0.003409 | 0.000524 | 0.0007665 | 0.007748012 | CBX5 |
| chr4 | 8079499 | 8079757 | 259 | 3 | 0.0002068 | 0.000996 | 0.002648 | 0.0007668 | 0.009448299 | ABLIM2 |
| chr1 | 53196571 | 53196848 | 278 | 4 | 0.00041126 | 0.001463 | 0.005667 | 0.0007682 | -0.00174282 | CPT2 |
| chr3 | 138115183 | 138115749 | 567 | 8 | 0.000266809 | 0.000389 | 0.010195 | 0.000769 | 0.001935381 | DZIP1L |
| chr1 | 160080485 | 160080591 | 107 | 2 | 0.000369087 | 0.015254 | 0.00031 | 0.0007721 | -0.01756158 | AL121987.1 |
| chr1 | 231282021 | 231282310 | 290 | 3 | 0.000178928 | 0.001327 | 0.001196 | 0.0007766 | 0.003990836 | NA |
| chr19 | 55302775 | 55302850 | 76 | 2 | 0.000353877 | 0.006346 | 0.000409 | 0.0007871 | -0.00262688 | BRSK1 |
| chr15 | 79433139 | 79433300 | 162 | 2 | 0.000285711 | 0.004611 | 0.000469 | 0.0007899 | 0.01162572 | MINAR1 |
| chr17 | 81082261 | 81082317 | 57 | 2 | 0.000306256 | 0.001762 | 0.000845 | 0.0007957 | -0.00322346 | BAIAP2 |
| chr13 | 25180580 | 25180713 | 134 | 2 | 0.000213331 | 0.000682 | 0.002601 | 0.0007965 | -0.01144784 | LINC01053 |
| chr6 | 99425773 | 99426067 | 295 | 3 | 0.000380876 | 0.000812 | 0.005627 | 0.0007976 | 0.000536739 | AL513550.1 |
| chr17 | 75846512 | 75847148 | 637 | 3 | 0.000174028 | 0.002961 | 0.000746 | 0.000798 | 0.002534811 | WBP2 |
| chr18 | 4106506 | 4107003 | 498 | 2 | 0.000276777 | 0.001296 | 0.001117 | 0.0007983 | 0.006288505 | DLGAP1 |
| chr22 | 50286415 | 50286420 | 6 | 2 | 0.000350423 | 0.000747 | 0.002277 | 0.0008053 | 0.006382204 | PLXNB2 |
| chr13 | 30617687 | 30618371 | 685 | 5 | 3.06E-05 | 0.002353 | 0.007712 | 0.0008071 | -0.00144361 | USPL1 |
| chr21 | 45748625 | 45749309 | 685 | 3 | 0.000469498 | 0.000637 | 0.008079 | 0.0008071 | 0.012058238 | PCBP3 |
| chr6 | 36196880 | 36197026 | 147 | 2 | 0.000204354 | 0.001115 | 0.001348 | 0.0008123 | 0.005344392 | BRPF3, Z84485.1 |
| chr8 | 143573277 | 143573383 | 107 | 3 | 0.000370366 | 0.000435 | 0.007857 | 0.0008155 | 0.008916086 | NA |
| chr13 | 80339205 | 80339313 | 109 | 2 | 0.00017883 | 0.005491 | 0.000453 | 0.0008166 | 0.006586909 | SPRY2 |
| chr1 | 32221135 | 32222317 | 1183 | 10 | 6.93E-05 | 0.018964 | 0.000751 | 0.0008217 | 0.001833496 | EIF3I, TMEM234 |
| chr20 | 18138005 | 18138068 | 64 | 3 | 0.000470758 | 0.001403 | 0.001614 | 0.0008308 | 0.001573825 | KAT14, PET117 |
| chr20 | 38032284 | 38032836 | 553 | 2 | 0.00023125 | 0.001531 | 0.001021 | 0.0008314 | 0.006126712 | TTI1 |
| chr9 | 93826268 | 93826548 | 281 | 3 | 0.000316362 | 0.000337 | 0.013614 | 0.0008326 | -0.00884314 | AL442224.1 |
| chr1 | 1144181 | 1144499 | 319 | 3 | 7.80E-05 | 0.058426 | 0.000151 | 0.0008351 | 0.002849625 | NA |
| chr17 | 48564142 | 48564346 | 205 | 2 | 0.000242258 | 0.000688 | 0.002916 | 0.0008394 | -0.01461472 | HOXB-AS3, HOXB3 |
| chr22 | 43173784 | 43174193 | 410 | 3 | 4.31E-05 | 0.023974 | 0.00017 | 0.0008444 | 0.002102554 | TTLL12 |
| chr4 | 67546196 | 67546247 | 52 | 2 | 0.000206655 | 0.021674 | 0.000319 | 0.0008507 | -0.01703279 | NA |
| chr20 | 61902599 | 61902761 | 163 | 2 | 0.000335484 | 0.002415 | 0.000744 | 0.0008546 | 0.003814558 | CDH4 |
| chr1 | 151717111 | 151717599 | 489 | 5 | 0.000198431 | 0.004828 | 0.001889 | 0.0008567 | 0.01117404 | RIIAD1 |
| chr7 | 100428723 | 100428861 | 139 | 5 | 0.000349988 | 0.000182 | 0.040157 | 0.0008593 | 0.000210261 | MEPCE, ZCWPW1 |
| chr19 | 46022191 | 46022308 | 118 | 2 | 0.000199526 | 0.000475 | 0.008926 | 0.0008597 | -0.0140453 | PGLYRP1 |
| chr5 | 16942726 | 16943301 | 576 | 2 | 0.000143148 | 0.007304 | 0.000431 | 0.0008625 | 0.000389208 | NA |
| chr19 | 12141078 | 12141296 | 219 | 2 | 0.000355528 | 0.000566 | 0.00494 | 0.0008627 | -0.01309898 | ZNF625-ZNF20 |
| chr2 | 235851610 | 235852296 | 687 | 4 | 0.000190483 | 0.003465 | 0.000558 | 0.000865 | 0.001793006 | AGAP1 |
| chr9 | 97468690 | 97469405 | 716 | 2 | 0.000197125 | 0.000869 | 0.002121 | 0.000868 | 0.005687725 | TDRD7 |
| chr9 | 137011862 | 137011930 | 69 | 2 | 0.000354403 | 0.003478 | 0.00061 | 0.0008714 | 0.00356649 | ABCA2 |
| chr7 | 150372488 | 150372801 | 314 | 5 | 0.00031479 | 0.247529 | 5.32E-05 | 0.0008761 | 0.001507699 | REPIN1, ZNF775, REPIN1-AS1 |
| chr14 | 67620163 | 67620622 | 460 | 2 | 2.71E-05 | 0.007962 | 0.000426 | 0.0008761 | 0.009216972 | ARG2 |
| chr19 | 10565510 | 10565642 | 133 | 2 | 0.00032054 | 0.001113 | 0.001557 | 0.0008774 | 0.003671694 | KRI1 |
| chr11 | 125112242 | 125112262 | 21 | 2 | 0.000398707 | 0.000482 | 0.009252 | 0.0008778 | 0.009921136 | NA |
| chr20 | 381718 | 382067 | 350 | 2 | 0.000109596 | 0.000672 | 0.003459 | 0.0008783 | 0.004032031 | TRIB3 |
| chr10 | 133376868 | 133377160 | 293 | 4 | 5.03E-05 | 0.008603 | 0.000251 | 0.000891 | 0.000726288 | NA |
| chr6 | 170265835 | 170266295 | 461 | 3 | 3.50E-05 | 0.021141 | 0.000184 | 0.0008923 | 0.004815528 | NA |
| chr5 | 75511126 | 75512004 | 879 | 8 | 2.50E-05 | 0.026222 | 0.002841 | 0.0008932 | 0.001056135 | POLK, CERT1 |
| chr10 | 101227051 | 101228337 | 1287 | 7 | 2.03E-05 | 0.056241 | 0.000159 | 0.000896 | 0.001308919 | LBX1 |
| chr8 | 22693207 | 22693444 | 238 | 5 | 0.000201249 | 0.004126 | 0.001166 | 0.0009053 | -0.00166419 | AC055854.1, EGR3 |
| chr5 | 3854351 | 3854479 | 129 | 3 | 0.00031523 | 0.001873 | 0.00499 | 0.0009068 | 0.005158119 | NA |
| chr9 | 136371841 | 136372475 | 635 | 3 | 0.000117955 | 0.035059 | 0.000159 | 0.0009188 | -0.00067751 | CARD9 |
| chr2 | 218658659 | 218659257 | 599 | 4 | 0.00016086 | 0.000829 | 0.003917 | 0.0009188 | 0.001041659 | BCS1L, ZNF142 |
| chr4 | 7067549 | 7067573 | 25 | 3 | 0.00037825 | 0.001216 | 0.006242 | 0.0009221 | 0.00254019 | GRPEL1 |
| chr16 | 70725192 | 70725731 | 540 | 3 | 0.000222242 | 0.000668 | 0.005128 | 0.0009256 | 0.006863576 | VAC14 |
| chr3 | 188901731 | 188901875 | 145 | 3 | 0.000314719 | 0.011205 | 0.000299 | 0.0009322 | 0.00287437 | NA |
| chr12 | 53972976 | 53973075 | 100 | 4 | 0.000344044 | 0.015081 | 0.000483 | 0.0009322 | 0.005025123 | HOTAIR |
| chr7 | 15562217 | 15562360 | 144 | 2 | 0.000346328 | 0.000521 | 0.008915 | 0.0009334 | 0.012217083 | NA |
| chr1 | 35577751 | 35578584 | 834 | 5 | 5.38E-05 | 0.166872 | 0.00049 | 0.0009355 | 0.003898276 | TFAP2E |
| chr1 | 145858961 | 145859318 | 358 | 8 | 0.000428269 | 0.001488 | 0.005073 | 0.000936 | 0.001702183 | PIAS3 |
| chr4 | 1494607 | 1495496 | 890 | 7 | 5.36E-05 | 0.037916 | 0.007092 | 0.0009407 | -0.00031028 | NA |
| chr1 | 47433239 | 47433558 | 320 | 3 | 0.000147746 | 0.001512 | 0.0059 | 0.0009417 | 0.009420917 | FOXD2-AS1 |
| chr4 | 498586 | 498672 | 87 | 2 | 0.000353204 | 0.000877 | 0.002532 | 0.0009455 | 0.014744704 | ZNF721 |
| chr16 | 1977570 | 1977628 | 59 | 2 | 0.000321951 | 0.001097 | 0.001863 | 0.000954 | 0.003314892 | TBL3 |
| chr1 | 46543774 | 46544137 | 364 | 3 | 0.000160146 | 0.006485 | 0.005405 | 0.0009541 | 0.010978537 | MKNK1-AS1, TMEM275 |
| chr10 | 132794176 | 132794375 | 200 | 2 | 0.000407678 | 0.008669 | 0.000458 | 0.0009597 | 0.010517945 | NA |
| chr8 | 27515284 | 27515631 | 348 | 2 | 0.000246058 | 0.00327 | 0.000723 | 0.0009639 | 0.00675049 | EPHX2 |
| chr15 | 88894479 | 88894977 | 499 | 3 | 5.60E-05 | 0.015641 | 0.000303 | 0.000964 | 0.002826707 | HAPLN3 |
| chr15 | 74606987 | 74607043 | 57 | 2 | 0.000404584 | 0.000661 | 0.00484 | 0.0009725 | 0.007161033 | CLK3 |
| chr3 | 183254837 | 183255244 | 408 | 3 | 0.000373688 | 0.000378 | 0.016676 | 0.0009734 | 0.010575912 | B3GNT5, MCF2L2 |
| chr17 | 48544650 | 48544792 | 143 | 4 | 0.000178475 | 0.00069 | 0.004557 | 0.0009738 | 0.006075625 | HOXB-AS1, HOXB2 |
| chr12 | 62794213 | 62794626 | 414 | 2 | 0.00011157 | 0.01631 | 0.000391 | 0.0009791 | -0.00697388 | PPM1H |
| chr1 | 153727545 | 153727670 | 126 | 4 | 0.000193898 | 0.004276 | 0.000763 | 0.0009819 | 0.002185477 | NA |
| chr9 | 21975024 | 21975623 | 600 | 8 | 0.000179411 | 0.073655 | 2.77E-05 | 0.0009875 | 0.001213823 | AL359922.1, CDKN2A |
| chr3 | 48446674 | 48447232 | 559 | 5 | 0.00017568 | 0.004011 | 0.005493 | 0.0009892 | 0.000690218 | ATRIP |
| chr6 | 32127693 | 32128582 | 890 | 22 | 0.000325559 | 0.001712 | 0.015488 | 0.0009918 | 0.0005038 | ATF6B |
| chr10 | 100972566 | 100972989 | 424 | 4 | 0.000120264 | 0.004038 | 0.001718 | 0.0010034 | 0.001824369 | SEMA4G, MRPL43 |
| chr11 | 121451526 | 121451530 | 5 | 2 | 0.000427626 | 0.000607 | 0.006899 | 0.0010043 | 0.007341874 | AP000977.1 |
| chr9 | 5089944 | 5090163 | 220 | 2 | 0.000401792 | 0.025323 | 0.000367 | 0.0010044 | 0.003287292 | JAK2 |
| chr1 | 924339 | 924642 | 304 | 4 | 0.000442015 | 0.00468 | 0.001549 | 0.0010051 | 0.001614898 | SAMD11 |
| chr7 | 142939373 | 142939743 | 371 | 5 | 8.45E-05 | 0.002669 | 0.001567 | 0.0010054 | 0.004030663 | LLCFC1 |
| chr19 | 4639227 | 4639832 | 606 | 8 | 0.000229873 | 0.006208 | 0.000539 | 0.0010059 | 0.001403831 | TNFAIP8L1 |
| chr1 | 238486528 | 238486590 | 63 | 2 | 0.000257816 | 0.000554 | 0.010001 | 0.0010093 | 0.012274142 | AL359551.1 |
| chr21 | 43734130 | 43734393 | 264 | 3 | 0.000466977 | 0.001884 | 0.003349 | 0.0010103 | -0.00959046 | PDXK |
| chr12 | 56223792 | 56224212 | 421 | 6 | 0.000144101 | 0.002104 | 0.000818 | 0.0010203 | -0.00531442 | NABP2 |
| chr6 | 99343084 | 99343187 | 104 | 2 | 0.00035596 | 0.001753 | 0.001255 | 0.0010221 | 0.007338799 | FAXC |
| chr4 | 75725167 | 75725445 | 279 | 3 | 0.000348236 | 0.004921 | 0.000547 | 0.0010223 | 0.001638733 | USO1 |
| chr6 | 43262820 | 43263087 | 268 | 3 | 0.00047772 | 0.000617 | 0.010884 | 0.0010253 | 0.00350455 | TTBK1 |
| chr2 | 55419294 | 55419486 | 193 | 4 | 0.000369079 | 0.046758 | 0.000344 | 0.0010271 | 0.002359766 | CCDC88A |
| chr19 | 384036 | 384251 | 216 | 2 | 0.000224625 | 0.000892 | 0.003006 | 0.0010297 | 0.01378847 | AC010641.2 |
| chr11 | 34916411 | 34916596 | 186 | 5 | 0.000431776 | 0.036084 | 0.000387 | 0.0010326 | 0.0016352 | PDHX |
| chr2 | 109613537 | 109614055 | 519 | 7 | 0.000267986 | 0.001586 | 0.000944 | 0.0010352 | -0.00018547 | SEPTIN10 |
| chr11 | 117098577 | 117098786 | 210 | 2 | 0.00035831 | 0.000573 | 0.010073 | 0.0010418 | 0.003178355 | NA |
| chr17 | 75088168 | 75088399 | 232 | 5 | 0.000324369 | 0.000481 | 0.01236 | 0.0010419 | 0.005944379 | SLC16A5 |
| chr11 | 134393149 | 134393797 | 649 | 6 | 0.000278231 | 0.002471 | 0.001589 | 0.0010421 | 0.005921164 | B3GAT1 |

| | | | | | | | | | | |
|---|---|---|---|---|---|---|---|---|---|---|
| chr1 | 197901948 | 197902496 | 549 | 7 | 0.000340893 | 0.002552 | 0.012692 | 0.0010491 | 0.004306911 | NA |
| chr1 | 180940563 | 180941072 | 510 | 3 | 0.000117936 | 0.004888 | 0.006537 | 0.0010546 | 0.00019239 | KIAA1614 |
| chr1 | 21824105 | 21824361 | 257 | 2 | 0.000281781 | 0.017285 | 0.000419 | 0.0010605 | 0.004501653 | LDLRAD2, HSPG2 |
| chr4 | 42399197 | 42399225 | 29 | 2 | 0.000474856 | 0.000806 | 0.003957 | 0.0010615 | 0.004565374 | SHISA3 |
| chr17 | 56885167 | 56885194 | 28 | 2 | 0.000475382 | 0.014517 | 0.000439 | 0.0010638 | 0.003469902 | TRIM25 |
| chr11 | 19241544 | 19241747 | 204 | 5 | 0.000226738 | 0.003283 | 0.013693 | 0.0010642 | -0.00125411 | CSRP3-AS1, E2F8 |
| chr19 | 51926727 | 51927011 | 285 | 3 | 0.000113564 | 0.013693 | 0.003014 | 0.0010735 | 0.001457581 | NA |
| chr1 | 17120050 | 17120273 | 224 | 2 | 0.00013039 | 0.00419 | 0.000723 | 0.0010756 | 0.007156004 | NA |
| chr13 | 108217916 | 108218101 | 186 | 4 | 0.000318389 | 0.001056 | 0.005191 | 0.0010816 | 0.000579452 | LIG4 |
| chr11 | 121024402 | 121024564 | 163 | 2 | 0.000255981 | 0.001302 | 0.001922 | 0.0010857 | -0.00072995 | TBCEL, TBCEL-TECTA |
| chr19 | 34677333 | 34677761 | 429 | 9 | 0.000392024 | 0.022973 | 0.001127 | 0.0010861 | 4.09E-05 | ZNF302, AC020910.6 |
| chr12 | 3492140 | 3492348 | 209 | 3 | 0.000321436 | 0.006276 | 0.000494 | 0.0010889 | 0.004414139 | PRMT8, THCAT155 |
| chr12 | 54033509 | 54034387 | 879 | 7 | 0.000127157 | 0.001292 | 0.005904 | 0.0010952 | 0.000466995 | AC012531.3, HOXC4, AC012531.2, HOXC5, MIR615 |
| chr8 | 123541248 | 123541963 | 716 | 6 | 0.000165004 | 0.001489 | 0.009095 | 0.0010988 | 0.000944505 | NA |
| chr7 | 99453842 | 99453924 | 83 | 3 | 0.000306164 | 0.005959 | 0.000822 | 0.0011019 | 0.005709758 | CPSF4, PTCD1, ATP5MF-PTCD1, ATP5MF |
| chr6 | 168872831 | 168872954 | 124 | 2 | 0.000374453 | 0.000606 | 0.010399 | 0.0011024 | 0.005264099 | NA |
| chr1 | 247331126 | 247331624 | 499 | 3 | 0.000221824 | 0.003318 | 0.002324 | 0.0011075 | -0.00119727 | ZNF496 |
| chr6 | 148272177 | 148272782 | 606 | 4 | 0.000257502 | 0.000476 | 0.018303 | 0.0011133 | 0.006296195 | SASH1, AL513164.1 |
| chr2 | 73269949 | 73270522 | 574 | 3 | 0.000262129 | 0.010663 | 0.001437 | 0.0011201 | 0.002517576 | FBXO41 |
| chr11 | 74468009 | 74468504 | 496 | 3 | 0.000250055 | 0.008656 | 0.000387 | 0.0011221 | 0.003418817 | AP001372.1 |
| chr4 | 7592374 | 7592397 | 24 | 2 | 0.000367318 | 0.000638 | 0.009217 | 0.0011232 | -0.01055438 | SORCS2 |
| chr4 | 47031552 | 47031617 | 66 | 3 | 0.000308657 | 0.000446 | 0.019063 | 0.0011248 | 0.001251116 | GABRB1 |
| chr2 | 216632820 | 216632894 | 75 | 2 | 0.000376831 | 0.002641 | 0.001037 | 0.0011252 | 0.006801452 | IGFBP2 |
| chr3 | 50234795 | 50235004 | 210 | 2 | 0.000140644 | 0.003581 | 0.000847 | 0.0011316 | 0.006773899 | GNAI2 |
| chr2 | 230824025 | 230824208 | 184 | 2 | 0.000245724 | 0.003048 | 0.000945 | 0.0011341 | 0.007045239 | NA |
| chr8 | 94638871 | 94638990 | 120 | 2 | 0.000245633 | 0.00287 | 0.000989 | 0.0011367 | 0.011905786 | AC108860.2 |
| chr17 | 28041572 | 28042023 | 452 | 6 | 0.000289467 | 0.001728 | 0.010536 | 0.0011374 | -0.00024188 | NLK |
| chr17 | 44383682 | 44383974 | 293 | 2 | 0.000267128 | 0.000832 | 0.004496 | 0.0011379 | 0.004597483 | ITGA2B |
| chr3 | 109316534 | 109316622 | 89 | 5 | 0.000432441 | 0.504355 | 8.42E-05 | 0.0011442 | -0.00124751 | NA |
| chr2 | 69012053 | 69012544 | 492 | 2 | 0.000115787 | 0.001181 | 0.002458 | 0.0011483 | -0.00691833 | NA |
| chr2 | 113156639 | 113157246 | 608 | 6 | 7.95E-05 | 0.010747 | 0.006487 | 0.001167 | 0.003484465 | NA |
| chr1 | 3141957 | 3142573 | 617 | 5 | 0.000259654 | 0.001577 | 0.009376 | 0.0011737 | 0.006000008 | PRDM16 |
| chr5 | 40797688 | 40798207 | 520 | 4 | 0.000330106 | 0.003126 | 0.002382 | 0.0011762 | 6.88E-05 | PRKAA1 |
| chr8 | 142292267 | 142292374 | 108 | 2 | 0.000332588 | 0.079238 | 0.000376 | 0.0011786 | 0.00251515 | TSNARE1 |
| chr4 | 122732368 | 122732441 | 74 | 3 | 0.000427703 | 0.000503 | 0.018397 | 0.0011908 | -0.00223781 | CETN4P |
| chr5 | 52799871 | 52800266 | 396 | 5 | 0.000382147 | 0.001384 | 0.003683 | 0.0011961 | -0.00100984 | ITGA1, PELO, AC022126.1 |
| chr7 | 1080805 | 1081488 | 684 | 6 | 3.86E-05 | 0.051183 | 0.000189 | 0.0011971 | 0.002308649 | AC091729.2, C7orf50 |
| chr17 | 1934970 | 1935145 | 176 | 2 | 0.000120948 | 0.001479 | 0.002008 | 0.0012055 | 0.004615245 | RTN4RL1 |
| chr1 | 210328592 | 210329230 | 639 | 9 | 0.000286137 | 0.00111 | 0.023263 | 0.0012095 | -0.00063661 | HHAT |
| chr17 | 81552165 | 81552262 | 98 | 3 | 0.000322659 | 0.000639 | 0.01437 | 0.0012118 | -0.00154134 | FAAP100 |
| chr2 | 74498683 | 74498798 | 116 | 2 | 0.00024136 | 0.000807 | 0.005871 | 0.0012125 | 0.000883912 | LBX2 |
| chr3 | 48664091 | 48664337 | 247 | 5 | 0.000334428 | 0.048512 | 0.000735 | 0.0012163 | 0.002907251 | LINC02585 |
| chr17 | 74273430 | 74274062 | 633 | 6 | 0.000280893 | 0.076633 | 0.000225 | 0.0012369 | -0.00292821 | NA |
| chr1 | 179085152 | 179085164 | 13 | 2 | 0.00029185 | 0.001424 | 0.00224 | 0.0012447 | 0.007858163 | TOR3A |
| chr2 | 26244876 | 26245587 | 712 | 11 | 0.000136758 | 0.008104 | 0.001769 | 0.0012474 | 0.0011541 | HADHB |
| chr3 | 13074343 | 13074526 | 184 | 2 | 0.00042989 | 0.000711 | 0.009725 | 0.0012486 | 0.012396892 | IQSEC1 |
| chr15 | 99976155 | 99976425 | 271 | 2 | 0.000383767 | 0.017951 | 0.000494 | 0.0012494 | 0.005718337 | ADAMTS17 |
| chr20 | 56628148 | 56628383 | 236 | 3 | 0.000454248 | 0.001263 | 0.00284 | 0.0012544 | 0.004325586 | NA |
| chr5 | 132812487 | 132813014 | 528 | 5 | 1.01E-05 | 0.047942 | 0.003292 | 0.0012588 | 0.007135155 | NA |
| chr15 | 95326689 | 95326825 | 137 | 2 | 0.000461301 | 0.000708 | 0.011423 | 0.0012894 | -0.00045963 | LINC00924, LINC01197 |
| chr2 | 199457021 | 199457311 | 291 | 2 | 0.000255104 | 0.001734 | 0.001892 | 0.0012905 | 0.001061491 | SATB2 |
| chr14 | 67411425 | 67411818 | 394 | 2 | 0.000197629 | 0.001319 | 0.002749 | 0.0013068 | 0.01060682 | PLEK2 |
| chr16 | 737902 | 738184 | 283 | 4 | 0.000454987 | 0.000351 | 0.035244 | 0.0013261 | 0.02060989 | CIAO3 |
| chr1 | 153977682 | 153978391 | 710 | 10 | 0.000264503 | 0.018563 | 0.002005 | 0.0013429 | -0.00084394 | AL358472.2, AL358472.6 |
| chr11 | 67605986 | 67606506 | 521 | 7 | 0.000183452 | 0.000951 | 0.010344 | 0.0013453 | 0.003746889 | AP003385.3, NDUFV1, C11orf72 |
| chr7 | 135686011 | 135686609 | 599 | 2 | 0.000451653 | 0.001217 | 0.003386 | 0.0013551 | 0.001080259 | STMP1, SLC13A4 |
| chr19 | 7348670 | 7348706 | 37 | 2 | 0.000428824 | 0.00077 | 0.010542 | 0.0013641 | 0.006944661 | NA |
| chr14 | 22494301 | 22494828 | 528 | 2 | 4.36E-05 | 0.031762 | 0.00048 | 0.0013643 | 0.007492604 | TRAJ44 |
| chr16 | 19522227 | 19522692 | 466 | 5 | 9.70E-05 | 0.005802 | 0.000648 | 0.00138 | 0.009242837 | NA |
| chr15 | 83285246 | 83286097 | 852 | 8 | 0.000248325 | 0.000639 | 0.031981 | 0.001393 | -0.00092662 | AC103876.1 |
| chr15 | 72168946 | 72169268 | 323 | 2 | 0.000121902 | 0.003844 | 0.001083 | 0.0013973 | 0.005595594 | GRAMD2A |
| chr1 | 206865061 | 206865151 | 91 | 3 | 0.000297003 | 0.003644 | 0.001034 | 0.0014012 | 0.005893258 | NA |
| chr1 | 70205236 | 70205503 | 268 | 4 | 0.000128842 | 0.003474 | 0.002249 | 0.0014063 | -0.00164782 | LRRC40 |
| chr1 | 19250427 | 19250542 | 116 | 2 | 9.53E-05 | 0.001099 | 0.004548 | 0.0014149 | 0.004941073 | EMC1 |
| chr11 | 47641986 | 47642114 | 129 | 2 | 0.000239101 | 0.05134 | 0.000466 | 0.0014179 | -0.00178317 | MTCH2 |
| chr20 | 44038490 | 44038819 | 330 | 2 | 0.00048301 | 0.000782 | 0.011866 | 0.0014188 | 0.009003951 | TOX2 |
| chr1 | 24744173 | 24744181 | 9 | 2 | 7.99E-05 | 0.004876 | 0.000958 | 0.0014244 | 0.002467595 | NA |
| chr1 | 32206231 | 32206283 | 53 | 3 | 0.000216851 | 0.000544 | 0.022472 | 0.0014286 | 0.011290279 | IQCC, AL049795.1 |
| chr12 | 107319272 | 107319639 | 368 | 2 | 8.58E-05 | 0.003318 | 0.001249 | 0.0014368 | 0.002329207 | BTBD11 |
| chr6 | 57089879 | 57090128 | 250 | 7 | 0.000340988 | 0.017099 | 0.003562 | 0.0014418 | -0.00097828 | ZNF451 |
| chr14 | 24428827 | 24429668 | 842 | 6 | 9.42E-05 | 0.006253 | 0.001498 | 0.0014419 | -0.00068791 | KHNYN, CBLN3 |
| chr10 | 99534543 | 99534695 | 153 | 3 | 0.000296195 | 0.004197 | 0.006082 | 0.0014506 | 0.006911882 | NKX2-3 |
| chr7 | 156950202 | 156950379 | 178 | 2 | 0.000233579 | 0.003296 | 0.001287 | 0.0014618 | -0.00152453 | NOM1 |
| chr5 | 1865387 | 1865548 | 162 | 2 | 0.000159 | 0.001517 | 0.003012 | 0.0014999 | 0.006277308 | NA |
| chr8 | 116756226 | 116756647 | 422 | 2 | 0.000181513 | 0.000972 | 0.007475 | 0.0015182 | 0.014068937 | EIF3H |
| chr19 | 13054777 | 13055470 | 694 | 3 | 0.000122712 | 0.008867 | 0.002401 | 0.0015379 | 0.005372998 | NFIX |
| chr7 | 86644369 | 86645127 | 759 | 6 | 7.20E-05 | 0.100752 | 0.001225 | 0.0015613 | 0.004152731 | GRM3 |
| chr6 | 33425335 | 33425406 | 72 | 3 | 0.000328838 | 0.006775 | 0.001112 | 0.0015659 | 0.00410672 | SYNGAP1 |
| chr19 | 55376714 | 55376778 | 65 | 2 | 0.000473396 | 0.000873 | 0.012337 | 0.0015762 | 0.015653872 | NA |
| chr4 | 186190687 | 186190991 | 305 | 5 | 0.000331265 | 0.010699 | 0.003426 | 0.0015796 | -0.00479343 | FLJ38576 |
| chr16 | 1471009 | 1471035 | 27 | 2 | 0.000283523 | 0.130222 | 0.000494 | 0.0016169 | 0.005867461 | CLCN7, AL031600.2 |
| chr7 | 1237468 | 1237885 | 418 | 4 | 0.000257999 | 0.001006 | 0.01053 | 0.0016289 | 0.004771841 | NA |
| chr12 | 122953066 | 122953093 | 28 | 2 | 0.000411717 | 0.006819 | 0.000964 | 0.0016496 | 0.002922656 | ABCB9 |
| chr2 | 42764203 | 42764419 | 217 | 4 | 0.000210002 | 0.029529 | 0.00459 | 0.0016607 | 0.0058328 | OXER1 |
| chr5 | 149960295 | 149960591 | 297 | 7 | 0.000342453 | 0.002441 | 0.002403 | 0.001663 | -0.00105371 | NA |
| chr2 | 6866496 | 6866655 | 160 | 2 | 0.000170171 | 0.001036 | 0.00883 | 0.0016768 | 0.008120279 | RSAD2, CMPK2 |
| chr2 | 86441110 | 86441613 | 504 | 10 | 0.00023831 | 0.001207 | 0.031594 | 0.0016793 | 0.000132763 | KDM3A |
| chr17 | 32444817 | 32444976 | 160 | 2 | 0.000318478 | 0.043287 | 0.000563 | 0.0016793 | 0.003150458 | PSMD11, AC005899.8 |
| chr2 | 230410909 | 230411358 | 450 | 3 | 0.000199602 | 0.010978 | 0.001022 | 0.0016816 | 0.00238182 | AC009949.1 |
| chr10 | 122879358 | 122879475 | 118 | 4 | 0.000472792 | 0.002101 | 0.006907 | 0.001693 | -0.01449298 | AC073585.1, FAM24B |
| chr3 | 52771305 | 52771622 | 318 | 4 | 0.000115201 | 0.017463 | 0.00055 | 0.0017082 | -0.00165938 | NA |
| chr1 | 224851974 | 224852146 | 173 | 2 | 0.000428898 | 0.04132 | 0.00058 | 0.001715 | 0.007430978 | AL391811.1 |
| chr19 | 7633679 | 7633697 | 19 | 2 | 0.000210135 | 0.003008 | 0.001775 | 0.0017152 | 0.003258071 | AC008763.2, PCP2 |
| chr17 | 77558040 | 77558064 | 25 | 2 | 0.000209941 | 0.001704 | 0.003459 | 0.001732 | -0.007873 | AC021683.2 |
| chr1 | 2069491 | 2069882 | 392 | 2 | 0.000303391 | 0.005931 | 0.001109 | 0.0017322 | 0.005126923 | PRKCZ |
| chr4 | 24913307 | 24913348 | 42 | 3 | 0.000426139 | 0.002361 | 0.008144 | 0.0017364 | 0.011602046 | CCDC149 |
| chr3 | 187746246 | 187746318 | 73 | 2 | 0.000394376 | 0.001107 | 0.008633 | 0.0017609 | -0.00018567 | NA |
| chr10 | 102394393 | 102395054 | 662 | 10 | 0.00012249 | 0.018787 | 0.000322 | 0.0017742 | 0.000994371 | NFKB2 |
| chr20 | 33644365 | 33644541 | 177 | 3 | 0.000238083 | 0.036728 | 0.000686 | 0.0018012 | 0.003126326 | CBFA2T2 |
| chr1 | 203626681 | 203627812 | 1132 | 7 | 0.000301734 | 0.014701 | 0.000368 | 0.0018079 | 0.001022242 | ATP2B4 |
| chr1 | 28648071 | 28648426 | 356 | 6 | 0.000367868 | 0.018536 | 0.001328 | 0.0018177 | 0.001972577 | LINC01715 |
| chr1 | 23368854 | 23369237 | 384 | 7 | 0.000268391 | 0.001998 | 0.01231 | 0.0018241 | 9.41E-05 | ZNF436-AS1, ZNF436 |
| chr3 | 179148907 | 179149216 | 310 | 2 | 0.000451093 | 0.001112 | 0.009817 | 0.0018318 | -0.00395661 | PIK3CA |
| chr7 | 1123913 | 1124427 | 515 | 2 | 0.000216409 | 0.002704 | 0.002199 | 0.0018359 | 0.005740281 | C7orf50 |
| chr19 | 33080735 | 33080801 | 67 | 3 | 0.000432315 | 0.121169 | 0.004885 | 0.0018605 | 0.007537535 | NA |
| chr12 | 8913043 | 8913057 | 15 | 2 | 0.000461643 | 0.001185 | 0.008696 | 0.0018642 | 0.013641217 | AC006581.2 |
| chr11 | 66466157 | 66466481 | 325 | 7 | 0.000372643 | 0.014199 | 0.022525 | 0.001867 | 0.005868914 | PELI3, MRPL11 |
| chr1 | 1337018 | 1337179 | 162 | 4 | 0.000410967 | 0.018817 | 0.003977 | 0.0018886 | -0.00090257 | DVL1 |
| chr10 | 100534797 | 100535174 | 378 | 2 | 8.16E-05 | 0.014627 | 0.000826 | 0.0018965 | 0.007847337 | HIF1AN |
| chr22 | 31107660 | 31107884 | 225 | 3 | 0.000441081 | 0.000925 | 0.015038 | 0.0019047 | -0.00020181 | SELENOM |
| chr5 | 177022841 | 177023178 | 338 | 4 | 0.000350434 | 0.000951 | 0.010547 | 0.0019127 | -0.00031217 | ZNF346 |
| chr1 | 36361582 | 36362146 | 565 | 2 | 0.000358039 | 0.066225 | 0.00061 | 0.0019211 | 0.003324015 | STK40 |
| chr2 | 95121289 | 95121851 | 563 | 4 | 4.90E-05 | 0.023839 | 0.00029 | 0.0019358 | 0.001316675 | MRPS5 |
| chr15 | 52783224 | 52783336 | 113 | 3 | 0.000303681 | 0.023347 | 0.001136 | 0.0019468 | 0.007903195 | ONECUT1 |
| chr21 | 36705373 | 36705671 | 299 | 2 | 0.000147734 | 0.010449 | 0.00098 | 0.0019654 | 0.005802076 | SIM2 |
| chr11 | 2133704 | 2133722 | 19 | 2 | 0.000398731 | 0.001191 | 0.010374 | 0.001969 | 0.010037748 | IGF2, AC132217.2, INS-IGF2 |
| chr14 | 103712758 | 103712823 | 66 | 3 | 0.000463516 | 0.000852 | 0.021101 | 0.0020346 | 0.002451272 | KLC1, XRCC3 |
| chr4 | 2793378 | 2793428 | 51 | 2 | 0.000316249 | 0.001406 | 0.007486 | 0.0020441 | 0.004733725 | SH3BP2 |
| chr14 | 79862107 | 79862252 | 146 | 2 | 0.000291152 | 0.029395 | 0.000742 | 0.0020487 | 0.003181624 | NRXN3 |
| chr6 | 84773337 | 84773616 | 280 | 2 | 0.000410125 | 0.001292 | 0.009452 | 0.0020539 | 0.004662119 | NA |
| chr4 | 109912652 | 109912788 | 137 | 3 | 0.000206454 | 0.006995 | 0.001214 | 0.0020559 | 0.003508578 | NA |
| chr16 | 3716891 | 3717167 | 277 | 3 | 9.72E-05 | 0.012127 | 0.00073 | 0.0020593 | 0.001898566 | TRAP1 |
| chr11 | 65636934 | 65636988 | 55 | 2 | 0.000268653 | 0.02997 | 0.000744 | 0.0020622 | 0.001815699 | PCNX3 |
| chr16 | 896408 | 896419 | 12 | 2 | 0.000424079 | 0.01282 | 0.00096 | 0.0020717 | 0.003638977 | LMF1 |
| chr10 | 75401371 | 75401767 | 397 | 4 | 0.000380311 | 0.004903 | 0.008328 | 0.002074 | 0.003069616 | ZNF503-AS2, ZNF503 |
| chr16 | 66919046 | 66919922 | 877 | 5 | 0.000103276 | 0.000538 | 0.045645 | 0.0020754 | 0.006721195 | NA |
| chr6 | 27238724 | 27238753 | 30 | 2 | 0.000476659 | 0.001424 | 0.007736 | 0.0020863 | 0.004103293 | NA |
| chr2 | 202036705 | 202036747 | 43 | 2 | 0.000497525 | 0.001164 | 0.014806 | 0.0021071 | 0.036800499 | FZD7 |
| chr1 | 213051607 | 213051638 | 32 | 2 | 0.000398818 | 0.001982 | 0.004313 | 0.0021239 | -0.00493564 | RPS6KC1 |
| chr2 | 221571429 | 221572227 | 799 | 3 | 3.85E-05 | 0.012147 | 0.001051 | 0.0021328 | 0.001645743 | EPHA4 |
| chr5 | 59893093 | 59893293 | 201 | 2 | 3.74E-05 | 0.004194 | 0.001913 | 0.0021537 | 0.009040759 | PDE4D |
| chr10 | 102231227 | 102232020 | 794 | 4 | 0.000102254 | 0.005697 | 0.001711 | 0.0021596 | -0.00129441 | PITX3 |
| chr6 | 43060028 | 43060266 | 239 | 3 | 0.000315389 | 0.018026 | 0.006743 | 0.00216 | 0.002390723 | KLC4 |
| chr6 | 16762237 | 16762347 | 111 | 2 | 0.000474812 | 0.008528 | 0.001245 | 0.0022057 | 0.002268542 | ATXN1-AS1 |
| chr6 | 133243328 | 133243440 | 113 | 3 | 0.000435295 | 0.231826 | 0.000227 | 0.002209 | 0.007828444 | EYA4 |
| chr2 | 3600938 | 3600964 | 27 | 3 | 0.000262296 | 0.041208 | 0.000391 | 0.0022137 | 0.004848959 | COLEC11 |
| chr11 | 76656881 | 76657034 | 154 | 2 | 0.000392436 | 0.001247 | 0.014431 | 0.0022255 | 0.017992342 | NA |
| chr8 | 96262465 | 96262769 | 305 | 2 | 5.14E-05 | 0.007594 | 0.001342 | 0.0022275 | -0.00074492 | PTDSS1 |
| chr1 | 225911946 | 225911949 | 4 | 2 | 0.000472352 | 0.001244 | 0.014628 | 0.002228 | 0.005988315 | AL117348.2 |
| chr4 | 150078099 | 150078128 | 30 | 5 | 0.000351606 | 0.006716 | 0.007721 | 0.0022293 | 0.001824186 | NA |
| chr19 | 56538320 | 56538488 | 169 | 3 | 0.000367912 | 0.000867 | 0.025182 | 0.0022302 | 0.000918013 | AC005498.2 |
| chr12 | 62603655 | 62603851 | 197 | 4 | 0.000245745 | 0.046214 | 0.001583 | 0.002276 | 0.002022674 | AC048341.1, MIRLET7I, LINC01465 |
| chr1 | 47416593 | 47416642 | 50 | 2 | 0.000273587 | 0.011028 | 0.001157 | 0.0022945 | 0.002276701 | FOXE3 |
| chr4 | 119628008 | 119628101 | 94 | 2 | 0.00040818 | 0.001775 | 0.006321 | 0.0022983 | -0.00127831 | PDE5A |
| chr3 | 48595290 | 48595711 | 422 | 6 | 0.000328838 | 0.075326 | 0.000259 | 0.0023042 | 0.003997149 | NA |
| chr5 | 1883100 | 1883826 | 727 | 4 | 4.34E-05 | 0.002944 | 0.010556 | 0.0023255 | 3.58E-05 | IRX4 |
| chr5 | 179257651 | 179258056 | 406 | 3 | 0.00033498 | 0.003847 | 0.005274 | 0.0023386 | -0.00344139 | ADAMTS2 |
| chr20 | 2801432 | 2801702 | 271 | 3 | 0.000327779 | 0.153832 | 0.000268 | 0.0023403 | 0.004678863 | NA |
| chr5 | 132555982 | 132556550 | 569 | 3 | 2.36E-05 | 0.134546 | 0.000279 | 0.0024321 | -0.00706695 | AC116366.2, RAD50, IL5 |
| chr11 | 86244677 | 86245279 | 603 | 7 | 0.000163639 | 0.016465 | 0.000547 | 0.0024729 | -0.00054748 | EED |
| chr11 | 2901234 | 2901622 | 389 | 8 | 9.50E-05 | 0.020138 | 0.000254 | 0.0024783 | 0.001029579 | SLC22A18, SLC22A18AS |
| chr16 | 56192434 | 56192438 | 5 | 2 | 0.000428136 | 0.002896 | 0.003534 | 0.0024792 | 0.005595336 | GNAO1 |
| chr5 | 2757688 | 2757974 | 287 | 3 | 0.00019887 | 0.001941 | 0.013219 | 0.0024878 | 0.007060248 | NA |
| chr1 | 914067 | 914096 | 30 | 2 | 0.000417812 | 0.148148 | 0.000743 | 0.0024931 | 0.003956981 | AL645608.6, AL645608.2 |
| chr11 | 64081399 | 64081500 | 102 | 2 | 0.000224446 | 0.001384 | 0.016283 | 0.0025043 | -0.00030623 | AP000721.2, MACROD1 |
| chr16 | 72787667 | 72787766 | 100 | 2 | 0.000322136 | 0.015806 | 0.001117 | 0.0025177 | 0.002412734 | AC004943.2, ZFHX3 |
| chr10 | 133358060 | 133358987 | 928 | 7 | 9.84E-05 | 0.007667 | 0.009812 | 0.0025473 | 0.003923709 | NA |
| chr12 | 25251869 | 25252346 | 478 | 4 | 0.000214541 | 0.029891 | 0.006854 | 0.0025546 | -0.00010566 | NA |
| chr1 | 45011953 | 45011964 | 12 | 2 | 0.00039407 | 0.001717 | 0.009206 | 0.002558 | 0.003605773 | UROD |
| chr16 | 50711082 | 50711191 | 110 | 3 | 0.000460048 | 0.001588 | 0.01472 | 0.0025624 | -0.00030918 | NOD2 |
| chr19 | 44954416 | 44955159 | 744 | 13 | 0.000315928 | 0.221003 | 0.002073 | 0.0025662 | 0.000910836 | CLPTM1 |
| chr2 | 216498897 | 216499100 | 204 | 4 | 0.000239711 | 0.013376 | 0.005547 | 0.0025793 | 0.000321612 | RPL37A |

| | | | | | | | | | | |
|---|---|---|---|---|---|---|---|---|---|---|
| chr8 | 42179652 | 42179869 | 218 | 2 | 0.00033416 | 0.001449 | 0.016063 | 0.0025962 | 0.018060602 | PLAT |
| chr15 | 42428057 | 42428070 | 14 | 2 | 0.000429617 | 0.002047 | 0.006732 | 0.0026165 | 0.000132058 | ZNF106 |
| chr5 | 181246096 | 181246203 | 108 | 3 | 0.000399354 | 0.144213 | 0.000339 | 0.0026546 | 0.000822078 | RACK1 |
| chr4 | 112286199 | 112286262 | 64 | 3 | 0.000363625 | 0.009649 | 0.009568 | 0.0027289 | 0.002324975 | ALPK1 |
| chr10 | 31000263 | 31000279 | 17 | 2 | 0.000478346 | 0.01681 | 0.001211 | 0.0027525 | 0.007013782 | ZNF438 |
| chr7 | 155372600 | 155373028 | 429 | 4 | 0.000309758 | 0.001937 | 0.020286 | 0.0028158 | 0.000772899 | NA |
| chr6 | 31534638 | 31535013 | 376 | 6 | 0.000370809 | 0.016896 | 0.00319 | 0.0028204 | 0.000416243 | DDX39B, ATP6V1G2-DDX39B |
| chr4 | 73620363 | 73620453 | 91 | 3 | 0.000371248 | 0.009677 | 0.00432 | 0.0028727 | -0.00193594 | RASSF6 |
| chr3 | 71065620 | 71065895 | 276 | 2 | 0.000160887 | 0.054197 | 0.000956 | 0.0029229 | 0.006331264 | FOXP1, AC097634.4 |
| chr13 | 40665475 | 40665595 | 121 | 2 | 0.000426425 | 0.00482 | 0.002818 | 0.0029552 | -0.00356707 | FOXO1 |
| chr17 | 39706174 | 39706309 | 136 | 2 | 0.0004174 | 0.003462 | 0.004044 | 0.0029732 | 0.000695474 | ERBB2 |
| chr3 | 141387034 | 141387078 | 45 | 2 | 0.000462692 | 0.043062 | 0.001024 | 0.0029954 | 0.00340345 | ZBTB38 |
| chr15 | 67147659 | 67147884 | 226 | 2 | 0.000423281 | 0.002615 | 0.006359 | 0.0030498 | 0.008600762 | SMAD3 |
| chr5 | 95822729 | 95822819 | 91 | 3 | 0.000343493 | 0.017979 | 0.000965 | 0.0030604 | 0.001041218 | RHOBTB3 |
| chr12 | 7128898 | 7129187 | 290 | 4 | 0.000291234 | 0.001763 | 0.01668 | 0.0030651 | 0.004691866 | AC018653.3 |
| chr2 | 62196198 | 62196581 | 384 | 2 | 0.000161165 | 0.002857 | 0.005703 | 0.0030933 | 0.000430538 | B3GNT2 |
| chr7 | 27184411 | 27184841 | 431 | 7 | 0.000289332 | 0.010113 | 0.010714 | 0.0031455 | -0.00094389 | HOXA11-AS, HOXA11 |
| chr1 | 40257858 | 40257940 | 83 | 5 | 0.00043145 | 0.002368 | 0.007097 | 0.0031641 | 0.000971649 | AL050341.2 |
| chr14 | 36505318 | 36505595 | 278 | 2 | 0.000141742 | 0.005815 | 0.002764 | 0.0032328 | 0.004778601 | AL132857.1, SFTA3, SFTA3 |
| chr15 | 52569199 | 52569822 | 624 | 8 | 0.000120548 | 0.054458 | 0.014057 | 0.0032368 | 0.003869724 | ARPP19 |
| chr20 | 63658924 | 63659042 | 119 | 3 | 0.000279127 | 0.022897 | 0.004419 | 0.0032403 | 0.000267315 | RTEL1 |
| chr6 | 144064124 | 144064669 | 546 | 4 | 0.000198853 | 0.013052 | 0.000933 | 0.003297 | -0.0012932 | PLAGL1 |
| chr17 | 55511133 | 55511271 | 139 | 2 | 0.000337515 | 0.005483 | 0.003032 | 0.0033137 | 0.000899627 | SMIM36 |
| chr17 | 75588279 | 75588536 | 258 | 2 | 8.25E-05 | 0.02402 | 0.001332 | 0.0033253 | -0.00300141 | MYO15B |
| chr1 | 156740969 | 156741209 | 241 | 6 | 0.000309207 | 0.013373 | 0.010376 | 0.0033364 | 0.000345364 | MRPL24 |
| chr2 | 218781934 | 218782444 | 511 | 2 | 0.000135472 | 0.007143 | 0.002489 | 0.0033582 | 0.001339697 | CYP27A1 |
| chr1 | 212564760 | 212564962 | 203 | 2 | 0.000333063 | 0.111433 | 0.001015 | 0.0033675 | 0.006327172 | NA |
| chr6 | 170553143 | 170553157 | 15 | 2 | 0.000485274 | 0.001905 | 0.019493 | 0.003433 | -0.00146803 | PSMB1 |
| chr8 | 109334323 | 109334812 | 490 | 5 | 0.000263621 | 0.016375 | 0.000852 | 0.0034581 | 0.000833745 | ENY2 |
| chr1 | 213016299 | 213016481 | 183 | 4 | 0.000299925 | 0.012346 | 0.001927 | 0.003523 | 0.000668591 | NA |
| chr18 | 61490348 | 61490646 | 299 | 3 | 0.000356826 | 0.023952 | 0.001208 | 0.0035277 | 0.001122881 | CDH20 |
| chr11 | 59556994 | 59557049 | 56 | 2 | 0.000328828 | 0.004616 | 0.004045 | 0.0035476 | 0.010406371 | AP003778.1 |
| chr4 | 162163706 | 162164083 | 378 | 3 | 0.00037819 | 0.002574 | 0.009462 | 0.0035589 | 0.008694753 | FSTL5 |
| chr1 | 185157509 | 185157961 | 453 | 4 | 0.000272707 | 0.001778 | 0.031342 | 0.0035626 | 0.002432259 | SWT1 |
| chr1 | 150877329 | 150877488 | 160 | 2 | 0.000128791 | 0.002005 | 0.019462 | 0.0035847 | 0.014946642 | NA |
| chr7 | 151278000 | 151278345 | 346 | 2 | 0.000318322 | 0.003644 | 0.005674 | 0.0036326 | 0.004800825 | NA |
| chr1 | 166166720 | 166166839 | 120 | 4 | 0.000398002 | 0.007772 | 0.003765 | 0.0036947 | -0.00041187 | AL596087.2, FAM78B |
| chr7 | 100828108 | 100828205 | 98 | 2 | 0.000428909 | 0.005644 | 0.003576 | 0.0037135 | 0.002762229 | SLC12A9 |
| chr6 | 39048003 | 39048405 | 403 | 2 | 8.50E-05 | 0.023049 | 0.001541 | 0.0037195 | 0.007024417 | NA |
| chr2 | 231394720 | 231394912 | 193 | 3 | 0.00022742 | 0.005697 | 0.013203 | 0.0037294 | 0.007244051 | AC017104.1 |
| chr20 | 43915006 | 43915538 | 533 | 5 | 0.000124471 | 0.387317 | 0.000951 | 0.003806 | 0.003188292 | TOX2 |
| chr20 | 2860074 | 2860109 | 36 | 2 | 0.000264428 | 0.002513 | 0.012575 | 0.0038114 | 0.003988208 | VPS16 |
| chr4 | 173522091 | 173522215 | 125 | 2 | 0.000235657 | 0.003623 | 0.006392 | 0.0038248 | 0.006636116 | NA |
| chr7 | 27114643 | 27114943 | 301 | 5 | 0.000358298 | 0.02633 | 0.004903 | 0.0038442 | 0.004016802 | HOXA-AS2, HOXA3 |
| chr21 | 36696281 | 36696462 | 182 | 2 | 0.000380876 | 0.002532 | 0.013077 | 0.0038825 | 0.011264088 | NA |
| chr1 | 46005890 | 46006314 | 425 | 2 | 0.000294186 | 0.015499 | 0.001914 | 0.0038845 | 0.001290002 | MAST2 |
| chr20 | 31605588 | 31605610 | 23 | 3 | 0.000481942 | 0.004895 | 0.004509 | 0.0040208 | 4.10E-05 | ID1 |
| chr3 | 9648803 | 9648886 | 84 | 3 | 0.000357766 | 0.012249 | 0.014458 | 0.0040715 | 0.000458174 | NA |
| chr15 | 25169616 | 25170252 | 637 | 5 | 0.000300244 | 0.012049 | 0.003642 | 0.0041199 | -0.0002682 | SNHG14, DMAC1P1 |
| chr2 | 176187959 | 176188617 | 659 | 11 | 0.0001652 | 0.032983 | 0.002943 | 0.0041346 | 2.75E-05 | HAGLR |
| chr2 | 218671726 | 218671865 | 140 | 5 | 0.000435073 | 0.005781 | 0.019139 | 0.0041519 | 0.000585895 | RNF25 |
| chr15 | 45130355 | 45130651 | 297 | 2 | 0.000218524 | 0.002374 | 0.019912 | 0.0041531 | 0.002595013 | DUOX1 |
| chr13 | 24270011 | 24270546 | 536 | 4 | 0.000144994 | 0.081287 | 0.002621 | 0.0043268 | 0.00508832 | AL359736.1, SPATA13 |
| chr14 | 103128394 | 103128632 | 239 | 2 | 8.08E-05 | 0.017186 | 0.002115 | 0.0043603 | 0.003535711 | TNFAIP2 |
| chr7 | 558822 | 559092 | 271 | 2 | 0.000367242 | 0.007379 | 0.003709 | 0.0043948 | -5.38E-05 | PRKAR1B |
| chr5 | 119269248 | 119269651 | 404 | 2 | 0.000178238 | 0.004208 | 0.007007 | 0.0044097 | 0.010538733 | TNFAIP8 |
| chr1 | 153678476 | 153678533 | 58 | 5 | 0.000446572 | 0.053154 | 0.015 | 0.0044368 | 0.0002529 | NA |
| chr6 | 44313044 | 44313295 | 252 | 3 | 0.000338674 | 0.003748 | 0.019166 | 0.0044572 | 0.002373896 | AL353588.1, AARS2 |
| chr15 | 99105229 | 99105457 | 229 | 2 | 0.000164533 | 0.003928 | 0.008026 | 0.0044691 | 4.84E-05 | SYNM, AC036108.1 |
| chr16 | 85310028 | 85310044 | 17 | 2 | 0.000487838 | 0.002708 | 0.017683 | 0.00448 | 0.009556327 | GSE1 |
| chr3 | 184362901 | 184363214 | 314 | 6 | 0.000416102 | 0.005433 | 0.008528 | 0.0046499 | 0.000205713 | POLR2H |
| chr1 | 983820 | 984042 | 223 | 2 | 0.000160522 | 0.005319 | 0.005768 | 0.0046625 | 0.009311976 | NA |
| chr15 | 72783594 | 72783678 | 85 | 3 | 0.000353639 | 0.00516 | 0.009249 | 0.0047263 | -0.00045319 | ADPGK-AS1, ADPGK |
| chr17 | 29390906 | 29391243 | 338 | 4 | 0.000141842 | 0.018794 | 0.001336 | 0.0047595 | 0.002129942 | AC068025.2, TAOK1 |
| chr2 | 151711003 | 151711235 | 233 | 2 | 0.000430163 | 0.006244 | 0.005078 | 0.0048003 | -0.01073088 | NEB |
| chr7 | 135663045 | 135663070 | 26 | 2 | 0.000385898 | 0.070287 | 0.001553 | 0.0048294 | -0.00154786 | STMP1 |
| chr2 | 207624724 | 207625187 | 464 | 5 | 0.000232261 | 0.008645 | 0.009415 | 0.0048761 | 0.001772587 | METTL21A |
| chr3 | 138936985 | 138937200 | 216 | 2 | 0.000362206 | 0.004133 | 0.009153 | 0.0048976 | 0.009571597 | LINC01391 |
| chr16 | 2989445 | 2989664 | 220 | 2 | 0.000301442 | 0.006921 | 0.004767 | 0.0049183 | 0.004206698 | LINC00514 |
| chr13 | 100089548 | 100089550 | 3 | 2 | 0.000186506 | 0.003446 | 0.013955 | 0.0050304 | 0.001790742 | PCCA |
| chr8 | 11708101 | 11708587 | 487 | 4 | 9.71E-05 | 0.017193 | 0.002008 | 0.0051923 | 0.004483417 | GATA4 |
| chr6 | 24719760 | 24720197 | 438 | 5 | 0.000205677 | 0.008125 | 0.006289 | 0.0052849 | 0.002361003 | C6orf62 |
| chr4 | 186227276 | 186227294 | 19 | 2 | 0.000494989 | 0.003232 | 0.020081 | 0.0053563 | -0.00534897 | KLKB1 |
| chr6 | 32097882 | 32098113 | 232 | 3 | 3.31E-05 | 0.118011 | 0.015096 | 0.0054343 | 0.004746227 | TNXB |
| chr12 | 6891370 | 6891637 | 268 | 2 | 0.000243992 | 0.020866 | 0.002641 | 0.0055528 | 0.001779296 | LRRC23 |
| chr16 | 31202965 | 31203559 | 595 | 10 | 0.000227323 | 0.026052 | 0.005243 | 0.0056199 | 0.000775002 | PYCARD-AS1, PYCARD |
| chr10 | 129963788 | 129964198 | 411 | 3 | 0.000160232 | 0.007298 | 0.005289 | 0.0056417 | -0.00080561 | EBF3 |
| chr10 | 16817847 | 16818141 | 295 | 2 | 0.000221897 | 0.004596 | 0.011132 | 0.0057204 | -0.00249796 | AL365215.2 |
| chr2 | 191014784 | 191014815 | 32 | 2 | 0.000452437 | 0.004169 | 0.013752 | 0.0057786 | 0.013495821 | STAT1 |
| chr6 | 5004836 | 5005254 | 419 | 2 | 0.000182989 | 0.003598 | 0.020834 | 0.0059016 | 0.007641135 | LYRM4-AS1 |
| chr11 | 67419104 | 67419351 | 248 | 2 | 0.000384419 | 0.016978 | 0.003229 | 0.0059434 | 0.005271781 | CARNS1, PPP1CA |
| chr3 | 50273044 | 50273210 | 167 | 6 | 0.000335878 | 0.120055 | 0.001751 | 0.0059694 | 0.000548724 | SEMA3B |
| chr21 | 46097912 | 46098154 | 243 | 2 | 0.000433558 | 0.021259 | 0.00289 | 0.005987 | -0.00175136 | COL6A2 |
| chr20 | 21102456 | 21102873 | 418 | 2 | 0.000323314 | 0.012177 | 0.004063 | 0.0059974 | 0.003137927 | LINC00237 |
| chr16 | 2149119 | 2149701 | 583 | 2 | 9.22E-05 | 0.087118 | 0.001904 | 0.0060309 | 0.005841662 | RAB26 |
| chr20 | 33674966 | 33675032 | 67 | 2 | 0.000338299 | 0.004053 | 0.017016 | 0.0060857 | 0.009077599 | AL121906.1 |
| chr15 | 37101788 | 37101965 | 178 | 2 | 0.000247774 | 0.010164 | 0.005042 | 0.0062661 | 0.0047076 | NA |
| chr22 | 20394520 | 20394619 | 100 | 2 | 0.000460749 | 0.005559 | 0.010402 | 0.0063421 | 0.000122297 | ZNF74 |
| chr19 | 38402515 | 38402612 | 98 | 2 | 0.000329701 | 0.201493 | 0.001923 | 0.0066025 | 0.010928579 | NA |
| chr2 | 176121732 | 176121931 | 200 | 3 | 0.00015839 | 0.006984 | 0.008144 | 0.0066675 | 0.006631819 | HOXD-AS2 |
| chr2 | 161160708 | 161161078 | 371 | 2 | 0.000125909 | 0.011472 | 0.005085 | 0.0067087 | -0.00053977 | TANK |
| chr12 | 108880223 | 108880465 | 243 | 2 | 0.000191708 | 0.003903 | 0.026898 | 0.0067653 | 0.003483457 | DAO |
| chr1 | 228144562 | 228144801 | 240 | 4 | 0.000142109 | 0.037451 | 0.002005 | 0.0067753 | 0.001202176 | GUK1 |
| chr2 | 219333178 | 219333267 | 90 | 2 | 0.000418187 | 0.003972 | 0.028694 | 0.0069814 | 0.005355713 | NA |
| chr1 | 221737206 | 221737544 | 339 | 3 | 0.00016854 | 0.043201 | 0.00308 | 0.0070398 | 0.005201653 | DUSP10 |
| chr1 | 214187410 | 214187611 | 202 | 2 | 0.000439016 | 0.031206 | 0.003262 | 0.0075137 | -0.00668806 | LINC02775 |
| chr12 | 49623489 | 49623629 | 141 | 5 | 0.000474825 | 0.005685 | 0.032451 | 0.0075596 | -0.00166481 | PRPF40B |
| chr10 | 129031365 | 129031406 | 42 | 3 | 0.000359205 | 0.051125 | 0.004455 | 0.0078258 | -0.00272936 | NA |
| chr19 | 3964149 | 3964839 | 691 | 4 | 0.000266021 | 0.011668 | 0.012696 | 0.0078508 | 0.001533278 | DAPK3 |
| chr1 | 21783659 | 21783899 | 241 | 4 | 0.000374038 | 0.221555 | 0.000498 | 0.0079336 | -7.11E-06 | NA |
| chr17 | 16381364 | 16381442 | 79 | 3 | 0.000384477 | 0.003939 | 0.040174 | 0.0079979 | -0.00310678 | UBB |
| chr19 | 11383088 | 11383244 | 157 | 2 | 0.000459999 | 0.010607 | 0.007605 | 0.00813 | 0.000678469 | EPOR |
| chr19 | 42279643 | 42280063 | 421 | 3 | 0.000132709 | 0.009732 | 0.015215 | 0.0082672 | 0.004160407 | CIC |
| chr4 | 154781918 | 154782018 | 101 | 3 | 0.000410825 | 0.00538 | 0.021188 | 0.008331 | -0.01686898 | RBM46 |
| chr11 | 117232478 | 117233110 | 633 | 5 | 0.000141088 | 0.006179 | 0.032804 | 0.0085534 | 0.000769813 | RNF214, PCSK7 |
| chr16 | 30634790 | 30634957 | 168 | 2 | 0.000368009 | 0.110361 | 0.002733 | 0.0086826 | -0.00994507 | NA |
| chr2 | 162343592 | 162343966 | 375 | 3 | 0.00025899 | 0.007535 | 0.012067 | 0.0086997 | 0.00725254 | GCA |
| chr12 | 26125744 | 26125834 | 91 | 2 | 0.000395561 | 0.131682 | 0.002729 | 0.0088641 | 0.001386927 | SSPN, AC022509.3 |
| chr21 | 42878849 | 42879066 | 218 | 2 | 0.000435906 | 0.009247 | 0.011286 | 0.0091935 | -0.0007747 | WDR4 |
| chr15 | 85794222 | 85794580 | 359 | 2 | 0.000170295 | 0.0934 | 0.003003 | 0.0092052 | 0.002738458 | KLHL25 |
| chr16 | 1786721 | 1786812 | 92 | 2 | 0.000368667 | 0.03457 | 0.004171 | 0.0093865 | 0.001476342 | NUBP2, SPSB3 |
| chr7 | 20614578 | 20614602 | 25 | 2 | 0.000465793 | 0.005624 | 0.034498 | 0.0097436 | -0.00085446 | NA |
| chr6 | 33406201 | 33406220 | 20 | 2 | 0.000366354 | 0.005784 | 0.034346 | 0.009956 | 0.002305144 | KIFC1 |
| chr7 | 130153314 | 130153493 | 180 | 2 | 0.000280942 | 0.024786 | 0.005515 | 0.0101262 | 0.004211282 | NA |
| chr4 | 56530724 | 56530959 | 236 | 3 | 0.000325559 | 0.018461 | 0.021614 | 0.0103707 | 0.008303886 | THEGL |
| chr14 | 22835944 | 22836030 | 87 | 2 | 0.000355263 | 0.07112 | 0.003943 | 0.0110155 | 0.00392068 | NA |
| chr2 | 119367520 | 119367610 | 91 | 3 | 0.000433642 | 0.013842 | 0.010728 | 0.011251 | 0.00012753 | DBI |
| chr8 | 8823393 | 8823461 | 69 | 3 | 0.000357244 | 0.046136 | 0.006859 | 0.0113225 | 0.004170007 | MFHAS1 |
| chr9 | 37592202 | 37592651 | 450 | 2 | 0.000271667 | 0.006557 | 0.038892 | 0.0114028 | -0.00039297 | AL513165.2, TOMM5, AL138752.2 |
| chr9 | 71446533 | 71446598 | 66 | 2 | 0.000418818 | 0.006733 | 0.036615 | 0.0114539 | 0.005558065 | TRPM3 |
| chr5 | 138876014 | 138876220 | 207 | 2 | 0.000248611 | 0.186973 | 0.003581 | 0.0117561 | -0.00193885 | CTNNA1 |
| chr5 | 122777345 | 122777402 | 58 | 2 | 0.000498258 | 0.171305 | 0.003666 | 0.011916 | 0.009362323 | SNX2 |
| chr19 | 49581026 | 49581158 | 133 | 4 | 0.000429917 | 0.100686 | 0.013468 | 0.0119999 | -0.00926128 | PRRG2, NOSIP |
| chr16 | 88883148 | 88883228 | 81 | 3 | 0.000413879 | 0.023238 | 0.010349 | 0.0120985 | 0.002116801 | AC092384.2, CBFA2T3 |
| chr1 | 54623311 | 54623431 | 121 | 2 | 0.000481783 | 0.021747 | 0.00902 | 0.0131406 | -0.00131723 | ACOT11, FAM151A, AL590093.1 |
| chr19 | 36014997 | 36015665 | 669 | 3 | 0.000113889 | 0.249721 | 0.001827 | 0.013242 | 0.003210881 | AC002116.2, CLIP3 |
| chr7 | 149855693 | 149855851 | 159 | 2 | 0.000419393 | 0.008466 | 0.038997 | 0.0139974 | 0.002989601 | ZNF862 |
| chr9 | 136848127 | 136848343 | 217 | 2 | 0.000244094 | 0.008799 | 0.044192 | 0.0149671 | 0.001857221 | AJM1 |
| chr6 | 133242204 | 133242244 | 41 | 2 | 0.00024312 | 0.275523 | 0.004702 | 0.0154792 | 0.003729267 | EYA4 |
| chr1 | 51322398 | 51322428 | 31 | 2 | 0.000490898 | 0.012022 | 0.034869 | 0.0175974 | 0.000974155 | TTC39A |
| chr1 | 203795009 | 203795403 | 395 | 2 | 0.000157066 | 0.015453 | 0.026225 | 0.0187671 | 0.002531291 | NA |
| chr1 | 182603950 | 182604489 | 540 | 8 | 0.000313403 | 0.0477 | 0.009583 | 0.0187912 | 0.002162054 | RGS16 |
| chr12 | 89353305 | 89353364 | 60 | 3 | 0.000364444 | 0.008434 | 0.071568 | 0.0193465 | 0.006041649 | NA |
| chr5 | 181223516 | 181224008 | 493 | 5 | 0.000183468 | 0.023534 | 0.06007 | 0.0196061 | 0.001917049 | TRIM41 |
| chr1 | 23484449 | 23484781 | 333 | 2 | 0.000390466 | 0.014337 | 0.039114 | 0.02085 | 0.000824106 | ASAP3 |
| chr1 | 21345077 | 21345597 | 521 | 4 | 0.000311292 | 0.014561 | 0.03675 | 0.0223284 | 0.001566985 | ECE1 |
| chr20 | 63741762 | 63741980 | 219 | 2 | 0.000219908 | 0.016866 | 0.034868 | 0.0223903 | -0.00112354 | SLC2A4RG |
| chr6 | 30185025 | 30185311 | 287 | 2 | 0.00031923 | 0.014813 | 0.045071 | 0.022457 | 0.002095606 | TRIM26 |
| chr3 | 123585496 | 123585578 | 83 | 4 | 0.000357497 | 0.009968 | 0.075258 | 0.023378 | -0.00013746 | MYLK-AS1 |
| chr3 | 23806499 | 23807021 | 523 | 3 | 0.000301113 | 0.048434 | 0.0142 | 0.026256 | 0.000585546 | UBE2E1, UBE2E1-AS1 |
| chr18 | 54827850 | 54827919 | 70 | 3 | 0.000473316 | 0.129045 | 0.005549 | 0.0275082 | -0.00697466 | RAB27B |
| chr2 | 191679084 | 191679182 | 99 | 2 | 0.000311289 | 0.018171 | 0.06198 | 0.0291375 | 0.01044335 | NABP1 |
| chr10 | 1199321 | 1199541 | 221 | 3 | 0.000378193 | 0.020801 | 0.064816 | 0.0323839 | 0.014839691 | LINC00200, ADARB2 |
| chr4 | 15701983 | 15702383 | 401 | 3 | 0.000289099 | 0.108383 | 0.0119 | 0.0354406 | 0.001801925 | AC114744.2 |
| chr9 | 37905116 | 37905216 | 101 | 2 | 0.000241803 | 0.056554 | 0.02362 | 0.0371279 | -0.00356928 | AL138752.2 |
| chr7 | 73843168 | 73843409 | 242 | 2 | 0.000322222 | 0.080035 | 0.019881 | 0.0384504 | -0.0087036 | NA |
| chr17 | 1907887 | 1908060 | 174 | 3 | 0.000423263 | 0.027485 | 0.069486 | 0.0396298 | 0.008493221 | NA |
| chr9 | 122221970 | 122222470 | 501 | 2 | 0.000108434 | 0.195653 | 0.015058 | 0.0401464 | -0.0029472 | LHX6 |
| chr16 | 56781646 | 56781914 | 269 | 4 | 0.00042145 | 0.110974 | 0.016555 | 0.0408371 | 0.001335834 | NUP93 |
| chr16 | 84789384 | 84789509 | 126 | 2 | 0.00040419 | 0.156188 | 0.016353 | 0.0408965 | -0.00200485 | NA |
| chr5 | 174692654 | 174693075 | 422 | 2 | 0.000418802 | 0.029836 | 0.06599 | 0.0427132 | 0.004352266 | NA |
| chr16 | 2047350 | 2047505 | 156 | 4 | 0.00048748 | 0.015962 | 0.120745 | 0.0428701 | -0.00082315 | NTHL1 |
| chr1 | 89995240 | 89995390 | 151 | 3 | 0.0004752 | 0.044501 | 0.040232 | 0.0435207 | 0.001278961 | ZNF326 |
| chr9 | 106620767 | 106621201 | 435 | 3 | 0.00024905 | 0.099745 | 0.022172 | 0.0458718 | -0.00365324 | LINC01505 |
| chr17 | 67245835 | 67245980 | 146 | 2 | 0.000204217 | 0.034774 | 0.06541 | 0.0472775 | -0.00170083 | AC007448.5, HELZ |
| chr20 | 41137720 | 41137866 | 147 | 3 | 0.000426295 | 0.090539 | 0.041828 | 0.0487411 | 0.001673168 | PLCG1, PLCG1-AS1 |
| chr1 | 150235739 | 150236133 | 395 | 3 | 0.000409647 | 0.026827 | 0.113079 | 0.0533407 | 0.000378985 | ANP32E |
| chr3 | 52706216 | 52706467 | 252 | 6 | 0.000388199 | 0.049784 | 0.139061 | 0.058719 | 3.30E-05 | SPCS1 |
| chr7 | 150450448 | 150450465 | 18 | 2 | 0.000480726 | 0.121851 | 0.03013 | 0.0591116 | 0.001796809 | NA |
| chr14 | 101025927 | 101026069 | 143 | 3 | 0.000433628 | 0.061187 | 0.04967 | 0.0652584 | 0.003932077 | MEG8, MIR758 |
| chr6 | 109094183 | 109094513 | 331 | 5 | 0.000305702 | 0.026868 | 0.142288 | 0.0680098 | -0.00061075 | SESN1 |
| chr14 | 24094585 | 24094858 | 274 | 3 | 0.000355488 | 0.181246 | 0.035338 | 0.0682366 | 0.001102488 | PCK2, NRL |

| | | | | | | | | | | |
|---|---|---|---|---|---|---|---|---|---|---|
| chr20 | 51802494 | 51802543 | 50 | 2 | 0.000453325 | 0.057673 | 0.102354 | 0.0797636 | 0.012606553 | SALL4 |
| chr9 | 91362464 | 91362499 | 36 | 2 | 0.000420369 | 0.057032 | 0.104771 | 0.0799407 | 0.001906507 | NA |
| chr6 | 31806380 | 31806476 | 97 | 2 | 0.000375775 | 0.109323 | 0.058956 | 0.0865749 | 0.001725165 | LSM2 |
| chr19 | 17419510 | 17419531 | 22 | 2 | 0.000437769 | 0.083746 | 0.076047 | 0.0870912 | 0.004771008 | MVB12A, AC010319.4, AC010319.3 |
| chr4 | 129093214 | 129093282 | 69 | 2 | 0.000299104 | 0.066225 | 0.120172 | 0.093462 | 0.00172883 | SCLT1 |
| chr17 | 50546530 | 50547077 | 548 | 7 | 0.000331144 | 0.141672 | 0.091419 | 0.1107859 | -0.0004649 | SPATA20 |
| chr16 | 2720813 | 2720865 | 53 | 2 | 0.000425644 | 0.248594 | 0.058761 | 0.1170435 | 0.004807851 | NA |
| chr1 | 13584301 | 13584598 | 298 | 2 | 0.000348303 | 0.210789 | 0.066297 | 0.1210509 | 0.007157135 | PDPN |
| chr19 | 12668814 | 12669121 | 308 | 3 | 0.000214344 | 0.358287 | 0.037294 | 0.1277277 | 0.00024435 | WDR83, AC010422.5, AC010422.8, WDR83OS |
| chr16 | 2215521 | 2215527 | 7 | 2 | 0.000489621 | 0.090901 | 0.16158 | 0.1304347 | 0.001638585 | NA |
| chr13 | 67230153 | 67230207 | 55 | 2 | 0.000468495 | 0.206175 | 0.08165 | 0.1377613 | 0.001725994 | PCDH9 |
| chr11 | 7576252 | 7576583 | 332 | 4 | 0.000370297 | 0.161343 | 0.122651 | 0.1407627 | 0.002643492 | PPFIBP2 |
| chr5 | 142325786 | 142325791 | 6 | 2 | 0.000482903 | 0.158849 | 0.115708 | 0.1522822 | 0.00115603 | SPRY4-AS1, SPRY4 |
| chr3 | 122183714 | 122183775 | 62 | 3 | 0.000464366 | 0.092008 | 0.1889 | 0.1571735 | 0.004860267 | CASR |
| chr16 | 89971527 | 89972061 | 535 | 4 | 0.000315948 | 0.152537 | 0.157252 | 0.1687515 | -0.00333515 | CENPBD1 |
| chr9 | 137017816 | 137017840 | 25 | 2 | 0.000470772 | 0.218462 | 0.210845 | 0.2485985 | 0.00318863 | ABCA2 |
| chr10 | 132786480 | 132786585 | 106 | 2 | 0.000476079 | 0.276133 | 0.18111 | 0.2541907 | -0.00388278 | NA |
| chr8 | 20255172 | 20255252 | 81 | 2 | 0.000459778 | 0.733056 | 0.157256 | 0.2929036 | -0.00380703 | LZTS1 |
| chr6 | 43509806 | 43510013 | 208 | 2 | 0.000422348 | 0.591878 | 0.236488 | 0.3853654 | 0.001042141 | POLR1C, LRRC73 |
| chr1 | 112956139 | 112956180 | 42 | 3 | 0.00036484 | 0.283096 | 0.361876 | 0.4212112 | -0.0013875 | SLC16A1 |
| chr7 | 130734914 | 130735031 | 118 | 3 | 0.000438698 | 0.603445 | 0.208874 | 0.4489721 | 0.002795256 | NA |
| chr7 | 114922261 | 114922742 | 482 | 2 | 0.000404957 | 0.382359 | 0.388315 | 0.458439 | 0.001116748 | MDFIC |
| chr6 | 31970313 | 31970430 | 118 | 4 | 0.000418996 | 0.527539 | 0.346649 | 0.5120048 | -0.0001995 | DXO |
| chr11 | 59755444 | 59755647 | 204 | 2 | 0.000345751 | 0.55834 | 0.425768 | 0.5636816 | 0.000841208 | STX3 |
| chr20 | 13638452 | 13638607 | 156 | 2 | 0.000267478 | 0.572575 | 0.551361 | 0.6659344 | -0.00053583 | TASP1 |
| chr12 | 118980305 | 118980562 | 258 | 2 | 0.00043244 | 0.781462 | 0.4934 | 0.6772303 | 0.000729759 | NA |
| chr12 | 54053565 | 54053646 | 82 | 2 | 0.000472169 | 0.724332 | 0.511225 | 0.6824328 | 0.002048781 | HOXC4 |
| chr14 | 105490416 | 105491011 | 596 | 3 | 0.000400515 | 0.72047 | 0.529665 | 0.7600784 | -0.00030762 | AL928654.3, TEDC1 |
| chr4 | 26860834 | 26860983 | 150 | 3 | 0.000264049 | 0.879147 | 0.5811 | 0.8550461 | 0.000935927 | STIM2 |
