## Supplementary Table S6 for "Early Epigenetic Biomarkers for Perinatal Suicidal Ideation: DNA Methylation Signatures Across the Peripartum Period"

**Supplementary Table S6.1. GO Pathway Results in Perinatal SI v NC at 17w gestation.**

| GO ID | Ontology | Term | N | DE | P.DE | FDR |
|---|---|---|---|---|---|---|
| GO:1903231 | MF | mRNA base-pairing translational repr | 259 | 49 | 2.26E-06 | 0.050338233 |
| GO:0031982 | CC | vesicle | 3944 | 1036 | 1.01E-05 | 0.11255394 |
| GO:0016442 | CC | RISC complex | 364 | 66 | 5.30E-05 | 0.288399041 |
| GO:0030371 | MF | translation repressor activity | 292 | 57 | 5.51E-05 | 0.288399041 |
| GO:0031332 | CC | RNAi effector complex | 365 | 66 | 6.47E-05 | 0.288399041 |
| GO:2000045 | BP | regulation of G1/S transition of mitot | 181 | 69 | 8.46E-05 | 0.300725459 |
| GO:1902806 | BP | regulation of cell cycle G1/S phase tr | 206 | 77 | 9.54E-05 | 0.300725459 |
| GO:0051937 | BP | catecholamine transport | 64 | 30 | 0.000108 | 0.300725459 |
| GO:0007166 | BP | cell surface receptor signaling pathw | 2733 | 757 | 0.000185 | 0.458438203 |
| GO:0035194 | BP | regulatory ncRNA-mediated post-trai | 549 | 95 | 0.000248 | 0.534151212 |
| GO:0035195 | BP | miRNA-mediated post-transcriptiona | 534 | 90 | 0.000264 | 0.534151212 |
| GO:0016441 | BP | post-transcriptional gene silencing | 555 | 96 | 0.000365 | 0.547528154 |
| GO:0016841 | MF | ammonia-lyase activity | 5 | 5 | 0.000389 | 0.547528154 |
| GO:0051693 | BP | actin filament capping | 40 | 21 | 0.000405 | 0.547528154 |
| GO:0044843 | BP | cell cycle G1/S phase transition | 269 | 94 | 0.000427 | 0.547528154 |
| GO:0032272 | BP | negative regulation of protein polym | 75 | 34 | 0.000435 | 0.547528154 |
| GO:0051016 | BP | barbed-end actin filament capping | 25 | 15 | 0.000446 | 0.547528154 |
| GO:0015872 | BP | dopamine transport | 45 | 22 | 0.000456 | 0.547528154 |
| GO:0016525 | BP | negative regulation of angiogenesis | 146 | 49 | 0.000467 | 0.547528154 |
| GO:0042748 | BP | circadian sleep/wake cycle, non-REM | 6 | 6 | 0.00056 | 0.601020198 |

**Supplementary Table S6.2. GO Pathway Results in Perinatal SI v NC at 38w gestation.**

| GO ID | Ontology | Term | N | DE | P.DE | FDR |
|---|---|---|---|---|---|---|
| GO:0048856 | BP | anatomical structure development | 5814 | 3664 | 3.12E-22 | 6.96E-18 |
| GO:0005515 | MF | protein binding | 13818 | 8211 | 1.08E-21 | 1.20E-17 |
| GO:0032502 | BP | developmental process | 6319 | 3947 | 2.07E-21 | 1.53E-17 |
| GO:0007275 | BP | multicellular organism development | 4625 | 2968 | 1.68E-20 | 9.37E-17 |
| GO:0005488 | MF | binding | 15772 | 9192 | 3.40E-20 | 1.51E-16 |
| GO:0071840 | BP | cellular component organization or l | 6603 | 4096 | 1.41E-19 | 5.25E-16 |
| GO:0048731 | BP | system development | 3974 | 2572 | 3.92E-19 | 1.25E-15 |
| GO:0016043 | BP | cellular component organization | 6400 | 3970 | 3.30E-18 | 9.18E-15 |
| GO:0007399 | BP | nervous system development | 2501 | 1686 | 5.38E-18 | 1.33E-14 |
| GO:0030154 | BP | cell differentiation | 4302 | 2715 | 2.29E-16 | 5.10E-13 |
| GO:0048869 | BP | cellular developmental process | 4304 | 2715 | 3.00E-16 | 6.09E-13 |
| GO:0043226 | CC | organelle | 13857 | 8024 | 1.55E-15 | 2.88E-12 |
| GO:0048518 | BP | positive regulation of biological proc | 6104 | 3743 | 2.16E-15 | 3.70E-12 |
| GO:0048468 | BP | cell development | 2779 | 1811 | 4.26E-15 | 6.77E-12 |
| GO:0048522 | BP | positive regulation of cellular proces | 5479 | 3389 | 6.47E-15 | 9.60E-12 |
| GO:0005622 | CC | intracellular anatomical structure | 14487 | 8394 | 9.02E-15 | 1.26E-11 |
| GO:0003674 | MF | molecular_function | 17111 | 9802 | 1.39E-14 | 1.82E-11 |
| GO:0022008 | BP | neurogenesis | 1724 | 1176 | 2.22E-14 | 2.75E-11 |
| GO:0043227 | CC | membrane-bounded organelle | 13037 | 7544 | 2.61E-14 | 3.06E-11 |
| GO:0043229 | CC | intracellular organelle | 13017 | 7563 | 4.40E-14 | 4.90E-11 |
| GO:0051179 | BP | localization | 5157 | 3168 | 5.41E-14 | 5.55E-11 |
| GO:0005737 | CC | cytoplasm | 11536 | 6819 | 5.48E-14 | 5.55E-11 |
| GO:0050793 | BP | regulation of developmental proces | 2447 | 1584 | 1.32E-13 | 1.27E-10 |
| GO:0043231 | CC | intracellular membrane-bounded or | 11936 | 6942 | 3.97E-13 | 3.69E-10 |
| GO:0110165 | CC | cellular anatomical entity | 17682 | 9974 | 5.01E-13 | 4.31E-10 |
| GO:0005575 | CC | cellular_component | 18008 | 10107 | 5.03E-13 | 4.31E-10 |
| GO:0009893 | BP | positive regulation of metabolic pro | 3570 | 2234 | 8.09E-13 | 6.68E-10 |
| GO:0051234 | BP | establishment of localization | 4522 | 2775 | 1.08E-12 | 8.58E-10 |
| GO:0009987 | BP | cellular process | 16153 | 9116 | 1.65E-12 | 1.27E-09 |
| GO:0009653 | BP | anatomical structure morphogenesis | 2684 | 1731 | 1.72E-12 | 1.28E-09 |
| GO:0006810 | BP | transport | 4218 | 2594 | 2.38E-12 | 1.71E-09 |
| GO:0005829 | CC | cytosol | 5245 | 3196 | 4.37E-12 | 3.05E-09 |
| GO:0048699 | BP | generation of neurons | 1475 | 1004 | 6.01E-12 | 4.06E-09 |
| GO:0008150 | BP | biological_process | 17165 | 9642 | 1.02E-11 | 6.68E-09 |
| GO:0005634 | CC | nucleus | 7580 | 4444 | 1.62E-11 | 1.03E-08 |
| GO:0030182 | BP | neuron differentiation | 1392 | 948 | 1.94E-11 | 1.20E-08 |
| GO:0051128 | BP | regulation of cellular component org | 2389 | 1540 | 2.08E-11 | 1.25E-08 |
| GO:0044085 | BP | cellular component biogenesis | 3257 | 2027 | 2.46E-11 | 1.44E-08 |
| GO:0036094 | MF | small molecule binding | 5922 | 3580 | 2.73E-11 | 1.56E-08 |
| GO:0050789 | BP | regulation of biological process | 11427 | 6586 | 3.11E-11 | 1.73E-08 |
| GO:0032991 | CC | protein-containing complex | 5658 | 3354 | 3.87E-11 | 2.10E-08 |
| GO:0005654 | CC | nucleoplasm | 3981 | 2475 | 4.54E-11 | 2.41E-08 |
| GO:0065007 | BP | biological regulation | 11793 | 6782 | 5.25E-11 | 2.72E-08 |
| GO:0008152 | BP | metabolic process | 11382 | 6463 | 6.05E-11 | 3.06E-08 |
| GO:0080090 | BP | regulation of primary metabolic pro | 5280 | 3195 | 7.97E-11 | 3.95E-08 |
| GO:0032501 | BP | multicellular organismal process | 7076 | 4176 | 9.02E-11 | 4.37E-08 |
| GO:0051641 | BP | cellular localization | 3311 | 2066 | 1.01E-10 | 4.77E-08 |

| | | | | | | |
|---|---|---|---|---|---|---|
| GO:0043167 | MF | ion binding | 5741 | 3469 | 1.12E-10 | 5.06E-08 |
| GO:0010604 | BP | positive regulation of macromolecul | 3275 | 2038 | 1.13E-10 | 5.06E-08 |
| GO:0048523 | BP | negative regulation of cellular proce | 5143 | 3069 | 1.14E-10 | 5.06E-08 |
| GO:0050794 | BP | regulation of cellular process | 10811 | 6233 | 1.51E-10 | 6.58E-08 |
| GO:0045935 | BP | positive regulation of nucleobase-co | 2042 | 1310 | 1.90E-10 | 8.15E-08 |
| GO:0044237 | BP | cellular metabolic process | 9669 | 5494 | 2.15E-10 | 9.02E-08 |
| GO:0006796 | BP | phosphate-containing compound me | 2253 | 1430 | 2.28E-10 | 9.41E-08 |
| GO:0006793 | BP | phosphorus metabolic process | 2255 | 1431 | 2.50E-10 | 1.01E-07 |
| GO:0044238 | BP | primary metabolic process | 9922 | 5713 | 2.74E-10 | 1.09E-07 |
| GO:0048519 | BP | negative regulation of biological pro | 5556 | 3293 | 4.03E-10 | 1.58E-07 |
| GO:0022607 | BP | cellular component assembly | 2995 | 1865 | 4.54E-10 | 1.74E-07 |
| GO:0048666 | BP | neuron development | 1126 | 770 | 5.16E-10 | 1.95E-07 |
| GO:0007417 | BP | central nervous system developmen | 992 | 685 | 5.59E-10 | 2.08E-07 |
| GO:0051239 | BP | regulation of multicellular organisma | 2984 | 1843 | 6.62E-10 | 2.42E-07 |
| GO:0007169 | BP | cell surface receptor protein tyrosine | 639 | 445 | 7.07E-10 | 2.54E-07 |
| GO:0031974 | CC | membrane-enclosed lumen | 5827 | 3422 | 8.66E-10 | 2.97E-07 |
| GO:0043233 | CC | organelle lumen | 5827 | 3422 | 8.66E-10 | 2.97E-07 |
| GO:0070013 | CC | intracellular organelle lumen | 5827 | 3422 | 8.66E-10 | 2.97E-07 |
| GO:0033554 | BP | cellular response to stress | 1799 | 1156 | 1.08E-09 | 3.63E-07 |
| GO:0019222 | BP | regulation of metabolic process | 6600 | 3861 | 1.25E-09 | 4.10E-07 |
| GO:0048513 | BP | animal organ development | 2955 | 1860 | 1.25E-09 | 4.10E-07 |
| GO:0007167 | BP | enzyme-linked receptor protein sign | 1012 | 672 | 1.27E-09 | 4.10E-07 |
| GO:0033036 | BP | macromolecule localization | 2950 | 1831 | 1.31E-09 | 4.18E-07 |
| GO:0010646 | BP | regulation of cell communication | 3478 | 2157 | 1.49E-09 | 4.68E-07 |
| GO:0031323 | BP | regulation of cellular metabolic proc | 5889 | 3452 | 1.75E-09 | 5.43E-07 |
| GO:0019219 | BP | regulation of nucleobase-containing | 3786 | 2320 | 1.81E-09 | 5.51E-07 |
| GO:0048583 | BP | regulation of response to stimulus | 3989 | 2428 | 1.98E-09 | 5.97E-07 |
| GO:0060255 | BP | regulation of macromolecule metab | 6082 | 3560 | 2.17E-09 | 6.44E-07 |
| GO:0048646 | BP | anatomical structure formation invo | 1247 | 813 | 2.32E-09 | 6.81E-07 |
| GO:0006996 | BP | organelle organization | 3490 | 2153 | 2.51E-09 | 7.26E-07 |
| GO:0031325 | BP | positive regulation of cellular metab | 2968 | 1854 | 2.63E-09 | 7.50E-07 |
| GO:0060322 | BP | head development | 749 | 526 | 2.97E-09 | 8.37E-07 |
| GO:0023051 | BP | regulation of signaling | 3471 | 2149 | 3.01E-09 | 8.37E-07 |
| GO:0042995 | CC | cell projection | 2320 | 1476 | 3.33E-09 | 9.16E-07 |
| GO:0043412 | BP | macromolecule modification | 2749 | 1717 | 3.48E-09 | 9.44E-07 |
| GO:0019899 | MF | enzyme binding | 2001 | 1279 | 4.15E-09 | 1.11E-06 |
| GO:0097159 | MF | organic cyclic compound binding | 5961 | 3527 | 4.34E-09 | 1.15E-06 |
| GO:0009889 | BP | regulation of biosynthetic process | 5349 | 3132 | 4.67E-09 | 1.22E-06 |
| GO:0009888 | BP | tissue development | 1979 | 1260 | 6.03E-09 | 1.56E-06 |
| GO:0007049 | BP | cell cycle | 1622 | 1035 | 7.82E-09 | 2.00E-06 |
| GO:0009891 | BP | positive regulation of biosynthetic p | 2605 | 1632 | 7.99E-09 | 2.02E-06 |
| GO:0065008 | BP | regulation of biological quality | 2908 | 1791 | 9.07E-09 | 2.27E-06 |
| GO:0031981 | CC | nuclear lumen | 4735 | 2789 | 9.35E-09 | 2.32E-06 |
| GO:0017076 | MF | purine nucleotide binding | 1864 | 1191 | 1.01E-08 | 2.45E-06 |
| GO:0007420 | BP | brain development | 700 | 491 | 1.01E-08 | 2.45E-06 |
| GO:0120025 | CC | plasma membrane bounded cell pro | 2204 | 1400 | 1.15E-08 | 2.75E-06 |
| GO:0031326 | BP | regulation of cellular biosynthetic pr | 5278 | 3084 | 1.30E-08 | 3.08E-06 |
| GO:0043170 | BP | macromolecule metabolic process | 9452 | 5350 | 1.43E-08 | 3.35E-06 |
| GO:0043005 | CC | neuron projection | 1276 | 846 | 1.53E-08 | 3.54E-06 |

| | | | | | | |
|---|---|---|---|---|---|---|
| GO:0016310 | BP | phosphorylation | 1308 | 854 | 1.54E-08 | 3.54E-06 |
| GO:0009058 | BP | biosynthetic process | 8141 | 4598 | 1.66E-08 | 3.77E-06 |
| GO:0051094 | BP | positive regulation of developmenta | 1341 | 874 | 1.76E-08 | 3.95E-06 |
| GO:0045595 | BP | regulation of cell differentiation | 1567 | 1012 | 1.81E-08 | 4.02E-06 |
| GO:1901265 | MF | nucleoside phosphate binding | 2032 | 1289 | 1.91E-08 | 4.22E-06 |
| GO:0051240 | BP | positive regulation of multicellular o | 1656 | 1049 | 2.33E-08 | 5.10E-06 |
| GO:0000166 | MF | nucleotide binding | 2014 | 1277 | 2.43E-08 | 5.25E-06 |
| GO:0036477 | CC | somatodendritic compartment | 817 | 560 | 2.60E-08 | 5.58E-06 |
| GO:0032879 | BP | regulation of localization | 2003 | 1257 | 3.29E-08 | 6.98E-06 |
| GO:0045892 | BP | negative regulation of DNA-templat | 1168 | 778 | 3.53E-08 | 7.43E-06 |
| GO:0036211 | BP | protein modification process | 2576 | 1607 | 3.84E-08 | 8.00E-06 |
| GO:0051252 | BP | regulation of RNA metabolic process | 3499 | 2137 | 4.01E-08 | 8.27E-06 |
| GO:0043232 | CC | intracellular non-membrane-bounde | 5542 | 3232 | 4.17E-08 | 8.52E-06 |
| GO:0010556 | BP | regulation of macromolecule biosyn | 5190 | 3026 | 4.53E-08 | 9.16E-06 |
| GO:0043228 | CC | non-membrane-bounded organelle | 5543 | 3232 | 4.56E-08 | 9.16E-06 |
| GO:0031328 | BP | positive regulation of cellular biosyn | 2559 | 1597 | 4.73E-08 | 9.40E-06 |
| GO:0009966 | BP | regulation of signal transduction | 3047 | 1880 | 5.11E-08 | 1.01E-05 |
| GO:0031175 | BP | neuron projection development | 983 | 667 | 5.25E-08 | 1.03E-05 |
| GO:0030030 | BP | cell projection organization | 1571 | 1025 | 5.87E-08 | 1.14E-05 |
| GO:0120036 | BP | plasma membrane bounded cell pro | 1529 | 999 | 6.49E-08 | 1.25E-05 |
| GO:0030554 | MF | adenyl nucleotide binding | 1542 | 998 | 6.61E-08 | 1.26E-05 |
| GO:0006468 | BP | protein phosphorylation | 1108 | 727 | 6.66E-08 | 1.26E-05 |
| GO:0032555 | MF | purine ribonucleotide binding | 1771 | 1128 | 6.87E-08 | 1.29E-05 |
| GO:0003824 | MF | catalytic activity | 5203 | 3088 | 7.10E-08 | 1.31E-05 |
| GO:0006355 | BP | regulation of DNA-templated transc | 3159 | 1948 | 7.12E-08 | 1.31E-05 |
| GO:0097367 | MF | carbohydrate derivative binding | 2144 | 1340 | 8.93E-08 | 1.63E-05 |
| GO:0010468 | BP | regulation of gene expression | 5073 | 2954 | 9.19E-08 | 1.67E-05 |
| GO:0019538 | BP | protein metabolic process | 4584 | 2727 | 9.35E-08 | 1.67E-05 |
| GO:1902679 | BP | negative regulation of RNA biosynth | 1182 | 782 | 9.39E-08 | 1.67E-05 |
| GO:0006950 | BP | response to stress | 3836 | 2279 | 1.00E-07 | 1.77E-05 |
| GO:2001141 | BP | regulation of RNA biosynthetic proc | 3177 | 1955 | 1.02E-07 | 1.78E-05 |
| GO:0032553 | MF | ribonucleotide binding | 1788 | 1136 | 1.03E-07 | 1.79E-05 |
| GO:0010557 | BP | positive regulation of macromolecul | 2502 | 1559 | 1.06E-07 | 1.83E-05 |
| GO:2000026 | BP | regulation of multicellular organism | 1445 | 923 | 1.17E-07 | 2.01E-05 |
| GO:0043168 | MF | anion binding | 2288 | 1430 | 1.24E-07 | 2.12E-05 |
| GO:0051254 | BP | positive regulation of RNA metaboli | 1847 | 1169 | 1.27E-07 | 2.14E-05 |
| GO:0070062 | CC | extracellular exosome | 2075 | 1251 | 1.45E-07 | 2.42E-05 |
| GO:1903561 | CC | extracellular vesicle | 2162 | 1290 | 1.47E-07 | 2.44E-05 |
| GO:0043230 | CC | extracellular organelle | 2163 | 1290 | 1.70E-07 | 2.78E-05 |
| GO:0065010 | CC | extracellular membrane-bounded or | 2163 | 1290 | 1.70E-07 | 2.78E-05 |
| GO:0035639 | MF | purine ribonucleoside triphosphate | 1724 | 1096 | 1.71E-07 | 2.79E-05 |
| GO:0043933 | BP | protein-containing complex organiza | 1763 | 1095 | 1.79E-07 | 2.88E-05 |
| GO:0098590 | CC | plasma membrane region | 1278 | 828 | 1.81E-07 | 2.91E-05 |
| GO:0008283 | BP | cell population proliferation | 1990 | 1234 | 1.92E-07 | 3.06E-05 |
| GO:0065003 | BP | protein-containing complex assembl | 1573 | 978 | 2.27E-07 | 3.59E-05 |
| GO:1901363 | MF | heterocyclic compound binding | 2159 | 1353 | 2.35E-07 | 3.70E-05 |
| GO:1902680 | BP | positive regulation of RNA biosynthe | 1633 | 1054 | 2.52E-07 | 3.93E-05 |
| GO:0051253 | BP | negative regulation of RNA metabol | 1280 | 839 | 2.90E-07 | 4.49E-05 |
| GO:0043169 | MF | cation binding | 4161 | 2498 | 3.08E-07 | 4.73E-05 |

| | | | | | | |
|---|---|---|---|---|---|---|
| GO:0000902 | BP | cell morphogenesis | 949 | 644 | 3.45E-07 | 5.25E-05 |
| GO:0045893 | BP | positive regulation of DNA-template | 1630 | 1051 | 3.46E-07 | 5.25E-05 |
| GO:0032559 | MF | adenyl ribonucleotide binding | 1450 | 936 | 4.05E-07 | 6.10E-05 |
| GO:0046872 | MF | metal ion binding | 4071 | 2446 | 4.16E-07 | 6.23E-05 |
| GO:0045597 | BP | positive regulation of cell differentia | 878 | 582 | 4.57E-07 | 6.78E-05 |
| GO:0009790 | BP | embryo development | 1099 | 727 | 4.59E-07 | 6.78E-05 |
| GO:0009628 | BP | response to abiotic stimulus | 1103 | 715 | 4.64E-07 | 6.80E-05 |
| GO:0070727 | BP | cellular macromolecule localization | 2480 | 1535 | 4.87E-07 | 7.10E-05 |
| GO:0098660 | BP | inorganic ion transmembrane transp | 908 | 587 | 5.28E-07 | 7.64E-05 |
| GO:0016020 | CC | membrane | 8749 | 5042 | 5.35E-07 | 7.70E-05 |
| GO:0040007 | BP | growth | 900 | 596 | 5.44E-07 | 7.78E-05 |
| GO:0006351 | BP | DNA-templated transcription | 3273 | 2006 | 5.54E-07 | 7.87E-05 |
| GO:0008104 | BP | protein localization | 2468 | 1528 | 6.32E-07 | 8.91E-05 |
| GO:0007610 | BP | behavior | 647 | 439 | 6.43E-07 | 9.01E-05 |
| GO:0010467 | BP | gene expression | 6780 | 3804 | 6.51E-07 | 9.07E-05 |
| GO:0045934 | BP | negative regulation of nucleobase-co | 1398 | 908 | 7.47E-07 | 0.00010341 |
| GO:0030054 | CC | cell junction | 2281 | 1453 | 7.66E-07 | 0.000105374 |
| GO:0044249 | BP | cellular biosynthetic process | 7739 | 4353 | 9.68E-07 | 0.000132389 |
| GO:0060341 | BP | regulation of cellular localization | 989 | 641 | 1.07E-06 | 0.000145678 |
| GO:0006915 | BP | apoptotic process | 1867 | 1168 | 1.12E-06 | 0.000151202 |
| GO:0022402 | BP | cell cycle process | 1266 | 807 | 1.23E-06 | 0.000165178 |
| GO:0045202 | CC | synapse | 1578 | 1023 | 1.24E-06 | 0.000165594 |
| GO:0030097 | BP | hemopoiesis | 928 | 606 | 1.26E-06 | 0.000166679 |
| GO:0001944 | BP | vasculature development | 823 | 531 | 1.26E-06 | 0.000166679 |
| GO:0050896 | BP | response to stimulus | 8395 | 4828 | 1.34E-06 | 0.000175001 |
| GO:0051649 | BP | establishment of localization in cell | 1901 | 1188 | 1.36E-06 | 0.000176905 |
| GO:0005524 | MF | ATP binding | 1410 | 910 | 1.43E-06 | 0.00018572 |
| GO:0006139 | BP | nucleobase-containing compound m | 5767 | 3325 | 1.48E-06 | 0.000191076 |
| GO:0007166 | BP | cell surface receptor signaling pathw | 2732 | 1662 | 1.69E-06 | 0.000216186 |
| GO:0048585 | BP | negative regulation of response to st | 1731 | 1069 | 1.72E-06 | 0.000219629 |
| GO:0019900 | MF | kinase binding | 754 | 502 | 2.22E-06 | 0.000280508 |
| GO:0042063 | BP | gliogenesis | 345 | 246 | 2.26E-06 | 0.000284207 |
| GO:0051338 | BP | regulation of transferase activity | 503 | 337 | 2.31E-06 | 0.000289029 |
| GO:0044297 | CC | cell body | 540 | 369 | 2.76E-06 | 0.000343788 |
| GO:0023057 | BP | negative regulation of signaling | 1479 | 925 | 2.78E-06 | 0.000344465 |
| GO:0012501 | BP | programmed cell death | 1933 | 1204 | 2.84E-06 | 0.000349472 |
| GO:0010648 | BP | negative regulation of cell communi | 1479 | 925 | 2.87E-06 | 0.000350588 |
| GO:0016604 | CC | nuclear body | 818 | 534 | 2.88E-06 | 0.000350588 |
| GO:0008219 | BP | cell death | 1937 | 1206 | 3.14E-06 | 0.000380355 |
| GO:0035239 | BP | tube morphogenesis | 934 | 602 | 3.51E-06 | 0.000423024 |
| GO:0003712 | MF | transcription coregulator activity | 491 | 338 | 3.71E-06 | 0.000444152 |
| GO:0009059 | BP | macromolecule biosynthetic process | 7202 | 4038 | 3.75E-06 | 0.000447053 |
| GO:0051130 | BP | positive regulation of cellular compo | 1083 | 702 | 4.01E-06 | 0.000475702 |
| GO:0043025 | CC | neuronal cell body | 476 | 329 | 4.20E-06 | 0.000495071 |
| GO:0051174 | BP | regulation of phosphorus metabolic | 866 | 560 | 4.38E-06 | 0.000513448 |
| GO:0090304 | BP | nucleic acid metabolic process | 5244 | 3019 | 4.63E-06 | 0.000540434 |
| GO:0009968 | BP | negative regulation of signal transdu | 1374 | 860 | 4.85E-06 | 0.000562943 |
| GO:0005615 | CC | extracellular space | 3286 | 1874 | 5.25E-06 | 0.000605688 |
| GO:0098662 | BP | inorganic cation transmembrane tra | 821 | 529 | 5.29E-06 | 0.000607574 |

| | | | | | | |
|---|---|---|---|---|---|---|
| GO:0019220 | BP | regulation of phosphate metabolic p | 865 | 559 | 5.51E-06 | 0.000627379 |
| GO:0072359 | BP | circulatory system development | 1209 | 769 | 5.52E-06 | 0.000627379 |
| GO:0031982 | CC | vesicle | 3944 | 2315 | 6.66E-06 | 0.000753311 |
| GO:0040008 | BP | regulation of growth | 583 | 393 | 6.90E-06 | 0.000773853 |
| GO:0001568 | BP | blood vessel development | 790 | 507 | 6.91E-06 | 0.000773853 |
| GO:0043067 | BP | regulation of programmed cell deatl | 1471 | 921 | 7.19E-06 | 0.000798449 |
| GO:0009056 | BP | catabolic process | 2621 | 1572 | 7.20E-06 | 0.000798449 |
| GO:0033043 | BP | regulation of organelle organization | 1141 | 732 | 8.61E-06 | 0.000949461 |
| GO:0010001 | BP | glial cell differentiation | 261 | 190 | 8.90E-06 | 0.000976512 |
| GO:0051716 | BP | cellular response to stimulus | 7066 | 4084 | 9.22E-06 | 0.001007403 |
| GO:0050905 | BP | neuromuscular process | 174 | 130 | 9.33E-06 | 0.001014298 |
| GO:0048667 | BP | cell morphogenesis involved in neur | 559 | 390 | 9.71E-06 | 0.001049857 |
| GO:0065009 | BP | regulation of molecular function | 1721 | 1056 | 1.01E-05 | 0.001088765 |
| GO:0051049 | BP | regulation of transport | 1596 | 991 | 1.02E-05 | 0.001089267 |
| GO:0000122 | BP | negative regulation of transcription | 858 | 573 | 1.04E-05 | 0.001112728 |
| GO:0034220 | BP | monoatomic ion transmembrane tra | 998 | 635 | 1.05E-05 | 0.001112728 |
| GO:0035295 | BP | tube development | 1147 | 731 | 1.06E-05 | 0.001114633 |
| GO:0140096 | MF | catalytic activity, acting on a protein | 2145 | 1323 | 1.14E-05 | 0.001193335 |
| GO:0048858 | BP | cell projection morphogenesis | 644 | 445 | 1.25E-05 | 0.001309773 |
| GO:0034654 | BP | nucleobase-containing compound bi | 4830 | 2789 | 1.27E-05 | 0.001324479 |
| GO:0071705 | BP | nitrogen compound transport | 1822 | 1122 | 1.37E-05 | 0.001412661 |
| GO:0005576 | CC | extracellular region | 4118 | 2319 | 1.37E-05 | 0.001412661 |
| GO:0009792 | BP | embryo development ending in birtl | 666 | 449 | 1.42E-05 | 0.00146122 |
| GO:0042981 | BP | regulation of apoptotic process | 1424 | 890 | 1.49E-05 | 0.001522055 |
| GO:0032880 | BP | regulation of protein localization | 881 | 570 | 1.57E-05 | 0.001599604 |
| GO:0012505 | CC | endomembrane system | 4560 | 2702 | 1.66E-05 | 0.001679277 |
| GO:0042325 | BP | regulation of phosphorylation | 742 | 482 | 1.68E-05 | 0.001690309 |
| GO:0034330 | BP | cell junction organization | 787 | 528 | 1.71E-05 | 0.001711927 |
| GO:0044877 | MF | protein-containing complex binding | 1247 | 789 | 1.71E-05 | 0.001711927 |
| GO:0043009 | BP | chordate embryonic development | 646 | 436 | 1.72E-05 | 0.001712445 |
| GO:0003714 | MF | transcription corepressor activity | 190 | 140 | 1.79E-05 | 0.001777013 |
| GO:0015318 | MF | inorganic molecular entity transmen | 679 | 444 | 1.82E-05 | 0.001791113 |
| GO:0051726 | BP | regulation of cell cycle | 1084 | 689 | 1.89E-05 | 0.00186027 |
| GO:0031324 | BP | negative regulation of cellular metal | 2827 | 1642 | 2.12E-05 | 0.002066881 |
| GO:0048584 | BP | positive regulation of response to st | 2291 | 1391 | 2.12E-05 | 0.002066881 |
| GO:0098655 | BP | monoatomic cation transmembrane | 843 | 538 | 2.32E-05 | 0.002248422 |
| GO:0050790 | BP | regulation of catalytic activity | 1158 | 719 | 2.39E-05 | 0.002296628 |
| GO:0009892 | BP | negative regulation of metabolic prc | 3217 | 1862 | 2.39E-05 | 0.002296628 |
| GO:0120039 | BP | plasma membrane bounded cell pro | 639 | 440 | 2.44E-05 | 0.002329099 |
| GO:0045859 | BP | regulation of protein kinase activity | 403 | 272 | 2.45E-05 | 0.002330516 |
| GO:0051054 | BP | positive regulation of DNA metaboli | 286 | 199 | 2.76E-05 | 0.002620265 |
| GO:0001525 | BP | angiogenesis | 613 | 391 | 2.90E-05 | 0.00273764 |
| GO:0044087 | BP | regulation of cellular component bic | 992 | 642 | 3.03E-05 | 0.002853492 |
| GO:0030424 | CC | axon | 629 | 426 | 3.07E-05 | 0.002878854 |
| GO:0031399 | BP | regulation of protein modification p | 953 | 610 | 3.18E-05 | 0.002963918 |
| GO:0043549 | BP | regulation of kinase activity | 436 | 292 | 3.19E-05 | 0.002963918 |
| GO:0035966 | BP | response to topologically incorrect p | 156 | 114 | 3.30E-05 | 0.003044968 |
| GO:0060429 | BP | epithelium development | 1203 | 763 | 3.31E-05 | 0.003044968 |
| GO:0048589 | BP | developmental growth | 651 | 434 | 3.55E-05 | 0.00325458 |

| | | | | | | |
|---|---|---|---|---|---|---|
| GO:0010605 | BP | negative regulation of macromolecu | 2996 | 1731 | 3.60E-05 | 0.003276666 |
| GO:0006811 | BP | monoatomic ion transport | 1216 | 758 | 3.61E-05 | 0.003276666 |
| GO:0051246 | BP | regulation of protein metabolic proc | 1978 | 1198 | 3.62E-05 | 0.003276666 |
| GO:0042802 | MF | identical protein binding | 2104 | 1288 | 3.69E-05 | 0.003324974 |
| GO:0060284 | BP | regulation of cell development | 819 | 538 | 3.77E-05 | 0.003387926 |
| GO:0006357 | BP | regulation of transcription by RNA p | 2416 | 1490 | 4.74E-05 | 0.004242198 |
| GO:0055085 | BP | transmembrane transport | 1508 | 927 | 4.76E-05 | 0.004246014 |
| GO:0007154 | BP | cell communication | 6256 | 3628 | 4.86E-05 | 0.004317112 |
| GO:0019901 | MF | protein kinase binding | 676 | 446 | 4.89E-05 | 0.004321951 |
| GO:0009408 | BP | response to heat | 98 | 76 | 4.97E-05 | 0.004375479 |
| GO:0030900 | BP | forebrain development | 398 | 279 | 5.08E-05 | 0.004458574 |
| GO:0042127 | BP | regulation of cell population prolifer | 1693 | 1038 | 5.35E-05 | 0.004678098 |
| GO:0048812 | BP | neuron projection morphogenesis | 623 | 427 | 5.48E-05 | 0.004768178 |
| GO:0023052 | BP | signaling | 6230 | 3611 | 5.50E-05 | 0.004768178 |
| GO:0071900 | BP | regulation of protein serine/threoni | 240 | 167 | 5.53E-05 | 0.004774164 |
| GO:0005215 | MF | transporter activity | 1176 | 733 | 5.55E-05 | 0.004777004 |
| GO:0045944 | BP | positive regulation of transcription b | 1212 | 783 | 5.76E-05 | 0.004933127 |
| GO:0097190 | BP | apoptotic signaling pathway | 606 | 394 | 6.04E-05 | 0.005155033 |
| GO:0030425 | CC | dendrite | 597 | 404 | 6.28E-05 | 0.005337899 |
| GO:0044093 | BP | positive regulation of molecular fun | 954 | 606 | 6.37E-05 | 0.005400763 |
| GO:0007010 | BP | cytoskeleton organization | 1480 | 930 | 6.43E-05 | 0.005426471 |
| GO:0048754 | BP | branching morphogenesis of an epit | 153 | 116 | 7.05E-05 | 0.005903975 |
| GO:0005509 | MF | calcium ion binding | 672 | 432 | 7.06E-05 | 0.005903975 |
| GO:0009314 | BP | response to radiation | 424 | 284 | 7.09E-05 | 0.005903975 |
| GO:0016070 | BP | RNA metabolic process | 4723 | 2699 | 7.10E-05 | 0.005903975 |
| GO:0001932 | BP | regulation of protein phosphorylatic | 688 | 445 | 7.25E-05 | 0.006009658 |
| GO:1901700 | BP | response to oxygen-containing comp | 1632 | 1003 | 7.32E-05 | 0.00604287 |
| GO:0007626 | BP | locomotory behavior | 208 | 150 | 7.55E-05 | 0.006193409 |
| GO:0006325 | BP | chromatin organization | 993 | 633 | 7.56E-05 | 0.006193409 |
| GO:0045927 | BP | positive regulation of growth | 242 | 171 | 7.87E-05 | 0.006425616 |
| GO:0141187 | BP | nucleic acid biosynthetic process | 4556 | 2617 | 7.94E-05 | 0.006460346 |
| GO:0061564 | BP | axon development | 494 | 342 | 8.32E-05 | 0.006738053 |
| GO:0033555 | BP | multicellular organismal response tc | 94 | 74 | 8.35E-05 | 0.006738053 |
| GO:0003676 | MF | nucleic acid binding | 4041 | 2352 | 8.66E-05 | 0.006969081 |
| GO:0022857 | MF | transmembrane transporter activity | 1063 | 664 | 8.94E-05 | 0.007165643 |
| GO:0051052 | BP | regulation of DNA metabolic proces: | 484 | 319 | 9.00E-05 | 0.007190391 |
| GO:0035556 | BP | intracellular signal transduction | 2922 | 1784 | 9.22E-05 | 0.007340356 |
| GO:0097447 | CC | dendritic tree | 599 | 404 | 9.35E-05 | 0.007410939 |
| GO:0099572 | CC | postsynaptic specialization | 377 | 262 | 9.54E-05 | 0.007517112 |
| GO:0022890 | MF | inorganic cation transmembrane tra | 564 | 369 | 9.55E-05 | 0.007517112 |
| GO:0007409 | BP | axonogenesis | 431 | 302 | 9.81E-05 | 0.007700535 |
| GO:0002009 | BP | morphogenesis of an epithelium | 485 | 330 | 0.000102 | 0.007945442 |
| GO:0010657 | BP | muscle cell apoptotic process | 100 | 74 | 0.000104 | 0.008070537 |
| GO:0010660 | BP | regulation of muscle cell apoptotic p | 94 | 70 | 0.000104 | 0.008070537 |
| GO:0071944 | CC | cell periphery | 5696 | 3283 | 0.000107 | 0.008288941 |
| GO:0008630 | BP | intrinsic apoptotic signaling pathway | 105 | 81 | 0.000113 | 0.008697584 |
| GO:0022037 | BP | metencephalon development | 110 | 86 | 0.000113 | 0.008707463 |
| GO:0048514 | BP | blood vessel morphogenesis | 703 | 444 | 0.000114 | 0.008746344 |
| GO:1901701 | BP | cellular response to oxygen-containi | 1165 | 726 | 0.000115 | 0.008760056 |

| | | | | | | |
|---|---|---|---|---|---|---|
| GO:0002521 | BP | leukocyte differentiation | 606 | 395 | 0.000116 | 0.008848094 |
| GO:0005815 | CC | microtubule organizing center | 852 | 545 | 0.000117 | 0.008851443 |
| GO:0016049 | BP | cell growth | 466 | 315 | 0.000119 | 0.008983562 |
| GO:0006366 | BP | transcription by RNA polymerase II | 2525 | 1547 | 0.00012 | 0.009004481 |
| GO:0009890 | BP | negative regulation of biosynthetic p | 2585 | 1488 | 0.000123 | 0.009194055 |
| GO:0000278 | BP | mitotic cell cycle | 897 | 572 | 0.000123 | 0.009204875 |
| GO:0016192 | BP | vesicle-mediated transport | 1511 | 934 | 0.000125 | 0.009296719 |
| GO:0034329 | BP | cell junction assembly | 484 | 330 | 0.000125 | 0.009296719 |
| GO:0051247 | BP | positive regulation of protein metab | 1050 | 661 | 0.000129 | 0.009536106 |
| GO:0032774 | BP | RNA biosynthetic process | 4447 | 2549 | 0.000129 | 0.009550609 |
| GO:0008016 | BP | regulation of heart contraction | 203 | 143 | 0.000134 | 0.009889992 |
| GO:0046486 | BP | glycerolipid metabolic process | 367 | 243 | 0.000138 | 0.010121734 |
| GO:0021549 | BP | cerebellum development | 100 | 79 | 0.000139 | 0.010187699 |
| GO:0001933 | BP | negative regulation of protein phosp | 234 | 163 | 0.000141 | 0.010240771 |
| GO:0035967 | BP | cellular response to topologically inc | 110 | 82 | 0.00016 | 0.011601583 |
| GO:0071495 | BP | cellular response to endogenous stir | 1247 | 781 | 0.000164 | 0.011876874 |
| GO:0006974 | BP | DNA damage response | 873 | 558 | 0.00017 | 0.012241617 |
| GO:0015075 | MF | monoatomic ion transmembrane tra | 711 | 456 | 0.000176 | 0.012655953 |
| GO:0006259 | BP | DNA metabolic process | 949 | 598 | 0.000179 | 0.012779302 |
| GO:0070925 | BP | organelle assembly | 989 | 623 | 0.000179 | 0.012779302 |
| GO:0080134 | BP | regulation of response to stress | 1389 | 839 | 0.000181 | 0.012858528 |
| GO:0031327 | BP | negative regulation of cellular biosyı | 2561 | 1471 | 0.000184 | 0.013059384 |
| GO:0045177 | CC | apical part of cell | 456 | 297 | 0.000185 | 0.013096237 |
| GO:0005694 | CC | chromosome | 1876 | 1164 | 0.000187 | 0.013152972 |
| GO:1903047 | BP | mitotic cell cycle process | 757 | 488 | 0.000191 | 0.01345727 |
| GO:0006629 | BP | lipid metabolic process | 1328 | 798 | 0.000195 | 0.013647413 |
| GO:0051301 | BP | cell division | 623 | 406 | 0.000198 | 0.013819833 |
| GO:0098772 | MF | molecular function regulator activity | 2082 | 1245 | 0.000198 | 0.013819833 |
| GO:0042326 | BP | negative regulation of phosphorylati | 254 | 175 | 0.000203 | 0.014075707 |
| GO:0016787 | MF | hydrolase activity | 2260 | 1341 | 0.000209 | 0.014438766 |
| GO:0099537 | BP | trans-synaptic signaling | 758 | 496 | 0.000209 | 0.014438766 |
| GO:0044057 | BP | regulation of system process | 565 | 366 | 0.000211 | 0.014466258 |
| GO:0010558 | BP | negative regulation of macromolecu | 2520 | 1447 | 0.000211 | 0.014466258 |
| GO:0016301 | MF | kinase activity | 691 | 456 | 0.000216 | 0.014746229 |
| GO:0006812 | BP | monoatomic cation transport | 1024 | 636 | 0.000218 | 0.01483719 |
| GO:0016772 | MF | transferase activity, transferring pho | 846 | 548 | 0.000224 | 0.015209581 |
| GO:0034976 | BP | response to endoplasmic reticulum s | 267 | 182 | 0.000227 | 0.015347862 |
| GO:0070887 | BP | cellular response to chemical stimulı | 2139 | 1283 | 0.000254 | 0.017151256 |
| GO:0098794 | CC | postsynapse | 743 | 491 | 0.000256 | 0.017237295 |
| GO:0001701 | BP | in utero embryonic development | 390 | 263 | 0.000258 | 0.017335122 |
| GO:0007268 | BP | chemical synaptic transmission | 753 | 492 | 0.00026 | 0.017350634 |
| GO:0098916 | BP | anterograde trans-synaptic signaling | 753 | 492 | 0.00026 | 0.017350634 |
| GO:0051093 | BP | negative regulation of development | 960 | 598 | 0.000273 | 0.018126892 |
| GO:0008324 | MF | monoatomic cation transmembrane | 604 | 389 | 0.000274 | 0.018164039 |
| GO:0016477 | BP | cell migration | 1568 | 961 | 0.000278 | 0.018411486 |
| GO:0033365 | BP | protein localization to organelle | 972 | 609 | 0.000283 | 0.01863483 |
| GO:0010720 | BP | positive regulation of cell developm | 440 | 298 | 0.000285 | 0.018751868 |
| GO:0045184 | BP | establishment of protein localizatior | 1653 | 1009 | 0.000292 | 0.01914041 |
| GO:0007267 | BP | cell-cell signaling | 1287 | 806 | 0.000296 | 0.019330801 |

| | | | | | | |
|---|---|---|---|---|---|---|
| GO:0006986 | BP | response to unfolded protein | 141 | 101 | 0.000305 | 0.019873236 |
| GO:0016773 | MF | phosphotransferase activity, alcohol | 643 | 426 | 0.00031 | 0.020123089 |
| GO:0043555 | BP | regulation of translation in response | 20 | 19 | 0.000312 | 0.020197515 |
| GO:0007165 | BP | signal transduction | 5747 | 3309 | 0.000316 | 0.020396392 |
| GO:1903844 | BP | regulation of cellular response to tra | 175 | 120 | 0.000324 | 0.020884831 |
| GO:0070314 | BP | G1 to G0 transition | 20 | 19 | 0.000338 | 0.021668976 |
| GO:0009719 | BP | response to endogenous stimulus | 1508 | 931 | 0.000339 | 0.021668976 |
| GO:0042592 | BP | homeostatic process | 1699 | 1041 | 0.000339 | 0.021668976 |
| GO:0061138 | BP | morphogenesis of a branching epith | 185 | 135 | 0.000343 | 0.02181656 |
| GO:0030178 | BP | negative regulation of Wnt signaling | 175 | 125 | 0.000345 | 0.021898938 |
| GO:0021782 | BP | glial cell development | 132 | 98 | 0.00035 | 0.022147163 |
| GO:0001501 | BP | skeletal system development | 517 | 346 | 0.000356 | 0.022453686 |
| GO:0070848 | BP | response to growth factor | 761 | 483 | 0.000362 | 0.022798006 |
| GO:0097193 | BP | intrinsic apoptotic signaling pathway | 317 | 212 | 0.000365 | 0.022913662 |
| GO:0005813 | CC | centrosome | 695 | 446 | 0.000374 | 0.02343327 |
| GO:0001558 | BP | regulation of cell growth | 387 | 262 | 0.000377 | 0.02353592 |
| GO:0090407 | BP | organophosphate biosynthetic proce | 574 | 367 | 0.000391 | 0.024318493 |
| GO:0002040 | BP | sprouting angiogenesis | 189 | 122 | 0.000392 | 0.024337142 |
| GO:0014069 | CC | postsynaptic density | 342 | 236 | 0.000393 | 0.024340637 |
| GO:0048762 | BP | mesenchymal cell differentiation | 267 | 184 | 0.000398 | 0.024528494 |
| GO:0004672 | MF | protein kinase activity | 540 | 361 | 0.000398 | 0.024528494 |
| GO:0051797 | BP | regulation of hair follicle developme | 20 | 19 | 0.0004 | 0.024568163 |
| GO:0050808 | BP | synapse organization | 524 | 352 | 0.000405 | 0.024788461 |
| GO:0099536 | BP | synaptic signaling | 785 | 510 | 0.000408 | 0.024912816 |
| GO:0042634 | BP | regulation of hair cycle | 29 | 26 | 0.000425 | 0.025859686 |
| GO:0141124 | BP | intracellular signaling cassette | 1882 | 1156 | 0.000427 | 0.025915296 |
| GO:1903829 | BP | positive regulation of protein localiz | 483 | 318 | 0.000436 | 0.026373023 |
| GO:0009057 | BP | macromolecule catabolic process | 1469 | 886 | 0.000452 | 0.027317119 |
| GO:1903131 | BP | mononuclear cell differentiation | 504 | 329 | 0.000462 | 0.027845318 |
| GO:0048870 | BP | cell motility | 1817 | 1098 | 0.000491 | 0.029502522 |
| GO:0060045 | BP | positive regulation of cardiac muscle | 30 | 26 | 0.000501 | 0.029988883 |
| GO:0098796 | CC | membrane protein complex | 1188 | 728 | 0.000511 | 0.030512343 |
| GO:0003015 | BP | heart process | 251 | 171 | 0.000519 | 0.030945944 |
| GO:0018958 | BP | phenol-containing compound metab | 108 | 78 | 0.000524 | 0.031147809 |
| GO:0051348 | BP | negative regulation of transferase ac | 190 | 131 | 0.000529 | 0.031357679 |
| GO:0015031 | BP | protein transport | 1372 | 840 | 0.000547 | 0.03235745 |
| GO:0060562 | BP | epithelial tube morphogenesis | 323 | 224 | 0.00055 | 0.032426143 |
| GO:0062197 | BP | cellular response to chemical stress | 309 | 207 | 0.000552 | 0.032426143 |
| GO:0022603 | BP | regulation of anatomical structure m | 860 | 546 | 0.000553 | 0.032426143 |
| GO:0017015 | BP | regulation of transforming growth fa | 172 | 117 | 0.000576 | 0.033664072 |
| GO:0003677 | MF | DNA binding | 2363 | 1425 | 0.00058 | 0.033827019 |
| GO:0140297 | MF | DNA-binding transcription factor bin | 463 | 311 | 0.000593 | 0.034478819 |
| GO:0034599 | BP | cellular response to oxidative stress | 242 | 165 | 0.000595 | 0.034524018 |
| GO:0005886 | CC | plasma membrane | 5237 | 3007 | 0.000605 | 0.034992187 |
| GO:0021591 | BP | ventricular system development | 32 | 28 | 0.000611 | 0.035267524 |
| GO:0046907 | BP | intracellular transport | 1308 | 808 | 0.000614 | 0.035355618 |
| GO:0048471 | CC | perinuclear region of cytoplasm | 718 | 460 | 0.000624 | 0.035822319 |
| GO:0019904 | MF | protein domain specific binding | 610 | 399 | 0.000626 | 0.035865792 |
| GO:0097194 | BP | execution phase of apoptosis | 80 | 61 | 0.00063 | 0.035990349 |

| | | | | | | |
|---|---|---|---|---|---|---|
| GO:0008092 | MF | cytoskeletal protein binding | 957 | 612 | 0.000631 | 0.035990349 |
| GO:0051259 | BP | protein complex oligomerization | 267 | 179 | 0.000643 | 0.036567655 |
| GO:0060047 | BP | heart contraction | 242 | 165 | 0.000657 | 0.037130578 |
| GO:0030098 | BP | lymphocyte differentiation | 412 | 271 | 0.000658 | 0.037130578 |
| GO:0033673 | BP | negative regulation of kinase activity | 166 | 116 | 0.000658 | 0.037130578 |
| GO:0070593 | BP | dendrite self-avoidance | 14 | 14 | 0.00066 | 0.037130578 |
| GO:0015630 | CC | microtubule cytoskeleton | 1366 | 844 | 0.000668 | 0.037511576 |
| GO:0061448 | BP | connective tissue development | 283 | 194 | 0.000704 | 0.039363419 |
| GO:0071902 | BP | positive regulation of protein serine, | 120 | 87 | 0.000705 | 0.039363419 |
| GO:0031400 | BP | negative regulation of protein modi | 327 | 218 | 0.00071 | 0.039536396 |
| GO:0051241 | BP | negative regulation of multicellular | 1176 | 706 | 0.000717 | 0.039839805 |
| GO:1901888 | BP | regulation of cell junction assembly | 250 | 176 | 0.000722 | 0.039996035 |
| GO:0008610 | BP | lipid biosynthetic process | 702 | 439 | 0.000724 | 0.040053173 |
| GO:0010647 | BP | positive regulation of cell communic | 1763 | 1078 | 0.000743 | 0.040970498 |
| GO:0043085 | BP | positive regulation of catalytic activi | 644 | 408 | 0.000767 | 0.042218551 |
| GO:0007155 | BP | cell adhesion | 1485 | 913 | 0.000773 | 0.042452421 |
| GO:0070382 | CC | exocytic vesicle | 240 | 166 | 0.000799 | 0.043762666 |
| GO:0051493 | BP | regulation of cytoskeleton organizat | 509 | 332 | 0.000821 | 0.044797592 |
| GO:0099634 | CC | postsynaptic specialization membra | 150 | 110 | 0.000822 | 0.044797592 |
| GO:0019637 | BP | organophosphate metabolic process | 1007 | 619 | 0.000824 | 0.044806735 |
| GO:0097435 | BP | supramolecular fiber organization | 837 | 528 | 0.000855 | 0.04625569 |
| GO:0043065 | BP | positive regulation of apoptotic proc | 550 | 348 | 0.000858 | 0.04625569 |
| GO:0043204 | CC | perikaryon | 143 | 103 | 0.000862 | 0.04625569 |
| GO:0030511 | BP | positive regulation of transforming g | 32 | 28 | 0.000864 | 0.04625569 |
| GO:1903846 | BP | positive regulation of cellular respor | 32 | 28 | 0.000864 | 0.04625569 |
| GO:0008284 | BP | positive regulation of cell populatior | 943 | 584 | 0.000865 | 0.04625569 |
| GO:0051276 | BP | chromosome organization | 621 | 392 | 0.000866 | 0.04625569 |
| GO:0048639 | BP | positive regulation of developmenta | 160 | 115 | 0.000868 | 0.046277653 |
| GO:0140352 | BP | export from cell | 907 | 564 | 0.000889 | 0.047299463 |
| GO:0045165 | BP | cell fate commitment | 291 | 201 | 0.000909 | 0.048214185 |
| GO:0031253 | CC | cell projection membrane | 345 | 232 | 0.000915 | 0.048343819 |
| GO:0045599 | BP | negative regulation of fat cell differe | 61 | 47 | 0.000917 | 0.048343819 |
| GO:0048638 | BP | regulation of developmental growth | 307 | 209 | 0.000918 | 0.048343819 |
| GO:0010656 | BP | negative regulation of muscle cell ap | 59 | 45 | 0.000925 | 0.048638362 |
| GO:0099150 | BP | regulation of postsynaptic specializa | 20 | 19 | 0.000932 | 0.04888574 |
| GO:0050821 | BP | protein stabilization | 219 | 148 | 0.000936 | 0.048942563 |
| GO:0051129 | BP | negative regulation of cellular comp | 699 | 445 | 0.000956 | 0.04988892 |

**Supplementary Table S6.3 GO Pathway Results in Perinatal SI v NC at 8w post-partum.**

| GO ID | Ontology | Term | N | DE | P.DE | FDR |
|---|---|---|---|---|---|---|
| GO:0048856 | BP | anatomical structure development | 5815 | 2327 | 3.89E-11 | 8.66E-07 |
| GO:0032502 | BP | developmental process | 6320 | 2495 | 2.49E-10 | 2.78E-06 |
| GO:0007275 | BP | multicellular organism development | 4625 | 1881 | 6.80E-09 | 5.05E-05 |
| GO:0048731 | BP | system development | 3974 | 1635 | 2.09E-08 | 0.000116174 |
| GO:0009653 | BP | anatomical structure morphogenesis | 2684 | 1122 | 1.45E-07 | 0.000647725 |
| GO:0048518 | BP | positive regulation of biological proce | 6105 | 2345 | 3.32E-06 | 0.009674512 |
| GO:0009887 | BP | animal organ morphogenesis | 995 | 449 | 3.56E-06 | 0.009674512 |
| GO:0030154 | BP | cell differentiation | 4304 | 1706 | 3.98E-06 | 0.009674512 |
| GO:0048513 | BP | animal organ development | 2955 | 1207 | 4.08E-06 | 0.009674512 |
| GO:0048869 | BP | cellular developmental process | 4306 | 1706 | 4.34E-06 | 0.009674512 |
| GO:0009888 | BP | tissue development | 1979 | 823 | 6.14E-06 | 0.012428446 |
| GO:0030856 | BP | regulation of epithelial cell differentia | 170 | 90 | 1.19E-05 | 0.022163564 |
| GO:0007389 | BP | pattern specification process | 459 | 221 | 2.61E-05 | 0.042106276 |
| GO:0008283 | BP | cell population proliferation | 1990 | 801 | 2.65E-05 | 0.042106276 |
| GO:0048468 | BP | cell development | 2780 | 1140 | 3.40E-05 | 0.050457584 |
| GO:0040007 | BP | growth | 900 | 399 | 4.11E-05 | 0.057255022 |
| GO:0010604 | BP | positive regulation of macromolecule | 3276 | 1290 | 4.90E-05 | 0.060677871 |
| GO:0048870 | BP | cell motility | 1818 | 730 | 4.90E-05 | 0.060677871 |
| GO:0001763 | BP | morphogenesis of a branching structu | 200 | 108 | 5.57E-05 | 0.065367539 |
| GO:0061138 | BP | morphogenesis of a branching epithe | 185 | 100 | 8.81E-05 | 0.093451328 |
